# An atlas of genetic regulation and disease associations of microRNAs

**DOI:** 10.1101/2022.11.10.22282180

**Authors:** Rima Mustafa, Michelle M.J. Mens, Arno van Hilten, Jian Huang, Gennady Roshchupkin, Tianxiao Huan, Linda Broer, Paul Elliott, Daniel Levy, M. Arfan Ikram, Marina Evangelou, Abbas Dehghan, Mohsen Ghanbari

## Abstract

MicroRNAs (miRNAs) are small non-coding RNAs that post-transcriptionally regulate gene expression. Identification of genetic variants influencing the transcription of miRNAs can provide an understanding of their genetic regulation and implication in human disease. Here we present genome-wide association studies of 2,083 plasma circulating miRNAs measured by next-generation sequencing in 2,178 participants of the Rotterdam Study to identify miRNA-expression quantitative trait loci (miR-eQTLs). We report 4,310 cis- and trans-miR-eQTLs for 64 miRNAs that have been replicated across independent studies. Many of these miR-eQTLs overlap with gene expression, protein, and metabolite-QTLs and with disease-associated variants. The consequences of perturbation in miRNA transcription on a wide range of clinical conditions are systematically investigated in phenome-wide association studies, with their causality tested using Mendelian randomization. Integration of genomics and miRNAs enables interrogation of the genetic architecture of miRNAs, revealing their clinical importance, and providing valuable resources for future studies of miRNAs in human disease.

## Introduction

MicroRNAs (miRNAs) are small non-coding RNAs of approximately 22 nucleotides that regulate gene expression at the post-transcriptional level and play critical roles in determining whether genes are (in)active and how much a particular protein is translated (1,2). Over 2,500 miRNAs have been identified in humans (3), which altogether regulate more than half of protein-coding genes through cleavage or translation repression of messenger(m)-RNAs (4,5). miRNAs have shown their potential as disease biomarkers (6) and, to a lesser extent, therapeutic targets (7). Identification of the role of miRNAs in regulating the expression of specific genes and their effects in clinical conditions has been a subject of extensive work in recent years. However, the genetic regulation of miRNAs remains less well understood.

Circulatory miRNAs are released from cells into circulation via extracellular vesicles such as exosomes (8). Genetic variants are known to regulate the level of miRNAs in the circulation (9-11) or tissues and cells (12-14), referred to as miRNA expression quantitative trait loci (miR-eQTLs). Previous studies showed that each miR-eQTL contributed to a relatively small proportion of variation in miRNA levels (9,10), with a tiny proportion of miR-eQTLs replicated across studies thus far (9). The identified miR-eQTLs have been used to study the effect of perturbation of miRNA levels on disease risk (9,10,14). However, such an effect on a wide range of clinical conditions remains to be elucidated. Unravelling the genetic regulation of nearly all high confidence miRNAs can provide insights into their roles in affecting disease risk and discover candidates for therapeutic targets.

This study measured plasma levels of 2,083 circulatory miRNAs in the population-based Rotterdam Study cohort using a next-generation sequencing platform (HTG EdgeSeq miRNA Whole Transcriptome Assay). This assay allows simultaneous, quantitative detection of miRNAs with a sensitivity and specificity of 97% (15,16). Subsequently, genome-wide association studies (GWAS) were conducted for 2,083 miRNAs to identify miR-eQTLs (N=2,178), followed by replication in two independent cohorts (9,10). We conducted downstream analyses to elucidate functional characteristics of the findings through cis and trans mapping of miR-eQTLs, cross-phenotype, and multi-omics QTLs look-up. A systematic investigation of the effects of genetically determined miRNA levels on a wide range of clinical conditions was conducted using phenome-wide association studies (PheWAS) in the UK Biobank (N=423,419) (17,18). We performed Mendelian randomisation (MR) to assess causality between miRNAs and clinical conditions (19). Potential downstream target genes that might be involved in the disease processes were highlighted, generating testable hypotheses for further functional studies to dissect the underlying molecular pathways.

## Results

### Genome-wide identification and replication of miR-eQTLs

An overview of the study workflow is presented in Fig. 1. The list of 2,083 miRNAs characterised in this study is shown in Supplementary Table 1. The miRNA expression profiling was performed for 2,754 participants from three sub-cohorts (RS-I-4, RS-II-2, and RS-IV-1) in the Rotterdam Study (20). Genotype data were available for 2,435 of 2,754 participants. Participants of non-European ancestries and relatives based on kinship coefficient > 0.088 were excluded, resulting in 2,178 participants in the analysis (Extended Data Fig. 1). The clinical characteristics of participants are summarised in Supplementary Table 2.

**Figure 1.**
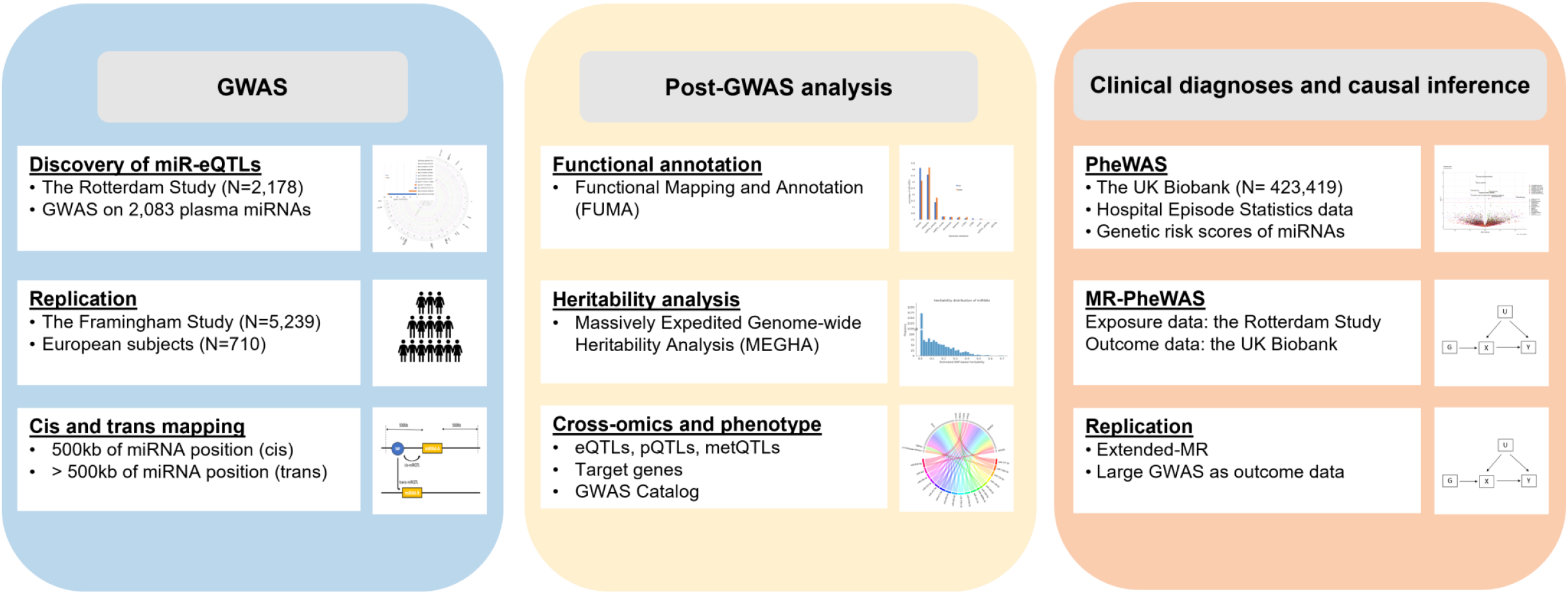
Overview of the study workflow.

We identified 3,292 associations between 1,289 SNPs and 63 miRNAs at P<2.4×10^-11^ (genome-wide threshold of P<5×10^-08^ and Bonferroni-corrected for 2,083 miRNAs) (Supplementary Table 3, Extended Data Fig. 2). After pruning with a series of linkage disequilibrium (LD) thresholds, the number of associations reduced to 75 (r^2^<0.1), 142 (r^2^<0.3), and 297 (r^2^<0.6). The 3,292 identified associations included 1,733 cis associations for 32 miRNAs and 1,559 trans associations for 33 miRNAs. The overall proportion of variance explained by each miR-eQTL ranged from 2% to 11% (median=2.9%). Eighteen miR-eQTLs (r^2^<0.6) were explaining more than 5% of the variation of corresponding miRNA levels (Supplementary Table 4). The highest proportion of variance explained was observed for miR-625-5p (11%) by rs2127868 (P=1.63×10^-60^), identified as cis-miR-eQTL.

We sought to replicate our findings in a previous study using the same HTG EdgeSeq platform by Nikpay et al. (9). Bonferroni correction was applied to address multiple testing (Online Methods). There were 2,254 associations for 57 miRNAs with identical SNPs available in the replication cohort by Nikpay et al. (9). Additionally, 27 associations for six miRNAs were tested for replication using proxy SNPs (Online Methods). There were 1,462 associations for 27 miRNAs replicated at Bonferroni-threshold (P<0.05/58) (Extended Data Fig. 2, Supplementary Tables 5-6). The effect estimates of replicated associations were strongly correlated (r=0.82, P<2.2×10^-16^) (Extended Data Fig. 3a). Additionally, 1,719 associations were nominally significant in Nikpay et al. (9), of which 1,685 (98%) were in a concordant direction (Supplementary Table 6).

### Replication of previously identified miR-eQTLs

In an alternative approach, we also attempted to replicate miR-eQTLs identified in previous studies (9,10). Associations reaching P<2.4×10^-11^ in Nikpay et al. (9) were tested for replication in our study (Extended Data Fig. 2), where 2,957 associations corresponding to 1,973 SNPs and 34 miRNAs were replicated (P<0.05/52) with a significant correlation between effect estimates (r=0.90, P<2.2×10^-16^) (Extended Data Fig. 3b). Additionally, we tested 8,820 cis and trans associations for 87 miRNAs that were previously reported by the Framingham Heart Study (10) in our data. Of those, 1,320 associations for 29 miRNAs were replicated (P<0.05/195) (Extended Data Fig. 3c). Collectively, 69% of associations in Nikpay et al. (20) and 15% of associations in the Framingham Heart Study (19) were replicated in our study. Altogether, our approaches successfully reported 4,310 associations pertaining to 64 miRNAs which have been replicated across studies (Extended Data Fig. 2, Supplementary Table 7).

### Functional annotation of miR-eQTLs

Using the Functional Mapping and Annotation (FUMA) (21), the identified and replicated miR-eQTLs were mapped into 22 loci (Fig. 2a). Over 70% of miR-eQTLs were located in the intronic and intergenic regions (Fig. 2b, Supplementary Table 8). Out of 22 loci, 11 loci were pleiotropic, i.e., linked to the level of multiple miRNAs. Fourteen loci regulated two miRNAs and eight loci regulated more than two miRNAs (Supplementary Table 9). Highly pleiotropic loci were identified in locus chr14:100655022-101244293, regulating 23 miRNAs (also known as 14q32 miRNA cluster), the majority of which were cis-miR-eQTLs (98%). This locus was mapped to *RP11-566J3*.*2, RP11-638I2*.*4, YY1, YY1:RP11-638I2*.*2, SLC25A29, WDR25, BEGAIN, DLK1, CTD-2644I21*.*1, LINC00523, RP11-566J3*.*4*, and *MEG3* (Extended Data Fig.4). The locus in chr9:136128546-136296530 was mapped to *ABO, ABO:RP11-430N14*.*4, Y_RNA*, and *LCN1P2* and regulated 18 miRNAs. This locus contained shared trans-miR-eQTLs for several well-known miRNAs, including miR-10, let-7, and miR-30 families (Supplementary Table 9).

**Figure 2.**
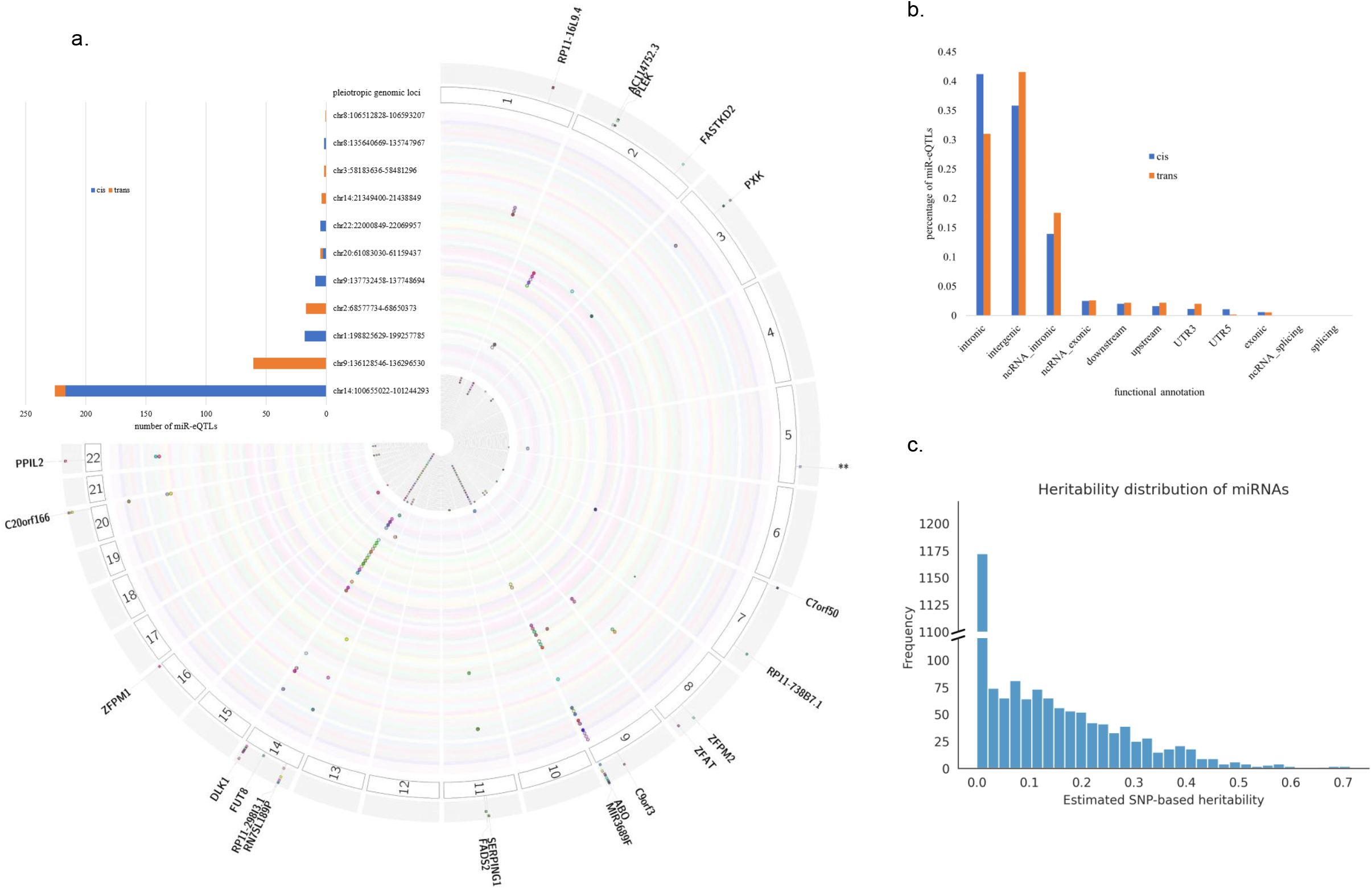
**a. Fujiplot of identified and replicated miR-eQTLs mapped into 22 genomic loci**. Each cular layer represents miRNA, and dots represent SNP-miRNA associations. The dots that form a radial ttern indicate loci associated with multiple miRNAs. The innermost histogram shows the number of miR-TLs identified in each locus. ** in chromosome 5 represents one locus mapped to *CTC-5298P*.*1*. The -left inset is a barplot to show the number of independent cis and trans-miR-eQTLs in pleiotropic loci, ch regulating the level of multiple miRNAs. Highly pleiotropic loci were identified in locus r14:100655022-101244293, regulating 23 miRNAs, the majority of which were cis-miR-eQTLs. The locus chr9:136128546-136296530 consisted of trans-miR-eQTLs and regulated 18 miRNAs. **b. Functional nsequences of identified miR-eQTLs on nearby and far genes. c. SNP-based heritability timates distribution for 2,083 miRNAs**.

Associations between 296 SNPs residing in seed, mature, or precursor genes of miRNAs and corresponding miRNAs were extracted from our GWAS results. Twelve associations were significant at Bonferroni-threshold considering the number of unique SNPs (P<0.05/296), consisting of three SNPs in the seed region of miR-4707-3p, miR-4482-5p, and miR-6891-3p, three SNPs in mature region of miR-3130-3p, miR-6891-3p, and miR-6839-5p, and six SNPs in precursor genes of six miRNAs. Additionally, 27 SNPs in 26 miRNAs were nominally significant (Supplementary Table 10).

### Heritability analysis

SNP-based heritability estimates for the plasma levels of 2,083 circulatory miRNAs were obtained using massively expedited genome-wide heritability analysis (MEGHA) (22). The distribution of heritability estimates is shown in Fig. 2c. Two miRNAs had a narrow-sense heritability estimate greater than 0.7, namely miR-30e-5p (0.72) and miR-6511a-5p (0.70). Twenty miRNAs had a narrow-sense heritability greater than 0.5, and 166 miRNAs had greater than 0.3. Heritability estimates for all miRNA are shown in Supplementary Table 11. Repeating the heritability analysis, including the first five principal components as covariates, resulted in SNP-based heritability estimates with a Pearson correlation of 0.99 with estimates without the principal components (Extended Data Fig. 5).

### Cross-phenotype and multi-omics QTLs look-up

As miRNAs dictate their role in biological processes by regulating the expression of their target genes, it is interesting to know whether miR-eQTLs are linked to the expression of other genes, including their host and target genes. To explore this, we sought overlaps between replicated miR-eQTLs and gene expression (eQTLs), protein (pQTLs), and metabolite-QTLs (met-QTLs). We further checked if any of the genes or proteins that shared QTLs were target genes of miRNAs.

Summary statistics for genetic variants that influence expression levels of mRNA transcripts (cis and trans-eQTLs) in whole blood (N=31,684) were used to identify miR-eQTLs that affect the expression of other genes (23). In this dataset, trans-eQTL analysis was conducted only for SNPs previously identified in GWAS (23). Cis-miR-eQTLs for 39 miRNAs overlapped with cis-eQTLs for 146 genes, 123 of which were protein-coding genes (Supplementary Table 12). Twelve intragenic miRNAs shared cis-miR-eQTLs with their host genes (Supplementary Table 13). In addition, miR-136-5p resides within the intronic region of *RTL1* but had overlapping cis-eQTLs with nearby genes, such as *DLK1, WARS, BEGAIN, MEG3*, and *SLC25A29*) (Supplementary Table 12).

Next, predicted target genes for miRNAs from TargetScan v7.2 (5) and experimentally validated target genes with robust validation methods (such as reporter assay, western blot or qRT-PCR) from miRTarBase (4) were retrieved to check whether miR-eQTLs overlap with eQTLs of their putative target genes. Twelve miRNAs shared either cis/trans-miR-eQTLs or cis/trans-eQTLs of their putative target genes (Supplementary Table 14). We identified shared trans-regulation by rs612169, located intronic to *ABO*, for miR-126-3p and its validated target *TCF4*.

Protein-QTLs (pQTLs) summary results were used to identify miR-eQTLs affecting protein levels in the blood (24,25). Cis-miR-eQTLs of 18 miRNAs overlapped with pQTLs for nine proteins. Cis-miR-eQTLs for 14q32 miRNA cluster were shared with pQTLs of *DLK1* located in the nearby genomic region and *SEMG2* in a distant region of chromosome 20 (Fig. 3a, Supplementary Table 15). These included shared miR-eQTLs of six intragenic miRNAs, of which four miRNAs (miR-127-3p, miR-136-5p, miR-431-5p, and miR-433-5p) reside in *RTL1*.

**Figure 3.**
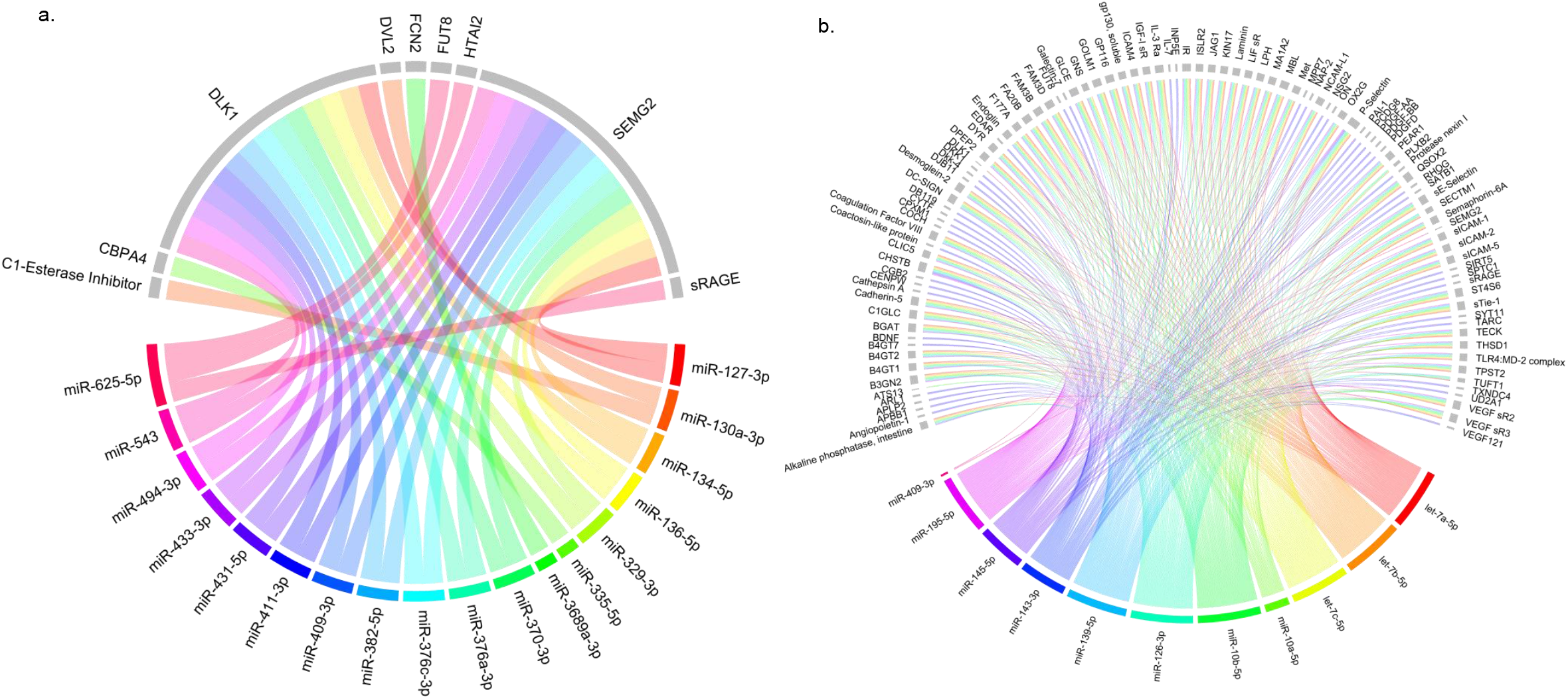
**a. Overlap between cis miR-eQTLs and proteins (pQTLs)**. The bottom half of the circle shows miRNAs in different colours, and the top half of the circle (grey coloured) shows the genes. **b. Overlap between trans-miR-eQTLs and proteins (pQTLs)**. Trans-miR-eQTLs are shown to be more pleiotropic than cis-miR-eQTLs.

Cis-miR-eQTLs of miR-625-5p overlapped with pQTLs for Alpha-(1,6)-fucosyltransferase, an enzyme encoded by *FUT8* gene where miR-625-5p resides. Cis-miR-eQTLs for miR-335-5p that resides in *MEST* overlapped with Carboxypeptidase A4, encoded by *CBPA4* in a nearby genomic region in chromosome 7. Trans-miR-eQTLs for 11 miRNAs overlapped with pQTLs for 103 proteins (Fig. 3b, Supplementary Table 16). Of these, *GNS* was identified as common predicted targets of let-7b-5p and let-7c-5p. Additionally, trans-miR-eQTLs of miR-145-5p and miR-195-5p overlapped with eQTLs of their predicted target genes. For cardiovascular proteins (25), cis-miR-eQTLs for miR-130a-3p overlapped with pQTLs of Pappalysin-1 (*PAPPA*). Overlap for trans-miR-QTLs was found with pQTLs of validated target genes of miR-126-3p (*TEK*) and miR-145-5p (*MMP1* and *VEGFA*) (Supplementary Table 17). The overlap of trans-miR-eQTLs between miR-143-3p and miR-145-5p with Dickkopf-related protein 1 (*DKK1*) were identified in both pQTLs datasets (24,25).

We used summary statistics from two common metabolomics platforms, Metabolon and Nightingale, to identify miR-eQTLs affecting metabolic pathways by investigating plasma levels of metabolites. Metabolon covers 529 metabolites (N=7,824) (26), while Nightingale covers 123 metabolites in Kettunen et al. (N>20,000) (27) and 249 metabolites in the UK Biobank (N=115,078) (28). Cis-miR-eQTLs for miR-1908-5p, miR-148a-3, miR-339-5p, and miR-130a-3p overlapped with met-QTLs for 218 metabolites in both platforms. For example, rs174561, located in the precursor gene of miR-1908-5p and intronic to *FADS1*, both known to be associated with lipid and obesity traits, was associated with metabolites in Nightingale that were also mainly lipid fractions. Trans-miR-eQTLs for nine miRNAs overlapped with four unnamed metabolites in Metabolon (M32740, M33801, M36115, M36230), and 146 metabolites in the Nightingale platform (Supplementary Table 18).

Finally, GWAS Catalog was used to identify miR-eQTLs associated with complex traits (29). At genome-wide significance, cis-miR-eQTLs were associated with GWAS traits, including mental health, haematological indices, cancers, anthropometric measures, lipid levels, and blood pressure (Supplementary Table 19). For example, cis-miR-eQTLs for miR-1908-5p were associated with multiple traits, mainly lipid through *FADS1, FADS2*, or *MYRF*, supporting the observed overlaps with eQTLs and pQTLs as described earlier. Trans-miR-eQTLs were associated with different traits and diseases, including haematological indices, cardiometabolic, cancer, and allergy. The pleiotropic regulatory region in chr9:136128546-136296530 consisted of trans-miR-eQTLs which were associated with protein and metabolite levels and complex traits. For example, rs687289, intronic to *ABO*, was identified as trans-miR-eQTL for six miRNAs and overlapped with pQTLs, met-QTLs, and associated with GWAS traits such as monocyte count, coagulation factor levels, and pancreatic cancer. Another variant in *ABO*, rs644234, was associated with 40 protein biomarkers, including five cardiovascular proteins (Supplementary Table 20). This observation might suggest the relevance of miR-eQTLs in *ABO* to cardiovascular traits.

### Phenome-wide association studies (PheWAS) in the UK-Biobank

We conducted phenome-wide association studies (PheWAS) in the UK Biobank (17) to investigate associations between genetically determined miRNA levels and a wide range of clinical conditions (Fig. 4a). The participants with genetic and hospital episode statistics data were used in the analysis. After excluding participants of non-European ancestries and one from each pair of related individuals, 423,419 individuals remained in the analysis (Extended Data Fig. 6). We tested the associations between genetically determined miRNA levels and 905 phecodes with at least 200 cases across 16 disease groups (Fig. 4b).

**Figure 4.**
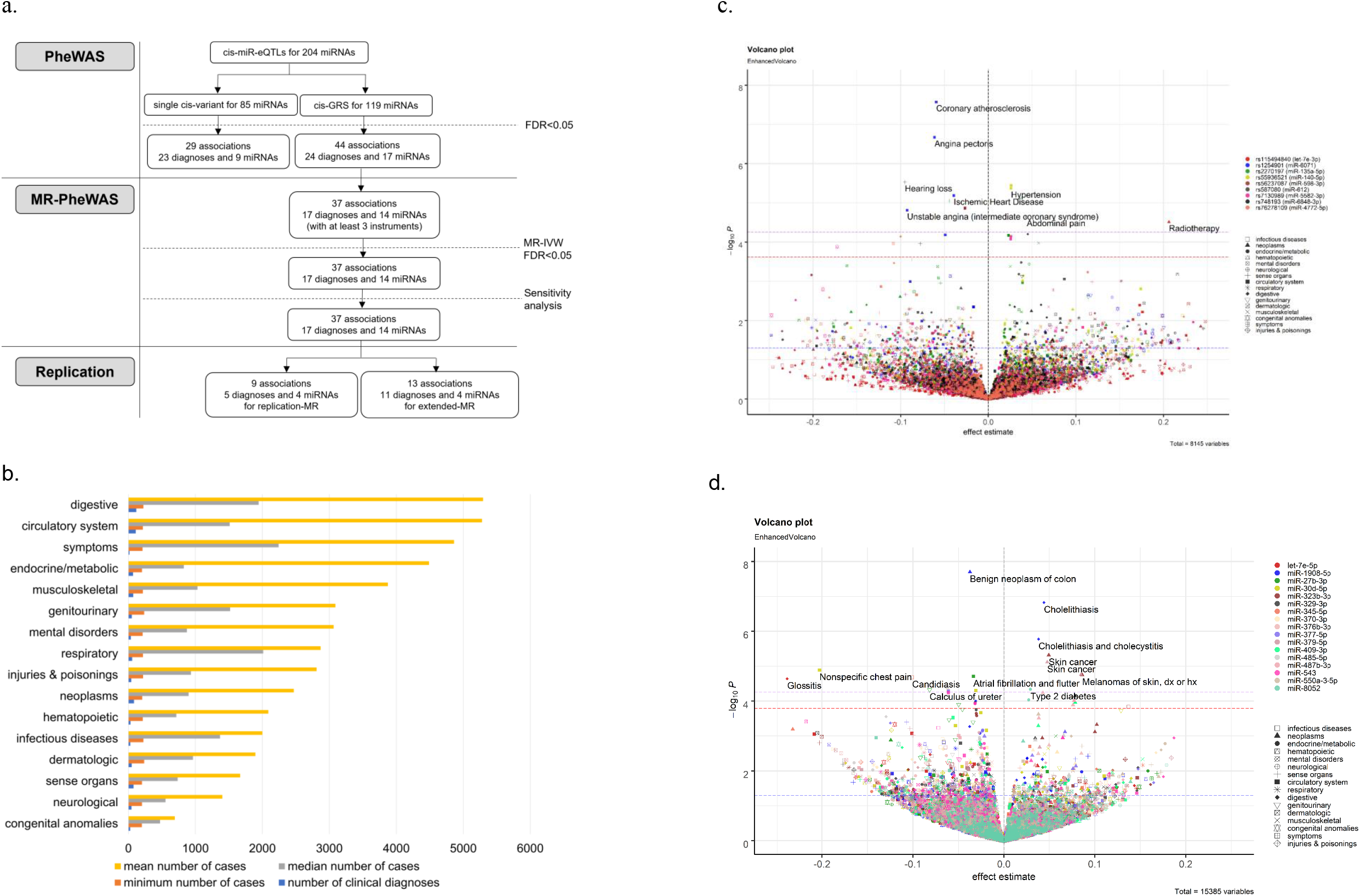
**a. Summary of PheWAS and MR. Cis-miR-eQTLs were used as proxies for miRNAs**. When ltiple cis-miR-eQTLs were available, cis-GRS was computed for PheWAS. Otherwise, single variant eWAS was conducted. MR were conducted for miRNA with at least three instruments. When available, ge GWAS data were used to replicate the findings. Otherwise, genome-wide trans-miR-eQTLs were ded in the extended-MR. **b**. Number of cases available within each disease group. The figure rresponds to clinical diagnoses with at least 200 cases.**c. Enhanced volcano plots for single variant eWAS and d. GRS PheWAS**. The X-axis denotes effect estimates for corresponding SNP or GRS. Y-s indicates -log10 of the association p-values between each SNP or GRS and clinical condition. Different ours of the dots represent different SNPs. Different shapes show different disease groups. Thresholds significance are indicated by dashed blue (nominal), red (FDR), and purple (Bonferroni) lines. Plots were ly created for SNP and GRS with at least one FDR-significant finding.

To extract genetic instruments for miRNAs, a threshold for cis-instruments at FDR<0.1 was calculated across all SNPs residing ±500 kb of each miRNA. This threshold was chosen to enable covering a higher number of miRNAs tested in PheWAS. At FDR<0.1, cis-instruments were identified for 204 miRNAs. Weak instruments were filtered out using F-statistics>10 (Online Methods). After LD clumping (r^2^<0.1), a single cis-instrument was available for 85 miRNAs for which a single variant PheWAS were conducted (Supplementary Table 21). Multiple cis-instruments were available to compute genetic risk scores (GRS) for 119 miRNAs and test in GRS-PheWAS (Fig. 4a). Since each clinical diagnosis is not entirely independent of the other, FDR correction was applied for each miRNA to account for multiple testing.

In the single variant PheWAS, 29 significant associations were identified between 9 SNPs and 23 clinical diagnoses at FDR<0.05 (Fig. 4a). Among these, rs2270197 (P=6.84×10^-05^), rs55936521 (P=4.27×10^-06^), rs5623708 (P=1.40×10^-05^) and rs7130989 (P=7.16×10^-05^) were associated with hypertension. rs1254901, located 2KB upstream of *VAMP5* and cis-miR-eQTL for miR-6701, was associated with ischemic heart disease-related conditions, including ischemic heart disease (P=6.51×10^-06^). Rs2270197, intronic to *ITIH1* and cis-miR-eQTL for miR-135a-5p, was associated with a range of clinical conditions, including osteoarthritis (P=4.22×10^-04^), hypertension (P=6.84×10^-05^), and bipolar disorders (P=4.02×10^-04^) (Fig. 4c, Supplementary Table 22).

In the genetic risk score (GRS) PheWAS, 44 associations between 17 cis-GRS and 24 diagnoses were identified at FDR<0.05 (Fig. 4a). The strongest association was identified between miR-1908-5p and benign neoplasm of colon (OR=0.96, P=1.99×10^-08^) and cholelithiasis (OR=1.04, P=1.51×10^-05^). Three miRNAs were associated with lower risk of obesity, namely miR-323b-3p (OR=0.97, P=2.28×10^-04^), miR-329-3p (OR=0.97, P=1.12×10^-04^), and miR-543 (OR=0.97, P=1.20×10^-04^). Three miRNAs were associated higher risk of skin cancer, including miR-323b-3p (OR=1.05, P=4.79×10^-06^), miR-376b-3p (OR= 1.04, P=5.81×10^-05^), and miR-379-5p (OR=1.05, P=7.68×10^-06^) (Fig. 4d, Supplementary Table 22). Out of 44 associations, extended-GRS was computed for 17 associations of five miRNAs with genome-wide significant trans-miR-eQTLs, where all remained statistically significant in a concordant direction (FDR<0.05) (Supplementary Table 23). Our PheWAS identified 73 associations between 45 clinical conditions and 26 miRNAs. Among those, 11 miRNAs were associated with circulatory disorders (Fig. 5a). Several miRNAs were associated with clinical diagnoses across different disease groups, indicating their pleiotropic properties, such as miR-323b-3p associated with endocrine/metabolic disease, infectious disease, and neoplasms (Fig. 5b).

**Figure 5.**
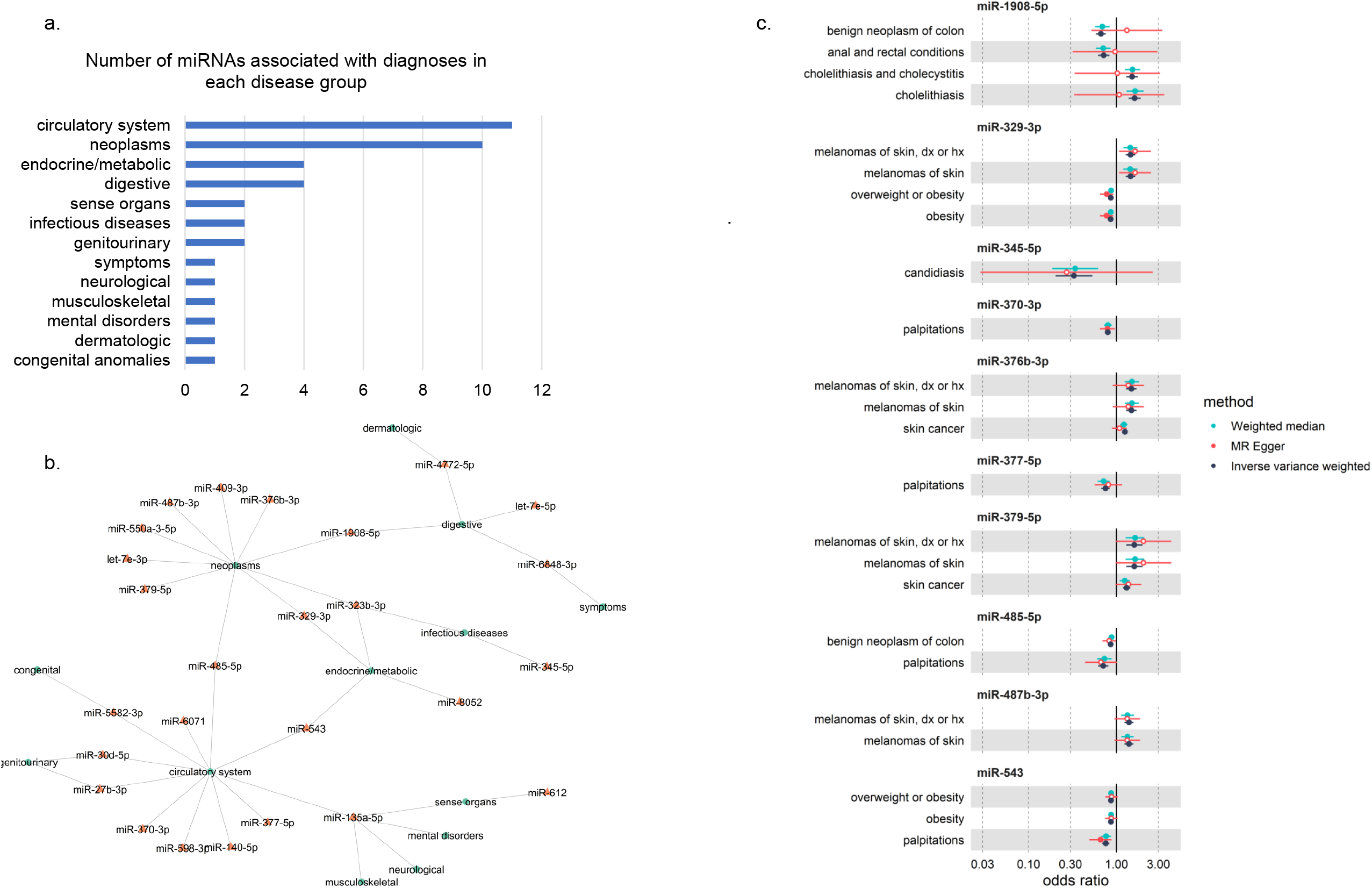
**a. Number of miRNAs associated with diagnoses in each disease group as identified in eWAS. b. Schematic network showing miRNAs and disease groups associations**. Each line corresponds to association between miRNA and clinical diagnosis that belong to a particular disease group. The colour of the circles indicates miRNA (orange) or disease groups (green). **c. Forest plots for 24 associations in MR-PheWAS with no genome-wide significant trans-miR-eQTLs**. Different colours correspond to different MR methods, as labelled.

### Mendelian randomisation

Mendelian randomisation (MR) was conducted for miRNAs with at least three independent instruments (Online Methods). Thirty-seven associations for 14 miRNAs with at least three instruments were tested in MR-PheWAS (Fig. 4a). The same cis or extended instruments for miRNAs were used in MR-PheWAS with effect estimates from the Rotterdam Study (N=2,178), whereas genetic associations with the outcomes were taken from the UK-Biobank (N=423,419). All 37 tested associations were significant in MR Inverse variance weighted (IVW) (FDR<0.05), had no indication of pleiotropy or heterogeneity and were in concordant direction with MR-Egger or Weighted Median (WM) (P<0.05) (Supplementary Table 24). Among those, the strongest association was observed between miR-1908-5p and the risk of benign neoplasm of the colon (MR-IVW estimate=-0.40, P=3.9×10^-10^). Six miRNAs were associated with a higher risk of melanoma, including miR-329-3p (MR-IVW estimate=0.37, P=5.4×10^-9^), miR-376b-3p (MR-IVW estimate=0.39, P=3.0×10^-8^), miR-323b-3p (MR-IVW estimate=0.44, P=1.4×10^-7^), and miR-379-5p (MR-IVW estimate=0.47, P=1.5×10^-5^). All 37 associations were significant when the correlation matrix between instruments was added, with no outliers detected by MRPRESSO. Of 37, 13 associations had genome-wide significant trans-miR-eQTLs to conduct extended-MR as a replication. For the remaining 24 associations, concordant direction across different MR methods was observed (Fig. 5c).

Extended-MR was conducted for 13 associations by adding genome-wide significant trans-miR-eQTLs. Twelve associations were significant (MR-IVW FDR<0.05) with no indication of pleiotropy and supported by MR-Egger or WM (P<0.05) (Supplementary Table 25). Nine associations from MR-PheWAS had genetic association data available in large GWAS for coronary artery disease, body mass index (BMI) and waist to hip ratio (WHR) (30-32) (Fig. 5a). Two associations were replicated between miR-543 and WHR (MR-IVW estimate=-0.02, P=1.72×10^-02^) and between miR-329-3p and BMI (MR-IVW estimate=-0.03, P=1.89×10^-02^), both in the same protective effects as in MR-PheWAS (Table 1, Extended Data Fig. 7, Supplementary Table 26). Reverse MR analyses were conducted for BMI and WHR as risk factors on miR-543 and miR-329-3p as the outcomes, where no significant effects in the opposite directions were observed (Supplementary Table 27).

**Table 1.** The results of Mendelian randomisation (MR-IVW) for replicated associations.

| exposure | MR-PheWAS |  |  |  |  |  |  | Replication |  |  |  |  |  |  |
| --- | --- | --- | --- | --- | --- | --- | --- | --- | --- | --- | --- | --- | --- | --- |
|  | outcome | n | beta | SE | P | P-het | Egger int P | outcome | n | beta | SE | P | P-het | Egger int P |
| **Replication in extended-MR** |  |  |  |  |  |  |  |  |  |  |  |  |  |  |
| miR-30d-5p | angina pectoris | 5 | -0.47 | 0.10 | 1.5×10<sup>-6</sup> | 3.4×10<sup>-1</sup> | 2.9×10<sup>-1</sup> | angina pectoris | 6 | -0.34 | 0.12 | 6.0×10<sup>-3</sup> | 3.1×10<sup>-2</sup> | 2.7×10<sup>-1</sup> |
|  | coronary atherosclerosis | 5 | -0.39 | 0.09 | 7.6×10<sup>-6</sup> | 3.6×10<sup>-1</sup> | 2.5×10<sup>-1</sup> | coronary atherosclerosis | 6 | -0.26 | 0.11 | 1.8×10<sup>-2</sup> | 2.8×10<sup>-2</sup> | 2.4×10<sup>-1</sup> |
|  | nephrotic syndrome | 5 | 1.91 | 0.41 | 3.9×10<sup>-6</sup> | 6.1×10<sup>-1</sup> | 5.7×10<sup>-1</sup> | nephrotic syndrome | 6 | 1.32 | 0.50 | 8.9×10<sup>-3</sup> | 7.0×10<sup>-2</sup> | 9.9×10<sup>-1</sup> |
|  | nonspecific chest pain | 5 | -3.07 | 0.56 | 4.9×10<sup>-8</sup> | 7.9×10<sup>-1</sup> | 9.2×10<sup>-1</sup> | nonspecific chest pain | 6 | -2.03 | 0.82 | 1.4×10<sup>-2</sup> | 1.2×10<sup>-2</sup> | 6.5×10<sup>-1</sup> |
| miR-323b-3p | melanomas of skin | 6 | 0.45 | 0.08 | 1.4×10<sup>-7</sup> | 8.8×10<sup>-1</sup> | 5.7×10<sup>-1</sup> | melanomas of skin | 7 | 0.31 | 0.11 | 4.7×10<sup>-3</sup> | 4.3×10<sup>-2</sup> | 8.1×10<sup>-1</sup> |
|  | obesity | 6 | -0.16 | 0.03 | 6.6×10<sup>-6</sup> | 7.7×10<sup>-1</sup> | 7.1×10<sup>-1</sup> | obesity | 7 | -0.11 | 0.05 | 1.9×10<sup>-2</sup> | 4.6×10<sup>-2</sup> | 8.7×10<sup>-1</sup> |
|  | overweight or obesity | 6 | -0.15 | 0.03 | 8.5×10<sup>-6</sup> | 7.1×10<sup>-1</sup> | 6.9×10<sup>-1</sup> | overweight or obesity | 7 | -0.11 | 0.04 | 1.7×10<sup>-2</sup> | 5.6×10<sup>-2</sup> | 8.6×10<sup>-1</sup> |
|  | skin cancer | 6 | 0.26 | 0.05 | 2.2×10<sup>-7</sup> | 2.9×10<sup>-1</sup> | 9.6×10<sup>-1</sup> | skin cancer | 7 | 0.21 | 0.06 | 3.4×10<sup>-4</sup> | 5.2×10<sup>-2</sup> | 9.0×10<sup>-1</sup> |
|  | viral enteritis | 6 | 0.72 | 0.15 | 1.7×10<sup>-6</sup> | 8.7×10<sup>-1</sup> | 7.3×10<sup>-1</sup> | viral enteritis | 7 | 0.61 | 0.13 | 3.2×10<sup>-6</sup> | 6.7×10<sup>-1</sup> | 6.6×10<sup>-1</sup> |
| miR-409-3p | melanomas of skin | 12 | 0.25 | 0.05 | 7.7×10<sup>-6</sup> | 1.1×10<sup>-1</sup> | 5.0×10<sup>-1</sup> | melanomas of skin | 13 | 0.24 | 0.05 | 3.3×10<sup>-6</sup> | 1.4×10<sup>-1</sup> | 4.7×10<sup>-1</sup> |
| **Replication using GWAS summary statistics** |  |  |  |  |  |  |  |  |  |  |  |  |  |  |
| miR-329-3p | overweight or obesity | 10 | -0.15 | 0.03 | 1.5×10<sup>-7</sup> | 3.2×10<sup>-1</sup> | 2.4×10<sup>-1</sup> | BMI | 6 | -0.03 | 0.01 | 1.9×10<sup>-2</sup> | 4.9×10<sup>-7</sup> | 9.9×10<sup>-2</sup> |
|  | obesity | 10 | -0.15 | 0.03 | 3.9×10<sup>-8</sup> | 3.5×10<sup>-1</sup> | 2.4×10<sup>-1</sup> |  |  |  |  |  |  |  |
| miR-543 | overweight or obesity | 8 | -0.15 | 0.03 | 9.5×10<sup>-9</sup> | 4.7×10<sup>-1</sup> | 8.4×10<sup>-1</sup> | WHR | 7 | -0.02 | 0.01 | 1.7×10<sup>-2</sup> | 7.7×10<sup>-1</sup> | 9.0×10<sup>-1</sup> |
|  | obesity | 8 | -0.15 | 0.03 | 1.1×10<sup>-8</sup> | 4.9×10<sup>-1</sup> | 8.9×10<sup>-1</sup> |  |  |  |  |  |  |  |

BMI: body mass index. WHR: waist-to-hip ratio. n is the number of genetic instruments used in the analysis. SE: standard error. P-het denotes P-value for heterogeneity of MR-IVW estimates. Egger int P: P-values for MR Egger intercept. The summary statistics presented are based on MR-IVW. Full results for other MR methods are presented in Supplementary Tables 25 and 26.

### Target genes and enrichment analysis

An in-silico search of target genes using TargetScan v7.2 and miRTarBase (5,33) identified eighty-two predicted and eighteen validated target genes of miR-543 associated with BMI or WHR. Forty-three predicted and fifty-eight validated target genes for miR-329-3p were also associated with BMI or WHR. Eighteen target genes were in common between miR-543 and miR-329-3p, where eleven genes were validated targets for at least one of them, including *BRCH1* and *TNRC6B* as validated targets of both miRNAs (Supplementary Table 28). Enrichment analysis was conducted as described in our previous work (34), resulting in a significant enrichment for BMI or WHR-related genes among validated targets of miR-543 (P=9.00×10^-03^) and predicted targets of miR-329-3p (P=3.18×10^-02^).

## Discussion

We present a genome-wide identification of miR-eQTLs using the next-generation sequencing method in 2,178 individuals in the population-based Rotterdam Study cohort. This study is currently the most extensive single-site analysis of 2,083 circulatory miRNA levels in a population of European ancestries. We discovered 3,292 genetic associations for 63 miRNAs. The highest proportion of variance explained was observed for miR-625-5p (11%) by rs2127868 (P=1.63×10^-60^), in perfect LD with rs2127870 associated with the level of miR-625-5p in plasma (P=2.9×10^-260^) and whole blood (P=2.98×10^-09^) (9,10). Altogether, 4,310 genetic associations for 64 miRNAs were replicated across different studies, including trans-miR-eQTLs, whose replication was previously minimal. Genetically proxied miRNAs were tested against a wide range of clinical conditions in the UK-Biobank, suggesting the pleiotropic properties of several miRNAs by being associated with clinical outcomes. Such observation is expected, given that miRNAs could potentially regulate many genes that are involved in different molecular pathways (4,5). Our MR analysis identified the potentially causal role of miRNAs in various complex traits and disorders.

We showed that miR-eQTLs overlap with gene expression QTLs and protein QTLs of their target genes, supporting their role in translational repression. Since target genes tend to be clustered to miRNAs according to their function (34,35), these shared miR-eQTLs might have biological relevance. Cis-miR-eQTLs that overlap with trans-mRNA-eQTLs might point to the downstream regulatory effect from miRNAs to their (direct or indirect) target genes. When cis-mRNA-eQTLs overlap with trans-miR-eQTLs, the effect might be going from the genes to miRNAs, pointing to bidirectional interaction between miRNAs and target genes as a feedback mechanism (36,37). However, when trans-miR-eQTLs overlap with trans-mRNA-eQTLs, a third factor may have contributed to simultaneous changes in miRNA and gene expression. As an example, a genetic variant could affect the regulatory region shared between miRNA and a gene that are co-expressed. We also hypothesise that cis- and trans-miR-eQTLs might have different clinical relevance. The magnitude of associations between miRNAs and complex traits appeared closer to the null when trans-miR-eQTLs were added as instruments. Trans-miR-eQTLs might affect the stability of mature miRNAs, whereas cis-miR-eQTLs influence the hairpin structure and regulate the expression of primary miRNAs (9).

The current analysis identified co-expression of miRNAs and host genes, which could occur through modification of promoter activity, chromatin accessibility, transcription factor binding, or DNA methylation (10). However, many miRNAs also have their own promoters (38), and the association could be independent of the host genes (10,39). This finding deepens our understanding that the relationship between miRNAs and gene expressions is more commonly driven by genetics (40). The genetic effect might be less strong for miRNAs than mRNAs (12,13), as shown by the small variation explained by miR-eQTLs, which could act as a mechanism to maintain biological function during evolution.

Given that each miRNA potentially regulates multiple target genes and pathways (1,2,5), even small changes in miRNA expression could result in considerable consequences. This concept aligns with the strong evolutionary constraint on miRNAs and their binding sites in gene 3-UTRs in humans and other species (5). Moreover, the seed, mature, and precursor regions of miRNA genes are known to have a lower density of genetic variation than the whole genome (41). Our study shows that the genetic variants in those regions could have functional importance, such as affecting miRNA expression. This functional consequence occurs by interfering with the processing of precursor to mature miRNA or the interaction between mature miRNA and target genes, resulting in gain and loss of function, which could deregulate biological pathways (42,43).

Human miRNAs can be categorised into families with similar functions due to their conserved structures in the mature or seed sequences (44) and clusters when they are encoded from the same region in our genome (3). Here, we showed that the 14q32 miRNA cluster shares cis-regulatory variants. We also showed that multiple miRNAs are regulated by shared miR-eQTLs (45), such as the pleiotropic trans-miR-eQTLs in the *ABO* gene. Several families sharing trans-regulatory variants in *ABO*, such as miR-10 family, miR-30 family, let-7 family, and miR-139-5p, were well-known in cardiometabolic traits and cancers (46-48). This finding agrees with the concept that several miRNAs can work in networks to control gene expression and pathways underlying diseases (49).

Several associations with complex traits highlighted in this study were reported in the literature. For example, miR-543 was released in plasma following a high-fat diet (50), which could be a physiological response to reduce the risk of obesity. Target genes of miR-329-3p were involved in lipid and glucose metabolism in rats (51). Low miR-329 expression was observed in melanoma cells, while miR-329 mimics could suppress the progression of melanoma (52). The effect in tumour tissue for miR-329 and miR-1908-5p (14,52) was opposite compared to our MR analysis which better captures the lifetime effect of miRNAs. This suggests the changes in the level of miRNAs in tumour tissue might be the consequence of disease processes and supports the hypothesis that the dysregulation of miRNA in diseased tissue might arise from negative feedback by downstream genes (36,37). It is also possible that the genetic effects have been buffered by canalisation (19), where people with a genetically higher level of miRNAs since the intra-uterine period might be resistant to the effect of higher miRNAs throughout life.

Here we would like to underline several aspects to be considered when attempting to replicate miR-eQTLs across studies. First, we found fewer trans were replicated than cis-miR-eQTLs, as observed in the large eQTL analysis (53). Trans-eQTLs are known to have weaker effects, are less replicable, and are more tissue-specific (54-56) than cis-eQTLs. Second, the concordant direction with those reported by Nikpay et al. (9) suggested that the type of biological sample and profiling method could have an effect. The lower replication rate in the Framingham Heart Study is likely due to differences in type of sample (whole blood vs plasma), as previously reported (57). Third, one should consider any systematic difference in participants’ characteristics across studies. This study came from a population-based cohort which makes the findings more generalisable. Other studies were in obese individuals (9) or enriched for a specific disease (11), making it particularly useful for investigating the relevant disease but not for a wide range of complex traits and disorders. Finally, since the overall proportion of variation explained by each miR-eQTL is relatively small, larger GWAS for miR-eQTLs identification will be a valuable resource to enrich the genetic studies on miRNAs. In particular, incorporating diverse ancestries could generate more transferrable findings for a wider population.

Collectively, the integration of genomics, molecular, and clinical data in this study has provided a better understanding of the genetic regulation of miRNAs and allowed us to perform a systematic investigation on the effect of perturbations of plasma miRNA levels on a wide range of clinical conditions. As an example, we highlight miR-543 and miR-329-3p associated with obesity-related traits with potential downstream targets. Although it is unlikely a single miRNA or its target genes will be entirely responsible for the disease mechanisms, it is plausible that the effect of identified miRNAs to be mediated at least in part through those target genes. Our approach allows generating testable hypotheses for further functional and clinical studies to dissect the underlying molecular and cellular pathways of various traits and diseases.

The summary statistics for miR-eQTLs identified in our study and their link to other omics layers and their associations with various clinical outcomes will be available through a web tool called miRNomics Atlas (www.mirnomicsatlas.com). This web tool allows the use of genetic association data of miR-eQTLs, serving as valuable resources for future research to decipher the association and causal role of miRNAs in human diseases and their regulatory pathways.

## Online Methods

### Cohort description

The Rotterdam Study (RS) is a large prospective population-based cohort study among middle-aged and elderly in the suburb Ommoord in Rotterdam, the Netherlands. In 1990, 7,983 inhabitants aged 55 years old and older were recruited to participate in the first cohort (RS-I). In 2000, the study was extended with a second cohort of 3,011 participants (RS-II) who became 55 years old or moved into the study district since the beginning of the study. In 2006, a further extension of the cohort (RS-III) was initiated, including 3,932 participants aged 45– 54 years. In 2016, the recruitment of another extension started (RS-IV), targeting participants aged 40 years and over, adding 3,005 new participants. A detailed description of the Rotterdam Study can be found elsewhere (20).

### Circulatory miRNA levels

Plasma miRNA levels were determined using the HTG EdgeSeq miRNA Whole Transcriptome Assay (WTA) to quantitatively detect the expression of 2,083 human miRNAs transcripts (HTG Molecular Diagnostics, Tuscon, AZ, USA) and using the Illumina NextSeq 500 sequencer (Illumina, San Diego, CA, USA). This method characterises miRNA expression patterns and measures the expression of 13 housekeeping genes to allow flexibility during data normalisation and analysis. Quantification of miRNA expression was based on counts per million (CPM). Log2 transformation of CPM was used as standardisation and adjustment for total reads within each sample. MiRNAs with log2 CPM <1.0 were indicated as not expressed in the samples.

### Genotype data

At baseline, blood was drawn for genotyping from 6,291 participants in RS-I, 2,157 in RS-II, and 2,654 in RS-IV. Genotyping in RS-I-II was performed using the HumanHap550 Duo BeadChip (Illumina, San Diego, California) for RS-I-II and the Global Screening Array (GSAMD-v3) Illumina array for RS-IV. Samples with a call rate below 97.5%, gender mismatches, excess autosomal heterozygosity, duplicates or family relations, and ethnic outliers were excluded. Variants with call rates below 95.0%, failing missingness test, Hardy‐Weinberg equilibrium P<10^-6^, and allele frequency below 1% were removed. Genotypes were imputed using the MaCH/minimac software to the 1000 Genomes phase I version 3 reference panel or phase 3 version 5 reference panels (for RS-IV). Genetic variants with minor allele frequency < 0.05 and imputation quality < 0.7 were filtered out after genotype imputation.

### Genome-wide association studies

Genome-wide association studies (GWAS) were conducted for 2,083 miRNAs in 2,178 participants randomly selected from three sub-cohorts of the Rotterdam Study to identify miRNA-expression quantitative trait loci (miR-eQTLs). Given the high number of miRNAs, GWAS was performed within the high-dimensional analysis framework (HASE) to reduce the computational burden and enable efficient implementation of GWAS on thousands of phenotypes (58). Multiple linear regression was used to test for association between each genetic variant and miRNA level, with miRNA level as the outcome and expected genotype count from imputation as predictors, with adjustment for age, sex, sub-cohort, and the first five principal components to account for population stratification.

We used the genome-wide threshold of P<5×10^-08^ and Bonferroni-corrected for 2,083 miRNAs (P<2.4×10^-11^) to identify significant associations. Associations reaching significance in the Rotterdam Study were taken forward for replication in a published miR-eQTLs study by Nikpay et al. (9) using the SNPs or their proxy SNPs (r^2^>0.7 within 500kb on either side of lead SNP position) obtained using LDlinkR (59). Linkage disequilibrium (LD) pruning was used to identify the number of independent SNPs for each miRNA (r^2^<0.01). Similarly, associations identified in previous GWAS by Nikpay et al. (at P<2.4×10^-11^) (9) and Huan et al. in the Framingham Heart Study (at FDR<0.1) (10) were also tested for replication. The Bonferroni threshold was used for replication (α<0.05/n, where n is the total number of SNP-miRNA pairs after pruning). Replication was defined when the associations between SNP and miRNA were Bonferroni-significant in an independent cohort with a concordant direction of effect.

### Functional annotation of miR-eQTLs

Genomic coordinates of miRNAs were extracted from miRBase v20 (ftp://mirbase.org/pub/mirbase/20/genomes/has.gff3) (3). Both SNPs and mature miRNA positions were based on Genome Reference Consortium Human Build 37 (GRCh37). The position of each miR-eQTL was mapped as cis or trans with respect to the miRNA position. SNPs located ±500kb upstream and downstream of the start position of mature miRNAs were identified as cis, and those located more than ±500kb away were identified as trans. To identify SNPs in seed, mature, or precursor genes of miRNA, the database was downloaded from http://bioinfo.life.hust.edu.cn/miRNASNP/#!/download (60).

The web-based tool Functional Mapping and Annotation (FUMA) was used to annotate identified miR-eQTLs. A detailed description of the FUMA workflow is described elsewhere (21). Independent significant miR-eQTLs were defined as those with P<5×10^-08^ in the discovery GWAS or those replicated in independent cohorts and moderate LD with each other at r^2^<0.6. LD calculation was referenced based on the 1000 Genomes phase 3 panel. These SNPs were further clumped to lead SNPs (r^2^ <0.1). Genomic risk loci were then defined based on the lead SNPs when they overlap with a maximum distance of 250kb between LD blocks. The major histocompatibility complex (MHC) region was excluded using the default region between *MOG* and *COL11A2* genes (21,61).

### Heritability analysis

The SNP-based heritability estimates for 2,083 circulatory miRNAs were obtained using massively expedited genome-wide heritability analysis (MEGHA) (22). A genetic relationship matrix was constructed from 1000 Genome imputed genotypes filtered on imputation quality (<0.5) and allele frequency (<0.1) using GCTA (62). After applying a stringent cut-off of 0.025 for genetic relatedness, 1,506 individuals were used for heritability estimation. Using MEGHA, the genetic relationship matrix, and age and sex as covariates, we computed the heritability and uncertainty (p-values based on 1000 permutations).

### Cross-phenotype and quantitative trait loci look-up

Cross-phenotype and quantitative trait loci look-up leveraged replicated miR-eQTLs with summary statistics for gene expression (eQTLs), protein-QTLs (pQTLs), metabolite-QTLs (met-QTLs), and complex traits. Summary for cis and trans-eQTLs analysis in whole blood were downloaded from https://www.eqtlgen.org/index.html (23). Summary statistics for pQTLs were from https://www.phpc.cam.ac.uk/ceu/proteins/ (24) and the SCALLOP consortium available through https://zenodo.org/record/2615265/ (25). Summary statistics for met-QTLs were from Metabolon (http://metabolomics.helmholtz-muenchen.de/gwas/) (26) and Nightingale from a published study (http://www.computationalmedicine.fi/data/NMR_GWAS/) (27) and in the UK Biobank available through OpenGWAS project (https://gwas.mrcieu.ac.uk) (28). The database for intragenic miRNAs was from https://bmi.ana.med.uni-muenchen.de/miriad/ (63). GWAS Catalog was downloaded from https://www.ebi.ac.uk/gwas/docs/file-downloads (29).

### Phenome-wide association studies

To investigate associations between genetically determined circulatory miRNA and a wide range of clinical diagnoses, a phenome-wide association study (PheWAS) was performed using hospital episode statistics data in the UK Biobank, a large prospective cohort study with over 500,000 individuals aged 40-69 years old recruited between 2006-2010 (17). In brief, participants with genotype and phenotype data were considered in the analysis. Quality control steps taken in the UK Biobank has been described elsewhere. Our analysis was restricted to participants who identified themselves as “White”. One from each pair of relatives and withdrawn individuals as of August 2021 were excluded (Extended Data Fig. 3). ICD (ninth and tenth editions) codes from the hospital episode statistics data were aligned into phecodes to identify clinically related phenotypes. The analysis was limited to phecodes with at least 200 cases to allow sufficient power for MR analysis (64). PheWAS was conducted using the PheWAS package in R (65).

For each miRNA, associations for genetic variants residing in 500kb on either side of the miRNA position (cis-SNPs) were extracted. The false discovery rate (FDR) was calculated across all cis-SNPs for each miRNA, where those with FDR<0.1 were selected as cis instruments (18). Trans-SNPs associated with circulatory miRNA at P<5×10^-08^ were added as trans instruments in the sensitivity analysis. Instruments were filtered for F-statistics> 10 to avoid weak instrument bias (66). Linkage disequilibrium (LD) clumping for the instruments was conducted using a threshold of r^2^ < 0.1 and a window of 10,000 kb.

For miRNA with single cis miR-eQTL satisfying the instrument criteria, cis-SNP was used as the proxy for corresponding miRNA in single-variant PheWAS. For miRNAs with multiple independent miR-eQTLs, weighted genetic risk scores (GRS) were computed for individuals in the UK Biobank as the sum score of miRNA-increasing alleles of miR-eQTLs identified in the Rotterdam Study using effect sizes as their weights as implemented using PLINK (36). The weighted GRS was rescaled by subtracting GRS from its mean and dividing by its standard deviation to express the association per-SD of the miRNA-increasing allele.

In the main analysis, GRS for each miRNA (miRNA-GRS) was computed from cis-miR-eQTLs (cis-GRS). Additionally, trans-miR-eQTLs at genome-wide significant (P<5×10^-08^) were added, in an extended analysis, to validate findings from cis-GRS. Multiple logistic regression was performed in the UK Biobank for each miRNA-GRS with adjustment for age, sex, genotyping array, and the first five principal components to account for population stratification. Given each phecode is not independent of the other, the false discovery rate (FDR) was calculated for each miRNA-GRS to account for multiple testing (67).

### Mendelian randomisation

Following PheWAS, two-sample Mendelian randomisation (MR) analysis was conducted to assess the causal relationship between candidate miRNAs and outcomes of interest identified from PheWAS. MR-PheWAS considered miRNAs with three or more independent instruments to enable performing robust MR methods as sensitivity analysis. The same set of genetic instruments used in PheWAS contributed to the exposure data in MR-PheWAS. The level of each candidate miRNA was rescaled by subtracting the value from its mean and dividing by its standard deviation (SD) to express the association per-SD increase of the miRNA level. The genetic association between the instruments and the outcome was taken from the UK Biobank. Associations that were significant at FDR<0.05 from MR-PheWAS were taken forward for validation using genetic association estimates (outcome data) from large GWAS consortia or by adding genome-wide significant trans-miR-eQTLs in an extended-MR.

The multiplicative random effect inverse variance weighted method (IVW) was used in the main analysis to combine the effect estimates of the genetic instruments assuming all instruments are valid (68). FDR adjustment was calculated for each miRNA based on p-values of MR-IVW since it was considered the most powerful method when all instruments are valid (68). Robust MR methods which allow the inclusion of pleiotropic variants were used as a sensitivity analysis, including weighted median (WM) or MR-Egger (69-71). WM estimate is valid if less than half of the weight of the genetic instrument is free from horizontal pleiotropy. MR-Egger does not force the regression line through an intercept of zero, making it statistically inefficient but provides a causal estimate corrected for directional horizontal pleiotropy. A non-null intercept in MR-Egger indicates evidence of pleiotropy (70). The agreement among different MR methods was examined to support a robust estimation of causal effects.

Since a liberal LD threshold (r^2^<0.1) was used for clumping, a further sensitivity analysis was conducted by incorporating the correlation matrix between genetic instruments in the fixed effect IVW method. MRPRESSO was used to detect outliers (72) and MR analysis was repeated after excluding outliers. Results for MR analysis using different MR methods were presented as forest plots. For replicated associations, reverse MR was conducted to assess the directionality of associations. Independent genetic instruments for complex traits (r^2^<0.001) were identified from large GWAS consortia for the outcomes of interest. Associations between the genetic instruments for complex traits with candidate miRNA levels were extracted from the Rotterdam Study.

### Target genes and enrichment analysis

To identify putative target genes for miRNAs, predicted and validated target genes of miRNAs were retrieved from TargetScan v7.2 and miRTarBase (4,5). For enrichment analysis, genes with predicted miRNA-target interaction (MTI) in TargetScan or validated MTI, including weak and strong validation methods in miRTaRBase, were considered. SNPs reaching genome-wide significance in large consortia GWAS for significant traits were mapped into protein-coding genes where they reside. Of those, predicted and validated target genes of miRNAs were identified. The enrichment analysis was conducted to test if the target genes of candidate miRNAs are enriched for the associated traits, as described in our previous work (34). Enrichment analysis was performed separately for predicted and validated target genes.

## Supporting information

All Supplementary Tables 1-28

## Data Availability

All data produced in the present study are available upon reasonable request to the authors. Requests should be directed towards the management team of the Rotterdam Study, which has a protocol for approving data requests.

## Acknowledgements

We would like to thank all participants of the Rotterdam Study and the UK Biobank. The Rotterdam Study is supported by the Erasmus MC University Medical Center and Erasmus University Rotterdam; The Netherlands Organisation for Scientific Research (NWO); The Netherlands Organisation for Health Research and Development (ZonMw); the Research Institute for Diseases in the Elderly (RIDE); The Netherlands Genomics Initiative (NGI); the Ministry of Education, Culture and Science; the Ministry of Health, Welfare and Sports; the European Commission (DG XII); and the Municipality of Rotterdam. The UK Biobank has approval from the North-West Multi-centre Research Ethics Committee (MREC) as a Research Tissue Bank (RTB) approval. Explicit informed consent was obtained from all participants when they enrolled in the UK Biobank. Access to the UK Biobank was provided through application 52569. This work was enabled by the computing resources and support from the Imperial College Research Computing Service and Erasmus MC. We thank Loukas Zagkos for helping with the visualisation of the results.

RM is supported by the President’s PhD Scholarship from Imperial College London. AD is funded by a Wellcome Trust seed award (206046/Z/17/Z). This project is partly supported by the Erasmus MC Fellowship (EMCF20213) and Alzheimer Nederland (WE.03-2021-10) grants of MG. PE acknowledges support from the Medical Research Council (MR/S019669/1) for the MRC Centre for Environment and Health, the British Heart Foundation (RE/18/4/34215) for the Imperial BHF Centre for Research Excellence, the UK Dementia Research Institute (MC_PC_17114) and the National Institute for Health Research Imperial College Biomedical Research Centre for infrastructure support.

## Code availability

For statistical analyses, we used the following software High-Dimensional Analysis Framework (HASE) (https://github.com/roshchupkin/hase), R version 3.6.3 (https://www.r-project.org), Plink v1.9 (https://www.cog-genomics.org/plink/), FUMA v1.4.1 (https://fuma.ctglab.nl/).

## Extended Data Figures

**Extended Data Fig. 1.**
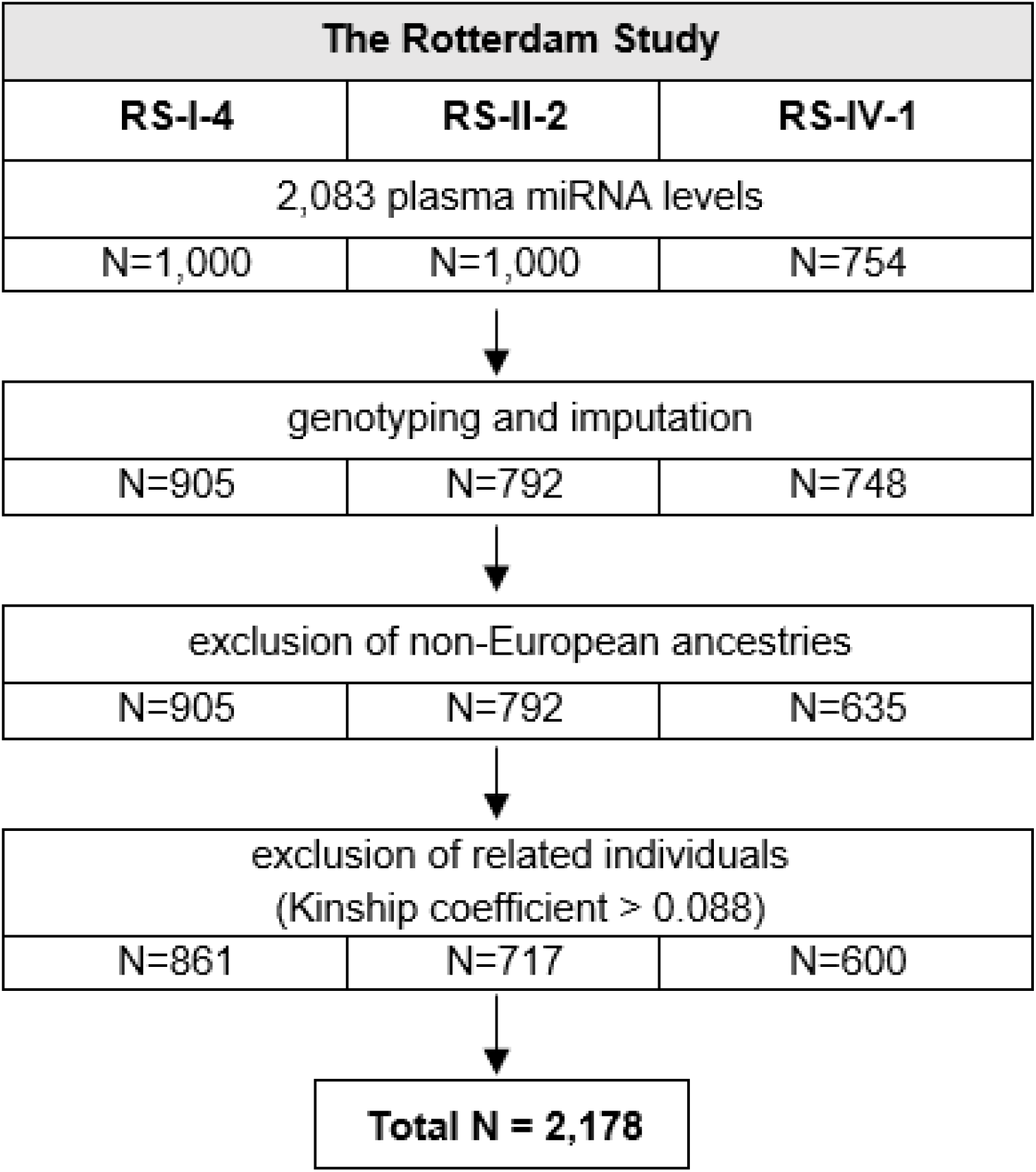
Selection of study participants in the Rotterdam Study.

**Extended Data Fig. 2.**
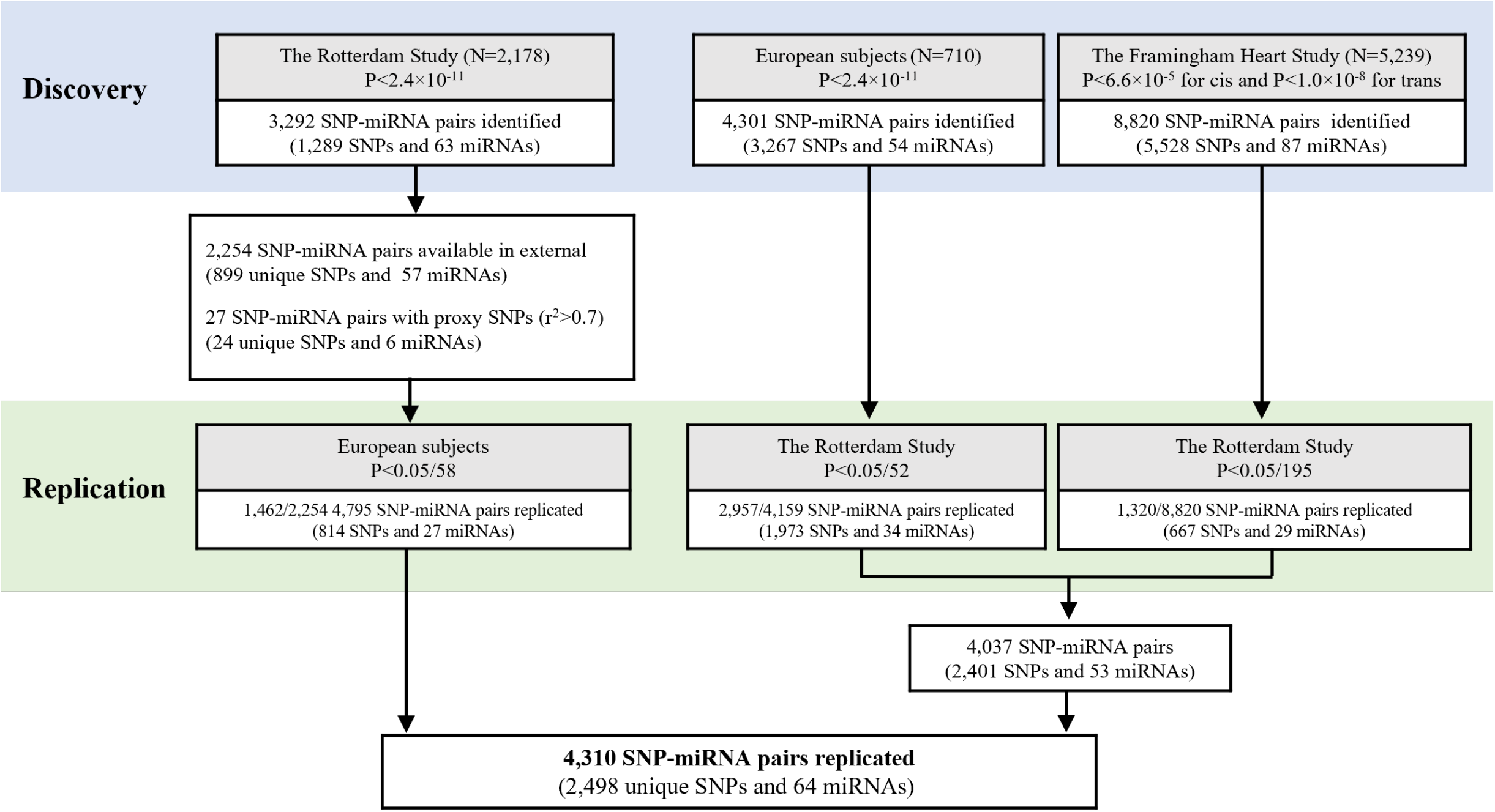
Identification of miR-eQTLs and replication in independent cohorts. In total, 3,292 significant associations were discovered for 63 miRNAs. Of those 1,462 out of 2,254 associations available in Nikpay et al. (9) were replicated for 27 miRNAs (P<0.05/58). On the other hand, 2,957 associations identified by Nikpay et al. (9) for 34 miRNAs were replicated (P<0.05/52) and 1,320 associations for 29 miRNAs identified in the Framingham Heart Study (10) were also replicated (P<0.05/195). Collectively, 4,310 associations for 64 miRNAs were successfully replicated across studies.

**Extended Data Fig. 3.**
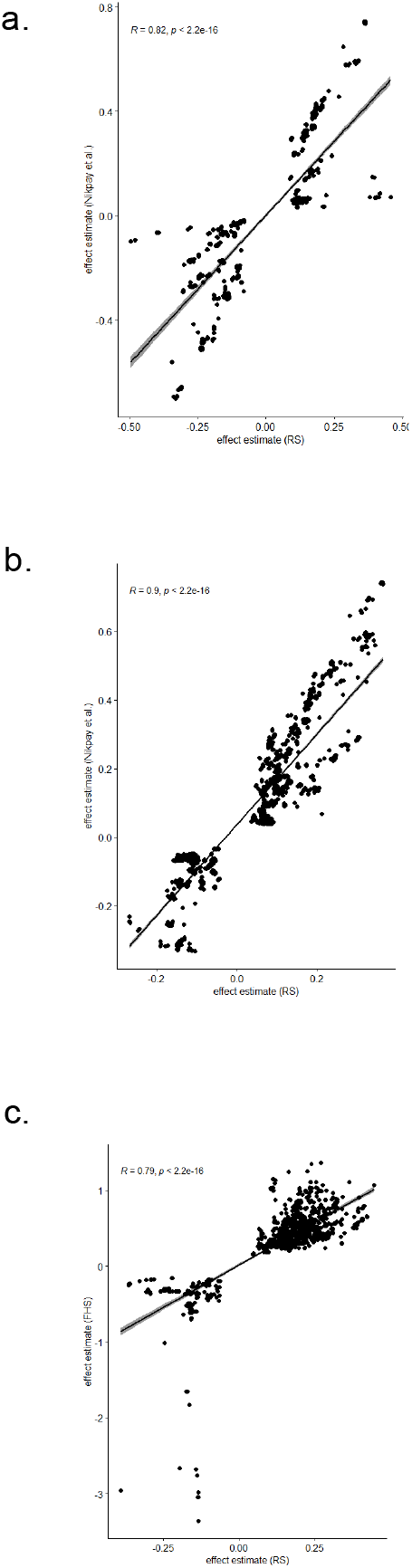
Correlation of effect estimates between discovery and replication of miR-eQTLs. a. Associations in the Rotterdam Study that were replicated in Nikpay et al.(9). b. Associations in Nikpay et al. (9) that were replicated in the Rotterdam Study. c. Associations in the Framingham Study that were replicated in the Rotterdam Study.

**Extended Data Fig. 4.**
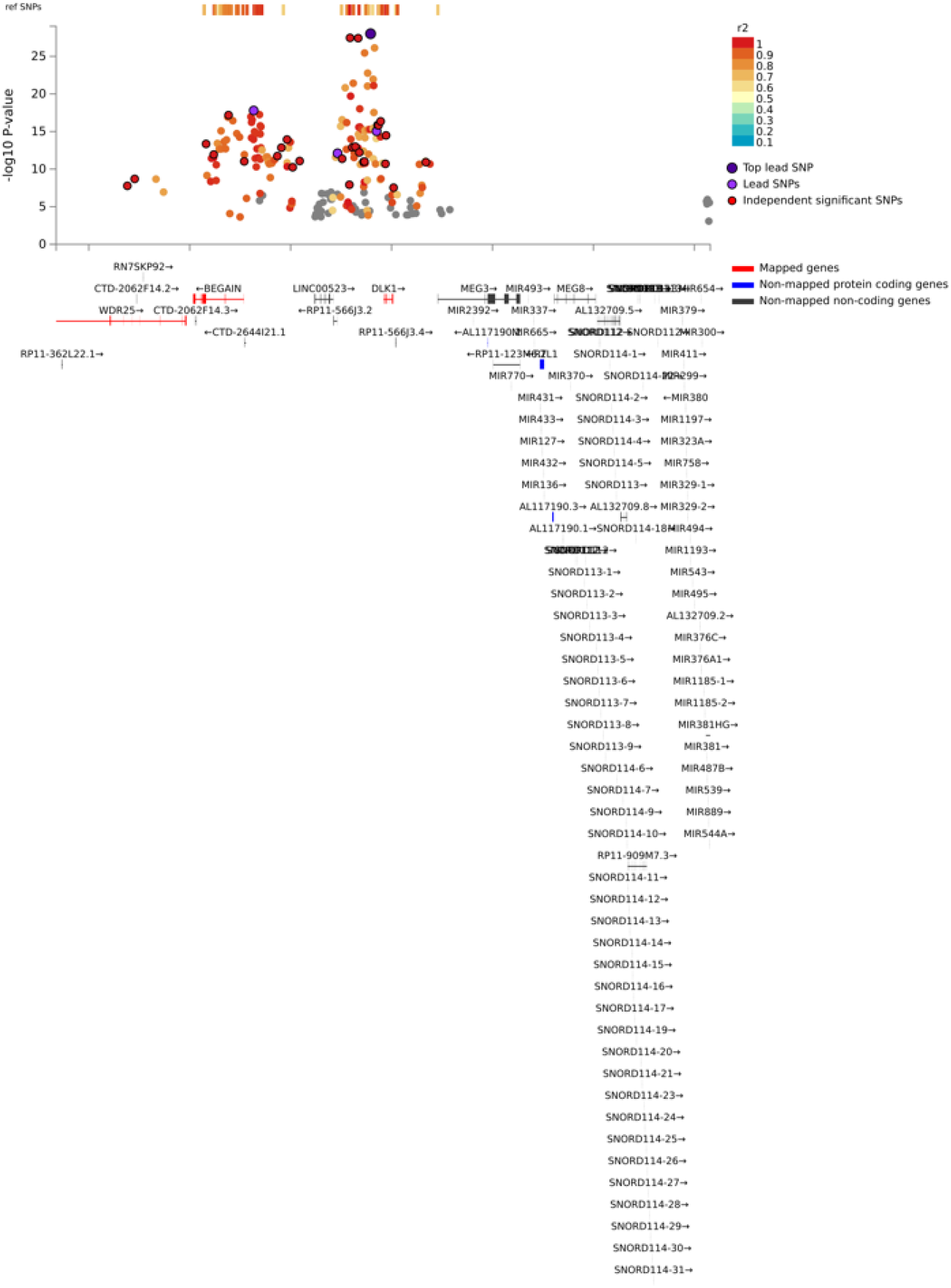
Regional plot for genomic risk loci in chr 14:100655022-101244293 harbouring cis-miR-eQTLs for 31 miRNAs that are clustered together. Plot was extracted from FUMA.

**Extended Data Fig. 5.**
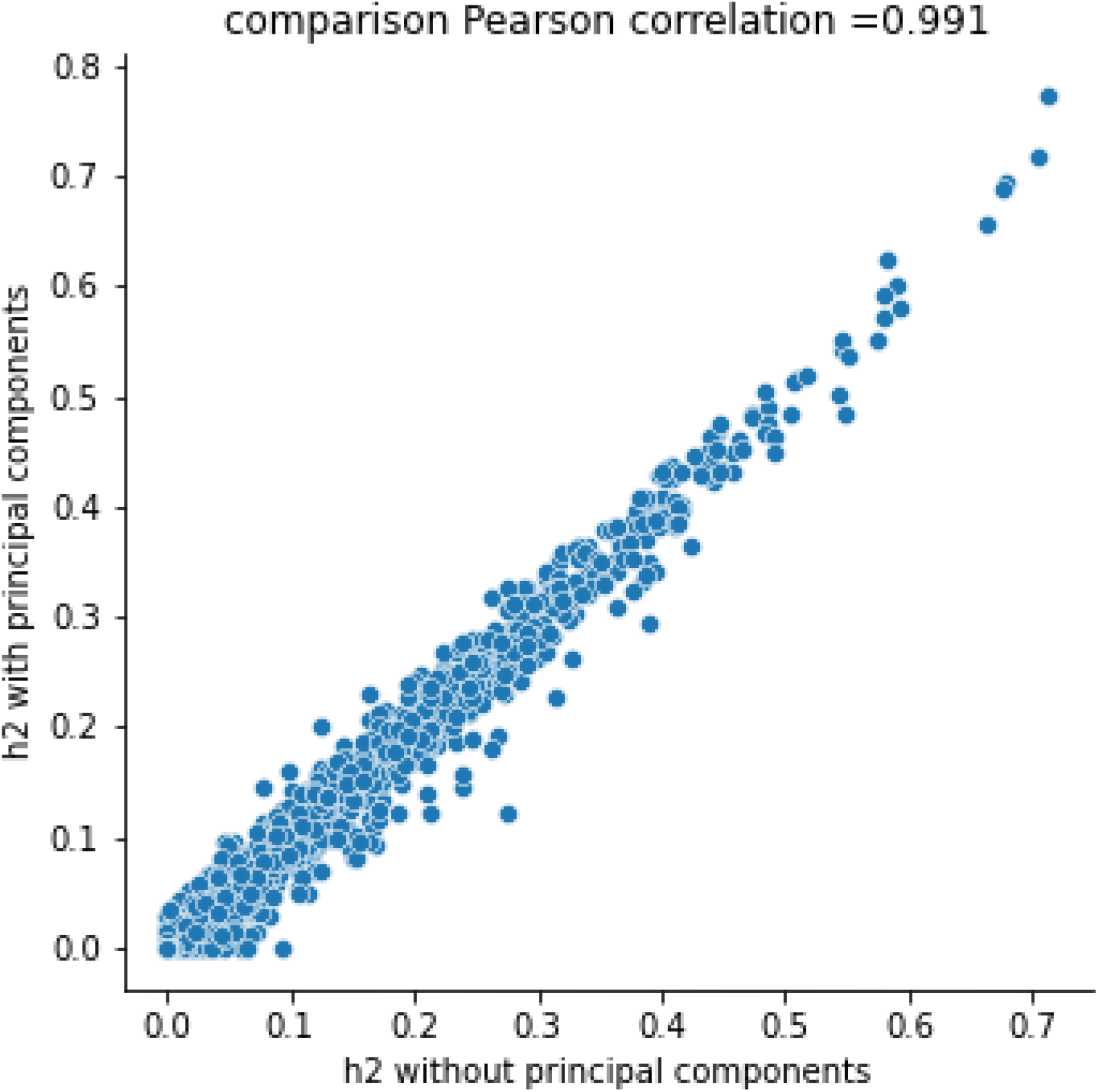
Pearson correlation of heritability estimates with and without principal components.

**Extended Data Fig. 6.**
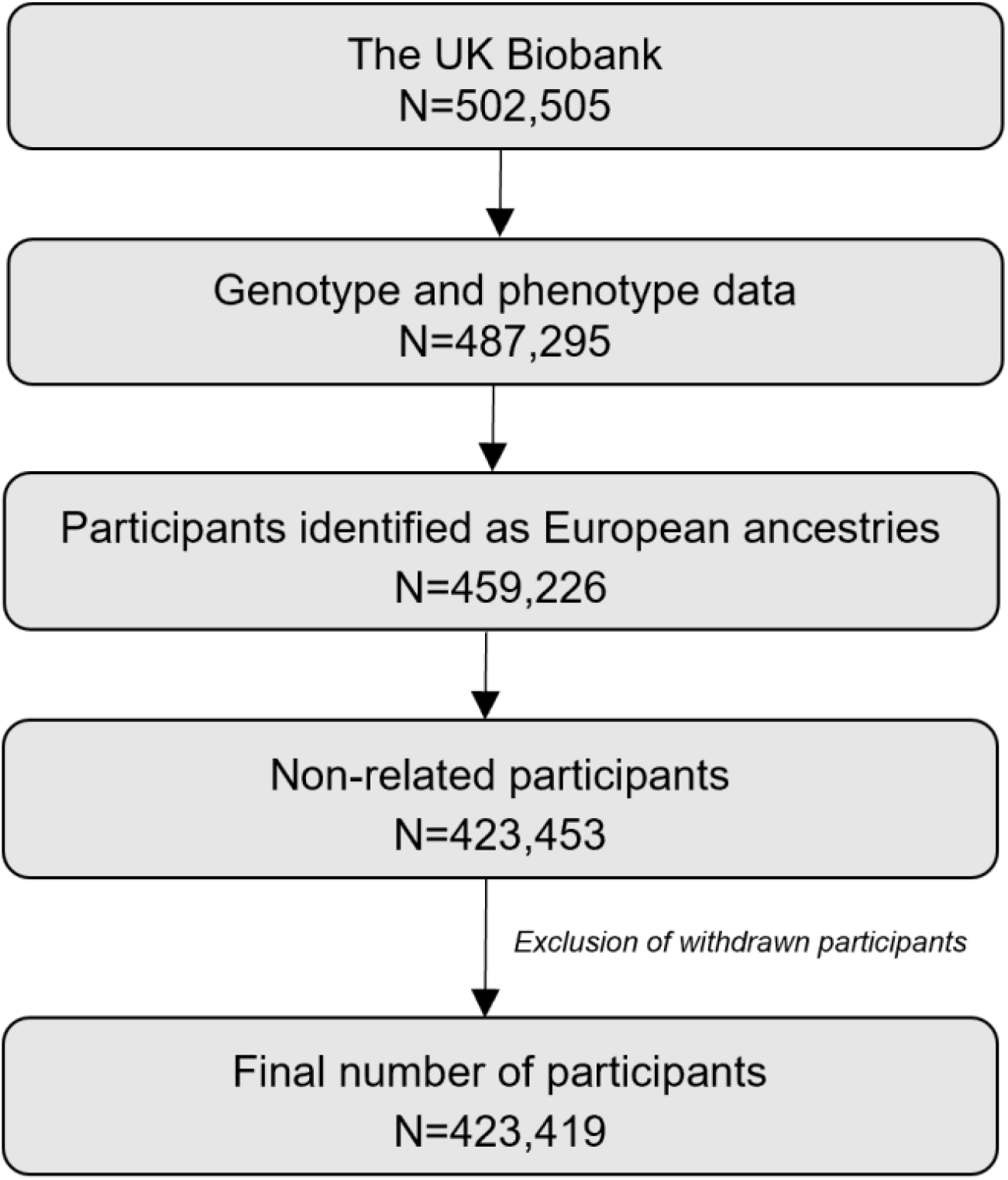
Selection of participants for PheWAS and MR-PheWAS in the UK Biobank.

**Extended Data Fig. 7.**
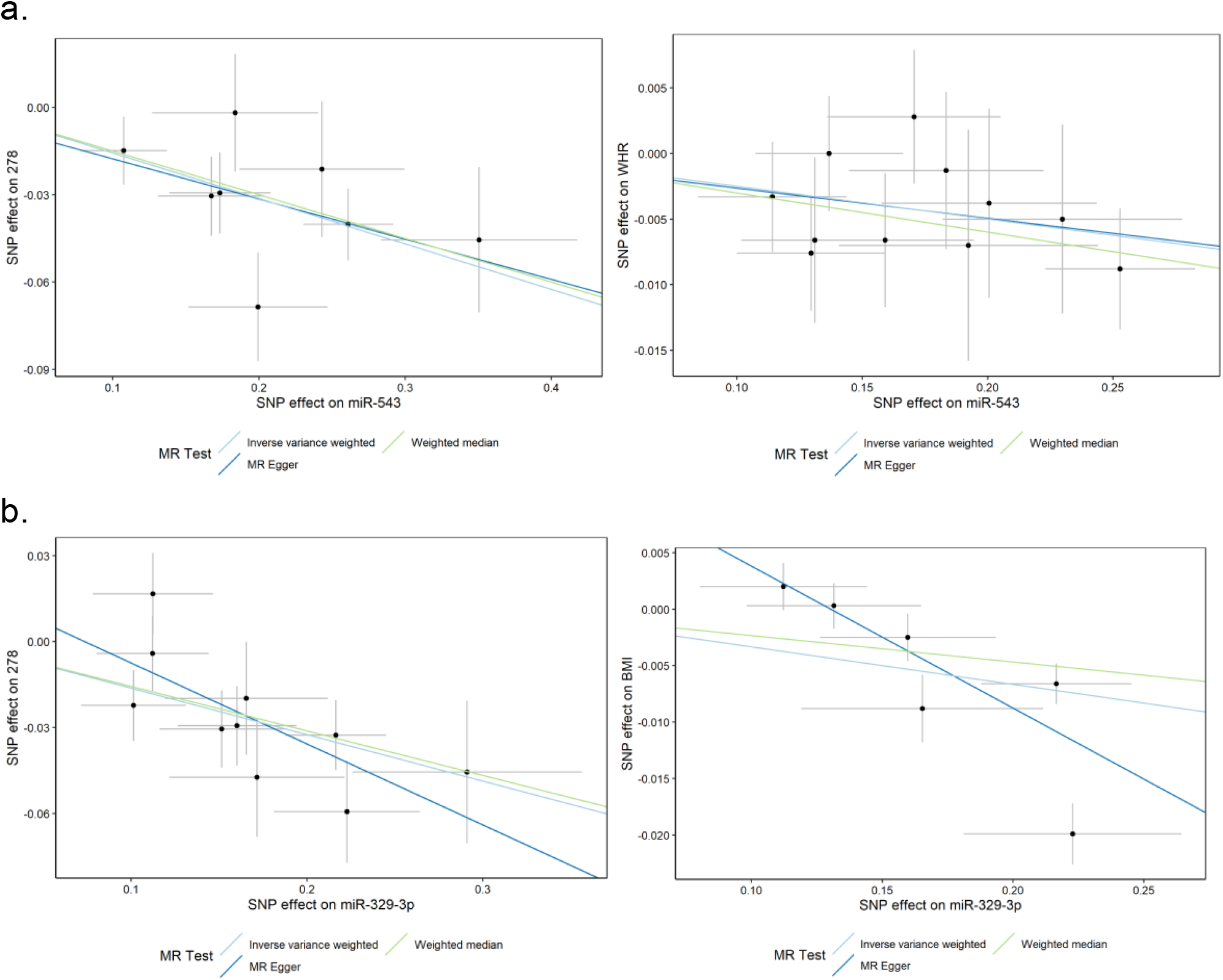
Scatter plots for MR-PheWAS and replication MR (bottom). a. miR-543 and obesity (left) and waist to hip ratio (right). b. miR-329-3p and obesity (left) and body mass index (right).

