## Supplementary material for "An atlas of genetic regulation and disease associations of microRNAs": All Supplementary Tables 1-28

| Tables | Title |
|---|---|
| **ST1** | List of 2083 miRNAs characterised in this study |
| **ST2** | Baseline characteristics of study participants |
| **ST3** | Genome-wide significant miR-eQTLs identified in this study |
| **ST4** | miR-eQTLs explaining high proportion of variation in miRNA level |
| **ST5** | Replicated miR-eQTLs in Nikpay et al |
| **ST6** | Summary of replication of miR-eQTLS |
| **ST7** | List of 4,310 replicated miR-eQTLs |
| **ST8** | Functional annotation of miR-eQTLs |
| **ST9** | List of 22 loci, miRNAs, and corresponding genes |
| **ST10** | SNPs in miRNA genes that also affected miRNA levels |
| **ST11** | SNP-based heritability estimates for 2,083 miRNAs |
| **ST12** | Overlap between cis-miR-eQTLs and gene expression QTLs (eQTLs) |
| **ST13** | miRNAs that shared cis-eQTLs with their host genes |
| **ST14** | miRNAs with shared eQTLs with their putative target genes |
| **ST15** | Overlap between cis-miR-eQTLs and pQTLs |
| **ST16** | Overlap between trans-miR-eQTLs and pQTLs |
| **ST17** | Overlap between trans-miR-eQTLs and pQTLs in Olink |
| **ST18** | Overlap between miR-eQTLs and met-QTLs |
| **ST19** | GWAS traits associated with miR-eQTLs |
| **ST20** | Association between miR-eQTLs in chr9:136128546-136296530 and other omics |
| **ST21** | Single cis-miR-eQTLs as proxy for 85 miRNAs in PheWAS |
| **ST22** | FDR-significant associations for PheWAS using single variant and cis-GRS |
| **ST23** | Cis and extended-GRS for 17 significant associations in MR-PheWAS |
| **ST24** | Full results for cis-MR analyses |
| **ST25** | Full results for extended-MR analyses |
| **ST26** | Full results for replication using large GWAS summary statistics |
| **ST27** | Reverse-MR to test association of obesity traits with miR-543 and miR-329-3p |
| **ST28** | Target genes of miR-543 and miR-329-3p associated with BMI or WHR |

**Table S1. List of 2083 miRNAs characterised in this study**

| | | | | | | | | | | |
|---|---|---|---|---|---|---|---|---|---|---|
| miRNA_ID | miR-1468-5p | miR-27b-3 | miR-3617- | miR-4288 | miR-4653- | miR-492 | miR-5585- | miR-660-3 | miR-6827- | miR-8060 |
| let-7a-2-3p | miR-146a-3p | miR-27b-5 | miR-3618 | miR-4289 | miR-4654 | miR-493-3 | miR-5586- | miR-660-5 | miR-6827- | miR-8061 |
| let-7a-3p | miR-146a-5p | miR-28-5p | miR-3619- | miR-429 | miR-4655- | miR-493-5 | miR-5586- | miR-661 | miR-6828- | miR-8062 |
| let-7a-5p | miR-146b-3p | miR-2861 | miR-3619- | miR-4290 | miR-4655- | miR-494-3 | miR-5587- | miR-662 | miR-6828- | miR-8063 |
| let-7b-5p | miR-146b-5p | miR-2909 | miR-362-3 | miR-4291 | miR-4656 | miR-494-5 | miR-5588- | miR-663b | miR-6829- | miR-8065 |
| let-7c-3p | miR-1471 | miR-296-3 | miR-362-5 | miR-4292 | miR-4657 | miR-495-3 | miR-5589- | miR-664a- | miR-6830- | miR-8067 |
| let-7c-5p | miR-147a | miR-296-5 | miR-3620- | miR-4293 | miR-4658 | miR-495-5 | miR-5589- | miR-664a- | miR-6830- | miR-8068 |
| let-7d-3p | miR-147b | miR-297 | miR-363-3 | miR-4294 | miR-4659a | miR-496 | miR-559 | miR-664b- | miR-6831- | miR-8069 |
| let-7d-5p | miR-148a-3p | miR-298 | miR-363-5 | miR-4295 | miR-4659b | miR-497-3 | miR-5590- | miR-664b- | miR-6831- | miR-8070 |
| let-7e-3p | miR-148b-3p | miR-299-3 | miR-3646 | miR-4296 | miR-466 | miR-498 | miR-561-3 | miR-668-3 | miR-6832- | miR-8071 |
| let-7e-5p | miR-149-3p | miR-299-5 | miR-3648 | miR-4297 | miR-4660 | miR-4999- | miR-561-5 | miR-668-5 | miR-6832- | miR-8074 |
| let-7f-1-3p | miR-149-5p | miR-29a-3 | miR-3649 | miR-4299 | miR-4661- | miR-499b- | miR-562 | miR-670-3 | miR-6833- | miR-8076 |
| let-7f-2-3p | miR-150-3p | miR-29a-5 | miR-3650 | miR-4300 | miR-4662a | miR-5000- | miR-563 | miR-670-5 | miR-6833- | miR-8077 |
| let-7f-5p | miR-150-5p | miR-29b-1 | miR-3652 | miR-4301 | miR-4663 | miR-5001- | miR-564 | miR-671-3 | miR-6834- | miR-8078 |
| let-7g-3p | miR-151a-3p | miR-29b-2 | miR-3653 | miR-4302 | miR-4664- | miR-5001- | miR-566 | miR-671-5 | miR-6834- | miR-8079 |
| let-7g-5p | miR-152-3p | miR-29b-3 | miR-3654 | miR-4303 | miR-4664- | miR-5002- | miR-567 | miR-6715b | miR-6835- | miR-8080 |
| let-7i-5p | miR-153-3p | miR-29c-3 | miR-3655 | miR-4304 | miR-4665- | miR-5002- | miR-568 | miR-6716- | miR-6835- | miR-8081 |
| miR-1 | miR-153-5p | miR-29c-5 | miR-3657 | miR-4305 | miR-4666a | miR-5003- | miR-5680 | miR-6716- | miR-6836- | miR-8082 |
| miR-100-3p | miR-1537-3p | miR-300 | miR-3658 | miR-4306 | miR-4666b | miR-5003- | miR-5682 | miR-6717- | miR-6837- | miR-8083 |
| miR-100-5p | miR-1537-5p | miR-301a- | miR-3659 | miR-4307 | miR-4667- | miR-5004- | miR-5684 | miR-6718- | miR-6837- | miR-8084 |
| miR-101-3p | miR-1538 | miR-301a- | miR-365a- | miR-4308 | miR-4667- | miR-5004- | miR-5685 | miR-6719- | miR-6838- | miR-8085 |
| miR-101-5p | miR-1539 | miR-301b | miR-365a- | miR-4309 | miR-4668- | miR-5006- | miR-5687 | miR-6720- | miR-6839- | miR-8086 |
| miR-103a-2-5p | miR-154-3p | miR-302a- | miR-365b- | miR-431-3 | miR-4668- | miR-5006- | miR-5688 | miR-6720- | miR-6840- | miR-8087 |
| miR-103a-3p | miR-154-5p | miR-302a- | miR-3660 | miR-431-5 | miR-4669 | miR-5007- | miR-5689 | miR-6721- | miR-6840- | miR-8088 |
| miR-105-3p | miR-155-3p | miR-302b- | miR-3661 | miR-4310 | miR-4670- | miR-5007- | miR-569 | miR-6722- | miR-6841- | miR-8089 |
| miR-105-5p | miR-155-5p | miR-302b- | miR-3662 | miR-4311 | miR-4671- | miR-5008- | miR-5690 | miR-6722- | miR-6841- | miR-873-3p |
| miR-106a-3p | miR-1587 | miR-302c- | miR-3663- | miR-4312 | miR-4671- | miR-5009- | miR-5691 | miR-6723- | miR-6842- | miR-873-5p |
| miR-106a-5p | miR-15a-3p | miR-302c- | miR-3664- | miR-4314 | miR-4672 | miR-5009- | miR-5692a | miR-6726- | miR-6842- | miR-874-3p |
| miR-106b-3p | miR-15a-5p | miR-302d- | miR-3666 | miR-4315 | miR-4673 | miR-500a- | miR-5692b | miR-6726- | miR-6843- | miR-874-5p |
| miR-106b-5p | miR-15b-3p | miR-302d- | miR-3667- | miR-4317 | miR-4675 | miR-500a- | miR-5692c | miR-6727- | miR-6844 | miR-875-3p |
| miR-107 | miR-15b-5p | miR-302e | miR-3667- | miR-4318 | miR-4676- | miR-500b- | miR-5693 | miR-6728- | miR-6845- | miR-875-5p |
| miR-10a-3p | miR-16-1-3p | miR-302f | miR-3668 | miR-4319 | miR-4677- | miR-500b- | miR-5694 | miR-6728- | miR-6846- | miR-876-3p |
| miR-10a-5p | miR-16-5p | miR-3064- | miR-367-3 | miR-432-3 | miR-4677- | miR-501-3 | miR-5695 | miR-6729- | miR-6846- | miR-876-5p |
| miR-10b-3p | miR-17-3p | miR-3064- | miR-367-5 | miR-432-5 | miR-4678 | miR-501-5 | miR-5696 | miR-6730- | miR-6847- | miR-877-3p |
| miR-10b-5p | miR-17-5p | miR-30a-3 | miR-3670 | miR-4320 | miR-4679 | miR-5010- | miR-5697 | miR-6731- | miR-6847- | miR-877-5p |
| miR-1178-3p | miR-181a-2-3p | miR-30a-5 | miR-3671 | miR-4322 | miR-4680- | miR-5010- | miR-5698 | miR-6731- | miR-6848- | miR-885-3p |
| miR-1178-5p | miR-181a-3p | miR-30b-3 | miR-3672 | miR-4324 | miR-4680- | miR-5011- | miR-5699- | miR-6733- | miR-6848- | miR-885-5p |
| miR-1179 | miR-181a-5p | miR-30b-5 | miR-3674 | miR-4325 | miR-4681 | miR-502-3 | miR-5700 | miR-6733- | miR-6849- | miR-887-3p |
| miR-1180-3p | miR-181b-3p | miR-30c-1 | miR-3675- | miR-4326 | miR-4682 | miR-502-5 | miR-5701 | miR-6734- | miR-6849- | miR-887-5p |
| miR-1180-5p | miR-181b-5p | miR-30c-2 | miR-3675- | miR-4327 | miR-4683 | miR-503-3 | miR-5702 | miR-6734- | miR-6851- | miR-888-3p |
| miR-1182 | miR-181c-3p | miR-30c-5 | miR-3677- | miR-4328 | miR-4684- | miR-503-5 | miR-5703 | miR-6735- | miR-6851- | miR-888-5p |
| miR-1183 | miR-181c-5p | miR-30d-3 | miR-3678- | miR-4329 | miR-4684- | miR-504-3 | miR-5704 | miR-6735- | miR-6852- | miR-889-3p |
| miR-1184 | miR-181d-3p | miR-30d-5 | miR-3679- | miR-433-3 | miR-4685- | miR-504-5 | miR-5705 | miR-6736- | miR-6852- | miR-889-5p |
| miR-1185-1-3p | miR-181d-5p | miR-30e-3 | miR-3679- | miR-433-5 | miR-4685- | miR-5047 | miR-5706 | miR-6736- | miR-6853- | miR-890 |
| miR-1185-2-3p | miR-182-3p | miR-30e-5 | miR-3680- | miR-4330 | miR-4686 | miR-505-3 | miR-5707 | miR-6737- | miR-6853- | miR-891a-3p |
| miR-1185-5p | miR-182-5p | miR-31-5p | miR-3680- | miR-4417 | miR-4687- | miR-505-5 | miR-571 | miR-6737- | miR-6854- | miR-891a-5p |
| miR-1193 | miR-183-3p | miR-3115 | miR-3681- | miR-4419a | miR-4688 | miR-506-3 | miR-572 | miR-6738- | miR-6854- | miR-891b |
| miR-1197 | miR-183-5p | miR-3116 | miR-3682- | miR-4419b | miR-4689 | miR-506-5 | miR-573 | miR-6738- | miR-6855- | miR-892a |
| miR-1199-3p | miR-184 | miR-3117- | miR-3682- | miR-4420 | miR-4690- | miR-507 | miR-5739 | miR-6739- | miR-6855- | miR-892b |
| miR-1200 | miR-185-3p | miR-3118 | miR-3683 | miR-4421 | miR-4690- | miR-508-3 | miR-574-3 | miR-6739- | miR-6856- | miR-892c-3p |
| miR-1202 | miR-185-5p | miR-3119 | miR-3684 | miR-4422 | miR-4691- | miR-508-5 | miR-574-5 | miR-6740- | miR-6856- | miR-892c-5p |
| miR-1204 | miR-186-3p | miR-3121- | miR-3685 | miR-4423- | miR-4691- | miR-5087 | miR-575 | miR-6740- | miR-6857- | miR-9-3p |
| miR-1205 | miR-186-5p | miR-3122 | miR-3686 | miR-4423- | miR-4692 | miR-5088- | miR-576-3 | miR-6741- | miR-6857- | miR-9-5p |
| miR-1206 | miR-187-3p | miR-3123 | miR-3687 | miR-4424 | miR-4693- | miR-5088- | miR-576-5 | miR-6741- | miR-6859- | miR-920 |
| miR-1207-3p | miR-187-5p | miR-3124- | miR-3688- | miR-4425 | miR-4693- | miR-5089- | miR-577 | miR-6742- | miR-6859- | miR-921 |
| miR-1207-5p | miR-188-3p | miR-3124- | miR-3689a | miR-4426 | miR-4694- | miR-509-3 | miR-578 | miR-6742- | miR-6861- | miR-922 |
| miR-1208 | miR-188-5p | miR-3125 | miR-3689a | miR-4427 | miR-4695- | miR-509-3 | miR-579-3 | miR-6744- | miR-6862- | miR-924 |
| miR-122-5p | miR-18a-3p | miR-3126- | miR-3689b | miR-4428 | miR-4695- | miR-509-5 | miR-579-5 | miR-6744- | miR-6862- | miR-92a-1-5p |
| miR-1224-5p | miR-18a-5p | miR-3126- | miR-3689d | miR-4429 | miR-4696 | miR-5090 | miR-580-3 | miR-6745 | miR-6863 | miR-92a-2-5p |
| miR-1225-3p | miR-18b-3p | miR-3127- | miR-3689f | miR-4430 | miR-4697- | miR-5091 | miR-580-5 | miR-6746- | miR-6864- | miR-92a-3p |
| miR-1225-5p | miR-18b-5p | miR-3127- | miR-369-3 | miR-4431 | miR-4697- | miR-5093 | miR-581 | miR-6747- | miR-6864- | miR-92b-3p |
| miR-1226-3p | miR-1908-5p | miR-3128 | miR-369-5 | miR-4432 | miR-4698 | miR-5094 | miR-582-5 | miR-6747- | miR-6865- | miR-92b-5p |
| miR-1227-3p | miR-1909-3p | miR-3129- | miR-3690 | miR-4433b | miR-4699- | miR-510-3 | miR-583 | miR-6748- | miR-6866- | miR-93-3p |
| miR-1228-3p | miR-1909-5p | miR-3130- | miR-3691- | miR-4434 | miR-4699- | miR-510-5 | miR-584-3 | miR-6748- | miR-6866- | miR-93-5p |
| miR-1229-3p | miR-190a-3p | miR-3131 | miR-3691- | miR-4435 | miR-4700- | miR-511-3 | miR-584-5 | miR-675-3 | miR-6867- | miR-933 |
| miR-1229-5p | miR-190a-5p | miR-3132 | miR-3692- | miR-4436a | miR-4701- | miR-511-5 | miR-585-3 | miR-675-5 | miR-6867- | miR-934 |

| | | | | | | | | | | |
|---|---|---|---|---|---|---|---|---|---|---|
| miR-1231 | miR-190b | miR-3133 | miR-3692- | miR-4436b | miR-4701- | miR-512-3 | miR-586 | miR-6750- | miR-6868- | miR-935 |
| miR-1233-3p | miR-191-3p | miR-3134 | miR-370-3 | miR-4437 | miR-4703- | miR-512-5 | miR-587 | miR-6750- | miR-6868- | miR-936 |
| miR-1234-3p | miR-191-5p | miR-3135a | miR-370-5 | miR-4438 | miR-4704- | miR-513a- | miR-588 | miR-6751- | miR-6869- | miR-937-3p |
| miR-1236-3p | miR-1910-3p | miR-3136- | miR-3713 | miR-4439 | miR-4704- | miR-513a- | miR-589-3 | miR-6751- | miR-6870- | miR-937-5p |
| miR-1236-5p | miR-1910-5p | miR-3137 | miR-3714 | miR-4440 | miR-4705 | miR-513b- | miR-589-5 | miR-6752- | miR-6870- | miR-938 |
| miR-1237-3p | miR-1911-3p | miR-3138 | miR-371b- | miR-4441 | miR-4706 | miR-513b- | miR-590-3 | miR-6753- | miR-6871- | miR-939-5p |
| miR-1237-5p | miR-1911-5p | miR-3140- | miR-371b- | miR-4442 | miR-4707- | miR-513c- | miR-590-5 | miR-6754- | miR-6872- | miR-941 |
| miR-1238-3p | miR-1912 | miR-3140- | miR-372-3 | miR-4444 | miR-4707- | miR-513c- | miR-591 | miR-6754- | miR-6872- | miR-942-3p |
| miR-1238-5p | miR-1913 | miR-3141 | miR-372-5 | miR-4446- | miR-4709- | miR-514a- | miR-592 | miR-6755- | miR-6873- | miR-943 |
| miR-124-3p | miR-1914-3p | miR-3142 | miR-373-3 | miR-4446- | miR-4709- | miR-514a- | miR-593-3 | miR-6755- | miR-6873- | miR-944 |
| miR-124-5p | miR-1914-5p | miR-3143 | miR-373-5 | miR-4447 | miR-4711- | miR-514b- | miR-593-5 | miR-6756- | miR-6874- | miR-95-3p |
| miR-1243 | miR-1915-3p | miR-3144- | miR-374a- | miR-4449 | miR-4712- | miR-514b- | miR-595 | miR-6757- | miR-6874- | miR-95-5p |
| miR-1244 | miR-1915-5p | miR-3144- | miR-374b- | miR-4450 | miR-4713- | miR-515-3 | miR-597-3 | miR-6757- | miR-6875- | miR-96-3p |
| miR-1245b-3p | miR-192-3p | miR-3145- | miR-374b- | miR-4451 | miR-4713- | miR-515-5 | miR-597-5 | miR-6758- | miR-6876- | miR-96-5p |
| miR-1247-3p | miR-192-5p | miR-3145- | miR-374c- | miR-4452 | miR-4714- | miR-516a- | miR-598-3 | miR-6758- | miR-6877- | miR-98-3p |
| miR-1247-5p | miR-193a-3p | miR-3146 | miR-375 | miR-4453 | miR-4718 | miR-516a- | miR-598-5 | miR-6759- | miR-6877- | miR-99a-5p |
| miR-1249 | miR-193a-5p | miR-3147 | miR-376a- | miR-4455 | miR-4719 | miR-516b- | miR-600 | miR-6759- | miR-6878- | miR-99b-3p |
| miR-1250-3p | miR-193b-3p | miR-3148 | miR-376a- | miR-4456 | miR-4720- | miR-517-5 | miR-601 | miR-676-3 | miR-6878- | miR-99b-5p |
| miR-1250-5p | miR-193b-5p | miR-3149 | miR-376a- | miR-4457 | miR-4720- | miR-517a- | miR-604 | miR-676-5 | miR-6880-5p | |
| miR-1251-3p | miR-194-3p | miR-3150b | miR-376b- | miR-4458 | miR-4721 | miR-517c- | miR-605-3 | miR-6760- | miR-6881-3p | |
| miR-1252-3p | miR-194-5p | miR-3152- | miR-376b- | miR-4459 | miR-4722- | miR-5186 | miR-605-5 | miR-6760- | miR-6881-5p | |
| miR-1252-5p | miR-195-3p | miR-3152- | miR-376c- | miR-4460 | miR-4722- | miR-5187- | miR-606 | miR-6761- | miR-6882-3p | |
| miR-1253 | miR-195-5p | miR-3153 | miR-376c- | miR-4461 | miR-4723- | miR-5187- | miR-6069 | miR-6761- | miR-6882-5p | |
| miR-1254 | miR-196a-3p | miR-3154 | miR-377-3 | miR-4462 | miR-4723- | miR-5188 | miR-607 | miR-6762- | miR-6883-3p | |
| miR-1255b-2-3p | miR-196a-5p | miR-3155a | miR-377-5 | miR-4463 | miR-4724- | miR-5189- | miR-6071 | miR-6764- | miR-6883-5p | |
| miR-1256 | miR-196b-3p | miR-3155b | miR-378a- | miR-4464 | miR-4725- | miR-5189- | miR-6072 | miR-6764- | miR-6884-3p | |
| miR-1257 | miR-196b-5p | miR-3156- | miR-378b | miR-4465 | miR-4725- | miR-518b | miR-6073 | miR-6765- | miR-6884-5p | |
| miR-1258 | miR-197-3p | miR-3157- | miR-378c | miR-4467 | miR-4726- | miR-518c- | miR-6075 | miR-6766- | miR-6885-3p | |
| miR-125a-3p | miR-197-5p | miR-3157- | miR-378d | miR-4468 | miR-4726- | miR-5190 | miR-6076 | miR-6766- | miR-6885-5p | |
| miR-125a-5p | miR-1976 | miR-3158- | miR-378e | miR-4470 | miR-4727- | miR-5191 | miR-6078 | miR-6767- | miR-6886-3p | |
| miR-125b-1-3p | miR-198 | miR-3159 | miR-378f | miR-4471 | miR-4727- | miR-5192 | miR-6079 | miR-6767- | miR-6886-5p | |
| miR-125b-2-3p | miR-199a-3p | miR-3161 | miR-378g | miR-4472 | miR-4728- | miR-5193 | miR-608 | miR-6768- | miR-6887-3p | |
| miR-125b-5p | miR-199a-5p | miR-3162- | miR-378h | miR-4473 | miR-4728- | miR-5194 | miR-6080 | miR-6768- | miR-6887-5p | |
| miR-126-3p | miR-199b-5p | miR-3163 | miR-378i | miR-4474- | miR-4729 | miR-5195- | miR-6081 | miR-6769a | miR-6888-3p | |
| miR-126-5p | miR-19a-3p | miR-3164 | miR-378j | miR-4475 | miR-4730 | miR-5195- | miR-6082 | miR-6769a | miR-6888-5p | |
| miR-1261 | miR-19a-5p | miR-3165 | miR-379-3 | miR-4476 | miR-4731- | miR-5196- | miR-6083 | miR-6769b | miR-6889-3p | |
| miR-1262 | miR-19b-1-5p | miR-3166 | miR-379-5 | miR-4477a | miR-4731- | miR-5196- | miR-6085 | miR-6770- | miR-6889-5p | |
| miR-1263 | miR-19b-2-5p | miR-3167 | miR-380-3 | miR-4477b | miR-4732- | miR-5197- | miR-6086 | miR-6770- | miR-6890-3p | |
| miR-1264 | miR-19b-3p | miR-3169 | miR-380-5 | miR-4478 | miR-4732- | miR-5197- | miR-6088 | miR-6771- | miR-6890-5p | |
| miR-1265 | miR-200a-3p | miR-3170 | miR-381-3 | miR-448 | miR-4733- | miR-519a- | miR-609 | miR-6771- | miR-6891-3p | |
| miR-1267 | miR-200a-5p | miR-3171 | miR-381-5 | miR-4480 | miR-4734 | miR-519b- | miR-610 | miR-6772- | miR-6892-3p | |
| miR-1269a | miR-200b-3p | miR-3174 | miR-382-3 | miR-4481 | miR-4735- | miR-519c- | miR-612 | miR-6772- | miR-6892-5p | |
| miR-1269b | miR-200b-5p | miR-3175 | miR-382-5 | miR-4482- | miR-4736 | miR-519d- | miR-6124 | miR-6773- | miR-6893-3p | |
| miR-127-3p | miR-200c-3p | miR-3176 | miR-383-3 | miR-4482- | miR-4737 | miR-519e- | miR-6126 | miR-6773- | miR-6893-5p | |
| miR-127-5p | miR-200c-5p | miR-3177- | miR-383-5 | miR-4483 | miR-4738- | miR-519e- | miR-6127 | miR-6774- | miR-6894-5p | |
| miR-1270 | miR-202-3p | miR-3177- | miR-384 | miR-4484 | miR-4738- | miR-520a- | miR-6128 | miR-6774- | miR-6895-3p | |
| miR-1271-5p | miR-202-5p | miR-3179 | miR-3907 | miR-4486 | miR-4742- | miR-520b | miR-6129 | miR-6775- | miR-6895-5p | |
| miR-1272 | miR-203b-3p | miR-3180 | miR-3908 | miR-4487 | miR-4742- | miR-520c- | miR-613 | miR-6775- | miR-7-1-3p | |
| miR-1273a | miR-203b-5p | miR-3180- | miR-3909 | miR-4489 | miR-4743- | miR-520d- | miR-6130 | miR-6776- | miR-7-2-3p | |
| miR-1273c | miR-204-3p | miR-3182 | miR-3910 | miR-4490 | miR-4743- | miR-520e | miR-6131 | miR-6776- | miR-7-5p | |
| miR-1273d | miR-204-5p | miR-3184- | miR-3912- | miR-4491 | miR-4744 | miR-520f- | miR-6132 | miR-6778- | miR-708-5p | |
| miR-1273e | miR-205-3p | miR-3185 | miR-3912- | miR-4493 | miR-4745- | miR-520g- | miR-6133 | miR-6778- | miR-7106-3p | |
| miR-1273g-5p | miR-205-5p | miR-3186- | miR-3913- | miR-4494 | miR-4746- | miR-521 | miR-6134 | miR-6779- | miR-7106-5p | |
| miR-1273h-5p | miR-2052 | miR-3186- | miR-3914 | miR-4495 | miR-4747- | miR-522-3 | miR-614 | miR-6779- | miR-7107-3p | |
| miR-1275 | miR-2053 | miR-3187- | miR-3915 | miR-4496 | miR-4747- | miR-523-3 | miR-615-3 | miR-6780a | miR-7107-5p | |
| miR-1276 | miR-2054 | miR-3187- | miR-3916 | miR-4498 | miR-4748 | miR-525-5 | miR-616-3 | miR-6780a | miR-7108-5p | |
| miR-1277-5p | miR-206 | miR-3188 | miR-3917 | miR-4499 | miR-4750- | miR-532-3 | miR-616-5 | miR-6780b | miR-7109-3p | |
| miR-1278 | miR-208a-3p | miR-3189- | miR-3919 | miR-449a | miR-4751 | miR-532-5 | miR-6165 | miR-6780b | miR-7109-5p | |
| miR-1279 | miR-208a-5p | miR-3189- | miR-3920 | miR-449b- | miR-4752 | miR-539-3 | miR-617 | miR-6781- | miR-711 | |
| miR-128-1-5p | miR-208b-3p | miR-3191- | miR-3921 | miR-449b- | miR-4753- | miR-539-5 | miR-618 | miR-6781- | miR-7111-3p | |
| miR-128-2-5p | miR-208b-5p | miR-3191- | miR-3922- | miR-449c- | miR-4753- | miR-541-3 | miR-619-3 | miR-6782- | miR-7111-5p | |
| miR-128-3p | miR-20a-3p | miR-3192- | miR-3922- | miR-449c- | miR-4756- | miR-541-5 | miR-620 | miR-6782- | miR-7112-3p | |
| miR-1282 | miR-20a-5p | miR-3192- | miR-3923 | miR-4500 | miR-4758- | miR-542-3 | miR-621 | miR-6783- | miR-7113-5p | |
| miR-1283 | miR-20b-3p | miR-3193 | miR-3924 | miR-4501 | miR-4759 | miR-542-5 | miR-622 | miR-6783- | miR-7114-3p | |
| miR-1285-5p | miR-20b-5p | miR-3197 | miR-3925- | miR-4502 | miR-4760- | miR-543 | miR-623 | miR-6784- | miR-7114-5p | |
| miR-1287-3p | miR-21-3p | miR-3198 | miR-3926 | miR-4503 | miR-4760- | miR-544a | miR-624-3 | miR-6785- | miR-7150 | |
| miR-1287-5p | miR-21-5p | miR-3199 | miR-3927- | miR-4504 | miR-4761- | miR-544b | miR-624-5 | miR-6786- | miR-7151-3p | |

| | | | | | | | | | |
|---|---|---|---|---|---|---|---|---|---|
| miR-1288-3p | miR-210-3p | miR-32-3p | miR-3928- | miR-4505 | miR-4761- | miR-545-3 | miR-625-5 | miR-6787- | miR-7151-5p |
| miR-1289 | miR-210-5p | miR-32-5p | miR-3928- | miR-4506 | miR-4762- | miR-545-5 | miR-626 | miR-6787- | miR-7152-3p |
| miR-129-1-3p | miR-211-3p | miR-3200- | miR-3929 | miR-4507 | miR-4763- | miR-548ab | miR-627-3 | miR-6788- | miR-7152-5p |
| miR-129-2-3p | miR-211-5p | miR-3201 | miR-3934- | miR-4509 | miR-4764- | miR-548ad | miR-628-5 | miR-6789- | miR-7153-3p |
| miR-129-5p | miR-2110 | miR-3202 | miR-3934- | miR-450a- | miR-4764- | miR-548ae | miR-629-5 | miR-6789- | miR-7153-5p |
| miR-1290 | miR-2113 | miR-320a | miR-3935 | miR-450a- | miR-4765 | miR-548ag | miR-630 | miR-6790- | miR-7154-3p |
| miR-1291 | miR-2114-3p | miR-320b | miR-3936 | miR-450a- | miR-4766- | miR-548ak | miR-631 | miR-6791- | miR-7154-5p |
| miR-1292-5p | miR-2114-5p | miR-320c | miR-3937 | miR-450b- | miR-4766- | miR-548al | miR-632 | miR-6791- | miR-7155-5p |
| miR-1293 | miR-2115-3p | miR-320d | miR-3938 | miR-450b- | miR-4768- | miR-548an | miR-633 | miR-6792- | miR-7156-3p |
| miR-1294 | miR-2115-5p | miR-320e | miR-3939 | miR-4510 | miR-4769- | miR-548ao | miR-634 | miR-6792- | miR-7156-5p |
| miR-1295b-3p | miR-2116-3p | miR-323a- | miR-3940- | miR-4511 | miR-4769- | miR-548ao | miR-635 | miR-6793- | miR-7157-3p |
| miR-1296-3p | miR-2116-5p | miR-323a- | miR-3941 | miR-4512 | miR-4770 | miR-548ar | miR-637 | miR-6794- | miR-7157-5p |
| miR-1296-5p | miR-2117 | miR-323b- | miR-3942- | miR-4513 | miR-4771 | miR-548as | miR-640 | miR-6794- | miR-7158-3p |
| miR-1297 | miR-212-3p | miR-323b- | miR-3943 | miR-4514 | miR-4772- | miR-548au | miR-641 | miR-6795- | miR-7159-3p |
| miR-1298-3p | miR-212-5p | miR-324-3 | miR-3944- | miR-4515 | miR-4773 | miR-548av | miR-642a- | miR-6796- | miR-7159-5p |
| miR-1298-5p | miR-214-5p | miR-324-5 | miR-3945 | miR-4517 | miR-4774- | miR-548aw | miR-642b- | miR-6796- | miR-7160-3p |
| miR-1299 | miR-215-3p | miR-325 | miR-3973 | miR-4518 | miR-4774- | miR-548ax | miR-643 | miR-6797- | miR-7160-5p |
| miR-1301-3p | miR-215-5p | miR-326 | miR-3974 | miR-4519 | miR-4775 | miR-548ay | miR-644a | miR-6797- | miR-7161-3p |
| miR-1301-5p | miR-216a-3p | miR-328-3 | miR-3975 | miR-451a | miR-4777- | miR-548ay | miR-645 | miR-6798- | miR-7161-5p |
| miR-1302 | miR-216a-5p | miR-329-3 | miR-3976 | miR-451b | miR-4777- | miR-548az | miR-646 | miR-6799- | miR-7162-3p |
| miR-1303 | miR-216b-3p | miR-329-5 | miR-3977 | miR-452-3 | miR-4778- | miR-548b- | miR-647 | miR-6799- | miR-7162-5p |
| miR-1304-3p | miR-216b-5p | miR-330-3 | miR-3978 | miR-452-5 | miR-4779 | miR-548b- | miR-648 | miR-6800- | miR-744-3p |
| miR-1304-5p | miR-217 | miR-330-5 | miR-409-3 | miR-4520a | miR-4780 | miR-548ba | miR-649 | miR-6800- | miR-744-5p |
| miR-1305 | miR-218-1-3p | miR-331-3 | miR-409-5 | miR-4521 | miR-4781- | miR-548d- | miR-6499- | miR-6801- | miR-7515 |
| miR-1306-3p | miR-218-2-3p | miR-331-5 | miR-410-3 | miR-4522 | miR-4781- | miR-548e- | miR-6499- | miR-6801- | miR-758-3p |
| miR-1306-5p | miR-218-5p | miR-335-3 | miR-410-5 | miR-4524a | miR-4782- | miR-548e- | miR-650 | miR-6802- | miR-758-5p |
| miR-1307-3p | miR-219a-1-3p | miR-335-5 | miR-411-3 | miR-4524 | miR-4783- | miR-548g- | miR-6500- | miR-6802- | miR-759 |
| miR-1307-5p | miR-219a-2-3p | miR-337-3 | miR-411-5 | miR-4525 | miR-4784 | miR-548h- | miR-6500- | miR-6803- | miR-760 |
| miR-130a-3p | miR-219b-3p | miR-337-5 | miR-412-3 | miR-4527 | miR-4786- | miR-548j-3 | miR-6501- | miR-6803- | miR-761 |
| miR-130a-5p | miR-22-3p | miR-338-3 | miR-412-5 | miR-4528 | miR-4786- | miR-548k | miR-6501- | miR-6804- | miR-764 |
| miR-130b-3p | miR-22-5p | miR-339-3 | miR-421 | miR-4530 | miR-4788 | miR-548l | miR-6502- | miR-6805- | miR-765 |
| miR-130b-5p | miR-221-3p | miR-339-5 | miR-422a | miR-4531 | miR-4789- | miR-548m | miR-6502- | miR-6806- | miR-766-3p |
| miR-132-3p | miR-221-5p | miR-33a-3 | miR-424-3 | miR-4533 | miR-4789- | miR-548s | miR-6503- | miR-6806- | miR-766-5p |
| miR-132-5p | miR-222-3p | miR-33a-5 | miR-424-5 | miR-4534 | miR-4790- | miR-548t- | miR-6503- | miR-6807- | miR-767-3p |
| miR-1321 | miR-222-5p | miR-33b-3 | miR-425-3 | miR-4535 | miR-4790- | miR-548v | miR-6504- | miR-6807- | miR-767-5p |
| miR-1322 | miR-223-3p | miR-33b-5 | miR-425-5 | miR-4536- | miR-4791 | miR-548w | miR-6505- | miR-6808- | miR-769-3p |
| miR-1323 | miR-223-5p | miR-340-3 | miR-4251 | miR-4537 | miR-4793- | miR-548x- | miR-6505- | miR-6809- | miR-769-5p |
| miR-1324 | miR-224-3p | miR-340-5 | miR-4252 | miR-4538 | miR-4793- | miR-548y | miR-6506- | miR-6809- | miR-770-5p |
| miR-133a-3p | miR-224-5p | miR-342-3 | miR-4253 | miR-4539 | miR-4794 | miR-549a | miR-6506- | miR-6810- | miR-7702 |
| miR-133a-5p | miR-2276-3p | miR-342-5 | miR-4254 | miR-454-3 | miR-4795- | miR-550a- | miR-6507- | miR-6810- | miR-7703 |
| miR-133b | miR-2276-5p | miR-345-3 | miR-4255 | miR-454-5 | miR-4795- | miR-550a- | miR-6508- | miR-6811- | miR-7705 |
| miR-134-3p | miR-2278 | miR-345-5 | miR-4256 | miR-4540 | miR-4796- | miR-550b- | miR-6509- | miR-6811- | miR-7706 |
| miR-134-5p | miR-2355-3p | miR-346 | miR-4257 | miR-455-3 | miR-4798- | miR-551a | miR-651-3 | miR-6812- | miR-7843-3p |
| miR-1343-5p | miR-2355-5p | miR-34a-3 | miR-4259 | miR-455-5 | miR-4798- | miR-551b- | miR-651-5 | miR-6813- | miR-7843-5p |
| miR-135a-3p | miR-2392 | miR-34a-5 | miR-4261 | miR-4632- | miR-4799- | miR-551b- | miR-6510- | miR-6813- | miR-7844-5p |
| miR-135a-5p | miR-23a-3p | miR-34b-3 | miR-4262 | miR-4632- | miR-4800- | miR-552-3 | miR-6511a | miR-6814- | miR-7845-5p |
| miR-135b-3p | miR-23a-5p | miR-34b-5 | miR-4263 | miR-4636 | miR-4800- | miR-552-5 | miR-6511a | miR-6814- | miR-7846-3p |
| miR-135b-5p | miR-23b-3p | miR-34c-3 | miR-4264 | miR-4637 | miR-4801 | miR-553 | miR-6511 | miR-6815- | miR-7848-3p |
| miR-136-3p | miR-23b-5p | miR-34c-5 | miR-4265 | miR-4638- | miR-4802- | miR-554 | miR-6513- | miR-6815- | miR-7850-5p |
| miR-136-5p | miR-23c | miR-3605- | miR-4266 | miR-4639- | miR-4802- | miR-555 | miR-6513- | miR-6816- | miR-7851-3p |
| miR-137 | miR-24-2-5p | miR-3605- | miR-4267 | miR-4639- | miR-4803 | miR-556-3 | miR-6514- | miR-6817- | miR-7852-3p |
| miR-138-1-3p | miR-24-3p | miR-3606- | miR-4268 | miR-4640- | miR-4804- | miR-556-5 | miR-6514- | miR-6817- | miR-7853-5p |
| miR-138-2-3p | miR-2467-3p | miR-3606- | miR-4269 | miR-4641 | miR-484 | miR-557 | miR-6515- | miR-6818- | miR-7854-3p |
| miR-138-5p | miR-2467-5p | miR-3607- | miR-4270 | miR-4642 | miR-485-3 | miR-5571- | miR-6515- | miR-6818- | miR-7856-5p |
| miR-139-3p | miR-25-3p | miR-3607- | miR-4271 | miR-4643 | miR-485-5 | miR-5571- | miR-6516- | miR-6819- | miR-7973 |
| miR-139-5p | miR-25-5p | miR-3609 | miR-4272 | miR-4644 | miR-486-5 | miR-5572 | miR-6516- | miR-6819- | miR-7974 |
| miR-140-5p | miR-2681-3p | miR-361-3 | miR-4273 | miR-4645- | miR-487a- | miR-5579- | miR-652-3 | miR-6820- | miR-7976 |
| miR-141-3p | miR-2681-5p | miR-361-5 | miR-4274 | miR-4646- | miR-487a- | miR-558 | miR-652-5 | miR-6821- | miR-7978 |
| miR-141-5p | miR-2682-3p | miR-3611 | miR-4275 | miR-4646- | miR-487b- | miR-5580- | miR-653-3 | miR-6821- | miR-802 |
| miR-142-5p | miR-2682-5p | miR-3612 | miR-4276 | miR-4647 | miR-487b- | miR-5581- | miR-654-3 | miR-6822- | miR-8052 |
| miR-143-3p | miR-26a-1-3p | miR-3613- | miR-4277 | miR-4648 | miR-488-3 | miR-5581- | miR-654-5 | miR-6822- | miR-8053 |
| miR-143-5p | miR-26a-2-3p | miR-3613- | miR-4278 | miR-4649- | miR-488-5 | miR-5582- | miR-655-3 | miR-6823- | miR-8054 |
| miR-144-3p | miR-26a-5p | miR-3614- | miR-4279 | miR-4649- | miR-489-3 | miR-5582- | miR-655-5 | miR-6823- | miR-8055 |
| miR-144-5p | miR-26b-3p | miR-3614- | miR-4280 | miR-4650- | miR-489-5 | miR-5583- | miR-656-3 | miR-6824- | miR-8056 |
| miR-145-3p | miR-26b-5p | miR-3616- | miR-4282 | miR-4652- | miR-490-5 | miR-5584- | miR-656-5 | miR-6825- | miR-8057 |
| miR-145-5p | miR-27a-3p | miR-3616- | miR-4283 | miR-4652- | miR-491-3 | miR-5584- | miR-657 | miR-6825- | miR-8058 |
| miR-1468-3p | miR-27a-5p | miR-3617- | miR-4287 | miR-4653- | miR-491-5 | miR-5585- | miR-658 | miR-6826- | miR-8059 |

**Table S2. Baseline characteristics of study participants**

| Characteristics | Sub-cohort | | |
|---|---|---|---|
| | **RS-I-4** | **RS-II-2** | **RS-IV-1** |
| Number of participants | 861 | 717 | 600 |
| Male (%) | 358 (41.6) | 323 (45.0) | 276 (46.0) |
| Age (years), mean ± SD | 75.2± 6.0 | 68.2±7.4 | 57.37±11.6 |
| Total cholesterol (mmol/L), mean ± SD | 5.6±1.0 | 5.8±1.0 | 5.4±1.0 |
| HDL (mmol/L), mean ± SD | 1.5±0.4 | 1.5±0.4 | 1.5±0.5 |
| SBP (mmHg), mean ± SD | 151.6±21.0 | 144.5±19.1 | 125.5±16.6 |
| DBP (mmHg), mean ± SD | 79.0±11.2 | 79.7±10.6 | 78.9±10.1 |
| Blood pressure lowering medication (%) | 424 (49.3) | 258 (36.3) | NA |
| Prevalent diabetes, N (%) | 109 (13.7) | 88 (12.3) | NA |
| Prevalent coronary artery disease (%) | 117 (13.9) | 48 (6.7) | NA |
| Prevalent of heart failure (%) | 66 (7.7) | 19 (2.6) | NA |
| Prevalent of dementia (%) | 5 (0.6) | 2 (0.3) | NA |
| BMI (kg/m$^2$), mean ± SD | 27.5±4.1 | 27.8±4.1 | 27.66±4.8 |
| Waist hip ratio, mean ± SD | 0.9 ±0.1 | 0.9±0.1 | 0.88±0.1 |
| Smoking status | | | |
| Current smoker (%) | 116 (13.5) | 108 (15.1) | 102 (17.0) |
| Former smoker (%) | 475 (55.2) | 390 (54.4) | 263 (43.8) |
| Never smoker (%) | 255 (29.6) | 211 (29.4) | 235 (39.2) |
| Education status | | | NA |
| Primary education (%) | 110 (12.8) | 48 (6.7) | 36 (6.0) |
| Lower/intermediate general education OR lower vocational education (%) | 377 (43.8) | 340 (47.4) | 173 (28.8) |
| Intermediate vocational education OR higher general education (%) | 270 (31.4) | 195 (27.2) | 211 (35.2) |
| Higher vocational education OR university (%) | 98 (11.4) | 127 (17.7) | 180 (30.0) |
| Alcohol consumption (median, IQR) | 6.0 (16.4) | 8.6 (18.6) | NA |

**Table S3. Genome-wide significant miR-eQTLs identified in this study**

| miRNA | SNP | position | snp_chr | snp_bp | effect_allel | other_allel | eaf | beta | se | pval | cismark |
|---|---|---|---|---|---|---|---|---|---|---|---|
| let-7b-5p | rs543040 | 9:1361430 | 9 | 1.36E+08 | A | T | 0.669196 | -0.10958 | 0.010251 | 1.13E-26 | trans |
| let-7b-5p | rs579459 | 9:1361541 | 9 | 1.36E+08 | T | C | 0.787293 | -0.11893 | 0.011614 | 1.30E-24 | trans |
| let-7b-5p | rs514659 | 9:1361422 | 9 | 1.36E+08 | A | C | 0.663927 | -0.10759 | 0.010066 | 1.15E-26 | trans |
| let-7b-5p | rs544873 | 9:1361432 | 9 | 1.36E+08 | G | A | 0.645742 | -0.10512 | 0.009877 | 1.88E-26 | trans |
| let-7b-5p | rs494242 | 9:1361451 | 9 | 1.36E+08 | C | T | 0.644977 | -0.10524 | 0.009882 | 1.74E-26 | trans |
| let-7b-5p | rs473533 | 9:1361480 | 9 | 1.36E+08 | T | C | 0.453861 | -0.06695 | 0.009927 | 1.54E-11 | trans |
| let-7b-5p | rs492488 | 9:1361449 | 9 | 1.36E+08 | G | A | 0.663579 | -0.10774 | 0.010074 | 1.07E-26 | trans |
| let-7b-5p | rs545971 | 9:1361433 | 9 | 1.36E+08 | C | T | 0.663766 | -0.10758 | 0.010061 | 1.11E-26 | trans |
| let-7b-5p | rs687621 | 9:1361370 | 9 | 1.36E+08 | A | G | 0.663896 | -0.10806 | 0.010063 | 6.72E-27 | trans |
| let-7b-5p | rs493246 | 9:1361449 | 9 | 1.36E+08 | G | A | 0.663234 | -0.10778 | 0.010079 | 1.09E-26 | trans |
| let-7b-5p | rs576125 | 9:1361443 | 9 | 1.36E+08 | G | A | 0.688677 | -0.11175 | 0.010799 | 4.29E-25 | trans |
| let-7b-5p | rs532436 | 9:1361498 | 9 | 1.36E+08 | G | A | 0.8067 | -0.12631 | 0.01205 | 1.04E-25 | trans |
| let-7b-5p | rs659104 | 9:1361478 | 9 | 1.36E+08 | T | G | 0.454107 | -0.06687 | 0.00992 | 1.57E-11 | trans |
| let-7b-5p | rs1165522 | 9:1361490 | 9 | 1.36E+08 | T | A | 0.653719 | -0.10753 | 0.01003 | 8.12E-27 | trans |
| let-7b-5p | rs657152 | 9:1361392 | 9 | 1.36E+08 | C | A | 0.645336 | -0.10511 | 0.009864 | 1.65E-26 | trans |
| let-7b-5p | rs687289 | 9:1361371 | 9 | 1.36E+08 | G | A | 0.668579 | -0.11129 | 0.010186 | 8.61E-28 | trans |
| let-7b-5p | rs2519093 | 9:1361418 | 9 | 1.36E+08 | C | T | 0.809892 | -0.12789 | 0.012263 | 1.82E-25 | trans |
| let-7b-5p | rs647800 | 9:1361480 | 9 | 1.36E+08 | A | G | 0.435519 | -0.06777 | 0.009886 | 7.08E-12 | trans |
| let-7b-5p | rs676457 | 9:1361462 | 9 | 1.36E+08 | A | T | 0.664166 | -0.10782 | 0.010067 | 9.10E-27 | trans |
| let-7b-5p | rs2769071 | 9:1361459 | 9 | 1.36E+08 | A | G | 0.663117 | -0.10792 | 0.010084 | 9.93E-27 | trans |
| let-7b-5p | rs630014 | 9:1361497 | 9 | 1.36E+08 | A | G | 0.494254 | -0.07974 | 0.009829 | 4.92E-16 | trans |
| let-7b-5p | rs643434 | 9:1361423 | 9 | 1.36E+08 | G | A | 0.645392 | -0.10516 | 0.009872 | 1.70E-26 | trans |
| let-7b-5p | rs616154 | 9:1361504 | 9 | 1.36E+08 | C | T | 0.501246 | -0.07964 | 0.009955 | 1.24E-15 | trans |
| let-7b-5p | rs491626 | 9:1361448 | 9 | 1.36E+08 | C | T | 0.664173 | -0.10815 | 0.01009 | 8.39E-27 | trans |
| let-7b-5p | rs649129 | 9:1361543 | 9 | 1.36E+08 | C | T | 0.787432 | -0.11902 | 0.011621 | 1.29E-24 | trans |
| let-7b-5p | rs1124406 | 9:1361539 | 9 | 1.36E+08 | C | T | 0.886198 | -0.12811 | 0.015849 | 6.32E-16 | trans |
| let-7b-5p | rs507666 | 9:1361493 | 9 | 1.36E+08 | G | A | 0.806573 | -0.12621 | 0.012042 | 1.06E-25 | trans |
| let-7b-5p | rs529565 | 9:1361495 | 9 | 1.36E+08 | T | C | 0.665911 | -0.11036 | 0.010051 | 4.77E-28 | trans |
| let-7b-5p | rs630510 | 9:1361495 | 9 | 1.36E+08 | A | G | 0.494403 | -0.07981 | 0.009835 | 4.85E-16 | trans |
| let-7b-5p | rs635634 | 9:1361550 | 9 | 1.36E+08 | C | T | 0.80599 | -0.12576 | 0.012018 | 1.26E-25 | trans |
| let-7b-5p | rs495828 | 9:1361548 | 9 | 1.36E+08 | G | T | 0.787276 | -0.11889 | 0.011613 | 1.35E-24 | trans |
| let-7b-5p | rs505922 | 9:1361492 | 9 | 1.36E+08 | T | C | 0.665748 | -0.11011 | 0.010045 | 5.90E-28 | trans |
| let-7b-5p | rs612169 | 9:1361434 | 9 | 1.36E+08 | A | G | 0.66434 | -0.10772 | 0.01008 | 1.17E-26 | trans |
| let-7b-5p | rs582094 | 9:1361454 | 9 | 1.36E+08 | A | T | 0.663709 | -0.10773 | 0.010063 | 9.59E-27 | trans |
| let-7b-5p | rs1401144 | 9:1361443 | 9 | 1.36E+08 | T | C | 0.690247 | -0.11075 | 0.010845 | 1.75E-24 | trans |
| let-7b-5p | rs8176663 | 9:1361444 | 9 | 1.36E+08 | T | C | 0.663959 | -0.10766 | 0.010066 | 1.07E-26 | trans |
| let-7b-5p | rs476410 | 9:1361483 | 9 | 1.36E+08 | G | C | 0.45414 | -0.06688 | 0.00992 | 1.57E-11 | trans |
| let-7b-5p | rs550057 | 9:1361465 | 9 | 1.36E+08 | C | T | 0.734085 | -0.09759 | 0.010867 | 2.70E-19 | trans |
| let-7b-5p | rs674302 | 9:1361466 | 9 | 1.36E+08 | T | A | 0.663989 | -0.10789 | 0.010058 | 7.63E-27 | trans |
| let-7b-5p | rs527210 | 9:1361464 | 9 | 1.36E+08 | C | T | 0.668712 | -0.10756 | 0.010189 | 4.75E-26 | trans |
| let-7b-5p | rs651007 | 9:1361538 | 9 | 1.36E+08 | C | T | 0.787179 | -0.11915 | 0.011615 | 1.09E-24 | trans |
| let-7b-5p | rs613534 | 9:1361431 | 9 | 1.36E+08 | A | G | 0.653693 | -0.106 | 0.01005 | 5.20E-26 | trans |
| let-7b-5p | rs660340 | 9:1361475 | 9 | 1.36E+08 | A | G | 0.454082 | -0.06686 | 0.009926 | 1.62E-11 | trans |
| let-7b-5p | rs554833 | 9:1361471 | 9 | 1.36E+08 | C | T | 0.664057 | -0.10782 | 0.01006 | 8.42E-27 | trans |
| let-7b-5p | rs645982 | 9:1361484 | 9 | 1.36E+08 | A | G | 0.454123 | -0.06688 | 0.009919 | 1.56E-11 | trans |
| let-7b-5p | rs600038 | 9:1361518 | 9 | 1.36E+08 | T | C | 0.787872 | -0.11951 | 0.011643 | 1.02E-24 | trans |
| let-7b-5p | rs582118 | 9:1361454 | 9 | 1.36E+08 | A | G | 0.663872 | -0.10778 | 0.010067 | 9.58E-27 | trans |
| let-7b-5p | rs543968 | 9:1361431 | 9 | 1.36E+08 | T | C | 0.653829 | -0.10593 | 0.010051 | 5.69E-26 | trans |
| let-7b-5p | rs633862 | 9:1361554 | 9 | 1.36E+08 | T | C | 0.460592 | -0.06788 | 0.00993 | 8.14E-12 | trans |
| let-7b-5p | rs495203 | 9:1361452 | 9 | 1.36E+08 | C | T | 0.663279 | -0.10772 | 0.010072 | 1.06E-26 | trans |
| let-7b-5p | rs559723 | 9:1361504 | 9 | 1.36E+08 | A | G | 0.508431 | -0.07846 | 0.010057 | 6.15E-15 | trans |
| let-7b-5p | rs676996 | 9:1361460 | 9 | 1.36E+08 | T | G | 0.667516 | -0.10802 | 0.010153 | 1.96E-26 | trans |
| let-7b-5p | rs581107 | 9:1361477 | 9 | 1.36E+08 | C | T | 0.452595 | -0.06708 | 0.009954 | 1.59E-11 | trans |
| let-7b-5p | rs475419 | 9:1361482 | 9 | 1.36E+08 | T | C | 0.454203 | -0.06691 | 0.00992 | 1.54E-11 | trans |
| let-7b-5p | rs597974 | 9:1361442 | 9 | 1.36E+08 | A | G | 0.687926 | -0.11232 | 0.01077 | 1.82E-25 | trans |
| let-7b-5p | rs677355 | 9:1361460 | 9 | 1.36E+08 | G | A | 0.670198 | -0.10865 | 0.010242 | 2.73E-26 | trans |
| let-7b-5p | rs644234 | 9:1361422 | 9 | 1.36E+08 | T | G | 0.645381 | -0.10519 | 0.009864 | 1.49E-26 | trans |
| let-7b-5p | rs597988 | 9:1361442 | 9 | 1.36E+08 | T | A | 0.686751 | -0.11225 | 0.010722 | 1.20E-25 | trans |
| let-7b-5p | rs9411378 | 9:1361454 | 9 | 1.36E+08 | C | A | 0.753652 | -0.09914 | 0.011778 | 3.83E-17 | trans |
| let-7c-5p | rs1165522 | 9:1361490 | 9 | 1.36E+08 | T | A | 0.653719 | -0.09078 | 0.00916 | 3.78E-23 | trans |
| let-7c-5p | rs635634 | 9:1361550 | 9 | 1.36E+08 | C | T | 0.80599 | -0.10652 | 0.010972 | 2.77E-22 | trans |

| | | | | | | | | | | |
|---|---|---|---|---|---|---|---|---|---|---|
| let-7c-5p | rs649129 | 9:1361543 | 9 | 1.36E+08 | C | T | 0.787432 | -0.10158 | 0.010605 | 9.83E-22 trans |
| let-7c-5p | rs491626 | 9:1361448 | 9 | 1.36E+08 | C | T | 0.664173 | -0.09187 | 0.009213 | 2.01E-23 trans |
| let-7c-5p | rs475419 | 9:1361482 | 9 | 1.36E+08 | T | C | 0.454203 | -0.06322 | 0.009019 | 2.39E-12 trans |
| let-7c-5p | rs529565 | 9:1361495 | 9 | 1.36E+08 | T | C | 0.665911 | -0.09316 | 0.009181 | 3.42E-24 trans |
| let-7c-5p | rs687621 | 9:1361370 | 9 | 1.36E+08 | A | G | 0.663896 | -0.09202 | 0.009187 | 1.28E-23 trans |
| let-7c-5p | rs579459 | 9:1361541 | 9 | 1.36E+08 | T | C | 0.787293 | -0.10147 | 0.010598 | 1.02E-21 trans |
| let-7c-5p | rs677355 | 9:1361460 | 9 | 1.36E+08 | G | A | 0.670198 | -0.09226 | 0.009351 | 5.81E-23 trans |
| let-7c-5p | rs597988 | 9:1361442 | 9 | 1.36E+08 | T | A | 0.686751 | -0.09546 | 0.009787 | 1.78E-22 trans |
| let-7c-5p | rs473533 | 9:1361480 | 9 | 1.36E+08 | T | C | 0.453861 | -0.06327 | 0.009025 | 2.38E-12 trans |
| let-7c-5p | rs687289 | 9:1361371 | 9 | 1.36E+08 | G | A | 0.668579 | -0.09391 | 0.009304 | 5.91E-24 trans |
| let-7c-5p | rs495828 | 9:1361548 | 9 | 1.36E+08 | G | T | 0.787276 | -0.10145 | 0.010598 | 1.04E-21 trans |
| let-7c-5p | rs532436 | 9:1361498 | 9 | 1.36E+08 | G | A | 0.8067 | -0.1071 | 0.011001 | 2.13E-22 trans |
| let-7c-5p | rs9411378 | 9:1361454 | 9 | 1.36E+08 | C | A | 0.753652 | -0.08394 | 0.01074 | 5.46E-15 trans |
| let-7c-5p | rs645982 | 9:1361484 | 9 | 1.36E+08 | A | G | 0.454123 | -0.0632 | 0.009018 | 2.43E-12 trans |
| let-7c-5p | rs600038 | 9:1361518 | 9 | 1.36E+08 | T | C | 0.787872 | -0.10202 | 0.010625 | 7.82E-22 trans |
| let-7c-5p | rs514659 | 9:1361422 | 9 | 1.36E+08 | A | C | 0.663927 | -0.09133 | 0.00919 | 2.86E-23 trans |
| let-7c-5p | rs616154 | 9:1361504 | 9 | 1.36E+08 | C | T | 0.501246 | -0.06284 | 0.009091 | 4.78E-12 trans |
| let-7c-5p | rs676457 | 9:1361462 | 9 | 1.36E+08 | A | T | 0.664166 | -0.09148 | 0.009192 | 2.46E-23 trans |
| let-7c-5p | rs582094 | 9:1361454 | 9 | 1.36E+08 | A | T | 0.663709 | -0.09135 | 0.009188 | 2.74E-23 trans |
| let-7c-5p | rs647800 | 9:1361480 | 9 | 1.36E+08 | A | G | 0.435519 | -0.06437 | 0.008986 | 7.87E-13 trans |
| let-7c-5p | rs559723 | 9:1361504 | 9 | 1.36E+08 | A | G | 0.508431 | -0.06193 | 0.009183 | 1.54E-11 trans |
| let-7c-5p | rs576125 | 9:1361443 | 9 | 1.36E+08 | G | A | 0.688677 | -0.09528 | 0.009856 | 4.16E-22 trans |
| let-7c-5p | rs633862 | 9:1361554 | 9 | 1.36E+08 | T | C | 0.460592 | -0.06289 | 0.009032 | 3.32E-12 trans |
| let-7c-5p | rs630014 | 9:1361497 | 9 | 1.36E+08 | A | G | 0.494254 | -0.06361 | 0.008975 | 1.37E-12 trans |
| let-7c-5p | rs476410 | 9:1361483 | 9 | 1.36E+08 | G | C | 0.45414 | -0.0632 | 0.009019 | 2.43E-12 trans |
| let-7c-5p | rs2519093 | 9:1361418 | 9 | 1.36E+08 | C | T | 0.809892 | -0.10804 | 0.011197 | 4.94E-22 trans |
| let-7c-5p | rs674302 | 9:1361466 | 9 | 1.36E+08 | T | A | 0.663989 | -0.09147 | 0.009184 | 2.28E-23 trans |
| let-7c-5p | rs527210 | 9:1361464 | 9 | 1.36E+08 | C | T | 0.668712 | -0.09077 | 0.009305 | 1.74E-22 trans |
| let-7c-5p | rs1401144 | 9:1361443 | 9 | 1.36E+08 | T | C | 0.690247 | -0.09443 | 0.009897 | 1.41E-21 trans |
| let-7c-5p | rs493246 | 9:1361449 | 9 | 1.36E+08 | G | A | 0.663234 | -0.0915 | 0.009202 | 2.70E-23 trans |
| let-7c-5p | rs544873 | 9:1361432 | 9 | 1.36E+08 | G | A | 0.645742 | -0.08952 | 0.009016 | 3.11E-23 trans |
| let-7c-5p | rs643434 | 9:1361423 | 9 | 1.36E+08 | G | A | 0.645392 | -0.08945 | 0.009012 | 3.20E-23 trans |
| let-7c-5p | rs581107 | 9:1361477 | 9 | 1.36E+08 | C | T | 0.452595 | -0.06337 | 0.00905 | 2.51E-12 trans |
| let-7c-5p | rs651007 | 9:1361538 | 9 | 1.36E+08 | C | T | 0.787179 | -0.10173 | 0.010599 | 8.16E-22 trans |
| let-7c-5p | rs500499 | 9:1361486 | 9 | 1.36E+08 | C | G | 0.465434 | -0.06256 | 0.009147 | 7.97E-12 trans |
| let-7c-5p | rs659104 | 9:1361478 | 9 | 1.36E+08 | T | G | 0.454107 | -0.0632 | 0.009019 | 2.43E-12 trans |
| let-7c-5p | rs676996 | 9:1361460 | 9 | 1.36E+08 | T | G | 0.667516 | -0.09165 | 0.00927 | 4.76E-23 trans |
| let-7c-5p | rs644234 | 9:1361422 | 9 | 1.36E+08 | T | G | 0.645381 | -0.08951 | 0.009005 | 2.80E-23 trans |
| let-7c-5p | rs554833 | 9:1361471 | 9 | 1.36E+08 | C | T | 0.664057 | -0.09142 | 0.009186 | 2.46E-23 trans |
| let-7c-5p | rs597974 | 9:1361442 | 9 | 1.36E+08 | A | G | 0.687926 | -0.09543 | 0.009831 | 2.81E-22 trans |
| let-7c-5p | rs550057 | 9:1361465 | 9 | 1.36E+08 | C | T | 0.734085 | -0.08406 | 0.009907 | 2.15E-17 trans |
| let-7c-5p | rs492488 | 9:1361449 | 9 | 1.36E+08 | G | A | 0.663579 | -0.09146 | 0.009197 | 2.67E-23 trans |
| let-7c-5p | rs630510 | 9:1361495 | 9 | 1.36E+08 | A | G | 0.494403 | -0.06368 | 0.008981 | 1.33E-12 trans |
| let-7c-5p | rs507666 | 9:1361493 | 9 | 1.36E+08 | G | A | 0.806573 | -0.10699 | 0.010994 | 2.21E-22 trans |
| let-7c-5p | rs543968 | 9:1361431 | 9 | 1.36E+08 | T | C | 0.653829 | -0.0907 | 0.009172 | 4.68E-23 trans |
| let-7c-5p | rs582118 | 9:1361454 | 9 | 1.36E+08 | A | G | 0.663872 | -0.09141 | 0.009192 | 2.66E-23 trans |
| let-7c-5p | rs8176663 | 9:1361444 | 9 | 1.36E+08 | T | C | 0.663959 | -0.09137 | 0.00919 | 2.74E-23 trans |
| let-7c-5p | rs657152 | 9:1361392 | 9 | 1.36E+08 | C | A | 0.645336 | -0.08946 | 0.009005 | 2.97E-23 trans |
| let-7c-5p | rs500498 | 9:1361486 | 9 | 1.36E+08 | C | T | 0.464233 | -0.0619 | 0.009136 | 1.24E-11 trans |
| let-7c-5p | rs494242 | 9:1361451 | 9 | 1.36E+08 | C | T | 0.644977 | -0.08968 | 0.00902 | 2.75E-23 trans |
| let-7c-5p | rs660340 | 9:1361475 | 9 | 1.36E+08 | A | G | 0.454082 | -0.06323 | 0.009024 | 2.44E-12 trans |
| let-7c-5p | rs612169 | 9:1361434 | 9 | 1.36E+08 | A | G | 0.66434 | -0.09149 | 0.009203 | 2.76E-23 trans |
| let-7c-5p | rs613534 | 9:1361431 | 9 | 1.36E+08 | A | G | 0.653693 | -0.09076 | 0.009171 | 4.30E-23 trans |
| let-7c-5p | rs545971 | 9:1361433 | 9 | 1.36E+08 | C | T | 0.663766 | -0.09129 | 0.009187 | 2.87E-23 trans |
| let-7c-5p | rs543040 | 9:1361430 | 9 | 1.36E+08 | A | T | 0.669196 | -0.0923 | 0.009362 | 6.26E-23 trans |
| let-7c-5p | rs495203 | 9:1361452 | 9 | 1.36E+08 | C | T | 0.663279 | -0.09143 | 0.009196 | 2.73E-23 trans |
| let-7c-5p | rs505922 | 9:1361492 | 9 | 1.36E+08 | T | C | 0.665748 | -0.09298 | 0.009176 | 3.95E-24 trans |
| let-7c-5p | rs1124406 | 9:1361539 | 9 | 1.36E+08 | C | T | 0.886198 | -0.10356 | 0.014468 | 8.20E-13 trans |
| let-7c-5p | rs2769071 | 9:1361459 | 9 | 1.36E+08 | A | G | 0.663117 | -0.09126 | 0.009208 | 3.76E-23 trans |
| let-7e-5p | rs657152 | 9:1361392 | 9 | 1.36E+08 | C | A | 0.645336 | -0.10995 | 0.014592 | 4.89E-14 trans |
| let-7e-5p | rs491626 | 9:1361448 | 9 | 1.36E+08 | C | T | 0.664173 | -0.10888 | 0.014943 | 3.20E-13 trans |
| let-7e-5p | rs576125 | 9:1361443 | 9 | 1.36E+08 | G | A | 0.688677 | -0.11147 | 0.015982 | 3.06E-12 trans |
| let-7e-5p | rs676996 | 9:1361460 | 9 | 1.36E+08 | T | G | 0.667516 | -0.10883 | 0.015033 | 4.51E-13 trans |

| | | | | | | | | | | | |
|---|---|---|---|---|---|---|---|---|---|---|---|
| let-7e-5p | rs514659 | 9:1361422( | 9 | 1.36E+08 | A | C | 0.663927 | -0.10821 | 0.014906 | 3.88E-13 | trans |
| let-7e-5p | rs493246 | 9:1361449! | 9 | 1.36E+08 | G | A | 0.663234 | -0.1085 | 0.014926 | 3.62E-13 | trans |
| let-7e-5p | rs544873 | 9:1361432 | 9 | 1.36E+08 | G | A | 0.645742 | -0.10985 | 0.014611 | 5.54E-14 | trans |
| let-7e-5p | rs651007 | 9:1361538 | 9 | 1.36E+08 | C | T | 0.787179 | -0.12976 | 0.017148 | 3.83E-14 | trans |
| let-7e-5p | rs635634 | 9:1361550( | 9 | 1.36E+08 | C | T | 0.80599 | -0.13032 | 0.017774 | 2.26E-13 | trans |
| let-7e-5p | rs613534 | 9:1361431 | 9 | 1.36E+08 | A | G | 0.653693 | -0.1123 | 0.014857 | 4.07E-14 | trans |
| let-7e-5p | rs529565 | 9:1361495( | 9 | 1.36E+08 | T | C | 0.665911 | -0.11071 | 0.014896 | 1.07E-13 | trans |
| let-7e-5p | rs494242 | 9:1361451 | 9 | 1.36E+08 | C | T | 0.644977 | -0.1103 | 0.014617 | 4.49E-14 | trans |
| let-7e-5p | rs527210 | 9:1361464 | 9 | 1.36E+08 | C | T | 0.668712 | -0.10694 | 0.015088 | 1.36E-12 | trans |
| let-7e-5p | rs579459 | 9:1361541( | 9 | 1.36E+08 | T | C | 0.787293 | -0.12954 | 0.017146 | 4.18E-14 | trans |
| let-7e-5p | rs676457 | 9:1361462 | 9 | 1.36E+08 | A | T | 0.664166 | -0.10811 | 0.01491 | 4.15E-13 | trans |
| let-7e-5p | rs597974 | 9:1361442! | 9 | 1.36E+08 | A | G | 0.687926 | -0.11249 | 0.01594 | 1.70E-12 | trans |
| let-7e-5p | rs582118 | 9:1361454 | 9 | 1.36E+08 | A | G | 0.663872 | -0.10829 | 0.014909 | 3.78E-13 | trans |
| let-7e-5p | rs612169 | 9:1361434 | 9 | 1.36E+08 | A | G | 0.66434 | -0.10831 | 0.014927 | 3.98E-13 | trans |
| let-7e-5p | rs649129 | 9:1361543( | 9 | 1.36E+08 | C | T | 0.787432 | -0.1297 | 0.017156 | 4.03E-14 | trans |
| let-7e-5p | rs550057 | 9:1361465! | 9 | 1.36E+08 | C | T | 0.734085 | -0.11064 | 0.015991 | 4.55E-12 | trans |
| let-7e-5p | rs597988 | 9:1361442 | 9 | 1.36E+08 | T | A | 0.686751 | -0.11271 | 0.01587 | 1.23E-12 | trans |
| let-7e-5p | rs2769071 | 9:1361459 | 9 | 1.36E+08 | A | G | 0.663117 | -0.10904 | 0.014931 | 2.82E-13 | trans |
| let-7e-5p | rs1401144! | 9:1361443( | 9 | 1.36E+08 | T | C | 0.690247 | -0.11076 | 0.016043 | 5.06E-12 | trans |
| let-7e-5p | rs687621 | 9:1361370( | 9 | 1.36E+08 | A | G | 0.663896 | -0.10775 | 0.014907 | 4.88E-13 | trans |
| let-7e-5p | rs543968 | 9:1361431 | 9 | 1.36E+08 | T | C | 0.653829 | -0.11239 | 0.014859 | 3.91E-14 | trans |
| let-7e-5p | rs492488 | 9:1361449( | 9 | 1.36E+08 | G | A | 0.663579 | -0.10842 | 0.014918 | 3.65E-13 | trans |
| let-7e-5p | rs582094 | 9:1361454 | 9 | 1.36E+08 | A | T | 0.663709 | -0.10819 | 0.014903 | 3.89E-13 | trans |
| let-7e-5p | rs1165522 | 9:1361490! | 9 | 1.36E+08 | T | A | 0.653719 | -0.11028 | 0.014847 | 1.11E-13 | trans |
| let-7e-5p | rs545971 | 9:1361433 | 9 | 1.36E+08 | C | T | 0.663766 | -0.1081 | 0.0149 | 4.01E-13 | trans |
| let-7e-5p | rs507666 | 9:1361493! | 9 | 1.36E+08 | G | A | 0.806573 | -0.1306 | 0.017811 | 2.26E-13 | trans |
| let-7e-5p | rs543040 | 9:1361430( | 9 | 1.36E+08 | A | T | 0.669196 | -0.11011 | 0.01518 | 4.05E-13 | trans |
| let-7e-5p | rs2519093 | 9:1361418 | 9 | 1.36E+08 | C | T | 0.809892 | -0.13199 | 0.018137 | 3.39E-13 | trans |
| let-7e-5p | rs495203 | 9:1361452 | 9 | 1.36E+08 | C | T | 0.663279 | -0.10839 | 0.014915 | 3.67E-13 | trans |
| let-7e-5p | rs687289 | 9:1361371( | 9 | 1.36E+08 | G | A | 0.668579 | -0.10974 | 0.0151 | 3.66E-13 | trans |
| let-7e-5p | rs644234 | 9:1361422 | 9 | 1.36E+08 | T | G | 0.645381 | -0.10992 | 0.014592 | 4.98E-14 | trans |
| let-7e-5p | rs495828 | 9:1361548( | 9 | 1.36E+08 | G | T | 0.787276 | -0.12951 | 0.017145 | 4.23E-14 | trans |
| let-7e-5p | rs532436 | 9:1361498 | 9 | 1.36E+08 | G | A | 0.8067 | -0.13076 | 0.017823 | 2.19E-13 | trans |
| let-7e-5p | rs643434 | 9:1361423 | 9 | 1.36E+08 | G | A | 0.645392 | -0.10992 | 0.014603 | 5.19E-14 | trans |
| let-7e-5p | rs674302 | 9:1361466( | 9 | 1.36E+08 | T | A | 0.663989 | -0.10806 | 0.014898 | 4.07E-13 | trans |
| let-7e-5p | rs8176663 | 9:1361444 | 9 | 1.36E+08 | T | C | 0.663959 | -0.1082 | 0.014906 | 3.90E-13 | trans |
| let-7e-5p | rs505922 | 9:1361492 | 9 | 1.36E+08 | T | C | 0.665748 | -0.11041 | 0.014887 | 1.21E-13 | trans |
| let-7e-5p | rs554833 | 9:1361471( | 9 | 1.36E+08 | C | T | 0.664057 | -0.10797 | 0.014901 | 4.28E-13 | trans |
| let-7e-5p | rs677355 | 9:1361460 | 9 | 1.36E+08 | G | A | 0.670198 | -0.10836 | 0.015167 | 9.03E-13 | trans |
| let-7e-5p | rs600038 | 9:1361518( | 9 | 1.36E+08 | T | C | 0.787872 | -0.13006 | 0.01719 | 3.86E-14 | trans |
| miR-10a-5p | rs633862 | 9:1361554 | 9 | 1.36E+08 | T | C | 0.460592 | -0.11826 | 0.01443 | 2.49E-16 | trans |
| miR-10a-5p | rs676457 | 9:1361462 | 9 | 1.36E+08 | A | T | 0.664166 | -0.18087 | 0.014572 | 2.24E-35 | trans |
| miR-10a-5p | rs1178913! | 9:1361853 | 9 | 1.36E+08 | C | G | 0.833223 | -0.13461 | 0.019822 | 1.11E-11 | trans |
| miR-10a-5p | rs687621 | 9:1361370( | 9 | 1.36E+08 | A | G | 0.663896 | -0.18219 | 0.01456 | 6.32E-36 | trans |
| miR-10a-5p | rs2073826 | 9:1361369( | 9 | 1.36E+08 | G | T | 0.60107 | 0.102208 | 0.014329 | 9.81E-13 | trans |
| miR-10a-5p | rs495203 | 9:1361452 | 9 | 1.36E+08 | C | T | 0.663279 | -0.18092 | 0.014577 | 2.29E-35 | trans |
| miR-10a-5p | rs8176691 | 9:1361382 | 9 | 1.36E+08 | C | T | 0.619638 | 0.105987 | 0.014449 | 2.21E-13 | trans |
| miR-10a-5p | rs473533 | 9:1361480 | 9 | 1.36E+08 | T | C | 0.453861 | -0.11837 | 0.014421 | 2.25E-16 | trans |
| miR-10a-5p | rs635634 | 9:1361550( | 9 | 1.36E+08 | C | T | 0.80599 | -0.21 | 0.017408 | 1.64E-33 | trans |
| miR-10a-5p | rs657152 | 9:1361392( | 9 | 1.36E+08 | C | A | 0.645336 | -0.17556 | 0.014284 | 1.02E-34 | trans |
| miR-10a-5p | rs544873 | 9:1361432 | 9 | 1.36E+08 | G | A | 0.645742 | -0.17561 | 0.014302 | 1.18E-34 | trans |
| miR-10a-5p | rs651007 | 9:1361538 | 9 | 1.36E+08 | C | T | 0.787179 | -0.19903 | 0.016829 | 2.84E-32 | trans |
| miR-10a-5p | rs505922 | 9:1361492 | 9 | 1.36E+08 | T | C | 0.665748 | -0.18379 | 0.01454 | 1.26E-36 | trans |
| miR-10a-5p | rs1124406 | 9:1361539 | 9 | 1.36E+08 | C | T | 0.886198 | -0.18406 | 0.023149 | 1.85E-15 | trans |
| miR-10a-5p | rs597974 | 9:1361442! | 9 | 1.36E+08 | A | G | 0.687926 | -0.19015 | 0.015586 | 3.11E-34 | trans |
| miR-10a-5p | rs495828 | 9:1361548( | 9 | 1.36E+08 | G | T | 0.787276 | -0.19862 | 0.016826 | 3.73E-32 | trans |
| miR-10a-5p | rs7857390 | 9:1361285 | 9 | 1.36E+08 | A | G | 0.377967 | -0.1041 | 0.015041 | 4.49E-12 | trans |
| miR-10a-5p | rs492488 | 9:1361449( | 9 | 1.36E+08 | G | A | 0.663579 | -0.18088 | 0.014581 | 2.44E-35 | trans |
| miR-10a-5p | rs559723 | 9:1361504 | 9 | 1.36E+08 | A | G | 0.508431 | -0.11333 | 0.014687 | 1.20E-14 | trans |
| miR-10a-5p | rs527210 | 9:1361464 | 9 | 1.36E+08 | C | T | 0.668712 | -0.17983 | 0.014756 | 3.64E-34 | trans |
| miR-10a-5p | rs8176715 | 9:1361331 | 9 | 1.36E+08 | T | C | 0.418056 | -0.09951 | 0.0147 | 1.29E-11 | trans |
| miR-10a-5p | rs7046674 | 9:1361470 | 9 | 1.36E+08 | C | T | 0.619013 | 0.107787 | 0.014473 | 9.50E-14 | trans |
| miR-10a-5p | rs660340 | 9:1361475 | 9 | 1.36E+08 | A | G | 0.454082 | -0.11823 | 0.014419 | 2.41E-16 | trans |

| | | | | | | | | | | | |
|---|---|---|---|---|---|---|---|---|---|---|---|
| miR-10a-5p | rs550057 | 9:1361465 | 9 | 1.36E+08 | C | T | 0.734085 | -0.16793 | 0.015749 | 1.52E-26 | trans |
| miR-10a-5p | rs600038 | 9:1361518 | 9 | 1.36E+08 | T | C | 0.787872 | -0.19936 | 0.01687 | 3.18E-32 | trans |
| miR-10a-5p | rs647800 | 9:1361480 | 9 | 1.36E+08 | A | G | 0.435519 | -0.11938 | 0.01436 | 9.33E-17 | trans |
| miR-10a-5p | rs500498 | 9:1361486 | 9 | 1.36E+08 | C | T | 0.464233 | -0.11512 | 0.014604 | 3.20E-15 | trans |
| miR-10a-5p | rs543968 | 9:1361431 | 9 | 1.36E+08 | T | C | 0.653829 | -0.17776 | 0.014552 | 2.57E-34 | trans |
| miR-10a-5p | rs476410 | 9:1361483 | 9 | 1.36E+08 | G | C | 0.45414 | -0.11814 | 0.014412 | 2.46E-16 | trans |
| miR-10a-5p | rs2073828 | 9:1361371 | 9 | 1.36E+08 | G | A | 0.601379 | 0.102564 | 0.014335 | 8.39E-13 | trans |
| miR-10a-5p | rs613534 | 9:1361431 | 9 | 1.36E+08 | A | G | 0.653693 | -0.17768 | 0.014551 | 2.72E-34 | trans |
| miR-10a-5p | rs176694 | 9:1361645 | 9 | 1.36E+08 | T | G | 0.900795 | -0.16574 | 0.024385 | 1.07E-11 | trans |
| miR-10a-5p | rs2519093 | 9:1361418 | 9 | 1.36E+08 | C | T | 0.809892 | -0.21091 | 0.017778 | 1.83E-32 | trans |
| miR-10a-5p | rs529565 | 9:1361495 | 9 | 1.36E+08 | T | C | 0.665911 | -0.18394 | 0.014549 | 1.22E-36 | trans |
| miR-10a-5p | rs7873635 | 9:1361320 | 9 | 1.36E+08 | T | C | 0.381256 | -0.10283 | 0.01466 | 2.32E-12 | trans |
| miR-10a-5p | rs543040 | 9:1361430 | 9 | 1.36E+08 | A | T | 0.669196 | -0.18259 | 0.014845 | 9.09E-35 | trans |
| miR-10a-5p | rs554833 | 9:1361471 | 9 | 1.36E+08 | C | T | 0.664057 | -0.1807 | 0.014563 | 2.35E-35 | trans |
| miR-10a-5p | rs8176690 | 9:1361383 | 9 | 1.36E+08 | A | G | 0.601023 | 0.101606 | 0.014332 | 1.35E-12 | trans |
| miR-10a-5p | rs630510 | 9:1361495 | 9 | 1.36E+08 | A | G | 0.494403 | -0.1169 | 0.014357 | 3.87E-16 | trans |
| miR-10a-5p | rs1401144 | 9:1361443 | 9 | 1.36E+08 | T | C | 0.690247 | -0.18845 | 0.015695 | 3.28E-33 | trans |
| miR-10a-5p | rs4962040 | 9:1361335 | 9 | 1.36E+08 | A | G | 0.619057 | 0.104354 | 0.014467 | 5.47E-13 | trans |
| miR-10a-5p | rs581107 | 9:1361477 | 9 | 1.36E+08 | C | T | 0.452595 | -0.11865 | 0.01446 | 2.30E-16 | trans |
| miR-10a-5p | rs9411378 | 9:1361454 | 9 | 1.36E+08 | C | A | 0.753652 | -0.17027 | 0.017086 | 2.16E-23 | trans |
| miR-10a-5p | rs2769071 | 9:1361459 | 9 | 1.36E+08 | A | G | 0.663117 | -0.18027 | 0.0146 | 5.06E-35 | trans |
| miR-10a-5p | rs644234 | 9:1361422 | 9 | 1.36E+08 | T | G | 0.645381 | -0.17563 | 0.014284 | 9.56E-35 | trans |
| miR-10a-5p | rs630014 | 9:1361497 | 9 | 1.36E+08 | A | G | 0.494254 | -0.11675 | 0.014347 | 4.05E-16 | trans |
| miR-10a-5p | rs579459 | 9:1361541 | 9 | 1.36E+08 | T | C | 0.787293 | -0.19863 | 0.016827 | 3.72E-32 | trans |
| miR-10a-5p | rs500499 | 9:1361486 | 9 | 1.36E+08 | C | G | 0.465434 | -0.11619 | 0.014621 | 1.91E-15 | trans |
| miR-10a-5p | rs532436 | 9:1361498 | 9 | 1.36E+08 | G | A | 0.8067 | -0.21073 | 0.017455 | 1.47E-33 | trans |
| miR-10a-5p | rs676996 | 9:1361460 | 9 | 1.36E+08 | T | G | 0.667516 | -0.18184 | 0.014695 | 3.60E-35 | trans |
| miR-10a-5p | rs507666 | 9:1361493 | 9 | 1.36E+08 | G | A | 0.806573 | -0.21053 | 0.017443 | 1.53E-33 | trans |
| miR-10a-5p | rs582094 | 9:1361454 | 9 | 1.36E+08 | A | T | 0.663709 | -0.18077 | 0.014566 | 2.28E-35 | trans |
| miR-10a-5p | rs8176663 | 9:1361444 | 9 | 1.36E+08 | T | C | 0.663959 | -0.18068 | 0.014569 | 2.58E-35 | trans |
| miR-10a-5p | rs2073827 | 9:1361371 | 9 | 1.36E+08 | G | C | 0.601379 | 0.102564 | 0.014335 | 8.39E-13 | trans |
| miR-10a-5p | rs612169 | 9:1361434 | 9 | 1.36E+08 | A | G | 0.66434 | -0.18095 | 0.014589 | 2.51E-35 | trans |
| miR-10a-5p | rs643434 | 9:1361423 | 9 | 1.36E+08 | G | A | 0.645392 | -0.17585 | 0.014293 | 8.76E-35 | trans |
| miR-10a-5p | rs649129 | 9:1361543 | 9 | 1.36E+08 | C | T | 0.787432 | -0.1988 | 0.016838 | 3.60E-32 | trans |
| miR-10a-5p | rs674302 | 9:1361466 | 9 | 1.36E+08 | T | A | 0.663989 | -0.18089 | 0.014559 | 1.93E-35 | trans |
| miR-10a-5p | rs8176668 | 9:1361440 | 9 | 1.36E+08 | A | T | 0.618711 | 0.106528 | 0.014474 | 1.84E-13 | trans |
| miR-10a-5p | rs1165522 | 9:1361490 | 9 | 1.36E+08 | T | A | 0.653719 | -0.17787 | 0.014532 | 1.91E-34 | trans |
| miR-10a-5p | rs494242 | 9:1361451 | 9 | 1.36E+08 | C | T | 0.644977 | -0.1761 | 0.014307 | 8.14E-35 | trans |
| miR-10a-5p | rs8176649 | 9:1361472 | 9 | 1.36E+08 | G | A | 0.618669 | 0.107644 | 0.014449 | 9.35E-14 | trans |
| miR-10a-5p | rs659104 | 9:1361478 | 9 | 1.36E+08 | T | G | 0.454107 | -0.11815 | 0.014412 | 2.44E-16 | trans |
| miR-10a-5p | rs8176681 | 9:1361397 | 9 | 1.36E+08 | T | C | 0.600667 | 0.102291 | 0.014344 | 9.94E-13 | trans |
| miR-10a-5p | rs616154 | 9:1361504 | 9 | 1.36E+08 | C | T | 0.501246 | -0.115 | 0.014537 | 2.55E-15 | trans |
| miR-10a-5p | rs8176682 | 9:1361392 | 9 | 1.36E+08 | C | T | 0.619011 | 0.105945 | 0.014446 | 2.24E-13 | trans |
| miR-10a-5p | rs475419 | 9:1361482 | 9 | 1.36E+08 | T | C | 0.454203 | -0.11819 | 0.014412 | 2.39E-16 | trans |
| miR-10a-5p | rs493246 | 9:1361449 | 9 | 1.36E+08 | G | A | 0.663234 | -0.18102 | 0.014588 | 2.34E-35 | trans |
| miR-10a-5p | rs576125 | 9:1361443 | 9 | 1.36E+08 | G | A | 0.688677 | -0.18959 | 0.015629 | 7.26E-34 | trans |
| miR-10a-5p | rs677355 | 9:1361460 | 9 | 1.36E+08 | G | A | 0.670198 | -0.18323 | 0.014822 | 4.18E-35 | trans |
| miR-10a-5p | rs1838531 | 9:1362190 | 9 | 1.36E+08 | T | A | 0.947229 | -0.24216 | 0.034161 | 1.35E-12 | trans |
| miR-10a-5p | rs597988 | 9:1361442 | 9 | 1.36E+08 | T | A | 0.686751 | -0.19028 | 0.015515 | 1.41E-34 | trans |
| miR-10a-5p | rs545971 | 9:1361433 | 9 | 1.36E+08 | C | T | 0.663766 | -0.18057 | 0.014563 | 2.65E-35 | trans |
| miR-10a-5p | rs582118 | 9:1361454 | 9 | 1.36E+08 | A | G | 0.663872 | -0.18085 | 0.014572 | 2.28E-35 | trans |
| miR-10a-5p | rs491626 | 9:1361448 | 9 | 1.36E+08 | C | T | 0.664173 | -0.18139 | 0.014606 | 2.05E-35 | trans |
| miR-10a-5p | rs687289 | 9:1361371 | 9 | 1.36E+08 | G | A | 0.668579 | -0.18573 | 0.014744 | 2.19E-36 | trans |
| miR-10a-5p | rs514659 | 9:1361422 | 9 | 1.36E+08 | A | C | 0.663927 | -0.18061 | 0.01457 | 2.75E-35 | trans |
| miR-10a-5p | rs7036642 | 9:1361446 | 9 | 1.36E+08 | G | A | 0.618956 | 0.106822 | 0.014464 | 1.52E-13 | trans |
| miR-10a-5p | rs645982 | 9:1361484 | 9 | 1.36E+08 | A | G | 0.454123 | -0.11814 | 0.014411 | 2.45E-16 | trans |
| miR-10a-5p | rs8176702 | 9:1361361 | 9 | 1.36E+08 | G | A | 0.6197 | 0.106041 | 0.014453 | 2.18E-13 | trans |
| miR-10b-5p | rs558240 | 9:1361571 | 9 | 1.36E+08 | G | A | 0.601396 | -0.09745 | 0.014384 | 1.24E-11 | trans |
| miR-10b-5p | rs579459 | 9:1361541 | 9 | 1.36E+08 | T | C | 0.787293 | -0.20136 | 0.016485 | 2.59E-34 | trans |
| miR-10b-5p | rs613534 | 9:1361431 | 9 | 1.36E+08 | A | G | 0.653693 | -0.17445 | 0.014285 | 2.68E-34 | trans |
| miR-10b-5p | rs597974 | 9:1361442 | 9 | 1.36E+08 | A | G | 0.687926 | -0.18704 | 0.0153 | 2.28E-34 | trans |
| miR-10b-5p | rs612169 | 9:1361434 | 9 | 1.36E+08 | A | G | 0.66434 | -0.17728 | 0.014325 | 3.54E-35 | trans |
| miR-10b-5p | rs657152 | 9:1361392 | 9 | 1.36E+08 | C | A | 0.645336 | -0.17187 | 0.014026 | 1.60E-34 | trans |

| | | | | | | | | | | | |
|---|---|---|---|---|---|---|---|---|---|---|---|
| miR-10b-5p | rs630510 | 9:1361495 | 9 | 1.36E+08 | A | G | 0.494403 | -0.11555 | 0.014092 | 2.40E-16 | trans |
| miR-10b-5p | rs550057 | 9:1361465 | 9 | 1.36E+08 | C | T | 0.734085 | -0.1679 | 0.015446 | 1.61E-27 | trans |
| miR-10b-5p | rs9411378 | 9:1361454 | 9 | 1.36E+08 | C | A | 0.753652 | -0.1719 | 0.016751 | 1.05E-24 | trans |
| miR-10b-5p | rs660340 | 9:1361475 | 9 | 1.36E+08 | A | G | 0.454082 | -0.12792 | 0.014109 | 1.23E-19 | trans |
| miR-10b-5p | rs674302 | 9:1361466 | 9 | 1.36E+08 | T | A | 0.663989 | -0.1772 | 0.014295 | 2.75E-35 | trans |
| miR-10b-5p | rs545971 | 9:1361433 | 9 | 1.36E+08 | C | T | 0.663766 | -0.177 | 0.014299 | 3.43E-35 | trans |
| miR-10b-5p | rs507666 | 9:1361493 | 9 | 1.36E+08 | G | A | 0.806573 | -0.21399 | 0.017083 | 5.37E-36 | trans |
| miR-10b-5p | rs649129 | 9:1361543 | 9 | 1.36E+08 | C | T | 0.787432 | -0.20151 | 0.016495 | 2.54E-34 | trans |
| miR-10b-5p | rs600038 | 9:1361518 | 9 | 1.36E+08 | T | C | 0.787872 | -0.20203 | 0.016527 | 2.32E-34 | trans |
| miR-10b-5p | rs643434 | 9:1361423 | 9 | 1.36E+08 | G | A | 0.645392 | -0.17197 | 0.014036 | 1.64E-34 | trans |
| miR-10b-5p | rs687289 | 9:1361371 | 9 | 1.36E+08 | G | A | 0.668579 | -0.18374 | 0.014466 | 5.80E-37 | trans |
| miR-10b-5p | rs645982 | 9:1361484 | 9 | 1.36E+08 | A | G | 0.454123 | -0.12777 | 0.0141 | 1.29E-19 | trans |
| miR-10b-5p | rs543968 | 9:1361431 | 9 | 1.36E+08 | T | C | 0.653829 | -0.17433 | 0.014287 | 3.05E-34 | trans |
| miR-10b-5p | rs495203 | 9:1361452 | 9 | 1.36E+08 | C | T | 0.663279 | -0.17727 | 0.014313 | 3.15E-35 | trans |
| miR-10b-5p | rs493246 | 9:1361449 | 9 | 1.36E+08 | G | A | 0.663234 | -0.17737 | 0.014324 | 3.23E-35 | trans |
| miR-10b-5p | rs492488 | 9:1361449 | 9 | 1.36E+08 | G | A | 0.663579 | -0.17726 | 0.014316 | 3.26E-35 | trans |
| miR-10b-5p | rs473533 | 9:1361480 | 9 | 1.36E+08 | T | C | 0.453861 | -0.12801 | 0.01411 | 1.17E-19 | trans |
| miR-10b-5p | rs2519093 | 9:1361418 | 9 | 1.36E+08 | C | T | 0.809892 | -0.21558 | 0.017406 | 3.13E-35 | trans |
| miR-10b-5p | rs505922 | 9:1361492 | 9 | 1.36E+08 | T | C | 0.665748 | -0.17903 | 0.014282 | 4.80E-36 | trans |
| miR-10b-5p | rs597988 | 9:1361442 | 9 | 1.36E+08 | T | A | 0.686751 | -0.18692 | 0.015231 | 1.27E-34 | trans |
| miR-10b-5p | rs1124406 | 9:1361539 | 9 | 1.36E+08 | C | T | 0.886198 | -0.20691 | 0.022623 | 5.93E-20 | trans |
| miR-10b-5p | rs475419 | 9:1361482 | 9 | 1.36E+08 | T | C | 0.454203 | -0.12782 | 0.014102 | 1.26E-19 | trans |
| miR-10b-5p | rs1165522 | 9:1361490 | 9 | 1.36E+08 | T | A | 0.653719 | -0.1745 | 0.014267 | 2.13E-34 | trans |
| miR-10b-5p | rs633862 | 9:1361554 | 9 | 1.36E+08 | T | C | 0.460592 | -0.12659 | 0.014125 | 3.19E-19 | trans |
| miR-10b-5p | rs635634 | 9:1361550 | 9 | 1.36E+08 | C | T | 0.80599 | -0.2135 | 0.017048 | 5.57E-36 | trans |
| miR-10b-5p | rs495828 | 9:1361548 | 9 | 1.36E+08 | G | T | 0.787276 | -0.20136 | 0.016484 | 2.56E-34 | trans |
| miR-10b-5p | rs687621 | 9:1361370 | 9 | 1.36E+08 | A | G | 0.663896 | -0.1787 | 0.014295 | 7.34E-36 | trans |
| miR-10b-5p | rs576125 | 9:1361443 | 9 | 1.36E+08 | G | A | 0.688677 | -0.18654 | 0.015341 | 5.11E-34 | trans |
| miR-10b-5p | rs559723 | 9:1361504 | 9 | 1.36E+08 | A | G | 0.508431 | -0.11196 | 0.014416 | 8.07E-15 | trans |
| miR-10b-5p | rs491626 | 9:1361448 | 9 | 1.36E+08 | C | T | 0.664173 | -0.17762 | 0.014341 | 3.14E-35 | trans |
| miR-10b-5p | rs676457 | 9:1361462 | 9 | 1.36E+08 | A | T | 0.664166 | -0.17724 | 0.014307 | 3.04E-35 | trans |
| miR-10b-5p | rs543040 | 9:1361430 | 9 | 1.36E+08 | A | T | 0.669196 | -0.17944 | 0.014573 | 7.68E-35 | trans |
| miR-10b-5p | rs529565 | 9:1361495 | 9 | 1.36E+08 | T | C | 0.665911 | -0.17932 | 0.01429 | 4.06E-36 | trans |
| miR-10b-5p | rs500499 | 9:1361486 | 9 | 1.36E+08 | C | G | 0.465434 | -0.12581 | 0.014309 | 1.46E-18 | trans |
| miR-10b-5p | rs582094 | 9:1361454 | 9 | 1.36E+08 | A | T | 0.663709 | -0.17709 | 0.014302 | 3.25E-35 | trans |
| miR-10b-5p | rs644234 | 9:1361422 | 9 | 1.36E+08 | T | G | 0.645381 | -0.17197 | 0.014025 | 1.46E-34 | trans |
| miR-10b-5p | rs582118 | 9:1361454 | 9 | 1.36E+08 | A | G | 0.663872 | -0.17715 | 0.014308 | 3.28E-35 | trans |
| miR-10b-5p | rs494242 | 9:1361451 | 9 | 1.36E+08 | C | T | 0.644977 | -0.17226 | 0.014049 | 1.47E-34 | trans |
| miR-10b-5p | rs544873 | 9:1361432 | 9 | 1.36E+08 | G | A | 0.645742 | -0.17201 | 0.014043 | 1.70E-34 | trans |
| miR-10b-5p | rs2769071 | 9:1361459 | 9 | 1.36E+08 | A | G | 0.663117 | -0.17738 | 0.014331 | 3.48E-35 | trans |
| miR-10b-5p | rs647800 | 9:1361480 | 9 | 1.36E+08 | A | G | 0.435519 | -0.12855 | 0.014052 | 5.81E-20 | trans |
| miR-10b-5p | rs554833 | 9:1361471 | 9 | 1.36E+08 | C | T | 0.664057 | -0.17706 | 0.014299 | 3.22E-35 | trans |
| miR-10b-5p | rs659104 | 9:1361478 | 9 | 1.36E+08 | T | G | 0.454107 | -0.12779 | 0.014102 | 1.28E-19 | trans |
| miR-10b-5p | rs500498 | 9:1361486 | 9 | 1.36E+08 | C | T | 0.464233 | -0.12501 | 0.014291 | 2.19E-18 | trans |
| miR-10b-5p | rs630014 | 9:1361497 | 9 | 1.36E+08 | A | G | 0.494254 | -0.11543 | 0.014082 | 2.46E-16 | trans |
| miR-10b-5p | rs581107 | 9:1361477 | 9 | 1.36E+08 | C | T | 0.452595 | -0.12844 | 0.014149 | 1.11E-19 | trans |
| miR-10b-5p | rs651007 | 9:1361538 | 9 | 1.36E+08 | C | T | 0.787179 | -0.2017 | 0.016486 | 2.03E-34 | trans |
| miR-10b-5p | rs1838531 | 9:1362190 | 9 | 1.36E+08 | T | A | 0.947229 | -0.22901 | 0.033565 | 8.93E-12 | trans |
| miR-10b-5p | rs1401144 | 9:1361443 | 9 | 1.36E+08 | T | C | 0.690247 | -0.18524 | 0.015407 | 2.70E-33 | trans |
| miR-10b-5p | rs8176663 | 9:1361444 | 9 | 1.36E+08 | T | C | 0.663959 | -0.17708 | 0.014305 | 3.40E-35 | trans |
| miR-10b-5p | rs676996 | 9:1361460 | 9 | 1.36E+08 | T | G | 0.667516 | -0.17752 | 0.014432 | 8.97E-35 | trans |
| miR-10b-5p | rs616154 | 9:1361504 | 9 | 1.36E+08 | C | T | 0.501246 | -0.11378 | 0.014268 | 1.53E-15 | trans |
| miR-10b-5p | rs514659 | 9:1361422 | 9 | 1.36E+08 | A | C | 0.663927 | -0.17705 | 0.014305 | 3.51E-35 | trans |
| miR-10b-5p | rs476410 | 9:1361483 | 9 | 1.36E+08 | G | C | 0.45414 | -0.12777 | 0.014101 | 1.29E-19 | trans |
| miR-10b-5p | rs532436 | 9:1361498 | 9 | 1.36E+08 | G | A | 0.8067 | -0.21417 | 0.017094 | 5.20E-36 | trans |
| miR-10b-5p | rs527210 | 9:1361464 | 9 | 1.36E+08 | C | T | 0.668712 | -0.17637 | 0.014487 | 4.26E-34 | trans |
| miR-10b-5p | rs677355 | 9:1361460 | 9 | 1.36E+08 | G | A | 0.670198 | -0.1789 | 0.014556 | 1.02E-34 | trans |
| miR-125a-5 | rs579459 | 9:1361541 | 9 | 1.36E+08 | T | C | 0.787293 | -0.1369 | 0.013561 | 5.80E-24 | trans |
| miR-125a-5 | rs597988 | 9:1361442 | 9 | 1.36E+08 | T | A | 0.686751 | -0.11935 | 0.012565 | 2.14E-21 | trans |
| miR-125a-5 | rs674302 | 9:1361466 | 9 | 1.36E+08 | T | A | 0.663989 | -0.11309 | 0.011797 | 9.15E-22 | trans |
| miR-125a-5 | rs9411378 | 9:1361454 | 9 | 1.36E+08 | C | A | 0.753652 | -0.11233 | 0.013756 | 3.19E-16 | trans |
| miR-125a-5 | rs507666 | 9:1361493 | 9 | 1.36E+08 | G | A | 0.806573 | -0.14538 | 0.014061 | 4.69E-25 | trans |
| miR-125a-5 | rs613534 | 9:1361431 | 9 | 1.36E+08 | A | G | 0.653693 | -0.11269 | 0.011778 | 1.09E-21 | trans |

| | | | | | | | | | | | |
|---|---|---|---|---|---|---|---|---|---|---|---|
| miR-125a-5 | rs657152 | 9:1361392 | 9 | 1.36E+08 | C | A | 0.645336 | -0.11014 | 0.011569 | 1.73E-21 | trans |
| miR-125a-5 | rs495203 | 9:1361452 | 9 | 1.36E+08 | C | T | 0.663279 | -0.11324 | 0.011811 | 9.04E-22 | trans |
| miR-125a-5 | rs582118 | 9:1361454 | 9 | 1.36E+08 | A | G | 0.663872 | -0.11318 | 0.011807 | 9.18E-22 | trans |
| miR-125a-5 | rs630014 | 9:1361497 | 9 | 1.36E+08 | A | G | 0.494254 | -0.07938 | 0.011516 | 5.49E-12 | trans |
| miR-125a-5 | rs597974 | 9:1361442 | 9 | 1.36E+08 | A | G | 0.687926 | -0.11925 | 0.012622 | 3.46E-21 | trans |
| miR-125a-5 | rs2519093 | 9:1361418 | 9 | 1.36E+08 | C | T | 0.809892 | -0.14661 | 0.014322 | 1.37E-24 | trans |
| miR-125a-5 | rs630510 | 9:1361495 | 9 | 1.36E+08 | A | G | 0.494403 | -0.07951 | 0.011524 | 5.21E-12 | trans |
| miR-125a-5 | rs687289 | 9:1361371 | 9 | 1.36E+08 | G | A | 0.668579 | -0.11659 | 0.011949 | 1.72E-22 | trans |
| miR-125a-5 | rs491626 | 9:1361448 | 9 | 1.36E+08 | C | T | 0.664173 | -0.11374 | 0.011833 | 7.10E-22 | trans |
| miR-125a-5 | rs651007 | 9:1361538 | 9 | 1.36E+08 | C | T | 0.787179 | -0.1372 | 0.013562 | 4.66E-24 | trans |
| miR-125a-5 | rs543968 | 9:1361431 | 9 | 1.36E+08 | T | C | 0.653829 | -0.11276 | 0.011779 | 1.03E-21 | trans |
| miR-125a-5 | rs1401144 | 9:1361443 | 9 | 1.36E+08 | T | C | 0.690247 | -0.11767 | 0.012707 | 2.04E-20 | trans |
| miR-125a-5 | rs600038 | 9:1361518 | 9 | 1.36E+08 | T | C | 0.787872 | -0.13758 | 0.013595 | 4.53E-24 | trans |
| miR-125a-5 | rs532436 | 9:1361498 | 9 | 1.36E+08 | G | A | 0.8067 | -0.14556 | 0.01407 | 4.40E-25 | trans |
| miR-125a-5 | rs1124406 | 9:1361539 | 9 | 1.36E+08 | C | T | 0.886198 | -0.13652 | 0.018541 | 1.80E-13 | trans |
| miR-125a-5 | rs643434 | 9:1361423 | 9 | 1.36E+08 | G | A | 0.645392 | -0.11011 | 0.011578 | 1.91E-21 | trans |
| miR-125a-5 | rs576125 | 9:1361443 | 9 | 1.36E+08 | G | A | 0.688677 | -0.11867 | 0.012655 | 6.79E-21 | trans |
| miR-125a-5 | rs676457 | 9:1361462 | 9 | 1.36E+08 | A | T | 0.664166 | -0.11313 | 0.011807 | 9.53E-22 | trans |
| miR-125a-5 | rs582094 | 9:1361454 | 9 | 1.36E+08 | A | T | 0.663709 | -0.11308 | 0.011802 | 9.54E-22 | trans |
| miR-125a-5 | rs644234 | 9:1361422 | 9 | 1.36E+08 | T | G | 0.645381 | -0.11013 | 0.011569 | 1.74E-21 | trans |
| miR-125a-5 | rs8176663 | 9:1361444 | 9 | 1.36E+08 | T | C | 0.663959 | -0.11308 | 0.011804 | 9.73E-22 | trans |
| miR-125a-5 | rs676996 | 9:1361460 | 9 | 1.36E+08 | T | G | 0.667516 | -0.1133 | 0.011908 | 1.82E-21 | trans |
| miR-125a-5 | rs514659 | 9:1361422 | 9 | 1.36E+08 | A | C | 0.663927 | -0.11306 | 0.011805 | 9.96E-22 | trans |
| miR-125a-5 | rs545971 | 9:1361433 | 9 | 1.36E+08 | C | T | 0.663766 | -0.11297 | 0.0118 | 1.03E-21 | trans |
| miR-125a-5 | rs505922 | 9:1361492 | 9 | 1.36E+08 | T | C | 0.665748 | -0.1157 | 0.011784 | 9.35E-23 | trans |
| miR-125a-5 | rs635634 | 9:1361550 | 9 | 1.36E+08 | C | T | 0.80599 | -0.14484 | 0.014033 | 5.63E-25 | trans |
| miR-125a-5 | rs492488 | 9:1361449 | 9 | 1.36E+08 | G | A | 0.663579 | -0.11329 | 0.011813 | 8.76E-22 | trans |
| miR-125a-5 | rs527210 | 9:1361464 | 9 | 1.36E+08 | C | T | 0.668712 | -0.11248 | 0.01195 | 4.84E-21 | trans |
| miR-125a-5 | rs612169 | 9:1361434 | 9 | 1.36E+08 | A | G | 0.66434 | -0.1132 | 0.011821 | 1.01E-21 | trans |
| miR-125a-5 | rs554833 | 9:1361471 | 9 | 1.36E+08 | C | T | 0.664057 | -0.11315 | 0.011799 | 8.84E-22 | trans |
| miR-125a-5 | rs495828 | 9:1361548 | 9 | 1.36E+08 | G | T | 0.787276 | -0.13688 | 0.01356 | 5.84E-24 | trans |
| miR-125a-5 | rs543040 | 9:1361430 | 9 | 1.36E+08 | A | T | 0.669196 | -0.1144 | 0.012024 | 1.83E-21 | trans |
| miR-125a-5 | rs616154 | 9:1361504 | 9 | 1.36E+08 | C | T | 0.501246 | -0.07884 | 0.011664 | 1.39E-11 | trans |
| miR-125a-5 | rs550057 | 9:1361465 | 9 | 1.36E+08 | C | T | 0.734085 | -0.11203 | 0.012688 | 1.05E-18 | trans |
| miR-125a-5 | rs529565 | 9:1361495 | 9 | 1.36E+08 | T | C | 0.665911 | -0.11591 | 0.01179 | 8.29E-23 | trans |
| miR-125a-5 | rs687621 | 9:1361370 | 9 | 1.36E+08 | A | G | 0.663896 | -0.11347 | 0.011802 | 6.91E-22 | trans |
| miR-125a-5 | rs493246 | 9:1361449 | 9 | 1.36E+08 | G | A | 0.663234 | -0.11332 | 0.01182 | 9.02E-22 | trans |
| miR-125a-5 | rs649129 | 9:1361543 | 9 | 1.36E+08 | C | T | 0.787432 | -0.13706 | 0.013569 | 5.46E-24 | trans |
| miR-125a-5 | rs544873 | 9:1361432 | 9 | 1.36E+08 | G | A | 0.645742 | -0.11013 | 0.011584 | 1.95E-21 | trans |
| miR-125a-5 | rs2769071 | 9:1361459 | 9 | 1.36E+08 | A | G | 0.663117 | -0.11313 | 0.011827 | 1.12E-21 | trans |
| miR-125a-5 | rs494242 | 9:1361451 | 9 | 1.36E+08 | C | T | 0.644977 | -0.11046 | 0.011588 | 1.54E-21 | trans |
| miR-125a-5 | rs1165522 | 9:1361490 | 9 | 1.36E+08 | T | A | 0.653719 | -0.11197 | 0.011767 | 1.81E-21 | trans |
| miR-125a-5 | rs677355 | 9:1361460 | 9 | 1.36E+08 | G | A | 0.670198 | -0.1132 | 0.012014 | 4.41E-21 | trans |
| miR-126-3p | rs8176681 | 9:1361397 | 9 | 1.36E+08 | T | C | 0.600667 | 0.112306 | 0.012838 | 2.18E-18 | trans |
| miR-126-3p | rs581107 | 9:1361477 | 9 | 1.36E+08 | C | T | 0.452595 | -0.09502 | 0.013059 | 3.43E-13 | trans |
| miR-126-3p | rs647800 | 9:1361480 | 9 | 1.36E+08 | A | G | 0.435519 | -0.09393 | 0.012976 | 4.52E-13 | trans |
| miR-126-3p | rs659104 | 9:1361478 | 9 | 1.36E+08 | T | G | 0.454107 | -0.09467 | 0.013015 | 3.50E-13 | trans |
| miR-126-3p | rs579459 | 9:1361541 | 9 | 1.36E+08 | T | C | 0.787293 | -0.19268 | 0.01507 | 1.96E-37 | trans |
| miR-126-3p | rs676457 | 9:1361462 | 9 | 1.36E+08 | A | T | 0.664166 | -0.17146 | 0.013067 | 2.46E-39 | trans |
| miR-126-3p | rs657152 | 9:1361392 | 9 | 1.36E+08 | C | A | 0.645336 | -0.1645 | 0.012821 | 1.11E-37 | trans |
| miR-126-3p | rs9411378 | 9:1361454 | 9 | 1.36E+08 | C | A | 0.753652 | -0.16555 | 0.015322 | 3.27E-27 | trans |
| miR-126-3p | rs2519093 | 9:1361418 | 9 | 1.36E+08 | C | T | 0.809892 | -0.21142 | 0.015879 | 1.90E-40 | trans |
| miR-126-3p | rs8176690 | 9:1361383 | 9 | 1.36E+08 | A | G | 0.601023 | 0.112118 | 0.012826 | 2.30E-18 | trans |
| miR-126-3p | rs576125 | 9:1361443 | 9 | 1.36E+08 | G | A | 0.688677 | -0.17847 | 0.014024 | 4.24E-37 | trans |
| miR-126-3p | rs494242 | 9:1361451 | 9 | 1.36E+08 | C | T | 0.644977 | -0.16489 | 0.012842 | 9.85E-38 | trans |
| miR-126-3p | rs651007 | 9:1361538 | 9 | 1.36E+08 | C | T | 0.787179 | -0.19282 | 0.015072 | 1.79E-37 | trans |
| miR-126-3p | rs514659 | 9:1361422 | 9 | 1.36E+08 | A | C | 0.663927 | -0.17127 | 0.013065 | 2.91E-39 | trans |
| miR-126-3p | rs630510 | 9:1361495 | 9 | 1.36E+08 | A | G | 0.494403 | -0.1155 | 0.012884 | 3.11E-19 | trans |
| miR-126-3p | rs7857390 | 9:1361285 | 9 | 1.36E+08 | A | G | 0.377967 | -0.11637 | 0.013459 | 5.31E-18 | trans |
| miR-126-3p | rs559723 | 9:1361504 | 9 | 1.36E+08 | A | G | 0.508431 | -0.11175 | 0.013185 | 2.33E-17 | trans |
| miR-126-3p | rs543040 | 9:1361430 | 9 | 1.36E+08 | A | T | 0.669196 | -0.1736 | 0.013309 | 6.96E-39 | trans |
| miR-126-3p | rs613534 | 9:1361431 | 9 | 1.36E+08 | A | G | 0.653693 | -0.16547 | 0.013067 | 9.46E-37 | trans |
| miR-126-3p | rs582094 | 9:1361454 | 9 | 1.36E+08 | A | T | 0.663709 | -0.17139 | 0.013061 | 2.46E-39 | trans |

| | | | | | | | | | | | |
|---|---|---|---|---|---|---|---|---|---|---|---|
| miR-126-3 | rs2073828 | 9:1361371 | 9 | 1.36E+08 | G | A | 0.601379 | 0.112127 | 0.012832 | 2.37E-18 | trans |
| miR-126-3 | rs493246 | 9:1361449 | 9 | 1.36E+08 | G | A | 0.663234 | -0.17162 | 0.013081 | 2.55E-39 | trans |
| miR-126-3 | rs8176702 | 9:1361361 | 9 | 1.36E+08 | G | A | 0.6197 | 0.114009 | 0.012941 | 1.25E-18 | trans |
| miR-126-3 | rs649129 | 9:1361543 | 9 | 1.36E+08 | C | T | 0.787432 | -0.19281 | 0.015079 | 1.95E-37 | trans |
| miR-126-3 | rs687621 | 9:1361370 | 9 | 1.36E+08 | A | G | 0.663896 | -0.17178 | 0.013061 | 1.64E-39 | trans |
| miR-126-3 | rs8176686 | 9:1361387 | 9 | 1.36E+08 | T | C | 0.595889 | 0.112828 | 0.013358 | 3.00E-17 | trans |
| miR-126-3 | rs612169 | 9:1361434 | 9 | 1.36E+08 | A | G | 0.66434 | -0.17151 | 0.013083 | 2.89E-39 | trans |
| miR-126-3 | rs674302 | 9:1361466 | 9 | 1.36E+08 | T | A | 0.663989 | -0.1715 | 0.013055 | 2.03E-39 | trans |
| miR-126-3 | rs7873635 | 9:1361320 | 9 | 1.36E+08 | T | C | 0.381256 | -0.11248 | 0.013126 | 1.04E-17 | trans |
| miR-126-3 | rs8176668 | 9:1361440 | 9 | 1.36E+08 | A | T | 0.618711 | 0.11469 | 0.012959 | 8.74E-19 | trans |
| miR-126-3 | rs676996 | 9:1361460 | 9 | 1.36E+08 | T | G | 0.667516 | -0.17228 | 0.013178 | 4.67E-39 | trans |
| miR-126-3 | rs597974 | 9:1361442 | 9 | 1.36E+08 | A | G | 0.687926 | -0.17932 | 0.013984 | 1.22E-37 | trans |
| miR-126-3 | rs491626 | 9:1361448 | 9 | 1.36E+08 | C | T | 0.664173 | -0.17201 | 0.013097 | 2.10E-39 | trans |
| miR-126-3 | rs616154 | 9:1361504 | 9 | 1.36E+08 | C | T | 0.501246 | -0.11412 | 0.013046 | 2.17E-18 | trans |
| miR-126-3 | rs8176649 | 9:1361472 | 9 | 1.36E+08 | G | A | 0.618669 | 0.114693 | 0.01294 | 7.78E-19 | trans |
| miR-126-3 | rs532436 | 9:1361498 | 9 | 1.36E+08 | G | A | 0.8067 | -0.20751 | 0.015609 | 2.49E-40 | trans |
| miR-126-3 | rs527210 | 9:1361464 | 9 | 1.36E+08 | C | T | 0.668712 | -0.17177 | 0.013226 | 1.44E-38 | trans |
| miR-126-3 | rs687289 | 9:1361371 | 9 | 1.36E+08 | G | A | 0.668579 | -0.17767 | 0.01321 | 3.06E-41 | trans |
| miR-126-3 | rs505922 | 9:1361492 | 9 | 1.36E+08 | T | C | 0.665748 | -0.17418 | 0.013036 | 1.02E-40 | trans |
| miR-126-3 | rs645982 | 9:1361484 | 9 | 1.36E+08 | A | G | 0.454123 | -0.09467 | 0.013014 | 3.48E-13 | trans |
| miR-126-3 | rs8176715 | 9:1361331 | 9 | 1.36E+08 | T | C | 0.418056 | -0.11649 | 0.013136 | 7.43E-19 | trans |
| miR-126-3 | rs8176682 | 9:1361392 | 9 | 1.36E+08 | C | T | 0.619011 | 0.113671 | 0.012936 | 1.54E-18 | trans |
| miR-126-3 | rs476410 | 9:1361483 | 9 | 1.36E+08 | G | C | 0.45414 | -0.09468 | 0.013015 | 3.48E-13 | trans |
| miR-126-3 | rs8176663 | 9:1361444 | 9 | 1.36E+08 | T | C | 0.663959 | -0.17133 | 0.013064 | 2.73E-39 | trans |
| miR-126-3 | rs495203 | 9:1361452 | 9 | 1.36E+08 | C | T | 0.663279 | -0.17152 | 0.013072 | 2.48E-39 | trans |
| miR-126-3 | rs644234 | 9:1361422 | 9 | 1.36E+08 | T | G | 0.645381 | -0.16456 | 0.012821 | 1.03E-37 | trans |
| miR-126-3 | rs8176691 | 9:1361382 | 9 | 1.36E+08 | C | T | 0.619638 | 0.114018 | 0.012938 | 1.22E-18 | trans |
| miR-126-3 | rs635634 | 9:1361550 | 9 | 1.36E+08 | C | T | 0.80599 | -0.20691 | 0.015566 | 2.57E-40 | trans |
| miR-126-3 | rs7036642 | 9:1361446 | 9 | 1.36E+08 | G | A | 0.618956 | 0.114538 | 0.012952 | 9.29E-19 | trans |
| miR-126-3 | rs495828 | 9:1361548 | 9 | 1.36E+08 | G | T | 0.787276 | -0.19264 | 0.015069 | 2.00E-37 | trans |
| miR-126-3 | rs600038 | 9:1361518 | 9 | 1.36E+08 | T | C | 0.787872 | -0.19328 | 0.015108 | 1.80E-37 | trans |
| miR-126-3 | rs550057 | 9:1361465 | 9 | 1.36E+08 | C | T | 0.734085 | -0.16349 | 0.014114 | 5.01E-31 | trans |
| miR-126-3 | rs529565 | 9:1361495 | 9 | 1.36E+08 | T | C | 0.665911 | -0.17441 | 0.013044 | 8.91E-41 | trans |
| miR-126-3 | rs633862 | 9:1361554 | 9 | 1.36E+08 | T | C | 0.460592 | -0.09673 | 0.013025 | 1.12E-13 | trans |
| miR-126-3 | rs473533 | 9:1361480 | 9 | 1.36E+08 | T | C | 0.453861 | -0.0947 | 0.013024 | 3.56E-13 | trans |
| miR-126-3 | rs2073826 | 9:1361369 | 9 | 1.36E+08 | G | T | 0.60107 | 0.112151 | 0.012825 | 2.24E-18 | trans |
| miR-126-3 | rs543968 | 9:1361431 | 9 | 1.36E+08 | T | C | 0.653829 | -0.16548 | 0.013068 | 9.52E-37 | trans |
| miR-126-3 | rs630014 | 9:1361497 | 9 | 1.36E+08 | A | G | 0.494254 | -0.1154 | 0.012875 | 3.15E-19 | trans |
| miR-126-3 | rs1165522 | 9:1361490 | 9 | 1.36E+08 | T | A | 0.653719 | -0.16755 | 0.013039 | 8.64E-38 | trans |
| miR-126-3 | rs677355 | 9:1361460 | 9 | 1.36E+08 | G | A | 0.670198 | -0.17272 | 0.013297 | 1.41E-38 | trans |
| miR-126-3 | rs660340 | 9:1361475 | 9 | 1.36E+08 | A | G | 0.454082 | -0.09478 | 0.013022 | 3.37E-13 | trans |
| miR-126-3 | rs597988 | 9:1361442 | 9 | 1.36E+08 | T | A | 0.686751 | -0.17928 | 0.01392 | 5.86E-38 | trans |
| miR-126-3 | rs545971 | 9:1361433 | 9 | 1.36E+08 | C | T | 0.663766 | -0.17125 | 0.013059 | 2.76E-39 | trans |
| miR-126-3 | rs500499 | 9:1361486 | 9 | 1.36E+08 | C | G | 0.465434 | -0.09086 | 0.013209 | 6.05E-12 | trans |
| miR-126-3 | rs643434 | 9:1361423 | 9 | 1.36E+08 | G | A | 0.645392 | -0.16456 | 0.012831 | 1.18E-37 | trans |
| miR-126-3 | rs1401144 | 9:1361443 | 9 | 1.36E+08 | T | C | 0.690247 | -0.17776 | 0.014083 | 1.60E-36 | trans |
| miR-126-3 | rs7046674 | 9:1361470 | 9 | 1.36E+08 | C | T | 0.619013 | 0.115101 | 0.01296 | 6.62E-19 | trans |
| miR-126-3 | rs1178913 | 9:1361853 | 9 | 1.36E+08 | C | G | 0.833223 | -0.12126 | 0.017843 | 1.08E-11 | trans |
| miR-126-3 | rs475419 | 9:1361482 | 9 | 1.36E+08 | T | C | 0.454203 | -0.09471 | 0.013015 | 3.41E-13 | trans |
| miR-126-3 | rs4962040 | 9:1361335 | 9 | 1.36E+08 | A | G | 0.619057 | 0.11337 | 0.012952 | 2.08E-18 | trans |
| miR-126-3 | rs492488 | 9:1361449 | 9 | 1.36E+08 | G | A | 0.663579 | -0.17151 | 0.013074 | 2.58E-39 | trans |
| miR-126-3 | rs582118 | 9:1361454 | 9 | 1.36E+08 | A | G | 0.663872 | -0.17144 | 0.013067 | 2.52E-39 | trans |
| miR-126-3 | rs554833 | 9:1361471 | 9 | 1.36E+08 | C | T | 0.664057 | -0.17137 | 0.013058 | 2.40E-39 | trans |
| miR-126-3 | rs500498 | 9:1361486 | 9 | 1.36E+08 | C | T | 0.464233 | -0.09029 | 0.013192 | 7.68E-12 | trans |
| miR-126-3 | rs2769071 | 9:1361459 | 9 | 1.36E+08 | A | G | 0.663117 | -0.17128 | 0.01309 | 4.06E-39 | trans |
| miR-126-3 | rs507666 | 9:1361493 | 9 | 1.36E+08 | G | A | 0.806573 | -0.20737 | 0.015598 | 2.50E-40 | trans |
| miR-126-3 | rs1124406 | 9:1361539 | 9 | 1.36E+08 | C | T | 0.886198 | -0.21457 | 0.020633 | 2.49E-25 | trans |
| miR-126-3 | rs2073827 | 9:1361371 | 9 | 1.36E+08 | G | C | 0.601379 | 0.112127 | 0.012832 | 2.37E-18 | trans |
| miR-126-3 | rs544873 | 9:1361432 | 9 | 1.36E+08 | G | A | 0.645742 | -0.16454 | 0.012837 | 1.31E-37 | trans |
| miR-126-5 | rs1014847 | 14:2143026 | 14 | 21430269 | C | A | 0.760451 | -0.15736 | 0.021765 | 4.84E-13 | trans |
| miR-126-5 | rs514659 | 9:1361422 | 9 | 1.36E+08 | A | C | 0.663927 | -0.12799 | 0.019019 | 1.70E-11 | trans |
| miR-126-5 | rs6197743 | 14:2143099 | 14 | 21430994 | G | A | 0.76048 | -0.15732 | 0.021768 | 4.93E-13 | trans |
| miR-126-5 | rs529565 | 9:1361495 | 9 | 1.36E+08 | T | C | 0.665911 | -0.12899 | 0.019013 | 1.17E-11 | trans |

| | | | | | | | | | | | |
|---|---|---|---|---|---|---|---|---|---|---|---|
| miR-126-5p | rs1289748 | 14:2143129 | 14 | 21431295 | C | T | 0.766905 | -0.15344 | 0.021953 | 2.76E-12 | trans |
| miR-126-5p | rs491626 | 9:1361448 | 9 | 1.36E+08 | C | T | 0.664173 | -0.1286 | 0.019067 | 1.54E-11 | trans |
| miR-126-5p | rs495203 | 9:1361452 | 9 | 1.36E+08 | C | T | 0.663279 | -0.12799 | 0.01903 | 1.75E-11 | trans |
| miR-126-5p | rs554833 | 9:1361471 | 9 | 1.36E+08 | C | T | 0.664057 | -0.12781 | 0.019011 | 1.78E-11 | trans |
| miR-126-5p | rs3790067 | 14:2142108 | 14 | 21421088 | C | T | 0.767067 | -0.15403 | 0.021945 | 2.24E-12 | trans |
| miR-126-5p | rs644234 | 9:1361422 | 9 | 1.36E+08 | T | G | 0.645381 | -0.13072 | 0.018618 | 2.21E-12 | trans |
| miR-126-5p | rs6197743 | 14:2143084 | 14 | 21430843 | G | A | 0.760478 | -0.15734 | 0.021767 | 4.90E-13 | trans |
| miR-126-5p | rs582094 | 9:1361454 | 9 | 1.36E+08 | A | T | 0.663709 | -0.12788 | 0.019015 | 1.75E-11 | trans |
| miR-126-5p | rs544873 | 9:1361432 | 9 | 1.36E+08 | G | A | 0.645742 | -0.13112 | 0.01864 | 2.00E-12 | trans |
| miR-126-5p | rs687289 | 9:1361371 | 9 | 1.36E+08 | G | A | 0.668579 | -0.13264 | 0.019257 | 5.66E-12 | trans |
| miR-126-5p | rs674302 | 9:1361466 | 9 | 1.36E+08 | T | A | 0.663989 | -0.1278 | 0.019008 | 1.77E-11 | trans |
| miR-126-5p | rs1288228 | 14:2143130 | 14 | 21431304 | T | C | 0.761865 | -0.15698 | 0.021787 | 5.79E-13 | trans |
| miR-126-5p | rs493246 | 9:1361449 | 9 | 1.36E+08 | G | A | 0.663234 | -0.12818 | 0.019044 | 1.69E-11 | trans |
| miR-126-5p | rs612169 | 9:1361434 | 9 | 1.36E+08 | A | G | 0.66434 | -0.12817 | 0.019045 | 1.70E-11 | trans |
| miR-126-5p | rs2769071 | 9:1361459 | 9 | 1.36E+08 | A | G | 0.663117 | -0.12801 | 0.019054 | 1.84E-11 | trans |
| miR-126-5p | rs543968 | 9:1361431 | 9 | 1.36E+08 | T | C | 0.653829 | -0.13102 | 0.018967 | 4.92E-12 | trans |
| miR-126-5p | rs1288230 | 14:2143134 | 14 | 21431349 | T | A | 0.766982 | -0.15321 | 0.021948 | 2.94E-12 | trans |
| miR-126-5p | rs6704901 | 14:2143110 | 14 | 21431105 | G | A | 0.761822 | -0.15709 | 0.021788 | 5.59E-13 | trans |
| miR-126-5p | rs613534 | 9:1361431 | 9 | 1.36E+08 | A | G | 0.653693 | -0.13103 | 0.018965 | 4.88E-12 | trans |
| miR-126-5p | rs505922 | 9:1361492 | 9 | 1.36E+08 | T | C | 0.665748 | -0.12889 | 0.019002 | 1.18E-11 | trans |
| miR-126-5p | rs643434 | 9:1361423 | 9 | 1.36E+08 | G | A | 0.645392 | -0.13124 | 0.01863 | 1.86E-12 | trans |
| miR-126-5p | rs1165522 | 9:1361490 | 9 | 1.36E+08 | T | A | 0.653719 | -0.13433 | 0.018932 | 1.29E-12 | trans |
| miR-126-5p | rs1289767 | 14:214314 | 14 | 21431413 | C | T | 0.760166 | -0.15737 | 0.021775 | 4.93E-13 | trans |
| miR-126-5p | rs8176663 | 9:1361444 | 9 | 1.36E+08 | T | C | 0.663959 | -0.12797 | 0.019019 | 1.72E-11 | trans |
| miR-126-5p | rs492488 | 9:1361449 | 9 | 1.36E+08 | G | A | 0.663579 | -0.12801 | 0.019034 | 1.75E-11 | trans |
| miR-126-5p | rs545971 | 9:1361433 | 9 | 1.36E+08 | C | T | 0.663766 | -0.12787 | 0.019011 | 1.74E-11 | trans |
| miR-126-5p | rs582118 | 9:1361454 | 9 | 1.36E+08 | A | G | 0.663872 | -0.12799 | 0.019023 | 1.72E-11 | trans |
| miR-126-5p | rs543040 | 9:1361430 | 9 | 1.36E+08 | A | T | 0.669196 | -0.12981 | 0.019369 | 2.06E-11 | trans |
| miR-126-5p | rs494242 | 9:1361451 | 9 | 1.36E+08 | C | T | 0.644977 | -0.13153 | 0.018648 | 1.75E-12 | trans |
| miR-126-5p | rs1289765 | 14:2143138 | 14 | 21431382 | C | T | 0.760157 | -0.15739 | 0.021775 | 4.91E-13 | trans |
| miR-126-5p | rs687621 | 9:1361370 | 9 | 1.36E+08 | A | G | 0.663896 | -0.12975 | 0.019012 | 8.81E-12 | trans |
| miR-126-5p | rs657152 | 9:1361392 | 9 | 1.36E+08 | C | A | 0.645336 | -0.13069 | 0.018619 | 2.23E-12 | trans |
| miR-126-5p | rs676457 | 9:1361462 | 9 | 1.36E+08 | A | T | 0.664166 | -0.128 | 0.019023 | 1.71E-11 | trans |
| miR-130a-3 | rs602861 | 11:576873 | 11 | 57687365 | G | C | 0.45465 | -0.15054 | 0.021197 | 1.23E-12 | cis |
| miR-130a-3 | rs1089666 | 11:576790 | 11 | 57679029 | C | A | 0.697394 | -0.21962 | 0.023047 | 1.58E-21 | cis |
| miR-130a-3 | rs2511993 | 11:573928 | 11 | 57392824 | C | T | 0.568694 | 0.154849 | 0.021586 | 7.31E-13 | cis |
| miR-130a-3 | rs7862440 | 11:573754 | 11 | 57375463 | G | C | 0.750533 | -0.29953 | 0.02326 | 6.00E-38 | cis |
| miR-130a-3 | rs1624389 | 11:574462 | 11 | 57446272 | A | G | 0.451986 | -0.16492 | 0.021048 | 4.67E-15 | cis |
| miR-130a-3 | rs1089664 | 11:575639 | 11 | 57563991 | C | T | 0.567891 | 0.160898 | 0.021008 | 1.88E-14 | cis |
| miR-130a-3 | rs1613592 | 11:574240 | 11 | 57424000 | G | T | 0.413386 | -0.1541 | 0.021359 | 5.40E-13 | cis |
| miR-130a-3 | rs7858720 | 11:576549 | 11 | 57654991 | T | G | 0.691717 | -0.22928 | 0.022913 | 1.43E-23 | cis |
| miR-130a-3 | rs588612 | 11:576128 | 11 | 57612891 | G | C | 0.439957 | -0.16246 | 0.021014 | 1.07E-14 | cis |
| miR-130a-3 | rs1122907 | 11:573990 | 11 | 57399072 | C | T | 0.749544 | -0.3014 | 0.023308 | 3.01E-38 | cis |
| miR-130a-3 | rs7102271 | 11:573968 | 11 | 57396825 | G | T | 0.749722 | -0.30147 | 0.023289 | 2.50E-38 | cis |
| miR-130a-3 | rs644883 | 11:576635 | 11 | 57663522 | C | G | 0.310308 | 0.226662 | 0.023122 | 1.10E-22 | cis |
| miR-130a-3 | rs1089664 | 11:575877 | 11 | 57587795 | A | G | 0.565548 | 0.160285 | 0.020992 | 2.25E-14 | cis |
| miR-130a-3 | rs1089665 | 11:576689 | 11 | 57668941 | C | G | 0.697383 | -0.22009 | 0.022989 | 1.03E-21 | cis |
| miR-130a-3 | rs1557522 | 11:573815 | 11 | 57381519 | T | C | 0.568447 | 0.152599 | 0.021513 | 1.31E-12 | cis |
| miR-130a-3 | rs639158 | 11:576020 | 11 | 57602039 | G | A | 0.441293 | -0.16365 | 0.021032 | 7.19E-15 | cis |
| miR-130a-3 | rs1122907 | 11:573910 | 11 | 57391023 | A | C | 0.750877 | -0.3019 | 0.023261 | 1.61E-38 | cis |
| miR-130a-3 | rs665058 | 11:575791 | 11 | 57579166 | T | C | 0.565581 | 0.160316 | 0.020993 | 2.23E-14 | cis |
| miR-130a-3 | rs929934 | 11:574112 | 11 | 57411252 | T | C | 0.447765 | -0.15731 | 0.021199 | 1.17E-13 | cis |
| miR-130a-3 | rs1160573 | 11:575240 | 11 | 57524089 | C | G | 0.683995 | -0.24275 | 0.022162 | 6.40E-28 | cis |
| miR-130a-3 | rs547891 | 11:574990 | 11 | 57499065 | C | T | 0.319553 | 0.241325 | 0.022213 | 1.70E-27 | cis |
| miR-130a-3 | rs9420 | 11:575102 | 11 | 57510294 | A | G | 0.309761 | 0.240756 | 0.0223 | 3.59E-27 | cis |
| miR-130a-3 | rs1278553 | 11:574866 | 11 | 57486651 | G | A | 0.685995 | -0.24279 | 0.022262 | 1.08E-27 | cis |
| miR-130a-3 | rs1160670 | 11:573835 | 11 | 57383575 | G | T | 0.750529 | -0.30167 | 0.023272 | 1.98E-38 | cis |
| miR-130a-3 | rs2511996 | 11:573913 | 11 | 57391395 | T | C | 0.568691 | 0.154709 | 0.021579 | 7.54E-13 | cis |
| miR-130a-3 | rs1160667 | 11:573833 | 11 | 57383377 | G | A | 0.750505 | -0.30163 | 0.023271 | 2.02E-38 | cis |
| miR-130a-3 | rs1160432 | 11:574970 | 11 | 57497011 | C | A | 0.688224 | -0.24209 | 0.022277 | 1.65E-27 | cis |
| miR-130a-3 | rs7836482 | 11:573707 | 11 | 57370742 | C | T | 0.752736 | -0.30697 | 0.023512 | 5.91E-39 | cis |
| miR-130a-3 | rs1798177 | 11:574121 | 11 | 57412150 | C | T | 0.4471 | -0.15357 | 0.021109 | 3.46E-13 | cis |
| miR-130a-3 | rs2081362 | 11:573938 | 11 | 57393884 | T | G | 0.567578 | 0.158761 | 0.021771 | 3.05E-13 | cis |

| | | | | | | | | | | | |
|---|---|---|---|---|---|---|---|---|---|---|---|
| miR-130a-3 | rs55920079 | 11:57410184 | 11 | 57410184 | T | C | 0.745296 | -0.29347 | 0.023524 | 1.02E-35 | cis |
| miR-130a-3 | rs11229067 | 11:57374946 | 11 | 57374946 | G | T | 0.750531 | -0.29953 | 0.02326 | 6.00E-38 | cis |
| miR-130a-3 | rs2508441 | 11:57375424 | 11 | 57375424 | G | A | 0.564809 | 0.14881 | 0.021438 | 3.88E-12 | cis |
| miR-130a-3 | rs641325 | 11:57681828 | 11 | 57681828 | T | G | 0.327793 | 0.198345 | 0.022691 | 2.30E-18 | cis |
| miR-130a-3 | rs2511994 | 11:57391691 | 11 | 57391691 | A | G | 0.56903 | 0.155056 | 0.021597 | 6.99E-13 | cis |
| miR-130a-3 | rs3781885 | 11:57570680 | 11 | 57570680 | C | T | 0.700878 | -0.24761 | 0.022418 | 2.31E-28 | cis |
| miR-130a-3 | rs35808061 | 11:57547000 | 11 | 57547000 | T | C | 0.684777 | -0.2434 | 0.022156 | 4.47E-28 | cis |
| miR-130a-3 | rs585761 | 11:57597367 | 11 | 57597367 | G | A | 0.43789 | -0.1607 | 0.021012 | 2.04E-14 | cis |
| miR-130a-3 | rs10792111 | 11:57565800 | 11 | 57565800 | C | T | 0.684897 | -0.24331 | 0.02216 | 4.77E-28 | cis |
| miR-130a-3 | rs11350477 | 11:57639926 | 11 | 57639926 | T | G | 0.651033 | 0.166552 | 0.024582 | 1.24E-11 | cis |
| miR-130a-3 | rs79564083 | 11:57361589 | 11 | 57361589 | T | C | 0.897878 | -0.29304 | 0.03481 | 3.83E-17 | cis |
| miR-130a-3 | rs11229060 | 11:57374871 | 11 | 57374871 | C | G | 0.750533 | -0.29953 | 0.02326 | 6.00E-38 | cis |
| miR-130a-3 | rs12270209 | 11:57537787 | 11 | 57537787 | G | A | 0.564726 | 0.161383 | 0.021024 | 1.64E-14 | cis |
| miR-130a-3 | rs631791 | 11:57675410 | 11 | 57675410 | A | G | 0.307763 | 0.220062 | 0.023047 | 1.32E-21 | cis |
| miR-130a-3 | rs11604331 | 11:57467035 | 11 | 57467035 | C | G | 0.686388 | -0.24468 | 0.022232 | 3.58E-28 | cis |
| miR-130a-3 | rs12274865 | 11:57557668 | 11 | 57557668 | C | T | 0.56562 | 0.161052 | 0.021005 | 1.76E-14 | cis |
| miR-130a-3 | rs708228 | 11:57585662 | 11 | 57585662 | C | T | 0.684575 | -0.24192 | 0.022159 | 9.51E-28 | cis |
| miR-130a-3 | rs1631684 | 11:57420885 | 11 | 57420885 | G | A | 0.412965 | -0.15412 | 0.021382 | 5.68E-13 | cis |
| miR-130a-3 | rs28362948 | 11:57369013 | 11 | 57369013 | A | C | 0.752624 | -0.30649 | 0.023509 | 7.52E-39 | cis |
| miR-130a-3 | rs1783824 | 11:57473012 | 11 | 57473012 | A | G | 0.452208 | -0.16406 | 0.020981 | 5.31E-15 | cis |
| miR-130a-3 | rs652908 | 11:57604009 | 11 | 57604009 | C | T | 0.441718 | -0.16383 | 0.021034 | 6.76E-15 | cis |
| miR-130a-3 | rs7120868 | 11:57530528 | 11 | 57530528 | T | G | 0.566777 | 0.161451 | 0.021059 | 1.77E-14 | cis |
| miR-130a-3 | rs686489 | 11:57659832 | 11 | 57659832 | A | G | 0.563137 | 0.159924 | 0.02141 | 8.05E-14 | cis |
| miR-130a-3 | rs1783828 | 11:57411521 | 11 | 57411521 | T | C | 0.446851 | -0.15659 | 0.021191 | 1.47E-13 | cis |
| miR-130a-3 | rs1783829 | 11:57419880 | 11 | 57419880 | T | C | 0.413999 | -0.15441 | 0.021345 | 4.68E-13 | cis |
| miR-130a-3 | rs11607127 | 11:57389737 | 11 | 57389737 | G | A | 0.743794 | -0.30287 | 0.023174 | 4.93E-39 | cis |
| miR-130a-3 | rs2511989 | 11:57378325 | 11 | 57378325 | C | T | 0.562864 | 0.148597 | 0.021337 | 3.30E-12 | cis |
| miR-130a-3 | rs1783818 | 11:57437551 | 11 | 57437551 | C | T | 0.454526 | -0.16598 | 0.021036 | 3.01E-15 | cis |
| miR-130a-3 | rs7105506 | 11:57497980 | 11 | 57497980 | G | A | 0.56028 | 0.172703 | 0.021219 | 3.98E-16 | cis |
| miR-130a-3 | rs71484468 | 11:57632799 | 11 | 57632799 | G | C | 0.683543 | -0.24161 | 0.02258 | 1.02E-26 | cis |
| miR-130a-3 | rs1798173 | 11:57432057 | 11 | 57432057 | T | C | 0.414473 | -0.1552 | 0.021395 | 4.05E-13 | cis |
| miR-130a-3 | rs2847308 | 11:57498232 | 11 | 57498232 | C | T | 0.319113 | 0.241055 | 0.022203 | 1.85E-27 | cis |
| miR-130a-3 | rs28362950 | 11:57371918 | 11 | 57371918 | G | A | 0.749181 | -0.30047 | 0.023454 | 1.43E-37 | cis |
| miR-130a-3 | rs2081361 | 11:57411742 | 11 | 57411742 | C | T | 0.745781 | -0.29375 | 0.023511 | 8.04E-36 | cis |
| miR-130a-3 | rs10896631 | 11:57424040 | 11 | 57424040 | T | C | 0.698591 | -0.25742 | 0.022716 | 9.10E-30 | cis |
| miR-130a-3 | rs2454660 | 11:57378640 | 11 | 57378640 | G | A | 0.564411 | 0.149045 | 0.021413 | 3.39E-12 | cis |
| miR-130a-3 | rs1786438 | 11:57597065 | 11 | 57597065 | T | G | 0.439556 | -0.16231 | 0.020983 | 1.03E-14 | cis |
| miR-130a-3 | rs1783984 | 11:57429480 | 11 | 57429480 | G | T | 0.423293 | -0.15785 | 0.021629 | 2.92E-13 | cis |
| miR-130a-3 | rs585337 | 11:57597274 | 11 | 57597274 | C | T | 0.439573 | -0.16233 | 0.020984 | 1.03E-14 | cis |
| miR-130a-3 | rs10792110 | 11:57536346 | 11 | 57536346 | C | T | 0.564735 | 0.161403 | 0.021024 | 1.63E-14 | cis |
| miR-130a-3 | rs4827 | 11:57507983 | 11 | 57507983 | C | T | 0.685347 | -0.24314 | 0.022155 | 5.06E-28 | cis |
| miR-130a-3 | rs633129 | 11:57685196 | 11 | 57685196 | G | A | 0.561364 | 0.141528 | 0.021176 | 2.33E-11 | cis |
| miR-130a-3 | rs11603020 | 11:57374332 | 11 | 57374332 | T | C | 0.750533 | -0.29953 | 0.02326 | 6.00E-38 | cis |
| miR-130a-3 | rs674094 | 11:57665336 | 11 | 57665336 | A | C | 0.289352 | 0.203736 | 0.02375 | 9.64E-18 | cis |
| miR-130a-3 | rs11606390 | 11:57435211 | 11 | 57435211 | C | T | 0.687897 | -0.24571 | 0.0223 | 3.12E-28 | cis |
| miR-130a-3 | rs2156638 | 11:57542998 | 11 | 57542998 | G | C | 0.565268 | 0.1611 | 0.021006 | 1.73E-14 | cis |
| miR-130a-3 | rs11229080 | 11:57399511 | 11 | 57399511 | T | C | 0.749539 | -0.3014 | 0.023308 | 3.01E-38 | cis |
| miR-130a-3 | rs2509808 | 11:57654605 | 11 | 57654605 | C | G | 0.317599 | 0.23097 | 0.022979 | 9.07E-24 | cis |
| miR-130a-3 | rs3824988 | 11:57377968 | 11 | 57377968 | A | G | 0.750533 | -0.29953 | 0.02326 | 6.00E-38 | cis |
| miR-130a-3 | rs79740710 | 11:57463155 | 11 | 57463155 | C | T | 0.686438 | -0.2449 | 0.022233 | 3.24E-28 | cis |
| miR-130a-3 | rs12362160 | 11:57453043 | 11 | 57453043 | C | T | 0.68725 | -0.24622 | 0.02227 | 2.05E-28 | cis |
| miR-130a-3 | rs682503 | 11:57640994 | 11 | 57640994 | C | G | 0.313309 | 0.230973 | 0.022934 | 7.40E-24 | cis |
| miR-130a-3 | rs7117878 | 11:57421457 | 11 | 57421457 | C | A | 0.697568 | -0.25888 | 0.022623 | 2.55E-30 | cis |
| miR-130a-3 | rs12364898 | 11:57691533 | 11 | 57691533 | T | C | 0.573821 | 0.146509 | 0.021518 | 9.86E-12 | cis |
| miR-130a-3 | rs12790190 | 11:57497847 | 11 | 57497847 | C | T | 0.686232 | -0.24408 | 0.022216 | 4.43E-28 | cis |
| miR-130a-3 | rs11607050 | 11:57496820 | 11 | 57496820 | C | T | 0.683884 | -0.2448 | 0.022215 | 3.08E-28 | cis |
| miR-130a-3 | rs11293607 | 11:57397244 | 11 | 57397244 | A | G | 0.749632 | -0.30124 | 0.023286 | 2.81E-38 | cis |
| miR-130a-3 | rs14425634 | 11:57371412 | 11 | 57371412 | C | T | 0.752772 | -0.30549 | 0.023522 | 1.44E-38 | cis |
| miR-130a-3 | rs12360939 | 11:57675925 | 11 | 57675925 | C | T | 0.697457 | -0.22014 | 0.023006 | 1.08E-21 | cis |
| miR-130a-3 | rs1783985 | 11:57431470 | 11 | 57431470 | G | A | 0.413264 | -0.15395 | 0.021402 | 6.32E-13 | cis |
| miR-130a-3 | rs12363024 | 11:57503266 | 11 | 57503266 | G | C | 0.556328 | 0.166301 | 0.021097 | 3.21E-15 | cis |
| miR-130a-3 | rs3758918 | 11:57363761 | 11 | 57363761 | T | C | 0.898325 | -0.29196 | 0.034803 | 4.91E-17 | cis |
| miR-130a-3 | rs34052104 | 11:57384519 | 11 | 57384519 | A | C | 0.750564 | -0.30174 | 0.023272 | 1.92E-38 | cis |

| | | | | | | | | | | | |
|---|---|---|---|---|---|---|---|---|---|---|---|
| miR-130a-3 | rs3758919 | 11:5736280 | 11 | 57362803 | G | A | 0.898333 | -0.29198 | 0.034801 | 4.86E-17 | cis |
| miR-130a-3 | rs10896630 | 11:5744803 | 11 | 57448032 | C | G | 0.68075 | -0.24314 | 0.022412 | 2.02E-27 | cis |
| miR-130a-3 | rs6591434 | 11:5764433 | 11 | 57644334 | A | G | 0.688831 | -0.22925 | 0.02291 | 1.42E-23 | cis |
| miR-130a-3 | rs600488 | 11:5768400 | 11 | 57684005 | C | T | 0.560573 | 0.142183 | 0.021211 | 2.04E-11 | cis |
| miR-130a-3 | rs11297668 | 11:5756483 | 11 | 57564836 | G | A | 0.566038 | 0.161154 | 0.021054 | 1.95E-14 | cis |
| miR-130a-3 | rs626043 | 11:5766944 | 11 | 57669443 | A | G | 0.447969 | -0.1488 | 0.021184 | 2.15E-12 | cis |
| miR-130a-3 | rs499188 | 11:5743412 | 11 | 57434122 | C | A | 0.686203 | -0.24159 | 0.022245 | 1.78E-27 | cis |
| miR-130a-3 | rs2509805 | 11:5765079 | 11 | 57650796 | T | C | 0.345913 | 0.241505 | 0.023207 | 2.31E-25 | cis |
| miR-130a-3 | rs71460549 | 11:5748413 | 11 | 57484130 | G | C | 0.661292 | -0.25716 | 0.022429 | 1.97E-30 | cis |
| miR-130a-3 | rs11602109 | 11:5743529 | 11 | 57435296 | G | A | 0.688626 | -0.24541 | 0.022271 | 3.09E-28 | cis |
| miR-130a-3 | rs2956981 | 11:5750118 | 11 | 57501182 | C | T | 0.554948 | 0.168947 | 0.02108 | 1.10E-15 | cis |
| miR-130a-3 | rs10896662 | 11:5767908 | 11 | 57679080 | A | G | 0.697393 | -0.21962 | 0.023047 | 1.58E-21 | cis |
| miR-130a-3 | rs11229113 | 11:5751082 | 11 | 57510823 | G | T | 0.692006 | -0.24347 | 0.022353 | 1.26E-27 | cis |
| miR-130a-3 | rs11600169 | 11:5746494 | 11 | 57464945 | T | A | 0.679868 | -0.24665 | 0.022456 | 4.59E-28 | cis |
| miR-130a-3 | rs11229130 | 11:5756216 | 11 | 57562169 | A | G | 0.688809 | -0.24307 | 0.022346 | 1.47E-27 | cis |
| miR-130a-3 | rs653889 | 11:5757043 | 11 | 57570430 | A | G | 0.567416 | 0.160942 | 0.021027 | 1.95E-14 | cis |
| miR-130a-3 | rs36036499 | 11:5749419 | 11 | 57494198 | T | C | 0.554138 | 0.164556 | 0.02107 | 5.72E-15 | cis |
| miR-130a-3 | rs1647403 | 11:5742642 | 11 | 57426428 | C | G | 0.41283 | -0.15409 | 0.021401 | 6.01E-13 | cis |
| miR-130a-3 | rs1785499 | 11:5746914 | 11 | 57469149 | T | G | 0.45303 | -0.16492 | 0.020975 | 3.76E-15 | cis |
| miR-130a-3 | rs80304923 | 11:5752282 | 11 | 57522823 | C | G | 0.684059 | -0.24278 | 0.022169 | 6.54E-28 | cis |
| miR-130a-3 | rs11229109 | 11:5749222 | 11 | 57492226 | A | G | 0.554137 | 0.164771 | 0.021077 | 5.39E-15 | cis |
| miR-130a-3 | rs77993059 | 11:5752284 | 11 | 57522840 | C | T | 0.684063 | -0.24278 | 0.022169 | 6.54E-28 | cis |
| miR-130a-3 | rs652191 | 11:5763504 | 11 | 57635048 | A | G | 0.568268 | 0.160772 | 0.021201 | 3.37E-14 | cis |
| miR-130a-3 | rs17455422 | 11:5753479 | 11 | 57534799 | C | T | 0.565004 | 0.15876 | 0.02111 | 5.46E-14 | cis |
| miR-130a-3 | rs606731 | 11:5768733 | 11 | 57687338 | G | A | 0.454605 | -0.15039 | 0.02119 | 1.27E-12 | cis |
| miR-130a-3 | rs2911727 | 11:5749137 | 11 | 57491372 | T | A | 0.57273 | 0.166825 | 0.022081 | 4.19E-14 | cis |
| miR-130a-3 | rs1783827 | 11:5740953 | 11 | 57409538 | G | A | 0.445411 | -0.15793 | 0.021234 | 1.03E-13 | cis |
| miR-130a-3 | rs645145 | 11:5760978 | 11 | 57609787 | G | T | 0.564908 | 0.160734 | 0.02104 | 2.18E-14 | cis |
| miR-130a-3 | rs7116341 | 11:5748115 | 11 | 57481155 | G | A | 0.685526 | -0.243 | 0.022225 | 7.92E-28 | cis |
| miR-130a-3 | rs543725 | 11:5748303 | 11 | 57483039 | A | T | 0.688868 | -0.23973 | 0.022465 | 1.39E-26 | cis |
| miR-130a-3 | rs28362934 | 11:5736354 | 11 | 57363543 | G | A | 0.898327 | -0.29196 | 0.034802 | 4.90E-17 | cis |
| miR-130a-3 | rs76525968 | 11:5737119 | 11 | 57371192 | G | T | 0.750432 | -0.31079 | 0.023869 | 9.35E-39 | cis |
| miR-130a-3 | rs1783983 | 11:5742078 | 11 | 57420780 | T | C | 0.412531 | -0.15385 | 0.021282 | 4.87E-13 | cis |
| miR-130a-3 | rs7936998 | 11:5763901 | 11 | 57639017 | G | A | 0.683653 | -0.24173 | 0.022605 | 1.09E-26 | cis |
| miR-130a-3 | rs731384 | 11:5740838 | 11 | 57408382 | G | A | 0.744474 | -0.29471 | 0.023568 | 7.02E-36 | cis |
| miR-130a-3 | rs12146541 | 11:5750867 | 11 | 57508678 | G | C | 0.685316 | -0.24321 | 0.022155 | 4.90E-28 | cis |
| miR-130a-3 | rs10896633 | 11:5739030 | 11 | 57390302 | G | A | 0.746727 | -0.30204 | 0.023122 | 5.37E-39 | cis |
| miR-130a-3 | rs9645687 | 11:5767625 | 11 | 57676251 | T | C | 0.697475 | -0.22012 | 0.023007 | 1.09E-21 | cis |
| miR-130a-3 | rs527528 | 11:5743332 | 11 | 57433327 | C | T | 0.681923 | -0.24347 | 0.022237 | 6.73E-28 | cis |
| miR-130a-3 | rs7115528 | 11:5765374 | 11 | 57653746 | T | C | 0.565776 | 0.161618 | 0.021398 | 4.26E-14 | cis |
| miR-130a-3 | rs10896647 | 11:5757188 | 11 | 57571883 | C | T | 0.685297 | -0.24254 | 0.022203 | 8.89E-28 | cis |
| miR-130a-3 | rs2508443 | 11:5737799 | 11 | 57377992 | G | T | 0.56737 | 0.151222 | 0.021429 | 1.70E-12 | cis |
| miR-130a-3 | rs7129727 | 11:5748466 | 11 | 57484660 | G | A | 0.701653 | -0.23686 | 0.023171 | 1.58E-24 | cis |
| miR-130a-3 | rs659165 | 11:5760899 | 11 | 57608992 | A | C | 0.571934 | 0.161004 | 0.021115 | 2.44E-14 | cis |
| miR-130a-3 | rs11601320 | 11:5764261 | 11 | 57642617 | T | C | 0.692075 | -0.23077 | 0.022881 | 6.38E-24 | cis |
| miR-130a-3 | rs1798172 | 11:5743013 | 11 | 57430136 | T | C | 0.413694 | -0.15428 | 0.021357 | 5.06E-13 | cis |
| miR-130a-3 | rs11229119 | 11:5753596 | 11 | 57535966 | C | T | 0.580179 | 0.160801 | 0.021726 | 1.35E-13 | cis |
| miR-130a-3 | rs11229131 | 11:5756252 | 11 | 57562525 | G | A | 0.684434 | -0.24273 | 0.022144 | 5.85E-28 | cis |
| miR-130a-3 | rs35196561 | 11:5739729 | 11 | 57397292 | A | T | 0.619582 | 0.163417 | 0.023051 | 1.35E-12 | cis |
| miR-130a-3 | rs11229160 | 11:5767604 | 11 | 57676044 | C | T | 0.697455 | -0.22014 | 0.023006 | 1.08E-21 | cis |
| miR-130a-3 | rs1268667 | 11:5757509 | 11 | 57575099 | A | G | 0.643725 | 0.178674 | 0.023634 | 4.02E-14 | cis |
| miR-130a-3 | rs71484461 | 11:5748889 | 11 | 57488890 | G | A | 0.689792 | -0.24297 | 0.022416 | 2.25E-27 | cis |
| miR-130a-3 | rs591322 | 11:5768468 | 11 | 57684680 | T | G | 0.560767 | 0.141968 | 0.021203 | 2.15E-11 | cis |
| miR-130a-3 | rs1647405 | 11:5747788 | 11 | 57477889 | C | T | 0.68315 | -0.23997 | 0.022171 | 2.67E-27 | cis |
| miR-130a-3 | rs2926664 | 11:5740049 | 11 | 57400495 | A | G | 0.564621 | 0.156233 | 0.021936 | 1.06E-12 | cis |
| miR-130a-3 | rs606499 | 11:5759384 | 11 | 57593840 | G | A | 0.439467 | -0.16225 | 0.020963 | 9.97E-15 | cis |
| miR-130a-3 | rs593239 | 11:5766711 | 11 | 57667111 | T | C | 0.560709 | 0.145744 | 0.021148 | 5.51E-12 | cis |
| miR-130a-3 | rs471398 | 11:5759361 | 11 | 57593617 | C | T | 0.438663 | -0.16039 | 0.020992 | 2.16E-14 | cis |
| miR-130a-3 | rs7117020 | 11:5749965 | 11 | 57499655 | T | G | 0.553432 | 0.16611 | 0.021055 | 3.04E-15 | cis |
| miR-130a-3 | rs11229063 | 11:5736973 | 11 | 57369730 | G | A | 0.752752 | -0.30693 | 0.023511 | 5.96E-39 | cis |
| miR-130a-3 | rs7117205 | 11:5764188 | 11 | 57641883 | T | C | 0.692088 | -0.23082 | 0.02288 | 6.23E-24 | cis |
| miR-130a-3 | rs4926 | 11:5738198 | 11 | 57381989 | G | A | 0.750066 | -0.30132 | 0.023228 | 1.75E-38 | cis |
| miR-130a-3 | rs11570190 | 11:5756045 | 11 | 57560452 | A | C | 0.684149 | -0.2427 | 0.022118 | 5.17E-28 | cis |

| | | | | | | | | | | | |
|---|---|---|---|---|---|---|---|---|---|---|---|
| miR-130a-3 | rs488769 | 11:574507 | 11 | 57450720 | A | C | 0.674485 | -0.23365 | 0.02225 | 8.54E-26 | cis |
| miR-130a-3 | rs1182004 | 11:573699 | 11 | 57369957 | T | C | 0.752731 | -0.30696 | 0.023511 | 5.90E-39 | cis |
| miR-130a-3 | rs1122907 | 11:573992 | 11 | 57399256 | T | C | 0.749405 | -0.30142 | 0.023313 | 3.09E-38 | cis |
| miR-130a-3 | rs2836294 | 11:573690 | 11 | 57369008 | C | G | 0.752625 | -0.30649 | 0.023509 | 7.51E-39 | cis |
| miR-130a-3 | rs8029706 | 11:577112 | 11 | 57711280 | T | C | 0.516195 | 0.156736 | 0.02328 | 1.67E-11 | cis |
| miR-130a-3 | rs7348056 | 11:574014 | 11 | 57401400 | C | T | 0.753813 | -0.30104 | 0.024024 | 5.07E-36 | cis |
| miR-130a-3 | rs7348055 | 11:574004 | 11 | 57400462 | G | C | 0.749419 | -0.30162 | 0.023319 | 2.87E-38 | cis |
| miR-130a-3 | rs2836295 | 11:573725 | 11 | 57372526 | A | G | 0.75263 | -0.30443 | 0.02343 | 1.34E-38 | cis |
| miR-130a-3 | rs1424753 | 11:574798 | 11 | 57479864 | T | C | 0.449285 | -0.16409 | 0.02102 | 5.87E-15 | cis |
| miR-130a-3 | rs1647396 | 11:574011 | 11 | 57401183 | G | A | 0.467844 | -0.18342 | 0.021728 | 3.13E-17 | cis |
| miR-130a-3 | rs1122914 | 11:576214 | 11 | 57621460 | A | T | 0.683461 | -0.2414 | 0.022336 | 3.17E-27 | cis |
| miR-130a-3 | rs1647395 | 11:574097 | 11 | 57409795 | C | G | 0.74253 | -0.29262 | 0.023526 | 1.62E-35 | cis |
| miR-130a-3 | rs1280725 | 11:576002 | 11 | 57600285 | C | T | 0.684858 | -0.24239 | 0.022231 | 1.11E-27 | cis |
| miR-130a-3 | rs1160144 | 11:575243 | 11 | 57524310 | G | A | 0.684 | -0.24276 | 0.022162 | 6.38E-28 | cis |
| miR-130a-3 | rs673344 | 11:576571 | 11 | 57657141 | G | A | 0.31116 | 0.230743 | 0.023075 | 1.53E-23 | cis |
| miR-130a-3 | rs1396209 | 11:577111 | 11 | 57711129 | C | G | 0.586505 | 0.150344 | 0.021913 | 6.84E-12 | cis |
| miR-130a-3 | rs5564951 | 11:574113 | 11 | 57411302 | C | G | 0.745279 | -0.29329 | 0.023516 | 1.06E-35 | cis |
| miR-130a-3 | rs681397 | 11:576820 | 11 | 57682011 | G | T | 0.558507 | 0.14526 | 0.021263 | 8.41E-12 | cis |
| miR-130a-3 | rs1745562 | 11:575444 | 11 | 57544484 | T | C | 0.567581 | 0.16129 | 0.021032 | 1.73E-14 | cis |
| miR-130a-3 | rs1122911 | 11:575345 | 11 | 57534542 | C | G | 0.683806 | -0.24446 | 0.022182 | 3.05E-28 | cis |
| miR-130a-3 | rs576859 | 11:574806 | 11 | 57480623 | C | A | 0.683179 | -0.24021 | 0.022172 | 2.37E-27 | cis |
| miR-130a-3 | rs1157018 | 11:575557 | 11 | 57555770 | T | C | 0.567672 | 0.161097 | 0.02102 | 1.80E-14 | cis |
| miR-130a-3 | rs589914 | 11:576610 | 11 | 57661032 | A | G | 0.311001 | 0.230655 | 0.02307 | 1.55E-23 | cis |
| miR-130a-3 | rs2956982 | 11:574907 | 11 | 57490750 | G | A | 0.552122 | 0.164782 | 0.021049 | 4.94E-15 | cis |
| miR-130a-3 | rs1783978 | 11:574635 | 11 | 57463500 | T | C | 0.45351 | -0.16484 | 0.020962 | 3.72E-15 | cis |
| miR-130a-3 | rs671508 | 11:576365 | 11 | 57636568 | A | T | 0.438934 | -0.16176 | 0.02122 | 2.48E-14 | cis |
| miR-130a-3 | rs682258 | 11:576371 | 11 | 57637197 | G | T | 0.438944 | -0.16177 | 0.021221 | 2.48E-14 | cis |
| miR-130a-3 | rs1127641 | 11:573858 | 11 | 57385856 | C | T | 0.7469 | -0.30227 | 0.023181 | 7.27E-39 | cis |
| miR-130a-3 | rs2836294 | 11:573719 | 11 | 57371911 | G | T | 0.749195 | -0.30047 | 0.023454 | 1.43E-37 | cis |
| miR-130a-3 | rs1745552 | 11:575404 | 11 | 57540458 | C | A | 0.684599 | -0.24348 | 0.022162 | 4.45E-28 | cis |
| miR-130a-3 | rs1647394 | 11:574185 | 11 | 57418588 | C | T | 0.408999 | -0.15747 | 0.021549 | 2.71E-13 | cis |
| miR-130a-3 | rs5639270 | 11:575183 | 11 | 57518392 | T | C | 0.557348 | 0.16563 | 0.021057 | 3.67E-15 | cis |
| miR-130a-3 | rs2926656 | 11:574653 | 11 | 57465389 | C | T | 0.683894 | -0.24208 | 0.022186 | 1.01E-27 | cis |
| miR-130a-3 | rs1783980 | 11:574483 | 11 | 57448325 | C | T | 0.4518 | -0.16507 | 0.021041 | 4.33E-15 | cis |
| miR-130a-3 | rs1783981 | 11:574510 | 11 | 57451090 | T | C | 0.454187 | -0.16544 | 0.021011 | 3.44E-15 | cis |
| miR-130a-3 | rs7938083 | 11:574936 | 11 | 57493622 | C | A | 0.686581 | -0.24555 | 0.022337 | 4.12E-28 | cis |
| miR-130a-3 | rs623541 | 11:575816 | 11 | 57581613 | C | A | 0.439413 | -0.16308 | 0.020971 | 7.45E-15 | cis |
| miR-130a-3 | rs615133 | 11:576849 | 11 | 57684949 | C | A | 0.561411 | 0.141502 | 0.021175 | 2.35E-11 | cis |
| miR-130a-3 | rs1157017 | 11:575294 | 11 | 57529429 | A | G | 0.566649 | 0.1615 | 0.021063 | 1.75E-14 | cis |
| miR-130a-3 | rs7949841 | 11:575247 | 11 | 57524700 | G | C | 0.684177 | -0.24311 | 0.022173 | 5.69E-28 | cis |
| miR-130a-3 | rs685149 | 11:576574 | 11 | 57657413 | A | G | 0.348389 | 0.209244 | 0.022731 | 3.41E-20 | cis |
| miR-130a-3 | rs1089662 | 11:573232 | 11 | 57323205 | A | G | 0.871328 | -0.21738 | 0.031621 | 6.22E-12 | cis |
| miR-130a-3 | rs1783976 | 11:574627 | 11 | 57462752 | T | C | 0.456765 | -0.16962 | 0.021185 | 1.18E-15 | cis |
| miR-130a-3 | rs547118 | 11:575950 | 11 | 57595062 | C | G | 0.438175 | -0.16318 | 0.020979 | 7.35E-15 | cis |
| miR-130a-3 | rs4611222 | 11:575522 | 11 | 57552265 | G | A | 0.56182 | 0.161086 | 0.020967 | 1.56E-14 | cis |
| miR-130a-3 | rs616911 | 11:576853 | 11 | 57685334 | G | A | 0.558798 | 0.145619 | 0.021196 | 6.42E-12 | cis |
| miR-130a-3 | rs636388 | 11:576795 | 11 | 57679524 | C | G | 0.562687 | 0.146202 | 0.021265 | 6.18E-12 | cis |
| miR-130a-3 | rs3017889 | 11:575208 | 11 | 57520849 | A | G | 0.556573 | 0.166392 | 0.021044 | 2.64E-15 | cis |
| miR-130a-3 | rs1075087 | 11:576238 | 11 | 57623815 | A | C | 0.437769 | -0.15918 | 0.021015 | 3.60E-14 | cis |
| miR-130a-3 | rs658936 | 11:575805 | 11 | 57580560 | T | G | 0.440007 | -0.16364 | 0.020988 | 6.35E-15 | cis |
| miR-130a-3 | rs644219 | 11:576100 | 11 | 57610017 | C | T | 0.56488 | 0.160737 | 0.02104 | 2.18E-14 | cis |
| miR-130a-3 | rs1280735 | 11:576569 | 11 | 57656902 | C | T | 0.69404 | -0.23074 | 0.023031 | 1.26E-23 | cis |
| miR-130a-3 | rs1783979 | 11:574681 | 11 | 57468129 | T | C | 0.451998 | -0.16499 | 0.021014 | 4.11E-15 | cis |
| miR-130a-3 | rs1089663 | 11:573812 | 11 | 57381263 | T | C | 0.750433 | -0.30148 | 0.023269 | 2.17E-38 | cis |
| miR-130a-3 | rs1409656 | 11:576761 | 11 | 57676139 | A | C | 0.571656 | 0.152375 | 0.021422 | 1.14E-12 | cis |
| miR-130a-3 | rs7932091 | 11:576180 | 11 | 57618068 | A | T | 0.683808 | -0.24188 | 0.022349 | 2.69E-27 | cis |
| miR-130a-3 | rs1785498 | 11:574489 | 11 | 57448932 | C | G | 0.451781 | -0.16507 | 0.021041 | 4.32E-15 | cis |
| miR-130a-3 | rs1280534 | 11:575247 | 11 | 57524724 | C | A | 0.566156 | 0.161834 | 0.021069 | 1.58E-14 | cis |
| miR-130a-3 | rs7947357 | 11:574143 | 11 | 57414387 | A | G | 0.450784 | -0.15575 | 0.021161 | 1.83E-13 | cis |
| miR-130a-3 | rs1279066 | 11:576672 | 11 | 57667222 | T | C | 0.697455 | -0.22008 | 0.022986 | 1.02E-21 | cis |
| miR-130a-3 | rs1280497 | 11:574637 | 11 | 57463767 | T | C | 0.686355 | -0.24483 | 0.022231 | 3.32E-28 | cis |
| miR-130a-3 | rs1387704 | 11:573761 | 11 | 57376131 | G | A | 0.750533 | -0.29953 | 0.02326 | 6.00E-38 | cis |
| miR-130a-3 | rs7123499 | 11:573526 | 11 | 57352603 | C | A | 0.871121 | -0.2237 | 0.031641 | 1.55E-12 | cis |

| | | | | | | | | | | | |
|---|---|---|---|---|---|---|---|---|---|---|---|
| miR-130a-3 | rs1783977 | 11:5746279 | 11 | 57462799 | T | C | 0.454153 | -0.16407 | 0.021046 | 6.40E-15 | cis |
| miR-130a-3 | rs7117896 | 11:5738855 | 11 | 57388551 | G | A | 0.746843 | -0.30234 | 0.023182 | 7.07E-39 | cis |
| miR-130a-3 | rs10750860 | 11:5740477 | 11 | 57404779 | A | G | 0.751752 | -0.30311 | 0.023739 | 2.46E-37 | cis |
| miR-130a-3 | rs12798200 | 11:5766847 | 11 | 57668479 | T | G | 0.697389 | -0.22009 | 0.022987 | 1.02E-21 | cis |
| miR-130a-3 | rs2911728 | 11:5749137 | 11 | 57491371 | T | A | 0.578254 | 0.16537 | 0.021815 | 3.44E-14 | cis |
| miR-130a-3 | rs79260745 | 11:5761296 | 11 | 57612960 | T | C | 0.687478 | -0.24572 | 0.022451 | 7.04E-28 | cis |
| miR-130a-3 | rs7479922 | 11:5747018 | 11 | 57470188 | A | G | 0.450674 | -0.16462 | 0.021018 | 4.78E-15 | cis |
| miR-130a-3 | rs12805841 | 11:5744384 | 11 | 57443842 | G | A | 0.687846 | -0.24589 | 0.022275 | 2.49E-28 | cis |
| miR-130a-3 | rs7121169 | 11:5745254 | 11 | 57452543 | G | A | 0.687601 | -0.24659 | 0.022277 | 1.77E-28 | cis |
| miR-130a-3 | rs627145 | 11:5760246 | 11 | 57602469 | G | A | 0.439595 | -0.16181 | 0.021052 | 1.52E-14 | cis |
| miR-130a-3 | rs1783826 | 11:5740296 | 11 | 57402964 | T | G | 0.465832 | -0.17354 | 0.021573 | 8.67E-16 | cis |
| miR-130b-3 | rs861814 | 22:2205800 | 22 | 22058000 | C | G | 0.470428 | -0.15596 | 0.023257 | 2.00E-11 | cis |
| miR-130b-3 | rs373001 | 22:2200742 | 22 | 22007426 | C | T | 0.563659 | 0.176513 | 0.022904 | 1.29E-14 | cis |
| miR-130b-3 | rs450469 | 22:2200562 | 22 | 22005629 | C | T | 0.550569 | 0.182711 | 0.023418 | 6.09E-15 | cis |
| miR-130b-3 | rs447001 | 22:2201091 | 22 | 22010918 | C | T | 0.558786 | 0.173303 | 0.02256 | 1.57E-14 | cis |
| miR-130b-3 | rs446142 | 22:2201017 | 22 | 22010173 | C | T | 0.564937 | 0.176373 | 0.022915 | 1.39E-14 | cis |
| miR-130b-3 | rs450129 | 22:2200299 | 22 | 22002999 | G | T | 0.646482 | 0.177549 | 0.025616 | 4.18E-12 | cis |
| miR-130b-3 | rs1210397 | 22:2200997 | 22 | 22009975 | C | T | 0.574327 | 0.176566 | 0.023175 | 2.56E-14 | cis |
| miR-130b-3 | rs449597 | 22:2200263 | 22 | 22002632 | A | C | 0.55856 | 0.184464 | 0.024055 | 1.74E-14 | cis |
| miR-130b-3 | rs5754754 | 22:2206067 | 22 | 22060673 | G | A | 0.59316 | 0.15629 | 0.023367 | 2.25E-11 | cis |
| miR-130b-3 | rs439760 | 22:2200305 | 22 | 22003050 | G | A | 0.560517 | 0.183713 | 0.023784 | 1.12E-14 | cis |
| miR-130b-3 | rs5754727 | 22:2205727 | 22 | 22057273 | G | A | 0.5885 | 0.157278 | 0.02306 | 9.08E-12 | cis |
| miR-133b | rs73918290 | 20:6115207 | 20 | 61152078 | C | T | 0.814283 | -0.26433 | 0.034449 | 1.68E-14 | trans |
| miR-133b | rs3934573 | 20:6114394 | 20 | 61143945 | T | G | 0.514746 | -0.1881 | 0.026782 | 2.17E-12 | trans |
| miR-133b | rs3934574 | 20:6114348 | 20 | 61143487 | T | C | 0.464792 | -0.17856 | 0.025429 | 2.19E-12 | trans |
| miR-133b | rs7271454 | 20:6114327 | 20 | 61143279 | T | C | 0.46691 | -0.17953 | 0.025676 | 2.71E-12 | trans |
| miR-133b | rs11635599 | 20:6115095 | 20 | 61150959 | G | A | 0.819024 | -0.26294 | 0.034375 | 2.02E-14 | trans |
| miR-133b | rs4305340 | 20:6115724 | 20 | 61157241 | C | T | 0.814576 | -0.25153 | 0.034149 | 1.76E-13 | trans |
| miR-133b | rs11785859 | 20:6114926 | 20 | 61149264 | G | A | 0.861146 | -0.3134 | 0.039142 | 1.18E-15 | trans |
| miR-133b | rs76213714 | 20:6115092 | 20 | 61150928 | A | G | 0.467251 | -0.19085 | 0.027215 | 2.34E-12 | trans |
| miR-133b | rs11906462 | 20:6115895 | 20 | 61158952 | C | T | 0.824779 | -0.24853 | 0.03256 | 2.29E-14 | trans |
| miR-133b | rs73920512 | 20:6115798 | 20 | 61157982 | C | T | 0.816102 | -0.2515 | 0.034264 | 2.13E-13 | trans |
| miR-133b | rs6062278 | 20:6114494 | 20 | 61144941 | A | G | 0.470455 | -0.17122 | 0.024967 | 6.98E-12 | trans |
| miR-133b | rs11741969 | 20:6115385 | 20 | 61153858 | G | A | 0.818584 | -0.2605 | 0.034321 | 3.20E-14 | trans |
| miR-133b | rs6062284 | 20:6114198 | 20 | 61141981 | T | G | 0.4602 | -0.17216 | 0.025716 | 2.16E-11 | trans |
| miR-133b | rs8122282 | 20:6115232 | 20 | 61152322 | G | A | 0.467131 | -0.18973 | 0.027217 | 3.15E-12 | trans |
| miR-133b | rs56703558 | 20:6115799 | 20 | 61157993 | C | T | 0.813578 | -0.24941 | 0.034128 | 2.71E-13 | trans |
| miR-133b | rs6062267 | 20:6115336 | 20 | 61153365 | T | C | 0.464907 | -0.1902 | 0.027172 | 2.56E-12 | trans |
| miR-133b | rs12624608 | 20:6115529 | 20 | 61155298 | C | A | 0.796375 | -0.26443 | 0.033362 | 2.26E-15 | trans |
| miR-133b | rs4637207 | 20:6114845 | 20 | 61148451 | A | G | 0.458537 | -0.19961 | 0.028232 | 1.55E-12 | trans |
| miR-133b | rs6062277 | 20:6114559 | 20 | 61145597 | A | G | 0.461496 | -0.1714 | 0.025325 | 1.30E-11 | trans |
| miR-134-5p | rs11847612 | 14:1010237 | 14 | 1.01E+08 | C | T | 0.753476 | 0.244494 | 0.034211 | 8.89E-13 | cis |
| miR-134-5p | rs12433699 | 14:1010355 | 14 | 1.01E+08 | C | G | 0.723403 | 0.241945 | 0.031754 | 2.55E-14 | cis |
| miR-134-5p | rs17099614 | 14:1011670 | 14 | 1.01E+08 | C | G | 0.668809 | 0.275723 | 0.030027 | 4.21E-20 | cis |
| miR-134-5p | rs879059 | 14:1010699 | 14 | 1.01E+08 | T | A | 0.31068 | -0.24629 | 0.03109 | 2.34E-15 | cis |
| miR-134-5p | rs8020868 | 14:1011669 | 14 | 1.01E+08 | A | G | 0.668343 | 0.274444 | 0.03002 | 6.13E-20 | cis |
| miR-134-5p | rs7154024 | 14:1010317 | 14 | 1.01E+08 | G | T | 0.725015 | 0.243619 | 0.031936 | 2.38E-14 | cis |
| miR-134-5p | rs12897601 | 14:1011822 | 14 | 1.01E+08 | C | T | 0.190452 | -0.33279 | 0.036531 | 8.25E-20 | cis |
| miR-134-5p | rs1190872 | 14:1010697 | 14 | 1.01E+08 | A | G | 0.310177 | -0.24621 | 0.031107 | 2.48E-15 | cis |
| miR-134-5p | rs7147982 | 14:1010460 | 14 | 1.01E+08 | C | T | 0.731753 | 0.252572 | 0.031775 | 1.88E-15 | cis |
| miR-134-5p | rs1190867 | 14:101069 | 14 | 1.01E+08 | G | T | 0.316788 | -0.24663 | 0.031332 | 3.50E-15 | cis |
| miR-134-5p | rs59939092 | 14:1011690 | 14 | 1.01E+08 | C | T | 0.892837 | 0.335736 | 0.04661 | 5.88E-13 | cis |
| miR-134-5p | rs4243729 | 14:1010660 | 14 | 1.01E+08 | A | C | 0.311439 | -0.24314 | 0.030911 | 3.66E-15 | cis |
| miR-134-5p | rs6575791 | 14:1010292 | 14 | 1.01E+08 | G | A | 0.719841 | 0.24412 | 0.03196 | 2.20E-14 | cis |
| miR-134-5p | rs7145337 | 14:1011555 | 14 | 1.01E+08 | C | T | 0.312091 | -0.26344 | 0.031513 | 6.30E-17 | cis |
| miR-134-5p | rs72631671 | 14:1011769 | 14 | 1.01E+08 | C | T | 0.889487 | 0.345579 | 0.045564 | 3.34E-14 | cis |
| miR-134-5p | rs10145513 | 14:1011685 | 14 | 1.01E+08 | A | G | 0.668173 | 0.274365 | 0.029985 | 5.68E-20 | cis |
| miR-134-5p | rs12434490 | 14:1011792 | 14 | 1.01E+08 | A | G | 0.18959 | -0.33712 | 0.036712 | 4.20E-20 | cis |
| miR-134-5p | rs12147008 | 14:101186 | 14 | 1.01E+08 | C | T | 0.223221 | -0.27445 | 0.034204 | 1.02E-15 | cis |
| miR-134-5p | rs4905999 | 14:101184 | 14 | 1.01E+08 | A | G | 0.181905 | -0.34229 | 0.037353 | 5.03E-20 | cis |
| miR-134-5p | rs1190862 | 14:1010677 | 14 | 1.01E+08 | T | G | 0.309628 | -0.24455 | 0.030926 | 2.62E-15 | cis |
| miR-134-5p | rs72631672 | 14:1011870 | 14 | 1.01E+08 | C | T | 0.881829 | 0.342201 | 0.045505 | 5.48E-14 | cis |
| miR-134-5p | rs7141210 | 14:1011824 | 14 | 1.01E+08 | T | C | 0.32902 | -0.29264 | 0.029875 | 1.18E-22 | cis |

| | | | | | | | | | | | |
|---|---|---|---|---|---|---|---|---|---|---|---|
| miR-134-5p | rs7149242 | 14:101159 | 14 | 1.01E+08 | T | G | 0.288371 | -0.26987 | 0.031974 | 3.17E-17 | cis |
| miR-134-5p | rs4905977 | 14:101061 | 14 | 1.01E+08 | C | G | 0.318473 | -0.2513 | 0.031262 | 9.09E-16 | cis |
| miR-134-5p | rs7493817 | 14:101038 | 14 | 1.01E+08 | G | A | 0.751723 | 0.27112 | 0.033102 | 2.60E-16 | cis |
| miR-134-5p | rs7155375 | 14:101188 | 14 | 1.01E+08 | C | T | 0.635995 | 0.219764 | 0.02937 | 7.29E-14 | cis |
| miR-134-5p | rs1288154 | 14:101176 | 14 | 1.01E+08 | G | C | 0.370047 | -0.29555 | 0.030832 | 9.16E-22 | cis |
| miR-134-5p | rs2183632 | 14:101063 | 14 | 1.01E+08 | C | T | 0.319851 | -0.24929 | 0.031275 | 1.57E-15 | cis |
| miR-134-5p | rs1288281 | 14:101183 | 14 | 1.01E+08 | C | G | 0.331398 | -0.29287 | 0.029975 | 1.51E-22 | cis |
| miR-134-5p | rs1888581 | 14:101042 | 14 | 1.01E+08 | A | C | 0.735363 | 0.256011 | 0.032003 | 1.25E-15 | cis |
| miR-134-5p | rs4905978 | 14:101061 | 14 | 1.01E+08 | C | T | 0.318452 | -0.25127 | 0.031262 | 9.15E-16 | cis |
| miR-134-5p | rs1184658 | 14:101017 | 14 | 1.01E+08 | C | G | 0.759077 | 0.254259 | 0.034744 | 2.52E-13 | trans |
| miR-134-5p | rs5982537 | 14:101168 | 14 | 1.01E+08 | G | T | 0.89241 | 0.336414 | 0.046621 | 5.36E-13 | cis |
| miR-134-5p | rs879060 | 14:101069 | 14 | 1.01E+08 | A | G | 0.310926 | -0.24678 | 0.031109 | 2.15E-15 | cis |
| miR-134-5p | rs873076 | 14:101070 | 14 | 1.01E+08 | A | G | 0.311961 | -0.24475 | 0.031043 | 3.16E-15 | cis |
| miR-134-5p | rs7142683 | 14:101158 | 14 | 1.01E+08 | A | G | 0.290748 | -0.27119 | 0.032064 | 2.73E-17 | cis |
| miR-134-5p | rs6199300 | 14:101030 | 14 | 1.01E+08 | G | A | 0.724196 | 0.242624 | 0.031867 | 2.66E-14 | cis |
| miR-134-5p | rs7143357 | 14:101054 | 14 | 1.01E+08 | T | C | 0.739636 | 0.27309 | 0.032349 | 3.12E-17 | cis |
| miR-134-5p | rs1289379 | 14:101178 | 14 | 1.01E+08 | C | G | 0.190813 | -0.33881 | 0.036736 | 2.89E-20 | cis |
| miR-134-5p | rs1116059 | 14:101049 | 14 | 1.01E+08 | C | T | 0.741662 | 0.279929 | 0.032294 | 4.40E-18 | cis |
| miR-134-5p | rs1258871 | 14:101149 | 14 | 1.01E+08 | G | C | 0.347269 | -0.23714 | 0.030193 | 4.03E-15 | cis |
| miR-134-5p | rs1555405 | 14:101176 | 14 | 1.01E+08 | G | A | 0.749075 | 0.256575 | 0.03237 | 2.26E-15 | cis |
| miR-134-5p | rs6001359 | 14:101021 | 14 | 1.01E+08 | C | T | 0.755543 | 0.243074 | 0.034341 | 1.46E-12 | cis |
| miR-134-5p | rs7140792 | 14:101172 | 14 | 1.01E+08 | C | T | 0.892517 | 0.337367 | 0.046442 | 3.75E-13 | cis |
| miR-134-5p | rs6199300 | 14:101034 | 14 | 1.01E+08 | G | A | 0.726044 | 0.24414 | 0.031964 | 2.21E-14 | cis |
| miR-134-5p | rs7492369 | 14:101020 | 14 | 1.01E+08 | C | T | 0.755698 | 0.241816 | 0.034313 | 1.82E-12 | trans |
| miR-134-5p | rs7155649 | 14:101181 | 14 | 1.01E+08 | C | A | 0.863608 | 0.287338 | 0.04258 | 1.50E-11 | cis |
| miR-134-5p | rs4905994 | 14:101154 | 14 | 1.01E+08 | T | C | 0.329146 | -0.24538 | 0.03112 | 3.15E-15 | cis |
| miR-134-5p | rs7140556 | 14:101036 | 14 | 1.01E+08 | C | A | 0.723818 | 0.244488 | 0.031891 | 1.77E-14 | cis |
| miR-134-5p | rs1162219 | 14:101016 | 14 | 1.01E+08 | G | A | 0.759852 | 0.260738 | 0.036289 | 6.72E-13 | trans |
| miR-134-5p | rs1162093 | 14:101151 | 14 | 1.01E+08 | C | G | 0.338205 | -0.23495 | 0.030459 | 1.23E-14 | cis |
| miR-134-5p | rs1288176 | 14:101176 | 14 | 1.01E+08 | G | C | 0.347791 | -0.29228 | 0.0304 | 6.93E-22 | cis |
| miR-134-5p | rs6130797 | 14:101177 | 14 | 1.01E+08 | C | T | 0.888723 | 0.3454 | 0.045621 | 3.70E-14 | cis |
| miR-134-5p | rs6030796 | 14:101027 | 14 | 1.01E+08 | C | T | 0.725784 | 0.250613 | 0.033118 | 3.81E-14 | cis |
| miR-134-5p | rs8021301 | 14:101166 | 14 | 1.01E+08 | G | C | 0.668324 | 0.274474 | 0.030023 | 6.12E-20 | cis |
| miR-134-5p | rs2837918 | 14:101168 | 14 | 1.01E+08 | G | T | 0.836434 | 0.329184 | 0.038556 | 1.37E-17 | cis |
| miR-134-5p | rs1014438 | 14:101173 | 14 | 1.01E+08 | C | T | 0.673446 | 0.288576 | 0.030143 | 1.03E-21 | cis |
| miR-134-5p | rs4905976 | 14:101060 | 14 | 1.01E+08 | G | A | 0.322862 | -0.24259 | 0.031329 | 9.67E-15 | cis |
| miR-134-5p | rs1467536 | 14:101061 | 14 | 1.01E+08 | A | G | 0.318958 | -0.25176 | 0.031214 | 7.29E-16 | cis |
| miR-134-5p | rs5870230 | 14:101167 | 14 | 1.01E+08 | G | A | 0.892706 | 0.3355 | 0.046665 | 6.50E-13 | cis |
| miR-134-5p | rs1184948 | 14:101037 | 14 | 1.01E+08 | C | G | 0.733365 | 0.258552 | 0.03208 | 7.65E-16 | cis |
| miR-134-5p | rs1190856 | 14:101065 | 14 | 1.01E+08 | T | C | 0.310244 | -0.24783 | 0.031135 | 1.72E-15 | cis |
| miR-134-5p | rs6762567 | 14:101173 | 14 | 1.01E+08 | A | G | 0.892632 | 0.33427 | 0.046381 | 5.72E-13 | cis |
| miR-134-5p | rs9324022 | 14:101172 | 14 | 1.01E+08 | T | C | 0.763739 | 0.242341 | 0.035125 | 5.23E-12 | cis |
| miR-134-5p | rs1014844 | 14:101026 | 14 | 1.01E+08 | G | A | 0.753241 | 0.244465 | 0.034117 | 7.75E-13 | cis |
| miR-134-5p | rs1243613 | 14:101031 | 14 | 1.01E+08 | C | T | 0.725015 | 0.243856 | 0.031964 | 2.36E-14 | cis |
| miR-134-5p | rs4905998 | 14:101181 | 14 | 1.01E+08 | A | G | 0.330615 | -0.29352 | 0.02995 | 1.12E-22 | cis |
| miR-134-5p | rs7141110 | 14:101172 | 14 | 1.01E+08 | G | C | 0.783235 | 0.248722 | 0.035855 | 4.01E-12 | cis |
| miR-134-5p | rs1185095 | 14:101056 | 14 | 1.01E+08 | G | T | 0.743981 | 0.273933 | 0.032153 | 1.60E-17 | cis |
| miR-134-5p | rs7142089 | 14:101172 | 14 | 1.01E+08 | T | A | 0.783402 | 0.253077 | 0.035888 | 1.77E-12 | cis |
| miR-134-5p | rs6575794 | 14:101039 | 14 | 1.01E+08 | C | T | 0.734189 | 0.259623 | 0.032124 | 6.37E-16 | cis |
| miR-134-5p | rs4900471 | 14:101162 | 14 | 1.01E+08 | G | A | 0.304173 | -0.25257 | 0.031587 | 1.29E-15 | cis |
| miR-134-5p | rs1184764 | 14:101023 | 14 | 1.01E+08 | C | T | 0.753952 | 0.2435 | 0.034249 | 1.16E-12 | cis |
| miR-134-5p | rs1190857 | 14:101066 | 14 | 1.01E+08 | G | T | 0.310273 | -0.24767 | 0.031142 | 1.82E-15 | cis |
| miR-134-5p | rs2150518 | 14:101063 | 14 | 1.01E+08 | C | T | 0.318506 | -0.25107 | 0.031307 | 1.06E-15 | cis |
| miR-134-5p | rs1190850 | 14:101064 | 14 | 1.01E+08 | A | G | 0.314302 | -0.25137 | 0.031341 | 1.05E-15 | cis |
| miR-134-5p | rs1190852 | 14:101064 | 14 | 1.01E+08 | A | G | 0.310295 | -0.24896 | 0.031157 | 1.34E-15 | cis |
| miR-134-5p | rs1014839 | 14:101026 | 14 | 1.01E+08 | A | G | 0.750738 | 0.242861 | 0.034134 | 1.12E-12 | cis |
| miR-134-5p | rs3470057 | 14:101173 | 14 | 1.01E+08 | C | T | 0.892725 | 0.334455 | 0.046405 | 5.71E-13 | cis |
| miR-134-5p | rs9671394 | 14:101159 | 14 | 1.01E+08 | C | T | 0.284232 | -0.27658 | 0.032415 | 1.43E-17 | cis |
| miR-134-5p | rs1243517 | 14:101155 | 14 | 1.01E+08 | G | A | 0.312183 | -0.26342 | 0.031521 | 6.44E-17 | cis |
| miR-134-5p | rs7145610 | 14:101049 | 14 | 1.01E+08 | T | C | 0.740391 | 0.280479 | 0.03238 | 4.63E-18 | cis |
| miR-134-5p | rs873077 | 14:101070 | 14 | 1.01E+08 | G | A | 0.312104 | -0.24465 | 0.031049 | 3.29E-15 | cis |
| miR-134-5p | rs879058 | 14:101069 | 14 | 1.01E+08 | G | T | 0.311807 | -0.24655 | 0.031107 | 2.26E-15 | cis |
| miR-134-5p | rs8012970 | 14:101168 | 14 | 1.01E+08 | T | C | 0.347902 | -0.26555 | 0.030584 | 3.87E-18 | cis |

| | | | | | | | | | | | |
|---|---|---|---|---|---|---|---|---|---|---|---|
| miR-134-5p | rs1014696 | 14:101170 | 14 | 1.01E+08 | T | C | 0.658875 | 0.277317 | 0.029485 | 5.18E-21 | cis |
| miR-136-5p | rs879059 | 14:101069 | 14 | 1.01E+08 | T | A | 0.31068 | -0.28748 | 0.038046 | 4.15E-14 | cis |
| miR-136-5p | rs1014438 | 14:101173 | 14 | 1.01E+08 | C | T | 0.673446 | 0.405272 | 0.036589 | 1.64E-28 | cis |
| miR-136-5p | rs1190872 | 14:101069 | 14 | 1.01E+08 | A | G | 0.310177 | -0.28736 | 0.038066 | 4.39E-14 | cis |
| miR-136-5p | rs4905998 | 14:101181 | 14 | 1.01E+08 | A | G | 0.330615 | -0.40992 | 0.036355 | 1.73E-29 | cis |
| miR-136-5p | rs8012970 | 14:101168 | 14 | 1.01E+08 | T | C | 0.347902 | -0.40152 | 0.037032 | 2.17E-27 | cis |
| miR-136-5p | rs1214700 | 14:101186 | 14 | 1.01E+08 | C | T | 0.223221 | -0.44024 | 0.041352 | 1.81E-26 | cis |
| miR-136-5p | rs1190856 | 14:101065 | 14 | 1.01E+08 | T | C | 0.310244 | -0.28863 | 0.038104 | 3.60E-14 | cis |
| miR-136-5p | rs6199300 | 14:101030 | 14 | 1.01E+08 | G | A | 0.724196 | 0.286266 | 0.038982 | 2.08E-13 | cis |
| miR-136-5p | rs879060 | 14:101069 | 14 | 1.01E+08 | A | G | 0.310926 | -0.28822 | 0.038068 | 3.70E-14 | cis |
| miR-136-5p | rs6199300 | 14:101034 | 14 | 1.01E+08 | G | A | 0.726044 | 0.287145 | 0.039104 | 2.09E-13 | cis |
| miR-136-5p | rs8008285 | 14:101146 | 14 | 1.01E+08 | T | C | 0.842905 | 0.378923 | 0.048707 | 7.27E-15 | cis |
| miR-136-5p | rs1190862 | 14:101067 | 14 | 1.01E+08 | T | G | 0.309628 | -0.28758 | 0.037837 | 2.95E-14 | cis |
| miR-136-5p | rs8021301 | 14:101166 | 14 | 1.01E+08 | G | C | 0.668324 | 0.400327 | 0.036391 | 3.80E-28 | cis |
| miR-136-5p | rs7145337 | 14:101155 | 14 | 1.01E+08 | C | T | 0.312091 | -0.38743 | 0.038235 | 3.96E-24 | cis |
| miR-136-5p | rs6030796 | 14:101027 | 14 | 1.01E+08 | C | T | 0.725784 | 0.30762 | 0.040471 | 2.94E-14 | cis |
| miR-136-5p | rs1243517 | 14:101155 | 14 | 1.01E+08 | G | A | 0.312183 | -0.38746 | 0.038245 | 4.02E-24 | cis |
| miR-136-5p | rs2837918 | 14:101168 | 14 | 1.01E+08 | G | T | 0.836434 | 0.456746 | 0.046892 | 2.03E-22 | cis |
| miR-136-5p | rs6575791 | 14:101029 | 14 | 1.01E+08 | G | A | 0.719841 | 0.286851 | 0.039101 | 2.20E-13 | cis |
| miR-136-5p | rs4905978 | 14:101061 | 14 | 1.01E+08 | C | T | 0.318452 | -0.29768 | 0.038241 | 7.01E-15 | cis |
| miR-136-5p | rs1258871 | 14:101149 | 14 | 1.01E+08 | G | C | 0.347269 | -0.3465 | 0.036675 | 3.46E-21 | cis |
| miR-136-5p | rs9671394 | 14:101159 | 14 | 1.01E+08 | C | T | 0.284232 | -0.39899 | 0.039355 | 3.73E-24 | cis |
| miR-136-5p | rs7493817 | 14:101038 | 14 | 1.01E+08 | G | A | 0.751723 | 0.297935 | 0.040576 | 2.09E-13 | cis |
| miR-136-5p | rs1190903 | 14:101096 | 14 | 1.01E+08 | C | T | 0.772566 | 0.29448 | 0.042363 | 3.62E-12 | cis |
| miR-136-5p | rs1014551 | 14:101168 | 14 | 1.01E+08 | A | G | 0.668173 | 0.399046 | 0.03635 | 4.89E-28 | cis |
| miR-136-5p | rs7140792 | 14:101172 | 14 | 1.01E+08 | C | T | 0.892517 | 0.447037 | 0.056638 | 2.95E-15 | cis |
| miR-136-5p | rs1162093 | 14:101151 | 14 | 1.01E+08 | C | G | 0.338205 | -0.3392 | 0.037024 | 5.11E-20 | cis |
| miR-136-5p | rs1259093 | 14:101163 | 14 | 1.01E+08 | A | T | 0.893649 | 0.419636 | 0.0585 | 7.32E-13 | cis |
| miR-136-5p | rs1190850 | 14:101064 | 14 | 1.01E+08 | A | G | 0.314302 | -0.29241 | 0.038357 | 2.47E-14 | cis |
| miR-136-5p | rs7142089 | 14:101172 | 14 | 1.01E+08 | T | A | 0.783402 | 0.330803 | 0.043789 | 4.21E-14 | cis |
| miR-136-5p | rs7147982 | 14:101046 | 14 | 1.01E+08 | C | T | 0.731753 | 0.279442 | 0.038937 | 7.13E-13 | cis |
| miR-136-5p | rs5993909 | 14:101169 | 14 | 1.01E+08 | C | T | 0.892837 | 0.451045 | 0.056823 | 2.06E-15 | cis |
| miR-136-5p | rs1190909 | 14:101101 | 14 | 1.01E+08 | G | A | 0.775531 | 0.292421 | 0.042859 | 8.92E-12 | cis |
| miR-136-5p | rs7154024 | 14:101031 | 14 | 1.01E+08 | G | T | 0.725015 | 0.287134 | 0.039067 | 1.99E-13 | cis |
| miR-136-5p | rs4243729 | 14:101066 | 14 | 1.01E+08 | A | C | 0.311439 | -0.28592 | 0.037818 | 4.02E-14 | cis |
| miR-136-5p | rs1243449 | 14:101179 | 14 | 1.01E+08 | A | G | 0.18959 | -0.49468 | 0.044481 | 9.88E-29 | cis |
| miR-136-5p | rs7145610 | 14:101049 | 14 | 1.01E+08 | T | C | 0.740391 | 0.310149 | 0.039698 | 5.59E-15 | cis |
| miR-136-5p | rs5910018 | 14:101184 | 14 | 1.01E+08 | G | A | 0.836014 | 0.39762 | 0.048235 | 1.67E-16 | cis |
| miR-136-5p | rs7141210 | 14:101182 | 14 | 1.01E+08 | T | C | 0.32902 | -0.40868 | 0.036265 | 1.86E-29 | cis |
| miR-136-5p | rs1190852 | 14:101064 | 14 | 1.01E+08 | A | G | 0.310295 | -0.28976 | 0.038132 | 2.99E-14 | cis |
| miR-136-5p | rs911330 | 14:101099 | 14 | 1.01E+08 | T | C | 0.774615 | 0.294165 | 0.042501 | 4.47E-12 | cis |
| miR-136-5p | rs1190867 | 14:101069 | 14 | 1.01E+08 | G | T | 0.316788 | -0.28574 | 0.038348 | 9.25E-14 | cis |
| miR-136-5p | rs4900471 | 14:101162 | 14 | 1.01E+08 | G | A | 0.304173 | -0.37862 | 0.038317 | 5.02E-23 | cis |
| miR-136-5p | rs1243369 | 14:101035 | 14 | 1.01E+08 | C | G | 0.723403 | 0.285505 | 0.038844 | 1.98E-13 | cis |
| miR-136-5p | rs1709961 | 14:101167 | 14 | 1.01E+08 | C | G | 0.668809 | 0.40362 | 0.036386 | 1.36E-28 | cis |
| miR-136-5p | rs4905999 | 14:101184 | 14 | 1.01E+08 | A | G | 0.181905 | -0.4793 | 0.045375 | 4.41E-26 | cis |
| miR-136-5p | rs7335877 | 14:101159 | 14 | 1.01E+08 | C | T | 0.891417 | 0.411586 | 0.058076 | 1.37E-12 | cis |
| miR-136-5p | rs1289379 | 14:101178 | 14 | 1.01E+08 | C | G | 0.190813 | -0.49409 | 0.044522 | 1.29E-28 | cis |
| miR-136-5p | rs1289760 | 14:101182 | 14 | 1.01E+08 | C | T | 0.190452 | -0.4956 | 0.044231 | 3.86E-29 | cis |
| miR-136-5p | rs7149242 | 14:101159 | 14 | 1.01E+08 | T | G | 0.288371 | -0.39204 | 0.038812 | 5.48E-24 | cis |
| miR-136-5p | rs8020868 | 14:101166 | 14 | 1.01E+08 | A | G | 0.668343 | 0.400258 | 0.036388 | 3.84E-28 | cis |
| miR-136-5p | rs2183632 | 14:101063 | 14 | 1.01E+08 | C | T | 0.319851 | -0.29421 | 0.038261 | 1.48E-14 | cis |
| miR-136-5p | rs5982537 | 14:101168 | 14 | 1.01E+08 | G | T | 0.89241 | 0.452013 | 0.056836 | 1.82E-15 | cis |
| miR-136-5p | rs6702222 | 14:101191 | 14 | 1.01E+08 | A | T | 0.731016 | 0.273571 | 0.040642 | 1.68E-11 | cis |
| miR-136-5p | rs7142683 | 14:101158 | 14 | 1.01E+08 | A | G | 0.290748 | -0.39275 | 0.038925 | 6.11E-24 | cis |
| miR-136-5p | rs1185095 | 14:101056 | 14 | 1.01E+08 | G | T | 0.743981 | 0.300419 | 0.039424 | 2.53E-14 | cis |
| miR-136-5p | rs6575794 | 14:101039 | 14 | 1.01E+08 | C | T | 0.734189 | 0.286049 | 0.039371 | 3.72E-13 | cis |
| miR-136-5p | rs6130797 | 14:101177 | 14 | 1.01E+08 | C | T | 0.888723 | 0.456524 | 0.055632 | 2.28E-16 | cis |
| miR-136-5p | rs7143357 | 14:101054 | 14 | 1.01E+08 | T | C | 0.739636 | 0.300872 | 0.039658 | 3.28E-14 | cis |
| miR-136-5p | rs879058 | 14:101069 | 14 | 1.01E+08 | G | T | 0.311807 | -0.28818 | 0.038065 | 3.71E-14 | cis |
| miR-136-5p | rs4905976 | 14:101060 | 14 | 1.01E+08 | G | A | 0.322862 | -0.28907 | 0.038315 | 4.54E-14 | cis |
| miR-136-5p | rs4905994 | 14:101154 | 14 | 1.01E+08 | T | C | 0.329146 | -0.3603 | 0.037792 | 1.52E-21 | cis |
| miR-136-5p | rs873077 | 14:101070 | 14 | 1.01E+08 | G | A | 0.312104 | -0.28533 | 0.037995 | 5.93E-14 | cis |

| | | | | | | | | | | | |
|---|---|---|---|---|---|---|---|---|---|---|---|
| miR-136-5p | rs1014696 | 14:101170 | 14 | 1.01E+08 | T | C | 0.658875 | 0.39858 | 0.035752 | 7.29E-29 | cis |
| miR-136-5p | rs7263167 | 14:101187 | 14 | 1.01E+08 | C | T | 0.881829 | 0.457921 | 0.05547 | 1.52E-16 | cis |
| miR-136-5p | rs5922882 | 14:101185 | 14 | 1.01E+08 | G | C | 0.745519 | 0.327292 | 0.040371 | 5.18E-16 | cis |
| miR-136-5p | rs9324022 | 14:101172 | 14 | 1.01E+08 | T | C | 0.763739 | 0.358128 | 0.042711 | 5.07E-17 | cis |
| miR-136-5p | rs1467536 | 14:101061 | 14 | 1.01E+08 | A | G | 0.318958 | -0.30086 | 0.038174 | 3.24E-15 | cis |
| miR-136-5p | rs1243613 | 14:101031 | 14 | 1.01E+08 | C | T | 0.725015 | 0.287507 | 0.039101 | 1.94E-13 | cis |
| miR-136-5p | rs1190904 | 14:101096 | 14 | 1.01E+08 | A | G | 0.772596 | 0.294548 | 0.042359 | 3.56E-12 | cis |
| miR-136-5p | rs1190857 | 14:101066 | 14 | 1.01E+08 | G | T | 0.310273 | -0.28852 | 0.038111 | 3.72E-14 | cis |
| miR-136-5p | rs1888581 | 14:101042 | 14 | 1.01E+08 | A | C | 0.735363 | 0.283234 | 0.039217 | 5.11E-13 | cis |
| miR-136-5p | rs3470057 | 14:101173 | 14 | 1.01E+08 | C | T | 0.892725 | 0.449012 | 0.056574 | 2.08E-15 | cis |
| miR-136-5p | rs7155649 | 14:101181 | 14 | 1.01E+08 | C | A | 0.863608 | 0.457293 | 0.051659 | 8.58E-19 | cis |
| miR-136-5p | rs6762567 | 14:101173 | 14 | 1.01E+08 | A | G | 0.892632 | 0.448685 | 0.056545 | 2.11E-15 | cis |
| miR-136-5p | rs1288176 | 14:101176 | 14 | 1.01E+08 | G | C | 0.347791 | -0.41693 | 0.036865 | 1.18E-29 | cis |
| miR-136-5p | rs1013940 | 14:101194 | 14 | 1.01E+08 | A | G | 0.323897 | -0.31112 | 0.037856 | 2.06E-16 | cis |
| miR-136-5p | rs1190907 | 14:101099 | 14 | 1.01E+08 | A | G | 0.772847 | 0.290916 | 0.042297 | 6.07E-12 | cis |
| miR-136-5p | rs7263167 | 14:101176 | 14 | 1.01E+08 | C | T | 0.889487 | 0.453054 | 0.055575 | 3.58E-16 | cis |
| miR-136-5p | rs5771269 | 14:101161 | 14 | 1.01E+08 | G | A | 0.894439 | 0.417877 | 0.058532 | 9.38E-13 | cis |
| miR-136-5p | rs1288281 | 14:101183 | 14 | 1.01E+08 | C | G | 0.331398 | -0.40977 | 0.036384 | 2.01E-29 | cis |
| miR-136-5p | rs5870230 | 14:101167 | 14 | 1.01E+08 | G | A | 0.892706 | 0.450591 | 0.056891 | 2.37E-15 | cis |
| miR-136-5p | rs7140556 | 14:101036 | 14 | 1.01E+08 | C | A | 0.723818 | 0.288841 | 0.039011 | 1.32E-13 | cis |
| miR-136-5p | rs2150518 | 14:101063 | 14 | 1.01E+08 | C | T | 0.318506 | -0.29583 | 0.038303 | 1.13E-14 | cis |
| miR-136-5p | rs7269872 | 14:101189 | 14 | 1.01E+08 | C | T | 0.825721 | 0.382143 | 0.045446 | 4.14E-17 | cis |
| miR-136-5p | rs1184948 | 14:101037 | 14 | 1.01E+08 | C | G | 0.733365 | 0.287472 | 0.039308 | 2.61E-13 | cis |
| miR-136-5p | rs1190906 | 14:101099 | 14 | 1.01E+08 | T | C | 0.772138 | 0.288893 | 0.042234 | 7.91E-12 | cis |
| miR-136-5p | rs7141110 | 14:101172 | 14 | 1.01E+08 | G | C | 0.783235 | 0.326991 | 0.043745 | 7.72E-14 | cis |
| miR-136-5p | rs4905977 | 14:101061 | 14 | 1.01E+08 | C | G | 0.318473 | -0.29773 | 0.038242 | 6.94E-15 | cis |
| miR-136-5p | rs1116059 | 14:101049 | 14 | 1.01E+08 | C | T | 0.741662 | 0.308914 | 0.039595 | 6.10E-15 | cis |
| miR-136-5p | rs873076 | 14:101070 | 14 | 1.01E+08 | A | G | 0.311961 | -0.28539 | 0.037988 | 5.80E-14 | cis |
| miR-136-5p | rs7155375 | 14:101188 | 14 | 1.01E+08 | C | T | 0.635995 | 0.320559 | 0.035698 | 2.71E-19 | cis |
| miR-136-5p | rs1288154 | 14:101176 | 14 | 1.01E+08 | G | C | 0.370047 | -0.42112 | 0.037394 | 2.03E-29 | cis |
| miR-136-5p | rs1214784 | 14:101144 | 14 | 1.01E+08 | C | T | 0.870989 | 0.380494 | 0.05243 | 3.95E-13 | cis |
| miR-136-5p | rs1258812 | 14:101162 | 14 | 1.01E+08 | T | C | 0.893709 | 0.419317 | 0.058496 | 7.59E-13 | cis |
| miR-136-5p | rs1757758 | 14:101189 | 14 | 1.01E+08 | C | T | 0.827094 | 0.384046 | 0.045714 | 4.42E-17 | cis |
| miR-136-5p | rs1555405 | 14:101176 | 14 | 1.01E+08 | G | A | 0.749075 | 0.322463 | 0.039528 | 3.41E-16 | cis |
| miR-139-5p | rs475419 | 9:1361482 | 9 | 1.36E+08 | T | C | 0.454203 | -0.09896 | 0.011534 | 9.49E-18 | trans |
| miR-139-5p | rs633862 | 9:1361554 | 9 | 1.36E+08 | T | C | 0.460592 | -0.09779 | 0.011554 | 2.58E-17 | trans |
| miR-139-5p | rs8176690 | 9:1361383 | 9 | 1.36E+08 | A | G | 0.601023 | 0.087564 | 0.011466 | 2.22E-14 | trans |
| miR-139-5p | rs8176682 | 9:1361392 | 9 | 1.36E+08 | C | T | 0.619011 | 0.089685 | 0.011561 | 8.67E-15 | trans |
| miR-139-5p | rs8176686 | 9:1361387 | 9 | 1.36E+08 | T | C | 0.595889 | 0.088317 | 0.011937 | 1.37E-13 | trans |
| miR-139-5p | rs687621 | 9:1361370 | 9 | 1.36E+08 | A | G | 0.663896 | -0.15832 | 0.011595 | 1.89E-42 | trans |
| miR-139-5p | rs576125 | 9:1361443 | 9 | 1.36E+08 | G | A | 0.688677 | -0.16349 | 0.012459 | 2.45E-39 | trans |
| miR-139-5p | rs7873635 | 9:1361320 | 9 | 1.36E+08 | T | C | 0.381256 | -0.08896 | 0.011728 | 3.31E-14 | trans |
| miR-139-5p | rs495203 | 9:1361452 | 9 | 1.36E+08 | C | T | 0.663279 | -0.15726 | 0.01161 | 8.37E-42 | trans |
| miR-139-5p | rs660340 | 9:1361475 | 9 | 1.36E+08 | A | G | 0.454082 | -0.09902 | 0.01154 | 9.44E-18 | trans |
| miR-139-5p | rs9411378 | 9:1361454 | 9 | 1.36E+08 | C | A | 0.753652 | -0.1514 | 0.013621 | 1.06E-28 | trans |
| miR-139-5p | rs1124406 | 9:1361539 | 9 | 1.36E+08 | C | T | 0.886198 | -0.17971 | 0.018422 | 1.75E-22 | trans |
| miR-139-5p | rs473533 | 9:1361480 | 9 | 1.36E+08 | T | C | 0.453861 | -0.09907 | 0.011541 | 9.15E-18 | trans |
| miR-139-5p | rs657152 | 9:1361392 | 9 | 1.36E+08 | C | A | 0.645336 | -0.15187 | 0.011382 | 1.30E-40 | trans |
| miR-139-5p | rs532436 | 9:1361498 | 9 | 1.36E+08 | G | A | 0.8067 | -0.17691 | 0.013943 | 6.95E-37 | trans |
| miR-139-5p | rs687289 | 9:1361371 | 9 | 1.36E+08 | G | A | 0.668579 | -0.16188 | 0.011737 | 2.87E-43 | trans |
| miR-139-5p | rs676457 | 9:1361462 | 9 | 1.36E+08 | A | T | 0.664166 | -0.15736 | 0.011604 | 6.87E-42 | trans |
| miR-139-5p | rs630510 | 9:1361495 | 9 | 1.36E+08 | A | G | 0.494403 | -0.10813 | 0.011448 | 3.54E-21 | trans |
| miR-139-5p | rs7046674 | 9:1361470 | 9 | 1.36E+08 | C | T | 0.619013 | 0.091154 | 0.011582 | 3.54E-15 | trans |
| miR-139-5p | rs676996 | 9:1361460 | 9 | 1.36E+08 | T | G | 0.667516 | -0.15735 | 0.011708 | 3.57E-41 | trans |
| miR-139-5p | rs492488 | 9:1361449 | 9 | 1.36E+08 | G | A | 0.663579 | -0.15731 | 0.011612 | 8.23E-42 | trans |
| miR-139-5p | rs647800 | 9:1361480 | 9 | 1.36E+08 | A | G | 0.435519 | -0.0992 | 0.011495 | 6.15E-18 | trans |
| miR-139-5p | rs8176691 | 9:1361382 | 9 | 1.36E+08 | C | T | 0.619638 | 0.090128 | 0.011562 | 6.44E-15 | trans |
| miR-139-5p | rs579459 | 9:1361541 | 9 | 1.36E+08 | T | C | 0.787293 | -0.16499 | 0.013454 | 1.43E-34 | trans |
| miR-139-5p | rs613534 | 9:1361431 | 9 | 1.36E+08 | A | G | 0.653693 | -0.15384 | 0.011595 | 3.54E-40 | trans |
| miR-139-5p | rs582094 | 9:1361454 | 9 | 1.36E+08 | A | T | 0.663709 | -0.1572 | 0.0116 | 7.69E-42 | trans |
| miR-139-5p | rs659104 | 9:1361478 | 9 | 1.36E+08 | T | G | 0.454107 | -0.09892 | 0.011534 | 9.80E-18 | trans |
| miR-139-5p | rs677355 | 9:1361460 | 9 | 1.36E+08 | G | A | 0.670198 | -0.15785 | 0.011814 | 1.01E-40 | trans |
| miR-139-5p | rs8176663 | 9:1361444 | 9 | 1.36E+08 | T | C | 0.663959 | -0.15722 | 0.011603 | 7.92E-42 | trans |

| | | | | | | | | | | | |
|---|---|---|---|---|---|---|---|---|---|---|---|
| miR-139-5p | rs1401144 | 9:1361443 | 9 | 1.36E+08 | T | C | 0.690247 | -0.16283 | 0.012511 | 1.01E-38 | trans |
| miR-139-5p | rs582118 | 9:1361454 | 9 | 1.36E+08 | A | G | 0.663872 | -0.1573 | 0.011605 | 7.44E-42 | trans |
| miR-139-5p | rs544873 | 9:1361432 | 9 | 1.36E+08 | G | A | 0.645742 | -0.15203 | 0.011396 | 1.34E-40 | trans |
| miR-139-5p | rs649129 | 9:1361543 | 9 | 1.36E+08 | C | T | 0.787432 | -0.16516 | 0.013462 | 1.34E-34 | trans |
| miR-139-5p | rs643434 | 9:1361423 | 9 | 1.36E+08 | G | A | 0.645392 | -0.15197 | 0.01139 | 1.31E-40 | trans |
| miR-139-5p | rs550057 | 9:1361465 | 9 | 1.36E+08 | C | T | 0.734085 | -0.14777 | 0.012554 | 5.52E-32 | trans |
| miR-139-5p | rs505922 | 9:1361492 | 9 | 1.36E+08 | T | C | 0.665748 | -0.15954 | 0.011579 | 3.41E-43 | trans |
| miR-139-5p | rs8176649 | 9:1361472 | 9 | 1.36E+08 | G | A | 0.618669 | 0.090781 | 0.011565 | 4.16E-15 | trans |
| miR-139-5p | rs491626 | 9:1361448 | 9 | 1.36E+08 | C | T | 0.664173 | -0.15758 | 0.011633 | 8.32E-42 | trans |
| miR-139-5p | rs2073828 | 9:1361371 | 9 | 1.36E+08 | G | A | 0.601379 | 0.088031 | 0.011469 | 1.65E-14 | trans |
| miR-139-5p | rs612169 | 9:1361434 | 9 | 1.36E+08 | A | G | 0.66434 | -0.15744 | 0.011618 | 7.83E-42 | trans |
| miR-139-5p | rs630014 | 9:1361497 | 9 | 1.36E+08 | A | G | 0.494254 | -0.10799 | 0.01144 | 3.74E-21 | trans |
| miR-139-5p | rs476410 | 9:1361483 | 9 | 1.36E+08 | G | C | 0.45414 | -0.09891 | 0.011534 | 9.87E-18 | trans |
| miR-139-5p | rs493246 | 9:1361449 | 9 | 1.36E+08 | G | A | 0.663234 | -0.15741 | 0.011618 | 8.06E-42 | trans |
| miR-139-5p | rs514659 | 9:1361422 | 9 | 1.36E+08 | A | C | 0.663927 | -0.15717 | 0.011603 | 8.33E-42 | trans |
| miR-139-5p | rs500499 | 9:1361486 | 9 | 1.36E+08 | C | G | 0.465434 | -0.0969 | 0.011704 | 1.24E-16 | trans |
| miR-139-5p | rs644234 | 9:1361422 | 9 | 1.36E+08 | T | G | 0.645381 | -0.15195 | 0.011382 | 1.17E-40 | trans |
| miR-139-5p | rs545971 | 9:1361433 | 9 | 1.36E+08 | C | T | 0.663766 | -0.1571 | 0.011598 | 8.40E-42 | trans |
| miR-139-5p | rs4962040 | 9:1361335 | 9 | 1.36E+08 | A | G | 0.619057 | 0.089968 | 0.011573 | 7.61E-15 | trans |
| miR-139-5p | rs554833 | 9:1361471 | 9 | 1.36E+08 | C | T | 0.664057 | -0.15734 | 0.011597 | 6.24E-42 | trans |
| miR-139-5p | rs543040 | 9:1361430 | 9 | 1.36E+08 | A | T | 0.669196 | -0.15905 | 0.011822 | 2.94E-41 | trans |
| miR-139-5p | rs494242 | 9:1361451 | 9 | 1.36E+08 | C | T | 0.644977 | -0.15225 | 0.011401 | 1.11E-40 | trans |
| miR-139-5p | rs559723 | 9:1361504 | 9 | 1.36E+08 | A | G | 0.508431 | -0.10594 | 0.011713 | 1.50E-19 | trans |
| miR-139-5p | rs543968 | 9:1361431 | 9 | 1.36E+08 | T | C | 0.653829 | -0.15371 | 0.011597 | 4.23E-40 | trans |
| miR-139-5p | rs527210 | 9:1361464 | 9 | 1.36E+08 | C | T | 0.668712 | -0.15683 | 0.011751 | 1.25E-40 | trans |
| miR-139-5p | rs500498 | 9:1361486 | 9 | 1.36E+08 | C | T | 0.464233 | -0.09645 | 0.011689 | 1.56E-16 | trans |
| miR-139-5p | rs7036642 | 9:1361446 | 9 | 1.36E+08 | G | A | 0.618956 | 0.090572 | 0.011575 | 5.09E-15 | trans |
| miR-139-5p | rs597988 | 9:1361442 | 9 | 1.36E+08 | T | A | 0.686751 | -0.16382 | 0.012368 | 4.80E-40 | trans |
| miR-139-5p | rs8176702 | 9:1361361 | 9 | 1.36E+08 | G | A | 0.6197 | 0.090201 | 0.011565 | 6.21E-15 | trans |
| miR-139-5p | rs2519093 | 9:1361418 | 9 | 1.36E+08 | C | T | 0.809892 | -0.1791 | 0.014192 | 1.63E-36 | trans |
| miR-139-5p | rs7857390 | 9:1361285 | 9 | 1.36E+08 | A | G | 0.377967 | -0.08896 | 0.012037 | 1.46E-13 | trans |
| miR-139-5p | rs2073827 | 9:1361371 | 9 | 1.36E+08 | G | C | 0.601379 | 0.088031 | 0.011469 | 1.65E-14 | trans |
| miR-139-5p | rs495828 | 9:1361548 | 9 | 1.36E+08 | G | T | 0.787276 | -0.16496 | 0.013454 | 1.46E-34 | trans |
| miR-139-5p | rs674302 | 9:1361466 | 9 | 1.36E+08 | T | A | 0.663989 | -0.15734 | 0.011595 | 6.05E-42 | trans |
| miR-139-5p | rs635634 | 9:1361550 | 9 | 1.36E+08 | C | T | 0.80599 | -0.1757 | 0.013909 | 1.42E-36 | trans |
| miR-139-5p | rs8176668 | 9:1361440 | 9 | 1.36E+08 | A | T | 0.618711 | 0.090485 | 0.011582 | 5.61E-15 | trans |
| miR-139-5p | rs645982 | 9:1361484 | 9 | 1.36E+08 | A | G | 0.454123 | -0.09891 | 0.011533 | 9.82E-18 | trans |
| miR-139-5p | rs8176715 | 9:1361331 | 9 | 1.36E+08 | T | C | 0.418056 | -0.08482 | 0.011765 | 5.63E-13 | trans |
| miR-139-5p | rs581107 | 9:1361477 | 9 | 1.36E+08 | C | T | 0.452595 | -0.09921 | 0.011573 | 1.01E-17 | trans |
| miR-139-5p | rs2073826 | 9:1361369 | 9 | 1.36E+08 | G | T | 0.60107 | 0.087903 | 0.011463 | 1.75E-14 | trans |
| miR-139-5p | rs529565 | 9:1361495 | 9 | 1.36E+08 | T | C | 0.665911 | -0.15979 | 0.011585 | 2.82E-43 | trans |
| miR-139-5p | rs597974 | 9:1361442 | 9 | 1.36E+08 | A | G | 0.687926 | -0.16395 | 0.012424 | 9.22E-40 | trans |
| miR-139-5p | rs1165522 | 9:1361490 | 9 | 1.36E+08 | T | A | 0.653719 | -0.15545 | 0.011571 | 3.77E-41 | trans |
| miR-139-5p | rs600038 | 9:1361518 | 9 | 1.36E+08 | T | C | 0.787872 | -0.16606 | 0.013486 | 7.68E-35 | trans |
| miR-139-5p | rs507666 | 9:1361493 | 9 | 1.36E+08 | G | A | 0.806573 | -0.17672 | 0.013934 | 7.43E-37 | trans |
| miR-139-5p | rs2769071 | 9:1361459 | 9 | 1.36E+08 | A | G | 0.663117 | -0.15721 | 0.011626 | 1.14E-41 | trans |
| miR-139-5p | rs1178913 | 9:1361853 | 9 | 1.36E+08 | C | G | 0.833223 | -0.10781 | 0.015887 | 1.15E-11 | trans |
| miR-139-5p | rs651007 | 9:1361538 | 9 | 1.36E+08 | C | T | 0.787179 | -0.16536 | 0.013455 | 1.03E-34 | trans |
| miR-139-5p | rs8176681 | 9:1361397 | 9 | 1.36E+08 | T | C | 0.600667 | 0.08773 | 0.011476 | 2.10E-14 | trans |
| miR-139-5p | rs616154 | 9:1361504 | 9 | 1.36E+08 | C | T | 0.501246 | -0.10645 | 0.011595 | 4.26E-20 | trans |
| miR-142-5p | rs6197743 | 14:2143099 | 14 | 21430994 | G | A | 0.76048 | -0.15496 | 0.018671 | 1.05E-16 | trans |
| miR-142-5p | rs6197743 | 14:2143084 | 14 | 21430843 | G | A | 0.760478 | -0.15497 | 0.018671 | 1.04E-16 | trans |
| miR-142-5p | rs1014847 | 14:2143026 | 14 | 21430269 | C | A | 0.760451 | -0.15506 | 0.018669 | 9.89E-17 | trans |
| miR-142-5p | rs4982384 | 14:2143243 | 14 | 21432436 | G | A | 0.308874 | 0.117524 | 0.01758 | 2.31E-11 | trans |
| miR-142-5p | rs1289748 | 14:2143129 | 14 | 21431295 | C | T | 0.766905 | -0.1519 | 0.018832 | 7.26E-16 | trans |
| miR-142-5p | rs2771329 | 14:2143545 | 14 | 21435450 | T | A | 0.641889 | -0.12408 | 0.016692 | 1.06E-13 | trans |
| miR-142-5p | rs1288230 | 14:2143134 | 14 | 21431349 | T | A | 0.766982 | -0.15158 | 0.018828 | 8.21E-16 | trans |
| miR-142-5p | rs1014269 | 14:2143810 | 14 | 21438108 | T | G | 0.398066 | 0.122952 | 0.016789 | 2.42E-13 | trans |
| miR-142-5p | rs1288228 | 14:2143130 | 14 | 21431304 | T | C | 0.761865 | -0.15555 | 0.018684 | 8.41E-17 | trans |
| miR-142-5p | rs2771327 | 14:2143437 | 14 | 21434371 | C | T | 0.643183 | -0.12534 | 0.016711 | 6.37E-14 | trans |
| miR-142-5p | rs6571588 | 14:2143884 | 14 | 21438849 | G | A | 0.424307 | 0.117914 | 0.016698 | 1.65E-12 | trans |
| miR-142-5p | rs1289765 | 14:2143138 | 14 | 21431382 | C | T | 0.760157 | -0.15514 | 0.018677 | 9.85E-17 | trans |
| miR-142-5p | rs1289767 | 14:2143141 | 14 | 21431413 | C | T | 0.760166 | -0.15514 | 0.018677 | 9.86E-17 | trans |

| | | | | | | | | | | | |
|---|---|---|---|---|---|---|---|---|---|---|---|
| miR-142-5p | rs2783794 | 14:214357 | 14 | 21435750 | T | C | 0.641653 | -0.12396 | 0.01668 | 1.07E-13 | trans |
| miR-142-5p | rs6704901 | 14:214311 | 14 | 21431105 | G | A | 0.761822 | -0.1557 | 0.018685 | 7.88E-17 | trans |
| miR-142-5p | rs3790067 | 14:214210 | 14 | 21421088 | C | T | 0.767067 | -0.15292 | 0.018823 | 4.49E-16 | trans |
| miR-143-3p | rs4734875 | 8:1065547 | 8 | 1.07E+08 | T | C | 0.78521 | -0.13589 | 0.020011 | 1.12E-11 | trans |
| miR-143-3p | rs2853985 | 8:1065467 | 8 | 1.07E+08 | C | G | 0.78472 | -0.13669 | 0.020019 | 8.63E-12 | trans |
| miR-143-3p | rs7267373 | 8:1065362 | 8 | 1.07E+08 | C | T | 0.788389 | -0.13654 | 0.020107 | 1.11E-11 | trans |
| miR-143-3p | rs1687334 | 8:1065494 | 8 | 1.07E+08 | G | C | 0.785426 | -0.13544 | 0.020034 | 1.37E-11 | trans |
| miR-154-5p | rs1014696 | 14:101170 | 14 | 1.01E+08 | T | C | 0.658875 | 0.196312 | 0.029305 | 2.10E-11 | cis |
| miR-193a-3 | rs687289 | 9:1361371 | 9 | 1.36E+08 | G | A | 0.668579 | -0.09395 | 0.013894 | 1.36E-11 | trans |
| miR-195-5p | rs527210 | 9:1361464 | 9 | 1.36E+08 | C | T | 0.668712 | -0.113 | 0.013578 | 8.63E-17 | trans |
| miR-195-5p | rs550057 | 9:1361465 | 9 | 1.36E+08 | C | T | 0.734085 | -0.1105 | 0.014416 | 1.79E-14 | trans |
| miR-195-5p | rs613534 | 9:1361431 | 9 | 1.36E+08 | A | G | 0.653693 | -0.11282 | 0.013386 | 3.51E-17 | trans |
| miR-195-5p | rs597988 | 9:1361442 | 9 | 1.36E+08 | T | A | 0.686751 | -0.1192 | 0.014281 | 7.00E-17 | trans |
| miR-195-5p | rs495203 | 9:1361452 | 9 | 1.36E+08 | C | T | 0.663279 | -0.11376 | 0.013422 | 2.34E-17 | trans |
| miR-195-5p | rs544873 | 9:1361432 | 9 | 1.36E+08 | G | A | 0.645742 | -0.11024 | 0.013164 | 5.54E-17 | trans |
| miR-195-5p | rs676457 | 9:1361462 | 9 | 1.36E+08 | A | T | 0.664166 | -0.11387 | 0.013416 | 2.12E-17 | trans |
| miR-195-5p | rs495828 | 9:1361548 | 9 | 1.36E+08 | G | T | 0.787276 | -0.13188 | 0.01544 | 1.33E-17 | trans |
| miR-195-5p | rs492488 | 9:1361449 | 9 | 1.36E+08 | G | A | 0.663579 | -0.11366 | 0.013425 | 2.54E-17 | trans |
| miR-195-5p | rs579459 | 9:1361541 | 9 | 1.36E+08 | T | C | 0.787293 | -0.13189 | 0.015441 | 1.32E-17 | trans |
| miR-195-5p | rs687621 | 9:1361370 | 9 | 1.36E+08 | A | G | 0.663896 | -0.11416 | 0.013411 | 1.70E-17 | trans |
| miR-195-5p | rs657152 | 9:1361392 | 9 | 1.36E+08 | C | A | 0.645336 | -0.11011 | 0.013148 | 5.54E-17 | trans |
| miR-195-5p | rs507666 | 9:1361493 | 9 | 1.36E+08 | G | A | 0.806573 | -0.13903 | 0.016019 | 3.98E-18 | trans |
| miR-195-5p | rs494242 | 9:1361451 | 9 | 1.36E+08 | C | T | 0.644977 | -0.11062 | 0.013169 | 4.48E-17 | trans |
| miR-195-5p | rs643434 | 9:1361423 | 9 | 1.36E+08 | G | A | 0.645392 | -0.11031 | 0.013157 | 5.10E-17 | trans |
| miR-195-5p | rs543040 | 9:1361430 | 9 | 1.36E+08 | A | T | 0.669196 | -0.11564 | 0.01366 | 2.56E-17 | trans |
| miR-195-5p | rs1165522 | 9:1361490 | 9 | 1.36E+08 | T | A | 0.653719 | -0.11298 | 0.013369 | 2.90E-17 | trans |
| miR-195-5p | rs491626 | 9:1361448 | 9 | 1.36E+08 | C | T | 0.664173 | -0.114 | 0.013448 | 2.32E-17 | trans |
| miR-195-5p | rs9411378 | 9:1361454 | 9 | 1.36E+08 | C | A | 0.753652 | -0.11231 | 0.015615 | 6.38E-13 | trans |
| miR-195-5p | rs677355 | 9:1361460 | 9 | 1.36E+08 | G | A | 0.670198 | -0.11434 | 0.013648 | 5.38E-17 | trans |
| miR-195-5p | rs1401144 | 9:1361443 | 9 | 1.36E+08 | T | C | 0.690247 | -0.11838 | 0.014435 | 2.38E-16 | trans |
| miR-195-5p | rs600038 | 9:1361518 | 9 | 1.36E+08 | T | C | 0.787872 | -0.13174 | 0.015483 | 1.76E-17 | trans |
| miR-195-5p | rs676996 | 9:1361460 | 9 | 1.36E+08 | T | G | 0.667516 | -0.11423 | 0.013529 | 3.08E-17 | trans |
| miR-195-5p | rs582094 | 9:1361454 | 9 | 1.36E+08 | A | T | 0.663709 | -0.11378 | 0.01341 | 2.17E-17 | trans |
| miR-195-5p | rs582118 | 9:1361454 | 9 | 1.36E+08 | A | G | 0.663872 | -0.11383 | 0.013416 | 2.17E-17 | trans |
| miR-195-5p | rs543968 | 9:1361431 | 9 | 1.36E+08 | T | C | 0.653829 | -0.11276 | 0.013387 | 3.66E-17 | trans |
| miR-195-5p | rs644234 | 9:1361422 | 9 | 1.36E+08 | T | G | 0.645381 | -0.11026 | 0.013148 | 5.01E-17 | trans |
| miR-195-5p | rs687289 | 9:1361371 | 9 | 1.36E+08 | G | A | 0.668579 | -0.11635 | 0.013584 | 1.08E-17 | trans |
| miR-195-5p | rs635634 | 9:1361550 | 9 | 1.36E+08 | C | T | 0.80599 | -0.13963 | 0.015982 | 2.40E-18 | trans |
| miR-195-5p | rs493246 | 9:1361449 | 9 | 1.36E+08 | G | A | 0.663234 | -0.11379 | 0.013432 | 2.42E-17 | trans |
| miR-195-5p | rs2769071 | 9:1361459 | 9 | 1.36E+08 | A | G | 0.663117 | -0.1134 | 0.01344 | 3.25E-17 | trans |
| miR-195-5p | rs651007 | 9:1361538 | 9 | 1.36E+08 | C | T | 0.787179 | -0.13229 | 0.015442 | 1.06E-17 | trans |
| miR-195-5p | rs532436 | 9:1361498 | 9 | 1.36E+08 | G | A | 0.8067 | -0.13917 | 0.01603 | 3.89E-18 | trans |
| miR-195-5p | rs545971 | 9:1361433 | 9 | 1.36E+08 | C | T | 0.663766 | -0.11352 | 0.013409 | 2.54E-17 | trans |
| miR-195-5p | rs554833 | 9:1361471 | 9 | 1.36E+08 | C | T | 0.664057 | -0.11379 | 0.013408 | 2.11E-17 | trans |
| miR-195-5p | rs529565 | 9:1361495 | 9 | 1.36E+08 | T | C | 0.665911 | -0.11648 | 0.013401 | 3.57E-18 | trans |
| miR-195-5p | rs612169 | 9:1361434 | 9 | 1.36E+08 | A | G | 0.66434 | -0.11371 | 0.013433 | 2.56E-17 | trans |
| miR-195-5p | rs514659 | 9:1361422 | 9 | 1.36E+08 | A | C | 0.663927 | -0.11351 | 0.013414 | 2.64E-17 | trans |
| miR-195-5p | rs8176663 | 9:1361444 | 9 | 1.36E+08 | T | C | 0.663959 | -0.1136 | 0.013414 | 2.48E-17 | trans |
| miR-195-5p | rs674302 | 9:1361466 | 9 | 1.36E+08 | T | A | 0.663989 | -0.11398 | 0.013405 | 1.85E-17 | trans |
| miR-195-5p | rs576125 | 9:1361443 | 9 | 1.36E+08 | G | A | 0.688677 | -0.11854 | 0.014381 | 1.68E-16 | trans |
| miR-195-5p | rs597974 | 9:1361442 | 9 | 1.36E+08 | A | G | 0.687926 | -0.11914 | 0.014344 | 9.92E-17 | trans |
| miR-195-5p | rs505922 | 9:1361492 | 9 | 1.36E+08 | T | C | 0.665748 | -0.11621 | 0.013393 | 4.07E-18 | trans |
| miR-195-5p | rs649129 | 9:1361543 | 9 | 1.36E+08 | C | T | 0.787432 | -0.13201 | 0.01545 | 1.29E-17 | trans |
| miR-195-5p | rs2519093 | 9:1361418 | 9 | 1.36E+08 | C | T | 0.809892 | -0.14041 | 0.016314 | 7.49E-18 | trans |
| miR-23b-3p | rs544873 | 9:1361432 | 9 | 1.36E+08 | G | A | 0.645742 | -0.07513 | 0.011225 | 2.18E-11 | trans |
| miR-23b-3p | rs643434 | 9:1361423 | 9 | 1.36E+08 | G | A | 0.645392 | -0.07518 | 0.01122 | 2.08E-11 | trans |
| miR-23b-3p | rs644234 | 9:1361422 | 9 | 1.36E+08 | T | G | 0.645381 | -0.07498 | 0.011212 | 2.27E-11 | trans |
| miR-23b-3p | rs494242 | 9:1361451 | 9 | 1.36E+08 | C | T | 0.644977 | -0.0754 | 0.01123 | 1.89E-11 | trans |
| miR-23b-3p | rs1165522 | 9:1361490 | 9 | 1.36E+08 | T | A | 0.653719 | -0.07861 | 0.011396 | 5.28E-12 | trans |
| miR-23b-3p | rs657152 | 9:1361392 | 9 | 1.36E+08 | C | A | 0.645336 | -0.07498 | 0.011212 | 2.28E-11 | trans |
| miR-26b-5p | rs6197743 | 14:214308 | 14 | 21430843 | G | A | 0.760478 | -0.12619 | 0.018249 | 4.67E-12 | trans |
| miR-26b-5p | rs1289748 | 14:214312 | 14 | 21431295 | C | T | 0.766905 | -0.12488 | 0.018397 | 1.14E-11 | trans |
| miR-26b-5p | rs1014847 | 14:214302 | 14 | 21430269 | C | A | 0.760451 | -0.12628 | 0.018247 | 4.50E-12 | trans |

| | | | | | | | | | | | |
|---|---|---|---|---|---|---|---|---|---|---|---|
| miR-26b-5p | rs1289767 | 14:214314 | 14 | 21431413 | C | T | 0.760166 | -0.12623 | 0.018255 | 4.69E-12 | trans |
| miR-26b-5p | rs6197743 | 14:214309 | 14 | 21430994 | G | A | 0.76048 | -0.12618 | 0.018249 | 4.70E-12 | trans |
| miR-26b-5p | rs1288230 | 14:214313 | 14 | 21431349 | T | A | 0.766982 | -0.12452 | 0.018394 | 1.29E-11 | trans |
| miR-26b-5p | rs3790067 | 14:214210 | 14 | 21421088 | C | T | 0.767067 | -0.12533 | 0.018391 | 9.44E-12 | trans |
| miR-26b-5p | rs6704901 | 14:214311 | 14 | 21431105 | G | A | 0.761822 | -0.12676 | 0.018263 | 3.91E-12 | trans |
| miR-26b-5p | rs1288228 | 14:214313 | 14 | 21431304 | T | C | 0.761865 | -0.12658 | 0.018263 | 4.18E-12 | trans |
| miR-26b-5p | rs1289765 | 14:214313 | 14 | 21431382 | C | T | 0.760157 | -0.12623 | 0.018255 | 4.68E-12 | trans |
| miR-27b-3p | rs1165522 | 9:1361490 | 9 | 1.36E+08 | T | A | 0.653719 | -0.10033 | 0.012871 | 6.47E-15 | trans |
| miR-27b-3p | rs493246 | 9:1361449 | 9 | 1.36E+08 | G | A | 0.663234 | -0.09559 | 0.012951 | 1.57E-13 | trans |
| miR-27b-3p | rs1401144 | 9:1361443 | 9 | 1.36E+08 | T | C | 0.690247 | -0.09649 | 0.013925 | 4.23E-12 | trans |
| miR-27b-3p | rs600038 | 9:1361518 | 9 | 1.36E+08 | T | C | 0.787872 | -0.10932 | 0.014934 | 2.48E-13 | trans |
| miR-27b-3p | rs2519093 | 9:1361418 | 9 | 1.36E+08 | C | T | 0.809892 | -0.10726 | 0.015767 | 1.03E-11 | trans |
| miR-27b-3p | rs613534 | 9:1361431 | 9 | 1.36E+08 | A | G | 0.653693 | -0.09948 | 0.01289 | 1.18E-14 | trans |
| miR-27b-3p | rs651007 | 9:1361538 | 9 | 1.36E+08 | C | T | 0.787179 | -0.10943 | 0.014896 | 2.04E-13 | trans |
| miR-27b-3p | rs576125 | 9:1361443 | 9 | 1.36E+08 | G | A | 0.688677 | -0.09697 | 0.013872 | 2.74E-12 | trans |
| miR-27b-3p | rs494242 | 9:1361451 | 9 | 1.36E+08 | C | T | 0.644977 | -0.0987 | 0.012677 | 6.93E-15 | trans |
| miR-27b-3p | rs687289 | 9:1361371 | 9 | 1.36E+08 | G | A | 0.668579 | -0.09793 | 0.013098 | 7.64E-14 | trans |
| miR-27b-3p | rs514659 | 9:1361422 | 9 | 1.36E+08 | A | C | 0.663927 | -0.09541 | 0.012934 | 1.62E-13 | trans |
| miR-27b-3p | rs554833 | 9:1361471 | 9 | 1.36E+08 | C | T | 0.664057 | -0.09544 | 0.012928 | 1.55E-13 | trans |
| miR-27b-3p | rs582094 | 9:1361454 | 9 | 1.36E+08 | A | T | 0.663709 | -0.09552 | 0.012931 | 1.50E-13 | trans |
| miR-27b-3p | rs687621 | 9:1361370 | 9 | 1.36E+08 | A | G | 0.663896 | -0.09623 | 0.01293 | 9.92E-14 | trans |
| miR-27b-3p | rs676457 | 9:1361462 | 9 | 1.36E+08 | A | T | 0.664166 | -0.09557 | 0.012936 | 1.49E-13 | trans |
| miR-27b-3p | rs657152 | 9:1361392 | 9 | 1.36E+08 | C | A | 0.645336 | -0.09824 | 0.012657 | 8.34E-15 | trans |
| miR-27b-3p | rs612169 | 9:1361434 | 9 | 1.36E+08 | A | G | 0.66434 | -0.09548 | 0.012952 | 1.68E-13 | trans |
| miR-27b-3p | rs597974 | 9:1361442 | 9 | 1.36E+08 | A | G | 0.687926 | -0.09727 | 0.013838 | 2.07E-12 | trans |
| miR-27b-3p | rs579459 | 9:1361541 | 9 | 1.36E+08 | T | C | 0.787293 | -0.10917 | 0.014894 | 2.31E-13 | trans |
| miR-27b-3p | rs529565 | 9:1361495 | 9 | 1.36E+08 | T | C | 0.665911 | -0.09807 | 0.012923 | 3.22E-14 | trans |
| miR-27b-3p | rs532436 | 9:1361498 | 9 | 1.36E+08 | G | A | 0.8067 | -0.10669 | 0.015493 | 5.73E-12 | trans |
| miR-27b-3p | rs8176663 | 9:1361444 | 9 | 1.36E+08 | T | C | 0.663959 | -0.09546 | 0.012934 | 1.57E-13 | trans |
| miR-27b-3p | rs543968 | 9:1361431 | 9 | 1.36E+08 | T | C | 0.653829 | -0.09936 | 0.012891 | 1.28E-14 | trans |
| miR-27b-3p | rs543040 | 9:1361430 | 9 | 1.36E+08 | A | T | 0.669196 | -0.09652 | 0.013173 | 2.36E-13 | trans |
| miR-27b-3p | rs492488 | 9:1361449 | 9 | 1.36E+08 | G | A | 0.663579 | -0.09549 | 0.012944 | 1.62E-13 | trans |
| miR-27b-3p | rs674302 | 9:1361466 | 9 | 1.36E+08 | T | A | 0.663989 | -0.09558 | 0.012926 | 1.42E-13 | trans |
| miR-27b-3p | rs550057 | 9:1361465 | 9 | 1.36E+08 | C | T | 0.734085 | -0.09358 | 0.013888 | 1.61E-11 | trans |
| miR-27b-3p | rs644234 | 9:1361422 | 9 | 1.36E+08 | T | G | 0.645381 | -0.09829 | 0.012656 | 8.09E-15 | trans |
| miR-27b-3p | rs582118 | 9:1361454 | 9 | 1.36E+08 | A | G | 0.663872 | -0.0956 | 0.012936 | 1.46E-13 | trans |
| miR-27b-3p | rs2769071 | 9:1361459 | 9 | 1.36E+08 | A | G | 0.663117 | -0.09506 | 0.012959 | 2.21E-13 | trans |
| miR-27b-3p | rs677355 | 9:1361460 | 9 | 1.36E+08 | G | A | 0.670198 | -0.09623 | 0.013158 | 2.60E-13 | trans |
| miR-27b-3p | rs649129 | 9:1361543 | 9 | 1.36E+08 | C | T | 0.787432 | -0.10932 | 0.014903 | 2.21E-13 | trans |
| miR-27b-3p | rs495203 | 9:1361452 | 9 | 1.36E+08 | C | T | 0.663279 | -0.09549 | 0.012942 | 1.60E-13 | trans |
| miR-27b-3p | rs597988 | 9:1361442 | 9 | 1.36E+08 | T | A | 0.686751 | -0.0974 | 0.013777 | 1.55E-12 | trans |
| miR-27b-3p | rs676996 | 9:1361460 | 9 | 1.36E+08 | T | G | 0.667516 | -0.09596 | 0.013044 | 1.89E-13 | trans |
| miR-27b-3p | rs544873 | 9:1361432 | 9 | 1.36E+08 | G | A | 0.645742 | -0.09838 | 0.012672 | 8.25E-15 | trans |
| miR-27b-3p | rs545971 | 9:1361433 | 9 | 1.36E+08 | C | T | 0.663766 | -0.09536 | 0.012929 | 1.63E-13 | trans |
| miR-27b-3p | rs495828 | 9:1361548 | 9 | 1.36E+08 | G | T | 0.787276 | -0.10918 | 0.014893 | 2.29E-13 | trans |
| miR-27b-3p | rs505922 | 9:1361492 | 9 | 1.36E+08 | T | C | 0.665748 | -0.09788 | 0.012916 | 3.49E-14 | trans |
| miR-27b-3p | rs527210 | 9:1361464 | 9 | 1.36E+08 | C | T | 0.668712 | -0.09445 | 0.013091 | 5.41E-13 | trans |
| miR-27b-3p | rs643434 | 9:1361423 | 9 | 1.36E+08 | G | A | 0.645392 | -0.09837 | 0.012665 | 8.06E-15 | trans |
| miR-27b-3p | rs507666 | 9:1361493 | 9 | 1.36E+08 | G | A | 0.806573 | -0.10654 | 0.015483 | 5.93E-12 | trans |
| miR-27b-3p | rs491626 | 9:1361448 | 9 | 1.36E+08 | C | T | 0.664173 | -0.09581 | 0.012967 | 1.48E-13 | trans |
| miR-27b-3p | rs635634 | 9:1361550 | 9 | 1.36E+08 | C | T | 0.80599 | -0.10667 | 0.01545 | 5.05E-12 | trans |
| miR-30a-5p | rs657152 | 9:1361392 | 9 | 1.36E+08 | C | A | 0.645336 | -0.06939 | 0.00818 | 2.20E-17 | trans |
| miR-30a-5p | rs649129 | 9:1361543 | 9 | 1.36E+08 | C | T | 0.787432 | -0.08661 | 0.009597 | 1.81E-19 | trans |
| miR-30a-5p | rs550057 | 9:1361465 | 9 | 1.36E+08 | C | T | 0.734085 | -0.06817 | 0.008974 | 3.04E-14 | trans |
| miR-30a-5p | rs1401144 | 9:1361443 | 9 | 1.36E+08 | T | C | 0.690247 | -0.0746 | 0.00898 | 9.83E-17 | trans |
| miR-30a-5p | rs505922 | 9:1361492 | 9 | 1.36E+08 | T | C | 0.665748 | -0.07284 | 0.008333 | 2.31E-18 | trans |
| miR-30a-5p | rs529565 | 9:1361495 | 9 | 1.36E+08 | T | C | 0.665911 | -0.07297 | 0.008338 | 2.11E-18 | trans |
| miR-30a-5p | rs651007 | 9:1361538 | 9 | 1.36E+08 | C | T | 0.787179 | -0.0867 | 0.009593 | 1.59E-19 | trans |
| miR-30a-5p | rs582118 | 9:1361454 | 9 | 1.36E+08 | A | G | 0.663872 | -0.07213 | 0.008345 | 5.44E-18 | trans |
| miR-30a-5p | rs493246 | 9:1361449 | 9 | 1.36E+08 | G | A | 0.663234 | -0.07218 | 0.008354 | 5.64E-18 | trans |
| miR-30a-5p | rs492488 | 9:1361449 | 9 | 1.36E+08 | G | A | 0.663579 | -0.0721 | 0.00835 | 5.86E-18 | trans |
| miR-30a-5p | rs643434 | 9:1361423 | 9 | 1.36E+08 | G | A | 0.645392 | -0.06946 | 0.008185 | 2.14E-17 | trans |
| miR-30a-5p | rs507666 | 9:1361493 | 9 | 1.36E+08 | G | A | 0.806573 | -0.09266 | 0.009944 | 1.19E-20 | trans |

| | | | | | | | | | | | |
|---|---|---|---|---|---|---|---|---|---|---|---|
| miR-30a-5p | rs9411378 | 9:1361454 | 9 | 1.36E+08 | C | A | 0.753652 | -0.06583 | 0.009731 | 1.33E-11 | trans |
| miR-30a-5p | rs544873 | 9:1361432 | 9 | 1.36E+08 | G | A | 0.645742 | -0.06958 | 0.008189 | 1.94E-17 | trans |
| miR-30a-5p | rs491626 | 9:1361448 | 9 | 1.36E+08 | C | T | 0.664173 | -0.07239 | 0.008364 | 4.95E-18 | trans |
| miR-30a-5p | rs597988 | 9:1361442 | 9 | 1.36E+08 | T | A | 0.686751 | -0.07517 | 0.008884 | 2.64E-17 | trans |
| miR-30a-5p | rs677355 | 9:1361460 | 9 | 1.36E+08 | G | A | 0.670198 | -0.07261 | 0.008488 | 1.18E-17 | trans |
| miR-30a-5p | rs687289 | 9:1361371 | 9 | 1.36E+08 | G | A | 0.668579 | -0.0759 | 0.00844 | 2.41E-19 | trans |
| miR-30a-5p | rs543040 | 9:1361430 | 9 | 1.36E+08 | A | T | 0.669196 | -0.07237 | 0.0085 | 1.68E-17 | trans |
| miR-30a-5p | rs597974 | 9:1361442 | 9 | 1.36E+08 | A | G | 0.687926 | -0.07518 | 0.008923 | 3.61E-17 | trans |
| miR-30a-5p | rs2519093 | 9:1361418 | 9 | 1.36E+08 | C | T | 0.809892 | -0.09249 | 0.010132 | 6.96E-20 | trans |
| miR-30a-5p | rs2769071 | 9:1361459 | 9 | 1.36E+08 | A | G | 0.663117 | -0.07198 | 0.008359 | 7.23E-18 | trans |
| miR-30a-5p | rs612169 | 9:1361434 | 9 | 1.36E+08 | A | G | 0.66434 | -0.07203 | 0.008355 | 6.64E-18 | trans |
| miR-30a-5p | rs495203 | 9:1361452 | 9 | 1.36E+08 | C | T | 0.663279 | -0.07211 | 0.008348 | 5.72E-18 | trans |
| miR-30a-5p | rs676457 | 9:1361462 | 9 | 1.36E+08 | A | T | 0.664166 | -0.07221 | 0.008344 | 4.99E-18 | trans |
| miR-30a-5p | rs635634 | 9:1361550 | 9 | 1.36E+08 | C | T | 0.80599 | -0.0922 | 0.009925 | 1.55E-20 | trans |
| miR-30a-5p | rs514659 | 9:1361422 | 9 | 1.36E+08 | A | C | 0.663927 | -0.07197 | 0.008343 | 6.32E-18 | trans |
| miR-30a-5p | rs1165522 | 9:1361490 | 9 | 1.36E+08 | T | A | 0.653719 | -0.06929 | 0.008324 | 8.57E-17 | trans |
| miR-30a-5p | rs582094 | 9:1361454 | 9 | 1.36E+08 | A | T | 0.663709 | -0.07209 | 0.008341 | 5.46E-18 | trans |
| miR-30a-5p | rs532436 | 9:1361498 | 9 | 1.36E+08 | G | A | 0.8067 | -0.09274 | 0.009951 | 1.16E-20 | trans |
| miR-30a-5p | rs644234 | 9:1361422 | 9 | 1.36E+08 | T | G | 0.645381 | -0.06948 | 0.008179 | 1.98E-17 | trans |
| miR-30a-5p | rs495828 | 9:1361548 | 9 | 1.36E+08 | G | T | 0.787276 | -0.08652 | 0.009591 | 1.87E-19 | trans |
| miR-30a-5p | rs8176663 | 9:1361444 | 9 | 1.36E+08 | T | C | 0.663959 | -0.07204 | 0.008343 | 5.92E-18 | trans |
| miR-30a-5p | rs613534 | 9:1361431 | 9 | 1.36E+08 | A | G | 0.653693 | -0.06926 | 0.008334 | 9.53E-17 | trans |
| miR-30a-5p | rs554833 | 9:1361471 | 9 | 1.36E+08 | C | T | 0.664057 | -0.07217 | 0.008339 | 4.97E-18 | trans |
| miR-30a-5p | rs676996 | 9:1361460 | 9 | 1.36E+08 | T | G | 0.667516 | -0.07246 | 0.008414 | 7.23E-18 | trans |
| miR-30a-5p | rs545971 | 9:1361433 | 9 | 1.36E+08 | C | T | 0.663766 | -0.07197 | 0.00834 | 6.12E-18 | trans |
| miR-30a-5p | rs1124406 | 9:1361539 | 9 | 1.36E+08 | C | T | 0.886198 | -0.09353 | 0.013063 | 8.07E-13 | trans |
| miR-30a-5p | rs543968 | 9:1361431 | 9 | 1.36E+08 | T | C | 0.653829 | -0.06916 | 0.008336 | 1.07E-16 | trans |
| miR-30a-5p | rs674302 | 9:1361466 | 9 | 1.36E+08 | T | A | 0.663989 | -0.07224 | 0.008337 | 4.51E-18 | trans |
| miR-30a-5p | rs576125 | 9:1361443 | 9 | 1.36E+08 | G | A | 0.688677 | -0.07502 | 0.008945 | 5.00E-17 | trans |
| miR-30a-5p | rs579459 | 9:1361541 | 9 | 1.36E+08 | T | C | 0.787293 | -0.08653 | 0.009592 | 1.86E-19 | trans |
| miR-30a-5p | rs600038 | 9:1361518 | 9 | 1.36E+08 | T | C | 0.787872 | -0.08701 | 0.009616 | 1.44E-19 | trans |
| miR-30a-5p | rs687621 | 9:1361370 | 9 | 1.36E+08 | A | G | 0.663896 | -0.07281 | 0.008339 | 2.52E-18 | trans |
| miR-30a-5p | rs494242 | 9:1361451 | 9 | 1.36E+08 | C | T | 0.644977 | -0.06974 | 0.008193 | 1.71E-17 | trans |
| miR-30a-5p | rs527210 | 9:1361464 | 9 | 1.36E+08 | C | T | 0.668712 | -0.07147 | 0.008446 | 2.63E-17 | trans |
| miR-30b-5p | rs505922 | 9:1361492 | 9 | 1.36E+08 | T | C | 0.665748 | -0.07123 | 0.01048 | 1.07E-11 | trans |
| miR-30b-5p | rs544873 | 9:1361432 | 9 | 1.36E+08 | G | A | 0.645742 | -0.07159 | 0.010283 | 3.37E-12 | trans |
| miR-30b-5p | rs643434 | 9:1361423 | 9 | 1.36E+08 | G | A | 0.645392 | -0.07145 | 0.010279 | 3.62E-12 | trans |
| miR-30b-5p | rs1165522 | 9:1361490 | 9 | 1.36E+08 | T | A | 0.653719 | -0.07402 | 0.010443 | 1.36E-12 | trans |
| miR-30b-5p | rs529565 | 9:1361495 | 9 | 1.36E+08 | T | C | 0.665911 | -0.0714 | 0.010486 | 9.85E-12 | trans |
| miR-30b-5p | rs494242 | 9:1361451 | 9 | 1.36E+08 | C | T | 0.644977 | -0.07179 | 0.010288 | 2.99E-12 | trans |
| miR-30b-5p | rs657152 | 9:1361392 | 9 | 1.36E+08 | C | A | 0.645336 | -0.07149 | 0.010271 | 3.40E-12 | trans |
| miR-30b-5p | rs644234 | 9:1361422 | 9 | 1.36E+08 | T | G | 0.645381 | -0.0715 | 0.010271 | 3.38E-12 | trans |
| miR-30b-5p | rs687621 | 9:1361370 | 9 | 1.36E+08 | A | G | 0.663896 | -0.07011 | 0.010491 | 2.34E-11 | trans |
| miR-30b-5p | rs543968 | 9:1361431 | 9 | 1.36E+08 | T | C | 0.653829 | -0.07287 | 0.010459 | 3.24E-12 | trans |
| miR-30b-5p | rs613534 | 9:1361431 | 9 | 1.36E+08 | A | G | 0.653693 | -0.07295 | 0.010458 | 3.04E-12 | trans |
| miR-30b-5p | rs687289 | 9:1361371 | 9 | 1.36E+08 | G | A | 0.668579 | -0.07173 | 0.010626 | 1.48E-11 | trans |
| miR-30d-5p | rs613534 | 9:1361431 | 9 | 1.36E+08 | A | G | 0.653693 | -0.05634 | 0.008318 | 1.25E-11 | trans |
| miR-30d-5p | rs676996 | 9:1361460 | 9 | 1.36E+08 | T | G | 0.667516 | -0.05668 | 0.008408 | 1.57E-11 | trans |
| miR-30d-5p | rs676457 | 9:1361462 | 9 | 1.36E+08 | A | T | 0.664166 | -0.05688 | 0.008337 | 8.92E-12 | trans |
| miR-30d-5p | rs543968 | 9:1361431 | 9 | 1.36E+08 | T | C | 0.653829 | -0.05635 | 0.008319 | 1.26E-11 | trans |
| miR-30d-5p | rs494242 | 9:1361451 | 9 | 1.36E+08 | C | T | 0.644977 | -0.05648 | 0.008179 | 5.03E-12 | trans |
| miR-30d-5p | rs579459 | 9:1361541 | 9 | 1.36E+08 | T | C | 0.787293 | -0.06698 | 0.009593 | 2.90E-12 | trans |
| miR-30d-5p | rs582094 | 9:1361454 | 9 | 1.36E+08 | A | T | 0.663709 | -0.05682 | 0.008334 | 9.24E-12 | trans |
| miR-30d-5p | rs545971 | 9:1361433 | 9 | 1.36E+08 | C | T | 0.663766 | -0.05673 | 0.008332 | 9.84E-12 | trans |
| miR-30d-5p | rs674302 | 9:1361466 | 9 | 1.36E+08 | T | A | 0.663989 | -0.0569 | 0.00833 | 8.48E-12 | trans |
| miR-30d-5p | rs677355 | 9:1361460 | 9 | 1.36E+08 | G | A | 0.670198 | -0.057 | 0.008481 | 1.81E-11 | trans |
| miR-30d-5p | rs657152 | 9:1361392 | 9 | 1.36E+08 | C | A | 0.645336 | -0.05633 | 0.008165 | 5.27E-12 | trans |
| miR-30d-5p | rs543040 | 9:1361430 | 9 | 1.36E+08 | A | T | 0.669196 | -0.05783 | 0.008489 | 9.58E-12 | trans |
| miR-30d-5p | rs492488 | 9:1361449 | 9 | 1.36E+08 | G | A | 0.663579 | -0.05688 | 0.008342 | 9.23E-12 | trans |
| miR-30d-5p | rs505922 | 9:1361492 | 9 | 1.36E+08 | T | C | 0.665748 | -0.05824 | 0.008325 | 2.63E-12 | trans |
| miR-30d-5p | rs554833 | 9:1361471 | 9 | 1.36E+08 | C | T | 0.664057 | -0.05682 | 0.008332 | 9.15E-12 | trans |
| miR-30d-5p | rs495203 | 9:1361452 | 9 | 1.36E+08 | C | T | 0.663279 | -0.05685 | 0.008341 | 9.36E-12 | trans |
| miR-30d-5p | rs544873 | 9:1361432 | 9 | 1.36E+08 | G | A | 0.645742 | -0.05636 | 0.008175 | 5.44E-12 | trans |

| | | | | | | | | | | | |
|---|---|---|---|---|---|---|---|---|---|---|---|
| miR-30d-5 | rs687621 | 9:1361370 | 9 | 1.36E+08 | A | G | 0.663896 | -0.05709 | 0.008334 | 7.39E-12 | trans |
| miR-30d-5 | rs687289 | 9:1361371 | 9 | 1.36E+08 | G | A | 0.668579 | -0.05939 | 0.008438 | 1.95E-12 | trans |
| miR-30d-5 | rs529565 | 9:1361495 | 9 | 1.36E+08 | T | C | 0.665911 | -0.05835 | 0.00833 | 2.46E-12 | trans |
| miR-30d-5 | rs493246 | 9:1361449 | 9 | 1.36E+08 | G | A | 0.663234 | -0.05687 | 0.008347 | 9.51E-12 | trans |
| miR-30d-5 | rs2769071 | 9:1361459 | 9 | 1.36E+08 | A | G | 0.663117 | -0.05697 | 0.008351 | 8.98E-12 | trans |
| miR-30d-5 | rs600038 | 9:1361518 | 9 | 1.36E+08 | T | C | 0.787872 | -0.0675 | 0.009617 | 2.23E-12 | trans |
| miR-30d-5 | rs643434 | 9:1361423 | 9 | 1.36E+08 | G | A | 0.645392 | -0.05626 | 0.008172 | 5.80E-12 | trans |
| miR-30d-5 | rs635634 | 9:1361550 | 9 | 1.36E+08 | C | T | 0.80599 | -0.0695 | 0.009936 | 2.67E-12 | trans |
| miR-30d-5 | rs495828 | 9:1361548 | 9 | 1.36E+08 | G | T | 0.787276 | -0.06694 | 0.009592 | 2.99E-12 | trans |
| miR-30d-5 | rs582118 | 9:1361454 | 9 | 1.36E+08 | A | G | 0.663872 | -0.05686 | 0.008337 | 9.11E-12 | trans |
| miR-30d-5 | rs651007 | 9:1361538 | 9 | 1.36E+08 | C | T | 0.787179 | -0.0671 | 0.009594 | 2.66E-12 | trans |
| miR-30d-5 | rs644234 | 9:1361422 | 9 | 1.36E+08 | T | G | 0.645381 | -0.05635 | 0.008165 | 5.16E-12 | trans |
| miR-30d-5 | rs507666 | 9:1361493 | 9 | 1.36E+08 | G | A | 0.806573 | -0.07002 | 0.009956 | 2.01E-12 | trans |
| miR-30d-5 | rs532436 | 9:1361498 | 9 | 1.36E+08 | G | A | 0.8067 | -0.07008 | 0.009962 | 2.00E-12 | trans |
| miR-30d-5 | rs514659 | 9:1361422 | 9 | 1.36E+08 | A | C | 0.663927 | -0.05676 | 0.008336 | 9.82E-12 | trans |
| miR-30d-5 | rs649129 | 9:1361543 | 9 | 1.36E+08 | C | T | 0.787432 | -0.06703 | 0.009598 | 2.89E-12 | trans |
| miR-30d-5 | rs491626 | 9:1361448 | 9 | 1.36E+08 | C | T | 0.664173 | -0.05706 | 0.008357 | 8.60E-12 | trans |
| miR-30d-5 | rs527210 | 9:1361464 | 9 | 1.36E+08 | C | T | 0.668712 | -0.05667 | 0.008435 | 1.83E-11 | trans |
| miR-30d-5 | rs2519093 | 9:1361418 | 9 | 1.36E+08 | C | T | 0.809892 | -0.07017 | 0.010139 | 4.51E-12 | trans |
| miR-30d-5 | rs8176663 | 9:1361444 | 9 | 1.36E+08 | T | C | 0.663959 | -0.05679 | 0.008336 | 9.57E-12 | trans |
| miR-30d-5 | rs612169 | 9:1361434 | 9 | 1.36E+08 | A | G | 0.66434 | -0.05689 | 0.008347 | 9.40E-12 | trans |
| miR-30d-5 | rs1165522 | 9:1361490 | 9 | 1.36E+08 | T | A | 0.653719 | -0.05677 | 0.008307 | 8.22E-12 | trans |
| miR-3135a | rs1809770 | 3:5831846 | 3 | 58318464 | A | C | 0.919174 | 0.222322 | 0.03309 | 1.83E-11 | trans |
| miR-3135a | rs3567747 | 3:5818363 | 3 | 58183636 | G | A | 0.912834 | 0.214933 | 0.03204 | 1.97E-11 | trans |
| miR-3135a | rs7308155 | 3:5830293 | 3 | 58302935 | C | T | 0.920004 | 0.223068 | 0.033101 | 1.60E-11 | trans |
| miR-320a | rs1300974 | 2:6858443 | 2 | 68584437 | C | A | 0.678986 | 0.125869 | 0.01732 | 3.67E-13 | trans |
| miR-320a | rs6546398 | 2:6863041 | 2 | 68630417 | G | C | 0.552757 | 0.152138 | 0.015759 | 4.72E-22 | trans |
| miR-320a | rs5618821 | 2:6861163 | 2 | 68611631 | T | C | 0.698075 | 0.139604 | 0.016858 | 1.22E-16 | trans |
| miR-320a | rs6546400 | 2:6863064 | 2 | 68630648 | G | A | 0.554382 | 0.152811 | 0.01582 | 4.47E-22 | trans |
| miR-320a | rs4610041 | 2:6862628 | 2 | 68626280 | C | T | 0.550745 | 0.155025 | 0.015748 | 7.25E-23 | trans |
| miR-320a | rs6729686 | 2:6863303 | 2 | 68633030 | C | T | 0.700171 | 0.145857 | 0.017461 | 6.63E-17 | trans |
| miR-320a | rs1164012 | 2:6862562 | 2 | 68625625 | C | T | 0.545777 | 0.151008 | 0.015571 | 3.08E-22 | trans |
| miR-320a | rs1017438 | 2:6859646 | 2 | 68596469 | C | T | 0.685243 | 0.144486 | 0.016618 | 3.47E-18 | trans |
| miR-320a | rs7420775 | 2:6862945 | 2 | 68629458 | G | T | 0.703122 | 0.143662 | 0.017427 | 1.67E-16 | trans |
| miR-320a | rs3419682 | 2:6862142 | 2 | 68621427 | G | A | 0.697518 | 0.139848 | 0.016851 | 1.05E-16 | trans |
| miR-320a | rs6214389 | 2:6860455 | 2 | 68604553 | C | T | 0.698307 | 0.139108 | 0.016881 | 1.71E-16 | trans |
| miR-320a | rs1301028 | 2:6862184 | 2 | 68621842 | G | T | 0.683506 | 0.145008 | 0.016554 | 1.96E-18 | trans |
| miR-320a | rs7572299 | 2:6858124 | 2 | 68581246 | T | A | 0.538033 | 0.140299 | 0.016042 | 2.22E-18 | trans |
| miR-320a | rs6744306 | 2:6863273 | 2 | 68632736 | T | C | 0.549956 | 0.155241 | 0.01588 | 1.43E-22 | trans |
| miR-320a | rs1486061 | 2:6862571 | 2 | 68625717 | T | G | 0.54743 | 0.151637 | 0.015624 | 2.86E-22 | trans |
| miR-320a | rs6546411 | 2:6863769 | 2 | 68637699 | A | G | 0.556248 | 0.157639 | 0.016098 | 1.21E-22 | trans |
| miR-320a | rs6214393 | 2:6864020 | 2 | 68640202 | A | G | 0.593308 | 0.142404 | 0.016446 | 4.76E-18 | trans |
| miR-320a | rs6546399 | 2:6863064 | 2 | 68630646 | C | T | 0.55394 | 0.15272 | 0.015805 | 4.34E-22 | trans |
| miR-320a | rs5888448 | 2:6864534 | 2 | 68645347 | G | A | 0.734214 | 0.144505 | 0.018417 | 4.28E-15 | trans |
| miR-320a | rs6546392 | 2:6862744 | 2 | 68627441 | A | G | 0.555235 | 0.153667 | 0.015829 | 2.79E-22 | trans |
| miR-320a | rs1049616 | 2:6862008 | 2 | 68620089 | C | T | 0.697496 | 0.139829 | 0.016852 | 1.06E-16 | trans |
| miR-320a | rs1300592 | 2:6862999 | 2 | 68629998 | T | A | 0.701261 | 0.142753 | 0.01735 | 1.90E-16 | trans |
| miR-320a | rs1300519 | 2:6863887 | 2 | 68638873 | A | G | 0.702186 | 0.147347 | 0.017627 | 6.31E-17 | trans |
| miR-320a | rs6214392 | 2:6863835 | 2 | 68638359 | C | G | 0.700378 | 0.148987 | 0.017611 | 2.68E-17 | trans |
| miR-320a | rs1158434 | 2:6863552 | 2 | 68635524 | G | C | 0.551072 | 0.155908 | 0.015994 | 1.88E-22 | trans |
| miR-320a | rs7598360 | 2:6863092 | 2 | 68630925 | A | G | 0.553166 | 0.155693 | 0.01579 | 6.20E-23 | trans |
| miR-320a | rs6715579 | 2:6858392 | 2 | 68583923 | C | T | 0.678794 | 0.125847 | 0.017319 | 3.69E-13 | trans |
| miR-320a | rs1484530 | 2:6863547 | 2 | 68635471 | A | G | 0.70445 | 0.148609 | 0.017695 | 4.53E-17 | trans |
| miR-320a | rs6546412 | 2:6863771 | 2 | 68637710 | T | C | 0.557086 | 0.15912 | 0.016127 | 5.79E-23 | trans |
| miR-320a | rs3471241 | 2:6857828 | 2 | 68578284 | G | A | 0.6787 | 0.124199 | 0.017366 | 8.55E-13 | trans |
| miR-320a | rs6546404 | 2:6863263 | 2 | 68632635 | G | T | 0.54995 | 0.15523 | 0.015879 | 1.43E-22 | trans |
| miR-320a | rs5999861 | 2:6864677 | 2 | 68646771 | G | T | 0.731295 | 0.13921 | 0.018406 | 3.93E-14 | trans |
| miR-320a | rs6546397 | 2:6863036 | 2 | 68630367 | G | A | 0.552756 | 0.152134 | 0.015759 | 4.74E-22 | trans |
| miR-320a | rs7608429 | 2:6858083 | 2 | 68580836 | G | T | 0.678719 | 0.125181 | 0.017336 | 5.16E-13 | trans |
| miR-320a | rs6214393 | 2:6864009 | 2 | 68640090 | T | C | 0.59325 | 0.14244 | 0.016445 | 4.65E-18 | trans |
| miR-320a | rs1050181 | 2:6862295 | 2 | 68622952 | G | C | 0.544992 | 0.150456 | 0.015516 | 3.10E-22 | trans |
| miR-320a | rs6714719 | 2:6859377 | 2 | 68593773 | G | C | 0.698404 | 0.139079 | 0.016929 | 2.11E-16 | trans |
| miR-320a | rs6709464 | 2:6862222 | 2 | 68622220 | C | T | 0.697279 | 0.139649 | 0.016836 | 1.09E-16 | trans |

| | | | | | | | | | | | |
|---|---|---|---|---|---|---|---|---|---|---|---|
| miR-320a | rs6546403 | 2:6863226 | 2 | 68632262 | C | T | 0.550801 | 0.155633 | 0.015897 | 1.24E-22 | trans |
| miR-320a | rs2241946 | 2:6861565 | 2 | 68615654 | T | C | 0.698076 | 0.139832 | 0.016864 | 1.12E-16 | trans |
| miR-320a | rs1442589 | 2:6863473 | 2 | 68634732 | G | T | 0.701896 | 0.146902 | 0.017592 | 6.80E-17 | trans |
| miR-320a | rs6742200 | 2:6862330 | 2 | 68623302 | A | G | 0.54552 | 0.150787 | 0.015557 | 3.25E-22 | trans |
| miR-320a | rs7451452 | 2:6863755 | 2 | 68637550 | T | G | 0.702552 | 0.14734 | 0.017642 | 6.74E-17 | trans |
| miR-320a | rs1760168 | 2:6861589 | 2 | 68615899 | G | A | 0.697578 | 0.139713 | 0.016864 | 1.18E-16 | trans |
| miR-320a | rs1390688 | 2:6862773 | 2 | 68627735 | C | T | 0.69721 | 0.142685 | 0.017183 | 1.01E-16 | trans |
| miR-320a | rs6546408 | 2:6863504 | 2 | 68635048 | T | C | 0.551062 | 0.155914 | 0.01599 | 1.84E-22 | trans |
| miR-320a | rs6546410 | 2:6863610 | 2 | 68636105 | C | G | 0.702434 | 0.147267 | 0.017635 | 6.78E-17 | trans |
| miR-320a | rs5628470 | 2:6863137 | 2 | 68631372 | G | A | 0.697508 | 0.14339 | 0.017241 | 9.03E-17 | trans |
| miR-320a | rs1542135 | 2:6862084 | 2 | 68620848 | C | T | 0.69752 | 0.139846 | 0.016851 | 1.05E-16 | trans |
| miR-320a | rs5633714 | 2:6864249 | 2 | 68642490 | G | A | 0.733417 | 0.145039 | 0.018352 | 2.72E-15 | trans |
| miR-320a | rs6757938 | 2:6863285 | 2 | 68632858 | A | G | 0.549958 | 0.155246 | 0.015882 | 1.44E-22 | trans |
| miR-320a | rs1188499 | 2:6864573 | 2 | 68645737 | T | C | 0.593791 | 0.142095 | 0.016485 | 6.73E-18 | trans |
| miR-320a | rs7597167 | 2:6861294 | 2 | 68612946 | C | T | 0.698051 | 0.13964 | 0.016858 | 1.20E-16 | trans |
| miR-320a | rs6546395 | 2:6862927 | 2 | 68629271 | A | G | 0.55448 | 0.152637 | 0.015805 | 4.56E-22 | trans |
| miR-320a | rs6546407 | 2:6863492 | 2 | 68634920 | A | C | 0.55091 | 0.155844 | 0.015974 | 1.74E-22 | trans |
| miR-320a | rs7424269 | 2:6862912 | 2 | 68629125 | A | G | 0.563426 | 0.155067 | 0.016005 | 3.36E-22 | trans |
| miR-320a | rs6546401 | 2:6863087 | 2 | 68630873 | A | T | 0.550127 | 0.153872 | 0.015747 | 1.49E-22 | trans |
| miR-320a | rs5933292 | 2:6863963 | 2 | 68639631 | G | A | 0.73529 | 0.147942 | 0.018524 | 1.39E-15 | trans |
| miR-320a | rs3611704 | 2:6857937 | 2 | 68579376 | A | C | 0.678545 | 0.124668 | 0.017351 | 6.71E-13 | trans |
| miR-320a | rs1752821 | 2:6857875 | 2 | 68578758 | G | T | 0.677689 | 0.124071 | 0.017388 | 9.65E-13 | trans |
| miR-320a | rs1455505 | 2:6862832 | 2 | 68628329 | G | A | 0.556221 | 0.155389 | 0.015875 | 1.26E-22 | trans |
| miR-320a | rs6729555 | 2:6863295 | 2 | 68632952 | C | A | 0.54995 | 0.155269 | 0.015883 | 1.43E-22 | trans |
| miR-320a | rs1302892 | 2:6862991 | 2 | 68629916 | C | T | 0.552648 | 0.152067 | 0.015754 | 4.78E-22 | trans |
| miR-320a | rs6546394 | 2:6862909 | 2 | 68629094 | T | C | 0.566701 | 0.154484 | 0.016052 | 6.33E-22 | trans |
| miR-320a | rs7590433 | 2:6862850 | 2 | 68628507 | A | G | 0.554708 | 0.156931 | 0.01586 | 4.38E-23 | trans |
| miR-320a | rs6708536 | 2:6862726 | 2 | 68627268 | G | A | 0.547249 | 0.152906 | 0.015663 | 1.63E-22 | trans |
| miR-320a | rs7578358 | 2:6862872 | 2 | 68628724 | T | C | 0.561556 | 0.15522 | 0.015987 | 2.75E-22 | trans |
| miR-320a | rs6419644 | 2:6863182 | 2 | 68631823 | A | G | 0.549898 | 0.155194 | 0.015873 | 1.41E-22 | trans |
| miR-320a | rs7800275 | 2:6862543 | 2 | 68625436 | A | G | 0.546309 | 0.150787 | 0.015553 | 3.17E-22 | trans |
| miR-320a | rs7605387 | 2:6863255 | 2 | 68632550 | A | G | 0.703499 | 0.147092 | 0.017558 | 5.40E-17 | trans |
| miR-320a | rs7581872 | 2:6858098 | 2 | 68580981 | A | C | 0.537959 | 0.14022 | 0.016042 | 2.31E-18 | trans |
| miR-320a | rs1760209 | 2:6862041 | 2 | 68620410 | T | A | 0.697529 | 0.139841 | 0.016851 | 1.05E-16 | trans |
| miR-320a | rs6743970 | 2:6858390 | 2 | 68583907 | A | T | 0.660237 | 0.133352 | 0.017104 | 6.37E-15 | trans |
| miR-320a | rs2165920 | 2:6862223 | 2 | 68622239 | T | C | 0.552012 | 0.148931 | 0.01552 | 8.32E-22 | trans |
| miR-320a | rs1474969 | 2:6862568 | 2 | 68625686 | G | A | 0.546544 | 0.151244 | 0.015595 | 3.06E-22 | trans |
| miR-320a | rs6747545 | 2:6863306 | 2 | 68633069 | T | A | 0.549964 | 0.155257 | 0.015884 | 1.45E-22 | trans |
| miR-320a | rs2241947 | 2:6861539 | 2 | 68615395 | C | T | 0.697988 | 0.13966 | 0.016857 | 1.18E-16 | trans |
| miR-320a | rs1301077 | 2:6863908 | 2 | 68639081 | C | T | 0.702339 | 0.147255 | 0.017632 | 6.73E-17 | trans |
| miR-320a | rs8761 | 2:6862426 | 2 | 68624260 | G | A | 0.545692 | 0.150887 | 0.015566 | 3.21E-22 | trans |
| miR-320a | rs1542136 | 2:6862093 | 2 | 68620930 | C | A | 0.697594 | 0.139843 | 0.016855 | 1.07E-16 | trans |
| miR-320a | rs9653552 | 2:6864396 | 2 | 68643966 | G | A | 0.593458 | 0.14288 | 0.016454 | 3.83E-18 | trans |
| miR-320a | rs6214393 | 2:6864062 | 2 | 68640621 | T | C | 0.733261 | 0.145064 | 0.018341 | 2.58E-15 | trans |
| miR-320a | rs3485223 | 2:6863859 | 2 | 68638599 | C | G | 0.550658 | 0.155791 | 0.015997 | 2.06E-22 | trans |
| miR-320a | rs1422402 | 2:6862752 | 2 | 68627521 | A | G | 0.698498 | 0.143247 | 0.017227 | 9.15E-17 | trans |
| miR-320a | rs1303482 | 2:6863029 | 2 | 68630297 | C | G | 0.552715 | 0.152114 | 0.015757 | 4.74E-22 | trans |
| miR-320a | rs1301107 | 2:6863931 | 2 | 68639313 | A | C | 0.700844 | 0.148254 | 0.017659 | 4.64E-17 | trans |
| miR-320a | rs7562439 | 2:6858256 | 2 | 68582569 | C | T | 0.6646 | 0.131508 | 0.017025 | 1.12E-14 | trans |
| miR-320a | rs7421228 | 2:6863012 | 2 | 68630124 | G | T | 0.55268 | 0.152092 | 0.015756 | 4.77E-22 | trans |
| miR-320a | rs5808991 | 2:6863966 | 2 | 68639660 | G | C | 0.734634 | 0.147894 | 0.018473 | 1.19E-15 | trans |
| miR-320a | rs1343065 | 2:6858771 | 2 | 68587710 | C | G | 0.699397 | 0.1397 | 0.017062 | 2.66E-16 | trans |
| miR-320a | rs7578602 | 2:6863786 | 2 | 68637868 | G | C | 0.556806 | 0.159076 | 0.016134 | 6.23E-23 | trans |
| miR-320a | rs2070171 | 2:6861371 | 2 | 68613710 | G | A | 0.698024 | 0.139616 | 0.016857 | 1.21E-16 | trans |
| miR-320a | rs1301143 | 2:6863937 | 2 | 68639379 | C | T | 0.702382 | 0.147277 | 0.017634 | 6.72E-17 | trans |
| miR-320a | rs1301157 | 2:6863927 | 2 | 68639273 | G | A | 0.702349 | 0.147267 | 0.017633 | 6.72E-17 | trans |
| miR-320a | rs1760149 | 2:6861171 | 2 | 68611712 | A | G | 0.69807 | 0.139607 | 0.016858 | 1.22E-16 | trans |
| miR-320a | rs7564214 | 2:6862869 | 2 | 68628696 | C | T | 0.569906 | 0.160165 | 0.016208 | 5.00E-23 | trans |
| miR-320a | rs1156629 | 2:6862663 | 2 | 68626631 | T | G | 0.547487 | 0.152478 | 0.015652 | 2.01E-22 | trans |
| miR-320a | rs1373200 | 2:6862265 | 2 | 68622658 | C | T | 0.547142 | 0.149223 | 0.015409 | 3.51E-22 | trans |
| miR-320a | rs6546393 | 2:6862757 | 2 | 68627575 | T | C | 0.554017 | 0.154967 | 0.015786 | 9.52E-23 | trans |
| miR-320a | rs1753019 | 2:6862133 | 2 | 68621338 | G | T | 0.683745 | 0.145203 | 0.016568 | 1.88E-18 | trans |
| miR-320a | rs6419643 | 2:6863126 | 2 | 68631260 | T | G | 0.548826 | 0.153407 | 0.015714 | 1.63E-22 | trans |

| | | | | | | | | | | | |
|---|---|---|---|---|---|---|---|---|---|---|---|
| miR-320a | rs1302897 | 2:6863003 | 2 | 68630038 | A | G | 0.54849 | 0.153419 | 0.015702 | 1.51E-22 | trans |
| miR-320a | rs6214393 | 2:6864041 | 2 | 68640415 | A | G | 0.593599 | 0.143051 | 0.01644 | 3.28E-18 | trans |
| miR-320a | rs8028813 | 2:6862547 | 2 | 68625478 | A | G | 0.548418 | 0.153418 | 0.015656 | 1.13E-22 | trans |
| miR-320a | rs7592526 | 2:6858352 | 2 | 68583528 | A | C | 0.538007 | 0.140533 | 0.01604 | 1.93E-18 | trans |
| miR-320a | rs6715909 | 2:6858399 | 2 | 68583995 | G | A | 0.664879 | 0.131663 | 0.01702 | 1.03E-14 | trans |
| miR-320a | rs3732044 | 2:6862391 | 2 | 68623915 | T | C | 0.545562 | 0.150811 | 0.01556 | 3.25E-22 | trans |
| miR-320a | rs1397159 | 2:6863425 | 2 | 68634258 | T | G | 0.705552 | 0.148814 | 0.017799 | 6.24E-17 | trans |
| miR-320a | rs6546409 | 2:6863587 | 2 | 68635879 | A | G | 0.551093 | 0.15591 | 0.015998 | 1.93E-22 | trans |
| miR-320a | rs6214393 | 2:6864038 | 2 | 68640381 | G | C | 0.593598 | 0.143062 | 0.01644 | 3.25E-18 | trans |
| miR-320a | rs7590450 | 2:6862856 | 2 | 68628563 | A | G | 0.549532 | 0.153227 | 0.015701 | 1.68E-22 | trans |
| miR-320a | rs1303372 | 2:6857805 | 2 | 68578058 | C | G | 0.670257 | 0.116708 | 0.017236 | 1.28E-11 | trans |
| miR-320a | rs6000716 | 2:6863963 | 2 | 68639636 | C | T | 0.735163 | 0.14802 | 0.018521 | 1.33E-15 | trans |
| miR-320a | rs6087844 | 2:6858211 | 2 | 68582115 | A | G | 0.538067 | 0.140648 | 0.016039 | 1.80E-18 | trans |
| miR-320a | rs1382899 | 2:6863782 | 2 | 68637829 | A | G | 0.702614 | 0.147363 | 0.017647 | 6.79E-17 | trans |
| miR-320a | rs6715706 | 2:6858399 | 2 | 68583996 | C | A | 0.664887 | 0.131667 | 0.01702 | 1.03E-14 | trans |
| miR-320a | rs5690424 | 2:6863847 | 2 | 68638477 | T | C | 0.550669 | 0.155791 | 0.015996 | 2.05E-22 | trans |
| miR-320e | rs6546399 | 2:6863064 | 2 | 68630646 | C | T | 0.55394 | 0.111105 | 0.015037 | 1.48E-13 | trans |
| miR-320e | rs6214393 | 2:6864020 | 2 | 68640202 | A | G | 0.593308 | 0.113862 | 0.015587 | 2.78E-13 | trans |
| miR-320e | rs6546398 | 2:6863041 | 2 | 68630417 | G | C | 0.552757 | 0.110975 | 0.014992 | 1.34E-13 | trans |
| miR-320e | rs1164012 | 2:6862562 | 2 | 68625625 | C | T | 0.545777 | 0.111024 | 0.014812 | 6.59E-14 | trans |
| miR-320e | rs7572299 | 2:6858124 | 2 | 68581246 | T | A | 0.538033 | 0.108792 | 0.015217 | 8.73E-13 | trans |
| miR-320e | rs6546407 | 2:6863492 | 2 | 68634920 | A | C | 0.55091 | 0.113961 | 0.015198 | 6.46E-14 | trans |
| miR-320e | rs1188499 | 2:6864573 | 2 | 68645737 | T | C | 0.593791 | 0.112826 | 0.015627 | 5.19E-13 | trans |
| miR-320e | rs1455505 | 2:6862832 | 2 | 68628329 | G | A | 0.556221 | 0.114653 | 0.015101 | 3.14E-14 | trans |
| miR-320e | rs6214393 | 2:6864009 | 2 | 68640090 | T | C | 0.59325 | 0.113891 | 0.015587 | 2.73E-13 | trans |
| miR-320e | rs7598360 | 2:6863092 | 2 | 68630925 | A | G | 0.553166 | 0.116479 | 0.015017 | 8.73E-15 | trans |
| miR-320e | rs6546397 | 2:6863036 | 2 | 68630367 | G | A | 0.552756 | 0.110969 | 0.014992 | 1.34E-13 | trans |
| miR-320e | rs6546411 | 2:6863769 | 2 | 68637699 | A | G | 0.556248 | 0.116536 | 0.015313 | 2.74E-14 | trans |
| miR-320e | rs6546408 | 2:6863504 | 2 | 68635048 | T | C | 0.551062 | 0.11385 | 0.015214 | 7.26E-14 | trans |
| miR-320e | rs2165920 | 2:6862223 | 2 | 68622239 | T | C | 0.552012 | 0.111604 | 0.014753 | 3.89E-14 | trans |
| miR-320e | rs6419644 | 2:6863182 | 2 | 68631823 | A | G | 0.549898 | 0.11406 | 0.015101 | 4.25E-14 | trans |
| miR-320e | rs7590433 | 2:6862850 | 2 | 68628507 | A | G | 0.554708 | 0.11533 | 0.015091 | 2.14E-14 | trans |
| miR-320e | rs1156629 | 2:6862663 | 2 | 68626631 | T | G | 0.547487 | 0.112242 | 0.014889 | 4.76E-14 | trans |
| miR-320e | rs6546412 | 2:6863771 | 2 | 68637710 | T | C | 0.557086 | 0.117526 | 0.015342 | 1.86E-14 | trans |
| miR-320e | rs6546400 | 2:6863064 | 2 | 68630648 | G | A | 0.554382 | 0.111156 | 0.015051 | 1.52E-13 | trans |
| miR-320e | rs8761 | 2:6862426 | 2 | 68624260 | G | A | 0.545692 | 0.110918 | 0.014806 | 6.81E-14 | trans |
| miR-320e | rs7564214 | 2:6862869 | 2 | 68628696 | C | T | 0.569906 | 0.113608 | 0.015437 | 1.85E-13 | trans |
| miR-320e | rs6546395 | 2:6862927 | 2 | 68629271 | A | G | 0.55448 | 0.11111 | 0.015036 | 1.47E-13 | trans |
| miR-320e | rs1373200 | 2:6862265 | 2 | 68622658 | C | T | 0.547142 | 0.110349 | 0.014654 | 5.07E-14 | trans |
| miR-320e | rs1158434 | 2:6863552 | 2 | 68635524 | G | C | 0.551072 | 0.11383 | 0.015217 | 7.41E-14 | trans |
| miR-320e | rs1486061 | 2:6862571 | 2 | 68625717 | T | G | 0.54743 | 0.111501 | 0.014862 | 6.26E-14 | trans |
| miR-320e | rs6546404 | 2:6863263 | 2 | 68632635 | G | T | 0.54995 | 0.114042 | 0.015107 | 4.39E-14 | trans |
| miR-320e | rs6747545 | 2:6863306 | 2 | 68633069 | T | A | 0.549964 | 0.114028 | 0.015112 | 4.50E-14 | trans |
| miR-320e | rs6546401 | 2:6863087 | 2 | 68630873 | A | T | 0.550127 | 0.113171 | 0.014981 | 4.20E-14 | trans |
| miR-320e | rs6546394 | 2:6862909 | 2 | 68629094 | T | C | 0.566701 | 0.112054 | 0.015272 | 2.18E-13 | trans |
| miR-320e | rs1050181 | 2:6862295 | 2 | 68622952 | G | C | 0.544992 | 0.110579 | 0.014759 | 6.76E-14 | trans |
| miR-320e | rs6744306 | 2:6863273 | 2 | 68632736 | T | C | 0.549956 | 0.114047 | 0.015107 | 4.38E-14 | trans |
| miR-320e | rs6546403 | 2:6863226 | 2 | 68632262 | C | T | 0.550801 | 0.114043 | 0.015125 | 4.70E-14 | trans |
| miR-320e | rs6757938 | 2:6863285 | 2 | 68632858 | A | G | 0.549958 | 0.114041 | 0.015109 | 4.43E-14 | trans |
| miR-320e | rs6214393 | 2:6864038 | 2 | 68640381 | G | C | 0.593598 | 0.114275 | 0.015583 | 2.24E-13 | trans |
| miR-320e | rs7800275 | 2:6862543 | 2 | 68625436 | A | G | 0.546309 | 0.110955 | 0.014794 | 6.38E-14 | trans |
| miR-320e | rs7424269 | 2:6862912 | 2 | 68629125 | A | G | 0.563426 | 0.113582 | 0.015225 | 8.63E-14 | trans |
| miR-320e | rs3732044 | 2:6862391 | 2 | 68623915 | T | C | 0.545562 | 0.11085 | 0.014801 | 6.91E-14 | trans |
| miR-320e | rs7421228 | 2:6863012 | 2 | 68630124 | G | T | 0.55268 | 0.110936 | 0.014989 | 1.35E-13 | trans |
| miR-320e | rs7590450 | 2:6862856 | 2 | 68628563 | A | G | 0.549532 | 0.11308 | 0.014935 | 3.68E-14 | trans |
| miR-320e | rs7578602 | 2:6863786 | 2 | 68637868 | G | C | 0.556806 | 0.117559 | 0.015349 | 1.87E-14 | trans |
| miR-320e | rs4610041 | 2:6862628 | 2 | 68626280 | C | T | 0.550745 | 0.11333 | 0.014985 | 3.95E-14 | trans |
| miR-320e | rs9653552 | 2:6864396 | 2 | 68643966 | G | A | 0.593458 | 0.113705 | 0.015597 | 3.10E-13 | trans |
| miR-320e | rs6087844 | 2:6858211 | 2 | 68582115 | A | G | 0.538067 | 0.108754 | 0.015216 | 8.84E-13 | trans |
| miR-320e | rs1474969 | 2:6862568 | 2 | 68625686 | G | A | 0.546544 | 0.111176 | 0.014834 | 6.65E-14 | trans |
| miR-320e | rs7578358 | 2:6862872 | 2 | 68628724 | T | C | 0.561556 | 0.114476 | 0.015206 | 5.13E-14 | trans |
| miR-320e | rs6214393 | 2:6864041 | 2 | 68640415 | A | G | 0.593599 | 0.114256 | 0.015583 | 2.27E-13 | trans |
| miR-320e | rs7581872 | 2:6858098 | 2 | 68580981 | A | C | 0.537959 | 0.108694 | 0.015217 | 9.15E-13 | trans |

| | | | | | | | | | | | |
|---|---|---|---|---|---|---|---|---|---|---|---|
| miR-320e | rs6729555 | 2:6863295: | 2 | 68632952 | C | A | 0.54995 | 0.114033 | 0.015111 | 4.47E-14 | trans |
| miR-320e | rs1303482! | 2:6863029 | 2 | 68630297 | C | G | 0.552715 | 0.110948 | 0.01499 | 1.35E-13 | trans |
| miR-320e | rs6546393 | 2:6862757 | 2 | 68627575 | T | C | 0.554017 | 0.114706 | 0.015016 | 2.19E-14 | trans |
| miR-320e | rs3485223 | 2:6863859 | 2 | 68638599 | C | G | 0.550658 | 0.113462 | 0.015221 | 9.04E-14 | trans |
| miR-320e | rs6546409 | 2:6863587 | 2 | 68635879 | A | G | 0.551093 | 0.113771 | 0.015222 | 7.77E-14 | trans |
| miR-320e | rs7592526 | 2:6858352 | 2 | 68583528 | A | C | 0.538007 | 0.109013 | 0.015216 | 7.81E-13 | trans |
| miR-320e | rs6419643 | 2:6863126 | 2 | 68631260 | T | G | 0.548826 | 0.113114 | 0.014948 | 3.81E-14 | trans |
| miR-320e | rs6742200 | 2:6862330 | 2 | 68623302 | A | G | 0.54552 | 0.110827 | 0.014798 | 6.92E-14 | trans |
| miR-320e | rs8028813 | 2:6862547 | 2 | 68625478 | A | G | 0.548418 | 0.112315 | 0.014897 | 4.71E-14 | trans |
| miR-320e | rs6546392 | 2:6862744 | 2 | 68627441 | A | G | 0.555235 | 0.113418 | 0.015055 | 4.94E-14 | trans |
| miR-320e | rs1302897 | 2:6863003 | 2 | 68630038 | A | G | 0.54849 | 0.112919 | 0.014938 | 4.05E-14 | trans |
| miR-320e | rs1302892 | 2:6862991 | 2 | 68629916 | C | T | 0.552648 | 0.110906 | 0.014987 | 1.36E-13 | trans |
| miR-320e | rs5690424 | 2:6863847 | 2 | 68638477 | T | C | 0.550669 | 0.113465 | 0.01522 | 9.00E-14 | trans |
| miR-320e | rs6708536 | 2:6862726 | 2 | 68627268 | G | A | 0.547249 | 0.112067 | 0.014901 | 5.45E-14 | trans |
| miR-337-5 | rs8021301 | 14:101166 | 14 | 1.01E+08 | G | C | 0.668324 | 0.200231 | 0.024692 | 5.09E-16 | cis |
| miR-337-5 | rs7142683 | 14:101158 | 14 | 1.01E+08 | A | G | 0.290748 | -0.19669 | 0.026359 | 8.53E-14 | cis |
| miR-337-5 | rs1289760 | 14:101182 | 14 | 1.01E+08 | C | T | 0.190452 | -0.25526 | 0.029996 | 1.74E-17 | cis |
| miR-337-5 | rs7155649 | 14:101181 | 14 | 1.01E+08 | C | A | 0.863608 | 0.235619 | 0.03488 | 1.43E-11 | cis |
| miR-337-5 | rs1014696 | 14:101170 | 14 | 1.01E+08 | T | C | 0.658875 | 0.199099 | 0.024267 | 2.31E-16 | cis |
| miR-337-5 | rs8012970 | 14:101168 | 14 | 1.01E+08 | T | C | 0.347902 | -0.20593 | 0.025099 | 2.31E-16 | cis |
| miR-337-5 | rs1709961 | 14:101167 | 14 | 1.01E+08 | C | G | 0.668809 | 0.202143 | 0.024692 | 2.69E-16 | cis |
| miR-337-5 | rs9671394 | 14:101159 | 14 | 1.01E+08 | C | T | 0.284232 | -0.20588 | 0.026631 | 1.07E-14 | cis |
| miR-337-5 | rs7145337 | 14:101155 | 14 | 1.01E+08 | C | T | 0.312091 | -0.18996 | 0.025908 | 2.27E-13 | cis |
| miR-337-5 | rs1288176 | 14:101176 | 14 | 1.01E+08 | G | C | 0.347791 | -0.21498 | 0.025005 | 8.14E-18 | cis |
| miR-337-5 | rs1289379 | 14:101178 | 14 | 1.01E+08 | C | G | 0.190813 | -0.25206 | 0.030196 | 6.97E-17 | cis |
| miR-337-5 | rs1214700 | 14:101186 | 14 | 1.01E+08 | C | T | 0.223221 | -0.21774 | 0.028045 | 8.23E-15 | cis |
| miR-337-5 | rs1243449 | 14:101179 | 14 | 1.01E+08 | A | G | 0.18959 | -0.25144 | 0.030173 | 7.85E-17 | cis |
| miR-337-5 | rs4905994 | 14:101154 | 14 | 1.01E+08 | T | C | 0.329146 | -0.18301 | 0.025555 | 8.00E-13 | cis |
| miR-337-5 | rs1288154 | 14:101176 | 14 | 1.01E+08 | G | C | 0.370047 | -0.22551 | 0.025327 | 5.39E-19 | cis |
| miR-337-5 | rs7149242 | 14:101159 | 14 | 1.01E+08 | T | G | 0.288371 | -0.19745 | 0.026279 | 5.76E-14 | cis |
| miR-337-5 | rs1243517 | 14:101155 | 14 | 1.01E+08 | G | A | 0.312183 | -0.18988 | 0.025915 | 2.35E-13 | cis |
| miR-337-5 | rs2837918 | 14:101168 | 14 | 1.01E+08 | G | T | 0.836434 | 0.216 | 0.031773 | 1.06E-11 | cis |
| miR-337-5 | rs4905999 | 14:101184 | 14 | 1.01E+08 | A | G | 0.181905 | -0.25453 | 0.030703 | 1.13E-16 | cis |
| miR-337-5 | rs1288281 | 14:101183 | 14 | 1.01E+08 | C | G | 0.331398 | -0.2046 | 0.024702 | 1.20E-16 | cis |
| miR-337-5 | rs7141210 | 14:101182 | 14 | 1.01E+08 | T | C | 0.32902 | -0.20498 | 0.024618 | 8.34E-17 | cis |
| miR-337-5 | rs8020868 | 14:101166 | 14 | 1.01E+08 | A | G | 0.668343 | 0.200186 | 0.02469 | 5.14E-16 | cis |
| miR-337-5 | rs4905998 | 14:101181 | 14 | 1.01E+08 | A | G | 0.330615 | -0.20503 | 0.024682 | 9.81E-17 | cis |
| miR-337-5 | rs4900471 | 14:101162 | 14 | 1.01E+08 | G | A | 0.304173 | -0.18924 | 0.025938 | 2.97E-13 | cis |
| miR-337-5 | rs1014438 | 14:101173 | 14 | 1.01E+08 | C | T | 0.673446 | 0.206362 | 0.024816 | 9.13E-17 | cis |
| miR-337-5 | rs1014551 | 14:101168 | 14 | 1.01E+08 | A | G | 0.668173 | 0.199673 | 0.024662 | 5.67E-16 | cis |
| miR-345-5 | rs1013476 | 14:100763 | 14 | 1.01E+08 | G | T | 0.661819 | -0.07961 | 0.011343 | 2.24E-12 | cis |
| miR-345-5 | rs1059264 | 14:100769 | 14 | 1.01E+08 | C | T | 0.659003 | -0.07799 | 0.011155 | 2.71E-12 | cis |
| miR-3689a | rs7846761 | 9:1377324 | 9 | 1.38E+08 | T | G | 0.293002 | 0.364447 | 0.032527 | 3.88E-29 | cis |
| miR-3689a | rs6257144 | 9:1377421 | 9 | 1.38E+08 | G | A | 0.461564 | 0.238583 | 0.026179 | 7.97E-20 | cis |
| miR-3689a | rs3124946 | 9:1377473 | 9 | 1.38E+08 | A | G | 0.206437 | 0.368571 | 0.034383 | 8.25E-27 | cis |
| miR-3689a | rs6257143 | 9:1377408 | 9 | 1.38E+08 | T | C | 0.360943 | 0.368427 | 0.029133 | 1.17E-36 | cis |
| miR-3689a | rs3128574 | 9:1377369 | 9 | 1.38E+08 | T | C | 0.256468 | 0.342667 | 0.02993 | 2.38E-30 | cis |
| miR-3689a | rs9410006 | 9:1377481 | 9 | 1.38E+08 | C | T | 0.413955 | 0.279702 | 0.026037 | 6.44E-27 | cis |
| miR-3689a | rs7250271 | 9:1377425 | 9 | 1.38E+08 | A | G | 0.844401 | -0.37583 | 0.037919 | 3.72E-23 | cis |
| miR-3689a | rs4625109 | 9:1377404 | 9 | 1.38E+08 | G | A | 0.461399 | 0.194923 | 0.028679 | 1.07E-11 | cis |
| miR-3689a | rs3128573 | 9:1377383 | 9 | 1.38E+08 | A | G | 0.256902 | 0.350572 | 0.030075 | 2.13E-31 | cis |
| miR-3689a | rs3128562 | 9:1377452 | 9 | 1.38E+08 | A | G | 0.300754 | 0.396962 | 0.02741 | 1.56E-47 | cis |
| miR-3689a | rs4504708 | 9:1377365 | 9 | 1.38E+08 | G | T | 0.258348 | 0.341103 | 0.029815 | 2.62E-30 | cis |
| miR-3689a | rs4446799 | 9:1377373 | 9 | 1.38E+08 | T | C | 0.252242 | 0.360583 | 0.030261 | 9.80E-33 | cis |
| miR-3689a | rs7041584 | 9:1377486 | 9 | 1.38E+08 | A | G | 0.156981 | 0.333081 | 0.039746 | 5.28E-17 | cis |
| miR-3689a | rs3443543 | 9:1377406 | 9 | 1.38E+08 | A | G | 0.453182 | 0.214849 | 0.029449 | 2.97E-13 | cis |
| miR-3689a | rs1134170 | 9:1377352 | 9 | 1.38E+08 | A | T | 0.299409 | 0.349032 | 0.030319 | 1.15E-30 | cis |
| miR-3689a | rs1110354 | 9:1377399 | 9 | 1.38E+08 | G | T | 0.285208 | 0.337768 | 0.029217 | 6.52E-31 | cis |
| miR-3689a | rs1110354 | 9:1377478 | 9 | 1.38E+08 | A | G | 0.844437 | -0.37771 | 0.037965 | 2.55E-23 | cis |
| miR-3689a | rs3128575 | 9:1377366 | 9 | 1.38E+08 | T | C | 0.295131 | 0.3169 | 0.028619 | 1.69E-28 | cis |
| miR-3689a | rs7250271 | 9:1377426 | 9 | 1.38E+08 | T | C | 0.844363 | -0.37586 | 0.037887 | 3.39E-23 | cis |
| miR-3689a | rs13946 | 9:1377342 | 9 | 1.38E+08 | C | T | 0.257382 | 0.3805 | 0.032828 | 4.59E-31 | cis |
| miR-3689a | rs3128570 | 9:1377433 | 9 | 1.38E+08 | G | A | 0.298028 | 0.390362 | 0.02716 | 7.70E-47 | cis |

| | | | | | | | | | | | |
|---|---|---|---|---|---|---|---|---|---|---|---|
| miR-3689a | rs3124944 | 9:1377452[illegible] | 9 | 1.38E+08 | A | G | 0.303119 | 0.395178 | 0.027506 | 8.32E-47 | cis |
| miR-3689a | rs3128572 | 9:1377383[illegible] | 9 | 1.38E+08 | G | A | 0.435312 | 0.182053 | 0.026952 | 1.43E-11 | cis |
| miR-3689a | rs3124945 | 9:1377453[illegible] | 9 | 1.38E+08 | A | T | 0.144081 | 0.386175 | 0.040266 | 8.76E-22 | cis |
| miR-3689a | rs9410005 | 9:1377478[illegible] | 9 | 1.38E+08 | A | G | 0.46835 | 0.220517 | 0.025857 | 1.49E-17 | cis |
| miR-3689a | rs4625108 | 9:1377404[illegible] | 9 | 1.38E+08 | G | A | 0.408685 | 0.262258 | 0.029007 | 1.55E-19 | cis |
| miR-3689a | rs1128254[illegible] | 9:1377412[illegible] | 9 | 1.38E+08 | T | C | 0.162721 | 0.352221 | 0.040698 | 4.95E-18 | cis |
| miR-3689a | rs3124937 | 9:1377381 | 9 | 1.38E+08 | G | C | 0.256872 | 0.34885 | 0.030055 | 3.80E-31 | cis |
| miR-3689a | rs9410004 | 9:1377477[illegible] | 9 | 1.38E+08 | C | T | 0.470272 | 0.220078 | 0.02582 | 1.55E-17 | cis |
| miR-3689a | rs3128571 | 9:1377394[illegible] | 9 | 1.38E+08 | T | A | 0.285728 | 0.325174 | 0.031338 | 3.18E-25 | cis |
| miR-3689a | rs3124938 | 9:1377381[illegible] | 9 | 1.38E+08 | G | A | 0.434499 | 0.184576 | 0.026943 | 7.35E-12 | cis |
| miR-3689a | rs3124939 | 9:1377381[illegible] | 9 | 1.38E+08 | T | C | 0.261117 | 0.342697 | 0.029997 | 3.17E-30 | cis |
| miR-3689a | rs9410006 | 9:1377481[illegible] | 9 | 1.38E+08 | C | T | 0.413955 | 0.243667 | 0.033543 | 3.75E-13 | cis |
| miR-370-3[illegible] | rs1289760[illegible] | 14:101182[illegible] | 14 | 1.01E+08 | C | T | 0.190452 | -0.28477 | 0.029658 | 7.86E-22 | cis |
| miR-370-3[illegible] | rs1190852 | 14:101064[illegible] | 14 | 1.01E+08 | A | G | 0.310295 | -0.20965 | 0.02532 | 1.23E-16 | cis |
| miR-370-3[illegible] | rs1162219[illegible] | 14:101016 | 14 | 1.01E+08 | G | A | 0.759852 | 0.199826 | 0.029561 | 1.38E-11 | cis |
| miR-370-3[illegible] | rs7145337 | 14:101155[illegible] | 14 | 1.01E+08 | C | T | 0.312091 | -0.25436 | 0.025467 | 1.72E-23 | cis |
| miR-370-3[illegible] | rs1709961[illegible] | 14:101167[illegible] | 14 | 1.01E+08 | C | G | 0.668809 | 0.252492 | 0.0243 | 2.73E-25 | cis |
| miR-370-3[illegible] | rs4905998 | 14:101181[illegible] | 14 | 1.01E+08 | A | G | 0.330615 | -0.26011 | 0.024263 | 8.18E-27 | cis |
| miR-370-3[illegible] | rs873076 | 14:101070[illegible] | 14 | 1.01E+08 | A | G | 0.311961 | -0.207 | 0.025224 | 2.28E-16 | cis |
| miR-370-3[illegible] | rs1190903 | 14:101096[illegible] | 14 | 1.01E+08 | C | T | 0.772566 | 0.217133 | 0.028126 | 1.16E-14 | cis |
| miR-370-3[illegible] | rs6199300[illegible] | 14:101034 | 14 | 1.01E+08 | G | A | 0.726044 | 0.200976 | 0.025994 | 1.06E-14 | cis |
| miR-370-3[illegible] | rs4905976 | 14:101060[illegible] | 14 | 1.01E+08 | G | A | 0.322862 | -0.20634 | 0.025453 | 5.20E-16 | cis |
| miR-370-3[illegible] | rs5922882[illegible] | 14:101185 | 14 | 1.01E+08 | G | C | 0.745519 | 0.205443 | 0.026917 | 2.30E-14 | cis |
| miR-370-3[illegible] | rs1289379[illegible] | 14:101178[illegible] | 14 | 1.01E+08 | C | G | 0.190813 | -0.28295 | 0.029853 | 2.59E-21 | cis |
| miR-370-3[illegible] | rs1243517[illegible] | 14:101155[illegible] | 14 | 1.01E+08 | G | A | 0.312183 | -0.25433 | 0.025473 | 1.79E-23 | cis |
| miR-370-3[illegible] | rs1013940[illegible] | 14:101194[illegible] | 14 | 1.01E+08 | A | G | 0.323897 | -0.171 | 0.025323 | 1.45E-11 | cis |
| miR-370-3[illegible] | rs1288281[illegible] | 14:101183 | 14 | 1.01E+08 | C | G | 0.331398 | -0.26046 | 0.02428 | 7.57E-27 | cis |
| miR-370-3[illegible] | rs8008285 | 14:101146[illegible] | 14 | 1.01E+08 | T | C | 0.842905 | 0.238019 | 0.03247 | 2.29E-13 | cis |
| miR-370-3[illegible] | rs7143357 | 14:101054 | 14 | 1.01E+08 | T | C | 0.739636 | 0.215159 | 0.026343 | 3.15E-16 | cis |
| miR-370-3[illegible] | rs4905977 | 14:101061[illegible] | 14 | 1.01E+08 | C | G | 0.318473 | -0.21246 | 0.025402 | 6.06E-17 | cis |
| miR-370-3[illegible] | rs7155649 | 14:101181[illegible] | 14 | 1.01E+08 | C | A | 0.863608 | 0.254793 | 0.034571 | 1.70E-13 | cis |
| miR-370-3[illegible] | rs879059 | 14:101069[illegible] | 14 | 1.01E+08 | T | A | 0.31068 | -0.20805 | 0.025263 | 1.79E-16 | cis |
| miR-370-3[illegible] | rs6575791 | 14:101029[illegible] | 14 | 1.01E+08 | G | A | 0.719841 | 0.19897 | 0.025998 | 1.96E-14 | cis |
| miR-370-3[illegible] | rs1116059[illegible] | 14:101049[illegible] | 14 | 1.01E+08 | C | T | 0.741662 | 0.216786 | 0.026314 | 1.75E-16 | cis |
| miR-370-3[illegible] | rs8012970 | 14:101168[illegible] | 14 | 1.01E+08 | T | C | 0.347902 | -0.24877 | 0.024739 | 8.67E-24 | cis |
| miR-370-3[illegible] | rs7141210 | 14:101182[illegible] | 14 | 1.01E+08 | T | C | 0.32902 | -0.2597 | 0.024201 | 7.28E-27 | cis |
| miR-370-3[illegible] | rs873077 | 14:101070[illegible] | 14 | 1.01E+08 | G | A | 0.312104 | -0.20691 | 0.025229 | 2.38E-16 | cis |
| miR-370-3[illegible] | rs7493817 | 14:101038[illegible] | 14 | 1.01E+08 | G | A | 0.751723 | 0.213483 | 0.026955 | 2.38E-15 | cis |
| miR-370-3[illegible] | rs7140556 | 14:101036[illegible] | 14 | 1.01E+08 | C | A | 0.723818 | 0.199916 | 0.02594 | 1.29E-14 | cis |
| miR-370-3[illegible] | rs9671394 | 14:101159[illegible] | 14 | 1.01E+08 | C | T | 0.284232 | -0.26639 | 0.026192 | 2.68E-24 | cis |
| miR-370-3[illegible] | rs7149242 | 14:101159[illegible] | 14 | 1.01E+08 | T | G | 0.288371 | -0.26026 | 0.025837 | 7.28E-24 | cis |
| miR-370-3[illegible] | rs1190862 | 14:101067[illegible] | 14 | 1.01E+08 | T | G | 0.309628 | -0.2062 | 0.025131 | 2.30E-16 | cis |
| miR-370-3[illegible] | rs6199300[illegible] | 14:101030[illegible] | 14 | 1.01E+08 | G | A | 0.724196 | 0.199112 | 0.025918 | 1.56E-14 | cis |
| miR-370-3[illegible] | rs1243369[illegible] | 14:101035[illegible] | 14 | 1.01E+08 | C | G | 0.723403 | 0.197119 | 0.025831 | 2.33E-14 | cis |
| miR-370-3[illegible] | rs1185095[illegible] | 14:101056[illegible] | 14 | 1.01E+08 | G | T | 0.743981 | 0.217094 | 0.026179 | 1.11E-16 | cis |
| miR-370-3[illegible] | rs1190909 | 14:101101[illegible] | 14 | 1.01E+08 | G | A | 0.775531 | 0.213631 | 0.028465 | 6.14E-14 | cis |
| miR-370-3[illegible] | rs1243449[illegible] | 14:101179[illegible] | 14 | 1.01E+08 | A | G | 0.18959 | -0.28265 | 0.029829 | 2.65E-21 | cis |
| miR-370-3[illegible] | rs1015151[illegible] | 14:101175[illegible] | 14 | 1.01E+08 | G | A | 0.806743 | 0.194814 | 0.029099 | 2.16E-11 | cis |
| miR-370-3[illegible] | rs1467536 | 14:101061[illegible] | 14 | 1.01E+08 | A | G | 0.318958 | -0.2145 | 0.025356 | 2.69E-17 | cis |
| miR-370-3[illegible] | rs1190904 | 14:101096[illegible] | 14 | 1.01E+08 | A | G | 0.772596 | 0.21724 | 0.028123 | 1.12E-14 | cis |
| miR-370-3[illegible] | rs7263167[illegible] | 14:101187[illegible] | 14 | 1.01E+08 | C | T | 0.881829 | 0.261606 | 0.037075 | 1.71E-12 | cis |
| miR-370-3[illegible] | rs4900471 | 14:101162[illegible] | 14 | 1.01E+08 | G | A | 0.304173 | -0.24701 | 0.025527 | 3.81E-22 | cis |
| miR-370-3[illegible] | rs5993909[illegible] | 14:101169[illegible] | 14 | 1.01E+08 | C | T | 0.892837 | 0.253904 | 0.037979 | 2.30E-11 | cis |
| miR-370-3[illegible] | rs1162093[illegible] | 14:101151[illegible] | 14 | 1.01E+08 | C | G | 0.338205 | -0.22338 | 0.024654 | 1.30E-19 | cis |
| miR-370-3[illegible] | rs5910018[illegible] | 14:101184[illegible] | 14 | 1.01E+08 | G | A | 0.836014 | 0.234711 | 0.032214 | 3.19E-13 | cis |
| miR-370-3[illegible] | rs1190907 | 14:101099[illegible] | 14 | 1.01E+08 | A | G | 0.772847 | 0.214936 | 0.028082 | 1.95E-14 | cis |
| miR-370-3[illegible] | rs1190857 | 14:101066[illegible] | 14 | 1.01E+08 | G | T | 0.310273 | -0.20891 | 0.025306 | 1.52E-16 | cis |
| miR-370-3[illegible] | rs911330 | 14:101099[illegible] | 14 | 1.01E+08 | T | C | 0.774615 | 0.215056 | 0.028225 | 2.55E-14 | cis |
| miR-370-3[illegible] | rs1014696[illegible] | 14:101170[illegible] | 14 | 1.01E+08 | T | C | 0.658875 | 0.258505 | 0.023832 | 2.06E-27 | cis |
| miR-370-3[illegible] | rs4905999 | 14:101184 | 14 | 1.01E+08 | A | G | 0.181905 | -0.27581 | 0.030398 | 1.16E-19 | cis |
| miR-370-3[illegible] | rs1190905 | 14:101097[illegible] | 14 | 1.01E+08 | G | T | 0.817561 | 0.21427 | 0.031146 | 6.01E-12 | cis |
| miR-370-3[illegible] | rs1214700[illegible] | 14:101186 | 14 | 1.01E+08 | C | T | 0.223221 | -0.26391 | 0.02766 | 1.41E-21 | cis |
| miR-370-3[illegible] | rs2183632 | 14:101063[illegible] | 14 | 1.01E+08 | C | T | 0.319851 | -0.21038 | 0.025414 | 1.25E-16 | cis |

| | | | | | | | | | | | |
|---|---|---|---|---|---|---|---|---|---|---|---|
| miR-370-3p | rs1190850 | 14:101064 | 14 | 1.01E+08 | A | G | 0.314302 | -0.21173 | 0.025468 | 9.29E-17 | cis |
| miR-370-3p | rs6130797 | 14:101177 | 14 | 1.01E+08 | C | T | 0.888723 | 0.264666 | 0.037169 | 1.07E-12 | cis |
| miR-370-3p | rs7154024 | 14:101031 | 14 | 1.01E+08 | G | T | 0.725015 | 0.199663 | 0.025974 | 1.51E-14 | cis |
| miR-370-3p | rs1184948 | 14:101037 | 14 | 1.01E+08 | C | G | 0.733365 | 0.198989 | 0.026138 | 2.68E-14 | cis |
| miR-370-3p | rs7271582 | 14:101096 | 14 | 1.01E+08 | C | T | 0.817473 | 0.214821 | 0.031245 | 6.18E-12 | cis |
| miR-370-3p | rs1190867 | 14:101069 | 14 | 1.01E+08 | G | T | 0.316788 | -0.21014 | 0.025453 | 1.51E-16 | cis |
| miR-370-3p | rs1757758 | 14:101189 | 14 | 1.01E+08 | C | T | 0.827094 | 0.237826 | 0.030495 | 6.25E-15 | cis |
| miR-370-3p | rs7142089 | 14:101172 | 14 | 1.01E+08 | T | A | 0.783402 | 0.215863 | 0.029162 | 1.34E-13 | cis |
| miR-370-3p | rs1709962 | 14:101173 | 14 | 1.01E+08 | A | C | 0.783449 | 0.191629 | 0.028093 | 9.02E-12 | cis |
| miR-370-3p | rs1288154 | 14:101176 | 14 | 1.01E+08 | G | C | 0.370047 | -0.2711 | 0.024937 | 1.58E-27 | cis |
| miR-370-3p | rs7140792 | 14:101172 | 14 | 1.01E+08 | C | T | 0.892517 | 0.253004 | 0.03785 | 2.32E-11 | cis |
| miR-370-3p | rs1888581 | 14:101042 | 14 | 1.01E+08 | A | C | 0.735363 | 0.196476 | 0.026077 | 4.90E-14 | cis |
| miR-370-3p | rs8021301 | 14:101166 | 14 | 1.01E+08 | G | C | 0.668324 | 0.250089 | 0.024303 | 7.80E-25 | cis |
| miR-370-3p | rs1190872 | 14:101069 | 14 | 1.01E+08 | A | G | 0.310177 | -0.20817 | 0.025276 | 1.79E-16 | cis |
| miR-370-3p | rs879058 | 14:101069 | 14 | 1.01E+08 | G | T | 0.311807 | -0.20823 | 0.025277 | 1.75E-16 | cis |
| miR-370-3p | rs6575794 | 14:101039 | 14 | 1.01E+08 | C | T | 0.734189 | 0.198744 | 0.026178 | 3.15E-14 | cis |
| miR-370-3p | rs3470057 | 14:101173 | 14 | 1.01E+08 | C | T | 0.892725 | 0.253411 | 0.03781 | 2.05E-11 | cis |
| miR-370-3p | rs879060 | 14:101069 | 14 | 1.01E+08 | A | G | 0.310926 | -0.2088 | 0.025277 | 1.45E-16 | cis |
| miR-370-3p | rs1258871 | 14:101149 | 14 | 1.01E+08 | G | C | 0.347269 | -0.22767 | 0.024425 | 1.15E-20 | cis |
| miR-370-3p | rs1288176 | 14:101176 | 14 | 1.01E+08 | G | C | 0.347791 | -0.2649 | 0.024603 | 4.91E-27 | cis |
| miR-370-3p | rs7141110 | 14:101172 | 14 | 1.01E+08 | G | C | 0.783235 | 0.212732 | 0.029134 | 2.84E-13 | cis |
| miR-370-3p | rs7269871 | 14:101173 | 14 | 1.01E+08 | G | T | 0.783221 | 0.191924 | 0.028096 | 8.44E-12 | cis |
| miR-370-3p | rs6030796 | 14:101027 | 14 | 1.01E+08 | C | T | 0.725784 | 0.207963 | 0.026927 | 1.14E-14 | cis |
| miR-370-3p | rs4905978 | 14:101061 | 14 | 1.01E+08 | C | T | 0.318452 | -0.21246 | 0.025401 | 6.05E-17 | cis |
| miR-370-3p | rs1014551 | 14:101168 | 14 | 1.01E+08 | A | G | 0.668173 | 0.249333 | 0.024276 | 9.53E-25 | cis |
| miR-370-3p | rs7142683 | 14:101158 | 14 | 1.01E+08 | A | G | 0.290748 | -0.26127 | 0.02591 | 6.52E-24 | cis |
| miR-370-3p | rs7155375 | 14:101188 | 14 | 1.01E+08 | C | T | 0.635995 | 0.206766 | 0.023788 | 3.56E-18 | cis |
| miR-370-3p | rs7147982 | 14:101046 | 14 | 1.01E+08 | C | T | 0.731753 | 0.191267 | 0.025899 | 1.52E-13 | cis |
| miR-370-3p | rs1555405 | 14:101176 | 14 | 1.01E+08 | G | A | 0.749075 | 0.215585 | 0.026307 | 2.51E-16 | cis |
| miR-370-3p | rs1014438 | 14:101173 | 14 | 1.01E+08 | C | T | 0.673446 | 0.258353 | 0.024412 | 3.57E-26 | cis |
| miR-370-3p | rs7269872 | 14:101189 | 14 | 1.01E+08 | C | T | 0.825721 | 0.239819 | 0.030305 | 2.50E-15 | cis |
| miR-370-3p | rs1243613 | 14:101031 | 14 | 1.01E+08 | C | T | 0.725015 | 0.199847 | 0.025997 | 1.50E-14 | cis |
| miR-370-3p | rs4243729 | 14:101066 | 14 | 1.01E+08 | A | C | 0.311439 | -0.20514 | 0.025119 | 3.17E-16 | cis |
| miR-370-3p | rs2837918 | 14:101168 | 14 | 1.01E+08 | G | T | 0.836434 | 0.269753 | 0.031357 | 7.80E-18 | cis |
| miR-370-3p | rs2150518 | 14:101063 | 14 | 1.01E+08 | C | T | 0.318506 | -0.21179 | 0.02544 | 8.44E-17 | cis |
| miR-370-3p | rs7263167 | 14:101176 | 14 | 1.01E+08 | C | T | 0.889487 | 0.260349 | 0.037136 | 2.37E-12 | cis |
| miR-370-3p | rs1190856 | 14:101065 | 14 | 1.01E+08 | T | C | 0.310244 | -0.20896 | 0.025301 | 1.47E-16 | cis |
| miR-370-3p | rs1190876 | 14:101071 | 14 | 1.01E+08 | A | T | 0.353354 | -0.16716 | 0.024812 | 1.61E-11 | cis |
| miR-370-3p | rs5982537 | 14:101168 | 14 | 1.01E+08 | G | T | 0.89241 | 0.254027 | 0.03799 | 2.28E-11 | cis |
| miR-370-3p | rs8020868 | 14:101166 | 14 | 1.01E+08 | A | G | 0.668343 | 0.250042 | 0.024301 | 7.89E-25 | cis |
| miR-370-3p | rs4905994 | 14:101154 | 14 | 1.01E+08 | T | C | 0.329146 | -0.24049 | 0.025152 | 1.16E-21 | cis |
| miR-370-3p | rs7145610 | 14:101049 | 14 | 1.01E+08 | T | C | 0.740391 | 0.21785 | 0.026381 | 1.49E-16 | cis |
| miR-370-3p | rs6762567 | 14:101173 | 14 | 1.01E+08 | A | G | 0.892632 | 0.253285 | 0.037791 | 2.05E-11 | cis |
| miR-370-3p | rs5892242 | 14:101097 | 14 | 1.01E+08 | C | G | 0.817892 | 0.215058 | 0.031189 | 5.37E-12 | cis |
| miR-370-3p | rs9324022 | 14:101172 | 14 | 1.01E+08 | T | C | 0.763739 | 0.21966 | 0.028499 | 1.28E-14 | cis |
| miR-370-3p | rs1190906 | 14:101099 | 14 | 1.01E+08 | T | C | 0.772138 | 0.214026 | 0.02804 | 2.29E-14 | cis |
| miR-376b-3 | rs9671394 | 14:101159 | 14 | 1.01E+08 | C | T | 0.284232 | -0.22098 | 0.032082 | 5.65E-12 | cis |
| miR-376c-3 | rs7143357 | 14:101054 | 14 | 1.01E+08 | T | C | 0.739636 | 0.179146 | 0.023568 | 2.94E-14 | cis |
| miR-376c-3 | rs1214700 | 14:101186 | 14 | 1.01E+08 | C | T | 0.223221 | -0.24082 | 0.024674 | 1.67E-22 | cis |
| miR-376c-3 | rs4905998 | 14:101181 | 14 | 1.01E+08 | A | G | 0.330615 | -0.22897 | 0.02168 | 4.51E-26 | cis |
| miR-376c-3 | rs1190867 | 14:101069 | 14 | 1.01E+08 | G | T | 0.316788 | -0.15714 | 0.022833 | 5.90E-12 | cis |
| miR-376c-3 | rs4905999 | 14:101184 | 14 | 1.01E+08 | A | G | 0.181905 | -0.27701 | 0.027005 | 1.09E-24 | cis |
| miR-376c-3 | rs1289760 | 14:101182 | 14 | 1.01E+08 | C | T | 0.190452 | -0.28274 | 0.026347 | 7.26E-27 | cis |
| miR-376c-3 | rs2183632 | 14:101063 | 14 | 1.01E+08 | C | T | 0.319851 | -0.16084 | 0.022787 | 1.69E-12 | cis |
| miR-376c-3 | rs1709961 | 14:101167 | 14 | 1.01E+08 | C | G | 0.668809 | 0.211678 | 0.021761 | 2.30E-22 | cis |
| miR-376c-3 | rs8008285 | 14:101146 | 14 | 1.01E+08 | T | C | 0.842905 | 0.213959 | 0.028987 | 1.57E-13 | cis |
| miR-376c-3 | rs5922882 | 14:101185 | 14 | 1.01E+08 | G | C | 0.745519 | 0.170194 | 0.024078 | 1.57E-12 | cis |
| miR-376c-3 | rs1190850 | 14:101064 | 14 | 1.01E+08 | A | G | 0.314302 | -0.15914 | 0.022846 | 3.27E-12 | cis |
| miR-376c-3 | rs1288281 | 14:101183 | 14 | 1.01E+08 | C | G | 0.331398 | -0.2289 | 0.021697 | 5.09E-26 | cis |
| miR-376c-3 | rs7263167 | 14:101187 | 14 | 1.01E+08 | C | T | 0.881829 | 0.278575 | 0.032942 | 2.76E-17 | cis |
| miR-376c-3 | rs7155649 | 14:101181 | 14 | 1.01E+08 | C | A | 0.863608 | 0.250619 | 0.030785 | 3.92E-16 | cis |
| miR-376c-3 | rs6575791 | 14:101029 | 14 | 1.01E+08 | G | A | 0.719841 | 0.163883 | 0.02326 | 1.85E-12 | cis |
| miR-376c-3 | rs7141210 | 14:101182 | 14 | 1.01E+08 | T | C | 0.32902 | -0.2286 | 0.021625 | 4.07E-26 | cis |

| | | | | | | | | | | | |
|---|---|---|---|---|---|---|---|---|---|---|---|
| miR-376c-3 | rs5993909 | 14:101169 | 14 | 1.01E+08 | C | T | 0.892837 | 0.263176 | 0.033789 | 6.76E-15 | cis |
| miR-376c-3 | rs6575794 | 14:101039 | 14 | 1.01E+08 | C | T | 0.734189 | 0.167266 | 0.023408 | 8.96E-13 | cis |
| miR-376c-3 | rs9671394 | 14:101159 | 14 | 1.01E+08 | C | T | 0.284232 | -0.22229 | 0.023456 | 2.62E-21 | cis |
| miR-376c-3 | rs1116059 | 14:101049 | 14 | 1.01E+08 | C | T | 0.741662 | 0.179342 | 0.023547 | 2.61E-14 | cis |
| miR-376c-3 | rs1289379 | 14:101178 | 14 | 1.01E+08 | C | G | 0.190813 | -0.28504 | 0.026504 | 5.65E-27 | cis |
| miR-376c-3 | rs7142683 | 14:101158 | 14 | 1.01E+08 | A | G | 0.290748 | -0.21451 | 0.023218 | 2.49E-20 | cis |
| miR-376c-3 | rs879059 | 14:101069 | 14 | 1.01E+08 | T | A | 0.31068 | -0.15658 | 0.022659 | 4.84E-12 | cis |
| miR-376c-3 | rs4900471 | 14:101162 | 14 | 1.01E+08 | G | A | 0.304173 | -0.2042 | 0.022863 | 4.20E-19 | cis |
| miR-376c-3 | rs8021301 | 14:101166 | 14 | 1.01E+08 | G | C | 0.668324 | 0.209581 | 0.021763 | 5.97E-22 | cis |
| miR-376c-3 | rs1888581 | 14:101042 | 14 | 1.01E+08 | A | C | 0.735363 | 0.165096 | 0.023318 | 1.44E-12 | cis |
| miR-376c-3 | rs1190852 | 14:101064 | 14 | 1.01E+08 | A | G | 0.310295 | -0.15773 | 0.022711 | 3.78E-12 | cis |
| miR-376c-3 | rs2837918 | 14:101168 | 14 | 1.01E+08 | G | T | 0.836434 | 0.247403 | 0.027972 | 9.18E-19 | cis |
| miR-376c-3 | rs7269872 | 14:101189 | 14 | 1.01E+08 | C | T | 0.825721 | 0.21176 | 0.027067 | 5.13E-15 | cis |
| miR-376c-3 | rs6702222 | 14:101191 | 14 | 1.01E+08 | A | T | 0.731016 | 0.169894 | 0.024131 | 1.92E-12 | cis |
| miR-376c-3 | rs7742021 | 14:101151 | 14 | 1.01E+08 | C | T | 0.945071 | 0.338224 | 0.048804 | 4.20E-12 | cis |
| miR-376c-3 | rs4243729 | 14:101066 | 14 | 1.01E+08 | A | C | 0.311439 | -0.1564 | 0.022522 | 3.80E-12 | cis |
| miR-376c-3 | rs3470057 | 14:101173 | 14 | 1.01E+08 | C | T | 0.892725 | 0.262038 | 0.033641 | 6.73E-15 | cis |
| miR-376c-3 | rs873076 | 14:101070 | 14 | 1.01E+08 | A | G | 0.311961 | -0.15498 | 0.022626 | 7.40E-12 | cis |
| miR-376c-3 | rs5870230 | 14:101167 | 14 | 1.01E+08 | G | A | 0.892706 | 0.263093 | 0.033829 | 7.41E-15 | cis |
| miR-376c-3 | rs7142089 | 14:101172 | 14 | 1.01E+08 | T | A | 0.783402 | 0.184603 | 0.026065 | 1.42E-12 | cis |
| miR-376c-3 | rs6199300 | 14:101034 | 14 | 1.01E+08 | G | A | 0.726044 | 0.163337 | 0.023265 | 2.21E-12 | cis |
| miR-376c-3 | rs1190862 | 14:101067 | 14 | 1.01E+08 | T | G | 0.309628 | -0.15724 | 0.022533 | 2.99E-12 | cis |
| miR-376c-3 | rs8012970 | 14:101168 | 14 | 1.01E+08 | T | C | 0.347902 | -0.2104 | 0.022142 | 2.05E-21 | cis |
| miR-376c-3 | rs1243449 | 14:101179 | 14 | 1.01E+08 | A | G | 0.18959 | -0.28495 | 0.026482 | 5.29E-27 | cis |
| miR-376c-3 | rs1014438 | 14:101173 | 14 | 1.01E+08 | C | T | 0.673446 | 0.223473 | 0.021831 | 1.36E-24 | cis |
| miR-376c-3 | rs1288154 | 14:101176 | 14 | 1.01E+08 | G | C | 0.370047 | -0.23937 | 0.022279 | 6.31E-27 | cis |
| miR-376c-3 | rs7493817 | 14:101038 | 14 | 1.01E+08 | G | A | 0.751723 | 0.178843 | 0.024109 | 1.19E-13 | cis |
| miR-376c-3 | rs1243369 | 14:101035 | 14 | 1.01E+08 | C | G | 0.723403 | 0.161295 | 0.023114 | 2.99E-12 | cis |
| miR-376c-3 | rs7154024 | 14:101031 | 14 | 1.01E+08 | G | T | 0.725015 | 0.162759 | 0.023245 | 2.52E-12 | cis |
| miR-376c-3 | rs8020868 | 14:101166 | 14 | 1.01E+08 | A | G | 0.668343 | 0.20953 | 0.021761 | 6.06E-22 | cis |
| miR-376c-3 | rs1288176 | 14:101176 | 14 | 1.01E+08 | G | C | 0.347791 | -0.23493 | 0.021975 | 1.12E-26 | cis |
| miR-376c-3 | rs6030796 | 14:101027 | 14 | 1.01E+08 | C | T | 0.725784 | 0.176828 | 0.024074 | 2.05E-13 | cis |
| miR-376c-3 | rs1190872 | 14:101069 | 14 | 1.01E+08 | A | G | 0.310177 | -0.15611 | 0.022672 | 5.76E-12 | cis |
| miR-376c-3 | rs7140556 | 14:101036 | 14 | 1.01E+08 | C | A | 0.723818 | 0.163817 | 0.023211 | 1.69E-12 | cis |
| miR-376c-3 | rs1555405 | 14:101176 | 14 | 1.01E+08 | G | A | 0.749075 | 0.187916 | 0.023506 | 1.30E-15 | cis |
| miR-376c-3 | rs7263167 | 14:101176 | 14 | 1.01E+08 | C | T | 0.889487 | 0.275601 | 0.033005 | 6.81E-17 | cis |
| miR-376c-3 | rs5910018 | 14:101184 | 14 | 1.01E+08 | G | A | 0.836014 | 0.21075 | 0.028759 | 2.33E-13 | cis |
| miR-376c-3 | rs7145337 | 14:101155 | 14 | 1.01E+08 | C | T | 0.312091 | -0.2074 | 0.022825 | 1.03E-19 | cis |
| miR-376c-3 | rs6199300 | 14:101030 | 14 | 1.01E+08 | G | A | 0.724196 | 0.162405 | 0.023194 | 2.52E-12 | cis |
| miR-376c-3 | rs1014551 | 14:101168 | 14 | 1.01E+08 | A | G | 0.668173 | 0.209202 | 0.021737 | 6.31E-22 | cis |
| miR-376c-3 | rs1467536 | 14:101061 | 14 | 1.01E+08 | A | G | 0.318958 | -0.16362 | 0.02274 | 6.24E-13 | cis |
| miR-376c-3 | rs1190857 | 14:101066 | 14 | 1.01E+08 | G | T | 0.310273 | -0.15701 | 0.022699 | 4.61E-12 | cis |
| miR-376c-3 | rs1243517 | 14:101155 | 14 | 1.01E+08 | G | A | 0.312183 | -0.20721 | 0.022832 | 1.13E-19 | cis |
| miR-376c-3 | rs879058 | 14:101069 | 14 | 1.01E+08 | G | T | 0.311807 | -0.15693 | 0.022671 | 4.44E-12 | cis |
| miR-376c-3 | rs1258871 | 14:101149 | 14 | 1.01E+08 | G | C | 0.347269 | -0.1894 | 0.021866 | 4.63E-18 | cis |
| miR-376c-3 | rs4905994 | 14:101154 | 14 | 1.01E+08 | T | C | 0.329146 | -0.19711 | 0.022532 | 2.18E-18 | cis |
| miR-376c-3 | rs873077 | 14:101070 | 14 | 1.01E+08 | G | A | 0.312104 | -0.15489 | 0.02263 | 7.67E-12 | cis |
| miR-376c-3 | rs7149242 | 14:101159 | 14 | 1.01E+08 | T | G | 0.288371 | -0.21449 | 0.02315 | 1.94E-20 | cis |
| miR-376c-3 | rs4905978 | 14:101061 | 14 | 1.01E+08 | C | T | 0.318452 | -0.16338 | 0.022774 | 7.29E-13 | cis |
| miR-376c-3 | rs1162093 | 14:101151 | 14 | 1.01E+08 | C | G | 0.338205 | -0.18492 | 0.022072 | 5.37E-17 | cis |
| miR-376c-3 | rs6762567 | 14:101173 | 14 | 1.01E+08 | A | G | 0.892632 | 0.261904 | 0.033623 | 6.74E-15 | cis |
| miR-376c-3 | rs1185095 | 14:101056 | 14 | 1.01E+08 | G | T | 0.743981 | 0.176232 | 0.023439 | 5.53E-14 | cis |
| miR-376c-3 | rs7155375 | 14:101188 | 14 | 1.01E+08 | C | T | 0.635995 | 0.184125 | 0.021242 | 4.40E-18 | cis |
| miR-376c-3 | rs9324022 | 14:101172 | 14 | 1.01E+08 | T | C | 0.763739 | 0.179459 | 0.025503 | 1.97E-12 | cis |
| miR-376c-3 | rs1757758 | 14:101189 | 14 | 1.01E+08 | C | T | 0.827094 | 0.214382 | 0.027221 | 3.39E-15 | cis |
| miR-376c-3 | rs5982537 | 14:101168 | 14 | 1.01E+08 | G | T | 0.89241 | 0.264334 | 0.033795 | 5.21E-15 | cis |
| miR-376c-3 | rs4905976 | 14:101060 | 14 | 1.01E+08 | G | A | 0.322862 | -0.16174 | 0.022805 | 1.32E-12 | cis |
| miR-376c-3 | rs6130797 | 14:101177 | 14 | 1.01E+08 | C | T | 0.888723 | 0.276405 | 0.033044 | 6.02E-17 | cis |
| miR-376c-3 | rs2150518 | 14:101063 | 14 | 1.01E+08 | C | T | 0.318506 | -0.16163 | 0.022813 | 1.39E-12 | cis |
| miR-376c-3 | rs7145610 | 14:101049 | 14 | 1.01E+08 | T | C | 0.740391 | 0.180314 | 0.023607 | 2.21E-14 | cis |
| miR-376c-3 | rs7147982 | 14:101046 | 14 | 1.01E+08 | C | T | 0.731753 | 0.161861 | 0.023155 | 2.74E-12 | cis |
| miR-376c-3 | rs1190856 | 14:101065 | 14 | 1.01E+08 | T | C | 0.310244 | -0.15713 | 0.022694 | 4.40E-12 | cis |
| miR-376c-3 | rs1013940 | 14:101194 | 14 | 1.01E+08 | A | G | 0.323897 | -0.19045 | 0.022477 | 2.40E-17 | cis |

| | | | | | | | | | | | |
|---|---|---|---|---|---|---|---|---|---|---|---|
| miR-376c-3 | rs879060 | 14:101069 | 14 | 1.01E+08 | A | G | 0.310926 | -0.1567 | 0.022673 | 4.81E-12 | cis |
| miR-376c-3 | rs7140792 | 14:101172 | 14 | 1.01E+08 | C | T | 0.892517 | 0.262816 | 0.033672 | 5.94E-15 | cis |
| miR-376c-3 | rs4905977 | 14:101061 | 14 | 1.01E+08 | C | G | 0.318473 | -0.16341 | 0.022775 | 7.23E-13 | cis |
| miR-376c-3 | rs1184948 | 14:101037 | 14 | 1.01E+08 | C | G | 0.733365 | 0.168474 | 0.02337 | 5.63E-13 | cis |
| miR-376c-3 | rs1214784 | 14:101144 | 14 | 1.01E+08 | C | T | 0.870989 | 0.218241 | 0.031186 | 2.60E-12 | cis |
| miR-376c-3 | rs1243613 | 14:101031 | 14 | 1.01E+08 | C | T | 0.725015 | 0.162954 | 0.023265 | 2.48E-12 | cis |
| miR-376c-3 | rs1014696 | 14:101170 | 14 | 1.01E+08 | T | C | 0.658875 | 0.213122 | 0.021364 | 1.95E-23 | cis |
| miR-376c-3 | rs7141110 | 14:101172 | 14 | 1.01E+08 | G | C | 0.783235 | 0.181998 | 0.02604 | 2.76E-12 | cis |
| miR-377-3p | rs4905998 | 14:101181 | 14 | 1.01E+08 | A | G | 0.330615 | -0.1687 | 0.021696 | 7.50E-15 | cis |
| miR-377-3p | rs7141210 | 14:101182 | 14 | 1.01E+08 | T | C | 0.32902 | -0.16828 | 0.021642 | 7.49E-15 | cis |
| miR-377-3p | rs1289379 | 14:101178 | 14 | 1.01E+08 | C | G | 0.190813 | -0.18378 | 0.026624 | 5.09E-12 | cis |
| miR-377-3p | rs1288154 | 14:101176 | 14 | 1.01E+08 | G | C | 0.370047 | -0.17673 | 0.022303 | 2.30E-15 | cis |
| miR-377-3p | rs1014438 | 14:101173 | 14 | 1.01E+08 | C | T | 0.673446 | 0.167537 | 0.021822 | 1.62E-14 | cis |
| miR-377-3p | rs1289760 | 14:101182 | 14 | 1.01E+08 | C | T | 0.190452 | -0.18937 | 0.026442 | 7.96E-13 | cis |
| miR-377-3p | rs1243449 | 14:101179 | 14 | 1.01E+08 | A | G | 0.18959 | -0.18371 | 0.026602 | 4.99E-12 | cis |
| miR-377-3p | rs1288176 | 14:101176 | 14 | 1.01E+08 | G | C | 0.347791 | -0.17185 | 0.022002 | 5.68E-15 | cis |
| miR-377-3p | rs1288281 | 14:101183 | 14 | 1.01E+08 | C | G | 0.331398 | -0.16888 | 0.021712 | 7.36E-15 | cis |
| miR-377-3p | rs4905999 | 14:101184 | 14 | 1.01E+08 | A | G | 0.181905 | -0.20793 | 0.026994 | 1.33E-14 | cis |
| miR-377-3p | rs1014696 | 14:101170 | 14 | 1.01E+08 | T | C | 0.658875 | 0.14783 | 0.021384 | 4.74E-12 | cis |
| miR-377-5p | rs1288281 | 14:101183 | 14 | 1.01E+08 | C | G | 0.331398 | -0.2352 | 0.03366 | 2.80E-12 | cis |
| miR-377-5p | rs1288154 | 14:101176 | 14 | 1.01E+08 | G | C | 0.370047 | -0.23953 | 0.034602 | 4.44E-12 | cis |
| miR-377-5p | rs1014438 | 14:101173 | 14 | 1.01E+08 | C | T | 0.673446 | 0.237087 | 0.033818 | 2.37E-12 | cis |
| miR-377-5p | rs1288176 | 14:101176 | 14 | 1.01E+08 | G | C | 0.347791 | -0.23731 | 0.034117 | 3.51E-12 | cis |
| miR-377-5p | rs4905998 | 14:101181 | 14 | 1.01E+08 | A | G | 0.330615 | -0.23455 | 0.033637 | 3.10E-12 | cis |
| miR-377-5p | rs1014696 | 14:101170 | 14 | 1.01E+08 | T | C | 0.658875 | 0.232313 | 0.033054 | 2.09E-12 | cis |
| miR-377-5p | rs7141210 | 14:101182 | 14 | 1.01E+08 | T | C | 0.32902 | -0.23425 | 0.033552 | 2.92E-12 | cis |
| miR-382-5p | rs879059 | 14:101069 | 14 | 1.01E+08 | T | A | 0.31068 | -0.25966 | 0.034839 | 9.11E-14 | cis |
| miR-382-5p | rs8012970 | 14:101168 | 14 | 1.01E+08 | T | C | 0.347902 | -0.32253 | 0.034109 | 3.21E-21 | cis |
| miR-382-5p | rs1243517 | 14:101155 | 14 | 1.01E+08 | G | A | 0.312183 | -0.32286 | 0.03515 | 4.11E-20 | cis |
| miR-382-5p | rs1709961 | 14:101167 | 14 | 1.01E+08 | C | G | 0.668809 | 0.326346 | 0.033515 | 2.09E-22 | cis |
| miR-382-5p | rs1190903 | 14:101096 | 14 | 1.01E+08 | C | T | 0.772566 | 0.268909 | 0.03878 | 4.08E-12 | cis |
| miR-382-5p | rs8021301 | 14:101166 | 14 | 1.01E+08 | G | C | 0.668324 | 0.324506 | 0.033512 | 3.55E-22 | cis |
| miR-382-5p | rs1288281 | 14:101183 | 14 | 1.01E+08 | C | G | 0.331398 | -0.33198 | 0.033515 | 3.95E-23 | cis |
| miR-382-5p | rs4905976 | 14:101060 | 14 | 1.01E+08 | G | A | 0.322862 | -0.253 | 0.035112 | 5.79E-13 | cis |
| miR-382-5p | rs4905977 | 14:101061 | 14 | 1.01E+08 | C | G | 0.318473 | -0.26323 | 0.035039 | 5.80E-14 | cis |
| miR-382-5p | rs1014696 | 14:101170 | 14 | 1.01E+08 | T | C | 0.658875 | 0.328407 | 0.032905 | 1.85E-23 | cis |
| miR-382-5p | rs5993909 | 14:101169 | 14 | 1.01E+08 | C | T | 0.892837 | 0.414387 | 0.052009 | 1.62E-15 | cis |
| miR-382-5p | rs7142089 | 14:101172 | 14 | 1.01E+08 | T | A | 0.783402 | 0.292345 | 0.04012 | 3.17E-13 | cis |
| miR-382-5p | rs1214700 | 14:101186 | 14 | 1.01E+08 | C | T | 0.223221 | -0.31605 | 0.038232 | 1.38E-16 | cis |
| miR-382-5p | rs7141210 | 14:101182 | 14 | 1.01E+08 | T | C | 0.32902 | -0.33035 | 0.03341 | 4.70E-23 | cis |
| miR-382-5p | rs7263167 | 14:101187 | 14 | 1.01E+08 | C | T | 0.881829 | 0.408715 | 0.050816 | 8.76E-16 | cis |
| miR-382-5p | rs7155649 | 14:101181 | 14 | 1.01E+08 | C | A | 0.863608 | 0.322021 | 0.047635 | 1.38E-11 | cis |
| miR-382-5p | rs1190904 | 14:101096 | 14 | 1.01E+08 | A | G | 0.772596 | 0.268729 | 0.038777 | 4.21E-12 | cis |
| miR-382-5p | rs7263167 | 14:101176 | 14 | 1.01E+08 | C | T | 0.889487 | 0.412186 | 0.050882 | 5.46E-16 | cis |
| miR-382-5p | rs2837918 | 14:101168 | 14 | 1.01E+08 | G | T | 0.836434 | 0.391528 | 0.043039 | 9.29E-20 | cis |
| miR-382-5p | rs1185095 | 14:101056 | 14 | 1.01E+08 | G | T | 0.743981 | 0.286172 | 0.036048 | 2.04E-15 | cis |
| miR-382-5p | rs1190905 | 14:101097 | 14 | 1.01E+08 | G | T | 0.817561 | 0.288861 | 0.042851 | 1.57E-11 | cis |
| miR-382-5p | rs6575794 | 14:101039 | 14 | 1.01E+08 | C | T | 0.734189 | 0.247015 | 0.036088 | 7.66E-12 | cis |
| miR-382-5p | rs1190906 | 14:101099 | 14 | 1.01E+08 | T | C | 0.772138 | 0.263048 | 0.038665 | 1.02E-11 | cis |
| miR-382-5p | rs1184948 | 14:101037 | 14 | 1.01E+08 | C | G | 0.733365 | 0.247145 | 0.036034 | 6.95E-12 | cis |
| miR-382-5p | rs7145610 | 14:101049 | 14 | 1.01E+08 | T | C | 0.740391 | 0.28563 | 0.036332 | 3.79E-15 | cis |
| miR-382-5p | rs1190867 | 14:101069 | 14 | 1.01E+08 | G | T | 0.316788 | -0.25992 | 0.035109 | 1.33E-13 | cis |
| miR-382-5p | rs4905978 | 14:101061 | 14 | 1.01E+08 | C | T | 0.318452 | -0.26323 | 0.035038 | 5.79E-14 | cis |
| miR-382-5p | rs1888581 | 14:101042 | 14 | 1.01E+08 | A | C | 0.735363 | 0.243774 | 0.035949 | 1.19E-11 | cis |
| miR-382-5p | rs879060 | 14:101069 | 14 | 1.01E+08 | A | G | 0.310926 | -0.25984 | 0.034861 | 9.08E-14 | cis |
| miR-382-5p | rs1190852 | 14:101064 | 14 | 1.01E+08 | A | G | 0.310295 | -0.26177 | 0.034917 | 6.54E-14 | cis |
| miR-382-5p | rs1757758 | 14:101189 | 14 | 1.01E+08 | C | T | 0.827094 | 0.286006 | 0.042075 | 1.06E-11 | cis |
| miR-382-5p | rs7147982 | 14:101046 | 14 | 1.01E+08 | C | T | 0.731753 | 0.239351 | 0.035694 | 2.01E-11 | cis |
| miR-382-5p | rs7335877 | 14:101159 | 14 | 1.01E+08 | C | T | 0.891417 | 0.368384 | 0.053189 | 4.33E-12 | cis |
| miR-382-5p | rs4905998 | 14:101181 | 14 | 1.01E+08 | A | G | 0.330615 | -0.33232 | 0.033488 | 3.29E-23 | cis |
| miR-382-5p | rs1190909 | 14:101101 | 14 | 1.01E+08 | G | A | 0.775531 | 0.26705 | 0.039234 | 9.99E-12 | cis |
| miR-382-5p | rs1288176 | 14:101176 | 14 | 1.01E+08 | G | C | 0.347791 | -0.33384 | 0.033979 | 8.79E-23 | cis |
| miR-382-5p | rs1013940 | 14:101194 | 14 | 1.01E+08 | A | G | 0.323897 | -0.238 | 0.034815 | 8.14E-12 | cis |

| | | | | | | | | | | | |
|---|---|---|---|---|---|---|---|---|---|---|---|
| miR-382-5p | rs1190850 | 14:101064 | 14 | 1.01E+08 | A | G | 0.314302 | -0.26488 | 0.035121 | 4.62E-14 | cis |
| miR-382-5p | rs5892242 | 14:101097 | 14 | 1.01E+08 | C | G | 0.817892 | 0.289588 | 0.042911 | 1.49E-11 | cis |
| miR-382-5p | rs873077 | 14:101070 | 14 | 1.01E+08 | G | A | 0.312104 | -0.25723 | 0.034793 | 1.43E-13 | cis |
| miR-382-5p | rs7143357 | 14:101054 | 14 | 1.01E+08 | T | C | 0.739636 | 0.283285 | 0.036274 | 5.74E-15 | cis |
| miR-382-5p | rs2150518 | 14:101063 | 14 | 1.01E+08 | C | T | 0.318506 | -0.26102 | 0.035096 | 1.03E-13 | cis |
| miR-382-5p | rs7493817 | 14:101038 | 14 | 1.01E+08 | G | A | 0.751723 | 0.269122 | 0.037155 | 4.38E-13 | cis |
| miR-382-5p | rs1258871 | 14:101149 | 14 | 1.01E+08 | G | C | 0.347269 | -0.29285 | 0.033673 | 3.41E-18 | cis |
| miR-382-5p | rs7145337 | 14:101155 | 14 | 1.01E+08 | C | T | 0.312091 | -0.32283 | 0.035142 | 4.06E-20 | cis |
| miR-382-5p | rs2183632 | 14:101063 | 14 | 1.01E+08 | C | T | 0.319851 | -0.26048 | 0.035054 | 1.08E-13 | cis |
| miR-382-5p | rs1288154 | 14:101176 | 14 | 1.01E+08 | G | C | 0.370047 | -0.33648 | 0.034467 | 1.63E-22 | cis |
| miR-382-5p | rs4905999 | 14:101184 | 14 | 1.01E+08 | A | G | 0.181905 | -0.39288 | 0.041746 | 4.92E-21 | cis |
| miR-382-5p | rs1014438 | 14:101173 | 14 | 1.01E+08 | C | T | 0.673446 | 0.332591 | 0.033679 | 5.32E-23 | cis |
| miR-382-5p | rs873076 | 14:101070 | 14 | 1.01E+08 | A | G | 0.311961 | -0.25736 | 0.034787 | 1.38E-13 | cis |
| miR-382-5p | rs1014551 | 14:101168 | 14 | 1.01E+08 | A | G | 0.668173 | 0.323644 | 0.033473 | 4.09E-22 | cis |
| miR-382-5p | rs1190872 | 14:101069 | 14 | 1.01E+08 | A | G | 0.310177 | -0.25962 | 0.034857 | 9.45E-14 | cis |
| miR-382-5p | rs1289379 | 14:101178 | 14 | 1.01E+08 | C | G | 0.190813 | -0.39242 | 0.04104 | 1.16E-21 | cis |
| miR-382-5p | rs7742021 | 14:101151 | 14 | 1.01E+08 | C | T | 0.945071 | 0.522734 | 0.075163 | 3.53E-12 | cis |
| miR-382-5p | rs4900471 | 14:101162 | 14 | 1.01E+08 | G | A | 0.304173 | -0.31319 | 0.035219 | 5.97E-19 | cis |
| miR-382-5p | rs7149242 | 14:101159 | 14 | 1.01E+08 | T | G | 0.288371 | -0.33031 | 0.035655 | 1.97E-20 | cis |
| miR-382-5p | rs1467536 | 14:101061 | 14 | 1.01E+08 | A | G | 0.318958 | -0.26409 | 0.034984 | 4.39E-14 | cis |
| miR-382-5p | rs1190862 | 14:101067 | 14 | 1.01E+08 | T | G | 0.309628 | -0.25649 | 0.034658 | 1.36E-13 | cis |
| miR-382-5p | rs4243729 | 14:101066 | 14 | 1.01E+08 | A | C | 0.311439 | -0.25456 | 0.034643 | 2.01E-13 | cis |
| miR-382-5p | rs5870230 | 14:101167 | 14 | 1.01E+08 | G | A | 0.892706 | 0.414331 | 0.05207 | 1.76E-15 | cis |
| miR-382-5p | rs8020868 | 14:101166 | 14 | 1.01E+08 | A | G | 0.668343 | 0.32446 | 0.033509 | 3.57E-22 | cis |
| miR-382-5p | rs1190856 | 14:101065 | 14 | 1.01E+08 | T | C | 0.310244 | -0.26085 | 0.034891 | 7.66E-14 | cis |
| miR-382-5p | rs1259093 | 14:101163 | 14 | 1.01E+08 | A | T | 0.893649 | 0.374255 | 0.053582 | 2.85E-12 | cis |
| miR-382-5p | rs1555405 | 14:101176 | 14 | 1.01E+08 | G | A | 0.749075 | 0.295258 | 0.036183 | 3.35E-16 | cis |
| miR-382-5p | rs7140792 | 14:101172 | 14 | 1.01E+08 | C | T | 0.892517 | 0.409611 | 0.051844 | 2.77E-15 | cis |
| miR-382-5p | rs5982537 | 14:101168 | 14 | 1.01E+08 | G | T | 0.89241 | 0.415931 | 0.052019 | 1.29E-15 | cis |
| miR-382-5p | rs4905994 | 14:101154 | 14 | 1.01E+08 | T | C | 0.329146 | -0.30272 | 0.034708 | 2.73E-18 | cis |
| miR-382-5p | rs1243449 | 14:101179 | 14 | 1.01E+08 | A | G | 0.18959 | -0.39163 | 0.041008 | 1.30E-21 | cis |
| miR-382-5p | rs7269872 | 14:101189 | 14 | 1.01E+08 | C | T | 0.825721 | 0.281693 | 0.041838 | 1.66E-11 | cis |
| miR-382-5p | rs879058 | 14:101069 | 14 | 1.01E+08 | G | T | 0.311807 | -0.25975 | 0.034858 | 9.21E-14 | cis |
| miR-382-5p | rs1258812 | 14:101162 | 14 | 1.01E+08 | T | C | 0.893709 | 0.374109 | 0.053578 | 2.90E-12 | cis |
| miR-382-5p | rs9671394 | 14:101159 | 14 | 1.01E+08 | C | T | 0.284232 | -0.33906 | 0.036142 | 6.52E-21 | cis |
| miR-382-5p | rs6130797 | 14:101177 | 14 | 1.01E+08 | C | T | 0.888723 | 0.410347 | 0.050953 | 8.05E-16 | cis |
| miR-382-5p | rs1190857 | 14:101066 | 14 | 1.01E+08 | G | T | 0.310273 | -0.26079 | 0.034898 | 7.85E-14 | cis |
| miR-382-5p | rs6762567 | 14:101173 | 14 | 1.01E+08 | A | G | 0.892632 | 0.410917 | 0.05176 | 2.04E-15 | cis |
| miR-382-5p | rs1190907 | 14:101099 | 14 | 1.01E+08 | A | G | 0.772847 | 0.264877 | 0.038722 | 7.89E-12 | cis |
| miR-382-5p | rs911330 | 14:101099 | 14 | 1.01E+08 | T | C | 0.774615 | 0.268715 | 0.038906 | 4.96E-12 | cis |
| miR-382-5p | rs1289760 | 14:101182 | 14 | 1.01E+08 | C | T | 0.190452 | -0.39113 | 0.040788 | 8.88E-22 | cis |
| miR-382-5p | rs3470057 | 14:101173 | 14 | 1.01E+08 | C | T | 0.892725 | 0.411051 | 0.051786 | 2.06E-15 | cis |
| miR-382-5p | rs1116059 | 14:101049 | 14 | 1.01E+08 | C | T | 0.741662 | 0.284332 | 0.036239 | 4.30E-15 | cis |
| miR-382-5p | rs7155375 | 14:101188 | 14 | 1.01E+08 | C | T | 0.635995 | 0.262309 | 0.032799 | 1.27E-15 | cis |
| miR-382-5p | rs7271582 | 14:101096 | 14 | 1.01E+08 | C | T | 0.817473 | 0.29028 | 0.042985 | 1.45E-11 | cis |
| miR-382-5p | rs1162093 | 14:101151 | 14 | 1.01E+08 | C | G | 0.338205 | -0.28742 | 0.033985 | 2.74E-17 | cis |
| miR-382-5p | rs7142683 | 14:101158 | 14 | 1.01E+08 | A | G | 0.290748 | -0.33134 | 0.035757 | 1.92E-20 | cis |
| miR-382-5p | rs5771269 | 14:101161 | 14 | 1.01E+08 | G | A | 0.894439 | 0.373589 | 0.053608 | 3.19E-12 | cis |
| miR-382-5p | rs7141110 | 14:101172 | 14 | 1.01E+08 | G | C | 0.783235 | 0.286492 | 0.040086 | 8.88E-13 | cis |
| miR-409-3p | rs1190876 | 14:101071 | 14 | 1.01E+08 | A | T | 0.353354 | -0.24103 | 0.033015 | 2.86E-13 | cis |
| miR-409-3p | rs1184761 | 14:101023 | 14 | 1.01E+08 | C | T | 0.753476 | 0.258867 | 0.037119 | 3.08E-12 | trans |
| miR-409-3p | rs1757758 | 14:101189 | 14 | 1.01E+08 | C | T | 0.827094 | 0.340337 | 0.040563 | 4.84E-17 | cis |
| miR-409-3p | rs873076 | 14:101070 | 14 | 1.01E+08 | A | G | 0.311961 | -0.28991 | 0.03357 | 5.81E-18 | cis |
| miR-409-3p | rs7147982 | 14:101046 | 14 | 1.01E+08 | C | T | 0.731753 | 0.28735 | 0.034406 | 6.73E-17 | cis |
| miR-409-3p | rs5993909 | 14:101169 | 14 | 1.01E+08 | C | T | 0.892837 | 0.406883 | 0.050393 | 6.79E-16 | cis |
| miR-409-3p | rs1190862 | 14:101067 | 14 | 1.01E+08 | T | G | 0.309628 | -0.29078 | 0.033438 | 3.43E-18 | cis |
| miR-409-3p | rs1288281 | 14:101183 | 14 | 1.01E+08 | C | G | 0.331398 | -0.36268 | 0.032287 | 2.80E-29 | cis |
| miR-409-3p | rs4905998 | 14:101181 | 14 | 1.01E+08 | A | G | 0.330615 | -0.36337 | 0.032258 | 1.97E-29 | cis |
| miR-409-3p | rs879059 | 14:101069 | 14 | 1.01E+08 | T | A | 0.31068 | -0.29175 | 0.03362 | 4.03E-18 | cis |
| miR-409-3p | rs8008285 | 14:101146 | 14 | 1.01E+08 | T | C | 0.842905 | 0.332888 | 0.043228 | 1.35E-14 | cis |
| miR-409-3p | rs7154024 | 14:101031 | 14 | 1.01E+08 | G | T | 0.725015 | 0.278312 | 0.03458 | 8.39E-16 | cis |
| miR-409-3p | rs7271582 | 14:101090 | 14 | 1.01E+08 | C | T | 0.93848 | 0.501542 | 0.067817 | 1.41E-13 | cis |
| miR-409-3p | rs7263167 | 14:101176 | 14 | 1.01E+08 | C | T | 0.889487 | 0.408556 | 0.049286 | 1.14E-16 | cis |

| | | | | | | | | | | | |
|---|---|---|---|---|---|---|---|---|---|---|---|
| miR-409-3p | rs1555407 | 14:101086 | 14 | 1.01E+08 | A | G | 0.331383 | -0.2408 | 0.034173 | 1.84E-12 | cis |
| miR-409-3p | rs1116059 | 14:101049 | 14 | 1.01E+08 | C | T | 0.741662 | 0.307143 | 0.035006 | 1.72E-18 | cis |
| miR-409-3p | rs1214700 | 14:101186 | 14 | 1.01E+08 | C | T | 0.223221 | -0.35214 | 0.03687 | 1.28E-21 | cis |
| miR-409-3p | rs1190903 | 14:101096 | 14 | 1.01E+08 | C | T | 0.772566 | 0.293624 | 0.037477 | 4.70E-15 | cis |
| miR-409-3p | rs1243369 | 14:101035 | 14 | 1.01E+08 | C | G | 0.723403 | 0.275821 | 0.034386 | 1.05E-15 | cis |
| miR-409-3p | rs1190867 | 14:101069 | 14 | 1.01E+08 | G | T | 0.316788 | -0.29269 | 0.03388 | 5.68E-18 | cis |
| miR-409-3p | rs2837918 | 14:101168 | 14 | 1.01E+08 | G | T | 0.836434 | 0.389638 | 0.041675 | 8.81E-21 | cis |
| miR-409-3p | rs7435633 | 14:101233 | 14 | 1.01E+08 | G | A | 0.942651 | 0.479819 | 0.070389 | 9.31E-12 | cis |
| miR-409-3p | rs6575791 | 14:101029 | 14 | 1.01E+08 | G | A | 0.719841 | 0.277286 | 0.034612 | 1.14E-15 | trans |
| miR-409-3p | rs1288263 | 14:101079 | 14 | 1.01E+08 | C | T | 0.354196 | -0.22096 | 0.032861 | 1.77E-11 | cis |
| miR-409-3p | rs1243517 | 14:101155 | 14 | 1.01E+08 | G | A | 0.312183 | -0.3695 | 0.033809 | 8.37E-28 | cis |
| miR-409-3p | rs1190850 | 14:101064 | 14 | 1.01E+08 | A | G | 0.314302 | -0.2972 | 0.03389 | 1.79E-18 | cis |
| miR-409-3p | rs7271583 | 14:101108 | 14 | 1.01E+08 | A | G | 0.929368 | 0.436852 | 0.063942 | 8.37E-12 | cis |
| miR-409-3p | rs1888581 | 14:101042 | 14 | 1.01E+08 | A | C | 0.735363 | 0.291849 | 0.03465 | 3.68E-17 | cis |
| miR-409-3p | rs7143357 | 14:101054 | 14 | 1.01E+08 | T | C | 0.739636 | 0.303185 | 0.035052 | 5.17E-18 | cis |
| miR-409-3p | rs1190904 | 14:101096 | 14 | 1.01E+08 | A | G | 0.772596 | 0.293462 | 0.037475 | 4.84E-15 | cis |
| miR-409-3p | rs1467536 | 14:101061 | 14 | 1.01E+08 | A | G | 0.318958 | -0.2991 | 0.033747 | 7.78E-19 | cis |
| miR-409-3p | rs2150518 | 14:101063 | 14 | 1.01E+08 | C | T | 0.318506 | -0.29761 | 0.033852 | 1.48E-18 | cis |
| miR-409-3p | rs7493817 | 14:101038 | 14 | 1.01E+08 | G | A | 0.751723 | 0.308992 | 0.035838 | 6.58E-18 | cis |
| miR-409-3p | rs7145337 | 14:101155 | 14 | 1.01E+08 | C | T | 0.312091 | -0.36953 | 0.0338 | 8.04E-28 | cis |
| miR-409-3p | rs1190872 | 14:101069 | 14 | 1.01E+08 | A | G | 0.310177 | -0.29146 | 0.033639 | 4.54E-18 | cis |
| miR-409-3p | rs9671394 | 14:101159 | 14 | 1.01E+08 | C | T | 0.284232 | -0.38828 | 0.03475 | 5.50E-29 | cis |
| miR-409-3p | rs879060 | 14:101069 | 14 | 1.01E+08 | A | G | 0.310926 | -0.29271 | 0.033638 | 3.27E-18 | cis |
| miR-409-3p | rs7155649 | 14:101181 | 14 | 1.01E+08 | C | A | 0.863608 | 0.338672 | 0.046086 | 2.00E-13 | cis |
| miR-409-3p | rs5910018 | 14:101184 | 14 | 1.01E+08 | G | A | 0.836014 | 0.332589 | 0.042873 | 8.66E-15 | cis |
| miR-409-3p | rs1190909 | 14:101101 | 14 | 1.01E+08 | G | A | 0.775531 | 0.287211 | 0.037936 | 3.70E-14 | cis |
| miR-409-3p | rs1190906 | 14:101099 | 14 | 1.01E+08 | T | C | 0.772138 | 0.287716 | 0.037368 | 1.37E-14 | cis |
| miR-409-3p | rs7153876 | 14:101077 | 14 | 1.01E+08 | G | A | 0.344769 | -0.23064 | 0.033203 | 3.75E-12 | cis |
| miR-409-3p | rs1014438 | 14:101173 | 14 | 1.01E+08 | C | T | 0.673446 | 0.364068 | 0.032442 | 3.17E-29 | cis |
| miR-409-3p | rs1190852 | 14:101064 | 14 | 1.01E+08 | A | G | 0.310295 | -0.2936 | 0.033696 | 2.95E-18 | cis |
| miR-409-3p | rs6702222 | 14:101191 | 14 | 1.01E+08 | A | T | 0.731016 | 0.243642 | 0.036058 | 1.41E-11 | cis |
| miR-409-3p | rs7269872 | 14:101189 | 14 | 1.01E+08 | C | T | 0.825721 | 0.335891 | 0.040335 | 8.26E-17 | cis |
| miR-409-3p | rs2183632 | 14:101063 | 14 | 1.01E+08 | C | T | 0.319851 | -0.29696 | 0.033812 | 1.60E-18 | cis |
| miR-409-3p | rs1190856 | 14:101065 | 14 | 1.01E+08 | T | C | 0.310244 | -0.29269 | 0.033671 | 3.54E-18 | cis |
| miR-409-3p | rs1288176 | 14:101176 | 14 | 1.01E+08 | G | C | 0.347791 | -0.37661 | 0.032675 | 9.76E-31 | cis |
| miR-409-3p | rs4905977 | 14:101061 | 14 | 1.01E+08 | C | G | 0.318473 | -0.29913 | 0.033797 | 8.69E-19 | cis |
| miR-409-3p | rs7269872 | 14:101193 | 14 | 1.01E+08 | T | A | 0.879081 | 0.326878 | 0.048744 | 2.00E-11 | cis |
| miR-409-3p | rs879058 | 14:101069 | 14 | 1.01E+08 | G | T | 0.311807 | -0.29189 | 0.033638 | 4.05E-18 | cis |
| miR-409-3p | rs873077 | 14:101070 | 14 | 1.01E+08 | G | A | 0.312104 | -0.28983 | 0.033576 | 6.03E-18 | cis |
| miR-409-3p | rs4905978 | 14:101061 | 14 | 1.01E+08 | C | T | 0.318452 | -0.2991 | 0.033797 | 8.76E-19 | cis |
| miR-409-3p | rs1184948 | 14:101037 | 14 | 1.01E+08 | C | G | 0.733365 | 0.296938 | 0.034724 | 1.22E-17 | cis |
| miR-409-3p | rs7154135 | 14:101077 | 14 | 1.01E+08 | C | A | 0.353569 | -0.22545 | 0.033188 | 1.10E-11 | cis |
| miR-409-3p | rs1184658 | 14:101017 | 14 | 1.01E+08 | C | G | 0.759077 | 0.275362 | 0.037678 | 2.71E-13 | trans |
| miR-409-3p | rs1185095 | 14:101056 | 14 | 1.01E+08 | G | T | 0.743981 | 0.304632 | 0.034836 | 2.24E-18 | cis |
| miR-409-3p | rs1289818 | 14:101078 | 14 | 1.01E+08 | T | G | 0.352585 | -0.22064 | 0.032918 | 2.05E-11 | cis |
| miR-409-3p | rs5922882 | 14:101185 | 14 | 1.01E+08 | G | C | 0.745519 | 0.287992 | 0.035829 | 9.14E-16 | cis |
| miR-409-3p | rs1014551 | 14:101168 | 14 | 1.01E+08 | A | G | 0.668173 | 0.355771 | 0.032244 | 2.63E-28 | cis |
| miR-409-3p | rs7142089 | 14:101172 | 14 | 1.01E+08 | T | A | 0.783402 | 0.301888 | 0.038824 | 7.49E-15 | cis |
| miR-409-3p | rs7140792 | 14:101172 | 14 | 1.01E+08 | C | T | 0.892517 | 0.404584 | 0.050225 | 7.92E-16 | cis |
| miR-409-3p | rs1184764 | 14:101023 | 14 | 1.01E+08 | C | T | 0.753952 | 0.257884 | 0.03716 | 3.93E-12 | trans |
| miR-409-3p | rs7141210 | 14:101182 | 14 | 1.01E+08 | T | C | 0.32902 | -0.36183 | 0.032181 | 2.49E-29 | cis |
| miR-409-3p | rs1014844 | 14:101026 | 14 | 1.01E+08 | G | A | 0.753241 | 0.259752 | 0.037014 | 2.26E-12 | trans |
| miR-409-3p | rs4243729 | 14:101066 | 14 | 1.01E+08 | A | C | 0.311439 | -0.28953 | 0.033421 | 4.59E-18 | cis |
| miR-409-3p | rs5892242 | 14:101097 | 14 | 1.01E+08 | C | G | 0.817892 | 0.307036 | 0.041508 | 1.39E-13 | cis |
| miR-409-3p | rs1555405 | 14:101176 | 14 | 1.01E+08 | G | A | 0.749075 | 0.306581 | 0.034993 | 1.93E-18 | cis |
| miR-409-3p | rs1289379 | 14:101178 | 14 | 1.01E+08 | C | G | 0.190813 | -0.42036 | 0.039594 | 2.49E-26 | cis |
| miR-409-3p | rs7141110 | 14:101172 | 14 | 1.01E+08 | G | C | 0.783235 | 0.296095 | 0.038793 | 2.30E-14 | cis |
| miR-409-3p | rs4905981 | 14:101081 | 14 | 1.01E+08 | C | T | 0.354168 | -0.22133 | 0.032864 | 1.64E-11 | cis |
| miR-409-3p | rs1015151 | 14:101175 | 14 | 1.01E+08 | G | A | 0.806743 | 0.282968 | 0.038714 | 2.69E-13 | cis |
| miR-409-3p | rs7271582 | 14:101096 | 14 | 1.01E+08 | C | T | 0.817473 | 0.307921 | 0.041579 | 1.30E-13 | cis |
| miR-409-3p | rs1162093 | 14:101151 | 14 | 1.01E+08 | C | G | 0.338205 | -0.32778 | 0.032732 | 1.32E-23 | cis |
| miR-409-3p | rs1014839 | 14:101026 | 14 | 1.01E+08 | A | G | 0.750738 | 0.259716 | 0.037027 | 2.31E-12 | trans |
| miR-409-3p | rs1190857 | 14:101066 | 14 | 1.01E+08 | G | T | 0.310273 | -0.29255 | 0.033678 | 3.74E-18 | cis |

| | | | | | | | | | | | |
|---|---|---|---|---|---|---|---|---|---|---|---|
| miR-409-3p | rs61993004 | 14:1010303 | 14 | 1.01E+08 | G | A | 0.724196 | 0.277295 | 0.034505 | 9.26E-16 | trans |
| miR-409-3p | rs12588120 | 14:1011623 | 14 | 1.01E+08 | T | C | 0.893709 | 0.386003 | 0.051855 | 9.78E-14 | cis |
| miR-409-3p | rs66695430 | 14:1010751 | 14 | 1.01E+08 | C | A | 0.349961 | -0.23216 | 0.033129 | 2.42E-12 | cis |
| miR-409-3p | rs8021301 | 14:1011669 | 14 | 1.01E+08 | G | C | 0.668324 | 0.356893 | 0.032281 | 2.05E-28 | cis |
| miR-409-3p | rs4905976 | 14:1010600 | 14 | 1.01E+08 | G | A | 0.322862 | -0.29011 | 0.033871 | 1.08E-17 | cis |
| miR-409-3p | rs57712693 | 14:1011613 | 14 | 1.01E+08 | G | A | 0.894439 | 0.384606 | 0.051887 | 1.24E-13 | cis |
| miR-409-3p | rs6575794 | 14:1010397 | 14 | 1.01E+08 | C | T | 0.734189 | 0.296437 | 0.034778 | 1.54E-17 | cis |
| miR-409-3p | rs4905999 | 14:101184 | 14 | 1.01E+08 | A | G | 0.181905 | -0.40763 | 0.040344 | 5.31E-24 | cis |
| miR-409-3p | rs8012970 | 14:1011684 | 14 | 1.01E+08 | T | C | 0.347902 | -0.35854 | 0.032846 | 9.69E-28 | cis |
| miR-409-3p | rs1190905 | 14:1010970 | 14 | 1.01E+08 | G | T | 0.817561 | 0.306166 | 0.041451 | 1.51E-13 | cis |
| miR-409-3p | rs77420215 | 14:1011510 | 14 | 1.01E+08 | C | T | 0.945071 | 0.583367 | 0.07259 | 9.25E-16 | cis |
| miR-409-3p | rs12436130 | 14:1010310 | 14 | 1.01E+08 | C | T | 0.725015 | 0.278583 | 0.03461 | 8.34E-16 | trans |
| miR-409-3p | rs8020868 | 14:1011669 | 14 | 1.01E+08 | A | G | 0.668343 | 0.356811 | 0.032278 | 2.09E-28 | cis |
| miR-409-3p | rs61993005 | 14:101034 | 14 | 1.01E+08 | G | A | 0.726044 | 0.279189 | 0.034609 | 7.21E-16 | cis |
| miR-409-3p | rs10136153 | 14:1010771 | 14 | 1.01E+08 | C | A | 0.344768 | -0.23064 | 0.033205 | 3.76E-12 | cis |
| miR-409-3p | rs58702304 | 14:1011671 | 14 | 1.01E+08 | G | A | 0.892706 | 0.406822 | 0.050452 | 7.42E-16 | cis |
| miR-409-3p | rs61307974 | 14:1011771 | 14 | 1.01E+08 | C | T | 0.888723 | 0.408882 | 0.049346 | 1.17E-16 | cis |
| miR-409-3p | rs4905994 | 14:1011540 | 14 | 1.01E+08 | T | C | 0.329146 | -0.34586 | 0.033412 | 4.12E-25 | cis |
| miR-409-3p | rs2776544 | 14:1010540 | 14 | 1.01E+08 | T | G | 0.396251 | -0.22709 | 0.033322 | 9.42E-12 | cis |
| miR-409-3p | rs7492369 | 14:1010208 | 14 | 1.01E+08 | C | T | 0.755698 | 0.257176 | 0.037225 | 4.89E-12 | trans |
| miR-409-3p | rs911324 | 14:1010839 | 14 | 1.01E+08 | C | A | 0.342046 | -0.22504 | 0.032695 | 5.86E-12 | cis |
| miR-409-3p | rs34700570 | 14:1011734 | 14 | 1.01E+08 | C | T | 0.892725 | 0.404351 | 0.050175 | 7.70E-16 | cis |
| miR-409-3p | rs67625670 | 14:1011730 | 14 | 1.01E+08 | A | G | 0.892632 | 0.404168 | 0.050149 | 7.67E-16 | cis |
| miR-409-3p | rs9324022 | 14:1011724 | 14 | 1.01E+08 | T | C | 0.763739 | 0.282318 | 0.038026 | 1.13E-13 | cis |
| miR-409-3p | rs17099614 | 14:1011670 | 14 | 1.01E+08 | C | G | 0.668809 | 0.358671 | 0.032282 | 1.11E-28 | cis |
| miR-409-3p | rs10139403 | 14:1011945 | 14 | 1.01E+08 | A | G | 0.323897 | -0.26497 | 0.03363 | 3.30E-15 | cis |
| miR-409-3p | rs10146962 | 14:1011705 | 14 | 1.01E+08 | T | C | 0.658875 | 0.356863 | 0.031706 | 2.18E-29 | cis |
| miR-409-3p | rs12588713 | 14:1011490 | 14 | 1.01E+08 | G | C | 0.347269 | -0.32898 | 0.032443 | 3.67E-24 | cis |
| miR-409-3p | rs7140556 | 14:1010364 | 14 | 1.01E+08 | C | A | 0.723818 | 0.280094 | 0.034528 | 4.98E-16 | cis |
| miR-409-3p | rs4900471 | 14:1011620 | 14 | 1.01E+08 | G | A | 0.304173 | -0.36033 | 0.033882 | 2.05E-26 | cis |
| miR-409-3p | rs72631673 | 14:1011870 | 14 | 1.01E+08 | C | T | 0.881829 | 0.407022 | 0.049215 | 1.34E-16 | cis |
| miR-409-3p | rs732565 | 14:1010768 | 14 | 1.01E+08 | G | A | 0.349911 | -0.2314 | 0.033 | 2.35E-12 | cis |
| miR-409-3p | rs1190907 | 14:1010993 | 14 | 1.01E+08 | A | G | 0.772847 | 0.289277 | 0.037424 | 1.08E-14 | cis |
| miR-409-3p | rs59825373 | 14:1011680 | 14 | 1.01E+08 | G | T | 0.89241 | 0.407638 | 0.050405 | 6.10E-16 | cis |
| miR-409-3p | rs11622193 | 14:101016 | 14 | 1.01E+08 | G | A | 0.759852 | 0.296621 | 0.039304 | 4.46E-14 | trans |
| miR-409-3p | rs7145610 | 14:1010494 | 14 | 1.01E+08 | T | C | 0.740391 | 0.307972 | 0.035097 | 1.71E-18 | cis |
| miR-409-3p | rs12434490 | 14:1011793 | 14 | 1.01E+08 | A | G | 0.18959 | -0.41952 | 0.039564 | 2.87E-26 | cis |
| miR-409-3p | rs7155375 | 14:1011887 | 14 | 1.01E+08 | C | T | 0.635995 | 0.276943 | 0.031705 | 2.43E-18 | cis |
| miR-409-3p | rs12897601 | 14:1011823 | 14 | 1.01E+08 | C | T | 0.190452 | -0.41904 | 0.03935 | 1.76E-26 | cis |
| miR-409-3p | rs7142683 | 14:1011588 | 14 | 1.01E+08 | A | G | 0.290748 | -0.37859 | 0.034391 | 3.48E-28 | cis |
| miR-409-3p | rs7149242 | 14:1011594 | 14 | 1.01E+08 | T | G | 0.288371 | -0.37747 | 0.034293 | 3.53E-28 | cis |
| miR-409-3p | rs12590933 | 14:1011630 | 14 | 1.01E+08 | A | T | 0.893649 | 0.38628 | 0.051858 | 9.42E-14 | cis |
| miR-409-3p | rs72698723 | 14:1011813 | 14 | 1.01E+08 | C | T | 0.808936 | 0.271522 | 0.039203 | 4.33E-12 | cis |
| miR-409-3p | rs1190881 | 14:1010798 | 14 | 1.01E+08 | T | C | 0.346494 | -0.22274 | 0.032893 | 1.27E-11 | cis |
| miR-409-3p | rs10873520 | 14:1011759 | 14 | 1.01E+08 | G | A | 0.809826 | 0.284259 | 0.039175 | 3.98E-13 | cis |
| miR-409-3p | rs911330 | 14:1010997 | 14 | 1.01E+08 | T | C | 0.774615 | 0.292431 | 0.037604 | 7.45E-15 | cis |
| miR-409-3p | rs60307965 | 14:1010274 | 14 | 1.01E+08 | C | T | 0.725784 | 0.29191 | 0.035841 | 3.80E-16 | trans |
| miR-409-3p | rs73358778 | 14:1011599 | 14 | 1.01E+08 | C | T | 0.891417 | 0.380225 | 0.051479 | 1.51E-13 | cis |
| miR-409-3p | rs60013593 | 14:1010215 | 14 | 1.01E+08 | C | T | 0.755543 | 0.258793 | 0.037255 | 3.74E-12 | trans |
| miR-409-3p | rs12881545 | 14:1011763 | 14 | 1.01E+08 | G | C | 0.370047 | -0.37648 | 0.033164 | 7.24E-30 | cis |
| miR-431-5p | rs4900471 | 14:1011620 | 14 | 1.01E+08 | G | A | 0.304173 | -0.25201 | 0.033359 | 4.21E-14 | cis |
| miR-431-5p | rs10144381 | 14:1011733 | 14 | 1.01E+08 | C | T | 0.673446 | 0.277728 | 0.031897 | 3.12E-18 | cis |
| miR-431-5p | rs12882815 | 14:101183 | 14 | 1.01E+08 | C | G | 0.331398 | -0.27383 | 0.031756 | 6.53E-18 | cis |
| miR-431-5p | rs8020868 | 14:1011669 | 14 | 1.01E+08 | A | G | 0.668343 | 0.235066 | 0.031862 | 1.61E-13 | cis |
| miR-431-5p | rs10145513 | 14:1011683 | 14 | 1.01E+08 | A | G | 0.668173 | 0.234361 | 0.031827 | 1.79E-13 | cis |
| miR-431-5p | rs7149242 | 14:1011594 | 14 | 1.01E+08 | T | G | 0.288371 | -0.25024 | 0.033841 | 1.42E-13 | cis |
| miR-431-5p | rs9671394 | 14:1011590 | 14 | 1.01E+08 | C | T | 0.284232 | -0.25976 | 0.034299 | 3.63E-14 | cis |
| miR-431-5p | rs12435171 | 14:1011551 | 14 | 1.01E+08 | G | A | 0.312183 | -0.25451 | 0.033323 | 2.22E-14 | cis |
| miR-431-5p | rs12881545 | 14:1011763 | 14 | 1.01E+08 | G | C | 0.370047 | -0.29185 | 0.032596 | 3.44E-19 | cis |
| miR-431-5p | rs8012970 | 14:1011684 | 14 | 1.01E+08 | T | C | 0.347902 | -0.23584 | 0.032412 | 3.44E-13 | cis |
| miR-431-5p | rs12434490 | 14:1011793 | 14 | 1.01E+08 | A | G | 0.18959 | -0.26938 | 0.03903 | 5.13E-12 | cis |
| miR-431-5p | rs12588713 | 14:1011490 | 14 | 1.01E+08 | G | C | 0.347269 | -0.21392 | 0.031959 | 2.18E-11 | cis |
| miR-431-5p | rs7142683 | 14:1011588 | 14 | 1.01E+08 | A | G | 0.290748 | -0.2502 | 0.03394 | 1.68E-13 | cis |

| | | | | | | | | | | | |
|---|---|---|---|---|---|---|---|---|---|---|---|
| miR-431-5p | rs8021301 | 14:101166 | 14 | 1.01E+08 | G | C | 0.668324 | 0.235108 | 0.031865 | 1.60E-13 | cis |
| miR-431-5p | rs1289379 | 14:101178 | 14 | 1.01E+08 | C | G | 0.190813 | -0.27089 | 0.039058 | 4.05E-12 | cis |
| miR-431-5p | rs4905999 | 14:101184 | 14 | 1.01E+08 | A | G | 0.181905 | -0.27513 | 0.039706 | 4.23E-12 | cis |
| miR-431-5p | rs7141210 | 14:101182 | 14 | 1.01E+08 | T | C | 0.32902 | -0.27413 | 0.031649 | 4.65E-18 | cis |
| miR-431-5p | rs1289760 | 14:101182 | 14 | 1.01E+08 | C | T | 0.190452 | -0.27629 | 0.0388 | 1.07E-12 | cis |
| miR-431-5p | rs1288176 | 14:101176 | 14 | 1.01E+08 | G | C | 0.347791 | -0.27611 | 0.03219 | 9.69E-18 | cis |
| miR-431-5p | rs1709961 | 14:101167 | 14 | 1.01E+08 | C | G | 0.668809 | 0.238542 | 0.031863 | 7.07E-14 | cis |
| miR-431-5p | rs4905998 | 14:101181 | 14 | 1.01E+08 | A | G | 0.330615 | -0.27423 | 0.031731 | 5.51E-18 | cis |
| miR-431-5p | rs7145337 | 14:101155 | 14 | 1.01E+08 | C | T | 0.312091 | -0.25478 | 0.033314 | 2.05E-14 | cis |
| miR-431-5p | rs4905994 | 14:101154 | 14 | 1.01E+08 | T | C | 0.329146 | -0.23067 | 0.032912 | 2.40E-12 | cis |
| miR-431-5p | rs1014696 | 14:101170 | 14 | 1.01E+08 | T | C | 0.658875 | 0.258244 | 0.03123 | 1.35E-16 | cis |
| miR-432-5p | rs7141210 | 14:101182 | 14 | 1.01E+08 | T | C | 0.32902 | -0.23621 | 0.027837 | 2.15E-17 | cis |
| miR-432-5p | rs1289379 | 14:101178 | 14 | 1.01E+08 | C | G | 0.190813 | -0.24936 | 0.034294 | 3.56E-13 | cis |
| miR-432-5p | rs7145337 | 14:101155 | 14 | 1.01E+08 | C | T | 0.312091 | -0.21893 | 0.0293 | 7.89E-14 | cis |
| miR-432-5p | rs1289760 | 14:101182 | 14 | 1.01E+08 | C | T | 0.190452 | -0.24735 | 0.034089 | 3.98E-13 | cis |
| miR-432-5p | rs4905999 | 14:101184 | 14 | 1.01E+08 | A | G | 0.181905 | -0.23808 | 0.034912 | 9.13E-12 | cis |
| miR-432-5p | rs9671394 | 14:101159 | 14 | 1.01E+08 | C | T | 0.284232 | -0.21504 | 0.030193 | 1.06E-12 | cis |
| miR-432-5p | rs1258871 | 14:101149 | 14 | 1.01E+08 | G | C | 0.347269 | -0.20907 | 0.028022 | 8.58E-14 | cis |
| miR-432-5p | rs9324022 | 14:101172 | 14 | 1.01E+08 | T | C | 0.763739 | 0.218021 | 0.032576 | 2.19E-11 | cis |
| miR-432-5p | rs7269871 | 14:101173 | 14 | 1.01E+08 | G | T | 0.783221 | 0.217853 | 0.032013 | 1.01E-11 | cis |
| miR-432-5p | rs1014438 | 14:101173 | 14 | 1.01E+08 | C | T | 0.673446 | 0.24004 | 0.028052 | 1.16E-17 | cis |
| miR-432-5p | rs1014696 | 14:101170 | 14 | 1.01E+08 | T | C | 0.658875 | 0.222658 | 0.027466 | 5.20E-16 | cis |
| miR-432-5p | rs8021301 | 14:101166 | 14 | 1.01E+08 | G | C | 0.668324 | 0.221507 | 0.027955 | 2.31E-15 | cis |
| miR-432-5p | rs1709961 | 14:101167 | 14 | 1.01E+08 | C | G | 0.668809 | 0.222003 | 0.027962 | 2.03E-15 | cis |
| miR-432-5p | rs4905994 | 14:101154 | 14 | 1.01E+08 | T | C | 0.329146 | -0.21402 | 0.028891 | 1.28E-13 | cis |
| miR-432-5p | rs1709962 | 14:101173 | 14 | 1.01E+08 | A | C | 0.783449 | 0.217512 | 0.032009 | 1.08E-11 | cis |
| miR-432-5p | rs1288176 | 14:101176 | 14 | 1.01E+08 | G | C | 0.347791 | -0.2438 | 0.028289 | 6.82E-18 | cis |
| miR-432-5p | rs1243449 | 14:101179 | 14 | 1.01E+08 | A | G | 0.18959 | -0.24767 | 0.034271 | 4.94E-13 | cis |
| miR-432-5p | rs7142683 | 14:101158 | 14 | 1.01E+08 | A | G | 0.290748 | -0.21673 | 0.029843 | 3.80E-13 | cis |
| miR-432-5p | rs7148436 | 14:101173 | 14 | 1.01E+08 | G | C | 0.786324 | 0.217699 | 0.032331 | 1.66E-11 | cis |
| miR-432-5p | rs4905998 | 14:101181 | 14 | 1.01E+08 | A | G | 0.330615 | -0.23608 | 0.02791 | 2.71E-17 | cis |
| miR-432-5p | rs1162093 | 14:101151 | 14 | 1.01E+08 | C | G | 0.338205 | -0.20425 | 0.028277 | 5.07E-13 | cis |
| miR-432-5p | rs4900471 | 14:101162 | 14 | 1.01E+08 | G | A | 0.304173 | -0.21884 | 0.029331 | 8.57E-14 | cis |
| miR-432-5p | rs7155375 | 14:101188 | 14 | 1.01E+08 | C | T | 0.635995 | 0.186573 | 0.027278 | 7.93E-12 | cis |
| miR-432-5p | rs7149242 | 14:101159 | 14 | 1.01E+08 | T | G | 0.288371 | -0.21645 | 0.029757 | 3.49E-13 | cis |
| miR-432-5p | rs1288154 | 14:101176 | 14 | 1.01E+08 | G | C | 0.370047 | -0.24744 | 0.028687 | 6.38E-18 | cis |
| miR-432-5p | rs1288281 | 14:101183 | 14 | 1.01E+08 | C | G | 0.331398 | -0.23608 | 0.027931 | 2.86E-17 | cis |
| miR-432-5p | rs8012970 | 14:101168 | 14 | 1.01E+08 | T | C | 0.347902 | -0.22299 | 0.028434 | 4.42E-15 | cis |
| miR-432-5p | rs8020868 | 14:101166 | 14 | 1.01E+08 | A | G | 0.668343 | 0.221473 | 0.027953 | 2.32E-15 | cis |
| miR-432-5p | rs1243517 | 14:101155 | 14 | 1.01E+08 | G | A | 0.312183 | -0.21856 | 0.029308 | 8.83E-14 | cis |
| miR-432-5p | rs1014551 | 14:101168 | 14 | 1.01E+08 | A | G | 0.668173 | 0.221036 | 0.027922 | 2.45E-15 | cis |
| miR-433-3p | rs1709961 | 14:101167 | 14 | 1.01E+08 | C | G | 0.668809 | 0.201794 | 0.028951 | 3.17E-12 | cis |
| miR-433-3p | rs1014438 | 14:101173 | 14 | 1.01E+08 | C | T | 0.673446 | 0.219739 | 0.029054 | 3.93E-14 | cis |
| miR-433-3p | rs8020868 | 14:101166 | 14 | 1.01E+08 | A | G | 0.668343 | 0.199579 | 0.028947 | 5.40E-12 | cis |
| miR-433-3p | rs1014551 | 14:101168 | 14 | 1.01E+08 | A | G | 0.668173 | 0.19846 | 0.028916 | 6.73E-12 | cis |
| miR-433-3p | rs1214700 | 14:101186 | 14 | 1.01E+08 | C | T | 0.223221 | -0.24514 | 0.032778 | 7.50E-14 | cis |
| miR-433-3p | rs8021301 | 14:101166 | 14 | 1.01E+08 | G | C | 0.668324 | 0.199652 | 0.028949 | 5.32E-12 | cis |
| miR-433-3p | rs7155375 | 14:101188 | 14 | 1.01E+08 | C | T | 0.635995 | 0.197083 | 0.028136 | 2.48E-12 | cis |
| miR-433-3p | rs4905999 | 14:101184 | 14 | 1.01E+08 | A | G | 0.181905 | -0.24163 | 0.036042 | 2.03E-11 | cis |
| miR-433-3p | rs7155649 | 14:101181 | 14 | 1.01E+08 | C | A | 0.863608 | 0.275895 | 0.040724 | 1.25E-11 | cis |
| miR-433-3p | rs8012970 | 14:101168 | 14 | 1.01E+08 | T | C | 0.347902 | -0.20011 | 0.029446 | 1.08E-11 | cis |
| miR-433-3p | rs1288154 | 14:101176 | 14 | 1.01E+08 | G | C | 0.370047 | -0.23417 | 0.029686 | 3.06E-15 | cis |
| miR-433-3p | rs7141210 | 14:101182 | 14 | 1.01E+08 | T | C | 0.32902 | -0.21412 | 0.028837 | 1.13E-13 | cis |
| miR-433-3p | rs1288281 | 14:101183 | 14 | 1.01E+08 | C | G | 0.331398 | -0.21423 | 0.028932 | 1.32E-13 | cis |
| miR-433-3p | rs1014696 | 14:101170 | 14 | 1.01E+08 | T | C | 0.658875 | 0.208041 | 0.028422 | 2.49E-13 | cis |
| miR-433-3p | rs1288176 | 14:101176 | 14 | 1.01E+08 | G | C | 0.347791 | -0.22114 | 0.029308 | 4.51E-14 | cis |
| miR-433-3p | rs1289379 | 14:101178 | 14 | 1.01E+08 | C | G | 0.190813 | -0.2534 | 0.035405 | 8.23E-13 | cis |
| miR-433-3p | rs4905998 | 14:101181 | 14 | 1.01E+08 | A | G | 0.330615 | -0.21372 | 0.028912 | 1.45E-13 | cis |
| miR-433-3p | rs1289760 | 14:101182 | 14 | 1.01E+08 | C | T | 0.190452 | -0.24967 | 0.035198 | 1.31E-12 | cis |
| miR-433-3p | rs1243449 | 14:101179 | 14 | 1.01E+08 | A | G | 0.18959 | -0.25164 | 0.03538 | 1.14E-12 | cis |
| miR-4429 | rs6214393 | 2:6864038 | 2 | 68640381 | G | C | 0.593598 | 0.109647 | 0.013593 | 7.24E-16 | trans |
| miR-4429 | rs7572299 | 2:6858124 | 2 | 68581246 | T | A | 0.538033 | 0.107773 | 0.013264 | 4.46E-16 | trans |
| miR-4429 | rs1164012 | 2:6862562 | 2 | 68625625 | C | T | 0.545777 | 0.114323 | 0.012888 | 7.30E-19 | trans |

| | | | | | | | | | | | |
|---|---|---|---|---|---|---|---|---|---|---|---|
| miR-4429 | rs1158434 | 2:6863552 | 2 | 68635524 | G | C | 0.551072 | 0.118342 | 0.013237 | 3.88E-19 | trans |
| miR-4429 | rs1486061 | 2:6862571 | 2 | 68625717 | T | G | 0.54743 | 0.115007 | 0.012931 | 5.91E-19 | trans |
| miR-4429 | rs6214393 | 2:6864020 | 2 | 68640202 | A | G | 0.593308 | 0.109092 | 0.013598 | 1.04E-15 | trans |
| miR-4429 | rs6546395 | 2:6862927 | 2 | 68629271 | A | G | 0.55448 | 0.116327 | 0.013078 | 5.85E-19 | trans |
| miR-4429 | rs7590433 | 2:6862850 | 2 | 68628507 | A | G | 0.554708 | 0.118314 | 0.013131 | 2.05E-19 | trans |
| miR-4429 | rs1301028 | 2:6862184 | 2 | 68621842 | G | T | 0.683506 | 0.105886 | 0.013707 | 1.12E-14 | trans |
| miR-4429 | rs1300974 | 2:6858443 | 2 | 68584437 | C | A | 0.678986 | 0.096595 | 0.01431 | 1.48E-11 | trans |
| miR-4429 | rs6546410 | 2:6863610 | 2 | 68636105 | C | G | 0.702434 | 0.109252 | 0.014591 | 7.02E-14 | trans |
| miR-4429 | rs1049616 | 2:6862008 | 2 | 68620089 | C | T | 0.697496 | 0.104328 | 0.013941 | 7.23E-14 | trans |
| miR-4429 | rs6214393 | 2:6864009 | 2 | 68640090 | T | C | 0.59325 | 0.109114 | 0.013597 | 1.02E-15 | trans |
| miR-4429 | rs6546407 | 2:6863492 | 2 | 68634920 | A | C | 0.55091 | 0.118316 | 0.013221 | 3.58E-19 | trans |
| miR-4429 | rs6546400 | 2:6863064 | 2 | 68630648 | G | A | 0.554382 | 0.116519 | 0.01309 | 5.51E-19 | trans |
| miR-4429 | rs5933292 | 2:6863963 | 2 | 68639631 | G | A | 0.73529 | 0.107748 | 0.015329 | 2.08E-12 | trans |
| miR-4429 | rs6714719 | 2:6859377 | 2 | 68593773 | G | C | 0.698404 | 0.104022 | 0.014003 | 1.10E-13 | trans |
| miR-4429 | rs1017438 | 2:6859646 | 2 | 68596469 | C | T | 0.685243 | 0.105793 | 0.013758 | 1.48E-14 | trans |
| miR-4429 | rs6546399 | 2:6863064 | 2 | 68630646 | C | T | 0.55394 | 0.116432 | 0.013078 | 5.44E-19 | trans |
| miR-4429 | rs6546404 | 2:6863263 | 2 | 68632635 | G | T | 0.54995 | 0.117873 | 0.013142 | 3.00E-19 | trans |
| miR-4429 | rs6546398 | 2:6863041 | 2 | 68630417 | G | C | 0.552757 | 0.116069 | 0.013039 | 5.51E-19 | trans |
| miR-4429 | rs5618821 | 2:6861163 | 2 | 68611631 | T | C | 0.698075 | 0.104494 | 0.013944 | 6.70E-14 | trans |
| miR-4429 | rs1760168 | 2:6861589 | 2 | 68615899 | G | A | 0.697578 | 0.104385 | 0.01395 | 7.27E-14 | trans |
| miR-4429 | rs1300519 | 2:6863887 | 2 | 68638873 | A | G | 0.702186 | 0.109146 | 0.014585 | 7.25E-14 | trans |
| miR-4429 | rs1484530 | 2:6863547 | 2 | 68635471 | A | G | 0.70445 | 0.109845 | 0.014643 | 6.31E-14 | trans |
| miR-4429 | rs6757938 | 2:6863285 | 2 | 68632858 | A | G | 0.549958 | 0.117883 | 0.013145 | 3.02E-19 | trans |
| miR-4429 | rs5690424 | 2:6863847 | 2 | 68638477 | T | C | 0.550669 | 0.118152 | 0.013239 | 4.48E-19 | trans |
| miR-4429 | rs6546411 | 2:6863769 | 2 | 68637699 | A | G | 0.556248 | 0.120734 | 0.013319 | 1.25E-19 | trans |
| miR-4429 | rs7608429 | 2:6858083 | 2 | 68580836 | G | T | 0.678719 | 0.09611 | 0.014323 | 1.94E-11 | trans |
| miR-4429 | rs7598360 | 2:6863092 | 2 | 68630925 | A | G | 0.553166 | 0.11928 | 0.013065 | 6.86E-20 | trans |
| miR-4429 | rs6546392 | 2:6862744 | 2 | 68627441 | A | G | 0.555235 | 0.116707 | 0.0131 | 5.15E-19 | trans |
| miR-4429 | rs1373200 | 2:6862265 | 2 | 68622658 | C | T | 0.547142 | 0.112939 | 0.012754 | 8.34E-19 | trans |
| miR-4429 | rs6546408 | 2:6863504 | 2 | 68635048 | T | C | 0.551062 | 0.118342 | 0.013234 | 3.82E-19 | trans |
| miR-4429 | rs6419643 | 2:6863126 | 2 | 68631260 | T | G | 0.548826 | 0.116437 | 0.013006 | 3.47E-19 | trans |
| miR-4429 | rs1302892 | 2:6862991 | 2 | 68629916 | C | T | 0.552648 | 0.115995 | 0.013035 | 5.65E-19 | trans |
| miR-4429 | rs6546412 | 2:6863771 | 2 | 68637710 | T | C | 0.557086 | 0.121849 | 0.013344 | 6.76E-20 | trans |
| miR-4429 | rs1188499 | 2:6864573 | 2 | 68645737 | T | C | 0.593791 | 0.108839 | 0.01363 | 1.41E-15 | trans |
| miR-4429 | rs6214392 | 2:6863835 | 2 | 68638359 | C | G | 0.700378 | 0.109847 | 0.014575 | 4.82E-14 | trans |
| miR-4429 | rs1300592 | 2:6862999 | 2 | 68629998 | T | A | 0.701261 | 0.106555 | 0.014352 | 1.13E-13 | trans |
| miR-4429 | rs6546397 | 2:6863036 | 2 | 68630367 | G | A | 0.552756 | 0.116061 | 0.01304 | 5.55E-19 | trans |
| miR-4429 | rs1760209 | 2:6862041 | 2 | 68620410 | T | A | 0.697529 | 0.104351 | 0.01394 | 7.12E-14 | trans |
| miR-4429 | rs1455505 | 2:6862832 | 2 | 68628329 | G | A | 0.556221 | 0.118259 | 0.013138 | 2.23E-19 | trans |
| miR-4429 | rs6546393 | 2:6862757 | 2 | 68627575 | T | C | 0.554017 | 0.117707 | 0.013065 | 2.08E-19 | trans |
| miR-4429 | rs2241947 | 2:6861539 | 2 | 68615395 | C | T | 0.697988 | 0.104503 | 0.013944 | 6.65E-14 | trans |
| miR-4429 | rs5888448 | 2:6864534 | 2 | 68645347 | G | A | 0.734214 | 0.105803 | 0.015237 | 3.82E-12 | trans |
| miR-4429 | rs6709464 | 2:6862222 | 2 | 68622220 | C | T | 0.697279 | 0.10404 | 0.013928 | 8.03E-14 | trans |
| miR-4429 | rs1050181 | 2:6862295 | 2 | 68622952 | G | C | 0.544992 | 0.113852 | 0.012842 | 7.63E-19 | trans |
| miR-4429 | rs6744306 | 2:6863273 | 2 | 68632736 | T | C | 0.549956 | 0.117881 | 0.013143 | 2.99E-19 | trans |
| miR-4429 | rs1301157 | 2:6863927 | 2 | 68639273 | G | A | 0.702349 | 0.109115 | 0.01459 | 7.50E-14 | trans |
| miR-4429 | rs6214389 | 2:6860455 | 2 | 68604553 | C | T | 0.698307 | 0.103959 | 0.013964 | 9.70E-14 | trans |
| miR-4429 | rs5999861 | 2:6864677 | 2 | 68646771 | G | T | 0.731295 | 0.101746 | 0.015226 | 2.35E-11 | trans |
| miR-4429 | rs1442589 | 2:6863473 | 2 | 68634732 | G | T | 0.701896 | 0.108994 | 0.014556 | 6.99E-14 | trans |
| miR-4429 | rs1422402 | 2:6862752 | 2 | 68627521 | A | G | 0.698498 | 0.105646 | 0.014255 | 1.25E-13 | trans |
| miR-4429 | rs1474969 | 2:6862568 | 2 | 68625686 | G | A | 0.546544 | 0.114556 | 0.012908 | 6.98E-19 | trans |
| miR-4429 | rs7592526 | 2:6858352 | 2 | 68583528 | A | C | 0.538007 | 0.108083 | 0.013262 | 3.64E-16 | trans |
| miR-4429 | rs6214393 | 2:6864062 | 2 | 68640621 | T | C | 0.733261 | 0.106445 | 0.015174 | 2.30E-12 | trans |
| miR-4429 | rs4610041 | 2:6862628 | 2 | 68626280 | C | T | 0.550745 | 0.116935 | 0.013037 | 2.99E-19 | trans |
| miR-4429 | rs1397159 | 2:6863425 | 2 | 68634258 | T | G | 0.705552 | 0.110694 | 0.014727 | 5.62E-14 | trans |
| miR-4429 | rs6743970 | 2:6858390 | 2 | 68583907 | A | T | 0.660237 | 0.101973 | 0.014137 | 5.46E-13 | trans |
| miR-4429 | rs6729686 | 2:6863303 | 2 | 68633030 | C | T | 0.700171 | 0.108074 | 0.014448 | 7.41E-14 | trans |
| miR-4429 | rs6729555 | 2:6863295 | 2 | 68632952 | C | A | 0.54995 | 0.117896 | 0.013146 | 3.01E-19 | trans |
| miR-4429 | rs7597167 | 2:6861294 | 2 | 68612946 | C | T | 0.698051 | 0.104514 | 0.013944 | 6.62E-14 | trans |
| miR-4429 | rs3485223 | 2:6863859 | 2 | 68638599 | C | G | 0.550658 | 0.118152 | 0.01324 | 4.50E-19 | trans |
| miR-4429 | rs7424269 | 2:6862912 | 2 | 68629125 | A | G | 0.563426 | 0.118034 | 0.013244 | 5.01E-19 | trans |
| miR-4429 | rs1303482 | 2:6863029 | 2 | 68630297 | C | G | 0.552715 | 0.116045 | 0.013038 | 5.56E-19 | trans |
| miR-4429 | rs6715579 | 2:6858392 | 2 | 68583923 | C | T | 0.678794 | 0.096668 | 0.014309 | 1.42E-11 | trans |

| | | | | | | | | | | | |
|---|---|---|---|---|---|---|---|---|---|---|---|
| miR-4429 | rs1542135 | 2:6862084 | 2 | 68620848 | C | T | 0.69752 | 0.104351 | 0.01394 | 7.11E-14 | trans |
| miR-4429 | rs1542136 | 2:6862093 | 2 | 68620930 | C | A | 0.697594 | 0.104377 | 0.013944 | 7.12E-14 | trans |
| miR-4429 | rs7581872 | 2:6858098 | 2 | 68580981 | A | C | 0.537959 | 0.107684 | 0.013264 | 4.71E-16 | trans |
| miR-4429 | rs2165920 | 2:6862223 | 2 | 68622239 | T | C | 0.552012 | 0.112932 | 0.012844 | 1.47E-18 | trans |
| miR-4429 | rs7605387 | 2:6863255 | 2 | 68632550 | A | G | 0.703499 | 0.108179 | 0.014531 | 9.72E-14 | trans |
| miR-4429 | rs7451452 | 2:6863755 | 2 | 68637550 | T | G | 0.702552 | 0.109286 | 0.014597 | 7.07E-14 | trans |
| miR-4429 | rs6546401 | 2:6863087 | 2 | 68630873 | A | T | 0.550127 | 0.116694 | 0.013034 | 3.45E-19 | trans |
| miR-4429 | rs3419682 | 2:6862142 | 2 | 68621427 | G | A | 0.697518 | 0.104352 | 0.01394 | 7.10E-14 | trans |
| miR-4429 | rs1760149 | 2:6861171 | 2 | 68611712 | A | G | 0.69807 | 0.104495 | 0.013944 | 6.70E-14 | trans |
| miR-4429 | rs2070171 | 2:6861371 | 2 | 68613710 | G | A | 0.698024 | 0.104489 | 0.013944 | 6.71E-14 | trans |
| miR-4429 | rs5628470 | 2:6863137 | 2 | 68631372 | G | A | 0.697508 | 0.106313 | 0.014265 | 9.15E-14 | trans |
| miR-4429 | rs6546403 | 2:6863226 | 2 | 68632262 | C | T | 0.550801 | 0.118031 | 0.013158 | 2.95E-19 | trans |
| miR-4429 | rs2241946 | 2:6861565 | 2 | 68615654 | T | C | 0.698076 | 0.104588 | 0.01395 | 6.51E-14 | trans |
| miR-4429 | rs7800275 | 2:6862543 | 2 | 68625436 | A | G | 0.546309 | 0.114214 | 0.012873 | 7.16E-19 | trans |
| miR-4429 | rs1156629 | 2:6862663 | 2 | 68626631 | T | G | 0.547487 | 0.115419 | 0.012956 | 5.17E-19 | trans |
| miR-4429 | rs6546394 | 2:6862909 | 2 | 68629094 | T | C | 0.566701 | 0.11724 | 0.013284 | 1.09E-18 | trans |
| miR-4429 | rs6546409 | 2:6863587 | 2 | 68635879 | A | G | 0.551093 | 0.118334 | 0.013241 | 4.00E-19 | trans |
| miR-4429 | rs7421228 | 2:6863012 | 2 | 68630124 | G | T | 0.55268 | 0.116024 | 0.013037 | 5.60E-19 | trans |
| miR-4429 | rs6715909 | 2:6858399 | 2 | 68583995 | G | A | 0.664879 | 0.098726 | 0.014073 | 2.30E-12 | trans |
| miR-4429 | rs9653552 | 2:6864396 | 2 | 68643966 | G | A | 0.593458 | 0.109389 | 0.013605 | 8.96E-16 | trans |
| miR-4429 | rs8761 | 2:6862426 | 2 | 68624260 | G | A | 0.545692 | 0.114212 | 0.012884 | 7.65E-19 | trans |
| miR-4429 | rs1302897 | 2:6863003 | 2 | 68630038 | A | G | 0.54849 | 0.116303 | 0.012997 | 3.60E-19 | trans |
| miR-4429 | rs3732044 | 2:6862391 | 2 | 68623915 | T | C | 0.545562 | 0.114139 | 0.012879 | 7.82E-19 | trans |
| miR-4429 | rs6747545 | 2:6863306 | 2 | 68633069 | T | A | 0.549964 | 0.11789 | 0.013147 | 3.04E-19 | trans |
| miR-4429 | rs6000716 | 2:6863963 | 2 | 68639636 | C | T | 0.735163 | 0.107772 | 0.015327 | 2.04E-12 | trans |
| miR-4429 | rs7562439 | 2:6858256 | 2 | 68582569 | C | T | 0.6646 | 0.098629 | 0.014077 | 2.44E-12 | trans |
| miR-4429 | rs6708536 | 2:6862726 | 2 | 68627268 | G | A | 0.547249 | 0.115405 | 0.012966 | 5.55E-19 | trans |
| miR-4429 | rs6419644 | 2:6863182 | 2 | 68631823 | A | G | 0.549898 | 0.117848 | 0.013138 | 2.96E-19 | trans |
| miR-4429 | rs1301143 | 2:6863937 | 2 | 68639379 | C | T | 0.702382 | 0.109113 | 0.014591 | 7.54E-14 | trans |
| miR-4429 | rs6742200 | 2:6862330 | 2 | 68623302 | A | G | 0.54552 | 0.114117 | 0.012877 | 7.84E-19 | trans |
| miR-4429 | rs1382899 | 2:6863782 | 2 | 68637829 | A | G | 0.702614 | 0.109295 | 0.014601 | 7.13E-14 | trans |
| miR-4429 | rs6087844 | 2:6858211 | 2 | 68582115 | A | G | 0.538067 | 0.108084 | 0.013261 | 3.62E-16 | trans |
| miR-4429 | rs7564214 | 2:6862869 | 2 | 68628696 | C | T | 0.569906 | 0.117322 | 0.013433 | 2.47E-18 | trans |
| miR-4429 | rs1390688 | 2:6862773 | 2 | 68627735 | C | T | 0.69721 | 0.105551 | 0.014218 | 1.14E-13 | trans |
| miR-4429 | rs1301077 | 2:6863908 | 2 | 68639081 | C | T | 0.702339 | 0.109105 | 0.014589 | 7.52E-14 | trans |
| miR-4429 | rs5633714 | 2:6864249 | 2 | 68642490 | G | A | 0.733417 | 0.106385 | 0.015183 | 2.44E-12 | trans |
| miR-4429 | rs7578602 | 2:6863786 | 2 | 68637868 | G | C | 0.556806 | 0.121802 | 0.01335 | 7.26E-20 | trans |
| miR-4429 | rs5808991 | 2:6863966 | 2 | 68639660 | G | C | 0.734634 | 0.10805 | 0.015286 | 1.57E-12 | trans |
| miR-4429 | rs1753019 | 2:6862133 | 2 | 68621338 | G | T | 0.683745 | 0.106181 | 0.013718 | 9.92E-15 | trans |
| miR-4429 | rs8028813 | 2:6862547 | 2 | 68625478 | A | G | 0.548418 | 0.11582 | 0.012961 | 4.03E-19 | trans |
| miR-4429 | rs7590450 | 2:6862856 | 2 | 68628563 | A | G | 0.549532 | 0.115985 | 0.012996 | 4.47E-19 | trans |
| miR-4429 | rs7578358 | 2:6862872 | 2 | 68628724 | T | C | 0.561556 | 0.117418 | 0.013232 | 7.09E-19 | trans |
| miR-4429 | rs1343065 | 2:6858771 | 2 | 68587710 | C | G | 0.699397 | 0.103919 | 0.014114 | 1.80E-13 | trans |
| miR-4429 | rs7420775 | 2:6862945 | 2 | 68629458 | G | T | 0.703122 | 0.107093 | 0.014416 | 1.10E-13 | trans |
| miR-4429 | rs1301107 | 2:6863931 | 2 | 68639313 | A | C | 0.700844 | 0.109516 | 0.014613 | 6.66E-14 | trans |
| miR-4429 | rs6715706 | 2:6858399 | 2 | 68583996 | C | A | 0.664887 | 0.098723 | 0.014073 | 2.30E-12 | trans |
| miR-4429 | rs6214393 | 2:6864041 | 2 | 68640415 | A | G | 0.593599 | 0.10964 | 0.013593 | 7.28E-16 | trans |
| miR-4433b | rs1426700 | 2:6456529 | 2 | 64565294 | G | A | 0.694437 | -0.08662 | 0.012902 | 1.89E-11 | cis |
| miR-4433b | rs1247741 | 2:6456768 | 2 | 64567680 | T | G | 0.733039 | 0.092647 | 0.013438 | 5.40E-12 | cis |
| miR-4433b | rs1300822 | 2:6457412 | 2 | 64574128 | G | A | 0.716086 | -0.08885 | 0.013143 | 1.38E-11 | cis |
| miR-4433b | rs1271350 | 2:6456984 | 2 | 64569840 | C | T | 0.714645 | -0.09125 | 0.013166 | 4.19E-12 | cis |
| miR-4433b | rs7598238 | 2:6455647 | 2 | 64556476 | G | A | 0.733848 | 0.095046 | 0.013564 | 2.43E-12 | cis |
| miR-4433b | rs2059631 | 2:6457468 | 2 | 64574682 | G | A | 0.560261 | -0.08164 | 0.012057 | 1.28E-11 | cis |
| miR-4433b | rs1247320 | 2:6456791 | 2 | 64567916 | C | G | 0.734857 | 0.09263 | 0.01353 | 7.58E-12 | cis |
| miR-4433b | rs7591530 | 2:6456755 | 2 | 64567550 | T | G | 0.68461 | -0.08729 | 0.01269 | 6.06E-12 | cis |
| miR-4707- | rs5782274 | 14:2340996 | 14 | 23409967 | G | A | 0.440759 | 0.093412 | 0.013618 | 6.91E-12 | cis |
| miR-4707- | rs6197774 | 14:2340097 | 14 | 23400979 | G | C | 0.440733 | 0.091819 | 0.013701 | 2.06E-11 | cis |
| miR-4707- | rs4982714 | 14:2341333 | 14 | 23413333 | C | T | 0.441229 | 0.093545 | 0.013601 | 6.08E-12 | cis |
| miR-4707- | rs5589413 | 14:2340883 | 14 | 23408837 | C | G | 0.443456 | 0.093971 | 0.013657 | 5.95E-12 | cis |
| miR-4707- | rs1258953 | 14:2339668 | 14 | 23396680 | T | C | 0.440305 | 0.091556 | 0.013703 | 2.37E-11 | cis |
| miR-4707- | rs3561518 | 14:2340438 | 14 | 23404389 | T | C | 0.441508 | 0.093065 | 0.013697 | 1.09E-11 | cis |
| miR-4707- | rs1243715 | 14:233729 | 14 | 23372913 | G | A | 0.437486 | 0.091279 | 0.013659 | 2.35E-11 | cis |
| miR-4707- | rs5618074 | 14:2337724 | 14 | 23377246 | T | C | 0.438286 | 0.091634 | 0.013678 | 2.10E-11 | cis |

| | | | | | | | | | | | |
|---|---|---|---|---|---|---|---|---|---|---|---|
| miR-4707-3 | rs4981451 | 14:2341028 | 14 | 23410281 | T | C | 0.441163 | 0.093487 | 0.013604 | 6.33E-12 | cis |
| miR-4707-3 | rs712486 | 14:2342504 | 14 | 23425041 | T | C | 0.441215 | 0.094678 | 0.013749 | 5.73E-12 | cis |
| miR-4707-3 | rs1923 | 14:2344117 | 14 | 23441179 | G | A | 0.569614 | -0.09073 | 0.013467 | 1.61E-11 | cis |
| miR-4707-3 | rs35085068 | 14:2340990 | 14 | 23409909 | C | A | 0.441051 | 0.09349 | 0.01361 | 6.45E-12 | cis |
| miR-4707-3 | rs4982713 | 14:2340410 | 14 | 23404102 | C | T | 0.441538 | 0.092987 | 0.013695 | 1.12E-11 | cis |
| miR-4707-3 | rs4981452 | 14:2341080 | 14 | 23410809 | A | G | 0.440913 | 0.093512 | 0.013614 | 6.47E-12 | cis |
| miR-4707-3 | rs8016027 | 14:2342015 | 14 | 23420153 | T | C | 0.441165 | 0.093615 | 0.013591 | 5.67E-12 | cis |
| miR-4707-3 | rs34496309 | 14:2340074 | 14 | 23400744 | G | C | 0.43983 | 0.093304 | 0.013786 | 1.30E-11 | cis |
| miR-4707-3 | rs34584578 | 14:2341169 | 14 | 23411690 | C | T | 0.44118 | 0.093538 | 0.013603 | 6.14E-12 | cis |
| miR-4707-3 | rs4536383 | 14:2341092 | 14 | 23410926 | T | C | 0.436129 | 0.091252 | 0.013624 | 2.11E-11 | cis |
| miR-4707-3 | rs2295682 | 14:2337486 | 14 | 23374862 | C | T | 0.43795 | 0.091553 | 0.013662 | 2.07E-11 | cis |
| miR-4707-3 | rs12884614 | 14:2340943 | 14 | 23409431 | A | G | 0.441135 | 0.093412 | 0.013605 | 6.60E-12 | cis |
| miR-4707-3 | rs2295680 | 14:2337443 | 14 | 23374435 | G | A | 0.437854 | 0.091485 | 0.013662 | 2.13E-11 | cis |
| miR-487b-3 | rs10145513 | 14:1011683 | 14 | 1.01E+08 | A | G | 0.668173 | 0.250766 | 0.031862 | 3.54E-15 | cis |
| miR-487b-3 | rs7155375 | 14:1011887 | 14 | 1.01E+08 | C | T | 0.635995 | 0.213013 | 0.031122 | 7.67E-12 | cis |
| miR-487b-3 | rs7142683 | 14:1011588 | 14 | 1.01E+08 | A | G | 0.290748 | -0.25189 | 0.034033 | 1.35E-13 | cis |
| miR-487b-3 | rs17577580 | 14:1011893 | 14 | 1.01E+08 | C | T | 0.827094 | 0.317512 | 0.039609 | 1.09E-15 | cis |
| miR-487b-3 | rs12881760 | 14:1011762 | 14 | 1.01E+08 | G | C | 0.347791 | -0.26701 | 0.03232 | 1.44E-16 | cis |
| miR-487b-3 | rs7145337 | 14:1011555 | 14 | 1.01E+08 | C | T | 0.312091 | -0.24879 | 0.033433 | 9.96E-14 | cis |
| miR-487b-3 | rs4905999 | 14:101184 | 14 | 1.01E+08 | A | G | 0.181905 | -0.33881 | 0.039594 | 1.16E-17 | cis |
| miR-487b-3 | rs8020868 | 14:1011669 | 14 | 1.01E+08 | A | G | 0.668343 | 0.251944 | 0.031895 | 2.81E-15 | cis |
| miR-487b-3 | rs1190814 | 14:1012275 | 14 | 1.01E+08 | T | C | 0.819011 | 0.259289 | 0.038786 | 2.31E-11 | cis |
| miR-487b-3 | rs12893790 | 14:1011788 | 14 | 1.01E+08 | C | G | 0.190813 | -0.31617 | 0.039015 | 5.33E-16 | cis |
| miR-487b-3 | rs12588718 | 14:1011490 | 14 | 1.01E+08 | G | C | 0.347269 | -0.22157 | 0.032027 | 4.58E-12 | cis |
| miR-487b-3 | rs9671394 | 14:1011590 | 14 | 1.01E+08 | C | T | 0.284232 | -0.25972 | 0.034399 | 4.34E-14 | cis |
| miR-487b-3 | rs8012970 | 14:1011684 | 14 | 1.01E+08 | T | C | 0.347902 | -0.24978 | 0.032458 | 1.41E-14 | cis |
| miR-487b-3 | rs28379180 | 14:1011680 | 14 | 1.01E+08 | G | T | 0.836434 | 0.294888 | 0.040964 | 6.08E-13 | cis |
| miR-487b-3 | rs12147008 | 14:101186 | 14 | 1.01E+08 | C | T | 0.223221 | -0.29654 | 0.036145 | 2.32E-16 | cis |
| miR-487b-3 | rs10144381 | 14:1011732 | 14 | 1.01E+08 | C | T | 0.673446 | 0.263683 | 0.032045 | 1.90E-16 | cis |
| miR-487b-3 | rs58540528 | 14:1012340 | 14 | 1.01E+08 | G | A | 0.822954 | 0.267534 | 0.039457 | 1.20E-11 | cis |
| miR-487b-3 | rs77420215 | 14:1011510 | 14 | 1.01E+08 | C | T | 0.945071 | 0.486945 | 0.071068 | 7.29E-12 | cis |
| miR-487b-3 | rs12435171 | 14:1011552 | 14 | 1.01E+08 | G | A | 0.312183 | -0.24856 | 0.033441 | 1.06E-13 | cis |
| miR-487b-3 | rs8021301 | 14:1011669 | 14 | 1.01E+08 | G | C | 0.668324 | 0.252033 | 0.031898 | 2.76E-15 | cis |
| miR-487b-3 | rs78894391 | 14:1012380 | 14 | 1.01E+08 | G | A | 0.827329 | 0.27051 | 0.040379 | 2.09E-11 | cis |
| miR-487b-3 | rs12882815 | 14:101183 | 14 | 1.01E+08 | C | G | 0.331398 | -0.26503 | 0.031884 | 9.38E-17 | cis |
| miR-487b-3 | rs4905998 | 14:1011817 | 14 | 1.01E+08 | A | G | 0.330615 | -0.26502 | 0.03186 | 8.93E-17 | cis |
| miR-487b-3 | rs1190815 | 14:1012283 | 14 | 1.01E+08 | G | A | 0.798481 | 0.253014 | 0.037527 | 1.56E-11 | cis |
| miR-487b-3 | rs12881545 | 14:1011762 | 14 | 1.01E+08 | G | C | 0.370047 | -0.28051 | 0.032737 | 1.05E-17 | cis |
| miR-487b-3 | rs10139403 | 14:1011943 | 14 | 1.01E+08 | A | G | 0.323897 | -0.2277 | 0.032899 | 4.48E-12 | cis |
| miR-487b-3 | rs4905994 | 14:1011540 | 14 | 1.01E+08 | T | C | 0.329146 | -0.23528 | 0.032992 | 9.94E-13 | cis |
| miR-487b-3 | rs10146962 | 14:1011705 | 14 | 1.01E+08 | T | C | 0.658875 | 0.253837 | 0.031338 | 5.49E-16 | cis |
| miR-487b-3 | rs12434490 | 14:1011793 | 14 | 1.01E+08 | A | G | 0.18959 | -0.31605 | 0.038982 | 5.16E-16 | cis |
| miR-487b-3 | rs72698727 | 14:1011890 | 14 | 1.01E+08 | C | T | 0.825721 | 0.314239 | 0.039383 | 1.47E-15 | cis |
| miR-487b-3 | rs11620937 | 14:1011510 | 14 | 1.01E+08 | C | G | 0.338205 | -0.22063 | 0.032303 | 8.49E-12 | cis |
| miR-487b-3 | rs7149242 | 14:1011594 | 14 | 1.01E+08 | T | G | 0.288371 | -0.25135 | 0.033936 | 1.30E-13 | cis |
| miR-487b-3 | rs7141210 | 14:1011824 | 14 | 1.01E+08 | T | C | 0.32902 | -0.26494 | 0.031778 | 7.62E-17 | cis |
| miR-487b-3 | rs17099614 | 14:1011670 | 14 | 1.01E+08 | C | G | 0.668809 | 0.254897 | 0.031897 | 1.34E-15 | cis |
| miR-487b-3 | rs12897601 | 14:1011822 | 14 | 1.01E+08 | C | T | 0.190452 | -0.31881 | 0.038763 | 1.96E-16 | cis |
| miR-487b-3 | rs4900471 | 14:1011620 | 14 | 1.01E+08 | G | A | 0.304173 | -0.24277 | 0.033488 | 4.19E-13 | cis |
| miR-494-3p | rs12882815 | 14:101183 | 14 | 1.01E+08 | C | G | 0.331398 | -0.17124 | 0.0256 | 2.25E-11 | cis |
| miR-494-3p | rs10144381 | 14:1011732 | 14 | 1.01E+08 | C | T | 0.673446 | 0.171904 | 0.025722 | 2.34E-11 | cis |
| miR-494-3p | rs12881545 | 14:1011762 | 14 | 1.01E+08 | G | C | 0.370047 | -0.18609 | 0.026279 | 1.43E-12 | cis |
| miR-494-3p | rs12881760 | 14:1011762 | 14 | 1.01E+08 | G | C | 0.347791 | -0.18034 | 0.025924 | 3.49E-12 | cis |
| miR-494-3p | rs7141210 | 14:1011824 | 14 | 1.01E+08 | T | C | 0.32902 | -0.17124 | 0.025516 | 1.93E-11 | cis |
| miR-494-3p | rs8008285 | 14:1011463 | 14 | 1.01E+08 | T | C | 0.842905 | 0.227618 | 0.033767 | 1.57E-11 | cis |
| miR-495-3p | rs10144381 | 14:1011732 | 14 | 1.01E+08 | C | T | 0.673446 | 0.235869 | 0.027016 | 2.53E-18 | cis |
| miR-495-3p | rs7149242 | 14:1011594 | 14 | 1.01E+08 | T | G | 0.288371 | -0.24075 | 0.028561 | 3.48E-17 | cis |
| miR-495-3p | rs12897601 | 14:1011822 | 14 | 1.01E+08 | C | T | 0.190452 | -0.28866 | 0.032665 | 9.85E-19 | cis |
| miR-495-3p | rs12588718 | 14:1011490 | 14 | 1.01E+08 | G | C | 0.347269 | -0.2102 | 0.026974 | 6.56E-15 | cis |
| miR-495-3p | rs12147008 | 14:101186 | 14 | 1.01E+08 | C | T | 0.223221 | -0.23682 | 0.030581 | 9.62E-15 | cis |
| miR-495-3p | rs7141210 | 14:1011824 | 14 | 1.01E+08 | T | C | 0.32902 | -0.23354 | 0.026803 | 2.96E-18 | cis |
| miR-495-3p | rs7155649 | 14:1011810 | 14 | 1.01E+08 | C | A | 0.863608 | 0.255264 | 0.038037 | 1.93E-11 | cis |
| miR-495-3p | rs7155375 | 14:1011887 | 14 | 1.01E+08 | C | T | 0.635995 | 0.178454 | 0.026292 | 1.14E-11 | cis |

| | | | | | | | | | | | |
|---|---|---|---|---|---|---|---|---|---|---|---|
| miR-495-3 | rs4905998 | 14:101181 | 14 | 1.01E+08 | A | G | 0.330615 | -0.23362 | 0.026873 | 3.51E-18 | cis |
| miR-495-3 | rs1243449 | 14:101179 | 14 | 1.01E+08 | A | G | 0.18959 | -0.29311 | 0.032824 | 4.27E-19 | cis |
| miR-495-3 | rs4905999 | 14:101184 | 14 | 1.01E+08 | A | G | 0.181905 | -0.2863 | 0.033443 | 1.12E-17 | cis |
| miR-495-3 | rs8012970 | 14:101168 | 14 | 1.01E+08 | T | C | 0.347902 | -0.21992 | 0.027384 | 9.66E-16 | cis |
| miR-495-3 | rs4900471 | 14:101162 | 14 | 1.01E+08 | G | A | 0.304173 | -0.23123 | 0.028193 | 2.37E-16 | cis |
| miR-495-3 | rs2837918 | 14:101168 | 14 | 1.01E+08 | G | T | 0.836434 | 0.239555 | 0.034632 | 4.61E-12 | cis |
| miR-495-3 | rs1288154 | 14:101176 | 14 | 1.01E+08 | G | C | 0.370047 | -0.24905 | 0.027603 | 1.84E-19 | cis |
| miR-495-3 | rs1243517 | 14:101155 | 14 | 1.01E+08 | G | A | 0.312183 | -0.23482 | 0.028156 | 7.43E-17 | cis |
| miR-495-3 | rs8008285 | 14:101146 | 14 | 1.01E+08 | T | C | 0.842905 | 0.238978 | 0.035723 | 2.24E-11 | cis |
| miR-495-3 | rs1014696 | 14:101170 | 14 | 1.01E+08 | T | C | 0.658875 | 0.222781 | 0.026438 | 3.56E-17 | cis |
| miR-495-3 | rs1014551 | 14:101168 | 14 | 1.01E+08 | A | G | 0.668173 | 0.221365 | 0.026877 | 1.78E-16 | cis |
| miR-495-3 | rs1709961 | 14:101167 | 14 | 1.01E+08 | C | G | 0.668809 | 0.224192 | 0.026909 | 7.99E-17 | cis |
| miR-495-3 | rs9671394 | 14:101159 | 14 | 1.01E+08 | C | T | 0.284232 | -0.25067 | 0.028939 | 4.63E-18 | cis |
| miR-495-3 | rs8021301 | 14:101166 | 14 | 1.01E+08 | G | C | 0.668324 | 0.221851 | 0.02691 | 1.66E-16 | cis |
| miR-495-3 | rs1289379 | 14:101178 | 14 | 1.01E+08 | C | G | 0.190813 | -0.29312 | 0.032852 | 4.56E-19 | cis |
| miR-495-3 | rs7145337 | 14:101155 | 14 | 1.01E+08 | C | T | 0.312091 | -0.23516 | 0.028148 | 6.58E-17 | cis |
| miR-495-3 | rs1162093 | 14:101151 | 14 | 1.01E+08 | C | G | 0.338205 | -0.20636 | 0.027219 | 3.41E-14 | cis |
| miR-495-3 | rs7142683 | 14:101158 | 14 | 1.01E+08 | A | G | 0.290748 | -0.24039 | 0.028646 | 4.79E-17 | cis |
| miR-495-3 | rs1757758 | 14:101189 | 14 | 1.01E+08 | C | T | 0.827094 | 0.232608 | 0.033579 | 4.29E-12 | cis |
| miR-495-3 | rs1288281 | 14:101183 | 14 | 1.01E+08 | C | G | 0.331398 | -0.23359 | 0.026893 | 3.76E-18 | cis |
| miR-495-3 | rs7269872 | 14:101189 | 14 | 1.01E+08 | C | T | 0.825721 | 0.229308 | 0.033389 | 6.52E-12 | cis |
| miR-495-3 | rs4905994 | 14:101154 | 14 | 1.01E+08 | T | C | 0.329146 | -0.21961 | 0.027795 | 2.76E-15 | cis |
| miR-495-3 | rs1288176 | 14:101176 | 14 | 1.01E+08 | G | C | 0.347791 | -0.24714 | 0.027213 | 1.07E-19 | cis |
| miR-495-3 | rs8020868 | 14:101166 | 14 | 1.01E+08 | A | G | 0.668343 | 0.22178 | 0.026908 | 1.69E-16 | cis |
| miR-543 | rs1014438 | 14:101173 | 14 | 1.01E+08 | C | T | 0.673446 | 0.250596 | 0.030086 | 8.14E-17 | cis |
| miR-543 | rs7149242 | 14:101159 | 14 | 1.01E+08 | T | G | 0.288371 | -0.24654 | 0.031837 | 9.65E-15 | cis |
| miR-543 | rs7141210 | 14:101182 | 14 | 1.01E+08 | T | C | 0.32902 | -0.25283 | 0.029831 | 2.35E-17 | cis |
| miR-543 | rs4900471 | 14:101162 | 14 | 1.01E+08 | G | A | 0.304173 | -0.25162 | 0.031369 | 1.05E-15 | cis |
| miR-543 | rs1288281 | 14:101183 | 14 | 1.01E+08 | C | G | 0.331398 | -0.25364 | 0.029928 | 2.35E-17 | cis |
| miR-543 | rs8012970 | 14:101168 | 14 | 1.01E+08 | T | C | 0.347902 | -0.2434 | 0.030454 | 1.32E-15 | cis |
| miR-543 | rs1709961 | 14:101167 | 14 | 1.01E+08 | C | G | 0.668809 | 0.247917 | 0.029927 | 1.19E-16 | cis |
| miR-543 | rs1258871 | 14:101149 | 14 | 1.01E+08 | G | C | 0.347269 | -0.20817 | 0.030081 | 4.50E-12 | cis |
| miR-543 | rs1014551 | 14:101168 | 14 | 1.01E+08 | A | G | 0.668173 | 0.24499 | 0.02989 | 2.48E-16 | cis |
| miR-543 | rs7142683 | 14:101158 | 14 | 1.01E+08 | A | G | 0.290748 | -0.24586 | 0.031933 | 1.37E-14 | cis |
| miR-543 | rs7145337 | 14:101155 | 14 | 1.01E+08 | C | T | 0.312091 | -0.24449 | 0.031363 | 6.41E-15 | cis |
| miR-543 | rs1288154 | 14:101176 | 14 | 1.01E+08 | G | C | 0.370047 | -0.26113 | 0.030757 | 2.07E-17 | cis |
| miR-543 | rs1288176 | 14:101176 | 14 | 1.01E+08 | G | C | 0.347791 | -0.25328 | 0.030346 | 7.03E-17 | cis |
| miR-543 | rs8020868 | 14:101166 | 14 | 1.01E+08 | A | G | 0.668343 | 0.245668 | 0.029923 | 2.21E-16 | cis |
| miR-543 | rs8021301 | 14:101166 | 14 | 1.01E+08 | G | C | 0.668324 | 0.245723 | 0.029926 | 2.19E-16 | cis |
| miR-543 | rs1014696 | 14:101170 | 14 | 1.01E+08 | T | C | 0.658875 | 0.244882 | 0.029409 | 8.30E-17 | cis |
| miR-543 | rs1555405 | 14:101176 | 14 | 1.01E+08 | G | A | 0.749075 | 0.22184 | 0.032262 | 6.15E-12 | cis |
| miR-543 | rs9671394 | 14:101159 | 14 | 1.01E+08 | C | T | 0.284232 | -0.25091 | 0.032284 | 7.74E-15 | cis |
| miR-543 | rs4905998 | 14:101181 | 14 | 1.01E+08 | A | G | 0.330615 | -0.25351 | 0.029906 | 2.31E-17 | cis |
| miR-543 | rs1243517 | 14:101155 | 14 | 1.01E+08 | G | A | 0.312183 | -0.24377 | 0.031373 | 7.85E-15 | cis |
| miR-543 | rs4905994 | 14:101154 | 14 | 1.01E+08 | T | C | 0.329146 | -0.22836 | 0.030962 | 1.64E-13 | cis |
| miR-543 | rs7155375 | 14:101188 | 14 | 1.01E+08 | C | T | 0.635995 | 0.202466 | 0.029223 | 4.26E-12 | cis |
| miR-543 | rs1162093 | 14:101151 | 14 | 1.01E+08 | C | G | 0.338205 | -0.20662 | 0.030341 | 9.77E-12 | cis |
| miR-584-5 | rs407696 | 5:1484225 | 5 | 1.48E+08 | A | G | 0.538877 | -0.17143 | 0.025051 | 7.73E-12 | cis |
| miR-584-5 | rs1779527 | 5:1484220 | 5 | 1.48E+08 | G | A | 0.854308 | -0.27333 | 0.035771 | 2.15E-14 | cis |
| miR-584-5 | rs6238010 | 5:1485004 | 5 | 1.49E+08 | G | T | 0.8464 | -0.27664 | 0.035415 | 5.66E-15 | cis |
| miR-584-5 | rs1173937 | 5:1485201 | 5 | 1.49E+08 | C | T | 0.400157 | 0.196896 | 0.026435 | 9.44E-14 | cis |
| miR-584-5 | rs1779544 | 5:1485160 | 5 | 1.49E+08 | G | A | 0.854008 | -0.28011 | 0.036799 | 2.70E-14 | cis |
| miR-584-5 | rs7697787 | 5:1484473 | 5 | 1.48E+08 | A | G | 0.844049 | -0.2714 | 0.034876 | 7.15E-15 | cis |
| miR-584-5 | rs5756988 | 5:1484501 | 5 | 1.48E+08 | A | G | 0.843968 | -0.2715 | 0.034894 | 7.21E-15 | cis |
| miR-584-5 | rs1318269 | 5:1484854 | 5 | 1.48E+08 | A | G | 0.474875 | 0.188851 | 0.025387 | 1.02E-13 | cis |
| miR-584-5 | rs2304034 | 5:1484109 | 5 | 1.48E+08 | A | C | 0.860163 | -0.2696 | 0.03781 | 1.00E-12 | cis |
| miR-584-5 | rs1116808 | 5:1484841 | 5 | 1.48E+08 | T | C | 0.408107 | 0.199694 | 0.02537 | 3.51E-15 | cis |
| miR-584-5 | rs6238010 | 5:1484921 | 5 | 1.48E+08 | C | A | 0.846193 | -0.27661 | 0.035432 | 5.86E-15 | cis |
| miR-584-5 | rs2915806 | 5:1484509 | 5 | 1.48E+08 | C | G | 0.5867 | -0.20673 | 0.025394 | 3.92E-16 | cis |
| miR-584-5 | rs1317416 | 5:1484883 | 5 | 1.48E+08 | T | C | 0.474697 | 0.188995 | 0.025386 | 9.70E-14 | cis |
| miR-584-5 | rs4705317 | 5:1485005 | 5 | 1.49E+08 | G | A | 0.408249 | 0.196678 | 0.025966 | 3.61E-14 | cis |
| miR-584-5 | rs4705305 | 5:1484708 | 5 | 1.48E+08 | G | T | 0.473411 | 0.187444 | 0.025456 | 1.79E-13 | cis |
| miR-584-5 | rs1779537 | 5:1484633 | 5 | 1.48E+08 | G | A | 0.845418 | -0.27472 | 0.035751 | 1.54E-14 | cis |

| | | | | | | | | | | | |
|---|---|---|---|---|---|---|---|---|---|---|---|
| miR-584-5p | rs1779525! | 5:1484169: | 5 | 1.48E+08 | T | C | 0.842839 | -0.26317 | 0.034503 | 2.40E-14 | cis |
| miR-584-5p | rs4705318 | 5:1485006: | 5 | 1.49E+08 | C | T | 0.464335 | 0.181964 | 0.02529 | 6.24E-13 | cis |
| miR-584-5p | rs6879243 | 5:1484732: | 5 | 1.48E+08 | G | C | 0.882379 | -0.30299 | 0.042731 | 1.33E-12 | cis |
| miR-584-5p | rs4705322 | 5:1485089: | 5 | 1.49E+08 | A | T | 0.473913 | 0.187238 | 0.025565 | 2.41E-13 | cis |
| miR-584-5p | rs1265507( | 5:1484855 | 5 | 1.48E+08 | A | G | 0.410692 | 0.201351 | 0.025491 | 2.81E-15 | cis |
| miR-584-5p | rs1044723: | 5:1485009( | 5 | 1.49E+08 | A | C | 0.474342 | 0.186354 | 0.02543 | 2.33E-13 | cis |
| miR-584-5p | rs1422577 | 5:1484863 | 5 | 1.48E+08 | G | T | 0.474749 | 0.18891 | 0.025385 | 9.93E-14 | cis |
| miR-584-5p | rs6861404 | 5:1484158: | 5 | 1.48E+08 | A | C | 0.536756 | -0.17368 | 0.02523 | 5.82E-12 | cis |
| miR-584-5p | rs6873489 | 5:1484926: | 5 | 1.48E+08 | T | C | 0.880859 | -0.31451 | 0.042076 | 7.73E-14 | cis |
| miR-584-5p | rs6873011 | 5:1484141 | 5 | 1.48E+08 | G | A | 0.842515 | -0.25933 | 0.034393 | 4.70E-14 | cis |
| miR-584-5p | rs1432792 | 5:1484149: | 5 | 1.48E+08 | C | T | 0.537202 | -0.17313 | 0.025206 | 6.47E-12 | cis |
| miR-584-5p | rs1173805! | 5:1485017: | 5 | 1.49E+08 | G | A | 0.473692 | 0.186237 | 0.02546 | 2.57E-13 | cis |
| miR-584-5p | rs7705960 | 5:1484430: | 5 | 1.48E+08 | T | C | 0.452092 | 0.207512 | 0.025894 | 1.11E-15 | cis |
| miR-584-5p | rs36081 | 5:1484290( | 5 | 1.48E+08 | A | G | 0.540101 | -0.17308 | 0.024956 | 4.05E-12 | cis |
| miR-584-5p | rs1422579 | 5:1484847: | 5 | 1.48E+08 | C | A | 0.509261 | 0.191642 | 0.026152 | 2.34E-13 | cis |
| miR-584-5p | rs1112745 | 5:1484718( | 5 | 1.48E+08 | G | A | 0.474305 | 0.187933 | 0.025488 | 1.66E-13 | cis |
| miR-584-5p | rs7846146( | 5:1484304! | 5 | 1.48E+08 | C | T | 0.843889 | -0.26844 | 0.034455 | 6.64E-15 | cis |
| miR-584-5p | rs6580593 | 5:1484636( | 5 | 1.48E+08 | G | C | 0.474061 | 0.188578 | 0.025475 | 1.34E-13 | cis |
| miR-584-5p | rs36047 | 5:1484413: | 5 | 1.48E+08 | C | T | 0.650304 | -0.23018 | 0.025932 | 6.90E-19 | cis |
| miR-584-5p | rs6874343 | 5:1484478( | 5 | 1.48E+08 | C | T | 0.844049 | -0.27141 | 0.034877 | 7.14E-15 | cis |
| miR-584-5p | rs1174699( | 5:1484994( | 5 | 1.48E+08 | T | C | 0.435388 | 0.185153 | 0.025551 | 4.28E-13 | cis |
| miR-584-5p | rs36077 | 5:1484319( | 5 | 1.48E+08 | G | A | 0.53902 | -0.17273 | 0.024829 | 3.48E-12 | cis |
| miR-584-5p | rs1046341: | 5:1484550: | 5 | 1.48E+08 | A | G | 0.844169 | -0.27268 | 0.034992 | 6.56E-15 | cis |
| miR-584-5p | rs6238006( | 5:1484422! | 5 | 1.48E+08 | C | T | 0.844027 | -0.27096 | 0.034834 | 7.34E-15 | cis |
| miR-584-5p | rs1174620( | 5:1484993: | 5 | 1.48E+08 | A | G | 0.473634 | 0.184438 | 0.025138 | 2.18E-13 | cis |
| miR-584-5p | rs7442677 | 5:1484293( | 5 | 1.48E+08 | C | T | 0.856689 | -0.27882 | 0.036018 | 9.87E-15 | cis |
| miR-584-5p | rs2915808 | 5:1484517! | 5 | 1.48E+08 | A | G | 0.586438 | -0.20555 | 0.025332 | 4.90E-16 | cis |
| miR-584-5p | rs6866101 | 5:1484159: | 5 | 1.48E+08 | T | C | 0.537249 | -0.17328 | 0.025199 | 6.14E-12 | cis |
| miR-625-5p | rs1003401 | 14:662377: | 14 | 66237720 | G | A | 0.386875 | 0.147199 | 0.019801 | 1.05E-13 | cis |
| miR-625-5p | rs1004787! | 14:657605 | 14 | 65760543 | C | A | 0.650175 | -0.21369 | 0.019842 | 4.80E-27 | cis |
| miR-625-5p | rs7733938! | 14:657580! | 14 | 65758097 | G | A | 0.84644 | -0.30998 | 0.02677 | 5.23E-31 | cis |
| miR-625-5p | rs7271246 | 14:657562: | 14 | 65756281 | A | T | 0.847244 | -0.32205 | 0.027281 | 3.68E-32 | cis |
| miR-625-5p | rs7271247: | 14:657771: | 14 | 65777124 | G | A | 0.846937 | -0.32457 | 0.026357 | 7.60E-35 | cis |
| miR-625-5p | rs7271248! | 14:657895: | 14 | 65789584 | A | T | 0.845779 | -0.32668 | 0.026156 | 8.48E-36 | cis |
| miR-625-5p | rs7271249! | 14:658023: | 14 | 65802338 | T | G | 0.845101 | -0.33788 | 0.026191 | 4.47E-38 | cis |
| miR-625-5p | rs7271250( | 14:658043: | 14 | 65804328 | G | A | 0.810519 | -0.23397 | 0.024476 | 1.19E-21 | cis |
| miR-625-5p | rs4073415 | 14:662595: | 14 | 66259530 | G | A | 0.538747 | -0.13833 | 0.019261 | 6.88E-13 | cis |
| miR-625-5p | rs1382362: | 14:658470: | 14 | 65847058 | C | T | 0.888755 | -0.35922 | 0.031703 | 9.26E-30 | cis |
| miR-625-5p | rs1815654 | 14:658552: | 14 | 65855232 | C | T | 0.393509 | 0.198121 | 0.019588 | 4.77E-24 | cis |
| miR-625-5p | rs1262272 | 14:657417: | 14 | 65741733 | C | G | 0.647443 | -0.19082 | 0.020345 | 6.64E-21 | cis |
| miR-625-5p | rs7271443! | 14:659169( | 14 | 65916969 | C | T | 0.81551 | -0.23085 | 0.024476 | 4.03E-21 | cis |
| miR-625-5p | rs7271444( | 14:659185! | 14 | 65918591 | C | T | 0.81551 | -0.23085 | 0.024476 | 4.03E-21 | cis |
| miR-625-5p | rs7271444: | 14:659190 | 14 | 65919046 | T | C | 0.81551 | -0.23085 | 0.024476 | 4.03E-21 | cis |
| miR-625-5p | rs7271444 | 14:659421 | 14 | 65942142 | C | A | 0.816196 | -0.22863 | 0.024764 | 2.64E-20 | cis |
| miR-625-5p | rs1121885 | 14:661569 | 14 | 66156914 | A | G | 0.34818 | 0.177649 | 0.020897 | 1.88E-17 | cis |
| miR-625-5p | rs7271446: | 14:659681: | 14 | 65968132 | A | G | 0.816433 | -0.23136 | 0.024716 | 7.92E-21 | cis |
| miR-625-5p | rs7271446: | 14:659782 | 14 | 65978214 | A | C | 0.819836 | -0.23334 | 0.024931 | 7.99E-21 | cis |
| miR-625-5p | rs7271447: | 14:659979: | 14 | 65997931 | G | A | 0.815051 | -0.23431 | 0.024801 | 3.47E-21 | cis |
| miR-625-5p | rs1392518( | 14:658295: | 14 | 65829552 | T | C | 0.813544 | -0.23719 | 0.02469 | 7.48E-22 | cis |
| miR-625-5p | rs7271448: | 14:660126: | 14 | 66012624 | G | A | 0.820577 | -0.23277 | 0.025061 | 1.57E-20 | cis |
| miR-625-5p | rs4080329 | 14:662334! | 14 | 66233490 | T | C | 0.372147 | 0.135947 | 0.020036 | 1.16E-11 | cis |
| miR-625-5p | rs7271449! | 14:660424: | 14 | 66042459 | C | T | 0.816154 | -0.23186 | 0.024844 | 1.03E-20 | cis |
| miR-625-5p | rs1162112: | 14:658224! | 14 | 65822493 | C | T | 0.388213 | 0.196561 | 0.019252 | 1.79E-24 | cis |
| miR-625-5p | rs1013171! | 14:661573 | 14 | 66157373 | T | C | 0.385228 | 0.195601 | 0.020646 | 2.69E-21 | cis |
| miR-625-5p | rs1014852! | 14:661047( | 14 | 66104703 | A | T | 0.345818 | 0.173355 | 0.020883 | 1.03E-16 | cis |
| miR-625-5p | rs1014890 | 14:658333 | 14 | 65833372 | C | G | 0.739103 | -0.14955 | 0.021844 | 7.58E-12 | cis |
| miR-625-5p | rs1015041: | 14:659167! | 14 | 65916791 | C | T | 0.358422 | 0.173561 | 0.020711 | 5.29E-17 | cis |
| miR-625-5p | rs7158153 | 14:660345: | 14 | 66034586 | C | A | 0.345985 | 0.173233 | 0.02063 | 4.57E-17 | cis |
| miR-625-5p | rs4143898 | 14:661888: | 14 | 66188882 | T | A | 0.450813 | 0.145864 | 0.01999 | 2.94E-13 | cis |
| miR-625-5p | rs7141536 | 14:659873 | 14 | 65987312 | T | A | 0.349283 | 0.174077 | 0.020739 | 4.71E-17 | cis |
| miR-625-5p | rs7141874 | 14:658219! | 14 | 65821994 | G | A | 0.071326 | 0.265544 | 0.038518 | 5.42E-12 | cis |
| miR-625-5p | rs1013805: | 14:658334 | 14 | 65833440 | T | C | 0.739224 | -0.14936 | 0.021863 | 8.39E-12 | cis |
| miR-625-5p | rs1013857( | 14:658600: | 14 | 65860038 | A | G | 0.470451 | 0.132126 | 0.019756 | 2.26E-11 | cis |

| | | | | | | | | | | | |
|---|---|---|---|---|---|---|---|---|---|---|---|
| miR-625-5p | rs1013867 | 14:658600 | 14 | 65860092 | A | G | 0.398463 | 0.171449 | 0.020209 | 2.18E-17 | cis |
| miR-625-5p | rs1269068 | 14:657673 | 14 | 65767333 | C | T | 0.643163 | -0.22338 | 0.019494 | 2.13E-30 | cis |
| miR-625-5p | rs1015200 | 14:660474 | 14 | 66047434 | A | T | 0.343755 | 0.172178 | 0.020581 | 5.97E-17 | cis |
| miR-625-5p | rs883082 | 14:658809 | 14 | 65880940 | A | G | 0.493786 | 0.147935 | 0.019805 | 8.05E-14 | cis |
| miR-625-5p | rs5672046 | 14:662563 | 14 | 66256336 | C | T | 0.539663 | -0.13926 | 0.019323 | 5.72E-13 | cis |
| miR-625-5p | rs8012278 | 14:660825 | 14 | 66082573 | A | G | 0.499774 | 0.145255 | 0.019787 | 2.12E-13 | cis |
| miR-625-5p | rs7271640 | 14:660652 | 14 | 66065231 | G | T | 0.820366 | -0.23312 | 0.025147 | 1.85E-20 | cis |
| miR-625-5p | rs7823084 | 14:660163 | 14 | 66016384 | G | A | 0.507675 | 0.145122 | 0.019792 | 2.26E-13 | cis |
| miR-625-5p | rs7142219 | 14:662417 | 14 | 66241781 | G | A | 0.452285 | 0.14296 | 0.019477 | 2.13E-13 | cis |
| miR-625-5p | rs7271642 | 14:661322 | 14 | 66132280 | C | T | 0.817792 | -0.23401 | 0.025168 | 1.43E-20 | cis |
| miR-625-5p | rs7271642 | 14:661345 | 14 | 66134551 | T | C | 0.817679 | -0.23419 | 0.025168 | 1.34E-20 | cis |
| miR-625-5p | rs7142651 | 14:661327 | 14 | 66132721 | C | G | 0.502009 | 0.147599 | 0.019835 | 9.98E-14 | cis |
| miR-625-5p | rs1889731 | 14:659427 | 14 | 65942711 | G | C | 0.34602 | 0.173026 | 0.020834 | 9.98E-17 | cis |
| miR-625-5p | rs1162740 | 14:658413 | 14 | 65841322 | T | G | 0.411248 | 0.199702 | 0.019998 | 1.75E-23 | cis |
| miR-625-5p | rs1162757 | 14:662344 | 14 | 66234448 | C | T | 0.452075 | 0.142993 | 0.019339 | 1.42E-13 | cis |
| miR-625-5p | rs1288338 | 14:661453 | 14 | 66145396 | C | T | 0.500344 | 0.147926 | 0.019848 | 9.13E-14 | cis |
| miR-625-5p | rs1162760 | 14:662343 | 14 | 66234313 | G | A | 0.452186 | 0.143034 | 0.019344 | 1.42E-13 | cis |
| miR-625-5p | rs2792213 | 14:657713 | 14 | 65771339 | T | C | 0.947488 | -0.34429 | 0.044198 | 6.72E-15 | cis |
| miR-625-5p | rs1014228 | 14:660441 | 14 | 66044129 | G | A | 0.346742 | 0.172676 | 0.02057 | 4.67E-17 | cis |
| miR-625-5p | rs6134870 | 14:661110 | 14 | 66111019 | T | A | 0.792537 | -0.17841 | 0.025227 | 1.53E-12 | cis |
| miR-625-5p | rs1288600 | 14:658092 | 14 | 65809247 | C | T | 0.390429 | 0.198498 | 0.019263 | 6.72E-25 | cis |
| miR-625-5p | rs1014320 | 14:661206 | 14 | 66120675 | C | T | 0.349542 | 0.175451 | 0.020878 | 4.33E-17 | cis |
| miR-625-5p | rs1287887 | 14:658046 | 14 | 65804665 | G | A | 0.386023 | 0.203653 | 0.019372 | 7.56E-26 | cis |
| miR-625-5p | rs1287920 | 14:659771 | 14 | 65977182 | C | T | 0.511623 | 0.146601 | 0.019927 | 1.88E-13 | cis |
| miR-625-5p | rs1287980 | 14:660477 | 14 | 66047795 | T | C | 0.498218 | 0.145168 | 0.019735 | 1.90E-13 | cis |
| miR-625-5p | rs1287997 | 14:659016 | 14 | 65901604 | G | A | 0.503137 | 0.145861 | 0.019778 | 1.65E-13 | cis |
| miR-625-5p | rs1131317 | 14:660641 | 14 | 66064120 | G | A | 0.501439 | 0.145533 | 0.019821 | 2.10E-13 | cis |
| miR-625-5p | rs1953416 | 14:658788 | 14 | 65878807 | C | T | 0.504724 | 0.147883 | 0.01976 | 7.20E-14 | cis |
| miR-625-5p | rs1953418 | 14:659152 | 14 | 65915226 | G | A | 0.355213 | 0.172927 | 0.020733 | 7.38E-17 | cis |
| miR-625-5p | rs1954052 | 14:658057 | 14 | 65805709 | T | A | 0.387387 | 0.206368 | 0.019447 | 2.63E-26 | cis |
| miR-625-5p | rs1956010 | 14:661510 | 14 | 66151090 | G | A | 0.382297 | 0.18771 | 0.020495 | 5.24E-20 | cis |
| miR-625-5p | rs7142906 | 14:659564 | 14 | 65956456 | A | T | 0.360633 | 0.183028 | 0.020527 | 4.82E-19 | cis |
| miR-625-5p | rs1288072 | 14:657998 | 14 | 65799805 | G | A | 0.386052 | 0.21297 | 0.019217 | 1.53E-28 | cis |
| miR-625-5p | rs7143109 | 14:658010 | 14 | 65801024 | G | A | 0.269639 | 0.305978 | 0.021066 | 8.46E-48 | cis |
| miR-625-5p | rs7143318 | 14:659406 | 14 | 65940689 | G | A | 0.35859 | 0.169987 | 0.020679 | 2.03E-16 | cis |
| miR-625-5p | rs1958559 | 14:660520 | 14 | 66052055 | C | T | 0.40803 | 0.166544 | 0.019995 | 8.14E-17 | cis |
| miR-625-5p | rs1958560 | 14:660367 | 14 | 66036795 | G | A | 0.400142 | 0.165944 | 0.019808 | 5.40E-17 | cis |
| miR-625-5p | rs1958561 | 14:660367 | 14 | 66036761 | T | G | 0.498978 | 0.146336 | 0.019735 | 1.22E-13 | cis |
| miR-625-5p | rs1959144 | 14:659460 | 14 | 65946051 | T | G | 0.346477 | 0.172772 | 0.020841 | 1.13E-16 | cis |
| miR-625-5p | rs883081 | 14:658806 | 14 | 65880621 | C | A | 0.497085 | 0.149391 | 0.019841 | 5.10E-14 | cis |
| miR-625-5p | rs1184844 | 14:662574 | 14 | 66257467 | A | G | 0.5414 | -0.13923 | 0.019364 | 6.47E-13 | cis |
| miR-625-5p | rs1984855 | 14:662392 | 14 | 66239257 | G | C | 0.387843 | 0.147118 | 0.019829 | 1.18E-13 | cis |
| miR-625-5p | rs2184601 | 14:658973 | 14 | 65897368 | T | A | 0.354646 | 0.174532 | 0.020745 | 3.99E-17 | cis |
| miR-625-5p | rs1014497 | 14:659976 | 14 | 65997638 | G | A | 0.355198 | 0.170483 | 0.02058 | 1.19E-16 | cis |
| miR-625-5p | rs1014514 | 14:660750 | 14 | 66075004 | C | T | 0.349632 | 0.180781 | 0.020698 | 2.45E-18 | cis |
| miR-625-5p | rs899961 | 14:658003 | 14 | 65800321 | A | C | 0.290881 | 0.291121 | 0.020561 | 1.65E-45 | cis |
| miR-625-5p | rs7143666 | 14:658099 | 14 | 65809977 | A | C | 0.348575 | 0.20683 | 0.019826 | 1.77E-25 | cis |
| miR-625-5p | rs7918480 | 14:659083 | 14 | 65908314 | A | C | 0.813031 | -0.23021 | 0.024633 | 9.11E-21 | cis |
| miR-625-5p | rs2845901 | 14:662585 | 14 | 66258528 | T | C | 0.538033 | -0.13862 | 0.019252 | 6.02E-13 | cis |
| miR-625-5p | rs7923073 | 14:657424 | 14 | 65742463 | C | T | 0.705814 | -0.1918 | 0.021535 | 5.28E-19 | cis |
| miR-625-5p | rs1289403 | 14:662387 | 14 | 66238763 | T | C | 0.453073 | 0.14422 | 0.019372 | 9.72E-14 | cis |
| miR-625-5p | rs7144415 | 14:659930 | 14 | 65993090 | G | C | 0.367432 | 0.176219 | 0.020741 | 1.96E-17 | cis |
| miR-625-5p | rs7144530 | 14:658405 | 14 | 65840566 | A | G | 0.053251 | 0.327275 | 0.045719 | 8.16E-13 | cis |
| miR-625-5p | rs7144971 | 14:661206 | 14 | 66120650 | G | A | 0.349544 | 0.175451 | 0.020879 | 4.34E-17 | cis |
| miR-625-5p | rs1289533 | 14:658074 | 14 | 65807442 | C | T | 0.348231 | 0.206644 | 0.019831 | 2.01E-25 | cis |
| miR-625-5p | rs1289540 | 14:660648 | 14 | 66064827 | G | A | 0.409413 | 0.168122 | 0.020142 | 7.00E-17 | cis |
| miR-625-5p | rs1075566 | 14:662394 | 14 | 66239457 | C | T | 0.454013 | 0.144041 | 0.019394 | 1.11E-13 | cis |
| miR-625-5p | rs4400971 | 14:662545 | 14 | 66254578 | A | C | 0.539825 | -0.13887 | 0.019316 | 6.51E-13 | cis |
| miR-625-5p | rs9635249 | 14:657977 | 14 | 65797784 | A | G | 0.22061 | 0.364734 | 0.022232 | 1.73E-60 | cis |
| miR-625-5p | rs1115859 | 14:659237 | 14 | 65923738 | A | G | 0.389613 | 0.1839 | 0.020429 | 2.22E-19 | cis |
| miR-625-5p | rs1115860 | 14:660253 | 14 | 66025363 | A | G | 0.499831 | 0.146827 | 0.019777 | 1.14E-13 | cis |
| miR-625-5p | rs1115860 | 14:660720 | 14 | 66072040 | C | T | 0.349651 | 0.180755 | 0.020698 | 2.48E-18 | cis |
| miR-625-5p | rs1115860 | 14:660944 | 14 | 66094450 | G | A | 0.346475 | 0.17345 | 0.020801 | 7.51E-17 | cis |

| | | | | | | | | | | | |
|---|---|---|---|---|---|---|---|---|---|---|---|
| miR-625-5p | rs1288175 | 14:658018 | 14 | 65801811 | A | G | 0.320867 | 0.23032 | 0.020035 | 1.38E-30 | cis |
| miR-625-5p | rs7969176 | 14:661562 | 14 | 66156264 | T | G | 0.889044 | -0.34407 | 0.032384 | 2.29E-26 | cis |
| miR-625-5p | rs4456409 | 14:658979 | 14 | 65897967 | C | T | 0.510659 | 0.147924 | 0.020252 | 2.79E-13 | cis |
| miR-625-5p | rs1205018 | 14:662430 | 14 | 66243080 | C | T | 0.455215 | 0.144476 | 0.019432 | 1.05E-13 | cis |
| miR-625-5p | rs2861669 | 14:662586 | 14 | 66258683 | C | T | 0.538909 | -0.13835 | 0.019264 | 6.87E-13 | cis |
| miR-625-5p | rs2866601 | 14:657581 | 14 | 65758148 | C | T | 0.775896 | -0.19508 | 0.022869 | 1.46E-17 | cis |
| miR-625-5p | rs7327068 | 14:662673 | 14 | 66267360 | C | T | 0.936339 | -0.29555 | 0.039548 | 7.82E-14 | cis |
| miR-625-5p | rs2064694 | 14:661482 | 14 | 66148246 | G | T | 0.501114 | 0.147758 | 0.019816 | 8.87E-14 | cis |
| miR-625-5p | rs2874089 | 14:657835 | 14 | 65783523 | G | A | 0.672319 | -0.15331 | 0.02031 | 4.39E-14 | cis |
| miR-625-5p | rs8003464 | 14:659082 | 14 | 65908235 | A | G | 0.35496 | 0.173425 | 0.020754 | 6.48E-17 | cis |
| miR-625-5p | rs8018278 | 14:661800 | 14 | 66180088 | A | G | 0.498198 | 0.145521 | 0.019847 | 2.26E-13 | cis |
| miR-625-5p | rs1184846 | 14:662572 | 14 | 66257294 | G | A | 0.541416 | -0.13927 | 0.01937 | 6.48E-13 | cis |
| miR-625-5p | rs4899174 | 14:658114 | 14 | 65811400 | A | T | 0.390371 | 0.198564 | 0.019268 | 6.68E-25 | cis |
| miR-625-5p | rs2898814 | 14:660239 | 14 | 66023961 | T | C | 0.347238 | 0.173957 | 0.020661 | 3.77E-17 | cis |
| miR-625-5p | rs2898818 | 14:659725 | 14 | 65972584 | G | C | 0.35329 | 0.170444 | 0.020622 | 1.40E-16 | cis |
| miR-625-5p | rs2898820 | 14:658793 | 14 | 65879354 | T | G | 0.35021 | 0.175123 | 0.020725 | 2.92E-17 | cis |
| miR-625-5p | rs8005710 | 14:660656 | 14 | 66065666 | G | A | 0.502323 | 0.147754 | 0.019868 | 1.03E-13 | cis |
| miR-625-5p | rs8022094 | 14:658468 | 14 | 65846841 | G | T | 0.396412 | 0.196013 | 0.019727 | 2.89E-23 | cis |
| miR-625-5p | rs8023016 | 14:658468 | 14 | 65846808 | T | A | 0.053493 | 0.326462 | 0.045712 | 9.22E-13 | cis |
| miR-625-5p | rs8023161 | 14:658468 | 14 | 65846865 | T | C | 0.063557 | 0.305586 | 0.045079 | 1.21E-11 | cis |
| miR-625-5p | rs1288191 | 14:658312 | 14 | 65831221 | T | C | 0.388492 | 0.19722 | 0.019447 | 3.62E-24 | cis |
| miR-625-5p | rs1288220 | 14:661578 | 14 | 66157861 | C | A | 0.503123 | 0.148195 | 0.019873 | 8.84E-14 | cis |
| miR-625-5p | rs1288226 | 14:658471 | 14 | 65847144 | T | G | 0.394348 | 0.195407 | 0.019702 | 3.47E-23 | cis |
| miR-625-5p | rs1184468 | 14:659108 | 14 | 65910844 | G | C | 0.356705 | 0.17288 | 0.02073 | 7.44E-17 | cis |
| miR-625-5p | rs7145759 | 14:658845 | 14 | 65884537 | C | A | 0.353283 | 0.175197 | 0.0207 | 2.60E-17 | cis |
| miR-625-5p | rs8029794 | 14:661006 | 14 | 66100698 | A | G | 0.542877 | 0.153421 | 0.021121 | 3.76E-13 | cis |
| miR-625-5p | rs7146742 | 14:660329 | 14 | 66032934 | G | A | 0.435936 | 0.139288 | 0.020069 | 3.91E-12 | cis |
| miR-625-5p | rs8007497 | 14:660444 | 14 | 66044476 | T | C | 0.338774 | 0.16985 | 0.02037 | 7.53E-17 | cis |
| miR-625-5p | rs8007846 | 14:662629 | 14 | 66262963 | A | G | 0.566372 | -0.15422 | 0.019109 | 7.00E-16 | cis |
| miR-625-5p | rs4587890 | 14:658637 | 14 | 65863784 | T | A | 0.391319 | 0.17464 | 0.020418 | 1.20E-17 | cis |
| miR-625-5p | rs7147536 | 14:658746 | 14 | 65874698 | C | T | 0.367002 | 0.185223 | 0.020461 | 1.39E-19 | cis |
| miR-625-5p | rs7147002 | 14:660325 | 14 | 66032546 | T | C | 0.342789 | 0.172498 | 0.020658 | 6.82E-17 | cis |
| miR-625-5p | rs1162160 | 14:658407 | 14 | 65840774 | A | G | 0.393883 | 0.198198 | 0.019616 | 5.32E-24 | cis |
| miR-625-5p | rs1288276 | 14:660780 | 14 | 66078025 | C | T | 0.395967 | 0.16261 | 0.021215 | 1.79E-14 | cis |
| miR-625-5p | rs1162201 | 14:660416 | 14 | 66041645 | C | T | 0.502262 | 0.14792 | 0.019779 | 7.51E-14 | cis |
| miR-625-5p | rs7157006 | 14:658282 | 14 | 65828207 | C | T | 0.387351 | 0.198515 | 0.019331 | 9.71E-25 | cis |
| miR-625-5p | rs7157449 | 14:662401 | 14 | 66240137 | T | G | 0.454044 | 0.144106 | 0.019397 | 1.09E-13 | cis |
| miR-625-5p | rs1289097 | 14:659828 | 14 | 65982890 | C | T | 0.504299 | 0.148214 | 0.019979 | 1.18E-13 | cis |
| miR-625-5p | rs7158347 | 14:658289 | 14 | 65828955 | C | T | 0.388449 | 0.198782 | 0.019394 | 1.19E-24 | cis |
| miR-625-5p | rs7158556 | 14:662407 | 14 | 66240729 | T | A | 0.373139 | 0.137414 | 0.020058 | 7.35E-12 | cis |
| miR-625-5p | rs1162227 | 14:661490 | 14 | 66149065 | A | T | 0.504871 | 0.146606 | 0.019892 | 1.71E-13 | cis |
| miR-625-5p | rs1162282 | 14:661917 | 14 | 66191782 | T | A | 0.496725 | 0.144626 | 0.019819 | 2.93E-13 | cis |
| miR-625-5p | rs8009991 | 14:662798 | 14 | 66279803 | C | T | 0.861309 | -0.25774 | 0.031501 | 2.79E-16 | cis |
| miR-625-5p | rs8010726 | 14:659325 | 14 | 65932597 | G | A | 0.358211 | 0.172184 | 0.020663 | 7.89E-17 | cis |
| miR-625-5p | rs1184844 | 14:662574 | 14 | 66257419 | C | T | 0.54144 | -0.13924 | 0.019367 | 6.49E-13 | cis |
| miR-625-5p | rs2268962 | 14:661472 | 14 | 66147273 | A | G | 0.501276 | 0.147836 | 0.01982 | 8.72E-14 | cis |
| miR-625-5p | rs8018379 | 14:662619 | 14 | 66261937 | C | G | 0.603691 | -0.16915 | 0.019661 | 7.76E-18 | cis |
| miR-625-5p | rs1256525 | 14:657385 | 14 | 65738550 | G | C | 0.646861 | -0.19091 | 0.020383 | 7.52E-21 | cis |
| miR-625-5p | rs4899175 | 14:658264 | 14 | 65826433 | A | G | 0.051502 | 0.327813 | 0.045295 | 4.58E-13 | cis |
| miR-625-5p | rs8012834 | 14:659665 | 14 | 65966555 | C | T | 0.362666 | 0.181923 | 0.020554 | 8.68E-19 | cis |
| miR-625-5p | rs1162588 | 14:662451 | 14 | 66245199 | G | A | 0.454755 | 0.141922 | 0.019347 | 2.21E-13 | cis |
| miR-625-5p | rs1162706 | 14:661829 | 14 | 66182953 | T | G | 0.488866 | 0.13317 | 0.01984 | 1.92E-11 | cis |
| miR-625-5p | rs1162708 | 14:659788 | 14 | 65978836 | T | G | 0.501066 | 0.146483 | 0.019949 | 2.09E-13 | cis |
| miR-625-5p | rs1162718 | 14:661214 | 14 | 66121443 | C | T | 0.501637 | 0.146864 | 0.019888 | 1.53E-13 | cis |
| miR-625-5p | rs5692256 | 14:662564 | 14 | 66256472 | A | T | 0.541356 | -0.13923 | 0.019369 | 6.57E-13 | cis |
| miR-625-5p | rs1162718 | 14:661214 | 14 | 66121492 | A | G | 0.501636 | 0.146867 | 0.019888 | 1.53E-13 | cis |
| miR-625-5p | rs2411351 | 14:661715 | 14 | 66171541 | T | C | 0.498384 | 0.147075 | 0.019765 | 9.99E-14 | cis |
| miR-625-5p | rs2411356 | 14:660531 | 14 | 66053161 | G | A | 0.388335 | 0.168635 | 0.020188 | 6.65E-17 | cis |
| miR-625-5p | rs1288381 | 14:659233 | 14 | 65923367 | C | T | 0.510962 | 0.147038 | 0.019928 | 1.60E-13 | cis |
| miR-625-5p | rs1162876 | 14:661684 | 14 | 66168449 | T | C | 0.500166 | 0.147858 | 0.019781 | 7.73E-14 | cis |
| miR-625-5p | rs1288584 | 14:659186 | 14 | 65918652 | C | T | 0.373139 | 0.182228 | 0.020451 | 5.08E-19 | cis |
| miR-625-5p | rs2411815 | 14:660200 | 14 | 66020071 | A | T | 0.354996 | 0.169079 | 0.020582 | 2.12E-16 | cis |
| miR-625-5p | rs1760978 | 14:657710 | 14 | 65771047 | T | G | 0.623429 | -0.19618 | 0.01985 | 4.92E-23 | cis |

| | | | | | | | | | | | |
|---|---|---|---|---|---|---|---|---|---|---|---|
| miR-625-5p | rs7435176! | 14:6580840 | 14 | 65808468 | C | T | 0.810279 | -0.2335 | 0.024456 | 1.33E-21 | cis |
| miR-625-5p | rs1288616 | 14:6580928 | 14 | 65809286 | C | A | 0.393656 | 0.204136 | 0.019134 | 1.43E-26 | cis |
| miR-625-5p | rs1288633 | 14:6609467 | 14 | 66094671 | T | G | 0.500184 | 0.146151 | 0.01985 | 1.80E-13 | cis |
| miR-625-5p | rs2411915 | 14:6579469 | 14 | 65794692 | C | T | 0.053213 | 0.337729 | 0.044685 | 4.09E-14 | cis |
| miR-625-5p | rs1288713 | 14:6604554 | 14 | 66045543 | G | C | 0.502159 | 0.147999 | 0.019774 | 7.18E-14 | cis |
| miR-625-5p | rs1288767 | 14:6622177 | 14 | 66221771 | T | C | 0.487094 | 0.131927 | 0.019729 | 2.28E-11 | cis |
| miR-625-5p | rs1288795 | 14:6624659 | 14 | 66246592 | G | A | 0.454723 | 0.142379 | 0.019362 | 1.93E-13 | cis |
| miR-625-5p | rs6198799 | 14:6607073 | 14 | 66070732 | C | G | 0.351584 | 0.180835 | 0.020721 | 2.61E-18 | cis |
| miR-625-5p | rs1288900 | 14:6606358 | 14 | 66063582 | C | G | 0.499455 | 0.145384 | 0.019766 | 1.91E-13 | cis |
| miR-625-5p | rs1289012 | 14:6599806 | 14 | 65998069 | C | T | 0.511668 | 0.146815 | 0.019882 | 1.53E-13 | cis |
| miR-625-5p | rs1289090 | 14:6611662 | 14 | 66116622 | T | A | 0.498435 | 0.144652 | 0.019829 | 2.98E-13 | cis |
| miR-625-5p | rs1865011 | 14:6615940 | 14 | 66159405 | G | A | 0.780937 | -0.19034 | 0.024831 | 1.78E-14 | cis |
| miR-625-5p | rs1289205 | 14:6590344 | 14 | 65903441 | T | C | 0.51011 | 0.146773 | 0.019879 | 1.54E-13 | cis |
| miR-625-5p | rs1048377 | 14:6591486 | 14 | 65914867 | A | G | 0.817942 | -0.23223 | 0.024543 | 3.02E-21 | cis |
| miR-625-5p | rs1048378 | 14:6598017 | 14 | 65980170 | T | C | 0.501165 | 0.146505 | 0.019946 | 2.05E-13 | cis |
| miR-625-5p | rs7151846 | 14:6625578 | 14 | 66255781 | C | G | 0.539502 | -0.13871 | 0.019301 | 6.65E-13 | cis |
| miR-625-5p | rs8016322 | 14:6605153 | 14 | 66051536 | G | A | 0.386004 | 0.167316 | 0.020156 | 1.03E-16 | cis |
| miR-625-5p | rs8017880 | 14:6594292 | 14 | 65942924 | G | C | 0.346067 | 0.172935 | 0.020833 | 1.03E-16 | cis |
| miR-625-5p | rs7152441 | 14:6582790 | 14 | 65827903 | A | G | 0.506387 | 0.156688 | 0.02018 | 8.19E-15 | cis |
| miR-625-5p | rs1162366 | 14:6602119 | 14 | 66021192 | T | G | 0.390012 | 0.168768 | 0.020334 | 1.04E-16 | cis |
| miR-625-5p | rs1162392 | 14:658193 | 14 | 65819314 | A | C | 0.389577 | 0.198029 | 0.019291 | 1.01E-24 | cis |
| miR-625-5p | rs1162404 | 14:6610801 | 14 | 66108018 | C | T | 0.498929 | 0.145121 | 0.019847 | 2.63E-13 | cis |
| miR-625-5p | rs8019762 | 14:6608717 | 14 | 66087178 | A | G | 0.346646 | 0.172489 | 0.020796 | 1.09E-16 | cis |
| miR-625-5p | rs8020305 | 14:6583750 | 14 | 65837503 | C | T | 0.066561 | 0.301123 | 0.040379 | 8.82E-14 | cis |
| miR-625-5p | rs3464038 | 14:6605300 | 14 | 66053006 | G | A | 0.819714 | -0.23302 | 0.025045 | 1.35E-20 | cis |
| miR-625-5p | rs7482562 | 14:6580778 | 14 | 65807781 | G | A | 0.810459 | -0.23324 | 0.024454 | 1.46E-21 | cis |
| miR-625-5p | rs7489777 | 14:6590900 | 14 | 65909005 | T | G | 0.81551 | -0.2309 | 0.024476 | 3.97E-21 | cis |
| miR-625-5p | rs1258705 | 14:6606637 | 14 | 66066375 | C | T | 0.345738 | 0.172451 | 0.020704 | 8.14E-17 | cis |
| miR-625-5p | rs6649234 | 14:6583948 | 14 | 65839489 | A | C | 0.739 | -0.14985 | 0.021839 | 6.80E-12 | cis |
| miR-625-5p | rs8025374 | 14:6610069 | 14 | 66100692 | G | A | 0.462994 | 0.179831 | 0.022323 | 7.89E-16 | cis |
| miR-625-5p | rs1289446 | 14:6624076 | 14 | 66240767 | G | A | 0.454589 | 0.14417 | 0.0194 | 1.07E-13 | cis |
| miR-625-5p | rs1289490 | 14:6616179 | 14 | 66161799 | C | T | 0.501572 | 0.148481 | 0.019827 | 6.95E-14 | cis |
| miR-625-5p | rs1289507 | 14:6597679 | 14 | 65976793 | G | T | 0.520727 | 0.14878 | 0.020225 | 1.89E-13 | cis |
| miR-625-5p | rs1115859 | 14:6585576 | 14 | 65855762 | C | T | 0.390273 | 0.199381 | 0.019678 | 3.98E-24 | cis |
| miR-625-5p | rs1115859 | 14:6585998 | 14 | 65859984 | G | A | 0.466718 | 0.134472 | 0.019789 | 1.08E-11 | cis |
| miR-625-5p | rs1289666 | 14:6610496 | 14 | 66104969 | G | A | 0.499 | 0.145192 | 0.01985 | 2.59E-13 | cis |
| miR-625-5p | rs1115859 | 14:6591197 | 14 | 65911975 | T | A | 0.507581 | 0.145384 | 0.01988 | 2.61E-13 | cis |
| miR-625-5p | rs1243359 | 14:6602884 | 14 | 66028847 | T | C | 0.343187 | 0.172769 | 0.020668 | 6.31E-17 | cis |
| miR-625-5p | rs2300864 | 14:6611980 | 14 | 66119800 | A | G | 0.349692 | 0.175475 | 0.020879 | 4.30E-17 | cis |
| miR-625-5p | rs3477200 | 14:6605309 | 14 | 66053091 | G | A | 0.819572 | -0.23305 | 0.02505 | 1.36E-20 | cis |
| miR-625-5p | rs7140341 | 14:660704 | 14 | 66070417 | C | G | 0.3503 | 0.181513 | 0.020699 | 1.80E-18 | cis |
| miR-625-5p | rs7682482 | 14:6575823 | 14 | 65758236 | G | A | 0.778334 | -0.19516 | 0.022955 | 1.87E-17 | cis |
| miR-625-5p | rs6198799 | 14:6607073 | 14 | 66070731 | C | T | 0.351584 | 0.180835 | 0.020721 | 2.61E-18 | cis |
| miR-625-5p | rs7156078 | 14:6623870 | 14 | 66238701 | G | A | 0.386926 | 0.147416 | 0.019811 | 9.98E-14 | cis |
| miR-625-5p | rs6198799 | 14:6607073 | 14 | 66070733 | A | G | 0.352209 | 0.180424 | 0.020737 | 3.30E-18 | cis |
| miR-625-5p | rs6198897 | 14:6625803 | 14 | 66258032 | C | A | 0.537113 | -0.13735 | 0.019389 | 1.40E-12 | cis |
| miR-625-5p | rs7154536 | 14:6599173 | 14 | 65991737 | A | G | 0.461219 | 0.147581 | 0.020129 | 2.27E-13 | cis |
| miR-625-5p | rs9323461 | 14:6603140 | 14 | 66031405 | T | C | 0.342775 | 0.172498 | 0.020658 | 6.81E-17 | cis |
| miR-625-5p | rs7327067 | 14:6626340 | 14 | 66263407 | C | T | 0.935874 | -0.29018 | 0.038987 | 9.84E-14 | cis |
| miR-625-5p | rs2268955 | 14:6609000 | 14 | 66090009 | T | A | 0.411712 | 0.169021 | 0.020476 | 1.52E-16 | cis |
| miR-625-5p | rs2268956 | 14:6609014 | 14 | 66090145 | G | A | 0.349005 | 0.172896 | 0.020779 | 8.74E-17 | cis |
| miR-625-5p | rs2268959 | 14:661453 | 14 | 66145318 | C | A | 0.818382 | -0.23441 | 0.02521 | 1.43E-20 | cis |
| miR-625-5p | rs1775350 | 14:6605745 | 14 | 66057452 | A | G | 0.821177 | -0.23294 | 0.025154 | 2.03E-20 | cis |
| miR-625-5p | rs1256523 | 14:6573812 | 14 | 65738126 | A | G | 0.647737 | -0.19099 | 0.020354 | 6.39E-21 | cis |
| miR-625-5p | rs1256524 | 14:6573843 | 14 | 65738435 | T | C | 0.647718 | -0.19092 | 0.020352 | 6.55E-21 | cis |
| miR-625-5p | rs8011224 | 14:6596967 | 14 | 65969670 | T | A | 0.357678 | 0.169952 | 0.020594 | 1.55E-16 | cis |
| miR-625-5p | rs1087319 | 14:6598143 | 14 | 65981439 | G | T | 0.361158 | 0.177 | 0.020825 | 1.91E-17 | cis |
| miR-625-5p | rs1087319 | 14:6611062 | 14 | 66110628 | T | C | 0.346549 | 0.173321 | 0.020847 | 9.27E-17 | cis |
| miR-625-5p | rs8022053 | 14:6586794 | 14 | 65867947 | T | G | 0.350181 | 0.176409 | 0.020719 | 1.68E-17 | cis |
| miR-625-5p | rs8022080 | 14:6599908 | 14 | 65999088 | T | C | 0.358857 | 0.182558 | 0.020463 | 4.61E-19 | cis |
| miR-625-5p | rs6573611 | 14:6586525 | 14 | 65865255 | G | T | 0.39802 | 0.170507 | 0.020229 | 3.49E-17 | cis |
| miR-625-5p | rs7271442 | 14:6589292 | 14 | 65892925 | A | C | 0.815998 | -0.23265 | 0.024632 | 3.55E-21 | cis |
| miR-625-5p | rs4073416 | 14:6625939 | 14 | 66259394 | T | C | 0.585483 | -0.13599 | 0.019383 | 2.29E-12 | cis |

| | | | | | | | | | | | |
|---|---|---|---|---|---|---|---|---|---|---|---|
| miR-625-5p | rs2002692 | 14:6589945 | 14 | 65899454 | A | G | 0.502596 | 0.145491 | 0.019792 | 1.97E-13 | cis |
| miR-625-5p | rs6573618 | 14:6605645 | 14 | 66056452 | A | G | 0.344637 | 0.172132 | 0.020646 | 7.60E-17 | cis |
| miR-625-5p | rs6573619 | 14:6606156 | 14 | 66061560 | T | C | 0.344872 | 0.172013 | 0.020629 | 7.53E-17 | cis |
| miR-625-5p | rs1162759 | 14:6579776 | 14 | 65797766 | G | T | 0.38607 | 0.212976 | 0.019217 | 1.52E-28 | cis |
| miR-625-5p | rs9783622 | 14:6574904 | 14 | 65749045 | C | G | 0.724193 | -0.2196 | 0.021928 | 1.31E-23 | cis |
| miR-625-5p | rs7271446 | 14:6596208 | 14 | 65962089 | A | G | 0.819561 | -0.23327 | 0.024859 | 6.37E-21 | cis |
| miR-625-5p | rs9796385 | 14:661012 | 14 | 66101219 | A | G | 0.518045 | 0.140926 | 0.020373 | 4.61E-12 | cis |
| miR-625-5p | rs6573627 | 14:6625232 | 14 | 66252326 | C | T | 0.542293 | -0.14076 | 0.019318 | 3.19E-13 | cis |
| miR-625-5p | rs1214799 | 14:6575162 | 14 | 65751627 | C | A | 0.729928 | -0.22238 | 0.022038 | 6.07E-24 | cis |
| miR-625-5p | rs7271448 | 14:6600660 | 14 | 66006602 | T | G | 0.815267 | -0.23397 | 0.024821 | 4.24E-21 | cis |
| miR-625-5p | rs2411822 | 14:6587839 | 14 | 65878395 | A | G | 0.501454 | 0.146326 | 0.019719 | 1.16E-13 | cis |
| miR-625-5p | rs2300865 | 14:661200 | 14 | 66120015 | G | C | 0.502657 | 0.147148 | 0.019872 | 1.31E-13 | cis |
| miR-625-5p | rs2411823 | 14:6586506 | 14 | 65865066 | C | T | 0.39822 | 0.170762 | 0.020236 | 3.22E-17 | cis |
| miR-625-5p | rs7660191 | 14:6608734 | 14 | 66087341 | G | C | 0.820834 | -0.23231 | 0.025252 | 3.59E-20 | cis |
| miR-625-5p | rs3594901 | 14:6613616 | 14 | 66136163 | C | A | 0.826447 | -0.23684 | 0.026022 | 8.90E-20 | cis |
| miR-625-5p | rs1185157 | 14:6590028 | 14 | 65900283 | T | C | 0.454741 | 0.146928 | 0.01999 | 1.98E-13 | cis |
| miR-625-5p | rs4902388 | 14:6579285 | 14 | 65792852 | A | G | 0.216378 | 0.360268 | 0.022447 | 5.75E-58 | cis |
| miR-625-5p | rs7147636 | 14:6601118 | 14 | 66011184 | T | C | 0.36432 | 0.172044 | 0.020591 | 6.53E-17 | cis |
| miR-625-5p | rs7157109 | 14:6590043 | 14 | 65900433 | C | G | 0.354575 | 0.174565 | 0.020746 | 3.94E-17 | cis |
| miR-625-5p | rs4902387 | 14:6579283 | 14 | 65792839 | C | T | 0.21638 | 0.360265 | 0.022447 | 5.75E-58 | cis |
| miR-625-5p | rs4902393 | 14:658395 | 14 | 65839514 | G | C | 0.390223 | 0.1982 | 0.019437 | 2.04E-24 | cis |
| miR-625-5p | rs4902399 | 14:6595689 | 14 | 65956897 | A | G | 0.345315 | 0.174461 | 0.020811 | 5.16E-17 | cis |
| miR-625-5p | rs3783709 | 14:6605866 | 14 | 66058664 | T | A | 0.499275 | 0.145547 | 0.01973 | 1.62E-13 | cis |
| miR-625-5p | rs3783711 | 14:6602844 | 14 | 66028446 | A | G | 0.343187 | 0.17277 | 0.020668 | 6.31E-17 | cis |
| miR-625-5p | rs2127868 | 14:6579432 | 14 | 65794327 | G | A | 0.220849 | 0.364388 | 0.022206 | 1.63E-60 | cis |
| miR-625-5p | rs7160780 | 14:660527 | 14 | 66052713 | G | T | 0.386614 | 0.16716 | 0.020127 | 9.96E-17 | cis |
| miR-625-5p | rs7161123 | 14:6605290 | 14 | 66052901 | G | A | 0.498649 | 0.145335 | 0.01972 | 1.71E-13 | cis |
| miR-625-5p | rs4902416 | 14:6623809 | 14 | 66238090 | G | C | 0.453194 | 0.143892 | 0.019371 | 1.10E-13 | cis |
| miR-625-5p | rs6048801 | 14:6582585 | 14 | 65825854 | C | A | 0.738599 | -0.14894 | 0.021849 | 9.30E-12 | cis |
| miR-625-5p | rs1243646 | 14:6580687 | 14 | 65806877 | T | C | 0.255814 | 0.28174 | 0.021604 | 7.15E-39 | cis |
| miR-625-5p | rs1959145 | 14:6595474 | 14 | 65954740 | A | G | 0.347293 | 0.172809 | 0.020786 | 9.28E-17 | cis |
| miR-625-5p | rs4899173 | 14:6579316 | 14 | 65793168 | T | C | 0.216374 | 0.360282 | 0.022447 | 5.72E-58 | cis |
| miR-625-5p | rs4899178 | 14:6586298 | 14 | 65862987 | C | T | 0.391583 | 0.174834 | 0.020417 | 1.10E-17 | cis |
| miR-625-5p | rs4899179 | 14:6592674 | 14 | 65926748 | T | G | 0.508099 | 0.145114 | 0.019871 | 2.81E-13 | cis |
| miR-625-5p | rs7142165 | 14:6625023 | 14 | 66250232 | G | A | 0.550826 | -0.14146 | 0.019388 | 2.95E-13 | cis |
| miR-625-5p | rs7271641 | 14:6612104 | 14 | 66121045 | G | A | 0.819456 | -0.23354 | 0.025291 | 2.60E-20 | cis |
| miR-625-5p | rs2149839 | 14:6598344 | 14 | 65983448 | G | A | 0.353418 | 0.170444 | 0.020619 | 1.38E-16 | cis |
| miR-625-5p | rs2149841 | 14:660103 | 14 | 66010319 | C | T | 0.503888 | 0.148409 | 0.019912 | 9.11E-14 | cis |
| miR-625-5p | rs7271643 | 14:661929 | 14 | 66192915 | G | T | 0.829643 | -0.21683 | 0.026052 | 8.58E-17 | cis |
| miR-625-5p | rs6573615 | 14:6604653 | 14 | 66046534 | G | A | 0.381045 | 0.166016 | 0.019917 | 7.73E-17 | cis |
| miR-625-5p | rs7271443 | 14:6590747 | 14 | 65907478 | A | G | 0.815498 | -0.23092 | 0.024477 | 3.95E-21 | cis |
| miR-625-5p | rs1115858 | 14:6579987 | 14 | 65799876 | G | A | 0.221744 | 0.35927 | 0.022176 | 5.00E-59 | cis |
| miR-625-5p | rs2411404 | 14:6623940 | 14 | 66239401 | C | G | 0.454146 | 0.144127 | 0.019397 | 1.08E-13 | cis |
| miR-625-5p | rs5830703 | 14:6582419 | 14 | 65824195 | A | T | 0.051498 | 0.327775 | 0.045296 | 4.61E-13 | cis |
| miR-625-5p | rs1162884 | 14:6623564 | 14 | 66235642 | A | G | 0.451742 | 0.143194 | 0.01935 | 1.36E-13 | cis |
| miR-625-5p | rs1115859 | 14:6590636 | 14 | 65906369 | A | G | 0.34826 | 0.175746 | 0.020926 | 4.52E-17 | cis |
| miR-625-5p | rs2411820 | 14:6594674 | 14 | 65946744 | C | T | 0.35728 | 0.171685 | 0.020672 | 9.97E-17 | cis |
| miR-625-5p | rs7146829 | 14:6619453 | 14 | 66194531 | T | C | 0.488699 | 0.132266 | 0.019792 | 2.34E-11 | cis |
| miR-625-5p | rs9972106 | 14:6602313 | 14 | 66023131 | A | T | 0.388305 | 0.167818 | 0.020231 | 1.08E-16 | cis |
| miR-625-5p | rs1115860 | 14:6607173 | 14 | 66071733 | T | G | 0.491419 | 0.14246 | 0.01987 | 7.53E-13 | cis |
| miR-625-5p | rs7146993 | 14:6604887 | 14 | 66048872 | C | T | 0.343625 | 0.171579 | 0.020581 | 7.63E-17 | cis |
| miR-625-5p | rs1258969 | 14:6592043 | 14 | 65920435 | G | A | 0.510971 | 0.147065 | 0.019929 | 1.59E-13 | cis |
| miR-625-5p | rs1115860 | 14:6610792 | 14 | 66107920 | T | A | 0.365947 | 0.173317 | 0.020738 | 6.42E-17 | cis |
| miR-625-5p | rs1115860 | 14:6611015 | 14 | 66110150 | G | A | 0.365256 | 0.176396 | 0.021218 | 9.29E-17 | cis |
| miR-625-5p | rs4902389 | 14:6579322 | 14 | 65793224 | A | G | 0.216372 | 0.360287 | 0.022448 | 5.70E-58 | cis |
| miR-625-5p | rs4902391 | 14:6581597 | 14 | 65815979 | G | T | 0.397869 | 0.205832 | 0.019183 | 7.36E-27 | cis |
| miR-625-5p | rs1256526 | 14:6573990 | 14 | 65739905 | T | G | 0.647195 | -0.19055 | 0.020338 | 7.31E-21 | cis |
| miR-625-5p | rs2092914 | 14:660791 | 14 | 66079115 | C | T | 0.36669 | 0.173049 | 0.020649 | 5.27E-17 | cis |
| miR-625-5p | rs7140695 | 14:6592913 | 14 | 65929132 | T | C | 0.350881 | 0.17535 | 0.020807 | 3.53E-17 | cis |
| miR-625-5p | rs7150448 | 14:6587150 | 14 | 65871502 | C | T | 0.345581 | 0.17931 | 0.020872 | 8.63E-18 | cis |
| miR-625-5p | rs1205023 | 14:662350 | 14 | 66235016 | T | C | 0.451755 | 0.143138 | 0.019345 | 1.37E-13 | cis |
| miR-625-5p | rs4902407 | 14:6607463 | 14 | 66074635 | T | G | 0.492329 | 0.142628 | 0.01991 | 7.85E-13 | cis |
| miR-625-5p | rs4902412 | 14:6618706 | 14 | 66187067 | T | C | 0.443 | 0.135201 | 0.019984 | 1.33E-11 | cis |

| | | | | | | | | | | | |
|---|---|---|---|---|---|---|---|---|---|---|---|
| miR-625-5p | rs4902415 | 14:6623569 | 14 | 66235690 | T | C | 0.451839 | 0.143041 | 0.019354 | 1.46E-13 | cis |
| miR-625-5p | rs899959 | 14:6579192 | 14 | 65791928 | A | G | 0.383745 | 0.208186 | 0.01953 | 1.57E-26 | cis |
| miR-625-5p | rs7152023 | 14:6616082 | 14 | 66160827 | C | A | 0.425722 | 0.183153 | 0.02014 | 9.56E-20 | cis |
| miR-625-5p | rs6137847 | 14:6625343 | 14 | 66253437 | G | A | 0.541244 | -0.14021 | 0.019376 | 4.62E-13 | cis |
| miR-625-5p | rs1256522 | 14:6573802 | 14 | 65738029 | C | T | 0.647567 | -0.19109 | 0.020355 | 6.13E-21 | cis |
| miR-625-5p | rs7608716 | 14:6606700 | 14 | 66067006 | T | C | 0.820364 | -0.23313 | 0.025147 | 1.85E-20 | cis |
| miR-625-5p | rs6573624 | 14:6622688 | 14 | 66226885 | G | A | 0.485559 | 0.132056 | 0.019672 | 1.91E-11 | cis |
| miR-625-5p | rs761831 | 14:6623937 | 14 | 66239370 | T | C | 0.454673 | 0.144038 | 0.019399 | 1.13E-13 | cis |
| miR-625-5p | rs1113962 | 14:6600579 | 14 | 66005798 | C | T | 0.355246 | 0.170317 | 0.02059 | 1.32E-16 | cis |
| miR-625-5p | rs3825639 | 14:6602850 | 14 | 66028505 | T | C | 0.343187 | 0.17277 | 0.020668 | 6.31E-17 | cis |
| miR-625-5p | rs3825640 | 14:6596120 | 14 | 65961204 | C | G | 0.503966 | 0.147669 | 0.019981 | 1.46E-13 | cis |
| miR-625-5p | rs1115689 | 14:6589789 | 14 | 65897898 | T | A | 0.523523 | 0.15347 | 0.020485 | 6.79E-14 | cis |
| miR-625-5p | rs1258684 | 14:6614502 | 14 | 66145022 | T | G | 0.500228 | 0.147394 | 0.019836 | 1.08E-13 | cis |
| miR-625-5p | rs7144345 | 14:6611184 | 14 | 66111841 | G | C | 0.345782 | 0.173329 | 0.020876 | 1.02E-16 | cis |
| miR-625-5p | rs1258883 | 14:6616263 | 14 | 66162638 | G | T | 0.500385 | 0.147861 | 0.019792 | 7.98E-14 | cis |
| miR-625-5p | rs747541 | 14:6580541 | 14 | 65805410 | C | A | 0.395726 | 0.206479 | 0.019239 | 7.18E-27 | cis |
| miR-625-5p | rs7155541 | 14:6604624 | 14 | 66046242 | G | C | 0.501962 | 0.146856 | 0.019792 | 1.17E-13 | cis |
| miR-625-5p | rs6573617 | 14:6605640 | 14 | 66056408 | C | T | 0.358534 | 0.178362 | 0.020632 | 5.39E-18 | cis |
| miR-625-5p | rs1115861 | 14:6623507 | 14 | 66235074 | G | A | 0.451752 | 0.143146 | 0.019346 | 1.37E-13 | cis |
| miR-625-5p | rs2236067 | 14:6624801 | 14 | 66248012 | A | G | 0.386784 | 0.145441 | 0.019837 | 2.27E-13 | cis |
| miR-625-5p | rs7493004 | 14:6612410 | 14 | 66124104 | A | C | 0.50236 | 0.147113 | 0.01987 | 1.32E-13 | cis |
| miR-625-5p | rs6573626 | 14:6624069 | 14 | 66240695 | G | C | 0.454572 | 0.145113 | 0.019413 | 7.72E-14 | cis |
| miR-625-5p | rs1710260 | 14:6578957 | 14 | 65789571 | A | C | 0.845741 | -0.32668 | 0.026149 | 8.16E-36 | cis |
| miR-625-5p | rs1162943 | 14:6589213 | 14 | 65892139 | C | G | 0.352083 | 0.17854 | 0.021058 | 2.28E-17 | cis |
| miR-625-5p | rs4581615 | 14:6605594 | 14 | 66055943 | C | A | 0.499128 | 0.145272 | 0.019735 | 1.82E-13 | cis |
| miR-625-5p | rs7271448 | 14:6601624 | 14 | 66016247 | A | G | 0.820587 | -0.23284 | 0.025077 | 1.62E-20 | cis |
| miR-625-5p | rs6573614 | 14:6595870 | 14 | 65958703 | G | A | 0.356282 | 0.171846 | 0.02064 | 8.37E-17 | cis |
| miR-625-5p | rs2127870 | 14:6579684 | 14 | 65796846 | G | C | 0.220153 | 0.363782 | 0.02224 | 3.86E-60 | cis |
| miR-625-5p | rs867972 | 14:6589576 | 14 | 65895761 | T | C | 0.50459 | 0.14613 | 0.019867 | 1.90E-13 | cis |
| miR-625-5p | rs1115860 | 14:6611079 | 14 | 66110797 | T | G | 0.365883 | 0.173324 | 0.020734 | 6.31E-17 | cis |
| miR-625-5p | rs2411355 | 14:6606070 | 14 | 66060707 | C | A | 0.49877 | 0.145602 | 0.019744 | 1.65E-13 | cis |
| miR-625-5p | rs4899172 | 14:6577990 | 14 | 65779904 | T | C | 0.213235 | 0.359928 | 0.022679 | 1.01E-56 | cis |
| miR-625-5p | rs7149301 | 14:6623041 | 14 | 66230414 | A | G | 0.452987 | 0.142786 | 0.019331 | 1.51E-13 | cis |
| miR-625-5p | rs7150094 | 14:6593636 | 14 | 65936361 | A | G | 0.346839 | 0.175864 | 0.020851 | 3.33E-17 | cis |
| miR-625-5p | rs8014475 | 14:6581153 | 14 | 65811537 | T | C | 0.34592 | 0.204093 | 0.019825 | 7.44E-25 | cis |
| miR-625-5p | rs7606377 | 14:6576777 | 14 | 65767779 | G | A | 0.843419 | -0.30851 | 0.02639 | 1.43E-31 | cis |
| miR-625-5p | rs1457728 | 14:6610281 | 14 | 66102816 | C | T | 0.821198 | -0.23415 | 0.02536 | 2.63E-20 | cis |
| miR-625-5p | rs3986780 | 14:6579476 | 14 | 65794763 | G | T | 0.053217 | 0.337746 | 0.044685 | 4.08E-14 | cis |
| miR-625-5p | rs1999725 | 14:6590423 | 14 | 65904237 | G | A | 0.373979 | 0.181949 | 0.020557 | 8.67E-19 | cis |
| miR-625-5p | rs2064695 | 14:6606708 | 14 | 66067088 | T | G | 0.40849 | 0.168086 | 0.020171 | 7.88E-17 | cis |
| miR-625-5p | rs1256520 | 14:6573719 | 14 | 65737193 | C | A | 0.64779 | -0.19132 | 0.020361 | 5.66E-21 | cis |
| miR-625-5p | rs2268961 | 14:6614676 | 14 | 66146765 | T | C | 0.501133 | 0.147737 | 0.019817 | 8.97E-14 | cis |
| miR-625-5p | rs1953415 | 14:6598803 | 14 | 65988034 | A | G | 0.355003 | 0.172319 | 0.020622 | 6.49E-17 | cis |
| miR-625-5p | rs7637563 | 14:6575394 | 14 | 65753942 | G | A | 0.888368 | -0.35516 | 0.032567 | 1.09E-27 | cis |
| miR-625-5p | rs1162063 | 14:6589217 | 14 | 65892171 | C | T | 0.417733 | 0.200817 | 0.021592 | 1.40E-20 | cis |
| miR-625-5p | rs1162074 | 14:6589980 | 14 | 65899804 | A | G | 0.502681 | 0.145468 | 0.019793 | 1.99E-13 | cis |
| miR-625-5p | rs6070872 | 14:6611679 | 14 | 66116790 | G | A | 0.819646 | -0.23961 | 0.025764 | 1.40E-20 | cis |
| miR-625-5p | rs8021946 | 14:6584689 | 14 | 65846895 | C | T | 0.054518 | 0.325363 | 0.045724 | 1.11E-12 | cis |
| miR-625-5p | rs7271442 | 14:6586356 | 14 | 65863561 | G | A | 0.814071 | -0.23348 | 0.024581 | 2.13E-21 | cis |
| miR-625-5p | rs730807 | 14:6623929 | 14 | 66239290 | C | T | 0.45402 | 0.143991 | 0.019392 | 1.12E-13 | cis |
| miR-625-5p | rs7142018 | 14:6624167 | 14 | 66241673 | G | T | 0.466429 | 0.152812 | 0.01959 | 6.16E-15 | cis |
| miR-625-5p | rs7465586 | 14:6583114 | 14 | 65831141 | G | A | 0.815043 | -0.23903 | 0.024818 | 5.91E-22 | cis |
| miR-625-5p | rs968540 | 14:6624906 | 14 | 66249064 | G | T | 0.633781 | -0.13512 | 0.020129 | 1.91E-11 | cis |
| miR-625-5p | rs7153297 | 14:6600346 | 14 | 66003466 | T | A | 0.51171 | 0.14732 | 0.019892 | 1.30E-13 | cis |
| miR-625-5p | rs1256521 | 14:6573760 | 14 | 65737608 | T | C | 0.646581 | -0.19102 | 0.020326 | 5.57E-21 | cis |
| miR-625-5p | rs8013442 | 14:6587186 | 14 | 65871861 | A | G | 0.342325 | 0.17882 | 0.020898 | 1.16E-17 | cis |
| miR-625-5p | rs6573625 | 14:6624063 | 14 | 66240634 | T | C | 0.37315 | 0.137387 | 0.020057 | 7.39E-12 | cis |
| miR-625-5p | rs743084 | 14:6623260 | 14 | 66232602 | T | C | 0.452662 | 0.142879 | 0.019322 | 1.42E-13 | cis |
| miR-625-5p | rs2411405 | 14:6623208 | 14 | 66232086 | A | G | 0.448097 | 0.141959 | 0.019234 | 1.57E-13 | cis |
| miR-625-5p | rs743085 | 14:6606813 | 14 | 66068133 | A | G | 0.499418 | 0.145261 | 0.019744 | 1.88E-13 | cis |
| miR-625-5p | rs6795753 | 14:6612458 | 14 | 66124585 | T | G | 0.50137 | 0.146801 | 0.01989 | 1.58E-13 | cis |
| miR-625-5p | rs2152374 | 14:6587344 | 14 | 65873447 | A | G | 0.349733 | 0.174624 | 0.021037 | 1.03E-16 | cis |
| miR-625-5p | rs2411816 | 14:6599488 | 14 | 65994880 | G | A | 0.50839 | 0.14508 | 0.019834 | 2.58E-13 | cis |

| | | | | | | | | | | | |
|---|---|---|---|---|---|---|---|---|---|---|---|
| miR-625-5 | rs7148737 | 14:662300 | 14 | 66230032 | C | A | 0.386987 | 0.146174 | 0.019789 | 1.51E-13 | cis |
| miR-625-5 | rs8015222 | 14:658360 | 14 | 65836095 | T | A | 0.052734 | 0.328492 | 0.045567 | 5.63E-13 | cis |
| miR-625-5 | rs7692276 | 14:657375 | 14 | 65737580 | C | G | 0.705647 | -0.19194 | 0.021474 | 3.95E-19 | cis |
| miR-625-5 | rs7271443 | 14:659028 | 14 | 65902814 | A | G | 0.815449 | -0.23104 | 0.024484 | 3.86E-21 | cis |
| miR-625-5 | rs2210805 | 14:658927 | 14 | 65892724 | C | G | 0.354478 | 0.174717 | 0.020743 | 3.67E-17 | cis |
| miR-625-5 | rs3935480 | 14:662879 | 14 | 66287921 | C | A | 0.881484 | -0.25286 | 0.031611 | 1.25E-15 | cis |
| miR-625-5 | rs1162819 | 14:658912 | 14 | 65891241 | A | G | 0.495088 | 0.14713 | 0.019857 | 1.27E-13 | cis |
| miR-625-5 | rs2411357 | 14:660511 | 14 | 66051100 | G | T | 0.499896 | 0.145832 | 0.019747 | 1.53E-13 | cis |
| miR-625-5 | rs1162536 | 14:662328 | 14 | 66232869 | G | A | 0.45263 | 0.142912 | 0.019323 | 1.40E-13 | cis |
| miR-625-5 | rs1214747 | 14:657950 | 14 | 65795081 | C | T | 0.845789 | -0.32979 | 0.02651 | 1.58E-35 | cis |
| miR-625-5 | rs1214777 | 14:657976 | 14 | 65797693 | A | T | 0.846077 | -0.32849 | 0.026194 | 4.47E-36 | cis |
| miR-625-5 | rs899960 | 14:657899 | 14 | 65789991 | C | G | 0.053722 | 0.334306 | 0.044654 | 7.07E-14 | cis |
| miR-625-5 | rs4902400 | 14:659571 | 14 | 65957115 | A | G | 0.508016 | 0.144622 | 0.019874 | 3.41E-13 | cis |
| miR-625-5 | rs1998036 | 14:661381 | 14 | 66138199 | G | C | 0.498979 | 0.148436 | 0.019879 | 8.20E-14 | cis |
| miR-625-5 | rs2127869 | 14:657943 | 14 | 65794352 | T | C | 0.291045 | 0.290832 | 0.020564 | 2.08E-45 | cis |
| miR-625-5 | rs1289309 | 14:657890 | 14 | 65789077 | G | A | 0.385962 | 0.21328 | 0.019213 | 1.24E-28 | cis |
| miR-625-5 | rs5963675 | 14:662564 | 14 | 66256489 | T | C | 0.541442 | -0.13934 | 0.019364 | 6.20E-13 | cis |
| miR-625-5 | rs6573616 | 14:660468 | 14 | 66046816 | A | G | 0.343484 | 0.172142 | 0.020598 | 6.41E-17 | cis |
| miR-625-5 | rs1177595 | 14:657590 | 14 | 65759023 | G | C | 0.845965 | -0.31163 | 0.026756 | 2.37E-31 | cis |
| miR-625-5 | rs7424586 | 14:660940 | 14 | 66094063 | A | G | 0.820602 | -0.23379 | 0.025222 | 1.87E-20 | cis |
| miR-625-5 | rs7151561 | 14:658906 | 14 | 65890603 | T | C | 0.350627 | 0.175974 | 0.020911 | 3.92E-17 | cis |
| miR-625-5 | rs4359350 | 14:658637 | 14 | 65863748 | T | C | 0.404776 | 0.184939 | 0.02022 | 5.88E-20 | cis |
| miR-625-5 | rs2183277 | 14:660354 | 14 | 66035429 | T | C | 0.342723 | 0.172432 | 0.020657 | 6.99E-17 | cis |
| miR-625-5 | rs7145500 | 14:658842 | 14 | 65884212 | G | A | 0.354536 | 0.173427 | 0.020668 | 4.81E-17 | cis |
| miR-625-5 | rs8022764 | 14:662446 | 14 | 66244655 | T | G | 0.464914 | 0.144871 | 0.019587 | 1.40E-13 | cis |
| miR-625-5 | rs9323460 | 14:659331 | 14 | 65933199 | T | G | 0.357542 | 0.174201 | 0.02067 | 3.53E-17 | cis |
| miR-625-5 | rs6573612 | 14:659128 | 14 | 65912829 | G | A | 0.355177 | 0.173057 | 0.020731 | 6.96E-17 | cis |
| miR-625-5 | rs4902404 | 14:660470 | 14 | 66047022 | T | C | 0.344394 | 0.172189 | 0.02055 | 5.34E-17 | cis |
| miR-625-5 | rs6573609 | 14:658614 | 14 | 65861402 | A | C | 0.390167 | 0.175181 | 0.020391 | 8.61E-18 | cis |
| miR-625-5 | rs1184726 | 14:657756 | 14 | 65775695 | T | G | 0.67304 | -0.15042 | 0.020266 | 1.15E-13 | cis |
| miR-625-5 | rs7271640 | 14:660660 | 14 | 66066001 | A | G | 0.820429 | -0.2331 | 0.025151 | 1.90E-20 | cis |
| miR-625-5 | rs1256518 | 14:657359 | 14 | 65735932 | T | G | 0.682112 | -0.20756 | 0.020966 | 4.17E-23 | cis |
| miR-625-5 | rs1256540 | 14:657640 | 14 | 65764069 | T | C | 0.645897 | -0.22306 | 0.019669 | 8.26E-30 | cis |
| miR-625-5 | rs8005243 | 14:660656 | 14 | 66065634 | C | G | 0.346908 | 0.172472 | 0.02067 | 7.17E-17 | cis |
| miR-625-5 | rs2185454 | 14:658705 | 14 | 65870594 | G | T | 0.352958 | 0.175668 | 0.020698 | 2.11E-17 | cis |
| miR-625-5 | rs7150648 | 14:658901 | 14 | 65890130 | T | C | 0.347708 | 0.175934 | 0.020902 | 3.87E-17 | cis |
| miR-625-5 | rs1214723 | 14:659949 | 14 | 65994949 | C | T | 0.931906 | -0.41769 | 0.042578 | 1.02E-22 | cis |
| miR-625-5 | rs2184603 | 14:659306 | 14 | 65930670 | A | C | 0.500695 | 0.146773 | 0.019945 | 1.86E-13 | cis |
| miR-625-5 | rs1256536 | 14:657624 | 14 | 65762476 | A | G | 0.656085 | -0.2135 | 0.020061 | 1.89E-26 | cis |
| miR-625-5 | rs1256544 | 14:657764 | 14 | 65776415 | G | C | 0.053147 | 0.335592 | 0.044505 | 4.68E-14 | cis |
| miR-6803- | rs871716 | 16:886212 | 16 | 88621200 | G | A | 0.806439 | -0.19619 | 0.02091 | 6.41E-21 | trans |
| miR-6803- | rs1164930 | 16:886168 | 16 | 88616811 | G | C | 0.653135 | 0.132491 | 0.019106 | 4.07E-12 | trans |
| miR-6803- | rs9935772 | 16:886117 | 16 | 88611754 | T | C | 0.689161 | -0.20718 | 0.01873 | 1.93E-28 | trans |
| miR-6803- | rs3598059 | 16:886173 | 16 | 88617310 | G | A | 0.7068 | -0.20629 | 0.017743 | 3.01E-31 | trans |
| miR-6803- | rs1164749 | 16:886200 | 16 | 88620037 | C | T | 0.42548 | 0.165562 | 0.016502 | 1.10E-23 | trans |
| miR-6803- | rs6204905 | 16:886195 | 16 | 88619579 | C | T | 0.652359 | 0.13183 | 0.019051 | 4.52E-12 | trans |
| miR-6803- | rs5865062 | 16:886161 | 16 | 88616153 | G | A | 0.621185 | 0.132207 | 0.018567 | 1.07E-12 | trans |
| miR-6803- | rs6749382 | 16:886078 | 16 | 88607889 | T | C | 0.671206 | -0.21255 | 0.018944 | 3.25E-29 | trans |
| miR-6803- | rs4782373 | 16:886067 | 16 | 88606707 | G | A | 0.774365 | -0.17766 | 0.019706 | 1.96E-19 | trans |
| miR-6803- | rs6204901 | 16:886143 | 16 | 88614351 | G | A | 0.753109 | -0.2025 | 0.019183 | 4.75E-26 | trans |
| miR-6803- | rs7195402 | 16:886222 | 16 | 88622286 | T | C | 0.628701 | 0.12964 | 0.01854 | 2.70E-12 | trans |
| miR-6803- | rs8062370 | 16:886187 | 16 | 88618757 | T | C | 0.511583 | -0.13481 | 0.017808 | 3.72E-14 | trans |
| miR-6803- | rs9928508 | 16:886120 | 16 | 88612013 | G | A | 0.738454 | -0.19327 | 0.019199 | 7.75E-24 | trans |
| miR-6803- | rs6204901 | 16:886098 | 16 | 88609891 | G | A | 0.692923 | -0.20254 | 0.018683 | 2.21E-27 | trans |
| miR-6803- | rs8062396 | 16:886187 | 16 | 88618795 | T | A | 0.511294 | -0.13476 | 0.017802 | 3.73E-14 | trans |
| miR-6803- | rs9935767 | 16:886117 | 16 | 88611740 | T | C | 0.687423 | -0.20665 | 0.018611 | 1.21E-28 | trans |
| miR-6803- | rs4782376 | 16:886125 | 16 | 88612565 | T | C | 0.73543 | -0.19561 | 0.019369 | 5.58E-24 | trans |
| miR-6803- | rs2086392 | 16:886198 | 16 | 88619835 | T | C | 0.448031 | 0.168637 | 0.016863 | 1.52E-23 | trans |
| miR-6803- | rs899739 | 16:886206 | 16 | 88620628 | A | G | 0.428159 | 0.165678 | 0.0166 | 1.85E-23 | trans |
| miR-6803- | rs2870365 | 16:886206 | 16 | 88620640 | G | A | 0.70835 | -0.20403 | 0.017584 | 3.97E-31 | trans |
| miR-6803- | rs1107666 | 16:886218 | 16 | 88621873 | T | C | 0.43175 | 0.16926 | 0.016724 | 4.46E-24 | trans |
| miR-6803- | rs9931930 | 16:886052 | 16 | 88605294 | C | T | 0.809345 | -0.20159 | 0.023199 | 3.64E-18 | trans |
| miR-6803- | rs1333440 | 16:886172 | 16 | 88617225 | C | T | 0.708025 | -0.20625 | 0.017751 | 3.30E-31 | trans |

| | | | | | | | | | | | |
|---|---|---|---|---|---|---|---|---|---|---|---|
| miR-6803-3 | rs9937571 | 16:886183 | 16 | 88618382 | A | G | 0.479994 | -0.13601 | 0.018358 | 1.27E-13 | trans |
| miR-6803-3 | rs1016341 | 16:886192 | 16 | 88619289 | C | A | 0.70803 | -0.20591 | 0.017701 | 2.81E-31 | trans |
| miR-6803-3 | rs1107666 | 16:886106 | 16 | 88610614 | C | G | 0.689457 | -0.20609 | 0.018446 | 5.57E-29 | trans |
| miR-6803-3 | rs3430110 | 16:886143 | 16 | 88614310 | T | C | 0.426741 | 0.169755 | 0.016876 | 8.37E-24 | trans |
| miR-6803-3 | rs6204905 | 16:886187 | 16 | 88618788 | T | C | 0.652329 | 0.132169 | 0.019049 | 3.97E-12 | trans |
| miR-6803-3 | rs9796884 | 16:886111 | 16 | 88611166 | C | G | 0.697275 | -0.20239 | 0.0187 | 2.68E-27 | trans |
| miR-6803-3 | rs1016341 | 16:886194 | 16 | 88619493 | C | T | 0.708147 | -0.20622 | 0.017703 | 2.33E-31 | trans |
| miR-6803-3 | rs9928428 | 16:886119 | 16 | 88611943 | G | C | 0.685418 | -0.20505 | 0.018497 | 1.48E-28 | trans |
| miR-6803-3 | rs2340959 | 16:886200 | 16 | 88620002 | G | A | 0.428157 | 0.167349 | 0.01656 | 5.23E-24 | trans |
| miR-6803-3 | rs2340960 | 16:886199 | 16 | 88619944 | T | C | 0.451698 | 0.160177 | 0.016877 | 2.30E-21 | trans |
| miR-6803-3 | rs7195401 | 16:886222 | 16 | 88622285 | T | C | 0.656089 | 0.133272 | 0.019131 | 3.25E-12 | trans |
| miR-6803-3 | rs2879897 | 16:886201 | 16 | 88620195 | T | G | 0.424016 | 0.16539 | 0.016455 | 9.09E-24 | trans |
| miR-6803-3 | rs3443273 | 16:886076 | 16 | 88607664 | G | A | 0.69548 | -0.20824 | 0.019111 | 1.21E-27 | trans |
| miR-6803-3 | rs3445792 | 16:886142 | 16 | 88614286 | A | G | 0.694459 | -0.20393 | 0.018086 | 1.74E-29 | trans |
| miR-99b-5p | rs579459 | 9:1361541 | 9 | 1.36E+08 | T | C | 0.787293 | -0.12665 | 0.015805 | 1.12E-15 | trans |
| miR-99b-5p | rs676457 | 9:1361462 | 9 | 1.36E+08 | A | T | 0.664166 | -0.10724 | 0.01374 | 5.96E-15 | trans |
| miR-99b-5p | rs677355 | 9:1361460 | 9 | 1.36E+08 | G | A | 0.670198 | -0.10743 | 0.013978 | 1.52E-14 | trans |
| miR-99b-5p | rs2769071 | 9:1361459 | 9 | 1.36E+08 | A | G | 0.663117 | -0.10759 | 0.013762 | 5.37E-15 | trans |
| miR-99b-5p | rs543968 | 9:1361431 | 9 | 1.36E+08 | T | C | 0.653829 | -0.10627 | 0.01371 | 9.07E-15 | trans |
| miR-99b-5p | rs649129 | 9:1361543 | 9 | 1.36E+08 | C | T | 0.787432 | -0.1268 | 0.015815 | 1.08E-15 | trans |
| miR-99b-5p | rs613534 | 9:1361431 | 9 | 1.36E+08 | A | G | 0.653693 | -0.10617 | 0.013709 | 9.58E-15 | trans |
| miR-99b-5p | rs597988 | 9:1361442 | 9 | 1.36E+08 | T | A | 0.686751 | -0.11166 | 0.014627 | 2.28E-14 | trans |
| miR-99b-5p | rs635634 | 9:1361550 | 9 | 1.36E+08 | C | T | 0.80599 | -0.13206 | 0.016369 | 7.16E-16 | trans |
| miR-99b-5p | rs550057 | 9:1361465 | 9 | 1.36E+08 | C | T | 0.734085 | -0.10212 | 0.014764 | 4.61E-12 | trans |
| miR-99b-5p | rs657152 | 9:1361392 | 9 | 1.36E+08 | C | A | 0.645336 | -0.10576 | 0.013458 | 3.88E-15 | trans |
| miR-99b-5p | rs494242 | 9:1361451 | 9 | 1.36E+08 | C | T | 0.644977 | -0.1059 | 0.013481 | 3.97E-15 | trans |
| miR-99b-5p | rs2519093 | 9:1361418 | 9 | 1.36E+08 | C | T | 0.809892 | -0.13188 | 0.01671 | 2.97E-15 | trans |
| miR-99b-5p | rs1401144 | 9:1361443 | 9 | 1.36E+08 | T | C | 0.690247 | -0.1104 | 0.014785 | 8.22E-14 | trans |
| miR-99b-5p | rs8176663 | 9:1361444 | 9 | 1.36E+08 | T | C | 0.663959 | -0.10722 | 0.013737 | 5.95E-15 | trans |
| miR-99b-5p | rs643434 | 9:1361423 | 9 | 1.36E+08 | G | A | 0.645392 | -0.10574 | 0.013468 | 4.11E-15 | trans |
| miR-99b-5p | rs612169 | 9:1361434 | 9 | 1.36E+08 | A | G | 0.66434 | -0.1073 | 0.013756 | 6.19E-15 | trans |
| miR-99b-5p | rs544873 | 9:1361432 | 9 | 1.36E+08 | G | A | 0.645742 | -0.10568 | 0.013475 | 4.40E-15 | trans |
| miR-99b-5p | rs651007 | 9:1361538 | 9 | 1.36E+08 | C | T | 0.787179 | -0.12698 | 0.015807 | 9.49E-16 | trans |
| miR-99b-5p | rs514659 | 9:1361422 | 9 | 1.36E+08 | A | C | 0.663927 | -0.10721 | 0.013737 | 5.98E-15 | trans |
| miR-99b-5p | rs1165522 | 9:1361490 | 9 | 1.36E+08 | T | A | 0.653719 | -0.10695 | 0.01369 | 5.62E-15 | trans |
| miR-99b-5p | rs597974 | 9:1361442 | 9 | 1.36E+08 | A | G | 0.687926 | -0.11159 | 0.014691 | 3.06E-14 | trans |
| miR-99b-5p | rs493246 | 9:1361449 | 9 | 1.36E+08 | G | A | 0.663234 | -0.10731 | 0.013756 | 6.12E-15 | trans |
| miR-99b-5p | rs543040 | 9:1361430 | 9 | 1.36E+08 | A | T | 0.669196 | -0.10913 | 0.013989 | 6.14E-15 | trans |
| miR-99b-5p | rs554833 | 9:1361471 | 9 | 1.36E+08 | C | T | 0.664057 | -0.10709 | 0.013732 | 6.26E-15 | trans |
| miR-99b-5p | rs491626 | 9:1361448 | 9 | 1.36E+08 | C | T | 0.664173 | -0.10777 | 0.013772 | 5.04E-15 | trans |
| miR-99b-5p | rs527210 | 9:1361464 | 9 | 1.36E+08 | C | T | 0.668712 | -0.10658 | 0.013904 | 1.78E-14 | trans |
| miR-99b-5p | rs582118 | 9:1361454 | 9 | 1.36E+08 | A | G | 0.663872 | -0.10735 | 0.01374 | 5.57E-15 | trans |
| miR-99b-5p | rs676996 | 9:1361460 | 9 | 1.36E+08 | T | G | 0.667516 | -0.10646 | 0.013859 | 1.58E-14 | trans |
| miR-99b-5p | rs545971 | 9:1361433 | 9 | 1.36E+08 | C | T | 0.663766 | -0.10713 | 0.013732 | 6.10E-15 | trans |
| miR-99b-5p | rs505922 | 9:1361492 | 9 | 1.36E+08 | T | C | 0.665748 | -0.10886 | 0.013721 | 2.12E-15 | trans |
| miR-99b-5p | rs495828 | 9:1361548 | 9 | 1.36E+08 | G | T | 0.787276 | -0.12664 | 0.015804 | 1.12E-15 | trans |
| miR-99b-5p | rs507666 | 9:1361493 | 9 | 1.36E+08 | G | A | 0.806573 | -0.13244 | 0.016402 | 6.77E-16 | trans |
| miR-99b-5p | rs532436 | 9:1361498 | 9 | 1.36E+08 | G | A | 0.8067 | -0.13261 | 0.016413 | 6.49E-16 | trans |
| miR-99b-5p | rs576125 | 9:1361443 | 9 | 1.36E+08 | G | A | 0.688677 | -0.11108 | 0.014728 | 4.64E-14 | trans |
| miR-99b-5p | rs687289 | 9:1361371 | 9 | 1.36E+08 | G | A | 0.668579 | -0.11056 | 0.013909 | 1.89E-15 | trans |
| miR-99b-5p | rs674302 | 9:1361466 | 9 | 1.36E+08 | T | A | 0.663989 | -0.10717 | 0.013729 | 5.92E-15 | trans |
| miR-99b-5p | rs687621 | 9:1361370 | 9 | 1.36E+08 | A | G | 0.663896 | -0.10767 | 0.013735 | 4.52E-15 | trans |
| miR-99b-5p | rs582094 | 9:1361454 | 9 | 1.36E+08 | A | T | 0.663709 | -0.10726 | 0.013734 | 5.72E-15 | trans |
| miR-99b-5p | rs600038 | 9:1361518 | 9 | 1.36E+08 | T | C | 0.787872 | -0.12718 | 0.015846 | 1.00E-15 | trans |
| miR-99b-5p | rs644234 | 9:1361422 | 9 | 1.36E+08 | T | G | 0.645381 | -0.10572 | 0.013458 | 3.98E-15 | trans |
| miR-99b-5p | rs492488 | 9:1361449 | 9 | 1.36E+08 | G | A | 0.663579 | -0.10726 | 0.013748 | 6.09E-15 | trans |
| miR-99b-5p | rs529565 | 9:1361495 | 9 | 1.36E+08 | T | C | 0.665911 | -0.10902 | 0.013729 | 2.00E-15 | trans |
| miR-99b-5p | rs495203 | 9:1361452 | 9 | 1.36E+08 | C | T | 0.663279 | -0.10728 | 0.013745 | 5.96E-15 | trans |
| let-7a-5p | rs1165522 | 9:1361490 | 9 | 1.36E+08 | T | A | 0.653719 | -0.08457 | 0.011334 | 8.59E-14 | trans |
| let-7a-5p | rs2519093 | 9:1361418 | 9 | 1.36E+08 | C | T | 0.809892 | -0.09389 | 0.01387 | 1.29E-11 | trans |
| let-7a-5p | rs494242 | 9:1361451 | 9 | 1.36E+08 | C | T | 0.644977 | -0.08006 | 0.011174 | 7.78E-13 | trans |
| let-7a-5p | rs495828 | 9:1361548 | 9 | 1.36E+08 | G | T | 0.787276 | -0.09272 | 0.013111 | 1.53E-12 | trans |
| let-7a-5p | rs505922 | 9:1361492 | 9 | 1.36E+08 | T | C | 0.665748 | -0.07803 | 0.011387 | 7.26E-12 | trans |

| | | | | | | | | | | | |
|---|---|---|---|---|---|---|---|---|---|---|---|
| let-7a-5p | rs529565 | 9:1361495 | 9 | 1.36E+08 | T | C | 0.665911 | -0.07822 | 0.011394 | 6.63E-12 | trans |
| let-7a-5p | rs543040 | 9:1361430 | 9 | 1.36E+08 | A | T | 0.669196 | -0.07765 | 0.011611 | 2.26E-11 | trans |
| let-7a-5p | rs543968 | 9:1361431 | 9 | 1.36E+08 | T | C | 0.653829 | -0.08143 | 0.011359 | 7.59E-13 | trans |
| let-7a-5p | rs544873 | 9:1361432 | 9 | 1.36E+08 | G | A | 0.645742 | -0.07991 | 0.011169 | 8.38E-13 | trans |
| let-7a-5p | rs579459 | 9:1361541 | 9 | 1.36E+08 | T | C | 0.787293 | -0.09273 | 0.013112 | 1.52E-12 | trans |
| let-7a-5p | rs600038 | 9:1361518 | 9 | 1.36E+08 | T | C | 0.787872 | -0.09303 | 0.013146 | 1.48E-12 | trans |
| let-7a-5p | rs613534 | 9:1361431 | 9 | 1.36E+08 | A | G | 0.653693 | -0.0815 | 0.011358 | 7.20E-13 | trans |
| let-7a-5p | rs643434 | 9:1361423 | 9 | 1.36E+08 | G | A | 0.645392 | -0.07973 | 0.011164 | 9.18E-13 | trans |
| let-7a-5p | rs644234 | 9:1361422 | 9 | 1.36E+08 | T | G | 0.645381 | -0.07981 | 0.011155 | 8.38E-13 | trans |
| let-7a-5p | rs649129 | 9:1361543 | 9 | 1.36E+08 | C | T | 0.787432 | -0.09284 | 0.01312 | 1.48E-12 | trans |
| let-7a-5p | rs651007 | 9:1361538 | 9 | 1.36E+08 | C | T | 0.787179 | -0.0928 | 0.013114 | 1.48E-12 | trans |
| let-7a-5p | rs657152 | 9:1361392 | 9 | 1.36E+08 | C | A | 0.645336 | -0.07978 | 0.011155 | 8.57E-13 | trans |
| let-7a-5p | rs687289 | 9:1361371 | 9 | 1.36E+08 | G | A | 0.668579 | -0.07734 | 0.01155 | 2.14E-11 | trans |
| miR-1254 | rs1382152 | 3:5837215 | 3 | 58372157 | G | A | 0.933752 | 0.253214 | 0.037565 | 1.58E-11 | trans |
| miR-1254 | rs1809770 | 3:5831846 | 3 | 58318464 | A | C | 0.919174 | 0.244344 | 0.034085 | 7.58E-13 | trans |
| miR-1254 | rs1854079 | 3:5831847 | 3 | 58318477 | G | A | 0.914351 | 0.234038 | 0.033995 | 5.80E-12 | trans |
| miR-1254 | rs3567747 | 3:5818363 | 3 | 58183636 | G | A | 0.912834 | 0.225624 | 0.033038 | 8.54E-12 | trans |
| miR-1254 | rs6772228 | 3:5837601 | 3 | 58376019 | T | A | 0.934269 | 0.254259 | 0.037613 | 1.38E-11 | trans |
| miR-1254 | rs7308155 | 3:5830293 | 3 | 58302935 | C | T | 0.920004 | 0.245075 | 0.034097 | 6.60E-13 | trans |
| miR-1254 | rs7748001 | 3:5833950 | 3 | 58339505 | G | A | 0.933847 | 0.256242 | 0.037593 | 9.35E-12 | trans |
| miR-133a-3 | rs1163559 | 20:611509 | 20 | 61150959 | G | A | 0.819024 | -0.17691 | 0.025998 | 1.01E-11 | cis |
| miR-133a-3 | rs1174196 | 20:611538 | 20 | 61153858 | G | A | 0.818584 | -0.17513 | 0.025957 | 1.51E-11 | cis |
| miR-133a-3 | rs1178585 | 20:611492 | 20 | 61149264 | G | A | 0.861146 | -0.20537 | 0.02963 | 4.17E-12 | cis |
| miR-133a-3 | rs1262460 | 20:611552 | 20 | 61155298 | C | A | 0.796375 | -0.17491 | 0.025247 | 4.27E-12 | cis |
| miR-133a-3 | rs7391829 | 20:611520 | 20 | 61152078 | C | T | 0.814283 | -0.17615 | 0.026061 | 1.39E-11 | cis |
| miR-181a-5 | rs1036332 | 1:1990124 | 1 | 1.99E+08 | A | C | 0.260189 | 0.172538 | 0.013837 | 1.10E-35 | cis |
| miR-181a-5 | rs1040959 | 1:1988958 | 1 | 1.99E+08 | A | C | 0.766999 | -0.10003 | 0.014691 | 9.81E-12 | cis |
| miR-181a-5 | rs1040960 | 1:1988959 | 1 | 1.99E+08 | G | T | 0.766066 | -0.1002 | 0.014695 | 9.21E-12 | cis |
| miR-181a-5 | rs1073755 | 1:1991209 | 1 | 1.99E+08 | T | C | 0.508916 | -0.09853 | 0.013264 | 1.10E-13 | cis |
| miR-181a-5 | rs1080059 | 1:1988708 | 1 | 1.99E+08 | T | C | 0.760087 | -0.10817 | 0.0147 | 1.86E-13 | cis |
| miR-181a-5 | rs1080060 | 1:1989145 | 1 | 1.99E+08 | A | G | 0.244491 | 0.102312 | 0.014406 | 1.23E-12 | cis |
| miR-181a-5 | rs1080060 | 1:1989196 | 1 | 1.99E+08 | A | G | 0.238849 | 0.106727 | 0.01456 | 2.30E-13 | cis |
| miR-181a-5 | rs1080060 | 1:1990049 | 1 | 1.99E+08 | C | G | 0.663982 | 0.087564 | 0.01307 | 2.09E-11 | cis |
| miR-181a-5 | rs1091960 | 1:1988826 | 1 | 1.99E+08 | G | A | 0.764985 | -0.10147 | 0.014749 | 6.00E-12 | cis |
| miR-181a-5 | rs1091961 | 1:1989749 | 1 | 1.99E+08 | C | T | 0.243033 | 0.166882 | 0.014556 | 1.99E-30 | cis |
| miR-181a-5 | rs1091961 | 1:1990380 | 1 | 1.99E+08 | T | C | 0.841859 | -0.15217 | 0.016799 | 1.33E-19 | cis |
| miR-181a-5 | rs1091965 | 1:1992486 | 1 | 1.99E+08 | A | G | 0.667251 | -0.10637 | 0.013119 | 5.15E-16 | cis |
| miR-181a-5 | rs1124025 | 1:1989798 | 1 | 1.99E+08 | C | G | 0.238162 | 0.170617 | 0.014497 | 5.62E-32 | cis |
| miR-181a-5 | rs1204800 | 1:1989153 | 1 | 1.99E+08 | T | C | 0.235546 | 0.100871 | 0.014669 | 6.14E-12 | cis |
| miR-181a-5 | rs1209687 | 1:1989144 | 1 | 1.99E+08 | G | A | 0.244151 | 0.101854 | 0.014401 | 1.52E-12 | cis |
| miR-181a-5 | rs1212331 | 1:1988726 | 1 | 1.99E+08 | G | T | 0.764104 | -0.10254 | 0.014805 | 4.33E-12 | cis |
| miR-181a-5 | rs1212520 | 1:1989920 | 1 | 1.99E+08 | A | G | 0.260715 | 0.177971 | 0.013845 | 8.14E-38 | cis |
| miR-181a-5 | rs1213024 | 1:1988841 | 1 | 1.99E+08 | G | C | 0.765569 | -0.10198 | 0.014754 | 4.79E-12 | cis |
| miR-181a-5 | rs1213201 | 1:1988820 | 1 | 1.99E+08 | T | C | 0.755495 | -0.10683 | 0.014472 | 1.55E-13 | cis |
| miR-181a-5 | rs1214202 | 1:1988815 | 1 | 1.99E+08 | C | T | 0.766397 | -0.10192 | 0.014796 | 5.65E-12 | cis |
| miR-181a-5 | rs1223917 | 1:1989764 | 1 | 1.99E+08 | C | T | 0.245157 | 0.163804 | 0.014397 | 5.38E-30 | cis |
| miR-181a-5 | rs1273277 | 1:1990687 | 1 | 1.99E+08 | C | A | 0.518257 | -0.12018 | 0.014272 | 3.73E-17 | cis |
| miR-181a-5 | rs1275009 | 1:1990708 | 1 | 1.99E+08 | C | T | 0.540441 | -0.10242 | 0.013128 | 6.10E-15 | cis |
| miR-181a-5 | rs1275163 | 1:1988916 | 1 | 1.99E+08 | C | T | 0.765073 | -0.1014 | 0.014694 | 5.18E-12 | cis |
| miR-181a-5 | rs1275164 | 1:1988944 | 1 | 1.99E+08 | A | G | 0.754678 | -0.10709 | 0.014304 | 7.08E-14 | cis |
| miR-181a-5 | rs1347323 | 1:1989704 | 1 | 1.99E+08 | A | G | 0.257803 | 0.11992 | 0.014395 | 8.03E-17 | cis |
| miR-181a-5 | rs1434282 | 1:1990107 | 1 | 1.99E+08 | C | T | 0.271104 | 0.177906 | 0.013618 | 5.30E-39 | cis |
| miR-181a-5 | rs1454632 | 1:1988420 | 1 | 1.99E+08 | A | G | 0.886807 | -0.13368 | 0.019816 | 1.52E-11 | cis |
| miR-181a-5 | rs1482660 | 1:1991115 | 1 | 1.99E+08 | A | G | 0.581738 | -0.09433 | 0.013553 | 3.39E-12 | cis |
| miR-181a-5 | rs167735 | 1:1990643 | 1 | 1.99E+08 | G | A | 0.505181 | -0.09148 | 0.01296 | 1.68E-12 | cis |
| miR-181a-5 | rs1684410 | 1:1990115 | 1 | 1.99E+08 | G | T | 0.842513 | -0.14945 | 0.016947 | 1.16E-18 | cis |
| miR-181a-5 | rs1762558 | 1:1989904 | 1 | 1.99E+08 | G | A | 0.74365 | -0.17886 | 0.013895 | 6.49E-38 | cis |
| miR-181a-5 | rs1763147 | 1:1992522 | 1 | 1.99E+08 | T | A | 0.668759 | -0.10637 | 0.013247 | 9.73E-16 | cis |
| miR-181a-5 | rs188926 | 1:1990672 | 1 | 1.99E+08 | T | C | 0.53384 | -0.10559 | 0.012882 | 2.47E-16 | cis |
| miR-181a-5 | rs1892432 | 1:1989406 | 1 | 1.99E+08 | T | C | 0.234225 | 0.101149 | 0.014723 | 6.41E-12 | cis |
| miR-181a-5 | rs1938038 | 1:1989248 | 1 | 1.99E+08 | A | G | 0.250919 | 0.099347 | 0.014575 | 9.35E-12 | cis |
| miR-181a-5 | rs2033565 | 1:1990150 | 1 | 1.99E+08 | A | T | 0.16327 | 0.149375 | 0.016865 | 8.21E-19 | cis |
| miR-181a-5 | rs2033566 | 1:1990148 | 1 | 1.99E+08 | T | G | 0.211961 | 0.117053 | 0.015337 | 2.31E-14 | cis |

| | | | | | | | | | | | |
|---|---|---|---|---|---|---|---|---|---|---|---|
| miR-181a-5 | rs2060581 | 1:1992502 | 1 | 1.99E+08 | T | C | 0.66838 | -0.1073 | 0.013201 | 4.36E-16 | cis |
| miR-181a-5 | rs2360962 | 1:1990047 | 1 | 1.99E+08 | A | T | 0.261757 | 0.178107 | 0.013794 | 3.87E-38 | cis |
| miR-181a-5 | rs2624882 | 1:1991077 | 1 | 1.99E+08 | A | G | 0.514062 | -0.10486 | 0.013065 | 1.01E-15 | cis |
| miR-181a-5 | rs2624884 | 1:1991177 | 1 | 1.99E+08 | G | C | 0.504555 | -0.10147 | 0.013174 | 1.34E-14 | cis |
| miR-181a-5 | rs2648770 | 1:1991092 | 1 | 1.99E+08 | T | G | 0.507982 | -0.09732 | 0.013186 | 1.57E-13 | cis |
| miR-181a-5 | rs2648772 | 1:1991147 | 1 | 1.99E+08 | C | T | 0.519051 | -0.10108 | 0.013173 | 1.67E-14 | cis |
| miR-181a-5 | rs2648773 | 1:1991157 | 1 | 1.99E+08 | G | A | 0.50507 | -0.10129 | 0.013172 | 1.47E-14 | cis |
| miR-181a-5 | rs2648774 | 1:1991161 | 1 | 1.99E+08 | C | G | 0.504916 | -0.10132 | 0.013175 | 1.47E-14 | cis |
| miR-181a-5 | rs322873 | 1:1991047 | 1 | 1.99E+08 | T | G | 0.538344 | -0.09694 | 0.013612 | 1.06E-12 | cis |
| miR-181a-5 | rs322874 | 1:1991034 | 1 | 1.99E+08 | C | A | 0.504259 | -0.09847 | 0.013128 | 6.34E-14 | cis |
| miR-181a-5 | rs322875 | 1:1991026 | 1 | 1.99E+08 | C | T | 0.518914 | -0.09953 | 0.013126 | 3.38E-14 | cis |
| miR-181a-5 | rs322876 | 1:1991020 | 1 | 1.99E+08 | C | A | 0.503793 | -0.09984 | 0.013118 | 2.72E-14 | cis |
| miR-181a-5 | rs322877 | 1:1990988 | 1 | 1.99E+08 | C | T | 0.508326 | -0.09461 | 0.013088 | 4.87E-13 | cis |
| miR-181a-5 | rs322879 | 1:1990382 | 1 | 1.99E+08 | C | T | 0.510352 | -0.09755 | 0.012799 | 2.50E-14 | cis |
| miR-181a-5 | rs322881 | 1:1990353 | 1 | 1.99E+08 | T | C | 0.506195 | -0.10244 | 0.01275 | 9.38E-16 | cis |
| miR-181a-5 | rs322887 | 1:1990263 | 1 | 1.99E+08 | A | G | 0.834392 | -0.15642 | 0.016492 | 2.43E-21 | cis |
| miR-181a-5 | rs322889 | 1:1990255 | 1 | 1.99E+08 | C | G | 0.786955 | -0.12461 | 0.015018 | 1.07E-16 | cis |
| miR-181a-5 | rs322890 | 1:1990627 | 1 | 1.99E+08 | C | T | 0.503457 | -0.09172 | 0.012934 | 1.33E-12 | cis |
| miR-181a-5 | rs322891 | 1:1990625 | 1 | 1.99E+08 | C | A | 0.4987 | -0.09256 | 0.013 | 1.08E-12 | cis |
| miR-181a-5 | rs322892 | 1:1990625 | 1 | 1.99E+08 | C | T | 0.503793 | -0.09161 | 0.012939 | 1.44E-12 | cis |
| miR-181a-5 | rs322893 | 1:1990616 | 1 | 1.99E+08 | T | C | 0.503696 | -0.09155 | 0.012937 | 1.48E-12 | cis |
| miR-181a-5 | rs322894 | 1:1990611 | 1 | 1.99E+08 | C | T | 0.516952 | -0.09237 | 0.013176 | 2.37E-12 | cis |
| miR-181a-5 | rs322895 | 1:1990938 | 1 | 1.99E+08 | C | T | 0.502544 | -0.09969 | 0.013068 | 2.37E-14 | cis |
| miR-181a-5 | rs322898 | 1:1990913 | 1 | 1.99E+08 | G | A | 0.514892 | -0.10346 | 0.013022 | 1.94E-15 | cis |
| miR-181a-5 | rs322901 | 1:1990894 | 1 | 1.99E+08 | G | C | 0.503019 | -0.09975 | 0.013055 | 2.16E-14 | cis |
| miR-181a-5 | rs322906 | 1:1990837 | 1 | 1.99E+08 | G | T | 0.499842 | -0.09783 | 0.013111 | 8.54E-14 | cis |
| miR-181a-5 | rs322907 | 1:1990825 | 1 | 1.99E+08 | G | T | 0.500608 | -0.09894 | 0.013128 | 4.84E-14 | cis |
| miR-181a-5 | rs322911 | 1:1990763 | 1 | 1.99E+08 | G | C | 0.504333 | -0.09998 | 0.013086 | 2.17E-14 | cis |
| miR-181a-5 | rs322912 | 1:1990743 | 1 | 1.99E+08 | C | T | 0.52926 | -0.09777 | 0.013089 | 8.01E-14 | cis |
| miR-181a-5 | rs322914 | 1:1990719 | 1 | 1.99E+08 | A | T | 0.513191 | -0.10012 | 0.013215 | 3.55E-14 | cis |
| miR-181a-5 | rs322915 | 1:1990707 | 1 | 1.99E+08 | G | A | 0.518776 | -0.09967 | 0.013107 | 2.86E-14 | cis |
| miR-181a-5 | rs322916 | 1:1990691 | 1 | 1.99E+08 | C | T | 0.50214 | -0.09929 | 0.013085 | 3.25E-14 | cis |
| miR-181a-5 | rs322918 | 1:1990686 | 1 | 1.99E+08 | A | G | 0.534816 | -0.10538 | 0.01288 | 2.81E-16 | cis |
| miR-181a-5 | rs322921 | 1:1990588 | 1 | 1.99E+08 | T | A | 0.498422 | -0.09055 | 0.013056 | 4.05E-12 | cis |
| miR-181a-5 | rs322927 | 1:1990218 | 1 | 1.99E+08 | T | G | 0.834224 | -0.15676 | 0.016443 | 1.52E-21 | cis |
| miR-181a-5 | rs322928 | 1:1990218 | 1 | 1.99E+08 | G | A | 0.833953 | -0.15677 | 0.016435 | 1.45E-21 | cis |
| miR-181a-5 | rs322929 | 1:1990209 | 1 | 1.99E+08 | T | C | 0.78491 | -0.12464 | 0.015004 | 9.80E-17 | cis |
| miR-181a-5 | rs322930 | 1:1990205 | 1 | 1.99E+08 | T | A | 0.790845 | -0.11926 | 0.015118 | 3.05E-15 | cis |
| miR-181a-5 | rs322931 | 1:1990198 | 1 | 1.99E+08 | G | A | 0.790484 | -0.1192 | 0.01509 | 2.81E-15 | cis |
| miR-181a-5 | rs322932 | 1:1990193 | 1 | 1.99E+08 | A | T | 0.789147 | -0.1191 | 0.01515 | 3.79E-15 | cis |
| miR-181a-5 | rs322933 | 1:1990180 | 1 | 1.99E+08 | C | T | 0.506039 | -0.10117 | 0.012628 | 1.13E-15 | cis |
| miR-181a-5 | rs3488967 | 1:1992490 | 1 | 1.99E+08 | A | G | 0.66157 | -0.10751 | 0.013292 | 6.03E-16 | cis |
| miR-181a-5 | rs3850622 | 1:1991204 | 1 | 1.99E+08 | G | T | 0.514795 | -0.09657 | 0.013338 | 4.47E-13 | cis |
| miR-181a-5 | rs3850623 | 1:1991205 | 1 | 1.99E+08 | G | A | 0.518649 | -0.10172 | 0.013222 | 1.43E-14 | cis |
| miR-181a-5 | rs427790 | 1:1990972 | 1 | 1.99E+08 | C | T | 0.519007 | -0.09882 | 0.01307 | 4.00E-14 | cis |
| miR-181a-5 | rs4915165 | 1:1989707 | 1 | 1.99E+08 | T | C | 0.253161 | 0.115248 | 0.01452 | 2.07E-15 | cis |
| miR-181a-5 | rs4915331 | 1:1989185 | 1 | 1.99E+08 | T | C | 0.235006 | 0.10124 | 0.014684 | 5.40E-12 | cis |
| miR-181a-5 | rs4915332 | 1:1989185 | 1 | 1.99E+08 | A | G | 0.234929 | 0.101276 | 0.014684 | 5.31E-12 | cis |
| miR-181a-5 | rs4915335 | 1:1989452 | 1 | 1.99E+08 | A | G | 0.238713 | 0.109407 | 0.01468 | 9.14E-14 | cis |
| miR-181a-5 | rs4915347 | 1:1991227 | 1 | 1.99E+08 | C | T | 0.556979 | -0.10226 | 0.014486 | 1.67E-12 | cis |
| miR-181a-5 | rs6427756 | 1:1990007 | 1 | 1.99E+08 | A | G | 0.263331 | 0.176572 | 0.013797 | 1.69E-37 | cis |
| miR-181a-5 | rs6703089 | 1:1989200 | 1 | 1.99E+08 | C | A | 0.236139 | 0.106716 | 0.014528 | 2.05E-13 | cis |
| miR-181a-5 | rs7273975 | 1:1988434 | 1 | 1.99E+08 | T | A | 0.886864 | -0.13378 | 0.019821 | 1.49E-11 | cis |
| miR-181a-5 | rs7273976 | 1:1988640 | 1 | 1.99E+08 | G | T | 0.765001 | -0.10288 | 0.014899 | 5.02E-12 | cis |
| miR-181a-5 | rs7524232 | 1:1990057 | 1 | 1.99E+08 | G | A | 0.670143 | 0.091241 | 0.013136 | 3.77E-12 | cis |
| miR-181a-5 | rs7524630 | 1:1991183 | 1 | 1.99E+08 | A | G | 0.514782 | -0.10413 | 0.01313 | 2.18E-15 | cis |
| miR-181a-5 | rs7529925 | 1:1990072 | 1 | 1.99E+08 | C | T | 0.255718 | 0.172939 | 0.013816 | 6.01E-36 | cis |
| miR-181a-5 | rs7536036 | 1:1990056 | 1 | 1.99E+08 | T | C | 0.669114 | 0.091188 | 0.013143 | 3.97E-12 | cis |
| miR-181a-5 | rs7539100 | 1:1990065 | 1 | 1.99E+08 | T | C | 0.473487 | 0.108623 | 0.012742 | 1.53E-17 | cis |
| miR-181a-5 | rs7539177 | 1:1989464 | 1 | 1.99E+08 | T | G | 0.238596 | 0.109677 | 0.014687 | 8.17E-14 | cis |
| miR-181a-5 | rs7547363 | 1:1991183 | 1 | 1.99E+08 | G | A | 0.519602 | -0.10696 | 0.013176 | 4.75E-16 | cis |
| miR-181a-5 | rs7550394 | 1:1990152 | 1 | 1.99E+08 | T | C | 0.216244 | 0.121548 | 0.015569 | 5.86E-15 | cis |
| miR-181a-5 | rs7890714 | 1:1989878 | 1 | 1.99E+08 | G | A | 0.904551 | -0.15289 | 0.021249 | 6.24E-13 | cis |

| | | | | | | | | | | | |
|---|---|---|---|---|---|---|---|---|---|---|---|
| miR-181a- | rs878254 | 1:1992571 | 1 | 1.99E+08 | T | A | 0.450913 | -0.09101 | 0.013036 | 2.93E-12 | cis |
| miR-181a- | rs881576 | 1:1992577 | 1 | 1.99E+08 | A | C | 0.44377 | -0.09064 | 0.013036 | 3.56E-12 | cis |
| miR-3130- | rs1019976 | 2:2076418 | 2 | 2.08E+08 | G | A | 0.842574 | -0.26684 | 0.032905 | 5.09E-16 | cis |
| miR-3130- | rs1048783 | 2:2076571 | 2 | 2.08E+08 | T | C | 0.762912 | -0.20853 | 0.030173 | 4.81E-12 | cis |
| miR-3130- | rs1093216 | 2:2076163 | 2 | 2.08E+08 | A | G | 0.855067 | -0.27942 | 0.034267 | 3.52E-16 | cis |
| miR-3130- | rs1155295 | 2:2076756 | 2 | 2.08E+08 | C | G | 0.874904 | -0.24728 | 0.036911 | 2.09E-11 | cis |
| miR-3130- | rs1189416 | 2:2076244 | 2 | 2.08E+08 | T | C | 0.859947 | -0.28432 | 0.034753 | 2.81E-16 | cis |
| miR-3130- | rs1189537 | 2:2076547 | 2 | 2.08E+08 | T | C | 0.842391 | -0.26485 | 0.033305 | 1.83E-15 | cis |
| miR-3130- | rs1246816 | 2:2076463 | 2 | 2.08E+08 | T | C | 0.842809 | -0.26718 | 0.032925 | 4.87E-16 | cis |
| miR-3130- | rs1247096 | 2:2076395 | 2 | 2.08E+08 | A | G | 0.866897 | -0.29906 | 0.035691 | 5.33E-17 | cis |
| miR-3130- | rs1247100 | 2:2076397 | 2 | 2.08E+08 | A | G | 0.842907 | -0.27172 | 0.032916 | 1.52E-16 | cis |
| miR-3130- | rs1247846 | 2:2076710 | 2 | 2.08E+08 | T | C | 0.846612 | -0.26339 | 0.033397 | 3.10E-15 | cis |
| miR-3130- | rs1342700 | 2:2076436 | 2 | 2.08E+08 | C | G | 0.84329 | -0.27067 | 0.032943 | 2.10E-16 | cis |
| miR-3130- | rs1683883 | 2:2076297 | 2 | 2.08E+08 | T | A | 0.84321 | -0.26971 | 0.032981 | 2.89E-16 | cis |
| miR-3130- | rs1862087 | 2:2076255 | 2 | 2.08E+08 | T | C | 0.859956 | -0.28439 | 0.034753 | 2.76E-16 | cis |
| miR-3130- | rs1862089 | 2:2076205 | 2 | 2.08E+08 | A | T | 0.854806 | -0.28145 | 0.034374 | 2.66E-16 | cis |
| miR-3130- | rs2059022 | 2:2076782 | 2 | 2.08E+08 | T | C | 0.876031 | -0.24747 | 0.036929 | 2.07E-11 | cis |
| miR-3130- | rs2241347 | 2:2076479 | 2 | 2.08E+08 | C | T | 0.841853 | -0.26645 | 0.032894 | 5.49E-16 | cis |
| miR-3130- | rs2287628 | 2:2076505 | 2 | 2.08E+08 | A | G | 0.843162 | -0.26632 | 0.032968 | 6.58E-16 | cis |
| miR-3130- | rs2287629 | 2:2076504 | 2 | 2.08E+08 | C | A | 0.842588 | -0.26634 | 0.032985 | 6.78E-16 | cis |
| miR-3130- | rs2836579 | 2:2076298 | 2 | 2.08E+08 | A | T | 0.842753 | -0.26995 | 0.032925 | 2.42E-16 | cis |
| miR-3130- | rs3815686 | 2:2076330 | 2 | 2.08E+08 | A | G | 0.843111 | -0.26891 | 0.03292 | 3.12E-16 | cis |
| miR-3130- | rs4673363 | 2:2076694 | 2 | 2.08E+08 | G | A | 0.846328 | -0.26326 | 0.033357 | 2.97E-15 | cis |
| miR-3130- | rs5760097 | 2:2076701 | 2 | 2.08E+08 | G | C | 0.846329 | -0.26298 | 0.033355 | 3.17E-15 | cis |
| miR-3130- | rs6001989 | 2:2076777 | 2 | 2.08E+08 | C | T | 0.87642 | -0.24753 | 0.03705 | 2.37E-11 | cis |
| miR-3130- | rs725980 | 2:2076385 | 2 | 2.08E+08 | C | G | 0.843297 | -0.27114 | 0.032939 | 1.85E-16 | cis |
| miR-3130- | rs7576591 | 2:2076492 | 2 | 2.08E+08 | G | C | 0.843106 | -0.26623 | 0.032952 | 6.52E-16 | cis |
| miR-3130- | rs7580138 | 2:2076152 | 2 | 2.08E+08 | C | A | 0.854791 | -0.27756 | 0.034201 | 4.84E-16 | cis |
| miR-3130- | rs7627046 | 2:2076536 | 2 | 2.08E+08 | G | A | 0.902503 | -0.27656 | 0.04049 | 8.47E-12 | cis |
| miR-3130- | rs768533 | 2:2076408 | 2 | 2.08E+08 | G | A | 0.842932 | -0.27166 | 0.032915 | 1.54E-16 | cis |
| miR-3130- | rs7949904 | 2:2076409 | 2 | 2.08E+08 | A | G | 0.902079 | -0.27782 | 0.04038 | 5.98E-12 | cis |
| miR-3130- | rs888621 | 2:2076493 | 2 | 2.08E+08 | T | G | 0.842681 | -0.26632 | 0.032863 | 5.32E-16 | cis |
| miR-3130- | rs957414 | 2:2076655 | 2 | 2.08E+08 | T | A | 0.84423 | -0.26426 | 0.033198 | 1.72E-15 | cis |
| miR-320b | rs1017438 | 2:6859646 | 2 | 68596469 | C | T | 0.685243 | 0.129366 | 0.014889 | 3.67E-18 | trans |
| miR-320b | rs1049616 | 2:6862008 | 2 | 68620089 | C | T | 0.697496 | 0.127045 | 0.015092 | 3.83E-17 | trans |
| miR-320b | rs1050181 | 2:6862295 | 2 | 68622952 | G | C | 0.544992 | 0.136202 | 0.013895 | 1.10E-22 | trans |
| miR-320b | rs1156629 | 2:6862663 | 2 | 68626631 | T | G | 0.547487 | 0.138094 | 0.014017 | 6.74E-23 | trans |
| miR-320b | rs1158434 | 2:6863552 | 2 | 68635524 | G | C | 0.551072 | 0.14116 | 0.014323 | 6.49E-23 | trans |
| miR-320b | rs1164012 | 2:6862562 | 2 | 68625625 | C | T | 0.545777 | 0.136712 | 0.013945 | 1.09E-22 | trans |
| miR-320b | rs1188499 | 2:6864573 | 2 | 68645737 | T | C | 0.593791 | 0.13089 | 0.014756 | 7.29E-19 | trans |
| miR-320b | rs1300519 | 2:6863887 | 2 | 68638873 | A | G | 0.702186 | 0.133519 | 0.015787 | 2.73E-17 | trans |
| miR-320b | rs1300592 | 2:6862999 | 2 | 68629998 | T | A | 0.701261 | 0.129992 | 0.015537 | 5.92E-17 | trans |
| miR-320b | rs1300974 | 2:6858443 | 2 | 68584437 | C | A | 0.678986 | 0.117096 | 0.015503 | 4.25E-14 | trans |
| miR-320b | rs1301028 | 2:6862184 | 2 | 68621842 | G | T | 0.683506 | 0.129676 | 0.014833 | 2.28E-18 | trans |
| miR-320b | rs1301077 | 2:6863908 | 2 | 68639081 | C | T | 0.702339 | 0.133415 | 0.015791 | 2.95E-17 | trans |
| miR-320b | rs1301107 | 2:6863931 | 2 | 68639313 | A | C | 0.700844 | 0.134034 | 0.015817 | 2.37E-17 | trans |
| miR-320b | rs1301143 | 2:6863937 | 2 | 68639379 | C | T | 0.702382 | 0.133434 | 0.015794 | 2.95E-17 | trans |
| miR-320b | rs1301157 | 2:6863927 | 2 | 68639273 | G | A | 0.702349 | 0.133427 | 0.015792 | 2.94E-17 | trans |
| miR-320b | rs1302892 | 2:6862991 | 2 | 68629916 | C | T | 0.552648 | 0.137763 | 0.014108 | 1.59E-22 | trans |
| miR-320b | rs1302897 | 2:6863003 | 2 | 68630038 | A | G | 0.54849 | 0.138955 | 0.014062 | 5.00E-23 | trans |
| miR-320b | rs1303372 | 2:6857805 | 2 | 68578058 | C | G | 0.670257 | 0.109332 | 0.015428 | 1.37E-12 | trans |
| miR-320b | rs1303482 | 2:6863029 | 2 | 68630297 | C | G | 0.552715 | 0.137808 | 0.014111 | 1.57E-22 | trans |
| miR-320b | rs1343065 | 2:6858771 | 2 | 68587710 | C | G | 0.699397 | 0.126957 | 0.01528 | 9.66E-17 | trans |
| miR-320b | rs1373200 | 2:6862265 | 2 | 68622658 | C | T | 0.547142 | 0.134711 | 0.013801 | 1.66E-22 | trans |
| miR-320b | rs1382899 | 2:6863782 | 2 | 68637829 | A | G | 0.702614 | 0.133506 | 0.015805 | 2.99E-17 | trans |
| miR-320b | rs1390688 | 2:6862773 | 2 | 68627735 | C | T | 0.69721 | 0.12989 | 0.015387 | 3.14E-17 | trans |
| miR-320b | rs1397159 | 2:6863425 | 2 | 68634258 | T | G | 0.705552 | 0.134497 | 0.015943 | 3.28E-17 | trans |
| miR-320b | rs1422402 | 2:6862752 | 2 | 68627521 | A | G | 0.698498 | 0.130556 | 0.015426 | 2.60E-17 | trans |
| miR-320b | rs1442589 | 2:6863473 | 2 | 68634732 | G | T | 0.701896 | 0.133113 | 0.015756 | 2.95E-17 | trans |
| miR-320b | rs1455505 | 2:6862832 | 2 | 68628329 | G | A | 0.556221 | 0.141184 | 0.014214 | 3.00E-23 | trans |
| miR-320b | rs1474969 | 2:6862568 | 2 | 68625686 | G | A | 0.546544 | 0.136939 | 0.013966 | 1.07E-22 | trans |
| miR-320b | rs1484530 | 2:6863547 | 2 | 68635471 | A | G | 0.70445 | 0.134492 | 0.015849 | 2.14E-17 | trans |
| miR-320b | rs1486061 | 2:6862571 | 2 | 68625717 | T | G | 0.54743 | 0.137325 | 0.013992 | 9.74E-23 | trans |

| | | | | | | | | | | | |
|---|---|---|---|---|---|---|---|---|---|---|---|
| miR-320b | rs1542135 | 2:6862084 | 2 | 68620848 | C | T | 0.69752 | 0.127051 | 0.015091 | 3.79E-17 | trans |
| miR-320b | rs1542136 | 2:6862093 | 2 | 68620930 | C | A | 0.697594 | 0.127041 | 0.015095 | 3.89E-17 | trans |
| miR-320b | rs1752821 | 2:6857875 | 2 | 68578758 | G | T | 0.677689 | 0.115156 | 0.015565 | 1.38E-13 | trans |
| miR-320b | rs1753019 | 2:6862133 | 2 | 68621338 | G | T | 0.683745 | 0.129844 | 0.014845 | 2.20E-18 | trans |
| miR-320b | rs1760149 | 2:6861171 | 2 | 68611712 | A | G | 0.69807 | 0.126937 | 0.015097 | 4.16E-17 | trans |
| miR-320b | rs1760168 | 2:6861589 | 2 | 68615899 | G | A | 0.697578 | 0.126954 | 0.015102 | 4.23E-17 | trans |
| miR-320b | rs1760209 | 2:6862041 | 2 | 68620410 | T | A | 0.697529 | 0.127053 | 0.015091 | 3.80E-17 | trans |
| miR-320b | rs2070171 | 2:6861371 | 2 | 68613710 | G | A | 0.698024 | 0.126917 | 0.015096 | 4.20E-17 | trans |
| miR-320b | rs2165920 | 2:6862223 | 2 | 68622239 | T | C | 0.552012 | 0.135016 | 0.013899 | 2.62E-22 | trans |
| miR-320b | rs2241946 | 2:6861565 | 2 | 68615654 | T | C | 0.698076 | 0.127092 | 0.015103 | 3.92E-17 | trans |
| miR-320b | rs2241947 | 2:6861539 | 2 | 68615395 | C | T | 0.697988 | 0.126954 | 0.015096 | 4.11E-17 | trans |
| miR-320b | rs3419682 | 2:6862142 | 2 | 68621427 | G | A | 0.697518 | 0.127052 | 0.015091 | 3.79E-17 | trans |
| miR-320b | rs3471241 | 2:6857828 | 2 | 68578284 | G | A | 0.6787 | 0.115519 | 0.015545 | 1.07E-13 | trans |
| miR-320b | rs3485223 | 2:6863859 | 2 | 68638599 | C | G | 0.550658 | 0.140992 | 0.014326 | 7.45E-23 | trans |
| miR-320b | rs3611704 | 2:6857937 | 2 | 68579376 | A | C | 0.678545 | 0.115935 | 0.015531 | 8.35E-14 | trans |
| miR-320b | rs3732044 | 2:6862391 | 2 | 68623915 | T | C | 0.545562 | 0.136529 | 0.013935 | 1.15E-22 | trans |
| miR-320b | rs4610041 | 2:6862628 | 2 | 68626280 | C | T | 0.550745 | 0.140566 | 0.014102 | 2.10E-23 | trans |
| miR-320b | rs5618821 | 2:6861163 | 2 | 68611631 | T | C | 0.698075 | 0.126937 | 0.015097 | 4.16E-17 | trans |
| miR-320b | rs5628470 | 2:6863137 | 2 | 68631372 | G | A | 0.697508 | 0.130591 | 0.015439 | 2.71E-17 | trans |
| miR-320b | rs5633714 | 2:6864249 | 2 | 68642490 | G | A | 0.733417 | 0.132351 | 0.016434 | 8.03E-16 | trans |
| miR-320b | rs5690424 | 2:6863847 | 2 | 68638477 | T | C | 0.550669 | 0.140995 | 0.014325 | 7.40E-23 | trans |
| miR-320b | rs5808991 | 2:6863966 | 2 | 68639660 | G | C | 0.734634 | 0.134981 | 0.016542 | 3.35E-16 | trans |
| miR-320b | rs5888448 | 2:6864534 | 2 | 68645347 | G | A | 0.734214 | 0.131847 | 0.016492 | 1.30E-15 | trans |
| miR-320b | rs5933292 | 2:6863963 | 2 | 68639631 | G | A | 0.73529 | 0.135103 | 0.016587 | 3.79E-16 | trans |
| miR-320b | rs5999861 | 2:6864677 | 2 | 68646771 | G | T | 0.731295 | 0.125973 | 0.016487 | 2.16E-14 | trans |
| miR-320b | rs6000716 | 2:6863963 | 2 | 68639636 | C | T | 0.735163 | 0.135202 | 0.016585 | 3.57E-16 | trans |
| miR-320b | rs6087844 | 2:6858211 | 2 | 68582115 | A | G | 0.538067 | 0.126281 | 0.014369 | 1.52E-18 | trans |
| miR-320b | rs6214389 | 2:6860455 | 2 | 68604553 | C | T | 0.698307 | 0.126569 | 0.015117 | 5.64E-17 | trans |
| miR-320b | rs6214392 | 2:6863835 | 2 | 68638359 | C | G | 0.700378 | 0.134816 | 0.015773 | 1.26E-17 | trans |
| miR-320b | rs6214393 | 2:6864009 | 2 | 68640090 | T | C | 0.59325 | 0.131146 | 0.01472 | 5.13E-19 | trans |
| miR-320b | rs6214393 | 2:6864020 | 2 | 68640202 | A | G | 0.593308 | 0.131089 | 0.014721 | 5.33E-19 | trans |
| miR-320b | rs6214393 | 2:6864038 | 2 | 68640381 | G | C | 0.593598 | 0.131783 | 0.014715 | 3.37E-19 | trans |
| miR-320b | rs6214393 | 2:6864041 | 2 | 68640415 | A | G | 0.593599 | 0.131772 | 0.014715 | 3.40E-19 | trans |
| miR-320b | rs6214393 | 2:6864062 | 2 | 68640621 | T | C | 0.733261 | 0.132408 | 0.016423 | 7.50E-16 | trans |
| miR-320b | rs6419643 | 2:6863126 | 2 | 68631260 | T | G | 0.548826 | 0.138972 | 0.014072 | 5.32E-23 | trans |
| miR-320b | rs6419644 | 2:6863182 | 2 | 68631823 | A | G | 0.549898 | 0.140681 | 0.014215 | 4.29E-23 | trans |
| miR-320b | rs6546392 | 2:6862744 | 2 | 68627441 | A | G | 0.555235 | 0.139524 | 0.014174 | 7.29E-23 | trans |
| miR-320b | rs6546393 | 2:6862757 | 2 | 68627575 | T | C | 0.554017 | 0.140713 | 0.014135 | 2.40E-23 | trans |
| miR-320b | rs6546394 | 2:6862909 | 2 | 68629094 | T | C | 0.566701 | 0.138978 | 0.014379 | 4.24E-22 | trans |
| miR-320b | rs6546395 | 2:6862927 | 2 | 68629271 | A | G | 0.55448 | 0.138124 | 0.014154 | 1.70E-22 | trans |
| miR-320b | rs6546397 | 2:6863036 | 2 | 68630367 | G | A | 0.552756 | 0.13783 | 0.014113 | 1.57E-22 | trans |
| miR-320b | rs6546398 | 2:6863041 | 2 | 68630417 | G | C | 0.552757 | 0.137831 | 0.014112 | 1.56E-22 | trans |
| miR-320b | rs6546399 | 2:6863064 | 2 | 68630646 | C | T | 0.55394 | 0.138212 | 0.014154 | 1.60E-22 | trans |
| miR-320b | rs6546400 | 2:6863064 | 2 | 68630648 | G | A | 0.554382 | 0.138315 | 0.014167 | 1.62E-22 | trans |
| miR-320b | rs6546401 | 2:6863087 | 2 | 68630873 | A | T | 0.550127 | 0.139225 | 0.014103 | 5.49E-23 | trans |
| miR-320b | rs6546403 | 2:6863226 | 2 | 68632262 | C | T | 0.550801 | 0.140899 | 0.014236 | 4.28E-23 | trans |
| miR-320b | rs6546404 | 2:6863263 | 2 | 68632635 | G | T | 0.54995 | 0.140703 | 0.01422 | 4.38E-23 | trans |
| miR-320b | rs6546407 | 2:6863492 | 2 | 68634920 | A | C | 0.55091 | 0.141146 | 0.014305 | 5.80E-23 | trans |
| miR-320b | rs6546408 | 2:6863504 | 2 | 68635048 | T | C | 0.551062 | 0.141172 | 0.01432 | 6.31E-23 | trans |
| miR-320b | rs6546409 | 2:6863587 | 2 | 68635879 | A | G | 0.551093 | 0.141154 | 0.014327 | 6.70E-23 | trans |
| miR-320b | rs6546410 | 2:6863610 | 2 | 68636105 | C | G | 0.702434 | 0.133424 | 0.015795 | 2.98E-17 | trans |
| miR-320b | rs6546411 | 2:6863769 | 2 | 68637699 | A | G | 0.556248 | 0.143009 | 0.014415 | 3.38E-23 | trans |
| miR-320b | rs6546412 | 2:6863771 | 2 | 68637710 | T | C | 0.557086 | 0.144247 | 0.014441 | 1.71E-23 | trans |
| miR-320b | rs6708536 | 2:6862726 | 2 | 68627268 | G | A | 0.547249 | 0.138343 | 0.014027 | 6.04E-23 | trans |
| miR-320b | rs6709464 | 2:6862222 | 2 | 68622220 | C | T | 0.697279 | 0.126869 | 0.015077 | 3.94E-17 | trans |
| miR-320b | rs6714719 | 2:6859377 | 2 | 68593773 | G | C | 0.698404 | 0.12667 | 0.015159 | 6.50E-17 | trans |
| miR-320b | rs6715579 | 2:6858392 | 2 | 68583923 | C | T | 0.678794 | 0.117162 | 0.015502 | 4.09E-14 | trans |
| miR-320b | rs6715706 | 2:6858399 | 2 | 68583996 | C | A | 0.664887 | 0.120184 | 0.015241 | 3.13E-15 | trans |
| miR-320b | rs6715909 | 2:6858399 | 2 | 68583995 | G | A | 0.664879 | 0.120181 | 0.015242 | 3.14E-15 | trans |
| miR-320b | rs6729555 | 2:6863295 | 2 | 68632952 | C | A | 0.54995 | 0.140731 | 0.014223 | 4.41E-23 | trans |
| miR-320b | rs6729686 | 2:6863303 | 2 | 68633030 | C | T | 0.700171 | 0.132218 | 0.015638 | 2.80E-17 | trans |
| miR-320b | rs6742200 | 2:6862330 | 2 | 68623302 | A | G | 0.54552 | 0.136507 | 0.013932 | 1.15E-22 | trans |
| miR-320b | rs6743970 | 2:6858390 | 2 | 68583907 | A | T | 0.660237 | 0.122904 | 0.015312 | 1.00E-15 | trans |

| | | | | | | | | | | | |
|---|---|---|---|---|---|---|---|---|---|---|---|
| miR-320b | rs6744306 | 2:6863273 | 2 | 68632736 | T | C | 0.549956 | 0.140712 | 0.01422 | 4.37E-23 | trans |
| miR-320b | rs6747545 | 2:6863306 | 2 | 68633069 | T | A | 0.549964 | 0.140721 | 0.014224 | 4.46E-23 | trans |
| miR-320b | rs6757938 | 2:6863285 | 2 | 68632858 | A | G | 0.549958 | 0.140714 | 0.014222 | 4.42E-23 | trans |
| miR-320b | rs7420775 | 2:6862945 | 2 | 68629458 | G | T | 0.703122 | 0.130665 | 0.015606 | 5.63E-17 | trans |
| miR-320b | rs7421228 | 2:6863012 | 2 | 68630124 | G | T | 0.55268 | 0.137787 | 0.01411 | 1.58E-22 | trans |
| miR-320b | rs7424269 | 2:6862912 | 2 | 68629125 | A | G | 0.563426 | 0.139819 | 0.014336 | 1.78E-22 | trans |
| miR-320b | rs7451452 | 2:6863755 | 2 | 68637550 | T | G | 0.702552 | 0.133486 | 0.015801 | 2.97E-17 | trans |
| miR-320b | rs7562439 | 2:6858256 | 2 | 68582569 | C | T | 0.6646 | 0.119946 | 0.015246 | 3.62E-15 | trans |
| miR-320b | rs7564214 | 2:6862869 | 2 | 68628696 | C | T | 0.569906 | 0.143898 | 0.01452 | 3.76E-23 | trans |
| miR-320b | rs7572299 | 2:6858124 | 2 | 68581246 | T | A | 0.538033 | 0.125959 | 0.014372 | 1.88E-18 | trans |
| miR-320b | rs7578358 | 2:6862872 | 2 | 68628724 | T | C | 0.561556 | 0.141146 | 0.014314 | 6.16E-23 | trans |
| miR-320b | rs7578602 | 2:6863786 | 2 | 68637868 | G | C | 0.556806 | 0.144256 | 0.014448 | 1.78E-23 | trans |
| miR-320b | rs7581872 | 2:6858098 | 2 | 68580981 | A | C | 0.537959 | 0.12589 | 0.014372 | 1.96E-18 | trans |
| miR-320b | rs7590433 | 2:6862850 | 2 | 68628507 | A | G | 0.554708 | 0.142374 | 0.014201 | 1.18E-23 | trans |
| miR-320b | rs7590450 | 2:6862856 | 2 | 68628563 | A | G | 0.549532 | 0.138968 | 0.014059 | 4.87E-23 | trans |
| miR-320b | rs7592526 | 2:6858352 | 2 | 68583528 | A | C | 0.538007 | 0.126303 | 0.01437 | 1.50E-18 | trans |
| miR-320b | rs7597167 | 2:6861294 | 2 | 68612946 | C | T | 0.698051 | 0.126955 | 0.015097 | 4.12E-17 | trans |
| miR-320b | rs7598360 | 2:6863092 | 2 | 68630925 | A | G | 0.553166 | 0.141628 | 0.014138 | 1.27E-23 | trans |
| miR-320b | rs7605387 | 2:6863255 | 2 | 68632550 | A | G | 0.703499 | 0.132498 | 0.015728 | 3.63E-17 | trans |
| miR-320b | rs7608429 | 2:6858083 | 2 | 68580836 | G | T | 0.678719 | 0.116442 | 0.015517 | 6.19E-14 | trans |
| miR-320b | rs7800275 | 2:6862543 | 2 | 68625436 | A | G | 0.546309 | 0.136492 | 0.013929 | 1.13E-22 | trans |
| miR-320b | rs8028813 | 2:6862547 | 2 | 68625478 | A | G | 0.548418 | 0.139006 | 0.01402 | 3.59E-23 | trans |
| miR-320b | rs8761 | 2:6862426 | 2 | 68624260 | G | A | 0.545692 | 0.136595 | 0.01394 | 1.14E-22 | trans |
| miR-320b | rs9653552 | 2:6864396 | 2 | 68643966 | G | A | 0.593458 | 0.13159 | 0.014728 | 4.08E-19 | trans |
| miR-320c | rs1017438 | 2:6859646 | 2 | 68596469 | C | T | 0.685243 | 0.117036 | 0.01396 | 5.12E-17 | trans |
| miR-320c | rs1049616 | 2:6862008 | 2 | 68620089 | C | T | 0.697496 | 0.113968 | 0.014152 | 8.08E-16 | trans |
| miR-320c | rs1050181 | 2:6862295 | 2 | 68622952 | G | C | 0.544992 | 0.123229 | 0.013032 | 3.20E-21 | trans |
| miR-320c | rs1156629 | 2:6862663 | 2 | 68626631 | T | G | 0.547487 | 0.12504 | 0.013146 | 1.88E-21 | trans |
| miR-320c | rs1158434 | 2:6863552 | 2 | 68635524 | G | C | 0.551072 | 0.128841 | 0.013428 | 8.41E-22 | trans |
| miR-320c | rs1164012 | 2:6862562 | 2 | 68625625 | C | T | 0.545777 | 0.123739 | 0.013078 | 3.04E-21 | trans |
| miR-320c | rs1188499 | 2:6864573 | 2 | 68645737 | T | C | 0.593791 | 0.119389 | 0.013832 | 6.05E-18 | trans |
| miR-320c | rs1300519 | 2:6863887 | 2 | 68638873 | A | G | 0.702186 | 0.120365 | 0.014802 | 4.24E-16 | trans |
| miR-320c | rs1300592 | 2:6862999 | 2 | 68629998 | T | A | 0.701261 | 0.117101 | 0.014568 | 9.09E-16 | trans |
| miR-320c | rs1300974 | 2:6858443 | 2 | 68584437 | C | A | 0.678986 | 0.104041 | 0.014538 | 8.27E-13 | trans |
| miR-320c | rs1301028 | 2:6862184 | 2 | 68621842 | G | T | 0.683506 | 0.11756 | 0.013906 | 2.82E-17 | trans |
| miR-320c | rs1301077 | 2:6863908 | 2 | 68639081 | C | T | 0.702339 | 0.120275 | 0.014806 | 4.54E-16 | trans |
| miR-320c | rs1301107 | 2:6863931 | 2 | 68639313 | A | C | 0.700844 | 0.120705 | 0.014831 | 3.99E-16 | trans |
| miR-320c | rs1301143 | 2:6863937 | 2 | 68639379 | C | T | 0.702382 | 0.12029 | 0.014808 | 4.55E-16 | trans |
| miR-320c | rs1301157 | 2:6863927 | 2 | 68639273 | G | A | 0.702349 | 0.120281 | 0.014807 | 4.55E-16 | trans |
| miR-320c | rs1302892 | 2:6862991 | 2 | 68629916 | C | T | 0.552648 | 0.125024 | 0.013229 | 3.37E-21 | trans |
| miR-320c | rs1302897 | 2:6863003 | 2 | 68630038 | A | G | 0.54849 | 0.125793 | 0.013188 | 1.45E-21 | trans |
| miR-320c | rs1303372 | 2:6857805 | 2 | 68578058 | C | G | 0.670257 | 0.09737 | 0.014464 | 1.67E-11 | trans |
| miR-320c | rs1303482 | 2:6863029 | 2 | 68630297 | C | G | 0.552715 | 0.125074 | 0.013232 | 3.32E-21 | trans |
| miR-320c | rs1343065 | 2:6858771 | 2 | 68587710 | C | G | 0.699397 | 0.114048 | 0.014328 | 1.72E-15 | trans |
| miR-320c | rs1373200 | 2:6862265 | 2 | 68622658 | C | T | 0.547142 | 0.122067 | 0.012943 | 4.05E-21 | trans |
| miR-320c | rs1382899 | 2:6863782 | 2 | 68637829 | A | G | 0.702614 | 0.12037 | 0.014819 | 4.56E-16 | trans |
| miR-320c | rs1390688 | 2:6862773 | 2 | 68627735 | C | T | 0.69721 | 0.116486 | 0.01443 | 6.88E-16 | trans |
| miR-320c | rs1397159 | 2:6863425 | 2 | 68634258 | T | G | 0.705552 | 0.121472 | 0.014948 | 4.42E-16 | trans |
| miR-320c | rs1422402 | 2:6862752 | 2 | 68627521 | A | G | 0.698498 | 0.116782 | 0.014467 | 6.91E-16 | trans |
| miR-320c | rs1442589 | 2:6863473 | 2 | 68634732 | G | T | 0.701896 | 0.119955 | 0.014773 | 4.67E-16 | trans |
| miR-320c | rs1455505 | 2:6862832 | 2 | 68628329 | G | A | 0.556221 | 0.127266 | 0.013334 | 1.36E-21 | trans |
| miR-320c | rs1474969 | 2:6862568 | 2 | 68625686 | G | A | 0.546544 | 0.123927 | 0.013098 | 3.04E-21 | trans |
| miR-320c | rs1484530 | 2:6863547 | 2 | 68635471 | A | G | 0.70445 | 0.121181 | 0.014861 | 3.51E-16 | trans |
| miR-320c | rs1486061 | 2:6862571 | 2 | 68625717 | T | G | 0.54743 | 0.124223 | 0.013123 | 2.90E-21 | trans |
| miR-320c | rs1542135 | 2:6862084 | 2 | 68620848 | C | T | 0.69752 | 0.113978 | 0.014151 | 8.00E-16 | trans |
| miR-320c | rs1542136 | 2:6862093 | 2 | 68620930 | C | A | 0.697594 | 0.113984 | 0.014155 | 8.12E-16 | trans |
| miR-320c | rs1752821 | 2:6857875 | 2 | 68578758 | G | T | 0.677689 | 0.102312 | 0.014595 | 2.38E-12 | trans |
| miR-320c | rs1753019 | 2:6862133 | 2 | 68621338 | G | T | 0.683745 | 0.117647 | 0.013918 | 2.84E-17 | trans |
| miR-320c | rs1760149 | 2:6861171 | 2 | 68611712 | A | G | 0.69807 | 0.113768 | 0.014157 | 9.29E-16 | trans |
| miR-320c | rs1760168 | 2:6861589 | 2 | 68615899 | G | A | 0.697578 | 0.113768 | 0.014163 | 9.51E-16 | trans |
| miR-320c | rs1760209 | 2:6862041 | 2 | 68620410 | T | A | 0.697529 | 0.113981 | 0.014152 | 8.00E-16 | trans |
| miR-320c | rs2070171 | 2:6861371 | 2 | 68613710 | G | A | 0.698024 | 0.113757 | 0.014157 | 9.33E-16 | trans |
| miR-320c | rs2165920 | 2:6862223 | 2 | 68622239 | T | C | 0.552012 | 0.121089 | 0.013039 | 1.60E-20 | trans |

| | | | | | | | | | | | |
|---|---|---|---|---|---|---|---|---|---|---|---|
| miR-320c | rs2241946 | 2:6861565 | 2 | 68615654 | T | C | 0.698076 | 0.113933 | 0.014163 | 8.66E-16 | trans |
| miR-320c | rs2241947 | 2:6861539 | 2 | 68615395 | C | T | 0.697988 | 0.113809 | 0.014157 | 9.05E-16 | trans |
| miR-320c | rs3419682 | 2:6862142 | 2 | 68621427 | G | A | 0.697518 | 0.113978 | 0.014151 | 7.99E-16 | trans |
| miR-320c | rs3471241 | 2:6857828 | 2 | 68578284 | G | A | 0.6787 | 0.102633 | 0.014576 | 1.91E-12 | trans |
| miR-320c | rs3485223 | 2:6863859 | 2 | 68638599 | C | G | 0.550658 | 0.128801 | 0.013431 | 8.81E-22 | trans |
| miR-320c | rs3611704 | 2:6857937 | 2 | 68579376 | A | C | 0.678545 | 0.102984 | 0.014564 | 1.53E-12 | trans |
| miR-320c | rs3732044 | 2:6862391 | 2 | 68623915 | T | C | 0.545562 | 0.123558 | 0.013069 | 3.24E-21 | trans |
| miR-320c | rs4610041 | 2:6862628 | 2 | 68626280 | C | T | 0.550745 | 0.127235 | 0.013226 | 6.58E-22 | trans |
| miR-320c | rs5618821 | 2:6861163 | 2 | 68611631 | T | C | 0.698075 | 0.113768 | 0.014157 | 9.29E-16 | trans |
| miR-320c | rs5628470 | 2:6863137 | 2 | 68631372 | G | A | 0.697508 | 0.117453 | 0.014477 | 4.93E-16 | trans |
| miR-320c | rs5633714 | 2:6864249 | 2 | 68642490 | G | A | 0.733417 | 0.119147 | 0.015407 | 1.05E-14 | trans |
| miR-320c | rs5690424 | 2:6863847 | 2 | 68638477 | T | C | 0.550669 | 0.128797 | 0.01343 | 8.80E-22 | trans |
| miR-320c | rs5808991 | 2:6863966 | 2 | 68639660 | G | C | 0.734634 | 0.121723 | 0.015509 | 4.20E-15 | trans |
| miR-320c | rs5888448 | 2:6864534 | 2 | 68645347 | G | A | 0.734214 | 0.118672 | 0.015462 | 1.65E-14 | trans |
| miR-320c | rs5933292 | 2:6863963 | 2 | 68639631 | G | A | 0.73529 | 0.1218 | 0.015551 | 4.79E-15 | trans |
| miR-320c | rs5999861 | 2:6864677 | 2 | 68646771 | G | T | 0.731295 | 0.112933 | 0.015457 | 2.75E-13 | trans |
| miR-320c | rs6000716 | 2:6863963 | 2 | 68639636 | C | T | 0.735163 | 0.121878 | 0.015549 | 4.56E-15 | trans |
| miR-320c | rs6087844 | 2:6858211 | 2 | 68582115 | A | G | 0.538067 | 0.11583 | 0.013466 | 7.86E-18 | trans |
| miR-320c | rs6214389 | 2:6860455 | 2 | 68604553 | C | T | 0.698307 | 0.113676 | 0.014176 | 1.06E-15 | trans |
| miR-320c | rs6214392 | 2:6863835 | 2 | 68638359 | C | G | 0.700378 | 0.121403 | 0.01479 | 2.25E-16 | trans |
| miR-320c | rs6214393 | 2:6864009 | 2 | 68640090 | T | C | 0.59325 | 0.11934 | 0.013799 | 5.22E-18 | trans |
| miR-320c | rs6214393 | 2:6864020 | 2 | 68640202 | A | G | 0.593308 | 0.119306 | 0.0138 | 5.36E-18 | trans |
| miR-320c | rs6214393 | 2:6864038 | 2 | 68640381 | G | C | 0.593598 | 0.119834 | 0.013795 | 3.72E-18 | trans |
| miR-320c | rs6214393 | 2:6864041 | 2 | 68640415 | A | G | 0.593599 | 0.11983 | 0.013795 | 3.74E-18 | trans |
| miR-320c | rs6214393 | 2:6864062 | 2 | 68640621 | T | C | 0.733261 | 0.119179 | 0.015398 | 9.95E-15 | trans |
| miR-320c | rs6419643 | 2:6863126 | 2 | 68631260 | T | G | 0.548826 | 0.12595 | 0.013197 | 1.38E-21 | trans |
| miR-320c | rs6419644 | 2:6863182 | 2 | 68631823 | A | G | 0.549898 | 0.128192 | 0.013328 | 6.69E-22 | trans |
| miR-320c | rs6546392 | 2:6862744 | 2 | 68627441 | A | G | 0.555235 | 0.125889 | 0.013295 | 2.82E-21 | trans |
| miR-320c | rs6546393 | 2:6862757 | 2 | 68627575 | T | C | 0.554017 | 0.126897 | 0.013259 | 1.06E-21 | trans |
| miR-320c | rs6546394 | 2:6862909 | 2 | 68629094 | T | C | 0.566701 | 0.125796 | 0.013485 | 1.07E-20 | trans |
| miR-320c | rs6546395 | 2:6862927 | 2 | 68629271 | A | G | 0.55448 | 0.125279 | 0.013273 | 3.79E-21 | trans |
| miR-320c | rs6546397 | 2:6863036 | 2 | 68630367 | G | A | 0.552756 | 0.125081 | 0.013234 | 3.34E-21 | trans |
| miR-320c | rs6546398 | 2:6863041 | 2 | 68630417 | G | C | 0.552757 | 0.125094 | 0.013234 | 3.30E-21 | trans |
| miR-320c | rs6546399 | 2:6863064 | 2 | 68630646 | C | T | 0.55394 | 0.125397 | 0.013273 | 3.48E-21 | trans |
| miR-320c | rs6546400 | 2:6863064 | 2 | 68630648 | G | A | 0.554382 | 0.125493 | 0.013286 | 3.52E-21 | trans |
| miR-320c | rs6546401 | 2:6863087 | 2 | 68630873 | A | T | 0.550127 | 0.126126 | 0.013226 | 1.48E-21 | trans |
| miR-320c | rs6546403 | 2:6863226 | 2 | 68632262 | C | T | 0.550801 | 0.128349 | 0.013348 | 6.89E-22 | trans |
| miR-320c | rs6546404 | 2:6863263 | 2 | 68632635 | G | T | 0.54995 | 0.128238 | 0.013333 | 6.69E-22 | trans |
| miR-320c | rs6546407 | 2:6863492 | 2 | 68634920 | A | C | 0.55091 | 0.128789 | 0.013412 | 7.79E-22 | trans |
| miR-320c | rs6546408 | 2:6863504 | 2 | 68635048 | T | C | 0.551062 | 0.128831 | 0.013426 | 8.32E-22 | trans |
| miR-320c | rs6546409 | 2:6863587 | 2 | 68635879 | A | G | 0.551093 | 0.128861 | 0.013432 | 8.51E-22 | trans |
| miR-320c | rs6546410 | 2:6863610 | 2 | 68636105 | C | G | 0.702434 | 0.120283 | 0.014809 | 4.58E-16 | trans |
| miR-320c | rs6546411 | 2:6863769 | 2 | 68637699 | A | G | 0.556248 | 0.130222 | 0.013516 | 5.73E-22 | trans |
| miR-320c | rs6546412 | 2:6863771 | 2 | 68637710 | T | C | 0.557086 | 0.131394 | 0.013541 | 2.91E-22 | trans |
| miR-320c | rs6708536 | 2:6862726 | 2 | 68627268 | G | A | 0.547249 | 0.125242 | 0.013155 | 1.73E-21 | trans |
| miR-320c | rs6709464 | 2:6862222 | 2 | 68622220 | C | T | 0.697279 | 0.113882 | 0.014138 | 7.96E-16 | trans |
| miR-320c | rs6714719 | 2:6859377 | 2 | 68593773 | G | C | 0.698404 | 0.113511 | 0.014216 | 1.41E-15 | trans |
| miR-320c | rs6715579 | 2:6858392 | 2 | 68583923 | C | T | 0.678794 | 0.10413 | 0.014536 | 7.87E-13 | trans |
| miR-320c | rs6715706 | 2:6858399 | 2 | 68583996 | C | A | 0.664887 | 0.10796 | 0.01429 | 4.18E-14 | trans |
| miR-320c | rs6715909 | 2:6858399 | 2 | 68583995 | G | A | 0.664879 | 0.107953 | 0.01429 | 4.21E-14 | trans |
| miR-320c | rs6729555 | 2:6863295 | 2 | 68632952 | C | A | 0.54995 | 0.128279 | 0.013336 | 6.64E-22 | trans |
| miR-320c | rs6729686 | 2:6863303 | 2 | 68633030 | C | T | 0.700171 | 0.119056 | 0.014663 | 4.69E-16 | trans |
| miR-320c | rs6742200 | 2:6862330 | 2 | 68623302 | A | G | 0.54552 | 0.123535 | 0.013066 | 3.25E-21 | trans |
| miR-320c | rs6743970 | 2:6858390 | 2 | 68583907 | A | T | 0.660237 | 0.110494 | 0.014356 | 1.40E-14 | trans |
| miR-320c | rs6744306 | 2:6863273 | 2 | 68632736 | T | C | 0.549956 | 0.128249 | 0.013333 | 6.66E-22 | trans |
| miR-320c | rs6747545 | 2:6863306 | 2 | 68633069 | T | A | 0.549964 | 0.128276 | 0.013337 | 6.70E-22 | trans |
| miR-320c | rs6757938 | 2:6863285 | 2 | 68632858 | A | G | 0.549958 | 0.128259 | 0.013335 | 6.69E-22 | trans |
| miR-320c | rs7420775 | 2:6862945 | 2 | 68629458 | G | T | 0.703122 | 0.117641 | 0.014633 | 9.02E-16 | trans |
| miR-320c | rs7421228 | 2:6863012 | 2 | 68630124 | G | T | 0.55268 | 0.125054 | 0.013231 | 3.34E-21 | trans |
| miR-320c | rs7424269 | 2:6862912 | 2 | 68629125 | A | G | 0.563426 | 0.126516 | 0.013445 | 4.95E-21 | trans |
| miR-320c | rs7451452 | 2:6863755 | 2 | 68637550 | T | G | 0.702552 | 0.120344 | 0.014815 | 4.55E-16 | trans |
| miR-320c | rs7562439 | 2:6858256 | 2 | 68582569 | C | T | 0.6646 | 0.107793 | 0.014294 | 4.66E-14 | trans |
| miR-320c | rs7564214 | 2:6862869 | 2 | 68628696 | C | T | 0.569906 | 0.129827 | 0.01362 | 1.54E-21 | trans |

| | | | | | | | | | | |
|---|---|---|---|---|---|---|---|---|---|---|
| miR-320c | rs7572299 | 2:6858124 | 2 | 68581246 | T | A | 0.538033 | 0.115531 | 0.013469 | 9.70E-18 trans |
| miR-320c | rs7578358 | 2:6862872 | 2 | 68628724 | T | C | 0.561556 | 0.127311 | 0.013427 | 2.50E-21 trans |
| miR-320c | rs7578602 | 2:6863786 | 2 | 68637868 | G | C | 0.556806 | 0.131453 | 0.013547 | 2.91E-22 trans |
| miR-320c | rs7581872 | 2:6858098 | 2 | 68580981 | A | C | 0.537959 | 0.115444 | 0.013469 | 1.02E-17 trans |
| miR-320c | rs7590433 | 2:6862850 | 2 | 68628507 | A | G | 0.554708 | 0.128831 | 0.01332 | 3.96E-22 trans |
| miR-320c | rs7590450 | 2:6862856 | 2 | 68628563 | A | G | 0.549532 | 0.12574 | 0.013186 | 1.49E-21 trans |
| miR-320c | rs7592526 | 2:6858352 | 2 | 68583528 | A | C | 0.538007 | 0.115839 | 0.013467 | 7.86E-18 trans |
| miR-320c | rs7597167 | 2:6861294 | 2 | 68612946 | C | T | 0.698051 | 0.11378 | 0.014157 | 9.21E-16 trans |
| miR-320c | rs7598360 | 2:6863092 | 2 | 68630925 | A | G | 0.553166 | 0.128065 | 0.01326 | 4.56E-22 trans |
| miR-320c | rs7605387 | 2:6863255 | 2 | 68632550 | A | G | 0.703499 | 0.119101 | 0.014748 | 6.72E-16 trans |
| miR-320c | rs7608429 | 2:6858083 | 2 | 68580836 | G | T | 0.678719 | 0.103457 | 0.014551 | 1.16E-12 trans |
| miR-320c | rs7800275 | 2:6862543 | 2 | 68625436 | A | G | 0.546309 | 0.123534 | 0.013063 | 3.18E-21 trans |
| miR-320c | rs8028813 | 2:6862547 | 2 | 68625478 | A | G | 0.548418 | 0.12585 | 0.013149 | 1.06E-21 trans |
| miR-320c | rs8761 | 2:6862426 | 2 | 68624260 | G | A | 0.545692 | 0.123625 | 0.013074 | 3.20E-21 trans |
| miR-320c | rs9653552 | 2:6864396 | 2 | 68643966 | G | A | 0.593458 | 0.11996 | 0.013806 | 3.65E-18 trans |
| miR-320d | rs1017438 | 2:6859646 | 2 | 68596469 | C | T | 0.685243 | 0.116538 | 0.014293 | 3.54E-16 trans |
| miR-320d | rs1049616 | 2:6862008 | 2 | 68620089 | C | T | 0.697496 | 0.116733 | 0.014478 | 7.45E-16 trans |
| miR-320d | rs1050181 | 2:6862295 | 2 | 68622952 | G | C | 0.544992 | 0.121982 | 0.01335 | 6.39E-20 trans |
| miR-320d | rs1156629 | 2:6862663 | 2 | 68626631 | T | G | 0.547487 | 0.123554 | 0.013468 | 4.55E-20 trans |
| miR-320d | rs1158434 | 2:6863552 | 2 | 68635524 | G | C | 0.551072 | 0.126979 | 0.013759 | 2.73E-20 trans |
| miR-320d | rs1164012 | 2:6862562 | 2 | 68625625 | C | T | 0.545777 | 0.122475 | 0.013397 | 6.14E-20 trans |
| miR-320d | rs1188499 | 2:6864573 | 2 | 68645737 | T | C | 0.593791 | 0.118599 | 0.014164 | 5.60E-17 trans |
| miR-320d | rs1300519 | 2:6863887 | 2 | 68638873 | A | G | 0.702186 | 0.121079 | 0.015151 | 1.33E-15 trans |
| miR-320d | rs1300592 | 2:6862999 | 2 | 68629998 | T | A | 0.701261 | 0.118515 | 0.014908 | 1.87E-15 trans |
| miR-320d | rs1300974 | 2:6858443 | 2 | 68584437 | C | A | 0.678986 | 0.108867 | 0.014864 | 2.41E-13 trans |
| miR-320d | rs1301028 | 2:6862184 | 2 | 68621842 | G | T | 0.683506 | 0.116353 | 0.014241 | 3.08E-16 trans |
| miR-320d | rs1301077 | 2:6863908 | 2 | 68639081 | C | T | 0.702339 | 0.12101 | 0.015155 | 1.41E-15 trans |
| miR-320d | rs1301107 | 2:6863931 | 2 | 68639313 | A | C | 0.700844 | 0.12145 | 0.01518 | 1.24E-15 trans |
| miR-320d | rs1301143 | 2:6863937 | 2 | 68639379 | C | T | 0.702382 | 0.121014 | 0.015157 | 1.42E-15 trans |
| miR-320d | rs1301157 | 2:6863927 | 2 | 68639273 | G | A | 0.702349 | 0.121015 | 0.015156 | 1.41E-15 trans |
| miR-320d | rs1302892 | 2:6862991 | 2 | 68629916 | C | T | 0.552648 | 0.124158 | 0.01355 | 5.05E-20 trans |
| miR-320d | rs1302897 | 2:6863003 | 2 | 68630038 | A | G | 0.54849 | 0.124504 | 0.01351 | 3.10E-20 trans |
| miR-320d | rs1303372 | 2:6857805 | 2 | 68578058 | C | G | 0.670257 | 0.10071 | 0.014794 | 9.92E-12 trans |
| miR-320d | rs1303482 | 2:6863029 | 2 | 68630297 | C | G | 0.552715 | 0.124211 | 0.013553 | 4.97E-20 trans |
| miR-320d | rs1343065 | 2:6858771 | 2 | 68587710 | C | G | 0.699397 | 0.117257 | 0.014655 | 1.23E-15 trans |
| miR-320d | rs1373200 | 2:6862265 | 2 | 68622658 | C | T | 0.547142 | 0.121474 | 0.013255 | 4.99E-20 trans |
| miR-320d | rs1382899 | 2:6863782 | 2 | 68637829 | A | G | 0.702614 | 0.121213 | 0.015168 | 1.33E-15 trans |
| miR-320d | rs1390688 | 2:6862773 | 2 | 68627735 | C | T | 0.69721 | 0.117637 | 0.014768 | 1.64E-15 trans |
| miR-320d | rs1397159 | 2:6863425 | 2 | 68634258 | T | G | 0.705552 | 0.122919 | 0.015297 | 9.32E-16 trans |
| miR-320d | rs1422402 | 2:6862752 | 2 | 68627521 | A | G | 0.698498 | 0.118434 | 0.014804 | 1.24E-15 trans |
| miR-320d | rs1442589 | 2:6863473 | 2 | 68634732 | G | T | 0.701896 | 0.120911 | 0.01512 | 1.28E-15 trans |
| miR-320d | rs1455505 | 2:6862832 | 2 | 68628329 | G | A | 0.556221 | 0.126528 | 0.013657 | 1.95E-20 trans |
| miR-320d | rs1474969 | 2:6862568 | 2 | 68625686 | G | A | 0.546544 | 0.122663 | 0.013417 | 6.13E-20 trans |
| miR-320d | rs1484530 | 2:6863547 | 2 | 68635471 | A | G | 0.70445 | 0.121983 | 0.01521 | 1.06E-15 trans |
| miR-320d | rs1486061 | 2:6862571 | 2 | 68625717 | T | G | 0.54743 | 0.123039 | 0.013442 | 5.53E-20 trans |
| miR-320d | rs1542135 | 2:6862084 | 2 | 68620848 | C | T | 0.69752 | 0.116748 | 0.014477 | 7.35E-16 trans |
| miR-320d | rs1542136 | 2:6862093 | 2 | 68620930 | C | A | 0.697594 | 0.116763 | 0.014481 | 7.42E-16 trans |
| miR-320d | rs1752821 | 2:6857875 | 2 | 68578758 | G | T | 0.677689 | 0.107228 | 0.014923 | 6.70E-13 trans |
| miR-320d | rs1753019 | 2:6862133 | 2 | 68621338 | G | T | 0.683745 | 0.116731 | 0.014252 | 2.61E-16 trans |
| miR-320d | rs1760149 | 2:6861171 | 2 | 68611712 | A | G | 0.69807 | 0.117074 | 0.014481 | 6.23E-16 trans |
| miR-320d | rs1760168 | 2:6861589 | 2 | 68615899 | G | A | 0.697578 | 0.11699 | 0.014486 | 6.71E-16 trans |
| miR-320d | rs1760209 | 2:6862041 | 2 | 68620410 | T | A | 0.697529 | 0.11675 | 0.014477 | 7.35E-16 trans |
| miR-320d | rs2070171 | 2:6861371 | 2 | 68613710 | G | A | 0.698024 | 0.117049 | 0.014481 | 6.31E-16 trans |
| miR-320d | rs2165920 | 2:6862223 | 2 | 68622239 | T | C | 0.552012 | 0.122188 | 0.013347 | 5.44E-20 trans |
| miR-320d | rs2241946 | 2:6861565 | 2 | 68615654 | T | C | 0.698076 | 0.117177 | 0.014487 | 6.03E-16 trans |
| miR-320d | rs2241947 | 2:6861539 | 2 | 68615395 | C | T | 0.697988 | 0.117037 | 0.014481 | 6.35E-16 trans |
| miR-320d | rs3419682 | 2:6862142 | 2 | 68621427 | G | A | 0.697518 | 0.116748 | 0.014477 | 7.35E-16 trans |
| miR-320d | rs3471241 | 2:6857828 | 2 | 68578284 | G | A | 0.6787 | 0.107313 | 0.014904 | 6.01E-13 trans |
| miR-320d | rs3485223 | 2:6863859 | 2 | 68638599 | C | G | 0.550658 | 0.126936 | 0.013761 | 2.86E-20 trans |
| miR-320d | rs3611704 | 2:6857937 | 2 | 68579376 | A | C | 0.678545 | 0.107728 | 0.014891 | 4.68E-13 trans |
| miR-320d | rs3732044 | 2:6862391 | 2 | 68623915 | T | C | 0.545562 | 0.122287 | 0.013387 | 6.57E-20 trans |
| miR-320d | rs4610041 | 2:6862628 | 2 | 68626280 | C | T | 0.550745 | 0.125806 | 0.01355 | 1.62E-20 trans |
| miR-320d | rs5618821 | 2:6861163 | 2 | 68611631 | T | C | 0.698075 | 0.117075 | 0.014481 | 6.23E-16 trans |

| | | | | | | | | | | | |
|---|---|---|---|---|---|---|---|---|---|---|---|
| miR-320d | rs5628470 | 2:6863137 | 2 | 68631372 | G | A | 0.697508 | 0.118242 | 0.014817 | 1.46E-15 | trans |
| miR-320d | rs5633714 | 2:6864249 | 2 | 68642490 | G | A | 0.733417 | 0.118238 | 0.015775 | 6.62E-14 | trans |
| miR-320d | rs5690424 | 2:6863847 | 2 | 68638477 | T | C | 0.550669 | 0.126931 | 0.01376 | 2.85E-20 | trans |
| miR-320d | rs5808991 | 2:6863966 | 2 | 68639660 | G | C | 0.734634 | 0.120732 | 0.015879 | 2.89E-14 | trans |
| miR-320d | rs5888448 | 2:6864534 | 2 | 68645347 | G | A | 0.734214 | 0.117789 | 0.01583 | 1.00E-13 | trans |
| miR-320d | rs5933292 | 2:6863963 | 2 | 68639631 | G | A | 0.73529 | 0.120638 | 0.015923 | 3.56E-14 | trans |
| miR-320d | rs5999861 | 2:6864677 | 2 | 68646771 | G | T | 0.731295 | 0.111989 | 0.015825 | 1.47E-12 | trans |
| miR-320d | rs6000716 | 2:6863963 | 2 | 68639636 | C | T | 0.735163 | 0.12067 | 0.015921 | 3.48E-14 | trans |
| miR-320d | rs6087844 | 2:6858211 | 2 | 68582115 | A | G | 0.538067 | 0.116986 | 0.013782 | 2.10E-17 | trans |
| miR-320d | rs6214389 | 2:6860455 | 2 | 68604553 | C | T | 0.698307 | 0.116699 | 0.014501 | 8.42E-16 | trans |
| miR-320d | rs6214392 | 2:6863835 | 2 | 68638359 | C | G | 0.700378 | 0.121704 | 0.01514 | 9.10E-16 | trans |
| miR-320d | rs6214393 | 2:6864009 | 2 | 68640090 | T | C | 0.59325 | 0.118698 | 0.01413 | 4.45E-17 | trans |
| miR-320d | rs6214393 | 2:6864020 | 2 | 68640202 | A | G | 0.593308 | 0.118688 | 0.014131 | 4.49E-17 | trans |
| miR-320d | rs6214393 | 2:6864038 | 2 | 68640381 | G | C | 0.593598 | 0.119156 | 0.014126 | 3.30E-17 | trans |
| miR-320d | rs6214393 | 2:6864041 | 2 | 68640415 | A | G | 0.593599 | 0.11915 | 0.014126 | 3.32E-17 | trans |
| miR-320d | rs6214393 | 2:6864062 | 2 | 68640621 | T | C | 0.733261 | 0.118334 | 0.015765 | 6.09E-14 | trans |
| miR-320d | rs6419643 | 2:6863126 | 2 | 68631260 | T | G | 0.548826 | 0.124619 | 0.01352 | 3.04E-20 | trans |
| miR-320d | rs6419644 | 2:6863182 | 2 | 68631823 | A | G | 0.549898 | 0.126235 | 0.013656 | 2.38E-20 | trans |
| miR-320d | rs6546392 | 2:6862744 | 2 | 68627441 | A | G | 0.555235 | 0.125564 | 0.013615 | 2.90E-20 | trans |
| miR-320d | rs6546393 | 2:6862757 | 2 | 68627575 | T | C | 0.554017 | 0.126078 | 0.013581 | 1.64E-20 | trans |
| miR-320d | rs6546394 | 2:6862909 | 2 | 68629094 | T | C | 0.566701 | 0.125273 | 0.01381 | 1.18E-19 | trans |
| miR-320d | rs6546395 | 2:6862927 | 2 | 68629271 | A | G | 0.55448 | 0.124433 | 0.013595 | 5.55E-20 | trans |
| miR-320d | rs6546397 | 2:6863036 | 2 | 68630367 | G | A | 0.552756 | 0.124221 | 0.013555 | 4.99E-20 | trans |
| miR-320d | rs6546398 | 2:6863041 | 2 | 68630417 | G | C | 0.552757 | 0.124235 | 0.013555 | 4.93E-20 | trans |
| miR-320d | rs6546399 | 2:6863064 | 2 | 68630646 | C | T | 0.55394 | 0.124567 | 0.013595 | 5.06E-20 | trans |
| miR-320d | rs6546400 | 2:6863064 | 2 | 68630648 | G | A | 0.554382 | 0.124654 | 0.013607 | 5.16E-20 | trans |
| miR-320d | rs6546401 | 2:6863087 | 2 | 68630873 | A | T | 0.550127 | 0.124835 | 0.013549 | 3.15E-20 | trans |
| miR-320d | rs6546403 | 2:6863226 | 2 | 68632262 | C | T | 0.550801 | 0.126407 | 0.013677 | 2.41E-20 | trans |
| miR-320d | rs6546404 | 2:6863263 | 2 | 68632635 | G | T | 0.54995 | 0.126284 | 0.013661 | 2.37E-20 | trans |
| miR-320d | rs6546407 | 2:6863492 | 2 | 68634920 | A | C | 0.55091 | 0.126901 | 0.013742 | 2.59E-20 | trans |
| miR-320d | rs6546408 | 2:6863504 | 2 | 68635048 | T | C | 0.551062 | 0.126965 | 0.013756 | 2.71E-20 | trans |
| miR-320d | rs6546409 | 2:6863587 | 2 | 68635879 | A | G | 0.551093 | 0.126997 | 0.013762 | 2.76E-20 | trans |
| miR-320d | rs6546410 | 2:6863610 | 2 | 68636105 | C | G | 0.702434 | 0.12118 | 0.015157 | 1.30E-15 | trans |
| miR-320d | rs6546411 | 2:6863769 | 2 | 68637699 | A | G | 0.556248 | 0.128784 | 0.013847 | 1.40E-20 | trans |
| miR-320d | rs6546412 | 2:6863771 | 2 | 68637710 | T | C | 0.557086 | 0.129709 | 0.013873 | 8.80E-21 | trans |
| miR-320d | rs6708536 | 2:6862726 | 2 | 68627268 | G | A | 0.547249 | 0.123687 | 0.013477 | 4.42E-20 | trans |
| miR-320d | rs6709464 | 2:6862222 | 2 | 68622220 | C | T | 0.697279 | 0.11635 | 0.014465 | 8.71E-16 | trans |
| miR-320d | rs6714719 | 2:6859377 | 2 | 68593773 | G | C | 0.698404 | 0.117036 | 0.01454 | 8.33E-16 | trans |
| miR-320d | rs6715579 | 2:6858392 | 2 | 68583923 | C | T | 0.678794 | 0.108857 | 0.014863 | 2.41E-13 | trans |
| miR-320d | rs6715706 | 2:6858399 | 2 | 68583996 | C | A | 0.664887 | 0.108852 | 0.014624 | 9.82E-14 | trans |
| miR-320d | rs6715909 | 2:6858399 | 2 | 68583995 | G | A | 0.664879 | 0.108849 | 0.014625 | 9.85E-14 | trans |
| miR-320d | rs6729555 | 2:6863295 | 2 | 68632952 | C | A | 0.54995 | 0.126323 | 0.013664 | 2.36E-20 | trans |
| miR-320d | rs6729686 | 2:6863303 | 2 | 68633030 | C | T | 0.700171 | 0.119939 | 0.015008 | 1.33E-15 | trans |
| miR-320d | rs6742200 | 2:6862330 | 2 | 68623302 | A | G | 0.54552 | 0.122264 | 0.013385 | 6.58E-20 | trans |
| miR-320d | rs6743970 | 2:6858390 | 2 | 68583907 | A | T | 0.660237 | 0.111589 | 0.014692 | 3.07E-14 | trans |
| miR-320d | rs6744306 | 2:6863273 | 2 | 68632736 | T | C | 0.549956 | 0.126294 | 0.013662 | 2.36E-20 | trans |
| miR-320d | rs6747545 | 2:6863306 | 2 | 68633069 | T | A | 0.549964 | 0.126319 | 0.013665 | 2.38E-20 | trans |
| miR-320d | rs6757938 | 2:6863285 | 2 | 68632858 | A | G | 0.549958 | 0.126303 | 0.013663 | 2.38E-20 | trans |
| miR-320d | rs7420775 | 2:6862945 | 2 | 68629458 | G | T | 0.703122 | 0.119074 | 0.014975 | 1.84E-15 | trans |
| miR-320d | rs7421228 | 2:6863012 | 2 | 68630124 | G | T | 0.55268 | 0.124188 | 0.013552 | 5.01E-20 | trans |
| miR-320d | rs7424269 | 2:6862912 | 2 | 68629125 | A | G | 0.563426 | 0.125995 | 0.013769 | 5.66E-20 | trans |
| miR-320d | rs7451452 | 2:6863755 | 2 | 68637550 | T | G | 0.702552 | 0.121207 | 0.015164 | 1.31E-15 | trans |
| miR-320d | rs7562439 | 2:6858256 | 2 | 68582569 | C | T | 0.6646 | 0.108725 | 0.014628 | 1.07E-13 | trans |
| miR-320d | rs7564214 | 2:6862869 | 2 | 68628696 | C | T | 0.569906 | 0.127732 | 0.013956 | 5.56E-20 | trans |
| miR-320d | rs7572299 | 2:6858124 | 2 | 68581246 | T | A | 0.538033 | 0.116665 | 0.013785 | 2.60E-17 | trans |
| miR-320d | rs7578358 | 2:6862872 | 2 | 68628724 | T | C | 0.561556 | 0.125497 | 0.013756 | 7.31E-20 | trans |
| miR-320d | rs7578602 | 2:6863786 | 2 | 68637868 | G | C | 0.556806 | 0.129683 | 0.01388 | 9.33E-21 | trans |
| miR-320d | rs7581872 | 2:6858098 | 2 | 68580981 | A | C | 0.537959 | 0.116574 | 0.013785 | 2.75E-17 | trans |
| miR-320d | rs7590433 | 2:6862850 | 2 | 68628507 | A | G | 0.554708 | 0.127536 | 0.013645 | 9.07E-21 | trans |
| miR-320d | rs7590450 | 2:6862856 | 2 | 68628563 | A | G | 0.549532 | 0.124555 | 0.013508 | 2.94E-20 | trans |
| miR-320d | rs7592526 | 2:6858352 | 2 | 68583528 | A | C | 0.538007 | 0.116969 | 0.013783 | 2.14E-17 | trans |
| miR-320d | rs7597167 | 2:6861294 | 2 | 68612946 | C | T | 0.698051 | 0.117082 | 0.014481 | 6.20E-16 | trans |
| miR-320d | rs7598360 | 2:6863092 | 2 | 68630925 | A | G | 0.553166 | 0.126722 | 0.013585 | 1.08E-20 | trans |

| | | | | | | | | | | | |
|---|---|---|---|---|---|---|---|---|---|---|---|
| miR-320d | rs7605387 | 2:6863255( | 2 | 68632550 | A | G | 0.703499 | 0.120593 | 0.015093 | 1.35E-15 | trans |
| miR-320d | rs7608429 | 2:6858083( | 2 | 68580836 | G | T | 0.678719 | 0.10826 | 0.014878 | 3.42E-13 | trans |
| miR-320d | rs7800275: | 2:6862543( | 2 | 68625436 | A | G | 0.546309 | 0.122332 | 0.013381 | 6.14E-20 | trans |
| miR-320d | rs8028813! | 2:6862547! | 2 | 68625478 | A | G | 0.548418 | 0.124394 | 0.013471 | 2.60E-20 | trans |
| miR-320d | rs8761 | 2:6862426( | 2 | 68624260 | G | A | 0.545692 | 0.12236 | 0.013392 | 6.45E-20 | trans |
| miR-320d | rs9653552 | 2:6864396( | 2 | 68643966 | G | A | 0.593458 | 0.119124 | 0.014138 | 3.58E-17 | trans |
| miR-329-3p | rs1014438: | 14:101173: | 14 | 1.01E+08 | C | T | 0.673446 | 0.215334 | 0.029343 | 2.16E-13 | cis |
| miR-329-3p | rs1014551: | 14:101168: | 14 | 1.01E+08 | A | G | 0.668173 | 0.20569 | 0.029165 | 1.76E-12 | cis |
| miR-329-3p | rs1014696: | 14:101170! | 14 | 1.01E+08 | T | C | 0.658875 | 0.21653 | 0.028661 | 4.20E-14 | cis |
| miR-329-3p | rs1243517: | 14:101155: | 14 | 1.01E+08 | G | A | 0.312183 | -0.20451 | 0.0306 | 2.34E-11 | cis |
| miR-329-3p | rs1288154! | 14:101176: | 14 | 1.01E+08 | G | C | 0.370047 | -0.21165 | 0.030044 | 1.86E-12 | cis |
| miR-329-3p | rs1288176( | 14:101176: | 14 | 1.01E+08 | G | C | 0.347791 | -0.21254 | 0.029614 | 7.12E-13 | cis |
| miR-329-3p | rs1288281! | 14:101183 | 14 | 1.01E+08 | C | G | 0.331398 | -0.21017 | 0.029219 | 6.33E-13 | cis |
| miR-329-3p | rs1709961( | 14:101167( | 14 | 1.01E+08 | C | G | 0.668809 | 0.207081 | 0.029207 | 1.34E-12 | cis |
| miR-329-3p | rs4905998 | 14:101181 | 14 | 1.01E+08 | A | G | 0.330615 | -0.21011 | 0.029197 | 6.18E-13 | cis |
| miR-329-3p | rs7141210 | 14:101182( | 14 | 1.01E+08 | T | C | 0.32902 | -0.21042 | 0.029121 | 4.98E-13 | cis |
| miR-329-3p | rs7142683 | 14:101158: | 14 | 1.01E+08 | A | G | 0.290748 | -0.21515 | 0.031116 | 4.69E-12 | cis |
| miR-329-3p | rs7145337 | 14:101155: | 14 | 1.01E+08 | C | T | 0.312091 | -0.20446 | 0.030593 | 2.34E-11 | cis |
| miR-329-3p | rs7149242 | 14:101159( | 14 | 1.01E+08 | T | G | 0.288371 | -0.21408 | 0.031029 | 5.23E-12 | cis |
| miR-329-3p | rs8012970 | 14:101168( | 14 | 1.01E+08 | T | C | 0.347902 | -0.20621 | 0.029703 | 3.85E-12 | cis |
| miR-329-3p | rs8020868 | 14:101166! | 14 | 1.01E+08 | A | G | 0.668343 | 0.205384 | 0.0292 | 2.01E-12 | cis |
| miR-329-3p | rs8021301 | 14:101166! | 14 | 1.01E+08 | G | C | 0.668324 | 0.205414 | 0.029203 | 2.01E-12 | cis |
| miR-329-3p | rs9671394 | 14:101159( | 14 | 1.01E+08 | C | T | 0.284232 | -0.216 | 0.031471 | 6.72E-12 | cis |
| miR-376a-3 | rs1013940: | 14:101194: | 14 | 1.01E+08 | A | G | 0.323897 | -0.21657 | 0.027713 | 5.51E-15 | cis |
| miR-376a-3 | rs1014438: | 14:101173: | 14 | 1.01E+08 | C | T | 0.673446 | 0.260434 | 0.026919 | 3.86E-22 | cis |
| miR-376a-3 | rs1014551: | 14:101168: | 14 | 1.01E+08 | A | G | 0.668173 | 0.239935 | 0.02681 | 3.58E-19 | cis |
| miR-376a-3 | rs1014696: | 14:101170! | 14 | 1.01E+08 | T | C | 0.658875 | 0.2434 | 0.026361 | 2.62E-20 | cis |
| miR-376a-3 | rs1116059( | 14:101049! | 14 | 1.01E+08 | C | T | 0.741662 | 0.208575 | 0.029004 | 6.42E-13 | cis |
| miR-376a-3 | rs1162093 | 14:101151( | 14 | 1.01E+08 | C | G | 0.338205 | -0.21614 | 0.02719 | 1.88E-15 | cis |
| miR-376a-3 | rs1184948! | 14:101037 | 14 | 1.01E+08 | C | G | 0.733365 | 0.196249 | 0.02878 | 9.17E-12 | cis |
| miR-376a-3 | rs1185095: | 14:101056: | 14 | 1.01E+08 | G | T | 0.743981 | 0.204733 | 0.02887 | 1.33E-12 | cis |
| miR-376a-3 | rs1214700: | 14:101186 | 14 | 1.01E+08 | C | T | 0.223221 | -0.26357 | 0.030488 | 5.37E-18 | cis |
| miR-376a-3 | rs1243449( | 14:101179: | 14 | 1.01E+08 | A | G | 0.18959 | -0.31532 | 0.032737 | 5.87E-22 | cis |
| miR-376a-3 | rs1243517: | 14:101155: | 14 | 1.01E+08 | G | A | 0.312183 | -0.24831 | 0.02811 | 1.01E-18 | cis |
| miR-376a-3 | rs1258871: | 14:101149( | 14 | 1.01E+08 | G | C | 0.347269 | -0.22149 | 0.026939 | 2.00E-16 | cis |
| miR-376a-3 | rs1288154! | 14:101176: | 14 | 1.01E+08 | G | C | 0.370047 | -0.27013 | 0.027519 | 9.60E-23 | cis |
| miR-376a-3 | rs1288176( | 14:101176: | 14 | 1.01E+08 | G | C | 0.347791 | -0.27085 | 0.027116 | 1.71E-23 | cis |
| miR-376a-3 | rs1288281! | 14:101183 | 14 | 1.01E+08 | C | G | 0.331398 | -0.26238 | 0.026777 | 1.14E-22 | cis |
| miR-376a-3 | rs1289379( | 14:101178: | 14 | 1.01E+08 | C | G | 0.190813 | -0.31572 | 0.032764 | 5.61E-22 | cis |
| miR-376a-3 | rs1289760: | 14:101182: | 14 | 1.01E+08 | C | T | 0.190452 | -0.31292 | 0.03257 | 7.42E-22 | cis |
| miR-376a-3 | rs1467536 | 14:101061: | 14 | 1.01E+08 | A | G | 0.318958 | -0.19165 | 0.028002 | 7.68E-12 | cis |
| miR-376a-3 | rs1555405 | 14:101176 | 14 | 1.01E+08 | G | A | 0.749075 | 0.214226 | 0.028973 | 1.42E-13 | cis |
| miR-376a-3 | rs1709961( | 14:101167( | 14 | 1.01E+08 | C | G | 0.668809 | 0.242595 | 0.026842 | 1.60E-19 | cis |
| miR-376a-3 | rs1757758( | 14:101189: | 14 | 1.01E+08 | C | T | 0.827094 | 0.245969 | 0.033544 | 2.25E-13 | cis |
| miR-376a-3 | rs1888581 | 14:101042 | 14 | 1.01E+08 | A | C | 0.735363 | 0.193551 | 0.028712 | 1.57E-11 | cis |
| miR-376a-3 | rs2837918( | 14:101168( | 14 | 1.01E+08 | G | T | 0.836434 | 0.286071 | 0.034477 | 1.06E-16 | cis |
| miR-376a-3 | rs3470057( | 14:101173( | 14 | 1.01E+08 | C | T | 0.892725 | 0.299403 | 0.041457 | 5.12E-13 | cis |
| miR-376a-3 | rs4900471 | 14:101162( | 14 | 1.01E+08 | G | A | 0.304173 | -0.24723 | 0.028137 | 1.54E-18 | cis |
| miR-376a-3 | rs4905977 | 14:101061: | 14 | 1.01E+08 | C | G | 0.318473 | -0.18833 | 0.028053 | 1.90E-11 | cis |
| miR-376a-3 | rs4905978 | 14:101061: | 14 | 1.01E+08 | C | T | 0.318452 | -0.18832 | 0.028053 | 1.91E-11 | cis |
| miR-376a-3 | rs4905994 | 14:101154( | 14 | 1.01E+08 | T | C | 0.329146 | -0.22658 | 0.027776 | 3.42E-16 | cis |
| miR-376a-3 | rs4905998 | 14:101181 | 14 | 1.01E+08 | A | G | 0.330615 | -0.26187 | 0.026759 | 1.29E-22 | cis |
| miR-376a-3 | rs4905999 | 14:101184 | 14 | 1.01E+08 | A | G | 0.181905 | -0.32006 | 0.03331 | 7.36E-22 | cis |
| miR-376a-3 | rs5870230( | 14:101167: | 14 | 1.01E+08 | G | A | 0.892706 | 0.30069 | 0.041688 | 5.48E-13 | cis |
| miR-376a-3 | rs5982537: | 14:101168( | 14 | 1.01E+08 | G | T | 0.89241 | 0.302137 | 0.041647 | 4.03E-13 | cis |
| miR-376a-3 | rs5993909: | 14:101169( | 14 | 1.01E+08 | C | T | 0.892837 | 0.301057 | 0.041638 | 4.82E-13 | cis |
| miR-376a-3 | rs6130797( | 14:101177: | 14 | 1.01E+08 | C | T | 0.888723 | 0.304021 | 0.040775 | 8.91E-14 | cis |
| miR-376a-3 | rs6575794 | 14:101039 | 14 | 1.01E+08 | C | T | 0.734189 | 0.193128 | 0.028832 | 2.11E-11 | cis |
| miR-376a-3 | rs6762567( | 14:101173( | 14 | 1.01E+08 | A | G | 0.892632 | 0.299289 | 0.041436 | 5.09E-13 | cis |
| miR-376a-3 | rs7140792 | 14:101172 | 14 | 1.01E+08 | C | T | 0.892517 | 0.30083 | 0.041494 | 4.17E-13 | cis |
| miR-376a-3 | rs7141110 | 14:101172: | 14 | 1.01E+08 | G | C | 0.783235 | 0.215415 | 0.032055 | 1.82E-11 | cis |
| miR-376a-3 | rs7141210 | 14:101182( | 14 | 1.01E+08 | T | C | 0.32902 | -0.26142 | 0.026691 | 1.19E-22 | cis |
| miR-376a-3 | rs7142089 | 14:101172: | 14 | 1.01E+08 | T | A | 0.783402 | 0.218184 | 0.032089 | 1.05E-11 | cis |

| | | | | | | | | | | | |
|---|---|---|---|---|---|---|---|---|---|---|---|
| miR-376a-3 | rs7142683 | 14:1011158 | 14 | 1.01E+08 | A | G | 0.290748 | -0.2537 | 0.0286 | 7.27E-19 | cis |
| miR-376a-3 | rs7143357 | 14:101054 | 14 | 1.01E+08 | T | C | 0.739636 | 0.206342 | 0.029036 | 1.19E-12 | cis |
| miR-376a-3 | rs7145337 | 14:1011155 | 14 | 1.01E+08 | C | T | 0.312091 | -0.24843 | 0.028103 | 9.58E-19 | cis |
| miR-376a-3 | rs7145610 | 14:1010494 | 14 | 1.01E+08 | T | C | 0.740391 | 0.209451 | 0.029079 | 5.90E-13 | cis |
| miR-376a-3 | rs7147982 | 14:1010460 | 14 | 1.01E+08 | C | T | 0.731753 | 0.190634 | 0.028507 | 2.27E-11 | cis |
| miR-376a-3 | rs7149242 | 14:1011594 | 14 | 1.01E+08 | T | G | 0.288371 | -0.25302 | 0.028519 | 7.17E-19 | cis |
| miR-376a-3 | rs7155375 | 14:1011887 | 14 | 1.01E+08 | C | T | 0.635995 | 0.215803 | 0.026169 | 1.63E-16 | cis |
| miR-376a-3 | rs7155649 | 14:1011810 | 14 | 1.01E+08 | C | A | 0.863608 | 0.270181 | 0.037999 | 1.16E-12 | cis |
| miR-376a-3 | rs7263167 | 14:1011769 | 14 | 1.01E+08 | C | T | 0.889487 | 0.302852 | 0.040728 | 1.04E-13 | cis |
| miR-376a-3 | rs7263167 | 14:1011870 | 14 | 1.01E+08 | C | T | 0.881829 | 0.303373 | 0.040663 | 8.61E-14 | cis |
| miR-376a-3 | rs7269872 | 14:1011890 | 14 | 1.01E+08 | C | T | 0.825721 | 0.243641 | 0.033351 | 2.76E-13 | cis |
| miR-376a-3 | rs7493817 | 14:1010384 | 14 | 1.01E+08 | G | A | 0.751723 | 0.205201 | 0.029703 | 4.90E-12 | cis |
| miR-376a-3 | rs8008285 | 14:1011463 | 14 | 1.01E+08 | T | C | 0.842905 | 0.255629 | 0.035679 | 7.79E-13 | cis |
| miR-376a-3 | rs8012970 | 14:1011684 | 14 | 1.01E+08 | T | C | 0.347902 | -0.24139 | 0.027308 | 9.60E-19 | cis |
| miR-376a-3 | rs8020868 | 14:1011669 | 14 | 1.01E+08 | A | G | 0.668343 | 0.240386 | 0.026841 | 3.37E-19 | cis |
| miR-376a-3 | rs8021301 | 14:1011669 | 14 | 1.01E+08 | G | C | 0.668324 | 0.240447 | 0.026843 | 3.32E-19 | cis |
| miR-376a-3 | rs9324022 | 14:1011724 | 14 | 1.01E+08 | T | C | 0.763739 | 0.221104 | 0.031367 | 1.80E-12 | cis |
| miR-376a-3 | rs9671394 | 14:1011590 | 14 | 1.01E+08 | C | T | 0.284232 | -0.26024 | 0.028907 | 2.20E-19 | cis |

**Table S4. miR-eQTLs explaining high proportion of variation in miRNA level**

| miRNA | SNP | position | effect_alle | other_alle | eaf | beta | se | pval | cismark | maf | R2 | F |
|---|---|---|---|---|---|---|---|---|---|---|---|---|
| let-7b-5p | rs529565 | 9:136149500 | T | C | 0.665911 | -0.11036 | 0.010051 | 4.77E-28 | trans | 0.334089 | 0.05245 | 120.4492 |
| miR-10a-5p | rs2519093 | 9:136141870 | C | T | 0.809892 | -0.21091 | 0.017778 | 1.83E-32 | trans | 0.190108 | 0.0607 | 140.6181 |
| miR-10a-5p | rs529565 | 9:136149500 | T | C | 0.665911 | -0.18394 | 0.014549 | 1.22E-36 | trans | 0.334089 | 0.068374 | 159.701 |
| miR-10b-5p | rs2519093 | 9:136141870 | C | T | 0.809892 | -0.21558 | 0.017406 | 3.13E-35 | trans | 0.190108 | 0.065798 | 153.2606 |
| miR-10b-5p | rs529565 | 9:136149500 | T | C | 0.665911 | -0.17932 | 0.01429 | 4.06E-36 | trans | 0.334089 | 0.067422 | 157.316 |
| miR-126-3p | rs2519093 | 9:136141870 | C | T | 0.809892 | -0.21142 | 0.015879 | 1.90E-40 | trans | 0.190108 | 0.075271 | 177.1213 |
| miR-126-3p | rs529565 | 9:136149500 | T | C | 0.665911 | -0.17441 | 0.013044 | 8.91E-41 | trans | 0.334089 | 0.075861 | 178.6248 |
| miR-130a-3p | rs11607122 | 11:57389737 | G | A | 0.743794 | -0.30287 | 0.023174 | 4.93E-39 | cis | 0.256207 | 0.072721 | 170.6505 |
| miR-130a-3p | rs71460549 | 11:57484130 | G | C | 0.661292 | -0.25716 | 0.022429 | 1.97E-30 | cis | 0.338708 | 0.056919 | 131.3321 |
| miR-136-5p | rs12897601 | 14:101182284 | C | T | 0.190452 | -0.4956 | 0.044231 | 3.86E-29 | cis | 0.190452 | 0.054502 | 125.4327 |
| miR-136-5p | rs12881760 | 14:101176335 | G | C | 0.347791 | -0.41693 | 0.036865 | 1.18E-29 | cis | 0.347791 | 0.05547 | 127.7916 |
| miR-139-5p | rs2519093 | 9:136141870 | C | T | 0.809892 | -0.1791 | 0.014192 | 1.63E-36 | trans | 0.190108 | 0.068146 | 159.1288 |
| miR-139-5p | rs529565 | 9:136149500 | T | C | 0.665911 | -0.15979 | 0.011585 | 2.82E-43 | trans | 0.334089 | 0.080329 | 190.0647 |
| miR-3689a-3p | rs9410006 | 9:137748168 | C | T | 0.413955 | 0.279702 | 0.026037 | 6.44E-27 | cis | 0.413955 | 0.050318 | 115.2921 |
| miR-3689a-3p | rs3128562 | 9:137745210 | A | G | 0.300754 | 0.396962 | 0.02741 | 1.56E-47 | cis | 0.300754 | 0.087841 | 209.5488 |
| miR-3689a-3p | rs11103545 | 9:137739980 | G | T | 0.285208 | 0.337768 | 0.029217 | 6.52E-31 | cis | 0.285208 | 0.057816 | 133.5271 |
| miR-3689a-3p | rs13946 | 9:137734232 | C | T | 0.257382 | 0.3805 | 0.032828 | 4.59E-31 | cis | 0.257382 | 0.0581 | 134.2239 |
| miR-370-3p | rs12881760 | 14:101176335 | G | C | 0.347791 | -0.2649 | 0.024603 | 4.91E-27 | cis | 0.347791 | 0.05054 | 115.8288 |
| miR-376c-3p | rs12897601 | 14:101182284 | C | T | 0.190452 | -0.28274 | 0.026347 | 7.26E-27 | cis | 0.190452 | 0.050219 | 115.0546 |
| miR-409-3p | rs9671394 | 14:101159674 | C | T | 0.284232 | -0.38828 | 0.03475 | 5.50E-29 | cis | 0.284232 | 0.054214 | 124.7325 |
| miR-409-3p | rs12881760 | 14:101176335 | G | C | 0.347791 | -0.37661 | 0.032675 | 9.76E-31 | cis | 0.347791 | 0.057489 | 132.7271 |
| miR-625-5p | rs1269068 | 14:65767333 | C | T | 0.643163 | -0.22338 | 0.019494 | 2.13E-30 | cis | 0.356837 | 0.056857 | 131.1792 |
| miR-625-5p | rs12881755 | 14:65801811 | A | G | 0.320867 | 0.23032 | 0.020035 | 1.38E-30 | cis | 0.320867 | 0.057207 | 132.0365 |
| miR-625-5p | rs2127868 | 14:65794327 | G | A | 0.220849 | 0.364388 | 0.022206 | 1.63E-60 | cis | 0.220849 | 0.110032 | 269.0331 |
| miR-625-5p | rs76375630 | 14:65753942 | G | A | 0.888368 | -0.35516 | 0.032567 | 1.09E-27 | cis | 0.111632 | 0.051776 | 118.8154 |
| miR-625-5p | rs12147473 | 14:65795081 | C | T | 0.845789 | -0.32979 | 0.02651 | 1.58E-35 | cis | 0.154211 | 0.066344 | 154.6231 |
| miR-6803-3p | rs10163414 | 16:88619493 | C | T | 0.708147 | -0.20622 | 0.017703 | 2.33E-31 | trans | 0.291853 | 0.058649 | 135.5703 |
| miR-181a-5p | rs1434282 | 1:199010721 | C | T | 0.271104 | 0.177906 | 0.013618 | 5.30E-39 | cis | 0.271104 | 0.072665 | 170.5082 |

**Table S5. Replicated miR-eQTLs in Nikpay et al**

| miRNA | SNP | position | The Rotterdam Study | | | | | | Nikpay et al | | | | | | cismark |
|---|---|---|---|---|---|---|---|---|---|---|---|---|---|---|---|
| | | | effect_allele | other_allele | eaf | beta | se.x | pval | A1 | A2 | freq | b | se.y | p | |
| miR-625-5p | rs1003401 | 14:66237772 | G | A | 0.386875 | 0.147199 | 0.019801 | 1.05E-13 | G | A | 0.3919 | 0.348 | 0.02598 | 1.43E-36 | cis |
| miR-625-5p | rs10047879 | 14:65760544 | C | A | 0.650175 | -0.213685 | 0.019842 | 4.80E-27 | A | C | 0.3948 | 0.4703 | 0.02249 | 2.33E-75 | cis |
| miR-625-5p | rs10131715 | 14:66157377 | T | C | 0.385228 | 0.195601 | 0.020646 | 2.69E-21 | T | C | 0.3719 | 0.4064 | 0.02475 | 2.52E-51 | cis |
| miR-625-5p | rs10138052 | 14:65833344 | T | C | 0.739224 | -0.149361 | 0.021863 | 8.39E-12 | C | T | 0.2372 | 0.2771 | 0.03091 | 3.05E-18 | cis |
| miR-625-5p | rs10138570 | 14:65860033 | A | G | 0.470451 | 0.132126 | 0.019756 | 2.26E-11 | A | G | 0.4731 | 0.2482 | 0.02659 | 1.33E-19 | cis |
| miR-625-5p | rs10138671 | 14:65860099 | A | G | 0.398463 | 0.171449 | 0.020209 | 2.18E-17 | A | G | 0.3924 | 0.3325 | 0.02618 | 2.51E-33 | cis |
| miR-625-5p | rs10142283 | 14:66044123 | G | A | 0.346742 | 0.172676 | 0.02057 | 4.67E-17 | G | A | 0.3139 | 0.3906 | 0.02665 | 1.42E-42 | cis |
| miR-625-5p | rs10143206 | 14:66120677 | C | T | 0.349542 | 0.175451 | 0.020878 | 4.33E-17 | C | T | 0.3135 | 0.3916 | 0.02678 | 2.14E-42 | cis |
| miR-136-5p | rs10144381 | 14:10117372 | C | T | 0.673446 | 0.405272 | 0.036589 | 1.64E-28 | T | C | 0.3153 | -0.07165 | 0.01346 | 1.39E-07 | cis |
| miR-625-5p | rs10144979 | 14:65997633 | G | A | 0.355198 | 0.170483 | 0.02058 | 1.19E-16 | G | A | 0.3201 | 0.3763 | 0.02678 | 1.17E-39 | cis |
| miR-625-5p | rs10145146 | 14:66075006 | C | T | 0.349632 | 0.180781 | 0.020698 | 2.45E-18 | C | T | 0.3251 | 0.4125 | 0.02633 | 2.37E-47 | cis |
| miR-136-5p | rs10145513 | 14:10116859 | A | G | 0.668173 | 0.399046 | 0.03635 | 4.89E-28 | G | A | 0.3257 | -0.06929 | 0.01344 | 3.31E-07 | cis |
| miR-376c-3 | rs10145513 | 14:10116859 | A | G | 0.668173 | 0.209202 | 0.021737 | 6.31E-22 | G | A | 0.3257 | -0.03484 | 0.01023 | 0.000697 | cis |
| miR-136-5p | rs10146962 | 14:10117059 | T | C | 0.658875 | 0.39858 | 0.035752 | 7.29E-29 | C | T | 0.3327 | -0.07021 | 0.01308 | 1.08E-07 | cis |
| miR-376c-3 | rs10146962 | 14:10117059 | T | C | 0.658875 | 0.213122 | 0.021364 | 1.95E-23 | C | T | 0.3327 | -0.03492 | 0.009912 | 0.000455 | cis |
| miR-625-5p | rs10148529 | 14:66104706 | A | T | 0.345818 | 0.173355 | 0.020883 | 1.03E-16 | A | T | 0.3131 | 0.3917 | 0.02666 | 9.02E-43 | cis |
| miR-625-5p | rs10148907 | 14:65833377 | C | G | 0.739103 | -0.14955 | 0.021844 | 7.58E-12 | G | C | 0.2372 | 0.2771 | 0.03091 | 3.05E-18 | cis |
| miR-625-5p | rs10150412 | 14:65916799 | C | T | 0.358422 | 0.173561 | 0.020711 | 5.29E-17 | C | T | 0.3197 | 0.3772 | 0.02676 | 7.19E-40 | cis |
| miR-625-5p | rs10152007 | 14:66047433 | A | T | 0.343755 | 0.172178 | 0.020581 | 5.97E-17 | A | T | 0.3133 | 0.393 | 0.0267 | 6.99E-43 | cis |
| miR-320a | rs10174380 | 2:68596469 | C | T | 0.685243 | 0.144486 | 0.016618 | 3.47E-18 | T | C | 0.3299 | -0.05389 | 0.006202 | 2.59E-17 | trans |
| miR-320b | rs10174380 | 2:68596469 | C | T | 0.685243 | 0.129366 | 0.014889 | 3.67E-18 | T | C | 0.3299 | -0.06117 | 0.005135 | 6.57E-30 | trans |
| miR-320c | rs10174380 | 2:68596469 | C | T | 0.685243 | 0.117036 | 0.01396 | 5.12E-17 | T | C | 0.3299 | -0.05389 | 0.005098 | 2.55E-24 | trans |
| miR-320d | rs10174380 | 2:68596469 | C | T | 0.685243 | 0.116538 | 0.014293 | 3.54E-16 | T | C | 0.3299 | -0.04934 | 0.005003 | 1.47E-21 | trans |
| miR-4429 | rs10174380 | 2:68596469 | C | T | 0.685243 | 0.105793 | 0.013758 | 1.48E-14 | T | C | 0.3299 | -0.06042 | 0.004863 | 3.81E-32 | trans |
| miR-3130-3 | rs10199763 | 2:20764180 | G | A | 0.842574 | -0.266836 | 0.032905 | 5.09E-16 | A | G | 0.1794 | 0.1673 | 0.03129 | 1.21E-07 | cis |
| miR-181a-5 | rs1036332 | 1:19901247 | A | C | 0.260189 | 0.172538 | 0.013837 | 1.10E-35 | A | C | 0.2553 | 0.3408 | 0.01322 | 2.71E-102 | cis |
| miR-181a-5 | rs1040959 | 1:19889584 | A | C | 0.766999 | -0.100032 | 0.014691 | 9.81E-12 | C | A | 0.2375 | 0.2323 | 0.01682 | 1.64E-38 | cis |
| miR-181a-5 | rs1040960 | 1:19889598 | G | T | 0.766066 | -0.100198 | 0.014695 | 9.21E-12 | T | G | 0.2388 | 0.2323 | 0.01682 | 1.64E-38 | cis |
| miR-584-5p | rs10447231 | 5:14850090 | A | C | 0.474342 | 0.186354 | 0.02543 | 2.33E-13 | A | C | 0.4498 | 0.1752 | 0.01324 | 9.98E-36 | cis |
| miR-584-5p | rs10463413 | 5:14845508 | A | G | 0.844169 | -0.272685 | 0.034992 | 6.56E-15 | G | A | 0.1546 | 0.2727 | 0.01754 | 5.95E-47 | cis |
| miR-625-5p | rs10483780 | 14:65980117 | T | C | 0.501165 | 0.146505 | 0.019946 | 2.05E-13 | C | T | 0.4775 | -0.3207 | 0.02578 | 3.39E-32 | cis |
| miR-320a | rs10496165 | 2:68620089 | C | T | 0.697496 | 0.139829 | 0.016852 | 1.06E-16 | T | C | 0.3198 | -0.05845 | 0.006246 | 1.12E-19 | trans |
| miR-320b | rs10496165 | 2:68620089 | C | T | 0.697496 | 0.127045 | 0.015092 | 3.83E-17 | T | C | 0.3198 | -0.06409 | 0.00518 | 6.68E-32 | trans |
| miR-320c | rs10496165 | 2:68620089 | C | T | 0.697496 | 0.113968 | 0.014152 | 8.08E-16 | T | C | 0.3198 | -0.05714 | 0.00516 | 2.40E-26 | trans |
| miR-320d | rs10496165 | 2:68620089 | C | T | 0.697496 | 0.116733 | 0.014478 | 7.45E-16 | T | C | 0.3198 | -0.05191 | 0.005054 | 3.97E-23 | trans |
| miR-4429 | rs10496165 | 2:68620089 | C | T | 0.697496 | 0.104328 | 0.013941 | 7.23E-14 | T | C | 0.3198 | -0.06411 | 0.004896 | 3.81E-35 | trans |
| miR-320a | rs1050181 | 2:68622952 | G | C | 0.544992 | 0.150456 | 0.015516 | 3.10E-22 | C | G | 0.4541 | -0.06727 | 0.005522 | 4.35E-31 | trans |
| miR-320b | rs1050181 | 2:68622952 | G | C | 0.544992 | 0.136202 | 0.013895 | 1.10E-22 | C | G | 0.4541 | -0.0665 | 0.004602 | 1.38E-41 | trans |
| miR-320c | rs1050181 | 2:68622952 | G | C | 0.544992 | 0.123229 | 0.013032 | 3.20E-21 | C | G | 0.4541 | -0.06396 | 0.004529 | 5.28E-40 | trans |
| miR-320d | rs1050181 | 2:68622952 | G | C | 0.544992 | 0.121982 | 0.01335 | 6.39E-20 | C | G | 0.4541 | -0.06049 | 0.00443 | 8.65E-38 | trans |
| miR-320e | rs1050181 | 2:68622952 | G | C | 0.544992 | 0.110579 | 0.014759 | 6.76E-14 | C | G | 0.4541 | -0.09639 | 0.006121 | 5.01E-48 | trans |
| miR-4429 | rs1050181 | 2:68622952 | G | C | 0.544992 | 0.113852 | 0.012842 | 7.63E-19 | C | G | 0.4541 | -0.06893 | 0.004297 | 1.62E-49 | trans |
| miR-130a-3 | rs10750870 | 11:5762381 | A | C | 0.437769 | -0.15918 | 0.021015 | 3.60E-14 | A | C | 0.4311 | -0.1707 | 0.01071 | 6.64E-49 | cis |
| miR-625-5p | rs1075566 | 14:66239459 | C | T | 0.454013 | 0.144041 | 0.019394 | 1.11E-13 | C | T | 0.4909 | 0.3186 | 0.02562 | 3.53E-32 | cis |
| miR-130a-3 | rs10792110 | 11:5753634 | C | T | 0.564735 | 0.161403 | 0.021024 | 1.63E-14 | T | C | 0.434 | -0.1713 | 0.01078 | 1.04E-48 | cis |
| miR-130a-3 | rs10792112 | 11:5756580 | C | T | 0.684897 | -0.243313 | 0.02216 | 4.77E-28 | T | C | 0.3019 | 0.2352 | 0.01037 | 1.76E-85 | cis |
| miR-181a-5 | rs10800599 | 1:19887083 | T | C | 0.760087 | -0.108167 | 0.0147 | 1.86E-13 | C | T | 0.2401 | 0.236 | 0.01655 | 1.52E-40 | cis |
| miR-181a-5 | rs10800602 | 1:19891451 | A | G | 0.244491 | 0.102312 | 0.014406 | 1.23E-12 | A | G | 0.2426 | 0.2416 | 0.01666 | 8.82E-42 | cis |
| miR-181a-5 | rs10800603 | 1:19891965 | A | G | 0.238849 | 0.106727 | 0.01456 | 2.30E-13 | A | G | 0.2446 | 0.237 | 0.01655 | 5.97E-41 | cis |
| miR-181a-5 | rs10800606 | 1:19900499 | C | G | 0.663982 | 0.087564 | 0.01307 | 2.09E-11 | G | C | 0.3635 | -0.141 | 0.01617 | 2.18E-17 | cis |
| miR-625-5p | rs10873191 | 14:66110627 | T | C | 0.346549 | 0.173321 | 0.020847 | 9.27E-17 | T | C | 0.3139 | 0.3929 | 0.02667 | 5.86E-43 | cis |
| miR-130a-3 | rs10896626 | 11:5732320 | A | G | 0.871328 | -0.217382 | 0.031621 | 6.22E-12 | G | A | 0.1333 | 0.2303 | 0.01607 | 8.75E-41 | cis |
| miR-130a-3 | rs10896631 | 11:5738120 | T | C | 0.750433 | -0.30148 | 0.023269 | 2.17E-38 | C | T | 0.2603 | 0.285 | 0.009157 | 5.49E-132 | cis |
| miR-130a-3 | rs10896632 | 11:5739030 | G | A | 0.746727 | -0.302037 | 0.023122 | 5.37E-39 | A | G | 0.2672 | 0.2894 | 0.008928 | 2.41E-141 | cis |
| miR-130a-3 | rs10896636 | 11:5744803 | C | G | 0.68075 | -0.243141 | 0.022412 | 2.02E-27 | G | C | 0.3016 | 0.2339 | 0.01072 | 4.89E-80 | cis |
| miR-130a-3 | rs10896644 | 11:5756399 | C | T | 0.567891 | 0.160898 | 0.021008 | 1.88E-14 | T | C | 0.4305 | -0.1701 | 0.0108 | 5.61E-48 | cis |
| miR-130a-3 | rs10896647 | 11:5757188 | C | T | 0.685297 | -0.242537 | 0.022203 | 8.89E-28 | T | C | 0.3032 | 0.2278 | 0.01056 | 2.53E-79 | cis |
| miR-130a-3 | rs10896649 | 11:5758779 | A | G | 0.565548 | 0.160285 | 0.020992 | 2.25E-14 | G | A | 0.4338 | -0.1712 | 0.01072 | 3.31E-49 | cis |
| miR-181a-5 | rs10919602 | 1:19888260 | G | A | 0.764985 | -0.101467 | 0.014749 | 6.00E-12 | A | G | 0.2414 | 0.2289 | 0.01699 | 8.39E-37 | cis |
| miR-181a-5 | rs10919619 | 1:19903800 | T | C | 0.841859 | -0.152167 | 0.016799 | 1.33E-19 | C | T | 0.1601 | 0.2939 | 0.01823 | 1.02E-49 | cis |
| miR-181a-5 | rs10919655 | 1:19924860 | A | G | 0.667251 | -0.106368 | 0.013119 | 5.15E-16 | G | A | 0.3384 | 0.1924 | 0.01485 | 1.47E-34 | cis |
| miR-3130-3 | rs10932164 | 2:20761637 | A | G | 0.855067 | -0.279415 | 0.034267 | 3.52E-16 | G | A | 0.1556 | 0.1715 | 0.03336 | 3.56E-07 | cis |
| miR-584-5p | rs1112745 | 5:14847180 | G | A | 0.474305 | 0.187933 | 0.025488 | 1.66E-13 | G | A | 0.4449 | 0.179 | 0.0132 | 2.64E-37 | cis |
| miR-625-5p | rs1113962 | 14:66005795 | C | T | 0.355246 | 0.170317 | 0.02059 | 1.32E-16 | C | T | 0.3201 | 0.3763 | 0.02678 | 1.17E-39 | cis |
| miR-625-5p | rs11158588 | 14:65799870 | G | A | 0.221744 | 0.35927 | 0.022176 | 5.00E-59 | G | A | 0.2468 | 0.7398 | 0.01684 | 8.19E-203 | cis |
| miR-625-5p | rs11158593 | 14:65859990 | G | A | 0.466718 | 0.134472 | 0.019789 | 1.08E-11 | G | A | 0.4664 | 0.2561 | 0.02686 | 2.56E-20 | cis |
| miR-625-5p | rs11158595 | 14:65906360 | A | G | 0.34826 | 0.175746 | 0.020926 | 4.52E-17 | A | G | 0.3135 | 0.3925 | 0.02667 | 6.98E-43 | cis |
| miR-625-5p | rs11158596 | 14:65911970 | T | A | 0.507581 | 0.145384 | 0.01988 | 2.61E-13 | A | T | 0.4699 | -0.314 | 0.02597 | 1.13E-30 | cis |
| miR-625-5p | rs11158598 | 14:65923770 | A | G | 0.389613 | 0.1839 | 0.020429 | 2.22E-19 | A | G | 0.3774 | 0.4052 | 0.02468 | 2.04E-51 | cis |
| miR-625-5p | rs11158601 | 14:66025300 | A | G | 0.499831 | 0.146827 | 0.019777 | 1.14E-13 | G | A | 0.4777 | -0.3183 | 0.02572 | 6.13E-32 | cis |
| miR-625-5p | rs11158602 | 14:66071730 | T | G | 0.491419 | 0.14246 | 0.01987 | 7.53E-13 | G | T | 0.4888 | -0.3007 | 0.02632 | 9.62E-28 | cis |
| miR-625-5p | rs11158603 | 14:66072040 | C | T | 0.349651 | 0.180755 | 0.020698 | 2.48E-18 | C | T | 0.3228 | 0.4153 | 0.02626 | 3.98E-48 | cis |
| miR-625-5p | rs11158604 | 14:66094450 | G | A | 0.346475 | 0.17345 | 0.020801 | 7.51E-17 | G | A | 0.3134 | 0.3927 | 0.02668 | 6.50E-43 | cis |
| miR-625-5p | rs11158605 | 14:66107920 | T | A | 0.365947 | 0.173317 | 0.020738 | 6.42E-17 | T | A | 0.3408 | 0.3788 | 0.02585 | 1.35E-42 | cis |
| miR-625-5p | rs11158607 | 14:66110790 | T | G | 0.365883 | 0.173324 | 0.020734 | 6.31E-17 | T | G | 0.3402 | 0.3798 | 0.02586 | 9.63E-43 | cis |
| miR-625-5p | rs11158611 | 14:66235070 | G | A | 0.451752 | 0.143146 | 0.019346 | 1.37E-13 | G | A | 0.4866 | 0.3222 | 0.02559 | 6.78E-33 | cis |
| miR-584-5p | rs11168080 | 5:14848418 | T | C | 0.408107 | 0.199694 | 0.02537 | 3.51E-15 | T | C | 0.3944 | 0.2109 | 0.0127 | 2.08E-52 | cis |
| miR-625-5p | rs1121885 | 14:66156910 | A | G | 0.34818 | 0.177649 | 0.020897 | 1.88E-17 | A | G | 0.3142 | 0.3915 | 0.02659 | 6.45E-43 | cis |
| miR-130a-3 | rs11229075 | 11:5739107 | A | C | 0.750877 | -0.301901 | 0.023261 | 1.61E-38 | C | A | 0.2601 | 0.2922 | 0.008899 | 1.23E-142 | cis |
| miR-130a-3 | rs11229109 | 11:5749222 | A | G | 0.554137 | 0.164771 | 0.021077 | 5.39E-15 | G | A | 0.4387 | -0.1757 | 0.01069 | 1.56E-51 | cis |
| miR-130a-3 | rs11229113 | 11:5751080 | G | T | 0.692006 | -0.243466 | 0.022353 | 1.26E-27 | T | G | 0.3003 | 0.2309 | 0.01061 | 2.52E-80 | cis |
| miR-130a-3 | rs11229116 | 11:5753454 | C | G | 0.683806 | -0.244457 | 0.022182 | 3.05E-28 | G | C | 0.3069 | 0.2287 | 0.01051 | 1.86E-80 | cis |
| miR-130a-3 | rs11229130 | 11:5756210 | A | G | 0.688809 | -0.243075 | 0.022346 | 1.47E-27 | G | A | 0.2915 | 0.225 | 0.01101 | 1.04E-72 | cis |

| | | | | | | | | | | | | | | | |
|---|---|---|---|---|---|---|---|---|---|---|---|---|---|---|---|
| miR-130a-3 | rs1122913 | 11:575625 | G | A | 0.684434 | -0.242732 | 0.022144 | 5.85E-28 | A | G | 0.3061 | 0.229 | 0.01048 | 5.57E-81 | cis |
| miR-130a-3 | rs1122914 | 11:576214 | A | T | 0.683461 | -0.2414 | 0.022336 | 3.17E-27 | T | A | 0.3018 | 0.2307 | 0.01054 | 5.47E-81 | cis |
| miR-10b-5p | rs1124406 | 9:1361539 | C | T | 0.886198 | -0.206905 | 0.022623 | 5.93E-20 | T | C | 0.1087 | 0.1079 | 0.02473 | 1.50E-05 | trans |
| miR-130a-3 | rs1127641 | 11:573858 | C | T | 0.7469 | -0.302271 | 0.023181 | 7.27E-39 | T | C | 0.2696 | 0.2862 | 0.009083 | 8.62E-135 | cis |
| miR-130a-3 | rs1129766 | 11:575648 | G | A | 0.566038 | 0.161154 | 0.021054 | 1.95E-14 | A | G | 0.4336 | -0.1731 | 0.01082 | 2.88E-49 | cis |
| miR-625-5p | rs1131317 | 14:660641 | G | A | 0.501439 | 0.145533 | 0.019821 | 2.10E-13 | A | G | 0.4723 | -0.318 | 0.02613 | 5.34E-31 | cis |
| miR-3130-3 | rs1155295 | 2:2076756 | C | G | 0.874904 | -0.247276 | 0.036911 | 2.09E-11 | G | C | 0.1428 | 0.1496 | 0.03406 | 1.30E-05 | cis |
| miR-320a | rs1156629 | 2:6862663 | T | G | 0.547487 | 0.152478 | 0.015652 | 2.01E-22 | G | T | 0.4541 | -0.06727 | 0.005522 | 4.35E-31 | trans |
| miR-320b | rs1156629 | 2:6862663 | T | G | 0.547487 | 0.138094 | 0.014017 | 6.74E-23 | G | T | 0.4541 | -0.0665 | 0.004602 | 1.38E-41 | trans |
| miR-320c | rs1156629 | 2:6862663 | T | G | 0.547487 | 0.12504 | 0.013146 | 1.88E-21 | G | T | 0.4541 | -0.06396 | 0.004529 | 5.28E-40 | trans |
| miR-320d | rs1156629 | 2:6862663 | T | G | 0.547487 | 0.123554 | 0.013468 | 4.55E-20 | G | T | 0.4541 | -0.06049 | 0.00443 | 8.65E-38 | trans |
| miR-320e | rs1156629 | 2:6862663 | T | G | 0.547487 | 0.112242 | 0.014889 | 4.76E-14 | G | T | 0.4541 | -0.09639 | 0.006121 | 5.01E-48 | trans |
| miR-4429 | rs1156629 | 2:6862663 | T | G | 0.547487 | 0.115419 | 0.012956 | 5.17E-19 | G | T | 0.4541 | -0.06893 | 0.004297 | 1.62E-49 | trans |
| miR-130a-3 | rs1157017 | 11:575294 | A | G | 0.566649 | 0.1615 | 0.021063 | 1.75E-14 | G | A | 0.4321 | -0.1711 | 0.01085 | 4.11E-48 | cis |
| miR-130a-3 | rs1157018 | 11:575557 | T | C | 0.567672 | 0.161097 | 0.02102 | 1.80E-14 | C | T | 0.4316 | -0.1706 | 0.01083 | 4.82E-48 | cis |
| miR-130a-3 | rs1157019 | 11:575604 | A | C | 0.684149 | -0.242699 | 0.022118 | 5.17E-28 | C | A | 0.3057 | 0.2293 | 0.01048 | 3.27E-81 | cis |
| miR-320a | rs1158434 | 2:6863552 | G | C | 0.551072 | 0.155908 | 0.015994 | 1.88E-22 | C | G | 0.4525 | -0.06653 | 0.005537 | 2.43E-30 | trans |
| miR-320b | rs1158434 | 2:6863552 | G | C | 0.551072 | 0.14116 | 0.014323 | 6.49E-23 | C | G | 0.4525 | -0.06512 | 0.004625 | 8.91E-40 | trans |
| miR-320c | rs1158434 | 2:6863552 | G | C | 0.551072 | 0.128841 | 0.013428 | 8.41E-22 | C | G | 0.4525 | -0.06344 | 0.004544 | 3.27E-39 | trans |
| miR-320d | rs1158434 | 2:6863552 | G | C | 0.551072 | 0.126979 | 0.013759 | 2.73E-20 | C | G | 0.4525 | -0.06088 | 0.00445 | 6.91E-38 | trans |
| miR-320e | rs1158434 | 2:6863552 | G | C | 0.551072 | 0.11383 | 0.015217 | 7.41E-14 | C | G | 0.4525 | -0.09579 | 0.006144 | 3.51E-47 | trans |
| miR-4429 | rs1158434 | 2:6863552 | G | C | 0.551072 | 0.118342 | 0.013237 | 3.88E-19 | C | G | 0.4525 | -0.06848 | 0.004328 | 2.36E-48 | trans |
| miR-130a-3 | rs1160144 | 11:575243 | G | A | 0.684 | -0.242757 | 0.022162 | 6.38E-28 | A | G | 0.3059 | 0.2296 | 0.01048 | 2.41E-81 | cis |
| miR-130a-3 | rs1160210 | 11:574352 | G | A | 0.688626 | -0.24541 | 0.022271 | 3.09E-28 | A | G | 0.303 | 0.2326 | 0.0105 | 1.56E-82 | cis |
| miR-130a-3 | rs1160432 | 11:574970 | C | A | 0.688224 | -0.242092 | 0.022277 | 1.65E-27 | A | C | 0.2997 | 0.2333 | 0.01058 | 5.22E-82 | cis |
| miR-130a-3 | rs1160433 | 11:574670 | C | G | 0.686388 | -0.244681 | 0.022232 | 3.58E-28 | G | C | 0.303 | 0.2326 | 0.0105 | 1.56E-82 | cis |
| miR-130a-3 | rs1160573 | 11:575240 | C | G | 0.683995 | -0.242751 | 0.022162 | 6.40E-28 | G | C | 0.3059 | 0.2296 | 0.01048 | 2.41E-81 | cis |
| miR-130a-3 | rs1160639 | 11:574352 | C | T | 0.687897 | -0.245707 | 0.0223 | 3.12E-28 | T | C | 0.303 | 0.2326 | 0.0105 | 1.56E-82 | cis |
| miR-130a-3 | rs1160667 | 11:573833 | G | A | 0.750505 | -0.301631 | 0.023271 | 2.02E-38 | A | G | 0.2603 | 0.285 | 0.009157 | 5.49E-132 | cis |
| miR-130a-3 | rs1160670 | 11:573835 | G | T | 0.750529 | -0.301671 | 0.023272 | 1.98E-38 | T | G | 0.2603 | 0.285 | 0.009157 | 5.49E-132 | cis |
| miR-130a-3 | rs1160705 | 11:574968 | C | T | 0.683884 | -0.244801 | 0.022215 | 3.08E-28 | T | C | 0.2995 | 0.2346 | 0.0107 | 3.31E-81 | cis |
| miR-130a-3 | rs1160712 | 11:573897 | G | A | 0.743794 | -0.302872 | 0.023174 | 4.93E-39 | A | G | 0.2723 | 0.2891 | 0.008958 | 5.30E-139 | cis |
| miR-625-5p | rs1162074 | 14:658998 | A | G | 0.502681 | 0.145468 | 0.019793 | 1.99E-13 | G | A | 0.4706 | -0.3144 | 0.02585 | 5.67E-31 | cis |
| miR-625-5p | rs1162112 | 14:658224 | C | T | 0.388213 | 0.196561 | 0.019252 | 1.79E-24 | C | T | 0.4309 | 0.4129 | 0.02387 | 5.00E-56 | cis |
| miR-625-5p | rs1162201 | 14:660416 | C | T | 0.502262 | 0.14792 | 0.019779 | 7.51E-14 | T | C | 0.4771 | -0.317 | 0.02573 | 1.07E-31 | cis |
| miR-625-5p | rs1162227 | 14:661490 | A | T | 0.504871 | 0.146606 | 0.019892 | 1.71E-13 | T | A | 0.473 | -0.3134 | 0.0259 | 1.08E-30 | cis |
| miR-625-5p | rs1162282 | 14:661917 | T | A | 0.496725 | 0.144626 | 0.019819 | 2.93E-13 | A | T | 0.477 | -0.3144 | 0.02593 | 8.02E-31 | cis |
| miR-625-5p | rs1162366 | 14:660211 | T | G | 0.390012 | 0.168768 | 0.020334 | 1.04E-16 | T | G | 0.3883 | 0.3408 | 0.02656 | 6.52E-34 | cis |
| miR-625-5p | rs1162392 | 14:658193 | A | C | 0.389577 | 0.198029 | 0.019291 | 1.01E-24 | A | C | 0.4298 | 0.4085 | 0.02394 | 8.73E-55 | cis |
| miR-625-5p | rs1162404 | 14:661080 | C | T | 0.498929 | 0.145121 | 0.019847 | 2.63E-13 | T | C | 0.4783 | -0.3166 | 0.02581 | 1.87E-31 | cis |
| miR-625-5p | rs1162536 | 14:662328 | G | A | 0.45263 | 0.142912 | 0.019323 | 1.40E-13 | G | A | 0.4905 | 0.32 | 0.02558 | 1.55E-32 | cis |
| miR-625-5p | rs1162588 | 14:662451 | G | A | 0.454755 | 0.141922 | 0.019347 | 2.21E-13 | G | A | 0.4935 | 0.3186 | 0.02562 | 3.53E-32 | cis |
| miR-625-5p | rs1162706 | 14:661829 | T | G | 0.488866 | 0.13317 | 0.01984 | 1.92E-11 | G | T | 0.4815 | -0.2981 | 0.02602 | 6.00E-28 | cis |
| miR-625-5p | rs1162708 | 14:659788 | T | G | 0.501066 | 0.146483 | 0.019949 | 2.09E-13 | G | T | 0.4775 | -0.3207 | 0.02578 | 3.39E-32 | cis |
| miR-625-5p | rs1162718 | 14:661214 | C | T | 0.501637 | 0.146864 | 0.019888 | 1.53E-13 | T | C | 0.4782 | -0.3154 | 0.02589 | 4.59E-31 | cis |
| miR-625-5p | rs1162718 | 14:661214 | A | G | 0.501636 | 0.146867 | 0.019888 | 1.53E-13 | G | A | 0.4782 | -0.3154 | 0.02589 | 4.59E-31 | cis |
| miR-625-5p | rs1162757 | 14:662344 | C | T | 0.452075 | 0.142993 | 0.019339 | 1.42E-13 | C | T | 0.4885 | 0.3222 | 0.02559 | 6.78E-33 | cis |
| miR-625-5p | rs1162759 | 14:657977 | G | T | 0.38607 | 0.212976 | 0.019217 | 1.52E-28 | G | T | 0.4342 | 0.4495 | 0.02264 | 1.39E-69 | cis |
| miR-625-5p | rs1162760 | 14:662343 | G | A | 0.452186 | 0.143034 | 0.019344 | 1.42E-13 | G | A | 0.4905 | 0.3222 | 0.02559 | 6.78E-33 | cis |
| miR-625-5p | rs1162819 | 14:658912 | A | G | 0.495088 | 0.14713 | 0.019857 | 1.27E-13 | G | A | 0.4781 | -0.3238 | 0.02562 | 4.51E-33 | cis |
| miR-625-5p | rs1162876 | 14:661684 | T | C | 0.500166 | 0.147858 | 0.019781 | 7.73E-14 | C | T | 0.4758 | -0.3151 | 0.0258 | 3.17E-31 | cis |
| miR-625-5p | rs1162884 | 14:662356 | A | G | 0.451742 | 0.143194 | 0.01935 | 1.36E-13 | A | G | 0.4873 | 0.3222 | 0.02559 | 6.78E-33 | cis |
| miR-625-5p | rs1162943 | 14:658921 | C | G | 0.352083 | 0.17854 | 0.021058 | 2.28E-17 | C | G | 0.3216 | 0.3754 | 0.02703 | 1.04E-38 | cis |
| miR-320a | rs1164012 | 2:6862562 | C | T | 0.545777 | 0.151008 | 0.015571 | 3.08E-22 | T | C | 0.4541 | -0.06727 | 0.005522 | 4.35E-31 | trans |
| miR-320b | rs1164012 | 2:6862562 | C | T | 0.545777 | 0.136712 | 0.013945 | 1.09E-22 | T | C | 0.4541 | -0.0665 | 0.004602 | 1.38E-41 | trans |
| miR-320c | rs1164012 | 2:6862562 | C | T | 0.545777 | 0.123739 | 0.013078 | 3.04E-21 | T | C | 0.4541 | -0.06396 | 0.004529 | 5.28E-40 | trans |
| miR-320d | rs1164012 | 2:6862562 | C | T | 0.545777 | 0.122475 | 0.013397 | 6.14E-20 | T | C | 0.4541 | -0.06049 | 0.00443 | 8.65E-38 | trans |
| miR-320e | rs1164012 | 2:6862562 | C | T | 0.545777 | 0.111024 | 0.014812 | 6.59E-14 | T | C | 0.4541 | -0.09639 | 0.006121 | 5.01E-48 | trans |
| miR-4429 | rs1164012 | 2:6862562 | C | T | 0.545777 | 0.114323 | 0.012888 | 7.30E-19 | T | C | 0.4541 | -0.06893 | 0.004297 | 1.62E-49 | trans |
| let-7a-5p | rs1165522 | 9:1361490 | T | A | 0.653719 | -0.084566 | 0.011334 | 8.59E-14 | A | T | 0.3436 | 0.01765 | 0.005048 | 0.0005 | trans |
| let-7b-5p | rs1165522 | 9:1361490 | T | A | 0.653719 | -0.107531 | 0.01003 | 8.12E-27 | A | T | 0.3436 | 0.02789 | 0.00657 | 2.48E-05 | trans |
| let-7c-5p | rs1165522 | 9:1361490 | T | A | 0.653719 | -0.090775 | 0.00916 | 3.78E-23 | A | T | 0.3436 | 0.02288 | 0.006292 | 0.000297 | trans |
| miR-10b-5p | rs1165522 | 9:1361490 | T | A | 0.653719 | -0.174502 | 0.014267 | 2.13E-34 | A | T | 0.3436 | 0.1072 | 0.0166 | 2.02E-10 | trans |
| miR-126-3p | rs1165522 | 9:1361490 | T | A | 0.653719 | -0.167546 | 0.013039 | 8.64E-38 | A | T | 0.3436 | 0.04986 | 0.008167 | 1.71E-09 | trans |
| miR-139-5p | rs1165522 | 9:1361490 | T | A | 0.653719 | -0.15545 | 0.011571 | 3.77E-41 | A | T | 0.3436 | 0.06164 | 0.01612 | 0.000143 | trans |
| miR-584-5p | rs1174620 | 5:1484993 | A | G | 0.473634 | 0.184438 | 0.025138 | 2.18E-13 | A | G | 0.4516 | 0.1762 | 0.01322 | 3.53E-36 | cis |
| miR-625-5p | rs1177595 | 14:657590 | G | C | 0.845965 | -0.311633 | 0.026756 | 2.37E-31 | C | G | 0.1549 | 0.6562 | 0.02918 | 4.54E-84 | cis |
| miR-625-5p | rs1184468 | 14:659108 | G | C | 0.356705 | 0.17288 | 0.02073 | 7.44E-17 | G | C | 0.3196 | 0.3803 | 0.02687 | 4.07E-40 | cis |
| miR-625-5p | rs1184726 | 14:657756 | T | G | 0.67304 | -0.150419 | 0.020266 | 1.15E-13 | G | T | 0.3597 | 0.298 | 0.02672 | 1.17E-26 | cis |
| miR-625-5p | rs1184844 | 14:662574 | C | T | 0.54144 | -0.139242 | 0.019367 | 6.49E-13 | C | T | 0.5 | -0.313 | 0.02632 | 9.16E-30 | cis |
| miR-625-5p | rs1184844 | 14:662574 | A | G | 0.5414 | -0.139233 | 0.019364 | 6.47E-13 | A | G | 0.5 | -0.313 | 0.02632 | 9.16E-30 | cis |
| miR-625-5p | rs1184846 | 14:662572 | G | A | 0.541416 | -0.139268 | 0.01937 | 6.48E-13 | A | G | 0.4993 | 0.3134 | 0.02627 | 6.16E-30 | cis |
| miR-625-5p | rs1185157 | 14:659002 | T | C | 0.454741 | 0.146928 | 0.01999 | 1.98E-13 | T | C | 0.4506 | 0.3265 | 0.02545 | 6.01E-34 | cis |
| miR-3130-3 | rs1189416 | 2:2076244 | T | C | 0.859947 | -0.284323 | 0.034753 | 2.81E-16 | C | T | 0.1528 | 0.1739 | 0.03373 | 3.30E-07 | cis |
| miR-3130-3 | rs1189537 | 2:2076547 | T | C | 0.842391 | -0.264848 | 0.033305 | 1.83E-15 | C | T | 0.1719 | 0.1741 | 0.03232 | 9.85E-08 | cis |
| miR-133b | rs1190646 | 20:611589 | C | T | 0.824779 | -0.248529 | 0.03256 | 2.29E-14 | T | C | 0.2036 | 0.4466 | 0.03799 | 3.06E-29 | trans |
| miR-181a-5 | rs1204800 | 1:1989153 | T | C | 0.235546 | 0.100871 | 0.014669 | 6.14E-12 | T | C | 0.2425 | 0.2315 | 0.01703 | 2.16E-37 | cis |
| miR-625-5p | rs1205018 | 14:662430 | C | T | 0.455215 | 0.144476 | 0.019432 | 1.05E-13 | C | T | 0.4942 | 0.3187 | 0.02561 | 3.38E-32 | cis |
| miR-625-5p | rs1205023 | 14:662350 | T | C | 0.451755 | 0.143138 | 0.019345 | 1.37E-13 | T | C | 0.4873 | 0.3222 | 0.02559 | 6.78E-33 | cis |
| miR-181a-5 | rs1209687 | 1:1989144 | G | A | 0.244151 | 0.101854 | 0.014401 | 1.52E-12 | G | A | 0.2426 | 0.2408 | 0.0167 | 2.06E-41 | cis |
| miR-181a-5 | rs1212331 | 1:1988726 | G | T | 0.764104 | -0.102538 | 0.014805 | 4.33E-12 | T | G | 0.2336 | 0.2327 | 0.01692 | 3.84E-38 | cis |
| miR-181a-5 | rs1212520 | 1:1989920 | A | G | 0.260715 | 0.177971 | 0.013845 | 8.14E-38 | A | G | 0.2659 | 0.3404 | 0.01251 | 1.13E-111 | cis |
| miR-181a-5 | rs1213024 | 1:1988841 | G | C | 0.765569 | -0.101979 | 0.014754 | 4.79E-12 | C | G | 0.2367 | 0.231 | 0.01681 | 3.99E-38 | cis |
| miR-181a-5 | rs1213201 | 1:1988820 | T | C | 0.755495 | -0.106835 | 0.014472 | 1.55E-13 | C | T | 0.2445 | 0.234 | 0.01639 | 1.17E-40 | cis |
| miR-181a-5 | rs1214202 | 1:1988815 | C | T | 0.766397 | -0.101919 | 0.014796 | 5.65E-12 | T | C | 0.2359 | 0.2324 | 0.01687 | 2.84E-38 | cis |
| miR-130a-3 | rs1214654 | 11:575086 | G | C | 0.685316 | -0.243208 | 0.022155 | 4.90E-28 | C | G | 0.3056 | 0.2297 | 0.01049 | 3.07E-81 | cis |

| | | | | | | | | | | | | | | | |
|---|---|---|---|---|---|---|---|---|---|---|---|---|---|---|---|
| miR-625-5p | rs1214747 | 14:6579508 | C | T | 0.845789 | -0.329795 | 0.02651 | 1.58E-35 | T | C | 0.1552 | 0.6992 | 0.02832 | 7.80E-97 | cis |
| miR-625-5p | rs1214777 | 14:6579769 | A | T | 0.846077 | -0.328488 | 0.026194 | 4.47E-36 | T | A | 0.1567 | 0.6934 | 0.02802 | 1.72E-97 | cis |
| miR-136-5p | rs1214784 | 14:101144 | C | T | 0.870989 | 0.380494 | 0.05243 | 3.95E-13 | T | C | 0.09763 | -0.07267 | 0.02081 | 0.00051 | cis |
| miR-130a-3 | rs1227020 | 11:575377 | G | A | 0.564726 | 0.161383 | 0.021024 | 1.64E-14 | A | G | 0.4358 | -0.17 | 0.01075 | 2.50E-48 | cis |
| miR-130a-3 | rs1227486 | 11:575576 | C | T | 0.56562 | 0.161052 | 0.021005 | 1.76E-14 | T | C | 0.4348 | -0.1718 | 0.01075 | 3.64E-49 | cis |
| miR-130a-3 | rs1236216 | 11:574530 | C | T | 0.68725 | -0.246218 | 0.02227 | 2.05E-28 | T | C | 0.3027 | 0.2325 | 0.01051 | 2.51E-82 | cis |
| miR-130a-3 | rs1236302 | 11:575032 | G | C | 0.556328 | 0.166301 | 0.021097 | 3.21E-15 | C | G | 0.4385 | -0.1756 | 0.01069 | 1.89E-51 | cis |
| miR-625-5p | rs1243359 | 14:660288 | T | C | 0.343187 | 0.172769 | 0.020668 | 6.31E-17 | T | C | 0.3115 | 0.3943 | 0.0267 | 4.19E-43 | cis |
| miR-136-5p | rs1243449 | 14:101179 | A | G | 0.18959 | -0.494684 | 0.044481 | 9.88E-29 | A | G | 0.1809 | -0.09697 | 0.01616 | 3.23E-09 | cis |
| miR-376c-3 | rs1243449 | 14:101179 | A | G | 0.18959 | -0.284953 | 0.026482 | 5.29E-27 | A | G | 0.1809 | -0.05216 | 0.01228 | 2.46E-05 | cis |
| miR-625-5p | rs1243646 | 14:658068 | T | C | 0.255814 | 0.28174 | 0.021604 | 7.15E-39 | T | C | 0.2575 | 0.6469 | 0.02164 | 2.09E-126 | cis |
| miR-4707-3 | rs1243715 | 14:233729 | G | A | 0.437486 | 0.091279 | 0.013659 | 2.35E-11 | G | A | 0.431 | 0.2952 | 0.01684 | 5.94E-57 | cis |
| miR-3130-3 | rs1246816 | 2:2076463 | T | C | 0.842809 | -0.267176 | 0.032925 | 4.87E-16 | C | T | 0.1783 | 0.1694 | 0.03125 | 8.17E-08 | cis |
| miR-3130-3 | rs1247096 | 2:2076395 | A | G | 0.866897 | -0.29906 | 0.035691 | 5.33E-17 | G | A | 0.1343 | 0.1868 | 0.03567 | 2.18E-07 | cis |
| miR-3130-3 | rs1247100 | 2:2076397 | A | G | 0.842907 | -0.27172 | 0.032916 | 1.52E-16 | G | A | 0.1794 | 0.1673 | 0.03129 | 1.21E-07 | cis |
| miR-3130-3 | rs1247846 | 2:2076710 | T | C | 0.846612 | -0.263395 | 0.033397 | 3.10E-15 | C | T | 0.1722 | 0.1649 | 0.03168 | 2.55E-07 | cis |
| miR-625-5p | rs1256520 | 14:657371 | C | A | 0.64779 | -0.191315 | 0.020361 | 5.66E-21 | A | C | 0.3665 | 0.4252 | 0.02444 | 2.22E-56 | cis |
| miR-625-5p | rs1256521 | 14:657376 | T | C | 0.646581 | -0.191015 | 0.020326 | 5.57E-21 | C | T | 0.3691 | 0.4338 | 0.02435 | 1.33E-58 | cis |
| miR-625-5p | rs1256522 | 14:657380 | C | T | 0.647567 | -0.191086 | 0.020355 | 6.13E-21 | T | C | 0.3658 | 0.4274 | 0.02445 | 7.33E-57 | cis |
| miR-625-5p | rs1256523 | 14:657381 | A | G | 0.647737 | -0.190991 | 0.020354 | 6.39E-21 | G | A | 0.3659 | 0.4274 | 0.02445 | 7.33E-57 | cis |
| miR-625-5p | rs1256524 | 14:657384 | T | C | 0.647718 | -0.190917 | 0.020352 | 6.55E-21 | C | T | 0.3674 | 0.4302 | 0.02436 | 7.26E-58 | cis |
| miR-625-5p | rs1256525 | 14:657385 | G | C | 0.646861 | -0.190909 | 0.020383 | 7.52E-21 | C | G | 0.3684 | 0.431 | 0.02445 | 1.21E-57 | cis |
| miR-625-5p | rs1256526 | 14:657399 | T | G | 0.647195 | -0.190548 | 0.020338 | 7.31E-21 | G | T | 0.3696 | 0.4319 | 0.02414 | 3.30E-59 | cis |
| miR-625-5p | rs1256536 | 14:657624 | A | G | 0.656085 | -0.2135 | 0.020061 | 1.89E-26 | G | A | 0.3906 | 0.4654 | 0.02258 | 1.43E-73 | cis |
| miR-625-5p | rs1256540 | 14:657640 | T | C | 0.645897 | -0.223056 | 0.019669 | 8.26E-30 | C | T | 0.4009 | 0.4761 | 0.02207 | 2.70E-79 | cis |
| miR-625-5p | rs1256544 | 14:657764 | G | C | 0.053147 | 0.335592 | 0.044505 | 4.68E-14 | G | C | 0.06346 | 0.5905 | 0.05286 | 9.40E-27 | cis |
| miR-625-5p | rs1258684 | 14:661450 | T | G | 0.500228 | 0.147394 | 0.019836 | 1.08E-13 | G | T | 0.4777 | -0.3158 | 0.02572 | 1.63E-31 | cis |
| miR-625-5p | rs1258705 | 14:660663 | C | T | 0.345738 | 0.172451 | 0.020704 | 8.14E-17 | C | T | 0.3136 | 0.3927 | 0.02668 | 6.50E-43 | cis |
| miR-625-5p | rs1258883 | 14:661626 | G | T | 0.500385 | 0.147861 | 0.019792 | 7.98E-14 | T | G | 0.4765 | -0.3158 | 0.02572 | 1.63E-31 | cis |
| miR-4707-3 | rs1258953 | 14:233966 | T | C | 0.440305 | 0.091556 | 0.013703 | 2.37E-11 | T | C | 0.4332 | 0.292 | 0.01658 | 1.81E-57 | cis |
| miR-625-5p | rs1258969 | 14:659204 | G | A | 0.510971 | 0.147065 | 0.019929 | 1.59E-13 | A | G | 0.4686 | -0.3129 | 0.02594 | 1.47E-30 | cis |
| miR-625-5p | rs1262272 | 14:657417 | C | G | 0.647443 | -0.190823 | 0.020345 | 6.64E-21 | G | C | 0.3709 | 0.4491 | 0.02392 | 1.49E-63 | cis |
| miR-133a-3 | rs1262460 | 20:611552 | C | A | 0.796375 | -0.174907 | 0.025247 | 4.27E-12 | A | C | 0.2288 | 0.394 | 0.03343 | 2.84E-29 | cis |
| miR-133b | rs1262460 | 20:611552 | C | A | 0.796375 | -0.264427 | 0.033362 | 2.26E-15 | A | C | 0.2288 | 0.4157 | 0.03565 | 9.61E-29 | trans |
| miR-584-5p | rs1265507 | 5:1484855 | A | G | 0.410692 | 0.201351 | 0.025491 | 2.81E-15 | A | G | 0.395 | 0.211 | 0.01272 | 2.48E-52 | cis |
| miR-625-5p | rs1269068 | 14:657673 | C | T | 0.643163 | -0.223379 | 0.019494 | 2.13E-30 | T | C | 0.4008 | 0.4756 | 0.02212 | 4.71E-79 | cis |
| miR-4433b | rs1271350 | 2:6456984 | C | T | 0.714645 | -0.091252 | 0.013166 | 4.19E-12 | T | C | 0.2913 | 0.2554 | 0.01953 | 4.12E-35 | cis |
| miR-181a-5 | rs1275009 | 1:1990708 | C | T | 0.540441 | -0.102424 | 0.013128 | 6.10E-15 | T | C | 0.465 | 0.2114 | 0.01453 | 5.44E-42 | cis |
| miR-181a-5 | rs1275163 | 1:1988916 | C | T | 0.765073 | -0.101395 | 0.014694 | 5.18E-12 | T | C | 0.2384 | 0.2355 | 0.01675 | 1.16E-39 | cis |
| miR-181a-5 | rs1275164 | 1:1988944 | A | G | 0.754678 | -0.107087 | 0.014304 | 7.08E-14 | G | A | 0.2494 | 0.2399 | 0.01628 | 5.86E-43 | cis |
| miR-130a-3 | rs1278553 | 11:574866 | G | A | 0.685995 | -0.242795 | 0.022262 | 1.08E-27 | A | G | 0.3018 | 0.2321 | 0.01051 | 2.87E-82 | cis |
| miR-130a-3 | rs1279019 | 11:574978 | C | T | 0.686232 | -0.244075 | 0.022216 | 4.43E-28 | T | C | 0.2979 | 0.2378 | 0.0104 | 2.25E-86 | cis |
| miR-130a-3 | rs1280497 | 11:574637 | T | C | 0.686355 | -0.244827 | 0.022231 | 3.32E-28 | C | T | 0.3036 | 0.2331 | 0.01051 | 8.36E-83 | cis |
| miR-130a-3 | rs1280534 | 11:575247 | C | A | 0.566156 | 0.161834 | 0.021069 | 1.58E-14 | A | C | 0.4346 | -0.1757 | 0.01082 | 1.98E-50 | cis |
| miR-130a-3 | rs1280584 | 11:574438 | G | A | 0.687846 | -0.245886 | 0.022275 | 2.49E-28 | A | G | 0.303 | 0.2326 | 0.0105 | 1.56E-82 | cis |
| miR-130a-3 | rs1280725 | 11:576002 | C | T | 0.684858 | -0.242393 | 0.022231 | 1.11E-27 | T | C | 0.3051 | 0.2285 | 0.01052 | 2.46E-80 | cis |
| miR-625-5p | rs1287887 | 14:658046 | G | A | 0.386023 | 0.203653 | 0.019372 | 7.56E-26 | G | A | 0.4197 | 0.4206 | 0.02362 | 1.36E-58 | cis |
| miR-625-5p | rs1287920 | 14:659771 | C | T | 0.511623 | 0.146601 | 0.019927 | 1.88E-13 | T | C | 0.4692 | -0.3114 | 0.02595 | 2.87E-30 | cis |
| miR-625-5p | rs1287980 | 14:660477 | T | C | 0.498218 | 0.145168 | 0.019735 | 1.90E-13 | C | T | 0.4826 | -0.3064 | 0.02594 | 1.88E-29 | cis |
| miR-625-5p | rs1287997 | 14:659016 | G | A | 0.503137 | 0.145861 | 0.019778 | 1.65E-13 | A | G | 0.4706 | -0.3144 | 0.02585 | 5.67E-31 | cis |
| miR-625-5p | rs1288072 | 14:657998 | G | A | 0.386052 | 0.21297 | 0.019217 | 1.53E-28 | G | A | 0.4307 | 0.4512 | 0.02279 | 3.08E-69 | cis |
| miR-625-5p | rs1288175 | 14:658018 | A | G | 0.320867 | 0.23032 | 0.020035 | 1.38E-30 | A | G | 0.3477 | 0.4793 | 0.02283 | 6.87E-76 | cis |
| miR-625-5p | rs1288191 | 14:658312 | T | C | 0.388492 | 0.19722 | 0.019447 | 3.62E-24 | T | C | 0.4303 | 0.4088 | 0.02388 | 5.73E-55 | cis |
| miR-625-5p | rs1288220 | 14:661578 | C | A | 0.503123 | 0.148195 | 0.019873 | 8.84E-14 | A | C | 0.4757 | -0.3148 | 0.02573 | 2.57E-31 | cis |
| miR-625-5p | rs1288338 | 14:661453 | C | T | 0.500344 | 0.147926 | 0.019848 | 9.13E-14 | T | C | 0.4776 | -0.3157 | 0.02578 | 2.31E-31 | cis |
| miR-625-5p | rs1288381 | 14:659233 | C | T | 0.510962 | 0.147038 | 0.019928 | 1.60E-13 | T | C | 0.4686 | -0.3129 | 0.02594 | 1.47E-30 | cis |
| miR-4707-3 | rs1288461 | 14:234094 | A | G | 0.441135 | 0.093412 | 0.013605 | 6.60E-12 | A | G | 0.4344 | 0.297 | 0.01632 | 1.13E-60 | cis |
| miR-625-5p | rs1288584 | 14:659186 | C | T | 0.373139 | 0.182228 | 0.020451 | 5.08E-19 | C | T | 0.3489 | 0.4095 | 0.02529 | 2.87E-50 | cis |
| miR-625-5p | rs1288600 | 14:658092 | C | T | 0.390429 | 0.198498 | 0.019263 | 6.72E-25 | C | T | 0.4305 | 0.4096 | 0.02389 | 2.98E-55 | cis |
| miR-625-5p | rs1288616 | 14:658092 | C | A | 0.393656 | 0.204136 | 0.019134 | 1.43E-26 | C | A | 0.4364 | 0.4216 | 0.02345 | 1.12E-59 | cis |
| miR-625-5p | rs1288633 | 14:660946 | T | G | 0.500184 | 0.146151 | 0.01985 | 1.80E-13 | G | T | 0.4777 | -0.3182 | 0.02577 | 8.25E-32 | cis |
| miR-625-5p | rs1288713 | 14:660455 | G | C | 0.502159 | 0.147999 | 0.019774 | 7.18E-14 | C | G | 0.4771 | -0.317 | 0.02573 | 1.07E-31 | cis |
| miR-625-5p | rs1288767 | 14:662217 | T | C | 0.487094 | 0.131927 | 0.019729 | 2.28E-11 | C | T | 0.4833 | -0.2973 | 0.02622 | 1.94E-27 | cis |
| miR-625-5p | rs1288795 | 14:662465 | G | A | 0.454723 | 0.142379 | 0.019362 | 1.93E-13 | G | A | 0.4935 | 0.3186 | 0.02562 | 3.53E-32 | cis |
| miR-625-5p | rs1288900 | 14:660635 | C | G | 0.499455 | 0.145384 | 0.019766 | 1.91E-13 | G | C | 0.4777 | -0.3181 | 0.02576 | 8.09E-32 | cis |
| miR-625-5p | rs1289012 | 14:659980 | C | T | 0.511668 | 0.146815 | 0.019882 | 1.53E-13 | T | C | 0.4699 | -0.3119 | 0.02587 | 1.57E-30 | cis |
| miR-625-5p | rs1289090 | 14:661166 | T | A | 0.498435 | 0.144652 | 0.019829 | 2.98E-13 | A | T | 0.4783 | -0.3166 | 0.02581 | 1.87E-31 | cis |
| miR-625-5p | rs1289097 | 14:659828 | C | T | 0.504299 | 0.148214 | 0.019979 | 1.18E-13 | T | C | 0.4775 | -0.3201 | 0.02569 | 2.70E-32 | cis |
| miR-625-5p | rs1289205 | 14:659034 | T | C | 0.51011 | 0.146773 | 0.019879 | 1.54E-13 | C | T | 0.4699 | -0.3133 | 0.02583 | 7.47E-31 | cis |
| miR-625-5p | rs1289309 | 14:657890 | G | A | 0.385962 | 0.21328 | 0.019213 | 1.24E-28 | G | A | 0.4348 | 0.4474 | 0.02273 | 1.14E-68 | cis |
| miR-625-5p | rs1289403 | 14:662387 | T | C | 0.453073 | 0.14422 | 0.019372 | 9.72E-14 | T | C | 0.4892 | 0.3195 | 0.02563 | 2.38E-32 | cis |
| miR-625-5p | rs1289446 | 14:662407 | G | A | 0.454589 | 0.14417 | 0.0194 | 1.07E-13 | G | A | 0.4942 | 0.3186 | 0.02562 | 3.53E-32 | cis |
| miR-625-5p | rs1289490 | 14:661617 | C | T | 0.501572 | 0.148481 | 0.019827 | 6.95E-14 | T | C | 0.4709 | -0.3174 | 0.02615 | 7.52E-31 | cis |
| miR-625-5p | rs1289507 | 14:659767 | G | T | 0.520727 | 0.14878 | 0.020225 | 1.89E-13 | T | G | 0.4642 | -0.3124 | 0.02624 | 7.63E-30 | cis |
| miR-625-5p | rs1289533 | 14:658074 | C | T | 0.348231 | 0.206644 | 0.019831 | 2.01E-25 | C | T | 0.3654 | 0.4442 | 0.02392 | 9.04E-63 | cis |
| miR-625-5p | rs1289540 | 14:660648 | G | A | 0.409413 | 0.168122 | 0.020142 | 7.00E-17 | G | A | 0.4135 | 0.34 | 0.02563 | 5.54E-36 | cis |
| miR-625-5p | rs1289666 | 14:661049 | G | A | 0.499 | 0.145192 | 0.01985 | 2.59E-13 | A | G | 0.4783 | -0.3166 | 0.02581 | 1.87E-31 | cis |
| miR-320a | rs1300519 | 2:6863887 | A | G | 0.702186 | 0.147347 | 0.017627 | 6.31E-17 | G | A | 0.325 | -0.05848 | 0.006216 | 7.45E-20 | trans |
| miR-320b | rs1300519 | 2:6863887 | A | G | 0.702186 | 0.133519 | 0.015787 | 2.73E-17 | G | A | 0.325 | -0.06318 | 0.005177 | 3.79E-31 | trans |
| miR-320c | rs1300519 | 2:6863887 | A | G | 0.702186 | 0.120365 | 0.014802 | 4.24E-16 | G | A | 0.325 | -0.05753 | 0.005144 | 8.48E-27 | trans |
| miR-320d | rs1300519 | 2:6863887 | A | G | 0.702186 | 0.121079 | 0.015151 | 1.33E-15 | G | A | 0.325 | -0.05234 | 0.005056 | 1.94E-23 | trans |
| miR-4429 | rs1300519 | 2:6863887 | A | G | 0.702186 | 0.109146 | 0.014585 | 7.25E-14 | G | A | 0.325 | -0.06464 | 0.004873 | 6.30E-36 | trans |
| miR-320a | rs1300592 | 2:6862999 | T | A | 0.701261 | 0.142753 | 0.01735 | 1.90E-16 | A | T | 0.3214 | -0.05868 | 0.006218 | 5.69E-20 | trans |
| miR-320b | rs1300592 | 2:6862999 | T | A | 0.701261 | 0.129992 | 0.015537 | 5.92E-17 | A | T | 0.3214 | -0.06358 | 0.005158 | 1.04E-31 | trans |
| miR-320c | rs1300592 | 2:6862999 | T | A | 0.701261 | 0.117101 | 0.014568 | 9.09E-16 | A | T | 0.3214 | -0.05548 | 0.005163 | 5.12E-25 | trans |
| miR-320d | rs1300592 | 2:6862999 | T | A | 0.701261 | 0.118515 | 0.014908 | 1.87E-15 | A | T | 0.3214 | -0.04991 | 0.005071 | 1.75E-21 | trans |

| | | | | | | | | | | | | | | | |
|---|---|---|---|---|---|---|---|---|---|---|---|---|---|---|---|
| miR-4429 | rs13005922 | 2:68629998 | T | A | 0.701261 | 0.106555 | 0.014352 | 1.13E-13 | A | T | 0.3214 | -0.06417 | 0.00487 | 1.55E-35 | trans |
| miR-4433b | rs13008221 | 2:64574128 | G | A | 0.716086 | -0.088848 | 0.013143 | 1.38E-11 | A | G | 0.2913 | 0.2554 | 0.01953 | 4.12E-35 | cis |
| miR-320a | rs13009741 | 2:68584437 | C | A | 0.678986 | 0.125869 | 0.01732 | 3.67E-13 | A | C | 0.3364 | -0.05219 | 0.006304 | 6.49E-16 | trans |
| miR-320b | rs13009741 | 2:68584437 | C | A | 0.678986 | 0.117096 | 0.015503 | 4.25E-14 | A | C | 0.3364 | -0.06047 | 0.005225 | 2.03E-28 | trans |
| miR-320c | rs13009741 | 2:68584437 | C | A | 0.678986 | 0.104041 | 0.014538 | 8.27E-13 | A | C | 0.3364 | -0.0516 | 0.005217 | 1.20E-21 | trans |
| miR-320d | rs13009741 | 2:68584437 | C | A | 0.678986 | 0.108867 | 0.014864 | 2.41E-13 | A | C | 0.3364 | -0.04835 | 0.005095 | 3.74E-20 | trans |
| miR-4429 | rs13009741 | 2:68584437 | C | A | 0.678986 | 0.096595 | 0.01431 | 1.48E-11 | A | C | 0.3364 | -0.05885 | 0.004948 | 8.62E-30 | trans |
| miR-320a | rs13010285 | 2:68621842 | G | T | 0.683506 | 0.145008 | 0.016554 | 1.96E-18 | T | G | 0.3323 | -0.05547 | 0.006196 | 3.24E-18 | trans |
| miR-320b | rs13010285 | 2:68621842 | G | T | 0.683506 | 0.129676 | 0.014833 | 2.28E-18 | T | G | 0.3323 | -0.06219 | 0.005128 | 8.32E-31 | trans |
| miR-320c | rs13010285 | 2:68621842 | G | T | 0.683506 | 0.11756 | 0.013906 | 2.82E-17 | T | G | 0.3323 | -0.05475 | 0.005116 | 7.90E-25 | trans |
| miR-320d | rs13010285 | 2:68621842 | G | T | 0.683506 | 0.116353 | 0.014241 | 3.08E-16 | T | G | 0.3323 | -0.05002 | 0.005005 | 4.78E-22 | trans |
| miR-4429 | rs13010285 | 2:68621842 | G | T | 0.683506 | 0.105886 | 0.013707 | 1.12E-14 | T | G | 0.3323 | -0.06218 | 0.004852 | 7.18E-34 | trans |
| miR-320a | rs13010774 | 2:68639081 | C | T | 0.702339 | 0.147255 | 0.017632 | 6.73E-17 | T | C | 0.325 | -0.05848 | 0.006216 | 7.45E-20 | trans |
| miR-320b | rs13010774 | 2:68639081 | C | T | 0.702339 | 0.133415 | 0.015791 | 2.95E-17 | T | C | 0.325 | -0.06318 | 0.005177 | 3.79E-31 | trans |
| miR-320c | rs13010774 | 2:68639081 | C | T | 0.702339 | 0.120275 | 0.014806 | 4.54E-16 | T | C | 0.325 | -0.05753 | 0.005144 | 8.48E-27 | trans |
| miR-320d | rs13010774 | 2:68639081 | C | T | 0.702339 | 0.12101 | 0.015155 | 1.41E-15 | T | C | 0.325 | -0.05234 | 0.005056 | 1.94E-23 | trans |
| miR-4429 | rs13010774 | 2:68639081 | C | T | 0.702339 | 0.109105 | 0.014589 | 7.52E-14 | T | C | 0.325 | -0.06464 | 0.004873 | 6.30E-36 | trans |
| miR-320a | rs13011075 | 2:68639313 | A | C | 0.700844 | 0.148254 | 0.017659 | 4.64E-17 | C | A | 0.3256 | -0.05836 | 0.006227 | 1.01E-19 | trans |
| miR-320b | rs13011075 | 2:68639313 | A | C | 0.700844 | 0.134034 | 0.015817 | 2.37E-17 | C | A | 0.3256 | -0.06327 | 0.005186 | 3.98E-31 | trans |
| miR-320c | rs13011075 | 2:68639313 | A | C | 0.700844 | 0.120705 | 0.014831 | 3.99E-16 | C | A | 0.3256 | -0.05739 | 0.005148 | 1.21E-26 | trans |
| miR-320d | rs13011075 | 2:68639313 | A | C | 0.700844 | 0.12145 | 0.01518 | 1.24E-15 | C | A | 0.3256 | -0.05247 | 0.005064 | 1.79E-23 | trans |
| miR-4429 | rs13011075 | 2:68639313 | A | C | 0.700844 | 0.109516 | 0.014613 | 6.66E-14 | C | A | 0.3256 | -0.06482 | 0.00488 | 5.44E-36 | trans |
| miR-320a | rs13011430 | 2:68639379 | C | T | 0.702382 | 0.147277 | 0.017634 | 6.72E-17 | T | C | 0.325 | -0.05848 | 0.006216 | 7.45E-20 | trans |
| miR-320b | rs13011430 | 2:68639379 | C | T | 0.702382 | 0.133434 | 0.015794 | 2.95E-17 | T | C | 0.325 | -0.06318 | 0.005177 | 3.79E-31 | trans |
| miR-320c | rs13011430 | 2:68639379 | C | T | 0.702382 | 0.12029 | 0.014808 | 4.55E-16 | T | C | 0.325 | -0.05753 | 0.005144 | 8.48E-27 | trans |
| miR-320d | rs13011430 | 2:68639379 | C | T | 0.702382 | 0.121014 | 0.015157 | 1.42E-15 | T | C | 0.325 | -0.05234 | 0.005056 | 1.94E-23 | trans |
| miR-4429 | rs13011430 | 2:68639379 | C | T | 0.702382 | 0.109113 | 0.014591 | 7.54E-14 | T | C | 0.325 | -0.06464 | 0.004873 | 6.30E-36 | trans |
| miR-320a | rs13011571 | 2:68639273 | G | A | 0.702349 | 0.147267 | 0.017633 | 6.72E-17 | A | G | 0.325 | -0.05848 | 0.006216 | 7.45E-20 | trans |
| miR-320b | rs13011571 | 2:68639273 | G | A | 0.702349 | 0.133427 | 0.015792 | 2.94E-17 | A | G | 0.325 | -0.06318 | 0.005177 | 3.79E-31 | trans |
| miR-320c | rs13011571 | 2:68639273 | G | A | 0.702349 | 0.120281 | 0.014807 | 4.55E-16 | A | G | 0.325 | -0.05753 | 0.005144 | 8.48E-27 | trans |
| miR-320d | rs13011571 | 2:68639273 | G | A | 0.702349 | 0.121015 | 0.015156 | 1.41E-15 | A | G | 0.325 | -0.05234 | 0.005056 | 1.94E-23 | trans |
| miR-4429 | rs13011571 | 2:68639273 | G | A | 0.702349 | 0.109115 | 0.01459 | 7.50E-14 | A | G | 0.325 | -0.06464 | 0.004873 | 6.30E-36 | trans |
| miR-320a | rs13028920 | 2:68629910 | C | T | 0.552648 | 0.152067 | 0.015754 | 4.78E-22 | T | C | 0.4488 | -0.06658 | 0.005541 | 2.34E-30 | trans |
| miR-320b | rs13028920 | 2:68629910 | C | T | 0.552648 | 0.137763 | 0.014108 | 1.59E-22 | T | C | 0.4488 | -0.06591 | 0.004617 | 9.87E-41 | trans |
| miR-320c | rs13028920 | 2:68629910 | C | T | 0.552648 | 0.125024 | 0.013229 | 3.37E-21 | T | C | 0.4488 | -0.06191 | 0.004572 | 3.06E-37 | trans |
| miR-320d | rs13028920 | 2:68629910 | C | T | 0.552648 | 0.124158 | 0.01355 | 5.05E-20 | T | C | 0.4488 | -0.05868 | 0.004467 | 2.30E-35 | trans |
| miR-320e | rs13028920 | 2:68629910 | C | T | 0.552648 | 0.110906 | 0.014987 | 1.36E-13 | T | C | 0.4488 | -0.09469 | 0.006167 | 5.01E-46 | trans |
| miR-4429 | rs13028920 | 2:68629910 | C | T | 0.552648 | 0.115995 | 0.013035 | 5.65E-19 | T | C | 0.4488 | -0.06805 | 0.004311 | 3.44E-48 | trans |
| miR-320a | rs13028971 | 2:68630038 | A | G | 0.54849 | 0.153419 | 0.015702 | 1.51E-22 | G | A | 0.4547 | -0.06692 | 0.005516 | 7.17E-31 | trans |
| miR-320b | rs13028971 | 2:68630038 | A | G | 0.54849 | 0.138955 | 0.014062 | 5.00E-23 | G | A | 0.4547 | -0.06612 | 0.004599 | 3.10E-41 | trans |
| miR-320c | rs13028971 | 2:68630038 | A | G | 0.54849 | 0.125793 | 0.013188 | 1.45E-21 | G | A | 0.4547 | -0.06386 | 0.004521 | 4.93E-40 | trans |
| miR-320d | rs13028971 | 2:68630038 | A | G | 0.54849 | 0.124504 | 0.01351 | 3.10E-20 | G | A | 0.4547 | -0.06054 | 0.004418 | 5.22E-38 | trans |
| miR-320e | rs13028971 | 2:68630038 | A | G | 0.54849 | 0.112919 | 0.014938 | 4.05E-14 | G | A | 0.4547 | -0.09626 | 0.006109 | 4.55E-48 | trans |
| miR-4429 | rs13028971 | 2:68630038 | A | G | 0.54849 | 0.116303 | 0.012997 | 3.60E-19 | G | A | 0.4547 | -0.06846 | 0.004297 | 5.96E-49 | trans |
| miR-320a | rs13034825 | 2:68630297 | C | G | 0.552715 | 0.152114 | 0.015757 | 4.74E-22 | G | C | 0.4488 | -0.06658 | 0.005541 | 2.34E-30 | trans |
| miR-320b | rs13034825 | 2:68630297 | C | G | 0.552715 | 0.137808 | 0.014111 | 1.57E-22 | G | C | 0.4488 | -0.06591 | 0.004617 | 9.87E-41 | trans |
| miR-320c | rs13034825 | 2:68630297 | C | G | 0.552715 | 0.125074 | 0.013232 | 3.32E-21 | G | C | 0.4488 | -0.06191 | 0.004572 | 3.06E-37 | trans |
| miR-320d | rs13034825 | 2:68630297 | C | G | 0.552715 | 0.124211 | 0.013553 | 4.97E-20 | G | C | 0.4488 | -0.05868 | 0.004467 | 2.30E-35 | trans |
| miR-320e | rs13034825 | 2:68630297 | C | G | 0.552715 | 0.110948 | 0.01499 | 1.35E-13 | G | C | 0.4488 | -0.09469 | 0.006167 | 5.01E-46 | trans |
| miR-4429 | rs13034825 | 2:68630297 | C | G | 0.552715 | 0.116045 | 0.013038 | 5.56E-19 | G | C | 0.4488 | -0.06805 | 0.004311 | 3.44E-48 | trans |
| miR-584-5p | rs13174164 | 5:14848831 | T | C | 0.474697 | 0.188995 | 0.025386 | 9.70E-14 | T | C | 0.4478 | 0.1802 | 0.01307 | 2.14E-38 | cis |
| miR-584-5p | rs13182697 | 5:14848544 | A | G | 0.474875 | 0.188851 | 0.025387 | 1.02E-13 | A | G | 0.4478 | 0.1802 | 0.01307 | 2.14E-38 | cis |
| miR-3130-3 | rs13427002 | 2:20764367 | C | G | 0.84329 | -0.270673 | 0.032943 | 2.10E-16 | G | C | 0.1794 | 0.1673 | 0.03129 | 1.21E-07 | cis |
| miR-320a | rs13430654 | 2:68587710 | C | G | 0.699397 | 0.1397 | 0.017062 | 2.66E-16 | G | C | 0.3167 | -0.05683 | 0.006272 | 1.29E-18 | trans |
| miR-320b | rs13430654 | 2:68587710 | C | G | 0.699397 | 0.126957 | 0.01528 | 9.66E-17 | G | C | 0.3167 | -0.06308 | 0.005196 | 6.74E-31 | trans |
| miR-320c | rs13430654 | 2:68587710 | C | G | 0.699397 | 0.114048 | 0.014328 | 1.72E-15 | G | C | 0.3167 | -0.0559 | 0.00516 | 2.24E-25 | trans |
| miR-320d | rs13430654 | 2:68587710 | C | G | 0.699397 | 0.117257 | 0.014655 | 1.23E-15 | G | C | 0.3167 | -0.05121 | 0.005062 | 1.57E-22 | trans |
| miR-4429 | rs13430654 | 2:68587710 | C | G | 0.699397 | 0.103919 | 0.014114 | 1.80E-13 | G | C | 0.3167 | -0.06231 | 0.004916 | 2.88E-33 | trans |
| miR-320a | rs1373200 | 2:68622658 | C | T | 0.547142 | 0.149223 | 0.015409 | 3.51E-22 | T | C | 0.4482 | -0.06706 | 0.00555 | 1.21E-30 | trans |
| miR-320b | rs1373200 | 2:68622658 | C | T | 0.547142 | 0.134711 | 0.013801 | 1.66E-22 | T | C | 0.4482 | -0.06678 | 0.004615 | 1.15E-41 | trans |
| miR-320c | rs1373200 | 2:68622658 | C | T | 0.547142 | 0.122067 | 0.012943 | 4.05E-21 | T | C | 0.4482 | -0.063 | 0.004562 | 1.64E-38 | trans |
| miR-320d | rs1373200 | 2:68622658 | C | T | 0.547142 | 0.121474 | 0.013255 | 4.99E-20 | T | C | 0.4482 | -0.05992 | 0.004447 | 6.41E-37 | trans |
| miR-320e | rs1373200 | 2:68622658 | C | T | 0.547142 | 0.110349 | 0.014654 | 5.07E-14 | T | C | 0.4482 | -0.09507 | 0.006173 | 2.96E-46 | trans |
| miR-4429 | rs1373200 | 2:68622658 | C | T | 0.547142 | 0.112939 | 0.012754 | 8.34E-19 | T | C | 0.4482 | -0.06807 | 0.004323 | 5.40E-48 | trans |
| miR-320a | rs13828992 | 2:68637829 | A | G | 0.702614 | 0.147363 | 0.017647 | 6.79E-17 | G | A | 0.323 | -0.05866 | 0.006255 | 9.61E-20 | trans |
| miR-320b | rs13828992 | 2:68637829 | A | G | 0.702614 | 0.133506 | 0.015805 | 2.99E-17 | G | A | 0.323 | -0.06221 | 0.00521 | 5.51E-30 | trans |
| miR-320c | rs13828992 | 2:68637829 | A | G | 0.702614 | 0.12037 | 0.014819 | 4.56E-16 | G | A | 0.323 | -0.05714 | 0.005174 | 3.22E-26 | trans |
| miR-320d | rs13828992 | 2:68637829 | A | G | 0.702614 | 0.121213 | 0.015168 | 1.33E-15 | G | A | 0.323 | -0.0528 | 0.005092 | 1.70E-23 | trans |
| miR-4429 | rs13828992 | 2:68637829 | A | G | 0.702614 | 0.109295 | 0.014601 | 7.13E-14 | G | A | 0.323 | -0.06486 | 0.00491 | 1.17E-35 | trans |
| miR-320a | rs13906881 | 2:68627735 | C | T | 0.69721 | 0.142685 | 0.017183 | 1.01E-16 | T | C | 0.3276 | -0.05868 | 0.006169 | 3.00E-20 | trans |
| miR-320b | rs13906881 | 2:68627735 | C | T | 0.69721 | 0.12989 | 0.015387 | 3.14E-17 | T | C | 0.3276 | -0.06324 | 0.005124 | 8.82E-32 | trans |
| miR-320c | rs13906881 | 2:68627735 | C | T | 0.69721 | 0.116486 | 0.01443 | 6.88E-16 | T | C | 0.3276 | -0.05743 | 0.005094 | 3.42E-27 | trans |
| miR-320d | rs13906881 | 2:68627735 | C | T | 0.69721 | 0.117637 | 0.014768 | 1.64E-15 | T | C | 0.3276 | -0.05189 | 0.005006 | 1.67E-23 | trans |
| miR-4429 | rs13906881 | 2:68627735 | C | T | 0.69721 | 0.105551 | 0.014218 | 1.14E-13 | T | C | 0.3276 | -0.06445 | 0.004835 | 2.92E-36 | trans |
| miR-625-5p | rs13925180 | 14:6582955 | T | C | 0.813544 | -0.23719 | 0.02469 | 7.48E-22 | C | T | 0.1561 | 0.5131 | 0.03307 | 1.17E-46 | cis |
| miR-320a | rs13971597 | 2:68634258 | T | G | 0.705552 | 0.148814 | 0.017799 | 6.24E-17 | G | T | 0.3178 | -0.0582 | 0.006295 | 3.00E-19 | trans |
| miR-320b | rs13971597 | 2:68634258 | T | G | 0.705552 | 0.134497 | 0.015943 | 3.28E-17 | G | T | 0.3178 | -0.0606 | 0.005246 | 2.69E-28 | trans |
| miR-320c | rs13971597 | 2:68634258 | T | G | 0.705552 | 0.121472 | 0.014948 | 4.42E-16 | G | T | 0.3178 | -0.05588 | 0.00524 | 1.20E-24 | trans |
| miR-320d | rs13971597 | 2:68634258 | T | G | 0.705552 | 0.122919 | 0.015297 | 9.32E-16 | G | T | 0.3178 | -0.05274 | 0.005167 | 7.58E-23 | trans |
| miR-4429 | rs13971597 | 2:68634258 | T | G | 0.705552 | 0.110694 | 0.014727 | 5.62E-14 | G | T | 0.3178 | -0.06442 | 0.004987 | 2.75E-34 | trans |
| miR-320a | rs14224022 | 2:68627521 | A | G | 0.698498 | 0.143247 | 0.017227 | 9.15E-17 | G | A | 0.3265 | -0.05936 | 0.006191 | 1.56E-20 | trans |
| miR-320b | rs14224022 | 2:68627521 | A | G | 0.698498 | 0.130556 | 0.015426 | 2.60E-17 | G | A | 0.3265 | -0.0637 | 0.00514 | 5.30E-32 | trans |
| miR-320c | rs14224022 | 2:68627521 | A | G | 0.698498 | 0.116782 | 0.014467 | 6.91E-16 | G | A | 0.3265 | -0.05738 | 0.005118 | 6.35E-27 | trans |
| miR-320d | rs14224022 | 2:68627521 | A | G | 0.698498 | 0.118434 | 0.014804 | 1.24E-15 | G | A | 0.3265 | -0.05236 | 0.005024 | 1.01E-23 | trans |
| miR-4429 | rs14224022 | 2:68627521 | A | G | 0.698498 | 0.105646 | 0.014255 | 1.25E-13 | G | A | 0.3265 | -0.06491 | 0.004851 | 1.75E-36 | trans |
| miR-584-5p | rs1422577 | 5:14848631 | G | T | 0.474749 | 0.18891 | 0.025385 | 9.93E-14 | G | T | 0.4478 | 0.1802 | 0.01307 | 2.14E-38 | cis |

| | | | | | | | | | | | | | | | |
|---|---|---|---|---|---|---|---|---|---|---|---|---|---|---|---|
| miR-584-5p | rs1422579 | 5:1484847 | C | A | 0.509261 | 0.191642 | 0.026152 | 2.34E-13 | C | A | 0.4833 | 0.1664 | 0.01344 | 7.95E-32 | cis |
| miR-130a-3 | rs1424753 | 11:574798 | T | C | 0.449285 | -0.164093 | 0.02102 | 5.87E-15 | T | C | 0.438 | -0.1758 | 0.01072 | 2.46E-51 | cis |
| miR-584-5p | rs1432792 | 5:1484149 | C | T | 0.537202 | -0.173133 | 0.025206 | 6.47E-12 | T | C | 0.4519 | 0.2084 | 0.0128 | 1.15E-50 | cis |
| miR-181a-5 | rs1434282 | 1:1990107 | C | T | 0.271104 | 0.177906 | 0.013618 | 5.30E-39 | C | T | 0.2656 | 0.3435 | 0.01264 | 1.87E-110 | cis |
| miR-130a-3 | rs1442563 | 11:573714 | C | T | 0.752772 | -0.305493 | 0.023522 | 1.44E-38 | T | C | 0.2483 | 0.2915 | 0.009325 | 7.04E-133 | cis |
| miR-320a | rs1442589 | 2:6863473 | G | T | 0.701896 | 0.146902 | 0.017592 | 6.80E-17 | T | G | 0.3233 | -0.05825 | 0.006222 | 1.08E-19 | trans |
| miR-320b | rs1442589 | 2:6863473 | G | T | 0.701896 | 0.133113 | 0.015756 | 2.95E-17 | T | G | 0.3233 | -0.062 | 0.00518 | 3.92E-30 | trans |
| miR-320c | rs1442589 | 2:6863473 | G | T | 0.701896 | 0.119955 | 0.014773 | 4.67E-16 | T | G | 0.3233 | -0.05689 | 0.005146 | 2.89E-26 | trans |
| miR-320d | rs1442589 | 2:6863473 | G | T | 0.701896 | 0.120911 | 0.01512 | 1.28E-15 | T | G | 0.3233 | -0.05241 | 0.005065 | 1.99E-23 | trans |
| miR-4429 | rs1442589 | 2:6863473 | G | T | 0.701896 | 0.108994 | 0.014556 | 6.99E-14 | T | G | 0.3233 | -0.06458 | 0.004894 | 1.28E-35 | trans |
| miR-320a | rs1455505 | 2:6862832 | G | A | 0.556221 | 0.155389 | 0.015875 | 1.26E-22 | A | G | 0.4401 | -0.06796 | 0.00565 | 2.76E-30 | trans |
| miR-320b | rs1455505 | 2:6862832 | G | A | 0.556221 | 0.141184 | 0.014214 | 3.00E-23 | A | G | 0.4401 | -0.06714 | 0.00473 | 3.85E-40 | trans |
| miR-320c | rs1455505 | 2:6862832 | G | A | 0.556221 | 0.127266 | 0.013334 | 1.36E-21 | A | G | 0.4401 | -0.06513 | 0.004656 | 3.69E-39 | trans |
| miR-320d | rs1455505 | 2:6862832 | G | A | 0.556221 | 0.126528 | 0.013657 | 1.95E-20 | A | G | 0.4401 | -0.06214 | 0.004585 | 3.99E-37 | trans |
| miR-320e | rs1455505 | 2:6862832 | G | A | 0.556221 | 0.114653 | 0.015101 | 3.14E-14 | A | G | 0.4401 | -0.09896 | 0.006215 | 1.35E-48 | trans |
| miR-4429 | rs1455505 | 2:6862832 | G | A | 0.556221 | 0.118259 | 0.013138 | 2.23E-19 | A | G | 0.4401 | -0.06832 | 0.004449 | 9.16E-46 | trans |
| miR-625-5p | rs1457728 | 14:661028 | C | T | 0.821198 | -0.234147 | 0.02536 | 2.63E-20 | T | C | 0.165 | 0.4832 | 0.03302 | 1.74E-42 | cis |
| miR-320a | rs1474969 | 2:6862568 | G | A | 0.546544 | 0.151244 | 0.015595 | 3.06E-22 | A | G | 0.4526 | -0.06639 | 0.005513 | 1.83E-30 | trans |
| miR-320b | rs1474969 | 2:6862568 | G | A | 0.546544 | 0.136939 | 0.013966 | 1.07E-22 | A | G | 0.4526 | -0.06577 | 0.004608 | 1.06E-40 | trans |
| miR-320c | rs1474969 | 2:6862568 | G | A | 0.546544 | 0.123927 | 0.013098 | 3.04E-21 | A | G | 0.4526 | -0.06362 | 0.004543 | 2.09E-39 | trans |
| miR-320d | rs1474969 | 2:6862568 | G | A | 0.546544 | 0.122663 | 0.013417 | 6.13E-20 | A | G | 0.4526 | -0.05982 | 0.004444 | 7.22E-37 | trans |
| miR-320e | rs1474969 | 2:6862568 | G | A | 0.546544 | 0.111176 | 0.014834 | 6.65E-14 | A | G | 0.4526 | -0.09636 | 0.006154 | 1.54E-47 | trans |
| miR-4429 | rs1474969 | 2:6862568 | G | A | 0.546544 | 0.114556 | 0.012908 | 6.98E-19 | A | G | 0.4526 | -0.06856 | 0.004297 | 4.73E-49 | trans |
| miR-320a | rs1484530 | 2:6863547 | A | G | 0.70445 | 0.148609 | 0.017695 | 4.53E-17 | G | A | 0.321 | -0.05858 | 0.006254 | 1.07E-19 | trans |
| miR-320b | rs1484530 | 2:6863547 | A | G | 0.70445 | 0.134492 | 0.015849 | 2.14E-17 | G | A | 0.321 | -0.06183 | 0.005223 | 1.52E-29 | trans |
| miR-320c | rs1484530 | 2:6863547 | A | G | 0.70445 | 0.121181 | 0.014861 | 3.51E-16 | G | A | 0.321 | -0.05652 | 0.005177 | 1.08E-25 | trans |
| miR-320d | rs1484530 | 2:6863547 | A | G | 0.70445 | 0.121983 | 0.01521 | 1.06E-15 | G | A | 0.321 | -0.05207 | 0.005101 | 7.23E-23 | trans |
| miR-4429 | rs1484530 | 2:6863547 | A | G | 0.70445 | 0.109845 | 0.014643 | 6.31E-14 | G | A | 0.321 | -0.0648 | 0.004922 | 1.92E-35 | trans |
| miR-320a | rs1486061 | 2:6862571 | T | G | 0.54743 | 0.151637 | 0.015624 | 2.86E-22 | G | T | 0.4525 | -0.0664 | 0.005491 | 1.15E-30 | trans |
| miR-320b | rs1486061 | 2:6862571 | T | G | 0.54743 | 0.137325 | 0.013992 | 9.74E-23 | G | T | 0.4525 | -0.06577 | 0.004596 | 7.08E-41 | trans |
| miR-320c | rs1486061 | 2:6862571 | T | G | 0.54743 | 0.124223 | 0.013123 | 2.90E-21 | G | T | 0.4525 | -0.06361 | 0.004549 | 2.64E-39 | trans |
| miR-320d | rs1486061 | 2:6862571 | T | G | 0.54743 | 0.123039 | 0.013442 | 5.53E-20 | G | T | 0.4525 | -0.05981 | 0.004448 | 8.81E-37 | trans |
| miR-320e | rs1486061 | 2:6862571 | T | G | 0.54743 | 0.111501 | 0.014862 | 6.26E-14 | G | T | 0.4525 | -0.09636 | 0.006163 | 2.08E-47 | trans |
| miR-4429 | rs1486061 | 2:6862571 | T | G | 0.54743 | 0.115007 | 0.012931 | 5.91E-19 | G | T | 0.4525 | -0.06854 | 0.004293 | 4.53E-49 | trans |
| miR-320a | rs1542135 | 2:6862084 | C | T | 0.69752 | 0.139846 | 0.016851 | 1.05E-16 | T | C | 0.3177 | -0.05845 | 0.006251 | 1.20E-19 | trans |
| miR-320b | rs1542135 | 2:6862084 | C | T | 0.69752 | 0.127051 | 0.015091 | 3.79E-17 | T | C | 0.3177 | -0.06404 | 0.005183 | 8.06E-32 | trans |
| miR-320c | rs1542135 | 2:6862084 | C | T | 0.69752 | 0.113978 | 0.014151 | 8.00E-16 | T | C | 0.3177 | -0.05706 | 0.005161 | 2.86E-26 | trans |
| miR-320d | rs1542135 | 2:6862084 | C | T | 0.69752 | 0.116748 | 0.014477 | 7.35E-16 | T | C | 0.3177 | -0.05188 | 0.005057 | 4.52E-23 | trans |
| miR-4429 | rs1542135 | 2:6862084 | C | T | 0.69752 | 0.104351 | 0.01394 | 7.11E-14 | T | C | 0.3177 | -0.06416 | 0.004899 | 3.68E-35 | trans |
| miR-320a | rs1542136 | 2:6862093 | C | A | 0.697594 | 0.139843 | 0.016855 | 1.07E-16 | A | C | 0.3183 | -0.05833 | 0.006257 | 1.51E-19 | trans |
| miR-320b | rs1542136 | 2:6862093 | C | A | 0.697594 | 0.127041 | 0.015095 | 3.89E-17 | A | C | 0.3183 | -0.06398 | 0.005187 | 9.99E-32 | trans |
| miR-320c | rs1542136 | 2:6862093 | C | A | 0.697594 | 0.113984 | 0.014155 | 8.12E-16 | A | C | 0.3183 | -0.05711 | 0.005164 | 2.74E-26 | trans |
| miR-320d | rs1542136 | 2:6862093 | C | A | 0.697594 | 0.116763 | 0.014481 | 7.42E-16 | A | C | 0.3183 | -0.05185 | 0.005061 | 5.05E-23 | trans |
| miR-4429 | rs1542136 | 2:6862093 | C | A | 0.697594 | 0.104377 | 0.013944 | 7.12E-14 | A | C | 0.3183 | -0.06421 | 0.004901 | 3.61E-35 | trans |
| miR-130a-3 | rs1624389 | 11:574462 | A | G | 0.451986 | -0.164916 | 0.021048 | 4.67E-15 | A | G | 0.4393 | -0.1739 | 0.01066 | 6.80E-51 | cis |
| miR-130a-3 | rs1631684 | 11:574208 | G | A | 0.412965 | -0.154123 | 0.021382 | 5.68E-13 | G | A | 0.4109 | -0.1796 | 0.01091 | 2.54E-51 | cis |
| miR-130a-3 | rs1647394 | 11:574185 | C | T | 0.408999 | -0.157474 | 0.021549 | 2.71E-13 | C | T | 0.4088 | -0.1798 | 0.011 | 1.25E-50 | cis |
| miR-130a-3 | rs1647405 | 11:574778 | C | T | 0.68315 | -0.239969 | 0.022171 | 2.67E-27 | T | C | 0.3103 | 0.2304 | 0.01049 | 1.58E-81 | cis |
| miR-3130-3 | rs1683883 | 2:2076297 | T | A | 0.84321 | -0.269712 | 0.032981 | 2.89E-16 | A | T | 0.1782 | 0.1711 | 0.03131 | 6.50E-08 | cis |
| miR-181a-5 | rs1684410 | 1:1990115 | G | T | 0.842513 | -0.14945 | 0.016947 | 1.16E-18 | T | G | 0.1636 | 0.3069 | 0.01767 | 3.69E-56 | cis |
| miR-136-5p | rs1709961 | 14:101167 | C | G | 0.668809 | 0.40362 | 0.036386 | 1.36E-28 | G | C | 0.3234 | -0.06964 | 0.01332 | 2.28E-07 | cis |
| miR-625-5p | rs1710260 | 14:657895 | A | C | 0.845741 | -0.326684 | 0.026149 | 8.16E-36 | C | A | 0.1565 | 0.6921 | 0.02811 | 7.93E-97 | cis |
| miR-130a-3 | rs1745542 | 11:575347 | C | T | 0.565004 | 0.15876 | 0.02111 | 5.46E-14 | T | C | 0.4314 | -0.1704 | 0.01083 | 6.29E-48 | cis |
| miR-130a-3 | rs1745552 | 11:575404 | C | A | 0.684599 | -0.243484 | 0.022162 | 4.45E-28 | A | C | 0.3065 | 0.2288 | 0.0105 | 1.04E-80 | cis |
| miR-130a-3 | rs1745562 | 11:575444 | T | C | 0.567581 | 0.16129 | 0.021032 | 1.73E-14 | C | T | 0.4316 | -0.1706 | 0.01083 | 4.82E-48 | cis |
| miR-320a | rs1753019 | 2:6862133 | G | T | 0.683745 | 0.145203 | 0.016568 | 1.88E-18 | T | G | 0.3323 | -0.05547 | 0.006196 | 3.24E-18 | trans |
| miR-320b | rs1753019 | 2:6862133 | G | T | 0.683745 | 0.129844 | 0.014845 | 2.20E-18 | T | G | 0.3323 | -0.06219 | 0.005128 | 8.32E-31 | trans |
| miR-320c | rs1753019 | 2:6862133 | G | T | 0.683745 | 0.117647 | 0.013918 | 2.84E-17 | T | G | 0.3323 | -0.05475 | 0.005116 | 7.90E-25 | trans |
| miR-320d | rs1753019 | 2:6862133 | G | T | 0.683745 | 0.116731 | 0.014252 | 2.61E-16 | T | G | 0.3323 | -0.05002 | 0.005005 | 4.78E-22 | trans |
| miR-4429 | rs1753019 | 2:6862133 | G | T | 0.683745 | 0.106181 | 0.013718 | 9.92E-15 | T | G | 0.3323 | -0.06218 | 0.004852 | 7.18E-34 | trans |
| miR-320a | rs1760149 | 2:6861171 | A | G | 0.69807 | 0.139607 | 0.016858 | 1.22E-16 | G | A | 0.3166 | -0.05774 | 0.006213 | 1.86E-19 | trans |
| miR-320b | rs1760149 | 2:6861171 | A | G | 0.69807 | 0.126937 | 0.015097 | 4.16E-17 | G | A | 0.3166 | -0.06338 | 0.005147 | 1.14E-31 | trans |
| miR-320c | rs1760149 | 2:6861171 | A | G | 0.69807 | 0.113768 | 0.014157 | 9.29E-16 | G | A | 0.3166 | -0.05691 | 0.005129 | 1.83E-26 | trans |
| miR-320d | rs1760149 | 2:6861171 | A | G | 0.69807 | 0.117074 | 0.014481 | 6.23E-16 | G | A | 0.3166 | -0.05224 | 0.005025 | 1.25E-23 | trans |
| miR-4429 | rs1760149 | 2:6861171 | A | G | 0.69807 | 0.104495 | 0.013944 | 6.70E-14 | G | A | 0.3166 | -0.06364 | 0.004888 | 7.60E-35 | trans |
| miR-320a | rs1760168 | 2:6861589 | G | A | 0.697578 | 0.139713 | 0.016864 | 1.18E-16 | A | G | 0.3168 | -0.05774 | 0.006213 | 1.86E-19 | trans |
| miR-320b | rs1760168 | 2:6861589 | G | A | 0.697578 | 0.126954 | 0.015102 | 4.23E-17 | A | G | 0.3168 | -0.06338 | 0.005147 | 1.14E-31 | trans |
| miR-320c | rs1760168 | 2:6861589 | G | A | 0.697578 | 0.113768 | 0.014163 | 9.51E-16 | A | G | 0.3168 | -0.05691 | 0.005129 | 1.83E-26 | trans |
| miR-320d | rs1760168 | 2:6861589 | G | A | 0.697578 | 0.11699 | 0.014486 | 6.71E-16 | A | G | 0.3168 | -0.05224 | 0.005025 | 1.25E-23 | trans |
| miR-4429 | rs1760168 | 2:6861589 | G | A | 0.697578 | 0.104385 | 0.01395 | 7.27E-14 | A | G | 0.3168 | -0.06364 | 0.004888 | 7.60E-35 | trans |
| miR-320a | rs1760209 | 2:6862041 | T | A | 0.697529 | 0.139841 | 0.016851 | 1.05E-16 | A | T | 0.3177 | -0.05845 | 0.006251 | 1.20E-19 | trans |
| miR-320b | rs1760209 | 2:6862041 | T | A | 0.697529 | 0.127053 | 0.015091 | 3.80E-17 | A | T | 0.3177 | -0.06404 | 0.005183 | 8.06E-32 | trans |
| miR-320c | rs1760209 | 2:6862041 | T | A | 0.697529 | 0.113981 | 0.014152 | 8.00E-16 | A | T | 0.3177 | -0.05706 | 0.005161 | 2.86E-26 | trans |
| miR-320d | rs1760209 | 2:6862041 | T | A | 0.697529 | 0.11675 | 0.014477 | 7.35E-16 | A | T | 0.3177 | -0.05188 | 0.005057 | 4.52E-23 | trans |
| miR-4429 | rs1760209 | 2:6862041 | T | A | 0.697529 | 0.104351 | 0.01394 | 7.12E-14 | A | T | 0.3177 | -0.06416 | 0.004899 | 3.68E-35 | trans |
| miR-181a-5 | rs1762558 | 1:1989904 | G | A | 0.74365 | -0.178857 | 0.013895 | 6.49E-38 | A | G | 0.265 | 0.3405 | 0.01253 | 2.18E-111 | cis |
| miR-181a-5 | rs1763147 | 1:1992522 | T | A | 0.668759 | -0.106374 | 0.013247 | 9.73E-16 | A | T | 0.3379 | 0.1982 | 0.01485 | 2.63E-36 | cis |
| miR-625-5p | rs1775350 | 14:660574 | A | G | 0.821177 | -0.232942 | 0.025154 | 2.03E-20 | G | A | 0.1641 | 0.485 | 0.033 | 8.28E-43 | cis |
| miR-584-5p | rs1779525 | 5:1484169 | T | C | 0.842839 | -0.263167 | 0.034503 | 2.40E-14 | C | T | 0.1546 | 0.2678 | 0.01753 | 1.35E-45 | cis |
| miR-584-5p | rs1779527 | 5:1484220 | G | A | 0.854308 | -0.273328 | 0.035771 | 2.15E-14 | A | G | 0.1469 | 0.2679 | 0.01784 | 2.84E-44 | cis |
| miR-584-5p | rs1779537 | 5:1484633 | G | A | 0.845418 | -0.274724 | 0.035751 | 1.54E-14 | A | G | 0.1503 | 0.2664 | 0.01776 | 3.34E-44 | cis |
| miR-130a-3 | rs1783818 | 11:574375 | C | T | 0.454526 | -0.165981 | 0.021036 | 3.01E-15 | C | T | 0.4434 | -0.1781 | 0.01062 | 3.59E-53 | cis |
| miR-130a-3 | rs1783824 | 11:574730 | A | G | 0.452208 | -0.164056 | 0.020981 | 5.31E-15 | A | G | 0.4419 | -0.1776 | 0.01062 | 5.11E-53 | cis |
| miR-130a-3 | rs1783829 | 11:574198 | T | C | 0.413999 | -0.154412 | 0.021345 | 4.68E-13 | T | C | 0.4137 | -0.1795 | 0.01083 | 7.32E-52 | cis |
| miR-130a-3 | rs1783976 | 11:574627 | T | C | 0.456765 | -0.169619 | 0.021185 | 1.18E-15 | T | C | 0.4487 | -0.1731 | 0.01083 | 5.53E-49 | cis |
| miR-130a-3 | rs1783977 | 11:574627 | T | C | 0.454153 | -0.16407 | 0.021046 | 6.40E-15 | T | C | 0.4385 | -0.1756 | 0.01069 | 1.62E-51 | cis |

| | | | | | | | | | | | | | | | |
|---|---|---|---|---|---|---|---|---|---|---|---|---|---|---|---|
| miR-130a-3 | rs1783978 | 11:574635( | T | C | 0.45351 | -0.164845 | 0.020962 | 3.72E-15 | T | C | 0.442 | -0.1776 | 0.01057 | 1.90E-53 | cis |
| miR-130a-3 | rs1783979 | 11:574681 | T | C | 0.451998 | -0.164991 | 0.021014 | 4.11E-15 | T | C | 0.4381 | -0.1757 | 0.01066 | 9.35E-52 | cis |
| miR-130a-3 | rs1783980 | 11:574483 | C | T | 0.4518 | -0.165066 | 0.021041 | 4.33E-15 | C | T | 0.4381 | -0.1757 | 0.01066 | 9.35E-52 | cis |
| miR-130a-3 | rs1783981 | 11:574510 | T | C | 0.454187 | -0.165439 | 0.021011 | 3.44E-15 | T | C | 0.4434 | -0.1781 | 0.01063 | 4.22E-53 | cis |
| miR-130a-3 | rs1783983 | 11:574207 | T | C | 0.412531 | -0.153845 | 0.021282 | 4.87E-13 | T | C | 0.4146 | -0.1783 | 0.01081 | 1.83E-51 | cis |
| miR-130a-3 | rs1785498 | 11:574489 | C | G | 0.451781 | -0.165074 | 0.021041 | 4.32E-15 | C | G | 0.4381 | -0.1757 | 0.01066 | 9.35E-52 | cis |
| miR-130a-3 | rs1785499 | 11:574691 | T | G | 0.45303 | -0.16492 | 0.020975 | 3.76E-15 | T | G | 0.442 | -0.1776 | 0.01057 | 1.90E-53 | cis |
| miR-130a-3 | rs1786438 | 11:575970( | T | G | 0.439556 | -0.162306 | 0.020983 | 1.03E-14 | T | G | 0.4377 | -0.1726 | 0.01072 | 6.79E-50 | cis |
| miR-3130-3 | rs1862087 | 2:2076255( | T | C | 0.859956 | -0.284389 | 0.034753 | 2.76E-16 | C | T | 0.1528 | 0.1739 | 0.03373 | 3.30E-07 | cis |
| miR-3130-3 | rs1862089 | 2:2076205 | A | T | 0.854806 | -0.281451 | 0.034374 | 2.66E-16 | T | A | 0.1556 | 0.17 | 0.03332 | 4.38E-07 | cis |
| miR-181a-5 | rs188926 | 1:1990672 | T | C | 0.53384 | -0.105587 | 0.012882 | 2.47E-16 | C | T | 0.4695 | 0.213 | 0.0144 | 3.71E-43 | cis |
| miR-625-5p | rs1889731 | 14:659427 | G | C | 0.34602 | 0.173026 | 0.020834 | 9.98E-17 | G | C | 0.3119 | 0.3939 | 0.02667 | 3.91E-43 | cis |
| miR-181a-5 | rs1892432 | 1:1989406 | T | C | 0.234225 | 0.101149 | 0.014723 | 6.41E-12 | T | C | 0.2378 | 0.2386 | 0.01691 | 7.19E-40 | cis |
| miR-625-5p | rs1953415 | 14:659880 | A | G | 0.355003 | 0.172319 | 0.020622 | 6.49E-17 | A | G | 0.3207 | 0.3763 | 0.02686 | 2.05E-39 | cis |
| miR-625-5p | rs1953416 | 14:658788( | C | T | 0.504724 | 0.147883 | 0.01976 | 7.20E-14 | T | C | 0.4718 | -0.3224 | 0.02588 | 3.23E-32 | cis |
| miR-625-5p | rs1953418 | 14:659152 | G | A | 0.355213 | 0.172927 | 0.020733 | 7.38E-17 | G | A | 0.3191 | 0.3795 | 0.02681 | 3.78E-40 | cis |
| miR-625-5p | rs1954052 | 14:658057( | T | A | 0.387387 | 0.206368 | 0.019447 | 2.63E-26 | T | A | 0.4249 | 0.4156 | 0.02384 | 1.25E-56 | cis |
| miR-625-5p | rs1956010 | 14:661510 | G | A | 0.382297 | 0.18771 | 0.020495 | 5.24E-20 | G | A | 0.369 | 0.4119 | 0.02437 | 5.49E-54 | cis |
| miR-625-5p | rs1958559 | 14:660520 | C | T | 0.40803 | 0.166544 | 0.019995 | 8.14E-17 | C | T | 0.4165 | 0.3333 | 0.02567 | 1.14E-34 | cis |
| miR-625-5p | rs1958560 | 14:660367 | G | A | 0.400142 | 0.165944 | 0.019808 | 5.40E-17 | G | A | 0.4096 | 0.3429 | 0.02566 | 1.97E-36 | cis |
| miR-625-5p | rs1958561 | 14:660367( | T | G | 0.498978 | 0.146336 | 0.019735 | 1.22E-13 | G | T | 0.4777 | -0.3181 | 0.02576 | 8.09E-32 | cis |
| miR-625-5p | rs1959144 | 14:659460 | T | G | 0.346477 | 0.172772 | 0.020841 | 1.13E-16 | T | G | 0.3119 | 0.3939 | 0.02667 | 3.91E-43 | cis |
| miR-625-5p | rs1959145 | 14:659547 | A | G | 0.347293 | 0.172809 | 0.020786 | 9.28E-17 | A | G | 0.3167 | 0.3882 | 0.02671 | 5.65E-42 | cis |
| miR-625-5p | rs1984855 | 14:662392 | G | C | 0.387843 | 0.147118 | 0.019829 | 1.18E-13 | G | C | 0.3939 | 0.3491 | 0.02598 | 1.00E-36 | cis |
| miR-625-5p | rs1998036 | 14:661381 | G | C | 0.498979 | 0.148436 | 0.019879 | 8.20E-14 | C | G | 0.4795 | -0.3146 | 0.02575 | 3.03E-31 | cis |
| miR-625-5p | rs1999725 | 14:659042 | G | A | 0.373979 | 0.181949 | 0.020557 | 8.67E-19 | G | A | 0.352 | 0.4122 | 0.02509 | 2.48E-51 | cis |
| miR-625-5p | rs2002692 | 14:658994 | A | G | 0.502596 | 0.145491 | 0.019792 | 1.97E-13 | G | A | 0.4706 | -0.3144 | 0.02585 | 5.67E-31 | cis |
| miR-181a-5 | rs2033565 | 1:1990150 | A | T | 0.16327 | 0.149375 | 0.016865 | 8.21E-19 | A | T | 0.1682 | 0.3054 | 0.01757 | 2.05E-56 | cis |
| miR-181a-5 | rs2033566 | 1:1990148 | T | G | 0.211961 | 0.117053 | 0.015337 | 2.31E-14 | T | G | 0.2125 | 0.2369 | 0.01676 | 4.59E-40 | cis |
| miR-3130-3 | rs2059022 | 2:2076782 | T | C | 0.876031 | -0.247467 | 0.036929 | 2.07E-11 | C | T | 0.1431 | 0.1551 | 0.03409 | 6.37E-06 | cis |
| miR-4433b | rs2059631 | 2:6457468 | G | A | 0.560261 | -0.081642 | 0.012057 | 1.28E-11 | A | G | 0.4444 | 0.2891 | 0.01658 | 1.57E-56 | cis |
| miR-181a-5 | rs2060581 | 1:1992502 | T | C | 0.66838 | -0.107303 | 0.013201 | 4.36E-16 | C | T | 0.3387 | 0.1965 | 0.01488 | 1.14E-35 | cis |
| miR-625-5p | rs2064694 | 14:661482 | G | T | 0.501114 | 0.147758 | 0.019816 | 8.87E-14 | T | G | 0.4765 | -0.3158 | 0.02572 | 1.63E-31 | cis |
| miR-625-5p | rs2064695 | 14:660670 | T | G | 0.40849 | 0.168086 | 0.020171 | 7.88E-17 | T | G | 0.4103 | 0.3449 | 0.02554 | 4.35E-37 | cis |
| miR-320a | rs2070171 | 2:6861371( | G | A | 0.698024 | 0.139616 | 0.016857 | 1.21E-16 | A | G | 0.3174 | -0.05771 | 0.006194 | 1.52E-19 | trans |
| miR-320b | rs2070171 | 2:6861371( | G | A | 0.698024 | 0.126917 | 0.015096 | 4.20E-17 | A | G | 0.3174 | -0.06341 | 0.005129 | 6.90E-32 | trans |
| miR-320c | rs2070171 | 2:6861371( | G | A | 0.698024 | 0.113757 | 0.014157 | 9.33E-16 | A | G | 0.3174 | -0.05623 | 0.005123 | 5.77E-26 | trans |
| miR-320d | rs2070171 | 2:6861371( | G | A | 0.698024 | 0.117049 | 0.014481 | 6.31E-16 | A | G | 0.3174 | -0.05172 | 0.005017 | 2.72E-23 | trans |
| miR-4429 | rs2070171 | 2:6861371( | G | A | 0.698024 | 0.104489 | 0.013944 | 6.71E-14 | A | G | 0.3174 | -0.06296 | 0.004884 | 2.90E-34 | trans |
| miR-10a-5p | rs2073826 | 9:1361369( | G | T | 0.60107 | 0.102208 | 0.014329 | 9.81E-13 | T | G | 0.4189 | -0.04958 | 0.0131 | 0.000167 | trans |
| miR-126-3p | rs2073826 | 9:1361369( | G | T | 0.60107 | 0.112151 | 0.012825 | 2.24E-18 | T | G | 0.4189 | -0.03375 | 0.007708 | 1.38E-05 | trans |
| miR-10a-5p | rs2073827 | 9:1361371 | G | C | 0.601379 | 0.102564 | 0.014335 | 8.39E-13 | C | G | 0.4192 | -0.04976 | 0.0131 | 0.000157 | trans |
| miR-126-3p | rs2073827 | 9:1361371 | G | C | 0.601379 | 0.112127 | 0.012832 | 2.37E-18 | C | G | 0.4192 | -0.03372 | 0.007709 | 1.40E-05 | trans |
| miR-10a-5p | rs2073828 | 9:1361371 | G | A | 0.601379 | 0.102564 | 0.014335 | 8.39E-13 | A | G | 0.4192 | -0.04976 | 0.0131 | 0.000157 | trans |
| miR-126-3p | rs2073828 | 9:1361371 | G | A | 0.601379 | 0.112127 | 0.012832 | 2.37E-18 | A | G | 0.4192 | -0.03372 | 0.007709 | 1.40E-05 | trans |
| miR-625-5p | rs2092914 | 14:660791 | C | T | 0.36669 | 0.173049 | 0.020649 | 5.27E-17 | C | T | 0.341 | 0.3796 | 0.02587 | 1.09E-42 | cis |
| miR-625-5p | rs2127868 | 14:657943 | G | A | 0.220849 | 0.364388 | 0.022206 | 1.63E-60 | G | A | 0.2432 | 0.7383 | 0.01673 | 1.94E-203 | cis |
| miR-625-5p | rs2127869 | 14:657943 | T | C | 0.291045 | 0.290832 | 0.020564 | 2.08E-45 | T | C | 0.3362 | 0.5809 | 0.0199 | 2.36E-122 | cis |
| miR-625-5p | rs2127870 | 14:657968 | G | C | 0.220153 | 0.363782 | 0.02224 | 3.86E-60 | G | C | 0.2358 | 0.7437 | 0.01664 | 2.87E-206 | cis |
| miR-625-5p | rs2149839 | 14:659834 | G | A | 0.353418 | 0.170444 | 0.020619 | 1.38E-16 | G | A | 0.3234 | 0.376 | 0.02699 | 4.97E-39 | cis |
| miR-625-5p | rs2149841 | 14:660103 | C | T | 0.503888 | 0.148409 | 0.019912 | 9.11E-14 | T | C | 0.477 | -0.3172 | 0.0257 | 8.88E-32 | cis |
| miR-130a-3 | rs2156638 | 11:575429 | G | C | 0.565268 | 0.1611 | 0.021006 | 1.73E-14 | C | G | 0.4341 | -0.1714 | 0.01078 | 8.62E-49 | cis |
| miR-320a | rs2165920 | 2:6862223 | T | C | 0.552012 | 0.148931 | 0.01552 | 8.32E-22 | C | T | 0.4436 | -0.06681 | 0.005599 | 5.57E-30 | trans |
| miR-320b | rs2165920 | 2:6862223 | T | C | 0.552012 | 0.135016 | 0.013899 | 2.62E-22 | C | T | 0.4436 | -0.06657 | 0.004657 | 8.15E-41 | trans |
| miR-320c | rs2165920 | 2:6862223 | T | C | 0.552012 | 0.121089 | 0.013039 | 1.60E-20 | C | T | 0.4436 | -0.06342 | 0.004593 | 1.75E-38 | trans |
| miR-320d | rs2165920 | 2:6862223 | T | C | 0.552012 | 0.122188 | 0.013347 | 5.44E-20 | C | T | 0.4436 | -0.05979 | 0.004484 | 2.88E-36 | trans |
| miR-320e | rs2165920 | 2:6862223 | T | C | 0.552012 | 0.111604 | 0.014753 | 3.89E-14 | C | T | 0.4436 | -0.09474 | 0.006229 | 2.79E-45 | trans |
| miR-4429 | rs2165920 | 2:6862223 | T | C | 0.552012 | 0.112932 | 0.012844 | 1.47E-18 | C | T | 0.4436 | -0.06834 | 0.004355 | 1.05E-47 | trans |
| miR-625-5p | rs2183277 | 14:660354 | T | C | 0.342723 | 0.172432 | 0.020657 | 6.99E-17 | T | C | 0.312 | 0.393 | 0.0267 | 6.99E-43 | cis |
| miR-625-5p | rs2184601 | 14:658973( | T | A | 0.354646 | 0.174532 | 0.020745 | 3.99E-17 | T | A | 0.3194 | 0.3752 | 0.02679 | 2.03E-39 | cis |
| miR-625-5p | rs2184603 | 14:659306 | A | C | 0.500695 | 0.146773 | 0.019945 | 1.86E-13 | C | A | 0.4775 | -0.3209 | 0.0258 | 3.38E-32 | cis |
| miR-625-5p | rs2185454 | 14:658705 | G | T | 0.352958 | 0.175668 | 0.020698 | 2.11E-17 | G | T | 0.319 | 0.3835 | 0.02664 | 3.25E-41 | cis |
| miR-625-5p | rs2210805 | 14:658927 | C | G | 0.354478 | 0.174717 | 0.020743 | 3.67E-17 | C | G | 0.3196 | 0.3777 | 0.02668 | 3.99E-40 | cis |
| miR-625-5p | rs2236067 | 14:662480 | A | G | 0.386784 | 0.145441 | 0.019837 | 2.27E-13 | A | G | 0.3925 | 0.3491 | 0.02598 | 1.00E-36 | cis |
| miR-3130-3 | rs2241347 | 2:2076479 | C | T | 0.841853 | -0.266447 | 0.032894 | 5.49E-16 | T | C | 0.1805 | 0.1677 | 0.03133 | 1.18E-07 | cis |
| miR-320a | rs2241946 | 2:6861565 | T | C | 0.698076 | 0.139832 | 0.016864 | 1.12E-16 | C | T | 0.3162 | -0.05774 | 0.006213 | 1.86E-19 | trans |
| miR-320b | rs2241946 | 2:6861565 | T | C | 0.698076 | 0.127092 | 0.015103 | 3.92E-17 | C | T | 0.3162 | -0.06338 | 0.005147 | 1.14E-31 | trans |
| miR-320c | rs2241946 | 2:6861565 | T | C | 0.698076 | 0.113933 | 0.014163 | 8.66E-16 | C | T | 0.3162 | -0.05691 | 0.005129 | 1.83E-26 | trans |
| miR-320d | rs2241946 | 2:6861565 | T | C | 0.698076 | 0.117177 | 0.014487 | 6.03E-16 | C | T | 0.3162 | -0.05224 | 0.005025 | 1.25E-23 | trans |
| miR-4429 | rs2241946 | 2:6861565 | T | C | 0.698076 | 0.104588 | 0.01395 | 6.51E-14 | C | T | 0.3162 | -0.06364 | 0.004888 | 7.60E-35 | trans |
| miR-320a | rs2241947 | 2:6861539 | C | T | 0.697988 | 0.13966 | 0.016857 | 1.18E-16 | T | C | 0.3162 | -0.05774 | 0.006213 | 1.86E-19 | trans |
| miR-320b | rs2241947 | 2:6861539 | C | T | 0.697988 | 0.126954 | 0.015096 | 4.11E-17 | T | C | 0.3162 | -0.06338 | 0.005147 | 1.14E-31 | trans |
| miR-320c | rs2241947 | 2:6861539 | C | T | 0.697988 | 0.113809 | 0.014157 | 9.05E-16 | T | C | 0.3162 | -0.05691 | 0.005129 | 1.83E-26 | trans |
| miR-320d | rs2241947 | 2:6861539 | C | T | 0.697988 | 0.117037 | 0.014481 | 6.35E-16 | T | C | 0.3162 | -0.05224 | 0.005025 | 1.25E-23 | trans |
| miR-4429 | rs2241947 | 2:6861539 | C | T | 0.697988 | 0.104503 | 0.013944 | 6.65E-14 | T | C | 0.3162 | -0.06364 | 0.004888 | 7.60E-35 | trans |
| miR-625-5p | rs2268955 | 14:660900( | T | A | 0.411712 | 0.169021 | 0.020476 | 1.52E-16 | T | A | 0.4089 | 0.336 | 0.02625 | 9.31E-34 | cis |
| miR-625-5p | rs2268956 | 14:660901 | G | A | 0.349005 | 0.172896 | 0.020779 | 8.74E-17 | G | A | 0.314 | 0.3928 | 0.02688 | 2.47E-42 | cis |
| miR-625-5p | rs2268959 | 14:661453 | C | A | 0.818382 | -0.234409 | 0.02521 | 1.43E-20 | A | C | 0.166 | 0.4814 | 0.03283 | 1.23E-42 | cis |
| miR-625-5p | rs2268961 | 14:661467( | T | C | 0.501133 | 0.147737 | 0.019817 | 8.97E-14 | C | T | 0.4765 | -0.3158 | 0.02572 | 1.63E-31 | cis |
| miR-625-5p | rs2268962 | 14:661472 | A | G | 0.501276 | 0.147836 | 0.01982 | 8.72E-14 | G | A | 0.4765 | -0.3158 | 0.02572 | 1.63E-31 | cis |
| miR-3130-3 | rs2287628 | 2:2076505 | A | G | 0.843162 | -0.266318 | 0.032968 | 6.58E-16 | G | A | 0.1794 | 0.1673 | 0.03129 | 1.21E-07 | cis |
| miR-3130-3 | rs2287629 | 2:2076504 | C | A | 0.842588 | -0.266338 | 0.032985 | 6.78E-16 | A | C | 0.1796 | 0.1677 | 0.03131 | 1.16E-07 | cis |
| miR-4707-3 | rs2295680 | 14:233744 | G | A | 0.437854 | 0.091485 | 0.013662 | 2.13E-11 | G | A | 0.4336 | 0.2965 | 0.01669 | 2.99E-58 | cis |
| miR-4707-3 | rs2295682 | 14:233748( | C | T | 0.43795 | 0.091553 | 0.013662 | 2.07E-11 | C | T | 0.4342 | 0.2947 | 0.01669 | 1.15E-57 | cis |
| miR-625-5p | rs2300864 | 14:661198( | A | G | 0.349692 | 0.175475 | 0.020879 | 4.30E-17 | A | G | 0.3135 | 0.3916 | 0.02678 | 2.14E-42 | cis |
| miR-625-5p | rs2300865 | 14:661200 | G | C | 0.502657 | 0.147148 | 0.019872 | 1.31E-13 | C | G | 0.4769 | -0.3154 | 0.02589 | 4.59E-31 | cis |

| | | | | | | | | | | | | | | | |
|---|---|---|---|---|---|---|---|---|---|---|---|---|---|---|---|
| miR-181a-5 | rs2360962 | 1:1990047( | A | T | 0.261757 | 0.178107 | 0.013794 | 3.87E-38 | A | T | 0.2629 | 0.3389 | 0.01261 | 2.85E-109 | cis |
| miR-625-5p | rs2411351 | 14:6617154 | T | C | 0.498384 | 0.147075 | 0.019765 | 9.99E-14 | C | T | 0.4784 | -0.3155 | 0.0257 | 1.67E-31 | cis |
| miR-625-5p | rs2411355 | 14:660607( | C | A | 0.49877 | 0.145602 | 0.019744 | 1.65E-13 | A | C | 0.4764 | -0.3128 | 0.02576 | 6.67E-31 | cis |
| miR-625-5p | rs2411356 | 14:660531( | G | A | 0.388335 | 0.168635 | 0.020188 | 6.65E-17 | G | A | 0.3845 | 0.3423 | 0.02588 | 9.26E-36 | cis |
| miR-625-5p | rs2411357 | 14:660511( | G | T | 0.499896 | 0.145832 | 0.019747 | 1.53E-13 | T | G | 0.475 | -0.3126 | 0.02588 | 1.25E-30 | cis |
| miR-625-5p | rs2411404 | 14:662394( | C | G | 0.454146 | 0.144127 | 0.019397 | 1.08E-13 | C | G | 0.4929 | 0.3186 | 0.02562 | 3.53E-32 | cis |
| miR-625-5p | rs2411405 | 14:6623208 | A | G | 0.448097 | 0.141959 | 0.019234 | 1.57E-13 | A | G | 0.486 | 0.3247 | 0.02565 | 3.39E-33 | cis |
| miR-625-5p | rs2411815 | 14:6602007 | A | T | 0.354996 | 0.169079 | 0.020582 | 2.12E-16 | A | T | 0.3227 | 0.3757 | 0.02704 | 7.97E-39 | cis |
| miR-625-5p | rs2411816 | 14:6599488 | G | A | 0.50839 | 0.14508 | 0.019834 | 2.58E-13 | A | G | 0.4706 | -0.3129 | 0.02589 | 1.21E-30 | cis |
| miR-625-5p | rs2411820 | 14:6594674 | C | T | 0.35728 | 0.171685 | 0.020672 | 9.97E-17 | C | T | 0.3231 | 0.3841 | 0.02679 | 6.16E-41 | cis |
| miR-625-5p | rs2411822 | 14:6587839 | A | G | 0.501454 | 0.146326 | 0.019719 | 1.16E-13 | G | A | 0.4701 | -0.3215 | 0.02578 | 2.66E-32 | cis |
| miR-625-5p | rs2411823 | 14:6586506 | C | T | 0.39822 | 0.170762 | 0.020236 | 3.22E-17 | C | T | 0.39 | 0.3333 | 0.02613 | 1.44E-33 | cis |
| miR-625-5p | rs2411915 | 14:6579469 | C | T | 0.053213 | 0.337729 | 0.044685 | 4.09E-14 | C | T | 0.06425 | 0.5932 | 0.05251 | 2.71E-27 | cis |
| let-7a-5p | rs2519093 | 9:1361418 | C | T | 0.809892 | -0.093892 | 0.01387 | 1.29E-11 | T | C | 0.1794 | 0.0216 | 0.006196 | 0.00052 | trans |
| let-7b-5p | rs2519093 | 9:1361418 | C | T | 0.809892 | -0.127895 | 0.012263 | 1.82E-25 | T | C | 0.1794 | 0.03811 | 0.008041 | 2.60E-06 | trans |
| let-7c-5p | rs2519093 | 9:1361418 | C | T | 0.809892 | -0.108043 | 0.011197 | 4.94E-22 | T | C | 0.1794 | 0.02875 | 0.007714 | 0.00021 | trans |
| miR-10b-5p | rs2519093 | 9:1361418 | C | T | 0.809892 | -0.215578 | 0.017406 | 3.13E-35 | T | C | 0.1794 | 0.1299 | 0.02037 | 3.31E-10 | trans |
| miR-126-3p | rs2519093 | 9:1361418 | C | T | 0.809892 | -0.211422 | 0.015879 | 1.90E-40 | T | C | 0.1794 | 0.06845 | 0.009937 | 1.26E-11 | trans |
| miR-139-5p | rs2519093 | 9:1361418 | C | T | 0.809892 | -0.179103 | 0.014192 | 1.63E-36 | T | C | 0.1794 | 0.08191 | 0.01981 | 3.99E-05 | trans |
| miR-195-5p | rs2519093 | 9:1361418 | C | T | 0.809892 | -0.140415 | 0.016314 | 7.49E-18 | T | C | 0.1794 | 0.07506 | 0.02204 | 0.000696 | trans |
| miR-181a-5 | rs2624882 | 1:1991077( | A | G | 0.514062 | -0.10486 | 0.013065 | 1.01E-15 | G | A | 0.4877 | 0.1931 | 0.0147 | 2.66E-35 | cis |
| miR-181a-5 | rs2648772 | 1:1991147 | C | T | 0.519051 | -0.10108 | 0.013173 | 1.67E-14 | T | C | 0.4832 | 0.1928 | 0.01493 | 2.79E-34 | cis |
| let-7b-5p | rs2769071 | 9:1361459 | A | G | 0.663117 | -0.107918 | 0.010084 | 9.93E-27 | G | A | 0.3208 | 0.03393 | 0.006656 | 4.44E-07 | trans |
| let-7c-5p | rs2769071 | 9:1361459 | A | G | 0.663117 | -0.091257 | 0.009208 | 3.76E-23 | G | A | 0.3208 | 0.02485 | 0.006368 | 0.000105 | trans |
| miR-10b-5p | rs2769071 | 9:1361459 | A | G | 0.663117 | -0.177378 | 0.014331 | 3.48E-35 | G | A | 0.3208 | 0.1132 | 0.01685 | 3.77E-11 | trans |
| miR-126-3p | rs2769071 | 9:1361459 | A | G | 0.663117 | -0.171277 | 0.01309 | 4.06E-39 | G | A | 0.3208 | 0.05308 | 0.008284 | 2.72E-10 | trans |
| miR-139-5p | rs2769071 | 9:1361459 | A | G | 0.663117 | -0.157213 | 0.011626 | 1.14E-41 | G | A | 0.3208 | 0.07071 | 0.01643 | 1.92E-05 | trans |
| miR-195-5p | rs2769071 | 9:1361459 | A | G | 0.663117 | -0.113399 | 0.01344 | 3.25E-17 | G | A | 0.3208 | 0.06681 | 0.01827 | 0.000275 | trans |
| miR-625-5p | rs2792213 | 14:6577133 | T | C | 0.947488 | -0.344288 | 0.044198 | 6.72E-15 | C | T | 0.06069 | 0.5597 | 0.05185 | 3.53E-25 | cis |
| miR-130a-3 | rs2836295 | 11:5737252 | A | G | 0.75263 | -0.304426 | 0.02343 | 1.34E-38 | G | A | 0.2473 | 0.292 | 0.009299 | 7.21E-134 | cis |
| miR-3130-3 | rs2836579 | 2:2076298 | A | T | 0.842753 | -0.269953 | 0.032925 | 2.42E-16 | T | A | 0.1782 | 0.1711 | 0.03131 | 6.50E-08 | cis |
| miR-136-5p | rs2837918( | 14:101168( | G | T | 0.836434 | 0.456746 | 0.046892 | 2.03E-22 | T | G | 0.1501 | -0.07182 | 0.01706 | 2.89E-05 | cis |
| miR-625-5p | rs2845901 | 14:6625852 | T | C | 0.538033 | -0.138619 | 0.019252 | 6.02E-13 | T | C | 0.4962 | -0.2971 | 0.02619 | 1.80E-27 | cis |
| miR-130a-3 | rs2847308 | 11:5749823 | C | T | 0.319113 | 0.241055 | 0.022203 | 1.85E-27 | C | T | 0.3102 | 0.2293 | 0.01051 | 8.15E-81 | cis |
| miR-625-5p | rs2861669 | 14:6625868 | C | T | 0.538909 | -0.138355 | 0.019264 | 6.87E-13 | C | T | 0.4962 | -0.2971 | 0.02619 | 1.80E-27 | cis |
| miR-625-5p | rs2866601( | 14:6575814 | C | T | 0.775896 | -0.195083 | 0.022869 | 1.46E-17 | T | C | 0.2187 | 0.478 | 0.02917 | 3.85E-51 | cis |
| miR-625-5p | rs2874089 | 14:6578352 | G | A | 0.672319 | -0.153314 | 0.02031 | 4.39E-14 | A | G | 0.358 | 0.3143 | 0.02707 | 1.57E-28 | cis |
| miR-625-5p | rs2898814 | 14:660239( | T | C | 0.347238 | 0.173957 | 0.020661 | 3.77E-17 | T | C | 0.3122 | 0.3918 | 0.02665 | 8.56E-43 | cis |
| miR-625-5p | rs2898818 | 14:6597258 | G | C | 0.35329 | 0.170444 | 0.020622 | 1.40E-16 | G | C | 0.3207 | 0.3786 | 0.02683 | 6.19E-40 | cis |
| miR-625-5p | rs2898820 | 14:6587935 | T | G | 0.35021 | 0.175123 | 0.020725 | 2.92E-17 | T | G | 0.3175 | 0.3875 | 0.02667 | 7.33E-42 | cis |
| miR-584-5p | rs2915806 | 5:1484509 | C | G | 0.5867 | -0.20673 | 0.025394 | 3.92E-16 | G | C | 0.4163 | 0.2245 | 0.01254 | 4.97E-59 | cis |
| miR-584-5p | rs2915808 | 5:1484517 | A | G | 0.586438 | -0.205548 | 0.025332 | 4.90E-16 | G | A | 0.417 | 0.2248 | 0.01251 | 1.67E-59 | cis |
| miR-130a-3 | rs2926656 | 11:5746538 | C | T | 0.683894 | -0.242084 | 0.022186 | 1.01E-27 | T | C | 0.3106 | 0.2311 | 0.01047 | 3.25E-82 | cis |
| miR-130a-3 | rs2956981 | 11:5750118 | C | T | 0.554948 | 0.168947 | 0.02108 | 1.10E-15 | T | C | 0.4404 | -0.1757 | 0.01064 | 6.86E-52 | cis |
| miR-130a-3 | rs2956982 | 11:5749077 | G | A | 0.552122 | 0.164782 | 0.021049 | 4.94E-15 | A | G | 0.4423 | -0.1774 | 0.01061 | 5.59E-53 | cis |
| miR-130a-3 | rs3017889 | 11:5752084 | A | G | 0.556573 | 0.166392 | 0.021044 | 2.64E-15 | G | A | 0.4401 | -0.1771 | 0.01067 | 2.92E-52 | cis |
| miR-3689a | rs3124944 | 9:1377452 | A | G | 0.303119 | 0.395178 | 0.027506 | 8.32E-47 | A | G | 0.3048 | 0.146 | 0.02319 | 5.42E-10 | cis |
| miR-3689a | rs3128562 | 9:1377452 | A | G | 0.300754 | 0.396962 | 0.02741 | 1.56E-47 | A | G | 0.304 | 0.1467 | 0.02319 | 4.54E-10 | cis |
| miR-3689a | rs3128570 | 9:1377433( | G | A | 0.298028 | 0.390362 | 0.02716 | 7.70E-47 | G | A | 0.3126 | 0.1479 | 0.02317 | 3.27E-10 | cis |
| miR-181a-5 | rs322875 | 1:1991026 | C | T | 0.518914 | -0.099532 | 0.013126 | 3.38E-14 | T | C | 0.4854 | 0.2017 | 0.01467 | 3.08E-38 | cis |
| miR-181a-5 | rs322877 | 1:1990988 | C | T | 0.508326 | -0.094607 | 0.013088 | 4.87E-13 | T | C | 0.4974 | 0.1901 | 0.01469 | 1.77E-34 | cis |
| miR-181a-5 | rs322887 | 1:1990263 | A | G | 0.834392 | -0.156419 | 0.016492 | 2.43E-21 | G | A | 0.1718 | 0.3048 | 0.01741 | 5.05E-57 | cis |
| miR-181a-5 | rs322889 | 1:1990255 | C | G | 0.786955 | -0.124607 | 0.015018 | 1.07E-16 | G | C | 0.2157 | 0.2444 | 0.01655 | 4.08E-43 | cis |
| miR-181a-5 | rs322898 | 1:1990913 | G | A | 0.514892 | -0.103458 | 0.013022 | 1.94E-15 | A | G | 0.4878 | 0.2046 | 0.01453 | 8.33E-40 | cis |
| miR-181a-5 | rs322912 | 1:1990743 | C | T | 0.52926 | -0.097774 | 0.013089 | 8.01E-14 | T | C | 0.4786 | 0.1974 | 0.01454 | 2.48E-37 | cis |
| miR-181a-5 | rs322915 | 1:1990707 | G | A | 0.518776 | -0.099672 | 0.013107 | 2.86E-14 | A | G | 0.4839 | 0.2033 | 0.01462 | 6.41E-39 | cis |
| miR-181a-5 | rs322918 | 1:1990686 | A | G | 0.534816 | -0.105378 | 0.01288 | 2.81E-16 | G | A | 0.4695 | 0.213 | 0.01441 | 3.93E-43 | cis |
| miR-181a-5 | rs322927 | 1:1990218 | T | G | 0.834224 | -0.156756 | 0.016443 | 1.52E-21 | G | T | 0.1739 | 0.3053 | 0.01718 | 1.66E-58 | cis |
| miR-181a-5 | rs322928 | 1:1990218 | G | A | 0.833953 | -0.15677 | 0.016435 | 1.45E-21 | A | G | 0.1744 | 0.3065 | 0.01721 | 1.14E-58 | cis |
| miR-181a-5 | rs322929 | 1:1990209 | T | C | 0.78491 | -0.124643 | 0.015004 | 9.80E-17 | C | T | 0.2197 | 0.24 | 0.01651 | 5.16E-42 | cis |
| miR-181a-5 | rs322930 | 1:1990205 | T | A | 0.790845 | -0.119262 | 0.015118 | 3.05E-15 | A | T | 0.2121 | 0.237 | 0.01663 | 1.25E-40 | cis |
| miR-181a-5 | rs322931 | 1:1990198 | G | A | 0.790484 | -0.1192 | 0.01509 | 2.81E-15 | A | G | 0.2125 | 0.237 | 0.01663 | 1.25E-40 | cis |
| miR-181a-5 | rs322932 | 1:1990193 | A | T | 0.789147 | -0.119104 | 0.01515 | 3.79E-15 | T | A | 0.2141 | 0.2356 | 0.0167 | 6.36E-40 | cis |
| miR-130a-3 | rs3405210 | 11:5738451 | A | C | 0.750564 | -0.301736 | 0.023272 | 1.92E-38 | C | A | 0.2603 | 0.285 | 0.009157 | 5.49E-132 | cis |
| miR-320a | rs3419682 | 2:6862142 | G | A | 0.697518 | 0.139848 | 0.016851 | 1.05E-16 | A | G | 0.3177 | -0.05845 | 0.006251 | 1.20E-19 | trans |
| miR-320b | rs3419682 | 2:6862142 | G | A | 0.697518 | 0.127052 | 0.015091 | 3.79E-17 | A | G | 0.3177 | -0.06404 | 0.005183 | 8.06E-32 | trans |
| miR-320c | rs3419682 | 2:6862142 | G | A | 0.697518 | 0.113978 | 0.014151 | 7.99E-16 | A | G | 0.3177 | -0.05706 | 0.005161 | 2.86E-26 | trans |
| miR-320d | rs3419682 | 2:6862142 | G | A | 0.697518 | 0.116748 | 0.014477 | 7.35E-16 | A | G | 0.3177 | -0.05188 | 0.005057 | 4.52E-23 | trans |
| miR-4429 | rs3419682 | 2:6862142 | G | A | 0.697518 | 0.104352 | 0.01394 | 7.10E-14 | A | G | 0.3177 | -0.06416 | 0.004899 | 3.68E-35 | trans |
| miR-4707-3 | rs3449630 | 14:2340074 | G | C | 0.43983 | 0.093304 | 0.013786 | 1.30E-11 | G | C | 0.4278 | 0.294 | 0.01664 | 1.04E-57 | cis |
| miR-4707-3 | rs3458457 | 14:2341169 | C | T | 0.44118 | 0.093538 | 0.013603 | 6.14E-12 | C | T | 0.4344 | 0.297 | 0.01632 | 1.13E-60 | cis |
| miR-625-5p | rs3464038 | 14:660530( | G | A | 0.819714 | -0.233017 | 0.025045 | 1.35E-20 | A | G | 0.1669 | 0.4712 | 0.03296 | 7.74E-41 | cis |
| miR-625-5p | rs3477200 | 14:660530 | G | A | 0.819572 | -0.233049 | 0.02505 | 1.36E-20 | A | G | 0.1671 | 0.4711 | 0.03305 | 1.28E-40 | cis |
| miR-320a | rs3485223 | 2:6863859 | C | G | 0.550658 | 0.155791 | 0.015997 | 2.06E-22 | G | C | 0.4536 | -0.06689 | 0.005545 | 1.54E-30 | trans |
| miR-320b | rs3485223 | 2:6863859 | C | G | 0.550658 | 0.140992 | 0.014326 | 7.45E-23 | G | C | 0.4536 | -0.06601 | 0.004631 | 1.40E-40 | trans |
| miR-320c | rs3485223 | 2:6863859 | C | G | 0.550658 | 0.128801 | 0.013431 | 8.81E-22 | G | C | 0.4536 | -0.06342 | 0.004547 | 4.08E-39 | trans |
| miR-320d | rs3485223 | 2:6863859 | C | G | 0.550658 | 0.126936 | 0.013761 | 2.86E-20 | G | C | 0.4536 | -0.06056 | 0.004448 | 1.47E-37 | trans |
| miR-320e | rs3485223 | 2:6863859 | C | G | 0.550658 | 0.113462 | 0.015221 | 9.04E-14 | G | C | 0.4536 | -0.0967 | 0.006138 | 5.48E-48 | trans |
| miR-4429 | rs3485223 | 2:6863859 | C | G | 0.550658 | 0.118152 | 0.01324 | 4.50E-19 | G | C | 0.4536 | -0.06832 | 0.004317 | 2.45E-48 | trans |
| miR-181a-5 | rs3488967 | 1:1992490( | A | G | 0.66157 | -0.107512 | 0.013292 | 6.03E-16 | G | A | 0.345 | 0.196 | 0.01479 | 7.30E-36 | cis |
| miR-4707-3 | rs3508506 | 14:234099( | C | A | 0.441051 | 0.09349 | 0.01361 | 6.45E-12 | C | A | 0.4337 | 0.297 | 0.01632 | 1.13E-60 | cis |
| miR-4707-3 | rs3561518( | 14:2340438 | T | C | 0.441508 | 0.093065 | 0.013697 | 1.09E-11 | T | C | 0.4344 | 0.2972 | 0.01634 | 1.25E-60 | cis |
| miR-130a-3 | rs3580806 | 11:575470( | T | C | 0.684777 | -0.2434 | 0.022156 | 4.47E-28 | C | T | 0.3061 | 0.2291 | 0.01049 | 6.10E-81 | cis |
| miR-130a-3 | rs3603649 | 11:5749419 | T | C | 0.554138 | 0.164556 | 0.02107 | 5.72E-15 | C | T | 0.4387 | -0.1757 | 0.01069 | 1.56E-51 | cis |
| miR-584-5p | rs36047 | 5:1484413 | C | T | 0.650304 | -0.230184 | 0.025932 | 6.90E-19 | T | C | 0.3655 | 0.2541 | 0.01183 | 9.35E-79 | cis |
| miR-584-5p | rs36077 | 5:1484319( | G | A | 0.53902 | -0.172731 | 0.024829 | 3.48E-12 | A | G | 0.4516 | 0.2118 | 0.01269 | 7.44E-53 | cis |

| | | | | | | | | | | | | | | | |
|---|---|---|---|---|---|---|---|---|---|---|---|---|---|---|---|
| miR-584-5p | rs36081 | 5:1484290( | A | G | 0.540101 | -0.173081 | 0.024956 | 4.05E-12 | G | A | 0.4507 | 0.2121 | 0.01262 | 1.87E-53 | cis |
| miR-320a | rs3732044 | 2:6862391! | T | C | 0.545562 | 0.150811 | 0.01556 | 3.25E-22 | C | T | 0.4541 | -0.06727 | 0.005522 | 4.35E-31 | trans |
| miR-320b | rs3732044 | 2:6862391! | T | C | 0.545562 | 0.136529 | 0.013935 | 1.15E-22 | C | T | 0.4541 | -0.0665 | 0.004602 | 1.38E-41 | trans |
| miR-320c | rs3732044 | 2:6862391! | T | C | 0.545562 | 0.123558 | 0.013069 | 3.24E-21 | C | T | 0.4541 | -0.06396 | 0.004529 | 5.28E-40 | trans |
| miR-320d | rs3732044 | 2:6862391! | T | C | 0.545562 | 0.122287 | 0.013387 | 6.57E-20 | C | T | 0.4541 | -0.06049 | 0.00443 | 8.65E-38 | trans |
| miR-320e | rs3732044 | 2:6862391! | T | C | 0.545562 | 0.11085 | 0.014801 | 6.91E-14 | C | T | 0.4541 | -0.09639 | 0.006121 | 5.01E-48 | trans |
| miR-4429 | rs3732044 | 2:6862391! | T | C | 0.545562 | 0.114139 | 0.012879 | 7.82E-19 | C | T | 0.4541 | -0.06893 | 0.004297 | 1.62E-49 | trans |
| miR-130a-3 | rs3781885 | 11:5757068 | C | T | 0.700878 | -0.247612 | 0.022418 | 2.31E-28 | T | C | 0.2906 | 0.2387 | 0.01056 | 9.74E-85 | cis |
| miR-625-5p | rs3783709 | 14:660586( | T | A | 0.499275 | 0.145547 | 0.01973 | 1.62E-13 | A | T | 0.4764 | -0.3128 | 0.02576 | 6.67E-31 | cis |
| miR-625-5p | rs3783711 | 14:660284 | A | G | 0.343187 | 0.17277 | 0.020668 | 6.31E-17 | A | G | 0.3115 | 0.3943 | 0.0267 | 4.19E-43 | cis |
| miR-3130-3 | rs3815686 | 2:2076330( | A | G | 0.843111 | -0.268909 | 0.03292 | 3.12E-16 | G | A | 0.1783 | 0.1694 | 0.03125 | 8.17E-08 | cis |
| miR-625-5p | rs3825639 | 14:660285( | T | C | 0.343187 | 0.17277 | 0.020668 | 6.31E-17 | T | C | 0.3115 | 0.3943 | 0.0267 | 4.19E-43 | cis |
| miR-625-5p | rs3825640 | 14:659612( | C | G | 0.503966 | 0.147669 | 0.019981 | 1.46E-13 | G | C | 0.4769 | -0.3197 | 0.02576 | 4.51E-32 | cis |
| miR-181a-5 | rs3850623 | 1:1991205 | G | A | 0.518649 | -0.10172 | 0.013222 | 1.43E-14 | A | G | 0.4794 | 0.1942 | 0.01515 | 8.63E-34 | cis |
| miR-133b | rs3934574 | 20:611434 | T | C | 0.464792 | -0.178563 | 0.025429 | 2.19E-12 | T | C | 0.4391 | -0.3184 | 0.03178 | 4.27E-22 | trans |
| miR-625-5p | rs3986780 | 14:657947( | G | T | 0.053217 | 0.337746 | 0.044685 | 4.08E-14 | G | T | 0.06425 | 0.5932 | 0.05251 | 2.71E-27 | cis |
| miR-625-5p | rs4073415 | 14:662595 | G | A | 0.538747 | -0.138325 | 0.019261 | 6.88E-13 | G | A | 0.4962 | -0.2971 | 0.02619 | 1.80E-27 | cis |
| miR-625-5p | rs4073416 | 14:662593 | T | C | 0.585483 | -0.135986 | 0.019383 | 2.29E-12 | C | T | 0.4338 | 0.3163 | 0.02607 | 7.34E-31 | cis |
| miR-584-5p | rs407696 | 5:1484225 | A | G | 0.538877 | -0.171433 | 0.025051 | 7.73E-12 | G | A | 0.452 | 0.211 | 0.01263 | 6.02E-53 | cis |
| miR-625-5p | rs4080329 | 14:662334 | T | C | 0.372147 | 0.135947 | 0.020036 | 1.16E-11 | T | C | 0.3605 | 0.3055 | 0.02699 | 2.20E-27 | cis |
| miR-625-5p | rs4143898 | 14:661888 | T | A | 0.450813 | 0.145864 | 0.01999 | 2.94E-13 | T | A | 0.4425 | 0.3291 | 0.02556 | 3.55E-34 | cis |
| miR-181a-5 | rs427790 | 1:1990972 | C | T | 0.519007 | -0.09882 | 0.01307 | 4.00E-14 | T | C | 0.4827 | 0.2019 | 0.01467 | 2.69E-38 | cis |
| miR-625-5p | rs4359350 | 14:658637 | T | C | 0.404776 | 0.184939 | 0.02022 | 5.88E-20 | T | C | 0.409 | 0.3904 | 0.02486 | 1.21E-47 | cis |
| miR-625-5p | rs4400971 | 14:662545 | A | C | 0.539825 | -0.138868 | 0.019316 | 6.51E-13 | A | C | 0.4967 | -0.3135 | 0.02631 | 7.51E-30 | cis |
| miR-130b-3 | rs447001 | 22:220109 | C | T | 0.558786 | 0.173303 | 0.02256 | 1.57E-14 | T | C | 0.4485 | -0.1546 | 0.008831 | 3.85E-57 | cis |
| miR-4707-3 | rs4536383 | 14:234109 | T | C | 0.436129 | 0.091252 | 0.013624 | 2.11E-11 | T | C | 0.4294 | 0.2942 | 0.01654 | 2.11E-58 | cis |
| miR-625-5p | rs4581615 | 14:660559 | C | A | 0.499128 | 0.145272 | 0.019735 | 1.82E-13 | A | C | 0.4758 | -0.3102 | 0.02578 | 1.97E-30 | cis |
| miR-625-5p | rs4587890 | 14:658637 | T | A | 0.391319 | 0.17464 | 0.020418 | 1.20E-17 | T | A | 0.3822 | 0.3451 | 0.02607 | 9.09E-36 | cis |
| miR-320a | rs4610041 | 2:6862628( | C | T | 0.550745 | 0.155025 | 0.015748 | 7.25E-23 | T | C | 0.4492 | -0.06777 | 0.005509 | 1.52E-31 | trans |
| miR-320b | rs4610041 | 2:6862628( | C | T | 0.550745 | 0.140566 | 0.014102 | 2.10E-23 | T | C | 0.4492 | -0.0662 | 0.004644 | 1.58E-40 | trans |
| miR-320c | rs4610041 | 2:6862628( | C | T | 0.550745 | 0.127235 | 0.013226 | 6.58E-22 | T | C | 0.4492 | -0.06484 | 0.004549 | 1.56E-40 | trans |
| miR-320d | rs4610041 | 2:6862628( | C | T | 0.550745 | 0.125806 | 0.01355 | 1.62E-20 | T | C | 0.4492 | -0.06085 | 0.004479 | 2.23E-37 | trans |
| miR-320e | rs4610041 | 2:6862628( | C | T | 0.550745 | 0.11333 | 0.014985 | 3.95E-14 | T | C | 0.4492 | -0.09655 | 0.006177 | 2.78E-47 | trans |
| miR-4429 | rs4610041 | 2:6862628( | C | T | 0.550745 | 0.116935 | 0.013037 | 2.99E-19 | T | C | 0.4492 | -0.06941 | 0.004316 | 1.45E-49 | trans |
| miR-130a-3 | rs4611222 | 11:575522( | G | A | 0.56182 | 0.161086 | 0.020967 | 1.56E-14 | A | G | 0.4337 | -0.1714 | 0.0108 | 1.58E-48 | cis |
| miR-3130-3 | rs4673363 | 2:2076694 | G | A | 0.846328 | -0.263262 | 0.033357 | 2.97E-15 | A | G | 0.1722 | 0.1649 | 0.03168 | 2.55E-07 | cis |
| miR-584-5p | rs4705305 | 5:1484708 | G | T | 0.473411 | 0.187444 | 0.025456 | 1.79E-13 | G | T | 0.4449 | 0.179 | 0.0132 | 2.64E-37 | cis |
| miR-130a-3 | rs471398 | 11:575936 | C | T | 0.438663 | -0.160389 | 0.020992 | 2.16E-14 | C | T | 0.4377 | -0.1726 | 0.01072 | 6.79E-50 | cis |
| miR-10b-5p | rs473533 | 9:1361480 | T | C | 0.453861 | -0.128006 | 0.01411 | 1.17E-19 | T | C | 0.4688 | -0.05911 | 0.01638 | 0.00033 | trans |
| miR-10b-5p | rs476410 | 9:1361483( | G | C | 0.45414 | -0.127771 | 0.014101 | 1.29E-19 | G | C | 0.4688 | -0.05911 | 0.01638 | 0.00033 | trans |
| miR-130a-3 | rs4827 | 11:575079 | C | T | 0.685347 | -0.243141 | 0.022155 | 5.06E-28 | T | C | 0.3056 | 0.2297 | 0.01049 | 3.07E-81 | cis |
| miR-130a-3 | rs488769 | 11:574507 | A | C | 0.674485 | -0.23365 | 0.02225 | 8.54E-26 | C | A | 0.3127 | 0.2284 | 0.01063 | 1.06E-78 | cis |
| miR-625-5p | rs4899172 | 14:657799( | T | C | 0.213235 | 0.359928 | 0.022679 | 1.01E-56 | T | C | 0.2277 | 0.7398 | 0.01773 | 3.74E-190 | cis |
| miR-625-5p | rs4899173 | 14:657931( | T | C | 0.216374 | 0.360282 | 0.022447 | 5.72E-58 | T | C | 0.2378 | 0.7437 | 0.01749 | 2.25E-195 | cis |
| miR-625-5p | rs4899174 | 14:658114( | A | T | 0.390371 | 0.198564 | 0.019268 | 6.68E-25 | A | T | 0.4305 | 0.4096 | 0.02389 | 2.98E-55 | cis |
| miR-625-5p | rs4899175 | 14:658264 | A | G | 0.051502 | 0.327813 | 0.045295 | 4.58E-13 | A | G | 0.05897 | 0.5849 | 0.05458 | 6.77E-25 | cis |
| miR-625-5p | rs4899178 | 14:658629 | C | T | 0.391583 | 0.174834 | 0.020417 | 1.10E-17 | C | T | 0.3835 | 0.3451 | 0.02607 | 9.09E-36 | cis |
| miR-625-5p | rs4899179 | 14:659267 | T | G | 0.508099 | 0.145114 | 0.019871 | 2.81E-13 | G | T | 0.4693 | -0.314 | 0.02597 | 1.13E-30 | cis |
| miR-625-5p | rs4902387 | 14:657928 | C | T | 0.21638 | 0.360265 | 0.022447 | 5.75E-58 | C | T | 0.2378 | 0.7437 | 0.01749 | 2.25E-195 | cis |
| miR-625-5p | rs4902388 | 14:657928 | A | G | 0.216378 | 0.360268 | 0.022447 | 5.75E-58 | A | G | 0.2378 | 0.7437 | 0.01749 | 2.25E-195 | cis |
| miR-625-5p | rs4902389 | 14:657932 | A | G | 0.216372 | 0.360287 | 0.022448 | 5.70E-58 | A | G | 0.2378 | 0.7437 | 0.01749 | 2.25E-195 | cis |
| miR-625-5p | rs4902391 | 14:658159 | G | T | 0.397869 | 0.205832 | 0.019183 | 7.36E-27 | G | T | 0.4365 | 0.4214 | 0.02339 | 8.48E-60 | cis |
| miR-625-5p | rs4902393 | 14:658395 | G | C | 0.390223 | 0.1982 | 0.019437 | 2.04E-24 | G | C | 0.4313 | 0.4045 | 0.0239 | 5.84E-54 | cis |
| miR-625-5p | rs4902399 | 14:659568 | A | G | 0.345315 | 0.174461 | 0.020811 | 5.16E-17 | A | G | 0.3119 | 0.3939 | 0.02667 | 3.91E-43 | cis |
| miR-625-5p | rs4902400 | 14:659571 | A | G | 0.508016 | 0.144622 | 0.019874 | 3.41E-13 | G | A | 0.4704 | -0.3124 | 0.02598 | 2.21E-30 | cis |
| miR-625-5p | rs4902404 | 14:660470 | T | C | 0.344394 | 0.172189 | 0.02055 | 5.34E-17 | T | C | 0.3146 | 0.393 | 0.0267 | 6.99E-43 | cis |
| miR-625-5p | rs4902412 | 14:661870( | T | C | 0.443 | 0.135201 | 0.019984 | 1.33E-11 | T | C | 0.4444 | 0.3181 | 0.02567 | 5.20E-32 | cis |
| miR-625-5p | rs4902415 | 14:662356 | T | C | 0.451839 | 0.143041 | 0.019354 | 1.46E-13 | T | C | 0.4873 | 0.3222 | 0.02559 | 6.78E-33 | cis |
| miR-625-5p | rs4902416 | 14:662380 | G | C | 0.453194 | 0.143892 | 0.019371 | 1.10E-13 | G | C | 0.4905 | 0.3195 | 0.02563 | 2.38E-32 | cis |
| miR-136-5p | rs4905999 | 14:101184 | A | G | 0.181905 | -0.479304 | 0.045375 | 4.41E-26 | A | G | 0.1828 | -0.09271 | 0.01623 | 1.67E-08 | cis |
| miR-376c-3 | rs4905999 | 14:101184 | A | G | 0.181905 | -0.277015 | 0.027005 | 1.09E-24 | A | G | 0.1828 | -0.04478 | 0.0123 | 0.000295 | cis |
| miR-181a-5 | rs4915331 | 1:1989185( | T | C | 0.235006 | 0.10124 | 0.014684 | 5.40E-12 | T | C | 0.2419 | 0.2311 | 0.01699 | 1.81E-37 | cis |
| miR-181a-5 | rs4915332 | 1:1989185 | A | G | 0.234929 | 0.101276 | 0.014684 | 5.31E-12 | A | G | 0.2422 | 0.2315 | 0.01697 | 1.22E-37 | cis |
| miR-181a-5 | rs4915335 | 1:1989452 | A | G | 0.238713 | 0.109407 | 0.01468 | 9.14E-14 | A | G | 0.2409 | 0.2413 | 0.01676 | 2.89E-41 | cis |
| let-7b-5p | rs491626 | 9:1361448 | C | T | 0.664173 | -0.108148 | 0.01009 | 8.39E-27 | T | C | 0.3217 | 0.03365 | 0.006656 | 5.49E-07 | trans |
| let-7c-5p | rs491626 | 9:1361448 | C | T | 0.664173 | -0.091873 | 0.009213 | 2.01E-23 | T | C | 0.3217 | 0.02438 | 0.006399 | 0.000151 | trans |
| miR-10b-5p | rs491626 | 9:1361448 | C | T | 0.664173 | -0.177622 | 0.014341 | 3.14E-35 | T | C | 0.3217 | 0.1137 | 0.01685 | 3.13E-11 | trans |
| miR-126-3p | rs491626 | 9:1361448 | C | T | 0.664173 | -0.172011 | 0.013097 | 2.10E-39 | T | C | 0.3217 | 0.05284 | 0.008284 | 3.25E-10 | trans |
| miR-139-5p | rs491626 | 9:1361448 | C | T | 0.664173 | -0.157582 | 0.011633 | 8.32E-42 | T | C | 0.3217 | 0.07078 | 0.01643 | 1.88E-05 | trans |
| miR-195-5p | rs491626 | 9:1361448 | C | T | 0.664173 | -0.113996 | 0.013448 | 2.32E-17 | T | C | 0.3217 | 0.06716 | 0.01826 | 0.000254 | trans |
| let-7b-5p | rs492488 | 9:1361449( | G | A | 0.663579 | -0.107737 | 0.010074 | 1.07E-26 | A | G | 0.3219 | 0.03377 | 0.006655 | 4.99E-07 | trans |
| let-7c-5p | rs492488 | 9:1361449( | G | A | 0.663579 | -0.091462 | 0.009197 | 2.67E-23 | A | G | 0.3219 | 0.02445 | 0.006395 | 0.000144 | trans |
| miR-10b-5p | rs492488 | 9:1361449( | G | A | 0.663579 | -0.177263 | 0.014316 | 3.26E-35 | A | G | 0.3219 | 0.1135 | 0.01684 | 3.31E-11 | trans |
| miR-126-3p | rs492488 | 9:1361449( | G | A | 0.663579 | -0.171514 | 0.013074 | 2.58E-39 | A | G | 0.3219 | 0.05294 | 0.008279 | 2.95E-10 | trans |
| miR-139-5p | rs492488 | 9:1361449( | G | A | 0.663579 | -0.157308 | 0.011612 | 8.23E-42 | A | G | 0.3219 | 0.0709 | 0.01641 | 1.79E-05 | trans |
| miR-195-5p | rs492488 | 9:1361449( | G | A | 0.663579 | -0.113656 | 0.013425 | 2.54E-17 | A | G | 0.3219 | 0.06692 | 0.01825 | 0.000265 | trans |
| miR-130a-3 | rs4926 | 11:573819 | G | A | 0.750066 | -0.301324 | 0.023228 | 1.75E-38 | A | G | 0.2603 | 0.285 | 0.009157 | 5.49E-132 | cis |
| let-7b-5p | rs493246 | 9:1361449 | G | A | 0.663234 | -0.107785 | 0.010079 | 1.09E-26 | A | G | 0.3219 | 0.03377 | 0.006655 | 4.99E-07 | trans |
| let-7c-5p | rs493246 | 9:1361449 | G | A | 0.663234 | -0.091502 | 0.009202 | 2.70E-23 | A | G | 0.3219 | 0.02445 | 0.006395 | 0.000144 | trans |
| miR-10b-5p | rs493246 | 9:1361449 | G | A | 0.663234 | -0.177369 | 0.014324 | 3.23E-35 | A | G | 0.3219 | 0.1135 | 0.01684 | 3.31E-11 | trans |
| miR-126-3p | rs493246 | 9:1361449 | G | A | 0.663234 | -0.171618 | 0.013081 | 2.55E-39 | A | G | 0.3219 | 0.05294 | 0.008279 | 2.95E-10 | trans |
| miR-139-5p | rs493246 | 9:1361449 | G | A | 0.663234 | -0.15741 | 0.011618 | 8.06E-42 | A | G | 0.3219 | 0.0709 | 0.01641 | 1.79E-05 | trans |
| miR-195-5p | rs493246 | 9:1361449 | G | A | 0.663234 | -0.11379 | 0.013432 | 2.42E-17 | A | G | 0.3219 | 0.06692 | 0.01825 | 0.000265 | trans |
| let-7a-5p | rs494242 | 9:1361451 | C | T | 0.644977 | -0.080059 | 0.011174 | 7.78E-13 | T | C | 0.3467 | 0.0169 | 0.005043 | 0.000849 | trans |
| let-7b-5p | rs494242 | 9:1361451 | C | T | 0.644977 | -0.105243 | 0.009882 | 1.74E-26 | T | C | 0.3467 | 0.02792 | 0.00656 | 2.36E-05 | trans |
| let-7c-5p | rs494242 | 9:1361451 | C | T | 0.644977 | -0.089675 | 0.00902 | 2.75E-23 | T | C | 0.3467 | 0.02285 | 0.006277 | 0.000292 | trans |

| | | | | | | | | | | | | | | | |
|---|---|---|---|---|---|---|---|---|---|---|---|---|---|---|---|
| miR-10b-5 | rs494242 | 9:1361451 | C | T | 0.644977 | -0.172256 | 0.014049 | 1.47E-34 | T | C | 0.3467 | 0.1083 | 0.01654 | 1.14E-10 | trans |
| miR-126-3 | rs494242 | 9:1361451 | C | T | 0.644977 | -0.16489 | 0.012842 | 9.85E-38 | T | C | 0.3467 | 0.04895 | 0.008145 | 2.98E-09 | trans |
| miR-139-5 | rs494242 | 9:1361451 | C | T | 0.644977 | -0.152252 | 0.011401 | 1.11E-40 | T | C | 0.3467 | 0.06736 | 0.01611 | 3.26E-05 | trans |
| let-7b-5p | rs495203 | 9:1361452 | C | T | 0.663279 | -0.107724 | 0.010072 | 1.06E-26 | T | C | 0.3219 | 0.03377 | 0.006655 | 4.99E-07 | trans |
| let-7c-5p | rs495203 | 9:1361452 | C | T | 0.663279 | -0.091426 | 0.009196 | 2.73E-23 | T | C | 0.3219 | 0.02445 | 0.006395 | 0.000144 | trans |
| miR-10b-5 | rs495203 | 9:1361452 | C | T | 0.663279 | -0.177267 | 0.014313 | 3.15E-35 | T | C | 0.3219 | 0.1135 | 0.01684 | 3.31E-11 | trans |
| miR-126-3 | rs495203 | 9:1361452 | C | T | 0.663279 | -0.17152 | 0.013072 | 2.48E-39 | T | C | 0.3219 | 0.05294 | 0.008279 | 2.95E-10 | trans |
| miR-139-5 | rs495203 | 9:1361452 | C | T | 0.663279 | -0.157265 | 0.01161 | 8.37E-42 | T | C | 0.3219 | 0.0709 | 0.01641 | 1.79E-05 | trans |
| miR-195-5 | rs495203 | 9:1361452 | C | T | 0.663279 | -0.113756 | 0.013422 | 2.34E-17 | T | C | 0.3219 | 0.06692 | 0.01825 | 0.000265 | trans |
| let-7a-5p | rs495828 | 9:1361548 | G | T | 0.787276 | -0.092717 | 0.013111 | 1.53E-12 | T | G | 0.2061 | 0.01976 | 0.005744 | 0.000616 | trans |
| miR-10b-5 | rs495828 | 9:1361548 | G | T | 0.787276 | -0.201363 | 0.016484 | 2.56E-34 | T | G | 0.2061 | 0.1077 | 0.01899 | 2.10E-08 | trans |
| miR-126-3 | rs495828 | 9:1361548 | G | T | 0.787276 | -0.192644 | 0.015069 | 2.00E-37 | T | G | 0.2061 | 0.05239 | 0.009309 | 2.64E-08 | trans |
| miR-139-5 | rs495828 | 9:1361548 | G | T | 0.787276 | -0.164962 | 0.013454 | 1.46E-34 | T | G | 0.2061 | 0.06801 | 0.01841 | 0.000237 | trans |
| miR-10a-5 | rs4962040 | 9:1361335 | A | G | 0.619057 | 0.104354 | 0.014467 | 5.47E-13 | G | A | 0.394 | -0.05033 | 0.01332 | 0.000172 | trans |
| miR-126-3 | rs4962040 | 9:1361335 | A | G | 0.619057 | 0.11337 | 0.012952 | 2.08E-18 | G | A | 0.394 | -0.03308 | 0.007848 | 2.83E-05 | trans |
| miR-4707-3 | rs4981451 | 14:234102 | T | C | 0.441163 | 0.093487 | 0.013604 | 6.33E-12 | T | C | 0.4344 | 0.297 | 0.01632 | 1.13E-60 | cis |
| miR-4707-3 | rs4981452 | 14:234108 | A | G | 0.440913 | 0.093512 | 0.013614 | 6.47E-12 | A | G | 0.4328 | 0.2974 | 0.0164 | 3.04E-60 | cis |
| miR-4707-3 | rs4982713 | 14:234041 | C | T | 0.441538 | 0.092987 | 0.013695 | 1.12E-11 | C | T | 0.4339 | 0.2948 | 0.01644 | 2.83E-59 | cis |
| miR-4707-3 | rs4982714 | 14:234133 | C | T | 0.441229 | 0.093545 | 0.013601 | 6.08E-12 | C | T | 0.4348 | 0.2974 | 0.01629 | 6.26E-61 | cis |
| miR-130a-3 | rs499188 | 11:574341 | C | A | 0.686203 | -0.241592 | 0.022245 | 1.78E-27 | A | C | 0.3097 | 0.2313 | 0.01051 | 6.67E-82 | cis |
| miR-130a-3 | rs527528 | 11:574333 | C | T | 0.681923 | -0.243471 | 0.022237 | 6.73E-28 | T | C | 0.3127 | 0.2299 | 0.01056 | 2.86E-80 | cis |
| let-7b-5p | rs529565 | 9:1361495 | T | C | 0.665911 | -0.110358 | 0.010051 | 4.77E-28 | C | T | 0.3195 | 0.03225 | 0.006664 | 1.61E-06 | trans |
| let-7c-5p | rs529565 | 9:1361495 | T | C | 0.665911 | -0.093158 | 0.009181 | 3.42E-24 | C | T | 0.3195 | 0.02299 | 0.006401 | 0.000352 | trans |
| miR-10b-5 | rs529565 | 9:1361495 | T | C | 0.665911 | -0.179319 | 0.01429 | 4.06E-36 | C | T | 0.3195 | 0.1101 | 0.01686 | 1.28E-10 | trans |
| miR-126-3 | rs529565 | 9:1361495 | T | C | 0.665911 | -0.174412 | 0.013044 | 8.91E-41 | C | T | 0.3195 | 0.0528 | 0.008276 | 3.23E-10 | trans |
| miR-139-5 | rs529565 | 9:1361495 | T | C | 0.665911 | -0.159786 | 0.011585 | 2.82E-43 | C | T | 0.3195 | 0.07001 | 0.01642 | 2.28E-05 | trans |
| miR-195-5 | rs529565 | 9:1361495 | T | C | 0.665911 | -0.116479 | 0.013401 | 3.57E-18 | C | T | 0.3195 | 0.06734 | 0.01825 | 0.000241 | trans |
| let-7b-5p | rs532436 | 9:1361498 | G | A | 0.8067 | -0.126313 | 0.01205 | 1.04E-25 | A | G | 0.1794 | 0.03659 | 0.008052 | 6.50E-06 | trans |
| let-7c-5p | rs532436 | 9:1361498 | G | A | 0.8067 | -0.107102 | 0.011001 | 2.13E-22 | A | G | 0.1794 | 0.02762 | 0.007721 | 0.000371 | trans |
| miR-10b-5 | rs532436 | 9:1361498 | G | A | 0.8067 | -0.214173 | 0.017094 | 5.20E-36 | A | G | 0.1794 | 0.128 | 0.02039 | 6.03E-10 | trans |
| miR-126-3 | rs532436 | 9:1361498 | G | A | 0.8067 | -0.207511 | 0.015609 | 2.49E-40 | A | G | 0.1794 | 0.06693 | 0.009953 | 3.67E-11 | trans |
| miR-139-5 | rs532436 | 9:1361498 | G | A | 0.8067 | -0.176906 | 0.013943 | 6.95E-37 | A | G | 0.1794 | 0.08337 | 0.0198 | 2.89E-05 | trans |
| miR-195-5 | rs532436 | 9:1361498 | G | A | 0.8067 | -0.13917 | 0.01603 | 3.89E-18 | A | G | 0.1794 | 0.07457 | 0.02204 | 0.000756 | trans |
| let-7b-5p | rs543040 | 9:1361430 | A | T | 0.669196 | -0.109582 | 0.010251 | 1.13E-26 | T | A | 0.322 | 0.03463 | 0.006797 | 4.57E-07 | trans |
| let-7c-5p | rs543040 | 9:1361430 | A | T | 0.669196 | -0.092304 | 0.009362 | 6.26E-23 | T | A | 0.322 | 0.02645 | 0.006486 | 5.10E-05 | trans |
| miR-10b-5 | rs543040 | 9:1361430 | A | T | 0.669196 | -0.179437 | 0.014573 | 7.68E-35 | T | A | 0.322 | 0.1135 | 0.01725 | 9.53E-11 | trans |
| miR-126-3 | rs543040 | 9:1361430 | A | T | 0.669196 | -0.173596 | 0.013309 | 6.96E-39 | T | A | 0.322 | 0.05485 | 0.008552 | 2.72E-10 | trans |
| miR-139-5 | rs543040 | 9:1361430 | A | T | 0.669196 | -0.159045 | 0.011822 | 2.94E-41 | T | A | 0.322 | 0.07256 | 0.01668 | 1.58E-05 | trans |
| miR-195-5 | rs543040 | 9:1361430 | A | T | 0.669196 | -0.115636 | 0.01366 | 2.56E-17 | T | A | 0.322 | 0.06796 | 0.01886 | 0.000339 | trans |
| miR-130a-3 | rs543725 | 11:574830 | A | T | 0.688868 | -0.239725 | 0.022465 | 1.39E-26 | T | A | 0.2988 | 0.2233 | 0.01072 | 7.29E-75 | cis |
| let-7b-5p | rs543968 | 9:1361431 | T | C | 0.653829 | -0.10593 | 0.010051 | 5.69E-26 | C | T | 0.3457 | 0.02922 | 0.006685 | 1.43E-05 | trans |
| let-7c-5p | rs543968 | 9:1361431 | T | C | 0.653829 | -0.090699 | 0.009172 | 4.68E-23 | C | T | 0.3457 | 0.02465 | 0.006356 | 0.000116 | trans |
| miR-10b-5 | rs543968 | 9:1361431 | T | C | 0.653829 | -0.174326 | 0.014287 | 3.05E-34 | C | T | 0.3457 | 0.1111 | 0.01688 | 9.54E-11 | trans |
| miR-126-3 | rs543968 | 9:1361431 | T | C | 0.653829 | -0.165481 | 0.013068 | 9.52E-37 | C | T | 0.3457 | 0.05053 | 0.008385 | 2.78E-09 | trans |
| miR-139-5 | rs543968 | 9:1361431 | T | C | 0.653829 | -0.153713 | 0.011597 | 4.23E-40 | C | T | 0.3457 | 0.06895 | 0.01639 | 2.94E-05 | trans |
| let-7a-5p | rs544873 | 9:1361432 | G | A | 0.645742 | -0.079908 | 0.011169 | 8.38E-13 | A | G | 0.3465 | 0.0169 | 0.005043 | 0.000849 | trans |
| let-7b-5p | rs544873 | 9:1361432 | G | A | 0.645742 | -0.105117 | 0.009877 | 1.88E-26 | A | G | 0.3465 | 0.02792 | 0.00656 | 2.36E-05 | trans |
| let-7c-5p | rs544873 | 9:1361432 | G | A | 0.645742 | -0.089522 | 0.009016 | 3.11E-23 | A | G | 0.3465 | 0.02285 | 0.006277 | 0.000292 | trans |
| miR-10b-5 | rs544873 | 9:1361432 | G | A | 0.645742 | -0.172011 | 0.014043 | 1.70E-34 | A | G | 0.3465 | 0.1083 | 0.01654 | 1.14E-10 | trans |
| miR-126-3 | rs544873 | 9:1361432 | G | A | 0.645742 | -0.16454 | 0.012837 | 1.31E-37 | A | G | 0.3465 | 0.04895 | 0.008145 | 2.98E-09 | trans |
| miR-139-5 | rs544873 | 9:1361432 | G | A | 0.645742 | -0.152029 | 0.011396 | 1.34E-40 | A | G | 0.3465 | 0.06736 | 0.01611 | 3.26E-05 | trans |
| let-7b-5p | rs545971 | 9:1361433 | C | T | 0.663766 | -0.107579 | 0.010061 | 1.11E-26 | T | C | 0.3219 | 0.03377 | 0.006655 | 4.99E-07 | trans |
| let-7c-5p | rs545971 | 9:1361433 | C | T | 0.663766 | -0.091289 | 0.009187 | 2.87E-23 | T | C | 0.3219 | 0.02445 | 0.006395 | 0.000144 | trans |
| miR-10b-5 | rs545971 | 9:1361433 | C | T | 0.663766 | -0.176997 | 0.014299 | 3.43E-35 | T | C | 0.3219 | 0.1135 | 0.01684 | 3.31E-11 | trans |
| miR-126-3 | rs545971 | 9:1361433 | C | T | 0.663766 | -0.171246 | 0.013059 | 2.76E-39 | T | C | 0.3219 | 0.05294 | 0.008279 | 2.95E-10 | trans |
| miR-139-5 | rs545971 | 9:1361433 | C | T | 0.663766 | -0.157102 | 0.011598 | 8.40E-42 | T | C | 0.3219 | 0.0709 | 0.01641 | 1.79E-05 | trans |
| miR-195-5 | rs545971 | 9:1361433 | C | T | 0.663766 | -0.113518 | 0.013409 | 2.54E-17 | T | C | 0.3219 | 0.06692 | 0.01825 | 0.000265 | trans |
| miR-130a-3 | rs547118 | 11:575950 | C | G | 0.438175 | -0.163182 | 0.020979 | 7.35E-15 | C | G | 0.4369 | -0.1723 | 0.01072 | 1.09E-49 | cis |
| miR-130a-3 | rs547891 | 11:574990 | C | T | 0.319553 | 0.241325 | 0.022213 | 1.70E-27 | C | T | 0.3103 | 0.2294 | 0.01052 | 9.88E-81 | cis |
| let-7b-5p | rs550057 | 9:1361465 | C | T | 0.734085 | -0.097587 | 0.010867 | 2.70E-19 | T | C | 0.2366 | 0.03111 | 0.007276 | 2.17E-05 | trans |
| miR-10b-5 | rs550057 | 9:1361465 | C | T | 0.734085 | -0.167896 | 0.015446 | 1.61E-27 | T | C | 0.2366 | 0.1028 | 0.01855 | 4.28E-08 | trans |
| miR-126-3 | rs550057 | 9:1361465 | C | T | 0.734085 | -0.163489 | 0.014114 | 5.01E-31 | T | C | 0.2366 | 0.04229 | 0.009146 | 4.50E-06 | trans |
| miR-139-5 | rs550057 | 9:1361465 | C | T | 0.734085 | -0.147772 | 0.012554 | 5.52E-32 | T | C | 0.2366 | 0.06132 | 0.01784 | 0.000625 | trans |
| let-7b-5p | rs554833 | 9:1361471 | C | T | 0.664057 | -0.107822 | 0.01006 | 8.42E-27 | T | C | 0.3217 | 0.03411 | 0.006647 | 3.72E-07 | trans |
| let-7c-5p | rs554833 | 9:1361471 | C | T | 0.664057 | -0.091422 | 0.009186 | 2.46E-23 | T | C | 0.3217 | 0.02468 | 0.006395 | 0.000124 | trans |
| miR-10b-5 | rs554833 | 9:1361471 | C | T | 0.664057 | -0.177063 | 0.014299 | 3.22E-35 | T | C | 0.3217 | 0.1139 | 0.01685 | 2.90E-11 | trans |
| miR-126-3 | rs554833 | 9:1361471 | C | T | 0.664057 | -0.171374 | 0.013058 | 2.40E-39 | T | C | 0.3217 | 0.05315 | 0.008284 | 2.59E-10 | trans |
| miR-139-5 | rs554833 | 9:1361471 | C | T | 0.664057 | -0.157337 | 0.011597 | 6.24E-42 | T | C | 0.3217 | 0.07034 | 0.01642 | 2.09E-05 | trans |
| miR-195-5 | rs554833 | 9:1361471 | C | T | 0.664057 | -0.113794 | 0.013408 | 2.11E-17 | T | C | 0.3217 | 0.06669 | 0.01827 | 0.000283 | trans |
| miR-4707-3 | rs5589413 | 14:234088 | C | G | 0.443456 | 0.093971 | 0.013657 | 5.95E-12 | C | G | 0.4384 | 0.2974 | 0.01642 | 3.74E-60 | cis |
| miR-4707-3 | rs5618074 | 14:233772 | T | C | 0.438286 | 0.091634 | 0.013678 | 2.10E-11 | T | C | 0.4336 | 0.2939 | 0.01667 | 1.58E-57 | cis |
| miR-320a | rs5618821 | 2:6861163 | T | C | 0.698075 | 0.139604 | 0.016858 | 1.22E-16 | C | T | 0.3166 | -0.05774 | 0.006213 | 1.86E-19 | trans |
| miR-320b | rs5618821 | 2:6861163 | T | C | 0.698075 | 0.126937 | 0.015097 | 4.16E-17 | C | T | 0.3166 | -0.06338 | 0.005147 | 1.14E-31 | trans |
| miR-320c | rs5618821 | 2:6861163 | T | C | 0.698075 | 0.113768 | 0.014157 | 9.29E-16 | C | T | 0.3166 | -0.05691 | 0.005129 | 1.83E-26 | trans |
| miR-320d | rs5618821 | 2:6861163 | T | C | 0.698075 | 0.117075 | 0.014481 | 6.23E-16 | C | T | 0.3166 | -0.05224 | 0.005025 | 1.25E-23 | trans |
| miR-4429 | rs5618821 | 2:6861163 | T | C | 0.698075 | 0.104494 | 0.013944 | 6.70E-14 | C | T | 0.3166 | -0.06364 | 0.004888 | 7.60E-35 | trans |
| miR-320a | rs5628470 | 2:6863137 | G | A | 0.697508 | 0.14339 | 0.017241 | 9.03E-17 | A | G | 0.3276 | -0.05868 | 0.006169 | 3.00E-20 | trans |
| miR-320b | rs5628470 | 2:6863137 | G | A | 0.697508 | 0.130591 | 0.015439 | 2.71E-17 | A | G | 0.3276 | -0.06324 | 0.005124 | 8.82E-32 | trans |
| miR-320c | rs5628470 | 2:6863137 | G | A | 0.697508 | 0.117453 | 0.014477 | 4.93E-16 | A | G | 0.3276 | -0.05743 | 0.005094 | 3.42E-27 | trans |
| miR-320d | rs5628470 | 2:6863137 | G | A | 0.697508 | 0.118242 | 0.014817 | 1.46E-15 | A | G | 0.3276 | -0.05189 | 0.005006 | 1.67E-23 | trans |
| miR-4429 | rs5628470 | 2:6863137 | G | A | 0.697508 | 0.106313 | 0.014265 | 9.15E-14 | A | G | 0.3276 | -0.06445 | 0.004835 | 2.92E-36 | trans |
| miR-320a | rs5633714 | 2:6864249 | G | A | 0.733417 | 0.145039 | 0.018352 | 2.72E-15 | A | G | 0.2867 | -0.05201 | 0.006605 | 1.35E-14 | trans |
| miR-320b | rs5633714 | 2:6864249 | G | A | 0.733417 | 0.132351 | 0.016434 | 8.03E-16 | A | G | 0.2867 | -0.05866 | 0.005526 | 1.82E-24 | trans |
| miR-320c | rs5633714 | 2:6864249 | G | A | 0.733417 | 0.119147 | 0.015407 | 1.05E-14 | A | G | 0.2867 | -0.05076 | 0.005512 | 4.03E-19 | trans |
| miR-320d | rs5633714 | 2:6864249 | G | A | 0.733417 | 0.118238 | 0.015775 | 6.62E-14 | A | G | 0.2867 | -0.05024 | 0.00533 | 6.51E-20 | trans |
| miR-4429 | rs5633714 | 2:6864249 | G | A | 0.733417 | 0.106385 | 0.015183 | 2.44E-12 | A | G | 0.2867 | -0.06032 | 0.005222 | 2.62E-28 | trans |

| | | | | | | | | | | | | | | |
|---|---|---|---|---|---|---|---|---|---|---|---|---|---|---|
| miR-130a-3 | rs5639270 | 11:575183 | T | C | 0.557348 | 0.16563 | 0.021057 | 3.67E-15 | C | T | 0.4404 | -0.1766 | 0.01066 | 3.57E-52 cis |
| miR-625-5p | rs5672046 | 14:662563 | C | T | 0.539663 | -0.139261 | 0.019323 | 5.72E-13 | C | T | 0.4967 | -0.3135 | 0.02627 | 6.33E-30 cis |
| miR-320a | rs5690424 | 2:6863847 | T | C | 0.550669 | 0.155791 | 0.015996 | 2.05E-22 | C | T | 0.4536 | -0.06689 | 0.005545 | 1.54E-30 trans |
| miR-320b | rs5690424 | 2:6863847 | T | C | 0.550669 | 0.140995 | 0.014325 | 7.40E-23 | C | T | 0.4536 | -0.06601 | 0.004631 | 1.40E-40 trans |
| miR-320c | rs5690424 | 2:6863847 | T | C | 0.550669 | 0.128797 | 0.01343 | 8.80E-22 | C | T | 0.4536 | -0.06342 | 0.004547 | 4.08E-39 trans |
| miR-320d | rs5690424 | 2:6863847 | T | C | 0.550669 | 0.126931 | 0.01376 | 2.85E-20 | C | T | 0.4536 | -0.06056 | 0.004448 | 1.47E-37 trans |
| miR-320e | rs5690424 | 2:6863847 | T | C | 0.550669 | 0.113465 | 0.01522 | 9.00E-14 | C | T | 0.4536 | -0.0967 | 0.006138 | 5.48E-48 trans |
| miR-4429 | rs5690424 | 2:6863847 | T | C | 0.550669 | 0.118152 | 0.013239 | 4.48E-19 | C | T | 0.4536 | -0.06832 | 0.004317 | 2.45E-48 trans |
| miR-625-5p | rs5692256 | 14:662564 | A | T | 0.541356 | -0.139228 | 0.019369 | 6.57E-13 | T | A | 0.4993 | 0.3135 | 0.02625 | 5.60E-30 cis |
| miR-130b-3 | rs5754727 | 22:220572 | G | A | 0.5885 | 0.157278 | 0.02306 | 9.08E-12 | A | G | 0.4292 | -0.1492 | 0.009083 | 1.97E-51 cis |
| miR-584-5p | rs5756988 | 5:1484501 | A | G | 0.843968 | -0.271505 | 0.034894 | 7.21E-15 | G | A | 0.1546 | 0.2727 | 0.01754 | 5.95E-47 cis |
| miR-3130-3 | rs5760097 | 2:2076701 | G | C | 0.846329 | -0.262977 | 0.033355 | 3.17E-15 | C | G | 0.1722 | 0.1649 | 0.03168 | 2.55E-07 cis |
| miR-130a-3 | rs576859 | 11:574806 | C | A | 0.683179 | -0.240212 | 0.022172 | 2.37E-27 | A | C | 0.3099 | 0.2301 | 0.01055 | 8.92E-81 cis |
| miR-136-5p | rs5771269 | 14:101161 | G | A | 0.894439 | 0.417877 | 0.058532 | 9.38E-13 | A | G | 0.09103 | -0.08607 | 0.02164 | 7.70E-05 cis |
| miR-4707-3 | rs5782274 | 14:234099 | G | A | 0.440759 | 0.093412 | 0.013618 | 6.91E-12 | G | A | 0.4344 | 0.297 | 0.01632 | 1.13E-60 cis |
| miR-320a | rs5808991 | 2:6863966 | G | C | 0.734634 | 0.147894 | 0.018473 | 1.19E-15 | C | G | 0.2812 | -0.05196 | 0.006637 | 1.92E-14 trans |
| miR-320b | rs5808991 | 2:6863966 | G | C | 0.734634 | 0.134981 | 0.016542 | 3.35E-16 | C | G | 0.2812 | -0.05892 | 0.005603 | 4.88E-24 trans |
| miR-320c | rs5808991 | 2:6863966 | G | C | 0.734634 | 0.121723 | 0.015509 | 4.20E-15 | C | G | 0.2812 | -0.05142 | 0.005585 | 4.24E-19 trans |
| miR-320d | rs5808991 | 2:6863966 | G | C | 0.734634 | 0.120732 | 0.015879 | 2.89E-14 | C | G | 0.2812 | -0.05108 | 0.005415 | 6.46E-20 trans |
| miR-4429 | rs5808991 | 2:6863966 | G | C | 0.734634 | 0.10805 | 0.015286 | 1.57E-12 | C | G | 0.2812 | -0.06162 | 0.005282 | 9.22E-29 trans |
| miR-10b-5p | rs581107 | 9:1361477 | C | T | 0.452595 | -0.128436 | 0.014149 | 1.11E-19 | C | T | 0.4679 | -0.058 | 0.01644 | 0.000445 trans |
| let-7b-5p | rs582094 | 9:1361454 | A | T | 0.663709 | -0.107729 | 0.010063 | 9.59E-27 | T | A | 0.3219 | 0.03377 | 0.006655 | 4.99E-07 trans |
| let-7c-5p | rs582094 | 9:1361454 | A | T | 0.663709 | -0.091349 | 0.009188 | 2.74E-23 | T | A | 0.3219 | 0.02445 | 0.006395 | 0.000144 trans |
| miR-10b-5p | rs582094 | 9:1361454 | A | T | 0.663709 | -0.17709 | 0.014302 | 3.25E-35 | T | A | 0.3219 | 0.1135 | 0.01684 | 3.31E-11 trans |
| miR-126-3p | rs582094 | 9:1361454 | A | T | 0.663709 | -0.171388 | 0.013061 | 2.46E-39 | T | A | 0.3219 | 0.05294 | 0.008279 | 2.95E-10 trans |
| miR-139-5p | rs582094 | 9:1361454 | A | T | 0.663709 | -0.157205 | 0.0116 | 7.69E-42 | T | A | 0.3219 | 0.0709 | 0.01641 | 1.79E-05 trans |
| miR-195-5p | rs582094 | 9:1361454 | A | T | 0.663709 | -0.113779 | 0.01341 | 2.17E-17 | T | A | 0.3219 | 0.06692 | 0.01825 | 0.000265 trans |
| let-7b-5p | rs582118 | 9:1361454 | A | G | 0.663872 | -0.107776 | 0.010067 | 9.58E-27 | G | A | 0.3219 | 0.03377 | 0.006655 | 4.99E-07 trans |
| let-7c-5p | rs582118 | 9:1361454 | A | G | 0.663872 | -0.091413 | 0.009192 | 2.66E-23 | G | A | 0.3219 | 0.02445 | 0.006395 | 0.000144 trans |
| miR-10b-5p | rs582118 | 9:1361454 | A | G | 0.663872 | -0.177155 | 0.014308 | 3.28E-35 | G | A | 0.3219 | 0.1135 | 0.01684 | 3.31E-11 trans |
| miR-126-3p | rs582118 | 9:1361454 | A | G | 0.663872 | -0.171439 | 0.013067 | 2.52E-39 | G | A | 0.3219 | 0.05294 | 0.008279 | 2.95E-10 trans |
| miR-139-5p | rs582118 | 9:1361454 | A | G | 0.663872 | -0.157298 | 0.011605 | 7.44E-42 | G | A | 0.3219 | 0.0709 | 0.01641 | 1.79E-05 trans |
| miR-195-5p | rs582118 | 9:1361454 | A | G | 0.663872 | -0.11383 | 0.013416 | 2.17E-17 | G | A | 0.3219 | 0.06692 | 0.01825 | 0.000265 trans |
| miR-625-5p | rs5830703 | 14:658241 | A | T | 0.051498 | 0.327775 | 0.045296 | 4.61E-13 | A | T | 0.05897 | 0.5849 | 0.05458 | 6.77E-25 cis |
| miR-130a-3 | rs585337 | 11:575972 | C | T | 0.439573 | -0.162328 | 0.020984 | 1.03E-14 | C | T | 0.4377 | -0.1726 | 0.01072 | 6.79E-50 cis |
| miR-130a-3 | rs585761 | 11:575973 | G | A | 0.43789 | -0.160703 | 0.021012 | 2.04E-14 | G | A | 0.433 | -0.1713 | 0.01074 | 4.63E-49 cis |
| miR-130a-3 | rs588612 | 11:576128 | G | C | 0.439957 | -0.162455 | 0.021014 | 1.07E-14 | G | C | 0.4344 | -0.1717 | 0.01074 | 3.15E-49 cis |
| miR-320a | rs5888448 | 2:6864534 | G | A | 0.734214 | 0.144505 | 0.018417 | 4.28E-15 | A | G | 0.2839 | -0.05177 | 0.006639 | 2.39E-14 trans |
| miR-320b | rs5888448 | 2:6864534 | G | A | 0.734214 | 0.131847 | 0.016492 | 1.30E-15 | A | G | 0.2839 | -0.05842 | 0.005551 | 4.31E-24 trans |
| miR-320c | rs5888448 | 2:6864534 | G | A | 0.734214 | 0.118672 | 0.015462 | 1.65E-14 | A | G | 0.2839 | -0.05075 | 0.005528 | 5.17E-19 trans |
| miR-320d | rs5888448 | 2:6864534 | G | A | 0.734214 | 0.117789 | 0.01583 | 1.00E-13 | A | G | 0.2839 | -0.04998 | 0.005333 | 1.05E-19 trans |
| miR-4429 | rs5888448 | 2:6864534 | G | A | 0.734214 | 0.105803 | 0.015237 | 3.82E-12 | A | G | 0.2839 | -0.0608 | 0.005229 | 1.29E-28 trans |
| miR-130a-3 | rs591322 | 11:576846 | T | G | 0.560767 | 0.141968 | 0.021203 | 2.15E-11 | G | T | 0.4439 | -0.138 | 0.01086 | 1.99E-33 cis |
| miR-320a | rs5933292 | 2:6863963 | G | A | 0.73529 | 0.147942 | 0.018524 | 1.39E-15 | A | G | 0.2773 | -0.05128 | 0.006664 | 5.14E-14 trans |
| miR-320b | rs5933292 | 2:6863963 | G | A | 0.73529 | 0.135103 | 0.016587 | 3.79E-16 | A | G | 0.2773 | -0.05912 | 0.005637 | 6.37E-24 trans |
| miR-320c | rs5933292 | 2:6863963 | G | A | 0.73529 | 0.1218 | 0.015551 | 4.79E-15 | A | G | 0.2773 | -0.05138 | 0.005634 | 8.87E-19 trans |
| miR-320d | rs5933292 | 2:6863963 | G | A | 0.73529 | 0.120638 | 0.015923 | 3.56E-14 | A | G | 0.2773 | -0.05067 | 0.005459 | 2.32E-19 trans |
| miR-4429 | rs5933292 | 2:6863963 | G | A | 0.73529 | 0.107748 | 0.015329 | 2.08E-12 | A | G | 0.2773 | -0.0615 | 0.005331 | 3.37E-28 trans |
| miR-625-5p | rs5963675 | 14:662564 | T | C | 0.541442 | -0.139344 | 0.019364 | 6.20E-13 | C | T | 0.4993 | 0.3135 | 0.02625 | 5.60E-30 cis |
| miR-320a | rs5999861 | 2:6864677 | G | T | 0.731295 | 0.13921 | 0.018406 | 3.93E-14 | T | G | 0.28 | -0.05365 | 0.006614 | 2.34E-15 trans |
| miR-320b | rs5999861 | 2:6864677 | G | T | 0.731295 | 0.125973 | 0.016487 | 2.16E-14 | T | G | 0.28 | -0.05887 | 0.005548 | 1.92E-24 trans |
| miR-320c | rs5999861 | 2:6864677 | G | T | 0.731295 | 0.112933 | 0.015457 | 2.75E-13 | T | G | 0.28 | -0.05065 | 0.005516 | 5.04E-19 trans |
| miR-320d | rs5999861 | 2:6864677 | G | T | 0.731295 | 0.111989 | 0.015825 | 1.47E-12 | T | G | 0.28 | -0.04846 | 0.005318 | 9.01E-19 trans |
| miR-4429 | rs5999861 | 2:6864677 | G | T | 0.731295 | 0.101746 | 0.015226 | 2.35E-11 | T | G | 0.28 | -0.0613 | 0.005228 | 4.78E-29 trans |
| let-7a-5p | rs600038 | 9:1361518 | T | C | 0.787872 | -0.093029 | 0.013146 | 1.48E-12 | C | T | 0.205 | 0.02068 | 0.005781 | 0.000372 trans |
| let-7b-5p | rs600038 | 9:1361518 | T | C | 0.787872 | -0.119508 | 0.011643 | 1.02E-24 | C | T | 0.205 | 0.02564 | 0.007562 | 0.000738 trans |
| miR-10b-5p | rs600038 | 9:1361518 | T | C | 0.787872 | -0.202026 | 0.016527 | 2.32E-34 | C | T | 0.205 | 0.1099 | 0.01912 | 1.37E-08 trans |
| miR-126-3p | rs600038 | 9:1361518 | T | C | 0.787872 | -0.193279 | 0.015108 | 1.80E-37 | C | T | 0.205 | 0.05476 | 0.009295 | 5.95E-09 trans |
| miR-139-5p | rs600038 | 9:1361518 | T | C | 0.787872 | -0.166058 | 0.013486 | 7.68E-35 | C | T | 0.205 | 0.07053 | 0.01847 | 0.000147 trans |
| miR-320a | rs6000716 | 2:6863963 | C | T | 0.735163 | 0.14802 | 0.018521 | 1.33E-15 | T | C | 0.2787 | -0.05128 | 0.006664 | 5.14E-14 trans |
| miR-320b | rs6000716 | 2:6863963 | C | T | 0.735163 | 0.135202 | 0.016585 | 3.57E-16 | T | C | 0.2787 | -0.05912 | 0.005637 | 6.37E-24 trans |
| miR-320c | rs6000716 | 2:6863963 | C | T | 0.735163 | 0.121878 | 0.015549 | 4.56E-15 | T | C | 0.2787 | -0.05138 | 0.005634 | 8.87E-19 trans |
| miR-320d | rs6000716 | 2:6863963 | C | T | 0.735163 | 0.12067 | 0.015921 | 3.48E-14 | T | C | 0.2787 | -0.05067 | 0.005459 | 2.32E-19 trans |
| miR-4429 | rs6000716 | 2:6863963 | C | T | 0.735163 | 0.107772 | 0.015327 | 2.04E-12 | T | C | 0.2787 | -0.0615 | 0.005331 | 3.37E-28 trans |
| miR-3130-3 | rs6001989 | 2:2076777 | C | T | 0.87642 | -0.247533 | 0.03705 | 2.37E-11 | T | C | 0.1431 | 0.1551 | 0.03409 | 6.37E-06 cis |
| miR-130a-3 | rs600488 | 11:576840 | C | T | 0.560573 | 0.142183 | 0.021211 | 2.04E-11 | T | C | 0.4432 | -0.1387 | 0.01082 | 6.28E-34 cis |
| miR-625-5p | rs6048801 | 14:658258 | C | A | 0.738599 | -0.148941 | 0.021849 | 9.30E-12 | A | C | 0.2368 | 0.2806 | 0.03084 | 9.88E-19 cis |
| miR-133b | rs6062267 | 20:611533 | T | C | 0.464907 | -0.190202 | 0.027172 | 2.56E-12 | T | C | 0.4472 | -0.3034 | 0.03057 | 8.33E-22 trans |
| miR-133b | rs6062284 | 20:611419 | T | G | 0.4602 | -0.17216 | 0.025716 | 2.16E-11 | T | G | 0.4391 | -0.3155 | 0.03191 | 1.40E-21 trans |
| miR-130a-3 | rs606499 | 11:575938 | G | A | 0.439467 | -0.162246 | 0.020963 | 9.97E-15 | G | A | 0.4377 | -0.1726 | 0.01072 | 6.79E-50 cis |
| miR-625-5p | rs6070872 | 14:661167 | G | A | 0.819646 | -0.239612 | 0.025764 | 1.40E-20 | A | G | 0.1658 | 0.5034 | 0.03337 | 1.71E-44 cis |
| miR-320a | rs6087844 | 2:6858211 | A | G | 0.538067 | 0.140648 | 0.016039 | 1.80E-18 | G | A | 0.4694 | -0.0584 | 0.005679 | 3.45E-23 trans |
| miR-320b | rs6087844 | 2:6858211 | A | G | 0.538067 | 0.126281 | 0.014369 | 1.52E-18 | G | A | 0.4694 | -0.06104 | 0.00472 | 1.91E-34 trans |
| miR-320c | rs6087844 | 2:6858211 | A | G | 0.538067 | 0.11583 | 0.013466 | 7.86E-18 | G | A | 0.4694 | -0.05563 | 0.004697 | 1.30E-29 trans |
| miR-320d | rs6087844 | 2:6858211 | A | G | 0.538067 | 0.116986 | 0.013782 | 2.10E-17 | G | A | 0.4694 | -0.05366 | 0.004566 | 3.18E-29 trans |
| miR-320e | rs6087844 | 2:6858211 | A | G | 0.538067 | 0.108754 | 0.015216 | 8.84E-13 | G | A | 0.4694 | -0.08422 | 0.006395 | 1.56E-35 trans |
| miR-4429 | rs6087844 | 2:6858211 | A | G | 0.538067 | 0.108084 | 0.013261 | 3.62E-16 | G | A | 0.4694 | -0.06028 | 0.004497 | 1.26E-36 trans |
| let-7b-5p | rs612169 | 9:1361434 | A | G | 0.66434 | -0.107722 | 0.01008 | 1.17E-26 | G | A | 0.3219 | 0.03377 | 0.006655 | 4.99E-07 trans |
| let-7c-5p | rs612169 | 9:1361434 | A | G | 0.66434 | -0.091485 | 0.009203 | 2.76E-23 | G | A | 0.3219 | 0.02445 | 0.006395 | 0.000144 trans |
| miR-10b-5p | rs612169 | 9:1361434 | A | G | 0.66434 | -0.177278 | 0.014325 | 3.54E-35 | G | A | 0.3219 | 0.1135 | 0.01684 | 3.31E-11 trans |
| miR-126-3p | rs612169 | 9:1361434 | A | G | 0.66434 | -0.171512 | 0.013083 | 2.89E-39 | G | A | 0.3219 | 0.05294 | 0.008279 | 2.95E-10 trans |
| miR-139-5p | rs612169 | 9:1361434 | A | G | 0.66434 | -0.157438 | 0.011618 | 7.83E-42 | G | A | 0.3219 | 0.0709 | 0.01641 | 1.79E-05 trans |
| miR-195-5p | rs612169 | 9:1361434 | A | G | 0.66434 | -0.113707 | 0.013433 | 2.56E-17 | G | A | 0.3219 | 0.06692 | 0.01825 | 0.000265 trans |
| let-7b-5p | rs613534 | 9:1361431 | A | G | 0.653693 | -0.106 | 0.01005 | 5.20E-26 | G | A | 0.3457 | 0.02922 | 0.006685 | 1.43E-05 trans |
| let-7c-5p | rs613534 | 9:1361431 | A | G | 0.653693 | -0.090765 | 0.009171 | 4.30E-23 | G | A | 0.3457 | 0.02465 | 0.006356 | 0.000116 trans |
| miR-10b-5p | rs613534 | 9:1361431 | A | G | 0.653693 | -0.174451 | 0.014285 | 2.68E-34 | G | A | 0.3457 | 0.1111 | 0.01688 | 9.54E-11 trans |

| | | | | | | | | | | | | | | | |
|---|---|---|---|---|---|---|---|---|---|---|---|---|---|---|---|
| miR-126-3p | rs613534 | 9:1361431 | A | G | 0.653693 | -0.165472 | 0.013067 | 9.46E-37 | G | A | 0.3457 | 0.05053 | 0.008385 | 2.78E-09 | trans |
| miR-139-5p | rs613534 | 9:1361431 | A | G | 0.653693 | -0.153841 | 0.011595 | 3.54E-40 | G | A | 0.3457 | 0.06895 | 0.01639 | 2.94E-05 | trans |
| miR-625-5p | rs6137847 | 14:662534 | G | A | 0.541244 | -0.140207 | 0.019376 | 4.62E-13 | G | A | 0.4987 | -0.3109 | 0.0263 | 1.90E-29 | cis |
| miR-130a-3 | rs615133 | 11:576849 | C | A | 0.561411 | 0.141502 | 0.021175 | 2.35E-11 | A | C | 0.4432 | -0.1387 | 0.01082 | 6.28E-34 | cis |
| miR-130a-3 | rs616911 | 11:576853 | G | A | 0.558798 | 0.145619 | 0.021196 | 6.42E-12 | A | G | 0.4432 | -0.1387 | 0.01081 | 5.62E-34 | cis |
| miR-4707-3 | rs6197774 | 14:234009 | G | C | 0.440733 | 0.091819 | 0.013701 | 2.06E-11 | G | C | 0.4328 | 0.2919 | 0.01651 | 6.26E-58 | cis |
| miR-625-5p | rs6198799 | 14:660707 | C | T | 0.351584 | 0.180835 | 0.020721 | 2.61E-18 | C | T | 0.3275 | 0.4141 | 0.0265 | 4.23E-47 | cis |
| miR-625-5p | rs6198799 | 14:660707 | C | G | 0.351584 | 0.180835 | 0.020721 | 2.61E-18 | C | G | 0.3275 | 0.4141 | 0.0265 | 4.23E-47 | cis |
| miR-625-5p | rs6198897 | 14:662580 | C | A | 0.537113 | -0.137354 | 0.019389 | 1.40E-12 | C | A | 0.4948 | -0.3009 | 0.02634 | 8.67E-28 | cis |
| miR-320a | rs6214389 | 2:6860455 | C | T | 0.698307 | 0.139108 | 0.016881 | 1.71E-16 | T | C | 0.3167 | -0.05686 | 0.006263 | 1.12E-18 | trans |
| miR-320b | rs6214389 | 2:6860455 | C | T | 0.698307 | 0.126569 | 0.015117 | 5.64E-17 | T | C | 0.3167 | -0.06312 | 0.005188 | 5.19E-31 | trans |
| miR-320c | rs6214389 | 2:6860455 | C | T | 0.698307 | 0.113676 | 0.014176 | 1.06E-15 | T | C | 0.3167 | -0.05612 | 0.005157 | 1.41E-25 | trans |
| miR-320d | rs6214389 | 2:6860455 | C | T | 0.698307 | 0.116699 | 0.014501 | 8.42E-16 | T | C | 0.3167 | -0.05128 | 0.005055 | 1.22E-22 | trans |
| miR-4429 | rs6214389 | 2:6860455 | C | T | 0.698307 | 0.103959 | 0.013964 | 9.70E-14 | T | C | 0.3167 | -0.06242 | 0.00491 | 1.92E-33 | trans |
| miR-320a | rs6214392 | 2:6863835 | C | G | 0.700378 | 0.148987 | 0.017611 | 2.68E-17 | G | C | 0.3284 | -0.05748 | 0.006199 | 2.34E-19 | trans |
| miR-320b | rs6214392 | 2:6863835 | C | G | 0.700378 | 0.134816 | 0.015773 | 1.26E-17 | G | C | 0.3284 | -0.06265 | 0.005152 | 6.09E-31 | trans |
| miR-320c | rs6214392 | 2:6863835 | C | G | 0.700378 | 0.121403 | 0.01479 | 2.25E-16 | G | C | 0.3284 | -0.05654 | 0.005072 | 1.24E-26 | trans |
| miR-320d | rs6214392 | 2:6863835 | C | G | 0.700378 | 0.121704 | 0.01514 | 9.10E-16 | G | C | 0.3284 | -0.05102 | 0.004972 | 4.43E-23 | trans |
| miR-4429 | rs6214392 | 2:6863835 | C | G | 0.700378 | 0.109847 | 0.014575 | 4.82E-14 | G | C | 0.3284 | -0.06392 | 0.004832 | 1.00E-35 | trans |
| miR-320a | rs6214393 | 2:6864009 | T | C | 0.59325 | 0.14244 | 0.016445 | 4.65E-18 | C | T | 0.4035 | -0.06073 | 0.005815 | 8.98E-24 | trans |
| miR-320b | rs6214393 | 2:6864009 | T | C | 0.59325 | 0.131146 | 0.01472 | 5.13E-19 | C | T | 0.4035 | -0.06085 | 0.004895 | 4.34E-32 | trans |
| miR-320c | rs6214393 | 2:6864009 | T | C | 0.59325 | 0.11934 | 0.013799 | 5.22E-18 | C | T | 0.4035 | -0.05706 | 0.004838 | 2.55E-29 | trans |
| miR-320d | rs6214393 | 2:6864009 | T | C | 0.59325 | 0.118698 | 0.01413 | 4.45E-17 | C | T | 0.4035 | -0.05807 | 0.004646 | 2.10E-32 | trans |
| miR-320e | rs6214393 | 2:6864009 | T | C | 0.59325 | 0.113891 | 0.015587 | 2.73E-13 | C | T | 0.4035 | -0.09326 | 0.006449 | 1.70E-41 | trans |
| miR-4429 | rs6214393 | 2:6864009 | T | C | 0.59325 | 0.109114 | 0.013597 | 1.02E-15 | C | T | 0.4035 | -0.0632 | 0.004591 | 3.46E-38 | trans |
| miR-320a | rs6214393 | 2:6864020 | A | G | 0.593308 | 0.142404 | 0.016446 | 4.76E-18 | G | A | 0.4026 | -0.06104 | 0.005832 | 7.44E-24 | trans |
| miR-320b | rs6214393 | 2:6864020 | A | G | 0.593308 | 0.131089 | 0.014721 | 5.33E-19 | G | A | 0.4026 | -0.06103 | 0.004911 | 4.55E-32 | trans |
| miR-320c | rs6214393 | 2:6864020 | A | G | 0.593308 | 0.119306 | 0.0138 | 5.36E-18 | G | A | 0.4026 | -0.0574 | 0.004851 | 1.72E-29 | trans |
| miR-320d | rs6214393 | 2:6864020 | A | G | 0.593308 | 0.118688 | 0.014131 | 4.49E-17 | G | A | 0.4026 | -0.05836 | 0.004659 | 1.63E-32 | trans |
| miR-320e | rs6214393 | 2:6864020 | A | G | 0.593308 | 0.113862 | 0.015587 | 2.78E-13 | G | A | 0.4026 | -0.09305 | 0.006461 | 3.41E-41 | trans |
| miR-4429 | rs6214393 | 2:6864020 | A | G | 0.593308 | 0.109092 | 0.013598 | 1.04E-15 | G | A | 0.4026 | -0.06283 | 0.004601 | 1.16E-37 | trans |
| miR-320a | rs6214393 | 2:6864038 | G | C | 0.593598 | 0.143062 | 0.01644 | 3.25E-18 | C | G | 0.4026 | -0.06104 | 0.005832 | 7.44E-24 | trans |
| miR-320b | rs6214393 | 2:6864038 | G | C | 0.593598 | 0.131783 | 0.014715 | 3.37E-19 | C | G | 0.4026 | -0.06103 | 0.004911 | 4.55E-32 | trans |
| miR-320c | rs6214393 | 2:6864038 | G | C | 0.593598 | 0.119834 | 0.013795 | 3.72E-18 | C | G | 0.4026 | -0.0574 | 0.004851 | 1.72E-29 | trans |
| miR-320d | rs6214393 | 2:6864038 | G | C | 0.593598 | 0.119156 | 0.014126 | 3.30E-17 | C | G | 0.4026 | -0.05836 | 0.004659 | 1.63E-32 | trans |
| miR-320e | rs6214393 | 2:6864038 | G | C | 0.593598 | 0.114275 | 0.015583 | 2.24E-13 | C | G | 0.4026 | -0.09305 | 0.006461 | 3.41E-41 | trans |
| miR-4429 | rs6214393 | 2:6864038 | G | C | 0.593598 | 0.109647 | 0.013593 | 7.24E-16 | C | G | 0.4026 | -0.06283 | 0.004601 | 1.16E-37 | trans |
| miR-320a | rs6214393 | 2:6864041 | A | G | 0.593599 | 0.143051 | 0.01644 | 3.28E-18 | G | A | 0.4026 | -0.06104 | 0.005832 | 7.44E-24 | trans |
| miR-320b | rs6214393 | 2:6864041 | A | G | 0.593599 | 0.131772 | 0.014715 | 3.40E-19 | G | A | 0.4026 | -0.06103 | 0.004911 | 4.55E-32 | trans |
| miR-320c | rs6214393 | 2:6864041 | A | G | 0.593599 | 0.11983 | 0.013795 | 3.74E-18 | G | A | 0.4026 | -0.0574 | 0.004851 | 1.72E-29 | trans |
| miR-320d | rs6214393 | 2:6864041 | A | G | 0.593599 | 0.11915 | 0.014126 | 3.32E-17 | G | A | 0.4026 | -0.05836 | 0.004659 | 1.63E-32 | trans |
| miR-320e | rs6214393 | 2:6864041 | A | G | 0.593599 | 0.114256 | 0.015583 | 2.27E-13 | G | A | 0.4026 | -0.09305 | 0.006461 | 3.41E-41 | trans |
| miR-4429 | rs6214393 | 2:6864041 | A | G | 0.593599 | 0.10964 | 0.013593 | 7.28E-16 | G | A | 0.4026 | -0.06283 | 0.004601 | 1.16E-37 | trans |
| miR-320a | rs6214393 | 2:6864062 | T | C | 0.733261 | 0.145064 | 0.018341 | 2.58E-15 | C | T | 0.2867 | -0.05181 | 0.006609 | 1.74E-14 | trans |
| miR-320b | rs6214393 | 2:6864062 | T | C | 0.733261 | 0.132408 | 0.016423 | 7.50E-16 | C | T | 0.2867 | -0.05861 | 0.005521 | 1.79E-24 | trans |
| miR-320c | rs6214393 | 2:6864062 | T | C | 0.733261 | 0.119179 | 0.015398 | 9.95E-15 | C | T | 0.2867 | -0.05092 | 0.005515 | 3.25E-19 | trans |
| miR-320d | rs6214393 | 2:6864062 | T | C | 0.733261 | 0.118334 | 0.015765 | 6.09E-14 | C | T | 0.2867 | -0.05033 | 0.005327 | 5.40E-20 | trans |
| miR-4429 | rs6214393 | 2:6864062 | T | C | 0.733261 | 0.106445 | 0.015174 | 2.30E-12 | C | T | 0.2867 | -0.06044 | 0.005222 | 2.05E-28 | trans |
| miR-130a-3 | rs623541 | 11:575816 | C | A | 0.439413 | -0.163085 | 0.020971 | 7.45E-15 | C | A | 0.4377 | -0.1726 | 0.01072 | 6.79E-50 | cis |
| miR-584-5p | rs6238006 | 5:1484422 | C | T | 0.844027 | -0.270959 | 0.034834 | 7.34E-15 | T | C | 0.1546 | 0.2727 | 0.01754 | 5.95E-47 | cis |
| miR-584-5p | rs6238010 | 5:1484921 | C | A | 0.846193 | -0.276613 | 0.035432 | 5.86E-15 | A | C | 0.1543 | 0.2561 | 0.01756 | 3.85E-42 | cis |
| miR-584-5p | rs6238010 | 5:1485004 | G | T | 0.8464 | -0.276639 | 0.035415 | 5.66E-15 | T | G | 0.1529 | 0.2553 | 0.01773 | 3.20E-41 | cis |
| miR-130a-3 | rs627145 | 11:576024 | G | A | 0.439595 | -0.161806 | 0.021052 | 1.52E-14 | G | A | 0.4323 | -0.1713 | 0.01074 | 4.63E-49 | cis |
| let-7b-5p | rs630014 | 9:1361497 | A | G | 0.494254 | -0.079742 | 0.009829 | 4.92E-16 | A | G | 0.4968 | -0.0219 | 0.006094 | 0.00035 | trans |
| miR-10b-5p | rs630014 | 9:1361497 | A | G | 0.494254 | -0.115432 | 0.014082 | 2.46E-16 | A | G | 0.4968 | -0.07768 | 0.0155 | 6.81E-07 | trans |
| miR-126-3p | rs630014 | 9:1361497 | A | G | 0.494254 | -0.115403 | 0.012875 | 3.15E-19 | A | G | 0.4968 | -0.03396 | 0.007623 | 9.79E-06 | trans |
| let-7b-5p | rs630510 | 9:1361495 | A | G | 0.494403 | -0.079814 | 0.009835 | 4.85E-16 | A | G | 0.4962 | -0.0219 | 0.006094 | 0.00035 | trans |
| miR-10b-5p | rs630510 | 9:1361495 | A | G | 0.494403 | -0.115552 | 0.014092 | 2.40E-16 | A | G | 0.4962 | -0.07768 | 0.0155 | 6.81E-07 | trans |
| miR-126-3p | rs630510 | 9:1361495 | A | G | 0.494403 | -0.115498 | 0.012884 | 3.11E-19 | A | G | 0.4962 | -0.03396 | 0.007623 | 9.79E-06 | trans |
| miR-130a-3 | rs633129 | 11:576851 | G | A | 0.561364 | 0.141528 | 0.021176 | 2.33E-11 | A | G | 0.4432 | -0.1387 | 0.01082 | 6.28E-34 | cis |
| miR-130a-3 | rs639158 | 11:576020 | G | A | 0.441293 | -0.163651 | 0.021032 | 7.19E-15 | G | A | 0.4362 | -0.1727 | 0.01074 | 9.50E-50 | cis |
| miR-320a | rs6419643 | 2:6863126 | T | G | 0.548826 | 0.153407 | 0.015714 | 1.63E-22 | G | T | 0.4541 | -0.06692 | 0.005508 | 6.09E-31 | trans |
| miR-320b | rs6419643 | 2:6863126 | T | G | 0.548826 | 0.138972 | 0.014072 | 5.32E-23 | G | T | 0.4541 | -0.06607 | 0.004593 | 2.87E-41 | trans |
| miR-320c | rs6419643 | 2:6863126 | T | G | 0.548826 | 0.12595 | 0.013197 | 1.38E-21 | G | T | 0.4541 | -0.06373 | 0.004516 | 6.01E-40 | trans |
| miR-320d | rs6419643 | 2:6863126 | T | G | 0.548826 | 0.124619 | 0.01352 | 3.04E-20 | G | T | 0.4541 | -0.06046 | 0.004413 | 5.28E-38 | trans |
| miR-320e | rs6419643 | 2:6863126 | T | G | 0.548826 | 0.113114 | 0.014948 | 3.81E-14 | G | T | 0.4541 | -0.09582 | 0.006109 | 1.04E-47 | trans |
| miR-4429 | rs6419643 | 2:6863126 | T | G | 0.548826 | 0.116437 | 0.013006 | 3.47E-19 | G | T | 0.4541 | -0.06832 | 0.004294 | 7.35E-49 | trans |
| miR-320a | rs6419644 | 2:6863182 | A | G | 0.549898 | 0.155194 | 0.015873 | 1.41E-22 | G | A | 0.4547 | -0.06692 | 0.005516 | 7.17E-31 | trans |
| miR-320b | rs6419644 | 2:6863182 | A | G | 0.549898 | 0.140681 | 0.014215 | 4.29E-23 | G | A | 0.4547 | -0.06612 | 0.004599 | 3.10E-41 | trans |
| miR-320c | rs6419644 | 2:6863182 | A | G | 0.549898 | 0.128192 | 0.013328 | 6.69E-22 | G | A | 0.4547 | -0.06386 | 0.004521 | 4.93E-40 | trans |
| miR-320d | rs6419644 | 2:6863182 | A | G | 0.549898 | 0.126235 | 0.013656 | 2.38E-20 | G | A | 0.4547 | -0.06054 | 0.004418 | 5.22E-38 | trans |
| miR-320e | rs6419644 | 2:6863182 | A | G | 0.549898 | 0.11406 | 0.015101 | 4.25E-14 | G | A | 0.4547 | -0.09626 | 0.006109 | 4.55E-48 | trans |
| miR-4429 | rs6419644 | 2:6863182 | A | G | 0.549898 | 0.117848 | 0.013138 | 2.96E-19 | G | A | 0.4547 | -0.06846 | 0.004297 | 5.96E-49 | trans |
| miR-181a-5 | rs6427756 | 1:1990007 | A | G | 0.263331 | 0.176572 | 0.013797 | 1.69E-37 | A | G | 0.2667 | 0.3404 | 0.01263 | 4.33E-110 | cis |
| let-7a-5p | rs643434 | 9:1361423 | G | A | 0.645392 | -0.079734 | 0.011164 | 9.18E-13 | A | G | 0.3467 | 0.0169 | 0.005043 | 0.000849 | trans |
| let-7b-5p | rs643434 | 9:1361423 | G | A | 0.645392 | -0.105157 | 0.009872 | 1.70E-26 | A | G | 0.3467 | 0.02792 | 0.00656 | 2.36E-05 | trans |
| let-7c-5p | rs643434 | 9:1361423 | G | A | 0.645392 | -0.089455 | 0.009012 | 3.20E-23 | A | G | 0.3467 | 0.02285 | 0.006277 | 0.000292 | trans |
| miR-10b-5p | rs643434 | 9:1361423 | G | A | 0.645392 | -0.171966 | 0.014036 | 1.64E-34 | A | G | 0.3467 | 0.1083 | 0.01654 | 1.14E-10 | trans |
| miR-126-3p | rs643434 | 9:1361423 | G | A | 0.645392 | -0.164561 | 0.012831 | 1.18E-37 | A | G | 0.3467 | 0.04895 | 0.008145 | 2.98E-09 | trans |
| miR-139-5p | rs643434 | 9:1361423 | G | A | 0.645392 | -0.151972 | 0.01139 | 1.31E-40 | A | G | 0.3467 | 0.06736 | 0.01611 | 3.26E-05 | trans |
| miR-130a-3 | rs644219 | 11:576100 | C | T | 0.56488 | 0.160737 | 0.02104 | 2.18E-14 | T | C | 0.4283 | -0.1693 | 0.01083 | 1.97E-47 | cis |
| let-7a-5p | rs644234 | 9:1361422 | T | G | 0.645381 | -0.079814 | 0.011155 | 8.38E-13 | G | T | 0.3467 | 0.0169 | 0.005043 | 0.000849 | trans |
| let-7b-5p | rs644234 | 9:1361422 | T | G | 0.645381 | -0.105194 | 0.009864 | 1.49E-26 | G | T | 0.3467 | 0.02792 | 0.00656 | 2.36E-05 | trans |
| let-7c-5p | rs644234 | 9:1361422 | T | G | 0.645381 | -0.089506 | 0.009005 | 2.80E-23 | G | T | 0.3467 | 0.02285 | 0.006277 | 0.000292 | trans |
| miR-10b-5p | rs644234 | 9:1361422 | T | G | 0.645381 | -0.171966 | 0.014025 | 1.46E-34 | G | T | 0.3467 | 0.1083 | 0.01654 | 1.14E-10 | trans |
| miR-126-3p | rs644234 | 9:1361422 | T | G | 0.645381 | -0.164565 | 0.012821 | 1.03E-37 | G | T | 0.3467 | 0.04895 | 0.008145 | 2.98E-09 | trans |

| | | | | | | | | | | | | | | | |
|---|---|---|---|---|---|---|---|---|---|---|---|---|---|---|---|
| miR-139-5p | rs644234 | 9:1361422 | T | G | 0.645381 | -0.151953 | 0.011382 | 1.17E-40 | G | T | 0.3467 | 0.06736 | 0.01611 | 3.26E-05 | trans |
| miR-130a-3 | rs645145 | 11:576097 | G | T | 0.564908 | 0.160734 | 0.02104 | 2.18E-14 | T | G | 0.4283 | -0.1693 | 0.01083 | 1.97E-47 | cis |
| miR-10b-5p | rs645982 | 9:1361484 | A | G | 0.454123 | -0.127768 | 0.0141 | 1.29E-19 | A | G | 0.4688 | -0.05911 | 0.01638 | 0.00033 | trans |
| miR-10b-5p | rs647800 | 9:1361480 | A | G | 0.435519 | -0.128548 | 0.014052 | 5.81E-20 | A | G | 0.4439 | -0.05815 | 0.01636 | 0.000405 | trans |
| let-7a-5p | rs649129 | 9:1361543 | C | T | 0.787432 | -0.092844 | 0.01312 | 1.48E-12 | T | C | 0.2055 | 0.02032 | 0.005769 | 0.000455 | trans |
| miR-10b-5p | rs649129 | 9:1361543 | C | T | 0.787432 | -0.201509 | 0.016495 | 2.54E-34 | T | C | 0.2055 | 0.1089 | 0.01907 | 1.68E-08 | trans |
| miR-126-3p | rs649129 | 9:1361543 | C | T | 0.787432 | -0.192807 | 0.015079 | 1.95E-37 | T | C | 0.2055 | 0.05301 | 0.00935 | 2.09E-08 | trans |
| miR-139-5p | rs649129 | 9:1361543 | C | T | 0.787432 | -0.165163 | 0.013462 | 1.34E-34 | T | C | 0.2055 | 0.06866 | 0.01849 | 0.000221 | trans |
| miR-130a-3 | rs652191 | 11:576350 | A | G | 0.568268 | 0.160772 | 0.021201 | 3.37E-14 | G | A | 0.4277 | -0.1692 | 0.01078 | 1.11E-47 | cis |
| miR-130a-3 | rs652908 | 11:576040 | C | T | 0.441718 | -0.163828 | 0.021034 | 6.76E-15 | C | T | 0.4362 | -0.1727 | 0.01074 | 9.50E-50 | cis |
| miR-130a-3 | rs653889 | 11:575704 | A | G | 0.567416 | 0.160942 | 0.021027 | 1.95E-14 | G | A | 0.43 | -0.17 | 0.01079 | 4.44E-48 | cis |
| miR-320a | rs6546393 | 2:6862757 | T | C | 0.554017 | 0.154967 | 0.015786 | 9.52E-23 | C | T | 0.4469 | -0.06776 | 0.005615 | 1.66E-30 | trans |
| miR-320b | rs6546393 | 2:6862757 | T | C | 0.554017 | 0.140713 | 0.014135 | 2.40E-23 | C | T | 0.4469 | -0.0667 | 0.004701 | 3.52E-40 | trans |
| miR-320c | rs6546393 | 2:6862757 | T | C | 0.554017 | 0.126897 | 0.013259 | 1.06E-21 | C | T | 0.4469 | -0.06463 | 0.004622 | 3.30E-39 | trans |
| miR-320d | rs6546393 | 2:6862757 | T | C | 0.554017 | 0.126078 | 0.013581 | 1.64E-20 | C | T | 0.4469 | -0.062 | 0.004533 | 9.11E-38 | trans |
| miR-320e | rs6546393 | 2:6862757 | T | C | 0.554017 | 0.114706 | 0.015016 | 2.19E-14 | C | T | 0.4469 | -0.0979 | 0.006212 | 6.84E-48 | trans |
| miR-4429 | rs6546393 | 2:6862757 | T | C | 0.554017 | 0.117707 | 0.013065 | 2.08E-19 | C | T | 0.4469 | -0.06899 | 0.004391 | 1.20E-47 | trans |
| miR-320a | rs6546395 | 2:6862927 | A | G | 0.55448 | 0.152637 | 0.015805 | 4.56E-22 | G | A | 0.4431 | -0.06708 | 0.005598 | 3.74E-30 | trans |
| miR-320b | rs6546395 | 2:6862927 | A | G | 0.55448 | 0.138124 | 0.014154 | 1.70E-22 | G | A | 0.4431 | -0.06519 | 0.004684 | 6.24E-39 | trans |
| miR-320c | rs6546395 | 2:6862927 | A | G | 0.55448 | 0.125279 | 0.013273 | 3.79E-21 | G | A | 0.4431 | -0.06082 | 0.0046 | 1.11E-35 | trans |
| miR-320d | rs6546395 | 2:6862927 | A | G | 0.55448 | 0.124433 | 0.013595 | 5.55E-20 | G | A | 0.4431 | -0.05871 | 0.004528 | 1.63E-34 | trans |
| miR-320e | rs6546395 | 2:6862927 | A | G | 0.55448 | 0.11111 | 0.015036 | 1.47E-13 | G | A | 0.4431 | -0.09397 | 0.006231 | 1.50E-44 | trans |
| miR-4429 | rs6546395 | 2:6862927 | A | G | 0.55448 | 0.116327 | 0.013078 | 5.85E-19 | G | A | 0.4431 | -0.06719 | 0.004354 | 2.81E-46 | trans |
| miR-320a | rs6546397 | 2:6863036 | G | A | 0.552756 | 0.152134 | 0.015759 | 4.74E-22 | A | G | 0.4488 | -0.06658 | 0.005541 | 2.34E-30 | trans |
| miR-320b | rs6546397 | 2:6863036 | G | A | 0.552756 | 0.13783 | 0.014113 | 1.57E-22 | A | G | 0.4488 | -0.06591 | 0.004617 | 9.87E-41 | trans |
| miR-320c | rs6546397 | 2:6863036 | G | A | 0.552756 | 0.125081 | 0.013234 | 3.34E-21 | A | G | 0.4488 | -0.06191 | 0.004572 | 3.06E-37 | trans |
| miR-320d | rs6546397 | 2:6863036 | G | A | 0.552756 | 0.124221 | 0.013555 | 4.99E-20 | A | G | 0.4488 | -0.05868 | 0.004467 | 2.30E-35 | trans |
| miR-320e | rs6546397 | 2:6863036 | G | A | 0.552756 | 0.110969 | 0.014992 | 1.34E-13 | A | G | 0.4488 | -0.09469 | 0.006167 | 5.01E-46 | trans |
| miR-4429 | rs6546397 | 2:6863036 | G | A | 0.552756 | 0.116061 | 0.01304 | 5.55E-19 | A | G | 0.4488 | -0.06805 | 0.004311 | 3.44E-48 | trans |
| miR-320a | rs6546398 | 2:6863041 | G | C | 0.552757 | 0.152138 | 0.015759 | 4.72E-22 | C | G | 0.4488 | -0.06658 | 0.005541 | 2.34E-30 | trans |
| miR-320b | rs6546398 | 2:6863041 | G | C | 0.552757 | 0.137831 | 0.014112 | 1.56E-22 | C | G | 0.4488 | -0.06591 | 0.004617 | 9.87E-41 | trans |
| miR-320c | rs6546398 | 2:6863041 | G | C | 0.552757 | 0.125094 | 0.013234 | 3.30E-21 | C | G | 0.4488 | -0.06191 | 0.004572 | 3.06E-37 | trans |
| miR-320d | rs6546398 | 2:6863041 | G | C | 0.552757 | 0.124235 | 0.013555 | 4.93E-20 | C | G | 0.4488 | -0.05868 | 0.004467 | 2.30E-35 | trans |
| miR-320e | rs6546398 | 2:6863041 | G | C | 0.552757 | 0.110975 | 0.014992 | 1.34E-13 | C | G | 0.4488 | -0.09469 | 0.006167 | 5.01E-46 | trans |
| miR-4429 | rs6546398 | 2:6863041 | G | C | 0.552757 | 0.116069 | 0.013039 | 5.51E-19 | C | G | 0.4488 | -0.06805 | 0.004311 | 3.44E-48 | trans |
| miR-320a | rs6546399 | 2:6863064 | C | T | 0.55394 | 0.15272 | 0.015805 | 4.34E-22 | T | C | 0.4462 | -0.06633 | 0.005564 | 6.53E-30 | trans |
| miR-320b | rs6546399 | 2:6863064 | C | T | 0.55394 | 0.138212 | 0.014154 | 1.60E-22 | T | C | 0.4462 | -0.06498 | 0.004653 | 3.42E-39 | trans |
| miR-320c | rs6546399 | 2:6863064 | C | T | 0.55394 | 0.125397 | 0.013273 | 3.48E-21 | T | C | 0.4462 | -0.06146 | 0.004609 | 3.13E-36 | trans |
| miR-320d | rs6546399 | 2:6863064 | C | T | 0.55394 | 0.124567 | 0.013595 | 5.06E-20 | T | C | 0.4462 | -0.05905 | 0.004508 | 3.82E-35 | trans |
| miR-320e | rs6546399 | 2:6863064 | C | T | 0.55394 | 0.111105 | 0.015037 | 1.48E-13 | T | C | 0.4462 | -0.09422 | 0.006221 | 6.44E-45 | trans |
| miR-4429 | rs6546399 | 2:6863064 | C | T | 0.55394 | 0.116432 | 0.013078 | 5.44E-19 | T | C | 0.4462 | -0.06799 | 0.004348 | 2.33E-47 | trans |
| miR-320a | rs6546400 | 2:6863064 | G | A | 0.554382 | 0.152811 | 0.01582 | 4.47E-22 | A | G | 0.4452 | -0.06638 | 0.005584 | 9.41E-30 | trans |
| miR-320b | rs6546400 | 2:6863064 | G | A | 0.554382 | 0.138315 | 0.014167 | 1.62E-22 | A | G | 0.4452 | -0.06509 | 0.004661 | 3.65E-39 | trans |
| miR-320c | rs6546400 | 2:6863064 | G | A | 0.554382 | 0.125493 | 0.013286 | 3.52E-21 | A | G | 0.4452 | -0.06162 | 0.004627 | 3.86E-36 | trans |
| miR-320d | rs6546400 | 2:6863064 | G | A | 0.554382 | 0.124654 | 0.013607 | 5.16E-20 | A | G | 0.4452 | -0.05905 | 0.004525 | 6.43E-35 | trans |
| miR-320e | rs6546400 | 2:6863064 | G | A | 0.554382 | 0.111156 | 0.015051 | 1.52E-13 | A | G | 0.4452 | -0.09441 | 0.006245 | 9.35E-45 | trans |
| miR-4429 | rs6546400 | 2:6863064 | G | A | 0.554382 | 0.116519 | 0.01309 | 5.51E-19 | A | G | 0.4452 | -0.06782 | 0.004365 | 7.59E-47 | trans |
| miR-320a | rs6546401 | 2:6863087 | A | T | 0.550127 | 0.153872 | 0.015747 | 1.49E-22 | T | A | 0.4522 | -0.06668 | 0.005538 | 1.95E-30 | trans |
| miR-320b | rs6546401 | 2:6863087 | A | T | 0.550127 | 0.139225 | 0.014103 | 5.49E-23 | T | A | 0.4522 | -0.0652 | 0.004634 | 1.07E-39 | trans |
| miR-320c | rs6546401 | 2:6863087 | A | T | 0.550127 | 0.126126 | 0.013226 | 1.48E-21 | T | A | 0.4522 | -0.06343 | 0.004557 | 5.38E-39 | trans |
| miR-320d | rs6546401 | 2:6863087 | A | T | 0.550127 | 0.124835 | 0.013549 | 3.15E-20 | T | A | 0.4522 | -0.06092 | 0.004458 | 8.88E-38 | trans |
| miR-320e | rs6546401 | 2:6863087 | A | T | 0.550127 | 0.113171 | 0.014981 | 4.20E-14 | T | A | 0.4522 | -0.09579 | 0.006161 | 6.20E-47 | trans |
| miR-4429 | rs6546401 | 2:6863087 | A | T | 0.550127 | 0.116694 | 0.013034 | 3.45E-19 | T | A | 0.4522 | -0.0684 | 0.004334 | 4.01E-48 | trans |
| miR-320a | rs6546403 | 2:6863226 | C | T | 0.550801 | 0.155633 | 0.015897 | 1.24E-22 | T | C | 0.4545 | -0.06698 | 0.005519 | 7.05E-31 | trans |
| miR-320b | rs6546403 | 2:6863226 | C | T | 0.550801 | 0.140899 | 0.014236 | 4.28E-23 | T | C | 0.4545 | -0.06616 | 0.004607 | 3.82E-41 | trans |
| miR-320c | rs6546403 | 2:6863226 | C | T | 0.550801 | 0.128349 | 0.013348 | 6.89E-22 | T | C | 0.4545 | -0.06388 | 0.00453 | 6.86E-40 | trans |
| miR-320d | rs6546403 | 2:6863226 | C | T | 0.550801 | 0.126407 | 0.013677 | 2.41E-20 | T | C | 0.4545 | -0.06058 | 0.004424 | 5.85E-38 | trans |
| miR-320e | rs6546403 | 2:6863226 | C | T | 0.550801 | 0.114043 | 0.015125 | 4.70E-14 | T | C | 0.4545 | -0.09624 | 0.006122 | 7.17E-48 | trans |
| miR-4429 | rs6546403 | 2:6863226 | C | T | 0.550801 | 0.118031 | 0.013158 | 2.95E-19 | T | C | 0.4545 | -0.06842 | 0.004302 | 8.46E-49 | trans |
| miR-320a | rs6546404 | 2:6863263 | G | T | 0.54995 | 0.15523 | 0.015879 | 1.43E-22 | T | G | 0.4547 | -0.06692 | 0.005516 | 7.17E-31 | trans |
| miR-320b | rs6546404 | 2:6863263 | G | T | 0.54995 | 0.140703 | 0.01422 | 4.38E-23 | T | G | 0.4547 | -0.06612 | 0.004599 | 3.10E-41 | trans |
| miR-320c | rs6546404 | 2:6863263 | G | T | 0.54995 | 0.128238 | 0.013333 | 6.69E-22 | T | G | 0.4547 | -0.06386 | 0.004521 | 4.93E-40 | trans |
| miR-320d | rs6546404 | 2:6863263 | G | T | 0.54995 | 0.126284 | 0.013661 | 2.37E-20 | T | G | 0.4547 | -0.06054 | 0.004418 | 5.22E-38 | trans |
| miR-320e | rs6546404 | 2:6863263 | G | T | 0.54995 | 0.114042 | 0.015107 | 4.39E-14 | T | G | 0.4547 | -0.09626 | 0.006109 | 4.55E-48 | trans |
| miR-4429 | rs6546404 | 2:6863263 | G | T | 0.54995 | 0.117873 | 0.013142 | 3.00E-19 | T | G | 0.4547 | -0.06846 | 0.004297 | 5.96E-49 | trans |
| miR-320a | rs6546407 | 2:6863492 | A | C | 0.55091 | 0.155844 | 0.015974 | 1.74E-22 | C | A | 0.4525 | -0.06653 | 0.005537 | 2.43E-30 | trans |
| miR-320b | rs6546407 | 2:6863492 | A | C | 0.55091 | 0.141146 | 0.014305 | 5.80E-23 | C | A | 0.4525 | -0.06512 | 0.004625 | 8.91E-40 | trans |
| miR-320c | rs6546407 | 2:6863492 | A | C | 0.55091 | 0.128789 | 0.013412 | 7.79E-22 | C | A | 0.4525 | -0.06344 | 0.004544 | 3.27E-39 | trans |
| miR-320d | rs6546407 | 2:6863492 | A | C | 0.55091 | 0.126901 | 0.013742 | 2.59E-20 | C | A | 0.4525 | -0.06088 | 0.00445 | 6.91E-38 | trans |
| miR-320e | rs6546407 | 2:6863492 | A | C | 0.55091 | 0.113961 | 0.015198 | 6.46E-14 | C | A | 0.4525 | -0.09579 | 0.006144 | 3.51E-47 | trans |
| miR-4429 | rs6546407 | 2:6863492 | A | C | 0.55091 | 0.118316 | 0.013221 | 3.58E-19 | C | A | 0.4525 | -0.06848 | 0.004328 | 2.36E-48 | trans |
| miR-320a | rs6546408 | 2:6863504 | T | C | 0.551062 | 0.155914 | 0.01599 | 1.84E-22 | C | T | 0.4525 | -0.06653 | 0.005537 | 2.43E-30 | trans |
| miR-320b | rs6546408 | 2:6863504 | T | C | 0.551062 | 0.141172 | 0.01432 | 6.31E-23 | C | T | 0.4525 | -0.06512 | 0.004625 | 8.91E-40 | trans |
| miR-320c | rs6546408 | 2:6863504 | T | C | 0.551062 | 0.128831 | 0.013426 | 8.32E-22 | C | T | 0.4525 | -0.06344 | 0.004544 | 3.27E-39 | trans |
| miR-320d | rs6546408 | 2:6863504 | T | C | 0.551062 | 0.126965 | 0.013756 | 2.71E-20 | C | T | 0.4525 | -0.06088 | 0.00445 | 6.91E-38 | trans |
| miR-320e | rs6546408 | 2:6863504 | T | C | 0.551062 | 0.11385 | 0.015214 | 7.26E-14 | C | T | 0.4525 | -0.09579 | 0.006144 | 3.51E-47 | trans |
| miR-4429 | rs6546408 | 2:6863504 | T | C | 0.551062 | 0.118342 | 0.013234 | 3.82E-19 | C | T | 0.4525 | -0.06848 | 0.004328 | 2.36E-48 | trans |
| miR-320a | rs6546409 | 2:6863587 | A | G | 0.551093 | 0.15591 | 0.015998 | 1.93E-22 | G | A | 0.4523 | -0.06634 | 0.005546 | 4.25E-30 | trans |
| miR-320b | rs6546409 | 2:6863587 | A | G | 0.551093 | 0.141154 | 0.014327 | 6.70E-23 | G | A | 0.4523 | -0.06498 | 0.004646 | 2.64E-39 | trans |
| miR-320c | rs6546409 | 2:6863587 | A | G | 0.551093 | 0.128861 | 0.013432 | 8.51E-22 | G | A | 0.4523 | -0.06295 | 0.004553 | 1.48E-38 | trans |
| miR-320d | rs6546409 | 2:6863587 | A | G | 0.551093 | 0.126997 | 0.013762 | 2.76E-20 | G | A | 0.4523 | -0.06074 | 0.004462 | 1.54E-37 | trans |
| miR-320e | rs6546409 | 2:6863587 | A | G | 0.551093 | 0.113771 | 0.015222 | 7.77E-14 | G | A | 0.4523 | -0.09552 | 0.006172 | 1.38E-46 | trans |
| miR-4429 | rs6546409 | 2:6863587 | A | G | 0.551093 | 0.118334 | 0.013241 | 4.00E-19 | G | A | 0.4523 | -0.06836 | 0.004337 | 5.00E-48 | trans |
| miR-320a | rs6546410 | 2:6863610 | C | G | 0.702434 | 0.147267 | 0.017635 | 6.78E-17 | G | C | 0.3239 | -0.05841 | 0.006234 | 1.04E-19 | trans |
| miR-320b | rs6546410 | 2:6863610 | C | G | 0.702434 | 0.133424 | 0.015795 | 2.98E-17 | G | C | 0.3239 | -0.06221 | 0.005192 | 3.57E-30 | trans |
| miR-320c | rs6546410 | 2:6863610 | C | G | 0.702434 | 0.120283 | 0.014809 | 4.58E-16 | G | C | 0.3239 | -0.05716 | 0.005155 | 2.14E-26 | trans |

| | | | | | | | | | | | | | | | |
|---|---|---|---|---|---|---|---|---|---|---|---|---|---|---|---|
| miR-320d | rs6546410 | 2:6863610! | C | G | 0.702434 | 0.12118 | 0.015157 | 1.30E-15 | G | C | 0.3239 | -0.05257 | 0.005075 | 1.85E-23 | trans |
| miR-4429 | rs6546410 | 2:6863610! | C | G | 0.702434 | 0.109252 | 0.014591 | 7.02E-14 | G | C | 0.3239 | -0.065 | 0.004893 | 5.25E-36 | trans |
| miR-320a | rs6546411 | 2:6863769! | A | G | 0.556248 | 0.157639 | 0.016098 | 1.21E-22 | G | A | 0.4439 | -0.06719 | 0.00569 | 2.30E-29 | trans |
| miR-320b | rs6546411 | 2:6863769! | A | G | 0.556248 | 0.143009 | 0.014415 | 3.38E-23 | G | A | 0.4439 | -0.06641 | 0.004763 | 5.49E-39 | trans |
| miR-320c | rs6546411 | 2:6863769! | A | G | 0.556248 | 0.130222 | 0.013516 | 5.73E-22 | G | A | 0.4439 | -0.06409 | 0.004665 | 5.01E-38 | trans |
| miR-320d | rs6546411 | 2:6863769! | A | G | 0.556248 | 0.128784 | 0.013847 | 1.40E-20 | G | A | 0.4439 | -0.06196 | 0.004585 | 5.74E-37 | trans |
| miR-320e | rs6546411 | 2:6863769! | A | G | 0.556248 | 0.116536 | 0.015313 | 2.74E-14 | G | A | 0.4439 | -0.09855 | 0.006247 | 6.61E-48 | trans |
| miR-4429 | rs6546411 | 2:6863769! | A | G | 0.556248 | 0.120734 | 0.013319 | 1.25E-19 | G | A | 0.4439 | -0.06938 | 0.00443 | 2.47E-47 | trans |
| miR-320a | rs6546412 | 2:6863771( | T | C | 0.557086 | 0.15912 | 0.016127 | 5.79E-23 | C | T | 0.4415 | -0.06695 | 0.005722 | 6.80E-29 | trans |
| miR-320b | rs6546412 | 2:6863771( | T | C | 0.557086 | 0.144247 | 0.014441 | 1.71E-23 | C | T | 0.4415 | -0.06656 | 0.004774 | 6.04E-39 | trans |
| miR-320c | rs6546412 | 2:6863771( | T | C | 0.557086 | 0.131394 | 0.013541 | 2.91E-22 | C | T | 0.4415 | -0.06397 | 0.004688 | 1.48E-37 | trans |
| miR-320d | rs6546412 | 2:6863771( | T | C | 0.557086 | 0.129709 | 0.013873 | 8.80E-21 | C | T | 0.4415 | -0.06184 | 0.004601 | 1.34E-36 | trans |
| miR-320e | rs6546412 | 2:6863771( | T | C | 0.557086 | 0.117526 | 0.015342 | 1.86E-14 | C | T | 0.4415 | -0.09844 | 0.006275 | 1.99E-47 | trans |
| miR-4429 | rs6546412 | 2:6863771( | T | C | 0.557086 | 0.121849 | 0.013344 | 6.76E-20 | C | T | 0.4415 | -0.06922 | 0.004432 | 4.48E-47 | trans |
| miR-625-5p | rs6573609 | 14:658614( | A | C | 0.390167 | 0.175181 | 0.020391 | 8.61E-18 | A | C | 0.3817 | 0.3462 | 0.02604 | 5.01E-36 | cis |
| miR-625-5p | rs6573611 | 14:658652! | G | T | 0.39802 | 0.170507 | 0.020229 | 3.49E-17 | G | T | 0.39 | 0.3333 | 0.02613 | 1.44E-33 | cis |
| miR-625-5p | rs6573612 | 14:659128. | G | A | 0.355177 | 0.173057 | 0.020731 | 6.96E-17 | G | A | 0.3191 | 0.3795 | 0.02681 | 3.78E-40 | cis |
| miR-625-5p | rs6573614 | 14:659587( | G | A | 0.356282 | 0.171846 | 0.02064 | 8.37E-17 | G | A | 0.3201 | 0.3772 | 0.02676 | 7.19E-40 | cis |
| miR-625-5p | rs6573615 | 14:660465. | G | A | 0.381045 | 0.166016 | 0.019917 | 7.73E-17 | G | A | 0.3865 | 0.3446 | 0.02619 | 1.79E-35 | cis |
| miR-625-5p | rs6573616 | 14:660468. | A | G | 0.343484 | 0.172142 | 0.020598 | 6.41E-17 | A | G | 0.3133 | 0.393 | 0.0267 | 6.99E-43 | cis |
| miR-625-5p | rs6573618 | 14:660564! | A | G | 0.344637 | 0.172132 | 0.020646 | 7.60E-17 | A | G | 0.3133 | 0.393 | 0.0267 | 6.99E-43 | cis |
| miR-625-5p | rs6573619 | 14:660615( | T | C | 0.344872 | 0.172013 | 0.020629 | 7.53E-17 | T | C | 0.3146 | 0.3844 | 0.02668 | 2.21E-41 | cis |
| miR-625-5p | rs6573624 | 14:662268( | G | A | 0.485559 | 0.132056 | 0.019672 | 1.91E-11 | A | G | 0.4833 | -0.2973 | 0.02622 | 1.94E-27 | cis |
| miR-625-5p | rs6573625 | 14:662406. | T | C | 0.37315 | 0.137387 | 0.020057 | 7.39E-12 | T | C | 0.3664 | 0.3054 | 0.02695 | 1.94E-27 | cis |
| miR-625-5p | rs6573626 | 14:662406! | G | C | 0.454572 | 0.145113 | 0.019413 | 7.72E-14 | G | C | 0.4908 | 0.3212 | 0.02595 | 7.69E-32 | cis |
| miR-625-5p | rs6573627 | 14:662523. | C | T | 0.542293 | -0.140757 | 0.019318 | 3.19E-13 | C | T | 0.5 | -0.3125 | 0.02632 | 1.21E-29 | cis |
| miR-584-5p | rs6580593 | 5:1484636( | G | C | 0.474061 | 0.188578 | 0.025475 | 1.34E-13 | G | C | 0.4437 | 0.1789 | 0.01316 | 2.04E-37 | cis |
| miR-130a-3 | rs658936 | 11:575805( | T | G | 0.440007 | -0.163641 | 0.020988 | 6.35E-15 | T | G | 0.4406 | -0.1743 | 0.01068 | 8.13E-51 | cis |
| miR-10b-5p | rs659104 | 9:1361478. | T | G | 0.454107 | -0.127786 | 0.014102 | 1.28E-19 | T | G | 0.4688 | -0.05911 | 0.01638 | 0.00033 | trans |
| miR-130a-3 | rs659165 | 11:576089! | A | C | 0.571934 | 0.161004 | 0.021115 | 2.44E-14 | C | A | 0.4192 | -0.1638 | 0.011 | 9.68E-44 | cis |
| miR-10b-5p | rs660340 | 9:1361475! | A | G | 0.454082 | -0.127916 | 0.014109 | 1.23E-19 | A | G | 0.4694 | -0.05888 | 0.01641 | 0.000358 | trans |
| miR-625-5p | rs6649234! | 14:658394( | A | C | 0.739 | -0.149853 | 0.021839 | 6.80E-12 | C | A | 0.2371 | 0.2715 | 0.03104 | 1.77E-17 | cis |
| miR-130a-3 | rs665058 | 11:575791( | T | C | 0.565581 | 0.160316 | 0.020993 | 2.23E-14 | C | T | 0.4338 | -0.1712 | 0.01072 | 3.31E-49 | cis |
| miR-181a-5 | rs6703089 | 1:1989200. | C | A | 0.236139 | 0.106716 | 0.014528 | 2.05E-13 | C | A | 0.2436 | 0.2373 | 0.01653 | 3.98E-41 | cis |
| miR-320a | rs6708536 | 2:6862726( | G | A | 0.547249 | 0.152906 | 0.015663 | 1.63E-22 | A | G | 0.4541 | -0.06727 | 0.005522 | 4.35E-31 | trans |
| miR-320b | rs6708536 | 2:6862726( | G | A | 0.547249 | 0.138343 | 0.014027 | 6.04E-23 | A | G | 0.4541 | -0.0665 | 0.004602 | 1.38E-41 | trans |
| miR-320c | rs6708536 | 2:6862726( | G | A | 0.547249 | 0.125242 | 0.013155 | 1.73E-21 | A | G | 0.4541 | -0.06396 | 0.004529 | 5.28E-40 | trans |
| miR-320d | rs6708536 | 2:6862726( | G | A | 0.547249 | 0.123687 | 0.013477 | 4.42E-20 | A | G | 0.4541 | -0.06049 | 0.00443 | 8.65E-38 | trans |
| miR-320e | rs6708536 | 2:6862726( | G | A | 0.547249 | 0.112067 | 0.014901 | 5.45E-14 | A | G | 0.4541 | -0.09639 | 0.006121 | 5.01E-48 | trans |
| miR-4429 | rs6708536 | 2:6862726( | G | A | 0.547249 | 0.115405 | 0.012966 | 5.55E-19 | A | G | 0.4541 | -0.06893 | 0.004297 | 1.62E-49 | trans |
| miR-320a | rs6709464 | 2:6862222( | C | T | 0.697279 | 0.139649 | 0.016836 | 1.09E-16 | T | C | 0.3197 | -0.05866 | 0.006214 | 5.57E-20 | trans |
| miR-320b | rs6709464 | 2:6862222( | C | T | 0.697279 | 0.126869 | 0.015077 | 3.94E-17 | T | C | 0.3197 | -0.06389 | 0.00515 | 4.61E-32 | trans |
| miR-320c | rs6709464 | 2:6862222( | C | T | 0.697279 | 0.113882 | 0.014138 | 7.96E-16 | T | C | 0.3197 | -0.05645 | 0.005139 | 5.47E-26 | trans |
| miR-320d | rs6709464 | 2:6862222( | C | T | 0.697279 | 0.11635 | 0.014465 | 8.71E-16 | T | C | 0.3197 | -0.05142 | 0.005035 | 6.62E-23 | trans |
| miR-4429 | rs6709464 | 2:6862222( | C | T | 0.697279 | 0.10404 | 0.013928 | 8.03E-14 | T | C | 0.3197 | -0.06381 | 0.004877 | 4.00E-35 | trans |
| miR-320a | rs6714719 | 2:6859377. | G | C | 0.698404 | 0.139079 | 0.016929 | 2.11E-16 | C | G | 0.3167 | -0.05686 | 0.006263 | 1.12E-18 | trans |
| miR-320b | rs6714719 | 2:6859377. | G | C | 0.698404 | 0.12667 | 0.015159 | 6.50E-17 | C | G | 0.3167 | -0.06312 | 0.005188 | 5.19E-31 | trans |
| miR-320c | rs6714719 | 2:6859377. | G | C | 0.698404 | 0.113511 | 0.014216 | 1.41E-15 | C | G | 0.3167 | -0.05612 | 0.005157 | 1.41E-25 | trans |
| miR-320d | rs6714719 | 2:6859377. | G | C | 0.698404 | 0.117036 | 0.01454 | 8.33E-16 | C | G | 0.3167 | -0.05128 | 0.005055 | 1.22E-22 | trans |
| miR-4429 | rs6714719 | 2:6859377. | G | C | 0.698404 | 0.104022 | 0.014003 | 1.10E-13 | C | G | 0.3167 | -0.06242 | 0.00491 | 1.92E-33 | trans |
| miR-130a-3 | rs671508 | 11:576365( | A | T | 0.438934 | -0.161761 | 0.02122 | 2.48E-14 | A | T | 0.4305 | -0.1703 | 0.01074 | 1.59E-48 | cis |
| miR-320a | rs6715579 | 2:6858392. | C | T | 0.678794 | 0.125847 | 0.017319 | 3.69E-13 | T | C | 0.3379 | -0.05223 | 0.006301 | 5.98E-16 | trans |
| miR-320b | rs6715579 | 2:6858392. | C | T | 0.678794 | 0.117162 | 0.015502 | 4.09E-14 | T | C | 0.3379 | -0.0602 | 0.00522 | 2.98E-28 | trans |
| miR-320c | rs6715579 | 2:6858392. | C | T | 0.678794 | 0.10413 | 0.014536 | 7.87E-13 | T | C | 0.3379 | -0.0518 | 0.005227 | 9.89E-22 | trans |
| miR-320d | rs6715579 | 2:6858392. | C | T | 0.678794 | 0.108857 | 0.014863 | 2.41E-13 | T | C | 0.3379 | -0.04869 | 0.005104 | 2.42E-20 | trans |
| miR-4429 | rs6715579 | 2:6858392. | C | T | 0.678794 | 0.096668 | 0.014309 | 1.42E-11 | T | C | 0.3379 | -0.05925 | 0.004971 | 6.66E-30 | trans |
| miR-320a | rs6715706 | 2:6858399( | C | A | 0.664887 | 0.131667 | 0.01702 | 1.03E-14 | A | C | 0.3513 | -0.04948 | 0.006255 | 1.02E-14 | trans |
| miR-320b | rs6715706 | 2:6858399( | C | A | 0.664887 | 0.120184 | 0.015241 | 3.13E-15 | A | C | 0.3513 | -0.05865 | 0.005177 | 2.18E-27 | trans |
| miR-320c | rs6715706 | 2:6858399( | C | A | 0.664887 | 0.10796 | 0.01429 | 4.18E-14 | A | C | 0.3513 | -0.04956 | 0.005176 | 1.81E-20 | trans |
| miR-320d | rs6715706 | 2:6858399( | C | A | 0.664887 | 0.108852 | 0.014624 | 9.82E-14 | A | C | 0.3513 | -0.04678 | 0.00505 | 2.50E-19 | trans |
| miR-4429 | rs6715706 | 2:6858399( | C | A | 0.664887 | 0.098723 | 0.014073 | 2.30E-12 | A | C | 0.3513 | -0.05726 | 0.004911 | 8.75E-29 | trans |
| miR-320a | rs6715909 | 2:6858399! | G | A | 0.664879 | 0.131663 | 0.01702 | 1.03E-14 | A | G | 0.3513 | -0.04948 | 0.006255 | 1.02E-14 | trans |
| miR-320b | rs6715909 | 2:6858399! | G | A | 0.664879 | 0.120181 | 0.015242 | 3.14E-15 | A | G | 0.3513 | -0.05865 | 0.005177 | 2.18E-27 | trans |
| miR-320c | rs6715909 | 2:6858399! | G | A | 0.664879 | 0.107953 | 0.01429 | 4.21E-14 | A | G | 0.3513 | -0.04956 | 0.005176 | 1.81E-20 | trans |
| miR-320d | rs6715909 | 2:6858399! | G | A | 0.664879 | 0.108849 | 0.014625 | 9.85E-14 | A | G | 0.3513 | -0.04678 | 0.00505 | 2.50E-19 | trans |
| miR-4429 | rs6715909 | 2:6858399! | G | A | 0.664879 | 0.098726 | 0.014073 | 2.30E-12 | A | G | 0.3513 | -0.05726 | 0.004911 | 8.75E-29 | trans |
| miR-320a | rs6729555 | 2:6863295. | C | A | 0.54995 | 0.155269 | 0.015883 | 1.43E-22 | A | C | 0.4547 | -0.06692 | 0.005516 | 7.17E-31 | trans |
| miR-320b | rs6729555 | 2:6863295. | C | A | 0.54995 | 0.140731 | 0.014223 | 4.41E-23 | A | C | 0.4547 | -0.06612 | 0.004599 | 3.10E-41 | trans |
| miR-320c | rs6729555 | 2:6863295. | C | A | 0.54995 | 0.128279 | 0.013336 | 6.64E-22 | A | C | 0.4547 | -0.06386 | 0.004521 | 4.93E-40 | trans |
| miR-320d | rs6729555 | 2:6863295. | C | A | 0.54995 | 0.126323 | 0.013664 | 2.36E-20 | A | C | 0.4547 | -0.06054 | 0.004418 | 5.22E-38 | trans |
| miR-320e | rs6729555 | 2:6863295. | C | A | 0.54995 | 0.114033 | 0.015111 | 4.47E-14 | A | C | 0.4547 | -0.09626 | 0.006109 | 4.55E-48 | trans |
| miR-4429 | rs6729555 | 2:6863295. | C | A | 0.54995 | 0.117896 | 0.013146 | 3.01E-19 | A | C | 0.4547 | -0.06846 | 0.004297 | 5.96E-49 | trans |
| miR-320a | rs6729686 | 2:6863303( | C | T | 0.700171 | 0.145857 | 0.017461 | 6.63E-17 | T | C | 0.3276 | -0.05868 | 0.006169 | 3.00E-20 | trans |
| miR-320b | rs6729686 | 2:6863303( | C | T | 0.700171 | 0.132218 | 0.015638 | 2.80E-17 | T | C | 0.3276 | -0.06324 | 0.005124 | 8.82E-32 | trans |
| miR-320c | rs6729686 | 2:6863303( | C | T | 0.700171 | 0.119056 | 0.014663 | 4.69E-16 | T | C | 0.3276 | -0.05743 | 0.005094 | 3.42E-27 | trans |
| miR-320d | rs6729686 | 2:6863303( | C | T | 0.700171 | 0.119939 | 0.015008 | 1.33E-15 | T | C | 0.3276 | -0.05189 | 0.005006 | 1.67E-23 | trans |
| miR-4429 | rs6729686 | 2:6863303( | C | T | 0.700171 | 0.108074 | 0.014448 | 7.41E-14 | T | C | 0.3276 | -0.06445 | 0.004835 | 2.92E-36 | trans |
| miR-320a | rs6742200 | 2:6862330. | A | G | 0.54552 | 0.150787 | 0.015557 | 3.25E-22 | G | A | 0.4541 | -0.06727 | 0.005522 | 4.35E-31 | trans |
| miR-320b | rs6742200 | 2:6862330. | A | G | 0.54552 | 0.136507 | 0.013932 | 1.15E-22 | G | A | 0.4541 | -0.0665 | 0.004602 | 1.38E-41 | trans |
| miR-320c | rs6742200 | 2:6862330. | A | G | 0.54552 | 0.123535 | 0.013066 | 3.25E-21 | G | A | 0.4541 | -0.06396 | 0.004529 | 5.28E-40 | trans |
| miR-320d | rs6742200 | 2:6862330. | A | G | 0.54552 | 0.122264 | 0.013385 | 6.58E-20 | G | A | 0.4541 | -0.06049 | 0.00443 | 8.65E-38 | trans |
| miR-320e | rs6742200 | 2:6862330. | A | G | 0.54552 | 0.110827 | 0.014798 | 6.92E-14 | G | A | 0.4541 | -0.09639 | 0.006121 | 5.01E-48 | trans |
| miR-4429 | rs6742200 | 2:6862330. | A | G | 0.54552 | 0.114117 | 0.012877 | 7.84E-19 | G | A | 0.4541 | -0.06893 | 0.004297 | 1.62E-49 | trans |
| let-7b-5p | rs674302 | 9:1361466( | T | A | 0.663989 | -0.107892 | 0.010058 | 7.63E-27 | A | T | 0.3219 | 0.03377 | 0.006655 | 4.99E-07 | trans |
| let-7c-5p | rs674302 | 9:1361466( | T | A | 0.663989 | -0.091475 | 0.009184 | 2.28E-23 | A | T | 0.3219 | 0.02445 | 0.006395 | 0.000144 | trans |
| miR-10b-5p | rs674302 | 9:1361466( | T | A | 0.663989 | -0.177203 | 0.014295 | 2.75E-35 | A | T | 0.3219 | 0.1135 | 0.01684 | 3.31E-11 | trans |

| | | | | | | | | | | | | | | | |
|---|---|---|---|---|---|---|---|---|---|---|---|---|---|---|---|
| miR-126-3p | rs674302 | 9:1361466( | T | A | 0.663989 | -0.171502 | 0.013055 | 2.03E-39 | A | T | 0.3219 | 0.05294 | 0.008279 | 2.95E-10 | trans |
| miR-139-5p | rs674302 | 9:1361466( | T | A | 0.663989 | -0.157335 | 0.011595 | 6.05E-42 | A | T | 0.3219 | 0.0709 | 0.01641 | 1.79E-05 | trans |
| miR-195-5p | rs674302 | 9:1361466( | T | A | 0.663989 | -0.113981 | 0.013405 | 1.85E-17 | A | T | 0.3219 | 0.06692 | 0.01825 | 0.000265 | trans |
| miR-320a | rs6743970 | 2:6858390 | A | T | 0.660237 | 0.133352 | 0.017104 | 6.37E-15 | T | A | 0.3589 | -0.05102 | 0.006281 | 2.18E-15 | trans |
| miR-320b | rs6743970 | 2:6858390 | A | T | 0.660237 | 0.122904 | 0.015312 | 1.00E-15 | T | A | 0.3589 | -0.0592 | 0.005228 | 2.50E-27 | trans |
| miR-320c | rs6743970 | 2:6858390 | A | T | 0.660237 | 0.110494 | 0.014356 | 1.40E-14 | T | A | 0.3589 | -0.05156 | 0.005245 | 2.13E-21 | trans |
| miR-320d | rs6743970 | 2:6858390 | A | T | 0.660237 | 0.111589 | 0.014692 | 3.07E-14 | T | A | 0.3589 | -0.0474 | 0.00512 | 2.78E-19 | trans |
| miR-4429 | rs6743970 | 2:6858390 | A | T | 0.660237 | 0.101973 | 0.014137 | 5.46E-13 | T | A | 0.3589 | -0.0589 | 0.004965 | 1.32E-29 | trans |
| miR-320a | rs6744306 | 2:6863273( | T | C | 0.549956 | 0.155241 | 0.01588 | 1.43E-22 | C | T | 0.4547 | -0.06692 | 0.005516 | 7.17E-31 | trans |
| miR-320b | rs6744306 | 2:6863273( | T | C | 0.549956 | 0.140712 | 0.01422 | 4.37E-23 | C | T | 0.4547 | -0.06612 | 0.004599 | 3.10E-41 | trans |
| miR-320c | rs6744306 | 2:6863273( | T | C | 0.549956 | 0.128249 | 0.013333 | 6.66E-22 | C | T | 0.4547 | -0.06386 | 0.004521 | 4.93E-40 | trans |
| miR-320d | rs6744306 | 2:6863273( | T | C | 0.549956 | 0.126294 | 0.013662 | 2.36E-20 | C | T | 0.4547 | -0.06054 | 0.004418 | 5.22E-38 | trans |
| miR-320e | rs6744306 | 2:6863273( | T | C | 0.549956 | 0.114047 | 0.015107 | 4.38E-14 | C | T | 0.4547 | -0.09626 | 0.006109 | 4.55E-48 | trans |
| miR-4429 | rs6744306 | 2:6863273( | T | C | 0.549956 | 0.117881 | 0.013143 | 2.99E-19 | C | T | 0.4547 | -0.06846 | 0.004297 | 5.96E-49 | trans |
| miR-320a | rs6747545 | 2:6863306 | T | A | 0.549964 | 0.155257 | 0.015884 | 1.45E-22 | A | T | 0.4547 | -0.06692 | 0.005516 | 7.17E-31 | trans |
| miR-320b | rs6747545 | 2:6863306 | T | A | 0.549964 | 0.140721 | 0.014224 | 4.46E-23 | A | T | 0.4547 | -0.06612 | 0.004599 | 3.10E-41 | trans |
| miR-320c | rs6747545 | 2:6863306 | T | A | 0.549964 | 0.128276 | 0.013337 | 6.70E-22 | A | T | 0.4547 | -0.06386 | 0.004521 | 4.93E-40 | trans |
| miR-320d | rs6747545 | 2:6863306 | T | A | 0.549964 | 0.126319 | 0.013665 | 2.38E-20 | A | T | 0.4547 | -0.06054 | 0.004418 | 5.22E-38 | trans |
| miR-320e | rs6747545 | 2:6863306 | T | A | 0.549964 | 0.114028 | 0.015112 | 4.50E-14 | A | T | 0.4547 | -0.09626 | 0.006109 | 4.55E-48 | trans |
| miR-4429 | rs6747545 | 2:6863306 | T | A | 0.549964 | 0.11789 | 0.013147 | 3.04E-19 | A | T | 0.4547 | -0.06846 | 0.004297 | 5.96E-49 | trans |
| miR-320a | rs6757938 | 2:6863285 | A | G | 0.549958 | 0.155246 | 0.015882 | 1.44E-22 | G | A | 0.4547 | -0.06692 | 0.005516 | 7.17E-31 | trans |
| miR-320b | rs6757938 | 2:6863285 | A | G | 0.549958 | 0.140714 | 0.014222 | 4.42E-23 | G | A | 0.4547 | -0.06612 | 0.004599 | 3.10E-41 | trans |
| miR-320c | rs6757938 | 2:6863285 | A | G | 0.549958 | 0.128259 | 0.013335 | 6.69E-22 | G | A | 0.4547 | -0.06386 | 0.004521 | 4.93E-40 | trans |
| miR-320d | rs6757938 | 2:6863285 | A | G | 0.549958 | 0.126303 | 0.013663 | 2.38E-20 | G | A | 0.4547 | -0.06054 | 0.004418 | 5.22E-38 | trans |
| miR-320e | rs6757938 | 2:6863285 | A | G | 0.549958 | 0.114041 | 0.015109 | 4.43E-14 | G | A | 0.4547 | -0.09626 | 0.006109 | 4.55E-48 | trans |
| miR-4429 | rs6757938 | 2:6863285 | A | G | 0.549958 | 0.117883 | 0.013145 | 3.02E-19 | G | A | 0.4547 | -0.06846 | 0.004297 | 5.96E-49 | trans |
| let-7b-5p | rs676457 | 9:1361462 | A | T | 0.664166 | -0.107822 | 0.010067 | 9.10E-27 | T | A | 0.3219 | 0.03377 | 0.006655 | 4.99E-07 | trans |
| let-7c-5p | rs676457 | 9:1361462 | A | T | 0.664166 | -0.091483 | 0.009192 | 2.46E-23 | T | A | 0.3219 | 0.02445 | 0.006395 | 0.000144 | trans |
| miR-10b-5p | rs676457 | 9:1361462 | A | T | 0.664166 | -0.177236 | 0.014307 | 3.04E-35 | T | A | 0.3219 | 0.1135 | 0.01684 | 3.31E-11 | trans |
| miR-126-3p | rs676457 | 9:1361462 | A | T | 0.664166 | -0.171461 | 0.013067 | 2.46E-39 | T | A | 0.3219 | 0.05294 | 0.008279 | 2.95E-10 | trans |
| miR-139-5p | rs676457 | 9:1361462 | A | T | 0.664166 | -0.15736 | 0.011604 | 6.87E-42 | T | A | 0.3219 | 0.0709 | 0.01641 | 1.79E-05 | trans |
| miR-195-5p | rs676457 | 9:1361462 | A | T | 0.664166 | -0.113865 | 0.013416 | 2.12E-17 | T | A | 0.3219 | 0.06692 | 0.01825 | 0.000265 | trans |
| let-7b-5p | rs676996 | 9:1361460 | T | G | 0.667516 | -0.108022 | 0.010153 | 1.96E-26 | G | T | 0.3114 | 0.03245 | 0.006779 | 2.08E-06 | trans |
| let-7c-5p | rs676996 | 9:1361460 | T | G | 0.667516 | -0.091652 | 0.00927 | 4.76E-23 | G | T | 0.3114 | 0.02428 | 0.006521 | 0.000213 | trans |
| miR-10b-5p | rs676996 | 9:1361460 | T | G | 0.667516 | -0.177525 | 0.014432 | 8.97E-35 | G | T | 0.3114 | 0.1107 | 0.01725 | 2.59E-10 | trans |
| miR-126-3p | rs676996 | 9:1361460 | T | G | 0.667516 | -0.172281 | 0.013178 | 4.67E-39 | G | T | 0.3114 | 0.05122 | 0.008368 | 1.58E-09 | trans |
| miR-139-5p | rs676996 | 9:1361460 | T | G | 0.667516 | -0.157346 | 0.011708 | 3.57E-41 | G | T | 0.3114 | 0.065 | 0.01689 | 0.00013 | trans |
| miR-195-5p | rs676996 | 9:1361460 | T | G | 0.667516 | -0.114229 | 0.013529 | 3.08E-17 | G | T | 0.3114 | 0.07026 | 0.01873 | 0.000191 | trans |
| let-7b-5p | rs677355 | 9:1361460 | G | A | 0.670198 | -0.108649 | 0.010242 | 2.73E-26 | A | G | 0.3097 | 0.03356 | 0.006813 | 1.06E-06 | trans |
| let-7c-5p | rs677355 | 9:1361460 | G | A | 0.670198 | -0.092257 | 0.009351 | 5.81E-23 | A | G | 0.3097 | 0.02433 | 0.006498 | 0.000197 | trans |
| miR-10b-5p | rs677355 | 9:1361460 | G | A | 0.670198 | -0.178902 | 0.014556 | 1.02E-34 | A | G | 0.3097 | 0.1127 | 0.01734 | 1.56E-10 | trans |
| miR-126-3p | rs677355 | 9:1361460 | G | A | 0.670198 | -0.172718 | 0.013297 | 1.41E-38 | A | G | 0.3097 | 0.05274 | 0.008451 | 7.73E-10 | trans |
| miR-139-5p | rs677355 | 9:1361460 | G | A | 0.670198 | -0.157852 | 0.011814 | 1.01E-40 | A | G | 0.3097 | 0.0686 | 0.01694 | 5.74E-05 | trans |
| miR-195-5p | rs677355 | 9:1361460 | G | A | 0.670198 | -0.114344 | 0.013648 | 5.38E-17 | A | G | 0.3097 | 0.07017 | 0.01887 | 0.000217 | trans |
| miR-625-5p | rs6795753 | 14:6612458 | T | G | 0.50137 | 0.146801 | 0.01989 | 1.58E-13 | G | T | 0.4782 | -0.3154 | 0.02589 | 4.59E-31 | cis |
| miR-130a-3 | rs681397 | 11:5768201 | G | T | 0.558507 | 0.14526 | 0.021263 | 8.41E-12 | T | G | 0.4416 | -0.1444 | 0.01097 | 2.44E-35 | cis |
| miR-130a-3 | rs682258 | 11:5763719 | G | T | 0.438944 | -0.161767 | 0.021221 | 2.48E-14 | G | T | 0.4305 | -0.1703 | 0.01074 | 1.59E-48 | cis |
| miR-584-5p | rs6861404 | 5:1484158 | A | C | 0.536756 | -0.173683 | 0.02523 | 5.82E-12 | C | A | 0.4542 | 0.2102 | 0.01287 | 7.21E-51 | cis |
| miR-584-5p | rs6866101 | 5:1484159 | T | C | 0.537249 | -0.173276 | 0.025199 | 6.14E-12 | C | T | 0.4519 | 0.2084 | 0.0128 | 1.15E-50 | cis |
| let-7b-5p | rs687289 | 9:1361371( | G | A | 0.668579 | -0.111294 | 0.010186 | 8.61E-28 | A | G | 0.321 | 0.03407 | 0.006661 | 4.05E-07 | trans |
| let-7c-5p | rs687289 | 9:1361371( | G | A | 0.668579 | -0.093907 | 0.009304 | 5.91E-24 | A | G | 0.321 | 0.02518 | 0.006394 | 9.04E-05 | trans |
| miR-10b-5p | rs687289 | 9:1361371( | G | A | 0.668579 | -0.183745 | 0.014466 | 5.80E-37 | A | G | 0.321 | 0.1139 | 0.01686 | 2.97E-11 | trans |
| miR-126-3p | rs687289 | 9:1361371( | G | A | 0.668579 | -0.177674 | 0.01321 | 3.06E-41 | A | G | 0.321 | 0.05269 | 0.008289 | 3.73E-10 | trans |
| miR-139-5p | rs687289 | 9:1361371( | G | A | 0.668579 | -0.161876 | 0.011737 | 2.87E-43 | A | G | 0.321 | 0.06887 | 0.01644 | 3.16E-05 | trans |
| miR-195-5p | rs687289 | 9:1361371( | G | A | 0.668579 | -0.116352 | 0.013584 | 1.08E-17 | A | G | 0.321 | 0.06877 | 0.01825 | 0.000179 | trans |
| miR-584-5p | rs6873011 | 5:1484141 | G | A | 0.842515 | -0.259329 | 0.034393 | 4.70E-14 | A | G | 0.1546 | 0.2678 | 0.01753 | 1.35E-45 | cis |
| miR-584-5p | rs6874343 | 5:1484478( | C | T | 0.844049 | -0.271414 | 0.034877 | 7.14E-15 | T | C | 0.1546 | 0.2727 | 0.01754 | 5.95E-47 | cis |
| miR-10a-5p | rs7036642 | 9:1361446 | G | A | 0.618956 | 0.106822 | 0.014464 | 1.52E-13 | A | G | 0.3931 | -0.05078 | 0.0133 | 0.000147 | trans |
| miR-126-3p | rs7036642 | 9:1361446 | G | A | 0.618956 | 0.114538 | 0.012952 | 9.29E-19 | A | G | 0.3931 | -0.03251 | 0.007841 | 3.80E-05 | trans |
| miR-10a-5p | rs7046674 | 9:1361470 | C | T | 0.619013 | 0.107787 | 0.014473 | 9.50E-14 | T | C | 0.391 | -0.04984 | 0.01333 | 0.0002 | trans |
| miR-126-3p | rs7046674 | 9:1361470 | C | T | 0.619013 | 0.115101 | 0.01296 | 6.62E-19 | T | C | 0.391 | -0.03305 | 0.007872 | 3.04E-05 | trans |
| miR-130a-3 | rs708228 | 11:5758560 | C | T | 0.684575 | -0.241919 | 0.022159 | 9.51E-28 | T | C | 0.3051 | 0.2285 | 0.01052 | 2.46E-80 | cis |
| miR-130a-3 | rs7105506 | 11:5749798 | G | A | 0.56028 | 0.172703 | 0.021219 | 3.98E-16 | A | G | 0.4304 | -0.1709 | 0.01079 | 2.09E-48 | cis |
| miR-130a-3 | rs7116341 | 11:5748115 | G | A | 0.685526 | -0.243004 | 0.022225 | 7.92E-28 | A | G | 0.303 | 0.232 | 0.01052 | 4.91E-82 | cis |
| miR-130a-3 | rs7117020 | 11:5749965 | T | G | 0.553432 | 0.16611 | 0.021055 | 3.04E-15 | G | T | 0.4418 | -0.1775 | 0.01062 | 5.65E-53 | cis |
| miR-130a-3 | rs7117896 | 11:5738855 | G | A | 0.746843 | -0.302337 | 0.023182 | 7.07E-39 | A | G | 0.2712 | 0.2872 | 0.009048 | 3.81E-136 | cis |
| miR-130a-3 | rs7120868 | 11:5753057 | T | G | 0.566777 | 0.161451 | 0.021059 | 1.77E-14 | G | T | 0.4321 | -0.1711 | 0.01085 | 4.11E-48 | cis |
| miR-130a-3 | rs7121169 | 11:5745254 | G | A | 0.687601 | -0.246593 | 0.022277 | 1.77E-28 | A | G | 0.301 | 0.2348 | 0.01043 | 1.80E-84 | cis |
| miR-4707-3 | rs712486 | 14:2342504 | T | C | 0.441215 | 0.094678 | 0.013749 | 5.73E-12 | T | C | 0.4387 | 0.3003 | 0.01619 | 2.39E-62 | cis |
| miR-625-5p | rs7140341 | 14:6607041 | C | G | 0.3503 | 0.181513 | 0.020699 | 1.80E-18 | C | G | 0.3238 | 0.4146 | 0.0263 | 7.15E-48 | cis |
| miR-625-5p | rs7140695 | 14:6592913 | T | C | 0.350881 | 0.17535 | 0.020807 | 3.53E-17 | T | C | 0.313 | 0.392 | 0.02655 | 3.79E-43 | cis |
| miR-625-5p | rs7141536 | 14:6598737 | T | A | 0.349283 | 0.174077 | 0.020739 | 4.71E-17 | T | A | 0.3147 | 0.3905 | 0.02672 | 2.37E-42 | cis |
| miR-625-5p | rs7141874 | 14:6582199 | G | A | 0.071326 | 0.265544 | 0.038518 | 5.42E-12 | G | A | 0.07874 | 0.4573 | 0.04728 | 8.08E-21 | cis |
| miR-625-5p | rs7142165 | 14:6625023 | G | A | 0.550826 | -0.141461 | 0.019388 | 2.95E-13 | A | G | 0.4927 | 0.321 | 0.026 | 1.04E-31 | cis |
| miR-625-5p | rs7142219 | 14:6624178 | G | A | 0.452285 | 0.14296 | 0.019477 | 2.13E-13 | G | A | 0.4868 | 0.3022 | 0.02599 | 1.29E-28 | cis |
| miR-625-5p | rs7142651 | 14:6613277 | C | G | 0.502009 | 0.147599 | 0.019835 | 9.98E-14 | G | C | 0.4765 | -0.3158 | 0.02572 | 1.63E-31 | cis |
| miR-136-5p | rs7142683 | 14:1011588 | A | G | 0.290748 | -0.392753 | 0.038925 | 6.11E-24 | A | G | 0.2819 | -0.065 | 0.01387 | 3.38E-06 | cis |
| miR-625-5p | rs7142906 | 14:6595645 | A | T | 0.360633 | 0.183028 | 0.020527 | 4.82E-19 | A | T | 0.3408 | 0.4237 | 0.02516 | 1.33E-53 | cis |
| miR-625-5p | rs7143109 | 14:6580107 | G | A | 0.269639 | 0.305978 | 0.021066 | 8.46E-48 | G | A | 0.3135 | 0.5818 | 0.02044 | 1.30E-117 | cis |
| miR-625-5p | rs7143318 | 14:6594068 | G | A | 0.35859 | 0.169987 | 0.020679 | 2.03E-16 | G | A | 0.321 | 0.3772 | 0.02676 | 7.19E-40 | cis |
| miR-625-5p | rs7143666 | 14:6580997 | A | C | 0.348575 | 0.20683 | 0.019826 | 1.77E-25 | A | C | 0.3695 | 0.4405 | 0.02421 | 1.22E-60 | cis |
| miR-625-5p | rs7144345 | 14:6611184 | G | C | 0.345782 | 0.173329 | 0.020876 | 1.02E-16 | G | C | 0.3134 | 0.3917 | 0.02666 | 9.02E-43 | cis |
| miR-625-5p | rs7144530 | 14:6584050 | A | G | 0.053251 | 0.327275 | 0.045719 | 8.16E-13 | A | G | 0.05727 | 0.5836 | 0.05495 | 1.66E-24 | cis |
| miR-625-5p | rs7144971 | 14:6612065 | G | A | 0.349544 | 0.175451 | 0.020879 | 4.34E-17 | G | A | 0.3135 | 0.3916 | 0.02678 | 2.14E-42 | cis |
| miR-625-5p | rs7145500 | 14:6588421 | G | A | 0.354536 | 0.173427 | 0.020668 | 4.81E-17 | G | A | 0.322 | 0.3773 | 0.02656 | 2.65E-40 | cis |
| miR-625-5p | rs7145759 | 14:6588453 | C | A | 0.353283 | 0.175197 | 0.0207 | 2.60E-17 | C | A | 0.3175 | 0.3822 | 0.02655 | 3.20E-41 | cis |

| | | | | | | | | | | | | | | | |
|---|---|---|---|---|---|---|---|---|---|---|---|---|---|---|---|
| miR-625-5p | rs7146742 | 14:6603293 | G | A | 0.435936 | 0.139288 | 0.020069 | 3.91E-12 | G | A | 0.4211 | 0.2881 | 0.02646 | 1.47E-25 | cis |
| miR-625-5p | rs7146829 | 14:6619453 | T | C | 0.488699 | 0.132266 | 0.019792 | 2.34E-11 | C | T | 0.4815 | -0.2981 | 0.02602 | 6.00E-28 | cis |
| miR-625-5p | rs7146993 | 14:6604887 | C | T | 0.343625 | 0.171579 | 0.020581 | 7.63E-17 | C | T | 0.3155 | 0.3822 | 0.0267 | 6.22E-41 | cis |
| miR-625-5p | rs7147002 | 14:6603254 | T | C | 0.342789 | 0.172498 | 0.020658 | 6.82E-17 | T | C | 0.312 | 0.393 | 0.0267 | 6.99E-43 | cis |
| miR-625-5p | rs7147536 | 14:6587469 | C | T | 0.367002 | 0.185223 | 0.020461 | 1.39E-19 | C | T | 0.3473 | 0.4223 | 0.02506 | 1.68E-53 | cis |
| miR-625-5p | rs7147636 | 14:6601118 | T | C | 0.36432 | 0.172044 | 0.020591 | 6.53E-17 | T | C | 0.3406 | 0.3792 | 0.02597 | 2.83E-42 | cis |
| miR-130a-3 | rs7148461 | 11:5748889 | G | A | 0.689792 | -0.242969 | 0.022416 | 2.25E-27 | A | G | 0.289 | 0.2275 | 0.01095 | 1.45E-74 | cis |
| miR-130a-3 | rs7148468 | 11:5763279 | G | C | 0.683543 | -0.241608 | 0.02258 | 1.02E-26 | C | G | 0.3019 | 0.2304 | 0.01054 | 9.27E-81 | cis |
| miR-625-5p | rs7148737 | 14:6623003 | C | A | 0.386987 | 0.146174 | 0.019789 | 1.51E-13 | C | A | 0.3929 | 0.3496 | 0.02609 | 1.52E-36 | cis |
| miR-136-5p | rs7149242 | 14:1011594 | T | G | 0.288371 | -0.392035 | 0.038812 | 5.48E-24 | T | G | 0.2817 | -0.06438 | 0.01382 | 3.82E-06 | cis |
| miR-625-5p | rs7149301 | 14:6623041 | A | G | 0.452987 | 0.142786 | 0.019331 | 1.51E-13 | A | G | 0.4923 | 0.3222 | 0.02571 | 1.26E-32 | cis |
| miR-625-5p | rs7150094 | 14:6593636 | A | G | 0.346839 | 0.175864 | 0.020851 | 3.33E-17 | A | G | 0.3119 | 0.3939 | 0.02667 | 3.91E-43 | cis |
| miR-625-5p | rs7150448 | 14:6587150 | C | T | 0.345581 | 0.17931 | 0.020872 | 8.63E-18 | C | T | 0.3121 | 0.4006 | 0.02648 | 8.63E-45 | cis |
| miR-625-5p | rs7150648 | 14:6589013 | T | C | 0.347708 | 0.175934 | 0.020902 | 3.87E-17 | T | C | 0.3144 | 0.3917 | 0.02647 | 3.07E-43 | cis |
| miR-625-5p | rs7151561 | 14:6589060 | T | C | 0.350627 | 0.175974 | 0.020911 | 3.92E-17 | T | C | 0.3159 | 0.3947 | 0.02649 | 1.03E-43 | cis |
| miR-625-5p | rs7151846 | 14:6625578 | C | G | 0.539502 | -0.138708 | 0.019301 | 6.65E-13 | C | G | 0.4967 | -0.3135 | 0.02629 | 6.96E-30 | cis |
| miR-625-5p | rs7152023 | 14:6616082 | C | A | 0.425722 | 0.183153 | 0.02014 | 9.56E-20 | C | A | 0.4383 | 0.3855 | 0.02437 | 2.14E-48 | cis |
| miR-625-5p | rs7153297 | 14:6600340 | T | A | 0.51171 | 0.14732 | 0.019892 | 1.30E-13 | A | T | 0.4693 | -0.309 | 0.02589 | 5.40E-30 | cis |
| miR-625-5p | rs7155541 | 14:6604624 | G | C | 0.501962 | 0.146856 | 0.019792 | 1.17E-13 | C | G | 0.4771 | -0.317 | 0.02573 | 1.07E-31 | cis |
| miR-625-5p | rs7156078 | 14:6623870 | G | A | 0.386926 | 0.147416 | 0.019811 | 9.98E-14 | G | A | 0.3919 | 0.348 | 0.02598 | 1.43E-36 | cis |
| miR-625-5p | rs7157006 | 14:6582820 | C | T | 0.387351 | 0.198515 | 0.019331 | 9.71E-25 | C | T | 0.4303 | 0.4102 | 0.02403 | 8.04E-55 | cis |
| miR-625-5p | rs7157109 | 14:6590043 | C | G | 0.354575 | 0.174565 | 0.020746 | 3.94E-17 | C | G | 0.3199 | 0.3758 | 0.02675 | 1.29E-39 | cis |
| miR-625-5p | rs7157449 | 14:6624013 | T | G | 0.454044 | 0.144106 | 0.019397 | 1.09E-13 | T | G | 0.4909 | 0.3186 | 0.02562 | 3.53E-32 | cis |
| miR-625-5p | rs7158153 | 14:6603458 | C | A | 0.345985 | 0.173233 | 0.02063 | 4.57E-17 | C | A | 0.3127 | 0.3906 | 0.02665 | 1.42E-42 | cis |
| miR-625-5p | rs7158347 | 14:6582895 | C | T | 0.388449 | 0.198782 | 0.019394 | 1.19E-24 | C | T | 0.4303 | 0.4102 | 0.02403 | 8.04E-55 | cis |
| miR-625-5p | rs7158556 | 14:6624072 | T | A | 0.373139 | 0.137414 | 0.020058 | 7.35E-12 | T | A | 0.3664 | 0.3054 | 0.02695 | 1.94E-27 | cis |
| miR-625-5p | rs7160780 | 14:6605271 | G | T | 0.386614 | 0.16716 | 0.020127 | 9.96E-17 | G | T | 0.3839 | 0.3378 | 0.02617 | 2.46E-34 | cis |
| miR-625-5p | rs7161123 | 14:6605290 | G | A | 0.498649 | 0.145335 | 0.01972 | 1.71E-13 | A | G | 0.4764 | -0.3128 | 0.02576 | 6.67E-31 | cis |
| miR-3130-3 | rs725980 | 2:20763858 | C | G | 0.843297 | -0.271139 | 0.032939 | 1.85E-16 | G | C | 0.1794 | 0.1673 | 0.03129 | 1.21E-07 | cis |
| miR-625-5p | rs72712464 | 14:6575628 | A | T | 0.847244 | -0.322051 | 0.027281 | 3.68E-32 | T | A | 0.1491 | 0.6661 | 0.02971 | 1.26E-83 | cis |
| miR-625-5p | rs72712473 | 14:6577712 | G | A | 0.846937 | -0.32457 | 0.026357 | 7.60E-35 | A | G | 0.1553 | 0.6917 | 0.0281 | 1.55E-96 | cis |
| miR-625-5p | rs72712485 | 14:6578958 | A | T | 0.845779 | -0.326679 | 0.026156 | 8.48E-36 | T | A | 0.1565 | 0.6921 | 0.02811 | 7.93E-97 | cis |
| miR-625-5p | rs72712499 | 14:6580233 | T | G | 0.845101 | -0.337884 | 0.026191 | 4.47E-38 | G | T | 0.1556 | 0.6938 | 0.02839 | 2.22E-95 | cis |
| miR-625-5p | rs72712500 | 14:6580432 | G | A | 0.810519 | -0.233969 | 0.024476 | 1.19E-21 | A | G | 0.1587 | 0.5062 | 0.03257 | 9.00E-47 | cis |
| miR-625-5p | rs72714423 | 14:6586350 | G | A | 0.814071 | -0.233481 | 0.024581 | 2.13E-21 | A | G | 0.1613 | 0.4694 | 0.033 | 1.93E-40 | cis |
| miR-625-5p | rs72714428 | 14:6589297 | A | C | 0.815998 | -0.232651 | 0.024632 | 3.55E-21 | C | A | 0.1643 | 0.4832 | 0.03272 | 3.85E-43 | cis |
| miR-625-5p | rs72714433 | 14:6590281 | A | G | 0.815449 | -0.231041 | 0.024484 | 3.86E-21 | G | A | 0.1648 | 0.4792 | 0.03287 | 3.15E-42 | cis |
| miR-625-5p | rs72714436 | 14:6590747 | A | G | 0.815498 | -0.230917 | 0.024477 | 3.95E-21 | G | A | 0.1648 | 0.4792 | 0.03287 | 3.15E-42 | cis |
| miR-625-5p | rs72714439 | 14:6591690 | C | T | 0.81551 | -0.230852 | 0.024476 | 4.03E-21 | T | C | 0.1648 | 0.4792 | 0.03287 | 3.15E-42 | cis |
| miR-625-5p | rs72714440 | 14:6591859 | C | T | 0.81551 | -0.230854 | 0.024476 | 4.03E-21 | T | C | 0.1648 | 0.4792 | 0.03287 | 3.15E-42 | cis |
| miR-625-5p | rs72714442 | 14:6591904 | T | C | 0.81551 | -0.230854 | 0.024476 | 4.03E-21 | C | T | 0.1648 | 0.4792 | 0.03287 | 3.15E-42 | cis |
| miR-625-5p | rs72714447 | 14:6594214 | C | A | 0.816196 | -0.22863 | 0.024764 | 2.64E-20 | A | C | 0.1648 | 0.4814 | 0.0331 | 5.08E-42 | cis |
| miR-625-5p | rs72714460 | 14:6596208 | A | G | 0.819561 | -0.233271 | 0.024859 | 6.37E-21 | G | A | 0.1641 | 0.485 | 0.033 | 8.28E-43 | cis |
| miR-625-5p | rs72714461 | 14:6596813 | A | G | 0.816433 | -0.231357 | 0.024716 | 7.92E-21 | G | A | 0.1648 | 0.4792 | 0.03287 | 3.15E-42 | cis |
| miR-625-5p | rs72714468 | 14:6597827 | A | C | 0.819836 | -0.233344 | 0.024931 | 7.99E-21 | C | A | 0.1641 | 0.485 | 0.033 | 8.28E-43 | cis |
| miR-625-5p | rs72714478 | 14:6599793 | G | A | 0.815051 | -0.234308 | 0.024801 | 3.47E-21 | A | G | 0.1662 | 0.4871 | 0.03269 | 9.29E-44 | cis |
| miR-625-5p | rs72714482 | 14:6600660 | T | G | 0.815267 | -0.233974 | 0.024821 | 4.24E-21 | G | T | 0.1662 | 0.4871 | 0.03269 | 9.29E-44 | cis |
| miR-625-5p | rs72714483 | 14:6601262 | G | A | 0.820577 | -0.232772 | 0.025061 | 1.57E-20 | A | G | 0.1641 | 0.485 | 0.033 | 8.28E-43 | cis |
| miR-625-5p | rs72714484 | 14:6601624 | A | G | 0.820587 | -0.232841 | 0.025077 | 1.62E-20 | G | A | 0.1641 | 0.485 | 0.033 | 8.28E-43 | cis |
| miR-625-5p | rs72714499 | 14:6604245 | C | T | 0.816154 | -0.231865 | 0.024844 | 1.03E-20 | T | C | 0.1662 | 0.4784 | 0.03287 | 4.33E-42 | cis |
| miR-625-5p | rs72716407 | 14:6606523 | G | T | 0.820366 | -0.233124 | 0.025147 | 1.85E-20 | T | G | 0.1654 | 0.485 | 0.033 | 8.28E-43 | cis |
| miR-625-5p | rs72716408 | 14:6606600 | A | G | 0.820429 | -0.233098 | 0.025151 | 1.90E-20 | G | A | 0.1654 | 0.485 | 0.033 | 8.28E-43 | cis |
| miR-625-5p | rs72716418 | 14:6612104 | G | A | 0.819456 | -0.233541 | 0.025291 | 2.60E-20 | A | G | 0.1635 | 0.4828 | 0.03322 | 5.98E-42 | cis |
| miR-625-5p | rs72716420 | 14:6613228 | C | T | 0.817792 | -0.234012 | 0.025168 | 1.43E-20 | T | C | 0.166 | 0.4814 | 0.03283 | 1.23E-42 | cis |
| miR-625-5p | rs72716421 | 14:6613455 | T | C | 0.817679 | -0.234194 | 0.025168 | 1.34E-20 | C | T | 0.166 | 0.4814 | 0.03283 | 1.23E-42 | cis |
| miR-625-5p | rs72716430 | 14:6619291 | G | T | 0.829643 | -0.216826 | 0.026052 | 8.58E-17 | T | G | 0.1594 | 0.4703 | 0.03369 | 3.17E-39 | cis |
| miR-181a-5 | rs72739764 | 1:19886402 | G | T | 0.765001 | -0.102876 | 0.014899 | 5.02E-12 | T | G | 0.2322 | 0.2337 | 0.01691 | 1.94E-38 | cis |
| miR-625-5p | rs730807 | 14:6623929 | C | T | 0.45402 | 0.143991 | 0.019392 | 1.12E-13 | C | T | 0.4909 | 0.3186 | 0.02562 | 3.53E-32 | cis |
| miR-136-5p | rs73358778 | 14:1011599 | C | T | 0.891417 | 0.411586 | 0.058076 | 1.37E-12 | T | C | 0.1 | -0.07117 | 0.02078 | 0.000651 | cis |
| miR-320a | rs7420775 | 2:68629458 | G | T | 0.703122 | 0.143662 | 0.017427 | 1.67E-16 | T | G | 0.3142 | -0.05966 | 0.006303 | 4.65E-20 | trans |
| miR-320b | rs7420775 | 2:68629458 | G | T | 0.703122 | 0.130665 | 0.015606 | 5.63E-17 | T | G | 0.3142 | -0.063 | 0.00525 | 3.11E-30 | trans |
| miR-320c | rs7420775 | 2:68629458 | G | T | 0.703122 | 0.117641 | 0.014633 | 9.02E-16 | T | G | 0.3142 | -0.05454 | 0.005259 | 1.69E-23 | trans |
| miR-320d | rs7420775 | 2:68629458 | G | T | 0.703122 | 0.119074 | 0.014975 | 1.84E-15 | T | G | 0.3142 | -0.05067 | 0.00515 | 1.88E-21 | trans |
| miR-4429 | rs7420775 | 2:68629458 | G | T | 0.703122 | 0.107093 | 0.014416 | 1.10E-13 | T | G | 0.3142 | -0.06449 | 0.004958 | 1.05E-34 | trans |
| miR-320a | rs7421228 | 2:68630124 | G | T | 0.55268 | 0.152092 | 0.015756 | 4.77E-22 | T | G | 0.4488 | -0.06658 | 0.005541 | 2.34E-30 | trans |
| miR-320b | rs7421228 | 2:68630124 | G | T | 0.55268 | 0.137787 | 0.01411 | 1.58E-22 | T | G | 0.4488 | -0.06591 | 0.004617 | 9.87E-41 | trans |
| miR-320c | rs7421228 | 2:68630124 | G | T | 0.55268 | 0.125054 | 0.013231 | 3.34E-21 | T | G | 0.4488 | -0.06191 | 0.004572 | 3.06E-37 | trans |
| miR-320d | rs7421228 | 2:68630124 | G | T | 0.55268 | 0.124188 | 0.013552 | 5.01E-20 | T | G | 0.4488 | -0.05868 | 0.004467 | 2.30E-35 | trans |
| miR-320e | rs7421228 | 2:68630124 | G | T | 0.55268 | 0.110936 | 0.014989 | 1.35E-13 | T | G | 0.4488 | -0.09469 | 0.006167 | 5.01E-46 | trans |
| miR-4429 | rs7421228 | 2:68630124 | G | T | 0.55268 | 0.116024 | 0.013037 | 5.60E-19 | T | G | 0.4488 | -0.06805 | 0.004311 | 3.44E-48 | trans |
| miR-625-5p | rs74245865 | 14:6609400 | A | G | 0.820602 | -0.233791 | 0.025222 | 1.87E-20 | G | A | 0.1654 | 0.485 | 0.033 | 8.28E-43 | cis |
| miR-625-5p | rs743084 | 14:6623260 | T | C | 0.452662 | 0.142879 | 0.019322 | 1.42E-13 | T | C | 0.4911 | 0.3195 | 0.02563 | 2.38E-32 | cis |
| miR-625-5p | rs743085 | 14:6606813 | A | G | 0.499418 | 0.145261 | 0.019744 | 1.88E-13 | G | A | 0.4777 | -0.3182 | 0.02577 | 8.06E-32 | cis |
| miR-625-5p | rs74351765 | 14:6580840 | C | T | 0.810279 | -0.2335 | 0.024456 | 1.33E-21 | T | C | 0.1615 | 0.506 | 0.03309 | 1.68E-45 | cis |
| miR-584-5p | rs7442677 | 5:14842930 | C | T | 0.856689 | -0.278817 | 0.036018 | 9.87E-15 | T | C | 0.1426 | 0.269 | 0.01797 | 5.23E-44 | cis |
| miR-320a | rs74514522 | 2:68637550 | T | G | 0.702552 | 0.14734 | 0.017642 | 6.74E-17 | G | T | 0.323 | -0.05866 | 0.006255 | 9.61E-20 | trans |
| miR-320b | rs74514522 | 2:68637550 | T | G | 0.702552 | 0.133486 | 0.015801 | 2.97E-17 | G | T | 0.323 | -0.06221 | 0.00521 | 5.51E-30 | trans |
| miR-320c | rs74514522 | 2:68637550 | T | G | 0.702552 | 0.120344 | 0.014815 | 4.55E-16 | G | T | 0.323 | -0.05714 | 0.005174 | 3.22E-26 | trans |
| miR-320d | rs74514522 | 2:68637550 | T | G | 0.702552 | 0.121207 | 0.015164 | 1.31E-15 | G | T | 0.323 | -0.0528 | 0.005092 | 1.70E-23 | trans |
| miR-4429 | rs74514522 | 2:68637550 | T | G | 0.702552 | 0.109286 | 0.014597 | 7.07E-14 | G | T | 0.323 | -0.06486 | 0.00491 | 1.17E-35 | trans |
| miR-625-5p | rs74655860 | 14:6583114 | G | A | 0.815043 | -0.239026 | 0.024818 | 5.91E-22 | A | G | 0.1533 | 0.5104 | 0.03342 | 2.12E-45 | cis |
| miR-625-5p | rs747541 | 14:6580541 | C | A | 0.395726 | 0.206479 | 0.019239 | 7.18E-27 | C | A | 0.4357 | 0.4216 | 0.02345 | 1.12E-59 | cis |
| miR-130a-3 | rs7479922 | 11:5747018 | A | G | 0.450674 | -0.164621 | 0.021018 | 4.78E-15 | A | G | 0.4381 | -0.1757 | 0.01066 | 9.35E-52 | cis |
| miR-625-5p | rs74825628 | 14:6580778 | G | A | 0.810459 | -0.23324 | 0.024454 | 1.46E-21 | A | G | 0.1596 | 0.5107 | 0.03278 | 5.33E-47 | cis |
| miR-625-5p | rs74897779 | 14:6590900 | T | G | 0.81551 | -0.230899 | 0.024476 | 3.97E-21 | G | T | 0.1648 | 0.4792 | 0.03287 | 3.15E-42 | cis |
| miR-625-5p | rs7493004 | 14:6612410 | A | C | 0.50236 | 0.147113 | 0.01987 | 1.32E-13 | C | A | 0.4769 | -0.3154 | 0.02589 | 4.59E-31 | cis |

| | | | | | | | | | | | | | | | |
|---|---|---|---|---|---|---|---|---|---|---|---|---|---|---|---|
| miR-181a-5 | rs7524232 | 1:1990057( | G | A | 0.670143 | 0.091241 | 0.013136 | 3.77E-12 | A | G | 0.3562 | -0.1484 | 0.01625 | 7.58E-19 | cis |
| miR-181a-5 | rs7524630 | 1:1991183: | A | G | 0.514782 | -0.104128 | 0.01313 | 2.18E-15 | G | A | 0.4883 | 0.1946 | 0.01476 | 1.55E-35 | cis |
| miR-181a-5 | rs7529925 | 1:1990072( | C | T | 0.255718 | 0.172939 | 0.013816 | 6.01E-36 | C | T | 0.2549 | 0.3413 | 0.01293 | 2.77E-106 | cis |
| miR-181a-5 | rs7536036 | 1:1990056: | T | C | 0.669114 | 0.091188 | 0.013143 | 3.97E-12 | C | T | 0.3573 | -0.1483 | 0.01624 | 7.81E-19 | cis |
| miR-181a-5 | rs7539177 | 1:1989464( | T | G | 0.238596 | 0.109677 | 0.014687 | 8.17E-14 | T | G | 0.2389 | 0.2392 | 0.01688 | 3.90E-40 | cis |
| miR-181a-5 | rs7547363 | 1:1991183( | G | A | 0.519602 | -0.106958 | 0.013176 | 4.75E-16 | A | G | 0.4834 | 0.1982 | 0.01499 | 1.25E-35 | cis |
| miR-320a | rs7562439 | 2:6858256: | C | T | 0.6646 | 0.131508 | 0.017025 | 1.12E-14 | T | C | 0.3534 | -0.04893 | 0.006235 | 1.62E-14 | trans |
| miR-320b | rs7562439 | 2:6858256: | C | T | 0.6646 | 0.119946 | 0.015246 | 3.62E-15 | T | C | 0.3534 | -0.05779 | 0.005169 | 8.87E-27 | trans |
| miR-320c | rs7562439 | 2:6858256: | C | T | 0.6646 | 0.107793 | 0.014294 | 4.66E-14 | T | C | 0.3534 | -0.04957 | 0.005173 | 1.69E-20 | trans |
| miR-320d | rs7562439 | 2:6858256: | C | T | 0.6646 | 0.108725 | 0.014628 | 1.07E-13 | T | C | 0.3534 | -0.04665 | 0.005045 | 2.82E-19 | trans |
| miR-4429 | rs7562439 | 2:6858256: | C | T | 0.6646 | 0.098629 | 0.014077 | 2.44E-12 | T | C | 0.3534 | -0.05744 | 0.004909 | 5.65E-29 | trans |
| miR-320a | rs7572299 | 2:6858124( | T | A | 0.538033 | 0.140299 | 0.016042 | 2.22E-18 | A | T | 0.4694 | -0.0584 | 0.005679 | 3.45E-23 | trans |
| miR-320b | rs7572299 | 2:6858124( | T | A | 0.538033 | 0.125959 | 0.014372 | 1.88E-18 | A | T | 0.4694 | -0.06104 | 0.00472 | 1.91E-34 | trans |
| miR-320c | rs7572299 | 2:6858124( | T | A | 0.538033 | 0.115531 | 0.013469 | 9.70E-18 | A | T | 0.4694 | -0.05563 | 0.004697 | 1.30E-29 | trans |
| miR-320d | rs7572299 | 2:6858124( | T | A | 0.538033 | 0.116665 | 0.013785 | 2.60E-17 | A | T | 0.4694 | -0.05366 | 0.004566 | 3.18E-29 | trans |
| miR-320e | rs7572299 | 2:6858124( | T | A | 0.538033 | 0.108792 | 0.015217 | 8.73E-13 | A | T | 0.4694 | -0.08422 | 0.006395 | 1.56E-35 | trans |
| miR-4429 | rs7572299 | 2:6858124( | T | A | 0.538033 | 0.107773 | 0.013264 | 4.46E-16 | A | T | 0.4694 | -0.06028 | 0.004497 | 1.26E-36 | trans |
| miR-3130-3 | rs7576591 | 2:2076492: | G | C | 0.843106 | -0.266231 | 0.032952 | 6.52E-16 | C | G | 0.1794 | 0.1673 | 0.03129 | 1.21E-07 | cis |
| miR-320a | rs7578602 | 2:6863786( | G | C | 0.556806 | 0.159076 | 0.016134 | 6.23E-23 | C | G | 0.443 | -0.06677 | 0.005704 | 6.39E-29 | trans |
| miR-320b | rs7578602 | 2:6863786( | G | C | 0.556806 | 0.144256 | 0.014448 | 1.78E-23 | C | G | 0.443 | -0.06656 | 0.00476 | 3.81E-39 | trans |
| miR-320c | rs7578602 | 2:6863786( | G | C | 0.556806 | 0.131453 | 0.013547 | 2.91E-22 | C | G | 0.443 | -0.06408 | 0.004673 | 7.00E-38 | trans |
| miR-320d | rs7578602 | 2:6863786( | G | C | 0.556806 | 0.129683 | 0.01388 | 9.33E-21 | C | G | 0.443 | -0.06177 | 0.004586 | 9.60E-37 | trans |
| miR-320e | rs7578602 | 2:6863786( | G | C | 0.556806 | 0.117559 | 0.015349 | 1.87E-14 | C | G | 0.443 | -0.09763 | 0.006286 | 1.16E-46 | trans |
| miR-4429 | rs7578602 | 2:6863786( | G | C | 0.556806 | 0.121802 | 0.01335 | 7.26E-20 | C | G | 0.443 | -0.06887 | 0.004424 | 7.53E-47 | trans |
| miR-3130-3 | rs7580138 | 2:2076152( | C | A | 0.854791 | -0.277559 | 0.034201 | 4.84E-16 | A | C | 0.1566 | 0.1707 | 0.03333 | 3.95E-07 | cis |
| miR-320a | rs7581872 | 2:6858098: | A | C | 0.537959 | 0.14022 | 0.016042 | 2.31E-18 | C | A | 0.4707 | -0.0584 | 0.005679 | 3.45E-23 | trans |
| miR-320b | rs7581872 | 2:6858098: | A | C | 0.537959 | 0.12589 | 0.014372 | 1.96E-18 | C | A | 0.4707 | -0.06104 | 0.00472 | 1.91E-34 | trans |
| miR-320c | rs7581872 | 2:6858098: | A | C | 0.537959 | 0.115444 | 0.013469 | 1.02E-17 | C | A | 0.4707 | -0.05563 | 0.004697 | 1.30E-29 | trans |
| miR-320d | rs7581872 | 2:6858098: | A | C | 0.537959 | 0.116574 | 0.013785 | 2.75E-17 | C | A | 0.4707 | -0.05366 | 0.004566 | 3.18E-29 | trans |
| miR-320e | rs7581872 | 2:6858098: | A | C | 0.537959 | 0.108694 | 0.015217 | 9.15E-13 | C | A | 0.4707 | -0.08422 | 0.006395 | 1.56E-35 | trans |
| miR-4429 | rs7581872 | 2:6858098: | A | C | 0.537959 | 0.107684 | 0.013264 | 4.71E-16 | C | A | 0.4707 | -0.06028 | 0.004497 | 1.26E-36 | trans |
| miR-320a | rs7590450 | 2:6862856: | A | G | 0.549532 | 0.153227 | 0.015701 | 1.68E-22 | G | A | 0.4512 | -0.06743 | 0.00555 | 6.35E-31 | trans |
| miR-320b | rs7590450 | 2:6862856: | A | G | 0.549532 | 0.138968 | 0.014059 | 4.87E-23 | G | A | 0.4512 | -0.06687 | 0.004615 | 9.60E-42 | trans |
| miR-320c | rs7590450 | 2:6862856: | A | G | 0.549532 | 0.12574 | 0.013186 | 1.49E-21 | G | A | 0.4512 | -0.06399 | 0.004548 | 9.94E-40 | trans |
| miR-320d | rs7590450 | 2:6862856: | A | G | 0.549532 | 0.124555 | 0.013508 | 2.94E-20 | G | A | 0.4512 | -0.06054 | 0.004456 | 1.92E-37 | trans |
| miR-320e | rs7590450 | 2:6862856: | A | G | 0.549532 | 0.11308 | 0.014935 | 3.68E-14 | G | A | 0.4512 | -0.09706 | 0.006143 | 3.03E-48 | trans |
| miR-4429 | rs7590450 | 2:6862856: | A | G | 0.549532 | 0.115985 | 0.012996 | 4.47E-19 | G | A | 0.4512 | -0.06902 | 0.004319 | 3.60E-49 | trans |
| miR-320a | rs7592526 | 2:6858352( | A | C | 0.538007 | 0.140533 | 0.01604 | 1.93E-18 | C | A | 0.4688 | -0.05862 | 0.005688 | 2.77E-23 | trans |
| miR-320b | rs7592526 | 2:6858352( | A | C | 0.538007 | 0.126303 | 0.01437 | 1.50E-18 | C | A | 0.4688 | -0.06122 | 0.004727 | 1.56E-34 | trans |
| miR-320c | rs7592526 | 2:6858352( | A | C | 0.538007 | 0.115839 | 0.013467 | 7.86E-18 | C | A | 0.4688 | -0.05564 | 0.004707 | 1.64E-29 | trans |
| miR-320d | rs7592526 | 2:6858352( | A | C | 0.538007 | 0.116969 | 0.013783 | 2.14E-17 | C | A | 0.4688 | -0.05353 | 0.004578 | 5.62E-29 | trans |
| miR-320e | rs7592526 | 2:6858352( | A | C | 0.538007 | 0.109013 | 0.015216 | 7.81E-13 | C | A | 0.4688 | -0.08431 | 0.006408 | 1.75E-35 | trans |
| miR-4429 | rs7592526 | 2:6858352( | A | C | 0.538007 | 0.108083 | 0.013262 | 3.64E-16 | C | A | 0.4688 | -0.06014 | 0.00451 | 2.63E-36 | trans |
| miR-320a | rs7597167 | 2:6861294( | C | T | 0.698051 | 0.13964 | 0.016858 | 1.20E-16 | T | C | 0.3166 | -0.05774 | 0.006213 | 1.86E-19 | trans |
| miR-320b | rs7597167 | 2:6861294( | C | T | 0.698051 | 0.126955 | 0.015097 | 4.12E-17 | T | C | 0.3166 | -0.06338 | 0.005147 | 1.14E-31 | trans |
| miR-320c | rs7597167 | 2:6861294( | C | T | 0.698051 | 0.11378 | 0.014157 | 9.21E-16 | T | C | 0.3166 | -0.05691 | 0.005129 | 1.83E-26 | trans |
| miR-320d | rs7597167 | 2:6861294( | C | T | 0.698051 | 0.117082 | 0.014481 | 6.20E-16 | T | C | 0.3166 | -0.05224 | 0.005025 | 1.25E-23 | trans |
| miR-4429 | rs7597167 | 2:6861294( | C | T | 0.698051 | 0.104514 | 0.013944 | 6.62E-14 | T | C | 0.3166 | -0.06364 | 0.004888 | 7.60E-35 | trans |
| miR-320a | rs7598360 | 2:6863092: | A | G | 0.553166 | 0.155693 | 0.01579 | 6.20E-23 | G | A | 0.4479 | -0.06742 | 0.005625 | 3.60E-30 | trans |
| miR-320b | rs7598360 | 2:6863092: | A | G | 0.553166 | 0.141628 | 0.014138 | 1.27E-23 | G | A | 0.4479 | -0.06756 | 0.004688 | 2.77E-41 | trans |
| miR-320c | rs7598360 | 2:6863092: | A | G | 0.553166 | 0.128065 | 0.01326 | 4.56E-22 | G | A | 0.4479 | -0.06528 | 0.004599 | 2.96E-40 | trans |
| miR-320d | rs7598360 | 2:6863092: | A | G | 0.553166 | 0.126722 | 0.013585 | 1.08E-20 | G | A | 0.4479 | -0.06147 | 0.0045 | 1.01E-37 | trans |
| miR-320e | rs7598360 | 2:6863092: | A | G | 0.553166 | 0.116479 | 0.015017 | 8.73E-15 | G | A | 0.4479 | -0.09778 | 0.006193 | 4.38E-48 | trans |
| miR-4429 | rs7598360 | 2:6863092: | A | G | 0.553166 | 0.11928 | 0.013065 | 6.86E-20 | G | A | 0.4479 | -0.06897 | 0.004348 | 1.90E-48 | trans |
| miR-320a | rs7605387 | 2:6863255( | A | G | 0.703499 | 0.147092 | 0.017558 | 5.40E-17 | G | A | 0.3247 | -0.05906 | 0.006192 | 2.43E-20 | trans |
| miR-320b | rs7605387 | 2:6863255( | A | G | 0.703499 | 0.132498 | 0.015728 | 3.63E-17 | G | A | 0.3247 | -0.06283 | 0.005134 | 2.69E-31 | trans |
| miR-320c | rs7605387 | 2:6863255( | A | G | 0.703499 | 0.119101 | 0.014748 | 6.72E-16 | G | A | 0.3247 | -0.05686 | 0.005136 | 2.45E-26 | trans |
| miR-320d | rs7605387 | 2:6863255( | A | G | 0.703499 | 0.120593 | 0.015093 | 1.35E-15 | G | A | 0.3247 | -0.05182 | 0.005044 | 3.91E-23 | trans |
| miR-4429 | rs7605387 | 2:6863255( | A | G | 0.703499 | 0.108179 | 0.014531 | 9.72E-14 | G | A | 0.3247 | -0.06403 | 0.004862 | 1.70E-35 | trans |
| miR-625-5p | rs7606377: | 14:657677: | G | A | 0.843419 | -0.308513 | 0.02639 | 1.43E-31 | A | G | 0.1591 | 0.6606 | 0.02865 | 1.26E-87 | cis |
| miR-320a | rs7608429 | 2:6858083( | G | T | 0.678719 | 0.125181 | 0.017336 | 5.16E-13 | T | G | 0.3396 | -0.05155 | 0.00629 | 1.21E-15 | trans |
| miR-320b | rs7608429 | 2:6858083( | G | T | 0.678719 | 0.116442 | 0.015517 | 6.19E-14 | T | G | 0.3396 | -0.05924 | 0.00522 | 1.75E-27 | trans |
| miR-320c | rs7608429 | 2:6858083( | G | T | 0.678719 | 0.103457 | 0.014551 | 1.16E-12 | T | G | 0.3396 | -0.05158 | 0.005218 | 1.23E-21 | trans |
| miR-320d | rs7608429 | 2:6858083( | G | T | 0.678719 | 0.10826 | 0.014878 | 3.42E-13 | T | G | 0.3396 | -0.04812 | 0.005089 | 4.88E-20 | trans |
| miR-4429 | rs7608429 | 2:6858083( | G | T | 0.678719 | 0.09611 | 0.014323 | 1.94E-11 | T | G | 0.3396 | -0.05907 | 0.004953 | 6.18E-30 | trans |
| miR-625-5p | rs7608716: | 14:660670( | T | C | 0.820364 | -0.233129 | 0.025147 | 1.85E-20 | C | T | 0.1654 | 0.485 | 0.033 | 8.28E-43 | cis |
| miR-625-5p | rs761831 | 14:662393: | T | C | 0.454673 | 0.144038 | 0.019399 | 1.13E-13 | T | C | 0.4942 | 0.3186 | 0.02562 | 3.53E-32 | cis |
| miR-133b | rs7621371: | 20:611509: | A | G | 0.467251 | -0.190849 | 0.027215 | 2.34E-12 | A | G | 0.4453 | -0.3171 | 0.03154 | 3.11E-22 | trans |
| miR-3130-3 | rs7627046: | 2:2076536: | G | A | 0.902503 | -0.276562 | 0.04049 | 8.47E-12 | A | G | 0.1013 | 0.1781 | 0.03919 | 6.51E-06 | cis |
| miR-625-5p | rs7660191( | 14:660873: | G | C | 0.820834 | -0.232309 | 0.025252 | 3.59E-20 | C | G | 0.1605 | 0.4746 | 0.03356 | 4.45E-40 | cis |
| miR-625-5p | rs7682482: | 14:657582: | G | A | 0.778334 | -0.195159 | 0.022955 | 1.87E-17 | A | G | 0.2158 | 0.4738 | 0.0293 | 4.90E-50 | cis |
| miR-3130-3 | rs768533 | 2:2076408: | G | A | 0.842932 | -0.271664 | 0.032915 | 1.54E-16 | A | G | 0.1794 | 0.1673 | 0.03129 | 1.21E-07 | cis |
| miR-625-5p | rs7692276: | 14:657375: | C | G | 0.705647 | -0.191943 | 0.021474 | 3.95E-19 | G | C | 0.3095 | 0.4704 | 0.02467 | 4.33E-65 | cis |
| miR-584-5p | rs7697787: | 5:1484473: | A | G | 0.844049 | -0.271396 | 0.034876 | 7.15E-15 | G | A | 0.1546 | 0.2727 | 0.01754 | 5.95E-47 | cis |
| miR-625-5p | rs7733938: | 14:657580: | G | A | 0.84644 | -0.309982 | 0.02677 | 5.23E-31 | A | G | 0.1542 | 0.6553 | 0.02911 | 2.73E-84 | cis |
| miR-130a-3 | rs7799305: | 11:575228: | C | T | 0.684063 | -0.24278 | 0.022169 | 6.54E-28 | T | C | 0.3059 | 0.2296 | 0.01048 | 2.41E-81 | cis |
| miR-320a | rs7800275: | 2:6862543( | A | G | 0.546309 | 0.150787 | 0.015553 | 3.17E-22 | G | A | 0.4488 | -0.06695 | 0.005536 | 1.10E-30 | trans |
| miR-320b | rs7800275: | 2:6862543( | A | G | 0.546309 | 0.136492 | 0.013929 | 1.13E-22 | G | A | 0.4488 | -0.06675 | 0.004599 | 7.04E-42 | trans |
| miR-320c | rs7800275: | 2:6862543( | A | G | 0.546309 | 0.123534 | 0.013063 | 3.18E-21 | G | A | 0.4488 | -0.06332 | 0.004553 | 5.89E-39 | trans |
| miR-320d | rs7800275: | 2:6862543( | A | G | 0.546309 | 0.122332 | 0.013381 | 6.14E-20 | G | A | 0.4488 | -0.05995 | 0.004447 | 6.06E-37 | trans |
| miR-320e | rs7800275: | 2:6862543( | A | G | 0.546309 | 0.110955 | 0.014794 | 6.38E-14 | G | A | 0.4488 | -0.09482 | 0.006169 | 4.26E-46 | trans |
| miR-4429 | rs7800275: | 2:6862543( | A | G | 0.546309 | 0.114214 | 0.012873 | 7.16E-19 | G | A | 0.4488 | -0.06798 | 0.004325 | 7.62E-48 | trans |
| miR-625-5p | rs7823084: | 14:660163: | G | A | 0.507675 | 0.145122 | 0.019792 | 2.26E-13 | A | G | 0.47 | -0.31 | 0.02592 | 4.19E-30 | cis |
| miR-584-5p | rs7846146( | 5:1484304: | C | T | 0.843889 | -0.26844 | 0.034455 | 6.64E-15 | T | C | 0.1562 | 0.2726 | 0.0173 | 4.95E-48 | cis |
| miR-10a-5p | rs7873635 | 9:1361320: | T | C | 0.381256 | -0.102828 | 0.01466 | 2.32E-12 | T | C | 0.3918 | -0.04805 | 0.01337 | 0.000349 | trans |
| miR-126-3p | rs7873635 | 9:1361320: | T | C | 0.381256 | -0.112479 | 0.013126 | 1.04E-17 | T | C | 0.3918 | -0.03298 | 0.007845 | 2.97E-05 | trans |

| | | | | | | | | | | | | | | | |
|---|---|---|---|---|---|---|---|---|---|---|---|---|---|---|---|
| miR-181a-5 | rs7890714 | 1:1989878( | G | A | 0.904551 | -0.152889 | 0.021249 | 6.24E-13 | A | G | 0.08475 | 0.2492 | 0.02665 | 1.22E-19 | cis |
| miR-625-5p | rs7918480 | 14:659083 | A | C | 0.813031 | -0.230215 | 0.024633 | 9.11E-21 | C | A | 0.1639 | 0.4871 | 0.03314 | 9.87E-43 | cis |
| miR-130a-3 | rs7926074 | 11:576129( | T | C | 0.687478 | -0.245723 | 0.022451 | 7.04E-28 | C | T | 0.2967 | 0.2395 | 0.01053 | 1.83E-85 | cis |
| miR-130a-3 | rs7932091 | 11:576180( | A | T | 0.683808 | -0.241876 | 0.022349 | 2.69E-27 | T | A | 0.2997 | 0.2348 | 0.0105 | 1.48E-83 | cis |
| miR-130a-3 | rs7936998 | 11:576390 | G | A | 0.683653 | -0.241729 | 0.022605 | 1.09E-26 | A | G | 0.3009 | 0.23 | 0.01057 | 4.44E-80 | cis |
| miR-130a-3 | rs7938083 | 11:574936 | C | A | 0.686581 | -0.245553 | 0.022337 | 4.12E-28 | A | C | 0.2929 | 0.2301 | 0.01066 | 4.48E-79 | cis |
| miR-130a-3 | rs7949841 | 11:575247( | G | C | 0.684177 | -0.243105 | 0.022173 | 5.69E-28 | C | G | 0.3054 | 0.2296 | 0.01046 | 1.75E-81 | cis |
| miR-3130-3 | rs7949904( | 2:20764093 | A | G | 0.902079 | -0.277819 | 0.04038 | 5.98E-12 | G | A | 0.1014 | 0.1779 | 0.03922 | 6.77E-06 | cis |
| miR-130a-3 | rs7974071( | 11:574631 | C | T | 0.686438 | -0.2449 | 0.022233 | 3.24E-28 | T | C | 0.303 | 0.2326 | 0.0105 | 1.56E-82 | cis |
| miR-625-5p | rs8003464 | 14:659082 | A | G | 0.35496 | 0.173425 | 0.020754 | 6.48E-17 | A | G | 0.3192 | 0.3811 | 0.02682 | 2.05E-40 | cis |
| miR-625-5p | rs8005243 | 14:660656 | C | G | 0.346908 | 0.172472 | 0.02067 | 7.17E-17 | C | G | 0.3162 | 0.3927 | 0.02668 | 6.50E-43 | cis |
| miR-625-5p | rs8005710 | 14:660656( | G | A | 0.502323 | 0.147754 | 0.019868 | 1.03E-13 | A | G | 0.4757 | -0.3165 | 0.02593 | 3.41E-31 | cis |
| miR-625-5p | rs8007497 | 14:660444 | T | C | 0.338774 | 0.16985 | 0.02037 | 7.53E-17 | T | C | 0.3178 | 0.393 | 0.0267 | 6.99E-43 | cis |
| miR-625-5p | rs8010726 | 14:659325 | G | A | 0.358211 | 0.172184 | 0.020663 | 7.89E-17 | G | A | 0.321 | 0.3772 | 0.02676 | 7.19E-40 | cis |
| miR-625-5p | rs8011224 | 14:659696 | T | A | 0.357678 | 0.169952 | 0.020594 | 1.55E-16 | T | A | 0.3269 | 0.3708 | 0.02681 | 1.38E-38 | cis |
| miR-625-5p | rs8012278 | 14:660825 | A | G | 0.499774 | 0.145255 | 0.019787 | 2.12E-13 | G | A | 0.4777 | -0.3182 | 0.02577 | 8.06E-32 | cis |
| miR-625-5p | rs8012834 | 14:659665 | C | T | 0.362666 | 0.181923 | 0.020554 | 8.68E-19 | C | T | 0.34 | 0.4159 | 0.02522 | 8.70E-52 | cis |
| miR-625-5p | rs8013442 | 14:658718( | A | G | 0.342325 | 0.17882 | 0.020898 | 1.16E-17 | A | G | 0.3127 | 0.4031 | 0.02654 | 4.13E-45 | cis |
| miR-625-5p | rs8014475 | 14:658115 | T | C | 0.34592 | 0.204093 | 0.019825 | 7.44E-25 | T | C | 0.3645 | 0.4429 | 0.02384 | 6.31E-63 | cis |
| miR-625-5p | rs8015222 | 14:658360 | T | A | 0.052734 | 0.328492 | 0.045567 | 5.63E-13 | T | A | 0.0572 | 0.5834 | 0.05492 | 1.60E-24 | cis |
| miR-4707-3 | rs8016027 | 14:234201 | T | C | 0.441165 | 0.093615 | 0.013591 | 5.67E-12 | T | C | 0.4364 | 0.2998 | 0.01624 | 4.76E-62 | cis |
| miR-625-5p | rs8016322 | 14:660515 | G | A | 0.386004 | 0.167316 | 0.020156 | 1.03E-16 | G | A | 0.3827 | 0.3378 | 0.02617 | 2.46E-34 | cis |
| miR-625-5p | rs8017880 | 14:659429 | G | C | 0.346067 | 0.172935 | 0.020833 | 1.03E-16 | G | C | 0.3119 | 0.3939 | 0.02667 | 3.91E-43 | cis |
| miR-625-5p | rs8018278 | 14:661800 | A | G | 0.498198 | 0.145521 | 0.019847 | 2.26E-13 | G | A | 0.4758 | -0.3144 | 0.02593 | 8.02E-31 | cis |
| miR-625-5p | rs8019762 | 14:660871 | A | G | 0.346646 | 0.172489 | 0.020796 | 1.09E-16 | A | G | 0.31 | 0.3841 | 0.02674 | 4.01E-41 | cis |
| miR-625-5p | rs8020305 | 14:658375( | C | T | 0.066561 | 0.301123 | 0.040379 | 8.82E-14 | C | T | 0.07898 | 0.5774 | 0.04507 | 7.98E-34 | cis |
| miR-136-5p | rs8020868 | 14:101166 | A | G | 0.668343 | 0.400258 | 0.036388 | 3.84E-28 | G | A | 0.3247 | -0.06964 | 0.01332 | 2.28E-07 | cis |
| miR-136-5p | rs8021301 | 14:101166 | G | C | 0.668324 | 0.400327 | 0.036391 | 3.80E-28 | C | G | 0.3247 | -0.06964 | 0.01332 | 2.28E-07 | cis |
| miR-625-5p | rs8021946 | 14:658468 | C | T | 0.054518 | 0.325363 | 0.045724 | 1.11E-12 | C | T | 0.06 | 0.591 | 0.05521 | 7.63E-25 | cis |
| miR-625-5p | rs8022053 | 14:658679 | T | G | 0.350181 | 0.176409 | 0.020719 | 1.68E-17 | T | G | 0.3215 | 0.3874 | 0.02652 | 3.42E-42 | cis |
| miR-625-5p | rs8022080 | 14:659990 | T | C | 0.358857 | 0.182558 | 0.020463 | 4.61E-19 | T | C | 0.3382 | 0.4297 | 0.02506 | 3.10E-55 | cis |
| miR-625-5p | rs8023016 | 14:658468( | T | A | 0.053493 | 0.326462 | 0.045712 | 9.22E-13 | T | A | 0.05742 | 0.591 | 0.05521 | 7.63E-25 | cis |
| miR-320a | rs8028813 | 2:6862547 | A | G | 0.548418 | 0.153418 | 0.015656 | 1.13E-22 | G | A | 0.4477 | -0.06844 | 0.005511 | 4.76E-32 | trans |
| miR-320b | rs8028813 | 2:6862547 | A | G | 0.548418 | 0.139006 | 0.01402 | 3.59E-23 | G | A | 0.4477 | -0.06693 | 0.004656 | 4.27E-41 | trans |
| miR-320c | rs8028813 | 2:6862547 | A | G | 0.548418 | 0.12585 | 0.013149 | 1.06E-21 | G | A | 0.4477 | -0.06552 | 0.004533 | 1.79E-41 | trans |
| miR-320d | rs8028813 | 2:6862547 | A | G | 0.548418 | 0.124394 | 0.013471 | 2.60E-20 | G | A | 0.4477 | -0.06131 | 0.004481 | 8.31E-38 | trans |
| miR-320e | rs8028813 | 2:6862547 | A | G | 0.548418 | 0.112315 | 0.014897 | 4.71E-14 | G | A | 0.4477 | -0.09729 | 0.006188 | 1.01E-47 | trans |
| miR-4429 | rs8028813 | 2:6862547 | A | G | 0.548418 | 0.11582 | 0.012961 | 4.03E-19 | G | A | 0.4477 | -0.07004 | 0.004315 | 2.69E-50 | trans |
| miR-130a-3 | rs8030492 | 11:575228 | C | G | 0.684059 | -0.24278 | 0.022169 | 6.54E-28 | G | C | 0.3059 | 0.2296 | 0.01048 | 2.41E-81 | cis |
| miR-133b | rs8122282 | 20:611523 | G | A | 0.467131 | -0.189725 | 0.027217 | 3.15E-12 | G | A | 0.4476 | -0.3127 | 0.03111 | 2.92E-22 | trans |
| miR-10a-5p | rs8176649 | 9:1361472 | G | A | 0.618669 | 0.107644 | 0.014449 | 9.35E-14 | A | G | 0.393 | -0.05095 | 0.0133 | 0.000139 | trans |
| miR-126-3p | rs8176649 | 9:1361472 | G | A | 0.618669 | 0.114693 | 0.01294 | 7.78E-19 | A | G | 0.393 | -0.03245 | 0.007845 | 3.96E-05 | trans |
| let-7b-5p | rs8176663 | 9:1361444 | T | C | 0.663959 | -0.107659 | 0.010066 | 1.07E-26 | C | T | 0.3219 | 0.03377 | 0.006655 | 4.99E-07 | trans |
| let-7c-5p | rs8176663 | 9:1361444 | T | C | 0.663959 | -0.091368 | 0.00919 | 2.74E-23 | C | T | 0.3219 | 0.02445 | 0.006395 | 0.000144 | trans |
| miR-10b-5p | rs8176663 | 9:1361444 | T | C | 0.663959 | -0.17708 | 0.014305 | 3.40E-35 | C | T | 0.3219 | 0.1135 | 0.01684 | 3.31E-11 | trans |
| miR-126-3p | rs8176663 | 9:1361444 | T | C | 0.663959 | -0.17133 | 0.013064 | 2.73E-39 | C | T | 0.3219 | 0.05294 | 0.008279 | 2.95E-10 | trans |
| miR-139-5p | rs8176663 | 9:1361444 | T | C | 0.663959 | -0.157215 | 0.011603 | 7.92E-42 | C | T | 0.3219 | 0.0709 | 0.01641 | 1.79E-05 | trans |
| miR-195-5p | rs8176663 | 9:1361444 | T | C | 0.663959 | -0.113603 | 0.013414 | 2.48E-17 | C | T | 0.3219 | 0.06692 | 0.01825 | 0.000265 | trans |
| miR-10a-5p | rs8176668 | 9:1361440 | A | T | 0.618711 | 0.106528 | 0.014474 | 1.84E-13 | T | A | 0.3931 | -0.05078 | 0.0133 | 0.000147 | trans |
| miR-126-3p | rs8176668 | 9:1361440 | A | T | 0.618711 | 0.11469 | 0.012959 | 8.74E-19 | T | A | 0.3931 | -0.03251 | 0.007841 | 3.80E-05 | trans |
| miR-10a-5p | rs8176681 | 9:1361397 | T | C | 0.600667 | 0.102291 | 0.014344 | 9.94E-13 | C | T | 0.4191 | -0.0496 | 0.01311 | 0.000168 | trans |
| miR-126-3p | rs8176681 | 9:1361397 | T | C | 0.600667 | 0.112306 | 0.012838 | 2.18E-18 | C | T | 0.4191 | -0.03292 | 0.007724 | 2.30E-05 | trans |
| miR-10a-5p | rs8176682 | 9:1361392 | C | T | 0.619011 | 0.105945 | 0.014446 | 2.24E-13 | T | C | 0.3944 | -0.04998 | 0.01332 | 0.00019 | trans |
| miR-126-3p | rs8176682 | 9:1361392 | C | T | 0.619011 | 0.113671 | 0.012936 | 1.54E-18 | T | C | 0.3944 | -0.03211 | 0.007851 | 4.82E-05 | trans |
| miR-10a-5p | rs8176690 | 9:1361383 | A | G | 0.601023 | 0.101606 | 0.014332 | 1.35E-12 | G | A | 0.4189 | -0.04846 | 0.01314 | 0.000243 | trans |
| miR-126-3p | rs8176690 | 9:1361383 | A | G | 0.601023 | 0.112118 | 0.012826 | 2.30E-18 | G | A | 0.4189 | -0.03316 | 0.007732 | 2.05E-05 | trans |
| miR-10a-5p | rs8176691 | 9:1361382 | C | T | 0.619638 | 0.105987 | 0.014449 | 2.21E-13 | T | C | 0.394 | -0.04887 | 0.01335 | 0.00027 | trans |
| miR-126-3p | rs8176691 | 9:1361382 | C | T | 0.619638 | 0.114018 | 0.012938 | 1.22E-18 | T | C | 0.394 | -0.03253 | 0.007861 | 3.93E-05 | trans |
| miR-10a-5p | rs8176702 | 9:1361361 | G | A | 0.6197 | 0.106041 | 0.014453 | 2.18E-13 | A | G | 0.3944 | -0.05032 | 0.0133 | 0.000168 | trans |
| miR-126-3p | rs8176702 | 9:1361361 | G | A | 0.6197 | 0.114009 | 0.012941 | 1.25E-18 | A | G | 0.3944 | -0.03303 | 0.007837 | 2.83E-05 | trans |
| miR-625-5p | rs867972 | 14:658957( | T | C | 0.50459 | 0.14613 | 0.019867 | 1.90E-13 | C | T | 0.4722 | -0.3116 | 0.0259 | 2.21E-30 | cis |
| miR-320a | rs8761 | 2:6862426( | G | A | 0.545692 | 0.150887 | 0.015566 | 3.21E-22 | A | G | 0.4548 | -0.06677 | 0.00552 | 1.05E-30 | trans |
| miR-320b | rs8761 | 2:6862426( | G | A | 0.545692 | 0.136595 | 0.01394 | 1.14E-22 | A | G | 0.4548 | -0.06621 | 0.004597 | 2.26E-41 | trans |
| miR-320c | rs8761 | 2:6862426( | G | A | 0.545692 | 0.123625 | 0.013074 | 3.20E-21 | A | G | 0.4548 | -0.06403 | 0.00452 | 3.13E-40 | trans |
| miR-320d | rs8761 | 2:6862426( | G | A | 0.545692 | 0.12236 | 0.013392 | 6.45E-20 | A | G | 0.4548 | -0.06052 | 0.00442 | 5.66E-38 | trans |
| miR-320e | rs8761 | 2:6862426( | G | A | 0.545692 | 0.110918 | 0.014806 | 6.81E-14 | A | G | 0.4548 | -0.09657 | 0.006109 | 2.43E-48 | trans |
| miR-4429 | rs8761 | 2:6862426( | G | A | 0.545692 | 0.114212 | 0.012884 | 7.65E-19 | A | G | 0.4548 | -0.0691 | 0.004289 | 7.07E-50 | trans |
| miR-181a-5 | rs881576 | 1:1992577 | A | C | 0.44377 | -0.090643 | 0.013036 | 3.56E-12 | A | C | 0.4742 | -0.1305 | 0.01538 | 1.39E-16 | cis |
| miR-625-5p | rs883081 | 14:658806 | C | A | 0.497085 | 0.149391 | 0.019841 | 5.10E-14 | A | C | 0.4778 | -0.3275 | 0.02557 | 8.61E-34 | cis |
| miR-625-5p | rs883082 | 14:658809 | A | G | 0.493786 | 0.147935 | 0.019805 | 8.05E-14 | G | A | 0.4785 | -0.3285 | 0.0256 | 6.47E-34 | cis |
| miR-3130-3 | rs888621 | 2:2076493 | T | G | 0.842681 | -0.266321 | 0.032863 | 5.32E-16 | G | T | 0.1794 | 0.1673 | 0.03129 | 1.21E-07 | cis |
| miR-625-5p | rs899960 | 14:657899 | C | G | 0.053722 | 0.334306 | 0.044654 | 7.07E-14 | C | G | 0.06491 | 0.5907 | 0.05263 | 5.74E-27 | cis |
| miR-625-5p | rs899961 | 14:658003 | A | C | 0.290881 | 0.291121 | 0.020561 | 1.65E-45 | A | C | 0.3327 | 0.5779 | 0.02006 | 6.86E-120 | cis |
| miR-625-5p | rs9323460 | 14:659331 | T | G | 0.357542 | 0.174201 | 0.02067 | 3.53E-17 | T | G | 0.3194 | 0.3778 | 0.02692 | 1.61E-39 | cis |
| miR-625-5p | rs9323461 | 14:660314( | T | C | 0.342775 | 0.172498 | 0.020658 | 6.81E-17 | T | C | 0.312 | 0.393 | 0.0267 | 6.99E-43 | cis |
| miR-3689a | rs9410004 | 9:1377477( | C | T | 0.470272 | 0.220078 | 0.02582 | 1.55E-17 | C | T | 0.4808 | 0.07998 | 0.02236 | 0.000372 | cis |
| miR-3689a | rs9410005 | 9:1377478 | A | G | 0.46835 | 0.220517 | 0.025857 | 1.49E-17 | A | G | 0.478 | 0.08011 | 0.02239 | 0.000372 | cis |
| miR-130a-3 | rs9420 | 11:575102 | A | G | 0.309761 | 0.240756 | 0.0223 | 3.59E-27 | A | G | 0.3054 | 0.2308 | 0.01061 | 2.36E-80 | cis |
| miR-3130-3 | rs957414 | 2:2076655 | T | A | 0.84423 | -0.26426 | 0.033198 | 1.72E-15 | A | T | 0.1779 | 0.1701 | 0.03121 | 7.06E-08 | cis |
| miR-625-5p | rs9635249 | 14:657977 | A | G | 0.22061 | 0.364734 | 0.022232 | 1.73E-60 | A | G | 0.2432 | 0.7383 | 0.01673 | 1.94E-203 | cis |
| miR-320a | rs9653552 | 2:6864396( | G | A | 0.593458 | 0.14288 | 0.016454 | 3.83E-18 | A | G | 0.4012 | -0.06118 | 0.005851 | 8.17E-24 | trans |
| miR-320b | rs9653552 | 2:6864396( | G | A | 0.593458 | 0.13159 | 0.014728 | 4.08E-19 | A | G | 0.4012 | -0.06116 | 0.00492 | 4.39E-32 | trans |
| miR-320c | rs9653552 | 2:6864396( | G | A | 0.593458 | 0.11996 | 0.013806 | 3.65E-18 | A | G | 0.4012 | -0.05748 | 0.004859 | 1.82E-29 | trans |
| miR-320d | rs9653552 | 2:6864396( | G | A | 0.593458 | 0.119124 | 0.014138 | 3.58E-17 | A | G | 0.4012 | -0.05833 | 0.00466 | 1.85E-32 | trans |
| miR-320e | rs9653552 | 2:6864396( | G | A | 0.593458 | 0.113705 | 0.015597 | 3.10E-13 | A | G | 0.4012 | -0.09302 | 0.006465 | 4.04E-41 | trans |
| miR-4429 | rs9653552 | 2:6864396( | G | A | 0.593458 | 0.109389 | 0.013605 | 8.96E-16 | A | G | 0.4012 | -0.06291 | 0.004609 | 1.26E-37 | trans |

| | | | | | | | | | | | | | | |
|---|---|---|---|---|---|---|---|---|---|---|---|---|---|---|
| miR-136-5p | rs9671394 | 14:101159( | C | T | 0.284232 | -0.398989 | 0.039355 | 3.73E-24 | C | T | 0.2799 | -0.0646 | 0.01382 | 3.52E-06 cis |
| miR-625-5p | rs968540 | 14:662490( | G | T | 0.633781 | -0.135121 | 0.020129 | 1.91E-11 | T | G | 0.366 | 0.3094 | 0.02698 | 5.55E-28 cis |
| miR-625-5p | rs9972106 | 14:660231: | A | T | 0.388305 | 0.167818 | 0.020231 | 1.08E-16 | A | T | 0.3863 | 0.3407 | 0.02618 | 8.45E-35 cis |

**Table S6. Summary of replication of miR-eQTLS**

| | Rotterdam Study (discovery) | | | Nikpay e | |
|---|---|---|---|---|---|
| | All | Unique SNPs | n miRNAs | Replication threshold | All |
| **all** | 2254 | 899 | 57 | P<0.05/N | 1462 |
| | | | | P<0.05 | 1685 |
| **cis** | 1068 | 720 | 30 | P<0.05/N | 651 |
| | | | | P<0.05 | 740 |
| **trans** | 1186 | 185 | 28 | P<0.05/N | 811 |
| | | | | P<0.05 | 945 |

| t al (replication) | | |
|---|---|---|
| % replication | Unique SNPs | n miRNAs |
| 64.86 | 814 | 27 |
| 74.76 | 861 | 37 |
| 60.96 | 647 | 12 |
| 69.29 | 687 | 17 |
| 68.38 | 168 | 15 |
| 79.68 | 175 | 20 |

**Table S7. List of 4,310 replicated miR-eQTLs**

| SNP | miRNA | position | effect_allel | other_allel | eaf | beta | se | pval | cismark |
|---|---|---|---|---|---|---|---|---|---|
| rs543040 | let-7b-5p | 9:1361430( | A | T | 0.669196 | -0.109582 | 0.010251 | 1.13E-26 | trans |
| rs544873 | let-7b-5p | 9:1361432: | G | A | 0.645742 | -0.105117 | 0.009877 | 1.88E-26 | trans |
| rs494242 | let-7b-5p | 9:1361451: | C | T | 0.644977 | -0.105243 | 0.009882 | 1.74E-26 | trans |
| rs492488 | let-7b-5p | 9:1361449( | G | A | 0.663579 | -0.107737 | 0.010074 | 1.07E-26 | trans |
| rs545971 | let-7b-5p | 9:1361433: | C | T | 0.663766 | -0.107579 | 0.010061 | 1.11E-26 | trans |
| rs493246 | let-7b-5p | 9:1361449! | G | A | 0.663234 | -0.107785 | 0.010079 | 1.09E-26 | trans |
| rs532436 | let-7b-5p | 9:1361498: | G | A | 0.8067 | -0.126313 | 0.01205 | 1.04E-25 | trans |
| rs1165522 | let-7b-5p | 9:1361490! | T | A | 0.653719 | -0.107531 | 0.01003 | 8.12E-27 | trans |
| rs687289 | let-7b-5p | 9:1361371( | G | A | 0.668579 | -0.111294 | 0.010186 | 8.61E-28 | trans |
| rs2519093 | let-7b-5p | 9:1361418: | C | T | 0.809892 | -0.127895 | 0.012263 | 1.82E-25 | trans |
| rs676457 | let-7b-5p | 9:1361462: | A | T | 0.664166 | -0.107822 | 0.010067 | 9.10E-27 | trans |
| rs2769071 | let-7b-5p | 9:1361459: | A | G | 0.663117 | -0.107918 | 0.010084 | 9.93E-27 | trans |
| rs630014 | let-7b-5p | 9:1361497: | A | G | 0.494254 | -0.079742 | 0.009829 | 4.92E-16 | trans |
| rs643434 | let-7b-5p | 9:1361423! | G | A | 0.645392 | -0.105157 | 0.009872 | 1.70E-26 | trans |
| rs491626 | let-7b-5p | 9:1361448: | C | T | 0.664173 | -0.108148 | 0.01009 | 8.39E-27 | trans |
| rs529565 | let-7b-5p | 9:1361495( | T | C | 0.665911 | -0.110358 | 0.010051 | 4.77E-28 | trans |
| rs630510 | let-7b-5p | 9:1361495! | A | G | 0.494403 | -0.079814 | 0.009835 | 4.85E-16 | trans |
| rs612169 | let-7b-5p | 9:1361434 | A | G | 0.66434 | -0.107722 | 0.01008 | 1.17E-26 | trans |
| rs582094 | let-7b-5p | 9:1361454! | A | T | 0.663709 | -0.107729 | 0.010063 | 9.59E-27 | trans |
| rs8176663 | let-7b-5p | 9:1361444: | T | C | 0.663959 | -0.107659 | 0.010066 | 1.07E-26 | trans |
| rs550057 | let-7b-5p | 9:1361465! | C | T | 0.734085 | -0.097587 | 0.010867 | 2.70E-19 | trans |
| rs674302 | let-7b-5p | 9:1361466( | T | A | 0.663989 | -0.107892 | 0.010058 | 7.63E-27 | trans |
| rs613534 | let-7b-5p | 9:1361431: | A | G | 0.653693 | -0.106 | 0.01005 | 5.20E-26 | trans |
| rs554833 | let-7b-5p | 9:1361471( | C | T | 0.664057 | -0.107822 | 0.01006 | 8.42E-27 | trans |
| rs600038 | let-7b-5p | 9:1361518( | T | C | 0.787872 | -0.119508 | 0.011643 | 1.02E-24 | trans |
| rs582118 | let-7b-5p | 9:1361454: | A | G | 0.663872 | -0.107776 | 0.010067 | 9.58E-27 | trans |
| rs543968 | let-7b-5p | 9:1361431: | T | C | 0.653829 | -0.10593 | 0.010051 | 5.69E-26 | trans |
| rs495203 | let-7b-5p | 9:1361452 | C | T | 0.663279 | -0.107724 | 0.010072 | 1.06E-26 | trans |
| rs676996 | let-7b-5p | 9:1361460: | T | G | 0.667516 | -0.108022 | 0.010153 | 1.96E-26 | trans |
| rs677355 | let-7b-5p | 9:1361460 | G | A | 0.670198 | -0.108649 | 0.010242 | 2.73E-26 | trans |
| rs644234 | let-7b-5p | 9:1361422: | T | G | 0.645381 | -0.105194 | 0.009864 | 1.49E-26 | trans |
| rs1165522 | let-7c-5p | 9:1361490! | T | A | 0.653719 | -0.090775 | 0.00916 | 3.78E-23 | trans |
| rs491626 | let-7c-5p | 9:1361448: | C | T | 0.664173 | -0.091873 | 0.009213 | 2.01E-23 | trans |
| rs529565 | let-7c-5p | 9:1361495( | T | C | 0.665911 | -0.093158 | 0.009181 | 3.42E-24 | trans |
| rs677355 | let-7c-5p | 9:1361460 | G | A | 0.670198 | -0.092257 | 0.009351 | 5.81E-23 | trans |
| rs687289 | let-7c-5p | 9:1361371( | G | A | 0.668579 | -0.093907 | 0.009304 | 5.91E-24 | trans |
| rs532436 | let-7c-5p | 9:1361498: | G | A | 0.8067 | -0.107102 | 0.011001 | 2.13E-22 | trans |
| rs676457 | let-7c-5p | 9:1361462: | A | T | 0.664166 | -0.091483 | 0.009192 | 2.46E-23 | trans |
| rs582094 | let-7c-5p | 9:1361454! | A | T | 0.663709 | -0.091349 | 0.009188 | 2.74E-23 | trans |
| rs2519093 | let-7c-5p | 9:1361418: | C | T | 0.809892 | -0.108043 | 0.011197 | 4.94E-22 | trans |
| rs674302 | let-7c-5p | 9:1361466( | T | A | 0.663989 | -0.091475 | 0.009184 | 2.28E-23 | trans |
| rs493246 | let-7c-5p | 9:1361449! | G | A | 0.663234 | -0.091502 | 0.009202 | 2.70E-23 | trans |
| rs544873 | let-7c-5p | 9:1361432: | G | A | 0.645742 | -0.089522 | 0.009016 | 3.11E-23 | trans |
| rs643434 | let-7c-5p | 9:1361423! | G | A | 0.645392 | -0.089455 | 0.009012 | 3.20E-23 | trans |
| rs676996 | let-7c-5p | 9:1361460: | T | G | 0.667516 | -0.091652 | 0.00927 | 4.76E-23 | trans |
| rs644234 | let-7c-5p | 9:1361422: | T | G | 0.645381 | -0.089506 | 0.009005 | 2.80E-23 | trans |
| rs554833 | let-7c-5p | 9:1361471( | C | T | 0.664057 | -0.091422 | 0.009186 | 2.46E-23 | trans |
| rs492488 | let-7c-5p | 9:1361449( | G | A | 0.663579 | -0.091462 | 0.009197 | 2.67E-23 | trans |
| rs543968 | let-7c-5p | 9:1361431: | T | C | 0.653829 | -0.090699 | 0.009172 | 4.68E-23 | trans |
| rs582118 | let-7c-5p | 9:1361454: | A | G | 0.663872 | -0.091413 | 0.009192 | 2.66E-23 | trans |
| rs8176663 | let-7c-5p | 9:1361444: | T | C | 0.663959 | -0.091368 | 0.00919 | 2.74E-23 | trans |

| | | | | | | | | | |
|---|---|---|---|---|---|---|---|---|---|
| rs494242 | let-7c-5p | 9:1361451: | C | T | 0.644977 | -0.089675 | 0.00902 | 2.75E-23 | trans |
| rs612169 | let-7c-5p | 9:1361434 | A | G | 0.66434 | -0.091485 | 0.009203 | 2.76E-23 | trans |
| rs613534 | let-7c-5p | 9:1361431 | A | G | 0.653693 | -0.090765 | 0.009171 | 4.30E-23 | trans |
| rs545971 | let-7c-5p | 9:1361433 | C | T | 0.663766 | -0.091289 | 0.009187 | 2.87E-23 | trans |
| rs543040 | let-7c-5p | 9:1361430 | A | T | 0.669196 | -0.092304 | 0.009362 | 6.26E-23 | trans |
| rs495203 | let-7c-5p | 9:1361452 | C | T | 0.663279 | -0.091426 | 0.009196 | 2.73E-23 | trans |
| rs2769071 | let-7c-5p | 9:1361459 | A | G | 0.663117 | -0.091257 | 0.009208 | 3.76E-23 | trans |
| rs2073826 | miR-10a-5p | 9:1361369 | G | T | 0.60107 | 0.102208 | 0.014329 | 9.81E-13 | trans |
| rs8176691 | miR-10a-5p | 9:1361382 | C | T | 0.619638 | 0.105987 | 0.014449 | 2.21E-13 | trans |
| rs7046674 | miR-10a-5p | 9:1361470 | C | T | 0.619013 | 0.107787 | 0.014473 | 9.50E-14 | trans |
| rs2073828 | miR-10a-5p | 9:1361371 | G | A | 0.601379 | 0.102564 | 0.014335 | 8.39E-13 | trans |
| rs7873635 | miR-10a-5p | 9:1361320 | T | C | 0.381256 | -0.102828 | 0.01466 | 2.32E-12 | trans |
| rs8176690 | miR-10a-5p | 9:1361383 | A | G | 0.601023 | 0.101606 | 0.014332 | 1.35E-12 | trans |
| rs4962040 | miR-10a-5p | 9:1361335 | A | G | 0.619057 | 0.104354 | 0.014467 | 5.47E-13 | trans |
| rs2073827 | miR-10a-5p | 9:1361371 | G | C | 0.601379 | 0.102564 | 0.014335 | 8.39E-13 | trans |
| rs8176668 | miR-10a-5p | 9:1361440 | A | T | 0.618711 | 0.106528 | 0.014474 | 1.84E-13 | trans |
| rs8176649 | miR-10a-5p | 9:1361472 | G | A | 0.618669 | 0.107644 | 0.014449 | 9.35E-14 | trans |
| rs8176681 | miR-10a-5p | 9:1361397 | T | C | 0.600667 | 0.102291 | 0.014344 | 9.94E-13 | trans |
| rs8176682 | miR-10a-5p | 9:1361392 | C | T | 0.619011 | 0.105945 | 0.014446 | 2.24E-13 | trans |
| rs7036642 | miR-10a-5p | 9:1361446 | G | A | 0.618956 | 0.106822 | 0.014464 | 1.52E-13 | trans |
| rs8176702 | miR-10a-5p | 9:1361361 | G | A | 0.6197 | 0.106041 | 0.014453 | 2.18E-13 | trans |
| rs613534 | miR-10b-5p | 9:1361431 | A | G | 0.653693 | -0.174451 | 0.014285 | 2.68E-34 | trans |
| rs612169 | miR-10b-5p | 9:1361434 | A | G | 0.66434 | -0.177278 | 0.014325 | 3.54E-35 | trans |
| rs630510 | miR-10b-5p | 9:1361495 | A | G | 0.494403 | -0.115552 | 0.014092 | 2.40E-16 | trans |
| rs550057 | miR-10b-5p | 9:1361465 | C | T | 0.734085 | -0.167896 | 0.015446 | 1.61E-27 | trans |
| rs660340 | miR-10b-5p | 9:1361475 | A | G | 0.454082 | -0.127916 | 0.014109 | 1.23E-19 | trans |
| rs674302 | miR-10b-5p | 9:1361466 | T | A | 0.663989 | -0.177203 | 0.014295 | 2.75E-35 | trans |
| rs545971 | miR-10b-5p | 9:1361433 | C | T | 0.663766 | -0.176997 | 0.014299 | 3.43E-35 | trans |
| rs649129 | miR-10b-5p | 9:1361543 | C | T | 0.787432 | -0.201509 | 0.016495 | 2.54E-34 | trans |
| rs600038 | miR-10b-5p | 9:1361518 | T | C | 0.787872 | -0.202026 | 0.016527 | 2.32E-34 | trans |
| rs643434 | miR-10b-5p | 9:1361423 | G | A | 0.645392 | -0.171966 | 0.014036 | 1.64E-34 | trans |
| rs687289 | miR-10b-5p | 9:1361371 | G | A | 0.668579 | -0.183745 | 0.014466 | 5.80E-37 | trans |
| rs645982 | miR-10b-5p | 9:1361484 | A | G | 0.454123 | -0.127768 | 0.0141 | 1.29E-19 | trans |
| rs543968 | miR-10b-5p | 9:1361431 | T | C | 0.653829 | -0.174326 | 0.014287 | 3.05E-34 | trans |
| rs495203 | miR-10b-5p | 9:1361452 | C | T | 0.663279 | -0.177267 | 0.014313 | 3.15E-35 | trans |
| rs493246 | miR-10b-5p | 9:1361449 | G | A | 0.663234 | -0.177369 | 0.014324 | 3.23E-35 | trans |
| rs492488 | miR-10b-5p | 9:1361449 | G | A | 0.663579 | -0.177263 | 0.014316 | 3.26E-35 | trans |
| rs473533 | miR-10b-5p | 9:1361480 | T | C | 0.453861 | -0.128006 | 0.01411 | 1.17E-19 | trans |
| rs2519093 | miR-10b-5p | 9:1361418 | C | T | 0.809892 | -0.215578 | 0.017406 | 3.13E-35 | trans |
| rs1124406 | miR-10b-5p | 9:1361539 | C | T | 0.886198 | -0.206905 | 0.022623 | 5.93E-20 | trans |
| rs1165522 | miR-10b-5p | 9:1361490 | T | A | 0.653719 | -0.174502 | 0.014267 | 2.13E-34 | trans |
| rs495828 | miR-10b-5p | 9:1361548 | G | T | 0.787276 | -0.201363 | 0.016484 | 2.56E-34 | trans |
| rs491626 | miR-10b-5p | 9:1361448 | C | T | 0.664173 | -0.177622 | 0.014341 | 3.14E-35 | trans |
| rs676457 | miR-10b-5p | 9:1361462 | A | T | 0.664166 | -0.177236 | 0.014307 | 3.04E-35 | trans |
| rs543040 | miR-10b-5p | 9:1361430 | A | T | 0.669196 | -0.179437 | 0.014573 | 7.68E-35 | trans |
| rs529565 | miR-10b-5p | 9:1361495 | T | C | 0.665911 | -0.179319 | 0.01429 | 4.06E-36 | trans |
| rs582094 | miR-10b-5p | 9:1361454 | A | T | 0.663709 | -0.17709 | 0.014302 | 3.25E-35 | trans |
| rs644234 | miR-10b-5p | 9:1361422 | T | G | 0.645381 | -0.171966 | 0.014025 | 1.46E-34 | trans |
| rs582118 | miR-10b-5p | 9:1361454 | A | G | 0.663872 | -0.177155 | 0.014308 | 3.28E-35 | trans |
| rs494242 | miR-10b-5p | 9:1361451 | C | T | 0.644977 | -0.172256 | 0.014049 | 1.47E-34 | trans |
| rs544873 | miR-10b-5p | 9:1361432 | G | A | 0.645742 | -0.172011 | 0.014043 | 1.70E-34 | trans |
| rs2769071 | miR-10b-5p | 9:1361459 | A | G | 0.663117 | -0.177378 | 0.014331 | 3.48E-35 | trans |
| rs647800 | miR-10b-5p | 9:1361480 | A | G | 0.435519 | -0.128548 | 0.014052 | 5.81E-20 | trans |

| | | | | | | | | | |
|---|---|---|---|---|---|---|---|---|---|
| rs554833 | miR-10b-5p | 9:1361471 | C | T | 0.664057 | -0.177063 | 0.014299 | 3.22E-35 | trans |
| rs659104 | miR-10b-5p | 9:1361478 | T | G | 0.454107 | -0.127786 | 0.014102 | 1.28E-19 | trans |
| rs630014 | miR-10b-5p | 9:1361497 | A | G | 0.494254 | -0.115432 | 0.014082 | 2.46E-16 | trans |
| rs581107 | miR-10b-5p | 9:1361477 | C | T | 0.452595 | -0.128436 | 0.014149 | 1.11E-19 | trans |
| rs8176663 | miR-10b-5p | 9:1361444 | T | C | 0.663959 | -0.17708 | 0.014305 | 3.40E-35 | trans |
| rs676996 | miR-10b-5p | 9:1361460 | T | G | 0.667516 | -0.177525 | 0.014432 | 8.97E-35 | trans |
| rs476410 | miR-10b-5p | 9:1361483 | G | C | 0.45414 | -0.127771 | 0.014101 | 1.29E-19 | trans |
| rs532436 | miR-10b-5p | 9:1361498 | G | A | 0.8067 | -0.214173 | 0.017094 | 5.20E-36 | trans |
| rs677355 | miR-10b-5p | 9:1361460 | G | A | 0.670198 | -0.178902 | 0.014556 | 1.02E-34 | trans |
| rs8176681 | miR-126-3p | 9:1361397 | T | C | 0.600667 | 0.112306 | 0.012838 | 2.18E-18 | trans |
| rs676457 | miR-126-3p | 9:1361462 | A | T | 0.664166 | -0.171461 | 0.013067 | 2.46E-39 | trans |
| rs2519093 | miR-126-3p | 9:1361418 | C | T | 0.809892 | -0.211422 | 0.015879 | 1.90E-40 | trans |
| rs8176690 | miR-126-3p | 9:1361383 | A | G | 0.601023 | 0.112118 | 0.012826 | 2.30E-18 | trans |
| rs494242 | miR-126-3p | 9:1361451 | C | T | 0.644977 | -0.16489 | 0.012842 | 9.85E-38 | trans |
| rs630510 | miR-126-3p | 9:1361495 | A | G | 0.494403 | -0.115498 | 0.012884 | 3.11E-19 | trans |
| rs543040 | miR-126-3p | 9:1361430 | A | T | 0.669196 | -0.173596 | 0.013309 | 6.96E-39 | trans |
| rs613534 | miR-126-3p | 9:1361431 | A | G | 0.653693 | -0.165472 | 0.013067 | 9.46E-37 | trans |
| rs582094 | miR-126-3p | 9:1361454 | A | T | 0.663709 | -0.171388 | 0.013061 | 2.46E-39 | trans |
| rs2073828 | miR-126-3p | 9:1361371 | G | A | 0.601379 | 0.112127 | 0.012832 | 2.37E-18 | trans |
| rs493246 | miR-126-3p | 9:1361449 | G | A | 0.663234 | -0.171618 | 0.013081 | 2.55E-39 | trans |
| rs8176702 | miR-126-3p | 9:1361361 | G | A | 0.6197 | 0.114009 | 0.012941 | 1.25E-18 | trans |
| rs649129 | miR-126-3p | 9:1361543 | C | T | 0.787432 | -0.192807 | 0.015079 | 1.95E-37 | trans |
| rs612169 | miR-126-3p | 9:1361434 | A | G | 0.66434 | -0.171512 | 0.013083 | 2.89E-39 | trans |
| rs674302 | miR-126-3p | 9:1361466 | T | A | 0.663989 | -0.171502 | 0.013055 | 2.03E-39 | trans |
| rs7873635 | miR-126-3p | 9:1361320 | T | C | 0.381256 | -0.112479 | 0.013126 | 1.04E-17 | trans |
| rs8176668 | miR-126-3p | 9:1361440 | A | T | 0.618711 | 0.11469 | 0.012959 | 8.74E-19 | trans |
| rs676996 | miR-126-3p | 9:1361460 | T | G | 0.667516 | -0.172281 | 0.013178 | 4.67E-39 | trans |
| rs491626 | miR-126-3p | 9:1361448 | C | T | 0.664173 | -0.172011 | 0.013097 | 2.10E-39 | trans |
| rs8176649 | miR-126-3p | 9:1361472 | G | A | 0.618669 | 0.114693 | 0.01294 | 7.78E-19 | trans |
| rs532436 | miR-126-3p | 9:1361498 | G | A | 0.8067 | -0.207511 | 0.015609 | 2.49E-40 | trans |
| rs687289 | miR-126-3p | 9:1361371 | G | A | 0.668579 | -0.177674 | 0.01321 | 3.06E-41 | trans |
| rs8176682 | miR-126-3p | 9:1361392 | C | T | 0.619011 | 0.113671 | 0.012936 | 1.54E-18 | trans |
| rs8176663 | miR-126-3p | 9:1361444 | T | C | 0.663959 | -0.17133 | 0.013064 | 2.73E-39 | trans |
| rs495203 | miR-126-3p | 9:1361452 | C | T | 0.663279 | -0.17152 | 0.013072 | 2.48E-39 | trans |
| rs644234 | miR-126-3p | 9:1361422 | T | G | 0.645381 | -0.164565 | 0.012821 | 1.03E-37 | trans |
| rs8176691 | miR-126-3p | 9:1361382 | C | T | 0.619638 | 0.114018 | 0.012938 | 1.22E-18 | trans |
| rs7036642 | miR-126-3p | 9:1361446 | G | A | 0.618956 | 0.114538 | 0.012952 | 9.29E-19 | trans |
| rs495828 | miR-126-3p | 9:1361548 | G | T | 0.787276 | -0.192644 | 0.015069 | 2.00E-37 | trans |
| rs600038 | miR-126-3p | 9:1361518 | T | C | 0.787872 | -0.193279 | 0.015108 | 1.80E-37 | trans |
| rs550057 | miR-126-3p | 9:1361465 | C | T | 0.734085 | -0.163489 | 0.014114 | 5.01E-31 | trans |
| rs529565 | miR-126-3p | 9:1361495 | T | C | 0.665911 | -0.174412 | 0.013044 | 8.91E-41 | trans |
| rs2073826 | miR-126-3p | 9:1361369 | G | T | 0.60107 | 0.112151 | 0.012825 | 2.24E-18 | trans |
| rs543968 | miR-126-3p | 9:1361431 | T | C | 0.653829 | -0.165481 | 0.013068 | 9.52E-37 | trans |
| rs630014 | miR-126-3p | 9:1361497 | A | G | 0.494254 | -0.115403 | 0.012875 | 3.15E-19 | trans |
| rs1165522 | miR-126-3p | 9:1361490 | T | A | 0.653719 | -0.167546 | 0.013039 | 8.64E-38 | trans |
| rs677355 | miR-126-3p | 9:1361460 | G | A | 0.670198 | -0.172718 | 0.013297 | 1.41E-38 | trans |
| rs545971 | miR-126-3p | 9:1361433 | C | T | 0.663766 | -0.171246 | 0.013059 | 2.76E-39 | trans |
| rs643434 | miR-126-3p | 9:1361423 | G | A | 0.645392 | -0.164561 | 0.012831 | 1.18E-37 | trans |
| rs7046674 | miR-126-3p | 9:1361470 | C | T | 0.619013 | 0.115101 | 0.01296 | 6.62E-19 | trans |
| rs4962040 | miR-126-3p | 9:1361335 | A | G | 0.619057 | 0.11337 | 0.012952 | 2.08E-18 | trans |
| rs492488 | miR-126-3p | 9:1361449 | G | A | 0.663579 | -0.171514 | 0.013074 | 2.58E-39 | trans |
| rs582118 | miR-126-3p | 9:1361454 | A | G | 0.663872 | -0.171439 | 0.013067 | 2.52E-39 | trans |
| rs554833 | miR-126-3p | 9:1361471 | C | T | 0.664057 | -0.171374 | 0.013058 | 2.40E-39 | trans |

| | | | | | | | | | |
|---|---|---|---|---|---|---|---|---|---|
| rs2769071 | miR-126-3p | 9:1361459 | A | G | 0.663117 | -0.171277 | 0.01309 | 4.06E-39 | trans |
| rs2073827 | miR-126-3p | 9:1361371 | G | C | 0.601379 | 0.112127 | 0.012832 | 2.37E-18 | trans |
| rs544873 | miR-126-3p | 9:1361432 | G | A | 0.645742 | -0.16454 | 0.012837 | 1.31E-37 | trans |
| rs1089666 | miR-130a-3 | 11:576790 | C | A | 0.697394 | -0.219619 | 0.023047 | 1.58E-21 | cis |
| rs1624389 | miR-130a-3 | 11:574462 | A | G | 0.451986 | -0.164916 | 0.021048 | 4.67E-15 | cis |
| rs1089664 | miR-130a-3 | 11:575639 | C | T | 0.567891 | 0.160898 | 0.021008 | 1.88E-14 | cis |
| rs588612 | miR-130a-3 | 11:576128 | G | C | 0.439957 | -0.162455 | 0.021014 | 1.07E-14 | cis |
| rs1122907 | miR-130a-3 | 11:573990 | C | T | 0.749544 | -0.3014 | 0.023308 | 3.01E-38 | cis |
| rs7102271 | miR-130a-3 | 11:573968 | G | T | 0.749722 | -0.301475 | 0.023289 | 2.50E-38 | cis |
| rs644883 | miR-130a-3 | 11:576635 | C | G | 0.310308 | 0.226662 | 0.023122 | 1.10E-22 | cis |
| rs1089664 | miR-130a-3 | 11:575877 | A | G | 0.565548 | 0.160285 | 0.020992 | 2.25E-14 | cis |
| rs1089665 | miR-130a-3 | 11:576689 | C | G | 0.697383 | -0.220092 | 0.022989 | 1.03E-21 | cis |
| rs639158 | miR-130a-3 | 11:576020 | G | A | 0.441293 | -0.163651 | 0.021032 | 7.19E-15 | cis |
| rs1122907 | miR-130a-3 | 11:573910 | A | C | 0.750877 | -0.301901 | 0.023261 | 1.61E-38 | cis |
| rs665058 | miR-130a-3 | 11:575791 | T | C | 0.565581 | 0.160316 | 0.020993 | 2.23E-14 | cis |
| rs1160573 | miR-130a-3 | 11:575240 | C | G | 0.683995 | -0.242751 | 0.022162 | 6.40E-28 | cis |
| rs547891 | miR-130a-3 | 11:574990 | C | T | 0.319553 | 0.241325 | 0.022213 | 1.70E-27 | cis |
| rs9420 | miR-130a-3 | 11:575102 | A | G | 0.309761 | 0.240756 | 0.0223 | 3.59E-27 | cis |
| rs1278553 | miR-130a-3 | 11:574866 | G | A | 0.685995 | -0.242795 | 0.022262 | 1.08E-27 | cis |
| rs6591443 | miR-130a-3 | 11:577199 | T | A | 0.44212 | -0.137702 | 0.021102 | 6.78E-11 | cis |
| rs1160670 | miR-130a-3 | 11:573835 | G | T | 0.750529 | -0.301671 | 0.023272 | 1.98E-38 | cis |
| rs1160667 | miR-130a-3 | 11:573833 | G | A | 0.750505 | -0.301631 | 0.023271 | 2.02E-38 | cis |
| rs1160432 | miR-130a-3 | 11:574970 | C | A | 0.688224 | -0.242092 | 0.022277 | 1.65E-27 | cis |
| rs4272798 | miR-130a-3 | 11:577436 | T | C | 0.602524 | 0.121564 | 0.021328 | 1.20E-08 | cis |
| rs1122906 | miR-130a-3 | 11:573749 | G | T | 0.750531 | -0.299533 | 0.02326 | 6.00E-38 | cis |
| rs641325 | miR-130a-3 | 11:576818 | T | G | 0.327793 | 0.198345 | 0.022691 | 2.30E-18 | cis |
| rs3781885 | miR-130a-3 | 11:575706 | C | T | 0.700878 | -0.247612 | 0.022418 | 2.31E-28 | cis |
| rs3580806 | miR-130a-3 | 11:575470 | T | C | 0.684777 | -0.2434 | 0.022156 | 4.47E-28 | cis |
| rs585761 | miR-130a-3 | 11:575973 | G | A | 0.43789 | -0.160703 | 0.021012 | 2.04E-14 | cis |
| rs1236606 | miR-130a-3 | 11:577350 | A | G | 0.560221 | 0.135284 | 0.02102 | 1.23E-10 | cis |
| rs1079211 | miR-130a-3 | 11:575658 | C | T | 0.684897 | -0.243313 | 0.02216 | 4.77E-28 | cis |
| rs1122906 | miR-130a-3 | 11:573748 | C | G | 0.750533 | -0.299532 | 0.02326 | 6.00E-38 | cis |
| rs632052 | miR-130a-3 | 11:576943 | A | T | 0.559771 | 0.139297 | 0.021184 | 4.84E-11 | cis |
| rs1227020 | miR-130a-3 | 11:575377 | G | A | 0.564726 | 0.161383 | 0.021024 | 1.64E-14 | cis |
| rs631791 | miR-130a-3 | 11:576754 | A | G | 0.307763 | 0.220062 | 0.023047 | 1.32E-21 | cis |
| rs1160433 | miR-130a-3 | 11:574670 | C | G | 0.686388 | -0.244681 | 0.022232 | 3.58E-28 | cis |
| rs1227486 | miR-130a-3 | 11:575576 | C | T | 0.56562 | 0.161052 | 0.021005 | 1.76E-14 | cis |
| rs708228 | miR-130a-3 | 11:575856 | C | T | 0.684575 | -0.241919 | 0.022159 | 9.51E-28 | cis |
| rs1631684 | miR-130a-3 | 11:574208 | G | A | 0.412965 | -0.154123 | 0.021382 | 5.68E-13 | cis |
| rs2836294 | miR-130a-3 | 11:573690 | A | C | 0.752624 | -0.306487 | 0.023509 | 7.52E-39 | cis |
| rs1783824 | miR-130a-3 | 11:574730 | A | G | 0.452208 | -0.164056 | 0.020981 | 5.31E-15 | cis |
| rs652908 | miR-130a-3 | 11:576040 | C | T | 0.441718 | -0.163828 | 0.021034 | 6.76E-15 | cis |
| rs1151273 | miR-130a-3 | 11:577565 | G | A | 0.603493 | 0.12256 | 0.021354 | 9.50E-09 | cis |
| rs7120868 | miR-130a-3 | 11:575305 | T | G | 0.566777 | 0.161451 | 0.021059 | 1.77E-14 | cis |
| rs1783829 | miR-130a-3 | 11:574198 | T | C | 0.413999 | -0.154412 | 0.021345 | 4.68E-13 | cis |
| rs1160712 | miR-130a-3 | 11:573897 | G | A | 0.743794 | -0.302872 | 0.023174 | 4.93E-39 | cis |
| rs1783818 | miR-130a-3 | 11:574375 | C | T | 0.454526 | -0.165981 | 0.021036 | 3.01E-15 | cis |
| rs7105506 | miR-130a-3 | 11:574979 | G | A | 0.56028 | 0.172703 | 0.021219 | 3.98E-16 | cis |
| rs7148446 | miR-130a-3 | 11:576327 | G | C | 0.683543 | -0.241608 | 0.02258 | 1.02E-26 | cis |
| rs2847308 | miR-130a-3 | 11:574982 | C | T | 0.319113 | 0.241055 | 0.022203 | 1.85E-27 | cis |
| rs1151273 | miR-130a-3 | 11:577564 | A | G | 0.602716 | 0.122135 | 0.021321 | 1.01E-08 | cis |
| rs2836295 | miR-130a-3 | 11:573719 | G | A | 0.749181 | -0.300466 | 0.023454 | 1.43E-37 | cis |
| rs2081361 | miR-130a-3 | 11:574117 | C | T | 0.745781 | -0.293753 | 0.023511 | 8.04E-36 | cis |

| | | | | | | | | | |
|---|---|---|---|---|---|---|---|---|---|
| rs1089663 | miR-130a-3 | 11:574240 | T | C | 0.698591 | -0.257416 | 0.022716 | 9.10E-30 | cis |
| rs1786438 | miR-130a-3 | 11:575970 | T | G | 0.439556 | -0.162306 | 0.020983 | 1.03E-14 | cis |
| rs585337 | miR-130a-3 | 11:575972 | C | T | 0.439573 | -0.162328 | 0.020984 | 1.03E-14 | cis |
| rs659756 | miR-130a-3 | 11:576918 | A | T | 0.559827 | 0.140626 | 0.021214 | 3.38E-11 | cis |
| rs1079211 | miR-130a-3 | 11:575363 | C | T | 0.564735 | 0.161403 | 0.021024 | 1.63E-14 | cis |
| rs4827 | miR-130a-3 | 11:575079 | C | T | 0.685347 | -0.243141 | 0.022155 | 5.06E-28 | cis |
| rs633129 | miR-130a-3 | 11:576851 | G | A | 0.561364 | 0.141528 | 0.021176 | 2.33E-11 | cis |
| rs1160302 | miR-130a-3 | 11:573743 | T | C | 0.750533 | -0.299532 | 0.02326 | 6.00E-38 | cis |
| rs674094 | miR-130a-3 | 11:576653 | A | C | 0.289352 | 0.203736 | 0.02375 | 9.64E-18 | cis |
| rs1160639 | miR-130a-3 | 11:574352 | C | T | 0.687897 | -0.245707 | 0.0223 | 3.12E-28 | cis |
| rs2156638 | miR-130a-3 | 11:575429 | G | C | 0.565268 | 0.1611 | 0.021006 | 1.73E-14 | cis |
| rs1122908 | miR-130a-3 | 11:573995 | T | C | 0.749539 | -0.3014 | 0.023308 | 3.01E-38 | cis |
| rs1122917 | miR-130a-3 | 11:577157 | A | G | 0.559886 | 0.137581 | 0.021098 | 6.98E-11 | cis |
| rs1079211 | miR-130a-3 | 11:577091 | A | G | 0.442386 | -0.138381 | 0.021145 | 5.97E-11 | cis |
| rs3824988 | miR-130a-3 | 11:573779 | A | G | 0.750533 | -0.299532 | 0.02326 | 6.00E-38 | cis |
| rs7974071 | miR-130a-3 | 11:574631 | C | T | 0.686438 | -0.2449 | 0.022233 | 3.24E-28 | cis |
| rs1236216 | miR-130a-3 | 11:574530 | C | T | 0.68725 | -0.246218 | 0.02227 | 2.05E-28 | cis |
| rs682503 | miR-130a-3 | 11:576409 | C | G | 0.313309 | 0.230973 | 0.022934 | 7.40E-24 | cis |
| rs7148448 | miR-130a-3 | 11:577530 | G | A | 0.606036 | 0.12257 | 0.021395 | 1.01E-08 | cis |
| rs7117878 | miR-130a-3 | 11:574214 | C | A | 0.697568 | -0.258876 | 0.022623 | 2.55E-30 | cis |
| rs1279019 | miR-130a-3 | 11:574978 | C | T | 0.686232 | -0.244075 | 0.022216 | 4.43E-28 | cis |
| rs1160705 | miR-130a-3 | 11:574968 | C | T | 0.683884 | -0.244801 | 0.022215 | 3.08E-28 | cis |
| rs1442563 | miR-130a-3 | 11:573714 | C | T | 0.752772 | -0.305493 | 0.023522 | 1.44E-38 | cis |
| rs1122918 | miR-130a-3 | 11:577354 | G | A | 0.560619 | 0.13566 | 0.021026 | 1.10E-10 | cis |
| rs1236302 | miR-130a-3 | 11:575032 | G | C | 0.556328 | 0.166301 | 0.021097 | 3.21E-15 | cis |
| rs3405210 | miR-130a-3 | 11:573845 | A | C | 0.750564 | -0.301736 | 0.023272 | 1.92E-38 | cis |
| rs1089663 | miR-130a-3 | 11:574480 | C | G | 0.68075 | -0.243141 | 0.022412 | 2.02E-27 | cis |
| rs600488 | miR-130a-3 | 11:576840 | C | T | 0.560573 | 0.142183 | 0.021211 | 2.04E-11 | cis |
| rs1129766 | miR-130a-3 | 11:575648 | G | A | 0.566038 | 0.161154 | 0.021054 | 1.95E-14 | cis |
| rs499188 | miR-130a-3 | 11:574341 | C | A | 0.686203 | -0.241592 | 0.022245 | 1.78E-27 | cis |
| rs1160210 | miR-130a-3 | 11:574352 | G | A | 0.688626 | -0.24541 | 0.022271 | 3.09E-28 | cis |
| rs2956981 | miR-130a-3 | 11:575011 | C | T | 0.554948 | 0.168947 | 0.02108 | 1.10E-15 | cis |
| rs1122911 | miR-130a-3 | 11:575108 | G | T | 0.692006 | -0.243466 | 0.022353 | 1.26E-27 | cis |
| rs1160016 | miR-130a-3 | 11:574649 | T | A | 0.679868 | -0.246647 | 0.022456 | 4.59E-28 | cis |
| rs1122913 | miR-130a-3 | 11:575621 | A | G | 0.688809 | -0.243075 | 0.022346 | 1.47E-27 | cis |
| rs653889 | miR-130a-3 | 11:575704 | A | G | 0.567416 | 0.160942 | 0.021027 | 1.95E-14 | cis |
| rs3603649 | miR-130a-3 | 11:574941 | T | C | 0.554138 | 0.164556 | 0.02107 | 5.72E-15 | cis |
| rs1785499 | miR-130a-3 | 11:574691 | T | G | 0.45303 | -0.16492 | 0.020975 | 3.76E-15 | cis |
| rs8030492 | miR-130a-3 | 11:575228 | C | G | 0.684059 | -0.24278 | 0.022169 | 6.54E-28 | cis |
| rs1122910 | miR-130a-3 | 11:574922 | A | G | 0.554137 | 0.164771 | 0.021077 | 5.39E-15 | cis |
| rs7799305 | miR-130a-3 | 11:575228 | C | T | 0.684063 | -0.24278 | 0.022169 | 6.54E-28 | cis |
| rs652191 | miR-130a-3 | 11:576350 | A | G | 0.568268 | 0.160772 | 0.021201 | 3.37E-14 | cis |
| rs1122917 | miR-130a-3 | 11:577076 | C | T | 0.560118 | 0.137979 | 0.021119 | 6.43E-11 | cis |
| rs1745542 | miR-130a-3 | 11:575347 | C | T | 0.565004 | 0.15876 | 0.02111 | 5.46E-14 | cis |
| rs645145 | miR-130a-3 | 11:576097 | G | T | 0.564908 | 0.160734 | 0.02104 | 2.18E-14 | cis |
| rs7116341 | miR-130a-3 | 11:574811 | G | A | 0.685526 | -0.243004 | 0.022225 | 7.92E-28 | cis |
| rs543725 | miR-130a-3 | 11:574830 | A | T | 0.688868 | -0.239725 | 0.022465 | 1.39E-26 | cis |
| rs1783983 | miR-130a-3 | 11:574207 | T | C | 0.412531 | -0.153845 | 0.021282 | 4.87E-13 | cis |
| rs7936998 | miR-130a-3 | 11:576390 | G | A | 0.683653 | -0.241729 | 0.022605 | 1.09E-26 | cis |
| rs731384 | miR-130a-3 | 11:574083 | G | A | 0.744474 | -0.294708 | 0.023568 | 7.02E-36 | cis |
| rs1214654 | miR-130a-3 | 11:575086 | G | C | 0.685316 | -0.243208 | 0.022155 | 4.90E-28 | cis |
| rs1089663 | miR-130a-3 | 11:573903 | G | A | 0.746727 | -0.302037 | 0.023122 | 5.37E-39 | cis |
| rs527528 | miR-130a-3 | 11:574333 | C | T | 0.681923 | -0.243471 | 0.022237 | 6.73E-28 | cis |

| | | | | | | | | | |
|---|---|---|---|---|---|---|---|---|---|
| rs1089664 | miR-130a-3 | 11:5757188 | C | T | 0.685297 | -0.242537 | 0.022203 | 8.89E-28 | cis |
| rs659165 | miR-130a-3 | 11:5760899 | A | C | 0.571934 | 0.161004 | 0.021115 | 2.44E-14 | cis |
| rs1160132 | miR-130a-3 | 11:5764261 | T | C | 0.692075 | -0.230773 | 0.022881 | 6.38E-24 | cis |
| rs1236464 | miR-130a-3 | 11:5770068 | C | A | 0.560601 | 0.139571 | 0.021186 | 4.46E-11 | cis |
| rs1122913 | miR-130a-3 | 11:5756252 | G | A | 0.684434 | -0.242732 | 0.022144 | 5.85E-28 | cis |
| rs7148446 | miR-130a-3 | 11:5748889 | G | A | 0.689792 | -0.242969 | 0.022416 | 2.25E-27 | cis |
| rs591322 | miR-130a-3 | 11:5768468 | T | G | 0.560767 | 0.141968 | 0.021203 | 2.15E-11 | cis |
| rs1647405 | miR-130a-3 | 11:5747788 | C | T | 0.68315 | -0.239969 | 0.022171 | 2.67E-27 | cis |
| rs606499 | miR-130a-3 | 11:5759384 | G | A | 0.439467 | -0.162246 | 0.020963 | 9.97E-15 | cis |
| rs471398 | miR-130a-3 | 11:5759361 | C | T | 0.438663 | -0.160389 | 0.020992 | 2.16E-14 | cis |
| rs7117020 | miR-130a-3 | 11:5749965 | T | G | 0.553432 | 0.16611 | 0.021055 | 3.04E-15 | cis |
| rs1122906 | miR-130a-3 | 11:5736973 | G | A | 0.752752 | -0.306934 | 0.023511 | 5.96E-39 | cis |
| rs7117205 | miR-130a-3 | 11:5764188 | T | C | 0.692088 | -0.23082 | 0.02288 | 6.23E-24 | cis |
| rs4926 | miR-130a-3 | 11:5738198 | G | A | 0.750066 | -0.301324 | 0.023228 | 1.75E-38 | cis |
| rs1157019 | miR-130a-3 | 11:5756045 | A | C | 0.684149 | -0.242699 | 0.022118 | 5.17E-28 | cis |
| rs4245215 | miR-130a-3 | 11:5771378 | G | A | 0.560778 | 0.137398 | 0.02113 | 7.90E-11 | cis |
| rs488769 | miR-130a-3 | 11:5745072 | A | C | 0.674485 | -0.23365 | 0.02225 | 8.54E-26 | cis |
| rs2836294 | miR-130a-3 | 11:5736900 | C | G | 0.752625 | -0.306489 | 0.023509 | 7.51E-39 | cis |
| rs631350 | miR-130a-3 | 11:5769094 | C | T | 0.560263 | 0.140189 | 0.021186 | 3.66E-11 | cis |
| rs2836295 | miR-130a-3 | 11:5737252 | A | G | 0.75263 | -0.304426 | 0.02343 | 1.34E-38 | cis |
| rs1424753 | miR-130a-3 | 11:5747986 | T | C | 0.449285 | -0.164093 | 0.02102 | 5.87E-15 | cis |
| rs1122914 | miR-130a-3 | 11:5762146 | A | T | 0.683461 | -0.2414 | 0.022336 | 3.17E-27 | cis |
| rs1122917 | miR-130a-3 | 11:5770593 | T | C | 0.560155 | 0.137989 | 0.021119 | 6.40E-11 | cis |
| rs1280725 | miR-130a-3 | 11:5760028 | C | T | 0.684858 | -0.242393 | 0.022231 | 1.11E-27 | cis |
| rs1160144 | miR-130a-3 | 11:5752431 | G | A | 0.684 | -0.242757 | 0.022162 | 6.38E-28 | cis |
| rs673344 | miR-130a-3 | 11:5765714 | G | A | 0.31116 | 0.230743 | 0.023075 | 1.53E-23 | cis |
| rs681397 | miR-130a-3 | 11:5768201 | G | T | 0.558507 | 0.14526 | 0.021263 | 8.41E-12 | cis |
| rs1745562 | miR-130a-3 | 11:5754448 | T | C | 0.567581 | 0.16129 | 0.021032 | 1.73E-14 | cis |
| rs1122911 | miR-130a-3 | 11:5753454 | C | G | 0.683806 | -0.244457 | 0.022182 | 3.05E-28 | cis |
| rs576859 | miR-130a-3 | 11:5748062 | C | A | 0.683179 | -0.240212 | 0.022172 | 2.37E-27 | cis |
| rs1157018 | miR-130a-3 | 11:5755577 | T | C | 0.567672 | 0.161097 | 0.02102 | 1.80E-14 | cis |
| rs662638 | miR-130a-3 | 11:5768676 | T | C | 0.437741 | -0.141722 | 0.021308 | 2.91E-11 | cis |
| rs589914 | miR-130a-3 | 11:5766103 | A | G | 0.311001 | 0.230655 | 0.02307 | 1.55E-23 | cis |
| rs2956982 | miR-130a-3 | 11:5749075 | G | A | 0.552122 | 0.164782 | 0.021049 | 4.94E-15 | cis |
| rs1783978 | miR-130a-3 | 11:5746350 | T | C | 0.45351 | -0.164845 | 0.020962 | 3.72E-15 | cis |
| rs1122917 | miR-130a-3 | 11:5771004 | A | G | 0.560212 | 0.137904 | 0.021115 | 6.54E-11 | cis |
| rs671508 | miR-130a-3 | 11:5763656 | A | T | 0.438934 | -0.161761 | 0.02122 | 2.48E-14 | cis |
| rs682258 | miR-130a-3 | 11:5763719 | G | T | 0.438944 | -0.161767 | 0.021221 | 2.48E-14 | cis |
| rs1127641 | miR-130a-3 | 11:5738585 | C | T | 0.7469 | -0.302271 | 0.023181 | 7.27E-39 | cis |
| rs2836294 | miR-130a-3 | 11:5737191 | G | T | 0.749195 | -0.300469 | 0.023454 | 1.43E-37 | cis |
| rs1745552 | miR-130a-3 | 11:5754045 | C | A | 0.684599 | -0.243484 | 0.022162 | 4.45E-28 | cis |
| rs7943793 | miR-130a-3 | 11:5770848 | T | C | 0.560165 | 0.137902 | 0.021114 | 6.52E-11 | cis |
| rs1647394 | miR-130a-3 | 11:5741858 | C | T | 0.408999 | -0.157474 | 0.021549 | 2.71E-13 | cis |
| rs7106195 | miR-130a-3 | 11:5775234 | T | C | 0.60381 | 0.122514 | 0.021291 | 8.70E-09 | cis |
| rs5639270 | miR-130a-3 | 11:5751839 | T | C | 0.557348 | 0.16563 | 0.021057 | 3.67E-15 | cis |
| rs2926656 | miR-130a-3 | 11:5746538 | C | T | 0.683894 | -0.242084 | 0.022186 | 1.01E-27 | cis |
| rs1783980 | miR-130a-3 | 11:5744832 | C | T | 0.4518 | -0.165066 | 0.021041 | 4.33E-15 | cis |
| rs1783981 | miR-130a-3 | 11:5745109 | T | C | 0.454187 | -0.165439 | 0.021011 | 3.44E-15 | cis |
| rs7938083 | miR-130a-3 | 11:5749362 | C | A | 0.686581 | -0.245553 | 0.022337 | 4.12E-28 | cis |
| rs623541 | miR-130a-3 | 11:5758161 | C | A | 0.439413 | -0.163085 | 0.020971 | 7.45E-15 | cis |
| rs615133 | miR-130a-3 | 11:5768494 | C | A | 0.561411 | 0.141502 | 0.021175 | 2.35E-11 | cis |
| rs1157017 | miR-130a-3 | 11:5752942 | A | G | 0.566649 | 0.1615 | 0.021063 | 1.75E-14 | cis |
| rs7949841 | miR-130a-3 | 11:5752470 | G | C | 0.684177 | -0.243105 | 0.022173 | 5.69E-28 | cis |

| | | | | | | | | | |
|---|---|---|---|---|---|---|---|---|---|
| rs1236442 | miR-130a-3 | 11:577311 | A | G | 0.560157 | 0.135455 | 0.021001 | 1.12E-10 | cis |
| rs685149 | miR-130a-3 | 11:576574 | A | G | 0.348389 | 0.209244 | 0.022731 | 3.41E-20 | cis |
| rs1089662 | miR-130a-3 | 11:573232 | A | G | 0.871328 | -0.217382 | 0.031621 | 6.22E-12 | cis |
| rs7926999 | miR-130a-3 | 11:577037 | T | C | 0.529289 | 0.133541 | 0.021102 | 2.48E-10 | cis |
| rs7123523 | miR-130a-3 | 11:577288 | G | T | 0.560828 | 0.136116 | 0.021036 | 9.76E-11 | cis |
| rs1783976 | miR-130a-3 | 11:574627 | T | C | 0.456765 | -0.169619 | 0.021185 | 1.18E-15 | cis |
| rs547118 | miR-130a-3 | 11:575950 | C | G | 0.438175 | -0.163182 | 0.020979 | 7.35E-15 | cis |
| rs4611222 | miR-130a-3 | 11:575522 | G | A | 0.56182 | 0.161086 | 0.020967 | 1.56E-14 | cis |
| rs7927640 | miR-130a-3 | 11:577264 | C | G | 0.442342 | -0.136876 | 0.021059 | 8.05E-11 | cis |
| rs616911 | miR-130a-3 | 11:576853 | G | A | 0.558798 | 0.145619 | 0.021196 | 6.42E-12 | cis |
| rs6190527 | miR-130a-3 | 11:577381 | A | G | 0.610228 | 0.124472 | 0.021419 | 6.20E-09 | cis |
| rs3017889 | miR-130a-3 | 11:575208 | A | G | 0.556573 | 0.166392 | 0.021044 | 2.64E-15 | cis |
| rs1075087 | miR-130a-3 | 11:576238 | A | C | 0.437769 | -0.15918 | 0.021015 | 3.60E-14 | cis |
| rs7119973 | miR-130a-3 | 11:577188 | A | T | 0.442122 | -0.137741 | 0.021104 | 6.72E-11 | cis |
| rs1397275 | miR-130a-3 | 11:577304 | C | A | 0.560097 | 0.135454 | 0.020999 | 1.11E-10 | cis |
| rs658936 | miR-130a-3 | 11:575805 | T | G | 0.440007 | -0.163641 | 0.020988 | 6.35E-15 | cis |
| rs644219 | miR-130a-3 | 11:576100 | C | T | 0.56488 | 0.160737 | 0.02104 | 2.18E-14 | cis |
| rs1280735 | miR-130a-3 | 11:576569 | C | T | 0.69404 | -0.230744 | 0.023031 | 1.26E-23 | cis |
| rs1783979 | miR-130a-3 | 11:574681 | T | C | 0.451998 | -0.164991 | 0.021014 | 4.11E-15 | cis |
| rs1089663 | miR-130a-3 | 11:573812 | T | C | 0.750433 | -0.30148 | 0.023269 | 2.17E-38 | cis |
| rs7932091 | miR-130a-3 | 11:576180 | A | T | 0.683808 | -0.241876 | 0.022349 | 2.69E-27 | cis |
| rs1785498 | miR-130a-3 | 11:574489 | C | G | 0.451781 | -0.165074 | 0.021041 | 4.32E-15 | cis |
| rs1280534 | miR-130a-3 | 11:575247 | C | A | 0.566156 | 0.161834 | 0.021069 | 1.58E-14 | cis |
| rs1279066 | miR-130a-3 | 11:576672 | T | C | 0.697455 | -0.220083 | 0.022986 | 1.02E-21 | cis |
| rs672537 | miR-130a-3 | 11:576932 | C | T | 0.559778 | 0.13944 | 0.02119 | 4.69E-11 | cis |
| rs1280497 | miR-130a-3 | 11:574637 | T | C | 0.686355 | -0.244827 | 0.022231 | 3.32E-28 | cis |
| rs7123782 | miR-130a-3 | 11:577289 | G | A | 0.559575 | 0.13451 | 0.021009 | 1.53E-10 | cis |
| rs1783977 | miR-130a-3 | 11:574627 | T | C | 0.454153 | -0.16407 | 0.021046 | 6.40E-15 | cis |
| rs7117896 | miR-130a-3 | 11:573885 | G | A | 0.746843 | -0.302337 | 0.023182 | 7.07E-39 | cis |
| rs1279820 | miR-130a-3 | 11:576684 | T | G | 0.697389 | -0.220092 | 0.022987 | 1.02E-21 | cis |
| rs7926074 | miR-130a-3 | 11:576129 | T | C | 0.687478 | -0.245723 | 0.022451 | 7.04E-28 | cis |
| rs1242022 | miR-130a-3 | 11:577145 | T | C | 0.560173 | 0.137818 | 0.021111 | 6.66E-11 | cis |
| rs7479922 | miR-130a-3 | 11:574701 | A | G | 0.450674 | -0.164621 | 0.021018 | 4.78E-15 | cis |
| rs1280584 | miR-130a-3 | 11:574438 | G | A | 0.687846 | -0.245886 | 0.022275 | 2.49E-28 | cis |
| rs7121169 | miR-130a-3 | 11:574525 | G | A | 0.687601 | -0.246593 | 0.022277 | 1.77E-28 | cis |
| rs627145 | miR-130a-3 | 11:576024 | G | A | 0.439595 | -0.161806 | 0.021052 | 1.52E-14 | cis |
| rs397422 | miR-130b-3 | 22:220276 | C | T | 0.642779 | 0.153977 | 0.024292 | 2.32E-10 | cis |
| rs3512262 | miR-130b-3 | 22:220468 | T | C | 0.644899 | 0.151204 | 0.02421 | 4.22E-10 | cis |
| rs861840 | miR-130b-3 | 22:220106 | G | A | 0.846604 | -0.194908 | 0.03122 | 4.29E-10 | cis |
| rs412596 | miR-130b-3 | 22:220057 | C | G | 0.183539 | 0.19505 | 0.030002 | 7.97E-11 | cis |
| rs1647705 | miR-130b-3 | 22:220254 | A | G | 0.834396 | -0.184583 | 0.030355 | 1.20E-09 | cis |
| rs373001 | miR-130b-3 | 22:220074 | C | T | 0.563659 | 0.176513 | 0.022904 | 1.29E-14 | cis |
| rs396913 | miR-130b-3 | 22:220398 | A | C | 0.64615 | 0.150807 | 0.024263 | 5.11E-10 | cis |
| rs1386471 | miR-130b-3 | 22:220311 | C | T | 0.844265 | -0.195246 | 0.030952 | 2.83E-10 | cis |
| rs861833 | miR-130b-3 | 22:220391 | C | T | 0.842381 | -0.19536 | 0.030506 | 1.51E-10 | cis |
| rs861832 | miR-130b-3 | 22:220403 | G | A | 0.841186 | -0.19377 | 0.030432 | 1.92E-10 | cis |
| rs369463 | miR-130b-3 | 22:220152 | C | T | 0.646159 | 0.161918 | 0.024635 | 4.94E-11 | cis |
| rs412360 | miR-130b-3 | 22:220197 | C | A | 0.666254 | 0.153878 | 0.024885 | 6.26E-10 | cis |
| rs861818 | miR-130b-3 | 22:220515 | G | T | 0.838891 | -0.190695 | 0.03043 | 3.69E-10 | cis |
| rs5749696 | miR-130b-3 | 22:220633 | T | C | 0.480386 | 0.138975 | 0.022812 | 1.11E-09 | cis |
| rs1248406 | miR-130b-3 | 22:220497 | C | T | 0.669142 | 0.147292 | 0.024624 | 2.21E-09 | cis |
| rs710176 | miR-130b-3 | 22:220044 | G | A | 0.844524 | -0.196467 | 0.031255 | 3.26E-10 | cis |
| rs861829 | miR-130b-3 | 22:220423 | C | T | 0.842272 | -0.195375 | 0.030524 | 1.55E-10 | cis |

| | | | | | | | | | |
|---|---|---|---|---|---|---|---|---|---|
| rs861826 | miR-130b-3 | 22:220456( | C | T | 0.382818 | -0.138417 | 0.023878 | 6.76E-09 | cis |
| rs2845458 | miR-130b-3 | 22:220652! | G | A | 0.833428 | -0.177547 | 0.030147 | 3.88E-09 | cis |
| rs450469 | miR-130b-3 | 22:220056. | C | T | 0.550569 | 0.182711 | 0.023418 | 6.09E-15 | cis |
| rs447001 | miR-130b-3 | 22:220109. | C | T | 0.558786 | 0.173303 | 0.02256 | 1.57E-14 | cis |
| rs861821 | miR-130b-3 | 22:220472! | G | A | 0.840506 | -0.193056 | 0.030471 | 2.36E-10 | cis |
| rs452614 | miR-130b-3 | 22:220404. | A | G | 0.636803 | 0.151915 | 0.0244 | 4.79E-10 | cis |
| rs5999098 | miR-130b-3 | 22:220488( | C | T | 0.667896 | 0.146662 | 0.024584 | 2.43E-09 | cis |
| rs2599423 | miR-130b-3 | 22:220329! | C | T | 0.384714 | -0.142462 | 0.023951 | 2.71E-09 | cis |
| rs368351 | miR-130b-3 | 22:220025! | T | C | 0.184797 | 0.202758 | 0.030783 | 4.50E-11 | cis |
| rs374659 | miR-130b-3 | 22:220056! | G | C | 0.183374 | 0.195102 | 0.029997 | 7.82E-11 | cis |
| rs1647711 | miR-130b-3 | 22:220371! | T | C | 0.841321 | -0.193677 | 0.030406 | 1.90E-10 | cis |
| rs7349039 | miR-130b-3 | 22:220681. | C | T | 0.461667 | 0.133934 | 0.022881 | 4.81E-09 | cis |
| rs436496 | miR-130b-3 | 22:220414. | C | T | 0.6462 | 0.150798 | 0.024265 | 5.14E-10 | cis |
| rs861820 | miR-130b-3 | 22:220493( | G | T | 0.841549 | -0.194398 | 0.030554 | 1.99E-10 | cis |
| rs1647710 | miR-130b-3 | 22:220341! | A | G | 0.386466 | -0.143744 | 0.023958 | 1.97E-09 | cis |
| rs370039 | miR-130b-3 | 22:220443! | G | A | 0.646238 | 0.150806 | 0.024267 | 5.15E-10 | cis |
| rs9607194 | miR-130b-3 | 22:220683. | G | A | 0.835071 | -0.177992 | 0.030306 | 4.28E-09 | cis |
| rs1103229 | miR-130b-3 | 22:220220( | G | A | 0.843078 | -0.195571 | 0.030515 | 1.46E-10 | cis |
| rs861813 | miR-130b-3 | 22:220585! | T | C | 0.568644 | -0.141856 | 0.022931 | 6.16E-10 | cis |
| rs881091 | miR-130b-3 | 22:220365. | C | T | 0.841213 | -0.193724 | 0.030408 | 1.88E-10 | cis |
| rs1669122 | miR-130b-3 | 22:220287. | A | G | 0.383988 | -0.140974 | 0.023886 | 3.59E-09 | cis |
| rs710171 | miR-130b-3 | 22:220644. | C | T | 0.835765 | -0.179973 | 0.03025 | 2.69E-09 | cis |
| rs2019746 | miR-130b-3 | 22:220654. | T | C | 0.194666 | 0.178022 | 0.029211 | 1.10E-09 | cis |
| rs1669116 | miR-130b-3 | 22:220346! | T | C | 0.402304 | -0.152351 | 0.023745 | 1.40E-10 | cis |
| rs861844 | miR-130b-3 | 22:220062. | G | T | 0.844522 | -0.193827 | 0.031096 | 4.57E-10 | cis |
| rs1669112 | miR-130b-3 | 22:220368. | T | C | 0.386517 | -0.14338 | 0.023868 | 1.89E-09 | cis |
| rs1669123 | miR-130b-3 | 22:220283. | G | A | 0.842806 | -0.195362 | 0.030525 | 1.55E-10 | cis |
| rs861831 | miR-130b-3 | 22:220407. | G | A | 0.842014 | -0.195551 | 0.030536 | 1.51E-10 | cis |
| rs372150 | miR-130b-3 | 22:220073. | T | C | 0.183322 | 0.193202 | 0.02973 | 8.11E-11 | cis |
| rs1669119 | miR-130b-3 | 22:220313. | C | T | 0.839997 | -0.184875 | 0.030889 | 2.16E-09 | cis |
| rs861811 | miR-130b-3 | 22:220604. | C | T | 0.835157 | -0.180714 | 0.030138 | 2.02E-09 | cis |
| rs861846 | miR-130b-3 | 22:220025! | A | G | 0.847398 | -0.205514 | 0.032039 | 1.41E-10 | cis |
| rs861843 | miR-130b-3 | 22:220075. | C | G | 0.84396 | -0.193491 | 0.030949 | 4.06E-10 | cis |
| rs449597 | miR-130b-3 | 22:220026. | A | C | 0.55856 | 0.184464 | 0.024055 | 1.74E-14 | cis |
| rs1782132. | miR-130b-3 | 22:220685. | G | A | 0.835489 | -0.177974 | 0.030285 | 4.19E-09 | cis |
| rs1669124 | miR-130b-3 | 22:220277. | A | G | 0.385747 | -0.142313 | 0.023882 | 2.54E-09 | cis |
| rs861825 | miR-130b-3 | 22:220463. | C | T | 0.822993 | -0.160586 | 0.029361 | 4.52E-08 | cis |
| rs108815 | miR-130b-3 | 22:220254. | A | G | 0.665524 | 0.151687 | 0.024717 | 8.41E-10 | cis |
| rs861827 | miR-130b-3 | 22:220450. | T | C | 0.382437 | -0.137602 | 0.023877 | 8.26E-09 | cis |
| rs2845457 | miR-130b-3 | 22:220652! | G | A | 0.834927 | -0.176858 | 0.030241 | 4.97E-09 | cis |
| rs439760 | miR-130b-3 | 22:220030! | G | A | 0.560517 | 0.183713 | 0.023784 | 1.12E-14 | cis |
| rs861841 | miR-130b-3 | 22:220101! | G | C | 0.844687 | -0.194394 | 0.030924 | 3.26E-10 | cis |
| rs861828 | miR-130b-3 | 22:220424. | C | T | 0.84079 | -0.193036 | 0.030403 | 2.16E-10 | cis |
| rs861845 | miR-130b-3 | 22:220039. | C | G | 0.847962 | -0.20262 | 0.031888 | 2.10E-10 | cis |
| rs5754727 | miR-130b-3 | 22:220572. | G | A | 0.5885 | 0.157278 | 0.02306 | 9.08E-12 | cis |
| rs3934574 | miR-133b | 20:611434! | T | C | 0.464792 | -0.178563 | 0.025429 | 2.19E-12 | trans |
| rs7621371. | miR-133b | 20:611509. | A | G | 0.467251 | -0.190849 | 0.027215 | 2.34E-12 | trans |
| rs1190646. | miR-133b | 20:611589! | C | T | 0.824779 | -0.248529 | 0.03256 | 2.29E-14 | trans |
| rs6062284 | miR-133b | 20:611419! | T | G | 0.4602 | -0.17216 | 0.025716 | 2.16E-11 | trans |
| rs8122282 | miR-133b | 20:611523. | G | A | 0.467131 | -0.189725 | 0.027217 | 3.15E-12 | trans |
| rs6062267 | miR-133b | 20:611533( | T | C | 0.464907 | -0.190202 | 0.027172 | 2.56E-12 | trans |
| rs1262460! | miR-133b | 20:611552! | C | A | 0.796375 | -0.264427 | 0.033362 | 2.26E-15 | trans |
| rs1243369! | miR-134-5p | 14:101035! | C | G | 0.723403 | 0.241945 | 0.031754 | 2.55E-14 | cis |

| | | | | | | | | | |
|---|---|---|---|---|---|---|---|---|---|
| rs879059 | miR-134-5ƿ | 14:101069! | T | A | 0.31068 | -0.24629 | 0.03109 | 2.34E-15 | cis |
| rs7154024 | miR-134-5ƿ | 14:101031: | G | T | 0.725015 | 0.243619 | 0.031936 | 2.38E-14 | cis |
| rs1190872 | miR-134-5ƿ | 14:101069: | A | G | 0.310177 | -0.246205 | 0.031107 | 2.48E-15 | cis |
| rs7147982 | miR-134-5ƿ | 14:101046( | C | T | 0.731753 | 0.252572 | 0.031775 | 1.88E-15 | cis |
| rs8008285 | miR-134-5ƿ | 14:101146! | T | C | 0.842905 | 0.251791 | 0.040042 | 3.21E-10 | cis |
| rs1190867 | miR-134-5ƿ | 14:101069: | G | T | 0.316788 | -0.246632 | 0.031332 | 3.50E-15 | cis |
| rs6575791 | miR-134-5ƿ | 14:101029: | G | A | 0.719841 | 0.24412 | 0.03196 | 2.20E-14 | cis |
| rs1243449( | miR-134-5ƿ | 14:101179: | A | G | 0.18959 | -0.337119 | 0.036712 | 4.20E-20 | cis |
| rs1214700ℰ | miR-134-5ƿ | 14:101186: | C | T | 0.223221 | -0.274446 | 0.034204 | 1.02E-15 | cis |
| rs1190862 | miR-134-5ƿ | 14:101067: | T | G | 0.309628 | -0.244552 | 0.030926 | 2.62E-15 | cis |
| rs7149242 | miR-134-5ƿ | 14:101159⁴ | T | G | 0.288371 | -0.269873 | 0.031974 | 3.17E-17 | cis |
| rs7493817 | miR-134-5ƿ | 14:101038⁴ | G | A | 0.751723 | 0.27112 | 0.033102 | 2.60E-16 | cis |
| rs7155375 | miR-134-5ƿ | 14:101188: | C | T | 0.635995 | 0.219764 | 0.02937 | 7.29E-14 | cis |
| rs1888581 | miR-134-5ƿ | 14:101042: | A | C | 0.735363 | 0.256011 | 0.032003 | 1.25E-15 | cis |
| rs879060 | miR-134-5ƿ | 14:101069ℰ | A | G | 0.310926 | -0.246775 | 0.031109 | 2.15E-15 | cis |
| rs7142683 | miR-134-5ƿ | 14:101158ℰ | A | G | 0.290748 | -0.271186 | 0.032064 | 2.73E-17 | cis |
| rs7143357 | miR-134-5ƿ | 14:101054: | T | C | 0.739636 | 0.27309 | 0.032349 | 3.12E-17 | cis |
| rs1289379( | miR-134-5ƿ | 14:101178ℰ | C | G | 0.190813 | -0.338808 | 0.036736 | 2.89E-20 | cis |
| rs1116059⁴ | miR-134-5ƿ | 14:101049! | C | T | 0.741662 | 0.279929 | 0.032294 | 4.40E-18 | cis |
| rs1555405 | miR-134-5ƿ | 14:101176: | G | A | 0.749075 | 0.256575 | 0.03237 | 2.26E-15 | cis |
| rs4905994 | miR-134-5ƿ | 14:101154( | T | C | 0.329146 | -0.245377 | 0.03112 | 3.15E-15 | cis |
| rs1015151! | miR-134-5ƿ | 14:101175: | G | A | 0.806743 | 0.212177 | 0.03585 | 3.25E-09 | cis |
| rs7140556 | miR-134-5ƿ | 14:101036⁴ | C | A | 0.723818 | 0.244488 | 0.031891 | 1.77E-14 | cis |
| rs1757758( | miR-134-5ƿ | 14:101189: | C | T | 0.827094 | 0.24034 | 0.037656 | 1.74E-10 | cis |
| rs1709962: | miR-134-5ƿ | 14:101173: | A | C | 0.783449 | 0.189368 | 0.034664 | 4.68E-08 | cis |
| rs1184948! | miR-134-5ƿ | 14:101037: | C | G | 0.733365 | 0.258552 | 0.03208 | 7.65E-16 | cis |
| rs1190856 | miR-134-5ƿ | 14:101065! | T | C | 0.310244 | -0.24783 | 0.031135 | 1.72E-15 | cis |
| rs9324022 | miR-134-5ƿ | 14:101172⁴ | T | C | 0.763739 | 0.242341 | 0.035125 | 5.23E-12 | cis |
| rs1243613( | miR-134-5ƿ | 14:101031( | C | T | 0.725015 | 0.243856 | 0.031964 | 2.36E-14 | cis |
| rs1190881 | miR-134-5ƿ | 14:101079ℰ | T | C | 0.346494 | -0.184288 | 0.030396 | 1.34E-09 | cis |
| rs1185095: | miR-134-5ƿ | 14:101056: | G | T | 0.743981 | 0.273933 | 0.032153 | 1.60E-17 | cis |
| rs6575794 | miR-134-5ƿ | 14:101039: | C | T | 0.734189 | 0.259623 | 0.032124 | 6.37E-16 | cis |
| rs1190857 | miR-134-5ƿ | 14:101066: | G | T | 0.310273 | -0.247675 | 0.031142 | 1.82E-15 | cis |
| rs1190850 | miR-134-5ƿ | 14:101064: | A | G | 0.314302 | -0.251371 | 0.031341 | 1.05E-15 | cis |
| rs1190852 | miR-134-5ƿ | 14:101064! | A | G | 0.310295 | -0.24896 | 0.031157 | 1.34E-15 | cis |
| rs1087352( | miR-134-5ƿ | 14:101175! | G | A | 0.809826 | 0.215086 | 0.036269 | 3.02E-09 | cis |
| rs9671394 | miR-134-5ƿ | 14:101159( | C | T | 0.284232 | -0.276585 | 0.032415 | 1.43E-17 | cis |
| rs7145610 | miR-134-5ƿ | 14:101049⁴ | T | C | 0.740391 | 0.280479 | 0.03238 | 4.63E-18 | cis |
| rs879058 | miR-134-5ƿ | 14:101069! | G | T | 0.311807 | -0.246554 | 0.031107 | 2.26E-15 | cis |
| rs1014438: | miR-136-5ƿ | 14:101173: | C | T | 0.673446 | 0.405272 | 0.036589 | 1.64E-28 | cis |
| rs8021301 | miR-136-5ƿ | 14:101166! | G | C | 0.668324 | 0.400327 | 0.036391 | 3.80E-28 | cis |
| rs2837918( | miR-136-5ƿ | 14:101168( | G | T | 0.836434 | 0.456746 | 0.046892 | 2.03E-22 | cis |
| rs9671394 | miR-136-5ƿ | 14:101159( | C | T | 0.284232 | -0.398989 | 0.039355 | 3.73E-24 | cis |
| rs1014551: | miR-136-5ƿ | 14:101168! | A | G | 0.668173 | 0.399046 | 0.03635 | 4.89E-28 | cis |
| rs1243449( | miR-136-5ƿ | 14:101179: | A | G | 0.18959 | -0.494684 | 0.044481 | 9.88E-29 | cis |
| rs1709961⁴ | miR-136-5ƿ | 14:101167( | C | G | 0.668809 | 0.40362 | 0.036386 | 1.36E-28 | cis |
| rs4905999 | miR-136-5ƿ | 14:101184: | A | G | 0.181905 | -0.479304 | 0.045375 | 4.41E-26 | cis |
| rs7335877ℰ | miR-136-5ƿ | 14:101159! | C | T | 0.891417 | 0.411586 | 0.058076 | 1.37E-12 | cis |
| rs7149242 | miR-136-5ƿ | 14:101159⁴ | T | G | 0.288371 | -0.392035 | 0.038812 | 5.48E-24 | cis |
| rs8020868 | miR-136-5ƿ | 14:101166! | A | G | 0.668343 | 0.400258 | 0.036388 | 3.84E-28 | cis |
| rs7142683 | miR-136-5ƿ | 14:101158ℰ | A | G | 0.290748 | -0.392753 | 0.038925 | 6.11E-24 | cis |
| rs1014696: | miR-136-5ƿ | 14:101170! | T | C | 0.658875 | 0.39858 | 0.035752 | 7.29E-29 | cis |
| rs5771269: | miR-136-5ƿ | 14:101161: | G | A | 0.894439 | 0.417877 | 0.058532 | 9.38E-13 | cis |

| | | | | | | | | | |
|---|---|---|---|---|---|---|---|---|---|
| rs1214784! | miR-136-5p | 14:101144! | C | T | 0.870989 | 0.380494 | 0.05243 | 3.95E-13 | cis |
| rs495203 | miR-139-5p | 9:1361452 | C | T | 0.663279 | -0.157265 | 0.01161 | 8.37E-42 | trans |
| rs532436 | miR-139-5p | 9:1361498 | G | A | 0.8067 | -0.176906 | 0.013943 | 6.95E-37 | trans |
| rs687289 | miR-139-5p | 9:1361371( | G | A | 0.668579 | -0.161876 | 0.011737 | 2.87E-43 | trans |
| rs676457 | miR-139-5p | 9:1361462 | A | T | 0.664166 | -0.15736 | 0.011604 | 6.87E-42 | trans |
| rs676996 | miR-139-5p | 9:1361460 | T | G | 0.667516 | -0.157346 | 0.011708 | 3.57E-41 | trans |
| rs492488 | miR-139-5p | 9:1361449 | G | A | 0.663579 | -0.157308 | 0.011612 | 8.23E-42 | trans |
| rs613534 | miR-139-5p | 9:1361431 | A | G | 0.653693 | -0.153841 | 0.011595 | 3.54E-40 | trans |
| rs582094 | miR-139-5p | 9:1361454 | A | T | 0.663709 | -0.157205 | 0.0116 | 7.69E-42 | trans |
| rs677355 | miR-139-5p | 9:1361460 | G | A | 0.670198 | -0.157852 | 0.011814 | 1.01E-40 | trans |
| rs8176663 | miR-139-5p | 9:1361444 | T | C | 0.663959 | -0.157215 | 0.011603 | 7.92E-42 | trans |
| rs582118 | miR-139-5p | 9:1361454 | A | G | 0.663872 | -0.157298 | 0.011605 | 7.44E-42 | trans |
| rs544873 | miR-139-5p | 9:1361432 | G | A | 0.645742 | -0.152029 | 0.011396 | 1.34E-40 | trans |
| rs649129 | miR-139-5p | 9:1361543( | C | T | 0.787432 | -0.165163 | 0.013462 | 1.34E-34 | trans |
| rs643434 | miR-139-5p | 9:1361423 | G | A | 0.645392 | -0.151972 | 0.01139 | 1.31E-40 | trans |
| rs550057 | miR-139-5p | 9:1361465 | C | T | 0.734085 | -0.147772 | 0.012554 | 5.52E-32 | trans |
| rs491626 | miR-139-5p | 9:1361448 | C | T | 0.664173 | -0.157582 | 0.011633 | 8.32E-42 | trans |
| rs612169 | miR-139-5p | 9:1361434 | A | G | 0.66434 | -0.157438 | 0.011618 | 7.83E-42 | trans |
| rs493246 | miR-139-5p | 9:1361449 | G | A | 0.663234 | -0.15741 | 0.011618 | 8.06E-42 | trans |
| rs644234 | miR-139-5p | 9:1361422 | T | G | 0.645381 | -0.151953 | 0.011382 | 1.17E-40 | trans |
| rs545971 | miR-139-5p | 9:1361433 | C | T | 0.663766 | -0.157102 | 0.011598 | 8.40E-42 | trans |
| rs554833 | miR-139-5p | 9:1361471 | C | T | 0.664057 | -0.157337 | 0.011597 | 6.24E-42 | trans |
| rs543040 | miR-139-5p | 9:1361430( | A | T | 0.669196 | -0.159045 | 0.011822 | 2.94E-41 | trans |
| rs494242 | miR-139-5p | 9:1361451 | C | T | 0.644977 | -0.152252 | 0.011401 | 1.11E-40 | trans |
| rs543968 | miR-139-5p | 9:1361431 | T | C | 0.653829 | -0.153713 | 0.011597 | 4.23E-40 | trans |
| rs2519093 | miR-139-5p | 9:1361418 | C | T | 0.809892 | -0.179103 | 0.014192 | 1.63E-36 | trans |
| rs495828 | miR-139-5p | 9:1361548 | G | T | 0.787276 | -0.164962 | 0.013454 | 1.46E-34 | trans |
| rs674302 | miR-139-5p | 9:1361466 | T | A | 0.663989 | -0.157335 | 0.011595 | 6.05E-42 | trans |
| rs529565 | miR-139-5p | 9:1361495( | T | C | 0.665911 | -0.159786 | 0.011585 | 2.82E-43 | trans |
| rs1165522 | miR-139-5p | 9:1361490 | T | A | 0.653719 | -0.15545 | 0.011571 | 3.77E-41 | trans |
| rs600038 | miR-139-5p | 9:1361518( | T | C | 0.787872 | -0.166058 | 0.013486 | 7.68E-35 | trans |
| rs2769071 | miR-139-5p | 9:1361459 | A | G | 0.663117 | -0.157213 | 0.011626 | 1.14E-41 | trans |
| rs3482677 | miR-143-3p | 8:1065709 | G | T | 0.749028 | -0.123023 | 0.018989 | 9.26E-11 | trans |
| rs2343592 | miR-143-3p | 8:1065722 | A | G | 0.747196 | -0.12391 | 0.018929 | 5.91E-11 | trans |
| rs4734879 | miR-143-3p | 8:1065831 | A | G | 0.731553 | -0.120528 | 0.018559 | 8.35E-11 | trans |
| rs7832219 | miR-143-3p | 8:1065789 | T | C | 0.740634 | -0.116485 | 0.018685 | 4.55E-10 | trans |
| rs4734879 | miR-145-5p | 8:1065831 | A | G | 0.731553 | -0.123659 | 0.022221 | 2.62E-08 | trans |
| rs3482677 | miR-145-5p | 8:1065709 | G | T | 0.749028 | -0.12525 | 0.022738 | 3.62E-08 | trans |
| rs2343592 | miR-145-5p | 8:1065722 | A | G | 0.747196 | -0.127334 | 0.022664 | 1.93E-08 | trans |
| rs3488967 | miR-181a-3 | 1:1992490( | A | G | 0.66157 | -0.153098 | 0.027876 | 3.97E-08 | cis |
| rs2060581 | miR-181a-3 | 1:1992502 | T | C | 0.66838 | -0.159056 | 0.027673 | 9.04E-09 | cis |
| rs1091965 | miR-181a-3 | 1:1992486 | A | G | 0.667251 | -0.151535 | 0.027515 | 3.64E-08 | cis |
| rs1763147 | miR-181a-3 | 1:1992522 | T | A | 0.668759 | -0.158031 | 0.027762 | 1.25E-08 | cis |
| rs1270690 | miR-182-5p | 7:1294318 | C | T | 0.353301 | 0.159476 | 0.025996 | 8.54E-10 | cis |
| rs174536 | miR-1908-5 | 11:615519 | A | C | 0.662082 | -0.09869 | 0.017362 | 1.31E-08 | cis |
| rs174592 | miR-1908-5 | 11:616186( | A | G | 0.64461 | -0.10446 | 0.017728 | 3.81E-09 | cis |
| rs174533 | miR-1908-5 | 11:615490 | G | A | 0.662238 | -0.098905 | 0.017379 | 1.26E-08 | cis |
| rs174548 | miR-1908-5 | 11:615713 | C | G | 0.697686 | -0.098291 | 0.01774 | 3.01E-08 | cis |
| rs174535 | miR-1908-5 | 11:615513 | T | C | 0.660499 | -0.100543 | 0.017361 | 6.98E-09 | cis |
| rs102274 | miR-1908-5 | 11:615578 | T | C | 0.665065 | -0.099704 | 0.017367 | 9.41E-09 | cis |
| rs174578 | miR-1908-5 | 11:616054 | T | A | 0.662872 | -0.095972 | 0.01723 | 2.55E-08 | cis |
| rs174566 | miR-1908-5 | 11:615923 | A | G | 0.662731 | -0.097547 | 0.017257 | 1.58E-08 | cis |
| rs174584 | miR-1908-5 | 11:616107 | G | A | 0.661198 | -0.099164 | 0.017473 | 1.39E-08 | cis |

| | | | | | | | | | |
|---|---|---|---|---|---|---|---|---|---|
| rs174577 | miR-1908-5 | 11:616048 | C | A | 0.662612 | -0.09691 | 0.017239 | 1.89E-08 | cis |
| rs174560 | miR-1908-5 | 11:615817 | T | C | 0.698363 | -0.096727 | 0.017734 | 4.91E-08 | cis |
| rs174553 | miR-1908-5 | 11:615751 | A | G | 0.666576 | -0.096501 | 0.017338 | 2.61E-08 | cis |
| rs174537 | miR-1908-5 | 11:615526 | G | T | 0.664974 | -0.099577 | 0.017345 | 9.41E-09 | cis |
| rs174530 | miR-1908-5 | 11:615465 | A | G | 0.637539 | -0.098809 | 0.017301 | 1.12E-08 | cis |
| rs99780 | miR-1908-5 | 11:615966 | C | T | 0.663955 | -0.097377 | 0.017279 | 1.74E-08 | cis |
| rs174545 | miR-1908-5 | 11:615693 | C | G | 0.66597 | -0.096772 | 0.017315 | 2.29E-08 | cis |
| rs174574 | miR-1908-5 | 11:616003 | A | C | 0.339789 | 0.099367 | 0.017256 | 8.50E-09 | cis |
| rs174564 | miR-1908-5 | 11:615883 | A | G | 0.667624 | -0.098529 | 0.017441 | 1.61E-08 | cis |
| rs174544 | miR-1908-5 | 11:615677 | C | A | 0.704181 | -0.100055 | 0.017907 | 2.30E-08 | cis |
| rs174568 | miR-1908-5 | 11:615938 | C | T | 0.669005 | -0.095364 | 0.017334 | 3.76E-08 | cis |
| rs174580 | miR-1908-5 | 11:616066 | A | G | 0.66143 | -0.097149 | 0.017258 | 1.81E-08 | cis |
| rs174581 | miR-1908-5 | 11:616066 | G | A | 0.661436 | -0.097359 | 0.017254 | 1.68E-08 | cis |
| rs174549 | miR-1908-5 | 11:615713 | G | A | 0.701463 | -0.097531 | 0.017769 | 4.04E-08 | cis |
| rs174562 | miR-1908-5 | 11:615851 | A | G | 0.667546 | -0.094474 | 0.017322 | 4.92E-08 | cis |
| rs174601 | miR-1908-5 | 11:616231 | C | T | 0.641558 | -0.103584 | 0.017806 | 5.98E-09 | cis |
| rs174554 | miR-1908-5 | 11:615794 | A | G | 0.666614 | -0.09578 | 0.017315 | 3.17E-08 | cis |
| rs174567 | miR-1908-5 | 11:615930 | A | G | 0.663295 | -0.098237 | 0.017285 | 1.32E-08 | cis |
| rs174576 | miR-1908-5 | 11:616035 | C | A | 0.663911 | -0.095346 | 0.01723 | 3.13E-08 | cis |
| rs495203 | miR-195-5 | 9:1361452 | C | T | 0.663279 | -0.113756 | 0.013422 | 2.34E-17 | trans |
| rs676457 | miR-195-5 | 9:1361462 | A | T | 0.664166 | -0.113865 | 0.013416 | 2.12E-17 | trans |
| rs492488 | miR-195-5 | 9:1361449 | G | A | 0.663579 | -0.113656 | 0.013425 | 2.54E-17 | trans |
| rs543040 | miR-195-5 | 9:1361430 | A | T | 0.669196 | -0.115636 | 0.01366 | 2.56E-17 | trans |
| rs491626 | miR-195-5 | 9:1361448 | C | T | 0.664173 | -0.113996 | 0.013448 | 2.32E-17 | trans |
| rs677355 | miR-195-5 | 9:1361460 | G | A | 0.670198 | -0.114344 | 0.013648 | 5.38E-17 | trans |
| rs676996 | miR-195-5 | 9:1361460 | T | G | 0.667516 | -0.114229 | 0.013529 | 3.08E-17 | trans |
| rs582094 | miR-195-5 | 9:1361454 | A | T | 0.663709 | -0.113779 | 0.01341 | 2.17E-17 | trans |
| rs582118 | miR-195-5 | 9:1361454 | A | G | 0.663872 | -0.11383 | 0.013416 | 2.17E-17 | trans |
| rs687289 | miR-195-5 | 9:1361371 | G | A | 0.668579 | -0.116352 | 0.013584 | 1.08E-17 | trans |
| rs493246 | miR-195-5 | 9:1361449 | G | A | 0.663234 | -0.11379 | 0.013432 | 2.42E-17 | trans |
| rs2769071 | miR-195-5 | 9:1361459 | A | G | 0.663117 | -0.113399 | 0.01344 | 3.25E-17 | trans |
| rs532436 | miR-195-5 | 9:1361498 | G | A | 0.8067 | -0.13917 | 0.01603 | 3.89E-18 | trans |
| rs545971 | miR-195-5 | 9:1361433 | C | T | 0.663766 | -0.113518 | 0.013409 | 2.54E-17 | trans |
| rs554833 | miR-195-5 | 9:1361471 | C | T | 0.664057 | -0.113794 | 0.013408 | 2.11E-17 | trans |
| rs529565 | miR-195-5 | 9:1361495 | T | C | 0.665911 | -0.116479 | 0.013401 | 3.57E-18 | trans |
| rs612169 | miR-195-5 | 9:1361434 | A | G | 0.66434 | -0.113707 | 0.013433 | 2.56E-17 | trans |
| rs8176663 | miR-195-5 | 9:1361444 | T | C | 0.663959 | -0.113603 | 0.013414 | 2.48E-17 | trans |
| rs674302 | miR-195-5 | 9:1361466 | T | A | 0.663989 | -0.113981 | 0.013405 | 1.85E-17 | trans |
| rs2519093 | miR-195-5 | 9:1361418 | C | T | 0.809892 | -0.140415 | 0.016314 | 7.49E-18 | trans |
| rs1076137 | miR-27b-3 | 9:9780708 | G | C | 0.193434 | 0.098613 | 0.016783 | 4.21E-09 | cis |
| rs4744428 | miR-27b-3 | 9:9781336 | T | C | 0.126668 | 0.114369 | 0.019986 | 1.05E-08 | cis |
| rs1506890 | miR-30d-5 | 8:1356803 | G | C | 0.822188 | -0.068614 | 0.010687 | 1.36E-10 | cis |
| rs1170105 | miR-30d-5 | 8:1356878 | C | T | 0.822163 | -0.068814 | 0.010681 | 1.17E-10 | cis |
| rs6776176 | miR-30d-5 | 8:1357189 | G | A | 0.822824 | -0.068631 | 0.010687 | 1.35E-10 | cis |
| rs5627761 | miR-30d-5 | 8:1357265 | G | A | 0.825158 | -0.066968 | 0.01076 | 4.85E-10 | cis |
| rs3426346 | miR-30d-5 | 8:1357211 | G | C | 0.822725 | -0.068469 | 0.010687 | 1.49E-10 | cis |
| rs1327100 | miR-30d-5 | 8:1356746 | T | G | 0.821675 | -0.069939 | 0.010683 | 5.88E-11 | cis |
| rs6728071 | miR-30d-5 | 8:1357056 | C | T | 0.822568 | -0.068179 | 0.010676 | 1.70E-10 | cis |
| rs9969668 | miR-30d-5 | 8:1357315 | G | A | 0.82631 | -0.066501 | 0.010765 | 6.50E-10 | cis |
| rs1328270 | miR-30d-5 | 8:1357321 | C | T | 0.826672 | -0.066772 | 0.010778 | 5.83E-10 | cis |
| rs7818560 | miR-30d-5 | 8:1357319 | T | C | 0.826469 | -0.066555 | 0.010774 | 6.51E-10 | cis |
| rs7152837 | miR-30d-5 | 8:1356846 | C | T | 0.828156 | -0.069795 | 0.010965 | 1.95E-10 | cis |
| rs3508255 | miR-30d-5 | 8:1357059 | C | T | 0.822769 | -0.068412 | 0.010683 | 1.51E-10 | cis |

| | | | | | | | | | |
|---|---|---|---|---|---|---|---|---|---|
| rs3403474! | miR-30d-5p | 8:1357380 | C | A | 0.827279 | -0.066767 | 0.010802 | 6.37E-10 | cis |
| rs1325501! | miR-30d-5p | 8:1357321 | G | A | 0.826669 | -0.06677 | 0.010778 | 5.84E-10 | cis |
| rs3544864 | miR-30d-5p | 8:1357024 | T | C | 0.822565 | -0.06818 | 0.010676 | 1.70E-10 | cis |
| rs6788020 | miR-30d-5p | 8:1357329 | C | T | 0.826501 | -0.066584 | 0.010773 | 6.38E-10 | cis |
| rs1325672! | miR-30d-5p | 8:1356676 | G | A | 0.820973 | -0.069604 | 0.010662 | 6.65E-11 | cis |
| rs6765498 | miR-30d-5p | 8:1357079 | G | A | 0.822544 | -0.068162 | 0.010675 | 1.71E-10 | cis |
| rs3524176 | miR-30d-5p | 8:1356754 | C | T | 0.821656 | -0.069884 | 0.010682 | 6.07E-11 | cis |
| rs5641351 | miR-30d-5p | 8:1356406 | G | A | 0.830335 | -0.066899 | 0.010897 | 8.30E-10 | cis |
| rs1470055 | miR-30d-5p | 8:1356906 | A | T | 0.822194 | -0.068799 | 0.010682 | 1.19E-10 | cis |
| rs1326279 | miR-30d-5p | 8:1357299 | G | C | 0.83353 | -0.065943 | 0.010999 | 2.03E-09 | cis |
| rs1198507 | miR-30d-5p | 8:1356717 | C | T | 0.82138 | -0.069704 | 0.010674 | 6.57E-11 | cis |
| rs1326623 | miR-30d-5p | 8:1357262 | G | T | 0.825149 | -0.06697 | 0.01076 | 4.84E-10 | cis |
| rs1326853 | miR-30d-5p | 8:1357271 | C | T | 0.825953 | -0.066189 | 0.010776 | 8.13E-10 | cis |
| rs1483166 | miR-30d-5p | 8:1356887 | C | A | 0.821815 | -0.068358 | 0.010678 | 1.54E-10 | cis |
| rs6650281 | miR-30d-5p | 8:1357189 | A | C | 0.822819 | -0.068628 | 0.010687 | 1.35E-10 | cis |
| rs5606523 | miR-30d-5p | 8:1357133 | T | C | 0.823842 | -0.070715 | 0.01072 | 4.22E-11 | cis |
| rs7272192 | miR-30d-5p | 8:1357276 | G | A | 0.825962 | -0.066185 | 0.010776 | 8.15E-10 | cis |
| rs1328246 | miR-30d-5p | 8:1357079 | C | T | 0.822381 | -0.068238 | 0.010677 | 1.65E-10 | cis |
| rs7271979 | miR-30d-5p | 8:1356585 | C | G | 0.820724 | -0.069577 | 0.010653 | 6.52E-11 | cis |
| rs6717003 | miR-30d-5p | 8:1356737 | G | C | 0.820817 | -0.069166 | 0.010679 | 9.38E-11 | cis |
| rs3407487 | miR-30d-5p | 8:1356525 | G | A | 0.822224 | -0.070798 | 0.010711 | 3.85E-11 | cis |
| rs6687637 | miR-30d-5p | 8:1357161 | A | G | 0.822526 | -0.068348 | 0.010674 | 1.52E-10 | cis |
| rs1326732 | miR-30d-5p | 8:1357351 | G | A | 0.827212 | -0.066557 | 0.010789 | 6.87E-10 | cis |
| rs1300974 | miR-320a | 2:6858443 | C | A | 0.678986 | 0.125869 | 0.01732 | 3.67E-13 | trans |
| rs6546398 | miR-320a | 2:6863041 | G | C | 0.552757 | 0.152138 | 0.015759 | 4.72E-22 | trans |
| rs5618821 | miR-320a | 2:6861163 | T | C | 0.698075 | 0.139604 | 0.016858 | 1.22E-16 | trans |
| rs6546400 | miR-320a | 2:6863064 | G | A | 0.554382 | 0.152811 | 0.01582 | 4.47E-22 | trans |
| rs4610041 | miR-320a | 2:6862628 | C | T | 0.550745 | 0.155025 | 0.015748 | 7.25E-23 | trans |
| rs6729686 | miR-320a | 2:6863303 | C | T | 0.700171 | 0.145857 | 0.017461 | 6.63E-17 | trans |
| rs1164012 | miR-320a | 2:6862562 | C | T | 0.545777 | 0.151008 | 0.015571 | 3.08E-22 | trans |
| rs1017438 | miR-320a | 2:6859646 | C | T | 0.685243 | 0.144486 | 0.016618 | 3.47E-18 | trans |
| rs7420775 | miR-320a | 2:6862945 | G | T | 0.703122 | 0.143662 | 0.017427 | 1.67E-16 | trans |
| rs3419682 | miR-320a | 2:6862142 | G | A | 0.697518 | 0.139848 | 0.016851 | 1.05E-16 | trans |
| rs6214389 | miR-320a | 2:6860455 | C | T | 0.698307 | 0.139108 | 0.016881 | 1.71E-16 | trans |
| rs1301028 | miR-320a | 2:6862184 | G | T | 0.683506 | 0.145008 | 0.016554 | 1.96E-18 | trans |
| rs7572299 | miR-320a | 2:6858124 | T | A | 0.538033 | 0.140299 | 0.016042 | 2.22E-18 | trans |
| rs6744306 | miR-320a | 2:6863273 | T | C | 0.549956 | 0.155241 | 0.01588 | 1.43E-22 | trans |
| rs1486061 | miR-320a | 2:6862571 | T | G | 0.54743 | 0.151637 | 0.015624 | 2.86E-22 | trans |
| rs6546411 | miR-320a | 2:6863769 | A | G | 0.556248 | 0.157639 | 0.016098 | 1.21E-22 | trans |
| rs6214393 | miR-320a | 2:6864020 | A | G | 0.593308 | 0.142404 | 0.016446 | 4.76E-18 | trans |
| rs6546399 | miR-320a | 2:6863064 | C | T | 0.55394 | 0.15272 | 0.015805 | 4.34E-22 | trans |
| rs5888448 | miR-320a | 2:6864534 | G | A | 0.734214 | 0.144505 | 0.018417 | 4.28E-15 | trans |
| rs1049616 | miR-320a | 2:6862008 | C | T | 0.697496 | 0.139829 | 0.016852 | 1.06E-16 | trans |
| rs1300592 | miR-320a | 2:6862999 | T | A | 0.701261 | 0.142753 | 0.01735 | 1.90E-16 | trans |
| rs1300519 | miR-320a | 2:6863887 | A | G | 0.702186 | 0.147347 | 0.017627 | 6.31E-17 | trans |
| rs6214392 | miR-320a | 2:6863835 | C | G | 0.700378 | 0.148987 | 0.017611 | 2.68E-17 | trans |
| rs1158434 | miR-320a | 2:6863552 | G | C | 0.551072 | 0.155908 | 0.015994 | 1.88E-22 | trans |
| rs7598360 | miR-320a | 2:6863092 | A | G | 0.553166 | 0.155693 | 0.01579 | 6.20E-23 | trans |
| rs6715579 | miR-320a | 2:6858392 | C | T | 0.678794 | 0.125847 | 0.017319 | 3.69E-13 | trans |
| rs1484530 | miR-320a | 2:6863547 | A | G | 0.70445 | 0.148609 | 0.017695 | 4.53E-17 | trans |
| rs6546412 | miR-320a | 2:6863771 | T | C | 0.557086 | 0.15912 | 0.016127 | 5.79E-23 | trans |
| rs6546404 | miR-320a | 2:6863263 | G | T | 0.54995 | 0.15523 | 0.015879 | 1.43E-22 | trans |
| rs5999861 | miR-320a | 2:6864677 | G | T | 0.731295 | 0.13921 | 0.018406 | 3.93E-14 | trans |

| | | | | | | | | | |
|---|---|---|---|---|---|---|---|---|---|
| rs6546397 | miR-320a | 2:6863036 | G | A | 0.552756 | 0.152134 | 0.015759 | 4.74E-22 | trans |
| rs7608429 | miR-320a | 2:6858083 | G | T | 0.678719 | 0.125181 | 0.017336 | 5.16E-13 | trans |
| rs6214393 | miR-320a | 2:6864009 | T | C | 0.59325 | 0.14244 | 0.016445 | 4.65E-18 | trans |
| rs1050181 | miR-320a | 2:6862295 | G | C | 0.544992 | 0.150456 | 0.015516 | 3.10E-22 | trans |
| rs6714719 | miR-320a | 2:6859377 | G | C | 0.698404 | 0.139079 | 0.016929 | 2.11E-16 | trans |
| rs6709464 | miR-320a | 2:6862222 | C | T | 0.697279 | 0.139649 | 0.016836 | 1.09E-16 | trans |
| rs6546403 | miR-320a | 2:6863226 | C | T | 0.550801 | 0.155633 | 0.015897 | 1.24E-22 | trans |
| rs2241946 | miR-320a | 2:6861565 | T | C | 0.698076 | 0.139832 | 0.016864 | 1.12E-16 | trans |
| rs1442589 | miR-320a | 2:6863473 | G | T | 0.701896 | 0.146902 | 0.017592 | 6.80E-17 | trans |
| rs6742200 | miR-320a | 2:6862330 | A | G | 0.54552 | 0.150787 | 0.015557 | 3.25E-22 | trans |
| rs7451452 | miR-320a | 2:6863755 | T | G | 0.702552 | 0.14734 | 0.017642 | 6.74E-17 | trans |
| rs1760168 | miR-320a | 2:6861589 | G | A | 0.697578 | 0.139713 | 0.016864 | 1.18E-16 | trans |
| rs1390688 | miR-320a | 2:6862773 | C | T | 0.69721 | 0.142685 | 0.017183 | 1.01E-16 | trans |
| rs6546408 | miR-320a | 2:6863504 | T | C | 0.551062 | 0.155914 | 0.01599 | 1.84E-22 | trans |
| rs6546410 | miR-320a | 2:6863610 | C | G | 0.702434 | 0.147267 | 0.017635 | 6.78E-17 | trans |
| rs5628470 | miR-320a | 2:6863137 | G | A | 0.697508 | 0.14339 | 0.017241 | 9.03E-17 | trans |
| rs1542135 | miR-320a | 2:6862084 | C | T | 0.69752 | 0.139846 | 0.016851 | 1.05E-16 | trans |
| rs5633714 | miR-320a | 2:6864249 | G | A | 0.733417 | 0.145039 | 0.018352 | 2.72E-15 | trans |
| rs6757938 | miR-320a | 2:6863285 | A | G | 0.549958 | 0.155246 | 0.015882 | 1.44E-22 | trans |
| rs7597167 | miR-320a | 2:6861294 | C | T | 0.698051 | 0.13964 | 0.016858 | 1.20E-16 | trans |
| rs6546395 | miR-320a | 2:6862927 | A | G | 0.55448 | 0.152637 | 0.015805 | 4.56E-22 | trans |
| rs6546407 | miR-320a | 2:6863492 | A | C | 0.55091 | 0.155844 | 0.015974 | 1.74E-22 | trans |
| rs6546401 | miR-320a | 2:6863087 | A | T | 0.550127 | 0.153872 | 0.015747 | 1.49E-22 | trans |
| rs5933292 | miR-320a | 2:6863963 | G | A | 0.73529 | 0.147942 | 0.018524 | 1.39E-15 | trans |
| rs1455505 | miR-320a | 2:6862832 | G | A | 0.556221 | 0.155389 | 0.015875 | 1.26E-22 | trans |
| rs6729555 | miR-320a | 2:6863295 | C | A | 0.54995 | 0.155269 | 0.015883 | 1.43E-22 | trans |
| rs1302892 | miR-320a | 2:6862991 | C | T | 0.552648 | 0.152067 | 0.015754 | 4.78E-22 | trans |
| rs6708536 | miR-320a | 2:6862726 | G | A | 0.547249 | 0.152906 | 0.015663 | 1.63E-22 | trans |
| rs6419644 | miR-320a | 2:6863182 | A | G | 0.549898 | 0.155194 | 0.015873 | 1.41E-22 | trans |
| rs7800275 | miR-320a | 2:6862543 | A | G | 0.546309 | 0.150787 | 0.015553 | 3.17E-22 | trans |
| rs7605387 | miR-320a | 2:6863255 | A | G | 0.703499 | 0.147092 | 0.017558 | 5.40E-17 | trans |
| rs7581872 | miR-320a | 2:6858098 | A | C | 0.537959 | 0.14022 | 0.016042 | 2.31E-18 | trans |
| rs1760209 | miR-320a | 2:6862041 | T | A | 0.697529 | 0.139841 | 0.016851 | 1.05E-16 | trans |
| rs6743970 | miR-320a | 2:6858390 | A | T | 0.660237 | 0.133352 | 0.017104 | 6.37E-15 | trans |
| rs2165920 | miR-320a | 2:6862223 | T | C | 0.552012 | 0.148931 | 0.01552 | 8.32E-22 | trans |
| rs1474969 | miR-320a | 2:6862568 | G | A | 0.546544 | 0.151244 | 0.015595 | 3.06E-22 | trans |
| rs6747545 | miR-320a | 2:6863306 | T | A | 0.549964 | 0.155257 | 0.015884 | 1.45E-22 | trans |
| rs2241947 | miR-320a | 2:6861539 | C | T | 0.697988 | 0.13966 | 0.016857 | 1.18E-16 | trans |
| rs1301077 | miR-320a | 2:6863908 | C | T | 0.702339 | 0.147255 | 0.017632 | 6.73E-17 | trans |
| rs8761 | miR-320a | 2:6862426 | G | A | 0.545692 | 0.150887 | 0.015566 | 3.21E-22 | trans |
| rs1542136 | miR-320a | 2:6862093 | C | A | 0.697594 | 0.139843 | 0.016855 | 1.07E-16 | trans |
| rs9653552 | miR-320a | 2:6864396 | G | A | 0.593458 | 0.14288 | 0.016454 | 3.83E-18 | trans |
| rs6214393 | miR-320a | 2:6864062 | T | C | 0.733261 | 0.145064 | 0.018341 | 2.58E-15 | trans |
| rs3485223 | miR-320a | 2:6863859 | C | G | 0.550658 | 0.155791 | 0.015997 | 2.06E-22 | trans |
| rs1422402 | miR-320a | 2:6862752 | A | G | 0.698498 | 0.143247 | 0.017227 | 9.15E-17 | trans |
| rs1303482 | miR-320a | 2:6863029 | C | G | 0.552715 | 0.152114 | 0.015757 | 4.74E-22 | trans |
| rs1301107 | miR-320a | 2:6863931 | A | C | 0.700844 | 0.148254 | 0.017659 | 4.64E-17 | trans |
| rs7562439 | miR-320a | 2:6858256 | C | T | 0.6646 | 0.131508 | 0.017025 | 1.12E-14 | trans |
| rs7421228 | miR-320a | 2:6863012 | G | T | 0.55268 | 0.152092 | 0.015756 | 4.77E-22 | trans |
| rs5808991 | miR-320a | 2:6863966 | G | C | 0.734634 | 0.147894 | 0.018473 | 1.19E-15 | trans |
| rs1343065 | miR-320a | 2:6858771 | C | G | 0.699397 | 0.1397 | 0.017062 | 2.66E-16 | trans |
| rs7578602 | miR-320a | 2:6863786 | G | C | 0.556806 | 0.159076 | 0.016134 | 6.23E-23 | trans |
| rs2070171 | miR-320a | 2:6861371 | G | A | 0.698024 | 0.139616 | 0.016857 | 1.21E-16 | trans |

| | | | | | | | | | |
|---|---|---|---|---|---|---|---|---|---|
| rs1301143 | miR-320a | 2:6863937 | C | T | 0.702382 | 0.147277 | 0.017634 | 6.72E-17 | trans |
| rs1301157 | miR-320a | 2:6863927 | G | A | 0.702349 | 0.147267 | 0.017633 | 6.72E-17 | trans |
| rs1760149 | miR-320a | 2:6861171 | A | G | 0.69807 | 0.139607 | 0.016858 | 1.22E-16 | trans |
| rs1156629 | miR-320a | 2:6862663 | T | G | 0.547487 | 0.152478 | 0.015652 | 2.01E-22 | trans |
| rs1373200 | miR-320a | 2:6862265 | C | T | 0.547142 | 0.149223 | 0.015409 | 3.51E-22 | trans |
| rs6546393 | miR-320a | 2:6862757 | T | C | 0.554017 | 0.154967 | 0.015786 | 9.52E-23 | trans |
| rs1753019 | miR-320a | 2:6862133 | G | T | 0.683745 | 0.145203 | 0.016568 | 1.88E-18 | trans |
| rs6419643 | miR-320a | 2:6863126 | T | G | 0.548826 | 0.153407 | 0.015714 | 1.63E-22 | trans |
| rs1302897 | miR-320a | 2:6863003 | A | G | 0.54849 | 0.153419 | 0.015702 | 1.51E-22 | trans |
| rs6214393 | miR-320a | 2:6864041 | A | G | 0.593599 | 0.143051 | 0.01644 | 3.28E-18 | trans |
| rs8028813 | miR-320a | 2:6862547 | A | G | 0.548418 | 0.153418 | 0.015656 | 1.13E-22 | trans |
| rs7592526 | miR-320a | 2:6858352 | A | C | 0.538007 | 0.140533 | 0.01604 | 1.93E-18 | trans |
| rs6715909 | miR-320a | 2:6858399 | G | A | 0.664879 | 0.131663 | 0.01702 | 1.03E-14 | trans |
| rs3732044 | miR-320a | 2:6862391 | T | C | 0.545562 | 0.150811 | 0.01556 | 3.25E-22 | trans |
| rs1397159 | miR-320a | 2:6863425 | T | G | 0.705552 | 0.148814 | 0.017799 | 6.24E-17 | trans |
| rs6546409 | miR-320a | 2:6863587 | A | G | 0.551093 | 0.15591 | 0.015998 | 1.93E-22 | trans |
| rs6214393 | miR-320a | 2:6864038 | G | C | 0.593598 | 0.143062 | 0.01644 | 3.25E-18 | trans |
| rs7590450 | miR-320a | 2:6862856 | A | G | 0.549532 | 0.153227 | 0.015701 | 1.68E-22 | trans |
| rs6000716 | miR-320a | 2:6863963 | C | T | 0.735163 | 0.14802 | 0.018521 | 1.33E-15 | trans |
| rs6087844 | miR-320a | 2:6858211 | A | G | 0.538067 | 0.140648 | 0.016039 | 1.80E-18 | trans |
| rs1382899 | miR-320a | 2:6863782 | A | G | 0.702614 | 0.147363 | 0.017647 | 6.79E-17 | trans |
| rs6715706 | miR-320a | 2:6858399 | C | A | 0.664887 | 0.131667 | 0.01702 | 1.03E-14 | trans |
| rs5690424 | miR-320a | 2:6863847 | T | C | 0.550669 | 0.155791 | 0.015996 | 2.05E-22 | trans |
| rs6546399 | miR-320e | 2:6863064 | C | T | 0.55394 | 0.111105 | 0.015037 | 1.48E-13 | trans |
| rs7605387 | miR-320e | 2:6863255 | A | G | 0.703499 | 0.104536 | 0.016676 | 3.64E-10 | trans |
| rs6214392 | miR-320e | 2:6863835 | C | G | 0.700378 | 0.10723 | 0.016725 | 1.44E-10 | trans |
| rs6214393 | miR-320e | 2:6864020 | A | G | 0.593308 | 0.113862 | 0.015587 | 2.78E-13 | trans |
| rs6546398 | miR-320e | 2:6863041 | G | C | 0.552757 | 0.110975 | 0.014992 | 1.34E-13 | trans |
| rs1164012 | miR-320e | 2:6862562 | C | T | 0.545777 | 0.111024 | 0.014812 | 6.59E-14 | trans |
| rs7572299 | miR-320e | 2:6858124 | T | A | 0.538033 | 0.108792 | 0.015217 | 8.73E-13 | trans |
| rs1301028 | miR-320e | 2:6862184 | G | T | 0.683506 | 0.098324 | 0.015747 | 4.26E-10 | trans |
| rs6546407 | miR-320e | 2:6863492 | A | C | 0.55091 | 0.113961 | 0.015198 | 6.46E-14 | trans |
| rs6729686 | miR-320e | 2:6863303 | C | T | 0.700171 | 0.105258 | 0.016579 | 2.17E-10 | trans |
| rs1017438 | miR-320e | 2:6859646 | C | T | 0.685243 | 0.09762 | 0.015806 | 6.58E-10 | trans |
| rs1484530 | miR-320e | 2:6863547 | A | G | 0.70445 | 0.107896 | 0.0168 | 1.34E-10 | trans |
| rs7451452 | miR-320e | 2:6863755 | T | G | 0.702552 | 0.107162 | 0.016749 | 1.57E-10 | trans |
| rs6214389 | miR-320e | 2:6860455 | C | T | 0.698307 | 0.099525 | 0.016028 | 5.32E-10 | trans |
| rs1455505 | miR-320e | 2:6862832 | G | A | 0.556221 | 0.114653 | 0.015101 | 3.14E-14 | trans |
| rs6214393 | miR-320e | 2:6864009 | T | C | 0.59325 | 0.113891 | 0.015587 | 2.73E-13 | trans |
| rs1442589 | miR-320e | 2:6863473 | G | T | 0.701896 | 0.106528 | 0.016702 | 1.79E-10 | trans |
| rs7598360 | miR-320e | 2:6863092 | A | G | 0.553166 | 0.116479 | 0.015017 | 8.73E-15 | trans |
| rs6546397 | miR-320e | 2:6863036 | G | A | 0.552756 | 0.110969 | 0.014992 | 1.34E-13 | trans |
| rs6546411 | miR-320e | 2:6863769 | A | G | 0.556248 | 0.116536 | 0.015313 | 2.74E-14 | trans |
| rs6546408 | miR-320e | 2:6863504 | T | C | 0.551062 | 0.11385 | 0.015214 | 7.26E-14 | trans |
| rs2165920 | miR-320e | 2:6862223 | T | C | 0.552012 | 0.111604 | 0.014753 | 3.89E-14 | trans |
| rs6419644 | miR-320e | 2:6863182 | A | G | 0.549898 | 0.11406 | 0.015101 | 4.25E-14 | trans |
| rs1300974 | miR-320e | 2:6858443 | C | A | 0.678986 | 0.097526 | 0.016401 | 2.74E-09 | trans |
| rs1301107 | miR-320e | 2:6863931 | A | C | 0.700844 | 0.107477 | 0.016766 | 1.45E-10 | trans |
| rs1156629 | miR-320e | 2:6862663 | T | G | 0.547487 | 0.112242 | 0.014889 | 4.76E-14 | trans |
| rs1300519 | miR-320e | 2:6863887 | A | G | 0.702186 | 0.107204 | 0.016734 | 1.49E-10 | trans |
| rs1753019 | miR-320e | 2:6862133 | G | T | 0.683745 | 0.098483 | 0.01576 | 4.13E-10 | trans |
| rs6546412 | miR-320e | 2:6863771 | T | C | 0.557086 | 0.117526 | 0.015342 | 1.86E-14 | trans |
| rs6546400 | miR-320e | 2:6863064 | G | A | 0.554382 | 0.111156 | 0.015051 | 1.52E-13 | trans |

| | | | | | | | | | |
|---|---|---|---|---|---|---|---|---|---|
| rs6546410 | miR-320e | 2:6863610 | C | G | 0.702434 | 0.107067 | 0.016742 | 1.60E-10 | trans |
| rs5888448 | miR-320e | 2:6864534 | G | A | 0.734214 | 0.111576 | 0.017452 | 1.62E-10 | trans |
| rs7608429 | miR-320e | 2:6858083 | G | T | 0.678719 | 0.097622 | 0.016413 | 2.72E-09 | trans |
| rs8761 | miR-320e | 2:6862426 | G | A | 0.545692 | 0.110918 | 0.014806 | 6.81E-14 | trans |
| rs5999861 | miR-320e | 2:6864677 | G | T | 0.731295 | 0.109875 | 0.017429 | 2.90E-10 | trans |
| rs6214393 | miR-320e | 2:6864062 | T | C | 0.733261 | 0.111997 | 0.017381 | 1.17E-10 | trans |
| rs6546395 | miR-320e | 2:6862927 | A | G | 0.55448 | 0.11111 | 0.015036 | 1.47E-13 | trans |
| rs3419682 | miR-320e | 2:6862142 | G | A | 0.697518 | 0.100578 | 0.015999 | 3.25E-10 | trans |
| rs1301077 | miR-320e | 2:6863908 | C | T | 0.702339 | 0.107187 | 0.016738 | 1.52E-10 | trans |
| rs1373200 | miR-320e | 2:6862265 | C | T | 0.547142 | 0.110349 | 0.014654 | 5.07E-14 | trans |
| rs5933292 | miR-320e | 2:6863963 | G | A | 0.73529 | 0.113856 | 0.017558 | 8.89E-11 | trans |
| rs5618821 | miR-320e | 2:6861163 | T | C | 0.698075 | 0.10022 | 0.016006 | 3.82E-10 | trans |
| rs1158434 | miR-320e | 2:6863552 | G | C | 0.551072 | 0.11383 | 0.015217 | 7.41E-14 | trans |
| rs1486061 | miR-320e | 2:6862571 | T | G | 0.54743 | 0.111501 | 0.014862 | 6.26E-14 | trans |
| rs6546404 | miR-320e | 2:6863263 | G | T | 0.54995 | 0.114042 | 0.015107 | 4.39E-14 | trans |
| rs2241946 | miR-320e | 2:6861565 | T | C | 0.698076 | 0.100416 | 0.016012 | 3.58E-10 | trans |
| rs1301157 | miR-320e | 2:6863927 | G | A | 0.702349 | 0.107197 | 0.016739 | 1.51E-10 | trans |
| rs6747545 | miR-320e | 2:6863306 | T | A | 0.549964 | 0.114028 | 0.015112 | 4.50E-14 | trans |
| rs7597167 | miR-320e | 2:6861294 | C | T | 0.698051 | 0.100272 | 0.016006 | 3.73E-10 | trans |
| rs6546401 | miR-320e | 2:6863087 | A | T | 0.550127 | 0.113171 | 0.014981 | 4.20E-14 | trans |
| rs1050181 | miR-320e | 2:6862295 | G | C | 0.544992 | 0.110579 | 0.014759 | 6.76E-14 | trans |
| rs1422402 | miR-320e | 2:6862752 | A | G | 0.698498 | 0.102502 | 0.016358 | 3.70E-10 | trans |
| rs1760168 | miR-320e | 2:6861589 | G | A | 0.697578 | 0.100273 | 0.016012 | 3.79E-10 | trans |
| rs6714719 | miR-320e | 2:6859377 | G | C | 0.698404 | 0.099991 | 0.016071 | 4.91E-10 | trans |
| rs6744306 | miR-320e | 2:6863273 | T | C | 0.549956 | 0.114047 | 0.015107 | 4.38E-14 | trans |
| rs1542136 | miR-320e | 2:6862093 | C | A | 0.697594 | 0.100615 | 0.016004 | 3.24E-10 | trans |
| rs6546403 | miR-320e | 2:6863226 | C | T | 0.550801 | 0.114043 | 0.015125 | 4.70E-14 | trans |
| rs6757938 | miR-320e | 2:6863285 | A | G | 0.549958 | 0.114041 | 0.015109 | 4.43E-14 | trans |
| rs6214393 | miR-320e | 2:6864038 | G | C | 0.593598 | 0.114275 | 0.015583 | 2.24E-13 | trans |
| rs7800275 | miR-320e | 2:6862543 | A | G | 0.546309 | 0.110955 | 0.014794 | 6.38E-14 | trans |
| rs1300592 | miR-320e | 2:6862999 | T | A | 0.701261 | 0.100993 | 0.016476 | 8.80E-10 | trans |
| rs1390688 | miR-320e | 2:6862773 | C | T | 0.69721 | 0.102067 | 0.016316 | 3.96E-10 | trans |
| rs3732044 | miR-320e | 2:6862391 | T | C | 0.545562 | 0.11085 | 0.014801 | 6.91E-14 | trans |
| rs7421228 | miR-320e | 2:6863012 | G | T | 0.55268 | 0.110936 | 0.014989 | 1.35E-13 | trans |
| rs6715579 | miR-320e | 2:6858392 | C | T | 0.678794 | 0.097675 | 0.016399 | 2.58E-09 | trans |
| rs7590450 | miR-320e | 2:6862856 | A | G | 0.549532 | 0.11308 | 0.014935 | 3.68E-14 | trans |
| rs7578602 | miR-320e | 2:6863786 | G | C | 0.556806 | 0.117559 | 0.015349 | 1.87E-14 | trans |
| rs5633714 | miR-320e | 2:6864249 | G | A | 0.733417 | 0.112098 | 0.017391 | 1.15E-10 | trans |
| rs6709464 | miR-320e | 2:6862222 | C | T | 0.697279 | 0.100407 | 0.015985 | 3.35E-10 | trans |
| rs7420775 | miR-320e | 2:6862945 | G | T | 0.703122 | 0.101277 | 0.01655 | 9.40E-10 | trans |
| rs4610041 | miR-320e | 2:6862628 | C | T | 0.550745 | 0.11333 | 0.014985 | 3.95E-14 | trans |
| rs1397159 | miR-320e | 2:6863425 | T | G | 0.705552 | 0.107312 | 0.016901 | 2.16E-10 | trans |
| rs9653552 | miR-320e | 2:6864396 | G | A | 0.593458 | 0.113705 | 0.015597 | 3.10E-13 | trans |
| rs6743970 | miR-320e | 2:6858390 | A | T | 0.660237 | 0.098626 | 0.01622 | 1.20E-09 | trans |
| rs6087844 | miR-320e | 2:6858211 | A | G | 0.538067 | 0.108754 | 0.015216 | 8.84E-13 | trans |
| rs1474969 | miR-320e | 2:6862568 | G | A | 0.546544 | 0.111176 | 0.014834 | 6.65E-14 | trans |
| rs6214393 | miR-320e | 2:6864041 | A | G | 0.593599 | 0.114256 | 0.015583 | 2.27E-13 | trans |
| rs1542135 | miR-320e | 2:6862084 | C | T | 0.69752 | 0.100575 | 0.015999 | 3.25E-10 | trans |
| rs1343065 | miR-320e | 2:6858771 | C | G | 0.699397 | 0.099802 | 0.016199 | 7.22E-10 | trans |
| rs7581872 | miR-320e | 2:6858098 | A | C | 0.537959 | 0.108694 | 0.015217 | 9.15E-13 | trans |
| rs1049616 | miR-320e | 2:6862008 | C | T | 0.697496 | 0.100537 | 0.016 | 3.31E-10 | trans |
| rs2241947 | miR-320e | 2:6861539 | C | T | 0.697988 | 0.100361 | 0.016005 | 3.60E-10 | trans |
| rs6729555 | miR-320e | 2:6863295 | C | A | 0.54995 | 0.114033 | 0.015111 | 4.47E-14 | trans |

| | | | | | | | | | |
|---|---|---|---|---|---|---|---|---|---|
| rs1760209 | miR-320e | 2:6862041( | T | A | 0.697529 | 0.100567 | 0.016 | 3.27E-10 | trans |
| rs1303482 | miR-320e | 2:6863029 | C | G | 0.552715 | 0.110948 | 0.01499 | 1.35E-13 | trans |
| rs6546393 | miR-320e | 2:6862757 | T | C | 0.554017 | 0.114706 | 0.015016 | 2.19E-14 | trans |
| rs3485223 | miR-320e | 2:6863859 | C | G | 0.550658 | 0.113462 | 0.015221 | 9.04E-14 | trans |
| rs6546409 | miR-320e | 2:6863587 | A | G | 0.551093 | 0.113771 | 0.015222 | 7.77E-14 | trans |
| rs7592526 | miR-320e | 2:6858352 | A | C | 0.538007 | 0.109013 | 0.015216 | 7.81E-13 | trans |
| rs6000716 | miR-320e | 2:6863963 | C | T | 0.735163 | 0.113878 | 0.017555 | 8.77E-11 | trans |
| rs6419643 | miR-320e | 2:6863126( | T | G | 0.548826 | 0.113114 | 0.014948 | 3.81E-14 | trans |
| rs1301143 | miR-320e | 2:6863937 | C | T | 0.702382 | 0.107216 | 0.016741 | 1.51E-10 | trans |
| rs6742200 | miR-320e | 2:6862330 | A | G | 0.54552 | 0.110827 | 0.014798 | 6.92E-14 | trans |
| rs2070171 | miR-320e | 2:6861371( | G | A | 0.698024 | 0.100279 | 0.016005 | 3.72E-10 | trans |
| rs1760149 | miR-320e | 2:6861171 | A | G | 0.69807 | 0.100226 | 0.016006 | 3.81E-10 | trans |
| rs8028813 | miR-320e | 2:6862547 | A | G | 0.548418 | 0.112315 | 0.014897 | 4.71E-14 | trans |
| rs6715909 | miR-320e | 2:6858399 | G | A | 0.664879 | 0.095462 | 0.016144 | 3.36E-09 | trans |
| rs1382899 | miR-320e | 2:6863782 | A | G | 0.702614 | 0.107213 | 0.016753 | 1.56E-10 | trans |
| rs7562439 | miR-320e | 2:6858256 | C | T | 0.6646 | 0.095513 | 0.016148 | 3.32E-09 | trans |
| rs6715706 | miR-320e | 2:6858399( | C | A | 0.664887 | 0.095464 | 0.016144 | 3.35E-09 | trans |
| rs1302897 | miR-320e | 2:6863003 | A | G | 0.54849 | 0.112919 | 0.014938 | 4.05E-14 | trans |
| rs1302892 | miR-320e | 2:6862991 | C | T | 0.552648 | 0.110906 | 0.014987 | 1.36E-13 | trans |
| rs5628470 | miR-320e | 2:6863137 | G | A | 0.697508 | 0.102907 | 0.016371 | 3.26E-10 | trans |
| rs5690424 | miR-320e | 2:6863847 | T | C | 0.550669 | 0.113465 | 0.01522 | 9.00E-14 | trans |
| rs5808991 | miR-320e | 2:6863966( | G | C | 0.734634 | 0.113408 | 0.017511 | 9.40E-11 | trans |
| rs6708536 | miR-320e | 2:6862726 | G | A | 0.547249 | 0.112067 | 0.014901 | 5.45E-14 | trans |
| rs2280725 | miR-339-5p | 7:1087426 | G | T | 0.421125 | -0.091865 | 0.016116 | 1.20E-08 | cis |
| rs1043514 | miR-339-5p | 7:1065988 | C | T | 0.42274 | -0.090218 | 0.016007 | 1.74E-08 | cis |
| rs7809203 | miR-339-5p | 7:1087454 | T | C | 0.42124 | -0.091841 | 0.016114 | 1.20E-08 | cis |
| rs1095159 | miR-339-5p | 7:1088607 | C | G | 0.420678 | -0.091986 | 0.01614 | 1.20E-08 | cis |
| rs2363285 | miR-339-5p | 7:1091644 | C | T | 0.421201 | -0.091691 | 0.016062 | 1.14E-08 | cis |
| rs4468532 | miR-345-5p | 14:100730 | G | C | 0.753211 | -0.067254 | 0.012258 | 4.10E-08 | cis |
| rs3783340 | miR-345-5p | 14:100734 | A | G | 0.753729 | -0.067149 | 0.012256 | 4.29E-08 | cis |
| rs8013532 | miR-345-5p | 14:100728 | G | T | 0.752838 | -0.067417 | 0.012265 | 3.87E-08 | cis |
| rs8021068 | miR-345-5p | 14:100738 | G | A | 0.753098 | -0.067265 | 0.012225 | 3.75E-08 | cis |
| rs8012853 | miR-345-5p | 14:100689 | A | C | 0.751989 | -0.06813 | 0.012337 | 3.34E-08 | cis |
| rs1986964 | miR-345-5p | 14:100708 | A | G | 0.753258 | -0.068083 | 0.012342 | 3.46E-08 | cis |
| rs1013176 | miR-345-5p | 14:100727 | C | T | 0.753383 | -0.06735 | 0.012259 | 3.94E-08 | cis |
| rs7159187 | miR-345-5p | 14:100701 | C | T | 0.752079 | -0.067809 | 0.012243 | 3.05E-08 | cis |
| rs9333573 | miR-345-5p | 14:100704 | C | T | 0.753293 | -0.067954 | 0.012311 | 3.39E-08 | cis |
| rs1288366 | miR-345-5p | 14:100736 | G | C | 0.752973 | -0.067235 | 0.012239 | 3.94E-08 | cis |
| rs1012920 | miR-345-5p | 14:100717 | T | C | 0.753859 | -0.068499 | 0.012404 | 3.35E-08 | cis |
| rs1013881 | miR-345-5p | 14:100692 | G | A | 0.754823 | -0.06922 | 0.012339 | 2.03E-08 | cis |
| rs1232381 | miR-345-5p | 14:100737 | C | A | 0.752905 | -0.06748 | 0.012236 | 3.49E-08 | cis |
| rs1162565 | miR-345-5p | 14:100723 | C | A | 0.753779 | -0.067756 | 0.012319 | 3.79E-08 | cis |
| rs3462948 | miR-345-5p | 14:100700 | C | T | 0.753899 | -0.068099 | 0.01234 | 3.41E-08 | cis |
| rs2895858 | miR-345-5p | 14:100730 | T | C | 0.753207 | -0.067248 | 0.012257 | 4.10E-08 | cis |
| rs7142599 | miR-345-5p | 14:100693 | G | A | 0.753695 | -0.067753 | 0.012311 | 3.72E-08 | cis |
| rs3128562 | miR-3689a | 9:1377452 | A | G | 0.300754 | 0.396962 | 0.02741 | 1.56E-47 | cis |
| rs3128570 | miR-3689a | 9:1377433( | G | A | 0.298028 | 0.390362 | 0.02716 | 7.70E-47 | cis |
| rs3124944 | miR-3689a | 9:1377452 | A | G | 0.303119 | 0.395178 | 0.027506 | 8.32E-47 | cis |
| rs9410005 | miR-3689a | 9:1377478 | A | G | 0.46835 | 0.220517 | 0.025857 | 1.49E-17 | cis |
| rs9410004 | miR-3689a | 9:1377477( | C | T | 0.470272 | 0.220078 | 0.02582 | 1.55E-17 | cis |
| rs1289760 | miR-370-3p | 14:101182 | C | T | 0.190452 | -0.284768 | 0.029658 | 7.86E-22 | cis |
| rs1190852 | miR-370-3p | 14:101064 | A | G | 0.310295 | -0.209646 | 0.02532 | 1.23E-16 | cis |
| rs2295659 | miR-370-3p | 14:101141 | G | T | 0.857995 | 0.18175 | 0.033334 | 4.97E-08 | cis |

| | | | | | | | | | |
|---|---|---|---|---|---|---|---|---|---|
| rs1162219 | miR-370-3p | 14:101016 | G | A | 0.759852 | 0.199826 | 0.029561 | 1.38E-11 | cis |
| rs3579985 | miR-370-3p | 14:101156 | C | T | 0.904015 | 0.238309 | 0.04161 | 1.02E-08 | cis |
| rs1190903 | miR-370-3p | 14:101096 | C | T | 0.772566 | 0.217133 | 0.028126 | 1.16E-14 | cis |
| rs1259093 | miR-370-3p | 14:101163 | A | T | 0.893649 | 0.250435 | 0.039029 | 1.39E-10 | cis |
| rs1289379 | miR-370-3p | 14:101178 | C | G | 0.190813 | -0.282952 | 0.029853 | 2.59E-21 | cis |
| rs1288162 | miR-370-3p | 14:101146 | A | G | 0.912196 | 0.256057 | 0.043325 | 3.42E-09 | cis |
| rs1184761 | miR-370-3p | 14:101023 | C | T | 0.753476 | 0.184975 | 0.027875 | 3.23E-11 | cis |
| rs8008285 | miR-370-3p | 14:101146 | T | C | 0.842905 | 0.238019 | 0.03247 | 2.29E-13 | cis |
| rs7143357 | miR-370-3p | 14:101054 | T | C | 0.739636 | 0.215159 | 0.026343 | 3.15E-16 | cis |
| rs7155649 | miR-370-3p | 14:101181 | C | A | 0.863608 | 0.254793 | 0.034571 | 1.70E-13 | cis |
| rs879059 | miR-370-3p | 14:101069 | T | A | 0.31068 | -0.208048 | 0.025263 | 1.79E-16 | cis |
| rs6575791 | miR-370-3p | 14:101029 | G | A | 0.719841 | 0.19897 | 0.025998 | 1.96E-14 | cis |
| rs1014844 | miR-370-3p | 14:101026 | G | A | 0.753241 | 0.184811 | 0.027799 | 2.97E-11 | cis |
| rs1190818 | miR-370-3p | 14:101230 | T | G | 0.818683 | 0.168083 | 0.03025 | 2.75E-08 | cis |
| rs1116059 | miR-370-3p | 14:101049 | C | T | 0.741662 | 0.216786 | 0.026314 | 1.75E-16 | cis |
| rs7493817 | miR-370-3p | 14:101038 | G | A | 0.751723 | 0.213483 | 0.026955 | 2.38E-15 | cis |
| rs7140556 | miR-370-3p | 14:101036 | C | A | 0.723818 | 0.199916 | 0.02594 | 1.29E-14 | cis |
| rs9671394 | miR-370-3p | 14:101159 | C | T | 0.284232 | -0.266385 | 0.026192 | 2.68E-24 | cis |
| rs7149242 | miR-370-3p | 14:101159 | T | G | 0.288371 | -0.260257 | 0.025837 | 7.28E-24 | cis |
| rs1190862 | miR-370-3p | 14:101067 | T | G | 0.309628 | -0.206203 | 0.025131 | 2.30E-16 | cis |
| rs1243369 | miR-370-3p | 14:101035 | C | G | 0.723403 | 0.197119 | 0.025831 | 2.33E-14 | cis |
| rs1185095 | miR-370-3p | 14:101056 | G | T | 0.743981 | 0.217094 | 0.026179 | 1.11E-16 | cis |
| rs1190909 | miR-370-3p | 14:101101 | G | A | 0.775531 | 0.213631 | 0.028465 | 6.14E-14 | cis |
| rs745797 | miR-370-3p | 14:101141 | G | A | 0.858727 | 0.183928 | 0.033436 | 3.78E-08 | cis |
| rs1243449 | miR-370-3p | 14:101179 | A | G | 0.18959 | -0.282653 | 0.029829 | 2.65E-21 | cis |
| rs1015151 | miR-370-3p | 14:101175 | G | A | 0.806743 | 0.194814 | 0.029099 | 2.16E-11 | cis |
| rs1190904 | miR-370-3p | 14:101096 | A | G | 0.772596 | 0.21724 | 0.028123 | 1.12E-14 | cis |
| rs7492369 | miR-370-3p | 14:101020 | C | T | 0.755698 | 0.183918 | 0.027954 | 4.73E-11 | cis |
| rs1184764 | miR-370-3p | 14:101023 | C | T | 0.753952 | 0.184275 | 0.027906 | 4.02E-11 | cis |
| rs1190907 | miR-370-3p | 14:101099 | A | G | 0.772847 | 0.214936 | 0.028082 | 1.95E-14 | cis |
| rs1190857 | miR-370-3p | 14:101066 | G | T | 0.310273 | -0.20891 | 0.025306 | 1.52E-16 | cis |
| rs911330 | miR-370-3p | 14:101099 | T | C | 0.774615 | 0.215056 | 0.028225 | 2.55E-14 | cis |
| rs1185164 | miR-370-3p | 14:100974 | G | A | 0.742035 | 0.170984 | 0.028421 | 1.79E-09 | cis |
| rs4905999 | miR-370-3p | 14:101184 | A | G | 0.181905 | -0.275806 | 0.030398 | 1.16E-19 | cis |
| rs1087352 | miR-370-3p | 14:101175 | G | A | 0.809826 | 0.19551 | 0.029445 | 3.14E-11 | cis |
| rs1214700 | miR-370-3p | 14:101186 | C | T | 0.223221 | -0.263911 | 0.02766 | 1.41E-21 | cis |
| rs2842154 | miR-370-3p | 14:101158 | C | T | 0.904763 | 0.237405 | 0.041658 | 1.21E-08 | cis |
| rs1190850 | miR-370-3p | 14:101064 | A | G | 0.314302 | -0.211731 | 0.025468 | 9.29E-17 | cis |
| rs1184658 | miR-370-3p | 14:101017 | C | G | 0.759077 | 0.183862 | 0.028338 | 8.69E-11 | cis |
| rs7144997 | miR-370-3p | 14:101155 | A | G | 0.899872 | 0.23041 | 0.0415 | 2.82E-08 | cis |
| rs7154024 | miR-370-3p | 14:101031 | G | T | 0.725015 | 0.199663 | 0.025974 | 1.51E-14 | cis |
| rs1184948 | miR-370-3p | 14:101037 | C | G | 0.733365 | 0.198989 | 0.026138 | 2.68E-14 | cis |
| rs1190867 | miR-370-3p | 14:101069 | G | T | 0.316788 | -0.210145 | 0.025453 | 1.51E-16 | cis |
| rs1757758 | miR-370-3p | 14:101189 | C | T | 0.827094 | 0.237826 | 0.030495 | 6.25E-15 | cis |
| rs1709962 | miR-370-3p | 14:101173 | A | C | 0.783449 | 0.191629 | 0.028093 | 9.02E-12 | cis |
| rs1116059 | miR-370-3p | 14:100965 | C | T | 0.745134 | 0.170875 | 0.028543 | 2.14E-09 | cis |
| rs1014839 | miR-370-3p | 14:101026 | A | G | 0.750738 | 0.185252 | 0.027807 | 2.70E-11 | cis |
| rs7140792 | miR-370-3p | 14:101172 | C | T | 0.892517 | 0.253004 | 0.03785 | 2.32E-11 | cis |
| rs1888581 | miR-370-3p | 14:101042 | A | C | 0.735363 | 0.196476 | 0.026077 | 4.90E-14 | cis |
| rs1190872 | miR-370-3p | 14:101069 | A | G | 0.310177 | -0.208165 | 0.025276 | 1.79E-16 | cis |
| rs879058 | miR-370-3p | 14:101069 | G | T | 0.311807 | -0.208233 | 0.025277 | 1.75E-16 | cis |
| rs1258812 | miR-370-3p | 14:101162 | T | C | 0.893709 | 0.250183 | 0.039027 | 1.45E-10 | cis |
| rs1288035 | miR-370-3p | 14:101158 | C | G | 0.904391 | 0.238821 | 0.041609 | 9.49E-09 | cis |

| | | | | | | | | | |
|---|---|---|---|---|---|---|---|---|---|
| rs6575794 | miR-370-3p | 14:101039 | C | T | 0.734189 | 0.198744 | 0.026178 | 3.15E-14 | cis |
| rs1214784 | miR-370-3p | 14:101144 | C | T | 0.870989 | 0.232326 | 0.034966 | 3.04E-11 | cis |
| rs1190881 | miR-370-3p | 14:101079 | T | C | 0.346494 | -0.15907 | 0.024701 | 1.20E-10 | cis |
| rs3470057 | miR-370-3p | 14:101173 | C | T | 0.892725 | 0.253411 | 0.03781 | 2.05E-11 | cis |
| rs879060 | miR-370-3p | 14:101069 | A | G | 0.310926 | -0.208801 | 0.025277 | 1.45E-16 | cis |
| rs7142683 | miR-370-3p | 14:101158 | A | G | 0.290748 | -0.26127 | 0.02591 | 6.52E-24 | cis |
| rs7155375 | miR-370-3p | 14:101188 | C | T | 0.635995 | 0.206766 | 0.023788 | 3.56E-18 | cis |
| rs7147982 | miR-370-3p | 14:101046 | C | T | 0.731753 | 0.191267 | 0.025899 | 1.52E-13 | cis |
| rs1555405 | miR-370-3p | 14:101176 | G | A | 0.749075 | 0.215585 | 0.026307 | 2.51E-16 | cis |
| rs2400940 | miR-370-3p | 14:101224 | G | A | 0.185552 | -0.163352 | 0.029621 | 3.49E-08 | cis |
| rs1243613 | miR-370-3p | 14:101031 | C | T | 0.725015 | 0.199847 | 0.025997 | 1.50E-14 | cis |
| rs1190878 | miR-370-3p | 14:101072 | C | A | 0.859763 | 0.19362 | 0.034305 | 1.66E-08 | cis |
| rs1243295 | miR-370-3p | 14:101205 | G | A | 0.231663 | -0.177159 | 0.027854 | 2.02E-10 | cis |
| rs2837918 | miR-370-3p | 14:101168 | G | T | 0.836434 | 0.269753 | 0.031357 | 7.80E-18 | cis |
| rs1190856 | miR-370-3p | 14:101065 | T | C | 0.310244 | -0.208965 | 0.025301 | 1.47E-16 | cis |
| rs4905994 | miR-370-3p | 14:101154 | T | C | 0.329146 | -0.240491 | 0.025152 | 1.16E-21 | cis |
| rs7145610 | miR-370-3p | 14:101049 | T | C | 0.740391 | 0.21785 | 0.026381 | 1.49E-16 | cis |
| rs1013035 | miR-370-3p | 14:101149 | T | C | 0.112665 | -0.227472 | 0.038907 | 5.02E-09 | cis |
| rs9324022 | miR-370-3p | 14:101172 | T | C | 0.763739 | 0.21966 | 0.028499 | 1.28E-14 | cis |
| rs1190906 | miR-370-3p | 14:101099 | T | C | 0.772138 | 0.214026 | 0.02804 | 2.29E-14 | cis |
| rs4905999 | miR-376c-3 | 14:101184 | A | G | 0.181905 | -0.277015 | 0.027005 | 1.09E-24 | cis |
| rs1243449 | miR-376c-3 | 14:101179 | A | G | 0.18959 | -0.284953 | 0.026482 | 5.29E-27 | cis |
| rs1014551 | miR-376c-3 | 14:101168 | A | G | 0.668173 | 0.209202 | 0.021737 | 6.31E-22 | cis |
| rs1014696 | miR-376c-3 | 14:101170 | T | C | 0.658875 | 0.213122 | 0.021364 | 1.95E-23 | cis |
| rs879059 | miR-382-5p | 14:101069 | T | A | 0.31068 | -0.25966 | 0.034839 | 9.11E-14 | cis |
| rs1190903 | miR-382-5p | 14:101096 | C | T | 0.772566 | 0.268909 | 0.03878 | 4.08E-12 | cis |
| rs1184761 | miR-382-5p | 14:101023 | C | T | 0.753476 | 0.225669 | 0.038417 | 4.25E-09 | cis |
| rs1184658 | miR-382-5p | 14:101017 | C | G | 0.759077 | 0.240563 | 0.039007 | 6.95E-10 | trans |
| rs8008285 | miR-382-5p | 14:101146 | T | C | 0.842905 | 0.286055 | 0.044784 | 1.69E-10 | cis |
| rs1162219 | miR-382-5p | 14:101016 | G | A | 0.759852 | 0.257157 | 0.040705 | 2.66E-10 | trans |
| rs7154024 | miR-382-5p | 14:101031 | G | T | 0.725015 | 0.229738 | 0.035866 | 1.50E-10 | cis |
| rs1214700 | miR-382-5p | 14:101186 | C | T | 0.223221 | -0.316047 | 0.038232 | 1.38E-16 | cis |
| rs7155649 | miR-382-5p | 14:101181 | C | A | 0.863608 | 0.322021 | 0.047635 | 1.38E-11 | cis |
| rs1190904 | miR-382-5p | 14:101096 | A | G | 0.772596 | 0.268729 | 0.038777 | 4.21E-12 | cis |
| rs1015151 | miR-382-5p | 14:101175 | G | A | 0.806743 | 0.24727 | 0.040079 | 6.85E-10 | cis |
| rs1243369 | miR-382-5p | 14:101035 | C | G | 0.723403 | 0.227165 | 0.035665 | 1.90E-10 | cis |
| rs2837918 | miR-382-5p | 14:101168 | G | T | 0.836434 | 0.391528 | 0.043039 | 9.29E-20 | cis |
| rs1185095 | miR-382-5p | 14:101056 | G | T | 0.743981 | 0.286172 | 0.036048 | 2.04E-15 | cis |
| rs1184764 | miR-382-5p | 14:101023 | C | T | 0.753952 | 0.225364 | 0.038457 | 4.62E-09 | cis |
| rs6575794 | miR-382-5p | 14:101039 | C | T | 0.734189 | 0.247015 | 0.036088 | 7.66E-12 | cis |
| rs1190906 | miR-382-5p | 14:101099 | T | C | 0.772138 | 0.263048 | 0.038665 | 1.02E-11 | cis |
| rs1184948 | miR-382-5p | 14:101037 | C | G | 0.733365 | 0.247145 | 0.036034 | 6.95E-12 | cis |
| rs1243613 | miR-382-5p | 14:101031 | C | T | 0.725015 | 0.22988 | 0.035898 | 1.52E-10 | cis |
| rs6575791 | miR-382-5p | 14:101029 | G | A | 0.719841 | 0.229519 | 0.035897 | 1.62E-10 | cis |
| rs7145610 | miR-382-5p | 14:101049 | T | C | 0.740391 | 0.28563 | 0.036332 | 3.79E-15 | cis |
| rs1190867 | miR-382-5p | 14:101069 | G | T | 0.316788 | -0.259921 | 0.035109 | 1.33E-13 | cis |
| rs1888581 | miR-382-5p | 14:101042 | A | C | 0.735363 | 0.243774 | 0.035949 | 1.19E-11 | cis |
| rs879060 | miR-382-5p | 14:101069 | A | G | 0.310926 | -0.259838 | 0.034861 | 9.08E-14 | cis |
| rs1190852 | miR-382-5p | 14:101064 | A | G | 0.310295 | -0.261768 | 0.034917 | 6.54E-14 | cis |
| rs1757758 | miR-382-5p | 14:101189 | C | T | 0.827094 | 0.286006 | 0.042075 | 1.06E-11 | cis |
| rs7147982 | miR-382-5p | 14:101046 | C | T | 0.731753 | 0.239351 | 0.035694 | 2.01E-11 | cis |
| rs1190909 | miR-382-5p | 14:101101 | G | A | 0.775531 | 0.26705 | 0.039234 | 9.99E-12 | cis |
| rs1190850 | miR-382-5p | 14:101064 | A | G | 0.314302 | -0.264885 | 0.035121 | 4.62E-14 | cis |

| | | | | | | | | | |
|---|---|---|---|---|---|---|---|---|---|
| rs7143357 | miR-382-5p | 14:101054 | T | C | 0.739636 | 0.283285 | 0.036274 | 5.74E-15 | cis |
| rs7493817 | miR-382-5p | 14:101038 | G | A | 0.751723 | 0.269122 | 0.037155 | 4.38E-13 | cis |
| rs1014844 | miR-382-5p | 14:101026 | G | A | 0.753241 | 0.226698 | 0.038309 | 3.27E-09 | cis |
| rs1214784 | miR-382-5p | 14:101144 | C | T | 0.870989 | 0.269987 | 0.048225 | 2.16E-08 | cis |
| rs4905999 | miR-382-5p | 14:101184 | A | G | 0.181905 | -0.392875 | 0.041746 | 4.92E-21 | cis |
| rs1014839 | miR-382-5p | 14:101026 | A | G | 0.750738 | 0.225819 | 0.038325 | 3.81E-09 | cis |
| rs1190872 | miR-382-5p | 14:101069 | A | G | 0.310177 | -0.259623 | 0.034857 | 9.45E-14 | cis |
| rs1289379 | miR-382-5p | 14:101178 | C | G | 0.190813 | -0.392419 | 0.04104 | 1.16E-21 | cis |
| rs7149242 | miR-382-5p | 14:101159 | T | G | 0.288371 | -0.330312 | 0.035655 | 1.97E-20 | cis |
| rs1190862 | miR-382-5p | 14:101067 | T | G | 0.309628 | -0.256488 | 0.034658 | 1.36E-13 | cis |
| rs1087352 | miR-382-5p | 14:101175 | G | A | 0.809826 | 0.248409 | 0.040554 | 9.05E-10 | cis |
| rs1190856 | miR-382-5p | 14:101065 | T | C | 0.310244 | -0.260849 | 0.034891 | 7.66E-14 | cis |
| rs1259093 | miR-382-5p | 14:101163 | A | T | 0.893649 | 0.374255 | 0.053582 | 2.85E-12 | cis |
| rs1555405 | miR-382-5p | 14:101176 | G | A | 0.749075 | 0.295258 | 0.036183 | 3.35E-16 | cis |
| rs7140792 | miR-382-5p | 14:101172 | C | T | 0.892517 | 0.409611 | 0.051844 | 2.77E-15 | cis |
| rs9324022 | miR-382-5p | 14:101172 | T | C | 0.763739 | 0.255537 | 0.039345 | 8.31E-11 | cis |
| rs4905994 | miR-382-5p | 14:101154 | T | C | 0.329146 | -0.302721 | 0.034708 | 2.73E-18 | cis |
| rs1243449 | miR-382-5p | 14:101179 | A | G | 0.18959 | -0.391633 | 0.041008 | 1.30E-21 | cis |
| rs879058 | miR-382-5p | 14:101069 | G | T | 0.311807 | -0.259751 | 0.034858 | 9.21E-14 | cis |
| rs1258812 | miR-382-5p | 14:101162 | T | C | 0.893709 | 0.374109 | 0.053578 | 2.90E-12 | cis |
| rs9671394 | miR-382-5p | 14:101159 | C | T | 0.284232 | -0.339058 | 0.036142 | 6.52E-21 | cis |
| rs7140556 | miR-382-5p | 14:101036 | C | A | 0.723818 | 0.230964 | 0.035817 | 1.13E-10 | cis |
| rs1190857 | miR-382-5p | 14:101066 | G | T | 0.310273 | -0.260786 | 0.034898 | 7.85E-14 | cis |
| rs1190907 | miR-382-5p | 14:101099 | A | G | 0.772847 | 0.264877 | 0.038722 | 7.89E-12 | cis |
| rs911330 | miR-382-5p | 14:101099 | T | C | 0.774615 | 0.268715 | 0.038906 | 4.96E-12 | cis |
| rs1289760 | miR-382-5p | 14:101182 | C | T | 0.190452 | -0.391127 | 0.040788 | 8.88E-22 | cis |
| rs3470057 | miR-382-5p | 14:101173 | C | T | 0.892725 | 0.411051 | 0.051786 | 2.06E-15 | cis |
| rs1116059 | miR-382-5p | 14:101049 | C | T | 0.741662 | 0.284332 | 0.036239 | 4.30E-15 | cis |
| rs7155375 | miR-382-5p | 14:101188 | C | T | 0.635995 | 0.262309 | 0.032799 | 1.27E-15 | cis |
| rs7492369 | miR-382-5p | 14:101020 | C | T | 0.755698 | 0.225958 | 0.03852 | 4.47E-09 | cis |
| rs7142683 | miR-382-5p | 14:101158 | A | G | 0.290748 | -0.331339 | 0.035757 | 1.92E-20 | cis |
| rs1185164 | miR-409-3p | 14:100974 | G | A | 0.742035 | 0.24402 | 0.037839 | 1.13E-10 | trans |
| rs1184761 | miR-409-3p | 14:101023 | C | T | 0.753476 | 0.258867 | 0.037119 | 3.08E-12 | trans |
| rs1757758 | miR-409-3p | 14:101189 | C | T | 0.827094 | 0.340337 | 0.040563 | 4.84E-17 | cis |
| rs7147982 | miR-409-3p | 14:101046 | C | T | 0.731753 | 0.28735 | 0.034406 | 6.73E-17 | cis |
| rs1765383 | miR-409-3p | 14:101101 | T | C | 0.877829 | 0.323085 | 0.048905 | 3.94E-11 | cis |
| rs1190862 | miR-409-3p | 14:101067 | T | G | 0.309628 | -0.290778 | 0.033438 | 3.43E-18 | cis |
| rs1288035 | miR-409-3p | 14:101158 | C | G | 0.904391 | 0.315671 | 0.055472 | 1.27E-08 | cis |
| rs745797 | miR-409-3p | 14:101141 | G | A | 0.858727 | 0.248489 | 0.044561 | 2.46E-08 | cis |
| rs1288162 | miR-409-3p | 14:101146 | A | G | 0.912196 | 0.31744 | 0.057815 | 4.01E-08 | cis |
| rs879059 | miR-409-3p | 14:101069 | T | A | 0.31068 | -0.291751 | 0.03362 | 4.03E-18 | cis |
| rs8008285 | miR-409-3p | 14:101146 | T | C | 0.842905 | 0.332888 | 0.043228 | 1.35E-14 | cis |
| rs7154024 | miR-409-3p | 14:101031 | G | T | 0.725015 | 0.278312 | 0.03458 | 8.39E-16 | cis |
| rs1116059 | miR-409-3p | 14:101049 | C | T | 0.741662 | 0.307143 | 0.035006 | 1.72E-18 | cis |
| rs1214700 | miR-409-3p | 14:101186 | C | T | 0.223221 | -0.352144 | 0.03687 | 1.28E-21 | cis |
| rs1190903 | miR-409-3p | 14:101096 | C | T | 0.772566 | 0.293624 | 0.037477 | 4.70E-15 | cis |
| rs1243369 | miR-409-3p | 14:101035 | C | G | 0.723403 | 0.275821 | 0.034386 | 1.05E-15 | cis |
| rs1190867 | miR-409-3p | 14:101069 | G | T | 0.316788 | -0.29269 | 0.03388 | 5.68E-18 | cis |
| rs2837918 | miR-409-3p | 14:101168 | G | T | 0.836434 | 0.389638 | 0.041675 | 8.81E-21 | cis |
| rs6575791 | miR-409-3p | 14:101029 | G | A | 0.719841 | 0.277286 | 0.034612 | 1.14E-15 | trans |
| rs1190850 | miR-409-3p | 14:101064 | A | G | 0.314302 | -0.297205 | 0.03389 | 1.79E-18 | cis |
| rs1190818 | miR-409-3p | 14:101230 | T | G | 0.818683 | 0.239831 | 0.040281 | 2.62E-09 | cis |
| rs1888581 | miR-409-3p | 14:101042 | A | C | 0.735363 | 0.291849 | 0.03465 | 3.68E-17 | cis |

| | | | | | | | | | |
|---|---|---|---|---|---|---|---|---|---|
| rs7143357 | miR-409-3p | 14:101054 | T | C | 0.739636 | 0.303185 | 0.035052 | 5.17E-18 | cis |
| rs1190904 | miR-409-3p | 14:101096 | A | G | 0.772596 | 0.293462 | 0.037475 | 4.84E-15 | cis |
| rs6575796 | miR-409-3p | 14:101160 | A | G | 0.094049 | -0.310509 | 0.056885 | 4.80E-08 | cis |
| rs7493817 | miR-409-3p | 14:101038 | G | A | 0.751723 | 0.308992 | 0.035838 | 6.58E-18 | cis |
| rs1190872 | miR-409-3p | 14:101069 | A | G | 0.310177 | -0.291458 | 0.033639 | 4.54E-18 | cis |
| rs1765384 | miR-409-3p | 14:101102 | A | T | 0.870972 | 0.321056 | 0.049009 | 5.72E-11 | cis |
| rs9671394 | miR-409-3p | 14:101159 | C | T | 0.284232 | -0.388278 | 0.03475 | 5.50E-29 | cis |
| rs879060 | miR-409-3p | 14:101069 | A | G | 0.310926 | -0.292711 | 0.033638 | 3.27E-18 | cis |
| rs7155649 | miR-409-3p | 14:101181 | C | A | 0.863608 | 0.338672 | 0.046086 | 2.00E-13 | cis |
| rs1190817 | miR-409-3p | 14:101229 | T | C | 0.798148 | 0.222068 | 0.038884 | 1.12E-08 | cis |
| rs1190909 | miR-409-3p | 14:101101 | G | A | 0.775531 | 0.287211 | 0.037936 | 3.70E-14 | cis |
| rs1243295 | miR-409-3p | 14:101205 | G | A | 0.231663 | -0.232771 | 0.03714 | 3.67E-10 | cis |
| rs1190906 | miR-409-3p | 14:101099 | T | C | 0.772138 | 0.287716 | 0.037368 | 1.37E-14 | cis |
| rs1190852 | miR-409-3p | 14:101064 | A | G | 0.310295 | -0.293602 | 0.033696 | 2.95E-18 | cis |
| rs1190856 | miR-409-3p | 14:101065 | T | C | 0.310244 | -0.292692 | 0.033671 | 3.54E-18 | cis |
| rs879058 | miR-409-3p | 14:101069 | G | T | 0.311807 | -0.29189 | 0.033638 | 4.05E-18 | cis |
| rs1184948 | miR-409-3p | 14:101037 | C | G | 0.733365 | 0.296938 | 0.034724 | 1.22E-17 | cis |
| rs1184658 | miR-409-3p | 14:101017 | C | G | 0.759077 | 0.275362 | 0.037678 | 2.71E-13 | trans |
| rs3579985 | miR-409-3p | 14:101156 | C | T | 0.904015 | 0.316092 | 0.05547 | 1.21E-08 | cis |
| rs1185095 | miR-409-3p | 14:101056 | G | T | 0.743981 | 0.304632 | 0.034836 | 2.24E-18 | cis |
| rs7144997 | miR-409-3p | 14:101155 | A | G | 0.899872 | 0.306471 | 0.055322 | 3.03E-08 | cis |
| rs7140792 | miR-409-3p | 14:101172 | C | T | 0.892517 | 0.404584 | 0.050225 | 7.92E-16 | cis |
| rs1184764 | miR-409-3p | 14:101023 | C | T | 0.753952 | 0.257884 | 0.03716 | 3.93E-12 | trans |
| rs1014844 | miR-409-3p | 14:101026 | G | A | 0.753241 | 0.259752 | 0.037014 | 2.26E-12 | trans |
| rs1555405 | miR-409-3p | 14:101176 | G | A | 0.749075 | 0.306581 | 0.034993 | 1.93E-18 | cis |
| rs1289379 | miR-409-3p | 14:101178 | C | G | 0.190813 | -0.420362 | 0.039594 | 2.49E-26 | cis |
| rs1190878 | miR-409-3p | 14:101072 | C | A | 0.859763 | 0.29174 | 0.045635 | 1.63E-10 | cis |
| rs1015151 | miR-409-3p | 14:101175 | G | A | 0.806743 | 0.282968 | 0.038714 | 2.69E-13 | cis |
| rs1014839 | miR-409-3p | 14:101026 | A | G | 0.750738 | 0.259716 | 0.037027 | 2.31E-12 | trans |
| rs1190857 | miR-409-3p | 14:101066 | G | T | 0.310273 | -0.292547 | 0.033678 | 3.74E-18 | cis |
| rs2295659 | miR-409-3p | 14:101141 | G | T | 0.857995 | 0.245953 | 0.044425 | 3.09E-08 | cis |
| rs1258812 | miR-409-3p | 14:101162 | T | C | 0.893709 | 0.386003 | 0.051855 | 9.78E-14 | cis |
| rs1116059 | miR-409-3p | 14:100966 | C | T | 0.745134 | 0.244078 | 0.038002 | 1.34E-10 | trans |
| rs2400940 | miR-409-3p | 14:101224 | G | A | 0.185552 | -0.233373 | 0.039444 | 3.29E-09 | cis |
| rs1214784 | miR-409-3p | 14:101144 | C | T | 0.870989 | 0.309359 | 0.04661 | 3.20E-11 | cis |
| rs6575794 | miR-409-3p | 14:101039 | C | T | 0.734189 | 0.296437 | 0.034778 | 1.54E-17 | cis |
| rs4905999 | miR-409-3p | 14:101184 | A | G | 0.181905 | -0.407631 | 0.040344 | 5.31E-24 | cis |
| rs1087351 | miR-409-3p | 14:100945 | G | T | 0.730696 | 0.230198 | 0.038387 | 2.01E-09 | trans |
| rs1243613 | miR-409-3p | 14:101031 | C | T | 0.725015 | 0.278583 | 0.03461 | 8.34E-16 | trans |
| rs4905994 | miR-409-3p | 14:101154 | T | C | 0.329146 | -0.345859 | 0.033412 | 4.12E-25 | cis |
| rs1190815 | miR-409-3p | 14:101228 | G | A | 0.798481 | 0.223251 | 0.038588 | 7.23E-09 | cis |
| rs7492369 | miR-409-3p | 14:101020 | C | T | 0.755698 | 0.257176 | 0.037225 | 4.89E-12 | trans |
| rs3470057 | miR-409-3p | 14:101173 | C | T | 0.892725 | 0.404351 | 0.050175 | 7.70E-16 | cis |
| rs9324022 | miR-409-3p | 14:101172 | T | C | 0.763739 | 0.282318 | 0.038026 | 1.13E-13 | cis |
| rs7140556 | miR-409-3p | 14:101036 | C | A | 0.723818 | 0.280094 | 0.034528 | 4.98E-16 | cis |
| rs1190907 | miR-409-3p | 14:101099 | A | G | 0.772847 | 0.289277 | 0.037424 | 1.08E-14 | cis |
| rs1162219 | miR-409-3p | 14:101016 | G | A | 0.759852 | 0.296621 | 0.039304 | 4.46E-14 | trans |
| rs7145610 | miR-409-3p | 14:101049 | T | C | 0.740391 | 0.307972 | 0.035097 | 1.71E-18 | cis |
| rs1243449 | miR-409-3p | 14:101179 | A | G | 0.18959 | -0.419523 | 0.039564 | 2.87E-26 | cis |
| rs1709962 | miR-409-3p | 14:101173 | A | C | 0.783449 | 0.248095 | 0.037471 | 3.57E-11 | cis |
| rs7155375 | miR-409-3p | 14:101188 | C | T | 0.635995 | 0.276943 | 0.031705 | 2.43E-18 | cis |
| rs1289760 | miR-409-3p | 14:101182 | C | T | 0.190452 | -0.419035 | 0.03935 | 1.76E-26 | cis |
| rs7142683 | miR-409-3p | 14:101158 | A | G | 0.290748 | -0.378587 | 0.034391 | 3.48E-28 | cis |

| | | | | | | | | | |
|---|---|---|---|---|---|---|---|---|---|
| rs7149242 | miR-409-3p | 14:101159 | T | G | 0.288371 | -0.377465 | 0.034293 | 3.53E-28 | cis |
| rs1259093 | miR-409-3p | 14:101163 | A | T | 0.893649 | 0.38628 | 0.051858 | 9.42E-14 | cis |
| rs1190881 | miR-409-3p | 14:101079 | T | C | 0.346494 | -0.222745 | 0.032893 | 1.27E-11 | cis |
| rs1087352 | miR-409-3p | 14:101175 | G | A | 0.809826 | 0.284259 | 0.039175 | 3.98E-13 | cis |
| rs2842154 | miR-409-3p | 14:101158 | C | T | 0.904763 | 0.314864 | 0.055534 | 1.43E-08 | cis |
| rs911330 | miR-409-3p | 14:101099 | T | C | 0.774615 | 0.292431 | 0.037604 | 7.45E-15 | cis |
| rs7152943 | miR-409-3p | 14:101174 | C | T | 0.924188 | 0.322446 | 0.058951 | 4.51E-08 | cis |
| rs1013035 | miR-409-3p | 14:101149 | T | C | 0.112665 | -0.2959 | 0.051883 | 1.18E-08 | cis |
| rs3579985 | miR-411-3p | 14:101156 | C | T | 0.904015 | 0.405334 | 0.061456 | 4.24E-11 | cis |
| rs2842154 | miR-411-3p | 14:101158 | C | T | 0.904763 | 0.402229 | 0.061534 | 6.29E-11 | cis |
| rs7149242 | miR-411-3p | 14:101159 | T | G | 0.288371 | -0.237007 | 0.038806 | 1.01E-09 | cis |
| rs6575796 | miR-411-3p | 14:101160 | A | G | 0.094049 | -0.405253 | 0.063016 | 1.27E-10 | cis |
| rs1288035 | miR-411-3p | 14:101158 | C | G | 0.904391 | 0.403679 | 0.061463 | 5.10E-11 | cis |
| rs1013035 | miR-411-3p | 14:101149 | T | C | 0.112665 | -0.336898 | 0.057604 | 4.96E-09 | cis |
| rs7142683 | miR-411-3p | 14:101158 | A | G | 0.290748 | -0.23718 | 0.038919 | 1.10E-09 | cis |
| rs7144997 | miR-411-3p | 14:101155 | A | G | 0.899872 | 0.391854 | 0.061304 | 1.64E-10 | cis |
| rs9671394 | miR-411-3p | 14:101159 | C | T | 0.284232 | -0.245899 | 0.03934 | 4.09E-10 | cis |
| rs7155375 | miR-431-5p | 14:101188 | C | T | 0.635995 | 0.189527 | 0.031102 | 1.10E-09 | cis |
| rs1243449 | miR-431-5p | 14:101179 | A | G | 0.18959 | -0.269382 | 0.03903 | 5.13E-12 | cis |
| rs1289379 | miR-431-5p | 14:101178 | C | G | 0.190813 | -0.270891 | 0.039058 | 4.05E-12 | cis |
| rs1214700 | miR-431-5p | 14:101186 | C | T | 0.223221 | -0.223582 | 0.036282 | 7.17E-10 | cis |
| rs1289760 | miR-431-5p | 14:101182 | C | T | 0.190452 | -0.276286 | 0.0388 | 1.07E-12 | cis |
| rs7149242 | miR-433-3p | 14:101159 | T | G | 0.288371 | -0.186034 | 0.030824 | 1.59E-09 | cis |
| rs1214700 | miR-433-3p | 14:101186 | C | T | 0.223221 | -0.245136 | 0.032778 | 7.50E-14 | cis |
| rs7142683 | miR-433-3p | 14:101158 | A | G | 0.290748 | -0.18614 | 0.030913 | 1.73E-09 | cis |
| rs7155375 | miR-433-3p | 14:101188 | C | T | 0.635995 | 0.197083 | 0.028136 | 2.48E-12 | cis |
| rs4905999 | miR-433-3p | 14:101184 | A | G | 0.181905 | -0.241627 | 0.036042 | 2.03E-11 | cis |
| rs4905994 | miR-433-3p | 14:101154 | T | C | 0.329146 | -0.183078 | 0.029933 | 9.58E-10 | cis |
| rs7155649 | miR-433-3p | 14:101181 | C | A | 0.863608 | 0.275895 | 0.040724 | 1.25E-11 | cis |
| rs1289379 | miR-433-3p | 14:101178 | C | G | 0.190813 | -0.253398 | 0.035405 | 8.23E-13 | cis |
| rs1289760 | miR-433-3p | 14:101182 | C | T | 0.190452 | -0.249669 | 0.035198 | 1.31E-12 | cis |
| rs1243449 | miR-433-3p | 14:101179 | A | G | 0.18959 | -0.251636 | 0.03538 | 1.14E-12 | cis |
| rs9324022 | miR-433-3p | 14:101172 | T | C | 0.763739 | 0.188894 | 0.033721 | 2.12E-08 | cis |
| rs9671394 | miR-433-3p | 14:101159 | C | T | 0.284232 | -0.188991 | 0.031259 | 1.49E-09 | cis |
| rs6214393 | miR-4429 | 2:6864038 | G | C | 0.593598 | 0.109647 | 0.013593 | 7.24E-16 | trans |
| rs7572299 | miR-4429 | 2:6858124 | T | A | 0.538033 | 0.107773 | 0.013264 | 4.46E-16 | trans |
| rs1164012 | miR-4429 | 2:6862562 | C | T | 0.545777 | 0.114323 | 0.012888 | 7.30E-19 | trans |
| rs1158434 | miR-4429 | 2:6863552 | G | C | 0.551072 | 0.118342 | 0.013237 | 3.88E-19 | trans |
| rs1486061 | miR-4429 | 2:6862571 | T | G | 0.54743 | 0.115007 | 0.012931 | 5.91E-19 | trans |
| rs1271364 | miR-4429 | 2:6860665 | G | A | 0.282933 | 0.082273 | 0.014215 | 7.13E-09 | trans |
| rs6214393 | miR-4429 | 2:6864020 | A | G | 0.593308 | 0.109092 | 0.013598 | 1.04E-15 | trans |
| rs6546395 | miR-4429 | 2:6862927 | A | G | 0.55448 | 0.116327 | 0.013078 | 5.85E-19 | trans |
| rs1301028 | miR-4429 | 2:6862184 | G | T | 0.683506 | 0.105886 | 0.013707 | 1.12E-14 | trans |
| rs7574592 | miR-4429 | 2:6859766 | T | G | 0.281934 | 0.080766 | 0.014188 | 1.25E-08 | trans |
| rs1300974 | miR-4429 | 2:6858443 | C | A | 0.678986 | 0.096595 | 0.01431 | 1.48E-11 | trans |
| rs6546410 | miR-4429 | 2:6863610 | C | G | 0.702434 | 0.109252 | 0.014591 | 7.02E-14 | trans |
| rs880668 | miR-4429 | 2:6859258 | T | C | 0.280228 | 0.079964 | 0.014273 | 2.11E-08 | trans |
| rs1049616 | miR-4429 | 2:6862008 | C | T | 0.697496 | 0.104328 | 0.013941 | 7.23E-14 | trans |
| rs6214393 | miR-4429 | 2:6864009 | T | C | 0.59325 | 0.109114 | 0.013597 | 1.02E-15 | trans |
| rs6546407 | miR-4429 | 2:6863492 | A | C | 0.55091 | 0.118316 | 0.013221 | 3.58E-19 | trans |
| rs6546400 | miR-4429 | 2:6863064 | G | A | 0.554382 | 0.116519 | 0.01309 | 5.51E-19 | trans |
| rs7600986 | miR-4429 | 2:6863692 | A | T | 0.285029 | 0.083204 | 0.014209 | 4.75E-09 | trans |
| rs6546396 | miR-4429 | 2:6862930 | A | T | 0.283762 | 0.082443 | 0.014187 | 6.21E-09 | trans |

| | | | | | | | | | |
|---|---|---|---|---|---|---|---|---|---|
| rs5933292 | miR-4429 | 2:6863963 | G | A | 0.73529 | 0.107748 | 0.015329 | 2.08E-12 | trans |
| rs6714719 | miR-4429 | 2:6859377 | G | C | 0.698404 | 0.104022 | 0.014003 | 1.10E-13 | trans |
| rs1017438 | miR-4429 | 2:6859646 | C | T | 0.685243 | 0.105793 | 0.013758 | 1.48E-14 | trans |
| rs6546399 | miR-4429 | 2:6863064 | C | T | 0.55394 | 0.116432 | 0.013078 | 5.44E-19 | trans |
| rs6546404 | miR-4429 | 2:6863263 | G | T | 0.54995 | 0.117873 | 0.013142 | 3.00E-19 | trans |
| rs6546398 | miR-4429 | 2:6863041 | G | C | 0.552757 | 0.116069 | 0.013039 | 5.51E-19 | trans |
| rs6546405 | miR-4429 | 2:6863311 | G | A | 0.28401 | 0.082999 | 0.014224 | 5.38E-09 | trans |
| rs5618821 | miR-4429 | 2:6861163 | T | C | 0.698075 | 0.104494 | 0.013944 | 6.70E-14 | trans |
| rs1760168 | miR-4429 | 2:6861589 | G | A | 0.697578 | 0.104385 | 0.01395 | 7.27E-14 | trans |
| rs6744216 | miR-4429 | 2:6863463 | A | G | 0.284465 | 0.083182 | 0.014224 | 4.97E-09 | trans |
| rs1300519 | miR-4429 | 2:6863887 | A | G | 0.702186 | 0.109146 | 0.014585 | 7.25E-14 | trans |
| rs1484530 | miR-4429 | 2:6863547 | A | G | 0.70445 | 0.109845 | 0.014643 | 6.31E-14 | trans |
| rs6757938 | miR-4429 | 2:6863285 | A | G | 0.549958 | 0.117883 | 0.013145 | 3.02E-19 | trans |
| rs5690424 | miR-4429 | 2:6863847 | T | C | 0.550669 | 0.118152 | 0.013239 | 4.48E-19 | trans |
| rs6546411 | miR-4429 | 2:6863769 | A | G | 0.556248 | 0.120734 | 0.013319 | 1.25E-19 | trans |
| rs7608429 | miR-4429 | 2:6858083 | G | T | 0.678719 | 0.09611 | 0.014323 | 1.94E-11 | trans |
| rs7598360 | miR-4429 | 2:6863092 | A | G | 0.553166 | 0.11928 | 0.013065 | 6.86E-20 | trans |
| rs2165919 | miR-4429 | 2:6861413 | C | T | 0.281408 | 0.082508 | 0.01418 | 5.94E-09 | trans |
| rs7420773 | miR-4429 | 2:6862945 | G | A | 0.28377 | 0.082444 | 0.014187 | 6.21E-09 | trans |
| rs1373200 | miR-4429 | 2:6862265 | C | T | 0.547142 | 0.112939 | 0.012754 | 8.34E-19 | trans |
| rs3426707 | miR-4429 | 2:6859915 | T | C | 0.282465 | 0.081328 | 0.014197 | 1.01E-08 | trans |
| rs6546408 | miR-4429 | 2:6863504 | T | C | 0.551062 | 0.118342 | 0.013234 | 3.82E-19 | trans |
| rs6419643 | miR-4429 | 2:6863126 | T | G | 0.548826 | 0.116437 | 0.013006 | 3.47E-19 | trans |
| rs1302892 | miR-4429 | 2:6862991 | C | T | 0.552648 | 0.115995 | 0.013035 | 5.65E-19 | trans |
| rs6546412 | miR-4429 | 2:6863771 | T | C | 0.557086 | 0.121849 | 0.013344 | 6.76E-20 | trans |
| rs6730974 | miR-4429 | 2:6863452 | T | A | 0.287755 | 0.085228 | 0.014255 | 2.25E-09 | trans |
| rs6546387 | miR-4429 | 2:6858758 | G | A | 0.281573 | 0.079049 | 0.014305 | 3.28E-08 | trans |
| rs964935 | miR-4429 | 2:6861228 | G | A | 0.281126 | 0.081712 | 0.014183 | 8.35E-09 | trans |
| rs6214392 | miR-4429 | 2:6863835 | C | G | 0.700378 | 0.109847 | 0.014575 | 4.82E-14 | trans |
| rs1300592 | miR-4429 | 2:6862999 | T | A | 0.701261 | 0.106555 | 0.014352 | 1.13E-13 | trans |
| rs6546397 | miR-4429 | 2:6863036 | G | A | 0.552756 | 0.116061 | 0.01304 | 5.55E-19 | trans |
| rs1760209 | miR-4429 | 2:6862041 | T | A | 0.697529 | 0.104351 | 0.01394 | 7.12E-14 | trans |
| rs1455505 | miR-4429 | 2:6862832 | G | A | 0.556221 | 0.118259 | 0.013138 | 2.23E-19 | trans |
| rs6546393 | miR-4429 | 2:6862757 | T | C | 0.554017 | 0.117707 | 0.013065 | 2.08E-19 | trans |
| rs2241947 | miR-4429 | 2:6861539 | C | T | 0.697988 | 0.104503 | 0.013944 | 6.65E-14 | trans |
| rs5888448 | miR-4429 | 2:6864534 | G | A | 0.734214 | 0.105803 | 0.015237 | 3.82E-12 | trans |
| rs6709464 | miR-4429 | 2:6862222 | C | T | 0.697279 | 0.10404 | 0.013928 | 8.03E-14 | trans |
| rs1050181 | miR-4429 | 2:6862295 | G | C | 0.544992 | 0.113852 | 0.012842 | 7.63E-19 | trans |
| rs6744306 | miR-4429 | 2:6863273 | T | C | 0.549956 | 0.117881 | 0.013143 | 2.99E-19 | trans |
| rs1301157 | miR-4429 | 2:6863927 | G | A | 0.702349 | 0.109115 | 0.01459 | 7.50E-14 | trans |
| rs6214389 | miR-4429 | 2:6860455 | C | T | 0.698307 | 0.103959 | 0.013964 | 9.70E-14 | trans |
| rs6546388 | miR-4429 | 2:6858948 | C | T | 0.281691 | 0.079334 | 0.014263 | 2.67E-08 | trans |
| rs5999861 | miR-4429 | 2:6864677 | G | T | 0.731295 | 0.101746 | 0.015226 | 2.35E-11 | trans |
| rs1442589 | miR-4429 | 2:6863473 | G | T | 0.701896 | 0.108994 | 0.014556 | 6.99E-14 | trans |
| rs1422402 | miR-4429 | 2:6862752 | A | G | 0.698498 | 0.105646 | 0.014255 | 1.25E-13 | trans |
| rs1474969 | miR-4429 | 2:6862568 | G | A | 0.546544 | 0.114556 | 0.012908 | 6.98E-19 | trans |
| rs7592526 | miR-4429 | 2:6858352 | A | C | 0.538007 | 0.108083 | 0.013262 | 3.64E-16 | trans |
| rs6546402 | miR-4429 | 2:6863130 | A | G | 0.283216 | 0.082643 | 0.014193 | 5.79E-09 | trans |
| rs1189593 | miR-4429 | 2:6861908 | A | G | 0.282175 | 0.0818 | 0.01416 | 7.61E-09 | trans |
| rs6214393 | miR-4429 | 2:6864062 | T | C | 0.733261 | 0.106445 | 0.015174 | 2.30E-12 | trans |
| rs7588432 | miR-4429 | 2:6858548 | G | A | 0.282438 | 0.079352 | 0.014543 | 4.86E-08 | trans |
| rs4610041 | miR-4429 | 2:6862628 | C | T | 0.550745 | 0.116935 | 0.013037 | 2.99E-19 | trans |
| rs1397159 | miR-4429 | 2:6863425 | T | G | 0.705552 | 0.110694 | 0.014727 | 5.62E-14 | trans |

| | | | | | | | | | |
|---|---|---|---|---|---|---|---|---|---|
| rs6743970 | miR-4429 | 2:68583907 | A | T | 0.660237 | 0.101973 | 0.014137 | 5.46E-13 | trans |
| rs6729686 | miR-4429 | 2:68633030 | C | T | 0.700171 | 0.108074 | 0.014448 | 7.41E-14 | trans |
| rs6729555 | miR-4429 | 2:68632952 | C | A | 0.54995 | 0.117896 | 0.013146 | 3.01E-19 | trans |
| rs7597167 | miR-4429 | 2:68612940 | C | T | 0.698051 | 0.104514 | 0.013944 | 6.62E-14 | trans |
| rs34852232 | miR-4429 | 2:68638599 | C | G | 0.550658 | 0.118152 | 0.01324 | 4.50E-19 | trans |
| rs13034825 | miR-4429 | 2:68630297 | C | G | 0.552715 | 0.116045 | 0.013038 | 5.56E-19 | trans |
| rs6715579 | miR-4429 | 2:68583923 | C | T | 0.678794 | 0.096668 | 0.014309 | 1.42E-11 | trans |
| rs1542135 | miR-4429 | 2:68620848 | C | T | 0.69752 | 0.104351 | 0.01394 | 7.11E-14 | trans |
| rs1542136 | miR-4429 | 2:68620930 | C | A | 0.697594 | 0.104377 | 0.013944 | 7.12E-14 | trans |
| rs7581872 | miR-4429 | 2:68580981 | A | C | 0.537959 | 0.107684 | 0.013264 | 4.71E-16 | trans |
| rs2165920 | miR-4429 | 2:68622239 | T | C | 0.552012 | 0.112932 | 0.012844 | 1.47E-18 | trans |
| rs7605387 | miR-4429 | 2:68632550 | A | G | 0.703499 | 0.108179 | 0.014531 | 9.72E-14 | trans |
| rs74514522 | miR-4429 | 2:68637550 | T | G | 0.702552 | 0.109286 | 0.014597 | 7.07E-14 | trans |
| rs6546401 | miR-4429 | 2:68630873 | A | T | 0.550127 | 0.116694 | 0.013034 | 3.45E-19 | trans |
| rs34196827 | miR-4429 | 2:68621427 | G | A | 0.697518 | 0.104352 | 0.01394 | 7.10E-14 | trans |
| rs4671208 | miR-4429 | 2:68600798 | A | G | 0.281916 | 0.080827 | 0.014188 | 1.22E-08 | trans |
| rs11126183 | miR-4429 | 2:68619138 | A | G | 0.282025 | 0.081766 | 0.014155 | 7.62E-09 | trans |
| rs17601498 | miR-4429 | 2:68611712 | A | G | 0.69807 | 0.104495 | 0.013944 | 6.70E-14 | trans |
| rs2070171 | miR-4429 | 2:68613710 | G | A | 0.698024 | 0.104489 | 0.013944 | 6.71E-14 | trans |
| rs56284706 | miR-4429 | 2:68631372 | G | A | 0.697508 | 0.106313 | 0.014265 | 9.15E-14 | trans |
| rs6546403 | miR-4429 | 2:68632262 | C | T | 0.550801 | 0.118031 | 0.013158 | 2.95E-19 | trans |
| rs2241946 | miR-4429 | 2:68615654 | T | C | 0.698076 | 0.104588 | 0.01395 | 6.51E-14 | trans |
| rs78002751 | miR-4429 | 2:68625430 | A | G | 0.546309 | 0.114214 | 0.012873 | 7.16E-19 | trans |
| rs75628896 | miR-4429 | 2:68625471 | T | C | 0.285916 | 0.080656 | 0.014206 | 1.36E-08 | trans |
| rs11566292 | miR-4429 | 2:68626631 | T | G | 0.547487 | 0.115419 | 0.012956 | 5.17E-19 | trans |
| rs6546409 | miR-4429 | 2:68635879 | A | G | 0.551093 | 0.118334 | 0.013241 | 4.00E-19 | trans |
| rs7421228 | miR-4429 | 2:68630124 | G | T | 0.55268 | 0.116024 | 0.013037 | 5.60E-19 | trans |
| rs10189903 | miR-4429 | 2:68606193 | C | G | 0.281778 | 0.081053 | 0.014164 | 1.05E-08 | trans |
| rs6715909 | miR-4429 | 2:68583995 | G | A | 0.664879 | 0.098726 | 0.014073 | 2.30E-12 | trans |
| rs7420782 | miR-4429 | 2:68629558 | G | T | 0.283742 | 0.082649 | 0.01419 | 5.73E-09 | trans |
| rs9653552 | miR-4429 | 2:68643960 | G | A | 0.593458 | 0.109389 | 0.013605 | 8.96E-16 | trans |
| rs8761 | miR-4429 | 2:68624260 | G | A | 0.545692 | 0.114212 | 0.012884 | 7.65E-19 | trans |
| rs4854536 | miR-4429 | 2:68631865 | T | A | 0.283938 | 0.082964 | 0.014224 | 5.45E-09 | trans |
| rs1373198 | miR-4429 | 2:68587983 | A | T | 0.281586 | 0.079054 | 0.014303 | 3.26E-08 | trans |
| rs13028971 | miR-4429 | 2:68630038 | A | G | 0.54849 | 0.116303 | 0.012997 | 3.60E-19 | trans |
| rs964934 | miR-4429 | 2:68611877 | G | A | 0.281023 | 0.081689 | 0.014181 | 8.38E-09 | trans |
| rs3732044 | miR-4429 | 2:68623915 | T | C | 0.545562 | 0.114139 | 0.012879 | 7.82E-19 | trans |
| rs1155426 | miR-4429 | 2:68621824 | T | C | 0.282037 | 0.081341 | 0.014154 | 9.08E-09 | trans |
| rs6747545 | miR-4429 | 2:68633069 | T | A | 0.549964 | 0.11789 | 0.013147 | 3.04E-19 | trans |
| rs60007165 | miR-4429 | 2:68639630 | C | T | 0.735163 | 0.107772 | 0.015327 | 2.04E-12 | trans |
| rs7562439 | miR-4429 | 2:68582569 | C | T | 0.6646 | 0.098629 | 0.014077 | 2.44E-12 | trans |
| rs6708536 | miR-4429 | 2:68627268 | G | A | 0.547249 | 0.115405 | 0.012966 | 5.55E-19 | trans |
| rs6419644 | miR-4429 | 2:68631823 | A | G | 0.549898 | 0.117848 | 0.013138 | 2.96E-19 | trans |
| rs13011436 | miR-4429 | 2:68639379 | C | T | 0.702382 | 0.109113 | 0.014591 | 7.54E-14 | trans |
| rs6742200 | miR-4429 | 2:68623302 | A | G | 0.54552 | 0.114117 | 0.012877 | 7.84E-19 | trans |
| rs13828992 | miR-4429 | 2:68637829 | A | G | 0.702614 | 0.109295 | 0.014601 | 7.13E-14 | trans |
| rs60878442 | miR-4429 | 2:68582115 | A | G | 0.538067 | 0.108084 | 0.013261 | 3.62E-16 | trans |
| rs13906881 | miR-4429 | 2:68627735 | C | T | 0.69721 | 0.105551 | 0.014218 | 1.14E-13 | trans |
| rs1867313 | miR-4429 | 2:68623652 | T | C | 0.282087 | 0.081215 | 0.014152 | 9.53E-09 | trans |
| rs13010774 | miR-4429 | 2:68639081 | C | T | 0.702339 | 0.109105 | 0.014589 | 7.52E-14 | trans |
| rs56337143 | miR-4429 | 2:68642490 | G | A | 0.733417 | 0.106385 | 0.015183 | 2.44E-12 | trans |
| rs7578602 | miR-4429 | 2:68637868 | G | C | 0.556806 | 0.121802 | 0.01335 | 7.26E-20 | trans |
| rs58089918 | miR-4429 | 2:68639660 | G | C | 0.734634 | 0.10805 | 0.015286 | 1.57E-12 | trans |

| | | | | | | | | | |
|---|---|---|---|---|---|---|---|---|---|
| rs1753019: | miR-4429 | 2:6862133: | G | T | 0.683745 | 0.106181 | 0.013718 | 9.92E-15 | trans |
| rs8028813! | miR-4429 | 2:6862547! | A | G | 0.548418 | 0.11582 | 0.012961 | 4.03E-19 | trans |
| rs7590450 | miR-4429 | 2:6862856: | A | G | 0.549532 | 0.115985 | 0.012996 | 4.47E-19 | trans |
| rs1343065 | miR-4429 | 2:6858771( | C | G | 0.699397 | 0.103919 | 0.014114 | 1.80E-13 | trans |
| rs7420775 | miR-4429 | 2:6862945: | G | T | 0.703122 | 0.107093 | 0.014416 | 1.10E-13 | trans |
| rs1301107! | miR-4429 | 2:6863931: | A | C | 0.700844 | 0.109516 | 0.014613 | 6.66E-14 | trans |
| rs6715706 | miR-4429 | 2:6858399( | C | A | 0.664887 | 0.098723 | 0.014073 | 2.30E-12 | trans |
| rs1018497! | miR-4429 | 2:6859988 | G | A | 0.286787 | 0.081077 | 0.014257 | 1.30E-08 | trans |
| rs6214393: | miR-4429 | 2:6864041! | A | G | 0.593599 | 0.10964 | 0.013593 | 7.28E-16 | trans |
| rs1300822: | miR-4433b | 2:6457412: | G | A | 0.716086 | -0.088848 | 0.013143 | 1.38E-11 | cis |
| rs1271350( | miR-4433b | 2:6456984( | C | T | 0.714645 | -0.091252 | 0.013166 | 4.19E-12 | cis |
| rs2059631 | miR-4433b | 2:6457468: | G | A | 0.560261 | -0.081642 | 0.012057 | 1.28E-11 | cis |
| rs1043209 | miR-4707-: | 14:233739: | G | A | 0.403581 | 0.087395 | 0.013671 | 1.63E-10 | cis |
| rs4982706 | miR-4707-: | 14:233816! | C | T | 0.404104 | 0.086894 | 0.013657 | 1.98E-10 | cis |
| rs4982711 | miR-4707-: | 14:233989! | T | G | 0.442327 | 0.091257 | 0.013718 | 2.89E-11 | cis |
| rs8005677 | miR-4707-: | 14:233766: | T | C | 0.403905 | 0.087174 | 0.013666 | 1.78E-10 | cis |
| rs9989230 | miR-4707-: | 14:233819: | C | T | 0.404111 | 0.086877 | 0.013656 | 2.00E-10 | cis |
| rs1956881 | miR-4707-: | 14:233940( | A | C | 0.406379 | 0.087141 | 0.013695 | 1.98E-10 | cis |
| rs5782274! | miR-4707-: | 14:234099( | G | A | 0.440759 | 0.093412 | 0.013618 | 6.91E-12 | cis |
| rs6197774: | miR-4707-: | 14:234009: | G | C | 0.440733 | 0.091819 | 0.013701 | 2.06E-11 | cis |
| rs4982707 | miR-4707-: | 14:233910: | C | T | 0.440299 | 0.091442 | 0.0137 | 2.48E-11 | cis |
| rs4982714 | miR-4707-: | 14:234133: | C | T | 0.441229 | 0.093545 | 0.013601 | 6.08E-12 | cis |
| rs1043675 | miR-4707-: | 14:234563: | T | C | 0.594711 | -0.081309 | 0.013445 | 1.47E-09 | cis |
| rs1115793( | miR-4707-: | 14:234031: | T | G | 0.407904 | 0.087831 | 0.013635 | 1.18E-10 | cis |
| rs5589413: | miR-4707-: | 14:234088: | C | G | 0.443456 | 0.093971 | 0.013657 | 5.95E-12 | cis |
| rs4982709 | miR-4707-: | 14:233946! | C | T | 0.407036 | 0.086929 | 0.013668 | 2.02E-10 | cis |
| rs1014821! | miR-4707-: | 14:233823 | C | T | 0.405428 | 0.085593 | 0.013692 | 4.07E-10 | cis |
| rs8012568 | miR-4707-: | 14:234528: | T | C | 0.587048 | -0.083383 | 0.013518 | 6.91E-10 | cis |
| rs3434488: | miR-4707-: | 14:233875: | A | G | 0.40749 | 0.087014 | 0.013677 | 1.99E-10 | cis |
| rs7145494 | miR-4707-: | 14:233997! | T | C | 0.407171 | 0.087186 | 0.013669 | 1.79E-10 | cis |
| rs3794453 | miR-4707-: | 14:233868: | A | T | 0.412393 | 0.089491 | 0.013682 | 6.13E-11 | cis |
| rs6197774( | miR-4707-: | 14:234008: | G | A | 0.408544 | 0.087112 | 0.013691 | 1.98E-10 | cis |
| rs1271742: | miR-4707-: | 14:234177! | C | T | 0.479709 | 0.088552 | 0.013614 | 7.79E-11 | cis |
| rs1258953! | miR-4707-: | 14:233966: | T | C | 0.440305 | 0.091556 | 0.013703 | 2.37E-11 | cis |
| rs3491719: | miR-4707-: | 14:233876: | T | C | 0.406925 | 0.086866 | 0.013671 | 2.10E-10 | cis |
| rs3561518( | miR-4707-: | 14:234043: | T | C | 0.441508 | 0.093065 | 0.013697 | 1.09E-11 | cis |
| rs1243715: | miR-4707-: | 14:233729: | G | A | 0.437486 | 0.091279 | 0.013659 | 2.35E-11 | cis |
| rs2145542 | miR-4707-: | 14:234190: | C | T | 0.480462 | 0.088607 | 0.013609 | 7.47E-11 | cis |
| rs5618074: | miR-4707-: | 14:233772 | T | C | 0.438286 | 0.091634 | 0.013678 | 2.10E-11 | cis |
| rs4981451 | miR-4707-: | 14:234102: | T | C | 0.441163 | 0.093487 | 0.013604 | 6.33E-12 | cis |
| rs712486 | miR-4707-: | 14:234250 | T | C | 0.441215 | 0.094678 | 0.013749 | 5.73E-12 | cis |
| rs3794452 | miR-4707-: | 14:233866: | T | G | 0.440231 | 0.091353 | 0.013699 | 2.59E-11 | cis |
| rs8016061 | miR-4707-: | 14:233758( | A | G | 0.403869 | 0.087223 | 0.013666 | 1.74E-10 | cis |
| rs7155857 | miR-4707-: | 14:234000! | C | A | 0.407819 | 0.087207 | 0.013677 | 1.81E-10 | cis |
| rs3508506: | miR-4707-: | 14:234099( | C | A | 0.441051 | 0.09349 | 0.01361 | 6.45E-12 | cis |
| rs4982710 | miR-4707-: | 14:233989( | C | T | 0.407129 | 0.087153 | 0.013676 | 1.86E-10 | cis |
| rs3538240 | miR-4707-: | 14:233798 | G | A | 0.43846 | 0.091125 | 0.013691 | 2.82E-11 | cis |
| rs8006396 | miR-4707-: | 14:233808! | A | G | 0.404084 | 0.086924 | 0.013658 | 1.96E-10 | cis |
| rs4982713 | miR-4707-: | 14:234041( | C | T | 0.441538 | 0.092987 | 0.013695 | 1.12E-11 | cis |
| rs4981449 | miR-4707-: | 14:233888: | C | G | 0.406993 | 0.08687 | 0.013668 | 2.08E-10 | cis |
| rs4981452 | miR-4707-: | 14:234108( | A | G | 0.440913 | 0.093512 | 0.013614 | 6.47E-12 | cis |
| rs1951119 | miR-4707-: | 14:233883: | A | C | 0.406974 | 0.086867 | 0.013669 | 2.09E-10 | cis |
| rs2295685 | miR-4707-: | 14:233981! | A | G | 0.40713 | 0.087125 | 0.013669 | 1.84E-10 | cis |

| | | | | | | | | | |
|---|---|---|---|---|---|---|---|---|---|
| rs8006875 | miR-4707- | 14:234052 | A | G | 0.407111 | 0.087858 | 0.013669 | 1.30E-10 | cis |
| rs8016027 | miR-4707- | 14:234201 | T | C | 0.441165 | 0.093615 | 0.013591 | 5.67E-12 | cis |
| rs2295687 | miR-4707- | 14:234210 | A | G | 0.48047 | 0.088553 | 0.013606 | 7.60E-11 | cis |
| rs1956880 | miR-4707- | 14:233940 | T | C | 0.406587 | 0.087253 | 0.013698 | 1.89E-10 | cis |
| rs8014936 | miR-4707- | 14:233751 | A | G | 0.40386 | 0.087238 | 0.013667 | 1.73E-10 | cis |
| rs4981450 | miR-4707- | 14:233893 | A | G | 0.440221 | 0.09141 | 0.013701 | 2.53E-11 | cis |
| rs1115792 | miR-4707- | 14:233862 | T | C | 0.407468 | 0.087604 | 0.013722 | 1.73E-10 | cis |
| rs3449630 | miR-4707- | 14:234007 | G | C | 0.43983 | 0.093304 | 0.013786 | 1.30E-11 | cis |
| rs3458457 | miR-4707- | 14:234116 | C | T | 0.44118 | 0.093538 | 0.013603 | 6.14E-12 | cis |
| rs8003934 | miR-4707- | 14:234011 | A | G | 0.407307 | 0.08715 | 0.013662 | 1.78E-10 | cis |
| rs8006409 | miR-4707- | 14:233965 | A | G | 0.408092 | 0.08703 | 0.013689 | 2.05E-10 | cis |
| rs8019902 | miR-4707- | 14:233760 | C | T | 0.402747 | 0.087636 | 0.013704 | 1.61E-10 | cis |
| rs1013075 | miR-4707- | 14:233966 | C | G | 0.407058 | 0.086976 | 0.013668 | 1.97E-10 | cis |
| rs4536383 | miR-4707- | 14:234109 | T | C | 0.436129 | 0.091252 | 0.013624 | 2.11E-11 | cis |
| rs2295683 | miR-4707- | 14:233883 | A | C | 0.406975 | 0.086865 | 0.013669 | 2.09E-10 | cis |
| rs1115793 | miR-4707- | 14:234031 | A | C | 0.407885 | 0.087845 | 0.013635 | 1.18E-10 | cis |
| rs2295682 | miR-4707- | 14:233748 | C | T | 0.43795 | 0.091553 | 0.013662 | 2.07E-11 | cis |
| rs1288461 | miR-4707- | 14:234094 | A | G | 0.441135 | 0.093412 | 0.013605 | 6.60E-12 | cis |
| rs4981446 | miR-4707- | 14:233814 | A | G | 0.404093 | 0.086907 | 0.013657 | 1.97E-10 | cis |
| rs2295680 | miR-4707- | 14:233744 | G | A | 0.437854 | 0.091485 | 0.013662 | 2.13E-11 | cis |
| rs1757758 | miR-494-3 | 14:101189 | C | T | 0.827094 | 0.186422 | 0.031844 | 4.79E-09 | cis |
| rs1243449 | miR-494-3 | 14:101179 | A | G | 0.18959 | -0.176854 | 0.031368 | 1.72E-08 | cis |
| rs1289760 | miR-494-3 | 14:101182 | C | T | 0.190452 | -0.17975 | 0.031195 | 8.30E-09 | cis |
| rs1013035 | miR-494-3 | 14:101149 | T | C | 0.112665 | -0.222111 | 0.040422 | 3.91E-08 | cis |
| rs7155375 | miR-494-3 | 14:101188 | C | T | 0.635995 | 0.147301 | 0.024919 | 3.39E-09 | cis |
| rs1289379 | miR-494-3 | 14:101178 | C | G | 0.190813 | -0.177745 | 0.031392 | 1.49E-08 | cis |
| rs1214784 | miR-494-3 | 14:101144 | C | T | 0.870989 | 0.232267 | 0.036321 | 1.61E-10 | cis |
| rs9671394 | miR-494-3 | 14:101159 | C | T | 0.284232 | -0.172553 | 0.02758 | 3.94E-10 | cis |
| rs7142683 | miR-494-3 | 14:101158 | A | G | 0.290748 | -0.169059 | 0.027277 | 5.73E-10 | cis |
| rs745797 | miR-494-3 | 14:101141 | G | A | 0.858727 | 0.198556 | 0.034686 | 1.04E-08 | cis |
| rs1214700 | miR-494-3 | 14:101186 | C | T | 0.223221 | -0.166887 | 0.029088 | 9.62E-09 | cis |
| rs7149242 | miR-494-3 | 14:101159 | T | G | 0.288371 | -0.170843 | 0.027193 | 3.33E-10 | cis |
| rs2295659 | miR-494-3 | 14:101141 | G | T | 0.857995 | 0.197342 | 0.034578 | 1.15E-08 | cis |
| rs8008285 | miR-494-3 | 14:101146 | T | C | 0.842905 | 0.227618 | 0.033767 | 1.57E-11 | cis |
| rs9324022 | miR-494-3 | 14:101172 | T | C | 0.763739 | 0.174811 | 0.029748 | 4.19E-09 | cis |
| rs4905994 | miR-494-3 | 14:101154 | T | C | 0.329146 | -0.164594 | 0.026417 | 4.65E-10 | cis |
| rs1709962 | miR-543 | 14:101173 | A | C | 0.783449 | 0.200842 | 0.034395 | 5.24E-09 | cis |
| rs7149242 | miR-543 | 14:101159 | T | G | 0.288371 | -0.246539 | 0.031837 | 9.65E-15 | cis |
| rs1214700 | miR-543 | 14:101186 | C | T | 0.223221 | -0.195442 | 0.034215 | 1.12E-08 | cis |
| rs7142683 | miR-543 | 14:101158 | A | G | 0.290748 | -0.24586 | 0.031933 | 1.37E-14 | cis |
| rs1243449 | miR-543 | 14:101179 | A | G | 0.18959 | -0.241613 | 0.0368 | 5.18E-11 | cis |
| rs4905999 | miR-543 | 14:101184 | A | G | 0.181905 | -0.237607 | 0.037465 | 2.27E-10 | cis |
| rs1087352 | miR-543 | 14:101175 | G | A | 0.809826 | 0.222745 | 0.035996 | 6.09E-10 | cis |
| rs1015151 | miR-543 | 14:101175 | G | A | 0.806743 | 0.223374 | 0.03557 | 3.39E-10 | cis |
| rs2837918 | miR-543 | 14:101168 | G | T | 0.836434 | 0.217924 | 0.038649 | 1.72E-08 | cis |
| rs1289379 | miR-543 | 14:101178 | C | G | 0.190813 | -0.242585 | 0.036828 | 4.49E-11 | cis |
| rs1555405 | miR-543 | 14:101176 | G | A | 0.749075 | 0.22184 | 0.032262 | 6.15E-12 | cis |
| rs9671394 | miR-543 | 14:101159 | C | T | 0.284232 | -0.250906 | 0.032284 | 7.74E-15 | cis |
| rs1757758 | miR-543 | 14:101189 | C | T | 0.827094 | 0.217425 | 0.037459 | 6.46E-09 | cis |
| rs1289760 | miR-543 | 14:101182 | C | T | 0.190452 | -0.240043 | 0.036609 | 5.50E-11 | cis |
| rs9324022 | miR-543 | 14:101172 | T | C | 0.763739 | 0.218125 | 0.034955 | 4.37E-10 | cis |
| rs4905994 | miR-543 | 14:101154 | T | C | 0.329146 | -0.228361 | 0.030962 | 1.64E-13 | cis |
| rs7155375 | miR-543 | 14:101188 | C | T | 0.635995 | 0.202466 | 0.029223 | 4.26E-12 | cis |

| | | | | | | | | | |
|---|---|---|---|---|---|---|---|---|---|
| rs1318719 | miR-584-5 | 5:1485254 | G | A | 0.518797 | 0.146142 | 0.025512 | 1.01E-08 | cis |
| rs407696 | miR-584-5 | 5:1484225 | A | G | 0.538877 | -0.171433 | 0.025051 | 7.73E-12 | cis |
| rs1779527 | miR-584-5 | 5:1484220 | G | A | 0.854308 | -0.273328 | 0.035771 | 2.15E-14 | cis |
| rs6238010 | miR-584-5 | 5:1485004 | G | T | 0.8464 | -0.276639 | 0.035415 | 5.66E-15 | cis |
| rs7697787 | miR-584-5 | 5:1484473 | A | G | 0.844049 | -0.271396 | 0.034876 | 7.15E-15 | cis |
| rs5756988 | miR-584-5 | 5:1484501 | A | G | 0.843968 | -0.271505 | 0.034894 | 7.21E-15 | cis |
| rs1318269 | miR-584-5 | 5:1484854 | A | G | 0.474875 | 0.188851 | 0.025387 | 1.02E-13 | cis |
| rs1116808 | miR-584-5 | 5:1484841 | T | C | 0.408107 | 0.199694 | 0.02537 | 3.51E-15 | cis |
| rs6238010 | miR-584-5 | 5:1484921 | C | A | 0.846193 | -0.276613 | 0.035432 | 5.86E-15 | cis |
| rs2915806 | miR-584-5 | 5:1484509 | C | G | 0.5867 | -0.20673 | 0.025394 | 3.92E-16 | cis |
| rs4077368 | miR-584-5 | 5:1485425 | G | A | 0.446252 | 0.15926 | 0.025527 | 4.41E-10 | cis |
| rs4077085 | miR-584-5 | 5:1485440 | A | G | 0.521708 | 0.146774 | 0.025794 | 1.27E-08 | cis |
| rs1317416 | miR-584-5 | 5:1484883 | T | C | 0.474697 | 0.188995 | 0.025386 | 9.70E-14 | cis |
| rs4705305 | miR-584-5 | 5:1484708 | G | T | 0.473411 | 0.187444 | 0.025456 | 1.79E-13 | cis |
| rs4314386 | miR-584-5 | 5:1485448 | A | G | 0.520646 | 0.145871 | 0.025585 | 1.19E-08 | cis |
| rs1779537 | miR-584-5 | 5:1484633 | G | A | 0.845418 | -0.274724 | 0.035751 | 1.54E-14 | cis |
| rs1779525 | miR-584-5 | 5:1484169 | T | C | 0.842839 | -0.263167 | 0.034503 | 2.40E-14 | cis |
| rs1265507 | miR-584-5 | 5:1484855 | A | G | 0.410692 | 0.201351 | 0.025491 | 2.81E-15 | cis |
| rs1044723 | miR-584-5 | 5:1485009 | A | C | 0.474342 | 0.186354 | 0.02543 | 2.33E-13 | cis |
| rs1422577 | miR-584-5 | 5:1484863 | G | T | 0.474749 | 0.18891 | 0.025385 | 9.93E-14 | cis |
| rs6861404 | miR-584-5 | 5:1484158 | A | C | 0.536756 | -0.173683 | 0.02523 | 5.82E-12 | cis |
| rs6873011 | miR-584-5 | 5:1484141 | G | A | 0.842515 | -0.259329 | 0.034393 | 4.70E-14 | cis |
| rs1432792 | miR-584-5 | 5:1484149 | C | T | 0.537202 | -0.173133 | 0.025206 | 6.47E-12 | cis |
| rs36081 | miR-584-5 | 5:1484290 | A | G | 0.540101 | -0.173081 | 0.024956 | 4.05E-12 | cis |
| rs7715362 | miR-584-5 | 5:1485453 | T | C | 0.523497 | 0.147445 | 0.025759 | 1.04E-08 | cis |
| rs4516842 | miR-584-5 | 5:1485463 | T | G | 0.44539 | 0.159287 | 0.02544 | 3.82E-10 | cis |
| rs4512126 | miR-584-5 | 5:1485457 | T | A | 0.446725 | 0.159635 | 0.02537 | 3.13E-10 | cis |
| rs1422579 | miR-584-5 | 5:1484847 | C | A | 0.509261 | 0.191642 | 0.026152 | 2.34E-13 | cis |
| rs1112745 | miR-584-5 | 5:1484718 | G | A | 0.474305 | 0.187933 | 0.025488 | 1.66E-13 | cis |
| rs7846146 | miR-584-5 | 5:1484304 | C | T | 0.843889 | -0.26844 | 0.034455 | 6.64E-15 | cis |
| rs6580593 | miR-584-5 | 5:1484636 | G | C | 0.474061 | 0.188578 | 0.025475 | 1.34E-13 | cis |
| rs36047 | miR-584-5 | 5:1484413 | C | T | 0.650304 | -0.230184 | 0.025932 | 6.90E-19 | cis |
| rs6874343 | miR-584-5 | 5:1484478 | C | T | 0.844049 | -0.271414 | 0.034877 | 7.14E-15 | cis |
| rs36077 | miR-584-5 | 5:1484319 | G | A | 0.53902 | -0.172731 | 0.024829 | 3.48E-12 | cis |
| rs1046341 | miR-584-5 | 5:1484550 | A | G | 0.844169 | -0.272685 | 0.034992 | 6.56E-15 | cis |
| rs4546368 | miR-584-5 | 5:1485391 | C | T | 0.448453 | 0.157063 | 0.025275 | 5.16E-10 | cis |
| rs4076805 | miR-584-5 | 5:1485434 | G | A | 0.445639 | 0.158921 | 0.025424 | 4.08E-10 | cis |
| rs6238006 | miR-584-5 | 5:1484422 | C | T | 0.844027 | -0.270959 | 0.034834 | 7.34E-15 | cis |
| rs1174620 | miR-584-5 | 5:1484993 | A | G | 0.473634 | 0.184438 | 0.025138 | 2.18E-13 | cis |
| rs4147470 | miR-584-5 | 5:1485281 | C | T | 0.518813 | 0.14546 | 0.025465 | 1.12E-08 | cis |
| rs7442677 | miR-584-5 | 5:1484293 | C | T | 0.856689 | -0.278817 | 0.036018 | 9.87E-15 | cis |
| rs2915808 | miR-584-5 | 5:1484517 | A | G | 0.586438 | -0.205548 | 0.025332 | 4.90E-16 | cis |
| rs6866101 | miR-584-5 | 5:1484159 | T | C | 0.537249 | -0.173276 | 0.025199 | 6.14E-12 | cis |
| rs6895500 | miR-584-5 | 5:1485261 | T | C | 0.518723 | 0.146035 | 0.025499 | 1.02E-08 | cis |
| rs1003401 | miR-625-5 | 14:662377 | G | A | 0.386875 | 0.147199 | 0.019801 | 1.05E-13 | cis |
| rs1004787 | miR-625-5 | 14:657605 | C | A | 0.650175 | -0.213685 | 0.019842 | 4.80E-27 | cis |
| rs7733938 | miR-625-5 | 14:657580 | G | A | 0.84644 | -0.309982 | 0.02677 | 5.23E-31 | cis |
| rs8003811 | miR-625-5 | 14:662112 | C | T | 0.407369 | 0.120565 | 0.020188 | 2.34E-09 | cis |
| rs1162410 | miR-625-5 | 14:661961 | A | G | 0.487531 | 0.131664 | 0.019816 | 3.05E-11 | cis |
| rs7271246 | miR-625-5 | 14:657562 | A | T | 0.847244 | -0.322051 | 0.027281 | 3.68E-32 | cis |
| rs7271247 | miR-625-5 | 14:657771 | G | A | 0.846937 | -0.32457 | 0.026357 | 7.60E-35 | cis |
| rs7271248 | miR-625-5 | 14:657895 | A | T | 0.845779 | -0.326679 | 0.026156 | 8.48E-36 | cis |
| rs7271249 | miR-625-5 | 14:658023 | T | G | 0.845101 | -0.337884 | 0.026191 | 4.47E-38 | cis |

| | | | | | | | | |
|---|---|---|---|---|---|---|---|---|
| rs7271250( | miR-625-5p | 14:658043; | G | A | 0.810519 | -0.233969 | 0.024476 | 1.19E-21 cis |
| rs4073415 | miR-625-5p | 14:662595: | G | A | 0.538747 | -0.138325 | 0.019261 | 6.88E-13 cis |
| rs1262272 | miR-625-5p | 14:657417: | C | G | 0.647443 | -0.190823 | 0.020345 | 6.64E-21 cis |
| rs7271443! | miR-625-5p | 14:659169( | C | T | 0.81551 | -0.230852 | 0.024476 | 4.03E-21 cis |
| rs7271444( | miR-625-5p | 14:659185! | C | T | 0.81551 | -0.230854 | 0.024476 | 4.03E-21 cis |
| rs7271444; | miR-625-5p | 14:659190ʻ | T | C | 0.81551 | -0.230854 | 0.024476 | 4.03E-21 cis |
| rs7271444; | miR-625-5p | 14:659421ʻ | C | A | 0.816196 | -0.22863 | 0.024764 | 2.64E-20 cis |
| rs1121885 | miR-625-5p | 14:661569: | A | G | 0.34818 | 0.177649 | 0.020897 | 1.88E-17 cis |
| rs7271446: | miR-625-5p | 14:659681: | A | G | 0.816433 | -0.231357 | 0.024716 | 7.92E-21 cis |
| rs7271446! | miR-625-5p | 14:659782: | A | C | 0.819836 | -0.233344 | 0.024931 | 7.99E-21 cis |
| rs7271447! | miR-625-5p | 14:659979: | G | A | 0.815051 | -0.234308 | 0.024801 | 3.47E-21 cis |
| rs1392518( | miR-625-5p | 14:658295! | T | C | 0.813544 | -0.23719 | 0.02469 | 7.48E-22 cis |
| rs7271448: | miR-625-5p | 14:660126; | G | A | 0.820577 | -0.232772 | 0.025061 | 1.57E-20 cis |
| rs4080329 | miR-625-5p | 14:662334! | T | C | 0.372147 | 0.135947 | 0.020036 | 1.16E-11 cis |
| rs7271449! | miR-625-5p | 14:660424! | C | T | 0.816154 | -0.231865 | 0.024844 | 1.03E-20 cis |
| rs1162112: | miR-625-5p | 14:658224! | C | T | 0.388213 | 0.196561 | 0.019252 | 1.79E-24 cis |
| rs1013171! | miR-625-5p | 14:661573: | T | C | 0.385228 | 0.195601 | 0.020646 | 2.69E-21 cis |
| rs1014852! | miR-625-5p | 14:661047( | A | T | 0.345818 | 0.173355 | 0.020883 | 1.03E-16 cis |
| rs1014890; | miR-625-5p | 14:658333: | C | G | 0.739103 | -0.14955 | 0.021844 | 7.58E-12 cis |
| rs4131095 | miR-625-5p | 14:662594: | G | A | 0.604284 | -0.125188 | 0.019816 | 2.66E-10 cis |
| rs1015041; | miR-625-5p | 14:659167! | C | T | 0.358422 | 0.173561 | 0.020711 | 5.29E-17 cis |
| rs7158153 | miR-625-5p | 14:660345! | C | A | 0.345985 | 0.173233 | 0.02063 | 4.57E-17 cis |
| rs4143898 | miR-625-5p | 14:661888! | T | A | 0.450813 | 0.145864 | 0.01999 | 2.94E-13 cis |
| rs7141536 | miR-625-5p | 14:659873: | T | A | 0.349283 | 0.174077 | 0.020739 | 4.71E-17 cis |
| rs7141874 | miR-625-5p | 14:658219! | G | A | 0.071326 | 0.265544 | 0.038518 | 5.42E-12 cis |
| rs1013805; | miR-625-5p | 14:658334ʻ | T | C | 0.739224 | -0.149361 | 0.021863 | 8.39E-12 cis |
| rs1013857( | miR-625-5p | 14:658600: | A | G | 0.470451 | 0.132126 | 0.019756 | 2.26E-11 cis |
| rs1013867: | miR-625-5p | 14:658600! | A | G | 0.398463 | 0.171449 | 0.020209 | 2.18E-17 cis |
| rs1269068 | miR-625-5p | 14:657673: | C | T | 0.643163 | -0.223379 | 0.019494 | 2.13E-30 cis |
| rs8010876 | miR-625-5p | 14:662069: | A | G | 0.487956 | 0.131312 | 0.019735 | 2.86E-11 cis |
| rs1015200; | miR-625-5p | 14:660474: | A | T | 0.343755 | 0.172178 | 0.020581 | 5.97E-17 cis |
| rs883082 | miR-625-5p | 14:658809ʻ | A | G | 0.493786 | 0.147935 | 0.019805 | 8.05E-14 cis |
| rs5672046( | miR-625-5p | 14:662563: | C | T | 0.539663 | -0.139261 | 0.019323 | 5.72E-13 cis |
| rs8012278 | miR-625-5p | 14:660825: | A | G | 0.499774 | 0.145255 | 0.019787 | 2.12E-13 cis |
| rs7271640; | miR-625-5p | 14:660652: | G | T | 0.820366 | -0.233124 | 0.025147 | 1.85E-20 cis |
| rs7823084! | miR-625-5p | 14:660163! | G | A | 0.507675 | 0.145122 | 0.019792 | 2.26E-13 cis |
| rs7142219 | miR-625-5p | 14:662417! | G | A | 0.452285 | 0.14296 | 0.019477 | 2.13E-13 cis |
| rs7271642( | miR-625-5p | 14:661322! | C | T | 0.817792 | -0.234012 | 0.025168 | 1.43E-20 cis |
| rs7271642: | miR-625-5p | 14:661345! | T | C | 0.817679 | -0.234194 | 0.025168 | 1.34E-20 cis |
| rs7142651 | miR-625-5p | 14:661327; | C | G | 0.502009 | 0.147599 | 0.019835 | 9.98E-14 cis |
| rs1889731 | miR-625-5p | 14:659427: | G | C | 0.34602 | 0.173026 | 0.020834 | 9.98E-17 cis |
| rs1162757! | miR-625-5p | 14:662344ʻ | C | T | 0.452075 | 0.142993 | 0.019339 | 1.42E-13 cis |
| rs1288338; | miR-625-5p | 14:661453! | C | T | 0.500344 | 0.147926 | 0.019848 | 9.13E-14 cis |
| rs1162760! | miR-625-5p | 14:662343: | G | A | 0.452186 | 0.143034 | 0.019344 | 1.42E-13 cis |
| rs2792213 | miR-625-5p | 14:657713: | T | C | 0.947488 | -0.344288 | 0.044198 | 6.72E-15 cis |
| rs1014228: | miR-625-5p | 14:660441; | G | A | 0.346742 | 0.172676 | 0.02057 | 4.67E-17 cis |
| rs1288600! | miR-625-5p | 14:658092ʻ | C | T | 0.390429 | 0.198498 | 0.019263 | 6.72E-25 cis |
| rs1014320( | miR-625-5p | 14:661206: | C | T | 0.349542 | 0.175451 | 0.020878 | 4.33E-17 cis |
| rs1950557 | miR-625-5p | 14:662017! | C | T | 0.757804 | -0.130219 | 0.022662 | 9.13E-09 cis |
| rs1287887; | miR-625-5p | 14:658046( | G | A | 0.386023 | 0.203653 | 0.019372 | 7.56E-26 cis |
| rs1287920; | miR-625-5p | 14:659771! | C | T | 0.511623 | 0.146601 | 0.019927 | 1.88E-13 cis |
| rs1287980! | miR-625-5p | 14:660477! | T | C | 0.498218 | 0.145168 | 0.019735 | 1.90E-13 cis |
| rs1287997: | miR-625-5p | 14:659016( | G | A | 0.503137 | 0.145861 | 0.019778 | 1.65E-13 cis |

| | | | | | | | | | |
|---|---|---|---|---|---|---|---|---|---|
| rs1131317: | miR-625-5p | 14:6606412 | G | A | 0.501439 | 0.145533 | 0.019821 | 2.10E-13 | cis |
| rs1953416 | miR-625-5p | 14:6587880 | C | T | 0.504724 | 0.147883 | 0.01976 | 7.20E-14 | cis |
| rs1953418 | miR-625-5p | 14:6591522 | G | A | 0.355213 | 0.172927 | 0.020733 | 7.38E-17 | cis |
| rs1954052 | miR-625-5p | 14:6580570 | T | A | 0.387387 | 0.206368 | 0.019447 | 2.63E-26 | cis |
| rs1956010 | miR-625-5p | 14:6615109 | G | A | 0.382297 | 0.18771 | 0.020495 | 5.24E-20 | cis |
| rs7142906 | miR-625-5p | 14:6595645 | A | T | 0.360633 | 0.183028 | 0.020527 | 4.82E-19 | cis |
| rs1288072! | miR-625-5p | 14:6579980 | G | A | 0.386052 | 0.21297 | 0.019217 | 1.53E-28 | cis |
| rs7143109 | miR-625-5p | 14:6580102 | G | A | 0.269639 | 0.305978 | 0.021066 | 8.46E-48 | cis |
| rs7143318 | miR-625-5p | 14:6594068 | G | A | 0.35859 | 0.169987 | 0.020679 | 2.03E-16 | cis |
| rs1958559 | miR-625-5p | 14:6605205 | C | T | 0.40803 | 0.166544 | 0.019995 | 8.14E-17 | cis |
| rs1958560 | miR-625-5p | 14:6603679 | G | A | 0.400142 | 0.165944 | 0.019808 | 5.40E-17 | cis |
| rs1958561 | miR-625-5p | 14:6603676 | T | G | 0.498978 | 0.146336 | 0.019735 | 1.22E-13 | cis |
| rs1959144 | miR-625-5p | 14:6594605 | T | G | 0.346477 | 0.172772 | 0.020841 | 1.13E-16 | cis |
| rs883081 | miR-625-5p | 14:6588062 | C | A | 0.497085 | 0.149391 | 0.019841 | 5.10E-14 | cis |
| rs1184844 | miR-625-5p | 14:6625746 | A | G | 0.5414 | -0.139233 | 0.019364 | 6.47E-13 | cis |
| rs946917 | miR-625-5p | 14:6581718 | A | G | 0.068596 | 0.260031 | 0.039377 | 4.01E-11 | cis |
| rs1984855 | miR-625-5p | 14:6623925 | G | C | 0.387843 | 0.147118 | 0.019829 | 1.18E-13 | cis |
| rs2184601 | miR-625-5p | 14:6589736 | T | A | 0.354646 | 0.174532 | 0.020745 | 3.99E-17 | cis |
| rs1014497! | miR-625-5p | 14:6599763 | G | A | 0.355198 | 0.170483 | 0.02058 | 1.19E-16 | cis |
| rs1014514( | miR-625-5p | 14:6607500 | C | T | 0.349632 | 0.180781 | 0.020698 | 2.45E-18 | cis |
| rs899961 | miR-625-5p | 14:6580032 | A | C | 0.290881 | 0.291121 | 0.020561 | 1.65E-45 | cis |
| rs7143666 | miR-625-5p | 14:6580997 | A | C | 0.348575 | 0.20683 | 0.019826 | 1.77E-25 | cis |
| rs7918480! | miR-625-5p | 14:6590831 | A | C | 0.813031 | -0.230215 | 0.024633 | 9.11E-21 | cis |
| rs2845901 | miR-625-5p | 14:6625852 | T | C | 0.538033 | -0.138619 | 0.019252 | 6.02E-13 | cis |
| rs1289403 | miR-625-5p | 14:6623876 | T | C | 0.453073 | 0.14422 | 0.019372 | 9.72E-14 | cis |
| rs7144530 | miR-625-5p | 14:6584056 | A | G | 0.053251 | 0.327275 | 0.045719 | 8.16E-13 | cis |
| rs7144971 | miR-625-5p | 14:6612065 | G | A | 0.349544 | 0.175451 | 0.020879 | 4.34E-17 | cis |
| rs1289533 | miR-625-5p | 14:6580744 | C | T | 0.348231 | 0.206644 | 0.019831 | 2.01E-25 | cis |
| rs1289540 | miR-625-5p | 14:6606482 | G | A | 0.409413 | 0.168122 | 0.020142 | 7.00E-17 | cis |
| rs1075566 | miR-625-5p | 14:6623945 | C | T | 0.454013 | 0.144041 | 0.019394 | 1.11E-13 | cis |
| rs4400971 | miR-625-5p | 14:6625457 | A | C | 0.539825 | -0.138868 | 0.019316 | 6.51E-13 | cis |
| rs9635249 | miR-625-5p | 14:6579778 | A | G | 0.22061 | 0.364734 | 0.022232 | 1.73E-60 | cis |
| rs1115859 | miR-625-5p | 14:6592373 | A | G | 0.389613 | 0.1839 | 0.020429 | 2.22E-19 | cis |
| rs1115860 | miR-625-5p | 14:6602536 | A | G | 0.499831 | 0.146827 | 0.019777 | 1.14E-13 | cis |
| rs927004 | miR-625-5p | 14:6620091 | G | C | 0.488311 | 0.130808 | 0.019782 | 3.78E-11 | cis |
| rs1115860 | miR-625-5p | 14:6607204 | C | T | 0.349651 | 0.180755 | 0.020698 | 2.48E-18 | cis |
| rs1115860 | miR-625-5p | 14:6609445 | G | A | 0.346475 | 0.17345 | 0.020801 | 7.51E-17 | cis |
| rs1288175! | miR-625-5p | 14:6580181 | A | G | 0.320867 | 0.23032 | 0.020035 | 1.38E-30 | cis |
| rs6027309 | miR-625-5p | 14:6621485 | C | T | 0.757774 | -0.130259 | 0.022657 | 8.96E-09 | cis |
| rs1205018 | miR-625-5p | 14:6624308 | C | T | 0.455215 | 0.144476 | 0.019432 | 1.05E-13 | cis |
| rs2861669 | miR-625-5p | 14:6625868 | C | T | 0.538909 | -0.138355 | 0.019264 | 6.87E-13 | cis |
| rs2866601( | miR-625-5p | 14:6575814 | C | T | 0.775896 | -0.195083 | 0.022869 | 1.46E-17 | cis |
| rs2064694 | miR-625-5p | 14:6614824 | G | T | 0.501114 | 0.147758 | 0.019816 | 8.87E-14 | cis |
| rs2874089! | miR-625-5p | 14:6578352 | G | A | 0.672319 | -0.153314 | 0.02031 | 4.39E-14 | cis |
| rs5584550 | miR-625-5p | 14:6625696 | C | T | 0.624387 | -0.130736 | 0.020069 | 7.30E-11 | cis |
| rs8003464 | miR-625-5p | 14:6590823 | A | G | 0.35496 | 0.173425 | 0.020754 | 6.48E-17 | cis |
| rs8018278 | miR-625-5p | 14:6618008 | A | G | 0.498198 | 0.145521 | 0.019847 | 2.26E-13 | cis |
| rs1184846 | miR-625-5p | 14:6625729 | G | A | 0.541416 | -0.139268 | 0.01937 | 6.48E-13 | cis |
| rs4899174 | miR-625-5p | 14:6581140 | A | T | 0.390371 | 0.198564 | 0.019268 | 6.68E-25 | cis |
| rs2898814 | miR-625-5p | 14:6602396 | T | C | 0.347238 | 0.173957 | 0.020661 | 3.77E-17 | cis |
| rs2898818 | miR-625-5p | 14:6597258 | G | C | 0.35329 | 0.170444 | 0.020622 | 1.40E-16 | cis |
| rs2898820 | miR-625-5p | 14:6587935 | T | G | 0.35021 | 0.175123 | 0.020725 | 2.92E-17 | cis |
| rs8005710 | miR-625-5p | 14:6606566 | G | A | 0.502323 | 0.147754 | 0.019868 | 1.03E-13 | cis |

| | | | | | | | | | |
|---|---|---|---|---|---|---|---|---|---|
| rs8023016 | miR-625-5p | 14:6584680 | T | A | 0.053493 | 0.326462 | 0.045712 | 9.22E-13 | cis |
| rs1288191 | miR-625-5p | 14:6583122 | T | C | 0.388492 | 0.19722 | 0.019447 | 3.62E-24 | cis |
| rs1288220 | miR-625-5p | 14:6615786 | C | A | 0.503123 | 0.148195 | 0.019873 | 8.84E-14 | cis |
| rs1184468 | miR-625-5p | 14:6591084 | G | C | 0.356705 | 0.17288 | 0.02073 | 7.44E-17 | cis |
| rs7145759 | miR-625-5p | 14:6588453 | C | A | 0.353283 | 0.175197 | 0.0207 | 2.60E-17 | cis |
| rs1184583 | miR-625-5p | 14:6626060 | C | A | 0.602996 | -0.124937 | 0.01984 | 3.03E-10 | cis |
| rs7146742 | miR-625-5p | 14:6603293 | G | A | 0.435936 | 0.139288 | 0.020069 | 3.91E-12 | cis |
| rs8007497 | miR-625-5p | 14:6604447 | T | C | 0.338774 | 0.16985 | 0.02037 | 7.53E-17 | cis |
| rs4587890 | miR-625-5p | 14:6586378 | T | A | 0.391319 | 0.17464 | 0.020418 | 1.20E-17 | cis |
| rs7147536 | miR-625-5p | 14:6587469 | C | T | 0.367002 | 0.185223 | 0.020461 | 1.39E-19 | cis |
| rs7147002 | miR-625-5p | 14:6603254 | T | C | 0.342789 | 0.172498 | 0.020658 | 6.82E-17 | cis |
| rs1162201 | miR-625-5p | 14:6604164 | C | T | 0.502262 | 0.14792 | 0.019779 | 7.51E-14 | cis |
| rs1288821 | miR-625-5p | 14:6620045 | T | C | 0.48838 | 0.130873 | 0.019792 | 3.78E-11 | cis |
| rs7157006 | miR-625-5p | 14:6582820 | C | T | 0.387351 | 0.198515 | 0.019331 | 9.71E-25 | cis |
| rs1014900 | miR-625-5p | 14:6583349 | C | T | 0.12871 | 0.175846 | 0.02978 | 3.53E-09 | cis |
| rs7157449 | miR-625-5p | 14:6624013 | T | G | 0.454044 | 0.144106 | 0.019397 | 1.09E-13 | cis |
| rs1289097 | miR-625-5p | 14:6598289 | C | T | 0.504299 | 0.148214 | 0.019979 | 1.18E-13 | cis |
| rs7158347 | miR-625-5p | 14:6582895 | C | T | 0.388449 | 0.198782 | 0.019394 | 1.19E-24 | cis |
| rs7158556 | miR-625-5p | 14:6624072 | T | A | 0.373139 | 0.137414 | 0.020058 | 7.35E-12 | cis |
| rs1162227 | miR-625-5p | 14:6614900 | A | T | 0.504871 | 0.146606 | 0.019892 | 1.71E-13 | cis |
| rs1162282 | miR-625-5p | 14:6619178 | T | A | 0.496725 | 0.144626 | 0.019819 | 2.93E-13 | cis |
| rs5637923 | miR-625-5p | 14:6625539 | A | C | 0.623894 | -0.1306 | 0.020071 | 7.68E-11 | cis |
| rs8010726 | miR-625-5p | 14:6593259 | G | A | 0.358211 | 0.172184 | 0.020663 | 7.89E-17 | cis |
| rs1184844 | miR-625-5p | 14:6625741 | C | T | 0.54144 | -0.139242 | 0.019367 | 6.49E-13 | cis |
| rs2268962 | miR-625-5p | 14:6614727 | A | G | 0.501276 | 0.147836 | 0.01982 | 8.72E-14 | cis |
| rs1256525 | miR-625-5p | 14:6573855 | G | C | 0.646861 | -0.190909 | 0.020383 | 7.52E-21 | cis |
| rs4899175 | miR-625-5p | 14:6582643 | A | G | 0.051502 | 0.327813 | 0.045295 | 4.58E-13 | cis |
| rs8012834 | miR-625-5p | 14:6596655 | C | T | 0.362666 | 0.181923 | 0.020554 | 8.68E-19 | cis |
| rs6129890 | miR-625-5p | 14:6621859 | C | T | 0.7576 | -0.130485 | 0.022666 | 8.57E-09 | cis |
| rs1162588 | miR-625-5p | 14:6624519 | G | A | 0.454755 | 0.141922 | 0.019347 | 2.21E-13 | cis |
| rs1162706 | miR-625-5p | 14:6618295 | T | G | 0.488866 | 0.13317 | 0.01984 | 1.92E-11 | cis |
| rs1162708 | miR-625-5p | 14:6597883 | T | G | 0.501066 | 0.146483 | 0.019949 | 2.09E-13 | cis |
| rs1162718 | miR-625-5p | 14:6612144 | C | T | 0.501637 | 0.146864 | 0.019888 | 1.53E-13 | cis |
| rs5692256 | miR-625-5p | 14:6625647 | A | T | 0.541356 | -0.139228 | 0.019369 | 6.57E-13 | cis |
| rs1162718 | miR-625-5p | 14:6612149 | A | G | 0.501636 | 0.146867 | 0.019888 | 1.53E-13 | cis |
| rs2411351 | miR-625-5p | 14:6617154 | T | C | 0.498384 | 0.147075 | 0.019765 | 9.99E-14 | cis |
| rs1185012 | miR-625-5p | 14:6626037 | T | C | 0.602946 | -0.125112 | 0.019839 | 2.86E-10 | cis |
| rs2411356 | miR-625-5p | 14:6605316 | G | A | 0.388335 | 0.168635 | 0.020188 | 6.65E-17 | cis |
| rs1288381 | miR-625-5p | 14:6592336 | C | T | 0.510962 | 0.147038 | 0.019928 | 1.60E-13 | cis |
| rs1162876 | miR-625-5p | 14:6616844 | T | C | 0.500166 | 0.147858 | 0.019781 | 7.73E-14 | cis |
| rs1288584 | miR-625-5p | 14:6591865 | C | T | 0.373139 | 0.182228 | 0.020451 | 5.08E-19 | cis |
| rs2411815 | miR-625-5p | 14:6602007 | A | T | 0.354996 | 0.169079 | 0.020582 | 2.12E-16 | cis |
| rs7435176 | miR-625-5p | 14:6580846 | C | T | 0.810279 | -0.2335 | 0.024456 | 1.33E-21 | cis |
| rs1288616 | miR-625-5p | 14:6580928 | C | A | 0.393656 | 0.204136 | 0.019134 | 1.43E-26 | cis |
| rs1288633 | miR-625-5p | 14:6609467 | T | G | 0.500184 | 0.146151 | 0.01985 | 1.80E-13 | cis |
| rs2411915 | miR-625-5p | 14:6579469 | C | T | 0.053213 | 0.337729 | 0.044685 | 4.09E-14 | cis |
| rs1288713 | miR-625-5p | 14:6604554 | G | C | 0.502159 | 0.147999 | 0.019774 | 7.18E-14 | cis |
| rs1288767 | miR-625-5p | 14:6622177 | T | C | 0.487094 | 0.131927 | 0.019729 | 2.28E-11 | cis |
| rs1288795 | miR-625-5p | 14:6624659 | G | A | 0.454723 | 0.142379 | 0.019362 | 1.93E-13 | cis |
| rs6198799 | miR-625-5p | 14:6607073 | C | G | 0.351584 | 0.180835 | 0.020721 | 2.61E-18 | cis |
| rs1288900 | miR-625-5p | 14:6606358 | C | G | 0.499455 | 0.145384 | 0.019766 | 1.91E-13 | cis |
| rs1289012 | miR-625-5p | 14:6599800 | C | T | 0.511668 | 0.146815 | 0.019882 | 1.53E-13 | cis |
| rs1289090 | miR-625-5p | 14:6611662 | T | A | 0.498435 | 0.144652 | 0.019829 | 2.98E-13 | cis |

| | | | | | | | | | |
|---|---|---|---|---|---|---|---|---|---|
| rs1289205 | miR-625-5p | 14:659034 | T | C | 0.51011 | 0.146773 | 0.019879 | 1.54E-13 | cis |
| rs1048378 | miR-625-5p | 14:659801 | T | C | 0.501165 | 0.146505 | 0.019946 | 2.05E-13 | cis |
| rs1048378 | miR-625-5p | 14:662195 | G | A | 0.486262 | 0.131301 | 0.01973 | 2.84E-11 | cis |
| rs7151846 | miR-625-5p | 14:662557 | C | G | 0.539502 | -0.138708 | 0.019301 | 6.65E-13 | cis |
| rs8016322 | miR-625-5p | 14:660515 | G | A | 0.386004 | 0.167316 | 0.020156 | 1.03E-16 | cis |
| rs8017880 | miR-625-5p | 14:659429 | G | C | 0.346067 | 0.172935 | 0.020833 | 1.03E-16 | cis |
| rs1162366 | miR-625-5p | 14:660211 | T | G | 0.390012 | 0.168768 | 0.020334 | 1.04E-16 | cis |
| rs1162392 | miR-625-5p | 14:658193 | A | C | 0.389577 | 0.198029 | 0.019291 | 1.01E-24 | cis |
| rs1162404 | miR-625-5p | 14:661080 | C | T | 0.498929 | 0.145121 | 0.019847 | 2.63E-13 | cis |
| rs8019762 | miR-625-5p | 14:660871 | A | G | 0.346646 | 0.172489 | 0.020796 | 1.09E-16 | cis |
| rs8020305 | miR-625-5p | 14:658375 | C | T | 0.066561 | 0.301123 | 0.040379 | 8.82E-14 | cis |
| rs3464038 | miR-625-5p | 14:660530 | G | A | 0.819714 | -0.233017 | 0.025045 | 1.35E-20 | cis |
| rs7482562 | miR-625-5p | 14:658077 | G | A | 0.810459 | -0.23324 | 0.024454 | 1.46E-21 | cis |
| rs1054218 | miR-625-5p | 14:662091 | C | G | 0.408277 | 0.120859 | 0.020155 | 2.02E-09 | cis |
| rs7489777 | miR-625-5p | 14:659090 | T | G | 0.81551 | -0.230899 | 0.024476 | 3.97E-21 | cis |
| rs1258705 | miR-625-5p | 14:660663 | C | T | 0.345738 | 0.172451 | 0.020704 | 8.14E-17 | cis |
| rs6649234 | miR-625-5p | 14:658394 | A | C | 0.739 | -0.149853 | 0.021839 | 6.80E-12 | cis |
| rs1289446 | miR-625-5p | 14:662407 | G | A | 0.454589 | 0.14417 | 0.0194 | 1.07E-13 | cis |
| rs1289490 | miR-625-5p | 14:661617 | C | T | 0.501572 | 0.148481 | 0.019827 | 6.95E-14 | cis |
| rs1289507 | miR-625-5p | 14:659767 | G | T | 0.520727 | 0.14878 | 0.020225 | 1.89E-13 | cis |
| rs1115859 | miR-625-5p | 14:658599 | G | T | 0.470037 | 0.131894 | 0.019761 | 2.48E-11 | cis |
| rs1115859 | miR-625-5p | 14:658599 | G | A | 0.466718 | 0.134472 | 0.019789 | 1.08E-11 | cis |
| rs1289666 | miR-625-5p | 14:661049 | G | A | 0.499 | 0.145192 | 0.01985 | 2.59E-13 | cis |
| rs1115859 | miR-625-5p | 14:659119 | T | A | 0.507581 | 0.145384 | 0.01988 | 2.61E-13 | cis |
| rs1243359 | miR-625-5p | 14:660288 | T | C | 0.343187 | 0.172769 | 0.020668 | 6.31E-17 | cis |
| rs2300864 | miR-625-5p | 14:661198 | A | G | 0.349692 | 0.175475 | 0.020879 | 4.30E-17 | cis |
| rs3477200 | miR-625-5p | 14:660530 | G | A | 0.819572 | -0.233049 | 0.02505 | 1.36E-20 | cis |
| rs7140341 | miR-625-5p | 14:660704 | C | G | 0.3503 | 0.181513 | 0.020699 | 1.80E-18 | cis |
| rs7682482 | miR-625-5p | 14:657582 | G | A | 0.778334 | -0.195159 | 0.022955 | 1.87E-17 | cis |
| rs6198799 | miR-625-5p | 14:660707 | C | T | 0.351584 | 0.180835 | 0.020721 | 2.61E-18 | cis |
| rs7156078 | miR-625-5p | 14:662387 | G | A | 0.386926 | 0.147416 | 0.019811 | 9.98E-14 | cis |
| rs6198897 | miR-625-5p | 14:662580 | C | A | 0.537113 | -0.137354 | 0.019389 | 1.40E-12 | cis |
| rs6198897 | miR-625-5p | 14:662581 | T | C | 0.550188 | -0.12846 | 0.019697 | 6.95E-11 | cis |
| rs1535173 | miR-625-5p | 14:661991 | C | G | 0.487949 | 0.131475 | 0.019789 | 3.05E-11 | cis |
| rs9323461 | miR-625-5p | 14:660314 | T | C | 0.342775 | 0.172498 | 0.020658 | 6.81E-17 | cis |
| rs2268955 | miR-625-5p | 14:660900 | T | A | 0.411712 | 0.169021 | 0.020476 | 1.52E-16 | cis |
| rs2268956 | miR-625-5p | 14:660901 | G | A | 0.349005 | 0.172896 | 0.020779 | 8.74E-17 | cis |
| rs2268959 | miR-625-5p | 14:661453 | C | A | 0.818382 | -0.234409 | 0.02521 | 1.43E-20 | cis |
| rs3520647 | miR-625-5p | 14:662232 | A | G | 0.757483 | -0.131248 | 0.022709 | 7.49E-09 | cis |
| rs1775350 | miR-625-5p | 14:660574 | A | G | 0.821177 | -0.232942 | 0.025154 | 2.03E-20 | cis |
| rs1256523 | miR-625-5p | 14:657381 | A | G | 0.647737 | -0.190991 | 0.020354 | 6.39E-21 | cis |
| rs1256524 | miR-625-5p | 14:657384 | T | C | 0.647718 | -0.190917 | 0.020352 | 6.55E-21 | cis |
| rs8011224 | miR-625-5p | 14:659696 | T | A | 0.357678 | 0.169952 | 0.020594 | 1.55E-16 | cis |
| rs1087319 | miR-625-5p | 14:661106 | T | C | 0.346549 | 0.173321 | 0.020847 | 9.27E-17 | cis |
| rs8022053 | miR-625-5p | 14:658679 | T | G | 0.350181 | 0.176409 | 0.020719 | 1.68E-17 | cis |
| rs8022080 | miR-625-5p | 14:659990 | T | C | 0.358857 | 0.182558 | 0.020463 | 4.61E-19 | cis |
| rs6573611 | miR-625-5p | 14:658652 | G | T | 0.39802 | 0.170507 | 0.020229 | 3.49E-17 | cis |
| rs7271442 | miR-625-5p | 14:658929 | A | C | 0.815998 | -0.232651 | 0.024632 | 3.55E-21 | cis |
| rs4073416 | miR-625-5p | 14:662593 | T | C | 0.585483 | -0.135986 | 0.019383 | 2.29E-12 | cis |
| rs2002692 | miR-625-5p | 14:658994 | A | G | 0.502596 | 0.145491 | 0.019792 | 1.97E-13 | cis |
| rs6573618 | miR-625-5p | 14:660564 | A | G | 0.344637 | 0.172132 | 0.020646 | 7.60E-17 | cis |
| rs6573619 | miR-625-5p | 14:660615 | T | C | 0.344872 | 0.172013 | 0.020629 | 7.53E-17 | cis |
| rs1162759 | miR-625-5p | 14:657977 | G | T | 0.38607 | 0.212976 | 0.019217 | 1.52E-28 | cis |

| | | | | | | | | | |
|---|---|---|---|---|---|---|---|---|---|
| rs9783622 | miR-625-5p | 14:6574904 | C | G | 0.724193 | -0.219602 | 0.021928 | 1.31E-23 | cis |
| rs7271446( | miR-625-5p | 14:6596208 | A | G | 0.819561 | -0.233271 | 0.024859 | 6.37E-21 | cis |
| rs6573627 | miR-625-5p | 14:6625232 | C | T | 0.542293 | -0.140757 | 0.019318 | 3.19E-13 | cis |
| rs1214799 | miR-625-5p | 14:6575162 | C | A | 0.729928 | -0.222383 | 0.022038 | 6.07E-24 | cis |
| rs7271448 | miR-625-5p | 14:6600660 | T | G | 0.815267 | -0.233974 | 0.024821 | 4.24E-21 | cis |
| rs2411822 | miR-625-5p | 14:6587839 | A | G | 0.501454 | 0.146326 | 0.019719 | 1.16E-13 | cis |
| rs2300865 | miR-625-5p | 14:6612001 | G | C | 0.502657 | 0.147148 | 0.019872 | 1.31E-13 | cis |
| rs2411823 | miR-625-5p | 14:6586506 | C | T | 0.39822 | 0.170762 | 0.020236 | 3.22E-17 | cis |
| rs7660191( | miR-625-5p | 14:6608734 | G | C | 0.820834 | -0.232309 | 0.025252 | 3.59E-20 | cis |
| rs1185157( | miR-625-5p | 14:6590028 | T | C | 0.454741 | 0.146928 | 0.01999 | 1.98E-13 | cis |
| rs4902388 | miR-625-5p | 14:6579285 | A | G | 0.216378 | 0.360268 | 0.022447 | 5.75E-58 | cis |
| rs7147636 | miR-625-5p | 14:6601118 | T | C | 0.36432 | 0.172044 | 0.020591 | 6.53E-17 | cis |
| rs7157109 | miR-625-5p | 14:6590043 | C | G | 0.354575 | 0.174565 | 0.020746 | 3.94E-17 | cis |
| rs4902387 | miR-625-5p | 14:6579283 | C | T | 0.21638 | 0.360265 | 0.022447 | 5.75E-58 | cis |
| rs4902393 | miR-625-5p | 14:6583951 | G | C | 0.390223 | 0.1982 | 0.019437 | 2.04E-24 | cis |
| rs4902399 | miR-625-5p | 14:6595689 | A | G | 0.345315 | 0.174461 | 0.020811 | 5.16E-17 | cis |
| rs3783709 | miR-625-5p | 14:6605860 | T | A | 0.499275 | 0.145547 | 0.01973 | 1.62E-13 | cis |
| rs3783711 | miR-625-5p | 14:6602844 | A | G | 0.343187 | 0.17277 | 0.020668 | 6.31E-17 | cis |
| rs2127868 | miR-625-5p | 14:6579432 | G | A | 0.220849 | 0.364388 | 0.022206 | 1.63E-60 | cis |
| rs7160780 | miR-625-5p | 14:6605271 | G | T | 0.386614 | 0.16716 | 0.020127 | 9.96E-17 | cis |
| rs7161123 | miR-625-5p | 14:6605290 | G | A | 0.498649 | 0.145335 | 0.01972 | 1.71E-13 | cis |
| rs4902416 | miR-625-5p | 14:6623809 | G | C | 0.453194 | 0.143892 | 0.019371 | 1.10E-13 | cis |
| rs6048801 | miR-625-5p | 14:6582585 | C | A | 0.738599 | -0.148941 | 0.021849 | 9.30E-12 | cis |
| rs1243646 | miR-625-5p | 14:6580687 | T | C | 0.255814 | 0.28174 | 0.021604 | 7.15E-39 | cis |
| rs1959145 | miR-625-5p | 14:6595474 | A | G | 0.347293 | 0.172809 | 0.020786 | 9.28E-17 | cis |
| rs4899173 | miR-625-5p | 14:6579316 | T | C | 0.216374 | 0.360282 | 0.022447 | 5.72E-58 | cis |
| rs4899178 | miR-625-5p | 14:6586298 | C | T | 0.391583 | 0.174834 | 0.020417 | 1.10E-17 | cis |
| rs4899179 | miR-625-5p | 14:6592674 | T | G | 0.508099 | 0.145114 | 0.019871 | 2.81E-13 | cis |
| rs7142165 | miR-625-5p | 14:6625023 | G | A | 0.550826 | -0.141461 | 0.019388 | 2.95E-13 | cis |
| rs7271641 | miR-625-5p | 14:6612104 | G | A | 0.819456 | -0.233541 | 0.025291 | 2.60E-20 | cis |
| rs2149839 | miR-625-5p | 14:6598344 | G | A | 0.353418 | 0.170444 | 0.020619 | 1.38E-16 | cis |
| rs2149841 | miR-625-5p | 14:6601031 | C | T | 0.503888 | 0.148409 | 0.019912 | 9.11E-14 | cis |
| rs7271643( | miR-625-5p | 14:6619291 | G | T | 0.829643 | -0.216826 | 0.026052 | 8.58E-17 | cis |
| rs6573615 | miR-625-5p | 14:6604653 | G | A | 0.381045 | 0.166016 | 0.019917 | 7.73E-17 | cis |
| rs7271443 | miR-625-5p | 14:6590747 | A | G | 0.815498 | -0.230917 | 0.024477 | 3.95E-21 | cis |
| rs1115858 | miR-625-5p | 14:6579987 | G | A | 0.221744 | 0.35927 | 0.022176 | 5.00E-59 | cis |
| rs3742597 | miR-625-5p | 14:6620017 | A | G | 0.757816 | -0.130189 | 0.022661 | 9.19E-09 | cis |
| rs2411404 | miR-625-5p | 14:6623940 | C | G | 0.454146 | 0.144127 | 0.019397 | 1.08E-13 | cis |
| rs5830703 | miR-625-5p | 14:6582419 | A | T | 0.051498 | 0.327775 | 0.045296 | 4.61E-13 | cis |
| rs1162884( | miR-625-5p | 14:6623564 | A | G | 0.451742 | 0.143194 | 0.01935 | 1.36E-13 | cis |
| rs1115859 | miR-625-5p | 14:6590636 | A | G | 0.34826 | 0.175746 | 0.020926 | 4.52E-17 | cis |
| rs2411820 | miR-625-5p | 14:6594674 | C | T | 0.35728 | 0.171685 | 0.020672 | 9.97E-17 | cis |
| rs7146829 | miR-625-5p | 14:6619453 | T | C | 0.488699 | 0.132266 | 0.019792 | 2.34E-11 | cis |
| rs9972106 | miR-625-5p | 14:6602313 | A | T | 0.388305 | 0.167818 | 0.020231 | 1.08E-16 | cis |
| rs1115860 | miR-625-5p | 14:6607173 | T | G | 0.491419 | 0.14246 | 0.01987 | 7.53E-13 | cis |
| rs7146993 | miR-625-5p | 14:6604887 | C | T | 0.343625 | 0.171579 | 0.020581 | 7.63E-17 | cis |
| rs1258969 | miR-625-5p | 14:6592043 | G | A | 0.510971 | 0.147065 | 0.019929 | 1.59E-13 | cis |
| rs1115860 | miR-625-5p | 14:6610792 | T | A | 0.365947 | 0.173317 | 0.020738 | 6.42E-17 | cis |
| rs4902389 | miR-625-5p | 14:6579322 | A | G | 0.216372 | 0.360287 | 0.022448 | 5.70E-58 | cis |
| rs4902391 | miR-625-5p | 14:6581597 | G | T | 0.397869 | 0.205832 | 0.019183 | 7.36E-27 | cis |
| rs1256526 | miR-625-5p | 14:6573990 | T | G | 0.647195 | -0.190548 | 0.020338 | 7.31E-21 | cis |
| rs2092914 | miR-625-5p | 14:6607911 | C | T | 0.36669 | 0.173049 | 0.020649 | 5.27E-17 | cis |
| rs7140695 | miR-625-5p | 14:6592913 | T | C | 0.350881 | 0.17535 | 0.020807 | 3.53E-17 | cis |

| | | | | | | | | | |
|---|---|---|---|---|---|---|---|---|---|
| rs7150448 | miR-625-5p | 14:658715 | C | T | 0.345581 | 0.17931 | 0.020872 | 8.63E-18 | cis |
| rs1205023 | miR-625-5p | 14:662350 | T | C | 0.451755 | 0.143138 | 0.019345 | 1.37E-13 | cis |
| rs4902412 | miR-625-5p | 14:661870 | T | C | 0.443 | 0.135201 | 0.019984 | 1.33E-11 | cis |
| rs4902415 | miR-625-5p | 14:662356 | T | C | 0.451839 | 0.143041 | 0.019354 | 1.46E-13 | cis |
| rs7152023 | miR-625-5p | 14:661608 | C | A | 0.425722 | 0.183153 | 0.02014 | 9.56E-20 | cis |
| rs6137847 | miR-625-5p | 14:662534 | G | A | 0.541244 | -0.140207 | 0.019376 | 4.62E-13 | cis |
| rs1256522 | miR-625-5p | 14:657380 | C | T | 0.647567 | -0.191086 | 0.020355 | 6.13E-21 | cis |
| rs7608716 | miR-625-5p | 14:660670 | T | C | 0.820364 | -0.233129 | 0.025147 | 1.85E-20 | cis |
| rs6573624 | miR-625-5p | 14:662268 | G | A | 0.485559 | 0.132056 | 0.019672 | 1.91E-11 | cis |
| rs761831 | miR-625-5p | 14:662393 | T | C | 0.454673 | 0.144038 | 0.019399 | 1.13E-13 | cis |
| rs1113962 | miR-625-5p | 14:660057 | C | T | 0.355246 | 0.170317 | 0.02059 | 1.32E-16 | cis |
| rs3825639 | miR-625-5p | 14:660285 | T | C | 0.343187 | 0.17277 | 0.020668 | 6.31E-17 | cis |
| rs3825640 | miR-625-5p | 14:659612 | C | G | 0.503966 | 0.147669 | 0.019981 | 1.46E-13 | cis |
| rs1258684 | miR-625-5p | 14:661450 | T | G | 0.500228 | 0.147394 | 0.019836 | 1.08E-13 | cis |
| rs7144345 | miR-625-5p | 14:661118 | G | C | 0.345782 | 0.173329 | 0.020876 | 1.02E-16 | cis |
| rs1258883 | miR-625-5p | 14:661626 | G | T | 0.500385 | 0.147861 | 0.019792 | 7.98E-14 | cis |
| rs747541 | miR-625-5p | 14:658054 | C | A | 0.395726 | 0.206479 | 0.019239 | 7.18E-27 | cis |
| rs7155541 | miR-625-5p | 14:660462 | G | C | 0.501962 | 0.146856 | 0.019792 | 1.17E-13 | cis |
| rs1115859 | miR-625-5p | 14:658211 | G | T | 0.068524 | 0.258694 | 0.039318 | 4.72E-11 | cis |
| rs1115861 | miR-625-5p | 14:662350 | G | A | 0.451752 | 0.143146 | 0.019346 | 1.37E-13 | cis |
| rs2236067 | miR-625-5p | 14:662480 | A | G | 0.386784 | 0.145441 | 0.019837 | 2.27E-13 | cis |
| rs7493004 | miR-625-5p | 14:661241 | A | C | 0.50236 | 0.147113 | 0.01987 | 1.32E-13 | cis |
| rs6573626 | miR-625-5p | 14:662406 | G | C | 0.454572 | 0.145113 | 0.019413 | 7.72E-14 | cis |
| rs1710260 | miR-625-5p | 14:657895 | A | C | 0.845741 | -0.326684 | 0.026149 | 8.16E-36 | cis |
| rs1162943 | miR-625-5p | 14:658921 | C | G | 0.352083 | 0.17854 | 0.021058 | 2.28E-17 | cis |
| rs4581615 | miR-625-5p | 14:660559 | C | A | 0.499128 | 0.145272 | 0.019735 | 1.82E-13 | cis |
| rs7271448 | miR-625-5p | 14:660162 | A | G | 0.820587 | -0.232841 | 0.025077 | 1.62E-20 | cis |
| rs6573614 | miR-625-5p | 14:659587 | G | A | 0.356282 | 0.171846 | 0.02064 | 8.37E-17 | cis |
| rs2127870 | miR-625-5p | 14:657968 | G | C | 0.220153 | 0.363782 | 0.02224 | 3.86E-60 | cis |
| rs867972 | miR-625-5p | 14:658957 | T | C | 0.50459 | 0.14613 | 0.019867 | 1.90E-13 | cis |
| rs1162168 | miR-625-5p | 14:660146 | T | G | 0.492955 | 0.13229 | 0.019812 | 2.44E-11 | cis |
| rs1115860 | miR-625-5p | 14:661107 | T | G | 0.365883 | 0.173324 | 0.020734 | 6.31E-17 | cis |
| rs2411355 | miR-625-5p | 14:660607 | C | A | 0.49877 | 0.145602 | 0.019744 | 1.65E-13 | cis |
| rs4899172 | miR-625-5p | 14:657799 | T | C | 0.213235 | 0.359928 | 0.022679 | 1.01E-56 | cis |
| rs7149301 | miR-625-5p | 14:662304 | A | G | 0.452987 | 0.142786 | 0.019331 | 1.51E-13 | cis |
| rs7150094 | miR-625-5p | 14:659363 | A | G | 0.346839 | 0.175864 | 0.020851 | 3.33E-17 | cis |
| rs8014475 | miR-625-5p | 14:658115 | T | C | 0.34592 | 0.204093 | 0.019825 | 7.44E-25 | cis |
| rs7606377 | miR-625-5p | 14:657677 | G | A | 0.843419 | -0.308513 | 0.02639 | 1.43E-31 | cis |
| rs1457728 | miR-625-5p | 14:661028 | C | T | 0.821198 | -0.234147 | 0.02536 | 2.63E-20 | cis |
| rs3986780 | miR-625-5p | 14:657947 | G | T | 0.053217 | 0.337746 | 0.044685 | 4.08E-14 | cis |
| rs1999725 | miR-625-5p | 14:659042 | G | A | 0.373979 | 0.181949 | 0.020557 | 8.67E-19 | cis |
| rs2064695 | miR-625-5p | 14:660670 | T | G | 0.40849 | 0.168086 | 0.020171 | 7.88E-17 | cis |
| rs1256520 | miR-625-5p | 14:657371 | C | A | 0.64779 | -0.191315 | 0.020361 | 5.66E-21 | cis |
| rs2268961 | miR-625-5p | 14:661467 | T | C | 0.501133 | 0.147737 | 0.019817 | 8.97E-14 | cis |
| rs1953415 | miR-625-5p | 14:659880 | A | G | 0.355003 | 0.172319 | 0.020622 | 6.49E-17 | cis |
| rs761830 | miR-625-5p | 14:662129 | G | A | 0.403845 | 0.115156 | 0.020216 | 1.23E-08 | cis |
| rs1162074 | miR-625-5p | 14:658998 | A | G | 0.502681 | 0.145468 | 0.019793 | 1.99E-13 | cis |
| rs6070872 | miR-625-5p | 14:661167 | G | A | 0.819646 | -0.239612 | 0.025764 | 1.40E-20 | cis |
| rs8021946 | miR-625-5p | 14:658468 | C | T | 0.054518 | 0.325363 | 0.045724 | 1.11E-12 | cis |
| rs7271442 | miR-625-5p | 14:658635 | G | A | 0.814071 | -0.233481 | 0.024581 | 2.13E-21 | cis |
| rs730807 | miR-625-5p | 14:662392 | C | T | 0.45402 | 0.143991 | 0.019392 | 1.12E-13 | cis |
| rs7465586 | miR-625-5p | 14:658311 | G | A | 0.815043 | -0.239026 | 0.024818 | 5.91E-22 | cis |
| rs968540 | miR-625-5p | 14:662490 | G | T | 0.633781 | -0.135121 | 0.020129 | 1.91E-11 | cis |

| | | | | | | | | | |
|---|---|---|---|---|---|---|---|---|---|
| rs7153297 | miR-625-5p | 14:660034( | T | A | 0.51171 | 0.14732 | 0.019892 | 1.30E-13 | cis |
| rs1256521 | miR-625-5p | 14:657376( | T | C | 0.646581 | -0.191015 | 0.020326 | 5.57E-21 | cis |
| rs8013442 | miR-625-5p | 14:658718( | A | G | 0.342325 | 0.17882 | 0.020898 | 1.16E-17 | cis |
| rs6573625 | miR-625-5p | 14:662406: | T | C | 0.37315 | 0.137387 | 0.020057 | 7.39E-12 | cis |
| rs743084 | miR-625-5p | 14:662326( | T | C | 0.452662 | 0.142879 | 0.019322 | 1.42E-13 | cis |
| rs2411405 | miR-625-5p | 14:662320{ | A | G | 0.448097 | 0.141959 | 0.019234 | 1.57E-13 | cis |
| rs743085 | miR-625-5p | 14:660681: | A | G | 0.499418 | 0.145261 | 0.019744 | 1.88E-13 | cis |
| rs6795753 | miR-625-5p | 14:661245{ | T | G | 0.50137 | 0.146801 | 0.01989 | 1.58E-13 | cis |
| rs2411816 | miR-625-5p | 14:659948{ | G | A | 0.50839 | 0.14508 | 0.019834 | 2.58E-13 | cis |
| rs7148737 | miR-625-5p | 14:662300: | C | A | 0.386987 | 0.146174 | 0.019789 | 1.51E-13 | cis |
| rs8015222 | miR-625-5p | 14:658360( | T | A | 0.052734 | 0.328492 | 0.045567 | 5.63E-13 | cis |
| rs7692276 | miR-625-5p | 14:657375{ | C | G | 0.705647 | -0.191943 | 0.021474 | 3.95E-19 | cis |
| rs7271443 | miR-625-5p | 14:659028: | A | G | 0.815449 | -0.231041 | 0.024484 | 3.86E-21 | cis |
| rs2210805 | miR-625-5p | 14:658927: | C | G | 0.354478 | 0.174717 | 0.020743 | 3.67E-17 | cis |
| rs1162819 | miR-625-5p | 14:658912 | A | G | 0.495088 | 0.14713 | 0.019857 | 1.27E-13 | cis |
| rs2411357 | miR-625-5p | 14:660511( | G | T | 0.499896 | 0.145832 | 0.019747 | 1.53E-13 | cis |
| rs1162536 | miR-625-5p | 14:662328( | G | A | 0.45263 | 0.142912 | 0.019323 | 1.40E-13 | cis |
| rs1214747 | miR-625-5p | 14:657950{ | C | T | 0.845789 | -0.329795 | 0.02651 | 1.58E-35 | cis |
| rs1214777 | miR-625-5p | 14:657976 | A | T | 0.846077 | -0.328488 | 0.026194 | 4.47E-36 | cis |
| rs899960 | miR-625-5p | 14:657899 | C | G | 0.053722 | 0.334306 | 0.044654 | 7.07E-14 | cis |
| rs4902400 | miR-625-5p | 14:659571 | A | G | 0.508016 | 0.144622 | 0.019874 | 3.41E-13 | cis |
| rs1998036 | miR-625-5p | 14:661381 | G | C | 0.498979 | 0.148436 | 0.019879 | 8.20E-14 | cis |
| rs2127869 | miR-625-5p | 14:657943 | T | C | 0.291045 | 0.290832 | 0.020564 | 2.08E-45 | cis |
| rs1289309 | miR-625-5p | 14:657890 | G | A | 0.385962 | 0.21328 | 0.019213 | 1.24E-28 | cis |
| rs4439682 | miR-625-5p | 14:662594 | A | G | 0.55661 | -0.127661 | 0.019553 | 6.63E-11 | cis |
| rs5963675 | miR-625-5p | 14:662564{ | T | C | 0.541442 | -0.139344 | 0.019364 | 6.20E-13 | cis |
| rs6573616 | miR-625-5p | 14:660468 | A | G | 0.343484 | 0.172142 | 0.020598 | 6.41E-17 | cis |
| rs1177595 | miR-625-5p | 14:657590 | G | C | 0.845965 | -0.311633 | 0.026756 | 2.37E-31 | cis |
| rs7424586 | miR-625-5p | 14:660940( | A | G | 0.820602 | -0.233791 | 0.025222 | 1.87E-20 | cis |
| rs7151561 | miR-625-5p | 14:658906( | T | C | 0.350627 | 0.175974 | 0.020911 | 3.92E-17 | cis |
| rs4359350 | miR-625-5p | 14:658637 | T | C | 0.404776 | 0.184939 | 0.02022 | 5.88E-20 | cis |
| rs2183277 | miR-625-5p | 14:660354 | T | C | 0.342723 | 0.172432 | 0.020657 | 6.99E-17 | cis |
| rs7145500 | miR-625-5p | 14:658842 | G | A | 0.354536 | 0.173427 | 0.020668 | 4.81E-17 | cis |
| rs9323460 | miR-625-5p | 14:659331 | T | G | 0.357542 | 0.174201 | 0.02067 | 3.53E-17 | cis |
| rs6573612 | miR-625-5p | 14:659128 | G | A | 0.355177 | 0.173057 | 0.020731 | 6.96E-17 | cis |
| rs4902404 | miR-625-5p | 14:660470 | T | C | 0.344394 | 0.172189 | 0.02055 | 5.34E-17 | cis |
| rs6573609 | miR-625-5p | 14:658614( | A | C | 0.390167 | 0.175181 | 0.020391 | 8.61E-18 | cis |
| rs1184726 | miR-625-5p | 14:657756 | T | G | 0.67304 | -0.150419 | 0.020266 | 1.15E-13 | cis |
| rs7271640 | miR-625-5p | 14:660660( | A | G | 0.820429 | -0.233098 | 0.025151 | 1.90E-20 | cis |
| rs1256540 | miR-625-5p | 14:657640( | T | C | 0.645897 | -0.223056 | 0.019669 | 8.26E-30 | cis |
| rs8005243 | miR-625-5p | 14:660656 | C | G | 0.346908 | 0.172472 | 0.02067 | 7.17E-17 | cis |
| rs2185454 | miR-625-5p | 14:658705 | G | T | 0.352958 | 0.175668 | 0.020698 | 2.11E-17 | cis |
| rs7150648 | miR-625-5p | 14:658901 | T | C | 0.347708 | 0.175934 | 0.020902 | 3.87E-17 | cis |
| rs6573606 | miR-625-5p | 14:658335 | T | C | 0.109568 | 0.205552 | 0.032428 | 2.32E-10 | cis |
| rs2184603 | miR-625-5p | 14:659306 | A | C | 0.500695 | 0.146773 | 0.019945 | 1.86E-13 | cis |
| rs1256536 | miR-625-5p | 14:657624 | A | G | 0.656085 | -0.2135 | 0.020061 | 1.89E-26 | cis |
| rs1256544 | miR-625-5p | 14:657764 | G | C | 0.053147 | 0.335592 | 0.044505 | 4.68E-14 | cis |
| rs1165522 | let-7a-5p | 9:1361490 | T | A | 0.653719 | -0.084566 | 0.011334 | 8.59E-14 | trans |
| rs2519093 | let-7a-5p | 9:1361418 | C | T | 0.809892 | -0.093892 | 0.01387 | 1.29E-11 | trans |
| rs494242 | let-7a-5p | 9:1361451 | C | T | 0.644977 | -0.080059 | 0.011174 | 7.78E-13 | trans |
| rs495828 | let-7a-5p | 9:1361548( | G | T | 0.787276 | -0.092717 | 0.013111 | 1.53E-12 | trans |
| rs544873 | let-7a-5p | 9:1361432 | G | A | 0.645742 | -0.079908 | 0.011169 | 8.38E-13 | trans |
| rs600038 | let-7a-5p | 9:1361518( | T | C | 0.787872 | -0.093029 | 0.013146 | 1.48E-12 | trans |

| | | | | | | | | | |
|---|---|---|---|---|---|---|---|---|---|
| rs643434 | let-7a-5p | 9:1361423! | G | A | 0.645392 | -0.079734 | 0.011164 | 9.18E-13 | trans |
| rs644234 | let-7a-5p | 9:1361422: | T | G | 0.645381 | -0.079814 | 0.011155 | 8.38E-13 | trans |
| rs649129 | let-7a-5p | 9:1361543( | C | T | 0.787432 | -0.092844 | 0.01312 | 1.48E-12 | trans |
| rs1190646: | miR-1 | 20:611589! | C | T | 0.824779 | -0.278766 | 0.050462 | 3.31E-08 | cis |
| rs1262460! | miR-1 | 20:611552! | C | A | 0.796375 | -0.297193 | 0.051729 | 9.18E-09 | cis |
| rs3934574 | miR-1 | 20:611434! | T | C | 0.464792 | -0.246928 | 0.03925 | 3.15E-10 | cis |
| rs6062267 | miR-1 | 20:611533( | T | C | 0.464907 | -0.267804 | 0.041926 | 1.69E-10 | cis |
| rs6062284 | miR-1 | 20:611419! | T | G | 0.4602 | -0.243553 | 0.03967 | 8.28E-10 | cis |
| rs762137 | miR-1 | 20:611509: | A | G | 0.467251 | -0.267207 | 0.041998 | 1.99E-10 | cis |
| rs8122282 | miR-1 | 20:611523: | G | A | 0.467131 | -0.267453 | 0.041995 | 1.91E-10 | cis |
| rs1190646: | miR-133a-3 | 20:611589! | C | T | 0.824779 | -0.163689 | 0.024636 | 3.05E-11 | cis |
| rs1262460! | miR-133a-3 | 20:611552! | C | A | 0.796375 | -0.174907 | 0.025247 | 4.27E-12 | cis |
| rs3934574 | miR-133a-3 | 20:611434! | T | C | 0.464792 | -0.111489 | 0.019248 | 6.95E-09 | cis |
| rs4305340 | miR-133a-3 | 20:611572 | C | T | 0.814576 | -0.170191 | 0.02582 | 4.35E-11 | cis |
| rs6062267 | miR-133a-3 | 20:611533( | T | C | 0.464907 | -0.118178 | 0.020569 | 9.16E-09 | cis |
| rs7271454 | miR-133a-3 | 20:611432: | T | C | 0.46691 | -0.112505 | 0.019434 | 7.07E-09 | cis |
| rs762137 | miR-133a-3 | 20:611509: | A | G | 0.467251 | -0.119241 | 0.0206 | 7.11E-09 | cis |
| rs8122282 | miR-133a-3 | 20:611523: | G | A | 0.467131 | -0.118017 | 0.020602 | 1.01E-08 | cis |
| rs1036332 | miR-181a-5 | 1:1990124: | A | C | 0.260189 | 0.172538 | 0.013837 | 1.10E-35 | cis |
| rs1040959 | miR-181a-5 | 1:1988958 | A | C | 0.766999 | -0.100032 | 0.014691 | 9.81E-12 | cis |
| rs1040960 | miR-181a-5 | 1:1988959! | G | T | 0.766066 | -0.100198 | 0.014695 | 9.21E-12 | cis |
| rs1075384: | miR-181a-5 | 1:1990930! | A | G | 0.671332 | -0.08257 | 0.013598 | 1.26E-09 | cis |
| rs1080059! | miR-181a-5 | 1:1988708: | T | C | 0.760087 | -0.108167 | 0.0147 | 1.86E-13 | cis |
| rs1080060: | miR-181a-5 | 1:1989145: | A | G | 0.244491 | 0.102312 | 0.014406 | 1.23E-12 | cis |
| rs1080060: | miR-181a-5 | 1:1989196! | A | G | 0.238849 | 0.106727 | 0.01456 | 2.30E-13 | cis |
| rs1080060( | miR-181a-5 | 1:1990049! | C | G | 0.663982 | 0.087564 | 0.01307 | 2.09E-11 | cis |
| rs1080060! | miR-181a-5 | 1:1990843: | A | C | 0.66159 | -0.075951 | 0.013497 | 1.83E-08 | cis |
| rs1080060! | miR-181a-5 | 1:1990916! | T | G | 0.671708 | -0.082217 | 0.013599 | 1.49E-09 | cis |
| rs1091960: | miR-181a-5 | 1:1988826( | G | A | 0.764985 | -0.101467 | 0.014749 | 6.00E-12 | cis |
| rs1091961! | miR-181a-5 | 1:1989749( | C | T | 0.243033 | 0.166882 | 0.014556 | 1.99E-30 | cis |
| rs1091961! | miR-181a-5 | 1:1990380 | T | C | 0.841859 | -0.152167 | 0.016799 | 1.33E-19 | cis |
| rs1091962: | miR-181a-5 | 1:1990912 | G | T | 0.671976 | -0.082295 | 0.013607 | 1.47E-09 | cis |
| rs1091965! | miR-181a-5 | 1:1992486( | A | G | 0.667251 | -0.106368 | 0.013119 | 5.15E-16 | cis |
| rs1124025 | miR-181a-5 | 1:1989798: | C | G | 0.238162 | 0.170617 | 0.014497 | 5.62E-32 | cis |
| rs1204800 | miR-181a-5 | 1:1989153( | T | C | 0.235546 | 0.100871 | 0.014669 | 6.14E-12 | cis |
| rs1209687( | miR-181a-5 | 1:1989144: | G | A | 0.244151 | 0.101854 | 0.014401 | 1.52E-12 | cis |
| rs1212331 | miR-181a-5 | 1:1988726! | G | T | 0.764104 | -0.102538 | 0.014805 | 4.33E-12 | cis |
| rs1212520( | miR-181a-5 | 1:1989920 | A | G | 0.260715 | 0.177971 | 0.013845 | 8.14E-38 | cis |
| rs1213024 | miR-181a-5 | 1:1988841: | G | C | 0.765569 | -0.101979 | 0.014754 | 4.79E-12 | cis |
| rs1213038: | miR-181a-5 | 1:1989376: | A | C | 0.845264 | -0.107981 | 0.017189 | 3.34E-10 | cis |
| rs1213201( | miR-181a-5 | 1:1988820: | T | C | 0.755495 | -0.106835 | 0.014472 | 1.55E-13 | cis |
| rs1214202! | miR-181a-5 | 1:1988815! | C | T | 0.766397 | -0.101919 | 0.014796 | 5.65E-12 | cis |
| rs1223917! | miR-181a-5 | 1:1989764! | C | T | 0.245157 | 0.163804 | 0.014397 | 5.38E-30 | cis |
| rs1275009 | miR-181a-5 | 1:1990708! | C | T | 0.540441 | -0.102424 | 0.013128 | 6.10E-15 | cis |
| rs1275163 | miR-181a-5 | 1:1988916: | C | T | 0.765073 | -0.101395 | 0.014694 | 5.18E-12 | cis |
| rs1275164 | miR-181a-5 | 1:1988944! | A | G | 0.754678 | -0.107087 | 0.014304 | 7.08E-14 | cis |
| rs1347323 | miR-181a-5 | 1:1989704: | A | G | 0.257803 | 0.11992 | 0.014395 | 8.03E-17 | cis |
| rs1434282 | miR-181a-5 | 1:1990107: | C | T | 0.271104 | 0.177906 | 0.013618 | 5.30E-39 | cis |
| rs167735 | miR-181a-5 | 1:1990643! | G | A | 0.505181 | -0.09148 | 0.01296 | 1.68E-12 | cis |
| rs1684410: | miR-181a-5 | 1:1990115: | G | T | 0.842513 | -0.14945 | 0.016947 | 1.16E-18 | cis |
| rs1762558: | miR-181a-5 | 1:1989904! | G | A | 0.74365 | -0.178857 | 0.013895 | 6.49E-38 | cis |
| rs1763147: | miR-181a-5 | 1:1992522! | T | A | 0.668759 | -0.106374 | 0.013247 | 9.73E-16 | cis |
| rs1806813 | miR-181a-5 | 1:1989266: | T | C | 0.844646 | -0.10745 | 0.017134 | 3.59E-10 | cis |

| | | | | | | | | | |
|---|---|---|---|---|---|---|---|---|---|
| rs188926 | miR-181a-5 | 1:1990672 | T | C | 0.53384 | -0.105587 | 0.012882 | 2.47E-16 | cis |
| rs1892432 | miR-181a-5 | 1:1989406 | T | C | 0.234225 | 0.101149 | 0.014723 | 6.41E-12 | cis |
| rs1938038 | miR-181a-5 | 1:1989248 | A | G | 0.250919 | 0.099347 | 0.014575 | 9.35E-12 | cis |
| rs2033565 | miR-181a-5 | 1:1990150 | A | T | 0.16327 | 0.149375 | 0.016865 | 8.21E-19 | cis |
| rs2033566 | miR-181a-5 | 1:1990148 | T | G | 0.211961 | 0.117053 | 0.015337 | 2.31E-14 | cis |
| rs2060581 | miR-181a-5 | 1:1992502 | T | C | 0.66838 | -0.107303 | 0.013201 | 4.36E-16 | cis |
| rs2360961 | miR-181a-5 | 1:1990002 | T | C | 0.615121 | 0.082632 | 0.012723 | 8.32E-11 | cis |
| rs2360962 | miR-181a-5 | 1:1990047 | A | T | 0.261757 | 0.178107 | 0.013794 | 3.87E-38 | cis |
| rs2360964 | miR-181a-5 | 1:1990541 | T | G | 0.669668 | -0.075828 | 0.013274 | 1.11E-08 | cis |
| rs2624882 | miR-181a-5 | 1:1991077 | A | G | 0.514062 | -0.10486 | 0.013065 | 1.01E-15 | cis |
| rs2648772 | miR-181a-5 | 1:1991147 | C | T | 0.519051 | -0.10108 | 0.013173 | 1.67E-14 | cis |
| rs2648773 | miR-181a-5 | 1:1991157 | G | A | 0.50507 | -0.101287 | 0.013172 | 1.47E-14 | cis |
| rs322875 | miR-181a-5 | 1:1991026 | C | T | 0.518914 | -0.099532 | 0.013126 | 3.38E-14 | cis |
| rs322877 | miR-181a-5 | 1:1990988 | C | T | 0.508326 | -0.094607 | 0.013088 | 4.87E-13 | cis |
| rs322887 | miR-181a-5 | 1:1990263 | A | G | 0.834392 | -0.156419 | 0.016492 | 2.43E-21 | cis |
| rs322889 | miR-181a-5 | 1:1990255 | C | G | 0.786955 | -0.124607 | 0.015018 | 1.07E-16 | cis |
| rs322898 | miR-181a-5 | 1:1990913 | G | A | 0.514892 | -0.103458 | 0.013022 | 1.94E-15 | cis |
| rs322907 | miR-181a-5 | 1:1990825 | G | T | 0.500608 | -0.098937 | 0.013128 | 4.84E-14 | cis |
| rs322912 | miR-181a-5 | 1:1990743 | C | T | 0.52926 | -0.097774 | 0.013089 | 8.01E-14 | cis |
| rs322915 | miR-181a-5 | 1:1990707 | G | A | 0.518776 | -0.099672 | 0.013107 | 2.86E-14 | cis |
| rs322916 | miR-181a-5 | 1:1990691 | C | T | 0.50214 | -0.099286 | 0.013085 | 3.25E-14 | cis |
| rs322918 | miR-181a-5 | 1:1990686 | A | G | 0.534816 | -0.105378 | 0.01288 | 2.81E-16 | cis |
| rs322927 | miR-181a-5 | 1:1990218 | T | G | 0.834224 | -0.156756 | 0.016443 | 1.52E-21 | cis |
| rs322928 | miR-181a-5 | 1:1990218 | G | A | 0.833953 | -0.15677 | 0.016435 | 1.45E-21 | cis |
| rs322929 | miR-181a-5 | 1:1990209 | T | C | 0.78491 | -0.124643 | 0.015004 | 9.80E-17 | cis |
| rs322930 | miR-181a-5 | 1:1990205 | T | A | 0.790845 | -0.119262 | 0.015118 | 3.05E-15 | cis |
| rs322931 | miR-181a-5 | 1:1990198 | G | A | 0.790484 | -0.1192 | 0.01509 | 2.81E-15 | cis |
| rs322932 | miR-181a-5 | 1:1990193 | A | T | 0.789147 | -0.119104 | 0.01515 | 3.79E-15 | cis |
| rs322933 | miR-181a-5 | 1:1990180 | C | T | 0.506039 | -0.101174 | 0.012628 | 1.13E-15 | cis |
| rs3488967 | miR-181a-5 | 1:1992490 | A | G | 0.66157 | -0.107512 | 0.013292 | 6.03E-16 | cis |
| rs3850623 | miR-181a-5 | 1:1991205 | G | A | 0.518649 | -0.10172 | 0.013222 | 1.43E-14 | cis |
| rs427790 | miR-181a-5 | 1:1990972 | C | T | 0.519007 | -0.09882 | 0.01307 | 4.00E-14 | cis |
| rs4915165 | miR-181a-5 | 1:1989707 | T | C | 0.253161 | 0.115248 | 0.01452 | 2.07E-15 | cis |
| rs4915331 | miR-181a-5 | 1:1989185 | T | C | 0.235006 | 0.10124 | 0.014684 | 5.40E-12 | cis |
| rs4915332 | miR-181a-5 | 1:1989185 | A | G | 0.234929 | 0.101276 | 0.014684 | 5.31E-12 | cis |
| rs4915335 | miR-181a-5 | 1:1989452 | A | G | 0.238713 | 0.109407 | 0.01468 | 9.14E-14 | cis |
| rs4915343 | miR-181a-5 | 1:1990778 | A | G | 0.661038 | -0.075465 | 0.013511 | 2.33E-08 | cis |
| rs6427756 | miR-181a-5 | 1:1990007 | A | G | 0.263331 | 0.176572 | 0.013797 | 1.69E-37 | cis |
| rs6679487 | miR-181a-5 | 1:1990963 | A | G | 0.676468 | -0.077739 | 0.013698 | 1.39E-08 | cis |
| rs6703089 | miR-181a-5 | 1:1989200 | C | A | 0.236139 | 0.106716 | 0.014528 | 2.05E-13 | cis |
| rs6703633 | miR-181a-5 | 1:1990870 | T | G | 0.661365 | -0.07571 | 0.013497 | 2.03E-08 | cis |
| rs7273976 | miR-181a-5 | 1:1988640 | G | T | 0.765001 | -0.102876 | 0.014899 | 5.02E-12 | cis |
| rs7273977 | miR-181a-5 | 1:1988868 | A | T | 0.849527 | -0.100557 | 0.017598 | 1.10E-08 | cis |
| rs7273977 | miR-181a-5 | 1:1988914 | C | G | 0.84758 | -0.098441 | 0.017385 | 1.49E-08 | cis |
| rs7273980 | miR-181a-5 | 1:1989245 | C | T | 0.893228 | -0.130536 | 0.020111 | 8.54E-11 | cis |
| rs7524232 | miR-181a-5 | 1:1990057 | G | A | 0.670143 | 0.091241 | 0.013136 | 3.77E-12 | cis |
| rs7524630 | miR-181a-5 | 1:1991183 | A | G | 0.514782 | -0.104128 | 0.01313 | 2.18E-15 | cis |
| rs7529925 | miR-181a-5 | 1:1990072 | C | T | 0.255718 | 0.172939 | 0.013816 | 6.01E-36 | cis |
| rs7536036 | miR-181a-5 | 1:1990056 | T | C | 0.669114 | 0.091188 | 0.013143 | 3.97E-12 | cis |
| rs7538077 | miR-181a-5 | 1:1990764 | T | A | 0.661658 | -0.075447 | 0.013513 | 2.36E-08 | cis |
| rs7538550 | miR-181a-5 | 1:1990801 | C | A | 0.676636 | -0.077086 | 0.013649 | 1.63E-08 | cis |
| rs7539177 | miR-181a-5 | 1:1989464 | T | G | 0.238596 | 0.109677 | 0.014687 | 8.17E-14 | cis |
| rs7544236 | miR-181a-5 | 1:1991084 | T | C | 0.641676 | -0.073938 | 0.013391 | 3.36E-08 | cis |

| | | | | | | | | | |
|---|---|---|---|---|---|---|---|---|---|
| rs7547363 | miR-181a-5 | 1:1991183( | G | A | 0.519602 | -0.106958 | 0.013176 | 4.75E-16 | cis |
| rs7890714 | miR-181a-5 | 1:1989878( | G | A | 0.904551 | -0.152889 | 0.021249 | 6.24E-13 | cis |
| rs881576 | miR-181a-5 | 1:1992577 | A | C | 0.44377 | -0.090643 | 0.013036 | 3.56E-12 | cis |
| rs1019976 | miR-3130-3 | 2:2076418( | G | A | 0.842574 | -0.266836 | 0.032905 | 5.09E-16 | cis |
| rs1093216 | miR-3130-3 | 2:2076163 | A | G | 0.855067 | -0.279415 | 0.034267 | 3.52E-16 | cis |
| rs1155295 | miR-3130-3 | 2:2076756( | C | G | 0.874904 | -0.247276 | 0.036911 | 2.09E-11 | cis |
| rs1189416 | miR-3130-3 | 2:2076244 | T | C | 0.859947 | -0.284323 | 0.034753 | 2.81E-16 | cis |
| rs1189537( | miR-3130-3 | 2:2076547 | T | C | 0.842391 | -0.264848 | 0.033305 | 1.83E-15 | cis |
| rs1246816( | miR-3130-3 | 2:2076463( | T | C | 0.842809 | -0.267176 | 0.032925 | 4.87E-16 | cis |
| rs1247096 | miR-3130-3 | 2:2076395 | A | G | 0.866897 | -0.29906 | 0.035691 | 5.33E-17 | cis |
| rs1247100 | miR-3130-3 | 2:2076397 | A | G | 0.842907 | -0.27172 | 0.032916 | 1.52E-16 | cis |
| rs1247846 | miR-3130-3 | 2:2076710 | T | C | 0.846612 | -0.263395 | 0.033397 | 3.10E-15 | cis |
| rs1342700 | miR-3130-3 | 2:2076436 | C | G | 0.84329 | -0.270673 | 0.032943 | 2.10E-16 | cis |
| rs1683883 | miR-3130-3 | 2:2076297 | T | A | 0.84321 | -0.269712 | 0.032981 | 2.89E-16 | cis |
| rs1862087 | miR-3130-3 | 2:2076255 | T | C | 0.859956 | -0.284389 | 0.034753 | 2.76E-16 | cis |
| rs1862089 | miR-3130-3 | 2:2076205 | A | T | 0.854806 | -0.281451 | 0.034374 | 2.66E-16 | cis |
| rs2059022 | miR-3130-3 | 2:2076782 | T | C | 0.876031 | -0.247467 | 0.036929 | 2.07E-11 | cis |
| rs2241347 | miR-3130-3 | 2:2076479 | C | T | 0.841853 | -0.266447 | 0.032894 | 5.49E-16 | cis |
| rs2287628 | miR-3130-3 | 2:2076505 | A | G | 0.843162 | -0.266318 | 0.032968 | 6.58E-16 | cis |
| rs2287629 | miR-3130-3 | 2:2076504 | C | A | 0.842588 | -0.266338 | 0.032985 | 6.78E-16 | cis |
| rs2836579 | miR-3130-3 | 2:2076298 | A | T | 0.842753 | -0.269953 | 0.032925 | 2.42E-16 | cis |
| rs3815686 | miR-3130-3 | 2:2076330( | A | G | 0.843111 | -0.268909 | 0.03292 | 3.12E-16 | cis |
| rs4673363 | miR-3130-3 | 2:2076694 | G | A | 0.846328 | -0.263262 | 0.033357 | 2.97E-15 | cis |
| rs5760097 | miR-3130-3 | 2:2076701 | G | C | 0.846329 | -0.262977 | 0.033355 | 3.17E-15 | cis |
| rs6001989 | miR-3130-3 | 2:2076777 | C | T | 0.87642 | -0.247533 | 0.03705 | 2.37E-11 | cis |
| rs725980 | miR-3130-3 | 2:2076385 | C | G | 0.843297 | -0.271139 | 0.032939 | 1.85E-16 | cis |
| rs7576591 | miR-3130-3 | 2:2076492 | G | C | 0.843106 | -0.266231 | 0.032952 | 6.52E-16 | cis |
| rs7580138 | miR-3130-3 | 2:2076152 | C | A | 0.854791 | -0.277559 | 0.034201 | 4.84E-16 | cis |
| rs7627046 | miR-3130-3 | 2:2076536 | G | A | 0.902503 | -0.276562 | 0.04049 | 8.47E-12 | cis |
| rs768533 | miR-3130-3 | 2:2076408 | G | A | 0.842932 | -0.271664 | 0.032915 | 1.54E-16 | cis |
| rs7949904( | miR-3130-3 | 2:2076409 | A | G | 0.902079 | -0.277819 | 0.04038 | 5.98E-12 | cis |
| rs888621 | miR-3130-3 | 2:2076493 | T | G | 0.842681 | -0.266321 | 0.032863 | 5.32E-16 | cis |
| rs957414 | miR-3130-3 | 2:2076655 | T | A | 0.84423 | -0.26426 | 0.033198 | 1.72E-15 | cis |
| rs1017438( | miR-320b | 2:6859646 | C | T | 0.685243 | 0.129366 | 0.014889 | 3.67E-18 | trans |
| rs1018497 | miR-320b | 2:6859988 | G | A | 0.286787 | 0.087293 | 0.015488 | 1.74E-08 | trans |
| rs1018990 | miR-320b | 2:6860619 | C | G | 0.281778 | 0.087109 | 0.015387 | 1.50E-08 | trans |
| rs1049616 | miR-320b | 2:6862008 | C | T | 0.697496 | 0.127045 | 0.015092 | 3.83E-17 | trans |
| rs1050181 | miR-320b | 2:6862295 | G | C | 0.544992 | 0.136202 | 0.013895 | 1.10E-22 | trans |
| rs1112618 | miR-320b | 2:6861913 | A | G | 0.282025 | 0.088026 | 0.015376 | 1.04E-08 | trans |
| rs1155426 | miR-320b | 2:6862182 | T | C | 0.282037 | 0.087768 | 0.015375 | 1.14E-08 | trans |
| rs1156629 | miR-320b | 2:6862663 | T | G | 0.547487 | 0.138094 | 0.014017 | 6.74E-23 | trans |
| rs1158434 | miR-320b | 2:6863552 | G | C | 0.551072 | 0.14116 | 0.014323 | 6.49E-23 | trans |
| rs1164012 | miR-320b | 2:6862562 | C | T | 0.545777 | 0.136712 | 0.013945 | 1.09E-22 | trans |
| rs1189593 | miR-320b | 2:6861908 | A | G | 0.282175 | 0.08814 | 0.015382 | 1.00E-08 | trans |
| rs1271364 | miR-320b | 2:6860665( | G | A | 0.282933 | 0.089045 | 0.01544 | 8.07E-09 | trans |
| rs1300495 | miR-320b | 2:6858081 | G | A | 0.306559 | 0.086658 | 0.015376 | 1.74E-08 | trans |
| rs1300519 | miR-320b | 2:6863887 | A | G | 0.702186 | 0.133519 | 0.015787 | 2.73E-17 | trans |
| rs1300592 | miR-320b | 2:6862999 | T | A | 0.701261 | 0.129992 | 0.015537 | 5.92E-17 | trans |
| rs1300974 | miR-320b | 2:6858443 | C | A | 0.678986 | 0.117096 | 0.015503 | 4.25E-14 | trans |
| rs1301028 | miR-320b | 2:6862184 | G | T | 0.683506 | 0.129676 | 0.014833 | 2.28E-18 | trans |
| rs1301077 | miR-320b | 2:6863908 | C | T | 0.702339 | 0.133415 | 0.015791 | 2.95E-17 | trans |
| rs1301107 | miR-320b | 2:6863931 | A | C | 0.700844 | 0.134034 | 0.015817 | 2.37E-17 | trans |
| rs1301143( | miR-320b | 2:6863937 | C | T | 0.702382 | 0.133434 | 0.015794 | 2.95E-17 | trans |

| | | | | | | | | | |
|---|---|---|---|---|---|---|---|---|---|
| rs1301157 | miR-320b | 2:6863927 | G | A | 0.702349 | 0.133427 | 0.015792 | 2.94E-17 | trans |
| rs1302892 | miR-320b | 2:6862991 | C | T | 0.552648 | 0.137763 | 0.014108 | 1.59E-22 | trans |
| rs1302897 | miR-320b | 2:6863003 | A | G | 0.54849 | 0.138955 | 0.014062 | 5.00E-23 | trans |
| rs1303482 | miR-320b | 2:6863029 | C | G | 0.552715 | 0.137808 | 0.014111 | 1.57E-22 | trans |
| rs1343065 | miR-320b | 2:6858771 | C | G | 0.699397 | 0.126957 | 0.01528 | 9.66E-17 | trans |
| rs1373198 | miR-320b | 2:6858798 | A | T | 0.281586 | 0.085614 | 0.015537 | 3.58E-08 | trans |
| rs1373200 | miR-320b | 2:6862265 | C | T | 0.547142 | 0.134711 | 0.013801 | 1.66E-22 | trans |
| rs1382899 | miR-320b | 2:6863782 | A | G | 0.702614 | 0.133506 | 0.015805 | 2.99E-17 | trans |
| rs1390688 | miR-320b | 2:6862773 | C | T | 0.69721 | 0.12989 | 0.015387 | 3.14E-17 | trans |
| rs1397159 | miR-320b | 2:6863425 | T | G | 0.705552 | 0.134497 | 0.015943 | 3.28E-17 | trans |
| rs1422402 | miR-320b | 2:6862752 | A | G | 0.698498 | 0.130556 | 0.015426 | 2.60E-17 | trans |
| rs1442589 | miR-320b | 2:6863473 | G | T | 0.701896 | 0.133113 | 0.015756 | 2.95E-17 | trans |
| rs1455505 | miR-320b | 2:6862832 | G | A | 0.556221 | 0.141184 | 0.014214 | 3.00E-23 | trans |
| rs1474969 | miR-320b | 2:6862568 | G | A | 0.546544 | 0.136939 | 0.013966 | 1.07E-22 | trans |
| rs1484530 | miR-320b | 2:6863547 | A | G | 0.70445 | 0.134492 | 0.015849 | 2.14E-17 | trans |
| rs1486061 | miR-320b | 2:6862571 | T | G | 0.54743 | 0.137325 | 0.013992 | 9.74E-23 | trans |
| rs1542135 | miR-320b | 2:6862084 | C | T | 0.69752 | 0.127051 | 0.015091 | 3.79E-17 | trans |
| rs1542136 | miR-320b | 2:6862093 | C | A | 0.697594 | 0.127041 | 0.015095 | 3.89E-17 | trans |
| rs1753019 | miR-320b | 2:6862133 | G | T | 0.683745 | 0.129844 | 0.014845 | 2.20E-18 | trans |
| rs1760149 | miR-320b | 2:6861171 | A | G | 0.69807 | 0.126937 | 0.015097 | 4.16E-17 | trans |
| rs1760168 | miR-320b | 2:6861589 | G | A | 0.697578 | 0.126954 | 0.015102 | 4.23E-17 | trans |
| rs1760209 | miR-320b | 2:6862041 | T | A | 0.697529 | 0.127053 | 0.015091 | 3.80E-17 | trans |
| rs1867313 | miR-320b | 2:6862365 | T | C | 0.282087 | 0.087699 | 0.015373 | 1.16E-08 | trans |
| rs2070171 | miR-320b | 2:6861371 | G | A | 0.698024 | 0.126917 | 0.015096 | 4.20E-17 | trans |
| rs2165919 | miR-320b | 2:6861413 | C | T | 0.281408 | 0.088625 | 0.015405 | 8.77E-09 | trans |
| rs2165920 | miR-320b | 2:6862223 | T | C | 0.552012 | 0.135016 | 0.013899 | 2.62E-22 | trans |
| rs2241946 | miR-320b | 2:6861565 | T | C | 0.698076 | 0.127092 | 0.015103 | 3.92E-17 | trans |
| rs2241947 | miR-320b | 2:6861539 | C | T | 0.697988 | 0.126954 | 0.015096 | 4.11E-17 | trans |
| rs3419682 | miR-320b | 2:6862142 | G | A | 0.697518 | 0.127052 | 0.015091 | 3.79E-17 | trans |
| rs3426707 | miR-320b | 2:6859915 | T | C | 0.282465 | 0.087895 | 0.015422 | 1.20E-08 | trans |
| rs3485223 | miR-320b | 2:6863859 | C | G | 0.550658 | 0.140992 | 0.014326 | 7.45E-23 | trans |
| rs3732044 | miR-320b | 2:6862391 | T | C | 0.545562 | 0.136529 | 0.013935 | 1.15E-22 | trans |
| rs4610041 | miR-320b | 2:6862628 | C | T | 0.550745 | 0.140566 | 0.014102 | 2.10E-23 | trans |
| rs4671208 | miR-320b | 2:6860079 | A | G | 0.281916 | 0.08725 | 0.015412 | 1.50E-08 | trans |
| rs4854536 | miR-320b | 2:6863186 | T | A | 0.283938 | 0.08972 | 0.015451 | 6.37E-09 | trans |
| rs5618821 | miR-320b | 2:6861163 | T | C | 0.698075 | 0.126937 | 0.015097 | 4.16E-17 | trans |
| rs5628470 | miR-320b | 2:6863137 | G | A | 0.697508 | 0.130591 | 0.015439 | 2.71E-17 | trans |
| rs5633714 | miR-320b | 2:6864249 | G | A | 0.733417 | 0.132351 | 0.016434 | 8.03E-16 | trans |
| rs5690424 | miR-320b | 2:6863847 | T | C | 0.550669 | 0.140995 | 0.014325 | 7.40E-23 | trans |
| rs5808991 | miR-320b | 2:6863966 | G | C | 0.734634 | 0.134981 | 0.016542 | 3.35E-16 | trans |
| rs5888448 | miR-320b | 2:6864534 | G | A | 0.734214 | 0.131847 | 0.016492 | 1.30E-15 | trans |
| rs5933292 | miR-320b | 2:6863963 | G | A | 0.73529 | 0.135103 | 0.016587 | 3.79E-16 | trans |
| rs5999861 | miR-320b | 2:6864677 | G | T | 0.731295 | 0.125973 | 0.016487 | 2.16E-14 | trans |
| rs6000716 | miR-320b | 2:6863963 | C | T | 0.735163 | 0.135202 | 0.016585 | 3.57E-16 | trans |
| rs6087844 | miR-320b | 2:6858211 | A | G | 0.538067 | 0.126281 | 0.014369 | 1.52E-18 | trans |
| rs6214389 | miR-320b | 2:6860455 | C | T | 0.698307 | 0.126569 | 0.015117 | 5.64E-17 | trans |
| rs6214392 | miR-320b | 2:6863835 | C | G | 0.700378 | 0.134816 | 0.015773 | 1.26E-17 | trans |
| rs6214393 | miR-320b | 2:6864009 | T | C | 0.59325 | 0.131146 | 0.01472 | 5.13E-19 | trans |
| rs6214393 | miR-320b | 2:6864020 | A | G | 0.593308 | 0.131089 | 0.014721 | 5.33E-19 | trans |
| rs6214393 | miR-320b | 2:6864038 | G | C | 0.593598 | 0.131783 | 0.014715 | 3.37E-19 | trans |
| rs6214393 | miR-320b | 2:6864041 | A | G | 0.593599 | 0.131772 | 0.014715 | 3.40E-19 | trans |
| rs6214393 | miR-320b | 2:6864062 | T | C | 0.733261 | 0.132408 | 0.016423 | 7.50E-16 | trans |
| rs6419643 | miR-320b | 2:6863126 | T | G | 0.548826 | 0.138972 | 0.014072 | 5.32E-23 | trans |

| | | | | | | | | | |
|---|---|---|---|---|---|---|---|---|---|
| rs6419644 | miR-320b | 2:6863182 | A | G | 0.549898 | 0.140681 | 0.014215 | 4.29E-23 | trans |
| rs6546387 | miR-320b | 2:6858758 | G | A | 0.281573 | 0.085611 | 0.015538 | 3.60E-08 | trans |
| rs6546388 | miR-320b | 2:6858948 | C | T | 0.281691 | 0.085982 | 0.015493 | 2.86E-08 | trans |
| rs6546393 | miR-320b | 2:6862757 | T | C | 0.554017 | 0.140713 | 0.014135 | 2.40E-23 | trans |
| rs6546395 | miR-320b | 2:6862927 | A | G | 0.55448 | 0.138124 | 0.014154 | 1.70E-22 | trans |
| rs6546396 | miR-320b | 2:6862930 | A | T | 0.283762 | 0.089047 | 0.015411 | 7.56E-09 | trans |
| rs6546397 | miR-320b | 2:6863036 | G | A | 0.552756 | 0.13783 | 0.014113 | 1.57E-22 | trans |
| rs6546398 | miR-320b | 2:6863041 | G | C | 0.552757 | 0.137831 | 0.014112 | 1.56E-22 | trans |
| rs6546399 | miR-320b | 2:6863064 | C | T | 0.55394 | 0.138212 | 0.014154 | 1.60E-22 | trans |
| rs6546400 | miR-320b | 2:6863064 | G | A | 0.554382 | 0.138315 | 0.014167 | 1.62E-22 | trans |
| rs6546401 | miR-320b | 2:6863087 | A | T | 0.550127 | 0.139225 | 0.014103 | 5.49E-23 | trans |
| rs6546402 | miR-320b | 2:6863130 | A | G | 0.283216 | 0.089228 | 0.015417 | 7.15E-09 | trans |
| rs6546403 | miR-320b | 2:6863226 | C | T | 0.550801 | 0.140899 | 0.014236 | 4.28E-23 | trans |
| rs6546404 | miR-320b | 2:6863263 | G | T | 0.54995 | 0.140703 | 0.01422 | 4.38E-23 | trans |
| rs6546405 | miR-320b | 2:6863311 | G | A | 0.28401 | 0.089745 | 0.015451 | 6.31E-09 | trans |
| rs6546407 | miR-320b | 2:6863492 | A | C | 0.55091 | 0.141146 | 0.014305 | 5.80E-23 | trans |
| rs6546408 | miR-320b | 2:6863504 | T | C | 0.551062 | 0.141172 | 0.01432 | 6.31E-23 | trans |
| rs6546409 | miR-320b | 2:6863587 | A | G | 0.551093 | 0.141154 | 0.014327 | 6.70E-23 | trans |
| rs6546410 | miR-320b | 2:6863610 | C | G | 0.702434 | 0.133424 | 0.015795 | 2.98E-17 | trans |
| rs6546411 | miR-320b | 2:6863769 | A | G | 0.556248 | 0.143009 | 0.014415 | 3.38E-23 | trans |
| rs6546412 | miR-320b | 2:6863771 | T | C | 0.557086 | 0.144247 | 0.014441 | 1.71E-23 | trans |
| rs6708536 | miR-320b | 2:6862726 | G | A | 0.547249 | 0.138343 | 0.014027 | 6.04E-23 | trans |
| rs6709464 | miR-320b | 2:6862222 | C | T | 0.697279 | 0.126869 | 0.015077 | 3.94E-17 | trans |
| rs6714719 | miR-320b | 2:6859377 | G | C | 0.698404 | 0.12667 | 0.015159 | 6.50E-17 | trans |
| rs6715579 | miR-320b | 2:6858392 | C | T | 0.678794 | 0.117162 | 0.015502 | 4.09E-14 | trans |
| rs6715706 | miR-320b | 2:6858399 | C | A | 0.664887 | 0.120184 | 0.015241 | 3.13E-15 | trans |
| rs6715909 | miR-320b | 2:6858399 | G | A | 0.664879 | 0.120181 | 0.015242 | 3.14E-15 | trans |
| rs6729555 | miR-320b | 2:6863295 | C | A | 0.54995 | 0.140731 | 0.014223 | 4.41E-23 | trans |
| rs6729686 | miR-320b | 2:6863303 | C | T | 0.700171 | 0.132218 | 0.015638 | 2.80E-17 | trans |
| rs6730974 | miR-320b | 2:6863452 | T | A | 0.287755 | 0.092003 | 0.015485 | 2.83E-09 | trans |
| rs6742200 | miR-320b | 2:6862330 | A | G | 0.54552 | 0.136507 | 0.013932 | 1.15E-22 | trans |
| rs6743970 | miR-320b | 2:6858390 | A | T | 0.660237 | 0.122904 | 0.015312 | 1.00E-15 | trans |
| rs6744216 | miR-320b | 2:6863463 | A | G | 0.284465 | 0.08985 | 0.015451 | 6.06E-09 | trans |
| rs6744306 | miR-320b | 2:6863273 | T | C | 0.549956 | 0.140712 | 0.01422 | 4.37E-23 | trans |
| rs6747545 | miR-320b | 2:6863306 | T | A | 0.549964 | 0.140721 | 0.014224 | 4.46E-23 | trans |
| rs6757938 | miR-320b | 2:6863285 | A | G | 0.549958 | 0.140714 | 0.014222 | 4.42E-23 | trans |
| rs7420773 | miR-320b | 2:6862945 | G | A | 0.28377 | 0.089047 | 0.015411 | 7.56E-09 | trans |
| rs7420775 | miR-320b | 2:6862945 | G | T | 0.703122 | 0.130665 | 0.015606 | 5.63E-17 | trans |
| rs7420782 | miR-320b | 2:6862955 | G | T | 0.283742 | 0.089257 | 0.015414 | 7.02E-09 | trans |
| rs7421228 | miR-320b | 2:6863012 | G | T | 0.55268 | 0.137787 | 0.01411 | 1.58E-22 | trans |
| rs7451452 | miR-320b | 2:6863755 | T | G | 0.702552 | 0.133486 | 0.015801 | 2.97E-17 | trans |
| rs7562439 | miR-320b | 2:6858256 | C | T | 0.6646 | 0.119946 | 0.015246 | 3.62E-15 | trans |
| rs7562889 | miR-320b | 2:6862547 | T | C | 0.285916 | 0.088205 | 0.015428 | 1.08E-08 | trans |
| rs7572299 | miR-320b | 2:6858124 | T | A | 0.538033 | 0.125959 | 0.014372 | 1.88E-18 | trans |
| rs7574592 | miR-320b | 2:6859766 | T | G | 0.281934 | 0.087165 | 0.015412 | 1.55E-08 | trans |
| rs7578602 | miR-320b | 2:6863786 | G | C | 0.556806 | 0.144256 | 0.014448 | 1.78E-23 | trans |
| rs7581872 | miR-320b | 2:6858098 | A | C | 0.537959 | 0.12589 | 0.014372 | 1.96E-18 | trans |
| rs7590450 | miR-320b | 2:6862856 | A | G | 0.549532 | 0.138968 | 0.014059 | 4.87E-23 | trans |
| rs7592526 | miR-320b | 2:6858352 | A | C | 0.538007 | 0.126303 | 0.01437 | 1.50E-18 | trans |
| rs7597167 | miR-320b | 2:6861294 | C | T | 0.698051 | 0.126955 | 0.015097 | 4.12E-17 | trans |
| rs7598360 | miR-320b | 2:6863092 | A | G | 0.553166 | 0.141628 | 0.014138 | 1.27E-23 | trans |
| rs7600986 | miR-320b | 2:6863692 | A | T | 0.285029 | 0.089812 | 0.015435 | 5.93E-09 | trans |
| rs7605387 | miR-320b | 2:6863255 | A | G | 0.703499 | 0.132498 | 0.015728 | 3.63E-17 | trans |

| | | | | | | | | | |
|---|---|---|---|---|---|---|---|---|---|
| rs7608429 | miR-320b | 2:6858083( | G | T | 0.678719 | 0.116442 | 0.015517 | 6.19E-14 | trans |
| rs7800275: | miR-320b | 2:6862543( | A | G | 0.546309 | 0.136492 | 0.013929 | 1.13E-22 | trans |
| rs8028813! | miR-320b | 2:6862547( | A | G | 0.548418 | 0.139006 | 0.01402 | 3.59E-23 | trans |
| rs8761 | miR-320b | 2:6862426( | G | A | 0.545692 | 0.136595 | 0.01394 | 1.14E-22 | trans |
| rs880668 | miR-320b | 2:6859258/ | T | C | 0.280228 | 0.086317 | 0.015504 | 2.59E-08 | trans |
| rs964934 | miR-320b | 2:6861187: | G | A | 0.281023 | 0.087809 | 0.015405 | 1.20E-08 | trans |
| rs964935 | miR-320b | 2:6861228/ | G | A | 0.281126 | 0.087853 | 0.015408 | 1.18E-08 | trans |
| rs9653552 | miR-320b | 2:6864396( | G | A | 0.593458 | 0.13159 | 0.014728 | 4.08E-19 | trans |
| rs1017438( | miR-320c | 2:6859646! | C | T | 0.685243 | 0.117036 | 0.01396 | 5.12E-17 | trans |
| rs1018990: | miR-320c | 2:6860619: | C | G | 0.281778 | 0.08316 | 0.014406 | 7.81E-09 | trans |
| rs1049616! | miR-320c | 2:6862008! | C | T | 0.697496 | 0.113968 | 0.014152 | 8.08E-16 | trans |
| rs1050181 | miR-320c | 2:6862295: | G | C | 0.544992 | 0.123229 | 0.013032 | 3.20E-21 | trans |
| rs1112618: | miR-320c | 2:6861913( | A | G | 0.282025 | 0.084201 | 0.014395 | 4.94E-09 | trans |
| rs1155426 | miR-320c | 2:6862182/ | T | C | 0.282037 | 0.083994 | 0.014394 | 5.36E-09 | trans |
| rs1156629: | miR-320c | 2:6862663: | T | G | 0.547487 | 0.12504 | 0.013146 | 1.88E-21 | trans |
| rs1158434: | miR-320c | 2:6863552/ | G | C | 0.551072 | 0.128841 | 0.013428 | 8.41E-22 | trans |
| rs1164012! | miR-320c | 2:6862562! | C | T | 0.545777 | 0.123739 | 0.013078 | 3.04E-21 | trans |
| rs1189593( | miR-320c | 2:6861908: | A | G | 0.282175 | 0.084324 | 0.0144 | 4.75E-09 | trans |
| rs1271364: | miR-320c | 2:6860665( | G | A | 0.282933 | 0.084921 | 0.014456 | 4.24E-09 | trans |
| rs1300495: | miR-320c | 2:6858081! | G | A | 0.306559 | 0.0811 | 0.0144 | 1.78E-08 | trans |
| rs1300519( | miR-320c | 2:6863887: | A | G | 0.702186 | 0.120365 | 0.014802 | 4.24E-16 | trans |
| rs1300592: | miR-320c | 2:6862999( | T | A | 0.701261 | 0.117101 | 0.014568 | 9.09E-16 | trans |
| rs1300974: | miR-320c | 2:6858443: | C | A | 0.678986 | 0.104041 | 0.014538 | 8.27E-13 | trans |
| rs1301028! | miR-320c | 2:6862184: | G | T | 0.683506 | 0.11756 | 0.013906 | 2.82E-17 | trans |
| rs1301077/ | miR-320c | 2:6863908: | C | T | 0.702339 | 0.120275 | 0.014806 | 4.54E-16 | trans |
| rs1301107! | miR-320c | 2:6863931: | A | C | 0.700844 | 0.120705 | 0.014831 | 3.99E-16 | trans |
| rs1301143( | miR-320c | 2:6863937! | C | T | 0.702382 | 0.12029 | 0.014808 | 4.55E-16 | trans |
| rs1301157: | miR-320c | 2:6863927: | G | A | 0.702349 | 0.120281 | 0.014807 | 4.55E-16 | trans |
| rs1302892( | miR-320c | 2:6862991( | C | T | 0.552648 | 0.125024 | 0.013229 | 3.37E-21 | trans |
| rs1302897: | miR-320c | 2:6863003( | A | G | 0.54849 | 0.125793 | 0.013188 | 1.45E-21 | trans |
| rs1303482! | miR-320c | 2:6863029: | C | G | 0.552715 | 0.125074 | 0.013232 | 3.32E-21 | trans |
| rs1343065/ | miR-320c | 2:6858771( | C | G | 0.699397 | 0.114048 | 0.014328 | 1.72E-15 | trans |
| rs1373198 | miR-320c | 2:6858798: | A | T | 0.281586 | 0.081867 | 0.014546 | 1.82E-08 | trans |
| rs1373200 | miR-320c | 2:6862265( | C | T | 0.547142 | 0.122067 | 0.012943 | 4.05E-21 | trans |
| rs1382899: | miR-320c | 2:6863782! | A | G | 0.702614 | 0.12037 | 0.014819 | 4.56E-16 | trans |
| rs1390688: | miR-320c | 2:6862773! | C | T | 0.69721 | 0.116486 | 0.01443 | 6.88E-16 | trans |
| rs1397159: | miR-320c | 2:6863425( | T | G | 0.705552 | 0.121472 | 0.014948 | 4.42E-16 | trans |
| rs1422402: | miR-320c | 2:6862752: | A | G | 0.698498 | 0.116782 | 0.014467 | 6.91E-16 | trans |
| rs1442589( | miR-320c | 2:6863473: | G | T | 0.701896 | 0.119955 | 0.014773 | 4.67E-16 | trans |
| rs1455505! | miR-320c | 2:6862832! | G | A | 0.556221 | 0.127266 | 0.013334 | 1.36E-21 | trans |
| rs1474969( | miR-320c | 2:6862568( | G | A | 0.546544 | 0.123927 | 0.013098 | 3.04E-21 | trans |
| rs1484530! | miR-320c | 2:6863547: | A | G | 0.70445 | 0.121181 | 0.014861 | 3.51E-16 | trans |
| rs1486061: | miR-320c | 2:6862571: | T | G | 0.54743 | 0.124223 | 0.013123 | 2.90E-21 | trans |
| rs1542135 | miR-320c | 2:6862084( | C | T | 0.69752 | 0.113978 | 0.014151 | 8.00E-16 | trans |
| rs1542136 | miR-320c | 2:6862093( | C | A | 0.697594 | 0.113984 | 0.014155 | 8.12E-16 | trans |
| rs1753019: | miR-320c | 2:6862133( | G | T | 0.683745 | 0.117647 | 0.013918 | 2.84E-17 | trans |
| rs1760149( | miR-320c | 2:6861171: | A | G | 0.69807 | 0.113768 | 0.014157 | 9.29E-16 | trans |
| rs1760168: | miR-320c | 2:6861589! | G | A | 0.697578 | 0.113768 | 0.014163 | 9.51E-16 | trans |
| rs1760209: | miR-320c | 2:6862041( | T | A | 0.697529 | 0.113981 | 0.014152 | 8.00E-16 | trans |
| rs1867313 | miR-320c | 2:6862365: | T | C | 0.282087 | 0.083946 | 0.014392 | 5.44E-09 | trans |
| rs2070171 | miR-320c | 2:6861371( | G | A | 0.698024 | 0.113757 | 0.014157 | 9.33E-16 | trans |
| rs2165919 | miR-320c | 2:6861413: | C | T | 0.281408 | 0.08472 | 0.014422 | 4.25E-09 | trans |
| rs2165920 | miR-320c | 2:6862223! | T | C | 0.552012 | 0.121089 | 0.013039 | 1.60E-20 | trans |

| | | | | | | | | | |
|---|---|---|---|---|---|---|---|---|---|
| rs2241946 | miR-320c | 2:6861565⁴ | T | C | 0.698076 | 0.113933 | 0.014163 | 8.66E-16 | trans |
| rs2241947 | miR-320c | 2:6861539! | C | T | 0.697988 | 0.113809 | 0.014157 | 9.05E-16 | trans |
| rs3419682; | miR-320c | 2:6862142; | G | A | 0.697518 | 0.113978 | 0.014151 | 7.99E-16 | trans |
| rs3426707; | miR-320c | 2:6859915⁴ | T | C | 0.282465 | 0.083992 | 0.014438 | 5.98E-09 | trans |
| rs3485223; | miR-320c | 2:6863859! | C | G | 0.550658 | 0.128801 | 0.013431 | 8.81E-22 | trans |
| rs3732044 | miR-320c | 2:6862391! | T | C | 0.545562 | 0.123558 | 0.013069 | 3.24E-21 | trans |
| rs4610041 | miR-320c | 2:6862628( | C | T | 0.550745 | 0.127235 | 0.013226 | 6.58E-22 | trans |
| rs4671208 | miR-320c | 2:6860079! | A | G | 0.281916 | 0.083393 | 0.014429 | 7.48E-09 | trans |
| rs4854536 | miR-320c | 2:6863186! | T | A | 0.283938 | 0.086515 | 0.014463 | 2.20E-09 | trans |
| rs5618821; | miR-320c | 2:6861163; | T | C | 0.698075 | 0.113768 | 0.014157 | 9.29E-16 | trans |
| rs5628470( | miR-320c | 2:6863137; | G | A | 0.697508 | 0.117453 | 0.014477 | 4.93E-16 | trans |
| rs5633714; | miR-320c | 2:6864249( | G | A | 0.733417 | 0.119147 | 0.015407 | 1.05E-14 | trans |
| rs5690424! | miR-320c | 2:6863847; | T | C | 0.550669 | 0.128797 | 0.01343 | 8.80E-22 | trans |
| rs5808991! | miR-320c | 2:6863966( | G | C | 0.734634 | 0.121723 | 0.015509 | 4.20E-15 | trans |
| rs5888448( | miR-320c | 2:6864534; | G | A | 0.734214 | 0.118672 | 0.015462 | 1.65E-14 | trans |
| rs5933292( | miR-320c | 2:6863963; | G | A | 0.73529 | 0.1218 | 0.015551 | 4.79E-15 | trans |
| rs5999861( | miR-320c | 2:6864677; | G | T | 0.731295 | 0.112933 | 0.015457 | 2.75E-13 | trans |
| rs6000716! | miR-320c | 2:6863963( | C | T | 0.735163 | 0.121878 | 0.015549 | 4.56E-15 | trans |
| rs6087844; | miR-320c | 2:6858211! | A | G | 0.538067 | 0.11583 | 0.013466 | 7.86E-18 | trans |
| rs6214389! | miR-320c | 2:6860455; | C | T | 0.698307 | 0.113676 | 0.014176 | 1.06E-15 | trans |
| rs6214392! | miR-320c | 2:6863835! | C | G | 0.700378 | 0.121403 | 0.01479 | 2.25E-16 | trans |
| rs6214393! | miR-320c | 2:6864009( | T | C | 0.59325 | 0.11934 | 0.013799 | 5.22E-18 | trans |
| rs6214393( | miR-320c | 2:6864020; | A | G | 0.593308 | 0.119306 | 0.0138 | 5.36E-18 | trans |
| rs6214393; | miR-320c | 2:6864038; | G | C | 0.593598 | 0.119834 | 0.013795 | 3.72E-18 | trans |
| rs6214393! | miR-320c | 2:6864041! | A | G | 0.593599 | 0.11983 | 0.013795 | 3.74E-18 | trans |
| rs6214393! | miR-320c | 2:6864062; | T | C | 0.733261 | 0.119179 | 0.015398 | 9.95E-15 | trans |
| rs6419643 | miR-320c | 2:6863126( | T | G | 0.548826 | 0.12595 | 0.013197 | 1.38E-21 | trans |
| rs6419644 | miR-320c | 2:6863182; | A | G | 0.549898 | 0.128192 | 0.013328 | 6.69E-22 | trans |
| rs6546387 | miR-320c | 2:6858758! | G | A | 0.281573 | 0.081863 | 0.014547 | 1.83E-08 | trans |
| rs6546388 | miR-320c | 2:6858948; | C | T | 0.281691 | 0.08214 | 0.014505 | 1.49E-08 | trans |
| rs6546393 | miR-320c | 2:6862757! | T | C | 0.554017 | 0.126897 | 0.013259 | 1.06E-21 | trans |
| rs6546395 | miR-320c | 2:6862927; | A | G | 0.55448 | 0.125279 | 0.013273 | 3.79E-21 | trans |
| rs6546396 | miR-320c | 2:6862930; | A | T | 0.283762 | 0.085197 | 0.014428 | 3.52E-09 | trans |
| rs6546397 | miR-320c | 2:6863036; | G | A | 0.552756 | 0.125081 | 0.013234 | 3.34E-21 | trans |
| rs6546398 | miR-320c | 2:6863041; | G | C | 0.552757 | 0.125094 | 0.013234 | 3.30E-21 | trans |
| rs6546399 | miR-320c | 2:6863064( | C | T | 0.55394 | 0.125397 | 0.013273 | 3.48E-21 | trans |
| rs6546400 | miR-320c | 2:6863064! | G | A | 0.554382 | 0.125493 | 0.013286 | 3.52E-21 | trans |
| rs6546401 | miR-320c | 2:6863087; | A | T | 0.550127 | 0.126126 | 0.013226 | 1.48E-21 | trans |
| rs6546402 | miR-320c | 2:6863130( | A | G | 0.283216 | 0.08541 | 0.014433 | 3.27E-09 | trans |
| rs6546403 | miR-320c | 2:6863226; | C | T | 0.550801 | 0.128349 | 0.013348 | 6.89E-22 | trans |
| rs6546404 | miR-320c | 2:6863263! | G | T | 0.54995 | 0.128238 | 0.013333 | 6.69E-22 | trans |
| rs6546405 | miR-320c | 2:6863311( | G | A | 0.28401 | 0.08655 | 0.014463 | 2.17E-09 | trans |
| rs6546407 | miR-320c | 2:6863492( | A | C | 0.55091 | 0.128789 | 0.013412 | 7.79E-22 | trans |
| rs6546408 | miR-320c | 2:6863504! | T | C | 0.551062 | 0.128831 | 0.013426 | 8.32E-22 | trans |
| rs6546409 | miR-320c | 2:6863587! | A | G | 0.551093 | 0.128861 | 0.013432 | 8.51E-22 | trans |
| rs6546410 | miR-320c | 2:6863610! | C | G | 0.702434 | 0.120283 | 0.014809 | 4.58E-16 | trans |
| rs6546411 | miR-320c | 2:6863769! | A | G | 0.556248 | 0.130222 | 0.013516 | 5.73E-22 | trans |
| rs6546412 | miR-320c | 2:6863771( | T | C | 0.557086 | 0.131394 | 0.013541 | 2.91E-22 | trans |
| rs6708536 | miR-320c | 2:6862726! | G | A | 0.547249 | 0.125242 | 0.013155 | 1.73E-21 | trans |
| rs6709464 | miR-320c | 2:6862222( | C | T | 0.697279 | 0.113882 | 0.014138 | 7.96E-16 | trans |
| rs6714719 | miR-320c | 2:6859377; | G | C | 0.698404 | 0.113511 | 0.014216 | 1.41E-15 | trans |
| rs6715579 | miR-320c | 2:6858392; | C | T | 0.678794 | 0.10413 | 0.014536 | 7.87E-13 | trans |
| rs6715706 | miR-320c | 2:6858399( | C | A | 0.664887 | 0.10796 | 0.01429 | 4.18E-14 | trans |

| | | | | | | | | | |
|---|---|---|---|---|---|---|---|---|---|
| rs6715909 | miR-320c | 2:6858399! | G | A | 0.664879 | 0.107953 | 0.01429 | 4.21E-14 | trans |
| rs6729555 | miR-320c | 2:6863295. | C | A | 0.54995 | 0.128279 | 0.013336 | 6.64E-22 | trans |
| rs6729686 | miR-320c | 2:6863303( | C | T | 0.700171 | 0.119056 | 0.014663 | 4.69E-16 | trans |
| rs6730974 | miR-320c | 2:6863452. | T | A | 0.287755 | 0.08875 | 0.014495 | 9.18E-10 | trans |
| rs6742200 | miR-320c | 2:6862330. | A | G | 0.54552 | 0.123535 | 0.013066 | 3.25E-21 | trans |
| rs6743970 | miR-320c | 2:6858390. | A | T | 0.660237 | 0.110494 | 0.014356 | 1.40E-14 | trans |
| rs6744216 | miR-320c | 2:6863463! | A | G | 0.284465 | 0.086696 | 0.014463 | 2.04E-09 | trans |
| rs6744306 | miR-320c | 2:6863273( | T | C | 0.549956 | 0.128249 | 0.013333 | 6.66E-22 | trans |
| rs6747545 | miR-320c | 2:6863306! | T | A | 0.549964 | 0.128276 | 0.013337 | 6.70E-22 | trans |
| rs6757938 | miR-320c | 2:6863285! | A | G | 0.549958 | 0.128259 | 0.013335 | 6.69E-22 | trans |
| rs7420773 | miR-320c | 2:6862945. | G | A | 0.28377 | 0.085197 | 0.014428 | 3.52E-09 | trans |
| rs7420775 | miR-320c | 2:6862945! | G | T | 0.703122 | 0.117641 | 0.014633 | 9.02E-16 | trans |
| rs7420782 | miR-320c | 2:6862955! | G | T | 0.283742 | 0.085395 | 0.014431 | 3.27E-09 | trans |
| rs7421228 | miR-320c | 2:6863012. | G | T | 0.55268 | 0.125054 | 0.013231 | 3.34E-21 | trans |
| rs7451452. | miR-320c | 2:6863755( | T | G | 0.702552 | 0.120344 | 0.014815 | 4.55E-16 | trans |
| rs7562439 | miR-320c | 2:6858256! | C | T | 0.6646 | 0.107793 | 0.014294 | 4.66E-14 | trans |
| rs7562889( | miR-320c | 2:6862547. | T | C | 0.285916 | 0.084406 | 0.014444 | 5.10E-09 | trans |
| rs7572299 | miR-320c | 2:6858124( | T | A | 0.538033 | 0.115531 | 0.013469 | 9.70E-18 | trans |
| rs7574592 | miR-320c | 2:6859766. | T | G | 0.281934 | 0.083326 | 0.014428 | 7.69E-09 | trans |
| rs7578602 | miR-320c | 2:6863786! | G | C | 0.556806 | 0.131453 | 0.013547 | 2.91E-22 | trans |
| rs7581872 | miR-320c | 2:6858098. | A | C | 0.537959 | 0.115444 | 0.013469 | 1.02E-17 | trans |
| rs7590450 | miR-320c | 2:6862856. | A | G | 0.549532 | 0.12574 | 0.013186 | 1.49E-21 | trans |
| rs7592526 | miR-320c | 2:6858352! | A | C | 0.538007 | 0.115839 | 0.013467 | 7.86E-18 | trans |
| rs7597167 | miR-320c | 2:6861294( | C | T | 0.698051 | 0.11378 | 0.014157 | 9.21E-16 | trans |
| rs7598360 | miR-320c | 2:6863092! | A | G | 0.553166 | 0.128065 | 0.01326 | 4.56E-22 | trans |
| rs7605387 | miR-320c | 2:6863255( | A | G | 0.703499 | 0.119101 | 0.014748 | 6.72E-16 | trans |
| rs7608429 | miR-320c | 2:6858083( | G | T | 0.678719 | 0.103457 | 0.014551 | 1.16E-12 | trans |
| rs7800275. | miR-320c | 2:6862543( | A | G | 0.546309 | 0.123534 | 0.013063 | 3.18E-21 | trans |
| rs8028813! | miR-320c | 2:6862547! | A | G | 0.548418 | 0.12585 | 0.013149 | 1.06E-21 | trans |
| rs8761 | miR-320c | 2:6862426( | G | A | 0.545692 | 0.123625 | 0.013074 | 3.20E-21 | trans |
| rs880668 | miR-320c | 2:6859258. | T | C | 0.280228 | 0.082308 | 0.014516 | 1.43E-08 | trans |
| rs964934 | miR-320c | 2:6861187. | G | A | 0.281023 | 0.083883 | 0.014422 | 6.02E-09 | trans |
| rs964935 | miR-320c | 2:6861228. | G | A | 0.281126 | 0.083917 | 0.014425 | 5.97E-09 | trans |
| rs9653552 | miR-320c | 2:6864396( | G | A | 0.593458 | 0.11996 | 0.013806 | 3.65E-18 | trans |
| rs1017438( | miR-320d | 2:6859646! | C | T | 0.685243 | 0.116538 | 0.014293 | 3.54E-16 | trans |
| rs1049616! | miR-320d | 2:6862008! | C | T | 0.697496 | 0.116733 | 0.014478 | 7.45E-16 | trans |
| rs1050181 | miR-320d | 2:6862295. | G | C | 0.544992 | 0.121982 | 0.01335 | 6.39E-20 | trans |
| rs1156629. | miR-320d | 2:6862663. | T | G | 0.547487 | 0.123554 | 0.013468 | 4.55E-20 | trans |
| rs1158434. | miR-320d | 2:6863552. | G | C | 0.551072 | 0.126979 | 0.013759 | 2.73E-20 | trans |
| rs1164012! | miR-320d | 2:6862562! | C | T | 0.545777 | 0.122475 | 0.013397 | 6.14E-20 | trans |
| rs1271364. | miR-320d | 2:6860665( | G | A | 0.282933 | 0.080783 | 0.014805 | 4.85E-08 | trans |
| rs1300519! | miR-320d | 2:6863887. | A | G | 0.702186 | 0.121079 | 0.015151 | 1.33E-15 | trans |
| rs1300592. | miR-320d | 2:6862999! | T | A | 0.701261 | 0.118515 | 0.014908 | 1.87E-15 | trans |
| rs1300974. | miR-320d | 2:6858443. | C | A | 0.678986 | 0.108867 | 0.014864 | 2.41E-13 | trans |
| rs1301028! | miR-320d | 2:6862184. | G | T | 0.683506 | 0.116353 | 0.014241 | 3.08E-16 | trans |
| rs1301077. | miR-320d | 2:6863908. | C | T | 0.702339 | 0.12101 | 0.015155 | 1.41E-15 | trans |
| rs1301107! | miR-320d | 2:6863931. | A | C | 0.700844 | 0.12145 | 0.01518 | 1.24E-15 | trans |
| rs1301143( | miR-320d | 2:6863937! | C | T | 0.702382 | 0.121014 | 0.015157 | 1.42E-15 | trans |
| rs1301157. | miR-320d | 2:6863927. | G | A | 0.702349 | 0.121015 | 0.015156 | 1.41E-15 | trans |
| rs1302892( | miR-320d | 2:6862991( | C | T | 0.552648 | 0.124158 | 0.01355 | 5.05E-20 | trans |
| rs1302897. | miR-320d | 2:6863003! | A | G | 0.54849 | 0.124504 | 0.01351 | 3.10E-20 | trans |
| rs1303482! | miR-320d | 2:6863029. | C | G | 0.552715 | 0.124211 | 0.013553 | 4.97E-20 | trans |
| rs1343065. | miR-320d | 2:6858771( | C | G | 0.699397 | 0.117257 | 0.014655 | 1.23E-15 | trans |

| | | | | | | | | | |
|---|---|---|---|---|---|---|---|---|---|
| rs1373200 | miR-320d | 2:6862265 | C | T | 0.547142 | 0.121474 | 0.013255 | 4.99E-20 | trans |
| rs1382899 | miR-320d | 2:6863782 | A | G | 0.702614 | 0.121213 | 0.015168 | 1.33E-15 | trans |
| rs1390688 | miR-320d | 2:6862773 | C | T | 0.69721 | 0.117637 | 0.014768 | 1.64E-15 | trans |
| rs1397159 | miR-320d | 2:6863425 | T | G | 0.705552 | 0.122919 | 0.015297 | 9.32E-16 | trans |
| rs1422402 | miR-320d | 2:6862752 | A | G | 0.698498 | 0.118434 | 0.014804 | 1.24E-15 | trans |
| rs1442589 | miR-320d | 2:6863473 | G | T | 0.701896 | 0.120911 | 0.01512 | 1.28E-15 | trans |
| rs1455505 | miR-320d | 2:6862832 | G | A | 0.556221 | 0.126528 | 0.013657 | 1.95E-20 | trans |
| rs1474969 | miR-320d | 2:6862568 | G | A | 0.546544 | 0.122663 | 0.013417 | 6.13E-20 | trans |
| rs1484530 | miR-320d | 2:6863547 | A | G | 0.70445 | 0.121983 | 0.01521 | 1.06E-15 | trans |
| rs1486061 | miR-320d | 2:6862571 | T | G | 0.54743 | 0.123039 | 0.013442 | 5.53E-20 | trans |
| rs1542135 | miR-320d | 2:6862084 | C | T | 0.69752 | 0.116748 | 0.014477 | 7.35E-16 | trans |
| rs1542136 | miR-320d | 2:6862093 | C | A | 0.697594 | 0.116763 | 0.014481 | 7.42E-16 | trans |
| rs1753019 | miR-320d | 2:6862133 | G | T | 0.683745 | 0.116731 | 0.014252 | 2.61E-16 | trans |
| rs1760149 | miR-320d | 2:6861171 | A | G | 0.69807 | 0.117074 | 0.014481 | 6.23E-16 | trans |
| rs1760168 | miR-320d | 2:6861589 | G | A | 0.697578 | 0.11699 | 0.014486 | 6.71E-16 | trans |
| rs1760209 | miR-320d | 2:6862041 | T | A | 0.697529 | 0.11675 | 0.014477 | 7.35E-16 | trans |
| rs2070171 | miR-320d | 2:6861371 | G | A | 0.698024 | 0.117049 | 0.014481 | 6.31E-16 | trans |
| rs2165919 | miR-320d | 2:6861413 | C | T | 0.281408 | 0.080688 | 0.01477 | 4.68E-08 | trans |
| rs2165920 | miR-320d | 2:6862223 | T | C | 0.552012 | 0.122188 | 0.013347 | 5.44E-20 | trans |
| rs2241946 | miR-320d | 2:6861565 | T | C | 0.698076 | 0.117177 | 0.014487 | 6.03E-16 | trans |
| rs2241947 | miR-320d | 2:6861539 | C | T | 0.697988 | 0.117037 | 0.014481 | 6.35E-16 | trans |
| rs3419682 | miR-320d | 2:6862142 | G | A | 0.697518 | 0.116748 | 0.014477 | 7.35E-16 | trans |
| rs3485223 | miR-320d | 2:6863859 | C | G | 0.550658 | 0.126936 | 0.013761 | 2.86E-20 | trans |
| rs3732044 | miR-320d | 2:6862391 | T | C | 0.545562 | 0.122287 | 0.013387 | 6.57E-20 | trans |
| rs4610041 | miR-320d | 2:6862628 | C | T | 0.550745 | 0.125806 | 0.01355 | 1.62E-20 | trans |
| rs4854536 | miR-320d | 2:6863186 | T | A | 0.283938 | 0.080854 | 0.014816 | 4.84E-08 | trans |
| rs5618821 | miR-320d | 2:6861163 | T | C | 0.698075 | 0.117075 | 0.014481 | 6.23E-16 | trans |
| rs5628470 | miR-320d | 2:6863137 | G | A | 0.697508 | 0.118242 | 0.014817 | 1.46E-15 | trans |
| rs5633714 | miR-320d | 2:6864249 | G | A | 0.733417 | 0.118238 | 0.015775 | 6.62E-14 | trans |
| rs5690424 | miR-320d | 2:6863847 | T | C | 0.550669 | 0.126931 | 0.01376 | 2.85E-20 | trans |
| rs5808991 | miR-320d | 2:6863966 | G | C | 0.734634 | 0.120732 | 0.015879 | 2.89E-14 | trans |
| rs5888448 | miR-320d | 2:6864534 | G | A | 0.734214 | 0.117789 | 0.01583 | 1.00E-13 | trans |
| rs5933292 | miR-320d | 2:6863963 | G | A | 0.73529 | 0.120638 | 0.015923 | 3.56E-14 | trans |
| rs5999861 | miR-320d | 2:6864677 | G | T | 0.731295 | 0.111989 | 0.015825 | 1.47E-12 | trans |
| rs6000716 | miR-320d | 2:6863963 | C | T | 0.735163 | 0.12067 | 0.015921 | 3.48E-14 | trans |
| rs6087844 | miR-320d | 2:6858211 | A | G | 0.538067 | 0.116986 | 0.013782 | 2.10E-17 | trans |
| rs6214389 | miR-320d | 2:6860455 | C | T | 0.698307 | 0.116699 | 0.014501 | 8.42E-16 | trans |
| rs6214392 | miR-320d | 2:6863835 | C | G | 0.700378 | 0.121704 | 0.01514 | 9.10E-16 | trans |
| rs6214393 | miR-320d | 2:6864009 | T | C | 0.59325 | 0.118698 | 0.01413 | 4.45E-17 | trans |
| rs6214393 | miR-320d | 2:6864020 | A | G | 0.593308 | 0.118688 | 0.014131 | 4.49E-17 | trans |
| rs6214393 | miR-320d | 2:6864038 | G | C | 0.593598 | 0.119156 | 0.014126 | 3.30E-17 | trans |
| rs6214393 | miR-320d | 2:6864041 | A | G | 0.593599 | 0.11915 | 0.014126 | 3.32E-17 | trans |
| rs6214393 | miR-320d | 2:6864062 | T | C | 0.733261 | 0.118334 | 0.015765 | 6.09E-14 | trans |
| rs6419643 | miR-320d | 2:6863126 | T | G | 0.548826 | 0.124619 | 0.01352 | 3.04E-20 | trans |
| rs6419644 | miR-320d | 2:6863182 | A | G | 0.549898 | 0.126235 | 0.013656 | 2.38E-20 | trans |
| rs6546393 | miR-320d | 2:6862757 | T | C | 0.554017 | 0.126078 | 0.013581 | 1.64E-20 | trans |
| rs6546395 | miR-320d | 2:6862927 | A | G | 0.55448 | 0.124433 | 0.013595 | 5.55E-20 | trans |
| rs6546397 | miR-320d | 2:6863036 | G | A | 0.552756 | 0.124221 | 0.013555 | 4.99E-20 | trans |
| rs6546398 | miR-320d | 2:6863041 | G | C | 0.552757 | 0.124235 | 0.013555 | 4.93E-20 | trans |
| rs6546399 | miR-320d | 2:6863064 | C | T | 0.55394 | 0.124567 | 0.013595 | 5.06E-20 | trans |
| rs6546400 | miR-320d | 2:6863064 | G | A | 0.554382 | 0.124654 | 0.013607 | 5.16E-20 | trans |
| rs6546401 | miR-320d | 2:6863087 | A | T | 0.550127 | 0.124835 | 0.013549 | 3.15E-20 | trans |
| rs6546403 | miR-320d | 2:6863226 | C | T | 0.550801 | 0.126407 | 0.013677 | 2.41E-20 | trans |

| | | | | | | | | | |
|---|---|---|---|---|---|---|---|---|---|
| rs6546404 | miR-320d | 2:6863263! | G | T | 0.54995 | 0.126284 | 0.013661 | 2.37E-20 | trans |
| rs6546405 | miR-320d | 2:6863311( | G | A | 0.28401 | 0.080885 | 0.014816 | 4.78E-08 | trans |
| rs6546407 | miR-320d | 2:6863492( | A | C | 0.55091 | 0.126901 | 0.013742 | 2.59E-20 | trans |
| rs6546408 | miR-320d | 2:6863504{ | T | C | 0.551062 | 0.126965 | 0.013756 | 2.71E-20 | trans |
| rs6546409 | miR-320d | 2:6863587! | A | G | 0.551093 | 0.126997 | 0.013762 | 2.76E-20 | trans |
| rs6546410 | miR-320d | 2:6863610! | C | G | 0.702434 | 0.12118 | 0.015157 | 1.30E-15 | trans |
| rs6546411 | miR-320d | 2:6863769! | A | G | 0.556248 | 0.128784 | 0.013847 | 1.40E-20 | trans |
| rs6546412 | miR-320d | 2:6863771( | T | C | 0.557086 | 0.129709 | 0.013873 | 8.80E-21 | trans |
| rs6708536 | miR-320d | 2:6862726{ | G | A | 0.547249 | 0.123687 | 0.013477 | 4.42E-20 | trans |
| rs6709464 | miR-320d | 2:6862222( | C | T | 0.697279 | 0.11635 | 0.014465 | 8.71E-16 | trans |
| rs6714719 | miR-320d | 2:6859377: | G | C | 0.698404 | 0.117036 | 0.01454 | 8.33E-16 | trans |
| rs6715579 | miR-320d | 2:6858392: | C | T | 0.678794 | 0.108857 | 0.014863 | 2.41E-13 | trans |
| rs6715706 | miR-320d | 2:6858399( | C | A | 0.664887 | 0.108852 | 0.014624 | 9.82E-14 | trans |
| rs6715909 | miR-320d | 2:6858399! | G | A | 0.664879 | 0.108849 | 0.014625 | 9.85E-14 | trans |
| rs6729555 | miR-320d | 2:6863295: | C | A | 0.54995 | 0.126323 | 0.013664 | 2.36E-20 | trans |
| rs6729686 | miR-320d | 2:6863303( | C | T | 0.700171 | 0.119939 | 0.015008 | 1.33E-15 | trans |
| rs6730974 | miR-320d | 2:6863452: | T | A | 0.287755 | 0.083307 | 0.014849 | 2.02E-08 | trans |
| rs6742200 | miR-320d | 2:6862330: | A | G | 0.54552 | 0.122264 | 0.013385 | 6.58E-20 | trans |
| rs6743970 | miR-320d | 2:6858390: | A | T | 0.660237 | 0.111589 | 0.014692 | 3.07E-14 | trans |
| rs6744216 | miR-320d | 2:6863463{ | A | G | 0.284465 | 0.081135 | 0.014816 | 4.34E-08 | trans |
| rs6744306 | miR-320d | 2:6863273( | T | C | 0.549956 | 0.126294 | 0.013662 | 2.36E-20 | trans |
| rs6747545 | miR-320d | 2:6863306! | T | A | 0.549964 | 0.126319 | 0.013665 | 2.38E-20 | trans |
| rs6757938 | miR-320d | 2:6863285{ | A | G | 0.549958 | 0.126303 | 0.013663 | 2.38E-20 | trans |
| rs7420775 | miR-320d | 2:6862945{ | G | T | 0.703122 | 0.119074 | 0.014975 | 1.84E-15 | trans |
| rs7421228 | miR-320d | 2:6863012( | G | T | 0.55268 | 0.124188 | 0.013552 | 5.01E-20 | trans |
| rs7451452: | miR-320d | 2:6863755( | T | G | 0.702552 | 0.121207 | 0.015164 | 1.31E-15 | trans |
| rs7562439 | miR-320d | 2:6858256! | C | T | 0.6646 | 0.108725 | 0.014628 | 1.07E-13 | trans |
| rs7572299 | miR-320d | 2:6858124( | T | A | 0.538033 | 0.116665 | 0.013785 | 2.60E-17 | trans |
| rs7578602 | miR-320d | 2:6863786{ | G | C | 0.556806 | 0.129683 | 0.01388 | 9.33E-21 | trans |
| rs7581872 | miR-320d | 2:6858098: | A | C | 0.537959 | 0.116574 | 0.013785 | 2.75E-17 | trans |
| rs7590450 | miR-320d | 2:6862856: | A | G | 0.549532 | 0.124555 | 0.013508 | 2.94E-20 | trans |
| rs7592526 | miR-320d | 2:6858352{ | A | C | 0.538007 | 0.116969 | 0.013783 | 2.14E-17 | trans |
| rs7597167 | miR-320d | 2:6861294( | C | T | 0.698051 | 0.117082 | 0.014481 | 6.20E-16 | trans |
| rs7598360 | miR-320d | 2:6863092! | A | G | 0.553166 | 0.126722 | 0.013585 | 1.08E-20 | trans |
| rs7600986 | miR-320d | 2:6863692: | A | T | 0.285029 | 0.081147 | 0.014801 | 4.19E-08 | trans |
| rs7605387 | miR-320d | 2:6863255( | A | G | 0.703499 | 0.120593 | 0.015093 | 1.35E-15 | trans |
| rs7608429 | miR-320d | 2:6858083( | G | T | 0.678719 | 0.10826 | 0.014878 | 3.42E-13 | trans |
| rs7800275: | miR-320d | 2:6862543( | A | G | 0.546309 | 0.122332 | 0.013381 | 6.14E-20 | trans |
| rs8028813! | miR-320d | 2:6862547{ | A | G | 0.548418 | 0.124394 | 0.013471 | 2.60E-20 | trans |
| rs8761 | miR-320d | 2:6862426( | G | A | 0.545692 | 0.12236 | 0.013392 | 6.45E-20 | trans |
| rs9653552 | miR-320d | 2:6864396( | G | A | 0.593458 | 0.119124 | 0.014138 | 3.58E-17 | trans |
| rs1214700{ | miR-329-3p | 14:101186: | C | T | 0.223221 | -0.211263 | 0.033195 | 1.96E-10 | cis |
| rs4905994 | miR-329-3p | 14:101154( | T | C | 0.329146 | -0.196223 | 0.030175 | 7.88E-11 | cis |
| rs7142683 | miR-329-3p | 14:101158{ | A | G | 0.290748 | -0.215153 | 0.031116 | 4.69E-12 | cis |
| rs7149242 | miR-329-3p | 14:101159( | T | G | 0.288371 | -0.214077 | 0.031029 | 5.23E-12 | cis |
| rs7155375 | miR-329-3p | 14:101188: | C | T | 0.635995 | 0.169832 | 0.028482 | 2.48E-09 | cis |
| rs9671394 | miR-329-3p | 14:101159( | C | T | 0.284232 | -0.216002 | 0.031471 | 6.72E-12 | cis |
| rs1013035( | miR-376a-3 | 14:101149! | T | C | 0.112665 | -0.248038 | 0.042734 | 6.46E-09 | cis |
| rs1013687( | miR-376a-3 | 14:101202( | G | T | 0.493366 | 0.146822 | 0.02648 | 2.94E-08 | cis |
| rs1015151! | miR-376a-3 | 14:101175: | G | A | 0.806743 | 0.185623 | 0.03204 | 6.89E-09 | cis |
| rs1087352( | miR-376a-3 | 14:101175! | G | A | 0.809826 | 0.1875 | 0.032416 | 7.29E-09 | cis |
| rs1116059( | miR-376a-3 | 14:101049! | C | T | 0.741662 | 0.208575 | 0.029004 | 6.42E-13 | cis |
| rs1184948! | miR-376a-3 | 14:101037: | C | G | 0.733365 | 0.196249 | 0.02878 | 9.17E-12 | cis |

| | | | | | | | | | |
|---|---|---|---|---|---|---|---|---|---|
| rs1185095 | miR-376a- | 14:101056 | G | T | 0.743981 | 0.204733 | 0.02887 | 1.33E-12 | cis |
| rs1190815 | miR-376a- | 14:101228 | G | A | 0.798481 | 0.181604 | 0.031798 | 1.12E-08 | cis |
| rs1190817 | miR-376a- | 14:101229 | T | C | 0.798148 | 0.181782 | 0.032039 | 1.40E-08 | cis |
| rs1190818 | miR-376a- | 14:101230 | T | G | 0.818683 | 0.197147 | 0.033188 | 2.84E-09 | cis |
| rs1190850 | miR-376a- | 14:101064 | A | G | 0.314302 | -0.185854 | 0.028131 | 3.93E-11 | cis |
| rs1190852 | miR-376a- | 14:101064 | A | G | 0.310295 | -0.184111 | 0.027965 | 4.59E-11 | cis |
| rs1190856 | miR-376a- | 14:101065 | T | C | 0.310244 | -0.183617 | 0.027944 | 5.00E-11 | cis |
| rs1190857 | miR-376a- | 14:101066 | G | T | 0.310273 | -0.183328 | 0.02795 | 5.41E-11 | cis |
| rs1190862 | miR-376a- | 14:101067 | T | G | 0.309628 | -0.182853 | 0.027749 | 4.41E-11 | cis |
| rs1190867 | miR-376a- | 14:101069 | G | T | 0.316788 | -0.182857 | 0.028117 | 7.85E-11 | cis |
| rs1190872 | miR-376a- | 14:101069 | A | G | 0.310177 | -0.182553 | 0.027916 | 6.18E-11 | cis |
| rs1190903 | miR-376a- | 14:101096 | C | T | 0.772566 | 0.204071 | 0.031002 | 4.63E-11 | cis |
| rs1190904 | miR-376a- | 14:101096 | A | G | 0.772596 | 0.20411 | 0.031 | 4.57E-11 | cis |
| rs1190906 | miR-376a- | 14:101099 | T | C | 0.772138 | 0.199604 | 0.030909 | 1.06E-10 | cis |
| rs1190907 | miR-376a- | 14:101099 | A | G | 0.772847 | 0.200134 | 0.030958 | 1.01E-10 | cis |
| rs1190909 | miR-376a- | 14:101101 | G | A | 0.775531 | 0.198097 | 0.031378 | 2.73E-10 | cis |
| rs1214700 | miR-376a- | 14:101186 | C | T | 0.223221 | -0.26357 | 0.030488 | 5.37E-18 | cis |
| rs1214784 | miR-376a- | 14:101144 | C | T | 0.870989 | 0.250029 | 0.038416 | 7.59E-11 | cis |
| rs1243295 | miR-376a- | 14:101205 | G | A | 0.231663 | -0.187686 | 0.03061 | 8.71E-10 | cis |
| rs1243449 | miR-376a- | 14:101179 | A | G | 0.18959 | -0.315322 | 0.032737 | 5.87E-22 | cis |
| rs1243613 | miR-376a- | 14:101031 | C | T | 0.725015 | 0.184446 | 0.028665 | 1.24E-10 | cis |
| rs1258812 | miR-376a- | 14:101162 | T | C | 0.893709 | 0.247901 | 0.042936 | 7.75E-09 | cis |
| rs1259093 | miR-376a- | 14:101163 | A | T | 0.893649 | 0.248131 | 0.042939 | 7.53E-09 | cis |
| rs1288162 | miR-376a- | 14:101146 | A | G | 0.912196 | 0.263957 | 0.047627 | 2.99E-08 | cis |
| rs1289379 | miR-376a- | 14:101178 | C | G | 0.190813 | -0.315723 | 0.032764 | 5.61E-22 | cis |
| rs1289760 | miR-376a- | 14:101182 | C | T | 0.190452 | -0.312925 | 0.03257 | 7.42E-22 | cis |
| rs1555405 | miR-376a- | 14:101176 | G | A | 0.749075 | 0.214226 | 0.028973 | 1.42E-13 | cis |
| rs1709962 | miR-376a- | 14:101173 | A | C | 0.783449 | 0.171531 | 0.030963 | 3.03E-08 | cis |
| rs1757758 | miR-376a- | 14:101189 | C | T | 0.827094 | 0.245969 | 0.033544 | 2.25E-13 | cis |
| rs1802710 | miR-376a- | 14:101200 | C | T | 0.493406 | 0.147537 | 0.026533 | 2.69E-08 | cis |
| rs1888581 | miR-376a- | 14:101042 | A | C | 0.735363 | 0.193551 | 0.028712 | 1.57E-11 | cis |
| rs2295659 | miR-376a- | 14:101141 | G | T | 0.857995 | 0.212033 | 0.036577 | 6.75E-09 | cis |
| rs2400940 | miR-376a- | 14:101224 | G | A | 0.185552 | -0.192104 | 0.032498 | 3.39E-09 | cis |
| rs2837918 | miR-376a- | 14:101168 | G | T | 0.836434 | 0.286071 | 0.034477 | 1.06E-16 | cis |
| rs3470057 | miR-376a- | 14:101173 | C | T | 0.892725 | 0.299403 | 0.041457 | 5.12E-13 | cis |
| rs4905994 | miR-376a- | 14:101154 | T | C | 0.329146 | -0.226581 | 0.027776 | 3.42E-16 | cis |
| rs4905999 | miR-376a- | 14:101184 | A | G | 0.181905 | -0.320062 | 0.03331 | 7.36E-22 | cis |
| rs6575791 | miR-376a- | 14:101029 | G | A | 0.719841 | 0.185562 | 0.02866 | 9.50E-11 | cis |
| rs6575794 | miR-376a- | 14:101039 | C | T | 0.734189 | 0.193128 | 0.028832 | 2.11E-11 | cis |
| rs7140556 | miR-376a- | 14:101036 | C | A | 0.723818 | 0.184412 | 0.028603 | 1.14E-10 | cis |
| rs7140792 | miR-376a- | 14:101172 | C | T | 0.892517 | 0.30083 | 0.041494 | 4.17E-13 | cis |
| rs7142683 | miR-376a- | 14:101158 | A | G | 0.290748 | -0.253704 | 0.0286 | 7.27E-19 | cis |
| rs7143357 | miR-376a- | 14:101054 | T | C | 0.739636 | 0.206342 | 0.029036 | 1.19E-12 | cis |
| rs7144375 | miR-376a- | 14:101137 | T | C | 0.866639 | 0.203834 | 0.037273 | 4.53E-08 | cis |
| rs7145610 | miR-376a- | 14:101049 | T | C | 0.740391 | 0.209451 | 0.029079 | 5.90E-13 | cis |
| rs7147982 | miR-376a- | 14:101046 | C | T | 0.731753 | 0.190634 | 0.028507 | 2.27E-11 | cis |
| rs7149242 | miR-376a- | 14:101159 | T | G | 0.288371 | -0.253025 | 0.028519 | 7.17E-19 | cis |
| rs7154024 | miR-376a- | 14:101031 | G | T | 0.725015 | 0.184202 | 0.02864 | 1.26E-10 | cis |
| rs7155375 | miR-376a- | 14:101188 | C | T | 0.635995 | 0.215803 | 0.026169 | 1.63E-16 | cis |
| rs7155649 | miR-376a- | 14:101181 | C | A | 0.863608 | 0.270181 | 0.037999 | 1.16E-12 | cis |
| rs745797 | miR-376a- | 14:101141 | G | A | 0.858727 | 0.214097 | 0.036689 | 5.36E-09 | cis |
| rs7493817 | miR-376a- | 14:101038 | G | A | 0.751723 | 0.205201 | 0.029703 | 4.90E-12 | cis |
| rs8008285 | miR-376a- | 14:101146 | T | C | 0.842905 | 0.255629 | 0.035679 | 7.79E-13 | cis |

| | | | | | | | | | |
|---|---|---|---|---|---|---|---|---|---|
| rs879058 | miR-376a-3 | 14:1010699 | G | T | 0.311807 | -0.183571 | 0.027914 | 4.82E-11 | cis |
| rs879059 | miR-376a-3 | 14:1010699 | T | A | 0.31068 | -0.183077 | 0.0279 | 5.31E-11 | cis |
| rs879060 | miR-376a-3 | 14:1010698 | A | G | 0.310926 | -0.183185 | 0.027918 | 5.32E-11 | cis |
| rs911330 | miR-376a-3 | 14:1010997 | T | C | 0.774615 | 0.204929 | 0.0311 | 4.42E-11 | cis |
| rs9671394 | miR-376a-3 | 14:1011596 | C | T | 0.284232 | -0.26024 | 0.028907 | 2.20E-19 | cis |
| rs1297485 | miR-125a-5 | 19:5218915 | G | A | 0.863745 | -0.067442 | 0.01778 | 0.000149 | cis |
| rs1190904 | miR-127-3p | 14:1010960 | A | G | 0.772596 | 0.38532 | 0.088025 | 1.20E-05 | cis |
| rs1190903 | miR-127-3p | 14:1010964 | C | T | 0.772566 | 0.38519 | 0.088032 | 1.21E-05 | cis |
| rs1555405 | miR-127-3p | 14:1011767 | G | A | 0.749075 | 0.314428 | 0.082567 | 0.00014 | cis |
| rs1015151 | miR-127-3p | 14:1011757 | G | A | 0.806743 | 0.377226 | 0.090815 | 3.27E-05 | cis |
| rs1190906 | miR-127-3p | 14:1010991 | T | C | 0.772138 | 0.38048 | 0.087742 | 1.45E-05 | cis |
| rs1190909 | miR-127-3p | 14:1011012 | G | A | 0.775531 | 0.386819 | 0.089033 | 1.39E-05 | cis |
| rs911330 | miR-127-3p | 14:1010997 | T | C | 0.774615 | 0.387391 | 0.088309 | 1.15E-05 | cis |
| rs9324022 | miR-127-3p | 14:1011724 | T | C | 0.763739 | 0.333537 | 0.089301 | 0.000188 | cis |
| rs1190907 | miR-127-3p | 14:1010993 | A | G | 0.772847 | 0.381185 | 0.087881 | 1.44E-05 | cis |
| rs1087352 | miR-127-3p | 14:1011759 | G | A | 0.809826 | 0.388538 | 0.091866 | 2.34E-05 | cis |
| rs4939170 | miR-130a-3 | 11:5772139 | C | T | 0.762033 | -0.093228 | 0.024594 | 0.00015 | cis |
| rs5624584 | miR-130a-3 | 11:5777643 | A | C | 0.784107 | 0.105795 | 0.025315 | 2.93E-05 | cis |
| rs7925491 | miR-130a-3 | 11:5776804 | C | T | 0.783849 | 0.107119 | 0.02524 | 2.20E-05 | cis |
| rs938011 | miR-130a-3 | 11:5770999 | A | C | 0.716098 | -0.081902 | 0.024073 | 0.000668 | cis |
| rs1745806 | miR-130a-3 | 11:5778607 | A | G | 0.792414 | 0.115423 | 0.025287 | 5.01E-06 | cis |
| rs5602368 | miR-130a-3 | 11:5780368 | G | A | 0.792377 | 0.114756 | 0.025291 | 5.69E-06 | cis |
| rs6681376 | miR-130a-3 | 11:5769825 | G | T | 0.719099 | -0.088551 | 0.024389 | 0.000283 | cis |
| rs7924376 | miR-130a-3 | 11:5774306 | C | A | 0.239323 | 0.094754 | 0.024494 | 0.00011 | cis |
| rs1169543 | miR-130a-3 | 11:5717172 | C | T | 0.892299 | -0.112242 | 0.033953 | 0.000947 | cis |
| rs1122917 | miR-130a-3 | 11:5772779 | G | A | 0.727084 | -0.09057 | 0.02324 | 9.73E-05 | cis |
| rs1089667 | miR-130a-3 | 11:5774112 | C | T | 0.761539 | -0.095681 | 0.024567 | 9.83E-05 | cis |
| rs1241917 | miR-130a-3 | 11:5770671 | T | C | 0.708383 | -0.088323 | 0.024102 | 0.000248 | cis |
| rs6190276 | miR-130a-3 | 11:5788075 | C | T | 0.783701 | 0.119494 | 0.02497 | 1.71E-06 | cis |
| rs6190396 | miR-130a-3 | 11:5780459 | T | C | 0.791272 | 0.113434 | 0.025291 | 7.28E-06 | cis |
| rs1160108 | miR-130a-3 | 11:5772708 | C | T | 0.71663 | -0.080692 | 0.023819 | 0.000705 | cis |
| rs3456326 | miR-130a-3 | 11:5769860 | A | T | 0.717362 | -0.086798 | 0.024348 | 0.000364 | cis |
| rs1241934 | miR-130a-3 | 11:5776760 | C | G | 0.78841 | 0.107197 | 0.025729 | 3.10E-05 | cis |
| rs5566520 | miR-130a-3 | 11:5780007 | T | C | 0.792454 | 0.115009 | 0.025275 | 5.36E-06 | cis |
| rs6190534 | miR-130a-3 | 11:5776992 | G | A | 0.784724 | 0.105792 | 0.025297 | 2.89E-05 | cis |
| rs1446103 | miR-130a-3 | 11:5761880 | C | T | 0.805715 | 0.12532 | 0.02664 | 2.55E-06 | cis |
| rs7927022 | miR-130a-3 | 11:5773392 | T | C | 0.682683 | -0.078113 | 0.023148 | 0.00074 | cis |
| rs2649663 | miR-130a-3 | 11:5735730 | G | A | 0.238562 | -0.084116 | 0.025109 | 0.000808 | cis |
| rs5592002 | miR-130a-3 | 11:5775589 | T | C | 0.785351 | 0.10476 | 0.025482 | 3.94E-05 | cis |
| rs6190401 | miR-130a-3 | 11:5781410 | G | A | 0.792325 | 0.114109 | 0.025483 | 7.54E-06 | cis |
| rs6190402 | miR-130a-3 | 11:5782355 | C | T | 0.784253 | 0.118804 | 0.025041 | 2.09E-06 | cis |
| rs3459466 | miR-130a-3 | 11:5770351 | C | T | 0.717404 | -0.085585 | 0.024309 | 0.00043 | cis |
| rs7291785 | miR-130a-3 | 11:5779612 | C | T | 0.792473 | 0.11513 | 0.025274 | 5.23E-06 | cis |
| rs7944888 | miR-130a-3 | 11:5777923 | T | C | 0.785845 | 0.107344 | 0.02537 | 2.33E-05 | cis |
| rs5605537 | miR-130a-3 | 11:5770561 | A | G | 0.71673 | -0.082212 | 0.024165 | 0.000669 | cis |
| rs2089748 | miR-130a-3 | 11:5774613 | C | T | 0.76272 | -0.093828 | 0.024577 | 0.000135 | cis |
| rs1079210 | miR-130a-3 | 11:5732815 | G | A | 0.771844 | -0.083559 | 0.024808 | 0.000756 | cis |
| rs1089667 | miR-130a-3 | 11:5773704 | A | C | 0.765249 | -0.09565 | 0.024737 | 0.00011 | cis |
| rs6190401 | miR-130a-3 | 11:5781144 | G | A | 0.792423 | 0.114142 | 0.025482 | 7.49E-06 | cis |
| rs1089667 | miR-130a-3 | 11:5773990 | T | C | 0.762488 | -0.094584 | 0.024564 | 0.000118 | cis |
| rs7291574 | miR-130a-3 | 11:5777961 | G | A | 0.785617 | 0.107444 | 0.025379 | 2.30E-05 | cis |
| rs7119218 | miR-130a-3 | 11:5771742 | C | T | 0.716158 | -0.080812 | 0.023993 | 0.000757 | cis |
| rs1160017 | miR-130a-3 | 11:5770070 | T | C | 0.718964 | -0.088369 | 0.024383 | 0.00029 | cis |

| | | | | | | | | | |
|---|---|---|---|---|---|---|---|---|---|
| rs1241733 | miR-130a- | 11:577772 | T | A | 0.785656 | 0.106606 | 0.02537 | 2.65E-05 | cis |
| rs7930234 | miR-130a- | 11:577198 | G | A | 0.717385 | -0.082458 | 0.023993 | 0.000589 | cis |
| rs1160244 | miR-130a- | 11:577194 | C | A | 0.717394 | -0.082471 | 0.023994 | 0.000588 | cis |
| rs602061 | miR-130a- | 11:575986 | A | G | 0.80593 | 0.126149 | 0.026629 | 2.17E-06 | cis |
| rs6190534 | miR-130a- | 11:577749 | A | G | 0.785126 | 0.106211 | 0.025296 | 2.68E-05 | cis |
| rs1160526 | miR-130a- | 11:576994 | C | T | 0.717302 | -0.086228 | 0.024352 | 0.000399 | cis |
| rs631015 | miR-130a- | 11:576908 | A | G | 0.758898 | -0.093682 | 0.025248 | 0.000207 | cis |
| rs5586744 | miR-130a- | 11:577604 | C | T | 0.78121 | 0.105106 | 0.02512 | 2.86E-05 | cis |
| rs3571343 | miR-130a- | 11:577084 | C | T | 0.717968 | -0.084389 | 0.024114 | 0.000466 | cis |
| rs7946863 | miR-130a- | 11:576995 | C | T | 0.717299 | -0.086214 | 0.024352 | 0.0004 | cis |
| rs1395903 | miR-130a- | 11:577662 | A | G | 0.782818 | 0.103888 | 0.025404 | 4.33E-05 | cis |
| rs1089661 | miR-130a- | 11:572089 | C | T | 0.888821 | -0.124809 | 0.034926 | 0.000352 | cis |
| rs1079212 | miR-130a- | 11:577133 | A | G | 0.761436 | -0.093448 | 0.024761 | 0.000161 | cis |
| rs1079212 | miR-130a- | 11:577474 | C | T | 0.762353 | -0.09585 | 0.024532 | 9.34E-05 | cis |
| rs3453471 | miR-130a- | 11:577008 | C | A | 0.71896 | -0.088372 | 0.024383 | 0.00029 | cis |
| rs4938870 | miR-130a- | 11:573066 | T | C | 0.656777 | -0.10311 | 0.021892 | 2.48E-06 | cis |
| rs1783819 | miR-130a- | 11:574475 | C | T | 0.191207 | -0.122196 | 0.026633 | 4.47E-06 | cis |
| rs1510419 | miR-130a- | 11:577519 | A | C | 0.23962 | 0.094898 | 0.024457 | 0.000104 | cis |
| rs7781441 | miR-130a- | 11:578331 | A | G | 0.783246 | 0.119027 | 0.024974 | 1.88E-06 | cis |
| rs2903399 | miR-130a- | 11:577232 | G | A | 0.762061 | -0.093204 | 0.024586 | 0.00015 | cis |
| rs6190396 | miR-130a- | 11:577788 | A | G | 0.786776 | 0.109145 | 0.025401 | 1.73E-05 | cis |
| rs6190396 | miR-130a- | 11:577800 | C | T | 0.785184 | 0.107096 | 0.025295 | 2.30E-05 | cis |
| rs7121756 | miR-130a- | 11:577478 | G | A | 0.702227 | 0.100233 | 0.022819 | 1.12E-05 | cis |
| rs6190401 | miR-130a- | 11:578078 | C | T | 0.79246 | 0.114441 | 0.025483 | 7.09E-06 | cis |
| rs7123570 | miR-130a- | 11:577059 | G | A | 0.716613 | -0.082581 | 0.024096 | 0.00061 | cis |
| rs1241861 | miR-130a- | 11:577754 | T | A | 0.785045 | 0.106107 | 0.025289 | 2.72E-05 | cis |
| rs1241935 | miR-130a- | 11:577676 | C | T | 0.783805 | 0.107092 | 0.025241 | 2.21E-05 | cis |
| rs714942 | miR-130a- | 11:576966 | T | C | 0.719031 | -0.088916 | 0.024411 | 0.00027 | cis |
| rs1242028 | miR-130a- | 11:577825 | G | A | 0.785127 | 0.10736 | 0.025302 | 2.20E-05 | cis |
| rs6190531 | miR-130a- | 11:577598 | G | A | 0.781448 | 0.104958 | 0.02511 | 2.92E-05 | cis |
| rs6190534 | miR-130a- | 11:577696 | G | A | 0.785556 | 0.10725 | 0.02532 | 2.28E-05 | cis |
| rs938012 | miR-130a- | 11:577455 | G | C | 0.239529 | 0.094677 | 0.024468 | 0.000109 | cis |
| rs1615585 | miR-130a- | 11:574736 | T | C | 0.193487 | -0.120457 | 0.026635 | 6.11E-06 | cis |
| rs1160584 | miR-130a- | 11:577004 | C | T | 0.760466 | -0.09462 | 0.025018 | 0.000156 | cis |
| rs7940223 | miR-130a- | 11:573509 | C | T | 0.771464 | -0.084549 | 0.024813 | 0.000656 | cis |
| rs1848883 | miR-130a- | 11:577411 | G | A | 0.762482 | -0.094641 | 0.024565 | 0.000117 | cis |
| rs1089667 | miR-130a- | 11:577370 | C | A | 0.765869 | -0.096347 | 0.024762 | 9.99E-05 | cis |
| rs7122921 | miR-130a- | 11:577481 | C | G | 0.702221 | 0.100228 | 0.022819 | 1.12E-05 | cis |
| rs7945120 | miR-130a- | 11:577794 | T | C | 0.785215 | 0.107003 | 0.025293 | 2.33E-05 | cis |
| rs1280169 | miR-130a- | 11:574602 | C | T | 0.742621 | 0.118275 | 0.024668 | 1.63E-06 | cis |
| rs1079210 | miR-130a- | 11:572966 | G | A | 0.774375 | -0.087451 | 0.02504 | 0.000479 | cis |
| rs7108167 | miR-130a- | 11:577576 | G | T | 0.781108 | 0.104558 | 0.025063 | 3.02E-05 | cis |
| rs1397273 | miR-130a- | 11:577053 | A | G | 0.714386 | -0.082838 | 0.024107 | 0.00059 | cis |
| rs1089666 | miR-130a- | 11:577058 | T | C | 0.76134 | -0.093725 | 0.024805 | 0.000158 | cis |
| rs7112126 | miR-130a- | 11:573526 | T | G | 0.771309 | -0.084824 | 0.024794 | 0.000624 | cis |
| rs7112942 | miR-130a- | 11:577889 | T | C | 0.792344 | 0.115453 | 0.025273 | 4.92E-06 | cis |
| rs2729354 | miR-130a- | 11:573559 | A | G | 0.238503 | -0.084099 | 0.025105 | 0.000809 | cis |
| rs6591444 | miR-130a- | 11:577240 | G | A | 0.68458 | -0.08083 | 0.023272 | 0.000514 | cis |
| rs2055855 | miR-130a- | 11:577228 | T | C | 0.761931 | -0.093124 | 0.024577 | 0.000151 | cis |
| rs2649650 | miR-130a- | 11:573275 | C | T | 0.239918 | -0.087759 | 0.025007 | 0.000449 | cis |
| rs2175276 | miR-130a- | 11:577392 | A | G | 0.219083 | -0.10639 | 0.025075 | 2.21E-05 | cis |
| rs7291983 | miR-130a- | 11:578331 | A | G | 0.784268 | 0.118804 | 0.024999 | 2.01E-06 | cis |
| rs687432 | miR-130a- | 11:576942 | A | C | 0.28675 | 0.087374 | 0.024332 | 0.00033 | cis |

| | | | | | | | | | |
|---|---|---|---|---|---|---|---|---|---|
| rs1510421 | miR-130a-3 | 11:577415: | G | A | 0.762319 | -0.094731 | 0.024574 | 0.000116 | cis |
| rs6190534: | miR-130a-3 | 11:577697: | T | C | 0.784718 | 0.105788 | 0.025297 | 2.89E-05 | cis |
| rs6190534 | miR-130a-3 | 11:577748 | A | G | 0.78511 | 0.106188 | 0.025294 | 2.69E-05 | cis |
| rs7128781 | miR-130a-3 | 11:577884: | A | G | 0.792351 | 0.115444 | 0.025274 | 4.93E-06 | cis |
| rs6190396: | miR-130a-3 | 11:577845: | T | C | 0.780053 | 0.108641 | 0.025019 | 1.41E-05 | cis |
| rs6190396 | miR-130a-3 | 11:577853: | G | A | 0.779995 | 0.108655 | 0.025013 | 1.40E-05 | cis |
| rs1510420 | miR-130a-3 | 11:577011 | T | C | 0.718951 | -0.088305 | 0.02438 | 0.000292 | cis |
| rs1242177: | miR-130a-3 | 11:577031 | C | T | 0.715081 | -0.085746 | 0.02429 | 0.000415 | cis |
| rs2866708 | miR-130a-3 | 11:577814: | C | T | 0.785175 | 0.107195 | 0.025299 | 2.26E-05 | cis |
| rs2729355 | miR-130a-3 | 11:573539: | T | G | 0.246615 | -0.090329 | 0.0251 | 0.00032 | cis |
| rs7950126 | miR-130a-3 | 11:573491: | T | G | 0.771081 | -0.084692 | 0.024833 | 0.000648 | cis |
| rs2848630 | miR-130a-3 | 11:573569 | T | C | 0.238482 | -0.083822 | 0.025099 | 0.000839 | cis |
| rs1280752: | miR-130a-3 | 11:577256 | C | T | 0.715049 | -0.082512 | 0.023859 | 0.000544 | cis |
| rs2649662 | miR-130a-3 | 11:573525 | A | C | 0.238716 | -0.08511 | 0.025121 | 0.000704 | cis |
| rs7126334 | miR-130a-3 | 11:577154: | A | G | 0.716228 | -0.080544 | 0.024007 | 0.000793 | cis |
| rs2866831 | miR-130a-3 | 11:577908: | G | A | 0.792831 | 0.11597 | 0.025293 | 4.54E-06 | cis |
| rs1280409: | miR-130a-3 | 11:577340: | T | C | 0.682355 | -0.077983 | 0.023126 | 0.000746 | cis |
| rs7109912 | miR-130a-3 | 11:577061 | A | G | 0.714528 | -0.082837 | 0.024086 | 0.000583 | cis |
| rs1647400 | miR-130a-3 | 11:574729 | G | C | 0.189881 | -0.121466 | 0.026792 | 5.80E-06 | cis |
| rs1278883 | miR-130a-3 | 11:576892: | G | A | 0.720363 | -0.09584 | 0.024669 | 0.000102 | cis |
| rs7101425 | miR-130a-3 | 11:573020: | T | G | 0.774814 | -0.08776 | 0.025052 | 0.00046 | cis |
| rs1848882 | miR-130a-3 | 11:577412: | C | G | 0.762513 | -0.094816 | 0.024591 | 0.000115 | cis |
| rs489858 | miR-130a-3 | 11:573475: | A | C | 0.239931 | -0.087229 | 0.025155 | 0.000525 | cis |
| rs7126553 | miR-130a-3 | 11:575566: | T | A | 0.810335 | 0.123684 | 0.026707 | 3.64E-06 | cis |
| rs7942126 | miR-130a-3 | 11:576915 | A | G | 0.717817 | -0.092753 | 0.024616 | 0.000165 | cis |
| rs6591447 | miR-130a-3 | 11:577421: | G | C | 0.763965 | -0.096619 | 0.024658 | 8.92E-05 | cis |
| rs7936212 | miR-130a-3 | 11:577165: | G | T | 0.717696 | -0.083176 | 0.024039 | 0.00054 | cis |
| rs3788331 | miR-130b-3 | 22:220592: | C | T | 0.556796 | 0.122908 | 0.023441 | 1.58E-07 | cis |
| rs2236643 | miR-130b-3 | 22:220583: | A | G | 0.55933 | 0.121474 | 0.023453 | 2.23E-07 | cis |
| rs861852 | miR-130b-3 | 22:219910: | T | C | 0.768782 | -0.131564 | 0.029281 | 7.02E-06 | cis |
| rs5994846 | miR-130b-3 | 22:220829: | G | A | 0.566035 | 0.099138 | 0.022988 | 1.61E-05 | cis |
| rs861855 | miR-130b-3 | 22:219868: | T | G | 0.802972 | -0.144464 | 0.031088 | 3.37E-06 | cis |
| rs861851 | miR-130b-3 | 22:219928 | T | C | 0.819735 | -0.138847 | 0.030269 | 4.50E-06 | cis |
| rs861853 | miR-130b-3 | 22:219886 | C | T | 0.817124 | -0.140064 | 0.030412 | 4.11E-06 | cis |
| rs861854 | miR-130b-3 | 22:219885: | C | T | 0.818581 | -0.139907 | 0.030378 | 4.12E-06 | cis |
| rs861850 | miR-130b-3 | 22:219930 | T | G | 0.820782 | -0.138475 | 0.030204 | 4.55E-06 | cis |
| rs8135758 | miR-130b-3 | 22:220764 | G | A | 0.822754 | -0.149916 | 0.029378 | 3.34E-07 | cis |
| rs743407 | miR-130b-3 | 22:220935: | G | A | 0.581852 | 0.095261 | 0.022556 | 2.41E-05 | cis |
| rs439760 | miR-130b-5 | 22:220030: | G | A | 0.560517 | 0.094329 | 0.019323 | 1.05E-06 | cis |
| rs369463 | miR-130b-5 | 22:220152: | C | T | 0.646159 | 0.080386 | 0.019975 | 5.71E-05 | cis |
| rs447001 | miR-130b-5 | 22:220109: | C | T | 0.558786 | 0.089871 | 0.018325 | 9.38E-07 | cis |
| rs5754727 | miR-130b-5 | 22:220572: | G | A | 0.5885 | 0.076464 | 0.01871 | 4.37E-05 | cis |
| rs373001 | miR-130b-5 | 22:220074: | C | T | 0.563659 | 0.090243 | 0.018609 | 1.24E-06 | cis |
| rs450469 | miR-130b-5 | 22:220056: | C | T | 0.550569 | 0.0945 | 0.019027 | 6.82E-07 | cis |
| rs449597 | miR-130b-5 | 22:220026: | A | C | 0.55856 | 0.096433 | 0.019538 | 7.98E-07 | cis |
| rs1765384: | miR-134-5p | 14:101102 | A | T | 0.870972 | 0.20167 | 0.045434 | 9.05E-06 | cis |
| rs1765383 | miR-134-5p | 14:101101: | T | C | 0.877829 | 0.208604 | 0.045329 | 4.19E-06 | cis |
| rs1190878 | miR-134-5p | 14:101072: | C | A | 0.859763 | 0.204493 | 0.042251 | 1.30E-06 | cis |
| rs1190868 | miR-134-5p | 14:101069: | G | A | 0.180475 | -0.194393 | 0.040042 | 1.21E-06 | cis |
| rs8022367 | miR-134-5p | 14:101189 | A | T | 0.295876 | -0.214093 | 0.038364 | 2.40E-08 | cis |
| rs1123554: | miR-139-3p | 11:723188: | G | A | 0.866946 | -0.162406 | 0.031243 | 2.01E-07 | cis |
| rs3781931 | miR-139-3p | 11:723272 | C | T | 0.853265 | -0.146586 | 0.030098 | 1.11E-06 | cis |
| rs1123555 | miR-139-3p | 11:723310: | T | C | 0.948914 | -0.245753 | 0.049184 | 5.84E-07 | cis |

| | | | | | | | | | |
|---|---|---|---|---|---|---|---|---|---|
| rs904469 | miR-139-3p | 11:7232403 | G | A | 0.853139 | -0.143613 | 0.029997 | 1.69E-06 | cis |
| rs754042 | miR-139-3p | 11:7232627 | C | T | 0.853191 | -0.145747 | 0.030075 | 1.26E-06 | cis |
| rs1123554 | miR-139-3p | 11:7228544 | G | A | 0.948453 | -0.195792 | 0.053322 | 0.000241 | cis |
| rs1513780 | miR-139-3p | 11:7232050 | G | T | 0.852078 | -0.142028 | 0.029951 | 2.12E-06 | cis |
| rs4944566 | miR-139-3p | 11:7232187 | G | A | 0.856071 | -0.142932 | 0.03032 | 2.43E-06 | cis |
| rs3781934 | miR-139-3p | 11:7232507 | G | A | 0.852984 | -0.144615 | 0.030038 | 1.48E-06 | cis |
| rs755933 | miR-139-3p | 11:7232548 | G | A | 0.853056 | -0.14474 | 0.030034 | 1.44E-06 | cis |
| rs4944563 | miR-139-3p | 11:7231688 | A | C | 0.852871 | -0.140987 | 0.029969 | 2.55E-06 | cis |
| rs7832219 | miR-145-5p | 8:1065789 | T | C | 0.740634 | -0.120482 | 0.022365 | 7.16E-08 | trans |
| rs4719841 | miR-148a-3 | 7:2599753 | A | G | 0.590618 | 0.063531 | 0.017296 | 0.000239 | cis |
| rs7806251 | miR-148a-3 | 7:2598349 | T | C | 0.267441 | -0.074391 | 0.019206 | 0.000107 | cis |
| rs7789194 | miR-148a-3 | 7:2599251 | T | C | 0.653481 | 0.074138 | 0.018257 | 4.89E-05 | cis |
| rs4574742 | miR-148a-3 | 7:2599506 | A | G | 0.685742 | 0.072873 | 0.018384 | 7.37E-05 | cis |
| rs2391174 | miR-148a-3 | 7:2597933 | A | G | 0.265442 | -0.072191 | 0.019156 | 0.000164 | cis |
| rs9639523 | miR-148a-3 | 7:2599197 | G | T | 0.657583 | 0.076903 | 0.018398 | 2.92E-05 | cis |
| rs1253258 | miR-148a-3 | 7:2598643 | C | T | 0.734918 | 0.07366 | 0.019212 | 0.000126 | cis |
| rs1267250 | miR-148a-3 | 7:2598744 | A | C | 0.7347 | 0.073976 | 0.019182 | 0.000115 | cis |
| rs6980357 | miR-148a-3 | 7:2599400 | G | A | 0.685623 | 0.073536 | 0.018337 | 6.07E-05 | cis |
| rs1253856 | miR-148a-3 | 7:2598591 | T | C | 0.734419 | 0.073635 | 0.019198 | 0.000125 | cis |
| rs6951827 | miR-148a-3 | 7:2597115 | G | A | 0.268174 | -0.07296 | 0.019118 | 0.000135 | cis |
| rs6968046 | miR-148a-3 | 7:2602304 | T | C | 0.699406 | -0.070774 | 0.019027 | 0.0002 | cis |
| rs1212520 | miR-181a-3 | 1:1989920 | A | G | 0.260715 | 0.151277 | 0.029708 | 3.54E-07 | cis |
| rs6703089 | miR-181a-3 | 1:1989200 | C | A | 0.236139 | 0.129439 | 0.030474 | 2.16E-05 | cis |
| rs1080060 | miR-181a-3 | 1:1989145 | A | G | 0.244491 | 0.128044 | 0.030195 | 2.23E-05 | cis |
| rs1080059 | miR-181a-3 | 1:1988708 | T | C | 0.760087 | -0.130762 | 0.030838 | 2.23E-05 | cis |
| rs4915335 | miR-181a-3 | 1:1989452 | A | G | 0.238713 | 0.129319 | 0.030808 | 2.70E-05 | cis |
| rs1080060 | miR-181a-3 | 1:1989196 | A | G | 0.238849 | 0.129199 | 0.030542 | 2.33E-05 | cis |
| rs4915331 | miR-181a-3 | 1:1989185 | T | C | 0.235006 | 0.132427 | 0.030754 | 1.66E-05 | cis |
| rs1762558 | miR-181a-3 | 1:1989904 | G | A | 0.74365 | -0.155588 | 0.029809 | 1.79E-07 | cis |
| rs4915332 | miR-181a-3 | 1:1989185 | A | G | 0.234929 | 0.132583 | 0.030754 | 1.62E-05 | cis |
| rs1209687 | miR-181a-3 | 1:1989144 | G | A | 0.244151 | 0.127137 | 0.030185 | 2.53E-05 | cis |
| rs7539177 | miR-181a-3 | 1:1989464 | T | G | 0.238596 | 0.129425 | 0.030825 | 2.68E-05 | cis |
| rs1214202 | miR-181a-3 | 1:1988815 | C | T | 0.766397 | -0.134455 | 0.030987 | 1.43E-05 | cis |
| rs7273976 | miR-181a-3 | 1:1988640 | G | T | 0.765001 | -0.134131 | 0.031205 | 1.72E-05 | cis |
| rs322887 | miR-181a-3 | 1:1990263 | A | G | 0.834392 | -0.118873 | 0.034925 | 0.000665 | cis |
| rs1212331 | miR-181a-3 | 1:1988726 | G | T | 0.764104 | -0.13272 | 0.031012 | 1.87E-05 | cis |
| rs1684410 | miR-181a-3 | 1:1990115 | G | T | 0.842513 | -0.124914 | 0.035788 | 0.000482 | cis |
| rs322928 | miR-181a-3 | 1:1990218 | G | A | 0.833953 | -0.119115 | 0.034813 | 0.000623 | cis |
| rs2360962 | miR-181a-3 | 1:1990047 | A | T | 0.261757 | 0.149666 | 0.029611 | 4.32E-07 | cis |
| rs1040959 | miR-181a-3 | 1:1988958 | A | C | 0.766999 | -0.133294 | 0.030758 | 1.47E-05 | cis |
| rs1036332 | miR-181a-3 | 1:1990124 | A | C | 0.260189 | 0.152488 | 0.029624 | 2.64E-07 | cis |
| rs6427756 | miR-181a-3 | 1:1990007 | A | G | 0.263331 | 0.150216 | 0.029597 | 3.87E-07 | cis |
| rs1213201 | miR-181a-3 | 1:1988820 | T | C | 0.755495 | -0.124851 | 0.030368 | 3.94E-05 | cis |
| rs7529925 | miR-181a-3 | 1:1990072 | C | T | 0.255718 | 0.152502 | 0.029587 | 2.54E-07 | cis |
| rs1275164 | miR-181a-3 | 1:1988944 | A | G | 0.754678 | -0.127135 | 0.030021 | 2.29E-05 | cis |
| rs1040960 | miR-181a-3 | 1:1988959 | G | T | 0.766066 | -0.132152 | 0.030772 | 1.75E-05 | cis |
| rs1434282 | miR-181a-3 | 1:1990107 | C | T | 0.271104 | 0.152077 | 0.029247 | 2.00E-07 | cis |
| rs1275163 | miR-181a-3 | 1:1988916 | C | T | 0.765073 | -0.132034 | 0.030777 | 1.79E-05 | cis |
| rs1213024 | miR-181a-3 | 1:1988841 | G | C | 0.765569 | -0.132247 | 0.030905 | 1.88E-05 | cis |
| rs1204800 | miR-181a-3 | 1:1989153 | T | C | 0.235546 | 0.131093 | 0.030724 | 1.98E-05 | cis |
| rs2033565 | miR-181a-3 | 1:1990150 | A | T | 0.16327 | 0.127034 | 0.035615 | 0.000361 | cis |
| rs1892432 | miR-181a-3 | 1:1989406 | T | C | 0.234225 | 0.13037 | 0.030838 | 2.36E-05 | cis |
| rs322927 | miR-181a-3 | 1:1990218 | T | G | 0.834224 | -0.118468 | 0.034829 | 0.00067 | cis |

| | | | | | | | | | |
|---|---|---|---|---|---|---|---|---|---|
| rs4731638 | miR-182-5p | 7:1295858 | T | C | 0.829083 | -0.139909 | 0.030981 | 6.30E-06 | cis |
| rs6975927 | miR-182-5p | 7:1292310 | C | T | 0.603702 | 0.098752 | 0.024043 | 4.00E-05 | cis |
| rs1157036 | miR-182-5p | 7:1292510 | T | C | 0.600503 | 0.103586 | 0.024305 | 2.03E-05 | cis |
| rs1755622 | miR-182-5p | 7:1293300 | C | G | 0.598776 | 0.09993 | 0.023709 | 2.50E-05 | cis |
| rs2466983 | miR-182-5p | 7:1294355 | T | A | 0.556113 | 0.101469 | 0.025762 | 8.19E-05 | cis |
| rs3817530 | miR-182-5p | 7:1295872 | T | G | 0.829033 | -0.136389 | 0.030774 | 9.34E-06 | cis |
| rs6976516 | miR-182-5p | 7:1292314 | C | T | 0.603765 | 0.099082 | 0.024057 | 3.81E-05 | cis |
| rs1197597 | miR-182-5p | 7:1292645 | G | C | 0.601924 | 0.110212 | 0.024231 | 5.41E-06 | cis |
| rs868056 | miR-182-5p | 7:1294351 | C | T | 0.556068 | 0.101649 | 0.025764 | 7.97E-05 | cis |
| rs4626538 | miR-182-5p | 7:1294131 | G | T | 0.482006 | 0.113353 | 0.023617 | 1.59E-06 | cis |
| rs1095424 | miR-182-5p | 7:1292682 | C | T | 0.600067 | 0.110449 | 0.024121 | 4.67E-06 | cis |
| rs3449871 | miR-182-5p | 7:1292460 | T | A | 0.599558 | 0.10107 | 0.024154 | 2.86E-05 | cis |
| rs1215434 | miR-182-5p | 7:1293569 | T | G | 0.599436 | 0.09985 | 0.023597 | 2.32E-05 | cis |
| rs1323153 | miR-182-5p | 7:1292671 | A | G | 0.600217 | 0.110484 | 0.024129 | 4.68E-06 | cis |
| rs4335057 | miR-182-5p | 7:1294160 | A | G | 0.485897 | 0.115383 | 0.023813 | 1.26E-06 | cis |
| rs7809195 | miR-182-5p | 7:1294097 | G | A | 0.445049 | 0.121354 | 0.024139 | 4.98E-07 | cis |
| rs1755643 | miR-182-5p | 7:1293462 | C | T | 0.599053 | 0.099394 | 0.023585 | 2.51E-05 | cis |
| rs2402967 | miR-182-5p | 7:1292981 | C | T | 0.589777 | 0.104461 | 0.023714 | 1.06E-05 | cis |
| rs2466984 | miR-182-5p | 7:1294356 | C | T | 0.558997 | 0.105535 | 0.025858 | 4.48E-05 | cis |
| rs1322627 | miR-182-5p | 7:1293082 | A | G | 0.592329 | 0.106125 | 0.023812 | 8.32E-06 | cis |
| rs1176914 | miR-182-5p | 7:1293491 | G | A | 0.597642 | 0.100085 | 0.02361 | 2.24E-05 | cis |
| rs1253346 | miR-182-5p | 7:1295694 | G | C | 0.829496 | -0.140414 | 0.030727 | 4.88E-06 | cis |
| rs1323622 | miR-182-5p | 7:1293335 | G | A | 0.599127 | 0.096829 | 0.023641 | 4.21E-05 | cis |
| rs4421293 | miR-182-5p | 7:1294142 | G | A | 0.486604 | 0.115293 | 0.023725 | 1.18E-06 | cis |
| rs3601778 | miR-182-5p | 7:1292591 | A | G | 0.600482 | 0.107399 | 0.024328 | 1.01E-05 | cis |
| rs2727471 | miR-182-5p | 7:1294358 | G | A | 0.555759 | 0.101783 | 0.025773 | 7.84E-05 | cis |
| rs3501994 | miR-182-5p | 7:1294095 | C | G | 0.516428 | 0.10927 | 0.023786 | 4.35E-06 | cis |
| rs2402975 | miR-182-5p | 7:1293345 | T | C | 0.598179 | 0.099176 | 0.023631 | 2.71E-05 | cis |
| rs1323901 | miR-182-5p | 7:1293639 | C | T | 0.599898 | 0.099277 | 0.023633 | 2.66E-05 | cis |
| rs6953987 | miR-182-5p | 7:1292939 | G | A | 0.592994 | 0.105061 | 0.023704 | 9.33E-06 | cis |
| rs1253858 | miR-182-5p | 7:1294163 | G | A | 0.519308 | 0.109949 | 0.024075 | 4.95E-06 | cis |
| rs6947908 | miR-182-5p | 7:1294167 | C | T | 0.487263 | 0.113755 | 0.023931 | 2.00E-06 | cis |
| rs1324463 | miR-182-5p | 7:1292582 | C | G | 0.602916 | 0.107133 | 0.024242 | 9.90E-06 | cis |
| rs3403823 | miR-182-5p | 7:1292080 | G | T | 0.6024 | 0.09312 | 0.023934 | 9.99E-05 | cis |
| rs2727474 | miR-182-5p | 7:1294245 | C | T | 0.580167 | 0.12335 | 0.025264 | 1.05E-06 | cis |
| rs3800599 | miR-182-5p | 7:1293546 | A | C | 0.599291 | 0.099086 | 0.023601 | 2.69E-05 | cis |
| rs1177302 | miR-182-5p | 7:1293439 | T | A | 0.597762 | 0.100044 | 0.023618 | 2.28E-05 | cis |
| rs2293629 | miR-182-5p | 7:1293572 | T | C | 0.599438 | 0.099859 | 0.023597 | 2.32E-05 | cis |
| rs1962038 | miR-182-5p | 7:1293514 | C | G | 0.597635 | 0.100089 | 0.02361 | 2.24E-05 | cis |
| rs1323174 | miR-182-5p | 7:1294153 | A | C | 0.516748 | 0.10884 | 0.023909 | 5.31E-06 | cis |
| rs3578790 | miR-182-5p | 7:1294057 | A | G | 0.515425 | 0.094554 | 0.023387 | 5.28E-05 | cis |
| rs4440554 | miR-182-5p | 7:1293381 | G | C | 0.600138 | 0.096421 | 0.023777 | 5.01E-05 | cis |
| rs1095424 | miR-182-5p | 7:1292297 | G | A | 0.602609 | 0.099532 | 0.024041 | 3.47E-05 | cis |
| rs1176143 | miR-182-5p | 7:1292905 | G | A | 0.592916 | 0.105012 | 0.023712 | 9.49E-06 | cis |
| rs1323067 | miR-182-5p | 7:1293371 | G | A | 0.60658 | 0.099524 | 0.023882 | 3.08E-05 | cis |
| rs1176058 | miR-182-5p | 7:1293070 | A | C | 0.591378 | 0.104231 | 0.023658 | 1.05E-05 | cis |
| rs1095425 | miR-182-5p | 7:1293294 | C | T | 0.598686 | 0.100006 | 0.023705 | 2.46E-05 | cis |
| rs3736626 | miR-182-5p | 7:1293501 | A | C | 0.596638 | 0.100288 | 0.023551 | 2.06E-05 | cis |
| rs1176060 | miR-182-5p | 7:1294355 | G | A | 0.557229 | 0.101801 | 0.025811 | 8.01E-05 | cis |
| rs1322562 | miR-182-5p | 7:1293206 | G | A | 0.599899 | 0.101112 | 0.023663 | 1.93E-05 | cis |
| rs174555 | miR-1908-5 | 11:615797 | T | C | 0.701955 | -0.095501 | 0.01775 | 7.43E-08 | cis |
| rs174556 | miR-1908-5 | 11:615806 | C | T | 0.701956 | -0.095381 | 0.017746 | 7.67E-08 | cis |
| rs2072113 | miR-1908-5 | 11:616049 | C | T | 0.881208 | -0.10584 | 0.025513 | 3.35E-05 | cis |

| | | | | | | | | | |
|---|---|---|---|---|---|---|---|---|---|
| rs2072114 | miR-1908-5 | 11:616052 | A | G | 0.875115 | -0.107941 | 0.024794 | 1.34E-05 | cis |
| rs174561 | miR-1908-5 | 11:615827 | T | C | 0.70223 | -0.095468 | 0.017762 | 7.66E-08 | cis |
| rs174572 | miR-1908-5 | 11:615982 | C | T | 0.743634 | -0.07722 | 0.01868 | 3.57E-05 | cis |
| rs174573 | miR-1908-5 | 11:616003 | G | A | 0.744012 | -0.077086 | 0.018695 | 3.73E-05 | cis |
| rs174575 | miR-1908-5 | 11:616020 | C | G | 0.74143 | -0.075351 | 0.018673 | 5.45E-05 | cis |
| rs28456 | miR-1908-5 | 11:615894 | A | G | 0.704555 | -0.09661 | 0.017907 | 6.85E-08 | cis |
| rs2524299 | miR-1908-5 | 11:616047 | A | T | 0.878659 | -0.102616 | 0.02525 | 4.82E-05 | cis |
| rs174559 | miR-1908-5 | 11:615816 | G | A | 0.729758 | -0.091142 | 0.018205 | 5.55E-07 | cis |
| rs174448 | miR-1908-5 | 11:616395 | G | A | 0.32712 | 0.075051 | 0.017575 | 1.95E-05 | cis |
| rs174449 | miR-1908-5 | 11:616403 | G | A | 0.336231 | 0.072481 | 0.017194 | 2.49E-05 | cis |
| rs2727271 | miR-1908-5 | 11:616033 | A | T | 0.881925 | -0.104784 | 0.025591 | 4.23E-05 | cis |
| rs174538 | miR-1908-5 | 11:615600 | G | A | 0.693741 | -0.093447 | 0.017724 | 1.35E-07 | cis |
| rs2727270 | miR-1908-5 | 11:616032 | C | T | 0.881959 | -0.104512 | 0.025597 | 4.45E-05 | cis |
| rs1012638 | miR-193a-5 | 17:301045 | G | A | 0.758199 | -0.083757 | 0.021036 | 6.85E-05 | cis |
| rs1165139 | miR-193a-5 | 17:301053 | G | A | 0.828639 | -0.100113 | 0.026705 | 0.000178 | cis |
| rs6682956 | miR-197-3p | 1:1101302 | G | A | 0.831458 | 0.065477 | 0.017658 | 0.000209 | cis |
| rs2301229 | miR-197-3p | 1:1101366 | C | T | 0.830979 | 0.066343 | 0.01762 | 0.000166 | cis |
| rs1203337 | miR-197-3p | 1:1101306 | A | T | 0.818337 | 0.059149 | 0.017117 | 0.000549 | cis |
| rs3525 | miR-197-3p | 1:1101368 | T | A | 0.830977 | 0.066348 | 0.01762 | 0.000166 | cis |
| rs1203797 | miR-197-3p | 1:1101248 | C | T | 0.831227 | 0.065756 | 0.01767 | 0.000198 | cis |
| rs7355073 | miR-197-3p | 1:1101297 | C | T | 0.818417 | 0.059162 | 0.017121 | 0.000549 | cis |
| rs6658855 | miR-197-3p | 1:1101233 | G | A | 0.816651 | 0.058965 | 0.017114 | 0.00057 | cis |
| rs1114278 | miR-204-5p | 9:7390148 | C | A | 0.804964 | -0.160259 | 0.033907 | 2.28E-06 | cis |
| rs1086899 | miR-204-5p | 9:7384828 | C | A | 0.805299 | -0.168174 | 0.033531 | 5.29E-07 | cis |
| rs2871376 | miR-204-5p | 9:7390198 | T | C | 0.808192 | -0.162322 | 0.0344 | 2.37E-06 | cis |
| rs1200513 | miR-204-5p | 9:7381117 | G | C | 0.806575 | -0.167101 | 0.033731 | 7.27E-07 | cis |
| rs1705662 | miR-204-5p | 9:7387369 | T | C | 0.805024 | -0.169575 | 0.033489 | 4.11E-07 | cis |
| rs1114276 | miR-204-5p | 9:7387750 | A | G | 0.80544 | -0.169387 | 0.033551 | 4.45E-07 | cis |
| rs1114277 | miR-204-5p | 9:7390064 | G | T | 0.810426 | -0.15697 | 0.034358 | 4.91E-06 | cis |
| rs1504392 | miR-204-5p | 9:7390334 | C | T | 0.804776 | -0.160523 | 0.033907 | 2.20E-06 | cis |
| rs1761679 | miR-204-5p | 9:7388065 | A | G | 0.805381 | -0.169167 | 0.033535 | 4.55E-07 | cis |
| rs1200502 | miR-204-5p | 9:7386908 | G | A | 0.798191 | -0.166606 | 0.03355 | 6.84E-07 | cis |
| rs1504404 | miR-204-5p | 9:7387082 | A | G | 0.805458 | -0.169842 | 0.033563 | 4.18E-07 | cis |
| rs1705648 | miR-204-5p | 9:7383267 | A | C | 0.804418 | -0.165743 | 0.033572 | 7.94E-07 | cis |
| rs7018911 | miR-204-5p | 9:7382828 | C | T | 0.219637 | 0.14593 | 0.032762 | 8.42E-06 | cis |
| rs7039341 | miR-204-5p | 9:7383302 | G | A | 0.804558 | -0.165891 | 0.033584 | 7.83E-07 | cis |
| rs1200394 | miR-204-5p | 9:7384427 | C | T | 0.805169 | -0.168037 | 0.033542 | 5.45E-07 | cis |
| rs1114277 | miR-204-5p | 9:7389541 | G | A | 0.79097 | -0.155258 | 0.033144 | 2.81E-06 | cis |
| rs1705642 | miR-204-5p | 9:7379436 | G | T | 0.799585 | -0.174775 | 0.033638 | 2.04E-07 | cis |
| rs1114277 | miR-204-5p | 9:7390065 | T | C | 0.810587 | -0.156814 | 0.034374 | 5.07E-06 | cis |
| rs1114277 | miR-204-5p | 9:7389504 | T | C | 0.804816 | -0.162197 | 0.033778 | 1.57E-06 | cis |
| rs1504393 | miR-204-5p | 9:7382038 | C | T | 0.806016 | -0.167463 | 0.033717 | 6.81E-07 | cis |
| rs1237860 | miR-204-5p | 9:7386553 | G | A | 0.792685 | -0.163209 | 0.032868 | 6.85E-07 | cis |
| rs1086899 | miR-204-5p | 9:7386785 | C | T | 0.79267 | -0.163241 | 0.032872 | 6.84E-07 | cis |
| rs1705660 | miR-204-5p | 9:7387164 | G | C | 0.805399 | -0.169881 | 0.033554 | 4.13E-07 | cis |
| rs1504388 | miR-204-5p | 9:7390247 | A | G | 0.804808 | -0.160547 | 0.033888 | 2.16E-06 | cis |
| rs497934 | miR-204-5p | 9:7363878 | C | G | 0.148725 | 0.15774 | 0.039102 | 5.48E-05 | cis |
| rs1504390 | miR-204-5p | 9:7390292 | A | T | 0.80471 | -0.160881 | 0.033891 | 2.06E-06 | cis |
| rs1114276 | miR-204-5p | 9:7388206 | C | T | 0.804988 | -0.162595 | 0.033671 | 1.37E-06 | cis |
| rs1238058 | miR-204-5p | 9:7387116 | G | A | 0.805323 | -0.170175 | 0.033545 | 3.92E-07 | cis |
| rs1114275 | miR-204-5p | 9:7386169 | C | T | 0.792851 | -0.162726 | 0.032877 | 7.44E-07 | cis |
| rs7027219 | miR-204-5p | 9:7386640 | T | A | 0.792485 | -0.163383 | 0.032877 | 6.71E-07 | cis |
| rs7021834 | miR-204-5p | 9:7389408 | C | T | 0.804857 | -0.162634 | 0.033746 | 1.44E-06 | cis |

| | | | | | | | | | |
|---|---|---|---|---|---|---|---|---|---|
| rs1532804 | miR-204-5p | 9:7390380 | T | C | 0.804424 | -0.161104 | 0.033898 | 2.01E-06 | cis |
| rs2309916 | miR-204-5p | 9:7390226 | C | A | 0.804969 | -0.160227 | 0.033898 | 2.28E-06 | cis |
| rs7854181 | miR-204-5p | 9:7390300 | T | G | 0.804525 | -0.161144 | 0.0339 | 2.00E-06 | cis |
| rs763359 | miR-204-5p | 9:7386513 | A | G | 0.805611 | -0.170222 | 0.033549 | 3.90E-07 | cis |
| rs2174308 | miR-204-5p | 9:7378395 | G | C | 0.807198 | -0.170845 | 0.034019 | 5.11E-07 | cis |
| rs7043726 | miR-204-5p | 9:7389196 | C | T | 0.804849 | -0.162886 | 0.033744 | 1.39E-06 | cis |
| rs985654 | miR-204-5p | 9:7380339 | C | A | 0.806843 | -0.166571 | 0.03374 | 7.94E-07 | cis |
| rs7848661 | miR-204-5p | 9:7389373 | G | A | 0.804859 | -0.162639 | 0.033746 | 1.44E-06 | cis |
| rs1504391 | miR-204-5p | 9:7390329 | T | C | 0.804627 | -0.161042 | 0.033894 | 2.02E-06 | cis |
| rs7854330 | miR-204-5p | 9:7390313 | T | G | 0.804591 | -0.161004 | 0.033895 | 2.03E-06 | cis |
| rs2309917 | miR-204-5p | 9:7390203 | G | A | 0.807914 | -0.159544 | 0.034489 | 3.73E-06 | cis |
| rs1114275 | miR-204-5p | 9:7385683 | T | G | 0.806038 | -0.169849 | 0.033595 | 4.29E-07 | cis |
| rs7870153 | miR-204-5p | 9:7379377 | T | C | 0.80642 | -0.168422 | 0.033818 | 6.35E-07 | cis |
| rs7040929 | miR-204-5p | 9:7386394 | T | C | 0.792736 | -0.163029 | 0.032863 | 7.02E-07 | cis |
| rs3437250 | miR-204-5p | 9:7384127 | A | T | 0.805216 | -0.167755 | 0.033533 | 5.65E-07 | cis |
| rs2309918 | miR-204-5p | 9:7390202 | G | A | 0.853031 | -0.183352 | 0.041034 | 7.88E-06 | cis |
| rs4079469 | miR-204-5p | 9:7385141 | T | A | 0.805239 | -0.1688 | 0.033539 | 4.83E-07 | cis |
| rs1237939 | miR-204-5p | 9:7386532 | T | C | 0.792689 | -0.163207 | 0.032869 | 6.86E-07 | cis |
| rs9785312 | miR-204-5p | 9:7385908 | C | A | 0.792465 | -0.162788 | 0.03281 | 6.99E-07 | cis |
| rs1504389 | miR-204-5p | 9:7390267 | C | A | 0.804889 | -0.160368 | 0.033903 | 2.24E-06 | cis |
| rs1532801 | miR-204-5p | 9:7390340 | C | T | 0.804766 | -0.160539 | 0.033907 | 2.19E-06 | cis |
| rs495259 | miR-204-5p | 9:7363246 | A | G | 0.149513 | 0.15665 | 0.039013 | 5.94E-05 | cis |
| rs1114274 | miR-204-5p | 9:7384351 | C | T | 0.805245 | -0.167919 | 0.033538 | 5.53E-07 | cis |
| rs1114273 | miR-204-5p | 9:7382778 | T | C | 0.805772 | -0.168315 | 0.033667 | 5.75E-07 | cis |
| rs4370599 | miR-204-5p | 9:7390233 | C | T | 0.804855 | -0.160694 | 0.033886 | 2.11E-06 | cis |
| rs1114271 | miR-204-5p | 9:7380275 | T | C | 0.80636 | -0.165262 | 0.03374 | 9.67E-07 | cis |
| rs1532802 | miR-204-5p | 9:7390341 | C | T | 0.804724 | -0.160474 | 0.033909 | 2.22E-06 | cis |
| rs523734 | miR-204-5p | 9:7368023 | C | T | 0.147447 | 0.157418 | 0.038979 | 5.38E-05 | cis |
| rs1114278 | miR-204-5p | 9:7390165 | C | T | 0.804871 | -0.160333 | 0.033899 | 2.25E-06 | cis |
| rs9785216 | miR-204-5p | 9:7385880 | A | G | 0.805704 | -0.169935 | 0.033541 | 4.05E-07 | cis |
| rs2430871 | miR-204-5p | 9:7362482 | G | C | 0.149621 | 0.156657 | 0.038981 | 5.85E-05 | cis |
| rs1009053 | miR-204-5p | 9:7387257 | T | C | 0.805257 | -0.16898 | 0.033549 | 4.73E-07 | cis |
| rs1114277 | miR-204-5p | 9:7390110 | C | G | 0.804851 | -0.160373 | 0.033901 | 2.24E-06 | cis |
| rs3487867 | miR-222-3p | 15:399501 | A | G | 0.837779 | 0.053783 | 0.01477 | 0.000271 | trans |
| rs2618156 | miR-222-3p | 15:398552 | C | T | 0.83701 | 0.062098 | 0.01481 | 2.75E-05 | trans |
| rs1052014 | miR-222-3p | 15:399307 | C | T | 0.83585 | 0.055171 | 0.014622 | 0.000161 | trans |
| rs1696954 | miR-222-3p | 15:399648 | G | A | 0.843008 | 0.058759 | 0.015081 | 9.77E-05 | trans |
| rs5818899 | miR-222-3p | 15:399069 | C | T | 0.835466 | 0.055399 | 0.014599 | 0.000148 | trans |
| rs2618165 | miR-222-3p | 15:398628 | C | A | 0.837166 | 0.060438 | 0.014715 | 4.00E-05 | trans |
| rs5957839 | miR-222-3p | 15:398938 | T | C | 0.83511 | 0.055152 | 0.014589 | 0.000157 | trans |
| rs2664123 | miR-222-3p | 15:398382 | T | A | 0.833481 | 0.061486 | 0.014768 | 3.14E-05 | trans |
| rs1813493 | miR-222-3p | 15:398594 | T | C | 0.837079 | 0.062088 | 0.014794 | 2.71E-05 | trans |
| rs2664137 | miR-222-3p | 15:398497 | C | T | 0.835996 | 0.061153 | 0.014808 | 3.63E-05 | trans |
| rs1770608 | miR-222-3p | 15:399110 | C | T | 0.834845 | 0.055278 | 0.014578 | 0.00015 | trans |
| rs2618167 | miR-222-3p | 15:398702 | G | C | 0.837517 | 0.060035 | 0.014707 | 4.46E-05 | trans |
| rs1052014 | miR-222-3p | 15:399594 | A | G | 0.841203 | 0.05951 | 0.014988 | 7.17E-05 | trans |
| rs1038097 | miR-222-3p | 15:398648 | G | A | 0.838144 | 0.060037 | 0.014747 | 4.68E-05 | trans |
| rs7400864 | miR-222-3p | 15:399179 | G | C | 0.83494 | 0.056167 | 0.01457 | 0.000116 | trans |
| rs7546542 | miR-222-3p | 15:398918 | C | T | 0.835687 | 0.055572 | 0.014612 | 0.000143 | trans |
| rs1770617 | miR-222-3p | 15:399151 | T | G | 0.835485 | 0.055536 | 0.014594 | 0.000142 | trans |
| rs1763321 | miR-222-3p | 15:399005 | C | A | 0.835558 | 0.055529 | 0.014608 | 0.000144 | trans |
| rs1600215 | miR-222-3p | 15:398375 | A | G | 0.833162 | 0.061584 | 0.014775 | 3.07E-05 | trans |
| rs1696948 | miR-222-3p | 15:399350 | C | T | 0.835975 | 0.05509 | 0.014634 | 0.000167 | trans |

| | | | | | | | | | |
|---|---|---|---|---|---|---|---|---|---|
| rs1994380 | miR-222-3p | 15:399861 | T | C | 0.840617 | 0.059606 | 0.01495 | 6.69E-05 | trans |
| rs4503768 | miR-222-3p | 15:399559 | G | C | 0.837595 | 0.054165 | 0.014828 | 0.000259 | trans |
| rs2664136 | miR-222-3p | 15:398491 | G | A | 0.834632 | 0.061388 | 0.01479 | 3.32E-05 | trans |
| rs1696949 | miR-222-3p | 15:399395 | A | T | 0.836829 | 0.055338 | 0.014713 | 0.000169 | trans |
| rs2861928 | miR-222-3p | 15:399182 | G | A | 0.835235 | 0.05643 | 0.014564 | 0.000107 | trans |
| rs2618161 | miR-222-3p | 15:398677 | G | T | 0.837831 | 0.060205 | 0.014712 | 4.27E-05 | trans |
| rs1770580 | miR-222-3p | 15:399004 | G | A | 0.83556 | 0.055533 | 0.014608 | 0.000144 | trans |
| rs1382444 | miR-222-3p | 15:398704 | G | C | 0.837505 | 0.060018 | 0.014707 | 4.48E-05 | trans |
| rs2618164 | miR-222-3p | 15:398638 | C | T | 0.83703 | 0.060222 | 0.01469 | 4.14E-05 | trans |
| rs1051442 | miR-222-3p | 15:398876 | C | T | 0.835802 | 0.055705 | 0.014608 | 0.000137 | trans |
| rs2837778 | miR-222-3p | 15:399182 | G | A | 0.834959 | 0.056534 | 0.014539 | 0.000101 | trans |
| rs5962857 | miR-222-3p | 15:399492 | C | A | 0.838352 | 0.052795 | 0.014803 | 0.000362 | trans |
| rs1988555 | miR-222-3p | 15:398252 | A | G | 0.174597 | -0.057112 | 0.014565 | 8.82E-05 | trans |
| rs7444792 | miR-222-3p | 15:399555 | T | G | 0.838179 | 0.054397 | 0.014823 | 0.000243 | trans |
| rs2618160 | miR-222-3p | 15:398333 | G | A | 0.832179 | 0.060202 | 0.014674 | 4.08E-05 | trans |
| rs1382443 | miR-222-3p | 15:398706 | A | T | 0.837271 | 0.060124 | 0.01471 | 4.37E-05 | trans |
| rs2664140 | miR-222-3p | 15:398321 | G | A | 0.827933 | 0.061279 | 0.014602 | 2.71E-05 | trans |
| rs7911559 | miR-222-3p | 15:398954 | G | A | 0.835478 | 0.055406 | 0.014593 | 0.000147 | trans |
| rs7653667 | miR-222-3p | 15:398590 | G | A | 0.837127 | 0.06206 | 0.014804 | 2.76E-05 | trans |
| rs7400864 | miR-222-3p | 15:399095 | T | C | 0.835363 | 0.055492 | 0.014588 | 0.000142 | trans |
| rs2664138 | miR-222-3p | 15:398299 | T | C | 0.82799 | 0.061465 | 0.014592 | 2.53E-05 | trans |
| rs6200238 | miR-222-3p | 15:398205 | C | T | 0.830418 | 0.058164 | 0.014554 | 6.43E-05 | trans |
| rs4131506 | miR-222-3p | 15:398882 | C | T | 0.835674 | 0.055587 | 0.014611 | 0.000142 | trans |
| rs7584210 | miR-222-3p | 15:399034 | G | C | 0.835367 | 0.055578 | 0.014591 | 0.000139 | trans |
| rs4546129 | miR-222-3p | 15:398846 | G | A | 0.835259 | 0.055939 | 0.0146 | 0.000127 | trans |
| rs4923835 | miR-222-3p | 15:398260 | G | A | 0.174023 | -0.057253 | 0.01457 | 8.51E-05 | trans |
| rs2228263 | miR-222-3p | 15:398853 | T | C | 0.835199 | 0.056028 | 0.014589 | 0.000123 | trans |
| rs2664142 | miR-222-3p | 15:398329 | C | T | 0.827964 | 0.061373 | 0.014606 | 2.65E-05 | trans |
| rs7592523 | miR-222-3p | 15:398956 | T | C | 0.83504 | 0.055047 | 0.014587 | 0.000161 | trans |
| rs4085744 | miR-222-3p | 15:399890 | G | A | 0.841832 | 0.060956 | 0.015055 | 5.15E-05 | trans |
| rs1763291 | miR-222-3p | 15:398855 | A | G | 0.835526 | 0.056239 | 0.014593 | 0.000116 | trans |
| rs880270 | miR-222-3p | 15:399188 | T | C | 0.835362 | 0.056388 | 0.014563 | 0.000108 | trans |
| rs6492905 | miR-222-3p | 15:398924 | C | A | 0.835596 | 0.055355 | 0.01463 | 0.000155 | trans |
| rs1763310 | miR-222-3p | 15:398882 | A | C | 0.835637 | 0.055589 | 0.014612 | 0.000142 | trans |
| rs2854932 | miR-222-3p | 15:399182 | T | C | 0.835233 | 0.05643 | 0.014564 | 0.000107 | trans |
| rs4415979 | miR-222-3p | 15:398944 | C | T | 0.83544 | 0.055397 | 0.014593 | 0.000147 | trans |
| rs2129457 | miR-222-3p | 15:399836 | G | T | 0.840479 | 0.059662 | 0.014926 | 6.41E-05 | trans |
| rs954152 | miR-222-3p | 15:398912 | C | T | 0.837263 | 0.054985 | 0.014758 | 0.000195 | trans |
| rs1052014 | miR-222-3p | 15:399419 | C | T | 0.836928 | 0.055015 | 0.014695 | 0.000181 | trans |
| rs9972350 | miR-222-3p | 15:399295 | T | C | 0.835787 | 0.055184 | 0.014618 | 0.00016 | trans |
| rs2865301 | miR-222-3p | 15:399284 | A | G | 0.835849 | 0.055233 | 0.014623 | 0.000159 | trans |
| rs2411316 | miR-222-3p | 15:399285 | C | A | 0.835814 | 0.055243 | 0.01462 | 0.000158 | trans |
| rs1052014 | miR-222-3p | 15:399790 | T | G | 0.838604 | 0.05853 | 0.014773 | 7.43E-05 | trans |
| rs1190167 | miR-26b-5p | 2:2196314 | A | G | 0.525732 | 0.056553 | 0.015965 | 0.000397 | cis |
| rs6745700 | miR-26b-5p | 2:2196398 | T | C | 0.521354 | 0.053615 | 0.015916 | 0.000756 | cis |
| rs7577018 | miR-26b-5p | 2:2196318 | A | T | 0.522028 | 0.055644 | 0.015814 | 0.000434 | cis |
| rs933994 | miR-26b-5p | 2:2196506 | C | T | 0.520846 | 0.053076 | 0.015931 | 0.000863 | cis |
| rs4672901 | miR-26b-5p | 2:2196367 | T | C | 0.52741 | 0.054501 | 0.016066 | 0.000693 | cis |
| rs1340830 | miR-26b-5p | 2:2196281 | G | A | 0.521729 | 0.055489 | 0.015765 | 0.000432 | cis |
| rs7559490 | miR-26b-5p | 2:2196568 | A | T | 0.579126 | 0.05679 | 0.01614 | 0.000434 | cis |
| rs1554622 | miR-26b-5p | 2:2196062 | T | G | 0.521083 | 0.055291 | 0.015771 | 0.000455 | cis |
| rs4672897 | miR-26b-5p | 2:2195881 | A | G | 0.525199 | 0.054051 | 0.01593 | 0.000691 | cis |
| rs6726695 | miR-26b-5p | 2:2196297 | T | G | 0.526361 | 0.056444 | 0.015915 | 0.00039 | cis |

| | | | | | | | | | |
|---|---|---|---|---|---|---|---|---|---|
| rs1341232 | miR-26b-5p | 2:2196205 | T | C | 0.521793 | 0.055497 | 0.015762 | 0.00043 | cis |
| rs1342890 | miR-26b-5p | 2:2196302 | T | C | 0.521913 | 0.055655 | 0.015807 | 0.00043 | cis |
| rs1554623 | miR-26b-5p | 2:2196061 | C | T | 0.521101 | 0.055281 | 0.015771 | 0.000456 | cis |
| rs1340842 | miR-26b-5p | 2:2196283 | C | T | 0.521233 | 0.055311 | 0.01578 | 0.000456 | cis |
| rs7355239 | miR-26b-5p | 2:2196338 | C | T | 0.526885 | 0.054074 | 0.016045 | 0.000751 | cis |
| rs6717107 | miR-26b-5p | 2:2195886 | A | G | 0.525172 | 0.054049 | 0.01593 | 0.000691 | cis |
| rs2556391 | miR-26b-5p | 2:2195563 | T | C | 0.613495 | 0.053869 | 0.016222 | 0.000898 | cis |
| rs1269444 | miR-26b-5p | 2:2196249 | G | A | 0.521946 | 0.055645 | 0.015769 | 0.000418 | cis |
| rs6710529 | miR-26b-5p | 2:2196455 | G | A | 0.521849 | 0.053713 | 0.015941 | 0.000753 | cis |
| rs6436074 | miR-26b-5p | 2:2196312 | G | A | 0.522006 | 0.055636 | 0.015813 | 0.000434 | cis |
| rs1757372 | miR-26b-5p | 2:2196521 | T | A | 0.579864 | 0.056971 | 0.016081 | 0.000396 | cis |
| rs3731877 | miR-26b-5p | 2:2196024 | G | C | 0.521743 | 0.055462 | 0.015764 | 0.000434 | cis |
| rs1757373 | miR-26b-5p | 2:2196521 | T | C | 0.579489 | 0.057165 | 0.016075 | 0.000376 | cis |
| rs5599679 | miR-26b-5p | 2:2195581 | C | G | 0.615245 | 0.054156 | 0.016194 | 0.000825 | cis |
| rs7562426 | miR-26b-5p | 2:2196051 | G | A | 0.521668 | 0.055455 | 0.015759 | 0.000433 | cis |
| rs7580147 | miR-26b-5p | 2:2196430 | A | G | 0.52136 | 0.053656 | 0.015917 | 0.000749 | cis |
| rs608047 | miR-26b-5p | 2:2196159 | G | T | 0.51862 | 0.054927 | 0.015863 | 0.000535 | cis |
| rs1269443 | miR-26b-5p | 2:2195908 | A | G | 0.52511 | 0.054081 | 0.015929 | 0.000686 | cis |
| rs690882 | miR-26b-5p | 2:2196406 | T | C | 0.517871 | 0.052841 | 0.015994 | 0.000954 | cis |
| rs1189620 | miR-26b-5p | 2:2195992 | G | A | 0.523861 | 0.055208 | 0.015899 | 0.000516 | cis |
| rs6729341 | miR-26b-5p | 2:2196114 | T | G | 0.521808 | 0.055493 | 0.015762 | 0.000431 | cis |
| rs5692347 | miR-26b-5p | 2:2196385 | A | G | 0.52138 | 0.053616 | 0.015916 | 0.000756 | cis |
| rs2891667 | miR-26b-5p | 2:2195951 | A | G | 0.522513 | 0.055386 | 0.015915 | 0.000501 | cis |
| rs7603709 | miR-26b-5p | 2:2196426 | G | A | 0.521505 | 0.053698 | 0.015922 | 0.000745 | cis |
| rs933995 | miR-26b-5p | 2:2196507 | G | A | 0.520745 | 0.052993 | 0.015929 | 0.000878 | cis |
| rs1344643 | miR-26b-5p | 2:2195849 | G | C | 0.582688 | 0.057592 | 0.016129 | 0.000356 | cis |
| rs7599568 | miR-26b-5p | 2:2196078 | A | G | 0.521626 | 0.055469 | 0.015756 | 0.000431 | cis |
| rs2037622 | miR-26b-5p | 2:2196094 | T | C | 0.521531 | 0.05546 | 0.015759 | 0.000433 | cis |
| rs3770219 | miR-26b-5p | 2:2196197 | T | C | 0.521657 | 0.055581 | 0.01576 | 0.000421 | cis |
| rs3770220 | miR-26b-5p | 2:2196195 | C | T | 0.521793 | 0.055497 | 0.015762 | 0.00043 | cis |
| rs6708650 | miR-26b-5p | 2:2196166 | A | G | 0.524507 | 0.05578 | 0.01585 | 0.000433 | cis |
| rs7559629 | miR-26b-5p | 2:2196570 | A | T | 0.578597 | 0.055428 | 0.016143 | 0.000596 | cis |
| rs7573999 | miR-26b-5p | 2:2196013 | A | G | 0.521472 | 0.055384 | 0.015826 | 0.000466 | cis |
| rs6436070 | miR-26b-5p | 2:2195979 | A | G | 0.523011 | 0.055308 | 0.015879 | 0.000496 | cis |
| rs6746040 | miR-26b-5p | 2:2195888 | G | T | 0.524516 | 0.05387 | 0.015949 | 0.000731 | cis |
| rs6436073 | miR-26b-5p | 2:2196182 | A | C | 0.521309 | 0.055309 | 0.015776 | 0.000455 | cis |
| rs6436081 | miR-26b-5p | 2:2196378 | A | G | 0.520597 | 0.053321 | 0.015911 | 0.000804 | cis |
| rs6436086 | miR-26b-5p | 2:2196401 | T | C | 0.521694 | 0.054689 | 0.015968 | 0.000615 | cis |
| rs6757772 | miR-26b-5p | 2:2196184 | T | G | 0.521818 | 0.055501 | 0.015762 | 0.00043 | cis |
| rs6761437 | miR-26b-5p | 2:2195980 | G | A | 0.522999 | 0.055317 | 0.015878 | 0.000494 | cis |
| rs3770211 | miR-26b-5p | 2:2192697 | G | T | 0.627162 | 0.071601 | 0.019396 | 0.000223 | cis |
| rs1541777 | miR-26b-5p | 2:2195872 | A | G | 0.523855 | 0.054074 | 0.015883 | 0.000663 | cis |
| rs6436072 | miR-26b-5p | 2:2195980 | C | T | 0.522979 | 0.055309 | 0.015878 | 0.000495 | cis |
| rs1747027 | miR-26b-5p | 2:2196519 | A | T | 0.579867 | 0.056971 | 0.016081 | 0.000396 | cis |
| rs7605055 | miR-26b-5p | 2:2196270 | G | C | 0.521751 | 0.055503 | 0.015763 | 0.00043 | cis |
| rs4674328 | miR-26b-5p | 2:2195932 | T | C | 0.523569 | 0.054302 | 0.015934 | 0.000655 | cis |
| rs6436084 | miR-26b-5p | 2:2196397 | A | G | 0.521376 | 0.053599 | 0.015917 | 0.000759 | cis |
| rs4674330 | miR-26b-5p | 2:2195957 | T | C | 0.523024 | 0.055309 | 0.015881 | 0.000496 | cis |
| rs921968 | miR-26b-5p | 2:2192722 | G | T | 0.418427 | -0.066112 | 0.016413 | 5.63E-05 | cis |
| rs6723334 | miR-26b-5p | 2:2196293 | T | C | 0.521746 | 0.055495 | 0.015767 | 0.000432 | cis |
| rs7586384 | miR-26b-5p | 2:2195828 | T | C | 0.524739 | 0.053776 | 0.015952 | 0.000748 | cis |
| rs6725488 | miR-26b-5p | 2:2196454 | T | C | 0.520854 | 0.053505 | 0.015998 | 0.000825 | cis |
| rs7573075 | miR-26b-5p | 2:2196306 | A | G | 0.522144 | 0.056373 | 0.015876 | 0.000384 | cis |

| | | | | | | | | | |
|---|---|---|---|---|---|---|---|---|---|
| rs1261354 | miR-26b-5p | 2:2196235 | C | T | 0.521807 | 0.055512 | 0.015761 | 0.000428 | cis |
| rs2059717 | miR-26b-5p | 2:2196003 | A | T | 0.522281 | 0.055193 | 0.015889 | 0.000513 | cis |
| rs5598365 | miR-26b-5p | 2:2196549 | T | C | 0.579795 | 0.057419 | 0.016117 | 0.000367 | cis |
| rs7594289 | miR-26b-5p | 2:2196522 | A | G | 0.579931 | 0.056914 | 0.016084 | 0.000403 | cis |
| rs6436069 | miR-26b-5p | 2:2195971 | T | C | 0.524437 | 0.055337 | 0.015916 | 0.000507 | cis |
| rs6436083 | miR-26b-5p | 2:2196397 | C | T | 0.521378 | 0.0536 | 0.015917 | 0.000758 | cis |
| rs4674334 | miR-26b-5p | 2:2196228 | T | C | 0.581424 | 0.06085 | 0.016081 | 0.000154 | cis |
| rs1261163 | miR-26b-5p | 2:2196212 | C | T | 0.521783 | 0.055489 | 0.015761 | 0.000431 | cis |
| rs4674338 | miR-26b-5p | 2:2196474 | G | A | 0.580172 | 0.057266 | 0.016054 | 0.000361 | cis |
| rs7593544 | miR-26b-5p | 2:2196237 | G | A | 0.521657 | 0.055646 | 0.015814 | 0.000434 | cis |
| rs1529382 | miR-26b-5p | 2:2196264 | C | T | 0.521787 | 0.055502 | 0.015764 | 0.00043 | cis |
| rs7595901 | miR-26b-5p | 2:2196069 | A | G | 0.521083 | 0.055291 | 0.015771 | 0.000455 | cis |
| rs7594046 | miR-26b-5p | 2:2196241 | G | A | 0.521286 | 0.055302 | 0.015776 | 0.000456 | cis |
| rs3569517 | miR-26b-5p | 2:2195891 | C | T | 0.525178 | 0.054057 | 0.01593 | 0.00069 | cis |
| rs4674332 | miR-26b-5p | 2:2196043 | T | C | 0.581884 | 0.060006 | 0.016042 | 0.000184 | cis |
| rs3518951 | miR-26b-5p | 2:2196287 | T | C | 0.521226 | 0.055313 | 0.01578 | 0.000456 | cis |
| rs1298700 | miR-26b-5p | 2:2196581 | A | T | 0.568859 | 0.054823 | 0.016169 | 0.000697 | cis |
| rs6436071 | miR-26b-5p | 2:2195980 | A | T | 0.522989 | 0.055303 | 0.015878 | 0.000496 | cis |
| rs1078803 | miR-26b-5p | 2:2196065 | C | A | 0.521605 | 0.055467 | 0.015756 | 0.000431 | cis |
| rs6219165 | miR-26b-5p | 2:2196305 | C | T | 0.521355 | 0.055593 | 0.015801 | 0.000434 | cis |
| rs3535851 | miR-26b-5p | 2:2196257 | A | C | 0.521781 | 0.055485 | 0.015763 | 0.000431 | cis |
| rs6436082 | miR-26b-5p | 2:2196394 | A | G | 0.521381 | 0.053601 | 0.015917 | 0.000758 | cis |
| rs755112 | miR-26b-5p | 2:2196060 | C | T | 0.521613 | 0.055468 | 0.015756 | 0.000431 | cis |
| rs7599365 | miR-26b-5p | 2:2196205 | G | A | 0.521803 | 0.055486 | 0.015762 | 0.000431 | cis |
| rs6219329 | miR-26b-5p | 2:2196552 | T | G | 0.578885 | 0.057091 | 0.016108 | 0.000394 | cis |
| rs1261239 | miR-26b-5p | 2:2196216 | G | C | 0.521291 | 0.055302 | 0.015776 | 0.000456 | cis |
| rs6722216 | miR-26b-5p | 2:2196132 | G | A | 0.522459 | 0.055597 | 0.015782 | 0.000427 | cis |
| rs1339019 | miR-26b-5p | 2:2196004 | A | G | 0.523808 | 0.055516 | 0.015896 | 0.000478 | cis |
| rs6436085 | miR-26b-5p | 2:2196400 | G | A | 0.525615 | 0.054308 | 0.016021 | 0.0007 | cis |
| rs2272188 | miR-26b-5p | 2:2196122 | A | G | 0.521792 | 0.055507 | 0.015762 | 0.000429 | cis |
| rs7599945 | miR-26b-5p | 2:2196415 | G | T | 0.521359 | 0.053605 | 0.015916 | 0.000757 | cis |
| rs7599984 | miR-26b-5p | 2:2196567 | T | G | 0.579124 | 0.056797 | 0.016139 | 0.000433 | cis |
| rs3731876 | miR-26b-5p | 2:2196028 | G | A | 0.519667 | 0.055064 | 0.015827 | 0.000503 | cis |
| rs3605995 | miR-26b-5p | 2:2196432 | G | T | 0.523691 | 0.053453 | 0.016012 | 0.000843 | cis |
| rs1338265 | miR-26b-5p | 2:2196281 | A | G | 0.521733 | 0.055487 | 0.015766 | 0.000432 | cis |
| rs6219165 | miR-26b-5p | 2:2196302 | A | G | 0.528749 | 0.055748 | 0.015991 | 0.00049 | cis |
| rs7574007 | miR-26b-5p | 2:2196315 | A | G | 0.522534 | 0.055797 | 0.015832 | 0.000425 | cis |
| rs3542263 | miR-26b-5p | 2:2196433 | C | T | 0.518267 | 0.054346 | 0.016078 | 0.000724 | cis |
| rs2272190 | miR-26b-5p | 2:2196099 | G | C | 0.521793 | 0.055507 | 0.015762 | 0.000429 | cis |
| rs7567622 | miR-26b-5p | 2:2196245 | A | T | 0.521781 | 0.055493 | 0.015761 | 0.00043 | cis |
| rs4674329 | miR-26b-5p | 2:2195955 | A | G | 0.523073 | 0.055309 | 0.015883 | 0.000497 | cis |
| rs7582329 | miR-26b-5p | 2:2196209 | C | T | 0.52178 | 0.055495 | 0.015762 | 0.00043 | cis |
| rs6436078 | miR-26b-5p | 2:2196361 | G | C | 0.522062 | 0.053471 | 0.015924 | 0.000786 | cis |
| rs3821035 | miR-26b-5p | 2:2196202 | T | G | 0.521793 | 0.055497 | 0.015762 | 0.00043 | cis |
| rs6436087 | miR-26b-5p | 2:2196454 | G | A | 0.521784 | 0.053366 | 0.015963 | 0.000829 | cis |
| rs2272191 | miR-26b-5p | 2:2196097 | G | A | 0.521718 | 0.055612 | 0.015766 | 0.00042 | cis |
| rs6733892 | miR-26b-5p | 2:2196160 | C | A | 0.524696 | 0.055999 | 0.015843 | 0.000409 | cis |
| rs3559467 | miR-26b-5p | 2:2195892 | T | A | 0.524995 | 0.054059 | 0.015935 | 0.000692 | cis |
| rs3755045 | miR-26b-5p | 2:2196091 | T | G | 0.521596 | 0.055469 | 0.015755 | 0.000431 | cis |
| rs692049 | miR-26b-5p | 2:2196227 | G | A | 0.51895 | 0.054854 | 0.015853 | 0.00054 | cis |
| rs1302258 | miR-26b-5p | 2:2195891 | T | C | 0.525156 | 0.054052 | 0.015929 | 0.000691 | cis |
| rs6219326 | miR-26b-5p | 2:2196305 | C | T | 0.521559 | 0.055568 | 0.015796 | 0.000435 | cis |
| rs586194 | miR-26b-5p | 2:2196104 | A | G | 0.519004 | 0.054873 | 0.015852 | 0.000537 | cis |

| | | | | | | | | | |
|---|---|---|---|---|---|---|---|---|---|
| rs5590024 | miR-26b-5p | 2:2196187( | T | C | 0.582356 | 0.060797 | 0.016062 | 0.000154 | cis |
| rs5767696 | miR-26b-5p | 2:2196230 | A | G | 0.520208 | 0.055483 | 0.01571 | 0.000413 | cis |
| rs7296602( | miR-26b-5p | 2:2196426 | C | G | 0.58315 | 0.058956 | 0.016161 | 0.000264 | cis |
| rs6752977 | miR-26b-5p | 2:2196260( | C | T | 0.521776 | 0.055485 | 0.015763 | 0.000432 | cis |
| rs6751527 | miR-26b-5p | 2:2196309 | C | G | 0.582826 | 0.060732 | 0.01608 | 0.000159 | cis |
| rs1079177 | miR-26b-5p | 2:2196059 | G | A | 0.521498 | 0.055552 | 0.01576 | 0.000423 | cis |
| rs6704575 | miR-26b-5p | 2:2195895 | T | C | 0.525144 | 0.054058 | 0.015929 | 0.00069 | cis |
| rs7582193 | miR-26b-5p | 2:2196525 | T | C | 0.579834 | 0.056853 | 0.016091 | 0.000411 | cis |
| rs7030617 | miR-27b-3p | 9:9749365( | T | C | 0.873326 | -0.066233 | 0.019125 | 0.000534 | cis |
| rs1233927 | miR-27b-3p | 9:9749091( | G | C | 0.873256 | -0.066391 | 0.019137 | 0.000522 | cis |
| rs1412488 | miR-27b-3p | 9:9778036 | C | G | 0.111617 | 0.104678 | 0.020257 | 2.37E-07 | cis |
| rs7853126 | miR-27b-3p | 9:9750525 | A | G | 0.873229 | -0.065517 | 0.019067 | 0.00059 | cis |
| rs1233770( | miR-27b-3p | 9:9752315 | A | G | 0.936811 | -0.086193 | 0.025972 | 0.000904 | cis |
| rs4537361 | miR-27b-3p | 9:9758524 | G | T | 0.133945 | 0.065405 | 0.019079 | 0.000608 | cis |
| rs3802454 | miR-27b-3p | 9:9778105 | T | G | 0.114317 | 0.103042 | 0.020091 | 2.91E-07 | cis |
| rs1082143 | miR-27b-3p | 9:9777271 | T | C | 0.920968 | -0.10743 | 0.023551 | 5.08E-06 | cis |
| rs7851173 | miR-27b-3p | 9:9777753 | T | C | 0.114158 | 0.102729 | 0.020171 | 3.53E-07 | cis |
| rs3433563( | miR-27b-3p | 9:9751721( | T | C | 0.936836 | -0.086267 | 0.025983 | 0.0009 | cis |
| rs1233799 | miR-27b-3p | 9:9752016 | G | A | 0.936702 | -0.086471 | 0.02598 | 0.000873 | cis |
| rs7873479 | miR-27b-3p | 9:9776947 | A | G | 0.111038 | 0.102358 | 0.020502 | 5.96E-07 | cis |
| rs7888717 | miR-27b-3p | 9:9776943 | G | C | 0.930373 | -0.085514 | 0.025025 | 0.000633 | cis |
| rs3802456 | miR-27b-3p | 9:9777185 | T | C | 0.114212 | 0.100854 | 0.020278 | 6.57E-07 | cis |
| rs396538 | miR-27b-3p | 9:9748770 | A | G | 0.87316 | -0.066557 | 0.019146 | 0.000509 | cis |
| rs7458754 | miR-27b-3p | 9:9750120 | T | C | 0.936873 | -0.086529 | 0.025993 | 0.000872 | cis |
| rs3739749 | miR-27b-3p | 9:9782456( | C | T | 0.928911 | -0.089342 | 0.024755 | 0.000307 | cis |
| rs962229 | miR-27b-3p | 9:9779216 | A | G | 0.113764 | 0.101241 | 0.020085 | 4.64E-07 | cis |
| rs4744436 | miR-27b-3p | 9:9783158 | G | A | 0.12894 | 0.094766 | 0.020146 | 2.55E-06 | cis |
| rs4744435 | miR-27b-3p | 9:9783147 | C | T | 0.117477 | 0.100143 | 0.020029 | 5.74E-07 | cis |
| rs4743981 | miR-27b-3p | 9:9781200 | G | A | 0.113585 | 0.101701 | 0.020083 | 4.11E-07 | cis |
| rs380205 | miR-27b-3p | 9:9753155 | C | T | 0.8732 | -0.065526 | 0.019068 | 0.00059 | cis |
| rs1125997( | miR-27b-3p | 9:9756002 | G | A | 0.875642 | -0.06709 | 0.01916 | 0.000462 | cis |
| rs193292 | miR-27b-3p | 9:9754708 | T | A | 0.872905 | -0.065177 | 0.019023 | 0.000612 | cis |
| rs1235227( | miR-27b-3p | 9:9749031 | A | G | 0.873236 | -0.066413 | 0.019138 | 0.00052 | cis |
| rs4744433 | miR-27b-3p | 9:9782702 | G | C | 0.114688 | 0.10334 | 0.020149 | 2.92E-07 | cis |
| rs7262141 | miR-27b-3p | 9:9777228 | C | T | 0.921642 | -0.105984 | 0.023642 | 7.36E-06 | cis |
| rs1113102 | miR-27b-3p | 9:9778016 | A | G | 0.114669 | 0.102364 | 0.020079 | 3.43E-07 | cis |
| rs418764 | miR-27b-3p | 9:9753133 | T | C | 0.8732 | -0.065526 | 0.019068 | 0.00059 | cis |
| rs384039 | miR-27b-3p | 9:9754856 | A | G | 0.87619 | -0.066336 | 0.019163 | 0.000537 | cis |
| rs1328534 | miR-27b-3p | 9:9759209( | C | G | 0.133952 | 0.065384 | 0.019079 | 0.00061 | cis |
| rs471161 | miR-27b-3p | 9:9750058( | T | G | 0.873518 | -0.065622 | 0.019085 | 0.000585 | cis |
| rs1011951 | miR-27b-3p | 9:9780838 | G | A | 0.114004 | 0.101477 | 0.020069 | 4.27E-07 | cis |
| rs7025234 | miR-27b-3p | 9:9760264 | G | A | 0.134081 | 0.06557 | 0.019089 | 0.000593 | cis |
| rs1810772 | miR-27b-3p | 9:9747717 | G | A | 0.867382 | -0.065043 | 0.01891 | 0.000582 | cis |
| rs1020963 | miR-27b-3p | 9:9778974 | A | G | 0.111091 | 0.10258 | 0.020233 | 3.98E-07 | cis |
| rs1234926 | miR-27b-3p | 9:9780503( | C | G | 0.928719 | -0.088585 | 0.024592 | 0.000316 | cis |
| rs672012 | miR-27b-3p | 9:9751005 | G | A | 0.873203 | -0.06554 | 0.019068 | 0.000588 | cis |
| rs2119954 | miR-27b-3p | 9:9758251 | C | T | 0.130584 | 0.066588 | 0.019219 | 0.000531 | cis |
| rs1073240( | miR-27b-3p | 9:9760007 | C | T | 0.134085 | 0.065572 | 0.019089 | 0.000593 | cis |
| rs1011737( | miR-27b-3p | 9:9781452 | A | G | 0.116136 | 0.09896 | 0.019994 | 7.44E-07 | cis |
| rs356134 | miR-27b-3p | 9:9755192 | C | T | 0.872901 | -0.06518 | 0.019023 | 0.000612 | cis |
| rs1051225 | miR-27b-3p | 9:9782835 | C | A | 0.114926 | 0.103523 | 0.020148 | 2.78E-07 | cis |
| rs1691166 | miR-27b-3p | 9:9749526 | A | G | 0.873186 | -0.066195 | 0.019124 | 0.000538 | cis |
| rs1099335( | miR-27b-3p | 9:9755951 | C | T | 0.93991 | -0.09471 | 0.026916 | 0.000434 | cis |

| | | | | | | | | | |
|---|---|---|---|---|---|---|---|---|---|
| rs4743979 | miR-27b-3p | 9:97805094 | A | C | 0.11362 | 0.100493 | 0.019985 | 4.94E-07 | cis |
| rs400231 | miR-27b-3p | 9:97533176 | T | G | 0.873187 | -0.065533 | 0.019068 | 0.000589 | cis |
| rs4437717 | miR-27b-3p | 9:97508281 | G | A | 0.8732 | -0.065547 | 0.019068 | 0.000587 | cis |
| rs2767404 | miR-27b-3p | 9:97573597 | A | G | 0.872614 | -0.064729 | 0.018999 | 0.000657 | cis |
| rs356124 | miR-27b-3p | 9:97561882 | A | T | 0.87288 | -0.065172 | 0.019024 | 0.000613 | cis |
| rs7857679 | miR-27b-3p | 9:97803860 | C | T | 0.114154 | 0.101292 | 0.020021 | 4.21E-07 | cis |
| rs7021351 | miR-27b-3p | 9:97793139 | A | T | 0.113411 | 0.101405 | 0.020024 | 4.10E-07 | cis |
| rs10732405 | miR-27b-3p | 9:97586542 | G | T | 0.12436 | 0.065691 | 0.019109 | 0.000587 | cis |
| rs4743980 | miR-27b-3p | 9:97809853 | C | T | 0.11354 | 0.10205 | 0.020111 | 3.89E-07 | cis |
| rs12346261 | miR-27b-3p | 9:97785778 | G | C | 0.920623 | -0.105495 | 0.023307 | 6.00E-06 | cis |
| rs4744425 | miR-27b-3p | 9:97792692 | C | T | 0.113909 | 0.101263 | 0.020071 | 4.53E-07 | cis |
| rs356135 | miR-27b-3p | 9:97550495 | G | A | 0.876227 | -0.066318 | 0.019168 | 0.000541 | cis |
| rs10993467 | miR-27b-3p | 9:97826698 | C | T | 0.114683 | 0.103292 | 0.020145 | 2.94E-07 | cis |
| rs10993456 | miR-27b-3p | 9:97783630 | T | C | 0.920166 | -0.107248 | 0.023249 | 3.97E-06 | cis |
| rs374708 | miR-27b-3p | 9:97528636 | A | C | 0.873196 | -0.06553 | 0.019068 | 0.000589 | cis |
| rs7022452 | miR-27b-3p | 9:97788002 | A | G | 0.114405 | 0.100534 | 0.020049 | 5.32E-07 | cis |
| rs4537362 | miR-27b-3p | 9:97798170 | C | T | 0.114017 | 0.100717 | 0.020008 | 4.81E-07 | cis |
| rs397617 | miR-27b-3p | 9:97523578 | T | C | 0.873178 | -0.065534 | 0.019066 | 0.000588 | cis |
| rs10993348 | miR-27b-3p | 9:97556168 | T | C | 0.876181 | -0.066316 | 0.019164 | 0.000539 | cis |
| rs12336308 | miR-27b-3p | 9:97538595 | G | T | 0.936745 | -0.086204 | 0.025954 | 0.000896 | cis |
| rs60514919 | miR-27b-3p | 9:97553468 | G | A | 0.9366 | -0.085759 | 0.025892 | 0.000926 | cis |
| rs4744426 | miR-27b-3p | 9:97805062 | G | A | 0.113621 | 0.100493 | 0.019985 | 4.94E-07 | cis |
| rs10733755 | miR-27b-3p | 9:97600063 | A | G | 0.134083 | 0.065571 | 0.019089 | 0.000593 | cis |
| rs55926255 | miR-27b-3p | 9:97606688 | A | G | 0.133755 | 0.066217 | 0.019133 | 0.000538 | cis |
| rs4744432 | miR-27b-3p | 9:97826939 | A | G | 0.114688 | 0.103331 | 0.020149 | 2.92E-07 | cis |
| rs7873567 | miR-27b-3p | 9:97807958 | G | A | 0.113558 | 0.101499 | 0.020066 | 4.23E-07 | cis |
| rs10821405 | miR-27b-3p | 9:97606591 | A | G | 0.131039 | 0.068284 | 0.0193 | 0.000403 | cis |
| rs2304884 | miR-27b-3p | 9:97587093 | G | A | 0.1306 | 0.06654 | 0.019217 | 0.000535 | cis |
| rs439052 | miR-27b-3p | 9:97569038 | C | T | 0.872645 | -0.064747 | 0.018999 | 0.000655 | cis |
| rs55795862 | miR-27b-3p | 9:97504650 | G | A | 0.873239 | -0.065509 | 0.019067 | 0.000591 | cis |
| rs1564234 | miR-27b-3p | 9:97781594 | G | A | 0.114214 | 0.103085 | 0.020085 | 2.86E-07 | cis |
| rs2061800 | miR-27b-3p | 9:97797501 | G | A | 0.114009 | 0.100718 | 0.020007 | 4.80E-07 | cis |
| rs2767403 | miR-27b-3p | 9:97574120 | C | G | 0.872633 | -0.064716 | 0.018999 | 0.000659 | cis |
| rs10761366 | miR-27b-3p | 9:97774879 | C | T | 0.114522 | 0.102412 | 0.020199 | 3.97E-07 | cis |
| rs11371161 | miR-27b-3p | 9:97544084 | C | T | 0.876232 | -0.066378 | 0.019164 | 0.000533 | cis |
| rs12336437 | miR-27b-3p | 9:97531079 | C | T | 0.936788 | -0.086126 | 0.025959 | 0.000907 | cis |
| rs75314995 | miR-27b-3p | 9:97804641 | A | C | 0.917394 | -0.102365 | 0.022938 | 8.10E-06 | cis |
| rs10993427 | miR-27b-3p | 9:97738137 | G | C | 0.902585 | -0.093633 | 0.021423 | 1.24E-05 | cis |
| rs12339639 | miR-27b-3p | 9:97786816 | C | T | 0.920633 | -0.104914 | 0.023284 | 6.61E-06 | cis |
| rs1004373 | miR-27b-3p | 9:97814770 | A | G | 0.113842 | 0.101763 | 0.020093 | 4.09E-07 | cis |
| rs12340251 | miR-27b-3p | 9:97494598 | T | C | 0.873328 | -0.066221 | 0.019123 | 0.000534 | cis |
| rs18885671 | miR-27b-3p | 9:97582244 | T | G | 0.941796 | -0.097397 | 0.027633 | 0.000424 | cis |
| rs10122127 | miR-27b-3p | 9:97603383 | G | C | 0.130745 | 0.066691 | 0.019229 | 0.000524 | cis |
| rs376902 | miR-27b-3p | 9:97554227 | T | C | 0.876186 | -0.066317 | 0.019164 | 0.000539 | cis |
| rs356129 | miR-27b-3p | 9:97570715 | A | T | 0.874739 | -0.065864 | 0.019132 | 0.000576 | cis |
| rs356137 | miR-27b-3p | 9:97549635 | C | T | 0.872882 | -0.065183 | 0.019023 | 0.000611 | cis |
| rs242259 | miR-27b-3p | 9:97552788 | C | A | 0.875522 | -0.066789 | 0.019157 | 0.00049 | cis |
| rs356140 | miR-27b-3p | 9:97547963 | T | C | 0.872889 | -0.065191 | 0.019023 | 0.000611 | cis |
| rs67625382 | miR-27b-3p | 9:97494761 | G | T | 0.873337 | -0.066201 | 0.019122 | 0.000536 | cis |
| rs57504746 | miR-27b-3p | 9:97506732 | T | C | 0.873135 | -0.065636 | 0.019159 | 0.000613 | cis |
| rs7031172 | miR-27b-3p | 9:97598180 | C | T | 0.130702 | 0.067077 | 0.019236 | 0.000489 | cis |
| rs2404456 | miR-27b-3p | 9:97823913 | A | G | 0.116933 | 0.100602 | 0.020131 | 5.82E-07 | cis |
| rs10993464 | miR-27b-3p | 9:97808086 | A | G | 0.113764 | 0.101414 | 0.020066 | 4.33E-07 | cis |

| | | | | | | | | | |
|---|---|---|---|---|---|---|---|---|---|
| rs356138 | miR-27b-3p | 9:9754896 | A | G | 0.872674 | -0.065384 | 0.019029 | 0.00059 | cis |
| rs1268470 | miR-27b-3p | 9:9749512 | G | A | 0.873265 | -0.066368 | 0.01912 | 0.000518 | cis |
| rs1228614 | miR-27b-3p | 9:9755639 | C | T | 0.872866 | -0.065151 | 0.019024 | 0.000616 | cis |
| rs1051223 | miR-27b-3p | 9:9750990 | T | G | 0.93686 | -0.08632 | 0.025994 | 0.000898 | cis |
| rs1076136 | miR-27b-3p | 9:9779398 | G | A | 0.88813 | -0.104152 | 0.02048 | 3.66E-07 | cis |
| rs1012176 | miR-27b-3p | 9:9782898 | T | C | 0.116919 | 0.103589 | 0.020066 | 2.44E-07 | cis |
| rs181488 | miR-27b-3p | 9:9754142 | A | G | 0.876233 | -0.06638 | 0.019164 | 0.000533 | cis |
| rs7520233 | miR-27b-3p | 9:9782169 | A | G | 0.928884 | -0.089303 | 0.024745 | 0.000307 | cis |
| rs6479565 | miR-27b-3p | 9:9747686 | G | A | 0.867528 | -0.065301 | 0.018861 | 0.000536 | cis |
| rs1012381 | miR-27b-3p | 9:9782887 | G | A | 0.11496 | 0.103546 | 0.020147 | 2.76E-07 | cis |
| rs4744431 | miR-27b-3p | 9:9782691 | A | G | 0.114511 | 0.103424 | 0.02015 | 2.86E-07 | cis |
| rs356123 | miR-27b-3p | 9:9756531 | A | G | 0.872612 | -0.06476 | 0.018999 | 0.000653 | cis |
| rs2584804 | miR-27b-3p | 9:9756586 | G | A | 0.875958 | -0.065888 | 0.019138 | 0.000576 | cis |
| rs1076136 | miR-27b-3p | 9:9776828 | T | C | 0.114089 | 0.100629 | 0.020328 | 7.41E-07 | cis |
| rs1234453 | miR-27b-3p | 9:9749878 | A | G | 0.873483 | -0.065703 | 0.019093 | 0.000579 | cis |
| rs7726965 | miR-27b-3p | 9:9777368 | C | A | 0.929611 | -0.088231 | 0.024872 | 0.000389 | cis |
| rs1012186 | miR-27b-3p | 9:9760612 | A | G | 0.133778 | 0.066212 | 0.019132 | 0.000539 | cis |
| rs2864728 | miR-27b-3p | 9:9750090 | C | A | 0.873228 | -0.06493 | 0.019089 | 0.000671 | cis |
| rs7045387 | miR-27b-3p | 9:9760278 | T | C | 0.130738 | 0.066693 | 0.019229 | 0.000524 | cis |
| rs4743984 | miR-27b-3p | 9:9781315 | G | A | 0.114329 | 0.101778 | 0.02009 | 4.06E-07 | cis |
| rs1099345 | miR-27b-3p | 9:9778487 | C | T | 0.92033 | -0.106072 | 0.023285 | 5.23E-06 | cis |
| rs356143 | miR-27b-3p | 9:9754581 | T | C | 0.87291 | -0.065177 | 0.019023 | 0.000612 | cis |
| rs4744430 | miR-27b-3p | 9:9781573 | T | C | 0.114335 | 0.101883 | 0.020098 | 3.99E-07 | cis |
| rs7681458 | miR-27b-3p | 9:9749552 | T | C | 0.93664 | -0.087311 | 0.026045 | 0.000801 | cis |
| rs356146 | miR-27b-3p | 9:9754270 | C | T | 0.87293 | -0.065192 | 0.019022 | 0.00061 | cis |
| rs1076135 | miR-27b-3p | 9:9759023 | T | C | 0.133968 | 0.065385 | 0.019079 | 0.00061 | cis |
| rs405110 | miR-27b-3p | 9:9754994 | T | C | 0.872904 | -0.065176 | 0.019023 | 0.000612 | cis |
| rs4744383 | miR-27b-3p | 9:9760437 | T | C | 0.131453 | 0.066313 | 0.01921 | 0.000556 | cis |
| rs356126 | miR-27b-3p | 9:9757214 | C | A | 0.872865 | -0.065152 | 0.019025 | 0.000616 | cis |
| rs415016 | miR-27b-3p | 9:9757580 | C | T | 0.875822 | -0.065841 | 0.019139 | 0.000581 | cis |
| rs7020709 | miR-27b-3p | 9:9783221 | G | A | 0.120586 | 0.105568 | 0.020195 | 1.72E-07 | cis |
| rs1099338 | miR-27b-3p | 9:9763946 | C | T | 0.933929 | -0.084609 | 0.025605 | 0.000952 | cis |
| rs7862700 | miR-27b-3p | 9:9758275 | T | C | 0.133852 | 0.065387 | 0.019079 | 0.00061 | cis |
| rs2404295 | miR-27b-3p | 9:9772386 | T | C | 0.889705 | -0.074988 | 0.020386 | 0.000235 | cis |
| rs1233954 | miR-27b-3p | 9:9763892 | C | A | 0.933932 | -0.084602 | 0.025606 | 0.000953 | cis |
| rs7865144 | miR-27b-3p | 9:9777109 | G | A | 0.113861 | 0.101143 | 0.020302 | 6.29E-07 | cis |
| rs4744424 | miR-27b-3p | 9:9779145 | T | A | 0.113866 | 0.101247 | 0.020069 | 4.54E-07 | cis |
| rs356130 | miR-27b-3p | 9:9756788 | G | A | 0.872648 | -0.064752 | 0.018999 | 0.000654 | cis |
| rs1099346 | miR-27b-3p | 9:9779517 | T | C | 0.9176 | -0.103035 | 0.022935 | 7.04E-06 | cis |
| rs2584803 | miR-27b-3p | 9:9756096 | C | T | 0.874243 | -0.065254 | 0.019155 | 0.000658 | cis |
| rs356122 | miR-27b-3p | 9:9756546 | T | C | 0.872312 | -0.064377 | 0.019003 | 0.000705 | cis |
| rs7871218 | miR-27b-3p | 9:9780779 | C | T | 0.114428 | 0.10167 | 0.020065 | 4.04E-07 | cis |
| rs1255369 | miR-27b-3p | 9:9750782 | C | G | 0.873201 | -0.065543 | 0.019068 | 0.000587 | cis |
| rs3494057 | miR-27b-3p | 9:9753050 | C | T | 0.873197 | -0.06552 | 0.019068 | 0.00059 | cis |
| rs356142 | miR-27b-3p | 9:9754614 | G | A | 0.875157 | -0.06639 | 0.019097 | 0.000508 | cis |
| rs1073998 | miR-27b-3p | 9:9760029 | T | C | 0.131269 | 0.066971 | 0.019238 | 0.000499 | cis |
| rs2406899 | miR-27b-3p | 9:9759301 | C | T | 0.130621 | 0.06653 | 0.019217 | 0.000536 | cis |
| rs3858087 | miR-27b-3p | 9:9758307 | C | A | 0.133907 | 0.065411 | 0.01908 | 0.000607 | cis |
| rs1021742 | miR-27b-3p | 9:9749435 | G | A | 0.873325 | -0.066223 | 0.019123 | 0.000534 | cis |
| rs7048941 | miR-27b-3p | 9:9780967 | T | C | 0.113775 | 0.101541 | 0.020075 | 4.23E-07 | cis |
| rs7043705 | miR-27b-3p | 9:9779129 | A | C | 0.113865 | 0.101249 | 0.020069 | 4.54E-07 | cis |
| rs1099335 | miR-27b-3p | 9:9756048 | T | C | 0.87288 | -0.065174 | 0.019024 | 0.000613 | cis |
| rs7862156 | miR-27b-3p | 9:9780516 | C | T | 0.113877 | 0.100546 | 0.019986 | 4.89E-07 | cis |

| | | | | | | | | | |
|---|---|---|---|---|---|---|---|---|---|
| rs7047864 | miR-27b-3p | 9:97497588 | T | C | 0.880682 | -0.067493 | 0.019792 | 0.000649 | cis |
| rs7863291 | miR-27b-3p | 9:97807983 | T | C | 0.113827 | 0.101501 | 0.020067 | 4.23E-07 | cis |
| rs356127 | miR-27b-3p | 9:97571455 | A | G | 0.872824 | -0.065125 | 0.019027 | 0.00062 | cis |
| rs79654060 | miR-27b-3p | 9:97509663 | G | A | 0.93686 | -0.086322 | 0.025994 | 0.000898 | cis |
| rs9409806 | miR-27b-3p | 9:97560276 | A | G | 0.872839 | -0.065214 | 0.019024 | 0.000608 | cis |
| rs1530332 | miR-27b-3p | 9:97595143 | C | G | 0.131402 | 0.068322 | 0.0192 | 0.000373 | cis |
| rs7869666 | miR-27b-3p | 9:97597137 | A | T | 0.159601 | 0.076886 | 0.019059 | 5.48E-05 | cis |
| rs58042095 | miR-27b-3p | 9:97527535 | G | C | 0.873201 | -0.065651 | 0.019071 | 0.000576 | cis |
| rs4744377 | miR-27b-3p | 9:97578933 | G | T | 0.134037 | 0.06538 | 0.01908 | 0.000611 | cis |
| rs76326161 | miR-27b-3p | 9:97812838 | T | A | 0.928889 | -0.089091 | 0.024713 | 0.000312 | cis |
| rs1571895 | miR-27b-3p | 9:97788956 | A | G | 0.113867 | 0.101252 | 0.02007 | 4.53E-07 | cis |
| rs6479591 | miR-27b-3p | 9:97795421 | G | A | 0.113637 | 0.10118 | 0.02001 | 4.27E-07 | cis |
| rs7861358 | miR-27b-3p | 9:97579712 | C | G | 0.134147 | 0.065267 | 0.019076 | 0.000623 | cis |
| rs4744422 | miR-27b-3p | 9:97770527 | C | G | 0.111129 | 0.102412 | 0.020496 | 5.83E-07 | cis |
| rs10993436 | miR-27b-3p | 9:97744927 | T | C | 0.89544 | -0.08946 | 0.021045 | 2.13E-05 | cis |
| rs6479592 | miR-27b-3p | 9:97795579 | C | T | 0.113637 | 0.101181 | 0.02001 | 4.27E-07 | cis |
| rs913182 | miR-27b-3p | 9:97478309 | T | C | 0.867892 | -0.065472 | 0.018898 | 0.000531 | cis |
| rs389365 | miR-27b-3p | 9:97516919 | T | C | 0.873214 | -0.065532 | 0.019067 | 0.000589 | cis |
| rs3858088 | miR-27b-3p | 9:97583063 | G | C | 0.133853 | 0.065388 | 0.019079 | 0.00061 | cis |
| rs7048786 | miR-27b-3p | 9:97809500 | T | C | 0.113775 | 0.101515 | 0.020073 | 4.25E-07 | cis |
| rs1571894 | miR-27b-3p | 9:97789107 | G | T | 0.114427 | 0.100509 | 0.02005 | 5.36E-07 | cis |
| rs55914113 | miR-27b-3p | 9:97606606 | A | G | 0.134885 | 0.068521 | 0.019094 | 0.000333 | cis |
| rs12343530 | miR-27b-3p | 9:97532193 | T | C | 0.936784 | -0.086113 | 0.025957 | 0.000908 | cis |
| rs6479572 | miR-27b-3p | 9:97579538 | T | C | 0.130519 | 0.066563 | 0.019218 | 0.000533 | cis |
| rs2165716 | miR-27b-3p | 9:97582102 | G | A | 0.133875 | 0.065416 | 0.019079 | 0.000607 | cis |
| rs2404305 | miR-27b-3p | 9:97801706 | G | T | 0.114115 | 0.10078 | 0.020009 | 4.74E-07 | cis |
| rs10761359 | miR-27b-3p | 9:97608113 | G | A | 0.139347 | 0.07069 | 0.019068 | 0.00021 | cis |
| rs6479588 | miR-27b-3p | 9:97775868 | T | C | 0.111048 | 0.104431 | 0.020373 | 2.96E-07 | cis |
| rs6479570 | miR-27b-3p | 9:97579455 | C | T | 0.130519 | 0.066562 | 0.019218 | 0.000533 | cis |
| rs10761368 | miR-27b-3p | 9:97788696 | G | T | 0.111439 | 0.102485 | 0.020226 | 4.04E-07 | cis |
| rs4744380 | miR-27b-3p | 9:97597317 | T | A | 0.130738 | 0.066706 | 0.01923 | 0.000523 | cis |
| rs4744423 | miR-27b-3p | 9:97770682 | C | T | 0.114254 | 0.100802 | 0.020295 | 6.80E-07 | cis |
| rs4744379 | miR-27b-3p | 9:97592569 | G | A | 0.130654 | 0.066561 | 0.019217 | 0.000533 | cis |
| rs12344584 | miR-27b-3p | 9:97787336 | A | G | 0.920633 | -0.104914 | 0.023284 | 6.61E-06 | cis |
| rs6479574 | miR-27b-3p | 9:97587520 | G | A | 0.130601 | 0.06654 | 0.019217 | 0.000535 | cis |
| rs6479571 | miR-27b-3p | 9:97579505 | G | T | 0.130525 | 0.066586 | 0.019218 | 0.000531 | cis |
| rs187163 | miR-27b-3p | 9:97553048 | G | C | 0.872884 | -0.065179 | 0.019024 | 0.000612 | cis |
| rs4743982 | miR-27b-3p | 9:97812340 | A | G | 0.113777 | 0.101656 | 0.020083 | 4.15E-07 | cis |
| rs56292909 | miR-30b-5p | 8:13564058 | A | G | 0.75197 | -0.041717 | 0.011851 | 0.000431 | cis |
| rs13282704 | miR-30b-5p | 8:13573212 | C | T | 0.826672 | -0.06902 | 0.0136 | 3.87E-07 | cis |
| rs34034745 | miR-30b-5p | 8:13573807 | C | A | 0.827279 | -0.06888 | 0.01363 | 4.33E-07 | cis |
| rs7008400 | miR-30b-5p | 8:13559163 | G | A | 0.758238 | -0.041768 | 0.011932 | 0.000464 | cis |
| rs9987094 | miR-30b-5p | 8:13558669 | C | T | 0.758034 | -0.041417 | 0.01191 | 0.000506 | cis |
| rs67170031 | miR-30b-5p | 8:13567378 | G | C | 0.820817 | -0.069425 | 0.013483 | 2.62E-07 | cis |
| rs66876375 | miR-30b-5p | 8:13571612 | A | G | 0.822526 | -0.066977 | 0.01348 | 6.74E-07 | cis |
| rs13256725 | miR-30b-5p | 8:13566763 | G | A | 0.820973 | -0.069691 | 0.013462 | 2.26E-07 | cis |
| rs6992521 | miR-30b-5p | 8:13557333 | G | A | 0.758587 | -0.040996 | 0.011945 | 0.000599 | cis |
| rs11701054 | miR-30b-5p | 8:13568786 | C | T | 0.822163 | -0.067629 | 0.013488 | 5.33E-07 | cis |
| rs13266238 | miR-30b-5p | 8:13572629 | G | T | 0.825149 | -0.06764 | 0.01358 | 6.33E-07 | cis |
| rs14831660 | miR-30b-5p | 8:13568875 | C | A | 0.821815 | -0.067196 | 0.013483 | 6.24E-07 | cis |
| rs35082558 | miR-30b-5p | 8:13570595 | C | T | 0.822769 | -0.066939 | 0.01349 | 6.98E-07 | cis |
| rs74648705 | miR-30b-5p | 8:13559222 | G | A | 0.776861 | -0.044578 | 0.012311 | 0.000294 | cis |
| rs35448643 | miR-30b-5p | 8:13570244 | T | C | 0.822565 | -0.066896 | 0.013481 | 6.97E-07 | cis |

| | | | | | | | | | |
|---|---|---|---|---|---|---|---|---|---|
| rs28450354 | miR-30b-5p | 8:13560507 | C | T | 0.750067 | -0.043288 | 0.011832 | 0.000254 | cis |
| rs66502815 | miR-30b-5p | 8:13571895 | A | C | 0.822819 | -0.067149 | 0.013496 | 6.51E-07 | cis |
| rs72719790 | miR-30b-5p | 8:13564208 | T | C | 0.748139 | -0.040128 | 0.011801 | 0.000673 | cis |
| rs4909608 | miR-30b-5p | 8:13570624 | C | G | 0.288481 | 0.044162 | 0.01134 | 9.85E-05 | cis |
| rs7829476 | miR-30b-5p | 8:13560748 | G | A | 0.761443 | -0.04426 | 0.012023 | 0.000232 | cis |
| rs13267329 | miR-30b-5p | 8:13573511 | G | A | 0.827212 | -0.068829 | 0.013612 | 4.27E-07 | cis |
| rs7829427 | miR-30b-5p | 8:13569795 | A | G | 0.281683 | 0.045917 | 0.011319 | 4.98E-05 | cis |
| rs15068900 | miR-30b-5p | 8:13568032 | G | C | 0.822188 | -0.067307 | 0.013496 | 6.13E-07 | cis |
| rs67654984 | miR-30b-5p | 8:13570798 | G | A | 0.822544 | -0.066882 | 0.01348 | 7.00E-07 | cis |
| rs72719787 | miR-30b-5p | 8:13563800 | C | T | 0.752768 | -0.042267 | 0.011844 | 0.000359 | cis |
| rs35241762 | miR-30b-5p | 8:13567547 | C | T | 0.821656 | -0.069682 | 0.013489 | 2.39E-07 | cis |
| rs34074875 | miR-30b-5p | 8:13565257 | G | A | 0.822224 | -0.06968 | 0.013529 | 2.60E-07 | cis |
| rs11166624 | miR-30b-5p | 8:13562528 | C | T | 0.752461 | -0.042004 | 0.011831 | 0.000385 | cis |
| rs67880203 | miR-30b-5p | 8:13573290 | C | T | 0.826501 | -0.069011 | 0.013592 | 3.83E-07 | cis |
| rs17771076 | miR-30b-5p | 8:13563740 | A | C | 0.752384 | -0.041934 | 0.011838 | 0.000397 | cis |
| rs56065230 | miR-30b-5p | 8:13571333 | T | C | 0.823842 | -0.070241 | 0.013538 | 2.12E-07 | cis |
| rs17769126 | miR-30b-5p | 8:13559723 | C | T | 0.749994 | -0.043476 | 0.011826 | 0.000237 | cis |
| rs1867058 | miR-30b-5p | 8:13564693 | G | C | 0.748581 | -0.041449 | 0.011807 | 0.000447 | cis |
| rs13271002 | miR-30b-5p | 8:13567460 | T | G | 0.821675 | -0.06975 | 0.01349 | 2.33E-07 | cis |
| rs4398863 | miR-30b-5p | 8:13569511 | G | C | 0.285279 | 0.046228 | 0.011376 | 4.83E-05 | cis |
| rs4573225 | miR-30b-5p | 8:13572045 | C | G | 0.284717 | 0.045274 | 0.011389 | 7.03E-05 | cis |
| rs58948487 | miR-30b-5p | 8:13563729 | A | G | 0.752382 | -0.041933 | 0.011838 | 0.000397 | cis |
| rs71528371 | miR-30b-5p | 8:13568463 | C | T | 0.828156 | -0.069591 | 0.013843 | 4.98E-07 | cis |
| rs13262791 | miR-30b-5p | 8:13572991 | G | C | 0.83353 | -0.067281 | 0.013877 | 1.25E-06 | cis |
| rs13255015 | miR-30b-5p | 8:13573211 | G | A | 0.826669 | -0.06902 | 0.013599 | 3.87E-07 | cis |
| rs7821751 | miR-30b-5p | 8:13571067 | C | A | 0.284025 | 0.045986 | 0.011359 | 5.15E-05 | cis |
| rs13268530 | miR-30b-5p | 8:13572719 | C | T | 0.825953 | -0.068472 | 0.013595 | 4.74E-07 | cis |
| rs72719796 | miR-30b-5p | 8:13565857 | C | G | 0.820724 | -0.069701 | 0.013451 | 2.20E-07 | cis |
| rs56413514 | miR-30b-5p | 8:13564066 | G | A | 0.830335 | -0.067757 | 0.013752 | 8.35E-07 | cis |
| rs56277619 | miR-30b-5p | 8:13572651 | G | A | 0.825158 | -0.067644 | 0.01358 | 6.32E-07 | cis |
| rs34263466 | miR-30b-5p | 8:13572112 | G | C | 0.822725 | -0.067257 | 0.013496 | 6.24E-07 | cis |
| rs17697948 | miR-30b-5p | 8:13562893 | G | A | 0.752507 | -0.041966 | 0.011829 | 0.000388 | cis |
| rs7839162 | miR-30b-5p | 8:13570003 | A | C | 0.284188 | 0.045952 | 0.011356 | 5.20E-05 | cis |
| rs17770304 | miR-30b-5p | 8:13562410 | G | A | 0.752786 | -0.041864 | 0.011843 | 0.000408 | cis |
| rs17697655 | miR-30b-5p | 8:13562467 | A | T | 0.752044 | -0.042617 | 0.011829 | 0.000315 | cis |
| rs13282464 | miR-30b-5p | 8:13570792 | C | T | 0.822381 | -0.067006 | 0.013483 | 6.70E-07 | cis |
| rs9969668 | miR-30b-5p | 8:13573157 | G | A | 0.82631 | -0.068997 | 0.013581 | 3.77E-07 | cis |
| rs11985072 | miR-30b-5p | 8:13567173 | C | T | 0.82138 | -0.069741 | 0.013478 | 2.28E-07 | cis |
| rs72721921 | miR-30b-5p | 8:13572768 | G | A | 0.825962 | -0.068474 | 0.013595 | 4.74E-07 | cis |
| rs67280714 | miR-30b-5p | 8:13570561 | C | T | 0.822568 | -0.066898 | 0.013481 | 6.97E-07 | cis |
| rs7003440 | miR-30b-5p | 8:13558627 | A | G | 0.758238 | -0.041422 | 0.011919 | 0.00051 | cis |
| rs17771094 | miR-30b-5p | 8:13563762 | G | A | 0.752835 | -0.04227 | 0.011848 | 0.00036 | cis |
| rs7818560 | miR-30b-5p | 8:13573195 | T | C | 0.826469 | -0.069013 | 0.013593 | 3.83E-07 | cis |
| rs67761769 | miR-30b-5p | 8:13571895 | G | A | 0.822824 | -0.067154 | 0.013496 | 6.50E-07 | cis |
| rs17771292 | miR-30b-5p | 8:13564073 | G | A | 0.752307 | -0.041952 | 0.011851 | 0.0004 | cis |
| rs14700551 | miR-30b-5p | 8:13569066 | A | T | 0.822194 | -0.067611 | 0.013489 | 5.38E-07 | cis |
| rs72719749 | miR-30b-5p | 8:13557691 | G | A | 0.777521 | -0.043881 | 0.012336 | 0.000375 | cis |
| rs72719790 | miR-30d-5p | 8:13564208 | T | C | 0.748139 | -0.042207 | 0.009361 | 6.52E-06 | cis |
| rs17771094 | miR-30d-5p | 8:13563762 | G | A | 0.752835 | -0.043341 | 0.009399 | 4.00E-06 | cis |
| rs7821751 | miR-30d-5p | 8:13571067 | C | A | 0.284025 | 0.04523 | 0.00901 | 5.17E-07 | cis |
| rs1867058 | miR-30d-5p | 8:13564693 | G | C | 0.748581 | -0.043202 | 0.009366 | 3.97E-06 | cis |
| rs72719749 | miR-30d-5p | 8:13557691 | G | A | 0.777521 | -0.038525 | 0.009799 | 8.44E-05 | cis |
| rs7829476 | miR-30d-5p | 8:13560748 | G | A | 0.761443 | -0.041629 | 0.009544 | 1.29E-05 | cis |

| | | | | | | | | | |
|---|---|---|---|---|---|---|---|---|---|
| rs1116662 | miR-30d-5p | 8:1356252 | C | T | 0.752461 | -0.043189 | 0.009385 | 4.19E-06 | cis |
| rs7464870 | miR-30d-5p | 8:1355922 | G | A | 0.776861 | -0.038198 | 0.009781 | 9.41E-05 | cis |
| rs2845035 | miR-30d-5p | 8:1356050 | C | T | 0.750067 | -0.035525 | 0.009403 | 0.000158 | cis |
| rs1777129 | miR-30d-5p | 8:1356407 | G | A | 0.752307 | -0.043277 | 0.009401 | 4.16E-06 | cis |
| rs4573225 | miR-30d-5p | 8:1357204 | C | G | 0.284717 | 0.044976 | 0.009033 | 6.40E-07 | cis |
| rs7271978 | miR-30d-5p | 8:1356380 | C | T | 0.752768 | -0.043325 | 0.009396 | 4.00E-06 | cis |
| rs4909608 | miR-30d-5p | 8:1357062 | C | G | 0.288481 | 0.044527 | 0.008994 | 7.40E-07 | cis |
| rs7829427 | miR-30d-5p | 8:1356979 | A | G | 0.281683 | 0.044876 | 0.00898 | 5.81E-07 | cis |
| rs1777107 | miR-30d-5p | 8:1356374 | A | C | 0.752384 | -0.04302 | 0.009391 | 4.63E-06 | cis |
| rs5894848 | miR-30d-5p | 8:1356372 | A | G | 0.752382 | -0.043018 | 0.009391 | 4.63E-06 | cis |
| rs4398863 | miR-30d-5p | 8:1356951 | G | C | 0.285279 | 0.045224 | 0.009024 | 5.40E-07 | cis |
| rs7839162 | miR-30d-5p | 8:1357000 | A | C | 0.284188 | 0.045033 | 0.009008 | 5.76E-07 | cis |
| rs5629290 | miR-30d-5p | 8:1356405 | A | G | 0.75197 | -0.043024 | 0.009401 | 4.73E-06 | cis |
| rs1769765 | miR-30d-5p | 8:1356246 | A | T | 0.752044 | -0.04342 | 0.009384 | 3.71E-06 | cis |
| rs1777030 | miR-30d-5p | 8:1356241 | G | A | 0.752786 | -0.043347 | 0.009395 | 3.95E-06 | cis |
| rs1769794 | miR-30d-5p | 8:1356289 | G | A | 0.752507 | -0.043205 | 0.009383 | 4.14E-06 | cis |
| rs1018397 | miR-320a | 2:6860491 | G | A | 0.720472 | -0.073544 | 0.017423 | 2.43E-05 | trans |
| rs7572216 | miR-320a | 2:6859194 | T | A | 0.719839 | -0.073193 | 0.017436 | 2.70E-05 | trans |
| rs4671914 | miR-320a | 2:6861229 | G | A | 0.751284 | -0.076921 | 0.01831 | 2.66E-05 | trans |
| rs7422209 | miR-320a | 2:6863221 | C | G | 0.75328 | -0.077601 | 0.019085 | 4.78E-05 | trans |
| rs1703537 | miR-320a | 2:6859650 | G | A | 0.754443 | -0.075732 | 0.018533 | 4.38E-05 | trans |
| rs1703537 | miR-320a | 2:6859890 | C | T | 0.720098 | -0.073047 | 0.017421 | 2.75E-05 | trans |
| rs2120334 | miR-320a | 2:6861608 | C | T | 0.751935 | -0.076359 | 0.018335 | 3.12E-05 | trans |
| rs4854474 | miR-320a | 2:6863328 | A | T | 0.734157 | -0.087601 | 0.018362 | 1.84E-06 | trans |
| rs7422735 | miR-320a | 2:6863318 | C | A | 0.752797 | -0.07763 | 0.019077 | 4.72E-05 | trans |
| rs7424172 | miR-320a | 2:6863238 | T | C | 0.734273 | -0.087565 | 0.018349 | 1.82E-06 | trans |
| rs4671912 | miR-320a | 2:6860225 | C | T | 0.720119 | -0.073069 | 0.017422 | 2.74E-05 | trans |
| rs7557983 | miR-320a | 2:6859171 | C | T | 0.724325 | -0.07488 | 0.017709 | 2.36E-05 | trans |
| rs3770650 | miR-320a | 2:6861643 | A | G | 0.751964 | -0.076379 | 0.018336 | 3.11E-05 | trans |
| rs1016765 | miR-320a | 2:6864556 | T | G | 0.735237 | -0.083927 | 0.018634 | 6.67E-06 | trans |
| rs5581986 | miR-320a | 2:6864591 | G | A | 0.722126 | -0.070833 | 0.018327 | 0.000111 | trans |
| rs1420691 | miR-320a | 2:6862748 | G | C | 0.737384 | -0.085431 | 0.018225 | 2.77E-06 | trans |
| rs2289689 | miR-320a | 2:6862015 | C | T | 0.752314 | -0.076664 | 0.018348 | 2.94E-05 | trans |
| rs7355584 | miR-320a | 2:6859828 | A | G | 0.720028 | -0.073134 | 0.017423 | 2.70E-05 | trans |
| rs1501855 | miR-320a | 2:6863467 | G | A | 0.734283 | -0.087188 | 0.018456 | 2.31E-06 | trans |
| rs4854435 | miR-320a | 2:6863971 | G | A | 0.753618 | -0.075977 | 0.019238 | 7.84E-05 | trans |
| rs1168809 | miR-320a | 2:6859604 | A | T | 0.751013 | -0.076026 | 0.01833 | 3.36E-05 | trans |
| rs1049616 | miR-320a | 2:6861012 | A | G | 0.750777 | -0.077152 | 0.018286 | 2.45E-05 | trans |
| rs9309415 | miR-320a | 2:6860533 | C | A | 0.72047 | -0.073548 | 0.017423 | 2.43E-05 | trans |
| rs2289688 | miR-320a | 2:6862046 | G | A | 0.752268 | -0.076663 | 0.018344 | 2.93E-05 | trans |
| rs6745671 | miR-320a | 2:6864349 | G | A | 0.754299 | -0.074859 | 0.019306 | 0.000106 | trans |
| rs1373199 | miR-320a | 2:6859568 | G | A | 0.750699 | -0.076216 | 0.018305 | 3.13E-05 | trans |
| rs1246827 | miR-320a | 2:6864587 | A | G | 0.735366 | -0.083936 | 0.018642 | 6.71E-06 | trans |
| rs5568893 | miR-320a | 2:6859897 | C | A | 0.720121 | -0.073016 | 0.01742 | 2.77E-05 | trans |
| rs1703535 | miR-320a | 2:6858932 | C | A | 0.750845 | -0.076307 | 0.018335 | 3.16E-05 | trans |
| rs1031117 | miR-320a | 2:6860115 | G | C | 0.721971 | -0.074534 | 0.017499 | 2.05E-05 | trans |
| rs6691649 | miR-320a | 2:6864346 | T | C | 0.72213 | -0.072066 | 0.018281 | 8.08E-05 | trans |
| rs3770648 | miR-320a | 2:6861939 | C | T | 0.752286 | -0.076612 | 0.018348 | 2.97E-05 | trans |
| rs1021019 | miR-320a | 2:6860353 | T | C | 0.720439 | -0.073546 | 0.017421 | 2.43E-05 | trans |
| rs5986226 | miR-320a | 2:6864491 | G | A | 0.720952 | -0.071998 | 0.018319 | 8.49E-05 | trans |
| rs1020987 | miR-320a | 2:6860519 | A | C | 0.72047 | -0.073546 | 0.017423 | 2.43E-05 | trans |
| rs4277504 | miR-320e | 2:6856956 | C | A | 0.559711 | 0.0633 | 0.01494 | 2.27E-05 | trans |
| rs964586 | miR-320e | 2:6857289 | C | T | 0.560066 | 0.063673 | 0.015001 | 2.19E-05 | trans |

| | | | | | | | | | |
|---|---|---|---|---|---|---|---|---|---|
| rs6546388 | miR-320e | 2:6858948: | C | T | 0.281691 | 0.071324 | 0.016354 | 1.29E-05 | trans |
| rs7355584 | miR-320e | 2:6859828! | A | G | 0.720028 | -0.060985 | 0.016448 | 0.000209 | trans |
| rs6546396 | miR-320e | 2:6862930: | A | T | 0.283762 | 0.075038 | 0.016269 | 3.98E-06 | trans |
| rs1975482 | miR-320e | 2:6857418: | A | G | 0.560651 | 0.064257 | 0.015027 | 1.90E-05 | trans |
| rs964585 | miR-320e | 2:6857243 | A | G | 0.559777 | 0.063611 | 0.015004 | 2.24E-05 | trans |
| rs2289689 | miR-320e | 2:6862015: | C | T | 0.752314 | -0.061155 | 0.017326 | 0.000416 | trans |
| rs7420773 | miR-320e | 2:6862945: | G | A | 0.28377 | 0.07504 | 0.016269 | 3.98E-06 | trans |
| rs7420782 | miR-320e | 2:6862955! | G | T | 0.283742 | 0.075243 | 0.016272 | 3.76E-06 | trans |
| rs1021019: | miR-320e | 2:6860353( | T | C | 0.720439 | -0.06082 | 0.016448 | 0.000217 | trans |
| rs1975481 | miR-320e | 2:6857399: | C | A | 0.560307 | 0.063774 | 0.015017 | 2.17E-05 | trans |
| rs7574580 | miR-320e | 2:6857324: | A | G | 0.561141 | 0.064211 | 0.015047 | 1.98E-05 | trans |
| rs9309415 | miR-320e | 2:6860533! | C | A | 0.72047 | -0.06082 | 0.016449 | 0.000218 | trans |
| rs7574592 | miR-320e | 2:6859766: | T | G | 0.281934 | 0.072213 | 0.01627 | 9.06E-06 | trans |
| rs1155426 | miR-320e | 2:6862182 | T | C | 0.282037 | 0.073783 | 0.01623 | 5.46E-06 | trans |
| rs1018497! | miR-320e | 2:6859988 | G | A | 0.286787 | 0.072876 | 0.016349 | 8.29E-06 | trans |
| rs6546387 | miR-320e | 2:6858758! | G | A | 0.281573 | 0.070982 | 0.016401 | 1.51E-05 | trans |
| rs964934 | miR-320e | 2:6861187: | G | A | 0.281023 | 0.073454 | 0.016262 | 6.28E-06 | trans |
| rs2289688 | miR-320e | 2:6862046: | G | A | 0.752268 | -0.061173 | 0.017322 | 0.000413 | trans |
| rs7562889( | miR-320e | 2:6862547: | T | C | 0.285916 | 0.072762 | 0.016289 | 7.93E-06 | trans |
| rs1703537! | miR-320e | 2:6859890: | C | T | 0.720098 | -0.060699 | 0.016447 | 0.000224 | trans |
| rs6546378 | miR-320e | 2:6857344: | C | G | 0.56041 | 0.064631 | 0.015054 | 1.76E-05 | trans |
| rs4671912 | miR-320e | 2:6860225! | C | T | 0.720119 | -0.060619 | 0.016448 | 0.000228 | trans |
| rs1262148! | miR-320e | 2:6859777! | A | G | 0.333832 | 0.060747 | 0.016015 | 0.000149 | trans |
| rs6546405 | miR-320e | 2:6863311( | G | A | 0.28401 | 0.074952 | 0.016313 | 4.33E-06 | trans |
| rs2120334 | miR-320e | 2:6861608! | C | T | 0.751935 | -0.060807 | 0.017314 | 0.000445 | trans |
| rs1501855( | miR-320e | 2:6863467: | G | A | 0.734283 | -0.061828 | 0.017446 | 0.000394 | trans |
| rs880668 | miR-320e | 2:6859258 | T | C | 0.280228 | 0.071999 | 0.016365 | 1.09E-05 | trans |
| rs964935 | miR-320e | 2:6861228 | G | A | 0.281126 | 0.073522 | 0.016265 | 6.18E-06 | trans |
| rs1112618: | miR-320e | 2:6861913! | A | G | 0.282025 | 0.074016 | 0.016232 | 5.12E-06 | trans |
| rs1703537 | miR-320e | 2:6859650: | G | A | 0.754443 | -0.062443 | 0.017496 | 0.000358 | trans |
| rs964587 | miR-320e | 2:6857291: | G | A | 0.560071 | 0.063677 | 0.015001 | 2.19E-05 | trans |
| rs1373198 | miR-320e | 2:6858798: | A | T | 0.281586 | 0.070999 | 0.016399 | 1.50E-05 | trans |
| rs6546379 | miR-320e | 2:6857362: | G | A | 0.559288 | 0.064067 | 0.015025 | 2.01E-05 | trans |
| rs7589952 | miR-320e | 2:6857009: | G | A | 0.559677 | 0.063225 | 0.014945 | 2.33E-05 | trans |
| rs6759864 | miR-320e | 2:6857115: | T | C | 0.558332 | 0.062767 | 0.014899 | 2.52E-05 | trans |
| rs3770648 | miR-320e | 2:6861939: | C | T | 0.752286 | -0.061049 | 0.017326 | 0.000426 | trans |
| rs3770650 | miR-320e | 2:6861643! | A | G | 0.751964 | -0.060849 | 0.017315 | 0.000441 | trans |
| rs1031117 | miR-320e | 2:6860115: | G | C | 0.721971 | -0.062654 | 0.01652 | 0.000149 | trans |
| rs7422735 | miR-320e | 2:6863318( | C | A | 0.752797 | -0.060972 | 0.018015 | 0.000713 | trans |
| rs7424172 | miR-320e | 2:6863238: | T | C | 0.734273 | -0.063045 | 0.017345 | 0.000278 | trans |
| rs1443651 | miR-320e | 2:6856931( | T | G | 0.559583 | 0.063298 | 0.014939 | 2.26E-05 | trans |
| rs9309412 | miR-320e | 2:6857056 | C | T | 0.559665 | 0.063159 | 0.014947 | 2.39E-05 | trans |
| rs9309411 | miR-320e | 2:6857046: | A | G | 0.566779 | 0.062751 | 0.015073 | 3.14E-05 | trans |
| rs1018990: | miR-320e | 2:6860619: | C | G | 0.281778 | 0.072591 | 0.016244 | 7.86E-06 | trans |
| rs1703535! | miR-320e | 2:6858932: | C | A | 0.750845 | -0.063074 | 0.017309 | 0.000269 | trans |
| rs7588432 | miR-320e | 2:6858548 | G | A | 0.282438 | 0.071179 | 0.016674 | 1.96E-05 | trans |
| rs4854474 | miR-320e | 2:6863328 | A | T | 0.734157 | -0.062892 | 0.017358 | 0.000291 | trans |
| rs7572216 | miR-320e | 2:6859194( | T | A | 0.719839 | -0.061514 | 0.01646 | 0.000186 | trans |
| rs7422209 | miR-320e | 2:6863221: | C | G | 0.75328 | -0.060993 | 0.018023 | 0.000714 | trans |
| rs7600829 | miR-320e | 2:6856977: | T | C | 0.56021 | 0.063307 | 0.01496 | 2.32E-05 | trans |
| rs1168809( | miR-320e | 2:6859604: | A | T | 0.751013 | -0.062559 | 0.017306 | 0.0003 | trans |
| rs1189593! | miR-320e | 2:6861908: | A | G | 0.282175 | 0.07403 | 0.016237 | 5.13E-06 | trans |
| rs1443652 | miR-320e | 2:6856965! | C | G | 0.559695 | 0.063305 | 0.014942 | 2.27E-05 | trans |

| | | | | | | | | | |
|---|---|---|---|---|---|---|---|---|---|
| rs1975480 | miR-320e | 2:6857386: | T | C | 0.559926 | 0.063662 | 0.015018 | 2.24E-05 | trans |
| rs4671914 | miR-320e | 2:6861229: | G | A | 0.751284 | -0.061605 | 0.01729 | 0.000366 | trans |
| rs1271364: | miR-320e | 2:6860665( | G | A | 0.282933 | 0.073329 | 0.016303 | 6.87E-06 | trans |
| rs1373199 | miR-320e | 2:6859568( | G | A | 0.750699 | -0.062697 | 0.017282 | 0.000286 | trans |
| rs6546402 | miR-320e | 2:6863130( | A | G | 0.283216 | 0.075201 | 0.016275 | 3.83E-06 | trans |
| rs4671208 | miR-320e | 2:6860079: | A | G | 0.281916 | 0.072274 | 0.01627 | 8.91E-06 | trans |
| rs5568893: | miR-320e | 2:6859897: | C | A | 0.720121 | -0.060601 | 0.016446 | 0.000229 | trans |
| rs7557983 | miR-320e | 2:6859171: | C | T | 0.724325 | -0.061992 | 0.01672 | 0.000209 | trans |
| rs1049616: | miR-320e | 2:6861012( | A | G | 0.750777 | -0.061821 | 0.017267 | 0.000343 | trans |
| rs6744216 | miR-320e | 2:6863463: | A | G | 0.284465 | 0.075207 | 0.016312 | 4.02E-06 | trans |
| rs6741866 | miR-320e | 2:6857097( | G | A | 0.565872 | 0.062147 | 0.015026 | 3.53E-05 | trans |
| rs6752922 | miR-320e | 2:6857308: | G | A | 0.559358 | 0.064094 | 0.015008 | 1.95E-05 | trans |
| rs6546377 | miR-320e | 2:6857022: | G | A | 0.559672 | 0.06341 | 0.014952 | 2.23E-05 | trans |
| rs1420691: | miR-320e | 2:6862748( | G | C | 0.737384 | -0.061462 | 0.017227 | 0.00036 | trans |
| rs7560374 | miR-320e | 2:6856998: | A | G | 0.559683 | 0.063255 | 0.014943 | 2.30E-05 | trans |
| rs4854536 | miR-320e | 2:6863186: | T | A | 0.283938 | 0.074906 | 0.016312 | 4.39E-06 | trans |
| rs3426707: | miR-320e | 2:6859915: | T | C | 0.282465 | 0.07261 | 0.016282 | 8.21E-06 | trans |
| rs2165919 | miR-320e | 2:6861413: | C | T | 0.281408 | 0.074349 | 0.016263 | 4.84E-06 | trans |
| rs1018397: | miR-320e | 2:6860491: | G | A | 0.720472 | -0.060822 | 0.016449 | 0.000218 | trans |
| rs1867313 | miR-320e | 2:6862365: | T | C | 0.282087 | 0.073705 | 0.016227 | 5.57E-06 | trans |
| rs6730974 | miR-320e | 2:6863452: | T | A | 0.287755 | 0.077158 | 0.01635 | 2.37E-06 | trans |
| rs1020987: | miR-320e | 2:6860519: | A | C | 0.72047 | -0.06082 | 0.016449 | 0.000218 | trans |
| rs7600986 | miR-320e | 2:6863692: | A | T | 0.285029 | 0.075299 | 0.016296 | 3.82E-06 | trans |
| rs736779 | miR-320e | 2:6857191: | T | A | 0.56004 | 0.063652 | 0.014997 | 2.19E-05 | trans |
| rs6546380 | miR-320e | 2:6857362( | G | A | 0.559433 | 0.064075 | 0.015022 | 1.99E-05 | trans |
| rs1300495: | miR-320e | 2:6858081: | G | A | 0.306559 | 0.067295 | 0.016241 | 3.42E-05 | trans |
| rs1014005: | miR-323b-: | 14:101512: | A | G | 0.78072 | 0.163006 | 0.034644 | 2.54E-06 | cis |
| rs1288271: | miR-323b-: | 14:101513: | C | T | 0.792278 | 0.1635 | 0.034869 | 2.75E-06 | cis |
| rs1210086: | miR-323b-: | 14:101513( | C | T | 0.7907 | 0.168405 | 0.034615 | 1.14E-06 | cis |
| rs3584427( | miR-323b-: | 14:101518( | T | C | 0.78988 | 0.16564 | 0.034885 | 2.05E-06 | cis |
| rs1116061: | miR-323b-: | 14:101512: | C | T | 0.790978 | 0.168873 | 0.034662 | 1.11E-06 | cis |
| rs1951032 | miR-323b-: | 14:101518: | G | A | 0.78919 | 0.16669 | 0.034903 | 1.79E-06 | cis |
| rs4900481 | miR-323b-: | 14:101519: | C | A | 0.783041 | 0.17001 | 0.034732 | 9.84E-07 | cis |
| rs1008340( | miR-323b-: | 14:101512: | A | C | 0.777007 | 0.159417 | 0.034686 | 4.31E-06 | cis |
| rs5610383: | miR-323b-: | 14:101522: | T | C | 0.788906 | 0.172218 | 0.034483 | 5.90E-07 | cis |
| rs1014419: | miR-323b-: | 14:101514: | A | T | 0.780379 | 0.161785 | 0.03459 | 2.91E-06 | cis |
| rs2875208: | miR-323b-: | 14:101520: | A | C | 0.785056 | 0.171466 | 0.034748 | 8.04E-07 | cis |
| rs2839883: | miR-323b-: | 14:101526: | C | G | 0.785451 | 0.173243 | 0.034679 | 5.87E-07 | cis |
| rs4143957 | miR-323b-: | 14:101520: | T | C | 0.785181 | 0.171518 | 0.034754 | 8.01E-07 | cis |
| rs1013291( | miR-323b-: | 14:101514: | T | C | 0.710292 | 0.104002 | 0.031146 | 0.00084 | cis |
| rs8003403 | miR-323b-: | 14:101529: | G | A | 0.703214 | 0.135086 | 0.030647 | 1.04E-05 | cis |
| rs7342570 | miR-323b-: | 14:101525: | G | A | 0.785229 | 0.17301 | 0.034577 | 5.63E-07 | cis |
| rs3403869: | miR-323b-: | 14:101512( | G | A | 0.785547 | 0.166581 | 0.034988 | 1.93E-06 | cis |
| rs1323913( | miR-335-5p | 7:1303317: | G | A | 0.453188 | -0.072567 | 0.018918 | 0.000125 | cis |
| rs1005171 | miR-335-5p | 7:1301444( | C | G | 0.448513 | -0.069713 | 0.018725 | 0.000197 | cis |
| rs3792318 | miR-335-5p | 7:1303002: | T | C | 0.453318 | -0.071787 | 0.018934 | 0.00015 | cis |
| rs1424417 | miR-335-5p | 7:1303155: | T | A | 0.453144 | -0.072184 | 0.018883 | 0.000132 | cis |
| rs9641881 | miR-335-5p | 7:1303360: | T | C | 0.452961 | -0.072486 | 0.018918 | 0.000127 | cis |
| rs2301335 | miR-335-5p | 7:1301328: | G | A | 0.447305 | -0.069249 | 0.018735 | 0.000219 | cis |
| rs3807348 | miR-335-5p | 7:1301361( | G | A | 0.447238 | -0.069356 | 0.018739 | 0.000215 | cis |
| rs3778859 | miR-335-5p | 7:1301331: | T | C | 0.448051 | -0.069321 | 0.018763 | 0.00022 | cis |
| rs1756239: | miR-335-5p | 7:1303293: | T | C | 0.453182 | -0.072558 | 0.018918 | 0.000125 | cis |
| rs2312789 | miR-335-5p | 7:1303275: | C | T | 0.453168 | -0.072519 | 0.01892 | 0.000127 | cis |

| | | | | | | | | | |
|---|---|---|---|---|---|---|---|---|---|
| rs6977338 | miR-335-5p | 7:1301430 | T | C | 0.448196 | -0.069744 | 0.01871 | 0.000193 | cis |
| rs3541601 | miR-335-5p | 7:1301375 | G | A | 0.448245 | -0.069754 | 0.018708 | 0.000193 | cis |
| rs2072573 | miR-335-5p | 7:1301369 | G | T | 0.447774 | -0.069566 | 0.018745 | 0.000206 | cis |
| rs1267017 | miR-335-5p | 7:1303043 | C | T | 0.452392 | -0.071857 | 0.018903 | 0.000144 | cis |
| rs1267429 | miR-335-5p | 7:1303412 | C | T | 0.452215 | -0.072009 | 0.01889 | 0.000138 | cis |
| rs1324192 | miR-335-5p | 7:1303152 | A | G | 0.453144 | -0.072184 | 0.018883 | 0.000132 | cis |
| rs2302826 | miR-335-5p | 7:1301473 | T | G | 0.448422 | -0.06981 | 0.018727 | 0.000193 | cis |
| rs3615339 | miR-335-5p | 7:1302559 | A | G | 0.451727 | -0.071299 | 0.018905 | 0.000162 | cis |
| rs3616459 | miR-335-5p | 7:1301528 | A | G | 0.447022 | -0.069076 | 0.018815 | 0.000241 | cis |
| rs1398210 | miR-335-5p | 7:1301545 | G | A | 0.450431 | -0.069795 | 0.019016 | 0.000242 | cis |
| rs1421141 | miR-335-5p | 7:1301249 | A | G | 0.448534 | -0.070053 | 0.01879 | 0.000193 | cis |
| rs992859 | miR-335-5p | 7:1301201 | C | T | 0.451559 | -0.071127 | 0.018793 | 0.000154 | cis |
| rs1324313 | miR-335-5p | 7:1303239 | T | C | 0.453734 | -0.072406 | 0.018912 | 0.000129 | cis |
| rs3616848 | miR-335-5p | 7:1302730 | G | A | 0.453452 | -0.071438 | 0.018961 | 0.000165 | cis |
| rs1270694 | miR-335-5p | 7:1301493 | G | A | 0.447937 | -0.06883 | 0.018773 | 0.000246 | cis |
| rs1270694 | miR-335-5p | 7:1302925 | T | C | 0.448372 | -0.072535 | 0.018977 | 0.000132 | cis |
| rs2072575 | miR-335-5p | 7:1301485 | G | A | 0.450703 | -0.069627 | 0.018803 | 0.000213 | cis |
| rs2129905 | miR-335-5p | 7:1303054 | A | G | 0.45218 | -0.072104 | 0.018909 | 0.000137 | cis |
| rs3614139 | miR-335-5p | 7:1302616 | G | A | 0.455181 | -0.071607 | 0.018999 | 0.000164 | cis |
| rs1266733 | miR-335-5p | 7:1303189 | C | T | 0.453823 | -0.072416 | 0.018914 | 0.000129 | cis |
| rs1270694 | miR-335-5p | 7:1301507 | G | A | 0.449057 | -0.069628 | 0.018769 | 0.000207 | cis |
| rs1270694 | miR-335-5p | 7:1301510 | A | G | 0.448056 | -0.068823 | 0.018781 | 0.000248 | cis |
| rs1038638 | miR-335-5p | 7:1303610 | G | A | 0.451503 | -0.07157 | 0.018852 | 0.000147 | cis |
| rs1270694 | miR-335-5p | 7:1303074 | C | T | 0.453614 | -0.071955 | 0.018949 | 0.000146 | cis |
| rs2072574 | miR-335-5p | 7:1301376 | A | G | 0.448229 | -0.069761 | 0.018708 | 0.000192 | cis |
| rs765205 | miR-335-5p | 7:1301407 | C | A | 0.446513 | -0.069497 | 0.018654 | 0.000195 | cis |
| rs3612720 | miR-335-5p | 7:1302741 | C | T | 0.452415 | -0.071304 | 0.018926 | 0.000165 | cis |
| rs1324663 | miR-335-5p | 7:1303110 | A | G | 0.453103 | -0.072154 | 0.018878 | 0.000132 | cis |
| rs6973680 | miR-339-5p | 7:1199632 | T | C | 0.394697 | -0.065422 | 0.016672 | 8.70E-05 | cis |
| rs1322141 | miR-339-5p | 7:1055619 | A | G | 0.694934 | -0.067789 | 0.017601 | 0.000117 | cis |
| rs7799917 | miR-339-5p | 7:1140650 | C | G | 0.504766 | -0.056358 | 0.015695 | 0.00033 | cis |
| rs7303336 | miR-339-5p | 7:1203631 | T | G | 0.390167 | -0.062948 | 0.016445 | 0.000129 | cis |
| rs1270212 | miR-339-5p | 7:1150028 | C | T | 0.493549 | -0.055584 | 0.016199 | 0.000601 | cis |
| rs7784502 | miR-339-5p | 7:1210898 | G | A | 0.390245 | -0.062816 | 0.016467 | 0.000136 | cis |
| rs1270201 | miR-339-5p | 7:1129392 | C | T | 0.359204 | -0.066932 | 0.016389 | 4.43E-05 | cis |
| rs7784334 | miR-339-5p | 7:1210760 | G | A | 0.382396 | -0.066969 | 0.016516 | 5.02E-05 | cis |
| rs2363279 | miR-339-5p | 7:1096143 | A | G | 0.40508 | -0.082272 | 0.016257 | 4.17E-07 | cis |
| rs6967431 | miR-339-5p | 7:1208882 | T | A | 0.380734 | -0.065584 | 0.016382 | 6.25E-05 | cis |
| rs2949181 | miR-339-5p | 7:1209368 | A | G | 0.38431 | -0.066004 | 0.016519 | 6.46E-05 | cis |
| rs7784607 | miR-339-5p | 7:1178694 | A | G | 0.344664 | -0.065247 | 0.017383 | 0.000174 | cis |
| rs7784202 | miR-339-5p | 7:1210688 | G | C | 0.389965 | -0.062814 | 0.016479 | 0.000138 | cis |
| rs1270243 | miR-339-5p | 7:1197994 | T | C | 0.390258 | -0.066554 | 0.016733 | 6.97E-05 | cis |
| rs2949198 | miR-339-5p | 7:1202422 | T | C | 0.377848 | -0.068238 | 0.016562 | 3.79E-05 | cis |
| rs7799391 | miR-339-5p | 7:1082566 | C | G | 0.39831 | -0.081465 | 0.016226 | 5.15E-07 | cis |
| rs2949199 | miR-339-5p | 7:1201843 | G | A | 0.380653 | -0.06648 | 0.016445 | 5.29E-05 | cis |
| rs6957733 | miR-339-5p | 7:1106327 | T | G | 0.464939 | -0.081158 | 0.015583 | 1.91E-07 | cis |
| rs2949203 | miR-339-5p | 7:1201208 | C | T | 0.380476 | -0.066636 | 0.01645 | 5.10E-05 | cis |
| rs1270185 | miR-339-5p | 7:1107707 | C | T | 0.401835 | -0.078191 | 0.015922 | 9.06E-07 | cis |
| rs2960856 | miR-339-5p | 7:1206638 | C | T | 0.405125 | -0.069291 | 0.017107 | 5.11E-05 | cis |
| rs6972326 | miR-339-5p | 7:1201736 | C | T | 0.383739 | -0.066578 | 0.016531 | 5.64E-05 | cis |
| rs1233429 | miR-339-5p | 7:1054517 | T | C | 0.684683 | -0.06611 | 0.017029 | 0.000104 | cis |
| rs1026366 | miR-339-5p | 7:1110628 | A | G | 0.453807 | -0.07515 | 0.016039 | 2.79E-06 | cis |
| rs7784647 | miR-339-5p | 7:1210987 | G | A | 0.38083 | -0.066127 | 0.016452 | 5.84E-05 | cis |

| | | | | | | | | | |
|---|---|---|---|---|---|---|---|---|---|
| rs2949178 | miR-339-5p | 7:1209925 | T | C | 0.379896 | -0.066333 | 0.01646 | 5.58E-05 | cis |
| rs1270243 | miR-339-5p | 7:1204368 | G | A | 0.380656 | -0.066097 | 0.016399 | 5.56E-05 | cis |
| rs2960838 | miR-339-5p | 7:1204225 | C | T | 0.409759 | -0.071002 | 0.017137 | 3.43E-05 | cis |
| rs4236381 | miR-339-5p | 7:1165723 | A | C | 0.345918 | -0.065225 | 0.016927 | 0.000117 | cis |
| rs1270211 | miR-339-5p | 7:1148989 | C | T | 0.496383 | -0.056168 | 0.016124 | 0.000495 | cis |
| rs9655470 | miR-339-5p | 7:1151713 | G | T | 0.496515 | -0.056309 | 0.016145 | 0.000487 | cis |
| rs3802141 | miR-339-5p | 7:1131356 | T | C | 0.35273 | -0.066189 | 0.016401 | 5.44E-05 | cis |
| rs2960841 | miR-339-5p | 7:1204894 | A | G | 0.380205 | -0.066074 | 0.0164 | 5.61E-05 | cis |
| rs1330773 | miR-339-5p | 7:1203745 | C | A | 0.380443 | -0.066236 | 0.016408 | 5.42E-05 | cis |
| rs6956265 | miR-339-5p | 7:1204504 | G | A | 0.389746 | -0.062757 | 0.016424 | 0.000133 | cis |
| rs1330894 | miR-339-5p | 7:1203716 | G | A | 0.383474 | -0.06692 | 0.016494 | 4.97E-05 | cis |
| rs2949187 | miR-339-5p | 7:1204932 | G | T | 0.380131 | -0.066079 | 0.016402 | 5.61E-05 | cis |
| rs7781991 | miR-339-5p | 7:1157436 | G | A | 0.488532 | -0.05873 | 0.016431 | 0.000351 | cis |
| rs1270196 | miR-339-5p | 7:1121849 | A | G | 0.34676 | -0.068169 | 0.016466 | 3.47E-05 | cis |
| rs6979076 | miR-339-5p | 7:1116247 | T | C | 0.354429 | -0.069998 | 0.016376 | 1.92E-05 | cis |
| rs1330893 | miR-339-5p | 7:1203642 | G | C | 0.392085 | -0.062829 | 0.016463 | 0.000135 | cis |
| rs7782651 | miR-339-5p | 7:1159895 | T | G | 0.345899 | -0.065584 | 0.016812 | 9.58E-05 | cis |
| rs1270208 | miR-339-5p | 7:1141245 | C | T | 0.504115 | -0.056346 | 0.015707 | 0.000334 | cis |
| rs6463504 | miR-339-5p | 7:1208598 | A | G | 0.381281 | -0.065421 | 0.016401 | 6.64E-05 | cis |
| rs868833 | miR-339-5p | 7:1054973 | C | G | 0.684744 | -0.066165 | 0.017033 | 0.000102 | cis |
| rs1331153 | miR-339-5p | 7:1205578 | C | A | 0.380577 | -0.065842 | 0.016384 | 5.85E-05 | cis |
| rs1270243 | miR-339-5p | 7:1204442 | C | T | 0.380481 | -0.066037 | 0.016389 | 5.59E-05 | cis |
| rs2949182 | miR-339-5p | 7:1209343 | G | A | 0.399032 | -0.068811 | 0.016928 | 4.80E-05 | cis |
| rs6978842 | miR-339-5p | 7:1204599 | A | C | 0.369392 | -0.064433 | 0.01675 | 0.00012 | cis |
| rs1270208 | miR-339-5p | 7:1141528 | T | C | 0.357805 | -0.065052 | 0.016094 | 5.30E-05 | cis |
| rs1270201 | miR-339-5p | 7:1129413 | C | T | 0.35658 | -0.066084 | 0.016333 | 5.21E-05 | cis |
| rs1043502 | miR-339-5p | 7:1065948 | A | G | 0.398064 | -0.081469 | 0.01617 | 4.70E-07 | cis |
| rs4075937 | miR-339-5p | 7:1198743 | C | A | 0.391469 | -0.067172 | 0.016729 | 5.93E-05 | cis |
| rs1028096 | miR-339-5p | 7:1103697 | T | C | 0.468617 | -0.079563 | 0.015528 | 2.99E-07 | cis |
| rs6975972 | miR-339-5p | 7:1070468 | A | G | 0.40294 | -0.080743 | 0.016139 | 5.64E-07 | cis |
| rs1324069 | miR-339-5p | 7:1079944 | C | T | 0.39913 | -0.081162 | 0.016181 | 5.28E-07 | cis |
| rs4285396 | miR-339-5p | 7:1123738 | T | C | 0.349641 | -0.066754 | 0.016311 | 4.27E-05 | cis |
| rs4720249 | miR-339-5p | 7:1072563 | C | G | 0.400053 | -0.080999 | 0.016103 | 4.90E-07 | cis |
| rs1323240 | miR-339-5p | 7:1055598 | G | A | 0.693869 | -0.067966 | 0.017507 | 0.000104 | cis |
| rs2949201 | miR-339-5p | 7:1201307 | T | C | 0.380379 | -0.066748 | 0.016454 | 4.98E-05 | cis |
| rs2960849 | miR-339-5p | 7:1211978 | C | G | 0.378585 | -0.066818 | 0.016575 | 5.55E-05 | cis |
| rs7497669 | miR-339-5p | 7:1160505 | C | T | 0.806544 | -0.067032 | 0.020303 | 0.000961 | cis |
| rs7800060 | miR-339-5p | 7:1206786 | G | A | 0.380583 | -0.065628 | 0.016388 | 6.21E-05 | cis |
| rs6968988 | miR-339-5p | 7:1169619 | A | G | 0.342514 | -0.065977 | 0.017125 | 0.000117 | cis |
| rs2949197 | miR-339-5p | 7:1202454 | C | T | 0.393091 | -0.067323 | 0.016874 | 6.61E-05 | cis |
| rs3735686 | miR-339-5p | 7:1062527 | G | A | 0.403542 | -0.080431 | 0.016083 | 5.70E-07 | cis |
| rs1881125 | miR-339-5p | 7:1096918 | A | G | 0.410917 | -0.084843 | 0.017057 | 6.56E-07 | cis |
| rs1197417 | miR-339-5p | 7:1065569 | C | T | 0.632265 | -0.066169 | 0.016445 | 5.73E-05 | cis |
| rs2867132 | miR-339-5p | 7:1174973 | G | A | 0.343823 | -0.06557 | 0.017151 | 0.000132 | cis |
| rs7798572 | miR-339-5p | 7:1210755 | C | T | 0.381866 | -0.067036 | 0.016503 | 4.86E-05 | cis |
| rs6463001 | miR-339-5p | 7:1108272 | A | C | 0.442797 | -0.069613 | 0.01569 | 9.13E-06 | cis |
| rs2960845 | miR-339-5p | 7:1205338 | A | G | 0.38161 | -0.066757 | 0.016411 | 4.74E-05 | cis |
| rs1197927 | miR-339-5p | 7:1069335 | A | T | 0.398971 | -0.080981 | 0.016118 | 5.06E-07 | cis |
| rs1270197 | miR-339-5p | 7:1121958 | T | C | 0.350538 | -0.066315 | 0.016309 | 4.78E-05 | cis |
| rs2949183 | miR-339-5p | 7:1209342 | T | A | 0.399553 | -0.068662 | 0.016944 | 5.07E-05 | cis |
| rs7799900 | miR-339-5p | 7:1206699 | G | A | 0.380547 | -0.06561 | 0.016388 | 6.24E-05 | cis |
| rs7784059 | miR-339-5p | 7:1210575 | G | A | 0.390412 | -0.062752 | 0.016459 | 0.000138 | cis |
| rs7784043 | miR-339-5p | 7:1210554 | G | T | 0.3904 | -0.062752 | 0.01646 | 0.000138 | cis |

| | | | | | | | | | |
|---|---|---|---|---|---|---|---|---|---|
| rs2030959 | miR-339-5p | 7:1075906 | T | C | 0.399923 | -0.081313 | 0.016186 | 5.07E-07 | cis |
| rs1323250 | miR-339-5p | 7:1055632 | G | C | 0.697628 | -0.068462 | 0.017712 | 0.000111 | cis |
| rs2960832 | miR-339-5p | 7:1202134 | A | G | 0.380632 | -0.066605 | 0.016443 | 5.11E-05 | cis |
| rs4723649 | miR-339-5p | 7:1071905 | T | C | 0.403365 | -0.080861 | 0.016157 | 5.60E-07 | cis |
| rs2280724 | miR-339-5p | 7:1064720 | A | G | 0.400293 | -0.080733 | 0.016016 | 4.64E-07 | cis |
| rs2960840 | miR-339-5p | 7:1204700 | C | T | 0.380448 | -0.066015 | 0.016391 | 5.64E-05 | cis |
| rs1025400 | miR-339-5p | 7:1207937 | T | C | 0.38071 | -0.065616 | 0.016384 | 6.21E-05 | cis |
| rs7806746 | miR-339-5p | 7:1075573 | A | C | 0.412128 | -0.080752 | 0.016136 | 5.61E-07 | cis |
| rs2949194 | miR-339-5p | 7:1203034 | C | G | 0.380544 | -0.066275 | 0.016416 | 5.41E-05 | cis |
| rs4596558 | miR-339-5p | 7:1108530 | T | C | 0.539806 | -0.073948 | 0.015843 | 3.05E-06 | cis |
| rs2960843 | miR-339-5p | 7:1205210 | A | T | 0.380548 | -0.065866 | 0.016385 | 5.82E-05 | cis |
| rs2949179 | miR-339-5p | 7:1209885 | C | G | 0.37949 | -0.066182 | 0.016449 | 5.73E-05 | cis |
| rs1270244 | miR-339-5p | 7:1205666 | C | A | 0.380606 | -0.065817 | 0.016396 | 5.96E-05 | cis |
| rs2960839 | miR-339-5p | 7:1204295 | A | G | 0.38047 | -0.066057 | 0.016391 | 5.58E-05 | cis |
| rs2960842 | miR-339-5p | 7:1205166 | G | T | 0.380622 | -0.065865 | 0.016389 | 5.85E-05 | cis |
| rs2960833 | miR-339-5p | 7:1202693 | A | C | 0.38564 | -0.066532 | 0.016534 | 5.72E-05 | cis |
| rs6947257 | miR-339-5p | 7:1046906 | T | C | 0.373285 | -0.082245 | 0.017053 | 1.41E-06 | cis |
| rs4724039 | miR-339-5p | 7:1111633 | T | C | 0.40621 | -0.080902 | 0.016028 | 4.47E-07 | cis |
| rs2949186 | miR-339-5p | 7:1205146 | T | C | 0.38089 | -0.065967 | 0.016396 | 5.74E-05 | cis |
| rs2949180 | miR-339-5p | 7:1209688 | T | A | 0.379089 | -0.066256 | 0.016479 | 5.81E-05 | cis |
| rs6463505 | miR-339-5p | 7:1208710 | C | G | 0.381871 | -0.065559 | 0.016414 | 6.49E-05 | cis |
| rs6950319 | miR-339-5p | 7:1161324 | A | G | 0.346351 | -0.065627 | 0.016815 | 9.51E-05 | cis |
| rs1270243 | miR-339-5p | 7:1204354 | T | A | 0.380439 | -0.06604 | 0.016392 | 5.61E-05 | cis |
| rs1574108 | miR-339-5p | 7:1105805 | T | C | 0.402737 | -0.078282 | 0.015935 | 8.99E-07 | cis |
| rs2960853 | miR-339-5p | 7:1207438 | T | A | 0.381537 | -0.065644 | 0.016411 | 6.33E-05 | cis |
| rs2949191 | miR-339-5p | 7:1203889 | C | G | 0.380477 | -0.066199 | 0.016405 | 5.46E-05 | cis |
| rs1022968 | miR-339-5p | 7:1158565 | C | T | 0.347824 | -0.065707 | 0.016838 | 9.53E-05 | cis |
| rs9801444 | miR-339-5p | 7:1080444 | G | T | 0.399147 | -0.081166 | 0.016182 | 5.28E-07 | cis |
| rs4998392 | miR-339-5p | 7:1113344 | A | C | 0.422412 | -0.084323 | 0.016627 | 3.95E-07 | cis |
| rs2960844 | miR-339-5p | 7:1205282 | A | G | 0.380698 | -0.065843 | 0.01639 | 5.89E-05 | cis |
| rs2960829 | miR-339-5p | 7:1200568 | C | T | 0.380647 | -0.067197 | 0.016457 | 4.44E-05 | cis |
| rs4585652 | miR-339-5p | 7:1145396 | A | G | 0.353725 | -0.063941 | 0.01616 | 7.60E-05 | cis |
| rs1198008 | miR-339-5p | 7:1069372 | T | C | 0.399655 | -0.081266 | 0.016201 | 5.27E-07 | cis |
| rs2949193 | miR-339-5p | 7:1203801 | T | C | 0.380208 | -0.065781 | 0.016407 | 6.09E-05 | cis |
| rs1205605 | miR-339-5p | 7:1197632 | A | C | 0.380419 | -0.070383 | 0.016763 | 2.69E-05 | cis |
| rs1270145 | miR-339-5p | 7:1055409 | A | G | 0.675744 | -0.069989 | 0.017196 | 4.70E-05 | cis |
| rs2960854 | miR-339-5p | 7:1207404 | A | G | 0.387301 | -0.066287 | 0.01657 | 6.32E-05 | cis |
| rs2949202 | miR-339-5p | 7:1201266 | G | A | 0.38048 | -0.066618 | 0.016448 | 5.12E-05 | cis |
| rs2949188 | miR-339-5p | 7:1204926 | C | G | 0.380134 | -0.066084 | 0.016402 | 5.60E-05 | cis |
| rs1270244 | miR-339-5p | 7:1205483 | G | A | 0.380563 | -0.065844 | 0.016384 | 5.85E-05 | cis |
| rs4724424 | miR-339-5p | 7:1154206 | T | C | 0.33352 | -0.06659 | 0.017392 | 0.000129 | cis |
| rs1095149 | miR-339-5p | 7:1065746 | A | G | 0.612938 | -0.065288 | 0.016243 | 5.83E-05 | cis |
| rs2949184 | miR-339-5p | 7:1209027 | T | G | 0.382303 | -0.066208 | 0.016443 | 5.66E-05 | cis |
| rs6945636 | miR-339-5p | 7:1204647 | T | A | 0.38988 | -0.062747 | 0.016419 | 0.000133 | cis |
| rs9655467 | miR-339-5p | 7:1150542 | T | A | 0.494917 | -0.056533 | 0.016173 | 0.000473 | cis |
| rs2949189 | miR-339-5p | 7:1204226 | A | G | 0.40961 | -0.071008 | 0.017129 | 3.39E-05 | cis |
| rs7805362 | miR-339-5p | 7:1182997 | A | G | 0.343701 | -0.065043 | 0.017343 | 0.000177 | cis |
| rs7809667 | miR-339-5p | 7:1208440 | G | C | 0.381092 | -0.065539 | 0.016396 | 6.41E-05 | cis |
| rs2949200 | miR-339-5p | 7:1201345 | T | C | 0.380481 | -0.06661 | 0.016448 | 5.13E-05 | cis |
| rs6945202 | miR-339-5p | 7:1116558 | T | C | 0.400481 | -0.064512 | 0.016245 | 7.15E-05 | cis |
| rs2140578 | miR-339-5p | 7:1214003 | T | C | 0.352201 | -0.057642 | 0.016795 | 0.000599 | cis |
| rs2960852 | miR-339-5p | 7:1208968 | G | A | 0.380737 | -0.06558 | 0.016382 | 6.25E-05 | cis |
| rs2949205 | miR-339-5p | 7:1200757 | C | T | 0.380827 | -0.067036 | 0.0165 | 4.85E-05 | cis |

| | | | | | | | | | |
|---|---|---|---|---|---|---|---|---|---|
| rs1133043 | miR-339-5p | 7:1133305 | C | G | 0.356957 | -0.065446 | 0.016176 | 5.21E-05 | cis |
| rs1881116 | miR-339-5p | 7:1053958 | C | T | 0.686633 | -0.064091 | 0.01699 | 0.000162 | cis |
| rs2949190 | miR-339-5p | 7:1203904 | T | A | 0.380455 | -0.066084 | 0.016396 | 5.57E-05 | cis |
| rs10276204 | miR-339-5p | 7:1047932 | T | C | 0.371855 | -0.083245 | 0.017188 | 1.28E-06 | cis |
| rs13235747 | miR-339-5p | 7:1129123 | T | C | 0.360607 | -0.067461 | 0.016461 | 4.16E-05 | cis |
| rs11767061 | miR-339-5p | 7:1199804 | C | T | 0.401612 | -0.065993 | 0.016847 | 8.96E-05 | cis |
| rs11766651 | miR-339-5p | 7:1161646 | T | C | 0.339979 | -0.065108 | 0.016767 | 0.000103 | cis |
| rs2960836 | miR-339-5p | 7:1202980 | T | G | 0.380547 | -0.066286 | 0.016416 | 5.40E-05 | cis |
| rs2766691 | miR-345-5p | 14:1006822 | T | C | 0.546649 | 0.045338 | 0.010652 | 2.08E-05 | cis |
| rs11627407 | miR-345-5p | 14:1006792 | A | G | 0.554108 | 0.046924 | 0.010811 | 1.42E-05 | cis |
| rs2757535 | miR-345-5p | 14:1006805 | G | A | 0.549851 | 0.047093 | 0.010744 | 1.17E-05 | cis |
| rs2757539 | miR-345-5p | 14:1006825 | A | T | 0.546641 | 0.045309 | 0.01065 | 2.10E-05 | cis |
| rs2757540 | miR-345-5p | 14:1006849 | C | T | 0.54653 | 0.045248 | 0.010639 | 2.11E-05 | cis |
| rs2757545 | miR-345-5p | 14:1006953 | T | C | 0.59028 | 0.044679 | 0.010684 | 2.89E-05 | cis |
| rs1004573 | miR-370-3p | 14:1011782 | G | C | 0.860589 | 0.12663 | 0.03365 | 0.000168 | cis |
| rs7160857 | miR-370-3p | 14:1012496 | A | G | 0.755456 | 0.118532 | 0.030136 | 8.38E-05 | cis |
| rs28478065 | miR-370-3p | 14:1011396 | C | A | 0.868556 | 0.167353 | 0.034208 | 9.97E-07 | cis |
| rs17576305 | miR-370-3p | 14:1011275 | G | C | 0.854847 | 0.143699 | 0.033439 | 1.73E-05 | cis |
| rs9635184 | miR-370-3p | 14:1011730 | C | T | 0.859552 | 0.132726 | 0.033238 | 6.52E-05 | cis |
| rs8017361 | miR-370-3p | 14:1012180 | G | A | 0.898047 | 0.183916 | 0.03882 | 2.16E-06 | cis |
| rs17576249 | miR-370-3p | 14:1011268 | G | C | 0.854853 | 0.143729 | 0.033458 | 1.74E-05 | cis |
| rs1190815 | miR-370-3p | 14:1012283 | G | A | 0.798481 | 0.151862 | 0.028988 | 1.62E-07 | cis |
| rs1555406 | miR-370-3p | 14:1012042 | C | T | 0.876511 | 0.131451 | 0.035483 | 0.000212 | cis |
| rs10136874 | miR-370-3p | 14:1012020 | G | T | 0.493366 | 0.123375 | 0.024136 | 3.19E-07 | cis |
| rs7156735 | miR-370-3p | 14:1011322 | T | C | 0.867137 | 0.161998 | 0.03401 | 1.91E-06 | cis |
| rs7152943 | miR-370-3p | 14:1011741 | C | T | 0.924188 | 0.235204 | 0.044241 | 1.06E-07 | cis |
| rs8017555 | miR-370-3p | 14:1012189 | C | T | 0.89727 | 0.187407 | 0.038807 | 1.37E-06 | cis |
| rs17576123 | miR-370-3p | 14:1011262 | C | T | 0.856231 | 0.141946 | 0.033588 | 2.38E-05 | cis |
| rs7150520 | miR-370-3p | 14:1011967 | G | A | 0.196892 | -0.139241 | 0.03089 | 6.55E-06 | cis |
| rs1190868 | miR-370-3p | 14:1010692 | G | A | 0.180475 | -0.156447 | 0.032578 | 1.57E-06 | cis |
| rs8014970 | miR-370-3p | 14:1011407 | T | G | 0.868329 | 0.167377 | 0.034241 | 1.02E-06 | cis |
| rs17653830 | miR-370-3p | 14:1011011 | T | C | 0.877829 | 0.197235 | 0.036812 | 8.42E-08 | cis |
| rs17099617 | miR-370-3p | 14:1011704 | A | G | 0.858798 | 0.127054 | 0.033014 | 0.000119 | cis |
| rs10220322 | miR-370-3p | 14:1011303 | T | G | 0.868038 | 0.160373 | 0.034061 | 2.50E-06 | cis |
| rs12888043 | miR-370-3p | 14:1011704 | G | A | 0.858051 | 0.126663 | 0.033109 | 0.00013 | cis |
| rs17653848 | miR-370-3p | 14:1011020 | A | T | 0.870972 | 0.196846 | 0.036887 | 9.48E-08 | cis |
| rs4905991 | miR-370-3p | 14:1011400 | T | C | 0.271223 | -0.140782 | 0.026547 | 1.14E-07 | cis |
| rs3783350 | miR-370-3p | 14:1011990 | G | A | 0.22096 | -0.140083 | 0.029086 | 1.46E-06 | cis |
| rs7144375 | miR-370-3p | 14:1011373 | T | C | 0.866639 | 0.167 | 0.033983 | 8.92E-07 | cis |
| rs1004572 | miR-370-3p | 14:1011783 | G | A | 0.86193 | 0.12884 | 0.033863 | 0.000142 | cis |
| rs6575796 | miR-370-3p | 14:1011603 | A | G | 0.094049 | -0.23082 | 0.042679 | 6.36E-08 | cis |
| rs1190817 | miR-370-3p | 14:1012299 | T | C | 0.798148 | 0.150156 | 0.029212 | 2.74E-07 | cis |
| rs28473022 | miR-370-3p | 14:1011618 | G | A | 0.949972 | 0.263653 | 0.058988 | 7.84E-06 | cis |
| rs10873517 | miR-370-3p | 14:1009454 | G | T | 0.730696 | 0.156152 | 0.028841 | 6.15E-08 | cis |
| rs10141715 | miR-370-3p | 14:1011726 | G | T | 0.923932 | 0.232858 | 0.044142 | 1.33E-07 | cis |
| rs1802710 | miR-370-3p | 14:1012000 | C | T | 0.493406 | 0.126102 | 0.02418 | 1.84E-07 | cis |
| rs35161451 | miR-370-3p | 14:1011641 | A | G | 0.624259 | 0.18375 | 0.028411 | 9.96E-11 | cis |
| rs7142298 | miR-370-3p | 14:1011285 | C | T | 0.854052 | 0.143809 | 0.033256 | 1.53E-05 | cis |
| rs17660210 | miR-370-3p | 14:1011314 | A | G | 0.868541 | 0.162812 | 0.034217 | 1.95E-06 | cis |
| rs8007901 | miR-370-3p | 14:1011305 | T | C | 0.867321 | 0.160815 | 0.033926 | 2.14E-06 | cis |
| rs9635298 | miR-370-3p | 14:1011725 | C | T | 0.859368 | 0.133159 | 0.033234 | 6.16E-05 | cis |
| rs17577779 | miR-370-3p | 14:1012004 | C | T | 0.495315 | 0.122279 | 0.0242 | 4.35E-07 | cis |
| rs17660362 | miR-370-3p | 14:1011361 | A | G | 0.868731 | 0.163014 | 0.034276 | 1.98E-06 | cis |

| | | | | | | | | | |
|---|---|---|---|---|---|---|---|---|---|
| rs1004574 | miR-370-3p | 14:101178( | G | C | 0.86063 | 0.126662 | 0.033649 | 0.000167 | cis |
| rs2852739: | miR-370-3p | 14:101130( | T | C | 0.868049 | 0.16039 | 0.034066 | 2.50E-06 | cis |
| rs8022367 | miR-370-3p | 14:101189⁄ | A | T | 0.295876 | -0.224145 | 0.031061 | 5.35E-13 | cis |
| rs3494675⁄ | miR-370-3p | 14:100938: | A | G | 0.904426 | 0.269181 | 0.046655 | 7.95E-09 | cis |
| rs1190815 | miR-382-5p | 14:101228: | G | A | 0.798481 | 0.177845 | 0.039934 | 8.45E-06 | cis |
| rs2847806! | miR-382-5p | 14:101139( | C | A | 0.868556 | 0.198176 | 0.047111 | 2.59E-05 | cis |
| rs1013687⁄ | miR-382-5p | 14:101202( | G | T | 0.493366 | 0.152509 | 0.033231 | 4.45E-06 | cis |
| rs1288162! | miR-382-5p | 14:101146⁄ | A | G | 0.912196 | 0.309333 | 0.059691 | 2.19E-07 | cis |
| rs1766036: | miR-382-5p | 14:101136: | A | G | 0.868731 | 0.194792 | 0.047197 | 3.67E-05 | cis |
| rs1013035⁄ | miR-382-5p | 14:101149! | T | C | 0.112665 | -0.231794 | 0.053696 | 1.58E-05 | cis |
| rs7156735 | miR-382-5p | 14:101132: | T | C | 0.867137 | 0.195148 | 0.046829 | 3.08E-05 | cis |
| rs1766021( | miR-382-5p | 14:101131⁄ | A | G | 0.868541 | 0.194646 | 0.047116 | 3.61E-05 | cis |
| rs7152943 | miR-382-5p | 14:101174: | C | T | 0.924188 | 0.257085 | 0.060987 | 2.49E-05 | cis |
| rs1288035! | miR-382-5p | 14:101158! | C | G | 0.904391 | 0.228236 | 0.057447 | 7.10E-05 | cis |
| rs1243295! | miR-382-5p | 14:101205! | G | A | 0.231663 | -0.191932 | 0.038441 | 5.95E-07 | cis |
| rs8022367 | miR-382-5p | 14:101189⁄ | A | T | 0.295876 | -0.266386 | 0.042846 | 5.06E-10 | cis |
| rs1087351: | miR-382-5p | 14:100945⁄ | G | T | 0.730696 | 0.189943 | 0.039721 | 1.74E-06 | trans |
| rs1765383( | miR-382-5p | 14:101101: | T | C | 0.877829 | 0.275327 | 0.050615 | 5.34E-08 | cis |
| rs7150520 | miR-382-5p | 14:101196: | G | A | 0.196892 | -0.201435 | 0.042459 | 2.09E-06 | cis |
| rs1190881 | miR-382-5p | 14:101079! | T | C | 0.346494 | -0.183687 | 0.034064 | 6.96E-08 | cis |
| rs1185164: | miR-382-5p | 14:100974( | G | A | 0.742035 | 0.207629 | 0.039157 | 1.14E-07 | trans |
| rs1190878 | miR-382-5p | 14:101072: | C | A | 0.859763 | 0.241638 | 0.047238 | 3.13E-07 | cis |
| rs1802710 | miR-382-5p | 14:101200( | C | T | 0.493406 | 0.152288 | 0.0333 | 4.80E-06 | cis |
| rs1014171! | miR-382-5p | 14:101172( | G | T | 0.923932 | 0.253423 | 0.06085 | 3.12E-05 | cis |
| rs6575796 | miR-382-5p | 14:101160: | A | G | 0.094049 | -0.227052 | 0.058887 | 0.000115 | cis |
| rs1022032: | miR-382-5p | 14:101130: | T | G | 0.868038 | 0.192922 | 0.046898 | 3.89E-05 | cis |
| rs1765384! | miR-382-5p | 14:101102( | A | T | 0.870972 | 0.268059 | 0.050734 | 1.27E-07 | cis |
| rs4905991 | miR-382-5p | 14:101140( | T | C | 0.271223 | -0.16005 | 0.036583 | 1.21E-05 | cis |
| rs8014970 | miR-382-5p | 14:101140: | T | G | 0.868329 | 0.198248 | 0.047156 | 2.62E-05 | cis |
| rs1757777! | miR-382-5p | 14:101200⁄ | C | T | 0.495315 | 0.145897 | 0.033328 | 1.20E-05 | cis |
| rs3516145: | miR-382-5p | 14:101164: | A | G | 0.624259 | 0.247236 | 0.039088 | 2.53E-10 | cis |
| rs2295659 | miR-382-5p | 14:101141! | G | T | 0.857995 | 0.207489 | 0.04594 | 6.29E-06 | cis |
| rs2842154! | miR-382-5p | 14:101158: | C | T | 0.904763 | 0.227689 | 0.057509 | 7.52E-05 | cis |
| rs1190818 | miR-382-5p | 14:101230! | T | G | 0.818683 | 0.192053 | 0.041692 | 4.10E-06 | cis |
| rs7144375 | miR-382-5p | 14:101137: | T | C | 0.866639 | 0.199253 | 0.046799 | 2.07E-05 | cis |
| rs2852739: | miR-382-5p | 14:101130( | T | C | 0.868049 | 0.192966 | 0.046904 | 3.89E-05 | cis |
| rs1190817 | miR-382-5p | 14:101229! | T | C | 0.798148 | 0.175807 | 0.04024 | 1.25E-05 | cis |
| rs7144997 | miR-382-5p | 14:101155⁄ | A | G | 0.899872 | 0.221597 | 0.057279 | 0.000109 | cis |
| rs3494675⁄ | miR-382-5p | 14:100938: | A | G | 0.904426 | 0.362073 | 0.064182 | 1.69E-08 | trans |
| rs1190868 | miR-382-5p | 14:101069: | G | A | 0.180475 | -0.181554 | 0.044871 | 5.21E-05 | cis |
| rs745797 | miR-382-5p | 14:101141! | G | A | 0.858727 | 0.20991 | 0.046081 | 5.23E-06 | cis |
| rs1116059( | miR-382-5p | 14:100965! | C | T | 0.745134 | 0.205347 | 0.03933 | 1.78E-07 | trans |
| rs2400940 | miR-382-5p | 14:101224! | G | A | 0.185552 | -0.18241 | 0.040833 | 7.93E-06 | cis |
| rs8007901 | miR-382-5p | 14:101130! | T | C | 0.867321 | 0.193567 | 0.046712 | 3.42E-05 | cis |
| rs3579985( | miR-382-5p | 14:101156! | C | T | 0.904015 | 0.229385 | 0.057443 | 6.52E-05 | cis |
| rs3783350 | miR-382-5p | 14:101199( | G | A | 0.22096 | -0.207582 | 0.039965 | 2.06E-07 | cis |
| rs1709962: | miR-382-5p | 14:101173: | A | C | 0.783449 | 0.203593 | 0.038801 | 1.54E-07 | cis |
| rs2847806! | miR-409-3p | 14:101139( | C | A | 0.868556 | 0.239726 | 0.04556 | 1.43E-07 | cis |
| rs8017361 | miR-409-3p | 14:101218( | G | A | 0.898047 | 0.248792 | 0.051739 | 1.52E-06 | cis |
| rs1214798: | miR-409-3p | 14:101224( | T | C | 0.65298 | -0.13239 | 0.033471 | 7.64E-05 | cis |
| rs1013687⁄ | miR-409-3p | 14:101202( | G | T | 0.493366 | 0.17103 | 0.032158 | 1.05E-07 | cis |
| rs8022367 | miR-409-3p | 14:101189⁄ | A | T | 0.295876 | -0.304169 | 0.041387 | 1.99E-13 | cis |
| rs1022032: | miR-409-3p | 14:101130: | T | G | 0.868038 | 0.235349 | 0.045355 | 2.11E-07 | cis |

| | | | | | | | | | |
|---|---|---|---|---|---|---|---|---|---|
| rs1766036[illegible] | miR-409-3[illegible] | 14:101136[illegible] | A | G | 0.868731 | 0.237507 | 0.045644 | 1.96E-07 | cis |
| rs1757624[illegible] | miR-409-3[illegible] | 14:101126[illegible] | G | C | 0.854853 | 0.209224 | 0.044563 | 2.67E-06 | cis |
| rs3783350 | miR-409-3[illegible] | 14:101199[illegible] | G | A | 0.22096 | -0.196382 | 0.03875 | 4.02E-07 | cis |
| rs2852739[illegible] | miR-409-3[illegible] | 14:101130[illegible] | T | C | 0.868049 | 0.235364 | 0.045361 | 2.12E-07 | cis |
| rs7160857 | miR-409-3[illegible] | 14:101249[illegible] | A | G | 0.755456 | 0.190724 | 0.040105 | 1.98E-06 | cis |
| rs6575803 | miR-409-3[illegible] | 14:101257[illegible] | C | T | 0.905637 | 0.260895 | 0.061917 | 2.51E-05 | cis |
| rs1757612[illegible] | miR-409-3[illegible] | 14:101126[illegible] | C | T | 0.856231 | 0.204385 | 0.044742 | 4.92E-06 | cis |
| rs4900472 | miR-409-3[illegible] | 14:101248[illegible] | G | A | 0.715172 | 0.150884 | 0.035897 | 2.63E-05 | cis |
| rs1766021[illegible] | miR-409-3[illegible] | 14:101131[illegible] | A | G | 0.868541 | 0.23757 | 0.045564 | 1.85E-07 | cis |
| rs1757777[illegible] | miR-409-3[illegible] | 14:101200[illegible] | C | T | 0.495315 | 0.1659 | 0.032251 | 2.69E-07 | cis |
| rs1757630[illegible] | miR-409-3[illegible] | 14:101127[illegible] | G | C | 0.854847 | 0.209235 | 0.044537 | 2.63E-06 | cis |
| rs7144375 | miR-409-3[illegible] | 14:101137[illegible] | T | C | 0.866639 | 0.240714 | 0.045257 | 1.04E-07 | cis |
| rs1802710 | miR-409-3[illegible] | 14:101200[illegible] | C | T | 0.493406 | 0.171774 | 0.032223 | 9.78E-08 | cis |
| rs4905991 | miR-409-3[illegible] | 14:101140[illegible] | T | C | 0.271223 | -0.170337 | 0.035428 | 1.52E-06 | cis |
| rs7156735 | miR-409-3[illegible] | 14:101132[illegible] | T | C | 0.867137 | 0.237202 | 0.045287 | 1.63E-07 | cis |
| rs7150520 | miR-409-3[illegible] | 14:101196[illegible] | G | A | 0.196892 | -0.205189 | 0.041133 | 6.09E-07 | cis |
| rs8014970 | miR-409-3[illegible] | 14:101140[illegible] | T | G | 0.868329 | 0.239334 | 0.045606 | 1.54E-07 | cis |
| rs8017555 | miR-409-3[illegible] | 14:101218[illegible] | C | T | 0.89727 | 0.254674 | 0.051719 | 8.47E-07 | cis |
| rs3494675[illegible] | miR-409-3[illegible] | 14:100938[illegible] | A | G | 0.904426 | 0.443923 | 0.061937 | 7.65E-13 | trans |
| rs7142298 | miR-409-3[illegible] | 14:101128[illegible] | C | T | 0.854052 | 0.209978 | 0.044292 | 2.13E-06 | cis |
| rs3516145[illegible] | miR-409-3[illegible] | 14:101164[illegible] | A | G | 0.624259 | 0.28071 | 0.037757 | 1.05E-13 | cis |
| rs1014171[illegible] | miR-409-3[illegible] | 14:101172[illegible] | G | T | 0.923932 | 0.319049 | 0.05882 | 5.82E-08 | cis |
| rs1190868 | miR-409-3[illegible] | 14:101069[illegible] | G | A | 0.180475 | -0.198823 | 0.043448 | 4.74E-06 | cis |
| rs8007901 | miR-409-3[illegible] | 14:101130[illegible] | T | C | 0.867321 | 0.235761 | 0.045174 | 1.80E-07 | cis |
| rs8008669 | miR-409-3[illegible] | 14:101164[illegible] | C | T | 0.943842 | -0.390254 | 0.091682 | 2.08E-05 | cis |
| rs9324022 | miR-411-3[illegible] | 14:101172[illegible] | T | C | 0.763739 | 0.168182 | 0.042616 | 7.93E-05 | cis |
| rs1766036[illegible] | miR-411-3[illegible] | 14:101136[illegible] | A | G | 0.868731 | 0.233278 | 0.050765 | 4.32E-06 | cis |
| rs4905994 | miR-411-3[illegible] | 14:101154[illegible] | T | C | 0.329146 | -0.205149 | 0.03776 | 5.54E-08 | cis |
| rs1022032[illegible] | miR-411-3[illegible] | 14:101130[illegible] | T | G | 0.868038 | 0.237074 | 0.050431 | 2.59E-06 | cis |
| rs1757624[illegible] | miR-411-3[illegible] | 14:101126[illegible] | G | C | 0.854853 | 0.209398 | 0.049544 | 2.37E-05 | cis |
| rs1243613[illegible] | miR-411-3[illegible] | 14:101031[illegible] | C | T | 0.725015 | 0.167784 | 0.038845 | 1.57E-05 | cis |
| rs1243369[illegible] | miR-411-3[illegible] | 14:101035[illegible] | C | G | 0.723403 | 0.163421 | 0.038595 | 2.29E-05 | cis |
| rs2847806[illegible] | miR-411-3[illegible] | 14:101139[illegible] | C | A | 0.868556 | 0.233812 | 0.050677 | 3.95E-06 | cis |
| rs7140556 | miR-411-3[illegible] | 14:101036[illegible] | C | A | 0.723818 | 0.162927 | 0.03877 | 2.64E-05 | cis |
| rs7144375 | miR-411-3[illegible] | 14:101137[illegible] | T | C | 0.866639 | 0.238331 | 0.050334 | 2.19E-06 | cis |
| rs1116059[illegible] | miR-411-3[illegible] | 14:101049[illegible] | C | T | 0.741662 | 0.145168 | 0.039442 | 0.000233 | cis |
| rs1214784[illegible] | miR-411-3[illegible] | 14:101144[illegible] | C | T | 0.870989 | 0.266017 | 0.05198 | 3.09E-07 | cis |
| rs8008285 | miR-411-3[illegible] | 14:101146[illegible] | T | C | 0.842905 | 0.239857 | 0.048392 | 7.18E-07 | cis |
| rs7147982 | miR-411-3[illegible] | 14:101046[illegible] | C | T | 0.731753 | 0.155976 | 0.03868 | 5.52E-05 | cis |
| rs1015151[illegible] | miR-411-3[illegible] | 14:101175[illegible] | G | A | 0.806743 | 0.220171 | 0.043269 | 3.61E-07 | cis |
| rs1757630[illegible] | miR-411-3[illegible] | 14:101127[illegible] | G | C | 0.854847 | 0.208991 | 0.049515 | 2.44E-05 | cis |
| rs1757612[illegible] | miR-411-3[illegible] | 14:101126[illegible] | C | T | 0.856231 | 0.215475 | 0.049719 | 1.46E-05 | cis |
| rs7142298 | miR-411-3[illegible] | 14:101128[illegible] | C | T | 0.854052 | 0.215361 | 0.049233 | 1.22E-05 | cis |
| rs1555405 | miR-411-3[illegible] | 14:101176[illegible] | G | A | 0.749075 | 0.152663 | 0.039412 | 0.000107 | cis |
| rs8022367 | miR-411-3[illegible] | 14:101189[illegible] | A | T | 0.295876 | -0.183868 | 0.046369 | 7.33E-05 | cis |
| rs2295659 | miR-411-3[illegible] | 14:101141[illegible] | G | T | 0.857995 | 0.244607 | 0.049412 | 7.41E-07 | cis |
| rs7156735 | miR-411-3[illegible] | 14:101132[illegible] | T | C | 0.867137 | 0.236976 | 0.050361 | 2.53E-06 | cis |
| rs2852739[illegible] | miR-411-3[illegible] | 14:101130[illegible] | T | C | 0.868049 | 0.237143 | 0.050437 | 2.58E-06 | cis |
| rs755837 | miR-411-3[illegible] | 14:101125[illegible] | G | A | 0.178884 | -0.178865 | 0.045949 | 9.92E-05 | cis |
| rs1184948[illegible] | miR-411-3[illegible] | 14:101037[illegible] | C | G | 0.733365 | 0.164515 | 0.039053 | 2.53E-05 | cis |
| rs7154024 | miR-411-3[illegible] | 14:101031[illegible] | G | T | 0.725015 | 0.167561 | 0.038811 | 1.58E-05 | cis |
| rs8014970 | miR-411-3[illegible] | 14:101140[illegible] | T | G | 0.868329 | 0.233938 | 0.050727 | 3.99E-06 | cis |
| rs1288162[illegible] | miR-411-3[illegible] | 14:101146[illegible] | A | G | 0.912196 | 0.283483 | 0.064373 | 1.06E-05 | cis |

| | | | | | | | | | |
|---|---|---|---|---|---|---|---|---|---|
| rs1888581 | miR-411-3p | 14:101042 | A | C | 0.735363 | 0.161186 | 0.038957 | 3.51E-05 | cis |
| rs745797 | miR-411-3p | 14:101141 | G | A | 0.858727 | 0.247351 | 0.049564 | 6.02E-07 | cis |
| rs1766021 | miR-411-3p | 14:101131 | A | G | 0.868541 | 0.231641 | 0.050681 | 4.86E-06 | cis |
| rs4905989 | miR-411-3p | 14:101124 | G | A | 0.176861 | -0.17014 | 0.04601 | 0.000217 | cis |
| rs7493817 | miR-411-3p | 14:101038 | G | A | 0.751723 | 0.176706 | 0.040303 | 1.16E-05 | cis |
| rs6575791 | miR-411-3p | 14:101029 | G | A | 0.719841 | 0.169886 | 0.038838 | 1.22E-05 | cis |
| rs6575794 | miR-411-3p | 14:101039 | C | T | 0.734189 | 0.166504 | 0.039106 | 2.07E-05 | cis |
| rs8007901 | miR-411-3p | 14:101130 | T | C | 0.867321 | 0.235487 | 0.050235 | 2.76E-06 | cis |
| rs1709962 | miR-411-3p | 14:101173 | A | C | 0.783449 | 0.192432 | 0.041833 | 4.23E-06 | cis |
| rs1087352 | miR-411-3p | 14:101175 | G | A | 0.809826 | 0.223681 | 0.043774 | 3.22E-07 | cis |
| rs8022367 | miR-431-5p | 14:101189 | A | T | 0.295876 | -0.215569 | 0.040478 | 1.01E-07 | cis |
| rs1757758 | miR-431-5p | 14:101189 | C | T | 0.827094 | 0.214406 | 0.039813 | 7.23E-08 | cis |
| rs1288035 | miR-433-3p | 14:101158 | C | G | 0.904391 | 0.215109 | 0.049076 | 1.17E-05 | cis |
| rs1757777 | miR-433-3p | 14:101200 | C | T | 0.495315 | 0.118673 | 0.028507 | 3.14E-05 | cis |
| rs1214784 | miR-433-3p | 14:101144 | C | T | 0.870989 | 0.163408 | 0.04138 | 7.85E-05 | cis |
| rs6575796 | miR-433-3p | 14:101160 | A | G | 0.094049 | -0.213175 | 0.05031 | 2.26E-05 | cis |
| rs8008285 | miR-433-3p | 14:101146 | T | C | 0.842905 | 0.173692 | 0.038467 | 6.32E-06 | cis |
| rs8022367 | miR-433-3p | 14:101189 | A | T | 0.295876 | -0.18311 | 0.036747 | 6.26E-07 | cis |
| rs7152943 | miR-433-3p | 14:101174 | C | T | 0.924188 | 0.244411 | 0.052092 | 2.71E-06 | cis |
| rs3579985 | miR-433-3p | 14:101156 | C | T | 0.904015 | 0.216203 | 0.049073 | 1.05E-05 | cis |
| rs1013687 | miR-433-3p | 14:101202 | G | T | 0.493366 | 0.118863 | 0.028435 | 2.91E-05 | cis |
| rs1013035 | miR-433-3p | 14:101149 | T | C | 0.112665 | -0.197967 | 0.045908 | 1.62E-05 | cis |
| rs1802710 | miR-433-3p | 14:101200 | C | T | 0.493406 | 0.121669 | 0.028488 | 1.95E-05 | cis |
| rs2842154 | miR-433-3p | 14:101158 | C | T | 0.904763 | 0.215895 | 0.049127 | 1.11E-05 | cis |
| rs1757758 | miR-433-3p | 14:101189 | C | T | 0.827094 | 0.180028 | 0.036148 | 6.35E-07 | cis |
| rs7144997 | miR-433-3p | 14:101155 | A | G | 0.899872 | 0.202488 | 0.048948 | 3.52E-05 | cis |
| rs1555405 | miR-433-3p | 14:101176 | G | A | 0.749075 | 0.157457 | 0.031224 | 4.59E-07 | cis |
| rs1709962 | miR-433-3p | 14:101173 | A | C | 0.783449 | 0.166518 | 0.033191 | 5.25E-07 | cis |
| rs1014171 | miR-433-3p | 14:101172 | G | T | 0.923932 | 0.242963 | 0.051971 | 2.94E-06 | cis |
| rs3783350 | miR-433-3p | 14:101199 | G | A | 0.22096 | -0.148382 | 0.034233 | 1.46E-05 | cis |
| rs7572216 | miR-4429 | 2:6859194 | T | A | 0.719839 | -0.051211 | 0.014399 | 0.000376 | trans |
| rs1016765 | miR-4429 | 2:6864556 | T | G | 0.735237 | -0.055175 | 0.015397 | 0.000339 | trans |
| rs1373199 | miR-4429 | 2:6859568 | G | A | 0.750699 | -0.055668 | 0.015112 | 0.00023 | trans |
| rs7355584 | miR-4429 | 2:6859828 | A | G | 0.720028 | -0.050909 | 0.014389 | 0.000403 | trans |
| rs7422209 | miR-4429 | 2:6863221 | C | G | 0.75328 | -0.056102 | 0.015757 | 0.00037 | trans |
| rs1501855 | miR-4429 | 2:6863467 | G | A | 0.734283 | -0.058275 | 0.015251 | 0.000133 | trans |
| rs1018397 | miR-4429 | 2:6860491 | G | A | 0.720472 | -0.051129 | 0.014389 | 0.00038 | trans |
| rs964586 | miR-4429 | 2:6857289 | C | T | 0.560066 | 0.047019 | 0.013134 | 0.000344 | trans |
| rs2120334 | miR-4429 | 2:6861608 | C | T | 0.751935 | -0.055192 | 0.015138 | 0.000266 | trans |
| rs7422735 | miR-4429 | 2:6863318 | C | A | 0.752797 | -0.056136 | 0.01575 | 0.000365 | trans |
| rs5568893 | miR-4429 | 2:6859897 | C | A | 0.720121 | -0.050682 | 0.014386 | 0.000427 | trans |
| rs1703537 | miR-4429 | 2:6859890 | C | T | 0.720098 | -0.05074 | 0.014387 | 0.000421 | trans |
| rs964587 | miR-4429 | 2:6857291 | G | A | 0.560071 | 0.047023 | 0.013135 | 0.000344 | trans |
| rs7574580 | miR-4429 | 2:6857324 | A | G | 0.561141 | 0.047236 | 0.013175 | 0.000337 | trans |
| rs4671912 | miR-4429 | 2:6860225 | C | T | 0.720119 | -0.050724 | 0.014388 | 0.000423 | trans |
| rs4854474 | miR-4429 | 2:6863328 | A | T | 0.734157 | -0.058856 | 0.015173 | 0.000105 | trans |
| rs4854435 | miR-4429 | 2:6863971 | G | A | 0.753618 | -0.054401 | 0.015883 | 0.000615 | trans |
| rs7557983 | miR-4429 | 2:6859171 | C | T | 0.724325 | -0.052062 | 0.014625 | 0.000371 | trans |
| rs1420691 | miR-4429 | 2:6862748 | G | C | 0.737384 | -0.057495 | 0.015059 | 0.000135 | trans |
| rs3770650 | miR-4429 | 2:6861643 | A | G | 0.751964 | -0.055236 | 0.015139 | 0.000264 | trans |
| rs2289689 | miR-4429 | 2:6862015 | C | T | 0.752314 | -0.055664 | 0.015148 | 0.000238 | trans |
| rs7600829 | miR-4429 | 2:6856977 | T | C | 0.56021 | 0.046462 | 0.013099 | 0.00039 | trans |
| rs1975482 | miR-4429 | 2:6857418 | A | G | 0.560651 | 0.04758 | 0.013158 | 0.000299 | trans |

| | | | | | | | | | |
|---|---|---|---|---|---|---|---|---|---|
| rs7589952 | miR-4429 | 2:6857009: | G | A | 0.559677 | 0.046373 | 0.013086 | 0.000394 | trans |
| rs1049616 | miR-4429 | 2:6861012( | A | G | 0.750777 | -0.055744 | 0.015098 | 0.000222 | trans |
| rs1703537 | miR-4429 | 2:6859650: | G | A | 0.754443 | -0.055427 | 0.0153 | 0.000292 | trans |
| rs4277504 | miR-4429 | 2:6856956( | C | A | 0.559711 | 0.046394 | 0.013082 | 0.00039 | trans |
| rs964585 | miR-4429 | 2:6857243 | A | G | 0.559777 | 0.047007 | 0.013137 | 0.000346 | trans |
| rs1300495 | miR-4429 | 2:6858081 | G | A | 0.306559 | 0.075363 | 0.014167 | 1.04E-07 | trans |
| rs6546378 | miR-4429 | 2:6857344 | C | G | 0.56041 | 0.047172 | 0.013182 | 0.000346 | trans |
| rs9309412 | miR-4429 | 2:6857056 | C | T | 0.559665 | 0.046357 | 0.013088 | 0.000397 | trans |
| rs7424172 | miR-4429 | 2:6863238 | T | C | 0.734273 | -0.058865 | 0.015162 | 0.000103 | trans |
| rs1031117 | miR-4429 | 2:6860115 | G | C | 0.721971 | -0.052564 | 0.014451 | 0.000275 | trans |
| rs1168809 | miR-4429 | 2:6859604 | A | T | 0.751013 | -0.055525 | 0.015133 | 0.000243 | trans |
| rs6741866 | miR-4429 | 2:6857097( | G | A | 0.565872 | 0.045908 | 0.013156 | 0.000484 | trans |
| rs6745671 | miR-4429 | 2:6864349 | G | A | 0.754299 | -0.053185 | 0.015939 | 0.000848 | trans |
| rs6546380 | miR-4429 | 2:6857362( | G | A | 0.559433 | 0.04694 | 0.013153 | 0.000359 | trans |
| rs7560374 | miR-4429 | 2:6856998 | A | G | 0.559683 | 0.046378 | 0.013084 | 0.000393 | trans |
| rs9309415 | miR-4429 | 2:6860533 | C | A | 0.72047 | -0.051133 | 0.014389 | 0.00038 | trans |
| rs1443652 | miR-4429 | 2:6856965 | C | G | 0.559695 | 0.046403 | 0.013084 | 0.00039 | trans |
| rs3770648 | miR-4429 | 2:6861939 | C | T | 0.752286 | -0.055576 | 0.015149 | 0.000244 | trans |
| rs4671914 | miR-4429 | 2:6861229 | G | A | 0.751284 | -0.055888 | 0.015117 | 0.000218 | trans |
| rs2289688 | miR-4429 | 2:6862046 | G | A | 0.752268 | -0.055686 | 0.015145 | 0.000236 | trans |
| rs1975480 | miR-4429 | 2:6857386 | T | C | 0.559926 | 0.04706 | 0.013149 | 0.000345 | trans |
| rs736779 | miR-4429 | 2:6857191 | T | A | 0.56004 | 0.047002 | 0.013131 | 0.000344 | trans |
| rs9309411 | miR-4429 | 2:6857046 | A | G | 0.566779 | 0.045206 | 0.013199 | 0.000615 | trans |
| rs1443651 | miR-4429 | 2:6856931( | T | G | 0.559583 | 0.046245 | 0.013081 | 0.000407 | trans |
| rs6759864 | miR-4429 | 2:6857115 | T | C | 0.558332 | 0.046099 | 0.013045 | 0.00041 | trans |
| rs1262148 | miR-4429 | 2:6859777 | A | G | 0.333832 | 0.068624 | 0.013975 | 9.08E-07 | trans |
| rs6752922 | miR-4429 | 2:6857308 | G | A | 0.559358 | 0.046966 | 0.013142 | 0.000352 | trans |
| rs1021019 | miR-4429 | 2:6860353( | T | C | 0.720439 | -0.051124 | 0.014388 | 0.00038 | trans |
| rs1246827 | miR-4429 | 2:6864587( | A | G | 0.735366 | -0.055164 | 0.015403 | 0.000342 | trans |
| rs6546377 | miR-4429 | 2:6857022 | G | A | 0.559672 | 0.046616 | 0.013092 | 0.00037 | trans |
| rs1703535 | miR-4429 | 2:6858932 | C | A | 0.750845 | -0.055971 | 0.015136 | 0.000217 | trans |
| rs1020987 | miR-4429 | 2:6860519 | A | C | 0.72047 | -0.051131 | 0.014389 | 0.00038 | trans |
| rs1975481 | miR-4429 | 2:6857399 | C | A | 0.560307 | 0.047114 | 0.013149 | 0.00034 | trans |
| rs6546379 | miR-4429 | 2:6857362 | G | A | 0.559288 | 0.046944 | 0.013156 | 0.000359 | trans |
| rs1162818 | miR-4707-3 | 14:234672 | C | T | 0.795396 | -0.072222 | 0.016523 | 1.24E-05 | cis |
| rs1127361 | miR-4707-3 | 14:233607 | A | G | 0.816648 | -0.058523 | 0.017418 | 0.00078 | cis |
| rs4981454 | miR-4707-3 | 14:234221 | G | A | 0.206909 | 0.057398 | 0.016723 | 0.000598 | cis |
| rs1162424 | miR-4707-3 | 14:234005 | A | C | 0.800309 | -0.075503 | 0.016989 | 8.82E-06 | cis |
| rs8021195 | miR-4707-3 | 14:234870 | G | A | 0.523769 | -0.065089 | 0.013304 | 9.97E-07 | cis |
| rs2295686 | miR-4707-3 | 14:234166 | G | A | 0.800269 | -0.07534 | 0.016774 | 7.07E-06 | cis |
| rs7147394 | miR-4707-3 | 14:234110 | C | G | 0.206653 | 0.057684 | 0.016733 | 0.000566 | cis |
| rs1123380 | miR-4707-3 | 14:234004( | T | C | 0.800101 | -0.075147 | 0.016912 | 8.86E-06 | cis |
| rs8015600 | miR-4707-3 | 14:234897 | A | G | 0.526169 | -0.064811 | 0.013382 | 1.28E-06 | cis |
| rs4982704 | miR-4707-3 | 14:233635 | C | A | 0.458169 | 0.046144 | 0.013499 | 0.00063 | cis |
| rs7350749 | miR-4707-3 | 14:233853 | T | C | 0.799345 | -0.074782 | 0.016904 | 9.69E-06 | cis |
| rs5623442 | miR-4707-3 | 14:234030( | C | T | 0.799056 | -0.075452 | 0.016862 | 7.66E-06 | cis |
| rs1014005 | miR-4707-3 | 14:233589 | G | A | 0.45857 | 0.047653 | 0.013696 | 0.000503 | cis |
| rs2295904 | miR-4707-3 | 14:233622( | T | C | 0.817393 | -0.060273 | 0.017397 | 0.000531 | cis |
| rs4982701 | miR-4707-3 | 14:233596( | A | G | 0.458563 | 0.047637 | 0.013697 | 0.000505 | cis |
| rs3751490 | miR-4707-3 | 14:233555 | G | A | 0.818869 | -0.060055 | 0.017507 | 0.000603 | cis |
| rs1243663 | miR-4707-3 | 14:234301 | C | T | 0.80047 | -0.079189 | 0.016842 | 2.58E-06 | cis |
| rs2016392 | miR-4707-3 | 14:234241 | A | G | 0.205834 | 0.056726 | 0.016839 | 0.000755 | cis |
| rs1185095 | miR-494-3p | 14:101056 | G | T | 0.743981 | 0.135604 | 0.027447 | 7.79E-07 | cis |

| | | | | | | | | | |
|---|---|---|---|---|---|---|---|---|---|
| rs1013687 | miR-494-3p | 14:101202 | G | T | 0.493366 | 0.115887 | 0.025081 | 3.83E-06 | cis |
| rs1190852 | miR-494-3p | 14:101064 | A | G | 0.310295 | -0.138992 | 0.026527 | 1.61E-07 | cis |
| rs7145610 | miR-494-3p | 14:101049 | T | C | 0.740391 | 0.139109 | 0.02765 | 4.88E-07 | cis |
| rs1190872 | miR-494-3p | 14:101069 | A | G | 0.310177 | -0.138348 | 0.026478 | 1.74E-07 | cis |
| rs1802710 | miR-494-3p | 14:101200 | C | T | 0.493406 | 0.117835 | 0.025128 | 2.74E-06 | cis |
| rs7147982 | miR-494-3p | 14:101046 | C | T | 0.731753 | 0.130731 | 0.027074 | 1.38E-06 | cis |
| rs1288035 | miR-494-3p | 14:101158 | C | G | 0.904391 | 0.20168 | 0.043301 | 3.20E-06 | cis |
| rs1087352 | miR-494-3p | 14:101175 | G | A | 0.809826 | 0.138199 | 0.03073 | 6.89E-06 | cis |
| rs7152943 | miR-494-3p | 14:101174 | C | T | 0.924188 | 0.181551 | 0.046056 | 8.08E-05 | cis |
| rs1888581 | miR-494-3p | 14:101042 | A | C | 0.735363 | 0.134125 | 0.027268 | 8.71E-07 | cis |
| rs1184948 | miR-494-3p | 14:101037 | C | G | 0.733365 | 0.135236 | 0.027338 | 7.54E-07 | cis |
| rs3579985 | miR-494-3p | 14:101156 | C | T | 0.904015 | 0.203734 | 0.043296 | 2.53E-06 | cis |
| rs1190856 | miR-494-3p | 14:101065 | T | C | 0.310244 | -0.138788 | 0.026506 | 1.64E-07 | cis |
| rs1555405 | miR-494-3p | 14:101176 | G | A | 0.749075 | 0.132345 | 0.027581 | 1.60E-06 | cis |
| rs1015151 | miR-494-3p | 14:101175 | G | A | 0.806743 | 0.138237 | 0.030371 | 5.32E-06 | cis |
| rs7143357 | miR-494-3p | 14:101054 | T | C | 0.739636 | 0.1381 | 0.027603 | 5.64E-07 | cis |
| rs2842154 | miR-494-3p | 14:101158 | C | T | 0.904763 | 0.201769 | 0.043348 | 3.24E-06 | cis |
| rs1190862 | miR-494-3p | 14:101067 | T | G | 0.309628 | -0.139587 | 0.026318 | 1.13E-07 | cis |
| rs2400940 | miR-494-3p | 14:101224 | G | A | 0.185552 | -0.137576 | 0.030821 | 8.05E-06 | cis |
| rs7144997 | miR-494-3p | 14:101155 | A | G | 0.899872 | 0.208627 | 0.04315 | 1.33E-06 | cis |
| rs879059 | miR-494-3p | 14:101069 | T | A | 0.31068 | -0.138705 | 0.026463 | 1.59E-07 | cis |
| rs1190867 | miR-494-3p | 14:101069 | G | T | 0.316788 | -0.142273 | 0.026658 | 9.45E-08 | cis |
| rs7493817 | miR-494-3p | 14:101038 | G | A | 0.751723 | 0.141855 | 0.028217 | 4.97E-07 | cis |
| rs6575796 | miR-494-3p | 14:101160 | A | G | 0.094049 | -0.194681 | 0.044402 | 1.16E-05 | cis |
| rs1243295 | miR-494-3p | 14:101205 | G | A | 0.231663 | -0.147521 | 0.029009 | 3.67E-07 | cis |
| rs1709962 | miR-494-3p | 14:101173 | A | C | 0.783449 | 0.150105 | 0.029295 | 2.99E-07 | cis |
| rs7155649 | miR-494-3p | 14:101181 | C | A | 0.863608 | 0.160841 | 0.036167 | 8.70E-06 | cis |
| rs1190857 | miR-494-3p | 14:101066 | G | T | 0.310273 | -0.138684 | 0.026511 | 1.68E-07 | cis |
| rs1190818 | miR-494-3p | 14:101230 | T | G | 0.818683 | 0.146906 | 0.031464 | 3.03E-06 | cis |
| rs2837918 | miR-494-3p | 14:101168 | G | T | 0.836434 | 0.134835 | 0.032973 | 4.33E-05 | cis |
| rs1757777 | miR-494-3p | 14:101200 | C | T | 0.495315 | 0.114634 | 0.025146 | 5.15E-06 | cis |
| rs879060 | miR-494-3p | 14:101069 | A | G | 0.310926 | -0.138556 | 0.026481 | 1.67E-07 | cis |
| rs879058 | miR-494-3p | 14:101069 | G | T | 0.311807 | -0.138979 | 0.026477 | 1.53E-07 | cis |
| rs6575794 | miR-494-3p | 14:101039 | C | T | 0.734189 | 0.134485 | 0.027379 | 9.02E-07 | cis |
| rs8022367 | miR-494-3p | 14:101189 | A | T | 0.295876 | -0.194219 | 0.032359 | 1.95E-09 | cis |
| rs1116059 | miR-494-3p | 14:101049 | C | T | 0.741662 | 0.13822 | 0.027579 | 5.39E-07 | cis |
| rs4905999 | miR-494-3p | 14:101184 | A | G | 0.181905 | -0.17147 | 0.031935 | 7.91E-08 | cis |
| rs1190850 | miR-494-3p | 14:101064 | A | G | 0.314302 | -0.139242 | 0.026687 | 1.81E-07 | cis |
| rs1014171 | miR-494-3p | 14:101172 | G | T | 0.923932 | 0.180477 | 0.045949 | 8.57E-05 | cis |
| rs7152943 | miR-543 | 14:101174 | C | T | 0.924188 | 0.198333 | 0.054197 | 0.000253 | cis |
| rs7145610 | miR-543 | 14:101049 | T | C | 0.740391 | 0.137491 | 0.032578 | 2.44E-05 | cis |
| rs7156735 | miR-543 | 14:101132 | T | C | 0.867137 | 0.158004 | 0.041601 | 0.000146 | cis |
| rs1190904 | miR-543 | 14:101096 | A | G | 0.772596 | 0.170559 | 0.034611 | 8.31E-07 | cis |
| rs1190852 | miR-543 | 14:101064 | A | G | 0.310295 | -0.136671 | 0.03126 | 1.23E-05 | cis |
| rs1190903 | miR-543 | 14:101096 | C | T | 0.772566 | 0.170636 | 0.034614 | 8.24E-07 | cis |
| rs1555406 | miR-543 | 14:101204 | C | T | 0.876511 | 0.167026 | 0.043309 | 0.000115 | cis |
| rs1022032 | miR-543 | 14:101130 | T | G | 0.868038 | 0.153843 | 0.041666 | 0.000222 | cis |
| rs1190815 | miR-543 | 14:101228 | G | A | 0.798481 | 0.158461 | 0.035451 | 7.83E-06 | cis |
| rs7140556 | miR-543 | 14:101036 | C | A | 0.723818 | 0.138492 | 0.031962 | 1.47E-05 | cis |
| rs2400940 | miR-543 | 14:101224 | G | A | 0.185552 | -0.171114 | 0.036231 | 2.33E-06 | cis |
| rs7492369 | miR-543 | 14:101020 | C | T | 0.755698 | 0.137229 | 0.034341 | 6.44E-05 | cis |
| rs879060 | miR-543 | 14:101069 | A | G | 0.310926 | -0.136148 | 0.031206 | 1.28E-05 | cis |
| rs1190872 | miR-543 | 14:101069 | A | G | 0.310177 | -0.135387 | 0.031203 | 1.43E-05 | cis |

| | | | | | | | | | |
|---|---|---|---|---|---|---|---|---|---|
| rs1288804 | miR-543 | 14:101170 | G | A | 0.858051 | 0.162733 | 0.040407 | 5.64E-05 | cis |
| rs911330 | miR-543 | 14:101099 | T | C | 0.774615 | 0.173473 | 0.034718 | 5.84E-07 | cis |
| rs1214784 | miR-543 | 14:101144 | C | T | 0.870989 | 0.200601 | 0.042906 | 2.93E-06 | cis |
| rs1190906 | miR-543 | 14:101099 | T | C | 0.772138 | 0.165929 | 0.034506 | 1.52E-06 | cis |
| rs1190817 | miR-543 | 14:101229 | T | C | 0.798148 | 0.158017 | 0.03572 | 9.70E-06 | cis |
| rs1190862 | miR-543 | 14:101067 | T | G | 0.309628 | -0.13507 | 0.031019 | 1.33E-05 | cis |
| rs1765384 | miR-543 | 14:101102 | A | T | 0.870972 | 0.175105 | 0.045173 | 0.000106 | cis |
| rs1116059 | miR-543 | 14:101049 | C | T | 0.741662 | 0.137411 | 0.032492 | 2.35E-05 | cis |
| rs7147982 | miR-543 | 14:101046 | C | T | 0.731753 | 0.129675 | 0.031894 | 4.79E-05 | cis |
| rs6575796 | miR-543 | 14:101160 | A | G | 0.094049 | -0.194343 | 0.05229 | 0.000202 | cis |
| rs1190857 | miR-543 | 14:101066 | G | T | 0.310273 | -0.136062 | 0.031242 | 1.33E-05 | cis |
| rs1709961 | miR-543 | 14:101170 | A | G | 0.858798 | 0.161565 | 0.040294 | 6.08E-05 | cis |
| rs2842154 | miR-543 | 14:101158 | C | T | 0.904763 | 0.201606 | 0.051056 | 7.86E-05 | cis |
| rs1014839 | miR-543 | 14:101026 | A | G | 0.750738 | 0.139869 | 0.034163 | 4.24E-05 | cis |
| rs1185095 | miR-543 | 14:101056 | G | T | 0.743981 | 0.132254 | 0.03234 | 4.32E-05 | cis |
| rs1765383 | miR-543 | 14:101101 | T | C | 0.877829 | 0.174275 | 0.045085 | 0.000111 | cis |
| rs1243613 | miR-543 | 14:101031 | C | T | 0.725015 | 0.139522 | 0.03203 | 1.32E-05 | cis |
| rs1243369 | miR-543 | 14:101035 | C | G | 0.723403 | 0.136924 | 0.031822 | 1.69E-05 | cis |
| rs2104070 | miR-543 | 14:101214 | G | A | 0.129596 | -0.164585 | 0.04292 | 0.000126 | cis |
| rs1190909 | miR-543 | 14:101101 | G | A | 0.775531 | 0.166989 | 0.035018 | 1.85E-06 | cis |
| rs3783350 | miR-543 | 14:101199 | G | A | 0.22096 | -0.131682 | 0.035588 | 0.000215 | cis |
| rs2847806 | miR-543 | 14:101139 | C | A | 0.868556 | 0.165494 | 0.041843 | 7.65E-05 | cis |
| rs1766021 | miR-543 | 14:101131 | A | G | 0.868541 | 0.159833 | 0.041852 | 0.000134 | cis |
| rs1802710 | miR-543 | 14:101200 | C | T | 0.493406 | 0.112858 | 0.029606 | 0.000138 | cis |
| rs1766036 | miR-543 | 14:101136 | A | G | 0.868731 | 0.159242 | 0.041925 | 0.000146 | cis |
| rs8008285 | miR-543 | 14:101146 | T | C | 0.842905 | 0.209707 | 0.039876 | 1.45E-07 | cis |
| rs3579985 | miR-543 | 14:101156 | C | T | 0.904015 | 0.202995 | 0.050997 | 6.88E-05 | cis |
| rs7493817 | miR-543 | 14:101038 | G | A | 0.751723 | 0.147012 | 0.033232 | 9.70E-06 | cis |
| rs7154024 | miR-543 | 14:101031 | G | T | 0.725015 | 0.139249 | 0.032002 | 1.35E-05 | cis |
| rs1709961 | miR-543 | 14:101164 | T | A | 0.903821 | 0.198975 | 0.049243 | 5.33E-05 | cis |
| rs1243295 | miR-543 | 14:101205 | G | A | 0.231663 | -0.175807 | 0.034114 | 2.56E-07 | cis |
| rs1184948 | miR-543 | 14:101037 | C | G | 0.733365 | 0.135753 | 0.032204 | 2.49E-05 | cis |
| rs1757777 | miR-543 | 14:101200 | C | T | 0.495315 | 0.109785 | 0.029625 | 0.000211 | cis |
| rs1288035 | miR-543 | 14:101158 | C | G | 0.904391 | 0.201455 | 0.051001 | 7.82E-05 | cis |
| rs1184764 | miR-543 | 14:101023 | C | T | 0.753952 | 0.137502 | 0.034283 | 6.05E-05 | cis |
| rs1013687 | miR-543 | 14:101202 | G | T | 0.493366 | 0.113444 | 0.029544 | 0.000123 | cis |
| rs879058 | miR-543 | 14:101069 | G | T | 0.311807 | -0.135882 | 0.031203 | 1.33E-05 | cis |
| rs4905991 | miR-543 | 14:101140 | T | C | 0.271223 | -0.129714 | 0.032501 | 6.58E-05 | cis |
| rs1190818 | miR-543 | 14:101230 | T | G | 0.818683 | 0.178149 | 0.036996 | 1.47E-06 | cis |
| rs2852739 | miR-543 | 14:101130 | T | C | 0.868049 | 0.153847 | 0.041672 | 0.000223 | cis |
| rs3516145 | miR-543 | 14:101164 | A | G | 0.624259 | 0.158704 | 0.034853 | 5.28E-06 | cis |
| rs1013035 | miR-543 | 14:101149 | T | C | 0.112665 | -0.199338 | 0.047682 | 2.91E-05 | cis |
| rs7155649 | miR-543 | 14:101181 | C | A | 0.863608 | 0.188511 | 0.04254 | 9.36E-06 | cis |
| rs8005989 | miR-543 | 14:101216 | G | A | 0.129787 | -0.164884 | 0.042922 | 0.000122 | cis |
| rs1258812 | miR-543 | 14:101162 | T | C | 0.893709 | 0.229501 | 0.047843 | 1.61E-06 | cis |
| rs3494675 | miR-543 | 14:100938 | A | G | 0.904426 | 0.235475 | 0.057172 | 3.81E-05 | trans |
| rs1004574 | miR-543 | 14:101178 | G | C | 0.86063 | 0.169602 | 0.041055 | 3.61E-05 | cis |
| rs7143357 | miR-543 | 14:101054 | T | C | 0.739636 | 0.13266 | 0.032528 | 4.54E-05 | cis |
| rs8014970 | miR-543 | 14:101140 | T | G | 0.868329 | 0.16553 | 0.041884 | 7.75E-05 | cis |
| rs7156824 | miR-543 | 14:101171 | C | A | 0.870119 | 0.16649 | 0.042069 | 7.57E-05 | cis |
| rs1116060 | miR-543 | 14:101220 | T | C | 0.874258 | 0.169116 | 0.043015 | 8.44E-05 | cis |
| rs6575791 | miR-543 | 14:101029 | G | A | 0.719841 | 0.141709 | 0.032023 | 9.63E-06 | cis |
| rs1190856 | miR-543 | 14:101065 | T | C | 0.310244 | -0.136302 | 0.031235 | 1.28E-05 | cis |

| | | | | | | | | | |
|---|---|---|---|---|---|---|---|---|---|
| rs1190850 | miR-543 | 14:101064 | A | G | 0.314302 | -0.138852 | 0.031445 | 1.01E-05 | cis |
| rs1190907 | miR-543 | 14:101099 | A | G | 0.772847 | 0.169157 | 0.034555 | 9.81E-07 | cis |
| rs2295659 | miR-543 | 14:101141 | G | T | 0.857995 | 0.169991 | 0.040812 | 3.11E-05 | cis |
| rs1004572 | miR-543 | 14:101178 | G | A | 0.86193 | 0.170058 | 0.041319 | 3.86E-05 | cis |
| rs9635298 | miR-543 | 14:101172 | C | T | 0.859368 | 0.166461 | 0.040567 | 4.07E-05 | cis |
| rs9635184 | miR-543 | 14:101173 | C | T | 0.859552 | 0.16671 | 0.040571 | 3.97E-05 | cis |
| rs3470057 | miR-543 | 14:101173 | C | T | 0.892725 | 0.222227 | 0.046393 | 1.67E-06 | cis |
| rs8007901 | miR-543 | 14:101130 | T | C | 0.867321 | 0.157146 | 0.041496 | 0.000152 | cis |
| rs1888581 | miR-543 | 14:101042 | A | C | 0.735363 | 0.133402 | 0.032123 | 3.28E-05 | cis |
| rs7144375 | miR-543 | 14:101137 | T | C | 0.866639 | 0.164708 | 0.04157 | 7.43E-05 | cis |
| rs6575794 | miR-543 | 14:101039 | C | T | 0.734189 | 0.133846 | 0.032254 | 3.33E-05 | cis |
| rs745797 | miR-543 | 14:101141 | G | A | 0.858727 | 0.171086 | 0.04094 | 2.93E-05 | cis |
| rs879059 | miR-543 | 14:101069 | T | A | 0.31068 | -0.135728 | 0.031187 | 1.35E-05 | cis |
| rs1162219 | miR-543 | 14:101016 | G | A | 0.759852 | 0.167489 | 0.03629 | 3.93E-06 | cis |
| rs1014844 | miR-543 | 14:101026 | G | A | 0.753241 | 0.141733 | 0.034148 | 3.32E-05 | cis |
| rs1184658 | miR-543 | 14:101017 | C | G | 0.759077 | 0.148572 | 0.034785 | 1.95E-05 | cis |
| rs1184761 | miR-543 | 14:101023 | C | T | 0.753476 | 0.137069 | 0.03425 | 6.28E-05 | cis |
| rs7144997 | miR-543 | 14:101155 | A | G | 0.899872 | 0.196244 | 0.050851 | 0.000114 | cis |
| rs1004573 | miR-543 | 14:101178 | G | C | 0.860589 | 0.16958 | 0.041055 | 3.62E-05 | cis |
| rs7140792 | miR-543 | 14:101172 | C | T | 0.892517 | 0.220987 | 0.046442 | 1.95E-06 | cis |
| rs1259093 | miR-543 | 14:101163 | A | T | 0.893649 | 0.229805 | 0.047847 | 1.56E-06 | cis |
| rs8022367 | miR-543 | 14:101189 | A | T | 0.295876 | -0.217458 | 0.03809 | 1.14E-08 | cis |
| rs1190878 | miR-543 | 14:101072 | C | A | 0.859763 | 0.153889 | 0.042059 | 0.000253 | cis |
| rs1190867 | miR-543 | 14:101069 | G | T | 0.316788 | -0.133999 | 0.031429 | 2.01E-05 | cis |
| rs1025475 | miR-584-5 | 5:1483896 | G | C | 0.59051 | -0.114539 | 0.025454 | 6.80E-06 | cis |
| rs1045942 | miR-584-5 | 5:1483811 | G | A | 0.567667 | -0.11594 | 0.025328 | 4.70E-06 | cis |
| rs250673 | miR-584-5 | 5:1484457 | C | T | 0.807036 | -0.113973 | 0.031638 | 0.000315 | cis |
| rs250672 | miR-584-5 | 5:1484461 | A | G | 0.807037 | -0.113973 | 0.031638 | 0.000315 | cis |
| rs250679 | miR-584-5 | 5:1484349 | T | A | 0.805009 | -0.113425 | 0.03168 | 0.000343 | cis |
| rs8030177 | miR-584-5 | 5:1484573 | T | A | 0.806138 | -0.113282 | 0.031811 | 0.000369 | cis |
| rs4620015 | miR-584-5 | 5:1485331 | T | A | 0.21298 | 0.16908 | 0.032046 | 1.32E-07 | cis |
| rs1004481 | miR-584-5 | 5:1485422 | T | C | 0.211057 | 0.167499 | 0.032112 | 1.83E-07 | cis |
| rs1025476 | miR-584-5 | 5:1483897 | G | A | 0.590572 | -0.114582 | 0.025455 | 6.75E-06 | cis |
| rs384979 | miR-584-5 | 5:1484592 | T | C | 0.804599 | -0.116367 | 0.031724 | 0.000244 | cis |
| rs36044 | miR-584-5 | 5:1484411 | T | C | 0.807043 | -0.113923 | 0.031636 | 0.000317 | cis |
| rs3213854 | miR-584-5 | 5:1483728 | T | C | 0.568146 | -0.115308 | 0.025333 | 5.32E-06 | cis |
| rs2963485 | miR-584-5 | 5:1484520 | C | T | 0.806974 | -0.112926 | 0.031698 | 0.000367 | cis |
| rs250674 | miR-584-5 | 5:1484385 | C | T | 0.805352 | -0.113373 | 0.031669 | 0.000344 | cis |
| rs733032 | miR-584-5 | 5:1483576 | T | G | 0.433976 | 0.117536 | 0.025261 | 3.27E-06 | cis |
| rs6871030 | miR-584-5 | 5:1483865 | T | G | 0.583882 | -0.113999 | 0.025536 | 8.03E-06 | cis |
| rs1432795 | miR-584-5 | 5:1483787 | C | T | 0.567724 | -0.11591 | 0.02533 | 4.74E-06 | cis |
| rs250671 | miR-584-5 | 5:1484465 | T | C | 0.806694 | -0.113726 | 0.031581 | 0.000317 | cis |
| rs36048 | miR-584-5 | 5:1484417 | G | C | 0.807038 | -0.113939 | 0.031636 | 0.000316 | cis |
| rs6580591 | miR-584-5 | 5:1484038 | A | G | 0.589871 | -0.115718 | 0.02567 | 6.55E-06 | cis |
| rs41181 | miR-584-5 | 5:1484442 | G | C | 0.807036 | -0.113956 | 0.031637 | 0.000316 | cis |
| rs3111150 | miR-584-5 | 5:1484483 | G | A | 0.810208 | -0.118293 | 0.032041 | 0.000223 | cis |
| rs4235739 | miR-584-5 | 5:1483665 | T | C | 0.413758 | 0.110563 | 0.025527 | 1.48E-05 | cis |
| rs250667 | miR-584-5 | 5:1484551 | G | A | 0.807242 | -0.112189 | 0.031723 | 0.000405 | cis |
| rs919733 | miR-584-5 | 5:1483493 | A | G | 0.432977 | 0.118117 | 0.02536 | 3.20E-06 | cis |
| rs1347130 | miR-584-5 | 5:1483769 | G | A | 0.568652 | -0.11677 | 0.025304 | 3.94E-06 | cis |
| rs4705067 | miR-584-5 | 5:1485424 | C | G | 0.212676 | 0.16803 | 0.032085 | 1.63E-07 | cis |
| rs36043 | miR-584-5 | 5:1484409 | G | A | 0.807028 | -0.113937 | 0.031636 | 0.000316 | cis |
| rs42364 | miR-584-5 | 5:1484388 | T | A | 0.804673 | -0.112774 | 0.031697 | 0.000374 | cis |

| | | | | | | | | | |
|---|---|---|---|---|---|---|---|---|---|
| rs4361503 | miR-584-5p | 5:1485448( | T | C | 0.208707 | 0.166154 | 0.031662 | 1.54E-07 | cis |
| rs36046 | miR-584-5p | 5:1484412! | A | C | 0.80704 | -0.113941 | 0.031636 | 0.000316 | cis |
| rs250670 | miR-584-5p | 5:1484466 | G | T | 0.806962 | -0.113866 | 0.031643 | 0.00032 | cis |
| rs40522 | miR-584-5p | 5:1484422! | G | A | 0.80695 | -0.113977 | 0.031633 | 0.000314 | cis |
| rs1265749 | miR-584-5p | 5:1483613( | G | C | 0.434856 | 0.115401 | 0.025312 | 5.14E-06 | cis |
| rs4134361 | miR-584-5p | 5:1485271( | G | C | 0.212728 | 0.170237 | 0.032157 | 1.20E-07 | cis |
| rs3095947 | miR-584-5p | 5:1484367 | T | C | 0.805269 | -0.113584 | 0.031694 | 0.000339 | cis |
| rs2116756 | miR-584-5p | 5:1483567 | G | T | 0.434475 | 0.115303 | 0.025271 | 5.05E-06 | cis |
| rs930205 | miR-584-5p | 5:1483652 | T | C | 0.435273 | 0.117656 | 0.02536 | 3.49E-06 | cis |
| rs36045 | miR-584-5p | 5:1484411 | G | A | 0.825102 | -0.108715 | 0.032812 | 0.000922 | cis |
| rs2915809 | miR-584-5p | 5:1484521 | A | T | 0.807189 | -0.112955 | 0.031694 | 0.000365 | cis |
| rs4418085 | miR-584-5p | 5:1484569 | C | A | 0.813279 | -0.114762 | 0.032597 | 0.00043 | cis |
| rs1821252 | miR-584-5p | 5:1483574( | T | A | 0.433491 | 0.116914 | 0.025243 | 3.63E-06 | cis |
| rs4705303 | miR-584-5p | 5:1483676 | C | T | 0.466328 | 0.114427 | 0.02579 | 9.13E-06 | cis |
| rs40523 | miR-584-5p | 5:1484441 | A | T | 0.80704 | -0.113959 | 0.031638 | 0.000316 | cis |
| rs4560536 | miR-584-5p | 5:1483647( | C | T | 0.433829 | 0.118853 | 0.025467 | 3.06E-06 | cis |
| rs168750 | miR-584-5p | 5:1484467! | A | G | 0.807145 | -0.114145 | 0.031657 | 0.000311 | cis |
| rs40149 | miR-584-5p | 5:1484442 | A | T | 0.807037 | -0.113962 | 0.031637 | 0.000316 | cis |
| rs4543254 | miR-584-5p | 5:1483682! | T | G | 0.413952 | 0.110431 | 0.025528 | 1.52E-05 | cis |
| rs2963484 | miR-584-5p | 5:1484511 | A | T | 0.807181 | -0.113055 | 0.031687 | 0.00036 | cis |
| rs185155 | miR-584-5p | 5:1484558! | T | C | 0.80724 | -0.11213 | 0.031725 | 0.000409 | cis |
| rs250663 | miR-584-5p | 5:1484531 | T | C | 0.807196 | -0.112783 | 0.031703 | 0.000374 | cis |
| rs1422581 | miR-584-5p | 5:1484373 | A | G | 0.805225 | -0.113393 | 0.031673 | 0.000343 | cis |
| rs1004062 | miR-584-5p | 5:1484375 | A | G | 0.80525 | -0.113391 | 0.031672 | 0.000343 | cis |
| rs1126599 | miR-584-5p | 5:1484578 | A | T | 0.808119 | -0.110561 | 0.031882 | 0.000525 | cis |
| rs250668 | miR-584-5p | 5:1484565 | T | C | 0.807366 | -0.111823 | 0.031773 | 0.000433 | cis |
| rs250680 | miR-584-5p | 5:1484360 | T | C | 0.805213 | -0.113587 | 0.031695 | 0.000339 | cis |
| rs166106 | miR-584-5p | 5:1484470( | T | C | 0.825108 | -0.108721 | 0.032813 | 0.000922 | cis |
| rs1549906 | miR-584-5p | 5:1483939 | A | G | 0.569385 | -0.122043 | 0.02545 | 1.62E-06 | cis |
| rs6893374 | miR-584-5p | 5:1483962 | A | G | 0.569809 | -0.124255 | 0.025543 | 1.15E-06 | cis |
| rs250678 | miR-584-5p | 5:1484348 | C | T | 0.804998 | -0.113427 | 0.03168 | 0.000343 | cis |
| rs36042 | miR-584-5p | 5:1484397 | G | T | 0.825023 | -0.108506 | 0.032835 | 0.000951 | cis |
| rs1006434 | miR-584-5p | 5:1484586 | G | A | 0.80727 | -0.111414 | 0.031766 | 0.000453 | cis |
| rs250664 | miR-584-5p | 5:1484538 | C | T | 0.807224 | -0.112395 | 0.031709 | 0.000393 | cis |
| rs250666 | miR-584-5p | 5:1484546 | A | G | 0.807229 | -0.112303 | 0.031714 | 0.000399 | cis |
| rs891919 | miR-584-5p | 5:1483695( | A | T | 0.568865 | -0.117038 | 0.025305 | 3.75E-06 | cis |
| rs6580598 | miR-584-5p | 5:1485337! | A | G | 0.209214 | 0.168006 | 0.031895 | 1.38E-07 | cis |
| rs168749 | miR-584-5p | 5:1484560 | A | G | 0.807287 | -0.111801 | 0.031779 | 0.000435 | cis |
| rs1036191 | miR-584-5p | 5:1483888 | C | T | 0.604903 | -0.124317 | 0.025774 | 1.41E-06 | cis |
| rs4705065 | miR-584-5p | 5:1484002 | A | G | 0.467652 | 0.123164 | 0.026182 | 2.55E-06 | cis |
| rs919732 | miR-584-5p | 5:1483495 | A | C | 0.431899 | 0.116519 | 0.025379 | 4.41E-06 | cis |
| rs1036190 | miR-584-5p | 5:1483876 | A | G | 0.589465 | -0.111569 | 0.025447 | 1.16E-05 | cis |
| rs1777940 | miR-625-5p | 14:662046 | G | A | 0.954809 | -0.321813 | 0.047741 | 1.57E-11 | cis |
| rs1718138 | miR-625-5p | 14:661180 | C | T | 0.955489 | -0.324084 | 0.048819 | 3.17E-11 | cis |
| rs7817768 | miR-625-5p | 14:658969 | A | C | 0.958266 | -0.312388 | 0.049535 | 2.86E-10 | cis |
| rs1256541 | miR-625-5p | 14:657692 | T | A | 0.951048 | -0.330826 | 0.045089 | 2.18E-13 | cis |
| rs1014624 | miR-625-5p | 14:657090 | G | A | 0.953383 | -0.248377 | 0.046554 | 9.54E-08 | cis |
| rs7888817( | miR-625-5p | 14:657356 | A | G | 0.959262 | -0.322794 | 0.05074 | 2.00E-10 | cis |
| rs1014633 | miR-625-5p | 14:657090 | G | A | 0.953216 | -0.248145 | 0.046558 | 9.83E-08 | cis |
| rs1048378 | miR-625-5p | 14:660765 | T | C | 0.956227 | -0.320765 | 0.048795 | 4.91E-11 | cis |
| rs1256542 | miR-625-5p | 14:657694 | A | T | 0.950471 | -0.331231 | 0.044931 | 1.68E-13 | cis |
| rs5739174 | miR-625-5p | 14:657066 | C | G | 0.952618 | -0.24363 | 0.045759 | 1.01E-07 | cis |
| rs5742565( | miR-625-5p | 14:656838 | A | T | 0.756453 | -0.078872 | 0.022688 | 0.000508 | cis |

| | | | | | | | | | |
|---|---|---|---|---|---|---|---|---|---|
| rs1407138 | miR-625-5p | 14:658468 | G | A | 0.959059 | -0.320584 | 0.050447 | 2.09E-10 | cis |
| rs1256537 | miR-625-5p | 14:657629 | T | C | 0.952007 | -0.321913 | 0.045372 | 1.29E-12 | cis |
| rs7327612 | miR-625-5p | 14:657141 | C | T | 0.919217 | -0.127804 | 0.036084 | 0.000397 | cis |
| rs7159220 | miR-625-5p | 14:656910 | C | T | 0.75928 | -0.08477 | 0.022799 | 0.000201 | cis |
| rs1038239 | miR-625-5p | 14:657679 | A | G | 0.950718 | -0.323316 | 0.045039 | 7.05E-13 | cis |
| rs1718099 | miR-625-5p | 14:658521 | A | T | 0.958988 | -0.318902 | 0.050362 | 2.42E-10 | cis |
| rs2101084 | miR-625-5p | 14:657196 | A | C | 0.951835 | -0.270858 | 0.046338 | 5.06E-09 | cis |
| rs766906 | miR-625-5p | 14:656603 | C | T | 0.753031 | -0.078085 | 0.022566 | 0.00054 | cis |
| rs7673402 | miR-625-5p | 14:657205 | G | A | 0.92213 | -0.135242 | 0.036972 | 0.000254 | cis |
| rs7583926 | miR-625-5p | 14:657998 | G | T | 0.961653 | -0.321471 | 0.052521 | 9.31E-10 | cis |
| rs1760984 | miR-625-5p | 14:656600 | A | G | 0.751725 | -0.078912 | 0.022496 | 0.000452 | cis |
| rs7785126 | miR-625-5p | 14:662702 | G | A | 0.951842 | -0.327317 | 0.046217 | 1.42E-12 | cis |
| rs7518509 | miR-625-5p | 14:662233 | C | G | 0.954314 | -0.324296 | 0.047597 | 9.53E-12 | cis |
| rs1014359 | miR-625-5p | 14:657013 | G | T | 0.950079 | -0.242547 | 0.04433 | 4.47E-08 | cis |
| rs7969543 | miR-625-5p | 14:657144 | G | A | 0.955272 | -0.257709 | 0.04803 | 8.07E-08 | cis |
| rs4131097 | miR-625-5p | 14:662591 | G | C | 0.754914 | -0.12278 | 0.022687 | 6.23E-08 | cis |
| rs8000324 | miR-625-5p | 14:661648 | G | A | 0.955494 | -0.324411 | 0.048471 | 2.19E-11 | cis |
| rs4299072 | miR-625-5p | 14:657052 | G | A | 0.952605 | -0.243207 | 0.045733 | 1.05E-07 | cis |
| rs7827466 | miR-625-5p | 14:659775 | A | G | 0.957792 | -0.314572 | 0.049335 | 1.82E-10 | cis |
| rs1391351 | miR-625-5p | 14:657211 | A | G | 0.952473 | -0.283045 | 0.046656 | 1.31E-09 | cis |
| rs1159809 | miR-625-5p | 14:658343 | A | G | 0.960304 | -0.324418 | 0.051353 | 2.66E-10 | cis |
| rs7757627 | miR-625-5p | 14:658320 | A | G | 0.964091 | -0.340905 | 0.055529 | 8.30E-10 | cis |
| rs1760983 | miR-625-5p | 14:656597 | A | G | 0.751685 | -0.079005 | 0.022495 | 0.000444 | cis |
| rs8034920 | miR-625-5p | 14:656968 | C | A | 0.952874 | -0.258333 | 0.046871 | 3.56E-08 | cis |
| rs7684188 | miR-625-5p | 14:658396 | A | G | 0.960013 | -0.323742 | 0.051185 | 2.53E-10 | cis |
| rs1171475 | miR-625-5p | 14:659124 | G | A | 0.957872 | -0.311447 | 0.049433 | 2.97E-10 | cis |
| rs7865452 | miR-625-5p | 14:657744 | C | G | 0.962554 | -0.319353 | 0.052879 | 1.55E-09 | cis |
| rs7565995 | miR-625-5p | 14:660328 | A | C | 0.956792 | -0.319133 | 0.048977 | 7.22E-11 | cis |
| rs1679879 | miR-625-5p | 14:656596 | C | T | 0.75012 | -0.078028 | 0.022332 | 0.000476 | cis |
| rs1679872 | miR-625-5p | 14:656693 | C | G | 0.751765 | -0.07773 | 0.02259 | 0.00058 | cis |
| rs1679873 | miR-625-5p | 14:656693 | A | G | 0.750818 | -0.078556 | 0.022529 | 0.000489 | cis |
| rs7964308 | miR-625-5p | 14:657299 | A | C | 0.953965 | -0.301059 | 0.046424 | 8.87E-11 | cis |
| rs1760985 | miR-625-5p | 14:656621 | A | G | 0.751842 | -0.078662 | 0.022498 | 0.000472 | cis |
| rs1679875 | miR-625-5p | 14:656633 | A | G | 0.75279 | -0.077783 | 0.02259 | 0.000575 | cis |
| rs1167701 | miR-625-5p | 14:659093 | A | G | 0.957923 | -0.312295 | 0.049431 | 2.65E-10 | cis |
| rs7951427 | miR-625-5p | 14:657031 | G | T | 0.952599 | -0.243778 | 0.045771 | 1.00E-07 | cis |
| rs7281925 | miR-744-5p | 17:119307 | A | T | 0.761846 | -0.117535 | 0.035178 | 0.000834 | cis |
| rs7282126 | miR-744-5p | 17:119879 | C | T | 0.763548 | -0.119524 | 0.035175 | 0.000679 | cis |
| rs1165372 | miR-744-5p | 17:119889 | G | T | 0.76356 | -0.119524 | 0.035175 | 0.000679 | cis |
| rs5945456 | miR-744-5p | 17:119705 | C | G | 0.761535 | -0.120492 | 0.035088 | 0.000595 | cis |
| rs1293688 | miR-744-5p | 17:120106 | A | G | 0.262735 | 0.118255 | 0.034722 | 0.00066 | cis |
| rs1205176 | miR-744-5p | 17:119779 | G | A | 0.252044 | 0.119363 | 0.034865 | 0.000618 | cis |
| rs2322123 | miR-744-5p | 17:119535 | G | A | 0.248462 | 0.118469 | 0.035056 | 0.000726 | cis |
| rs1165572 | miR-744-5p | 17:119586 | T | C | 0.7631 | -0.119056 | 0.035129 | 0.000701 | cis |
| rs7282126 | miR-744-5p | 17:119953 | G | C | 0.76315 | -0.120405 | 0.035149 | 0.000613 | cis |
| rs7210799 | miR-744-5p | 17:119666 | C | G | 0.24936 | 0.120452 | 0.035143 | 0.000609 | cis |
| rs1165446 | miR-744-5p | 17:119855 | T | C | 0.763551 | -0.119321 | 0.035173 | 0.000693 | cis |
| rs757594 | miR-744-5p | 17:120006 | G | A | 0.240611 | 0.119067 | 0.034631 | 0.000586 | cis |
| rs5593978 | miR-744-5p | 17:119336 | A | G | 0.761961 | -0.11771 | 0.035164 | 0.000816 | cis |
| rs8082185 | miR-744-5p | 17:119339 | A | T | 0.249147 | 0.117158 | 0.035214 | 0.000878 | cis |
| rs1232587 | miR-744-5p | 17:119951 | A | G | 0.76342 | -0.120725 | 0.035141 | 0.000592 | cis |
| rs3414245 | miR-133a- | 20:610884 | C | T | 0.707746 | -0.139283 | 0.026124 | 9.73E-08 | cis |
| rs4429540 | miR-133a- | 20:610870 | C | T | 0.687203 | -0.136631 | 0.026004 | 1.49E-07 | cis |

| | | | | | | | | | |
|---|---|---|---|---|---|---|---|---|---|
| rs4508677 | miR-133a-3 | 20:611483: | A | G | 0.412643 | -0.116625 | 0.021971 | 1.11E-07 | cis |
| rs6061259 | miR-133a-3 | 20:610877 | G | A | 0.682582 | -0.135364 | 0.02604 | 2.01E-07 | cis |
| rs6061571 | miR-133a-3 | 20:610862 | A | G | 0.677911 | -0.134668 | 0.026078 | 2.42E-07 | cis |
| rs6061572 | miR-133a-3 | 20:610867 | G | C | 0.677483 | -0.134673 | 0.02607 | 2.39E-07 | cis |
| rs6061576 | miR-133a-3 | 20:610888 | G | C | 0.698154 | -0.143014 | 0.026079 | 4.16E-08 | cis |
| rs6062277 | miR-133a-3 | 20:611455 | A | G | 0.461496 | -0.104082 | 0.019172 | 5.67E-08 | cis |
| rs6062278 | miR-133a-3 | 20:611449 | A | G | 0.470455 | -0.102881 | 0.018906 | 5.28E-08 | cis |
| rs6062284 | miR-133a-3 | 20:611419 | T | G | 0.4602 | -0.105041 | 0.019465 | 6.80E-08 | cis |
| rs6089750 | miR-133a-3 | 20:611556 | C | T | 0.58104 | 0.111273 | 0.021759 | 3.16E-07 | cis |
| rs6122011 | miR-133a-3 | 20:611569 | G | T | 0.578584 | 0.111915 | 0.021838 | 2.98E-07 | cis |
| rs6143048 | miR-133a-3 | 20:611594 | C | T | 0.62443 | 0.162891 | 0.027868 | 5.07E-09 | cis |
| rs7269123 | miR-133a-3 | 20:611579 | C | T | 0.579224 | 0.111666 | 0.022805 | 9.75E-07 | cis |
| rs1049478 | miR-181a-5 | 1:1991617 | T | C | 0.694823 | -0.066336 | 0.013463 | 8.33E-07 | cis |
| rs1073755 | miR-181a-5 | 1:1990298 | T | G | 0.68782 | -0.070755 | 0.013487 | 1.55E-07 | cis |
| rs1075384 | miR-181a-5 | 1:1990517 | C | A | 0.65941 | -0.067258 | 0.013226 | 3.67E-07 | cis |
| rs1075384 | miR-181a-5 | 1:1990518 | C | T | 0.634887 | -0.059074 | 0.013043 | 5.93E-06 | cis |
| rs1080059 | miR-181a-5 | 1:1988680 | G | A | 0.373371 | -0.046359 | 0.012918 | 0.000332 | cis |
| rs1080059 | miR-181a-5 | 1:1988695 | T | C | 0.372444 | -0.046008 | 0.012901 | 0.000362 | cis |
| rs1080060 | miR-181a-5 | 1:1990295 | T | C | 0.691896 | -0.065566 | 0.013563 | 1.34E-06 | cis |
| rs1080061 | miR-181a-5 | 1:1991394 | C | T | 0.734662 | -0.066308 | 0.014124 | 2.67E-06 | cis |
| rs1080061 | miR-181a-5 | 1:1991582 | C | A | 0.750461 | -0.074457 | 0.014445 | 2.54E-07 | cis |
| rs1080061 | miR-181a-5 | 1:1991678 | G | T | 0.734306 | -0.067614 | 0.014296 | 2.25E-06 | cis |
| rs1091962 | miR-181a-5 | 1:1990539 | T | C | 0.658927 | -0.069731 | 0.013207 | 1.29E-07 | cis |
| rs1091963 | miR-181a-5 | 1:1991674 | T | C | 0.679446 | -0.060814 | 0.013389 | 5.57E-06 | cis |
| rs1180362 | miR-181a-5 | 1:1987805 | C | T | 0.884045 | -0.079819 | 0.019853 | 5.81E-05 | cis |
| rs1180498 | miR-181a-5 | 1:1987836 | C | T | 0.884595 | -0.08016 | 0.019805 | 5.18E-05 | cis |
| rs1180776 | miR-181a-5 | 1:1988058 | C | G | 0.821168 | -0.062706 | 0.016863 | 0.0002 | cis |
| rs1180840 | miR-181a-5 | 1:1988020 | C | G | 0.886021 | -0.084055 | 0.019702 | 1.99E-05 | cis |
| rs1207100 | miR-181a-5 | 1:1988011 | T | C | 0.884463 | -0.08179 | 0.019507 | 2.75E-05 | cis |
| rs1212146 | miR-181a-5 | 1:1990624 | T | C | 0.663581 | -0.066298 | 0.013345 | 6.76E-07 | cis |
| rs1213074 | miR-181a-5 | 1:1988076 | A | G | 0.883608 | -0.094515 | 0.019597 | 1.42E-06 | cis |
| rs1213415 | miR-181a-5 | 1:1988631 | C | T | 0.375349 | -0.046858 | 0.01292 | 0.000287 | cis |
| rs1213722 | miR-181a-5 | 1:1987769 | G | A | 0.881396 | -0.090827 | 0.019727 | 4.14E-06 | cis |
| rs1214000 | miR-181a-5 | 1:1988091 | C | G | 0.819834 | -0.061848 | 0.016841 | 0.00024 | cis |
| rs1272976 | miR-181a-5 | 1:1989920 | G | C | 0.644616 | 0.056081 | 0.013043 | 1.71E-05 | cis |
| rs1273650 | miR-181a-5 | 1:1989866 | G | A | 0.645541 | 0.05572 | 0.013025 | 1.89E-05 | cis |
| rs1273729 | miR-181a-5 | 1:1989813 | A | G | 0.647996 | 0.054741 | 0.013058 | 2.76E-05 | cis |
| rs1274639 | miR-181a-5 | 1:1989881 | C | T | 0.644011 | 0.055092 | 0.013048 | 2.42E-05 | cis |
| rs1274829 | miR-181a-5 | 1:1989833 | C | G | 0.646513 | 0.055369 | 0.013032 | 2.15E-05 | cis |
| rs1275160 | miR-181a-5 | 1:1988856 | G | T | 0.370157 | -0.04621 | 0.012938 | 0.000355 | cis |
| rs1275161 | miR-181a-5 | 1:1988864 | A | G | 0.372 | -0.045008 | 0.012815 | 0.000445 | cis |
| rs1275165 | miR-181a-5 | 1:1988947 | C | T | 0.37301 | -0.043694 | 0.012803 | 0.000643 | cis |
| rs1275166 | miR-181a-5 | 1:1988999 | G | A | 0.373025 | -0.043351 | 0.012807 | 0.000712 | cis |
| rs1275167 | miR-181a-5 | 1:1989013 | G | A | 0.373683 | -0.042972 | 0.012821 | 0.000803 | cis |
| rs1275169 | miR-181a-5 | 1:1989016 | T | G | 0.372822 | -0.043254 | 0.012815 | 0.000737 | cis |
| rs1275667 | miR-181a-5 | 1:1989955 | G | T | 0.641183 | 0.054797 | 0.012988 | 2.45E-05 | cis |
| rs1275768 | miR-181a-5 | 1:1989812 | T | C | 0.64568 | 0.054862 | 0.013025 | 2.53E-05 | cis |
| rs1322913 | miR-181a-5 | 1:1988530 | G | A | 0.372443 | -0.045407 | 0.013021 | 0.000488 | cis |
| rs1322914 | miR-181a-5 | 1:1988532 | C | T | 0.372444 | -0.0454 | 0.013021 | 0.000489 | cis |
| rs1337454 | miR-181a-5 | 1:1991098 | C | T | 0.816536 | -0.06665 | 0.016914 | 8.13E-05 | cis |
| rs1407683 | miR-181a-5 | 1:1988528 | A | G | 0.372404 | -0.045422 | 0.01302 | 0.000485 | cis |
| rs1684415 | miR-181a-5 | 1:1990546 | C | T | 0.643092 | -0.069517 | 0.013032 | 9.60E-08 | cis |
| rs1684416 | miR-181a-5 | 1:1990546 | T | C | 0.642911 | -0.069224 | 0.013029 | 1.08E-07 | cis |

| | | | | | | | | | |
|---|---|---|---|---|---|---|---|---|---|
| rs2068269 | miR-181a-5 | 1:1991588: | G | A | 0.750438 | -0.074467 | 0.014443 | 2.53E-07 | cis |
| rs2117334 | miR-181a-5 | 1:1990744: | C | G | 0.665573 | -0.071151 | 0.013574 | 1.59E-07 | cis |
| rs2117335 | miR-181a-5 | 1:1990721: | T | C | 0.663711 | -0.071277 | 0.013595 | 1.58E-07 | cis |
| rs2117336 | miR-181a-5 | 1:1990621: | T | C | 0.661347 | -0.068865 | 0.013265 | 2.09E-07 | cis |
| rs2360965 | miR-181a-5 | 1:1990543: | G | A | 0.641186 | -0.068611 | 0.012972 | 1.23E-07 | cis |
| rs3861924 | miR-181a-5 | 1:1991213: | G | A | 0.818633 | -0.066924 | 0.017032 | 8.52E-05 | cis |
| rs4915342 | miR-181a-5 | 1:1990750: | A | G | 0.639545 | -0.070608 | 0.013386 | 1.33E-07 | cis |
| rs6063971 | miR-181a-5 | 1:1988676: | G | T | 0.371658 | -0.046193 | 0.012876 | 0.000334 | cis |
| rs6662894 | miR-181a-5 | 1:1988802 | A | G | 0.372178 | -0.045197 | 0.012854 | 0.000438 | cis |
| rs6678208 | miR-181a-5 | 1:1988652: | A | G | 0.372039 | -0.046116 | 0.012928 | 0.000361 | cis |
| rs6684332 | miR-181a-5 | 1:1991504: | C | T | 0.696891 | -0.064831 | 0.013356 | 1.21E-06 | cis |
| rs7273972 | miR-181a-5 | 1:1987791 | T | A | 0.880889 | -0.089435 | 0.019697 | 5.61E-06 | cis |
| rs7273972 | miR-181a-5 | 1:1987801: | G | A | 0.884616 | -0.079686 | 0.019869 | 6.06E-05 | cis |
| rs7273972 | miR-181a-5 | 1:1987872 | A | G | 0.881438 | -0.089783 | 0.019614 | 4.71E-06 | cis |
| rs7273972 | miR-181a-5 | 1:1987884: | G | C | 0.88146 | -0.08985 | 0.01961 | 4.61E-06 | cis |
| rs7273973 | miR-181a-5 | 1:1987886 | T | A | 0.88146 | -0.089856 | 0.01961 | 4.60E-06 | cis |
| rs7273973 | miR-181a-5 | 1:1988013 | T | C | 0.885103 | -0.082087 | 0.019587 | 2.78E-05 | cis |
| rs7273973 | miR-181a-5 | 1:1988066: | G | T | 0.885639 | -0.084616 | 0.019785 | 1.90E-05 | cis |
| rs7474686 | miR-181a-5 | 1:1990232 | A | G | 0.87922 | -0.097557 | 0.019349 | 4.61E-07 | cis |
| rs7521271 | miR-181a-5 | 1:1989960 | T | A | 0.644767 | 0.056137 | 0.013045 | 1.68E-05 | cis |
| rs7527688 | miR-181a-5 | 1:1989920: | T | C | 0.644613 | 0.056087 | 0.013043 | 1.71E-05 | cis |
| rs7537080 | miR-181a-5 | 1:1987911 | A | G | 0.880842 | -0.090022 | 0.019593 | 4.33E-06 | cis |
| rs7544504 | miR-181a-5 | 1:1988518: | C | T | 0.372062 | -0.045571 | 0.013023 | 0.000467 | cis |
| rs7550980 | miR-181a-5 | 1:1988710: | A | G | 0.372407 | -0.045949 | 0.0129 | 0.000368 | cis |
| rs962793 | miR-181a-5 | 1:1990349 | A | G | 0.686838 | -0.070028 | 0.013428 | 1.84E-07 | cis |
| rs1434282 | miR-181b-5 | 1:1990107: | C | T | 0.271104 | 0.0533 | 0.015963 | 0.000841 | cis |
| rs1762558 | miR-181b-5 | 1:1989904: | G | A | 0.74365 | -0.054289 | 0.016271 | 0.000848 | cis |
| rs1016765 | miR-320b | 2:6864556 | T | G | 0.735237 | -0.077325 | 0.016691 | 3.61E-06 | trans |
| rs1018397 | miR-320b | 2:6860491: | G | A | 0.720472 | -0.070064 | 0.015602 | 7.10E-06 | trans |
| rs1020987 | miR-320b | 2:6860519 | A | C | 0.72047 | -0.070065 | 0.015602 | 7.09E-06 | trans |
| rs1021019 | miR-320b | 2:6860353 | T | C | 0.720439 | -0.070071 | 0.0156 | 7.07E-06 | trans |
| rs1031117 | miR-320b | 2:6860115: | G | C | 0.721971 | -0.070805 | 0.01567 | 6.23E-06 | trans |
| rs1049616 | miR-320b | 2:6861012 | A | G | 0.750777 | -0.073154 | 0.016376 | 7.92E-06 | trans |
| rs1168809 | miR-320b | 2:6859604: | A | T | 0.751013 | -0.072542 | 0.016414 | 9.90E-06 | trans |
| rs1246827 | miR-320b | 2:6864587 | A | G | 0.735366 | -0.077326 | 0.016698 | 3.64E-06 | trans |
| rs1262148 | miR-320b | 2:6859777: | A | G | 0.333832 | 0.076988 | 0.015173 | 3.90E-07 | trans |
| rs1373199 | miR-320b | 2:6859568 | G | A | 0.750699 | -0.072682 | 0.016392 | 9.25E-06 | trans |
| rs1420691 | miR-320b | 2:6862748 | G | C | 0.737384 | -0.079722 | 0.016322 | 1.04E-06 | trans |
| rs1501855 | miR-320b | 2:6863467: | G | A | 0.734283 | -0.080357 | 0.016531 | 1.17E-06 | trans |
| rs1703535 | miR-320b | 2:6858932: | C | A | 0.750845 | -0.072626 | 0.016419 | 9.72E-06 | trans |
| rs1703537 | miR-320b | 2:6859650: | G | A | 0.754443 | -0.072895 | 0.016595 | 1.12E-05 | trans |
| rs1703537 | miR-320b | 2:6859890: | C | T | 0.720098 | -0.069748 | 0.0156 | 7.79E-06 | trans |
| rs1975480 | miR-320b | 2:6857386: | T | C | 0.559926 | 0.055177 | 0.014276 | 0.000111 | trans |
| rs1975481 | miR-320b | 2:6857399: | C | A | 0.560307 | 0.055261 | 0.014275 | 0.000108 | trans |
| rs1975482 | miR-320b | 2:6857418: | A | G | 0.560651 | 0.055579 | 0.014285 | 9.99E-05 | trans |
| rs2120334 | miR-320b | 2:6861608: | C | T | 0.751935 | -0.072685 | 0.016419 | 9.56E-06 | trans |
| rs2289688 | miR-320b | 2:6862046: | G | A | 0.752268 | -0.072997 | 0.016427 | 8.84E-06 | trans |
| rs2289689 | miR-320b | 2:6862015: | C | T | 0.752314 | -0.072996 | 0.01643 | 8.88E-06 | trans |
| rs2857967 | miR-320b | 2:6858911: | G | A | 0.743089 | -0.065173 | 0.016357 | 6.77E-05 | trans |
| rs3770648 | miR-320b | 2:6861939: | C | T | 0.752286 | -0.072964 | 0.016431 | 8.97E-06 | trans |
| rs3770650 | miR-320b | 2:6861643: | A | G | 0.751964 | -0.072703 | 0.01642 | 9.53E-06 | trans |
| rs4671912 | miR-320b | 2:6860225: | C | T | 0.720119 | -0.069752 | 0.015601 | 7.78E-06 | trans |
| rs4671914 | miR-320b | 2:6861229: | G | A | 0.751284 | -0.07332 | 0.016396 | 7.76E-06 | trans |

| | | | | | | | | | |
|---|---|---|---|---|---|---|---|---|---|
| rs4854435 | miR-320b | 2:6863971 | G | A | 0.753618 | -0.073063 | 0.017227 | 2.22E-05 | trans |
| rs4854474 | miR-320b | 2:6863328 | A | T | 0.734157 | -0.080831 | 0.016446 | 8.89E-07 | trans |
| rs5568893 | miR-320b | 2:6859897 | C | A | 0.720121 | -0.069711 | 0.015599 | 7.86E-06 | trans |
| rs5581986 | miR-320b | 2:6864591 | G | A | 0.722126 | -0.067175 | 0.016413 | 4.26E-05 | trans |
| rs5986226 | miR-320b | 2:6864491 | G | A | 0.720952 | -0.068285 | 0.016406 | 3.15E-05 | trans |
| rs6546379 | miR-320b | 2:6857362 | G | A | 0.559288 | 0.055078 | 0.014283 | 0.000115 | trans |
| rs6546380 | miR-320b | 2:6857362 | G | A | 0.559433 | 0.055047 | 0.01428 | 0.000116 | trans |
| rs6691649 | miR-320b | 2:6864346 | T | C | 0.72213 | -0.068325 | 0.016372 | 3.00E-05 | trans |
| rs6741866 | miR-320b | 2:6857097 | G | A | 0.565872 | 0.054225 | 0.014282 | 0.000147 | trans |
| rs6745671 | miR-320b | 2:6864349 | G | A | 0.754299 | -0.072078 | 0.017288 | 3.06E-05 | trans |
| rs6752922 | miR-320b | 2:6857308 | G | A | 0.559358 | 0.05504 | 0.014267 | 0.000114 | trans |
| rs6759864 | miR-320b | 2:6857115 | T | C | 0.558332 | 0.05433 | 0.014162 | 0.000125 | trans |
| rs7355584 | miR-320b | 2:6859828 | A | G | 0.720028 | -0.069849 | 0.015602 | 7.57E-06 | trans |
| rs736779 | miR-320b | 2:6857191 | T | A | 0.56004 | 0.055109 | 0.014256 | 0.000111 | trans |
| rs7422209 | miR-320b | 2:6863221 | C | G | 0.75328 | -0.074715 | 0.01709 | 1.23E-05 | trans |
| rs7422735 | miR-320b | 2:6863318 | C | A | 0.752797 | -0.074618 | 0.017082 | 1.25E-05 | trans |
| rs7424172 | miR-320b | 2:6863238 | T | C | 0.734273 | -0.080816 | 0.016434 | 8.76E-07 | trans |
| rs7557983 | miR-320b | 2:6859171 | C | T | 0.724325 | -0.071371 | 0.015858 | 6.78E-06 | trans |
| rs7572216 | miR-320b | 2:6859194 | T | A | 0.719839 | -0.069718 | 0.015614 | 8.00E-06 | trans |
| rs7588432 | miR-320b | 2:6858548 | G | A | 0.282438 | 0.085949 | 0.015797 | 5.30E-08 | trans |
| rs7595717 | miR-320b | 2:6858747 | C | T | 0.738999 | -0.063962 | 0.016113 | 7.20E-05 | trans |
| rs9309411 | miR-320b | 2:6857046 | A | G | 0.566779 | 0.052162 | 0.014331 | 0.000273 | trans |
| rs9309412 | miR-320b | 2:6857056 | C | T | 0.559665 | 0.054506 | 0.014209 | 0.000125 | trans |
| rs9309415 | miR-320b | 2:6860533 | C | A | 0.72047 | -0.070067 | 0.015602 | 7.09E-06 | trans |
| rs964585 | miR-320b | 2:6857243 | A | G | 0.559777 | 0.055091 | 0.014262 | 0.000112 | trans |
| rs964586 | miR-320b | 2:6857289 | C | T | 0.560066 | 0.055128 | 0.014259 | 0.000111 | trans |
| rs964587 | miR-320b | 2:6857291 | G | A | 0.560071 | 0.055134 | 0.01426 | 0.00011 | trans |
| rs1016765 | miR-320c | 2:6864556 | T | G | 0.735237 | -0.065378 | 0.015645 | 2.93E-05 | trans |
| rs1018397 | miR-320c | 2:6860491 | G | A | 0.720472 | -0.058335 | 0.014625 | 6.64E-05 | trans |
| rs1020987 | miR-320c | 2:6860519 | A | C | 0.72047 | -0.058335 | 0.014625 | 6.64E-05 | trans |
| rs1021019 | miR-320c | 2:6860353 | T | C | 0.720439 | -0.058342 | 0.014624 | 6.62E-05 | trans |
| rs1031117 | miR-320c | 2:6860115 | G | C | 0.721971 | -0.058896 | 0.01469 | 6.09E-05 | trans |
| rs1049616 | miR-320c | 2:6861012 | A | G | 0.750777 | -0.064468 | 0.015344 | 2.65E-05 | trans |
| rs1168809 | miR-320c | 2:6859604 | A | T | 0.751013 | -0.063623 | 0.01538 | 3.52E-05 | trans |
| rs1246827 | miR-320c | 2:6864587 | A | G | 0.735366 | -0.065387 | 0.015651 | 2.94E-05 | trans |
| rs1262148 | miR-320c | 2:6859777 | A | G | 0.333832 | 0.073868 | 0.014206 | 1.99E-07 | trans |
| rs1373199 | miR-320c | 2:6859568 | G | A | 0.750699 | -0.063801 | 0.015359 | 3.27E-05 | trans |
| rs1420691 | miR-320c | 2:6862748 | G | C | 0.737384 | -0.066897 | 0.015302 | 1.23E-05 | trans |
| rs1501855 | miR-320c | 2:6863467 | G | A | 0.734283 | -0.067972 | 0.015496 | 1.15E-05 | trans |
| rs1703535 | miR-320c | 2:6858932 | C | A | 0.750845 | -0.063922 | 0.015384 | 3.25E-05 | trans |
| rs1703537 | miR-320c | 2:6859650 | G | A | 0.754443 | -0.063855 | 0.015549 | 4.02E-05 | trans |
| rs1703537 | miR-320c | 2:6859890 | C | T | 0.720098 | -0.057772 | 0.014624 | 7.80E-05 | trans |
| rs2120334 | miR-320c | 2:6861608 | C | T | 0.751935 | -0.063735 | 0.015385 | 3.43E-05 | trans |
| rs2289688 | miR-320c | 2:6862046 | G | A | 0.752268 | -0.063974 | 0.015392 | 3.24E-05 | trans |
| rs2289689 | miR-320c | 2:6862015 | C | T | 0.752314 | -0.06397 | 0.015395 | 3.25E-05 | trans |
| rs3770648 | miR-320c | 2:6861939 | C | T | 0.752286 | -0.06394 | 0.015396 | 3.28E-05 | trans |
| rs3770650 | miR-320c | 2:6861643 | A | G | 0.751964 | -0.06375 | 0.015386 | 3.42E-05 | trans |
| rs4671912 | miR-320c | 2:6860225 | C | T | 0.720119 | -0.057775 | 0.014625 | 7.80E-05 | trans |
| rs4671914 | miR-320c | 2:6861229 | G | A | 0.751284 | -0.064402 | 0.015363 | 2.77E-05 | trans |
| rs4854435 | miR-320c | 2:6863971 | G | A | 0.753618 | -0.064153 | 0.016141 | 7.05E-05 | trans |
| rs4854474 | miR-320c | 2:6863328 | A | T | 0.734157 | -0.068344 | 0.015418 | 9.30E-06 | trans |
| rs5568893 | miR-320c | 2:6859897 | C | A | 0.720121 | -0.057755 | 0.014623 | 7.83E-05 | trans |
| rs5581986 | miR-320c | 2:6864591 | G | A | 0.722126 | -0.055858 | 0.015383 | 0.000282 | trans |

| | | | | | | | | | |
|---|---|---|---|---|---|---|---|---|---|
| rs5986226 | miR-320c | 2:6864491 | G | A | 0.720952 | -0.056798 | 0.015377 | 0.000221 | trans |
| rs6691649 | miR-320c | 2:6864346 | T | C | 0.72213 | -0.056818 | 0.015345 | 0.000213 | trans |
| rs6745671 | miR-320c | 2:6864349 | G | A | 0.754299 | -0.06316 | 0.016198 | 9.65E-05 | trans |
| rs7355584 | miR-320c | 2:6859828 | A | G | 0.720028 | -0.057818 | 0.014626 | 7.71E-05 | trans |
| rs7422209 | miR-320c | 2:6863221 | C | G | 0.75328 | -0.065232 | 0.016014 | 4.63E-05 | trans |
| rs7422735 | miR-320c | 2:6863318 | C | A | 0.752797 | -0.065301 | 0.016006 | 4.51E-05 | trans |
| rs7424172 | miR-320c | 2:6863238 | T | C | 0.734273 | -0.068349 | 0.015406 | 9.15E-06 | trans |
| rs7557983 | miR-320c | 2:6859171 | C | T | 0.724325 | -0.059058 | 0.014866 | 7.11E-05 | trans |
| rs7572216 | miR-320c | 2:6859194 | T | A | 0.719839 | -0.057776 | 0.014637 | 7.90E-05 | trans |
| rs9309415 | miR-320c | 2:6860533 | C | A | 0.72047 | -0.058336 | 0.014625 | 6.64E-05 | trans |
| rs1016765 | miR-320d | 2:6864556 | T | G | 0.735237 | -0.069456 | 0.016 | 1.42E-05 | trans |
| rs1018397 | miR-320d | 2:6860491 | G | A | 0.720472 | -0.065468 | 0.014951 | 1.19E-05 | trans |
| rs1018497 | miR-320d | 2:6859988 | G | A | 0.286787 | 0.078742 | 0.014851 | 1.14E-07 | trans |
| rs1018990 | miR-320d | 2:6860619 | C | G | 0.281778 | 0.079179 | 0.014753 | 8.01E-08 | trans |
| rs1020987 | miR-320d | 2:6860519 | A | C | 0.72047 | -0.065468 | 0.014951 | 1.19E-05 | trans |
| rs1021019 | miR-320d | 2:6860353 | T | C | 0.720439 | -0.065488 | 0.014949 | 1.18E-05 | trans |
| rs1031117 | miR-320d | 2:6860115 | G | C | 0.721971 | -0.066439 | 0.015016 | 9.66E-06 | trans |
| rs1112618 | miR-320d | 2:6861913 | A | G | 0.282025 | 0.079944 | 0.014743 | 5.87E-08 | trans |
| rs1155426 | miR-320d | 2:6862182 | T | C | 0.282037 | 0.079522 | 0.014742 | 6.88E-08 | trans |
| rs1168809 | miR-320d | 2:6859604 | A | T | 0.751013 | -0.068165 | 0.015729 | 1.47E-05 | trans |
| rs1189593 | miR-320d | 2:6861908 | A | G | 0.282175 | 0.080028 | 0.014748 | 5.75E-08 | trans |
| rs1246827 | miR-320d | 2:6864587 | A | G | 0.735366 | -0.06947 | 0.016007 | 1.42E-05 | trans |
| rs1262148 | miR-320d | 2:6859777 | A | G | 0.333832 | 0.067958 | 0.01455 | 3.00E-06 | trans |
| rs1300495 | miR-320d | 2:6858081 | G | A | 0.306559 | 0.078977 | 0.014742 | 8.44E-08 | trans |
| rs1373198 | miR-320d | 2:6858798 | A | T | 0.281586 | 0.077628 | 0.014896 | 1.87E-07 | trans |
| rs1373199 | miR-320d | 2:6859568 | G | A | 0.750699 | -0.068322 | 0.015707 | 1.36E-05 | trans |
| rs1420691 | miR-320d | 2:6862748 | G | C | 0.737384 | -0.070321 | 0.015651 | 7.02E-06 | trans |
| rs1443651 | miR-320d | 2:6856931 | T | G | 0.559583 | 0.056215 | 0.013598 | 3.57E-05 | trans |
| rs1443652 | miR-320d | 2:6856965 | C | G | 0.559695 | 0.05627 | 0.013601 | 3.51E-05 | trans |
| rs1501855 | miR-320d | 2:6863467 | G | A | 0.734283 | -0.071823 | 0.015848 | 5.85E-06 | trans |
| rs1703537 | miR-320d | 2:6859650 | G | A | 0.754443 | -0.068244 | 0.015902 | 1.77E-05 | trans |
| rs1703537 | miR-320d | 2:6859890 | C | T | 0.720098 | -0.065018 | 0.014949 | 1.37E-05 | trans |
| rs1867313 | miR-320d | 2:6862365 | T | C | 0.282087 | 0.079407 | 0.01474 | 7.15E-08 | trans |
| rs1975480 | miR-320d | 2:6857386 | T | C | 0.559926 | 0.056909 | 0.013669 | 3.14E-05 | trans |
| rs1975481 | miR-320d | 2:6857399 | C | A | 0.560307 | 0.056955 | 0.013669 | 3.09E-05 | trans |
| rs1975482 | miR-320d | 2:6857418 | A | G | 0.560651 | 0.057272 | 0.013678 | 2.83E-05 | trans |
| rs2120334 | miR-320d | 2:6861608 | C | T | 0.751935 | -0.068022 | 0.015734 | 1.54E-05 | trans |
| rs2289688 | miR-320d | 2:6862046 | G | A | 0.752268 | -0.068404 | 0.015741 | 1.39E-05 | trans |
| rs2289689 | miR-320d | 2:6862015 | C | T | 0.752314 | -0.06839 | 0.015744 | 1.40E-05 | trans |
| rs3426707 | miR-320d | 2:6859915 | T | C | 0.282465 | 0.07967 | 0.014787 | 7.13E-08 | trans |
| rs3770648 | miR-320d | 2:6861939 | C | T | 0.752286 | -0.068321 | 0.015745 | 1.43E-05 | trans |
| rs3770650 | miR-320d | 2:6861643 | A | G | 0.751964 | -0.068057 | 0.015735 | 1.52E-05 | trans |
| rs4277504 | miR-320d | 2:6856956 | C | A | 0.559711 | 0.056267 | 0.013599 | 3.51E-05 | trans |
| rs4671208 | miR-320d | 2:6860079 | A | G | 0.281916 | 0.079072 | 0.014777 | 8.74E-08 | trans |
| rs4671912 | miR-320d | 2:6860225 | C | T | 0.720119 | -0.064987 | 0.01495 | 1.38E-05 | trans |
| rs4671914 | miR-320d | 2:6861229 | G | A | 0.751284 | -0.068595 | 0.015712 | 1.27E-05 | trans |
| rs4854435 | miR-320d | 2:6863971 | G | A | 0.753618 | -0.068279 | 0.016508 | 3.53E-05 | trans |
| rs4854474 | miR-320d | 2:6863328 | A | T | 0.734157 | -0.072183 | 0.015768 | 4.70E-06 | trans |
| rs5568893 | miR-320d | 2:6859897 | C | A | 0.720121 | -0.064969 | 0.014949 | 1.39E-05 | trans |
| rs5581986 | miR-320d | 2:6864591 | G | A | 0.722126 | -0.062787 | 0.015728 | 6.55E-05 | trans |
| rs5986226 | miR-320d | 2:6864491 | G | A | 0.720952 | -0.063997 | 0.015721 | 4.69E-05 | trans |
| rs6546377 | miR-320d | 2:6857022 | G | A | 0.559672 | 0.05651 | 0.01361 | 3.29E-05 | trans |
| rs6546378 | miR-320d | 2:6857344 | C | G | 0.56041 | 0.056781 | 0.013704 | 3.42E-05 | trans |

| | | | | | | | | | |
|---|---|---|---|---|---|---|---|---|---|
| rs6546379 | miR-320d | 2:6857362 | G | A | 0.559288 | 0.056625 | 0.013677 | 3.47E-05 | trans |
| rs6546380 | miR-320d | 2:6857362 | G | A | 0.559433 | 0.056612 | 0.013674 | 3.47E-05 | trans |
| rs6546387 | miR-320d | 2:6858758 | G | A | 0.281573 | 0.077623 | 0.014898 | 1.88E-07 | trans |
| rs6546388 | miR-320d | 2:6858948 | C | T | 0.281691 | 0.07798 | 0.014854 | 1.52E-07 | trans |
| rs6546396 | miR-320d | 2:6862930 | A | T | 0.283762 | 0.08027 | 0.014778 | 5.58E-08 | trans |
| rs6546402 | miR-320d | 2:6863130 | A | G | 0.283216 | 0.080485 | 0.014784 | 5.20E-08 | trans |
| rs6691649 | miR-320d | 2:6864346 | T | C | 0.72213 | -0.063634 | 0.015689 | 4.99E-05 | trans |
| rs6741866 | miR-320d | 2:6857097 | G | A | 0.565872 | 0.055583 | 0.013676 | 4.82E-05 | trans |
| rs6745671 | miR-320d | 2:6864349 | G | A | 0.754299 | -0.067304 | 0.016566 | 4.85E-05 | trans |
| rs6752922 | miR-320d | 2:6857308 | G | A | 0.559358 | 0.056626 | 0.013662 | 3.40E-05 | trans |
| rs6759864 | miR-320d | 2:6857115 | T | C | 0.558332 | 0.055856 | 0.013561 | 3.81E-05 | trans |
| rs7355584 | miR-320d | 2:6859828 | A | G | 0.720028 | -0.065155 | 0.014951 | 1.31E-05 | trans |
| rs736779 | miR-320d | 2:6857191 | T | A | 0.56004 | 0.056818 | 0.01365 | 3.15E-05 | trans |
| rs7420773 | miR-320d | 2:6862945 | G | A | 0.28377 | 0.080269 | 0.014778 | 5.58E-08 | trans |
| rs7420782 | miR-320d | 2:6862955 | G | T | 0.283742 | 0.080513 | 0.014781 | 5.12E-08 | trans |
| rs7422209 | miR-320d | 2:6863221 | C | G | 0.75328 | -0.069442 | 0.016377 | 2.23E-05 | trans |
| rs7422735 | miR-320d | 2:6863318 | C | A | 0.752797 | -0.069494 | 0.01637 | 2.18E-05 | trans |
| rs7424172 | miR-320d | 2:6863238 | T | C | 0.734273 | -0.072174 | 0.015757 | 4.64E-06 | trans |
| rs7560374 | miR-320d | 2:6856998 | A | G | 0.559683 | 0.056244 | 0.013601 | 3.55E-05 | trans |
| rs7562889 | miR-320d | 2:6862547 | T | C | 0.285916 | 0.079614 | 0.014794 | 7.38E-08 | trans |
| rs7572216 | miR-320d | 2:6859194 | T | A | 0.719839 | -0.065375 | 0.014962 | 1.25E-05 | trans |
| rs7574580 | miR-320d | 2:6857324 | A | G | 0.561141 | 0.057005 | 0.013696 | 3.15E-05 | trans |
| rs7574592 | miR-320d | 2:6859766 | T | G | 0.281934 | 0.078996 | 0.014777 | 8.99E-08 | trans |
| rs7588432 | miR-320d | 2:6858548 | G | A | 0.282438 | 0.077732 | 0.015146 | 2.86E-07 | trans |
| rs7589952 | miR-320d | 2:6857009 | G | A | 0.559677 | 0.05623 | 0.013603 | 3.57E-05 | trans |
| rs7600829 | miR-320d | 2:6856977 | T | C | 0.56021 | 0.056403 | 0.013617 | 3.44E-05 | trans |
| rs880668 | miR-320d | 2:6859258 | T | C | 0.280228 | 0.078264 | 0.014865 | 1.40E-07 | trans |
| rs9309411 | miR-320d | 2:6857046 | A | G | 0.566779 | 0.054713 | 0.013722 | 6.68E-05 | trans |
| rs9309412 | miR-320d | 2:6857056 | C | T | 0.559665 | 0.056194 | 0.013606 | 3.62E-05 | trans |
| rs9309415 | miR-320d | 2:6860533 | C | A | 0.72047 | -0.065469 | 0.014951 | 1.19E-05 | trans |
| rs964585 | miR-320d | 2:6857243 | A | G | 0.559777 | 0.056836 | 0.013656 | 3.16E-05 | trans |
| rs964586 | miR-320d | 2:6857289 | C | T | 0.560066 | 0.056836 | 0.013654 | 3.15E-05 | trans |
| rs964587 | miR-320d | 2:6857291 | G | A | 0.560071 | 0.056842 | 0.013654 | 3.14E-05 | trans |
| rs964934 | miR-320d | 2:6861187 | G | A | 0.281023 | 0.07994 | 0.01477 | 6.22E-08 | trans |
| rs964935 | miR-320d | 2:6861228 | G | A | 0.281126 | 0.079972 | 0.014772 | 6.17E-08 | trans |
| rs1013035 | miR-329-3p | 14:101149 | T | C | 0.112665 | -0.23242 | 0.046261 | 5.06E-07 | cis |
| rs1014839 | miR-329-3p | 14:101026 | A | G | 0.750738 | 0.138159 | 0.033199 | 3.16E-05 | cis |
| rs1014844 | miR-329-3p | 14:101026 | G | A | 0.753241 | 0.137679 | 0.033189 | 3.35E-05 | cis |
| rs1116059 | miR-329-3p | 14:101049 | C | T | 0.741662 | 0.127724 | 0.03159 | 5.27E-05 | cis |
| rs1162219 | miR-329-3p | 14:101016 | G | A | 0.759852 | 0.151508 | 0.035294 | 1.77E-05 | cis |
| rs1184658 | miR-329-3p | 14:101017 | C | G | 0.759077 | 0.133188 | 0.03383 | 8.25E-05 | cis |
| rs1184948 | miR-329-3p | 14:101037 | C | G | 0.733365 | 0.12599 | 0.03131 | 5.72E-05 | cis |
| rs1185095 | miR-329-3p | 14:101056 | G | T | 0.743981 | 0.129015 | 0.03143 | 4.05E-05 | cis |
| rs1190850 | miR-329-3p | 14:101064 | A | G | 0.314302 | -0.125559 | 0.03058 | 4.03E-05 | cis |
| rs1190852 | miR-329-3p | 14:101064 | A | G | 0.310295 | -0.124566 | 0.030398 | 4.17E-05 | cis |
| rs1190856 | miR-329-3p | 14:101065 | T | C | 0.310244 | -0.124194 | 0.030374 | 4.34E-05 | cis |
| rs1190857 | miR-329-3p | 14:101066 | G | T | 0.310273 | -0.124033 | 0.03038 | 4.45E-05 | cis |
| rs1190862 | miR-329-3p | 14:101067 | T | G | 0.309628 | -0.124061 | 0.030162 | 3.90E-05 | cis |
| rs1190867 | miR-329-3p | 14:101069 | G | T | 0.316788 | -0.124028 | 0.030558 | 4.93E-05 | cis |
| rs1190872 | miR-329-3p | 14:101069 | A | G | 0.310177 | -0.122862 | 0.030343 | 5.14E-05 | cis |
| rs1214784 | miR-329-3p | 14:101144 | C | T | 0.870989 | 0.222748 | 0.041636 | 8.80E-08 | cis |
| rs1243369 | miR-329-3p | 14:101035 | C | G | 0.723403 | 0.129329 | 0.030935 | 2.91E-05 | cis |
| rs1243613 | miR-329-3p | 14:101031 | C | T | 0.725015 | 0.130653 | 0.03114 | 2.72E-05 | cis |

| | | | | | | | | | |
|---|---|---|---|---|---|---|---|---|---|
| rs1288035{ | miR-329-3p | 14:1011588 | C | G | 0.904391 | 0.221489 | 0.049519 | 7.72E-06 | cis |
| rs1709962: | miR-329-3p | 14:101173: | A | C | 0.783449 | 0.130329 | 0.033574 | 0.000104 | cis |
| rs1757624( | miR-329-3p | 14:101126{ | G | C | 0.854853 | 0.153892 | 0.039733 | 0.000107 | cis |
| rs1888581 | miR-329-3p | 14:101042: | A | C | 0.735363 | 0.122166 | 0.031235 | 9.18E-05 | cis |
| rs2842154( | miR-329-3p | 14:101158: | C | T | 0.904763 | 0.221821 | 0.049571 | 7.65E-06 | cis |
| rs3579985( | miR-329-3p | 14:101156{ | C | T | 0.904015 | 0.222695 | 0.049515 | 6.88E-06 | cis |
| rs6575791 | miR-329-3p | 14:101029: | G | A | 0.719841 | 0.133429 | 0.031132 | 1.82E-05 | cis |
| rs6575794 | miR-329-3p | 14:101039: | C | T | 0.734189 | 0.123387 | 0.031361 | 8.34E-05 | cis |
| rs6575796 | miR-329-3p | 14:101160: | A | G | 0.094049 | -0.220221 | 0.050763 | 1.44E-05 | cis |
| rs7140556 | miR-329-3p | 14:101036 | C | A | 0.723818 | 0.130763 | 0.031072 | 2.57E-05 | cis |
| rs7143357 | miR-329-3p | 14:101054: | T | C | 0.739636 | 0.131999 | 0.031609 | 2.97E-05 | cis |
| rs7144997 | miR-329-3p | 14:101155 | A | G | 0.899872 | 0.221634 | 0.049364 | 7.13E-06 | cis |
| rs7145610 | miR-329-3p | 14:101049 | T | C | 0.740391 | 0.128339 | 0.031673 | 5.08E-05 | cis |
| rs7147982 | miR-329-3p | 14:101046( | C | T | 0.731753 | 0.120992 | 0.031008 | 9.54E-05 | cis |
| rs7154024 | miR-329-3p | 14:101031: | G | T | 0.725015 | 0.130349 | 0.031113 | 2.79E-05 | cis |
| rs745797 | miR-329-3p | 14:101141( | G | A | 0.858727 | 0.193655 | 0.039733 | 1.09E-06 | cis |
| rs7493817 | miR-329-3p | 14:101038 | G | A | 0.751723 | 0.132187 | 0.032319 | 4.31E-05 | cis |
| rs8008285 | miR-329-3p | 14:101146! | T | C | 0.842905 | 0.19737 | 0.038772 | 3.57E-07 | cis |
| rs879058 | miR-329-3p | 14:101069( | G | T | 0.311807 | -0.123579 | 0.030343 | 4.65E-05 | cis |
| rs879059 | miR-329-3p | 14:101069( | T | A | 0.31068 | -0.123062 | 0.030328 | 4.95E-05 | cis |
| rs879060 | miR-329-3p | 14:101069{ | A | G | 0.310926 | -0.124113 | 0.030345 | 4.31E-05 | cis |
| rs1014171! | miR-376a-3 | 14:101172( | G | T | 0.923932 | 0.240171 | 0.048515 | 7.40E-07 | cis |
| rs1022032: | miR-376a-3 | 14:101130: | T | G | 0.868038 | 0.193611 | 0.037367 | 2.20E-07 | cis |
| rs1190878 | miR-376a-3 | 14:101072: | C | A | 0.859763 | 0.175367 | 0.037764 | 3.42E-06 | cis |
| rs1190881 | miR-376a-3 | 14:101079{ | T | C | 0.346494 | -0.141208 | 0.027217 | 2.12E-07 | cis |
| rs1288035{ | miR-376a-3 | 14:1011588 | C | G | 0.904391 | 0.237148 | 0.04576 | 2.19E-07 | cis |
| rs1757612: | miR-376a-3 | 14:101126: | C | T | 0.856231 | 0.159721 | 0.03688 | 1.49E-05 | cis |
| rs1757624( | miR-376a-3 | 14:101126{ | G | C | 0.854853 | 0.165717 | 0.036729 | 6.43E-06 | cis |
| rs1757630! | miR-376a-3 | 14:101127! | G | C | 0.854847 | 0.16583 | 0.036707 | 6.25E-06 | cis |
| rs1757777( | miR-376a-3 | 14:101200 | C | T | 0.495315 | 0.143807 | 0.026554 | 6.11E-08 | cis |
| rs1765383( | miR-376a-3 | 14:101101: | T | C | 0.877829 | 0.197681 | 0.040473 | 1.04E-06 | cis |
| rs1765384{ | miR-376a-3 | 14:101102( | A | T | 0.870972 | 0.202208 | 0.040544 | 6.12E-07 | cis |
| rs1766021( | miR-376a-3 | 14:101131 | A | G | 0.868541 | 0.19568 | 0.03754 | 1.86E-07 | cis |
| rs1766036: | miR-376a-3 | 14:101136: | A | G | 0.868731 | 0.195684 | 0.037605 | 1.95E-07 | cis |
| rs2842154( | miR-376a-3 | 14:101158: | C | T | 0.904763 | 0.237472 | 0.045809 | 2.17E-07 | cis |
| rs2847806! | miR-376a-3 | 14:101139( | C | A | 0.868556 | 0.200998 | 0.037528 | 8.51E-08 | cis |
| rs2852739: | miR-376a-3 | 14:101130( | T | C | 0.868049 | 0.193618 | 0.037373 | 2.21E-07 | cis |
| rs3516145: | miR-376a-3 | 14:101164: | A | G | 0.624259 | 0.193792 | 0.031225 | 5.43E-10 | cis |
| rs3579985( | miR-376a-3 | 14:101156{ | C | T | 0.904015 | 0.238541 | 0.045756 | 1.85E-07 | cis |
| rs4905991 | miR-376a-3 | 14:101140( | T | C | 0.271223 | -0.118159 | 0.029234 | 5.30E-05 | cis |
| rs6575796 | miR-376a-3 | 14:101160: | A | G | 0.094049 | -0.238421 | 0.046908 | 3.72E-07 | cis |
| rs7142298 | miR-376a-3 | 14:101128! | C | T | 0.854052 | 0.167239 | 0.036504 | 4.62E-06 | cis |
| rs7144997 | miR-376a-3 | 14:101155 | A | G | 0.899872 | 0.232316 | 0.045628 | 3.55E-07 | cis |
| rs7152943 | miR-376a-3 | 14:101174: | C | T | 0.924188 | 0.241748 | 0.048626 | 6.64E-07 | cis |
| rs7156735 | miR-376a-3 | 14:101132: | T | C | 0.867137 | 0.196655 | 0.037308 | 1.36E-07 | cis |
| rs8007901 | miR-376a-3 | 14:101130! | T | C | 0.867321 | 0.194601 | 0.037217 | 1.71E-07 | cis |
| rs8014970 | miR-376a-3 | 14:101140: | T | G | 0.868329 | 0.201058 | 0.037564 | 8.68E-08 | cis |
| rs8017361 | miR-376a-3 | 14:101218( | G | A | 0.898047 | 0.181152 | 0.042677 | 2.19E-05 | cis |
| rs8017555 | miR-376a-3 | 14:101218( | C | T | 0.89727 | 0.186206 | 0.04266 | 1.27E-05 | cis |
| rs8022367 | miR-376a-3 | 14:101189 | A | T | 0.295876 | -0.239525 | 0.034134 | 2.26E-12 | cis |
| rs2247381 | miR-941 | 20:625466: | A | G | 0.789223 | 0.113853 | 0.030315 | 0.000173 | cis |
| rs2427546 | miR-941 | 20:625470: | T | C | 0.789295 | 0.113908 | 0.030322 | 0.000172 | cis |
| rs2427550 | miR-941 | 20:625475: | C | A | 0.789276 | 0.113806 | 0.030312 | 0.000174 | cis |

| | | | | | | | | | |
|---|---|---|---|---|---|---|---|---|---|
| rs4809244 | miR-941 | 20:625307[illegible] | C | T | 0.950318 | 0.212612 | 0.056743 | 0.000179 | cis |
| rs4809249 | miR-941 | 20:625506[illegible] | C | T | 0.949005 | 0.278933 | 0.05743 | 1.19E-06 | cis |
| rs4809382 | miR-941 | 20:625447[illegible] | A | G | 0.949663 | 0.282925 | 0.056457 | 5.41E-07 | cis |
| rs6011222 | miR-941 | 20:625460[illegible] | T | G | 0.789234 | 0.113903 | 0.030324 | 0.000172 | cis |
| rs6062330 | miR-941 | 20:625306[illegible] | C | T | 0.950314 | 0.21261 | 0.05674 | 0.000179 | cis |
| rs6062331 | miR-941 | 20:625320[illegible] | A | G | 0.95036 | 0.217584 | 0.05704 | 0.000136 | cis |
| rs6062332 | miR-941 | 20:625321[illegible] | C | T | 0.950204 | 0.217499 | 0.056894 | 0.000132 | cis |
| rs6062573 | miR-941 | 20:625309[illegible] | G | A | 0.948917 | 0.212851 | 0.056764 | 0.000177 | cis |
| rs6062576 | miR-941 | 20:625442[illegible] | C | T | 0.949659 | 0.282851 | 0.056455 | 5.44E-07 | cis |
| rs6062578 | miR-941 | 20:625450[illegible] | G | C | 0.789137 | 0.113944 | 0.030322 | 0.000171 | cis |
| rs6062581 | miR-941 | 20:625460[illegible] | G | T | 0.94965 | 0.283136 | 0.056466 | 5.32E-07 | cis |
| rs6062583 | miR-941 | 20:625473[illegible] | G | A | 0.949677 | 0.282944 | 0.056468 | 5.42E-07 | cis |
| rs6062584 | miR-941 | 20:625477[illegible] | C | T | 0.949531 | 0.28305 | 0.056477 | 5.39E-07 | cis |
| rs6062587 | miR-941 | 20:625517[illegible] | C | T | 0.949069 | 0.278155 | 0.057289 | 1.20E-06 | cis |
| rs7409760 | miR-941 | 20:625485[illegible] | T | C | 0.949686 | 0.282991 | 0.056473 | 5.41E-07 | cis |
| rs817371 | miR-941 | 20:625329[illegible] | G | C | 0.787683 | 0.115284 | 0.030443 | 0.000153 | cis |
| rs817372 | miR-941 | 20:625331[illegible] | G | C | 0.787659 | 0.115629 | 0.030429 | 0.000145 | cis |
| rs817374 | miR-941 | 20:625334[illegible] | T | G | 0.787389 | 0.115765 | 0.030462 | 0.000145 | cis |
| rs817379 | miR-941 | 20:625392[illegible] | C | A | 0.788135 | 0.114758 | 0.030378 | 0.000158 | cis |

**Table S8. Functional annotation of miR-eQTLs**

Functional annotation of cis-miR-eQTLs

| annot | ref.count | ref.prop | count | prop | enrichmen | fisher.P |
|---|---|---|---|---|---|---|
| UTR3 | 233824 | 0.009322 | 23 | 0.011154 | 1.196484 | 0.358725 |
| UTR5 | 71546 | 0.002853 | 22 | 0.010669 | 3.740292 | 2.65E-07 |
| downstrea | 284177 | 0.01133 | 42 | 0.020369 | 1.797747 | 0.000369 |
| exonic | 254736 | 0.010156 | 12 | 0.00582 | 0.573006 | 0.047482 |
| intergenic | 11684523 | 0.465859 | 739 | 0.35839 | 0.76931 | 6.16E-23 |
| intronic | 9137749 | 0.36432 | 850 | 0.412221 | 1.131482 | 7.99E-06 |
| ncRNA_exc | 259951 | 0.010364 | 52 | 0.025218 | 2.433212 | 1.26E-08 |
| ncRNA_intı | 2884355 | 0.114998 | 288 | 0.13967 | 1.21454 | 0.000625 |
| ncRNA_spli | 1313 | 5.23E-05 | 1 | 0.000485 | 9.264104 | 0.102328 |
| splicing | 2830 | 0.000113 | 0 | 0 | 0 | 1 |
| upstream | 266686 | 0.010633 | 33 | 0.016004 | 1.505157 | 0.023403 |

Functional annotation of trans-miR-eQTLs

| annot | ref.count | ref.prop | count | prop | enrichmen | fisher.P |
|---|---|---|---|---|---|---|
| UTR3 | 233824 | 0.009322 | 11 | 0.020333 | 2.181037 | 0.020364 |
| UTR5 | 71546 | 0.002853 | 1 | 0.001848 | 0.647999 | 1 |
| downstrea | 284177 | 0.01133 | 12 | 0.022181 | 1.957726 | 0.024583 |
| exonic | 254736 | 0.010156 | 3 | 0.005545 | 0.545997 | 0.38976 |
| intergenic | 11684523 | 0.465859 | 225 | 0.415896 | 0.892752 | 0.019965 |
| intronic | 9137749 | 0.36432 | 168 | 0.310536 | 0.852373 | 0.009519 |
| ncRNA_exc | 259951 | 0.010364 | 14 | 0.025878 | 2.496871 | 0.001883 |
| ncRNA_intı | 2884355 | 0.114998 | 95 | 0.175601 | 1.526984 | 3.54E-05 |
| ncRNA_spli | 1313 | 5.23E-05 | 0 | 0 | 0 | 1 |
| splicing | 2830 | 0.000113 | 0 | 0 | 0 | 1 |
| upstream | 266686 | 0.010633 | 12 | 0.022181 | 2.086126 | 0.017638 |

**Table S9. List of 22 loci, miRNAs, and corresponding genes**

| Genomic Loci | n_miRNAs | miRNA | gene |
|---|---|---|---|
| 14:100655022-101244293 | 23 | miR-382-5p | RP11-566J3.2,RP11-638I2.4,YY1,YY |
| 9:136128546-136296530 | 18 | miR-195-5p | ABO,ABO:RP11-430N14.4,Y_RNA,L |
| 2:68577734-68650373 | 6 | miR-320e,r | PLEK,AC015969.3,AC015969.3:PLE |
| 1:198825629-199257785 | 3 | miR-181a-5 | MIR181A1HG,RP11-16L9.1,RP11-1 |
| 14:21349400-21438849 | 3 | miR-142-5p | RN7SL189P,RNASE2 |
| 20:61083030-61159437 | 3 | miR-133a-3 | RP13-379O24.2,C20orf166,C20orf |
| 3:58183636-58481296 | 2 | miR-1254,r | PXK,RPP14,DNASE1L3 |
| 8:106512828-106593207 | 2 | miR-143-3p | ZFPM2 |
| 8:135640669-135747967 | 2 | miR-30b-5p | ZFAT,CTD-2342N23.3 |
| 9:137732458-137748694 | 2 | miR-3689a- | MIR3689F,MIR3689D2,AL603650.1 |
| 22:22000849-22069957 | 2 | miR-130b-3 | PPIL2,YPEL1 |
| 2:64556476-64577491 | 1 | miR-4433b | AC114752.3:MIR4433,AC114752.3 |
| 2:207615284-207685904 | 1 | miR-3130-3 | FASTKD2,RP11-95H11.1,MDH1B,F/ |
| 5:148410920-148546347 | 1 | miR-584-5p | CTC-529P8.1,SH3TC2,RN7SKP145,/ |
| 7:1015407-1161324 | 1 | miR-339-5p | C7orf50,C7orf50:AC073957.15,C7c |
| 7:129431052-129432094 | 1 | miR-182-5p | RP11-738B7.1 |
| 9:97723868-97828989 | 1 | miR-27b-3p | C9orf3 |
| 11:56767507-57756568 | 1 | miR-130a-3 | OR5BD1P,TMX2-CTNND1:RP11-69 |
| 11:61542006-61624181 | 1 | miR-1908-5 | MYRF:TMEM258,FADS2,FADS2:FA |
| 14:23372913-23478860 | 1 | miR-4707-3 | RBM23,RP11-298I3.1,PRMT5-AS1, |
| 14:65665376-66287921 | 1 | miR-625-5p | FUT8,CTD-2014B16.1,MIR4708,CT |
| 16:88602910-88622286 | 1 | miR-6803-3 | ZFPM1,ZC3H18 |

/1:RP11-638I2.2,SLC25A29,WDR25,BEGAIN,DLK1,CTD-2644I21.1,LINC00523,RP11-566J3.4,MEG3
.CN1P2,SURF1
:K
.6L9.2,RP11-16L9.3,RP11-16L9.3:RP11-16L9.4,RP11-16L9.4,RP11-382E9.1

166-AS1,C20orf166-AS1:C20orf166

2,MIR3689C,COL5A1

},AC114752.2,AC114752.1
ASTKD2:MIR3130-1
ABLIM3,ABLIM3:RP11-331K21.1
orf50:GPR146,C7orf50:GPR146:RP11-449P15.1,C7orf50:GPER1,C7orf50:AC091729.7

1N7.6:CTNND1,TMX2-CTNND1:RP11-691N7.6:BTBD18,RP11-691N7.6:CTNND1,CTNND1,OR5BA1P,RP11-734C1
DS1,FADS2:FADS1:MIR1908,TMEM258
RP11-298I3.1:HAUS4:RP11-298I3.5,HAUS4:RP11-298I3.5,RP11-298I3.5:AJUBA,RP11-298I3.4,RP11-298I3.4:C14
D-2509G16.5,CTD-2509G16.3,CTD-2509G16.5:CTD-2509G16.1

.4.2,OR5AZ1P,UBE2L6,RPS4XP13,SERPING1,CYCSP26,AP000662.4,AP000662.4:YPEL4,AP000662.4:CLP1,

orf93,PRMT5-AS1:PRMT5,PRMT5

CLP1,ZDHHC5,TMX2:TMX2-CTNND1,TMX2-CTNND1,TMX2-CTNND1:C11orf31:RP11-691N7.6,MED19

**Table S10. SNPs in miRNA genes that also affected miRNA levels**

| miRNA | SNP | position | location | effect_alle | other_allel | eaf | beta | se | pval | signif |
|---|---|---|---|---|---|---|---|---|---|---|
| miR-3130- | rs2241347 | 2:2076479 | Mature | C | T | 0.841853 | -0.26645 | 0.032894 | 5.49E-16 | Bonferroni-sig |
| miR-4707- | rs2273626 | 14:234261 | Seed | A | C | 0.488021 | 0.091058 | 0.013739 | 3.41E-11 | Bonferroni-sig |
| miR-1908- | rs174561 | 11:615827 | pre-miRNA | T | C | 0.70223 | -0.09547 | 0.017762 | 7.66E-08 | Bonferroni-sig |
| miR-323b- | rs5610383 | 14:101522 | pre-miRNA | T | C | 0.788906 | 0.172218 | 0.034483 | 5.90E-07 | Bonferroni-sig |
| miR-2110 | rs1709140 | 10:115933 | pre-miRNA | C | T | 0.884998 | 0.137732 | 0.027697 | 6.60E-07 | Bonferroni-sig |
| miR-4482- | rs4559684 | 10:106028 | Seed | G | A | 0.690649 | 0.206064 | 0.042009 | 9.33E-07 | Bonferroni-sig |
| miR-6891- | rs2276448 | 6:3132302 | Seed | T | C | 0.748517 | 0.127075 | 0.026937 | 2.39E-06 | Bonferroni-sig |
| miR-6891- | rs2854001 | 6:3132301 | Mature | G | A | 0.85292 | 0.17272 | 0.039316 | 1.12E-05 | Bonferroni-sig |
| miR-618 | rs2682818 | 12:813295 | pre-miRNA | A | C | 0.128889 | -0.2331 | 0.053556 | 1.35E-05 | Bonferroni-sig |
| miR-27a-5 | rs895819 | 19:139472 | pre-miRNA | T | C | 0.666223 | 0.118003 | 0.027699 | 2.04E-05 | Bonferroni-sig |
| miR-6839- | rs7804972 | 7:6413946 | Mature | G | A | 0.597858 | 0.14158 | 0.035113 | 5.53E-05 | Bonferroni-sig |
| miR-4482- | rs641071 | 10:106028 | pre-miRNA | G | T | 0.642509 | 0.155901 | 0.040595 | 0.000123 | Bonferroni-sig |
| miR-6868- | rs7208391 | 17:740941 | Mature | C | G | 0.57323 | 0.149522 | 0.040036 | 0.000188 | nominally-sig |
| miR-629-5 | rs7821277 | 15:703717 | Mature | C | G | 0.988445 | 0.364687 | 0.10252 | 0.000375 | nominally-sig |
| miR-6841- | rs7634784 | 8:2481132 | Mature | A | G | 0.940856 | 0.163006 | 0.047526 | 0.000604 | nominally-sig |
| miR-4326 | rs6062431 | 20:619181 | pre-miRNA | C | G | 0.317658 | -0.10837 | 0.033748 | 0.001322 | nominally-sig |
| miR-7157- | rs5614856 | 2:1413442 | Mature | T | C | 0.778924 | 0.144622 | 0.046368 | 0.001815 | nominally-sig |
| miR-3977 | rs7951280 | 5:8213602 | pre-miRNA | T | G | 0.992036 | 0.974143 | 0.315052 | 0.001988 | nominally-sig |
| miR-5707 | rs8012858 | 7:1583843 | pre-miRNA | G | A | 0.959124 | -0.352 | 0.116558 | 0.002528 | nominally-sig |
| miR-4772- | rs6215497 | 2:1030487 | Mature | C | T | 0.904965 | 0.193118 | 0.064191 | 0.002625 | nominally-sig |
| miR-8060 | rs1514422 | 3:9607886 | Mature | G | A | 0.949442 | 0.213778 | 0.077067 | 0.005539 | nominally-sig |
| miR-216a- | rs4129117 | 2:5621609 | pre-miRNA | A | T | 0.953417 | -0.15069 | 0.055721 | 0.006844 | nominally-sig |
| miR-182-5 | rs7648177 | 7:1294102 | pre-miRNA | C | T | 0.894847 | -0.102 | 0.038433 | 0.007953 | nominally-sig |
| miR-6717- | rs1176501 | 14:214915 | Seed | G | A | 0.971572 | 0.26135 | 0.103455 | 0.01153 | nominally-sig |
| miR-7150 | rs5860547 | 9:1262478 | pre-miRNA | A | G | 0.783001 | 0.044897 | 0.01792 | 0.012233 | nominally-sig |
| miR-4679 | rs7281095 | 10:908231 | Mature | G | A | 0.98284 | -0.38579 | 0.154193 | 0.012349 | nominally-sig |
| miR-1307- | rs7911488 | 10:105154 | pre-miRNA | A | G | 0.676679 | 0.041295 | 0.017191 | 0.016299 | nominally-sig |
| miR-3689f | rs7250271 | 9:1377425 | pre-miRNA | A | G | 0.844401 | -0.11187 | 0.048342 | 0.020658 | nominally-sig |
| miR-4274 | rs1251266 | 4:7461769 | pre-miRNA | A | G | 0.575801 | -0.02168 | 0.00944 | 0.021663 | nominally-sig |
| miR-8070 | rs1385521 | 11:118047 | Seed | C | T | 0.975041 | 0.273628 | 0.120744 | 0.02344 | nominally-sig |
| miR-4657 | rs1129089 | 7:4492138 | Mature | C | A | 0.988128 | 0.542813 | 0.244984 | 0.026712 | nominally-sig |
| miR-612 | rs550894 | 11:652119 | pre-miRNA | C | A | 0.907567 | 0.106676 | 0.04828 | 0.027138 | nominally-sig |
| miR-938 | rs1241660 | 10:298912 | Seed | C | T | 0.740841 | 0.111801 | 0.050837 | 0.027865 | nominally-sig |
| miR-1206 | rs2114358 | 8:1290211 | pre-miRNA | G | A | 0.380682 | -0.09545 | 0.043794 | 0.029301 | nominally-sig |
| miR-4636 | rs257095 | 5:9053945 | pre-miRNA | C | T | 0.126973 | 0.128531 | 0.060357 | 0.033212 | nominally-sig |
| miR-612 | rs1280391 | 11:652119 | pre-miRNA | G | A | 0.768095 | 0.070547 | 0.03413 | 0.038736 | nominally-sig |
| miR-2117 | rs7207008 | 17:415222 | pre-miRNA | A | T | 0.5367 | 0.063948 | 0.031343 | 0.041325 | nominally-sig |
| miR-4513 | rs2168518 | 15:750810 | Seed | G | A | 0.318808 | 0.079313 | 0.039296 | 0.043557 | nominally-sig |
| miR-1269a | rs7323913 | 4:6714262 | Mature | G | A | 0.764952 | 0.071193 | 0.036118 | 0.048708 | nominally-sig |

**Table S11. SNP-based heritability estimates for 2083 miRNAs**

| miRNA | h2 | Pval | PermPval | PermFWEcPval |
|---|---|---|---|---|
| miR-30e-5p | 0.71358344 | 0.00058216 | 0.002 | 0.716 |
| miR-6511a-5p | 0.70359235 | 0.00068236 | 0.002 | 0.773 |
| let-7i-5p | 0.67812368 | 0.00101398 | 0.002 | 0.869 |
| miR-8078 | 0.67512863 | 0.00106146 | 0.001 | 0.877 |
| miR-505-3p | 0.6641229 | 0.00125394 | 0.005 | 0.91 |
| miR-6814-5p | 0.59373968 | 0.00344537 | 0.007 | 0.997 |
| miR-3687 | 0.59115907 | 0.00356902 | 0.005 | 0.997 |
| miR-4786-5p | 0.58182282 | 0.00405033 | 0.003 | 0.998 |
| miR-4287 | 0.58065969 | 0.00411419 | 0.003 | 0.999 |
| miR-4259 | 0.58028349 | 0.00413504 | 0.005 | 0.999 |
| miR-5571-5p | 0.57347432 | 0.00452907 | 0.009 | 0.999 |
| miR-625-5p | 0.55207703 | 0.0059944 | 0.004 | 1 |
| miR-548b-3p | 0.54832035 | 0.00629116 | 0.004 | 1 |
| miR-16-1-3p | 0.54651653 | 0.00643822 | 0.007 | 1 |
| miR-1273h-5p | 0.54472565 | 0.00658723 | 0.006 | 1 |
| miR-6832-3p | 0.54205919 | 0.00681474 | 0.01 | 1 |
| miR-6827-5p | 0.51638888 | 0.00938522 | 0.008 | 1 |
| miR-1185-1-3p | 0.50827975 | 0.01035706 | 0.016 | 1 |
| miR-561-3p | 0.50775835 | 0.01042245 | 0.012 | 1 |
| miR-6883-5p | 0.50478923 | 0.0108017 | 0.017 | 1 |
| miR-614 | 0.49105855 | 0.01271583 | 0.012 | 1 |
| miR-944 | 0.4900853 | 0.01286201 | 0.014 | 1 |
| miR-1273d | 0.48564767 | 0.01354712 | 0.012 | 1 |
| miR-374a-3p | 0.48542794 | 0.01358185 | 0.016 | 1 |
| miR-5698 | 0.48432443 | 0.01375741 | 0.007 | 1 |
| miR-3197 | 0.48386396 | 0.01383125 | 0.015 | 1 |
| miR-370-5p | 0.47248644 | 0.01576727 | 0.011 | 1 |
| miR-10a-5p | 0.4716158 | 0.01592456 | 0.022 | 1 |
| miR-6782-5p | 0.46418489 | 0.01732259 | 0.013 | 1 |
| miR-136-5p | 0.46250866 | 0.01765205 | 0.021 | 1 |
| miR-548au-3p | 0.45728936 | 0.01871237 | 0.018 | 1 |
| miR-493-3p | 0.45618021 | 0.01894455 | 0.024 | 1 |
| miR-4679 | 0.44786506 | 0.02076443 | 0.027 | 1 |
| miR-7856-5p | 0.44703673 | 0.02095358 | 0.023 | 1 |
| miR-6843-3p | 0.44551078 | 0.02130583 | 0.019 | 1 |
| miR-566 | 0.44424544 | 0.02160171 | 0.03 | 1 |
| miR-136-3p | 0.44352232 | 0.02177236 | 0.022 | 1 |
| miR-139-5p | 0.44258877 | 0.02199435 | 0.012 | 1 |
| miR-1247-5p | 0.44256364 | 0.02200035 | 0.018 | 1 |
| miR-299-3p | 0.43949226 | 0.02274438 | 0.023 | 1 |
| miR-4699-3p | 0.43914351 | 0.02283019 | 0.025 | 1 |
| miR-4426 | 0.43635246 | 0.0235268 | 0.021 | 1 |
| miR-302e | 0.43593347 | 0.0236329 | 0.031 | 1 |
| miR-513c-5p | 0.43164827 | 0.02474134 | 0.03 | 1 |
| miR-4461 | 0.43026396 | 0.02510861 | 0.033 | 1 |
| miR-525-5p | 0.43006973 | 0.02516051 | 0.029 | 1 |
| miR-5590-5p | 0.42704643 | 0.02597993 | 0.026 | 1 |
| miR-892a | 0.42299394 | 0.02711321 | 0.026 | 1 |

| | | | | |
|---|---|---|---|---|
| miR-5007-5p | 0.41631931 | 0.02906955 | 0.04 | 1 |
| miR-6862-3p | 0.41601649 | 0.02916102 | 0.026 | 1 |
| miR-6731-3p | 0.41423564 | 0.02970379 | 0.028 | 1 |
| miR-875-3p | 0.41276491 | 0.03015833 | 0.035 | 1 |
| miR-675-3p | 0.41253089 | 0.03023119 | 0.038 | 1 |
| miR-7106-3p | 0.41134888 | 0.0306014 | 0.035 | 1 |
| miR-1321 | 0.41047348 | 0.030878 | 0.034 | 1 |
| miR-2117 | 0.41010143 | 0.03099618 | 0.031 | 1 |
| miR-656-5p | 0.4090795 | 0.03132272 | 0.04 | 1 |
| miR-147b | 0.40751174 | 0.0318292 | 0.034 | 1 |
| miR-378h | 0.40739627 | 0.03186677 | 0.029 | 1 |
| miR-548b-5p | 0.40304432 | 0.03330965 | 0.036 | 1 |
| miR-3688-5p | 0.40247187 | 0.03350338 | 0.031 | 1 |
| miR-7851-3p | 0.40047003 | 0.03418819 | 0.041 | 1 |
| miR-4437 | 0.40022331 | 0.03427338 | 0.029 | 1 |
| miR-3937 | 0.39971461 | 0.03444958 | 0.039 | 1 |
| miR-593-5p | 0.39957969 | 0.03449643 | 0.024 | 1 |
| miR-4484 | 0.39925129 | 0.03461071 | 0.034 | 1 |
| miR-1255b-2-: | 0.39828322 | 0.03494937 | 0.036 | 1 |
| miR-548av-5p | 0.3979389 | 0.03507048 | 0.036 | 1 |
| miR-4801 | 0.39703065 | 0.03539159 | 0.036 | 1 |
| miR-323a-5p | 0.39695771 | 0.03541749 | 0.04 | 1 |
| miR-1254 | 0.39537314 | 0.03598383 | 0.036 | 1 |
| miR-6723-5p | 0.39531007 | 0.03600652 | 0.043 | 1 |
| miR-1299 | 0.3949031 | 0.03615324 | 0.052 | 1 |
| miR-342-5p | 0.39371653 | 0.03658382 | 0.035 | 1 |
| miR-3117-5p | 0.38886625 | 0.03838772 | 0.042 | 1 |
| miR-708-5p | 0.38885226 | 0.03839302 | 0.037 | 1 |
| miR-106a-3p | 0.38856998 | 0.03850022 | 0.038 | 1 |
| miR-449b-5p | 0.38808149 | 0.03868629 | 0.039 | 1 |
| miR-6866-5p | 0.38686474 | 0.03915295 | 0.028 | 1 |
| miR-4639-3p | 0.38628805 | 0.03937572 | 0.033 | 1 |
| miR-595 | 0.38613362 | 0.03943555 | 0.043 | 1 |
| miR-4782-5p | 0.38470505 | 0.03999253 | 0.033 | 1 |
| miR-5007-3p | 0.38406636 | 0.04024361 | 0.057 | 1 |
| miR-6126 | 0.38249622 | 0.04086629 | 0.041 | 1 |
| miR-337-5p | 0.37971614 | 0.04198797 | 0.038 | 1 |
| miR-1273c | 0.37957064 | 0.04204736 | 0.048 | 1 |
| miR-195-3p | 0.37865526 | 0.04242252 | 0.032 | 1 |
| miR-873-3p | 0.37737083 | 0.0429535 | 0.028 | 1 |
| miR-582-5p | 0.37679356 | 0.04319389 | 0.046 | 1 |
| miR-138-5p | 0.37619391 | 0.04344474 | 0.052 | 1 |
| miR-542-5p | 0.37445949 | 0.04417693 | 0.042 | 1 |
| miR-5584-3p | 0.36978011 | 0.04620206 | 0.053 | 1 |
| miR-4688 | 0.36821006 | 0.046898 | 0.041 | 1 |
| miR-4999-5p | 0.36517929 | 0.04826521 | 0.048 | 1 |
| miR-1908-5p | 0.36456357 | 0.04854682 | 0.049 | 1 |
| miR-4436b-3p | 0.36452037 | 0.04856663 | 0.037 | 1 |
| miR-4436a | 0.36380233 | 0.04889681 | 0.05 | 1 |
| miR-6863 | 0.36368752 | 0.04894977 | 0.055 | 1 |

| | | | | |
|---|---|---|---|---|
| miR-1224-5p | 0.36282357 | 0.04934976 | 0.056 | 1 |
| miR-6837-5p | 0.36240493 | 0.04954452 | 0.055 | 1 |
| miR-4712-5p | 0.36210419 | 0.04968481 | 0.058 | 1 |
| miR-4425 | 0.36142513 | 0.05000274 | 0.045 | 1 |
| miR-1914-5p | 0.36081245 | 0.05029098 | 0.048 | 1 |
| miR-4684-5p | 0.36013045 | 0.0506134 | 0.049 | 1 |
| miR-574-5p | 0.35909605 | 0.05110554 | 0.055 | 1 |
| miR-4524b-5p | 0.3531029 | 0.05403234 | 0.052 | 1 |
| miR-6856-5p | 0.35277461 | 0.05419642 | 0.049 | 1 |
| miR-593-3p | 0.35191758 | 0.05462662 | 0.051 | 1 |
| miR-5586-5p | 0.35147765 | 0.05484849 | 0.061 | 1 |
| miR-4539 | 0.35085551 | 0.05516348 | 0.054 | 1 |
| miR-335-3p | 0.35022901 | 0.05548212 | 0.045 | 1 |
| miR-5585-3p | 0.34682523 | 0.05723875 | 0.063 | 1 |
| miR-4677-5p | 0.34556568 | 0.05789977 | 0.056 | 1 |
| miR-369-5p | 0.34215537 | 0.05971965 | 0.07 | 1 |
| miR-6733-3p | 0.34188792 | 0.05986425 | 0.051 | 1 |
| miR-4439 | 0.34128577 | 0.0601908 | 0.071 | 1 |
| miR-3674 | 0.34090383 | 0.06039866 | 0.074 | 1 |
| let-7c-5p | 0.33887085 | 0.06151448 | 0.054 | 1 |
| miR-1273a | 0.33754109 | 0.062253 | 0.067 | 1 |
| miR-519d-3p | 0.33703273 | 0.06253716 | 0.084 | 1 |
| miR-6780a-5p | 0.33477714 | 0.06381015 | 0.056 | 1 |
| miR-20a-5p | 0.3327939 | 0.06494601 | 0.075 | 1 |
| miR-4696 | 0.33241616 | 0.06516413 | 0.071 | 1 |
| miR-3607-5p | 0.33164162 | 0.06561313 | 0.06 | 1 |
| miR-2113 | 0.32941294 | 0.0669185 | 0.071 | 1 |
| miR-412-5p | 0.32860053 | 0.06739931 | 0.066 | 1 |
| miR-3605-5p | 0.32848259 | 0.06746933 | 0.083 | 1 |
| miR-511-3p | 0.32624568 | 0.06880805 | 0.065 | 1 |
| miR-513c-3p | 0.32553368 | 0.06923843 | 0.074 | 1 |
| miR-1244 | 0.32490818 | 0.06961824 | 0.066 | 1 |
| miR-3921 | 0.32439478 | 0.06993117 | 0.08 | 1 |
| miR-4713-3p | 0.32434042 | 0.06996437 | 0.087 | 1 |
| miR-4795-3p | 0.32320429 | 0.07066098 | 0.073 | 1 |
| miR-4657 | 0.32226872 | 0.0712386 | 0.074 | 1 |
| miR-1275 | 0.32216147 | 0.07130505 | 0.077 | 1 |
| miR-615-3p | 0.32153855 | 0.07169193 | 0.073 | 1 |
| miR-601 | 0.32046606 | 0.07236179 | 0.086 | 1 |
| miR-20b-5p | 0.31932156 | 0.07308191 | 0.069 | 1 |
| miR-7706 | 0.31904458 | 0.07325701 | 0.074 | 1 |
| miR-345-5p | 0.31834165 | 0.07370282 | 0.074 | 1 |
| miR-548e-3p | 0.31825309 | 0.07375913 | 0.082 | 1 |
| miR-2116-5p | 0.31800887 | 0.07391459 | 0.064 | 1 |
| miR-4737 | 0.31755094 | 0.07420677 | 0.09 | 1 |
| let-7d-5p | 0.31752656 | 0.07422235 | 0.069 | 1 |
| miR-7160-3p | 0.31715949 | 0.07445723 | 0.081 | 1 |
| miR-3142 | 0.31654638 | 0.07485081 | 0.072 | 1 |
| miR-10b-5p | 0.31592115 | 0.07525381 | 0.078 | 1 |
| miR-6804-3p | 0.31549319 | 0.0755306 | 0.074 | 1 |

| | | | | |
|---|---|---|---|---|
| miR-4302 | 0.31423676 | 0.07634772 | 0.08 | 1 |
| miR-651-5p | 0.31290323 | 0.07722231 | 0.064 | 1 |
| miR-6833-5p | 0.31263916 | 0.0773964 | 0.089 | 1 |
| miR-6716-3p | 0.31231959 | 0.07760747 | 0.064 | 1 |
| miR-320e | 0.31184478 | 0.0779219 | 0.079 | 1 |
| miR-6731-5p | 0.31063595 | 0.07872675 | 0.086 | 1 |
| miR-513a-3p | 0.31059323 | 0.07875531 | 0.065 | 1 |
| miR-7848-3p | 0.30988741 | 0.07922828 | 0.078 | 1 |
| miR-5187-3p | 0.30918875 | 0.07969858 | 0.083 | 1 |
| miR-1228-3p | 0.30876638 | 0.07998391 | 0.068 | 1 |
| let-7b-5p | 0.30715049 | 0.08108266 | 0.069 | 1 |
| miR-5583-5p | 0.30641064 | 0.08158953 | 0.095 | 1 |
| miR-3140-3p | 0.30558699 | 0.0821566 | 0.084 | 1 |
| miR-125b-2-3 | 0.30459405 | 0.08284418 | 0.083 | 1 |
| miR-3614-5p | 0.30313265 | 0.08386401 | 0.09 | 1 |
| miR-363-5p | 0.30288797 | 0.08403567 | 0.097 | 1 |
| miR-6793-5p | 0.30187875 | 0.08474653 | 0.081 | 1 |
| miR-6891-3p | 0.30174565 | 0.08484061 | 0.073 | 1 |
| miR-1909-3p | 0.29959422 | 0.08637229 | 0.084 | 1 |
| miR-30c-2-3p | 0.29956878 | 0.08639052 | 0.071 | 1 |
| miR-6765-5p | 0.29611928 | 0.08888975 | 0.097 | 1 |
| miR-6073 | 0.29520105 | 0.08956401 | 0.104 | 1 |
| miR-6866-3p | 0.29463909 | 0.08997853 | 0.08 | 1 |
| miR-4695-5p | 0.29418635 | 0.09031353 | 0.085 | 1 |
| miR-4293 | 0.29393964 | 0.09049646 | 0.092 | 1 |
| miR-4764-3p | 0.29184201 | 0.09206298 | 0.106 | 1 |
| miR-922 | 0.29156112 | 0.09227426 | 0.089 | 1 |
| miR-125a-5p | 0.29127496 | 0.09248988 | 0.101 | 1 |
| miR-3123 | 0.29102749 | 0.09267665 | 0.077 | 1 |
| miR-7703 | 0.29074781 | 0.09288806 | 0.1 | 1 |
| miR-877-3p | 0.28927815 | 0.09400484 | 0.11 | 1 |
| miR-376c-5p | 0.28920786 | 0.0940585 | 0.088 | 1 |
| miR-431-5p | 0.28911181 | 0.09413186 | 0.11 | 1 |
| miR-4483 | 0.2885579 | 0.09455575 | 0.083 | 1 |
| miR-4790-5p | 0.28840281 | 0.09467469 | 0.084 | 1 |
| miR-4540 | 0.28811278 | 0.09489741 | 0.1 | 1 |
| miR-3613-3p | 0.28783127 | 0.09511396 | 0.101 | 1 |
| miR-4732-5p | 0.28770271 | 0.09521297 | 0.096 | 1 |
| miR-4779 | 0.28607737 | 0.09647131 | 0.097 | 1 |
| miR-548ay-5p | 0.28607336 | 0.09647443 | 0.094 | 1 |
| miR-6878-5p | 0.28479746 | 0.09747078 | 0.112 | 1 |
| miR-4495 | 0.28413867 | 0.09798818 | 0.076 | 1 |
| miR-378j | 0.28354389 | 0.09845702 | 0.123 | 1 |
| miR-107 | 0.28314364 | 0.09877345 | 0.124 | 1 |
| miR-574-3p | 0.28167071 | 0.09994434 | 0.1 | 1 |
| miR-1265 | 0.28125622 | 0.10027564 | 0.087 | 1 |
| miR-4788 | 0.28123738 | 0.10029072 | 0.106 | 1 |
| miR-6795-3p | 0.28084202 | 0.10060753 | 0.096 | 1 |
| miR-4652-5p | 0.28080702 | 0.10063561 | 0.093 | 1 |
| miR-208a-3p | 0.28032433 | 0.10102346 | 0.1 | 1 |

| | | | | |
|---|---|---|---|---|
| miR-7154-5p | 0.27927724 | 0.10186855 | 0.087 | 1 |
| miR-764 | 0.27908743 | 0.10202229 | 0.113 | 1 |
| miR-200b-3p | 0.27819844 | 0.1027446 | 0.102 | 1 |
| miR-4465 | 0.2771041 | 0.10363886 | 0.1 | 1 |
| miR-503-5p | 0.27676723 | 0.10391527 | 0.095 | 1 |
| miR-4299 | 0.27626275 | 0.10433021 | 0.109 | 1 |
| miR-4800-3p | 0.27602449 | 0.1045266 | 0.101 | 1 |
| miR-539-5p | 0.27549136 | 0.10496701 | 0.109 | 1 |
| miR-6791-3p | 0.27497882 | 0.10539167 | 0.093 | 1 |
| miR-6792-5p | 0.27455879 | 0.10574061 | 0.118 | 1 |
| miR-140-5p | 0.27446316 | 0.10582017 | 0.109 | 1 |
| miR-4430 | 0.27443673 | 0.10584216 | 0.118 | 1 |
| miR-135a-3p | 0.27411772 | 0.10610792 | 0.091 | 1 |
| miR-548d-5p | 0.27300492 | 0.10703875 | 0.104 | 1 |
| miR-7846-3p | 0.27225975 | 0.10766534 | 0.092 | 1 |
| miR-29b-2-5p | 0.27220203 | 0.10771398 | 0.12 | 1 |
| miR-208b-3p | 0.27213153 | 0.10777342 | 0.088 | 1 |
| miR-548ao-5p | 0.27104471 | 0.10869269 | 0.102 | 1 |
| miR-6745 | 0.27055727 | 0.10910681 | 0.083 | 1 |
| miR-8082 | 0.27016815 | 0.10943822 | 0.104 | 1 |
| miR-4481 | 0.26853123 | 0.11084025 | 0.11 | 1 |
| miR-4537 | 0.26794904 | 0.111342 | 0.097 | 1 |
| miR-5196-3p | 0.26784343 | 0.11143318 | 0.102 | 1 |
| miR-517c-3p | 0.26745899 | 0.11176559 | 0.101 | 1 |
| miR-515-3p | 0.26742645 | 0.11179375 | 0.113 | 1 |
| miR-4763-3p | 0.26686086 | 0.11228416 | 0.116 | 1 |
| miR-4421 | 0.26621986 | 0.1128418 | 0.115 | 1 |
| miR-6075 | 0.2652049 | 0.11372883 | 0.11 | 1 |
| miR-590-5p | 0.26388267 | 0.11489182 | 0.1 | 1 |
| miR-4665-5p | 0.26294845 | 0.11571862 | 0.119 | 1 |
| miR-589-3p | 0.26284017 | 0.11581472 | 0.122 | 1 |
| miR-451a | 0.26263355 | 0.11599826 | 0.118 | 1 |
| miR-4273 | 0.26260198 | 0.11602632 | 0.121 | 1 |
| miR-4261 | 0.2621197 | 0.1164556 | 0.109 | 1 |
| miR-181c-5p | 0.26101499 | 0.11744316 | 0.13 | 1 |
| miR-6828-5p | 0.26026132 | 0.1181203 | 0.118 | 1 |
| miR-449a | 0.25917942 | 0.11909716 | 0.127 | 1 |
| miR-605-3p | 0.25788157 | 0.12027651 | 0.124 | 1 |
| miR-6499-5p | 0.25596517 | 0.12203292 | 0.122 | 1 |
| miR-6809-3p | 0.25508282 | 0.12284765 | 0.13 | 1 |
| miR-6764-5p | 0.25472307 | 0.12318091 | 0.131 | 1 |
| miR-4753-3p | 0.25468043 | 0.12322046 | 0.114 | 1 |
| miR-6127 | 0.2546404 | 0.12325759 | 0.142 | 1 |
| miR-4278 | 0.25451973 | 0.12336957 | 0.118 | 1 |
| miR-218-5p | 0.25404226 | 0.12381334 | 0.124 | 1 |
| miR-409-5p | 0.25351495 | 0.12430474 | 0.114 | 1 |
| miR-212-5p | 0.25321418 | 0.12458564 | 0.135 | 1 |
| miR-6888-3p | 0.251033 | 0.12663597 | 0.116 | 1 |
| miR-5190 | 0.25073406 | 0.12691879 | 0.13 | 1 |
| miR-3620-3p | 0.24986642 | 0.12774215 | 0.138 | 1 |

| | | | | |
|---|---|---|---|---|
| miR-4521 | 0.249601 | 0.12799477 | 0.123 | 1 |
| miR-629-5p | 0.24909011 | 0.12848199 | 0.12 | 1 |
| miR-1234-3p | 0.24906672 | 0.12850432 | 0.128 | 1 |
| miR-129-2-3p | 0.24784215 | 0.12967753 | 0.119 | 1 |
| miR-7153-5p | 0.24728587 | 0.13021293 | 0.137 | 1 |
| miR-6832-5p | 0.24711809 | 0.1303747 | 0.13 | 1 |
| miR-202-5p | 0.2469728 | 0.13051491 | 0.113 | 1 |
| miR-206 | 0.24665957 | 0.13081753 | 0.114 | 1 |
| miR-210-5p | 0.24651559 | 0.1309568 | 0.149 | 1 |
| miR-6796-5p | 0.24569792 | 0.13174965 | 0.132 | 1 |
| miR-6735-3p | 0.24492775 | 0.13249947 | 0.123 | 1 |
| miR-6830-5p | 0.24449646 | 0.13292064 | 0.14 | 1 |
| miR-5196-5p | 0.24429802 | 0.13311473 | 0.13 | 1 |
| miR-1285-5p | 0.24417802 | 0.1332322 | 0.126 | 1 |
| miR-130b-5p | 0.24416648 | 0.1332435 | 0.147 | 1 |
| miR-1257 | 0.2439211 | 0.13348394 | 0.146 | 1 |
| miR-4307 | 0.24327889 | 0.13411462 | 0.144 | 1 |
| miR-376a-5p | 0.24232342 | 0.13505673 | 0.125 | 1 |
| miR-488-3p | 0.24139353 | 0.13597796 | 0.136 | 1 |
| miR-4534 | 0.24139268 | 0.1359788 | 0.152 | 1 |
| miR-1322 | 0.24070944 | 0.13665841 | 0.147 | 1 |
| miR-28-5p | 0.24059564 | 0.13677183 | 0.136 | 1 |
| miR-181a-2-3p | 0.24045835 | 0.13690875 | 0.126 | 1 |
| miR-1250-3p | 0.24022795 | 0.13713874 | 0.139 | 1 |
| miR-135b-3p | 0.24013423 | 0.13723237 | 0.157 | 1 |
| miR-5587-3p | 0.23912735 | 0.13824101 | 0.115 | 1 |
| miR-4266 | 0.23855126 | 0.13882038 | 0.126 | 1 |
| miR-513b-3p | 0.23791784 | 0.13945931 | 0.151 | 1 |
| miR-509-3p | 0.23705175 | 0.14033617 | 0.139 | 1 |
| miR-6757-3p | 0.23674934 | 0.14064321 | 0.134 | 1 |
| miR-3124-5p | 0.23659362 | 0.1408015 | 0.143 | 1 |
| miR-8070 | 0.23452625 | 0.1429144 | 0.143 | 1 |
| miR-126-5p | 0.23424482 | 0.14320367 | 0.159 | 1 |
| miR-198 | 0.23389899 | 0.14355969 | 0.137 | 1 |
| miR-3920 | 0.23358634 | 0.14388205 | 0.151 | 1 |
| miR-5188 | 0.23284069 | 0.14465286 | 0.138 | 1 |
| miR-450b-3p | 0.23170775 | 0.14582932 | 0.154 | 1 |
| miR-6728-3p | 0.23133793 | 0.14621475 | 0.149 | 1 |
| miR-4493 | 0.22981652 | 0.14780751 | 0.158 | 1 |
| miR-4254 | 0.22856337 | 0.14912812 | 0.151 | 1 |
| miR-4781-5p | 0.22792673 | 0.14980205 | 0.168 | 1 |
| miR-219b-3p | 0.22746267 | 0.15029457 | 0.153 | 1 |
| miR-4455 | 0.22627671 | 0.15155815 | 0.162 | 1 |
| miR-6877-5p | 0.2256632 | 0.15221458 | 0.139 | 1 |
| miR-3132 | 0.22511489 | 0.15280284 | 0.161 | 1 |
| miR-199a-5p | 0.22467469 | 0.15327621 | 0.155 | 1 |
| miR-1252-3p | 0.22462662 | 0.15332796 | 0.164 | 1 |
| miR-103a-3p | 0.22353891 | 0.15450204 | 0.164 | 1 |
| miR-511-5p | 0.22332572 | 0.15473285 | 0.165 | 1 |
| miR-340-3p | 0.22228335 | 0.15586467 | 0.128 | 1 |

| | | | | |
|---|---|---|---|---|
| miR-7153-3p | 0.22212401 | 0.15603817 | 0.17 | 1 |
| miR-320a | 0.22191957 | 0.15626095 | 0.167 | 1 |
| miR-1273e | 0.22144234 | 0.15678181 | 0.158 | 1 |
| miR-4750-5p | 0.22085065 | 0.1574292 | 0.181 | 1 |
| miR-5197-3p | 0.22046445 | 0.15785269 | 0.158 | 1 |
| miR-4503 | 0.21989709 | 0.1584762 | 0.152 | 1 |
| miR-188-3p | 0.21956705 | 0.15883964 | 0.144 | 1 |
| miR-222-3p | 0.21929281 | 0.15914205 | 0.159 | 1 |
| miR-6761-5p | 0.21890904 | 0.15956588 | 0.175 | 1 |
| miR-5010-5p | 0.21857622 | 0.15993403 | 0.148 | 1 |
| miR-550a-3-5p | 0.21643167 | 0.16231958 | 0.155 | 1 |
| miR-329-5p | 0.21638468 | 0.1623721 | 0.165 | 1 |
| miR-1298-3p | 0.21534492 | 0.16353724 | 0.161 | 1 |
| miR-1231 | 0.21476612 | 0.16418818 | 0.195 | 1 |
| miR-4304 | 0.21471857 | 0.16424173 | 0.174 | 1 |
| miR-3174 | 0.21441005 | 0.16458947 | 0.157 | 1 |
| miR-6797-5p | 0.21361031 | 0.16549307 | 0.176 | 1 |
| miR-6508-5p | 0.21329924 | 0.16584541 | 0.16 | 1 |
| miR-508-3p | 0.21288028 | 0.16632071 | 0.17 | 1 |
| miR-8079 | 0.21265005 | 0.16658228 | 0.152 | 1 |
| miR-4644 | 0.21258446 | 0.16665685 | 0.178 | 1 |
| miR-885-5p | 0.21215841 | 0.16714173 | 0.158 | 1 |
| miR-937-5p | 0.21208276 | 0.16722792 | 0.179 | 1 |
| miR-548ar-3p | 0.21187175 | 0.16746848 | 0.173 | 1 |
| miR-3683 | 0.21141507 | 0.16798988 | 0.156 | 1 |
| miR-600 | 0.2112894 | 0.16813355 | 0.151 | 1 |
| miR-4429 | 0.21117025 | 0.16826983 | 0.162 | 1 |
| miR-4506 | 0.21072112 | 0.16878418 | 0.183 | 1 |
| miR-7162-3p | 0.21065296 | 0.16886232 | 0.16 | 1 |
| miR-6514-3p | 0.21057045 | 0.16895696 | 0.161 | 1 |
| miR-8067 | 0.20969811 | 0.16995951 | 0.176 | 1 |
| miR-2052 | 0.20968889 | 0.16997014 | 0.173 | 1 |
| miR-647 | 0.20968245 | 0.16997755 | 0.188 | 1 |
| miR-1538 | 0.20967439 | 0.16998683 | 0.167 | 1 |
| miR-3185 | 0.20930281 | 0.17041508 | 0.168 | 1 |
| miR-299-5p | 0.20871147 | 0.17109802 | 0.169 | 1 |
| miR-6894-5p | 0.20846426 | 0.17138405 | 0.189 | 1 |
| miR-588 | 0.20842341 | 0.17143134 | 0.166 | 1 |
| miR-3179 | 0.20818444 | 0.17170818 | 0.163 | 1 |
| miR-32-3p | 0.20811724 | 0.17178608 | 0.192 | 1 |
| miR-199a-3p | 0.20580165 | 0.17448407 | 0.192 | 1 |
| miR-4463 | 0.20576076 | 0.17453195 | 0.199 | 1 |
| miR-8061 | 0.20531958 | 0.17504911 | 0.175 | 1 |
| miR-6874-3p | 0.20514538 | 0.17525359 | 0.181 | 1 |
| miR-4294 | 0.20467832 | 0.17580256 | 0.164 | 1 |
| miR-6729-3p | 0.20362367 | 0.17704615 | 0.174 | 1 |
| miR-892c-5p | 0.20348342 | 0.17721194 | 0.199 | 1 |
| miR-497-3p | 0.2027784 | 0.17804686 | 0.172 | 1 |
| miR-2681-5p | 0.20236902 | 0.17853281 | 0.184 | 1 |
| miR-378d | 0.20006449 | 0.18128389 | 0.178 | 1 |

| | | | | |
|---|---|---|---|---|
| miR-513a-5p | 0.19991181 | 0.18146709 | 0.199 | 1 |
| miR-3689f | 0.19981159 | 0.1815874 | 0.204 | 1 |
| miR-517a-3p | 0.19946947 | 0.18199849 | 0.192 | 1 |
| miR-4290 | 0.19943371 | 0.18204149 | 0.192 | 1 |
| miR-554 | 0.19895172 | 0.18262173 | 0.178 | 1 |
| miR-409-3p | 0.19876569 | 0.18284599 | 0.171 | 1 |
| miR-4800-5p | 0.19726384 | 0.18466275 | 0.191 | 1 |
| miR-135a-5p | 0.19720777 | 0.1847308 | 0.166 | 1 |
| miR-4446-5p | 0.19700922 | 0.18497187 | 0.186 | 1 |
| miR-3612 | 0.19694599 | 0.18504869 | 0.183 | 1 |
| miR-6790-5p | 0.19679918 | 0.1852271 | 0.186 | 1 |
| miR-4417 | 0.19676337 | 0.18527064 | 0.183 | 1 |
| miR-1910-5p | 0.19614512 | 0.18602331 | 0.188 | 1 |
| miR-17-5p | 0.19600825 | 0.1861902 | 0.195 | 1 |
| miR-8076 | 0.19589843 | 0.18632416 | 0.183 | 1 |
| miR-6744-3p | 0.19579469 | 0.18645077 | 0.169 | 1 |
| miR-96-3p | 0.19503415 | 0.18738058 | 0.178 | 1 |
| miR-520g-5p | 0.19464411 | 0.18785854 | 0.171 | 1 |
| miR-650 | 0.19440711 | 0.18814933 | 0.176 | 1 |
| miR-6877-3p | 0.19364075 | 0.18909153 | 0.169 | 1 |
| miR-6082 | 0.19346583 | 0.18930698 | 0.201 | 1 |
| miR-5093 | 0.1934265 | 0.18935545 | 0.18 | 1 |
| miR-664a-5p | 0.19336891 | 0.18942644 | 0.212 | 1 |
| miR-4784 | 0.19301539 | 0.18986252 | 0.2 | 1 |
| miR-6873-3p | 0.19213793 | 0.19094757 | 0.184 | 1 |
| miR-7853-5p | 0.19185675 | 0.19129607 | 0.203 | 1 |
| miR-6719-3p | 0.19172488 | 0.19145965 | 0.197 | 1 |
| miR-449c-5p | 0.19141146 | 0.19184878 | 0.187 | 1 |
| miR-205-3p | 0.19118159 | 0.19213449 | 0.176 | 1 |
| miR-320c | 0.19035766 | 0.19316068 | 0.203 | 1 |
| miR-548t-5p | 0.19001816 | 0.19358449 | 0.189 | 1 |
| miR-548k | 0.18993355 | 0.1936902 | 0.235 | 1 |
| miR-148a-3p | 0.18887557 | 0.195015 | 0.184 | 1 |
| miR-223-3p | 0.1888023 | 0.19510696 | 0.198 | 1 |
| miR-4675 | 0.18828721 | 0.19575413 | 0.208 | 1 |
| miR-6870-3p | 0.18813013 | 0.19595175 | 0.195 | 1 |
| miR-6768-3p | 0.18783065 | 0.19632884 | 0.209 | 1 |
| miR-887-5p | 0.18727258 | 0.19703273 | 0.178 | 1 |
| miR-301a-3p | 0.18671708 | 0.1977349 | 0.196 | 1 |
| miR-4464 | 0.18631133 | 0.19824872 | 0.195 | 1 |
| miR-6129 | 0.18620169 | 0.19838771 | 0.195 | 1 |
| miR-4428 | 0.18569468 | 0.19903117 | 0.199 | 1 |
| miR-6833-3p | 0.1844221 | 0.20065178 | 0.176 | 1 |
| miR-610 | 0.18407587 | 0.20109406 | 0.224 | 1 |
| miR-8088 | 0.18399963 | 0.20119153 | 0.181 | 1 |
| miR-4723-3p | 0.18288558 | 0.20261902 | 0.173 | 1 |
| miR-4736 | 0.18282936 | 0.20269123 | 0.206 | 1 |
| miR-6884-3p | 0.18214643 | 0.20356948 | 0.222 | 1 |
| miR-1185-5p | 0.18198156 | 0.20378184 | 0.205 | 1 |
| miR-4280 | 0.18053431 | 0.20565164 | 0.203 | 1 |

| | | | | |
|---|---|---|---|---|
| miR-3177-3p | 0.18034995 | 0.20589056 | 0.188 | 1 |
| miR-7157-5p | 0.1790124 | 0.20762885 | 0.205 | 1 |
| miR-3938 | 0.17857562 | 0.20819837 | 0.197 | 1 |
| miR-579-3p | 0.17825156 | 0.20862151 | 0.214 | 1 |
| miR-3199 | 0.17812555 | 0.20878618 | 0.202 | 1 |
| miR-3912-5p | 0.17698489 | 0.21028029 | 0.196 | 1 |
| miR-32-5p | 0.17692492 | 0.21035902 | 0.215 | 1 |
| miR-1307-3p | 0.17684505 | 0.21046389 | 0.2 | 1 |
| miR-500a-5p | 0.17641179 | 0.21103335 | 0.191 | 1 |
| miR-137 | 0.17534002 | 0.21244588 | 0.236 | 1 |
| miR-6844 | 0.17528416 | 0.21251966 | 0.195 | 1 |
| miR-4315 | 0.17497654 | 0.21292618 | 0.199 | 1 |
| miR-7-1-3p | 0.17489988 | 0.21302756 | 0.218 | 1 |
| miR-411-5p | 0.17445432 | 0.21361735 | 0.187 | 1 |
| miR-1236-3p | 0.1740645 | 0.21413413 | 0.217 | 1 |
| miR-3679-3p | 0.1736314 | 0.21470913 | 0.198 | 1 |
| miR-31-5p | 0.17348696 | 0.2149011 | 0.214 | 1 |
| miR-6727-5p | 0.1731829 | 0.21530553 | 0.245 | 1 |
| miR-6736-3p | 0.17264228 | 0.21602572 | 0.204 | 1 |
| miR-1296-3p | 0.17234427 | 0.21642329 | 0.211 | 1 |
| miR-4758-5p | 0.1720199 | 0.21685653 | 0.213 | 1 |
| miR-200c-5p | 0.17181392 | 0.2171319 | 0.198 | 1 |
| miR-6854-3p | 0.17139693 | 0.21768997 | 0.208 | 1 |
| miR-4726-5p | 0.17108854 | 0.21810325 | 0.197 | 1 |
| miR-6819-5p | 0.17103732 | 0.21817193 | 0.207 | 1 |
| miR-98-3p | 0.17093474 | 0.21830951 | 0.18 | 1 |
| miR-541-3p | 0.17069083 | 0.21863688 | 0.219 | 1 |
| let-7c-3p | 0.17062685 | 0.21872279 | 0.223 | 1 |
| miR-27b-3p | 0.16995307 | 0.21962876 | 0.228 | 1 |
| miR-6829-5p | 0.16953003 | 0.22019869 | 0.206 | 1 |
| miR-5094 | 0.16952254 | 0.22020878 | 0.218 | 1 |
| miR-548as-3p | 0.16948664 | 0.22025719 | 0.217 | 1 |
| miR-501-3p | 0.16946377 | 0.22028803 | 0.23 | 1 |
| miR-552-5p | 0.16924408 | 0.2205844 | 0.231 | 1 |
| miR-1304-3p | 0.16790635 | 0.22239397 | 0.246 | 1 |
| miR-3122 | 0.16748533 | 0.22296523 | 0.229 | 1 |
| miR-2861 | 0.16745939 | 0.22300046 | 0.213 | 1 |
| miR-532-3p | 0.16651067 | 0.22429092 | 0.234 | 1 |
| miR-4525 | 0.16558976 | 0.2255476 | 0.215 | 1 |
| miR-670-5p | 0.16525976 | 0.22599889 | 0.242 | 1 |
| miR-320b | 0.16515988 | 0.22613558 | 0.223 | 1 |
| miR-514b-3p | 0.16476438 | 0.2266773 | 0.24 | 1 |
| miR-4431 | 0.16473498 | 0.22671759 | 0.241 | 1 |
| miR-3154 | 0.16440969 | 0.22716374 | 0.213 | 1 |
| miR-4419b | 0.16429862 | 0.22731619 | 0.222 | 1 |
| miR-6823-3p | 0.16422522 | 0.22741697 | 0.231 | 1 |
| miR-5586-3p | 0.16379804 | 0.22800398 | 0.214 | 1 |
| miR-4672 | 0.1633594 | 0.22860762 | 0.22 | 1 |
| miR-561-5p | 0.16295748 | 0.22916151 | 0.242 | 1 |
| miR-1909-5p | 0.16282285 | 0.22934722 | 0.217 | 1 |

| | | | | |
|---|---|---|---|---|
| miR-4276 | 0.16272407 | 0.22948353 | 0.256 | 1 |
| miR-5009-5p | 0.16261186 | 0.22963843 | 0.238 | 1 |
| miR-6500-3p | 0.16189994 | 0.23062253 | 0.222 | 1 |
| miR-548az-5p | 0.16164504 | 0.23097545 | 0.2 | 1 |
| miR-6760-5p | 0.16063532 | 0.23237644 | 0.239 | 1 |
| miR-3133 | 0.16003616 | 0.23321 | 0.239 | 1 |
| miR-4289 | 0.15943078 | 0.2340539 | 0.24 | 1 |
| miR-185-3p | 0.1591346 | 0.23446739 | 0.236 | 1 |
| miR-320d | 0.15904007 | 0.23459945 | 0.235 | 1 |
| miR-891a-5p | 0.15881231 | 0.2349178 | 0.207 | 1 |
| miR-485-3p | 0.15855148 | 0.23528266 | 0.273 | 1 |
| miR-8074 | 0.15805154 | 0.23598286 | 0.246 | 1 |
| miR-6834-3p | 0.15758026 | 0.23664398 | 0.217 | 1 |
| miR-125b-1-3 | 0.15753556 | 0.23670675 | 0.245 | 1 |
| miR-6853-5p | 0.15732229 | 0.2370063 | 0.242 | 1 |
| miR-4646-5p | 0.15694587 | 0.23753552 | 0.252 | 1 |
| miR-363-3p | 0.1566486 | 0.23795392 | 0.226 | 1 |
| miR-933 | 0.15596811 | 0.23891322 | 0.208 | 1 |
| miR-301a-5p | 0.15507176 | 0.24018004 | 0.25 | 1 |
| miR-484 | 0.15396178 | 0.24175384 | 0.24 | 1 |
| miR-585-3p | 0.15347906 | 0.24244001 | 0.24 | 1 |
| miR-6747-5p | 0.15329937 | 0.24269571 | 0.247 | 1 |
| miR-769-3p | 0.15322592 | 0.24280027 | 0.223 | 1 |
| miR-8055 | 0.15261745 | 0.24366738 | 0.268 | 1 |
| miR-3975 | 0.15227902 | 0.2441504 | 0.245 | 1 |
| miR-7161-3p | 0.15189968 | 0.24469242 | 0.257 | 1 |
| miR-568 | 0.15188737 | 0.24471002 | 0.257 | 1 |
| miR-3926 | 0.15142967 | 0.24536488 | 0.237 | 1 |
| miR-6516-5p | 0.15119913 | 0.24569509 | 0.213 | 1 |
| miR-183-5p | 0.15041928 | 0.24681386 | 0.241 | 1 |
| miR-4762-5p | 0.15036434 | 0.24689277 | 0.239 | 1 |
| miR-18b-3p | 0.15008009 | 0.24730129 | 0.231 | 1 |
| miR-6134 | 0.1497372 | 0.24779459 | 0.265 | 1 |
| miR-4709-3p | 0.14922157 | 0.24853737 | 0.257 | 1 |
| miR-6855-3p | 0.14904257 | 0.2487955 | 0.232 | 1 |
| miR-6890-5p | 0.14840868 | 0.24971077 | 0.236 | 1 |
| miR-1324 | 0.14822096 | 0.24998216 | 0.25 | 1 |
| miR-4522 | 0.14820888 | 0.24999962 | 0.256 | 1 |
| miR-1282 | 0.14787493 | 0.25048283 | 0.232 | 1 |
| miR-518c-3p | 0.14785173 | 0.25051641 | 0.259 | 1 |
| miR-29a-5p | 0.14710369 | 0.25160064 | 0.26 | 1 |
| miR-876-3p | 0.14681956 | 0.25201311 | 0.225 | 1 |
| miR-4256 | 0.14660112 | 0.25233047 | 0.239 | 1 |
| miR-8071 | 0.14646038 | 0.25253505 | 0.278 | 1 |
| miR-664b-5p | 0.14605718 | 0.25312162 | 0.271 | 1 |
| miR-543 | 0.14571575 | 0.2536189 | 0.258 | 1 |
| miR-3672 | 0.14538716 | 0.25409796 | 0.25 | 1 |
| miR-8085 | 0.14535431 | 0.25414588 | 0.226 | 1 |
| miR-578 | 0.14518983 | 0.25438588 | 0.226 | 1 |
| miR-890 | 0.14513745 | 0.25446234 | 0.259 | 1 |

| | | | | |
|---|---|---|---|---|
| miR-3177-5p | 0.1444213 | 0.25550885 | 0.262 | 1 |
| miR-548e-5p | 0.14434276 | 0.25562376 | 0.275 | 1 |
| miR-24-2-5p | 0.14370889 | 0.25655213 | 0.288 | 1 |
| miR-150-3p | 0.1436445 | 0.25664654 | 0.228 | 1 |
| miR-4733-3p | 0.14347162 | 0.25690008 | 0.264 | 1 |
| miR-379-5p | 0.1423435 | 0.25855784 | 0.259 | 1 |
| miR-6515-5p | 0.142157 | 0.25883243 | 0.253 | 1 |
| miR-769-5p | 0.14117016 | 0.2602879 | 0.27 | 1 |
| miR-5187-5p | 0.1405095 | 0.26126464 | 0.278 | 1 |
| miR-489-3p | 0.14038076 | 0.2614552 | 0.233 | 1 |
| miR-4311 | 0.14017934 | 0.26175347 | 0.247 | 1 |
| miR-6820-3p | 0.13994341 | 0.26210306 | 0.267 | 1 |
| miR-18b-5p | 0.13993788 | 0.26211127 | 0.261 | 1 |
| miR-18a-3p | 0.13985823 | 0.26222935 | 0.244 | 1 |
| miR-495-3p | 0.13983435 | 0.26226474 | 0.25 | 1 |
| miR-134-5p | 0.13969796 | 0.26246703 | 0.24 | 1 |
| miR-7705 | 0.1391215 | 0.26332282 | 0.255 | 1 |
| miR-548ax | 0.1383771 | 0.26443006 | 0.268 | 1 |
| miR-8080 | 0.13802791 | 0.26495026 | 0.289 | 1 |
| miR-589-5p | 0.13751105 | 0.2657212 | 0.271 | 1 |
| miR-6849-5p | 0.13731128 | 0.26601949 | 0.285 | 1 |
| miR-30e-3p | 0.13726481 | 0.2660889 | 0.262 | 1 |
| miR-552-3p | 0.1370771 | 0.26636936 | 0.272 | 1 |
| miR-150-5p | 0.13683462 | 0.26673189 | 0.255 | 1 |
| miR-4768-5p | 0.13635632 | 0.26744768 | 0.261 | 1 |
| miR-99b-3p | 0.13622753 | 0.2676406 | 0.279 | 1 |
| miR-5582-5p | 0.13611266 | 0.26781271 | 0.278 | 1 |
| miR-196b-5p | 0.13598811 | 0.26799941 | 0.285 | 1 |
| miR-500a-3p | 0.13592676 | 0.26809139 | 0.248 | 1 |
| miR-3129-3p | 0.13504458 | 0.26941578 | 0.253 | 1 |
| miR-556-3p | 0.13467271 | 0.26997502 | 0.261 | 1 |
| miR-100-5p | 0.13435061 | 0.27045991 | 0.268 | 1 |
| miR-181b-5p | 0.13434364 | 0.2704704 | 0.248 | 1 |
| miR-548g-3p | 0.13432332 | 0.27050101 | 0.29 | 1 |
| miR-375 | 0.13431744 | 0.27050986 | 0.269 | 1 |
| miR-766-3p | 0.13403749 | 0.27093167 | 0.297 | 1 |
| miR-181a-5p | 0.13358633 | 0.27161215 | 0.288 | 1 |
| miR-4669 | 0.13287354 | 0.27268897 | 0.271 | 1 |
| miR-1471 | 0.13263093 | 0.27305598 | 0.26 | 1 |
| miR-6873-5p | 0.13243714 | 0.27334929 | 0.264 | 1 |
| miR-195-5p | 0.13192366 | 0.27412726 | 0.286 | 1 |
| miR-411-3p | 0.13093019 | 0.27563555 | 0.257 | 1 |
| miR-6130 | 0.13037884 | 0.27647436 | 0.296 | 1 |
| miR-937-3p | 0.13002081 | 0.27701973 | 0.314 | 1 |
| miR-6794-3p | 0.12993206 | 0.277155 | 0.264 | 1 |
| miR-1272 | 0.12978127 | 0.27738491 | 0.27 | 1 |
| miR-1238-3p | 0.12957568 | 0.27769851 | 0.297 | 1 |
| miR-3116 | 0.12932776 | 0.27807692 | 0.241 | 1 |
| miR-676-3p | 0.12895572 | 0.27864524 | 0.293 | 1 |
| miR-4778-3p | 0.12884519 | 0.2788142 | 0.274 | 1 |

| | | | | |
|---|---|---|---|---|
| miR-938 | 0.12859254 | 0.27920057 | 0.3 | 1 |
| miR-598-3p | 0.12800825 | 0.28009513 | 0.275 | 1 |
| let-7f-5p | 0.12792164 | 0.28022786 | 0.282 | 1 |
| miR-591 | 0.12698895 | 0.28165902 | 0.3 | 1 |
| miR-5739 | 0.12654712 | 0.28233821 | 0.281 | 1 |
| miR-30c-1-3p | 0.12601483 | 0.28315751 | 0.263 | 1 |
| miR-6812-5p | 0.12584843 | 0.28341386 | 0.276 | 1 |
| miR-105-5p | 0.12565884 | 0.28370609 | 0.305 | 1 |
| miR-15b-3p | 0.12550016 | 0.28395078 | 0.287 | 1 |
| miR-6791-5p | 0.12542429 | 0.28406779 | 0.256 | 1 |
| miR-4474-3p | 0.12508727 | 0.28458793 | 0.282 | 1 |
| miR-616-5p | 0.12501398 | 0.28470109 | 0.296 | 1 |
| miR-143-3p | 0.12486605 | 0.28492958 | 0.295 | 1 |
| miR-7515 | 0.12477111 | 0.28507627 | 0.279 | 1 |
| miR-6802-5p | 0.12454751 | 0.28542189 | 0.292 | 1 |
| miR-6852-5p | 0.1243336 | 0.28575272 | 0.273 | 1 |
| miR-6085 | 0.12397645 | 0.28630547 | 0.282 | 1 |
| miR-6774-5p | 0.12364562 | 0.28681796 | 0.3 | 1 |
| miR-6772-5p | 0.12362709 | 0.28684667 | 0.283 | 1 |
| miR-506-3p | 0.12180514 | 0.2896769 | 0.298 | 1 |
| miR-145-5p | 0.12146171 | 0.29021184 | 0.296 | 1 |
| miR-1252-5p | 0.12069571 | 0.29140668 | 0.289 | 1 |
| miR-8056 | 0.12014697 | 0.29226404 | 0.301 | 1 |
| miR-3692-5p | 0.11989333 | 0.29266072 | 0.283 | 1 |
| miR-199b-5p | 0.1196213 | 0.29308644 | 0.304 | 1 |
| miR-4467 | 0.11944306 | 0.29336555 | 0.278 | 1 |
| miR-92a-1-5p | 0.11939889 | 0.29343472 | 0.286 | 1 |
| miR-888-3p | 0.11931631 | 0.2935641 | 0.311 | 1 |
| miR-5687 | 0.11902426 | 0.29402181 | 0.28 | 1 |
| miR-3131 | 0.11884142 | 0.29430855 | 0.284 | 1 |
| miR-6752-5p | 0.11878824 | 0.29439197 | 0.291 | 1 |
| miR-7107-3p | 0.11841428 | 0.29497887 | 0.293 | 1 |
| miR-6133 | 0.11804039 | 0.29556622 | 0.294 | 1 |
| miR-3149 | 0.11748375 | 0.29644165 | 0.319 | 1 |
| miR-372-3p | 0.11713521 | 0.29699039 | 0.312 | 1 |
| miR-6849-3p | 0.11697715 | 0.29723939 | 0.305 | 1 |
| miR-4664-3p | 0.11685781 | 0.29742747 | 0.284 | 1 |
| miR-6846-3p | 0.11683124 | 0.29746935 | 0.282 | 1 |
| miR-4691-5p | 0.11620538 | 0.2984566 | 0.29 | 1 |
| miR-151a-3p | 0.11584593 | 0.29902429 | 0.294 | 1 |
| miR-2909 | 0.11522045 | 0.30001329 | 0.301 | 1 |
| miR-3135a | 0.11444696 | 0.30123836 | 0.333 | 1 |
| miR-128-2-5p | 0.11399322 | 0.30195805 | 0.289 | 1 |
| miR-4711-3p | 0.11383327 | 0.30221192 | 0.309 | 1 |
| miR-4639-5p | 0.11375103 | 0.30234251 | 0.322 | 1 |
| miR-3928-5p | 0.11361588 | 0.30255714 | 0.314 | 1 |
| miR-6758-5p | 0.11344485 | 0.30282885 | 0.295 | 1 |
| miR-3944-5p | 0.11343007 | 0.30285235 | 0.282 | 1 |
| miR-4468 | 0.11338496 | 0.30292403 | 0.286 | 1 |
| miR-6124 | 0.11288786 | 0.30371451 | 0.321 | 1 |

| | | | | |
|---|---|---|---|---|
| miR-4308 | 0.11239359 | 0.3045014 | 0.3 | 1 |
| miR-4632-5p | 0.11169817 | 0.30561006 | 0.292 | 1 |
| miR-3978 | 0.11140621 | 0.30607604 | 0.287 | 1 |
| miR-4505 | 0.11083406 | 0.30699013 | 0.314 | 1 |
| miR-4794 | 0.11075179 | 0.30712168 | 0.306 | 1 |
| miR-3153 | 0.11065159 | 0.30728191 | 0.337 | 1 |
| miR-29a-3p | 0.1103818 | 0.30771354 | 0.311 | 1 |
| miR-5685 | 0.11033637 | 0.30778625 | 0.299 | 1 |
| miR-2467-3p | 0.11027682 | 0.30788157 | 0.31 | 1 |
| miR-4519 | 0.11017444 | 0.30804546 | 0.303 | 1 |
| miR-6859-5p | 0.10981365 | 0.30862337 | 0.301 | 1 |
| miR-331-3p | 0.10975335 | 0.30872 | 0.327 | 1 |
| miR-6881-5p | 0.10947042 | 0.30917358 | 0.283 | 1 |
| miR-935 | 0.10911474 | 0.30974421 | 0.29 | 1 |
| miR-4330 | 0.10851883 | 0.31070127 | 0.305 | 1 |
| miR-649 | 0.10825014 | 0.31113321 | 0.309 | 1 |
| miR-6746-5p | 0.10807016 | 0.31142269 | 0.322 | 1 |
| miR-2682-3p | 0.10781601 | 0.31183168 | 0.318 | 1 |
| miR-1256 | 0.10755544 | 0.31225122 | 0.331 | 1 |
| miR-5189-5p | 0.1072259 | 0.31278218 | 0.289 | 1 |
| miR-376b-5p | 0.10709291 | 0.31299655 | 0.313 | 1 |
| miR-5690 | 0.10691365 | 0.31328563 | 0.289 | 1 |
| miR-7111-5p | 0.10657983 | 0.31382424 | 0.305 | 1 |
| miR-4668-3p | 0.10652097 | 0.31391926 | 0.297 | 1 |
| miR-770-5p | 0.10648441 | 0.31397828 | 0.344 | 1 |
| miR-6854-5p | 0.10617543 | 0.31447726 | 0.293 | 1 |
| miR-5682 | 0.10559305 | 0.31541871 | 0.323 | 1 |
| miR-586 | 0.10552168 | 0.31553416 | 0.322 | 1 |
| miR-421 | 0.10524417 | 0.31598327 | 0.3 | 1 |
| miR-4704-5p | 0.10482758 | 0.31665795 | 0.303 | 1 |
| miR-127-3p | 0.10480924 | 0.31668767 | 0.318 | 1 |
| miR-889-3p | 0.10475884 | 0.31676935 | 0.319 | 1 |
| miR-129-5p | 0.1041484 | 0.31775924 | 0.318 | 1 |
| miR-3936 | 0.10397174 | 0.31804596 | 0.316 | 1 |
| miR-4447 | 0.10341132 | 0.31895623 | 0.304 | 1 |
| miR-6852-3p | 0.10327443 | 0.31917873 | 0.324 | 1 |
| miR-3137 | 0.10310832 | 0.31944882 | 0.323 | 1 |
| miR-6722-5p | 0.10299737 | 0.31962928 | 0.335 | 1 |
| miR-3919 | 0.10207597 | 0.32112957 | 0.319 | 1 |
| miR-548al | 0.10205397 | 0.32116544 | 0.353 | 1 |
| miR-514a-5p | 0.10167499 | 0.3217834 | 0.334 | 1 |
| miR-6851-3p | 0.10077977 | 0.32324509 | 0.317 | 1 |
| miR-4446-3p | 0.10043309 | 0.32381188 | 0.332 | 1 |
| miR-146b-5p | 0.10020669 | 0.32418224 | 0.324 | 1 |
| miR-4728-3p | 0.1000957 | 0.32436387 | 0.305 | 1 |
| miR-450a-2-3p | 0.09981659 | 0.3248208 | 0.314 | 1 |
| miR-1915-3p | 0.09880499 | 0.32647912 | 0.345 | 1 |
| miR-4650-3p | 0.09870863 | 0.32663725 | 0.318 | 1 |
| miR-92b-3p | 0.09859067 | 0.32683088 | 0.351 | 1 |
| miR-361-5p | 0.09856931 | 0.32686595 | 0.312 | 1 |

| | | | | |
|---|---|---|---|---|
| miR-2116-3p | 0.09839516 | 0.32715191 | 0.334 | 1 |
| miR-4735-5p | 0.09773824 | 0.32823155 | 0.294 | 1 |
| miR-486-5p | 0.09740994 | 0.32877164 | 0.305 | 1 |
| miR-5195-3p | 0.09717093 | 0.32916506 | 0.327 | 1 |
| miR-515-5p | 0.09698007 | 0.32947935 | 0.346 | 1 |
| miR-6875-5p | 0.0969427 | 0.32954091 | 0.318 | 1 |
| miR-7151-5p | 0.09625004 | 0.33068265 | 0.328 | 1 |
| miR-4517 | 0.09595048 | 0.33117691 | 0.335 | 1 |
| miR-4670-5p | 0.09514552 | 0.33250654 | 0.32 | 1 |
| miR-223-5p | 0.0946491 | 0.33332758 | 0.316 | 1 |
| miR-548l | 0.09420368 | 0.33406493 | 0.341 | 1 |
| miR-4274 | 0.0935508 | 0.33514688 | 0.316 | 1 |
| miR-106b-5p | 0.09340115 | 0.33539509 | 0.321 | 1 |
| miR-3689a-3p | 0.09330464 | 0.33555519 | 0.335 | 1 |
| miR-200a-5p | 0.09296997 | 0.33611059 | 0.315 | 1 |
| miR-15a-5p | 0.09282478 | 0.33635167 | 0.33 | 1 |
| miR-664b-3p | 0.09280458 | 0.33638522 | 0.33 | 1 |
| miR-5688 | 0.09213785 | 0.33749311 | 0.326 | 1 |
| miR-196a-3p | 0.09204263 | 0.33765146 | 0.339 | 1 |
| miR-6824-5p | 0.09151468 | 0.33852993 | 0.318 | 1 |
| miR-7160-5p | 0.09121752 | 0.33902476 | 0.339 | 1 |
| miR-6131 | 0.09046599 | 0.34027746 | 0.328 | 1 |
| miR-4432 | 0.09015565 | 0.34079526 | 0.327 | 1 |
| miR-6775-3p | 0.08949659 | 0.3418959 | 0.326 | 1 |
| miR-4489 | 0.08949483 | 0.34189884 | 0.355 | 1 |
| miR-3613-5p | 0.08910122 | 0.34255684 | 0.324 | 1 |
| miR-6893-3p | 0.08890291 | 0.34288852 | 0.347 | 1 |
| miR-6840-5p | 0.0886424 | 0.34332444 | 0.34 | 1 |
| miR-487b-3p | 0.08855822 | 0.34346533 | 0.328 | 1 |
| miR-3689a-5p | 0.08808146 | 0.34426375 | 0.326 | 1 |
| miR-6506-3p | 0.08763556 | 0.34501111 | 0.349 | 1 |
| miR-3945 | 0.08745471 | 0.3453144 | 0.345 | 1 |
| miR-892b | 0.08720291 | 0.34573686 | 0.357 | 1 |
| miR-4473 | 0.08708043 | 0.3459424 | 0.352 | 1 |
| miR-22-5p | 0.08527998 | 0.34896926 | 0.365 | 1 |
| miR-4668-5p | 0.08503811 | 0.34937661 | 0.325 | 1 |
| miR-21-3p | 0.08489312 | 0.34962089 | 0.332 | 1 |
| miR-297 | 0.08474944 | 0.34986302 | 0.377 | 1 |
| miR-6872-3p | 0.08471491 | 0.34992122 | 0.342 | 1 |
| miR-6825-3p | 0.08456512 | 0.35017373 | 0.378 | 1 |
| miR-4472 | 0.08440335 | 0.35044651 | 0.357 | 1 |
| miR-7844-5p | 0.08435249 | 0.35053227 | 0.338 | 1 |
| miR-2355-5p | 0.08428251 | 0.35065032 | 0.324 | 1 |
| miR-3126-3p | 0.08384807 | 0.3513834 | 0.356 | 1 |
| miR-6783-5p | 0.08349311 | 0.35198279 | 0.376 | 1 |
| miR-487b-5p | 0.08301404 | 0.35279233 | 0.351 | 1 |
| miR-4705 | 0.0828654 | 0.35304364 | 0.325 | 1 |
| miR-6827-3p | 0.08273394 | 0.35326596 | 0.346 | 1 |
| miR-4303 | 0.08184912 | 0.35476362 | 0.337 | 1 |
| miR-4728-5p | 0.08080353 | 0.35653629 | 0.342 | 1 |

| | | | | |
|---|---|---|---|---|
| miR-4686 | 0.08068845 | 0.35673159 | 0.364 | 1 |
| miR-508-5p | 0.08049475 | 0.35706038 | 0.362 | 1 |
| miR-141-5p | 0.08028481 | 0.35741687 | 0.37 | 1 |
| miR-5589-3p | 0.08018411 | 0.3575879 | 0.338 | 1 |
| miR-2467-5p | 0.07966747 | 0.35846585 | 0.346 | 1 |
| miR-15b-5p | 0.07951939 | 0.35871764 | 0.355 | 1 |
| miR-5192 | 0.07934358 | 0.35901664 | 0.36 | 1 |
| miR-4761-3p | 0.07929044 | 0.35910703 | 0.385 | 1 |
| miR-4498 | 0.07916501 | 0.35932042 | 0.354 | 1 |
| miR-410-3p | 0.07912641 | 0.3593861 | 0.366 | 1 |
| miR-342-3p | 0.07898668 | 0.35962389 | 0.34 | 1 |
| miR-660-5p | 0.07854427 | 0.36037713 | 0.37 | 1 |
| miR-3680-5p | 0.07827398 | 0.3608376 | 0.354 | 1 |
| miR-4277 | 0.07750976 | 0.3621406 | 0.327 | 1 |
| miR-376a-2-5 | 0.07708548 | 0.36286469 | 0.375 | 1 |
| miR-1249 | 0.07700325 | 0.36300508 | 0.351 | 1 |
| miR-6789-3p | 0.07694922 | 0.36309733 | 0.377 | 1 |
| miR-3684 | 0.07685298 | 0.3632617 | 0.369 | 1 |
| miR-4714-5p | 0.07677121 | 0.36340134 | 0.359 | 1 |
| miR-623 | 0.07665762 | 0.36359538 | 0.36 | 1 |
| miR-20b-3p | 0.07638966 | 0.36405326 | 0.336 | 1 |
| miR-1305 | 0.07636542 | 0.36409468 | 0.354 | 1 |
| miR-8068 | 0.07627813 | 0.36424389 | 0.372 | 1 |
| miR-101-3p | 0.07625142 | 0.36428955 | 0.364 | 1 |
| miR-6503-5p | 0.07566889 | 0.36528584 | 0.373 | 1 |
| miR-300 | 0.0756502 | 0.36531781 | 0.387 | 1 |
| miR-425-3p | 0.07534969 | 0.36583213 | 0.359 | 1 |
| miR-556-5p | 0.07530491 | 0.36590879 | 0.373 | 1 |
| miR-4535 | 0.0750373 | 0.36636705 | 0.345 | 1 |
| miR-6778-5p | 0.07498755 | 0.36645226 | 0.356 | 1 |
| miR-6821-3p | 0.07426557 | 0.36768959 | 0.365 | 1 |
| miR-618 | 0.07417416 | 0.36784635 | 0.345 | 1 |
| miR-6837-3p | 0.07402454 | 0.36810298 | 0.337 | 1 |
| miR-6818-3p | 0.0738973 | 0.36832127 | 0.369 | 1 |
| miR-4440 | 0.07386459 | 0.3683774 | 0.348 | 1 |
| miR-3155a | 0.07378822 | 0.36850844 | 0.352 | 1 |
| miR-558 | 0.07354489 | 0.36892608 | 0.375 | 1 |
| miR-3940-5p | 0.0735167 | 0.36897447 | 0.396 | 1 |
| miR-6815-3p | 0.07325218 | 0.36942868 | 0.371 | 1 |
| miR-181d-5p | 0.07316254 | 0.36958264 | 0.384 | 1 |
| miR-148b-3p | 0.0730674 | 0.36974608 | 0.385 | 1 |
| miR-6831-3p | 0.07287673 | 0.37007367 | 0.368 | 1 |
| miR-920 | 0.07238216 | 0.37092386 | 0.357 | 1 |
| miR-6773-3p | 0.07219462 | 0.37124642 | 0.369 | 1 |
| miR-3159 | 0.07214951 | 0.37132402 | 0.349 | 1 |
| miR-5707 | 0.07207584 | 0.37145076 | 0.378 | 1 |
| miR-3660 | 0.07207018 | 0.3714605 | 0.358 | 1 |
| miR-7976 | 0.07206031 | 0.37147747 | 0.364 | 1 |
| miR-4798-3p | 0.07131182 | 0.37276599 | 0.378 | 1 |
| miR-492 | 0.07100741 | 0.37329043 | 0.364 | 1 |

| | | | | |
|---|---|---|---|---|
| miR-6857-3p | 0.07080149 | 0.37364531 | 0.358 | 1 |
| miR-617 | 0.0707062 | 0.37380958 | 0.369 | 1 |
| miR-153-5p | 0.07055919 | 0.37406305 | 0.392 | 1 |
| miR-1269a | 0.07016311 | 0.37474623 | 0.383 | 1 |
| miR-583 | 0.07009496 | 0.37486381 | 0.406 | 1 |
| miR-6810-5p | 0.06992488 | 0.37515733 | 0.357 | 1 |
| miR-4774-3p | 0.06949518 | 0.37589918 | 0.354 | 1 |
| miR-519b-3p | 0.06907615 | 0.37662307 | 0.372 | 1 |
| miR-5693 | 0.06892618 | 0.37688225 | 0.365 | 1 |
| miR-6737-3p | 0.06855843 | 0.37751803 | 0.361 | 1 |
| miR-6887-5p | 0.06835102 | 0.37787675 | 0.347 | 1 |
| miR-6867-3p | 0.0676503 | 0.37908949 | 0.364 | 1 |
| miR-4271 | 0.06721439 | 0.3798445 | 0.374 | 1 |
| miR-6842-3p | 0.06688309 | 0.38041864 | 0.38 | 1 |
| miR-941 | 0.06643207 | 0.38120068 | 0.385 | 1 |
| miR-6740-3p | 0.06636104 | 0.38132387 | 0.381 | 1 |
| miR-6822-5p | 0.06617068 | 0.38165412 | 0.378 | 1 |
| miR-6720-3p | 0.06589103 | 0.38213943 | 0.402 | 1 |
| miR-4660 | 0.06588607 | 0.38214805 | 0.364 | 1 |
| miR-6848-3p | 0.06536591 | 0.38305123 | 0.362 | 1 |
| miR-6088 | 0.06429933 | 0.38490519 | 0.389 | 1 |
| miR-3161 | 0.06371281 | 0.38592583 | 0.387 | 1 |
| miR-138-1-3p | 0.06368838 | 0.38596837 | 0.398 | 1 |
| miR-744-3p | 0.06347828 | 0.38633417 | 0.375 | 1 |
| miR-362-5p | 0.0628137 | 0.38749195 | 0.394 | 1 |
| miR-432-3p | 0.06262351 | 0.38782347 | 0.375 | 1 |
| miR-490-5p | 0.06222359 | 0.38852084 | 0.405 | 1 |
| miR-1237-5p | 0.06191189 | 0.38906461 | 0.374 | 1 |
| miR-767-3p | 0.06030604 | 0.39186954 | 0.394 | 1 |
| miR-193a-5p | 0.06013229 | 0.39217337 | 0.387 | 1 |
| miR-449b-3p | 0.05961731 | 0.39307428 | 0.382 | 1 |
| miR-6733-5p | 0.05935509 | 0.39353323 | 0.384 | 1 |
| miR-7107-5p | 0.05909328 | 0.39399159 | 0.379 | 1 |
| miR-3200-5p | 0.05897865 | 0.39419234 | 0.382 | 1 |
| miR-563 | 0.0587864 | 0.39452908 | 0.408 | 1 |
| miR-620 | 0.05801373 | 0.39588322 | 0.413 | 1 |
| miR-943 | 0.05789058 | 0.39609917 | 0.388 | 1 |
| miR-501-5p | 0.05772844 | 0.39638353 | 0.392 | 1 |
| miR-4692 | 0.05753235 | 0.39672751 | 0.38 | 1 |
| miR-6785-3p | 0.05691754 | 0.39780651 | 0.39 | 1 |
| miR-3649 | 0.05656167 | 0.39843144 | 0.383 | 1 |
| miR-668-3p | 0.05616246 | 0.39913277 | 0.384 | 1 |
| miR-1912 | 0.05575518 | 0.39984863 | 0.385 | 1 |
| miR-2355-3p | 0.05526872 | 0.40070409 | 0.377 | 1 |
| miR-517-5p | 0.05518898 | 0.40084435 | 0.369 | 1 |
| miR-373-3p | 0.05469919 | 0.40170623 | 0.402 | 1 |
| miR-4637 | 0.05461616 | 0.40185239 | 0.383 | 1 |
| miR-548s | 0.05458236 | 0.4019119 | 0.384 | 1 |
| miR-621 | 0.05319997 | 0.40434734 | 0.408 | 1 |
| miR-10b-3p | 0.05308725 | 0.40454611 | 0.435 | 1 |

| | | | | |
|---|---|---|---|---|
| miR-204-3p | 0.05298723 | 0.40472247 | 0.413 | 1 |
| miR-641 | 0.05294015 | 0.40480551 | 0.397 | 1 |
| miR-4263 | 0.05273267 | 0.40517146 | 0.383 | 1 |
| miR-5580-3p | 0.05259274 | 0.40541833 | 0.37 | 1 |
| miR-106a-5p | 0.05254399 | 0.40550434 | 0.437 | 1 |
| miR-6878-3p | 0.05249566 | 0.40558961 | 0.412 | 1 |
| miR-1304-5p | 0.05202884 | 0.40641352 | 0.415 | 1 |
| miR-191-3p | 0.05187773 | 0.4066803 | 0.415 | 1 |
| miR-6502-3p | 0.05137757 | 0.40756365 | 0.429 | 1 |
| miR-204-5p | 0.051338 | 0.40763356 | 0.417 | 1 |
| miR-29c-3p | 0.05074494 | 0.40868164 | 0.426 | 1 |
| miR-302c-5p | 0.05049131 | 0.40913007 | 0.404 | 1 |
| miR-302a-5p | 0.05015712 | 0.4097211 | 0.408 | 1 |
| miR-133a-3p | 0.05001756 | 0.409968 | 0.396 | 1 |
| miR-520e | 0.04968922 | 0.41054896 | 0.413 | 1 |
| miR-4745-3p | 0.04965272 | 0.41061356 | 0.407 | 1 |
| miR-6757-5p | 0.04957246 | 0.41075561 | 0.413 | 1 |
| miR-6730-5p | 0.0494479 | 0.41097608 | 0.435 | 1 |
| miR-433-5p | 0.04907799 | 0.41163102 | 0.411 | 1 |
| miR-3128 | 0.04851831 | 0.4126224 | 0.426 | 1 |
| miR-4326 | 0.04817564 | 0.41322966 | 0.416 | 1 |
| miR-3655 | 0.04695937 | 0.41538673 | 0.416 | 1 |
| miR-371b-5p | 0.0467257 | 0.41580146 | 0.426 | 1 |
| miR-485-5p | 0.04668096 | 0.41588087 | 0.418 | 1 |
| miR-6811-3p | 0.04666774 | 0.41590433 | 0.422 | 1 |
| miR-6780b-5p | 0.04646474 | 0.41626472 | 0.436 | 1 |
| miR-1293 | 0.04571628 | 0.41759401 | 0.431 | 1 |
| miR-5579-5p | 0.04533564 | 0.41827042 | 0.44 | 1 |
| miR-19a-5p | 0.04522283 | 0.41847093 | 0.394 | 1 |
| miR-26b-3p | 0.04482289 | 0.41918197 | 0.396 | 1 |
| miR-6726-3p | 0.04479031 | 0.4192399 | 0.412 | 1 |
| miR-5003-3p | 0.0446329 | 0.41951984 | 0.403 | 1 |
| miR-4318 | 0.04440953 | 0.41991713 | 0.418 | 1 |
| miR-652-3p | 0.04349133 | 0.42155117 | 0.424 | 1 |
| miR-1306-3p | 0.04341429 | 0.42168833 | 0.402 | 1 |
| miR-4538 | 0.04339395 | 0.42172456 | 0.417 | 1 |
| miR-6165 | 0.04319218 | 0.42208384 | 0.438 | 1 |
| miR-1323 | 0.0431516 | 0.4221561 | 0.448 | 1 |
| miR-7112-3p | 0.04299949 | 0.42242701 | 0.424 | 1 |
| miR-6764-3p | 0.04193588 | 0.42432234 | 0.426 | 1 |
| miR-4433b-5p | 0.04186703 | 0.42444508 | 0.426 | 1 |
| miR-8053 | 0.04176343 | 0.42462979 | 0.398 | 1 |
| miR-3652 | 0.04134421 | 0.42537744 | 0.413 | 1 |
| miR-346 | 0.04101428 | 0.42596601 | 0.413 | 1 |
| miR-381-5p | 0.04053339 | 0.42682421 | 0.407 | 1 |
| miR-181d-3p | 0.04040275 | 0.4270574 | 0.412 | 1 |
| miR-8084 | 0.04012444 | 0.42755428 | 0.406 | 1 |
| miR-4478 | 0.03988536 | 0.42798121 | 0.435 | 1 |
| miR-378b | 0.03958758 | 0.42851308 | 0.457 | 1 |
| miR-3119 | 0.03912798 | 0.42933423 | 0.42 | 1 |

| | | | | |
|---|---|---|---|---|
| miR-6755-5p | 0.03881677 | 0.42989044 | 0.403 | 1 |
| miR-550b-2-5 | 0.03867281 | 0.43014778 | 0.433 | 1 |
| miR-6716-5p | 0.03774742 | 0.43180266 | 0.432 | 1 |
| miR-4771 | 0.03772107 | 0.43184981 | 0.406 | 1 |
| miR-3115 | 0.03757765 | 0.4321064 | 0.449 | 1 |
| miR-4727-3p | 0.03747115 | 0.43229696 | 0.426 | 1 |
| miR-4721 | 0.03740915 | 0.4324079 | 0.4 | 1 |
| miR-4641 | 0.0373048 | 0.43259463 | 0.43 | 1 |
| miR-4729 | 0.03721529 | 0.43275483 | 0.455 | 1 |
| miR-3152-3p | 0.03713521 | 0.43289817 | 0.418 | 1 |
| miR-6779-5p | 0.03665329 | 0.43376087 | 0.42 | 1 |
| miR-33a-5p | 0.03651076 | 0.43401608 | 0.443 | 1 |
| miR-4722-3p | 0.03649684 | 0.43404101 | 0.426 | 1 |
| miR-4482-5p | 0.03636712 | 0.43427331 | 0.413 | 1 |
| miR-6750-3p | 0.0363635 | 0.43427979 | 0.44 | 1 |
| miR-613 | 0.03582624 | 0.43524214 | 0.43 | 1 |
| miR-4744 | 0.03566919 | 0.43552353 | 0.425 | 1 |
| let-7f-1-3p | 0.03561917 | 0.43561315 | 0.434 | 1 |
| miR-5006-3p | 0.03518683 | 0.43638799 | 0.448 | 1 |
| miR-4482-3p | 0.0349792 | 0.43676016 | 0.434 | 1 |
| miR-616-3p | 0.0348438 | 0.43700292 | 0.443 | 1 |
| miR-1911-5p | 0.03410863 | 0.43832133 | 0.434 | 1 |
| miR-4662a-3p | 0.03361304 | 0.43921048 | 0.452 | 1 |
| miR-4760-3p | 0.03244974 | 0.44129881 | 0.453 | 1 |
| miR-6766-5p | 0.03176765 | 0.44252404 | 0.458 | 1 |
| miR-4459 | 0.0315305 | 0.44295016 | 0.428 | 1 |
| miR-4654 | 0.03121051 | 0.44352522 | 0.439 | 1 |
| miR-4689 | 0.03119565 | 0.44355193 | 0.445 | 1 |
| miR-4777-3p | 0.03093162 | 0.44402654 | 0.427 | 1 |
| miR-338-3p | 0.02968214 | 0.44627357 | 0.435 | 1 |
| miR-6798-5p | 0.02960217 | 0.44641744 | 0.452 | 1 |
| miR-7978 | 0.02948861 | 0.44662176 | 0.432 | 1 |
| miR-1976 | 0.02916579 | 0.44720267 | 0.444 | 1 |
| miR-6794-5p | 0.02864781 | 0.44813499 | 0.445 | 1 |
| miR-3682-3p | 0.02847331 | 0.44844915 | 0.415 | 1 |
| miR-154-3p | 0.02816318 | 0.44900754 | 0.437 | 1 |
| miR-4502 | 0.02806412 | 0.44918593 | 0.444 | 1 |
| miR-584-3p | 0.02788838 | 0.44950243 | 0.415 | 1 |
| miR-410-5p | 0.0277926 | 0.44967493 | 0.42 | 1 |
| miR-6734-5p | 0.02777611 | 0.44970463 | 0.456 | 1 |
| miR-4766-5p | 0.02772983 | 0.44978798 | 0.439 | 1 |
| miR-6509-5p | 0.02752902 | 0.45014971 | 0.461 | 1 |
| miR-3141 | 0.02736244 | 0.45044978 | 0.468 | 1 |
| miR-5694 | 0.02723511 | 0.45067918 | 0.436 | 1 |
| miR-377-3p | 0.02691496 | 0.45125605 | 0.454 | 1 |
| miR-3668 | 0.0268899 | 0.45130121 | 0.452 | 1 |
| miR-4319 | 0.02647174 | 0.45205483 | 0.449 | 1 |
| miR-425-5p | 0.02646316 | 0.45207028 | 0.445 | 1 |
| miR-5088-3p | 0.02631891 | 0.45233031 | 0.441 | 1 |
| miR-4666b | 0.02593008 | 0.45303128 | 0.44 | 1 |

| | | | | |
|---|---|---|---|---|
| miR-1296-5p | 0.02508498 | 0.4545553 | 0.451 | 1 |
| miR-6886-5p | 0.02491279 | 0.45486592 | 0.44 | 1 |
| miR-6748-3p | 0.02472326 | 0.45520783 | 0.465 | 1 |
| miR-4636 | 0.02463088 | 0.45537449 | 0.457 | 1 |
| miR-6862-5p | 0.02441933 | 0.4557562 | 0.455 | 1 |
| miR-518b | 0.02428268 | 0.45600277 | 0.456 | 1 |
| miR-7159-3p | 0.02335422 | 0.45767857 | 0.439 | 1 |
| miR-328-3p | 0.02318382 | 0.45798623 | 0.472 | 1 |
| miR-761 | 0.02316243 | 0.45802484 | 0.475 | 1 |
| miR-3686 | 0.02215941 | 0.45983623 | 0.46 | 1 |
| miR-4720-5p | 0.02201026 | 0.46010565 | 0.476 | 1 |
| miR-378a-3p | 0.02162803 | 0.4607962 | 0.488 | 1 |
| miR-5684 | 0.02143447 | 0.46114595 | 0.467 | 1 |
| miR-5696 | 0.02098307 | 0.46196167 | 0.466 | 1 |
| miR-4656 | 0.0204716 | 0.46288617 | 0.472 | 1 |
| miR-3973 | 0.0204548 | 0.46291654 | 0.434 | 1 |
| miR-633 | 0.02015515 | 0.46345826 | 0.49 | 1 |
| miR-6722-3p | 0.02011703 | 0.46352718 | 0.458 | 1 |
| miR-4297 | 0.02009461 | 0.46356771 | 0.482 | 1 |
| miR-6838-3p | 0.01988415 | 0.46394824 | 0.488 | 1 |
| miR-6513-5p | 0.01976234 | 0.46416848 | 0.464 | 1 |
| miR-7109-3p | 0.01974889 | 0.4641928 | 0.442 | 1 |
| miR-4444 | 0.0197145 | 0.46425499 | 0.466 | 1 |
| miR-4730 | 0.01887139 | 0.46577983 | 0.458 | 1 |
| miR-30d-5p | 0.0182556 | 0.46689386 | 0.462 | 1 |
| miR-3138 | 0.01825454 | 0.46689576 | 0.465 | 1 |
| miR-509-3-5p | 0.01817651 | 0.46703695 | 0.464 | 1 |
| miR-18a-5p | 0.01810056 | 0.46717437 | 0.475 | 1 |
| miR-219a-2-3p | 0.01769264 | 0.46791252 | 0.462 | 1 |
| miR-125b-5p | 0.01716172 | 0.46887341 | 0.465 | 1 |
| miR-4707-3p | 0.01712607 | 0.46893794 | 0.459 | 1 |
| miR-6726-5p | 0.01686109 | 0.46941759 | 0.504 | 1 |
| miR-3150b-5p | 0.01669338 | 0.4697212 | 0.477 | 1 |
| miR-6888-5p | 0.01607341 | 0.47084367 | 0.472 | 1 |
| miR-4743-3p | 0.01552484 | 0.47183707 | 0.481 | 1 |
| miR-33a-3p | 0.01544531 | 0.4719811 | 0.465 | 1 |
| miR-758-5p | 0.01483195 | 0.47309205 | 0.484 | 1 |
| miR-6754-5p | 0.0147374 | 0.47326333 | 0.492 | 1 |
| miR-5582-3p | 0.01442437 | 0.47383041 | 0.457 | 1 |
| miR-7152-3p | 0.01424482 | 0.4741557 | 0.489 | 1 |
| miR-520c-3p | 0.01419907 | 0.47423858 | 0.466 | 1 |
| miR-4272 | 0.01359552 | 0.47533218 | 0.482 | 1 |
| miR-2110 | 0.01355979 | 0.47539693 | 0.466 | 1 |
| miR-337-3p | 0.01332621 | 0.47582022 | 0.45 | 1 |
| miR-5589-5p | 0.01312748 | 0.47618036 | 0.463 | 1 |
| miR-6864-5p | 0.01213235 | 0.47798411 | 0.483 | 1 |
| miR-3156-5p | 0.01188701 | 0.47842888 | 0.45 | 1 |
| miR-196a-5p | 0.01153177 | 0.47907293 | 0.48 | 1 |
| miR-4509 | 0.01098741 | 0.48005994 | 0.49 | 1 |
| miR-4527 | 0.01086031 | 0.48029042 | 0.482 | 1 |

| | | | | |
|---|---|---|---|---|
| miR-6768-5p | 0.01028061 | 0.48134169 | 0.484 | 1 |
| miR-191-5p | 0.01023582 | 0.48142291 | 0.504 | 1 |
| miR-147a | 0.00999229 | 0.48186459 | 0.458 | 1 |
| miR-624-3p | 0.00936106 | 0.48300953 | 0.476 | 1 |
| miR-3152-5p | 0.00911983 | 0.48344709 | 0.487 | 1 |
| miR-6807-5p | 0.00910736 | 0.48346972 | 0.467 | 1 |
| miR-4301 | 0.00906799 | 0.48354115 | 0.455 | 1 |
| miR-581 | 0.00900741 | 0.48365103 | 0.463 | 1 |
| miR-5692b | 0.00889224 | 0.48385996 | 0.504 | 1 |
| miR-3175 | 0.0088501 | 0.48393641 | 0.463 | 1 |
| miR-1180-5p | 0.00873575 | 0.48414385 | 0.491 | 1 |
| miR-6825-5p | 0.00774466 | 0.48594198 | 0.467 | 1 |
| miR-4647 | 0.00767129 | 0.48607511 | 0.507 | 1 |
| miR-4723-5p | 0.00759318 | 0.48621682 | 0.495 | 1 |
| miR-5088-5p | 0.00737253 | 0.48661719 | 0.482 | 1 |
| miR-5571-3p | 0.00674116 | 0.4877629 | 0.497 | 1 |
| miR-6839-5p | 0.00656076 | 0.48809027 | 0.485 | 1 |
| miR-4731-3p | 0.00582592 | 0.4894239 | 0.498 | 1 |
| miR-6841-5p | 0.00568411 | 0.48968128 | 0.484 | 1 |
| miR-6769b-3p | 0.00527556 | 0.49042279 | 0.464 | 1 |
| miR-194-3p | 0.00506027 | 0.49081356 | 0.502 | 1 |
| miR-218-2-3p | 0.00495418 | 0.49100612 | 0.494 | 1 |
| miR-572 | 0.00493302 | 0.49104452 | 0.451 | 1 |
| miR-130a-3p | 0.00493158 | 0.49104715 | 0.497 | 1 |
| miR-500b-5p | 0.00487697 | 0.49114626 | 0.492 | 1 |
| miR-580-5p | 0.0047374 | 0.49139961 | 0.495 | 1 |
| miR-6892-5p | 0.00426669 | 0.49225403 | 0.468 | 1 |
| miR-30b-5p | 0.00406315 | 0.4926235 | 0.502 | 1 |
| miR-30a-5p | 0.00404335 | 0.49265944 | 0.499 | 1 |
| miR-5002-5p | 0.00381318 | 0.49307726 | 0.49 | 1 |
| miR-4520a-5p | 0.00359388 | 0.49347537 | 0.483 | 1 |
| miR-23c | 0.0033617 | 0.49389686 | 0.475 | 1 |
| miR-523-3p | 0.00321432 | 0.4941644 | 0.48 | 1 |
| miR-7843-5p | 0.00319813 | 0.49419379 | 0.494 | 1 |
| miR-194-5p | 0.00211269 | 0.49616433 | 0.492 | 1 |
| miR-4504 | 0.00185252 | 0.49663666 | 0.493 | 1 |
| miR-4512 | 0.00148235 | 0.49730872 | 0.528 | 1 |
| miR-95-3p | 0.00144497 | 0.49737657 | 0.536 | 1 |
| miR-30c-5p | 0.0013916 | 0.49747347 | 0.504 | 1 |
| miR-7152-5p | 0.00060376 | 0.49890384 | 0.486 | 1 |
| miR-4257 | 0.00024477 | 0.4995556 | 0.504 | 1 |
| miR-4309 | 0.00017083 | 0.49968984 | 0.488 | 1 |
| miR-378i | 0.00010559 | 0.4998083 | 0.492 | 1 |
| miR-553 | 0 | 0.83864671 | 0.851 | 1 |
| miR-889-5p | 0 | 0.7836474 | 0.781 | 1 |
| miR-92a-3p | 0 | 0.89765668 | 0.902 | 1 |
| miR-766-5p | 0 | 0.65857637 | 0.653 | 1 |
| miR-590-3p | 0 | 0.94106891 | 0.939 | 1 |
| miR-892c-3p | 0 | 0.87727648 | 0.868 | 1 |
| miR-767-5p | 0 | 0.87142553 | 0.872 | 1 |

| | | | | |
|---|---|---|---|---|
| miR-92a-2-5p | 0 | 0.81421436 | 0.815 | 1 |
| miR-557 | 0 | 0.50544391 | 0.514 | 1 |
| miR-891a-3p | 0 | 0.63563664 | 0.623 | 1 |
| miR-7702 | 0 | 0.9731817 | 0.972 | 1 |
| miR-891b | 0 | 0.56488792 | 0.569 | 1 |
| miR-555 | 0 | 0.63250611 | 0.653 | 1 |
| miR-924 | 0 | 0.86044553 | 0.869 | 1 |
| miR-5572 | 0 | 0.71701499 | 0.757 | 1 |
| miR-921 | 0 | 0.78847854 | 0.786 | 1 |
| miR-5706 | 0 | 0.56255789 | 0.558 | 1 |
| miR-551b-5p | 0 | 0.86716432 | 0.889 | 1 |
| miR-9-3p | 0 | 0.66462839 | 0.656 | 1 |
| miR-597-5p | 0 | 0.54803571 | 0.534 | 1 |
| miR-939-5p | 0 | 0.52768139 | 0.512 | 1 |
| miR-548m | 0 | 0.67304681 | 0.709 | 1 |
| miR-598-5p | 0 | 0.80693242 | 0.811 | 1 |
| miR-548j-3p | 0 | 0.57511643 | 0.556 | 1 |
| miR-548h-5p | 0 | 0.58936444 | 0.589 | 1 |
| miR-942-3p | 0 | 0.92624018 | 0.936 | 1 |
| miR-597-3p | 0 | 0.64564398 | 0.651 | 1 |
| miR-758-3p | 0 | 0.6609364 | 0.652 | 1 |
| miR-95-5p | 0 | 0.89199503 | 0.89 | 1 |
| miR-744-5p | 0 | 0.9261648 | 0.938 | 1 |
| miR-9-5p | 0 | 0.61581707 | 0.605 | 1 |
| miR-96-5p | 0 | 0.73346173 | 0.739 | 1 |
| miR-99a-5p | 0 | 0.57756187 | 0.58 | 1 |
| miR-548v | 0 | 0.97049137 | 0.969 | 1 |
| miR-759 | 0 | 0.52025359 | 0.514 | 1 |
| miR-92b-5p | 0 | 0.7265142 | 0.736 | 1 |
| miR-592 | 0 | 0.62813316 | 0.639 | 1 |
| miR-551b-3p | 0 | 0.51516503 | 0.504 | 1 |
| miR-551a | 0 | 0.69430908 | 0.683 | 1 |
| miR-93-3p | 0 | 0.51665627 | 0.554 | 1 |
| miR-765 | 0 | 0.54678404 | 0.544 | 1 |
| miR-888-5p | 0 | 0.76157472 | 0.765 | 1 |
| miR-550a-5p | 0 | 0.67236865 | 0.676 | 1 |
| miR-93-5p | 0 | 0.92258536 | 0.914 | 1 |
| miR-548w | 0 | 0.64265441 | 0.65 | 1 |
| miR-760 | 0 | 0.72963839 | 0.73 | 1 |
| miR-7-5p | 0 | 0.89702604 | 0.888 | 1 |
| miR-549a | 0 | 0.52846136 | 0.539 | 1 |
| miR-936 | 0 | 0.54266152 | 0.557 | 1 |
| miR-548y | 0 | 0.78730638 | 0.771 | 1 |
| miR-548x-3p | 0 | 0.58687213 | 0.583 | 1 |
| miR-934 | 0 | 0.51309347 | 0.501 | 1 |
| miR-5584-5p | 0 | 0.68298714 | 0.703 | 1 |
| miR-5581-3p | 0 | 0.53614438 | 0.515 | 1 |
| miR-7973 | 0 | 0.93951306 | 0.93 | 1 |
| miR-7850-5p | 0 | 0.97396048 | 0.977 | 1 |
| miR-8059 | 0 | 0.8822902 | 0.893 | 1 |

| | | | | |
|---|---|---|---|---|
| miR-7852-3p | 0 | 0.59697772 | 0.61 | 1 |
| miR-577 | 0 | 0.76230979 | 0.759 | 1 |
| miR-8058 | 0 | 0.9918853 | 0.991 | 1 |
| miR-8057 | 0 | 0.99156846 | 0.991 | 1 |
| miR-576-5p | 0 | 0.88609283 | 0.879 | 1 |
| miR-5689 | 0 | 0.93936199 | 0.935 | 1 |
| miR-576-3p | 0 | 0.68549243 | 0.685 | 1 |
| miR-569 | 0 | 0.73810071 | 0.729 | 1 |
| miR-5691 | 0 | 0.85563549 | 0.865 | 1 |
| miR-5692a | 0 | 0.94023328 | 0.933 | 1 |
| miR-575 | 0 | 0.86909856 | 0.871 | 1 |
| miR-7854-3p | 0 | 0.82813457 | 0.84 | 1 |
| miR-5692c | 0 | 0.71790544 | 0.717 | 1 |
| miR-5581-5p | 0 | 0.65066013 | 0.655 | 1 |
| miR-5695 | 0 | 0.51837656 | 0.507 | 1 |
| miR-5697 | 0 | 0.50685823 | 0.504 | 1 |
| miR-8054 | 0 | 0.7553057 | 0.728 | 1 |
| miR-5699-5p | 0 | 0.88964776 | 0.882 | 1 |
| miR-5700 | 0 | 0.77061652 | 0.766 | 1 |
| miR-7974 | 0 | 0.53753863 | 0.532 | 1 |
| miR-5701 | 0 | 0.91383883 | 0.92 | 1 |
| miR-5702 | 0 | 0.87297711 | 0.878 | 1 |
| miR-5703 | 0 | 0.89531367 | 0.909 | 1 |
| miR-573 | 0 | 0.93249988 | 0.931 | 1 |
| miR-5704 | 0 | 0.86775306 | 0.871 | 1 |
| miR-8052 | 0 | 0.50557489 | 0.501 | 1 |
| miR-802 | 0 | 0.72470959 | 0.709 | 1 |
| miR-571 | 0 | 0.75086754 | 0.751 | 1 |
| miR-8060 | 0 | 0.7910282 | 0.797 | 1 |
| miR-7845-5p | 0 | 0.81407788 | 0.828 | 1 |
| miR-5680 | 0 | 0.79995913 | 0.782 | 1 |
| miR-8062 | 0 | 0.94412995 | 0.96 | 1 |
| miR-887-3p | 0 | 0.96443671 | 0.973 | 1 |
| miR-885-3p | 0 | 0.77457021 | 0.759 | 1 |
| miR-877-5p | 0 | 0.52219968 | 0.527 | 1 |
| miR-876-5p | 0 | 0.97837591 | 0.979 | 1 |
| miR-875-5p | 0 | 0.64056282 | 0.641 | 1 |
| miR-874-5p | 0 | 0.7234966 | 0.703 | 1 |
| miR-587 | 0 | 0.90735307 | 0.905 | 1 |
| miR-5705 | 0 | 0.6954153 | 0.67 | 1 |
| miR-874-3p | 0 | 0.88595329 | 0.884 | 1 |
| miR-873-5p | 0 | 0.70150411 | 0.677 | 1 |
| miR-8089 | 0 | 0.69668344 | 0.696 | 1 |
| miR-5585-5p | 0 | 0.8529943 | 0.829 | 1 |
| miR-8087 | 0 | 0.52208159 | 0.492 | 1 |
| miR-8086 | 0 | 0.65498621 | 0.676 | 1 |
| miR-584-5p | 0 | 0.62254821 | 0.627 | 1 |
| miR-8083 | 0 | 0.69675074 | 0.665 | 1 |
| miR-5588-5p | 0 | 0.76763435 | 0.744 | 1 |
| miR-8081 | 0 | 0.64211391 | 0.651 | 1 |

| | | | | |
|---|---|---|---|---|
| miR-559 | 0 | 0.80987989 | 0.811 | 1 |
| miR-8077 | 0 | 0.51920198 | 0.506 | 1 |
| miR-8069 | 0 | 0.93445058 | 0.927 | 1 |
| miR-562 | 0 | 0.52239349 | 0.536 | 1 |
| miR-7843-3p | 0 | 0.72876973 | 0.71 | 1 |
| miR-8065 | 0 | 0.69823067 | 0.708 | 1 |
| miR-580-3p | 0 | 0.86392449 | 0.857 | 1 |
| miR-564 | 0 | 0.68488994 | 0.708 | 1 |
| miR-8063 | 0 | 0.5758989 | 0.593 | 1 |
| miR-567 | 0 | 0.9367243 | 0.941 | 1 |
| miR-579-5p | 0 | 0.61744556 | 0.614 | 1 |
| miR-604 | 0 | 0.69882348 | 0.69 | 1 |
| miR-6815-5p | 0 | 0.77418398 | 0.776 | 1 |
| miR-605-5p | 0 | 0.94122972 | 0.936 | 1 |
| miR-6751-3p | 0 | 0.6975865 | 0.704 | 1 |
| miR-6753-3p | 0 | 0.71940136 | 0.729 | 1 |
| miR-6848-5p | 0 | 0.90372526 | 0.905 | 1 |
| miR-6754-3p | 0 | 0.98979681 | 0.99 | 1 |
| miR-6755-3p | 0 | 0.6841257 | 0.662 | 1 |
| miR-675-5p | 0 | 0.50404727 | 0.509 | 1 |
| miR-6756-5p | 0 | 0.68680732 | 0.693 | 1 |
| miR-6847-5p | 0 | 0.50637574 | 0.484 | 1 |
| miR-6758-3p | 0 | 0.57673882 | 0.597 | 1 |
| miR-6759-3p | 0 | 0.91394288 | 0.912 | 1 |
| miR-6847-3p | 0 | 0.84132037 | 0.842 | 1 |
| miR-6846-5p | 0 | 0.84473819 | 0.875 | 1 |
| miR-6759-5p | 0 | 0.68297679 | 0.697 | 1 |
| miR-6760-3p | 0 | 0.52143421 | 0.49 | 1 |
| miR-6761-3p | 0 | 0.96005691 | 0.967 | 1 |
| miR-6845-5p | 0 | 0.52361213 | 0.533 | 1 |
| miR-6762-3p | 0 | 0.5141247 | 0.508 | 1 |
| miR-676-5p | 0 | 0.52474294 | 0.518 | 1 |
| miR-6766-3p | 0 | 0.58352519 | 0.582 | 1 |
| miR-6767-3p | 0 | 0.64325007 | 0.644 | 1 |
| miR-6767-5p | 0 | 0.82972336 | 0.842 | 1 |
| miR-6769a-3p | 0 | 0.65647503 | 0.667 | 1 |
| miR-6769a-5p | 0 | 0.54607404 | 0.538 | 1 |
| miR-6842-5p | 0 | 0.65692486 | 0.667 | 1 |
| miR-6770-3p | 0 | 0.93337499 | 0.93 | 1 |
| miR-6770-5p | 0 | 0.98351935 | 0.985 | 1 |
| miR-6771-3p | 0 | 0.91035745 | 0.915 | 1 |
| miR-6771-5p | 0 | 0.63953814 | 0.634 | 1 |
| miR-6751-5p | 0 | 0.99216445 | 0.993 | 1 |
| miR-6750-5p | 0 | 0.75835682 | 0.772 | 1 |
| miR-606 | 0 | 0.91490683 | 0.901 | 1 |
| miR-6748-5p | 0 | 0.63733266 | 0.632 | 1 |
| miR-6868-3p | 0 | 0.6071504 | 0.599 | 1 |
| miR-6728-5p | 0 | 0.62751698 | 0.646 | 1 |
| miR-6867-5p | 0 | 0.84651486 | 0.84 | 1 |
| miR-6865-5p | 0 | 0.69742832 | 0.715 | 1 |

| | | | | |
|---|---|---|---|---|
| miR-6864-3p | 0 | 0.94627661 | 0.953 | 1 |
| miR-6734-3p | 0 | 0.69165614 | 0.714 | 1 |
| miR-6861-3p | 0 | 0.64925495 | 0.645 | 1 |
| miR-6735-5p | 0 | 0.90181296 | 0.91 | 1 |
| miR-6859-3p | 0 | 0.60636012 | 0.591 | 1 |
| miR-6857-5p | 0 | 0.65065176 | 0.636 | 1 |
| miR-6736-5p | 0 | 0.59920112 | 0.568 | 1 |
| miR-6737-5p | 0 | 0.80299841 | 0.831 | 1 |
| miR-6738-3p | 0 | 0.78740078 | 0.797 | 1 |
| miR-6856-3p | 0 | 0.64282818 | 0.646 | 1 |
| miR-6738-5p | 0 | 0.81962845 | 0.815 | 1 |
| miR-6739-3p | 0 | 0.72545414 | 0.749 | 1 |
| miR-6855-5p | 0 | 0.59804842 | 0.615 | 1 |
| miR-6739-5p | 0 | 0.91001784 | 0.902 | 1 |
| miR-6740-5p | 0 | 0.60905869 | 0.577 | 1 |
| miR-6741-3p | 0 | 0.87597008 | 0.866 | 1 |
| miR-6741-5p | 0 | 0.94069564 | 0.935 | 1 |
| miR-6742-3p | 0 | 0.66822598 | 0.673 | 1 |
| miR-6742-5p | 0 | 0.59720711 | 0.6 | 1 |
| miR-6853-3p | 0 | 0.93561616 | 0.928 | 1 |
| miR-6744-5p | 0 | 0.95501385 | 0.964 | 1 |
| miR-6747-3p | 0 | 0.78901728 | 0.778 | 1 |
| miR-6851-5p | 0 | 0.99342683 | 0.995 | 1 |
| miR-6772-3p | 0 | 0.89772468 | 0.889 | 1 |
| miR-6773-5p | 0 | 0.80721663 | 0.822 | 1 |
| miR-6774-3p | 0 | 0.700905 | 0.709 | 1 |
| miR-6841-3p | 0 | 0.61982896 | 0.628 | 1 |
| miR-6799-3p | 0 | 0.84450705 | 0.856 | 1 |
| miR-6799-5p | 0 | 0.65382471 | 0.658 | 1 |
| miR-6800-3p | 0 | 0.90639756 | 0.903 | 1 |
| miR-6800-5p | 0 | 0.66400117 | 0.678 | 1 |
| miR-6822-3p | 0 | 0.64326133 | 0.622 | 1 |
| miR-6801-3p | 0 | 0.88073961 | 0.883 | 1 |
| miR-6821-5p | 0 | 0.57237146 | 0.556 | 1 |
| miR-6801-5p | 0 | 0.85127441 | 0.849 | 1 |
| miR-6802-3p | 0 | 0.93486771 | 0.953 | 1 |
| miR-6803-3p | 0 | 0.85997529 | 0.862 | 1 |
| miR-6803-5p | 0 | 0.90754731 | 0.915 | 1 |
| miR-6805-5p | 0 | 0.65562884 | 0.636 | 1 |
| miR-6819-3p | 0 | 0.52005287 | 0.492 | 1 |
| miR-6806-3p | 0 | 0.71199648 | 0.696 | 1 |
| miR-6806-5p | 0 | 0.82203698 | 0.819 | 1 |
| miR-6818-5p | 0 | 0.5233943 | 0.499 | 1 |
| miR-6807-3p | 0 | 0.97215937 | 0.965 | 1 |
| miR-6808-5p | 0 | 0.9563589 | 0.954 | 1 |
| miR-6817-5p | 0 | 0.8903676 | 0.891 | 1 |
| miR-6809-5p | 0 | 0.65843262 | 0.635 | 1 |
| miR-6810-3p | 0 | 0.63319544 | 0.629 | 1 |
| miR-6811-5p | 0 | 0.96523353 | 0.967 | 1 |
| miR-6813-3p | 0 | 0.81330664 | 0.801 | 1 |

| | | | | |
|---|---|---|---|---|
| miR-6813-5p | 0 | 0.80069083 | 0.815 | 1 |
| miR-6817-3p | 0 | 0.63234284 | 0.629 | 1 |
| miR-6814-3p | 0 | 0.87384903 | 0.883 | 1 |
| miR-6816-3p | 0 | 0.76248715 | 0.776 | 1 |
| miR-6823-5p | 0 | 0.76605517 | 0.775 | 1 |
| miR-6797-3p | 0 | 0.72960414 | 0.727 | 1 |
| miR-6796-3p | 0 | 0.90604519 | 0.907 | 1 |
| miR-6782-3p | 0 | 0.9647426 | 0.968 | 1 |
| miR-6775-5p | 0 | 0.5382715 | 0.528 | 1 |
| miR-6776-3p | 0 | 0.82996875 | 0.82 | 1 |
| miR-6776-5p | 0 | 0.63325443 | 0.629 | 1 |
| miR-6778-3p | 0 | 0.56075425 | 0.542 | 1 |
| miR-6840-3p | 0 | 0.75589305 | 0.741 | 1 |
| miR-6779-3p | 0 | 0.98019796 | 0.982 | 1 |
| miR-6780a-3p | 0 | 0.77899861 | 0.79 | 1 |
| miR-6780b-3p | 0 | 0.95657068 | 0.939 | 1 |
| miR-6781-3p | 0 | 0.75161391 | 0.738 | 1 |
| miR-6836-5p | 0 | 0.55275073 | 0.577 | 1 |
| miR-6835-5p | 0 | 0.92392665 | 0.94 | 1 |
| miR-6781-5p | 0 | 0.88644734 | 0.88 | 1 |
| miR-6835-3p | 0 | 0.86848435 | 0.872 | 1 |
| miR-6826-5p | 0 | 0.60584169 | 0.583 | 1 |
| miR-6783-3p | 0 | 0.72160344 | 0.705 | 1 |
| miR-6834-5p | 0 | 0.81200136 | 0.831 | 1 |
| miR-6784-5p | 0 | 0.89187884 | 0.903 | 1 |
| miR-6786-3p | 0 | 0.77167109 | 0.765 | 1 |
| miR-6787-3p | 0 | 0.82772558 | 0.827 | 1 |
| miR-6787-5p | 0 | 0.84661669 | 0.841 | 1 |
| miR-6831-5p | 0 | 0.96837638 | 0.974 | 1 |
| miR-6788-3p | 0 | 0.55720317 | 0.545 | 1 |
| miR-6789-5p | 0 | 0.51890783 | 0.508 | 1 |
| miR-6830-3p | 0 | 0.5336789 | 0.545 | 1 |
| miR-6828-3p | 0 | 0.77633419 | 0.771 | 1 |
| miR-6792-3p | 0 | 0.97756854 | 0.979 | 1 |
| miR-6868-5p | 0 | 0.63717567 | 0.642 | 1 |
| miR-6721-5p | 0 | 0.82147102 | 0.819 | 1 |
| miR-6720-5p | 0 | 0.74397891 | 0.728 | 1 |
| miR-6501-5p | 0 | 0.57835296 | 0.559 | 1 |
| miR-624-5p | 0 | 0.50400471 | 0.496 | 1 |
| miR-7114-5p | 0 | 0.68151696 | 0.702 | 1 |
| miR-626 | 0 | 0.5923003 | 0.6 | 1 |
| miR-627-3p | 0 | 0.83929669 | 0.852 | 1 |
| miR-628-5p | 0 | 0.66743365 | 0.678 | 1 |
| miR-7114-3p | 0 | 0.51084175 | 0.515 | 1 |
| miR-630 | 0 | 0.98453612 | 0.992 | 1 |
| miR-631 | 0 | 0.63903557 | 0.63 | 1 |
| miR-632 | 0 | 0.82834042 | 0.823 | 1 |
| miR-634 | 0 | 0.60547184 | 0.586 | 1 |
| miR-635 | 0 | 0.57026428 | 0.574 | 1 |
| miR-7113-5p | 0 | 0.84785662 | 0.852 | 1 |

| | | | | |
|---|---|---|---|---|
| miR-637 | 0 | 0.95881563 | 0.961 | 1 |
| miR-640 | 0 | 0.51465464 | 0.503 | 1 |
| miR-642a-5p | 0 | 0.69928613 | 0.696 | 1 |
| miR-642b-5p | 0 | 0.7654618 | 0.759 | 1 |
| miR-643 | 0 | 0.97643643 | 0.982 | 1 |
| miR-644a | 0 | 0.86519589 | 0.876 | 1 |
| miR-645 | 0 | 0.65740345 | 0.644 | 1 |
| miR-646 | 0 | 0.97109337 | 0.966 | 1 |
| miR-7111-3p | 0 | 0.95489627 | 0.957 | 1 |
| miR-648 | 0 | 0.97810355 | 0.983 | 1 |
| miR-6499-3p | 0 | 0.51988882 | 0.517 | 1 |
| miR-711 | 0 | 0.99124477 | 0.99 | 1 |
| miR-7109-5p | 0 | 0.67116011 | 0.651 | 1 |
| miR-7108-5p | 0 | 0.89305328 | 0.875 | 1 |
| miR-6500-5p | 0 | 0.95286776 | 0.958 | 1 |
| miR-7150 | 0 | 0.62459311 | 0.613 | 1 |
| miR-622 | 0 | 0.55420337 | 0.576 | 1 |
| miR-7151-3p | 0 | 0.6685468 | 0.671 | 1 |
| miR-6080 | 0 | 0.93105904 | 0.927 | 1 |
| miR-7-2-3p | 0 | 0.73096527 | 0.718 | 1 |
| miR-7162-5p | 0 | 0.90509552 | 0.902 | 1 |
| miR-6069 | 0 | 0.71825831 | 0.692 | 1 |
| miR-607 | 0 | 0.87810786 | 0.874 | 1 |
| miR-6071 | 0 | 0.84794618 | 0.835 | 1 |
| miR-7161-5p | 0 | 0.8158851 | 0.805 | 1 |
| miR-6072 | 0 | 0.83325221 | 0.84 | 1 |
| miR-7159-5p | 0 | 0.70516294 | 0.708 | 1 |
| miR-6076 | 0 | 0.83784917 | 0.829 | 1 |
| miR-6078 | 0 | 0.52462135 | 0.509 | 1 |
| miR-6079 | 0 | 0.84996185 | 0.864 | 1 |
| miR-608 | 0 | 0.5078565 | 0.494 | 1 |
| miR-6081 | 0 | 0.61831427 | 0.585 | 1 |
| miR-619-3p | 0 | 0.9890609 | 0.996 | 1 |
| miR-7158-3p | 0 | 0.64637623 | 0.642 | 1 |
| miR-7157-3p | 0 | 0.55507615 | 0.527 | 1 |
| miR-7156-5p | 0 | 0.8761892 | 0.869 | 1 |
| miR-6083 | 0 | 0.76084394 | 0.753 | 1 |
| miR-7156-3p | 0 | 0.76819334 | 0.77 | 1 |
| miR-6086 | 0 | 0.68256011 | 0.659 | 1 |
| miR-609 | 0 | 0.64114587 | 0.611 | 1 |
| miR-612 | 0 | 0.74306345 | 0.723 | 1 |
| miR-7155-5p | 0 | 0.90388506 | 0.907 | 1 |
| miR-6128 | 0 | 0.78600572 | 0.777 | 1 |
| miR-7154-3p | 0 | 0.82642687 | 0.825 | 1 |
| miR-6132 | 0 | 0.63813429 | 0.623 | 1 |
| miR-6501-3p | 0 | 0.56799081 | 0.576 | 1 |
| miR-6502-5p | 0 | 0.67841863 | 0.674 | 1 |
| miR-6869-5p | 0 | 0.70537998 | 0.702 | 1 |
| miR-6503-3p | 0 | 0.89756363 | 0.899 | 1 |
| miR-655-5p | 0 | 0.70595964 | 0.686 | 1 |

| | | | | |
|---|---|---|---|---|
| miR-656-3p | 0 | 0.70456511 | 0.696 | 1 |
| miR-6884-5p | 0 | 0.519671 | 0.519 | 1 |
| miR-657 | 0 | 0.65767153 | 0.655 | 1 |
| miR-658 | 0 | 0.95819583 | 0.969 | 1 |
| miR-6883-3p | 0 | 0.85031974 | 0.855 | 1 |
| miR-6882-5p | 0 | 0.78016106 | 0.784 | 1 |
| miR-660-3p | 0 | 0.60208827 | 0.579 | 1 |
| miR-6882-3p | 0 | 0.95631585 | 0.956 | 1 |
| miR-661 | 0 | 0.76523646 | 0.748 | 1 |
| miR-662 | 0 | 0.69647646 | 0.693 | 1 |
| miR-663b | 0 | 0.51209826 | 0.475 | 1 |
| miR-664a-3p | 0 | 0.63389676 | 0.663 | 1 |
| miR-6881-3p | 0 | 0.61705286 | 0.636 | 1 |
| miR-6880-5p | 0 | 0.89820775 | 0.901 | 1 |
| miR-668-5p | 0 | 0.71672836 | 0.689 | 1 |
| miR-670-3p | 0 | 0.55999702 | 0.547 | 1 |
| miR-6876-3p | 0 | 0.50575783 | 0.501 | 1 |
| miR-671-3p | 0 | 0.60603018 | 0.609 | 1 |
| miR-6715b-3p | 0 | 0.51905531 | 0.507 | 1 |
| miR-6874-5p | 0 | 0.76373148 | 0.74 | 1 |
| miR-671-5p | 0 | 0.9263925 | 0.916 | 1 |
| miR-6872-5p | 0 | 0.91330367 | 0.914 | 1 |
| miR-6717-5p | 0 | 0.81358789 | 0.801 | 1 |
| miR-6718-5p | 0 | 0.63229622 | 0.644 | 1 |
| miR-6871-5p | 0 | 0.57988072 | 0.612 | 1 |
| miR-6870-5p | 0 | 0.67373038 | 0.676 | 1 |
| miR-655-3p | 0 | 0.81383892 | 0.818 | 1 |
| miR-654-5p | 0 | 0.87121617 | 0.877 | 1 |
| miR-654-3p | 0 | 0.5042216 | 0.508 | 1 |
| miR-6513-3p | 0 | 0.51749411 | 0.525 | 1 |
| miR-6504-5p | 0 | 0.6954093 | 0.721 | 1 |
| miR-6505-3p | 0 | 0.74662465 | 0.742 | 1 |
| miR-7106-5p | 0 | 0.85208819 | 0.849 | 1 |
| miR-6895-5p | 0 | 0.93530555 | 0.93 | 1 |
| miR-6505-5p | 0 | 0.73321072 | 0.693 | 1 |
| miR-6895-3p | 0 | 0.68651725 | 0.66 | 1 |
| miR-6506-5p | 0 | 0.52115968 | 0.553 | 1 |
| miR-6507-5p | 0 | 0.7928179 | 0.798 | 1 |
| miR-6893-5p | 0 | 0.56247505 | 0.565 | 1 |
| miR-6510-3p | 0 | 0.8426354 | 0.854 | 1 |
| miR-6511a-3p | 0 | 0.78167546 | 0.772 | 1 |
| miR-6511b-3p | 0 | 0.55924027 | 0.56 | 1 |
| miR-651-3p | 0 | 0.81043973 | 0.819 | 1 |
| miR-6885-3p | 0 | 0.86706079 | 0.858 | 1 |
| miR-6892-3p | 0 | 0.64003523 | 0.651 | 1 |
| miR-6514-5p | 0 | 0.75744057 | 0.771 | 1 |
| miR-6890-3p | 0 | 0.96075826 | 0.955 | 1 |
| miR-6515-3p | 0 | 0.91497993 | 0.917 | 1 |
| miR-6889-5p | 0 | 0.68560161 | 0.684 | 1 |
| miR-6889-3p | 0 | 0.70183576 | 0.711 | 1 |

| | | | | |
|---|---|---|---|---|
| miR-6887-3p | 0 | 0.84695188 | 0.843 | 1 |
| miR-6516-3p | 0 | 0.54462534 | 0.527 | 1 |
| miR-6886-3p | 0 | 0.70683948 | 0.706 | 1 |
| miR-652-5p | 0 | 0.62112883 | 0.619 | 1 |
| miR-653-3p | 0 | 0.83579592 | 0.847 | 1 |
| miR-6885-5p | 0 | 0.64362879 | 0.626 | 1 |
| let-7a-2-3p | 0 | 0.70938786 | 0.71 | 1 |
| miR-4685-3p | 0 | 0.99230913 | 0.992 | 1 |
| miR-548ba | 0 | 0.75937722 | 0.76 | 1 |
| miR-3144-3p | 0 | 0.71457897 | 0.697 | 1 |
| miR-3140-5p | 0 | 0.73933527 | 0.727 | 1 |
| miR-3136-5p | 0 | 0.74874427 | 0.755 | 1 |
| miR-3134 | 0 | 0.80207543 | 0.787 | 1 |
| miR-3130-3p | 0 | 0.97781712 | 0.984 | 1 |
| miR-3127-5p | 0 | 0.78434675 | 0.771 | 1 |
| miR-3127-3p | 0 | 0.74873394 | 0.742 | 1 |
| miR-3126-5p | 0 | 0.97586393 | 0.979 | 1 |
| miR-3125 | 0 | 0.92082881 | 0.935 | 1 |
| miR-3124-3p | 0 | 0.94203431 | 0.939 | 1 |
| miR-3121-5p | 0 | 0.65843866 | 0.661 | 1 |
| miR-3118 | 0 | 0.85240193 | 0.853 | 1 |
| miR-30d-3p | 0 | 0.51205309 | 0.481 | 1 |
| miR-30b-3p | 0 | 0.92672967 | 0.919 | 1 |
| miR-30a-3p | 0 | 0.55743118 | 0.53 | 1 |
| miR-3064-5p | 0 | 0.8348687 | 0.836 | 1 |
| miR-3064-3p | 0 | 0.73447336 | 0.759 | 1 |
| miR-302f | 0 | 0.9632915 | 0.971 | 1 |
| miR-302d-5p | 0 | 0.57711833 | 0.575 | 1 |
| miR-302d-3p | 0 | 0.80325076 | 0.811 | 1 |
| miR-3143 | 0 | 0.73131995 | 0.737 | 1 |
| miR-3144-5p | 0 | 0.86362062 | 0.86 | 1 |
| miR-3182 | 0 | 0.76133529 | 0.786 | 1 |
| miR-3145-3p | 0 | 0.73862741 | 0.728 | 1 |
| miR-3180 | 0 | 0.97538671 | 0.979 | 1 |
| miR-3176 | 0 | 0.86860828 | 0.87 | 1 |
| miR-3171 | 0 | 0.80822116 | 0.806 | 1 |
| miR-3170 | 0 | 0.65695682 | 0.621 | 1 |
| miR-3169 | 0 | 0.5376411 | 0.553 | 1 |
| miR-3167 | 0 | 0.67959668 | 0.691 | 1 |
| miR-3166 | 0 | 0.55367023 | 0.562 | 1 |
| miR-3165 | 0 | 0.97777338 | 0.978 | 1 |
| miR-3164 | 0 | 0.71415267 | 0.732 | 1 |
| miR-3163 | 0 | 0.73010056 | 0.734 | 1 |
| miR-3162-5p | 0 | 0.64478977 | 0.657 | 1 |
| miR-3158-5p | 0 | 0.84904201 | 0.858 | 1 |
| miR-3157-5p | 0 | 0.72824257 | 0.732 | 1 |
| miR-3157-3p | 0 | 0.56059363 | 0.538 | 1 |
| miR-3155b | 0 | 0.63116547 | 0.633 | 1 |
| miR-3148 | 0 | 0.9335975 | 0.935 | 1 |
| miR-3147 | 0 | 0.51355182 | 0.487 | 1 |

| | | | | |
|---|---|---|---|---|
| miR-3146 | 0 | 0.70398931 | 0.714 | 1 |
| miR-3145-5p | 0 | 0.89540009 | 0.906 | 1 |
| miR-302c-3p | 0 | 0.95033567 | 0.953 | 1 |
| miR-302b-5p | 0 | 0.87587747 | 0.876 | 1 |
| miR-302b-3p | 0 | 0.89575456 | 0.9 | 1 |
| miR-302a-3p | 0 | 0.90945858 | 0.912 | 1 |
| miR-23a-3p | 0 | 0.83215979 | 0.845 | 1 |
| miR-2392 | 0 | 0.67946798 | 0.679 | 1 |
| miR-2278 | 0 | 0.78988006 | 0.782 | 1 |
| miR-2276-5p | 0 | 0.87742138 | 0.87 | 1 |
| miR-2276-3p | 0 | 0.83686504 | 0.831 | 1 |
| miR-224-5p | 0 | 0.98166703 | 0.98 | 1 |
| miR-224-3p | 0 | 0.90355517 | 0.897 | 1 |
| miR-22-3p | 0 | 0.74493097 | 0.755 | 1 |
| miR-222-5p | 0 | 0.87139208 | 0.87 | 1 |
| miR-221-5p | 0 | 0.64717867 | 0.667 | 1 |
| miR-221-3p | 0 | 0.59002581 | 0.589 | 1 |
| miR-219a-1-3p | 0 | 0.71340733 | 0.693 | 1 |
| miR-218-1-3p | 0 | 0.55764311 | 0.55 | 1 |
| miR-217 | 0 | 0.80494098 | 0.81 | 1 |
| miR-216b-5p | 0 | 0.99330166 | 0.991 | 1 |
| miR-216b-3p | 0 | 0.53543458 | 0.55 | 1 |
| miR-216a-5p | 0 | 0.67516331 | 0.646 | 1 |
| miR-216a-3p | 0 | 0.53836651 | 0.521 | 1 |
| miR-21-5p | 0 | 0.62948994 | 0.627 | 1 |
| miR-23a-5p | 0 | 0.54340037 | 0.553 | 1 |
| miR-23b-3p | 0 | 0.78291162 | 0.791 | 1 |
| miR-23b-5p | 0 | 0.95484395 | 0.964 | 1 |
| miR-27a-5p | 0 | 0.68451263 | 0.712 | 1 |
| miR-301b | 0 | 0.94731835 | 0.946 | 1 |
| miR-29c-5p | 0 | 0.62844707 | 0.635 | 1 |
| miR-29b-3p | 0 | 0.66620378 | 0.666 | 1 |
| miR-29b-1-5p | 0 | 0.77446641 | 0.765 | 1 |
| miR-298 | 0 | 0.77864387 | 0.777 | 1 |
| miR-296-5p | 0 | 0.58638139 | 0.596 | 1 |
| miR-296-3p | 0 | 0.90086847 | 0.895 | 1 |
| miR-27b-5p | 0 | 0.61345806 | 0.629 | 1 |
| miR-27a-3p | 0 | 0.82380604 | 0.845 | 1 |
| miR-24-3p | 0 | 0.60556773 | 0.636 | 1 |
| miR-26b-5p | 0 | 0.66200938 | 0.655 | 1 |
| miR-26a-5p | 0 | 0.72711297 | 0.744 | 1 |
| miR-26a-2-3p | 0 | 0.88089726 | 0.905 | 1 |
| miR-26a-1-3p | 0 | 0.57089666 | 0.552 | 1 |
| miR-2682-5p | 0 | 0.88892973 | 0.871 | 1 |
| miR-2681-3p | 0 | 0.68634788 | 0.661 | 1 |
| miR-25-5p | 0 | 0.60398769 | 0.601 | 1 |
| miR-25-3p | 0 | 0.59257373 | 0.574 | 1 |
| miR-3180-3p | 0 | 0.96583007 | 0.975 | 1 |
| miR-3184-3p | 0 | 0.57310593 | 0.563 | 1 |
| miR-215-3p | 0 | 0.61212662 | 0.612 | 1 |

| | | | | |
|---|---|---|---|---|
| miR-3661 | 0 | 0.61978466 | 0.61 | 1 |
| miR-365a-5p | 0 | 0.92745091 | 0.909 | 1 |
| miR-365a-3p | 0 | 0.74081904 | 0.759 | 1 |
| miR-3659 | 0 | 0.96229428 | 0.961 | 1 |
| miR-3658 | 0 | 0.70289615 | 0.722 | 1 |
| miR-3657 | 0 | 0.81536706 | 0.813 | 1 |
| miR-3654 | 0 | 0.69301315 | 0.695 | 1 |
| miR-3653 | 0 | 0.79546563 | 0.816 | 1 |
| miR-3650 | 0 | 0.9839083 | 0.985 | 1 |
| miR-3648 | 0 | 0.73439031 | 0.754 | 1 |
| miR-3646 | 0 | 0.51950161 | 0.537 | 1 |
| miR-362-3p | 0 | 0.79058676 | 0.783 | 1 |
| miR-3619-5p | 0 | 0.74038697 | 0.761 | 1 |
| miR-3619-3p | 0 | 0.6323732 | 0.634 | 1 |
| miR-3618 | 0 | 0.55921766 | 0.535 | 1 |
| miR-3617-5p | 0 | 0.92586049 | 0.932 | 1 |
| miR-3617-3p | 0 | 0.75088462 | 0.744 | 1 |
| miR-3616-5p | 0 | 0.75892927 | 0.746 | 1 |
| miR-3616-3p | 0 | 0.62860259 | 0.615 | 1 |
| miR-3614-3p | 0 | 0.84945621 | 0.86 | 1 |
| miR-365b-5p | 0 | 0.90057515 | 0.892 | 1 |
| miR-3662 | 0 | 0.64737434 | 0.663 | 1 |
| miR-3186-3p | 0 | 0.84282195 | 0.854 | 1 |
| miR-3663-5p | 0 | 0.58201307 | 0.567 | 1 |
| miR-3689d | 0 | 0.78474425 | 0.78 | 1 |
| miR-3689b-3p | 0 | 0.83021326 | 0.833 | 1 |
| miR-3685 | 0 | 0.76431669 | 0.761 | 1 |
| miR-3682-5p | 0 | 0.69490895 | 0.692 | 1 |
| miR-3681-5p | 0 | 0.63589361 | 0.649 | 1 |
| miR-3680-3p | 0 | 0.60124166 | 0.626 | 1 |
| miR-3679-5p | 0 | 0.98042796 | 0.98 | 1 |
| miR-3678-3p | 0 | 0.56268214 | 0.553 | 1 |
| miR-3677-5p | 0 | 0.71359072 | 0.737 | 1 |
| miR-367-5p | 0 | 0.76200417 | 0.758 | 1 |
| miR-3675-5p | 0 | 0.62798071 | 0.647 | 1 |
| miR-3675-3p | 0 | 0.95562325 | 0.936 | 1 |
| miR-367-3p | 0 | 0.67184731 | 0.68 | 1 |
| miR-3671 | 0 | 0.57533262 | 0.59 | 1 |
| miR-3670 | 0 | 0.8164356 | 0.81 | 1 |
| miR-3667-5p | 0 | 0.57134254 | 0.562 | 1 |
| miR-3667-3p | 0 | 0.71263741 | 0.683 | 1 |
| miR-3666 | 0 | 0.88555845 | 0.884 | 1 |
| miR-3664-5p | 0 | 0.78784041 | 0.79 | 1 |
| miR-361-3p | 0 | 0.95413996 | 0.969 | 1 |
| miR-3611 | 0 | 0.98738423 | 0.985 | 1 |
| miR-3609 | 0 | 0.89181292 | 0.889 | 1 |
| miR-3607-3p | 0 | 0.87135071 | 0.886 | 1 |
| miR-324-5p | 0 | 0.97324807 | 0.985 | 1 |
| miR-324-3p | 0 | 0.68860321 | 0.694 | 1 |
| miR-323b-5p | 0 | 0.68478559 | 0.685 | 1 |

| | | | | |
|---|---|---|---|---|
| miR-323b-3p | 0 | 0.95946004 | 0.964 | 1 |
| miR-323a-3p | 0 | 0.61616198 | 0.608 | 1 |
| miR-3202 | 0 | 0.97025154 | 0.961 | 1 |
| miR-3201 | 0 | 0.88786632 | 0.887 | 1 |
| miR-3198 | 0 | 0.95708914 | 0.958 | 1 |
| miR-3193 | 0 | 0.72742802 | 0.738 | 1 |
| miR-3192-5p | 0 | 0.7155434 | 0.711 | 1 |
| miR-3192-3p | 0 | 0.87436187 | 0.88 | 1 |
| miR-3191-5p | 0 | 0.87737163 | 0.883 | 1 |
| miR-3191-3p | 0 | 0.91108275 | 0.923 | 1 |
| miR-3189-5p | 0 | 0.80464029 | 0.799 | 1 |
| miR-3189-3p | 0 | 0.92980161 | 0.928 | 1 |
| miR-3188 | 0 | 0.7740931 | 0.772 | 1 |
| miR-3187-5p | 0 | 0.80749094 | 0.802 | 1 |
| miR-3187-3p | 0 | 0.72257566 | 0.745 | 1 |
| miR-3186-5p | 0 | 0.65215354 | 0.643 | 1 |
| miR-325 | 0 | 0.9070842 | 0.909 | 1 |
| miR-326 | 0 | 0.59449656 | 0.597 | 1 |
| miR-329-3p | 0 | 0.99759774 | 1 | 1 |
| miR-34a-3p | 0 | 0.52988473 | 0.546 | 1 |
| miR-3606-5p | 0 | 0.86550224 | 0.894 | 1 |
| miR-3606-3p | 0 | 0.68399147 | 0.696 | 1 |
| miR-3605-3p | 0 | 0.99200693 | 0.991 | 1 |
| miR-34c-5p | 0 | 0.94809498 | 0.951 | 1 |
| miR-34c-3p | 0 | 0.92685307 | 0.93 | 1 |
| miR-34b-5p | 0 | 0.86023581 | 0.868 | 1 |
| miR-34b-3p | 0 | 0.54482602 | 0.532 | 1 |
| miR-34a-5p | 0 | 0.75505015 | 0.768 | 1 |
| miR-345-3p | 0 | 0.86313832 | 0.863 | 1 |
| miR-330-3p | 0 | 0.98713688 | 0.989 | 1 |
| miR-340-5p | 0 | 0.96641716 | 0.963 | 1 |
| miR-33b-5p | 0 | 0.78244022 | 0.788 | 1 |
| miR-33b-3p | 0 | 0.85155568 | 0.862 | 1 |
| miR-339-5p | 0 | 0.65960513 | 0.661 | 1 |
| miR-339-3p | 0 | 0.87843648 | 0.865 | 1 |
| miR-335-5p | 0 | 0.94743222 | 0.955 | 1 |
| miR-331-5p | 0 | 0.51196116 | 0.529 | 1 |
| miR-330-5p | 0 | 0.81348047 | 0.827 | 1 |
| miR-215-5p | 0 | 0.72724816 | 0.73 | 1 |
| miR-214-5p | 0 | 0.54595048 | 0.554 | 1 |
| miR-3691-3p | 0 | 0.9964654 | 0.997 | 1 |
| miR-1276 | 0 | 0.50819514 | 0.478 | 1 |
| miR-1273g-5p | 0 | 0.56780622 | 0.574 | 1 |
| miR-1271-5p | 0 | 0.9790201 | 0.986 | 1 |
| miR-1270 | 0 | 0.83585825 | 0.845 | 1 |
| miR-1269b | 0 | 0.74313137 | 0.753 | 1 |
| miR-1267 | 0 | 0.89041247 | 0.911 | 1 |
| miR-1264 | 0 | 0.70526386 | 0.692 | 1 |
| miR-126-3p | 0 | 0.53696154 | 0.578 | 1 |
| miR-1263 | 0 | 0.77151996 | 0.763 | 1 |

| | | | | |
|---|---|---|---|---|
| miR-1262 | 0 | 0.64242312 | 0.635 | 1 |
| miR-1261 | 0 | 0.99572241 | 0.992 | 1 |
| miR-125a-3p | 0 | 0.533708 | 0.528 | 1 |
| miR-1258 | 0 | 0.59315028 | 0.589 | 1 |
| miR-1253 | 0 | 0.85315558 | 0.861 | 1 |
| miR-1251-3p | 0 | 0.91442599 | 0.93 | 1 |
| miR-1250-5p | 0 | 0.6582261 | 0.671 | 1 |
| miR-1247-3p | 0 | 0.50117796 | 0.502 | 1 |
| miR-124-5p | 0 | 0.88195712 | 0.888 | 1 |
| miR-1245b-3p | 0 | 0.60512587 | 0.618 | 1 |
| miR-124-3p | 0 | 0.54613378 | 0.543 | 1 |
| miR-127-5p | 0 | 0.89164496 | 0.876 | 1 |
| miR-1277-5p | 0 | 0.68175543 | 0.663 | 1 |
| miR-1306-5p | 0 | 0.82793048 | 0.829 | 1 |
| miR-1278 | 0 | 0.53804057 | 0.546 | 1 |
| miR-1302 | 0 | 0.75031689 | 0.754 | 1 |
| miR-1301-5p | 0 | 0.79321478 | 0.76 | 1 |
| miR-1301-3p | 0 | 0.6374388 | 0.653 | 1 |
| miR-1298-5p | 0 | 0.82389319 | 0.84 | 1 |
| miR-1297 | 0 | 0.54450566 | 0.534 | 1 |
| miR-1295b-3p | 0 | 0.87660684 | 0.89 | 1 |
| miR-1294 | 0 | 0.78500614 | 0.778 | 1 |
| miR-1292-5p | 0 | 0.80791857 | 0.798 | 1 |
| miR-129-1-3p | 0 | 0.83227583 | 0.816 | 1 |
| miR-1291 | 0 | 0.95490788 | 0.964 | 1 |
| miR-1290 | 0 | 0.99506933 | 0.993 | 1 |
| miR-1289 | 0 | 0.57116626 | 0.59 | 1 |
| miR-1288-3p | 0 | 0.54783258 | 0.549 | 1 |
| miR-1287-5p | 0 | 0.690908 | 0.7 | 1 |
| miR-1287-3p | 0 | 0.61949754 | 0.622 | 1 |
| miR-128-3p | 0 | 0.56292911 | 0.551 | 1 |
| miR-1283 | 0 | 0.88342282 | 0.889 | 1 |
| miR-128-1-5p | 0 | 0.80963829 | 0.813 | 1 |
| miR-1279 | 0 | 0.86278966 | 0.856 | 1 |
| miR-1243 | 0 | 0.85580521 | 0.845 | 1 |
| miR-1238-5p | 0 | 0.74101212 | 0.748 | 1 |
| miR-1237-3p | 0 | 0.55994758 | 0.547 | 1 |
| miR-1236-5p | 0 | 0.93225853 | 0.939 | 1 |
| miR-1182 | 0 | 0.52130153 | 0.531 | 1 |
| miR-1180-3p | 0 | 0.79098541 | 0.785 | 1 |
| miR-1179 | 0 | 0.83615742 | 0.812 | 1 |
| miR-1178-5p | 0 | 0.53032702 | 0.493 | 1 |
| miR-1178-3p | 0 | 0.90963428 | 0.901 | 1 |
| miR-10a-3p | 0 | 0.56932035 | 0.562 | 1 |
| miR-106b-3p | 0 | 0.82284823 | 0.845 | 1 |
| miR-105-3p | 0 | 0.6393976 | 0.626 | 1 |
| miR-103a-2-5p | 0 | 0.64509264 | 0.627 | 1 |
| miR-101-5p | 0 | 0.91892346 | 0.923 | 1 |
| miR-100-3p | 0 | 0.60768598 | 0.589 | 1 |
| miR-1 | 0 | 0.87018973 | 0.867 | 1 |

| | | | | |
|---|---|---|---|---|
| let-7g-5p | 0 | 0.69815504 | 0.678 | 1 |
| let-7g-3p | 0 | 0.8379443 | 0.833 | 1 |
| let-7f-2-3p | 0 | 0.76767304 | 0.779 | 1 |
| let-7e-5p | 0 | 0.58347071 | 0.572 | 1 |
| let-7e-3p | 0 | 0.58017647 | 0.567 | 1 |
| let-7d-3p | 0 | 0.57980399 | 0.597 | 1 |
| let-7a-5p | 0 | 0.69607448 | 0.683 | 1 |
| miR-1183 | 0 | 0.55407668 | 0.541 | 1 |
| miR-1184 | 0 | 0.75295333 | 0.762 | 1 |
| miR-1185-2-3 | 0 | 0.68648456 | 0.673 | 1 |
| miR-1208 | 0 | 0.68261719 | 0.679 | 1 |
| miR-1233-3p | 0 | 0.50266646 | 0.507 | 1 |
| miR-1229-5p | 0 | 0.73324137 | 0.732 | 1 |
| miR-1229-3p | 0 | 0.67645139 | 0.675 | 1 |
| miR-1227-3p | 0 | 0.90318326 | 0.911 | 1 |
| miR-1226-3p | 0 | 0.86856957 | 0.875 | 1 |
| miR-122-5p | 0 | 0.83167303 | 0.82 | 1 |
| miR-1225-5p | 0 | 0.74855695 | 0.743 | 1 |
| miR-1225-3p | 0 | 0.54358494 | 0.528 | 1 |
| miR-1207-5p | 0 | 0.59660138 | 0.606 | 1 |
| miR-1193 | 0 | 0.89970044 | 0.897 | 1 |
| miR-1207-3p | 0 | 0.62559189 | 0.63 | 1 |
| miR-1206 | 0 | 0.8097061 | 0.798 | 1 |
| miR-1205 | 0 | 0.74088759 | 0.745 | 1 |
| miR-1204 | 0 | 0.72594972 | 0.701 | 1 |
| miR-1202 | 0 | 0.89343955 | 0.9 | 1 |
| miR-1200 | 0 | 0.88557228 | 0.899 | 1 |
| miR-1199-3p | 0 | 0.91010366 | 0.923 | 1 |
| miR-1197 | 0 | 0.8042317 | 0.804 | 1 |
| miR-1303 | 0 | 0.51677433 | 0.513 | 1 |
| miR-1307-5p | 0 | 0.59687848 | 0.611 | 1 |
| miR-212-3p | 0 | 0.81537994 | 0.788 | 1 |
| miR-19b-1-5p | 0 | 0.85620451 | 0.855 | 1 |
| miR-197-5p | 0 | 0.81335475 | 0.815 | 1 |
| miR-197-3p | 0 | 0.87315237 | 0.883 | 1 |
| miR-196b-3p | 0 | 0.68256473 | 0.667 | 1 |
| miR-193b-5p | 0 | 0.55246915 | 0.547 | 1 |
| miR-193b-3p | 0 | 0.60345004 | 0.617 | 1 |
| miR-193a-3p | 0 | 0.89452144 | 0.898 | 1 |
| miR-192-5p | 0 | 0.86784552 | 0.86 | 1 |
| miR-192-3p | 0 | 0.54258929 | 0.533 | 1 |
| miR-1915-5p | 0 | 0.57066878 | 0.564 | 1 |
| miR-1914-3p | 0 | 0.53374604 | 0.516 | 1 |
| miR-1913 | 0 | 0.53454187 | 0.54 | 1 |
| miR-1911-3p | 0 | 0.94027439 | 0.947 | 1 |
| miR-1910-3p | 0 | 0.83211446 | 0.834 | 1 |
| miR-190b | 0 | 0.53367416 | 0.504 | 1 |
| miR-190a-5p | 0 | 0.98816332 | 0.989 | 1 |
| miR-190a-3p | 0 | 0.84273676 | 0.849 | 1 |
| miR-188-5p | 0 | 0.6909533 | 0.671 | 1 |

| | | | | |
|---|---|---|---|---|
| miR-187-5p | 0 | 0.75581718 | 0.739 | 1 |
| miR-187-3p | 0 | 0.8347456 | 0.811 | 1 |
| miR-19a-3p | 0 | 0.74341268 | 0.725 | 1 |
| miR-19b-2-5p | 0 | 0.6078378 | 0.657 | 1 |
| miR-130a-5p | 0 | 0.6751409 | 0.67 | 1 |
| miR-19b-3p | 0 | 0.79293847 | 0.783 | 1 |
| miR-211-5p | 0 | 0.86353028 | 0.868 | 1 |
| miR-2115-5p | 0 | 0.50571687 | 0.467 | 1 |
| miR-2115-3p | 0 | 0.75283465 | 0.747 | 1 |
| miR-2114-5p | 0 | 0.81984634 | 0.812 | 1 |
| miR-2114-3p | 0 | 0.79992224 | 0.777 | 1 |
| miR-211-3p | 0 | 0.76194484 | 0.768 | 1 |
| miR-210-3p | 0 | 0.71066671 | 0.714 | 1 |
| miR-20a-3p | 0 | 0.91744525 | 0.919 | 1 |
| miR-208b-5p | 0 | 0.51762856 | 0.525 | 1 |
| miR-208a-5p | 0 | 0.63247266 | 0.609 | 1 |
| miR-205-5p | 0 | 0.86751325 | 0.862 | 1 |
| miR-2054 | 0 | 0.75560319 | 0.746 | 1 |
| miR-2053 | 0 | 0.73754589 | 0.727 | 1 |
| miR-203b-5p | 0 | 0.60364652 | 0.6 | 1 |
| miR-203b-3p | 0 | 0.76061466 | 0.763 | 1 |
| miR-202-3p | 0 | 0.75087333 | 0.751 | 1 |
| miR-200c-3p | 0 | 0.80832497 | 0.799 | 1 |
| miR-200b-5p | 0 | 0.78232726 | 0.799 | 1 |
| miR-200a-3p | 0 | 0.91621893 | 0.921 | 1 |
| miR-186-5p | 0 | 0.65388208 | 0.684 | 1 |
| miR-186-3p | 0 | 0.66262012 | 0.64 | 1 |
| miR-185-5p | 0 | 0.87877644 | 0.854 | 1 |
| miR-184 | 0 | 0.91783224 | 0.917 | 1 |
| miR-146a-3p | 0 | 0.97112934 | 0.963 | 1 |
| miR-1468-5p | 0 | 0.96556118 | 0.966 | 1 |
| miR-1468-3p | 0 | 0.85076439 | 0.849 | 1 |
| miR-145-3p | 0 | 0.89824525 | 0.891 | 1 |
| miR-144-5p | 0 | 0.6887942 | 0.681 | 1 |
| miR-144-3p | 0 | 0.58794532 | 0.592 | 1 |
| miR-143-5p | 0 | 0.77988267 | 0.798 | 1 |
| miR-142-5p | 0 | 0.69409868 | 0.691 | 1 |
| miR-141-3p | 0 | 0.98130624 | 0.987 | 1 |
| miR-139-3p | 0 | 0.74892885 | 0.744 | 1 |
| miR-138-2-3p | 0 | 0.87780469 | 0.873 | 1 |
| miR-135b-5p | 0 | 0.5801692 | 0.61 | 1 |
| miR-134-3p | 0 | 0.71197745 | 0.708 | 1 |
| miR-1343-5p | 0 | 0.8205097 | 0.811 | 1 |
| miR-133b | 0 | 0.70898208 | 0.722 | 1 |
| miR-133a-5p | 0 | 0.92614554 | 0.922 | 1 |
| miR-132-5p | 0 | 0.9804571 | 0.982 | 1 |
| miR-132-3p | 0 | 0.50396053 | 0.498 | 1 |
| miR-130b-3p | 0 | 0.50153401 | 0.514 | 1 |
| miR-146a-5p | 0 | 0.86655892 | 0.858 | 1 |
| miR-146b-3p | 0 | 0.64238523 | 0.67 | 1 |

| | | | | |
|---|---|---|---|---|
| miR-149-3p | 0 | 0.60508832 | 0.633 | 1 |
| miR-15a-3p | 0 | 0.59635211 | 0.603 | 1 |
| miR-183-3p | 0 | 0.53398616 | 0.52 | 1 |
| miR-182-5p | 0 | 0.882128 | 0.878 | 1 |
| miR-182-3p | 0 | 0.78293104 | 0.767 | 1 |
| miR-181c-3p | 0 | 0.9141885 | 0.923 | 1 |
| miR-181b-3p | 0 | 0.82094799 | 0.809 | 1 |
| miR-181a-3p | 0 | 0.52946552 | 0.533 | 1 |
| miR-17-3p | 0 | 0.70923158 | 0.717 | 1 |
| miR-16-5p | 0 | 0.64719665 | 0.66 | 1 |
| miR-1587 | 0 | 0.66159309 | 0.674 | 1 |
| miR-149-5p | 0 | 0.9994125 | 1 | 1 |
| miR-155-5p | 0 | 0.96174595 | 0.96 | 1 |
| miR-155-3p | 0 | 0.96682685 | 0.964 | 1 |
| miR-154-5p | 0 | 0.50316345 | 0.497 | 1 |
| miR-1539 | 0 | 0.967744 | 0.956 | 1 |
| miR-1537-5p | 0 | 0.86232 | 0.865 | 1 |
| miR-1537-3p | 0 | 0.68018391 | 0.668 | 1 |
| miR-153-3p | 0 | 0.80926775 | 0.805 | 1 |
| miR-152-3p | 0 | 0.5686394 | 0.58 | 1 |
| miR-3690 | 0 | 0.98224277 | 0.976 | 1 |
| miR-3691-5p | 0 | 0.98978709 | 0.994 | 1 |
| miR-548ay-3p | 0 | 0.78866999 | 0.787 | 1 |
| miR-4761-5p | 0 | 0.85606173 | 0.848 | 1 |
| miR-4759 | 0 | 0.64722382 | 0.652 | 1 |
| miR-4756-5p | 0 | 0.98218463 | 0.982 | 1 |
| miR-4753-5p | 0 | 0.84106341 | 0.848 | 1 |
| miR-4752 | 0 | 0.89127223 | 0.903 | 1 |
| miR-4751 | 0 | 0.88612673 | 0.887 | 1 |
| miR-4748 | 0 | 0.97333586 | 0.978 | 1 |
| miR-4747-5p | 0 | 0.58179309 | 0.551 | 1 |
| miR-4747-3p | 0 | 0.76949279 | 0.758 | 1 |
| miR-4746-5p | 0 | 0.9773819 | 0.974 | 1 |
| miR-4743-5p | 0 | 0.78668695 | 0.765 | 1 |
| miR-4742-5p | 0 | 0.53063688 | 0.544 | 1 |
| miR-4742-3p | 0 | 0.82947844 | 0.829 | 1 |
| miR-4738-5p | 0 | 0.59001934 | 0.582 | 1 |
| miR-4738-3p | 0 | 0.58544894 | 0.588 | 1 |
| miR-4734 | 0 | 0.55445552 | 0.556 | 1 |
| miR-4732-3p | 0 | 0.64891978 | 0.668 | 1 |
| miR-4731-5p | 0 | 0.93728144 | 0.944 | 1 |
| miR-4727-5p | 0 | 0.53974085 | 0.533 | 1 |
| miR-4726-3p | 0 | 0.65436368 | 0.638 | 1 |
| miR-4760-5p | 0 | 0.90517899 | 0.897 | 1 |
| miR-4764-5p | 0 | 0.53294404 | 0.554 | 1 |
| miR-4796-5p | 0 | 0.93235138 | 0.921 | 1 |
| miR-4765 | 0 | 0.84419656 | 0.849 | 1 |
| miR-4793-5p | 0 | 0.76128925 | 0.753 | 1 |
| miR-4793-3p | 0 | 0.86587399 | 0.872 | 1 |
| miR-4791 | 0 | 0.94681148 | 0.952 | 1 |

| | | | | |
|---|---|---|---|---|
| miR-4790-3p | 0 | 0.57639948 | 0.583 | 1 |
| miR-4789-5p | 0 | 0.93252551 | 0.948 | 1 |
| miR-4789-3p | 0 | 0.89304043 | 0.903 | 1 |
| miR-4786-3p | 0 | 0.66052946 | 0.677 | 1 |
| miR-4783-3p | 0 | 0.87589634 | 0.874 | 1 |
| miR-4781-3p | 0 | 0.66852572 | 0.652 | 1 |
| miR-4780 | 0 | 0.82765834 | 0.817 | 1 |
| miR-4777-5p | 0 | 0.84929442 | 0.846 | 1 |
| miR-4775 | 0 | 0.81063593 | 0.807 | 1 |
| miR-4774-5p | 0 | 0.71232797 | 0.695 | 1 |
| miR-4773 | 0 | 0.85354066 | 0.844 | 1 |
| miR-4772-5p | 0 | 0.51850769 | 0.503 | 1 |
| miR-4770 | 0 | 0.61328322 | 0.57 | 1 |
| miR-4769-5p | 0 | 0.7534865 | 0.766 | 1 |
| miR-4769-3p | 0 | 0.78306832 | 0.776 | 1 |
| miR-4766-3p | 0 | 0.580444 | 0.588 | 1 |
| miR-4725-5p | 0 | 0.78017454 | 0.778 | 1 |
| miR-4725-3p | 0 | 0.51448015 | 0.487 | 1 |
| miR-4724-3p | 0 | 0.51974045 | 0.514 | 1 |
| miR-4722-5p | 0 | 0.97233624 | 0.972 | 1 |
| miR-4690-3p | 0 | 0.87989173 | 0.86 | 1 |
| miR-4687-3p | 0 | 0.88188973 | 0.881 | 1 |
| miR-4685-5p | 0 | 0.84539381 | 0.843 | 1 |
| let-7a-3p | 0 | 0.71531555 | 0.705 | 1 |
| miR-4684-3p | 0 | 0.75093178 | 0.755 | 1 |
| miR-4683 | 0 | 0.67586315 | 0.668 | 1 |
| miR-4682 | 0 | 0.90874759 | 0.912 | 1 |
| miR-4681 | 0 | 0.79130483 | 0.793 | 1 |
| miR-4680-5p | 0 | 0.60455026 | 0.596 | 1 |
| miR-4680-3p | 0 | 0.64242871 | 0.62 | 1 |
| miR-4678 | 0 | 0.98844943 | 0.991 | 1 |
| miR-4677-3p | 0 | 0.51597845 | 0.488 | 1 |
| miR-4676-3p | 0 | 0.85279644 | 0.836 | 1 |
| miR-4673 | 0 | 0.94457133 | 0.941 | 1 |
| miR-4671-5p | 0 | 0.50789882 | 0.497 | 1 |
| miR-4671-3p | 0 | 0.51164572 | 0.504 | 1 |
| miR-4667-5p | 0 | 0.88861019 | 0.877 | 1 |
| miR-4667-3p | 0 | 0.54220026 | 0.533 | 1 |
| miR-4666a-5p | 0 | 0.80265483 | 0.817 | 1 |
| miR-4690-5p | 0 | 0.67063114 | 0.672 | 1 |
| miR-4691-3p | 0 | 0.93396675 | 0.933 | 1 |
| miR-4693-3p | 0 | 0.99065837 | 0.996 | 1 |
| miR-4703-5p | 0 | 0.89474134 | 0.881 | 1 |
| miR-4720-3p | 0 | 0.86359237 | 0.861 | 1 |
| miR-4719 | 0 | 0.78899597 | 0.779 | 1 |
| miR-4718 | 0 | 0.71310247 | 0.715 | 1 |
| miR-4713-5p | 0 | 0.91533942 | 0.906 | 1 |
| miR-4709-5p | 0 | 0.94798062 | 0.95 | 1 |
| miR-4707-5p | 0 | 0.64980651 | 0.619 | 1 |
| miR-4706 | 0 | 0.67646602 | 0.66 | 1 |

| | | | | |
|---|---|---|---|---|
| miR-4704-3p | 0 | 0.77412753 | 0.786 | 1 |
| miR-4701-5p | 0 | 0.6815382 | 0.687 | 1 |
| miR-4693-5p | 0 | 0.83860359 | 0.845 | 1 |
| miR-4701-3p | 0 | 0.58049327 | 0.562 | 1 |
| miR-4700-5p | 0 | 0.80768593 | 0.789 | 1 |
| miR-4699-5p | 0 | 0.76359804 | 0.758 | 1 |
| miR-4698 | 0 | 0.81170335 | 0.797 | 1 |
| miR-4697-5p | 0 | 0.7421525 | 0.717 | 1 |
| miR-4697-3p | 0 | 0.86131497 | 0.854 | 1 |
| miR-4695-3p | 0 | 0.81283585 | 0.812 | 1 |
| miR-4694-5p | 0 | 0.86929373 | 0.853 | 1 |
| miR-4795-5p | 0 | 0.8505142 | 0.842 | 1 |
| miR-4798-5p | 0 | 0.90207237 | 0.897 | 1 |
| miR-4663 | 0 | 0.71882886 | 0.713 | 1 |
| miR-520a-5p | 0 | 0.84439343 | 0.854 | 1 |
| miR-519e-3p | 0 | 0.57398036 | 0.587 | 1 |
| miR-519c-3p | 0 | 0.84525046 | 0.848 | 1 |
| miR-519a-3p | 0 | 0.57585799 | 0.6 | 1 |
| miR-5197-5p | 0 | 0.57695232 | 0.597 | 1 |
| miR-5195-5p | 0 | 0.92059714 | 0.915 | 1 |
| miR-5194 | 0 | 0.80157338 | 0.79 | 1 |
| miR-5193 | 0 | 0.56749008 | 0.556 | 1 |
| miR-5191 | 0 | 0.57691607 | 0.583 | 1 |
| miR-5189-3p | 0 | 0.88918044 | 0.893 | 1 |
| miR-5186 | 0 | 0.75298018 | 0.746 | 1 |
| miR-516b-5p | 0 | 0.94458945 | 0.944 | 1 |
| miR-516a-5p | 0 | 0.5793023 | 0.578 | 1 |
| miR-516a-3p | 0 | 0.93324662 | 0.941 | 1 |
| miR-514b-5p | 0 | 0.66484698 | 0.636 | 1 |
| miR-514a-3p | 0 | 0.90665013 | 0.902 | 1 |
| miR-513b-5p | 0 | 0.72937161 | 0.74 | 1 |
| miR-512-5p | 0 | 0.95427325 | 0.944 | 1 |
| miR-512-3p | 0 | 0.54416412 | 0.556 | 1 |
| miR-510-5p | 0 | 0.53415274 | 0.531 | 1 |
| miR-519e-5p | 0 | 0.99308575 | 0.997 | 1 |
| miR-520b | 0 | 0.89648566 | 0.892 | 1 |
| miR-4799-5p | 0 | 0.71586849 | 0.728 | 1 |
| miR-520d-3p | 0 | 0.56385742 | 0.544 | 1 |
| miR-548aw | 0 | 0.70746687 | 0.673 | 1 |
| miR-548ao-3p | 0 | 0.93156969 | 0.925 | 1 |
| miR-548an | 0 | 0.64342055 | 0.638 | 1 |
| miR-548ak | 0 | 0.5728496 | 0.562 | 1 |
| miR-548ag | 0 | 0.95243216 | 0.945 | 1 |
| miR-548ae | 0 | 0.65700025 | 0.638 | 1 |
| miR-548ad | 0 | 0.72195884 | 0.723 | 1 |
| miR-548ab | 0 | 0.72138285 | 0.746 | 1 |
| miR-545-5p | 0 | 0.96885787 | 0.975 | 1 |
| miR-545-3p | 0 | 0.97597882 | 0.985 | 1 |
| miR-544b | 0 | 0.68753514 | 0.671 | 1 |
| miR-544a | 0 | 0.51686303 | 0.511 | 1 |

| | | | | |
|---|---|---|---|---|
| miR-542-3p | 0 | 0.91837291 | 0.921 | 1 |
| miR-541-5p | 0 | 0.53571772 | 0.539 | 1 |
| miR-539-3p | 0 | 0.79084332 | 0.783 | 1 |
| miR-532-5p | 0 | 0.61061441 | 0.597 | 1 |
| miR-522-3p | 0 | 0.61331518 | 0.591 | 1 |
| miR-521 | 0 | 0.96389758 | 0.968 | 1 |
| miR-520f-3p | 0 | 0.81317983 | 0.799 | 1 |
| miR-510-3p | 0 | 0.78929651 | 0.802 | 1 |
| miR-509-5p | 0 | 0.93971885 | 0.942 | 1 |
| miR-5091 | 0 | 0.99787937 | 0.998 | 1 |
| miR-5090 | 0 | 0.76006112 | 0.745 | 1 |
| miR-5001-3p | 0 | 0.58296794 | 0.571 | 1 |
| miR-5000-5p | 0 | 0.59978213 | 0.608 | 1 |
| miR-499b-3p | 0 | 0.55151805 | 0.552 | 1 |
| miR-498 | 0 | 0.64292637 | 0.644 | 1 |
| miR-496 | 0 | 0.9210157 | 0.924 | 1 |
| miR-495-5p | 0 | 0.68531845 | 0.683 | 1 |
| miR-494-5p | 0 | 0.55862372 | 0.546 | 1 |
| miR-494-3p | 0 | 0.85441095 | 0.849 | 1 |
| miR-493-5p | 0 | 0.83365382 | 0.854 | 1 |
| miR-491-5p | 0 | 0.64253706 | 0.645 | 1 |
| miR-491-3p | 0 | 0.98893373 | 0.993 | 1 |
| miR-489-5p | 0 | 0.51260658 | 0.501 | 1 |
| miR-488-5p | 0 | 0.57570391 | 0.574 | 1 |
| miR-487a-5p | 0 | 0.80273222 | 0.79 | 1 |
| miR-487a-3p | 0 | 0.91123461 | 0.909 | 1 |
| miR-4804-5p | 0 | 0.58591127 | 0.595 | 1 |
| miR-4803 | 0 | 0.75618696 | 0.782 | 1 |
| miR-4802-5p | 0 | 0.70040661 | 0.698 | 1 |
| miR-4802-3p | 0 | 0.68683343 | 0.687 | 1 |
| miR-5001-5p | 0 | 0.88452067 | 0.897 | 1 |
| miR-5002-3p | 0 | 0.97989015 | 0.984 | 1 |
| miR-5003-5p | 0 | 0.98764323 | 0.991 | 1 |
| miR-503-3p | 0 | 0.59085645 | 0.61 | 1 |
| miR-5089-3p | 0 | 0.99363948 | 0.996 | 1 |
| miR-5087 | 0 | 0.94785604 | 0.943 | 1 |
| miR-507 | 0 | 0.97819691 | 0.976 | 1 |
| miR-506-5p | 0 | 0.71820444 | 0.718 | 1 |
| miR-505-5p | 0 | 0.63675917 | 0.639 | 1 |
| miR-5047 | 0 | 0.6214201 | 0.649 | 1 |
| miR-504-5p | 0 | 0.93621344 | 0.93 | 1 |
| miR-504-3p | 0 | 0.61296222 | 0.623 | 1 |
| miR-502-5p | 0 | 0.74212373 | 0.767 | 1 |
| miR-5004-3p | 0 | 0.79041256 | 0.789 | 1 |
| miR-502-3p | 0 | 0.55559378 | 0.555 | 1 |
| miR-5011-5p | 0 | 0.64746009 | 0.626 | 1 |
| miR-5010-3p | 0 | 0.54135872 | 0.542 | 1 |
| miR-500b-3p | 0 | 0.92692995 | 0.937 | 1 |
| miR-5009-3p | 0 | 0.83457385 | 0.828 | 1 |
| miR-5008-5p | 0 | 0.65480677 | 0.662 | 1 |

| | | | | |
|---|---|---|---|---|
| miR-5006-5p | 0 | 0.58986827 | 0.607 | 1 |
| miR-5004-5p | 0 | 0.58883768 | 0.579 | 1 |
| miR-4664-5p | 0 | 0.82345288 | 0.844 | 1 |
| miR-4661-5p | 0 | 0.99174466 | 0.992 | 1 |
| miR-3692-3p | 0 | 0.54865534 | 0.539 | 1 |
| miR-4265 | 0 | 0.81962394 | 0.817 | 1 |
| miR-4262 | 0 | 0.81436567 | 0.82 | 1 |
| miR-4255 | 0 | 0.63357775 | 0.632 | 1 |
| miR-4253 | 0 | 0.83591382 | 0.83 | 1 |
| miR-4252 | 0 | 0.94239481 | 0.947 | 1 |
| miR-4251 | 0 | 0.65738789 | 0.653 | 1 |
| miR-424-5p | 0 | 0.87326438 | 0.876 | 1 |
| miR-424-3p | 0 | 0.61517726 | 0.601 | 1 |
| miR-422a | 0 | 0.98510411 | 0.989 | 1 |
| miR-412-3p | 0 | 0.91026826 | 0.911 | 1 |
| miR-3977 | 0 | 0.94579076 | 0.944 | 1 |
| miR-3976 | 0 | 0.92509389 | 0.935 | 1 |
| miR-3974 | 0 | 0.64959711 | 0.639 | 1 |
| miR-3943 | 0 | 0.76790099 | 0.774 | 1 |
| miR-3942-5p | 0 | 0.771393 | 0.756 | 1 |
| miR-3941 | 0 | 0.99581751 | 0.993 | 1 |
| miR-3939 | 0 | 0.8806849 | 0.873 | 1 |
| miR-3935 | 0 | 0.91034352 | 0.916 | 1 |
| miR-3934-5p | 0 | 0.66789104 | 0.678 | 1 |
| miR-3934-3p | 0 | 0.78567028 | 0.784 | 1 |
| miR-4264 | 0 | 0.63660004 | 0.631 | 1 |
| miR-4267 | 0 | 0.62012119 | 0.62 | 1 |
| miR-4320 | 0 | 0.9297955 | 0.932 | 1 |
| miR-4268 | 0 | 0.65943562 | 0.675 | 1 |
| miR-4314 | 0 | 0.85632134 | 0.866 | 1 |
| miR-431-3p | 0 | 0.72472878 | 0.717 | 1 |
| miR-4312 | 0 | 0.84605821 | 0.829 | 1 |
| miR-4310 | 0 | 0.89393386 | 0.896 | 1 |
| miR-4306 | 0 | 0.85565074 | 0.835 | 1 |
| miR-4305 | 0 | 0.51359876 | 0.526 | 1 |
| miR-4300 | 0 | 0.73574376 | 0.732 | 1 |
| miR-4296 | 0 | 0.62892493 | 0.627 | 1 |
| miR-4295 | 0 | 0.76328413 | 0.771 | 1 |
| miR-4292 | 0 | 0.85776594 | 0.867 | 1 |
| miR-4291 | 0 | 0.73674951 | 0.733 | 1 |
| miR-429 | 0 | 0.58178783 | 0.568 | 1 |
| miR-4288 | 0 | 0.96646156 | 0.962 | 1 |
| miR-4283 | 0 | 0.6923597 | 0.702 | 1 |
| miR-4282 | 0 | 0.50030878 | 0.496 | 1 |
| miR-4279 | 0 | 0.66202788 | 0.659 | 1 |
| miR-4275 | 0 | 0.87128066 | 0.889 | 1 |
| miR-4270 | 0 | 0.74503291 | 0.745 | 1 |
| miR-4269 | 0 | 0.50898108 | 0.511 | 1 |
| miR-3929 | 0 | 0.89605287 | 0.907 | 1 |
| miR-3928-3p | 0 | 0.64388241 | 0.633 | 1 |

| | | | | |
|---|---|---|---|---|
| miR-3927-5p | 0 | 0.57904079 | 0.565 | 1 |
| miR-3925-5p | 0 | 0.55265983 | 0.547 | 1 |
| miR-379-3p | 0 | 0.5892093 | 0.572 | 1 |
| miR-378g | 0 | 0.84881743 | 0.846 | 1 |
| miR-378f | 0 | 0.57837378 | 0.575 | 1 |
| miR-378e | 0 | 0.90763124 | 0.915 | 1 |
| miR-378c | 0 | 0.50108996 | 0.511 | 1 |
| miR-377-5p | 0 | 0.60193085 | 0.615 | 1 |
| miR-376c-3p | 0 | 0.6285955 | 0.639 | 1 |
| miR-376b-3p | 0 | 0.97549051 | 0.985 | 1 |
| miR-376a-3p | 0 | 0.63884985 | 0.658 | 1 |
| miR-374c-5p | 0 | 0.51033993 | 0.507 | 1 |
| miR-374b-5p | 0 | 0.95906585 | 0.955 | 1 |
| miR-374b-3p | 0 | 0.92310552 | 0.927 | 1 |
| miR-373-5p | 0 | 0.82550947 | 0.796 | 1 |
| miR-372-5p | 0 | 0.64097168 | 0.668 | 1 |
| miR-371b-3p | 0 | 0.85116511 | 0.87 | 1 |
| miR-3714 | 0 | 0.74256599 | 0.743 | 1 |
| miR-3713 | 0 | 0.68920412 | 0.707 | 1 |
| miR-370-3p | 0 | 0.55609725 | 0.566 | 1 |
| miR-369-3p | 0 | 0.50093936 | 0.477 | 1 |
| miR-380-3p | 0 | 0.87959941 | 0.878 | 1 |
| miR-380-5p | 0 | 0.56852411 | 0.591 | 1 |
| miR-381-3p | 0 | 0.62750198 | 0.66 | 1 |
| miR-3913-5p | 0 | 0.81334023 | 0.812 | 1 |
| miR-3924 | 0 | 0.90928943 | 0.918 | 1 |
| miR-3923 | 0 | 0.69901404 | 0.701 | 1 |
| miR-3922-5p | 0 | 0.66007945 | 0.651 | 1 |
| miR-3922-3p | 0 | 0.97062998 | 0.974 | 1 |
| miR-3917 | 0 | 0.74994068 | 0.721 | 1 |
| miR-3916 | 0 | 0.87303027 | 0.879 | 1 |
| miR-3915 | 0 | 0.94459312 | 0.946 | 1 |
| miR-3914 | 0 | 0.97669268 | 0.967 | 1 |
| miR-3912-3p | 0 | 0.53026161 | 0.498 | 1 |
| miR-382-3p | 0 | 0.85863493 | 0.863 | 1 |
| miR-3910 | 0 | 0.79281962 | 0.785 | 1 |
| miR-3909 | 0 | 0.90480246 | 0.916 | 1 |
| miR-3908 | 0 | 0.5870272 | 0.613 | 1 |
| miR-3907 | 0 | 0.78319586 | 0.774 | 1 |
| miR-384 | 0 | 0.91254423 | 0.925 | 1 |
| miR-383-5p | 0 | 0.80909641 | 0.809 | 1 |
| miR-383-3p | 0 | 0.85037266 | 0.852 | 1 |
| miR-382-5p | 0 | 0.84220688 | 0.826 | 1 |
| miR-4317 | 0 | 0.50874126 | 0.503 | 1 |
| miR-4322 | 0 | 0.7113109 | 0.713 | 1 |
| miR-466 | 0 | 0.50628958 | 0.501 | 1 |
| miR-454-3p | 0 | 0.54652409 | 0.547 | 1 |
| miR-4533 | 0 | 0.79561817 | 0.812 | 1 |
| miR-4531 | 0 | 0.88916759 | 0.883 | 1 |
| miR-4530 | 0 | 0.67095809 | 0.65 | 1 |

| | | | | |
|---|---|---|---|---|
| miR-4528 | 0 | 0.97316233 | 0.979 | 1 |
| miR-452-5p | 0 | 0.81838496 | 0.826 | 1 |
| miR-4524a-5p | 0 | 0.6395877 | 0.638 | 1 |
| miR-452-3p | 0 | 0.96029779 | 0.959 | 1 |
| miR-451b | 0 | 0.82316578 | 0.838 | 1 |
| miR-4518 | 0 | 0.64559518 | 0.646 | 1 |
| miR-4515 | 0 | 0.76099679 | 0.768 | 1 |
| miR-4514 | 0 | 0.93975923 | 0.95 | 1 |
| miR-4513 | 0 | 0.85787223 | 0.851 | 1 |
| miR-4511 | 0 | 0.51666119 | 0.53 | 1 |
| miR-4510 | 0 | 0.76946364 | 0.757 | 1 |
| miR-450b-5p | 0 | 0.68295191 | 0.688 | 1 |
| miR-450a-5p | 0 | 0.75413683 | 0.754 | 1 |
| miR-450a-1-3p | 0 | 0.77402854 | 0.777 | 1 |
| miR-4507 | 0 | 0.52705911 | 0.512 | 1 |
| miR-4501 | 0 | 0.97895203 | 0.977 | 1 |
| miR-4536-5p | 0 | 0.72750821 | 0.743 | 1 |
| miR-454-5p | 0 | 0.53734769 | 0.544 | 1 |
| miR-4324 | 0 | 0.92309287 | 0.92 | 1 |
| miR-455-3p | 0 | 0.99125179 | 0.995 | 1 |
| miR-4659b-5p | 0 | 0.56685477 | 0.542 | 1 |
| miR-4659a-5p | 0 | 0.56690503 | 0.607 | 1 |
| miR-4658 | 0 | 0.76863915 | 0.758 | 1 |
| miR-4655-5p | 0 | 0.66918429 | 0.659 | 1 |
| miR-4655-3p | 0 | 0.89159085 | 0.91 | 1 |
| miR-4653-5p | 0 | 0.96770468 | 0.965 | 1 |
| miR-4653-3p | 0 | 0.60461328 | 0.606 | 1 |
| miR-4652-3p | 0 | 0.50742345 | 0.51 | 1 |
| miR-4649-5p | 0 | 0.76774406 | 0.773 | 1 |
| miR-4649-3p | 0 | 0.88019756 | 0.858 | 1 |
| miR-4648 | 0 | 0.50634525 | 0.499 | 1 |
| miR-4646-3p | 0 | 0.68750395 | 0.683 | 1 |
| miR-4645-3p | 0 | 0.8309048 | 0.832 | 1 |
| miR-4643 | 0 | 0.81002375 | 0.817 | 1 |
| miR-4642 | 0 | 0.95286947 | 0.958 | 1 |
| miR-4640-3p | 0 | 0.6606236 | 0.648 | 1 |
| miR-4638-5p | 0 | 0.87311092 | 0.888 | 1 |
| miR-4632-3p | 0 | 0.92144024 | 0.924 | 1 |
| miR-455-5p | 0 | 0.94262101 | 0.948 | 1 |
| miR-4500 | 0 | 0.82468613 | 0.824 | 1 |
| miR-449c-3p | 0 | 0.72397084 | 0.72 | 1 |
| miR-4499 | 0 | 0.57957433 | 0.58 | 1 |
| miR-4496 | 0 | 0.83615117 | 0.829 | 1 |
| miR-4449 | 0 | 0.88707686 | 0.904 | 1 |
| miR-4442 | 0 | 0.68751005 | 0.674 | 1 |
| miR-4441 | 0 | 0.61856715 | 0.629 | 1 |
| miR-4438 | 0 | 0.86250058 | 0.849 | 1 |
| miR-4435 | 0 | 0.59949297 | 0.604 | 1 |
| miR-4434 | 0 | 0.63242354 | 0.605 | 1 |
| miR-4427 | 0 | 0.96245738 | 0.959 | 1 |

| | | | | |
|---|---|---|---|---|
| miR-4424 | 0 | 0.94134917 | 0.927 | 1 |
| miR-4423-5p | 0 | 0.6183625 | 0.614 | 1 |
| miR-4423-3p | 0 | 0.67232552 | 0.674 | 1 |
| miR-4422 | 0 | 0.7499736 | 0.735 | 1 |
| miR-4420 | 0 | 0.52801817 | 0.532 | 1 |
| miR-4419a | 0 | 0.51871399 | 0.53 | 1 |
| miR-433-3p | 0 | 0.77300404 | 0.743 | 1 |
| miR-4329 | 0 | 0.87213038 | 0.886 | 1 |
| miR-4328 | 0 | 0.80414015 | 0.809 | 1 |
| miR-4327 | 0 | 0.58233284 | 0.568 | 1 |
| miR-432-5p | 0 | 0.97398186 | 0.969 | 1 |
| miR-4325 | 0 | 0.55805108 | 0.546 | 1 |
| miR-4450 | 0 | 0.81071249 | 0.806 | 1 |
| miR-4451 | 0 | 0.92601071 | 0.934 | 1 |
| miR-4452 | 0 | 0.54653745 | 0.506 | 1 |
| miR-4477a | 0 | 0.70890196 | 0.699 | 1 |
| miR-4494 | 0 | 0.84645296 | 0.85 | 1 |
| miR-4491 | 0 | 0.67740767 | 0.669 | 1 |
| miR-4490 | 0 | 0.52858856 | 0.507 | 1 |
| miR-4487 | 0 | 0.63374696 | 0.656 | 1 |
| miR-4486 | 0 | 0.95287201 | 0.951 | 1 |
| miR-4480 | 0 | 0.86372845 | 0.858 | 1 |
| miR-448 | 0 | 0.95632222 | 0.954 | 1 |
| miR-4477b | 0 | 0.8236321 | 0.813 | 1 |
| miR-4476 | 0 | 0.94505763 | 0.947 | 1 |
| miR-4453 | 0 | 0.64932934 | 0.649 | 1 |
| miR-4475 | 0 | 0.68684536 | 0.67 | 1 |
| miR-4471 | 0 | 0.86600379 | 0.864 | 1 |
| miR-4470 | 0 | 0.75182623 | 0.761 | 1 |
| miR-4462 | 0 | 0.97048348 | 0.966 | 1 |
| miR-4460 | 0 | 0.83839875 | 0.841 | 1 |
| miR-4458 | 0 | 0.80351377 | 0.819 | 1 |
| miR-4457 | 0 | 0.5469392 | 0.558 | 1 |
| miR-4456 | 0 | 0.62489458 | 0.624 | 1 |
| miR-99b-5p | 0 | 0.63586561 | 0.643 | 1 |

**S12. Overlap between cis-miR-eQTLs and gene expression QTLs (eQTLs)**

| | | | miR-eQTLs (The Rotterdam Study) | | | | | | | | | | |
|---|---|---|---|---|---|---|---|---|---|---|---|---|---|
| SNP | miRNA | position | effect_alle | other_allel | eaf | beta | se | pval | mir_chr | mir_start | mir_end | cismark | Pvalue |
| rs1003401 | miR-625-5 | 14:662377 | G | A | 0.386875 | 0.147199 | 0.019801 | 1.05E-13 | 14 | 65937834 | 65937854 | cis | 2.27E-159 |
| rs1004373 | miR-27b-3 | 9:9781477 | A | G | 0.113842 | 0.101763 | 0.020093 | 4.09E-07 | 9 | 97847787 | 97847807 | cis | 9.89E-83 |
| rs1004481 | miR-584-5 | 5:1485422 | T | C | 0.211057 | 0.167499 | 0.032112 | 1.83E-07 | 5 | 148441936 | 148441957 | cis | 1.20E-05 |
| rs1004481 | miR-584-5 | 5:1485422 | T | C | 0.211057 | 0.167499 | 0.032112 | 1.83E-07 | 5 | 148441936 | 148441957 | cis | 6.38E-09 |
| rs1004481 | miR-584-5 | 5:1485422 | T | C | 0.211057 | 0.167499 | 0.032112 | 1.83E-07 | 5 | 148441936 | 148441957 | cis | 1.93E-13 |
| rs1004572 | miR-370-3 | 14:101178 | G | A | 0.86193 | 0.12884 | 0.033863 | 0.0001419 | 14 | 101377523 | 101377544 | cis | 9.73E-203 |
| rs1004572 | miR-370-3 | 14:101178 | G | A | 0.86193 | 0.12884 | 0.033863 | 0.0001419 | 14 | 101377523 | 101377544 | cis | 4.37E-10 |
| rs1004572 | miR-370-3 | 14:101178 | G | A | 0.86193 | 0.12884 | 0.033863 | 0.0001419 | 14 | 101377523 | 101377544 | cis | 1.24E-07 |
| rs1004572 | miR-543 | 14:101178 | G | A | 0.86193 | 0.170058 | 0.041319 | 3.86E-05 | 14 | 101498370 | 101498391 | cis | 9.73E-203 |
| rs1004572 | miR-543 | 14:101178 | G | A | 0.86193 | 0.170058 | 0.041319 | 3.86E-05 | 14 | 101498370 | 101498391 | cis | 4.37E-10 |
| rs1004572 | miR-543 | 14:101178 | G | A | 0.86193 | 0.170058 | 0.041319 | 3.86E-05 | 14 | 101498370 | 101498391 | cis | 1.24E-07 |
| rs1005171 | miR-335-5 | 7:1301444 | C | G | 0.448513 | -0.069713 | 0.018725 | 0.0001968 | 7 | 130135967 | 130135989 | cis | 1.04E-12 |
| rs1005171 | miR-335-5 | 7:1301444 | C | G | 0.448513 | -0.069713 | 0.018725 | 0.0001968 | 7 | 130135967 | 130135989 | cis | 6.08E-65 |
| rs1005171 | miR-335-5 | 7:1301444 | C | G | 0.448513 | -0.069713 | 0.018725 | 0.0001968 | 7 | 130135967 | 130135989 | cis | 2.22E-112 |
| rs1005171 | miR-335-5 | 7:1301444 | C | G | 0.448513 | -0.069713 | 0.018725 | 0.0001968 | 7 | 130135967 | 130135989 | cis | 7.40E-20 |
| rs1012638 | miR-193a- | 17:301045 | G | A | 0.758199 | -0.083757 | 0.021036 | 6.85E-05 | 17 | 29887035 | 29887056 | cis | 2.86E-17 |
| rs1012920 | miR-345-5 | 14:100717 | T | C | 0.753859 | -0.068499 | 0.012404 | 3.35E-08 | 14 | 100774213 | 100774234 | cis | 6.30E-09 |
| rs1012920 | miR-345-5 | 14:100717 | T | C | 0.753859 | -0.068499 | 0.012404 | 3.35E-08 | 14 | 100774213 | 100774234 | cis | 3.2717000000 |
| rs1012920 | miR-345-5 | 14:100717 | T | C | 0.753859 | -0.068499 | 0.012404 | 3.35E-08 | 14 | 100774213 | 100774234 | cis | 2.43E-65 |
| rs1012920 | miR-345-5 | 14:100717 | T | C | 0.753859 | -0.068499 | 0.012404 | 3.35E-08 | 14 | 100774213 | 100774234 | cis | 8.07E-14 |
| rs1012920 | miR-345-5 | 14:100717 | T | C | 0.753859 | -0.068499 | 0.012404 | 3.35E-08 | 14 | 100774213 | 100774234 | cis | 1.07E-05 |
| rs1013035 | miR-382-5 | 14:101149 | T | C | 0.112665 | -0.231794 | 0.053696 | 1.58E-05 | 14 | 101520653 | 101520674 | cis | 1.93E-12 |
| rs1013035 | miR-382-5 | 14:101149 | T | C | 0.112665 | -0.231794 | 0.053696 | 1.58E-05 | 14 | 101520653 | 101520674 | cis | 9.99E-10 |
| rs1013035 | miR-382-5 | 14:101149 | T | C | 0.112665 | -0.231794 | 0.053696 | 1.58E-05 | 14 | 101520653 | 101520674 | cis | 4.51E-13 |
| rs1013035 | miR-382-5 | 14:101149 | T | C | 0.112665 | -0.231794 | 0.053696 | 1.58E-05 | 14 | 101520653 | 101520674 | cis | 7.80E-73 |
| rs1013035 | miR-382-5 | 14:101149 | T | C | 0.112665 | -0.231794 | 0.053696 | 1.58E-05 | 14 | 101520653 | 101520674 | cis | 3.34E-10 |
| rs1013035 | miR-494-3 | 14:101149 | T | C | 0.112665 | -0.222111 | 0.040422 | 3.91E-08 | 14 | 101496018 | 101496039 | cis | 1.93E-12 |
| rs1013035 | miR-494-3 | 14:101149 | T | C | 0.112665 | -0.222111 | 0.040422 | 3.91E-08 | 14 | 101496018 | 101496039 | cis | 9.99E-10 |
| rs1013035 | miR-494-3 | 14:101149 | T | C | 0.112665 | -0.222111 | 0.040422 | 3.91E-08 | 14 | 101496018 | 101496039 | cis | 4.51E-13 |
| rs1013035 | miR-494-3 | 14:101149 | T | C | 0.112665 | -0.222111 | 0.040422 | 3.91E-08 | 14 | 101496018 | 101496039 | cis | 7.80E-73 |
| rs1013035 | miR-494-3 | 14:101149 | T | C | 0.112665 | -0.222111 | 0.040422 | 3.91E-08 | 14 | 101496018 | 101496039 | cis | 3.34E-10 |
| rs1013035 | miR-433-3 | 14:101149 | T | C | 0.112665 | -0.197967 | 0.045908 | 1.62E-05 | 14 | 101348286 | 101348307 | cis | 1.93E-12 |
| rs1013035 | miR-433-3 | 14:101149 | T | C | 0.112665 | -0.197967 | 0.045908 | 1.62E-05 | 14 | 101348286 | 101348307 | cis | 9.99E-10 |
| rs1013035 | miR-433-3 | 14:101149 | T | C | 0.112665 | -0.197967 | 0.045908 | 1.62E-05 | 14 | 101348286 | 101348307 | cis | 4.51E-13 |
| rs1013035 | miR-433-3 | 14:101149 | T | C | 0.112665 | -0.197967 | 0.045908 | 1.62E-05 | 14 | 101348286 | 101348307 | cis | 7.80E-73 |
| rs1013035 | miR-433-3 | 14:101149 | T | C | 0.112665 | -0.197967 | 0.045908 | 1.62E-05 | 14 | 101348286 | 101348307 | cis | 3.34E-10 |
| rs1013035 | miR-329-3 | 14:101149 | T | C | 0.112665 | -0.23242 | 0.046261 | 5.06E-07 | 14 | 101493171 | 101493192 | cis | 1.93E-12 |
| rs1013035 | miR-329-3 | 14:101149 | T | C | 0.112665 | -0.23242 | 0.046261 | 5.06E-07 | 14 | 101493171 | 101493192 | cis | 9.99E-10 |
| rs1013035 | miR-329-3 | 14:101149 | T | C | 0.112665 | -0.23242 | 0.046261 | 5.06E-07 | 14 | 101493171 | 101493192 | cis | 4.51E-13 |
| rs1013035 | miR-329-3 | 14:101149 | T | C | 0.112665 | -0.23242 | 0.046261 | 5.06E-07 | 14 | 101493171 | 101493192 | cis | 7.80E-73 |
| rs1013035 | miR-329-3 | 14:101149 | T | C | 0.112665 | -0.23242 | 0.046261 | 5.06E-07 | 14 | 101493171 | 101493192 | cis | 3.34E-10 |
| rs1013035 | miR-409-3 | 14:101149 | T | C | 0.112665 | -0.2959 | 0.051883 | 1.18E-08 | 14 | 101531683 | 101531704 | cis | 1.93E-12 |
| rs1013035 | miR-409-3 | 14:101149 | T | C | 0.112665 | -0.2959 | 0.051883 | 1.18E-08 | 14 | 101531683 | 101531704 | cis | 9.99E-10 |
| rs1013035 | miR-409-3 | 14:101149 | T | C | 0.112665 | -0.2959 | 0.051883 | 1.18E-08 | 14 | 101531683 | 101531704 | cis | 4.51E-13 |
| rs1013035 | miR-409-3 | 14:101149 | T | C | 0.112665 | -0.2959 | 0.051883 | 1.18E-08 | 14 | 101531683 | 101531704 | cis | 7.80E-73 |
| rs1013035 | miR-409-3 | 14:101149 | T | C | 0.112665 | -0.2959 | 0.051883 | 1.18E-08 | 14 | 101531683 | 101531704 | cis | 3.34E-10 |
| rs1013035 | miR-370-3 | 14:101149 | T | C | 0.112665 | -0.227472 | 0.038907 | 5.02E-09 | 14 | 101377523 | 101377544 | cis | 1.93E-12 |
| rs1013035 | miR-370-3 | 14:101149 | T | C | 0.112665 | -0.227472 | 0.038907 | 5.02E-09 | 14 | 101377523 | 101377544 | cis | 4.51E-13 |
| rs1013035 | miR-543 | 14:101149 | T | C | 0.112665 | -0.199338 | 0.047682 | 2.91E-05 | 14 | 101498370 | 101498391 | cis | 1.93E-12 |
| rs1013035 | miR-543 | 14:101149 | T | C | 0.112665 | -0.199338 | 0.047682 | 2.91E-05 | 14 | 101498370 | 101498391 | cis | 4.51E-13 |
| rs1013035 | miR-376a- | 14:101149 | T | C | 0.112665 | -0.248038 | 0.042734 | 6.46E-09 | 14 | 101506455 | 101506475 | cis | 1.93E-12 |
| rs1013035 | miR-376a- | 14:101149 | T | C | 0.112665 | -0.248038 | 0.042734 | 6.46E-09 | 14 | 101506455 | 101506475 | cis | 9.99E-10 |
| rs1013035 | miR-376a- | 14:101149 | T | C | 0.112665 | -0.248038 | 0.042734 | 6.46E-09 | 14 | 101506455 | 101506475 | cis | 4.51E-13 |
| rs1013035 | miR-376a- | 14:101149 | T | C | 0.112665 | -0.248038 | 0.042734 | 6.46E-09 | 14 | 101506455 | 101506475 | cis | 7.80E-73 |
| rs1013035 | miR-376a- | 14:101149 | T | C | 0.112665 | -0.248038 | 0.042734 | 6.46E-09 | 14 | 101506455 | 101506475 | cis | 3.34E-10 |
| rs1013035 | miR-411-3 | 14:101149 | T | C | 0.112665 | -0.336898 | 0.057604 | 4.96E-09 | 14 | 101489712 | 101489733 | cis | 1.93E-12 |
| rs1013035 | miR-411-3 | 14:101149 | T | C | 0.112665 | -0.336898 | 0.057604 | 4.96E-09 | 14 | 101489712 | 101489733 | cis | 9.99E-10 |
| rs1013035 | miR-411-3 | 14:101149 | T | C | 0.112665 | -0.336898 | 0.057604 | 4.96E-09 | 14 | 101489712 | 101489733 | cis | 4.51E-13 |
| rs1013035 | miR-411-3 | 14:101149 | T | C | 0.112665 | -0.336898 | 0.057604 | 4.96E-09 | 14 | 101489712 | 101489733 | cis | 7.80E-73 |
| rs1013035 | miR-411-3 | 14:101149 | T | C | 0.112665 | -0.336898 | 0.057604 | 4.96E-09 | 14 | 101489712 | 101489733 | cis | 3.34E-10 |
| rs1013075 | miR-4707- | 14:233966 | C | G | 0.407058 | 0.086976 | 0.013668 | 1.97E-10 | 14 | 23426166 | 23426187 | cis | 4.51E-49 |
| rs1013075 | miR-4707- | 14:233966 | C | G | 0.407058 | 0.086976 | 0.013668 | 1.97E-10 | 14 | 23426166 | 23426187 | cis | 1.52E-45 |
| rs1013075 | miR-4707- | 14:233966 | C | G | 0.407058 | 0.086976 | 0.013668 | 1.97E-10 | 14 | 23426166 | 23426187 | cis | 1.03E-10 |
| rs1013075 | miR-4707- | 14:233966 | C | G | 0.407058 | 0.086976 | 0.013668 | 1.97E-10 | 14 | 23426166 | 23426187 | cis | 1.41E-30 |
| rs1013075 | miR-4707- | 14:233966 | C | G | 0.407058 | 0.086976 | 0.013668 | 1.97E-10 | 14 | 23426166 | 23426187 | cis | 1.15E-09 |
| rs1013075 | miR-4707- | 14:233966 | C | G | 0.407058 | 0.086976 | 0.013668 | 1.97E-10 | 14 | 23426166 | 23426187 | cis | 3.2717000000 |
| rs1013075 | miR-4707- | 14:233966 | C | G | 0.407058 | 0.086976 | 0.013668 | 1.97E-10 | 14 | 23426166 | 23426187 | cis | 2.09E-07 |
| rs1013075 | miR-4707- | 14:233966 | C | G | 0.407058 | 0.086976 | 0.013668 | 1.97E-10 | 14 | 23426166 | 23426187 | cis | 1.12E-163 |
| rs1014005 | miR-4707- | 14:233589 | G | A | 0.45857 | 0.047653 | 0.013696 | 0.0005028 | 14 | 23426166 | 23426187 | cis | 1.28E-05 |
| rs1014005 | miR-4707- | 14:233589 | G | A | 0.45857 | 0.047653 | 0.013696 | 0.0005028 | 14 | 23426166 | 23426187 | cis | 9.35E-85 |
| rs1014438 | miR-136-5 | 14:101173 | C | T | 0.673446 | 0.405272 | 0.036589 | 1.64E-28 | 14 | 101351053 | 101351075 | cis | 4.83E-46 |
| rs1014438 | miR-136-5 | 14:101173 | C | T | 0.673446 | 0.405272 | 0.036589 | 1.64E-28 | 14 | 101351053 | 101351075 | cis | 1.80E-14 |
| rs1014438 | miR-136-5 | 14:101173 | C | T | 0.673446 | 0.405272 | 0.036589 | 1.64E-28 | 14 | 101351053 | 101351075 | cis | 6.89E-09 |
| rs1014438 | miR-136-5 | 14:101173 | C | T | 0.673446 | 0.405272 | 0.036589 | 1.64E-28 | 14 | 101351053 | 101351075 | cis | 3.2717000000 |
| rs1014551 | miR-376c- | 14:101168 | A | G | 0.668173 | 0.209202 | 0.021737 | 6.31E-22 | 14 | 101506069 | 101506089 | cis | 3.67E-12 |
| rs1014551 | miR-376c- | 14:101168 | A | G | 0.668173 | 0.209202 | 0.021737 | 6.31E-22 | 14 | 101506069 | 101506089 | cis | 3.2717000000 |
| rs1014551 | miR-376c- | 14:101168 | A | G | 0.668173 | 0.209202 | 0.021737 | 6.31E-22 | 14 | 101506069 | 101506089 | cis | 2.39E-09 |
| rs1014551 | miR-376c- | 14:101168 | A | G | 0.668173 | 0.209202 | 0.021737 | 6.31E-22 | 14 | 101506069 | 101506089 | cis | 1.28E-48 |
| rs1014839 | miR-543 | 14:101026 | A | G | 0.750738 | 0.139869 | 0.034163 | 4.24E-05 | 14 | 101498370 | 101498391 | cis | 1.63E-61 |
| rs1014839 | miR-543 | 14:101026 | A | G | 0.750738 | 0.139869 | 0.034163 | 4.24E-05 | 14 | 101498370 | 101498391 | cis | 2.15E-12 |
| rs1014839 | miR-543 | 14:101026 | A | G | 0.750738 | 0.139869 | 0.034163 | 4.24E-05 | 14 | 101498370 | 101498391 | cis | 5.97E-32 |
| rs1014839 | miR-382-5 | 14:101026 | A | G | 0.750738 | 0.225819 | 0.038325 | 3.81E-09 | 14 | 101520653 | 101520674 | cis | 1.63E-61 |
| rs1014839 | miR-382-5 | 14:101026 | A | G | 0.750738 | 0.225819 | 0.038325 | 3.81E-09 | 14 | 101520653 | 101520674 | cis | 2.15E-12 |
| rs1014839 | miR-382-5 | 14:101026 | A | G | 0.750738 | 0.225819 | 0.038325 | 3.81E-09 | 14 | 101520653 | 101520674 | cis | 5.97E-32 |
| rs1014839 | miR-329-3 | 14:101026 | A | G | 0.750738 | 0.138159 | 0.033199 | 3.16E-05 | 14 | 101493171 | 101493192 | cis | 1.63E-61 |
| rs1014839 | miR-329-3 | 14:101026 | A | G | 0.750738 | 0.138159 | 0.033199 | 3.16E-05 | 14 | 101493171 | 101493192 | cis | 2.15E-12 |
| rs1014839 | miR-329-3 | 14:101026 | A | G | 0.750738 | 0.138159 | 0.033199 | 3.16E-05 | 14 | 101493171 | 101493192 | cis | 5.97E-32 |
| rs1014839 | miR-370-3 | 14:101026 | A | G | 0.750738 | 0.185252 | 0.027807 | 2.70E-11 | 14 | 101377523 | 101377544 | cis | 1.63E-61 |
| rs1014839 | miR-370-3 | 14:101026 | A | G | 0.750738 | 0.185252 | 0.027807 | 2.70E-11 | 14 | 101377523 | 101377544 | cis | 2.15E-12 |
| rs1014839 | miR-370-3 | 14:101026 | A | G | 0.750738 | 0.185252 | 0.027807 | 2.70E-11 | 14 | 101377523 | 101377544 | cis | 5.97E-32 |
| rs1015151 | miR-127-3 | 14:101175 | G | A | 0.806743 | 0.377226 | 0.090815 | 3.27E-05 | 14 | 101349372 | 101349393 | cis | 1.57E-22 |
| rs1015151 | miR-127-3 | 14:101175 | G | A | 0.806743 | 0.377226 | 0.090815 | 3.27E-05 | 14 | 101349372 | 101349393 | cis | 1.12E-35 |
| rs1015151 | miR-127-3 | 14:101175 | G | A | 0.806743 | 0.377226 | 0.090815 | 3.27E-05 | 14 | 101349372 | 101349393 | cis | 5.55E-13 |
| rs1015151 | miR-127-3 | 14:101175 | G | A | 0.806743 | 0.377226 | 0.090815 | 3.27E-05 | 14 | 101349372 | 101349393 | cis | 3.2717000000 |
| rs1015151 | miR-127-3 | 14:101175 | G | A | 0.806743 | 0.377226 | 0.090815 | 3.27E-05 | 14 | 101349372 | 101349393 | cis | 3.10E-07 |
| rs1015151 | miR-134-5 | 14:101175 | G | A | 0.806743 | 0.212177 | 0.03585 | 3.25E-09 | 14 | 101521031 | 101521052 | cis | 1.57E-22 |
| rs1015151 | miR-134-5 | 14:101175 | G | A | 0.806743 | 0.212177 | 0.03585 | 3.25E-09 | 14 | 101521031 | 101521052 | cis | 1.12E-35 |
| rs1015151 | miR-134-5 | 14:101175 | G | A | 0.806743 | 0.212177 | 0.03585 | 3.25E-09 | 14 | 101521031 | 101521052 | cis | 5.55E-13 |
| rs1015151 | miR-134-5 | 14:101175 | G | A | 0.806743 | 0.212177 | 0.03585 | 3.25E-09 | 14 | 101521031 | 101521052 | cis | 3.2717000000 |
| rs1015151 | miR-134-5 | 14:101175 | G | A | 0.806743 | 0.212177 | 0.03585 | 3.25E-09 | 14 | 101521031 | 101521052 | cis | 3.10E-07 |
| rs1019976 | miR-3130- | 2:2076418 | G | A | 0.842574 | -0.266836 | 0.032905 | 5.09E-16 | 2 | 207647969 | 207647989 | cis | 2.92E-09 |
| rs102274 | miR-1908- | 11:615578 | T | C | 0.665065 | -0.099704 | 0.017367 | 9.41E-09 | 11 | 61582681 | 61582701 | cis | 1.44E-239 |
| rs102274 | miR-1908- | 11:615578 | T | C | 0.665065 | -0.099704 | 0.017367 | 9.41E-09 | 11 | 61582681 | 61582701 | cis | 1.54E-07 |
| rs102274 | miR-1908- | 11:615578 | T | C | 0.665065 | -0.099704 | 0.017367 | 9.41E-09 | 11 | 61582681 | 61582701 | cis | 3.2717000000 |
| rs102274 | miR-1908- | 11:615578 | T | C | 0.665065 | -0.099704 | 0.017367 | 9.41E-09 | 11 | 61582681 | 61582701 | cis | 1.61E-18 |
| rs102274 | miR-1908- | 11:615578 | T | C | 0.665065 | -0.099704 | 0.017367 | 9.41E-09 | 11 | 61582681 | 61582701 | cis | 3.2717000000 |

| | | | | | | | | | | | | | |
|---|---|---|---|---|---|---|---|---|---|---|---|---|---|
| rs102274 | miR-1908-! | 11:615578: | T | C | 0.665065 | -0.099704 | 0.017367 | 9.41E-09 | 11 | 61582681 | 61582701 | cis | 1.36E-23 |
| rs1022968( | miR-339-5| | 7:1158565 | C | T | 0.347824 | -0.065707 | 0.016838 | 9.53E-05 | 7 | 1062626 | 1062648 | cis | 4.07E-55 |
| rs1022968( | miR-339-5| | 7:1158565 | C | T | 0.347824 | -0.065707 | 0.016838 | 9.53E-05 | 7 | 1062626 | 1062648 | cis | 3.2717000000 |
| rs1022968( | miR-339-5| | 7:1158565 | C | T | 0.347824 | -0.065707 | 0.016838 | 9.53E-05 | 7 | 1062626 | 1062648 | cis | 1.05E-07 |
| rs1022968( | miR-339-5| | 7:1158565 | C | T | 0.347824 | -0.065707 | 0.016838 | 9.53E-05 | 7 | 1062626 | 1062648 | cis | 7.05E-11 |
| rs1022968( | miR-339-5| | 7:1158565 | C | T | 0.347824 | -0.065707 | 0.016838 | 9.53E-05 | 7 | 1062626 | 1062648 | cis | 1.97E-06 |
| rs1022968( | miR-339-5| | 7:1158565 | C | T | 0.347824 | -0.065707 | 0.016838 | 9.53E-05 | 7 | 1062626 | 1062648 | cis | 3.79E-07 |
| rs1022968( | miR-339-5| | 7:1158565 | C | T | 0.347824 | -0.065707 | 0.016838 | 9.53E-05 | 7 | 1062626 | 1062648 | cis | 2.25E-14 |
| rs1022968( | miR-339-5| | 7:1158565 | C | T | 0.347824 | -0.065707 | 0.016838 | 9.53E-05 | 7 | 1062626 | 1062648 | cis | 3.2717000000 |
| rs1028096( | miR-339-5| | 7:1103697 | T | C | 0.468617 | -0.079563 | 0.015528 | 2.99E-07 | 7 | 1062626 | 1062648 | cis | 6.42E-06 |
| rs1051223 | miR-27b-3| | 9:9750990! | T | G | 0.93686 | -0.08632 | 0.025994 | 0.0008978 | 9 | 97847787 | 97847807 | cis | 8.19E-07 |
| rs1075087( | miR-130a-: | 11:576238: | A | C | 0.437769 | -0.15918 | 0.021015 | 3.60E-14 | 11 | 57408725 | 57408746 | cis | 2.41E-123 |
| rs1075087( | miR-130a-: | 11:576238: | A | C | 0.437769 | -0.15918 | 0.021015 | 3.60E-14 | 11 | 57408725 | 57408746 | cis | 5.04E-47 |
| rs1075087( | miR-130a-: | 11:576238: | A | C | 0.437769 | -0.15918 | 0.021015 | 3.60E-14 | 11 | 57408725 | 57408746 | cis | 6.01E-20 |
| rs1075087( | miR-130a-: | 11:576238: | A | C | 0.437769 | -0.15918 | 0.021015 | 3.60E-14 | 11 | 57408725 | 57408746 | cis | 4.80E-47 |
| rs1075087( | miR-130a-: | 11:576238: | A | C | 0.437769 | -0.15918 | 0.021015 | 3.60E-14 | 11 | 57408725 | 57408746 | cis | 1.42E-20 |
| rs1075087( | miR-130a-: | 11:576238: | A | C | 0.437769 | -0.15918 | 0.021015 | 3.60E-14 | 11 | 57408725 | 57408746 | cis | 5.21E-138 |
| rs1075087( | miR-130a-: | 11:576238: | A | C | 0.437769 | -0.15918 | 0.021015 | 3.60E-14 | 11 | 57408725 | 57408746 | cis | 2.07E-09 |
| rs1075087( | miR-130a-: | 11:576238: | A | C | 0.437769 | -0.15918 | 0.021015 | 3.60E-14 | 11 | 57408725 | 57408746 | cis | 3.16E-117 |
| rs1075087( | miR-130a-: | 11:576238: | A | C | 0.437769 | -0.15918 | 0.021015 | 3.60E-14 | 11 | 57408725 | 57408746 | cis | 4.65E-07 |
| rs1075087( | miR-130a-: | 11:576238: | A | C | 0.437769 | -0.15918 | 0.021015 | 3.60E-14 | 11 | 57408725 | 57408746 | cis | 3.61E-22 |
| rs1075087( | miR-130a-: | 11:576238: | A | C | 0.437769 | -0.15918 | 0.021015 | 3.60E-14 | 11 | 57408725 | 57408746 | cis | 4.91E-12 |
| rs1078803 | miR-26b-5| | 2:2196065: | C | A | 0.521605 | 0.055467 | 0.015756 | 0.0004308 | 2 | 219267380 | 219267400 | cis | 7.98E-53 |
| rs1078803 | miR-26b-5| | 2:2196065: | C | A | 0.521605 | 0.055467 | 0.015756 | 0.0004308 | 2 | 219267380 | 219267400 | cis | 6.04E-12 |
| rs1078803 | miR-26b-5| | 2:2196065: | C | A | 0.521605 | 0.055467 | 0.015756 | 0.0004308 | 2 | 219267380 | 219267400 | cis | 3.2717000000 |
| rs1078803 | miR-26b-5| | 2:2196065: | C | A | 0.521605 | 0.055467 | 0.015756 | 0.0004308 | 2 | 219267380 | 219267400 | cis | 3.2717000000 |
| rs1078803 | miR-26b-5| | 2:2196065: | C | A | 0.521605 | 0.055467 | 0.015756 | 0.0004308 | 2 | 219267380 | 219267400 | cis | 3.12E-56 |
| rs1078803 | miR-26b-5| | 2:2196065: | C | A | 0.521605 | 0.055467 | 0.015756 | 0.0004308 | 2 | 219267380 | 219267400 | cis | 1.29E-05 |
| rs1078803 | miR-26b-5| | 2:2196065: | C | A | 0.521605 | 0.055467 | 0.015756 | 0.0004308 | 2 | 219267380 | 219267400 | cis | 2.15E-64 |
| rs1078803 | miR-26b-5| | 2:2196065: | C | A | 0.521605 | 0.055467 | 0.015756 | 0.0004308 | 2 | 219267380 | 219267400 | cis | 5.43E-26 |
| rs1078803 | miR-26b-5| | 2:2196065: | C | A | 0.521605 | 0.055467 | 0.015756 | 0.0004308 | 2 | 219267380 | 219267400 | cis | 4.61E-106 |
| rs1078803 | miR-26b-5| | 2:2196065: | C | A | 0.521605 | 0.055467 | 0.015756 | 0.0004308 | 2 | 219267380 | 219267400 | cis | 5.46E-80 |
| rs1078803 | miR-26b-5| | 2:2196065: | C | A | 0.521605 | 0.055467 | 0.015756 | 0.0004308 | 2 | 219267380 | 219267400 | cis | 3.00E-149 |
| rs1079210: | miR-130a-: | 11:572966! | G | A | 0.774375 | -0.087451 | 0.02504 | 0.0004785 | 11 | 57408725 | 57408746 | cis | 1.02E-08 |
| rs1079210: | miR-130a-: | 11:572966! | G | A | 0.774375 | -0.087451 | 0.02504 | 0.0004785 | 11 | 57408725 | 57408746 | cis | 5.23E-59 |
| rs108815 | miR-130b-: | 22:220254 | A | G | 0.665524 | 0.151687 | 0.024717 | 8.41E-10 | 22 | 22007643 | 22007664 | cis | 3.20E-89 |
| rs108815 | miR-130b-: | 22:220254 | A | G | 0.665524 | 0.151687 | 0.024717 | 8.41E-10 | 22 | 22007643 | 22007664 | cis | 5.74E-33 |
| rs108815 | miR-130b-: | 22:220254 | A | G | 0.665524 | 0.151687 | 0.024717 | 8.41E-10 | 22 | 22007643 | 22007664 | cis | 5.46E-12 |
| rs108815 | miR-130b-: | 22:220254 | A | G | 0.665524 | 0.151687 | 0.024717 | 8.41E-10 | 22 | 22007643 | 22007664 | cis | 2.38E-06 |
| rs108815 | miR-130b-: | 22:220254 | A | G | 0.665524 | 0.151687 | 0.024717 | 8.41E-10 | 22 | 22007643 | 22007664 | cis | 6.33E-18 |
| rs108815 | miR-130b-: | 22:220254 | A | G | 0.665524 | 0.151687 | 0.024717 | 8.41E-10 | 22 | 22007643 | 22007664 | cis | 2.72E-09 |
| rs1089661 | miR-130a-: | 11:572089 | C | T | 0.888821 | -0.124809 | 0.034926 | 0.0003522 | 11 | 57408725 | 57408746 | cis | 9.24E-09 |
| rs1095149! | miR-339-5| | 7:1065746 | A | G | 0.612938 | -0.065288 | 0.016243 | 5.83E-05 | 7 | 1062626 | 1062648 | cis | 1.29E-10 |
| rs1103229 | miR-130b-: | 22:220220( | G | A | 0.843078 | -0.195571 | 0.030515 | 1.46E-10 | 22 | 22007643 | 22007664 | cis | 2.23E-10 |
| rs1103229 | miR-130b-: | 22:220220( | G | A | 0.843078 | -0.195571 | 0.030515 | 1.46E-10 | 22 | 22007643 | 22007664 | cis | 6.36E-18 |
| rs1103229 | miR-130b-: | 22:220220( | G | A | 0.843078 | -0.195571 | 0.030515 | 1.46E-10 | 22 | 22007643 | 22007664 | cis | 1.65E-12 |
| rs1115859( | miR-625-5| | 14:659119 | T | A | 0.507581 | 0.145384 | 0.01988 | 2.61E-13 | 14 | 65937834 | 65937854 | cis | 3.51E-06 |
| rs1116059 | miR-376a-: | 14:101049! | C | T | 0.741662 | 0.208575 | 0.029004 | 6.42E-13 | 14 | 101506455 | 101506475 | cis | 5.04E-15 |
| rs1116059 | miR-376a-: | 14:101049! | C | T | 0.741662 | 0.208575 | 0.029004 | 6.42E-13 | 14 | 101506455 | 101506475 | cis | 2.20E-27 |
| rs1116059 | miR-376a-: | 14:101049! | C | T | 0.741662 | 0.208575 | 0.029004 | 6.42E-13 | 14 | 101506455 | 101506475 | cis | 1.80E-06 |
| rs1116059 | miR-494-3| | 14:101049! | C | T | 0.741662 | 0.13822 | 0.027579 | 5.39E-07 | 14 | 101496018 | 101496039 | cis | 5.04E-15 |
| rs1116059 | miR-494-3| | 14:101049! | C | T | 0.741662 | 0.13822 | 0.027579 | 5.39E-07 | 14 | 101496018 | 101496039 | cis | 2.20E-27 |
| rs1116059 | miR-494-3| | 14:101049! | C | T | 0.741662 | 0.13822 | 0.027579 | 5.39E-07 | 14 | 101496018 | 101496039 | cis | 1.80E-06 |
| rs1116059 | miR-409-3| | 14:101049! | C | T | 0.741662 | 0.307143 | 0.035006 | 1.72E-18 | 14 | 101531683 | 101531704 | cis | 5.04E-15 |
| rs1116059 | miR-409-3| | 14:101049! | C | T | 0.741662 | 0.307143 | 0.035006 | 1.72E-18 | 14 | 101531683 | 101531704 | cis | 2.20E-27 |
| rs1116059 | miR-409-3| | 14:101049! | C | T | 0.741662 | 0.307143 | 0.035006 | 1.72E-18 | 14 | 101531683 | 101531704 | cis | 1.80E-06 |
| rs1116059 | miR-411-3| | 14:101049! | C | T | 0.741662 | 0.145168 | 0.039442 | 0.0002327 | 14 | 101489712 | 101489733 | cis | 5.04E-15 |
| rs1116059 | miR-411-3| | 14:101049! | C | T | 0.741662 | 0.145168 | 0.039442 | 0.0002327 | 14 | 101489712 | 101489733 | cis | 2.20E-27 |
| rs1116059 | miR-411-3| | 14:101049! | C | T | 0.741662 | 0.145168 | 0.039442 | 0.0002327 | 14 | 101489712 | 101489733 | cis | 1.80E-06 |
| rs1116059 | miR-134-5| | 14:101049! | C | T | 0.741662 | 0.279929 | 0.032294 | 4.40E-18 | 14 | 101521031 | 101521052 | cis | 5.04E-15 |
| rs1116059 | miR-134-5| | 14:101049! | C | T | 0.741662 | 0.279929 | 0.032294 | 4.40E-18 | 14 | 101521031 | 101521052 | cis | 2.20E-27 |
| rs1116059 | miR-134-5| | 14:101049! | C | T | 0.741662 | 0.279929 | 0.032294 | 4.40E-18 | 14 | 101521031 | 101521052 | cis | 1.80E-06 |
| rs1116662 | miR-30b-5| | 8:1356252: | C | T | 0.752461 | -0.042004 | 0.011831 | 0.0003845 | 8 | 135812813 | 135812834 | cis | 4.29E-14 |
| rs1116662 | miR-30b-5| | 8:1356252: | C | T | 0.752461 | -0.042004 | 0.011831 | 0.0003845 | 8 | 135812813 | 135812834 | cis | 1.77E-20 |
| rs1116662 | miR-30d-5| | 8:1356252: | C | T | 0.752461 | -0.043189 | 0.009385 | 4.19E-06 | 8 | 135817162 | 135817183 | cis | 4.29E-14 |
| rs1116662 | miR-30d-5| | 8:1356252: | C | T | 0.752461 | -0.043189 | 0.009385 | 4.19E-06 | 8 | 135817162 | 135817183 | cis | 1.77E-20 |
| rs1123554: | miR-139-3| | 11:722854 | G | A | 0.948453 | -0.195792 | 0.053322 | 0.0002408 | 11 | 72326110 | 72326132 | cis | 2.40E-07 |
| rs1123554: | miR-139-3| | 11:722854 | G | A | 0.948453 | -0.195792 | 0.053322 | 0.0002408 | 11 | 72326110 | 72326132 | cis | 4.00E-17 |
| rs1123554: | miR-139-3| | 11:722854 | G | A | 0.948453 | -0.195792 | 0.053322 | 0.0002408 | 11 | 72326110 | 72326132 | cis | 1.87E-13 |
| rs1123554: | miR-139-3| | 11:722854 | G | A | 0.948453 | -0.195792 | 0.053322 | 0.0002408 | 11 | 72326110 | 72326132 | cis | 1.19E-14 |
| rs1162740 | miR-345-5| | 14:100679: | A | G | 0.554108 | 0.046924 | 0.010811 | 1.42E-05 | 14 | 100774213 | 100774234 | cis | 1.21E-05 |
| rs1162759! | miR-625-5| | 14:657977( | G | T | 0.38607 | 0.212976 | 0.019217 | 1.52E-28 | 14 | 65937834 | 65937854 | cis | 4.45E-07 |
| rs1165139! | miR-193a-! | 17:301053: | G | A | 0.828639 | -0.100113 | 0.026705 | 0.0001776 | 17 | 29887035 | 29887056 | cis | 2.09E-10 |
| rs1165372 | miR-744-5| | 17:119889: | G | T | 0.76356 | -0.119524 | 0.035175 | 0.0006789 | 17 | 11985226 | 11985247 | cis | 2.08E-08 |
| rs1165372 | miR-744-5| | 17:119889: | G | T | 0.76356 | -0.119524 | 0.035175 | 0.0006789 | 17 | 11985226 | 11985247 | cis | 4.43E-13 |
| rs1177595: | miR-625-5| | 14:657590: | G | C | 0.845965 | -0.311633 | 0.026756 | 2.37E-31 | 14 | 65937834 | 65937854 | cis | 5.18E-07 |
| rs1184726: | miR-625-5| | 14:657756! | T | G | 0.67304 | -0.150419 | 0.020266 | 1.15E-13 | 14 | 65937834 | 65937854 | cis | 9.73E-06 |
| rs1190903 | miR-127-3| | 14:101096: | C | T | 0.772566 | 0.38519 | 0.088032 | 1.21E-05 | 14 | 101349372 | 101349393 | cis | 2.36E-13 |
| rs1203337( | miR-197-3| | 1:1101306 | A | T | 0.818337 | 0.059149 | 0.017117 | 0.000549 | 1 | 110141562 | 110141583 | cis | 3.47E-08 |
| rs1203337( | miR-197-3| | 1:1101306 | A | T | 0.818337 | 0.059149 | 0.017117 | 0.000549 | 1 | 110141562 | 110141583 | cis | 6.20E-08 |
| rs1203337( | miR-197-3| | 1:1101306 | A | T | 0.818337 | 0.059149 | 0.017117 | 0.000549 | 1 | 110141562 | 110141583 | cis | 9.17E-08 |
| rs1203337( | miR-197-3| | 1:1101306 | A | T | 0.818337 | 0.059149 | 0.017117 | 0.000549 | 1 | 110141562 | 110141583 | cis | 2.63E-57 |
| rs1203337( | miR-197-3| | 1:1101306 | A | T | 0.818337 | 0.059149 | 0.017117 | 0.000549 | 1 | 110141562 | 110141583 | cis | 2.08E-07 |
| rs1203337( | miR-197-3| | 1:1101306 | A | T | 0.818337 | 0.059149 | 0.017117 | 0.000549 | 1 | 110141562 | 110141583 | cis | 3.2717000000 |
| rs1203337( | miR-197-3| | 1:1101306 | A | T | 0.818337 | 0.059149 | 0.017117 | 0.000549 | 1 | 110141562 | 110141583 | cis | 1.52E-06 |
| rs1203337( | miR-197-3| | 1:1101306 | A | T | 0.818337 | 0.059149 | 0.017117 | 0.000549 | 1 | 110141562 | 110141583 | cis | 1.33E-16 |
| rs1203337( | miR-197-3| | 1:1101306 | A | T | 0.818337 | 0.059149 | 0.017117 | 0.000549 | 1 | 110141562 | 110141583 | cis | 1.68E-08 |
| rs1203337( | miR-197-3| | 1:1101306 | A | T | 0.818337 | 0.059149 | 0.017117 | 0.000549 | 1 | 110141562 | 110141583 | cis | 7.20E-07 |
| rs1203337( | miR-197-3| | 1:1101306 | A | T | 0.818337 | 0.059149 | 0.017117 | 0.000549 | 1 | 110141562 | 110141583 | cis | 7.45E-09 |
| rs1214700! | miR-433-3| | 14:101186: | C | T | 0.223221 | -0.245136 | 0.032778 | 7.50E-14 | 14 | 101348286 | 101348307 | cis | 1.05E-09 |
| rs1214700! | miR-431-5| | 14:101186: | C | T | 0.223221 | -0.223582 | 0.036282 | 7.17E-10 | 14 | 101347363 | 101347383 | cis | 1.68E-295 |
| rs1214700! | miR-431-5| | 14:101186: | C | T | 0.223221 | -0.223582 | 0.036282 | 7.17E-10 | 14 | 101347363 | 101347383 | cis | 4.22E-08 |
| rs1214700! | miR-431-5| | 14:101186: | C | T | 0.223221 | -0.223582 | 0.036282 | 7.17E-10 | 14 | 101347363 | 101347383 | cis | 2.22E-16 |
| rs1214700! | miR-431-5| | 14:101186: | C | T | 0.223221 | -0.223582 | 0.036282 | 7.17E-10 | 14 | 101347363 | 101347383 | cis | 2.83E-38 |
| rs1214700! | miR-431-5| | 14:101186: | C | T | 0.223221 | -0.223582 | 0.036282 | 7.17E-10 | 14 | 101347363 | 101347383 | cis | 9.54E-35 |
| rs1214700! | miR-431-5| | 14:101186: | C | T | 0.223221 | -0.223582 | 0.036282 | 7.17E-10 | 14 | 101347363 | 101347383 | cis | 1.05E-09 |
| rs1214784! | miR-136-5| | 14:101144! | C | T | 0.870989 | 0.380494 | 0.05243 | 3.95E-13 | 14 | 101351053 | 101351075 | cis | 5.94E-06 |
| rs1243449( | miR-376c-: | 14:101179: | A | G | 0.18959 | -0.284953 | 0.026482 | 5.29E-27 | 14 | 101506069 | 101506089 | cis | 1.70E-09 |
| rs1243449( | miR-136-5| | 14:101179: | A | G | 0.18959 | -0.494684 | 0.044481 | 9.88E-29 | 14 | 101351053 | 101351075 | cis | 1.70E-09 |
| rs1243715: | miR-4707-: | 14:233729: | G | A | 0.437486 | 0.091279 | 0.013659 | 2.35E-11 | 14 | 23426166 | 23426187 | cis | 2.09E-09 |
| rs1253346! | miR-182-5| | 7:1295694: | G | C | 0.829496 | -0.140414 | 0.030727 | 4.88E-06 | 7 | 129410287 | 129410310 | cis | 7.95E-06 |
| rs1253346! | miR-182-5| | 7:1295694: | G | C | 0.829496 | -0.140414 | 0.030727 | 4.88E-06 | 7 | 129410287 | 129410310 | cis | 1.88E-07 |
| rs1253346! | miR-182-5| | 7:1295694: | G | C | 0.829496 | -0.140414 | 0.030727 | 4.88E-06 | 7 | 129410287 | 129410310 | cis | 7.41E-06 |
| rs1261354! | miR-26b-5| | 2:2196235( | C | T | 0.521807 | 0.055512 | 0.015761 | 0.0004283 | 2 | 219267380 | 219267400 | cis | 1.76E-05 |
| rs1271350( | miR-4433b | 2:6456984( | C | T | 0.714645 | -0.091252 | 0.013166 | 4.19E-12 | 2 | 64567943 | 64567963 | cis | 1.86E-18 |
| rs1271350( | miR-4433b | 2:6456984( | C | T | 0.714645 | -0.091252 | 0.013166 | 4.19E-12 | 2 | 64567943 | 64567963 | cis | 3.00E-06 |
| rs1271350( | miR-4433b | 2:6456984( | C | T | 0.714645 | -0.091252 | 0.013166 | 4.19E-12 | 2 | 64567943 | 64567963 | cis | 7.20E-11 |
| rs1271350( | miR-4433b | 2:6456984( | C | T | 0.714645 | -0.091252 | 0.013166 | 4.19E-12 | 2 | 64567943 | 64567963 | cis | 1.32E-09 |
| rs1297485! | miR-125a-! | 19:521891! | G | A | 0.863745 | -0.067442 | 0.01778 | 0.0001488 | 19 | 52196521 | 52196544 | cis | 1.13E-15 |

| | | | | | | | | | | | | | |
|---|---|---|---|---|---|---|---|---|---|---|---|---|---|
| rs1297485 | miR-125a- | 19:521891 | G | A | 0.863745 | -0.067442 | 0.01778 | 0.0001488 | 19 | 52196521 | 52196544 | cis | 2.41E-21 |
| rs1297485 | miR-125a- | 19:521891 | G | A | 0.863745 | -0.067442 | 0.01778 | 0.0001488 | 19 | 52196521 | 52196544 | cis | 2.81E-67 |
| rs1297485 | miR-125a- | 19:521891 | G | A | 0.863745 | -0.067442 | 0.01778 | 0.0001488 | 19 | 52196521 | 52196544 | cis | 1.47E-27 |
| rs1297485 | miR-125a- | 19:521891 | G | A | 0.863745 | -0.067442 | 0.01778 | 0.0001488 | 19 | 52196521 | 52196544 | cis | 2.29E-23 |
| rs1318719 | miR-584-5 | 5:1485254 | G | A | 0.518797 | 0.146142 | 0.025512 | 1.01E-08 | 5 | 148441936 | 148441957 | cis | 2.91E-06 |
| rs1647710 | miR-130b- | 22:220341 | A | G | 0.386466 | -0.143744 | 0.023958 | 1.97E-09 | 22 | 22007643 | 22007664 | cis | 1.71E-08 |
| rs1669119 | miR-130b- | 22:220313 | C | T | 0.839997 | -0.184875 | 0.030889 | 2.16E-09 | 22 | 22007643 | 22007664 | cis | 1.36E-05 |
| rs174572 | miR-1908- | 11:615982 | C | T | 0.743634 | -0.07722 | 0.01868 | 3.57E-05 | 11 | 61582681 | 61582701 | cis | 9.57E-07 |
| rs2016392 | miR-4707- | 14:234241 | A | G | 0.205834 | 0.056726 | 0.016839 | 0.0007552 | 14 | 23426166 | 23426187 | cis | 5.33E-06 |
| rs2016392 | miR-4707- | 14:234241 | A | G | 0.205834 | 0.056726 | 0.016839 | 0.0007552 | 14 | 23426166 | 23426187 | cis | 2.58E-13 |
| rs2019746 | miR-130b- | 22:220654 | T | C | 0.194666 | 0.178022 | 0.029211 | 1.10E-09 | 22 | 22007643 | 22007664 | cis | 2.65E-06 |
| rs2019746 | miR-130b- | 22:220654 | T | C | 0.194666 | 0.178022 | 0.029211 | 1.10E-09 | 22 | 22007643 | 22007664 | cis | 1.76E-06 |
| rs2236643 | miR-130b- | 22:220583 | A | G | 0.55933 | 0.121474 | 0.023453 | 2.23E-07 | 22 | 22007643 | 22007664 | cis | 3.71E-15 |
| rs2247381 | miR-941 | 20:625466 | A | G | 0.789223 | 0.113853 | 0.030315 | 0.0001729 | 20 | 62550848 | 62550870 | cis | 1.50E-103 |
| rs2247381 | miR-941 | 20:625466 | A | G | 0.789223 | 0.113853 | 0.030315 | 0.0001729 | 20 | 62550848 | 62550870 | cis | 1.85E-05 |
| rs2247381 | miR-941 | 20:625466 | A | G | 0.789223 | 0.113853 | 0.030315 | 0.0001729 | 20 | 62550848 | 62550870 | cis | 1.63E-05 |
| rs2247381 | miR-941 | 20:625466 | A | G | 0.789223 | 0.113853 | 0.030315 | 0.0001729 | 20 | 62550848 | 62550870 | cis | 1.52E-10 |
| rs2247381 | miR-941 | 20:625466 | A | G | 0.789223 | 0.113853 | 0.030315 | 0.0001729 | 20 | 62550848 | 62550870 | cis | 8.74E-30 |
| rs2247381 | miR-941 | 20:625466 | A | G | 0.789223 | 0.113853 | 0.030315 | 0.0001729 | 20 | 62550848 | 62550870 | cis | 4.02E-06 |
| rs2247381 | miR-941 | 20:625466 | A | G | 0.789223 | 0.113853 | 0.030315 | 0.0001729 | 20 | 62550848 | 62550870 | cis | 9.20E-13 |
| rs2556391 | miR-26b-5 | 2:2195563 | T | C | 0.613495 | 0.053869 | 0.016222 | 0.0008977 | 2 | 219267380 | 219267400 | cis | 2.55E-06 |
| rs3124944 | miR-3689a | 9:1377452 | A | G | 0.303119 | 0.395178 | 0.027506 | 8.32E-47 | 9 | 137741340 | 137741361 | cis | 8.81E-11 |
| rs369463 | miR-130b- | 22:220152 | C | T | 0.646159 | 0.080386 | 0.019975 | 5.71E-05 | 22 | 22007605 | 22007625 | cis | 8.44E-61 |
| rs369463 | miR-130b- | 22:220152 | C | T | 0.646159 | 0.080386 | 0.019975 | 5.71E-05 | 22 | 22007605 | 22007625 | cis | 4.55E-15 |
| rs369463 | miR-130b- | 22:220152 | C | T | 0.646159 | 0.080386 | 0.019975 | 5.71E-05 | 22 | 22007605 | 22007625 | cis | 4.88E-36 |
| rs369463 | miR-130b- | 22:220152 | C | T | 0.646159 | 0.080386 | 0.019975 | 5.71E-05 | 22 | 22007605 | 22007625 | cis | 1.05E-07 |
| rs369463 | miR-130b- | 22:220152 | C | T | 0.646159 | 0.080386 | 0.019975 | 5.71E-05 | 22 | 22007605 | 22007625 | cis | 3.36E-07 |
| rs369463 | miR-130b- | 22:220152 | C | T | 0.646159 | 0.080386 | 0.019975 | 5.71E-05 | 22 | 22007605 | 22007625 | cis | 4.42E-20 |
| rs3770211 | miR-26b-5 | 2:2192697 | G | T | 0.627162 | 0.071601 | 0.019396 | 0.0002229 | 2 | 219267380 | 219267400 | cis | 1.49E-06 |
| rs4719841 | miR-148a- | 7:2599753 | A | G | 0.590618 | 0.063531 | 0.017296 | 0.0002395 | 7 | 25989542 | 25989563 | cis | 1.54E-06 |
| rs4809244 | miR-941 | 20:625307 | C | T | 0.950318 | 0.212612 | 0.056743 | 0.000179 | 20 | 62550848 | 62550870 | cis | 1.50E-14 |
| rs4809244 | miR-941 | 20:625307 | C | T | 0.950318 | 0.212612 | 0.056743 | 0.000179 | 20 | 62550848 | 62550870 | cis | 3.47E-07 |
| rs8015600 | miR-4707- | 14:234897 | A | G | 0.526169 | -0.064811 | 0.013382 | 1.28E-06 | 14 | 23426166 | 23426187 | cis | 1.86E-09 |
| rs8034920 | miR-625-5 | 14:656968 | C | A | 0.952874 | -0.258333 | 0.046871 | 3.56E-08 | 14 | 65937834 | 65937854 | cis | 8.97E-06 |
| rs921968 | miR-26b-5 | 2:2192722 | G | T | 0.418427 | -0.066112 | 0.016413 | 5.63E-05 | 2 | 219267380 | 219267400 | cis | 8.16E-08 |
| rs921968 | miR-26b-5 | 2:2192722 | G | T | 0.418427 | -0.066112 | 0.016413 | 5.63E-05 | 2 | 219267380 | 219267400 | cis | 2.56E-07 |
| rs921968 | miR-26b-5 | 2:2192722 | G | T | 0.418427 | -0.066112 | 0.016413 | 5.63E-05 | 2 | 219267380 | 219267400 | cis | 4.69E-09 |
| rs921968 | miR-26b-5 | 2:2192722 | G | T | 0.418427 | -0.066112 | 0.016413 | 5.63E-05 | 2 | 219267380 | 219267400 | cis | 5.20E-11 |

| eQTLs (Võsa et al, 2021) | | | | | | | | | | | | |
|---|---|---|---|---|---|---|---|---|---|---|---|---|
| SNPChr | SNPPos | AssessedAl | OtherAllele | Zscore | Gene | GeneSymb | GeneChr | GenePos | NrCohorts | NrSamples | FDR | BonferroniP |
| 14 | 66237720 | G | A | -26.8986 | ENSG0000( | FUT8 | 14 | 66044074 | 35 | 31060 | 0 | 2.89E-151 |
| 9 | 97814770 | A | G | 19.2685 | ENSG0000( | C9orf3 | 9 | 97669212 | 27 | 26080 | 0 | 1.26E-74 |
| 5 | 148542270 | T | C | -4.3769 | ENSG0000( | ADRB2 | 5 | 148207176 | 36 | 31567 | 0.0311791 | 1 |
| 5 | 148542270 | T | C | -5.8064 | ENSG0000( | PCYOX1L | 5 | 148743393 | 36 | 31567 | 4.52E-05 | 0.811854899 |
| 5 | 148542270 | T | C | 7.3537 | ENSG0000( | SH3TC2 | 5 | 148372964 | 36 | 31567 | 0 | 2.46E-05 |
| 14 | 101178330 | A | G | -30.381 | ENSG0000( | MEG3 | 14 | 101286557 | 33 | 23509 | 0 | 1.24E-194 |
| 14 | 101178330 | A | G | -6.2401 | ENSG0000( | DLK1 | 14 | 101196790 | 36 | 31565 | 0 | 0.055649639 |
| 14 | 101178330 | A | G | -5.288 | ENSG0000( | BEGAIN | 14 | 101028618 | 35 | 31351 | 0.0003831 | 1 |
| 14 | 101178330 | A | G | -30.381 | ENSG0000( | MEG3 | 14 | 101286557 | 33 | 23509 | 0 | 1.24E-194 |
| 14 | 101178330 | A | G | -6.2401 | ENSG0000( | DLK1 | 14 | 101196790 | 36 | 31565 | 0 | 0.055649639 |
| 14 | 101178330 | A | G | -5.288 | ENSG0000( | BEGAIN | 14 | 101028618 | 35 | 31351 | 0.0003831 | 1 |
| 7 | 130144402 | C | G | -7.1256 | ENSG0000( | MEST | 7 | 130136089 | 36 | 31568 | 0 | 0.000131964 |
| 7 | 130144402 | C | G | -17.0174 | ENSG0000( | COPG2 | 7 | 130249843 | 34 | 26279 | 0 | 7.74E-57 |
| 7 | 130144402 | C | G | 22.5278 | ENSG0000( | CPA5 | 7 | 129996600 | 16 | 14262 | 0 | 2.82E-104 |
| 7 | 130144402 | C | G | 9.1218 | ENSG0000( | CEP41 | 7 | 130059324 | 33 | 25360 | 0 | 9.43E-12 |
| 17 | 30104598 | A | G | 8.4521 | ENSG0000( | COPRS | 17 | 30182619 | 33 | 25237 | 0 | 3.65E-09 |
| 14 | 100717821 | C | T | -5.8086 | ENSG0000( | RP11-638I2 | 14 | 100833111 | 13 | 5500 | 4.52E-05 | 0.802215125 |
| 14 | 100717821 | C | T | 50.9075 | ENSG0000( | SLC25A29 | 14 | 100765166 | 34 | 29414 | 0 | 4.17E-302 |
| 14 | 100717821 | C | T | -17.0711 | ENSG0000( | WARS | 14 | 100821633 | 33 | 24874 | 0 | 3.10E-57 |
| 14 | 100717821 | C | T | 7.4692 | ENSG0000( | WDR25 | 14 | 100919697 | 35 | 30163 | 0 | 1.03E-05 |
| 14 | 100717821 | C | T | 4.4029 | ENSG0000( | RP11-638I2 | 14 | 100730930 | 9 | 3831 | 0.0277381 | 1 |
| 14 | 101149511 | T | C | -7.0392 | ENSG0000( | WARS | 14 | 100821633 | 30 | 24788 | 0 | 0.000246215 |
| 14 | 101149511 | T | C | 6.1097 | ENSG0000( | BEGAIN | 14 | 101028618 | 31 | 29863 | 1.32E-05 | 0.127190261 |
| 14 | 101149511 | T | C | 7.2394 | ENSG0000( | WDR25 | 14 | 100919697 | 32 | 30077 | 0 | 5.74E-05 |
| 14 | 101149511 | T | C | -18.0505 | ENSG0000( | MEG3 | 14 | 101286557 | 29 | 22021 | 0 | 9.94E-65 |
| 14 | 101149511 | T | C | -6.2821 | ENSG0000( | DLK1 | 14 | 101196790 | 32 | 30077 | 0 | 0.042509239 |
| 14 | 101149511 | T | C | -7.0392 | ENSG0000( | WARS | 14 | 100821633 | 30 | 24788 | 0 | 0.000246215 |
| 14 | 101149511 | T | C | 6.1097 | ENSG0000( | BEGAIN | 14 | 101028618 | 31 | 29863 | 1.32E-05 | 0.127190261 |
| 14 | 101149511 | T | C | 7.2394 | ENSG0000( | WDR25 | 14 | 100919697 | 32 | 30077 | 0 | 5.74E-05 |
| 14 | 101149511 | T | C | -18.0505 | ENSG0000( | MEG3 | 14 | 101286557 | 29 | 22021 | 0 | 9.94E-65 |
| 14 | 101149511 | T | C | -6.2821 | ENSG0000( | DLK1 | 14 | 101196790 | 32 | 30077 | 0 | 0.042509239 |
| 14 | 101149511 | T | C | -7.0392 | ENSG0000( | WARS | 14 | 100821633 | 30 | 24788 | 0 | 0.000246215 |
| 14 | 101149511 | T | C | 6.1097 | ENSG0000( | BEGAIN | 14 | 101028618 | 31 | 29863 | 1.32E-05 | 0.127190261 |
| 14 | 101149511 | T | C | 7.2394 | ENSG0000( | WDR25 | 14 | 100919697 | 32 | 30077 | 0 | 5.74E-05 |
| 14 | 101149511 | T | C | -18.0505 | ENSG0000( | MEG3 | 14 | 101286557 | 29 | 22021 | 0 | 9.94E-65 |
| 14 | 101149511 | T | C | -6.2821 | ENSG0000( | DLK1 | 14 | 101196790 | 32 | 30077 | 0 | 0.042509239 |
| 14 | 101149511 | T | C | -7.0392 | ENSG0000( | WARS | 14 | 100821633 | 30 | 24788 | 0 | 0.000246215 |
| 14 | 101149511 | T | C | 6.1097 | ENSG0000( | BEGAIN | 14 | 101028618 | 31 | 29863 | 1.32E-05 | 0.127190261 |
| 14 | 101149511 | T | C | 7.2394 | ENSG0000( | WDR25 | 14 | 100919697 | 32 | 30077 | 0 | 5.74E-05 |
| 14 | 101149511 | T | C | -18.0505 | ENSG0000( | MEG3 | 14 | 101286557 | 29 | 22021 | 0 | 9.94E-65 |
| 14 | 101149511 | T | C | -6.2821 | ENSG0000( | DLK1 | 14 | 101196790 | 32 | 30077 | 0 | 0.042509239 |
| 14 | 101149511 | T | C | -7.0392 | ENSG0000( | WARS | 14 | 100821633 | 30 | 24788 | 0 | 0.000246215 |
| 14 | 101149511 | T | C | 6.1097 | ENSG0000( | BEGAIN | 14 | 101028618 | 31 | 29863 | 1.32E-05 | 0.127190261 |
| 14 | 101149511 | T | C | 7.2394 | ENSG0000( | WDR25 | 14 | 100919697 | 32 | 30077 | 0 | 5.74E-05 |
| 14 | 101149511 | T | C | -18.0505 | ENSG0000( | MEG3 | 14 | 101286557 | 29 | 22021 | 0 | 9.94E-65 |
| 14 | 101149511 | T | C | -6.2821 | ENSG0000( | DLK1 | 14 | 101196790 | 32 | 30077 | 0 | 0.042509239 |
| 14 | 101149511 | T | C | -7.0392 | ENSG0000( | WARS | 14 | 100821633 | 30 | 24788 | 0 | 0.000246215 |
| 14 | 101149511 | T | C | 7.2394 | ENSG0000( | WDR25 | 14 | 100919697 | 32 | 30077 | 0 | 5.74E-05 |
| 14 | 101149511 | T | C | -7.0392 | ENSG0000( | WARS | 14 | 100821633 | 30 | 24788 | 0 | 0.000246215 |
| 14 | 101149511 | T | C | 7.2394 | ENSG0000( | WDR25 | 14 | 100919697 | 32 | 30077 | 0 | 5.74E-05 |
| 14 | 101149511 | T | C | -7.0392 | ENSG0000( | WARS | 14 | 100821633 | 30 | 24788 | 0 | 0.000246215 |
| 14 | 101149511 | T | C | 6.1097 | ENSG0000( | BEGAIN | 14 | 101028618 | 31 | 29863 | 1.32E-05 | 0.127190261 |
| 14 | 101149511 | T | C | 7.2394 | ENSG0000( | WDR25 | 14 | 100919697 | 32 | 30077 | 0 | 5.74E-05 |
| 14 | 101149511 | T | C | -18.0505 | ENSG0000( | MEG3 | 14 | 101286557 | 29 | 22021 | 0 | 9.94E-65 |
| 14 | 101149511 | T | C | -6.2821 | ENSG0000( | DLK1 | 14 | 101196790 | 32 | 30077 | 0 | 0.042509239 |
| 14 | 101149511 | T | C | -7.0392 | ENSG0000( | WARS | 14 | 100821633 | 30 | 24788 | 0 | 0.000246215 |
| 14 | 101149511 | T | C | 6.1097 | ENSG0000( | BEGAIN | 14 | 101028618 | 31 | 29863 | 1.32E-05 | 0.127190261 |
| 14 | 101149511 | T | C | 7.2394 | ENSG0000( | WDR25 | 14 | 100919697 | 32 | 30077 | 0 | 5.74E-05 |
| 14 | 101149511 | T | C | -18.0505 | ENSG0000( | MEG3 | 14 | 101286557 | 29 | 22021 | 0 | 9.94E-65 |
| 14 | 101149511 | T | C | -6.2821 | ENSG0000( | DLK1 | 14 | 101196790 | 32 | 30077 | 0 | 0.042509239 |
| 14 | 23396600 | C | G | -14.724 | ENSG0000( | RP11-298I3 | 14 | 23411794 | 13 | 5500 | 0 | 5.74E-41 |
| 14 | 23396600 | C | G | 14.1648 | ENSG0000( | HAUS4 | 14 | 23420903 | 35 | 30818 | 0 | 1.93E-37 |
| 14 | 23396600 | C | G | 6.463 | ENSG0000( | PSMB11 | 14 | 23512322 | 22 | 19334 | 0 | 0.013071636 |
| 14 | 23396600 | C | G | 11.4942 | ENSG0000( | REM2 | 14 | 23354634 | 36 | 31567 | 0 | 1.80E-22 |
| 14 | 23396600 | C | G | -6.0862 | ENSG0000( | PRMT5-AS1 | 14 | 23392385 | 11 | 4655 | 1.31E-05 | 0.147054308 |
| 14 | 23396600 | C | G | -41.0864 | ENSG0000( | PRMT5 | 14 | 23394257 | 34 | 30434 | 0 | 4.17E-302 |
| 14 | 23396600 | C | G | 5.1909 | ENSG0000( | C14orf93 | 14 | 23467742 | 36 | 31567 | 0.0006433 | 1 |
| 14 | 23396600 | C | G | -27.2643 | ENSG0000( | RBM23 | 14 | 23379123 | 36 | 31567 | 0 | 1.43E-155 |
| 14 | 23358922 | G | A | 4.3636 | ENSG0000( | ABHD4 | 14 | 23074205 | 35 | 30156 | 0.0330267 | 1 |
| 14 | 23358922 | G | A | 19.5083 | ENSG0000( | LRP10 | 14 | 23345805 | 35 | 30156 | 0 | 1.19E-76 |
| 14 | 101173297 | T | C | -14.2447 | ENSG0000( | DLK1 | 14 | 101196790 | 34 | 29642 | 0 | 6.15E-38 |
| 14 | 101173297 | T | C | -7.6644 | ENSG0000( | BEGAIN | 14 | 101028618 | 33 | 29428 | 0 | 2.29E-06 |
| 14 | 101173297 | T | C | 5.7938 | ENSG0000( | WARS | 14 | 100821633 | 32 | 24353 | 4.52E-05 | 0.877295849 |
| 14 | 101173297 | T | C | -52.4401 | ENSG0000( | MEG3 | 14 | 101286557 | 31 | 21586 | 0 | 4.17E-302 |
| 14 | 101168587 | G | A | -6.9494 | ENSG0000( | BEGAIN | 14 | 101028618 | 35 | 31345 | 0 | 0.000467141 |
| 14 | 101168587 | G | A | -51.5421 | ENSG0000( | MEG3 | 14 | 101286557 | 33 | 23503 | 0 | 4.17E-302 |
| 14 | 101168587 | G | A | 5.9688 | ENSG0000( | WARS | 14 | 100821633 | 34 | 26270 | 2.61E-05 | 0.304346897 |
| 14 | 101168587 | G | A | -14.6532 | ENSG0000( | DLK1 | 14 | 101196790 | 36 | 31559 | 0 | 1.63E-40 |
| 14 | 101026299 | G | A | 16.5488 | ENSG0000( | SLC25A29 | 14 | 100765166 | 34 | 28049 | 0 | 2.08E-53 |
| 14 | 101026299 | G | A | 7.0244 | ENSG0000( | RP11-638I2 | 14 | 100833111 | 13 | 5498 | 0 | 0.000273772 |
| 14 | 101026299 | G | A | -11.764 | ENSG0000( | CTD-2062F | 14 | 101006011 | 19 | 15590 | 0 | 7.60E-24 |
| 14 | 101026299 | G | A | 16.5488 | ENSG0000( | SLC25A29 | 14 | 100765166 | 34 | 28049 | 0 | 2.08E-53 |
| 14 | 101026299 | G | A | 7.0244 | ENSG0000( | RP11-638I2 | 14 | 100833111 | 13 | 5498 | 0 | 0.000273772 |
| 14 | 101026299 | G | A | -11.764 | ENSG0000( | CTD-2062F | 14 | 101006011 | 19 | 15590 | 0 | 7.60E-24 |
| 14 | 101026299 | G | A | 16.5488 | ENSG0000( | SLC25A29 | 14 | 100765166 | 34 | 28049 | 0 | 2.08E-53 |
| 14 | 101026299 | G | A | 7.0244 | ENSG0000( | RP11-638I2 | 14 | 100833111 | 13 | 5498 | 0 | 0.000273772 |
| 14 | 101026299 | G | A | -11.764 | ENSG0000( | CTD-2062F | 14 | 101006011 | 19 | 15590 | 0 | 7.60E-24 |
| 14 | 101026299 | G | A | 16.5488 | ENSG0000( | SLC25A29 | 14 | 100765166 | 34 | 28049 | 0 | 2.08E-53 |
| 14 | 101026299 | G | A | 7.0244 | ENSG0000( | RP11-638I2 | 14 | 100833111 | 13 | 5498 | 0 | 0.000273772 |
| 14 | 101026299 | G | A | -11.764 | ENSG0000( | CTD-2062F | 14 | 101006011 | 19 | 15590 | 0 | 7.60E-24 |
| 14 | 101175798 | A | G | -9.7661 | ENSG0000( | DLK1 | 14 | 101196790 | 34 | 30074 | 0 | 2.00E-14 |
| 14 | 101175798 | A | G | -12.4679 | ENSG0000( | BEGAIN | 14 | 101028618 | 33 | 29860 | 0 | 1.42E-27 |
| 14 | 101175798 | A | G | 7.2113 | ENSG0000( | WARS | 14 | 100821633 | 32 | 24785 | 0 | 7.06E-05 |
| 14 | 101175798 | A | G | -39.0538 | ENSG0000( | MEG3 | 14 | 101286557 | 31 | 22018 | 0 | 4.17E-302 |
| 14 | 101175798 | A | G | -5.117 | ENSG0000( | WDR25 | 14 | 100919697 | 34 | 30074 | 0.0009506 | 1 |
| 14 | 101175798 | A | G | -9.7661 | ENSG0000( | DLK1 | 14 | 101196790 | 34 | 30074 | 0 | 2.00E-14 |
| 14 | 101175798 | A | G | -12.4679 | ENSG0000( | BEGAIN | 14 | 101028618 | 33 | 29860 | 0 | 1.42E-27 |
| 14 | 101175798 | A | G | 7.2113 | ENSG0000( | WARS | 14 | 100821633 | 32 | 24785 | 0 | 7.06E-05 |
| 14 | 101175798 | A | G | -39.0538 | ENSG0000( | MEG3 | 14 | 101286557 | 31 | 22018 | 0 | 4.17E-302 |
| 14 | 101175798 | A | G | -5.117 | ENSG0000( | WDR25 | 14 | 100919697 | 34 | 30074 | 0.0009506 | 1 |
| 2 | 207641868 | A | G | 5.9362 | ENSG0000( | FASTKD2 | 2 | 207643657 | 36 | 31568 | 2.60E-05 | 0.371456025 |
| 11 | 61557826 | C | T | 33.0525 | ENSG0000( | TMEM258 | 11 | 61558354 | 34 | 25574 | 0 | 1.83E-231 |
| 11 | 61557826 | C | T | 5.248 | ENSG0000( | FTH1 | 11 | 61731161 | 34 | 31144 | 0.0004826 | 1 |
| 11 | 61557826 | C | T | 41.6904 | ENSG0000( | FADS1 | 11 | 61581944 | 36 | 31568 | 0 | 4.17E-302 |
| 11 | 61557826 | C | T | 8.7815 | ENSG0000( | BEST1 | 11 | 61725140 | 36 | 31568 | 0 | 2.06E-10 |
| 11 | 61557826 | C | T | 67.4729 | ENSG0000( | FADS2 | 11 | 61597639 | 35 | 31354 | 0 | 4.17E-302 |

| | | | | | | | | | | | | |
|---|---|---|---|---|---|---|---|---|---|---|---|---|
| 11 | 61557826 | C | T | 10.0113 | ENSG0000( | MYRF | 11 | 61538052 | 34 | 25574 | 0 | 1.73E-15 |
| 7 | 1158565 | C | T | -15.6369 | ENSG0000( | GPR146 | 7 | 1091554 | 37 | 31684 | 0 | 5.19E-47 |
| 7 | 1158565 | C | T | 61.5601 | ENSG0000( | AC091729. | 7 | 1202456 | 13 | 5502 | 0 | 4.17E-302 |
| 7 | 1158565 | C | T | 5.3178 | ENSG0000( | COX19 | 7 | 976825 | 36 | 26609 | 0.0003271 | 1 |
| 7 | 1158565 | C | T | 6.5196 | ENSG0000( | ADAP1 | 7 | 966291 | 29 | 26410 | 0 | 0.00897696 |
| 7 | 1158565 | C | T | -4.7566 | ENSG0000( | GPER | 7 | 1127647 | 36 | 26609 | 0.0053753 | 1 |
| 7 | 1158565 | C | T | -5.0793 | ENSG0000( | C7orf50 | 7 | 1107259 | 37 | 31684 | 0.0011159 | 1 |
| 7 | 1158565 | C | T | 7.6352 | ENSG0000( | GET4 | 7 | 926131 | 36 | 31470 | 0 | 2.87E-06 |
| 7 | 1158565 | C | T | 91.222 | ENSG0000( | ZFAND2A | 7 | 1196051 | 37 | 31684 | 0 | 4.17E-302 |
| 7 | 1103697 | T | C | 4.5121 | ENSG0000( | MIR339 | 7 | 1062615 | 19 | 11813 | 0.0170004 | 1 |
| 9 | 97509909 | G | T | 4.9309 | ENSG0000( | FBP1 | 9 | 97383973 | 35 | 30606 | 0.0024213 | 1 |
| 11 | 57623815 | A | C | -23.6197 | ENSG0000( | TIMM10 | 11 | 57297106 | 37 | 31682 | 0 | 3.07E-115 |
| 11 | 57623815 | A | C | -14.4016 | ENSG0000( | MED19 | 11 | 57475439 | 37 | 31682 | 0 | 6.42E-39 |
| 11 | 57623815 | A | C | -9.1441 | ENSG0000( | CLP1 | 11 | 57422902 | 37 | 31682 | 0 | 7.65E-12 |
| 11 | 57623815 | A | C | -14.4051 | ENSG0000( | TMX2 | 11 | 57494258 | 14 | 6419 | 0 | 6.11E-39 |
| 11 | 57623815 | A | C | -9.2989 | ENSG0000( | SMTNL1 | 11 | 57313543 | 31 | 25671 | 0 | 1.81E-12 |
| 11 | 57623815 | A | C | -25.0063 | ENSG0000( | ZDHHC5 | 11 | 57451939 | 36 | 30933 | 0 | 6.63E-130 |
| 11 | 57623815 | A | C | 5.9922 | ENSG0000( | LPXN | 11 | 58320018 | 37 | 31682 | 1.96E-05 | 0.263954079 |
| 11 | 57623815 | A | C | -23.0166 | ENSG0000( | SERPING1 | 11 | 57373593 | 37 | 31682 | 0 | 4.03E-109 |
| 11 | 57623815 | A | C | -5.0401 | ENSG0000( | ZFP91 | 11 | 58367549 | 37 | 31682 | 0.0014161 | 1 |
| 11 | 57623815 | A | C | -9.6815 | ENSG0000( | YPEL4 | 11 | 57414988 | 37 | 31682 | 0 | 4.60E-14 |
| 11 | 57623815 | A | C | -6.9081 | ENSG0000( | CTNND1 | 11 | 57553866 | 36 | 31468 | 0 | 0.000625044 |
| 2 | 219606538 | A | C | 15.2974 | ENSG0000( | PRKAG3 | 2 | 219691957 | 32 | 29317 | 0 | 1.02E-44 |
| 2 | 219606538 | A | C | -6.8786 | ENSG0000( | BCS1L | 2 | 219525826 | 35 | 26494 | 0 | 0.000769705 |
| 2 | 219606538 | A | C | -40.2315 | ENSG0000( | RQCD1 | 2 | 219447553 | 15 | 9188 | 0 | 4.17E-302 |
| 2 | 219606538 | A | C | 134.4068 | ENSG0000( | CYP27A1 | 2 | 219663244 | 36 | 31569 | 0 | 4.17E-302 |
| 2 | 219606538 | A | C | 15.8 | ENSG0000( | USP37 | 2 | 219374029 | 36 | 31569 | 0 | 3.97E-48 |
| 2 | 219606538 | A | C | -4.3611 | ENSG0000( | CTDSP1 | 2 | 219266821 | 36 | 31569 | 0.0333511 | 1 |
| 2 | 219606538 | A | C | 16.9437 | ENSG0000( | SLC11A1 | 2 | 219254184 | 36 | 31569 | 0 | 2.73E-56 |
| 2 | 219606538 | A | C | -10.5438 | ENSG0000( | STK36 | 2 | 219552094 | 36 | 31569 | 0 | 6.91E-18 |
| 2 | 219606538 | A | C | 21.874 | ENSG0000( | ZNF142 | 2 | 219513508 | 36 | 31569 | 0 | 5.88E-98 |
| 2 | 219606538 | A | C | -18.9387 | ENSG0000( | VIL1 | 2 | 219300916 | 16 | 14263 | 0 | 6.96E-72 |
| 2 | 219606538 | A | C | 26.0193 | ENSG0000( | TTLL4 | 2 | 219597853 | 36 | 31569 | 0 | 3.82E-141 |
| 11 | 57296659 | A | G | -5.7269 | ENSG0000( | SLC43A1 | 11 | 57267633 | 35 | 31086 | 5.80E-05 | 1 |
| 11 | 57296659 | A | G | 16.1978 | ENSG0000( | UBE2L6 | 11 | 57327443 | 37 | 31684 | 0 | 6.66E-51 |
| 22 | 22025476 | G | A | 20.0271 | ENSG0000( | PPIL2 | 22 | 22029562 | 37 | 31684 | 0 | 4.08E-81 |
| 22 | 22025476 | G | A | 11.9604 | ENSG0000( | UBE2L3 | 22 | 21941029 | 37 | 31684 | 0 | 7.31E-25 |
| 22 | 22025476 | G | A | 6.8931 | ENSG0000( | PPM1F | 22 | 22290501 | 36 | 31470 | 0 | 0.000695643 |
| 22 | 22025476 | G | A | -4.7178 | ENSG0000( | HIC2 | 22 | 21788722 | 37 | 31684 | 0.0064395 | 1 |
| 22 | 22025476 | G | A | 8.6265 | ENSG0000( | LL22NC03- | 22 | 22295204 | 14 | 8269 | 0 | 8.06E-10 |
| 22 | 22025476 | G | A | -5.9475 | ENSG0000( | IGLV8-61 | 22 | 22453366 | 30 | 24878 | 2.61E-05 | 0.346497032 |
| 11 | 57208915 | T | C | 5.7443 | ENSG0000( | TNKS1BP1 | 11 | 57079769 | 16 | 14248 | 5.80E-05 | 1 |
| 7 | 1065746 | G | A | 6.4281 | ENSG0000( | SUN1 | 7 | 895800 | 35 | 24939 | 0 | 0.016464021 |
| 22 | 22022005 | A | G | -6.3447 | ENSG0000( | PI4KAP2 | 22 | 21849555 | 14 | 6421 | 0 | 0.028360292 |
| 22 | 22022005 | A | G | -8.6258 | ENSG0000( | YDJC | 22 | 21983365 | 33 | 26395 | 0 | 8.10E-10 |
| 22 | 22022005 | A | G | -7.0611 | ENSG0000( | YPEL1 | 22 | 22070978 | 36 | 31569 | 0 | 0.000210203 |
| 14 | 65911975 | T | A | 4.6386 | ENSG0000( | ZBTB25 | 14 | 64943877 | 33 | 29613 | 0.0094069 | 1 |
| 14 | 101049906 | T | C | -7.8257 | ENSG0000( | CTD-2062F | 14 | 101006011 | 19 | 15590 | 0 | 6.42E-07 |
| 14 | 101049906 | T | C | 10.8412 | ENSG0000( | SLC25A29 | 14 | 100765166 | 35 | 30808 | 0 | 2.80E-19 |
| 14 | 101049906 | T | C | 4.7747 | ENSG0000( | RP11-638I2 | 14 | 100833111 | 13 | 5490 | 0.0049525 | 1 |
| 14 | 101049906 | T | C | -7.8257 | ENSG0000( | CTD-2062F | 14 | 101006011 | 19 | 15590 | 0 | 6.42E-07 |
| 14 | 101049906 | T | C | 10.8412 | ENSG0000( | SLC25A29 | 14 | 100765166 | 35 | 30808 | 0 | 2.80E-19 |
| 14 | 101049906 | T | C | 4.7747 | ENSG0000( | RP11-638I2 | 14 | 100833111 | 13 | 5490 | 0.0049525 | 1 |
| 14 | 101049906 | T | C | -7.8257 | ENSG0000( | CTD-2062F | 14 | 101006011 | 19 | 15590 | 0 | 6.42E-07 |
| 14 | 101049906 | T | C | 10.8412 | ENSG0000( | SLC25A29 | 14 | 100765166 | 35 | 30808 | 0 | 2.80E-19 |
| 14 | 101049906 | T | C | 4.7747 | ENSG0000( | RP11-638I2 | 14 | 100833111 | 13 | 5490 | 0.0049525 | 1 |
| 14 | 101049906 | T | C | -7.8257 | ENSG0000( | CTD-2062F | 14 | 101006011 | 19 | 15590 | 0 | 6.42E-07 |
| 14 | 101049906 | T | C | 10.8412 | ENSG0000( | SLC25A29 | 14 | 100765166 | 35 | 30808 | 0 | 2.80E-19 |
| 14 | 101049906 | T | C | 4.7747 | ENSG0000( | RP11-638I2 | 14 | 100833111 | 13 | 5490 | 0.0049525 | 1 |
| 14 | 101049906 | T | C | -7.8257 | ENSG0000( | CTD-2062F | 14 | 101006011 | 19 | 15590 | 0 | 6.42E-07 |
| 14 | 101049906 | T | C | 10.8412 | ENSG0000( | SLC25A29 | 14 | 100765166 | 35 | 30808 | 0 | 2.80E-19 |
| 14 | 101049906 | T | C | 4.7747 | ENSG0000( | RP11-638I2 | 14 | 100833111 | 13 | 5490 | 0.0049525 | 1 |
| 8 | 135625287 | T | C | 7.5522 | ENSG0000( | AC083843. | 8 | 135807389 | 13 | 5502 | 0 | 5.46E-06 |
| 8 | 135625287 | T | C | 9.2756 | ENSG0000( | ZFAT | 8 | 135607661 | 36 | 31569 | 0 | 2.25E-12 |
| 8 | 135625287 | T | C | 7.5522 | ENSG0000( | AC083843. | 8 | 135807389 | 13 | 5502 | 0 | 5.46E-06 |
| 8 | 135625287 | T | C | 9.2756 | ENSG0000( | ZFAT | 8 | 135607661 | 36 | 31569 | 0 | 2.25E-12 |
| 11 | 72285444 | A | G | 5.1655 | ENSG0000( | P2RY2 | 11 | 72938370 | 27 | 27766 | 0.0007422 | 1 |
| 11 | 72285444 | A | G | 8.4129 | ENSG0000( | PDE2A | 11 | 72336410 | 28 | 27980 | 0 | 5.10E-09 |
| 11 | 72285444 | A | G | 7.358 | ENSG0000( | STARD10 | 11 | 72485493 | 13 | 13100 | 0 | 2.38E-05 |
| 11 | 72285444 | A | G | 7.7177 | ENSG0000( | RP11-169D | 11 | 72297319 | 10 | 4339 | 0 | 1.51E-06 |
| 14 | 100679288 | G | A | 4.3766 | ENSG0000( | BEGAIN | 14 | 101028618 | 35 | 30066 | 0.0312248 | 1 |
| 14 | 65797766 | G | T | -5.0485 | ENSG0000( | FNTB | 14 | 65491403 | 33 | 24705 | 0.0013179 | 1 |
| 17 | 30105322 | A | G | 6.355 | ENSG0000( | CDK5R1 | 17 | 30815955 | 35 | 31315 | 0 | 0.026576233 |
| 17 | 11988916 | T | G | -5.6054 | ENSG0000( | ZNF18 | 17 | 11890794 | 34 | 30604 | 0.0001086 | 1 |
| 17 | 11988916 | T | G | 7.2419 | ENSG0000( | MAP2K4 | 17 | 11985644 | 16 | 14261 | 0 | 5.64E-05 |
| 14 | 65759023 | C | G | 5.0198 | ENSG0000( | CHURC1 | 14 | 65396194 | 34 | 29943 | 0.0015627 | 1 |
| 14 | 65775695 | G | T | -4.4231 | ENSG0000( | PTBP1P | 14 | 65746934 | 9 | 3831 | 0.0253383 | 1 |
| 14 | 101096414 | T | C | 7.3269 | ENSG0000( | SLC25A29 | 14 | 100765166 | 35 | 30814 | 0 | 3.00E-05 |
| 1 | 110130641 | T | A | 5.516 | ENSG0000( | AMIGO1 | 1 | 110049578 | 36 | 31567 | 0.0001336 | 1 |
| 1 | 110130641 | T | A | 5.4128 | ENSG0000( | GSTM3 | 1 | 110280469 | 35 | 31183 | 0.0002466 | 1 |
| 1 | 110130641 | T | A | -5.3422 | ENSG0000( | RP5-1160K | 1 | 110139625 | 13 | 5500 | 0.0003086 | 1 |
| 1 | 110130641 | T | A | 15.9551 | ENSG0000( | GSTM4 | 1 | 110203410 | 34 | 26278 | 0 | 3.35E-49 |
| 1 | 110130641 | T | A | -5.1922 | ENSG0000( | PSRC1 | 1 | 109823993 | 36 | 31567 | 0.0006309 | 1 |
| 1 | 110130641 | T | A | -65.6868 | ENSG0000( | GNAI3 | 1 | 110114104 | 36 | 31567 | 0 | 4.17E-302 |
| 1 | 110130641 | T | A | 4.8082 | ENSG0000( | ATXN7L2 | 1 | 110030763 | 16 | 14261 | 0.0042526 | 1 |
| 1 | 110130641 | T | A | 8.2708 | ENSG0000( | GSTM2 | 1 | 110231408 | 32 | 23626 | 0 | 1.69E-08 |
| 1 | 110130641 | T | A | -5.6415 | ENSG0000( | MIR197 | 1 | 110141552 | 21 | 16157 | 9.61E-05 | 1 |
| 1 | 110130641 | T | A | 4.956 | ENSG0000( | GSTM1 | 1 | 110240871 | 32 | 23626 | 0.0021239 | 1 |
| 1 | 110130641 | T | A | -5.7804 | ENSG0000( | KIAA1324 | 1 | 109702851 | 35 | 30818 | 4.52E-05 | 0.948989281 |
| 14 | 101186175 | C | T | 6.1023 | ENSG0000( | SLC25A29 | 14 | 100765166 | 35 | 29531 | 1.31E-05 | 0.133224989 |
| 14 | 101186175 | C | T | -36.7399 | ENSG0000( | MEG3 | 14 | 101286557 | 33 | 22224 | 0 | 2.14E-287 |
| 14 | 101186175 | C | T | -5.4813 | ENSG0000( | WDR25 | 14 | 100919697 | 36 | 30280 | 0.0001397 | 1 |
| 14 | 101186175 | C | T | -8.2097 | ENSG0000( | BEGAIN | 14 | 101028618 | 35 | 30066 | 0 | 2.82E-08 |
| 14 | 101186175 | C | T | 12.9359 | ENSG0000( | WARS | 14 | 100821633 | 34 | 24991 | 0 | 3.60E-30 |
| 14 | 101186175 | C | T | -12.2957 | ENSG0000( | DLK1 | 14 | 101196790 | 36 | 30280 | 0 | 1.21E-26 |
| 14 | 101186175 | C | T | 6.1023 | ENSG0000( | SLC25A29 | 14 | 100765166 | 35 | 29531 | 1.31E-05 | 0.133224989 |
| 14 | 101144596 | T | C | 4.5286 | ENSG0000( | WDR25 | 14 | 100919697 | 33 | 29751 | 0.0157705 | 1 |
| 14 | 101179222 | A | G | 6.0238 | ENSG0000( | SLC25A29 | 14 | 100765166 | 35 | 30817 | 1.31E-05 | 0.216952221 |
| 14 | 101179222 | A | G | 6.0238 | ENSG0000( | SLC25A29 | 14 | 100765166 | 35 | 30817 | 1.31E-05 | 0.216952221 |
| 14 | 23372913 | G | A | 5.9912 | ENSG0000( | MRPL52 | 14 | 23301667 | 35 | 31354 | 1.96E-05 | 0.26557132 |
| 7 | 129569411 | C | G | 4.4666 | ENSG0000( | UBE2H | 7 | 129531680 | 36 | 31569 | 0.0207773 | 1 |
| 7 | 129569411 | C | G | 5.2112 | ENSG0000( | CPA5 | 7 | 129996600 | 16 | 14263 | 0.0005877 | 1 |
| 7 | 129569411 | C | G | 4.4818 | ENSG0000( | TMEM209 | 7 | 129826082 | 36 | 31569 | 0.0195058 | 1 |
| 2 | 219623504 | T | C | 4.2935 | ENSG0000( | WNT6 | 2 | 219731749 | 37 | 31684 | 0.0442877 | 1 |
| 2 | 64569840 | T | C | 8.7652 | ENSG0000( | LGALSL | 2 | 64684809 | 33 | 25187 | 0 | 2.37E-10 |
| 2 | 64569840 | T | C | 4.6706 | ENSG0000( | AC008074. | 2 | 64677444 | 12 | 4990 | 0.0080492 | 1 |
| 2 | 64569840 | T | C | -6.5162 | ENSG0000( | AC114752. | 2 | 64566991 | 13 | 5498 | 0 | 0.00917052 |
| 2 | 64569840 | T | C | 6.0644 | ENSG0000( | AC008074. | 2 | 64651643 | 12 | 4990 | 1.31E-05 | 0.168664211 |
| 19 | 52189152 | A | G | 8.0124 | ENSG0000( | SIGLEC5 | 19 | 52132466 | 34 | 26165 | 0 | 1.43E-07 |

| | | | | | | | | | | | | |
|---|---|---|---|---|---|---|---|---|---|---|---|---|
| 19 | 52189152 | A | G | 9.4857 | ENSG0000( | ZNF175 | 19 | 52083771 | 35 | 31240 | 0 | 3.07E-13 |
| 19 | 52189152 | A | G | 17.3297 | ENSG0000( | SIGLEC14 | 19 | 52147930 | 25 | 20677 | 0 | 3.58E-59 |
| 19 | 52189152 | A | G | 10.8779 | ENSG0000( | HAS1 | 19 | 52221806 | 34 | 30732 | 0 | 1.87E-19 |
| 19 | 52189152 | A | G | 9.9597 | ENSG0000( | LINC00085 | 19 | 52206116 | 31 | 25147 | 0 | 2.92E-15 |
| 5 | 148525488 | A | G | 4.6773 | ENSG0000( | ABLIM3 | 5 | 148580575 | 36 | 31560 | 0.0078228 | 1 |
| 22 | 22034152 | A | G | -5.6386 | ENSG0000( | SDF2L1 | 22 | 21997568 | 36 | 31564 | 9.61E-05 | 1 |
| 22 | 22031373 | T | C | -4.3501 | ENSG0000( | TOP3BP1 | 22 | 22578269 | 11 | 4776 | 0.0349463 | 1 |
| 11 | 61598288 | T | C | -4.9 | ENSG0000( | FEN1 | 11 | 61562412 | 35 | 31353 | 0.0027667 | 1 |
| 14 | 23424186 | A | G | -4.551 | ENSG0000( | SLC7A8 | 14 | 23623693 | 36 | 31559 | 0.0143252 | 1 |
| 14 | 23424186 | A | G | 7.3148 | ENSG0000( | PSMB5 | 14 | 23495095 | 36 | 31559 | 0 | 3.29E-05 |
| 22 | 22065422 | T | C | 4.6964 | ENSG0000( | MAPK1 | 22 | 22165379 | 34 | 30605 | 0.0070107 | 1 |
| 22 | 22065422 | T | C | -4.7796 | ENSG0000( | RIMBP3C | 22 | 21902698 | 10 | 4338 | 0.0048266 | 1 |
| 22 | 22058355 | G | A | 7.8645 | ENSG0000( | TOP3B | 22 | 22324305 | 16 | 14262 | 0 | 4.72E-07 |
| 20 | 62546616 | G | A | -21.6083 | ENSG0000( | UCKL1 | 20 | 62579477 | 34 | 28618 | 0 | 1.91E-95 |
| 20 | 62546616 | G | A | -4.2824 | ENSG0000( | EEF1A2 | 20 | 62124935 | 34 | 28618 | 0.0462205 | 1 |
| 20 | 62546616 | G | A | -4.3105 | ENSG0000( | ZNF512B | 20 | 62634084 | 35 | 28832 | 0.0411501 | 1 |
| 20 | 62546616 | G | A | -6.4035 | ENSG0000( | PPDPF | 20 | 62152818 | 35 | 28832 | 0 | 0.019325391 |
| 20 | 62546616 | G | A | -11.3355 | ENSG0000( | TPD52L2 | 20 | 62509770 | 34 | 28618 | 0 | 1.11E-21 |
| 20 | 62546616 | G | A | -4.61 | ENSG0000( | ZBTB46 | 20 | 62418808 | 34 | 23757 | 0.0107906 | 1 |
| 20 | 62546616 | G | A | -7.1419 | ENSG0000( | DNAJC5 | 20 | 62546951 | 34 | 28618 | 0 | 0.000117133 |
| 2 | 219556346 | C | T | 4.7045 | ENSG0000( | SLC23A3 | 2 | 219987800 | 36 | 31567 | 0.0068265 | 1 |
| 9 | 137745249 | A | G | 6.4862 | ENSG0000( | FCN1 | 9 | 137805620 | 36 | 31568 | 0 | 0.011224543 |
| 22 | 22015234 | T | C | 16.4497 | ENSG0000( | PPIL2 | 22 | 22029562 | 37 | 31682 | 0 | 1.08E-52 |
| 22 | 22015234 | T | C | 7.839 | ENSG0000( | PPM1F | 22 | 22290501 | 36 | 31468 | 0 | 5.79E-07 |
| 22 | 22015234 | T | C | 12.5339 | ENSG0000( | UBE2L3 | 22 | 21941029 | 37 | 31682 | 0 | 6.21E-28 |
| 22 | 22015234 | T | C | -5.3185 | ENSG0000( | IGLV8-61 | 22 | 22453366 | 30 | 24876 | 0.0003271 | 1 |
| 22 | 22015234 | T | C | -5.1017 | ENSG0000( | HIC2 | 22 | 21788722 | 37 | 31682 | 0.0010302 | 1 |
| 22 | 22015234 | T | C | 9.1776 | ENSG0000( | LL22NC03- | 22 | 22295204 | 14 | 8267 | 0 | 5.62E-12 |
| 2 | 219269729 | T | G | -4.8129 | ENSG0000( | PNKD | 2 | 219173315 | 28 | 24031 | 0.0041496 | 1 |
| 7 | 25997536 | G | A | -4.8062 | ENSG0000( | NFE2L3 | 7 | 26209302 | 16 | 14263 | 0.0042948 | 1 |
| 20 | 62530746 | T | C | 7.6879 | ENSG0000( | OPRL1 | 20 | 62721761 | 31 | 27679 | 0 | 1.91E-06 |
| 20 | 62530746 | T | C | -5.0957 | ENSG0000( | SAMD10 | 20 | 62608413 | 32 | 27893 | 0.001042 | 1 |
| 14 | 23489788 | G | A | 6.0099 | ENSG0000( | CDH24 | 14 | 23521509 | 35 | 30162 | 1.96E-05 | 0.23653739 |
| 14 | 65696896 | A | C | 4.4407 | ENSG0000( | MAX | 14 | 65521152 | 34 | 29830 | 0.0234283 | 1 |
| 2 | 219272294 | G | T | -5.3634 | ENSG0000( | CXCR2 | 2 | 218995994 | 35 | 26389 | 0.0003026 | 1 |
| 2 | 219272294 | G | T | -5.1532 | ENSG0000( | PLCD4 | 2 | 219487197 | 37 | 31678 | 0.0007661 | 1 |
| 2 | 219272294 | G | T | -5.8577 | ENSG0000( | CXCR1 | 2 | 219029643 | 37 | 31678 | 3.24E-05 | 0.597283969 |
| 2 | 219272294 | G | T | -6.565 | ENSG0000( | RP11-378A | 2 | 219121064 | 13 | 5496 | 0 | 0.006617062 |

**Table S13. miRNAs that shared cis-eQTLs with their host genes**

| miRNA | miRNA context | SNP | SNP position | EA | P | |
|---|---|---|---|---|---|---|
| | | | | | miR-eQTL | eQTL |
| miR-625-5p | intragenic (*FUT8* ) | rs1003401 | 14:66237720 | G | 1.05E-13 | 2.27E-159 |
| miR-27b-3p | intragenic (*C9orf3* ) | rs1004373 | 9:97814770 | A | 4.09E-07 | 9.89E-83 |
| miR-584-5p | intragenic (*SH3TC2* ) | rs10044814 | 5:148542270 | T | 1.83E-07 | 1.93E-13 |
| miR-335-5p | intragenic (*MEST* ) | rs1005171 | 7:130144402 | C | 1.97E-04 | 1.04E-12 |
| miR-4707-3p | intragenic (*HAUS4* ) | rs10130756 | 14:23396600 | C | 1.97E-10 | 1.52E-45 |
| miR-3130-3p | intragenic (*FASTKD2* ) | rs10199766 | 2:207641868 | G | 5.09E-16 | 2.92E-09 |
| miR-1908-5p | intragenic (*FADS1* ) | rs102274 | 11:61557826 | T | 9.41E-09 | 3.27E-310 |
| miR-339-5p | intragenic (*C7orf50* ) | rs10229680 | 7:1158565 | C | 9.53E-05 | 3.79E-07 |
| miR-26b-5p | intragenic (*CTDSP1* ) | rs1078803 | 2:219606538 | C | 4.31E-04 | 1.29E-05 |
| miR-139-3p | intragenic (*PDE2A* ) | rs11235541 | 11:72285444 | G | 2.41E-04 | 4.00E-17 |
| miR-744-5p | intragenic (*MAP2K4)* | rs11653721 | 17:11988916 | G | 6.79E-04 | 4.43E-13 |
| miR-941 | intragenic (*DNAJC5* ) | rs2247381 | 20:62546616 | A | 1.73E-04 | 9.20E-13 |

EA: effect allele, chr: chromosome, bp: base pair

The table shows 12 miRNAs locted in intragenic region of protein-coding genes that share cis-eQTLs with their host genes.

**Table S14. miRNAs with shared eQTLs with their putative target genes**

| miRNA | Target gen | Gene chr | Gene pos | MTI | Type |
|---|---|---|---|---|---|
| miR-127-3p | *DLK1* | 14 | 101196790 | predicted | cis-miR-eQTLs overlap with cis-eQTLs |
| miR-130a-3p | *ZFP91* | 11 | 58367549 | predicted | cis-miR-eQTLs overlap with cis-eQTLs |
| miR-130b-3p | *MAPK1* | 22 | 22165379 | predicted | cis-miR-eQTLs overlap with cis-eQTLs |
| miR-26b-5p | *USP37* | 2 | 219374029 | predicted | cis-miR-eQTLs overlap with cis-eQTLs |
| miR-181a-5p | *CD4* | 12 | 6912999 | predicted | cis-miR-eQTLs overlap with trans-eQTLs |
| miR-181a-5p | *PBX1* | 1 | 164696677 | predicted | cis-miR-eQTLs overlap with trans-eQTLs |
| miR-181a-5p | *PDE5A* | 4 | 120482848 | predicted | cis-miR-eQTLs overlap with trans-eQTLs |
| miR-181a-5p | *SESN3* | 11 | 94932204 | predicted | cis-miR-eQTLs overlap with trans-eQTLs |
| miR-181b-5p | *CD4* | 12 | 6912999 | predicted | cis-miR-eQTLs overlap with trans-eQTLs |
| miR-181b-5p | *PBX1* | 1 | 164696677 | predicted | cis-miR-eQTLs overlap with trans-eQTLs |
| miR-181b-5p | *PDE5A* | 4 | 120482848 | predicted | cis-miR-eQTLs overlap with trans-eQTLs |
| miR-181b-5p | *SESN3* | 11 | 94932204 | predicted | cis-miR-eQTLs overlap with trans-eQTLs |
| let-7a-5p | *CACFD1* | 9 | 136330529 | predicted | trans-miR-eQTLs overlap with cis-eQTLs |
| let-7b-5p | *CACFD1* | 9 | 136330529 | predicted | trans-miR-eQTLs overlap with cis-eQTLs |
| let-7c-5p | *CACFD1* | 9 | 136330529 | predicted | trans-miR-eQTLs overlap with cis-eQTLs |
| miR-126-3p | *TCF4* | 18 | 53110790 | validated | trans-miR-eQTLs overlap with trans-eQTLs |
| miR-139-5p | *TCF4* | 18 | 53110790 | predicted | trans-miR-eQTLs overlap with trans-eQTLs |
| miR-320b | *YWHAH* | 22 | 32347018 | predicted | trans-miR-eQTLs overlap with trans-eQTLs |

MTI: miRNA-target interaction

The table shows miRNAs that share eQTLs with their some of their putative target genes extracted from TargetScan v7.2.

**Table S15. Overlap between cis-miR-eQTLs and pQTLs**

| miRNA | mir_chr | mir_start | mir_end | n_cismiRQTLs | UniProt | TargetFullName |
|---|---|---|---|---|---|---|
| miR-127-3p | 14 | 101349372 | 101349393 | 10 | Q02383 | Semenogelin-2 |
| miR-127-3p | 14 | 101349372 | 101349393 | 10 | P80370 | Protein delta homolog 1 |
| miR-130a-3 | 11 | 57408725 | 57408746 | 24 | O14641 | Segment polarity protein dishevelled hom |
| miR-130a-3 | 11 | 57408725 | 57408746 | 246 | P05155 | Plasma protease C1 inhibitor |
| miR-134-5p | 14 | 101521031 | 101521052 | 38 | P80370 | Protein delta homolog 1 |
| miR-134-5p | 14 | 101521031 | 101521052 | 42 | Q02383 | Semenogelin-2 |
| miR-136-5p | 14 | 101351053 | 101351075 | 15 | P80370 | Protein delta homolog 1 |
| miR-136-5p | 14 | 101351053 | 101351075 | 15 | Q02383 | Semenogelin-2 |
| miR-329-3p | 14 | 101493171 | 101493192 | 37 | P80370 | Protein delta homolog 1 |
| miR-329-3p | 14 | 101493488 | 101493509 | 37 | P80370 | Protein delta homolog 1 |
| miR-329-3p | 14 | 101493171 | 101493192 | 41 | Q02383 | Semenogelin-2 |
| miR-329-3p | 14 | 101493488 | 101493509 | 41 | Q02383 | Semenogelin-2 |
| miR-335-5p | 7 | 130135967 | 130135989 | 38 | Q9UI42 | Carboxypeptidase A4 |
| miR-3689a- | 9 | 137741340 | 137741361 | 3 | Q15485 | Ficolin-2 |
| miR-370-3p | 14 | 101377523 | 101377544 | 101 | P80370 | Protein delta homolog 1 |
| miR-370-3p | 14 | 101377523 | 101377544 | 105 | Q02383 | Semenogelin-2 |
| miR-376a-3 | 14 | 101507162 | 101507182 | 87 | P80370 | Protein delta homolog 1 |
| miR-376a-3 | 14 | 101506455 | 101506475 | 87 | P80370 | Protein delta homolog 1 |
| miR-376a-3 | 14 | 101507162 | 101507182 | 91 | Q02383 | Semenogelin-2 |
| miR-376a-3 | 14 | 101506455 | 101506475 | 91 | Q02383 | Semenogelin-2 |
| miR-376c-3 | 14 | 101506069 | 101506089 | 4 | P80370 | Protein delta homolog 1 |
| miR-376c-3 | 14 | 101506069 | 101506089 | 4 | Q02383 | Semenogelin-2 |
| miR-382-5p | 14 | 101520653 | 101520674 | 85 | P80370 | Protein delta homolog 1 |
| miR-382-5p | 14 | 101520653 | 101520674 | 89 | Q02383 | Semenogelin-2 |
| miR-409-3p | 14 | 101531683 | 101531704 | 93 | P80370 | Protein delta homolog 1 |
| miR-409-3p | 14 | 101531683 | 101531704 | 97 | Q02383 | Semenogelin-2 |
| miR-411-3p | 14 | 101489712 | 101489733 | 43 | P80370 | Protein delta homolog 1 |
| miR-411-3p | 14 | 101489712 | 101489733 | 47 | Q02383 | Semenogelin-2 |
| miR-431-5p | 14 | 101347363 | 101347383 | 6 | Q02383 | Semenogelin-2 |
| miR-431-5p | 14 | 101347363 | 101347383 | 6 | P80370 | Protein delta homolog 1 |
| miR-433-3p | 14 | 101348286 | 101348307 | 29 | P80370 | Protein delta homolog 1 |
| miR-433-3p | 14 | 101348286 | 101348307 | 29 | Q02383 | Semenogelin-2 |
| miR-494-3p | 14 | 101496018 | 101496039 | 53 | P80370 | Protein delta homolog 1 |
| miR-494-3p | 14 | 101496018 | 101496039 | 55 | Q02383 | Semenogelin-2 |
| miR-543 | 14 | 101498370 | 101498391 | 95 | P80370 | Protein delta homolog 1 |
| miR-543 | 14 | 101498370 | 101498391 | 99 | Q02383 | Semenogelin-2 |
| miR-625-5p | 14 | 65937834 | 65937854 | 409 | Q9BUP3 | Oxidoreductase HTATIP2 |
| miR-625-5p | 14 | 65937834 | 65937854 | 304 | Q15109 | Advanced glycosylation end product-speci |
| miR-625-5p | 14 | 65937834 | 65937854 | 414 | Q9BYC5 | Alpha-(1,6)-fucosyltransferase |

| Target | SOMAMER_ID | miRNA_context |
|---|---|---|
| SEMG2 | SEMG2.6373.54.3 | intragenic (RTL1) |
| DLK1 | DLK1.6496.60.3 | intragenic (RTL1) |
| DVL2 | DVL2.13575.40.3 | intergenic |
| C1-Esterase Inhibitor | SERPING1.4479.14.2 | intergenic |
| DLK1 | DLK1.6496.60.3 | intergenic |
| SEMG2 | SEMG2.6373.54.3 | intergenic |
| DLK1 | DLK1.6496.60.3 | intragenic (RTL1) |
| SEMG2 | SEMG2.6373.54.3 | intragenic (RTL1) |
| DLK1 | DLK1.6496.60.3 | intergenic |
| DLK1 | DLK1.6496.60.3 | intergenic |
| SEMG2 | SEMG2.6373.54.3 | intergenic |
| SEMG2 | SEMG2.6373.54.3 | intergenic |
| CBPA4 | CPA4.9267.2.3 | intragenic (MEST) |
| FCN2 | FCN2.3313.21.2 | intergenic |
| DLK1 | DLK1.6496.60.3 | intergenic |
| SEMG2 | SEMG2.6373.54.3 | intergenic |
| DLK1 | DLK1.6496.60.3 | intergenic |
| DLK1 | DLK1.6496.60.3 | intergenic |
| SEMG2 | SEMG2.6373.54.3 | intergenic |
| SEMG2 | SEMG2.6373.54.3 | intergenic |
| DLK1 | DLK1.6496.60.3 | intergenic |
| SEMG2 | SEMG2.6373.54.3 | intergenic |
| DLK1 | DLK1.6496.60.3 | intergenic |
| SEMG2 | SEMG2.6373.54.3 | intergenic |
| DLK1 | DLK1.6496.60.3 | intergenic |
| SEMG2 | SEMG2.6373.54.3 | intergenic |
| DLK1 | DLK1.6496.60.3 | intergenic |
| SEMG2 | SEMG2.6373.54.3 | intergenic |
| SEMG2 | SEMG2.6373.54.3 | intragenic (RTL1) |
| DLK1 | DLK1.6496.60.3 | intragenic (RTL1) |
| DLK1 | DLK1.6496.60.3 | intragenic (RTL1) |
| SEMG2 | SEMG2.6373.54.3 | intragenic (RTL1) |
| DLK1 | DLK1.6496.60.3 | intergenic |
| SEMG2 | SEMG2.6373.54.3 | intergenic |
| DLK1 | DLK1.6496.60.3 | intergenic |
| SEMG2 | SEMG2.6373.54.3 | intergenic |
| HTAI2 | HTATIP2.7753.21.3 | intragenic (FUT8) |
| sRAGE | AGER.4125.52.2 | intragenic (FUT8) |
| FUT8 | FUT8.8244.16.3 | intragenic (FUT8) |

**Table S16. Overlap between trans-miR-eQTLs and pQTLs**

| miRNA | n_transmiRQTLs | UniProt | TargetFullName | Target | SOMAMER_ID | MTI |
|---|---|---|---|---|---|---|
| let-7b-5p | 20 | P15586 | N-acetylglucosamine-6-sulfatase | GNS | GNS.3616.3.5 | predicted MTI |
| let-7c-5p | 19 | P15586 | N-acetylglucosamine-6-sulfatase | GNS | GNS.3616.3.5 | predicted MTI |
| miR-145-5p | 4 | Q9GZP0 | Platelet-derived growth factor D | PDGFD | PDGFD.9341.1.3 | predicted MTI |
| miR-195-5p | 20 | O94923 | D-glucuronyl C5-epimerase | GLCE | GLCE.7808.5.3 | predicted MTI |
| let-7a-5p | 8 | P16442 | Histo-blood group ABO system transferase | BGAT | ABO.9253.52.3 | none |
| let-7c-5p | 26 | P16442 | Histo-blood group ABO system transferase | BGAT | ABO.9253.52.3 | none |
| miR-10b-5p | 40 | P16442 | Histo-blood group ABO system transferase | BGAT | ABO.9253.52.3 | none |
| let-7b-5p | 30 | P16442 | Histo-blood group ABO system transferase | BGAT | ABO.9253.52.3 | none |
| miR-195-5p | 20 | P16442 | Histo-blood group ABO system transferase | BGAT | ABO.9253.52.3 | none |
| miR-139-5p | 30 | P16442 | Histo-blood group ABO system transferase | BGAT | ABO.9253.52.3 | none |
| miR-126-3p | 46 | P16442 | Histo-blood group ABO system transferase | BGAT | ABO.9253.52.3 | none |
| miR-10a-5p | 14 | P16442 | Histo-blood group ABO system transferase | BGAT | ABO.9253.52.3 | none |
| miR-10b-5p | 6 | Q76LX8 | A disintegrin and metalloproteinase with thrombospondin motifs 13 | ATS13 | ADAMTS13.3175.51.5 | none |
| miR-195-5p | 20 | Q8IZF2 | Adhesion G protein-coupled receptor F5 | GP116 | ADGRF5.6409.57.3 | none |
| miR-10b-5p | 40 | Q8IZF2 | Adhesion G protein-coupled receptor F5 | GP116 | ADGRF5.6409.57.3 | none |
| let-7b-5p | 30 | Q8IZF2 | Adhesion G protein-coupled receptor F5 | GP116 | ADGRF5.6409.57.3 | none |
| miR-10a-5p | 14 | Q8IZF2 | Adhesion G protein-coupled receptor F5 | GP116 | ADGRF5.6409.57.3 | none |
| miR-126-3p | 46 | Q8IZF2 | Adhesion G protein-coupled receptor F5 | GP116 | ADGRF5.6409.57.3 | none |
| let-7c-5p | 26 | Q8IZF2 | Adhesion G protein-coupled receptor F5 | GP116 | ADGRF5.6409.57.3 | none |
| miR-139-5p | 30 | Q8IZF2 | Adhesion G protein-coupled receptor F5 | GP116 | ADGRF5.6409.57.3 | none |
| let-7a-5p | 8 | Q8IZF2 | Adhesion G protein-coupled receptor F5 | GP116 | ADGRF5.6409.57.3 | none |
| miR-145-5p | 4 | Q15109 | Advanced glycosylation end product-specific receptor, soluble | sRAGE | AGER.4125.52.2 | none |
| miR-143-3p | 4 | Q15109 | Advanced glycosylation end product-specific receptor, soluble | sRAGE | AGER.4125.52.2 | none |
| miR-10b-5p | 35 | P09923 | Intestinal-type alkaline phosphatase | Alkaline ph | ALPI.10463.23.3 | none |
| miR-195-5p | 20 | P09923 | Intestinal-type alkaline phosphatase | Alkaline ph | ALPI.10463.23.3 | none |
| miR-126-3p | 30 | P09923 | Intestinal-type alkaline phosphatase | Alkaline ph | ALPI.10463.23.3 | none |
| miR-139-5p | 30 | P09923 | Intestinal-type alkaline phosphatase | Alkaline ph | ALPI.10463.23.3 | none |
| let-7c-5p | 26 | P09923 | Intestinal-type alkaline phosphatase | Alkaline ph | ALPI.10463.23.3 | none |
| let-7a-5p | 8 | P09923 | Intestinal-type alkaline phosphatase | Alkaline ph | ALPI.10463.23.3 | none |
| let-7b-5p | 28 | P09923 | Intestinal-type alkaline phosphatase | Alkaline ph | ALPI.10463.23.3 | none |
| miR-143-3p | 4 | Q15389 | Angiopoietin-1 | Angiopoiet | ANGPT1.2811.27.1 | none |
| miR-145-5p | 4 | Q15389 | Angiopoietin-1 | Angiopoiet | ANGPT1.2811.27.1 | none |
| miR-143-3p | 1 | O00213 | Amyloid beta A4 precursor protein-binding family B member 1 | APBB1 | APBB1.14206.28.3 | none |
| miR-145-5p | 1 | O00213 | Amyloid beta A4 precursor protein-binding family B member 1 | APBB1 | APBB1.14206.28.3 | none |
| miR-145-5p | 4 | Q06481 | Amyloid-like protein 2 | APLP2 | APLP2.10627.87.3 | none |
| miR-143-3p | 4 | Q06481 | Amyloid-like protein 2 | APLP2 | APLP2.10627.87.3 | none |
| miR-143-3p | 3 | P40616 | ADP-ribosylation factor-like protein 1 | ARL1 | ARL1.12392.30.3 | none |
| miR-145-5p | 3 | P40616 | ADP-ribosylation factor-like protein 1 | ARL1 | ARL1.12392.30.3 | none |
| miR-126-3p | 44 | Q9NY97 | N-acetyllactosaminide beta-1,3-N-acetylglucosaminyltransferase 2 | B3GN2 | B3GNT2.7980.72.3 | none |
| miR-195-5p | 20 | Q9NY97 | N-acetyllactosaminide beta-1,3-N-acetylglucosaminyltransferase 2 | B3GN2 | B3GNT2.7980.72.3 | none |
| let-7c-5p | 26 | Q9NY97 | N-acetyllactosaminide beta-1,3-N-acetylglucosaminyltransferase 2 | B3GN2 | B3GNT2.7980.72.3 | none |
| let-7b-5p | 28 | Q9NY97 | N-acetyllactosaminide beta-1,3-N-acetylglucosaminyltransferase 2 | B3GN2 | B3GNT2.7980.72.3 | none |
| miR-10a-5p | 14 | Q9NY97 | N-acetyllactosaminide beta-1,3-N-acetylglucosaminyltransferase 2 | B3GN2 | B3GNT2.7980.72.3 | none |
| let-7a-5p | 8 | Q9NY97 | N-acetyllactosaminide beta-1,3-N-acetylglucosaminyltransferase 2 | B3GN2 | B3GNT2.7980.72.3 | none |
| miR-10b-5p | 31 | Q9NY97 | N-acetyllactosaminide beta-1,3-N-acetylglucosaminyltransferase 2 | B3GN2 | B3GNT2.7980.72.3 | none |
| miR-139-5p | 30 | Q9NY97 | N-acetyllactosaminide beta-1,3-N-acetylglucosaminyltransferase 2 | B3GN2 | B3GNT2.7980.72.3 | none |
| miR-195-5p | 4 | P15291 | Beta-1,4-galactosyltransferase 1 | B4GT1 | B4GALT1.13381.49.3 | none |
| let-7a-5p | 8 | P15291 | Beta-1,4-galactosyltransferase 1 | B4GT1 | B4GALT1.13381.49.3 | none |
| miR-139-5p | 13 | P15291 | Beta-1,4-galactosyltransferase 1 | B4GT1 | B4GALT1.13381.49.3 | none |
| let-7b-5p | 11 | P15291 | Beta-1,4-galactosyltransferase 1 | B4GT1 | B4GALT1.13381.49.3 | none |
| miR-126-3p | 13 | P15291 | Beta-1,4-galactosyltransferase 1 | B4GT1 | B4GALT1.13381.49.3 | none |
| miR-10b-5p | 14 | P15291 | Beta-1,4-galactosyltransferase 1 | B4GT1 | B4GALT1.13381.49.3 | none |
| let-7c-5p | 9 | P15291 | Beta-1,4-galactosyltransferase 1 | B4GT1 | B4GALT1.13381.49.3 | none |
| let-7b-5p | 4 | O60909 | Beta-1,4-galactosyltransferase 2 | B4GT2 | B4GALT2.9595.11.3 | none |
| miR-195-5p | 2 | O60909 | Beta-1,4-galactosyltransferase 2 | B4GT2 | B4GALT2.9595.11.3 | none |
| miR-126-3p | 4 | O60909 | Beta-1,4-galactosyltransferase 2 | B4GT2 | B4GALT2.9595.11.3 | none |
| let-7a-5p | 2 | O60909 | Beta-1,4-galactosyltransferase 2 | B4GT2 | B4GALT2.9595.11.3 | none |
| let-7c-5p | 2 | O60909 | Beta-1,4-galactosyltransferase 2 | B4GT2 | B4GALT2.9595.11.3 | none |
| miR-10b-5p | 5 | O60909 | Beta-1,4-galactosyltransferase 2 | B4GT2 | B4GALT2.9595.11.3 | none |
| miR-139-5p | 4 | O60909 | Beta-1,4-galactosyltransferase 2 | B4GT2 | B4GALT2.9595.11.3 | none |
| miR-143-3p | 1 | Q9UBV7 | Beta-1,4-galactosyltransferase 7 | B4GT7 | B4GALT7.7806.33.3 | none |
| miR-145-5p | 1 | Q9UBV7 | Beta-1,4-galactosyltransferase 7 | B4GT7 | B4GALT7.7806.33.3 | none |
| miR-145-5p | 4 | P23560 | Brain-derived neurotrophic factor | BDNF | BDNF.2421.7.3 | none |
| miR-143-3p | 4 | P23560 | Brain-derived neurotrophic factor | BDNF | BDNF.2421.7.3 | none |
| miR-195-5p | 20 | Q96EU7 | C1GALT1-specific chaperone 1 | C1GLC | C1GALT1C1.5735.54.3 | none |
| miR-126-3p | 46 | Q96EU7 | C1GALT1-specific chaperone 1 | C1GLC | C1GALT1C1.5735.54.3 | none |
| let-7b-5p | 30 | Q96EU7 | C1GALT1-specific chaperone 1 | C1GLC | C1GALT1C1.5735.54.3 | none |
| miR-139-5p | 30 | Q96EU7 | C1GALT1-specific chaperone 1 | C1GLC | C1GALT1C1.5735.54.3 | none |
| let-7a-5p | 8 | Q96EU7 | C1GALT1-specific chaperone 1 | C1GLC | C1GALT1C1.5735.54.3 | none |
| miR-10b-5p | 40 | Q96EU7 | C1GALT1-specific chaperone 1 | C1GLC | C1GALT1C1.5735.54.3 | none |
| miR-10a-5p | 14 | Q96EU7 | C1GALT1-specific chaperone 1 | C1GLC | C1GALT1C1.5735.54.3 | none |
| let-7c-5p | 26 | Q96EU7 | C1GALT1-specific chaperone 1 | C1GLC | C1GALT1C1.5735.54.3 | none |
| miR-145-5p | 4 | Q92583 | C-C motif chemokine 17 | TARC | CCL17.3519.3.2 | none |
| miR-143-3p | 4 | Q92583 | C-C motif chemokine 17 | TARC | CCL17.3519.3.2 | none |
| miR-195-5p | 18 | O15444 | C-C motif chemokine 25 | TECK | CCL25.14068.29.3 | none |
| let-7b-5p | 25 | O15444 | C-C motif chemokine 25 | TECK | CCL25.14068.29.3 | none |
| miR-126-3p | 25 | O15444 | C-C motif chemokine 25 | TECK | CCL25.14068.29.3 | none |
| miR-139-5p | 25 | O15444 | C-C motif chemokine 25 | TECK | CCL25.14068.29.3 | none |
| let-7c-5p | 24 | O15444 | C-C motif chemokine 25 | TECK | CCL25.14068.29.3 | none |
| miR-10b-5p | 25 | O15444 | C-C motif chemokine 25 | TECK | CCL25.14068.29.3 | none |
| let-7a-5p | 4 | O15444 | C-C motif chemokine 25 | TECK | CCL25.14068.29.3 | none |
| miR-126-3p | 2 | P41217 | OX-2 membrane glycoprotein | OX2G | CD200.5112.73.3 | none |
| miR-139-5p | 2 | P41217 | OX-2 membrane glycoprotein | OX2G | CD200.5112.73.3 | none |
| miR-10b-5p | 2 | P41217 | OX-2 membrane glycoprotein | OX2G | CD200.5112.73.3 | none |
| let-7b-5p | 2 | P41217 | OX-2 membrane glycoprotein | OX2G | CD200.5112.73.3 | none |
| let-7c-5p | 2 | P41217 | OX-2 membrane glycoprotein | OX2G | CD200.5112.73.3 | none |
| let-7a-5p | 1 | P41217 | OX-2 membrane glycoprotein | OX2G | CD200.5112.73.3 | none |

| | | | | | | |
|---|---|---|---|---|---|---|
| miR-195-5p | 2 | P41217 | OX-2 membrane glycoprotein | OX2G | CD200.5112.73.3 | none |
| miR-139-5p | 30 | Q9NNX6 | CD209 antigen | DC-SIGN | CD209.3029.52.2 | none |
| miR-10b-5p | 40 | Q9NNX6 | CD209 antigen | DC-SIGN | CD209.3029.52.2 | none |
| miR-126-3p | 46 | Q9NNX6 | CD209 antigen | DC-SIGN | CD209.3029.52.2 | none |
| miR-195-5p | 20 | Q9NNX6 | CD209 antigen | DC-SIGN | CD209.3029.52.2 | none |
| let-7b-5p | 30 | Q9NNX6 | CD209 antigen | DC-SIGN | CD209.3029.52.2 | none |
| miR-10a-5p | 14 | Q9NNX6 | CD209 antigen | DC-SIGN | CD209.3029.52.2 | none |
| let-7a-5p | 8 | Q9NNX6 | CD209 antigen | DC-SIGN | CD209.3029.52.2 | none |
| let-7c-5p | 26 | Q9NNX6 | CD209 antigen | DC-SIGN | CD209.3029.52.2 | none |
| miR-139-5p | 30 | P33151 | Cadherin-5 | Cadherin-5 | CDH5.2819.23.2 | none |
| let-7b-5p | 28 | P33151 | Cadherin-5 | Cadherin-5 | CDH5.2819.23.2 | none |
| miR-10b-5p | 30 | P33151 | Cadherin-5 | Cadherin-5 | CDH5.2819.23.2 | none |
| miR-126-3p | 39 | P33151 | Cadherin-5 | Cadherin-5 | CDH5.2819.23.2 | none |
| let-7c-5p | 26 | P33151 | Cadherin-5 | Cadherin-5 | CDH5.2819.23.2 | none |
| miR-10a-5p | 9 | P33151 | Cadherin-5 | Cadherin-5 | CDH5.2819.23.2 | none |
| let-7a-5p | 8 | P33151 | Cadherin-5 | Cadherin-5 | CDH5.2819.23.2 | none |
| miR-195-5p | 20 | P33151 | Cadherin-5 | Cadherin-5 | CDH5.2819.23.2 | none |
| miR-143-3p | 2 | Q5EE01 | Centromere protein W | CENPW | CENPW.8864.59.3 | none |
| miR-145-5p | 2 | Q5EE01 | Centromere protein W | CENPW | CENPW.8864.59.3 | none |
| miR-145-5p | 2 | Q6NT52 | Choriogonadotropin subunit beta variant 2 | CGB2 | CGB2.6213.10.3 | none |
| miR-143-3p | 2 | Q6NT52 | Choriogonadotropin subunit beta variant 2 | CGB2 | CGB2.6213.10.3 | none |
| miR-10b-5p | 30 | Q9NPF2 | Carbohydrate sulfotransferase 11 | CHSTB | CHST11.7779.86.3 | none |
| miR-126-3p | 30 | Q9NPF2 | Carbohydrate sulfotransferase 11 | CHSTB | CHST11.7779.86.3 | none |
| miR-139-5p | 30 | Q9NPF2 | Carbohydrate sulfotransferase 11 | CHSTB | CHST11.7779.86.3 | none |
| let-7b-5p | 28 | Q9NPF2 | Carbohydrate sulfotransferase 11 | CHSTB | CHST11.7779.86.3 | none |
| let-7a-5p | 8 | Q9NPF2 | Carbohydrate sulfotransferase 11 | CHSTB | CHST11.7779.86.3 | none |
| miR-143-3p | 2 | Q9NPF2 | Carbohydrate sulfotransferase 11 | CHSTB | CHST11.7779.86.3 | none |
| miR-145-5p | 2 | Q9NPF2 | Carbohydrate sulfotransferase 11 | CHSTB | CHST11.7779.86.3 | none |
| let-7c-5p | 26 | Q9NPF2 | Carbohydrate sulfotransferase 11 | CHSTB | CHST11.7779.86.3 | none |
| miR-195-5p | 20 | Q9NPF2 | Carbohydrate sulfotransferase 11 | CHSTB | CHST11.7779.86.3 | none |
| let-7b-5p | 30 | Q7LFX5 | Carbohydrate sulfotransferase 15 | ST4S6 | CHST15.14097.86.3 | none |
| miR-10b-5p | 40 | Q7LFX5 | Carbohydrate sulfotransferase 15 | ST4S6 | CHST15.14097.86.3 | none |
| let-7c-5p | 26 | Q7LFX5 | Carbohydrate sulfotransferase 15 | ST4S6 | CHST15.14097.86.3 | none |
| let-7a-5p | 8 | Q7LFX5 | Carbohydrate sulfotransferase 15 | ST4S6 | CHST15.14097.86.3 | none |
| miR-195-5p | 20 | Q7LFX5 | Carbohydrate sulfotransferase 15 | ST4S6 | CHST15.14097.86.3 | none |
| miR-126-3p | 32 | Q7LFX5 | Carbohydrate sulfotransferase 15 | ST4S6 | CHST15.14097.86.3 | none |
| miR-139-5p | 30 | Q7LFX5 | Carbohydrate sulfotransferase 15 | ST4S6 | CHST15.14097.86.3 | none |
| miR-10a-5p | 10 | Q7LFX5 | Carbohydrate sulfotransferase 15 | ST4S6 | CHST15.4469.78.2 | none |
| let-7c-5p | 25 | Q9NZA1 | Chloride intracellular channel protein 5 | CLIC5 | CLIC5.12475.48.3 | none |
| miR-10b-5p | 25 | Q9NZA1 | Chloride intracellular channel protein 5 | CLIC5 | CLIC5.12475.48.3 | none |
| miR-126-3p | 25 | Q9NZA1 | Chloride intracellular channel protein 5 | CLIC5 | CLIC5.12475.48.3 | none |
| miR-139-5p | 25 | Q9NZA1 | Chloride intracellular channel protein 5 | CLIC5 | CLIC5.12475.48.3 | none |
| let-7b-5p | 25 | Q9NZA1 | Chloride intracellular channel protein 5 | CLIC5 | CLIC5.12475.48.3 | none |
| let-7a-5p | 4 | Q9NZA1 | Chloride intracellular channel protein 5 | CLIC5 | CLIC5.12475.48.3 | none |
| miR-195-5p | 20 | Q9NZA1 | Chloride intracellular channel protein 5 | CLIC5 | CLIC5.12475.48.3 | none |
| miR-145-5p | 4 | O43405 | Cochlin | COCH | COCH.7227.75.3 | none |
| miR-143-3p | 4 | O43405 | Cochlin | COCH | COCH.7227.75.3 | none |
| miR-145-5p | 4 | Q14019 | Coactosin-like protein | Coactosin-l | COTL1.4905.63.1 | none |
| miR-143-3p | 4 | Q14019 | Coactosin-like protein | Coactosin-l | COTL1.4905.63.1 | none |
| miR-143-3p | 4 | Q96SM3 | Probable carboxypeptidase X1 | CPXM1 | CPXM1.6255.74.3 | none |
| miR-145-5p | 4 | Q96SM3 | Probable carboxypeptidase X1 | CPXM1 | CPXM1.6255.74.3 | none |
| miR-143-3p | 4 | O76096 | Cystatin-F | CYTF | CST7.3302.58.1 | none |
| miR-145-5p | 4 | O76096 | Cystatin-F | CYTF | CST7.3302.58.1 | none |
| miR-145-5p | 4 | P10619 | Lysosomal protective protein | Cathepsin A | CTSA.3179.51.2 | none |
| miR-143-3p | 4 | P10619 | Lysosomal protective protein | Cathepsin A | CTSA.3179.51.2 | none |
| miR-145-5p | 2 | Q8N690 | Beta-defensin 119 | DB119 | DEFB119.8315.5.3 | none |
| miR-143-3p | 2 | Q8N690 | Beta-defensin 119 | DB119 | DEFB119.8315.5.3 | none |
| let-7c-5p | 24 | P00374 | Dihydrofolate reductase | DYR | DHFR.9823.2.3 | none |
| miR-10b-5p | 24 | P00374 | Dihydrofolate reductase | DYR | DHFR.9823.2.3 | none |
| let-7b-5p | 24 | P00374 | Dihydrofolate reductase | DYR | DHFR.9823.2.3 | none |
| miR-195-5p | 18 | P00374 | Dihydrofolate reductase | DYR | DHFR.9823.2.3 | none |
| miR-139-5p | 24 | P00374 | Dihydrofolate reductase | DYR | DHFR.9823.2.3 | none |
| let-7a-5p | 4 | P00374 | Dihydrofolate reductase | DYR | DHFR.9823.2.3 | none |
| miR-126-3p | 24 | P00374 | Dihydrofolate reductase | DYR | DHFR.9823.2.3 | none |
| miR-143-3p | 4 | O94907 | Dickkopf-related protein 1 | DKK1 | DKK1.3535.84.1 | none |
| miR-145-5p | 4 | O94907 | Dickkopf-related protein 1 | DKK1 | DKK1.3535.84.1 | none |
| miR-143-3p | 4 | Q9UBT3 | Dickkopf-related protein 4 | Dkk-4 | DKK4.3365.7.2 | none |
| miR-145-5p | 4 | Q9UBT3 | Dickkopf-related protein 4 | Dkk-4 | DKK4.3365.7.2 | none |
| miR-409-3p | 2 | P80370 | Protein delta homolog 1 | DLK1 | DLK1.6496.60.3 | none |
| miR-145-5p | 4 | Q9UBS4 | DnaJ homolog subfamily B member 11 | DJB11 | DNAJB11.7110.2.3 | none |
| miR-143-3p | 4 | Q9UBS4 | DnaJ homolog subfamily B member 11 | DJB11 | DNAJB11.7110.2.3 | none |
| miR-126-3p | 6 | Q9H4A9 | Dipeptidase 2 | DPEP2 | DPEP2.8327.26.3 | none |
| miR-195-5p | 2 | Q9H4A9 | Dipeptidase 2 | DPEP2 | DPEP2.8327.26.3 | none |
| miR-139-5p | 6 | Q9H4A9 | Dipeptidase 2 | DPEP2 | DPEP2.8327.26.3 | none |
| let-7c-5p | 2 | Q9H4A9 | Dipeptidase 2 | DPEP2 | DPEP2.8327.26.3 | none |
| let-7a-5p | 4 | Q9H4A9 | Dipeptidase 2 | DPEP2 | DPEP2.8327.26.3 | none |
| miR-10b-5p | 6 | Q9H4A9 | Dipeptidase 2 | DPEP2 | DPEP2.8327.26.3 | none |
| let-7b-5p | 4 | Q9H4A9 | Dipeptidase 2 | DPEP2 | DPEP2.8327.26.3 | none |
| miR-195-5p | 19 | Q14126 | Desmoglein-2 | Desmoglei | DSG2.9484.75.3 | none |
| miR-126-3p | 39 | Q14126 | Desmoglein-2 | Desmoglei | DSG2.9484.75.3 | none |
| let-7a-5p | 5 | Q14126 | Desmoglein-2 | Desmoglei | DSG2.9484.75.3 | none |
| let-7c-5p | 25 | Q14126 | Desmoglein-2 | Desmoglei | DSG2.9484.75.3 | none |
| miR-10b-5p | 25 | Q14126 | Desmoglein-2 | Desmoglei | DSG2.9484.75.3 | none |
| let-7b-5p | 25 | Q14126 | Desmoglein-2 | Desmoglei | DSG2.9484.75.3 | none |
| miR-10a-5p | 14 | Q14126 | Desmoglein-2 | Desmoglei | DSG2.9484.75.3 | none |
| miR-139-5p | 25 | Q14126 | Desmoglein-2 | Desmoglei | DSG2.9484.75.3 | none |
| miR-145-5p | 4 | Q9UNE0 | Tumor necrosis factor receptor superfamily member EDAR | EDAR | EDAR.2977.7.2 | none |
| miR-143-3p | 4 | Q9UNE0 | Tumor necrosis factor receptor superfamily member EDAR | EDAR | EDAR.2977.7.2 | none |
| miR-126-3p | 6 | P17813 | Endoglin | Endoglin | ENG.4908.6.1 | none |

| | | | | | | |
|---|---|---|---|---|---|---|
| miR-195-5p | 2 | P17813 | Endoglin | Endoglin | ENG.4908.6.1 | none |
| let-7c-5p | 2 | P17813 | Endoglin | Endoglin | ENG.4908.6.1 | none |
| let-7b-5p | 4 | P17813 | Endoglin | Endoglin | ENG.4908.6.1 | none |
| miR-10b-5p | 7 | P17813 | Endoglin | Endoglin | ENG.4908.6.1 | none |
| let-7a-5p | 4 | P17813 | Endoglin | Endoglin | ENG.4908.6.1 | none |
| miR-139-5p | 6 | P17813 | Endoglin | Endoglin | ENG.4908.6.1 | none |
| miR-145-5p | 4 | Q9BS26 | Endoplasmic reticulum resident protein 44 | TXNDC4 | ERP44.6064.4.3 | none |
| miR-143-3p | 4 | Q9BS26 | Endoplasmic reticulum resident protein 44 | TXNDC4 | ERP44.6064.4.3 | none |
| let-7b-5p | 30 | P00451 | Coagulation Factor VIII | Coagulation | F8.13499.30.3 | none |
| miR-139-5p | 30 | P00451 | Coagulation Factor VIII | Coagulation | F8.13499.30.3 | none |
| let-7c-5p | 26 | P00451 | Coagulation Factor VIII | Coagulation | F8.13499.30.3 | none |
| miR-195-5p | 20 | P00451 | Coagulation Factor VIII | Coagulation | F8.13499.30.3 | none |
| miR-10a-5p | 14 | P00451 | Coagulation Factor VIII | Coagulation | F8.13499.30.3 | none |
| let-7a-5p | 8 | P00451 | Coagulation Factor VIII | Coagulation | F8.13499.30.3 | none |
| miR-126-3p | 46 | P00451 | Coagulation Factor VIII | Coagulation | F8.13499.30.3 | none |
| miR-10b-5p | 40 | P00451 | Coagulation Factor VIII | Coagulation | F8.13499.30.3 | none |
| let-7b-5p | 28 | Q8N128 | Protein FAM177A1 | F177A | FAM177A1.8039.41.3 | none |
| miR-195-5p | 20 | Q8N128 | Protein FAM177A1 | F177A | FAM177A1.8039.41.3 | none |
| miR-139-5p | 30 | Q8N128 | Protein FAM177A1 | F177A | FAM177A1.8039.41.3 | none |
| let-7c-5p | 26 | Q8N128 | Protein FAM177A1 | F177A | FAM177A1.8039.41.3 | none |
| let-7a-5p | 8 | Q8N128 | Protein FAM177A1 | F177A | FAM177A1.8039.41.3 | none |
| miR-126-3p | 30 | Q8N128 | Protein FAM177A1 | F177A | FAM177A1.8039.41.3 | none |
| miR-10b-5p | 30 | Q8N128 | Protein FAM177A1 | F177A | FAM177A1.8039.41.3 | none |
| miR-10a-5p | 1 | O75063 | Glycosaminoglycan xylosylkinase | FA20B | FAM20B.7198.197.3 | none |
| let-7b-5p | 4 | O75063 | Glycosaminoglycan xylosylkinase | FA20B | FAM20B.7198.197.3 | none |
| miR-195-5p | 2 | O75063 | Glycosaminoglycan xylosylkinase | FA20B | FAM20B.7198.197.3 | none |
| let-7a-5p | 4 | O75063 | Glycosaminoglycan xylosylkinase | FA20B | FAM20B.7198.197.3 | none |
| miR-10b-5p | 7 | O75063 | Glycosaminoglycan xylosylkinase | FA20B | FAM20B.7198.197.3 | none |
| miR-126-3p | 7 | O75063 | Glycosaminoglycan xylosylkinase | FA20B | FAM20B.7198.197.3 | none |
| let-7c-5p | 2 | O75063 | Glycosaminoglycan xylosylkinase | FA20B | FAM20B.7198.197.3 | none |
| miR-139-5p | 6 | O75063 | Glycosaminoglycan xylosylkinase | FA20B | FAM20B.7198.197.3 | none |
| miR-195-5p | 2 | P58499 | Protein FAM3B | FAM3B | FAM3B.5618.50.3 | none |
| miR-126-3p | 5 | P58499 | Protein FAM3B | FAM3B | FAM3B.5618.50.3 | none |
| miR-10b-5p | 6 | P58499 | Protein FAM3B | FAM3B | FAM3B.5618.50.3 | none |
| let-7b-5p | 3 | P58499 | Protein FAM3B | FAM3B | FAM3B.5618.50.3 | none |
| let-7c-5p | 2 | P58499 | Protein FAM3B | FAM3B | FAM3B.5618.50.3 | none |
| miR-139-5p | 5 | P58499 | Protein FAM3B | FAM3B | FAM3B.5618.50.3 | none |
| let-7a-5p | 4 | P58499 | Protein FAM3B | FAM3B | FAM3B.5618.50.3 | none |
| miR-10a-5p | 14 | P58499 | Protein FAM3B | FAM3B | FAM3B.9177.6.3 | none |
| miR-10a-5p | 14 | Q96BQ1 | Protein FAM3D | FAM3D | FAM3D.13102.1.3 | none |
| miR-10b-5p | 40 | Q96BQ1 | Protein FAM3D | FAM3D | FAM3D.13102.1.3 | none |
| miR-195-5p | 20 | Q96BQ1 | Protein FAM3D | FAM3D | FAM3D.13102.1.3 | none |
| let-7b-5p | 30 | Q96BQ1 | Protein FAM3D | FAM3D | FAM3D.13102.1.3 | none |
| let-7a-5p | 8 | Q96BQ1 | Protein FAM3D | FAM3D | FAM3D.13102.1.3 | none |
| let-7c-5p | 26 | Q96BQ1 | Protein FAM3D | FAM3D | FAM3D.13102.1.3 | none |
| miR-139-5p | 30 | Q96BQ1 | Protein FAM3D | FAM3D | FAM3D.13102.1.3 | none |
| miR-126-3p | 46 | Q96BQ1 | Protein FAM3D | FAM3D | FAM3D.13102.1.3 | none |
| miR-139-5p | 30 | P35916 | Vascular endothelial growth factor receptor 3 | VEGF sR3 | FLT4.2358.19.2 | none |
| miR-10b-5p | 40 | P35916 | Vascular endothelial growth factor receptor 3 | VEGF sR3 | FLT4.2358.19.2 | none |
| let-7c-5p | 26 | P35916 | Vascular endothelial growth factor receptor 3 | VEGF sR3 | FLT4.2358.19.2 | none |
| let-7a-5p | 8 | P35916 | Vascular endothelial growth factor receptor 3 | VEGF sR3 | FLT4.2358.19.2 | none |
| miR-10a-5p | 14 | P35916 | Vascular endothelial growth factor receptor 3 | VEGF sR3 | FLT4.2358.19.2 | none |
| let-7b-5p | 30 | P35916 | Vascular endothelial growth factor receptor 3 | VEGF sR3 | FLT4.2358.19.2 | none |
| miR-195-5p | 20 | P35916 | Vascular endothelial growth factor receptor 3 | VEGF sR3 | FLT4.2358.19.2 | none |
| miR-126-3p | 46 | P35916 | Vascular endothelial growth factor receptor 3 | VEGF sR3 | FLT4.2358.19.2 | none |
| miR-143-3p | 2 | Q9BYC5 | Alpha-(1,6)-fucosyltransferase | FUT8 | FUT8.8244.16.3 | none |
| miR-145-5p | 2 | Q9BYC5 | Alpha-(1,6)-fucosyltransferase | FUT8 | FUT8.8244.16.3 | none |
| miR-10a-5p | 14 | O94923 | D-glucuronyl C5-epimerase | GLCE | GLCE.7808.5.3 | none |
| let-7c-5p | 26 | O94923 | D-glucuronyl C5-epimerase | GLCE | GLCE.7808.5.3 | none |
| miR-139-5p | 30 | O94923 | D-glucuronyl C5-epimerase | GLCE | GLCE.7808.5.3 | none |
| miR-10b-5p | 40 | O94923 | D-glucuronyl C5-epimerase | GLCE | GLCE.7808.5.3 | none |
| let-7a-5p | 8 | O94923 | D-glucuronyl C5-epimerase | GLCE | GLCE.7808.5.3 | none |
| miR-126-3p | 46 | O94923 | D-glucuronyl C5-epimerase | GLCE | GLCE.7808.5.3 | none |
| let-7b-5p | 30 | O94923 | D-glucuronyl C5-epimerase | GLCE | GLCE.7808.5.3 | none |
| miR-139-5p | 20 | P15586 | N-acetylglucosamine-6-sulfatase | GNS | GNS.3616.3.5 | none |
| miR-10b-5p | 20 | P15586 | N-acetylglucosamine-6-sulfatase | GNS | GNS.3616.3.5 | none |
| miR-126-3p | 20 | P15586 | N-acetylglucosamine-6-sulfatase | GNS | GNS.3616.3.5 | none |
| miR-195-5p | 18 | P15586 | N-acetylglucosamine-6-sulfatase | GNS | GNS.3616.3.5 | none |
| let-7c-5p | 26 | Q8NBJ4 | Golgi membrane protein 1 | GOLM1 | GOLM1.8983.7.3 | none |
| miR-139-5p | 30 | Q8NBJ4 | Golgi membrane protein 1 | GOLM1 | GOLM1.8983.7.3 | none |
| miR-126-3p | 46 | Q8NBJ4 | Golgi membrane protein 1 | GOLM1 | GOLM1.8983.7.3 | none |
| let-7b-5p | 30 | Q8NBJ4 | Golgi membrane protein 1 | GOLM1 | GOLM1.8983.7.3 | none |
| miR-195-5p | 20 | Q8NBJ4 | Golgi membrane protein 1 | GOLM1 | GOLM1.8983.7.3 | none |
| let-7a-5p | 8 | Q8NBJ4 | Golgi membrane protein 1 | GOLM1 | GOLM1.8983.7.3 | none |
| miR-10a-5p | 14 | Q8NBJ4 | Golgi membrane protein 1 | GOLM1 | GOLM1.8983.7.3 | none |
| miR-10b-5p | 40 | Q8NBJ4 | Golgi membrane protein 1 | GOLM1 | GOLM1.8983.7.3 | none |
| let-7a-5p | 8 | P05362 | Intercellular adhesion molecule 1 | sICAM-1 | ICAM1.4342.10.3 | none |
| let-7c-5p | 26 | P05362 | Intercellular adhesion molecule 1 | sICAM-1 | ICAM1.4342.10.3 | none |
| miR-195-5p | 20 | P05362 | Intercellular adhesion molecule 1 | sICAM-1 | ICAM1.4342.10.3 | none |
| miR-10b-5p | 31 | P05362 | Intercellular adhesion molecule 1 | sICAM-1 | ICAM1.4342.10.3 | none |
| miR-139-5p | 30 | P05362 | Intercellular adhesion molecule 1 | sICAM-1 | ICAM1.4342.10.3 | none |
| let-7b-5p | 28 | P05362 | Intercellular adhesion molecule 1 | sICAM-1 | ICAM1.4342.10.3 | none |
| miR-126-3p | 30 | P05362 | Intercellular adhesion molecule 1 | sICAM-1 | ICAM1.4342.10.3 | none |
| let-7c-5p | 26 | P13598 | Intercellular adhesion molecule 2 | sICAM-2 | ICAM2.5486.73.3 | none |
| miR-10b-5p | 31 | P13598 | Intercellular adhesion molecule 2 | sICAM-2 | ICAM2.5486.73.3 | none |
| let-7a-5p | 8 | P13598 | Intercellular adhesion molecule 2 | sICAM-2 | ICAM2.5486.73.3 | none |
| miR-195-5p | 20 | P13598 | Intercellular adhesion molecule 2 | sICAM-2 | ICAM2.5486.73.3 | none |
| miR-126-3p | 30 | P13598 | Intercellular adhesion molecule 2 | sICAM-2 | ICAM2.5486.73.3 | none |
| miR-139-5p | 30 | P13598 | Intercellular adhesion molecule 2 | sICAM-2 | ICAM2.5486.73.3 | none |

| | | | | | | |
|---|---|---|---|---|---|---|
| let-7b-5p | 28 | P13598 | Intercellular adhesion molecule 2 | sICAM-2 | ICAM2.5486.73.3 | none |
| miR-139-5p | 5 | Q14773 | Intercellular adhesion molecule 4 | ICAM4 | ICAM4.6550.4.3 | none |
| miR-195-5p | 2 | Q14773 | Intercellular adhesion molecule 4 | ICAM4 | ICAM4.6550.4.3 | none |
| miR-126-3p | 5 | Q14773 | Intercellular adhesion molecule 4 | ICAM4 | ICAM4.6550.4.3 | none |
| let-7a-5p | 4 | Q14773 | Intercellular adhesion molecule 4 | ICAM4 | ICAM4.6550.4.3 | none |
| miR-10b-5p | 5 | Q14773 | Intercellular adhesion molecule 4 | ICAM4 | ICAM4.6550.4.3 | none |
| let-7c-5p | 2 | Q14773 | Intercellular adhesion molecule 4 | ICAM4 | ICAM4.6550.4.3 | none |
| let-7b-5p | 3 | Q14773 | Intercellular adhesion molecule 4 | ICAM4 | ICAM4.6550.4.3 | none |
| let-7c-5p | 20 | Q9UMF0 | Intercellular adhesion molecule 5 | sICAM-5 | ICAM5.5124.62.3 | none |
| miR-195-5p | 20 | Q9UMF0 | Intercellular adhesion molecule 5 | sICAM-5 | ICAM5.5124.62.3 | none |
| miR-139-5p | 24 | Q9UMF0 | Intercellular adhesion molecule 5 | sICAM-5 | ICAM5.5124.62.3 | none |
| miR-126-3p | 24 | Q9UMF0 | Intercellular adhesion molecule 5 | sICAM-5 | ICAM5.5124.62.3 | none |
| let-7a-5p | 4 | Q9UMF0 | Intercellular adhesion molecule 5 | sICAM-5 | ICAM5.5124.62.3 | none |
| let-7b-5p | 22 | Q9UMF0 | Intercellular adhesion molecule 5 | sICAM-5 | ICAM5.5124.62.3 | none |
| miR-10b-5p | 25 | Q9UMF0 | Intercellular adhesion molecule 5 | sICAM-5 | ICAM5.5124.62.3 | none |
| miR-195-5p | 2 | P08069 | Insulin-like growth factor 1 receptor | IGF-I sR | IGF1R.4232.19.2 | none |
| let-7a-5p | 4 | P08069 | Insulin-like growth factor 1 receptor | IGF-I sR | IGF1R.4232.19.2 | none |
| let-7b-5p | 4 | P08069 | Insulin-like growth factor 1 receptor | IGF-I sR | IGF1R.4232.19.2 | none |
| miR-10b-5p | 6 | P08069 | Insulin-like growth factor 1 receptor | IGF-I sR | IGF1R.4232.19.2 | none |
| let-7c-5p | 2 | P08069 | Insulin-like growth factor 1 receptor | IGF-I sR | IGF1R.4232.19.2 | none |
| miR-126-3p | 6 | P08069 | Insulin-like growth factor 1 receptor | IGF-I sR | IGF1R.4232.19.2 | none |
| miR-139-5p | 6 | P08069 | Insulin-like growth factor 1 receptor | IGF-I sR | IGF1R.4232.19.2 | none |
| let-7b-5p | 30 | P26951 | Interleukin-3 receptor subunit alpha | IL-3 Ra | IL3RA.13744.37.3 | none |
| miR-195-5p | 20 | P26951 | Interleukin-3 receptor subunit alpha | IL-3 Ra | IL3RA.13744.37.3 | none |
| miR-10a-5p | 14 | P26951 | Interleukin-3 receptor subunit alpha | IL-3 Ra | IL3RA.13744.37.3 | none |
| let-7c-5p | 26 | P26951 | Interleukin-3 receptor subunit alpha | IL-3 Ra | IL3RA.13744.37.3 | none |
| miR-126-3p | 46 | P26951 | Interleukin-3 receptor subunit alpha | IL-3 Ra | IL3RA.13744.37.3 | none |
| let-7a-5p | 8 | P26951 | Interleukin-3 receptor subunit alpha | IL-3 Ra | IL3RA.13744.37.3 | none |
| miR-10b-5p | 40 | P26951 | Interleukin-3 receptor subunit alpha | IL-3 Ra | IL3RA.13744.37.3 | none |
| miR-139-5p | 30 | P26951 | Interleukin-3 receptor subunit alpha | IL-3 Ra | IL3RA.13744.37.3 | none |
| let-7b-5p | 4 | P40189 | Interleukin-6 receptor subunit beta | gp130, solu | IL6ST.2620.4.2 | none |
| let-7a-5p | 4 | P40189 | Interleukin-6 receptor subunit beta | gp130, solu | IL6ST.2620.4.2 | none |
| miR-126-3p | 6 | P40189 | Interleukin-6 receptor subunit beta | gp130, solu | IL6ST.2620.4.2 | none |
| miR-139-5p | 6 | P40189 | Interleukin-6 receptor subunit beta | gp130, solu | IL6ST.2620.4.2 | none |
| let-7c-5p | 2 | P40189 | Interleukin-6 receptor subunit beta | gp130, solu | IL6ST.2620.4.2 | none |
| miR-10b-5p | 6 | P40189 | Interleukin-6 receptor subunit beta | gp130, solu | IL6ST.2620.4.2 | none |
| miR-195-5p | 2 | P40189 | Interleukin-6 receptor subunit beta | gp130, solu | IL6ST.2620.4.2 | none |
| miR-143-3p | 2 | P13232 | Interleukin-7 | IL-7 | IL7.14049.17.3 | none |
| miR-145-5p | 2 | P13232 | Interleukin-7 | IL-7 | IL7.14049.17.3 | none |
| miR-145-5p | 4 | Q9NRR6 | 72 kDa inositol polyphosphate 5-phosphatase | INP5E | INPP5E.11370.20.3 | none |
| miR-143-3p | 4 | Q9NRR6 | 72 kDa inositol polyphosphate 5-phosphatase | INP5E | INPP5E.11370.20.3 | none |
| miR-139-5p | 30 | P06213 | Insulin receptor | IR | INSR.3448.13.2 | none |
| let-7c-5p | 26 | P06213 | Insulin receptor | IR | INSR.3448.13.2 | none |
| miR-10a-5p | 14 | P06213 | Insulin receptor | IR | INSR.3448.13.2 | none |
| miR-126-3p | 46 | P06213 | Insulin receptor | IR | INSR.3448.13.2 | none |
| miR-10b-5p | 40 | P06213 | Insulin receptor | IR | INSR.3448.13.2 | none |
| let-7a-5p | 8 | P06213 | Insulin receptor | IR | INSR.3448.13.2 | none |
| miR-195-5p | 20 | P06213 | Insulin receptor | IR | INSR.3448.13.2 | none |
| let-7b-5p | 30 | P06213 | Insulin receptor | IR | INSR.3448.13.2 | none |
| miR-139-5p | 24 | Q6UXK2 | Immunoglobulin superfamily containing leucine-rich repeat protein 2 | ISLR2 | ISLR2.13124.20.3 | none |
| miR-10b-5p | 25 | Q6UXK2 | Immunoglobulin superfamily containing leucine-rich repeat protein 2 | ISLR2 | ISLR2.13124.20.3 | none |
| miR-195-5p | 20 | Q6UXK2 | Immunoglobulin superfamily containing leucine-rich repeat protein 2 | ISLR2 | ISLR2.13124.20.3 | none |
| let-7c-5p | 20 | Q6UXK2 | Immunoglobulin superfamily containing leucine-rich repeat protein 2 | ISLR2 | ISLR2.13124.20.3 | none |
| let-7a-5p | 4 | Q6UXK2 | Immunoglobulin superfamily containing leucine-rich repeat protein 2 | ISLR2 | ISLR2.13124.20.3 | none |
| let-7b-5p | 22 | Q6UXK2 | Immunoglobulin superfamily containing leucine-rich repeat protein 2 | ISLR2 | ISLR2.13124.20.3 | none |
| miR-126-3p | 24 | Q6UXK2 | Immunoglobulin superfamily containing leucine-rich repeat protein 2 | ISLR2 | ISLR2.13124.20.3 | none |
| miR-10b-5p | 6 | P78504 | Protein jagged-1 | JAG1 | JAG1.5092.51.3 | none |
| miR-195-5p | 2 | P78504 | Protein jagged-1 | JAG1 | JAG1.5092.51.3 | none |
| let-7a-5p | 4 | P78504 | Protein jagged-1 | JAG1 | JAG1.5092.51.3 | none |
| miR-126-3p | 6 | P78504 | Protein jagged-1 | JAG1 | JAG1.5092.51.3 | none |
| let-7b-5p | 4 | P78504 | Protein jagged-1 | JAG1 | JAG1.5092.51.3 | none |
| let-7c-5p | 2 | P78504 | Protein jagged-1 | JAG1 | JAG1.5092.51.3 | none |
| miR-139-5p | 6 | P78504 | Protein jagged-1 | JAG1 | JAG1.5092.51.3 | none |
| miR-10a-5p | 14 | P35968 | Vascular endothelial growth factor receptor 2 | VEGF sR2 | KDR.3651.50.5 | none |
| let-7a-5p | 8 | P35968 | Vascular endothelial growth factor receptor 2 | VEGF sR2 | KDR.3651.50.5 | none |
| miR-195-5p | 20 | P35968 | Vascular endothelial growth factor receptor 2 | VEGF sR2 | KDR.3651.50.5 | none |
| let-7c-5p | 26 | P35968 | Vascular endothelial growth factor receptor 2 | VEGF sR2 | KDR.3651.50.5 | none |
| let-7b-5p | 30 | P35968 | Vascular endothelial growth factor receptor 2 | VEGF sR2 | KDR.3651.50.5 | none |
| miR-10b-5p | 40 | P35968 | Vascular endothelial growth factor receptor 2 | VEGF sR2 | KDR.3651.50.5 | none |
| miR-139-5p | 30 | P35968 | Vascular endothelial growth factor receptor 2 | VEGF sR2 | KDR.3651.50.5 | none |
| miR-126-3p | 46 | P35968 | Vascular endothelial growth factor receptor 2 | VEGF sR2 | KDR.3651.50.5 | none |
| miR-139-5p | 30 | O60870 | DNA/RNA-binding protein KIN17 | KIN17 | KIN.14643.27.3 | none |
| let-7b-5p | 28 | O60870 | DNA/RNA-binding protein KIN17 | KIN17 | KIN.14643.27.3 | none |
| let-7c-5p | 26 | O60870 | DNA/RNA-binding protein KIN17 | KIN17 | KIN.14643.27.3 | none |
| miR-195-5p | 20 | O60870 | DNA/RNA-binding protein KIN17 | KIN17 | KIN.14643.27.3 | none |
| miR-10b-5p | 37 | O60870 | DNA/RNA-binding protein KIN17 | KIN17 | KIN.14643.27.3 | none |
| let-7a-5p | 8 | O60870 | DNA/RNA-binding protein KIN17 | KIN17 | KIN.14643.27.3 | none |
| miR-126-3p | 30 | O60870 | DNA/RNA-binding protein KIN17 | KIN17 | KIN.14643.27.3 | none |
| miR-195-5p | 2 | P32004 | Neural cell adhesion molecule L1 | NCAM-L1 | L1CAM.4246.40.2 | none |
| let-7a-5p | 4 | P32004 | Neural cell adhesion molecule L1 | NCAM-L1 | L1CAM.4246.40.2 | none |
| let-7b-5p | 4 | P32004 | Neural cell adhesion molecule L1 | NCAM-L1 | L1CAM.4246.40.2 | none |
| miR-139-5p | 6 | P32004 | Neural cell adhesion molecule L1 | NCAM-L1 | L1CAM.4246.40.2 | none |
| let-7c-5p | 2 | P32004 | Neural cell adhesion molecule L1 | NCAM-L1 | L1CAM.4246.40.2 | none |
| miR-126-3p | 6 | P32004 | Neural cell adhesion molecule L1 | NCAM-L1 | L1CAM.4246.40.2 | none |
| miR-10b-5p | 6 | P32004 | Neural cell adhesion molecule L1 | NCAM-L1 | L1CAM.4246.40.2 | none |
| let-7b-5p | 2 | P07942,P1 | Laminin | Laminin | LAMA1.LAMB1.LAMC1.2728. | none |
| let-7a-5p | 1 | P07942,P1 | Laminin | Laminin | LAMA1.LAMB1.LAMC1.2728. | none |
| let-7c-5p | 2 | P07942,P1 | Laminin | Laminin | LAMA1.LAMB1.LAMC1.2728. | none |
| miR-139-5p | 2 | P07942,P1 | Laminin | Laminin | LAMA1.LAMB1.LAMC1.2728. | none |

| | | | | | | |
|---|---|---|---|---|---|---|
| miR-195-5p | 2 | P07942,P1 | Laminin | Laminin | LAMA1.LAMB1.LAMC1.2728. | none |
| miR-126-3p | 2 | P07942,P1 | Laminin | Laminin | LAMA1.LAMB1.LAMC1.2728. | none |
| miR-10b-5p | 2 | P07942,P1 | Laminin | Laminin | LAMA1.LAMB1.LAMC1.2728. | none |
| miR-126-3p | 3 | P09848 | Lactase-phlorizin hydrolase | LPH | LCT.9017.58.3 | none |
| miR-139-5p | 3 | P09848 | Lactase-phlorizin hydrolase | LPH | LCT.9017.58.3 | none |
| let-7a-5p | 1 | P09848 | Lactase-phlorizin hydrolase | LPH | LCT.9017.58.3 | none |
| miR-10b-5p | 3 | P09848 | Lactase-phlorizin hydrolase | LPH | LCT.9017.58.3 | none |
| miR-195-5p | 2 | P09848 | Lactase-phlorizin hydrolase | LPH | LCT.9017.58.3 | none |
| let-7b-5p | 3 | P09848 | Lactase-phlorizin hydrolase | LPH | LCT.9017.58.3 | none |
| let-7c-5p | 2 | P09848 | Lactase-phlorizin hydrolase | LPH | LCT.9017.58.3 | none |
| miR-145-5p | 4 | P47929 | Galectin-7 | Galectin-7 | LGALS7.9196.8.3 | none |
| miR-143-3p | 4 | P47929 | Galectin-7 | Galectin-7 | LGALS7.9196.8.3 | none |
| miR-139-5p | 16 | P42702 | Leukemia inhibitory factor receptor | LIF sR | LIFR.5837.49.3 | none |
| let-7a-5p | 4 | P42702 | Leukemia inhibitory factor receptor | LIF sR | LIFR.5837.49.3 | none |
| miR-126-3p | 16 | P42702 | Leukemia inhibitory factor receptor | LIF sR | LIFR.5837.49.3 | none |
| miR-10b-5p | 17 | P42702 | Leukemia inhibitory factor receptor | LIF sR | LIFR.5837.49.3 | none |
| let-7b-5p | 14 | P42702 | Leukemia inhibitory factor receptor | LIF sR | LIFR.5837.49.3 | none |
| let-7c-5p | 12 | P42702 | Leukemia inhibitory factor receptor | LIF sR | LIFR.5837.49.3 | none |
| miR-195-5p | 12 | P42702 | Leukemia inhibitory factor receptor | LIF sR | LIFR.5837.49.3 | none |
| let-7a-5p | 4 | O60476 | Mannosyl-oligosaccharide 1,2-alpha-mannosidase IB | MA1A2 | MAN1A2.9077.10.3 | none |
| miR-195-5p | 2 | O60476 | Mannosyl-oligosaccharide 1,2-alpha-mannosidase IB | MA1A2 | MAN1A2.9077.10.3 | none |
| miR-10b-5p | 5 | O60476 | Mannosyl-oligosaccharide 1,2-alpha-mannosidase IB | MA1A2 | MAN1A2.9077.10.3 | none |
| miR-139-5p | 5 | O60476 | Mannosyl-oligosaccharide 1,2-alpha-mannosidase IB | MA1A2 | MAN1A2.9077.10.3 | none |
| miR-126-3p | 5 | O60476 | Mannosyl-oligosaccharide 1,2-alpha-mannosidase IB | MA1A2 | MAN1A2.9077.10.3 | none |
| let-7b-5p | 3 | O60476 | Mannosyl-oligosaccharide 1,2-alpha-mannosidase IB | MA1A2 | MAN1A2.9077.10.3 | none |
| let-7c-5p | 2 | O60476 | Mannosyl-oligosaccharide 1,2-alpha-mannosidase IB | MA1A2 | MAN1A2.9077.10.3 | none |
| miR-195-5p | 20 | P11226 | Mannose-binding protein C | MBL | MBL2.3000.66.1 | none |
| let-7b-5p | 30 | P11226 | Mannose-binding protein C | MBL | MBL2.3000.66.1 | none |
| let-7c-5p | 26 | P11226 | Mannose-binding protein C | MBL | MBL2.3000.66.1 | none |
| miR-10b-5p | 33 | P11226 | Mannose-binding protein C | MBL | MBL2.3000.66.1 | none |
| miR-10a-5p | 5 | P11226 | Mannose-binding protein C | MBL | MBL2.3000.66.1 | none |
| miR-126-3p | 37 | P11226 | Mannose-binding protein C | MBL | MBL2.3000.66.1 | none |
| miR-139-5p | 30 | P11226 | Mannose-binding protein C | MBL | MBL2.3000.66.1 | none |
| let-7a-5p | 8 | P11226 | Mannose-binding protein C | MBL | MBL2.3000.66.1 | none |
| let-7b-5p | 30 | P08581 | Hepatocyte growth factor receptor | Met | MET.2837.3.2 | none |
| let-7c-5p | 26 | P08581 | Hepatocyte growth factor receptor | Met | MET.2837.3.2 | none |
| let-7a-5p | 8 | P08581 | Hepatocyte growth factor receptor | Met | MET.2837.3.2 | none |
| miR-126-3p | 32 | P08581 | Hepatocyte growth factor receptor | Met | MET.2837.3.2 | none |
| miR-139-5p | 30 | P08581 | Hepatocyte growth factor receptor | Met | MET.2837.3.2 | none |
| miR-195-5p | 20 | P08581 | Hepatocyte growth factor receptor | Met | MET.2837.3.2 | none |
| miR-10b-5p | 40 | P08581 | Hepatocyte growth factor receptor | Met | MET.2837.3.2 | none |
| miR-145-5p | 3 | Q5T2T1 | MAGUK p55 subfamily member 7 | MPP7 | MPP7.12732.13.3 | none |
| miR-143-3p | 3 | Q5T2T1 | MAGUK p55 subfamily member 7 | MPP7 | MPP7.12732.13.3 | none |
| miR-143-3p | 4 | Q9Y328 | Neuron-specific protein family member 2 | NSG2 | NSG2.13409.9.3 | none |
| miR-145-5p | 4 | Q9Y328 | Neuron-specific protein family member 2 | NSG2 | NSG2.13409.9.3 | none |
| miR-143-3p | 1 | Q9Y5G5 | Protocadherin gamma-A8 | PCDG8 | PCDHGA8.11259.71.3 | none |
| miR-145-5p | 1 | Q9Y5G5 | Protocadherin gamma-A8 | PCDG8 | PCDHGA8.11259.71.3 | none |
| miR-143-3p | 4 | P04085 | Platelet-derived growth factor subunit A | PDGF-AA | PDGFA.4499.21.1 | none |
| miR-145-5p | 4 | P04085 | Platelet-derived growth factor subunit A | PDGF-AA | PDGFA.4499.21.1 | none |
| miR-145-5p | 4 | P01127 | Platelet-derived growth factor subunit B | PDGF-BB | PDGFB.4149.8.2 | none |
| miR-143-3p | 4 | P01127 | Platelet-derived growth factor subunit B | PDGF-BB | PDGFB.4149.8.2 | none |
| miR-143-3p | 4 | Q9GZP0 | Platelet-derived growth factor D | PDGFD | PDGFD.9341.1.3 | none |
| let-7a-5p | 4 | Q5VY43 | Platelet endothelial aggregation receptor 1 | PEAR1 | PEAR1.8275.31.3 | none |
| let-7b-5p | 4 | Q5VY43 | Platelet endothelial aggregation receptor 1 | PEAR1 | PEAR1.8275.31.3 | none |
| miR-126-3p | 6 | Q5VY43 | Platelet endothelial aggregation receptor 1 | PEAR1 | PEAR1.8275.31.3 | none |
| miR-139-5p | 6 | Q5VY43 | Platelet endothelial aggregation receptor 1 | PEAR1 | PEAR1.8275.31.3 | none |
| let-7c-5p | 2 | Q5VY43 | Platelet endothelial aggregation receptor 1 | PEAR1 | PEAR1.8275.31.3 | none |
| miR-195-5p | 2 | Q5VY43 | Platelet endothelial aggregation receptor 1 | PEAR1 | PEAR1.8275.31.3 | none |
| miR-10b-5p | 6 | Q5VY43 | Platelet endothelial aggregation receptor 1 | PEAR1 | PEAR1.8275.31.3 | none |
| miR-10b-5p | 2 | O15031 | Plexin-B2 | PLXB2 | PLXNB2.9216.100.3 | none |
| miR-126-3p | 2 | O15031 | Plexin-B2 | PLXB2 | PLXNB2.9216.100.3 | none |
| let-7b-5p | 2 | O15031 | Plexin-B2 | PLXB2 | PLXNB2.9216.100.3 | none |
| miR-143-3p | 4 | P02775 | Neutrophil-activating peptide 2 | NAP-2 | PPBP.2790.54.2 | none |
| miR-145-5p | 4 | P02775 | Neutrophil-activating peptide 2 | NAP-2 | PPBP.2790.54.2 | none |
| miR-139-5p | 30 | Q6ZRP7 | Sulfhydryl oxidase 2 | QSOX2 | QSOX2.8397.147.3 | none |
| miR-10b-5p | 40 | Q6ZRP7 | Sulfhydryl oxidase 2 | QSOX2 | QSOX2.8397.147.3 | none |
| miR-10a-5p | 14 | Q6ZRP7 | Sulfhydryl oxidase 2 | QSOX2 | QSOX2.8397.147.3 | none |
| let-7b-5p | 30 | Q6ZRP7 | Sulfhydryl oxidase 2 | QSOX2 | QSOX2.8397.147.3 | none |
| let-7c-5p | 26 | Q6ZRP7 | Sulfhydryl oxidase 2 | QSOX2 | QSOX2.8397.147.3 | none |
| let-7a-5p | 8 | Q6ZRP7 | Sulfhydryl oxidase 2 | QSOX2 | QSOX2.8397.147.3 | none |
| miR-195-5p | 20 | Q6ZRP7 | Sulfhydryl oxidase 2 | QSOX2 | QSOX2.8397.147.3 | none |
| miR-126-3p | 46 | Q6ZRP7 | Sulfhydryl oxidase 2 | QSOX2 | QSOX2.8397.147.3 | none |
| miR-143-3p | 4 | P84095 | Rho-related GTP-binding protein RhoG | RHOG | RHOG.12540.25.3 | none |
| miR-145-5p | 4 | P84095 | Rho-related GTP-binding protein RhoG | RHOG | RHOG.12540.25.3 | none |
| miR-145-5p | 1 | Q01826 | DNA-binding protein SATB1 | SATB1 | SATB1.13511.29.3 | none |
| miR-143-3p | 1 | Q01826 | DNA-binding protein SATB1 | SATB1 | SATB1.13511.29.3 | none |
| miR-139-5p | 3 | Q8WVN6 | Secreted and transmembrane protein 1 | SECTM1 | SECTM1.13093.6.3 | none |
| miR-10b-5p | 3 | Q8WVN6 | Secreted and transmembrane protein 1 | SECTM1 | SECTM1.13093.6.3 | none |
| let-7a-5p | 3 | Q8WVN6 | Secreted and transmembrane protein 1 | SECTM1 | SECTM1.13093.6.3 | none |
| let-7b-5p | 1 | Q8WVN6 | Secreted and transmembrane protein 1 | SECTM1 | SECTM1.13093.6.3 | none |
| miR-126-3p | 3 | Q8WVN6 | Secreted and transmembrane protein 1 | SECTM1 | SECTM1.13093.6.3 | none |
| miR-139-5p | 30 | P16581 | E-selectin | sE-Selectin | SELE.3470.1.2 | none |
| let-7b-5p | 30 | P16581 | E-selectin | sE-Selectin | SELE.3470.1.2 | none |
| miR-126-3p | 46 | P16581 | E-selectin | sE-Selectin | SELE.3470.1.2 | none |
| let-7c-5p | 26 | P16581 | E-selectin | sE-Selectin | SELE.3470.1.2 | none |
| miR-10a-5p | 14 | P16581 | E-selectin | sE-Selectin | SELE.3470.1.2 | none |
| miR-10b-5p | 40 | P16581 | E-selectin | sE-Selectin | SELE.3470.1.2 | none |
| let-7a-5p | 8 | P16581 | E-selectin | sE-Selectin | SELE.3470.1.2 | none |
| miR-195-5p | 20 | P16581 | E-selectin | sE-Selectin | SELE.3470.1.2 | none |

| | | | | | | |
|---|---|---|---|---|---|---|
| miR-145-5p | 4 | P16109 | P-selectin | P-Selectin | SELP.4154.57.2 | none |
| miR-143-3p | 4 | P16109 | P-selectin | P-Selectin | SELP.4154.57.2 | none |
| miR-126-3p | 40 | P16109 | P-selectin | P-Selectin | SELP.4154.57.2 | none |
| miR-139-5p | 30 | P16109 | P-selectin | P-Selectin | SELP.4154.57.2 | none |
| miR-10a-5p | 8 | P16109 | P-selectin | P-Selectin | SELP.4154.57.2 | none |
| miR-195-5p | 20 | P16109 | P-selectin | P-Selectin | SELP.4154.57.2 | none |
| miR-10b-5p | 40 | P16109 | P-selectin | P-Selectin | SELP.4154.57.2 | none |
| let-7b-5p | 30 | P16109 | P-selectin | P-Selectin | SELP.4154.57.2 | none |
| let-7c-5p | 26 | P16109 | P-selectin | P-Selectin | SELP.4154.57.2 | none |
| let-7a-5p | 8 | P16109 | P-selectin | P-Selectin | SELP.4154.57.2 | none |
| miR-126-3p | 6 | Q9H2E6 | Semaphorin-6A | Semaphori | SEMA6A.7945.10.3 | none |
| let-7a-5p | 4 | Q9H2E6 | Semaphorin-6A | Semaphori | SEMA6A.7945.10.3 | none |
| miR-139-5p | 6 | Q9H2E6 | Semaphorin-6A | Semaphori | SEMA6A.7945.10.3 | none |
| miR-195-5p | 2 | Q9H2E6 | Semaphorin-6A | Semaphori | SEMA6A.7945.10.3 | none |
| let-7c-5p | 2 | Q9H2E6 | Semaphorin-6A | Semaphori | SEMA6A.7945.10.3 | none |
| miR-10b-5p | 7 | Q9H2E6 | Semaphorin-6A | Semaphori | SEMA6A.7945.10.3 | none |
| let-7b-5p | 4 | Q9H2E6 | Semaphorin-6A | Semaphori | SEMA6A.7945.10.3 | none |
| miR-409-3p | 2 | Q02383 | Semenogelin-2 | SEMG2 | SEMG2.6373.54.3 | none |
| miR-143-3p | 4 | P05121 | Plasminogen activator inhibitor 1 | PAI-1 | SERPINE1.2925.9.1 | none |
| miR-145-5p | 4 | P05121 | Plasminogen activator inhibitor 1 | PAI-1 | SERPINE1.2925.9.1 | none |
| miR-145-5p | 1 | P07093 | Glia-derived nexin | Protease n | SERPINE2.3217.74.2 | none |
| miR-143-3p | 1 | P07093 | Glia-derived nexin | Protease n | SERPINE2.3217.74.2 | none |
| miR-143-3p | 4 | Q9NXA8 | NAD-dependent protein deacylase sirtuin-5, mitochondrial | SIRT5 | SIRT5.12461.8.3 | none |
| miR-145-5p | 4 | Q9NXA8 | NAD-dependent protein deacylase sirtuin-5, mitochondrial | SIRT5 | SIRT5.12461.8.3 | none |
| miR-145-5p | 4 | P09486 | SPARC | ON | SPARC.3043.49.2 | none |
| miR-143-3p | 4 | P09486 | SPARC | ON | SPARC.3043.49.2 | none |
| miR-143-3p | 4 | O15269 | Serine palmitoyltransferase 1 | SPTC1 | SPTLC1.7886.26.3 | none |
| miR-145-5p | 4 | O15269 | Serine palmitoyltransferase 1 | SPTC1 | SPTLC1.7886.26.3 | none |
| miR-143-3p | 4 | Q9BT88 | Synaptotagmin-11 | SYT11 | SYT11.7089.42.3 | none |
| miR-145-5p | 4 | Q9BT88 | Synaptotagmin-11 | SYT11 | SYT11.7089.42.3 | none |
| let-7a-5p | 8 | Q9NS62 | Thrombospondin type-1 domain-containing protein 1 | THSD1 | THSD1.5621.64.3 | none |
| let-7c-5p | 26 | Q9NS62 | Thrombospondin type-1 domain-containing protein 1 | THSD1 | THSD1.5621.64.3 | none |
| miR-126-3p | 30 | Q9NS62 | Thrombospondin type-1 domain-containing protein 1 | THSD1 | THSD1.5621.64.3 | none |
| miR-10b-5p | 31 | Q9NS62 | Thrombospondin type-1 domain-containing protein 1 | THSD1 | THSD1.5621.64.3 | none |
| miR-195-5p | 20 | Q9NS62 | Thrombospondin type-1 domain-containing protein 1 | THSD1 | THSD1.5621.64.3 | none |
| miR-139-5p | 30 | Q9NS62 | Thrombospondin type-1 domain-containing protein 1 | THSD1 | THSD1.5621.64.3 | none |
| let-7b-5p | 28 | Q9NS62 | Thrombospondin type-1 domain-containing protein 1 | THSD1 | THSD1.5621.64.3 | none |
| miR-10a-5p | 14 | P35590 | Tyrosine-protein kinase receptor Tie-1, soluble | sTie-1 | TIE1.2844.53.2 | none |
| let-7a-5p | 8 | P35590 | Tyrosine-protein kinase receptor Tie-1, soluble | sTie-1 | TIE1.2844.53.2 | none |
| let-7c-5p | 26 | P35590 | Tyrosine-protein kinase receptor Tie-1, soluble | sTie-1 | TIE1.2844.53.2 | none |
| miR-126-3p | 46 | P35590 | Tyrosine-protein kinase receptor Tie-1, soluble | sTie-1 | TIE1.2844.53.2 | none |
| miR-10b-5p | 40 | P35590 | Tyrosine-protein kinase receptor Tie-1, soluble | sTie-1 | TIE1.2844.53.2 | none |
| let-7b-5p | 30 | P35590 | Tyrosine-protein kinase receptor Tie-1, soluble | sTie-1 | TIE1.2844.53.2 | none |
| miR-139-5p | 30 | P35590 | Tyrosine-protein kinase receptor Tie-1, soluble | sTie-1 | TIE1.2844.53.2 | none |
| miR-195-5p | 20 | P35590 | Tyrosine-protein kinase receptor Tie-1, soluble | sTie-1 | TIE1.2844.53.2 | none |
| let-7a-5p | 8 | O00206,Q9 | Toll-like receptor 4:Lymphocyte antigen 96 complex | TLR4:MD-2 | TLR4.LY96.3647.49.4 | none |
| let-7c-5p | 26 | O00206,Q9 | Toll-like receptor 4:Lymphocyte antigen 96 complex | TLR4:MD-2 | TLR4.LY96.3647.49.4 | none |
| let-7b-5p | 28 | O00206,Q9 | Toll-like receptor 4:Lymphocyte antigen 96 complex | TLR4:MD-2 | TLR4.LY96.3647.49.4 | none |
| miR-126-3p | 30 | O00206,Q9 | Toll-like receptor 4:Lymphocyte antigen 96 complex | TLR4:MD-2 | TLR4.LY96.3647.49.4 | none |
| miR-139-5p | 30 | O00206,Q9 | Toll-like receptor 4:Lymphocyte antigen 96 complex | TLR4:MD-2 | TLR4.LY96.3647.49.4 | none |
| miR-195-5p | 20 | O00206,Q9 | Toll-like receptor 4:Lymphocyte antigen 96 complex | TLR4:MD-2 | TLR4.LY96.3647.49.4 | none |
| miR-10b-5p | 31 | O00206,Q9 | Toll-like receptor 4:Lymphocyte antigen 96 complex | TLR4:MD-2 | TLR4.LY96.3647.49.4 | none |
| let-7c-5p | 26 | O60704 | Protein-tyrosine sulfotransferase 2 | TPST2 | TPST2.8024.64.3 | none |
| miR-126-3p | 32 | O60704 | Protein-tyrosine sulfotransferase 2 | TPST2 | TPST2.8024.64.3 | none |
| miR-195-5p | 20 | O60704 | Protein-tyrosine sulfotransferase 2 | TPST2 | TPST2.8024.64.3 | none |
| let-7a-5p | 8 | O60704 | Protein-tyrosine sulfotransferase 2 | TPST2 | TPST2.8024.64.3 | none |
| miR-10b-5p | 33 | O60704 | Protein-tyrosine sulfotransferase 2 | TPST2 | TPST2.8024.64.3 | none |
| miR-139-5p | 30 | O60704 | Protein-tyrosine sulfotransferase 2 | TPST2 | TPST2.8024.64.3 | none |
| let-7b-5p | 30 | O60704 | Protein-tyrosine sulfotransferase 2 | TPST2 | TPST2.8024.64.3 | none |
| let-7a-5p | 2 | Q9NNX1 | Tuftelin | TUFT1 | TUFT1.5690.49.3 | none |
| miR-10b-5p | 2 | Q9NNX1 | Tuftelin | TUFT1 | TUFT1.5690.49.3 | none |
| miR-126-3p | 2 | Q9NNX1 | Tuftelin | TUFT1 | TUFT1.5690.49.3 | none |
| miR-139-5p | 2 | Q9NNX1 | Tuftelin | TUFT1 | TUFT1.5690.49.3 | none |
| miR-143-3p | 4 | Q9Y4X1 | UDP-glucuronosyltransferase 2A1 | UD2A1 | UGT2A1.8907.11.3 | none |
| miR-145-5p | 4 | Q9Y4X1 | UDP-glucuronosyltransferase 2A1 | UD2A1 | UGT2A1.8907.11.3 | none |
| miR-143-3p | 4 | P15692 | Vascular endothelial growth factor A, isoform 121 | VEGF121 | VEGFA.14032.2.3 | none |
| miR-145-5p | 4 | P15692 | Vascular endothelial growth factor A, isoform 121 | VEGF121 | VEGFA.14032.2.3 | none |

**Table S17. Overlap between trans-miR-eQTLs and pQTLs in Olink**

| miRNA | n_transmiRQTLs | UniProt.ID | Gene | Olink_assay | annotation |
|---|---|---|---|---|---|
| miR-126-3p | 47 | Q02763 | TEK | 122_TIE2 | TIE2 |
| miR-145-5p | 4 | P03956 | MMP1 | 142_MMP-1 | MMP-1 |
| miR-145-5p | 4 | P15692 | VEGFA | 102_VEGF-A | VEGF-A |
| miR-195-5p | 2 | P78423 | CX3CL1 | 186_CX3CL1 | CX3CL1 |
| let-7a-5p | 4 | P78423 | CX3CL1 | 186_CX3CL1 | CX3CL1 |
| let-7c-5p | 2 | P78423 | CX3CL1 | 186_CX3CL1 | CX3CL1 |
| miR-139-5p | 5 | P78423 | CX3CL1 | 186_CX3CL1 | CX3CL1 |
| miR-10b-5p | 5 | P78423 | CX3CL1 | 186_CX3CL1 | CX3CL1 |
| let-7b-5p | 3 | P78423 | CX3CL1 | 186_CX3CL1 | CX3CL1 |
| miR-126-3p | 5 | P78423 | CX3CL1 | 186_CX3CL1 | CX3CL1 |
| miR-143-3p | 4 | O94907 | DKK1 | 171_Dkk-1 | Dkk-1 |
| miR-145-5p | 4 | O94907 | DKK1 | 171_Dkk-1 | Dkk-1 |
| let-7c-5p | 27 | Q9GZV9 | FGF23 | 131_FGF-23 | FGF-23 |
| miR-139-5p | 30 | Q9GZV9 | FGF23 | 131_FGF-23 | FGF-23 |
| let-7a-5p | 8 | Q9GZV9 | FGF23 | 131_FGF-23 | FGF-23 |
| let-7b-5p | 29 | Q9GZV9 | FGF23 | 131_FGF-23 | FGF-23 |
| miR-195-5p | 20 | Q9GZV9 | FGF23 | 131_FGF-23 | FGF-23 |
| miR-126-3p | 30 | Q9GZV9 | FGF23 | 131_FGF-23 | FGF-23 |
| miR-10b-5p | 30 | Q9GZV9 | FGF23 | 131_FGF-23 | FGF-23 |
| miR-143-3p | 4 | P03956 | MMP1 | 142_MMP-1 | MMP-1 |
| let-7c-5p | 27 | P16284 | PECAM1 | 194_PECAM-1 | PECAM-1 |
| let-7b-5p | 31 | P16284 | PECAM1 | 194_PECAM-1 | PECAM-1 |
| miR-195-5p | 20 | P16284 | PECAM1 | 194_PECAM-1 | PECAM-1 |
| miR-139-5p | 31 | P16284 | PECAM1 | 194_PECAM-1 | PECAM-1 |
| miR-126-3p | 47 | P16284 | PECAM1 | 194_PECAM-1 | PECAM-1 |
| miR-10b-5p | 41 | P16284 | PECAM1 | 194_PECAM-1 | PECAM-1 |
| let-7a-5p | 9 | P16284 | PECAM1 | 194_PECAM-1 | PECAM-1 |
| miR-10a-5p | 14 | P16284 | PECAM1 | 194_PECAM-1 | PECAM-1 |
| let-7b-5p | 1 | P21583 | KITLG | 132_SCF | SCF |
| miR-139-5p | 3 | P21583 | KITLG | 132_SCF | SCF |
| miR-126-3p | 3 | P21583 | KITLG | 132_SCF | SCF |
| miR-10b-5p | 3 | P21583 | KITLG | 132_SCF | SCF |
| let-7a-5p | 3 | P21583 | KITLG | 132_SCF | SCF |
| let-7c-5p | 27 | P16581 | SELE | 108_SELE | SELE |
| let-7a-5p | 9 | P16581 | SELE | 108_SELE | SELE |
| miR-126-3p | 47 | P16581 | SELE | 108_SELE | SELE |
| miR-10a-5p | 14 | P16581 | SELE | 108_SELE | SELE |
| miR-10b-5p | 41 | P16581 | SELE | 108_SELE | SELE |
| let-7b-5p | 31 | P16581 | SELE | 108_SELE | SELE |
| miR-195-5p | 20 | P16581 | SELE | 108_SELE | SELE |
| miR-139-5p | 31 | P16581 | SELE | 108_SELE | SELE |
| miR-139-5p | 5 | Q01638 | IL1RL1 | 151_ST2 | ST2 |
| let-7a-5p | 4 | Q01638 | IL1RL1 | 151_ST2 | ST2 |
| let-7c-5p | 2 | Q01638 | IL1RL1 | 151_ST2 | ST2 |
| miR-126-3p | 5 | Q01638 | IL1RL1 | 151_ST2 | ST2 |
| miR-195-5p | 2 | Q01638 | IL1RL1 | 151_ST2 | ST2 |
| miR-10b-5p | 5 | Q01638 | IL1RL1 | 151_ST2 | ST2 |
| let-7b-5p | 3 | Q01638 | IL1RL1 | 151_ST2 | ST2 |
| miR-139-5p | 6 | P13726 | F3 | 123_TF | TF |
| let-7a-5p | 4 | P13726 | F3 | 123_TF | TF |
| let-7b-5p | 4 | P13726 | F3 | 123_TF | TF |
| let-7c-5p | 2 | P13726 | F3 | 123_TF | TF |
| miR-126-3p | 6 | P13726 | F3 | 123_TF | TF |
| miR-10b-5p | 7 | P13726 | F3 | 123_TF | TF |
| miR-195-5p | 2 | P13726 | F3 | 123_TF | TF |
| let-7b-5p | 31 | Q02763 | TEK | 122_TIE2 | TIE2 |
| let-7a-5p | 9 | Q02763 | TEK | 122_TIE2 | TIE2 |
| miR-139-5p | 31 | Q02763 | TEK | 122_TIE2 | TIE2 |
| let-7c-5p | 27 | Q02763 | TEK | 122_TIE2 | TIE2 |
| miR-195-5p | 20 | Q02763 | TEK | 122_TIE2 | TIE2 |
| miR-10b-5p | 41 | Q02763 | TEK | 122_TIE2 | TIE2 |
| miR-10a-5p | 14 | Q02763 | TEK | 122_TIE2 | TIE2 |
| let-7b-5p | 31 | P07204 | THBD | 159_TM | TM |
| let-7a-5p | 9 | P07204 | THBD | 159_TM | TM |
| miR-10a-5p | 14 | P07204 | THBD | 159_TM | TM |
| let-7c-5p | 27 | P07204 | THBD | 159_TM | TM |
| miR-126-3p | 47 | P07204 | THBD | 159_TM | TM |
| miR-195-5p | 20 | P07204 | THBD | 159_TM | TM |
| miR-139-5p | 31 | P07204 | THBD | 159_TM | TM |
| miR-10b-5p | 41 | P07204 | THBD | 159_TM | TM |
| miR-143-3p | 4 | P15692 | VEGFA | 102_VEGF-A | VEGF-A |

| Long_name | MTI |
|---|---|
| Angiopoietin-1 receptor | validated MTI |
| Matrix metalloproteinase-1 | validated MTI |
| Vascular endothelial growth fa | validated MTI |
| Fractalkine | predicted MTI |
| Fractalkine | none |
| Fractalkine | none |
| Fractalkine | none |
| Fractalkine | none |
| Fractalkine | none |
| Fractalkine | none |
| Dickkopf-related protein 1 | none |
| Dickkopf-related protein 1 | none |
| Fibroblast growth factor 23 | none |
| Fibroblast growth factor 23 | none |
| Fibroblast growth factor 23 | none |
| Fibroblast growth factor 23 | none |
| Fibroblast growth factor 23 | none |
| Fibroblast growth factor 23 | none |
| Fibroblast growth factor 23 | none |
| Matrix metalloproteinase-1 | none |
| Platelet endothelial cell adhes | none |
| Platelet endothelial cell adhes | none |
| Platelet endothelial cell adhes | none |
| Platelet endothelial cell adhes | none |
| Platelet endothelial cell adhes | none |
| Platelet endothelial cell adhes | none |
| Platelet endothelial cell adhes | none |
| Platelet endothelial cell adhes | none |
| Stem cell factor | none |
| Stem cell factor | none |
| Stem cell factor | none |
| Stem cell factor | none |
| Stem cell factor | none |
| E-selectin | none |
| E-selectin | none |
| E-selectin | none |
| E-selectin | none |
| E-selectin | none |
| E-selectin | none |
| E-selectin | none |
| E-selectin | none |
| ST2 protein | none |
| ST2 protein | none |
| ST2 protein | none |
| ST2 protein | none |
| ST2 protein | none |
| ST2 protein | none |
| ST2 protein | none |
| Tissue factor | none |
| Tissue factor | none |
| Tissue factor | none |
| Tissue factor | none |
| Tissue factor | none |
| Tissue factor | none |
| Tissue factor | none |
| Angiopoietin-1 receptor | none |
| Angiopoietin-1 receptor | none |
| Angiopoietin-1 receptor | none |
| Angiopoietin-1 receptor | none |
| Angiopoietin-1 receptor | none |
| Angiopoietin-1 receptor | none |
| Angiopoietin-1 receptor | none |
| Thrombomodulin | none |
| Thrombomodulin | none |
| Thrombomodulin | none |
| Thrombomodulin | none |
| Thrombomodulin | none |
| Thrombomodulin | none |
| Thrombomodulin | none |
| Thrombomodulin | none |
| Vascular endothelial growth fa | none |

**Table S18. Overlap between miR-eQTLs and met-QTLs**

| SNP | miRNA | cismark | pval_RS | platform | metabolite | description | effect_allel | other_allel | effect | se | pval |
|---|---|---|---|---|---|---|---|---|---|---|---|
| rs174577 | miR-1908-! | cis | 1.89E-08 | Metabolon | M01110 | arachidona | a | c | -0.0458 | 0.0025 | 1.37E-75 |
| rs174548 | miR-1908-! | cis | 3.01E-08 | Metabolon | M01110 | arachidona | c | g | 0.0488 | 0.0025 | 1.43E-84 |
| rs174538 | miR-1908-! | cis | 1.35E-07 | Metabolon | M01110 | arachidona | a | g | -0.0464 | 0.0025 | 6.24E-76 |
| rs174555 | miR-1908-! | cis | 7.43E-08 | Metabolon | M01110 | arachidona | t | c | 0.0481 | 0.0025 | 6.58E-82 |
| rs174576 | miR-1908-! | cis | 3.13E-08 | Metabolon | M01110 | arachidona | a | c | -0.0461 | 0.0025 | 4.18E-77 |
| rs2727270 | miR-1908-! | cis | 4.45E-05 | Metabolon | M01110 | arachidona | t | c | -0.0506 | 0.0034 | 6.09E-50 |
| rs174578 | miR-1908-! | cis | 2.55E-08 | Metabolon | M01110 | arachidona | a | t | -0.0461 | 0.0025 | 5.68E-77 |
| rs174448 | miR-1908-! | cis | 1.95E-05 | Metabolon | M01110 | arachidona | a | g | 0.0278 | 0.0025 | 7.85E-29 |
| rs174574 | miR-1908-! | cis | 8.50E-09 | Metabolon | M01110 | arachidona | a | c | -0.0475 | 0.0025 | 1.23E-81 |
| rs174549 | miR-1908-! | cis | 4.04E-08 | Metabolon | M01110 | arachidona | a | g | -0.048 | 0.0025 | 8.18E-82 |
| rs174535 | miR-1908-! | cis | 6.98E-09 | Metabolon | M01110 | arachidona | t | c | 0.0475 | 0.0025 | 2.05E-81 |
| rs174545 | miR-1908-! | cis | 2.29E-08 | Metabolon | M01110 | arachidona | c | g | 0.0476 | 0.0025 | 8.31E-82 |
| rs174536 | miR-1908-! | cis | 1.31E-08 | Metabolon | M01110 | arachidona | a | c | 0.0475 | 0.0025 | 2.05E-81 |
| rs2727271 | miR-1908-! | cis | 4.23E-05 | Metabolon | M01110 | arachidona | a | t | 0.0506 | 0.0034 | 6.18E-50 |
| rs174449 | miR-1908-! | cis | 2.49E-05 | Metabolon | M01110 | arachidona | a | g | 0.0268 | 0.0025 | 3.89E-26 |
| rs2072114 | miR-1908-! | cis | 1.34E-05 | Metabolon | M01110 | arachidona | a | g | 0.0461 | 0.0034 | 6.91E-42 |
| rs174537 | miR-1908-! | cis | 9.41E-09 | Metabolon | M01110 | arachidona | t | g | -0.0475 | 0.0025 | 2.05E-81 |
| rs2524299 | miR-1908-! | cis | 4.82E-05 | Metabolon | M01110 | arachidona | a | t | 0.047 | 0.0034 | 2.10E-43 |
| rs174556 | miR-1908-! | cis | 7.67E-08 | Metabolon | M01110 | arachidona | t | c | -0.0488 | 0.0025 | 2.98E-84 |
| rs174601 | miR-1908-! | cis | 5.98E-09 | Metabolon | M01110 | arachidona | t | c | -0.0463 | 0.0025 | 4.99E-76 |
| rs174575 | miR-1908-! | cis | 5.45E-05 | Metabolon | M01110 | arachidona | c | g | 0.0359 | 0.0026 | 5.25E-44 |
| rs2727271 | miR-1908-! | cis | 4.23E-05 | Metabolon | M18467 | eicosapent | a | t | 0.0389 | 0.0054 | 4.44E-13 |
| rs174578 | miR-1908-! | cis | 2.55E-08 | Metabolon | M18467 | eicosapent | a | t | -0.0329 | 0.0036 | 7.91E-20 |
| rs174538 | miR-1908-! | cis | 1.35E-07 | Metabolon | M18467 | eicosapent | a | g | -0.0344 | 0.0037 | 5.20E-21 |
| rs174601 | miR-1908-! | cis | 5.98E-09 | Metabolon | M18467 | eicosapent | t | c | -0.0343 | 0.0042 | 5.68E-16 |
| rs174537 | miR-1908-! | cis | 9.41E-09 | Metabolon | M18467 | eicosapent | t | g | -0.0343 | 0.0036 | 2.55E-21 |
| rs174556 | miR-1908-! | cis | 7.67E-08 | Metabolon | M18467 | eicosapent | t | c | -0.0356 | 0.0037 | 1.97E-22 |
| rs174548 | miR-1908-! | cis | 3.01E-08 | Metabolon | M18467 | eicosapent | c | g | 0.0353 | 0.0036 | 3.71E-22 |
| rs174535 | miR-1908-! | cis | 6.98E-09 | Metabolon | M18467 | eicosapent | t | c | 0.0343 | 0.0036 | 2.55E-21 |
| rs2727270 | miR-1908-! | cis | 4.45E-05 | Metabolon | M18467 | eicosapent | t | c | -0.0389 | 0.0054 | 4.32E-13 |
| rs2072114 | miR-1908-! | cis | 1.34E-05 | Metabolon | M18467 | eicosapent | a | g | 0.038 | 0.0054 | 1.46E-12 |
| rs174555 | miR-1908-! | cis | 7.43E-08 | Metabolon | M18467 | eicosapent | t | c | 0.0356 | 0.0037 | 1.97E-22 |
| rs174574 | miR-1908-! | cis | 8.50E-09 | Metabolon | M18467 | eicosapent | a | c | -0.0341 | 0.0036 | 4.34E-21 |
| rs2524299 | miR-1908-! | cis | 4.82E-05 | Metabolon | M18467 | eicosapent | a | t | 0.038 | 0.0054 | 1.42E-12 |
| rs174577 | miR-1908-! | cis | 1.89E-08 | Metabolon | M18467 | eicosapent | a | c | -0.0331 | 0.0036 | 5.32E-20 |
| rs174536 | miR-1908-! | cis | 1.31E-08 | Metabolon | M18467 | eicosapent | a | c | 0.0343 | 0.0036 | 2.55E-21 |
| rs174576 | miR-1908-! | cis | 3.13E-08 | Metabolon | M18467 | eicosapent | a | c | -0.0323 | 0.0036 | 3.74E-19 |
| rs174545 | miR-1908-! | cis | 2.29E-08 | Metabolon | M18467 | eicosapent | c | g | 0.0342 | 0.0036 | 3.02E-21 |
| rs174575 | miR-1908-! | cis | 5.45E-05 | Metabolon | M18467 | eicosapent | c | g | 0.0216 | 0.0037 | 7.87E-09 |
| rs174549 | miR-1908-! | cis | 4.04E-08 | Metabolon | M18467 | eicosapent | a | g | -0.0356 | 0.0037 | 2.02E-22 |
| rs174577 | miR-1908-! | cis | 1.89E-08 | Metabolon | M32504 | docosapen | a | c | -0.0248 | 0.0035 | 1.35E-12 |
| rs174601 | miR-1908-! | cis | 5.98E-09 | Metabolon | M32504 | docosapen | t | c | -0.0247 | 0.0041 | 1.24E-09 |
| rs174538 | miR-1908-! | cis | 1.35E-07 | Metabolon | M32504 | docosapen | a | g | -0.0264 | 0.0035 | 8.90E-14 |
| rs174576 | miR-1908-! | cis | 3.13E-08 | Metabolon | M32504 | docosapen | a | c | -0.0234 | 0.0035 | 1.85E-11 |
| rs174536 | miR-1908-! | cis | 1.31E-08 | Metabolon | M32504 | docosapen | a | c | 0.0252 | 0.0035 | 5.07E-13 |
| rs174548 | miR-1908-! | cis | 3.01E-08 | Metabolon | M32504 | docosapen | c | g | 0.0256 | 0.0041 | 2.94E-10 |
| rs174556 | miR-1908-! | cis | 7.67E-08 | Metabolon | M32504 | docosapen | t | c | -0.0258 | 0.0041 | 2.66E-10 |
| rs174537 | miR-1908-! | cis | 9.41E-09 | Metabolon | M32504 | docosapen | t | g | -0.0252 | 0.0035 | 5.07E-13 |
| rs174545 | miR-1908-! | cis | 2.29E-08 | Metabolon | M32504 | docosapen | c | g | 0.0252 | 0.0035 | 4.87E-13 |
| rs174578 | miR-1908-! | cis | 2.55E-08 | Metabolon | M32504 | docosapen | a | t | -0.0241 | 0.0035 | 5.34E-12 |
| rs174535 | miR-1908-! | cis | 6.98E-09 | Metabolon | M32504 | docosapen | t | c | 0.0252 | 0.0035 | 5.06E-13 |
| rs174555 | miR-1908-! | cis | 7.43E-08 | Metabolon | M32504 | docosapen | t | c | 0.0258 | 0.0041 | 2.66E-10 |
| rs174549 | miR-1908-! | cis | 4.04E-08 | Metabolon | M32504 | docosapen | a | g | -0.0256 | 0.0041 | 3.45E-10 |
| rs174574 | miR-1908-! | cis | 8.50E-09 | Metabolon | M32504 | docosapen | a | c | -0.0245 | 0.0035 | 2.54E-12 |
| rs174575 | miR-1908-! | cis | 5.45E-05 | Metabolon | M32635 | 1-linoleoyl | c | g | -0.0314 | 0.0036 | 5.63E-18 |
| rs174577 | miR-1908-! | cis | 1.89E-08 | Metabolon | M32635 | 1-linoleoyl | a | c | 0.0413 | 0.0035 | 3.47E-32 |
| rs174538 | miR-1908-! | cis | 1.35E-07 | Metabolon | M32635 | 1-linoleoyl | a | g | 0.0424 | 0.0035 | 5.75E-33 |
| rs174555 | miR-1908-! | cis | 7.43E-08 | Metabolon | M32635 | 1-linoleoyl | t | c | -0.0423 | 0.0035 | 6.39E-33 |
| rs174578 | miR-1908-! | cis | 2.55E-08 | Metabolon | M32635 | 1-linoleoyl | a | t | 0.0407 | 0.0035 | 2.44E-31 |
| rs174448 | miR-1908-! | cis | 1.95E-05 | Metabolon | M32635 | 1-linoleoyl | a | g | -0.0236 | 0.0035 | 1.90E-11 |
| rs174535 | miR-1908-! | cis | 6.98E-09 | Metabolon | M32635 | 1-linoleoyl | t | c | -0.044 | 0.0035 | 2.82E-36 |

| | | | | | | | | | | | |
|---|---|---|---|---|---|---|---|---|---|---|---|
| rs174536 | miR-1908-5 | cis | 1.31E-08 | Metabolon | M32635 | 1-linoleoyl | a | c | -0.044 | 0.0035 | 2.82E-36 |
| rs174449 | miR-1908-5 | cis | 2.49E-05 | Metabolon | M32635 | 1-linoleoyl | a | g | -0.0223 | 0.0036 | 3.90E-10 |
| rs174545 | miR-1908-5 | cis | 2.29E-08 | Metabolon | M32635 | 1-linoleoyl | c | g | -0.0439 | 0.0035 | 3.03E-36 |
| rs174574 | miR-1908-5 | cis | 8.50E-09 | Metabolon | M32635 | 1-linoleoyl | a | c | 0.0432 | 0.0035 | 5.58E-35 |
| rs174601 | miR-1908-5 | cis | 5.98E-09 | Metabolon | M32635 | 1-linoleoyl | t | c | 0.0395 | 0.0035 | 5.36E-29 |
| rs174576 | miR-1908-5 | cis | 3.13E-08 | Metabolon | M32635 | 1-linoleoyl | a | c | 0.0415 | 0.0035 | 1.73E-32 |
| rs2524299 | miR-1908-5 | cis | 4.82E-05 | Metabolon | M32635 | 1-linoleoyl | a | t | -0.0432 | 0.0052 | 9.70E-17 |
| rs174548 | miR-1908-5 | cis | 3.01E-08 | Metabolon | M32635 | 1-linoleoyl | c | g | -0.0431 | 0.0035 | 3.16E-34 |
| rs174537 | miR-1908-5 | cis | 9.41E-09 | Metabolon | M32635 | 1-linoleoyl | t | g | 0.044 | 0.0035 | 2.82E-36 |
| rs2727271 | miR-1908-5 | cis | 4.23E-05 | Metabolon | M32635 | 1-linoleoyl | a | t | -0.044 | 0.0052 | 2.60E-17 |
| rs174556 | miR-1908-5 | cis | 7.67E-08 | Metabolon | M32635 | 1-linoleoyl | t | c | 0.0423 | 0.0035 | 6.37E-33 |
| rs174549 | miR-1908-5 | cis | 4.04E-08 | Metabolon | M32635 | 1-linoleoyl | a | g | 0.0421 | 0.0035 | 1.46E-32 |
| rs2727270 | miR-1908-5 | cis | 4.45E-05 | Metabolon | M32635 | 1-linoleoyl | t | c | 0.0441 | 0.0052 | 2.57E-17 |
| rs2072114 | miR-1908-5 | cis | 1.34E-05 | Metabolon | M32635 | 1-linoleoyl | a | g | -0.0433 | 0.0052 | 8.92E-17 |
| rs174556 | miR-1908-5 | cis | 7.67E-08 | Metabolon | M32980 | adrenate (2 | t | c | -0.0321 | 0.0035 | 4.08E-20 |
| rs174549 | miR-1908-5 | cis | 4.04E-08 | Metabolon | M32980 | adrenate (2 | a | g | -0.0319 | 0.0035 | 6.19E-20 |
| rs174577 | miR-1908-5 | cis | 1.89E-08 | Metabolon | M32980 | adrenate (2 | a | c | -0.0335 | 0.0035 | 3.19E-22 |
| rs174535 | miR-1908-5 | cis | 6.98E-09 | Metabolon | M32980 | adrenate (2 | t | c | 0.0341 | 0.0035 | 5.17E-23 |
| rs174538 | miR-1908-5 | cis | 1.35E-07 | Metabolon | M32980 | adrenate (2 | a | g | -0.0328 | 0.0035 | 5.60E-21 |
| rs174536 | miR-1908-5 | cis | 1.31E-08 | Metabolon | M32980 | adrenate (2 | a | c | 0.0341 | 0.0035 | 5.18E-23 |
| rs174578 | miR-1908-5 | cis | 2.55E-08 | Metabolon | M32980 | adrenate (2 | a | t | -0.0331 | 0.0034 | 8.12E-22 |
| rs2727270 | miR-1908-5 | cis | 4.45E-05 | Metabolon | M32980 | adrenate (2 | t | c | -0.0299 | 0.0051 | 5.41E-09 |
| rs174575 | miR-1908-5 | cis | 5.45E-05 | Metabolon | M32980 | adrenate (2 | c | g | 0.0302 | 0.0036 | 3.03E-17 |
| rs174537 | miR-1908-5 | cis | 9.41E-09 | Metabolon | M32980 | adrenate (2 | t | g | -0.0341 | 0.0035 | 5.19E-23 |
| rs174601 | miR-1908-5 | cis | 5.98E-09 | Metabolon | M32980 | adrenate (2 | t | c | -0.0339 | 0.0035 | 2.53E-22 |
| rs2727271 | miR-1908-5 | cis | 4.23E-05 | Metabolon | M32980 | adrenate (2 | a | t | 0.0299 | 0.0051 | 5.53E-09 |
| rs174555 | miR-1908-5 | cis | 7.43E-08 | Metabolon | M32980 | adrenate (2 | t | c | 0.0321 | 0.0035 | 4.08E-20 |
| rs2072114 | miR-1908-5 | cis | 1.34E-05 | Metabolon | M32980 | adrenate (2 | a | g | 0.0268 | 0.0045 | 2.75E-09 |
| rs174545 | miR-1908-5 | cis | 2.29E-08 | Metabolon | M32980 | adrenate (2 | c | g | 0.0341 | 0.0034 | 4.83E-23 |
| rs174548 | miR-1908-5 | cis | 3.01E-08 | Metabolon | M32980 | adrenate (2 | c | g | 0.0318 | 0.0035 | 6.81E-20 |
| rs174576 | miR-1908-5 | cis | 3.13E-08 | Metabolon | M32980 | adrenate (2 | a | c | -0.0331 | 0.0034 | 7.72E-22 |
| rs174574 | miR-1908-5 | cis | 8.50E-09 | Metabolon | M32980 | adrenate (2 | a | c | -0.034 | 0.0034 | 6.07E-23 |
| rs174448 | miR-1908-5 | cis | 1.95E-05 | Metabolon | M33228 | 1-arachido | a | g | 0.0336 | 0.0027 | 7.70E-35 |
| rs174535 | miR-1908-5 | cis | 6.98E-09 | Metabolon | M33228 | 1-arachido | t | c | 0.0561 | 0.0027 | 1.86E-94 |
| rs174576 | miR-1908-5 | cis | 3.13E-08 | Metabolon | M33228 | 1-arachido | a | c | -0.055 | 0.0027 | 7.46E-91 |
| rs174548 | miR-1908-5 | cis | 3.01E-08 | Metabolon | M33228 | 1-arachido | c | g | 0.0526 | 0.0027 | 3.08E-82 |
| rs174537 | miR-1908-5 | cis | 9.41E-09 | Metabolon | M33228 | 1-arachido | t | g | -0.0553 | 0.0027 | 9.18E-92 |
| rs174449 | miR-1908-5 | cis | 2.49E-05 | Metabolon | M33228 | 1-arachido | a | g | 0.0309 | 0.0028 | 2.71E-29 |
| rs2072114 | miR-1908-5 | cis | 1.34E-05 | Metabolon | M33228 | 1-arachido | a | g | 0.0513 | 0.0044 | 4.54E-31 |
| rs174538 | miR-1908-5 | cis | 1.35E-07 | Metabolon | M33228 | 1-arachido | a | g | -0.0518 | 0.0027 | 9.83E-80 |
| rs174545 | miR-1908-5 | cis | 2.29E-08 | Metabolon | M33228 | 1-arachido | c | g | 0.056 | 0.0027 | 3.72E-94 |
| rs174601 | miR-1908-5 | cis | 5.98E-09 | Metabolon | M33228 | 1-arachido | t | c | -0.0548 | 0.0034 | 1.53E-57 |
| rs174536 | miR-1908-5 | cis | 1.31E-08 | Metabolon | M33228 | 1-arachido | a | c | 0.0553 | 0.0027 | 9.18E-92 |
| rs174556 | miR-1908-5 | cis | 7.67E-08 | Metabolon | M33228 | 1-arachido | t | c | -0.0528 | 0.0027 | 9.51E-83 |
| rs174549 | miR-1908-5 | cis | 4.04E-08 | Metabolon | M33228 | 1-arachido | a | g | -0.0528 | 0.0027 | 9.36E-83 |
| rs2727270 | miR-1908-5 | cis | 4.45E-05 | Metabolon | M33228 | 1-arachido | t | c | -0.0546 | 0.0044 | 6.79E-35 |
| rs2727271 | miR-1908-5 | cis | 4.23E-05 | Metabolon | M33228 | 1-arachido | a | t | 0.0546 | 0.0044 | 7.02E-35 |
| rs174577 | miR-1908-5 | cis | 1.89E-08 | Metabolon | M33228 | 1-arachido | a | c | -0.0551 | 0.0027 | 4.61E-91 |
| rs174555 | miR-1908-5 | cis | 7.43E-08 | Metabolon | M33228 | 1-arachido | t | c | 0.0528 | 0.0027 | 9.53E-83 |
| rs174575 | miR-1908-5 | cis | 5.45E-05 | Metabolon | M33228 | 1-arachido | c | g | 0.0455 | 0.0035 | 3.04E-38 |
| rs174578 | miR-1908-5 | cis | 2.55E-08 | Metabolon | M33228 | 1-arachido | a | t | -0.0541 | 0.0027 | 3.38E-88 |
| rs174574 | miR-1908-5 | cis | 8.50E-09 | Metabolon | M33228 | 1-arachido | a | c | -0.0552 | 0.0027 | 1.97E-91 |
| rs2524299 | miR-1908-5 | cis | 4.82E-05 | Metabolon | M33228 | 1-arachido | a | t | 0.0536 | 0.0044 | 7.99E-34 |
| rs174555 | miR-1908-5 | cis | 7.43E-08 | Metabolon | M33821 | 1-eicosatri | t | c | -0.0271 | 0.0035 | 1.74E-14 |
| rs174535 | miR-1908-5 | cis | 6.98E-09 | Metabolon | M33821 | 1-eicosatri | t | c | -0.0204 | 0.0035 | 5.80E-09 |
| rs174538 | miR-1908-5 | cis | 1.35E-07 | Metabolon | M33821 | 1-eicosatri | a | g | 0.0217 | 0.0035 | 9.13E-10 |
| rs174536 | miR-1908-5 | cis | 1.31E-08 | Metabolon | M33821 | 1-eicosatri | a | c | -0.0204 | 0.0035 | 5.80E-09 |
| rs174576 | miR-1908-5 | cis | 3.13E-08 | Metabolon | M33821 | 1-eicosatri | a | c | 0.0204 | 0.0035 | 5.50E-09 |
| rs174556 | miR-1908-5 | cis | 7.67E-08 | Metabolon | M33821 | 1-eicosatri | t | c | 0.0271 | 0.0035 | 1.74E-14 |
| rs174578 | miR-1908-5 | cis | 2.55E-08 | Metabolon | M33821 | 1-eicosatri | a | t | 0.0211 | 0.0035 | 1.71E-09 |
| rs174449 | miR-1908-5 | cis | 2.49E-05 | Metabolon | M33821 | 1-eicosatri | a | g | -0.0196 | 0.0036 | 3.71E-08 |
| rs174549 | miR-1908-5 | cis | 4.04E-08 | Metabolon | M33821 | 1-eicosatri | a | g | 0.0271 | 0.0035 | 1.82E-14 |
| rs174545 | miR-1908-5 | cis | 2.29E-08 | Metabolon | M33821 | 1-eicosatri | c | g | -0.0207 | 0.0035 | 3.83E-09 |

| | | | | | | | | | | | |
|---|---|---|---|---|---|---|---|---|---|---|---|
| rs174601 | miR-1908-5 | cis | 5.98E-09 | Metabolon | M33821 | 1-eicosatri | t | c | 0.0214 | 0.0035 | 1.37E-09 |
| rs174448 | miR-1908-5 | cis | 1.95E-05 | Metabolon | M33821 | 1-eicosatri | a | g | -0.0203 | 0.0035 | 7.30E-09 |
| rs174577 | miR-1908-5 | cis | 1.89E-08 | Metabolon | M33821 | 1-eicosatri | a | c | 0.0205 | 0.0035 | 5.31E-09 |
| rs174537 | miR-1908-5 | cis | 9.41E-09 | Metabolon | M33821 | 1-eicosatri | t | g | 0.0204 | 0.0035 | 5.79E-09 |
| rs174574 | miR-1908-5 | cis | 8.50E-09 | Metabolon | M33821 | 1-eicosatri | a | c | 0.0201 | 0.0035 | 1.01E-08 |
| rs174548 | miR-1908-5 | cis | 3.01E-08 | Metabolon | M33821 | 1-eicosatri | c | g | -0.0272 | 0.0035 | 1.30E-14 |
| rs174575 | miR-1908-5 | cis | 5.45E-05 | Metabolon | M33821 | 1-eicosatri | c | g | -0.0237 | 0.0036 | 5.41E-11 |
| rs174536 | miR-1908-5 | cis | 1.31E-08 | Metabolon | M33871 | 1-eicosadie | a | c | -0.0194 | 0.0035 | 2.91E-08 |
| rs174537 | miR-1908-5 | cis | 9.41E-09 | Metabolon | M33871 | 1-eicosadie | t | g | 0.0194 | 0.0035 | 2.91E-08 |
| rs174535 | miR-1908-5 | cis | 6.98E-09 | Metabolon | M33871 | 1-eicosadie | t | c | -0.0194 | 0.0035 | 2.92E-08 |
| rs174545 | miR-1908-5 | cis | 2.29E-08 | Metabolon | M33871 | 1-eicosadie | c | g | -0.0195 | 0.0035 | 2.44E-08 |
| rs174601 | miR-1908-5 | cis | 5.98E-09 | Metabolon | M33969 | stearidona | t | c | -0.0336 | 0.0042 | 7.93E-16 |
| rs174538 | miR-1908-5 | cis | 1.35E-07 | Metabolon | M33969 | stearidona | a | g | -0.0305 | 0.0042 | 3.30E-13 |
| rs2727271 | miR-1908-5 | cis | 4.23E-05 | Metabolon | M33969 | stearidona | a | t | 0.0398 | 0.0059 | 1.11E-11 |
| rs174577 | miR-1908-5 | cis | 1.89E-08 | Metabolon | M33969 | stearidona | a | c | -0.0331 | 0.0041 | 1.11E-15 |
| rs174574 | miR-1908-5 | cis | 8.50E-09 | Metabolon | M33969 | stearidona | a | c | -0.0329 | 0.0041 | 1.67E-15 |
| rs2727270 | miR-1908-5 | cis | 4.45E-05 | Metabolon | M33969 | stearidona | t | c | -0.0398 | 0.0059 | 1.08E-11 |
| rs2524299 | miR-1908-5 | cis | 4.82E-05 | Metabolon | M33969 | stearidona | a | t | 0.0372 | 0.0059 | 1.96E-10 |
| rs174549 | miR-1908-5 | cis | 4.04E-08 | Metabolon | M33969 | stearidona | a | g | -0.0312 | 0.0042 | 8.04E-14 |
| rs174556 | miR-1908-5 | cis | 7.67E-08 | Metabolon | M33969 | stearidona | t | c | -0.0312 | 0.0042 | 9.13E-14 |
| rs174535 | miR-1908-5 | cis | 6.98E-09 | Metabolon | M33969 | stearidona | t | c | 0.0331 | 0.0041 | 1.07E-15 |
| rs174537 | miR-1908-5 | cis | 9.41E-09 | Metabolon | M33969 | stearidona | t | g | -0.0331 | 0.0041 | 1.07E-15 |
| rs174536 | miR-1908-5 | cis | 1.31E-08 | Metabolon | M33969 | stearidona | a | c | 0.0331 | 0.0041 | 1.07E-15 |
| rs174575 | miR-1908-5 | cis | 5.45E-05 | Metabolon | M33969 | stearidona | c | g | 0.0239 | 0.0043 | 2.40E-08 |
| rs174555 | miR-1908-5 | cis | 7.43E-08 | Metabolon | M33969 | stearidona | t | c | 0.0312 | 0.0042 | 9.13E-14 |
| rs174576 | miR-1908-5 | cis | 3.13E-08 | Metabolon | M33969 | stearidona | a | c | -0.0313 | 0.0041 | 3.24E-14 |
| rs174578 | miR-1908-5 | cis | 2.55E-08 | Metabolon | M33969 | stearidona | a | t | -0.032 | 0.0041 | 9.59E-15 |
| rs2072114 | miR-1908-5 | cis | 1.34E-05 | Metabolon | M33969 | stearidona | a | g | 0.0375 | 0.0059 | 1.46E-10 |
| rs174548 | miR-1908-5 | cis | 3.01E-08 | Metabolon | M33969 | stearidona | c | g | 0.0307 | 0.0042 | 1.97E-13 |
| rs174545 | miR-1908-5 | cis | 2.29E-08 | Metabolon | M33969 | stearidona | c | g | 0.033 | 0.0041 | 1.33E-15 |
| rs174556 | miR-1908-5 | cis | 7.67E-08 | Metabolon | M34214 | 1-arachido | t | c | -0.0263 | 0.0026 | 2.63E-23 |
| rs174536 | miR-1908-5 | cis | 1.31E-08 | Metabolon | M34214 | 1-arachido | a | c | 0.0257 | 0.0026 | 1.04E-22 |
| rs2524299 | miR-1908-5 | cis | 4.82E-05 | Metabolon | M34214 | 1-arachido | a | t | 0.0282 | 0.0042 | 2.17E-11 |
| rs174537 | miR-1908-5 | cis | 9.41E-09 | Metabolon | M34214 | 1-arachido | t | g | -0.0257 | 0.0026 | 1.04E-22 |
| rs174545 | miR-1908-5 | cis | 2.29E-08 | Metabolon | M34214 | 1-arachido | c | g | 0.0258 | 0.0026 | 6.60E-23 |
| rs174449 | miR-1908-5 | cis | 2.49E-05 | Metabolon | M34214 | 1-arachido | a | g | 0.0176 | 0.0027 | 2.95E-11 |
| rs174538 | miR-1908-5 | cis | 1.35E-07 | Metabolon | M34214 | 1-arachido | a | g | -0.0249 | 0.0026 | 4.59E-21 |
| rs174601 | miR-1908-5 | cis | 5.98E-09 | Metabolon | M34214 | 1-arachido | t | c | -0.0252 | 0.0032 | 7.89E-15 |
| rs174549 | miR-1908-5 | cis | 4.04E-08 | Metabolon | M34214 | 1-arachido | a | g | -0.0264 | 0.0026 | 1.98E-23 |
| rs2727270 | miR-1908-5 | cis | 4.45E-05 | Metabolon | M34214 | 1-arachido | t | c | -0.0294 | 0.0042 | 3.15E-12 |
| rs174576 | miR-1908-5 | cis | 3.13E-08 | Metabolon | M34214 | 1-arachido | a | c | -0.0259 | 0.0026 | 5.52E-23 |
| rs174448 | miR-1908-5 | cis | 1.95E-05 | Metabolon | M34214 | 1-arachido | a | g | 0.0195 | 0.0026 | 1.10E-13 |
| rs174555 | miR-1908-5 | cis | 7.43E-08 | Metabolon | M34214 | 1-arachido | t | c | 0.0263 | 0.0026 | 2.64E-23 |
| rs174575 | miR-1908-5 | cis | 5.45E-05 | Metabolon | M34214 | 1-arachido | c | g | 0.0186 | 0.0033 | 2.68E-08 |
| rs174548 | miR-1908-5 | cis | 3.01E-08 | Metabolon | M34214 | 1-arachido | c | g | 0.0262 | 0.0026 | 3.19E-23 |
| rs174578 | miR-1908-5 | cis | 2.55E-08 | Metabolon | M34214 | 1-arachido | a | t | -0.0266 | 0.0026 | 3.02E-24 |
| rs174574 | miR-1908-5 | cis | 8.50E-09 | Metabolon | M34214 | 1-arachido | a | c | -0.0259 | 0.0026 | 5.44E-23 |
| rs174535 | miR-1908-5 | cis | 6.98E-09 | Metabolon | M34214 | 1-arachido | t | c | 0.0257 | 0.0026 | 1.04E-22 |
| rs174577 | miR-1908-5 | cis | 1.89E-08 | Metabolon | M34214 | 1-arachido | a | c | -0.0263 | 0.0026 | 1.11E-23 |
| rs2727271 | miR-1908-5 | cis | 4.23E-05 | Metabolon | M34214 | 1-arachido | a | t | 0.0294 | 0.0042 | 3.15E-12 |
| rs2072114 | miR-1908-5 | cis | 1.34E-05 | Metabolon | M34214 | 1-arachido | a | g | 0.0272 | 0.0042 | 1.07E-10 |
| rs2727271 | miR-1908-5 | cis | 4.23E-05 | Metabolon | M34674 | X-12990 | a | t | 0.0551 | 0.0083 | 2.74E-11 |
| rs2524299 | miR-1908-5 | cis | 4.82E-05 | Metabolon | M34674 | X-12990 | a | t | 0.0475 | 0.0081 | 3.89E-09 |
| rs2072114 | miR-1908-5 | cis | 1.34E-05 | Metabolon | M34674 | X-12990 | a | g | 0.0482 | 0.008 | 2.12E-09 |
| rs2727270 | miR-1908-5 | cis | 4.45E-05 | Metabolon | M34674 | X-12990 | t | c | -0.0551 | 0.0083 | 2.76E-11 |
| rs174601 | miR-1908-5 | cis | 5.98E-09 | Metabolon | M35186 | 1-arachido | t | c | -0.0364 | 0.0032 | 2.13E-29 |
| rs174549 | miR-1908-5 | cis | 4.04E-08 | Metabolon | M35186 | 1-arachido | a | g | -0.0336 | 0.0026 | 2.57E-37 |
| rs174448 | miR-1908-5 | cis | 1.95E-05 | Metabolon | M35186 | 1-arachido | a | g | 0.0254 | 0.0026 | 2.27E-22 |
| rs174576 | miR-1908-5 | cis | 3.13E-08 | Metabolon | M35186 | 1-arachido | a | c | -0.0354 | 0.0026 | 7.35E-42 |
| rs174574 | miR-1908-5 | cis | 8.50E-09 | Metabolon | M35186 | 1-arachido | a | c | -0.035 | 0.0026 | 7.49E-41 |
| rs174449 | miR-1908-5 | cis | 2.49E-05 | Metabolon | M35186 | 1-arachido | a | g | 0.0241 | 0.0026 | 9.20E-20 |
| rs174545 | miR-1908-5 | cis | 2.29E-08 | Metabolon | M35186 | 1-arachido | c | g | 0.0349 | 0.0026 | 1.25E-40 |
| rs2072114 | miR-1908-5 | cis | 1.34E-05 | Metabolon | M35186 | 1-arachido | a | g | 0.0298 | 0.0042 | 1.43E-12 |

| | | | | | | | | | | | |
|---|---|---|---|---|---|---|---|---|---|---|---|
| rs174577 | miR-1908-5 | cis | 1.89E-08 | Metabolon | M35186 | 1-arachido | a | c | -0.0354 | 0.0026 | 8.89E-42 |
| rs2524299 | miR-1908-5 | cis | 4.82E-05 | Metabolon | M35186 | 1-arachido | a | t | 0.0311 | 0.0042 | 1.35E-13 |
| rs174535 | miR-1908-5 | cis | 6.98E-09 | Metabolon | M35186 | 1-arachido | t | c | 0.0341 | 0.0026 | 9.87E-39 |
| rs174578 | miR-1908-5 | cis | 2.55E-08 | Metabolon | M35186 | 1-arachido | a | t | -0.0354 | 0.0026 | 6.12E-42 |
| rs174536 | miR-1908-5 | cis | 1.31E-08 | Metabolon | M35186 | 1-arachido | a | c | 0.0341 | 0.0026 | 9.85E-39 |
| rs174575 | miR-1908-5 | cis | 5.45E-05 | Metabolon | M35186 | 1-arachido | c | g | 0.0305 | 0.0027 | 8.92E-30 |
| rs174555 | miR-1908-5 | cis | 7.43E-08 | Metabolon | M35186 | 1-arachido | t | c | 0.0337 | 0.0026 | 1.92E-37 |
| rs2727271 | miR-1908-5 | cis | 4.23E-05 | Metabolon | M35186 | 1-arachido | a | t | 0.0332 | 0.0042 | 2.69E-15 |
| rs174548 | miR-1908-5 | cis | 3.01E-08 | Metabolon | M35186 | 1-arachido | c | g | 0.0337 | 0.0026 | 1.30E-37 |
| rs2727270 | miR-1908-5 | cis | 4.45E-05 | Metabolon | M35186 | 1-arachido | t | c | -0.0332 | 0.0042 | 2.73E-15 |
| rs174537 | miR-1908-5 | cis | 9.41E-09 | Metabolon | M35186 | 1-arachido | t | g | -0.0341 | 0.0026 | 9.84E-39 |
| rs174538 | miR-1908-5 | cis | 1.35E-07 | Metabolon | M35186 | 1-arachido | a | g | -0.0325 | 0.0026 | 7.71E-35 |
| rs174556 | miR-1908-5 | cis | 7.67E-08 | Metabolon | M35186 | 1-arachido | t | c | -0.0337 | 0.0026 | 1.92E-37 |
| rs174578 | miR-1908-5 | cis | 2.55E-08 | Metabolon | M35718 | dihomo-lin | a | t | 0.0193 | 0.0026 | 1.02E-13 |
| rs174575 | miR-1908-5 | cis | 5.45E-05 | Metabolon | M35718 | dihomo-lin | c | g | -0.0219 | 0.0027 | 2.36E-16 |
| rs174601 | miR-1908-5 | cis | 5.98E-09 | Metabolon | M35718 | dihomo-lin | t | c | 0.0186 | 0.0026 | 1.33E-12 |
| rs174574 | miR-1908-5 | cis | 8.50E-09 | Metabolon | M35718 | dihomo-lin | a | c | 0.0192 | 0.0026 | 1.68E-13 |
| rs174537 | miR-1908-5 | cis | 9.41E-09 | Metabolon | M35718 | dihomo-lin | t | g | 0.0191 | 0.0026 | 2.32E-13 |
| rs174576 | miR-1908-5 | cis | 3.13E-08 | Metabolon | M35718 | dihomo-lin | a | c | 0.0186 | 0.0026 | 7.92E-13 |
| rs174549 | miR-1908-5 | cis | 4.04E-08 | Metabolon | M35718 | dihomo-lin | a | g | 0.0224 | 0.0026 | 1.33E-17 |
| rs174449 | miR-1908-5 | cis | 2.49E-05 | Metabolon | M35718 | dihomo-lin | a | g | -0.0168 | 0.0026 | 1.57E-10 |
| rs174545 | miR-1908-5 | cis | 2.29E-08 | Metabolon | M35718 | dihomo-lin | c | g | -0.0191 | 0.0026 | 2.04E-13 |
| rs174555 | miR-1908-5 | cis | 7.43E-08 | Metabolon | M35718 | dihomo-lin | t | c | -0.0224 | 0.0026 | 1.23E-17 |
| rs174448 | miR-1908-5 | cis | 1.95E-05 | Metabolon | M35718 | dihomo-lin | a | g | -0.0178 | 0.0026 | 6.36E-12 |
| rs174577 | miR-1908-5 | cis | 1.89E-08 | Metabolon | M35718 | dihomo-lin | a | c | 0.019 | 0.0026 | 2.81E-13 |
| rs174535 | miR-1908-5 | cis | 6.98E-09 | Metabolon | M35718 | dihomo-lin | t | c | -0.0191 | 0.0026 | 2.32E-13 |
| rs174548 | miR-1908-5 | cis | 3.01E-08 | Metabolon | M35718 | dihomo-lin | c | g | -0.0222 | 0.0026 | 2.19E-17 |
| rs174538 | miR-1908-5 | cis | 1.35E-07 | Metabolon | M35718 | dihomo-lin | a | g | 0.0193 | 0.0026 | 1.74E-13 |
| rs174536 | miR-1908-5 | cis | 1.31E-08 | Metabolon | M35718 | dihomo-lin | a | c | -0.0191 | 0.0026 | 2.32E-13 |
| rs174556 | miR-1908-5 | cis | 7.67E-08 | Metabolon | M35718 | dihomo-lin | t | c | 0.0216 | 0.0026 | 1.47E-16 |
| rs495828 | miR-10b-5p | trans | 2.56E-34 | Metabolon | M32740 | X-11423 | t | g | -0.0136 | 0.0024 | 1.46E-08 |
| rs649129 | let-7a-5p | trans | 1.48E-12 | Metabolon | M32740 | X-11423 | t | c | -0.0142 | 0.0024 | 3.02E-09 |
| rs649129 | miR-139-5p | trans | 1.34E-34 | Metabolon | M32740 | X-11423 | t | c | -0.0142 | 0.0024 | 3.02E-09 |
| rs495828 | miR-126-3p | trans | 2.00E-37 | Metabolon | M32740 | X-11423 | t | g | -0.0136 | 0.0024 | 1.46E-08 |
| rs649129 | miR-10b-5p | trans | 2.54E-34 | Metabolon | M32740 | X-11423 | t | c | -0.0142 | 0.0024 | 3.02E-09 |
| rs495828 | miR-139-5p | trans | 1.46E-34 | Metabolon | M32740 | X-11423 | t | g | -0.0136 | 0.0024 | 1.46E-08 |
| rs649129 | miR-126-3p | trans | 1.95E-37 | Metabolon | M32740 | X-11423 | t | c | -0.0142 | 0.0024 | 3.02E-09 |
| rs495828 | let-7a-5p | trans | 1.53E-12 | Metabolon | M32740 | X-11423 | t | g | -0.0136 | 0.0024 | 1.46E-08 |
| rs529565 | let-7c-5p | trans | 3.42E-24 | Metabolon | M33801 | ADpSGEGD | t | c | -0.0435 | 0.006 | 5.41E-13 |
| rs612169 | miR-139-5p | trans | 7.83E-42 | Metabolon | M33801 | ADpSGEGD | a | g | -0.0452 | 0.0061 | 9.66E-14 |
| rs529565 | miR-195-5p | trans | 3.57E-18 | Metabolon | M33801 | ADpSGEGD | t | c | -0.0435 | 0.006 | 5.41E-13 |
| rs529565 | let-7b-5p | trans | 4.77E-28 | Metabolon | M33801 | ADpSGEGD | t | c | -0.0435 | 0.006 | 5.41E-13 |
| rs545971 | miR-10b-5p | trans | 3.43E-35 | Metabolon | M33801 | ADpSGEGD | t | c | 0.0439 | 0.006 | 3.33E-13 |
| rs495828 | let-7a-5p | trans | 1.53E-12 | Metabolon | M33801 | ADpSGEGD | t | g | 0.0603 | 0.0069 | 2.72E-18 |
| rs495828 | miR-10b-5p | trans | 2.56E-34 | Metabolon | M33801 | ADpSGEGD | t | g | 0.0603 | 0.0069 | 2.72E-18 |
| rs643434 | let-7c-5p | trans | 3.20E-23 | Metabolon | M33801 | ADpSGEGD | a | g | 0.044 | 0.0059 | 1.45E-13 |
| rs612169 | miR-195-5p | trans | 2.56E-17 | Metabolon | M33801 | ADpSGEGD | a | g | -0.0452 | 0.0061 | 9.66E-14 |
| rs529565 | miR-10b-5p | trans | 4.06E-36 | Metabolon | M33801 | ADpSGEGD | t | c | -0.0435 | 0.006 | 5.41E-13 |
| rs495828 | miR-126-3p | trans | 2.00E-37 | Metabolon | M33801 | ADpSGEGD | t | g | 0.0603 | 0.0069 | 2.72E-18 |
| rs612169 | miR-10b-5p | trans | 3.54E-35 | Metabolon | M33801 | ADpSGEGD | a | g | -0.0452 | 0.0061 | 9.66E-14 |
| rs612169 | miR-126-3p | trans | 2.89E-39 | Metabolon | M33801 | ADpSGEGD | a | g | -0.0452 | 0.0061 | 9.66E-14 |
| rs529565 | miR-139-5p | trans | 2.82E-43 | Metabolon | M33801 | ADpSGEGD | t | c | -0.0435 | 0.006 | 5.41E-13 |
| rs495828 | miR-139-5p | trans | 1.46E-34 | Metabolon | M33801 | ADpSGEGD | t | g | 0.0603 | 0.0069 | 2.72E-18 |
| rs545971 | let-7b-5p | trans | 1.11E-26 | Metabolon | M33801 | ADpSGEGD | t | c | 0.0439 | 0.006 | 3.33E-13 |
| rs643434 | miR-139-5p | trans | 1.31E-40 | Metabolon | M33801 | ADpSGEGD | a | g | 0.044 | 0.0059 | 1.45E-13 |
| rs545971 | miR-195-5p | trans | 2.54E-17 | Metabolon | M33801 | ADpSGEGD | t | c | 0.0439 | 0.006 | 3.33E-13 |
| rs545971 | miR-126-3p | trans | 2.76E-39 | Metabolon | M33801 | ADpSGEGD | t | c | 0.0439 | 0.006 | 3.33E-13 |
| rs545971 | miR-139-5p | trans | 8.40E-42 | Metabolon | M33801 | ADpSGEGD | t | c | 0.0439 | 0.006 | 3.33E-13 |
| rs674302 | let-7c-5p | trans | 2.28E-23 | Metabolon | M33801 | ADpSGEGD | a | t | 0.0439 | 0.006 | 3.56E-13 |
| rs643434 | let-7a-5p | trans | 9.18E-13 | Metabolon | M33801 | ADpSGEGD | a | g | 0.044 | 0.0059 | 1.45E-13 |
| rs612169 | let-7b-5p | trans | 1.17E-26 | Metabolon | M33801 | ADpSGEGD | a | g | -0.0452 | 0.0061 | 9.66E-14 |
| rs529565 | miR-126-3p | trans | 8.91E-41 | Metabolon | M33801 | ADpSGEGD | t | c | -0.0435 | 0.006 | 5.41E-13 |
| rs644234 | miR-139-5p | trans | 1.17E-40 | Metabolon | M33801 | ADpSGEGD | t | g | -0.044 | 0.0059 | 1.43E-13 |

| | | | | | | | | | | | |
|---|---|---|---|---|---|---|---|---|---|---|---|
| rs644234 | miR-10b-5p | trans | 1.46E-34 | Metabolon | M33801 | ADpSGEGD | t | g | -0.044 | 0.0059 | 1.43E-13 |
| rs630014 | miR-126-3p | trans | 3.15E-19 | Metabolon | M33801 | ADpSGEGD | a | g | -0.0349 | 0.0059 | 3.53E-09 |
| rs545971 | let-7c-5p | trans | 2.87E-23 | Metabolon | M33801 | ADpSGEGD | t | c | 0.0439 | 0.006 | 3.33E-13 |
| rs630014 | miR-10b-5p | trans | 2.46E-16 | Metabolon | M33801 | ADpSGEGD | a | g | -0.0349 | 0.0059 | 3.53E-09 |
| rs687289 | let-7b-5p | trans | 8.61E-28 | Metabolon | M33801 | ADpSGEGD | a | g | 0.0443 | 0.006 | 2.30E-13 |
| rs643434 | miR-126-3p | trans | 1.18E-37 | Metabolon | M33801 | ADpSGEGD | a | g | 0.044 | 0.0059 | 1.45E-13 |
| rs643434 | miR-10b-5p | trans | 1.64E-34 | Metabolon | M33801 | ADpSGEGD | a | g | 0.044 | 0.0059 | 1.45E-13 |
| rs643434 | let-7b-5p | trans | 1.70E-26 | Metabolon | M33801 | ADpSGEGD | a | g | 0.044 | 0.0059 | 1.45E-13 |
| rs687289 | miR-195-5p | trans | 1.08E-17 | Metabolon | M33801 | ADpSGEGD | a | g | 0.0443 | 0.006 | 2.30E-13 |
| rs674302 | miR-195-5p | trans | 1.85E-17 | Metabolon | M33801 | ADpSGEGD | a | t | 0.0439 | 0.006 | 3.56E-13 |
| rs644234 | miR-126-3p | trans | 1.03E-37 | Metabolon | M33801 | ADpSGEGD | t | g | -0.044 | 0.0059 | 1.43E-13 |
| rs612169 | let-7c-5p | trans | 2.76E-23 | Metabolon | M33801 | ADpSGEGD | a | g | -0.0452 | 0.0061 | 9.66E-14 |
| rs687289 | let-7c-5p | trans | 5.91E-24 | Metabolon | M33801 | ADpSGEGD | a | g | 0.0443 | 0.006 | 2.30E-13 |
| rs687289 | miR-10b-5p | trans | 5.80E-37 | Metabolon | M33801 | ADpSGEGD | a | g | 0.0443 | 0.006 | 2.30E-13 |
| rs644234 | let-7c-5p | trans | 2.80E-23 | Metabolon | M33801 | ADpSGEGD | t | g | -0.044 | 0.0059 | 1.43E-13 |
| rs630014 | let-7b-5p | trans | 4.92E-16 | Metabolon | M33801 | ADpSGEGD | a | g | -0.0349 | 0.0059 | 3.53E-09 |
| rs674302 | miR-126-3p | trans | 2.03E-39 | Metabolon | M33801 | ADpSGEGD | a | t | 0.0439 | 0.006 | 3.56E-13 |
| rs674302 | let-7b-5p | trans | 7.63E-27 | Metabolon | M33801 | ADpSGEGD | a | t | 0.0439 | 0.006 | 3.56E-13 |
| rs649129 | miR-126-3p | trans | 1.95E-37 | Metabolon | M33801 | ADpSGEGD | t | c | 0.0623 | 0.0069 | 2.09E-19 |
| rs649129 | miR-139-5p | trans | 1.34E-34 | Metabolon | M33801 | ADpSGEGD | t | c | 0.0623 | 0.0069 | 2.09E-19 |
| rs644234 | let-7b-5p | trans | 1.49E-26 | Metabolon | M33801 | ADpSGEGD | t | g | -0.044 | 0.0059 | 1.43E-13 |
| rs687289 | miR-139-5p | trans | 2.87E-43 | Metabolon | M33801 | ADpSGEGD | a | g | 0.0443 | 0.006 | 2.30E-13 |
| rs649129 | let-7a-5p | trans | 1.48E-12 | Metabolon | M33801 | ADpSGEGD | t | c | 0.0623 | 0.0069 | 2.09E-19 |
| rs644234 | let-7a-5p | trans | 8.38E-13 | Metabolon | M33801 | ADpSGEGD | t | g | -0.044 | 0.0059 | 1.43E-13 |
| rs649129 | miR-10b-5p | trans | 2.54E-34 | Metabolon | M33801 | ADpSGEGD | t | c | 0.0623 | 0.0069 | 2.09E-19 |
| rs674302 | miR-10b-5p | trans | 2.75E-35 | Metabolon | M33801 | ADpSGEGD | a | t | 0.0439 | 0.006 | 3.56E-13 |
| rs674302 | miR-139-5p | trans | 6.05E-42 | Metabolon | M33801 | ADpSGEGD | a | t | 0.0439 | 0.006 | 3.56E-13 |
| rs687289 | miR-126-3p | trans | 3.06E-41 | Metabolon | M33801 | ADpSGEGD | a | g | 0.0443 | 0.006 | 2.30E-13 |
| rs649129 | miR-10b-5p | trans | 2.54E-34 | Metabolon | M36115 | X-14189 | t | c | -0.0437 | 0.0074 | 4.32E-09 |
| rs649129 | miR-139-5p | trans | 1.34E-34 | Metabolon | M36115 | X-14189 | t | c | -0.0437 | 0.0074 | 4.32E-09 |
| rs649129 | miR-126-3p | trans | 1.95E-37 | Metabolon | M36115 | X-14189 | t | c | -0.0437 | 0.0074 | 4.32E-09 |
| rs649129 | let-7a-5p | trans | 1.48E-12 | Metabolon | M36115 | X-14189 | t | c | -0.0437 | 0.0074 | 4.32E-09 |
| rs495828 | miR-126-3p | trans | 2.00E-37 | Metabolon | M36115 | X-14189 | t | g | -0.0435 | 0.0073 | 2.26E-09 |
| rs495828 | miR-139-5p | trans | 1.46E-34 | Metabolon | M36115 | X-14189 | t | g | -0.0435 | 0.0073 | 2.26E-09 |
| rs495828 | miR-10b-5p | trans | 2.56E-34 | Metabolon | M36115 | X-14189 | t | g | -0.0435 | 0.0073 | 2.26E-09 |
| rs495828 | let-7a-5p | trans | 1.53E-12 | Metabolon | M36115 | X-14189 | t | g | -0.0435 | 0.0073 | 2.26E-09 |
| rs495828 | miR-10b-5p | trans | 2.56E-34 | Metabolon | M36230 | X-14304 | t | g | -0.0431 | 0.0077 | 1.98E-08 |
| rs649129 | miR-139-5p | trans | 1.34E-34 | Metabolon | M36230 | X-14304 | t | c | -0.0437 | 0.0077 | 1.26E-08 |
| rs495828 | miR-126-3p | trans | 2.00E-37 | Metabolon | M36230 | X-14304 | t | g | -0.0431 | 0.0077 | 1.98E-08 |
| rs649129 | let-7a-5p | trans | 1.48E-12 | Metabolon | M36230 | X-14304 | t | c | -0.0437 | 0.0077 | 1.26E-08 |
| rs495828 | miR-139-5p | trans | 1.46E-34 | Metabolon | M36230 | X-14304 | t | g | -0.0431 | 0.0077 | 1.98E-08 |
| rs649129 | miR-10b-5p | trans | 2.54E-34 | Metabolon | M36230 | X-14304 | t | c | -0.0437 | 0.0077 | 1.26E-08 |
| rs495828 | let-7a-5p | trans | 1.53E-12 | Metabolon | M36230 | X-14304 | t | g | -0.0431 | 0.0077 | 1.98E-08 |
| rs649129 | miR-126-3p | trans | 1.95E-37 | Metabolon | M36230 | X-14304 | t | c | -0.0437 | 0.0077 | 1.26E-08 |
| rs174574 | miR-1908-5 | cis | 8.50E-09 | Nightingale | L.HDL.P | Concentrat | C | A | 0.087189 | 0.010488 | 2.27E-16 |
| rs28456 | miR-1908-5 | cis | 6.85E-08 | Nightingale | L.HDL.P | Concentrat | G | A | -0.08061 | 0.010672 | 8.92E-14 |
| rs174548 | miR-1908-5 | cis | 3.01E-08 | Nightingale | L.HDL.P | Concentrat | G | C | -0.08288 | 0.01056 | 9.34E-15 |
| rs174578 | miR-1908-5 | cis | 2.55E-08 | Nightingale | L.HDL.P | Concentrat | A | T | -0.08914 | 0.010489 | 4.89E-17 |
| rs2727270 | miR-1908-5 | cis | 4.45E-05 | Nightingale | L.HDL.P | Concentrat | T | C | -0.08852 | 0.011895 | 2.05E-13 |
| rs174533 | miR-1908-5 | cis | 1.26E-08 | Nightingale | L.HDL.P | Concentrat | A | G | -0.0866 | 0.01047 | 3.20E-16 |
| rs2072114 | miR-1908-5 | cis | 1.34E-05 | Nightingale | L.HDL.P | Concentrat | G | A | -0.09036 | 0.011889 | 6.26E-14 |
| rs102274 | miR-1908-5 | cis | 9.41E-09 | Nightingale | L.HDL.P | Concentrat | C | T | -0.0874 | 0.010461 | 1.62E-16 |
| rs174544 | miR-1908-5 | cis | 2.30E-08 | Nightingale | L.HDL.P | Concentrat | A | C | -0.08317 | 0.010619 | 1.07E-14 |
| rs174536 | miR-1908-5 | cis | 1.31E-08 | Nightingale | L.HDL.P | Concentrat | C | A | -0.08727 | 0.010465 | 1.84E-16 |
| rs174581 | miR-1908-5 | cis | 1.68E-08 | Nightingale | L.HDL.P | Concentrat | A | G | -0.08904 | 0.010506 | 5.92E-17 |
| rs174562 | miR-1908-5 | cis | 4.92E-08 | Nightingale | L.HDL.P | Concentrat | G | A | -0.0855 | 0.010473 | 7.72E-16 |
| rs174448 | miR-1908-5 | cis | 1.95E-05 | Nightingale | L.HDL.P | Concentrat | A | G | 0.076576 | 0.010717 | 1.74E-12 |
| rs174577 | miR-1908-5 | cis | 1.89E-08 | Nightingale | L.HDL.P | Concentrat | A | C | -0.08854 | 0.010489 | 7.90E-17 |
| rs174584 | miR-1908-5 | cis | 1.39E-08 | Nightingale | L.HDL.P | Concentrat | A | G | -0.08793 | 0.010622 | 3.04E-16 |
| rs174530 | miR-1908-5 | cis | 1.12E-08 | Nightingale | L.HDL.P | Concentrat | G | A | -0.08789 | 0.0105 | 1.43E-16 |
| rs2727271 | miR-1908-5 | cis | 4.23E-05 | Nightingale | L.HDL.P | Concentrat | T | A | -0.08891 | 0.011883 | 1.51E-13 |
| rs174564 | miR-1908-5 | cis | 1.61E-08 | Nightingale | L.HDL.P | Concentrat | G | A | -0.08593 | 0.010534 | 8.06E-16 |
| rs174555 | miR-1908-5 | cis | 7.43E-08 | Nightingale | L.HDL.P | Concentrat | C | T | -0.08171 | 0.010569 | 2.31E-14 |

| | | | | | | | | | | | |
|---|---|---|---|---|---|---|---|---|---|---|---|
| rs174537 | miR-1908-5 | cis | 9.41E-09 | Nightingale | L.HDL.P | Concentrat | T | G | -0.08676 | 0.010476 | 2.95E-16 |
| rs174568 | miR-1908-5 | cis | 3.76E-08 | Nightingale | L.HDL.P | Concentrat | T | C | -0.08617 | 0.01052 | 6.19E-16 |
| rs174592 | miR-1908-5 | cis | 3.81E-09 | Nightingale | L.HDL.P | Concentrat | G | A | -0.08639 | 0.010527 | 5.47E-16 |
| rs174553 | miR-1908-5 | cis | 2.61E-08 | Nightingale | L.HDL.P | Concentrat | G | A | -0.08742 | 0.010501 | 2.07E-16 |
| rs174566 | miR-1908-5 | cis | 1.58E-08 | Nightingale | L.HDL.P | Concentrat | G | A | -0.08597 | 0.010476 | 5.47E-16 |
| rs174580 | miR-1908-5 | cis | 1.81E-08 | Nightingale | L.HDL.P | Concentrat | G | A | -0.08905 | 0.010505 | 5.86E-17 |
| rs99780 | miR-1908-5 | cis | 1.74E-08 | Nightingale | L.HDL.P | Concentrat | T | C | -0.08565 | 0.010487 | 7.48E-16 |
| rs174549 | miR-1908-5 | cis | 4.04E-08 | Nightingale | L.HDL.P | Concentrat | A | G | -0.08225 | 0.010563 | 1.51E-14 |
| rs174449 | miR-1908-5 | cis | 2.49E-05 | Nightingale | L.HDL.P | Concentrat | A | G | 0.071026 | 0.010681 | 5.23E-11 |
| rs174601 | miR-1908-5 | cis | 5.98E-09 | Nightingale | L.HDL.P | Concentrat | T | C | -0.08682 | 0.010584 | 5.62E-16 |
| rs174556 | miR-1908-5 | cis | 7.67E-08 | Nightingale | L.HDL.P | Concentrat | T | C | -0.0808 | 0.010577 | 4.67E-14 |
| rs2524299 | miR-1908-5 | cis | 4.82E-05 | Nightingale | L.HDL.P | Concentrat | T | A | -0.09001 | 0.011866 | 7.00E-14 |
| rs174560 | miR-1908-5 | cis | 4.91E-08 | Nightingale | L.HDL.P | Concentrat | C | T | -0.08099 | 0.010567 | 3.84E-14 |
| rs174567 | miR-1908-5 | cis | 1.32E-08 | Nightingale | L.HDL.P | Concentrat | G | A | -0.08546 | 0.010487 | 8.67E-16 |
| rs174554 | miR-1908-5 | cis | 3.17E-08 | Nightingale | L.HDL.P | Concentrat | G | A | -0.08653 | 0.010465 | 3.29E-16 |
| rs174559 | miR-1908-5 | cis | 5.55E-07 | Nightingale | L.HDL.P | Concentrat | A | G | -0.07227 | 0.01094 | 6.99E-11 |
| rs174545 | miR-1908-5 | cis | 2.29E-08 | Nightingale | L.HDL.P | Concentrat | G | C | -0.08717 | 0.010465 | 1.99E-16 |
| rs174576 | miR-1908-5 | cis | 3.13E-08 | Nightingale | L.HDL.P | Concentrat | A | C | -0.0875 | 0.010494 | 1.86E-16 |
| rs2072113 | miR-1908-5 | cis | 3.35E-05 | Nightingale | L.HDL.P | Concentrat | T | C | -0.08933 | 0.011895 | 1.23E-13 |
| rs174535 | miR-1908-5 | cis | 6.98E-09 | Nightingale | L.HDL.P | Concentrat | C | T | -0.08704 | 0.010474 | 2.33E-16 |
| rs174561 | miR-1908-5 | cis | 7.66E-08 | Nightingale | L.HDL.P | Concentrat | C | T | -0.08071 | 0.010573 | 4.86E-14 |
| rs174538 | miR-1908-5 | cis | 1.35E-07 | Nightingale | L.HDL.P | Concentrat | A | G | -0.07256 | 0.010861 | 4.26E-11 |
| rs174556 | miR-1908-5 | cis | 7.67E-08 | Nightingale | XL.HDL.P | Concentrat | T | C | -0.08034 | 0.010587 | 5.47E-14 |
| rs174566 | miR-1908-5 | cis | 1.58E-08 | Nightingale | XL.HDL.P | Concentrat | G | A | -0.08182 | 0.010492 | 1.09E-14 |
| rs174548 | miR-1908-5 | cis | 3.01E-08 | Nightingale | XL.HDL.P | Concentrat | G | C | -0.0809 | 0.010575 | 3.42E-14 |
| rs174561 | miR-1908-5 | cis | 7.66E-08 | Nightingale | XL.HDL.P | Concentrat | C | T | -0.07999 | 0.010587 | 7.02E-14 |
| rs2727271 | miR-1908-5 | cis | 4.23E-05 | Nightingale | XL.HDL.P | Concentrat | T | A | -0.08982 | 0.011893 | 7.23E-14 |
| rs174568 | miR-1908-5 | cis | 3.76E-08 | Nightingale | XL.HDL.P | Concentrat | T | C | -0.0834 | 0.010535 | 4.33E-15 |
| rs174537 | miR-1908-5 | cis | 9.41E-09 | Nightingale | XL.HDL.P | Concentrat | T | G | -0.08289 | 0.010487 | 4.76E-15 |
| rs174562 | miR-1908-5 | cis | 4.92E-08 | Nightingale | XL.HDL.P | Concentrat | G | A | -0.08345 | 0.010488 | 3.14E-15 |
| rs174559 | miR-1908-5 | cis | 5.55E-07 | Nightingale | XL.HDL.P | Concentrat | A | G | -0.07218 | 0.010953 | 6.54E-11 |
| rs174581 | miR-1908-5 | cis | 1.68E-08 | Nightingale | XL.HDL.P | Concentrat | A | G | -0.08238 | 0.010522 | 8.57E-15 |
| rs174560 | miR-1908-5 | cis | 4.91E-08 | Nightingale | XL.HDL.P | Concentrat | C | T | -0.07955 | 0.010581 | 9.32E-14 |
| rs99780 | miR-1908-5 | cis | 1.74E-08 | Nightingale | XL.HDL.P | Concentrat | T | C | -0.08239 | 0.010501 | 7.52E-15 |
| rs174584 | miR-1908-5 | cis | 1.39E-08 | Nightingale | XL.HDL.P | Concentrat | A | G | -0.08091 | 0.010639 | 4.81E-14 |
| rs174538 | miR-1908-5 | cis | 1.35E-07 | Nightingale | XL.HDL.P | Concentrat | A | G | -0.07208 | 0.010869 | 4.97E-11 |
| rs174544 | miR-1908-5 | cis | 2.30E-08 | Nightingale | XL.HDL.P | Concentrat | A | C | -0.08238 | 0.010633 | 1.62E-14 |
| rs174535 | miR-1908-5 | cis | 6.98E-09 | Nightingale | XL.HDL.P | Concentrat | C | T | -0.08348 | 0.010483 | 2.97E-15 |
| rs174533 | miR-1908-5 | cis | 1.26E-08 | Nightingale | XL.HDL.P | Concentrat | A | G | -0.08294 | 0.010484 | 4.53E-15 |
| rs174578 | miR-1908-5 | cis | 2.55E-08 | Nightingale | XL.HDL.P | Concentrat | A | T | -0.08263 | 0.010505 | 6.46E-15 |
| rs174536 | miR-1908-5 | cis | 1.31E-08 | Nightingale | XL.HDL.P | Concentrat | C | A | -0.0837 | 0.01048 | 2.48E-15 |
| rs174448 | miR-1908-5 | cis | 1.95E-05 | Nightingale | XL.HDL.P | Concentrat | A | G | 0.062762 | 0.010728 | 6.73E-09 |
| rs2072114 | miR-1908-5 | cis | 1.34E-05 | Nightingale | XL.HDL.P | Concentrat | G | A | -0.09193 | 0.011896 | 1.88E-14 |
| rs174553 | miR-1908-5 | cis | 2.61E-08 | Nightingale | XL.HDL.P | Concentrat | G | A | -0.08496 | 0.010516 | 1.18E-15 |
| rs2072113 | miR-1908-5 | cis | 3.35E-05 | Nightingale | XL.HDL.P | Concentrat | T | C | -0.09053 | 0.011905 | 4.87E-14 |
| rs174592 | miR-1908-5 | cis | 3.81E-09 | Nightingale | XL.HDL.P | Concentrat | G | A | -0.08183 | 0.010542 | 1.44E-14 |
| rs174554 | miR-1908-5 | cis | 3.17E-08 | Nightingale | XL.HDL.P | Concentrat | G | A | -0.08458 | 0.010479 | 1.27E-15 |
| rs174545 | miR-1908-5 | cis | 2.29E-08 | Nightingale | XL.HDL.P | Concentrat | G | C | -0.0847 | 0.010479 | 1.15E-15 |
| rs2727270 | miR-1908-5 | cis | 4.45E-05 | Nightingale | XL.HDL.P | Concentrat | T | C | -0.08976 | 0.011899 | 7.73E-14 |
| rs174580 | miR-1908-5 | cis | 1.81E-08 | Nightingale | XL.HDL.P | Concentrat | G | A | -0.0824 | 0.010522 | 8.46E-15 |
| rs174574 | miR-1908-5 | cis | 8.50E-09 | Nightingale | XL.HDL.P | Concentrat | C | A | 0.082345 | 0.010504 | 7.91E-15 |
| rs102274 | miR-1908-5 | cis | 9.41E-09 | Nightingale | XL.HDL.P | Concentrat | C | T | -0.08356 | 0.010476 | 2.69E-15 |
| rs174567 | miR-1908-5 | cis | 1.32E-08 | Nightingale | XL.HDL.P | Concentrat | G | A | -0.0821 | 0.010502 | 9.36E-15 |
| rs28456 | miR-1908-5 | cis | 6.85E-08 | Nightingale | XL.HDL.P | Concentrat | G | A | -0.07909 | 0.010686 | 2.23E-13 |
| rs2524299 | miR-1908-5 | cis | 4.82E-05 | Nightingale | XL.HDL.P | Concentrat | T | A | -0.08927 | 0.011878 | 9.46E-14 |
| rs174576 | miR-1908-5 | cis | 3.13E-08 | Nightingale | XL.HDL.P | Concentrat | A | C | -0.08227 | 0.010508 | 8.60E-15 |
| rs174601 | miR-1908-5 | cis | 5.98E-09 | Nightingale | XL.HDL.P | Concentrat | T | C | -0.08241 | 0.010598 | 1.30E-14 |
| rs174549 | miR-1908-5 | cis | 4.04E-08 | Nightingale | XL.HDL.P | Concentrat | A | G | -0.08136 | 0.010577 | 2.48E-14 |
| rs174530 | miR-1908-5 | cis | 1.12E-08 | Nightingale | XL.HDL.P | Concentrat | G | A | -0.08347 | 0.010516 | 3.65E-15 |
| rs174564 | miR-1908-5 | cis | 1.61E-08 | Nightingale | XL.HDL.P | Concentrat | G | A | -0.0847 | 0.010547 | 1.74E-15 |
| rs174577 | miR-1908-5 | cis | 1.89E-08 | Nightingale | XL.HDL.P | Concentrat | A | C | -0.08219 | 0.010499 | 8.65E-15 |
| rs174555 | miR-1908-5 | cis | 7.43E-08 | Nightingale | XL.HDL.P | Concentrat | C | T | -0.08103 | 0.010583 | 3.27E-14 |
| rs102274 | miR-1908-5 | cis | 9.41E-09 | Nightingale | L.HDL.FC | Free choles | C | T | -0.08053 | 0.010094 | 2.85E-15 |

| | | | | | | | | | | | |
|---|---|---|---|---|---|---|---|---|---|---|---|
| rs174549 | miR-1908-5 | cis | 4.04E-08 | Nightingale | L.HDL.FC | Free choles | A | G | -0.07536 | 0.010225 | 2.98E-13 |
| rs174554 | miR-1908-5 | cis | 3.17E-08 | Nightingale | L.HDL.FC | Free choles | G | A | -0.08058 | 0.010107 | 2.98E-15 |
| rs174559 | miR-1908-5 | cis | 5.55E-07 | Nightingale | L.HDL.FC | Free choles | A | G | -0.06913 | 0.010608 | 1.12E-10 |
| rs28456 | miR-1908-5 | cis | 6.85E-08 | Nightingale | L.HDL.FC | Free choles | G | A | -0.07463 | 0.010321 | 8.22E-13 |
| rs174535 | miR-1908-5 | cis | 6.98E-09 | Nightingale | L.HDL.FC | Free choles | C | T | -0.07972 | 0.010084 | 5.06E-15 |
| rs174449 | miR-1908-5 | cis | 2.49E-05 | Nightingale | L.HDL.FC | Free choles | A | G | 0.065316 | 0.010202 | 2.34E-10 |
| rs174561 | miR-1908-5 | cis | 7.66E-08 | Nightingale | L.HDL.FC | Free choles | C | T | -0.07477 | 0.010237 | 4.85E-13 |
| rs174537 | miR-1908-5 | cis | 9.41E-09 | Nightingale | L.HDL.FC | Free choles | T | G | -0.07954 | 0.010088 | 5.97E-15 |
| rs174538 | miR-1908-5 | cis | 1.35E-07 | Nightingale | L.HDL.FC | Free choles | A | G | -0.06905 | 0.01047 | 6.64E-11 |
| rs2727271 | miR-1908-5 | cis | 4.23E-05 | Nightingale | L.HDL.FC | Free choles | T | A | -0.082 | 0.011579 | 2.39E-12 |
| rs174545 | miR-1908-5 | cis | 2.29E-08 | Nightingale | L.HDL.FC | Free choles | G | C | -0.08065 | 0.010105 | 2.78E-15 |
| rs174556 | miR-1908-5 | cis | 7.67E-08 | Nightingale | L.HDL.FC | Free choles | T | C | -0.07481 | 0.010242 | 4.84E-13 |
| rs174577 | miR-1908-5 | cis | 1.89E-08 | Nightingale | L.HDL.FC | Free choles | A | C | -0.08125 | 0.010036 | 1.11E-15 |
| rs99780 | miR-1908-5 | cis | 1.74E-08 | Nightingale | L.HDL.FC | Free choles | T | C | -0.0797 | 0.01007 | 4.71E-15 |
| rs174448 | miR-1908-5 | cis | 1.95E-05 | Nightingale | L.HDL.FC | Free choles | A | G | 0.071092 | 0.010322 | 9.27E-12 |
| rs174553 | miR-1908-5 | cis | 2.61E-08 | Nightingale | L.HDL.FC | Free choles | G | A | -0.08112 | 0.010142 | 2.43E-15 |
| rs2072114 | miR-1908-5 | cis | 1.34E-05 | Nightingale | L.HDL.FC | Free choles | G | A | -0.08301 | 0.01158 | 1.29E-12 |
| rs174568 | miR-1908-5 | cis | 3.76E-08 | Nightingale | L.HDL.FC | Free choles | T | C | -0.08 | 0.010121 | 5.14E-15 |
| rs174566 | miR-1908-5 | cis | 1.58E-08 | Nightingale | L.HDL.FC | Free choles | G | A | -0.07981 | 0.010058 | 4.03E-15 |
| rs2727270 | miR-1908-5 | cis | 4.45E-05 | Nightingale | L.HDL.FC | Free choles | T | C | -0.08188 | 0.011588 | 2.67E-12 |
| rs174578 | miR-1908-5 | cis | 2.55E-08 | Nightingale | L.HDL.FC | Free choles | A | T | -0.08168 | 0.010039 | 8.02E-16 |
| rs174601 | miR-1908-5 | cis | 5.98E-09 | Nightingale | L.HDL.FC | Free choles | T | C | -0.08182 | 0.01025 | 2.76E-15 |
| rs174574 | miR-1908-5 | cis | 8.50E-09 | Nightingale | L.HDL.FC | Free choles | C | A | 0.080463 | 0.01007 | 2.58E-15 |
| rs174530 | miR-1908-5 | cis | 1.12E-08 | Nightingale | L.HDL.FC | Free choles | G | A | -0.08053 | 0.010097 | 2.91E-15 |
| rs174567 | miR-1908-5 | cis | 1.32E-08 | Nightingale | L.HDL.FC | Free choles | G | A | -0.07949 | 0.010069 | 5.50E-15 |
| rs174560 | miR-1908-5 | cis | 4.91E-08 | Nightingale | L.HDL.FC | Free choles | C | T | -0.07457 | 0.010222 | 5.17E-13 |
| rs174576 | miR-1908-5 | cis | 3.13E-08 | Nightingale | L.HDL.FC | Free choles | A | C | -0.08048 | 0.010036 | 2.07E-15 |
| rs174581 | miR-1908-5 | cis | 1.68E-08 | Nightingale | L.HDL.FC | Free choles | A | G | -0.0815 | 0.010057 | 1.04E-15 |
| rs174580 | miR-1908-5 | cis | 1.81E-08 | Nightingale | L.HDL.FC | Free choles | G | A | -0.0815 | 0.010056 | 1.03E-15 |
| rs174536 | miR-1908-5 | cis | 1.31E-08 | Nightingale | L.HDL.FC | Free choles | C | A | -0.07995 | 0.010076 | 4.00E-15 |
| rs174544 | miR-1908-5 | cis | 2.30E-08 | Nightingale | L.HDL.FC | Free choles | A | C | -0.0764 | 0.01028 | 1.90E-13 |
| rs174562 | miR-1908-5 | cis | 4.92E-08 | Nightingale | L.HDL.FC | Free choles | G | A | -0.07995 | 0.010112 | 5.04E-15 |
| rs174533 | miR-1908-5 | cis | 1.26E-08 | Nightingale | L.HDL.FC | Free choles | A | G | -0.07939 | 0.010078 | 6.33E-15 |
| rs174592 | miR-1908-5 | cis | 3.81E-09 | Nightingale | L.HDL.FC | Free choles | G | A | -0.08109 | 0.010173 | 3.03E-15 |
| rs174555 | miR-1908-5 | cis | 7.43E-08 | Nightingale | L.HDL.FC | Free choles | C | T | -0.07543 | 0.010233 | 2.96E-13 |
| rs174584 | miR-1908-5 | cis | 1.39E-08 | Nightingale | L.HDL.FC | Free choles | A | G | -0.08173 | 0.01021 | 2.31E-15 |
| rs174564 | miR-1908-5 | cis | 1.61E-08 | Nightingale | L.HDL.FC | Free choles | G | A | -0.08058 | 0.010162 | 4.19E-15 |
| rs2524299 | miR-1908-5 | cis | 4.82E-05 | Nightingale | L.HDL.FC | Free choles | T | A | -0.0822 | 0.011559 | 1.93E-12 |
| rs174548 | miR-1908-5 | cis | 3.01E-08 | Nightingale | L.HDL.FC | Free choles | G | C | -0.07562 | 0.01022 | 2.40E-13 |
| rs2072113 | miR-1908-5 | cis | 3.35E-05 | Nightingale | L.HDL.FC | Free choles | T | C | -0.08242 | 0.011587 | 1.91E-12 |
| rs174535 | miR-1908-5 | cis | 6.98E-09 | Nightingale | XL.HDL.FC | Free choles | C | T | -0.08585 | 0.010093 | 2.12E-17 |
| rs174548 | miR-1908-5 | cis | 3.01E-08 | Nightingale | XL.HDL.FC | Free choles | G | C | -0.08125 | 0.010233 | 2.33E-15 |
| rs174448 | miR-1908-5 | cis | 1.95E-05 | Nightingale | XL.HDL.FC | Free choles | A | G | 0.062573 | 0.010332 | 1.52E-09 |
| rs174578 | miR-1908-5 | cis | 2.55E-08 | Nightingale | XL.HDL.FC | Free choles | A | T | -0.08771 | 0.010055 | 3.21E-18 |
| rs174545 | miR-1908-5 | cis | 2.29E-08 | Nightingale | XL.HDL.FC | Free choles | G | C | -0.08732 | 0.010118 | 7.19E-18 |
| rs174538 | miR-1908-5 | cis | 1.35E-07 | Nightingale | XL.HDL.FC | Free choles | A | G | -0.0771 | 0.010476 | 2.09E-13 |
| rs174567 | miR-1908-5 | cis | 1.32E-08 | Nightingale | XL.HDL.FC | Free choles | G | A | -0.08501 | 0.010082 | 3.98E-17 |
| rs2727271 | miR-1908-5 | cis | 4.23E-05 | Nightingale | XL.HDL.FC | Free choles | T | A | -0.08578 | 0.011596 | 1.57E-13 |
| rs174576 | miR-1908-5 | cis | 3.13E-08 | Nightingale | XL.HDL.FC | Free choles | A | C | -0.08684 | 0.010044 | 6.26E-18 |
| rs174561 | miR-1908-5 | cis | 7.66E-08 | Nightingale | XL.HDL.FC | Free choles | C | T | -0.08129 | 0.01025 | 2.50E-15 |
| rs174562 | miR-1908-5 | cis | 4.92E-08 | Nightingale | XL.HDL.FC | Free choles | G | A | -0.08672 | 0.010123 | 1.26E-17 |
| rs174554 | miR-1908-5 | cis | 3.17E-08 | Nightingale | XL.HDL.FC | Free choles | G | A | -0.08752 | 0.010118 | 6.11E-18 |
| rs174577 | miR-1908-5 | cis | 1.89E-08 | Nightingale | XL.HDL.FC | Free choles | A | C | -0.08747 | 0.010046 | 3.70E-18 |
| rs28456 | miR-1908-5 | cis | 6.85E-08 | Nightingale | XL.HDL.FC | Free choles | G | A | -0.08092 | 0.010334 | 5.55E-15 |
| rs174533 | miR-1908-5 | cis | 1.26E-08 | Nightingale | XL.HDL.FC | Free choles | A | G | -0.08585 | 0.010093 | 2.13E-17 |
| rs2072114 | miR-1908-5 | cis | 1.34E-05 | Nightingale | XL.HDL.FC | Free choles | G | A | -0.08695 | 0.011585 | 6.93E-14 |
| rs174584 | miR-1908-5 | cis | 1.39E-08 | Nightingale | XL.HDL.FC | Free choles | A | G | -0.08809 | 0.010224 | 8.13E-18 |
| rs174564 | miR-1908-5 | cis | 1.61E-08 | Nightingale | XL.HDL.FC | Free choles | G | A | -0.08723 | 0.010173 | 1.18E-17 |
| rs174553 | miR-1908-5 | cis | 2.61E-08 | Nightingale | XL.HDL.FC | Free choles | G | A | -0.08736 | 0.010154 | 9.12E-18 |
| rs174566 | miR-1908-5 | cis | 1.58E-08 | Nightingale | XL.HDL.FC | Free choles | G | A | -0.0851 | 0.010071 | 3.43E-17 |
| rs174556 | miR-1908-5 | cis | 7.67E-08 | Nightingale | XL.HDL.FC | Free choles | T | C | -0.08152 | 0.010245 | 2.03E-15 |
| rs174530 | miR-1908-5 | cis | 1.12E-08 | Nightingale | XL.HDL.FC | Free choles | G | A | -0.08696 | 0.010113 | 9.52E-18 |
| rs174449 | miR-1908-5 | cis | 2.49E-05 | Nightingale | XL.HDL.FC | Free choles | A | G | 0.056974 | 0.010205 | 2.54E-08 |

| | | | | | | | | | | | | |
|---|---|---|---|---|---|---|---|---|---|---|---|---|
| rs174568 | miR-1908-5 | cis | | 3.76E-08 | Nightingale | XL.HDL.FC | Free choles | T | C | -0.08691 | 0.010134 | 1.16E-17 |
| rs174537 | miR-1908-5 | cis | | 9.41E-09 | Nightingale | XL.HDL.FC | Free choles | T | G | -0.0854 | 0.010093 | 3.12E-17 |
| rs174592 | miR-1908-5 | cis | | 3.81E-09 | Nightingale | XL.HDL.FC | Free choles | G | A | -0.08692 | 0.010185 | 1.66E-17 |
| rs174544 | miR-1908-5 | cis | | 2.30E-08 | Nightingale | XL.HDL.FC | Free choles | A | C | -0.08304 | 0.010294 | 8.37E-16 |
| rs174601 | miR-1908-5 | cis | | 5.98E-09 | Nightingale | XL.HDL.FC | Free choles | T | C | -0.08698 | 0.010261 | 2.71E-17 |
| rs174560 | miR-1908-5 | cis | | 4.91E-08 | Nightingale | XL.HDL.FC | Free choles | C | T | -0.08053 | 0.010235 | 4.16E-15 |
| rs2524299 | miR-1908-5 | cis | | 4.82E-05 | Nightingale | XL.HDL.FC | Free choles | T | A | -0.08539 | 0.011576 | 1.84E-13 |
| rs174574 | miR-1908-5 | cis | | 8.50E-09 | Nightingale | XL.HDL.FC | Free choles | C | A | 0.085951 | 0.010083 | 1.81E-17 |
| rs174549 | miR-1908-5 | cis | | 4.04E-08 | Nightingale | XL.HDL.FC | Free choles | A | G | -0.08168 | 0.010239 | 1.73E-15 |
| rs174555 | miR-1908-5 | cis | | 7.43E-08 | Nightingale | XL.HDL.FC | Free choles | C | T | -0.08203 | 0.010246 | 1.37E-15 |
| rs2727270 | miR-1908-5 | cis | | 4.45E-05 | Nightingale | XL.HDL.FC | Free choles | T | C | -0.08568 | 0.011598 | 1.69E-13 |
| rs2072113 | miR-1908-5 | cis | | 3.35E-05 | Nightingale | XL.HDL.FC | Free choles | T | C | -0.08669 | 0.011604 | 9.05E-14 |
| rs174536 | miR-1908-5 | cis | | 1.31E-08 | Nightingale | XL.HDL.FC | Free choles | C | A | -0.08608 | 0.010091 | 1.71E-17 |
| rs102274 | miR-1908-5 | cis | | 9.41E-09 | Nightingale | XL.HDL.FC | Free choles | C | T | -0.08659 | 0.010107 | 1.25E-17 |
| rs174580 | miR-1908-5 | cis | | 1.81E-08 | Nightingale | XL.HDL.FC | Free choles | G | A | -0.0875 | 0.010072 | 4.39E-18 |
| rs174559 | miR-1908-5 | cis | | 5.55E-07 | Nightingale | XL.HDL.FC | Free choles | A | G | -0.07479 | 0.01062 | 2.11E-12 |
| rs174581 | miR-1908-5 | cis | | 1.68E-08 | Nightingale | XL.HDL.FC | Free choles | A | G | -0.0875 | 0.010073 | 4.43E-18 |
| rs99780 | miR-1908-5 | cis | | 1.74E-08 | Nightingale | XL.HDL.FC | Free choles | T | C | -0.08582 | 0.010083 | 2.00E-17 |
| rs174573 | miR-1908-5 | cis | | 3.73E-05 | Nightingale | FAw3 | Omega-3 fa | A | G | -0.10577 | 0.014827 | 1.40E-12 |
| rs174573 | miR-1908-5 | cis | | 3.73E-05 | Nightingale | FAw3 | Omega-3 fa | A | G | -0.10577 | 0.014827 | 1.40E-12 |
| rs174576 | miR-1908-5 | cis | | 3.13E-08 | Nightingale | FAw3 | Omega-3 fa | A | C | -0.1515 | 0.012457 | 1.34E-33 |
| rs174576 | miR-1908-5 | cis | | 3.13E-08 | Nightingale | FAw3 | Omega-3 fa | A | C | -0.1515 | 0.012457 | 1.34E-33 |
| rs174536 | miR-1908-5 | cis | | 1.31E-08 | Nightingale | FAw3 | Omega-3 fa | C | A | -0.1529 | 0.012432 | 2.55E-34 |
| rs174536 | miR-1908-5 | cis | | 1.31E-08 | Nightingale | FAw3 | Omega-3 fa | C | A | -0.1529 | 0.012432 | 2.55E-34 |
| rs174584 | miR-1908-5 | cis | | 1.39E-08 | Nightingale | FAw3 | Omega-3 fa | A | G | -0.15173 | 0.012639 | 8.87E-33 |
| rs174584 | miR-1908-5 | cis | | 1.39E-08 | Nightingale | FAw3 | Omega-3 fa | A | G | -0.15173 | 0.012639 | 8.87E-33 |
| rs174554 | miR-1908-5 | cis | | 3.17E-08 | Nightingale | FAw3 | Omega-3 fa | G | A | -0.15261 | 0.012432 | 3.39E-34 |
| rs174554 | miR-1908-5 | cis | | 3.17E-08 | Nightingale | FAw3 | Omega-3 fa | G | A | -0.15261 | 0.012432 | 3.39E-34 |
| rs174564 | miR-1908-5 | cis | | 1.61E-08 | Nightingale | FAw3 | Omega-3 fa | G | A | -0.1514 | 0.012503 | 2.56E-33 |
| rs174564 | miR-1908-5 | cis | | 1.61E-08 | Nightingale | FAw3 | Omega-3 fa | G | A | -0.1514 | 0.012503 | 2.56E-33 |
| rs2072113 | miR-1908-5 | cis | | 3.35E-05 | Nightingale | FAw3 | Omega-3 fa | T | C | -0.12704 | 0.014097 | 3.57E-19 |
| rs2072113 | miR-1908-5 | cis | | 3.35E-05 | Nightingale | FAw3 | Omega-3 fa | T | C | -0.12704 | 0.014097 | 3.57E-19 |
| rs174581 | miR-1908-5 | cis | | 1.68E-08 | Nightingale | FAw3 | Omega-3 fa | A | G | -0.15297 | 0.012479 | 4.18E-34 |
| rs174581 | miR-1908-5 | cis | | 1.68E-08 | Nightingale | FAw3 | Omega-3 fa | A | G | -0.15297 | 0.012479 | 4.18E-34 |
| rs174448 | miR-1908-5 | cis | | 1.95E-05 | Nightingale | FAw3 | Omega-3 fa | A | G | 0.117587 | 0.012808 | 7.70E-20 |
| rs174448 | miR-1908-5 | cis | | 1.95E-05 | Nightingale | FAw3 | Omega-3 fa | A | G | 0.117587 | 0.012808 | 7.70E-20 |
| rs2072114 | miR-1908-5 | cis | | 1.34E-05 | Nightingale | FAw3 | Omega-3 fa | G | A | -0.12815 | 0.014077 | 1.56E-19 |
| rs2072114 | miR-1908-5 | cis | | 1.34E-05 | Nightingale | FAw3 | Omega-3 fa | G | A | -0.12815 | 0.014077 | 1.56E-19 |
| rs174577 | miR-1908-5 | cis | | 1.89E-08 | Nightingale | FAw3 | Omega-3 fa | A | C | -0.15212 | 0.012455 | 7.25E-34 |
| rs174577 | miR-1908-5 | cis | | 1.89E-08 | Nightingale | FAw3 | Omega-3 fa | A | C | -0.15212 | 0.012455 | 7.25E-34 |
| rs2727271 | miR-1908-5 | cis | | 4.23E-05 | Nightingale | FAw3 | Omega-3 fa | T | A | -0.12748 | 0.014098 | 2.69E-19 |
| rs2727271 | miR-1908-5 | cis | | 4.23E-05 | Nightingale | FAw3 | Omega-3 fa | T | A | -0.12748 | 0.014098 | 2.69E-19 |
| rs174580 | miR-1908-5 | cis | | 1.81E-08 | Nightingale | FAw3 | Omega-3 fa | G | A | -0.15297 | 0.012478 | 4.17E-34 |
| rs174580 | miR-1908-5 | cis | | 1.81E-08 | Nightingale | FAw3 | Omega-3 fa | G | A | -0.15297 | 0.012478 | 4.17E-34 |
| rs174578 | miR-1908-5 | cis | | 2.55E-08 | Nightingale | FAw3 | Omega-3 fa | A | T | -0.15261 | 0.012458 | 4.66E-34 |
| rs174578 | miR-1908-5 | cis | | 2.55E-08 | Nightingale | FAw3 | Omega-3 fa | A | T | -0.15261 | 0.012458 | 4.66E-34 |
| rs174548 | miR-1908-5 | cis | | 3.01E-08 | Nightingale | FAw3 | Omega-3 fa | G | C | -0.1527 | 0.012538 | 1.09E-33 |
| rs174548 | miR-1908-5 | cis | | 3.01E-08 | Nightingale | FAw3 | Omega-3 fa | G | C | -0.1527 | 0.012538 | 1.09E-33 |
| rs174559 | miR-1908-5 | cis | | 5.55E-07 | Nightingale | FAw3 | Omega-3 fa | A | G | -0.14358 | 0.012992 | 4.95E-28 |
| rs174559 | miR-1908-5 | cis | | 5.55E-07 | Nightingale | FAw3 | Omega-3 fa | A | G | -0.14358 | 0.012992 | 4.95E-28 |
| rs174567 | miR-1908-5 | cis | | 1.32E-08 | Nightingale | FAw3 | Omega-3 fa | G | A | -0.15045 | 0.012454 | 3.60E-33 |
| rs174567 | miR-1908-5 | cis | | 1.32E-08 | Nightingale | FAw3 | Omega-3 fa | G | A | -0.15045 | 0.012454 | 3.60E-33 |
| rs28456 | miR-1908-5 | cis | | 6.85E-08 | Nightingale | FAw3 | Omega-3 fa | G | A | -0.15011 | 0.012671 | 5.83E-32 |
| rs28456 | miR-1908-5 | cis | | 6.85E-08 | Nightingale | FAw3 | Omega-3 fa | G | A | -0.15011 | 0.012671 | 5.83E-32 |
| rs174544 | miR-1908-5 | cis | | 2.30E-08 | Nightingale | FAw3 | Omega-3 fa | A | C | -0.15301 | 0.012611 | 1.91E-33 |
| rs174544 | miR-1908-5 | cis | | 2.30E-08 | Nightingale | FAw3 | Omega-3 fa | A | C | -0.15301 | 0.012611 | 1.91E-33 |
| rs174555 | miR-1908-5 | cis | | 7.43E-08 | Nightingale | FAw3 | Omega-3 fa | C | T | -0.1519 | 0.012552 | 2.79E-33 |
| rs174555 | miR-1908-5 | cis | | 7.43E-08 | Nightingale | FAw3 | Omega-3 fa | C | T | -0.1519 | 0.012552 | 2.79E-33 |
| rs174568 | miR-1908-5 | cis | | 3.76E-08 | Nightingale | FAw3 | Omega-3 fa | T | C | -0.14971 | 0.012496 | 1.19E-32 |
| rs174568 | miR-1908-5 | cis | | 3.76E-08 | Nightingale | FAw3 | Omega-3 fa | T | C | -0.14971 | 0.012496 | 1.19E-32 |
| rs174530 | miR-1908-5 | cis | | 1.12E-08 | Nightingale | FAw3 | Omega-3 fa | G | A | -0.14879 | 0.012484 | 2.50E-32 |
| rs174530 | miR-1908-5 | cis | | 1.12E-08 | Nightingale | FAw3 | Omega-3 fa | G | A | -0.14879 | 0.012484 | 2.50E-32 |
| rs174562 | miR-1908-5 | cis | | 4.92E-08 | Nightingale | FAw3 | Omega-3 fa | G | A | -0.15077 | 0.012437 | 2.18E-33 |

| | | | | | | | | | | | |
|---|---|---|---|---|---|---|---|---|---|---|---|
| rs174562 | miR-1908-5 | cis | 4.92E-08 | Nightingale | FAw3 | Omega-3 fa | G | A | -0.15077 | 0.012437 | 2.18E-33 |
| rs2727270 | miR-1908-5 | cis | 4.45E-05 | Nightingale | FAw3 | Omega-3 fa | T | C | -0.12645 | 0.01411 | 5.56E-19 |
| rs2727270 | miR-1908-5 | cis | 4.45E-05 | Nightingale | FAw3 | Omega-3 fa | T | C | -0.12645 | 0.01411 | 5.56E-19 |
| rs99780 | miR-1908-5 | cis | 1.74E-08 | Nightingale | FAw3 | Omega-3 fa | T | C | -0.1492 | 0.012463 | 1.31E-32 |
| rs99780 | miR-1908-5 | cis | 1.74E-08 | Nightingale | FAw3 | Omega-3 fa | T | C | -0.1492 | 0.012463 | 1.31E-32 |
| rs174592 | miR-1908-5 | cis | 3.81E-09 | Nightingale | FAw3 | Omega-3 fa | G | A | -0.15043 | 0.012554 | 1.16E-32 |
| rs174592 | miR-1908-5 | cis | 3.81E-09 | Nightingale | FAw3 | Omega-3 fa | G | A | -0.15043 | 0.012554 | 1.16E-32 |
| rs174533 | miR-1908-5 | cis | 1.26E-08 | Nightingale | FAw3 | Omega-3 fa | A | G | -0.15254 | 0.012436 | 3.81E-34 |
| rs174533 | miR-1908-5 | cis | 1.26E-08 | Nightingale | FAw3 | Omega-3 fa | A | G | -0.15254 | 0.012436 | 3.81E-34 |
| rs174566 | miR-1908-5 | cis | 1.58E-08 | Nightingale | FAw3 | Omega-3 fa | G | A | -0.15057 | 0.01244 | 2.74E-33 |
| rs174566 | miR-1908-5 | cis | 1.58E-08 | Nightingale | FAw3 | Omega-3 fa | G | A | -0.15057 | 0.01244 | 2.74E-33 |
| rs174549 | miR-1908-5 | cis | 4.04E-08 | Nightingale | FAw3 | Omega-3 fa | A | G | -0.15265 | 0.012543 | 1.22E-33 |
| rs174549 | miR-1908-5 | cis | 4.04E-08 | Nightingale | FAw3 | Omega-3 fa | A | G | -0.15265 | 0.012543 | 1.22E-33 |
| rs174560 | miR-1908-5 | cis | 4.91E-08 | Nightingale | FAw3 | Omega-3 fa | C | T | -0.15052 | 0.012544 | 9.48E-33 |
| rs174560 | miR-1908-5 | cis | 4.91E-08 | Nightingale | FAw3 | Omega-3 fa | C | T | -0.15052 | 0.012544 | 9.48E-33 |
| rs174561 | miR-1908-5 | cis | 7.66E-08 | Nightingale | FAw3 | Omega-3 fa | C | T | -0.15012 | 0.012554 | 1.57E-32 |
| rs174561 | miR-1908-5 | cis | 7.66E-08 | Nightingale | FAw3 | Omega-3 fa | C | T | -0.15012 | 0.012554 | 1.57E-32 |
| rs2524299 | miR-1908-5 | cis | 4.82E-05 | Nightingale | FAw3 | Omega-3 fa | T | A | -0.12688 | 0.014061 | 3.21E-19 |
| rs2524299 | miR-1908-5 | cis | 4.82E-05 | Nightingale | FAw3 | Omega-3 fa | T | A | -0.12688 | 0.014061 | 3.21E-19 |
| rs174575 | miR-1908-5 | cis | 5.45E-05 | Nightingale | FAw3 | Omega-3 fa | G | C | -0.10579 | 0.014804 | 1.28E-12 |
| rs174575 | miR-1908-5 | cis | 5.45E-05 | Nightingale | FAw3 | Omega-3 fa | G | C | -0.10579 | 0.014804 | 1.28E-12 |
| rs174601 | miR-1908-5 | cis | 5.98E-09 | Nightingale | FAw3 | Omega-3 fa | T | C | -0.1508 | 0.012611 | 1.55E-32 |
| rs174601 | miR-1908-5 | cis | 5.98E-09 | Nightingale | FAw3 | Omega-3 fa | T | C | -0.1508 | 0.012611 | 1.55E-32 |
| rs102274 | miR-1908-5 | cis | 9.41E-09 | Nightingale | FAw3 | Omega-3 fa | C | T | -0.15282 | 0.012428 | 2.64E-34 |
| rs102274 | miR-1908-5 | cis | 9.41E-09 | Nightingale | FAw3 | Omega-3 fa | C | T | -0.15282 | 0.012428 | 2.64E-34 |
| rs174574 | miR-1908-5 | cis | 8.50E-09 | Nightingale | FAw3 | Omega-3 fa | C | A | 0.152226 | 0.01246 | 6.93E-34 |
| rs174574 | miR-1908-5 | cis | 8.50E-09 | Nightingale | FAw3 | Omega-3 fa | C | A | 0.152226 | 0.01246 | 6.93E-34 |
| rs174556 | miR-1908-5 | cis | 7.67E-08 | Nightingale | FAw3 | Omega-3 fa | T | C | -0.15013 | 0.012563 | 1.71E-32 |
| rs174556 | miR-1908-5 | cis | 7.67E-08 | Nightingale | FAw3 | Omega-3 fa | T | C | -0.15013 | 0.012563 | 1.71E-32 |
| rs174545 | miR-1908-5 | cis | 2.29E-08 | Nightingale | FAw3 | Omega-3 fa | G | C | -0.15329 | 0.012432 | 1.75E-34 |
| rs174545 | miR-1908-5 | cis | 2.29E-08 | Nightingale | FAw3 | Omega-3 fa | G | C | -0.15329 | 0.012432 | 1.75E-34 |
| rs174572 | miR-1908-5 | cis | 3.57E-05 | Nightingale | FAw3 | Omega-3 fa | T | C | -0.1054 | 0.014809 | 1.57E-12 |
| rs174572 | miR-1908-5 | cis | 3.57E-05 | Nightingale | FAw3 | Omega-3 fa | T | C | -0.1054 | 0.014809 | 1.57E-12 |
| rs174538 | miR-1908-5 | cis | 1.35E-07 | Nightingale | FAw3 | Omega-3 fa | A | G | -0.13735 | 0.0129 | 3.91E-26 |
| rs174538 | miR-1908-5 | cis | 1.35E-07 | Nightingale | FAw3 | Omega-3 fa | A | G | -0.13735 | 0.0129 | 3.91E-26 |
| rs174553 | miR-1908-5 | cis | 2.61E-08 | Nightingale | FAw3 | Omega-3 fa | G | A | -0.15301 | 0.012473 | 3.75E-34 |
| rs174553 | miR-1908-5 | cis | 2.61E-08 | Nightingale | FAw3 | Omega-3 fa | G | A | -0.15301 | 0.012473 | 3.75E-34 |
| rs174449 | miR-1908-5 | cis | 2.49E-05 | Nightingale | FAw3 | Omega-3 fa | A | G | 0.113224 | 0.012723 | 9.79E-19 |
| rs174449 | miR-1908-5 | cis | 2.49E-05 | Nightingale | FAw3 | Omega-3 fa | A | G | 0.113224 | 0.012723 | 9.79E-19 |
| rs174535 | miR-1908-5 | cis | 6.98E-09 | Nightingale | FAw3 | Omega-3 fa | C | T | -0.15324 | 0.012435 | 1.89E-34 |
| rs174535 | miR-1908-5 | cis | 6.98E-09 | Nightingale | FAw3 | Omega-3 fa | C | T | -0.15324 | 0.012435 | 1.89E-34 |
| rs174537 | miR-1908-5 | cis | 9.41E-09 | Nightingale | FAw3 | Omega-3 fa | T | G | -0.15178 | 0.012446 | 9.02E-34 |
| rs174537 | miR-1908-5 | cis | 9.41E-09 | Nightingale | FAw3 | Omega-3 fa | T | G | -0.15178 | 0.012446 | 9.02E-34 |
| rs174449 | miR-1908-5 | cis | 2.49E-05 | Nightingale | L.HDL.PL | Phospholip | A | G | 0.070027 | 0.010678 | 9.97E-11 |
| rs174537 | miR-1908-5 | cis | 9.41E-09 | Nightingale | L.HDL.PL | Phospholip | T | G | -0.08365 | 0.010473 | 3.37E-15 |
| rs174538 | miR-1908-5 | cis | 1.35E-07 | Nightingale | L.HDL.PL | Phospholip | A | G | -0.0707 | 0.010856 | 1.34E-10 |
| rs2072114 | miR-1908-5 | cis | 1.34E-05 | Nightingale | L.HDL.PL | Phospholip | G | A | -0.08742 | 0.011886 | 4.06E-13 |
| rs174448 | miR-1908-5 | cis | 1.95E-05 | Nightingale | L.HDL.PL | Phospholip | A | G | 0.075367 | 0.010712 | 3.97E-12 |
| rs174561 | miR-1908-5 | cis | 7.66E-08 | Nightingale | L.HDL.PL | Phospholip | C | T | -0.0777 | 0.010569 | 4.15E-13 |
| rs174568 | miR-1908-5 | cis | 3.76E-08 | Nightingale | L.HDL.PL | Phospholip | T | C | -0.08313 | 0.010516 | 6.40E-15 |
| rs174556 | miR-1908-5 | cis | 7.67E-08 | Nightingale | L.HDL.PL | Phospholip | T | C | -0.07775 | 0.010574 | 4.12E-13 |
| rs174567 | miR-1908-5 | cis | 1.32E-08 | Nightingale | L.HDL.PL | Phospholip | G | A | -0.08236 | 0.010483 | 9.31E-15 |
| rs99780 | miR-1908-5 | cis | 1.74E-08 | Nightingale | L.HDL.PL | Phospholip | T | C | -0.08259 | 0.010483 | 7.85E-15 |
| rs174564 | miR-1908-5 | cis | 1.61E-08 | Nightingale | L.HDL.PL | Phospholip | G | A | -0.08263 | 0.01053 | 1.00E-14 |
| rs174566 | miR-1908-5 | cis | 1.58E-08 | Nightingale | L.HDL.PL | Phospholip | G | A | -0.08295 | 0.010472 | 5.65E-15 |
| rs174533 | miR-1908-5 | cis | 1.26E-08 | Nightingale | L.HDL.PL | Phospholip | A | G | -0.08348 | 0.010466 | 3.64E-15 |
| rs2727270 | miR-1908-5 | cis | 4.45E-05 | Nightingale | L.HDL.PL | Phospholip | T | C | -0.0856 | 0.01189 | 1.25E-12 |
| rs174580 | miR-1908-5 | cis | 1.81E-08 | Nightingale | L.HDL.PL | Phospholip | G | A | -0.08621 | 0.010501 | 5.67E-16 |
| rs174536 | miR-1908-5 | cis | 1.31E-08 | Nightingale | L.HDL.PL | Phospholip | C | A | -0.08413 | 0.010461 | 2.17E-15 |
| rs174601 | miR-1908-5 | cis | 5.98E-09 | Nightingale | L.HDL.PL | Phospholip | T | C | -0.08405 | 0.01058 | 4.71E-15 |
| rs2727271 | miR-1908-5 | cis | 4.23E-05 | Nightingale | L.HDL.PL | Phospholip | T | A | -0.08605 | 0.011877 | 9.00E-13 |
| rs174553 | miR-1908-5 | cis | 2.61E-08 | Nightingale | L.HDL.PL | Phospholip | G | A | -0.08412 | 0.010497 | 2.72E-15 |
| rs174560 | miR-1908-5 | cis | 4.91E-08 | Nightingale | L.HDL.PL | Phospholip | C | T | -0.07804 | 0.010562 | 3.18E-13 |

| | | | | | | | | | | | |
|---|---|---|---|---|---|---|---|---|---|---|---|
| rs174530 | miR-1908-5 | cis | 1.12E-08 | Nightingale | L.HDL.PL | Phospholip | G | A | -0.08474 | 0.010496 | 1.69E-15 |
| rs174577 | miR-1908-5 | cis | 1.89E-08 | Nightingale | L.HDL.PL | Phospholip | A | C | -0.0856 | 0.010486 | 8.22E-16 |
| rs174548 | miR-1908-5 | cis | 3.01E-08 | Nightingale | L.HDL.PL | Phospholip | G | C | -0.07995 | 0.010555 | 8.03E-14 |
| rs174545 | miR-1908-5 | cis | 2.29E-08 | Nightingale | L.HDL.PL | Phospholip | G | C | -0.0839 | 0.010461 | 2.59E-15 |
| rs28456 | miR-1908-5 | cis | 6.85E-08 | Nightingale | L.HDL.PL | Phospholip | G | A | -0.07758 | 0.010668 | 7.43E-13 |
| rs174574 | miR-1908-5 | cis | 8.50E-09 | Nightingale | L.HDL.PL | Phospholip | C | A | 0.084287 | 0.010484 | 2.21E-15 |
| rs174578 | miR-1908-5 | cis | 2.55E-08 | Nightingale | L.HDL.PL | Phospholip | A | T | -0.08625 | 0.010485 | 4.95E-16 |
| rs174592 | miR-1908-5 | cis | 3.81E-09 | Nightingale | L.HDL.PL | Phospholip | G | A | -0.08349 | 0.010523 | 5.09E-15 |
| rs2524299 | miR-1908-5 | cis | 4.82E-05 | Nightingale | L.HDL.PL | Phospholip | T | A | -0.08739 | 0.011861 | 3.70E-13 |
| rs174555 | miR-1908-5 | cis | 7.43E-08 | Nightingale | L.HDL.PL | Phospholip | C | T | -0.0787 | 0.010564 | 2.03E-13 |
| rs174554 | miR-1908-5 | cis | 3.17E-08 | Nightingale | L.HDL.PL | Phospholip | G | A | -0.08323 | 0.010461 | 4.29E-15 |
| rs2072113 | miR-1908-5 | cis | 3.35E-05 | Nightingale | L.HDL.PL | Phospholip | T | C | -0.0864 | 0.011889 | 7.71E-13 |
| rs102274 | miR-1908-5 | cis | 9.41E-09 | Nightingale | L.HDL.PL | Phospholip | C | T | -0.08428 | 0.010458 | 1.90E-15 |
| rs174584 | miR-1908-5 | cis | 1.39E-08 | Nightingale | L.HDL.PL | Phospholip | A | G | -0.08532 | 0.010618 | 2.28E-15 |
| rs174544 | miR-1908-5 | cis | 2.30E-08 | Nightingale | L.HDL.PL | Phospholip | A | C | -0.08014 | 0.010615 | 9.66E-14 |
| rs174581 | miR-1908-5 | cis | 1.68E-08 | Nightingale | L.HDL.PL | Phospholip | A | G | -0.0862 | 0.010502 | 5.71E-16 |
| rs174576 | miR-1908-5 | cis | 3.13E-08 | Nightingale | L.HDL.PL | Phospholip | A | C | -0.08458 | 0.01049 | 1.84E-15 |
| rs174559 | miR-1908-5 | cis | 5.55E-07 | Nightingale | L.HDL.PL | Phospholip | A | G | -0.07023 | 0.010935 | 2.39E-10 |
| rs174535 | miR-1908-5 | cis | 6.98E-09 | Nightingale | L.HDL.PL | Phospholip | C | T | -0.08386 | 0.01047 | 2.83E-15 |
| rs174562 | miR-1908-5 | cis | 4.92E-08 | Nightingale | L.HDL.PL | Phospholip | G | A | -0.08221 | 0.010469 | 9.66E-15 |
| rs174549 | miR-1908-5 | cis | 4.04E-08 | Nightingale | L.HDL.PL | Phospholip | A | G | -0.07926 | 0.010558 | 1.33E-13 |
| rs174530 | miR-1908-5 | cis | 1.12E-08 | Nightingale | XL.HDL.PL | Phospholip | G | A | -0.0931 | 0.0105 | 1.39E-18 |
| rs174549 | miR-1908-5 | cis | 4.04E-08 | Nightingale | XL.HDL.PL | Phospholip | A | G | -0.08682 | 0.010564 | 3.49E-16 |
| rs28456 | miR-1908-5 | cis | 6.85E-08 | Nightingale | XL.HDL.PL | Phospholip | G | A | -0.08449 | 0.010674 | 4.00E-15 |
| rs174580 | miR-1908-5 | cis | 1.81E-08 | Nightingale | XL.HDL.PL | Phospholip | G | A | -0.09202 | 0.010506 | 3.58E-18 |
| rs174559 | miR-1908-5 | cis | 5.55E-07 | Nightingale | XL.HDL.PL | Phospholip | A | G | -0.07851 | 0.01094 | 1.07E-12 |
| rs99780 | miR-1908-5 | cis | 1.74E-08 | Nightingale | XL.HDL.PL | Phospholip | T | C | -0.09024 | 0.010486 | 1.35E-17 |
| rs174592 | miR-1908-5 | cis | 3.81E-09 | Nightingale | XL.HDL.PL | Phospholip | G | A | -0.09072 | 0.010527 | 1.22E-17 |
| rs2727270 | miR-1908-5 | cis | 4.45E-05 | Nightingale | XL.HDL.PL | Phospholip | T | C | -0.09052 | 0.011894 | 4.31E-14 |
| rs2524299 | miR-1908-5 | cis | 4.82E-05 | Nightingale | XL.HDL.PL | Phospholip | T | A | -0.09031 | 0.011873 | 4.42E-14 |
| rs174561 | miR-1908-5 | cis | 7.66E-08 | Nightingale | XL.HDL.PL | Phospholip | C | T | -0.08558 | 0.010574 | 9.65E-16 |
| rs2072114 | miR-1908-5 | cis | 1.34E-05 | Nightingale | XL.HDL.PL | Phospholip | G | A | -0.09206 | 0.011897 | 1.61E-14 |
| rs174548 | miR-1908-5 | cis | 3.01E-08 | Nightingale | XL.HDL.PL | Phospholip | G | C | -0.08664 | 0.010561 | 3.94E-16 |
| rs2072113 | miR-1908-5 | cis | 3.35E-05 | Nightingale | XL.HDL.PL | Phospholip | T | C | -0.09123 | 0.011901 | 2.82E-14 |
| rs174584 | miR-1908-5 | cis | 1.39E-08 | Nightingale | XL.HDL.PL | Phospholip | A | G | -0.09114 | 0.010622 | 1.67E-17 |
| rs174562 | miR-1908-5 | cis | 4.92E-08 | Nightingale | XL.HDL.PL | Phospholip | G | A | -0.09118 | 0.010473 | 5.62E-18 |
| rs174567 | miR-1908-5 | cis | 1.32E-08 | Nightingale | XL.HDL.PL | Phospholip | G | A | -0.08976 | 0.010487 | 2.00E-17 |
| rs174554 | miR-1908-5 | cis | 3.17E-08 | Nightingale | XL.HDL.PL | Phospholip | G | A | -0.09212 | 0.010465 | 2.42E-18 |
| rs174535 | miR-1908-5 | cis | 6.98E-09 | Nightingale | XL.HDL.PL | Phospholip | C | T | -0.09248 | 0.010467 | 1.82E-18 |
| rs174566 | miR-1908-5 | cis | 1.58E-08 | Nightingale | XL.HDL.PL | Phospholip | G | A | -0.08995 | 0.010477 | 1.60E-17 |
| rs102274 | miR-1908-5 | cis | 9.41E-09 | Nightingale | XL.HDL.PL | Phospholip | C | T | -0.09254 | 0.010461 | 1.67E-18 |
| rs174564 | miR-1908-5 | cis | 1.61E-08 | Nightingale | XL.HDL.PL | Phospholip | G | A | -0.09168 | 0.010533 | 5.77E-18 |
| rs174449 | miR-1908-5 | cis | 2.49E-05 | Nightingale | XL.HDL.PL | Phospholip | A | G | 0.06373 | 0.010679 | 3.18E-09 |
| rs174545 | miR-1908-5 | cis | 2.29E-08 | Nightingale | XL.HDL.PL | Phospholip | G | C | -0.09264 | 0.010464 | 1.56E-18 |
| rs174578 | miR-1908-5 | cis | 2.55E-08 | Nightingale | XL.HDL.PL | Phospholip | A | T | -0.09226 | 0.010489 | 2.58E-18 |
| rs174577 | miR-1908-5 | cis | 1.89E-08 | Nightingale | XL.HDL.PL | Phospholip | A | C | -0.09178 | 0.010483 | 3.68E-18 |
| rs174536 | miR-1908-5 | cis | 1.31E-08 | Nightingale | XL.HDL.PL | Phospholip | C | A | -0.09261 | 0.010465 | 1.61E-18 |
| rs174576 | miR-1908-5 | cis | 3.13E-08 | Nightingale | XL.HDL.PL | Phospholip | A | C | -0.09097 | 0.010491 | 7.66E-18 |
| rs174574 | miR-1908-5 | cis | 8.50E-09 | Nightingale | XL.HDL.PL | Phospholip | C | A | 0.090636 | 0.010489 | 9.89E-18 |
| rs174601 | miR-1908-5 | cis | 5.98E-09 | Nightingale | XL.HDL.PL | Phospholip | T | C | -0.09138 | 0.010584 | 1.06E-17 |
| rs174553 | miR-1908-5 | cis | 2.61E-08 | Nightingale | XL.HDL.PL | Phospholip | G | A | -0.0926 | 0.010501 | 2.12E-18 |
| rs174568 | miR-1908-5 | cis | 3.76E-08 | Nightingale | XL.HDL.PL | Phospholip | T | C | -0.09077 | 0.01052 | 1.11E-17 |
| rs174544 | miR-1908-5 | cis | 2.30E-08 | Nightingale | XL.HDL.PL | Phospholip | A | C | -0.08816 | 0.010621 | 1.77E-16 |
| rs174537 | miR-1908-5 | cis | 9.41E-09 | Nightingale | XL.HDL.PL | Phospholip | T | G | -0.09197 | 0.010476 | 3.00E-18 |
| rs174581 | miR-1908-5 | cis | 1.68E-08 | Nightingale | XL.HDL.PL | Phospholip | A | G | -0.092 | 0.010506 | 3.63E-18 |
| rs174448 | miR-1908-5 | cis | 1.95E-05 | Nightingale | XL.HDL.PL | Phospholip | A | G | 0.070307 | 0.010714 | 7.43E-11 |
| rs2727271 | miR-1908-5 | cis | 4.23E-05 | Nightingale | XL.HDL.PL | Phospholip | T | A | -0.09051 | 0.011889 | 4.23E-14 |
| rs174538 | miR-1908-5 | cis | 1.35E-07 | Nightingale | XL.HDL.PL | Phospholip | A | G | -0.08253 | 0.010852 | 4.48E-14 |
| rs174556 | miR-1908-5 | cis | 7.67E-08 | Nightingale | XL.HDL.PL | Phospholip | T | C | -0.08587 | 0.010579 | 7.95E-16 |
| rs174533 | miR-1908-5 | cis | 1.26E-08 | Nightingale | XL.HDL.PL | Phospholip | A | G | -0.09198 | 0.010469 | 2.83E-18 |
| rs174555 | miR-1908-5 | cis | 7.43E-08 | Nightingale | XL.HDL.PL | Phospholip | C | T | -0.08643 | 0.01057 | 4.88E-16 |
| rs174560 | miR-1908-5 | cis | 4.91E-08 | Nightingale | XL.HDL.PL | Phospholip | C | T | -0.08508 | 0.010568 | 1.36E-15 |
| rs174449 | miR-1908-5 | cis | 2.49E-05 | Nightingale | L.HDL.L | Total lipids | A | G | 0.069487 | 0.010683 | 1.33E-10 |

| | | | | | | | | | | | |
|---|---|---|---|---|---|---|---|---|---|---|---|
| rs174538 | miR-1908-5 | cis | 1.35E-07 | Nightingale | L.HDL.L | Total lipids | A | G | -0.07097 | 0.010861 | 1.10E-10 |
| rs174559 | miR-1908-5 | cis | 5.55E-07 | Nightingale | L.HDL.L | Total lipids | A | G | -0.07098 | 0.010941 | 1.49E-10 |
| rs174561 | miR-1908-5 | cis | 7.66E-08 | Nightingale | L.HDL.L | Total lipids | C | T | -0.07956 | 0.010574 | 1.09E-13 |
| rs174577 | miR-1908-5 | cis | 1.89E-08 | Nightingale | L.HDL.L | Total lipids | A | C | -0.08724 | 0.01049 | 2.16E-16 |
| rs174566 | miR-1908-5 | cis | 1.58E-08 | Nightingale | L.HDL.L | Total lipids | G | A | -0.08473 | 0.010478 | 1.40E-15 |
| rs174537 | miR-1908-5 | cis | 9.41E-09 | Nightingale | L.HDL.L | Total lipids | T | G | -0.08557 | 0.010477 | 7.30E-16 |
| rs174545 | miR-1908-5 | cis | 2.29E-08 | Nightingale | L.HDL.L | Total lipids | G | C | -0.08603 | 0.010466 | 4.75E-16 |
| rs174548 | miR-1908-5 | cis | 3.01E-08 | Nightingale | L.HDL.L | Total lipids | G | C | -0.08165 | 0.010561 | 2.28E-14 |
| rs2072114 | miR-1908-5 | cis | 1.34E-05 | Nightingale | L.HDL.L | Total lipids | G | A | -0.08919 | 0.011891 | 1.30E-13 |
| rs174549 | miR-1908-5 | cis | 4.04E-08 | Nightingale | L.HDL.L | Total lipids | A | G | -0.08103 | 0.010564 | 3.61E-14 |
| rs174601 | miR-1908-5 | cis | 5.98E-09 | Nightingale | L.HDL.L | Total lipids | T | C | -0.0855 | 0.010586 | 1.51E-15 |
| rs174553 | miR-1908-5 | cis | 2.61E-08 | Nightingale | L.HDL.L | Total lipids | G | A | -0.08628 | 0.010502 | 4.97E-16 |
| rs102274 | miR-1908-5 | cis | 9.41E-09 | Nightingale | L.HDL.L | Total lipids | C | T | -0.08621 | 0.010463 | 4.06E-16 |
| rs174562 | miR-1908-5 | cis | 4.92E-08 | Nightingale | L.HDL.L | Total lipids | G | A | -0.08441 | 0.010474 | 1.75E-15 |
| rs174530 | miR-1908-5 | cis | 1.12E-08 | Nightingale | L.HDL.L | Total lipids | G | A | -0.08689 | 0.010501 | 3.05E-16 |
| rs174578 | miR-1908-5 | cis | 2.55E-08 | Nightingale | L.HDL.L | Total lipids | A | T | -0.08783 | 0.01049 | 1.36E-16 |
| rs174581 | miR-1908-5 | cis | 1.68E-08 | Nightingale | L.HDL.L | Total lipids | A | G | -0.08772 | 0.010507 | 1.67E-16 |
| rs174592 | miR-1908-5 | cis | 3.81E-09 | Nightingale | L.HDL.L | Total lipids | G | A | -0.08509 | 0.010529 | 1.45E-15 |
| rs174568 | miR-1908-5 | cis | 3.76E-08 | Nightingale | L.HDL.L | Total lipids | T | C | -0.08491 | 0.010521 | 1.60E-15 |
| rs2072113 | miR-1908-5 | cis | 3.35E-05 | Nightingale | L.HDL.L | Total lipids | T | C | -0.08813 | 0.011896 | 2.56E-13 |
| rs174556 | miR-1908-5 | cis | 7.67E-08 | Nightingale | L.HDL.L | Total lipids | T | C | -0.07964 | 0.010579 | 1.06E-13 |
| rs2727270 | miR-1908-5 | cis | 4.45E-05 | Nightingale | L.HDL.L | Total lipids | T | C | -0.08731 | 0.011896 | 4.24E-13 |
| rs174576 | miR-1908-5 | cis | 3.13E-08 | Nightingale | L.HDL.L | Total lipids | A | C | -0.08616 | 0.010497 | 5.23E-16 |
| rs174535 | miR-1908-5 | cis | 6.98E-09 | Nightingale | L.HDL.L | Total lipids | C | T | -0.08592 | 0.010474 | 5.48E-16 |
| rs174574 | miR-1908-5 | cis | 8.50E-09 | Nightingale | L.HDL.L | Total lipids | C | A | 0.085863 | 0.010489 | 6.28E-16 |
| rs174584 | miR-1908-5 | cis | 1.39E-08 | Nightingale | L.HDL.L | Total lipids | A | G | -0.08673 | 0.010623 | 7.50E-16 |
| rs174560 | miR-1908-5 | cis | 4.91E-08 | Nightingale | L.HDL.L | Total lipids | C | T | -0.0798 | 0.010568 | 8.87E-14 |
| rs174544 | miR-1908-5 | cis | 2.30E-08 | Nightingale | L.HDL.L | Total lipids | A | C | -0.082 | 0.01062 | 2.45E-14 |
| rs174567 | miR-1908-5 | cis | 1.32E-08 | Nightingale | L.HDL.L | Total lipids | G | A | -0.08423 | 0.010488 | 2.18E-15 |
| rs174448 | miR-1908-5 | cis | 1.95E-05 | Nightingale | L.HDL.L | Total lipids | A | G | 0.074987 | 0.010718 | 4.88E-12 |
| rs2524299 | miR-1908-5 | cis | 4.82E-05 | Nightingale | L.HDL.L | Total lipids | T | A | -0.08863 | 0.011867 | 1.64E-13 |
| rs174554 | miR-1908-5 | cis | 3.17E-08 | Nightingale | L.HDL.L | Total lipids | G | A | -0.08542 | 0.010466 | 7.66E-16 |
| rs174533 | miR-1908-5 | cis | 1.26E-08 | Nightingale | L.HDL.L | Total lipids | A | G | -0.08549 | 0.010471 | 7.46E-16 |
| rs2727271 | miR-1908-5 | cis | 4.23E-05 | Nightingale | L.HDL.L | Total lipids | T | A | -0.08765 | 0.011884 | 3.26E-13 |
| rs174564 | miR-1908-5 | cis | 1.61E-08 | Nightingale | L.HDL.L | Total lipids | G | A | -0.08486 | 0.010535 | 1.80E-15 |
| rs174536 | miR-1908-5 | cis | 1.31E-08 | Nightingale | L.HDL.L | Total lipids | C | A | -0.08611 | 0.010466 | 4.48E-16 |
| rs28456 | miR-1908-5 | cis | 6.85E-08 | Nightingale | L.HDL.L | Total lipids | G | A | -0.0795 | 0.010674 | 1.90E-13 |
| rs174555 | miR-1908-5 | cis | 7.43E-08 | Nightingale | L.HDL.L | Total lipids | C | T | -0.08052 | 0.01057 | 5.36E-14 |
| rs174580 | miR-1908-5 | cis | 1.81E-08 | Nightingale | L.HDL.L | Total lipids | G | A | -0.08772 | 0.010506 | 1.65E-16 |
| rs99780 | miR-1908-5 | cis | 1.74E-08 | Nightingale | L.HDL.L | Total lipids | T | C | -0.08447 | 0.010488 | 1.81E-15 |
| rs174560 | miR-1908-5 | cis | 4.91E-08 | Nightingale | XL.HDL.L | Total lipids | C | T | -0.08456 | 0.010561 | 1.59E-15 |
| rs174535 | miR-1908-5 | cis | 6.98E-09 | Nightingale | XL.HDL.L | Total lipids | C | T | -0.09188 | 0.010457 | 2.20E-18 |
| rs174538 | miR-1908-5 | cis | 1.35E-07 | Nightingale | XL.HDL.L | Total lipids | A | G | -0.08197 | 0.010843 | 5.28E-14 |
| rs2727270 | miR-1908-5 | cis | 4.45E-05 | Nightingale | XL.HDL.L | Total lipids | T | C | -0.08967 | 0.011884 | 5.88E-14 |
| rs174449 | miR-1908-5 | cis | 2.49E-05 | Nightingale | XL.HDL.L | Total lipids | A | G | 0.060122 | 0.010659 | 1.97E-08 |
| rs174548 | miR-1908-5 | cis | 3.01E-08 | Nightingale | XL.HDL.L | Total lipids | G | C | -0.08616 | 0.010554 | 4.41E-16 |
| rs174581 | miR-1908-5 | cis | 1.68E-08 | Nightingale | XL.HDL.L | Total lipids | A | G | -0.09106 | 0.010499 | 5.92E-18 |
| rs174559 | miR-1908-5 | cis | 5.55E-07 | Nightingale | XL.HDL.L | Total lipids | A | G | -0.07862 | 0.010932 | 8.18E-13 |
| rs28456 | miR-1908-5 | cis | 6.85E-08 | Nightingale | XL.HDL.L | Total lipids | G | A | -0.08443 | 0.010666 | 3.28E-15 |
| rs174554 | miR-1908-5 | cis | 3.17E-08 | Nightingale | XL.HDL.L | Total lipids | G | A | -0.09199 | 0.010457 | 2.00E-18 |
| rs174556 | miR-1908-5 | cis | 7.67E-08 | Nightingale | XL.HDL.L | Total lipids | T | C | -0.08573 | 0.010562 | 6.49E-16 |
| rs174567 | miR-1908-5 | cis | 1.32E-08 | Nightingale | XL.HDL.L | Total lipids | G | A | -0.08872 | 0.01048 | 3.53E-17 |
| rs174574 | miR-1908-5 | cis | 8.50E-09 | Nightingale | XL.HDL.L | Total lipids | C | A | 0.089468 | 0.010481 | 1.94E-17 |
| rs174566 | miR-1908-5 | cis | 1.58E-08 | Nightingale | XL.HDL.L | Total lipids | G | A | -0.08891 | 0.010469 | 2.81E-17 |
| rs174536 | miR-1908-5 | cis | 1.31E-08 | Nightingale | XL.HDL.L | Total lipids | C | A | -0.09201 | 0.010457 | 1.98E-18 |
| rs174578 | miR-1908-5 | cis | 2.55E-08 | Nightingale | XL.HDL.L | Total lipids | A | T | -0.0913 | 0.010482 | 4.29E-18 |
| rs174576 | miR-1908-5 | cis | 3.13E-08 | Nightingale | XL.HDL.L | Total lipids | A | C | -0.09006 | 0.010482 | 1.20E-17 |
| rs174533 | miR-1908-5 | cis | 1.26E-08 | Nightingale | XL.HDL.L | Total lipids | A | G | -0.09174 | 0.010461 | 2.55E-18 |
| rs174555 | miR-1908-5 | cis | 7.43E-08 | Nightingale | XL.HDL.L | Total lipids | C | T | -0.08643 | 0.010563 | 3.78E-16 |
| rs174562 | miR-1908-5 | cis | 4.92E-08 | Nightingale | XL.HDL.L | Total lipids | G | A | -0.09084 | 0.010465 | 5.56E-18 |
| rs174537 | miR-1908-5 | cis | 9.41E-09 | Nightingale | XL.HDL.L | Total lipids | T | G | -0.09114 | 0.01046 | 4.18E-18 |
| rs174568 | miR-1908-5 | cis | 3.76E-08 | Nightingale | XL.HDL.L | Total lipids | T | C | -0.09093 | 0.010512 | 7.23E-18 |
| rs174564 | miR-1908-5 | cis | 1.61E-08 | Nightingale | XL.HDL.L | Total lipids | G | A | -0.09094 | 0.010525 | 7.92E-18 |

| | | | | | | | | | | | |
|---|---|---|---|---|---|---|---|---|---|---|---|
| rs102274 | miR-1908-! | cis | 9.41E-09 | Nightingale | XL.HDL.L | Total lipids | C | T | -0.09196 | 0.010453 | 2.00E-18 |
| rs174448 | miR-1908-! | cis | 1.95E-05 | Nightingale | XL.HDL.L | Total lipids | A | G | 0.066085 | 0.010704 | 7.98E-10 |
| rs2524299 | miR-1908-! | cis | 4.82E-05 | Nightingale | XL.HDL.L | Total lipids | T | A | -0.08887 | 0.011865 | 8.90E-14 |
| rs174584 | miR-1908-! | cis | 1.39E-08 | Nightingale | XL.HDL.L | Total lipids | A | G | -0.09064 | 0.010614 | 1.89E-17 |
| rs174577 | miR-1908-! | cis | 1.89E-08 | Nightingale | XL.HDL.L | Total lipids | A | C | -0.09105 | 0.010473 | 4.94E-18 |
| rs174530 | miR-1908-! | cis | 1.12E-08 | Nightingale | XL.HDL.L | Total lipids | G | A | -0.09283 | 0.010492 | 1.29E-18 |
| rs99780 | miR-1908-! | cis | 1.74E-08 | Nightingale | XL.HDL.L | Total lipids | T | C | -0.0895 | 0.010479 | 1.86E-17 |
| rs174561 | miR-1908-! | cis | 7.66E-08 | Nightingale | XL.HDL.L | Total lipids | C | T | -0.08543 | 0.010567 | 8.44E-16 |
| rs2072114 | miR-1908-! | cis | 1.34E-05 | Nightingale | XL.HDL.L | Total lipids | G | A | -0.09106 | 0.011878 | 2.32E-14 |
| rs174592 | miR-1908-! | cis | 3.81E-09 | Nightingale | XL.HDL.L | Total lipids | G | A | -0.08979 | 0.010519 | 1.95E-17 |
| rs174553 | miR-1908-! | cis | 2.61E-08 | Nightingale | XL.HDL.L | Total lipids | G | A | -0.09232 | 0.010493 | 1.98E-18 |
| rs2072113 | miR-1908-! | cis | 3.35E-05 | Nightingale | XL.HDL.L | Total lipids | T | C | -0.09061 | 0.011893 | 3.34E-14 |
| rs174580 | miR-1908-! | cis | 1.81E-08 | Nightingale | XL.HDL.L | Total lipids | G | A | -0.09107 | 0.010498 | 5.85E-18 |
| rs174544 | miR-1908-! | cis | 2.30E-08 | Nightingale | XL.HDL.L | Total lipids | A | C | -0.08817 | 0.010613 | 1.34E-16 |
| rs174549 | miR-1908-! | cis | 4.04E-08 | Nightingale | XL.HDL.L | Total lipids | A | G | -0.08675 | 0.010557 | 2.84E-16 |
| rs174545 | miR-1908-! | cis | 2.29E-08 | Nightingale | XL.HDL.L | Total lipids | G | C | -0.09242 | 0.010457 | 1.39E-18 |
| rs2727271 | miR-1908-! | cis | 4.23E-05 | Nightingale | XL.HDL.L | Total lipids | T | A | -0.08976 | 0.011881 | 5.45E-14 |
| rs174601 | miR-1908-! | cis | 5.98E-09 | Nightingale | XL.HDL.L | Total lipids | T | C | -0.09026 | 0.010577 | 1.98E-17 |
| rs174545 | miR-1908-! | cis | 2.29E-08 | Nightingale | XL.HDL.TG | Triglyceride | G | C | -0.07943 | 0.01011 | 8.13E-15 |
| rs174561 | miR-1908-! | cis | 7.66E-08 | Nightingale | XL.HDL.TG | Triglyceride | C | T | -0.07126 | 0.010241 | 6.08E-12 |
| rs174544 | miR-1908-! | cis | 2.30E-08 | Nightingale | XL.HDL.TG | Triglyceride | A | C | -0.07244 | 0.010287 | 3.40E-12 |
| rs174562 | miR-1908-! | cis | 4.92E-08 | Nightingale | XL.HDL.TG | Triglyceride | G | A | -0.07874 | 0.010114 | 1.42E-14 |
| rs2072113 | miR-1908-! | cis | 3.35E-05 | Nightingale | XL.HDL.TG | Triglyceride | T | C | -0.0699 | 0.011594 | 2.54E-09 |
| rs174592 | miR-1908-! | cis | 3.81E-09 | Nightingale | XL.HDL.TG | Triglyceride | G | A | -0.08092 | 0.010178 | 3.91E-15 |
| rs174574 | miR-1908-! | cis | 8.50E-09 | Nightingale | XL.HDL.TG | Triglyceride | C | A | 0.079607 | 0.010074 | 5.68E-15 |
| rs174530 | miR-1908-! | cis | 1.12E-08 | Nightingale | XL.HDL.TG | Triglyceride | G | A | -0.07405 | 0.010116 | 4.65E-13 |
| rs174560 | miR-1908-! | cis | 4.91E-08 | Nightingale | XL.HDL.TG | Triglyceride | C | T | -0.06991 | 0.010227 | 1.41E-11 |
| rs28456 | miR-1908-! | cis | 6.85E-08 | Nightingale | XL.HDL.TG | Triglyceride | G | A | -0.07029 | 0.010325 | 1.71E-11 |
| rs174568 | miR-1908-! | cis | 3.76E-08 | Nightingale | XL.HDL.TG | Triglyceride | T | C | -0.08037 | 0.010124 | 4.28E-15 |
| rs174601 | miR-1908-! | cis | 5.98E-09 | Nightingale | XL.HDL.TG | Triglyceride | T | C | -0.08171 | 0.01025 | 3.28E-15 |
| rs174555 | miR-1908-! | cis | 7.43E-08 | Nightingale | XL.HDL.TG | Triglyceride | C | T | -0.07232 | 0.010237 | 2.91E-12 |
| rs174581 | miR-1908-! | cis | 1.68E-08 | Nightingale | XL.HDL.TG | Triglyceride | A | G | -0.07944 | 0.010071 | 6.37E-15 |
| rs102274 | miR-1908-! | cis | 9.41E-09 | Nightingale | XL.HDL.TG | Triglyceride | C | T | -0.07843 | 0.010103 | 1.68E-14 |
| rs174535 | miR-1908-! | cis | 6.98E-09 | Nightingale | XL.HDL.TG | Triglyceride | C | T | -0.07716 | 0.010096 | 4.24E-14 |
| rs2524299 | miR-1908-! | cis | 4.82E-05 | Nightingale | XL.HDL.TG | Triglyceride | T | A | -0.06965 | 0.011564 | 2.63E-09 |
| rs174548 | miR-1908-! | cis | 3.01E-08 | Nightingale | XL.HDL.TG | Triglyceride | G | C | -0.07055 | 0.010225 | 9.13E-12 |
| rs174538 | miR-1908-! | cis | 1.35E-07 | Nightingale | XL.HDL.TG | Triglyceride | A | G | -0.07271 | 0.010477 | 6.91E-12 |
| rs174449 | miR-1908-! | cis | 2.49E-05 | Nightingale | XL.HDL.TG | Triglyceride | A | G | 0.05764 | 0.010213 | 2.43E-08 |
| rs174536 | miR-1908-! | cis | 1.31E-08 | Nightingale | XL.HDL.TG | Triglyceride | C | A | -0.07722 | 0.01009 | 3.92E-14 |
| rs174559 | miR-1908-! | cis | 5.55E-07 | Nightingale | XL.HDL.TG | Triglyceride | A | G | -0.06718 | 0.010611 | 3.92E-10 |
| rs174533 | miR-1908-! | cis | 1.26E-08 | Nightingale | XL.HDL.TG | Triglyceride | A | G | -0.07651 | 0.010093 | 6.75E-14 |
| rs174549 | miR-1908-! | cis | 4.04E-08 | Nightingale | XL.HDL.TG | Triglyceride | A | G | -0.07206 | 0.010231 | 3.38E-12 |
| rs174576 | miR-1908-! | cis | 3.13E-08 | Nightingale | XL.HDL.TG | Triglyceride | A | C | -0.07928 | 0.01004 | 5.97E-15 |
| rs174567 | miR-1908-! | cis | 1.32E-08 | Nightingale | XL.HDL.TG | Triglyceride | G | A | -0.07818 | 0.010072 | 1.69E-14 |
| rs174553 | miR-1908-! | cis | 2.61E-08 | Nightingale | XL.HDL.TG | Triglyceride | G | A | -0.07963 | 0.010146 | 8.65E-15 |
| rs99780 | miR-1908-! | cis | 1.74E-08 | Nightingale | XL.HDL.TG | Triglyceride | T | C | -0.07799 | 0.010074 | 1.99E-14 |
| rs174448 | miR-1908-! | cis | 1.95E-05 | Nightingale | XL.HDL.TG | Triglyceride | A | G | 0.064465 | 0.010329 | 6.90E-10 |
| rs2727271 | miR-1908-! | cis | 4.23E-05 | Nightingale | XL.HDL.TG | Triglyceride | T | A | -0.07071 | 0.011585 | 1.61E-09 |
| rs174554 | miR-1908-! | cis | 3.17E-08 | Nightingale | XL.HDL.TG | Triglyceride | G | A | -0.07982 | 0.01011 | 6.02E-15 |
| rs174580 | miR-1908-! | cis | 1.81E-08 | Nightingale | XL.HDL.TG | Triglyceride | G | A | -0.07945 | 0.01007 | 6.29E-15 |
| rs2072114 | miR-1908-! | cis | 1.34E-05 | Nightingale | XL.HDL.TG | Triglyceride | G | A | -0.07069 | 0.011584 | 1.62E-09 |
| rs174584 | miR-1908-! | cis | 1.39E-08 | Nightingale | XL.HDL.TG | Triglyceride | A | G | -0.0786 | 0.01022 | 2.91E-14 |
| rs174556 | miR-1908-! | cis | 7.67E-08 | Nightingale | XL.HDL.TG | Triglyceride | T | C | -0.07148 | 0.010243 | 5.32E-12 |
| rs2727270 | miR-1908-! | cis | 4.45E-05 | Nightingale | XL.HDL.TG | Triglyceride | T | C | -0.06985 | 0.011592 | 2.59E-09 |
| rs174578 | miR-1908-! | cis | 2.55E-08 | Nightingale | XL.HDL.TG | Triglyceride | A | T | -0.0794 | 0.010053 | 5.90E-15 |
| rs174564 | miR-1908-! | cis | 1.61E-08 | Nightingale | XL.HDL.TG | Triglyceride | G | A | -0.07864 | 0.010164 | 2.06E-14 |
| rs174577 | miR-1908-! | cis | 1.89E-08 | Nightingale | XL.HDL.TG | Triglyceride | A | C | -0.07934 | 0.010048 | 5.98E-15 |
| rs174566 | miR-1908-! | cis | 1.58E-08 | Nightingale | XL.HDL.TG | Triglyceride | G | A | -0.07821 | 0.010062 | 1.55E-14 |
| rs174537 | miR-1908-! | cis | 9.41E-09 | Nightingale | XL.HDL.TG | Triglyceride | T | G | -0.07698 | 0.010099 | 4.92E-14 |
| rs550057 | miR-10b-5p | trans | 1.61E-27 | Nightingale | IDL.P | Concentrat | T | C | 0.068167 | 0.011288 | 2.74E-09 |
| rs2519093 | miR-139-5p | trans | 1.63E-36 | Nightingale | IDL.P | Concentrat | T | C | 0.082512 | 0.012886 | 2.87E-10 |
| rs2519093 | let-7b-5p | trans | 1.82E-25 | Nightingale | IDL.P | Concentrat | T | C | 0.082512 | 0.012886 | 2.87E-10 |
| rs600038 | let-7b-5p | trans | 1.02E-24 | Nightingale | IDL.P | Concentrat | C | T | 0.077109 | 0.012319 | 7.10E-10 |

| | | | | | | | | | | | |
|---|---|---|---|---|---|---|---|---|---|---|---|
| rs2519093 | miR-10b-5p | trans | 3.13E-35 | Nightingale | IDL.P | Concentrat | T | C | 0.082512 | 0.012886 | 2.87E-10 |
| rs550057 | let-7b-5p | trans | 2.70E-19 | Nightingale | IDL.P | Concentrat | T | C | 0.068167 | 0.011288 | 2.74E-09 |
| rs600038 | miR-10b-5p | trans | 2.32E-34 | Nightingale | IDL.P | Concentrat | C | T | 0.077109 | 0.012319 | 7.10E-10 |
| rs532436 | miR-10b-5p | trans | 5.20E-36 | Nightingale | IDL.P | Concentrat | A | G | 0.083816 | 0.01282 | 1.21E-10 |
| rs495828 | miR-126-3p | trans | 2.00E-37 | Nightingale | IDL.P | Concentrat | T | G | 0.077045 | 0.012167 | 4.50E-10 |
| rs550057 | miR-126-3p | trans | 5.01E-31 | Nightingale | IDL.P | Concentrat | T | C | 0.068167 | 0.011288 | 2.74E-09 |
| rs550057 | miR-139-5p | trans | 5.52E-32 | Nightingale | IDL.P | Concentrat | T | C | 0.068167 | 0.011288 | 2.74E-09 |
| rs649129 | let-7a-5p | trans | 1.48E-12 | Nightingale | IDL.P | Concentrat | T | C | 0.077615 | 0.012322 | 5.54E-10 |
| rs495828 | miR-10b-5p | trans | 2.56E-34 | Nightingale | IDL.P | Concentrat | T | G | 0.077045 | 0.012167 | 4.50E-10 |
| rs2519093 | miR-126-3p | trans | 1.90E-40 | Nightingale | IDL.P | Concentrat | T | C | 0.082512 | 0.012886 | 2.87E-10 |
| rs2519093 | let-7a-5p | trans | 1.29E-11 | Nightingale | IDL.P | Concentrat | T | C | 0.082512 | 0.012886 | 2.87E-10 |
| rs532436 | let-7c-5p | trans | 2.13E-22 | Nightingale | IDL.P | Concentrat | A | G | 0.083816 | 0.01282 | 1.21E-10 |
| rs495828 | miR-139-5p | trans | 1.46E-34 | Nightingale | IDL.P | Concentrat | T | G | 0.077045 | 0.012167 | 4.50E-10 |
| rs532436 | miR-195-5p | trans | 3.89E-18 | Nightingale | IDL.P | Concentrat | A | G | 0.083816 | 0.01282 | 1.21E-10 |
| rs649129 | miR-10b-5p | trans | 2.54E-34 | Nightingale | IDL.P | Concentrat | T | C | 0.077615 | 0.012322 | 5.54E-10 |
| rs649129 | miR-126-3p | trans | 1.95E-37 | Nightingale | IDL.P | Concentrat | T | C | 0.077615 | 0.012322 | 5.54E-10 |
| rs600038 | miR-139-5p | trans | 7.68E-35 | Nightingale | IDL.P | Concentrat | C | T | 0.077109 | 0.012319 | 7.10E-10 |
| rs532436 | miR-139-5p | trans | 6.95E-37 | Nightingale | IDL.P | Concentrat | A | G | 0.083816 | 0.01282 | 1.21E-10 |
| rs600038 | let-7a-5p | trans | 1.48E-12 | Nightingale | IDL.P | Concentrat | C | T | 0.077109 | 0.012319 | 7.10E-10 |
| rs649129 | miR-139-5p | trans | 1.34E-34 | Nightingale | IDL.P | Concentrat | T | C | 0.077615 | 0.012322 | 5.54E-10 |
| rs532436 | miR-126-3p | trans | 2.49E-40 | Nightingale | IDL.P | Concentrat | A | G | 0.083816 | 0.01282 | 1.21E-10 |
| rs2519093 | miR-195-5p | trans | 7.49E-18 | Nightingale | IDL.P | Concentrat | T | C | 0.082512 | 0.012886 | 2.87E-10 |
| rs2519093 | let-7c-5p | trans | 4.94E-22 | Nightingale | IDL.P | Concentrat | T | C | 0.082512 | 0.012886 | 2.87E-10 |
| rs532436 | let-7b-5p | trans | 1.04E-25 | Nightingale | IDL.P | Concentrat | A | G | 0.083816 | 0.01282 | 1.21E-10 |
| rs495828 | let-7a-5p | trans | 1.53E-12 | Nightingale | IDL.P | Concentrat | T | G | 0.077045 | 0.012167 | 4.50E-10 |
| rs600038 | miR-126-3p | trans | 1.80E-37 | Nightingale | IDL.P | Concentrat | C | T | 0.077109 | 0.012319 | 7.10E-10 |
| rs550057 | miR-10b-5p | trans | 1.61E-27 | Nightingale | L.LDL.P | Concentrat | T | C | 0.062556 | 0.01128 | 4.88E-08 |
| rs649129 | miR-10b-5p | trans | 2.54E-34 | Nightingale | L.LDL.P | Concentrat | T | C | 0.071735 | 0.012314 | 9.99E-09 |
| rs495828 | let-7a-5p | trans | 1.53E-12 | Nightingale | L.LDL.P | Concentrat | T | G | 0.071954 | 0.012162 | 5.88E-09 |
| rs495828 | miR-139-5p | trans | 1.46E-34 | Nightingale | L.LDL.P | Concentrat | T | G | 0.071954 | 0.012162 | 5.88E-09 |
| rs550057 | miR-139-5p | trans | 5.52E-32 | Nightingale | L.LDL.P | Concentrat | T | C | 0.062556 | 0.01128 | 4.88E-08 |
| rs532436 | let-7b-5p | trans | 1.04E-25 | Nightingale | L.LDL.P | Concentrat | A | G | 0.076685 | 0.012812 | 3.91E-09 |
| rs600038 | miR-139-5p | trans | 7.68E-35 | Nightingale | L.LDL.P | Concentrat | C | T | 0.071302 | 0.012311 | 1.22E-08 |
| rs2519093 | miR-126-3p | trans | 1.90E-40 | Nightingale | L.LDL.P | Concentrat | T | C | 0.076205 | 0.012878 | 5.84E-09 |
| rs532436 | miR-195-5p | trans | 3.89E-18 | Nightingale | L.LDL.P | Concentrat | A | G | 0.076685 | 0.012812 | 3.91E-09 |
| rs2519093 | let-7b-5p | trans | 1.82E-25 | Nightingale | L.LDL.P | Concentrat | T | C | 0.076205 | 0.012878 | 5.84E-09 |
| rs600038 | miR-126-3p | trans | 1.80E-37 | Nightingale | L.LDL.P | Concentrat | C | T | 0.071302 | 0.012311 | 1.22E-08 |
| rs2519093 | miR-195-5p | trans | 7.49E-18 | Nightingale | L.LDL.P | Concentrat | T | C | 0.076205 | 0.012878 | 5.84E-09 |
| rs600038 | let-7a-5p | trans | 1.48E-12 | Nightingale | L.LDL.P | Concentrat | C | T | 0.071302 | 0.012311 | 1.22E-08 |
| rs550057 | miR-126-3p | trans | 5.01E-31 | Nightingale | L.LDL.P | Concentrat | T | C | 0.062556 | 0.01128 | 4.88E-08 |
| rs649129 | miR-126-3p | trans | 1.95E-37 | Nightingale | L.LDL.P | Concentrat | T | C | 0.071735 | 0.012314 | 9.99E-09 |
| rs649129 | miR-139-5p | trans | 1.34E-34 | Nightingale | L.LDL.P | Concentrat | T | C | 0.071735 | 0.012314 | 9.99E-09 |
| rs649129 | let-7a-5p | trans | 1.48E-12 | Nightingale | L.LDL.P | Concentrat | T | C | 0.071735 | 0.012314 | 9.99E-09 |
| rs600038 | miR-10b-5p | trans | 2.32E-34 | Nightingale | L.LDL.P | Concentrat | C | T | 0.071302 | 0.012311 | 1.22E-08 |
| rs532436 | let-7c-5p | trans | 2.13E-22 | Nightingale | L.LDL.P | Concentrat | A | G | 0.076685 | 0.012812 | 3.91E-09 |
| rs2519093 | miR-139-5p | trans | 1.63E-36 | Nightingale | L.LDL.P | Concentrat | T | C | 0.076205 | 0.012878 | 5.84E-09 |
| rs532436 | miR-139-5p | trans | 6.95E-37 | Nightingale | L.LDL.P | Concentrat | A | G | 0.076685 | 0.012812 | 3.91E-09 |
| rs495828 | miR-126-3p | trans | 2.00E-37 | Nightingale | L.LDL.P | Concentrat | T | G | 0.071954 | 0.012162 | 5.88E-09 |
| rs2519093 | miR-10b-5p | trans | 3.13E-35 | Nightingale | L.LDL.P | Concentrat | T | C | 0.076205 | 0.012878 | 5.84E-09 |
| rs550057 | let-7b-5p | trans | 2.70E-19 | Nightingale | L.LDL.P | Concentrat | T | C | 0.062556 | 0.01128 | 4.88E-08 |
| rs2519093 | let-7c-5p | trans | 4.94E-22 | Nightingale | L.LDL.P | Concentrat | T | C | 0.076205 | 0.012878 | 5.84E-09 |
| rs532436 | miR-126-3p | trans | 2.49E-40 | Nightingale | L.LDL.P | Concentrat | A | G | 0.076685 | 0.012812 | 3.91E-09 |
| rs600038 | let-7b-5p | trans | 1.02E-24 | Nightingale | L.LDL.P | Concentrat | C | T | 0.071302 | 0.012311 | 1.22E-08 |
| rs495828 | miR-10b-5p | trans | 2.56E-34 | Nightingale | L.LDL.P | Concentrat | T | G | 0.071954 | 0.012162 | 5.88E-09 |
| rs532436 | miR-10b-5p | trans | 5.20E-36 | Nightingale | L.LDL.P | Concentrat | A | G | 0.076685 | 0.012812 | 3.91E-09 |
| rs2519093 | let-7a-5p | trans | 1.29E-11 | Nightingale | L.LDL.P | Concentrat | T | C | 0.076205 | 0.012878 | 5.84E-09 |
| rs550057 | miR-10b-5p | trans | 1.61E-27 | Nightingale | XS.VLDL.P | Concentrat | T | C | 0.065357 | 0.011295 | 1.22E-08 |
| rs495828 | miR-126-3p | trans | 2.00E-37 | Nightingale | XS.VLDL.P | Concentrat | T | G | 0.06784 | 0.012176 | 4.11E-08 |
| rs2519093 | miR-126-3p | trans | 1.90E-40 | Nightingale | XS.VLDL.P | Concentrat | T | C | 0.074723 | 0.012894 | 1.16E-08 |
| rs550057 | let-7b-5p | trans | 2.70E-19 | Nightingale | XS.VLDL.P | Concentrat | T | C | 0.065357 | 0.011295 | 1.22E-08 |
| rs532436 | miR-126-3p | trans | 2.49E-40 | Nightingale | XS.VLDL.P | Concentrat | A | G | 0.076221 | 0.012829 | 4.92E-09 |
| rs600038 | let-7a-5p | trans | 1.48E-12 | Nightingale | XS.VLDL.P | Concentrat | C | T | 0.068852 | 0.012329 | 3.82E-08 |
| rs600038 | miR-10b-5p | trans | 2.32E-34 | Nightingale | XS.VLDL.P | Concentrat | C | T | 0.068852 | 0.012329 | 3.82E-08 |

| | | | | | | | | | | | |
|---|---|---|---|---|---|---|---|---|---|---|---|
| rs600038 | let-7b-5p | trans | 1.02E-24 | Nightingale | XS.VLDL.P | Concentrat | C | T | 0.068852 | 0.012329 | 3.82E-08 |
| rs495828 | miR-139-5p | trans | 1.46E-34 | Nightingale | XS.VLDL.P | Concentrat | T | G | 0.06784 | 0.012176 | 4.11E-08 |
| rs2519093 | let-7b-5p | trans | 1.82E-25 | Nightingale | XS.VLDL.P | Concentrat | T | C | 0.074723 | 0.012894 | 1.16E-08 |
| rs532436 | miR-139-5p | trans | 6.95E-37 | Nightingale | XS.VLDL.P | Concentrat | A | G | 0.076221 | 0.012829 | 4.92E-09 |
| rs649129 | let-7a-5p | trans | 1.48E-12 | Nightingale | XS.VLDL.P | Concentrat | T | C | 0.069004 | 0.012332 | 3.59E-08 |
| rs532436 | miR-195-5p | trans | 3.89E-18 | Nightingale | XS.VLDL.P | Concentrat | A | G | 0.076221 | 0.012829 | 4.92E-09 |
| rs2519093 | miR-10b-5p | trans | 3.13E-35 | Nightingale | XS.VLDL.P | Concentrat | T | C | 0.074723 | 0.012894 | 1.16E-08 |
| rs600038 | miR-126-3p | trans | 1.80E-37 | Nightingale | XS.VLDL.P | Concentrat | C | T | 0.068852 | 0.012329 | 3.82E-08 |
| rs495828 | miR-10b-5p | trans | 2.56E-34 | Nightingale | XS.VLDL.P | Concentrat | T | G | 0.06784 | 0.012176 | 4.11E-08 |
| rs649129 | miR-126-3p | trans | 1.95E-37 | Nightingale | XS.VLDL.P | Concentrat | T | C | 0.069004 | 0.012332 | 3.59E-08 |
| rs532436 | miR-10b-5p | trans | 5.20E-36 | Nightingale | XS.VLDL.P | Concentrat | A | G | 0.076221 | 0.012829 | 4.92E-09 |
| rs550057 | miR-126-3p | trans | 5.01E-31 | Nightingale | XS.VLDL.P | Concentrat | T | C | 0.065357 | 0.011295 | 1.22E-08 |
| rs2519093 | let-7c-5p | trans | 4.94E-22 | Nightingale | XS.VLDL.P | Concentrat | T | C | 0.074723 | 0.012894 | 1.16E-08 |
| rs649129 | miR-139-5p | trans | 1.34E-34 | Nightingale | XS.VLDL.P | Concentrat | T | C | 0.069004 | 0.012332 | 3.59E-08 |
| rs649129 | miR-10b-5p | trans | 2.54E-34 | Nightingale | XS.VLDL.P | Concentrat | T | C | 0.069004 | 0.012332 | 3.59E-08 |
| rs2519093 | miR-139-5p | trans | 1.63E-36 | Nightingale | XS.VLDL.P | Concentrat | T | C | 0.074723 | 0.012894 | 1.16E-08 |
| rs550057 | miR-139-5p | trans | 5.52E-32 | Nightingale | XS.VLDL.P | Concentrat | T | C | 0.065357 | 0.011295 | 1.22E-08 |
| rs2519093 | let-7a-5p | trans | 1.29E-11 | Nightingale | XS.VLDL.P | Concentrat | T | C | 0.074723 | 0.012894 | 1.16E-08 |
| rs532436 | let-7c-5p | trans | 2.13E-22 | Nightingale | XS.VLDL.P | Concentrat | A | G | 0.076221 | 0.012829 | 4.92E-09 |
| rs532436 | let-7b-5p | trans | 1.04E-25 | Nightingale | XS.VLDL.P | Concentrat | A | G | 0.076221 | 0.012829 | 4.92E-09 |
| rs495828 | let-7a-5p | trans | 1.53E-12 | Nightingale | XS.VLDL.P | Concentrat | T | G | 0.06784 | 0.012176 | 4.11E-08 |
| rs2519093 | miR-195-5p | trans | 7.49E-18 | Nightingale | XS.VLDL.P | Concentrat | T | C | 0.074723 | 0.012894 | 1.16E-08 |
| rs600038 | miR-139-5p | trans | 7.68E-35 | Nightingale | XS.VLDL.P | Concentrat | C | T | 0.068852 | 0.012329 | 3.82E-08 |
| rs2519093 | miR-126-3p | trans | 1.90E-40 | Nightingale | IDL.FC | Free choles | T | C | 0.074858 | 0.012388 | 2.47E-09 |
| rs649129 | let-7a-5p | trans | 1.48E-12 | Nightingale | IDL.FC | Free choles | T | C | 0.070778 | 0.011901 | 4.36E-09 |
| rs532436 | let-7c-5p | trans | 2.13E-22 | Nightingale | IDL.FC | Free choles | A | G | 0.076427 | 0.012349 | 1.01E-09 |
| rs600038 | miR-126-3p | trans | 1.80E-37 | Nightingale | IDL.FC | Free choles | C | T | 0.070293 | 0.011892 | 5.42E-09 |
| rs649129 | miR-126-3p | trans | 1.95E-37 | Nightingale | IDL.FC | Free choles | T | C | 0.070778 | 0.011901 | 4.36E-09 |
| rs600038 | let-7a-5p | trans | 1.48E-12 | Nightingale | IDL.FC | Free choles | C | T | 0.070293 | 0.011892 | 5.42E-09 |
| rs600038 | miR-139-5p | trans | 7.68E-35 | Nightingale | IDL.FC | Free choles | C | T | 0.070293 | 0.011892 | 5.42E-09 |
| rs2519093 | miR-139-5p | trans | 1.63E-36 | Nightingale | IDL.FC | Free choles | T | C | 0.074858 | 0.012388 | 2.47E-09 |
| rs600038 | miR-10b-5p | trans | 2.32E-34 | Nightingale | IDL.FC | Free choles | C | T | 0.070293 | 0.011892 | 5.42E-09 |
| rs2519093 | miR-10b-5p | trans | 3.13E-35 | Nightingale | IDL.FC | Free choles | T | C | 0.074858 | 0.012388 | 2.47E-09 |
| rs495828 | miR-10b-5p | trans | 2.56E-34 | Nightingale | IDL.FC | Free choles | T | G | 0.070792 | 0.011761 | 2.84E-09 |
| rs532436 | miR-126-3p | trans | 2.49E-40 | Nightingale | IDL.FC | Free choles | A | G | 0.076427 | 0.012349 | 1.01E-09 |
| rs649129 | miR-139-5p | trans | 1.34E-34 | Nightingale | IDL.FC | Free choles | T | C | 0.070778 | 0.011901 | 4.36E-09 |
| rs532436 | miR-139-5p | trans | 6.95E-37 | Nightingale | IDL.FC | Free choles | A | G | 0.076427 | 0.012349 | 1.01E-09 |
| rs495828 | miR-139-5p | trans | 1.46E-34 | Nightingale | IDL.FC | Free choles | T | G | 0.070792 | 0.011761 | 2.84E-09 |
| rs532436 | miR-195-5p | trans | 3.89E-18 | Nightingale | IDL.FC | Free choles | A | G | 0.076427 | 0.012349 | 1.01E-09 |
| rs2519093 | let-7c-5p | trans | 4.94E-22 | Nightingale | IDL.FC | Free choles | T | C | 0.074858 | 0.012388 | 2.47E-09 |
| rs2519093 | let-7b-5p | trans | 1.82E-25 | Nightingale | IDL.FC | Free choles | T | C | 0.074858 | 0.012388 | 2.47E-09 |
| rs532436 | let-7b-5p | trans | 1.04E-25 | Nightingale | IDL.FC | Free choles | A | G | 0.076427 | 0.012349 | 1.01E-09 |
| rs600038 | let-7b-5p | trans | 1.02E-24 | Nightingale | IDL.FC | Free choles | C | T | 0.070293 | 0.011892 | 5.42E-09 |
| rs495828 | miR-126-3p | trans | 2.00E-37 | Nightingale | IDL.FC | Free choles | T | G | 0.070792 | 0.011761 | 2.84E-09 |
| rs532436 | miR-10b-5p | trans | 5.20E-36 | Nightingale | IDL.FC | Free choles | A | G | 0.076427 | 0.012349 | 1.01E-09 |
| rs2519093 | let-7a-5p | trans | 1.29E-11 | Nightingale | IDL.FC | Free choles | T | C | 0.074858 | 0.012388 | 2.47E-09 |
| rs2519093 | miR-195-5p | trans | 7.49E-18 | Nightingale | IDL.FC | Free choles | T | C | 0.074858 | 0.012388 | 2.47E-09 |
| rs495828 | let-7a-5p | trans | 1.53E-12 | Nightingale | IDL.FC | Free choles | T | G | 0.070792 | 0.011761 | 2.84E-09 |
| rs649129 | miR-10b-5p | trans | 2.54E-34 | Nightingale | IDL.FC | Free choles | T | C | 0.070778 | 0.011901 | 4.36E-09 |
| rs2519093 | let-7a-5p | trans | 1.29E-11 | Nightingale | L.LDL.FC | Free choles | T | C | 0.075458 | 0.012377 | 1.91E-09 |
| rs532436 | let-7b-5p | trans | 1.04E-25 | Nightingale | L.LDL.FC | Free choles | A | G | 0.076272 | 0.012338 | 1.14E-09 |
| rs649129 | miR-10b-5p | trans | 2.54E-34 | Nightingale | L.LDL.FC | Free choles | T | C | 0.073994 | 0.01189 | 8.80E-10 |
| rs532436 | miR-126-3p | trans | 2.49E-40 | Nightingale | L.LDL.FC | Free choles | A | G | 0.076272 | 0.012338 | 1.14E-09 |
| rs532436 | let-7c-5p | trans | 2.13E-22 | Nightingale | L.LDL.FC | Free choles | A | G | 0.076272 | 0.012338 | 1.14E-09 |
| rs600038 | miR-126-3p | trans | 1.80E-37 | Nightingale | L.LDL.FC | Free choles | C | T | 0.073478 | 0.011881 | 1.12E-09 |
| rs495828 | miR-139-5p | trans | 1.46E-34 | Nightingale | L.LDL.FC | Free choles | T | G | 0.074325 | 0.011751 | 4.67E-10 |
| rs532436 | miR-10b-5p | trans | 5.20E-36 | Nightingale | L.LDL.FC | Free choles | A | G | 0.076272 | 0.012338 | 1.14E-09 |
| rs600038 | let-7b-5p | trans | 1.02E-24 | Nightingale | L.LDL.FC | Free choles | C | T | 0.073478 | 0.011881 | 1.12E-09 |
| rs600038 | let-7a-5p | trans | 1.48E-12 | Nightingale | L.LDL.FC | Free choles | C | T | 0.073478 | 0.011881 | 1.12E-09 |
| rs550057 | miR-126-3p | trans | 5.01E-31 | Nightingale | L.LDL.FC | Free choles | T | C | 0.06219 | 0.010884 | 1.82E-08 |
| rs2519093 | let-7c-5p | trans | 4.94E-22 | Nightingale | L.LDL.FC | Free choles | T | C | 0.075458 | 0.012377 | 1.91E-09 |
| rs2519093 | miR-10b-5p | trans | 3.13E-35 | Nightingale | L.LDL.FC | Free choles | T | C | 0.075458 | 0.012377 | 1.91E-09 |
| rs495828 | miR-10b-5p | trans | 2.56E-34 | Nightingale | L.LDL.FC | Free choles | T | G | 0.074325 | 0.011751 | 4.67E-10 |

| | | | | | | | | | | |
|---|---|---|---|---|---|---|---|---|---|---|
| rs532436 | miR-195-5p | trans | 3.89E-18 | Nightingale | L.LDL.FC | Free choles | A | G | 0.076272 | 0.012338 | 1.14E-09 |
| rs495828 | miR-126-3p | trans | 2.00E-37 | Nightingale | L.LDL.FC | Free choles | T | G | 0.074325 | 0.011751 | 4.67E-10 |
| rs600038 | miR-139-5p | trans | 7.68E-35 | Nightingale | L.LDL.FC | Free choles | C | T | 0.073478 | 0.011881 | 1.12E-09 |
| rs495828 | let-7a-5p | trans | 1.53E-12 | Nightingale | L.LDL.FC | Free choles | T | G | 0.074325 | 0.011751 | 4.67E-10 |
| rs649129 | miR-126-3p | trans | 1.95E-37 | Nightingale | L.LDL.FC | Free choles | T | C | 0.073994 | 0.01189 | 8.80E-10 |
| rs2519093 | let-7b-5p | trans | 1.82E-25 | Nightingale | L.LDL.FC | Free choles | T | C | 0.075458 | 0.012377 | 1.91E-09 |
| rs550057 | miR-139-5p | trans | 5.52E-32 | Nightingale | L.LDL.FC | Free choles | T | C | 0.06219 | 0.010884 | 1.82E-08 |
| rs600038 | miR-10b-5p | trans | 2.32E-34 | Nightingale | L.LDL.FC | Free choles | C | T | 0.073478 | 0.011881 | 1.12E-09 |
| rs649129 | let-7a-5p | trans | 1.48E-12 | Nightingale | L.LDL.FC | Free choles | T | C | 0.073994 | 0.01189 | 8.80E-10 |
| rs550057 | let-7b-5p | trans | 2.70E-19 | Nightingale | L.LDL.FC | Free choles | T | C | 0.06219 | 0.010884 | 1.82E-08 |
| rs2519093 | miR-195-5p | trans | 7.49E-18 | Nightingale | L.LDL.FC | Free choles | T | C | 0.075458 | 0.012377 | 1.91E-09 |
| rs532436 | miR-139-5p | trans | 6.95E-37 | Nightingale | L.LDL.FC | Free choles | A | G | 0.076272 | 0.012338 | 1.14E-09 |
| rs2519093 | miR-126-3p | trans | 1.90E-40 | Nightingale | L.LDL.FC | Free choles | T | C | 0.075458 | 0.012377 | 1.91E-09 |
| rs550057 | miR-10b-5p | trans | 1.61E-27 | Nightingale | L.LDL.FC | Free choles | T | C | 0.06219 | 0.010884 | 1.82E-08 |
| rs2519093 | miR-139-5p | trans | 1.63E-36 | Nightingale | L.LDL.FC | Free choles | T | C | 0.075458 | 0.012377 | 1.91E-09 |
| rs649129 | miR-139-5p | trans | 1.34E-34 | Nightingale | L.LDL.FC | Free choles | T | C | 0.073994 | 0.01189 | 8.80E-10 |
| rs2519093 | let-7b-5p | trans | 1.82E-25 | Nightingale | IDL.PL | Phospholip | T | C | 0.074614 | 0.012391 | 2.78E-09 |
| rs2519093 | miR-10b-5p | trans | 3.13E-35 | Nightingale | IDL.PL | Phospholip | T | C | 0.074614 | 0.012391 | 2.78E-09 |
| rs2519093 | miR-139-5p | trans | 1.63E-36 | Nightingale | IDL.PL | Phospholip | T | C | 0.074614 | 0.012391 | 2.78E-09 |
| rs550057 | let-7b-5p | trans | 2.70E-19 | Nightingale | IDL.PL | Phospholip | T | C | 0.061516 | 0.010897 | 2.51E-08 |
| rs532436 | miR-139-5p | trans | 6.95E-37 | Nightingale | IDL.PL | Phospholip | A | G | 0.076039 | 0.012352 | 1.23E-09 |
| rs600038 | miR-126-3p | trans | 1.80E-37 | Nightingale | IDL.PL | Phospholip | C | T | 0.068903 | 0.011895 | 1.08E-08 |
| rs2519093 | let-7c-5p | trans | 4.94E-22 | Nightingale | IDL.PL | Phospholip | T | C | 0.074614 | 0.012391 | 2.78E-09 |
| rs495828 | miR-139-5p | trans | 1.46E-34 | Nightingale | IDL.PL | Phospholip | T | G | 0.069638 | 0.011765 | 5.14E-09 |
| rs532436 | miR-126-3p | trans | 2.49E-40 | Nightingale | IDL.PL | Phospholip | A | G | 0.076039 | 0.012352 | 1.23E-09 |
| rs495828 | let-7a-5p | trans | 1.53E-12 | Nightingale | IDL.PL | Phospholip | T | G | 0.069638 | 0.011765 | 5.14E-09 |
| rs649129 | let-7a-5p | trans | 1.48E-12 | Nightingale | IDL.PL | Phospholip | T | C | 0.069591 | 0.011904 | 7.90E-09 |
| rs600038 | miR-10b-5p | trans | 2.32E-34 | Nightingale | IDL.PL | Phospholip | C | T | 0.068903 | 0.011895 | 1.08E-08 |
| rs532436 | miR-10b-5p | trans | 5.20E-36 | Nightingale | IDL.PL | Phospholip | A | G | 0.076039 | 0.012352 | 1.23E-09 |
| rs600038 | let-7a-5p | trans | 1.48E-12 | Nightingale | IDL.PL | Phospholip | C | T | 0.068903 | 0.011895 | 1.08E-08 |
| rs2519093 | let-7a-5p | trans | 1.29E-11 | Nightingale | IDL.PL | Phospholip | T | C | 0.074614 | 0.012391 | 2.78E-09 |
| rs495828 | miR-126-3p | trans | 2.00E-37 | Nightingale | IDL.PL | Phospholip | T | G | 0.069638 | 0.011765 | 5.14E-09 |
| rs532436 | miR-195-5p | trans | 3.89E-18 | Nightingale | IDL.PL | Phospholip | A | G | 0.076039 | 0.012352 | 1.23E-09 |
| rs550057 | miR-139-5p | trans | 5.52E-32 | Nightingale | IDL.PL | Phospholip | T | C | 0.061516 | 0.010897 | 2.51E-08 |
| rs600038 | let-7b-5p | trans | 1.02E-24 | Nightingale | IDL.PL | Phospholip | C | T | 0.068903 | 0.011895 | 1.08E-08 |
| rs2519093 | miR-126-3p | trans | 1.90E-40 | Nightingale | IDL.PL | Phospholip | T | C | 0.074614 | 0.012391 | 2.78E-09 |
| rs600038 | miR-139-5p | trans | 7.68E-35 | Nightingale | IDL.PL | Phospholip | C | T | 0.068903 | 0.011895 | 1.08E-08 |
| rs550057 | miR-126-3p | trans | 5.01E-31 | Nightingale | IDL.PL | Phospholip | T | C | 0.061516 | 0.010897 | 2.51E-08 |
| rs649129 | miR-126-3p | trans | 1.95E-37 | Nightingale | IDL.PL | Phospholip | T | C | 0.069591 | 0.011904 | 7.90E-09 |
| rs532436 | let-7c-5p | trans | 2.13E-22 | Nightingale | IDL.PL | Phospholip | A | G | 0.076039 | 0.012352 | 1.23E-09 |
| rs550057 | miR-10b-5p | trans | 1.61E-27 | Nightingale | IDL.PL | Phospholip | T | C | 0.061516 | 0.010897 | 2.51E-08 |
| rs495828 | miR-10b-5p | trans | 2.56E-34 | Nightingale | IDL.PL | Phospholip | T | G | 0.069638 | 0.011765 | 5.14E-09 |
| rs532436 | let-7b-5p | trans | 1.04E-25 | Nightingale | IDL.PL | Phospholip | A | G | 0.076039 | 0.012352 | 1.23E-09 |
| rs2519093 | miR-195-5p | trans | 7.49E-18 | Nightingale | IDL.PL | Phospholip | T | C | 0.074614 | 0.012391 | 2.78E-09 |
| rs649129 | miR-10b-5p | trans | 2.54E-34 | Nightingale | IDL.PL | Phospholip | T | C | 0.069591 | 0.011904 | 7.90E-09 |
| rs649129 | miR-139-5p | trans | 1.34E-34 | Nightingale | IDL.PL | Phospholip | T | C | 0.069591 | 0.011904 | 7.90E-09 |
| rs2519093 | miR-195-5p | trans | 7.49E-18 | Nightingale | L.LDL.PL | Phospholip | T | C | 0.076902 | 0.012383 | 9.21E-10 |
| rs649129 | miR-126-3p | trans | 1.95E-37 | Nightingale | L.LDL.PL | Phospholip | T | C | 0.073743 | 0.011896 | 9.88E-10 |
| rs495828 | miR-139-5p | trans | 1.46E-34 | Nightingale | L.LDL.PL | Phospholip | T | G | 0.073952 | 0.011759 | 5.63E-10 |
| rs600038 | let-7a-5p | trans | 1.48E-12 | Nightingale | L.LDL.PL | Phospholip | C | T | 0.07316 | 0.011888 | 1.30E-09 |
| rs2519093 | let-7a-5p | trans | 1.29E-11 | Nightingale | L.LDL.PL | Phospholip | T | C | 0.076902 | 0.012383 | 9.21E-10 |
| rs600038 | miR-10b-5p | trans | 2.32E-34 | Nightingale | L.LDL.PL | Phospholip | C | T | 0.07316 | 0.011888 | 1.30E-09 |
| rs600038 | miR-139-5p | trans | 7.68E-35 | Nightingale | L.LDL.PL | Phospholip | C | T | 0.07316 | 0.011888 | 1.30E-09 |
| rs2519093 | miR-126-3p | trans | 1.90E-40 | Nightingale | L.LDL.PL | Phospholip | T | C | 0.076902 | 0.012383 | 9.21E-10 |
| rs532436 | miR-126-3p | trans | 2.49E-40 | Nightingale | L.LDL.PL | Phospholip | A | G | 0.077502 | 0.012344 | 6.03E-10 |
| rs600038 | let-7b-5p | trans | 1.02E-24 | Nightingale | L.LDL.PL | Phospholip | C | T | 0.07316 | 0.011888 | 1.30E-09 |
| rs550057 | let-7b-5p | trans | 2.70E-19 | Nightingale | L.LDL.PL | Phospholip | T | C | 0.064195 | 0.010889 | 6.16E-09 |
| rs649129 | miR-10b-5p | trans | 2.54E-34 | Nightingale | L.LDL.PL | Phospholip | T | C | 0.073743 | 0.011896 | 9.88E-10 |
| rs550057 | miR-126-3p | trans | 5.01E-31 | Nightingale | L.LDL.PL | Phospholip | T | C | 0.064195 | 0.010889 | 6.16E-09 |
| rs550057 | miR-139-5p | trans | 5.52E-32 | Nightingale | L.LDL.PL | Phospholip | T | C | 0.064195 | 0.010889 | 6.16E-09 |
| rs2519093 | miR-139-5p | trans | 1.63E-36 | Nightingale | L.LDL.PL | Phospholip | T | C | 0.076902 | 0.012383 | 9.21E-10 |
| rs532436 | miR-139-5p | trans | 6.95E-37 | Nightingale | L.LDL.PL | Phospholip | A | G | 0.077502 | 0.012344 | 6.03E-10 |
| rs532436 | miR-195-5p | trans | 3.89E-18 | Nightingale | L.LDL.PL | Phospholip | A | G | 0.077502 | 0.012344 | 6.03E-10 |

| | | | | | | | | | | | |
|---|---|---|---|---|---|---|---|---|---|---|---|
| rs532436 | let-7b-5p | trans | 1.04E-25 | Nightingale | L.LDL.PL | Phospholip | A | G | 0.077502 | 0.012344 | 6.03E-10 |
| rs495828 | let-7a-5p | trans | 1.53E-12 | Nightingale | L.LDL.PL | Phospholip | T | G | 0.073952 | 0.011759 | 5.63E-10 |
| rs649129 | miR-139-5p | trans | 1.34E-34 | Nightingale | L.LDL.PL | Phospholip | T | C | 0.073743 | 0.011896 | 9.88E-10 |
| rs532436 | let-7c-5p | trans | 2.13E-22 | Nightingale | L.LDL.PL | Phospholip | A | G | 0.077502 | 0.012344 | 6.03E-10 |
| rs2519093 | let-7c-5p | trans | 4.94E-22 | Nightingale | L.LDL.PL | Phospholip | T | C | 0.076902 | 0.012383 | 9.21E-10 |
| rs649129 | let-7a-5p | trans | 1.48E-12 | Nightingale | L.LDL.PL | Phospholip | T | C | 0.073743 | 0.011896 | 9.88E-10 |
| rs550057 | miR-10b-5p | trans | 1.61E-27 | Nightingale | L.LDL.PL | Phospholip | T | C | 0.064195 | 0.010889 | 6.16E-09 |
| rs2519093 | let-7b-5p | trans | 1.82E-25 | Nightingale | L.LDL.PL | Phospholip | T | C | 0.076902 | 0.012383 | 9.21E-10 |
| rs495828 | miR-10b-5p | trans | 2.56E-34 | Nightingale | L.LDL.PL | Phospholip | T | G | 0.073952 | 0.011759 | 5.63E-10 |
| rs495828 | miR-126-3p | trans | 2.00E-37 | Nightingale | L.LDL.PL | Phospholip | T | G | 0.073952 | 0.011759 | 5.63E-10 |
| rs532436 | miR-10b-5p | trans | 5.20E-36 | Nightingale | L.LDL.PL | Phospholip | A | G | 0.077502 | 0.012344 | 6.03E-10 |
| rs600038 | miR-126-3p | trans | 1.80E-37 | Nightingale | L.LDL.PL | Phospholip | C | T | 0.07316 | 0.011888 | 1.30E-09 |
| rs2519093 | miR-10b-5p | trans | 3.13E-35 | Nightingale | L.LDL.PL | Phospholip | T | C | 0.076902 | 0.012383 | 9.21E-10 |
| rs2519093 | miR-126-3p | trans | 1.90E-40 | Nightingale | XS.VLDL.PL | Phospholip | T | C | 0.080266 | 0.012895 | 8.39E-10 |
| rs532436 | let-7c-5p | trans | 2.13E-22 | Nightingale | XS.VLDL.PL | Phospholip | A | G | 0.082256 | 0.012829 | 2.59E-10 |
| rs649129 | let-7a-5p | trans | 1.48E-12 | Nightingale | XS.VLDL.PL | Phospholip | T | C | 0.074089 | 0.012331 | 3.14E-09 |
| rs600038 | miR-10b-5p | trans | 2.32E-34 | Nightingale | XS.VLDL.PL | Phospholip | C | T | 0.073833 | 0.012329 | 3.53E-09 |
| rs550057 | let-7b-5p | trans | 2.70E-19 | Nightingale | XS.VLDL.PL | Phospholip | T | C | 0.066376 | 0.011296 | 6.89E-09 |
| rs649129 | miR-126-3p | trans | 1.95E-37 | Nightingale | XS.VLDL.PL | Phospholip | T | C | 0.074089 | 0.012331 | 3.14E-09 |
| rs2519093 | miR-195-5p | trans | 7.49E-18 | Nightingale | XS.VLDL.PL | Phospholip | T | C | 0.080266 | 0.012895 | 8.39E-10 |
| rs495828 | miR-126-3p | trans | 2.00E-37 | Nightingale | XS.VLDL.PL | Phospholip | T | G | 0.073247 | 0.012179 | 3.03E-09 |
| rs2519093 | let-7a-5p | trans | 1.29E-11 | Nightingale | XS.VLDL.PL | Phospholip | T | C | 0.080266 | 0.012895 | 8.39E-10 |
| rs532436 | miR-10b-5p | trans | 5.20E-36 | Nightingale | XS.VLDL.PL | Phospholip | A | G | 0.082256 | 0.012829 | 2.59E-10 |
| rs495828 | miR-139-5p | trans | 1.46E-34 | Nightingale | XS.VLDL.PL | Phospholip | T | G | 0.073247 | 0.012179 | 3.03E-09 |
| rs2519093 | let-7c-5p | trans | 4.94E-22 | Nightingale | XS.VLDL.PL | Phospholip | T | C | 0.080266 | 0.012895 | 8.39E-10 |
| rs532436 | let-7b-5p | trans | 1.04E-25 | Nightingale | XS.VLDL.PL | Phospholip | A | G | 0.082256 | 0.012829 | 2.59E-10 |
| rs495828 | let-7a-5p | trans | 1.53E-12 | Nightingale | XS.VLDL.PL | Phospholip | T | G | 0.073247 | 0.012179 | 3.03E-09 |
| rs600038 | miR-126-3p | trans | 1.80E-37 | Nightingale | XS.VLDL.PL | Phospholip | C | T | 0.073833 | 0.012329 | 3.53E-09 |
| rs550057 | miR-139-5p | trans | 5.52E-32 | Nightingale | XS.VLDL.PL | Phospholip | T | C | 0.066376 | 0.011296 | 6.89E-09 |
| rs2519093 | let-7b-5p | trans | 1.82E-25 | Nightingale | XS.VLDL.PL | Phospholip | T | C | 0.080266 | 0.012895 | 8.39E-10 |
| rs532436 | miR-139-5p | trans | 6.95E-37 | Nightingale | XS.VLDL.PL | Phospholip | A | G | 0.082256 | 0.012829 | 2.59E-10 |
| rs600038 | miR-139-5p | trans | 7.68E-35 | Nightingale | XS.VLDL.PL | Phospholip | C | T | 0.073833 | 0.012329 | 3.53E-09 |
| rs649129 | miR-139-5p | trans | 1.34E-34 | Nightingale | XS.VLDL.PL | Phospholip | T | C | 0.074089 | 0.012331 | 3.14E-09 |
| rs532436 | miR-126-3p | trans | 2.49E-40 | Nightingale | XS.VLDL.PL | Phospholip | A | G | 0.082256 | 0.012829 | 2.59E-10 |
| rs550057 | miR-126-3p | trans | 5.01E-31 | Nightingale | XS.VLDL.PL | Phospholip | T | C | 0.066376 | 0.011296 | 6.89E-09 |
| rs649129 | miR-10b-5p | trans | 2.54E-34 | Nightingale | XS.VLDL.PL | Phospholip | T | C | 0.074089 | 0.012331 | 3.14E-09 |
| rs532436 | miR-195-5p | trans | 3.89E-18 | Nightingale | XS.VLDL.PL | Phospholip | A | G | 0.082256 | 0.012829 | 2.59E-10 |
| rs2519093 | miR-139-5p | trans | 1.63E-36 | Nightingale | XS.VLDL.PL | Phospholip | T | C | 0.080266 | 0.012895 | 8.39E-10 |
| rs600038 | let-7a-5p | trans | 1.48E-12 | Nightingale | XS.VLDL.PL | Phospholip | C | T | 0.073833 | 0.012329 | 3.53E-09 |
| rs550057 | miR-10b-5p | trans | 1.61E-27 | Nightingale | XS.VLDL.PL | Phospholip | T | C | 0.066376 | 0.011296 | 6.89E-09 |
| rs600038 | let-7b-5p | trans | 1.02E-24 | Nightingale | XS.VLDL.PL | Phospholip | C | T | 0.073833 | 0.012329 | 3.53E-09 |
| rs495828 | miR-10b-5p | trans | 2.56E-34 | Nightingale | XS.VLDL.PL | Phospholip | T | G | 0.073247 | 0.012179 | 3.03E-09 |
| rs2519093 | miR-10b-5p | trans | 3.13E-35 | Nightingale | XS.VLDL.PL | Phospholip | T | C | 0.080266 | 0.012895 | 8.39E-10 |
| rs532436 | miR-195-5p | trans | 3.89E-18 | Nightingale | IDL.L | Total lipids | A | G | 0.084425 | 0.01282 | 8.77E-11 |
| rs495828 | miR-10b-5p | trans | 2.56E-34 | Nightingale | IDL.L | Total lipids | T | G | 0.078164 | 0.01217 | 2.51E-10 |
| rs2519093 | miR-126-3p | trans | 1.90E-40 | Nightingale | IDL.L | Total lipids | T | C | 0.083299 | 0.012885 | 1.92E-10 |
| rs2519093 | miR-139-5p | trans | 1.63E-36 | Nightingale | IDL.L | Total lipids | T | C | 0.083299 | 0.012885 | 1.92E-10 |
| rs2519093 | let-7a-5p | trans | 1.29E-11 | Nightingale | IDL.L | Total lipids | T | C | 0.083299 | 0.012885 | 1.92E-10 |
| rs2519093 | let-7b-5p | trans | 1.82E-25 | Nightingale | IDL.L | Total lipids | T | C | 0.083299 | 0.012885 | 1.92E-10 |
| rs532436 | miR-139-5p | trans | 6.95E-37 | Nightingale | IDL.L | Total lipids | A | G | 0.084425 | 0.01282 | 8.77E-11 |
| rs649129 | miR-10b-5p | trans | 2.54E-34 | Nightingale | IDL.L | Total lipids | T | C | 0.078692 | 0.012321 | 3.15E-10 |
| rs600038 | miR-126-3p | trans | 1.80E-37 | Nightingale | IDL.L | Total lipids | C | T | 0.078247 | 0.012318 | 3.93E-10 |
| rs550057 | miR-139-5p | trans | 5.52E-32 | Nightingale | IDL.L | Total lipids | T | C | 0.068346 | 0.011288 | 2.46E-09 |
| rs550057 | miR-10b-5p | trans | 1.61E-27 | Nightingale | IDL.L | Total lipids | T | C | 0.068346 | 0.011288 | 2.46E-09 |
| rs550057 | let-7b-5p | trans | 2.70E-19 | Nightingale | IDL.L | Total lipids | T | C | 0.068346 | 0.011288 | 2.46E-09 |
| rs532436 | let-7c-5p | trans | 2.13E-22 | Nightingale | IDL.L | Total lipids | A | G | 0.084425 | 0.01282 | 8.77E-11 |
| rs550057 | miR-126-3p | trans | 5.01E-31 | Nightingale | IDL.L | Total lipids | T | C | 0.068346 | 0.011288 | 2.46E-09 |
| rs495828 | miR-126-3p | trans | 2.00E-37 | Nightingale | IDL.L | Total lipids | T | G | 0.078164 | 0.01217 | 2.51E-10 |
| rs532436 | miR-10b-5p | trans | 5.20E-36 | Nightingale | IDL.L | Total lipids | A | G | 0.084425 | 0.01282 | 8.77E-11 |
| rs2519093 | let-7c-5p | trans | 4.94E-22 | Nightingale | IDL.L | Total lipids | T | C | 0.083299 | 0.012885 | 1.92E-10 |
| rs495828 | miR-139-5p | trans | 1.46E-34 | Nightingale | IDL.L | Total lipids | T | G | 0.078164 | 0.01217 | 2.51E-10 |
| rs2519093 | miR-195-5p | trans | 7.49E-18 | Nightingale | IDL.L | Total lipids | T | C | 0.083299 | 0.012885 | 1.92E-10 |
| rs532436 | let-7b-5p | trans | 1.04E-25 | Nightingale | IDL.L | Total lipids | A | G | 0.084425 | 0.01282 | 8.77E-11 |

| | | | | | | | | | | |
|---|---|---|---|---|---|---|---|---|---|---|
| rs495828 | let-7a-5p | trans | 1.53E-12 | Nightingale | IDL.L | Total lipids T | G | 0.078164 | 0.01217 | 2.51E-10 |
| rs600038 | let-7a-5p | trans | 1.48E-12 | Nightingale | IDL.L | Total lipids C | T | 0.078247 | 0.012318 | 3.93E-10 |
| rs649129 | miR-126-3p | trans | 1.95E-37 | Nightingale | IDL.L | Total lipids T | C | 0.078692 | 0.012321 | 3.15E-10 |
| rs600038 | miR-10b-5p | trans | 2.32E-34 | Nightingale | IDL.L | Total lipids C | T | 0.078247 | 0.012318 | 3.93E-10 |
| rs600038 | miR-139-5p | trans | 7.68E-35 | Nightingale | IDL.L | Total lipids C | T | 0.078247 | 0.012318 | 3.93E-10 |
| rs649129 | let-7a-5p | trans | 1.48E-12 | Nightingale | IDL.L | Total lipids T | C | 0.078692 | 0.012321 | 3.15E-10 |
| rs532436 | miR-126-3p | trans | 2.49E-40 | Nightingale | IDL.L | Total lipids A | G | 0.084425 | 0.01282 | 8.77E-11 |
| rs2519093 | miR-10b-5p | trans | 3.13E-35 | Nightingale | IDL.L | Total lipids T | C | 0.083299 | 0.012885 | 1.92E-10 |
| rs649129 | miR-139-5p | trans | 1.34E-34 | Nightingale | IDL.L | Total lipids T | C | 0.078692 | 0.012321 | 3.15E-10 |
| rs600038 | let-7b-5p | trans | 1.02E-24 | Nightingale | IDL.L | Total lipids C | T | 0.078247 | 0.012318 | 3.93E-10 |
| rs2519093 | let-7a-5p | trans | 1.29E-11 | Nightingale | L.LDL.L | Total lipids T | C | 0.075633 | 0.012877 | 7.67E-09 |
| rs532436 | miR-195-5p | trans | 3.89E-18 | Nightingale | L.LDL.L | Total lipids A | G | 0.07602 | 0.012812 | 5.39E-09 |
| rs600038 | miR-10b-5p | trans | 2.32E-34 | Nightingale | L.LDL.L | Total lipids C | T | 0.07085 | 0.01231 | 1.52E-08 |
| rs2519093 | miR-195-5p | trans | 7.49E-18 | Nightingale | L.LDL.L | Total lipids T | C | 0.075633 | 0.012877 | 7.67E-09 |
| rs600038 | let-7a-5p | trans | 1.48E-12 | Nightingale | L.LDL.L | Total lipids C | T | 0.07085 | 0.01231 | 1.52E-08 |
| rs600038 | miR-139-5p | trans | 7.68E-35 | Nightingale | L.LDL.L | Total lipids C | T | 0.07085 | 0.01231 | 1.52E-08 |
| rs495828 | miR-126-3p | trans | 2.00E-37 | Nightingale | L.LDL.L | Total lipids T | G | 0.071589 | 0.012161 | 7.09E-09 |
| rs2519093 | miR-10b-5p | trans | 3.13E-35 | Nightingale | L.LDL.L | Total lipids T | C | 0.075633 | 0.012877 | 7.67E-09 |
| rs600038 | let-7b-5p | trans | 1.02E-24 | Nightingale | L.LDL.L | Total lipids C | T | 0.07085 | 0.01231 | 1.52E-08 |
| rs532436 | let-7c-5p | trans | 2.13E-22 | Nightingale | L.LDL.L | Total lipids A | G | 0.07602 | 0.012812 | 5.39E-09 |
| rs532436 | let-7b-5p | trans | 1.04E-25 | Nightingale | L.LDL.L | Total lipids A | G | 0.07602 | 0.012812 | 5.39E-09 |
| rs2519093 | miR-126-3p | trans | 1.90E-40 | Nightingale | L.LDL.L | Total lipids T | C | 0.075633 | 0.012877 | 7.67E-09 |
| rs649129 | miR-10b-5p | trans | 2.54E-34 | Nightingale | L.LDL.L | Total lipids T | C | 0.071226 | 0.012313 | 1.28E-08 |
| rs2519093 | miR-139-5p | trans | 1.63E-36 | Nightingale | L.LDL.L | Total lipids T | C | 0.075633 | 0.012877 | 7.67E-09 |
| rs2519093 | let-7c-5p | trans | 4.94E-22 | Nightingale | L.LDL.L | Total lipids T | C | 0.075633 | 0.012877 | 7.67E-09 |
| rs495828 | miR-10b-5p | trans | 2.56E-34 | Nightingale | L.LDL.L | Total lipids T | G | 0.071589 | 0.012161 | 7.09E-09 |
| rs649129 | let-7a-5p | trans | 1.48E-12 | Nightingale | L.LDL.L | Total lipids T | C | 0.071226 | 0.012313 | 1.28E-08 |
| rs495828 | let-7a-5p | trans | 1.53E-12 | Nightingale | L.LDL.L | Total lipids T | G | 0.071589 | 0.012161 | 7.09E-09 |
| rs649129 | miR-139-5p | trans | 1.34E-34 | Nightingale | L.LDL.L | Total lipids T | C | 0.071226 | 0.012313 | 1.28E-08 |
| rs495828 | miR-139-5p | trans | 1.46E-34 | Nightingale | L.LDL.L | Total lipids T | G | 0.071589 | 0.012161 | 7.09E-09 |
| rs532436 | miR-139-5p | trans | 6.95E-37 | Nightingale | L.LDL.L | Total lipids A | G | 0.07602 | 0.012812 | 5.39E-09 |
| rs600038 | miR-126-3p | trans | 1.80E-37 | Nightingale | L.LDL.L | Total lipids C | T | 0.07085 | 0.01231 | 1.52E-08 |
| rs532436 | miR-126-3p | trans | 2.49E-40 | Nightingale | L.LDL.L | Total lipids A | G | 0.07602 | 0.012812 | 5.39E-09 |
| rs532436 | miR-10b-5p | trans | 5.20E-36 | Nightingale | L.LDL.L | Total lipids A | G | 0.07602 | 0.012812 | 5.39E-09 |
| rs649129 | miR-126-3p | trans | 1.95E-37 | Nightingale | L.LDL.L | Total lipids T | C | 0.071226 | 0.012313 | 1.28E-08 |
| rs2519093 | let-7b-5p | trans | 1.82E-25 | Nightingale | L.LDL.L | Total lipids T | C | 0.075633 | 0.012877 | 7.67E-09 |
| rs649129 | miR-126-3p | trans | 1.95E-37 | Nightingale | XS.VLDL.L | Total lipids T | C | 0.073514 | 0.012332 | 4.24E-09 |
| rs532436 | let-7c-5p | trans | 2.13E-22 | Nightingale | XS.VLDL.L | Total lipids A | G | 0.080264 | 0.01283 | 7.07E-10 |
| rs2519093 | miR-126-3p | trans | 1.90E-40 | Nightingale | XS.VLDL.L | Total lipids T | C | 0.078795 | 0.012895 | 1.73E-09 |
| rs2519093 | let-7b-5p | trans | 1.82E-25 | Nightingale | XS.VLDL.L | Total lipids T | C | 0.078795 | 0.012895 | 1.73E-09 |
| rs2519093 | miR-195-5p | trans | 7.49E-18 | Nightingale | XS.VLDL.L | Total lipids T | C | 0.078795 | 0.012895 | 1.73E-09 |
| rs600038 | miR-10b-5p | trans | 2.32E-34 | Nightingale | XS.VLDL.L | Total lipids C | T | 0.073399 | 0.012329 | 4.45E-09 |
| rs550057 | let-7b-5p | trans | 2.70E-19 | Nightingale | XS.VLDL.L | Total lipids T | C | 0.067994 | 0.011296 | 3.00E-09 |
| rs2519093 | miR-10b-5p | trans | 3.13E-35 | Nightingale | XS.VLDL.L | Total lipids T | C | 0.078795 | 0.012895 | 1.73E-09 |
| rs532436 | let-7b-5p | trans | 1.04E-25 | Nightingale | XS.VLDL.L | Total lipids A | G | 0.080264 | 0.01283 | 7.07E-10 |
| rs2519093 | miR-139-5p | trans | 1.63E-36 | Nightingale | XS.VLDL.L | Total lipids T | C | 0.078795 | 0.012895 | 1.73E-09 |
| rs600038 | let-7b-5p | trans | 1.02E-24 | Nightingale | XS.VLDL.L | Total lipids C | T | 0.073399 | 0.012329 | 4.45E-09 |
| rs649129 | miR-10b-5p | trans | 2.54E-34 | Nightingale | XS.VLDL.L | Total lipids T | C | 0.073514 | 0.012332 | 4.24E-09 |
| rs600038 | let-7a-5p | trans | 1.48E-12 | Nightingale | XS.VLDL.L | Total lipids C | T | 0.073399 | 0.012329 | 4.45E-09 |
| rs495828 | miR-139-5p | trans | 1.46E-34 | Nightingale | XS.VLDL.L | Total lipids T | G | 0.072199 | 0.012178 | 5.15E-09 |
| rs495828 | miR-126-3p | trans | 2.00E-37 | Nightingale | XS.VLDL.L | Total lipids T | G | 0.072199 | 0.012178 | 5.15E-09 |
| rs532436 | miR-10b-5p | trans | 5.20E-36 | Nightingale | XS.VLDL.L | Total lipids A | G | 0.080264 | 0.01283 | 7.07E-10 |
| rs550057 | miR-126-3p | trans | 5.01E-31 | Nightingale | XS.VLDL.L | Total lipids T | C | 0.067994 | 0.011296 | 3.00E-09 |
| rs600038 | miR-126-3p | trans | 1.80E-37 | Nightingale | XS.VLDL.L | Total lipids C | T | 0.073399 | 0.012329 | 4.45E-09 |
| rs649129 | let-7a-5p | trans | 1.48E-12 | Nightingale | XS.VLDL.L | Total lipids T | C | 0.073514 | 0.012332 | 4.24E-09 |
| rs532436 | miR-195-5p | trans | 3.89E-18 | Nightingale | XS.VLDL.L | Total lipids A | G | 0.080264 | 0.01283 | 7.07E-10 |
| rs495828 | let-7a-5p | trans | 1.53E-12 | Nightingale | XS.VLDL.L | Total lipids T | G | 0.072199 | 0.012178 | 5.15E-09 |
| rs495828 | miR-10b-5p | trans | 2.56E-34 | Nightingale | XS.VLDL.L | Total lipids T | G | 0.072199 | 0.012178 | 5.15E-09 |
| rs649129 | miR-139-5p | trans | 1.34E-34 | Nightingale | XS.VLDL.L | Total lipids T | C | 0.073514 | 0.012332 | 4.24E-09 |
| rs550057 | miR-10b-5p | trans | 1.61E-27 | Nightingale | XS.VLDL.L | Total lipids T | C | 0.067994 | 0.011296 | 3.00E-09 |
| rs550057 | miR-139-5p | trans | 5.52E-32 | Nightingale | XS.VLDL.L | Total lipids T | C | 0.067994 | 0.011296 | 3.00E-09 |
| rs600038 | miR-139-5p | trans | 7.68E-35 | Nightingale | XS.VLDL.L | Total lipids C | T | 0.073399 | 0.012329 | 4.45E-09 |
| rs532436 | miR-139-5p | trans | 6.95E-37 | Nightingale | XS.VLDL.L | Total lipids A | G | 0.080264 | 0.01283 | 7.07E-10 |

| | | | | | | | | | | | |
|---|---|---|---|---|---|---|---|---|---|---|---|
| rs532436 | miR-126-3p | trans | 2.49E-40 | Nightingale | XS.VLDL.L | Total lipids | A | G | 0.080264 | 0.01283 | 7.07E-10 |
| rs2519093 | let-7c-5p | trans | 4.94E-22 | Nightingale | XS.VLDL.L | Total lipids | T | C | 0.078795 | 0.012895 | 1.73E-09 |
| rs2519093 | let-7a-5p | trans | 1.29E-11 | Nightingale | XS.VLDL.L | Total lipids | T | C | 0.078795 | 0.012895 | 1.73E-09 |
| rs102274 | miR-1908-5 | cis | 9.41E-09 | Nightingale | LA | NA | C | T | 0.082086 | 0.004257 | 4.30E-83 |
| rs102274 | miR-1908-5 | cis | 9.41E-09 | Nightingale | M_HDL_TG | NA | C | T | 0.049596 | 0.004287 | 1.00E-31 |
| rs102274 | miR-1908-5 | cis | 9.41E-09 | Nightingale | M_LDL_PL_ | NA | C | T | 0.045772 | 0.004338 | 4.20E-27 |
| rs102274 | miR-1908-5 | cis | 9.41E-09 | Nightingale | LDL_FC | NA | C | T | -0.06509 | 0.004294 | 1.10E-54 |
| rs102274 | miR-1908-5 | cis | 9.41E-09 | Nightingale | LDL_TG | NA | C | T | 0.029086 | 0.004293 | 2.60E-11 |
| rs102274 | miR-1908-5 | cis | 9.41E-09 | Nightingale | L_VLDL_PL_ | NA | C | T | 0.087012 | 0.004213 | 3.00E-98 |
| rs102274 | miR-1908-5 | cis | 9.41E-09 | Nightingale | XL_VLDL_T | NA | C | T | 0.056434 | 0.004152 | 9.80E-44 |
| rs102274 | miR-1908-5 | cis | 9.41E-09 | Nightingale | XL_VLDL_L | NA | C | T | 0.0509 | 0.004177 | 1.80E-35 |
| rs102274 | miR-1908-5 | cis | 9.41E-09 | Nightingale | Total_FA | NA | C | T | -0.02458 | 0.004306 | 3.50E-09 |
| rs102274 | miR-1908-5 | cis | 9.41E-09 | Nightingale | S_VLDL_PL | NA | C | T | -0.07884 | 0.004153 | 3.80E-84 |
| rs102274 | miR-1908-5 | cis | 9.41E-09 | Nightingale | M_HDL_TG | NA | C | T | 0.066073 | 0.004162 | 2.10E-59 |
| rs102274 | miR-1908-5 | cis | 9.41E-09 | Nightingale | L_VLDL_C | NA | C | T | 0.02821 | 0.004245 | 1.40E-11 |
| rs102274 | miR-1908-5 | cis | 9.41E-09 | Nightingale | XL_VLDL_F | NA | C | T | 0.042737 | 0.004204 | 4.10E-25 |
| rs102274 | miR-1908-5 | cis | 9.41E-09 | Nightingale | S_LDL_P | NA | C | T | -0.03339 | 0.004345 | 3.10E-15 |
| rs102274 | miR-1908-5 | cis | 9.41E-09 | Nightingale | VLDL_size | NA | C | T | 0.072288 | 0.004046 | 1.70E-75 |
| rs102274 | miR-1908-5 | cis | 9.41E-09 | Nightingale | XS_VLDL_F | NA | C | T | -0.05736 | 0.004265 | 5.30E-44 |
| rs102274 | miR-1908-5 | cis | 9.41E-09 | Nightingale | IDL_CE | NA | C | T | -0.07353 | 0.00419 | 5.70E-73 |
| rs102274 | miR-1908-5 | cis | 9.41E-09 | Nightingale | XS_VLDL_P | NA | C | T | -0.04492 | 0.004283 | 1.20E-27 |
| rs102274 | miR-1908-5 | cis | 9.41E-09 | Nightingale | HDL_size | NA | C | T | -0.07292 | 0.003846 | 1.10E-86 |
| rs102274 | miR-1908-5 | cis | 9.41E-09 | Nightingale | S_LDL_PL_ | NA | C | T | -0.06143 | 0.004332 | 9.90E-48 |
| rs102274 | miR-1908-5 | cis | 9.41E-09 | Nightingale | L_LDL_L | NA | C | T | -0.05506 | 0.004298 | 1.10E-39 |
| rs102274 | miR-1908-5 | cis | 9.41E-09 | Nightingale | L_LDL_TG_ | NA | C | T | 0.077854 | 0.004256 | 2.90E-76 |
| rs102274 | miR-1908-5 | cis | 9.41E-09 | Nightingale | S_VLDL_FC | NA | C | T | -0.02921 | 0.004342 | 5.40E-12 |
| rs102274 | miR-1908-5 | cis | 9.41E-09 | Nightingale | L_VLDL_CE | NA | C | T | -0.05936 | 0.004154 | 4.10E-49 |
| rs102274 | miR-1908-5 | cis | 9.41E-09 | Nightingale | XS_VLDL_P | NA | C | T | -0.03568 | 0.00428 | 2.20E-18 |
| rs102274 | miR-1908-5 | cis | 9.41E-09 | Nightingale | L_VLDL_TG | NA | C | T | 0.036878 | 0.004177 | 2.30E-19 |
| rs102274 | miR-1908-5 | cis | 9.41E-09 | Nightingale | L_HDL_FC_ | NA | C | T | -0.11319 | 0.004032 | 1.60E-188 |
| rs102274 | miR-1908-5 | cis | 9.41E-09 | Nightingale | Unsaturati | NA | C | T | -0.33829 | 0.004111 | 0 |
| rs102274 | miR-1908-5 | cis | 9.41E-09 | Nightingale | XL_VLDL_C | NA | C | T | 0.029603 | 0.004272 | 2.00E-12 |
| rs102274 | miR-1908-5 | cis | 9.41E-09 | Nightingale | MUFA_pct | NA | C | T | 0.196989 | 0.004154 | 0 |
| rs102274 | miR-1908-5 | cis | 9.41E-09 | Nightingale | IDL_TG_pct | NA | C | T | 0.088485 | 0.004193 | 1.70E-102 |
| rs102274 | miR-1908-5 | cis | 9.41E-09 | Nightingale | IDL_FC_pct | NA | C | T | -0.10349 | 0.004306 | 5.00E-129 |
| rs102274 | miR-1908-5 | cis | 9.41E-09 | Nightingale | M_LDL_TG | NA | C | T | 0.036088 | 0.004277 | 3.60E-17 |
| rs102274 | miR-1908-5 | cis | 9.41E-09 | Nightingale | M_VLDL_C | NA | C | T | -0.0763 | 0.004075 | 2.60E-81 |
| rs102274 | miR-1908-5 | cis | 9.41E-09 | Nightingale | S_LDL_CE_ | NA | C | T | 0.056462 | 0.004333 | 1.30E-40 |
| rs102274 | miR-1908-5 | cis | 9.41E-09 | Nightingale | XL_HDL_CE | NA | C | T | -0.02561 | 0.004338 | 8.80E-09 |
| rs102274 | miR-1908-5 | cis | 9.41E-09 | Nightingale | XXL_VLDL_ | NA | C | T | 0.044457 | 0.004364 | 2.80E-25 |
| rs102274 | miR-1908-5 | cis | 9.41E-09 | Nightingale | SFA | NA | C | T | -0.03223 | 0.004302 | 2.60E-14 |
| rs102274 | miR-1908-5 | cis | 9.41E-09 | Nightingale | non_HDL_( | NA | C | T | -0.04886 | 0.004317 | 3.70E-31 |
| rs102274 | miR-1908-5 | cis | 9.41E-09 | Nightingale | Phosphogly | NA | C | T | -0.0419 | 0.004112 | 3.50E-26 |
| rs102274 | miR-1908-5 | cis | 9.41E-09 | Nightingale | SFA_pct | NA | C | T | -0.04289 | 0.004282 | 1.30E-23 |
| rs102274 | miR-1908-5 | cis | 9.41E-09 | Nightingale | Total_CE | NA | C | T | -0.06799 | 0.004187 | 1.50E-62 |
| rs102274 | miR-1908-5 | cis | 9.41E-09 | Nightingale | S_LDL_TG | NA | C | T | 0.049764 | 0.004234 | 2.30E-32 |
| rs102274 | miR-1908-5 | cis | 9.41E-09 | Nightingale | Remnant_( | NA | C | T | -0.04536 | 0.004313 | 2.60E-27 |
| rs102274 | miR-1908-5 | cis | 9.41E-09 | Nightingale | DHA | NA | C | T | -0.28748 | 0.004183 | 0 |
| rs102274 | miR-1908-5 | cis | 9.41E-09 | Nightingale | IDL_C_pct | NA | C | T | -0.07528 | 0.004238 | 3.60E-72 |
| rs102274 | miR-1908-5 | cis | 9.41E-09 | Nightingale | XXL_VLDL_ | NA | C | T | -0.02632 | 0.004319 | 2.10E-10 |
| rs102274 | miR-1908-5 | cis | 9.41E-09 | Nightingale | S_LDL_PL | NA | C | T | -0.0522 | 0.00433 | 2.30E-35 |
| rs102274 | miR-1908-5 | cis | 9.41E-09 | Nightingale | M_LDL_C | NA | C | T | -0.03473 | 0.004349 | 3.50E-16 |
| rs102274 | miR-1908-5 | cis | 9.41E-09 | Nightingale | HDL_P | NA | C | T | -0.0358 | 0.004076 | 1.20E-19 |
| rs102274 | miR-1908-5 | cis | 9.41E-09 | Nightingale | LDL_L | NA | C | T | -0.04629 | 0.004326 | 4.40E-28 |
| rs102274 | miR-1908-5 | cis | 9.41E-09 | Nightingale | XL_VLDL_C | NA | C | T | -0.06394 | 0.00416 | 1.10E-54 |
| rs102274 | miR-1908-5 | cis | 9.41E-09 | Nightingale | XXL_VLDL_ | NA | C | T | 0.067188 | 0.004182 | 8.10E-61 |
| rs102274 | miR-1908-5 | cis | 9.41E-09 | Nightingale | XS_VLDL_T | NA | C | T | 0.097894 | 0.004123 | 2.40E-130 |
| rs102274 | miR-1908-5 | cis | 9.41E-09 | Nightingale | L_VLDL_P | NA | C | T | 0.041355 | 0.004191 | 7.90E-24 |
| rs102274 | miR-1908-5 | cis | 9.41E-09 | Nightingale | S_HDL_TG | NA | C | T | 0.068426 | 0.004209 | 3.70E-62 |
| rs102274 | miR-1908-5 | cis | 9.41E-09 | Nightingale | M_HDL_P | NA | C | T | -0.03452 | 0.003999 | 2.80E-19 |
| rs102274 | miR-1908-5 | cis | 9.41E-09 | Nightingale | M_LDL_CE | NA | C | T | -0.02702 | 0.004347 | 2.20E-10 |
| rs102274 | miR-1908-5 | cis | 9.41E-09 | Nightingale | M_HDL_C_ | NA | C | T | -0.06533 | 0.004132 | 3.20E-59 |
| rs102274 | miR-1908-5 | cis | 9.41E-09 | Nightingale | XL_HDL_PL | NA | C | T | -0.08959 | 0.003902 | 8.49E-127 |

| | | | | | | | | | | | |
|---|---|---|---|---|---|---|---|---|---|---|---|
| rs102274 | miR-1908-5 | cis | 9.41E-09 | Nightingale | M_VLDL_C | NA | C | T | -0.04502 | 0.004329 | 2.40E-26 |
| rs102274 | miR-1908-5 | cis | 9.41E-09 | Nightingale | M_LDL_L | NA | C | T | -0.02887 | 0.004347 | 1.20E-11 |
| rs102274 | miR-1908-5 | cis | 9.41E-09 | Nightingale | Tyr | NA | C | T | 0.033379 | 0.004335 | 1.40E-14 |
| rs102274 | miR-1908-5 | cis | 9.41E-09 | Nightingale | XXL_VLDL_ | NA | C | T | 0.065823 | 0.004181 | 2.90E-58 |
| rs102274 | miR-1908-5 | cis | 9.41E-09 | Nightingale | M_HDL_C | NA | C | T | -0.04088 | 0.003994 | 4.10E-26 |
| rs102274 | miR-1908-5 | cis | 9.41E-09 | Nightingale | XL_HDL_P | NA | C | T | -0.0902 | 0.003906 | 8.20E-129 |
| rs102274 | miR-1908-5 | cis | 9.41E-09 | Nightingale | IDL_P | NA | C | T | -0.04647 | 0.004295 | 9.90E-29 |
| rs102274 | miR-1908-5 | cis | 9.41E-09 | Nightingale | Clinical_LD | NA | C | T | -0.05606 | 0.004316 | 2.20E-40 |
| rs102274 | miR-1908-5 | cis | 9.41E-09 | Nightingale | XS_VLDL_C | NA | C | T | -0.09268 | 0.004092 | 1.40E-118 |
| rs102274 | miR-1908-5 | cis | 9.41E-09 | Nightingale | M_HDL_CE | NA | C | T | -0.04626 | 0.004209 | 6.90E-29 |
| rs102274 | miR-1908-5 | cis | 9.41E-09 | Nightingale | XL_HDL_TG | NA | C | T | 0.087129 | 0.004089 | 1.90E-106 |
| rs102274 | miR-1908-5 | cis | 9.41E-09 | Nightingale | S_VLDL_C_ | NA | C | T | -0.05695 | 0.004223 | 1.60E-42 |
| rs102274 | miR-1908-5 | cis | 9.41E-09 | Nightingale | M_HDL_CE | NA | C | T | -0.03787 | 0.004009 | 1.90E-22 |
| rs102274 | miR-1908-5 | cis | 9.41E-09 | Nightingale | M_LDL_FC | NA | C | T | -0.05302 | 0.004336 | 3.60E-36 |
| rs102274 | miR-1908-5 | cis | 9.41E-09 | Nightingale | ApoA1 | NA | C | T | -0.04152 | 0.003972 | 1.70E-27 |
| rs102274 | miR-1908-5 | cis | 9.41E-09 | Nightingale | M_HDL_PL | NA | C | T | 0.058061 | 0.004117 | 5.30E-47 |
| rs102274 | miR-1908-5 | cis | 9.41E-09 | Nightingale | L_HDL_PL | NA | C | T | -0.07635 | 0.00382 | 2.00E-96 |
| rs102274 | miR-1908-5 | cis | 9.41E-09 | Nightingale | Citrate | NA | C | T | -0.03313 | 0.004313 | 1.40E-15 |
| rs102274 | miR-1908-5 | cis | 9.41E-09 | Nightingale | Total_C | NA | C | T | -0.06623 | 0.004202 | 5.50E-59 |
| rs102274 | miR-1908-5 | cis | 9.41E-09 | Nightingale | XS_VLDL_C | NA | C | T | -0.09853 | 0.004113 | 2.70E-132 |
| rs102274 | miR-1908-5 | cis | 9.41E-09 | Nightingale | XL_VLDL_C | NA | C | T | -0.06081 | 0.004162 | 1.10E-49 |
| rs102274 | miR-1908-5 | cis | 9.41E-09 | Nightingale | Cholines | NA | C | T | -0.03806 | 0.004082 | 1.80E-22 |
| rs102274 | miR-1908-5 | cis | 9.41E-09 | Nightingale | M_LDL_FC_ | NA | C | T | -0.0715 | 0.004158 | 1.20E-68 |
| rs102274 | miR-1908-5 | cis | 9.41E-09 | Nightingale | XS_VLDL_T | NA | C | T | 0.030961 | 0.004274 | 1.60E-12 |
| rs102274 | miR-1908-5 | cis | 9.41E-09 | Nightingale | XXL_VLDL_ | NA | C | T | 0.071327 | 0.004166 | 1.20E-68 |
| rs102274 | miR-1908-5 | cis | 9.41E-09 | Nightingale | L_LDL_FC | NA | C | T | -0.06922 | 0.004262 | 1.10E-62 |
| rs102274 | miR-1908-5 | cis | 9.41E-09 | Nightingale | L_LDL_PL | NA | C | T | -0.05587 | 0.004308 | 3.10E-40 |
| rs102274 | miR-1908-5 | cis | 9.41E-09 | Nightingale | LA_pct | NA | C | T | 0.183862 | 0.004242 | 0 |
| rs102274 | miR-1908-5 | cis | 9.41E-09 | Nightingale | XL_HDL_PL | NA | C | T | -0.06758 | 0.003988 | 1.80E-69 |
| rs102274 | miR-1908-5 | cis | 9.41E-09 | Nightingale | TG_by_PG | NA | C | T | 0.068508 | 0.004028 | 5.10E-68 |
| rs102274 | miR-1908-5 | cis | 9.41E-09 | Nightingale | LDL_P | NA | C | T | -0.04116 | 0.004338 | 2.10E-22 |
| rs102274 | miR-1908-5 | cis | 9.41E-09 | Nightingale | Omega_6_ | NA | C | T | 0.023859 | 0.004225 | 8.10E-09 |
| rs102274 | miR-1908-5 | cis | 9.41E-09 | Nightingale | L_VLDL_FC | NA | C | T | 0.028298 | 0.0043 | 2.40E-11 |
| rs102274 | miR-1908-5 | cis | 9.41E-09 | Nightingale | M_VLDL_P | NA | C | T | -0.06323 | 0.004165 | 1.90E-55 |
| rs102274 | miR-1908-5 | cis | 9.41E-09 | Nightingale | S_LDL_FC | NA | C | T | -0.06337 | 0.004334 | 3.10E-51 |
| rs102274 | miR-1908-5 | cis | 9.41E-09 | Nightingale | L_HDL_C | NA | C | T | -0.0805 | 0.003822 | 7.80E-107 |
| rs102274 | miR-1908-5 | cis | 9.41E-09 | Nightingale | S_LDL_TG_ | NA | C | T | 0.085997 | 0.004206 | 6.80E-96 |
| rs102274 | miR-1908-5 | cis | 9.41E-09 | Nightingale | XL_VLDL_F | NA | C | T | -0.0687 | 0.004206 | 9.80E-63 |
| rs102274 | miR-1908-5 | cis | 9.41E-09 | Nightingale | S_HDL_TG_ | NA | C | T | 0.068624 | 0.004128 | 7.80E-65 |
| rs102274 | miR-1908-5 | cis | 9.41E-09 | Nightingale | XL_HDL_CE | NA | C | T | -0.09754 | 0.003887 | 1.20E-150 |
| rs102274 | miR-1908-5 | cis | 9.41E-09 | Nightingale | L_VLDL_C_ | NA | C | T | -0.03873 | 0.00424 | 4.20E-21 |
| rs102274 | miR-1908-5 | cis | 9.41E-09 | Nightingale | VLDL_TG | NA | C | T | 0.047165 | 0.004177 | 1.50E-30 |
| rs102274 | miR-1908-5 | cis | 9.41E-09 | Nightingale | Total_FC | NA | C | T | -0.06051 | 0.00424 | 8.10E-49 |
| rs102274 | miR-1908-5 | cis | 9.41E-09 | Nightingale | XL_VLDL_T | NA | C | T | 0.059613 | 0.004205 | 4.90E-47 |
| rs102274 | miR-1908-5 | cis | 9.41E-09 | Nightingale | MUFA | NA | C | T | 0.063137 | 0.004296 | 6.90E-50 |
| rs102274 | miR-1908-5 | cis | 9.41E-09 | Nightingale | M_HDL_FC | NA | C | T | -0.10364 | 0.003846 | 7.10E-174 |
| rs102274 | miR-1908-5 | cis | 9.41E-09 | Nightingale | M_VLDL_TG | NA | C | T | 0.030937 | 0.004217 | 9.40E-14 |
| rs102274 | miR-1908-5 | cis | 9.41E-09 | Nightingale | IDL_FC | NA | C | T | -0.0908 | 0.004201 | 3.70E-109 |
| rs102274 | miR-1908-5 | cis | 9.41E-09 | Nightingale | DHA_pct | NA | C | T | -0.27234 | 0.004214 | 0 |
| rs102274 | miR-1908-5 | cis | 9.41E-09 | Nightingale | HDL_CE | NA | C | T | -0.06163 | 0.003895 | 1.20E-60 |
| rs102274 | miR-1908-5 | cis | 9.41E-09 | Nightingale | L_VLDL_L | NA | C | T | 0.037681 | 0.004195 | 5.30E-20 |
| rs102274 | miR-1908-5 | cis | 9.41E-09 | Nightingale | M_HDL_FC | NA | C | T | -0.05179 | 0.00395 | 5.70E-42 |
| rs102274 | miR-1908-5 | cis | 9.41E-09 | Nightingale | HDL_FC | NA | C | T | -0.07438 | 0.003845 | 5.70E-90 |
| rs102274 | miR-1908-5 | cis | 9.41E-09 | Nightingale | XL_HDL_FC | NA | C | T | 0.061555 | 0.003848 | 1.30E-63 |
| rs102274 | miR-1908-5 | cis | 9.41E-09 | Nightingale | IDL_PL | NA | C | T | -0.07409 | 0.004188 | 6.50E-75 |
| rs102274 | miR-1908-5 | cis | 9.41E-09 | Nightingale | XL_HDL_C_ | NA | C | T | 0.024556 | 0.004147 | 1.30E-10 |
| rs102274 | miR-1908-5 | cis | 9.41E-09 | Nightingale | L_LDL_C_p | NA | C | T | -0.07728 | 0.004229 | 8.00E-77 |
| rs102274 | miR-1908-5 | cis | 9.41E-09 | Nightingale | Total_TG | NA | C | T | 0.046168 | 0.004202 | 5.00E-29 |
| rs102274 | miR-1908-5 | cis | 9.41E-09 | Nightingale | L_LDL_C | NA | C | T | -0.05866 | 0.004292 | 6.60E-45 |
| rs102274 | miR-1908-5 | cis | 9.41E-09 | Nightingale | Total_PL | NA | C | T | -0.05217 | 0.004133 | 8.80E-39 |
| rs102274 | miR-1908-5 | cis | 9.41E-09 | Nightingale | L_HDL_FC | NA | C | T | -0.08498 | 0.003804 | 6.50E-120 |
| rs102274 | miR-1908-5 | cis | 9.41E-09 | Nightingale | HDL_PL | NA | C | T | -0.04885 | 0.003924 | 4.60E-38 |
| rs102274 | miR-1908-5 | cis | 9.41E-09 | Nightingale | L_VLDL_FC | NA | C | T | 0.039588 | 0.004199 | 5.90E-22 |

| | | | | | | | | | | | |
|---|---|---|---|---|---|---|---|---|---|---|---|
| rs102274 | miR-1908-5 | cis | 9.41E-09 | Nightingale | XS_VLDL_L | NA | C | T | -0.0429 | 0.004267 | 8.80E-26 |
| rs102274 | miR-1908-5 | cis | 9.41E-09 | Nightingale | XL_HDL_L | NA | C | T | -0.09355 | 0.003901 | 4.00E-138 |
| rs102274 | miR-1908-5 | cis | 9.41E-09 | Nightingale | S_VLDL_TG | NA | C | T | 0.064833 | 0.0042 | 2.40E-55 |
| rs102274 | miR-1908-5 | cis | 9.41E-09 | Nightingale | L_LDL_FC_ | NA | C | T | -0.08314 | 0.004137 | 7.50E-93 |
| rs102274 | miR-1908-5 | cis | 9.41E-09 | Nightingale | IDL_CE_pct | NA | C | T | -0.0389 | 0.004259 | 7.70E-20 |
| rs102274 | miR-1908-5 | cis | 9.41E-09 | Nightingale | S_HDL_C_p | NA | C | T | -0.0416 | 0.00429 | 4.90E-22 |
| rs102274 | miR-1908-5 | cis | 9.41E-09 | Nightingale | M_VLDL_F | NA | C | T | -0.09179 | 0.004127 | 3.70E-115 |
| rs102274 | miR-1908-5 | cis | 9.41E-09 | Nightingale | L_LDL_CE | NA | C | T | -0.05413 | 0.004302 | 2.50E-38 |
| rs102274 | miR-1908-5 | cis | 9.41E-09 | Nightingale | S_LDL_C | NA | C | T | -0.03296 | 0.004345 | 6.80E-15 |
| rs102274 | miR-1908-5 | cis | 9.41E-09 | Nightingale | L_VLDL_PL | NA | C | T | 0.048178 | 0.004188 | 6.10E-32 |
| rs102274 | miR-1908-5 | cis | 9.41E-09 | Nightingale | Omega_3 | NA | C | T | -0.33468 | 0.004254 | 0 |
| rs102274 | miR-1908-5 | cis | 9.41E-09 | Nightingale | M_LDL_PL | NA | C | T | -0.02421 | 0.004347 | 1.20E-08 |
| rs102274 | miR-1908-5 | cis | 9.41E-09 | Nightingale | M_HDL_PL | NA | C | T | -0.02178 | 0.004059 | 2.40E-08 |
| rs102274 | miR-1908-5 | cis | 9.41E-09 | Nightingale | Omega_3_ | NA | C | T | -0.38811 | 0.004241 | 0 |
| rs102274 | miR-1908-5 | cis | 9.41E-09 | Nightingale | M_HDL_L | NA | C | T | -0.02791 | 0.00403 | 6.20E-13 |
| rs102274 | miR-1908-5 | cis | 9.41E-09 | Nightingale | LDL_C | NA | C | T | -0.05057 | 0.004322 | 3.20E-33 |
| rs102274 | miR-1908-5 | cis | 9.41E-09 | Nightingale | S_LDL_L | NA | C | T | -0.03324 | 0.00434 | 3.30E-15 |
| rs102274 | miR-1908-5 | cis | 9.41E-09 | Nightingale | S_LDL_FC_ | NA | C | T | -0.07935 | 0.004206 | 4.20E-82 |
| rs102274 | miR-1908-5 | cis | 9.41E-09 | Nightingale | XXL_VLDL_ | NA | C | T | 0.060478 | 0.004189 | 2.70E-49 |
| rs102274 | miR-1908-5 | cis | 9.41E-09 | Nightingale | LDL_CE | NA | C | T | -0.04439 | 0.00433 | 7.00E-26 |
| rs102274 | miR-1908-5 | cis | 9.41E-09 | Nightingale | VLDL_L | NA | C | T | 0.026578 | 0.004256 | 2.50E-10 |
| rs102274 | miR-1908-5 | cis | 9.41E-09 | Nightingale | L_HDL_L | NA | C | T | -0.07748 | 0.003809 | 5.30E-100 |
| rs102274 | miR-1908-5 | cis | 9.41E-09 | Nightingale | S_HDL_FC_ | NA | C | T | -0.10306 | 0.004117 | 2.50E-153 |
| rs102274 | miR-1908-5 | cis | 9.41E-09 | Nightingale | L_HDL_TG_ | NA | C | T | 0.078286 | 0.004096 | 1.80E-86 |
| rs102274 | miR-1908-5 | cis | 9.41E-09 | Nightingale | L_LDL_P | NA | C | T | -0.03835 | 0.004326 | 5.60E-20 |
| rs102274 | miR-1908-5 | cis | 9.41E-09 | Nightingale | PUFA_by_N | NA | C | T | -0.1702 | 0.004158 | 0 |
| rs102274 | miR-1908-5 | cis | 9.41E-09 | Nightingale | PUFA | NA | C | T | -0.09727 | 0.004233 | 7.29E-124 |
| rs102274 | miR-1908-5 | cis | 9.41E-09 | Nightingale | L_HDL_P | NA | C | T | -0.07815 | 0.003802 | 4.80E-102 |
| rs102274 | miR-1908-5 | cis | 9.41E-09 | Nightingale | XL_VLDL_P | NA | C | T | 0.052306 | 0.004172 | 2.00E-37 |
| rs102274 | miR-1908-5 | cis | 9.41E-09 | Nightingale | Total_L | NA | C | T | -0.03845 | 0.00427 | 1.30E-20 |
| rs102274 | miR-1908-5 | cis | 9.41E-09 | Nightingale | LDL_PL | NA | C | T | -0.04701 | 0.004331 | 1.10E-28 |
| rs102274 | miR-1908-5 | cis | 9.41E-09 | Nightingale | Total_P | NA | C | T | -0.0407 | 0.004085 | 5.30E-25 |
| rs102274 | miR-1908-5 | cis | 9.41E-09 | Nightingale | ApoB | NA | C | T | -0.03949 | 0.004335 | 7.70E-21 |
| rs102274 | miR-1908-5 | cis | 9.41E-09 | Nightingale | Sphingomy | NA | C | T | -0.11576 | 0.004056 | 3.40E-188 |
| rs102274 | miR-1908-5 | cis | 9.41E-09 | Nightingale | IDL_C | NA | C | T | -0.0785 | 0.004191 | 8.30E-83 |
| rs102274 | miR-1908-5 | cis | 9.41E-09 | Nightingale | M_VLDL_F | NA | C | T | -0.03039 | 0.00434 | 7.40E-13 |
| rs102274 | miR-1908-5 | cis | 9.41E-09 | Nightingale | L_HDL_C_p | NA | C | T | -0.06819 | 0.004091 | 6.60E-67 |
| rs102274 | miR-1908-5 | cis | 9.41E-09 | Nightingale | Omega_6_ | NA | C | T | 0.368122 | 0.004278 | 0 |
| rs102274 | miR-1908-5 | cis | 9.41E-09 | Nightingale | XL_HDL_C | NA | C | T | -0.0997 | 0.003914 | 4.70E-155 |
| rs102274 | miR-1908-5 | cis | 9.41E-09 | Nightingale | S_VLDL_TG | NA | C | T | 0.038447 | 0.004212 | 2.00E-20 |
| rs102274 | miR-1908-5 | cis | 9.41E-09 | Nightingale | XL_VLDL_P | NA | C | T | 0.047566 | 0.004189 | 5.40E-31 |
| rs102274 | miR-1908-5 | cis | 9.41E-09 | Nightingale | XL_VLDL_C | NA | C | T | 0.036736 | 0.004239 | 1.10E-18 |
| rs102274 | miR-1908-5 | cis | 9.41E-09 | Nightingale | L_HDL_CE | NA | C | T | -0.07888 | 0.003833 | 5.60E-102 |
| rs102274 | miR-1908-5 | cis | 9.41E-09 | Nightingale | XXL_VLDL_ | NA | C | T | 0.070897 | 0.004183 | 5.70E-67 |
| rs102274 | miR-1908-5 | cis | 9.41E-09 | Nightingale | XS_VLDL_C | NA | C | T | -0.06958 | 0.004213 | 4.70E-65 |
| rs102274 | miR-1908-5 | cis | 9.41E-09 | Nightingale | HDL_L | NA | C | T | -0.05373 | 0.003898 | 2.50E-46 |
| rs102274 | miR-1908-5 | cis | 9.41E-09 | Nightingale | XXL_VLDL_ | NA | C | T | 0.072839 | 0.004192 | 5.60E-70 |
| rs102274 | miR-1908-5 | cis | 9.41E-09 | Nightingale | M_VLDL_C | NA | C | T | -0.05367 | 0.004301 | 2.30E-37 |
| rs102274 | miR-1908-5 | cis | 9.41E-09 | Nightingale | PUFA_pct | NA | C | T | -0.121 | 0.004175 | 4.90E-186 |
| rs102274 | miR-1908-5 | cis | 9.41E-09 | Nightingale | S_VLDL_FC | NA | C | T | -0.08329 | 0.004145 | 1.10E-93 |
| rs102274 | miR-1908-5 | cis | 9.41E-09 | Nightingale | HDL_TG | NA | C | T | 0.045479 | 0.00428 | 7.90E-27 |
| rs102274 | miR-1908-5 | cis | 9.41E-09 | Nightingale | L_HDL_CE_ | NA | C | T | -0.04965 | 0.004153 | 4.20E-35 |
| rs102274 | miR-1908-5 | cis | 9.41E-09 | Nightingale | S_VLDL_CE | NA | C | T | -0.02928 | 0.004282 | 4.70E-12 |
| rs102274 | miR-1908-5 | cis | 9.41E-09 | Nightingale | M_LDL_P | NA | C | T | -0.0474 | 0.004353 | 1.90E-28 |
| rs102274 | miR-1908-5 | cis | 9.41E-09 | Nightingale | M_LDL_TG | NA | C | T | 0.075932 | 0.004286 | 1.50E-71 |
| rs102274 | miR-1908-5 | cis | 9.41E-09 | Nightingale | M_VLDL_T | NA | C | T | 0.078428 | 0.0041 | 2.50E-85 |
| rs102274 | miR-1908-5 | cis | 9.41E-09 | Nightingale | M_LDL_C_ | NA | C | T | -0.10058 | 0.004344 | 6.10E-119 |
| rs102274 | miR-1908-5 | cis | 9.41E-09 | Nightingale | XS_VLDL_P | NA | C | T | 0.0581 | 0.004235 | 1.60E-43 |
| rs102274 | miR-1908-5 | cis | 9.41E-09 | Nightingale | XS_VLDL_C | NA | C | T | -0.07337 | 0.004188 | 6.70E-73 |
| rs102274 | miR-1908-5 | cis | 9.41E-09 | Nightingale | XXL_VLDL_ | NA | C | T | 0.06536 | 0.004175 | 8.60E-58 |
| rs102274 | miR-1908-5 | cis | 9.41E-09 | Nightingale | XS_VLDL_F | NA | C | T | -0.11296 | 0.004308 | 1.30E-154 |
| rs102274 | miR-1908-5 | cis | 9.41E-09 | Nightingale | M_VLDL_C | NA | C | T | -0.08107 | 0.004086 | 3.30E-91 |
| rs102274 | miR-1908-5 | cis | 9.41E-09 | Nightingale | HDL_C | NA | C | T | -0.06498 | 0.003876 | 4.50E-68 |

| | | | | | | | | | | | | |
|---|---|---|---|---|---|---|---|---|---|---|---|---|
| rs102274 | miR-1908-5 | cis | | 9.41E-09 | Nightingale | IDL_L | NA | C | T | -0.07367 | 0.004196 | 1.90E-73 |
| rs102274 | miR-1908-5 | cis | | 9.41E-09 | Nightingale | XL_HDL_FC | NA | C | T | -0.10335 | 0.004044 | 3.00E-155 |
| rs102274 | miR-1908-5 | cis | | 9.41E-09 | Nightingale | L_HDL_PL_ | NA | C | T | 0.043326 | 0.004201 | 1.40E-26 |
| rs1022968( | miR-339-5p | cis | | 9.53E-05 | Nightingale | L_VLDL_C_ | NA | T | C | -0.02205 | 0.00418 | 2.70E-08 |
| rs1022968( | miR-339-5p | cis | | 9.53E-05 | Nightingale | L_VLDL_CE | NA | T | C | -0.02238 | 0.004094 | 2.20E-08 |
| rs1025400 | miR-339-5p | cis | | 6.21E-05 | Nightingale | L_VLDL_C_ | NA | C | T | -0.0258 | 0.004128 | 3.40E-11 |
| rs1025400 | miR-339-5p | cis | | 6.21E-05 | Nightingale | XL_VLDL_C | NA | C | T | -0.02201 | 0.004053 | 2.70E-08 |
| rs1025400 | miR-339-5p | cis | | 6.21E-05 | Nightingale | XL_VLDL_F | NA | C | T | -0.02464 | 0.004098 | 1.10E-09 |
| rs1025400 | miR-339-5p | cis | | 6.21E-05 | Nightingale | M_VLDL_F | NA | C | T | -0.02334 | 0.00402 | 2.10E-09 |
| rs1025400 | miR-339-5p | cis | | 6.21E-05 | Nightingale | M_VLDL_C | NA | C | T | -0.02174 | 0.003969 | 2.10E-08 |
| rs1025400 | miR-339-5p | cis | | 6.21E-05 | Nightingale | L_VLDL_CE | NA | C | T | -0.02574 | 0.004044 | 3.90E-11 |
| rs1025400 | miR-339-5p | cis | | 6.21E-05 | Nightingale | VLDL_size | NA | C | T | 0.023023 | 0.003941 | 1.40E-08 |
| rs1025400 | miR-339-5p | cis | | 6.21E-05 | Nightingale | M_VLDL_T | NA | C | T | 0.02314 | 0.003993 | 2.10E-09 |
| rs1025400 | miR-339-5p | cis | | 6.21E-05 | Nightingale | M_VLDL_P | NA | C | T | -0.02454 | 0.004056 | 2.70E-10 |
| rs1025400 | miR-339-5p | cis | | 6.21E-05 | Nightingale | M_VLDL_C | NA | C | T | -0.02239 | 0.00398 | 7.20E-09 |
| rs1025400 | miR-339-5p | cis | | 6.21E-05 | Nightingale | XL_VLDL_T | NA | C | T | 0.023816 | 0.004097 | 1.20E-09 |
| rs1027620 | miR-339-5p | cis | | 1.28E-06 | Nightingale | L_HDL_CE | NA | C | T | -0.02119 | 0.003813 | 5.00E-08 |
| rs1027620 | miR-339-5p | cis | | 1.28E-06 | Nightingale | L_HDL_PL | NA | C | T | -0.02175 | 0.0038 | 1.90E-08 |
| rs1027620 | miR-339-5p | cis | | 1.28E-06 | Nightingale | L_HDL_C | NA | C | T | -0.02154 | 0.003803 | 2.70E-08 |
| rs1027620 | miR-339-5p | cis | | 1.28E-06 | Nightingale | XL_HDL_FC | NA | C | T | 0.021578 | 0.003827 | 1.20E-08 |
| rs1027620 | miR-339-5p | cis | | 1.28E-06 | Nightingale | L_HDL_L | NA | C | T | -0.02206 | 0.003789 | 9.60E-09 |
| rs1027620 | miR-339-5p | cis | | 1.28E-06 | Nightingale | L_HDL_FC | NA | C | T | -0.02225 | 0.003784 | 6.90E-09 |
| rs1027620 | miR-339-5p | cis | | 1.28E-06 | Nightingale | HDL_size | NA | C | T | -0.02226 | 0.003826 | 1.20E-08 |
| rs1027620 | miR-339-5p | cis | | 1.28E-06 | Nightingale | L_HDL_P | NA | C | T | -0.02224 | 0.003782 | 6.90E-09 |
| rs1028096( | miR-339-5p | cis | | 2.99E-07 | Nightingale | L_VLDL_C_ | NA | C | T | -0.02322 | 0.004035 | 4.60E-09 |
| rs1028096( | miR-339-5p | cis | | 2.99E-07 | Nightingale | L_VLDL_CE | NA | C | T | -0.02247 | 0.003953 | 1.30E-08 |
| rs1028096( | miR-339-5p | cis | | 2.99E-07 | Nightingale | XL_VLDL_T | NA | C | T | 0.021895 | 0.004004 | 3.30E-08 |
| rs1043502( | miR-339-5p | cis | | 4.70E-07 | Nightingale | L_HDL_FC | NA | G | A | -0.02053 | 0.003679 | 4.70E-08 |
| rs1043514 | miR-339-5p | cis | | 1.74E-08 | Nightingale | L_VLDL_CE | NA | T | C | -0.02489 | 0.003983 | 3.90E-10 |
| rs1043514 | miR-339-5p | cis | | 1.74E-08 | Nightingale | L_HDL_FC | NA | T | C | -0.0202 | 0.00365 | 3.30E-08 |
| rs1043514 | miR-339-5p | cis | | 1.74E-08 | Nightingale | XL_VLDL_C | NA | T | C | -0.02195 | 0.003991 | 3.80E-08 |
| rs1043514 | miR-339-5p | cis | | 1.74E-08 | Nightingale | L_VLDL_C_ | NA | T | C | -0.0251 | 0.004066 | 2.90E-10 |
| rs1043514 | miR-339-5p | cis | | 1.74E-08 | Nightingale | M_VLDL_P | NA | T | C | -0.02408 | 0.003995 | 2.10E-09 |
| rs1043514 | miR-339-5p | cis | | 1.74E-08 | Nightingale | XL_VLDL_F | NA | T | C | -0.02292 | 0.004035 | 1.40E-08 |
| rs1043514 | miR-339-5p | cis | | 1.74E-08 | Nightingale | M_VLDL_F | NA | T | C | -0.02359 | 0.003959 | 4.20E-09 |
| rs1043514 | miR-339-5p | cis | | 1.74E-08 | Nightingale | M_VLDL_T | NA | T | C | 0.022801 | 0.003932 | 1.00E-08 |
| rs1043514 | miR-339-5p | cis | | 1.74E-08 | Nightingale | M_VLDL_C | NA | T | C | -0.02204 | 0.003919 | 2.60E-08 |
| rs1043514 | miR-339-5p | cis | | 1.74E-08 | Nightingale | XL_VLDL_T | NA | T | C | 0.023573 | 0.004034 | 3.10E-09 |
| rs1095159 | miR-339-5p | cis | | 1.20E-08 | Nightingale | M_VLDL_T | NA | G | C | 0.021754 | 0.003927 | 3.80E-08 |
| rs1095159 | miR-339-5p | cis | | 1.20E-08 | Nightingale | L_VLDL_C_ | NA | G | C | -0.02417 | 0.00406 | 9.00E-10 |
| rs1095159 | miR-339-5p | cis | | 1.20E-08 | Nightingale | L_VLDL_CE | NA | G | C | -0.02386 | 0.003978 | 1.60E-09 |
| rs1095159 | miR-339-5p | cis | | 1.20E-08 | Nightingale | XL_VLDL_T | NA | G | C | 0.022434 | 0.004029 | 1.40E-08 |
| rs1095159 | miR-339-5p | cis | | 1.20E-08 | Nightingale | M_VLDL_F | NA | G | C | -0.02235 | 0.003954 | 2.20E-08 |
| rs1095159 | miR-339-5p | cis | | 1.20E-08 | Nightingale | M_VLDL_P | NA | G | C | -0.02306 | 0.003989 | 8.10E-09 |
| rs1133043 | miR-339-5p | cis | | 5.21E-05 | Nightingale | L_HDL_L | NA | G | C | -0.02113 | 0.003737 | 2.20E-08 |
| rs1133043 | miR-339-5p | cis | | 5.21E-05 | Nightingale | L_VLDL_C_ | NA | G | C | -0.02387 | 0.004157 | 2.00E-09 |
| rs1133043 | miR-339-5p | cis | | 5.21E-05 | Nightingale | L_HDL_CE | NA | G | C | -0.02131 | 0.003761 | 2.50E-08 |
| rs1133043 | miR-339-5p | cis | | 5.21E-05 | Nightingale | HDL_size | NA | G | C | -0.02165 | 0.003774 | 1.80E-08 |
| rs1133043 | miR-339-5p | cis | | 5.21E-05 | Nightingale | L_HDL_FC | NA | G | C | -0.02152 | 0.003733 | 1.20E-08 |
| rs1133043 | miR-339-5p | cis | | 5.21E-05 | Nightingale | M_VLDL_P | NA | G | C | -0.0222 | 0.004085 | 3.00E-08 |
| rs1133043 | miR-339-5p | cis | | 5.21E-05 | Nightingale | L_VLDL_CE | NA | G | C | -0.0245 | 0.004073 | 7.60E-10 |
| rs1133043 | miR-339-5p | cis | | 5.21E-05 | Nightingale | VLDL_size | NA | G | C | 0.023904 | 0.003969 | 7.10E-09 |
| rs1133043 | miR-339-5p | cis | | 5.21E-05 | Nightingale | L_HDL_P | NA | G | C | -0.02139 | 0.003731 | 1.30E-08 |
| rs1133043 | miR-339-5p | cis | | 5.21E-05 | Nightingale | L_HDL_C | NA | G | C | -0.02144 | 0.003751 | 1.80E-08 |
| rs1176706 | miR-339-5p | cis | | 8.96E-05 | Nightingale | M_VLDL_T | NA | T | C | 0.022079 | 0.003977 | 1.00E-08 |
| rs1176706 | miR-339-5p | cis | | 8.96E-05 | Nightingale | M_VLDL_F | NA | T | C | -0.02233 | 0.004004 | 9.30E-09 |
| rs1176706 | miR-339-5p | cis | | 8.96E-05 | Nightingale | M_VLDL_P | NA | T | C | -0.02371 | 0.00404 | 9.10E-10 |
| rs1176706 | miR-339-5p | cis | | 8.96E-05 | Nightingale | M_VLDL_C | NA | T | C | -0.02128 | 0.003964 | 3.50E-08 |
| rs1176706 | miR-339-5p | cis | | 8.96E-05 | Nightingale | XL_VLDL_T | NA | T | C | 0.02262 | 0.004081 | 6.90E-09 |
| rs1176706 | miR-339-5p | cis | | 8.96E-05 | Nightingale | L_VLDL_CE | NA | T | C | -0.02444 | 0.004028 | 3.20E-10 |
| rs1176706 | miR-339-5p | cis | | 8.96E-05 | Nightingale | XL_VLDL_F | NA | T | C | -0.02309 | 0.004082 | 9.30E-09 |
| rs1176706 | miR-339-5p | cis | | 8.96E-05 | Nightingale | L_VLDL_C_ | NA | T | C | -0.02481 | 0.004112 | 1.70E-10 |
| rs1205605 | miR-339-5p | cis | | 2.69E-05 | Nightingale | M_VLDL_F | NA | C | A | -0.02415 | 0.004025 | 6.40E-10 |

| | | | | | | | | | | | |
|---|---|---|---|---|---|---|---|---|---|---|---|
| rs1205605: | miR-339-5p | cis | 2.69E-05 | Nightingale | VLDL_size | NA | C | A | 0.023549 | 0.003945 | 7.60E-09 |
| rs1205605: | miR-339-5p | cis | 2.69E-05 | Nightingale | L_VLDL_C_ | NA | C | A | -0.02666 | 0.004133 | 8.60E-12 |
| rs1205605: | miR-339-5p | cis | 2.69E-05 | Nightingale | XL_VLDL_F | NA | C | A | -0.02548 | 0.004102 | 3.10E-10 |
| rs1205605: | miR-339-5p | cis | 2.69E-05 | Nightingale | M_VLDL_T | NA | C | A | 0.023852 | 0.003997 | 7.70E-10 |
| rs1205605: | miR-339-5p | cis | 2.69E-05 | Nightingale | M_VLDL_C | NA | C | A | -0.02239 | 0.003974 | 9.20E-09 |
| rs1205605: | miR-339-5p | cis | 2.69E-05 | Nightingale | L_VLDL_CE | NA | C | A | -0.02653 | 0.004048 | 1.10E-11 |
| rs1205605: | miR-339-5p | cis | 2.69E-05 | Nightingale | XL_VLDL_C | NA | C | A | -0.02268 | 0.004057 | 1.10E-08 |
| rs1205605: | miR-339-5p | cis | 2.69E-05 | Nightingale | L_VLDL_TG | NA | C | A | 0.020897 | 0.004212 | 4.40E-08 |
| rs1205605: | miR-339-5p | cis | 2.69E-05 | Nightingale | M_VLDL_C | NA | C | A | -0.02308 | 0.003984 | 2.80E-09 |
| rs1205605: | miR-339-5p | cis | 2.69E-05 | Nightingale | M_VLDL_P | NA | C | A | -0.02527 | 0.004061 | 8.80E-11 |
| rs1205605: | miR-339-5p | cis | 2.69E-05 | Nightingale | XL_VLDL_T | NA | C | A | 0.024621 | 0.004102 | 3.70E-10 |
| rs1233429( | miR-339-5p | cis | 0.000104 | Nightingale | XS_VLDL_C | NA | C | T | -0.02338 | 0.004331 | 3.90E-08 |
| rs1233429( | miR-339-5p | cis | 0.000104 | Nightingale | XS_VLDL_C | NA | C | T | -0.02368 | 0.004306 | 2.10E-08 |
| rs1253258( | miR-148a-3 | cis | 0.000126 | Nightingale | S_VLDL_PL | NA | T | C | -0.0288 | 0.004492 | 2.20E-11 |
| rs1253258( | miR-148a-3 | cis | 0.000126 | Nightingale | S_LDL_TG_ | NA | T | C | 0.029568 | 0.00455 | 1.70E-11 |
| rs1253258( | miR-148a-3 | cis | 0.000126 | Nightingale | XS_VLDL_T | NA | T | C | 0.025105 | 0.00446 | 5.60E-09 |
| rs1253258( | miR-148a-3 | cis | 0.000126 | Nightingale | L_HDL_CE_ | NA | T | C | -0.02609 | 0.004492 | 2.20E-09 |
| rs1253258( | miR-148a-3 | cis | 0.000126 | Nightingale | M_HDL_TG | NA | T | C | 0.024664 | 0.004504 | 1.50E-08 |
| rs1253258( | miR-148a-3 | cis | 0.000126 | Nightingale | L_VLDL_L | NA | T | C | 0.025165 | 0.004537 | 1.50E-08 |
| rs1253258( | miR-148a-3 | cis | 0.000126 | Nightingale | L_HDL_TG_ | NA | T | C | 0.025088 | 0.004433 | 7.60E-09 |
| rs1253258( | miR-148a-3 | cis | 0.000126 | Nightingale | XL_VLDL_T | NA | T | C | 0.025963 | 0.004547 | 1.20E-09 |
| rs1253258( | miR-148a-3 | cis | 0.000126 | Nightingale | XL_VLDL_L | NA | T | C | 0.026084 | 0.004517 | 2.80E-09 |
| rs1253258( | miR-148a-3 | cis | 0.000126 | Nightingale | M_VLDL_C | NA | T | C | -0.03102 | 0.004419 | 1.00E-13 |
| rs1253258( | miR-148a-3 | cis | 0.000126 | Nightingale | M_VLDL_T | NA | T | C | 0.029961 | 0.004433 | 7.80E-13 |
| rs1253258( | miR-148a-3 | cis | 0.000126 | Nightingale | L_LDL_FC_ | NA | T | C | -0.02694 | 0.004476 | 7.50E-10 |
| rs1253258( | miR-148a-3 | cis | 0.000126 | Nightingale | M_HDL_CE | NA | T | C | -0.02593 | 0.004554 | 4.10E-09 |
| rs1253258( | miR-148a-3 | cis | 0.000126 | Nightingale | XXL_VLDL_ | NA | T | C | 0.028223 | 0.00453 | 6.50E-11 |
| rs1253258( | miR-148a-3 | cis | 0.000126 | Nightingale | S_LDL_FC_ | NA | T | C | -0.03108 | 0.004551 | 1.10E-12 |
| rs1253258( | miR-148a-3 | cis | 0.000126 | Nightingale | XL_VLDL_P | NA | T | C | 0.026073 | 0.004512 | 2.40E-09 |
| rs1253258( | miR-148a-3 | cis | 0.000126 | Nightingale | S_VLDL_TG | NA | T | C | 0.027269 | 0.004556 | 1.00E-09 |
| rs1253258( | miR-148a-3 | cis | 0.000126 | Nightingale | VLDL_TG | NA | T | C | 0.02801 | 0.004518 | 2.00E-10 |
| rs1253258( | miR-148a-3 | cis | 0.000126 | Nightingale | XL_VLDL_T | NA | T | C | 0.027975 | 0.00449 | 1.10E-10 |
| rs1253258( | miR-148a-3 | cis | 0.000126 | Nightingale | S_HDL_C_p | NA | T | C | -0.02824 | 0.00464 | 1.80E-10 |
| rs1253258( | miR-148a-3 | cis | 0.000126 | Nightingale | XXL_VLDL_ | NA | T | C | 0.025373 | 0.004535 | 4.80E-09 |
| rs1253258( | miR-148a-3 | cis | 0.000126 | Nightingale | XXL_VLDL_ | NA | T | C | 0.028231 | 0.004516 | 8.40E-11 |
| rs1253258( | miR-148a-3 | cis | 0.000126 | Nightingale | HDL_TG | NA | T | C | 0.02788 | 0.00463 | 6.10E-10 |
| rs1253258( | miR-148a-3 | cis | 0.000126 | Nightingale | XL_VLDL_F | NA | T | C | 0.024246 | 0.004547 | 3.70E-08 |
| rs1253258( | miR-148a-3 | cis | 0.000126 | Nightingale | XL_HDL_TG | NA | T | C | 0.02633 | 0.004424 | 6.20E-10 |
| rs1253258( | miR-148a-3 | cis | 0.000126 | Nightingale | L_VLDL_PL | NA | T | C | 0.024538 | 0.00453 | 2.50E-08 |
| rs1253258( | miR-148a-3 | cis | 0.000126 | Nightingale | L_VLDL_CE | NA | T | C | -0.02301 | 0.00449 | 3.30E-08 |
| rs1253258( | miR-148a-3 | cis | 0.000126 | Nightingale | XXL_VLDL_ | NA | T | C | 0.028233 | 0.004506 | 6.20E-11 |
| rs1253258( | miR-148a-3 | cis | 0.000126 | Nightingale | XL_VLDL_C | NA | T | C | -0.02736 | 0.004498 | 8.10E-11 |
| rs1253258( | miR-148a-3 | cis | 0.000126 | Nightingale | S_VLDL_TG | NA | T | C | 0.028659 | 0.004542 | 6.00E-11 |
| rs1253258( | miR-148a-3 | cis | 0.000126 | Nightingale | S_VLDL_FC | NA | T | C | -0.02783 | 0.004483 | 6.70E-11 |
| rs1253258( | miR-148a-3 | cis | 0.000126 | Nightingale | S_VLDL_C_ | NA | T | C | -0.02801 | 0.004566 | 1.90E-10 |
| rs1253258( | miR-148a-3 | cis | 0.000126 | Nightingale | M_VLDL_C | NA | T | C | -0.0319 | 0.004407 | 2.10E-14 |
| rs1253258( | miR-148a-3 | cis | 0.000126 | Nightingale | XXL_VLDL_ | NA | T | C | 0.028636 | 0.004522 | 4.00E-11 |
| rs1253258( | miR-148a-3 | cis | 0.000126 | Nightingale | M_VLDL_F | NA | T | C | -0.02688 | 0.004463 | 1.20E-10 |
| rs1253258( | miR-148a-3 | cis | 0.000126 | Nightingale | M_VLDL_P | NA | T | C | -0.02372 | 0.004503 | 2.30E-08 |
| rs1253258( | miR-148a-3 | cis | 0.000126 | Nightingale | TG_by_PG | NA | T | C | 0.025181 | 0.004357 | 3.10E-09 |
| rs1253258( | miR-148a-3 | cis | 0.000126 | Nightingale | S_VLDL_CE | NA | T | C | -0.02531 | 0.00463 | 1.90E-08 |
| rs1253258( | miR-148a-3 | cis | 0.000126 | Nightingale | XXL_VLDL_ | NA | T | C | 0.028171 | 0.004523 | 8.20E-11 |
| rs1253258( | miR-148a-3 | cis | 0.000126 | Nightingale | M_HDL_C_ | NA | T | C | -0.02427 | 0.004471 | 1.30E-08 |
| rs1253258( | miR-148a-3 | cis | 0.000126 | Nightingale | XL_VLDL_P | NA | T | C | 0.024762 | 0.004531 | 1.60E-08 |
| rs1253258( | miR-148a-3 | cis | 0.000126 | Nightingale | M_LDL_FC | NA | T | C | -0.03018 | 0.004498 | 1.70E-12 |
| rs1253258( | miR-148a-3 | cis | 0.000126 | Nightingale | Omega_6_ | NA | T | C | -0.02754 | 0.004568 | 1.40E-09 |
| rs1253258( | miR-148a-3 | cis | 0.000126 | Nightingale | L_HDL_C_p | NA | T | C | -0.02625 | 0.004426 | 7.60E-10 |
| rs1253258( | miR-148a-3 | cis | 0.000126 | Nightingale | L_VLDL_P | NA | T | C | 0.025019 | 0.004533 | 1.60E-08 |
| rs1253258( | miR-148a-3 | cis | 0.000126 | Nightingale | S_HDL_TG | NA | T | C | 0.026707 | 0.004554 | 1.30E-09 |
| rs1253258( | miR-148a-3 | cis | 0.000126 | Nightingale | M_VLDL_T | NA | T | C | 0.025577 | 0.004561 | 1.10E-08 |
| rs1253258( | miR-148a-3 | cis | 0.000126 | Nightingale | XXL_VLDL_ | NA | T | C | 0.027097 | 0.004525 | 4.40E-10 |
| rs1253258( | miR-148a-3 | cis | 0.000126 | Nightingale | M_HDL_TG | NA | T | C | 0.028832 | 0.004637 | 1.30E-10 |
| rs1253258( | miR-148a-3 | cis | 0.000126 | Nightingale | L_VLDL_TG | NA | T | C | 0.026953 | 0.004516 | 1.10E-09 |

| | | | | | | | | | | | |
|---|---|---|---|---|---|---|---|---|---|---|---|
| rs1253258( | miR-148a-3 | cis | 0.000126 | Nightingale | VLDL_size | NA | T | C | 0.026286 | 0.004376 | 2.30E-10 |
| rs1253258( | miR-148a-3 | cis | 0.000126 | Nightingale | Total_TG | NA | T | C | 0.027746 | 0.004544 | 3.60E-10 |
| rs1253258( | miR-148a-3 | cis | 0.000126 | Nightingale | XL_VLDL_C | NA | T | C | -0.02826 | 0.0045 | 1.90E-11 |
| rs1253856 | miR-148a-3 | cis | 0.000125 | Nightingale | M_HDL_TG | NA | C | T | 0.024734 | 0.004504 | 1.30E-08 |
| rs1253856 | miR-148a-3 | cis | 0.000125 | Nightingale | L_VLDL_CE | NA | C | T | -0.02314 | 0.004489 | 2.80E-08 |
| rs1253856 | miR-148a-3 | cis | 0.000125 | Nightingale | TG_by_PG | NA | C | T | 0.025275 | 0.004356 | 2.80E-09 |
| rs1253856 | miR-148a-3 | cis | 0.000125 | Nightingale | S_VLDL_TG | NA | C | T | 0.02743 | 0.004555 | 8.20E-10 |
| rs1253856 | miR-148a-3 | cis | 0.000125 | Nightingale | XL_VLDL_T | NA | C | T | 0.028105 | 0.004489 | 9.40E-11 |
| rs1253856 | miR-148a-3 | cis | 0.000125 | Nightingale | XS_VLDL_T | NA | C | T | 0.025233 | 0.004459 | 4.60E-09 |
| rs1253856 | miR-148a-3 | cis | 0.000125 | Nightingale | XL_VLDL_C | NA | C | T | -0.02749 | 0.004497 | 6.50E-11 |
| rs1253856 | miR-148a-3 | cis | 0.000125 | Nightingale | S_LDL_FC_ | NA | C | T | -0.03117 | 0.00455 | 9.10E-13 |
| rs1253856 | miR-148a-3 | cis | 0.000125 | Nightingale | XL_VLDL_P | NA | C | T | 0.026197 | 0.004512 | 2.00E-09 |
| rs1253856 | miR-148a-3 | cis | 0.000125 | Nightingale | M_HDL_C_ | NA | C | T | -0.02433 | 0.00447 | 1.20E-08 |
| rs1253856 | miR-148a-3 | cis | 0.000125 | Nightingale | S_VLDL_PL | NA | C | T | -0.02888 | 0.004492 | 1.90E-11 |
| rs1253856 | miR-148a-3 | cis | 0.000125 | Nightingale | M_VLDL_P | NA | C | T | -0.02376 | 0.004503 | 2.10E-08 |
| rs1253856 | miR-148a-3 | cis | 0.000125 | Nightingale | XL_VLDL_T | NA | C | T | 0.026061 | 0.004546 | 1.00E-09 |
| rs1253856 | miR-148a-3 | cis | 0.000125 | Nightingale | M_VLDL_C | NA | C | T | -0.03198 | 0.004406 | 1.80E-14 |
| rs1253856 | miR-148a-3 | cis | 0.000125 | Nightingale | M_VLDL_C | NA | C | T | -0.0311 | 0.004418 | 8.70E-14 |
| rs1253856 | miR-148a-3 | cis | 0.000125 | Nightingale | M_HDL_CE | NA | C | T | -0.026 | 0.004553 | 3.70E-09 |
| rs1253856 | miR-148a-3 | cis | 0.000125 | Nightingale | S_VLDL_C_ | NA | C | T | -0.02811 | 0.004565 | 1.60E-10 |
| rs1253856 | miR-148a-3 | cis | 0.000125 | Nightingale | VLDL_TG | NA | C | T | 0.02815 | 0.004517 | 1.70E-10 |
| rs1253856 | miR-148a-3 | cis | 0.000125 | Nightingale | L_VLDL_L | NA | C | T | 0.025278 | 0.004536 | 1.30E-08 |
| rs1253856 | miR-148a-3 | cis | 0.000125 | Nightingale | XXL_VLDL_ | NA | C | T | 0.02831 | 0.004522 | 6.80E-11 |
| rs1253856 | miR-148a-3 | cis | 0.000125 | Nightingale | M_VLDL_T | NA | C | T | 0.030035 | 0.004432 | 6.80E-13 |
| rs1253856 | miR-148a-3 | cis | 0.000125 | Nightingale | L_VLDL_PL | NA | C | T | 0.024655 | 0.004529 | 2.20E-08 |
| rs1253856 | miR-148a-3 | cis | 0.000125 | Nightingale | S_HDL_C_p | NA | C | T | -0.0283 | 0.004639 | 1.60E-10 |
| rs1253856 | miR-148a-3 | cis | 0.000125 | Nightingale | L_LDL_FC_ | NA | C | T | -0.02709 | 0.004475 | 6.10E-10 |
| rs1253856 | miR-148a-3 | cis | 0.000125 | Nightingale | M_HDL_TG | NA | C | T | 0.028946 | 0.004636 | 1.10E-10 |
| rs1253856 | miR-148a-3 | cis | 0.000125 | Nightingale | XS_VLDL_C | NA | C | T | -0.02362 | 0.004448 | 4.90E-08 |
| rs1253856 | miR-148a-3 | cis | 0.000125 | Nightingale | XL_VLDL_L | NA | C | T | 0.026208 | 0.004516 | 2.40E-09 |
| rs1253856 | miR-148a-3 | cis | 0.000125 | Nightingale | M_VLDL_T | NA | C | T | 0.025711 | 0.00456 | 9.40E-09 |
| rs1253856 | miR-148a-3 | cis | 0.000125 | Nightingale | XL_HDL_TG | NA | C | T | 0.026433 | 0.004423 | 5.40E-10 |
| rs1253856 | miR-148a-3 | cis | 0.000125 | Nightingale | VLDL_size | NA | C | T | 0.026367 | 0.004375 | 2.00E-10 |
| rs1253856 | miR-148a-3 | cis | 0.000125 | Nightingale | XXL_VLDL_ | NA | C | T | 0.025509 | 0.004534 | 4.00E-09 |
| rs1253856 | miR-148a-3 | cis | 0.000125 | Nightingale | L_VLDL_TG | NA | C | T | 0.027069 | 0.004516 | 9.00E-10 |
| rs1253856 | miR-148a-3 | cis | 0.000125 | Nightingale | S_VLDL_TG | NA | C | T | 0.02876 | 0.004541 | 5.10E-11 |
| rs1253856 | miR-148a-3 | cis | 0.000125 | Nightingale | XXL_VLDL_ | NA | C | T | 0.028768 | 0.004521 | 3.30E-11 |
| rs1253856 | miR-148a-3 | cis | 0.000125 | Nightingale | XXL_VLDL_ | NA | C | T | 0.028373 | 0.004505 | 5.10E-11 |
| rs1253856 | miR-148a-3 | cis | 0.000125 | Nightingale | XL_VLDL_P | NA | C | T | 0.024886 | 0.00453 | 1.40E-08 |
| rs1253856 | miR-148a-3 | cis | 0.000125 | Nightingale | XXL_VLDL_ | NA | C | T | 0.027236 | 0.004524 | 3.60E-10 |
| rs1253856 | miR-148a-3 | cis | 0.000125 | Nightingale | S_HDL_TG | NA | C | T | 0.02684 | 0.004553 | 1.10E-09 |
| rs1253856 | miR-148a-3 | cis | 0.000125 | Nightingale | S_LDL_TG_ | NA | C | T | 0.029697 | 0.004549 | 1.40E-11 |
| rs1253856 | miR-148a-3 | cis | 0.000125 | Nightingale | S_VLDL_FC | NA | C | T | -0.02792 | 0.004482 | 5.80E-11 |
| rs1253856 | miR-148a-3 | cis | 0.000125 | Nightingale | M_VLDL_F | NA | C | T | -0.02696 | 0.004463 | 1.00E-10 |
| rs1253856 | miR-148a-3 | cis | 0.000125 | Nightingale | S_VLDL_CE | NA | C | T | -0.02542 | 0.004629 | 1.60E-08 |
| rs1253856 | miR-148a-3 | cis | 0.000125 | Nightingale | XXL_VLDL_ | NA | C | T | 0.028344 | 0.004529 | 5.40E-11 |
| rs1253856 | miR-148a-3 | cis | 0.000125 | Nightingale | L_HDL_CE_ | NA | C | T | -0.02615 | 0.004491 | 2.00E-09 |
| rs1253856 | miR-148a-3 | cis | 0.000125 | Nightingale | L_HDL_C_p | NA | C | T | -0.0263 | 0.004425 | 6.80E-10 |
| rs1253856 | miR-148a-3 | cis | 0.000125 | Nightingale | XL_VLDL_C | NA | C | T | -0.0284 | 0.0045 | 1.50E-11 |
| rs1253856 | miR-148a-3 | cis | 0.000125 | Nightingale | L_VLDL_P | NA | C | T | 0.02514 | 0.004532 | 1.40E-08 |
| rs1253856 | miR-148a-3 | cis | 0.000125 | Nightingale | Omega_6_ | NA | C | T | -0.02765 | 0.004567 | 1.20E-09 |
| rs1253856 | miR-148a-3 | cis | 0.000125 | Nightingale | L_HDL_TG_ | NA | C | T | 0.025129 | 0.004432 | 7.20E-09 |
| rs1253856 | miR-148a-3 | cis | 0.000125 | Nightingale | HDL_TG | NA | C | T | 0.028004 | 0.004629 | 5.10E-10 |
| rs1253856 | miR-148a-3 | cis | 0.000125 | Nightingale | Total_TG | NA | C | T | 0.027896 | 0.004544 | 2.90E-10 |
| rs1253856 | miR-148a-3 | cis | 0.000125 | Nightingale | XL_VLDL_F | NA | C | T | 0.024364 | 0.004546 | 3.30E-08 |
| rs1253856 | miR-148a-3 | cis | 0.000125 | Nightingale | XXL_VLDL_ | NA | C | T | 0.028366 | 0.004515 | 6.90E-11 |
| rs1253856 | miR-148a-3 | cis | 0.000125 | Nightingale | M_LDL_FC_ | NA | C | T | -0.03024 | 0.004497 | 1.60E-12 |
| rs1267250 | miR-148a-3 | cis | 0.000115 | Nightingale | M_VLDL_F | NA | C | A | -0.02699 | 0.004462 | 9.70E-11 |
| rs1267250 | miR-148a-3 | cis | 0.000115 | Nightingale | XXL_VLDL_ | NA | C | A | 0.028286 | 0.004529 | 5.80E-11 |
| rs1267250 | miR-148a-3 | cis | 0.000115 | Nightingale | M_VLDL_C | NA | C | A | -0.032 | 0.004406 | 1.70E-14 |
| rs1267250 | miR-148a-3 | cis | 0.000115 | Nightingale | M_HDL_CE | NA | C | A | -0.02603 | 0.004553 | 3.60E-09 |
| rs1267250 | miR-148a-3 | cis | 0.000115 | Nightingale | XL_VLDL_L | NA | C | A | 0.026154 | 0.004516 | 2.50E-09 |
| rs1267250 | miR-148a-3 | cis | 0.000115 | Nightingale | S_VLDL_CE | NA | C | A | -0.02545 | 0.004628 | 1.50E-08 |

| | | | | | | | | | | | |
|---|---|---|---|---|---|---|---|---|---|---|---|
| rs1267250! | miR-148a-3 | cis | 0.000115 | Nightingale | S_VLDL_FC | NA | C | A | -0.02796 | 0.004482 | 5.30E-11 |
| rs1267250! | miR-148a-3 | cis | 0.000115 | Nightingale | M_LDL_FC_ | NA | C | A | -0.03028 | 0.004497 | 1.50E-12 |
| rs1267250! | miR-148a-3 | cis | 0.000115 | Nightingale | M_HDL_TG | NA | C | A | 0.024751 | 0.004503 | 1.30E-08 |
| rs1267250! | miR-148a-3 | cis | 0.000115 | Nightingale | HDL_TG | NA | C | A | 0.028015 | 0.004628 | 5.00E-10 |
| rs1267250! | miR-148a-3 | cis | 0.000115 | Nightingale | XL_VLDL_T | NA | C | A | 0.028054 | 0.004489 | 9.90E-11 |
| rs1267250! | miR-148a-3 | cis | 0.000115 | Nightingale | L_VLDL_L | NA | C | A | 0.025223 | 0.004536 | 1.40E-08 |
| rs1267250! | miR-148a-3 | cis | 0.000115 | Nightingale | XL_VLDL_P | NA | C | A | 0.02615 | 0.004511 | 2.10E-09 |
| rs1267250! | miR-148a-3 | cis | 0.000115 | Nightingale | Omega_6_ | NA | C | A | -0.02764 | 0.004566 | 1.20E-09 |
| rs1267250! | miR-148a-3 | cis | 0.000115 | Nightingale | M_VLDL_P | NA | C | A | -0.02379 | 0.004502 | 2.00E-08 |
| rs1267250! | miR-148a-3 | cis | 0.000115 | Nightingale | TG_by_PG | NA | C | A | 0.025242 | 0.004356 | 2.80E-09 |
| rs1267250! | miR-148a-3 | cis | 0.000115 | Nightingale | S_LDL_TG_ | NA | C | A | 0.029722 | 0.004549 | 1.30E-11 |
| rs1267250! | miR-148a-3 | cis | 0.000115 | Nightingale | XL_VLDL_T | NA | C | A | 0.026042 | 0.004546 | 1.00E-09 |
| rs1267250! | miR-148a-3 | cis | 0.000115 | Nightingale | S_HDL_C_p | NA | C | A | -0.02841 | 0.004639 | 1.40E-10 |
| rs1267250! | miR-148a-3 | cis | 0.000115 | Nightingale | L_VLDL_PL | NA | C | A | 0.02461 | 0.004529 | 2.30E-08 |
| rs1267250! | miR-148a-3 | cis | 0.000115 | Nightingale | S_VLDL_C_ | NA | C | A | -0.02815 | 0.004565 | 1.50E-10 |
| rs1267250! | miR-148a-3 | cis | 0.000115 | Nightingale | XXL_VLDL_ | NA | C | A | 0.028237 | 0.004522 | 7.40E-11 |
| rs1267250! | miR-148a-3 | cis | 0.000115 | Nightingale | S_VLDL_TG | NA | C | A | 0.027401 | 0.004555 | 8.20E-10 |
| rs1267250! | miR-148a-3 | cis | 0.000115 | Nightingale | S_LDL_FC_ | NA | C | A | -0.03117 | 0.00455 | 9.20E-13 |
| rs1267250! | miR-148a-3 | cis | 0.000115 | Nightingale | L_VLDL_TG | NA | C | A | 0.02701 | 0.004515 | 9.50E-10 |
| rs1267250! | miR-148a-3 | cis | 0.000115 | Nightingale | L_HDL_C_p | NA | C | A | -0.02631 | 0.004425 | 6.70E-10 |
| rs1267250! | miR-148a-3 | cis | 0.000115 | Nightingale | M_VLDL_C_ | NA | C | A | -0.03113 | 0.004418 | 8.10E-14 |
| rs1267250! | miR-148a-3 | cis | 0.000115 | Nightingale | Total_TG | NA | C | A | 0.027854 | 0.004543 | 3.00E-10 |
| rs1267250! | miR-148a-3 | cis | 0.000115 | Nightingale | XXL_VLDL_ | NA | C | A | 0.025454 | 0.004534 | 4.20E-09 |
| rs1267250! | miR-148a-3 | cis | 0.000115 | Nightingale | XL_VLDL_C | NA | C | A | -0.02833 | 0.004499 | 1.60E-11 |
| rs1267250! | miR-148a-3 | cis | 0.000115 | Nightingale | L_HDL_CE_ | NA | C | A | -0.02616 | 0.004491 | 2.00E-09 |
| rs1267250! | miR-148a-3 | cis | 0.000115 | Nightingale | VLDL_TG | NA | C | A | 0.028101 | 0.004517 | 1.80E-10 |
| rs1267250! | miR-148a-3 | cis | 0.000115 | Nightingale | XXL_VLDL_ | NA | C | A | 0.028292 | 0.004505 | 5.50E-11 |
| rs1267250! | miR-148a-3 | cis | 0.000115 | Nightingale | XXL_VLDL_ | NA | C | A | 0.02829 | 0.004515 | 7.50E-11 |
| rs1267250! | miR-148a-3 | cis | 0.000115 | Nightingale | XS_VLDL_T | NA | C | A | 0.025267 | 0.004459 | 4.30E-09 |
| rs1267250! | miR-148a-3 | cis | 0.000115 | Nightingale | M_VLDL_T | NA | C | A | 0.025653 | 0.00456 | 1.00E-08 |
| rs1267250! | miR-148a-3 | cis | 0.000115 | Nightingale | M_HDL_TG | NA | C | A | 0.028957 | 0.004636 | 1.10E-10 |
| rs1267250! | miR-148a-3 | cis | 0.000115 | Nightingale | S_VLDL_PL | NA | C | A | -0.02892 | 0.004491 | 1.80E-11 |
| rs1267250! | miR-148a-3 | cis | 0.000115 | Nightingale | XL_VLDL_F | NA | C | A | 0.024299 | 0.004546 | 3.50E-08 |
| rs1267250! | miR-148a-3 | cis | 0.000115 | Nightingale | VLDL_size | NA | C | A | 0.02631 | 0.004375 | 2.10E-10 |
| rs1267250! | miR-148a-3 | cis | 0.000115 | Nightingale | L_VLDL_CE | NA | C | A | -0.02313 | 0.004489 | 2.80E-08 |
| rs1267250! | miR-148a-3 | cis | 0.000115 | Nightingale | L_VLDL_P | NA | C | A | 0.025091 | 0.004532 | 1.50E-08 |
| rs1267250! | miR-148a-3 | cis | 0.000115 | Nightingale | L_HDL_TG_ | NA | C | A | 0.025106 | 0.004432 | 7.40E-09 |
| rs1267250! | miR-148a-3 | cis | 0.000115 | Nightingale | XS_VLDL_C | NA | C | A | -0.02366 | 0.004448 | 4.50E-08 |
| rs1267250! | miR-148a-3 | cis | 0.000115 | Nightingale | S_VLDL_TG | NA | C | A | 0.028796 | 0.00454 | 4.60E-11 |
| rs1267250! | miR-148a-3 | cis | 0.000115 | Nightingale | XL_VLDL_C | NA | C | A | -0.02744 | 0.004497 | 6.90E-11 |
| rs1267250! | miR-148a-3 | cis | 0.000115 | Nightingale | XXL_VLDL_ | NA | C | A | 0.028688 | 0.004521 | 3.60E-11 |
| rs1267250! | miR-148a-3 | cis | 0.000115 | Nightingale | M_HDL_C_ | NA | C | A | -0.02435 | 0.00447 | 1.10E-08 |
| rs1267250! | miR-148a-3 | cis | 0.000115 | Nightingale | XXL_VLDL_ | NA | C | A | 0.027178 | 0.004524 | 3.80E-10 |
| rs1267250! | miR-148a-3 | cis | 0.000115 | Nightingale | XL_VLDL_P | NA | C | A | 0.024824 | 0.00453 | 1.50E-08 |
| rs1267250! | miR-148a-3 | cis | 0.000115 | Nightingale | M_VLDL_T | NA | C | A | 0.030059 | 0.004432 | 6.30E-13 |
| rs1267250! | miR-148a-3 | cis | 0.000115 | Nightingale | XL_HDL_TG | NA | C | A | 0.026416 | 0.004423 | 5.40E-10 |
| rs1267250! | miR-148a-3 | cis | 0.000115 | Nightingale | S_HDL_TG | NA | C | A | 0.026836 | 0.004553 | 1.00E-09 |
| rs1267250! | miR-148a-3 | cis | 0.000115 | Nightingale | L_LDL_FC_ | NA | C | A | -0.0271 | 0.004475 | 5.90E-10 |
| rs1270145! | miR-339-5p | cis | 4.70E-05 | Nightingale | XS_VLDL_C | NA | G | A | -0.02541 | 0.004269 | 1.70E-09 |
| rs1270145! | miR-339-5p | cis | 4.70E-05 | Nightingale | XS_VLDL_L | NA | G | A | -0.02421 | 0.004324 | 1.10E-08 |
| rs1270145! | miR-339-5p | cis | 4.70E-05 | Nightingale | M_VLDL_P | NA | G | A | -0.02356 | 0.00422 | 4.30E-08 |
| rs1270145! | miR-339-5p | cis | 4.70E-05 | Nightingale | XS_VLDL_C | NA | G | A | -0.02568 | 0.004244 | 8.70E-10 |
| rs1270145! | miR-339-5p | cis | 4.70E-05 | Nightingale | XS_VLDL_P | NA | G | A | -0.02444 | 0.00434 | 9.70E-09 |
| rs1270145! | miR-339-5p | cis | 4.70E-05 | Nightingale | XS_VLDL_F | NA | G | A | -0.02379 | 0.004322 | 2.20E-08 |
| rs1270145! | miR-339-5p | cis | 4.70E-05 | Nightingale | IDL_PL | NA | G | A | -0.02409 | 0.004244 | 8.20E-09 |
| rs1270145! | miR-339-5p | cis | 4.70E-05 | Nightingale | IDL_L | NA | G | A | -0.02385 | 0.004253 | 1.60E-08 |
| rs1270185( | miR-339-5p | cis | 9.06E-07 | Nightingale | L_VLDL_C_ | NA | T | C | -0.02179 | 0.004086 | 5.00E-08 |
| rs1270185( | miR-339-5p | cis | 9.06E-07 | Nightingale | L_VLDL_CE | NA | T | C | -0.02217 | 0.004002 | 3.40E-08 |
| rs1270196! | miR-339-5p | cis | 3.47E-05 | Nightingale | L_HDL_FC | NA | G | A | -0.02175 | 0.003799 | 1.90E-08 |
| rs1270196! | miR-339-5p | cis | 3.47E-05 | Nightingale | L_VLDL_CE | NA | G | A | -0.02289 | 0.004145 | 1.30E-08 |
| rs1270196! | miR-339-5p | cis | 3.47E-05 | Nightingale | L_VLDL_C_ | NA | G | A | -0.0222 | 0.004231 | 2.60E-08 |
| rs1270196! | miR-339-5p | cis | 3.47E-05 | Nightingale | HDL_size | NA | G | A | -0.02181 | 0.003841 | 3.20E-08 |
| rs1270196! | miR-339-5p | cis | 3.47E-05 | Nightingale | L_HDL_L | NA | G | A | -0.02133 | 0.003804 | 3.70E-08 |

| | | | | | | | | | | | |
|---|---|---|---|---|---|---|---|---|---|---|---|
| rs1270196! | miR-339-5p | cis | 3.47E-05 | Nightingale | S_HDL_FC_ | NA | G | A | -0.02231 | 0.004112 | 4.80E-08 |
| rs1270196! | miR-339-5p | cis | 3.47E-05 | Nightingale | L_HDL_P | NA | G | A | -0.02155 | 0.003797 | 2.40E-08 |
| rs1270197( | miR-339-5p | cis | 4.78E-05 | Nightingale | XL_HDL_CE | NA | C | T | -0.02211 | 0.003845 | 4.80E-08 |
| rs1270197( | miR-339-5p | cis | 4.78E-05 | Nightingale | L_HDL_FC | NA | C | T | -0.0224 | 0.003762 | 5.00E-09 |
| rs1270197( | miR-339-5p | cis | 4.78E-05 | Nightingale | L_VLDL_CE | NA | C | T | -0.02456 | 0.004105 | 1.00E-09 |
| rs1270197( | miR-339-5p | cis | 4.78E-05 | Nightingale | L_HDL_P | NA | C | T | -0.022 | 0.00376 | 8.50E-09 |
| rs1270197( | miR-339-5p | cis | 4.78E-05 | Nightingale | L_VLDL_C_ | NA | C | T | -0.02396 | 0.00419 | 2.10E-09 |
| rs1270197( | miR-339-5p | cis | 4.78E-05 | Nightingale | M_VLDL_P | NA | C | T | -0.02314 | 0.004117 | 1.10E-08 |
| rs1270197( | miR-339-5p | cis | 4.78E-05 | Nightingale | L_HDL_CE | NA | C | T | -0.02156 | 0.003791 | 3.20E-08 |
| rs1270197( | miR-339-5p | cis | 4.78E-05 | Nightingale | L_HDL_L | NA | C | T | -0.02174 | 0.003766 | 1.40E-08 |
| rs1270197( | miR-339-5p | cis | 4.78E-05 | Nightingale | VLDL_size | NA | C | T | 0.023243 | 0.004 | 3.60E-08 |
| rs1270197( | miR-339-5p | cis | 4.78E-05 | Nightingale | XL_HDL_P | NA | C | T | -0.02219 | 0.003863 | 2.50E-08 |
| rs1270197( | miR-339-5p | cis | 4.78E-05 | Nightingale | L_HDL_C | NA | C | T | -0.02185 | 0.00378 | 1.80E-08 |
| rs1270197( | miR-339-5p | cis | 4.78E-05 | Nightingale | XL_HDL_L | NA | C | T | -0.02239 | 0.003858 | 3.70E-08 |
| rs1270197( | miR-339-5p | cis | 4.78E-05 | Nightingale | S_HDL_FC_ | NA | C | T | -0.02332 | 0.004072 | 1.30E-08 |
| rs1270197( | miR-339-5p | cis | 4.78E-05 | Nightingale | HDL_size | NA | C | T | -0.02245 | 0.003803 | 8.80E-09 |
| rs1270197( | miR-339-5p | cis | 4.78E-05 | Nightingale | M_VLDL_F | NA | C | T | -0.02242 | 0.00408 | 3.50E-08 |
| rs1270197( | miR-339-5p | cis | 4.78E-05 | Nightingale | XL_HDL_PL | NA | C | T | -0.02244 | 0.003859 | 4.30E-08 |
| rs1270201( | miR-339-5p | cis | 4.43E-05 | Nightingale | L_HDL_FC | NA | T | C | -0.02158 | 0.003736 | 1.30E-08 |
| rs1270201( | miR-339-5p | cis | 4.43E-05 | Nightingale | L_HDL_P | NA | T | C | -0.02138 | 0.003734 | 1.70E-08 |
| rs1270201( | miR-339-5p | cis | 4.43E-05 | Nightingale | L_HDL_C | NA | T | C | -0.02143 | 0.003754 | 2.40E-08 |
| rs1270201( | miR-339-5p | cis | 4.43E-05 | Nightingale | L_VLDL_CE | NA | T | C | -0.02404 | 0.004076 | 1.40E-09 |
| rs1270201( | miR-339-5p | cis | 4.43E-05 | Nightingale | VLDL_size | NA | T | C | 0.024058 | 0.003973 | 6.90E-09 |
| rs1270201( | miR-339-5p | cis | 4.43E-05 | Nightingale | L_HDL_L | NA | T | C | -0.0211 | 0.003741 | 2.80E-08 |
| rs1270201( | miR-339-5p | cis | 4.43E-05 | Nightingale | M_VLDL_T | NA | T | C | 0.021747 | 0.004025 | 4.80E-08 |
| rs1270201( | miR-339-5p | cis | 4.43E-05 | Nightingale | L_HDL_CE | NA | T | C | -0.02127 | 0.003765 | 3.40E-08 |
| rs1270201( | miR-339-5p | cis | 4.43E-05 | Nightingale | HDL_size | NA | T | C | -0.02167 | 0.003777 | 2.20E-08 |
| rs1270201( | miR-339-5p | cis | 4.43E-05 | Nightingale | M_VLDL_F | NA | T | C | -0.02193 | 0.004052 | 4.40E-08 |
| rs1270201( | miR-339-5p | cis | 4.43E-05 | Nightingale | M_VLDL_P | NA | T | C | -0.02242 | 0.004089 | 2.00E-08 |
| rs1270201( | miR-339-5p | cis | 4.43E-05 | Nightingale | L_VLDL_C_ | NA | T | C | -0.02332 | 0.004161 | 3.90E-09 |
| rs1270201( | miR-339-5p | cis | 4.43E-05 | Nightingale | S_HDL_FC_ | NA | T | C | -0.02239 | 0.004044 | 4.80E-08 |
| rs1270201 | miR-339-5p | cis | 5.21E-05 | Nightingale | L_HDL_FC | NA | T | C | -0.02168 | 0.003737 | 1.20E-08 |
| rs1270201 | miR-339-5p | cis | 5.21E-05 | Nightingale | L_VLDL_CE | NA | T | C | -0.02419 | 0.004077 | 1.10E-09 |
| rs1270201 | miR-339-5p | cis | 5.21E-05 | Nightingale | L_HDL_C | NA | T | C | -0.02152 | 0.003755 | 2.10E-08 |
| rs1270201 | miR-339-5p | cis | 5.21E-05 | Nightingale | L_HDL_P | NA | T | C | -0.02148 | 0.003734 | 1.40E-08 |
| rs1270201 | miR-339-5p | cis | 5.21E-05 | Nightingale | M_VLDL_P | NA | T | C | -0.02253 | 0.004089 | 1.80E-08 |
| rs1270201 | miR-339-5p | cis | 5.21E-05 | Nightingale | M_VLDL_T | NA | T | C | 0.021837 | 0.004025 | 4.30E-08 |
| rs1270201 | miR-339-5p | cis | 5.21E-05 | Nightingale | VLDL_size | NA | T | C | 0.024084 | 0.003973 | 6.90E-09 |
| rs1270201 | miR-339-5p | cis | 5.21E-05 | Nightingale | L_VLDL_C_ | NA | T | C | -0.02346 | 0.004162 | 3.20E-09 |
| rs1270201 | miR-339-5p | cis | 5.21E-05 | Nightingale | M_VLDL_F | NA | T | C | -0.02202 | 0.004053 | 3.90E-08 |
| rs1270201 | miR-339-5p | cis | 5.21E-05 | Nightingale | L_HDL_CE | NA | T | C | -0.02136 | 0.003765 | 3.00E-08 |
| rs1270201 | miR-339-5p | cis | 5.21E-05 | Nightingale | L_HDL_L | NA | T | C | -0.0212 | 0.003741 | 2.50E-08 |
| rs1270201 | miR-339-5p | cis | 5.21E-05 | Nightingale | HDL_size | NA | T | C | -0.02178 | 0.003777 | 1.90E-08 |
| rs1270208 | miR-339-5p | cis | 5.30E-05 | Nightingale | L_VLDL_CE | NA | C | T | -0.02294 | 0.004066 | 6.80E-09 |
| rs1270208 | miR-339-5p | cis | 5.30E-05 | Nightingale | L_HDL_L | NA | C | T | -0.02105 | 0.003731 | 2.90E-08 |
| rs1270208 | miR-339-5p | cis | 5.30E-05 | Nightingale | VLDL_size | NA | C | T | 0.023462 | 0.003962 | 1.50E-08 |
| rs1270208 | miR-339-5p | cis | 5.30E-05 | Nightingale | L_HDL_FC | NA | C | T | -0.02141 | 0.003727 | 1.60E-08 |
| rs1270208 | miR-339-5p | cis | 5.30E-05 | Nightingale | L_VLDL_C_ | NA | C | T | -0.02238 | 0.00415 | 1.40E-08 |
| rs1270208 | miR-339-5p | cis | 5.30E-05 | Nightingale | L_HDL_CE | NA | C | T | -0.02115 | 0.003755 | 4.10E-08 |
| rs1270208 | miR-339-5p | cis | 5.30E-05 | Nightingale | HDL_size | NA | C | T | -0.02157 | 0.003767 | 2.50E-08 |
| rs1270208 | miR-339-5p | cis | 5.30E-05 | Nightingale | L_HDL_P | NA | C | T | -0.02128 | 0.003724 | 1.90E-08 |
| rs1270208 | miR-339-5p | cis | 5.30E-05 | Nightingale | L_HDL_C | NA | C | T | -0.02129 | 0.003745 | 2.90E-08 |
| rs1270243! | miR-339-5p | cis | 6.97E-05 | Nightingale | M_VLDL_P | NA | C | T | -0.02409 | 0.004041 | 5.30E-10 |
| rs1270243! | miR-339-5p | cis | 6.97E-05 | Nightingale | L_VLDL_C_ | NA | C | T | -0.02521 | 0.004113 | 9.10E-11 |
| rs1270243! | miR-339-5p | cis | 6.97E-05 | Nightingale | XL_VLDL_T | NA | C | T | 0.022962 | 0.004082 | 4.40E-09 |
| rs1270243! | miR-339-5p | cis | 6.97E-05 | Nightingale | M_VLDL_F | NA | C | T | -0.0227 | 0.004006 | 5.70E-09 |
| rs1270243! | miR-339-5p | cis | 6.97E-05 | Nightingale | XL_VLDL_F | NA | C | T | -0.02335 | 0.004083 | 6.50E-09 |
| rs1270243! | miR-339-5p | cis | 6.97E-05 | Nightingale | M_VLDL_C_ | NA | C | T | -0.02163 | 0.003965 | 2.30E-08 |
| rs1270243! | miR-339-5p | cis | 6.97E-05 | Nightingale | L_VLDL_CE | NA | C | T | -0.02482 | 0.004029 | 1.80E-10 |
| rs1270243! | miR-339-5p | cis | 6.97E-05 | Nightingale | M_VLDL_T | NA | C | T | 0.022435 | 0.003978 | 6.40E-09 |
| rs1270243( | miR-339-5p | cis | 5.61E-05 | Nightingale | XL_VLDL_F | NA | A | T | -0.02476 | 0.004098 | 9.00E-10 |
| rs1270243( | miR-339-5p | cis | 5.61E-05 | Nightingale | XL_VLDL_T | NA | A | T | 0.024015 | 0.004097 | 9.10E-10 |
| rs1270243( | miR-339-5p | cis | 5.61E-05 | Nightingale | L_VLDL_C_ | NA | A | T | -0.02596 | 0.004128 | 2.50E-11 |

| | | | | | | | | | | | |
|---|---|---|---|---|---|---|---|---|---|---|---|
| rs1270243( | miR-339-5p | cis | 5.61E-05 | Nightingale | M_VLDL_P | NA | A | T | -0.02455 | 0.004056 | 2.60E-10 |
| rs1270243( | miR-339-5p | cis | 5.61E-05 | Nightingale | M_VLDL_C | NA | A | T | -0.02188 | 0.00397 | 1.80E-08 |
| rs1270243( | miR-339-5p | cis | 5.61E-05 | Nightingale | L_VLDL_CE | NA | A | T | -0.02592 | 0.004044 | 2.90E-11 |
| rs1270243( | miR-339-5p | cis | 5.61E-05 | Nightingale | M_VLDL_F( | NA | A | T | -0.02343 | 0.00402 | 1.80E-09 |
| rs1270243( | miR-339-5p | cis | 5.61E-05 | Nightingale | VLDL_size | NA | A | T | 0.023367 | 0.003941 | 9.10E-09 |
| rs1270243( | miR-339-5p | cis | 5.61E-05 | Nightingale | XL_VLDL_C | NA | A | T | -0.02223 | 0.004053 | 2.00E-08 |
| rs1270243( | miR-339-5p | cis | 5.61E-05 | Nightingale | M_VLDL_T( | NA | A | T | 0.023246 | 0.003993 | 1.80E-09 |
| rs1270243( | miR-339-5p | cis | 5.61E-05 | Nightingale | M_VLDL_C | NA | A | T | -0.02252 | 0.00398 | 6.10E-09 |
| rs1270243' | miR-339-5p | cis | 5.56E-05 | Nightingale | VLDL_size | NA | A | G | 0.023369 | 0.003941 | 9.10E-09 |
| rs1270243' | miR-339-5p | cis | 5.56E-05 | Nightingale | L_VLDL_C_ | NA | A | G | -0.02596 | 0.004128 | 2.60E-11 |
| rs1270243' | miR-339-5p | cis | 5.56E-05 | Nightingale | M_VLDL_T( | NA | A | G | 0.023243 | 0.003993 | 1.90E-09 |
| rs1270243' | miR-339-5p | cis | 5.56E-05 | Nightingale | XL_VLDL_C | NA | A | G | -0.02223 | 0.004053 | 2.00E-08 |
| rs1270243' | miR-339-5p | cis | 5.56E-05 | Nightingale | M_VLDL_P | NA | A | G | -0.02455 | 0.004056 | 2.60E-10 |
| rs1270243' | miR-339-5p | cis | 5.56E-05 | Nightingale | M_VLDL_C | NA | A | G | -0.02251 | 0.00398 | 6.20E-09 |
| rs1270243' | miR-339-5p | cis | 5.56E-05 | Nightingale | M_VLDL_F( | NA | A | G | -0.02342 | 0.00402 | 1.90E-09 |
| rs1270243' | miR-339-5p | cis | 5.56E-05 | Nightingale | M_VLDL_C | NA | A | G | -0.02188 | 0.003969 | 1.80E-08 |
| rs1270243' | miR-339-5p | cis | 5.56E-05 | Nightingale | XL_VLDL_F | NA | A | G | -0.02476 | 0.004098 | 9.00E-10 |
| rs1270243' | miR-339-5p | cis | 5.56E-05 | Nightingale | L_VLDL_CE | NA | A | G | -0.02592 | 0.004044 | 2.90E-11 |
| rs1270243' | miR-339-5p | cis | 5.56E-05 | Nightingale | XL_VLDL_T | NA | A | G | 0.024015 | 0.004097 | 9.10E-10 |
| rs1270243: | miR-339-5p | cis | 5.59E-05 | Nightingale | M_VLDL_P | NA | T | C | -0.02458 | 0.004057 | 2.60E-10 |
| rs1270243: | miR-339-5p | cis | 5.59E-05 | Nightingale | M_VLDL_C | NA | T | C | -0.02199 | 0.003971 | 1.60E-08 |
| rs1270243: | miR-339-5p | cis | 5.59E-05 | Nightingale | VLDL_size | NA | T | C | 0.023401 | 0.003942 | 8.40E-09 |
| rs1270243: | miR-339-5p | cis | 5.59E-05 | Nightingale | M_VLDL_F( | NA | T | C | -0.0235 | 0.004021 | 1.70E-09 |
| rs1270243: | miR-339-5p | cis | 5.59E-05 | Nightingale | M_VLDL_T( | NA | T | C | 0.023336 | 0.003994 | 1.70E-09 |
| rs1270243: | miR-339-5p | cis | 5.59E-05 | Nightingale | XL_VLDL_C | NA | T | C | -0.02225 | 0.004054 | 1.90E-08 |
| rs1270243: | miR-339-5p | cis | 5.59E-05 | Nightingale | XL_VLDL_F | NA | T | C | -0.02485 | 0.004099 | 8.00E-10 |
| rs1270243: | miR-339-5p | cis | 5.59E-05 | Nightingale | M_VLDL_C | NA | T | C | -0.02262 | 0.003981 | 5.40E-09 |
| rs1270243: | miR-339-5p | cis | 5.59E-05 | Nightingale | XL_VLDL_T | NA | T | C | 0.024035 | 0.004098 | 9.10E-10 |
| rs1270243: | miR-339-5p | cis | 5.59E-05 | Nightingale | L_VLDL_CE | NA | T | C | -0.02589 | 0.004045 | 3.10E-11 |
| rs1270243: | miR-339-5p | cis | 5.59E-05 | Nightingale | L_VLDL_C_ | NA | T | C | -0.0259 | 0.004129 | 3.00E-11 |
| rs1270244( | miR-339-5p | cis | 5.85E-05 | Nightingale | M_VLDL_T( | NA | A | G | 0.023113 | 0.003992 | 2.20E-09 |
| rs1270244( | miR-339-5p | cis | 5.85E-05 | Nightingale | M_VLDL_P | NA | A | G | -0.02447 | 0.004056 | 3.00E-10 |
| rs1270244( | miR-339-5p | cis | 5.85E-05 | Nightingale | M_VLDL_F( | NA | A | G | -0.02329 | 0.004019 | 2.20E-09 |
| rs1270244( | miR-339-5p | cis | 5.85E-05 | Nightingale | XL_VLDL_C | NA | A | G | -0.02202 | 0.004052 | 2.60E-08 |
| rs1270244( | miR-339-5p | cis | 5.85E-05 | Nightingale | M_VLDL_C | NA | A | G | -0.02174 | 0.003969 | 2.10E-08 |
| rs1270244( | miR-339-5p | cis | 5.85E-05 | Nightingale | M_VLDL_C | NA | A | G | -0.02238 | 0.003979 | 7.20E-09 |
| rs1270244( | miR-339-5p | cis | 5.85E-05 | Nightingale | XL_VLDL_F | NA | A | G | -0.02457 | 0.004097 | 1.20E-09 |
| rs1270244( | miR-339-5p | cis | 5.85E-05 | Nightingale | XL_VLDL_T | NA | A | G | 0.023767 | 0.004096 | 1.30E-09 |
| rs1270244( | miR-339-5p | cis | 5.85E-05 | Nightingale | VLDL_size | NA | A | G | 0.023113 | 0.00394 | 1.30E-08 |
| rs1270244( | miR-339-5p | cis | 5.85E-05 | Nightingale | L_VLDL_CE | NA | A | G | -0.02571 | 0.004043 | 4.10E-11 |
| rs1270244( | miR-339-5p | cis | 5.85E-05 | Nightingale | L_VLDL_C_ | NA | A | G | -0.02575 | 0.004128 | 3.70E-11 |
| rs1270244: | miR-339-5p | cis | 5.96E-05 | Nightingale | XL_VLDL_C | NA | A | C | -0.02199 | 0.004052 | 2.70E-08 |
| rs1270244: | miR-339-5p | cis | 5.96E-05 | Nightingale | XL_VLDL_F | NA | A | C | -0.02452 | 0.004097 | 1.30E-09 |
| rs1270244: | miR-339-5p | cis | 5.96E-05 | Nightingale | M_VLDL_F( | NA | A | C | -0.02325 | 0.00402 | 2.30E-09 |
| rs1270244: | miR-339-5p | cis | 5.96E-05 | Nightingale | VLDL_size | NA | A | C | 0.023089 | 0.003941 | 1.30E-08 |
| rs1270244: | miR-339-5p | cis | 5.96E-05 | Nightingale | L_VLDL_C_ | NA | A | C | -0.02574 | 0.004128 | 3.70E-11 |
| rs1270244: | miR-339-5p | cis | 5.96E-05 | Nightingale | L_VLDL_CE | NA | A | C | -0.02569 | 0.004044 | 4.20E-11 |
| rs1270244: | miR-339-5p | cis | 5.96E-05 | Nightingale | XL_VLDL_T | NA | A | C | 0.023741 | 0.004097 | 1.30E-09 |
| rs1270244: | miR-339-5p | cis | 5.96E-05 | Nightingale | M_VLDL_P | NA | A | C | -0.02444 | 0.004056 | 3.10E-10 |
| rs1270244: | miR-339-5p | cis | 5.96E-05 | Nightingale | M_VLDL_C | NA | A | C | -0.02235 | 0.003979 | 7.40E-09 |
| rs1270244: | miR-339-5p | cis | 5.96E-05 | Nightingale | M_VLDL_C | NA | A | C | -0.02171 | 0.003969 | 2.20E-08 |
| rs1270244: | miR-339-5p | cis | 5.96E-05 | Nightingale | M_VLDL_T( | NA | A | C | 0.023083 | 0.003993 | 2.30E-09 |
| rs1322141' | miR-339-5p | cis | 0.000117 | Nightingale | XS_VLDL_C | NA | G | A | -0.02394 | 0.004305 | 1.50E-08 |
| rs1322141' | miR-339-5p | cis | 0.000117 | Nightingale | XS_VLDL_C | NA | G | A | -0.02365 | 0.00433 | 2.80E-08 |
| rs1323240: | miR-339-5p | cis | 0.000104 | Nightingale | XS_VLDL_C | NA | A | G | -0.02406 | 0.004306 | 1.30E-08 |
| rs1323240: | miR-339-5p | cis | 0.000104 | Nightingale | XS_VLDL_C | NA | A | G | -0.02378 | 0.004332 | 2.30E-08 |
| rs1323250: | miR-339-5p | cis | 0.000111 | Nightingale | XS_VLDL_C | NA | C | G | -0.02377 | 0.004327 | 2.40E-08 |
| rs1323250: | miR-339-5p | cis | 0.000111 | Nightingale | XS_VLDL_C | NA | C | G | -0.0241 | 0.004302 | 1.30E-08 |
| rs1323574' | miR-339-5p | cis | 4.16E-05 | Nightingale | L_HDL_C | NA | C | T | -0.02158 | 0.003749 | 1.70E-08 |
| rs1323574' | miR-339-5p | cis | 4.16E-05 | Nightingale | L_VLDL_C_ | NA | C | T | -0.02311 | 0.004155 | 4.70E-09 |
| rs1323574' | miR-339-5p | cis | 4.16E-05 | Nightingale | VLDL_size | NA | C | T | 0.023954 | 0.003967 | 7.10E-09 |
| rs1323574' | miR-339-5p | cis | 4.16E-05 | Nightingale | L_VLDL_CE | NA | C | T | -0.02387 | 0.004071 | 1.50E-09 |
| rs1323574' | miR-339-5p | cis | 4.16E-05 | Nightingale | L_HDL_FC | NA | C | T | -0.02169 | 0.003731 | 1.00E-08 |

| | | | | | | | | | | | |
|---|---|---|---|---|---|---|---|---|---|---|---|
| rs1323574 | miR-339-5p | cis | 4.16E-05 | Nightingale | L_HDL_L | NA | C | T | -0.02126 | 0.003735 | 2.00E-08 |
| rs1323574 | miR-339-5p | cis | 4.16E-05 | Nightingale | M_VLDL_P | NA | C | T | -0.02217 | 0.004083 | 2.50E-08 |
| rs1323574 | miR-339-5p | cis | 4.16E-05 | Nightingale | L_HDL_P | NA | C | T | -0.02155 | 0.003729 | 1.20E-08 |
| rs1323574 | miR-339-5p | cis | 4.16E-05 | Nightingale | L_HDL_CE | NA | C | T | -0.02144 | 0.003759 | 2.40E-08 |
| rs1323574 | miR-339-5p | cis | 4.16E-05 | Nightingale | HDL_size | NA | C | T | -0.02181 | 0.003772 | 1.60E-08 |
| rs1330773 | miR-339-5p | cis | 5.42E-05 | Nightingale | L_VLDL_CE | NA | A | C | -0.02593 | 0.004044 | 2.90E-11 |
| rs1330773 | miR-339-5p | cis | 5.42E-05 | Nightingale | XL_VLDL_C | NA | A | C | -0.02221 | 0.004053 | 2.10E-08 |
| rs1330773 | miR-339-5p | cis | 5.42E-05 | Nightingale | M_VLDL_P | NA | A | C | -0.02458 | 0.004056 | 2.60E-10 |
| rs1330773 | miR-339-5p | cis | 5.42E-05 | Nightingale | M_VLDL_F | NA | A | C | -0.02342 | 0.00402 | 1.90E-09 |
| rs1330773 | miR-339-5p | cis | 5.42E-05 | Nightingale | M_VLDL_T | NA | A | C | 0.023247 | 0.003993 | 1.90E-09 |
| rs1330773 | miR-339-5p | cis | 5.42E-05 | Nightingale | XL_VLDL_T | NA | A | C | 0.023993 | 0.004097 | 9.80E-10 |
| rs1330773 | miR-339-5p | cis | 5.42E-05 | Nightingale | XL_VLDL_F | NA | A | C | -0.02468 | 0.004098 | 1.00E-09 |
| rs1330773 | miR-339-5p | cis | 5.42E-05 | Nightingale | M_VLDL_C | NA | A | C | -0.02251 | 0.00398 | 6.40E-09 |
| rs1330773 | miR-339-5p | cis | 5.42E-05 | Nightingale | M_VLDL_C | NA | A | C | -0.02188 | 0.00397 | 1.90E-08 |
| rs1330773 | miR-339-5p | cis | 5.42E-05 | Nightingale | VLDL_size | NA | A | C | 0.023346 | 0.003941 | 9.80E-09 |
| rs1330773 | miR-339-5p | cis | 5.42E-05 | Nightingale | L_VLDL_C_ | NA | A | C | -0.026 | 0.004128 | 2.50E-11 |
| rs1330893 | miR-339-5p | cis | 0.000135 | Nightingale | L_VLDL_C_ | NA | C | G | -0.02447 | 0.00411 | 2.80E-10 |
| rs1330893 | miR-339-5p | cis | 0.000135 | Nightingale | L_VLDL_CE | NA | C | G | -0.02419 | 0.004026 | 4.50E-10 |
| rs1330893 | miR-339-5p | cis | 0.000135 | Nightingale | XL_VLDL_T | NA | C | G | 0.022345 | 0.004079 | 1.00E-08 |
| rs1330893 | miR-339-5p | cis | 0.000135 | Nightingale | M_VLDL_P | NA | C | G | -0.02332 | 0.004038 | 1.60E-09 |
| rs1330893 | miR-339-5p | cis | 0.000135 | Nightingale | M_VLDL_C | NA | C | G | -0.02107 | 0.003962 | 4.80E-08 |
| rs1330893 | miR-339-5p | cis | 0.000135 | Nightingale | M_VLDL_F | NA | C | G | -0.02198 | 0.004002 | 1.50E-08 |
| rs1330893 | miR-339-5p | cis | 0.000135 | Nightingale | XL_VLDL_F | NA | C | G | -0.02266 | 0.00408 | 1.70E-08 |
| rs1330893 | miR-339-5p | cis | 0.000135 | Nightingale | M_VLDL_T | NA | C | G | 0.021821 | 0.003975 | 1.50E-08 |
| rs1330894 | miR-339-5p | cis | 4.97E-05 | Nightingale | XL_VLDL_C | NA | A | G | -0.0222 | 0.004053 | 2.10E-08 |
| rs1330894 | miR-339-5p | cis | 4.97E-05 | Nightingale | L_VLDL_CE | NA | A | G | -0.02587 | 0.004044 | 3.20E-11 |
| rs1330894 | miR-339-5p | cis | 4.97E-05 | Nightingale | M_VLDL_F | NA | A | G | -0.02339 | 0.00402 | 2.00E-09 |
| rs1330894 | miR-339-5p | cis | 4.97E-05 | Nightingale | M_VLDL_T | NA | A | G | 0.023213 | 0.003993 | 2.00E-09 |
| rs1330894 | miR-339-5p | cis | 4.97E-05 | Nightingale | M_VLDL_P | NA | A | G | -0.02453 | 0.004056 | 2.80E-10 |
| rs1330894 | miR-339-5p | cis | 4.97E-05 | Nightingale | L_VLDL_C_ | NA | A | G | -0.02592 | 0.004128 | 2.80E-11 |
| rs1330894 | miR-339-5p | cis | 4.97E-05 | Nightingale | VLDL_size | NA | A | G | 0.023328 | 0.003941 | 9.80E-09 |
| rs1330894 | miR-339-5p | cis | 4.97E-05 | Nightingale | M_VLDL_C | NA | A | G | -0.02185 | 0.003969 | 1.90E-08 |
| rs1330894 | miR-339-5p | cis | 4.97E-05 | Nightingale | XL_VLDL_F | NA | A | G | -0.0247 | 0.004098 | 9.80E-10 |
| rs1330894 | miR-339-5p | cis | 4.97E-05 | Nightingale | M_VLDL_C | NA | A | G | -0.02248 | 0.00398 | 6.50E-09 |
| rs1330894 | miR-339-5p | cis | 4.97E-05 | Nightingale | XL_VLDL_T | NA | A | G | 0.023973 | 0.004097 | 9.90E-10 |
| rs1331153 | miR-339-5p | cis | 5.85E-05 | Nightingale | L_VLDL_CE | NA | A | C | -0.02571 | 0.004044 | 4.10E-11 |
| rs1331153 | miR-339-5p | cis | 5.85E-05 | Nightingale | M_VLDL_P | NA | A | C | -0.02444 | 0.004056 | 3.00E-10 |
| rs1331153 | miR-339-5p | cis | 5.85E-05 | Nightingale | XL_VLDL_F | NA | A | C | -0.02454 | 0.004097 | 1.20E-09 |
| rs1331153 | miR-339-5p | cis | 5.85E-05 | Nightingale | M_VLDL_C | NA | A | C | -0.02235 | 0.003979 | 7.50E-09 |
| rs1331153 | miR-339-5p | cis | 5.85E-05 | Nightingale | XL_VLDL_T | NA | A | C | 0.02376 | 0.004097 | 1.30E-09 |
| rs1331153 | miR-339-5p | cis | 5.85E-05 | Nightingale | M_VLDL_F | NA | A | C | -0.02326 | 0.00402 | 2.30E-09 |
| rs1331153 | miR-339-5p | cis | 5.85E-05 | Nightingale | L_VLDL_C_ | NA | A | C | -0.02576 | 0.004128 | 3.60E-11 |
| rs1331153 | miR-339-5p | cis | 5.85E-05 | Nightingale | XL_VLDL_C | NA | A | C | -0.02201 | 0.004052 | 2.60E-08 |
| rs1331153 | miR-339-5p | cis | 5.85E-05 | Nightingale | VLDL_size | NA | A | C | 0.023103 | 0.003941 | 1.30E-08 |
| rs1331153 | miR-339-5p | cis | 5.85E-05 | Nightingale | M_VLDL_C | NA | A | C | -0.02171 | 0.003969 | 2.20E-08 |
| rs1331153 | miR-339-5p | cis | 5.85E-05 | Nightingale | M_VLDL_T | NA | A | C | 0.023082 | 0.003992 | 2.30E-09 |
| rs1574108 | miR-339-5p | cis | 8.99E-07 | Nightingale | L_VLDL_C_ | NA | C | T | -0.02256 | 0.004087 | 1.80E-08 |
| rs1574108 | miR-339-5p | cis | 8.99E-07 | Nightingale | L_VLDL_CE | NA | C | T | -0.02295 | 0.004004 | 1.20E-08 |
| rs1574108 | miR-339-5p | cis | 8.99E-07 | Nightingale | L_HDL_FC | NA | C | T | -0.02052 | 0.00367 | 4.50E-08 |
| rs174448 | miR-1908-5 | cis | 1.95E-05 | Nightingale | M_HDL_C_ | NA | A | G | 0.041705 | 0.004113 | 5.90E-26 |
| rs174448 | miR-1908-5 | cis | 1.95E-05 | Nightingale | HDL_size | NA | A | G | 0.048895 | 0.003828 | 4.40E-41 |
| rs174448 | miR-1908-5 | cis | 1.95E-05 | Nightingale | XXL_VLDL_ | NA | A | G | -0.04128 | 0.004161 | 2.30E-24 |
| rs174448 | miR-1908-5 | cis | 1.95E-05 | Nightingale | Total_L | NA | A | G | 0.028128 | 0.004251 | 2.80E-11 |
| rs174448 | miR-1908-5 | cis | 1.95E-05 | Nightingale | Total_C | NA | A | G | 0.046889 | 0.004183 | 9.70E-30 |
| rs174448 | miR-1908-5 | cis | 1.95E-05 | Nightingale | M_LDL_TG | NA | A | G | -0.02323 | 0.004257 | 3.40E-08 |
| rs174448 | miR-1908-5 | cis | 1.95E-05 | Nightingale | IDL_FC_pct | NA | A | G | 0.072558 | 0.004286 | 1.20E-65 |
| rs174448 | miR-1908-5 | cis | 1.95E-05 | Nightingale | S_LDL_PL | NA | A | G | 0.037482 | 0.00431 | 3.60E-18 |
| rs174448 | miR-1908-5 | cis | 1.95E-05 | Nightingale | L_VLDL_C_ | NA | A | G | 0.028566 | 0.004221 | 3.10E-12 |
| rs174448 | miR-1908-5 | cis | 1.95E-05 | Nightingale | ApoA1 | NA | A | G | 0.025691 | 0.003954 | 7.00E-12 |
| rs174448 | miR-1908-5 | cis | 1.95E-05 | Nightingale | LDL_FC | NA | A | G | 0.046719 | 0.004275 | 8.10E-28 |
| rs174448 | miR-1908-5 | cis | 1.95E-05 | Nightingale | M_HDL_CE | NA | A | G | 0.022431 | 0.00399 | 3.20E-09 |
| rs174448 | miR-1908-5 | cis | 1.95E-05 | Nightingale | VLDL_TG | NA | A | G | -0.02944 | 0.004158 | 3.60E-13 |
| rs174448 | miR-1908-5 | cis | 1.95E-05 | Nightingale | XS_VLDL_P | NA | A | G | 0.03303 | 0.004263 | 2.20E-15 |

| | | | | | | | | | | | |
|---|---|---|---|---|---|---|---|---|---|---|---|
| rs174448 | miR-1908-! | cis | 1.95E-05 | Nightingale | ApoB | NA | A | G | 0.030147 | 0.004315 | 3.00E-12 |
| rs174448 | miR-1908-! | cis | 1.95E-05 | Nightingale | M_VLDL_C | NA | A | G | 0.052788 | 0.004056 | 8.10E-40 |
| rs174448 | miR-1908-! | cis | 1.95E-05 | Nightingale | L_VLDL_L | NA | A | G | -0.02242 | 0.004175 | 2.10E-08 |
| rs174448 | miR-1908-! | cis | 1.95E-05 | Nightingale | Total_PL | NA | A | G | 0.036143 | 0.004114 | 2.80E-19 |
| rs174448 | miR-1908-! | cis | 1.95E-05 | Nightingale | LDL_P | NA | A | G | 0.031185 | 0.004318 | 5.90E-13 |
| rs174448 | miR-1908-! | cis | 1.95E-05 | Nightingale | Remnant_( | NA | A | G | 0.034197 | 0.004293 | 9.70E-16 |
| rs174448 | miR-1908-! | cis | 1.95E-05 | Nightingale | DHA_pct | NA | A | G | 0.179296 | 0.004194 | 0 |
| rs174448 | miR-1908-! | cis | 1.95E-05 | Nightingale | L_HDL_L | NA | A | G | 0.051379 | 0.003791 | 1.70E-46 |
| rs174448 | miR-1908-! | cis | 1.95E-05 | Nightingale | MUFA_pct | NA | A | G | -0.13005 | 0.004134 | 2.80E-224 |
| rs174448 | miR-1908-! | cis | 1.95E-05 | Nightingale | S_LDL_C | NA | A | G | 0.025591 | 0.004325 | 6.60E-09 |
| rs174448 | miR-1908-! | cis | 1.95E-05 | Nightingale | PUFA_by_I | NA | A | G | 0.111832 | 0.004138 | 1.90E-164 |
| rs174448 | miR-1908-! | cis | 1.95E-05 | Nightingale | L_LDL_FC | NA | A | G | 0.049461 | 0.004243 | 7.30E-32 |
| rs174448 | miR-1908-! | cis | 1.95E-05 | Nightingale | S_HDL_C_p | NA | A | G | 0.028178 | 0.004271 | 6.80E-11 |
| rs174448 | miR-1908-! | cis | 1.95E-05 | Nightingale | L_VLDL_PL | NA | A | G | -0.05588 | 0.004193 | 2.40E-42 |
| rs174448 | miR-1908-! | cis | 1.95E-05 | Nightingale | IDL_P | NA | A | G | 0.034744 | 0.004275 | 4.50E-16 |
| rs174448 | miR-1908-! | cis | 1.95E-05 | Nightingale | S_VLDL_FC | NA | A | G | 0.057021 | 0.004126 | 8.30E-45 |
| rs174448 | miR-1908-! | cis | 1.95E-05 | Nightingale | HDL_P | NA | A | G | 0.022252 | 0.004057 | 1.00E-08 |
| rs174448 | miR-1908-! | cis | 1.95E-05 | Nightingale | M_VLDL_F | NA | A | G | 0.063454 | 0.004108 | 3.80E-56 |
| rs174448 | miR-1908-! | cis | 1.95E-05 | Nightingale | L_VLDL_CE | NA | A | G | 0.041532 | 0.004135 | 3.00E-25 |
| rs174448 | miR-1908-! | cis | 1.95E-05 | Nightingale | MUFA | NA | A | G | -0.04069 | 0.004276 | 3.50E-22 |
| rs174448 | miR-1908-! | cis | 1.95E-05 | Nightingale | S_LDL_L | NA | A | G | 0.025545 | 0.00432 | 5.80E-09 |
| rs174448 | miR-1908-! | cis | 1.95E-05 | Nightingale | L_HDL_C | NA | A | G | 0.053637 | 0.003805 | 4.90E-50 |
| rs174448 | miR-1908-! | cis | 1.95E-05 | Nightingale | M_HDL_FC | NA | A | G | 0.032399 | 0.003932 | 5.10E-18 |
| rs174448 | miR-1908-! | cis | 1.95E-05 | Nightingale | SFA_pct | NA | A | G | 0.030173 | 0.004262 | 1.50E-12 |
| rs174448 | miR-1908-! | cis | 1.95E-05 | Nightingale | L_HDL_PL_ | NA | A | G | -0.03252 | 0.004182 | 4.00E-15 |
| rs174448 | miR-1908-! | cis | 1.95E-05 | Nightingale | S_VLDL_TG | NA | A | G | -0.04542 | 0.004181 | 1.50E-27 |
| rs174448 | miR-1908-! | cis | 1.95E-05 | Nightingale | XL_VLDL_P | NA | A | G | -0.03245 | 0.004153 | 7.10E-16 |
| rs174448 | miR-1908-! | cis | 1.95E-05 | Nightingale | L_HDL_P | NA | A | G | 0.05221 | 0.003784 | 4.10E-48 |
| rs174448 | miR-1908-! | cis | 1.95E-05 | Nightingale | VLDL_size | NA | A | G | -0.04556 | 0.004027 | 6.10E-32 |
| rs174448 | miR-1908-! | cis | 1.95E-05 | Nightingale | M_HDL_TG | NA | A | G | -0.04162 | 0.004143 | 2.60E-25 |
| rs174448 | miR-1908-! | cis | 1.95E-05 | Nightingale | XL_HDL_FC | NA | A | G | 0.071173 | 0.004025 | 4.30E-75 |
| rs174448 | miR-1908-! | cis | 1.95E-05 | Nightingale | XL_VLDL_C | NA | A | G | 0.040842 | 0.004142 | 1.90E-23 |
| rs174448 | miR-1908-! | cis | 1.95E-05 | Nightingale | IDL_C_pct | NA | A | G | 0.051872 | 0.004218 | 3.60E-35 |
| rs174448 | miR-1908-! | cis | 1.95E-05 | Nightingale | Omega_3_ | NA | A | G | 0.257166 | 0.004221 | 0 |
| rs174448 | miR-1908-! | cis | 1.95E-05 | Nightingale | LDL_PL | NA | A | G | 0.034755 | 0.004311 | 1.70E-15 |
| rs174448 | miR-1908-! | cis | 1.95E-05 | Nightingale | M_HDL_P | NA | A | G | 0.020639 | 0.00398 | 3.90E-08 |
| rs174448 | miR-1908-! | cis | 1.95E-05 | Nightingale | L_LDL_TG_ | NA | A | G | -0.05428 | 0.004237 | 6.80E-38 |
| rs174448 | miR-1908-! | cis | 1.95E-05 | Nightingale | XXL_VLDL_ | NA | A | G | -0.04562 | 0.004146 | 1.80E-29 |
| rs174448 | miR-1908-! | cis | 1.95E-05 | Nightingale | M_LDL_P | NA | A | G | 0.035535 | 0.004333 | 3.50E-16 |
| rs174448 | miR-1908-! | cis | 1.95E-05 | Nightingale | IDL_FC | NA | A | G | 0.064167 | 0.004182 | 9.20E-55 |
| rs174448 | miR-1908-! | cis | 1.95E-05 | Nightingale | LDL_CE | NA | A | G | 0.032938 | 0.00431 | 4.10E-14 |
| rs174448 | miR-1908-! | cis | 1.95E-05 | Nightingale | XL_VLDL_F | NA | A | G | 0.045906 | 0.004187 | 2.10E-29 |
| rs174448 | miR-1908-! | cis | 1.95E-05 | Nightingale | M_VLDL_C | NA | A | G | 0.039453 | 0.004282 | 3.10E-20 |
| rs174448 | miR-1908-! | cis | 1.95E-05 | Nightingale | M_VLDL_T | NA | A | G | -0.05451 | 0.004081 | 3.00E-42 |
| rs174448 | miR-1908-! | cis | 1.95E-05 | Nightingale | L_HDL_FC | NA | A | G | 0.057071 | 0.003787 | 1.40E-56 |
| rs174448 | miR-1908-! | cis | 1.95E-05 | Nightingale | S_HDL_TG_ | NA | A | G | -0.04394 | 0.004109 | 2.00E-28 |
| rs174448 | miR-1908-! | cis | 1.95E-05 | Nightingale | Sphingomy | NA | A | G | 0.079051 | 0.004037 | 3.20E-89 |
| rs174448 | miR-1908-! | cis | 1.95E-05 | Nightingale | IDL_L | NA | A | G | 0.052372 | 0.004177 | 3.10E-37 |
| rs174448 | miR-1908-! | cis | 1.95E-05 | Nightingale | S_LDL_FC_ | NA | A | G | 0.05346 | 0.004187 | 1.90E-38 |
| rs174448 | miR-1908-! | cis | 1.95E-05 | Nightingale | PUFA | NA | A | G | 0.065808 | 0.004213 | 9.70E-57 |
| rs174448 | miR-1908-! | cis | 1.95E-05 | Nightingale | Total_FC | NA | A | G | 0.04339 | 0.004221 | 4.10E-25 |
| rs174448 | miR-1908-! | cis | 1.95E-05 | Nightingale | XL_HDL_PL | NA | A | G | 0.061516 | 0.003884 | 1.40E-61 |
| rs174448 | miR-1908-! | cis | 1.95E-05 | Nightingale | L_VLDL_FC | NA | A | G | -0.02344 | 0.004179 | 4.00E-09 |
| rs174448 | miR-1908-! | cis | 1.95E-05 | Nightingale | XL_HDL_P | NA | A | G | 0.062641 | 0.003888 | 8.40E-64 |
| rs174448 | miR-1908-! | cis | 1.95E-05 | Nightingale | L_HDL_FC_ | NA | A | G | 0.079402 | 0.004014 | 8.90E-93 |
| rs174448 | miR-1908-! | cis | 1.95E-05 | Nightingale | HDL_TG | NA | A | G | -0.02937 | 0.00426 | 2.40E-12 |
| rs174448 | miR-1908-! | cis | 1.95E-05 | Nightingale | Omega_3 | NA | A | G | 0.222553 | 0.004235 | 0 |
| rs174448 | miR-1908-! | cis | 1.95E-05 | Nightingale | S_LDL_PL_ | NA | A | G | 0.037399 | 0.004312 | 4.60E-19 |
| rs174448 | miR-1908-! | cis | 1.95E-05 | Nightingale | IDL_CE_pct | NA | A | G | 0.026902 | 0.00424 | 3.50E-10 |
| rs174448 | miR-1908-! | cis | 1.95E-05 | Nightingale | M_LDL_FC | NA | A | G | 0.03872 | 0.004316 | 5.90E-19 |
| rs174448 | miR-1908-! | cis | 1.95E-05 | Nightingale | L_HDL_C_p | NA | A | G | 0.046763 | 0.004073 | 5.10E-33 |
| rs174448 | miR-1908-! | cis | 1.95E-05 | Nightingale | XL_HDL_L | NA | A | G | 0.064346 | 0.003883 | 3.40E-67 |
| rs174448 | miR-1908-! | cis | 1.95E-05 | Nightingale | M_VLDL_F | NA | A | G | 0.024097 | 0.00432 | 2.70E-08 |

| | | | | | | | | | | | |
|---|---|---|---|---|---|---|---|---|---|---|---|
| rs174448 | miR-1908-! | cis | 1.95E-05 | Nightingale | XL_HDL_PL | NA | A | G | 0.045149 | 0.003969 | 2.10E-32 |
| rs174448 | miR-1908-! | cis | 1.95E-05 | Nightingale | Omega_6_ | NA | A | G | -0.24423 | 0.004258 | 0 |
| rs174448 | miR-1908-! | cis | 1.95E-05 | Nightingale | L_VLDL_TG | NA | A | G | -0.02221 | 0.004157 | 2.40E-08 |
| rs174448 | miR-1908-! | cis | 1.95E-05 | Nightingale | M_LDL_PL_ | NA | A | G | -0.03166 | 0.004318 | 1.90E-14 |
| rs174448 | miR-1908-! | cis | 1.95E-05 | Nightingale | XS_VLDL_C | NA | A | G | 0.052382 | 0.004169 | 1.30E-37 |
| rs174448 | miR-1908-! | cis | 1.95E-05 | Nightingale | M_HDL_FC | NA | A | G | 0.068258 | 0.003829 | 4.20E-78 |
| rs174448 | miR-1908-! | cis | 1.95E-05 | Nightingale | Total_CE | NA | A | G | 0.047915 | 0.004168 | 2.40E-31 |
| rs174448 | miR-1908-! | cis | 1.95E-05 | Nightingale | S_VLDL_C_ | NA | A | G | 0.040489 | 0.004204 | 7.60E-22 |
| rs174448 | miR-1908-! | cis | 1.95E-05 | Nightingale | L_HDL_PL | NA | A | G | 0.050089 | 0.003803 | 6.10E-44 |
| rs174448 | miR-1908-! | cis | 1.95E-05 | Nightingale | S_LDL_TG_ | NA | A | G | -0.05844 | 0.004187 | 3.50E-45 |
| rs174448 | miR-1908-! | cis | 1.95E-05 | Nightingale | XXL_VLDL_ | NA | A | G | -0.03797 | 0.004169 | 9.40E-21 |
| rs174448 | miR-1908-! | cis | 1.95E-05 | Nightingale | IDL_TG_pc | NA | A | G | -0.06137 | 0.004174 | 3.40E-50 |
| rs174448 | miR-1908-! | cis | 1.95E-05 | Nightingale | M_LDL_C | NA | A | G | 0.026522 | 0.004329 | 2.00E-09 |
| rs174448 | miR-1908-! | cis | 1.95E-05 | Nightingale | L_HDL_CE | NA | A | G | 0.052392 | 0.003815 | 1.20E-47 |
| rs174448 | miR-1908-! | cis | 1.95E-05 | Nightingale | L_LDL_L | NA | A | G | 0.040006 | 0.004279 | 8.30E-21 |
| rs174448 | miR-1908-! | cis | 1.95E-05 | Nightingale | S_HDL_FC_ | NA | A | G | 0.0698 | 0.004098 | 1.30E-71 |
| rs174448 | miR-1908-! | cis | 1.95E-05 | Nightingale | XL_VLDL_T | NA | A | G | -0.0415 | 0.004186 | 1.10E-23 |
| rs174448 | miR-1908-! | cis | 1.95E-05 | Nightingale | M_VLDL_P | NA | A | G | 0.044608 | 0.004146 | 1.30E-28 |
| rs174448 | miR-1908-! | cis | 1.95E-05 | Nightingale | L_HDL_CE_ | NA | A | G | 0.033335 | 0.004134 | 2.80E-17 |
| rs174448 | miR-1908-! | cis | 1.95E-05 | Nightingale | LA_pct | NA | A | G | -0.12302 | 0.004222 | 3.40E-188 |
| rs174448 | miR-1908-! | cis | 1.95E-05 | Nightingale | XL_HDL_CE | NA | A | G | 0.066394 | 0.003869 | 4.10E-72 |
| rs174448 | miR-1908-! | cis | 1.95E-05 | Nightingale | M_HDL_C | NA | A | G | 0.024553 | 0.003975 | 8.50E-11 |
| rs174448 | miR-1908-! | cis | 1.95E-05 | Nightingale | L_LDL_CE | NA | A | G | 0.039348 | 0.004282 | 4.30E-20 |
| rs174448 | miR-1908-! | cis | 1.95E-05 | Nightingale | XS_VLDL_F | NA | A | G | 0.077604 | 0.004288 | 5.90E-74 |
| rs174448 | miR-1908-! | cis | 1.95E-05 | Nightingale | XS_VLDL_P | NA | A | G | -0.04166 | 0.004216 | 2.60E-23 |
| rs174448 | miR-1908-! | cis | 1.95E-05 | Nightingale | M_HDL_TG | NA | A | G | -0.03236 | 0.004267 | 1.10E-14 |
| rs174448 | miR-1908-! | cis | 1.95E-05 | Nightingale | XS_VLDL_L | NA | A | G | 0.031145 | 0.004247 | 4.20E-14 |
| rs174448 | miR-1908-! | cis | 1.95E-05 | Nightingale | M_HDL_CE | NA | A | G | 0.028923 | 0.00419 | 1.20E-12 |
| rs174448 | miR-1908-! | cis | 1.95E-05 | Nightingale | IDL_CE | NA | A | G | 0.052094 | 0.004171 | 1.20E-36 |
| rs174448 | miR-1908-! | cis | 1.95E-05 | Nightingale | HDL_PL | NA | A | G | 0.030429 | 0.003906 | 2.30E-16 |
| rs174448 | miR-1908-! | cis | 1.95E-05 | Nightingale | S_VLDL_PL | NA | A | G | 0.05395 | 0.004134 | 3.60E-40 |
| rs174448 | miR-1908-! | cis | 1.95E-05 | Nightingale | S_HDL_TG | NA | A | G | -0.04471 | 0.00419 | 5.00E-28 |
| rs174448 | miR-1908-! | cis | 1.95E-05 | Nightingale | M_LDL_FC_ | NA | A | G | 0.047779 | 0.004139 | 7.00E-32 |
| rs174448 | miR-1908-! | cis | 1.95E-05 | Nightingale | XL_VLDL_P | NA | A | G | -0.02888 | 0.00417 | 7.50E-13 |
| rs174448 | miR-1908-! | cis | 1.95E-05 | Nightingale | XL_VLDL_L | NA | A | G | -0.03129 | 0.004157 | 8.10E-15 |
| rs174448 | miR-1908-! | cis | 1.95E-05 | Nightingale | LDL_C | NA | A | G | 0.037122 | 0.004302 | 1.10E-17 |
| rs174448 | miR-1908-! | cis | 1.95E-05 | Nightingale | Total_P | NA | A | G | 0.026313 | 0.004066 | 1.70E-11 |
| rs174448 | miR-1908-! | cis | 1.95E-05 | Nightingale | Phosphogl | NA | A | G | 0.028174 | 0.004092 | 1.00E-12 |
| rs174448 | miR-1908-! | cis | 1.95E-05 | Nightingale | XS_VLDL_C | NA | A | G | 0.064378 | 0.004073 | 3.20E-58 |
| rs174448 | miR-1908-! | cis | 1.95E-05 | Nightingale | XL_HDL_TG | NA | A | G | -0.05788 | 0.00407 | 2.40E-48 |
| rs174448 | miR-1908-! | cis | 1.95E-05 | Nightingale | S_VLDL_TG | NA | A | G | -0.02445 | 0.004193 | 2.50E-09 |
| rs174448 | miR-1908-! | cis | 1.95E-05 | Nightingale | IDL_PL | NA | A | G | 0.052733 | 0.004169 | 3.90E-38 |
| rs174448 | miR-1908-! | cis | 1.95E-05 | Nightingale | L_LDL_C_p | NA | A | G | 0.053741 | 0.00421 | 8.70E-38 |
| rs174448 | miR-1908-! | cis | 1.95E-05 | Nightingale | TG_by_PG | NA | A | G | -0.04423 | 0.004009 | 2.80E-30 |
| rs174448 | miR-1908-! | cis | 1.95E-05 | Nightingale | XL_HDL_FC | NA | A | G | -0.04124 | 0.003831 | 1.00E-30 |
| rs174448 | miR-1908-! | cis | 1.95E-05 | Nightingale | M_LDL_C_ | NA | A | G | 0.069409 | 0.004324 | 1.20E-57 |
| rs174448 | miR-1908-! | cis | 1.95E-05 | Nightingale | L_HDL_TG_ | NA | A | G | -0.05084 | 0.004077 | 2.40E-38 |
| rs174448 | miR-1908-! | cis | 1.95E-05 | Nightingale | L_LDL_P | NA | A | G | 0.028928 | 0.004306 | 1.90E-11 |
| rs174448 | miR-1908-! | cis | 1.95E-05 | Nightingale | Unsaturati | NA | A | G | 0.223545 | 0.004092 | 0 |
| rs174448 | miR-1908-! | cis | 1.95E-05 | Nightingale | XS_VLDL_P | NA | A | G | 0.025975 | 0.00426 | 3.30E-10 |
| rs174448 | miR-1908-! | cis | 1.95E-05 | Nightingale | L_VLDL_P | NA | A | G | -0.02472 | 0.004172 | 6.90E-10 |
| rs174448 | miR-1908-! | cis | 1.95E-05 | Nightingale | Clinical_LD | NA | A | G | 0.040945 | 0.004296 | 2.70E-21 |
| rs174448 | miR-1908-! | cis | 1.95E-05 | Nightingale | HDL_CE | NA | A | G | 0.038948 | 0.003878 | 2.70E-26 |
| rs174448 | miR-1908-! | cis | 1.95E-05 | Nightingale | S_LDL_FC | NA | A | G | 0.045091 | 0.004314 | 2.00E-25 |
| rs174448 | miR-1908-! | cis | 1.95E-05 | Nightingale | M_LDL_TG | NA | A | G | -0.05299 | 0.004266 | 1.90E-35 |
| rs174448 | miR-1908-! | cis | 1.95E-05 | Nightingale | L_LDL_PL | NA | A | G | 0.040759 | 0.004288 | 3.40E-21 |
| rs174448 | miR-1908-! | cis | 1.95E-05 | Nightingale | HDL_L | NA | A | G | 0.033868 | 0.00388 | 4.50E-20 |
| rs174448 | miR-1908-! | cis | 1.95E-05 | Nightingale | M_VLDL_C | NA | A | G | 0.03395 | 0.004309 | 4.50E-15 |
| rs174448 | miR-1908-! | cis | 1.95E-05 | Nightingale | XXL_VLDL_ | NA | A | G | -0.02913 | 0.004347 | 2.60E-11 |
| rs174448 | miR-1908-! | cis | 1.95E-05 | Nightingale | M_HDL_PL | NA | A | G | -0.03816 | 0.004098 | 6.70E-22 |
| rs174448 | miR-1908-! | cis | 1.95E-05 | Nightingale | L_LDL_C | NA | A | G | 0.042424 | 0.004272 | 2.80E-23 |
| rs174448 | miR-1908-! | cis | 1.95E-05 | Nightingale | non_HDL_( | NA | A | G | 0.03628 | 0.004297 | 3.80E-17 |
| rs174448 | miR-1908-! | cis | 1.95E-05 | Nightingale | XS_VLDL_F | NA | A | G | 0.040921 | 0.004245 | 4.90E-23 |

| | | | | | | | | | | | |
|---|---|---|---|---|---|---|---|---|---|---|---|
| rs174448 | miR-1908-5 | cis | 1.95E-05 | Nightingale | XXL_VLDL_ | NA | A | G | -0.04271 | 0.004162 | 7.70E-26 |
| rs174448 | miR-1908-5 | cis | 1.95E-05 | Nightingale | XL_HDL_C | NA | A | G | 0.068133 | 0.003896 | 1.60E-74 |
| rs174448 | miR-1908-5 | cis | 1.95E-05 | Nightingale | HDL_FC | NA | A | G | 0.048517 | 0.003828 | 1.90E-40 |
| rs174448 | miR-1908-5 | cis | 1.95E-05 | Nightingale | XL_VLDL_F | NA | A | G | -0.02542 | 0.004185 | 2.70E-10 |
| rs174448 | miR-1908-5 | cis | 1.95E-05 | Nightingale | XXL_VLDL_ | NA | A | G | -0.04488 | 0.004173 | 3.20E-28 |
| rs174448 | miR-1908-5 | cis | 1.95E-05 | Nightingale | M_VLDL_C | NA | A | G | 0.056168 | 0.004067 | 1.10E-44 |
| rs174448 | miR-1908-5 | cis | 1.95E-05 | Nightingale | XXL_VLDL_ | NA | A | G | -0.04451 | 0.004164 | 6.70E-28 |
| rs174448 | miR-1908-5 | cis | 1.95E-05 | Nightingale | IDL_C | NA | A | G | 0.055559 | 0.004172 | 1.50E-41 |
| rs174448 | miR-1908-5 | cis | 1.95E-05 | Nightingale | S_LDL_TG | NA | A | G | -0.03163 | 0.004215 | 2.70E-14 |
| rs174448 | miR-1908-5 | cis | 1.95E-05 | Nightingale | XXL_VLDL_ | NA | A | G | -0.04146 | 0.004156 | 9.90E-25 |
| rs174448 | miR-1908-5 | cis | 1.95E-05 | Nightingale | S_LDL_P | NA | A | G | 0.026732 | 0.004325 | 6.80E-10 |
| rs174448 | miR-1908-5 | cis | 1.95E-05 | Nightingale | L_LDL_FC_ | NA | A | G | 0.056734 | 0.004118 | 8.90E-45 |
| rs174448 | miR-1908-5 | cis | 1.95E-05 | Nightingale | DHA | NA | A | G | 0.190196 | 0.004163 | 0 |
| rs174448 | miR-1908-5 | cis | 1.95E-05 | Nightingale | Cholines | NA | A | G | 0.025873 | 0.004063 | 3.10E-11 |
| rs174448 | miR-1908-5 | cis | 1.95E-05 | Nightingale | PUFA_pct | NA | A | G | 0.078813 | 0.004156 | 2.80E-81 |
| rs174448 | miR-1908-5 | cis | 1.95E-05 | Nightingale | HDL_C | NA | A | G | 0.04138 | 0.003858 | 1.00E-29 |
| rs174448 | miR-1908-5 | cis | 1.95E-05 | Nightingale | LDL_L | NA | A | G | 0.034184 | 0.004307 | 3.40E-15 |
| rs174448 | miR-1908-5 | cis | 1.95E-05 | Nightingale | XS_VLDL_C | NA | A | G | 0.068296 | 0.004094 | 1.60E-64 |
| rs174448 | miR-1908-5 | cis | 1.95E-05 | Nightingale | LA | NA | A | G | -0.0532 | 0.004237 | 2.50E-37 |
| rs174448 | miR-1908-5 | cis | 1.95E-05 | Nightingale | XS_VLDL_C | NA | A | G | 0.049679 | 0.004193 | 1.10E-33 |
| rs174448 | miR-1908-5 | cis | 1.95E-05 | Nightingale | L_VLDL_PL | NA | A | G | -0.02967 | 0.004169 | 1.40E-13 |
| rs174448 | miR-1908-5 | cis | 1.95E-05 | Nightingale | XS_VLDL_T | NA | A | G | -0.06779 | 0.004104 | 3.50E-63 |
| rs174448 | miR-1908-5 | cis | 1.95E-05 | Nightingale | S_LDL_CE_ | NA | A | G | -0.03436 | 0.004313 | 2.90E-16 |
| rs174448 | miR-1908-5 | cis | 1.95E-05 | Nightingale | XL_VLDL_T | NA | A | G | -0.03554 | 0.004133 | 9.80E-19 |
| rs174448 | miR-1908-5 | cis | 1.95E-05 | Nightingale | Total_TG | NA | A | G | -0.02901 | 0.004182 | 9.80E-13 |
| rs174448 | miR-1908-5 | cis | 1.95E-05 | Nightingale | XL_VLDL_C | NA | A | G | 0.042912 | 0.00414 | 1.10E-25 |
| rs174449 | miR-1908-5 | cis | 2.49E-05 | Nightingale | ApoA1 | NA | A | G | 0.025671 | 0.003961 | 7.50E-12 |
| rs174449 | miR-1908-5 | cis | 2.49E-05 | Nightingale | M_HDL_CE | NA | A | G | 0.022236 | 0.003997 | 4.50E-09 |
| rs174449 | miR-1908-5 | cis | 2.49E-05 | Nightingale | LDL_PL | NA | A | G | 0.035799 | 0.004319 | 4.50E-16 |
| rs174449 | miR-1908-5 | cis | 2.49E-05 | Nightingale | VLDL_TG | NA | A | G | -0.02795 | 0.004165 | 6.20E-12 |
| rs174449 | miR-1908-5 | cis | 2.49E-05 | Nightingale | Total_PL | NA | A | G | 0.036833 | 0.004122 | 8.10E-20 |
| rs174449 | miR-1908-5 | cis | 2.49E-05 | Nightingale | S_LDL_PL | NA | A | G | 0.038288 | 0.004318 | 1.40E-18 |
| rs174449 | miR-1908-5 | cis | 2.49E-05 | Nightingale | L_VLDL_PL | NA | A | G | -0.05413 | 0.004201 | 8.70E-40 |
| rs174449 | miR-1908-5 | cis | 2.49E-05 | Nightingale | SFA_pct | NA | A | G | 0.029909 | 0.00427 | 2.50E-12 |
| rs174449 | miR-1908-5 | cis | 2.49E-05 | Nightingale | L_HDL_PL | NA | A | G | 0.049036 | 0.003809 | 4.70E-42 |
| rs174449 | miR-1908-5 | cis | 2.49E-05 | Nightingale | VLDL_size | NA | A | G | -0.04408 | 0.004035 | 8.80E-30 |
| rs174449 | miR-1908-5 | cis | 2.49E-05 | Nightingale | M_HDL_C_ | NA | A | G | 0.040699 | 0.00412 | 1.30E-24 |
| rs174449 | miR-1908-5 | cis | 2.49E-05 | Nightingale | M_LDL_C_ | NA | A | G | 0.069779 | 0.004332 | 6.70E-58 |
| rs174449 | miR-1908-5 | cis | 2.49E-05 | Nightingale | L_HDL_L | NA | A | G | 0.050262 | 0.003798 | 1.90E-44 |
| rs174449 | miR-1908-5 | cis | 2.49E-05 | Nightingale | MUFA_pct | NA | A | G | -0.12794 | 0.004142 | 3.30E-216 |
| rs174449 | miR-1908-5 | cis | 2.49E-05 | Nightingale | L_LDL_PL | NA | A | G | 0.041698 | 0.004296 | 8.80E-22 |
| rs174449 | miR-1908-5 | cis | 2.49E-05 | Nightingale | M_VLDL_F | NA | A | G | 0.062998 | 0.004116 | 7.20E-55 |
| rs174449 | miR-1908-5 | cis | 2.49E-05 | Nightingale | HDL_size | NA | A | G | 0.047667 | 0.003835 | 4.70E-39 |
| rs174449 | miR-1908-5 | cis | 2.49E-05 | Nightingale | ApoB | NA | A | G | 0.03121 | 0.004323 | 8.70E-13 |
| rs174449 | miR-1908-5 | cis | 2.49E-05 | Nightingale | IDL_CE | NA | A | G | 0.052628 | 0.004178 | 6.50E-37 |
| rs174449 | miR-1908-5 | cis | 2.49E-05 | Nightingale | M_VLDL_T | NA | A | G | -0.05377 | 0.004088 | 9.70E-41 |
| rs174449 | miR-1908-5 | cis | 2.49E-05 | Nightingale | HDL_P | NA | A | G | 0.022569 | 0.004064 | 7.10E-09 |
| rs174449 | miR-1908-5 | cis | 2.49E-05 | Nightingale | M_VLDL_F | NA | A | G | 0.025259 | 0.004328 | 8.10E-09 |
| rs174449 | miR-1908-5 | cis | 2.49E-05 | Nightingale | S_VLDL_TG | NA | A | G | -0.0448 | 0.004188 | 1.70E-26 |
| rs174449 | miR-1908-5 | cis | 2.49E-05 | Nightingale | Sphingomy | NA | A | G | 0.079348 | 0.004044 | 3.90E-89 |
| rs174449 | miR-1908-5 | cis | 2.49E-05 | Nightingale | HDL_L | NA | A | G | 0.033551 | 0.003887 | 1.10E-19 |
| rs174449 | miR-1908-5 | cis | 2.49E-05 | Nightingale | Total_C | NA | A | G | 0.047574 | 0.00419 | 3.30E-30 |
| rs174449 | miR-1908-5 | cis | 2.49E-05 | Nightingale | L_HDL_FC_ | NA | A | G | 0.077986 | 0.004021 | 1.30E-89 |
| rs174449 | miR-1908-5 | cis | 2.49E-05 | Nightingale | L_HDL_P | NA | A | G | 0.051118 | 0.003791 | 4.30E-46 |
| rs174449 | miR-1908-5 | cis | 2.49E-05 | Nightingale | XXL_VLDL_ | NA | A | G | -0.03981 | 0.004169 | 1.50E-22 |
| rs174449 | miR-1908-5 | cis | 2.49E-05 | Nightingale | S_VLDL_FC | NA | A | G | 0.05614 | 0.004133 | 5.00E-43 |
| rs174449 | miR-1908-5 | cis | 2.49E-05 | Nightingale | M_HDL_FC | NA | A | G | 0.032215 | 0.003939 | 8.20E-18 |
| rs174449 | miR-1908-5 | cis | 2.49E-05 | Nightingale | XS_VLDL_C | NA | A | G | 0.05019 | 0.004201 | 8.80E-34 |
| rs174449 | miR-1908-5 | cis | 2.49E-05 | Nightingale | Total_P | NA | A | G | 0.026763 | 0.004073 | 8.30E-12 |
| rs174449 | miR-1908-5 | cis | 2.49E-05 | Nightingale | Unsaturati | NA | A | G | 0.222522 | 0.004099 | 0 |
| rs174449 | miR-1908-5 | cis | 2.49E-05 | Nightingale | M_LDL_P | NA | A | G | 0.036824 | 0.004341 | 5.00E-17 |
| rs174449 | miR-1908-5 | cis | 2.49E-05 | Nightingale | S_VLDL_PL | NA | A | G | 0.053036 | 0.004142 | 1.90E-38 |
| rs174449 | miR-1908-5 | cis | 2.49E-05 | Nightingale | XL_VLDL_P | NA | A | G | -0.03085 | 0.00416 | 2.00E-14 |

| | | | | | | | | | | | | |
|---|---|---|---|---|---|---|---|---|---|---|---|---|
| rs174449 | miR-1908-! | cis | | 2.49E-05 | Nightingale | L_HDL_C | NA | A | G | 0.05241 | 0.003812 | 1.00E-47 |
| rs174449 | miR-1908-! | cis | | 2.49E-05 | Nightingale | M_HDL_TG | NA | A | G | -0.04033 | 0.004151 | 9.80E-24 |
| rs174449 | miR-1908-! | cis | | 2.49E-05 | Nightingale | DHA_pct | NA | A | G | 0.178093 | 0.004201 | 0 |
| rs174449 | miR-1908-! | cis | | 2.49E-05 | Nightingale | S_LDL_C | NA | A | G | 0.026793 | 0.004333 | 1.90E-09 |
| rs174449 | miR-1908-! | cis | | 2.49E-05 | Nightingale | XL_VLDL_F | NA | A | G | 0.045426 | 0.004194 | 1.40E-28 |
| rs174449 | miR-1908-! | cis | | 2.49E-05 | Nightingale | M_HDL_C | NA | A | G | 0.024363 | 0.003983 | 1.20E-10 |
| rs174449 | miR-1908-! | cis | | 2.49E-05 | Nightingale | XL_HDL_C | NA | A | G | 0.066604 | 0.003903 | 3.60E-71 |
| rs174449 | miR-1908-! | cis | | 2.49E-05 | Nightingale | XL_HDL_PL | NA | A | G | 0.043647 | 0.003976 | 1.80E-30 |
| rs174449 | miR-1908-! | cis | | 2.49E-05 | Nightingale | M_HDL_FC | NA | A | G | 0.067635 | 0.003836 | 3.20E-76 |
| rs174449 | miR-1908-! | cis | | 2.49E-05 | Nightingale | Remnant_( | NA | A | G | 0.035195 | 0.004301 | 2.90E-16 |
| rs174449 | miR-1908-! | cis | | 2.49E-05 | Nightingale | L_LDL_TG_ | NA | A | G | -0.05408 | 0.004244 | 3.30E-37 |
| rs174449 | miR-1908-! | cis | | 2.49E-05 | Nightingale | IDL_C_pct | NA | A | G | 0.05139 | 0.004226 | 3.20E-34 |
| rs174449 | miR-1908-! | cis | | 2.49E-05 | Nightingale | L_HDL_TG_ | NA | A | G | -0.04929 | 0.004084 | 6.90E-36 |
| rs174449 | miR-1908-! | cis | | 2.49E-05 | Nightingale | XL_HDL_P | NA | A | G | 0.061319 | 0.003895 | 4.00E-61 |
| rs174449 | miR-1908-! | cis | | 2.49E-05 | Nightingale | IDL_FC | NA | A | G | 0.06455 | 0.004189 | 7.80E-55 |
| rs174449 | miR-1908-! | cis | | 2.49E-05 | Nightingale | LDL_P | NA | A | G | 0.032249 | 0.004325 | 1.70E-13 |
| rs174449 | miR-1908-! | cis | | 2.49E-05 | Nightingale | PUFA_by_N | NA | A | G | 0.110027 | 0.004146 | 1.70E-158 |
| rs174449 | miR-1908-! | cis | | 2.49E-05 | Nightingale | XS_VLDL_P | NA | A | G | 0.033929 | 0.004271 | 7.80E-16 |
| rs174449 | miR-1908-! | cis | | 2.49E-05 | Nightingale | IDL_L | NA | A | G | 0.052952 | 0.004184 | 1.60E-37 |
| rs174449 | miR-1908-! | cis | | 2.49E-05 | Nightingale | IDL_CE_pct | NA | A | G | 0.026698 | 0.004247 | 6.70E-10 |
| rs174449 | miR-1908-! | cis | | 2.49E-05 | Nightingale | LDL_L | NA | A | G | 0.035257 | 0.004314 | 8.70E-16 |
| rs174449 | miR-1908-! | cis | | 2.49E-05 | Nightingale | S_HDL_TG | NA | A | G | -0.04298 | 0.004197 | 6.30E-26 |
| rs174449 | miR-1908-! | cis | | 2.49E-05 | Nightingale | L_HDL_C_p | NA | A | G | 0.045541 | 0.00408 | 3.00E-31 |
| rs174449 | miR-1908-! | cis | | 2.49E-05 | Nightingale | M_HDL_P | NA | A | G | 0.020602 | 0.003988 | 3.90E-08 |
| rs174449 | miR-1908-! | cis | | 2.49E-05 | Nightingale | S_LDL_FC_ | NA | A | G | 0.052 | 0.004195 | 6.00E-36 |
| rs174449 | miR-1908-! | cis | | 2.49E-05 | Nightingale | TG_by_PG | NA | A | G | -0.04297 | 0.004017 | 1.90E-28 |
| rs174449 | miR-1908-! | cis | | 2.49E-05 | Nightingale | L_LDL_C_p | NA | A | G | 0.053393 | 0.004217 | 6.70E-37 |
| rs174449 | miR-1908-! | cis | | 2.49E-05 | Nightingale | M_VLDL_P | NA | A | G | 0.044492 | 0.004153 | 3.90E-28 |
| rs174449 | miR-1908-! | cis | | 2.49E-05 | Nightingale | M_VLDL_C | NA | A | G | 0.040114 | 0.004289 | 1.40E-20 |
| rs174449 | miR-1908-! | cis | | 2.49E-05 | Nightingale | Total_L | NA | A | G | 0.029165 | 0.004258 | 7.40E-12 |
| rs174449 | miR-1908-! | cis | | 2.49E-05 | Nightingale | SFA | NA | A | G | 0.024216 | 0.004289 | 1.50E-08 |
| rs174449 | miR-1908-! | cis | | 2.49E-05 | Nightingale | S_LDL_FC | NA | A | G | 0.045696 | 0.004321 | 1.10E-25 |
| rs174449 | miR-1908-! | cis | | 2.49E-05 | Nightingale | Clinical_LD | NA | A | G | 0.041845 | 0.004304 | 7.50E-22 |
| rs174449 | miR-1908-! | cis | | 2.49E-05 | Nightingale | L_LDL_P | NA | A | G | 0.029848 | 0.004314 | 7.70E-12 |
| rs174449 | miR-1908-! | cis | | 2.49E-05 | Nightingale | XS_VLDL_P | NA | A | G | -0.04122 | 0.004223 | 1.10E-22 |
| rs174449 | miR-1908-! | cis | | 2.49E-05 | Nightingale | XL_HDL_TG | NA | A | G | -0.05585 | 0.004077 | 6.90E-45 |
| rs174449 | miR-1908-! | cis | | 2.49E-05 | Nightingale | M_HDL_CE | NA | A | G | 0.027941 | 0.004197 | 8.20E-12 |
| rs174449 | miR-1908-! | cis | | 2.49E-05 | Nightingale | XL_VLDL_C | NA | A | G | 0.039494 | 0.00415 | 1.20E-21 |
| rs174449 | miR-1908-! | cis | | 2.49E-05 | Nightingale | L_VLDL_C_ | NA | A | G | 0.028408 | 0.004228 | 6.20E-12 |
| rs174449 | miR-1908-! | cis | | 2.49E-05 | Nightingale | L_HDL_FC | NA | A | G | 0.05585 | 0.003793 | 3.80E-54 |
| rs174449 | miR-1908-! | cis | | 2.49E-05 | Nightingale | S_HDL_TG_ | NA | A | G | -0.0426 | 0.004117 | 1.10E-26 |
| rs174449 | miR-1908-! | cis | | 2.49E-05 | Nightingale | Omega_3_ | NA | A | G | 0.256793 | 0.004229 | 0 |
| rs174449 | miR-1908-! | cis | | 2.49E-05 | Nightingale | IDL_P | NA | A | G | 0.035441 | 0.004283 | 2.30E-16 |
| rs174449 | miR-1908-! | cis | | 2.49E-05 | Nightingale | M_LDL_FC | NA | A | G | 0.039642 | 0.004324 | 1.70E-19 |
| rs174449 | miR-1908-! | cis | | 2.49E-05 | Nightingale | S_LDL_TG_ | NA | A | G | -0.05756 | 0.004194 | 2.50E-43 |
| rs174449 | miR-1908-! | cis | | 2.49E-05 | Nightingale | DHA | NA | A | G | 0.189859 | 0.004171 | 0 |
| rs174449 | miR-1908-! | cis | | 2.49E-05 | Nightingale | HDL_C | NA | A | G | 0.040862 | 0.003865 | 7.00E-29 |
| rs174449 | miR-1908-! | cis | | 2.49E-05 | Nightingale | IDL_TG_pc | NA | A | G | -0.06088 | 0.004182 | 5.80E-49 |
| rs174449 | miR-1908-! | cis | | 2.49E-05 | Nightingale | LA_pct | NA | A | G | -0.12338 | 0.00423 | 5.40E-189 |
| rs174449 | miR-1908-! | cis | | 2.49E-05 | Nightingale | S_LDL_CE_ | NA | A | G | -0.03249 | 0.00432 | 1.30E-14 |
| rs174449 | miR-1908-! | cis | | 2.49E-05 | Nightingale | S_LDL_PL_ | NA | A | G | 0.035736 | 0.00432 | 2.10E-17 |
| rs174449 | miR-1908-! | cis | | 2.49E-05 | Nightingale | M_LDL_C | NA | A | G | 0.027839 | 0.004337 | 4.70E-10 |
| rs174449 | miR-1908-! | cis | | 2.49E-05 | Nightingale | XL_HDL_FC | NA | A | G | 0.069626 | 0.004032 | 9.70E-72 |
| rs174449 | miR-1908-! | cis | | 2.49E-05 | Nightingale | HDL_FC | NA | A | G | 0.047985 | 0.003834 | 1.90E-39 |
| rs174449 | miR-1908-! | cis | | 2.49E-05 | Nightingale | IDL_FC_pct | NA | A | G | 0.071649 | 0.004294 | 1.20E-63 |
| rs174449 | miR-1908-! | cis | | 2.49E-05 | Nightingale | Omega_6_ | NA | A | G | -0.24416 | 0.004265 | 0 |
| rs174449 | miR-1908-! | cis | | 2.49E-05 | Nightingale | S_HDL_C_p | NA | A | G | 0.027701 | 0.004278 | 2.40E-10 |
| rs174449 | miR-1908-! | cis | | 2.49E-05 | Nightingale | LDL_CE | NA | A | G | 0.034082 | 0.004317 | 9.50E-15 |
| rs174449 | miR-1908-! | cis | | 2.49E-05 | Nightingale | S_LDL_P | NA | A | G | 0.027843 | 0.004333 | 2.00E-10 |
| rs174449 | miR-1908-! | cis | | 2.49E-05 | Nightingale | Omega_3 | NA | A | G | 0.222919 | 0.004242 | 0 |
| rs174449 | miR-1908-! | cis | | 2.49E-05 | Nightingale | L_VLDL_FC | NA | A | G | -0.02183 | 0.004187 | 4.00E-08 |
| rs174449 | miR-1908-! | cis | | 2.49E-05 | Nightingale | non_HDL_( | NA | A | G | 0.037301 | 0.004305 | 9.90E-18 |
| rs174449 | miR-1908-! | cis | | 2.49E-05 | Nightingale | IDL_C | NA | A | G | 0.05606 | 0.00418 | 8.80E-42 |

| | | | | | | | | | | | |
|---|---|---|---|---|---|---|---|---|---|---|---|
| rs174449 | miR-1908-5 | cis | 2.49E-05 | Nightingale | Phosphogly | NA | A | G | 0.028927 | 0.0041 | 3.10E-13 |
| rs174449 | miR-1908-5 | cis | 2.49E-05 | Nightingale | XXL_VLDL_ | NA | A | G | -0.044 | 0.004154 | 2.80E-27 |
| rs174449 | miR-1908-5 | cis | 2.49E-05 | Nightingale | XL_HDL_FC | NA | A | G | -0.04001 | 0.003838 | 4.80E-29 |
| rs174449 | miR-1908-5 | cis | 2.49E-05 | Nightingale | XL_HDL_PL | NA | A | G | 0.059931 | 0.003891 | 1.80E-58 |
| rs174449 | miR-1908-5 | cis | 2.49E-05 | Nightingale | IDL_PL | NA | A | G | 0.053315 | 0.004176 | 2.00E-38 |
| rs174449 | miR-1908-5 | cis | 2.49E-05 | Nightingale | L_LDL_FC | NA | A | G | 0.050122 | 0.00425 | 3.10E-32 |
| rs174449 | miR-1908-5 | cis | 2.49E-05 | Nightingale | L_VLDL_P | NA | A | G | -0.02321 | 0.004179 | 7.30E-09 |
| rs174449 | miR-1908-5 | cis | 2.49E-05 | Nightingale | XXL_VLDL_ | NA | A | G | -0.03988 | 0.004163 | 8.50E-23 |
| rs174449 | miR-1908-5 | cis | 2.49E-05 | Nightingale | M_VLDL_C | NA | A | G | 0.051765 | 0.004063 | 5.90E-38 |
| rs174449 | miR-1908-5 | cis | 2.49E-05 | Nightingale | M_LDL_PL_ | NA | A | G | -0.03243 | 0.004326 | 4.50E-15 |
| rs174449 | miR-1908-5 | cis | 2.49E-05 | Nightingale | HDL_CE | NA | A | G | 0.038435 | 0.003884 | 1.50E-25 |
| rs174449 | miR-1908-5 | cis | 2.49E-05 | Nightingale | S_VLDL_TG | NA | A | G | -0.02291 | 0.0042 | 2.60E-08 |
| rs174449 | miR-1908-5 | cis | 2.49E-05 | Nightingale | XL_VLDL_T | NA | A | G | -0.03408 | 0.00414 | 3.10E-17 |
| rs174449 | miR-1908-5 | cis | 2.49E-05 | Nightingale | XL_HDL_L | NA | A | G | 0.062813 | 0.00389 | 5.20E-64 |
| rs174449 | miR-1908-5 | cis | 2.49E-05 | Nightingale | XL_VLDL_T | NA | A | G | -0.0406 | 0.004193 | 1.90E-22 |
| rs174449 | miR-1908-5 | cis | 2.49E-05 | Nightingale | L_LDL_L | NA | A | G | 0.040929 | 0.004286 | 2.30E-21 |
| rs174449 | miR-1908-5 | cis | 2.49E-05 | Nightingale | PUFA | NA | A | G | 0.06687 | 0.004221 | 4.10E-58 |
| rs174449 | miR-1908-5 | cis | 2.49E-05 | Nightingale | L_LDL_FC_ | NA | A | G | 0.055693 | 0.004126 | 8.70E-43 |
| rs174449 | miR-1908-5 | cis | 2.49E-05 | Nightingale | LDL_FC | NA | A | G | 0.04745 | 0.004282 | 3.00E-28 |
| rs174449 | miR-1908-5 | cis | 2.49E-05 | Nightingale | L_LDL_CE | NA | A | G | 0.040297 | 0.00429 | 1.20E-20 |
| rs174449 | miR-1908-5 | cis | 2.49E-05 | Nightingale | S_VLDL_C_ | NA | A | G | 0.040018 | 0.004211 | 4.40E-21 |
| rs174449 | miR-1908-5 | cis | 2.49E-05 | Nightingale | XL_VLDL_L | NA | A | G | -0.02973 | 0.004165 | 1.90E-13 |
| rs174449 | miR-1908-5 | cis | 2.49E-05 | Nightingale | XXL_VLDL_ | NA | A | G | -0.04108 | 0.00417 | 8.60E-24 |
| rs174449 | miR-1908-5 | cis | 2.49E-05 | Nightingale | XS_VLDL_C | NA | A | G | 0.052821 | 0.004177 | 1.10E-37 |
| rs174449 | miR-1908-5 | cis | 2.49E-05 | Nightingale | L_VLDL_CE | NA | A | G | 0.04089 | 0.004142 | 2.70E-24 |
| rs174449 | miR-1908-5 | cis | 2.49E-05 | Nightingale | XXL_VLDL_ | NA | A | G | -0.028 | 0.004354 | 1.40E-10 |
| rs174449 | miR-1908-5 | cis | 2.49E-05 | Nightingale | Cholines | NA | A | G | 0.026504 | 0.004071 | 1.30E-11 |
| rs174449 | miR-1908-5 | cis | 2.49E-05 | Nightingale | LA | NA | A | G | -0.05196 | 0.004244 | 9.50E-36 |
| rs174449 | miR-1908-5 | cis | 2.49E-05 | Nightingale | XS_VLDL_F | NA | A | G | 0.076773 | 0.004296 | 6.30E-72 |
| rs174449 | miR-1908-5 | cis | 2.49E-05 | Nightingale | XS_VLDL_L | NA | A | G | 0.03205 | 0.004255 | 1.50E-14 |
| rs174449 | miR-1908-5 | cis | 2.49E-05 | Nightingale | HDL_TG | NA | A | G | -0.02785 | 0.004268 | 3.40E-11 |
| rs174449 | miR-1908-5 | cis | 2.49E-05 | Nightingale | M_LDL_TG | NA | A | G | -0.05288 | 0.004274 | 5.50E-35 |
| rs174449 | miR-1908-5 | cis | 2.49E-05 | Nightingale | HDL_PL | NA | A | G | 0.030157 | 0.003913 | 3.80E-16 |
| rs174449 | miR-1908-5 | cis | 2.49E-05 | Nightingale | Total_CE | NA | A | G | 0.048548 | 0.004175 | 9.00E-32 |
| rs174449 | miR-1908-5 | cis | 2.49E-05 | Nightingale | XS_VLDL_T | NA | A | G | -0.0669 | 0.004111 | 4.00E-61 |
| rs174449 | miR-1908-5 | cis | 2.49E-05 | Nightingale | L_HDL_PL_ | NA | A | G | -0.03193 | 0.004189 | 1.10E-14 |
| rs174449 | miR-1908-5 | cis | 2.49E-05 | Nightingale | LDL_C | NA | A | G | 0.038156 | 0.00431 | 2.60E-18 |
| rs174449 | miR-1908-5 | cis | 2.49E-05 | Nightingale | M_LDL_FC_ | NA | A | G | 0.046254 | 0.004147 | 1.40E-29 |
| rs174449 | miR-1908-5 | cis | 2.49E-05 | Nightingale | L_HDL_CE_ | NA | A | G | 0.032333 | 0.004141 | 3.20E-16 |
| rs174449 | miR-1908-5 | cis | 2.49E-05 | Nightingale | L_VLDL_PL | NA | A | G | -0.02814 | 0.004176 | 2.30E-12 |
| rs174449 | miR-1908-5 | cis | 2.49E-05 | Nightingale | XS_VLDL_C | NA | A | G | 0.067415 | 0.004101 | 1.90E-62 |
| rs174449 | miR-1908-5 | cis | 2.49E-05 | Nightingale | XS_VLDL_P | NA | A | G | 0.026936 | 0.004268 | 1.20E-10 |
| rs174449 | miR-1908-5 | cis | 2.49E-05 | Nightingale | M_HDL_PL | NA | A | G | -0.03762 | 0.004106 | 4.50E-21 |
| rs174449 | miR-1908-5 | cis | 2.49E-05 | Nightingale | XL_HDL_CE | NA | A | G | 0.064899 | 0.003876 | 7.40E-69 |
| rs174449 | miR-1908-5 | cis | 2.49E-05 | Nightingale | S_HDL_FC_ | NA | A | G | 0.069154 | 0.004106 | 7.70E-70 |
| rs174449 | miR-1908-5 | cis | 2.49E-05 | Nightingale | L_LDL_C | NA | A | G | 0.043294 | 0.00428 | 8.30E-24 |
| rs174449 | miR-1908-5 | cis | 2.49E-05 | Nightingale | PUFA_pct | NA | A | G | 0.077412 | 0.004163 | 5.40E-78 |
| rs174449 | miR-1908-5 | cis | 2.49E-05 | Nightingale | XL_VLDL_P | NA | A | G | -0.02732 | 0.004177 | 1.30E-11 |
| rs174449 | miR-1908-5 | cis | 2.49E-05 | Nightingale | MUFA | NA | A | G | -0.03877 | 0.004284 | 3.20E-20 |
| rs174449 | miR-1908-5 | cis | 2.49E-05 | Nightingale | S_LDL_L | NA | A | G | 0.026699 | 0.004328 | 1.80E-09 |
| rs174449 | miR-1908-5 | cis | 2.49E-05 | Nightingale | M_VLDL_C | NA | A | G | 0.03482 | 0.004317 | 1.70E-15 |
| rs174449 | miR-1908-5 | cis | 2.49E-05 | Nightingale | XXL_VLDL_ | NA | A | G | -0.03642 | 0.004177 | 5.20E-19 |
| rs174449 | miR-1908-5 | cis | 2.49E-05 | Nightingale | XXL_VLDL_ | NA | A | G | -0.04329 | 0.00418 | 3.70E-26 |
| rs174449 | miR-1908-5 | cis | 2.49E-05 | Nightingale | L_HDL_CE | NA | A | G | 0.051167 | 0.003822 | 2.10E-45 |
| rs174449 | miR-1908-5 | cis | 2.49E-05 | Nightingale | XXL_VLDL_ | NA | A | G | -0.04288 | 0.004171 | 8.50E-26 |
| rs174449 | miR-1908-5 | cis | 2.49E-05 | Nightingale | Total_TG | NA | A | G | -0.02748 | 0.00419 | 1.70E-11 |
| rs174449 | miR-1908-5 | cis | 2.49E-05 | Nightingale | XS_VLDL_C | NA | A | G | 0.063497 | 0.00408 | 3.30E-56 |
| rs174449 | miR-1908-5 | cis | 2.49E-05 | Nightingale | S_LDL_TG | NA | A | G | -0.0301 | 0.004222 | 4.90E-13 |
| rs174449 | miR-1908-5 | cis | 2.49E-05 | Nightingale | Total_FC | NA | A | G | 0.044201 | 0.004229 | 1.30E-25 |
| rs174449 | miR-1908-5 | cis | 2.49E-05 | Nightingale | M_VLDL_C_ | NA | A | G | 0.055278 | 0.004074 | 6.40E-43 |
| rs174449 | miR-1908-5 | cis | 2.49E-05 | Nightingale | XL_VLDL_C | NA | A | G | 0.0417 | 0.004148 | 5.10E-24 |
| rs174449 | miR-1908-5 | cis | 2.49E-05 | Nightingale | XS_VLDL_F | NA | A | G | 0.041632 | 0.004253 | 2.30E-23 |
| rs174449 | miR-1908-5 | cis | 2.49E-05 | Nightingale | XL_VLDL_F | NA | A | G | -0.02384 | 0.004192 | 3.20E-09 |

| | | | | | | | | | | | |
|---|---|---|---|---|---|---|---|---|---|---|---|
| rs174449 | miR-1908-5 | cis | 2.49E-05 | Nightingale | M_HDL_TG | NA | A | G | -0.03084 | 0.004275 | 2.30E-13 |
| rs174530 | miR-1908-5 | cis | 1.12E-08 | Nightingale | VLDL_size | NA | G | A | 0.066325 | 0.003981 | 1.60E-65 |
| rs174530 | miR-1908-5 | cis | 1.12E-08 | Nightingale | L_VLDL_FC | NA | G | A | 0.036071 | 0.004132 | 7.30E-19 |
| rs174530 | miR-1908-5 | cis | 1.12E-08 | Nightingale | S_VLDL_CE | NA | G | A | -0.02735 | 0.004214 | 6.70E-11 |
| rs174530 | miR-1908-5 | cis | 1.12E-08 | Nightingale | XXL_VLDL_ | NA | G | A | -0.0235 | 0.004251 | 1.00E-08 |
| rs174530 | miR-1908-5 | cis | 1.12E-08 | Nightingale | XL_HDL_TG | NA | G | A | 0.078592 | 0.004024 | 1.40E-89 |
| rs174530 | miR-1908-5 | cis | 1.12E-08 | Nightingale | M_HDL_TG | NA | G | A | 0.045752 | 0.004219 | 1.50E-27 |
| rs174530 | miR-1908-5 | cis | 1.12E-08 | Nightingale | L_LDL_TG_ | NA | G | A | 0.070034 | 0.004188 | 1.40E-63 |
| rs174530 | miR-1908-5 | cis | 1.12E-08 | Nightingale | LDL_PL | NA | G | A | -0.04299 | 0.004262 | 4.50E-25 |
| rs174530 | miR-1908-5 | cis | 1.12E-08 | Nightingale | XL_HDL_P | NA | G | A | -0.08161 | 0.003844 | 1.00E-110 |
| rs174530 | miR-1908-5 | cis | 1.12E-08 | Nightingale | PUFA_pct | NA | G | A | -0.10927 | 0.004108 | 4.10E-157 |
| rs174530 | miR-1908-5 | cis | 1.12E-08 | Nightingale | XS_VLDL_T | NA | G | A | 0.027598 | 0.004206 | 3.00E-10 |
| rs174530 | miR-1908-5 | cis | 1.12E-08 | Nightingale | XL_VLDL_C | NA | G | A | 0.033444 | 0.004172 | 4.70E-16 |
| rs174530 | miR-1908-5 | cis | 1.12E-08 | Nightingale | MUFA | NA | G | A | 0.057052 | 0.004228 | 3.40E-42 |
| rs174530 | miR-1908-5 | cis | 1.12E-08 | Nightingale | XS_VLDL_F | NA | G | A | -0.10343 | 0.004239 | 5.30E-134 |
| rs174530 | miR-1908-5 | cis | 1.12E-08 | Nightingale | S_LDL_TG | NA | G | A | 0.044968 | 0.004167 | 2.30E-27 |
| rs174530 | miR-1908-5 | cis | 1.12E-08 | Nightingale | M_HDL_PL | NA | G | A | 0.052838 | 0.004052 | 2.80E-40 |
| rs174530 | miR-1908-5 | cis | 1.12E-08 | Nightingale | M_LDL_PL_ | NA | G | A | 0.040645 | 0.004269 | 1.30E-22 |
| rs174530 | miR-1908-5 | cis | 1.12E-08 | Nightingale | M_HDL_C_ | NA | G | A | -0.05934 | 0.004066 | 3.30E-50 |
| rs174530 | miR-1908-5 | cis | 1.12E-08 | Nightingale | IDL_FC_pct | NA | G | A | -0.0956 | 0.004237 | 1.30E-113 |
| rs174530 | miR-1908-5 | cis | 1.12E-08 | Nightingale | Omega_3_ | NA | G | A | -0.35679 | 0.004173 | 0 |
| rs174530 | miR-1908-5 | cis | 1.12E-08 | Nightingale | TG_by_PG | NA | G | A | 0.062415 | 0.003964 | 3.20E-58 |
| rs174530 | miR-1908-5 | cis | 1.12E-08 | Nightingale | LDL_C | NA | G | A | -0.04607 | 0.004253 | 8.20E-29 |
| rs174530 | miR-1908-5 | cis | 1.12E-08 | Nightingale | XXL_VLDL_ | NA | G | A | 0.059711 | 0.004114 | 1.70E-49 |
| rs174530 | miR-1908-5 | cis | 1.12E-08 | Nightingale | M_VLDL_C | NA | G | A | -0.04084 | 0.00426 | 8.60E-23 |
| rs174530 | miR-1908-5 | cis | 1.12E-08 | Nightingale | PUFA | NA | G | A | -0.08837 | 0.004165 | 1.60E-106 |
| rs174530 | miR-1908-5 | cis | 1.12E-08 | Nightingale | Total_C | NA | G | A | -0.06008 | 0.004135 | 1.00E-50 |
| rs174530 | miR-1908-5 | cis | 1.12E-08 | Nightingale | IDL_C | NA | G | A | -0.07144 | 0.004124 | 2.30E-71 |
| rs174530 | miR-1908-5 | cis | 1.12E-08 | Nightingale | IDL_PL | NA | G | A | -0.06768 | 0.004121 | 3.10E-65 |
| rs174530 | miR-1908-5 | cis | 1.12E-08 | Nightingale | XS_VLDL_P | NA | G | A | 0.051598 | 0.004168 | 6.90E-36 |
| rs174530 | miR-1908-5 | cis | 1.12E-08 | Nightingale | SFA | NA | G | A | -0.02997 | 0.004233 | 5.30E-13 |
| rs174530 | miR-1908-5 | cis | 1.12E-08 | Nightingale | XL_HDL_PL | NA | G | A | -0.08076 | 0.003839 | 2.60E-108 |
| rs174530 | miR-1908-5 | cis | 1.12E-08 | Nightingale | DHA_pct | NA | G | A | -0.24962 | 0.004146 | 0 |
| rs174530 | miR-1908-5 | cis | 1.12E-08 | Nightingale | HDL_TG | NA | G | A | 0.041761 | 0.004212 | 4.50E-23 |
| rs174530 | miR-1908-5 | cis | 1.12E-08 | Nightingale | L_VLDL_PL | NA | G | A | 0.077293 | 0.004145 | 6.00E-80 |
| rs174530 | miR-1908-5 | cis | 1.12E-08 | Nightingale | LDL_FC | NA | G | A | -0.05943 | 0.004226 | 1.90E-47 |
| rs174530 | miR-1908-5 | cis | 1.12E-08 | Nightingale | Omega_3 | NA | G | A | -0.30784 | 0.004186 | 0 |
| rs174530 | miR-1908-5 | cis | 1.12E-08 | Nightingale | S_VLDL_PL | NA | G | A | -0.07157 | 0.004087 | 2.00E-71 |
| rs174530 | miR-1908-5 | cis | 1.12E-08 | Nightingale | L_VLDL_TG | NA | G | A | 0.034156 | 0.00411 | 4.30E-17 |
| rs174530 | miR-1908-5 | cis | 1.12E-08 | Nightingale | S_VLDL_C_ | NA | G | A | -0.05218 | 0.004156 | 5.90E-37 |
| rs174530 | miR-1908-5 | cis | 1.12E-08 | Nightingale | L_VLDL_FC | NA | G | A | 0.02368 | 0.004231 | 7.50E-09 |
| rs174530 | miR-1908-5 | cis | 1.12E-08 | Nightingale | XL_HDL_C_ | NA | G | A | 0.02244 | 0.00408 | 1.20E-09 |
| rs174530 | miR-1908-5 | cis | 1.12E-08 | Nightingale | L_VLDL_C | NA | G | A | 0.0258 | 0.004178 | 4.50E-10 |
| rs174530 | miR-1908-5 | cis | 1.12E-08 | Nightingale | M_VLDL_C | NA | G | A | -0.04882 | 0.004233 | 2.80E-32 |
| rs174530 | miR-1908-5 | cis | 1.12E-08 | Nightingale | S_HDL_TG_ | NA | G | A | 0.061852 | 0.004062 | 3.40E-54 |
| rs174530 | miR-1908-5 | cis | 1.12E-08 | Nightingale | XS_VLDL_P | NA | G | A | -0.04167 | 0.004214 | 3.00E-25 |
| rs174530 | miR-1908-5 | cis | 1.12E-08 | Nightingale | Tyr | NA | G | A | 0.03099 | 0.004266 | 3.70E-13 |
| rs174530 | miR-1908-5 | cis | 1.12E-08 | Nightingale | S_LDL_FC_ | NA | G | A | -0.07258 | 0.004139 | 1.10E-70 |
| rs174530 | miR-1908-5 | cis | 1.12E-08 | Nightingale | L_HDL_TG_ | NA | G | A | 0.071031 | 0.004031 | 1.50E-73 |
| rs174530 | miR-1908-5 | cis | 1.12E-08 | Nightingale | S_LDL_CE_ | NA | G | A | 0.052748 | 0.004263 | 1.90E-36 |
| rs174530 | miR-1908-5 | cis | 1.12E-08 | Nightingale | M_HDL_FC | NA | G | A | -0.04616 | 0.003887 | 4.70E-35 |
| rs174530 | miR-1908-5 | cis | 1.12E-08 | Nightingale | LDL_L | NA | G | A | -0.04228 | 0.004257 | 1.60E-24 |
| rs174530 | miR-1908-5 | cis | 1.12E-08 | Nightingale | PUFA_by_M | NA | G | A | -0.15418 | 0.004091 | 6.39734972 |
| rs174530 | miR-1908-5 | cis | 1.12E-08 | Nightingale | L_LDL_C | NA | G | A | -0.05345 | 0.004223 | 6.60E-39 |
| rs174530 | miR-1908-5 | cis | 1.12E-08 | Nightingale | XL_VLDL_T | NA | G | A | 0.051465 | 0.004085 | 1.80E-37 |
| rs174530 | miR-1908-5 | cis | 1.12E-08 | Nightingale | XS_VLDL_C | NA | G | A | -0.06687 | 0.004121 | 1.60E-63 |
| rs174530 | miR-1908-5 | cis | 1.12E-08 | Nightingale | L_HDL_CE_ | NA | G | A | -0.04515 | 0.004087 | 5.80E-30 |
| rs174530 | miR-1908-5 | cis | 1.12E-08 | Nightingale | LDL_P | NA | G | A | -0.03744 | 0.004268 | 1.60E-19 |
| rs174530 | miR-1908-5 | cis | 1.12E-08 | Nightingale | M_LDL_L | NA | G | A | -0.02642 | 0.004278 | 2.20E-10 |
| rs174530 | miR-1908-5 | cis | 1.12E-08 | Nightingale | L_VLDL_CE | NA | G | A | -0.05408 | 0.004087 | 2.20E-42 |
| rs174530 | miR-1908-5 | cis | 1.12E-08 | Nightingale | XL_VLDL_L | NA | G | A | 0.046352 | 0.00411 | 2.40E-30 |
| rs174530 | miR-1908-5 | cis | 1.12E-08 | Nightingale | S_VLDL_FC | NA | G | A | -0.02669 | 0.004272 | 1.10E-10 |
| rs174530 | miR-1908-5 | cis | 1.12E-08 | Nightingale | XL_HDL_L | NA | G | A | -0.08445 | 0.003838 | 3.60E-118 |

| | | | | | | | | | | | |
|---|---|---|---|---|---|---|---|---|---|---|---|
| rs174530 | miR-1908-5 | cis | 1.12E-08 | Nightingale | LDL_CE | NA | G | A | -0.0404 | 0.00426 | 1.70E-22 |
| rs174530 | miR-1908-5 | cis | 1.12E-08 | Nightingale | XXL_VLDL_ | NA | G | A | 0.059364 | 0.004108 | 3.50E-49 |
| rs174530 | miR-1908-5 | cis | 1.12E-08 | Nightingale | Sphingomy | NA | G | A | -0.10589 | 0.003991 | 1.10E-163 |
| rs174530 | miR-1908-5 | cis | 1.12E-08 | Nightingale | S_LDL_P | NA | G | A | -0.03068 | 0.004275 | 1.30E-13 |
| rs174530 | miR-1908-5 | cis | 1.12E-08 | Nightingale | DHA | NA | G | A | -0.26367 | 0.004116 | 0 |
| rs174530 | miR-1908-5 | cis | 1.12E-08 | Nightingale | S_HDL_FC_ | NA | G | A | -0.09492 | 0.004051 | 3.90E-136 |
| rs174530 | miR-1908-5 | cis | 1.12E-08 | Nightingale | S_VLDL_FC | NA | G | A | -0.07569 | 0.004079 | 1.10E-79 |
| rs174530 | miR-1908-5 | cis | 1.12E-08 | Nightingale | XL_VLDL_F | NA | G | A | -0.06242 | 0.004139 | 1.00E-53 |
| rs174530 | miR-1908-5 | cis | 1.12E-08 | Nightingale | L_HDL_L | NA | G | A | -0.06991 | 0.003748 | 1.90E-85 |
| rs174530 | miR-1908-5 | cis | 1.12E-08 | Nightingale | L_HDL_FC | NA | G | A | -0.07674 | 0.003743 | 1.50E-102 |
| rs174530 | miR-1908-5 | cis | 1.12E-08 | Nightingale | M_VLDL_T | NA | G | A | 0.071684 | 0.004034 | 1.60E-73 |
| rs174530 | miR-1908-5 | cis | 1.12E-08 | Nightingale | L_LDL_L | NA | G | A | -0.05028 | 0.00423 | 1.50E-34 |
| rs174530 | miR-1908-5 | cis | 1.12E-08 | Nightingale | Omega_6_ | NA | G | A | 0.024082 | 0.004157 | 3.30E-09 |
| rs174530 | miR-1908-5 | cis | 1.12E-08 | Nightingale | LA | NA | G | A | 0.07731 | 0.004189 | 8.40E-76 |
| rs174530 | miR-1908-5 | cis | 1.12E-08 | Nightingale | Citrate | NA | G | A | -0.02965 | 0.004244 | 5.60E-13 |
| rs174530 | miR-1908-5 | cis | 1.12E-08 | Nightingale | M_LDL_P | NA | G | A | -0.04332 | 0.004284 | 6.10E-25 |
| rs174530 | miR-1908-5 | cis | 1.12E-08 | Nightingale | XL_VLDL_C | NA | G | A | 0.026918 | 0.004204 | 1.10E-10 |
| rs174530 | miR-1908-5 | cis | 1.12E-08 | Nightingale | VLDL_TG | NA | G | A | 0.043146 | 0.00411 | 2.40E-26 |
| rs174530 | miR-1908-5 | cis | 1.12E-08 | Nightingale | L_HDL_FC_ | NA | G | A | -0.1037 | 0.003968 | 3.10E-165 |
| rs174530 | miR-1908-5 | cis | 1.12E-08 | Nightingale | Total_CE | NA | G | A | -0.06173 | 0.00412 | 7.60E-54 |
| rs174530 | miR-1908-5 | cis | 1.12E-08 | Nightingale | HDL_FC | NA | G | A | -0.06701 | 0.003784 | 1.70E-76 |
| rs174530 | miR-1908-5 | cis | 1.12E-08 | Nightingale | IDL_L | NA | G | A | -0.06721 | 0.004129 | 1.30E-63 |
| rs174530 | miR-1908-5 | cis | 1.12E-08 | Nightingale | XXL_VLDL_ | NA | G | A | 0.061027 | 0.004115 | 9.40E-52 |
| rs174530 | miR-1908-5 | cis | 1.12E-08 | Nightingale | L_LDL_PL | NA | G | A | -0.05099 | 0.004239 | 5.70E-35 |
| rs174530 | miR-1908-5 | cis | 1.12E-08 | Nightingale | L_LDL_CE | NA | G | A | -0.0493 | 0.004233 | 2.90E-33 |
| rs174530 | miR-1908-5 | cis | 1.12E-08 | Nightingale | L_VLDL_C_ | NA | G | A | -0.03584 | 0.004172 | 8.30E-19 |
| rs174530 | miR-1908-5 | cis | 1.12E-08 | Nightingale | L_VLDL_P | NA | G | A | 0.037801 | 0.004124 | 1.50E-20 |
| rs174530 | miR-1908-5 | cis | 1.12E-08 | Nightingale | Unsaturati | NA | G | A | -0.30927 | 0.004045 | 0 |
| rs174530 | miR-1908-5 | cis | 1.12E-08 | Nightingale | Phosphogly | NA | G | A | -0.03766 | 0.004046 | 2.00E-22 |
| rs174530 | miR-1908-5 | cis | 1.12E-08 | Nightingale | XS_VLDL_C | NA | G | A | -0.06353 | 0.004146 | 5.20E-57 |
| rs174530 | miR-1908-5 | cis | 1.12E-08 | Nightingale | Remnant_( | NA | G | A | -0.04128 | 0.004244 | 8.10E-24 |
| rs174530 | miR-1908-5 | cis | 1.12E-08 | Nightingale | Total_L | NA | G | A | -0.03469 | 0.004202 | 8.00E-18 |
| rs174530 | miR-1908-5 | cis | 1.12E-08 | Nightingale | XL_VLDL_C | NA | G | A | -0.05801 | 0.004093 | 2.60E-46 |
| rs174530 | miR-1908-5 | cis | 1.12E-08 | Nightingale | Cholines | NA | G | A | -0.03396 | 0.004017 | 4.50E-19 |
| rs174530 | miR-1908-5 | cis | 1.12E-08 | Nightingale | XXL_VLDL_ | NA | G | A | 0.066049 | 0.004125 | 2.80E-59 |
| rs174530 | miR-1908-5 | cis | 1.12E-08 | Nightingale | ApoB | NA | G | A | -0.03607 | 0.004266 | 2.60E-18 |
| rs174530 | miR-1908-5 | cis | 1.12E-08 | Nightingale | M_LDL_C_ | NA | G | A | -0.08981 | 0.004275 | 1.40E-98 |
| rs174530 | miR-1908-5 | cis | 1.12E-08 | Nightingale | XS_VLDL_P | NA | G | A | -0.03309 | 0.004211 | 5.30E-17 |
| rs174530 | miR-1908-5 | cis | 1.12E-08 | Nightingale | SFA_pct | NA | G | A | -0.04024 | 0.004214 | 1.30E-21 |
| rs174530 | miR-1908-5 | cis | 1.12E-08 | Nightingale | XL_HDL_CE | NA | G | A | -0.08794 | 0.003825 | 2.00E-128 |
| rs174530 | miR-1908-5 | cis | 1.12E-08 | Nightingale | Omega_6_ | NA | G | A | 0.339063 | 0.004209 | 0 |
| rs174530 | miR-1908-5 | cis | 1.12E-08 | Nightingale | XL_VLDL_P | NA | G | A | 0.047691 | 0.004105 | 4.40E-32 |
| rs174530 | miR-1908-5 | cis | 1.12E-08 | Nightingale | XL_VLDL_P | NA | G | A | 0.043321 | 0.004122 | 1.40E-26 |
| rs174530 | miR-1908-5 | cis | 1.12E-08 | Nightingale | L_HDL_PL | NA | G | A | -0.06884 | 0.003759 | 2.60E-82 |
| rs174530 | miR-1908-5 | cis | 1.12E-08 | Nightingale | M_HDL_FC | NA | G | A | -0.09392 | 0.003785 | 7.69E-149 |
| rs174530 | miR-1908-5 | cis | 1.12E-08 | Nightingale | M_LDL_FC_ | NA | G | A | -0.06556 | 0.004092 | 2.60E-59 |
| rs174530 | miR-1908-5 | cis | 1.12E-08 | Nightingale | M_HDL_CE | NA | G | A | -0.04208 | 0.004142 | 1.30E-24 |
| rs174530 | miR-1908-5 | cis | 1.12E-08 | Nightingale | L_LDL_FC_ | NA | G | A | -0.07526 | 0.004071 | 2.10E-78 |
| rs174530 | miR-1908-5 | cis | 1.12E-08 | Nightingale | XS_VLDL_L | NA | G | A | -0.03954 | 0.004198 | 2.20E-23 |
| rs174530 | miR-1908-5 | cis | 1.12E-08 | Nightingale | M_HDL_C | NA | G | A | -0.03613 | 0.00393 | 2.00E-21 |
| rs174530 | miR-1908-5 | cis | 1.12E-08 | Nightingale | M_LDL_CE | NA | G | A | -0.0245 | 0.004278 | 3.70E-09 |
| rs174530 | miR-1908-5 | cis | 1.12E-08 | Nightingale | M_LDL_TG | NA | G | A | 0.032043 | 0.004208 | 4.10E-14 |
| rs174530 | miR-1908-5 | cis | 1.12E-08 | Nightingale | MUFA_pct | NA | G | A | 0.178799 | 0.004087 | 0 |
| rs174530 | miR-1908-5 | cis | 1.12E-08 | Nightingale | M_VLDL_F | NA | G | A | -0.02754 | 0.004271 | 2.70E-11 |
| rs174530 | miR-1908-5 | cis | 1.12E-08 | Nightingale | M_LDL_TG | NA | G | A | 0.068366 | 0.004217 | 8.80E-60 |
| rs174530 | miR-1908-5 | cis | 1.12E-08 | Nightingale | XL_VLDL_T | NA | G | A | 0.054236 | 0.004138 | 3.20E-40 |
| rs174530 | miR-1908-5 | cis | 1.12E-08 | Nightingale | LDL_TG | NA | G | A | 0.025567 | 0.004224 | 4.10E-09 |
| rs174530 | miR-1908-5 | cis | 1.12E-08 | Nightingale | XXL_VLDL_ | NA | G | A | 0.054933 | 0.004122 | 6.10E-42 |
| rs174530 | miR-1908-5 | cis | 1.12E-08 | Nightingale | XL_VLDL_C | NA | G | A | -0.05534 | 0.004095 | 2.60E-42 |
| rs174530 | miR-1908-5 | cis | 1.12E-08 | Nightingale | S_LDL_PL | NA | G | A | -0.04803 | 0.004261 | 4.80E-31 |
| rs174530 | miR-1908-5 | cis | 1.12E-08 | Nightingale | M_VLDL_F | NA | G | A | -0.08405 | 0.004061 | 7.00E-100 |
| rs174530 | miR-1908-5 | cis | 1.12E-08 | Nightingale | IDL_C_pct | NA | G | A | -0.06741 | 0.00417 | 7.10E-60 |
| rs174530 | miR-1908-5 | cis | 1.12E-08 | Nightingale | XXL_VLDL_ | NA | G | A | 0.06472 | 0.004099 | 2.80E-58 |

| | | | | | | | | | | | | |
|---|---|---|---|---|---|---|---|---|---|---|---|---|
| rs174530 | miR-1908-5 | cis | | 1.12E-08 | Nightingale | XS_VLDL_F | NA | G | A | -0.05265 | 0.004197 | 3.40E-39 |
| rs174530 | miR-1908-5 | cis | | 1.12E-08 | Nightingale | L_HDL_PL_ | NA | G | A | 0.039804 | 0.004134 | 2.00E-23 |
| rs174530 | miR-1908-5 | cis | | 1.12E-08 | Nightingale | XL_HDL_FC | NA | G | A | 0.055249 | 0.003787 | 2.80E-54 |
| rs174530 | miR-1908-5 | cis | | 1.12E-08 | Nightingale | M_VLDL_C | NA | G | A | -0.07415 | 0.00402 | 1.40E-78 |
| rs174530 | miR-1908-5 | cis | | 1.12E-08 | Nightingale | M_VLDL_T | NA | G | A | 0.02863 | 0.00415 | 4.10E-12 |
| rs174530 | miR-1908-5 | cis | | 1.12E-08 | Nightingale | Total_FA | NA | G | A | -0.02274 | 0.004237 | 2.30E-08 |
| rs174530 | miR-1908-5 | cis | | 1.12E-08 | Nightingale | Total_TG | NA | G | A | 0.042223 | 0.004135 | 4.80E-25 |
| rs174530 | miR-1908-5 | cis | | 1.12E-08 | Nightingale | M_LDL_C | NA | G | A | -0.03162 | 0.00428 | 3.50E-14 |
| rs174530 | miR-1908-5 | cis | | 1.12E-08 | Nightingale | HDL_PL | NA | G | A | -0.04339 | 0.003861 | 1.30E-31 |
| rs174530 | miR-1908-5 | cis | | 1.12E-08 | Nightingale | IDL_FC | NA | G | A | -0.08307 | 0.004134 | 6.30E-95 |
| rs174530 | miR-1908-5 | cis | | 1.12E-08 | Nightingale | M_LDL_FC | NA | G | A | -0.04857 | 0.004266 | 1.10E-31 |
| rs174530 | miR-1908-5 | cis | | 1.12E-08 | Nightingale | S_LDL_PL_ | NA | G | A | -0.05665 | 0.004263 | 9.40E-42 |
| rs174530 | miR-1908-5 | cis | | 1.12E-08 | Nightingale | S_VLDL_TG | NA | G | A | 0.034894 | 0.004145 | 2.20E-17 |
| rs174530 | miR-1908-5 | cis | | 1.12E-08 | Nightingale | M_HDL_CE | NA | G | A | -0.03338 | 0.003945 | 2.30E-18 |
| rs174530 | miR-1908-5 | cis | | 1.12E-08 | Nightingale | XS_VLDL_C | NA | G | A | -0.08908 | 0.004047 | 4.90E-112 |
| rs174530 | miR-1908-5 | cis | | 1.12E-08 | Nightingale | non_HDL_( | NA | G | A | -0.04445 | 0.004248 | 4.80E-27 |
| rs174530 | miR-1908-5 | cis | | 1.12E-08 | Nightingale | HDL_CE | NA | G | A | -0.05516 | 0.003833 | 1.20E-50 |
| rs174530 | miR-1908-5 | cis | | 1.12E-08 | Nightingale | S_HDL_C_p | NA | G | A | -0.03885 | 0.004222 | 8.80E-20 |
| rs174530 | miR-1908-5 | cis | | 1.12E-08 | Nightingale | XL_HDL_FC | NA | G | A | -0.09383 | 0.003979 | 6.30E-134 |
| rs174530 | miR-1908-5 | cis | | 1.12E-08 | Nightingale | L_LDL_C_p | NA | G | A | -0.0694 | 0.004162 | 1.80E-64 |
| rs174530 | miR-1908-5 | cis | | 1.12E-08 | Nightingale | IDL_P | NA | G | A | -0.04283 | 0.004227 | 1.80E-25 |
| rs174530 | miR-1908-5 | cis | | 1.12E-08 | Nightingale | M_HDL_L | NA | G | A | -0.02428 | 0.003965 | 1.50E-10 |
| rs174530 | miR-1908-5 | cis | | 1.12E-08 | Nightingale | LA_pct | NA | G | A | 0.172207 | 0.004174 | 0 |
| rs174530 | miR-1908-5 | cis | | 1.12E-08 | Nightingale | L_VLDL_L | NA | G | A | 0.034639 | 0.004128 | 2.10E-17 |
| rs174530 | miR-1908-5 | cis | | 1.12E-08 | Nightingale | M_HDL_TG | NA | G | A | 0.059986 | 0.004096 | 3.70E-50 |
| rs174530 | miR-1908-5 | cis | | 1.12E-08 | Nightingale | M_HDL_P | NA | G | A | -0.03037 | 0.003935 | 7.30E-16 |
| rs174530 | miR-1908-5 | cis | | 1.12E-08 | Nightingale | HDL_L | NA | G | A | -0.04791 | 0.003836 | 1.10E-38 |
| rs174530 | miR-1908-5 | cis | | 1.12E-08 | Nightingale | HDL_C | NA | G | A | -0.05825 | 0.003814 | 4.10E-57 |
| rs174530 | miR-1908-5 | cis | | 1.12E-08 | Nightingale | IDL_CE_pct | NA | G | A | -0.03364 | 0.004191 | 1.70E-15 |
| rs174530 | miR-1908-5 | cis | | 1.12E-08 | Nightingale | S_HDL_TG | NA | G | A | 0.062032 | 0.004142 | 1.00E-52 |
| rs174530 | miR-1908-5 | cis | | 1.12E-08 | Nightingale | XL_VLDL_F | NA | G | A | 0.038941 | 0.004137 | 1.40E-21 |
| rs174530 | miR-1908-5 | cis | | 1.12E-08 | Nightingale | Total_FC | NA | G | A | -0.05475 | 0.004173 | 8.40E-42 |
| rs174530 | miR-1908-5 | cis | | 1.12E-08 | Nightingale | L_HDL_CE | NA | G | A | -0.07111 | 0.003772 | 8.60E-87 |
| rs174530 | miR-1908-5 | cis | | 1.12E-08 | Nightingale | XXL_VLDL_ | NA | G | A | 0.038961 | 0.004296 | 1.30E-20 |
| rs174530 | miR-1908-5 | cis | | 1.12E-08 | Nightingale | L_HDL_C_p | NA | G | A | -0.06218 | 0.004026 | 1.50E-57 |
| rs174530 | miR-1908-5 | cis | | 1.12E-08 | Nightingale | S_VLDL_TG | NA | G | A | 0.059206 | 0.004133 | 1.10E-47 |
| rs174530 | miR-1908-5 | cis | | 1.12E-08 | Nightingale | M_VLDL_P | NA | G | A | -0.05774 | 0.004099 | 3.40E-48 |
| rs174530 | miR-1908-5 | cis | | 1.12E-08 | Nightingale | Total_P | NA | G | A | -0.03637 | 0.00402 | 5.70E-21 |
| rs174530 | miR-1908-5 | cis | | 1.12E-08 | Nightingale | S_LDL_TG_ | NA | G | A | 0.078216 | 0.004139 | 9.40E-82 |
| rs174530 | miR-1908-5 | cis | | 1.12E-08 | Nightingale | L_VLDL_PL | NA | G | A | 0.043976 | 0.004121 | 1.80E-27 |
| rs174530 | miR-1908-5 | cis | | 1.12E-08 | Nightingale | XS_VLDL_T | NA | G | A | 0.088837 | 0.004057 | 4.00E-111 |
| rs174530 | miR-1908-5 | cis | | 1.12E-08 | Nightingale | L_HDL_P | NA | G | A | -0.07048 | 0.003741 | 4.50E-87 |
| rs174530 | miR-1908-5 | cis | | 1.12E-08 | Nightingale | XS_VLDL_C | NA | G | A | -0.08362 | 0.004026 | 4.30E-100 |
| rs174530 | miR-1908-5 | cis | | 1.12E-08 | Nightingale | L_LDL_FC | NA | G | A | -0.06309 | 0.004194 | 2.80E-54 |
| rs174530 | miR-1908-5 | cis | | 1.12E-08 | Nightingale | Clinical_LD | NA | G | A | -0.05111 | 0.004247 | 5.00E-35 |
| rs174530 | miR-1908-5 | cis | | 1.12E-08 | Nightingale | L_HDL_C | NA | G | A | -0.07261 | 0.003761 | 4.70E-91 |
| rs174530 | miR-1908-5 | cis | | 1.12E-08 | Nightingale | XL_HDL_PL | NA | G | A | -0.06076 | 0.003924 | 2.20E-59 |
| rs174530 | miR-1908-5 | cis | | 1.12E-08 | Nightingale | XXL_VLDL_ | NA | G | A | 0.064342 | 0.004116 | 7.20E-57 |
| rs174530 | miR-1908-5 | cis | | 1.12E-08 | Nightingale | IDL_CE | NA | G | A | -0.06676 | 0.004123 | 1.20E-62 |
| rs174530 | miR-1908-5 | cis | | 1.12E-08 | Nightingale | L_LDL_P | NA | G | A | -0.03472 | 0.004257 | 2.80E-17 |
| rs174530 | miR-1908-5 | cis | | 1.12E-08 | Nightingale | ApoA1 | NA | G | A | -0.03682 | 0.003909 | 7.50E-23 |
| rs174530 | miR-1908-5 | cis | | 1.12E-08 | Nightingale | S_LDL_C | NA | G | A | -0.03008 | 0.004276 | 4.40E-13 |
| rs174530 | miR-1908-5 | cis | | 1.12E-08 | Nightingale | VLDL_L | NA | G | A | 0.024226 | 0.004188 | 6.50E-09 |
| rs174530 | miR-1908-5 | cis | | 1.12E-08 | Nightingale | S_LDL_FC | NA | G | A | -0.05809 | 0.004264 | 7.60E-45 |
| rs174530 | miR-1908-5 | cis | | 1.12E-08 | Nightingale | S_LDL_L | NA | G | A | -0.03051 | 0.004271 | 1.80E-13 |
| rs174530 | miR-1908-5 | cis | | 1.12E-08 | Nightingale | M_VLDL_C | NA | G | A | -0.06978 | 0.00401 | 5.80E-70 |
| rs174530 | miR-1908-5 | cis | | 1.12E-08 | Nightingale | XL_HDL_C | NA | G | A | -0.09003 | 0.003851 | 1.50E-132 |
| rs174530 | miR-1908-5 | cis | | 1.12E-08 | Nightingale | Total_PL | NA | G | A | -0.04708 | 0.004067 | 3.20E-33 |
| rs174530 | miR-1908-5 | cis | | 1.12E-08 | Nightingale | HDL_size | NA | G | A | -0.06583 | 0.003785 | 1.50E-74 |
| rs174530 | miR-1908-5 | cis | | 1.12E-08 | Nightingale | IDL_TG_pct | NA | G | A | 0.079886 | 0.004126 | 3.60E-86 |
| rs174530 | miR-1908-5 | cis | | 1.12E-08 | Nightingale | HDL_P | NA | G | A | -0.03177 | 0.004011 | 2.40E-16 |
| rs174533 | miR-1908-5 | cis | | 1.26E-08 | Nightingale | Total_CE | NA | A | G | -0.06709 | 0.004187 | 6.90E-61 |
| rs174533 | miR-1908-5 | cis | | 1.26E-08 | Nightingale | LDL_PL | NA | A | G | -0.04631 | 0.004331 | 8.60E-28 |

| | | | | | | | | | | | |
|---|---|---|---|---|---|---|---|---|---|---|---|
| rs174533 | miR-1908-! | cis | 1.26E-08 | Nightingale | M_HDL_CE | NA | A | G | -0.04637 | 0.004209 | 5.80E-29 |
| rs174533 | miR-1908-! | cis | 1.26E-08 | Nightingale | S_LDL_TG | NA | A | G | 0.050058 | 0.004234 | 1.10E-32 |
| rs174533 | miR-1908-! | cis | 1.26E-08 | Nightingale | S_LDL_P | NA | A | G | -0.03262 | 0.004345 | 1.40E-14 |
| rs174533 | miR-1908-! | cis | 1.26E-08 | Nightingale | XL_VLDL_T | NA | A | G | 0.056709 | 0.004152 | 4.30E-44 |
| rs174533 | miR-1908-! | cis | 1.26E-08 | Nightingale | M_HDL_PL | NA | A | G | 0.057862 | 0.004117 | 1.00E-46 |
| rs174533 | miR-1908-! | cis | 1.26E-08 | Nightingale | XL_VLDL_C | NA | A | G | -0.06382 | 0.004159 | 2.30E-54 |
| rs174533 | miR-1908-! | cis | 1.26E-08 | Nightingale | Total_PL | NA | A | G | -0.05124 | 0.004133 | 1.40E-37 |
| rs174533 | miR-1908-! | cis | 1.26E-08 | Nightingale | Citrate | NA | A | G | -0.03295 | 0.004313 | 1.60E-15 |
| rs174533 | miR-1908-! | cis | 1.26E-08 | Nightingale | M_HDL_C_ | NA | A | G | -0.06526 | 0.004132 | 4.50E-59 |
| rs174533 | miR-1908-! | cis | 1.26E-08 | Nightingale | S_LDL_FC | NA | A | G | -0.06282 | 0.004333 | 3.20E-50 |
| rs174533 | miR-1908-! | cis | 1.26E-08 | Nightingale | XL_VLDL_P | NA | A | G | 0.047968 | 0.004189 | 1.80E-31 |
| rs174533 | miR-1908-! | cis | 1.26E-08 | Nightingale | XL_VLDL_C | NA | A | G | -0.06076 | 0.004161 | 1.80E-49 |
| rs174533 | miR-1908-! | cis | 1.26E-08 | Nightingale | XL_HDL_CE | NA | A | G | -0.02587 | 0.004338 | 6.80E-09 |
| rs174533 | miR-1908-! | cis | 1.26E-08 | Nightingale | S_LDL_CE_ | NA | A | G | 0.057274 | 0.004332 | 1.10E-41 |
| rs174533 | miR-1908-! | cis | 1.26E-08 | Nightingale | L_HDL_CE | NA | A | G | -0.07822 | 0.003833 | 1.20E-100 |
| rs174533 | miR-1908-! | cis | 1.26E-08 | Nightingale | S_VLDL_PL | NA | A | G | -0.07832 | 0.004153 | 7.20E-83 |
| rs174533 | miR-1908-! | cis | 1.26E-08 | Nightingale | L_VLDL_TG | NA | A | G | 0.037168 | 0.004176 | 1.20E-19 |
| rs174533 | miR-1908-! | cis | 1.26E-08 | Nightingale | MUFA | NA | A | G | 0.063693 | 0.004296 | 1.20E-50 |
| rs174533 | miR-1908-! | cis | 1.26E-08 | Nightingale | XXL_VLDL_ | NA | A | G | -0.02672 | 0.004319 | 1.30E-10 |
| rs174533 | miR-1908-! | cis | 1.26E-08 | Nightingale | L_LDL_CE | NA | A | G | -0.05339 | 0.004302 | 2.80E-37 |
| rs174533 | miR-1908-! | cis | 1.26E-08 | Nightingale | M_LDL_P | NA | A | G | -0.04664 | 0.004353 | 1.50E-27 |
| rs174533 | miR-1908-! | cis | 1.26E-08 | Nightingale | M_HDL_FC | NA | A | G | -0.05119 | 0.00395 | 2.90E-41 |
| rs174533 | miR-1908-! | cis | 1.26E-08 | Nightingale | LDL_TG | NA | A | G | 0.029201 | 0.004292 | 2.20E-11 |
| rs174533 | miR-1908-! | cis | 1.26E-08 | Nightingale | XS_VLDL_F | NA | A | G | -0.11258 | 0.004308 | 3.00E-153 |
| rs174533 | miR-1908-! | cis | 1.26E-08 | Nightingale | M_LDL_C_ | NA | A | G | -0.09951 | 0.004344 | 2.00E-116 |
| rs174533 | miR-1908-! | cis | 1.26E-08 | Nightingale | S_LDL_FC_ | NA | A | G | -0.07941 | 0.004206 | 5.20E-82 |
| rs174533 | miR-1908-! | cis | 1.26E-08 | Nightingale | XXL_VLDL_ | NA | A | G | 0.066399 | 0.00418 | 3.60E-59 |
| rs174533 | miR-1908-! | cis | 1.26E-08 | Nightingale | XS_VLDL_P | NA | A | G | -0.04424 | 0.004283 | 7.20E-27 |
| rs174533 | miR-1908-! | cis | 1.26E-08 | Nightingale | non_HDL_( | NA | A | G | -0.04801 | 0.004317 | 4.40E-30 |
| rs174533 | miR-1908-! | cis | 1.26E-08 | Nightingale | M_VLDL_C | NA | A | G | -0.04419 | 0.004329 | 2.10E-25 |
| rs174533 | miR-1908-! | cis | 1.26E-08 | Nightingale | IDL_CE | NA | A | G | -0.07259 | 0.00419 | 4.60E-71 |
| rs174533 | miR-1908-! | cis | 1.26E-08 | Nightingale | XL_VLDL_T | NA | A | G | 0.059387 | 0.004205 | 1.60E-46 |
| rs174533 | miR-1908-! | cis | 1.26E-08 | Nightingale | M_HDL_L | NA | A | G | -0.02741 | 0.004029 | 1.20E-12 |
| rs174533 | miR-1908-! | cis | 1.26E-08 | Nightingale | M_LDL_PL_ | NA | A | G | 0.045099 | 0.004338 | 1.90E-26 |
| rs174533 | miR-1908-! | cis | 1.26E-08 | Nightingale | M_HDL_TG | NA | A | G | 0.04991 | 0.004287 | 5.40E-32 |
| rs174533 | miR-1908-! | cis | 1.26E-08 | Nightingale | LDL_FC | NA | A | G | -0.06438 | 0.004294 | 2.20E-53 |
| rs174533 | miR-1908-! | cis | 1.26E-08 | Nightingale | XS_VLDL_P | NA | A | G | -0.03507 | 0.00428 | 7.90E-18 |
| rs174533 | miR-1908-! | cis | 1.26E-08 | Nightingale | DHA_pct | NA | A | G | -0.27299 | 0.004213 | 0 |
| rs174533 | miR-1908-! | cis | 1.26E-08 | Nightingale | Total_FA | NA | A | G | -0.02381 | 0.004306 | 9.60E-09 |
| rs174533 | miR-1908-! | cis | 1.26E-08 | Nightingale | M_HDL_CE | NA | A | G | -0.03743 | 0.004008 | 3.80E-22 |
| rs174533 | miR-1908-! | cis | 1.26E-08 | Nightingale | M_LDL_C | NA | A | G | -0.03408 | 0.004349 | 1.30E-15 |
| rs174533 | miR-1908-! | cis | 1.26E-08 | Nightingale | M_HDL_FC | NA | A | G | -0.10291 | 0.003846 | 1.00E-171 |
| rs174533 | miR-1908-! | cis | 1.26E-08 | Nightingale | SFA_pct | NA | A | G | -0.04238 | 0.004282 | 4.30E-23 |
| rs174533 | miR-1908-! | cis | 1.26E-08 | Nightingale | L_VLDL_FC | NA | A | G | 0.028226 | 0.004299 | 2.30E-11 |
| rs174533 | miR-1908-! | cis | 1.26E-08 | Nightingale | XL_VLDL_L | NA | A | G | 0.051251 | 0.004176 | 6.90E-36 |
| rs174533 | miR-1908-! | cis | 1.26E-08 | Nightingale | M_LDL_L | NA | A | G | -0.02825 | 0.004347 | 3.50E-11 |
| rs174533 | miR-1908-! | cis | 1.26E-08 | Nightingale | SFA | NA | A | G | -0.03143 | 0.004301 | 9.50E-14 |
| rs174533 | miR-1908-! | cis | 1.26E-08 | Nightingale | S_VLDL_FC | NA | A | G | -0.02859 | 0.004341 | 1.50E-11 |
| rs174533 | miR-1908-! | cis | 1.26E-08 | Nightingale | HDL_CE | NA | A | G | -0.061 | 0.003895 | 1.10E-59 |
| rs174533 | miR-1908-! | cis | 1.26E-08 | Nightingale | IDL_P | NA | A | G | -0.04561 | 0.004295 | 1.10E-27 |
| rs174533 | miR-1908-! | cis | 1.26E-08 | Nightingale | Remnant_( | NA | A | G | -0.04445 | 0.004312 | 2.80E-26 |
| rs174533 | miR-1908-! | cis | 1.26E-08 | Nightingale | M_LDL_TG | NA | A | G | 0.036201 | 0.004277 | 2.90E-17 |
| rs174533 | miR-1908-! | cis | 1.26E-08 | Nightingale | LDL_CE | NA | A | G | -0.04366 | 0.004329 | 4.80E-25 |
| rs174533 | miR-1908-! | cis | 1.26E-08 | Nightingale | Total_FC | NA | A | G | -0.05956 | 0.00424 | 2.50E-47 |
| rs174533 | miR-1908-! | cis | 1.26E-08 | Nightingale | Clinical_LD | NA | A | G | -0.05528 | 0.004316 | 3.30E-39 |
| rs174533 | miR-1908-! | cis | 1.26E-08 | Nightingale | XS_VLDL_P | NA | A | G | 0.05744 | 0.004235 | 1.10E-42 |
| rs174533 | miR-1908-! | cis | 1.26E-08 | Nightingale | XS_VLDL_T | NA | A | G | 0.09716 | 0.004123 | 3.60E-128 |
| rs174533 | miR-1908-! | cis | 1.26E-08 | Nightingale | Tyr | NA | A | G | 0.03318 | 0.004335 | 1.90E-14 |
| rs174533 | miR-1908-! | cis | 1.26E-08 | Nightingale | M_LDL_TG | NA | A | G | 0.075281 | 0.004286 | 3.40E-70 |
| rs174533 | miR-1908-! | cis | 1.26E-08 | Nightingale | XL_HDL_FC | NA | A | G | 0.060883 | 0.003848 | 1.70E-62 |
| rs174533 | miR-1908-! | cis | 1.26E-08 | Nightingale | L_LDL_C_p | NA | A | G | -0.07661 | 0.004229 | 1.90E-75 |
| rs174533 | miR-1908-! | cis | 1.26E-08 | Nightingale | XL_HDL_FC | NA | A | G | -0.10254 | 0.004043 | 8.59E-153 |
| rs174533 | miR-1908-! | cis | 1.26E-08 | Nightingale | L_VLDL_FC | NA | A | G | 0.03991 | 0.004198 | 2.60E-22 |

| | | | | | | | | | | | |
|---|---|---|---|---|---|---|---|---|---|---|---|
| rs174533 | miR-1908-! | cis | 1.26E-08 | Nightingale | TG_by_PG | NA | A | G | 0.068452 | 0.004028 | 6.30E-68 |
| rs174533 | miR-1908-! | cis | 1.26E-08 | Nightingale | M_VLDL_T | NA | A | G | 0.077865 | 0.004099 | 5.50E-84 |
| rs174533 | miR-1908-! | cis | 1.26E-08 | Nightingale | S_HDL_C_p | NA | A | G | -0.04186 | 0.00429 | 3.80E-22 |
| rs174533 | miR-1908-! | cis | 1.26E-08 | Nightingale | L_LDL_TG_ | NA | A | G | 0.07711 | 0.004256 | 1.10E-74 |
| rs174533 | miR-1908-! | cis | 1.26E-08 | Nightingale | XL_HDL_TC | NA | A | G | 0.086765 | 0.004089 | 1.50E-105 |
| rs174533 | miR-1908-! | cis | 1.26E-08 | Nightingale | XS_VLDL_C | NA | A | G | -0.09187 | 0.004091 | 2.00E-116 |
| rs174533 | miR-1908-! | cis | 1.26E-08 | Nightingale | XS_VLDL_F | NA | A | G | -0.05665 | 0.004265 | 6.10E-43 |
| rs174533 | miR-1908-! | cis | 1.26E-08 | Nightingale | XXL_VLDL_ | NA | A | G | 0.067729 | 0.004182 | 1.20E-61 |
| rs174533 | miR-1908-! | cis | 1.26E-08 | Nightingale | XXL_VLDL_ | NA | A | G | 0.043751 | 0.004364 | 1.30E-24 |
| rs174533 | miR-1908-! | cis | 1.26E-08 | Nightingale | IDL_C | NA | A | G | -0.07757 | 0.004191 | 8.20E-81 |
| rs174533 | miR-1908-! | cis | 1.26E-08 | Nightingale | LDL_L | NA | A | G | -0.04558 | 0.004326 | 3.20E-27 |
| rs174533 | miR-1908-! | cis | 1.26E-08 | Nightingale | XL_HDL_L | NA | A | G | -0.09272 | 0.0039 | 7.91E-136 |
| rs174533 | miR-1908-! | cis | 1.26E-08 | Nightingale | L_VLDL_C | NA | A | G | 0.028598 | 0.004245 | 6.70E-12 |
| rs174533 | miR-1908-! | cis | 1.26E-08 | Nightingale | HDL_C | NA | A | G | -0.06431 | 0.003876 | 5.40E-67 |
| rs174533 | miR-1908-! | cis | 1.26E-08 | Nightingale | PUFA | NA | A | G | -0.09646 | 0.004233 | 6.90E-122 |
| rs174533 | miR-1908-! | cis | 1.26E-08 | Nightingale | XS_VLDL_L | NA | A | G | -0.0422 | 0.004266 | 5.10E-25 |
| rs174533 | miR-1908-! | cis | 1.26E-08 | Nightingale | XL_HDL_P | NA | A | G | -0.08932 | 0.003906 | 1.90E-126 |
| rs174533 | miR-1908-! | cis | 1.26E-08 | Nightingale | L_VLDL_CE | NA | A | G | -0.05911 | 0.004154 | 1.30E-48 |
| rs174533 | miR-1908-! | cis | 1.26E-08 | Nightingale | L_LDL_L | NA | A | G | -0.05432 | 0.004298 | 1.40E-38 |
| rs174533 | miR-1908-! | cis | 1.26E-08 | Nightingale | PUFA_by_M | NA | A | G | -0.17028 | 0.004157 | 0 |
| rs174533 | miR-1908-! | cis | 1.26E-08 | Nightingale | LA | NA | A | G | 0.083038 | 0.004256 | 5.50E-85 |
| rs174533 | miR-1908-! | cis | 1.26E-08 | Nightingale | L_LDL_C | NA | A | G | -0.05791 | 0.004292 | 9.80E-44 |
| rs174533 | miR-1908-! | cis | 1.26E-08 | Nightingale | M_VLDL_F | NA | A | G | -0.02966 | 0.00434 | 2.60E-12 |
| rs174533 | miR-1908-! | cis | 1.26E-08 | Nightingale | L_HDL_P | NA | A | G | -0.07741 | 0.003801 | 1.60E-100 |
| rs174533 | miR-1908-! | cis | 1.26E-08 | Nightingale | XL_HDL_PL | NA | A | G | -0.08878 | 0.003902 | 1.00E-124 |
| rs174533 | miR-1908-! | cis | 1.26E-08 | Nightingale | MUFA_pct | NA | A | G | 0.196942 | 0.004154 | 0 |
| rs174533 | miR-1908-! | cis | 1.26E-08 | Nightingale | XL_HDL_CE | NA | A | G | -0.09675 | 0.003887 | 2.10E-148 |
| rs174533 | miR-1908-! | cis | 1.26E-08 | Nightingale | M_HDL_TG | NA | A | G | 0.066093 | 0.004162 | 2.20E-59 |
| rs174533 | miR-1908-! | cis | 1.26E-08 | Nightingale | LDL_P | NA | A | G | -0.04042 | 0.004337 | 1.30E-21 |
| rs174533 | miR-1908-! | cis | 1.26E-08 | Nightingale | S_LDL_C | NA | A | G | -0.03226 | 0.004345 | 2.70E-14 |
| rs174533 | miR-1908-! | cis | 1.26E-08 | Nightingale | L_VLDL_PL | NA | A | G | 0.087068 | 0.004213 | 2.30E-98 |
| rs174533 | miR-1908-! | cis | 1.26E-08 | Nightingale | S_VLDL_C_ | NA | A | G | -0.05651 | 0.004223 | 1.10E-41 |
| rs174533 | miR-1908-! | cis | 1.26E-08 | Nightingale | VLDL_size | NA | A | G | 0.072538 | 0.004046 | 6.00E-76 |
| rs174533 | miR-1908-! | cis | 1.26E-08 | Nightingale | Total_P | NA | A | G | -0.04011 | 0.004085 | 1.90E-24 |
| rs174533 | miR-1908-! | cis | 1.26E-08 | Nightingale | M_HDL_C | NA | A | G | -0.04041 | 0.003994 | 1.00E-25 |
| rs174533 | miR-1908-! | cis | 1.26E-08 | Nightingale | Total_C | NA | A | G | -0.06531 | 0.004202 | 2.30E-57 |
| rs174533 | miR-1908-! | cis | 1.26E-08 | Nightingale | HDL_P | NA | A | G | -0.03529 | 0.004076 | 3.00E-19 |
| rs174533 | miR-1908-! | cis | 1.26E-08 | Nightingale | XL_VLDL_P | NA | A | G | 0.052622 | 0.004172 | 8.00E-38 |
| rs174533 | miR-1908-! | cis | 1.26E-08 | Nightingale | ApoB | NA | A | G | -0.03873 | 0.004335 | 4.60E-20 |
| rs174533 | miR-1908-! | cis | 1.26E-08 | Nightingale | M_LDL_CE | NA | A | G | -0.02637 | 0.004347 | 6.10E-10 |
| rs174533 | miR-1908-! | cis | 1.26E-08 | Nightingale | XL_HDL_C | NA | A | G | -0.0989 | 0.003914 | 1.10E-152 |
| rs174533 | miR-1908-! | cis | 1.26E-08 | Nightingale | L_HDL_L | NA | A | G | -0.07677 | 0.003808 | 1.50E-98 |
| rs174533 | miR-1908-! | cis | 1.26E-08 | Nightingale | L_LDL_PL | NA | A | G | -0.05509 | 0.004308 | 4.60E-39 |
| rs174533 | miR-1908-! | cis | 1.26E-08 | Nightingale | HDL_TG | NA | A | G | 0.045774 | 0.00428 | 4.50E-27 |
| rs174533 | miR-1908-! | cis | 1.26E-08 | Nightingale | XL_VLDL_F | NA | A | G | -0.06833 | 0.004206 | 4.80E-62 |
| rs174533 | miR-1908-! | cis | 1.26E-08 | Nightingale | Total_L | NA | A | G | -0.03754 | 0.00427 | 9.60E-20 |
| rs174533 | miR-1908-! | cis | 1.26E-08 | Nightingale | M_LDL_PL | NA | A | G | -0.02365 | 0.004347 | 2.70E-08 |
| rs174533 | miR-1908-! | cis | 1.26E-08 | Nightingale | Total_TG | NA | A | G | 0.046498 | 0.004201 | 2.40E-29 |
| rs174533 | miR-1908-! | cis | 1.26E-08 | Nightingale | IDL_C_pct | NA | A | G | -0.07461 | 0.004237 | 8.40E-71 |
| rs174533 | miR-1908-! | cis | 1.26E-08 | Nightingale | Cholines | NA | A | G | -0.03722 | 0.004082 | 1.10E-21 |
| rs174533 | miR-1908-! | cis | 1.26E-08 | Nightingale | S_HDL_TG | NA | A | G | 0.06844 | 0.004209 | 4.30E-62 |
| rs174533 | miR-1908-! | cis | 1.26E-08 | Nightingale | Omega_6_ | NA | A | G | 0.023864 | 0.004224 | 7.70E-09 |
| rs174533 | miR-1908-! | cis | 1.26E-08 | Nightingale | HDL_PL | NA | A | G | -0.04816 | 0.003924 | 3.00E-37 |
| rs174533 | miR-1908-! | cis | 1.26E-08 | Nightingale | XS_VLDL_T | NA | A | G | 0.031054 | 0.004274 | 1.60E-12 |
| rs174533 | miR-1908-! | cis | 1.26E-08 | Nightingale | LA_pct | NA | A | G | 0.184284 | 0.004242 | 0 |
| rs174533 | miR-1908-! | cis | 1.26E-08 | Nightingale | XS_VLDL_C | NA | A | G | -0.09776 | 0.004113 | 4.20E-130 |
| rs174533 | miR-1908-! | cis | 1.26E-08 | Nightingale | L_HDL_TG_ | NA | A | G | 0.078011 | 0.004096 | 6.60E-86 |
| rs174533 | miR-1908-! | cis | 1.26E-08 | Nightingale | L_VLDL_P | NA | A | G | 0.04165 | 0.004191 | 3.80E-24 |
| rs174533 | miR-1908-! | cis | 1.26E-08 | Nightingale | Phosphogly | NA | A | G | -0.04109 | 0.004111 | 2.30E-25 |
| rs174533 | miR-1908-! | cis | 1.26E-08 | Nightingale | S_VLDL_FC | NA | A | G | -0.08275 | 0.004145 | 2.90E-92 |
| rs174533 | miR-1908-! | cis | 1.26E-08 | Nightingale | L_HDL_FC_ | NA | A | G | -0.11226 | 0.004032 | 2.10E-185 |
| rs174533 | miR-1908-! | cis | 1.26E-08 | Nightingale | DHA | NA | A | G | -0.28773 | 0.004183 | 0 |
| rs174533 | miR-1908-! | cis | 1.26E-08 | Nightingale | HDL_size | NA | A | G | -0.07222 | 0.003846 | 2.40E-85 |

| | | | | | | | | | | | |
|---|---|---|---|---|---|---|---|---|---|---|---|
| rs174533 | miR-1908-! | cis | 1.26E-08 | Nightingale | HDL_FC | NA | A | G | -0.07361 | 0.003845 | 2.00E-88 |
| rs174533 | miR-1908-! | cis | 1.26E-08 | Nightingale | XXL_VLDL_ | NA | A | G | 0.071435 | 0.004183 | 7.40E-68 |
| rs174533 | miR-1908-! | cis | 1.26E-08 | Nightingale | IDL_L | NA | A | G | -0.07276 | 0.004196 | 1.30E-71 |
| rs174533 | miR-1908-! | cis | 1.26E-08 | Nightingale | M_LDL_FC | NA | A | G | -0.05241 | 0.004335 | 2.80E-35 |
| rs174533 | miR-1908-! | cis | 1.26E-08 | Nightingale | L_LDL_FC | NA | A | G | -0.06845 | 0.004262 | 3.00E-61 |
| rs174533 | miR-1908-! | cis | 1.26E-08 | Nightingale | VLDL_L | NA | A | G | 0.027095 | 0.004256 | 1.10E-10 |
| rs174533 | miR-1908-! | cis | 1.26E-08 | Nightingale | L_VLDL_PL | NA | A | G | 0.048468 | 0.004188 | 2.60E-32 |
| rs174533 | miR-1908-! | cis | 1.26E-08 | Nightingale | XL_VLDL_F | NA | A | G | 0.043197 | 0.004204 | 1.20E-25 |
| rs174533 | miR-1908-! | cis | 1.26E-08 | Nightingale | M_VLDL_T | NA | A | G | 0.031224 | 0.004217 | 5.80E-14 |
| rs174533 | miR-1908-! | cis | 1.26E-08 | Nightingale | L_HDL_C | NA | A | G | -0.0798 | 0.003822 | 2.30E-105 |
| rs174533 | miR-1908-! | cis | 1.26E-08 | Nightingale | IDL_CE_pct | NA | A | G | -0.03817 | 0.004259 | 4.30E-19 |
| rs174533 | miR-1908-! | cis | 1.26E-08 | Nightingale | HDL_L | NA | A | G | -0.05302 | 0.003898 | 2.20E-45 |
| rs174533 | miR-1908-! | cis | 1.26E-08 | Nightingale | S_LDL_L | NA | A | G | -0.03259 | 0.00434 | 1.20E-14 |
| rs174533 | miR-1908-! | cis | 1.26E-08 | Nightingale | M_HDL_P | NA | A | G | -0.03397 | 0.003999 | 7.00E-19 |
| rs174533 | miR-1908-! | cis | 1.26E-08 | Nightingale | L_HDL_C_p | NA | A | G | -0.06774 | 0.004091 | 4.50E-66 |
| rs174533 | miR-1908-! | cis | 1.26E-08 | Nightingale | PUFA_pct | NA | A | G | -0.12123 | 0.004175 | 1.10E-186 |
| rs174533 | miR-1908-! | cis | 1.26E-08 | Nightingale | XS_VLDL_C | NA | A | G | -0.0724 | 0.004188 | 5.10E-71 |
| rs174533 | miR-1908-! | cis | 1.26E-08 | Nightingale | Omega_3_ | NA | A | G | -0.38858 | 0.004241 | 0 |
| rs174533 | miR-1908-! | cis | 1.26E-08 | Nightingale | XXL_VLDL_ | NA | A | G | 0.065837 | 0.004175 | 1.60E-58 |
| rs174533 | miR-1908-! | cis | 1.26E-08 | Nightingale | M_HDL_PL | NA | A | G | -0.02128 | 0.004059 | 3.90E-08 |
| rs174533 | miR-1908-! | cis | 1.26E-08 | Nightingale | L_HDL_CE_ | NA | A | G | -0.04935 | 0.004153 | 1.00E-34 |
| rs174533 | miR-1908-! | cis | 1.26E-08 | Nightingale | XL_HDL_C_ | NA | A | G | 0.023983 | 0.004146 | 2.70E-10 |
| rs174533 | miR-1908-! | cis | 1.26E-08 | Nightingale | L_HDL_PL_ | NA | A | G | 0.042694 | 0.004201 | 8.90E-26 |
| rs174533 | miR-1908-! | cis | 1.26E-08 | Nightingale | S_LDL_PL | NA | A | G | -0.05169 | 0.00433 | 1.30E-34 |
| rs174533 | miR-1908-! | cis | 1.26E-08 | Nightingale | S_HDL_TG_ | NA | A | G | 0.068512 | 0.004128 | 1.40E-64 |
| rs174533 | miR-1908-! | cis | 1.26E-08 | Nightingale | M_VLDL_C | NA | A | G | -0.07583 | 0.004075 | 3.40E-80 |
| rs174533 | miR-1908-! | cis | 1.26E-08 | Nightingale | L_HDL_PL | NA | A | G | -0.07568 | 0.00382 | 4.50E-95 |
| rs174533 | miR-1908-! | cis | 1.26E-08 | Nightingale | XXL_VLDL_ | NA | A | G | 0.071701 | 0.004165 | 3.20E-69 |
| rs174533 | miR-1908-! | cis | 1.26E-08 | Nightingale | L_HDL_FC | NA | A | G | -0.08419 | 0.003804 | 4.60E-118 |
| rs174533 | miR-1908-! | cis | 1.26E-08 | Nightingale | Omega_6_ | NA | A | G | 0.368576 | 0.004278 | 0 |
| rs174533 | miR-1908-! | cis | 1.26E-08 | Nightingale | L_VLDL_C_ | NA | A | G | -0.03859 | 0.00424 | 7.20E-21 |
| rs174533 | miR-1908-! | cis | 1.26E-08 | Nightingale | S_HDL_FC_ | NA | A | G | -0.10275 | 0.004117 | 2.40E-152 |
| rs174533 | miR-1908-! | cis | 1.26E-08 | Nightingale | VLDL_TG | NA | A | G | 0.047489 | 0.004177 | 7.00E-31 |
| rs174533 | miR-1908-! | cis | 1.26E-08 | Nightingale | S_LDL_TG_ | NA | A | G | 0.085808 | 0.004206 | 3.70E-95 |
| rs174533 | miR-1908-! | cis | 1.26E-08 | Nightingale | XXL_VLDL_ | NA | A | G | 0.061051 | 0.004188 | 4.20E-50 |
| rs174533 | miR-1908-! | cis | 1.26E-08 | Nightingale | XL_VLDL_C | NA | A | G | 0.037208 | 0.004239 | 3.70E-19 |
| rs174533 | miR-1908-! | cis | 1.26E-08 | Nightingale | XL_HDL_PL | NA | A | G | -0.06696 | 0.003987 | 2.10E-68 |
| rs174533 | miR-1908-! | cis | 1.26E-08 | Nightingale | M_VLDL_P | NA | A | G | -0.06252 | 0.004165 | 3.80E-54 |
| rs174533 | miR-1908-! | cis | 1.26E-08 | Nightingale | S_VLDL_TG | NA | A | G | 0.038551 | 0.004212 | 1.80E-20 |
| rs174533 | miR-1908-! | cis | 1.26E-08 | Nightingale | XS_VLDL_C | NA | A | G | -0.06867 | 0.004213 | 2.10E-63 |
| rs174533 | miR-1908-! | cis | 1.26E-08 | Nightingale | L_VLDL_L | NA | A | G | 0.038003 | 0.004195 | 2.50E-20 |
| rs174533 | miR-1908-! | cis | 1.26E-08 | Nightingale | IDL_TG_pc | NA | A | G | 0.087724 | 0.004193 | 1.50E-100 |
| rs174533 | miR-1908-! | cis | 1.26E-08 | Nightingale | Unsaturati | NA | A | G | -0.33836 | 0.004111 | 0 |
| rs174533 | miR-1908-! | cis | 1.26E-08 | Nightingale | M_VLDL_C | NA | A | G | -0.08056 | 0.004085 | 6.00E-90 |
| rs174533 | miR-1908-! | cis | 1.26E-08 | Nightingale | M_VLDL_F | NA | A | G | -0.09119 | 0.004127 | 1.50E-113 |
| rs174533 | miR-1908-! | cis | 1.26E-08 | Nightingale | S_VLDL_TG | NA | A | G | 0.064364 | 0.0042 | 2.20E-54 |
| rs174533 | miR-1908-! | cis | 1.26E-08 | Nightingale | XL_VLDL_C | NA | A | G | 0.030078 | 0.004272 | 8.10E-13 |
| rs174533 | miR-1908-! | cis | 1.26E-08 | Nightingale | Sphingomy | NA | A | G | -0.115 | 0.004056 | 1.00E-185 |
| rs174533 | miR-1908-! | cis | 1.26E-08 | Nightingale | XXL_VLDL_ | NA | A | G | 0.073344 | 0.004192 | 8.00E-71 |
| rs174533 | miR-1908-! | cis | 1.26E-08 | Nightingale | M_LDL_FC_ | NA | A | G | -0.07149 | 0.004158 | 1.90E-68 |
| rs174533 | miR-1908-! | cis | 1.26E-08 | Nightingale | Omega_3 | NA | A | G | -0.33473 | 0.004254 | 0 |
| rs174533 | miR-1908-! | cis | 1.26E-08 | Nightingale | S_LDL_PL_ | NA | A | G | -0.06215 | 0.004332 | 9.10E-49 |
| rs174533 | miR-1908-! | cis | 1.26E-08 | Nightingale | S_VLDL_CE | NA | A | G | -0.02898 | 0.004282 | 1.00E-11 |
| rs174533 | miR-1908-! | cis | 1.26E-08 | Nightingale | LDL_C | NA | A | G | -0.04984 | 0.004322 | 3.10E-32 |
| rs174533 | miR-1908-! | cis | 1.26E-08 | Nightingale | M_VLDL_C | NA | A | G | -0.05283 | 0.004301 | 3.70E-36 |
| rs174533 | miR-1908-! | cis | 1.26E-08 | Nightingale | L_LDL_P | NA | A | G | -0.03766 | 0.004326 | 2.70E-19 |
| rs174533 | miR-1908-! | cis | 1.26E-08 | Nightingale | IDL_PL | NA | A | G | -0.07315 | 0.004188 | 5.20E-73 |
| rs174533 | miR-1908-! | cis | 1.26E-08 | Nightingale | IDL_FC | NA | A | G | -0.08995 | 0.004201 | 5.00E-107 |
| rs174533 | miR-1908-! | cis | 1.26E-08 | Nightingale | IDL_FC_pct | NA | A | G | -0.10339 | 0.004306 | 1.10E-128 |
| rs174533 | miR-1908-! | cis | 1.26E-08 | Nightingale | L_LDL_FC_ | NA | A | G | -0.08273 | 0.004137 | 1.10E-91 |
| rs174533 | miR-1908-! | cis | 1.26E-08 | Nightingale | ApoA1 | NA | A | G | -0.0409 | 0.003972 | 6.70E-27 |
| rs174535 | miR-1908-! | cis | 6.98E-09 | Nightingale | M_LDL_P | NA | C | T | -0.0464 | 0.004341 | 1.20E-27 |
| rs174535 | miR-1908-! | cis | 6.98E-09 | Nightingale | M_HDL_PL | NA | C | T | 0.056818 | 0.004106 | 3.20E-45 |

| | | | | | | | | | | | |
|---|---|---|---|---|---|---|---|---|---|---|---|
| rs174535 | miR-1908-5 | cis | 6.98E-09 | Nightingale | M_HDL_TG | NA | C | T | 0.048949 | 0.004275 | 3.70E-31 |
| rs174535 | miR-1908-5 | cis | 6.98E-09 | Nightingale | XL_VLDL_P | NA | C | T | 0.047139 | 0.004177 | 1.40E-30 |
| rs174535 | miR-1908-5 | cis | 6.98E-09 | Nightingale | S_LDL_CE_ | NA | C | T | 0.05601 | 0.00432 | 4.10E-40 |
| rs174535 | miR-1908-5 | cis | 6.98E-09 | Nightingale | M_HDL_CE | NA | C | T | -0.04541 | 0.004197 | 5.70E-28 |
| rs174535 | miR-1908-5 | cis | 6.98E-09 | Nightingale | XS_VLDL_P | NA | C | T | -0.03513 | 0.004268 | 5.40E-18 |
| rs174535 | miR-1908-5 | cis | 6.98E-09 | Nightingale | DHA_pct | NA | C | T | -0.26894 | 0.004201 | 0 |
| rs174535 | miR-1908-5 | cis | 6.98E-09 | Nightingale | M_LDL_C | NA | C | T | -0.0336 | 0.004337 | 1.90E-15 |
| rs174535 | miR-1908-5 | cis | 6.98E-09 | Nightingale | L_VLDL_FC | NA | C | T | 0.026826 | 0.004287 | 1.90E-10 |
| rs174535 | miR-1908-5 | cis | 6.98E-09 | Nightingale | Total_FC | NA | C | T | -0.05898 | 0.004228 | 5.90E-47 |
| rs174535 | miR-1908-5 | cis | 6.98E-09 | Nightingale | L_HDL_TG_ | NA | C | T | 0.07721 | 0.004084 | 1.60E-84 |
| rs174535 | miR-1908-5 | cis | 6.98E-09 | Nightingale | XL_VLDL_F | NA | C | T | -0.06812 | 0.004194 | 4.70E-62 |
| rs174535 | miR-1908-5 | cis | 6.98E-09 | Nightingale | Phosphogly | NA | C | T | -0.04097 | 0.0041 | 2.60E-25 |
| rs174535 | miR-1908-5 | cis | 6.98E-09 | Nightingale | L_VLDL_PL | NA | C | T | 0.084977 | 0.004201 | 4.10E-94 |
| rs174535 | miR-1908-5 | cis | 6.98E-09 | Nightingale | SFA_pct | NA | C | T | -0.0428 | 0.00427 | 1.20E-23 |
| rs174535 | miR-1908-5 | cis | 6.98E-09 | Nightingale | XL_VLDL_L | NA | C | T | 0.050465 | 0.004165 | 5.50E-35 |
| rs174535 | miR-1908-5 | cis | 6.98E-09 | Nightingale | XS_VLDL_F | NA | C | T | -0.11137 | 0.004296 | 3.10E-151 |
| rs174535 | miR-1908-5 | cis | 6.98E-09 | Nightingale | XL_VLDL_C | NA | C | T | -0.05963 | 0.00415 | 3.50E-48 |
| rs174535 | miR-1908-5 | cis | 6.98E-09 | Nightingale | MUFA | NA | C | T | 0.062248 | 0.004284 | 9.00E-49 |
| rs174535 | miR-1908-5 | cis | 6.98E-09 | Nightingale | XL_VLDL_C | NA | C | T | -0.06282 | 0.004148 | 3.30E-53 |
| rs174535 | miR-1908-5 | cis | 6.98E-09 | Nightingale | LA | NA | C | T | 0.082304 | 0.004244 | 5.60E-84 |
| rs174535 | miR-1908-5 | cis | 6.98E-09 | Nightingale | XL_HDL_FC | NA | C | T | 0.060654 | 0.003837 | 5.20E-62 |
| rs174535 | miR-1908-5 | cis | 6.98E-09 | Nightingale | S_VLDL_FC | NA | C | T | -0.0284 | 0.004329 | 1.40E-11 |
| rs174535 | miR-1908-5 | cis | 6.98E-09 | Nightingale | XL_VLDL_T | NA | C | T | 0.055933 | 0.00414 | 3.60E-43 |
| rs174535 | miR-1908-5 | cis | 6.98E-09 | Nightingale | L_LDL_FC | NA | C | T | -0.06769 | 0.00425 | 1.30E-60 |
| rs174535 | miR-1908-5 | cis | 6.98E-09 | Nightingale | Remnant_( | NA | C | T | -0.04419 | 0.0043 | 2.60E-26 |
| rs174535 | miR-1908-5 | cis | 6.98E-09 | Nightingale | Tyr | NA | C | T | 0.033724 | 0.004323 | 6.10E-15 |
| rs174535 | miR-1908-5 | cis | 6.98E-09 | Nightingale | VLDL_size | NA | C | T | 0.071593 | 0.004035 | 2.30E-74 |
| rs174535 | miR-1908-5 | cis | 6.98E-09 | Nightingale | L_HDL_CE | NA | C | T | -0.07764 | 0.003822 | 5.50E-99 |
| rs174535 | miR-1908-5 | cis | 6.98E-09 | Nightingale | HDL_P | NA | C | T | -0.03489 | 0.004064 | 6.30E-19 |
| rs174535 | miR-1908-5 | cis | 6.98E-09 | Nightingale | S_VLDL_PL | NA | C | T | -0.07728 | 0.004142 | 1.90E-81 |
| rs174535 | miR-1908-5 | cis | 6.98E-09 | Nightingale | HDL_C | NA | C | T | -0.06373 | 0.003865 | 1.00E-65 |
| rs174535 | miR-1908-5 | cis | 6.98E-09 | Nightingale | XL_HDL_CE | NA | C | T | -0.02466 | 0.004326 | 2.40E-08 |
| rs174535 | miR-1908-5 | cis | 6.98E-09 | Nightingale | S_VLDL_C_ | NA | C | T | -0.05594 | 0.004211 | 2.20E-41 |
| rs174535 | miR-1908-5 | cis | 6.98E-09 | Nightingale | Total_CE | NA | C | T | -0.06643 | 0.004175 | 2.40E-60 |
| rs174535 | miR-1908-5 | cis | 6.98E-09 | Nightingale | non_HDL_( | NA | C | T | -0.04756 | 0.004305 | 6.60E-30 |
| rs174535 | miR-1908-5 | cis | 6.98E-09 | Nightingale | TG_by_PG | NA | C | T | 0.067713 | 0.004017 | 8.20E-67 |
| rs174535 | miR-1908-5 | cis | 6.98E-09 | Nightingale | Total_C | NA | C | T | -0.06467 | 0.00419 | 7.40E-57 |
| rs174535 | miR-1908-5 | cis | 6.98E-09 | Nightingale | S_LDL_FC | NA | C | T | -0.06188 | 0.004321 | 1.80E-49 |
| rs174535 | miR-1908-5 | cis | 6.98E-09 | Nightingale | XL_HDL_L | NA | C | T | -0.09207 | 0.003889 | 4.00E-134 |
| rs174535 | miR-1908-5 | cis | 6.98E-09 | Nightingale | L_VLDL_C | NA | C | T | 0.028107 | 0.004233 | 1.70E-11 |
| rs174535 | miR-1908-5 | cis | 6.98E-09 | Nightingale | M_HDL_C_ | NA | C | T | -0.06419 | 0.00412 | 2.80E-57 |
| rs174535 | miR-1908-5 | cis | 6.98E-09 | Nightingale | Total_FA | NA | C | T | -0.02397 | 0.004294 | 7.10E-09 |
| rs174535 | miR-1908-5 | cis | 6.98E-09 | Nightingale | S_LDL_P | NA | C | T | -0.0326 | 0.004332 | 9.00E-15 |
| rs174535 | miR-1908-5 | cis | 6.98E-09 | Nightingale | XL_HDL_CE | NA | C | T | -0.0959 | 0.003876 | 4.10E-146 |
| rs174535 | miR-1908-5 | cis | 6.98E-09 | Nightingale | XXL_VLDL_ | NA | C | T | -0.0262 | 0.004307 | 2.40E-10 |
| rs174535 | miR-1908-5 | cis | 6.98E-09 | Nightingale | XL_HDL_TG | NA | C | T | 0.086058 | 0.004077 | 3.20E-104 |
| rs174535 | miR-1908-5 | cis | 6.98E-09 | Nightingale | M_VLDL_C | NA | C | T | -0.0524 | 0.004289 | 5.90E-36 |
| rs174535 | miR-1908-5 | cis | 6.98E-09 | Nightingale | LDL_TG | NA | C | T | 0.028598 | 0.00428 | 4.90E-11 |
| rs174535 | miR-1908-5 | cis | 6.98E-09 | Nightingale | L_HDL_P | NA | C | T | -0.07687 | 0.003791 | 5.90E-99 |
| rs174535 | miR-1908-5 | cis | 6.98E-09 | Nightingale | M_HDL_C | NA | C | T | -0.04005 | 0.003982 | 2.80E-25 |
| rs174535 | miR-1908-5 | cis | 6.98E-09 | Nightingale | M_LDL_TG | NA | C | T | 0.035612 | 0.004265 | 7.60E-17 |
| rs174535 | miR-1908-5 | cis | 6.98E-09 | Nightingale | L_LDL_CE | NA | C | T | -0.05272 | 0.00429 | 7.80E-37 |
| rs174535 | miR-1908-5 | cis | 6.98E-09 | Nightingale | Clinical_LD | NA | C | T | -0.05468 | 0.004304 | 7.40E-39 |
| rs174535 | miR-1908-5 | cis | 6.98E-09 | Nightingale | M_HDL_CE | NA | C | T | -0.0371 | 0.003997 | 9.80E-22 |
| rs174535 | miR-1908-5 | cis | 6.98E-09 | Nightingale | S_LDL_PL_ | NA | C | T | -0.06084 | 0.00432 | 5.10E-47 |
| rs174535 | miR-1908-5 | cis | 6.98E-09 | Nightingale | L_VLDL_TG | NA | C | T | 0.036905 | 0.004165 | 1.90E-19 |
| rs174535 | miR-1908-5 | cis | 6.98E-09 | Nightingale | L_VLDL_P | NA | C | T | 0.041084 | 0.004179 | 1.40E-23 |
| rs174535 | miR-1908-5 | cis | 6.98E-09 | Nightingale | Omega_3_ | NA | C | T | -0.38338 | 0.004229 | 0 |
| rs174535 | miR-1908-5 | cis | 6.98E-09 | Nightingale | M_HDL_FC | NA | C | T | -0.10185 | 0.003835 | 2.00E-168 |
| rs174535 | miR-1908-5 | cis | 6.98E-09 | Nightingale | LDL_C | NA | C | T | -0.04921 | 0.00431 | 7.10E-32 |
| rs174535 | miR-1908-5 | cis | 6.98E-09 | Nightingale | DHA | NA | C | T | -0.28376 | 0.004171 | 0 |
| rs174535 | miR-1908-5 | cis | 6.98E-09 | Nightingale | L_HDL_L | NA | C | T | -0.07628 | 0.003798 | 3.70E-97 |
| rs174535 | miR-1908-5 | cis | 6.98E-09 | Nightingale | L_HDL_PL | NA | C | T | -0.07522 | 0.003809 | 8.50E-94 |

| | | | | | | | | | | | |
|---|---|---|---|---|---|---|---|---|---|---|---|
| rs174535 | miR-1908-5 | cis | 6.98E-09 | Nightingale | LDL_L | NA | C | T | -0.04499 | 0.004314 | 6.50E-27 |
| rs174535 | miR-1908-5 | cis | 6.98E-09 | Nightingale | L_LDL_L | NA | C | T | -0.05368 | 0.004286 | 3.60E-38 |
| rs174535 | miR-1908-5 | cis | 6.98E-09 | Nightingale | S_VLDL_TG | NA | C | T | 0.063644 | 0.004188 | 8.80E-54 |
| rs174535 | miR-1908-5 | cis | 6.98E-09 | Nightingale | IDL_CE | NA | C | T | -0.07187 | 0.004178 | 2.20E-70 |
| rs174535 | miR-1908-5 | cis | 6.98E-09 | Nightingale | PUFA | NA | C | T | -0.09528 | 0.004221 | 9.71E-120 |
| rs174535 | miR-1908-5 | cis | 6.98E-09 | Nightingale | M_LDL_TG | NA | C | T | 0.074065 | 0.004274 | 9.40E-69 |
| rs174535 | miR-1908-5 | cis | 6.98E-09 | Nightingale | S_LDL_TG | NA | C | T | 0.04928 | 0.004222 | 6.20E-32 |
| rs174535 | miR-1908-5 | cis | 6.98E-09 | Nightingale | S_LDL_FC_ | NA | C | T | -0.078 | 0.004194 | 7.50E-80 |
| rs174535 | miR-1908-5 | cis | 6.98E-09 | Nightingale | IDL_FC_pct | NA | C | T | -0.10265 | 0.004294 | 9.40E-128 |
| rs174535 | miR-1908-5 | cis | 6.98E-09 | Nightingale | L_HDL_C | NA | C | T | -0.07922 | 0.003811 | 1.10E-103 |
| rs174535 | miR-1908-5 | cis | 6.98E-09 | Nightingale | Total_P | NA | C | T | -0.03967 | 0.004073 | 4.20E-24 |
| rs174535 | miR-1908-5 | cis | 6.98E-09 | Nightingale | LDL_FC | NA | C | T | -0.0636 | 0.004282 | 9.00E-53 |
| rs174535 | miR-1908-5 | cis | 6.98E-09 | Nightingale | XXL_VLDL_ | NA | C | T | 0.059834 | 0.004177 | 1.90E-48 |
| rs174535 | miR-1908-5 | cis | 6.98E-09 | Nightingale | XS_VLDL_C | NA | C | T | -0.09102 | 0.00408 | 3.40E-115 |
| rs174535 | miR-1908-5 | cis | 6.98E-09 | Nightingale | M_VLDL_C | NA | C | T | -0.07492 | 0.004063 | 6.50E-79 |
| rs174535 | miR-1908-5 | cis | 6.98E-09 | Nightingale | HDL_CE | NA | C | T | -0.06044 | 0.003884 | 1.50E-58 |
| rs174535 | miR-1908-5 | cis | 6.98E-09 | Nightingale | M_LDL_CE | NA | C | T | -0.026 | 0.004335 | 7.20E-10 |
| rs174535 | miR-1908-5 | cis | 6.98E-09 | Nightingale | M_HDL_TG | NA | C | T | 0.065075 | 0.004151 | 8.90E-58 |
| rs174535 | miR-1908-5 | cis | 6.98E-09 | Nightingale | XL_HDL_P | NA | C | T | -0.08859 | 0.003895 | 1.30E-124 |
| rs174535 | miR-1908-5 | cis | 6.98E-09 | Nightingale | M_LDL_PL_ | NA | C | T | 0.045336 | 0.004326 | 1.10E-26 |
| rs174535 | miR-1908-5 | cis | 6.98E-09 | Nightingale | LDL_P | NA | C | T | -0.03997 | 0.004325 | 1.80E-21 |
| rs174535 | miR-1908-5 | cis | 6.98E-09 | Nightingale | M_LDL_L | NA | C | T | -0.02781 | 0.004335 | 4.50E-11 |
| rs174535 | miR-1908-5 | cis | 6.98E-09 | Nightingale | M_VLDL_C | NA | C | T | -0.04384 | 0.004316 | 2.30E-25 |
| rs174535 | miR-1908-5 | cis | 6.98E-09 | Nightingale | VLDL_L | NA | C | T | 0.026561 | 0.004244 | 2.70E-10 |
| rs174535 | miR-1908-5 | cis | 6.98E-09 | Nightingale | Total_TG | NA | C | T | 0.045817 | 0.00419 | 1.00E-28 |
| rs174535 | miR-1908-5 | cis | 6.98E-09 | Nightingale | XL_VLDL_C | NA | C | T | 0.029587 | 0.00426 | 2.10E-12 |
| rs174535 | miR-1908-5 | cis | 6.98E-09 | Nightingale | XS_VLDL_P | NA | C | T | -0.04422 | 0.004271 | 4.80E-27 |
| rs174535 | miR-1908-5 | cis | 6.98E-09 | Nightingale | Omega_3 | NA | C | T | -0.33048 | 0.004242 | 0 |
| rs174535 | miR-1908-5 | cis | 6.98E-09 | Nightingale | HDL_L | NA | C | T | -0.05265 | 0.003887 | 9.80E-45 |
| rs174535 | miR-1908-5 | cis | 6.98E-09 | Nightingale | XS_VLDL_F | NA | C | T | -0.05649 | 0.004253 | 5.20E-43 |
| rs174535 | miR-1908-5 | cis | 6.98E-09 | Nightingale | XS_VLDL_P | NA | C | T | 0.056813 | 0.004223 | 4.00E-42 |
| rs174535 | miR-1908-5 | cis | 6.98E-09 | Nightingale | M_LDL_C_ | NA | C | T | -0.0984 | 0.004332 | 6.61E-115 |
| rs174535 | miR-1908-5 | cis | 6.98E-09 | Nightingale | M_VLDL_T | NA | C | T | 0.077104 | 0.004088 | 5.30E-83 |
| rs174535 | miR-1908-5 | cis | 6.98E-09 | Nightingale | L_HDL_C_ | NA | C | T | -0.06686 | 0.00408 | 1.20E-64 |
| rs174535 | miR-1908-5 | cis | 6.98E-09 | Nightingale | M_VLDL_F | NA | C | T | -0.09051 | 0.004115 | 1.20E-112 |
| rs174535 | miR-1908-5 | cis | 6.98E-09 | Nightingale | LA_pct | NA | C | T | 0.183225 | 0.00423 | 0 |
| rs174535 | miR-1908-5 | cis | 6.98E-09 | Nightingale | HDL_TG | NA | C | T | 0.04493 | 0.004268 | 1.90E-26 |
| rs174535 | miR-1908-5 | cis | 6.98E-09 | Nightingale | IDL_PL | NA | C | T | -0.07265 | 0.004176 | 1.60E-72 |
| rs174535 | miR-1908-5 | cis | 6.98E-09 | Nightingale | S_VLDL_TG | NA | C | T | 0.03804 | 0.0042 | 4.30E-20 |
| rs174535 | miR-1908-5 | cis | 6.98E-09 | Nightingale | M_LDL_PL | NA | C | T | -0.02318 | 0.004335 | 3.60E-08 |
| rs174535 | miR-1908-5 | cis | 6.98E-09 | Nightingale | XL_VLDL_P | NA | C | T | 0.051843 | 0.00416 | 7.00E-37 |
| rs174535 | miR-1908-5 | cis | 6.98E-09 | Nightingale | M_VLDL_T | NA | C | T | 0.030978 | 0.004205 | 8.10E-14 |
| rs174535 | miR-1908-5 | cis | 6.98E-09 | Nightingale | L_HDL_CE_ | NA | C | T | -0.04849 | 0.004141 | 1.10E-33 |
| rs174535 | miR-1908-5 | cis | 6.98E-09 | Nightingale | S_HDL_C_ | NA | C | T | -0.04087 | 0.004278 | 1.70E-21 |
| rs174535 | miR-1908-5 | cis | 6.98E-09 | Nightingale | ApoB | NA | C | T | -0.03839 | 0.004323 | 5.30E-20 |
| rs174535 | miR-1908-5 | cis | 6.98E-09 | Nightingale | XXL_VLDL_ | NA | C | T | 0.064954 | 0.004169 | 5.20E-57 |
| rs174535 | miR-1908-5 | cis | 6.98E-09 | Nightingale | M_HDL_L | NA | C | T | -0.02729 | 0.004018 | 1.50E-12 |
| rs174535 | miR-1908-5 | cis | 6.98E-09 | Nightingale | IDL_L | NA | C | T | -0.07215 | 0.004184 | 5.30E-71 |
| rs174535 | miR-1908-5 | cis | 6.98E-09 | Nightingale | L_LDL_TG_ | NA | C | T | 0.075803 | 0.004244 | 5.10E-73 |
| rs174535 | miR-1908-5 | cis | 6.98E-09 | Nightingale | M_LDL_FC | NA | C | T | -0.05168 | 0.004323 | 8.20E-35 |
| rs174535 | miR-1908-5 | cis | 6.98E-09 | Nightingale | LDL_CE | NA | C | T | -0.0431 | 0.004317 | 8.70E-25 |
| rs174535 | miR-1908-5 | cis | 6.98E-09 | Nightingale | XXL_VLDL_ | NA | C | T | 0.066362 | 0.00417 | 1.40E-59 |
| rs174535 | miR-1908-5 | cis | 6.98E-09 | Nightingale | L_LDL_PL | NA | C | T | -0.0545 | 0.004296 | 9.90E-39 |
| rs174535 | miR-1908-5 | cis | 6.98E-09 | Nightingale | XS_VLDL_C | NA | C | T | -0.09683 | 0.004101 | 1.00E-128 |
| rs174535 | miR-1908-5 | cis | 6.98E-09 | Nightingale | S_LDL_C | NA | C | T | -0.03179 | 0.004333 | 3.90E-14 |
| rs174535 | miR-1908-5 | cis | 6.98E-09 | Nightingale | Cholines | NA | C | T | -0.03711 | 0.004071 | 1.20E-21 |
| rs174535 | miR-1908-5 | cis | 6.98E-09 | Nightingale | L_VLDL_FC | NA | C | T | 0.03928 | 0.004187 | 1.10E-21 |
| rs174535 | miR-1908-5 | cis | 6.98E-09 | Nightingale | XXL_VLDL_ | NA | C | T | 0.064516 | 0.004163 | 1.40E-56 |
| rs174535 | miR-1908-5 | cis | 6.98E-09 | Nightingale | M_HDL_FC | NA | C | T | -0.05077 | 0.003939 | 1.30E-40 |
| rs174535 | miR-1908-5 | cis | 6.98E-09 | Nightingale | IDL_CE_pct | NA | C | T | -0.03716 | 0.004247 | 2.50E-18 |
| rs174535 | miR-1908-5 | cis | 6.98E-09 | Nightingale | Citrate | NA | C | T | -0.03273 | 0.004301 | 2.60E-15 |
| rs174535 | miR-1908-5 | cis | 6.98E-09 | Nightingale | L_VLDL_C_ | NA | C | T | -0.03877 | 0.004228 | 3.40E-21 |
| rs174535 | miR-1908-5 | cis | 6.98E-09 | Nightingale | S_LDL_PL | NA | C | T | -0.05075 | 0.004318 | 6.30E-34 |

| | | | | | | | | | | | |
|---|---|---|---|---|---|---|---|---|---|---|---|
| rs174535 | miR-1908-5 | cis | 6.98E-09 | Nightingale | ApoA1 | NA | C | T | -0.04058 | 0.003961 | 1.70E-26 |
| rs174535 | miR-1908-5 | cis | 6.98E-09 | Nightingale | L_LDL_FC_ | NA | C | T | -0.08159 | 0.004125 | 7.80E-90 |
| rs174535 | miR-1908-5 | cis | 6.98E-09 | Nightingale | HDL_size | NA | C | T | -0.07186 | 0.003835 | 2.30E-84 |
| rs174535 | miR-1908-5 | cis | 6.98E-09 | Nightingale | L_HDL_FC | NA | C | T | -0.08364 | 0.003793 | 1.90E-116 |
| rs174535 | miR-1908-5 | cis | 6.98E-09 | Nightingale | XL_HDL_C | NA | C | T | -0.09804 | 0.003903 | 2.00E-150 |
| rs174535 | miR-1908-5 | cis | 6.98E-09 | Nightingale | XXL_VLDL_ | NA | C | T | 0.070431 | 0.004154 | 3.00E-67 |
| rs174535 | miR-1908-5 | cis | 6.98E-09 | Nightingale | S_LDL_TG_ | NA | C | T | 0.084449 | 0.004194 | 4.00E-93 |
| rs174535 | miR-1908-5 | cis | 6.98E-09 | Nightingale | Unsaturati | NA | C | T | -0.33356 | 0.004099 | 0 |
| rs174535 | miR-1908-5 | cis | 6.98E-09 | Nightingale | XS_VLDL_L | NA | C | T | -0.0422 | 0.004254 | 3.50E-25 |
| rs174535 | miR-1908-5 | cis | 6.98E-09 | Nightingale | L_VLDL_CE | NA | C | T | -0.05877 | 0.004142 | 1.90E-48 |
| rs174535 | miR-1908-5 | cis | 6.98E-09 | Nightingale | S_HDL_TG_ | NA | C | T | 0.067452 | 0.004117 | 6.90E-63 |
| rs174535 | miR-1908-5 | cis | 6.98E-09 | Nightingale | S_HDL_FC_ | NA | C | T | -0.10125 | 0.004105 | 1.40E-148 |
| rs174535 | miR-1908-5 | cis | 6.98E-09 | Nightingale | XXL_VLDL_ | NA | C | T | 0.043222 | 0.004352 | 3.60E-24 |
| rs174535 | miR-1908-5 | cis | 6.98E-09 | Nightingale | L_LDL_P | NA | C | T | -0.03704 | 0.004314 | 5.70E-19 |
| rs174535 | miR-1908-5 | cis | 6.98E-09 | Nightingale | L_VLDL_PL | NA | C | T | 0.047818 | 0.004176 | 1.50E-31 |
| rs174535 | miR-1908-5 | cis | 6.98E-09 | Nightingale | Sphingomy | NA | C | T | -0.11402 | 0.004045 | 6.61E-184 |
| rs174535 | miR-1908-5 | cis | 6.98E-09 | Nightingale | IDL_P | NA | C | T | -0.04542 | 0.004283 | 1.00E-27 |
| rs174535 | miR-1908-5 | cis | 6.98E-09 | Nightingale | IDL_C_pct | NA | C | T | -0.0735 | 0.004225 | 2.20E-69 |
| rs174535 | miR-1908-5 | cis | 6.98E-09 | Nightingale | M_HDL_PL | NA | C | T | -0.0213 | 0.004047 | 4.00E-08 |
| rs174535 | miR-1908-5 | cis | 6.98E-09 | Nightingale | XL_VLDL_T | NA | C | T | 0.058669 | 0.004193 | 7.60E-46 |
| rs174535 | miR-1908-5 | cis | 6.98E-09 | Nightingale | S_LDL_L | NA | C | T | -0.03205 | 0.004328 | 1.90E-14 |
| rs174535 | miR-1908-5 | cis | 6.98E-09 | Nightingale | HDL_PL | NA | C | T | -0.0479 | 0.003913 | 7.80E-37 |
| rs174535 | miR-1908-5 | cis | 6.98E-09 | Nightingale | L_LDL_C_p | NA | C | T | -0.07512 | 0.004217 | 2.10E-73 |
| rs174535 | miR-1908-5 | cis | 6.98E-09 | Nightingale | IDL_TG_pc | NA | C | T | 0.086519 | 0.004181 | 9.30E-99 |
| rs174535 | miR-1908-5 | cis | 6.98E-09 | Nightingale | XL_HDL_PL | NA | C | T | -0.08826 | 0.003891 | 2.30E-123 |
| rs174535 | miR-1908-5 | cis | 6.98E-09 | Nightingale | MUFA_pct | NA | C | T | 0.193683 | 0.004142 | 0 |
| rs174535 | miR-1908-5 | cis | 6.98E-09 | Nightingale | S_VLDL_CE | NA | C | T | -0.02885 | 0.00427 | 7.80E-12 |
| rs174535 | miR-1908-5 | cis | 6.98E-09 | Nightingale | L_VLDL_L | NA | C | T | 0.037569 | 0.004183 | 6.20E-20 |
| rs174535 | miR-1908-5 | cis | 6.98E-09 | Nightingale | XL_HDL_C_ | NA | C | T | 0.024645 | 0.004135 | 1.30E-10 |
| rs174535 | miR-1908-5 | cis | 6.98E-09 | Nightingale | M_LDL_FC_ | NA | C | T | -0.07022 | 0.004146 | 1.40E-66 |
| rs174535 | miR-1908-5 | cis | 6.98E-09 | Nightingale | M_HDL_P | NA | C | T | -0.03378 | 0.003987 | 1.20E-18 |
| rs174535 | miR-1908-5 | cis | 6.98E-09 | Nightingale | HDL_FC | NA | C | T | -0.07298 | 0.003834 | 5.80E-87 |
| rs174535 | miR-1908-5 | cis | 6.98E-09 | Nightingale | S_VLDL_FC | NA | C | T | -0.08171 | 0.004133 | 8.50E-91 |
| rs174535 | miR-1908-5 | cis | 6.98E-09 | Nightingale | XL_VLDL_C | NA | C | T | 0.036546 | 0.004227 | 1.60E-18 |
| rs174535 | miR-1908-5 | cis | 6.98E-09 | Nightingale | S_HDL_TG | NA | C | T | 0.067449 | 0.004197 | 1.30E-60 |
| rs174535 | miR-1908-5 | cis | 6.98E-09 | Nightingale | M_VLDL_C_ | NA | C | T | -0.0797 | 0.004074 | 9.70E-89 |
| rs174535 | miR-1908-5 | cis | 6.98E-09 | Nightingale | SFA | NA | C | T | -0.03169 | 0.004289 | 5.00E-14 |
| rs174535 | miR-1908-5 | cis | 6.98E-09 | Nightingale | L_LDL_C | NA | C | T | -0.05721 | 0.00428 | 3.20E-43 |
| rs174535 | miR-1908-5 | cis | 6.98E-09 | Nightingale | VLDL_TG | NA | C | T | 0.04683 | 0.004165 | 2.90E-30 |
| rs174535 | miR-1908-5 | cis | 6.98E-09 | Nightingale | XS_VLDL_T | NA | C | T | 0.030357 | 0.004262 | 4.00E-12 |
| rs174535 | miR-1908-5 | cis | 6.98E-09 | Nightingale | M_VLDL_P | NA | C | T | -0.06223 | 0.004153 | 5.00E-54 |
| rs174535 | miR-1908-5 | cis | 6.98E-09 | Nightingale | Total_L | NA | C | T | -0.03725 | 0.004258 | 1.10E-19 |
| rs174535 | miR-1908-5 | cis | 6.98E-09 | Nightingale | IDL_C | NA | C | T | -0.07686 | 0.004179 | 4.20E-80 |
| rs174535 | miR-1908-5 | cis | 6.98E-09 | Nightingale | Omega_6_ | NA | C | T | 0.024449 | 0.004213 | 3.20E-09 |
| rs174535 | miR-1908-5 | cis | 6.98E-09 | Nightingale | XL_HDL_PL | NA | C | T | -0.06691 | 0.003976 | 3.30E-68 |
| rs174535 | miR-1908-5 | cis | 6.98E-09 | Nightingale | PUFA_by_M | NA | C | T | -0.16722 | 0.004146 | 0 |
| rs174535 | miR-1908-5 | cis | 6.98E-09 | Nightingale | M_VLDL_F | NA | C | T | -0.02947 | 0.004328 | 2.40E-12 |
| rs174535 | miR-1908-5 | cis | 6.98E-09 | Nightingale | IDL_FC | NA | C | T | -0.08926 | 0.004189 | 2.50E-106 |
| rs174535 | miR-1908-5 | cis | 6.98E-09 | Nightingale | XS_VLDL_T | NA | C | T | 0.09618 | 0.004111 | 1.30E-126 |
| rs174535 | miR-1908-5 | cis | 6.98E-09 | Nightingale | L_HDL_PL_ | NA | C | T | 0.042021 | 0.004189 | 3.80E-25 |
| rs174535 | miR-1908-5 | cis | 6.98E-09 | Nightingale | XL_VLDL_F | NA | C | T | 0.042348 | 0.004192 | 9.50E-25 |
| rs174535 | miR-1908-5 | cis | 6.98E-09 | Nightingale | XS_VLDL_C | NA | C | T | -0.07219 | 0.004176 | 5.10E-71 |
| rs174535 | miR-1908-5 | cis | 6.98E-09 | Nightingale | Total_PL | NA | C | T | -0.05091 | 0.004121 | 2.40E-37 |
| rs174535 | miR-1908-5 | cis | 6.98E-09 | Nightingale | PUFA_pct | NA | C | T | -0.11862 | 0.004163 | 7.80E-180 |
| rs174535 | miR-1908-5 | cis | 6.98E-09 | Nightingale | XL_HDL_FC | NA | C | T | -0.10173 | 0.004032 | 7.69E-151 |
| rs174535 | miR-1908-5 | cis | 6.98E-09 | Nightingale | XS_VLDL_C | NA | C | T | -0.06847 | 0.004201 | 2.20E-63 |
| rs174535 | miR-1908-5 | cis | 6.98E-09 | Nightingale | LDL_PL | NA | C | T | -0.0457 | 0.004319 | 1.80E-27 |
| rs174535 | miR-1908-5 | cis | 6.98E-09 | Nightingale | Omega_6_ | NA | C | T | 0.363838 | 0.004266 | 0 |
| rs174535 | miR-1908-5 | cis | 6.98E-09 | Nightingale | XXL_VLDL_ | NA | C | T | 0.071872 | 0.00418 | 2.40E-68 |
| rs174535 | miR-1908-5 | cis | 6.98E-09 | Nightingale | XXL_VLDL_ | NA | C | T | 0.069988 | 0.004171 | 1.70E-65 |
| rs174535 | miR-1908-5 | cis | 6.98E-09 | Nightingale | L_HDL_FC_ | NA | C | T | -0.11153 | 0.004021 | 1.20E-183 |
| rs174536 | miR-1908-5 | cis | 1.31E-08 | Nightingale | M_HDL_CE | NA | C | A | -0.04569 | 0.004202 | 3.10E-28 |
| rs174536 | miR-1908-5 | cis | 1.31E-08 | Nightingale | XL_VLDL_C | NA | C | A | -0.05971 | 0.004154 | 4.40E-48 |

| | | | | | | | | | | | |
|---|---|---|---|---|---|---|---|---|---|---|---|
| rs174536 | miR-1908-! | cis | 1.31E-08 | Nightingale | LA | NA | C | A | 0.082331 | 0.004249 | 5.60E-84 |
| rs174536 | miR-1908-! | cis | 1.31E-08 | Nightingale | LDL_L | NA | C | A | -0.04552 | 0.004318 | 2.60E-27 |
| rs174536 | miR-1908-! | cis | 1.31E-08 | Nightingale | XL_VLDL_C | NA | C | A | -0.0628 | 0.004152 | 5.90E-53 |
| rs174536 | miR-1908-! | cis | 1.31E-08 | Nightingale | M_LDL_C | NA | C | A | -0.0341 | 0.004341 | 1.00E-15 |
| rs174536 | miR-1908-! | cis | 1.31E-08 | Nightingale | M_LDL_P | NA | C | A | -0.04675 | 0.004345 | 7.70E-28 |
| rs174536 | miR-1908-! | cis | 1.31E-08 | Nightingale | Remnant_( | NA | C | A | -0.04458 | 0.004305 | 1.60E-26 |
| rs174536 | miR-1908-! | cis | 1.31E-08 | Nightingale | XL_HDL_P | NA | C | A | -0.08899 | 0.003899 | 6.30E-126 |
| rs174536 | miR-1908-! | cis | 1.31E-08 | Nightingale | XL_HDL_FC | NA | C | A | 0.060834 | 0.003841 | 2.00E-62 |
| rs174536 | miR-1908-! | cis | 1.31E-08 | Nightingale | L_LDL_CE | NA | C | A | -0.05329 | 0.004294 | 2.40E-37 |
| rs174536 | miR-1908-! | cis | 1.31E-08 | Nightingale | XL_VLDL_T | NA | C | A | 0.055786 | 0.004144 | 7.40E-43 |
| rs174536 | miR-1908-! | cis | 1.31E-08 | Nightingale | TG_by_PG | NA | C | A | 0.067755 | 0.004021 | 8.90E-67 |
| rs174536 | miR-1908-! | cis | 1.31E-08 | Nightingale | MUFA | NA | C | A | 0.062474 | 0.004288 | 5.30E-49 |
| rs174536 | miR-1908-! | cis | 1.31E-08 | Nightingale | XL_VLDL_C | NA | C | A | 0.029406 | 0.004264 | 2.80E-12 |
| rs174536 | miR-1908-! | cis | 1.31E-08 | Nightingale | Citrate | NA | C | A | -0.03304 | 0.004305 | 1.40E-15 |
| rs174536 | miR-1908-! | cis | 1.31E-08 | Nightingale | Total_FA | NA | C | A | -0.02441 | 0.004298 | 4.00E-09 |
| rs174536 | miR-1908-! | cis | 1.31E-08 | Nightingale | IDL_CE | NA | C | A | -0.07228 | 0.004182 | 8.80E-71 |
| rs174536 | miR-1908-! | cis | 1.31E-08 | Nightingale | L_VLDL_TG | NA | C | A | 0.036519 | 0.004169 | 4.90E-19 |
| rs174536 | miR-1908-! | cis | 1.31E-08 | Nightingale | XS_VLDL_C | NA | C | A | -0.09119 | 0.004084 | 3.60E-115 |
| rs174536 | miR-1908-! | cis | 1.31E-08 | Nightingale | Clinical_LD | NA | C | A | -0.05514 | 0.004308 | 3.10E-39 |
| rs174536 | miR-1908-! | cis | 1.31E-08 | Nightingale | SFA | NA | C | A | -0.03202 | 0.004294 | 3.00E-14 |
| rs174536 | miR-1908-! | cis | 1.31E-08 | Nightingale | XL_HDL_PL | NA | C | A | -0.06681 | 0.00398 | 3.70E-68 |
| rs174536 | miR-1908-! | cis | 1.31E-08 | Nightingale | IDL_CE_pct | NA | C | A | -0.03754 | 0.004251 | 1.50E-18 |
| rs174536 | miR-1908-! | cis | 1.31E-08 | Nightingale | M_HDL_TG | NA | C | A | 0.049154 | 0.004279 | 2.50E-31 |
| rs174536 | miR-1908-! | cis | 1.31E-08 | Nightingale | L_VLDL_FC | NA | C | A | 0.027716 | 0.004292 | 5.00E-11 |
| rs174536 | miR-1908-! | cis | 1.31E-08 | Nightingale | S_LDL_FC | NA | C | A | -0.06239 | 0.004326 | 6.00E-50 |
| rs174536 | miR-1908-! | cis | 1.31E-08 | Nightingale | non_HDL_( | NA | C | A | -0.04804 | 0.004309 | 2.90E-30 |
| rs174536 | miR-1908-! | cis | 1.31E-08 | Nightingale | HDL_CE | NA | C | A | -0.06067 | 0.003888 | 5.50E-59 |
| rs174536 | miR-1908-! | cis | 1.31E-08 | Nightingale | Total_PL | NA | C | A | -0.05141 | 0.004126 | 7.20E-38 |
| rs174536 | miR-1908-! | cis | 1.31E-08 | Nightingale | XL_HDL_C_ | NA | C | A | 0.024411 | 0.004139 | 1.50E-10 |
| rs174536 | miR-1908-! | cis | 1.31E-08 | Nightingale | M_HDL_PL | NA | C | A | 0.057172 | 0.00411 | 1.20E-45 |
| rs174536 | miR-1908-! | cis | 1.31E-08 | Nightingale | S_VLDL_TG | NA | C | A | 0.063618 | 0.004192 | 1.60E-53 |
| rs174536 | miR-1908-! | cis | 1.31E-08 | Nightingale | LDL_FC | NA | C | A | -0.0641 | 0.004287 | 2.90E-53 |
| rs174536 | miR-1908-! | cis | 1.31E-08 | Nightingale | SFA_pct | NA | C | A | -0.04263 | 0.004274 | 2.00E-23 |
| rs174536 | miR-1908-! | cis | 1.31E-08 | Nightingale | M_HDL_C_ | NA | C | A | -0.06452 | 0.004124 | 7.80E-58 |
| rs174536 | miR-1908-! | cis | 1.31E-08 | Nightingale | S_VLDL_PL | NA | C | A | -0.07744 | 0.004146 | 1.70E-81 |
| rs174536 | miR-1908-! | cis | 1.31E-08 | Nightingale | Total_FC | NA | C | A | -0.05954 | 0.004233 | 1.70E-47 |
| rs174536 | miR-1908-! | cis | 1.31E-08 | Nightingale | S_LDL_P | NA | C | A | -0.0329 | 0.004337 | 6.80E-15 |
| rs174536 | miR-1908-! | cis | 1.31E-08 | Nightingale | S_LDL_CE_ | NA | C | A | 0.056256 | 0.004325 | 1.80E-40 |
| rs174536 | miR-1908-! | cis | 1.31E-08 | Nightingale | DHA_pct | NA | C | A | -0.27069 | 0.004206 | 0 |
| rs174536 | miR-1908-! | cis | 1.31E-08 | Nightingale | XS_VLDL_F | NA | C | A | -0.05674 | 0.004257 | 3.40E-43 |
| rs174536 | miR-1908-! | cis | 1.31E-08 | Nightingale | LDL_TG | NA | C | A | 0.028605 | 0.004285 | 5.10E-11 |
| rs174536 | miR-1908-! | cis | 1.31E-08 | Nightingale | Cholines | NA | C | A | -0.03745 | 0.004075 | 6.30E-22 |
| rs174536 | miR-1908-! | cis | 1.31E-08 | Nightingale | M_HDL_FC | NA | C | A | -0.10234 | 0.003839 | 5.30E-170 |
| rs174536 | miR-1908-! | cis | 1.31E-08 | Nightingale | Omega_3_ | NA | C | A | -0.38615 | 0.004233 | 0 |
| rs174536 | miR-1908-! | cis | 1.31E-08 | Nightingale | XS_VLDL_C | NA | C | A | -0.09702 | 0.004105 | 1.00E-128 |
| rs174536 | miR-1908-! | cis | 1.31E-08 | Nightingale | S_LDL_PL_ | NA | C | A | -0.06104 | 0.004324 | 2.40E-47 |
| rs174536 | miR-1908-! | cis | 1.31E-08 | Nightingale | M_HDL_TG | NA | C | A | 0.065362 | 0.004155 | 3.20E-58 |
| rs174536 | miR-1908-! | cis | 1.31E-08 | Nightingale | XS_VLDL_P | NA | C | A | 0.057182 | 0.004227 | 2.10E-42 |
| rs174536 | miR-1908-! | cis | 1.31E-08 | Nightingale | XXL_VLDL_ | NA | C | A | 0.065047 | 0.004173 | 4.80E-57 |
| rs174536 | miR-1908-! | cis | 1.31E-08 | Nightingale | L_LDL_TG_ | NA | C | A | 0.076412 | 0.004248 | 8.20E-74 |
| rs174536 | miR-1908-! | cis | 1.31E-08 | Nightingale | L_VLDL_PL | NA | C | A | 0.085731 | 0.004205 | 1.30E-95 |
| rs174536 | miR-1908-! | cis | 1.31E-08 | Nightingale | M_LDL_TG | NA | C | A | 0.035564 | 0.004269 | 9.30E-17 |
| rs174536 | miR-1908-! | cis | 1.31E-08 | Nightingale | L_VLDL_P | NA | C | A | 0.040891 | 0.004183 | 2.30E-23 |
| rs174536 | miR-1908-! | cis | 1.31E-08 | Nightingale | XL_VLDL_C | NA | C | A | 0.036411 | 0.004232 | 2.20E-18 |
| rs174536 | miR-1908-! | cis | 1.31E-08 | Nightingale | L_LDL_FC | NA | C | A | -0.06816 | 0.004254 | 4.50E-61 |
| rs174536 | miR-1908-! | cis | 1.31E-08 | Nightingale | XL_VLDL_T | NA | C | A | 0.058465 | 0.004198 | 2.30E-45 |
| rs174536 | miR-1908-! | cis | 1.31E-08 | Nightingale | L_HDL_C_p | NA | C | A | -0.0671 | 0.004084 | 4.50E-65 |
| rs174536 | miR-1908-! | cis | 1.31E-08 | Nightingale | M_HDL_FC | NA | C | A | -0.05104 | 0.003943 | 5.30E-41 |
| rs174536 | miR-1908-! | cis | 1.31E-08 | Nightingale | XS_VLDL_P | NA | C | A | -0.03531 | 0.004272 | 4.40E-18 |
| rs174536 | miR-1908-! | cis | 1.31E-08 | Nightingale | XL_VLDL_F | NA | C | A | -0.0677 | 0.004198 | 4.10E-61 |
| rs174536 | miR-1908-! | cis | 1.31E-08 | Nightingale | XL_HDL_FC | NA | C | A | -0.10201 | 0.004036 | 1.00E-151 |
| rs174536 | miR-1908-! | cis | 1.31E-08 | Nightingale | L_LDL_L | NA | C | A | -0.05421 | 0.004291 | 1.30E-38 |
| rs174536 | miR-1908-! | cis | 1.31E-08 | Nightingale | Total_CE | NA | C | A | -0.06695 | 0.004179 | 6.10E-61 |

| | | | | | | | | | | | | |
|---|---|---|---|---|---|---|---|---|---|---|---|---|
| rs174536 | miR-1908-! | cis | | 1.31E-08 | Nightingale | XS_VLDL_T | NA | C | A | 0.096366 | 0.004115 | 1.10E-126 |
| rs174536 | miR-1908-! | cis | | 1.31E-08 | Nightingale | MUFA_pct | NA | C | A | 0.194977 | 0.004146 | 0 |
| rs174536 | miR-1908-! | cis | | 1.31E-08 | Nightingale | Omega_6_ | NA | C | A | 0.024475 | 0.004217 | 3.10E-09 |
| rs174536 | miR-1908-! | cis | | 1.31E-08 | Nightingale | PUFA_pct | NA | C | A | -0.11968 | 0.004167 | 1.60E-182 |
| rs174536 | miR-1908-! | cis | | 1.31E-08 | Nightingale | IDL_P | NA | C | A | -0.04559 | 0.004287 | 9.80E-28 |
| rs174536 | miR-1908-! | cis | | 1.31E-08 | Nightingale | XS_VLDL_P | NA | C | A | -0.04443 | 0.004275 | 3.80E-27 |
| rs174536 | miR-1908-! | cis | | 1.31E-08 | Nightingale | Tyr | NA | C | A | 0.033752 | 0.004327 | 6.20E-15 |
| rs174536 | miR-1908-! | cis | | 1.31E-08 | Nightingale | M_HDL_CE | NA | C | A | -0.03726 | 0.004001 | 7.20E-22 |
| rs174536 | miR-1908-! | cis | | 1.31E-08 | Nightingale | L_HDL_CE | NA | C | A | -0.07772 | 0.003826 | 2.30E-99 |
| rs174536 | miR-1908-! | cis | | 1.31E-08 | Nightingale | S_VLDL_FC | NA | C | A | -0.02878 | 0.004334 | 9.80E-12 |
| rs174536 | miR-1908-! | cis | | 1.31E-08 | Nightingale | VLDL_size | NA | C | A | 0.071664 | 0.004039 | 2.00E-74 |
| rs174536 | miR-1908-! | cis | | 1.31E-08 | Nightingale | XL_VLDL_L | NA | C | A | 0.050332 | 0.004169 | 9.20E-35 |
| rs174536 | miR-1908-! | cis | | 1.31E-08 | Nightingale | L_VLDL_C | NA | C | A | 0.027896 | 0.004238 | 2.30E-11 |
| rs174536 | miR-1908-! | cis | | 1.31E-08 | Nightingale | L_LDL_PL | NA | C | A | -0.05496 | 0.0043 | 4.20E-39 |
| rs174536 | miR-1908-! | cis | | 1.31E-08 | Nightingale | Phosphogly | NA | C | A | -0.04135 | 0.004104 | 1.10E-25 |
| rs174536 | miR-1908-! | cis | | 1.31E-08 | Nightingale | M_VLDL_C | NA | C | A | -0.04423 | 0.004321 | 1.50E-25 |
| rs174536 | miR-1908-! | cis | | 1.31E-08 | Nightingale | M_LDL_PL_ | NA | C | A | 0.045362 | 0.00433 | 1.00E-26 |
| rs174536 | miR-1908-! | cis | | 1.31E-08 | Nightingale | M_VLDL_T | NA | C | A | 0.030591 | 0.004209 | 1.70E-13 |
| rs174536 | miR-1908-! | cis | | 1.31E-08 | Nightingale | XXL_VLDL_ | NA | C | A | 0.059767 | 0.004181 | 3.20E-48 |
| rs174536 | miR-1908-! | cis | | 1.31E-08 | Nightingale | Unsaturati | NA | C | A | -0.33597 | 0.004103 | 0 |
| rs174536 | miR-1908-! | cis | | 1.31E-08 | Nightingale | XXL_VLDL_ | NA | C | A | 0.064657 | 0.004167 | 1.00E-56 |
| rs174536 | miR-1908-! | cis | | 1.31E-08 | Nightingale | XL_HDL_L | NA | C | A | -0.09234 | 0.003893 | 4.50E-135 |
| rs174536 | miR-1908-! | cis | | 1.31E-08 | Nightingale | M_VLDL_C | NA | C | A | -0.07505 | 0.004067 | 7.30E-79 |
| rs174536 | miR-1908-! | cis | | 1.31E-08 | Nightingale | HDL_C | NA | C | A | -0.06399 | 0.003869 | 2.90E-66 |
| rs174536 | miR-1908-! | cis | | 1.31E-08 | Nightingale | LDL_CE | NA | C | A | -0.04365 | 0.004322 | 3.50E-25 |
| rs174536 | miR-1908-! | cis | | 1.31E-08 | Nightingale | IDL_L | NA | C | A | -0.07256 | 0.004189 | 1.90E-71 |
| rs174536 | miR-1908-! | cis | | 1.31E-08 | Nightingale | S_LDL_TG | NA | C | A | 0.049228 | 0.004227 | 9.10E-32 |
| rs174536 | miR-1908-! | cis | | 1.31E-08 | Nightingale | XXL_VLDL_ | NA | C | A | 0.066492 | 0.004174 | 1.00E-59 |
| rs174536 | miR-1908-! | cis | | 1.31E-08 | Nightingale | IDL_FC | NA | C | A | -0.08965 | 0.004194 | 9.40E-107 |
| rs174536 | miR-1908-! | cis | | 1.31E-08 | Nightingale | Total_TG | NA | C | A | 0.045692 | 0.004194 | 1.70E-28 |
| rs174536 | miR-1908-! | cis | | 1.31E-08 | Nightingale | XL_HDL_C | NA | C | A | -0.09837 | 0.003906 | 1.60E-151 |
| rs174536 | miR-1908-! | cis | | 1.31E-08 | Nightingale | M_HDL_P | NA | C | A | -0.03395 | 0.003991 | 8.90E-19 |
| rs174536 | miR-1908-! | cis | | 1.31E-08 | Nightingale | S_VLDL_C_ | NA | C | A | -0.05584 | 0.004215 | 4.20E-41 |
| rs174536 | miR-1908-! | cis | | 1.31E-08 | Nightingale | Total_C | NA | C | A | -0.06521 | 0.004194 | 1.90E-57 |
| rs174536 | miR-1908-! | cis | | 1.31E-08 | Nightingale | IDL_TG_pct | NA | C | A | 0.08697 | 0.004186 | 2.30E-99 |
| rs174536 | miR-1908-! | cis | | 1.31E-08 | Nightingale | L_HDL_CE_ | NA | C | A | -0.04864 | 0.004145 | 7.10E-34 |
| rs174536 | miR-1908-! | cis | | 1.31E-08 | Nightingale | L_VLDL_CE | NA | C | A | -0.05842 | 0.004146 | 8.80E-48 |
| rs174536 | miR-1908-! | cis | | 1.31E-08 | Nightingale | M_HDL_L | NA | C | A | -0.02744 | 0.004022 | 1.30E-12 |
| rs174536 | miR-1908-! | cis | | 1.31E-08 | Nightingale | S_LDL_FC_ | NA | C | A | -0.07839 | 0.004199 | 2.00E-80 |
| rs174536 | miR-1908-! | cis | | 1.31E-08 | Nightingale | M_LDL_CE | NA | C | A | -0.02647 | 0.004339 | 4.50E-10 |
| rs174536 | miR-1908-! | cis | | 1.31E-08 | Nightingale | LDL_P | NA | C | A | -0.04046 | 0.00433 | 8.90E-22 |
| rs174536 | miR-1908-! | cis | | 1.31E-08 | Nightingale | XL_VLDL_P | NA | C | A | 0.047076 | 0.004181 | 1.90E-30 |
| rs174536 | miR-1908-! | cis | | 1.31E-08 | Nightingale | L_LDL_C | NA | C | A | -0.05776 | 0.004284 | 9.70E-44 |
| rs174536 | miR-1908-! | cis | | 1.31E-08 | Nightingale | M_LDL_PL | NA | C | A | -0.02368 | 0.004339 | 2.30E-08 |
| rs174536 | miR-1908-! | cis | | 1.31E-08 | Nightingale | L_HDL_P | NA | C | A | -0.07701 | 0.003795 | 1.80E-99 |
| rs174536 | miR-1908-! | cis | | 1.31E-08 | Nightingale | S_HDL_TG | NA | C | A | 0.067545 | 0.004201 | 1.00E-60 |
| rs174536 | miR-1908-! | cis | | 1.31E-08 | Nightingale | IDL_C | NA | C | A | -0.07727 | 0.004184 | 1.60E-80 |
| rs174536 | miR-1908-! | cis | | 1.31E-08 | Nightingale | XS_VLDL_L | NA | C | A | -0.04242 | 0.004259 | 2.70E-25 |
| rs174536 | miR-1908-! | cis | | 1.31E-08 | Nightingale | XL_VLDL_P | NA | C | A | 0.051749 | 0.004164 | 1.00E-36 |
| rs174536 | miR-1908-! | cis | | 1.31E-08 | Nightingale | HDL_L | NA | C | A | -0.05288 | 0.003891 | 4.30E-45 |
| rs174536 | miR-1908-! | cis | | 1.31E-08 | Nightingale | ApoB | NA | C | A | -0.03881 | 0.004327 | 3.10E-20 |
| rs174536 | miR-1908-! | cis | | 1.31E-08 | Nightingale | XS_VLDL_T | NA | C | A | 0.030331 | 0.004266 | 4.30E-12 |
| rs174536 | miR-1908-! | cis | | 1.31E-08 | Nightingale | XXL_VLDL_ | NA | C | A | -0.02559 | 0.004311 | 6.30E-10 |
| rs174536 | miR-1908-! | cis | | 1.31E-08 | Nightingale | L_LDL_C_p | NA | C | A | -0.07603 | 0.004221 | 8.40E-75 |
| rs174536 | miR-1908-! | cis | | 1.31E-08 | Nightingale | XXL_VLDL_ | NA | C | A | 0.043838 | 0.004356 | 1.00E-24 |
| rs174536 | miR-1908-! | cis | | 1.31E-08 | Nightingale | XS_VLDL_F | NA | C | A | -0.11175 | 0.0043 | 8.49E-152 |
| rs174536 | miR-1908-! | cis | | 1.31E-08 | Nightingale | XL_HDL_TG | NA | C | A | 0.086099 | 0.004082 | 3.10E-104 |
| rs174536 | miR-1908-! | cis | | 1.31E-08 | Nightingale | PUFA | NA | C | A | -0.09634 | 0.004225 | 7.29E-122 |
| rs174536 | miR-1908-! | cis | | 1.31E-08 | Nightingale | Omega_3 | NA | C | A | -0.333 | 0.004246 | 0 |
| rs174536 | miR-1908-! | cis | | 1.31E-08 | Nightingale | XL_HDL_CE | NA | C | A | -0.09622 | 0.00388 | 3.30E-147 |
| rs174536 | miR-1908-! | cis | | 1.31E-08 | Nightingale | DHA | NA | C | A | -0.28576 | 0.004175 | 0 |
| rs174536 | miR-1908-! | cis | | 1.31E-08 | Nightingale | XL_HDL_CE | NA | C | A | -0.02505 | 0.00433 | 1.70E-08 |
| rs174536 | miR-1908-! | cis | | 1.31E-08 | Nightingale | XS_VLDL_C | NA | C | A | -0.07241 | 0.004181 | 3.60E-71 |

| | | | | | | | | | | | |
|---|---|---|---|---|---|---|---|---|---|---|---|
| rs174536 | miR-1908-5 | cis | 1.31E-08 | Nightingale | M_LDL_L | NA | C | A | -0.02831 | 0.004339 | 2.60E-11 |
| rs174536 | miR-1908-5 | cis | 1.31E-08 | Nightingale | M_VLDL_F | NA | C | A | -0.09053 | 0.00412 | 2.60E-112 |
| rs174536 | miR-1908-5 | cis | 1.31E-08 | Nightingale | XL_HDL_PL | NA | C | A | -0.08844 | 0.003895 | 4.70E-124 |
| rs174536 | miR-1908-5 | cis | 1.31E-08 | Nightingale | Sphingomy | NA | C | A | -0.11457 | 0.004049 | 5.40E-185 |
| rs174536 | miR-1908-5 | cis | 1.31E-08 | Nightingale | HDL_PL | NA | C | A | -0.04808 | 0.003917 | 4.40E-37 |
| rs174536 | miR-1908-5 | cis | 1.31E-08 | Nightingale | L_VLDL_PL | NA | C | A | 0.04768 | 0.00418 | 2.30E-31 |
| rs174536 | miR-1908-5 | cis | 1.31E-08 | Nightingale | S_VLDL_CE | NA | C | A | -0.02849 | 0.004275 | 1.50E-11 |
| rs174536 | miR-1908-5 | cis | 1.31E-08 | Nightingale | HDL_size | NA | C | A | -0.07182 | 0.003839 | 1.70E-84 |
| rs174536 | miR-1908-5 | cis | 1.31E-08 | Nightingale | XXL_VLDL_ | NA | C | A | 0.07052 | 0.004158 | 2.70E-67 |
| rs174536 | miR-1908-5 | cis | 1.31E-08 | Nightingale | Total_P | NA | C | A | -0.04009 | 0.004077 | 1.90E-24 |
| rs174536 | miR-1908-5 | cis | 1.31E-08 | Nightingale | XXL_VLDL_ | NA | C | A | 0.072063 | 0.004184 | 1.40E-68 |
| rs174536 | miR-1908-5 | cis | 1.31E-08 | Nightingale | S_HDL_C_p | NA | C | A | -0.04126 | 0.004282 | 8.60E-22 |
| rs174536 | miR-1908-5 | cis | 1.31E-08 | Nightingale | IDL_PL | NA | C | A | -0.07304 | 0.00418 | 5.40E-73 |
| rs174536 | miR-1908-5 | cis | 1.31E-08 | Nightingale | L_HDL_FC_ | NA | C | A | -0.11187 | 0.004025 | 1.10E-184 |
| rs174536 | miR-1908-5 | cis | 1.31E-08 | Nightingale | M_HDL_C | NA | C | A | -0.04024 | 0.003986 | 1.90E-25 |
| rs174536 | miR-1908-5 | cis | 1.31E-08 | Nightingale | L_VLDL_FC | NA | C | A | 0.039129 | 0.004191 | 1.60E-21 |
| rs174536 | miR-1908-5 | cis | 1.31E-08 | Nightingale | XS_VLDL_C | NA | C | A | -0.0687 | 0.004205 | 1.40E-63 |
| rs174536 | miR-1908-5 | cis | 1.31E-08 | Nightingale | L_HDL_C | NA | C | A | -0.07932 | 0.003815 | 4.00E-104 |
| rs174536 | miR-1908-5 | cis | 1.31E-08 | Nightingale | XL_VLDL_F | NA | C | A | 0.042284 | 0.004197 | 1.20E-24 |
| rs174536 | miR-1908-5 | cis | 1.31E-08 | Nightingale | L_HDL_FC | NA | C | A | -0.08379 | 0.003797 | 4.70E-117 |
| rs174536 | miR-1908-5 | cis | 1.31E-08 | Nightingale | L_HDL_PL_ | NA | C | A | 0.042296 | 0.004193 | 1.90E-25 |
| rs174536 | miR-1908-5 | cis | 1.31E-08 | Nightingale | S_LDL_L | NA | C | A | -0.03259 | 0.004332 | 9.70E-15 |
| rs174536 | miR-1908-5 | cis | 1.31E-08 | Nightingale | L_LDL_FC_ | NA | C | A | -0.08199 | 0.00413 | 2.30E-90 |
| rs174536 | miR-1908-5 | cis | 1.31E-08 | Nightingale | IDL_FC_pct | NA | C | A | -0.10289 | 0.004298 | 6.00E-128 |
| rs174536 | miR-1908-5 | cis | 1.31E-08 | Nightingale | M_VLDL_C | NA | C | A | -0.05273 | 0.004293 | 3.50E-36 |
| rs174536 | miR-1908-5 | cis | 1.31E-08 | Nightingale | M_LDL_C_p | NA | C | A | -0.09895 | 0.004336 | 1.40E-115 |
| rs174536 | miR-1908-5 | cis | 1.31E-08 | Nightingale | XXL_VLDL_ | NA | C | A | 0.070151 | 0.004175 | 1.00E-65 |
| rs174536 | miR-1908-5 | cis | 1.31E-08 | Nightingale | ApoA1 | NA | C | A | -0.04083 | 0.003965 | 9.50E-27 |
| rs174536 | miR-1908-5 | cis | 1.31E-08 | Nightingale | S_VLDL_FC | NA | C | A | -0.08192 | 0.004137 | 6.30E-91 |
| rs174536 | miR-1908-5 | cis | 1.31E-08 | Nightingale | M_LDL_FC_ | NA | C | A | -0.07072 | 0.004151 | 2.40E-67 |
| rs174536 | miR-1908-5 | cis | 1.31E-08 | Nightingale | IDL_C_pct | NA | C | A | -0.07387 | 0.00423 | 1.10E-69 |
| rs174536 | miR-1908-5 | cis | 1.31E-08 | Nightingale | L_HDL_TG_ | NA | C | A | 0.077274 | 0.004088 | 1.50E-84 |
| rs174536 | miR-1908-5 | cis | 1.31E-08 | Nightingale | M_LDL_TG | NA | C | A | 0.074524 | 0.004278 | 2.90E-69 |
| rs174536 | miR-1908-5 | cis | 1.31E-08 | Nightingale | Omega_6_ | NA | C | A | 0.366468 | 0.00427 | 0 |
| rs174536 | miR-1908-5 | cis | 1.31E-08 | Nightingale | PUFA_by_M | NA | C | A | -0.16846 | 0.00415 | 0 |
| rs174536 | miR-1908-5 | cis | 1.31E-08 | Nightingale | Total_L | NA | C | A | -0.03779 | 0.004262 | 4.60E-20 |
| rs174536 | miR-1908-5 | cis | 1.31E-08 | Nightingale | LDL_PL | NA | C | A | -0.0462 | 0.004323 | 7.60E-28 |
| rs174536 | miR-1908-5 | cis | 1.31E-08 | Nightingale | M_LDL_FC | NA | C | A | -0.05221 | 0.004328 | 2.90E-35 |
| rs174536 | miR-1908-5 | cis | 1.31E-08 | Nightingale | L_VLDL_C_ | NA | C | A | -0.03814 | 0.004232 | 1.60E-20 |
| rs174536 | miR-1908-5 | cis | 1.31E-08 | Nightingale | S_LDL_TG_ | NA | C | A | 0.08477 | 0.004198 | 1.60E-93 |
| rs174536 | miR-1908-5 | cis | 1.31E-08 | Nightingale | HDL_TG | NA | C | A | 0.045049 | 0.004272 | 1.70E-26 |
| rs174536 | miR-1908-5 | cis | 1.31E-08 | Nightingale | L_LDL_P | NA | C | A | -0.03761 | 0.004318 | 2.50E-19 |
| rs174536 | miR-1908-5 | cis | 1.31E-08 | Nightingale | M_VLDL_C_ | NA | C | A | -0.0798 | 0.004078 | 1.30E-88 |
| rs174536 | miR-1908-5 | cis | 1.31E-08 | Nightingale | S_HDL_FC_ | NA | C | A | -0.10206 | 0.00411 | 9.91E-151 |
| rs174536 | miR-1908-5 | cis | 1.31E-08 | Nightingale | HDL_P | NA | C | A | -0.03525 | 0.004068 | 3.40E-19 |
| rs174536 | miR-1908-5 | cis | 1.31E-08 | Nightingale | S_HDL_TG_ | NA | C | A | 0.067742 | 0.004121 | 2.60E-63 |
| rs174536 | miR-1908-5 | cis | 1.31E-08 | Nightingale | M_VLDL_F | NA | C | A | -0.02989 | 0.004332 | 1.50E-12 |
| rs174536 | miR-1908-5 | cis | 1.31E-08 | Nightingale | VLDL_TG | NA | C | A | 0.046647 | 0.004169 | 5.70E-30 |
| rs174536 | miR-1908-5 | cis | 1.31E-08 | Nightingale | M_VLDL_P | NA | C | A | -0.06203 | 0.004157 | 1.60E-53 |
| rs174536 | miR-1908-5 | cis | 1.31E-08 | Nightingale | L_VLDL_L | NA | C | A | 0.037293 | 0.004187 | 1.20E-19 |
| rs174536 | miR-1908-5 | cis | 1.31E-08 | Nightingale | M_HDL_PL | NA | C | A | -0.0214 | 0.004052 | 3.60E-08 |
| rs174536 | miR-1908-5 | cis | 1.31E-08 | Nightingale | HDL_FC | NA | C | A | -0.07333 | 0.003838 | 8.60E-88 |
| rs174536 | miR-1908-5 | cis | 1.31E-08 | Nightingale | L_HDL_L | NA | C | A | -0.07636 | 0.003802 | 1.70E-97 |
| rs174536 | miR-1908-5 | cis | 1.31E-08 | Nightingale | VLDL_L | NA | C | A | 0.026348 | 0.004248 | 3.60E-10 |
| rs174536 | miR-1908-5 | cis | 1.31E-08 | Nightingale | S_LDL_C | NA | C | A | -0.03228 | 0.004337 | 2.20E-14 |
| rs174536 | miR-1908-5 | cis | 1.31E-08 | Nightingale | S_LDL_PL | NA | C | A | -0.0514 | 0.004322 | 1.80E-34 |
| rs174536 | miR-1908-5 | cis | 1.31E-08 | Nightingale | M_VLDL_T | NA | C | A | 0.077138 | 0.004092 | 9.20E-83 |
| rs174536 | miR-1908-5 | cis | 1.31E-08 | Nightingale | L_HDL_PL | NA | C | A | -0.0753 | 0.003813 | 4.00E-94 |
| rs174536 | miR-1908-5 | cis | 1.31E-08 | Nightingale | S_VLDL_TG | NA | C | A | 0.037772 | 0.004204 | 8.00E-20 |
| rs174536 | miR-1908-5 | cis | 1.31E-08 | Nightingale | LDL_C | NA | C | A | -0.04975 | 0.004314 | 2.60E-32 |
| rs174536 | miR-1908-5 | cis | 1.31E-08 | Nightingale | LA_pct | NA | C | A | 0.184014 | 0.004234 | 0 |
| rs174537 | miR-1908-5 | cis | 9.41E-09 | Nightingale | L_LDL_CE | NA | T | G | -0.054 | 0.004302 | 4.50E-38 |
| rs174537 | miR-1908-5 | cis | 9.41E-09 | Nightingale | SFA_pct | NA | T | G | -0.04249 | 0.004282 | 3.30E-23 |

| | | | | | | | | | | | |
|---|---|---|---|---|---|---|---|---|---|---|---|
| rs174537 | miR-1908-5 | cis | 9.41E-09 | Nightingale | M_LDL_TG | NA | T | G | 0.03642 | 0.004277 | 2.00E-17 |
| rs174537 | miR-1908-5 | cis | 9.41E-09 | Nightingale | DHA | NA | T | G | -0.2883 | 0.004183 | 0 |
| rs174537 | miR-1908-5 | cis | 9.41E-09 | Nightingale | Total_C | NA | T | G | -0.06596 | 0.004202 | 2.10E-58 |
| rs174537 | miR-1908-5 | cis | 9.41E-09 | Nightingale | L_HDL_PL | NA | T | G | -0.07616 | 0.00382 | 6.00E-96 |
| rs174537 | miR-1908-5 | cis | 9.41E-09 | Nightingale | XS_VLDL_C | NA | T | G | -0.09276 | 0.004092 | 1.30E-118 |
| rs174537 | miR-1908-5 | cis | 9.41E-09 | Nightingale | Phosphogly | NA | T | G | -0.04151 | 0.004112 | 9.50E-26 |
| rs174537 | miR-1908-5 | cis | 9.41E-09 | Nightingale | MUFA_pct | NA | T | G | 0.197467 | 0.004154 | 0 |
| rs174537 | miR-1908-5 | cis | 9.41E-09 | Nightingale | SFA | NA | T | G | -0.03178 | 0.004302 | 5.40E-14 |
| rs174537 | miR-1908-5 | cis | 9.41E-09 | Nightingale | LDL_TG | NA | T | G | 0.029451 | 0.004293 | 1.50E-11 |
| rs174537 | miR-1908-5 | cis | 9.41E-09 | Nightingale | XL_VLDL_C | NA | T | G | 0.029907 | 0.004272 | 1.20E-12 |
| rs174537 | miR-1908-5 | cis | 9.41E-09 | Nightingale | M_HDL_C | NA | T | G | -0.04081 | 0.003994 | 4.70E-26 |
| rs174537 | miR-1908-5 | cis | 9.41E-09 | Nightingale | TG_by_PG | NA | T | G | 0.06879 | 0.004028 | 1.90E-68 |
| rs174537 | miR-1908-5 | cis | 9.41E-09 | Nightingale | M_HDL_FC | NA | T | G | -0.10357 | 0.003846 | 1.00E-173 |
| rs174537 | miR-1908-5 | cis | 9.41E-09 | Nightingale | XL_VLDL_C | NA | T | G | -0.06446 | 0.00416 | 1.80E-55 |
| rs174537 | miR-1908-5 | cis | 9.41E-09 | Nightingale | S_VLDL_FC | NA | T | G | -0.02904 | 0.004342 | 7.50E-12 |
| rs174537 | miR-1908-5 | cis | 9.41E-09 | Nightingale | L_HDL_PL_ | NA | T | G | 0.043288 | 0.004201 | 1.60E-26 |
| rs174537 | miR-1908-5 | cis | 9.41E-09 | Nightingale | S_VLDL_TG | NA | T | G | 0.065088 | 0.0042 | 1.10E-55 |
| rs174537 | miR-1908-5 | cis | 9.41E-09 | Nightingale | Unsaturati | NA | T | G | -0.33922 | 0.004111 | 0 |
| rs174537 | miR-1908-5 | cis | 9.41E-09 | Nightingale | XS_VLDL_F | NA | T | G | -0.05701 | 0.004265 | 2.10E-43 |
| rs174537 | miR-1908-5 | cis | 9.41E-09 | Nightingale | Clinical_LD | NA | T | G | -0.05594 | 0.004316 | 4.20E-40 |
| rs174537 | miR-1908-5 | cis | 9.41E-09 | Nightingale | XS_VLDL_P | NA | T | G | -0.03525 | 0.00428 | 5.80E-18 |
| rs174537 | miR-1908-5 | cis | 9.41E-09 | Nightingale | MUFA | NA | T | G | 0.063644 | 0.004296 | 1.40E-50 |
| rs174537 | miR-1908-5 | cis | 9.41E-09 | Nightingale | XS_VLDL_P | NA | T | G | -0.04454 | 0.004283 | 3.70E-27 |
| rs174537 | miR-1908-5 | cis | 9.41E-09 | Nightingale | S_LDL_CE_ | NA | T | G | 0.056991 | 0.004333 | 2.70E-41 |
| rs174537 | miR-1908-5 | cis | 9.41E-09 | Nightingale | L_HDL_P | NA | T | G | -0.07795 | 0.003802 | 1.20E-101 |
| rs174537 | miR-1908-5 | cis | 9.41E-09 | Nightingale | M_LDL_C | NA | T | G | -0.03464 | 0.004349 | 4.60E-16 |
| rs174537 | miR-1908-5 | cis | 9.41E-09 | Nightingale | XL_HDL_FC | NA | T | G | 0.061289 | 0.003849 | 4.20E-63 |
| rs174537 | miR-1908-5 | cis | 9.41E-09 | Nightingale | XS_VLDL_T | NA | T | G | 0.098015 | 0.004123 | 1.90E-130 |
| rs174537 | miR-1908-5 | cis | 9.41E-09 | Nightingale | M_HDL_C_ | NA | T | G | -0.0658 | 0.004132 | 5.80E-60 |
| rs174537 | miR-1908-5 | cis | 9.41E-09 | Nightingale | M_HDL_L | NA | T | G | -0.02771 | 0.00403 | 8.20E-13 |
| rs174537 | miR-1908-5 | cis | 9.41E-09 | Nightingale | Remnant_( | NA | T | G | -0.04501 | 0.004313 | 7.40E-27 |
| rs174537 | miR-1908-5 | cis | 9.41E-09 | Nightingale | L_LDL_FC | NA | T | G | -0.06913 | 0.004262 | 2.10E-62 |
| rs174537 | miR-1908-5 | cis | 9.41E-09 | Nightingale | XL_HDL_CE | NA | T | G | -0.02616 | 0.004338 | 4.50E-09 |
| rs174537 | miR-1908-5 | cis | 9.41E-09 | Nightingale | S_LDL_FC | NA | T | G | -0.06344 | 0.004334 | 3.10E-51 |
| rs174537 | miR-1908-5 | cis | 9.41E-09 | Nightingale | L_VLDL_C | NA | T | G | 0.028487 | 0.004246 | 9.40E-12 |
| rs174537 | miR-1908-5 | cis | 9.41E-09 | Nightingale | M_HDL_PL | NA | T | G | 0.058412 | 0.004117 | 1.60E-47 |
| rs174537 | miR-1908-5 | cis | 9.41E-09 | Nightingale | M_LDL_TG | NA | T | G | 0.076315 | 0.004286 | 3.60E-72 |
| rs174537 | miR-1908-5 | cis | 9.41E-09 | Nightingale | L_VLDL_C_ | NA | T | G | -0.03921 | 0.00424 | 1.50E-21 |
| rs174537 | miR-1908-5 | cis | 9.41E-09 | Nightingale | XS_VLDL_C | NA | T | G | -0.07296 | 0.004188 | 5.40E-72 |
| rs174537 | miR-1908-5 | cis | 9.41E-09 | Nightingale | LDL_FC | NA | T | G | -0.06505 | 0.004295 | 1.80E-54 |
| rs174537 | miR-1908-5 | cis | 9.41E-09 | Nightingale | XS_VLDL_C | NA | T | G | -0.09866 | 0.004113 | 1.70E-132 |
| rs174537 | miR-1908-5 | cis | 9.41E-09 | Nightingale | LDL_P | NA | T | G | -0.04099 | 0.004338 | 3.40E-22 |
| rs174537 | miR-1908-5 | cis | 9.41E-09 | Nightingale | IDL_FC_pct | NA | T | G | -0.10416 | 0.004306 | 1.10E-130 |
| rs174537 | miR-1908-5 | cis | 9.41E-09 | Nightingale | LDL_L | NA | T | G | -0.04615 | 0.004326 | 7.50E-28 |
| rs174537 | miR-1908-5 | cis | 9.41E-09 | Nightingale | L_VLDL_P | NA | T | G | 0.041652 | 0.004191 | 4.40E-24 |
| rs174537 | miR-1908-5 | cis | 9.41E-09 | Nightingale | non_HDL_( | NA | T | G | -0.04861 | 0.004317 | 8.80E-31 |
| rs174537 | miR-1908-5 | cis | 9.41E-09 | Nightingale | XL_VLDL_F | NA | T | G | -0.0691 | 0.004206 | 2.10E-63 |
| rs174537 | miR-1908-5 | cis | 9.41E-09 | Nightingale | S_LDL_P | NA | T | G | -0.03317 | 0.004345 | 4.90E-15 |
| rs174537 | miR-1908-5 | cis | 9.41E-09 | Nightingale | HDL_P | NA | T | G | -0.0357 | 0.004076 | 1.40E-19 |
| rs174537 | miR-1908-5 | cis | 9.41E-09 | Nightingale | Total_TG | NA | T | G | 0.046606 | 0.004202 | 1.80E-29 |
| rs174537 | miR-1908-5 | cis | 9.41E-09 | Nightingale | XL_VLDL_C | NA | T | G | 0.03707 | 0.00424 | 6.00E-19 |
| rs174537 | miR-1908-5 | cis | 9.41E-09 | Nightingale | L_HDL_C_p | NA | T | G | -0.06839 | 0.004091 | 2.70E-67 |
| rs174537 | miR-1908-5 | cis | 9.41E-09 | Nightingale | Total_FA | NA | T | G | -0.02416 | 0.004306 | 6.10E-09 |
| rs174537 | miR-1908-5 | cis | 9.41E-09 | Nightingale | XXL_VLDL_ | NA | T | G | 0.067715 | 0.004182 | 1.30E-61 |
| rs174537 | miR-1908-5 | cis | 9.41E-09 | Nightingale | XL_VLDL_F | NA | T | G | 0.0431 | 0.004204 | 1.90E-25 |
| rs174537 | miR-1908-5 | cis | 9.41E-09 | Nightingale | L_HDL_FC_ | NA | T | G | -0.11276 | 0.004033 | 5.11E-187 |
| rs174537 | miR-1908-5 | cis | 9.41E-09 | Nightingale | M_VLDL_T | NA | T | G | 0.031213 | 0.004217 | 6.30E-14 |
| rs174537 | miR-1908-5 | cis | 9.41E-09 | Nightingale | L_VLDL_FC | NA | T | G | 0.039859 | 0.004199 | 3.60E-22 |
| rs174537 | miR-1908-5 | cis | 9.41E-09 | Nightingale | IDL_P | NA | T | G | -0.04611 | 0.004295 | 3.30E-28 |
| rs174537 | miR-1908-5 | cis | 9.41E-09 | Nightingale | HDL_C | NA | T | G | -0.06486 | 0.003876 | 6.80E-68 |
| rs174537 | miR-1908-5 | cis | 9.41E-09 | Nightingale | IDL_CE_pct | NA | T | G | -0.03872 | 0.004259 | 1.30E-19 |
| rs174537 | miR-1908-5 | cis | 9.41E-09 | Nightingale | S_VLDL_PL | NA | T | G | -0.0791 | 0.004153 | 1.50E-84 |
| rs174537 | miR-1908-5 | cis | 9.41E-09 | Nightingale | L_HDL_C | NA | T | G | -0.08034 | 0.003822 | 1.80E-106 |

| | | | | | | | | | | | |
|---|---|---|---|---|---|---|---|---|---|---|---|
| rs174537 | miR-1908-5 | cis | 9.41E-09 | Nightingale | XL_HDL_PL | NA | T | G | -0.06719 | 0.003988 | 1.10E-68 |
| rs174537 | miR-1908-5 | cis | 9.41E-09 | Nightingale | XL_HDL_TG | NA | T | G | 0.087267 | 0.004089 | 1.20E-106 |
| rs174537 | miR-1908-5 | cis | 9.41E-09 | Nightingale | L_LDL_FC_ | NA | T | G | -0.08335 | 0.004137 | 3.90E-93 |
| rs174537 | miR-1908-5 | cis | 9.41E-09 | Nightingale | M_LDL_PL_ | NA | T | G | 0.045569 | 0.004338 | 5.80E-27 |
| rs174537 | miR-1908-5 | cis | 9.41E-09 | Nightingale | DHA_pct | NA | T | G | -0.2734 | 0.004214 | 0 |
| rs174537 | miR-1908-5 | cis | 9.41E-09 | Nightingale | M_LDL_P | NA | T | G | -0.04729 | 0.004353 | 2.80E-28 |
| rs174537 | miR-1908-5 | cis | 9.41E-09 | Nightingale | LDL_C | NA | T | G | -0.05046 | 0.004322 | 5.50E-33 |
| rs174537 | miR-1908-5 | cis | 9.41E-09 | Nightingale | XS_VLDL_L | NA | T | G | -0.04248 | 0.004267 | 2.90E-25 |
| rs174537 | miR-1908-5 | cis | 9.41E-09 | Nightingale | IDL_FC | NA | T | G | -0.09065 | 0.004201 | 1.30E-108 |
| rs174537 | miR-1908-5 | cis | 9.41E-09 | Nightingale | HDL_CE | NA | T | G | -0.06154 | 0.003896 | 1.60E-60 |
| rs174537 | miR-1908-5 | cis | 9.41E-09 | Nightingale | XL_HDL_L | NA | T | G | -0.09326 | 0.003901 | 3.20E-137 |
| rs174537 | miR-1908-5 | cis | 9.41E-09 | Nightingale | M_VLDL_C | NA | T | G | -0.07651 | 0.004075 | 1.20E-81 |
| rs174537 | miR-1908-5 | cis | 9.41E-09 | Nightingale | LA | NA | T | G | 0.082806 | 0.004257 | 1.30E-84 |
| rs174537 | miR-1908-5 | cis | 9.41E-09 | Nightingale | L_VLDL_FC | NA | T | G | 0.027965 | 0.0043 | 4.00E-11 |
| rs174537 | miR-1908-5 | cis | 9.41E-09 | Nightingale | XXL_VLDL_ | NA | T | G | 0.071761 | 0.004166 | 2.50E-69 |
| rs174537 | miR-1908-5 | cis | 9.41E-09 | Nightingale | Omega_6_ | NA | T | G | 0.369165 | 0.004278 | 0 |
| rs174537 | miR-1908-5 | cis | 9.41E-09 | Nightingale | M_HDL_P | NA | T | G | -0.03434 | 0.003999 | 3.90E-19 |
| rs174537 | miR-1908-5 | cis | 9.41E-09 | Nightingale | Cholines | NA | T | G | -0.03768 | 0.004082 | 4.60E-22 |
| rs174537 | miR-1908-5 | cis | 9.41E-09 | Nightingale | XL_HDL_PL | NA | T | G | -0.08929 | 0.003902 | 6.40E-126 |
| rs174537 | miR-1908-5 | cis | 9.41E-09 | Nightingale | S_LDL_L | NA | T | G | -0.03311 | 0.00434 | 4.40E-15 |
| rs174537 | miR-1908-5 | cis | 9.41E-09 | Nightingale | Omega_6_ | NA | T | G | 0.023759 | 0.004225 | 8.70E-09 |
| rs174537 | miR-1908-5 | cis | 9.41E-09 | Nightingale | S_LDL_PL_ | NA | T | G | -0.06187 | 0.004332 | 2.40E-48 |
| rs174537 | miR-1908-5 | cis | 9.41E-09 | Nightingale | Omega_3_ | NA | T | G | -0.38924 | 0.004241 | 0 |
| rs174537 | miR-1908-5 | cis | 9.41E-09 | Nightingale | IDL_TG_pct | NA | T | G | 0.088709 | 0.004193 | 7.80E-103 |
| rs174537 | miR-1908-5 | cis | 9.41E-09 | Nightingale | IDL_CE | NA | T | G | -0.07321 | 0.00419 | 3.30E-72 |
| rs174537 | miR-1908-5 | cis | 9.41E-09 | Nightingale | Total_FC | NA | T | G | -0.06019 | 0.004241 | 3.20E-48 |
| rs174537 | miR-1908-5 | cis | 9.41E-09 | Nightingale | XS_VLDL_C | NA | T | G | -0.06918 | 0.004213 | 3.20E-64 |
| rs174537 | miR-1908-5 | cis | 9.41E-09 | Nightingale | Total_PL | NA | T | G | -0.05173 | 0.004133 | 3.90E-38 |
| rs174537 | miR-1908-5 | cis | 9.41E-09 | Nightingale | XL_HDL_P | NA | T | G | -0.08987 | 0.003906 | 6.50E-128 |
| rs174537 | miR-1908-5 | cis | 9.41E-09 | Nightingale | L_HDL_CE | NA | T | G | -0.07875 | 0.003833 | 1.00E-101 |
| rs174537 | miR-1908-5 | cis | 9.41E-09 | Nightingale | XXL_VLDL_ | NA | T | G | 0.071417 | 0.004183 | 8.60E-68 |
| rs174537 | miR-1908-5 | cis | 9.41E-09 | Nightingale | L_LDL_C | NA | T | G | -0.05855 | 0.004292 | 1.30E-44 |
| rs174537 | miR-1908-5 | cis | 9.41E-09 | Nightingale | M_LDL_PL | NA | T | G | -0.02413 | 0.004347 | 1.40E-08 |
| rs174537 | miR-1908-5 | cis | 9.41E-09 | Nightingale | XL_VLDL_C | NA | T | G | -0.06134 | 0.004162 | 1.80E-50 |
| rs174537 | miR-1908-5 | cis | 9.41E-09 | Nightingale | IDL_C | NA | T | G | -0.07822 | 0.004191 | 4.50E-82 |
| rs174537 | miR-1908-5 | cis | 9.41E-09 | Nightingale | Omega_3 | NA | T | G | -0.33541 | 0.004254 | 0 |
| rs174537 | miR-1908-5 | cis | 9.41E-09 | Nightingale | S_VLDL_C_ | NA | T | G | -0.05719 | 0.004223 | 8.70E-43 |
| rs174537 | miR-1908-5 | cis | 9.41E-09 | Nightingale | ApoB | NA | T | G | -0.03926 | 0.004335 | 1.40E-20 |
| rs174537 | miR-1908-5 | cis | 9.41E-09 | Nightingale | M_HDL_CE | NA | T | G | -0.04682 | 0.004209 | 1.70E-29 |
| rs174537 | miR-1908-5 | cis | 9.41E-09 | Nightingale | L_LDL_L | NA | T | G | -0.05492 | 0.004299 | 2.20E-39 |
| rs174537 | miR-1908-5 | cis | 9.41E-09 | Nightingale | L_VLDL_PL_ | NA | T | G | 0.087451 | 0.004213 | 4.60E-99 |
| rs174537 | miR-1908-5 | cis | 9.41E-09 | Nightingale | L_VLDL_PL | NA | T | G | 0.048515 | 0.004188 | 2.80E-32 |
| rs174537 | miR-1908-5 | cis | 9.41E-09 | Nightingale | M_VLDL_C | NA | T | G | -0.0448 | 0.004329 | 4.90E-26 |
| rs174537 | miR-1908-5 | cis | 9.41E-09 | Nightingale | M_LDL_L | NA | T | G | -0.02876 | 0.004347 | 1.50E-11 |
| rs174537 | miR-1908-5 | cis | 9.41E-09 | Nightingale | XL_VLDL_P | NA | T | G | 0.047931 | 0.004189 | 2.30E-31 |
| rs174537 | miR-1908-5 | cis | 9.41E-09 | Nightingale | XL_VLDL_T | NA | T | G | 0.056841 | 0.004152 | 3.10E-44 |
| rs174537 | miR-1908-5 | cis | 9.41E-09 | Nightingale | HDL_L | NA | T | G | -0.05349 | 0.003898 | 5.40E-46 |
| rs174537 | miR-1908-5 | cis | 9.41E-09 | Nightingale | M_VLDL_F | NA | T | G | -0.03016 | 0.00434 | 1.10E-12 |
| rs174537 | miR-1908-5 | cis | 9.41E-09 | Nightingale | IDL_L | NA | T | G | -0.07335 | 0.004196 | 1.10E-72 |
| rs174537 | miR-1908-5 | cis | 9.41E-09 | Nightingale | Tyr | NA | T | G | 0.03371 | 0.004335 | 7.50E-15 |
| rs174537 | miR-1908-5 | cis | 9.41E-09 | Nightingale | M_HDL_TG | NA | T | G | 0.066571 | 0.004162 | 4.00E-60 |
| rs174537 | miR-1908-5 | cis | 9.41E-09 | Nightingale | HDL_FC | NA | T | G | -0.07413 | 0.003845 | 1.80E-89 |
| rs174537 | miR-1908-5 | cis | 9.41E-09 | Nightingale | LDL_CE | NA | T | G | -0.04425 | 0.00433 | 1.20E-25 |
| rs174537 | miR-1908-5 | cis | 9.41E-09 | Nightingale | M_LDL_C_ | NA | T | G | -0.10073 | 0.004344 | 3.20E-119 |
| rs174537 | miR-1908-5 | cis | 9.41E-09 | Nightingale | XL_HDL_FC | NA | T | G | -0.10305 | 0.004044 | 2.90E-154 |
| rs174537 | miR-1908-5 | cis | 9.41E-09 | Nightingale | S_VLDL_TG | NA | T | G | 0.038743 | 0.004212 | 1.20E-20 |
| rs174537 | miR-1908-5 | cis | 9.41E-09 | Nightingale | L_LDL_P | NA | T | G | -0.03817 | 0.004326 | 9.20E-20 |
| rs174537 | miR-1908-5 | cis | 9.41E-09 | Nightingale | HDL_size | NA | T | G | -0.07269 | 0.003846 | 3.80E-86 |
| rs174537 | miR-1908-5 | cis | 9.41E-09 | Nightingale | XL_HDL_C | NA | T | G | -0.09946 | 0.003914 | 2.90E-154 |
| rs174537 | miR-1908-5 | cis | 9.41E-09 | Nightingale | XXL_VLDL_ | NA | T | G | 0.061009 | 0.004189 | 5.10E-50 |
| rs174537 | miR-1908-5 | cis | 9.41E-09 | Nightingale | XXL_VLDL_ | NA | T | G | 0.073314 | 0.004192 | 1.00E-70 |
| rs174537 | miR-1908-5 | cis | 9.41E-09 | Nightingale | XL_HDL_C_ | NA | T | G | 0.024068 | 0.004147 | 2.70E-10 |
| rs174537 | miR-1908-5 | cis | 9.41E-09 | Nightingale | M_LDL_FC_ | NA | T | G | -0.07186 | 0.004158 | 3.50E-69 |

| | | | | | | | | | | | |
|---|---|---|---|---|---|---|---|---|---|---|---|
| rs174537 | miR-1908-5 | cis | 9.41E-09 | Nightingale | Total_CE | NA | T | G | -0.06774 | 0.004187 | 5.70E-62 |
| rs174537 | miR-1908-5 | cis | 9.41E-09 | Nightingale | M_HDL_CE | NA | T | G | -0.03782 | 0.004009 | 2.00E-22 |
| rs174537 | miR-1908-5 | cis | 9.41E-09 | Nightingale | M_HDL_TG | NA | T | G | 0.050212 | 0.004287 | 2.00E-32 |
| rs174537 | miR-1908-5 | cis | 9.41E-09 | Nightingale | L_LDL_PL | NA | T | G | -0.05572 | 0.004308 | 6.50E-40 |
| rs174537 | miR-1908-5 | cis | 9.41E-09 | Nightingale | XL_VLDL_L | NA | T | G | 0.051267 | 0.004177 | 7.50E-36 |
| rs174537 | miR-1908-5 | cis | 9.41E-09 | Nightingale | LDL_PL | NA | T | G | -0.04689 | 0.004331 | 1.90E-28 |
| rs174537 | miR-1908-5 | cis | 9.41E-09 | Nightingale | M_VLDL_C | NA | T | G | -0.08129 | 0.004086 | 1.40E-91 |
| rs174537 | miR-1908-5 | cis | 9.41E-09 | Nightingale | XL_VLDL_T | NA | T | G | 0.060136 | 0.004205 | 9.50E-48 |
| rs174537 | miR-1908-5 | cis | 9.41E-09 | Nightingale | L_HDL_TG_ | NA | T | G | 0.078598 | 0.004096 | 5.00E-87 |
| rs174537 | miR-1908-5 | cis | 9.41E-09 | Nightingale | XL_HDL_CE | NA | T | G | -0.09732 | 0.003887 | 5.40E-150 |
| rs174537 | miR-1908-5 | cis | 9.41E-09 | Nightingale | LA_pct | NA | T | G | 0.184364 | 0.004242 | 0 |
| rs174537 | miR-1908-5 | cis | 9.41E-09 | Nightingale | S_HDL_TG | NA | T | G | 0.068817 | 0.004209 | 1.10E-62 |
| rs174537 | miR-1908-5 | cis | 9.41E-09 | Nightingale | L_HDL_CE_ | NA | T | G | -0.04996 | 0.004153 | 1.60E-35 |
| rs174537 | miR-1908-5 | cis | 9.41E-09 | Nightingale | M_HDL_FC | NA | T | G | -0.05161 | 0.00395 | 9.60E-42 |
| rs174537 | miR-1908-5 | cis | 9.41E-09 | Nightingale | S_LDL_TG_ | NA | T | G | 0.086488 | 0.004206 | 8.90E-97 |
| rs174537 | miR-1908-5 | cis | 9.41E-09 | Nightingale | M_LDL_CE | NA | T | G | -0.0269 | 0.004347 | 2.80E-10 |
| rs174537 | miR-1908-5 | cis | 9.41E-09 | Nightingale | PUFA | NA | T | G | -0.09704 | 0.004233 | 3.40E-123 |
| rs174537 | miR-1908-5 | cis | 9.41E-09 | Nightingale | L_LDL_C_p | NA | T | G | -0.07752 | 0.004229 | 3.30E-77 |
| rs174537 | miR-1908-5 | cis | 9.41E-09 | Nightingale | S_LDL_TG | NA | T | G | 0.050199 | 0.004234 | 8.30E-33 |
| rs174537 | miR-1908-5 | cis | 9.41E-09 | Nightingale | S_LDL_C | NA | T | G | -0.03282 | 0.004345 | 9.60E-15 |
| rs174537 | miR-1908-5 | cis | 9.41E-09 | Nightingale | L_VLDL_CE | NA | T | G | -0.05976 | 0.004154 | 1.10E-49 |
| rs174537 | miR-1908-5 | cis | 9.41E-09 | Nightingale | S_LDL_FC_ | NA | T | G | -0.07995 | 0.004206 | 3.40E-83 |
| rs174537 | miR-1908-5 | cis | 9.41E-09 | Nightingale | VLDL_L | NA | T | G | 0.026948 | 0.004256 | 1.50E-10 |
| rs174537 | miR-1908-5 | cis | 9.41E-09 | Nightingale | XL_VLDL_P | NA | T | G | 0.05266 | 0.004172 | 8.10E-38 |
| rs174537 | miR-1908-5 | cis | 9.41E-09 | Nightingale | XXL_VLDL_ | NA | T | G | 0.065856 | 0.004175 | 1.60E-58 |
| rs174537 | miR-1908-5 | cis | 9.41E-09 | Nightingale | M_HDL_PL | NA | T | G | -0.02152 | 0.004059 | 3.20E-08 |
| rs174537 | miR-1908-5 | cis | 9.41E-09 | Nightingale | PUFA_pct | NA | T | G | -0.12156 | 0.004175 | 1.40E-187 |
| rs174537 | miR-1908-5 | cis | 9.41E-09 | Nightingale | Total_L | NA | T | G | -0.03802 | 0.00427 | 3.60E-20 |
| rs174537 | miR-1908-5 | cis | 9.41E-09 | Nightingale | HDL_TG | NA | T | G | 0.046075 | 0.00428 | 1.90E-27 |
| rs174537 | miR-1908-5 | cis | 9.41E-09 | Nightingale | S_HDL_FC_ | NA | T | G | -0.10306 | 0.004117 | 3.10E-153 |
| rs174537 | miR-1908-5 | cis | 9.41E-09 | Nightingale | Citrate | NA | T | G | -0.03304 | 0.004313 | 1.50E-15 |
| rs174537 | miR-1908-5 | cis | 9.41E-09 | Nightingale | XXL_VLDL_ | NA | T | G | 0.044374 | 0.004364 | 3.30E-25 |
| rs174537 | miR-1908-5 | cis | 9.41E-09 | Nightingale | PUFA_by_N | NA | T | G | -0.17072 | 0.004158 | 0 |
| rs174537 | miR-1908-5 | cis | 9.41E-09 | Nightingale | Sphingomy | NA | T | G | -0.11565 | 0.004056 | 1.40E-187 |
| rs174537 | miR-1908-5 | cis | 9.41E-09 | Nightingale | IDL_PL | NA | T | G | -0.07376 | 0.004188 | 4.00E-74 |
| rs174537 | miR-1908-5 | cis | 9.41E-09 | Nightingale | IDL_C_pct | NA | T | G | -0.07541 | 0.004238 | 2.80E-72 |
| rs174537 | miR-1908-5 | cis | 9.41E-09 | Nightingale | L_LDL_TG_ | NA | T | G | 0.078174 | 0.004256 | 8.50E-77 |
| rs174537 | miR-1908-5 | cis | 9.41E-09 | Nightingale | L_HDL_L | NA | T | G | -0.07728 | 0.003809 | 1.50E-99 |
| rs174537 | miR-1908-5 | cis | 9.41E-09 | Nightingale | XS_VLDL_F | NA | T | G | -0.11341 | 0.004308 | 1.20E-155 |
| rs174537 | miR-1908-5 | cis | 9.41E-09 | Nightingale | XXL_VLDL_ | NA | T | G | -0.02683 | 0.004319 | 1.00E-10 |
| rs174537 | miR-1908-5 | cis | 9.41E-09 | Nightingale | S_HDL_C_p | NA | T | G | -0.04236 | 0.00429 | 9.70E-23 |
| rs174537 | miR-1908-5 | cis | 9.41E-09 | Nightingale | M_VLDL_T | NA | T | G | 0.078609 | 0.0041 | 1.30E-85 |
| rs174537 | miR-1908-5 | cis | 9.41E-09 | Nightingale | VLDL_TG | NA | T | G | 0.047554 | 0.004177 | 6.00E-31 |
| rs174537 | miR-1908-5 | cis | 9.41E-09 | Nightingale | S_HDL_TG_ | NA | T | G | 0.069012 | 0.004128 | 2.10E-65 |
| rs174537 | miR-1908-5 | cis | 9.41E-09 | Nightingale | L_VLDL_TG | NA | T | G | 0.037178 | 0.004177 | 1.30E-19 |
| rs174537 | miR-1908-5 | cis | 9.41E-09 | Nightingale | Total_P | NA | T | G | -0.04056 | 0.004085 | 7.20E-25 |
| rs174537 | miR-1908-5 | cis | 9.41E-09 | Nightingale | XXL_VLDL_ | NA | T | G | 0.066394 | 0.004181 | 4.00E-59 |
| rs174537 | miR-1908-5 | cis | 9.41E-09 | Nightingale | M_LDL_FC | NA | T | G | -0.05304 | 0.004336 | 4.20E-36 |
| rs174537 | miR-1908-5 | cis | 9.41E-09 | Nightingale | L_HDL_FC | NA | T | G | -0.08474 | 0.003804 | 2.80E-119 |
| rs174537 | miR-1908-5 | cis | 9.41E-09 | Nightingale | HDL_PL | NA | T | G | -0.04856 | 0.003924 | 1.10E-37 |
| rs174537 | miR-1908-5 | cis | 9.41E-09 | Nightingale | ApoA1 | NA | T | G | -0.04131 | 0.003972 | 2.80E-27 |
| rs174537 | miR-1908-5 | cis | 9.41E-09 | Nightingale | M_VLDL_P | NA | T | G | -0.06325 | 0.004165 | 2.10E-55 |
| rs174537 | miR-1908-5 | cis | 9.41E-09 | Nightingale | S_VLDL_FC | NA | T | G | -0.08351 | 0.004145 | 5.10E-94 |
| rs174537 | miR-1908-5 | cis | 9.41E-09 | Nightingale | L_VLDL_L | NA | T | G | 0.03798 | 0.004195 | 3.00E-20 |
| rs174537 | miR-1908-5 | cis | 9.41E-09 | Nightingale | S_VLDL_CE | NA | T | G | -0.02948 | 0.004282 | 3.60E-12 |
| rs174537 | miR-1908-5 | cis | 9.41E-09 | Nightingale | XS_VLDL_T | NA | T | G | 0.031382 | 0.004274 | 8.70E-13 |
| rs174537 | miR-1908-5 | cis | 9.41E-09 | Nightingale | XS_VLDL_P | NA | T | G | 0.058226 | 0.004235 | 1.10E-43 |
| rs174537 | miR-1908-5 | cis | 9.41E-09 | Nightingale | S_LDL_PL | NA | T | G | -0.05216 | 0.00433 | 2.80E-35 |
| rs174537 | miR-1908-5 | cis | 9.41E-09 | Nightingale | VLDL_size | NA | T | G | 0.072479 | 0.004046 | 9.20E-76 |
| rs174537 | miR-1908-5 | cis | 9.41E-09 | Nightingale | M_VLDL_F | NA | T | G | -0.09201 | 0.004127 | 1.40E-115 |
| rs174537 | miR-1908-5 | cis | 9.41E-09 | Nightingale | M_VLDL_C | NA | T | G | -0.05347 | 0.004301 | 5.40E-37 |
| rs174538 | miR-1908-5 | cis | 1.35E-07 | Nightingale | XL_VLDL_T | NA | A | G | 0.053899 | 0.004243 | 1.40E-38 |
| rs174538 | miR-1908-5 | cis | 1.35E-07 | Nightingale | L_VLDL_FC | NA | A | G | 0.024998 | 0.004394 | 4.90E-09 |

| | | | | | | | | | | | |
|---|---|---|---|---|---|---|---|---|---|---|---|
| rs174538 | miR-1908-! | cis | 1.35E-07 | Nightingale | S_LDL_CE_ | NA | A | G | 0.05396 | 0.004428 | 3.40E-36 |
| rs174538 | miR-1908-! | cis | 1.35E-07 | Nightingale | PUFA | NA | A | G | -0.08911 | 0.004327 | 1.10E-99 |
| rs174538 | miR-1908-! | cis | 1.35E-07 | Nightingale | XL_VLDL_C | NA | A | G | -0.05855 | 0.004254 | 6.20E-44 |
| rs174538 | miR-1908-! | cis | 1.35E-07 | Nightingale | XXL_VLDL_ | NA | A | G | 0.062822 | 0.004273 | 3.20E-51 |
| rs174538 | miR-1908-! | cis | 1.35E-07 | Nightingale | XL_HDL_P | NA | A | G | -0.08436 | 0.003992 | 1.10E-108 |
| rs174538 | miR-1908-! | cis | 1.35E-07 | Nightingale | M_HDL_L | NA | A | G | -0.02457 | 0.004119 | 4.20E-10 |
| rs174538 | miR-1908-! | cis | 1.35E-07 | Nightingale | XXL_VLDL_ | NA | A | G | 0.04063 | 0.00446 | 1.90E-20 |
| rs174538 | miR-1908-! | cis | 1.35E-07 | Nightingale | XS_VLDL_T | NA | A | G | 0.029746 | 0.004368 | 4.40E-11 |
| rs174538 | miR-1908-! | cis | 1.35E-07 | Nightingale | XS_VLDL_F | NA | A | G | -0.10732 | 0.004403 | 2.50E-133 |
| rs174538 | miR-1908-! | cis | 1.35E-07 | Nightingale | HDL_C | NA | A | G | -0.05952 | 0.003961 | 2.90E-55 |
| rs174538 | miR-1908-! | cis | 1.35E-07 | Nightingale | ApoA1 | NA | A | G | -0.03732 | 0.00406 | 9.70E-22 |
| rs174538 | miR-1908-! | cis | 1.35E-07 | Nightingale | L_VLDL_TG | NA | A | G | 0.0356 | 0.004269 | 1.40E-17 |
| rs174538 | miR-1908-! | cis | 1.35E-07 | Nightingale | HDL_L | NA | A | G | -0.04882 | 0.003984 | 3.70E-37 |
| rs174538 | miR-1908-! | cis | 1.35E-07 | Nightingale | M_HDL_PL | NA | A | G | 0.053761 | 0.004208 | 1.40E-38 |
| rs174538 | miR-1908-! | cis | 1.35E-07 | Nightingale | LDL_CE | NA | A | G | -0.04052 | 0.004425 | 1.20E-20 |
| rs174538 | miR-1908-! | cis | 1.35E-07 | Nightingale | M_HDL_CE | NA | A | G | -0.04323 | 0.004302 | 2.70E-24 |
| rs174538 | miR-1908-! | cis | 1.35E-07 | Nightingale | LDL_TG | NA | A | G | 0.0284 | 0.004387 | 2.80E-10 |
| rs174538 | miR-1908-! | cis | 1.35E-07 | Nightingale | S_HDL_FC_ | NA | A | G | -0.09674 | 0.004208 | 1.30E-130 |
| rs174538 | miR-1908-! | cis | 1.35E-07 | Nightingale | DHA_pct | NA | A | G | -0.25738 | 0.004307 | 0 |
| rs174538 | miR-1908-! | cis | 1.35E-07 | Nightingale | M_HDL_P | NA | A | G | -0.03076 | 0.004087 | 3.90E-15 |
| rs174538 | miR-1908-! | cis | 1.35E-07 | Nightingale | XXL_VLDL_ | NA | A | G | 0.057876 | 0.004281 | 1.50E-43 |
| rs174538 | miR-1908-! | cis | 1.35E-07 | Nightingale | Citrate | NA | A | G | -0.03132 | 0.004408 | 1.40E-13 |
| rs174538 | miR-1908-! | cis | 1.35E-07 | Nightingale | XL_VLDL_F | NA | A | G | -0.06561 | 0.004299 | 8.20E-55 |
| rs174538 | miR-1908-! | cis | 1.35E-07 | Nightingale | M_LDL_C_ | NA | A | G | -0.09501 | 0.00444 | 6.81E-101 |
| rs174538 | miR-1908-! | cis | 1.35E-07 | Nightingale | Omega_3_ | NA | A | G | -0.36539 | 0.004335 | 0 |
| rs174538 | miR-1908-! | cis | 1.35E-07 | Nightingale | M_VLDL_C | NA | A | G | -0.04998 | 0.004396 | 5.60E-31 |
| rs174538 | miR-1908-! | cis | 1.35E-07 | Nightingale | L_LDL_FC_ | NA | A | G | -0.07872 | 0.004228 | 2.30E-79 |
| rs174538 | miR-1908-! | cis | 1.35E-07 | Nightingale | L_VLDL_CE | NA | A | G | -0.05826 | 0.004245 | 4.60E-45 |
| rs174538 | miR-1908-! | cis | 1.35E-07 | Nightingale | XL_VLDL_P | NA | A | G | 0.045208 | 0.004281 | 3.20E-27 |
| rs174538 | miR-1908-! | cis | 1.35E-07 | Nightingale | HDL_FC | NA | A | G | -0.06844 | 0.00393 | 1.10E-73 |
| rs174538 | miR-1908-! | cis | 1.35E-07 | Nightingale | IDL_CE | NA | A | G | -0.06848 | 0.004283 | 2.30E-60 |
| rs174538 | miR-1908-! | cis | 1.35E-07 | Nightingale | L_VLDL_C | NA | A | G | 0.026933 | 0.004339 | 2.10E-10 |
| rs174538 | miR-1908-! | cis | 1.35E-07 | Nightingale | IDL_CE_pct | NA | A | G | -0.03618 | 0.004353 | 2.30E-16 |
| rs174538 | miR-1908-! | cis | 1.35E-07 | Nightingale | L_HDL_CE | NA | A | G | -0.07333 | 0.003917 | 4.10E-85 |
| rs174538 | miR-1908-! | cis | 1.35E-07 | Nightingale | M_HDL_FC | NA | A | G | -0.04706 | 0.004037 | 9.90E-34 |
| rs174538 | miR-1908-! | cis | 1.35E-07 | Nightingale | L_HDL_PL_ | NA | A | G | 0.040806 | 0.004294 | 9.60E-23 |
| rs174538 | miR-1908-! | cis | 1.35E-07 | Nightingale | S_LDL_TG | NA | A | G | 0.047681 | 0.004328 | 1.70E-28 |
| rs174538 | miR-1908-! | cis | 1.35E-07 | Nightingale | HDL_TG | NA | A | G | 0.04352 | 0.004375 | 1.10E-23 |
| rs174538 | miR-1908-! | cis | 1.35E-07 | Nightingale | M_LDL_C | NA | A | G | -0.03148 | 0.004445 | 9.00E-13 |
| rs174538 | miR-1908-! | cis | 1.35E-07 | Nightingale | M_VLDL_P | NA | A | G | -0.06122 | 0.004257 | 1.20E-49 |
| rs174538 | miR-1908-! | cis | 1.35E-07 | Nightingale | Cholines | NA | A | G | -0.03397 | 0.004172 | 1.60E-17 |
| rs174538 | miR-1908-! | cis | 1.35E-07 | Nightingale | M_LDL_FC | NA | A | G | -0.04864 | 0.004431 | 5.40E-29 |
| rs174538 | miR-1908-! | cis | 1.35E-07 | Nightingale | L_HDL_PL | NA | A | G | -0.07078 | 0.003904 | 3.10E-80 |
| rs174538 | miR-1908-! | cis | 1.35E-07 | Nightingale | M_HDL_CE | NA | A | G | -0.03405 | 0.004097 | 7.30E-18 |
| rs174538 | miR-1908-! | cis | 1.35E-07 | Nightingale | XS_VLDL_P | NA | A | G | -0.04215 | 0.004377 | 1.70E-23 |
| rs174538 | miR-1908-! | cis | 1.35E-07 | Nightingale | MUFA | NA | A | G | 0.060356 | 0.004391 | 3.60E-44 |
| rs174538 | miR-1908-! | cis | 1.35E-07 | Nightingale | LA | NA | A | G | 0.080096 | 0.004351 | 2.70E-76 |
| rs174538 | miR-1908-! | cis | 1.35E-07 | Nightingale | DHA | NA | A | G | -0.2709 | 0.004275 | 0 |
| rs174538 | miR-1908-! | cis | 1.35E-07 | Nightingale | S_LDL_L | NA | A | G | -0.03013 | 0.004436 | 4.00E-12 |
| rs174538 | miR-1908-! | cis | 1.35E-07 | Nightingale | VLDL_L | NA | A | G | 0.025527 | 0.00435 | 2.40E-09 |
| rs174538 | miR-1908-! | cis | 1.35E-07 | Nightingale | S_VLDL_PL | NA | A | G | -0.0748 | 0.004245 | 2.60E-72 |
| rs174538 | miR-1908-! | cis | 1.35E-07 | Nightingale | XL_VLDL_C | NA | A | G | 0.034966 | 0.004333 | 1.50E-16 |
| rs174538 | miR-1908-! | cis | 1.35E-07 | Nightingale | M_HDL_C_ | NA | A | G | -0.06081 | 0.004223 | 3.20E-49 |
| rs174538 | miR-1908-! | cis | 1.35E-07 | Nightingale | IDL_C | NA | A | G | -0.07315 | 0.004284 | 1.50E-68 |
| rs174538 | miR-1908-! | cis | 1.35E-07 | Nightingale | S_LDL_TG_ | NA | A | G | 0.081466 | 0.004299 | 4.30E-82 |
| rs174538 | miR-1908-! | cis | 1.35E-07 | Nightingale | M_HDL_TG | NA | A | G | 0.06164 | 0.004254 | 8.40E-50 |
| rs174538 | miR-1908-! | cis | 1.35E-07 | Nightingale | SFA_pct | NA | A | G | -0.04072 | 0.004377 | 1.30E-20 |
| rs174538 | miR-1908-! | cis | 1.35E-07 | Nightingale | VLDL_size | NA | A | G | 0.068488 | 0.004135 | 2.30E-65 |
| rs174538 | miR-1908-! | cis | 1.35E-07 | Nightingale | IDL_L | NA | A | G | -0.06861 | 0.004289 | 7.50E-61 |
| rs174538 | miR-1908-! | cis | 1.35E-07 | Nightingale | S_LDL_PL_ | NA | A | G | -0.05908 | 0.004427 | 5.00E-43 |
| rs174538 | miR-1908-! | cis | 1.35E-07 | Nightingale | Total_FC | NA | A | G | -0.05571 | 0.004334 | 1.20E-39 |
| rs174538 | miR-1908-! | cis | 1.35E-07 | Nightingale | M_VLDL_T | NA | A | G | 0.075198 | 0.00419 | 6.20E-75 |
| rs174538 | miR-1908-! | cis | 1.35E-07 | Nightingale | Total_CE | NA | A | G | -0.06253 | 0.004279 | 1.60E-50 |

| | | | | | | | | | | | |
|---|---|---|---|---|---|---|---|---|---|---|---|
| rs174538 | miR-1908-! | cis | 1.35E-07 | Nightingale | XL_HDL_C | NA | A | G | -0.09346 | 0.004 | 4.70E-131 |
| rs174538 | miR-1908-! | cis | 1.35E-07 | Nightingale | S_HDL_TG | NA | A | G | 0.064959 | 0.004302 | 6.80E-54 |
| rs174538 | miR-1908-! | cis | 1.35E-07 | Nightingale | M_LDL_P | NA | A | G | -0.04367 | 0.004449 | 4.10E-23 |
| rs174538 | miR-1908-! | cis | 1.35E-07 | Nightingale | M_HDL_TG | NA | A | G | 0.047421 | 0.004382 | 4.80E-28 |
| rs174538 | miR-1908-! | cis | 1.35E-07 | Nightingale | HDL_PL | NA | A | G | -0.04423 | 0.004011 | 2.00E-30 |
| rs174538 | miR-1908-! | cis | 1.35E-07 | Nightingale | L_HDL_TG_ | NA | A | G | 0.072909 | 0.004186 | 4.00E-72 |
| rs174538 | miR-1908-! | cis | 1.35E-07 | Nightingale | IDL_TG_pc | NA | A | G | 0.083361 | 0.004286 | 1.90E-86 |
| rs174538 | miR-1908-! | cis | 1.35E-07 | Nightingale | XL_HDL_PL | NA | A | G | -0.0637 | 0.004075 | 1.60E-59 |
| rs174538 | miR-1908-! | cis | 1.35E-07 | Nightingale | Total_PL | NA | A | G | -0.0474 | 0.004224 | 7.10E-31 |
| rs174538 | miR-1908-! | cis | 1.35E-07 | Nightingale | M_LDL_FC_ | NA | A | G | -0.06715 | 0.00425 | 7.00E-58 |
| rs174538 | miR-1908-! | cis | 1.35E-07 | Nightingale | Omega_6_ | NA | A | G | 0.34687 | 0.004372 | 0 |
| rs174538 | miR-1908-! | cis | 1.35E-07 | Nightingale | L_LDL_C | NA | A | G | -0.05404 | 0.004387 | 2.60E-36 |
| rs174538 | miR-1908-! | cis | 1.35E-07 | Nightingale | PUFA_pct | NA | A | G | -0.11289 | 0.004267 | 3.90E-155 |
| rs174538 | miR-1908-! | cis | 1.35E-07 | Nightingale | L_HDL_C_p | NA | A | G | -0.06386 | 0.004182 | 2.90E-56 |
| rs174538 | miR-1908-! | cis | 1.35E-07 | Nightingale | S_LDL_PL | NA | A | G | -0.0482 | 0.004426 | 6.60E-29 |
| rs174538 | miR-1908-! | cis | 1.35E-07 | Nightingale | LDL_C | NA | A | G | -0.04638 | 0.004418 | 1.20E-26 |
| rs174538 | miR-1908-! | cis | 1.35E-07 | Nightingale | ApoB | NA | A | G | -0.03644 | 0.004431 | 4.50E-17 |
| rs174538 | miR-1908-! | cis | 1.35E-07 | Nightingale | XL_HDL_PL | NA | A | G | -0.08419 | 0.003988 | 6.61E-108 |
| rs174538 | miR-1908-! | cis | 1.35E-07 | Nightingale | S_VLDL_TG | NA | A | G | 0.06224 | 0.004293 | 1.40E-48 |
| rs174538 | miR-1908-! | cis | 1.35E-07 | Nightingale | Phosphogly | NA | A | G | -0.03762 | 0.004202 | 1.40E-20 |
| rs174538 | miR-1908-! | cis | 1.35E-07 | Nightingale | XXL_VLDL_ | NA | A | G | -0.02476 | 0.004414 | 4.50E-09 |
| rs174538 | miR-1908-! | cis | 1.35E-07 | Nightingale | M_LDL_PL_ | NA | A | G | 0.043824 | 0.004434 | 5.30E-24 |
| rs174538 | miR-1908-! | cis | 1.35E-07 | Nightingale | L_LDL_PL | NA | A | G | -0.05153 | 0.004403 | 1.30E-32 |
| rs174538 | miR-1908-! | cis | 1.35E-07 | Nightingale | M_LDL_L | NA | A | G | -0.02591 | 0.004443 | 4.10E-09 |
| rs174538 | miR-1908-! | cis | 1.35E-07 | Nightingale | XL_VLDL_L | NA | A | G | 0.048477 | 0.004269 | 3.50E-31 |
| rs174538 | miR-1908-! | cis | 1.35E-07 | Nightingale | L_VLDL_PL | NA | A | G | 0.04577 | 0.00428 | 5.50E-28 |
| rs174538 | miR-1908-! | cis | 1.35E-07 | Nightingale | PUFA_by_N | NA | A | G | -0.15922 | 0.004249 | 1.90107827 |
| rs174538 | miR-1908-! | cis | 1.35E-07 | Nightingale | XS_VLDL_C | NA | A | G | -0.0692 | 0.004281 | 6.80E-62 |
| rs174538 | miR-1908-! | cis | 1.35E-07 | Nightingale | M_LDL_CE | NA | A | G | -0.0243 | 0.004443 | 3.40E-08 |
| rs174538 | miR-1908-! | cis | 1.35E-07 | Nightingale | S_VLDL_C_ | NA | A | G | -0.05505 | 0.004316 | 8.20E-38 |
| rs174538 | miR-1908-! | cis | 1.35E-07 | Nightingale | Total_L | NA | A | G | -0.03454 | 0.004365 | 3.10E-16 |
| rs174538 | miR-1908-! | cis | 1.35E-07 | Nightingale | XXL_VLDL_ | NA | A | G | 0.062112 | 0.004267 | 2.20E-50 |
| rs174538 | miR-1908-! | cis | 1.35E-07 | Nightingale | Remnant_( | NA | A | G | -0.042 | 0.004408 | 1.80E-22 |
| rs174538 | miR-1908-! | cis | 1.35E-07 | Nightingale | XXL_VLDL_ | NA | A | G | 0.067899 | 0.004258 | 5.40E-60 |
| rs174538 | miR-1908-! | cis | 1.35E-07 | Nightingale | L_VLDL_P | NA | A | G | 0.039566 | 0.004284 | 3.30E-21 |
| rs174538 | miR-1908-! | cis | 1.35E-07 | Nightingale | S_HDL_C_p | NA | A | G | -0.0393 | 0.004385 | 6.40E-19 |
| rs174538 | miR-1908-! | cis | 1.35E-07 | Nightingale | XL_VLDL_P | NA | A | G | 0.049689 | 0.004264 | 9.50E-33 |
| rs174538 | miR-1908-! | cis | 1.35E-07 | Nightingale | M_VLDL_C | NA | A | G | -0.07287 | 0.004165 | 7.00E-71 |
| rs174538 | miR-1908-! | cis | 1.35E-07 | Nightingale | L_HDL_CE_ | NA | A | G | -0.04629 | 0.004245 | 3.70E-29 |
| rs174538 | miR-1908-! | cis | 1.35E-07 | Nightingale | Omega_6_ | NA | A | G | 0.023191 | 0.004318 | 3.20E-08 |
| rs174538 | miR-1908-! | cis | 1.35E-07 | Nightingale | S_HDL_TG_ | NA | A | G | 0.064255 | 0.004219 | 1.20E-54 |
| rs174538 | miR-1908-! | cis | 1.35E-07 | Nightingale | L_VLDL_C_ | NA | A | G | -0.03875 | 0.004334 | 5.70E-20 |
| rs174538 | miR-1908-! | cis | 1.35E-07 | Nightingale | XS_VLDL_C | NA | A | G | -0.06563 | 0.004306 | 2.00E-55 |
| rs174538 | miR-1908-! | cis | 1.35E-07 | Nightingale | M_VLDL_C_ | NA | A | G | -0.07753 | 0.004176 | 9.50E-80 |
| rs174538 | miR-1908-! | cis | 1.35E-07 | Nightingale | M_LDL_TG | NA | A | G | 0.035045 | 0.004371 | 1.90E-15 |
| rs174538 | miR-1908-! | cis | 1.35E-07 | Nightingale | XL_VLDL_C | NA | A | G | 0.028263 | 0.004367 | 3.70E-11 |
| rs174538 | miR-1908-! | cis | 1.35E-07 | Nightingale | M_VLDL_F | NA | A | G | -0.08788 | 0.004218 | 8.39E-101 |
| rs174538 | miR-1908-! | cis | 1.35E-07 | Nightingale | M_VLDL_C | NA | A | G | -0.04184 | 0.004424 | 1.00E-21 |
| rs174538 | miR-1908-! | cis | 1.35E-07 | Nightingale | L_LDL_C_p | NA | A | G | -0.07217 | 0.004322 | 1.50E-63 |
| rs174538 | miR-1908-! | cis | 1.35E-07 | Nightingale | Tyr | NA | A | G | 0.033362 | 0.004431 | 5.10E-14 |
| rs174538 | miR-1908-! | cis | 1.35E-07 | Nightingale | IDL_FC | NA | A | G | -0.08474 | 0.004294 | 1.20E-90 |
| rs174538 | miR-1908-! | cis | 1.35E-07 | Nightingale | IDL_C_pct | NA | A | G | -0.07034 | 0.004331 | 5.50E-60 |
| rs174538 | miR-1908-! | cis | 1.35E-07 | Nightingale | S_VLDL_TG | NA | A | G | 0.037096 | 0.004305 | 2.10E-18 |
| rs174538 | miR-1908-! | cis | 1.35E-07 | Nightingale | L_HDL_P | NA | A | G | -0.07255 | 0.003886 | 4.50E-85 |
| rs174538 | miR-1908-! | cis | 1.35E-07 | Nightingale | LDL_P | NA | A | G | -0.038 | 0.004433 | 2.30E-18 |
| rs174538 | miR-1908-! | cis | 1.35E-07 | Nightingale | M_LDL_TG | NA | A | G | 0.071789 | 0.004381 | 1.40E-60 |
| rs174538 | miR-1908-! | cis | 1.35E-07 | Nightingale | LDL_PL | NA | A | G | -0.04312 | 0.004426 | 4.50E-23 |
| rs174538 | miR-1908-! | cis | 1.35E-07 | Nightingale | XL_HDL_C_ | NA | A | G | 0.023009 | 0.004238 | 3.20E-09 |
| rs174538 | miR-1908-! | cis | 1.35E-07 | Nightingale | L_VLDL_FC | NA | A | G | 0.037727 | 0.004291 | 2.00E-19 |
| rs174538 | miR-1908-! | cis | 1.35E-07 | Nightingale | S_LDL_FC_ | NA | A | G | -0.07477 | 0.004299 | 6.10E-70 |
| rs174538 | miR-1908-! | cis | 1.35E-07 | Nightingale | LA_pct | NA | A | G | 0.174397 | 0.004336 | 0 |
| rs174538 | miR-1908-! | cis | 1.35E-07 | Nightingale | XL_HDL_TC | NA | A | G | 0.081771 | 0.004179 | 3.00E-90 |
| rs174538 | miR-1908-! | cis | 1.35E-07 | Nightingale | XL_HDL_CE | NA | A | G | -0.09128 | 0.003973 | 5.50E-127 |

| | | | | | | | | | | | |
|---|---|---|---|---|---|---|---|---|---|---|---|
| rs174538 | miR-1908-5 | cis | 1.35E-07 | Nightingale | HDL_size | NA | A | G | -0.0678 | 0.003931 | 1.50E-72 |
| rs174538 | miR-1908-5 | cis | 1.35E-07 | Nightingale | L_LDL_P | NA | A | G | -0.03551 | 0.004421 | 1.70E-16 |
| rs174538 | miR-1908-5 | cis | 1.35E-07 | Nightingale | LDL_FC | NA | A | G | -0.06023 | 0.004389 | 1.60E-44 |
| rs174538 | miR-1908-5 | cis | 1.35E-07 | Nightingale | IDL_PL | NA | A | G | -0.0692 | 0.00428 | 1.50E-62 |
| rs174538 | miR-1908-5 | cis | 1.35E-07 | Nightingale | Clinical_LD | NA | A | G | -0.0517 | 0.004411 | 9.70E-33 |
| rs174538 | miR-1908-5 | cis | 1.35E-07 | Nightingale | HDL_CE | NA | A | G | -0.05638 | 0.003981 | 3.30E-49 |
| rs174538 | miR-1908-5 | cis | 1.35E-07 | Nightingale | S_VLDL_CE | NA | A | G | -0.02946 | 0.004377 | 1.70E-11 |
| rs174538 | miR-1908-5 | cis | 1.35E-07 | Nightingale | XXL_VLDL_ | NA | A | G | 0.064019 | 0.004274 | 2.40E-53 |
| rs174538 | miR-1908-5 | cis | 1.35E-07 | Nightingale | HDL_P | NA | A | G | -0.0318 | 0.004166 | 3.00E-15 |
| rs174538 | miR-1908-5 | cis | 1.35E-07 | Nightingale | L_VLDL_L | NA | A | G | 0.036087 | 0.004287 | 7.00E-18 |
| rs174538 | miR-1908-5 | cis | 1.35E-07 | Nightingale | L_LDL_TG_ | NA | A | G | 0.073401 | 0.00435 | 3.20E-64 |
| rs174538 | miR-1908-5 | cis | 1.35E-07 | Nightingale | SFA | NA | A | G | -0.02887 | 0.004396 | 2.60E-11 |
| rs174538 | miR-1908-5 | cis | 1.35E-07 | Nightingale | XS_VLDL_C | NA | A | G | -0.09333 | 0.004204 | 5.30E-113 |
| rs174538 | miR-1908-5 | cis | 1.35E-07 | Nightingale | XS_VLDL_L | NA | A | G | -0.0404 | 0.004361 | 4.10E-22 |
| rs174538 | miR-1908-5 | cis | 1.35E-07 | Nightingale | L_LDL_CE | NA | A | G | -0.0497 | 0.004397 | 7.30E-31 |
| rs174538 | miR-1908-5 | cis | 1.35E-07 | Nightingale | MUFA_pct | NA | A | G | 0.184639 | 0.004246 | 0 |
| rs174538 | miR-1908-5 | cis | 1.35E-07 | Nightingale | Total_TG | NA | A | G | 0.044148 | 0.004294 | 1.00E-25 |
| rs174538 | miR-1908-5 | cis | 1.35E-07 | Nightingale | S_LDL_FC | NA | A | G | -0.05861 | 0.004429 | 9.00E-42 |
| rs174538 | miR-1908-5 | cis | 1.35E-07 | Nightingale | XXL_VLDL_ | NA | A | G | 0.06914 | 0.004284 | 7.60E-61 |
| rs174538 | miR-1908-5 | cis | 1.35E-07 | Nightingale | L_HDL_C | NA | A | G | -0.07486 | 0.003907 | 3.20E-89 |
| rs174538 | miR-1908-5 | cis | 1.35E-07 | Nightingale | XS_VLDL_P | NA | A | G | -0.0337 | 0.004374 | 6.10E-16 |
| rs174538 | miR-1908-5 | cis | 1.35E-07 | Nightingale | L_HDL_FC_ | NA | A | G | -0.10645 | 0.004121 | 1.90E-160 |
| rs174538 | miR-1908-5 | cis | 1.35E-07 | Nightingale | IDL_P | NA | A | G | -0.04301 | 0.00439 | 1.30E-23 |
| rs174538 | miR-1908-5 | cis | 1.35E-07 | Nightingale | IDL_FC_pct | NA | A | G | -0.09729 | 0.004401 | 2.80E-109 |
| rs174538 | miR-1908-5 | cis | 1.35E-07 | Nightingale | XL_VLDL_C | NA | A | G | -0.06145 | 0.004251 | 3.70E-48 |
| rs174538 | miR-1908-5 | cis | 1.35E-07 | Nightingale | L_HDL_FC | NA | A | G | -0.07914 | 0.003888 | 2.20E-100 |
| rs174538 | miR-1908-5 | cis | 1.35E-07 | Nightingale | S_LDL_P | NA | A | G | -0.03044 | 0.004441 | 2.70E-12 |
| rs174538 | miR-1908-5 | cis | 1.35E-07 | Nightingale | L_VLDL_PL | NA | A | G | 0.08155 | 0.004306 | 7.50E-83 |
| rs174538 | miR-1908-5 | cis | 1.35E-07 | Nightingale | XS_VLDL_T | NA | A | G | 0.093089 | 0.004214 | 3.20E-112 |
| rs174538 | miR-1908-5 | cis | 1.35E-07 | Nightingale | M_VLDL_T | NA | A | G | 0.030107 | 0.00431 | 1.00E-12 |
| rs174538 | miR-1908-5 | cis | 1.35E-07 | Nightingale | XL_HDL_FC | NA | A | G | -0.09743 | 0.004133 | 1.30E-132 |
| rs174538 | miR-1908-5 | cis | 1.35E-07 | Nightingale | XL_VLDL_F | NA | A | G | 0.040607 | 0.004297 | 4.50E-22 |
| rs174538 | miR-1908-5 | cis | 1.35E-07 | Nightingale | M_HDL_C | NA | A | G | -0.03685 | 0.004082 | 8.80E-21 |
| rs174538 | miR-1908-5 | cis | 1.35E-07 | Nightingale | XS_VLDL_C | NA | A | G | -0.08775 | 0.004182 | 4.00E-101 |
| rs174538 | miR-1908-5 | cis | 1.35E-07 | Nightingale | XL_HDL_L | NA | A | G | -0.08776 | 0.003987 | 5.00E-117 |
| rs174538 | miR-1908-5 | cis | 1.35E-07 | Nightingale | M_HDL_FC | NA | A | G | -0.096 | 0.003931 | 1.70E-143 |
| rs174538 | miR-1908-5 | cis | 1.35E-07 | Nightingale | L_LDL_FC | NA | A | G | -0.06422 | 0.004356 | 2.60E-51 |
| rs174538 | miR-1908-5 | cis | 1.35E-07 | Nightingale | L_LDL_L | NA | A | G | -0.05065 | 0.004393 | 4.70E-32 |
| rs174538 | miR-1908-5 | cis | 1.35E-07 | Nightingale | XL_VLDL_T | NA | A | G | 0.057682 | 0.004298 | 5.70E-42 |
| rs174538 | miR-1908-5 | cis | 1.35E-07 | Nightingale | TG_by_PG | NA | A | G | 0.064335 | 0.004117 | 9.70E-58 |
| rs174538 | miR-1908-5 | cis | 1.35E-07 | Nightingale | non_HDL_( | NA | A | G | -0.045 | 0.004413 | 2.90E-25 |
| rs174538 | miR-1908-5 | cis | 1.35E-07 | Nightingale | XL_HDL_FC | NA | A | G | 0.05703 | 0.003933 | 4.50E-53 |
| rs174538 | miR-1908-5 | cis | 1.35E-07 | Nightingale | S_LDL_C | NA | A | G | -0.02978 | 0.004441 | 9.20E-12 |
| rs174538 | miR-1908-5 | cis | 1.35E-07 | Nightingale | XS_VLDL_F | NA | A | G | -0.05413 | 0.004359 | 1.10E-37 |
| rs174538 | miR-1908-5 | cis | 1.35E-07 | Nightingale | M_VLDL_F | NA | A | G | -0.02804 | 0.004436 | 1.20E-10 |
| rs174538 | miR-1908-5 | cis | 1.35E-07 | Nightingale | Omega_3 | NA | A | G | -0.31438 | 0.004348 | 0 |
| rs174538 | miR-1908-5 | cis | 1.35E-07 | Nightingale | S_VLDL_FC | NA | A | G | -0.02691 | 0.004437 | 6.70E-10 |
| rs174538 | miR-1908-5 | cis | 1.35E-07 | Nightingale | XXL_VLDL_ | NA | A | G | 0.067401 | 0.004275 | 1.90E-58 |
| rs174538 | miR-1908-5 | cis | 1.35E-07 | Nightingale | Sphingomy | NA | A | G | -0.10796 | 0.004146 | 1.80E-156 |
| rs174538 | miR-1908-5 | cis | 1.35E-07 | Nightingale | LDL_L | NA | A | G | -0.04234 | 0.004422 | 1.70E-22 |
| rs174538 | miR-1908-5 | cis | 1.35E-07 | Nightingale | Unsaturati | NA | A | G | -0.31741 | 0.004202 | 0 |
| rs174538 | miR-1908-5 | cis | 1.35E-07 | Nightingale | Total_C | NA | A | G | -0.06092 | 0.004295 | 1.10E-47 |
| rs174538 | miR-1908-5 | cis | 1.35E-07 | Nightingale | L_HDL_L | NA | A | G | -0.07186 | 0.003893 | 2.80E-83 |
| rs174538 | miR-1908-5 | cis | 1.35E-07 | Nightingale | Total_P | NA | A | G | -0.03639 | 0.004175 | 1.80E-19 |
| rs174538 | miR-1908-5 | cis | 1.35E-07 | Nightingale | VLDL_TG | NA | A | G | 0.045154 | 0.004269 | 3.80E-27 |
| rs174538 | miR-1908-5 | cis | 1.35E-07 | Nightingale | S_VLDL_FC | NA | A | G | -0.07906 | 0.004236 | 1.30E-80 |
| rs174538 | miR-1908-5 | cis | 1.35E-07 | Nightingale | XS_VLDL_P | NA | A | G | 0.053641 | 0.004329 | 3.20E-35 |
| rs174544 | miR-1908-5 | cis | 2.30E-08 | Nightingale | XXL_VLDL_ | NA | A | C | 0.072279 | 0.004308 | 4.60E-66 |
| rs174544 | miR-1908-5 | cis | 2.30E-08 | Nightingale | S_VLDL_FC | NA | A | C | -0.08186 | 0.004269 | 2.30E-85 |
| rs174544 | miR-1908-5 | cis | 2.30E-08 | Nightingale | S_VLDL_TG | NA | A | C | 0.0395 | 0.004338 | 1.00E-20 |
| rs174544 | miR-1908-5 | cis | 2.30E-08 | Nightingale | IDL_PL | NA | A | C | -0.06975 | 0.004313 | 6.00E-63 |
| rs174544 | miR-1908-5 | cis | 2.30E-08 | Nightingale | S_LDL_PL_ | NA | A | C | -0.06149 | 0.004462 | 3.80E-45 |
| rs174544 | miR-1908-5 | cis | 2.30E-08 | Nightingale | S_VLDL_C_ | NA | A | C | -0.05601 | 0.00435 | 5.10E-39 |

| | | | | | | | | | | | |
|---|---|---|---|---|---|---|---|---|---|---|---|
| rs174544 | miR-1908-5 | cis | 2.30E-08 | Nightingale | Total_P | NA | A | C | -0.03895 | 0.004207 | 1.10E-21 |
| rs174544 | miR-1908-5 | cis | 2.30E-08 | Nightingale | XS_VLDL_L | NA | A | C | -0.03829 | 0.004394 | 7.20E-20 |
| rs174544 | miR-1908-5 | cis | 2.30E-08 | Nightingale | XS_VLDL_C | NA | A | C | -0.06855 | 0.004314 | 1.60E-60 |
| rs174544 | miR-1908-5 | cis | 2.30E-08 | Nightingale | HDL_P | NA | A | C | -0.03445 | 0.004198 | 2.70E-17 |
| rs174544 | miR-1908-5 | cis | 2.30E-08 | Nightingale | XS_VLDL_T | NA | A | C | 0.033228 | 0.004402 | 9.80E-14 |
| rs174544 | miR-1908-5 | cis | 2.30E-08 | Nightingale | L_VLDL_P | NA | A | C | 0.042991 | 0.004317 | 2.00E-24 |
| rs174544 | miR-1908-5 | cis | 2.30E-08 | Nightingale | M_HDL_CE | NA | A | C | -0.03712 | 0.004129 | 2.10E-20 |
| rs174544 | miR-1908-5 | cis | 2.30E-08 | Nightingale | L_HDL_FC | NA | A | C | -0.08204 | 0.003918 | 1.10E-105 |
| rs174544 | miR-1908-5 | cis | 2.30E-08 | Nightingale | Total_CE | NA | A | C | -0.06468 | 0.004312 | 1.80E-53 |
| rs174544 | miR-1908-5 | cis | 2.30E-08 | Nightingale | M_HDL_FC | NA | A | C | -0.04949 | 0.004068 | 1.90E-36 |
| rs174544 | miR-1908-5 | cis | 2.30E-08 | Nightingale | VLDL_TG | NA | A | C | 0.049003 | 0.004302 | 2.20E-31 |
| rs174544 | miR-1908-5 | cis | 2.30E-08 | Nightingale | M_LDL_FC | NA | A | C | -0.05146 | 0.004465 | 4.30E-32 |
| rs174544 | miR-1908-5 | cis | 2.30E-08 | Nightingale | Sphingomy | NA | A | C | -0.1121 | 0.004178 | 4.60E-166 |
| rs174544 | miR-1908-5 | cis | 2.30E-08 | Nightingale | L_HDL_C | NA | A | C | -0.07825 | 0.003937 | 2.20E-95 |
| rs174544 | miR-1908-5 | cis | 2.30E-08 | Nightingale | L_LDL_C_p | NA | A | C | -0.07629 | 0.004356 | 2.70E-70 |
| rs174544 | miR-1908-5 | cis | 2.30E-08 | Nightingale | L_HDL_TG_ | NA | A | C | 0.078507 | 0.004219 | 7.10E-82 |
| rs174544 | miR-1908-5 | cis | 2.30E-08 | Nightingale | XS_VLDL_P | NA | A | C | -0.04053 | 0.004411 | 1.20E-21 |
| rs174544 | miR-1908-5 | cis | 2.30E-08 | Nightingale | XL_VLDL_C | NA | A | C | -0.06476 | 0.004284 | 4.90E-53 |
| rs174544 | miR-1908-5 | cis | 2.30E-08 | Nightingale | XL_VLDL_C | NA | A | C | 0.031461 | 0.0044 | 3.40E-13 |
| rs174544 | miR-1908-5 | cis | 2.30E-08 | Nightingale | M_VLDL_P | NA | A | C | -0.06091 | 0.00429 | 7.40E-49 |
| rs174544 | miR-1908-5 | cis | 2.30E-08 | Nightingale | HDL_C | NA | A | C | -0.06327 | 0.003992 | 5.20E-61 |
| rs174544 | miR-1908-5 | cis | 2.30E-08 | Nightingale | IDL_TG_pct | NA | A | C | 0.087356 | 0.004319 | 2.60E-94 |
| rs174544 | miR-1908-5 | cis | 2.30E-08 | Nightingale | S_LDL_C | NA | A | C | -0.03094 | 0.004475 | 1.70E-12 |
| rs174544 | miR-1908-5 | cis | 2.30E-08 | Nightingale | LDL_P | NA | A | C | -0.0386 | 0.004468 | 1.00E-18 |
| rs174544 | miR-1908-5 | cis | 2.30E-08 | Nightingale | XXL_VLDL_ | NA | A | C | 0.067198 | 0.0043 | 6.00E-58 |
| rs174544 | miR-1908-5 | cis | 2.30E-08 | Nightingale | S_LDL_TG | NA | A | C | 0.05172 | 0.004361 | 2.10E-33 |
| rs174544 | miR-1908-5 | cis | 2.30E-08 | Nightingale | HDL_L | NA | A | C | -0.05136 | 0.004014 | 2.90E-40 |
| rs174544 | miR-1908-5 | cis | 2.30E-08 | Nightingale | M_VLDL_C | NA | A | C | -0.05102 | 0.00443 | 5.80E-32 |
| rs174544 | miR-1908-5 | cis | 2.30E-08 | Nightingale | S_HDL_FC_ | NA | A | C | -0.10003 | 0.00424 | 6.61E-136 |
| rs174544 | miR-1908-5 | cis | 2.30E-08 | Nightingale | S_LDL_FC | NA | A | C | -0.06169 | 0.004463 | 1.10E-45 |
| rs174544 | miR-1908-5 | cis | 2.30E-08 | Nightingale | S_HDL_TG_ | NA | A | C | 0.069217 | 0.004252 | 2.40E-62 |
| rs174544 | miR-1908-5 | cis | 2.30E-08 | Nightingale | XL_VLDL_P | NA | A | C | 0.049552 | 0.004314 | 7.10E-32 |
| rs174544 | miR-1908-5 | cis | 2.30E-08 | Nightingale | XL_VLDL_C | NA | A | C | 0.038742 | 0.004366 | 1.30E-19 |
| rs174544 | miR-1908-5 | cis | 2.30E-08 | Nightingale | XL_HDL_CE | NA | A | C | -0.02838 | 0.004468 | 9.20E-10 |
| rs174544 | miR-1908-5 | cis | 2.30E-08 | Nightingale | M_HDL_PL | NA | A | C | 0.059912 | 0.004241 | 7.90E-47 |
| rs174544 | miR-1908-5 | cis | 2.30E-08 | Nightingale | XL_HDL_C | NA | A | C | -0.09664 | 0.004031 | 1.60E-137 |
| rs174544 | miR-1908-5 | cis | 2.30E-08 | Nightingale | XS_VLDL_P | NA | A | C | 0.058185 | 0.004362 | 2.70E-41 |
| rs174544 | miR-1908-5 | cis | 2.30E-08 | Nightingale | IDL_CE_pct | NA | A | C | -0.03761 | 0.004387 | 1.80E-17 |
| rs174544 | miR-1908-5 | cis | 2.30E-08 | Nightingale | M_LDL_TG | NA | A | C | 0.076026 | 0.004414 | 1.00E-67 |
| rs174544 | miR-1908-5 | cis | 2.30E-08 | Nightingale | L_HDL_CE | NA | A | C | -0.07684 | 0.003947 | 2.50E-91 |
| rs174544 | miR-1908-5 | cis | 2.30E-08 | Nightingale | M_VLDL_F | NA | A | C | -0.02738 | 0.00447 | 3.80E-10 |
| rs174544 | miR-1908-5 | cis | 2.30E-08 | Nightingale | XL_HDL_PL | NA | A | C | -0.08626 | 0.004019 | 1.70E-111 |
| rs174544 | miR-1908-5 | cis | 2.30E-08 | Nightingale | S_VLDL_CE | NA | A | C | -0.02884 | 0.004411 | 2.30E-11 |
| rs174544 | miR-1908-5 | cis | 2.30E-08 | Nightingale | Total_TG | NA | A | C | 0.048398 | 0.004327 | 2.20E-30 |
| rs174544 | miR-1908-5 | cis | 2.30E-08 | Nightingale | M_VLDL_T | NA | A | C | 0.076974 | 0.004222 | 1.70E-77 |
| rs174544 | miR-1908-5 | cis | 2.30E-08 | Nightingale | S_HDL_C_p | NA | A | C | -0.04502 | 0.004419 | 8.00E-24 |
| rs174544 | miR-1908-5 | cis | 2.30E-08 | Nightingale | L_HDL_CE_ | NA | A | C | -0.05139 | 0.004277 | 3.00E-35 |
| rs174544 | miR-1908-5 | cis | 2.30E-08 | Nightingale | IDL_L | NA | A | C | -0.06946 | 0.004322 | 5.90E-62 |
| rs174544 | miR-1908-5 | cis | 2.30E-08 | Nightingale | S_LDL_FC_ | NA | A | C | -0.08097 | 0.004332 | 1.00E-80 |
| rs174544 | miR-1908-5 | cis | 2.30E-08 | Nightingale | L_LDL_FC | NA | A | C | -0.06657 | 0.00439 | 1.10E-54 |
| rs174544 | miR-1908-5 | cis | 2.30E-08 | Nightingale | XL_VLDL_T | NA | A | C | 0.05811 | 0.004276 | 5.50E-44 |
| rs174544 | miR-1908-5 | cis | 2.30E-08 | Nightingale | XL_VLDL_L | NA | A | C | 0.052683 | 0.004301 | 5.00E-36 |
| rs174544 | miR-1908-5 | cis | 2.30E-08 | Nightingale | SFA_pct | NA | A | C | -0.03913 | 0.004411 | 7.30E-19 |
| rs174544 | miR-1908-5 | cis | 2.30E-08 | Nightingale | IDL_FC_pct | NA | A | C | -0.10428 | 0.004435 | 1.90E-123 |
| rs174544 | miR-1908-5 | cis | 2.30E-08 | Nightingale | M_VLDL_C | NA | A | C | -0.07546 | 0.004197 | 6.20E-75 |
| rs174544 | miR-1908-5 | cis | 2.30E-08 | Nightingale | M_HDL_P | NA | A | C | -0.03287 | 0.004118 | 9.30E-17 |
| rs174544 | miR-1908-5 | cis | 2.30E-08 | Nightingale | M_HDL_TG | NA | A | C | 0.067505 | 0.004287 | 2.60E-58 |
| rs174544 | miR-1908-5 | cis | 2.30E-08 | Nightingale | XXL_VLDL_ | NA | A | C | 0.042944 | 0.004493 | 2.90E-22 |
| rs174544 | miR-1908-5 | cis | 2.30E-08 | Nightingale | HDL_TG | NA | A | C | 0.04792 | 0.004408 | 6.30E-28 |
| rs174544 | miR-1908-5 | cis | 2.30E-08 | Nightingale | L_LDL_C | NA | A | C | -0.05604 | 0.00442 | 1.10E-38 |
| rs174544 | miR-1908-5 | cis | 2.30E-08 | Nightingale | Total_L | NA | A | C | -0.03447 | 0.004398 | 5.40E-16 |
| rs174544 | miR-1908-5 | cis | 2.30E-08 | Nightingale | M_HDL_L | NA | A | C | -0.02621 | 0.00415 | 4.70E-11 |
| rs174544 | miR-1908-5 | cis | 2.30E-08 | Nightingale | LDL_FC | NA | A | C | -0.06281 | 0.004423 | 6.00E-48 |

| | | | | | | | | | | | |
|---|---|---|---|---|---|---|---|---|---|---|---|
| rs174544 | miR-1908-5 | cis | 2.30E-08 | Nightingale | Total_PL | NA | A | C | -0.04798 | 0.004257 | 2.40E-31 |
| rs174544 | miR-1908-5 | cis | 2.30E-08 | Nightingale | ApoB | NA | A | C | -0.03652 | 0.004465 | 5.00E-17 |
| rs174544 | miR-1908-5 | cis | 2.30E-08 | Nightingale | LA | NA | A | C | 0.083574 | 0.004384 | 2.80E-81 |
| rs174544 | miR-1908-5 | cis | 2.30E-08 | Nightingale | LDL_PL | NA | A | C | -0.04469 | 0.004461 | 1.60E-24 |
| rs174544 | miR-1908-5 | cis | 2.30E-08 | Nightingale | VLDL_L | NA | A | C | 0.029114 | 0.004383 | 1.30E-11 |
| rs174544 | miR-1908-5 | cis | 2.30E-08 | Nightingale | XXL_VLDL_ | NA | A | C | 0.072907 | 0.00429 | 4.80E-68 |
| rs174544 | miR-1908-5 | cis | 2.30E-08 | Nightingale | S_HDL_TG | NA | A | C | 0.069078 | 0.004335 | 3.00E-60 |
| rs174544 | miR-1908-5 | cis | 2.30E-08 | Nightingale | L_VLDL_C_ | NA | A | C | -0.03907 | 0.004367 | 1.80E-20 |
| rs174544 | miR-1908-5 | cis | 2.30E-08 | Nightingale | XL_VLDL_P | NA | A | C | 0.053915 | 0.004297 | 9.90E-38 |
| rs174544 | miR-1908-5 | cis | 2.30E-08 | Nightingale | L_VLDL_FC | NA | A | C | 0.027353 | 0.004428 | 6.20E-10 |
| rs174544 | miR-1908-5 | cis | 2.30E-08 | Nightingale | HDL_PL | NA | A | C | -0.04617 | 0.004042 | 1.80E-32 |
| rs174544 | miR-1908-5 | cis | 2.30E-08 | Nightingale | S_LDL_TG_ | NA | A | C | 0.086475 | 0.004332 | 1.20E-91 |
| rs174544 | miR-1908-5 | cis | 2.30E-08 | Nightingale | DHA | NA | A | C | -0.28195 | 0.004308 | 0 |
| rs174544 | miR-1908-5 | cis | 2.30E-08 | Nightingale | L_HDL_L | NA | A | C | -0.07492 | 0.003923 | 1.40E-88 |
| rs174544 | miR-1908-5 | cis | 2.30E-08 | Nightingale | L_VLDL_PL | NA | A | C | 0.087611 | 0.004339 | 7.90E-94 |
| rs174544 | miR-1908-5 | cis | 2.30E-08 | Nightingale | L_VLDL_C | NA | A | C | 0.030071 | 0.004373 | 2.20E-12 |
| rs174544 | miR-1908-5 | cis | 2.30E-08 | Nightingale | Omega_6_ | NA | A | C | 0.361721 | 0.004406 | 0 |
| rs174544 | miR-1908-5 | cis | 2.30E-08 | Nightingale | LDL_CE | NA | A | C | -0.04195 | 0.004459 | 6.30E-22 |
| rs174544 | miR-1908-5 | cis | 2.30E-08 | Nightingale | L_HDL_C_p | NA | A | C | -0.06852 | 0.004214 | 1.80E-63 |
| rs174544 | miR-1908-5 | cis | 2.30E-08 | Nightingale | IDL_P | NA | A | C | -0.0425 | 0.004424 | 4.80E-23 |
| rs174544 | miR-1908-5 | cis | 2.30E-08 | Nightingale | HDL_size | NA | A | C | -0.07024 | 0.003961 | 2.40E-76 |
| rs174544 | miR-1908-5 | cis | 2.30E-08 | Nightingale | Phosphogly | NA | A | C | -0.03807 | 0.004235 | 8.40E-21 |
| rs174544 | miR-1908-5 | cis | 2.30E-08 | Nightingale | ApoA1 | NA | A | C | -0.03951 | 0.004091 | 8.60E-24 |
| rs174544 | miR-1908-5 | cis | 2.30E-08 | Nightingale | LDL_TG | NA | A | C | 0.031647 | 0.004421 | 9.70E-13 |
| rs174544 | miR-1908-5 | cis | 2.30E-08 | Nightingale | XL_HDL_P | NA | A | C | -0.08686 | 0.004023 | 8.00E-113 |
| rs174544 | miR-1908-5 | cis | 2.30E-08 | Nightingale | non_HDL_( | NA | A | C | -0.04566 | 0.004447 | 6.20E-26 |
| rs174544 | miR-1908-5 | cis | 2.30E-08 | Nightingale | IDL_C_pct | NA | A | C | -0.07446 | 0.004364 | 1.00E-66 |
| rs174544 | miR-1908-5 | cis | 2.30E-08 | Nightingale | IDL_FC | NA | A | C | -0.08692 | 0.004327 | 1.40E-94 |
| rs174544 | miR-1908-5 | cis | 2.30E-08 | Nightingale | XS_VLDL_T | NA | A | C | 0.095587 | 0.004246 | 5.11E-118 |
| rs174544 | miR-1908-5 | cis | 2.30E-08 | Nightingale | S_LDL_L | NA | A | C | -0.03106 | 0.00447 | 1.20E-12 |
| rs174544 | miR-1908-5 | cis | 2.30E-08 | Nightingale | M_HDL_C_ | NA | A | C | -0.06695 | 0.004256 | 1.60E-58 |
| rs174544 | miR-1908-5 | cis | 2.30E-08 | Nightingale | M_HDL_CE | NA | A | C | -0.04938 | 0.004335 | 8.30E-31 |
| rs174544 | miR-1908-5 | cis | 2.30E-08 | Nightingale | XL_VLDL_C | NA | A | C | -0.06201 | 0.004286 | 9.80E-49 |
| rs174544 | miR-1908-5 | cis | 2.30E-08 | Nightingale | HDL_FC | NA | A | C | -0.07132 | 0.00396 | 2.70E-78 |
| rs174544 | miR-1908-5 | cis | 2.30E-08 | Nightingale | DHA_pct | NA | A | C | -0.26891 | 0.00434 | 0 |
| rs174544 | miR-1908-5 | cis | 2.30E-08 | Nightingale | LDL_L | NA | A | C | -0.04383 | 0.004456 | 7.50E-24 |
| rs174544 | miR-1908-5 | cis | 2.30E-08 | Nightingale | PUFA_by_M | NA | A | C | -0.16798 | 0.004282 | 0 |
| rs174544 | miR-1908-5 | cis | 2.30E-08 | Nightingale | XXL_VLDL_ | NA | A | C | 0.062494 | 0.004314 | 8.70E-50 |
| rs174544 | miR-1908-5 | cis | 2.30E-08 | Nightingale | LDL_C | NA | A | C | -0.04817 | 0.004452 | 1.70E-28 |
| rs174544 | miR-1908-5 | cis | 2.30E-08 | Nightingale | Citrate | NA | A | C | -0.03381 | 0.004442 | 2.80E-15 |
| rs174544 | miR-1908-5 | cis | 2.30E-08 | Nightingale | L_LDL_FC_ | NA | A | C | -0.08282 | 0.004261 | 4.50E-87 |
| rs174544 | miR-1908-5 | cis | 2.30E-08 | Nightingale | M_VLDL_C | NA | A | C | -0.04214 | 0.004458 | 5.10E-22 |
| rs174544 | miR-1908-5 | cis | 2.30E-08 | Nightingale | S_VLDL_PL | NA | A | C | -0.07785 | 0.004278 | 2.80E-77 |
| rs174544 | miR-1908-5 | cis | 2.30E-08 | Nightingale | L_HDL_PL_ | NA | A | C | 0.043686 | 0.004327 | 2.70E-25 |
| rs174544 | miR-1908-5 | cis | 2.30E-08 | Nightingale | Total_FC | NA | A | C | -0.05683 | 0.004367 | 5.60E-41 |
| rs174544 | miR-1908-5 | cis | 2.30E-08 | Nightingale | LA_pct | NA | A | C | 0.180125 | 0.004369 | 0 |
| rs174544 | miR-1908-5 | cis | 2.30E-08 | Nightingale | M_LDL_P | NA | A | C | -0.04484 | 0.004484 | 3.80E-24 |
| rs174544 | miR-1908-5 | cis | 2.30E-08 | Nightingale | L_LDL_P | NA | A | C | -0.03603 | 0.004455 | 8.50E-17 |
| rs174544 | miR-1908-5 | cis | 2.30E-08 | Nightingale | PUFA | NA | A | C | -0.09242 | 0.00436 | 8.49E-106 |
| rs174544 | miR-1908-5 | cis | 2.30E-08 | Nightingale | PUFA_pct | NA | A | C | -0.12052 | 0.0043 | 3.70E-174 |
| rs174544 | miR-1908-5 | cis | 2.30E-08 | Nightingale | Omega_3_ | NA | A | C | -0.38182 | 0.004368 | 0 |
| rs174544 | miR-1908-5 | cis | 2.30E-08 | Nightingale | IDL_C | NA | A | C | -0.07449 | 0.004317 | 1.40E-70 |
| rs174544 | miR-1908-5 | cis | 2.30E-08 | Nightingale | M_LDL_FC_ | NA | A | C | -0.07263 | 0.004283 | 5.90E-67 |
| rs174544 | miR-1908-5 | cis | 2.30E-08 | Nightingale | VLDL_size | NA | A | C | 0.072456 | 0.004167 | 7.60E-72 |
| rs174544 | miR-1908-5 | cis | 2.30E-08 | Nightingale | S_VLDL_TG | NA | A | C | 0.063861 | 0.004326 | 1.10E-50 |
| rs174544 | miR-1908-5 | cis | 2.30E-08 | Nightingale | Total_C | NA | A | C | -0.06279 | 0.004328 | 3.40E-50 |
| rs174544 | miR-1908-5 | cis | 2.30E-08 | Nightingale | M_HDL_C | NA | A | C | -0.03981 | 0.004113 | 1.80E-23 |
| rs174544 | miR-1908-5 | cis | 2.30E-08 | Nightingale | MUFA_pct | NA | A | C | 0.193755 | 0.004278 | 0 |
| rs174544 | miR-1908-5 | cis | 2.30E-08 | Nightingale | L_LDL_PL | NA | A | C | -0.05326 | 0.004437 | 1.30E-34 |
| rs174544 | miR-1908-5 | cis | 2.30E-08 | Nightingale | Tyr | NA | A | C | 0.032729 | 0.004465 | 2.30E-13 |
| rs174544 | miR-1908-5 | cis | 2.30E-08 | Nightingale | L_VLDL_FC | NA | A | C | 0.041208 | 0.004324 | 1.60E-22 |
| rs174544 | miR-1908-5 | cis | 2.30E-08 | Nightingale | M_LDL_CE | NA | A | C | -0.02506 | 0.004477 | 1.40E-08 |
| rs174544 | miR-1908-5 | cis | 2.30E-08 | Nightingale | XXL_VLDL_ | NA | A | C | 0.067636 | 0.004306 | 2.40E-58 |

| | | | | | | | | | | | | |
|---|---|---|---|---|---|---|---|---|---|---|---|---|
| rs174544 | miR-1908-5 | cis | | 2.30E-08 | Nightingale | L_VLDL_PL | NA | A | C | 0.049711 | 0.004313 | 3.40E-32 |
| rs174544 | miR-1908-5 | cis | | 2.30E-08 | Nightingale | L_VLDL_TG | NA | A | C | 0.038632 | 0.004302 | 3.80E-20 |
| rs174544 | miR-1908-5 | cis | | 2.30E-08 | Nightingale | IDL_CE | NA | A | C | -0.06952 | 0.004315 | 9.00E-62 |
| rs174544 | miR-1908-5 | cis | | 2.30E-08 | Nightingale | L_HDL_FC_ | NA | A | C | -0.10815 | 0.004153 | 1.00E-161 |
| rs174544 | miR-1908-5 | cis | | 2.30E-08 | Nightingale | XXL_VLDL_ | NA | A | C | 0.068838 | 0.004307 | 1.60E-60 |
| rs174544 | miR-1908-5 | cis | | 2.30E-08 | Nightingale | Remnant_( | NA | A | C | -0.04147 | 0.004442 | 6.40E-22 |
| rs174544 | miR-1908-5 | cis | | 2.30E-08 | Nightingale | L_LDL_TG_ | NA | A | C | 0.07763 | 0.004384 | 1.40E-71 |
| rs174544 | miR-1908-5 | cis | | 2.30E-08 | Nightingale | S_VLDL_FC | NA | A | C | -0.02666 | 0.004472 | 1.00E-09 |
| rs174544 | miR-1908-5 | cis | | 2.30E-08 | Nightingale | Clinical_LD | NA | A | C | -0.0535 | 0.004445 | 9.20E-35 |
| rs174544 | miR-1908-5 | cis | | 2.30E-08 | Nightingale | M_VLDL_C | NA | A | C | -0.07987 | 0.004208 | 1.80E-83 |
| rs174544 | miR-1908-5 | cis | | 2.30E-08 | Nightingale | M_LDL_C | NA | A | C | -0.03284 | 0.004479 | 9.50E-14 |
| rs174544 | miR-1908-5 | cis | | 2.30E-08 | Nightingale | L_HDL_PL | NA | A | C | -0.07365 | 0.003935 | 4.00E-85 |
| rs174544 | miR-1908-5 | cis | | 2.30E-08 | Nightingale | HDL_CE | NA | A | C | -0.06032 | 0.004012 | 6.20E-55 |
| rs174544 | miR-1908-5 | cis | | 2.30E-08 | Nightingale | XS_VLDL_P | NA | A | C | -0.03111 | 0.004408 | 9.50E-14 |
| rs174544 | miR-1908-5 | cis | | 2.30E-08 | Nightingale | XXL_VLDL_ | NA | A | C | 0.0739 | 0.004317 | 2.30E-68 |
| rs174544 | miR-1908-5 | cis | | 2.30E-08 | Nightingale | M_VLDL_T | NA | A | C | 0.032543 | 0.004343 | 1.90E-14 |
| rs174544 | miR-1908-5 | cis | | 2.30E-08 | Nightingale | XL_HDL_L | NA | A | C | -0.09027 | 0.004017 | 1.40E-121 |
| rs174544 | miR-1908-5 | cis | | 2.30E-08 | Nightingale | L_VLDL_CE | NA | A | C | -0.05914 | 0.004278 | 4.10E-46 |
| rs174544 | miR-1908-5 | cis | | 2.30E-08 | Nightingale | XL_HDL_FC | NA | A | C | -0.09929 | 0.004165 | 4.80E-135 |
| rs174544 | miR-1908-5 | cis | | 2.30E-08 | Nightingale | M_LDL_TG | NA | A | C | 0.038192 | 0.004405 | 2.20E-18 |
| rs174544 | miR-1908-5 | cis | | 2.30E-08 | Nightingale | M_HDL_TG | NA | A | C | 0.051991 | 0.004415 | 1.30E-32 |
| rs174544 | miR-1908-5 | cis | | 2.30E-08 | Nightingale | XL_HDL_FC | NA | A | C | 0.06016 | 0.003964 | 1.30E-57 |
| rs174544 | miR-1908-5 | cis | | 2.30E-08 | Nightingale | M_LDL_L | NA | A | C | -0.02701 | 0.004477 | 9.70E-10 |
| rs174544 | miR-1908-5 | cis | | 2.30E-08 | Nightingale | Cholines | NA | A | C | -0.03462 | 0.004205 | 6.50E-18 |
| rs174544 | miR-1908-5 | cis | | 2.30E-08 | Nightingale | XS_VLDL_F | NA | A | C | -0.1106 | 0.004437 | 6.70E-140 |
| rs174544 | miR-1908-5 | cis | | 2.30E-08 | Nightingale | XS_VLDL_C | NA | A | C | -0.06474 | 0.004339 | 1.20E-53 |
| rs174544 | miR-1908-5 | cis | | 2.30E-08 | Nightingale | L_LDL_CE | NA | A | C | -0.05153 | 0.004431 | 7.70E-33 |
| rs174544 | miR-1908-5 | cis | | 2.30E-08 | Nightingale | M_HDL_FC | NA | A | C | -0.10005 | 0.003961 | 1.00E-152 |
| rs174544 | miR-1908-5 | cis | | 2.30E-08 | Nightingale | XL_HDL_CE | NA | A | C | -0.09477 | 0.004003 | 4.00E-134 |
| rs174544 | miR-1908-5 | cis | | 2.30E-08 | Nightingale | XL_VLDL_F | NA | A | C | -0.06836 | 0.004332 | 1.50E-58 |
| rs174544 | miR-1908-5 | cis | | 2.30E-08 | Nightingale | S_LDL_PL | NA | A | C | -0.0498 | 0.00446 | 2.30E-30 |
| rs174544 | miR-1908-5 | cis | | 2.30E-08 | Nightingale | TG_by_PG | NA | A | C | 0.069046 | 0.004149 | 1.60E-65 |
| rs174544 | miR-1908-5 | cis | | 2.30E-08 | Nightingale | XXL_VLDL_ | NA | A | C | -0.02921 | 0.004446 | 8.00E-12 |
| rs174544 | miR-1908-5 | cis | | 2.30E-08 | Nightingale | XS_VLDL_C | NA | A | C | -0.0966 | 0.004236 | 1.60E-120 |
| rs174544 | miR-1908-5 | cis | | 2.30E-08 | Nightingale | XS_VLDL_C | NA | A | C | -0.09089 | 0.004214 | 3.80E-108 |
| rs174544 | miR-1908-5 | cis | | 2.30E-08 | Nightingale | M_LDL_C_ | NA | A | C | -0.09863 | 0.004474 | 7.19E-108 |
| rs174544 | miR-1908-5 | cis | | 2.30E-08 | Nightingale | S_LDL_CE_ | NA | A | C | 0.057424 | 0.004462 | 1.70E-39 |
| rs174544 | miR-1908-5 | cis | | 2.30E-08 | Nightingale | L_HDL_P | NA | A | C | -0.0757 | 0.003915 | 1.40E-90 |
| rs174544 | miR-1908-5 | cis | | 2.30E-08 | Nightingale | L_LDL_L | NA | A | C | -0.05236 | 0.004427 | 6.30E-34 |
| rs174544 | miR-1908-5 | cis | | 2.30E-08 | Nightingale | XL_HDL_C_ | NA | A | C | 0.021559 | 0.004271 | 2.20E-08 |
| rs174544 | miR-1908-5 | cis | | 2.30E-08 | Nightingale | XL_HDL_TG | NA | A | C | 0.086677 | 0.004211 | 7.60E-100 |
| rs174544 | miR-1908-5 | cis | | 2.30E-08 | Nightingale | XL_HDL_PL | NA | A | C | -0.06436 | 0.004107 | 3.40E-60 |
| rs174544 | miR-1908-5 | cis | | 2.30E-08 | Nightingale | S_LDL_P | NA | A | C | -0.03036 | 0.004475 | 4.10E-12 |
| rs174544 | miR-1908-5 | cis | | 2.30E-08 | Nightingale | M_LDL_PL_ | NA | A | C | 0.042685 | 0.004468 | 8.80E-23 |
| rs174544 | miR-1908-5 | cis | | 2.30E-08 | Nightingale | XS_VLDL_F | NA | A | C | -0.05262 | 0.004393 | 2.30E-35 |
| rs174544 | miR-1908-5 | cis | | 2.30E-08 | Nightingale | MUFA | NA | A | C | 0.06466 | 0.004425 | 8.00E-50 |
| rs174544 | miR-1908-5 | cis | | 2.30E-08 | Nightingale | XL_VLDL_F | NA | A | C | 0.044915 | 0.00433 | 2.80E-26 |
| rs174544 | miR-1908-5 | cis | | 2.30E-08 | Nightingale | L_VLDL_L | NA | A | C | 0.039417 | 0.00432 | 1.00E-20 |
| rs174544 | miR-1908-5 | cis | | 2.30E-08 | Nightingale | Omega_3 | NA | A | C | -0.32784 | 0.004382 | 0 |
| rs174544 | miR-1908-5 | cis | | 2.30E-08 | Nightingale | Unsaturati | NA | A | C | -0.33274 | 0.004234 | 0 |
| rs174544 | miR-1908-5 | cis | | 2.30E-08 | Nightingale | M_VLDL_F | NA | A | C | -0.08965 | 0.004251 | 1.20E-103 |
| rs174544 | miR-1908-5 | cis | | 2.30E-08 | Nightingale | SFA | NA | A | C | -0.02782 | 0.00443 | 2.30E-10 |
| rs174544 | miR-1908-5 | cis | | 2.30E-08 | Nightingale | XL_VLDL_T | NA | A | C | 0.060239 | 0.004331 | 1.80E-45 |
| rs174545 | miR-1908-5 | cis | | 2.29E-08 | Nightingale | S_HDL_C_p | NA | G | C | -0.04254 | 0.004291 | 5.20E-23 |
| rs174545 | miR-1908-5 | cis | | 2.29E-08 | Nightingale | IDL_L | NA | G | C | -0.07261 | 0.004197 | 1.80E-71 |
| rs174545 | miR-1908-5 | cis | | 2.29E-08 | Nightingale | XS_VLDL_P | NA | G | C | -0.0438 | 0.004283 | 2.10E-26 |
| rs174545 | miR-1908-5 | cis | | 2.29E-08 | Nightingale | L_HDL_CE_ | NA | G | C | -0.05032 | 0.004154 | 4.10E-36 |
| rs174545 | miR-1908-5 | cis | | 2.29E-08 | Nightingale | S_LDL_L | NA | G | C | -0.03253 | 0.004341 | 1.10E-14 |
| rs174545 | miR-1908-5 | cis | | 2.29E-08 | Nightingale | SFA_pct | NA | G | C | -0.04238 | 0.004283 | 4.40E-23 |
| rs174545 | miR-1908-5 | cis | | 2.29E-08 | Nightingale | XL_VLDL_C | NA | G | C | -0.06433 | 0.00416 | 2.30E-55 |
| rs174545 | miR-1908-5 | cis | | 2.29E-08 | Nightingale | M_HDL_CE | NA | G | C | -0.03774 | 0.004009 | 1.90E-22 |
| rs174545 | miR-1908-5 | cis | | 2.29E-08 | Nightingale | PUFA_pct | NA | G | C | -0.12181 | 0.004176 | 1.30E-188 |
| rs174545 | miR-1908-5 | cis | | 2.29E-08 | Nightingale | L_LDL_C | NA | G | C | -0.05793 | 0.004293 | 7.40E-44 |

| | | | | | | | | | | | |
|---|---|---|---|---|---|---|---|---|---|---|---|
| rs174545 | miR-1908-! | cis | 2.29E-08 | Nightingale | HDL_size | NA | G | C | -0.07278 | 0.003847 | 1.50E-86 |
| rs174545 | miR-1908-! | cis | 2.29E-08 | Nightingale | XL_HDL_C | NA | G | C | -0.09955 | 0.003914 | 8.30E-155 |
| rs174545 | miR-1908-! | cis | 2.29E-08 | Nightingale | L_VLDL_P | NA | G | C | 0.042177 | 0.004192 | 9.90E-25 |
| rs174545 | miR-1908-! | cis | 2.29E-08 | Nightingale | XL_VLDL_T | NA | G | C | 0.057209 | 0.004152 | 6.40E-45 |
| rs174545 | miR-1908-! | cis | 2.29E-08 | Nightingale | S_VLDL_CE | NA | G | C | -0.02901 | 0.004283 | 7.10E-12 |
| rs174545 | miR-1908-! | cis | 2.29E-08 | Nightingale | S_LDL_PL | NA | G | C | -0.05171 | 0.004331 | 9.10E-35 |
| rs174545 | miR-1908-! | cis | 2.29E-08 | Nightingale | VLDL_size | NA | G | C | 0.072814 | 0.004047 | 1.10E-76 |
| rs174545 | miR-1908-! | cis | 2.29E-08 | Nightingale | L_LDL_C_p | NA | G | C | -0.07724 | 0.00423 | 7.50E-77 |
| rs174545 | miR-1908-! | cis | 2.29E-08 | Nightingale | XS_VLDL_L | NA | G | C | -0.04176 | 0.004267 | 1.50E-24 |
| rs174545 | miR-1908-! | cis | 2.29E-08 | Nightingale | M_HDL_C_ | NA | G | C | -0.06611 | 0.004133 | 1.00E-60 |
| rs174545 | miR-1908-! | cis | 2.29E-08 | Nightingale | XS_VLDL_T | NA | G | C | 0.031731 | 0.004275 | 3.90E-13 |
| rs174545 | miR-1908-! | cis | 2.29E-08 | Nightingale | S_LDL_TG | NA | G | C | 0.050634 | 0.004235 | 1.70E-33 |
| rs174545 | miR-1908-! | cis | 2.29E-08 | Nightingale | IDL_P | NA | G | C | -0.04518 | 0.004296 | 2.90E-27 |
| rs174545 | miR-1908-! | cis | 2.29E-08 | Nightingale | M_VLDL_C | NA | G | C | -0.08099 | 0.004086 | 3.90E-91 |
| rs174545 | miR-1908-! | cis | 2.29E-08 | Nightingale | M_HDL_P | NA | G | C | -0.03413 | 0.003999 | 5.40E-19 |
| rs174545 | miR-1908-! | cis | 2.29E-08 | Nightingale | XXL_VLDL_ | NA | G | C | 0.068316 | 0.004182 | 7.50E-63 |
| rs174545 | miR-1908-! | cis | 2.29E-08 | Nightingale | S_LDL_FC | NA | G | C | -0.063 | 0.004334 | 1.10E-50 |
| rs174545 | miR-1908-! | cis | 2.29E-08 | Nightingale | L_VLDL_C_ | NA | G | C | -0.03879 | 0.004241 | 3.80E-21 |
| rs174545 | miR-1908-! | cis | 2.29E-08 | Nightingale | M_HDL_C | NA | G | C | -0.0407 | 0.003994 | 4.80E-26 |
| rs174545 | miR-1908-! | cis | 2.29E-08 | Nightingale | XL_HDL_CE | NA | G | C | -0.0263 | 0.004339 | 3.10E-09 |
| rs174545 | miR-1908-! | cis | 2.29E-08 | Nightingale | XXL_VLDL_ | NA | G | C | 0.044099 | 0.004364 | 5.70E-25 |
| rs174545 | miR-1908-! | cis | 2.29E-08 | Nightingale | XL_HDL_L | NA | G | C | -0.09334 | 0.003901 | 1.20E-137 |
| rs174545 | miR-1908-! | cis | 2.29E-08 | Nightingale | XS_VLDL_F | NA | G | C | -0.11317 | 0.004309 | 2.60E-155 |
| rs174545 | miR-1908-! | cis | 2.29E-08 | Nightingale | S_HDL_FC_ | NA | G | C | -0.10302 | 0.004118 | 2.90E-153 |
| rs174545 | miR-1908-! | cis | 2.29E-08 | Nightingale | M_HDL_TG | NA | G | C | 0.050759 | 0.004288 | 3.40E-33 |
| rs174545 | miR-1908-! | cis | 2.29E-08 | Nightingale | L_VLDL_FC | NA | G | C | 0.028536 | 0.0043 | 1.60E-11 |
| rs174545 | miR-1908-! | cis | 2.29E-08 | Nightingale | L_VLDL_TG | NA | G | C | 0.037587 | 0.004177 | 4.70E-20 |
| rs174545 | miR-1908-! | cis | 2.29E-08 | Nightingale | XL_VLDL_C | NA | G | C | 0.030529 | 0.004273 | 4.30E-13 |
| rs174545 | miR-1908-! | cis | 2.29E-08 | Nightingale | M_HDL_FC | NA | G | C | -0.0514 | 0.003951 | 1.50E-41 |
| rs174545 | miR-1908-! | cis | 2.29E-08 | Nightingale | M_VLDL_C | NA | G | C | -0.07625 | 0.004076 | 2.50E-81 |
| rs174545 | miR-1908-! | cis | 2.29E-08 | Nightingale | Omega_6_ | NA | G | C | 0.369846 | 0.004278 | 0 |
| rs174545 | miR-1908-! | cis | 2.29E-08 | Nightingale | LDL_TG | NA | G | C | 0.029823 | 0.004293 | 7.40E-12 |
| rs174545 | miR-1908-! | cis | 2.29E-08 | Nightingale | XL_HDL_C_ | NA | G | C | 0.024089 | 0.004147 | 2.90E-10 |
| rs174545 | miR-1908-! | cis | 2.29E-08 | Nightingale | M_LDL_C | NA | G | C | -0.03404 | 0.00435 | 1.20E-15 |
| rs174545 | miR-1908-! | cis | 2.29E-08 | Nightingale | Sphingomy | NA | G | C | -0.11547 | 0.004057 | 3.10E-187 |
| rs174545 | miR-1908-! | cis | 2.29E-08 | Nightingale | XXL_VLDL_ | NA | G | C | 0.066956 | 0.004181 | 3.10E-60 |
| rs174545 | miR-1908-! | cis | 2.29E-08 | Nightingale | DHA | NA | G | C | -0.28865 | 0.004184 | 0 |
| rs174545 | miR-1908-! | cis | 2.29E-08 | Nightingale | L_HDL_PL | NA | G | C | -0.07621 | 0.003821 | 2.40E-96 |
| rs174545 | miR-1908-! | cis | 2.29E-08 | Nightingale | XL_HDL_PL | NA | G | C | -0.0894 | 0.003902 | 2.40E-126 |
| rs174545 | miR-1908-! | cis | 2.29E-08 | Nightingale | S_LDL_TG_ | NA | G | C | 0.086527 | 0.004207 | 3.50E-97 |
| rs174545 | miR-1908-! | cis | 2.29E-08 | Nightingale | ApoA1 | NA | G | C | -0.04108 | 0.003973 | 4.60E-27 |
| rs174545 | miR-1908-! | cis | 2.29E-08 | Nightingale | XXL_VLDL_ | NA | G | C | 0.072368 | 0.004166 | 1.10E-70 |
| rs174545 | miR-1908-! | cis | 2.29E-08 | Nightingale | Phosphogl | NA | G | C | -0.04092 | 0.004112 | 4.20E-25 |
| rs174545 | miR-1908-! | cis | 2.29E-08 | Nightingale | Total_CE | NA | G | C | -0.06713 | 0.004187 | 4.60E-61 |
| rs174545 | miR-1908-! | cis | 2.29E-08 | Nightingale | M_LDL_CE | NA | G | C | -0.02629 | 0.004348 | 6.20E-10 |
| rs174545 | miR-1908-! | cis | 2.29E-08 | Nightingale | IDL_TG_pc | NA | G | C | 0.088349 | 0.004194 | 2.40E-102 |
| rs174545 | miR-1908-! | cis | 2.29E-08 | Nightingale | L_LDL_P | NA | G | C | -0.03756 | 0.004327 | 3.00E-19 |
| rs174545 | miR-1908-! | cis | 2.29E-08 | Nightingale | Tyr | NA | G | C | 0.033816 | 0.004336 | 6.20E-15 |
| rs174545 | miR-1908-! | cis | 2.29E-08 | Nightingale | L_VLDL_PL | NA | G | C | 0.049032 | 0.004189 | 5.10E-33 |
| rs174545 | miR-1908-! | cis | 2.29E-08 | Nightingale | Remnant_( | NA | G | C | -0.04417 | 0.004313 | 4.70E-26 |
| rs174545 | miR-1908-! | cis | 2.29E-08 | Nightingale | Total_FA | NA | G | C | -0.02363 | 0.004307 | 1.20E-08 |
| rs174545 | miR-1908-! | cis | 2.29E-08 | Nightingale | HDL_PL | NA | G | C | -0.04837 | 0.003925 | 1.70E-37 |
| rs174545 | miR-1908-! | cis | 2.29E-08 | Nightingale | IDL_FC | NA | G | C | -0.08998 | 0.004202 | 2.60E-107 |
| rs174545 | miR-1908-! | cis | 2.29E-08 | Nightingale | XXL_VLDL_ | NA | G | C | 0.061548 | 0.004189 | 5.80E-51 |
| rs174545 | miR-1908-! | cis | 2.29E-08 | Nightingale | PUFA_by_I | NA | G | C | -0.171 | 0.004158 | 0 |
| rs174545 | miR-1908-! | cis | 2.29E-08 | Nightingale | S_HDL_TG | NA | G | C | 0.069248 | 0.00421 | 1.20E-63 |
| rs174545 | miR-1908-! | cis | 2.29E-08 | Nightingale | ApoB | NA | G | C | -0.03856 | 0.004336 | 5.70E-20 |
| rs174545 | miR-1908-! | cis | 2.29E-08 | Nightingale | XS_VLDL_C | NA | G | C | -0.09824 | 0.004113 | 8.39E-132 |
| rs174545 | miR-1908-! | cis | 2.29E-08 | Nightingale | MUFA_pct | NA | G | C | 0.197718 | 0.004154 | 0 |
| rs174545 | miR-1908-! | cis | 2.29E-08 | Nightingale | Omega_3 | NA | G | C | -0.33578 | 0.004255 | 0 |
| rs174545 | miR-1908-! | cis | 2.29E-08 | Nightingale | HDL_CE | NA | G | C | -0.0615 | 0.003896 | 1.20E-60 |
| rs174545 | miR-1908-! | cis | 2.29E-08 | Nightingale | XL_HDL_FC | NA | G | C | 0.061385 | 0.003849 | 2.00E-63 |
| rs174545 | miR-1908-! | cis | 2.29E-08 | Nightingale | TG_by_PG | NA | G | C | 0.068976 | 0.004029 | 4.60E-69 |

| | | | | | | | | | | | |
|---|---|---|---|---|---|---|---|---|---|---|---|
| rs174545 | miR-1908-5 | cis | 2.29E-08 | Nightingale | LDL_P | NA | G | C | -0.04038 | 0.004338 | 1.20E-21 |
| rs174545 | miR-1908-5 | cis | 2.29E-08 | Nightingale | XL_HDL_TG | NA | G | C | 0.087632 | 0.00409 | 9.51E-108 |
| rs174545 | miR-1908-5 | cis | 2.29E-08 | Nightingale | M_HDL_CE | NA | G | C | -0.04725 | 0.00421 | 4.20E-30 |
| rs174545 | miR-1908-5 | cis | 2.29E-08 | Nightingale | L_HDL_FC_ | NA | G | C | -0.11275 | 0.004033 | 3.50E-187 |
| rs174545 | miR-1908-5 | cis | 2.29E-08 | Nightingale | IDL_C | NA | G | C | -0.07748 | 0.004192 | 8.40E-81 |
| rs174545 | miR-1908-5 | cis | 2.29E-08 | Nightingale | XL_HDL_P | NA | G | C | -0.08983 | 0.003907 | 5.11E-128 |
| rs174545 | miR-1908-5 | cis | 2.29E-08 | Nightingale | L_HDL_P | NA | G | C | -0.07799 | 0.003802 | 5.60E-102 |
| rs174545 | miR-1908-5 | cis | 2.29E-08 | Nightingale | IDL_CE | NA | G | C | -0.07246 | 0.004191 | 5.70E-71 |
| rs174545 | miR-1908-5 | cis | 2.29E-08 | Nightingale | M_LDL_P | NA | G | C | -0.04676 | 0.004354 | 9.50E-28 |
| rs174545 | miR-1908-5 | cis | 2.29E-08 | Nightingale | XL_VLDL_L | NA | G | C | 0.051764 | 0.004177 | 1.30E-36 |
| rs174545 | miR-1908-5 | cis | 2.29E-08 | Nightingale | IDL_PL | NA | G | C | -0.07308 | 0.004189 | 5.60E-73 |
| rs174545 | miR-1908-5 | cis | 2.29E-08 | Nightingale | L_HDL_L | NA | G | C | -0.07734 | 0.003809 | 6.00E-100 |
| rs174545 | miR-1908-5 | cis | 2.29E-08 | Nightingale | VLDL_TG | NA | G | C | 0.047977 | 0.004178 | 1.40E-31 |
| rs174545 | miR-1908-5 | cis | 2.29E-08 | Nightingale | L_LDL_PL | NA | G | C | -0.05506 | 0.004309 | 3.90E-39 |
| rs174545 | miR-1908-5 | cis | 2.29E-08 | Nightingale | DHA_pct | NA | G | C | -0.27405 | 0.004214 | 0 |
| rs174545 | miR-1908-5 | cis | 2.29E-08 | Nightingale | Total_L | NA | G | C | -0.0373 | 0.004271 | 1.50E-19 |
| rs174545 | miR-1908-5 | cis | 2.29E-08 | Nightingale | XS_VLDL_C | NA | G | C | -0.07218 | 0.004189 | 1.10E-70 |
| rs174545 | miR-1908-5 | cis | 2.29E-08 | Nightingale | Cholines | NA | G | C | -0.03708 | 0.004083 | 1.80E-21 |
| rs174545 | miR-1908-5 | cis | 2.29E-08 | Nightingale | HDL_FC | NA | G | C | -0.07395 | 0.003846 | 3.40E-89 |
| rs174545 | miR-1908-5 | cis | 2.29E-08 | Nightingale | XXL_VLDL_ | NA | G | C | 0.073931 | 0.004192 | 4.40E-72 |
| rs174545 | miR-1908-5 | cis | 2.29E-08 | Nightingale | S_LDL_PL_ | NA | G | C | -0.06255 | 0.004332 | 1.90E-49 |
| rs174545 | miR-1908-5 | cis | 2.29E-08 | Nightingale | XL_VLDL_P | NA | G | C | 0.053174 | 0.004173 | 1.20E-38 |
| rs174545 | miR-1908-5 | cis | 2.29E-08 | Nightingale | XL_VLDL_P | NA | G | C | 0.048543 | 0.004189 | 3.20E-32 |
| rs174545 | miR-1908-5 | cis | 2.29E-08 | Nightingale | Unsaturati | NA | G | C | -0.33982 | 0.004111 | 0 |
| rs174545 | miR-1908-5 | cis | 2.29E-08 | Nightingale | LDL_CE | NA | G | C | -0.04363 | 0.00433 | 4.40E-25 |
| rs174545 | miR-1908-5 | cis | 2.29E-08 | Nightingale | Total_TG | NA | G | C | 0.04706 | 0.004202 | 4.10E-30 |
| rs174545 | miR-1908-5 | cis | 2.29E-08 | Nightingale | LDL_PL | NA | G | C | -0.04626 | 0.004332 | 7.50E-28 |
| rs174545 | miR-1908-5 | cis | 2.29E-08 | Nightingale | XS_VLDL_C | NA | G | C | -0.06841 | 0.004214 | 5.10E-63 |
| rs174545 | miR-1908-5 | cis | 2.29E-08 | Nightingale | Clinical_LD | NA | G | C | -0.05529 | 0.004316 | 2.30E-39 |
| rs174545 | miR-1908-5 | cis | 2.29E-08 | Nightingale | M_VLDL_T | NA | G | C | 0.031664 | 0.004218 | 2.50E-14 |
| rs174545 | miR-1908-5 | cis | 2.29E-08 | Nightingale | S_LDL_CE_ | NA | G | C | 0.057686 | 0.004333 | 2.60E-42 |
| rs174545 | miR-1908-5 | cis | 2.29E-08 | Nightingale | Omega_6_ | NA | G | C | 0.023821 | 0.004225 | 9.30E-09 |
| rs174545 | miR-1908-5 | cis | 2.29E-08 | Nightingale | XS_VLDL_C | NA | G | C | -0.09231 | 0.004092 | 6.40E-118 |
| rs174545 | miR-1908-5 | cis | 2.29E-08 | Nightingale | LDL_FC | NA | G | C | -0.06447 | 0.004295 | 1.10E-53 |
| rs174545 | miR-1908-5 | cis | 2.29E-08 | Nightingale | L_LDL_CE | NA | G | C | -0.05339 | 0.004303 | 2.30E-37 |
| rs174545 | miR-1908-5 | cis | 2.29E-08 | Nightingale | IDL_C_pct | NA | G | C | -0.07492 | 0.004238 | 1.30E-71 |
| rs174545 | miR-1908-5 | cis | 2.29E-08 | Nightingale | S_VLDL_C_ | NA | G | C | -0.05676 | 0.004224 | 2.70E-42 |
| rs174545 | miR-1908-5 | cis | 2.29E-08 | Nightingale | XS_VLDL_T | NA | G | C | 0.097663 | 0.004124 | 5.90E-130 |
| rs174545 | miR-1908-5 | cis | 2.29E-08 | Nightingale | S_LDL_P | NA | G | C | -0.03254 | 0.004345 | 1.40E-14 |
| rs174545 | miR-1908-5 | cis | 2.29E-08 | Nightingale | LDL_L | NA | G | C | -0.04552 | 0.004327 | 3.10E-27 |
| rs174545 | miR-1908-5 | cis | 2.29E-08 | Nightingale | M_LDL_L | NA | G | C | -0.02816 | 0.004348 | 3.60E-11 |
| rs174545 | miR-1908-5 | cis | 2.29E-08 | Nightingale | M_HDL_PL | NA | G | C | 0.058621 | 0.004118 | 6.00E-48 |
| rs174545 | miR-1908-5 | cis | 2.29E-08 | Nightingale | S_VLDL_PL | NA | G | C | -0.07882 | 0.004154 | 3.30E-84 |
| rs174545 | miR-1908-5 | cis | 2.29E-08 | Nightingale | M_VLDL_F | NA | G | C | -0.02945 | 0.004341 | 3.40E-12 |
| rs174545 | miR-1908-5 | cis | 2.29E-08 | Nightingale | MUFA | NA | G | C | 0.06412 | 0.004297 | 2.20E-51 |
| rs174545 | miR-1908-5 | cis | 2.29E-08 | Nightingale | L_VLDL_FC | NA | G | C | 0.040416 | 0.004199 | 8.30E-23 |
| rs174545 | miR-1908-5 | cis | 2.29E-08 | Nightingale | HDL_TG | NA | G | C | 0.046586 | 0.004281 | 3.90E-28 |
| rs174545 | miR-1908-5 | cis | 2.29E-08 | Nightingale | M_VLDL_C | NA | G | C | -0.05289 | 0.004302 | 2.50E-36 |
| rs174545 | miR-1908-5 | cis | 2.29E-08 | Nightingale | M_LDL_FC | NA | G | C | -0.0525 | 0.004336 | 1.60E-35 |
| rs174545 | miR-1908-5 | cis | 2.29E-08 | Nightingale | non_HDL_( | NA | G | C | -0.04787 | 0.004318 | 5.30E-30 |
| rs174545 | miR-1908-5 | cis | 2.29E-08 | Nightingale | XL_HDL_PL | NA | G | C | -0.06735 | 0.003988 | 4.90E-69 |
| rs174545 | miR-1908-5 | cis | 2.29E-08 | Nightingale | S_VLDL_FC | NA | G | C | -0.0832 | 0.004145 | 1.30E-93 |
| rs174545 | miR-1908-5 | cis | 2.29E-08 | Nightingale | XS_VLDL_P | NA | G | C | 0.057714 | 0.004236 | 4.60E-43 |
| rs174545 | miR-1908-5 | cis | 2.29E-08 | Nightingale | VLDL_L | NA | G | C | 0.027585 | 0.004257 | 5.30E-11 |
| rs174545 | miR-1908-5 | cis | 2.29E-08 | Nightingale | HDL_L | NA | G | C | -0.05331 | 0.003898 | 7.70E-46 |
| rs174545 | miR-1908-5 | cis | 2.29E-08 | Nightingale | L_HDL_FC | NA | G | C | -0.08477 | 0.003805 | 1.30E-119 |
| rs174545 | miR-1908-5 | cis | 2.29E-08 | Nightingale | M_LDL_TG | NA | G | C | 0.076048 | 0.004287 | 6.10E-72 |
| rs174545 | miR-1908-5 | cis | 2.29E-08 | Nightingale | XXL_VLDL_ | NA | G | C | 0.066453 | 0.004176 | 1.00E-59 |
| rs174545 | miR-1908-5 | cis | 2.29E-08 | Nightingale | XL_VLDL_C | NA | G | C | -0.06136 | 0.004162 | 1.30E-50 |
| rs174545 | miR-1908-5 | cis | 2.29E-08 | Nightingale | S_LDL_FC_ | NA | G | C | -0.0801 | 0.004207 | 7.90E-84 |
| rs174545 | miR-1908-5 | cis | 2.29E-08 | Nightingale | L_LDL_FC_ | NA | G | C | -0.08303 | 0.004138 | 8.80E-93 |
| rs174545 | miR-1908-5 | cis | 2.29E-08 | Nightingale | L_HDL_C | NA | G | C | -0.08044 | 0.003823 | 5.30E-107 |
| rs174545 | miR-1908-5 | cis | 2.29E-08 | Nightingale | S_HDL_TG_ | NA | G | C | 0.069249 | 0.004129 | 4.10E-66 |

| | | | | | | | | | | | |
|---|---|---|---|---|---|---|---|---|---|---|---|
| rs174545 | miR-1908-5 | cis | 2.29E-08 | Nightingale | LA | NA | G | C | 0.083779 | 0.004257 | 2.00E-86 |
| rs174545 | miR-1908-5 | cis | 2.29E-08 | Nightingale | Total_C | NA | G | C | -0.06532 | 0.004203 | 1.70E-57 |
| rs174545 | miR-1908-5 | cis | 2.29E-08 | Nightingale | M_HDL_TG | NA | G | C | 0.066942 | 0.004163 | 5.30E-61 |
| rs174545 | miR-1908-5 | cis | 2.29E-08 | Nightingale | L_HDL_PL_ | NA | G | C | 0.043397 | 0.004202 | 9.90E-27 |
| rs174545 | miR-1908-5 | cis | 2.29E-08 | Nightingale | IDL_CE_pct | NA | G | C | -0.03814 | 0.00426 | 3.80E-19 |
| rs174545 | miR-1908-5 | cis | 2.29E-08 | Nightingale | M_LDL_C_ | NA | G | C | -0.10042 | 0.004345 | 8.49E-119 |
| rs174545 | miR-1908-5 | cis | 2.29E-08 | Nightingale | PUFA | NA | G | C | -0.09652 | 0.004234 | 4.60E-122 |
| rs174545 | miR-1908-5 | cis | 2.29E-08 | Nightingale | L_VLDL_CE | NA | G | C | -0.05941 | 0.004154 | 3.50E-49 |
| rs174545 | miR-1908-5 | cis | 2.29E-08 | Nightingale | Total_FC | NA | G | C | -0.0595 | 0.004241 | 2.40E-47 |
| rs174545 | miR-1908-5 | cis | 2.29E-08 | Nightingale | M_LDL_PL | NA | G | C | -0.02352 | 0.004348 | 2.80E-08 |
| rs174545 | miR-1908-5 | cis | 2.29E-08 | Nightingale | M_VLDL_F | NA | G | C | -0.09158 | 0.004128 | 9.20E-115 |
| rs174545 | miR-1908-5 | cis | 2.29E-08 | Nightingale | L_VLDL_PL | NA | G | C | 0.087889 | 0.004213 | 2.40E-100 |
| rs174545 | miR-1908-5 | cis | 2.29E-08 | Nightingale | IDL_FC_pct | NA | G | C | -0.1042 | 0.004306 | 5.90E-131 |
| rs174545 | miR-1908-5 | cis | 2.29E-08 | Nightingale | HDL_P | NA | G | C | -0.03542 | 0.004076 | 2.50E-19 |
| rs174545 | miR-1908-5 | cis | 2.29E-08 | Nightingale | S_VLDL_TG | NA | G | C | 0.064704 | 0.004201 | 3.50E-55 |
| rs174545 | miR-1908-5 | cis | 2.29E-08 | Nightingale | XXL_VLDL_ | NA | G | C | 0.072034 | 0.004184 | 4.00E-69 |
| rs174545 | miR-1908-5 | cis | 2.29E-08 | Nightingale | L_LDL_L | NA | G | C | -0.05429 | 0.004299 | 1.20E-38 |
| rs174545 | miR-1908-5 | cis | 2.29E-08 | Nightingale | XL_VLDL_F | NA | G | C | -0.06841 | 0.004206 | 3.30E-62 |
| rs174545 | miR-1908-5 | cis | 2.29E-08 | Nightingale | M_VLDL_C | NA | G | C | -0.04413 | 0.004329 | 2.10E-25 |
| rs174545 | miR-1908-5 | cis | 2.29E-08 | Nightingale | L_LDL_FC | NA | G | C | -0.06851 | 0.004263 | 1.70E-61 |
| rs174545 | miR-1908-5 | cis | 2.29E-08 | Nightingale | M_HDL_PL | NA | G | C | -0.02126 | 0.00406 | 4.30E-08 |
| rs174545 | miR-1908-5 | cis | 2.29E-08 | Nightingale | XL_HDL_CE | NA | G | C | -0.09742 | 0.003888 | 1.40E-150 |
| rs174545 | miR-1908-5 | cis | 2.29E-08 | Nightingale | Citrate | NA | G | C | -0.03355 | 0.004314 | 6.80E-16 |
| rs174545 | miR-1908-5 | cis | 2.29E-08 | Nightingale | L_HDL_C_p | NA | G | C | -0.06868 | 0.004092 | 5.20E-68 |
| rs174545 | miR-1908-5 | cis | 2.29E-08 | Nightingale | S_VLDL_TG | NA | G | C | 0.03904 | 0.004213 | 4.80E-21 |
| rs174545 | miR-1908-5 | cis | 2.29E-08 | Nightingale | XL_VLDL_C | NA | G | C | 0.0377 | 0.00424 | 1.40E-19 |
| rs174545 | miR-1908-5 | cis | 2.29E-08 | Nightingale | M_LDL_FC_ | NA | G | C | -0.07195 | 0.004159 | 1.30E-69 |
| rs174545 | miR-1908-5 | cis | 2.29E-08 | Nightingale | S_LDL_C | NA | G | C | -0.03223 | 0.004346 | 2.50E-14 |
| rs174545 | miR-1908-5 | cis | 2.29E-08 | Nightingale | M_VLDL_P | NA | G | C | -0.06269 | 0.004166 | 1.40E-54 |
| rs174545 | miR-1908-5 | cis | 2.29E-08 | Nightingale | XS_VLDL_P | NA | G | C | -0.03461 | 0.00428 | 2.00E-17 |
| rs174545 | miR-1908-5 | cis | 2.29E-08 | Nightingale | L_VLDL_C | NA | G | C | 0.029091 | 0.004246 | 3.30E-12 |
| rs174545 | miR-1908-5 | cis | 2.29E-08 | Nightingale | L_VLDL_L | NA | G | C | 0.038481 | 0.004195 | 8.50E-21 |
| rs174545 | miR-1908-5 | cis | 2.29E-08 | Nightingale | HDL_C | NA | G | C | -0.06478 | 0.003876 | 6.20E-68 |
| rs174545 | miR-1908-5 | cis | 2.29E-08 | Nightingale | M_LDL_TG | NA | G | C | 0.036768 | 0.004277 | 8.30E-18 |
| rs174545 | miR-1908-5 | cis | 2.29E-08 | Nightingale | LA_pct | NA | G | C | 0.185202 | 0.004243 | 0 |
| rs174545 | miR-1908-5 | cis | 2.29E-08 | Nightingale | M_HDL_L | NA | G | C | -0.02749 | 0.00403 | 1.10E-12 |
| rs174545 | miR-1908-5 | cis | 2.29E-08 | Nightingale | XL_VLDL_T | NA | G | C | 0.059718 | 0.004206 | 3.70E-47 |
| rs174545 | miR-1908-5 | cis | 2.29E-08 | Nightingale | XL_VLDL_F | NA | G | C | 0.043746 | 0.004205 | 3.10E-26 |
| rs174545 | miR-1908-5 | cis | 2.29E-08 | Nightingale | L_HDL_TG_ | NA | G | C | 0.079045 | 0.004097 | 3.10E-88 |
| rs174545 | miR-1908-5 | cis | 2.29E-08 | Nightingale | S_VLDL_FC | NA | G | C | -0.02837 | 0.004342 | 2.00E-11 |
| rs174545 | miR-1908-5 | cis | 2.29E-08 | Nightingale | M_VLDL_T | NA | G | C | 0.078248 | 0.0041 | 5.20E-85 |
| rs174545 | miR-1908-5 | cis | 2.29E-08 | Nightingale | M_LDL_PL_ | NA | G | C | 0.045461 | 0.004339 | 8.80E-27 |
| rs174545 | miR-1908-5 | cis | 2.29E-08 | Nightingale | LDL_C | NA | G | C | -0.04984 | 0.004323 | 2.50E-32 |
| rs174545 | miR-1908-5 | cis | 2.29E-08 | Nightingale | XXL_VLDL_ | NA | G | C | -0.02712 | 0.004319 | 7.00E-11 |
| rs174545 | miR-1908-5 | cis | 2.29E-08 | Nightingale | XL_HDL_FC | NA | G | C | -0.10313 | 0.004044 | 1.10E-154 |
| rs174545 | miR-1908-5 | cis | 2.29E-08 | Nightingale | XS_VLDL_F | NA | G | C | -0.05629 | 0.004266 | 1.80E-42 |
| rs174545 | miR-1908-5 | cis | 2.29E-08 | Nightingale | Total_P | NA | G | C | -0.04019 | 0.004086 | 1.80E-24 |
| rs174545 | miR-1908-5 | cis | 2.29E-08 | Nightingale | L_LDL_TG_ | NA | G | C | 0.077841 | 0.004257 | 2.00E-76 |
| rs174545 | miR-1908-5 | cis | 2.29E-08 | Nightingale | SFA | NA | G | C | -0.03129 | 0.004302 | 1.30E-13 |
| rs174545 | miR-1908-5 | cis | 2.29E-08 | Nightingale | Omega_3_ | NA | G | C | -0.38997 | 0.004242 | 0 |
| rs174545 | miR-1908-5 | cis | 2.29E-08 | Nightingale | L_HDL_CE | NA | G | C | -0.07888 | 0.003833 | 2.90E-102 |
| rs174545 | miR-1908-5 | cis | 2.29E-08 | Nightingale | M_HDL_FC | NA | G | C | -0.10339 | 0.003847 | 2.40E-173 |
| rs174545 | miR-1908-5 | cis | 2.29E-08 | Nightingale | Total_PL | NA | G | C | -0.05111 | 0.004134 | 2.30E-37 |
| rs174548 | miR-1908-5 | cis | 3.01E-08 | Nightingale | IDL_C_pct | NA | G | C | -0.07409 | 0.004338 | 2.00E-67 |
| rs174548 | miR-1908-5 | cis | 3.01E-08 | Nightingale | S_LDL_P | NA | G | C | -0.0304 | 0.004448 | 2.80E-12 |
| rs174548 | miR-1908-5 | cis | 3.01E-08 | Nightingale | L_LDL_PL | NA | G | C | -0.05311 | 0.004411 | 5.70E-35 |
| rs174548 | miR-1908-5 | cis | 3.01E-08 | Nightingale | XS_VLDL_F | NA | G | C | -0.05259 | 0.004366 | 1.30E-35 |
| rs174548 | miR-1908-5 | cis | 3.01E-08 | Nightingale | L_HDL_P | NA | G | C | -0.07493 | 0.003892 | 2.00E-89 |
| rs174548 | miR-1908-5 | cis | 3.01E-08 | Nightingale | HDL_CE | NA | G | C | -0.05993 | 0.003988 | 1.20E-54 |
| rs174548 | miR-1908-5 | cis | 3.01E-08 | Nightingale | L_VLDL_PL | NA | G | C | 0.087231 | 0.004313 | 2.50E-94 |
| rs174548 | miR-1908-5 | cis | 3.01E-08 | Nightingale | XS_VLDL_T | NA | G | C | 0.094882 | 0.004221 | 5.00E-118 |
| rs174548 | miR-1908-5 | cis | 3.01E-08 | Nightingale | XS_VLDL_C | NA | G | C | -0.0683 | 0.004288 | 8.10E-61 |
| rs174548 | miR-1908-5 | cis | 3.01E-08 | Nightingale | HDL_size | NA | G | C | -0.06952 | 0.003938 | 3.40E-75 |

| | | | | | | | | | | | | |
|---|---|---|---|---|---|---|---|---|---|---|---|---|
| rs174548 | miR-1908-5 | cis | | 3.01E-08 | Nightingale | LA | NA | G | C | 0.084052 | 0.004358 | 2.60E-83 |
| rs174548 | miR-1908-5 | cis | | 3.01E-08 | Nightingale | IDL_PL | NA | G | C | -0.06935 | 0.004288 | 5.20E-63 |
| rs174548 | miR-1908-5 | cis | | 3.01E-08 | Nightingale | XL_HDL_PL | NA | G | C | -0.08512 | 0.003995 | 3.20E-109 |
| rs174548 | miR-1908-5 | cis | | 3.01E-08 | Nightingale | Clinical_LD | NA | G | C | -0.05333 | 0.004418 | 4.10E-35 |
| rs174548 | miR-1908-5 | cis | | 3.01E-08 | Nightingale | M_HDL_C_ | NA | G | C | -0.06635 | 0.00423 | 2.90E-58 |
| rs174548 | miR-1908-5 | cis | | 3.01E-08 | Nightingale | M_HDL_C | NA | G | C | -0.03979 | 0.004089 | 1.20E-23 |
| rs174548 | miR-1908-5 | cis | | 3.01E-08 | Nightingale | XS_VLDL_C | NA | G | C | -0.09599 | 0.004211 | 8.00E-121 |
| rs174548 | miR-1908-5 | cis | | 3.01E-08 | Nightingale | S_LDL_CE_ | NA | G | C | 0.056734 | 0.004436 | 3.10E-39 |
| rs174548 | miR-1908-5 | cis | | 3.01E-08 | Nightingale | XL_VLDL_L | NA | G | C | 0.051979 | 0.004276 | 1.20E-35 |
| rs174548 | miR-1908-5 | cis | | 3.01E-08 | Nightingale | SFA | NA | G | C | -0.02846 | 0.004404 | 7.90E-11 |
| rs174548 | miR-1908-5 | cis | | 3.01E-08 | Nightingale | S_VLDL_FC | NA | G | C | -0.02672 | 0.004445 | 7.70E-10 |
| rs174548 | miR-1908-5 | cis | | 3.01E-08 | Nightingale | LDL_C | NA | G | C | -0.04799 | 0.004425 | 1.00E-28 |
| rs174548 | miR-1908-5 | cis | | 3.01E-08 | Nightingale | HDL_FC | NA | G | C | -0.07086 | 0.003937 | 7.80E-78 |
| rs174548 | miR-1908-5 | cis | | 3.01E-08 | Nightingale | L_VLDL_CE | NA | G | C | -0.05881 | 0.004253 | 3.50E-46 |
| rs174548 | miR-1908-5 | cis | | 3.01E-08 | Nightingale | M_HDL_TG | NA | G | C | 0.051458 | 0.004389 | 1.40E-32 |
| rs174548 | miR-1908-5 | cis | | 3.01E-08 | Nightingale | VLDL_size | NA | G | C | 0.071598 | 0.004142 | 4.80E-71 |
| rs174548 | miR-1908-5 | cis | | 3.01E-08 | Nightingale | L_LDL_P | NA | G | C | -0.03558 | 0.004429 | 1.20E-16 |
| rs174548 | miR-1908-5 | cis | | 3.01E-08 | Nightingale | Omega_3 | NA | G | C | -0.3287 | 0.004356 | 0 |
| rs174548 | miR-1908-5 | cis | | 3.01E-08 | Nightingale | XXL_VLDL_ | NA | G | C | 0.073204 | 0.004291 | 3.20E-68 |
| rs174548 | miR-1908-5 | cis | | 3.01E-08 | Nightingale | Unsaturati | NA | G | C | -0.33312 | 0.004209 | 0 |
| rs174548 | miR-1908-5 | cis | | 3.01E-08 | Nightingale | L_VLDL_FC | NA | G | C | 0.040617 | 0.004298 | 3.00E-22 |
| rs174548 | miR-1908-5 | cis | | 3.01E-08 | Nightingale | XS_VLDL_C | NA | G | C | -0.06455 | 0.004313 | 6.20E-54 |
| rs174548 | miR-1908-5 | cis | | 3.01E-08 | Nightingale | L_LDL_FC_ | NA | G | C | -0.08175 | 0.004236 | 2.20E-86 |
| rs174548 | miR-1908-5 | cis | | 3.01E-08 | Nightingale | IDL_FC | NA | G | C | -0.08664 | 0.004301 | 2.40E-95 |
| rs174548 | miR-1908-5 | cis | | 3.01E-08 | Nightingale | LDL_L | NA | G | C | -0.04368 | 0.004429 | 4.80E-24 |
| rs174548 | miR-1908-5 | cis | | 3.01E-08 | Nightingale | XL_HDL_FC | NA | G | C | -0.09791 | 0.00414 | 2.20E-132 |
| rs174548 | miR-1908-5 | cis | | 3.01E-08 | Nightingale | M_LDL_FC_ | NA | G | C | -0.07167 | 0.004257 | 1.50E-66 |
| rs174548 | miR-1908-5 | cis | | 3.01E-08 | Nightingale | Phosphogly | NA | G | C | -0.03813 | 0.004209 | 6.80E-21 |
| rs174548 | miR-1908-5 | cis | | 3.01E-08 | Nightingale | L_LDL_L | NA | G | C | -0.05209 | 0.004401 | 4.40E-34 |
| rs174548 | miR-1908-5 | cis | | 3.01E-08 | Nightingale | M_HDL_CE | NA | G | C | -0.04874 | 0.004309 | 1.50E-30 |
| rs174548 | miR-1908-5 | cis | | 3.01E-08 | Nightingale | LA_pct | NA | G | C | 0.181625 | 0.004343 | 0 |
| rs174548 | miR-1908-5 | cis | | 3.01E-08 | Nightingale | XXL_VLDL_ | NA | G | C | 0.0665 | 0.004274 | 1.30E-57 |
| rs174548 | miR-1908-5 | cis | | 3.01E-08 | Nightingale | M_VLDL_F | NA | G | C | -0.02745 | 0.004443 | 2.70E-10 |
| rs174548 | miR-1908-5 | cis | | 3.01E-08 | Nightingale | S_VLDL_C_ | NA | G | C | -0.05573 | 0.004324 | 3.00E-39 |
| rs174548 | miR-1908-5 | cis | | 3.01E-08 | Nightingale | L_LDL_CE | NA | G | C | -0.05125 | 0.004404 | 5.90E-33 |
| rs174548 | miR-1908-5 | cis | | 3.01E-08 | Nightingale | Total_TG | NA | G | C | 0.047803 | 0.004302 | 3.60E-30 |
| rs174548 | miR-1908-5 | cis | | 3.01E-08 | Nightingale | IDL_CE | NA | G | C | -0.06917 | 0.00429 | 3.90E-62 |
| rs174548 | miR-1908-5 | cis | | 3.01E-08 | Nightingale | SFA_pct | NA | G | C | -0.04002 | 0.004384 | 7.00E-20 |
| rs174548 | miR-1908-5 | cis | | 3.01E-08 | Nightingale | M_VLDL_C | NA | G | C | -0.05076 | 0.004404 | 4.30E-32 |
| rs174548 | miR-1908-5 | cis | | 3.01E-08 | Nightingale | Remnant_( | NA | G | C | -0.04146 | 0.004415 | 3.50E-22 |
| rs174548 | miR-1908-5 | cis | | 3.01E-08 | Nightingale | M_VLDL_C_ | NA | G | C | -0.07928 | 0.004183 | 1.60E-83 |
| rs174548 | miR-1908-5 | cis | | 3.01E-08 | Nightingale | PUFA_pct | NA | G | C | -0.12012 | 0.004274 | 1.90E-175 |
| rs174548 | miR-1908-5 | cis | | 3.01E-08 | Nightingale | Total_C | NA | G | C | -0.0625 | 0.004302 | 2.40E-50 |
| rs174548 | miR-1908-5 | cis | | 3.01E-08 | Nightingale | XL_VLDL_P | NA | G | C | 0.048935 | 0.004288 | 1.30E-31 |
| rs174548 | miR-1908-5 | cis | | 3.01E-08 | Nightingale | VLDL_L | NA | G | C | 0.028572 | 0.004357 | 1.90E-11 |
| rs174548 | miR-1908-5 | cis | | 3.01E-08 | Nightingale | S_LDL_FC | NA | G | C | -0.06157 | 0.004437 | 2.80E-46 |
| rs174548 | miR-1908-5 | cis | | 3.01E-08 | Nightingale | Omega_6_ | NA | G | C | 0.362759 | 0.004379 | 0 |
| rs174548 | miR-1908-5 | cis | | 3.01E-08 | Nightingale | IDL_P | NA | G | C | -0.04254 | 0.004397 | 1.80E-23 |
| rs174548 | miR-1908-5 | cis | | 3.01E-08 | Nightingale | XXL_VLDL_ | NA | G | C | -0.02906 | 0.00442 | 7.90E-12 |
| rs174548 | miR-1908-5 | cis | | 3.01E-08 | Nightingale | XL_HDL_P | NA | G | C | -0.08556 | 0.003999 | 2.60E-110 |
| rs174548 | miR-1908-5 | cis | | 3.01E-08 | Nightingale | M_LDL_FC | NA | G | C | -0.05133 | 0.004439 | 1.90E-32 |
| rs174548 | miR-1908-5 | cis | | 3.01E-08 | Nightingale | XL_HDL_CE | NA | G | C | -0.02834 | 0.004441 | 5.30E-10 |
| rs174548 | miR-1908-5 | cis | | 3.01E-08 | Nightingale | M_VLDL_T | NA | G | C | 0.032026 | 0.004317 | 3.00E-14 |
| rs174548 | miR-1908-5 | cis | | 3.01E-08 | Nightingale | MUFA | NA | G | C | 0.064351 | 0.004399 | 3.90E-50 |
| rs174548 | miR-1908-5 | cis | | 3.01E-08 | Nightingale | ApoA1 | NA | G | C | -0.03944 | 0.004067 | 8.10E-24 |
| rs174548 | miR-1908-5 | cis | | 3.01E-08 | Nightingale | L_VLDL_PL | NA | G | C | 0.049132 | 0.004288 | 6.00E-32 |
| rs174548 | miR-1908-5 | cis | | 3.01E-08 | Nightingale | DHA_pct | NA | G | C | -0.26936 | 0.004314 | 0 |
| rs174548 | miR-1908-5 | cis | | 3.01E-08 | Nightingale | S_LDL_TG | NA | G | C | 0.051258 | 0.004335 | 1.80E-33 |
| rs174548 | miR-1908-5 | cis | | 3.01E-08 | Nightingale | S_LDL_TG_ | NA | G | C | 0.085888 | 0.004306 | 4.20E-92 |
| rs174548 | miR-1908-5 | cis | | 3.01E-08 | Nightingale | XL_HDL_CE | NA | G | C | -0.09361 | 0.003979 | 6.00E-132 |
| rs174548 | miR-1908-5 | cis | | 3.01E-08 | Nightingale | XL_VLDL_C | NA | G | C | -0.06419 | 0.004258 | 6.60E-53 |
| rs174548 | miR-1908-5 | cis | | 3.01E-08 | Nightingale | Tyr | NA | G | C | 0.031897 | 0.004438 | 6.60E-13 |
| rs174548 | miR-1908-5 | cis | | 3.01E-08 | Nightingale | IDL_C | NA | G | C | -0.07416 | 0.004291 | 4.70E-71 |

| | | | | | | | | | | | |
|---|---|---|---|---|---|---|---|---|---|---|---|
| rs174548 | miR-1908-5 | cis | 3.01E-08 | Nightingale | M_HDL_FC | NA | G | C | -0.04943 | 0.004044 | 1.40E-36 |
| rs174548 | miR-1908-5 | cis | 3.01E-08 | Nightingale | S_HDL_FC_ | NA | G | C | -0.09878 | 0.004215 | 1.30E-133 |
| rs174548 | miR-1908-5 | cis | 3.01E-08 | Nightingale | L_VLDL_TG | NA | G | C | 0.038059 | 0.004276 | 7.50E-20 |
| rs174548 | miR-1908-5 | cis | 3.01E-08 | Nightingale | XXL_VLDL_ | NA | G | C | 0.04272 | 0.004467 | 2.10E-22 |
| rs174548 | miR-1908-5 | cis | 3.01E-08 | Nightingale | L_VLDL_P | NA | G | C | 0.042382 | 0.004291 | 3.90E-24 |
| rs174548 | miR-1908-5 | cis | 3.01E-08 | Nightingale | LDL_FC | NA | G | C | -0.06249 | 0.004397 | 3.20E-48 |
| rs174548 | miR-1908-5 | cis | 3.01E-08 | Nightingale | HDL_C | NA | G | C | -0.06287 | 0.003968 | 1.10E-60 |
| rs174548 | miR-1908-5 | cis | 3.01E-08 | Nightingale | HDL_PL | NA | G | C | -0.04608 | 0.004017 | 1.70E-32 |
| rs174548 | miR-1908-5 | cis | 3.01E-08 | Nightingale | XL_VLDL_C | NA | G | C | -0.0614 | 0.00426 | 1.50E-48 |
| rs174548 | miR-1908-5 | cis | 3.01E-08 | Nightingale | Total_CE | NA | G | C | -0.06438 | 0.004286 | 1.20E-53 |
| rs174548 | miR-1908-5 | cis | 3.01E-08 | Nightingale | HDL_TG | NA | G | C | 0.047543 | 0.004382 | 4.80E-28 |
| rs174548 | miR-1908-5 | cis | 3.01E-08 | Nightingale | XXL_VLDL_ | NA | G | C | 0.061975 | 0.004288 | 1.10E-49 |
| rs174548 | miR-1908-5 | cis | 3.01E-08 | Nightingale | non_HDL_( | NA | G | C | -0.04555 | 0.00442 | 3.50E-26 |
| rs174548 | miR-1908-5 | cis | 3.01E-08 | Nightingale | M_LDL_C_ | NA | G | C | -0.09865 | 0.004447 | 9.91E-110 |
| rs174548 | miR-1908-5 | cis | 3.01E-08 | Nightingale | Total_FC | NA | G | C | -0.05659 | 0.004341 | 4.10E-41 |
| rs174548 | miR-1908-5 | cis | 3.01E-08 | Nightingale | S_VLDL_PL | NA | G | C | -0.07752 | 0.004252 | 7.40E-78 |
| rs174548 | miR-1908-5 | cis | 3.01E-08 | Nightingale | S_VLDL_CE | NA | G | C | -0.02874 | 0.004384 | 1.70E-11 |
| rs174548 | miR-1908-5 | cis | 3.01E-08 | Nightingale | S_VLDL_FC | NA | G | C | -0.08136 | 0.004243 | 1.20E-85 |
| rs174548 | miR-1908-5 | cis | 3.01E-08 | Nightingale | S_LDL_C | NA | G | C | -0.0309 | 0.004449 | 1.10E-12 |
| rs174548 | miR-1908-5 | cis | 3.01E-08 | Nightingale | XXL_VLDL_ | NA | G | C | 0.068116 | 0.004281 | 3.20E-60 |
| rs174548 | miR-1908-5 | cis | 3.01E-08 | Nightingale | S_LDL_PL_ | NA | G | C | -0.06054 | 0.004435 | 1.70E-44 |
| rs174548 | miR-1908-5 | cis | 3.01E-08 | Nightingale | XXL_VLDL_ | NA | G | C | 0.072318 | 0.004265 | 5.60E-68 |
| rs174548 | miR-1908-5 | cis | 3.01E-08 | Nightingale | M_HDL_P | NA | G | C | -0.03294 | 0.004094 | 6.70E-17 |
| rs174548 | miR-1908-5 | cis | 3.01E-08 | Nightingale | XL_VLDL_T | NA | G | C | 0.060033 | 0.004305 | 7.90E-46 |
| rs174548 | miR-1908-5 | cis | 3.01E-08 | Nightingale | HDL_P | NA | G | C | -0.03458 | 0.004173 | 1.50E-17 |
| rs174548 | miR-1908-5 | cis | 3.01E-08 | Nightingale | L_LDL_C | NA | G | C | -0.05571 | 0.004394 | 7.90E-39 |
| rs174548 | miR-1908-5 | cis | 3.01E-08 | Nightingale | L_VLDL_L | NA | G | C | 0.03883 | 0.004295 | 1.90E-20 |
| rs174548 | miR-1908-5 | cis | 3.01E-08 | Nightingale | S_LDL_PL | NA | G | C | -0.04949 | 0.004433 | 1.60E-30 |
| rs174548 | miR-1908-5 | cis | 3.01E-08 | Nightingale | XS_VLDL_T | NA | G | C | 0.032782 | 0.004376 | 1.00E-13 |
| rs174548 | miR-1908-5 | cis | 3.01E-08 | Nightingale | XL_VLDL_T | NA | G | C | 0.05736 | 0.00425 | 1.90E-43 |
| rs174548 | miR-1908-5 | cis | 3.01E-08 | Nightingale | M_HDL_PL | NA | G | C | 0.059061 | 0.004215 | 3.50E-46 |
| rs174548 | miR-1908-5 | cis | 3.01E-08 | Nightingale | L_VLDL_FC | NA | G | C | 0.027025 | 0.004402 | 9.70E-10 |
| rs174548 | miR-1908-5 | cis | 3.01E-08 | Nightingale | HDL_L | NA | G | C | -0.05114 | 0.00399 | 3.90E-40 |
| rs174548 | miR-1908-5 | cis | 3.01E-08 | Nightingale | XL_VLDL_C | NA | G | C | 0.030951 | 0.004374 | 4.90E-13 |
| rs174548 | miR-1908-5 | cis | 3.01E-08 | Nightingale | XXL_VLDL_ | NA | G | C | 0.071566 | 0.004283 | 7.90E-66 |
| rs174548 | miR-1908-5 | cis | 3.01E-08 | Nightingale | M_LDL_P | NA | G | C | -0.04512 | 0.004457 | 9.60E-25 |
| rs174548 | miR-1908-5 | cis | 3.01E-08 | Nightingale | M_LDL_PL_ | NA | G | C | 0.043635 | 0.004441 | 9.10E-24 |
| rs174548 | miR-1908-5 | cis | 3.01E-08 | Nightingale | L_LDL_TG_ | NA | G | C | 0.07695 | 0.004357 | 1.30E-71 |
| rs174548 | miR-1908-5 | cis | 3.01E-08 | Nightingale | S_VLDL_TG | NA | G | C | 0.063558 | 0.0043 | 4.90E-51 |
| rs174548 | miR-1908-5 | cis | 3.01E-08 | Nightingale | LDL_PL | NA | G | C | -0.04455 | 0.004434 | 8.80E-25 |
| rs174548 | miR-1908-5 | cis | 3.01E-08 | Nightingale | L_LDL_C_p | NA | G | C | -0.07538 | 0.00433 | 9.00E-70 |
| rs174548 | miR-1908-5 | cis | 3.01E-08 | Nightingale | M_LDL_TG | NA | G | C | 0.075585 | 0.004388 | 3.20E-68 |
| rs174548 | miR-1908-5 | cis | 3.01E-08 | Nightingale | L_HDL_FC | NA | G | C | -0.08123 | 0.003894 | 1.70E-104 |
| rs174548 | miR-1908-5 | cis | 3.01E-08 | Nightingale | Total_L | NA | G | C | -0.03445 | 0.004372 | 4.10E-16 |
| rs174548 | miR-1908-5 | cis | 3.01E-08 | Nightingale | XL_VLDL_F | NA | G | C | 0.044244 | 0.004304 | 5.50E-26 |
| rs174548 | miR-1908-5 | cis | 3.01E-08 | Nightingale | L_HDL_PL | NA | G | C | -0.07309 | 0.003911 | 2.10E-84 |
| rs174548 | miR-1908-5 | cis | 3.01E-08 | Nightingale | VLDL_TG | NA | G | C | 0.048387 | 0.004276 | 4.00E-31 |
| rs174548 | miR-1908-5 | cis | 3.01E-08 | Nightingale | LDL_P | NA | G | C | -0.03841 | 0.004441 | 8.40E-19 |
| rs174548 | miR-1908-5 | cis | 3.01E-08 | Nightingale | XS_VLDL_P | NA | G | C | -0.04052 | 0.004385 | 8.60E-22 |
| rs174548 | miR-1908-5 | cis | 3.01E-08 | Nightingale | PUFA | NA | G | C | -0.09262 | 0.004334 | 3.20E-107 |
| rs174548 | miR-1908-5 | cis | 3.01E-08 | Nightingale | XL_HDL_TC | NA | G | C | 0.085883 | 0.004186 | 2.90E-99 |
| rs174548 | miR-1908-5 | cis | 3.01E-08 | Nightingale | L_HDL_C | NA | G | C | -0.07744 | 0.003913 | 3.70E-94 |
| rs174548 | miR-1908-5 | cis | 3.01E-08 | Nightingale | XS_VLDL_P | NA | G | C | -0.03105 | 0.004382 | 1.10E-13 |
| rs174548 | miR-1908-5 | cis | 3.01E-08 | Nightingale | LDL_CE | NA | G | C | -0.04183 | 0.004433 | 4.00E-22 |
| rs174548 | miR-1908-5 | cis | 3.01E-08 | Nightingale | DHA | NA | G | C | -0.28258 | 0.004282 | 0 |
| rs174548 | miR-1908-5 | cis | 3.01E-08 | Nightingale | M_VLDL_T | NA | G | C | 0.076426 | 0.004197 | 1.50E-77 |
| rs174548 | miR-1908-5 | cis | 3.01E-08 | Nightingale | L_HDL_CE | NA | G | C | -0.07602 | 0.003924 | 4.10E-90 |
| rs174548 | miR-1908-5 | cis | 3.01E-08 | Nightingale | Sphingomy | NA | G | C | -0.11192 | 0.004153 | 1.10E-167 |
| rs174548 | miR-1908-5 | cis | 3.01E-08 | Nightingale | M_HDL_L | NA | G | C | -0.02638 | 0.004125 | 3.20E-11 |
| rs174548 | miR-1908-5 | cis | 3.01E-08 | Nightingale | L_VLDL_C_ | NA | G | C | -0.03912 | 0.004341 | 8.30E-21 |
| rs174548 | miR-1908-5 | cis | 3.01E-08 | Nightingale | M_LDL_C | NA | G | C | -0.03297 | 0.004453 | 4.70E-14 |
| rs174548 | miR-1908-5 | cis | 3.01E-08 | Nightingale | L_LDL_FC | NA | G | C | -0.06615 | 0.004363 | 7.80E-55 |
| rs174548 | miR-1908-5 | cis | 3.01E-08 | Nightingale | XS_VLDL_C | NA | G | C | -0.09027 | 0.004189 | 2.40E-108 |

| | | | | | | | | | | | |
|---|---|---|---|---|---|---|---|---|---|---|---|
| rs174548 | miR-1908-! | cis | 3.01E-08 | Nightingale | LDL_TG | NA | G | C | 0.031248 | 0.004395 | 9.80E-13 |
| rs174548 | miR-1908-! | cis | 3.01E-08 | Nightingale | S_HDL_C_p | NA | G | C | -0.04451 | 0.004392 | 7.80E-24 |
| rs174548 | miR-1908-! | cis | 3.01E-08 | Nightingale | Cholines | NA | G | C | -0.03452 | 0.004179 | 7.20E-18 |
| rs174548 | miR-1908-! | cis | 3.01E-08 | Nightingale | Total_P | NA | G | C | -0.03903 | 0.004182 | 6.40E-22 |
| rs174548 | miR-1908-! | cis | 3.01E-08 | Nightingale | L_HDL_PL_ | NA | G | C | 0.042111 | 0.004301 | 1.10E-23 |
| rs174548 | miR-1908-! | cis | 3.01E-08 | Nightingale | Total_PL | NA | G | C | -0.04789 | 0.004232 | 2.00E-31 |
| rs174548 | miR-1908-! | cis | 3.01E-08 | Nightingale | S_VLDL_TG | NA | G | C | 0.0391 | 0.004312 | 1.00E-20 |
| rs174548 | miR-1908-! | cis | 3.01E-08 | Nightingale | L_HDL_CE_ | NA | G | C | -0.05028 | 0.004252 | 4.10E-34 |
| rs174548 | miR-1908-! | cis | 3.01E-08 | Nightingale | XL_HDL_L | NA | G | C | -0.08906 | 0.003993 | 3.40E-119 |
| rs174548 | miR-1908-! | cis | 3.01E-08 | Nightingale | XXL_VLDL_ | NA | G | C | 0.066931 | 0.00428 | 5.50E-58 |
| rs174548 | miR-1908-! | cis | 3.01E-08 | Nightingale | L_HDL_C_p | NA | G | C | -0.0674 | 0.004189 | 4.70E-62 |
| rs174548 | miR-1908-! | cis | 3.01E-08 | Nightingale | IDL_FC_pct | NA | G | C | -0.10417 | 0.004408 | 8.00E-125 |
| rs174548 | miR-1908-! | cis | 3.01E-08 | Nightingale | Omega_3_ | NA | G | C | -0.38268 | 0.004342 | 0 |
| rs174548 | miR-1908-! | cis | 3.01E-08 | Nightingale | Citrate | NA | G | C | -0.03355 | 0.004416 | 3.70E-15 |
| rs174548 | miR-1908-! | cis | 3.01E-08 | Nightingale | M_HDL_FC | NA | G | C | -0.09957 | 0.003938 | 7.19E-153 |
| rs174548 | miR-1908-! | cis | 3.01E-08 | Nightingale | M_VLDL_C | NA | G | C | -0.07487 | 0.004172 | 5.80E-75 |
| rs174548 | miR-1908-! | cis | 3.01E-08 | Nightingale | XL_VLDL_P | NA | G | C | 0.053221 | 0.004271 | 2.30E-37 |
| rs174548 | miR-1908-! | cis | 3.01E-08 | Nightingale | IDL_TG_pct | NA | G | C | 0.086656 | 0.004293 | 1.70E-94 |
| rs174548 | miR-1908-! | cis | 3.01E-08 | Nightingale | IDL_CE_pct | NA | G | C | -0.03719 | 0.004361 | 1.30E-17 |
| rs174548 | miR-1908-! | cis | 3.01E-08 | Nightingale | XL_HDL_C | NA | G | C | -0.09543 | 0.004007 | 3.30E-135 |
| rs174548 | miR-1908-! | cis | 3.01E-08 | Nightingale | S_HDL_TG | NA | G | C | 0.068446 | 0.004309 | 6.10E-60 |
| rs174548 | miR-1908-! | cis | 3.01E-08 | Nightingale | L_HDL_L | NA | G | C | -0.07425 | 0.003899 | 1.20E-87 |
| rs174548 | miR-1908-! | cis | 3.01E-08 | Nightingale | MUFA_pct | NA | G | C | 0.193804 | 0.004253 | 0 |
| rs174548 | miR-1908-! | cis | 3.01E-08 | Nightingale | XS_VLDL_F | NA | G | C | -0.11022 | 0.004411 | 8.71E-141 |
| rs174548 | miR-1908-! | cis | 3.01E-08 | Nightingale | L_VLDL_C | NA | G | C | 0.029596 | 0.004346 | 3.00E-12 |
| rs174548 | miR-1908-! | cis | 3.01E-08 | Nightingale | M_LDL_CE | NA | G | C | -0.02529 | 0.00445 | 8.00E-09 |
| rs174548 | miR-1908-! | cis | 3.01E-08 | Nightingale | XL_VLDL_F | NA | G | C | -0.06791 | 0.004306 | 1.50E-58 |
| rs174548 | miR-1908-! | cis | 3.01E-08 | Nightingale | S_LDL_L | NA | G | C | -0.03095 | 0.004443 | 9.00E-13 |
| rs174548 | miR-1908-! | cis | 3.01E-08 | Nightingale | XL_VLDL_C | NA | G | C | 0.038178 | 0.00434 | 2.00E-19 |
| rs174548 | miR-1908-! | cis | 3.01E-08 | Nightingale | PUFA_by_I | NA | G | C | -0.1678 | 0.004256 | 0 |
| rs174548 | miR-1908-! | cis | 3.01E-08 | Nightingale | ApoB | NA | G | C | -0.03643 | 0.004438 | 3.50E-17 |
| rs174548 | miR-1908-! | cis | 3.01E-08 | Nightingale | S_HDL_TG_ | NA | G | C | 0.068847 | 0.004226 | 1.10E-62 |
| rs174548 | miR-1908-! | cis | 3.01E-08 | Nightingale | L_HDL_TG_ | NA | G | C | 0.078121 | 0.004193 | 4.60E-82 |
| rs174548 | miR-1908-! | cis | 3.01E-08 | Nightingale | XS_VLDL_P | NA | G | C | 0.058312 | 0.004336 | 2.70E-42 |
| rs174548 | miR-1908-! | cis | 3.01E-08 | Nightingale | IDL_L | NA | G | C | -0.06914 | 0.004296 | 2.90E-62 |
| rs174548 | miR-1908-! | cis | 3.01E-08 | Nightingale | M_VLDL_F | NA | G | C | -0.08914 | 0.004225 | 6.70E-104 |
| rs174548 | miR-1908-! | cis | 3.01E-08 | Nightingale | M_VLDL_P | NA | G | C | -0.06056 | 0.004264 | 6.00E-49 |
| rs174548 | miR-1908-! | cis | 3.01E-08 | Nightingale | M_HDL_CE | NA | G | C | -0.03711 | 0.004104 | 1.40E-20 |
| rs174548 | miR-1908-! | cis | 3.01E-08 | Nightingale | XL_HDL_PL | NA | G | C | -0.06346 | 0.004082 | 9.60E-59 |
| rs174548 | miR-1908-! | cis | 3.01E-08 | Nightingale | M_HDL_TG | NA | G | C | 0.067099 | 0.004261 | 1.80E-58 |
| rs174548 | miR-1908-! | cis | 3.01E-08 | Nightingale | S_LDL_FC_ | NA | G | C | -0.08058 | 0.004306 | 1.70E-81 |
| rs174548 | miR-1908-! | cis | 3.01E-08 | Nightingale | M_LDL_L | NA | G | C | -0.02712 | 0.004451 | 6.00E-10 |
| rs174548 | miR-1908-! | cis | 3.01E-08 | Nightingale | XL_HDL_FC | NA | G | C | 0.059042 | 0.00394 | 1.20E-55 |
| rs174548 | miR-1908-! | cis | 3.01E-08 | Nightingale | TG_by_PG | NA | G | C | 0.068463 | 0.004124 | 1.80E-65 |
| rs174548 | miR-1908-! | cis | 3.01E-08 | Nightingale | M_LDL_TG | NA | G | C | 0.037781 | 0.004378 | 2.20E-18 |
| rs174548 | miR-1908-! | cis | 3.01E-08 | Nightingale | XS_VLDL_L | NA | G | C | -0.03826 | 0.004368 | 6.10E-20 |
| rs174548 | miR-1908-! | cis | 3.01E-08 | Nightingale | L_HDL_FC_ | NA | G | C | -0.1072 | 0.004128 | 4.90E-160 |
| rs174548 | miR-1908-! | cis | 3.01E-08 | Nightingale | M_VLDL_C | NA | G | C | -0.04203 | 0.004432 | 3.20E-22 |
| rs174549 | miR-1908-! | cis | 4.04E-08 | Nightingale | Sphingomy | NA | A | G | -0.11204 | 0.004173 | 2.80E-166 |
| rs174549 | miR-1908-! | cis | 4.04E-08 | Nightingale | S_VLDL_C_ | NA | A | G | -0.05578 | 0.004345 | 7.00E-39 |
| rs174549 | miR-1908-! | cis | 4.04E-08 | Nightingale | L_LDL_P | NA | A | G | -0.03576 | 0.004451 | 1.40E-16 |
| rs174549 | miR-1908-! | cis | 4.04E-08 | Nightingale | XL_VLDL_F | NA | A | G | -0.06809 | 0.004327 | 3.20E-58 |
| rs174549 | miR-1908-! | cis | 4.04E-08 | Nightingale | XL_VLDL_C | NA | A | G | 0.038632 | 0.004362 | 1.20E-19 |
| rs174549 | miR-1908-! | cis | 4.04E-08 | Nightingale | DHA | NA | A | G | -0.28236 | 0.004304 | 0 |
| rs174549 | miR-1908-! | cis | 4.04E-08 | Nightingale | L_VLDL_FC | NA | A | G | 0.02734 | 0.004424 | 6.00E-10 |
| rs174549 | miR-1908-! | cis | 4.04E-08 | Nightingale | S_LDL_C | NA | A | G | -0.03073 | 0.004471 | 2.30E-12 |
| rs174549 | miR-1908-! | cis | 4.04E-08 | Nightingale | Total_P | NA | A | G | -0.03889 | 0.004203 | 1.20E-21 |
| rs174549 | miR-1908-! | cis | 4.04E-08 | Nightingale | M_VLDL_F | NA | A | G | -0.02725 | 0.004465 | 4.90E-10 |
| rs174549 | miR-1908-! | cis | 4.04E-08 | Nightingale | Total_CE | NA | A | G | -0.06444 | 0.004308 | 3.20E-53 |
| rs174549 | miR-1908-! | cis | 4.04E-08 | Nightingale | XXL_VLDL_ | NA | A | G | 0.042873 | 0.004488 | 2.70E-22 |
| rs174549 | miR-1908-! | cis | 4.04E-08 | Nightingale | IDL_PL | NA | A | G | -0.06954 | 0.004309 | 1.10E-62 |
| rs174549 | miR-1908-! | cis | 4.04E-08 | Nightingale | XL_HDL_TG | NA | A | G | 0.086347 | 0.004207 | 1.80E-99 |
| rs174549 | miR-1908-! | cis | 4.04E-08 | Nightingale | XS_VLDL_T | NA | A | G | 0.032883 | 0.004397 | 1.50E-13 |

| | | | | | | | | | | | |
|---|---|---|---|---|---|---|---|---|---|---|---|
| rs174549 | miR-1908-! | cis | 4.04E-08 | Nightingale | Total_PL | NA | A | G | -0.04794 | 0.004253 | 2.40E-31 |
| rs174549 | miR-1908-! | cis | 4.04E-08 | Nightingale | L_HDL_L | NA | A | G | -0.07464 | 0.003919 | 4.20E-88 |
| rs174549 | miR-1908-! | cis | 4.04E-08 | Nightingale | S_LDL_PL | NA | A | G | -0.04961 | 0.004455 | 3.40E-30 |
| rs174549 | miR-1908-! | cis | 4.04E-08 | Nightingale | L_VLDL_L | NA | A | G | 0.039287 | 0.004316 | 9.80E-21 |
| rs174549 | miR-1908-! | cis | 4.04E-08 | Nightingale | XXL_VLDL_ | NA | A | G | 0.07209 | 0.004304 | 5.90E-66 |
| rs174549 | miR-1908-! | cis | 4.04E-08 | Nightingale | IDL_FC_pct | NA | A | G | -0.10399 | 0.00443 | 4.50E-123 |
| rs174549 | miR-1908-! | cis | 4.04E-08 | Nightingale | IDL_CE_pct | NA | A | G | -0.03708 | 0.004382 | 4.00E-17 |
| rs174549 | miR-1908-! | cis | 4.04E-08 | Nightingale | MUFA | NA | A | G | 0.064463 | 0.00442 | 1.00E-49 |
| rs174549 | miR-1908-! | cis | 4.04E-08 | Nightingale | M_HDL_TG | NA | A | G | 0.051741 | 0.004411 | 1.80E-32 |
| rs174549 | miR-1908-! | cis | 4.04E-08 | Nightingale | HDL_TG | NA | A | G | 0.047713 | 0.004404 | 7.70E-28 |
| rs174549 | miR-1908-! | cis | 4.04E-08 | Nightingale | VLDL_L | NA | A | G | 0.028974 | 0.004379 | 1.30E-11 |
| rs174549 | miR-1908-! | cis | 4.04E-08 | Nightingale | XL_HDL_C_ | NA | A | G | 0.021554 | 0.004266 | 2.40E-08 |
| rs174549 | miR-1908-! | cis | 4.04E-08 | Nightingale | M_LDL_TG | NA | A | G | 0.075569 | 0.00441 | 3.20E-67 |
| rs174549 | miR-1908-! | cis | 4.04E-08 | Nightingale | XL_HDL_P | NA | A | G | -0.08643 | 0.004019 | 7.69E-112 |
| rs174549 | miR-1908-! | cis | 4.04E-08 | Nightingale | IDL_L | NA | A | G | -0.06923 | 0.004317 | 1.10E-61 |
| rs174549 | miR-1908-! | cis | 4.04E-08 | Nightingale | M_LDL_FC | NA | A | G | -0.05118 | 0.004461 | 7.30E-32 |
| rs174549 | miR-1908-! | cis | 4.04E-08 | Nightingale | S_LDL_TG | NA | A | G | 0.051479 | 0.004357 | 2.90E-33 |
| rs174549 | miR-1908-! | cis | 4.04E-08 | Nightingale | HDL_L | NA | A | G | -0.05128 | 0.00401 | 3.50E-40 |
| rs174549 | miR-1908-! | cis | 4.04E-08 | Nightingale | XS_VLDL_C | NA | A | G | -0.0684 | 0.004309 | 2.60E-60 |
| rs174549 | miR-1908-! | cis | 4.04E-08 | Nightingale | XS_VLDL_P | NA | A | G | -0.03123 | 0.004403 | 9.10E-14 |
| rs174549 | miR-1908-! | cis | 4.04E-08 | Nightingale | XS_VLDL_F | NA | A | G | -0.05264 | 0.004388 | 2.40E-35 |
| rs174549 | miR-1908-! | cis | 4.04E-08 | Nightingale | L_VLDL_PL | NA | A | G | 0.049558 | 0.004309 | 3.40E-32 |
| rs174549 | miR-1908-! | cis | 4.04E-08 | Nightingale | S_HDL_FC_ | NA | A | G | -0.09968 | 0.004236 | 3.90E-135 |
| rs174549 | miR-1908-! | cis | 4.04E-08 | Nightingale | L_HDL_FC | NA | A | G | -0.08172 | 0.003914 | 3.90E-105 |
| rs174549 | miR-1908-! | cis | 4.04E-08 | Nightingale | LDL_TG | NA | A | G | 0.031368 | 0.004416 | 1.30E-12 |
| rs174549 | miR-1908-! | cis | 4.04E-08 | Nightingale | L_HDL_C_p | NA | A | G | -0.068 | 0.004209 | 8.50E-63 |
| rs174549 | miR-1908-! | cis | 4.04E-08 | Nightingale | M_LDL_P | NA | A | G | -0.04481 | 0.004479 | 4.00E-24 |
| rs174549 | miR-1908-! | cis | 4.04E-08 | Nightingale | M_LDL_PL_ | NA | A | G | 0.042919 | 0.004463 | 5.40E-23 |
| rs174549 | miR-1908-! | cis | 4.04E-08 | Nightingale | IDL_CE | NA | A | G | -0.06922 | 0.004311 | 1.90E-61 |
| rs174549 | miR-1908-! | cis | 4.04E-08 | Nightingale | XS_VLDL_L | NA | A | G | -0.03835 | 0.00439 | 7.30E-20 |
| rs174549 | miR-1908-! | cis | 4.04E-08 | Nightingale | S_LDL_PL_ | NA | A | G | -0.0615 | 0.004457 | 2.60E-45 |
| rs174549 | miR-1908-! | cis | 4.04E-08 | Nightingale | M_VLDL_F | NA | A | G | -0.08942 | 0.004246 | 2.00E-103 |
| rs174549 | miR-1908-! | cis | 4.04E-08 | Nightingale | M_HDL_L | NA | A | G | -0.02626 | 0.004146 | 4.10E-11 |
| rs174549 | miR-1908-! | cis | 4.04E-08 | Nightingale | XS_VLDL_C | NA | A | G | -0.09035 | 0.00421 | 2.20E-107 |
| rs174549 | miR-1908-! | cis | 4.04E-08 | Nightingale | XL_HDL_CE | NA | A | G | -0.02805 | 0.004463 | 1.20E-09 |
| rs174549 | miR-1908-! | cis | 4.04E-08 | Nightingale | Total_L | NA | A | G | -0.03439 | 0.004394 | 6.00E-16 |
| rs174549 | miR-1908-! | cis | 4.04E-08 | Nightingale | VLDL_TG | NA | A | G | 0.0488 | 0.004298 | 2.70E-31 |
| rs174549 | miR-1908-! | cis | 4.04E-08 | Nightingale | S_VLDL_TG | NA | A | G | 0.039305 | 0.004334 | 1.20E-20 |
| rs174549 | miR-1908-! | cis | 4.04E-08 | Nightingale | XXL_VLDL_ | NA | A | G | 0.067409 | 0.004301 | 3.80E-58 |
| rs174549 | miR-1908-! | cis | 4.04E-08 | Nightingale | XL_HDL_C | NA | A | G | -0.09622 | 0.004027 | 1.30E-136 |
| rs174549 | miR-1908-! | cis | 4.04E-08 | Nightingale | HDL_size | NA | A | G | -0.06993 | 0.003957 | 7.80E-76 |
| rs174549 | miR-1908-! | cis | 4.04E-08 | Nightingale | XL_VLDL_C | NA | A | G | -0.06172 | 0.004282 | 1.90E-48 |
| rs174549 | miR-1908-! | cis | 4.04E-08 | Nightingale | S_LDL_CE_ | NA | A | G | 0.057421 | 0.004458 | 1.20E-39 |
| rs174549 | miR-1908-! | cis | 4.04E-08 | Nightingale | XL_HDL_PL | NA | A | G | -0.06423 | 0.004102 | 4.80E-60 |
| rs174549 | miR-1908-! | cis | 4.04E-08 | Nightingale | LDL_PL | NA | A | G | -0.04446 | 0.004456 | 2.40E-24 |
| rs174549 | miR-1908-! | cis | 4.04E-08 | Nightingale | XS_VLDL_F | NA | A | G | -0.1104 | 0.004432 | 8.89E-140 |
| rs174549 | miR-1908-! | cis | 4.04E-08 | Nightingale | S_VLDL_FC | NA | A | G | -0.08153 | 0.004264 | 4.60E-85 |
| rs174549 | miR-1908-! | cis | 4.04E-08 | Nightingale | S_LDL_TG_ | NA | A | G | 0.086059 | 0.004327 | 3.60E-91 |
| rs174549 | miR-1908-! | cis | 4.04E-08 | Nightingale | PUFA_by_N | NA | A | G | -0.16777 | 0.004278 | 0 |
| rs174549 | miR-1908-! | cis | 4.04E-08 | Nightingale | HDL_C | NA | A | G | -0.06307 | 0.003988 | 9.30E-61 |
| rs174549 | miR-1908-! | cis | 4.04E-08 | Nightingale | M_HDL_PL | NA | A | G | 0.059427 | 0.004236 | 2.60E-46 |
| rs174549 | miR-1908-! | cis | 4.04E-08 | Nightingale | L_LDL_FC_ | NA | A | G | -0.08217 | 0.004257 | 3.50E-86 |
| rs174549 | miR-1908-! | cis | 4.04E-08 | Nightingale | M_LDL_L | NA | A | G | -0.02688 | 0.004473 | 1.20E-09 |
| rs174549 | miR-1908-! | cis | 4.04E-08 | Nightingale | M_LDL_C | NA | A | G | -0.03269 | 0.004475 | 1.20E-13 |
| rs174549 | miR-1908-! | cis | 4.04E-08 | Nightingale | M_LDL_TG | NA | A | G | 0.037939 | 0.0044 | 2.90E-18 |
| rs174549 | miR-1908-! | cis | 4.04E-08 | Nightingale | LDL_P | NA | A | G | -0.03841 | 0.004463 | 1.50E-18 |
| rs174549 | miR-1908-! | cis | 4.04E-08 | Nightingale | L_HDL_C | NA | A | G | -0.07789 | 0.003933 | 9.80E-95 |
| rs174549 | miR-1908-! | cis | 4.04E-08 | Nightingale | HDL_P | NA | A | G | -0.03442 | 0.004193 | 2.70E-17 |
| rs174549 | miR-1908-! | cis | 4.04E-08 | Nightingale | LDL_CE | NA | A | G | -0.04175 | 0.004455 | 9.00E-22 |
| rs174549 | miR-1908-! | cis | 4.04E-08 | Nightingale | L_VLDL_C_ | NA | A | G | -0.03895 | 0.004363 | 2.30E-20 |
| rs174549 | miR-1908-! | cis | 4.04E-08 | Nightingale | M_HDL_FC | NA | A | G | -0.04946 | 0.004064 | 1.80E-36 |
| rs174549 | miR-1908-! | cis | 4.04E-08 | Nightingale | ApoA1 | NA | A | G | -0.03947 | 0.004087 | 8.90E-24 |
| rs174549 | miR-1908-! | cis | 4.04E-08 | Nightingale | L_HDL_CE_ | NA | A | G | -0.05083 | 0.004273 | 1.10E-34 |

| | | | | | | | | | | | |
|---|---|---|---|---|---|---|---|---|---|---|---|
| rs174549 | miR-1908-5 | cis | 4.04E-08 | Nightingale | Remnant_( | NA | A | G | -0.0413 | 0.004437 | 9.50E-22 |
| rs174549 | miR-1908-5 | cis | 4.04E-08 | Nightingale | XL_VLDL_T | NA | A | G | 0.057863 | 0.004272 | 8.00E-44 |
| rs174549 | miR-1908-5 | cis | 4.04E-08 | Nightingale | M_LDL_FC_ | NA | A | G | -0.07212 | 0.004278 | 2.40E-66 |
| rs174549 | miR-1908-5 | cis | 4.04E-08 | Nightingale | L_VLDL_TG | NA | A | G | 0.0385 | 0.004297 | 4.00E-20 |
| rs174549 | miR-1908-5 | cis | 4.04E-08 | Nightingale | XXL_VLDL_ | NA | A | G | 0.066979 | 0.004295 | 8.90E-58 |
| rs174549 | miR-1908-5 | cis | 4.04E-08 | Nightingale | LDL_C | NA | A | G | -0.04793 | 0.004447 | 2.60E-28 |
| rs174549 | miR-1908-5 | cis | 4.04E-08 | Nightingale | Citrate | NA | A | G | -0.03418 | 0.004438 | 1.20E-15 |
| rs174549 | miR-1908-5 | cis | 4.04E-08 | Nightingale | S_HDL_C_p | NA | A | G | -0.04462 | 0.004414 | 1.60E-23 |
| rs174549 | miR-1908-5 | cis | 4.04E-08 | Nightingale | XL_VLDL_T | NA | A | G | 0.059995 | 0.004327 | 3.10E-45 |
| rs174549 | miR-1908-5 | cis | 4.04E-08 | Nightingale | Total_FC | NA | A | G | -0.05662 | 0.004363 | 9.40E-41 |
| rs174549 | miR-1908-5 | cis | 4.04E-08 | Nightingale | L_VLDL_C | NA | A | G | 0.029991 | 0.004368 | 2.00E-12 |
| rs174549 | miR-1908-5 | cis | 4.04E-08 | Nightingale | IDL_P | NA | A | G | -0.04226 | 0.004419 | 8.00E-23 |
| rs174549 | miR-1908-5 | cis | 4.04E-08 | Nightingale | XXL_VLDL_ | NA | A | G | 0.07269 | 0.004286 | 7.30E-68 |
| rs174549 | miR-1908-5 | cis | 4.04E-08 | Nightingale | M_LDL_CE | NA | A | G | -0.02496 | 0.004473 | 1.60E-08 |
| rs174549 | miR-1908-5 | cis | 4.04E-08 | Nightingale | XS_VLDL_C | NA | A | G | -0.06463 | 0.004334 | 1.70E-53 |
| rs174549 | miR-1908-5 | cis | 4.04E-08 | Nightingale | XL_VLDL_P | NA | A | G | 0.049368 | 0.00431 | 8.10E-32 |
| rs174549 | miR-1908-5 | cis | 4.04E-08 | Nightingale | XXL_VLDL_ | NA | A | G | 0.073759 | 0.004313 | 2.20E-68 |
| rs174549 | miR-1908-5 | cis | 4.04E-08 | Nightingale | VLDL_size | NA | A | G | 0.072305 | 0.004163 | 7.70E-72 |
| rs174549 | miR-1908-5 | cis | 4.04E-08 | Nightingale | S_VLDL_CE | NA | A | G | -0.0287 | 0.004406 | 2.60E-11 |
| rs174549 | miR-1908-5 | cis | 4.04E-08 | Nightingale | PUFA | NA | A | G | -0.09246 | 0.004355 | 4.60E-106 |
| rs174549 | miR-1908-5 | cis | 4.04E-08 | Nightingale | LA_pct | NA | A | G | 0.181254 | 0.004364 | 0 |
| rs174549 | miR-1908-5 | cis | 4.04E-08 | Nightingale | S_VLDL_TG | NA | A | G | 0.063611 | 0.004321 | 1.50E-50 |
| rs174549 | miR-1908-5 | cis | 4.04E-08 | Nightingale | IDL_TG_pct | NA | A | G | 0.08678 | 0.004314 | 1.70E-93 |
| rs174549 | miR-1908-5 | cis | 4.04E-08 | Nightingale | ApoB | NA | A | G | -0.03635 | 0.00446 | 7.00E-17 |
| rs174549 | miR-1908-5 | cis | 4.04E-08 | Nightingale | M_VLDL_T | NA | A | G | 0.0767 | 0.004218 | 3.10E-77 |
| rs174549 | miR-1908-5 | cis | 4.04E-08 | Nightingale | HDL_FC | NA | A | G | -0.07115 | 0.003956 | 5.10E-78 |
| rs174549 | miR-1908-5 | cis | 4.04E-08 | Nightingale | M_HDL_P | NA | A | G | -0.03287 | 0.004114 | 8.60E-17 |
| rs174549 | miR-1908-5 | cis | 4.04E-08 | Nightingale | L_VLDL_FC | NA | A | G | 0.041079 | 0.00432 | 1.60E-22 |
| rs174549 | miR-1908-5 | cis | 4.04E-08 | Nightingale | Cholines | NA | A | G | -0.03454 | 0.0042 | 7.50E-18 |
| rs174549 | miR-1908-5 | cis | 4.04E-08 | Nightingale | XL_HDL_CE | NA | A | G | -0.09435 | 0.003999 | 3.30E-133 |
| rs174549 | miR-1908-5 | cis | 4.04E-08 | Nightingale | XL_HDL_L | NA | A | G | -0.0899 | 0.004013 | 8.89E-121 |
| rs174549 | miR-1908-5 | cis | 4.04E-08 | Nightingale | M_HDL_CE | NA | A | G | -0.04904 | 0.004331 | 1.40E-30 |
| rs174549 | miR-1908-5 | cis | 4.04E-08 | Nightingale | M_LDL_C_ | NA | A | G | -0.09829 | 0.004469 | 1.60E-107 |
| rs174549 | miR-1908-5 | cis | 4.04E-08 | Nightingale | XS_VLDL_T | NA | A | G | 0.095148 | 0.004242 | 1.80E-117 |
| rs174549 | miR-1908-5 | cis | 4.04E-08 | Nightingale | XS_VLDL_P | NA | A | G | 0.057636 | 0.004357 | 9.70E-41 |
| rs174549 | miR-1908-5 | cis | 4.04E-08 | Nightingale | L_HDL_TG_ | NA | A | G | 0.078256 | 0.004214 | 1.00E-81 |
| rs174549 | miR-1908-5 | cis | 4.04E-08 | Nightingale | L_VLDL_CE | NA | A | G | -0.05891 | 0.004274 | 7.40E-46 |
| rs174549 | miR-1908-5 | cis | 4.04E-08 | Nightingale | M_HDL_TG | NA | A | G | 0.067269 | 0.004283 | 3.50E-58 |
| rs174549 | miR-1908-5 | cis | 4.04E-08 | Nightingale | XL_HDL_FC | NA | A | G | -0.09891 | 0.00416 | 2.70E-134 |
| rs174549 | miR-1908-5 | cis | 4.04E-08 | Nightingale | XL_VLDL_C | NA | A | G | 0.031397 | 0.004396 | 3.00E-13 |
| rs174549 | miR-1908-5 | cis | 4.04E-08 | Nightingale | Tyr | NA | A | G | 0.03287 | 0.00446 | 1.70E-13 |
| rs174549 | miR-1908-5 | cis | 4.04E-08 | Nightingale | S_VLDL_PL | NA | A | G | -0.07757 | 0.004273 | 4.10E-77 |
| rs174549 | miR-1908-5 | cis | 4.04E-08 | Nightingale | M_HDL_C_ | NA | A | G | -0.0666 | 0.004251 | 3.30E-58 |
| rs174549 | miR-1908-5 | cis | 4.04E-08 | Nightingale | XXL_VLDL_ | NA | A | G | -0.0291 | 0.004442 | 9.30E-12 |
| rs174549 | miR-1908-5 | cis | 4.04E-08 | Nightingale | S_LDL_P | NA | A | G | -0.03023 | 0.00447 | 5.20E-12 |
| rs174549 | miR-1908-5 | cis | 4.04E-08 | Nightingale | L_HDL_FC_ | NA | A | G | -0.10793 | 0.004149 | 2.40E-161 |
| rs174549 | miR-1908-5 | cis | 4.04E-08 | Nightingale | non_HDL_( | NA | A | G | -0.04545 | 0.004442 | 9.70E-26 |
| rs174549 | miR-1908-5 | cis | 4.04E-08 | Nightingale | DHA_pct | NA | A | G | -0.2692 | 0.004335 | 0 |
| rs174549 | miR-1908-5 | cis | 4.04E-08 | Nightingale | S_HDL_TG_ | NA | A | G | 0.068966 | 0.004247 | 3.40E-62 |
| rs174549 | miR-1908-5 | cis | 4.04E-08 | Nightingale | L_LDL_PL | NA | A | G | -0.05301 | 0.004432 | 2.20E-34 |
| rs174549 | miR-1908-5 | cis | 4.04E-08 | Nightingale | IDL_FC | NA | A | G | -0.08666 | 0.004323 | 2.70E-94 |
| rs174549 | miR-1908-5 | cis | 4.04E-08 | Nightingale | L_HDL_PL_ | NA | A | G | 0.042956 | 0.004322 | 1.40E-24 |
| rs174549 | miR-1908-5 | cis | 4.04E-08 | Nightingale | M_VLDL_C | NA | A | G | -0.05073 | 0.004425 | 1.10E-31 |
| rs174549 | miR-1908-5 | cis | 4.04E-08 | Nightingale | M_VLDL_C | NA | A | G | -0.07516 | 0.004193 | 1.10E-74 |
| rs174549 | miR-1908-5 | cis | 4.04E-08 | Nightingale | L_VLDL_P | NA | A | G | 0.042841 | 0.004312 | 2.10E-24 |
| rs174549 | miR-1908-5 | cis | 4.04E-08 | Nightingale | Clinical_LD | NA | A | G | -0.05322 | 0.00444 | 1.60E-34 |
| rs174549 | miR-1908-5 | cis | 4.04E-08 | Nightingale | M_VLDL_C | NA | A | G | -0.04192 | 0.004454 | 8.50E-22 |
| rs174549 | miR-1908-5 | cis | 4.04E-08 | Nightingale | TG_by_PG | NA | A | G | 0.068794 | 0.004144 | 2.30E-65 |
| rs174549 | miR-1908-5 | cis | 4.04E-08 | Nightingale | XS_VLDL_C | NA | A | G | -0.09608 | 0.004232 | 8.61E-120 |
| rs174549 | miR-1908-5 | cis | 4.04E-08 | Nightingale | XL_VLDL_L | NA | A | G | 0.052474 | 0.004297 | 6.10E-36 |
| rs174549 | miR-1908-5 | cis | 4.04E-08 | Nightingale | L_LDL_FC | NA | A | G | -0.06622 | 0.004385 | 2.60E-54 |
| rs174549 | miR-1908-5 | cis | 4.04E-08 | Nightingale | S_LDL_FC_ | NA | A | G | -0.08056 | 0.004328 | 3.00E-80 |
| rs174549 | miR-1908-5 | cis | 4.04E-08 | Nightingale | LDL_L | NA | A | G | -0.04363 | 0.004451 | 1.10E-23 |

| | | | | | | | | | | | |
|---|---|---|---|---|---|---|---|---|---|---|---|
| rs174549 | miR-1908-! | cis | 4.04E-08 | Nightingale | Omega_6_ | NA | A | G | 0.362558 | 0.004401 | 0 |
| rs174549 | miR-1908-! | cis | 4.04E-08 | Nightingale | M_HDL_CE | NA | A | G | -0.03707 | 0.004124 | 2.00E-20 |
| rs174549 | miR-1908-! | cis | 4.04E-08 | Nightingale | XL_VLDL_P | NA | A | G | 0.05373 | 0.004292 | 1.10E-37 |
| rs174549 | miR-1908-! | cis | 4.04E-08 | Nightingale | S_HDL_TG | NA | A | G | 0.068781 | 0.00433 | 5.50E-60 |
| rs174549 | miR-1908-! | cis | 4.04E-08 | Nightingale | L_HDL_CE | NA | A | G | -0.07646 | 0.003943 | 1.10E-90 |
| rs174549 | miR-1908-! | cis | 4.04E-08 | Nightingale | L_LDL_C | NA | A | G | -0.05576 | 0.004416 | 1.90E-38 |
| rs174549 | miR-1908-! | cis | 4.04E-08 | Nightingale | L_HDL_PL | NA | A | G | -0.07344 | 0.00393 | 8.30E-85 |
| rs174549 | miR-1908-! | cis | 4.04E-08 | Nightingale | IDL_C | NA | A | G | -0.0742 | 0.004312 | 2.90E-70 |
| rs174549 | miR-1908-! | cis | 4.04E-08 | Nightingale | MUFA_pct | NA | A | G | 0.193652 | 0.004274 | 0 |
| rs174549 | miR-1908-! | cis | 4.04E-08 | Nightingale | LA | NA | A | G | 0.084007 | 0.004379 | 2.50E-82 |
| rs174549 | miR-1908-! | cis | 4.04E-08 | Nightingale | XL_VLDL_F | NA | A | G | 0.044756 | 0.004326 | 2.90E-26 |
| rs174549 | miR-1908-! | cis | 4.04E-08 | Nightingale | L_LDL_TG_ | NA | A | G | 0.07705 | 0.004379 | 8.40E-71 |
| rs174549 | miR-1908-! | cis | 4.04E-08 | Nightingale | SFA | NA | A | G | -0.02811 | 0.004426 | 1.50E-10 |
| rs174549 | miR-1908-! | cis | 4.04E-08 | Nightingale | Unsaturati | NA | A | G | -0.33309 | 0.00423 | 0 |
| rs174549 | miR-1908-! | cis | 4.04E-08 | Nightingale | M_HDL_C | NA | A | G | -0.03976 | 0.004109 | 1.70E-23 |
| rs174549 | miR-1908-! | cis | 4.04E-08 | Nightingale | S_LDL_L | NA | A | G | -0.03088 | 0.004465 | 1.60E-12 |
| rs174549 | miR-1908-! | cis | 4.04E-08 | Nightingale | PUFA_pct | NA | A | G | -0.12023 | 0.004296 | 7.80E-174 |
| rs174549 | miR-1908-! | cis | 4.04E-08 | Nightingale | M_HDL_FC | NA | A | G | -0.09983 | 0.003957 | 2.80E-152 |
| rs174549 | miR-1908-! | cis | 4.04E-08 | Nightingale | IDL_C_pct | NA | A | G | -0.07392 | 0.00436 | 4.60E-66 |
| rs174549 | miR-1908-! | cis | 4.04E-08 | Nightingale | Phosphogly | NA | A | G | -0.03808 | 0.00423 | 8.10E-21 |
| rs174549 | miR-1908-! | cis | 4.04E-08 | Nightingale | XXL_VLDL_ | NA | A | G | 0.068629 | 0.004302 | 2.30E-60 |
| rs174549 | miR-1908-! | cis | 4.04E-08 | Nightingale | LDL_FC | NA | A | G | -0.06248 | 0.004418 | 1.30E-47 |
| rs174549 | miR-1908-! | cis | 4.04E-08 | Nightingale | Total_TG | NA | A | G | 0.048165 | 0.004323 | 3.00E-30 |
| rs174549 | miR-1908-! | cis | 4.04E-08 | Nightingale | Omega_3 | NA | A | G | -0.32854 | 0.004377 | 0 |
| rs174549 | miR-1908-! | cis | 4.04E-08 | Nightingale | XL_HDL_PL | NA | A | G | -0.08594 | 0.004014 | 7.50E-111 |
| rs174549 | miR-1908-! | cis | 4.04E-08 | Nightingale | XXL_VLDL_ | NA | A | G | 0.062276 | 0.004309 | 1.40E-49 |
| rs174549 | miR-1908-! | cis | 4.04E-08 | Nightingale | L_LDL_CE | NA | A | G | -0.05129 | 0.004426 | 1.20E-32 |
| rs174549 | miR-1908-! | cis | 4.04E-08 | Nightingale | M_VLDL_P | NA | A | G | -0.06069 | 0.004285 | 1.20E-48 |
| rs174549 | miR-1908-! | cis | 4.04E-08 | Nightingale | L_LDL_C_p | NA | A | G | -0.07573 | 0.004351 | 1.40E-69 |
| rs174549 | miR-1908-! | cis | 4.04E-08 | Nightingale | XS_VLDL_P | NA | A | G | -0.04057 | 0.004406 | 1.20E-21 |
| rs174549 | miR-1908-! | cis | 4.04E-08 | Nightingale | L_HDL_P | NA | A | G | -0.07536 | 0.003911 | 6.10E-90 |
| rs174549 | miR-1908-! | cis | 4.04E-08 | Nightingale | M_VLDL_C | NA | A | G | -0.07958 | 0.004204 | 3.10E-83 |
| rs174549 | miR-1908-! | cis | 4.04E-08 | Nightingale | M_VLDL_T | NA | A | G | 0.032416 | 0.004339 | 1.90E-14 |
| rs174549 | miR-1908-! | cis | 4.04E-08 | Nightingale | L_VLDL_PL | NA | A | G | 0.087356 | 0.004334 | 1.00E-93 |
| rs174549 | miR-1908-! | cis | 4.04E-08 | Nightingale | S_LDL_FC | NA | A | G | -0.06138 | 0.004459 | 2.20E-45 |
| rs174549 | miR-1908-! | cis | 4.04E-08 | Nightingale | HDL_CE | NA | A | G | -0.06011 | 0.004008 | 1.10E-54 |
| rs174549 | miR-1908-! | cis | 4.04E-08 | Nightingale | L_LDL_L | NA | A | G | -0.05212 | 0.004422 | 1.00E-33 |
| rs174549 | miR-1908-! | cis | 4.04E-08 | Nightingale | Omega_3_ | NA | A | G | -0.38255 | 0.004364 | 0 |
| rs174549 | miR-1908-! | cis | 4.04E-08 | Nightingale | S_VLDL_FC | NA | A | G | -0.02658 | 0.004467 | 1.30E-09 |
| rs174549 | miR-1908-! | cis | 4.04E-08 | Nightingale | XL_HDL_FC | NA | A | G | 0.059815 | 0.003959 | 5.00E-57 |
| rs174549 | miR-1908-! | cis | 4.04E-08 | Nightingale | XL_VLDL_C | NA | A | G | -0.06444 | 0.004279 | 1.10E-52 |
| rs174549 | miR-1908-! | cis | 4.04E-08 | Nightingale | Total_C | NA | A | G | -0.06256 | 0.004323 | 5.80E-50 |
| rs174549 | miR-1908-! | cis | 4.04E-08 | Nightingale | SFA_pct | NA | A | G | -0.0396 | 0.004406 | 2.50E-19 |
| rs174549 | miR-1908-! | cis | 4.04E-08 | Nightingale | HDL_PL | NA | A | G | -0.04617 | 0.004037 | 1.70E-32 |
| rs174553 | miR-1908-! | cis | 2.61E-08 | Nightingale | XXL_VLDL_ | NA | G | A | 0.044137 | 0.004365 | 5.10E-25 |
| rs174553 | miR-1908-! | cis | 2.61E-08 | Nightingale | S_VLDL_C_ | NA | G | A | -0.05676 | 0.004224 | 2.90E-42 |
| rs174553 | miR-1908-! | cis | 2.61E-08 | Nightingale | M_LDL_FC | NA | G | A | -0.05245 | 0.004337 | 2.00E-35 |
| rs174553 | miR-1908-! | cis | 2.61E-08 | Nightingale | XXL_VLDL_ | NA | G | A | 0.066528 | 0.004176 | 7.70E-60 |
| rs174553 | miR-1908-! | cis | 2.61E-08 | Nightingale | S_HDL_FC_ | NA | G | A | -0.10304 | 0.004118 | 3.30E-153 |
| rs174553 | miR-1908-! | cis | 2.61E-08 | Nightingale | XL_HDL_FC | NA | G | A | -0.10316 | 0.004044 | 1.00E-154 |
| rs174553 | miR-1908-! | cis | 2.61E-08 | Nightingale | XS_VLDL_C | NA | G | A | -0.06834 | 0.004214 | 7.70E-63 |
| rs174553 | miR-1908-! | cis | 2.61E-08 | Nightingale | L_LDL_FC_ | NA | G | A | -0.08307 | 0.004138 | 7.50E-93 |
| rs174553 | miR-1908-! | cis | 2.61E-08 | Nightingale | HDL_C | NA | G | A | -0.06483 | 0.003877 | 5.00E-68 |
| rs174553 | miR-1908-! | cis | 2.61E-08 | Nightingale | XL_VLDL_C | NA | G | A | -0.06141 | 0.004162 | 1.20E-50 |
| rs174553 | miR-1908-! | cis | 2.61E-08 | Nightingale | L_LDL_C | NA | G | A | -0.05789 | 0.004293 | 9.00E-44 |
| rs174553 | miR-1908-! | cis | 2.61E-08 | Nightingale | M_LDL_C_ | NA | G | A | -0.10027 | 0.004345 | 2.00E-118 |
| rs174553 | miR-1908-! | cis | 2.61E-08 | Nightingale | XXL_VLDL_ | NA | G | A | 0.072092 | 0.004184 | 3.20E-69 |
| rs174553 | miR-1908-! | cis | 2.61E-08 | Nightingale | Omega_6_ | NA | G | A | 0.369862 | 0.004279 | 0 |
| rs174553 | miR-1908-! | cis | 2.61E-08 | Nightingale | Omega_6_ | NA | G | A | 0.023774 | 0.004226 | 1.00E-08 |
| rs174553 | miR-1908-! | cis | 2.61E-08 | Nightingale | M_VLDL_F | NA | G | A | -0.02934 | 0.004341 | 4.10E-12 |
| rs174553 | miR-1908-! | cis | 2.61E-08 | Nightingale | XL_VLDL_L | NA | G | A | 0.05186 | 0.004177 | 9.40E-37 |
| rs174553 | miR-1908-! | cis | 2.61E-08 | Nightingale | M_LDL_P | NA | G | A | -0.04663 | 0.004354 | 1.40E-27 |
| rs174553 | miR-1908-! | cis | 2.61E-08 | Nightingale | M_HDL_PL | NA | G | A | 0.058675 | 0.004118 | 4.90E-48 |

| | | | | | | | | | | | |
|---|---|---|---|---|---|---|---|---|---|---|---|
| rs174553 | miR-1908-5 | cis | 2.61E-08 | Nightingale | M_VLDL_C | NA | G | A | -0.08101 | 0.004087 | 3.90E-91 |
| rs174553 | miR-1908-5 | cis | 2.61E-08 | Nightingale | XS_VLDL_L | NA | G | A | -0.04169 | 0.004267 | 1.90E-24 |
| rs174553 | miR-1908-5 | cis | 2.61E-08 | Nightingale | XXL_VLDL_ | NA | G | A | 0.061613 | 0.00419 | 4.80E-51 |
| rs174553 | miR-1908-5 | cis | 2.61E-08 | Nightingale | M_HDL_PL | NA | G | A | -0.0213 | 0.00406 | 4.00E-08 |
| rs174553 | miR-1908-5 | cis | 2.61E-08 | Nightingale | L_LDL_PL | NA | G | A | -0.055 | 0.004309 | 5.00E-39 |
| rs174553 | miR-1908-5 | cis | 2.61E-08 | Nightingale | XS_VLDL_F | NA | G | A | -0.11311 | 0.004309 | 4.30E-155 |
| rs174553 | miR-1908-5 | cis | 2.61E-08 | Nightingale | XXL_VLDL_ | NA | G | A | 0.068388 | 0.004183 | 5.80E-63 |
| rs174553 | miR-1908-5 | cis | 2.61E-08 | Nightingale | S_VLDL_FC | NA | G | A | -0.0832 | 0.004146 | 1.40E-93 |
| rs174553 | miR-1908-5 | cis | 2.61E-08 | Nightingale | L_VLDL_FC | NA | G | A | 0.040514 | 0.0042 | 6.40E-23 |
| rs174553 | miR-1908-5 | cis | 2.61E-08 | Nightingale | Unsaturati | NA | G | A | -0.33987 | 0.004112 | 0 |
| rs174553 | miR-1908-5 | cis | 2.61E-08 | Nightingale | M_VLDL_F | NA | G | A | -0.09158 | 0.004128 | 1.00E-114 |
| rs174553 | miR-1908-5 | cis | 2.61E-08 | Nightingale | L_HDL_CE | NA | G | A | -0.07894 | 0.003834 | 2.00E-102 |
| rs174553 | miR-1908-5 | cis | 2.61E-08 | Nightingale | HDL_PL | NA | G | A | -0.0484 | 0.003925 | 1.50E-37 |
| rs174553 | miR-1908-5 | cis | 2.61E-08 | Nightingale | Total_CE | NA | G | A | -0.06709 | 0.004188 | 5.70E-61 |
| rs174553 | miR-1908-5 | cis | 2.61E-08 | Nightingale | M_HDL_L | NA | G | A | -0.02753 | 0.004031 | 1.00E-12 |
| rs174553 | miR-1908-5 | cis | 2.61E-08 | Nightingale | VLDL_L | NA | G | A | 0.027686 | 0.004257 | 4.50E-11 |
| rs174553 | miR-1908-5 | cis | 2.61E-08 | Nightingale | XL_HDL_CE | NA | G | A | -0.02629 | 0.004339 | 3.10E-09 |
| rs174553 | miR-1908-5 | cis | 2.61E-08 | Nightingale | M_HDL_C | NA | G | A | -0.04075 | 0.003995 | 4.10E-26 |
| rs174553 | miR-1908-5 | cis | 2.61E-08 | Nightingale | Cholines | NA | G | A | -0.03707 | 0.004083 | 1.90E-21 |
| rs174553 | miR-1908-5 | cis | 2.61E-08 | Nightingale | HDL_TG | NA | G | A | 0.046634 | 0.004281 | 3.30E-28 |
| rs174553 | miR-1908-5 | cis | 2.61E-08 | Nightingale | Clinical_LD | NA | G | A | -0.05523 | 0.004317 | 3.00E-39 |
| rs174553 | miR-1908-5 | cis | 2.61E-08 | Nightingale | L_VLDL_TG | NA | G | A | 0.037691 | 0.004177 | 3.70E-20 |
| rs174553 | miR-1908-5 | cis | 2.61E-08 | Nightingale | M_HDL_TG | NA | G | A | 0.050802 | 0.004288 | 2.90E-33 |
| rs174553 | miR-1908-5 | cis | 2.61E-08 | Nightingale | XS_VLDL_T | NA | G | A | 0.0318 | 0.004275 | 3.40E-13 |
| rs174553 | miR-1908-5 | cis | 2.61E-08 | Nightingale | M_LDL_TG | NA | G | A | 0.076016 | 0.004287 | 7.10E-72 |
| rs174553 | miR-1908-5 | cis | 2.61E-08 | Nightingale | M_LDL_PL | NA | G | A | -0.02346 | 0.004348 | 3.20E-08 |
| rs174553 | miR-1908-5 | cis | 2.61E-08 | Nightingale | Total_L | NA | G | A | -0.03724 | 0.004271 | 1.70E-19 |
| rs174553 | miR-1908-5 | cis | 2.61E-08 | Nightingale | Total_FC | NA | G | A | -0.05943 | 0.004241 | 3.20E-47 |
| rs174553 | miR-1908-5 | cis | 2.61E-08 | Nightingale | S_LDL_CE_ | NA | G | A | 0.057783 | 0.004334 | 2.00E-42 |
| rs174553 | miR-1908-5 | cis | 2.61E-08 | Nightingale | Total_P | NA | G | A | -0.04021 | 0.004086 | 1.70E-24 |
| rs174553 | miR-1908-5 | cis | 2.61E-08 | Nightingale | L_HDL_L | NA | G | A | -0.07741 | 0.003809 | 4.40E-100 |
| rs174553 | miR-1908-5 | cis | 2.61E-08 | Nightingale | S_LDL_PL_ | NA | G | A | -0.06264 | 0.004333 | 1.50E-49 |
| rs174553 | miR-1908-5 | cis | 2.61E-08 | Nightingale | XL_HDL_PL | NA | G | A | -0.08945 | 0.003903 | 1.80E-126 |
| rs174553 | miR-1908-5 | cis | 2.61E-08 | Nightingale | Phosphogly | NA | G | A | -0.04091 | 0.004112 | 4.30E-25 |
| rs174553 | miR-1908-5 | cis | 2.61E-08 | Nightingale | LDL_CE | NA | G | A | -0.04357 | 0.004331 | 5.40E-25 |
| rs174553 | miR-1908-5 | cis | 2.61E-08 | Nightingale | S_LDL_TG_ | NA | G | A | 0.086547 | 0.004207 | 3.30E-97 |
| rs174553 | miR-1908-5 | cis | 2.61E-08 | Nightingale | XS_VLDL_T | NA | G | A | 0.097667 | 0.004124 | 6.30E-130 |
| rs174553 | miR-1908-5 | cis | 2.61E-08 | Nightingale | IDL_C_pct | NA | G | A | -0.07489 | 0.004238 | 1.50E-71 |
| rs174553 | miR-1908-5 | cis | 2.61E-08 | Nightingale | Omega_3 | NA | G | A | -0.33578 | 0.004255 | 0 |
| rs174553 | miR-1908-5 | cis | 2.61E-08 | Nightingale | S_VLDL_PL | NA | G | A | -0.07882 | 0.004154 | 3.50E-84 |
| rs174553 | miR-1908-5 | cis | 2.61E-08 | Nightingale | XL_HDL_L | NA | G | A | -0.09339 | 0.003901 | 9.71E-138 |
| rs174553 | miR-1908-5 | cis | 2.61E-08 | Nightingale | IDL_CE | NA | G | A | -0.0724 | 0.004191 | 8.00E-71 |
| rs174553 | miR-1908-5 | cis | 2.61E-08 | Nightingale | XXL_VLDL_ | NA | G | A | 0.073979 | 0.004193 | 3.70E-72 |
| rs174553 | miR-1908-5 | cis | 2.61E-08 | Nightingale | L_HDL_P | NA | G | A | -0.07805 | 0.003803 | 4.10E-102 |
| rs174553 | miR-1908-5 | cis | 2.61E-08 | Nightingale | PUFA_pct | NA | G | A | -0.12185 | 0.004176 | 9.59E-189 |
| rs174553 | miR-1908-5 | cis | 2.61E-08 | Nightingale | XL_HDL_C | NA | G | A | -0.0996 | 0.003915 | 6.50E-155 |
| rs174553 | miR-1908-5 | cis | 2.61E-08 | Nightingale | S_LDL_L | NA | G | A | -0.03246 | 0.004341 | 1.30E-14 |
| rs174553 | miR-1908-5 | cis | 2.61E-08 | Nightingale | XL_VLDL_C | NA | G | A | -0.06437 | 0.00416 | 2.10E-55 |
| rs174553 | miR-1908-5 | cis | 2.61E-08 | Nightingale | L_LDL_C_p | NA | G | A | -0.07728 | 0.00423 | 6.20E-77 |
| rs174553 | miR-1908-5 | cis | 2.61E-08 | Nightingale | XXL_VLDL_ | NA | G | A | -0.02714 | 0.004319 | 6.90E-11 |
| rs174553 | miR-1908-5 | cis | 2.61E-08 | Nightingale | M_VLDL_P | NA | G | A | -0.06268 | 0.004166 | 1.60E-54 |
| rs174553 | miR-1908-5 | cis | 2.61E-08 | Nightingale | Remnant_( | NA | G | A | -0.04408 | 0.004314 | 6.30E-26 |
| rs174553 | miR-1908-5 | cis | 2.61E-08 | Nightingale | PUFA | NA | G | A | -0.09648 | 0.004234 | 6.30E-122 |
| rs174553 | miR-1908-5 | cis | 2.61E-08 | Nightingale | XL_HDL_P | NA | G | A | -0.08988 | 0.003907 | 4.20E-128 |
| rs174553 | miR-1908-5 | cis | 2.61E-08 | Nightingale | L_HDL_CE_ | NA | G | A | -0.0504 | 0.004154 | 3.10E-36 |
| rs174553 | miR-1908-5 | cis | 2.61E-08 | Nightingale | Sphingomy | NA | G | A | -0.11544 | 0.004057 | 4.50E-187 |
| rs174553 | miR-1908-5 | cis | 2.61E-08 | Nightingale | LA | NA | G | A | 0.083839 | 0.004258 | 1.50E-86 |
| rs174553 | miR-1908-5 | cis | 2.61E-08 | Nightingale | S_LDL_C | NA | G | A | -0.03215 | 0.004346 | 3.00E-14 |
| rs174553 | miR-1908-5 | cis | 2.61E-08 | Nightingale | IDL_P | NA | G | A | -0.04511 | 0.004296 | 3.70E-27 |
| rs174553 | miR-1908-5 | cis | 2.61E-08 | Nightingale | S_LDL_FC | NA | G | A | -0.06294 | 0.004335 | 1.40E-50 |
| rs174553 | miR-1908-5 | cis | 2.61E-08 | Nightingale | M_HDL_C_ | NA | G | A | -0.06617 | 0.004133 | 7.50E-61 |
| rs174553 | miR-1908-5 | cis | 2.61E-08 | Nightingale | M_HDL_CE | NA | G | A | -0.04732 | 0.00421 | 3.20E-30 |
| rs174553 | miR-1908-5 | cis | 2.61E-08 | Nightingale | LDL_PL | NA | G | A | -0.0462 | 0.004332 | 9.40E-28 |

| | | | | | | | | | | | |
|---|---|---|---|---|---|---|---|---|---|---|---|
| rs174553 | miR-1908-5 | cis | 2.61E-08 | Nightingale | M_VLDL_T | NA | G | A | 0.078259 | 0.0041 | 5.40E-85 |
| rs174553 | miR-1908-5 | cis | 2.61E-08 | Nightingale | XL_VLDL_P | NA | G | A | 0.053267 | 0.004173 | 9.20E-39 |
| rs174553 | miR-1908-5 | cis | 2.61E-08 | Nightingale | M_HDL_FC | NA | G | A | -0.05144 | 0.003951 | 1.30E-41 |
| rs174553 | miR-1908-5 | cis | 2.61E-08 | Nightingale | L_HDL_PL_ | NA | G | A | 0.043403 | 0.004202 | 9.90E-27 |
| rs174553 | miR-1908-5 | cis | 2.61E-08 | Nightingale | LDL_L | NA | G | A | -0.04546 | 0.004327 | 3.70E-27 |
| rs174553 | miR-1908-5 | cis | 2.61E-08 | Nightingale | MUFA_pct | NA | G | A | 0.197756 | 0.004155 | 0 |
| rs174553 | miR-1908-5 | cis | 2.61E-08 | Nightingale | L_LDL_P | NA | G | A | -0.03751 | 0.004327 | 3.40E-19 |
| rs174553 | miR-1908-5 | cis | 2.61E-08 | Nightingale | XXL_VLDL_ | NA | G | A | 0.067018 | 0.004182 | 2.50E-60 |
| rs174553 | miR-1908-5 | cis | 2.61E-08 | Nightingale | IDL_C | NA | G | A | -0.07743 | 0.004192 | 1.20E-80 |
| rs174553 | miR-1908-5 | cis | 2.61E-08 | Nightingale | HDL_FC | NA | G | A | -0.07398 | 0.003846 | 2.90E-89 |
| rs174553 | miR-1908-5 | cis | 2.61E-08 | Nightingale | XXL_VLDL_ | NA | G | A | 0.072438 | 0.004167 | 8.60E-71 |
| rs174553 | miR-1908-5 | cis | 2.61E-08 | Nightingale | PUFA_by_I | NA | G | A | -0.17104 | 0.004159 | 0 |
| rs174553 | miR-1908-5 | cis | 2.61E-08 | Nightingale | L_VLDL_P | NA | G | A | 0.042273 | 0.004192 | 7.70E-25 |
| rs174553 | miR-1908-5 | cis | 2.61E-08 | Nightingale | S_VLDL_FC | NA | G | A | -0.02828 | 0.004343 | 2.40E-11 |
| rs174553 | miR-1908-5 | cis | 2.61E-08 | Nightingale | XS_VLDL_C | NA | G | A | -0.09823 | 0.004114 | 9.40E-132 |
| rs174553 | miR-1908-5 | cis | 2.61E-08 | Nightingale | L_LDL_L | NA | G | A | -0.05424 | 0.004299 | 1.50E-38 |
| rs174553 | miR-1908-5 | cis | 2.61E-08 | Nightingale | S_LDL_TG | NA | G | A | 0.050705 | 0.004235 | 1.40E-33 |
| rs174553 | miR-1908-5 | cis | 2.61E-08 | Nightingale | XL_VLDL_P | NA | G | A | 0.048644 | 0.00419 | 2.40E-32 |
| rs174553 | miR-1908-5 | cis | 2.61E-08 | Nightingale | XS_VLDL_C | NA | G | A | -0.07212 | 0.004189 | 1.60E-70 |
| rs174553 | miR-1908-5 | cis | 2.61E-08 | Nightingale | SFA | NA | G | A | -0.03124 | 0.004302 | 1.40E-13 |
| rs174553 | miR-1908-5 | cis | 2.61E-08 | Nightingale | XS_VLDL_P | NA | G | A | -0.03454 | 0.004281 | 2.50E-17 |
| rs174553 | miR-1908-5 | cis | 2.61E-08 | Nightingale | L_VLDL_CE | NA | G | A | -0.05944 | 0.004155 | 3.50E-49 |
| rs174553 | miR-1908-5 | cis | 2.61E-08 | Nightingale | SFA_pct | NA | G | A | -0.04235 | 0.004283 | 4.70E-23 |
| rs174553 | miR-1908-5 | cis | 2.61E-08 | Nightingale | XL_VLDL_C | NA | G | A | 0.030632 | 0.004273 | 3.60E-13 |
| rs174553 | miR-1908-5 | cis | 2.61E-08 | Nightingale | HDL_CE | NA | G | A | -0.06155 | 0.003896 | 9.60E-61 |
| rs174553 | miR-1908-5 | cis | 2.61E-08 | Nightingale | S_LDL_FC_ | NA | G | A | -0.08014 | 0.004207 | 6.90E-84 |
| rs174553 | miR-1908-5 | cis | 2.61E-08 | Nightingale | XS_VLDL_P | NA | G | A | 0.057685 | 0.004236 | 5.10E-43 |
| rs174553 | miR-1908-5 | cis | 2.61E-08 | Nightingale | XL_VLDL_T | NA | G | A | 0.059738 | 0.004206 | 3.60E-47 |
| rs174553 | miR-1908-5 | cis | 2.61E-08 | Nightingale | XL_HDL_TG | NA | G | A | 0.0877 | 0.00409 | 6.50E-108 |
| rs174553 | miR-1908-5 | cis | 2.61E-08 | Nightingale | L_VLDL_PL | NA | G | A | 0.049131 | 0.004189 | 3.80E-33 |
| rs174553 | miR-1908-5 | cis | 2.61E-08 | Nightingale | M_VLDL_C | NA | G | A | -0.07628 | 0.004076 | 2.40E-81 |
| rs174553 | miR-1908-5 | cis | 2.61E-08 | Nightingale | ApoB | NA | G | A | -0.03848 | 0.004336 | 7.10E-20 |
| rs174553 | miR-1908-5 | cis | 2.61E-08 | Nightingale | ApoA1 | NA | G | A | -0.04111 | 0.003973 | 4.20E-27 |
| rs174553 | miR-1908-5 | cis | 2.61E-08 | Nightingale | L_HDL_FC | NA | G | A | -0.08483 | 0.003805 | 9.51E-120 |
| rs174553 | miR-1908-5 | cis | 2.61E-08 | Nightingale | XL_HDL_FC | NA | G | A | 0.061459 | 0.003849 | 1.50E-63 |
| rs174553 | miR-1908-5 | cis | 2.61E-08 | Nightingale | M_VLDL_T | NA | G | A | 0.031761 | 0.004218 | 2.00E-14 |
| rs174553 | miR-1908-5 | cis | 2.61E-08 | Nightingale | XL_VLDL_F | NA | G | A | -0.06842 | 0.004207 | 3.20E-62 |
| rs174553 | miR-1908-5 | cis | 2.61E-08 | Nightingale | M_LDL_C | NA | G | A | -0.03396 | 0.00435 | 1.50E-15 |
| rs174553 | miR-1908-5 | cis | 2.61E-08 | Nightingale | S_HDL_TG | NA | G | A | 0.069309 | 0.00421 | 9.40E-64 |
| rs174553 | miR-1908-5 | cis | 2.61E-08 | Nightingale | LDL_C | NA | G | A | -0.04979 | 0.004323 | 3.10E-32 |
| rs174553 | miR-1908-5 | cis | 2.61E-08 | Nightingale | M_HDL_CE | NA | G | A | -0.03779 | 0.00401 | 1.60E-22 |
| rs174553 | miR-1908-5 | cis | 2.61E-08 | Nightingale | L_VLDL_C | NA | G | A | 0.029197 | 0.004246 | 2.70E-12 |
| rs174553 | miR-1908-5 | cis | 2.61E-08 | Nightingale | M_LDL_CE | NA | G | A | -0.0262 | 0.004348 | 7.40E-10 |
| rs174553 | miR-1908-5 | cis | 2.61E-08 | Nightingale | S_LDL_P | NA | G | A | -0.03244 | 0.004346 | 1.80E-14 |
| rs174553 | miR-1908-5 | cis | 2.61E-08 | Nightingale | LDL_FC | NA | G | A | -0.06443 | 0.004295 | 1.40E-53 |
| rs174553 | miR-1908-5 | cis | 2.61E-08 | Nightingale | L_LDL_CE | NA | G | A | -0.05335 | 0.004303 | 2.70E-37 |
| rs174553 | miR-1908-5 | cis | 2.61E-08 | Nightingale | Total_PL | NA | G | A | -0.05108 | 0.004134 | 2.70E-37 |
| rs174553 | miR-1908-5 | cis | 2.61E-08 | Nightingale | S_VLDL_CE | NA | G | A | -0.029 | 0.004283 | 7.50E-12 |
| rs174553 | miR-1908-5 | cis | 2.61E-08 | Nightingale | MUFA | NA | G | A | 0.064176 | 0.004297 | 1.80E-51 |
| rs174553 | miR-1908-5 | cis | 2.61E-08 | Nightingale | HDL_size | NA | G | A | -0.07284 | 0.003847 | 1.20E-86 |
| rs174553 | miR-1908-5 | cis | 2.61E-08 | Nightingale | IDL_PL | NA | G | A | -0.07303 | 0.004189 | 7.60E-73 |
| rs174553 | miR-1908-5 | cis | 2.61E-08 | Nightingale | L_VLDL_PL | NA | G | A | 0.087945 | 0.004214 | 1.70E-100 |
| rs174553 | miR-1908-5 | cis | 2.61E-08 | Nightingale | DHA | NA | G | A | -0.28866 | 0.004184 | 0 |
| rs174553 | miR-1908-5 | cis | 2.61E-08 | Nightingale | LDL_P | NA | G | A | -0.0403 | 0.004339 | 1.50E-21 |
| rs174553 | miR-1908-5 | cis | 2.61E-08 | Nightingale | DHA_pct | NA | G | A | -0.27409 | 0.004214 | 0 |
| rs174553 | miR-1908-5 | cis | 2.61E-08 | Nightingale | L_HDL_PL | NA | G | A | -0.07628 | 0.003821 | 1.80E-96 |
| rs174553 | miR-1908-5 | cis | 2.61E-08 | Nightingale | L_HDL_TG_ | NA | G | A | 0.079133 | 0.004097 | 2.00E-88 |
| rs174553 | miR-1908-5 | cis | 2.61E-08 | Nightingale | S_VLDL_TG | NA | G | A | 0.039116 | 0.004213 | 4.00E-21 |
| rs174553 | miR-1908-5 | cis | 2.61E-08 | Nightingale | VLDL_TG | NA | G | A | 0.048068 | 0.004178 | 1.10E-31 |
| rs174553 | miR-1908-5 | cis | 2.61E-08 | Nightingale | XL_VLDL_C | NA | G | A | 0.037803 | 0.00424 | 1.10E-19 |
| rs174553 | miR-1908-5 | cis | 2.61E-08 | Nightingale | L_LDL_FC | NA | G | A | -0.06848 | 0.004263 | 2.10E-61 |
| rs174553 | miR-1908-5 | cis | 2.61E-08 | Nightingale | M_LDL_TG | NA | G | A | 0.03683 | 0.004278 | 7.20E-18 |
| rs174553 | miR-1908-5 | cis | 2.61E-08 | Nightingale | Total_C | NA | G | A | -0.06528 | 0.004203 | 2.20E-57 |

| | | | | | | | | | | | |
|---|---|---|---|---|---|---|---|---|---|---|---|
| rs174553 | miR-1908-5 | cis | 2.61E-08 | Nightingale | IDL_CE_pct | NA | G | A | -0.03807 | 0.00426 | 4.50E-19 |
| rs174553 | miR-1908-5 | cis | 2.61E-08 | Nightingale | IDL_L | NA | G | A | -0.07256 | 0.004197 | 2.50E-71 |
| rs174553 | miR-1908-5 | cis | 2.61E-08 | Nightingale | XL_HDL_CE | NA | G | A | -0.09747 | 0.003888 | 1.10E-150 |
| rs174553 | miR-1908-5 | cis | 2.61E-08 | Nightingale | IDL_FC_pct | NA | G | A | -0.10426 | 0.004307 | 4.30E-131 |
| rs174553 | miR-1908-5 | cis | 2.61E-08 | Nightingale | Total_TG | NA | G | A | 0.047145 | 0.004203 | 3.20E-30 |
| rs174553 | miR-1908-5 | cis | 2.61E-08 | Nightingale | M_HDL_P | NA | G | A | -0.03417 | 0.004 | 4.90E-19 |
| rs174553 | miR-1908-5 | cis | 2.61E-08 | Nightingale | M_HDL_TG | NA | G | A | 0.067013 | 0.004163 | 3.80E-61 |
| rs174553 | miR-1908-5 | cis | 2.61E-08 | Nightingale | M_LDL_FC_ | NA | G | A | -0.07204 | 0.004159 | 9.20E-70 |
| rs174553 | miR-1908-5 | cis | 2.61E-08 | Nightingale | Citrate | NA | G | A | -0.03361 | 0.004314 | 6.20E-16 |
| rs174553 | miR-1908-5 | cis | 2.61E-08 | Nightingale | XL_VLDL_T | NA | G | A | 0.0573 | 0.004153 | 4.80E-45 |
| rs174553 | miR-1908-5 | cis | 2.61E-08 | Nightingale | VLDL_size | NA | G | A | 0.0729 | 0.004047 | 8.10E-77 |
| rs174553 | miR-1908-5 | cis | 2.61E-08 | Nightingale | XS_VLDL_F | NA | G | A | -0.05621 | 0.004266 | 2.60E-42 |
| rs174553 | miR-1908-5 | cis | 2.61E-08 | Nightingale | HDL_L | NA | G | A | -0.05335 | 0.003899 | 6.80E-46 |
| rs174553 | miR-1908-5 | cis | 2.61E-08 | Nightingale | XS_VLDL_P | NA | G | A | -0.04373 | 0.004284 | 2.70E-26 |
| rs174553 | miR-1908-5 | cis | 2.61E-08 | Nightingale | Tyr | NA | G | A | 0.033878 | 0.004336 | 5.60E-15 |
| rs174553 | miR-1908-5 | cis | 2.61E-08 | Nightingale | M_VLDL_C | NA | G | A | -0.0528 | 0.004302 | 3.40E-36 |
| rs174553 | miR-1908-5 | cis | 2.61E-08 | Nightingale | LA_pct | NA | G | A | 0.185226 | 0.004243 | 0 |
| rs174553 | miR-1908-5 | cis | 2.61E-08 | Nightingale | L_VLDL_L | NA | G | A | 0.038585 | 0.004196 | 6.70E-21 |
| rs174553 | miR-1908-5 | cis | 2.61E-08 | Nightingale | L_HDL_FC_ | NA | G | A | -0.11278 | 0.004033 | 3.40E-187 |
| rs174553 | miR-1908-5 | cis | 2.61E-08 | Nightingale | Total_FA | NA | G | A | -0.02358 | 0.004307 | 1.30E-08 |
| rs174553 | miR-1908-5 | cis | 2.61E-08 | Nightingale | L_VLDL_FC | NA | G | A | 0.028521 | 0.004301 | 1.60E-11 |
| rs174553 | miR-1908-5 | cis | 2.61E-08 | Nightingale | XS_VLDL_C | NA | G | A | -0.09232 | 0.004093 | 6.81E-118 |
| rs174553 | miR-1908-5 | cis | 2.61E-08 | Nightingale | S_VLDL_TG | NA | G | A | 0.0647 | 0.004201 | 3.80E-55 |
| rs174553 | miR-1908-5 | cis | 2.61E-08 | Nightingale | S_LDL_PL | NA | G | A | -0.05165 | 0.004331 | 1.10E-34 |
| rs174553 | miR-1908-5 | cis | 2.61E-08 | Nightingale | S_HDL_TG_ | NA | G | A | 0.069311 | 0.004129 | 3.00E-66 |
| rs174553 | miR-1908-5 | cis | 2.61E-08 | Nightingale | HDL_P | NA | G | A | -0.03544 | 0.004077 | 2.30E-19 |
| rs174553 | miR-1908-5 | cis | 2.61E-08 | Nightingale | S_HDL_C_p | NA | G | A | -0.04259 | 0.004291 | 4.70E-23 |
| rs174553 | miR-1908-5 | cis | 2.61E-08 | Nightingale | TG_by_PG | NA | G | A | 0.069065 | 0.004029 | 3.10E-69 |
| rs174553 | miR-1908-5 | cis | 2.61E-08 | Nightingale | M_HDL_FC | NA | G | A | -0.10341 | 0.003847 | 2.30E-173 |
| rs174553 | miR-1908-5 | cis | 2.61E-08 | Nightingale | IDL_TG_pct | NA | G | A | 0.088345 | 0.004194 | 2.50E-102 |
| rs174553 | miR-1908-5 | cis | 2.61E-08 | Nightingale | M_LDL_L | NA | G | A | -0.02808 | 0.004348 | 4.20E-11 |
| rs174553 | miR-1908-5 | cis | 2.61E-08 | Nightingale | XL_HDL_C_ | NA | G | A | 0.024137 | 0.004148 | 2.70E-10 |
| rs174553 | miR-1908-5 | cis | 2.61E-08 | Nightingale | Omega_3_ | NA | G | A | -0.39 | 0.004242 | 0 |
| rs174553 | miR-1908-5 | cis | 2.61E-08 | Nightingale | LDL_TG | NA | G | A | 0.029882 | 0.004293 | 6.50E-12 |
| rs174553 | miR-1908-5 | cis | 2.61E-08 | Nightingale | L_VLDL_C_ | NA | G | A | -0.03881 | 0.004241 | 3.80E-21 |
| rs174553 | miR-1908-5 | cis | 2.61E-08 | Nightingale | non_HDL_( | NA | G | A | -0.0478 | 0.004318 | 6.70E-30 |
| rs174553 | miR-1908-5 | cis | 2.61E-08 | Nightingale | M_LDL_PL_ | NA | G | A | 0.04534 | 0.004339 | 1.20E-26 |
| rs174553 | miR-1908-5 | cis | 2.61E-08 | Nightingale | L_HDL_C_p | NA | G | A | -0.06875 | 0.004092 | 3.90E-68 |
| rs174553 | miR-1908-5 | cis | 2.61E-08 | Nightingale | L_HDL_C | NA | G | A | -0.08051 | 0.003823 | 3.80E-107 |
| rs174553 | miR-1908-5 | cis | 2.61E-08 | Nightingale | XL_HDL_PL | NA | G | A | -0.0674 | 0.003988 | 4.20E-69 |
| rs174553 | miR-1908-5 | cis | 2.61E-08 | Nightingale | IDL_FC | NA | G | A | -0.08995 | 0.004202 | 3.50E-107 |
| rs174553 | miR-1908-5 | cis | 2.61E-08 | Nightingale | M_VLDL_C | NA | G | A | -0.04404 | 0.00433 | 2.80E-25 |
| rs174553 | miR-1908-5 | cis | 2.61E-08 | Nightingale | XL_VLDL_F | NA | G | A | 0.043847 | 0.004205 | 2.30E-26 |
| rs174553 | miR-1908-5 | cis | 2.61E-08 | Nightingale | L_LDL_TG_ | NA | G | A | 0.077837 | 0.004257 | 2.00E-76 |
| rs174554 | miR-1908-5 | cis | 3.17E-08 | Nightingale | Omega_3 | NA | G | A | -0.33493 | 0.004258 | 0 |
| rs174554 | miR-1908-5 | cis | 3.17E-08 | Nightingale | L_LDL_FC | NA | G | A | -0.06873 | 0.004265 | 8.10E-62 |
| rs174554 | miR-1908-5 | cis | 3.17E-08 | Nightingale | Unsaturati | NA | G | A | -0.33928 | 0.004114 | 0 |
| rs174554 | miR-1908-5 | cis | 3.17E-08 | Nightingale | IDL_C_pct | NA | G | A | -0.07526 | 0.004241 | 4.30E-72 |
| rs174554 | miR-1908-5 | cis | 3.17E-08 | Nightingale | L_VLDL_CE | NA | G | A | -0.05952 | 0.004157 | 2.80E-49 |
| rs174554 | miR-1908-5 | cis | 3.17E-08 | Nightingale | MUFA_pct | NA | G | A | 0.197653 | 0.004157 | 0 |
| rs174554 | miR-1908-5 | cis | 3.17E-08 | Nightingale | L_HDL_FC_ | NA | G | A | -0.11296 | 0.004035 | 8.71E-188 |
| rs174554 | miR-1908-5 | cis | 3.17E-08 | Nightingale | IDL_FC | NA | G | A | -0.09016 | 0.004204 | 1.30E-107 |
| rs174554 | miR-1908-5 | cis | 3.17E-08 | Nightingale | S_LDL_FC_ | NA | G | A | -0.08028 | 0.00421 | 5.20E-84 |
| rs174554 | miR-1908-5 | cis | 3.17E-08 | Nightingale | S_VLDL_TG | NA | G | A | 0.039125 | 0.004215 | 4.50E-21 |
| rs174554 | miR-1908-5 | cis | 3.17E-08 | Nightingale | XS_VLDL_P | NA | G | A | 0.057995 | 0.004238 | 2.40E-43 |
| rs174554 | miR-1908-5 | cis | 3.17E-08 | Nightingale | Total_FA | NA | G | A | -0.02354 | 0.00431 | 1.30E-08 |
| rs174554 | miR-1908-5 | cis | 3.17E-08 | Nightingale | XL_HDL_CE | NA | G | A | -0.09774 | 0.00389 | 2.10E-151 |
| rs174554 | miR-1908-5 | cis | 3.17E-08 | Nightingale | Omega_6_ | NA | G | A | 0.368815 | 0.004281 | 0 |
| rs174554 | miR-1908-5 | cis | 3.17E-08 | Nightingale | VLDL_TG | NA | G | A | 0.048071 | 0.00418 | 1.40E-31 |
| rs174554 | miR-1908-5 | cis | 3.17E-08 | Nightingale | XL_VLDL_F | NA | G | A | -0.0685 | 0.004209 | 2.50E-62 |
| rs174554 | miR-1908-5 | cis | 3.17E-08 | Nightingale | M_VLDL_F( | NA | G | A | -0.02954 | 0.004343 | 2.60E-12 |
| rs174554 | miR-1908-5 | cis | 3.17E-08 | Nightingale | M_HDL_CE | NA | G | A | -0.04738 | 0.004212 | 3.60E-30 |
| rs174554 | miR-1908-5 | cis | 3.17E-08 | Nightingale | M_HDL_PL | NA | G | A | 0.058887 | 0.00412 | 3.20E-48 |

| | | | | | | | | | | | |
|---|---|---|---|---|---|---|---|---|---|---|---|
| rs174554 | miR-1908-5 | cis | 3.17E-08 | Nightingale | L_HDL_FC | NA | G | A | -0.08502 | 0.003807 | 3.50E-120 |
| rs174554 | miR-1908-5 | cis | 3.17E-08 | Nightingale | Remnant_( | NA | G | A | -0.04431 | 0.004316 | 2.90E-26 |
| rs174554 | miR-1908-5 | cis | 3.17E-08 | Nightingale | S_LDL_P | NA | G | A | -0.0326 | 0.004348 | 1.10E-14 |
| rs174554 | miR-1908-5 | cis | 3.17E-08 | Nightingale | XS_VLDL_F | NA | G | A | -0.11323 | 0.004311 | 2.80E-155 |
| rs174554 | miR-1908-5 | cis | 3.17E-08 | Nightingale | Total_PL | NA | G | A | -0.05118 | 0.004136 | 2.00E-37 |
| rs174554 | miR-1908-5 | cis | 3.17E-08 | Nightingale | MUFA | NA | G | A | 0.064167 | 0.0043 | 2.50E-51 |
| rs174554 | miR-1908-5 | cis | 3.17E-08 | Nightingale | M_VLDL_C | NA | G | A | -0.05309 | 0.004305 | 1.30E-36 |
| rs174554 | miR-1908-5 | cis | 3.17E-08 | Nightingale | L_HDL_TG_ | NA | G | A | 0.07916 | 0.004099 | 2.70E-88 |
| rs174554 | miR-1908-5 | cis | 3.17E-08 | Nightingale | IDL_CE | NA | G | A | -0.07267 | 0.004193 | 2.60E-71 |
| rs174554 | miR-1908-5 | cis | 3.17E-08 | Nightingale | Omega_3_ | NA | G | A | -0.38901 | 0.004244 | 0 |
| rs174554 | miR-1908-5 | cis | 3.17E-08 | Nightingale | M_HDL_C_ | NA | G | A | -0.06624 | 0.004135 | 8.50E-61 |
| rs174554 | miR-1908-5 | cis | 3.17E-08 | Nightingale | LDL_TG | NA | G | A | 0.029986 | 0.004296 | 6.40E-12 |
| rs174554 | miR-1908-5 | cis | 3.17E-08 | Nightingale | XL_VLDL_C | NA | G | A | -0.06147 | 0.004165 | 1.10E-50 |
| rs174554 | miR-1908-5 | cis | 3.17E-08 | Nightingale | S_LDL_TG | NA | G | A | 0.050753 | 0.004238 | 1.50E-33 |
| rs174554 | miR-1908-5 | cis | 3.17E-08 | Nightingale | S_VLDL_PL | NA | G | A | -0.07893 | 0.004156 | 3.40E-84 |
| rs174554 | miR-1908-5 | cis | 3.17E-08 | Nightingale | S_VLDL_FC | NA | G | A | -0.08335 | 0.004148 | 9.80E-94 |
| rs174554 | miR-1908-5 | cis | 3.17E-08 | Nightingale | VLDL_L | NA | G | A | 0.027643 | 0.004259 | 5.70E-11 |
| rs174554 | miR-1908-5 | cis | 3.17E-08 | Nightingale | Citrate | NA | G | A | -0.03332 | 0.004316 | 1.20E-15 |
| rs174554 | miR-1908-5 | cis | 3.17E-08 | Nightingale | XL_VLDL_T | NA | G | A | 0.05977 | 0.004208 | 3.40E-47 |
| rs174554 | miR-1908-5 | cis | 3.17E-08 | Nightingale | L_VLDL_L | NA | G | A | 0.038548 | 0.004198 | 8.90E-21 |
| rs174554 | miR-1908-5 | cis | 3.17E-08 | Nightingale | L_VLDL_C_ | NA | G | A | -0.03881 | 0.004243 | 3.80E-21 |
| rs174554 | miR-1908-5 | cis | 3.17E-08 | Nightingale | S_LDL_C | NA | G | A | -0.03234 | 0.004349 | 1.90E-14 |
| rs174554 | miR-1908-5 | cis | 3.17E-08 | Nightingale | XL_VLDL_P | NA | G | A | 0.048633 | 0.004192 | 3.20E-32 |
| rs174554 | miR-1908-5 | cis | 3.17E-08 | Nightingale | M_LDL_C | NA | G | A | -0.0341 | 0.004352 | 1.10E-15 |
| rs174554 | miR-1908-5 | cis | 3.17E-08 | Nightingale | XL_VLDL_T | NA | G | A | 0.057327 | 0.004155 | 5.70E-45 |
| rs174554 | miR-1908-5 | cis | 3.17E-08 | Nightingale | S_LDL_L | NA | G | A | -0.03263 | 0.004343 | 8.90E-15 |
| rs174554 | miR-1908-5 | cis | 3.17E-08 | Nightingale | L_VLDL_TG | NA | G | A | 0.037662 | 0.00418 | 4.60E-20 |
| rs174554 | miR-1908-5 | cis | 3.17E-08 | Nightingale | HDL_L | NA | G | A | -0.05338 | 0.003901 | 6.50E-46 |
| rs174554 | miR-1908-5 | cis | 3.17E-08 | Nightingale | L_HDL_L | NA | G | A | -0.07757 | 0.003811 | 1.90E-100 |
| rs174554 | miR-1908-5 | cis | 3.17E-08 | Nightingale | M_HDL_TG | NA | G | A | 0.050819 | 0.00429 | 3.60E-33 |
| rs174554 | miR-1908-5 | cis | 3.17E-08 | Nightingale | XL_HDL_L | NA | G | A | -0.09364 | 0.003903 | 1.90E-138 |
| rs174554 | miR-1908-5 | cis | 3.17E-08 | Nightingale | XS_VLDL_C | NA | G | A | -0.09857 | 0.004116 | 2.30E-132 |
| rs174554 | miR-1908-5 | cis | 3.17E-08 | Nightingale | XS_VLDL_F | NA | G | A | -0.05631 | 0.004268 | 1.30E-42 |
| rs174554 | miR-1908-5 | cis | 3.17E-08 | Nightingale | M_VLDL_P | NA | G | A | -0.06279 | 0.004168 | 1.10E-54 |
| rs174554 | miR-1908-5 | cis | 3.17E-08 | Nightingale | S_LDL_PL | NA | G | A | -0.0518 | 0.004334 | 6.70E-35 |
| rs174554 | miR-1908-5 | cis | 3.17E-08 | Nightingale | XL_HDL_P | NA | G | A | -0.09018 | 0.003909 | 6.10E-129 |
| rs174554 | miR-1908-5 | cis | 3.17E-08 | Nightingale | LDL_CE | NA | G | A | -0.04373 | 0.004333 | 3.30E-25 |
| rs174554 | miR-1908-5 | cis | 3.17E-08 | Nightingale | M_VLDL_C | NA | G | A | -0.07641 | 0.004078 | 2.00E-81 |
| rs174554 | miR-1908-5 | cis | 3.17E-08 | Nightingale | PUFA | NA | G | A | -0.09649 | 0.004236 | 6.00E-122 |
| rs174554 | miR-1908-5 | cis | 3.17E-08 | Nightingale | XXL_VLDL_ | NA | G | A | 0.066996 | 0.004184 | 3.40E-60 |
| rs174554 | miR-1908-5 | cis | 3.17E-08 | Nightingale | LDL_C | NA | G | A | -0.04997 | 0.004325 | 1.70E-32 |
| rs174554 | miR-1908-5 | cis | 3.17E-08 | Nightingale | Total_P | NA | G | A | -0.04022 | 0.004088 | 1.70E-24 |
| rs174554 | miR-1908-5 | cis | 3.17E-08 | Nightingale | XXL_VLDL_ | NA | G | A | 0.072058 | 0.004186 | 5.00E-69 |
| rs174554 | miR-1908-5 | cis | 3.17E-08 | Nightingale | XS_VLDL_C | NA | G | A | -0.07233 | 0.004191 | 5.60E-71 |
| rs174554 | miR-1908-5 | cis | 3.17E-08 | Nightingale | XXL_VLDL_ | NA | G | A | -0.02707 | 0.004322 | 7.80E-11 |
| rs174554 | miR-1908-5 | cis | 3.17E-08 | Nightingale | XS_VLDL_P | NA | G | A | -0.03458 | 0.004283 | 1.80E-17 |
| rs174554 | miR-1908-5 | cis | 3.17E-08 | Nightingale | XL_VLDL_F | NA | G | A | 0.043816 | 0.004207 | 3.20E-26 |
| rs174554 | miR-1908-5 | cis | 3.17E-08 | Nightingale | L_HDL_C | NA | G | A | -0.08071 | 0.003825 | 1.40E-107 |
| rs174554 | miR-1908-5 | cis | 3.17E-08 | Nightingale | S_HDL_TG_ | NA | G | A | 0.069336 | 0.004131 | 4.10E-66 |
| rs174554 | miR-1908-5 | cis | 3.17E-08 | Nightingale | XS_VLDL_T | NA | G | A | 0.031924 | 0.004277 | 3.20E-13 |
| rs174554 | miR-1908-5 | cis | 3.17E-08 | Nightingale | S_HDL_FC_ | NA | G | A | -0.10322 | 0.00412 | 8.00E-154 |
| rs174554 | miR-1908-5 | cis | 3.17E-08 | Nightingale | XL_HDL_CE | NA | G | A | -0.02639 | 0.004341 | 3.00E-09 |
| rs174554 | miR-1908-5 | cis | 3.17E-08 | Nightingale | M_HDL_L | NA | G | A | -0.02746 | 0.004033 | 1.20E-12 |
| rs174554 | miR-1908-5 | cis | 3.17E-08 | Nightingale | L_VLDL_PL | NA | G | A | 0.049101 | 0.004191 | 5.50E-33 |
| rs174554 | miR-1908-5 | cis | 3.17E-08 | Nightingale | L_LDL_P | NA | G | A | -0.03775 | 0.004329 | 1.80E-19 |
| rs174554 | miR-1908-5 | cis | 3.17E-08 | Nightingale | HDL_C | NA | G | A | -0.06491 | 0.003879 | 4.10E-68 |
| rs174554 | miR-1908-5 | cis | 3.17E-08 | Nightingale | VLDL_size | NA | G | A | 0.072916 | 0.004049 | 1.10E-76 |
| rs174554 | miR-1908-5 | cis | 3.17E-08 | Nightingale | L_VLDL_FC | NA | G | A | 0.028567 | 0.004303 | 1.60E-11 |
| rs174554 | miR-1908-5 | cis | 3.17E-08 | Nightingale | L_LDL_PL | NA | G | A | -0.05521 | 0.004311 | 2.40E-39 |
| rs174554 | miR-1908-5 | cis | 3.17E-08 | Nightingale | non_HDL_( | NA | G | A | -0.04801 | 0.004321 | 3.40E-30 |
| rs174554 | miR-1908-5 | cis | 3.17E-08 | Nightingale | SFA | NA | G | A | -0.03112 | 0.004305 | 1.60E-13 |
| rs174554 | miR-1908-5 | cis | 3.17E-08 | Nightingale | M_VLDL_F | NA | G | A | -0.09168 | 0.00413 | 7.50E-115 |
| rs174554 | miR-1908-5 | cis | 3.17E-08 | Nightingale | L_HDL_C_p | NA | G | A | -0.06903 | 0.004094 | 1.60E-68 |

| | | | | | | | | | | | |
|---|---|---|---|---|---|---|---|---|---|---|---|
| rs174554 | miR-1908-5 | cis | 3.17E-08 | Nightingale | IDL_L | NA | G | A | -0.07278 | 0.004199 | 9.30E-72 |
| rs174554 | miR-1908-5 | cis | 3.17E-08 | Nightingale | S_VLDL_C_ | NA | G | A | -0.05685 | 0.004226 | 2.60E-42 |
| rs174554 | miR-1908-5 | cis | 3.17E-08 | Nightingale | IDL_PL | NA | G | A | -0.07323 | 0.004191 | 2.90E-73 |
| rs174554 | miR-1908-5 | cis | 3.17E-08 | Nightingale | ApoA1 | NA | G | A | -0.04111 | 0.003975 | 4.40E-27 |
| rs174554 | miR-1908-5 | cis | 3.17E-08 | Nightingale | L_HDL_PL_ | NA | G | A | 0.043957 | 0.004204 | 2.40E-27 |
| rs174554 | miR-1908-5 | cis | 3.17E-08 | Nightingale | M_LDL_CE | NA | G | A | -0.02631 | 0.00435 | 5.60E-10 |
| rs174554 | miR-1908-5 | cis | 3.17E-08 | Nightingale | Sphingomy | NA | G | A | -0.11549 | 0.004059 | 4.20E-187 |
| rs174554 | miR-1908-5 | cis | 3.17E-08 | Nightingale | L_LDL_TG_ | NA | G | A | 0.078167 | 0.004259 | 6.40E-77 |
| rs174554 | miR-1908-5 | cis | 3.17E-08 | Nightingale | XS_VLDL_C | NA | G | A | -0.09266 | 0.004095 | 1.60E-118 |
| rs174554 | miR-1908-5 | cis | 3.17E-08 | Nightingale | M_LDL_L | NA | G | A | -0.02821 | 0.004351 | 3.10E-11 |
| rs174554 | miR-1908-5 | cis | 3.17E-08 | Nightingale | TG_by_PG | NA | G | A | 0.069106 | 0.004031 | 3.90E-69 |
| rs174554 | miR-1908-5 | cis | 3.17E-08 | Nightingale | HDL_P | NA | G | A | -0.03542 | 0.004079 | 2.60E-19 |
| rs174554 | miR-1908-5 | cis | 3.17E-08 | Nightingale | XL_HDL_C_ | NA | G | A | 0.024214 | 0.00415 | 2.30E-10 |
| rs174554 | miR-1908-5 | cis | 3.17E-08 | Nightingale | M_LDL_FC | NA | G | A | -0.05265 | 0.004339 | 1.10E-35 |
| rs174554 | miR-1908-5 | cis | 3.17E-08 | Nightingale | XXL_VLDL_ | NA | G | A | 0.044175 | 0.004367 | 5.40E-25 |
| rs174554 | miR-1908-5 | cis | 3.17E-08 | Nightingale | L_LDL_FC_ | NA | G | A | -0.08348 | 0.00414 | 1.70E-93 |
| rs174554 | miR-1908-5 | cis | 3.17E-08 | Nightingale | L_HDL_CE | NA | G | A | -0.07915 | 0.003836 | 7.50E-103 |
| rs174554 | miR-1908-5 | cis | 3.17E-08 | Nightingale | M_VLDL_T | NA | G | A | 0.07838 | 0.004103 | 4.10E-85 |
| rs174554 | miR-1908-5 | cis | 3.17E-08 | Nightingale | S_HDL_C_p | NA | G | A | -0.04266 | 0.004294 | 4.50E-23 |
| rs174554 | miR-1908-5 | cis | 3.17E-08 | Nightingale | M_LDL_PL | NA | G | A | -0.02361 | 0.004351 | 2.40E-08 |
| rs174554 | miR-1908-5 | cis | 3.17E-08 | Nightingale | Total_FC | NA | G | A | -0.05965 | 0.004244 | 1.40E-47 |
| rs174554 | miR-1908-5 | cis | 3.17E-08 | Nightingale | M_LDL_C_p | NA | G | A | -0.10046 | 0.004347 | 1.30E-118 |
| rs174554 | miR-1908-5 | cis | 3.17E-08 | Nightingale | M_LDL_PL_ | NA | G | A | 0.045212 | 0.004341 | 1.70E-26 |
| rs174554 | miR-1908-5 | cis | 3.17E-08 | Nightingale | XS_VLDL_T | NA | G | A | 0.097958 | 0.004126 | 2.00E-130 |
| rs174554 | miR-1908-5 | cis | 3.17E-08 | Nightingale | LA | NA | G | A | 0.083285 | 0.00426 | 2.90E-85 |
| rs174554 | miR-1908-5 | cis | 3.17E-08 | Nightingale | M_LDL_TG | NA | G | A | 0.07627 | 0.004289 | 3.40E-72 |
| rs174554 | miR-1908-5 | cis | 3.17E-08 | Nightingale | XL_HDL_TG | NA | G | A | 0.087827 | 0.004092 | 5.50E-108 |
| rs174554 | miR-1908-5 | cis | 3.17E-08 | Nightingale | LDL_PL | NA | G | A | -0.04639 | 0.004334 | 5.30E-28 |
| rs174554 | miR-1908-5 | cis | 3.17E-08 | Nightingale | M_LDL_P | NA | G | A | -0.04672 | 0.004357 | 9.60E-28 |
| rs174554 | miR-1908-5 | cis | 3.17E-08 | Nightingale | LA_pct | NA | G | A | 0.184179 | 0.004245 | 0 |
| rs174554 | miR-1908-5 | cis | 3.17E-08 | Nightingale | IDL_TG_pct | NA | G | A | 0.088705 | 0.004196 | 5.90E-103 |
| rs174554 | miR-1908-5 | cis | 3.17E-08 | Nightingale | XXL_VLDL_ | NA | G | A | 0.072445 | 0.004169 | 1.10E-70 |
| rs174554 | miR-1908-5 | cis | 3.17E-08 | Nightingale | XL_HDL_C | NA | G | A | -0.09987 | 0.003917 | 1.20E-155 |
| rs174554 | miR-1908-5 | cis | 3.17E-08 | Nightingale | Omega_6_ | NA | G | A | 0.023273 | 0.004228 | 1.90E-08 |
| rs174554 | miR-1908-5 | cis | 3.17E-08 | Nightingale | S_LDL_CE_ | NA | G | A | 0.057593 | 0.004336 | 4.00E-42 |
| rs174554 | miR-1908-5 | cis | 3.17E-08 | Nightingale | IDL_CE_pct | NA | G | A | -0.0385 | 0.004263 | 1.90E-19 |
| rs174554 | miR-1908-5 | cis | 3.17E-08 | Nightingale | XL_HDL_FC | NA | G | A | 0.061676 | 0.003851 | 5.40E-64 |
| rs174554 | miR-1908-5 | cis | 3.17E-08 | Nightingale | M_VLDL_C | NA | G | A | -0.04429 | 0.004332 | 1.20E-25 |
| rs174554 | miR-1908-5 | cis | 3.17E-08 | Nightingale | XXL_VLDL_ | NA | G | A | 0.068373 | 0.004185 | 8.00E-63 |
| rs174554 | miR-1908-5 | cis | 3.17E-08 | Nightingale | S_VLDL_TG | NA | G | A | 0.064798 | 0.004203 | 3.60E-55 |
| rs174554 | miR-1908-5 | cis | 3.17E-08 | Nightingale | HDL_FC | NA | G | A | -0.07407 | 0.003848 | 1.90E-89 |
| rs174554 | miR-1908-5 | cis | 3.17E-08 | Nightingale | XS_VLDL_L | NA | G | A | -0.04176 | 0.00427 | 1.20E-24 |
| rs174554 | miR-1908-5 | cis | 3.17E-08 | Nightingale | M_HDL_PL | NA | G | A | -0.02119 | 0.004062 | 4.80E-08 |
| rs174554 | miR-1908-5 | cis | 3.17E-08 | Nightingale | Total_TG | NA | G | A | 0.047168 | 0.004205 | 3.80E-30 |
| rs174554 | miR-1908-5 | cis | 3.17E-08 | Nightingale | LDL_L | NA | G | A | -0.04563 | 0.00433 | 2.20E-27 |
| rs174554 | miR-1908-5 | cis | 3.17E-08 | Nightingale | ApoB | NA | G | A | -0.03868 | 0.004338 | 3.90E-20 |
| rs174554 | miR-1908-5 | cis | 3.17E-08 | Nightingale | Clinical_LD | NA | G | A | -0.05546 | 0.004319 | 1.40E-39 |
| rs174554 | miR-1908-5 | cis | 3.17E-08 | Nightingale | S_VLDL_FC | NA | G | A | -0.02843 | 0.004345 | 1.60E-11 |
| rs174554 | miR-1908-5 | cis | 3.17E-08 | Nightingale | M_HDL_FC | NA | G | A | -0.1035 | 0.003849 | 1.50E-173 |
| rs174554 | miR-1908-5 | cis | 3.17E-08 | Nightingale | XL_VLDL_L | NA | G | A | 0.05187 | 0.00418 | 1.20E-36 |
| rs174554 | miR-1908-5 | cis | 3.17E-08 | Nightingale | M_HDL_C | NA | G | A | -0.04071 | 0.003997 | 5.20E-26 |
| rs174554 | miR-1908-5 | cis | 3.17E-08 | Nightingale | L_VLDL_PL | NA | G | A | 0.087984 | 0.004216 | 2.70E-100 |
| rs174554 | miR-1908-5 | cis | 3.17E-08 | Nightingale | IDL_FC_pct | NA | G | A | -0.10434 | 0.004309 | 4.00E-131 |
| rs174554 | miR-1908-5 | cis | 3.17E-08 | Nightingale | XL_HDL_PL | NA | G | A | -0.0675 | 0.003991 | 2.80E-69 |
| rs174554 | miR-1908-5 | cis | 3.17E-08 | Nightingale | M_HDL_FC | NA | G | A | -0.05141 | 0.003953 | 1.60E-41 |
| rs174554 | miR-1908-5 | cis | 3.17E-08 | Nightingale | XL_VLDL_P | NA | G | A | 0.05326 | 0.004175 | 1.20E-38 |
| rs174554 | miR-1908-5 | cis | 3.17E-08 | Nightingale | S_LDL_PL_ | NA | G | A | -0.06245 | 0.004335 | 3.20E-49 |
| rs174554 | miR-1908-5 | cis | 3.17E-08 | Nightingale | M_HDL_P | NA | G | A | -0.03413 | 0.004002 | 5.70E-19 |
| rs174554 | miR-1908-5 | cis | 3.17E-08 | Nightingale | Cholines | NA | G | A | -0.03718 | 0.004085 | 1.40E-21 |
| rs174554 | miR-1908-5 | cis | 3.17E-08 | Nightingale | HDL_size | NA | G | A | -0.07305 | 0.003849 | 4.00E-87 |
| rs174554 | miR-1908-5 | cis | 3.17E-08 | Nightingale | Phosphogly | NA | G | A | -0.04096 | 0.004115 | 3.80E-25 |
| rs174554 | miR-1908-5 | cis | 3.17E-08 | Nightingale | L_VLDL_FC | NA | G | A | 0.040474 | 0.004202 | 8.90E-23 |
| rs174554 | miR-1908-5 | cis | 3.17E-08 | Nightingale | Total_CE | NA | G | A | -0.06729 | 0.00419 | 2.70E-61 |

| | | | | | | | | | | | |
|---|---|---|---|---|---|---|---|---|---|---|---|
| rs174554 | miR-1908-! | cis | 3.17E-08 | Nightingale | L_LDL_L | NA | G | A | -0.05442 | 0.004302 | 8.20E-39 |
| rs174554 | miR-1908-! | cis | 3.17E-08 | Nightingale | L_LDL_C | NA | G | A | -0.05809 | 0.004295 | 4.70E-44 |
| rs174554 | miR-1908-! | cis | 3.17E-08 | Nightingale | S_VLDL_CE | NA | G | A | -0.02906 | 0.004286 | 7.20E-12 |
| rs174554 | miR-1908-! | cis | 3.17E-08 | Nightingale | M_LDL_FC_ | NA | G | A | -0.07228 | 0.004161 | 4.70E-70 |
| rs174554 | miR-1908-! | cis | 3.17E-08 | Nightingale | S_LDL_TG_ | NA | G | A | 0.086719 | 0.004209 | 2.20E-97 |
| rs174554 | miR-1908-! | cis | 3.17E-08 | Nightingale | M_HDL_TG | NA | G | A | 0.066994 | 0.004166 | 6.00E-61 |
| rs174554 | miR-1908-! | cis | 3.17E-08 | Nightingale | HDL_CE | NA | G | A | -0.06162 | 0.003898 | 8.10E-61 |
| rs174554 | miR-1908-! | cis | 3.17E-08 | Nightingale | L_LDL_CE | NA | G | A | -0.05352 | 0.004305 | 1.60E-37 |
| rs174554 | miR-1908-! | cis | 3.17E-08 | Nightingale | PUFA_by_I | NA | G | A | -0.17103 | 0.004161 | 0 |
| rs174554 | miR-1908-! | cis | 3.17E-08 | Nightingale | DHA_pct | NA | G | A | -0.27352 | 0.004217 | 0 |
| rs174554 | miR-1908-! | cis | 3.17E-08 | Nightingale | Tyr | NA | G | A | 0.033498 | 0.004338 | 1.20E-14 |
| rs174554 | miR-1908-! | cis | 3.17E-08 | Nightingale | IDL_C | NA | G | A | -0.07769 | 0.004194 | 3.80E-81 |
| rs174554 | miR-1908-! | cis | 3.17E-08 | Nightingale | XL_VLDL_C | NA | G | A | 0.037732 | 0.004243 | 1.60E-19 |
| rs174554 | miR-1908-! | cis | 3.17E-08 | Nightingale | L_VLDL_C | NA | G | A | 0.02911 | 0.004249 | 3.70E-12 |
| rs174554 | miR-1908-! | cis | 3.17E-08 | Nightingale | M_VLDL_C | NA | G | A | -0.08113 | 0.004089 | 3.20E-91 |
| rs174554 | miR-1908-! | cis | 3.17E-08 | Nightingale | M_LDL_TG | NA | G | A | 0.036923 | 0.00428 | 7.00E-18 |
| rs174554 | miR-1908-! | cis | 3.17E-08 | Nightingale | PUFA_pct | NA | G | A | -0.12191 | 0.004178 | 1.20E-188 |
| rs174554 | miR-1908-! | cis | 3.17E-08 | Nightingale | L_HDL_P | NA | G | A | -0.07826 | 0.003805 | 1.40E-102 |
| rs174554 | miR-1908-! | cis | 3.17E-08 | Nightingale | M_VLDL_T | NA | G | A | 0.031735 | 0.00422 | 2.50E-14 |
| rs174554 | miR-1908-! | cis | 3.17E-08 | Nightingale | L_HDL_CE_ | NA | G | A | -0.05069 | 0.004156 | 1.70E-36 |
| rs174554 | miR-1908-! | cis | 3.17E-08 | Nightingale | L_VLDL_P | NA | G | A | 0.042245 | 0.004194 | 1.00E-24 |
| rs174554 | miR-1908-! | cis | 3.17E-08 | Nightingale | IDL_P | NA | G | A | -0.04537 | 0.004298 | 1.60E-27 |
| rs174554 | miR-1908-! | cis | 3.17E-08 | Nightingale | SFA_pct | NA | G | A | -0.04203 | 0.004286 | 1.00E-22 |
| rs174554 | miR-1908-! | cis | 3.17E-08 | Nightingale | XL_VLDL_C | NA | G | A | -0.06446 | 0.004162 | 1.70E-55 |
| rs174554 | miR-1908-! | cis | 3.17E-08 | Nightingale | XXL_VLDL_ | NA | G | A | 0.066524 | 0.004178 | 9.90E-60 |
| rs174554 | miR-1908-! | cis | 3.17E-08 | Nightingale | LDL_P | NA | G | A | -0.0405 | 0.004341 | 8.20E-22 |
| rs174554 | miR-1908-! | cis | 3.17E-08 | Nightingale | XXL_VLDL_ | NA | G | A | 0.061594 | 0.004192 | 5.90E-51 |
| rs174554 | miR-1908-! | cis | 3.17E-08 | Nightingale | M_HDL_CE | NA | G | A | -0.03775 | 0.004012 | 2.00E-22 |
| rs174554 | miR-1908-! | cis | 3.17E-08 | Nightingale | Total_C | NA | G | A | -0.06548 | 0.004205 | 1.00E-57 |
| rs174554 | miR-1908-! | cis | 3.17E-08 | Nightingale | HDL_PL | NA | G | A | -0.04839 | 0.003927 | 1.60E-37 |
| rs174554 | miR-1908-! | cis | 3.17E-08 | Nightingale | XL_HDL_PL | NA | G | A | -0.08968 | 0.003905 | 4.60E-127 |
| rs174554 | miR-1908-! | cis | 3.17E-08 | Nightingale | S_LDL_FC | NA | G | A | -0.06314 | 0.004337 | 7.10E-51 |
| rs174554 | miR-1908-! | cis | 3.17E-08 | Nightingale | LDL_FC | NA | G | A | -0.06466 | 0.004298 | 6.10E-54 |
| rs174554 | miR-1908-! | cis | 3.17E-08 | Nightingale | XS_VLDL_C | NA | G | A | -0.06852 | 0.004216 | 2.90E-63 |
| rs174554 | miR-1908-! | cis | 3.17E-08 | Nightingale | DHA | NA | G | A | -0.28804 | 0.004186 | 0 |
| rs174554 | miR-1908-! | cis | 3.17E-08 | Nightingale | L_LDL_C_p | NA | G | A | -0.07751 | 0.004232 | 3.00E-77 |
| rs174554 | miR-1908-! | cis | 3.17E-08 | Nightingale | XL_HDL_FC | NA | G | A | -0.10344 | 0.004047 | 1.80E-155 |
| rs174554 | miR-1908-! | cis | 3.17E-08 | Nightingale | Total_L | NA | G | A | -0.03739 | 0.004274 | 1.20E-19 |
| rs174554 | miR-1908-! | cis | 3.17E-08 | Nightingale | XL_VLDL_C | NA | G | A | 0.030528 | 0.004275 | 5.10E-13 |
| rs174554 | miR-1908-! | cis | 3.17E-08 | Nightingale | L_HDL_PL | NA | G | A | -0.07639 | 0.003823 | 1.10E-96 |
| rs174554 | miR-1908-! | cis | 3.17E-08 | Nightingale | S_HDL_TG | NA | G | A | 0.069387 | 0.004212 | 9.70E-64 |
| rs174554 | miR-1908-! | cis | 3.17E-08 | Nightingale | HDL_TG | NA | G | A | 0.046622 | 0.004283 | 4.30E-28 |
| rs174554 | miR-1908-! | cis | 3.17E-08 | Nightingale | XXL_VLDL_ | NA | G | A | 0.073914 | 0.004195 | 7.00E-72 |
| rs174554 | miR-1908-! | cis | 3.17E-08 | Nightingale | XS_VLDL_P | NA | G | A | -0.04382 | 0.004286 | 1.60E-26 |
| rs174555 | miR-1908-! | cis | 7.43E-08 | Nightingale | MUFA | NA | C | T | 0.064427 | 0.004421 | 1.20E-49 |
| rs174555 | miR-1908-! | cis | 7.43E-08 | Nightingale | M_HDL_TG | NA | C | T | 0.051789 | 0.004412 | 1.70E-32 |
| rs174555 | miR-1908-! | cis | 7.43E-08 | Nightingale | S_LDL_P | NA | C | T | -0.03021 | 0.004471 | 5.50E-12 |
| rs174555 | miR-1908-! | cis | 7.43E-08 | Nightingale | M_VLDL_C | NA | C | T | -0.04188 | 0.004455 | 9.80E-22 |
| rs174555 | miR-1908-! | cis | 7.43E-08 | Nightingale | VLDL_size | NA | C | T | 0.072294 | 0.004164 | 9.80E-72 |
| rs174555 | miR-1908-! | cis | 7.43E-08 | Nightingale | Omega_6_ | NA | C | T | 0.362818 | 0.004402 | 0 |
| rs174555 | miR-1908-! | cis | 7.43E-08 | Nightingale | XL_HDL_CE | NA | C | T | -0.02805 | 0.004464 | 1.30E-09 |
| rs174555 | miR-1908-! | cis | 7.43E-08 | Nightingale | S_VLDL_C_ | NA | C | T | -0.05576 | 0.004346 | 7.90E-39 |
| rs174555 | miR-1908-! | cis | 7.43E-08 | Nightingale | L_VLDL_PL | NA | C | T | 0.087444 | 0.004335 | 1.10E-93 |
| rs174555 | miR-1908-! | cis | 7.43E-08 | Nightingale | HDL_FC | NA | C | T | -0.07129 | 0.003957 | 3.30E-78 |
| rs174555 | miR-1908-! | cis | 7.43E-08 | Nightingale | M_LDL_PL_ | NA | C | T | 0.042739 | 0.004464 | 7.50E-23 |
| rs174555 | miR-1908-! | cis | 7.43E-08 | Nightingale | L_HDL_CE_ | NA | C | T | -0.0509 | 0.004274 | 1.10E-34 |
| rs174555 | miR-1908-! | cis | 7.43E-08 | Nightingale | M_HDL_FC | NA | C | T | -0.09999 | 0.003958 | 1.40E-152 |
| rs174555 | miR-1908-! | cis | 7.43E-08 | Nightingale | M_LDL_L | NA | C | T | -0.02687 | 0.004474 | 1.20E-09 |
| rs174555 | miR-1908-! | cis | 7.43E-08 | Nightingale | L_HDL_FC_ | NA | C | T | -0.10801 | 0.00415 | 2.30E-161 |
| rs174555 | miR-1908-! | cis | 7.43E-08 | Nightingale | XL_VLDL_F | NA | C | T | -0.06817 | 0.004328 | 2.40E-58 |
| rs174555 | miR-1908-! | cis | 7.43E-08 | Nightingale | L_HDL_C | NA | C | T | -0.07804 | 0.003933 | 6.90E-95 |
| rs174555 | miR-1908-! | cis | 7.43E-08 | Nightingale | XS_VLDL_C | NA | C | T | -0.09042 | 0.004211 | 2.40E-107 |
| rs174555 | miR-1908-! | cis | 7.43E-08 | Nightingale | XL_HDL_L | NA | C | T | -0.09002 | 0.004014 | 6.40E-121 |

| | | | | | | | | | | | |
|---|---|---|---|---|---|---|---|---|---|---|---|
| rs174555 | miR-1908-5 | cis | 7.43E-08 | Nightingale | TG_by_PG | NA | C | T | 0.068871 | 0.004145 | 2.20E-65 |
| rs174555 | miR-1908-5 | cis | 7.43E-08 | Nightingale | S_LDL_CE_ | NA | C | T | 0.057532 | 0.004459 | 1.00E-39 |
| rs174555 | miR-1908-5 | cis | 7.43E-08 | Nightingale | M_VLDL_C_ | NA | C | T | -0.07963 | 0.004205 | 3.40E-83 |
| rs174555 | miR-1908-5 | cis | 7.43E-08 | Nightingale | XL_VLDL_L | NA | C | T | 0.052516 | 0.004298 | 6.50E-36 |
| rs174555 | miR-1908-5 | cis | 7.43E-08 | Nightingale | L_LDL_TG_ | NA | C | T | 0.077087 | 0.00438 | 8.90E-71 |
| rs174555 | miR-1908-5 | cis | 7.43E-08 | Nightingale | XXL_VLDL_ | NA | C | T | 0.067482 | 0.004302 | 3.40E-58 |
| rs174555 | miR-1908-5 | cis | 7.43E-08 | Nightingale | L_HDL_PL_ | NA | C | T | 0.042938 | 0.004323 | 1.60E-24 |
| rs174555 | miR-1908-5 | cis | 7.43E-08 | Nightingale | HDL_size | NA | C | T | -0.07008 | 0.003958 | 5.60E-76 |
| rs174555 | miR-1908-5 | cis | 7.43E-08 | Nightingale | XS_VLDL_C | NA | C | T | -0.09613 | 0.004233 | 9.79E-120 |
| rs174555 | miR-1908-5 | cis | 7.43E-08 | Nightingale | XL_VLDL_T | NA | C | T | 0.060073 | 0.004328 | 2.50E-45 |
| rs174555 | miR-1908-5 | cis | 7.43E-08 | Nightingale | Total_FC | NA | C | T | -0.05664 | 0.004364 | 1.00E-40 |
| rs174555 | miR-1908-5 | cis | 7.43E-08 | Nightingale | XS_VLDL_P | NA | C | T | -0.03119 | 0.004404 | 9.90E-14 |
| rs174555 | miR-1908-5 | cis | 7.43E-08 | Nightingale | L_VLDL_TG | NA | C | T | 0.03852 | 0.004298 | 4.20E-20 |
| rs174555 | miR-1908-5 | cis | 7.43E-08 | Nightingale | LDL_PL | NA | C | T | -0.04447 | 0.004457 | 2.60E-24 |
| rs174555 | miR-1908-5 | cis | 7.43E-08 | Nightingale | IDL_CE | NA | C | T | -0.06927 | 0.004312 | 2.00E-61 |
| rs174555 | miR-1908-5 | cis | 7.43E-08 | Nightingale | M_LDL_P | NA | C | T | -0.04472 | 0.00448 | 5.10E-24 |
| rs174555 | miR-1908-5 | cis | 7.43E-08 | Nightingale | LA | NA | C | T | 0.083959 | 0.004381 | 3.10E-82 |
| rs174555 | miR-1908-5 | cis | 7.43E-08 | Nightingale | L_LDL_PL | NA | C | T | -0.05301 | 0.004433 | 2.50E-34 |
| rs174555 | miR-1908-5 | cis | 7.43E-08 | Nightingale | XXL_VLDL_ | NA | C | T | 0.07273 | 0.004287 | 7.60E-68 |
| rs174555 | miR-1908-5 | cis | 7.43E-08 | Nightingale | XXL_VLDL_ | NA | C | T | -0.02906 | 0.004443 | 9.50E-12 |
| rs174555 | miR-1908-5 | cis | 7.43E-08 | Nightingale | S_VLDL_TG | NA | C | T | 0.063616 | 0.004322 | 1.70E-50 |
| rs174555 | miR-1908-5 | cis | 7.43E-08 | Nightingale | M_VLDL_F | NA | C | T | -0.08943 | 0.004247 | 2.50E-103 |
| rs174555 | miR-1908-5 | cis | 7.43E-08 | Nightingale | M_HDL_PL | NA | C | T | 0.059542 | 0.004237 | 2.10E-46 |
| rs174555 | miR-1908-5 | cis | 7.43E-08 | Nightingale | S_VLDL_PL | NA | C | T | -0.07762 | 0.004274 | 4.40E-77 |
| rs174555 | miR-1908-5 | cis | 7.43E-08 | Nightingale | L_LDL_FC | NA | C | T | -0.06625 | 0.004386 | 2.80E-54 |
| rs174555 | miR-1908-5 | cis | 7.43E-08 | Nightingale | L_VLDL_CE | NA | C | T | -0.05891 | 0.004275 | 7.30E-46 |
| rs174555 | miR-1908-5 | cis | 7.43E-08 | Nightingale | LDL_TG | NA | C | T | 0.031334 | 0.004417 | 1.40E-12 |
| rs174555 | miR-1908-5 | cis | 7.43E-08 | Nightingale | Citrate | NA | C | T | -0.03409 | 0.004439 | 1.60E-15 |
| rs174555 | miR-1908-5 | cis | 7.43E-08 | Nightingale | IDL_C_pct | NA | C | T | -0.07401 | 0.004361 | 4.30E-66 |
| rs174555 | miR-1908-5 | cis | 7.43E-08 | Nightingale | XS_VLDL_T | NA | C | T | 0.032914 | 0.004398 | 1.50E-13 |
| rs174555 | miR-1908-5 | cis | 7.43E-08 | Nightingale | S_HDL_TG | NA | C | T | 0.068849 | 0.004331 | 5.50E-60 |
| rs174555 | miR-1908-5 | cis | 7.43E-08 | Nightingale | S_LDL_C | NA | C | T | -0.03073 | 0.004472 | 2.40E-12 |
| rs174555 | miR-1908-5 | cis | 7.43E-08 | Nightingale | XS_VLDL_F | NA | C | T | -0.11031 | 0.004433 | 1.90E-139 |
| rs174555 | miR-1908-5 | cis | 7.43E-08 | Nightingale | IDL_TG_pct | NA | C | T | 0.086846 | 0.004315 | 1.60E-93 |
| rs174555 | miR-1908-5 | cis | 7.43E-08 | Nightingale | XL_VLDL_P | NA | C | T | 0.049406 | 0.004311 | 8.70E-32 |
| rs174555 | miR-1908-5 | cis | 7.43E-08 | Nightingale | L_VLDL_FC | NA | C | T | 0.027375 | 0.004425 | 6.10E-10 |
| rs174555 | miR-1908-5 | cis | 7.43E-08 | Nightingale | M_VLDL_T | NA | C | T | 0.076741 | 0.004219 | 3.30E-77 |
| rs174555 | miR-1908-5 | cis | 7.43E-08 | Nightingale | Remnant_( | NA | C | T | -0.04129 | 0.004438 | 1.10E-21 |
| rs174555 | miR-1908-5 | cis | 7.43E-08 | Nightingale | M_HDL_CE | NA | C | T | -0.04914 | 0.004332 | 1.30E-30 |
| rs174555 | miR-1908-5 | cis | 7.43E-08 | Nightingale | XL_VLDL_F | NA | C | T | 0.044795 | 0.004327 | 3.20E-26 |
| rs174555 | miR-1908-5 | cis | 7.43E-08 | Nightingale | XL_HDL_CE | NA | C | T | -0.09447 | 0.004 | 2.50E-133 |
| rs174555 | miR-1908-5 | cis | 7.43E-08 | Nightingale | VLDL_L | NA | C | T | 0.029004 | 0.00438 | 1.30E-11 |
| rs174555 | miR-1908-5 | cis | 7.43E-08 | Nightingale | Total_PL | NA | C | T | -0.04799 | 0.004254 | 2.30E-31 |
| rs174555 | miR-1908-5 | cis | 7.43E-08 | Nightingale | Omega_3_ | NA | C | T | -0.38283 | 0.004365 | 0 |
| rs174555 | miR-1908-5 | cis | 7.43E-08 | Nightingale | M_VLDL_P | NA | C | T | -0.06073 | 0.004286 | 1.30E-48 |
| rs174555 | miR-1908-5 | cis | 7.43E-08 | Nightingale | XL_HDL_C | NA | C | T | -0.09634 | 0.004028 | 1.00E-136 |
| rs174555 | miR-1908-5 | cis | 7.43E-08 | Nightingale | PUFA_by_M | NA | C | T | -0.16782 | 0.004279 | 0 |
| rs174555 | miR-1908-5 | cis | 7.43E-08 | Nightingale | S_LDL_TG | NA | C | T | 0.051454 | 0.004358 | 3.50E-33 |
| rs174555 | miR-1908-5 | cis | 7.43E-08 | Nightingale | L_HDL_CE | NA | C | T | -0.07661 | 0.003944 | 7.80E-91 |
| rs174555 | miR-1908-5 | cis | 7.43E-08 | Nightingale | XL_VLDL_P | NA | C | T | 0.053769 | 0.004293 | 1.20E-37 |
| rs174555 | miR-1908-5 | cis | 7.43E-08 | Nightingale | LDL_CE | NA | C | T | -0.04176 | 0.004456 | 9.50E-22 |
| rs174555 | miR-1908-5 | cis | 7.43E-08 | Nightingale | M_HDL_C_ | NA | C | T | -0.06673 | 0.004252 | 2.60E-58 |
| rs174555 | miR-1908-5 | cis | 7.43E-08 | Nightingale | XL_HDL_P | NA | C | T | -0.08655 | 0.00402 | 5.00E-112 |
| rs174555 | miR-1908-5 | cis | 7.43E-08 | Nightingale | IDL_FC | NA | C | T | -0.08667 | 0.004324 | 3.60E-94 |
| rs174555 | miR-1908-5 | cis | 7.43E-08 | Nightingale | M_VLDL_T | NA | C | T | 0.032445 | 0.00434 | 2.00E-14 |
| rs174555 | miR-1908-5 | cis | 7.43E-08 | Nightingale | XL_VLDL_T | NA | C | T | 0.057923 | 0.004273 | 7.80E-44 |
| rs174555 | miR-1908-5 | cis | 7.43E-08 | Nightingale | XL_VLDL_C | NA | C | T | -0.06176 | 0.004283 | 1.80E-48 |
| rs174555 | miR-1908-5 | cis | 7.43E-08 | Nightingale | MUFA_pct | NA | C | T | 0.193649 | 0.004275 | 0 |
| rs174555 | miR-1908-5 | cis | 7.43E-08 | Nightingale | L_HDL_C_p | NA | C | T | -0.06809 | 0.00421 | 7.80E-63 |
| rs174555 | miR-1908-5 | cis | 7.43E-08 | Nightingale | S_VLDL_FC | NA | C | T | -0.08157 | 0.004265 | 5.10E-85 |
| rs174555 | miR-1908-5 | cis | 7.43E-08 | Nightingale | M_LDL_FC | NA | C | T | -0.05119 | 0.004462 | 8.00E-32 |
| rs174555 | miR-1908-5 | cis | 7.43E-08 | Nightingale | IDL_CE_pct | NA | C | T | -0.03719 | 0.004383 | 3.70E-17 |
| rs174555 | miR-1908-5 | cis | 7.43E-08 | Nightingale | M_HDL_TG | NA | C | T | 0.067391 | 0.004284 | 3.00E-58 |

| | | | | | | | | | | | |
|---|---|---|---|---|---|---|---|---|---|---|---|
| rs174555 | miR-1908-! | cis | 7.43E-08 | Nightingale | M_VLDL_F | NA | C | T | -0.02721 | 0.004466 | 5.20E-10 |
| rs174555 | miR-1908-! | cis | 7.43E-08 | Nightingale | M_LDL_C | NA | C | T | -0.03267 | 0.004476 | 1.30E-13 |
| rs174555 | miR-1908-! | cis | 7.43E-08 | Nightingale | Omega_3 | NA | C | T | -0.32878 | 0.004378 | 0 |
| rs174555 | miR-1908-! | cis | 7.43E-08 | Nightingale | XL_HDL_TC | NA | C | T | 0.086398 | 0.004208 | 2.00E-99 |
| rs174555 | miR-1908-! | cis | 7.43E-08 | Nightingale | S_VLDL_TG | NA | C | T | 0.039345 | 0.004335 | 1.30E-20 |
| rs174555 | miR-1908-! | cis | 7.43E-08 | Nightingale | Clinical_LD | NA | C | T | -0.05323 | 0.004441 | 1.80E-34 |
| rs174555 | miR-1908-! | cis | 7.43E-08 | Nightingale | L_VLDL_L | NA | C | T | 0.039321 | 0.004317 | 1.00E-20 |
| rs174555 | miR-1908-! | cis | 7.43E-08 | Nightingale | HDL_P | NA | C | T | -0.03452 | 0.004194 | 2.40E-17 |
| rs174555 | miR-1908-! | cis | 7.43E-08 | Nightingale | S_LDL_FC_ | NA | C | T | -0.08062 | 0.004329 | 2.90E-80 |
| rs174555 | miR-1908-! | cis | 7.43E-08 | Nightingale | S_LDL_PL | NA | C | T | -0.04964 | 0.004456 | 3.50E-30 |
| rs174555 | miR-1908-! | cis | 7.43E-08 | Nightingale | L_VLDL_C | NA | C | T | 0.03005 | 0.004369 | 2.00E-12 |
| rs174555 | miR-1908-! | cis | 7.43E-08 | Nightingale | M_HDL_FC | NA | C | T | -0.0496 | 0.004065 | 1.30E-36 |
| rs174555 | miR-1908-! | cis | 7.43E-08 | Nightingale | S_LDL_L | NA | C | T | -0.03088 | 0.004466 | 1.60E-12 |
| rs174555 | miR-1908-! | cis | 7.43E-08 | Nightingale | S_HDL_C_p | NA | C | T | -0.0447 | 0.004415 | 1.40E-23 |
| rs174555 | miR-1908-! | cis | 7.43E-08 | Nightingale | M_LDL_CE | NA | C | T | -0.02493 | 0.004474 | 1.70E-08 |
| rs174555 | miR-1908-! | cis | 7.43E-08 | Nightingale | M_LDL_C_ | NA | C | T | -0.09818 | 0.00447 | 3.50E-107 |
| rs174555 | miR-1908-! | cis | 7.43E-08 | Nightingale | M_LDL_TG | NA | C | T | 0.037897 | 0.004401 | 3.20E-18 |
| rs174555 | miR-1908-! | cis | 7.43E-08 | Nightingale | Total_C | NA | C | T | -0.06259 | 0.004324 | 6.10E-50 |
| rs174555 | miR-1908-! | cis | 7.43E-08 | Nightingale | IDL_PL | NA | C | T | -0.06955 | 0.00431 | 1.30E-62 |
| rs174555 | miR-1908-! | cis | 7.43E-08 | Nightingale | L_HDL_FC | NA | C | T | -0.08187 | 0.003915 | 2.60E-105 |
| rs174555 | miR-1908-! | cis | 7.43E-08 | Nightingale | L_VLDL_PL | NA | C | T | 0.049608 | 0.00431 | 3.50E-32 |
| rs174555 | miR-1908-! | cis | 7.43E-08 | Nightingale | IDL_FC_pct | NA | C | T | -0.10392 | 0.004431 | 7.91E-123 |
| rs174555 | miR-1908-! | cis | 7.43E-08 | Nightingale | HDL_TG | NA | C | T | 0.047743 | 0.004405 | 7.80E-28 |
| rs174555 | miR-1908-! | cis | 7.43E-08 | Nightingale | M_HDL_P | NA | C | T | -0.03299 | 0.004115 | 7.30E-17 |
| rs174555 | miR-1908-! | cis | 7.43E-08 | Nightingale | ApoB | NA | C | T | -0.03633 | 0.004461 | 7.60E-17 |
| rs174555 | miR-1908-! | cis | 7.43E-08 | Nightingale | LDL_FC | NA | C | T | -0.0625 | 0.004419 | 1.40E-47 |
| rs174555 | miR-1908-! | cis | 7.43E-08 | Nightingale | M_HDL_CE | NA | C | T | -0.0372 | 0.004125 | 1.70E-20 |
| rs174555 | miR-1908-! | cis | 7.43E-08 | Nightingale | XS_VLDL_P | NA | C | T | 0.057715 | 0.004358 | 9.10E-41 |
| rs174555 | miR-1908-! | cis | 7.43E-08 | Nightingale | M_HDL_L | NA | C | T | -0.02638 | 0.004147 | 3.60E-11 |
| rs174555 | miR-1908-! | cis | 7.43E-08 | Nightingale | Unsaturati | NA | C | T | -0.33325 | 0.00423 | 0 |
| rs174555 | miR-1908-! | cis | 7.43E-08 | Nightingale | L_LDL_FC_ | NA | C | T | -0.08223 | 0.004258 | 3.80E-86 |
| rs174555 | miR-1908-! | cis | 7.43E-08 | Nightingale | IDL_C | NA | C | T | -0.07424 | 0.004313 | 3.20E-70 |
| rs174555 | miR-1908-! | cis | 7.43E-08 | Nightingale | XL_HDL_PL | NA | C | T | -0.06431 | 0.004103 | 4.20E-60 |
| rs174555 | miR-1908-! | cis | 7.43E-08 | Nightingale | L_VLDL_C_ | NA | C | T | -0.03894 | 0.004364 | 2.20E-20 |
| rs174555 | miR-1908-! | cis | 7.43E-08 | Nightingale | XS_VLDL_C | NA | C | T | -0.06837 | 0.00431 | 3.40E-60 |
| rs174555 | miR-1908-! | cis | 7.43E-08 | Nightingale | Cholines | NA | C | T | -0.03462 | 0.004201 | 6.80E-18 |
| rs174555 | miR-1908-! | cis | 7.43E-08 | Nightingale | L_HDL_L | NA | C | T | -0.07479 | 0.003919 | 2.80E-88 |
| rs174555 | miR-1908-! | cis | 7.43E-08 | Nightingale | HDL_L | NA | C | T | -0.05141 | 0.004011 | 2.50E-40 |
| rs174555 | miR-1908-! | cis | 7.43E-08 | Nightingale | L_HDL_PL | NA | C | T | -0.0736 | 0.003931 | 5.30E-85 |
| rs174555 | miR-1908-! | cis | 7.43E-08 | Nightingale | IDL_P | NA | C | T | -0.04229 | 0.00442 | 8.40E-23 |
| rs174555 | miR-1908-! | cis | 7.43E-08 | Nightingale | S_LDL_TG_ | NA | C | T | 0.086078 | 0.004328 | 4.30E-91 |
| rs174555 | miR-1908-! | cis | 7.43E-08 | Nightingale | S_LDL_PL_ | NA | C | T | -0.06158 | 0.004458 | 2.40E-45 |
| rs174555 | miR-1908-! | cis | 7.43E-08 | Nightingale | M_VLDL_C | NA | C | T | -0.0752 | 0.004193 | 1.20E-74 |
| rs174555 | miR-1908-! | cis | 7.43E-08 | Nightingale | XS_VLDL_L | NA | C | T | -0.03832 | 0.004391 | 8.20E-20 |
| rs174555 | miR-1908-! | cis | 7.43E-08 | Nightingale | L_LDL_P | NA | C | T | -0.03578 | 0.004452 | 1.40E-16 |
| rs174555 | miR-1908-! | cis | 7.43E-08 | Nightingale | XXL_VLDL_ | NA | C | T | 0.07218 | 0.004305 | 5.20E-66 |
| rs174555 | miR-1908-! | cis | 7.43E-08 | Nightingale | M_LDL_TG | NA | C | T | 0.075559 | 0.004411 | 4.00E-67 |
| rs174555 | miR-1908-! | cis | 7.43E-08 | Nightingale | M_VLDL_C | NA | C | T | -0.0507 | 0.004426 | 1.40E-31 |
| rs174555 | miR-1908-! | cis | 7.43E-08 | Nightingale | L_VLDL_FC | NA | C | T | 0.041127 | 0.004321 | 1.60E-22 |
| rs174555 | miR-1908-! | cis | 7.43E-08 | Nightingale | XL_VLDL_C | NA | C | T | 0.038685 | 0.004363 | 1.20E-19 |
| rs174555 | miR-1908-! | cis | 7.43E-08 | Nightingale | HDL_C | NA | C | T | -0.06321 | 0.003989 | 6.60E-61 |
| rs174555 | miR-1908-! | cis | 7.43E-08 | Nightingale | XXL_VLDL_ | NA | C | T | 0.067025 | 0.004296 | 9.00E-58 |
| rs174555 | miR-1908-! | cis | 7.43E-08 | Nightingale | XL_HDL_PL | NA | C | T | -0.08606 | 0.004015 | 5.60E-111 |
| rs174555 | miR-1908-! | cis | 7.43E-08 | Nightingale | L_VLDL_P | NA | C | T | 0.042882 | 0.004313 | 2.20E-24 |
| rs174555 | miR-1908-! | cis | 7.43E-08 | Nightingale | L_HDL_TG_ | NA | C | T | 0.078354 | 0.004215 | 1.00E-81 |
| rs174555 | miR-1908-! | cis | 7.43E-08 | Nightingale | XS_VLDL_F | NA | C | T | -0.05259 | 0.004389 | 3.00E-35 |
| rs174555 | miR-1908-! | cis | 7.43E-08 | Nightingale | XS_VLDL_T | NA | C | T | 0.095168 | 0.004243 | 2.30E-117 |
| rs174555 | miR-1908-! | cis | 7.43E-08 | Nightingale | M_HDL_C | NA | C | T | -0.0399 | 0.00411 | 1.40E-23 |
| rs174555 | miR-1908-! | cis | 7.43E-08 | Nightingale | XL_HDL_C_ | NA | C | T | 0.021648 | 0.004267 | 2.10E-08 |
| rs174555 | miR-1908-! | cis | 7.43E-08 | Nightingale | S_HDL_TG_ | NA | C | T | 0.06906 | 0.004248 | 3.20E-62 |
| rs174555 | miR-1908-! | cis | 7.43E-08 | Nightingale | S_HDL_FC_ | NA | C | T | -0.0998 | 0.004237 | 2.90E-135 |
| rs174555 | miR-1908-! | cis | 7.43E-08 | Nightingale | L_LDL_L | NA | C | T | -0.05215 | 0.004424 | 1.10E-33 |
| rs174555 | miR-1908-! | cis | 7.43E-08 | Nightingale | Total_L | NA | C | T | -0.03442 | 0.004395 | 6.10E-16 |

| | | | | | | | | | | | |
|---|---|---|---|---|---|---|---|---|---|---|---|
| rs174555 | miR-1908-5 | cis | 7.43E-08 | Nightingale | IDL_L | NA | C | T | -0.06926 | 0.004318 | 1.20E-61 |
| rs174555 | miR-1908-5 | cis | 7.43E-08 | Nightingale | LA_pct | NA | C | T | 0.181223 | 0.004365 | 0 |
| rs174555 | miR-1908-5 | cis | 7.43E-08 | Nightingale | L_LDL_C | NA | C | T | -0.0558 | 0.004417 | 2.00E-38 |
| rs174555 | miR-1908-5 | cis | 7.43E-08 | Nightingale | L_LDL_CE | NA | C | T | -0.05132 | 0.004427 | 1.30E-32 |
| rs174555 | miR-1908-5 | cis | 7.43E-08 | Nightingale | ApoA1 | NA | C | T | -0.03959 | 0.004088 | 7.30E-24 |
| rs174555 | miR-1908-5 | cis | 7.43E-08 | Nightingale | Total_P | NA | C | T | -0.03898 | 0.004204 | 1.10E-21 |
| rs174555 | miR-1908-5 | cis | 7.43E-08 | Nightingale | L_HDL_P | NA | C | T | -0.0755 | 0.003912 | 4.00E-90 |
| rs174555 | miR-1908-5 | cis | 7.43E-08 | Nightingale | Sphingomy | NA | C | T | -0.11205 | 0.004174 | 4.10E-166 |
| rs174555 | miR-1908-5 | cis | 7.43E-08 | Nightingale | DHA | NA | C | T | -0.28262 | 0.004305 | 0 |
| rs174555 | miR-1908-5 | cis | 7.43E-08 | Nightingale | DHA_pct | NA | C | T | -0.26943 | 0.004336 | 0 |
| rs174555 | miR-1908-5 | cis | 7.43E-08 | Nightingale | S_VLDL_FC | NA | C | T | -0.02653 | 0.004468 | 1.30E-09 |
| rs174555 | miR-1908-5 | cis | 7.43E-08 | Nightingale | SFA | NA | C | T | -0.0281 | 0.004427 | 1.60E-10 |
| rs174555 | miR-1908-5 | cis | 7.43E-08 | Nightingale | XL_HDL_FC | NA | C | T | -0.099 | 0.004161 | 2.30E-134 |
| rs174555 | miR-1908-5 | cis | 7.43E-08 | Nightingale | LDL_L | NA | C | T | -0.04364 | 0.004452 | 1.10E-23 |
| rs174555 | miR-1908-5 | cis | 7.43E-08 | Nightingale | M_LDL_FC_ | NA | C | T | -0.07219 | 0.004279 | 2.30E-66 |
| rs174555 | miR-1908-5 | cis | 7.43E-08 | Nightingale | HDL_CE | NA | C | T | -0.06025 | 0.004009 | 8.10E-55 |
| rs174555 | miR-1908-5 | cis | 7.43E-08 | Nightingale | XL_HDL_FC | NA | C | T | 0.059926 | 0.00396 | 4.00E-57 |
| rs174555 | miR-1908-5 | cis | 7.43E-08 | Nightingale | XXL_VLDL_ | NA | C | T | 0.068691 | 0.004303 | 2.20E-60 |
| rs174555 | miR-1908-5 | cis | 7.43E-08 | Nightingale | HDL_PL | NA | C | T | -0.0463 | 0.004038 | 1.30E-32 |
| rs174555 | miR-1908-5 | cis | 7.43E-08 | Nightingale | XS_VLDL_P | NA | C | T | -0.04053 | 0.004407 | 1.40E-21 |
| rs174555 | miR-1908-5 | cis | 7.43E-08 | Nightingale | XL_VLDL_C | NA | C | T | 0.031467 | 0.004397 | 2.90E-13 |
| rs174555 | miR-1908-5 | cis | 7.43E-08 | Nightingale | LDL_P | NA | C | T | -0.03839 | 0.004464 | 1.60E-18 |
| rs174555 | miR-1908-5 | cis | 7.43E-08 | Nightingale | VLDL_TG | NA | C | T | 0.048835 | 0.004299 | 2.80E-31 |
| rs174555 | miR-1908-5 | cis | 7.43E-08 | Nightingale | XXL_VLDL_ | NA | C | T | 0.062303 | 0.00431 | 1.40E-49 |
| rs174555 | miR-1908-5 | cis | 7.43E-08 | Nightingale | PUFA_pct | NA | C | T | -0.12033 | 0.004297 | 5.40E-174 |
| rs174555 | miR-1908-5 | cis | 7.43E-08 | Nightingale | Tyr | NA | C | T | 0.03302 | 0.004461 | 1.40E-13 |
| rs174555 | miR-1908-5 | cis | 7.43E-08 | Nightingale | L_LDL_C_p | NA | C | T | -0.07583 | 0.004352 | 1.30E-69 |
| rs174555 | miR-1908-5 | cis | 7.43E-08 | Nightingale | Total_CE | NA | C | T | -0.06448 | 0.004309 | 3.30E-53 |
| rs174555 | miR-1908-5 | cis | 7.43E-08 | Nightingale | XXL_VLDL_ | NA | C | T | 0.042912 | 0.004489 | 2.80E-22 |
| rs174555 | miR-1908-5 | cis | 7.43E-08 | Nightingale | XXL_VLDL_ | NA | C | T | 0.073857 | 0.004314 | 1.90E-68 |
| rs174555 | miR-1908-5 | cis | 7.43E-08 | Nightingale | S_LDL_FC | NA | C | T | -0.06141 | 0.00446 | 2.40E-45 |
| rs174555 | miR-1908-5 | cis | 7.43E-08 | Nightingale | non_HDL_( | NA | C | T | -0.04545 | 0.004443 | 1.10E-25 |
| rs174555 | miR-1908-5 | cis | 7.43E-08 | Nightingale | S_VLDL_CE | NA | C | T | -0.02862 | 0.004407 | 2.90E-11 |
| rs174555 | miR-1908-5 | cis | 7.43E-08 | Nightingale | SFA_pct | NA | C | T | -0.03941 | 0.004407 | 3.80E-19 |
| rs174555 | miR-1908-5 | cis | 7.43E-08 | Nightingale | Phosphogly | NA | C | T | -0.03816 | 0.004231 | 7.00E-21 |
| rs174555 | miR-1908-5 | cis | 7.43E-08 | Nightingale | LDL_C | NA | C | T | -0.04794 | 0.004448 | 2.80E-28 |
| rs174555 | miR-1908-5 | cis | 7.43E-08 | Nightingale | PUFA | NA | C | T | -0.09257 | 0.004356 | 3.50E-106 |
| rs174555 | miR-1908-5 | cis | 7.43E-08 | Nightingale | Total_TG | NA | C | T | 0.048195 | 0.004324 | 3.20E-30 |
| rs174555 | miR-1908-5 | cis | 7.43E-08 | Nightingale | XS_VLDL_C | NA | C | T | -0.0646 | 0.004335 | 2.30E-53 |
| rs174555 | miR-1908-5 | cis | 7.43E-08 | Nightingale | XL_VLDL_C | NA | C | T | -0.0645 | 0.00428 | 9.90E-53 |
| rs174556 | miR-1908-5 | cis | 7.67E-08 | Nightingale | XL_VLDL_F | NA | T | C | -0.06832 | 0.004332 | 1.80E-58 |
| rs174556 | miR-1908-5 | cis | 7.67E-08 | Nightingale | L_LDL_TG_ | NA | T | C | 0.077518 | 0.004384 | 2.30E-71 |
| rs174556 | miR-1908-5 | cis | 7.67E-08 | Nightingale | XL_HDL_L | NA | T | C | -0.09031 | 0.004017 | 1.30E-121 |
| rs174556 | miR-1908-5 | cis | 7.67E-08 | Nightingale | XL_VLDL_F | NA | T | C | 0.044861 | 0.00433 | 3.40E-26 |
| rs174556 | miR-1908-5 | cis | 7.67E-08 | Nightingale | L_VLDL_CE | NA | T | C | -0.05908 | 0.004278 | 5.10E-46 |
| rs174556 | miR-1908-5 | cis | 7.67E-08 | Nightingale | LA | NA | T | C | 0.083492 | 0.004384 | 4.10E-81 |
| rs174556 | miR-1908-5 | cis | 7.67E-08 | Nightingale | XS_VLDL_C | NA | T | C | -0.09083 | 0.004214 | 5.40E-108 |
| rs174556 | miR-1908-5 | cis | 7.67E-08 | Nightingale | M_LDL_L | NA | T | C | -0.02701 | 0.004478 | 9.60E-10 |
| rs174556 | miR-1908-5 | cis | 7.67E-08 | Nightingale | XXL_VLDL_ | NA | T | C | -0.02908 | 0.004447 | 9.70E-12 |
| rs174556 | miR-1908-5 | cis | 7.67E-08 | Nightingale | XS_VLDL_P | NA | T | C | -0.03118 | 0.004408 | 8.20E-14 |
| rs174556 | miR-1908-5 | cis | 7.67E-08 | Nightingale | M_LDL_PL_ | NA | T | C | 0.042618 | 0.004468 | 1.00E-22 |
| rs174556 | miR-1908-5 | cis | 7.67E-08 | Nightingale | M_VLDL_C | NA | T | C | -0.04213 | 0.004459 | 5.10E-22 |
| rs174556 | miR-1908-5 | cis | 7.67E-08 | Nightingale | M_HDL_C | NA | T | C | -0.03985 | 0.004114 | 1.70E-23 |
| rs174556 | miR-1908-5 | cis | 7.67E-08 | Nightingale | XS_VLDL_P | NA | T | C | 0.058071 | 0.004362 | 4.10E-41 |
| rs174556 | miR-1908-5 | cis | 7.67E-08 | Nightingale | IDL_CE | NA | T | C | -0.06955 | 0.004316 | 8.00E-62 |
| rs174556 | miR-1908-5 | cis | 7.67E-08 | Nightingale | non_HDL_( | NA | T | C | -0.04567 | 0.004447 | 6.00E-26 |
| rs174556 | miR-1908-5 | cis | 7.67E-08 | Nightingale | S_VLDL_C_ | NA | T | C | -0.05595 | 0.00435 | 6.20E-39 |
| rs174556 | miR-1908-5 | cis | 7.67E-08 | Nightingale | L_VLDL_PL | NA | T | C | 0.087593 | 0.004339 | 1.00E-93 |
| rs174556 | miR-1908-5 | cis | 7.67E-08 | Nightingale | XS_VLDL_T | NA | T | C | 0.033135 | 0.004402 | 1.20E-13 |
| rs174556 | miR-1908-5 | cis | 7.67E-08 | Nightingale | L_VLDL_L | NA | T | C | 0.039369 | 0.004321 | 1.20E-20 |
| rs174556 | miR-1908-5 | cis | 7.67E-08 | Nightingale | M_VLDL_P | NA | T | C | -0.06089 | 0.00429 | 8.20E-49 |
| rs174556 | miR-1908-5 | cis | 7.67E-08 | Nightingale | XL_VLDL_C | NA | T | C | -0.06193 | 0.004286 | 1.30E-48 |
| rs174556 | miR-1908-5 | cis | 7.67E-08 | Nightingale | M_LDL_FC | NA | T | C | -0.05143 | 0.004466 | 4.50E-32 |

| | | | | | | | | | | | |
|---|---|---|---|---|---|---|---|---|---|---|---|
| rs174556 | miR-1908-! | cis | 7.67E-08 | Nightingale | IDL_FC | NA | T | C | -0.08693 | 0.004327 | 1.40E-94 |
| rs174556 | miR-1908-! | cis | 7.67E-08 | Nightingale | LDL_TG | NA | T | C | 0.03153 | 0.004421 | 1.20E-12 |
| rs174556 | miR-1908-! | cis | 7.67E-08 | Nightingale | XL_VLDL_P | NA | T | C | 0.049496 | 0.004314 | 8.90E-32 |
| rs174556 | miR-1908-! | cis | 7.67E-08 | Nightingale | XS_VLDL_C | NA | T | C | -0.09654 | 0.004236 | 2.20E-120 |
| rs174556 | miR-1908-! | cis | 7.67E-08 | Nightingale | TG_by_PG | NA | T | C | 0.069003 | 0.004149 | 2.00E-65 |
| rs174556 | miR-1908-! | cis | 7.67E-08 | Nightingale | XL_VLDL_T | NA | T | C | 0.058055 | 0.004276 | 7.10E-44 |
| rs174556 | miR-1908-! | cis | 7.67E-08 | Nightingale | XL_HDL_PL | NA | T | C | -0.0644 | 0.004107 | 3.20E-60 |
| rs174556 | miR-1908-! | cis | 7.67E-08 | Nightingale | L_HDL_C_p | NA | T | C | -0.06847 | 0.004214 | 2.50E-63 |
| rs174556 | miR-1908-! | cis | 7.67E-08 | Nightingale | M_LDL_TG | NA | T | C | 0.038084 | 0.004405 | 2.70E-18 |
| rs174556 | miR-1908-! | cis | 7.67E-08 | Nightingale | Unsaturati | NA | T | C | -0.33277 | 0.004234 | 0 |
| rs174556 | miR-1908-! | cis | 7.67E-08 | Nightingale | Citrate | NA | T | C | -0.03382 | 0.004442 | 2.90E-15 |
| rs174556 | miR-1908-! | cis | 7.67E-08 | Nightingale | S_LDL_CE_ | NA | T | C | 0.057432 | 0.004463 | 1.70E-39 |
| rs174556 | miR-1908-! | cis | 7.67E-08 | Nightingale | MUFA_pct | NA | T | C | 0.193669 | 0.004278 | 0 |
| rs174556 | miR-1908-! | cis | 7.67E-08 | Nightingale | XL_HDL_CE | NA | T | C | -0.02824 | 0.004468 | 1.10E-09 |
| rs174556 | miR-1908-! | cis | 7.67E-08 | Nightingale | XL_HDL_C_ | NA | T | C | 0.021667 | 0.004271 | 1.90E-08 |
| rs174556 | miR-1908-! | cis | 7.67E-08 | Nightingale | L_LDL_FC | NA | T | C | -0.06656 | 0.00439 | 1.10E-54 |
| rs174556 | miR-1908-! | cis | 7.67E-08 | Nightingale | M_LDL_P | NA | T | C | -0.0448 | 0.004484 | 4.00E-24 |
| rs174556 | miR-1908-! | cis | 7.67E-08 | Nightingale | IDL_FC_pct | NA | T | C | -0.10417 | 0.004435 | 3.50E-123 |
| rs174556 | miR-1908-! | cis | 7.67E-08 | Nightingale | L_HDL_C | NA | T | C | -0.07828 | 0.003937 | 2.30E-95 |
| rs174556 | miR-1908-! | cis | 7.67E-08 | Nightingale | L_VLDL_FC | NA | T | C | 0.027396 | 0.004428 | 6.10E-10 |
| rs174556 | miR-1908-! | cis | 7.67E-08 | Nightingale | M_HDL_CE | NA | T | C | -0.0493 | 0.004335 | 1.10E-30 |
| rs174556 | miR-1908-! | cis | 7.67E-08 | Nightingale | M_HDL_L | NA | T | C | -0.02628 | 0.00415 | 4.40E-11 |
| rs174556 | miR-1908-! | cis | 7.67E-08 | Nightingale | M_LDL_C_ | NA | T | C | -0.09845 | 0.004474 | 1.80E-107 |
| rs174556 | miR-1908-! | cis | 7.67E-08 | Nightingale | XS_VLDL_F | NA | T | C | -0.11052 | 0.004437 | 9.79E-140 |
| rs174556 | miR-1908-! | cis | 7.67E-08 | Nightingale | LDL_PL | NA | T | C | -0.04468 | 0.004461 | 1.60E-24 |
| rs174556 | miR-1908-! | cis | 7.67E-08 | Nightingale | M_LDL_C | NA | T | C | -0.03282 | 0.00448 | 9.50E-14 |
| rs174556 | miR-1908-! | cis | 7.67E-08 | Nightingale | L_LDL_PL | NA | T | C | -0.05325 | 0.004437 | 1.30E-34 |
| rs174556 | miR-1908-! | cis | 7.67E-08 | Nightingale | Remnant_( | NA | T | C | -0.0415 | 0.004442 | 5.90E-22 |
| rs174556 | miR-1908-! | cis | 7.67E-08 | Nightingale | XXL_VLDL_ | NA | T | C | 0.068785 | 0.004307 | 2.10E-60 |
| rs174556 | miR-1908-! | cis | 7.67E-08 | Nightingale | S_VLDL_TG | NA | T | C | 0.063801 | 0.004326 | 1.30E-50 |
| rs174556 | miR-1908-! | cis | 7.67E-08 | Nightingale | HDL_L | NA | T | C | -0.05143 | 0.004015 | 2.50E-40 |
| rs174556 | miR-1908-! | cis | 7.67E-08 | Nightingale | XL_HDL_P | NA | T | C | -0.0869 | 0.004023 | 7.80E-113 |
| rs174556 | miR-1908-! | cis | 7.67E-08 | Nightingale | M_VLDL_F | NA | T | C | -0.08961 | 0.004251 | 1.50E-103 |
| rs174556 | miR-1908-! | cis | 7.67E-08 | Nightingale | S_LDL_PL_ | NA | T | C | -0.0615 | 0.004462 | 3.90E-45 |
| rs174556 | miR-1908-! | cis | 7.67E-08 | Nightingale | S_LDL_TG_ | NA | T | C | 0.086374 | 0.004332 | 1.90E-91 |
| rs174556 | miR-1908-! | cis | 7.67E-08 | Nightingale | L_HDL_FC_ | NA | T | C | -0.10822 | 0.004153 | 6.70E-162 |
| rs174556 | miR-1908-! | cis | 7.67E-08 | Nightingale | HDL_P | NA | T | C | -0.03449 | 0.004198 | 2.60E-17 |
| rs174556 | miR-1908-! | cis | 7.67E-08 | Nightingale | XXL_VLDL_ | NA | T | C | 0.06757 | 0.004306 | 3.30E-58 |
| rs174556 | miR-1908-! | cis | 7.67E-08 | Nightingale | Total_C | NA | T | C | -0.06281 | 0.004328 | 3.20E-50 |
| rs174556 | miR-1908-! | cis | 7.67E-08 | Nightingale | M_HDL_TG | NA | T | C | 0.051911 | 0.004415 | 1.60E-32 |
| rs174556 | miR-1908-! | cis | 7.67E-08 | Nightingale | L_LDL_C | NA | T | C | -0.05603 | 0.004421 | 1.10E-38 |
| rs174556 | miR-1908-! | cis | 7.67E-08 | Nightingale | M_HDL_C_ | NA | T | C | -0.0669 | 0.004256 | 2.10E-58 |
| rs174556 | miR-1908-! | cis | 7.67E-08 | Nightingale | S_LDL_FC_ | NA | T | C | -0.08088 | 0.004332 | 1.50E-80 |
| rs174556 | miR-1908-! | cis | 7.67E-08 | Nightingale | S_LDL_PL | NA | T | C | -0.04982 | 0.00446 | 2.10E-30 |
| rs174556 | miR-1908-! | cis | 7.67E-08 | Nightingale | Omega_3 | NA | T | C | -0.32796 | 0.004382 | 0 |
| rs174556 | miR-1908-! | cis | 7.67E-08 | Nightingale | HDL_FC | NA | T | C | -0.07138 | 0.003961 | 2.30E-78 |
| rs174556 | miR-1908-! | cis | 7.67E-08 | Nightingale | XL_VLDL_T | NA | T | C | 0.060184 | 0.004331 | 2.10E-45 |
| rs174556 | miR-1908-! | cis | 7.67E-08 | Nightingale | M_HDL_FC | NA | T | C | -0.10011 | 0.003962 | 8.79E-153 |
| rs174556 | miR-1908-! | cis | 7.67E-08 | Nightingale | M_HDL_TG | NA | T | C | 0.067463 | 0.004287 | 3.40E-58 |
| rs174556 | miR-1908-! | cis | 7.67E-08 | Nightingale | L_HDL_CE | NA | T | C | -0.07686 | 0.003948 | 2.70E-91 |
| rs174556 | miR-1908-! | cis | 7.67E-08 | Nightingale | M_VLDL_T | NA | T | C | 0.07694 | 0.004222 | 2.20E-77 |
| rs174556 | miR-1908-! | cis | 7.67E-08 | Nightingale | MUFA | NA | T | C | 0.064531 | 0.004425 | 1.30E-49 |
| rs174556 | miR-1908-! | cis | 7.67E-08 | Nightingale | SFA | NA | T | C | -0.02792 | 0.00443 | 2.00E-10 |
| rs174556 | miR-1908-! | cis | 7.67E-08 | Nightingale | XS_VLDL_T | NA | T | C | 0.095539 | 0.004247 | 6.61E-118 |
| rs174556 | miR-1908-! | cis | 7.67E-08 | Nightingale | XL_VLDL_C | NA | T | C | 0.038698 | 0.004367 | 1.50E-19 |
| rs174556 | miR-1908-! | cis | 7.67E-08 | Nightingale | M_VLDL_T | NA | T | C | 0.0325 | 0.004344 | 2.10E-14 |
| rs174556 | miR-1908-! | cis | 7.67E-08 | Nightingale | L_VLDL_PL | NA | T | C | 0.049668 | 0.004314 | 4.10E-32 |
| rs174556 | miR-1908-! | cis | 7.67E-08 | Nightingale | VLDL_L | NA | T | C | 0.029052 | 0.004384 | 1.50E-11 |
| rs174556 | miR-1908-! | cis | 7.67E-08 | Nightingale | Omega_3_ | NA | T | C | -0.38191 | 0.004368 | 0 |
| rs174556 | miR-1908-! | cis | 7.67E-08 | Nightingale | S_VLDL_PL | NA | T | C | -0.07781 | 0.004278 | 3.50E-77 |
| rs174556 | miR-1908-! | cis | 7.67E-08 | Nightingale | XL_HDL_C | NA | T | C | -0.09667 | 0.004031 | 1.70E-137 |
| rs174556 | miR-1908-! | cis | 7.67E-08 | Nightingale | XL_VLDL_L | NA | T | C | 0.052626 | 0.004302 | 6.30E-36 |
| rs174556 | miR-1908-! | cis | 7.67E-08 | Nightingale | IDL_TG_pc | NA | T | C | 0.087289 | 0.004319 | 3.50E-94 |

| | | | | | | | | | | | |
|---|---|---|---|---|---|---|---|---|---|---|---|
| rs174556 | miR-1908-5 | cis | 7.67E-08 | Nightingale | M_VLDL_F | NA | T | C | -0.02739 | 0.00447 | 3.60E-10 |
| rs174556 | miR-1908-5 | cis | 7.67E-08 | Nightingale | L_HDL_PL_ | NA | T | C | 0.043583 | 0.004327 | 3.40E-25 |
| rs174556 | miR-1908-5 | cis | 7.67E-08 | Nightingale | XL_HDL_TG | NA | T | C | 0.086616 | 0.004212 | 1.20E-99 |
| rs174556 | miR-1908-5 | cis | 7.67E-08 | Nightingale | VLDL_size | NA | T | C | 0.07241 | 0.004167 | 9.70E-72 |
| rs174556 | miR-1908-5 | cis | 7.67E-08 | Nightingale | XL_HDL_CE | NA | T | C | -0.09479 | 0.004003 | 4.40E-134 |
| rs174556 | miR-1908-5 | cis | 7.67E-08 | Nightingale | M_HDL_PL | NA | T | C | 0.059851 | 0.004241 | 1.00E-46 |
| rs174556 | miR-1908-5 | cis | 7.67E-08 | Nightingale | L_VLDL_C_ | NA | T | C | -0.039 | 0.004367 | 2.00E-20 |
| rs174556 | miR-1908-5 | cis | 7.67E-08 | Nightingale | Total_L | NA | T | C | -0.03453 | 0.004398 | 4.90E-16 |
| rs174556 | miR-1908-5 | cis | 7.67E-08 | Nightingale | L_VLDL_TG | NA | T | C | 0.03858 | 0.004302 | 4.50E-20 |
| rs174556 | miR-1908-5 | cis | 7.67E-08 | Nightingale | S_LDL_TG | NA | T | C | 0.051614 | 0.004361 | 3.00E-33 |
| rs174556 | miR-1908-5 | cis | 7.67E-08 | Nightingale | ApoB | NA | T | C | -0.03653 | 0.004465 | 4.80E-17 |
| rs174556 | miR-1908-5 | cis | 7.67E-08 | Nightingale | HDL_size | NA | T | C | -0.07031 | 0.003961 | 2.10E-76 |
| rs174556 | miR-1908-5 | cis | 7.67E-08 | Nightingale | Total_PL | NA | T | C | -0.04806 | 0.004257 | 2.00E-31 |
| rs174556 | miR-1908-5 | cis | 7.67E-08 | Nightingale | IDL_L | NA | T | C | -0.06949 | 0.004322 | 5.20E-62 |
| rs174556 | miR-1908-5 | cis | 7.67E-08 | Nightingale | M_HDL_CE | NA | T | C | -0.03716 | 0.004129 | 2.00E-20 |
| rs174556 | miR-1908-5 | cis | 7.67E-08 | Nightingale | IDL_C_pct | NA | T | C | -0.07443 | 0.004365 | 1.20E-66 |
| rs174556 | miR-1908-5 | cis | 7.67E-08 | Nightingale | PUFA_pct | NA | T | C | -0.12047 | 0.0043 | 5.40E-174 |
| rs174556 | miR-1908-5 | cis | 7.67E-08 | Nightingale | LDL_L | NA | T | C | -0.04383 | 0.004456 | 7.30E-24 |
| rs174556 | miR-1908-5 | cis | 7.67E-08 | Nightingale | Sphingomy | NA | T | C | -0.11213 | 0.004178 | 4.00E-166 |
| rs174556 | miR-1908-5 | cis | 7.67E-08 | Nightingale | ApoA1 | NA | T | C | -0.03957 | 0.004091 | 7.90E-24 |
| rs174556 | miR-1908-5 | cis | 7.67E-08 | Nightingale | S_VLDL_TG | NA | T | C | 0.039444 | 0.004338 | 1.20E-20 |
| rs174556 | miR-1908-5 | cis | 7.67E-08 | Nightingale | Omega_6_ | NA | T | C | 0.36182 | 0.004406 | 0 |
| rs174556 | miR-1908-5 | cis | 7.67E-08 | Nightingale | XS_VLDL_C | NA | T | C | -0.06857 | 0.004314 | 1.50E-60 |
| rs174556 | miR-1908-5 | cis | 7.67E-08 | Nightingale | XXL_VLDL_ | NA | T | C | 0.072851 | 0.00429 | 6.60E-68 |
| rs174556 | miR-1908-5 | cis | 7.67E-08 | Nightingale | L_HDL_CE_ | NA | T | C | -0.0513 | 0.004278 | 4.30E-35 |
| rs174556 | miR-1908-5 | cis | 7.67E-08 | Nightingale | M_HDL_P | NA | T | C | -0.03293 | 0.004119 | 8.60E-17 |
| rs174556 | miR-1908-5 | cis | 7.67E-08 | Nightingale | S_LDL_L | NA | T | C | -0.03107 | 0.00447 | 1.10E-12 |
| rs174556 | miR-1908-5 | cis | 7.67E-08 | Nightingale | XS_VLDL_P | NA | T | C | -0.04058 | 0.004411 | 1.00E-21 |
| rs174556 | miR-1908-5 | cis | 7.67E-08 | Nightingale | VLDL_TG | NA | T | C | 0.04894 | 0.004302 | 2.80E-31 |
| rs174556 | miR-1908-5 | cis | 7.67E-08 | Nightingale | S_LDL_P | NA | T | C | -0.03036 | 0.004475 | 4.00E-12 |
| rs174556 | miR-1908-5 | cis | 7.67E-08 | Nightingale | XXL_VLDL_ | NA | T | C | 0.072239 | 0.004309 | 6.10E-66 |
| rs174556 | miR-1908-5 | cis | 7.67E-08 | Nightingale | M_LDL_CE | NA | T | C | -0.02505 | 0.004477 | 1.40E-08 |
| rs174556 | miR-1908-5 | cis | 7.67E-08 | Nightingale | M_LDL_TG | NA | T | C | 0.075908 | 0.004414 | 1.60E-67 |
| rs174556 | miR-1908-5 | cis | 7.67E-08 | Nightingale | L_HDL_L | NA | T | C | -0.07498 | 0.003923 | 1.20E-88 |
| rs174556 | miR-1908-5 | cis | 7.67E-08 | Nightingale | S_LDL_C | NA | T | C | -0.03094 | 0.004476 | 1.60E-12 |
| rs174556 | miR-1908-5 | cis | 7.67E-08 | Nightingale | L_LDL_P | NA | T | C | -0.03604 | 0.004456 | 8.20E-17 |
| rs174556 | miR-1908-5 | cis | 7.67E-08 | Nightingale | L_HDL_FC | NA | T | C | -0.08209 | 0.003918 | 9.71E-106 |
| rs174556 | miR-1908-5 | cis | 7.67E-08 | Nightingale | L_VLDL_C | NA | T | C | 0.030036 | 0.004373 | 2.40E-12 |
| rs174556 | miR-1908-5 | cis | 7.67E-08 | Nightingale | S_HDL_TG | NA | T | C | 0.069024 | 0.004335 | 4.20E-60 |
| rs174556 | miR-1908-5 | cis | 7.67E-08 | Nightingale | L_HDL_PL | NA | T | C | -0.07372 | 0.003935 | 3.30E-85 |
| rs174556 | miR-1908-5 | cis | 7.67E-08 | Nightingale | L_VLDL_P | NA | T | C | 0.04294 | 0.004317 | 2.40E-24 |
| rs174556 | miR-1908-5 | cis | 7.67E-08 | Nightingale | XL_VLDL_P | NA | T | C | 0.05386 | 0.004297 | 1.30E-37 |
| rs174556 | miR-1908-5 | cis | 7.67E-08 | Nightingale | HDL_C | NA | T | C | -0.06331 | 0.003992 | 5.10E-61 |
| rs174556 | miR-1908-5 | cis | 7.67E-08 | Nightingale | Total_FC | NA | T | C | -0.05686 | 0.004368 | 5.20E-41 |
| rs174556 | miR-1908-5 | cis | 7.67E-08 | Nightingale | L_VLDL_FC | NA | T | C | 0.041167 | 0.004324 | 1.90E-22 |
| rs174556 | miR-1908-5 | cis | 7.67E-08 | Nightingale | LA_pct | NA | T | C | 0.180196 | 0.004369 | 0 |
| rs174556 | miR-1908-5 | cis | 7.67E-08 | Nightingale | M_VLDL_C | NA | T | C | -0.07983 | 0.004208 | 2.30E-83 |
| rs174556 | miR-1908-5 | cis | 7.67E-08 | Nightingale | M_VLDL_C | NA | T | C | -0.051 | 0.00443 | 5.90E-32 |
| rs174556 | miR-1908-5 | cis | 7.67E-08 | Nightingale | Total_P | NA | T | C | -0.03898 | 0.004207 | 1.10E-21 |
| rs174556 | miR-1908-5 | cis | 7.67E-08 | Nightingale | IDL_PL | NA | T | C | -0.06977 | 0.004314 | 5.70E-63 |
| rs174556 | miR-1908-5 | cis | 7.67E-08 | Nightingale | S_VLDL_FC | NA | T | C | -0.08181 | 0.004269 | 3.00E-85 |
| rs174556 | miR-1908-5 | cis | 7.67E-08 | Nightingale | XXL_VLDL_ | NA | T | C | 0.042989 | 0.004493 | 2.70E-22 |
| rs174556 | miR-1908-5 | cis | 7.67E-08 | Nightingale | XL_HDL_FC | NA | T | C | -0.09933 | 0.004165 | 4.20E-135 |
| rs174556 | miR-1908-5 | cis | 7.67E-08 | Nightingale | DHA_pct | NA | T | C | -0.26895 | 0.00434 | 0 |
| rs174556 | miR-1908-5 | cis | 7.67E-08 | Nightingale | XL_VLDL_C | NA | T | C | 0.031431 | 0.0044 | 3.70E-13 |
| rs174556 | miR-1908-5 | cis | 7.67E-08 | Nightingale | XL_HDL_FC | NA | T | C | 0.060176 | 0.003964 | 1.40E-57 |
| rs174556 | miR-1908-5 | cis | 7.67E-08 | Nightingale | HDL_CE | NA | T | C | -0.06035 | 0.004012 | 6.30E-55 |
| rs174556 | miR-1908-5 | cis | 7.67E-08 | Nightingale | L_HDL_TG_ | NA | T | C | 0.078471 | 0.004219 | 9.70E-82 |
| rs174556 | miR-1908-5 | cis | 7.67E-08 | Nightingale | Cholines | NA | T | C | -0.03469 | 0.004205 | 5.70E-18 |
| rs174556 | miR-1908-5 | cis | 7.67E-08 | Nightingale | S_HDL_C_p | NA | T | C | -0.0449 | 0.004419 | 1.00E-23 |
| rs174556 | miR-1908-5 | cis | 7.67E-08 | Nightingale | L_LDL_FC_ | NA | T | C | -0.08276 | 0.004261 | 6.40E-87 |
| rs174556 | miR-1908-5 | cis | 7.67E-08 | Nightingale | LDL_CE | NA | T | C | -0.04194 | 0.004459 | 6.30E-22 |
| rs174556 | miR-1908-5 | cis | 7.67E-08 | Nightingale | M_HDL_FC | NA | T | C | -0.04955 | 0.004068 | 1.70E-36 |

| | | | | | | | | | | | |
|---|---|---|---|---|---|---|---|---|---|---|---|
| rs174556 | miR-1908-5 | cis | 7.67E-08 | Nightingale | XS_VLDL_C | NA | T | C | -0.06476 | 0.004339 | 1.10E-53 |
| rs174556 | miR-1908-5 | cis | 7.67E-08 | Nightingale | IDL_CE_pct | NA | T | C | -0.03761 | 0.004387 | 1.80E-17 |
| rs174556 | miR-1908-5 | cis | 7.67E-08 | Nightingale | S_HDL_FC_ | NA | T | C | -0.10005 | 0.004241 | 6.61E-136 |
| rs174556 | miR-1908-5 | cis | 7.67E-08 | Nightingale | PUFA | NA | T | C | -0.09253 | 0.00436 | 4.90E-106 |
| rs174556 | miR-1908-5 | cis | 7.67E-08 | Nightingale | HDL_PL | NA | T | C | -0.04625 | 0.004042 | 1.50E-32 |
| rs174556 | miR-1908-5 | cis | 7.67E-08 | Nightingale | XXL_VLDL_ | NA | T | C | 0.062408 | 0.004314 | 1.20E-49 |
| rs174556 | miR-1908-5 | cis | 7.67E-08 | Nightingale | PUFA_by_M | NA | T | C | -0.16791 | 0.004282 | 0 |
| rs174556 | miR-1908-5 | cis | 7.67E-08 | Nightingale | M_VLDL_C | NA | T | C | -0.07542 | 0.004197 | 7.80E-75 |
| rs174556 | miR-1908-5 | cis | 7.67E-08 | Nightingale | XS_VLDL_L | NA | T | C | -0.03835 | 0.004394 | 6.30E-20 |
| rs174556 | miR-1908-5 | cis | 7.67E-08 | Nightingale | LDL_C | NA | T | C | -0.04815 | 0.004452 | 1.70E-28 |
| rs174556 | miR-1908-5 | cis | 7.67E-08 | Nightingale | IDL_C | NA | T | C | -0.07452 | 0.004317 | 1.30E-70 |
| rs174556 | miR-1908-5 | cis | 7.67E-08 | Nightingale | DHA | NA | T | C | -0.28204 | 0.004308 | 0 |
| rs174556 | miR-1908-5 | cis | 7.67E-08 | Nightingale | HDL_TG | NA | T | C | 0.047834 | 0.004408 | 8.00E-28 |
| rs174556 | miR-1908-5 | cis | 7.67E-08 | Nightingale | LDL_P | NA | T | C | -0.0386 | 0.004468 | 9.90E-19 |
| rs174556 | miR-1908-5 | cis | 7.67E-08 | Nightingale | L_LDL_C_p | NA | T | C | -0.07618 | 0.004356 | 4.30E-70 |
| rs174556 | miR-1908-5 | cis | 7.67E-08 | Nightingale | XS_VLDL_F | NA | T | C | -0.05267 | 0.004393 | 2.00E-35 |
| rs174556 | miR-1908-5 | cis | 7.67E-08 | Nightingale | LDL_FC | NA | T | C | -0.06279 | 0.004423 | 6.20E-48 |
| rs174556 | miR-1908-5 | cis | 7.67E-08 | Nightingale | M_LDL_FC_ | NA | T | C | -0.07256 | 0.004283 | 8.30E-67 |
| rs174556 | miR-1908-5 | cis | 7.67E-08 | Nightingale | Total_TG | NA | T | C | 0.048323 | 0.004328 | 2.90E-30 |
| rs174556 | miR-1908-5 | cis | 7.67E-08 | Nightingale | Clinical_LD | NA | T | C | -0.05349 | 0.004445 | 9.20E-35 |
| rs174556 | miR-1908-5 | cis | 7.67E-08 | Nightingale | IDL_P | NA | T | C | -0.04253 | 0.004424 | 4.50E-23 |
| rs174556 | miR-1908-5 | cis | 7.67E-08 | Nightingale | L_HDL_P | NA | T | C | -0.07574 | 0.003916 | 1.30E-90 |
| rs174556 | miR-1908-5 | cis | 7.67E-08 | Nightingale | L_LDL_L | NA | T | C | -0.05236 | 0.004427 | 6.20E-34 |
| rs174556 | miR-1908-5 | cis | 7.67E-08 | Nightingale | XL_HDL_PL | NA | T | C | -0.08631 | 0.004019 | 1.50E-111 |
| rs174556 | miR-1908-5 | cis | 7.67E-08 | Nightingale | SFA_pct | NA | T | C | -0.03911 | 0.004411 | 7.60E-19 |
| rs174556 | miR-1908-5 | cis | 7.67E-08 | Nightingale | L_LDL_CE | NA | T | C | -0.05152 | 0.004431 | 7.70E-33 |
| rs174556 | miR-1908-5 | cis | 7.67E-08 | Nightingale | XXL_VLDL_ | NA | T | C | 0.067135 | 0.0043 | 8.20E-58 |
| rs174556 | miR-1908-5 | cis | 7.67E-08 | Nightingale | S_VLDL_CE | NA | T | C | -0.02876 | 0.004411 | 2.60E-11 |
| rs174556 | miR-1908-5 | cis | 7.67E-08 | Nightingale | Tyr | NA | T | C | 0.032685 | 0.004465 | 2.50E-13 |
| rs174556 | miR-1908-5 | cis | 7.67E-08 | Nightingale | Total_CE | NA | T | C | -0.0647 | 0.004312 | 1.70E-53 |
| rs174556 | miR-1908-5 | cis | 7.67E-08 | Nightingale | S_LDL_FC | NA | T | C | -0.06166 | 0.004463 | 1.20E-45 |
| rs174556 | miR-1908-5 | cis | 7.67E-08 | Nightingale | S_HDL_TG_ | NA | T | C | 0.069172 | 0.004252 | 3.10E-62 |
| rs174556 | miR-1908-5 | cis | 7.67E-08 | Nightingale | S_VLDL_FC | NA | T | C | -0.02667 | 0.004472 | 1.00E-09 |
| rs174556 | miR-1908-5 | cis | 7.67E-08 | Nightingale | Phosphogly | NA | T | C | -0.03816 | 0.004235 | 7.00E-21 |
| rs174556 | miR-1908-5 | cis | 7.67E-08 | Nightingale | XL_VLDL_C | NA | T | C | -0.06469 | 0.004284 | 6.40E-53 |
| rs174556 | miR-1908-5 | cis | 7.67E-08 | Nightingale | XXL_VLDL_ | NA | T | C | 0.073871 | 0.004317 | 2.80E-68 |
| rs174559 | miR-1908-5 | cis | 5.55E-07 | Nightingale | PUFA_pct | NA | A | G | -0.11225 | 0.004422 | 4.80E-143 |
| rs174559 | miR-1908-5 | cis | 5.55E-07 | Nightingale | M_HDL_C | NA | A | G | -0.03607 | 0.00423 | 1.20E-18 |
| rs174559 | miR-1908-5 | cis | 5.55E-07 | Nightingale | L_LDL_C | NA | A | G | -0.05162 | 0.004545 | 6.70E-31 |
| rs174559 | miR-1908-5 | cis | 5.55E-07 | Nightingale | M_VLDL_C | NA | A | G | -0.0393 | 0.004584 | 5.50E-18 |
| rs174559 | miR-1908-5 | cis | 5.55E-07 | Nightingale | XXL_VLDL_ | NA | A | G | 0.065504 | 0.004429 | 2.10E-52 |
| rs174559 | miR-1908-5 | cis | 5.55E-07 | Nightingale | SFA | NA | A | G | -0.02527 | 0.004556 | 2.70E-08 |
| rs174559 | miR-1908-5 | cis | 5.55E-07 | Nightingale | L_VLDL_CE | NA | A | G | -0.05847 | 0.004399 | 2.30E-42 |
| rs174559 | miR-1908-5 | cis | 5.55E-07 | Nightingale | XL_HDL_FC | NA | A | G | 0.056185 | 0.004075 | 3.50E-48 |
| rs174559 | miR-1908-5 | cis | 5.55E-07 | Nightingale | M_LDL_FC_ | NA | A | G | -0.06797 | 0.004404 | 1.70E-55 |
| rs174559 | miR-1908-5 | cis | 5.55E-07 | Nightingale | TG_by_PG | NA | A | G | 0.064988 | 0.004266 | 2.40E-55 |
| rs174559 | miR-1908-5 | cis | 5.55E-07 | Nightingale | IDL_FC | NA | A | G | -0.08151 | 0.004449 | 2.70E-78 |
| rs174559 | miR-1908-5 | cis | 5.55E-07 | Nightingale | L_LDL_P | NA | A | G | -0.03345 | 0.004581 | 9.40E-14 |
| rs174559 | miR-1908-5 | cis | 5.55E-07 | Nightingale | XL_VLDL_F | NA | A | G | -0.0657 | 0.004454 | 3.70E-51 |
| rs174559 | miR-1908-5 | cis | 5.55E-07 | Nightingale | LDL_P | NA | A | G | -0.0357 | 0.004594 | 3.60E-15 |
| rs174559 | miR-1908-5 | cis | 5.55E-07 | Nightingale | L_LDL_CE | NA | A | G | -0.04731 | 0.004556 | 4.30E-26 |
| rs174559 | miR-1908-5 | cis | 5.55E-07 | Nightingale | LDL_L | NA | A | G | -0.04008 | 0.004582 | 7.70E-19 |
| rs174559 | miR-1908-5 | cis | 5.55E-07 | Nightingale | Clinical_LD | NA | A | G | -0.04935 | 0.004571 | 8.00E-28 |
| rs174559 | miR-1908-5 | cis | 5.55E-07 | Nightingale | XL_HDL_PL | NA | A | G | -0.0817 | 0.004132 | 3.80E-95 |
| rs174559 | miR-1908-5 | cis | 5.55E-07 | Nightingale | L_LDL_TG_ | NA | A | G | 0.072736 | 0.004507 | 8.20E-59 |
| rs174559 | miR-1908-5 | cis | 5.55E-07 | Nightingale | XS_VLDL_T | NA | A | G | 0.09106 | 0.004366 | 6.30E-101 |
| rs174559 | miR-1908-5 | cis | 5.55E-07 | Nightingale | Total_C | NA | A | G | -0.05796 | 0.00445 | 2.60E-40 |
| rs174559 | miR-1908-5 | cis | 5.55E-07 | Nightingale | M_VLDL_T | NA | A | G | 0.03175 | 0.004466 | 2.50E-13 |
| rs174559 | miR-1908-5 | cis | 5.55E-07 | Nightingale | XS_VLDL_F | NA | A | G | -0.05042 | 0.004517 | 1.20E-30 |
| rs174559 | miR-1908-5 | cis | 5.55E-07 | Nightingale | M_LDL_PL_ | NA | A | G | 0.041306 | 0.004594 | 2.50E-20 |
| rs174559 | miR-1908-5 | cis | 5.55E-07 | Nightingale | HDL_CE | NA | A | G | -0.05537 | 0.004125 | 2.70E-44 |
| rs174559 | miR-1908-5 | cis | 5.55E-07 | Nightingale | XS_VLDL_P | NA | A | G | -0.03019 | 0.004532 | 3.00E-12 |
| rs174559 | miR-1908-5 | cis | 5.55E-07 | Nightingale | VLDL_L | NA | A | G | 0.027797 | 0.004507 | 2.30E-10 |

| | | | | | | | | | | | | |
|---|---|---|---|---|---|---|---|---|---|---|---|---|
| rs174559 | miR-1908-5 | cis | | 5.55E-07 | Nightingale | IDL_C | NA | A | G | -0.06978 | 0.004439 | 2.80E-58 |
| rs174559 | miR-1908-5 | cis | | 5.55E-07 | Nightingale | LDL_FC | NA | A | G | -0.05806 | 0.004548 | 1.80E-38 |
| rs174559 | miR-1908-5 | cis | | 5.55E-07 | Nightingale | VLDL_TG | NA | A | G | 0.04692 | 0.004424 | 1.10E-27 |
| rs174559 | miR-1908-5 | cis | | 5.55E-07 | Nightingale | L_HDL_P | NA | A | G | -0.07061 | 0.004026 | 3.50E-75 |
| rs174559 | miR-1908-5 | cis | | 5.55E-07 | Nightingale | L_HDL_PL | NA | A | G | -0.06872 | 0.004046 | 7.20E-71 |
| rs174559 | miR-1908-5 | cis | | 5.55E-07 | Nightingale | L_HDL_L | NA | A | G | -0.06988 | 0.004033 | 1.20E-73 |
| rs174559 | miR-1908-5 | cis | | 5.55E-07 | Nightingale | Phosphogly | NA | A | G | -0.03448 | 0.004354 | 2.00E-16 |
| rs174559 | miR-1908-5 | cis | | 5.55E-07 | Nightingale | Cholines | NA | A | G | -0.03112 | 0.004323 | 5.40E-14 |
| rs174559 | miR-1908-5 | cis | | 5.55E-07 | Nightingale | IDL_P | NA | A | G | -0.03968 | 0.004549 | 5.50E-19 |
| rs174559 | miR-1908-5 | cis | | 5.55E-07 | Nightingale | L_HDL_TG_ | NA | A | G | 0.073041 | 0.004338 | 1.90E-67 |
| rs174559 | miR-1908-5 | cis | | 5.55E-07 | Nightingale | L_LDL_FC_ | NA | A | G | -0.07826 | 0.004381 | 2.20E-73 |
| rs174559 | miR-1908-5 | cis | | 5.55E-07 | Nightingale | S_VLDL_TG | NA | A | G | 0.06155 | 0.004448 | 2.40E-44 |
| rs174559 | miR-1908-5 | cis | | 5.55E-07 | Nightingale | DHA_pct | NA | A | G | -0.25499 | 0.004462 | 0 |
| rs174559 | miR-1908-5 | cis | | 5.55E-07 | Nightingale | LA_pct | NA | A | G | 0.172119 | 0.004492 | 0 |
| rs174559 | miR-1908-5 | cis | | 5.55E-07 | Nightingale | ApoA1 | NA | A | G | -0.03576 | 0.004207 | 7.70E-19 |
| rs174559 | miR-1908-5 | cis | | 5.55E-07 | Nightingale | L_LDL_L | NA | A | G | -0.0482 | 0.004552 | 4.10E-27 |
| rs174559 | miR-1908-5 | cis | | 5.55E-07 | Nightingale | S_VLDL_TG | NA | A | G | 0.038047 | 0.004461 | 1.80E-18 |
| rs174559 | miR-1908-5 | cis | | 5.55E-07 | Nightingale | Total_TG | NA | A | G | 0.046172 | 0.00445 | 1.40E-26 |
| rs174559 | miR-1908-5 | cis | | 5.55E-07 | Nightingale | HDL_FC | NA | A | G | -0.06605 | 0.004072 | 5.20E-64 |
| rs174559 | miR-1908-5 | cis | | 5.55E-07 | Nightingale | HDL_PL | NA | A | G | -0.04222 | 0.004156 | 4.10E-26 |
| rs174559 | miR-1908-5 | cis | | 5.55E-07 | Nightingale | L_HDL_FC_ | NA | A | G | -0.1025 | 0.00427 | 8.20E-138 |
| rs174559 | miR-1908-5 | cis | | 5.55E-07 | Nightingale | XXL_VLDL_ | NA | A | G | -0.02752 | 0.004572 | 3.10E-10 |
| rs174559 | miR-1908-5 | cis | | 5.55E-07 | Nightingale | L_VLDL_L | NA | A | G | 0.037843 | 0.004442 | 1.70E-18 |
| rs174559 | miR-1908-5 | cis | | 5.55E-07 | Nightingale | M_LDL_TG | NA | A | G | 0.036791 | 0.004529 | 3.40E-16 |
| rs174559 | miR-1908-5 | cis | | 5.55E-07 | Nightingale | S_VLDL_FC | NA | A | G | -0.07781 | 0.004389 | 7.60E-73 |
| rs174559 | miR-1908-5 | cis | | 5.55E-07 | Nightingale | XL_HDL_FC | NA | A | G | -0.09419 | 0.004282 | 3.10E-115 |
| rs174559 | miR-1908-5 | cis | | 5.55E-07 | Nightingale | S_LDL_C | NA | A | G | -0.02784 | 0.004602 | 1.10E-09 |
| rs174559 | miR-1908-5 | cis | | 5.55E-07 | Nightingale | IDL_PL | NA | A | G | -0.06563 | 0.004435 | 1.30E-52 |
| rs174559 | miR-1908-5 | cis | | 5.55E-07 | Nightingale | L_HDL_C | NA | A | G | -0.07306 | 0.004048 | 3.10E-79 |
| rs174559 | miR-1908-5 | cis | | 5.55E-07 | Nightingale | Total_PL | NA | A | G | -0.04395 | 0.004377 | 5.10E-25 |
| rs174559 | miR-1908-5 | cis | | 5.55E-07 | Nightingale | Omega_3_ | NA | A | G | -0.36097 | 0.004492 | 0 |
| rs174559 | miR-1908-5 | cis | | 5.55E-07 | Nightingale | Total_P | NA | A | G | -0.03491 | 0.004326 | 9.70E-17 |
| rs174559 | miR-1908-5 | cis | | 5.55E-07 | Nightingale | L_LDL_C_p | NA | A | G | -0.07078 | 0.004479 | 1.00E-56 |
| rs174559 | miR-1908-5 | cis | | 5.55E-07 | Nightingale | non_HDL_( | NA | A | G | -0.04219 | 0.004572 | 7.30E-21 |
| rs174559 | miR-1908-5 | cis | | 5.55E-07 | Nightingale | XL_VLDL_F | NA | A | G | 0.042632 | 0.004452 | 6.80E-23 |
| rs174559 | miR-1908-5 | cis | | 5.55E-07 | Nightingale | Omega_6_ | NA | A | G | 0.342443 | 0.00453 | 0 |
| rs174559 | miR-1908-5 | cis | | 5.55E-07 | Nightingale | S_LDL_PL_ | NA | A | G | -0.05923 | 0.004588 | 2.30E-40 |
| rs174559 | miR-1908-5 | cis | | 5.55E-07 | Nightingale | M_VLDL_T | NA | A | G | 0.074297 | 0.004341 | 4.00E-68 |
| rs174559 | miR-1908-5 | cis | | 5.55E-07 | Nightingale | M_HDL_L | NA | A | G | -0.02334 | 0.004267 | 9.70E-09 |
| rs174559 | miR-1908-5 | cis | | 5.55E-07 | Nightingale | IDL_TG_pc | NA | A | G | 0.082036 | 0.004441 | 3.50E-78 |
| rs174559 | miR-1908-5 | cis | | 5.55E-07 | Nightingale | L_HDL_CE_ | NA | A | G | -0.04747 | 0.004398 | 2.00E-28 |
| rs174559 | miR-1908-5 | cis | | 5.55E-07 | Nightingale | M_LDL_FC | NA | A | G | -0.04705 | 0.004592 | 3.20E-25 |
| rs174559 | miR-1908-5 | cis | | 5.55E-07 | Nightingale | L_VLDL_PL | NA | A | G | 0.047279 | 0.004435 | 4.50E-28 |
| rs174559 | miR-1908-5 | cis | | 5.55E-07 | Nightingale | S_LDL_PL | NA | A | G | -0.04595 | 0.004586 | 1.90E-24 |
| rs174559 | miR-1908-5 | cis | | 5.55E-07 | Nightingale | M_VLDL_F | NA | A | G | -0.08638 | 0.004371 | 1.00E-90 |
| rs174559 | miR-1908-5 | cis | | 5.55E-07 | Nightingale | XS_VLDL_C | NA | A | G | -0.09161 | 0.004356 | 4.60E-102 |
| rs174559 | miR-1908-5 | cis | | 5.55E-07 | Nightingale | XXL_VLDL_ | NA | A | G | 0.059747 | 0.004436 | 2.00E-43 |
| rs174559 | miR-1908-5 | cis | | 5.55E-07 | Nightingale | IDL_C_pct | NA | A | G | -0.06939 | 0.004488 | 1.50E-54 |
| rs174559 | miR-1908-5 | cis | | 5.55E-07 | Nightingale | M_VLDL_F | NA | A | G | -0.02535 | 0.004596 | 2.60E-08 |
| rs174559 | miR-1908-5 | cis | | 5.55E-07 | Nightingale | L_VLDL_C | NA | A | G | 0.028721 | 0.004496 | 4.40E-11 |
| rs174559 | miR-1908-5 | cis | | 5.55E-07 | Nightingale | XL_VLDL_T | NA | A | G | 0.05554 | 0.004397 | 1.60E-38 |
| rs174559 | miR-1908-5 | cis | | 5.55E-07 | Nightingale | XXL_VLDL_ | NA | A | G | 0.070124 | 0.004439 | 7.80E-59 |
| rs174559 | miR-1908-5 | cis | | 5.55E-07 | Nightingale | L_VLDL_FC | NA | A | G | 0.039348 | 0.004446 | 7.10E-20 |
| rs174559 | miR-1908-5 | cis | | 5.55E-07 | Nightingale | M_VLDL_C | NA | A | G | -0.07682 | 0.004327 | 6.10E-73 |
| rs174559 | miR-1908-5 | cis | | 5.55E-07 | Nightingale | IDL_CE_pct | NA | A | G | -0.03477 | 0.004511 | 4.30E-14 |
| rs174559 | miR-1908-5 | cis | | 5.55E-07 | Nightingale | HDL_P | NA | A | G | -0.03068 | 0.004316 | 2.50E-13 |
| rs174559 | miR-1908-5 | cis | | 5.55E-07 | Nightingale | M_HDL_FC | NA | A | G | -0.09298 | 0.004073 | 4.80E-125 |
| rs174559 | miR-1908-5 | cis | | 5.55E-07 | Nightingale | L_LDL_FC | NA | A | G | -0.06177 | 0.004514 | 3.50E-44 |
| rs174559 | miR-1908-5 | cis | | 5.55E-07 | Nightingale | Citrate | NA | A | G | -0.03208 | 0.004568 | 2.90E-13 |
| rs174559 | miR-1908-5 | cis | | 5.55E-07 | Nightingale | XS_VLDL_C | NA | A | G | -0.0862 | 0.004333 | 1.60E-91 |
| rs174559 | miR-1908-5 | cis | | 5.55E-07 | Nightingale | M_HDL_TG | NA | A | G | 0.049216 | 0.00454 | 5.20E-28 |
| rs174559 | miR-1908-5 | cis | | 5.55E-07 | Nightingale | XS_VLDL_P | NA | A | G | 0.053626 | 0.004485 | 6.40E-33 |
| rs174559 | miR-1908-5 | cis | | 5.55E-07 | Nightingale | M_HDL_TG | NA | A | G | 0.062723 | 0.004408 | 4.80E-48 |

| | | | | | | | | | | | |
|---|---|---|---|---|---|---|---|---|---|---|---|
| rs174559 | miR-1908-5 | cis | 5.55E-07 | Nightingale | SFA_pct | NA | A | G | -0.03807 | 0.004535 | 4.70E-17 |
| rs174559 | miR-1908-5 | cis | 5.55E-07 | Nightingale | M_VLDL_C | NA | A | G | -0.07246 | 0.004315 | 2.80E-65 |
| rs174559 | miR-1908-5 | cis | 5.55E-07 | Nightingale | M_HDL_P | NA | A | G | -0.02954 | 0.004235 | 3.20E-13 |
| rs174559 | miR-1908-5 | cis | 5.55E-07 | Nightingale | L_VLDL_TG | NA | A | G | 0.037469 | 0.004423 | 2.70E-18 |
| rs174559 | miR-1908-5 | cis | 5.55E-07 | Nightingale | L_HDL_PL_ | NA | A | G | 0.040678 | 0.004449 | 5.90E-21 |
| rs174559 | miR-1908-5 | cis | 5.55E-07 | Nightingale | IDL_CE | NA | A | G | -0.06508 | 0.004437 | 5.90E-51 |
| rs174559 | miR-1908-5 | cis | 5.55E-07 | Nightingale | XL_VLDL_T | NA | A | G | 0.05855 | 0.004454 | 1.90E-40 |
| rs174559 | miR-1908-5 | cis | 5.55E-07 | Nightingale | IDL_FC_pct | NA | A | G | -0.09802 | 0.00456 | 3.80E-103 |
| rs174559 | miR-1908-5 | cis | 5.55E-07 | Nightingale | M_VLDL_P | NA | A | G | -0.05965 | 0.004411 | 4.00E-44 |
| rs174559 | miR-1908-5 | cis | 5.55E-07 | Nightingale | XS_VLDL_C | NA | A | G | -0.06543 | 0.004436 | 8.30E-52 |
| rs174559 | miR-1908-5 | cis | 5.55E-07 | Nightingale | M_LDL_TG | NA | A | G | 0.071475 | 0.004539 | 4.90E-56 |
| rs174559 | miR-1908-5 | cis | 5.55E-07 | Nightingale | XXL_VLDL_ | NA | A | G | 0.064428 | 0.004427 | 1.50E-50 |
| rs174559 | miR-1908-5 | cis | 5.55E-07 | Nightingale | XL_VLDL_L | NA | A | G | 0.050176 | 0.004423 | 2.00E-31 |
| rs174559 | miR-1908-5 | cis | 5.55E-07 | Nightingale | Remnant_( | NA | A | G | -0.03871 | 0.004567 | 5.20E-18 |
| rs174559 | miR-1908-5 | cis | 5.55E-07 | Nightingale | S_LDL_FC | NA | A | G | -0.0569 | 0.004589 | 1.50E-36 |
| rs174559 | miR-1908-5 | cis | 5.55E-07 | Nightingale | L_VLDL_P | NA | A | G | 0.04119 | 0.004439 | 1.20E-21 |
| rs174559 | miR-1908-5 | cis | 5.55E-07 | Nightingale | XL_VLDL_C | NA | A | G | -0.06226 | 0.004405 | 3.60E-46 |
| rs174559 | miR-1908-5 | cis | 5.55E-07 | Nightingale | M_VLDL_C | NA | A | G | -0.04767 | 0.004555 | 2.50E-26 |
| rs174559 | miR-1908-5 | cis | 5.55E-07 | Nightingale | XL_VLDL_C | NA | A | G | -0.05966 | 0.004407 | 1.60E-42 |
| rs174559 | miR-1908-5 | cis | 5.55E-07 | Nightingale | XL_HDL_PL | NA | A | G | -0.06146 | 0.004223 | 2.60E-52 |
| rs174559 | miR-1908-5 | cis | 5.55E-07 | Nightingale | S_LDL_L | NA | A | G | -0.02806 | 0.004596 | 6.90E-10 |
| rs174559 | miR-1908-5 | cis | 5.55E-07 | Nightingale | L_HDL_FC | NA | A | G | -0.07686 | 0.004029 | 2.30E-88 |
| rs174559 | miR-1908-5 | cis | 5.55E-07 | Nightingale | L_VLDL_C_ | NA | A | G | -0.03949 | 0.00449 | 1.70E-19 |
| rs174559 | miR-1908-5 | cis | 5.55E-07 | Nightingale | XL_HDL_L | NA | A | G | -0.08527 | 0.004131 | 4.80E-103 |
| rs174559 | miR-1908-5 | cis | 5.55E-07 | Nightingale | S_LDL_P | NA | A | G | -0.0277 | 0.004601 | 1.20E-09 |
| rs174559 | miR-1908-5 | cis | 5.55E-07 | Nightingale | S_LDL_CE_ | NA | A | G | 0.054896 | 0.004588 | 7.20E-35 |
| rs174559 | miR-1908-5 | cis | 5.55E-07 | Nightingale | XL_HDL_C | NA | A | G | -0.09109 | 0.004145 | 6.40E-116 |
| rs174559 | miR-1908-5 | cis | 5.55E-07 | Nightingale | M_HDL_CE | NA | A | G | -0.03354 | 0.004245 | 3.40E-16 |
| rs174559 | miR-1908-5 | cis | 5.55E-07 | Nightingale | IDL_L | NA | A | G | -0.06513 | 0.004444 | 2.50E-51 |
| rs174559 | miR-1908-5 | cis | 5.55E-07 | Nightingale | S_HDL_C_p | NA | A | G | -0.04179 | 0.004544 | 1.50E-19 |
| rs174559 | miR-1908-5 | cis | 5.55E-07 | Nightingale | XL_VLDL_P | NA | A | G | 0.051252 | 0.004418 | 8.70E-33 |
| rs174559 | miR-1908-5 | cis | 5.55E-07 | Nightingale | M_HDL_CE | NA | A | G | -0.0457 | 0.004458 | 3.30E-25 |
| rs174559 | miR-1908-5 | cis | 5.55E-07 | Nightingale | PUFA_by_M | NA | A | G | -0.15718 | 0.004403 | 6.10E-285 |
| rs174559 | miR-1908-5 | cis | 5.55E-07 | Nightingale | L_HDL_C_p | NA | A | G | -0.06394 | 0.004333 | 2.40E-52 |
| rs174559 | miR-1908-5 | cis | 5.55E-07 | Nightingale | M_LDL_P | NA | A | G | -0.04135 | 0.00461 | 2.20E-19 |
| rs174559 | miR-1908-5 | cis | 5.55E-07 | Nightingale | XXL_VLDL_ | NA | A | G | 0.068634 | 0.00443 | 6.20E-57 |
| rs174559 | miR-1908-5 | cis | 5.55E-07 | Nightingale | XS_VLDL_T | NA | A | G | 0.031331 | 0.004526 | 1.10E-11 |
| rs174559 | miR-1908-5 | cis | 5.55E-07 | Nightingale | XXL_VLDL_ | NA | A | G | 0.039417 | 0.00462 | 6.20E-18 |
| rs174559 | miR-1908-5 | cis | 5.55E-07 | Nightingale | LDL_TG | NA | A | G | 0.030441 | 0.004546 | 3.30E-11 |
| rs174559 | miR-1908-5 | cis | 5.55E-07 | Nightingale | XS_VLDL_C | NA | A | G | -0.06185 | 0.004462 | 4.00E-46 |
| rs174559 | miR-1908-5 | cis | 5.55E-07 | Nightingale | L_LDL_PL | NA | A | G | -0.04919 | 0.004562 | 9.30E-28 |
| rs174559 | miR-1908-5 | cis | 5.55E-07 | Nightingale | M_LDL_C_p | NA | A | G | -0.09309 | 0.0046 | 3.50E-90 |
| rs174559 | miR-1908-5 | cis | 5.55E-07 | Nightingale | S_HDL_FC_ | NA | A | G | -0.09426 | 0.00436 | 4.60E-115 |
| rs174559 | miR-1908-5 | cis | 5.55E-07 | Nightingale | Total_CE | NA | A | G | -0.05966 | 0.004434 | 7.10E-43 |
| rs174559 | miR-1908-5 | cis | 5.55E-07 | Nightingale | PUFA | NA | A | G | -0.08496 | 0.004483 | 1.20E-84 |
| rs174559 | miR-1908-5 | cis | 5.55E-07 | Nightingale | XL_HDL_P | NA | A | G | -0.08177 | 0.004137 | 4.60E-95 |
| rs174559 | miR-1908-5 | cis | 5.55E-07 | Nightingale | L_HDL_CE | NA | A | G | -0.07167 | 0.004059 | 1.00E-75 |
| rs174559 | miR-1908-5 | cis | 5.55E-07 | Nightingale | XL_HDL_CE | NA | A | G | -0.08915 | 0.004116 | 1.20E-112 |
| rs174559 | miR-1908-5 | cis | 5.55E-07 | Nightingale | S_LDL_TG_ | NA | A | G | 0.081647 | 0.004454 | 4.20E-77 |
| rs174559 | miR-1908-5 | cis | 5.55E-07 | Nightingale | S_LDL_FC_ | NA | A | G | -0.07597 | 0.004455 | 2.20E-67 |
| rs174559 | miR-1908-5 | cis | 5.55E-07 | Nightingale | Sphingomy | NA | A | G | -0.10503 | 0.004296 | 1.60E-137 |
| rs174559 | miR-1908-5 | cis | 5.55E-07 | Nightingale | XL_VLDL_P | NA | A | G | 0.047055 | 0.004436 | 9.80E-28 |
| rs174559 | miR-1908-5 | cis | 5.55E-07 | Nightingale | LDL_C | NA | A | G | -0.04413 | 0.004577 | 1.50E-22 |
| rs174559 | miR-1908-5 | cis | 5.55E-07 | Nightingale | XS_VLDL_P | NA | A | G | -0.03876 | 0.004536 | 1.00E-18 |
| rs174559 | miR-1908-5 | cis | 5.55E-07 | Nightingale | Unsaturati | NA | A | G | -0.31309 | 0.004354 | 0 |
| rs174559 | miR-1908-5 | cis | 5.55E-07 | Nightingale | LA | NA | A | G | 0.081802 | 0.004508 | 2.90E-74 |
| rs174559 | miR-1908-5 | cis | 5.55E-07 | Nightingale | VLDL_size | NA | A | G | 0.069091 | 0.004285 | 2.50E-62 |
| rs174559 | miR-1908-5 | cis | 5.55E-07 | Nightingale | MUFA_pct | NA | A | G | 0.181902 | 0.004399 | 0 |
| rs174559 | miR-1908-5 | cis | 5.55E-07 | Nightingale | S_LDL_TG | NA | A | G | 0.049303 | 0.004484 | 6.60E-29 |
| rs174559 | miR-1908-5 | cis | 5.55E-07 | Nightingale | S_VLDL_C_ | NA | A | G | -0.05444 | 0.004473 | 1.10E-34 |
| rs174559 | miR-1908-5 | cis | 5.55E-07 | Nightingale | XL_HDL_TG | NA | A | G | 0.081516 | 0.00433 | 3.80E-84 |
| rs174559 | miR-1908-5 | cis | 5.55E-07 | Nightingale | S_VLDL_PL | NA | A | G | -0.07401 | 0.004398 | 6.10E-66 |
| rs174559 | miR-1908-5 | cis | 5.55E-07 | Nightingale | LDL_CE | NA | A | G | -0.03825 | 0.004585 | 3.30E-17 |

| | | | | | | | | | | | |
|---|---|---|---|---|---|---|---|---|---|---|---|
| rs174559 | miR-1908-! | cis | 5.55E-07 | Nightingale | M_HDL_C_ | NA | A | G | -0.06199 | 0.004376 | 1.20E-47 |
| rs174559 | miR-1908-! | cis | 5.55E-07 | Nightingale | HDL_C | NA | A | G | -0.0582 | 0.004105 | 2.90E-49 |
| rs174559 | miR-1908-! | cis | 5.55E-07 | Nightingale | LDL_PL | NA | A | G | -0.04098 | 0.004587 | 1.90E-19 |
| rs174559 | miR-1908-! | cis | 5.55E-07 | Nightingale | S_HDL_TG_ | NA | A | G | 0.064623 | 0.004372 | 9.60E-52 |
| rs174559 | miR-1908-! | cis | 5.55E-07 | Nightingale | M_HDL_PL | NA | A | G | 0.055094 | 0.00436 | 1.60E-37 |
| rs174559 | miR-1908-! | cis | 5.55E-07 | Nightingale | XL_VLDL_C | NA | A | G | 0.030025 | 0.004524 | 8.40E-12 |
| rs174559 | miR-1908-! | cis | 5.55E-07 | Nightingale | DHA | NA | A | G | -0.2668 | 0.00443 | 0 |
| rs174559 | miR-1908-! | cis | 5.55E-07 | Nightingale | M_LDL_C | NA | A | G | -0.02965 | 0.004606 | 1.30E-10 |
| rs174559 | miR-1908-! | cis | 5.55E-07 | Nightingale | Omega_3 | NA | A | G | -0.30942 | 0.004506 | 0 |
| rs174559 | miR-1908-! | cis | 5.55E-07 | Nightingale | MUFA | NA | A | G | 0.061604 | 0.00455 | 1.80E-43 |
| rs174559 | miR-1908-! | cis | 5.55E-07 | Nightingale | S_HDL_TG | NA | A | G | 0.06541 | 0.004457 | 1.60E-51 |
| rs174559 | miR-1908-! | cis | 5.55E-07 | Nightingale | L_VLDL_PL | NA | A | G | 0.081877 | 0.004461 | 6.50E-78 |
| rs174559 | miR-1908-! | cis | 5.55E-07 | Nightingale | XS_VLDL_F | NA | A | G | -0.10504 | 0.004562 | 4.00E-119 |
| rs174559 | miR-1908-! | cis | 5.55E-07 | Nightingale | Tyr | NA | A | G | 0.032482 | 0.004591 | 1.50E-12 |
| rs174559 | miR-1908-! | cis | 5.55E-07 | Nightingale | HDL_L | NA | A | G | -0.04703 | 0.004128 | 2.50E-32 |
| rs174559 | miR-1908-! | cis | 5.55E-07 | Nightingale | HDL_TG | NA | A | G | 0.0454 | 0.004533 | 6.30E-24 |
| rs174559 | miR-1908-! | cis | 5.55E-07 | Nightingale | XS_VLDL_L | NA | A | G | -0.03685 | 0.004518 | 2.10E-17 |
| rs174559 | miR-1908-! | cis | 5.55E-07 | Nightingale | XL_VLDL_C | NA | A | G | 0.036854 | 0.00449 | 2.90E-17 |
| rs174559 | miR-1908-! | cis | 5.55E-07 | Nightingale | Total_FC | NA | A | G | -0.05258 | 0.004491 | 4.00E-33 |
| rs174559 | miR-1908-! | cis | 5.55E-07 | Nightingale | HDL_size | NA | A | G | -0.06575 | 0.004073 | 5.80E-64 |
| rs174559 | miR-1908-! | cis | 5.55E-07 | Nightingale | XXL_VLDL_ | NA | A | G | 0.063763 | 0.004421 | 7.50E-50 |
| rs174559 | miR-1908-! | cis | 5.55E-07 | Nightingale | M_HDL_FC | NA | A | G | -0.04529 | 0.004183 | 2.60E-29 |
| rs174559 | miR-1908-! | cis | 5.55E-07 | Nightingale | XXL_VLDL_ | NA | A | G | 0.069377 | 0.004411 | 8.70E-59 |
| rs174559 | miR-1908-! | cis | 5.55E-07 | Nightingale | S_VLDL_CE | NA | A | G | -0.02941 | 0.004535 | 5.90E-11 |
| rs174559 | miR-1908-! | cis | 5.55E-07 | Nightingale | Total_L | NA | A | G | -0.03112 | 0.004522 | 1.40E-12 |
| rs174559 | miR-1908-! | cis | 5.55E-07 | Nightingale | ApoB | NA | A | G | -0.03385 | 0.004591 | 7.40E-14 |
| rs174560 | miR-1908-! | cis | 4.91E-08 | Nightingale | XL_HDL_P | NA | C | T | -0.08569 | 0.004 | 1.70E-110 |
| rs174560 | miR-1908-! | cis | 4.91E-08 | Nightingale | M_LDL_FC | NA | C | T | -0.05141 | 0.00444 | 1.80E-32 |
| rs174560 | miR-1908-! | cis | 4.91E-08 | Nightingale | LDL_TG | NA | C | T | 0.031284 | 0.004396 | 9.50E-13 |
| rs174560 | miR-1908-! | cis | 4.91E-08 | Nightingale | L_HDL_PL_ | NA | C | T | 0.042071 | 0.004302 | 1.20E-23 |
| rs174560 | miR-1908-! | cis | 4.91E-08 | Nightingale | L_LDL_TG_ | NA | C | T | 0.077076 | 0.004359 | 1.00E-71 |
| rs174560 | miR-1908-! | cis | 4.91E-08 | Nightingale | Unsaturati | NA | C | T | -0.33342 | 0.00421 | 0 |
| rs174560 | miR-1908-! | cis | 4.91E-08 | Nightingale | Omega_6_ | NA | C | T | 0.363131 | 0.004381 | 0 |
| rs174560 | miR-1908-! | cis | 4.91E-08 | Nightingale | S_LDL_TG | NA | C | T | 0.051294 | 0.004337 | 1.90E-33 |
| rs174560 | miR-1908-! | cis | 4.91E-08 | Nightingale | L_LDL_FC | NA | C | T | -0.06623 | 0.004365 | 7.40E-55 |
| rs174560 | miR-1908-! | cis | 4.91E-08 | Nightingale | S_VLDL_PL | NA | C | T | -0.07758 | 0.004254 | 7.80E-78 |
| rs174560 | miR-1908-! | cis | 4.91E-08 | Nightingale | M_HDL_L | NA | C | T | -0.0266 | 0.004127 | 2.40E-11 |
| rs174560 | miR-1908-! | cis | 4.91E-08 | Nightingale | IDL_CE | NA | C | T | -0.06923 | 0.004291 | 4.10E-62 |
| rs174560 | miR-1908-! | cis | 4.91E-08 | Nightingale | L_HDL_FC_ | NA | C | T | -0.10722 | 0.00413 | 6.90E-160 |
| rs174560 | miR-1908-! | cis | 4.91E-08 | Nightingale | L_LDL_C | NA | C | T | -0.0558 | 0.004395 | 7.20E-39 |
| rs174560 | miR-1908-! | cis | 4.91E-08 | Nightingale | XL_VLDL_P | NA | C | T | 0.053312 | 0.004273 | 2.20E-37 |
| rs174560 | miR-1908-! | cis | 4.91E-08 | Nightingale | S_VLDL_C_ | NA | C | T | -0.05569 | 0.004325 | 3.90E-39 |
| rs174560 | miR-1908-! | cis | 4.91E-08 | Nightingale | M_VLDL_C | NA | C | T | -0.07491 | 0.004173 | 6.60E-75 |
| rs174560 | miR-1908-! | cis | 4.91E-08 | Nightingale | IDL_CE_pct | NA | C | T | -0.03734 | 0.004362 | 1.20E-17 |
| rs174560 | miR-1908-! | cis | 4.91E-08 | Nightingale | XL_VLDL_C | NA | C | T | -0.06147 | 0.004262 | 1.40E-48 |
| rs174560 | miR-1908-! | cis | 4.91E-08 | Nightingale | XL_HDL_L | NA | C | T | -0.08918 | 0.003995 | 2.60E-119 |
| rs174560 | miR-1908-! | cis | 4.91E-08 | Nightingale | M_LDL_PL | NA | C | T | 0.043517 | 0.004443 | 1.10E-23 |
| rs174560 | miR-1908-! | cis | 4.91E-08 | Nightingale | L_LDL_P | NA | C | T | -0.03559 | 0.00443 | 1.30E-16 |
| rs174560 | miR-1908-! | cis | 4.91E-08 | Nightingale | LA_pct | NA | C | T | 0.181674 | 0.004344 | 0 |
| rs174560 | miR-1908-! | cis | 4.91E-08 | Nightingale | M_VLDL_T | NA | C | T | 0.076455 | 0.004198 | 1.80E-77 |
| rs174560 | miR-1908-! | cis | 4.91E-08 | Nightingale | L_LDL_L | NA | C | T | -0.05216 | 0.004402 | 4.30E-34 |
| rs174560 | miR-1908-! | cis | 4.91E-08 | Nightingale | MUFA_pct | NA | C | T | 0.19389 | 0.004254 | 0 |
| rs174560 | miR-1908-! | cis | 4.91E-08 | Nightingale | L_HDL_CE | NA | C | T | -0.07622 | 0.003925 | 2.40E-90 |
| rs174560 | miR-1908-! | cis | 4.91E-08 | Nightingale | XL_VLDL_F | NA | C | T | -0.06796 | 0.004307 | 1.30E-58 |
| rs174560 | miR-1908-! | cis | 4.91E-08 | Nightingale | XS_VLDL_C | NA | C | T | -0.09035 | 0.00419 | 2.60E-108 |
| rs174560 | miR-1908-! | cis | 4.91E-08 | Nightingale | LDL_CE | NA | C | T | -0.04189 | 0.004434 | 4.00E-22 |
| rs174560 | miR-1908-! | cis | 4.91E-08 | Nightingale | M_VLDL_T | NA | C | T | 0.032089 | 0.004319 | 2.90E-14 |
| rs174560 | miR-1908-! | cis | 4.91E-08 | Nightingale | M_HDL_C | NA | C | T | -0.04005 | 0.00409 | 7.60E-24 |
| rs174560 | miR-1908-! | cis | 4.91E-08 | Nightingale | S_VLDL_FC | NA | C | T | -0.08141 | 0.004245 | 1.30E-85 |
| rs174560 | miR-1908-! | cis | 4.91E-08 | Nightingale | XS_VLDL_C | NA | C | T | -0.06823 | 0.004289 | 1.20E-60 |
| rs174560 | miR-1908-! | cis | 4.91E-08 | Nightingale | LDL_PL | NA | C | T | -0.0446 | 0.004435 | 9.20E-25 |
| rs174560 | miR-1908-! | cis | 4.91E-08 | Nightingale | HDL_FC | NA | C | T | -0.07107 | 0.003938 | 3.80E-78 |
| rs174560 | miR-1908-! | cis | 4.91E-08 | Nightingale | Total_FC | NA | C | T | -0.05663 | 0.004343 | 4.30E-41 |

| | | | | | | | | | | | |
|---|---|---|---|---|---|---|---|---|---|---|---|
| rs174560 | miR-1908-5 | cis | 4.91E-08 | Nightingale | S_LDL_PL_ | NA | C | T | -0.06058 | 0.004436 | 1.80E-44 |
| rs174560 | miR-1908-5 | cis | 4.91E-08 | Nightingale | S_LDL_TG_ | NA | C | T | 0.085958 | 0.004307 | 4.60E-92 |
| rs174560 | miR-1908-5 | cis | 4.91E-08 | Nightingale | XS_VLDL_C | NA | C | T | -0.09605 | 0.004212 | 9.51E-121 |
| rs174560 | miR-1908-5 | cis | 4.91E-08 | Nightingale | PUFA_by_M | NA | C | T | -0.16791 | 0.004258 | 0 |
| rs174560 | miR-1908-5 | cis | 4.91E-08 | Nightingale | XL_VLDL_P | NA | C | T | 0.049035 | 0.00429 | 1.20E-31 |
| rs174560 | miR-1908-5 | cis | 4.91E-08 | Nightingale | M_LDL_TG | NA | C | T | 0.037798 | 0.00438 | 2.20E-18 |
| rs174560 | miR-1908-5 | cis | 4.91E-08 | Nightingale | XL_VLDL_F | NA | C | T | 0.044346 | 0.004306 | 5.20E-26 |
| rs174560 | miR-1908-5 | cis | 4.91E-08 | Nightingale | XL_VLDL_L | NA | C | T | 0.052074 | 0.004277 | 1.10E-35 |
| rs174560 | miR-1908-5 | cis | 4.91E-08 | Nightingale | HDL_C | NA | C | T | -0.06311 | 0.003969 | 5.50E-61 |
| rs174560 | miR-1908-5 | cis | 4.91E-08 | Nightingale | L_HDL_P | NA | C | T | -0.07512 | 0.003893 | 1.10E-89 |
| rs174560 | miR-1908-5 | cis | 4.91E-08 | Nightingale | Tyr | NA | C | T | 0.032081 | 0.00444 | 5.00E-13 |
| rs174560 | miR-1908-5 | cis | 4.91E-08 | Nightingale | XXL_VLDL_ | NA | C | T | 0.062065 | 0.00429 | 9.50E-50 |
| rs174560 | miR-1908-5 | cis | 4.91E-08 | Nightingale | M_HDL_TG | NA | C | T | 0.067317 | 0.004263 | 1.10E-58 |
| rs174560 | miR-1908-5 | cis | 4.91E-08 | Nightingale | S_HDL_FC_ | NA | C | T | -0.09886 | 0.004216 | 1.20E-133 |
| rs174560 | miR-1908-5 | cis | 4.91E-08 | Nightingale | HDL_CE | NA | C | T | -0.06017 | 0.003989 | 6.10E-55 |
| rs174560 | miR-1908-5 | cis | 4.91E-08 | Nightingale | M_LDL_C | NA | C | T | -0.03299 | 0.004454 | 4.90E-14 |
| rs174560 | miR-1908-5 | cis | 4.91E-08 | Nightingale | Remnant_( | NA | C | T | -0.04142 | 0.004417 | 4.20E-22 |
| rs174560 | miR-1908-5 | cis | 4.91E-08 | Nightingale | PUFA_pct | NA | C | T | -0.12028 | 0.004276 | 9.59E-176 |
| rs174560 | miR-1908-5 | cis | 4.91E-08 | Nightingale | XL_VLDL_T | NA | C | T | 0.057462 | 0.004252 | 1.70E-43 |
| rs174560 | miR-1908-5 | cis | 4.91E-08 | Nightingale | S_VLDL_TG | NA | C | T | 0.039173 | 0.004314 | 1.00E-20 |
| rs174560 | miR-1908-5 | cis | 4.91E-08 | Nightingale | L_HDL_C | NA | C | T | -0.07763 | 0.003914 | 2.20E-94 |
| rs174560 | miR-1908-5 | cis | 4.91E-08 | Nightingale | Total_P | NA | C | T | -0.03925 | 0.004183 | 4.30E-22 |
| rs174560 | miR-1908-5 | cis | 4.91E-08 | Nightingale | Sphingomy | NA | C | T | -0.112 | 0.004154 | 1.20E-167 |
| rs174560 | miR-1908-5 | cis | 4.91E-08 | Nightingale | XS_VLDL_P | NA | C | T | -0.04039 | 0.004386 | 1.20E-21 |
| rs174560 | miR-1908-5 | cis | 4.91E-08 | Nightingale | ApoB | NA | C | T | -0.0364 | 0.00444 | 4.00E-17 |
| rs174560 | miR-1908-5 | cis | 4.91E-08 | Nightingale | Omega_3 | NA | C | T | -0.32904 | 0.004357 | 0 |
| rs174560 | miR-1908-5 | cis | 4.91E-08 | Nightingale | S_HDL_TG_ | NA | C | T | 0.069042 | 0.004228 | 7.20E-63 |
| rs174560 | miR-1908-5 | cis | 4.91E-08 | Nightingale | XXL_VLDL_ | NA | C | T | 0.072399 | 0.004266 | 5.40E-68 |
| rs174560 | miR-1908-5 | cis | 4.91E-08 | Nightingale | S_HDL_C_p | NA | C | T | -0.0447 | 0.004394 | 5.80E-24 |
| rs174560 | miR-1908-5 | cis | 4.91E-08 | Nightingale | M_LDL_CE | NA | C | T | -0.02529 | 0.004452 | 8.30E-09 |
| rs174560 | miR-1908-5 | cis | 4.91E-08 | Nightingale | MUFA | NA | C | T | 0.064347 | 0.0044 | 4.30E-50 |
| rs174560 | miR-1908-5 | cis | 4.91E-08 | Nightingale | IDL_TG_pct | NA | C | T | 0.086784 | 0.004294 | 1.30E-94 |
| rs174560 | miR-1908-5 | cis | 4.91E-08 | Nightingale | L_HDL_CE_ | NA | C | T | -0.0504 | 0.004253 | 3.40E-34 |
| rs174560 | miR-1908-5 | cis | 4.91E-08 | Nightingale | XS_VLDL_T | NA | C | T | 0.032891 | 0.004377 | 9.10E-14 |
| rs174560 | miR-1908-5 | cis | 4.91E-08 | Nightingale | XS_VLDL_C | NA | C | T | -0.06446 | 0.004314 | 9.60E-54 |
| rs174560 | miR-1908-5 | cis | 4.91E-08 | Nightingale | S_VLDL_FC | NA | C | T | -0.02665 | 0.004446 | 8.70E-10 |
| rs174560 | miR-1908-5 | cis | 4.91E-08 | Nightingale | ApoA1 | NA | C | T | -0.03967 | 0.004068 | 5.20E-24 |
| rs174560 | miR-1908-5 | cis | 4.91E-08 | Nightingale | M_VLDL_C | NA | C | T | -0.07932 | 0.004184 | 1.90E-83 |
| rs174560 | miR-1908-5 | cis | 4.91E-08 | Nightingale | SFA_pct | NA | C | T | -0.03986 | 0.004386 | 1.00E-19 |
| rs174560 | miR-1908-5 | cis | 4.91E-08 | Nightingale | HDL_size | NA | C | T | -0.06965 | 0.003939 | 2.70E-75 |
| rs174560 | miR-1908-5 | cis | 4.91E-08 | Nightingale | M_LDL_C_p | NA | C | T | -0.09865 | 0.004449 | 1.30E-109 |
| rs174560 | miR-1908-5 | cis | 4.91E-08 | Nightingale | XXL_VLDL_ | NA | C | T | 0.073331 | 0.004293 | 2.60E-68 |
| rs174560 | miR-1908-5 | cis | 4.91E-08 | Nightingale | S_LDL_CE_ | NA | C | T | 0.056821 | 0.004437 | 2.80E-39 |
| rs174560 | miR-1908-5 | cis | 4.91E-08 | Nightingale | M_HDL_FC | NA | C | T | -0.04967 | 0.004045 | 7.90E-37 |
| rs174560 | miR-1908-5 | cis | 4.91E-08 | Nightingale | Phosphogly | NA | C | T | -0.03827 | 0.004211 | 5.30E-21 |
| rs174560 | miR-1908-5 | cis | 4.91E-08 | Nightingale | IDL_C_pct | NA | C | T | -0.07426 | 0.00434 | 1.50E-67 |
| rs174560 | miR-1908-5 | cis | 4.91E-08 | Nightingale | L_LDL_C_p | NA | C | T | -0.0756 | 0.004331 | 5.20E-70 |
| rs174560 | miR-1908-5 | cis | 4.91E-08 | Nightingale | XS_VLDL_T | NA | C | T | 0.094896 | 0.004222 | 7.19E-118 |
| rs174560 | miR-1908-5 | cis | 4.91E-08 | Nightingale | L_VLDL_FC | NA | C | T | 0.027085 | 0.004403 | 9.60E-10 |
| rs174560 | miR-1908-5 | cis | 4.91E-08 | Nightingale | Clinical_LD | NA | C | T | -0.05338 | 0.00442 | 4.30E-35 |
| rs174560 | miR-1908-5 | cis | 4.91E-08 | Nightingale | XXL_VLDL_ | NA | C | T | -0.02914 | 0.004422 | 6.80E-12 |
| rs174560 | miR-1908-5 | cis | 4.91E-08 | Nightingale | M_HDL_FC | NA | C | T | -0.09978 | 0.003939 | 2.90E-153 |
| rs174560 | miR-1908-5 | cis | 4.91E-08 | Nightingale | IDL_C | NA | C | T | -0.07422 | 0.004292 | 5.10E-71 |
| rs174560 | miR-1908-5 | cis | 4.91E-08 | Nightingale | DHA | NA | C | T | -0.28293 | 0.004284 | 0 |
| rs174560 | miR-1908-5 | cis | 4.91E-08 | Nightingale | M_HDL_CE | NA | C | T | -0.04895 | 0.004311 | 1.10E-30 |
| rs174560 | miR-1908-5 | cis | 4.91E-08 | Nightingale | M_HDL_C_ | NA | C | T | -0.06658 | 0.004232 | 1.70E-58 |
| rs174560 | miR-1908-5 | cis | 4.91E-08 | Nightingale | XL_HDL_CE | NA | C | T | -0.02846 | 0.004443 | 5.00E-10 |
| rs174560 | miR-1908-5 | cis | 4.91E-08 | Nightingale | L_HDL_TG_ | NA | C | T | 0.078292 | 0.004195 | 3.40E-82 |
| rs174560 | miR-1908-5 | cis | 4.91E-08 | Nightingale | Citrate | NA | C | T | -0.03348 | 0.004417 | 4.50E-15 |
| rs174560 | miR-1908-5 | cis | 4.91E-08 | Nightingale | HDL_P | NA | C | T | -0.03482 | 0.004174 | 1.00E-17 |
| rs174560 | miR-1908-5 | cis | 4.91E-08 | Nightingale | Omega_3_ | NA | C | T | -0.38308 | 0.004344 | 0 |
| rs174560 | miR-1908-5 | cis | 4.91E-08 | Nightingale | M_LDL_TG | NA | C | T | 0.075652 | 0.004389 | 3.30E-68 |
| rs174560 | miR-1908-5 | cis | 4.91E-08 | Nightingale | S_LDL_C | NA | C | T | -0.03091 | 0.00445 | 1.20E-12 |

| | | | | | | | | | | | |
|---|---|---|---|---|---|---|---|---|---|---|---|
| rs174560 | miR-1908-! | cis | 4.91E-08 | Nightingale | Cholines | NA | C | T | -0.03466 | 0.004181 | 5.90E-18 |
| rs174560 | miR-1908-! | cis | 4.91E-08 | Nightingale | M_LDL_P | NA | C | T | -0.04505 | 0.004458 | 1.20E-24 |
| rs174560 | miR-1908-! | cis | 4.91E-08 | Nightingale | Total_C | NA | C | T | -0.06259 | 0.004303 | 2.20E-50 |
| rs174560 | miR-1908-! | cis | 4.91E-08 | Nightingale | M_LDL_L | NA | C | T | -0.02714 | 0.004452 | 6.20E-10 |
| rs174560 | miR-1908-! | cis | 4.91E-08 | Nightingale | PUFA | NA | C | T | -0.09276 | 0.004335 | 2.10E-107 |
| rs174560 | miR-1908-! | cis | 4.91E-08 | Nightingale | M_HDL_CE | NA | C | T | -0.03737 | 0.004105 | 9.00E-21 |
| rs174560 | miR-1908-! | cis | 4.91E-08 | Nightingale | M_HDL_P | NA | C | T | -0.03318 | 0.004095 | 4.50E-17 |
| rs174560 | miR-1908-! | cis | 4.91E-08 | Nightingale | IDL_L | NA | C | T | -0.06918 | 0.004297 | 3.20E-62 |
| rs174560 | miR-1908-! | cis | 4.91E-08 | Nightingale | XS_VLDL_L | NA | C | T | -0.03814 | 0.004369 | 8.10E-20 |
| rs174560 | miR-1908-! | cis | 4.91E-08 | Nightingale | Total_TG | NA | C | T | 0.047883 | 0.004303 | 3.50E-30 |
| rs174560 | miR-1908-! | cis | 4.91E-08 | Nightingale | XL_HDL_PL | NA | C | T | -0.08522 | 0.003996 | 2.70E-109 |
| rs174560 | miR-1908-! | cis | 4.91E-08 | Nightingale | XS_VLDL_F | NA | C | T | -0.05246 | 0.004368 | 1.90E-35 |
| rs174560 | miR-1908-! | cis | 4.91E-08 | Nightingale | XXL_VLDL_ | NA | C | T | 0.042781 | 0.004468 | 2.20E-22 |
| rs174560 | miR-1908-! | cis | 4.91E-08 | Nightingale | L_VLDL_PL | NA | C | T | 0.04924 | 0.004289 | 5.40E-32 |
| rs174560 | miR-1908-! | cis | 4.91E-08 | Nightingale | L_HDL_FC | NA | C | T | -0.08141 | 0.003896 | 1.00E-104 |
| rs174560 | miR-1908-! | cis | 4.91E-08 | Nightingale | VLDL_TG | NA | C | T | 0.048466 | 0.004278 | 3.80E-31 |
| rs174560 | miR-1908-! | cis | 4.91E-08 | Nightingale | L_VLDL_FC | NA | C | T | 0.040714 | 0.0043 | 2.80E-22 |
| rs174560 | miR-1908-! | cis | 4.91E-08 | Nightingale | XL_HDL_TG | NA | C | T | 0.08597 | 0.004188 | 2.90E-99 |
| rs174560 | miR-1908-! | cis | 4.91E-08 | Nightingale | LDL_P | NA | C | T | -0.03838 | 0.004442 | 9.50E-19 |
| rs174560 | miR-1908-! | cis | 4.91E-08 | Nightingale | XXL_VLDL_ | NA | C | T | 0.068211 | 0.004283 | 2.90E-60 |
| rs174560 | miR-1908-! | cis | 4.91E-08 | Nightingale | L_VLDL_PL | NA | C | T | 0.087443 | 0.004314 | 1.60E-94 |
| rs174560 | miR-1908-! | cis | 4.91E-08 | Nightingale | HDL_PL | NA | C | T | -0.04629 | 0.004019 | 1.00E-32 |
| rs174560 | miR-1908-! | cis | 4.91E-08 | Nightingale | XL_VLDL_T | NA | C | T | 0.060081 | 0.004307 | 7.40E-46 |
| rs174560 | miR-1908-! | cis | 4.91E-08 | Nightingale | XS_VLDL_P | NA | C | T | 0.058556 | 0.004337 | 1.60E-42 |
| rs174560 | miR-1908-! | cis | 4.91E-08 | Nightingale | Total_PL | NA | C | T | -0.04799 | 0.004233 | 1.80E-31 |
| rs174560 | miR-1908-! | cis | 4.91E-08 | Nightingale | VLDL_size | NA | C | T | 0.071617 | 0.004144 | 5.70E-71 |
| rs174560 | miR-1908-! | cis | 4.91E-08 | Nightingale | S_LDL_FC | NA | C | T | -0.06163 | 0.004438 | 2.90E-46 |
| rs174560 | miR-1908-! | cis | 4.91E-08 | Nightingale | XL_VLDL_C | NA | C | T | -0.06426 | 0.00426 | 5.90E-53 |
| rs174560 | miR-1908-! | cis | 4.91E-08 | Nightingale | HDL_TG | NA | C | T | 0.047615 | 0.004383 | 4.70E-28 |
| rs174560 | miR-1908-! | cis | 4.91E-08 | Nightingale | IDL_P | NA | C | T | -0.04255 | 0.004399 | 2.00E-23 |
| rs174560 | miR-1908-! | cis | 4.91E-08 | Nightingale | M_VLDL_P | NA | C | T | -0.06055 | 0.004266 | 7.30E-49 |
| rs174560 | miR-1908-! | cis | 4.91E-08 | Nightingale | S_VLDL_CE | NA | C | T | -0.02863 | 0.004386 | 2.10E-11 |
| rs174560 | miR-1908-! | cis | 4.91E-08 | Nightingale | M_VLDL_C | NA | C | T | -0.05071 | 0.004405 | 5.50E-32 |
| rs174560 | miR-1908-! | cis | 4.91E-08 | Nightingale | M_HDL_PL | NA | C | T | 0.059279 | 0.004217 | 2.10E-46 |
| rs174560 | miR-1908-! | cis | 4.91E-08 | Nightingale | M_VLDL_F | NA | C | T | -0.08914 | 0.004227 | 8.99E-104 |
| rs174560 | miR-1908-! | cis | 4.91E-08 | Nightingale | L_LDL_CE | NA | C | T | -0.05133 | 0.004406 | 5.50E-33 |
| rs174560 | miR-1908-! | cis | 4.91E-08 | Nightingale | LDL_FC | NA | C | T | -0.06256 | 0.004398 | 3.10E-48 |
| rs174560 | miR-1908-! | cis | 4.91E-08 | Nightingale | L_VLDL_P | NA | C | T | 0.042472 | 0.004292 | 3.70E-24 |
| rs174560 | miR-1908-! | cis | 4.91E-08 | Nightingale | XXL_VLDL_ | NA | C | T | 0.066593 | 0.004276 | 1.20E-57 |
| rs174560 | miR-1908-! | cis | 4.91E-08 | Nightingale | VLDL_L | NA | C | T | 0.028662 | 0.004359 | 1.80E-11 |
| rs174560 | miR-1908-! | cis | 4.91E-08 | Nightingale | Total_CE | NA | C | T | -0.06447 | 0.004288 | 1.10E-53 |
| rs174560 | miR-1908-! | cis | 4.91E-08 | Nightingale | M_VLDL_F | NA | C | T | -0.02738 | 0.004445 | 3.00E-10 |
| rs174560 | miR-1908-! | cis | 4.91E-08 | Nightingale | L_HDL_L | NA | C | T | -0.07444 | 0.0039 | 7.10E-88 |
| rs174560 | miR-1908-! | cis | 4.91E-08 | Nightingale | XL_HDL_C | NA | C | T | -0.09556 | 0.004008 | 2.50E-135 |
| rs174560 | miR-1908-! | cis | 4.91E-08 | Nightingale | L_VLDL_C | NA | C | T | 0.029711 | 0.004348 | 2.80E-12 |
| rs174560 | miR-1908-! | cis | 4.91E-08 | Nightingale | HDL_L | NA | C | T | -0.05135 | 0.003992 | 2.20E-40 |
| rs174560 | miR-1908-! | cis | 4.91E-08 | Nightingale | M_VLDL_C | NA | C | T | -0.04197 | 0.004433 | 3.90E-22 |
| rs174560 | miR-1908-! | cis | 4.91E-08 | Nightingale | S_LDL_L | NA | C | T | -0.03097 | 0.004445 | 9.40E-13 |
| rs174560 | miR-1908-! | cis | 4.91E-08 | Nightingale | L_VLDL_L | NA | C | T | 0.038916 | 0.004296 | 1.80E-20 |
| rs174560 | miR-1908-! | cis | 4.91E-08 | Nightingale | XL_VLDL_C | NA | C | T | 0.031077 | 0.004375 | 4.30E-13 |
| rs174560 | miR-1908-! | cis | 4.91E-08 | Nightingale | XS_VLDL_F | NA | C | T | -0.1101 | 0.004412 | 2.40E-140 |
| rs174560 | miR-1908-! | cis | 4.91E-08 | Nightingale | M_LDL_FC_ | NA | C | T | -0.07182 | 0.004259 | 1.20E-66 |
| rs174560 | miR-1908-! | cis | 4.91E-08 | Nightingale | non_HDL_( | NA | C | T | -0.04556 | 0.004421 | 3.80E-26 |
| rs174560 | miR-1908-! | cis | 4.91E-08 | Nightingale | L_LDL_PL | NA | C | T | -0.05316 | 0.004412 | 6.00E-35 |
| rs174560 | miR-1908-! | cis | 4.91E-08 | Nightingale | XL_HDL_CE | NA | C | T | -0.09375 | 0.003981 | 4.40E-132 |
| rs174560 | miR-1908-! | cis | 4.91E-08 | Nightingale | L_VLDL_TG | NA | C | T | 0.038122 | 0.004277 | 7.40E-20 |
| rs174560 | miR-1908-! | cis | 4.91E-08 | Nightingale | XL_HDL_PL | NA | C | T | -0.06349 | 0.004084 | 1.00E-58 |
| rs174560 | miR-1908-! | cis | 4.91E-08 | Nightingale | S_VLDL_TG | NA | C | T | 0.063553 | 0.004301 | 6.10E-51 |
| rs174560 | miR-1908-! | cis | 4.91E-08 | Nightingale | S_LDL_PL | NA | C | T | -0.04951 | 0.004435 | 1.70E-30 |
| rs174560 | miR-1908-! | cis | 4.91E-08 | Nightingale | XL_HDL_FC | NA | C | T | -0.09797 | 0.004141 | 2.20E-132 |
| rs174560 | miR-1908-! | cis | 4.91E-08 | Nightingale | XXL_VLDL_ | NA | C | T | 0.067055 | 0.004282 | 4.40E-58 |
| rs174560 | miR-1908-! | cis | 4.91E-08 | Nightingale | LDL_L | NA | C | T | -0.04373 | 0.004431 | 4.80E-24 |
| rs174560 | miR-1908-! | cis | 4.91E-08 | Nightingale | IDL_FC | NA | C | T | -0.08669 | 0.004303 | 2.60E-95 |

| | | | | | | | | | | | |
|---|---|---|---|---|---|---|---|---|---|---|---|
| rs174560 | miR-1908-5 | cis | 4.91E-08 | Nightingale | M_HDL_TG | NA | C | T | 0.051538 | 0.00439 | 1.40E-32 |
| rs174560 | miR-1908-5 | cis | 4.91E-08 | Nightingale | XXL_VLDL_ | NA | C | T | 0.071688 | 0.004284 | 6.60E-66 |
| rs174560 | miR-1908-5 | cis | 4.91E-08 | Nightingale | LA | NA | C | T | 0.08405 | 0.004359 | 2.80E-83 |
| rs174560 | miR-1908-5 | cis | 4.91E-08 | Nightingale | IDL_FC_pct | NA | C | T | -0.10423 | 0.00441 | 7.19E-125 |
| rs174560 | miR-1908-5 | cis | 4.91E-08 | Nightingale | L_VLDL_C_ | NA | C | T | -0.03908 | 0.004342 | 8.50E-21 |
| rs174560 | miR-1908-5 | cis | 4.91E-08 | Nightingale | S_HDL_TG | NA | C | T | 0.06855 | 0.004311 | 5.60E-60 |
| rs174560 | miR-1908-5 | cis | 4.91E-08 | Nightingale | IDL_PL | NA | C | T | -0.06936 | 0.004289 | 6.20E-63 |
| rs174560 | miR-1908-5 | cis | 4.91E-08 | Nightingale | XL_HDL_FC | NA | C | T | 0.059198 | 0.003941 | 8.30E-56 |
| rs174560 | miR-1908-5 | cis | 4.91E-08 | Nightingale | TG_by_PG | NA | C | T | 0.068612 | 0.004125 | 1.40E-65 |
| rs174560 | miR-1908-5 | cis | 4.91E-08 | Nightingale | L_VLDL_CE | NA | C | T | -0.05879 | 0.004254 | 3.70E-46 |
| rs174560 | miR-1908-5 | cis | 4.91E-08 | Nightingale | S_LDL_P | NA | C | T | -0.03033 | 0.00445 | 3.20E-12 |
| rs174560 | miR-1908-5 | cis | 4.91E-08 | Nightingale | S_LDL_FC_ | NA | C | T | -0.08071 | 0.004308 | 1.50E-81 |
| rs174560 | miR-1908-5 | cis | 4.91E-08 | Nightingale | Total_L | NA | C | T | -0.03451 | 0.004373 | 4.00E-16 |
| rs174560 | miR-1908-5 | cis | 4.91E-08 | Nightingale | L_LDL_FC_ | NA | C | T | -0.08185 | 0.004237 | 2.10E-86 |
| rs174560 | miR-1908-5 | cis | 4.91E-08 | Nightingale | L_HDL_PL | NA | C | T | -0.07329 | 0.003912 | 1.20E-84 |
| rs174560 | miR-1908-5 | cis | 4.91E-08 | Nightingale | XS_VLDL_P | NA | C | T | -0.03091 | 0.004383 | 1.30E-13 |
| rs174560 | miR-1908-5 | cis | 4.91E-08 | Nightingale | SFA | NA | C | T | -0.02846 | 0.004405 | 7.90E-11 |
| rs174560 | miR-1908-5 | cis | 4.91E-08 | Nightingale | DHA_pct | NA | C | T | -0.26967 | 0.004315 | 0 |
| rs174560 | miR-1908-5 | cis | 4.91E-08 | Nightingale | XL_VLDL_C | NA | C | T | 0.038291 | 0.004342 | 1.80E-19 |
| rs174560 | miR-1908-5 | cis | 4.91E-08 | Nightingale | L_HDL_C_p | NA | C | T | -0.06752 | 0.00419 | 4.00E-62 |
| rs174560 | miR-1908-5 | cis | 4.91E-08 | Nightingale | LDL_C | NA | C | T | -0.04805 | 0.004426 | 9.80E-29 |
| rs174561 | miR-1908-5 | cis | 7.66E-08 | Nightingale | PUFA_pct | NA | C | T | -0.1203 | 0.004297 | 8.09E-174 |
| rs174561 | miR-1908-5 | cis | 7.66E-08 | Nightingale | IDL_C_pct | NA | C | T | -0.07397 | 0.004362 | 5.50E-66 |
| rs174561 | miR-1908-5 | cis | 7.66E-08 | Nightingale | SFA | NA | C | T | -0.02815 | 0.004428 | 1.50E-10 |
| rs174561 | miR-1908-5 | cis | 7.66E-08 | Nightingale | Total_C | NA | C | T | -0.06261 | 0.004325 | 5.80E-50 |
| rs174561 | miR-1908-5 | cis | 7.66E-08 | Nightingale | non_HDL_( | NA | C | T | -0.04548 | 0.004444 | 1.00E-25 |
| rs174561 | miR-1908-5 | cis | 7.66E-08 | Nightingale | L_LDL_C | NA | C | T | -0.05582 | 0.004418 | 1.90E-38 |
| rs174561 | miR-1908-5 | cis | 7.66E-08 | Nightingale | L_VLDL_P | NA | C | T | 0.042857 | 0.004314 | 2.30E-24 |
| rs174561 | miR-1908-5 | cis | 7.66E-08 | Nightingale | M_LDL_TG | NA | C | T | 0.037847 | 0.004402 | 3.60E-18 |
| rs174561 | miR-1908-5 | cis | 7.66E-08 | Nightingale | XL_VLDL_F | NA | C | T | 0.04477 | 0.004328 | 3.40E-26 |
| rs174561 | miR-1908-5 | cis | 7.66E-08 | Nightingale | HDL_CE | NA | C | T | -0.06026 | 0.00401 | 8.20E-55 |
| rs174561 | miR-1908-5 | cis | 7.66E-08 | Nightingale | IDL_P | NA | C | T | -0.04231 | 0.004421 | 8.10E-23 |
| rs174561 | miR-1908-5 | cis | 7.66E-08 | Nightingale | XS_VLDL_F | NA | C | T | -0.05265 | 0.00439 | 2.50E-35 |
| rs174561 | miR-1908-5 | cis | 7.66E-08 | Nightingale | M_HDL_TG | NA | C | T | 0.051764 | 0.004413 | 1.90E-32 |
| rs174561 | miR-1908-5 | cis | 7.66E-08 | Nightingale | LDL_FC | NA | C | T | -0.06252 | 0.00442 | 1.40E-47 |
| rs174561 | miR-1908-5 | cis | 7.66E-08 | Nightingale | Unsaturati | NA | C | T | -0.3333 | 0.004231 | 0 |
| rs174561 | miR-1908-5 | cis | 7.66E-08 | Nightingale | M_VLDL_P | NA | C | T | -0.06074 | 0.004287 | 1.20E-48 |
| rs174561 | miR-1908-5 | cis | 7.66E-08 | Nightingale | L_VLDL_L | NA | C | T | 0.039299 | 0.004318 | 1.10E-20 |
| rs174561 | miR-1908-5 | cis | 7.66E-08 | Nightingale | LDL_L | NA | C | T | -0.04367 | 0.004453 | 1.10E-23 |
| rs174561 | miR-1908-5 | cis | 7.66E-08 | Nightingale | XL_HDL_PL | NA | C | T | -0.08606 | 0.004016 | 5.70E-111 |
| rs174561 | miR-1908-5 | cis | 7.66E-08 | Nightingale | XXL_VLDL_ | NA | C | T | 0.062295 | 0.004311 | 1.50E-49 |
| rs174561 | miR-1908-5 | cis | 7.66E-08 | Nightingale | L_HDL_P | NA | C | T | -0.07552 | 0.003913 | 3.90E-90 |
| rs174561 | miR-1908-5 | cis | 7.66E-08 | Nightingale | S_VLDL_FC | NA | C | T | -0.02657 | 0.004469 | 1.30E-09 |
| rs174561 | miR-1908-5 | cis | 7.66E-08 | Nightingale | XL_HDL_L | NA | C | T | -0.09003 | 0.004015 | 6.30E-121 |
| rs174561 | miR-1908-5 | cis | 7.66E-08 | Nightingale | IDL_C | NA | C | T | -0.07426 | 0.004314 | 3.20E-70 |
| rs174561 | miR-1908-5 | cis | 7.66E-08 | Nightingale | XXL_VLDL_ | NA | C | T | 0.067464 | 0.004303 | 3.80E-58 |
| rs174561 | miR-1908-5 | cis | 7.66E-08 | Nightingale | L_HDL_C_p | NA | C | T | -0.06808 | 0.004211 | 8.70E-63 |
| rs174561 | miR-1908-5 | cis | 7.66E-08 | Nightingale | XL_VLDL_T | NA | C | T | 0.060057 | 0.004328 | 2.80E-45 |
| rs174561 | miR-1908-5 | cis | 7.66E-08 | Nightingale | Total_L | NA | C | T | -0.03446 | 0.004395 | 5.80E-16 |
| rs174561 | miR-1908-5 | cis | 7.66E-08 | Nightingale | L_LDL_PL | NA | C | T | -0.05304 | 0.004434 | 2.40E-34 |
| rs174561 | miR-1908-5 | cis | 7.66E-08 | Nightingale | M_LDL_FC_ | NA | C | T | -0.07216 | 0.00428 | 2.80E-66 |
| rs174561 | miR-1908-5 | cis | 7.66E-08 | Nightingale | L_HDL_PL | NA | C | T | -0.07361 | 0.003932 | 5.10E-85 |
| rs174561 | miR-1908-5 | cis | 7.66E-08 | Nightingale | M_LDL_C | NA | C | T | -0.0327 | 0.004477 | 1.20E-13 |
| rs174561 | miR-1908-5 | cis | 7.66E-08 | Nightingale | IDL_L | NA | C | T | -0.06928 | 0.004319 | 1.10E-61 |
| rs174561 | miR-1908-5 | cis | 7.66E-08 | Nightingale | Total_TG | NA | C | T | 0.048172 | 0.004325 | 3.40E-30 |
| rs174561 | miR-1908-5 | cis | 7.66E-08 | Nightingale | M_HDL_C | NA | C | T | -0.03991 | 0.004111 | 1.40E-23 |
| rs174561 | miR-1908-5 | cis | 7.66E-08 | Nightingale | L_VLDL_CE | NA | C | T | -0.05891 | 0.004276 | 7.50E-46 |
| rs174561 | miR-1908-5 | cis | 7.66E-08 | Nightingale | Remnant_( | NA | C | T | -0.04132 | 0.004439 | 1.00E-21 |
| rs174561 | miR-1908-5 | cis | 7.66E-08 | Nightingale | L_LDL_TG_ | NA | C | T | 0.077041 | 0.004381 | 1.20E-70 |
| rs174561 | miR-1908-5 | cis | 7.66E-08 | Nightingale | IDL_FC | NA | C | T | -0.0867 | 0.004324 | 3.30E-94 |
| rs174561 | miR-1908-5 | cis | 7.66E-08 | Nightingale | XL_HDL_PL | NA | C | T | -0.06429 | 0.004104 | 4.40E-60 |
| rs174561 | miR-1908-5 | cis | 7.66E-08 | Nightingale | S_VLDL_TG | NA | C | T | 0.063625 | 0.004323 | 1.70E-50 |
| rs174561 | miR-1908-5 | cis | 7.66E-08 | Nightingale | XL_VLDL_T | NA | C | T | 0.057896 | 0.004273 | 8.80E-44 |

| | | | | | | | | | | | |
|---|---|---|---|---|---|---|---|---|---|---|---|
| rs174561 | miR-1908-5 | cis | 7.66E-08 | Nightingale | M_HDL_P | NA | C | T | -0.03301 | 0.004116 | 7.10E-17 |
| rs174561 | miR-1908-5 | cis | 7.66E-08 | Nightingale | M_LDL_FC | NA | C | T | -0.05121 | 0.004463 | 7.80E-32 |
| rs174561 | miR-1908-5 | cis | 7.66E-08 | Nightingale | XL_HDL_P | NA | C | T | -0.08657 | 0.004021 | 4.80E-112 |
| rs174561 | miR-1908-5 | cis | 7.66E-08 | Nightingale | DHA_pct | NA | C | T | -0.26949 | 0.004337 | 0 |
| rs174561 | miR-1908-5 | cis | 7.66E-08 | Nightingale | XL_VLDL_C | NA | C | T | 0.031447 | 0.004398 | 3.00E-13 |
| rs174561 | miR-1908-5 | cis | 7.66E-08 | Nightingale | L_VLDL_PL | NA | C | T | 0.087421 | 0.004336 | 1.30E-93 |
| rs174561 | miR-1908-5 | cis | 7.66E-08 | Nightingale | IDL_CE | NA | C | T | -0.06928 | 0.004313 | 2.00E-61 |
| rs174561 | miR-1908-5 | cis | 7.66E-08 | Nightingale | Citrate | NA | C | T | -0.03405 | 0.004439 | 1.70E-15 |
| rs174561 | miR-1908-5 | cis | 7.66E-08 | Nightingale | M_VLDL_C | NA | C | T | -0.0419 | 0.004456 | 9.50E-22 |
| rs174561 | miR-1908-5 | cis | 7.66E-08 | Nightingale | S_HDL_TG_ | NA | C | T | 0.069036 | 0.004249 | 3.70E-62 |
| rs174561 | miR-1908-5 | cis | 7.66E-08 | Nightingale | XL_HDL_C_ | NA | C | T | 0.021648 | 0.004268 | 2.10E-08 |
| rs174561 | miR-1908-5 | cis | 7.66E-08 | Nightingale | DHA | NA | C | T | -0.2827 | 0.004305 | 0 |
| rs174561 | miR-1908-5 | cis | 7.66E-08 | Nightingale | L_VLDL_C_ | NA | C | T | -0.03895 | 0.004364 | 2.20E-20 |
| rs174561 | miR-1908-5 | cis | 7.66E-08 | Nightingale | HDL_FC | NA | C | T | -0.07132 | 0.003958 | 3.00E-78 |
| rs174561 | miR-1908-5 | cis | 7.66E-08 | Nightingale | S_VLDL_C_ | NA | C | T | -0.05578 | 0.004347 | 7.80E-39 |
| rs174561 | miR-1908-5 | cis | 7.66E-08 | Nightingale | XS_VLDL_P | NA | C | T | -0.03125 | 0.004405 | 8.90E-14 |
| rs174561 | miR-1908-5 | cis | 7.66E-08 | Nightingale | TG_by_PG | NA | C | T | 0.068859 | 0.004146 | 2.40E-65 |
| rs174561 | miR-1908-5 | cis | 7.66E-08 | Nightingale | L_LDL_CE | NA | C | T | -0.05135 | 0.004428 | 1.20E-32 |
| rs174561 | miR-1908-5 | cis | 7.66E-08 | Nightingale | XS_VLDL_F | NA | C | T | -0.11032 | 0.004434 | 1.90E-139 |
| rs174561 | miR-1908-5 | cis | 7.66E-08 | Nightingale | Phosphogly | NA | C | T | -0.03819 | 0.004232 | 6.70E-21 |
| rs174561 | miR-1908-5 | cis | 7.66E-08 | Nightingale | L_LDL_FC | NA | C | T | -0.06627 | 0.004387 | 2.80E-54 |
| rs174561 | miR-1908-5 | cis | 7.66E-08 | Nightingale | XL_VLDL_C | NA | C | T | -0.06175 | 0.004283 | 2.00E-48 |
| rs174561 | miR-1908-5 | cis | 7.66E-08 | Nightingale | XS_VLDL_T | NA | C | T | 0.095143 | 0.004244 | 2.90E-117 |
| rs174561 | miR-1908-5 | cis | 7.66E-08 | Nightingale | XL_HDL_CE | NA | C | T | -0.02806 | 0.004465 | 1.30E-09 |
| rs174561 | miR-1908-5 | cis | 7.66E-08 | Nightingale | XL_VLDL_F | NA | C | T | -0.06818 | 0.004329 | 2.50E-58 |
| rs174561 | miR-1908-5 | cis | 7.66E-08 | Nightingale | LA | NA | C | T | 0.083992 | 0.004381 | 2.90E-82 |
| rs174561 | miR-1908-5 | cis | 7.66E-08 | Nightingale | L_VLDL_TG | NA | C | T | 0.038503 | 0.004299 | 4.40E-20 |
| rs174561 | miR-1908-5 | cis | 7.66E-08 | Nightingale | XL_HDL_FC | NA | C | T | 0.059934 | 0.003961 | 4.00E-57 |
| rs174561 | miR-1908-5 | cis | 7.66E-08 | Nightingale | MUFA_pct | NA | C | T | 0.193668 | 0.004276 | 0 |
| rs174561 | miR-1908-5 | cis | 7.66E-08 | Nightingale | S_LDL_PL_ | NA | C | T | -0.06158 | 0.004459 | 2.40E-45 |
| rs174561 | miR-1908-5 | cis | 7.66E-08 | Nightingale | L_HDL_CE_ | NA | C | T | -0.05088 | 0.004275 | 1.10E-34 |
| rs174561 | miR-1908-5 | cis | 7.66E-08 | Nightingale | Clinical_LD | NA | C | T | -0.05325 | 0.004442 | 1.70E-34 |
| rs174561 | miR-1908-5 | cis | 7.66E-08 | Nightingale | XL_HDL_C | NA | C | T | -0.09635 | 0.004028 | 1.00E-136 |
| rs174561 | miR-1908-5 | cis | 7.66E-08 | Nightingale | M_VLDL_C | NA | C | T | -0.0752 | 0.004194 | 1.20E-74 |
| rs174561 | miR-1908-5 | cis | 7.66E-08 | Nightingale | S_VLDL_TG | NA | C | T | 0.039318 | 0.004336 | 1.40E-20 |
| rs174561 | miR-1908-5 | cis | 7.66E-08 | Nightingale | L_LDL_L | NA | C | T | -0.05217 | 0.004424 | 1.00E-33 |
| rs174561 | miR-1908-5 | cis | 7.66E-08 | Nightingale | M_VLDL_C | NA | C | T | -0.05071 | 0.004427 | 1.40E-31 |
| rs174561 | miR-1908-5 | cis | 7.66E-08 | Nightingale | S_VLDL_CE | NA | C | T | -0.02865 | 0.004408 | 2.70E-11 |
| rs174561 | miR-1908-5 | cis | 7.66E-08 | Nightingale | XXL_VLDL_ | NA | C | T | -0.0291 | 0.004444 | 9.00E-12 |
| rs174561 | miR-1908-5 | cis | 7.66E-08 | Nightingale | S_LDL_CE_ | NA | C | T | 0.057531 | 0.00446 | 1.00E-39 |
| rs174561 | miR-1908-5 | cis | 7.66E-08 | Nightingale | Omega_3 | NA | C | T | -0.32888 | 0.004379 | 0 |
| rs174561 | miR-1908-5 | cis | 7.66E-08 | Nightingale | M_HDL_PL | NA | C | T | 0.059508 | 0.004238 | 2.40E-46 |
| rs174561 | miR-1908-5 | cis | 7.66E-08 | Nightingale | IDL_FC_pct | NA | C | T | -0.10395 | 0.004432 | 7.50E-123 |
| rs174561 | miR-1908-5 | cis | 7.66E-08 | Nightingale | L_LDL_C_p | NA | C | T | -0.0758 | 0.004353 | 1.50E-69 |
| rs174561 | miR-1908-5 | cis | 7.66E-08 | Nightingale | IDL_CE_pct | NA | C | T | -0.03712 | 0.004384 | 4.30E-17 |
| rs174561 | miR-1908-5 | cis | 7.66E-08 | Nightingale | XXL_VLDL_ | NA | C | T | 0.067 | 0.004297 | 1.00E-57 |
| rs174561 | miR-1908-5 | cis | 7.66E-08 | Nightingale | XS_VLDL_C | NA | C | T | -0.06464 | 0.004336 | 2.00E-53 |
| rs174561 | miR-1908-5 | cis | 7.66E-08 | Nightingale | XS_VLDL_T | NA | C | T | 0.032855 | 0.004399 | 1.70E-13 |
| rs174561 | miR-1908-5 | cis | 7.66E-08 | Nightingale | XXL_VLDL_ | NA | C | T | 0.068664 | 0.004304 | 2.50E-60 |
| rs174561 | miR-1908-5 | cis | 7.66E-08 | Nightingale | M_LDL_L | NA | C | T | -0.02691 | 0.004475 | 1.20E-09 |
| rs174561 | miR-1908-5 | cis | 7.66E-08 | Nightingale | VLDL_size | NA | C | T | 0.072303 | 0.004165 | 9.80E-72 |
| rs174561 | miR-1908-5 | cis | 7.66E-08 | Nightingale | HDL_PL | NA | C | T | -0.04632 | 0.004039 | 1.20E-32 |
| rs174561 | miR-1908-5 | cis | 7.66E-08 | Nightingale | M_HDL_C_ | NA | C | T | -0.06671 | 0.004253 | 3.00E-58 |
| rs174561 | miR-1908-5 | cis | 7.66E-08 | Nightingale | XL_VLDL_L | NA | C | T | 0.052488 | 0.004299 | 7.20E-36 |
| rs174561 | miR-1908-5 | cis | 7.66E-08 | Nightingale | L_HDL_FC_ | NA | C | T | -0.10803 | 0.004151 | 2.00E-161 |
| rs174561 | miR-1908-5 | cis | 7.66E-08 | Nightingale | M_HDL_CE | NA | C | T | -0.03721 | 0.004126 | 1.70E-20 |
| rs174561 | miR-1908-5 | cis | 7.66E-08 | Nightingale | Omega_3_ | NA | C | T | -0.38293 | 0.004366 | 0 |
| rs174561 | miR-1908-5 | cis | 7.66E-08 | Nightingale | XXL_VLDL_ | NA | C | T | 0.073841 | 0.004315 | 2.20E-68 |
| rs174561 | miR-1908-5 | cis | 7.66E-08 | Nightingale | Total_PL | NA | C | T | -0.04803 | 0.004254 | 2.20E-31 |
| rs174561 | miR-1908-5 | cis | 7.66E-08 | Nightingale | PUFA_by_I | NA | C | T | -0.16781 | 0.004279 | 0 |
| rs174561 | miR-1908-5 | cis | 7.66E-08 | Nightingale | M_VLDL_C | NA | C | T | -0.07963 | 0.004205 | 3.70E-83 |
| rs174561 | miR-1908-5 | cis | 7.66E-08 | Nightingale | M_VLDL_T | NA | C | T | 0.076744 | 0.00422 | 3.50E-77 |
| rs174561 | miR-1908-5 | cis | 7.66E-08 | Nightingale | LA_pct | NA | C | T | 0.181345 | 0.004366 | 0 |

| | | | | | | | | | | | |
|---|---|---|---|---|---|---|---|---|---|---|---|
| rs174561 | miR-1908-! | cis | 7.66E-08 | Nightingale | VLDL_TG | NA | C | T | 0.048814 | 0.004299 | 3.00E-31 |
| rs174561 | miR-1908-! | cis | 7.66E-08 | Nightingale | M_VLDL_F | NA | C | T | -0.02724 | 0.004467 | 5.10E-10 |
| rs174561 | miR-1908-! | cis | 7.66E-08 | Nightingale | L_VLDL_PL | NA | C | T | 0.049585 | 0.004311 | 3.80E-32 |
| rs174561 | miR-1908-! | cis | 7.66E-08 | Nightingale | S_VLDL_PL | NA | C | T | -0.07761 | 0.004275 | 4.90E-77 |
| rs174561 | miR-1908-! | cis | 7.66E-08 | Nightingale | SFA_pct | NA | C | T | -0.0395 | 0.004408 | 3.20E-19 |
| rs174561 | miR-1908-! | cis | 7.66E-08 | Nightingale | M_HDL_CE | NA | C | T | -0.04911 | 0.004333 | 1.40E-30 |
| rs174561 | miR-1908-! | cis | 7.66E-08 | Nightingale | M_HDL_L | NA | C | T | -0.0264 | 0.004148 | 3.50E-11 |
| rs174561 | miR-1908-! | cis | 7.66E-08 | Nightingale | L_VLDL_C | NA | C | T | 0.030026 | 0.00437 | 2.00E-12 |
| rs174561 | miR-1908-! | cis | 7.66E-08 | Nightingale | M_LDL_CE | NA | C | T | -0.02497 | 0.004475 | 1.70E-08 |
| rs174561 | miR-1908-! | cis | 7.66E-08 | Nightingale | M_VLDL_T | NA | C | T | 0.032426 | 0.004341 | 2.10E-14 |
| rs174561 | miR-1908-! | cis | 7.66E-08 | Nightingale | Total_P | NA | C | T | -0.039 | 0.004205 | 1.10E-21 |
| rs174561 | miR-1908-! | cis | 7.66E-08 | Nightingale | L_HDL_C | NA | C | T | -0.07805 | 0.003934 | 6.80E-95 |
| rs174561 | miR-1908-! | cis | 7.66E-08 | Nightingale | LDL_PL | NA | C | T | -0.0445 | 0.004458 | 2.50E-24 |
| rs174561 | miR-1908-! | cis | 7.66E-08 | Nightingale | IDL_PL | NA | C | T | -0.06959 | 0.004311 | 1.20E-62 |
| rs174561 | miR-1908-! | cis | 7.66E-08 | Nightingale | M_HDL_TG | NA | C | T | 0.067381 | 0.004284 | 3.30E-58 |
| rs174561 | miR-1908-! | cis | 7.66E-08 | Nightingale | LDL_CE | NA | C | T | -0.04179 | 0.004457 | 9.10E-22 |
| rs174561 | miR-1908-! | cis | 7.66E-08 | Nightingale | L_LDL_FC_ | NA | C | T | -0.0822 | 0.004258 | 4.70E-86 |
| rs174561 | miR-1908-! | cis | 7.66E-08 | Nightingale | MUFA | NA | C | T | 0.064413 | 0.004422 | 1.30E-49 |
| rs174561 | miR-1908-! | cis | 7.66E-08 | Nightingale | Total_CE | NA | C | T | -0.0645 | 0.004309 | 3.20E-53 |
| rs174561 | miR-1908-! | cis | 7.66E-08 | Nightingale | HDL_size | NA | C | T | -0.07009 | 0.003959 | 5.50E-76 |
| rs174561 | miR-1908-! | cis | 7.66E-08 | Nightingale | XL_VLDL_C | NA | C | T | -0.06449 | 0.004281 | 1.10E-52 |
| rs174561 | miR-1908-! | cis | 7.66E-08 | Nightingale | L_VLDL_FC | NA | C | T | 0.041103 | 0.004322 | 1.70E-22 |
| rs174561 | miR-1908-! | cis | 7.66E-08 | Nightingale | S_HDL_TG | NA | C | T | 0.068812 | 0.004332 | 6.70E-60 |
| rs174561 | miR-1908-! | cis | 7.66E-08 | Nightingale | HDL_L | NA | C | T | -0.05143 | 0.004012 | 2.40E-40 |
| rs174561 | miR-1908-! | cis | 7.66E-08 | Nightingale | M_HDL_FC | NA | C | T | -0.04962 | 0.004066 | 1.30E-36 |
| rs174561 | miR-1908-! | cis | 7.66E-08 | Nightingale | VLDL_L | NA | C | T | 0.028977 | 0.004381 | 1.40E-11 |
| rs174561 | miR-1908-! | cis | 7.66E-08 | Nightingale | L_LDL_P | NA | C | T | -0.03582 | 0.004453 | 1.40E-16 |
| rs174561 | miR-1908-! | cis | 7.66E-08 | Nightingale | S_VLDL_FC | NA | C | T | -0.08156 | 0.004266 | 5.60E-85 |
| rs174561 | miR-1908-! | cis | 7.66E-08 | Nightingale | S_LDL_C | NA | C | T | -0.03075 | 0.004473 | 2.30E-12 |
| rs174561 | miR-1908-! | cis | 7.66E-08 | Nightingale | Tyr | NA | C | T | 0.032968 | 0.004462 | 1.50E-13 |
| rs174561 | miR-1908-! | cis | 7.66E-08 | Nightingale | S_HDL_FC_ | NA | C | T | -0.09983 | 0.004238 | 2.50E-135 |
| rs174561 | miR-1908-! | cis | 7.66E-08 | Nightingale | XS_VLDL_C | NA | C | T | -0.0961 | 0.004233 | 1.30E-119 |
| rs174561 | miR-1908-! | cis | 7.66E-08 | Nightingale | S_LDL_TG | NA | C | T | 0.051424 | 0.004358 | 3.90E-33 |
| rs174561 | miR-1908-! | cis | 7.66E-08 | Nightingale | S_LDL_PL | NA | C | T | -0.04967 | 0.004457 | 3.30E-30 |
| rs174561 | miR-1908-! | cis | 7.66E-08 | Nightingale | XL_VLDL_P | NA | C | T | 0.053742 | 0.004294 | 1.40E-37 |
| rs174561 | miR-1908-! | cis | 7.66E-08 | Nightingale | M_LDL_TG | NA | C | T | 0.075522 | 0.004412 | 5.10E-67 |
| rs174561 | miR-1908-! | cis | 7.66E-08 | Nightingale | PUFA | NA | C | T | -0.09259 | 0.004357 | 3.50E-106 |
| rs174561 | miR-1908-! | cis | 7.66E-08 | Nightingale | M_LDL_C_ | NA | C | T | -0.09818 | 0.004471 | 3.80E-107 |
| rs174561 | miR-1908-! | cis | 7.66E-08 | Nightingale | LDL_TG | NA | C | T | 0.031276 | 0.004418 | 1.50E-12 |
| rs174561 | miR-1908-! | cis | 7.66E-08 | Nightingale | XS_VLDL_L | NA | C | T | -0.03837 | 0.004392 | 7.30E-20 |
| rs174561 | miR-1908-! | cis | 7.66E-08 | Nightingale | L_HDL_L | NA | C | T | -0.0748 | 0.00392 | 2.70E-88 |
| rs174561 | miR-1908-! | cis | 7.66E-08 | Nightingale | L_HDL_PL_ | NA | C | T | 0.042896 | 0.004324 | 1.70E-24 |
| rs174561 | miR-1908-! | cis | 7.66E-08 | Nightingale | M_LDL_P | NA | C | T | -0.04475 | 0.004481 | 4.80E-24 |
| rs174561 | miR-1908-! | cis | 7.66E-08 | Nightingale | S_LDL_FC_ | NA | C | T | -0.08061 | 0.00433 | 3.40E-80 |
| rs174561 | miR-1908-! | cis | 7.66E-08 | Nightingale | XS_VLDL_P | NA | C | T | -0.04058 | 0.004408 | 1.20E-21 |
| rs174561 | miR-1908-! | cis | 7.66E-08 | Nightingale | XXL_VLDL_ | NA | C | T | 0.042869 | 0.00449 | 3.20E-22 |
| rs174561 | miR-1908-! | cis | 7.66E-08 | Nightingale | S_LDL_TG_ | NA | C | T | 0.08606 | 0.004329 | 5.20E-91 |
| rs174561 | miR-1908-! | cis | 7.66E-08 | Nightingale | LDL_P | NA | C | T | -0.03843 | 0.004465 | 1.50E-18 |
| rs174561 | miR-1908-! | cis | 7.66E-08 | Nightingale | Sphingomy | NA | C | T | -0.11208 | 0.004175 | 3.80E-166 |
| rs174561 | miR-1908-! | cis | 7.66E-08 | Nightingale | M_HDL_FC | NA | C | T | -0.10001 | 0.003959 | 1.30E-152 |
| rs174561 | miR-1908-! | cis | 7.66E-08 | Nightingale | XL_VLDL_C | NA | C | T | 0.038661 | 0.004364 | 1.30E-19 |
| rs174561 | miR-1908-! | cis | 7.66E-08 | Nightingale | HDL_P | NA | C | T | -0.03454 | 0.004195 | 2.40E-17 |
| rs174561 | miR-1908-! | cis | 7.66E-08 | Nightingale | L_HDL_TG_ | NA | C | T | 0.078362 | 0.004216 | 1.10E-81 |
| rs174561 | miR-1908-! | cis | 7.66E-08 | Nightingale | M_LDL_PL_ | NA | C | T | 0.042803 | 0.004465 | 6.60E-23 |
| rs174561 | miR-1908-! | cis | 7.66E-08 | Nightingale | M_VLDL_F | NA | C | T | -0.08944 | 0.004248 | 2.50E-103 |
| rs174561 | miR-1908-! | cis | 7.66E-08 | Nightingale | XXL_VLDL_ | NA | C | T | 0.072159 | 0.004306 | 6.00E-66 |
| rs174561 | miR-1908-! | cis | 7.66E-08 | Nightingale | IDL_TG_pc | NA | C | T | 0.086803 | 0.004316 | 2.20E-93 |
| rs174561 | miR-1908-! | cis | 7.66E-08 | Nightingale | XL_HDL_FC | NA | C | T | -0.09901 | 0.004162 | 2.30E-134 |
| rs174561 | miR-1908-! | cis | 7.66E-08 | Nightingale | L_VLDL_FC | NA | C | T | 0.027337 | 0.004426 | 6.50E-10 |
| rs174561 | miR-1908-! | cis | 7.66E-08 | Nightingale | XXL_VLDL_ | NA | C | T | 0.072704 | 0.004288 | 8.90E-68 |
| rs174561 | miR-1908-! | cis | 7.66E-08 | Nightingale | S_LDL_FC | NA | C | T | -0.06142 | 0.004461 | 2.40E-45 |
| rs174561 | miR-1908-! | cis | 7.66E-08 | Nightingale | ApoB | NA | C | T | -0.03637 | 0.004462 | 7.20E-17 |
| rs174561 | miR-1908-! | cis | 7.66E-08 | Nightingale | Total_FC | NA | C | T | -0.05666 | 0.004365 | 9.80E-41 |

| | | | | | | | | | | | | |
|---|---|---|---|---|---|---|---|---|---|---|---|---|
| rs174561 | miR-1908-! | cis | | 7.66E-08 | Nightingale | L_HDL_FC | NA | C | T | -0.08189 | 0.003915 | 2.50E-105 |
| rs174561 | miR-1908-! | cis | | 7.66E-08 | Nightingale | S_HDL_C_p | NA | C | T | -0.04468 | 0.004416 | 1.50E-23 |
| rs174561 | miR-1908-! | cis | | 7.66E-08 | Nightingale | S_LDL_P | NA | C | T | -0.03024 | 0.004472 | 5.30E-12 |
| rs174561 | miR-1908-! | cis | | 7.66E-08 | Nightingale | Omega_6_ | NA | C | T | 0.362928 | 0.004403 | 0 |
| rs174561 | miR-1908-! | cis | | 7.66E-08 | Nightingale | XS_VLDL_P | NA | C | T | 0.057686 | 0.004359 | 1.00E-40 |
| rs174561 | miR-1908-! | cis | | 7.66E-08 | Nightingale | XS_VLDL_C | NA | C | T | -0.09038 | 0.004212 | 3.10E-107 |
| rs174561 | miR-1908-! | cis | | 7.66E-08 | Nightingale | L_HDL_CE | NA | C | T | -0.07662 | 0.003945 | 7.90E-91 |
| rs174561 | miR-1908-! | cis | | 7.66E-08 | Nightingale | XL_VLDL_P | NA | C | T | 0.04938 | 0.004312 | 9.60E-32 |
| rs174561 | miR-1908-! | cis | | 7.66E-08 | Nightingale | XL_HDL_TG | NA | C | T | 0.086385 | 0.004209 | 2.30E-99 |
| rs174561 | miR-1908-! | cis | | 7.66E-08 | Nightingale | LDL_C | NA | C | T | -0.04797 | 0.004449 | 2.70E-28 |
| rs174561 | miR-1908-! | cis | | 7.66E-08 | Nightingale | HDL_C | NA | C | T | -0.06322 | 0.00399 | 6.50E-61 |
| rs174561 | miR-1908-! | cis | | 7.66E-08 | Nightingale | HDL_TG | NA | C | T | 0.047718 | 0.004406 | 8.60E-28 |
| rs174561 | miR-1908-! | cis | | 7.66E-08 | Nightingale | Cholines | NA | C | T | -0.03464 | 0.004202 | 6.50E-18 |
| rs174561 | miR-1908-! | cis | | 7.66E-08 | Nightingale | S_LDL_L | NA | C | T | -0.03092 | 0.004467 | 1.60E-12 |
| rs174561 | miR-1908-! | cis | | 7.66E-08 | Nightingale | ApoA1 | NA | C | T | -0.0396 | 0.004089 | 7.10E-24 |
| rs174561 | miR-1908-! | cis | | 7.66E-08 | Nightingale | XS_VLDL_C | NA | C | T | -0.06841 | 0.004311 | 3.00E-60 |
| rs174561 | miR-1908-! | cis | | 7.66E-08 | Nightingale | XL_HDL_CE | NA | C | T | -0.09448 | 0.004001 | 2.60E-133 |
| rs174562 | miR-1908-! | cis | | 4.92E-08 | Nightingale | L_VLDL_L | NA | G | A | 0.03847 | 0.004197 | 8.70E-21 |
| rs174562 | miR-1908-! | cis | | 4.92E-08 | Nightingale | L_VLDL_TG | NA | G | A | 0.037588 | 0.004179 | 4.60E-20 |
| rs174562 | miR-1908-! | cis | | 4.92E-08 | Nightingale | VLDL_size | NA | G | A | 0.072817 | 0.004048 | 1.20E-76 |
| rs174562 | miR-1908-! | cis | | 4.92E-08 | Nightingale | Total_FA | NA | G | A | -0.02363 | 0.004309 | 1.30E-08 |
| rs174562 | miR-1908-! | cis | | 4.92E-08 | Nightingale | M_HDL_CE | NA | G | A | -0.04717 | 0.004211 | 5.40E-30 |
| rs174562 | miR-1908-! | cis | | 4.92E-08 | Nightingale | L_HDL_C_p | NA | G | A | -0.06862 | 0.004093 | 7.60E-68 |
| rs174562 | miR-1908-! | cis | | 4.92E-08 | Nightingale | XL_VLDL_F | NA | G | A | -0.06837 | 0.004208 | 3.60E-62 |
| rs174562 | miR-1908-! | cis | | 4.92E-08 | Nightingale | L_LDL_L | NA | G | A | -0.05418 | 0.004301 | 1.80E-38 |
| rs174562 | miR-1908-! | cis | | 4.92E-08 | Nightingale | MUFA | NA | G | A | 0.064111 | 0.004299 | 2.20E-51 |
| rs174562 | miR-1908-! | cis | | 4.92E-08 | Nightingale | M_LDL_L | NA | G | A | -0.02807 | 0.00435 | 4.20E-11 |
| rs174562 | miR-1908-! | cis | | 4.92E-08 | Nightingale | XS_VLDL_T | NA | G | A | 0.097547 | 0.004125 | 1.30E-129 |
| rs174562 | miR-1908-! | cis | | 4.92E-08 | Nightingale | S_LDL_TG | NA | G | A | 0.050582 | 0.004237 | 2.00E-33 |
| rs174562 | miR-1908-! | cis | | 4.92E-08 | Nightingale | non_HDL_( | NA | G | A | -0.04777 | 0.00432 | 7.30E-30 |
| rs174562 | miR-1908-! | cis | | 4.92E-08 | Nightingale | XS_VLDL_F | NA | G | A | -0.05629 | 0.004267 | 1.80E-42 |
| rs174562 | miR-1908-! | cis | | 4.92E-08 | Nightingale | XXL_VLDL_ | NA | G | A | 0.066896 | 0.004183 | 4.10E-60 |
| rs174562 | miR-1908-! | cis | | 4.92E-08 | Nightingale | MUFA_pct | NA | G | A | 0.197678 | 0.004156 | 0 |
| rs174562 | miR-1908-! | cis | | 4.92E-08 | Nightingale | S_VLDL_PL | NA | G | A | -0.07873 | 0.004156 | 5.40E-84 |
| rs174562 | miR-1908-! | cis | | 4.92E-08 | Nightingale | IDL_FC_pct | NA | G | A | -0.10416 | 0.004308 | 8.39E-131 |
| rs174562 | miR-1908-! | cis | | 4.92E-08 | Nightingale | IDL_FC | NA | G | A | -0.08987 | 0.004203 | 5.11E-107 |
| rs174562 | miR-1908-! | cis | | 4.92E-08 | Nightingale | XL_HDL_CE | NA | G | A | -0.02615 | 0.00434 | 3.90E-09 |
| rs174562 | miR-1908-! | cis | | 4.92E-08 | Nightingale | Total_C | NA | G | A | -0.06519 | 0.004204 | 3.20E-57 |
| rs174562 | miR-1908-! | cis | | 4.92E-08 | Nightingale | XS_VLDL_P | NA | G | A | -0.03466 | 0.004282 | 1.90E-17 |
| rs174562 | miR-1908-! | cis | | 4.92E-08 | Nightingale | Omega_3 | NA | G | A | -0.33588 | 0.004257 | 0 |
| rs174562 | miR-1908-! | cis | | 4.92E-08 | Nightingale | XL_VLDL_F | NA | G | A | 0.043711 | 0.004206 | 3.40E-26 |
| rs174562 | miR-1908-! | cis | | 4.92E-08 | Nightingale | HDL_P | NA | G | A | -0.03534 | 0.004078 | 3.10E-19 |
| rs174562 | miR-1908-! | cis | | 4.92E-08 | Nightingale | IDL_C_pct | NA | G | A | -0.07471 | 0.00424 | 3.40E-71 |
| rs174562 | miR-1908-! | cis | | 4.92E-08 | Nightingale | Total_PL | NA | G | A | -0.05104 | 0.004135 | 3.10E-37 |
| rs174562 | miR-1908-! | cis | | 4.92E-08 | Nightingale | L_VLDL_PL | NA | G | A | 0.0878 | 0.004215 | 4.80E-100 |
| rs174562 | miR-1908-! | cis | | 4.92E-08 | Nightingale | L_LDL_TG_ | NA | G | A | 0.077623 | 0.004258 | 5.40E-76 |
| rs174562 | miR-1908-! | cis | | 4.92E-08 | Nightingale | L_LDL_PL | NA | G | A | -0.05494 | 0.00431 | 5.80E-39 |
| rs174562 | miR-1908-! | cis | | 4.92E-08 | Nightingale | M_LDL_TG | NA | G | A | 0.036681 | 0.004279 | 9.90E-18 |
| rs174562 | miR-1908-! | cis | | 4.92E-08 | Nightingale | Citrate | NA | G | A | -0.03333 | 0.004315 | 1.10E-15 |
| rs174562 | miR-1908-! | cis | | 4.92E-08 | Nightingale | XXL_VLDL_ | NA | G | A | 0.061495 | 0.004191 | 7.30E-51 |
| rs174562 | miR-1908-! | cis | | 4.92E-08 | Nightingale | L_VLDL_CE | NA | G | A | -0.05936 | 0.004156 | 4.20E-49 |
| rs174562 | miR-1908-! | cis | | 4.92E-08 | Nightingale | L_HDL_FC_ | NA | G | A | -0.11281 | 0.004035 | 3.10E-187 |
| rs174562 | miR-1908-! | cis | | 4.92E-08 | Nightingale | S_LDL_CE_ | NA | G | A | 0.057782 | 0.004335 | 1.90E-42 |
| rs174562 | miR-1908-! | cis | | 4.92E-08 | Nightingale | Omega_3_ | NA | G | A | -0.3901 | 0.004243 | 0 |
| rs174562 | miR-1908-! | cis | | 4.92E-08 | Nightingale | S_LDL_PL | NA | G | A | -0.05165 | 0.004333 | 1.10E-34 |
| rs174562 | miR-1908-! | cis | | 4.92E-08 | Nightingale | XS_VLDL_C | NA | G | A | -0.09215 | 0.004094 | 1.80E-117 |
| rs174562 | miR-1908-! | cis | | 4.92E-08 | Nightingale | XL_HDL_FC | NA | G | A | 0.061367 | 0.00385 | 2.50E-63 |
| rs174562 | miR-1908-! | cis | | 4.92E-08 | Nightingale | LA | NA | G | A | 0.083973 | 0.004259 | 8.50E-87 |
| rs174562 | miR-1908-! | cis | | 4.92E-08 | Nightingale | XXL_VLDL_ | NA | G | A | -0.02708 | 0.004321 | 7.50E-11 |
| rs174562 | miR-1908-! | cis | | 4.92E-08 | Nightingale | M_LDL_PL_ | NA | G | A | 0.045375 | 0.00434 | 1.10E-26 |
| rs174562 | miR-1908-! | cis | | 4.92E-08 | Nightingale | XL_VLDL_C | NA | G | A | -0.06131 | 0.004164 | 1.70E-50 |
| rs174562 | miR-1908-! | cis | | 4.92E-08 | Nightingale | M_VLDL_F( | NA | G | A | -0.09151 | 0.004129 | 1.50E-114 |
| rs174562 | miR-1908-! | cis | | 4.92E-08 | Nightingale | M_LDL_C | NA | G | A | -0.03393 | 0.004351 | 1.60E-15 |

| | | | | | | | | | | | | |
|---|---|---|---|---|---|---|---|---|---|---|---|---|
| rs174562 | miR-1908-! | cis | | 4.92E-08 | Nightingale | M_LDL_FC_ | NA | G | A | -0.07187 | 0.00416 | 2.00E-69 |
| rs174562 | miR-1908-! | cis | | 4.92E-08 | Nightingale | TG_by_PG | NA | G | A | 0.068921 | 0.00403 | 6.10E-69 |
| rs174562 | miR-1908-! | cis | | 4.92E-08 | Nightingale | XL_HDL_TG | NA | G | A | 0.087589 | 0.004091 | 1.40E-107 |
| rs174562 | miR-1908-! | cis | | 4.92E-08 | Nightingale | M_VLDL_P | NA | G | A | -0.06263 | 0.004167 | 1.80E-54 |
| rs174562 | miR-1908-! | cis | | 4.92E-08 | Nightingale | S_HDL_TG_ | NA | G | A | 0.069155 | 0.00413 | 6.70E-66 |
| rs174562 | miR-1908-! | cis | | 4.92E-08 | Nightingale | Sphingomy | NA | G | A | -0.11535 | 0.004058 | 9.51E-187 |
| rs174562 | miR-1908-! | cis | | 4.92E-08 | Nightingale | S_LDL_TG_ | NA | G | A | 0.086423 | 0.004208 | 6.50E-97 |
| rs174562 | miR-1908-! | cis | | 4.92E-08 | Nightingale | XL_HDL_PL | NA | G | A | -0.06734 | 0.00399 | 5.50E-69 |
| rs174562 | miR-1908-! | cis | | 4.92E-08 | Nightingale | S_LDL_P | NA | G | A | -0.03246 | 0.004347 | 1.70E-14 |
| rs174562 | miR-1908-! | cis | | 4.92E-08 | Nightingale | L_HDL_C | NA | G | A | -0.08039 | 0.003824 | 8.99E-107 |
| rs174562 | miR-1908-! | cis | | 4.92E-08 | Nightingale | XL_VLDL_L | NA | G | A | 0.051714 | 0.004179 | 1.50E-36 |
| rs174562 | miR-1908-! | cis | | 4.92E-08 | Nightingale | M_HDL_CE | NA | G | A | -0.03768 | 0.004011 | 2.30E-22 |
| rs174562 | miR-1908-! | cis | | 4.92E-08 | Nightingale | XXL_VLDL_ | NA | G | A | 0.068259 | 0.004184 | 9.90E-63 |
| rs174562 | miR-1908-! | cis | | 4.92E-08 | Nightingale | SFA | NA | G | A | -0.03133 | 0.004304 | 1.20E-13 |
| rs174562 | miR-1908-! | cis | | 4.92E-08 | Nightingale | XXL_VLDL_ | NA | G | A | 0.073878 | 0.004194 | 5.90E-72 |
| rs174562 | miR-1908-! | cis | | 4.92E-08 | Nightingale | S_VLDL_CE | NA | G | A | -0.02904 | 0.004285 | 6.70E-12 |
| rs174562 | miR-1908-! | cis | | 4.92E-08 | Nightingale | XS_VLDL_F | NA | G | A | -0.11312 | 0.00431 | 4.40E-155 |
| rs174562 | miR-1908-! | cis | | 4.92E-08 | Nightingale | M_HDL_TG | NA | G | A | 0.050741 | 0.004289 | 3.70E-33 |
| rs174562 | miR-1908-! | cis | | 4.92E-08 | Nightingale | L_LDL_FC | NA | G | A | -0.06836 | 0.004264 | 3.20E-61 |
| rs174562 | miR-1908-! | cis | | 4.92E-08 | Nightingale | M_LDL_CE | NA | G | A | -0.02619 | 0.004349 | 7.30E-10 |
| rs174562 | miR-1908-! | cis | | 4.92E-08 | Nightingale | L_VLDL_C | NA | G | A | 0.029092 | 0.004248 | 3.30E-12 |
| rs174562 | miR-1908-! | cis | | 4.92E-08 | Nightingale | Total_P | NA | G | A | -0.0401 | 0.004087 | 2.40E-24 |
| rs174562 | miR-1908-! | cis | | 4.92E-08 | Nightingale | M_VLDL_F | NA | G | A | -0.02936 | 0.004342 | 4.00E-12 |
| rs174562 | miR-1908-! | cis | | 4.92E-08 | Nightingale | VLDL_TG | NA | G | A | 0.047956 | 0.004179 | 1.50E-31 |
| rs174562 | miR-1908-! | cis | | 4.92E-08 | Nightingale | XL_HDL_P | NA | G | A | -0.0898 | 0.003908 | 7.91E-128 |
| rs174562 | miR-1908-! | cis | | 4.92E-08 | Nightingale | XS_VLDL_C | NA | G | A | -0.09808 | 0.004115 | 2.40E-131 |
| rs174562 | miR-1908-! | cis | | 4.92E-08 | Nightingale | DHA_pct | NA | G | A | -0.27409 | 0.004216 | 0 |
| rs174562 | miR-1908-! | cis | | 4.92E-08 | Nightingale | L_LDL_C | NA | G | A | -0.05781 | 0.004294 | 1.20E-43 |
| rs174562 | miR-1908-! | cis | | 4.92E-08 | Nightingale | XXL_VLDL_ | NA | G | A | 0.072297 | 0.004168 | 1.60E-70 |
| rs174562 | miR-1908-! | cis | | 4.92E-08 | Nightingale | M_VLDL_C | NA | G | A | -0.05274 | 0.004304 | 4.00E-36 |
| rs174562 | miR-1908-! | cis | | 4.92E-08 | Nightingale | PUFA | NA | G | A | -0.09643 | 0.004235 | 9.91E-122 |
| rs174562 | miR-1908-! | cis | | 4.92E-08 | Nightingale | XL_VLDL_C | NA | G | A | 0.037679 | 0.004242 | 1.50E-19 |
| rs174562 | miR-1908-! | cis | | 4.92E-08 | Nightingale | S_LDL_PL_ | NA | G | A | -0.06261 | 0.004334 | 1.60E-49 |
| rs174562 | miR-1908-! | cis | | 4.92E-08 | Nightingale | XL_VLDL_T | NA | G | A | 0.057158 | 0.004154 | 7.90E-45 |
| rs174562 | miR-1908-! | cis | | 4.92E-08 | Nightingale | Unsaturati | NA | G | A | -0.33979 | 0.004113 | 0 |
| rs174562 | miR-1908-! | cis | | 4.92E-08 | Nightingale | S_VLDL_TG | NA | G | A | 0.039019 | 0.004214 | 5.00E-21 |
| rs174562 | miR-1908-! | cis | | 4.92E-08 | Nightingale | S_LDL_C | NA | G | A | -0.03213 | 0.004348 | 3.00E-14 |
| rs174562 | miR-1908-! | cis | | 4.92E-08 | Nightingale | HDL_size | NA | G | A | -0.07279 | 0.003848 | 1.70E-86 |
| rs174562 | miR-1908-! | cis | | 4.92E-08 | Nightingale | S_VLDL_C_ | NA | G | A | -0.05673 | 0.004225 | 3.00E-42 |
| rs174562 | miR-1908-! | cis | | 4.92E-08 | Nightingale | PUFA_by_I | NA | G | A | -0.17093 | 0.00416 | 0 |
| rs174562 | miR-1908-! | cis | | 4.92E-08 | Nightingale | M_VLDL_C | NA | G | A | -0.07618 | 0.004077 | 3.70E-81 |
| rs174562 | miR-1908-! | cis | | 4.92E-08 | Nightingale | PUFA_pct | NA | G | A | -0.12169 | 0.004177 | 3.50E-188 |
| rs174562 | miR-1908-! | cis | | 4.92E-08 | Nightingale | XL_HDL_FC | NA | G | A | -0.10309 | 0.004046 | 1.80E-154 |
| rs174562 | miR-1908-! | cis | | 4.92E-08 | Nightingale | L_VLDL_C_ | NA | G | A | -0.03878 | 0.004242 | 3.80E-21 |
| rs174562 | miR-1908-! | cis | | 4.92E-08 | Nightingale | DHA | NA | G | A | -0.28869 | 0.004185 | 0 |
| rs174562 | miR-1908-! | cis | | 4.92E-08 | Nightingale | IDL_PL | NA | G | A | -0.07299 | 0.00419 | 9.00E-73 |
| rs174562 | miR-1908-! | cis | | 4.92E-08 | Nightingale | S_HDL_TG | NA | G | A | 0.06917 | 0.004211 | 1.80E-63 |
| rs174562 | miR-1908-! | cis | | 4.92E-08 | Nightingale | L_VLDL_FC | NA | G | A | 0.028441 | 0.004302 | 1.90E-11 |
| rs174562 | miR-1908-! | cis | | 4.92E-08 | Nightingale | XL_VLDL_T | NA | G | A | 0.059683 | 0.004207 | 4.10E-47 |
| rs174562 | miR-1908-! | cis | | 4.92E-08 | Nightingale | L_HDL_TG_ | NA | G | A | 0.079066 | 0.004098 | 3.20E-88 |
| rs174562 | miR-1908-! | cis | | 4.92E-08 | Nightingale | L_VLDL_P | NA | G | A | 0.042152 | 0.004193 | 1.10E-24 |
| rs174562 | miR-1908-! | cis | | 4.92E-08 | Nightingale | M_HDL_P | NA | G | A | -0.03408 | 0.004001 | 6.20E-19 |
| rs174562 | miR-1908-! | cis | | 4.92E-08 | Nightingale | L_HDL_CE_ | NA | G | A | -0.05024 | 0.004155 | 5.40E-36 |
| rs174562 | miR-1908-! | cis | | 4.92E-08 | Nightingale | IDL_C | NA | G | A | -0.07735 | 0.004194 | 1.70E-80 |
| rs174562 | miR-1908-! | cis | | 4.92E-08 | Nightingale | XL_HDL_C_ | NA | G | A | 0.024211 | 0.004149 | 2.40E-10 |
| rs174562 | miR-1908-! | cis | | 4.92E-08 | Nightingale | S_VLDL_TG | NA | G | A | 0.064658 | 0.004202 | 4.30E-55 |
| rs174562 | miR-1908-! | cis | | 4.92E-08 | Nightingale | IDL_L | NA | G | A | -0.07251 | 0.004198 | 3.10E-71 |
| rs174562 | miR-1908-! | cis | | 4.92E-08 | Nightingale | HDL_TG | NA | G | A | 0.046575 | 0.004282 | 4.10E-28 |
| rs174562 | miR-1908-! | cis | | 4.92E-08 | Nightingale | L_LDL_P | NA | G | A | -0.03751 | 0.004328 | 3.40E-19 |
| rs174562 | miR-1908-! | cis | | 4.92E-08 | Nightingale | S_LDL_L | NA | G | A | -0.03246 | 0.004342 | 1.30E-14 |
| rs174562 | miR-1908-! | cis | | 4.92E-08 | Nightingale | XL_HDL_L | NA | G | A | -0.09331 | 0.003902 | 1.80E-137 |
| rs174562 | miR-1908-! | cis | | 4.92E-08 | Nightingale | M_LDL_FC | NA | G | A | -0.05237 | 0.004338 | 2.40E-35 |
| rs174562 | miR-1908-! | cis | | 4.92E-08 | Nightingale | XS_VLDL_P | NA | G | A | 0.0575 | 0.004237 | 9.90E-43 |

| | | | | | | | | | | | | |
|---|---|---|---|---|---|---|---|---|---|---|---|---|
| rs174562 | miR-1908-! | cis | | 4.92E-08 | Nightingale | HDL_L | NA | G | A | -0.05327 | 0.0039 | 9.90E-46 |
| rs174562 | miR-1908-! | cis | | 4.92E-08 | Nightingale | ApoB | NA | G | A | -0.03849 | 0.004337 | 6.80E-20 |
| rs174562 | miR-1908-! | cis | | 4.92E-08 | Nightingale | Remnant_( | NA | G | A | -0.04408 | 0.004315 | 6.10E-26 |
| rs174562 | miR-1908-! | cis | | 4.92E-08 | Nightingale | XS_VLDL_L | NA | G | A | -0.04178 | 0.004269 | 1.40E-24 |
| rs174562 | miR-1908-! | cis | | 4.92E-08 | Nightingale | Phosphogly | NA | G | A | -0.04087 | 0.004114 | 4.90E-25 |
| rs174562 | miR-1908-! | cis | | 4.92E-08 | Nightingale | M_HDL_C | NA | G | A | -0.04064 | 0.003996 | 5.90E-26 |
| rs174562 | miR-1908-! | cis | | 4.92E-08 | Nightingale | HDL_PL | NA | G | A | -0.04834 | 0.003926 | 1.90E-37 |
| rs174562 | miR-1908-! | cis | | 4.92E-08 | Nightingale | L_HDL_P | NA | G | A | -0.07795 | 0.003804 | 8.49E-102 |
| rs174562 | miR-1908-! | cis | | 4.92E-08 | Nightingale | Total_L | NA | G | A | -0.03723 | 0.004273 | 1.80E-19 |
| rs174562 | miR-1908-! | cis | | 4.92E-08 | Nightingale | L_HDL_L | NA | G | A | -0.07732 | 0.003811 | 8.50E-100 |
| rs174562 | miR-1908-! | cis | | 4.92E-08 | Nightingale | L_LDL_FC_ | NA | G | A | -0.08288 | 0.004139 | 1.90E-92 |
| rs174562 | miR-1908-! | cis | | 4.92E-08 | Nightingale | M_HDL_PL | NA | G | A | 0.058459 | 0.004119 | 1.10E-47 |
| rs174562 | miR-1908-! | cis | | 4.92E-08 | Nightingale | M_HDL_TG | NA | G | A | 0.066904 | 0.004165 | 6.90E-61 |
| rs174562 | miR-1908-! | cis | | 4.92E-08 | Nightingale | M_LDL_TG | NA | G | A | 0.075855 | 0.004288 | 1.50E-71 |
| rs174562 | miR-1908-! | cis | | 4.92E-08 | Nightingale | IDL_CE | NA | G | A | -0.07231 | 0.004192 | 1.10E-70 |
| rs174562 | miR-1908-! | cis | | 4.92E-08 | Nightingale | M_LDL_C_ | NA | G | A | -0.10011 | 0.004346 | 5.30E-118 |
| rs174562 | miR-1908-! | cis | | 4.92E-08 | Nightingale | XL_VLDL_P | NA | G | A | 0.053124 | 0.004174 | 1.50E-38 |
| rs174562 | miR-1908-! | cis | | 4.92E-08 | Nightingale | LDL_FC | NA | G | A | -0.06432 | 0.004297 | 2.00E-53 |
| rs174562 | miR-1908-! | cis | | 4.92E-08 | Nightingale | HDL_CE | NA | G | A | -0.06141 | 0.003897 | 2.00E-60 |
| rs174562 | miR-1908-! | cis | | 4.92E-08 | Nightingale | LDL_L | NA | G | A | -0.04542 | 0.004329 | 4.10E-27 |
| rs174562 | miR-1908-! | cis | | 4.92E-08 | Nightingale | L_LDL_C_p | NA | G | A | -0.07702 | 0.004231 | 2.20E-76 |
| rs174562 | miR-1908-! | cis | | 4.92E-08 | Nightingale | IDL_P | NA | G | A | -0.0451 | 0.004297 | 3.70E-27 |
| rs174562 | miR-1908-! | cis | | 4.92E-08 | Nightingale | L_LDL_CE | NA | G | A | -0.05328 | 0.004304 | 3.30E-37 |
| rs174562 | miR-1908-! | cis | | 4.92E-08 | Nightingale | Total_CE | NA | G | A | -0.06699 | 0.004189 | 8.80E-61 |
| rs174562 | miR-1908-! | cis | | 4.92E-08 | Nightingale | XL_HDL_C | NA | G | A | -0.0995 | 0.003916 | 1.60E-154 |
| rs174562 | miR-1908-! | cis | | 4.92E-08 | Nightingale | M_HDL_PL | NA | G | A | -0.02126 | 0.004061 | 4.40E-08 |
| rs174562 | miR-1908-! | cis | | 4.92E-08 | Nightingale | XXL_VLDL_ | NA | G | A | 0.071977 | 0.004185 | 5.30E-69 |
| rs174562 | miR-1908-! | cis | | 4.92E-08 | Nightingale | L_HDL_FC | NA | G | A | -0.08475 | 0.003806 | 1.80E-119 |
| rs174562 | miR-1908-! | cis | | 4.92E-08 | Nightingale | LDL_PL | NA | G | A | -0.04616 | 0.004333 | 1.00E-27 |
| rs174562 | miR-1908-! | cis | | 4.92E-08 | Nightingale | S_HDL_C_p | NA | G | A | -0.04246 | 0.004293 | 6.40E-23 |
| rs174562 | miR-1908-! | cis | | 4.92E-08 | Nightingale | M_VLDL_C | NA | G | A | -0.04401 | 0.004331 | 2.80E-25 |
| rs174562 | miR-1908-! | cis | | 4.92E-08 | Nightingale | Total_TG | NA | G | A | 0.047034 | 0.004204 | 4.40E-30 |
| rs174562 | miR-1908-! | cis | | 4.92E-08 | Nightingale | L_HDL_PL | NA | G | A | -0.0762 | 0.003822 | 3.00E-96 |
| rs174562 | miR-1908-! | cis | | 4.92E-08 | Nightingale | XL_HDL_PL | NA | G | A | -0.08937 | 0.003904 | 3.30E-126 |
| rs174562 | miR-1908-! | cis | | 4.92E-08 | Nightingale | S_VLDL_FC | NA | G | A | -0.0283 | 0.004344 | 2.30E-11 |
| rs174562 | miR-1908-! | cis | | 4.92E-08 | Nightingale | Clinical_LD | NA | G | A | -0.05516 | 0.004318 | 3.60E-39 |
| rs174562 | miR-1908-! | cis | | 4.92E-08 | Nightingale | XS_VLDL_C | NA | G | A | -0.06836 | 0.004215 | 6.50E-63 |
| rs174562 | miR-1908-! | cis | | 4.92E-08 | Nightingale | L_VLDL_PL | NA | G | A | 0.048997 | 0.00419 | 5.70E-33 |
| rs174562 | miR-1908-! | cis | | 4.92E-08 | Nightingale | M_HDL_L | NA | G | A | -0.02746 | 0.004032 | 1.20E-12 |
| rs174562 | miR-1908-! | cis | | 4.92E-08 | Nightingale | M_VLDL_T | NA | G | A | 0.031679 | 0.004219 | 2.40E-14 |
| rs174562 | miR-1908-! | cis | | 4.92E-08 | Nightingale | S_HDL_FC_ | NA | G | A | -0.10302 | 0.004119 | 3.90E-153 |
| rs174562 | miR-1908-! | cis | | 4.92E-08 | Nightingale | LDL_CE | NA | G | A | -0.04352 | 0.004332 | 5.90E-25 |
| rs174562 | miR-1908-! | cis | | 4.92E-08 | Nightingale | XL_VLDL_C | NA | G | A | 0.030529 | 0.004275 | 4.30E-13 |
| rs174562 | miR-1908-! | cis | | 4.92E-08 | Nightingale | LA_pct | NA | G | A | 0.185507 | 0.004244 | 0 |
| rs174562 | miR-1908-! | cis | | 4.92E-08 | Nightingale | Omega_6_ | NA | G | A | 0.023968 | 0.004227 | 7.90E-09 |
| rs174562 | miR-1908-! | cis | | 4.92E-08 | Nightingale | S_LDL_FC | NA | G | A | -0.06288 | 0.004336 | 1.80E-50 |
| rs174562 | miR-1908-! | cis | | 4.92E-08 | Nightingale | L_HDL_CE | NA | G | A | -0.07881 | 0.003835 | 5.00E-102 |
| rs174562 | miR-1908-! | cis | | 4.92E-08 | Nightingale | Total_FC | NA | G | A | -0.05938 | 0.004243 | 4.00E-47 |
| rs174562 | miR-1908-! | cis | | 4.92E-08 | Nightingale | XL_VLDL_C | NA | G | A | -0.06428 | 0.004162 | 2.90E-55 |
| rs174562 | miR-1908-! | cis | | 4.92E-08 | Nightingale | SFA_pct | NA | G | A | -0.04257 | 0.004285 | 2.90E-23 |
| rs174562 | miR-1908-! | cis | | 4.92E-08 | Nightingale | VLDL_L | NA | G | A | 0.027573 | 0.004258 | 5.40E-11 |
| rs174562 | miR-1908-! | cis | | 4.92E-08 | Nightingale | XS_VLDL_C | NA | G | A | -0.07212 | 0.00419 | 1.60E-70 |
| rs174562 | miR-1908-! | cis | | 4.92E-08 | Nightingale | LDL_P | NA | G | A | -0.0403 | 0.00434 | 1.40E-21 |
| rs174562 | miR-1908-! | cis | | 4.92E-08 | Nightingale | XL_HDL_CE | NA | G | A | -0.09736 | 0.003889 | 2.80E-150 |
| rs174562 | miR-1908-! | cis | | 4.92E-08 | Nightingale | Tyr | NA | G | A | 0.033726 | 0.004337 | 7.50E-15 |
| rs174562 | miR-1908-! | cis | | 4.92E-08 | Nightingale | Cholines | NA | G | A | -0.037 | 0.004084 | 2.20E-21 |
| rs174562 | miR-1908-! | cis | | 4.92E-08 | Nightingale | HDL_C | NA | G | A | -0.06471 | 0.003878 | 9.80E-68 |
| rs174562 | miR-1908-! | cis | | 4.92E-08 | Nightingale | XS_VLDL_P | NA | G | A | -0.04382 | 0.004285 | 2.00E-26 |
| rs174562 | miR-1908-! | cis | | 4.92E-08 | Nightingale | M_VLDL_C | NA | G | A | -0.08091 | 0.004088 | 5.90E-91 |
| rs174562 | miR-1908-! | cis | | 4.92E-08 | Nightingale | M_LDL_PL | NA | G | A | -0.02344 | 0.00435 | 3.20E-08 |
| rs174562 | miR-1908-! | cis | | 4.92E-08 | Nightingale | ApoA1 | NA | G | A | -0.04102 | 0.003974 | 5.70E-27 |
| rs174562 | miR-1908-! | cis | | 4.92E-08 | Nightingale | XXL_VLDL_ | NA | G | A | 0.066387 | 0.004177 | 1.40E-59 |
| rs174562 | miR-1908-! | cis | | 4.92E-08 | Nightingale | XXL_VLDL_ | NA | G | A | 0.044065 | 0.004366 | 6.60E-25 |

| | | | | | | | | | | | |
|---|---|---|---|---|---|---|---|---|---|---|---|
| rs174562 | miR-1908-! | cis | 4.92E-08 | Nightingale | IDL_TG_pc | NA | G | A | 0.088155 | 0.004195 | 6.81E-102 |
| rs174562 | miR-1908-! | cis | 4.92E-08 | Nightingale | L_VLDL_FC | NA | G | A | 0.040395 | 0.004201 | 8.60E-23 |
| rs174562 | miR-1908-! | cis | 4.92E-08 | Nightingale | S_VLDL_FC | NA | G | A | -0.08309 | 0.004147 | 2.30E-93 |
| rs174562 | miR-1908-! | cis | 4.92E-08 | Nightingale | M_LDL_P | NA | G | A | -0.04664 | 0.004356 | 1.30E-27 |
| rs174562 | miR-1908-! | cis | 4.92E-08 | Nightingale | M_VLDL_T | NA | G | A | 0.078178 | 0.004102 | 7.60E-85 |
| rs174562 | miR-1908-! | cis | 4.92E-08 | Nightingale | L_HDL_PL_ | NA | G | A | 0.043203 | 0.004203 | 1.60E-26 |
| rs174562 | miR-1908-! | cis | 4.92E-08 | Nightingale | XS_VLDL_T | NA | G | A | 0.031635 | 0.004276 | 4.60E-13 |
| rs174562 | miR-1908-! | cis | 4.92E-08 | Nightingale | HDL_FC | NA | G | A | -0.07391 | 0.003847 | 4.80E-89 |
| rs174562 | miR-1908-! | cis | 4.92E-08 | Nightingale | M_HDL_FC | NA | G | A | -0.05136 | 0.003952 | 1.80E-41 |
| rs174562 | miR-1908-! | cis | 4.92E-08 | Nightingale | S_LDL_FC_ | NA | G | A | -0.08004 | 0.004209 | 1.20E-83 |
| rs174562 | miR-1908-! | cis | 4.92E-08 | Nightingale | Omega_6_ | NA | G | A | 0.37002 | 0.00428 | 0 |
| rs174562 | miR-1908-! | cis | 4.92E-08 | Nightingale | M_HDL_FC | NA | G | A | -0.10334 | 0.003848 | 5.11E-173 |
| rs174562 | miR-1908-! | cis | 4.92E-08 | Nightingale | LDL_C | NA | G | A | -0.04972 | 0.004324 | 3.60E-32 |
| rs174562 | miR-1908-! | cis | 4.92E-08 | Nightingale | XL_VLDL_P | NA | G | A | 0.048497 | 0.004191 | 3.80E-32 |
| rs174562 | miR-1908-! | cis | 4.92E-08 | Nightingale | LDL_TG | NA | G | A | 0.029716 | 0.004295 | 8.70E-12 |
| rs174562 | miR-1908-! | cis | 4.92E-08 | Nightingale | M_HDL_C_ | NA | G | A | -0.06602 | 0.004134 | 1.60E-60 |
| rs174562 | miR-1908-! | cis | 4.92E-08 | Nightingale | IDL_CE_pct | NA | G | A | -0.03789 | 0.004262 | 6.80E-19 |
| rs174564 | miR-1908-! | cis | 1.61E-08 | Nightingale | Total_FA | NA | G | A | -0.02379 | 0.004294 | 9.50E-09 |
| rs174564 | miR-1908-! | cis | 1.61E-08 | Nightingale | L_LDL_L | NA | G | A | -0.05421 | 0.004286 | 7.20E-39 |
| rs174564 | miR-1908-! | cis | 1.61E-08 | Nightingale | XXL_VLDL_ | NA | G | A | 0.066609 | 0.004168 | 4.70E-60 |
| rs174564 | miR-1908-! | cis | 1.61E-08 | Nightingale | L_LDL_PL | NA | G | A | -0.05505 | 0.004295 | 1.60E-39 |
| rs174564 | miR-1908-! | cis | 1.61E-08 | Nightingale | XL_VLDL_L | NA | G | A | 0.051436 | 0.004164 | 2.00E-36 |
| rs174564 | miR-1908-! | cis | 1.61E-08 | Nightingale | S_LDL_P | NA | G | A | -0.03257 | 0.004332 | 9.50E-15 |
| rs174564 | miR-1908-! | cis | 1.61E-08 | Nightingale | XL_VLDL_T | NA | G | A | 0.056831 | 0.00414 | 1.40E-44 |
| rs174564 | miR-1908-! | cis | 1.61E-08 | Nightingale | L_HDL_FC_ | NA | G | A | -0.11275 | 0.004021 | 7.80E-188 |
| rs174564 | miR-1908-! | cis | 1.61E-08 | Nightingale | DHA | NA | G | A | -0.28971 | 0.004171 | 0 |
| rs174564 | miR-1908-! | cis | 1.61E-08 | Nightingale | L_VLDL_FC | NA | G | A | 0.028662 | 0.004287 | 1.40E-11 |
| rs174564 | miR-1908-! | cis | 1.61E-08 | Nightingale | MUFA | NA | G | A | 0.064379 | 0.004284 | 3.30E-52 |
| rs174564 | miR-1908-! | cis | 1.61E-08 | Nightingale | LA | NA | G | A | 0.084276 | 0.004244 | 5.10E-88 |
| rs174564 | miR-1908-! | cis | 1.61E-08 | Nightingale | M_HDL_TG | NA | G | A | 0.050625 | 0.004274 | 2.60E-33 |
| rs174564 | miR-1908-! | cis | 1.61E-08 | Nightingale | XL_HDL_FC | NA | G | A | 0.060882 | 0.003837 | 2.30E-62 |
| rs174564 | miR-1908-! | cis | 1.61E-08 | Nightingale | XL_HDL_CE | NA | G | A | -0.02609 | 0.004325 | 3.20E-09 |
| rs174564 | miR-1908-! | cis | 1.61E-08 | Nightingale | non_HDL_( | NA | G | A | -0.04789 | 0.004305 | 2.50E-30 |
| rs174564 | miR-1908-! | cis | 1.61E-08 | Nightingale | M_VLDL_C | NA | G | A | -0.04415 | 0.004316 | 1.00E-25 |
| rs174564 | miR-1908-! | cis | 1.61E-08 | Nightingale | LDL_C | NA | G | A | -0.04979 | 0.004309 | 1.50E-32 |
| rs174564 | miR-1908-! | cis | 1.61E-08 | Nightingale | VLDL_size | NA | G | A | 0.072538 | 0.004034 | 1.60E-76 |
| rs174564 | miR-1908-! | cis | 1.61E-08 | Nightingale | S_LDL_CE_ | NA | G | A | 0.057447 | 0.00432 | 2.10E-42 |
| rs174564 | miR-1908-! | cis | 1.61E-08 | Nightingale | XL_VLDL_F | NA | G | A | 0.04345 | 0.004192 | 4.20E-26 |
| rs174564 | miR-1908-! | cis | 1.61E-08 | Nightingale | IDL_C_pct | NA | G | A | -0.07502 | 0.004225 | 1.30E-72 |
| rs174564 | miR-1908-! | cis | 1.61E-08 | Nightingale | XS_VLDL_P | NA | G | A | -0.0346 | 0.004267 | 1.70E-17 |
| rs174564 | miR-1908-! | cis | 1.61E-08 | Nightingale | M_HDL_CE | NA | G | A | -0.04691 | 0.004197 | 6.50E-30 |
| rs174564 | miR-1908-! | cis | 1.61E-08 | Nightingale | ApoA1 | NA | G | A | -0.04095 | 0.003961 | 6.50E-27 |
| rs174564 | miR-1908-! | cis | 1.61E-08 | Nightingale | HDL_CE | NA | G | A | -0.06131 | 0.003884 | 1.90E-60 |
| rs174564 | miR-1908-! | cis | 1.61E-08 | Nightingale | L_HDL_PL | NA | G | A | -0.07607 | 0.003809 | 2.70E-96 |
| rs174564 | miR-1908-! | cis | 1.61E-08 | Nightingale | M_LDL_L | NA | G | A | -0.02816 | 0.004335 | 2.70E-11 |
| rs174564 | miR-1908-! | cis | 1.61E-08 | Nightingale | DHA_pct | NA | G | A | -0.2751 | 0.004201 | 0 |
| rs174564 | miR-1908-! | cis | 1.61E-08 | Nightingale | XXL_VLDL_ | NA | G | A | 0.06801 | 0.004169 | 8.00E-63 |
| rs174564 | miR-1908-! | cis | 1.61E-08 | Nightingale | PUFA | NA | G | A | -0.09678 | 0.004221 | 2.90E-123 |
| rs174564 | miR-1908-! | cis | 1.61E-08 | Nightingale | L_VLDL_CE | NA | G | A | -0.05914 | 0.004142 | 3.80E-49 |
| rs174564 | miR-1908-! | cis | 1.61E-08 | Nightingale | S_LDL_TG | NA | G | A | 0.05055 | 0.004222 | 1.10E-33 |
| rs174564 | miR-1908-! | cis | 1.61E-08 | Nightingale | XS_VLDL_T | NA | G | A | 0.097594 | 0.004111 | 9.71E-131 |
| rs174564 | miR-1908-! | cis | 1.61E-08 | Nightingale | L_LDL_FC | NA | G | A | -0.06841 | 0.00425 | 7.40E-62 |
| rs174564 | miR-1908-! | cis | 1.61E-08 | Nightingale | M_LDL_C | NA | G | A | -0.03407 | 0.004336 | 8.20E-16 |
| rs174564 | miR-1908-! | cis | 1.61E-08 | Nightingale | XXL_VLDL_ | NA | G | A | -0.0266 | 0.004306 | 1.40E-10 |
| rs174564 | miR-1908-! | cis | 1.61E-08 | Nightingale | PUFA_pct | NA | G | A | -0.12214 | 0.004163 | 4.60E-191 |
| rs174564 | miR-1908-! | cis | 1.61E-08 | Nightingale | S_VLDL_TG | NA | G | A | 0.038923 | 0.0042 | 4.00E-21 |
| rs174564 | miR-1908-! | cis | 1.61E-08 | Nightingale | VLDL_L | NA | G | A | 0.027358 | 0.004243 | 6.50E-11 |
| rs174564 | miR-1908-! | cis | 1.61E-08 | Nightingale | XXL_VLDL_ | NA | G | A | 0.061291 | 0.004176 | 6.80E-51 |
| rs174564 | miR-1908-! | cis | 1.61E-08 | Nightingale | XS_VLDL_C | NA | G | A | -0.09826 | 0.004101 | 8.79E-133 |
| rs174564 | miR-1908-! | cis | 1.61E-08 | Nightingale | L_LDL_P | NA | G | A | -0.03738 | 0.004313 | 2.90E-19 |
| rs174564 | miR-1908-! | cis | 1.61E-08 | Nightingale | HDL_C | NA | G | A | -0.06461 | 0.003865 | 9.20E-68 |
| rs174564 | miR-1908-! | cis | 1.61E-08 | Nightingale | XS_VLDL_F | NA | G | A | -0.11319 | 0.004295 | 1.90E-156 |
| rs174564 | miR-1908-! | cis | 1.61E-08 | Nightingale | S_HDL_TG_ | NA | G | A | 0.069142 | 0.004116 | 2.10E-66 |

| | | | | | | | | | | | |
|---|---|---|---|---|---|---|---|---|---|---|---|
| rs174564 | miR-1908-5 | cis | 1.61E-08 | Nightingale | L_VLDL_TG | NA | G | A | 0.037307 | 0.004164 | 7.00E-20 |
| rs174564 | miR-1908-5 | cis | 1.61E-08 | Nightingale | M_LDL_TG | NA | G | A | 0.036746 | 0.004264 | 6.00E-18 |
| rs174564 | miR-1908-5 | cis | 1.61E-08 | Nightingale | L_VLDL_PL | NA | G | A | 0.087913 | 0.0042 | 5.90E-101 |
| rs174564 | miR-1908-5 | cis | 1.61E-08 | Nightingale | XL_VLDL_F | NA | G | A | -0.06807 | 0.004193 | 4.10E-62 |
| rs174564 | miR-1908-5 | cis | 1.61E-08 | Nightingale | Citrate | NA | G | A | -0.03267 | 0.0043 | 3.10E-15 |
| rs174564 | miR-1908-5 | cis | 1.61E-08 | Nightingale | S_VLDL_C_ | NA | G | A | -0.05661 | 0.004211 | 1.90E-42 |
| rs174564 | miR-1908-5 | cis | 1.61E-08 | Nightingale | TG_by_PG | NA | G | A | 0.068765 | 0.004016 | 3.40E-69 |
| rs174564 | miR-1908-5 | cis | 1.61E-08 | Nightingale | Omega_3_ | NA | G | A | -0.39151 | 0.004229 | 0 |
| rs174564 | miR-1908-5 | cis | 1.61E-08 | Nightingale | L_HDL_TG_ | NA | G | A | 0.078974 | 0.004084 | 1.70E-88 |
| rs174564 | miR-1908-5 | cis | 1.61E-08 | Nightingale | HDL_PL | NA | G | A | -0.04828 | 0.003913 | 2.10E-37 |
| rs174564 | miR-1908-5 | cis | 1.61E-08 | Nightingale | XL_VLDL_C | NA | G | A | 0.037466 | 0.004227 | 1.70E-19 |
| rs174564 | miR-1908-5 | cis | 1.61E-08 | Nightingale | Tyr | NA | G | A | 0.032334 | 0.004322 | 7.40E-14 |
| rs174564 | miR-1908-5 | cis | 1.61E-08 | Nightingale | M_HDL_L | NA | G | A | -0.02743 | 0.004018 | 1.30E-12 |
| rs174564 | miR-1908-5 | cis | 1.61E-08 | Nightingale | Total_C | NA | G | A | -0.06524 | 0.00419 | 1.00E-57 |
| rs174564 | miR-1908-5 | cis | 1.61E-08 | Nightingale | M_HDL_TG | NA | G | A | 0.066806 | 0.00415 | 3.80E-61 |
| rs174564 | miR-1908-5 | cis | 1.61E-08 | Nightingale | XS_VLDL_F | NA | G | A | -0.05639 | 0.004252 | 5.80E-43 |
| rs174564 | miR-1908-5 | cis | 1.61E-08 | Nightingale | IDL_FC | NA | G | A | -0.09004 | 0.004189 | 2.20E-108 |
| rs174564 | miR-1908-5 | cis | 1.61E-08 | Nightingale | XL_VLDL_T | NA | G | A | 0.059545 | 0.004193 | 2.40E-47 |
| rs174564 | miR-1908-5 | cis | 1.61E-08 | Nightingale | XL_HDL_PL | NA | G | A | -0.08913 | 0.00389 | 6.10E-126 |
| rs174564 | miR-1908-5 | cis | 1.61E-08 | Nightingale | Total_P | NA | G | A | -0.0401 | 0.004073 | 2.10E-24 |
| rs174564 | miR-1908-5 | cis | 1.61E-08 | Nightingale | XL_HDL_FC | NA | G | A | -0.10284 | 0.004032 | 1.60E-154 |
| rs174564 | miR-1908-5 | cis | 1.61E-08 | Nightingale | SFA | NA | G | A | -0.0316 | 0.004289 | 6.60E-14 |
| rs174564 | miR-1908-5 | cis | 1.61E-08 | Nightingale | S_LDL_C | NA | G | A | -0.03219 | 0.004333 | 1.80E-14 |
| rs174564 | miR-1908-5 | cis | 1.61E-08 | Nightingale | VLDL_TG | NA | G | A | 0.047733 | 0.004165 | 1.70E-31 |
| rs174564 | miR-1908-5 | cis | 1.61E-08 | Nightingale | XL_HDL_C_ | NA | G | A | 0.02379 | 0.004134 | 5.60E-10 |
| rs174564 | miR-1908-5 | cis | 1.61E-08 | Nightingale | M_LDL_FC | NA | G | A | -0.0525 | 0.004323 | 7.60E-36 |
| rs174564 | miR-1908-5 | cis | 1.61E-08 | Nightingale | S_LDL_PL | NA | G | A | -0.05155 | 0.004318 | 6.20E-35 |
| rs174564 | miR-1908-5 | cis | 1.61E-08 | Nightingale | L_VLDL_L | NA | G | A | 0.038223 | 0.004182 | 1.20E-20 |
| rs174564 | miR-1908-5 | cis | 1.61E-08 | Nightingale | L_HDL_C | NA | G | A | -0.08026 | 0.003811 | 6.81E-107 |
| rs174564 | miR-1908-5 | cis | 1.61E-08 | Nightingale | M_LDL_P | NA | G | A | -0.04681 | 0.004341 | 4.60E-28 |
| rs174564 | miR-1908-5 | cis | 1.61E-08 | Nightingale | L_VLDL_C | NA | G | A | 0.028909 | 0.004233 | 3.80E-12 |
| rs174564 | miR-1908-5 | cis | 1.61E-08 | Nightingale | M_VLDL_F | NA | G | A | -0.02951 | 0.004327 | 2.30E-12 |
| rs174564 | miR-1908-5 | cis | 1.61E-08 | Nightingale | S_LDL_PL_ | NA | G | A | -0.06206 | 0.004319 | 3.60E-49 |
| rs174564 | miR-1908-5 | cis | 1.61E-08 | Nightingale | XL_HDL_C | NA | G | A | -0.0993 | 0.003902 | 1.10E-154 |
| rs174564 | miR-1908-5 | cis | 1.61E-08 | Nightingale | HDL_P | NA | G | A | -0.03533 | 0.004064 | 2.80E-19 |
| rs174564 | miR-1908-5 | cis | 1.61E-08 | Nightingale | L_LDL_CE | NA | G | A | -0.05332 | 0.00429 | 1.40E-37 |
| rs174564 | miR-1908-5 | cis | 1.61E-08 | Nightingale | M_LDL_C_ | NA | G | A | -0.10051 | 0.004331 | 5.40E-120 |
| rs174564 | miR-1908-5 | cis | 1.61E-08 | Nightingale | M_VLDL_C | NA | G | A | -0.08077 | 0.004074 | 2.00E-91 |
| rs174564 | miR-1908-5 | cis | 1.61E-08 | Nightingale | XL_HDL_CE | NA | G | A | -0.09716 | 0.003875 | 2.00E-150 |
| rs174564 | miR-1908-5 | cis | 1.61E-08 | Nightingale | L_VLDL_PL | NA | G | A | 0.048809 | 0.004176 | 6.30E-33 |
| rs174564 | miR-1908-5 | cis | 1.61E-08 | Nightingale | Remnant_( | NA | G | A | -0.04428 | 0.0043 | 2.00E-26 |
| rs174564 | miR-1908-5 | cis | 1.61E-08 | Nightingale | XL_VLDL_C | NA | G | A | -0.061 | 0.004149 | 1.60E-50 |
| rs174564 | miR-1908-5 | cis | 1.61E-08 | Nightingale | XS_VLDL_P | NA | G | A | 0.058267 | 0.004223 | 2.70E-44 |
| rs174564 | miR-1908-5 | cis | 1.61E-08 | Nightingale | IDL_FC_pct | NA | G | A | -0.10445 | 0.004293 | 1.70E-132 |
| rs174564 | miR-1908-5 | cis | 1.61E-08 | Nightingale | M_HDL_PL | NA | G | A | 0.058191 | 0.004105 | 1.50E-47 |
| rs174564 | miR-1908-5 | cis | 1.61E-08 | Nightingale | M_LDL_FC_ | NA | G | A | -0.07183 | 0.004146 | 3.60E-70 |
| rs174564 | miR-1908-5 | cis | 1.61E-08 | Nightingale | LDL_FC | NA | G | A | -0.0644 | 0.004282 | 4.60E-54 |
| rs174564 | miR-1908-5 | cis | 1.61E-08 | Nightingale | Total_FC | NA | G | A | -0.05945 | 0.004228 | 1.40E-47 |
| rs174564 | miR-1908-5 | cis | 1.61E-08 | Nightingale | M_VLDL_F | NA | G | A | -0.09134 | 0.004115 | 4.10E-115 |
| rs174564 | miR-1908-5 | cis | 1.61E-08 | Nightingale | S_VLDL_CE | NA | G | A | -0.02891 | 0.00427 | 6.10E-12 |
| rs174564 | miR-1908-5 | cis | 1.61E-08 | Nightingale | LDL_CE | NA | G | A | -0.04359 | 0.004317 | 2.90E-25 |
| rs174564 | miR-1908-5 | cis | 1.61E-08 | Nightingale | LDL_PL | NA | G | A | -0.04621 | 0.004318 | 4.50E-28 |
| rs174564 | miR-1908-5 | cis | 1.61E-08 | Nightingale | XXL_VLDL_ | NA | G | A | 0.071751 | 0.004171 | 3.50E-69 |
| rs174564 | miR-1908-5 | cis | 1.61E-08 | Nightingale | S_VLDL_FC | NA | G | A | -0.08303 | 0.004133 | 5.20E-94 |
| rs174564 | miR-1908-5 | cis | 1.61E-08 | Nightingale | M_VLDL_C | NA | G | A | -0.05285 | 0.004289 | 1.20E-36 |
| rs174564 | miR-1908-5 | cis | 1.61E-08 | Nightingale | S_HDL_TG | NA | G | A | 0.069102 | 0.004197 | 9.90E-64 |
| rs174564 | miR-1908-5 | cis | 1.61E-08 | Nightingale | M_HDL_CE | NA | G | A | -0.0376 | 0.003997 | 2.40E-22 |
| rs174564 | miR-1908-5 | cis | 1.61E-08 | Nightingale | L_HDL_C_p | NA | G | A | -0.06836 | 0.004079 | 1.50E-67 |
| rs174564 | miR-1908-5 | cis | 1.61E-08 | Nightingale | S_VLDL_TG | NA | G | A | 0.064551 | 0.004188 | 2.10E-55 |
| rs174564 | miR-1908-5 | cis | 1.61E-08 | Nightingale | S_VLDL_PL | NA | G | A | -0.07869 | 0.004141 | 1.30E-84 |
| rs174564 | miR-1908-5 | cis | 1.61E-08 | Nightingale | LDL_P | NA | G | A | -0.04029 | 0.004325 | 8.90E-22 |
| rs174564 | miR-1908-5 | cis | 1.61E-08 | Nightingale | L_VLDL_P | NA | G | A | 0.041921 | 0.004179 | 1.40E-24 |
| rs174564 | miR-1908-5 | cis | 1.61E-08 | Nightingale | Cholines | NA | G | A | -0.03695 | 0.00407 | 2.40E-21 |

| | | | | | | | | | | | |
|---|---|---|---|---|---|---|---|---|---|---|---|
| rs174564 | miR-1908-! | cis | 1.61E-08 | Nightingale | XL_HDL_P | NA | G | A | -0.08953 | 0.003895 | 9.20E-128 |
| rs174564 | miR-1908-! | cis | 1.61E-08 | Nightingale | L_VLDL_C_ | NA | G | A | -0.0386 | 0.004228 | 3.30E-21 |
| rs174564 | miR-1908-! | cis | 1.61E-08 | Nightingale | Omega_6_ | NA | G | A | 0.02425 | 0.004212 | 5.30E-09 |
| rs174564 | miR-1908-! | cis | 1.61E-08 | Nightingale | S_LDL_TG_ | NA | G | A | 0.08636 | 0.004194 | 8.60E-98 |
| rs174564 | miR-1908-! | cis | 1.61E-08 | Nightingale | LDL_TG | NA | G | A | 0.029788 | 0.00428 | 5.80E-12 |
| rs174564 | miR-1908-! | cis | 1.61E-08 | Nightingale | S_LDL_L | NA | G | A | -0.03246 | 0.004327 | 9.10E-15 |
| rs174564 | miR-1908-! | cis | 1.61E-08 | Nightingale | L_HDL_P | NA | G | A | -0.07782 | 0.003791 | 6.50E-102 |
| rs174564 | miR-1908-! | cis | 1.61E-08 | Nightingale | L_HDL_PL_ | NA | G | A | 0.042867 | 0.004189 | 3.90E-26 |
| rs174564 | miR-1908-! | cis | 1.61E-08 | Nightingale | PUFA_by_I | NA | G | A | -0.17165 | 0.004145 | 0 |
| rs174564 | miR-1908-! | cis | 1.61E-08 | Nightingale | LDL_L | NA | G | A | -0.04547 | 0.004314 | 2.00E-27 |
| rs174564 | miR-1908-! | cis | 1.61E-08 | Nightingale | M_HDL_C_ | NA | G | A | -0.06584 | 0.00412 | 1.50E-60 |
| rs174564 | miR-1908-! | cis | 1.61E-08 | Nightingale | M_VLDL_T | NA | G | A | 0.078021 | 0.004087 | 3.10E-85 |
| rs174564 | miR-1908-! | cis | 1.61E-08 | Nightingale | HDL_TG | NA | G | A | 0.046498 | 0.004268 | 2.80E-28 |
| rs174564 | miR-1908-! | cis | 1.61E-08 | Nightingale | XS_VLDL_C | NA | G | A | -0.06851 | 0.0042 | 8.80E-64 |
| rs174564 | miR-1908-! | cis | 1.61E-08 | Nightingale | SFA_pct | NA | G | A | -0.04301 | 0.00427 | 7.30E-24 |
| rs174564 | miR-1908-! | cis | 1.61E-08 | Nightingale | Total_L | NA | G | A | -0.03732 | 0.004258 | 1.20E-19 |
| rs174564 | miR-1908-! | cis | 1.61E-08 | Nightingale | L_HDL_CE | NA | G | A | -0.07868 | 0.003821 | 4.10E-102 |
| rs174564 | miR-1908-! | cis | 1.61E-08 | Nightingale | XL_HDL_L | NA | G | A | -0.09307 | 0.003889 | 2.30E-137 |
| rs174564 | miR-1908-! | cis | 1.61E-08 | Nightingale | L_LDL_C | NA | G | A | -0.05785 | 0.004279 | 4.10E-44 |
| rs174564 | miR-1908-! | cis | 1.61E-08 | Nightingale | Sphingomy | NA | G | A | -0.11543 | 0.004044 | 2.00E-188 |
| rs174564 | miR-1908-! | cis | 1.61E-08 | Nightingale | M_VLDL_P | NA | G | A | -0.06249 | 0.004153 | 1.00E-54 |
| rs174564 | miR-1908-! | cis | 1.61E-08 | Nightingale | XL_HDL_PL | NA | G | A | -0.0671 | 0.003976 | 1.40E-68 |
| rs174564 | miR-1908-! | cis | 1.61E-08 | Nightingale | LA_pct | NA | G | A | 0.186242 | 0.004229 | 0 |
| rs174564 | miR-1908-! | cis | 1.61E-08 | Nightingale | M_HDL_C | NA | G | A | -0.04057 | 0.003982 | 6.30E-26 |
| rs174564 | miR-1908-! | cis | 1.61E-08 | Nightingale | XXL_VLDL_ | NA | G | A | 0.073652 | 0.004179 | 3.50E-72 |
| rs174564 | miR-1908-! | cis | 1.61E-08 | Nightingale | L_HDL_CE_ | NA | G | A | -0.04991 | 0.004141 | 1.30E-35 |
| rs174564 | miR-1908-! | cis | 1.61E-08 | Nightingale | S_LDL_FC_ | NA | G | A | -0.0802 | 0.004194 | 6.00E-85 |
| rs174564 | miR-1908-! | cis | 1.61E-08 | Nightingale | HDL_size | NA | G | A | -0.07269 | 0.003835 | 1.60E-86 |
| rs174564 | miR-1908-! | cis | 1.61E-08 | Nightingale | Omega_6_ | NA | G | A | 0.371404 | 0.004265 | 0 |
| rs174564 | miR-1908-! | cis | 1.61E-08 | Nightingale | XL_HDL_TC | NA | G | A | 0.087406 | 0.004077 | 8.30E-108 |
| rs174564 | miR-1908-! | cis | 1.61E-08 | Nightingale | IDL_PL | NA | G | A | -0.07303 | 0.004176 | 1.80E-73 |
| rs174564 | miR-1908-! | cis | 1.61E-08 | Nightingale | L_HDL_L | NA | G | A | -0.0772 | 0.003797 | 6.90E-100 |
| rs174564 | miR-1908-! | cis | 1.61E-08 | Nightingale | XXL_VLDL_ | NA | G | A | 0.07217 | 0.004153 | 7.60E-71 |
| rs174564 | miR-1908-! | cis | 1.61E-08 | Nightingale | XS_VLDL_C | NA | G | A | -0.09234 | 0.00408 | 7.69E-119 |
| rs174564 | miR-1908-! | cis | 1.61E-08 | Nightingale | S_HDL_C_p | NA | G | A | -0.04229 | 0.004278 | 4.80E-23 |
| rs174564 | miR-1908-! | cis | 1.61E-08 | Nightingale | XXL_VLDL_ | NA | G | A | 0.066134 | 0.004163 | 1.30E-59 |
| rs174564 | miR-1908-! | cis | 1.61E-08 | Nightingale | IDL_CE_pct | NA | G | A | -0.03816 | 0.004247 | 1.80E-19 |
| rs174564 | miR-1908-! | cis | 1.61E-08 | Nightingale | XL_VLDL_P | NA | G | A | 0.052879 | 0.00416 | 1.70E-38 |
| rs174564 | miR-1908-! | cis | 1.61E-08 | Nightingale | L_LDL_C_p | NA | G | A | -0.07709 | 0.004217 | 3.70E-77 |
| rs174564 | miR-1908-! | cis | 1.61E-08 | Nightingale | IDL_P | NA | G | A | -0.04545 | 0.004283 | 6.50E-28 |
| rs174564 | miR-1908-! | cis | 1.61E-08 | Nightingale | L_LDL_TG_ | NA | G | A | 0.077727 | 0.004244 | 5.80E-77 |
| rs174564 | miR-1908-! | cis | 1.61E-08 | Nightingale | HDL_FC | NA | G | A | -0.07382 | 0.003834 | 3.50E-89 |
| rs174564 | miR-1908-! | cis | 1.61E-08 | Nightingale | M_VLDL_C | NA | G | A | -0.07606 | 0.004063 | 1.30E-81 |
| rs174564 | miR-1908-! | cis | 1.61E-08 | Nightingale | XXL_VLDL_ | NA | G | A | 0.044329 | 0.004351 | 2.00E-25 |
| rs174564 | miR-1908-! | cis | 1.61E-08 | Nightingale | IDL_L | NA | G | A | -0.07261 | 0.004184 | 4.40E-72 |
| rs174564 | miR-1908-! | cis | 1.61E-08 | Nightingale | M_VLDL_T | NA | G | A | 0.031469 | 0.004205 | 3.00E-14 |
| rs174564 | miR-1908-! | cis | 1.61E-08 | Nightingale | S_HDL_FC_ | NA | G | A | -0.10275 | 0.004105 | 3.50E-153 |
| rs174564 | miR-1908-! | cis | 1.61E-08 | Nightingale | Total_PL | NA | G | A | -0.05106 | 0.004121 | 2.30E-37 |
| rs174564 | miR-1908-! | cis | 1.61E-08 | Nightingale | HDL_L | NA | G | A | -0.05319 | 0.003886 | 1.00E-45 |
| rs174564 | miR-1908-! | cis | 1.61E-08 | Nightingale | Phosphogly | NA | G | A | -0.0409 | 0.004099 | 4.30E-25 |
| rs174564 | miR-1908-! | cis | 1.61E-08 | Nightingale | IDL_C | NA | G | A | -0.07749 | 0.004179 | 1.50E-81 |
| rs174564 | miR-1908-! | cis | 1.61E-08 | Nightingale | XS_VLDL_L | NA | G | A | -0.04182 | 0.004254 | 8.60E-25 |
| rs174564 | miR-1908-! | cis | 1.61E-08 | Nightingale | XS_VLDL_C | NA | G | A | -0.07228 | 0.004176 | 1.60E-71 |
| rs174564 | miR-1908-! | cis | 1.61E-08 | Nightingale | IDL_CE | NA | G | A | -0.07245 | 0.004178 | 1.30E-71 |
| rs174564 | miR-1908-! | cis | 1.61E-08 | Nightingale | M_LDL_PL | NA | G | A | -0.02346 | 0.004335 | 2.40E-08 |
| rs174564 | miR-1908-! | cis | 1.61E-08 | Nightingale | M_HDL_FC | NA | G | A | -0.05132 | 0.003938 | 1.70E-41 |
| rs174564 | miR-1908-! | cis | 1.61E-08 | Nightingale | Total_CE | NA | G | A | -0.06704 | 0.004174 | 2.70E-61 |
| rs174564 | miR-1908-! | cis | 1.61E-08 | Nightingale | L_VLDL_FC | NA | G | A | 0.040208 | 0.004186 | 1.00E-22 |
| rs174564 | miR-1908-! | cis | 1.61E-08 | Nightingale | L_HDL_FC | NA | G | A | -0.08461 | 0.003793 | 1.20E-119 |
| rs174564 | miR-1908-! | cis | 1.61E-08 | Nightingale | M_LDL_PL_ | NA | G | A | 0.045939 | 0.004325 | 2.40E-27 |
| rs174564 | miR-1908-! | cis | 1.61E-08 | Nightingale | M_HDL_P | NA | G | A | -0.03404 | 0.003987 | 6.80E-19 |
| rs174564 | miR-1908-! | cis | 1.61E-08 | Nightingale | XL_VLDL_C | NA | G | A | -0.06402 | 0.004147 | 2.30E-55 |
| rs174564 | miR-1908-! | cis | 1.61E-08 | Nightingale | M_LDL_TG | NA | G | A | 0.075938 | 0.004273 | 1.80E-72 |

| | | | | | | | | | | | |
|---|---|---|---|---|---|---|---|---|---|---|---|
| rs174564 | miR-1908-5 | cis | 1.61E-08 | Nightingale | M_HDL_PL | NA | G | A | -0.02127 | 0.004047 | 4.70E-08 |
| rs174564 | miR-1908-5 | cis | 1.61E-08 | Nightingale | XL_VLDL_C | NA | G | A | 0.030318 | 0.00426 | 5.20E-13 |
| rs174564 | miR-1908-5 | cis | 1.61E-08 | Nightingale | XL_VLDL_P | NA | G | A | 0.048285 | 0.004176 | 4.10E-32 |
| rs174564 | miR-1908-5 | cis | 1.61E-08 | Nightingale | M_HDL_FC | NA | G | A | -0.10333 | 0.003835 | 3.50E-174 |
| rs174564 | miR-1908-5 | cis | 1.61E-08 | Nightingale | XS_VLDL_T | NA | G | A | 0.031653 | 0.004261 | 3.10E-13 |
| rs174564 | miR-1908-5 | cis | 1.61E-08 | Nightingale | IDL_TG_pc | NA | G | A | 0.088271 | 0.004181 | 3.60E-103 |
| rs174564 | miR-1908-5 | cis | 1.61E-08 | Nightingale | Clinical_LD | NA | G | A | -0.05528 | 0.004303 | 9.80E-40 |
| rs174564 | miR-1908-5 | cis | 1.61E-08 | Nightingale | S_LDL_FC | NA | G | A | -0.06304 | 0.004321 | 3.10E-51 |
| rs174564 | miR-1908-5 | cis | 1.61E-08 | Nightingale | S_VLDL_FC | NA | G | A | -0.02837 | 0.004329 | 1.50E-11 |
| rs174564 | miR-1908-5 | cis | 1.61E-08 | Nightingale | XS_VLDL_P | NA | G | A | -0.04389 | 0.00427 | 9.70E-27 |
| rs174564 | miR-1908-5 | cis | 1.61E-08 | Nightingale | M_LDL_CE | NA | G | A | -0.02633 | 0.004334 | 4.70E-10 |
| rs174564 | miR-1908-5 | cis | 1.61E-08 | Nightingale | Omega_3 | NA | G | A | -0.33709 | 0.004242 | 0 |
| rs174564 | miR-1908-5 | cis | 1.61E-08 | Nightingale | MUFA_pct | NA | G | A | 0.198596 | 0.004142 | 0 |
| rs174564 | miR-1908-5 | cis | 1.61E-08 | Nightingale | ApoB | NA | G | A | -0.03857 | 0.004322 | 3.50E-20 |
| rs174564 | miR-1908-5 | cis | 1.61E-08 | Nightingale | L_LDL_FC_ | NA | G | A | -0.08283 | 0.004125 | 4.10E-93 |
| rs174564 | miR-1908-5 | cis | 1.61E-08 | Nightingale | Unsaturati | NA | G | A | -0.34088 | 0.004099 | 0 |
| rs174564 | miR-1908-5 | cis | 1.61E-08 | Nightingale | Total_TG | NA | G | A | 0.046855 | 0.004189 | 4.20E-30 |
| rs174566 | miR-1908-5 | cis | 1.58E-08 | Nightingale | M_HDL_TG | NA | G | A | 0.066902 | 0.004148 | 2.40E-61 |
| rs174566 | miR-1908-5 | cis | 1.58E-08 | Nightingale | XL_VLDL_L | NA | G | A | 0.051145 | 0.004162 | 4.20E-36 |
| rs174566 | miR-1908-5 | cis | 1.58E-08 | Nightingale | L_VLDL_PL | NA | G | A | 0.087963 | 0.004198 | 4.60E-101 |
| rs174566 | miR-1908-5 | cis | 1.58E-08 | Nightingale | XS_VLDL_P | NA | G | A | -0.03424 | 0.004265 | 3.30E-17 |
| rs174566 | miR-1908-5 | cis | 1.58E-08 | Nightingale | ApoB | NA | G | A | -0.03847 | 0.00432 | 4.20E-20 |
| rs174566 | miR-1908-5 | cis | 1.58E-08 | Nightingale | HDL_C | NA | G | A | -0.06501 | 0.003862 | 1.40E-68 |
| rs174566 | miR-1908-5 | cis | 1.58E-08 | Nightingale | Total_FC | NA | G | A | -0.05942 | 0.004226 | 1.30E-47 |
| rs174566 | miR-1908-5 | cis | 1.58E-08 | Nightingale | L_HDL_P | NA | G | A | -0.07784 | 0.003788 | 5.70E-102 |
| rs174566 | miR-1908-5 | cis | 1.58E-08 | Nightingale | M_LDL_TG | NA | G | A | 0.075697 | 0.004271 | 4.10E-72 |
| rs174566 | miR-1908-5 | cis | 1.58E-08 | Nightingale | VLDL_size | NA | G | A | 0.071883 | 0.004032 | 2.80E-75 |
| rs174566 | miR-1908-5 | cis | 1.58E-08 | Nightingale | L_VLDL_CE | NA | G | A | -0.05882 | 0.004139 | 9.30E-49 |
| rs174566 | miR-1908-5 | cis | 1.58E-08 | Nightingale | XS_VLDL_C | NA | G | A | -0.09778 | 0.004098 | 1.30E-131 |
| rs174566 | miR-1908-5 | cis | 1.58E-08 | Nightingale | S_LDL_CE_ | NA | G | A | 0.056978 | 0.004317 | 8.50E-42 |
| rs174566 | miR-1908-5 | cis | 1.58E-08 | Nightingale | XS_VLDL_F | NA | G | A | -0.05598 | 0.00425 | 1.90E-42 |
| rs174566 | miR-1908-5 | cis | 1.58E-08 | Nightingale | M_VLDL_C | NA | G | A | -0.08032 | 0.004071 | 1.70E-90 |
| rs174566 | miR-1908-5 | cis | 1.58E-08 | Nightingale | XXL_VLDL_ | NA | G | A | 0.073364 | 0.004177 | 9.10E-72 |
| rs174566 | miR-1908-5 | cis | 1.58E-08 | Nightingale | LDL_PL | NA | G | A | -0.04628 | 0.004316 | 3.50E-28 |
| rs174566 | miR-1908-5 | cis | 1.58E-08 | Nightingale | XL_VLDL_P | NA | G | A | 0.04802 | 0.004174 | 7.50E-32 |
| rs174566 | miR-1908-5 | cis | 1.58E-08 | Nightingale | M_VLDL_T | NA | G | A | 0.077585 | 0.004085 | 2.20E-84 |
| rs174566 | miR-1908-5 | cis | 1.58E-08 | Nightingale | L_HDL_CE_ | NA | G | A | -0.04958 | 0.004139 | 3.60E-35 |
| rs174566 | miR-1908-5 | cis | 1.58E-08 | Nightingale | LDL_CE | NA | G | A | -0.04371 | 0.004314 | 2.10E-25 |
| rs174566 | miR-1908-5 | cis | 1.58E-08 | Nightingale | L_VLDL_FC | NA | G | A | 0.039888 | 0.004184 | 1.90E-22 |
| rs174566 | miR-1908-5 | cis | 1.58E-08 | Nightingale | IDL_C | NA | G | A | -0.07729 | 0.004177 | 3.10E-81 |
| rs174566 | miR-1908-5 | cis | 1.58E-08 | Nightingale | TG_by_PG | NA | G | A | 0.068629 | 0.004014 | 5.40E-69 |
| rs174566 | miR-1908-5 | cis | 1.58E-08 | Nightingale | M_VLDL_C | NA | G | A | -0.07561 | 0.004061 | 1.10E-80 |
| rs174566 | miR-1908-5 | cis | 1.58E-08 | Nightingale | M_LDL_CE | NA | G | A | -0.0266 | 0.004332 | 3.10E-10 |
| rs174566 | miR-1908-5 | cis | 1.58E-08 | Nightingale | XS_VLDL_C | NA | G | A | -0.07174 | 0.004174 | 1.20E-70 |
| rs174566 | miR-1908-5 | cis | 1.58E-08 | Nightingale | L_LDL_PL | NA | G | A | -0.05505 | 0.004293 | 1.50E-39 |
| rs174566 | miR-1908-5 | cis | 1.58E-08 | Nightingale | Cholines | NA | G | A | -0.03741 | 0.004068 | 8.00E-22 |
| rs174566 | miR-1908-5 | cis | 1.58E-08 | Nightingale | XXL_VLDL_ | NA | G | A | 0.065828 | 0.00416 | 3.40E-59 |
| rs174566 | miR-1908-5 | cis | 1.58E-08 | Nightingale | XS_VLDL_C | NA | G | A | -0.06801 | 0.004198 | 5.30E-63 |
| rs174566 | miR-1908-5 | cis | 1.58E-08 | Nightingale | XXL_VLDL_ | NA | G | A | 0.061123 | 0.004174 | 9.50E-51 |
| rs174566 | miR-1908-5 | cis | 1.58E-08 | Nightingale | XL_HDL_C | NA | G | A | -0.09871 | 0.0039 | 6.81E-153 |
| rs174566 | miR-1908-5 | cis | 1.58E-08 | Nightingale | XS_VLDL_L | NA | G | A | -0.04151 | 0.004252 | 1.70E-24 |
| rs174566 | miR-1908-5 | cis | 1.58E-08 | Nightingale | IDL_TG_pc | NA | G | A | 0.08794 | 0.004179 | 1.80E-102 |
| rs174566 | miR-1908-5 | cis | 1.58E-08 | Nightingale | LA | NA | G | A | 0.083772 | 0.004242 | 3.30E-87 |
| rs174566 | miR-1908-5 | cis | 1.58E-08 | Nightingale | XS_VLDL_T | NA | G | A | 0.096917 | 0.004109 | 4.30E-129 |
| rs174566 | miR-1908-5 | cis | 1.58E-08 | Nightingale | PUFA_pct | NA | G | A | -0.12138 | 0.004161 | 5.00E-189 |
| rs174566 | miR-1908-5 | cis | 1.58E-08 | Nightingale | S_LDL_P | NA | G | A | -0.03254 | 0.00433 | 9.90E-15 |
| rs174566 | miR-1908-5 | cis | 1.58E-08 | Nightingale | M_VLDL_T | NA | G | A | 0.031089 | 0.004202 | 5.30E-14 |
| rs174566 | miR-1908-5 | cis | 1.58E-08 | Nightingale | S_HDL_C_p | NA | G | A | -0.04239 | 0.004275 | 3.70E-23 |
| rs174566 | miR-1908-5 | cis | 1.58E-08 | Nightingale | M_HDL_CE | NA | G | A | -0.03833 | 0.003995 | 4.00E-23 |
| rs174566 | miR-1908-5 | cis | 1.58E-08 | Nightingale | XS_VLDL_T | NA | G | A | 0.031411 | 0.004259 | 4.30E-13 |
| rs174566 | miR-1908-5 | cis | 1.58E-08 | Nightingale | non_HDL_C | NA | G | A | -0.04787 | 0.004302 | 2.50E-30 |
| rs174566 | miR-1908-5 | cis | 1.58E-08 | Nightingale | XL_HDL_PL | NA | G | A | -0.06649 | 0.003974 | 1.80E-67 |
| rs174566 | miR-1908-5 | cis | 1.58E-08 | Nightingale | HDL_CE | NA | G | A | -0.06173 | 0.003882 | 3.10E-61 |

| | | | | | | | | | | | | |
|---|---|---|---|---|---|---|---|---|---|---|---|---|
| rs174566 | miR-1908-5 | cis | | 1.58E-08 | Nightingale | PUFA_by_M | NA | G | A | -0.17068 | 0.004143 | 0 |
| rs174566 | miR-1908-5 | cis | | 1.58E-08 | Nightingale | L_VLDL_C | NA | G | A | 0.028722 | 0.004231 | 4.80E-12 |
| rs174566 | miR-1908-5 | cis | | 1.58E-08 | Nightingale | S_LDL_TG_ | NA | G | A | 0.08608 | 0.004191 | 2.90E-97 |
| rs174566 | miR-1908-5 | cis | | 1.58E-08 | Nightingale | Remnant_( | NA | G | A | -0.04416 | 0.004298 | 2.40E-26 |
| rs174566 | miR-1908-5 | cis | | 1.58E-08 | Nightingale | XL_HDL_CE | NA | G | A | -0.09667 | 0.003873 | 6.70E-149 |
| rs174566 | miR-1908-5 | cis | | 1.58E-08 | Nightingale | IDL_PL | NA | G | A | -0.07267 | 0.004173 | 7.10E-73 |
| rs174566 | miR-1908-5 | cis | | 1.58E-08 | Nightingale | Total_FA | NA | G | A | -0.02423 | 0.004291 | 5.80E-09 |
| rs174566 | miR-1908-5 | cis | | 1.58E-08 | Nightingale | IDL_C_pct | NA | G | A | -0.07501 | 0.004223 | 1.20E-72 |
| rs174566 | miR-1908-5 | cis | | 1.58E-08 | Nightingale | IDL_CE_pct | NA | G | A | -0.03818 | 0.004244 | 1.60E-19 |
| rs174566 | miR-1908-5 | cis | | 1.58E-08 | Nightingale | M_HDL_TG | NA | G | A | 0.050221 | 0.004272 | 6.80E-33 |
| rs174566 | miR-1908-5 | cis | | 1.58E-08 | Nightingale | M_HDL_P | NA | G | A | -0.03477 | 0.003985 | 1.30E-19 |
| rs174566 | miR-1908-5 | cis | | 1.58E-08 | Nightingale | M_HDL_PL | NA | G | A | 0.058369 | 0.004103 | 7.50E-48 |
| rs174566 | miR-1908-5 | cis | | 1.58E-08 | Nightingale | M_LDL_C_ | NA | G | A | -0.10032 | 0.004329 | 8.39E-120 |
| rs174566 | miR-1908-5 | cis | | 1.58E-08 | Nightingale | L_LDL_C | NA | G | A | -0.05784 | 0.004277 | 3.90E-44 |
| rs174566 | miR-1908-5 | cis | | 1.58E-08 | Nightingale | Omega_6_ | NA | G | A | 0.024545 | 0.00421 | 3.60E-09 |
| rs174566 | miR-1908-5 | cis | | 1.58E-08 | Nightingale | XL_VLDL_T | NA | G | A | 0.056534 | 0.004137 | 3.20E-44 |
| rs174566 | miR-1908-5 | cis | | 1.58E-08 | Nightingale | L_LDL_L | NA | G | A | -0.05423 | 0.004283 | 6.60E-39 |
| rs174566 | miR-1908-5 | cis | | 1.58E-08 | Nightingale | XL_VLDL_P | NA | G | A | 0.052536 | 0.004157 | 4.40E-38 |
| rs174566 | miR-1908-5 | cis | | 1.58E-08 | Nightingale | LDL_C | NA | G | A | -0.04986 | 0.004307 | 1.20E-32 |
| rs174566 | miR-1908-5 | cis | | 1.58E-08 | Nightingale | S_LDL_PL_ | NA | G | A | -0.06145 | 0.004317 | 2.60E-48 |
| rs174566 | miR-1908-5 | cis | | 1.58E-08 | Nightingale | Sphingomy | NA | G | A | -0.11535 | 0.004042 | 2.10E-188 |
| rs174566 | miR-1908-5 | cis | | 1.58E-08 | Nightingale | M_LDL_TG | NA | G | A | 0.036249 | 0.004262 | 1.40E-17 |
| rs174566 | miR-1908-5 | cis | | 1.58E-08 | Nightingale | L_VLDL_FC | NA | G | A | 0.028632 | 0.004285 | 1.70E-11 |
| rs174566 | miR-1908-5 | cis | | 1.58E-08 | Nightingale | M_HDL_C | NA | G | A | -0.04129 | 0.00398 | 9.20E-27 |
| rs174566 | miR-1908-5 | cis | | 1.58E-08 | Nightingale | Phosphogly | NA | G | A | -0.04139 | 0.004097 | 1.30E-25 |
| rs174566 | miR-1908-5 | cis | | 1.58E-08 | Nightingale | L_LDL_C_p | NA | G | A | -0.07685 | 0.004214 | 9.80E-77 |
| rs174566 | miR-1908-5 | cis | | 1.58E-08 | Nightingale | LDL_TG | NA | G | A | 0.029363 | 0.004277 | 9.90E-12 |
| rs174566 | miR-1908-5 | cis | | 1.58E-08 | Nightingale | HDL_FC | NA | G | A | -0.07412 | 0.003832 | 5.50E-90 |
| rs174566 | miR-1908-5 | cis | | 1.58E-08 | Nightingale | L_HDL_CE | NA | G | A | -0.07863 | 0.003819 | 5.90E-102 |
| rs174566 | miR-1908-5 | cis | | 1.58E-08 | Nightingale | L_VLDL_P | NA | G | A | 0.041602 | 0.004176 | 2.70E-24 |
| rs174566 | miR-1908-5 | cis | | 1.58E-08 | Nightingale | XL_HDL_TG | NA | G | A | 0.086896 | 0.004075 | 1.10E-106 |
| rs174566 | miR-1908-5 | cis | | 1.58E-08 | Nightingale | M_HDL_CE | NA | G | A | -0.04702 | 0.004195 | 4.60E-30 |
| rs174566 | miR-1908-5 | cis | | 1.58E-08 | Nightingale | L_HDL_PL | NA | G | A | -0.0762 | 0.003807 | 1.30E-96 |
| rs174566 | miR-1908-5 | cis | | 1.58E-08 | Nightingale | S_VLDL_PL | NA | G | A | -0.07841 | 0.004139 | 4.50E-84 |
| rs174566 | miR-1908-5 | cis | | 1.58E-08 | Nightingale | L_LDL_FC_ | NA | G | A | -0.08234 | 0.004123 | 3.80E-92 |
| rs174566 | miR-1908-5 | cis | | 1.58E-08 | Nightingale | M_LDL_PL | NA | G | A | -0.0237 | 0.004332 | 1.70E-08 |
| rs174566 | miR-1908-5 | cis | | 1.58E-08 | Nightingale | L_LDL_FC | NA | G | A | -0.06829 | 0.004247 | 1.00E-61 |
| rs174566 | miR-1908-5 | cis | | 1.58E-08 | Nightingale | L_HDL_C_p | NA | G | A | -0.06792 | 0.004077 | 1.10E-66 |
| rs174566 | miR-1908-5 | cis | | 1.58E-08 | Nightingale | L_VLDL_C_ | NA | G | A | -0.0384 | 0.004225 | 4.20E-21 |
| rs174566 | miR-1908-5 | cis | | 1.58E-08 | Nightingale | XL_VLDL_C | NA | G | A | -0.06374 | 0.004145 | 6.20E-55 |
| rs174566 | miR-1908-5 | cis | | 1.58E-08 | Nightingale | S_HDL_FC_ | NA | G | A | -0.1023 | 0.004103 | 6.40E-152 |
| rs174566 | miR-1908-5 | cis | | 1.58E-08 | Nightingale | XS_VLDL_P | NA | G | A | -0.04357 | 0.004268 | 2.00E-26 |
| rs174566 | miR-1908-5 | cis | | 1.58E-08 | Nightingale | Total_PL | NA | G | A | -0.05134 | 0.004119 | 9.50E-38 |
| rs174566 | miR-1908-5 | cis | | 1.58E-08 | Nightingale | Total_C | NA | G | A | -0.06528 | 0.004187 | 7.80E-58 |
| rs174566 | miR-1908-5 | cis | | 1.58E-08 | Nightingale | S_VLDL_C_ | NA | G | A | -0.0562 | 0.004209 | 6.10E-42 |
| rs174566 | miR-1908-5 | cis | | 1.58E-08 | Nightingale | SFA | NA | G | A | -0.03199 | 0.004286 | 3.70E-14 |
| rs174566 | miR-1908-5 | cis | | 1.58E-08 | Nightingale | IDL_P | NA | G | A | -0.04524 | 0.00428 | 9.80E-28 |
| rs174566 | miR-1908-5 | cis | | 1.58E-08 | Nightingale | XL_VLDL_C | NA | G | A | 0.037263 | 0.004225 | 2.40E-19 |
| rs174566 | miR-1908-5 | cis | | 1.58E-08 | Nightingale | XL_VLDL_F | NA | G | A | 0.043174 | 0.00419 | 7.60E-26 |
| rs174566 | miR-1908-5 | cis | | 1.58E-08 | Nightingale | HDL_size | NA | G | A | -0.07256 | 0.003833 | 3.40E-86 |
| rs174566 | miR-1908-5 | cis | | 1.58E-08 | Nightingale | S_HDL_TG | NA | G | A | 0.068688 | 0.004194 | 4.50E-63 |
| rs174566 | miR-1908-5 | cis | | 1.58E-08 | Nightingale | M_HDL_PL | NA | G | A | -0.02208 | 0.004045 | 1.50E-08 |
| rs174566 | miR-1908-5 | cis | | 1.58E-08 | Nightingale | S_HDL_TG_ | NA | G | A | 0.06918 | 0.004114 | 1.50E-66 |
| rs174566 | miR-1908-5 | cis | | 1.58E-08 | Nightingale | L_VLDL_L | NA | G | A | 0.037913 | 0.00418 | 2.20E-20 |
| rs174566 | miR-1908-5 | cis | | 1.58E-08 | Nightingale | L_HDL_C | NA | G | A | -0.0802 | 0.003809 | 1.10E-106 |
| rs174566 | miR-1908-5 | cis | | 1.58E-08 | Nightingale | XL_HDL_P | NA | G | A | -0.08906 | 0.003893 | 1.60E-126 |
| rs174566 | miR-1908-5 | cis | | 1.58E-08 | Nightingale | L_LDL_CE | NA | G | A | -0.05336 | 0.004287 | 1.20E-37 |
| rs174566 | miR-1908-5 | cis | | 1.58E-08 | Nightingale | XS_VLDL_F | NA | G | A | -0.11226 | 0.004293 | 3.40E-154 |
| rs174566 | miR-1908-5 | cis | | 1.58E-08 | Nightingale | M_HDL_FC | NA | G | A | -0.10348 | 0.003833 | 8.00E-175 |
| rs174566 | miR-1908-5 | cis | | 1.58E-08 | Nightingale | S_LDL_L | NA | G | A | -0.03258 | 0.004325 | 7.00E-15 |
| rs174566 | miR-1908-5 | cis | | 1.58E-08 | Nightingale | Omega_3_ | NA | G | A | -0.39026 | 0.004226 | 0 |
| rs174566 | miR-1908-5 | cis | | 1.58E-08 | Nightingale | LA_pct | NA | G | A | 0.185991 | 0.004227 | 0 |
| rs174566 | miR-1908-5 | cis | | 1.58E-08 | Nightingale | S_LDL_TG | NA | G | A | 0.050153 | 0.00422 | 2.60E-33 |

| | | | | | | | | | | | |
|---|---|---|---|---|---|---|---|---|---|---|---|
| rs174566 | miR-1908-5 | cis | 1.58E-08 | Nightingale | XXL_VLDL_ | NA | G | A | 0.067657 | 0.004167 | 2.60E-62 |
| rs174566 | miR-1908-5 | cis | 1.58E-08 | Nightingale | S_LDL_FC | NA | G | A | -0.06299 | 0.004318 | 3.10E-51 |
| rs174566 | miR-1908-5 | cis | 1.58E-08 | Nightingale | M_HDL_C_ | NA | G | A | -0.06599 | 0.004118 | 7.60E-61 |
| rs174566 | miR-1908-5 | cis | 1.58E-08 | Nightingale | M_LDL_FC | NA | G | A | -0.05256 | 0.004321 | 5.70E-36 |
| rs174566 | miR-1908-5 | cis | 1.58E-08 | Nightingale | L_VLDL_TG | NA | G | A | 0.036956 | 0.004162 | 1.40E-19 |
| rs174566 | miR-1908-5 | cis | 1.58E-08 | Nightingale | S_VLDL_FC | NA | G | A | -0.02829 | 0.004326 | 1.70E-11 |
| rs174566 | miR-1908-5 | cis | 1.58E-08 | Nightingale | S_VLDL_TG | NA | G | A | 0.038614 | 0.004197 | 7.00E-21 |
| rs174566 | miR-1908-5 | cis | 1.58E-08 | Nightingale | Total_P | NA | G | A | -0.0408 | 0.004071 | 3.20E-25 |
| rs174566 | miR-1908-5 | cis | 1.58E-08 | Nightingale | ApoA1 | NA | G | A | -0.04159 | 0.003958 | 1.00E-27 |
| rs174566 | miR-1908-5 | cis | 1.58E-08 | Nightingale | HDL_TG | NA | G | A | 0.046149 | 0.004265 | 5.70E-28 |
| rs174566 | miR-1908-5 | cis | 1.58E-08 | Nightingale | S_LDL_PL | NA | G | A | -0.05152 | 0.004315 | 5.70E-35 |
| rs174566 | miR-1908-5 | cis | 1.58E-08 | Nightingale | Clinical_LD | NA | G | A | -0.05524 | 0.004301 | 1.00E-39 |
| rs174566 | miR-1908-5 | cis | 1.58E-08 | Nightingale | XXL_VLDL_ | NA | G | A | 0.071876 | 0.004151 | 1.90E-70 |
| rs174566 | miR-1908-5 | cis | 1.58E-08 | Nightingale | Citrate | NA | G | A | -0.03297 | 0.004298 | 2.00E-15 |
| rs174566 | miR-1908-5 | cis | 1.58E-08 | Nightingale | XL_HDL_C_ | NA | G | A | 0.023455 | 0.004132 | 8.70E-10 |
| rs174566 | miR-1908-5 | cis | 1.58E-08 | Nightingale | LDL_FC | NA | G | A | -0.06433 | 0.004279 | 5.10E-54 |
| rs174566 | miR-1908-5 | cis | 1.58E-08 | Nightingale | MUFA_pct | NA | G | A | 0.197502 | 0.004139 | 0 |
| rs174566 | miR-1908-5 | cis | 1.58E-08 | Nightingale | Total_TG | NA | G | A | 0.046503 | 0.004187 | 9.30E-30 |
| rs174566 | miR-1908-5 | cis | 1.58E-08 | Nightingale | HDL_P | NA | G | A | -0.03611 | 0.004061 | 4.50E-20 |
| rs174566 | miR-1908-5 | cis | 1.58E-08 | Nightingale | M_LDL_P | NA | G | A | -0.04685 | 0.004338 | 3.80E-28 |
| rs174566 | miR-1908-5 | cis | 1.58E-08 | Nightingale | Total_L | NA | G | A | -0.03758 | 0.004255 | 7.00E-20 |
| rs174566 | miR-1908-5 | cis | 1.58E-08 | Nightingale | XL_HDL_L | NA | G | A | -0.09251 | 0.003887 | 8.30E-136 |
| rs174566 | miR-1908-5 | cis | 1.58E-08 | Nightingale | S_VLDL_FC | NA | G | A | -0.08269 | 0.00413 | 2.50E-93 |
| rs174566 | miR-1908-5 | cis | 1.58E-08 | Nightingale | XL_HDL_PL | NA | G | A | -0.08856 | 0.003888 | 2.00E-124 |
| rs174566 | miR-1908-5 | cis | 1.58E-08 | Nightingale | M_VLDL_F | NA | G | A | -0.02945 | 0.004325 | 2.50E-12 |
| rs174566 | miR-1908-5 | cis | 1.58E-08 | Nightingale | L_LDL_P | NA | G | A | -0.03724 | 0.004311 | 3.80E-19 |
| rs174566 | miR-1908-5 | cis | 1.58E-08 | Nightingale | XS_VLDL_C | NA | G | A | -0.09192 | 0.004077 | 7.29E-118 |
| rs174566 | miR-1908-5 | cis | 1.58E-08 | Nightingale | XXL_VLDL_ | NA | G | A | 0.066388 | 0.004166 | 8.40E-60 |
| rs174566 | miR-1908-5 | cis | 1.58E-08 | Nightingale | S_VLDL_CE | NA | G | A | -0.02843 | 0.004267 | 1.20E-11 |
| rs174566 | miR-1908-5 | cis | 1.58E-08 | Nightingale | PUFA | NA | G | A | -0.09687 | 0.004218 | 2.00E-123 |
| rs174566 | miR-1908-5 | cis | 1.58E-08 | Nightingale | L_HDL_PL_ | NA | G | A | 0.041998 | 0.004186 | 3.40E-25 |
| rs174566 | miR-1908-5 | cis | 1.58E-08 | Nightingale | IDL_L | NA | G | A | -0.07239 | 0.004182 | 9.50E-72 |
| rs174566 | miR-1908-5 | cis | 1.58E-08 | Nightingale | M_LDL_PL_ | NA | G | A | 0.046183 | 0.004323 | 1.00E-27 |
| rs174566 | miR-1908-5 | cis | 1.58E-08 | Nightingale | M_VLDL_F | NA | G | A | -0.09086 | 0.004113 | 4.30E-114 |
| rs174566 | miR-1908-5 | cis | 1.58E-08 | Nightingale | DHA | NA | G | A | -0.2891 | 0.004168 | 0 |
| rs174566 | miR-1908-5 | cis | 1.58E-08 | Nightingale | M_VLDL_C | NA | G | A | -0.0439 | 0.004314 | 1.70E-25 |
| rs174566 | miR-1908-5 | cis | 1.58E-08 | Nightingale | L_HDL_L | NA | G | A | -0.07725 | 0.003795 | 5.20E-100 |
| rs174566 | miR-1908-5 | cis | 1.58E-08 | Nightingale | XL_HDL_FC | NA | G | A | 0.060846 | 0.003835 | 2.40E-62 |
| rs174566 | miR-1908-5 | cis | 1.58E-08 | Nightingale | S_LDL_FC_ | NA | G | A | -0.07994 | 0.004192 | 1.80E-84 |
| rs174566 | miR-1908-5 | cis | 1.58E-08 | Nightingale | IDL_FC | NA | G | A | -0.08978 | 0.004187 | 6.70E-108 |
| rs174566 | miR-1908-5 | cis | 1.58E-08 | Nightingale | XL_HDL_FC | NA | G | A | -0.10192 | 0.004029 | 6.30E-152 |
| rs174566 | miR-1908-5 | cis | 1.58E-08 | Nightingale | XL_HDL_CE | NA | G | A | -0.02643 | 0.004323 | 2.00E-09 |
| rs174566 | miR-1908-5 | cis | 1.58E-08 | Nightingale | L_HDL_FC_ | NA | G | A | -0.11193 | 0.004018 | 2.70E-185 |
| rs174566 | miR-1908-5 | cis | 1.58E-08 | Nightingale | M_HDL_L | NA | G | A | -0.02821 | 0.004016 | 3.10E-13 |
| rs174566 | miR-1908-5 | cis | 1.58E-08 | Nightingale | XL_VLDL_C | NA | G | A | 0.030195 | 0.004257 | 6.00E-13 |
| rs174566 | miR-1908-5 | cis | 1.58E-08 | Nightingale | XS_VLDL_P | NA | G | A | 0.058669 | 0.00422 | 5.20E-45 |
| rs174566 | miR-1908-5 | cis | 1.58E-08 | Nightingale | M_HDL_FC | NA | G | A | -0.05194 | 0.003936 | 1.90E-42 |
| rs174566 | miR-1908-5 | cis | 1.58E-08 | Nightingale | DHA_pct | NA | G | A | -0.27421 | 0.004199 | 0 |
| rs174566 | miR-1908-5 | cis | 1.58E-08 | Nightingale | XL_VLDL_T | NA | G | A | 0.05932 | 0.00419 | 4.50E-47 |
| rs174566 | miR-1908-5 | cis | 1.58E-08 | Nightingale | S_VLDL_TG | NA | G | A | 0.064183 | 0.004185 | 7.20E-55 |
| rs174566 | miR-1908-5 | cis | 1.58E-08 | Nightingale | Tyr | NA | G | A | 0.032762 | 0.00432 | 3.40E-14 |
| rs174566 | miR-1908-5 | cis | 1.58E-08 | Nightingale | LDL_L | NA | G | A | -0.04556 | 0.004311 | 1.50E-27 |
| rs174566 | miR-1908-5 | cis | 1.58E-08 | Nightingale | VLDL_TG | NA | G | A | 0.047387 | 0.004162 | 3.90E-31 |
| rs174566 | miR-1908-5 | cis | 1.58E-08 | Nightingale | M_LDL_C | NA | G | A | -0.03429 | 0.004334 | 5.20E-16 |
| rs174566 | miR-1908-5 | cis | 1.58E-08 | Nightingale | MUFA | NA | G | A | 0.063599 | 0.004281 | 3.30E-51 |
| rs174566 | miR-1908-5 | cis | 1.58E-08 | Nightingale | Total_CE | NA | G | A | -0.0671 | 0.004172 | 1.80E-61 |
| rs174566 | miR-1908-5 | cis | 1.58E-08 | Nightingale | LDL_P | NA | G | A | -0.04021 | 0.004322 | 1.00E-21 |
| rs174566 | miR-1908-5 | cis | 1.58E-08 | Nightingale | S_LDL_C | NA | G | A | -0.03232 | 0.00433 | 1.40E-14 |
| rs174566 | miR-1908-5 | cis | 1.58E-08 | Nightingale | HDL_L | NA | G | A | -0.05368 | 0.003884 | 1.50E-46 |
| rs174566 | miR-1908-5 | cis | 1.58E-08 | Nightingale | Omega_3 | NA | G | A | -0.33624 | 0.004239 | 0 |
| rs174566 | miR-1908-5 | cis | 1.58E-08 | Nightingale | M_VLDL_P | NA | G | A | -0.06211 | 0.004151 | 3.20E-54 |
| rs174566 | miR-1908-5 | cis | 1.58E-08 | Nightingale | VLDL_L | NA | G | A | 0.0271 | 0.004241 | 8.80E-11 |
| rs174566 | miR-1908-5 | cis | 1.58E-08 | Nightingale | M_LDL_L | NA | G | A | -0.0284 | 0.004332 | 1.80E-11 |

| | | | | | | | | | | | |
|---|---|---|---|---|---|---|---|---|---|---|---|
| rs174566 | miR-1908-5 | cis | 1.58E-08 | Nightingale | M_VLDL_C | NA | G | A | -0.05245 | 0.004286 | 3.70E-36 |
| rs174566 | miR-1908-5 | cis | 1.58E-08 | Nightingale | XXL_VLDL_ | NA | G | A | 0.071456 | 0.004169 | 9.00E-69 |
| rs174566 | miR-1908-5 | cis | 1.58E-08 | Nightingale | Unsaturatio | NA | G | A | -0.33962 | 0.004096 | 0 |
| rs174566 | miR-1908-5 | cis | 1.58E-08 | Nightingale | L_VLDL_PL | NA | G | A | 0.048519 | 0.004173 | 1.30E-32 |
| rs174566 | miR-1908-5 | cis | 1.58E-08 | Nightingale | L_LDL_TG_ | NA | G | A | 0.077449 | 0.004241 | 1.60E-76 |
| rs174566 | miR-1908-5 | cis | 1.58E-08 | Nightingale | IDL_CE | NA | G | A | -0.07226 | 0.004175 | 2.40E-71 |
| rs174566 | miR-1908-5 | cis | 1.58E-08 | Nightingale | XL_VLDL_F | NA | G | A | -0.06787 | 0.004191 | 7.80E-62 |
| rs174566 | miR-1908-5 | cis | 1.58E-08 | Nightingale | IDL_FC_pct | NA | G | A | -0.10412 | 0.004291 | 6.70E-132 |
| rs174566 | miR-1908-5 | cis | 1.58E-08 | Nightingale | M_LDL_FC_ | NA | G | A | -0.07138 | 0.004144 | 2.50E-69 |
| rs174566 | miR-1908-5 | cis | 1.58E-08 | Nightingale | L_HDL_TG_ | NA | G | A | 0.078932 | 0.004082 | 2.10E-88 |
| rs174566 | miR-1908-5 | cis | 1.58E-08 | Nightingale | SFA_pct | NA | G | A | -0.04296 | 0.004267 | 7.80E-24 |
| rs174566 | miR-1908-5 | cis | 1.58E-08 | Nightingale | XL_VLDL_C | NA | G | A | -0.06073 | 0.004147 | 3.90E-50 |
| rs174566 | miR-1908-5 | cis | 1.58E-08 | Nightingale | XXL_VLDL_ | NA | G | A | -0.02695 | 0.004304 | 7.70E-11 |
| rs174566 | miR-1908-5 | cis | 1.58E-08 | Nightingale | HDL_PL | NA | G | A | -0.04881 | 0.00391 | 3.30E-38 |
| rs174566 | miR-1908-5 | cis | 1.58E-08 | Nightingale | L_HDL_FC | NA | G | A | -0.08452 | 0.003791 | 2.20E-119 |
| rs174566 | miR-1908-5 | cis | 1.58E-08 | Nightingale | Omega_6_ | NA | G | A | 0.370347 | 0.004263 | 0 |
| rs174566 | miR-1908-5 | cis | 1.58E-08 | Nightingale | XXL_VLDL_ | NA | G | A | 0.044165 | 0.004349 | 3.20E-25 |
| rs174567 | miR-1908-5 | cis | 1.32E-08 | Nightingale | M_VLDL_C | NA | G | A | -0.07542 | 0.00406 | 1.90E-80 |
| rs174567 | miR-1908-5 | cis | 1.32E-08 | Nightingale | S_LDL_PL | NA | G | A | -0.05139 | 0.004314 | 6.90E-35 |
| rs174567 | miR-1908-5 | cis | 1.32E-08 | Nightingale | XXL_VLDL_ | NA | G | A | 0.06101 | 0.004173 | 1.30E-50 |
| rs174567 | miR-1908-5 | cis | 1.32E-08 | Nightingale | L_VLDL_C_ | NA | G | A | -0.0383 | 0.004224 | 5.30E-21 |
| rs174567 | miR-1908-5 | cis | 1.32E-08 | Nightingale | XL_VLDL_C | NA | G | A | -0.06351 | 0.004144 | 1.30E-54 |
| rs174567 | miR-1908-5 | cis | 1.32E-08 | Nightingale | S_HDL_FC_ | NA | G | A | -0.10217 | 0.004102 | 9.29E-152 |
| rs174567 | miR-1908-5 | cis | 1.32E-08 | Nightingale | L_LDL_C | NA | G | A | -0.05773 | 0.004276 | 4.60E-44 |
| rs174567 | miR-1908-5 | cis | 1.32E-08 | Nightingale | L_LDL_TG_ | NA | G | A | 0.07717 | 0.00424 | 4.50E-76 |
| rs174567 | miR-1908-5 | cis | 1.32E-08 | Nightingale | L_VLDL_L | NA | G | A | 0.037844 | 0.004179 | 2.40E-20 |
| rs174567 | miR-1908-5 | cis | 1.32E-08 | Nightingale | HDL_CE | NA | G | A | -0.0616 | 0.003881 | 4.80E-61 |
| rs174567 | miR-1908-5 | cis | 1.32E-08 | Nightingale | XL_HDL_CE | NA | G | A | -0.09654 | 0.003872 | 1.20E-148 |
| rs174567 | miR-1908-5 | cis | 1.32E-08 | Nightingale | Total_TG | NA | G | A | 0.046364 | 0.004186 | 1.30E-29 |
| rs174567 | miR-1908-5 | cis | 1.32E-08 | Nightingale | Phosphogly | NA | G | A | -0.04139 | 0.004096 | 1.20E-25 |
| rs174567 | miR-1908-5 | cis | 1.32E-08 | Nightingale | XL_VLDL_C | NA | G | A | 0.037188 | 0.004224 | 2.70E-19 |
| rs174567 | miR-1908-5 | cis | 1.32E-08 | Nightingale | XS_VLDL_C | NA | G | A | -0.06804 | 0.004197 | 3.60E-63 |
| rs174567 | miR-1908-5 | cis | 1.32E-08 | Nightingale | HDL_TG | NA | G | A | 0.045935 | 0.004264 | 9.10E-28 |
| rs174567 | miR-1908-5 | cis | 1.32E-08 | Nightingale | XS_VLDL_F | NA | G | A | -0.11218 | 0.004292 | 4.30E-154 |
| rs174567 | miR-1908-5 | cis | 1.32E-08 | Nightingale | S_LDL_FC | NA | G | A | -0.06282 | 0.004317 | 4.50E-51 |
| rs174567 | miR-1908-5 | cis | 1.32E-08 | Nightingale | XXL_VLDL_ | NA | G | A | 0.067493 | 0.004166 | 4.30E-62 |
| rs174567 | miR-1908-5 | cis | 1.32E-08 | Nightingale | L_HDL_P | NA | G | A | -0.07773 | 0.003788 | 8.61E-102 |
| rs174567 | miR-1908-5 | cis | 1.32E-08 | Nightingale | M_VLDL_P | NA | G | A | -0.062 | 0.00415 | 3.80E-54 |
| rs174567 | miR-1908-5 | cis | 1.32E-08 | Nightingale | XS_VLDL_C | NA | G | A | -0.09755 | 0.004098 | 3.30E-131 |
| rs174567 | miR-1908-5 | cis | 1.32E-08 | Nightingale | XL_HDL_FC | NA | G | A | -0.10184 | 0.004028 | 8.39E-152 |
| rs174567 | miR-1908-5 | cis | 1.32E-08 | Nightingale | XS_VLDL_P | NA | G | A | -0.04366 | 0.004267 | 1.40E-26 |
| rs174567 | miR-1908-5 | cis | 1.32E-08 | Nightingale | XXL_VLDL_ | NA | G | A | 0.065682 | 0.004159 | 5.20E-59 |
| rs174567 | miR-1908-5 | cis | 1.32E-08 | Nightingale | M_VLDL_F | NA | G | A | -0.09071 | 0.004112 | 6.50E-114 |
| rs174567 | miR-1908-5 | cis | 1.32E-08 | Nightingale | S_LDL_FC_ | NA | G | A | -0.07968 | 0.004191 | 4.50E-84 |
| rs174567 | miR-1908-5 | cis | 1.32E-08 | Nightingale | L_HDL_C_p | NA | G | A | -0.06772 | 0.004076 | 2.10E-66 |
| rs174567 | miR-1908-5 | cis | 1.32E-08 | Nightingale | PUFA_pct | NA | G | A | -0.12111 | 0.00416 | 2.60E-188 |
| rs174567 | miR-1908-5 | cis | 1.32E-08 | Nightingale | M_LDL_TG | NA | G | A | 0.075454 | 0.00427 | 9.40E-72 |
| rs174567 | miR-1908-5 | cis | 1.32E-08 | Nightingale | XL_VLDL_L | NA | G | A | 0.051037 | 0.004161 | 5.20E-36 |
| rs174567 | miR-1908-5 | cis | 1.32E-08 | Nightingale | IDL_FC_pct | NA | G | A | -0.104 | 0.00429 | 1.10E-131 |
| rs174567 | miR-1908-5 | cis | 1.32E-08 | Nightingale | S_LDL_C | NA | G | A | -0.03222 | 0.004329 | 1.50E-14 |
| rs174567 | miR-1908-5 | cis | 1.32E-08 | Nightingale | HDL_P | NA | G | A | -0.03603 | 0.004061 | 5.30E-20 |
| rs174567 | miR-1908-5 | cis | 1.32E-08 | Nightingale | XL_VLDL_T | NA | G | A | 0.059068 | 0.004189 | 9.80E-47 |
| rs174567 | miR-1908-5 | cis | 1.32E-08 | Nightingale | IDL_P | NA | G | A | -0.04516 | 0.004279 | 1.00E-27 |
| rs174567 | miR-1908-5 | cis | 1.32E-08 | Nightingale | L_LDL_C_p | NA | G | A | -0.07656 | 0.004213 | 2.80E-76 |
| rs174567 | miR-1908-5 | cis | 1.32E-08 | Nightingale | SFA | NA | G | A | -0.03209 | 0.004285 | 3.00E-14 |
| rs174567 | miR-1908-5 | cis | 1.32E-08 | Nightingale | Total_PL | NA | G | A | -0.05133 | 0.004118 | 8.90E-38 |
| rs174567 | miR-1908-5 | cis | 1.32E-08 | Nightingale | Omega_6_ | NA | G | A | 0.370162 | 0.004262 | 0 |
| rs174567 | miR-1908-5 | cis | 1.32E-08 | Nightingale | M_VLDL_C | NA | G | A | -0.04384 | 0.004313 | 1.70E-25 |
| rs174567 | miR-1908-5 | cis | 1.32E-08 | Nightingale | S_LDL_CE_ | NA | G | A | 0.056868 | 0.004317 | 1.10E-41 |
| rs174567 | miR-1908-5 | cis | 1.32E-08 | Nightingale | non_HDL_( | NA | G | A | -0.0478 | 0.004301 | 2.60E-30 |
| rs174567 | miR-1908-5 | cis | 1.32E-08 | Nightingale | L_LDL_L | NA | G | A | -0.05413 | 0.004282 | 7.10E-39 |
| rs174567 | miR-1908-5 | cis | 1.32E-08 | Nightingale | M_LDL_FC_ | NA | G | A | -0.07115 | 0.004143 | 5.20E-69 |
| rs174567 | miR-1908-5 | cis | 1.32E-08 | Nightingale | L_HDL_C | NA | G | A | -0.08008 | 0.003808 | 1.70E-106 |

| | | | | | | | | | | | |
|---|---|---|---|---|---|---|---|---|---|---|---|
| rs174567 | miR-1908-! | cis | 1.32E-08 | Nightingale | LDL_L | NA | G | A | -0.04547 | 0.00431 | 1.60E-27 |
| rs174567 | miR-1908-! | cis | 1.32E-08 | Nightingale | XL_HDL_PL | NA | G | A | -0.0665 | 0.003973 | 1.70E-67 |
| rs174567 | miR-1908-! | cis | 1.32E-08 | Nightingale | XXL_VLDL_ | NA | G | A | 0.071724 | 0.00415 | 3.20E-70 |
| rs174567 | miR-1908-! | cis | 1.32E-08 | Nightingale | L_HDL_FC | NA | G | A | -0.08442 | 0.00379 | 3.20E-119 |
| rs174567 | miR-1908-! | cis | 1.32E-08 | Nightingale | L_LDL_CE | NA | G | A | -0.05325 | 0.004286 | 1.30E-37 |
| rs174567 | miR-1908-! | cis | 1.32E-08 | Nightingale | XXL_VLDL_ | NA | G | A | 0.073222 | 0.004176 | 1.40E-71 |
| rs174567 | miR-1908-! | cis | 1.32E-08 | Nightingale | L_VLDL_P | NA | G | A | 0.041519 | 0.004176 | 3.00E-24 |
| rs174567 | miR-1908-! | cis | 1.32E-08 | Nightingale | XL_HDL_PL | NA | G | A | -0.08848 | 0.003887 | 2.70E-124 |
| rs174567 | miR-1908-! | cis | 1.32E-08 | Nightingale | Citrate | NA | G | A | -0.03309 | 0.004297 | 1.50E-15 |
| rs174567 | miR-1908-! | cis | 1.32E-08 | Nightingale | L_VLDL_PL | NA | G | A | 0.087723 | 0.004197 | 1.20E-100 |
| rs174567 | miR-1908-! | cis | 1.32E-08 | Nightingale | S_LDL_P | NA | G | A | -0.03247 | 0.004329 | 1.00E-14 |
| rs174567 | miR-1908-! | cis | 1.32E-08 | Nightingale | Total_FA | NA | G | A | -0.02428 | 0.00429 | 5.30E-09 |
| rs174567 | miR-1908-! | cis | 1.32E-08 | Nightingale | M_HDL_C | NA | G | A | -0.0412 | 0.003979 | 1.10E-26 |
| rs174567 | miR-1908-! | cis | 1.32E-08 | Nightingale | L_HDL_PL | NA | G | A | -0.07612 | 0.003806 | 1.60E-96 |
| rs174567 | miR-1908-! | cis | 1.32E-08 | Nightingale | XS_VLDL_P | NA | G | A | 0.058414 | 0.004219 | 1.10E-44 |
| rs174567 | miR-1908-! | cis | 1.32E-08 | Nightingale | XL_HDL_C_ | NA | G | A | 0.023556 | 0.004131 | 7.70E-10 |
| rs174567 | miR-1908-! | cis | 1.32E-08 | Nightingale | VLDL_TG | NA | G | A | 0.047269 | 0.004162 | 4.90E-31 |
| rs174567 | miR-1908-! | cis | 1.32E-08 | Nightingale | HDL_PL | NA | G | A | -0.04877 | 0.00391 | 3.60E-38 |
| rs174567 | miR-1908-! | cis | 1.32E-08 | Nightingale | LDL_FC | NA | G | A | -0.06418 | 0.004278 | 6.60E-54 |
| rs174567 | miR-1908-! | cis | 1.32E-08 | Nightingale | LDL_P | NA | G | A | -0.04015 | 0.004322 | 1.00E-21 |
| rs174567 | miR-1908-! | cis | 1.32E-08 | Nightingale | S_VLDL_CE | NA | G | A | -0.02839 | 0.004266 | 1.30E-11 |
| rs174567 | miR-1908-! | cis | 1.32E-08 | Nightingale | XL_HDL_L | NA | G | A | -0.09241 | 0.003886 | 1.20E-135 |
| rs174567 | miR-1908-! | cis | 1.32E-08 | Nightingale | M_VLDL_C | NA | G | A | -0.05238 | 0.004285 | 3.80E-36 |
| rs174567 | miR-1908-! | cis | 1.32E-08 | Nightingale | L_HDL_FC_ | NA | G | A | -0.11188 | 0.004017 | 2.40E-185 |
| rs174567 | miR-1908-! | cis | 1.32E-08 | Nightingale | S_VLDL_TG | NA | G | A | 0.063997 | 0.004184 | 1.20E-54 |
| rs174567 | miR-1908-! | cis | 1.32E-08 | Nightingale | XL_VLDL_F | NA | G | A | 0.043085 | 0.004189 | 8.60E-26 |
| rs174567 | miR-1908-! | cis | 1.32E-08 | Nightingale | M_HDL_L | NA | G | A | -0.02818 | 0.004015 | 3.20E-13 |
| rs174567 | miR-1908-! | cis | 1.32E-08 | Nightingale | S_HDL_TG_ | NA | G | A | 0.068939 | 0.004113 | 3.50E-66 |
| rs174567 | miR-1908-! | cis | 1.32E-08 | Nightingale | XS_VLDL_C | NA | G | A | -0.07176 | 0.004173 | 8.70E-71 |
| rs174567 | miR-1908-! | cis | 1.32E-08 | Nightingale | XL_VLDL_P | NA | G | A | 0.052424 | 0.004156 | 5.50E-38 |
| rs174567 | miR-1908-! | cis | 1.32E-08 | Nightingale | L_LDL_FC | NA | G | A | -0.06815 | 0.004246 | 1.30E-61 |
| rs174567 | miR-1908-! | cis | 1.32E-08 | Nightingale | M_VLDL_T | NA | G | A | 0.031033 | 0.004202 | 5.60E-14 |
| rs174567 | miR-1908-! | cis | 1.32E-08 | Nightingale | Total_CE | NA | G | A | -0.067 | 0.004171 | 2.20E-61 |
| rs174567 | miR-1908-! | cis | 1.32E-08 | Nightingale | Clinical_LD | NA | G | A | -0.05513 | 0.0043 | 1.10E-39 |
| rs174567 | miR-1908-! | cis | 1.32E-08 | Nightingale | Tyr | NA | G | A | 0.032542 | 0.004319 | 4.90E-14 |
| rs174567 | miR-1908-! | cis | 1.32E-08 | Nightingale | LA | NA | G | A | 0.083821 | 0.004241 | 2.80E-87 |
| rs174567 | miR-1908-! | cis | 1.32E-08 | Nightingale | XS_VLDL_P | NA | G | A | -0.03438 | 0.004264 | 2.30E-17 |
| rs174567 | miR-1908-! | cis | 1.32E-08 | Nightingale | XS_VLDL_C | NA | G | A | -0.09168 | 0.004076 | 1.90E-117 |
| rs174567 | miR-1908-! | cis | 1.32E-08 | Nightingale | PUFA | NA | G | A | -0.09679 | 0.004217 | 2.30E-123 |
| rs174567 | miR-1908-! | cis | 1.32E-08 | Nightingale | HDL_L | NA | G | A | -0.05361 | 0.003883 | 1.80E-46 |
| rs174567 | miR-1908-! | cis | 1.32E-08 | Nightingale | S_LDL_L | NA | G | A | -0.03249 | 0.004324 | 7.50E-15 |
| rs174567 | miR-1908-! | cis | 1.32E-08 | Nightingale | M_VLDL_F | NA | G | A | -0.02943 | 0.004324 | 2.40E-12 |
| rs174567 | miR-1908-! | cis | 1.32E-08 | Nightingale | VLDL_L | NA | G | A | 0.027025 | 0.00424 | 9.60E-11 |
| rs174567 | miR-1908-! | cis | 1.32E-08 | Nightingale | LA_pct | NA | G | A | 0.186163 | 0.004226 | 0 |
| rs174567 | miR-1908-! | cis | 1.32E-08 | Nightingale | L_LDL_FC_ | NA | G | A | -0.08209 | 0.004122 | 9.30E-92 |
| rs174567 | miR-1908-! | cis | 1.32E-08 | Nightingale | IDL_C | NA | G | A | -0.07718 | 0.004176 | 3.50E-81 |
| rs174567 | miR-1908-! | cis | 1.32E-08 | Nightingale | Unsaturati | NA | G | A | -0.33936 | 0.004096 | 0 |
| rs174567 | miR-1908-! | cis | 1.32E-08 | Nightingale | SFA_pct | NA | G | A | -0.04321 | 0.004266 | 4.20E-24 |
| rs174567 | miR-1908-! | cis | 1.32E-08 | Nightingale | DHA | NA | G | A | -0.28887 | 0.004167 | 0 |
| rs174567 | miR-1908-! | cis | 1.32E-08 | Nightingale | XS_VLDL_T | NA | G | A | 0.096719 | 0.004108 | 9.40E-129 |
| rs174567 | miR-1908-! | cis | 1.32E-08 | Nightingale | Sphingomy | NA | G | A | -0.11532 | 0.004041 | 1.60E-188 |
| rs174567 | miR-1908-! | cis | 1.32E-08 | Nightingale | L_VLDL_CE | NA | G | A | -0.05869 | 0.004138 | 1.40E-48 |
| rs174567 | miR-1908-! | cis | 1.32E-08 | Nightingale | M_LDL_TG | NA | G | A | 0.036141 | 0.004261 | 1.70E-17 |
| rs174567 | miR-1908-! | cis | 1.32E-08 | Nightingale | Omega_3_ | NA | G | A | -0.39002 | 0.004225 | 0 |
| rs174567 | miR-1908-! | cis | 1.32E-08 | Nightingale | L_VLDL_PL | NA | G | A | 0.048414 | 0.004172 | 1.60E-32 |
| rs174567 | miR-1908-! | cis | 1.32E-08 | Nightingale | IDL_CE | NA | G | A | -0.07215 | 0.004174 | 2.80E-71 |
| rs174567 | miR-1908-! | cis | 1.32E-08 | Nightingale | HDL_C | NA | G | A | -0.06489 | 0.003861 | 2.20E-68 |
| rs174567 | miR-1908-! | cis | 1.32E-08 | Nightingale | S_VLDL_FC | NA | G | A | -0.02828 | 0.004325 | 1.60E-11 |
| rs174567 | miR-1908-! | cis | 1.32E-08 | Nightingale | Cholines | NA | G | A | -0.03739 | 0.004067 | 7.90E-22 |
| rs174567 | miR-1908-! | cis | 1.32E-08 | Nightingale | M_LDL_CE | NA | G | A | -0.02654 | 0.004331 | 3.10E-10 |
| rs174567 | miR-1908-! | cis | 1.32E-08 | Nightingale | M_VLDL_C | NA | G | A | -0.08014 | 0.00407 | 3.00E-90 |
| rs174567 | miR-1908-! | cis | 1.32E-08 | Nightingale | Total_L | NA | G | A | -0.03756 | 0.004254 | 6.60E-20 |
| rs174567 | miR-1908-! | cis | 1.32E-08 | Nightingale | M_LDL_PL_ | NA | G | A | 0.046321 | 0.004322 | 7.70E-28 |

| | | | | | | | | | | | |
|---|---|---|---|---|---|---|---|---|---|---|---|
| rs174567 | miR-1908-5 | cis | 1.32E-08 | Nightingale | IDL_FC | NA | G | A | -0.08968 | 0.004186 | 7.00E-108 |
| rs174567 | miR-1908-5 | cis | 1.32E-08 | Nightingale | XS_VLDL_F | NA | G | A | -0.05607 | 0.004249 | 1.20E-42 |
| rs174567 | miR-1908-5 | cis | 1.32E-08 | Nightingale | XL_VLDL_C | NA | G | A | -0.06051 | 0.004146 | 7.90E-50 |
| rs174567 | miR-1908-5 | cis | 1.32E-08 | Nightingale | L_HDL_L | NA | G | A | -0.07715 | 0.003794 | 7.10E-100 |
| rs174567 | miR-1908-5 | cis | 1.32E-08 | Nightingale | L_VLDL_C | NA | G | A | 0.02865 | 0.00423 | 5.20E-12 |
| rs174567 | miR-1908-5 | cis | 1.32E-08 | Nightingale | S_VLDL_TG | NA | G | A | 0.038458 | 0.004196 | 9.40E-21 |
| rs174567 | miR-1908-5 | cis | 1.32E-08 | Nightingale | L_VLDL_TG | NA | G | A | 0.036899 | 0.004161 | 1.40E-19 |
| rs174567 | miR-1908-5 | cis | 1.32E-08 | Nightingale | L_HDL_TG_ | NA | G | A | 0.078752 | 0.004081 | 3.90E-88 |
| rs174567 | miR-1908-5 | cis | 1.32E-08 | Nightingale | M_HDL_FC | NA | G | A | -0.05187 | 0.003935 | 2.30E-42 |
| rs174567 | miR-1908-5 | cis | 1.32E-08 | Nightingale | IDL_TG_pct | NA | G | A | 0.087661 | 0.004178 | 5.70E-102 |
| rs174567 | miR-1908-5 | cis | 1.32E-08 | Nightingale | Omega_3 | NA | G | A | -0.33607 | 0.004239 | 0 |
| rs174567 | miR-1908-5 | cis | 1.32E-08 | Nightingale | LDL_CE | NA | G | A | -0.04362 | 0.004314 | 2.30E-25 |
| rs174567 | miR-1908-5 | cis | 1.32E-08 | Nightingale | XL_HDL_TG | NA | G | A | 0.086743 | 0.004074 | 2.10E-106 |
| rs174567 | miR-1908-5 | cis | 1.32E-08 | Nightingale | IDL_C_pct | NA | G | A | -0.07472 | 0.004222 | 3.20E-72 |
| rs174567 | miR-1908-5 | cis | 1.32E-08 | Nightingale | L_LDL_P | NA | G | A | -0.03718 | 0.00431 | 3.80E-19 |
| rs174567 | miR-1908-5 | cis | 1.32E-08 | Nightingale | ApoA1 | NA | G | A | -0.04153 | 0.003958 | 1.20E-27 |
| rs174567 | miR-1908-5 | cis | 1.32E-08 | Nightingale | XL_VLDL_T | NA | G | A | 0.056392 | 0.004136 | 4.60E-44 |
| rs174567 | miR-1908-5 | cis | 1.32E-08 | Nightingale | M_LDL_C_ | NA | G | A | -0.10016 | 0.004328 | 1.70E-119 |
| rs174567 | miR-1908-5 | cis | 1.32E-08 | Nightingale | M_HDL_TG | NA | G | A | 0.066647 | 0.004147 | 5.70E-61 |
| rs174567 | miR-1908-5 | cis | 1.32E-08 | Nightingale | S_VLDL_FC | NA | G | A | -0.08245 | 0.004129 | 5.80E-93 |
| rs174567 | miR-1908-5 | cis | 1.32E-08 | Nightingale | S_LDL_TG_ | NA | G | A | 0.085841 | 0.00419 | 7.30E-97 |
| rs174567 | miR-1908-5 | cis | 1.32E-08 | Nightingale | L_HDL_CE | NA | G | A | -0.0785 | 0.003818 | 9.59E-102 |
| rs174567 | miR-1908-5 | cis | 1.32E-08 | Nightingale | XL_VLDL_C | NA | G | A | 0.030132 | 0.004256 | 6.60E-13 |
| rs174567 | miR-1908-5 | cis | 1.32E-08 | Nightingale | M_LDL_PL | NA | G | A | -0.02362 | 0.004331 | 1.80E-08 |
| rs174567 | miR-1908-5 | cis | 1.32E-08 | Nightingale | L_VLDL_FC | NA | G | A | 0.039808 | 0.004183 | 2.20E-22 |
| rs174567 | miR-1908-5 | cis | 1.32E-08 | Nightingale | XXL_VLDL_ | NA | G | A | -0.02699 | 0.004303 | 7.30E-11 |
| rs174567 | miR-1908-5 | cis | 1.32E-08 | Nightingale | Total_C | NA | G | A | -0.06518 | 0.004186 | 8.80E-58 |
| rs174567 | miR-1908-5 | cis | 1.32E-08 | Nightingale | LDL_TG | NA | G | A | 0.029232 | 0.004277 | 1.20E-11 |
| rs174567 | miR-1908-5 | cis | 1.32E-08 | Nightingale | MUFA_pct | NA | G | A | 0.197321 | 0.004138 | 0 |
| rs174567 | miR-1908-5 | cis | 1.32E-08 | Nightingale | Total_P | NA | G | A | -0.04073 | 0.00407 | 3.70E-25 |
| rs174567 | miR-1908-5 | cis | 1.32E-08 | Nightingale | M_LDL_FC | NA | G | A | -0.05241 | 0.00432 | 7.00E-36 |
| rs174567 | miR-1908-5 | cis | 1.32E-08 | Nightingale | S_LDL_TG | NA | G | A | 0.05004 | 0.004219 | 3.40E-33 |
| rs174567 | miR-1908-5 | cis | 1.32E-08 | Nightingale | Total_FC | NA | G | A | -0.05936 | 0.004225 | 1.30E-47 |
| rs174567 | miR-1908-5 | cis | 1.32E-08 | Nightingale | XS_VLDL_L | NA | G | A | -0.04162 | 0.004251 | 1.10E-24 |
| rs174567 | miR-1908-5 | cis | 1.32E-08 | Nightingale | M_HDL_C_ | NA | G | A | -0.06572 | 0.004117 | 1.90E-60 |
| rs174567 | miR-1908-5 | cis | 1.32E-08 | Nightingale | LDL_PL | NA | G | A | -0.04618 | 0.004315 | 3.80E-28 |
| rs174567 | miR-1908-5 | cis | 1.32E-08 | Nightingale | HDL_size | NA | G | A | -0.07248 | 0.003832 | 4.20E-86 |
| rs174567 | miR-1908-5 | cis | 1.32E-08 | Nightingale | LDL_C | NA | G | A | -0.04975 | 0.004306 | 1.30E-32 |
| rs174567 | miR-1908-5 | cis | 1.32E-08 | Nightingale | S_HDL_C_p | NA | G | A | -0.04208 | 0.004274 | 6.80E-23 |
| rs174567 | miR-1908-5 | cis | 1.32E-08 | Nightingale | M_LDL_C | NA | G | A | -0.03421 | 0.004333 | 5.40E-16 |
| rs174567 | miR-1908-5 | cis | 1.32E-08 | Nightingale | IDL_PL | NA | G | A | -0.07263 | 0.004172 | 6.10E-73 |
| rs174567 | miR-1908-5 | cis | 1.32E-08 | Nightingale | XXL_VLDL_ | NA | G | A | 0.043976 | 0.004348 | 4.40E-25 |
| rs174567 | miR-1908-5 | cis | 1.32E-08 | Nightingale | S_VLDL_PL | NA | G | A | -0.07818 | 0.004138 | 9.70E-84 |
| rs174567 | miR-1908-5 | cis | 1.32E-08 | Nightingale | XL_VLDL_P | NA | G | A | 0.047918 | 0.004173 | 9.10E-32 |
| rs174567 | miR-1908-5 | cis | 1.32E-08 | Nightingale | XL_HDL_P | NA | G | A | -0.08895 | 0.003892 | 2.60E-126 |
| rs174567 | miR-1908-5 | cis | 1.32E-08 | Nightingale | S_LDL_PL_ | NA | G | A | -0.06135 | 0.004316 | 3.20E-48 |
| rs174567 | miR-1908-5 | cis | 1.32E-08 | Nightingale | HDL_FC | NA | G | A | -0.07403 | 0.003831 | 7.70E-90 |
| rs174567 | miR-1908-5 | cis | 1.32E-08 | Nightingale | M_HDL_PL | NA | G | A | -0.02207 | 0.004044 | 1.50E-08 |
| rs174567 | miR-1908-5 | cis | 1.32E-08 | Nightingale | M_VLDL_T | NA | G | A | 0.077415 | 0.004084 | 3.50E-84 |
| rs174567 | miR-1908-5 | cis | 1.32E-08 | Nightingale | VLDL_size | NA | G | A | 0.071868 | 0.004031 | 2.50E-75 |
| rs174567 | miR-1908-5 | cis | 1.32E-08 | Nightingale | PUFA_by_N | NA | G | A | -0.17045 | 0.004142 | 0 |
| rs174567 | miR-1908-5 | cis | 1.32E-08 | Nightingale | Remnant_( | NA | G | A | -0.04413 | 0.004297 | 2.20E-26 |
| rs174567 | miR-1908-5 | cis | 1.32E-08 | Nightingale | XL_VLDL_F | NA | G | A | -0.06764 | 0.00419 | 1.70E-61 |
| rs174567 | miR-1908-5 | cis | 1.32E-08 | Nightingale | TG_by_PG | NA | G | A | 0.068471 | 0.004013 | 8.90E-69 |
| rs174567 | miR-1908-5 | cis | 1.32E-08 | Nightingale | ApoB | NA | G | A | -0.03842 | 0.004319 | 4.00E-20 |
| rs174567 | miR-1908-5 | cis | 1.32E-08 | Nightingale | XXL_VLDL_ | NA | G | A | 0.071295 | 0.004168 | 1.50E-68 |
| rs174567 | miR-1908-5 | cis | 1.32E-08 | Nightingale | XL_HDL_CE | NA | G | A | -0.02628 | 0.004322 | 2.30E-09 |
| rs174567 | miR-1908-5 | cis | 1.32E-08 | Nightingale | L_HDL_CE_ | NA | G | A | -0.04938 | 0.004138 | 6.00E-35 |
| rs174567 | miR-1908-5 | cis | 1.32E-08 | Nightingale | IDL_L | NA | G | A | -0.07232 | 0.004181 | 9.30E-72 |
| rs174567 | miR-1908-5 | cis | 1.32E-08 | Nightingale | L_HDL_PL_ | NA | G | A | 0.041891 | 0.004185 | 4.20E-25 |
| rs174567 | miR-1908-5 | cis | 1.32E-08 | Nightingale | L_LDL_PL | NA | G | A | -0.05495 | 0.004292 | 1.60E-39 |
| rs174567 | miR-1908-5 | cis | 1.32E-08 | Nightingale | Omega_6_ | NA | G | A | 0.024716 | 0.004209 | 2.80E-09 |
| rs174567 | miR-1908-5 | cis | 1.32E-08 | Nightingale | S_HDL_TG | NA | G | A | 0.068472 | 0.004193 | 9.40E-63 |

| | | | | | | | | | | | |
|---|---|---|---|---|---|---|---|---|---|---|---|
| rs174567 | miR-1908-! | cis | 1.32E-08 | Nightingale | M_HDL_CE | NA | G | A | -0.04676 | 0.004194 | 8.20E-30 |
| rs174567 | miR-1908-! | cis | 1.32E-08 | Nightingale | M_LDL_P | NA | G | A | -0.04682 | 0.004337 | 3.60E-28 |
| rs174567 | miR-1908-! | cis | 1.32E-08 | Nightingale | M_LDL_L | NA | G | A | -0.02833 | 0.004331 | 1.90E-11 |
| rs174567 | miR-1908-! | cis | 1.32E-08 | Nightingale | XXL_VLDL_ | NA | G | A | 0.066234 | 0.004165 | 1.30E-59 |
| rs174567 | miR-1908-! | cis | 1.32E-08 | Nightingale | XS_VLDL_T | NA | G | A | 0.031189 | 0.004258 | 6.10E-13 |
| rs174567 | miR-1908-! | cis | 1.32E-08 | Nightingale | S_VLDL_C_ | NA | G | A | -0.05604 | 0.004208 | 9.00E-42 |
| rs174567 | miR-1908-! | cis | 1.32E-08 | Nightingale | L_VLDL_FC | NA | G | A | 0.028565 | 0.004284 | 1.90E-11 |
| rs174567 | miR-1908-! | cis | 1.32E-08 | Nightingale | MUFA | NA | G | A | 0.063493 | 0.00428 | 4.70E-51 |
| rs174567 | miR-1908-! | cis | 1.32E-08 | Nightingale | IDL_CE_pct | NA | G | A | -0.03792 | 0.004243 | 2.60E-19 |
| rs174567 | miR-1908-! | cis | 1.32E-08 | Nightingale | M_HDL_TG | NA | G | A | 0.049984 | 0.004271 | 1.20E-32 |
| rs174567 | miR-1908-! | cis | 1.32E-08 | Nightingale | M_HDL_P | NA | G | A | -0.03471 | 0.003984 | 1.40E-19 |
| rs174567 | miR-1908-! | cis | 1.32E-08 | Nightingale | M_HDL_CE | NA | G | A | -0.03824 | 0.003994 | 4.70E-23 |
| rs174567 | miR-1908-! | cis | 1.32E-08 | Nightingale | M_HDL_FC | NA | G | A | -0.10331 | 0.003832 | 2.10E-174 |
| rs174567 | miR-1908-! | cis | 1.32E-08 | Nightingale | XL_HDL_FC | NA | G | A | 0.060781 | 0.003834 | 3.00E-62 |
| rs174567 | miR-1908-! | cis | 1.32E-08 | Nightingale | M_HDL_PL | NA | G | A | 0.058121 | 0.004102 | 1.60E-47 |
| rs174567 | miR-1908-! | cis | 1.32E-08 | Nightingale | DHA_pct | NA | G | A | -0.27396 | 0.004198 | 0 |
| rs174567 | miR-1908-! | cis | 1.32E-08 | Nightingale | XL_HDL_C | NA | G | A | -0.09859 | 0.003899 | 1.10E-152 |
| rs174568 | miR-1908-! | cis | 3.76E-08 | Nightingale | DHA_pct | NA | T | C | -0.27306 | 0.004217 | 0 |
| rs174568 | miR-1908-! | cis | 3.76E-08 | Nightingale | L_LDL_TG_ | NA | T | C | 0.077658 | 0.00426 | 6.10E-76 |
| rs174568 | miR-1908-! | cis | 3.76E-08 | Nightingale | M_HDL_FC | NA | T | C | -0.1035 | 0.00385 | 1.10E-173 |
| rs174568 | miR-1908-! | cis | 3.76E-08 | Nightingale | XL_HDL_L | NA | T | C | -0.0934 | 0.003904 | 7.69E-138 |
| rs174568 | miR-1908-! | cis | 3.76E-08 | Nightingale | LDL_C | NA | T | C | -0.04994 | 0.004326 | 2.00E-32 |
| rs174568 | miR-1908-! | cis | 3.76E-08 | Nightingale | M_VLDL_C | NA | T | C | -0.04407 | 0.004333 | 2.00E-25 |
| rs174568 | miR-1908-! | cis | 3.76E-08 | Nightingale | S_LDL_P | NA | T | C | -0.03257 | 0.004349 | 1.20E-14 |
| rs174568 | miR-1908-! | cis | 3.76E-08 | Nightingale | XS_VLDL_F | NA | T | C | -0.11263 | 0.004312 | 1.10E-153 |
| rs174568 | miR-1908-! | cis | 3.76E-08 | Nightingale | HDL_TG | NA | T | C | 0.046242 | 0.004284 | 1.00E-27 |
| rs174568 | miR-1908-! | cis | 3.76E-08 | Nightingale | XL_HDL_TG | NA | T | C | 0.087333 | 0.004093 | 7.69E-107 |
| rs174568 | miR-1908-! | cis | 3.76E-08 | Nightingale | S_LDL_FC_ | NA | T | C | -0.07996 | 0.00421 | 2.30E-83 |
| rs174568 | miR-1908-! | cis | 3.76E-08 | Nightingale | S_HDL_FC_ | NA | T | C | -0.10324 | 0.004121 | 5.50E-154 |
| rs174568 | miR-1908-! | cis | 3.76E-08 | Nightingale | IDL_FC | NA | T | C | -0.08996 | 0.004205 | 3.30E-107 |
| rs174568 | miR-1908-! | cis | 3.76E-08 | Nightingale | Total_TG | NA | T | C | 0.046858 | 0.004205 | 8.10E-30 |
| rs174568 | miR-1908-! | cis | 3.76E-08 | Nightingale | L_HDL_PL | NA | T | C | -0.07642 | 0.003824 | 8.00E-97 |
| rs174568 | miR-1908-! | cis | 3.76E-08 | Nightingale | L_LDL_L | NA | T | C | -0.05439 | 0.004302 | 9.60E-39 |
| rs174568 | miR-1908-! | cis | 3.76E-08 | Nightingale | LDL_P | NA | T | C | -0.04043 | 0.004341 | 9.60E-22 |
| rs174568 | miR-1908-! | cis | 3.76E-08 | Nightingale | Total_FA | NA | T | C | -0.02373 | 0.00431 | 1.10E-08 |
| rs174568 | miR-1908-! | cis | 3.76E-08 | Nightingale | L_HDL_P | NA | T | C | -0.07824 | 0.003805 | 1.40E-102 |
| rs174568 | miR-1908-! | cis | 3.76E-08 | Nightingale | XXL_VLDL_ | NA | T | C | 0.061459 | 0.004192 | 9.90E-51 |
| rs174568 | miR-1908-! | cis | 3.76E-08 | Nightingale | Total_PL | NA | T | C | -0.05126 | 0.004137 | 1.70E-37 |
| rs174568 | miR-1908-! | cis | 3.76E-08 | Nightingale | DHA | NA | T | C | -0.2877 | 0.004187 | 0 |
| rs174568 | miR-1908-! | cis | 3.76E-08 | Nightingale | Unsaturati | NA | T | C | -0.33839 | 0.004114 | 0 |
| rs174568 | miR-1908-! | cis | 3.76E-08 | Nightingale | XL_HDL_PL | NA | T | C | -0.08941 | 0.003905 | 2.10E-126 |
| rs174568 | miR-1908-! | cis | 3.76E-08 | Nightingale | M_VLDL_C | NA | T | C | -0.05274 | 0.004305 | 3.40E-36 |
| rs174568 | miR-1908-! | cis | 3.76E-08 | Nightingale | IDL_C | NA | T | C | -0.07747 | 0.004195 | 9.60E-81 |
| rs174568 | miR-1908-! | cis | 3.76E-08 | Nightingale | L_VLDL_CE | NA | T | C | -0.05919 | 0.004157 | 8.30E-49 |
| rs174568 | miR-1908-! | cis | 3.76E-08 | Nightingale | Total_L | NA | T | C | -0.0375 | 0.004274 | 1.00E-19 |
| rs174568 | miR-1908-! | cis | 3.76E-08 | Nightingale | IDL_C_pct | NA | T | C | -0.07482 | 0.004241 | 2.90E-71 |
| rs174568 | miR-1908-! | cis | 3.76E-08 | Nightingale | Omega_6_ | NA | T | C | 0.36837 | 0.004281 | 0 |
| rs174568 | miR-1908-! | cis | 3.76E-08 | Nightingale | XL_VLDL_T | NA | T | C | 0.059518 | 0.004209 | 7.90E-47 |
| rs174568 | miR-1908-! | cis | 3.76E-08 | Nightingale | XXL_VLDL_ | NA | T | C | 0.066812 | 0.004184 | 6.50E-60 |
| rs174568 | miR-1908-! | cis | 3.76E-08 | Nightingale | LDL_FC | NA | T | C | -0.06449 | 0.004298 | 1.00E-53 |
| rs174568 | miR-1908-! | cis | 3.76E-08 | Nightingale | M_HDL_C | NA | T | C | -0.04089 | 0.003997 | 3.40E-26 |
| rs174568 | miR-1908-! | cis | 3.76E-08 | Nightingale | S_VLDL_CE | NA | T | C | -0.02881 | 0.004286 | 9.90E-12 |
| rs174568 | miR-1908-! | cis | 3.76E-08 | Nightingale | S_VLDL_FC | NA | T | C | -0.02837 | 0.004345 | 1.80E-11 |
| rs174568 | miR-1908-! | cis | 3.76E-08 | Nightingale | S_HDL_TG_ | NA | T | C | 0.069055 | 0.004132 | 1.40E-65 |
| rs174568 | miR-1908-! | cis | 3.76E-08 | Nightingale | XS_VLDL_T | NA | T | C | 0.097365 | 0.004127 | 6.30E-129 |
| rs174568 | miR-1908-! | cis | 3.76E-08 | Nightingale | Phosphogly | NA | T | C | -0.04114 | 0.004115 | 2.50E-25 |
| rs174568 | miR-1908-! | cis | 3.76E-08 | Nightingale | Total_C | NA | T | C | -0.06539 | 0.004206 | 1.50E-57 |
| rs174568 | miR-1908-! | cis | 3.76E-08 | Nightingale | XL_VLDL_F | NA | T | C | 0.043568 | 0.004208 | 5.80E-26 |
| rs174568 | miR-1908-! | cis | 3.76E-08 | Nightingale | XS_VLDL_C | NA | T | C | -0.09805 | 0.004116 | 5.40E-131 |
| rs174568 | miR-1908-! | cis | 3.76E-08 | Nightingale | XS_VLDL_P | NA | T | C | 0.058044 | 0.004239 | 1.70E-43 |
| rs174568 | miR-1908-! | cis | 3.76E-08 | Nightingale | S_HDL_TG | NA | T | C | 0.06893 | 0.004213 | 5.70E-63 |
| rs174568 | miR-1908-! | cis | 3.76E-08 | Nightingale | XL_VLDL_C | NA | T | C | -0.0642 | 0.004163 | 4.90E-55 |
| rs174568 | miR-1908-! | cis | 3.76E-08 | Nightingale | L_LDL_C | NA | T | C | -0.05802 | 0.004296 | 6.00E-44 |

| | | | | | | | | | | | |
|---|---|---|---|---|---|---|---|---|---|---|---|
| rs174568 | miR-1908-5 | cis | 3.76E-08 | Nightingale | IDL_PL | NA | T | C | -0.07306 | 0.004192 | 5.20E-73 |
| rs174568 | miR-1908-5 | cis | 3.76E-08 | Nightingale | M_LDL_PL_ | NA | T | C | 0.045593 | 0.004342 | 5.60E-27 |
| rs174568 | miR-1908-5 | cis | 3.76E-08 | Nightingale | PUFA | NA | T | C | -0.09637 | 0.004237 | 2.00E-121 |
| rs174568 | miR-1908-5 | cis | 3.76E-08 | Nightingale | M_VLDL_T | NA | T | C | 0.031467 | 0.004221 | 3.80E-14 |
| rs174568 | miR-1908-5 | cis | 3.76E-08 | Nightingale | HDL_CE | NA | T | C | -0.06167 | 0.003899 | 6.70E-61 |
| rs174568 | miR-1908-5 | cis | 3.76E-08 | Nightingale | non_HDL_( | NA | T | C | -0.04795 | 0.004321 | 4.00E-30 |
| rs174568 | miR-1908-5 | cis | 3.76E-08 | Nightingale | XXL_VLDL_ | NA | T | C | 0.068074 | 0.004185 | 2.40E-62 |
| rs174568 | miR-1908-5 | cis | 3.76E-08 | Nightingale | SFA | NA | T | C | -0.03136 | 0.004305 | 1.20E-13 |
| rs174568 | miR-1908-5 | cis | 3.76E-08 | Nightingale | LDL_L | NA | T | C | -0.04563 | 0.00433 | 2.30E-27 |
| rs174568 | miR-1908-5 | cis | 3.76E-08 | Nightingale | IDL_P | NA | T | C | -0.04519 | 0.004299 | 2.30E-27 |
| rs174568 | miR-1908-5 | cis | 3.76E-08 | Nightingale | L_VLDL_P | NA | T | C | 0.041965 | 0.004195 | 2.00E-24 |
| rs174568 | miR-1908-5 | cis | 3.76E-08 | Nightingale | XL_HDL_C_ | NA | T | C | 0.024239 | 0.00415 | 2.10E-10 |
| rs174568 | miR-1908-5 | cis | 3.76E-08 | Nightingale | HDL_size | NA | T | C | -0.07301 | 0.003849 | 4.90E-87 |
| rs174568 | miR-1908-5 | cis | 3.76E-08 | Nightingale | M_VLDL_F | NA | T | C | -0.09127 | 0.004131 | 5.70E-114 |
| rs174568 | miR-1908-5 | cis | 3.76E-08 | Nightingale | L_VLDL_PL | NA | T | C | 0.048818 | 0.004192 | 1.10E-32 |
| rs174568 | miR-1908-5 | cis | 3.76E-08 | Nightingale | Citrate | NA | T | C | -0.03306 | 0.004317 | 1.80E-15 |
| rs174568 | miR-1908-5 | cis | 3.76E-08 | Nightingale | S_LDL_PL | NA | T | C | -0.05171 | 0.004334 | 8.50E-35 |
| rs174568 | miR-1908-5 | cis | 3.76E-08 | Nightingale | XL_VLDL_C | NA | T | C | 0.037546 | 0.004243 | 2.30E-19 |
| rs174568 | miR-1908-5 | cis | 3.76E-08 | Nightingale | PUFA_pct | NA | T | C | -0.12131 | 0.004179 | 8.79E-187 |
| rs174568 | miR-1908-5 | cis | 3.76E-08 | Nightingale | XL_VLDL_L | NA | T | C | 0.051569 | 0.00418 | 2.80E-36 |
| rs174568 | miR-1908-5 | cis | 3.76E-08 | Nightingale | M_LDL_TG | NA | T | C | 0.036449 | 0.00428 | 1.80E-17 |
| rs174568 | miR-1908-5 | cis | 3.76E-08 | Nightingale | M_LDL_FC_ | NA | T | C | -0.07187 | 0.004162 | 2.80E-69 |
| rs174568 | miR-1908-5 | cis | 3.76E-08 | Nightingale | L_HDL_C_p | NA | T | C | -0.06871 | 0.004095 | 6.60E-68 |
| rs174568 | miR-1908-5 | cis | 3.76E-08 | Nightingale | L_HDL_C | NA | T | C | -0.08064 | 0.003826 | 2.10E-107 |
| rs174568 | miR-1908-5 | cis | 3.76E-08 | Nightingale | M_HDL_C_ | NA | T | C | -0.06607 | 0.004136 | 1.60E-60 |
| rs174568 | miR-1908-5 | cis | 3.76E-08 | Nightingale | M_VLDL_C | NA | T | C | -0.076 | 0.004078 | 1.30E-80 |
| rs174568 | miR-1908-5 | cis | 3.76E-08 | Nightingale | M_VLDL_P | NA | T | C | -0.0625 | 0.004169 | 2.40E-54 |
| rs174568 | miR-1908-5 | cis | 3.76E-08 | Nightingale | IDL_FC_pct | NA | T | C | -0.10415 | 0.00431 | 1.00E-130 |
| rs174568 | miR-1908-5 | cis | 3.76E-08 | Nightingale | L_LDL_C_p | NA | T | C | -0.07718 | 0.004233 | 1.40E-76 |
| rs174568 | miR-1908-5 | cis | 3.76E-08 | Nightingale | L_LDL_P | NA | T | C | -0.03766 | 0.00433 | 2.30E-19 |
| rs174568 | miR-1908-5 | cis | 3.76E-08 | Nightingale | XS_VLDL_P | NA | T | C | -0.03463 | 0.004283 | 1.50E-17 |
| rs174568 | miR-1908-5 | cis | 3.76E-08 | Nightingale | XL_HDL_CE | NA | T | C | -0.09747 | 0.00389 | 1.20E-150 |
| rs174568 | miR-1908-5 | cis | 3.76E-08 | Nightingale | S_LDL_CE_ | NA | T | C | 0.057327 | 0.004336 | 8.00E-42 |
| rs174568 | miR-1908-5 | cis | 3.76E-08 | Nightingale | XS_VLDL_F | NA | T | C | -0.05627 | 0.004269 | 1.20E-42 |
| rs174568 | miR-1908-5 | cis | 3.76E-08 | Nightingale | M_HDL_CE | NA | T | C | -0.04719 | 0.004213 | 5.80E-30 |
| rs174568 | miR-1908-5 | cis | 3.76E-08 | Nightingale | XXL_VLDL_ | NA | T | C | 0.072148 | 0.004169 | 3.40E-70 |
| rs174568 | miR-1908-5 | cis | 3.76E-08 | Nightingale | M_HDL_P | NA | T | C | -0.03433 | 0.004002 | 3.80E-19 |
| rs174568 | miR-1908-5 | cis | 3.76E-08 | Nightingale | XS_VLDL_C | NA | T | C | -0.06836 | 0.004217 | 4.10E-63 |
| rs174568 | miR-1908-5 | cis | 3.76E-08 | Nightingale | XS_VLDL_C | NA | T | C | -0.07212 | 0.004192 | 1.00E-70 |
| rs174568 | miR-1908-5 | cis | 3.76E-08 | Nightingale | Total_CE | NA | T | C | -0.0672 | 0.00419 | 4.00E-61 |
| rs174568 | miR-1908-5 | cis | 3.76E-08 | Nightingale | S_LDL_TG | NA | T | C | 0.050396 | 0.004238 | 4.30E-33 |
| rs174568 | miR-1908-5 | cis | 3.76E-08 | Nightingale | XXL_VLDL_ | NA | T | C | 0.043826 | 0.004368 | 1.20E-24 |
| rs174568 | miR-1908-5 | cis | 3.76E-08 | Nightingale | Total_P | NA | T | C | -0.04036 | 0.004088 | 1.30E-24 |
| rs174568 | miR-1908-5 | cis | 3.76E-08 | Nightingale | XXL_VLDL_ | NA | T | C | 0.071821 | 0.004187 | 1.20E-68 |
| rs174568 | miR-1908-5 | cis | 3.76E-08 | Nightingale | LA | NA | T | C | 0.08322 | 0.00426 | 3.30E-85 |
| rs174568 | miR-1908-5 | cis | 3.76E-08 | Nightingale | L_VLDL_PL | NA | T | C | 0.087815 | 0.004216 | 8.70E-100 |
| rs174568 | miR-1908-5 | cis | 3.76E-08 | Nightingale | L_LDL_FC | NA | T | C | -0.06854 | 0.004266 | 1.60E-61 |
| rs174568 | miR-1908-5 | cis | 3.76E-08 | Nightingale | XL_HDL_PL | NA | T | C | -0.06722 | 0.003991 | 8.00E-69 |
| rs174568 | miR-1908-5 | cis | 3.76E-08 | Nightingale | XXL_VLDL_ | NA | T | C | 0.066241 | 0.004179 | 2.80E-59 |
| rs174568 | miR-1908-5 | cis | 3.76E-08 | Nightingale | S_LDL_FC | NA | T | C | -0.06296 | 0.004337 | 1.30E-50 |
| rs174568 | miR-1908-5 | cis | 3.76E-08 | Nightingale | L_VLDL_L | NA | T | C | 0.038279 | 0.004198 | 1.50E-20 |
| rs174568 | miR-1908-5 | cis | 3.76E-08 | Nightingale | HDL_PL | NA | T | C | -0.04855 | 0.003928 | 9.20E-38 |
| rs174568 | miR-1908-5 | cis | 3.76E-08 | Nightingale | XS_VLDL_C | NA | T | C | -0.09218 | 0.004095 | 2.80E-117 |
| rs174568 | miR-1908-5 | cis | 3.76E-08 | Nightingale | M_LDL_TG | NA | T | C | 0.075809 | 0.00429 | 2.40E-71 |
| rs174568 | miR-1908-5 | cis | 3.76E-08 | Nightingale | SFA_pct | NA | T | C | -0.04232 | 0.004286 | 5.40E-23 |
| rs174568 | miR-1908-5 | cis | 3.76E-08 | Nightingale | S_VLDL_PL | NA | T | C | -0.07849 | 0.004157 | 2.50E-83 |
| rs174568 | miR-1908-5 | cis | 3.76E-08 | Nightingale | L_VLDL_C | NA | T | C | 0.028944 | 0.004249 | 4.60E-12 |
| rs174568 | miR-1908-5 | cis | 3.76E-08 | Nightingale | L_HDL_L | NA | T | C | -0.07756 | 0.003812 | 1.90E-100 |
| rs174568 | miR-1908-5 | cis | 3.76E-08 | Nightingale | S_VLDL_FC | NA | T | C | -0.08291 | 0.004148 | 8.30E-93 |
| rs174568 | miR-1908-5 | cis | 3.76E-08 | Nightingale | XL_HDL_FC | NA | T | C | -0.10302 | 0.004047 | 1.80E-154 |
| rs174568 | miR-1908-5 | cis | 3.76E-08 | Nightingale | M_HDL_TG | NA | T | C | 0.050436 | 0.004291 | 9.00E-33 |
| rs174568 | miR-1908-5 | cis | 3.76E-08 | Nightingale | S_VLDL_TG | NA | T | C | 0.064421 | 0.004204 | 1.30E-54 |
| rs174568 | miR-1908-5 | cis | 3.76E-08 | Nightingale | L_VLDL_C_ | NA | T | C | -0.03854 | 0.004244 | 6.30E-21 |

| | | | | | | | | | | | |
|---|---|---|---|---|---|---|---|---|---|---|---|
| rs174568 | miR-1908-5 | cis | 3.76E-08 | Nightingale | S_LDL_L | NA | T | C | -0.03263 | 0.004344 | 8.80E-15 |
| rs174568 | miR-1908-5 | cis | 3.76E-08 | Nightingale | Omega_3_ | NA | T | C | -0.38839 | 0.004245 | 0 |
| rs174568 | miR-1908-5 | cis | 3.76E-08 | Nightingale | XS_VLDL_P | NA | T | C | -0.04385 | 0.004287 | 1.30E-26 |
| rs174568 | miR-1908-5 | cis | 3.76E-08 | Nightingale | LA_pct | NA | T | C | 0.184342 | 0.004246 | 0 |
| rs174568 | miR-1908-5 | cis | 3.76E-08 | Nightingale | L_HDL_TG_ | NA | T | C | 0.078962 | 0.0041 | 7.10E-88 |
| rs174568 | miR-1908-5 | cis | 3.76E-08 | Nightingale | XL_VLDL_F | NA | T | C | -0.0683 | 0.004209 | 6.20E-62 |
| rs174568 | miR-1908-5 | cis | 3.76E-08 | Nightingale | M_HDL_PL | NA | T | C | 0.058703 | 0.004121 | 5.90E-48 |
| rs174568 | miR-1908-5 | cis | 3.76E-08 | Nightingale | S_LDL_C | NA | T | C | -0.03232 | 0.004349 | 1.90E-14 |
| rs174568 | miR-1908-5 | cis | 3.76E-08 | Nightingale | HDL_P | NA | T | C | -0.0356 | 0.004079 | 1.80E-19 |
| rs174568 | miR-1908-5 | cis | 3.76E-08 | Nightingale | VLDL_TG | NA | T | C | 0.047783 | 0.004181 | 2.90E-31 |
| rs174568 | miR-1908-5 | cis | 3.76E-08 | Nightingale | XXL_VLDL_ | NA | T | C | -0.0272 | 0.004322 | 6.60E-11 |
| rs174568 | miR-1908-5 | cis | 3.76E-08 | Nightingale | VLDL_size | NA | T | C | 0.072611 | 0.00405 | 4.10E-76 |
| rs174568 | miR-1908-5 | cis | 3.76E-08 | Nightingale | M_LDL_CE | NA | T | C | -0.02641 | 0.004351 | 4.80E-10 |
| rs174568 | miR-1908-5 | cis | 3.76E-08 | Nightingale | XL_VLDL_C | NA | T | C | 0.030424 | 0.004276 | 5.80E-13 |
| rs174568 | miR-1908-5 | cis | 3.76E-08 | Nightingale | LDL_CE | NA | T | C | -0.04375 | 0.004333 | 3.30E-25 |
| rs174568 | miR-1908-5 | cis | 3.76E-08 | Nightingale | L_HDL_FC | NA | T | C | -0.08498 | 0.003807 | 4.40E-120 |
| rs174568 | miR-1908-5 | cis | 3.76E-08 | Nightingale | Clinical_LD | NA | T | C | -0.05532 | 0.00432 | 2.10E-39 |
| rs174568 | miR-1908-5 | cis | 3.76E-08 | Nightingale | XS_VLDL_L | NA | T | C | -0.04182 | 0.00427 | 9.60E-25 |
| rs174568 | miR-1908-5 | cis | 3.76E-08 | Nightingale | TG_by_PG | NA | T | C | 0.068851 | 0.004032 | 1.20E-68 |
| rs174568 | miR-1908-5 | cis | 3.76E-08 | Nightingale | M_LDL_FC | NA | T | C | -0.05255 | 0.004339 | 1.40E-35 |
| rs174568 | miR-1908-5 | cis | 3.76E-08 | Nightingale | M_LDL_C_ | NA | T | C | -0.10018 | 0.004348 | 5.79E-118 |
| rs174568 | miR-1908-5 | cis | 3.76E-08 | Nightingale | HDL_C | NA | T | C | -0.06497 | 0.003879 | 2.90E-68 |
| rs174568 | miR-1908-5 | cis | 3.76E-08 | Nightingale | VLDL_L | NA | T | C | 0.027418 | 0.00426 | 7.50E-11 |
| rs174568 | miR-1908-5 | cis | 3.76E-08 | Nightingale | L_VLDL_FC | NA | T | C | 0.040229 | 0.004202 | 1.50E-22 |
| rs174568 | miR-1908-5 | cis | 3.76E-08 | Nightingale | XXL_VLDL_ | NA | T | C | 0.073706 | 0.004195 | 1.50E-71 |
| rs174568 | miR-1908-5 | cis | 3.76E-08 | Nightingale | L_LDL_CE | NA | T | C | -0.0535 | 0.004306 | 1.80E-37 |
| rs174568 | miR-1908-5 | cis | 3.76E-08 | Nightingale | XL_HDL_FC | NA | T | C | 0.061628 | 0.003852 | 6.60E-64 |
| rs174568 | miR-1908-5 | cis | 3.76E-08 | Nightingale | XL_VLDL_C | NA | T | C | -0.0612 | 0.004165 | 2.90E-50 |
| rs174568 | miR-1908-5 | cis | 3.76E-08 | Nightingale | PUFA_by_M | NA | T | C | -0.17039 | 0.004161 | 0 |
| rs174568 | miR-1908-5 | cis | 3.76E-08 | Nightingale | HDL_L | NA | T | C | -0.05352 | 0.003901 | 3.70E-46 |
| rs174568 | miR-1908-5 | cis | 3.76E-08 | Nightingale | IDL_CE | NA | T | C | -0.07245 | 0.004194 | 6.40E-71 |
| rs174568 | miR-1908-5 | cis | 3.76E-08 | Nightingale | M_LDL_L | NA | T | C | -0.02829 | 0.004351 | 2.70E-11 |
| rs174568 | miR-1908-5 | cis | 3.76E-08 | Nightingale | M_LDL_P | NA | T | C | -0.0467 | 0.004357 | 9.50E-28 |
| rs174568 | miR-1908-5 | cis | 3.76E-08 | Nightingale | L_LDL_PL | NA | T | C | -0.05512 | 0.004312 | 3.20E-39 |
| rs174568 | miR-1908-5 | cis | 3.76E-08 | Nightingale | S_VLDL_TG | NA | T | C | 0.038782 | 0.004216 | 9.20E-21 |
| rs174568 | miR-1908-5 | cis | 3.76E-08 | Nightingale | M_HDL_PL | NA | T | C | -0.02149 | 0.004063 | 3.30E-08 |
| rs174568 | miR-1908-5 | cis | 3.76E-08 | Nightingale | M_VLDL_F | NA | T | C | -0.02947 | 0.004344 | 2.90E-12 |
| rs174568 | miR-1908-5 | cis | 3.76E-08 | Nightingale | Omega_3 | NA | T | C | -0.33452 | 0.004258 | 0 |
| rs174568 | miR-1908-5 | cis | 3.76E-08 | Nightingale | ApoA1 | NA | T | C | -0.04128 | 0.003976 | 2.70E-27 |
| rs174568 | miR-1908-5 | cis | 3.76E-08 | Nightingale | HDL_FC | NA | T | C | -0.07418 | 0.003849 | 8.60E-90 |
| rs174568 | miR-1908-5 | cis | 3.76E-08 | Nightingale | LDL_PL | NA | T | C | -0.04633 | 0.004335 | 6.10E-28 |
| rs174568 | miR-1908-5 | cis | 3.76E-08 | Nightingale | IDL_TG_pct | NA | T | C | 0.088155 | 0.004197 | 1.10E-101 |
| rs174568 | miR-1908-5 | cis | 3.76E-08 | Nightingale | L_VLDL_TG | NA | T | C | 0.037379 | 0.00418 | 8.10E-20 |
| rs174568 | miR-1908-5 | cis | 3.76E-08 | Nightingale | XS_VLDL_T | NA | T | C | 0.031463 | 0.004278 | 6.70E-13 |
| rs174568 | miR-1908-5 | cis | 3.76E-08 | Nightingale | S_HDL_C_p | NA | T | C | -0.04251 | 0.004294 | 5.90E-23 |
| rs174568 | miR-1908-5 | cis | 3.76E-08 | Nightingale | M_HDL_FC | NA | T | C | -0.05161 | 0.003953 | 7.80E-42 |
| rs174568 | miR-1908-5 | cis | 3.76E-08 | Nightingale | XL_VLDL_P | NA | T | C | 0.048357 | 0.004192 | 6.70E-32 |
| rs174568 | miR-1908-5 | cis | 3.76E-08 | Nightingale | Tyr | NA | T | C | 0.033189 | 0.004339 | 2.00E-14 |
| rs174568 | miR-1908-5 | cis | 3.76E-08 | Nightingale | IDL_L | NA | T | C | -0.07262 | 0.0042 | 1.70E-71 |
| rs174568 | miR-1908-5 | cis | 3.76E-08 | Nightingale | IDL_CE_pct | NA | T | C | -0.03804 | 0.004263 | 5.10E-19 |
| rs174568 | miR-1908-5 | cis | 3.76E-08 | Nightingale | Cholines | NA | T | C | -0.03732 | 0.004086 | 1.00E-21 |
| rs174568 | miR-1908-5 | cis | 3.76E-08 | Nightingale | LDL_TG | NA | T | C | 0.029496 | 0.004296 | 1.30E-11 |
| rs174568 | miR-1908-5 | cis | 3.76E-08 | Nightingale | MUFA_pct | NA | T | C | 0.197006 | 0.004157 | 0 |
| rs174568 | miR-1908-5 | cis | 3.76E-08 | Nightingale | Omega_6_ | NA | T | C | 0.023642 | 0.004228 | 1.20E-08 |
| rs174568 | miR-1908-5 | cis | 3.76E-08 | Nightingale | M_VLDL_C | NA | T | C | -0.08073 | 0.004089 | 2.10E-90 |
| rs174568 | miR-1908-5 | cis | 3.76E-08 | Nightingale | XL_HDL_CE | NA | T | C | -0.02622 | 0.004342 | 3.90E-09 |
| rs174568 | miR-1908-5 | cis | 3.76E-08 | Nightingale | M_HDL_CE | NA | T | C | -0.03793 | 0.004012 | 1.40E-22 |
| rs174568 | miR-1908-5 | cis | 3.76E-08 | Nightingale | S_VLDL_C_ | NA | T | C | -0.05651 | 0.004227 | 6.90E-42 |
| rs174568 | miR-1908-5 | cis | 3.76E-08 | Nightingale | Remnant_( | NA | T | C | -0.04421 | 0.004316 | 3.50E-26 |
| rs174568 | miR-1908-5 | cis | 3.76E-08 | Nightingale | M_LDL_PL | NA | T | C | -0.02365 | 0.004351 | 2.30E-08 |
| rs174568 | miR-1908-5 | cis | 3.76E-08 | Nightingale | S_LDL_TG_ | NA | T | C | 0.086337 | 0.00421 | 1.60E-96 |
| rs174568 | miR-1908-5 | cis | 3.76E-08 | Nightingale | XL_VLDL_T | NA | T | C | 0.057013 | 0.004155 | 1.60E-44 |
| rs174568 | miR-1908-5 | cis | 3.76E-08 | Nightingale | L_HDL_PL_ | NA | T | C | 0.043367 | 0.004205 | 9.40E-27 |

| | | | | | | | | | | | |
|---|---|---|---|---|---|---|---|---|---|---|---|
| rs174568 | miR-1908-5 | cis | 3.76E-08 | Nightingale | L_HDL_CE_ | NA | T | C | -0.05035 | 0.004157 | 4.60E-36 |
| rs174568 | miR-1908-5 | cis | 3.76E-08 | Nightingale | M_VLDL_T | NA | T | C | 0.077998 | 0.004103 | 2.20E-84 |
| rs174568 | miR-1908-5 | cis | 3.76E-08 | Nightingale | S_LDL_PL_ | NA | T | C | -0.06209 | 0.004336 | 9.30E-49 |
| rs174568 | miR-1908-5 | cis | 3.76E-08 | Nightingale | MUFA | NA | T | C | 0.063775 | 0.0043 | 8.20E-51 |
| rs174568 | miR-1908-5 | cis | 3.76E-08 | Nightingale | Sphingomy | NA | T | C | -0.11528 | 0.00406 | 1.60E-186 |
| rs174568 | miR-1908-5 | cis | 3.76E-08 | Nightingale | M_HDL_TG | NA | T | C | 0.066832 | 0.004166 | 1.20E-60 |
| rs174568 | miR-1908-5 | cis | 3.76E-08 | Nightingale | XL_VLDL_P | NA | T | C | 0.052945 | 0.004176 | 3.10E-38 |
| rs174568 | miR-1908-5 | cis | 3.76E-08 | Nightingale | XL_HDL_P | NA | T | C | -0.09001 | 0.00391 | 1.40E-128 |
| rs174568 | miR-1908-5 | cis | 3.76E-08 | Nightingale | L_LDL_FC_ | NA | T | C | -0.08299 | 0.004141 | 1.90E-92 |
| rs174568 | miR-1908-5 | cis | 3.76E-08 | Nightingale | L_HDL_CE | NA | T | C | -0.07905 | 0.003836 | 1.20E-102 |
| rs174568 | miR-1908-5 | cis | 3.76E-08 | Nightingale | ApoB | NA | T | C | -0.03861 | 0.004339 | 4.60E-20 |
| rs174568 | miR-1908-5 | cis | 3.76E-08 | Nightingale | Total_FC | NA | T | C | -0.05956 | 0.004244 | 1.90E-47 |
| rs174568 | miR-1908-5 | cis | 3.76E-08 | Nightingale | M_LDL_C | NA | T | C | -0.03415 | 0.004353 | 9.50E-16 |
| rs174568 | miR-1908-5 | cis | 3.76E-08 | Nightingale | M_HDL_L | NA | T | C | -0.02772 | 0.004033 | 8.00E-13 |
| rs174568 | miR-1908-5 | cis | 3.76E-08 | Nightingale | L_VLDL_FC | NA | T | C | 0.028483 | 0.004303 | 2.00E-11 |
| rs174568 | miR-1908-5 | cis | 3.76E-08 | Nightingale | L_HDL_FC_ | NA | T | C | -0.11278 | 0.004036 | 2.20E-187 |
| rs174568 | miR-1908-5 | cis | 3.76E-08 | Nightingale | XL_HDL_C | NA | T | C | -0.09957 | 0.003917 | 8.00E-155 |
| rs174572 | miR-1908-5 | cis | 3.57E-05 | Nightingale | IDL_TG_pc | NA | T | C | 0.065764 | 0.004495 | 1.10E-51 |
| rs174572 | miR-1908-5 | cis | 3.57E-05 | Nightingale | DHA_pct | NA | T | C | -0.21107 | 0.004517 | 0 |
| rs174572 | miR-1908-5 | cis | 3.57E-05 | Nightingale | HDL_C | NA | T | C | -0.04402 | 0.004155 | 1.70E-29 |
| rs174572 | miR-1908-5 | cis | 3.57E-05 | Nightingale | LDL_CE | NA | T | C | -0.03278 | 0.004641 | 2.40E-13 |
| rs174572 | miR-1908-5 | cis | 3.57E-05 | Nightingale | M_HDL_FC | NA | T | C | -0.07396 | 0.004123 | 3.00E-81 |
| rs174572 | miR-1908-5 | cis | 3.57E-05 | Nightingale | IDL_L | NA | T | C | -0.05293 | 0.004498 | 1.10E-34 |
| rs174572 | miR-1908-5 | cis | 3.57E-05 | Nightingale | S_LDL_TG_ | NA | T | C | 0.064769 | 0.004509 | 1.50E-49 |
| rs174572 | miR-1908-5 | cis | 3.57E-05 | Nightingale | XL_HDL_L | NA | T | C | -0.06959 | 0.004181 | 9.80E-70 |
| rs174572 | miR-1908-5 | cis | 3.57E-05 | Nightingale | LDL_C | NA | T | C | -0.03726 | 0.004633 | 7.30E-17 |
| rs174572 | miR-1908-5 | cis | 3.57E-05 | Nightingale | M_HDL_TG | NA | T | C | 0.047731 | 0.004462 | 8.20E-29 |
| rs174572 | miR-1908-5 | cis | 3.57E-05 | Nightingale | M_LDL_PL_ | NA | T | C | 0.038949 | 0.004651 | 2.50E-17 |
| rs174572 | miR-1908-5 | cis | 3.57E-05 | Nightingale | IDL_CE | NA | T | C | -0.0524 | 0.004492 | 4.30E-34 |
| rs174572 | miR-1908-5 | cis | 3.57E-05 | Nightingale | XXL_VLDL_ | NA | T | C | 0.050388 | 0.004482 | 5.00E-31 |
| rs174572 | miR-1908-5 | cis | 3.57E-05 | Nightingale | PUFA_by_I | NA | T | C | -0.12933 | 0.004457 | 4.30E-190 |
| rs174572 | miR-1908-5 | cis | 3.57E-05 | Nightingale | XL_HDL_P | NA | T | C | -0.06676 | 0.004188 | 6.10E-65 |
| rs174572 | miR-1908-5 | cis | 3.57E-05 | Nightingale | M_VLDL_T | NA | T | C | 0.056437 | 0.004395 | 4.10E-40 |
| rs174572 | miR-1908-5 | cis | 3.57E-05 | Nightingale | L_LDL_TG_ | NA | T | C | 0.058812 | 0.004563 | 9.40E-40 |
| rs174572 | miR-1908-5 | cis | 3.57E-05 | Nightingale | SFA_pct | NA | T | C | -0.03213 | 0.004591 | 2.60E-12 |
| rs174572 | miR-1908-5 | cis | 3.57E-05 | Nightingale | HDL_FC | NA | T | C | -0.05114 | 0.004122 | 1.20E-39 |
| rs174572 | miR-1908-5 | cis | 3.57E-05 | Nightingale | M_LDL_FC | NA | T | C | -0.039 | 0.004648 | 2.50E-18 |
| rs174572 | miR-1908-5 | cis | 3.57E-05 | Nightingale | XL_VLDL_T | NA | T | C | 0.04145 | 0.004451 | 1.10E-21 |
| rs174572 | miR-1908-5 | cis | 3.57E-05 | Nightingale | MUFA_pct | NA | T | C | 0.149644 | 0.004453 | 6.81E-256 |
| rs174572 | miR-1908-5 | cis | 3.57E-05 | Nightingale | XL_VLDL_C | NA | T | C | -0.0439 | 0.004461 | 7.70E-24 |
| rs174572 | miR-1908-5 | cis | 3.57E-05 | Nightingale | XL_HDL_C | NA | T | C | -0.07429 | 0.004196 | 1.20E-78 |
| rs174572 | miR-1908-5 | cis | 3.57E-05 | Nightingale | HDL_size | NA | T | C | -0.05181 | 0.004123 | 3.30E-40 |
| rs174572 | miR-1908-5 | cis | 3.57E-05 | Nightingale | Remnant_( | NA | T | C | -0.03295 | 0.004623 | 1.20E-13 |
| rs174572 | miR-1908-5 | cis | 3.57E-05 | Nightingale | M_LDL_TG | NA | T | C | 0.057856 | 0.004595 | 7.50E-38 |
| rs174572 | miR-1908-5 | cis | 3.57E-05 | Nightingale | Cholines | NA | T | C | -0.02409 | 0.004376 | 5.50E-09 |
| rs174572 | miR-1908-5 | cis | 3.57E-05 | Nightingale | L_VLDL_FC | NA | T | C | 0.028073 | 0.004501 | 9.00E-11 |
| rs174572 | miR-1908-5 | cis | 3.57E-05 | Nightingale | Omega_6_ | NA | T | C | 0.288612 | 0.004586 | 0 |
| rs174572 | miR-1908-5 | cis | 3.57E-05 | Nightingale | M_LDL_C | NA | T | C | -0.02591 | 0.004662 | 1.00E-08 |
| rs174572 | miR-1908-5 | cis | 3.57E-05 | Nightingale | M_HDL_TG | NA | T | C | 0.039022 | 0.004596 | 2.70E-18 |
| rs174572 | miR-1908-5 | cis | 3.57E-05 | Nightingale | XS_VLDL_F | NA | T | C | -0.08944 | 0.004618 | 1.70E-86 |
| rs174572 | miR-1908-5 | cis | 3.57E-05 | Nightingale | XXL_VLDL_ | NA | T | C | 0.029098 | 0.004679 | 8.60E-11 |
| rs174572 | miR-1908-5 | cis | 3.57E-05 | Nightingale | XL_VLDL_P | NA | T | C | 0.034655 | 0.00449 | 1.70E-15 |
| rs174572 | miR-1908-5 | cis | 3.57E-05 | Nightingale | L_HDL_CE_ | NA | T | C | -0.03767 | 0.004452 | 1.20E-19 |
| rs174572 | miR-1908-5 | cis | 3.57E-05 | Nightingale | M_VLDL_C | NA | T | C | -0.05468 | 0.004368 | 6.10E-38 |
| rs174572 | miR-1908-5 | cis | 3.57E-05 | Nightingale | L_LDL_P | NA | T | C | -0.02865 | 0.004637 | 1.10E-10 |
| rs174572 | miR-1908-5 | cis | 3.57E-05 | Nightingale | IDL_PL | NA | T | C | -0.05443 | 0.00449 | 6.20E-37 |
| rs174572 | miR-1908-5 | cis | 3.57E-05 | Nightingale | S_LDL_FC_ | NA | T | C | -0.05811 | 0.004509 | 4.00E-40 |
| rs174572 | miR-1908-5 | cis | 3.57E-05 | Nightingale | S_LDL_L | NA | T | C | -0.02463 | 0.004653 | 3.40E-08 |
| rs174572 | miR-1908-5 | cis | 3.57E-05 | Nightingale | XL_HDL_PL | NA | T | C | -0.05098 | 0.004275 | 7.40E-36 |
| rs174572 | miR-1908-5 | cis | 3.57E-05 | Nightingale | S_VLDL_FC | NA | T | C | -0.0606 | 0.004443 | 3.30E-45 |
| rs174572 | miR-1908-5 | cis | 3.57E-05 | Nightingale | M_VLDL_P | NA | T | C | -0.04472 | 0.004465 | 6.70E-26 |
| rs174572 | miR-1908-5 | cis | 3.57E-05 | Nightingale | XS_VLDL_P | NA | T | C | -0.03317 | 0.004591 | 3.70E-14 |
| rs174572 | miR-1908-5 | cis | 3.57E-05 | Nightingale | XXL_VLDL_ | NA | T | C | 0.055961 | 0.004494 | 1.40E-37 |

| | | | | | | | | | | | |
|---|---|---|---|---|---|---|---|---|---|---|---|
| rs174572 | miR-1908-5 | cis | 3.57E-05 | Nightingale | XS_VLDL_C | NA | T | C | -0.05118 | 0.004516 | 3.00E-32 |
| rs174572 | miR-1908-5 | cis | 3.57E-05 | Nightingale | L_HDL_PL_ | NA | T | C | 0.035227 | 0.004504 | 8.30E-17 |
| rs174572 | miR-1908-5 | cis | 3.57E-05 | Nightingale | IDL_CE_pct | NA | T | C | -0.02562 | 0.004566 | 1.40E-08 |
| rs174572 | miR-1908-5 | cis | 3.57E-05 | Nightingale | S_VLDL_TG | NA | T | C | 0.026732 | 0.004516 | 7.10E-10 |
| rs174572 | miR-1908-5 | cis | 3.57E-05 | Nightingale | S_HDL_C_p | NA | T | C | -0.0307 | 0.004599 | 1.80E-11 |
| rs174572 | miR-1908-5 | cis | 3.57E-05 | Nightingale | L_HDL_PL | NA | T | C | -0.05349 | 0.004095 | 1.40E-43 |
| rs174572 | miR-1908-5 | cis | 3.57E-05 | Nightingale | M_VLDL_C | NA | T | C | -0.03432 | 0.004641 | 1.80E-14 |
| rs174572 | miR-1908-5 | cis | 3.57E-05 | Nightingale | M_VLDL_C_ | NA | T | C | -0.05859 | 0.00438 | 4.20E-43 |
| rs174572 | miR-1908-5 | cis | 3.57E-05 | Nightingale | M_LDL_C_ | NA | T | C | -0.07882 | 0.004657 | 6.60E-65 |
| rs174572 | miR-1908-5 | cis | 3.57E-05 | Nightingale | S_VLDL_TG | NA | T | C | 0.04647 | 0.004502 | 3.10E-26 |
| rs174572 | miR-1908-5 | cis | 3.57E-05 | Nightingale | S_LDL_P | NA | T | C | -0.02526 | 0.004658 | 2.40E-08 |
| rs174572 | miR-1908-5 | cis | 3.57E-05 | Nightingale | IDL_FC | NA | T | C | -0.06601 | 0.004504 | 2.10E-52 |
| rs174572 | miR-1908-5 | cis | 3.57E-05 | Nightingale | XL_VLDL_C | NA | T | C | -0.04682 | 0.004459 | 8.10E-27 |
| rs174572 | miR-1908-5 | cis | 3.57E-05 | Nightingale | XL_HDL_FC | NA | T | C | -0.07833 | 0.004335 | 8.70E-81 |
| rs174572 | miR-1908-5 | cis | 3.57E-05 | Nightingale | Total_CE | NA | T | C | -0.04871 | 0.004488 | 1.40E-29 |
| rs174572 | miR-1908-5 | cis | 3.57E-05 | Nightingale | Total_L | NA | T | C | -0.02698 | 0.004578 | 1.20E-09 |
| rs174572 | miR-1908-5 | cis | 3.57E-05 | Nightingale | L_HDL_CE | NA | T | C | -0.05639 | 0.004109 | 4.10E-48 |
| rs174572 | miR-1908-5 | cis | 3.57E-05 | Nightingale | M_HDL_FC | NA | T | C | -0.03344 | 0.004234 | 9.20E-17 |
| rs174572 | miR-1908-5 | cis | 3.57E-05 | Nightingale | ApoA1 | NA | T | C | -0.02589 | 0.004258 | 1.30E-10 |
| rs174572 | miR-1908-5 | cis | 3.57E-05 | Nightingale | Citrate | NA | T | C | -0.02815 | 0.004624 | 3.70E-10 |
| rs174572 | miR-1908-5 | cis | 3.57E-05 | Nightingale | L_VLDL_P | NA | T | C | 0.02921 | 0.004493 | 1.60E-11 |
| rs174572 | miR-1908-5 | cis | 3.57E-05 | Nightingale | Omega_3_ | NA | T | C | -0.30369 | 0.004547 | 0 |
| rs174572 | miR-1908-5 | cis | 3.57E-05 | Nightingale | L_HDL_L | NA | T | C | -0.05465 | 0.004083 | 7.30E-46 |
| rs174572 | miR-1908-5 | cis | 3.57E-05 | Nightingale | S_HDL_TG_ | NA | T | C | 0.04953 | 0.004426 | 3.40E-31 |
| rs174572 | miR-1908-5 | cis | 3.57E-05 | Nightingale | L_HDL_C_p | NA | T | C | -0.0513 | 0.004386 | 1.20E-35 |
| rs174572 | miR-1908-5 | cis | 3.57E-05 | Nightingale | XL_VLDL_F | NA | T | C | -0.05279 | 0.004509 | 2.70E-33 |
| rs174572 | miR-1908-5 | cis | 3.57E-05 | Nightingale | M_LDL_P | NA | T | C | -0.03646 | 0.004667 | 1.90E-15 |
| rs174572 | miR-1908-5 | cis | 3.57E-05 | Nightingale | non_HDL_( | NA | T | C | -0.03568 | 0.004628 | 1.50E-15 |
| rs174572 | miR-1908-5 | cis | 3.57E-05 | Nightingale | XS_VLDL_P | NA | T | C | -0.02705 | 0.004588 | 4.10E-10 |
| rs174572 | miR-1908-5 | cis | 3.57E-05 | Nightingale | HDL_CE | NA | T | C | -0.04163 | 0.004176 | 3.00E-26 |
| rs174572 | miR-1908-5 | cis | 3.57E-05 | Nightingale | Total_FC | NA | T | C | -0.04391 | 0.004546 | 9.90E-24 |
| rs174572 | miR-1908-5 | cis | 3.57E-05 | Nightingale | L_VLDL_TG | NA | T | C | 0.02562 | 0.004477 | 3.80E-09 |
| rs174572 | miR-1908-5 | cis | 3.57E-05 | Nightingale | XL_HDL_FC | NA | T | C | 0.044273 | 0.004126 | 1.20E-30 |
| rs174572 | miR-1908-5 | cis | 3.57E-05 | Nightingale | PUFA | NA | T | C | -0.07246 | 0.004538 | 1.30E-60 |
| rs174572 | miR-1908-5 | cis | 3.57E-05 | Nightingale | L_VLDL_C_ | NA | T | C | -0.02907 | 0.004546 | 3.10E-11 |
| rs174572 | miR-1908-5 | cis | 3.57E-05 | Nightingale | Unsaturati | NA | T | C | -0.2645 | 0.004407 | 0 |
| rs174572 | miR-1908-5 | cis | 3.57E-05 | Nightingale | L_HDL_P | NA | T | C | -0.05537 | 0.004075 | 2.80E-47 |
| rs174572 | miR-1908-5 | cis | 3.57E-05 | Nightingale | L_VLDL_PL | NA | T | C | 0.034611 | 0.00449 | 1.40E-15 |
| rs174572 | miR-1908-5 | cis | 3.57E-05 | Nightingale | XXL_VLDL_ | NA | T | C | 0.053965 | 0.004484 | 2.10E-35 |
| rs174572 | miR-1908-5 | cis | 3.57E-05 | Nightingale | Omega_3 | NA | T | C | -0.26051 | 0.004561 | 0 |
| rs174572 | miR-1908-5 | cis | 3.57E-05 | Nightingale | Clinical_LD | NA | T | C | -0.041 | 0.004627 | 4.40E-20 |
| rs174572 | miR-1908-5 | cis | 3.57E-05 | Nightingale | XL_HDL_TG | NA | T | C | 0.064848 | 0.004383 | 1.10E-53 |
| rs174572 | miR-1908-5 | cis | 3.57E-05 | Nightingale | LDL_PL | NA | T | C | -0.03408 | 0.004643 | 3.20E-14 |
| rs174572 | miR-1908-5 | cis | 3.57E-05 | Nightingale | M_LDL_TG | NA | T | C | 0.027581 | 0.004585 | 9.40E-10 |
| rs174572 | miR-1908-5 | cis | 3.57E-05 | Nightingale | HDL_TG | NA | T | C | 0.03528 | 0.004588 | 3.60E-15 |
| rs174572 | miR-1908-5 | cis | 3.57E-05 | Nightingale | LDL_P | NA | T | C | -0.0311 | 0.00465 | 4.50E-12 |
| rs174572 | miR-1908-5 | cis | 3.57E-05 | Nightingale | L_LDL_CE | NA | T | C | -0.03982 | 0.004612 | 2.60E-19 |
| rs174572 | miR-1908-5 | cis | 3.57E-05 | Nightingale | HDL_L | NA | T | C | -0.03513 | 0.004179 | 6.00E-19 |
| rs174572 | miR-1908-5 | cis | 3.57E-05 | Nightingale | LDL_FC | NA | T | C | -0.04775 | 0.004604 | 2.70E-27 |
| rs174572 | miR-1908-5 | cis | 3.57E-05 | Nightingale | M_VLDL_C | NA | T | C | -0.04046 | 0.004611 | 1.10E-19 |
| rs174572 | miR-1908-5 | cis | 3.57E-05 | Nightingale | XL_VLDL_C | NA | T | C | 0.026601 | 0.004545 | 1.30E-09 |
| rs174572 | miR-1908-5 | cis | 3.57E-05 | Nightingale | L_LDL_L | NA | T | C | -0.04014 | 0.004608 | 1.30E-19 |
| rs174572 | miR-1908-5 | cis | 3.57E-05 | Nightingale | L_VLDL_CE | NA | T | C | -0.04478 | 0.004453 | 2.30E-25 |
| rs174572 | miR-1908-5 | cis | 3.57E-05 | Nightingale | L_LDL_C | NA | T | C | -0.04307 | 0.004601 | 1.80E-22 |
| rs174572 | miR-1908-5 | cis | 3.57E-05 | Nightingale | XXL_VLDL_ | NA | T | C | 0.049187 | 0.004476 | 6.20E-30 |
| rs174572 | miR-1908-5 | cis | 3.57E-05 | Nightingale | XS_VLDL_P | NA | T | C | 0.038229 | 0.00454 | 4.80E-18 |
| rs174572 | miR-1908-5 | cis | 3.57E-05 | Nightingale | M_HDL_C_ | NA | T | C | -0.04706 | 0.00443 | 1.70E-28 |
| rs174572 | miR-1908-5 | cis | 3.57E-05 | Nightingale | XS_VLDL_F | NA | T | C | -0.04311 | 0.004572 | 7.40E-23 |
| rs174572 | miR-1908-5 | cis | 3.57E-05 | Nightingale | S_LDL_PL_ | NA | T | C | -0.04812 | 0.004644 | 1.90E-27 |
| rs174572 | miR-1908-5 | cis | 3.57E-05 | Nightingale | XL_VLDL_P | NA | T | C | 0.038109 | 0.004472 | 1.60E-18 |
| rs174572 | miR-1908-5 | cis | 3.57E-05 | Nightingale | L_HDL_C | NA | T | C | -0.05745 | 0.004098 | 3.10E-50 |
| rs174572 | miR-1908-5 | cis | 3.57E-05 | Nightingale | XL_HDL_CE | NA | T | C | -0.0722 | 0.004167 | 1.40E-75 |
| rs174572 | miR-1908-5 | cis | 3.57E-05 | Nightingale | XL_HDL_PL | NA | T | C | -0.06671 | 0.004183 | 6.20E-64 |

| | | | | | | | | | | | |
|---|---|---|---|---|---|---|---|---|---|---|---|
| rs174572 | miR-1908-! | cis | 3.57E-05 | Nightingale | XS_VLDL_C | NA | T | C | -0.05361 | 0.00449 | 1.50E-35 |
| rs174572 | miR-1908-! | cis | 3.57E-05 | Nightingale | M_HDL_CE | NA | T | C | -0.0332 | 0.004512 | 2.90E-14 |
| rs174572 | miR-1908-! | cis | 3.57E-05 | Nightingale | LDL_L | NA | T | C | -0.03383 | 0.004638 | 3.70E-14 |
| rs174572 | miR-1908-! | cis | 3.57E-05 | Nightingale | M_HDL_PL | NA | T | C | 0.042028 | 0.004414 | 3.90E-23 |
| rs174572 | miR-1908-! | cis | 3.57E-05 | Nightingale | IDL_C | NA | T | C | -0.05627 | 0.004493 | 5.60E-39 |
| rs174572 | miR-1908-! | cis | 3.57E-05 | Nightingale | M_LDL_FC_ | NA | T | C | -0.05093 | 0.004458 | 4.80E-32 |
| rs174572 | miR-1908-! | cis | 3.57E-05 | Nightingale | IDL_C_pct | NA | T | C | -0.0534 | 0.004543 | 1.90E-33 |
| rs174572 | miR-1908-! | cis | 3.57E-05 | Nightingale | M_HDL_CE | NA | T | C | -0.02302 | 0.004297 | 1.70E-08 |
| rs174572 | miR-1908-! | cis | 3.57E-05 | Nightingale | L_HDL_TG_ | NA | T | C | 0.056685 | 0.004391 | 6.00E-42 |
| rs174572 | miR-1908-! | cis | 3.57E-05 | Nightingale | L_LDL_PL | NA | T | C | -0.04035 | 0.004618 | 1.60E-19 |
| rs174572 | miR-1908-! | cis | 3.57E-05 | Nightingale | L_VLDL_PL | NA | T | C | 0.065514 | 0.004516 | 2.10E-50 |
| rs174572 | miR-1908-! | cis | 3.57E-05 | Nightingale | L_HDL_FC | NA | T | C | -0.0605 | 0.004078 | 7.10E-56 |
| rs174572 | miR-1908-! | cis | 3.57E-05 | Nightingale | L_LDL_C_p | NA | T | C | -0.0591 | 0.004534 | 2.10E-41 |
| rs174572 | miR-1908-! | cis | 3.57E-05 | Nightingale | S_VLDL_PL | NA | T | C | -0.05649 | 0.004452 | 1.50E-39 |
| rs174572 | miR-1908-! | cis | 3.57E-05 | Nightingale | S_LDL_FC | NA | T | C | -0.0474 | 0.004646 | 1.70E-26 |
| rs174572 | miR-1908-! | cis | 3.57E-05 | Nightingale | VLDL_TG | NA | T | C | 0.034052 | 0.004478 | 5.10E-15 |
| rs174572 | miR-1908-! | cis | 3.57E-05 | Nightingale | S_LDL_PL | NA | T | C | -0.03937 | 0.004642 | 7.20E-19 |
| rs174572 | miR-1908-! | cis | 3.57E-05 | Nightingale | XXL_VLDL_ | NA | T | C | 0.05089 | 0.004483 | 6.10E-32 |
| rs174572 | miR-1908-! | cis | 3.57E-05 | Nightingale | XXL_VLDL_ | NA | T | C | 0.053375 | 0.004466 | 4.30E-35 |
| rs174572 | miR-1908-! | cis | 3.57E-05 | Nightingale | S_VLDL_C_ | NA | T | C | -0.0408 | 0.004527 | 2.60E-20 |
| rs174572 | miR-1908-! | cis | 3.57E-05 | Nightingale | XXL_VLDL_ | NA | T | C | 0.046335 | 0.00449 | 2.20E-26 |
| rs174572 | miR-1908-! | cis | 3.57E-05 | Nightingale | Sphingomy | NA | T | C | -0.08669 | 0.004348 | 1.50E-94 |
| rs174572 | miR-1908-! | cis | 3.57E-05 | Nightingale | HDL_PL | NA | T | C | -0.03144 | 0.004207 | 3.50E-15 |
| rs174572 | miR-1908-! | cis | 3.57E-05 | Nightingale | Phosphogly | NA | T | C | -0.02727 | 0.004407 | 1.20E-10 |
| rs174572 | miR-1908-! | cis | 3.57E-05 | Nightingale | LA_pct | NA | T | C | 0.148682 | 0.004547 | 3.20E-236 |
| rs174572 | miR-1908-! | cis | 3.57E-05 | Nightingale | XL_VLDL_T | NA | T | C | 0.044407 | 0.004508 | 8.40E-24 |
| rs174572 | miR-1908-! | cis | 3.57E-05 | Nightingale | Total_PL | NA | T | C | -0.03603 | 0.004431 | 3.80E-17 |
| rs174572 | miR-1908-! | cis | 3.57E-05 | Nightingale | LA | NA | T | C | 0.069196 | 0.004563 | 6.10E-52 |
| rs174572 | miR-1908-! | cis | 3.57E-05 | Nightingale | S_LDL_CE_ | NA | T | C | 0.043048 | 0.004645 | 2.80E-22 |
| rs174572 | miR-1908-! | cis | 3.57E-05 | Nightingale | DHA | NA | T | C | -0.22179 | 0.004484 | 0 |
| rs174572 | miR-1908-! | cis | 3.57E-05 | Nightingale | IDL_P | NA | T | C | -0.03217 | 0.004605 | 4.10E-13 |
| rs174572 | miR-1908-! | cis | 3.57E-05 | Nightingale | M_VLDL_F | NA | T | C | -0.06772 | 0.004424 | 1.50E-56 |
| rs174572 | miR-1908-! | cis | 3.57E-05 | Nightingale | VLDL_size | NA | T | C | 0.052183 | 0.004337 | 1.20E-35 |
| rs174572 | miR-1908-! | cis | 3.57E-05 | Nightingale | Total_C | NA | T | C | -0.04762 | 0.004505 | 4.90E-28 |
| rs174572 | miR-1908-! | cis | 3.57E-05 | Nightingale | XL_VLDL_F | NA | T | C | 0.030978 | 0.004507 | 1.20E-12 |
| rs174572 | miR-1908-! | cis | 3.57E-05 | Nightingale | S_LDL_TG | NA | T | C | 0.037563 | 0.004539 | 2.30E-17 |
| rs174572 | miR-1908-! | cis | 3.57E-05 | Nightingale | S_HDL_FC_ | NA | T | C | -0.07715 | 0.004414 | 9.70E-79 |
| rs174572 | miR-1908-! | cis | 3.57E-05 | Nightingale | XL_VLDL_L | NA | T | C | 0.037313 | 0.004477 | 9.30E-18 |
| rs174572 | miR-1908-! | cis | 3.57E-05 | Nightingale | S_HDL_TG | NA | T | C | 0.051704 | 0.004512 | 1.30E-32 |
| rs174572 | miR-1908-! | cis | 3.57E-05 | Nightingale | MUFA | NA | T | C | 0.048802 | 0.004606 | 3.80E-27 |
| rs174572 | miR-1908-! | cis | 3.57E-05 | Nightingale | XS_VLDL_L | NA | T | C | -0.03177 | 0.004574 | 2.30E-13 |
| rs174572 | miR-1908-! | cis | 3.57E-05 | Nightingale | TG_by_PG | NA | T | C | 0.048338 | 0.004318 | 3.80E-31 |
| rs174572 | miR-1908-! | cis | 3.57E-05 | Nightingale | PUFA_pct | NA | T | C | -0.09175 | 0.004476 | 9.20E-95 |
| rs174572 | miR-1908-! | cis | 3.57E-05 | Nightingale | L_LDL_FC_ | NA | T | C | -0.06034 | 0.004435 | 9.20E-45 |
| rs174572 | miR-1908-! | cis | 3.57E-05 | Nightingale | L_VLDL_L | NA | T | C | 0.026267 | 0.004497 | 1.40E-09 |
| rs174572 | miR-1908-! | cis | 3.57E-05 | Nightingale | L_HDL_FC_ | NA | T | C | -0.08413 | 0.004323 | 6.70E-94 |
| rs174572 | miR-1908-! | cis | 3.57E-05 | Nightingale | L_LDL_FC | NA | T | C | -0.0506 | 0.004569 | 5.00E-31 |
| rs174572 | miR-1908-! | cis | 3.57E-05 | Nightingale | M_HDL_C | NA | T | C | -0.02523 | 0.004281 | 5.50E-10 |
| rs174572 | miR-1908-! | cis | 3.57E-05 | Nightingale | Total_P | NA | T | C | -0.02589 | 0.004379 | 4.80E-10 |
| rs174572 | miR-1908-! | cis | 3.57E-05 | Nightingale | IDL_FC_pct | NA | T | C | -0.0775 | 0.004616 | 8.70E-65 |
| rs174572 | miR-1908-! | cis | 3.57E-05 | Nightingale | XS_VLDL_C | NA | T | C | -0.07216 | 0.004409 | 1.50E-64 |
| rs174572 | miR-1908-! | cis | 3.57E-05 | Nightingale | XS_VLDL_C | NA | T | C | -0.067 | 0.004386 | 1.00E-56 |
| rs174572 | miR-1908-! | cis | 3.57E-05 | Nightingale | XS_VLDL_T | NA | T | C | 0.072908 | 0.00442 | 1.30E-65 |
| rs174572 | miR-1908-! | cis | 3.57E-05 | Nightingale | ApoB | NA | T | C | -0.02934 | 0.004647 | 6.00E-11 |
| rs174572 | miR-1908-! | cis | 3.57E-05 | Nightingale | Total_TG | NA | T | C | 0.033554 | 0.004504 | 1.50E-14 |
| rs174573 | miR-1908-! | cis | 3.73E-05 | Nightingale | IDL_L | NA | A | G | -0.05257 | 0.004498 | 3.00E-34 |
| rs174573 | miR-1908-! | cis | 3.73E-05 | Nightingale | HDL_FC | NA | A | G | -0.05135 | 0.004122 | 4.50E-40 |
| rs174573 | miR-1908-! | cis | 3.73E-05 | Nightingale | SFA_pct | NA | A | G | -0.03202 | 0.00459 | 3.00E-12 |
| rs174573 | miR-1908-! | cis | 3.73E-05 | Nightingale | L_HDL_PL_ | NA | A | G | 0.035294 | 0.004504 | 7.00E-17 |
| rs174573 | miR-1908-! | cis | 3.73E-05 | Nightingale | Sphingomy | NA | A | G | -0.08661 | 0.004348 | 2.10E-94 |
| rs174573 | miR-1908-! | cis | 3.73E-05 | Nightingale | Remnant_( | NA | A | G | -0.0326 | 0.004623 | 2.00E-13 |
| rs174573 | miR-1908-! | cis | 3.73E-05 | Nightingale | PUFA_by_I | NA | A | G | -0.12927 | 0.004457 | 9.40E-190 |
| rs174573 | miR-1908-! | cis | 3.73E-05 | Nightingale | M_HDL_TG | NA | A | G | 0.048057 | 0.004462 | 4.50E-29 |

| | | | | | | | | | | | |
|---|---|---|---|---|---|---|---|---|---|---|---|
| rs174573 | miR-1908-5 | cis | 3.73E-05 | Nightingale | LDL_PL | NA | A | G | -0.03388 | 0.004643 | 4.90E-14 |
| rs174573 | miR-1908-5 | cis | 3.73E-05 | Nightingale | M_LDL_FC | NA | A | G | -0.03891 | 0.004648 | 3.50E-18 |
| rs174573 | miR-1908-5 | cis | 3.73E-05 | Nightingale | M_VLDL_C | NA | A | G | -0.05439 | 0.004368 | 1.70E-37 |
| rs174573 | miR-1908-5 | cis | 3.73E-05 | Nightingale | IDL_CE | NA | A | G | -0.05207 | 0.004492 | 1.20E-33 |
| rs174573 | miR-1908-5 | cis | 3.73E-05 | Nightingale | S_LDL_TG_ | NA | A | G | 0.06482 | 0.004509 | 1.90E-49 |
| rs174573 | miR-1908-5 | cis | 3.73E-05 | Nightingale | XL_VLDL_C | NA | A | G | -0.04389 | 0.004461 | 8.90E-24 |
| rs174573 | miR-1908-5 | cis | 3.73E-05 | Nightingale | Cholines | NA | A | G | -0.02411 | 0.004376 | 5.20E-09 |
| rs174573 | miR-1908-5 | cis | 3.73E-05 | Nightingale | DHA_pct | NA | A | G | -0.21077 | 0.004517 | 0 |
| rs174573 | miR-1908-5 | cis | 3.73E-05 | Nightingale | M_VLDL_C | NA | A | G | -0.03403 | 0.00464 | 3.10E-14 |
| rs174573 | miR-1908-5 | cis | 3.73E-05 | Nightingale | XL_HDL_PL | NA | A | G | -0.06661 | 0.004183 | 6.90E-64 |
| rs174573 | miR-1908-5 | cis | 3.73E-05 | Nightingale | XL_VLDL_P | NA | A | G | 0.034789 | 0.00449 | 1.60E-15 |
| rs174573 | miR-1908-5 | cis | 3.73E-05 | Nightingale | XL_HDL_C | NA | A | G | -0.07416 | 0.004195 | 1.50E-78 |
| rs174573 | miR-1908-5 | cis | 3.73E-05 | Nightingale | IDL_TG_pct | NA | A | G | 0.065813 | 0.004495 | 1.30E-51 |
| rs174573 | miR-1908-5 | cis | 3.73E-05 | Nightingale | LDL_CE | NA | A | G | -0.03264 | 0.004641 | 3.30E-13 |
| rs174573 | miR-1908-5 | cis | 3.73E-05 | Nightingale | L_LDL_TG_ | NA | A | G | 0.059029 | 0.004563 | 7.20E-40 |
| rs174573 | miR-1908-5 | cis | 3.73E-05 | Nightingale | XS_VLDL_C | NA | A | G | -0.06692 | 0.004386 | 1.80E-56 |
| rs174573 | miR-1908-5 | cis | 3.73E-05 | Nightingale | M_LDL_PL_ | NA | A | G | 0.039274 | 0.00465 | 1.20E-17 |
| rs174573 | miR-1908-5 | cis | 3.73E-05 | Nightingale | HDL_size | NA | A | G | -0.05197 | 0.004123 | 1.40E-40 |
| rs174573 | miR-1908-5 | cis | 3.73E-05 | Nightingale | S_LDL_TG | NA | A | G | 0.037831 | 0.004539 | 1.70E-17 |
| rs174573 | miR-1908-5 | cis | 3.73E-05 | Nightingale | LDL_C | NA | A | G | -0.03712 | 0.004633 | 1.10E-16 |
| rs174573 | miR-1908-5 | cis | 3.73E-05 | Nightingale | XXL_VLDL_ | NA | A | G | 0.028836 | 0.004679 | 1.50E-10 |
| rs174573 | miR-1908-5 | cis | 3.73E-05 | Nightingale | Clinical_LD | NA | A | G | -0.04083 | 0.004626 | 7.00E-20 |
| rs174573 | miR-1908-5 | cis | 3.73E-05 | Nightingale | M_HDL_CE | NA | A | G | -0.03367 | 0.004512 | 1.50E-14 |
| rs174573 | miR-1908-5 | cis | 3.73E-05 | Nightingale | L_VLDL_TG | NA | A | G | 0.025622 | 0.004477 | 4.20E-09 |
| rs174573 | miR-1908-5 | cis | 3.73E-05 | Nightingale | M_VLDL_C_ | NA | A | G | -0.05826 | 0.00438 | 1.40E-42 |
| rs174573 | miR-1908-5 | cis | 3.73E-05 | Nightingale | L_HDL_CE_ | NA | A | G | -0.03806 | 0.004452 | 4.90E-20 |
| rs174573 | miR-1908-5 | cis | 3.73E-05 | Nightingale | IDL_CE_pct | NA | A | G | -0.0256 | 0.004566 | 1.70E-08 |
| rs174573 | miR-1908-5 | cis | 3.73E-05 | Nightingale | XXL_VLDL_ | NA | A | G | 0.053897 | 0.004484 | 3.40E-35 |
| rs174573 | miR-1908-5 | cis | 3.73E-05 | Nightingale | M_VLDL_T | NA | A | G | 0.056046 | 0.004395 | 1.60E-39 |
| rs174573 | miR-1908-5 | cis | 3.73E-05 | Nightingale | S_VLDL_PL | NA | A | G | -0.05636 | 0.004452 | 3.20E-39 |
| rs174573 | miR-1908-5 | cis | 3.73E-05 | Nightingale | M_VLDL_C | NA | A | G | -0.04017 | 0.004611 | 2.00E-19 |
| rs174573 | miR-1908-5 | cis | 3.73E-05 | Nightingale | M_HDL_FC | NA | A | G | -0.03382 | 0.004234 | 3.80E-17 |
| rs174573 | miR-1908-5 | cis | 3.73E-05 | Nightingale | XL_HDL_P | NA | A | G | -0.0666 | 0.004187 | 8.60E-65 |
| rs174573 | miR-1908-5 | cis | 3.73E-05 | Nightingale | S_HDL_TG_ | NA | A | G | 0.049858 | 0.004426 | 1.90E-31 |
| rs174573 | miR-1908-5 | cis | 3.73E-05 | Nightingale | XL_VLDL_C | NA | A | G | -0.04676 | 0.004459 | 1.00E-26 |
| rs174573 | miR-1908-5 | cis | 3.73E-05 | Nightingale | S_VLDL_TG | NA | A | G | 0.046289 | 0.004502 | 6.30E-26 |
| rs174573 | miR-1908-5 | cis | 3.73E-05 | Nightingale | L_LDL_PL | NA | A | G | -0.0402 | 0.004618 | 2.50E-19 |
| rs174573 | miR-1908-5 | cis | 3.73E-05 | Nightingale | M_HDL_C_ | NA | A | G | -0.04746 | 0.00443 | 7.00E-29 |
| rs174573 | miR-1908-5 | cis | 3.73E-05 | Nightingale | Total_FC | NA | A | G | -0.04368 | 0.004546 | 1.60E-23 |
| rs174573 | miR-1908-5 | cis | 3.73E-05 | Nightingale | L_VLDL_PL | NA | A | G | 0.065841 | 0.004516 | 8.80E-51 |
| rs174573 | miR-1908-5 | cis | 3.73E-05 | Nightingale | XL_VLDL_C | NA | A | G | 0.02673 | 0.004545 | 1.20E-09 |
| rs174573 | miR-1908-5 | cis | 3.73E-05 | Nightingale | L_VLDL_FC | NA | A | G | 0.028148 | 0.004501 | 9.10E-11 |
| rs174573 | miR-1908-5 | cis | 3.73E-05 | Nightingale | XS_VLDL_C | NA | A | G | -0.05069 | 0.004516 | 1.00E-31 |
| rs174573 | miR-1908-5 | cis | 3.73E-05 | Nightingale | L_HDL_PL | NA | A | G | -0.05374 | 0.004095 | 4.00E-44 |
| rs174573 | miR-1908-5 | cis | 3.73E-05 | Nightingale | L_LDL_L | NA | A | G | -0.03997 | 0.004608 | 2.00E-19 |
| rs174573 | miR-1908-5 | cis | 3.73E-05 | Nightingale | S_VLDL_FC | NA | A | G | -0.0605 | 0.004443 | 6.50E-45 |
| rs174573 | miR-1908-5 | cis | 3.73E-05 | Nightingale | XS_VLDL_F | NA | A | G | -0.04256 | 0.004572 | 2.20E-22 |
| rs174573 | miR-1908-5 | cis | 3.73E-05 | Nightingale | IDL_PL | NA | A | G | -0.05405 | 0.00449 | 1.70E-36 |
| rs174573 | miR-1908-5 | cis | 3.73E-05 | Nightingale | L_VLDL_PL | NA | A | G | 0.03473 | 0.004489 | 1.30E-15 |
| rs174573 | miR-1908-5 | cis | 3.73E-05 | Nightingale | XL_VLDL_T | NA | A | G | 0.041466 | 0.004451 | 1.20E-21 |
| rs174573 | miR-1908-5 | cis | 3.73E-05 | Nightingale | XXL_VLDL_ | NA | A | G | 0.050388 | 0.004482 | 5.60E-31 |
| rs174573 | miR-1908-5 | cis | 3.73E-05 | Nightingale | Citrate | NA | A | G | -0.02883 | 0.004623 | 1.50E-10 |
| rs174573 | miR-1908-5 | cis | 3.73E-05 | Nightingale | L_HDL_FC_ | NA | A | G | -0.08363 | 0.004323 | 7.90E-93 |
| rs174573 | miR-1908-5 | cis | 3.73E-05 | Nightingale | S_HDL_C_p | NA | A | G | -0.031 | 0.004599 | 1.40E-11 |
| rs174573 | miR-1908-5 | cis | 3.73E-05 | Nightingale | M_VLDL_F | NA | A | G | -0.0673 | 0.004424 | 7.30E-56 |
| rs174573 | miR-1908-5 | cis | 3.73E-05 | Nightingale | IDL_C_pct | NA | A | G | -0.05342 | 0.004543 | 2.50E-33 |
| rs174573 | miR-1908-5 | cis | 3.73E-05 | Nightingale | HDL_C | NA | A | G | -0.0444 | 0.004155 | 4.90E-30 |
| rs174573 | miR-1908-5 | cis | 3.73E-05 | Nightingale | L_LDL_C_p | NA | A | G | -0.05928 | 0.004533 | 1.80E-41 |
| rs174573 | miR-1908-5 | cis | 3.73E-05 | Nightingale | XL_HDL_CE | NA | A | G | -0.07215 | 0.004167 | 1.20E-75 |
| rs174573 | miR-1908-5 | cis | 3.73E-05 | Nightingale | L_HDL_L | NA | A | G | -0.05491 | 0.004083 | 2.00E-46 |
| rs174573 | miR-1908-5 | cis | 3.73E-05 | Nightingale | XS_VLDL_P | NA | A | G | -0.02641 | 0.004588 | 9.00E-10 |
| rs174573 | miR-1908-5 | cis | 3.73E-05 | Nightingale | IDL_C | NA | A | G | -0.05596 | 0.004493 | 1.60E-38 |
| rs174573 | miR-1908-5 | cis | 3.73E-05 | Nightingale | L_VLDL_C_ | NA | A | G | -0.02885 | 0.004545 | 4.10E-11 |

| | | | | | | | | | | | |
|---|---|---|---|---|---|---|---|---|---|---|---|
| rs174573 | miR-1908-5 | cis | 3.73E-05 | Nightingale | M_HDL_CE | NA | A | G | -0.02355 | 0.004297 | 8.40E-09 |
| rs174573 | miR-1908-5 | cis | 3.73E-05 | Nightingale | M_HDL_TG | NA | A | G | 0.039044 | 0.004596 | 3.20E-18 |
| rs174573 | miR-1908-5 | cis | 3.73E-05 | Nightingale | XL_HDL_FC | NA | A | G | -0.07791 | 0.004335 | 4.00E-80 |
| rs174573 | miR-1908-5 | cis | 3.73E-05 | Nightingale | XL_HDL_TG | NA | A | G | 0.065034 | 0.004383 | 6.00E-54 |
| rs174573 | miR-1908-5 | cis | 3.73E-05 | Nightingale | S_VLDL_C_ | NA | A | G | -0.04061 | 0.004527 | 4.80E-20 |
| rs174573 | miR-1908-5 | cis | 3.73E-05 | Nightingale | MUFA | NA | A | G | 0.048942 | 0.004605 | 3.40E-27 |
| rs174573 | miR-1908-5 | cis | 3.73E-05 | Nightingale | XS_VLDL_C | NA | A | G | -0.05315 | 0.00449 | 5.00E-35 |
| rs174573 | miR-1908-5 | cis | 3.73E-05 | Nightingale | M_VLDL_P | NA | A | G | -0.04414 | 0.004465 | 2.70E-25 |
| rs174573 | miR-1908-5 | cis | 3.73E-05 | Nightingale | L_LDL_P | NA | A | G | -0.02835 | 0.004637 | 1.80E-10 |
| rs174573 | miR-1908-5 | cis | 3.73E-05 | Nightingale | Total_L | NA | A | G | -0.02681 | 0.004578 | 1.40E-09 |
| rs174573 | miR-1908-5 | cis | 3.73E-05 | Nightingale | IDL_P | NA | A | G | -0.03166 | 0.004604 | 9.80E-13 |
| rs174573 | miR-1908-5 | cis | 3.73E-05 | Nightingale | XL_HDL_PL | NA | A | G | -0.05087 | 0.004274 | 8.10E-36 |
| rs174573 | miR-1908-5 | cis | 3.73E-05 | Nightingale | M_LDL_TG | NA | A | G | 0.05805 | 0.004594 | 6.60E-38 |
| rs174573 | miR-1908-5 | cis | 3.73E-05 | Nightingale | M_LDL_C_ | NA | A | G | -0.07932 | 0.004657 | 1.30E-65 |
| rs174573 | miR-1908-5 | cis | 3.73E-05 | Nightingale | XS_VLDL_F | NA | A | G | -0.08943 | 0.004618 | 2.50E-86 |
| rs174573 | miR-1908-5 | cis | 3.73E-05 | Nightingale | XXL_VLDL_ | NA | A | G | 0.053434 | 0.004465 | 4.50E-35 |
| rs174573 | miR-1908-5 | cis | 3.73E-05 | Nightingale | Total_P | NA | A | G | -0.0263 | 0.004379 | 2.60E-10 |
| rs174573 | miR-1908-5 | cis | 3.73E-05 | Nightingale | S_VLDL_TG | NA | A | G | 0.026882 | 0.004515 | 6.50E-10 |
| rs174573 | miR-1908-5 | cis | 3.73E-05 | Nightingale | M_HDL_FC | NA | A | G | -0.07408 | 0.004123 | 1.30E-81 |
| rs174573 | miR-1908-5 | cis | 3.73E-05 | Nightingale | HDL_TG | NA | A | G | 0.03539 | 0.004588 | 3.70E-15 |
| rs174573 | miR-1908-5 | cis | 3.73E-05 | Nightingale | Total_C | NA | A | G | -0.04748 | 0.004504 | 7.00E-28 |
| rs174573 | miR-1908-5 | cis | 3.73E-05 | Nightingale | L_HDL_FC | NA | A | G | -0.06066 | 0.004078 | 2.70E-56 |
| rs174573 | miR-1908-5 | cis | 3.73E-05 | Nightingale | XXL_VLDL_ | NA | A | G | 0.046398 | 0.00449 | 2.00E-26 |
| rs174573 | miR-1908-5 | cis | 3.73E-05 | Nightingale | non_HDL_C | NA | A | G | -0.03543 | 0.004628 | 2.40E-15 |
| rs174573 | miR-1908-5 | cis | 3.73E-05 | Nightingale | M_LDL_TG | NA | A | G | 0.027888 | 0.004585 | 6.90E-10 |
| rs174573 | miR-1908-5 | cis | 3.73E-05 | Nightingale | L_HDL_CE | NA | A | G | -0.05668 | 0.004109 | 1.00E-48 |
| rs174573 | miR-1908-5 | cis | 3.73E-05 | Nightingale | S_LDL_FC | NA | A | G | -0.04717 | 0.004646 | 3.80E-26 |
| rs174573 | miR-1908-5 | cis | 3.73E-05 | Nightingale | Total_CE | NA | A | G | -0.04861 | 0.004488 | 1.80E-29 |
| rs174573 | miR-1908-5 | cis | 3.73E-05 | Nightingale | XL_HDL_L | NA | A | G | -0.06945 | 0.004181 | 1.20E-69 |
| rs174573 | miR-1908-5 | cis | 3.73E-05 | Nightingale | M_LDL_P | NA | A | G | -0.03626 | 0.004667 | 2.70E-15 |
| rs174573 | miR-1908-5 | cis | 3.73E-05 | Nightingale | S_LDL_PL_ | NA | A | G | -0.0475 | 0.004644 | 9.10E-27 |
| rs174573 | miR-1908-5 | cis | 3.73E-05 | Nightingale | XL_HDL_FC | NA | A | G | 0.044555 | 0.004125 | 4.40E-31 |
| rs174573 | miR-1908-5 | cis | 3.73E-05 | Nightingale | VLDL_TG | NA | A | G | 0.034153 | 0.004478 | 4.90E-15 |
| rs174573 | miR-1908-5 | cis | 3.73E-05 | Nightingale | L_LDL_CE | NA | A | G | -0.03967 | 0.004612 | 3.80E-19 |
| rs174573 | miR-1908-5 | cis | 3.73E-05 | Nightingale | Omega_3_ | NA | A | G | -0.30311 | 0.004546 | 0 |
| rs174573 | miR-1908-5 | cis | 3.73E-05 | Nightingale | L_VLDL_P | NA | A | G | 0.029295 | 0.004493 | 1.60E-11 |
| rs174573 | miR-1908-5 | cis | 3.73E-05 | Nightingale | XS_VLDL_P | NA | A | G | -0.03257 | 0.004591 | 9.30E-14 |
| rs174573 | miR-1908-5 | cis | 3.73E-05 | Nightingale | ApoA1 | NA | A | G | -0.02631 | 0.004258 | 6.00E-11 |
| rs174573 | miR-1908-5 | cis | 3.73E-05 | Nightingale | L_HDL_TG_ | NA | A | G | 0.057052 | 0.004391 | 2.20E-42 |
| rs174573 | miR-1908-5 | cis | 3.73E-05 | Nightingale | XXL_VLDL_ | NA | A | G | 0.055868 | 0.004494 | 2.40E-37 |
| rs174573 | miR-1908-5 | cis | 3.73E-05 | Nightingale | S_HDL_TG | NA | A | G | 0.051773 | 0.004512 | 1.30E-32 |
| rs174573 | miR-1908-5 | cis | 3.73E-05 | Nightingale | L_HDL_C | NA | A | G | -0.05772 | 0.004097 | 8.00E-51 |
| rs174573 | miR-1908-5 | cis | 3.73E-05 | Nightingale | L_LDL_C | NA | A | G | -0.04293 | 0.004601 | 2.70E-22 |
| rs174573 | miR-1908-5 | cis | 3.73E-05 | Nightingale | Total_TG | NA | A | G | 0.033697 | 0.004504 | 1.30E-14 |
| rs174573 | miR-1908-5 | cis | 3.73E-05 | Nightingale | M_HDL_C | NA | A | G | -0.02573 | 0.004281 | 2.50E-10 |
| rs174573 | miR-1908-5 | cis | 3.73E-05 | Nightingale | ApoB | NA | A | G | -0.02898 | 0.004647 | 1.00E-10 |
| rs174573 | miR-1908-5 | cis | 3.73E-05 | Nightingale | XS_VLDL_L | NA | A | G | -0.03118 | 0.004574 | 5.20E-13 |
| rs174573 | miR-1908-5 | cis | 3.73E-05 | Nightingale | LDL_P | NA | A | G | -0.03081 | 0.00465 | 7.20E-12 |
| rs174573 | miR-1908-5 | cis | 3.73E-05 | Nightingale | M_HDL_PL | NA | A | G | 0.042506 | 0.004414 | 1.40E-23 |
| rs174573 | miR-1908-5 | cis | 3.73E-05 | Nightingale | M_LDL_C | NA | A | G | -0.02582 | 0.004662 | 1.20E-08 |
| rs174573 | miR-1908-5 | cis | 3.73E-05 | Nightingale | M_LDL_FC_ | NA | A | G | -0.05097 | 0.004458 | 5.70E-32 |
| rs174573 | miR-1908-5 | cis | 3.73E-05 | Nightingale | IDL_FC_pct | NA | A | G | -0.0778 | 0.004616 | 3.80E-65 |
| rs174573 | miR-1908-5 | cis | 3.73E-05 | Nightingale | XL_VLDL_F | NA | A | G | 0.031101 | 0.004507 | 1.20E-12 |
| rs174573 | miR-1908-5 | cis | 3.73E-05 | Nightingale | S_LDL_PL | NA | A | G | -0.03891 | 0.004642 | 2.00E-18 |
| rs174573 | miR-1908-5 | cis | 3.73E-05 | Nightingale | S_LDL_FC_ | NA | A | G | -0.05816 | 0.004509 | 5.00E-40 |
| rs174573 | miR-1908-5 | cis | 3.73E-05 | Nightingale | LA_pct | NA | A | G | 0.148448 | 0.004547 | 1.80E-235 |
| rs174573 | miR-1908-5 | cis | 3.73E-05 | Nightingale | XXL_VLDL_ | NA | A | G | 0.049232 | 0.004475 | 6.50E-30 |
| rs174573 | miR-1908-5 | cis | 3.73E-05 | Nightingale | Omega_3 | NA | A | G | -0.25992 | 0.00456 | 0 |
| rs174573 | miR-1908-5 | cis | 3.73E-05 | Nightingale | L_VLDL_L | NA | A | G | 0.026337 | 0.004497 | 1.50E-09 |
| rs174573 | miR-1908-5 | cis | 3.73E-05 | Nightingale | XL_VLDL_T | NA | A | G | 0.044288 | 0.004508 | 1.20E-23 |
| rs174573 | miR-1908-5 | cis | 3.73E-05 | Nightingale | HDL_L | NA | A | G | -0.03547 | 0.004178 | 2.50E-19 |
| rs174573 | miR-1908-5 | cis | 3.73E-05 | Nightingale | XS_VLDL_P | NA | A | G | 0.038627 | 0.00454 | 2.50E-18 |
| rs174573 | miR-1908-5 | cis | 3.73E-05 | Nightingale | XL_VLDL_F | NA | A | G | -0.05261 | 0.004509 | 4.30E-33 |

| | | | | | | | | | | | |
|---|---|---|---|---|---|---|---|---|---|---|---|
| rs174573 | miR-1908-! | cis | 3.73E-05 | Nightingale | S_LDL_CE_ | NA | A | G | 0.042504 | 0.004645 | 9.40E-22 |
| rs174573 | miR-1908-! | cis | 3.73E-05 | Nightingale | LA | NA | A | G | 0.069288 | 0.004563 | 4.90E-52 |
| rs174573 | miR-1908-! | cis | 3.73E-05 | Nightingale | PUFA_pct | NA | A | G | -0.0917 | 0.004475 | 1.30E-94 |
| rs174573 | miR-1908-! | cis | 3.73E-05 | Nightingale | HDL_PL | NA | A | G | -0.03175 | 0.004207 | 1.70E-15 |
| rs174573 | miR-1908-! | cis | 3.73E-05 | Nightingale | Phosphogly | NA | A | G | -0.02725 | 0.004407 | 1.10E-10 |
| rs174573 | miR-1908-! | cis | 3.73E-05 | Nightingale | XL_VLDL_P | NA | A | G | 0.03819 | 0.004472 | 1.60E-18 |
| rs174573 | miR-1908-! | cis | 3.73E-05 | Nightingale | L_HDL_P | NA | A | G | -0.05564 | 0.004075 | 7.40E-48 |
| rs174573 | miR-1908-! | cis | 3.73E-05 | Nightingale | L_LDL_FC | NA | A | G | -0.05046 | 0.004569 | 8.70E-31 |
| rs174573 | miR-1908-! | cis | 3.73E-05 | Nightingale | DHA | NA | A | G | -0.22135 | 0.004484 | 0 |
| rs174573 | miR-1908-! | cis | 3.73E-05 | Nightingale | L_LDL_FC_ | NA | A | G | -0.06036 | 0.004435 | 1.00E-44 |
| rs174573 | miR-1908-! | cis | 3.73E-05 | Nightingale | MUFA_pct | NA | A | G | 0.149587 | 0.004453 | 1.40E-255 |
| rs174573 | miR-1908-! | cis | 3.73E-05 | Nightingale | XS_VLDL_T | NA | A | G | 0.072732 | 0.00442 | 3.30E-65 |
| rs174573 | miR-1908-! | cis | 3.73E-05 | Nightingale | S_HDL_FC_ | NA | A | G | -0.07664 | 0.004413 | 7.40E-78 |
| rs174573 | miR-1908-! | cis | 3.73E-05 | Nightingale | XL_VLDL_L | NA | A | G | 0.037381 | 0.004477 | 9.70E-18 |
| rs174573 | miR-1908-! | cis | 3.73E-05 | Nightingale | Total_PL | NA | A | G | -0.03595 | 0.004431 | 4.20E-17 |
| rs174573 | miR-1908-! | cis | 3.73E-05 | Nightingale | TG_by_PG | NA | A | G | 0.048518 | 0.004318 | 2.70E-31 |
| rs174573 | miR-1908-! | cis | 3.73E-05 | Nightingale | PUFA | NA | A | G | -0.07218 | 0.004538 | 3.30E-60 |
| rs174573 | miR-1908-! | cis | 3.73E-05 | Nightingale | LDL_FC | NA | A | G | -0.04761 | 0.004604 | 4.80E-27 |
| rs174573 | miR-1908-! | cis | 3.73E-05 | Nightingale | XXL_VLDL_ | NA | A | G | 0.050823 | 0.004483 | 9.20E-32 |
| rs174573 | miR-1908-! | cis | 3.73E-05 | Nightingale | XS_VLDL_C | NA | A | G | -0.07208 | 0.004409 | 2.70E-64 |
| rs174573 | miR-1908-! | cis | 3.73E-05 | Nightingale | L_HDL_C_p | NA | A | G | -0.05149 | 0.004386 | 6.70E-36 |
| rs174573 | miR-1908-! | cis | 3.73E-05 | Nightingale | IDL_FC | NA | A | G | -0.06573 | 0.004504 | 6.20E-52 |
| rs174573 | miR-1908-! | cis | 3.73E-05 | Nightingale | VLDL_size | NA | A | G | 0.052017 | 0.004337 | 2.00E-35 |
| rs174573 | miR-1908-! | cis | 3.73E-05 | Nightingale | LDL_L | NA | A | G | -0.03365 | 0.004638 | 5.40E-14 |
| rs174573 | miR-1908-! | cis | 3.73E-05 | Nightingale | Omega_6_ | NA | A | G | 0.288031 | 0.004585 | 0 |
| rs174573 | miR-1908-! | cis | 3.73E-05 | Nightingale | HDL_CE | NA | A | G | -0.04204 | 0.004176 | 8.60E-27 |
| rs174573 | miR-1908-! | cis | 3.73E-05 | Nightingale | Unsaturati | NA | A | G | -0.26415 | 0.004407 | 0 |
| rs174573 | miR-1908-! | cis | 3.73E-05 | Nightingale | L_VLDL_CE | NA | A | G | -0.0445 | 0.004453 | 4.40E-25 |
| rs174573 | miR-1908-! | cis | 3.73E-05 | Nightingale | S_LDL_P | NA | A | G | -0.02491 | 0.004658 | 3.60E-08 |
| rs174574 | miR-1908-! | cis | 8.50E-09 | Nightingale | XS_VLDL_P | NA | C | A | 0.043016 | 0.004263 | 5.40E-26 |
| rs174574 | miR-1908-! | cis | 8.50E-09 | Nightingale | XXL_VLDL_ | NA | C | A | -0.07286 | 0.004172 | 6.90E-71 |
| rs174574 | miR-1908-! | cis | 8.50E-09 | Nightingale | S_LDL_FC | NA | C | A | 0.062664 | 0.004313 | 1.10E-50 |
| rs174574 | miR-1908-! | cis | 8.50E-09 | Nightingale | M_HDL_C | NA | C | A | 0.041592 | 0.003975 | 2.90E-27 |
| rs174574 | miR-1908-! | cis | 8.50E-09 | Nightingale | PUFA | NA | C | A | 0.096131 | 0.004213 | 4.90E-122 |
| rs174574 | miR-1908-! | cis | 8.50E-09 | Nightingale | XXL_VLDL_ | NA | C | A | -0.06545 | 0.004155 | 1.60E-58 |
| rs174574 | miR-1908-! | cis | 8.50E-09 | Nightingale | S_LDL_TG_ | NA | C | A | -0.08589 | 0.004186 | 1.30E-96 |
| rs174574 | miR-1908-! | cis | 8.50E-09 | Nightingale | L_LDL_C | NA | C | A | 0.057729 | 0.004272 | 5.00E-44 |
| rs174574 | miR-1908-! | cis | 8.50E-09 | Nightingale | S_LDL_P | NA | C | A | 0.032272 | 0.004324 | 1.50E-14 |
| rs174574 | miR-1908-! | cis | 8.50E-09 | Nightingale | L_VLDL_L | NA | C | A | -0.03768 | 0.004175 | 3.30E-20 |
| rs174574 | miR-1908-! | cis | 8.50E-09 | Nightingale | S_LDL_L | NA | C | A | 0.032311 | 0.004319 | 1.20E-14 |
| rs174574 | miR-1908-! | cis | 8.50E-09 | Nightingale | M_VLDL_C | NA | C | A | 0.05213 | 0.004281 | 8.10E-36 |
| rs174574 | miR-1908-! | cis | 8.50E-09 | Nightingale | XL_HDL_FC | NA | C | A | 0.101306 | 0.004024 | 8.09E-151 |
| rs174574 | miR-1908-! | cis | 8.50E-09 | Nightingale | HDL_PL | NA | C | A | 0.049016 | 0.003906 | 8.60E-39 |
| rs174574 | miR-1908-! | cis | 8.50E-09 | Nightingale | XL_HDL_CE | NA | C | A | 0.096255 | 0.003869 | 2.00E-148 |
| rs174574 | miR-1908-! | cis | 8.50E-09 | Nightingale | L_VLDL_FC | NA | C | A | -0.03969 | 0.004179 | 2.60E-22 |
| rs174574 | miR-1908-! | cis | 8.50E-09 | Nightingale | XL_HDL_C_ | NA | C | A | -0.02321 | 0.004127 | 8.30E-10 |
| rs174574 | miR-1908-! | cis | 8.50E-09 | Nightingale | XS_VLDL_T | NA | C | A | -0.09671 | 0.004104 | 1.20E-128 |
| rs174574 | miR-1908-! | cis | 8.50E-09 | Nightingale | Clinical_LD | NA | C | A | 0.055104 | 0.004295 | 1.50E-39 |
| rs174574 | miR-1908-! | cis | 8.50E-09 | Nightingale | SFA_pct | NA | C | A | 0.04298 | 0.004262 | 6.50E-24 |
| rs174574 | miR-1908-! | cis | 8.50E-09 | Nightingale | XXL_VLDL_ | NA | C | A | -0.06075 | 0.004169 | 3.20E-50 |
| rs174574 | miR-1908-! | cis | 8.50E-09 | Nightingale | IDL_C | NA | C | A | 0.077004 | 0.004172 | 8.90E-81 |
| rs174574 | miR-1908-! | cis | 8.50E-09 | Nightingale | Total_L | NA | C | A | 0.037521 | 0.00425 | 6.60E-20 |
| rs174574 | miR-1908-! | cis | 8.50E-09 | Nightingale | XS_VLDL_F | NA | C | A | 0.11207 | 0.004288 | 9.29E-154 |
| rs174574 | miR-1908-! | cis | 8.50E-09 | Nightingale | LDL_C | NA | C | A | 0.049755 | 0.004302 | 1.50E-32 |
| rs174574 | miR-1908-! | cis | 8.50E-09 | Nightingale | XL_HDL_TG | NA | C | A | -0.08673 | 0.00407 | 2.30E-106 |
| rs174574 | miR-1908-! | cis | 8.50E-09 | Nightingale | L_VLDL_C_ | NA | C | A | 0.037766 | 0.00422 | 1.30E-20 |
| rs174574 | miR-1908-! | cis | 8.50E-09 | Nightingale | M_VLDL_T | NA | C | A | -0.03092 | 0.004197 | 6.80E-14 |
| rs174574 | miR-1908-! | cis | 8.50E-09 | Nightingale | Total_FA | NA | C | A | 0.024047 | 0.004286 | 6.60E-09 |
| rs174574 | miR-1908-! | cis | 8.50E-09 | Nightingale | HDL_C | NA | C | A | 0.065081 | 0.003858 | 4.70E-69 |
| rs174574 | miR-1908-! | cis | 8.50E-09 | Nightingale | L_LDL_C_p | NA | C | A | 0.076934 | 0.004209 | 7.70E-77 |
| rs174574 | miR-1908-! | cis | 8.50E-09 | Nightingale | S_VLDL_FC | NA | C | A | 0.028038 | 0.004321 | 2.40E-11 |
| rs174574 | miR-1908-! | cis | 8.50E-09 | Nightingale | M_VLDL_P | NA | C | A | 0.061498 | 0.004145 | 2.50E-53 |
| rs174574 | miR-1908-! | cis | 8.50E-09 | Nightingale | PUFA_by_M | NA | C | A | 0.169377 | 0.004138 | 0 |

| | | | | | | | | | | | |
|---|---|---|---|---|---|---|---|---|---|---|---|
| rs174574 | miR-1908-5 | cis | 8.50E-09 | Nightingale | L_VLDL_PL | NA | C | A | -0.04833 | 0.004168 | 1.90E-32 |
| rs174574 | miR-1908-5 | cis | 8.50E-09 | Nightingale | XXL_VLDL_ | NA | C | A | -0.06729 | 0.004162 | 1.20E-61 |
| rs174574 | miR-1908-5 | cis | 8.50E-09 | Nightingale | S_VLDL_FC | NA | C | A | 0.082416 | 0.004125 | 9.80E-93 |
| rs174574 | miR-1908-5 | cis | 8.50E-09 | Nightingale | XS_VLDL_F | NA | C | A | 0.055491 | 0.004245 | 5.10E-42 |
| rs174574 | miR-1908-5 | cis | 8.50E-09 | Nightingale | non_HDL_( | NA | C | A | 0.047701 | 0.004297 | 3.60E-30 |
| rs174574 | miR-1908-5 | cis | 8.50E-09 | Nightingale | Cholines | NA | C | A | 0.037513 | 0.004063 | 4.30E-22 |
| rs174574 | miR-1908-5 | cis | 8.50E-09 | Nightingale | S_VLDL_CE | NA | C | A | 0.028188 | 0.004262 | 1.90E-11 |
| rs174574 | miR-1908-5 | cis | 8.50E-09 | Nightingale | HDL_FC | NA | C | A | 0.074032 | 0.003827 | 2.40E-90 |
| rs174574 | miR-1908-5 | cis | 8.50E-09 | Nightingale | M_LDL_C_ | NA | C | A | 0.100453 | 0.004323 | 3.20E-120 |
| rs174574 | miR-1908-5 | cis | 8.50E-09 | Nightingale | Omega_6_ | NA | C | A | -0.02452 | 0.004205 | 3.20E-09 |
| rs174574 | miR-1908-5 | cis | 8.50E-09 | Nightingale | S_HDL_C_p | NA | C | A | 0.042132 | 0.00427 | 9.90E-23 |
| rs174574 | miR-1908-5 | cis | 8.50E-09 | Nightingale | L_LDL_FC_ | NA | C | A | 0.082361 | 0.004118 | 2.60E-92 |
| rs174574 | miR-1908-5 | cis | 8.50E-09 | Nightingale | Tyr | NA | C | A | -0.03212 | 0.004315 | 9.70E-14 |
| rs174574 | miR-1908-5 | cis | 8.50E-09 | Nightingale | XS_VLDL_C | NA | C | A | 0.067565 | 0.004193 | 1.60E-62 |
| rs174574 | miR-1908-5 | cis | 8.50E-09 | Nightingale | Sphingomy | NA | C | A | 0.114884 | 0.004037 | 1.90E-187 |
| rs174574 | miR-1908-5 | cis | 8.50E-09 | Nightingale | LDL_PL | NA | C | A | 0.046083 | 0.004311 | 5.90E-28 |
| rs174574 | miR-1908-5 | cis | 8.50E-09 | Nightingale | XL_HDL_C | NA | C | A | 0.098256 | 0.003895 | 2.60E-152 |
| rs174574 | miR-1908-5 | cis | 8.50E-09 | Nightingale | M_VLDL_C | NA | C | A | 0.074992 | 0.004056 | 1.70E-79 |
| rs174574 | miR-1908-5 | cis | 8.50E-09 | Nightingale | HDL_TG | NA | C | A | -0.04588 | 0.00426 | 1.80E-27 |
| rs174574 | miR-1908-5 | cis | 8.50E-09 | Nightingale | M_LDL_PL | NA | C | A | 0.023598 | 0.004327 | 2.00E-08 |
| rs174574 | miR-1908-5 | cis | 8.50E-09 | Nightingale | M_HDL_FC | NA | C | A | 0.103135 | 0.003828 | 1.80E-174 |
| rs174574 | miR-1908-5 | cis | 8.50E-09 | Nightingale | XL_VLDL_C | NA | C | A | -0.03008 | 0.004252 | 6.80E-13 |
| rs174574 | miR-1908-5 | cis | 8.50E-09 | Nightingale | Total_C | NA | C | A | 0.065152 | 0.004182 | 9.20E-58 |
| rs174574 | miR-1908-5 | cis | 8.50E-09 | Nightingale | L_HDL_PL | NA | C | A | 0.076155 | 0.003802 | 3.70E-97 |
| rs174574 | miR-1908-5 | cis | 8.50E-09 | Nightingale | Total_TG | NA | C | A | -0.04633 | 0.004182 | 1.50E-29 |
| rs174574 | miR-1908-5 | cis | 8.50E-09 | Nightingale | LDL_FC | NA | C | A | 0.064184 | 0.004274 | 7.90E-54 |
| rs174574 | miR-1908-5 | cis | 8.50E-09 | Nightingale | XL_HDL_L | NA | C | A | 0.092076 | 0.003882 | 2.10E-135 |
| rs174574 | miR-1908-5 | cis | 8.50E-09 | Nightingale | L_LDL_TG_ | NA | C | A | -0.07751 | 0.004236 | 1.90E-76 |
| rs174574 | miR-1908-5 | cis | 8.50E-09 | Nightingale | M_VLDL_C | NA | C | A | 0.043644 | 0.004308 | 2.80E-25 |
| rs174574 | miR-1908-5 | cis | 8.50E-09 | Nightingale | XXL_VLDL_ | NA | C | A | -0.071 | 0.004163 | 6.10E-68 |
| rs174574 | miR-1908-5 | cis | 8.50E-09 | Nightingale | VLDL_TG | NA | C | A | -0.04719 | 0.004157 | 6.50E-31 |
| rs174574 | miR-1908-5 | cis | 8.50E-09 | Nightingale | M_VLDL_F | NA | C | A | 0.090296 | 0.004108 | 5.50E-113 |
| rs174574 | miR-1908-5 | cis | 8.50E-09 | Nightingale | M_LDL_TG | NA | C | A | -0.07576 | 0.004266 | 5.50E-72 |
| rs174574 | miR-1908-5 | cis | 8.50E-09 | Nightingale | L_LDL_P | NA | C | A | 0.037004 | 0.004305 | 6.30E-19 |
| rs174574 | miR-1908-5 | cis | 8.50E-09 | Nightingale | Omega_3_ | NA | C | A | 0.386925 | 0.004221 | 0 |
| rs174574 | miR-1908-5 | cis | 8.50E-09 | Nightingale | XL_VLDL_C | NA | C | A | -0.03711 | 0.004219 | 3.10E-19 |
| rs174574 | miR-1908-5 | cis | 8.50E-09 | Nightingale | L_LDL_L | NA | C | A | 0.054083 | 0.004278 | 9.00E-39 |
| rs174574 | miR-1908-5 | cis | 8.50E-09 | Nightingale | XS_VLDL_P | NA | C | A | -0.05867 | 0.004215 | 4.90E-45 |
| rs174574 | miR-1908-5 | cis | 8.50E-09 | Nightingale | HDL_P | NA | C | A | 0.036447 | 0.004057 | 1.60E-20 |
| rs174574 | miR-1908-5 | cis | 8.50E-09 | Nightingale | XL_VLDL_P | NA | C | A | -0.04779 | 0.004169 | 1.30E-31 |
| rs174574 | miR-1908-5 | cis | 8.50E-09 | Nightingale | XL_HDL_FC | NA | C | A | -0.06069 | 0.00383 | 1.60E-62 |
| rs174574 | miR-1908-5 | cis | 8.50E-09 | Nightingale | XL_VLDL_F | NA | C | A | -0.04296 | 0.004184 | 1.20E-25 |
| rs174574 | miR-1908-5 | cis | 8.50E-09 | Nightingale | S_LDL_PL_ | NA | C | A | 0.060594 | 0.004311 | 4.70E-47 |
| rs174574 | miR-1908-5 | cis | 8.50E-09 | Nightingale | LDL_CE | NA | C | A | 0.043621 | 0.004309 | 2.50E-25 |
| rs174574 | miR-1908-5 | cis | 8.50E-09 | Nightingale | IDL_TG_pc | NA | C | A | -0.08789 | 0.004174 | 3.30E-102 |
| rs174574 | miR-1908-5 | cis | 8.50E-09 | Nightingale | S_LDL_FC_ | NA | C | A | 0.079562 | 0.004187 | 1.60E-83 |
| rs174574 | miR-1908-5 | cis | 8.50E-09 | Nightingale | XXL_VLDL_ | NA | C | A | -0.0444 | 0.004344 | 2.20E-25 |
| rs174574 | miR-1908-5 | cis | 8.50E-09 | Nightingale | M_LDL_FC | NA | C | A | 0.052399 | 0.004315 | 8.90E-36 |
| rs174574 | miR-1908-5 | cis | 8.50E-09 | Nightingale | L_HDL_TG_ | NA | C | A | -0.07883 | 0.004077 | 3.50E-88 |
| rs174574 | miR-1908-5 | cis | 8.50E-09 | Nightingale | IDL_FC | NA | C | A | 0.089425 | 0.004181 | 2.60E-107 |
| rs174574 | miR-1908-5 | cis | 8.50E-09 | Nightingale | VLDL_size | NA | C | A | -0.07133 | 0.004027 | 1.90E-74 |
| rs174574 | miR-1908-5 | cis | 8.50E-09 | Nightingale | L_VLDL_CE | NA | C | A | 0.058063 | 0.004134 | 8.50E-48 |
| rs174574 | miR-1908-5 | cis | 8.50E-09 | Nightingale | XL_HDL_PL | NA | C | A | 0.088154 | 0.003883 | 4.20E-124 |
| rs174574 | miR-1908-5 | cis | 8.50E-09 | Nightingale | M_HDL_TG | NA | C | A | -0.0498 | 0.004267 | 3.40E-32 |
| rs174574 | miR-1908-5 | cis | 8.50E-09 | Nightingale | M_HDL_PL | NA | C | A | -0.05816 | 0.004098 | 1.40E-47 |
| rs174574 | miR-1908-5 | cis | 8.50E-09 | Nightingale | S_VLDL_TG | NA | C | A | -0.03867 | 0.004192 | 6.60E-21 |
| rs174574 | miR-1908-5 | cis | 8.50E-09 | Nightingale | L_HDL_C | NA | C | A | 0.080093 | 0.003804 | 4.60E-107 |
| rs174574 | miR-1908-5 | cis | 8.50E-09 | Nightingale | M_VLDL_F | NA | C | A | 0.029298 | 0.00432 | 3.00E-12 |
| rs174574 | miR-1908-5 | cis | 8.50E-09 | Nightingale | Citrate | NA | C | A | 0.033304 | 0.004293 | 1.00E-15 |
| rs174574 | miR-1908-5 | cis | 8.50E-09 | Nightingale | ApoA1 | NA | C | A | 0.041861 | 0.003954 | 3.20E-28 |
| rs174574 | miR-1908-5 | cis | 8.50E-09 | Nightingale | M_LDL_CE | NA | C | A | 0.026583 | 0.004327 | 3.10E-10 |
| rs174574 | miR-1908-5 | cis | 8.50E-09 | Nightingale | M_VLDL_C | NA | C | A | 0.079725 | 0.004066 | 2.60E-89 |
| rs174574 | miR-1908-5 | cis | 8.50E-09 | Nightingale | M_LDL_PL_ | NA | C | A | -0.04643 | 0.004318 | 3.40E-28 |

| | | | | | | | | | | | |
|---|---|---|---|---|---|---|---|---|---|---|---|
| rs174574 | miR-1908-5 | cis | 8.50E-09 | Nightingale | SFA | NA | C | A | 0.031832 | 0.004281 | 3.90E-14 |
| rs174574 | miR-1908-5 | cis | 8.50E-09 | Nightingale | L_HDL_C_p | NA | C | A | 0.067696 | 0.004072 | 2.30E-66 |
| rs174574 | miR-1908-5 | cis | 8.50E-09 | Nightingale | XL_HDL_PL | NA | C | A | 0.066223 | 0.003969 | 2.00E-67 |
| rs174574 | miR-1908-5 | cis | 8.50E-09 | Nightingale | DHA_pct | NA | C | A | 0.27186 | 0.004194 | 0 |
| rs174574 | miR-1908-5 | cis | 8.50E-09 | Nightingale | XS_VLDL_P | NA | C | A | 0.033735 | 0.00426 | 6.40E-17 |
| rs174574 | miR-1908-5 | cis | 8.50E-09 | Nightingale | Total_FC | NA | C | A | 0.059221 | 0.00422 | 1.80E-47 |
| rs174574 | miR-1908-5 | cis | 8.50E-09 | Nightingale | LA | NA | C | A | -0.08307 | 0.004237 | 5.60E-86 |
| rs174574 | miR-1908-5 | cis | 8.50E-09 | Nightingale | IDL_CE_pct | NA | C | A | 0.038244 | 0.004239 | 1.80E-19 |
| rs174574 | miR-1908-5 | cis | 8.50E-09 | Nightingale | M_LDL_FC_ | NA | C | A | 0.071012 | 0.004139 | 1.40E-68 |
| rs174574 | miR-1908-5 | cis | 8.50E-09 | Nightingale | XL_HDL_P | NA | C | A | 0.088617 | 0.003888 | 3.50E-126 |
| rs174574 | miR-1908-5 | cis | 8.50E-09 | Nightingale | HDL_size | NA | C | A | 0.072426 | 0.003828 | 1.70E-86 |
| rs174574 | miR-1908-5 | cis | 8.50E-09 | Nightingale | ApoB | NA | C | A | 0.038206 | 0.004315 | 7.00E-20 |
| rs174574 | miR-1908-5 | cis | 8.50E-09 | Nightingale | XL_VLDL_P | NA | C | A | -0.05225 | 0.004152 | 9.60E-38 |
| rs174574 | miR-1908-5 | cis | 8.50E-09 | Nightingale | M_HDL_FC | NA | C | A | 0.052128 | 0.003931 | 5.50E-43 |
| rs174574 | miR-1908-5 | cis | 8.50E-09 | Nightingale | XS_VLDL_L | NA | C | A | 0.041006 | 0.004246 | 3.70E-24 |
| rs174574 | miR-1908-5 | cis | 8.50E-09 | Nightingale | M_HDL_PL | NA | C | A | 0.022494 | 0.00404 | 6.50E-09 |
| rs174574 | miR-1908-5 | cis | 8.50E-09 | Nightingale | HDL_CE | NA | C | A | 0.061849 | 0.003877 | 9.70E-62 |
| rs174574 | miR-1908-5 | cis | 8.50E-09 | Nightingale | M_HDL_CE | NA | C | A | 0.038667 | 0.00399 | 1.30E-23 |
| rs174574 | miR-1908-5 | cis | 8.50E-09 | Nightingale | M_LDL_TG | NA | C | A | -0.03648 | 0.004257 | 9.40E-18 |
| rs174574 | miR-1908-5 | cis | 8.50E-09 | Nightingale | IDL_FC_pct | NA | C | A | 0.103854 | 0.004286 | 2.60E-131 |
| rs174574 | miR-1908-5 | cis | 8.50E-09 | Nightingale | VLDL_L | NA | C | A | -0.02705 | 0.004236 | 9.40E-11 |
| rs174574 | miR-1908-5 | cis | 8.50E-09 | Nightingale | M_HDL_C_ | NA | C | A | 0.065801 | 0.004113 | 1.60E-60 |
| rs174574 | miR-1908-5 | cis | 8.50E-09 | Nightingale | Total_PL | NA | C | A | 0.051307 | 0.004114 | 6.40E-38 |
| rs174574 | miR-1908-5 | cis | 8.50E-09 | Nightingale | L_VLDL_P | NA | C | A | -0.04139 | 0.004171 | 4.10E-24 |
| rs174574 | miR-1908-5 | cis | 8.50E-09 | Nightingale | M_HDL_CE | NA | C | A | 0.046884 | 0.004189 | 7.80E-30 |
| rs174574 | miR-1908-5 | cis | 8.50E-09 | Nightingale | IDL_CE | NA | C | A | 0.072004 | 0.00417 | 6.20E-71 |
| rs174574 | miR-1908-5 | cis | 8.50E-09 | Nightingale | XXL_VLDL_ | NA | C | A | 0.026668 | 0.004299 | 9.40E-11 |
| rs174574 | miR-1908-5 | cis | 8.50E-09 | Nightingale | M_LDL_L | NA | C | A | 0.02832 | 0.004327 | 2.10E-11 |
| rs174574 | miR-1908-5 | cis | 8.50E-09 | Nightingale | IDL_C_pct | NA | C | A | 0.074953 | 0.004218 | 2.00E-72 |
| rs174574 | miR-1908-5 | cis | 8.50E-09 | Nightingale | L_LDL_PL | NA | C | A | 0.054889 | 0.004288 | 2.30E-39 |
| rs174574 | miR-1908-5 | cis | 8.50E-09 | Nightingale | S_LDL_C | NA | C | A | 0.032161 | 0.004325 | 1.90E-14 |
| rs174574 | miR-1908-5 | cis | 8.50E-09 | Nightingale | LDL_TG | NA | C | A | -0.02964 | 0.004272 | 7.10E-12 |
| rs174574 | miR-1908-5 | cis | 8.50E-09 | Nightingale | XS_VLDL_C | NA | C | A | 0.091735 | 0.004072 | 1.60E-117 |
| rs174574 | miR-1908-5 | cis | 8.50E-09 | Nightingale | XXL_VLDL_ | NA | C | A | -0.07153 | 0.004146 | 8.60E-70 |
| rs174574 | miR-1908-5 | cis | 8.50E-09 | Nightingale | DHA | NA | C | A | 0.286614 | 0.004163 | 0 |
| rs174574 | miR-1908-5 | cis | 8.50E-09 | Nightingale | L_HDL_CE | NA | C | A | 0.078539 | 0.003815 | 2.40E-102 |
| rs174574 | miR-1908-5 | cis | 8.50E-09 | Nightingale | L_HDL_CE_ | NA | C | A | 0.049562 | 0.004134 | 3.50E-35 |
| rs174574 | miR-1908-5 | cis | 8.50E-09 | Nightingale | LDL_L | NA | C | A | 0.045414 | 0.004306 | 2.10E-27 |
| rs174574 | miR-1908-5 | cis | 8.50E-09 | Nightingale | XL_VLDL_C | NA | C | A | 0.063044 | 0.00414 | 6.90E-54 |
| rs174574 | miR-1908-5 | cis | 8.50E-09 | Nightingale | L_LDL_FC | NA | C | A | 0.068184 | 0.004242 | 1.30E-61 |
| rs174574 | miR-1908-5 | cis | 8.50E-09 | Nightingale | L_HDL_FC_ | NA | C | A | 0.111119 | 0.004013 | 2.90E-183 |
| rs174574 | miR-1908-5 | cis | 8.50E-09 | Nightingale | TG_by_PG | NA | C | A | -0.0685 | 0.004009 | 8.20E-69 |
| rs174574 | miR-1908-5 | cis | 8.50E-09 | Nightingale | M_HDL_L | NA | C | A | 0.02859 | 0.004011 | 1.10E-13 |
| rs174574 | miR-1908-5 | cis | 8.50E-09 | Nightingale | L_VLDL_FC | NA | C | A | -0.02891 | 0.004279 | 1.10E-11 |
| rs174574 | miR-1908-5 | cis | 8.50E-09 | Nightingale | S_HDL_TG_ | NA | C | A | -0.06914 | 0.004109 | 1.70E-66 |
| rs174574 | miR-1908-5 | cis | 8.50E-09 | Nightingale | Phosphogly | NA | C | A | 0.041422 | 0.004092 | 7.50E-26 |
| rs174574 | miR-1908-5 | cis | 8.50E-09 | Nightingale | IDL_P | NA | C | A | 0.044942 | 0.004275 | 2.00E-27 |
| rs174574 | miR-1908-5 | cis | 8.50E-09 | Nightingale | M_LDL_P | NA | C | A | 0.046605 | 0.004333 | 6.10E-28 |
| rs174574 | miR-1908-5 | cis | 8.50E-09 | Nightingale | M_HDL_P | NA | C | A | 0.035118 | 0.00398 | 4.00E-20 |
| rs174574 | miR-1908-5 | cis | 8.50E-09 | Nightingale | Omega_6_ | NA | C | A | -0.36715 | 0.004258 | 0 |
| rs174574 | miR-1908-5 | cis | 8.50E-09 | Nightingale | IDL_L | NA | C | A | 0.072059 | 0.004176 | 2.90E-71 |
| rs174574 | miR-1908-5 | cis | 8.50E-09 | Nightingale | MUFA_pct | NA | C | A | -0.19611 | 0.004134 | 0 |
| rs174574 | miR-1908-5 | cis | 8.50E-09 | Nightingale | L_HDL_PL_ | NA | C | A | -0.04169 | 0.004181 | 6.00E-25 |
| rs174574 | miR-1908-5 | cis | 8.50E-09 | Nightingale | S_VLDL_C_ | NA | C | A | 0.055931 | 0.004203 | 1.70E-41 |
| rs174574 | miR-1908-5 | cis | 8.50E-09 | Nightingale | L_HDL_L | NA | C | A | 0.077177 | 0.003791 | 1.60E-100 |
| rs174574 | miR-1908-5 | cis | 8.50E-09 | Nightingale | M_LDL_C | NA | C | A | 0.034229 | 0.004329 | 5.60E-16 |
| rs174574 | miR-1908-5 | cis | 8.50E-09 | Nightingale | IDL_PL | NA | C | A | 0.072313 | 0.004168 | 2.20E-72 |
| rs174574 | miR-1908-5 | cis | 8.50E-09 | Nightingale | XS_VLDL_C | NA | C | A | 0.071331 | 0.004169 | 3.70E-70 |
| rs174574 | miR-1908-5 | cis | 8.50E-09 | Nightingale | L_VLDL_PL | NA | C | A | -0.08798 | 0.004193 | 2.70E-101 |
| rs174574 | miR-1908-5 | cis | 8.50E-09 | Nightingale | XS_VLDL_T | NA | C | A | -0.03172 | 0.004254 | 3.20E-13 |
| rs174574 | miR-1908-5 | cis | 8.50E-09 | Nightingale | Total_P | NA | C | A | 0.041083 | 0.004066 | 1.10E-25 |
| rs174574 | miR-1908-5 | cis | 8.50E-09 | Nightingale | L_LDL_CE | NA | C | A | 0.053246 | 0.004282 | 1.50E-37 |
| rs174574 | miR-1908-5 | cis | 8.50E-09 | Nightingale | MUFA | NA | C | A | -0.06316 | 0.004276 | 1.50E-50 |

| | | | | | | | | | | | |
|---|---|---|---|---|---|---|---|---|---|---|---|
| rs174574 | miR-1908-5 | cis | 8.50E-09 | Nightingale | M_VLDL_T | NA | C | A | -0.07697 | 0.00408 | 3.00E-83 |
| rs174574 | miR-1908-5 | cis | 8.50E-09 | Nightingale | XL_VLDL_C | NA | C | A | 0.060055 | 0.004142 | 3.80E-49 |
| rs174574 | miR-1908-5 | cis | 8.50E-09 | Nightingale | M_HDL_TG | NA | C | A | -0.06675 | 0.004143 | 5.10E-61 |
| rs174574 | miR-1908-5 | cis | 8.50E-09 | Nightingale | XXL_VLDL_ | NA | C | A | -0.06591 | 0.004161 | 5.40E-59 |
| rs174574 | miR-1908-5 | cis | 8.50E-09 | Nightingale | Total_CE | NA | C | A | 0.067008 | 0.004167 | 1.80E-61 |
| rs174574 | miR-1908-5 | cis | 8.50E-09 | Nightingale | S_HDL_FC_ | NA | C | A | 0.101212 | 0.004098 | 1.80E-149 |
| rs174574 | miR-1908-5 | cis | 8.50E-09 | Nightingale | Omega_3 | NA | C | A | 0.333382 | 0.004234 | 0 |
| rs174574 | miR-1908-5 | cis | 8.50E-09 | Nightingale | L_VLDL_C | NA | C | A | -0.02859 | 0.004225 | 5.80E-12 |
| rs174574 | miR-1908-5 | cis | 8.50E-09 | Nightingale | S_VLDL_TG | NA | C | A | -0.06387 | 0.00418 | 2.50E-54 |
| rs174574 | miR-1908-5 | cis | 8.50E-09 | Nightingale | PUFA_pct | NA | C | A | 0.120214 | 0.004155 | 8.20E-186 |
| rs174574 | miR-1908-5 | cis | 8.50E-09 | Nightingale | S_VLDL_PL | NA | C | A | 0.07802 | 0.004134 | 3.20E-83 |
| rs174574 | miR-1908-5 | cis | 8.50E-09 | Nightingale | L_HDL_P | NA | C | A | 0.077722 | 0.003784 | 2.00E-102 |
| rs174574 | miR-1908-5 | cis | 8.50E-09 | Nightingale | S_HDL_TG | NA | C | A | -0.06843 | 0.004189 | 1.40E-62 |
| rs174574 | miR-1908-5 | cis | 8.50E-09 | Nightingale | S_LDL_PL | NA | C | A | 0.050991 | 0.00431 | 2.90E-34 |
| rs174574 | miR-1908-5 | cis | 8.50E-09 | Nightingale | XL_VLDL_T | NA | C | A | -0.05618 | 0.004132 | 9.90E-44 |
| rs174574 | miR-1908-5 | cis | 8.50E-09 | Nightingale | Unsaturati | NA | C | A | 0.336838 | 0.004092 | 0 |
| rs174574 | miR-1908-5 | cis | 8.50E-09 | Nightingale | XL_VLDL_F | NA | C | A | 0.067256 | 0.004186 | 5.90E-61 |
| rs174574 | miR-1908-5 | cis | 8.50E-09 | Nightingale | XS_VLDL_C | NA | C | A | 0.097597 | 0.004093 | 3.00E-131 |
| rs174574 | miR-1908-5 | cis | 8.50E-09 | Nightingale | L_HDL_FC | NA | C | A | 0.084333 | 0.003786 | 1.20E-119 |
| rs174574 | miR-1908-5 | cis | 8.50E-09 | Nightingale | XL_VLDL_T | NA | C | A | -0.05866 | 0.004185 | 3.50E-46 |
| rs174574 | miR-1908-5 | cis | 8.50E-09 | Nightingale | XL_HDL_CE | NA | C | A | 0.026408 | 0.004318 | 2.50E-09 |
| rs174574 | miR-1908-5 | cis | 8.50E-09 | Nightingale | L_VLDL_TG | NA | C | A | -0.03662 | 0.004157 | 2.60E-19 |
| rs174574 | miR-1908-5 | cis | 8.50E-09 | Nightingale | LDL_P | NA | C | A | 0.039956 | 0.004317 | 1.70E-21 |
| rs174574 | miR-1908-5 | cis | 8.50E-09 | Nightingale | S_LDL_CE_ | NA | C | A | -0.0561 | 0.004312 | 1.20E-40 |
| rs174574 | miR-1908-5 | cis | 8.50E-09 | Nightingale | Remnant_( | NA | C | A | 0.043929 | 0.004292 | 3.60E-26 |
| rs174574 | miR-1908-5 | cis | 8.50E-09 | Nightingale | LA_pct | NA | C | A | -0.18463 | 0.004222 | 0 |
| rs174574 | miR-1908-5 | cis | 8.50E-09 | Nightingale | HDL_L | NA | C | A | 0.053833 | 0.003879 | 4.20E-47 |
| rs174574 | miR-1908-5 | cis | 8.50E-09 | Nightingale | S_LDL_TG | NA | C | A | -0.05024 | 0.004214 | 2.30E-33 |
| rs174574 | miR-1908-5 | cis | 8.50E-09 | Nightingale | XL_VLDL_L | NA | C | A | -0.05085 | 0.004157 | 9.40E-36 |
| rs174575 | miR-1908-5 | cis | 5.45E-05 | Nightingale | L_HDL_FC_ | NA | G | C | -0.08317 | 0.004297 | 1.50E-92 |
| rs174575 | miR-1908-5 | cis | 5.45E-05 | Nightingale | XL_VLDL_T | NA | G | C | 0.041188 | 0.004424 | 1.20E-21 |
| rs174575 | miR-1908-5 | cis | 5.45E-05 | Nightingale | M_VLDL_C | NA | G | C | -0.05422 | 0.004342 | 9.70E-38 |
| rs174575 | miR-1908-5 | cis | 5.45E-05 | Nightingale | Sphingomy | NA | G | C | -0.08653 | 0.004322 | 2.80E-95 |
| rs174575 | miR-1908-5 | cis | 5.45E-05 | Nightingale | M_HDL_TG | NA | G | C | 0.048056 | 0.004435 | 1.80E-29 |
| rs174575 | miR-1908-5 | cis | 5.45E-05 | Nightingale | M_HDL_CE | NA | G | C | -0.02379 | 0.004272 | 5.10E-09 |
| rs174575 | miR-1908-5 | cis | 5.45E-05 | Nightingale | L_VLDL_PL | NA | G | C | 0.066102 | 0.004489 | 8.40E-52 |
| rs174575 | miR-1908-5 | cis | 5.45E-05 | Nightingale | LDL_PL | NA | G | C | -0.03378 | 0.004615 | 4.10E-14 |
| rs174575 | miR-1908-5 | cis | 5.45E-05 | Nightingale | S_HDL_TG | NA | G | C | 0.051689 | 0.004485 | 6.50E-33 |
| rs174575 | miR-1908-5 | cis | 5.45E-05 | Nightingale | S_LDL_PL_ | NA | G | C | -0.04674 | 0.004616 | 2.00E-26 |
| rs174575 | miR-1908-5 | cis | 5.45E-05 | Nightingale | M_LDL_FC | NA | G | C | -0.03888 | 0.00462 | 2.40E-18 |
| rs174575 | miR-1908-5 | cis | 5.45E-05 | Nightingale | XL_HDL_P | NA | G | C | -0.0662 | 0.004162 | 9.50E-65 |
| rs174575 | miR-1908-5 | cis | 5.45E-05 | Nightingale | L_HDL_CE_ | NA | G | C | -0.03778 | 0.004426 | 6.80E-20 |
| rs174575 | miR-1908-5 | cis | 5.45E-05 | Nightingale | XXL_VLDL_ | NA | G | C | 0.029671 | 0.004651 | 3.10E-11 |
| rs174575 | miR-1908-5 | cis | 5.45E-05 | Nightingale | M_LDL_P | NA | G | C | -0.03619 | 0.004639 | 2.30E-15 |
| rs174575 | miR-1908-5 | cis | 5.45E-05 | Nightingale | M_VLDL_T | NA | G | C | 0.055769 | 0.004368 | 1.20E-39 |
| rs174575 | miR-1908-5 | cis | 5.45E-05 | Nightingale | L_LDL_PL | NA | G | C | -0.04015 | 0.004591 | 1.60E-19 |
| rs174575 | miR-1908-5 | cis | 5.45E-05 | Nightingale | S_LDL_TG | NA | G | C | 0.037878 | 0.004512 | 7.10E-18 |
| rs174575 | miR-1908-5 | cis | 5.45E-05 | Nightingale | LDL_CE | NA | G | C | -0.03253 | 0.004614 | 2.90E-13 |
| rs174575 | miR-1908-5 | cis | 5.45E-05 | Nightingale | S_VLDL_PL | NA | G | C | -0.05626 | 0.004426 | 1.50E-39 |
| rs174575 | miR-1908-5 | cis | 5.45E-05 | Nightingale | HDL_size | NA | G | C | -0.05189 | 0.004098 | 9.80E-41 |
| rs174575 | miR-1908-5 | cis | 5.45E-05 | Nightingale | IDL_TG_pc | NA | G | C | 0.065922 | 0.004468 | 1.50E-52 |
| rs174575 | miR-1908-5 | cis | 5.45E-05 | Nightingale | XS_VLDL_T | NA | G | C | 0.072631 | 0.004394 | 9.10E-66 |
| rs174575 | miR-1908-5 | cis | 5.45E-05 | Nightingale | XS_VLDL_C | NA | G | C | -0.06706 | 0.00436 | 2.20E-57 |
| rs174575 | miR-1908-5 | cis | 5.45E-05 | Nightingale | XS_VLDL_C | NA | G | C | -0.07217 | 0.004383 | 3.40E-65 |
| rs174575 | miR-1908-5 | cis | 5.45E-05 | Nightingale | L_HDL_PL_ | NA | G | C | 0.034841 | 0.004477 | 1.50E-16 |
| rs174575 | miR-1908-5 | cis | 5.45E-05 | Nightingale | M_HDL_CE | NA | G | C | -0.03353 | 0.004485 | 1.10E-14 |
| rs174575 | miR-1908-5 | cis | 5.45E-05 | Nightingale | S_VLDL_C_ | NA | G | C | -0.04028 | 0.0045 | 5.80E-20 |
| rs174575 | miR-1908-5 | cis | 5.45E-05 | Nightingale | L_VLDL_TG | NA | G | C | 0.025397 | 0.004451 | 4.40E-09 |
| rs174575 | miR-1908-5 | cis | 5.45E-05 | Nightingale | S_LDL_FC_ | NA | G | C | -0.05834 | 0.004482 | 5.30E-41 |
| rs174575 | miR-1908-5 | cis | 5.45E-05 | Nightingale | L_HDL_P | NA | G | C | -0.05555 | 0.004051 | 3.80E-48 |
| rs174575 | miR-1908-5 | cis | 5.45E-05 | Nightingale | Total_FC | NA | G | C | -0.04362 | 0.004519 | 1.20E-23 |
| rs174575 | miR-1908-5 | cis | 5.45E-05 | Nightingale | XL_HDL_PL | NA | G | C | -0.06621 | 0.004158 | 1.20E-63 |
| rs174575 | miR-1908-5 | cis | 5.45E-05 | Nightingale | IDL_L | NA | G | C | -0.05246 | 0.004472 | 1.60E-34 |

| | | | | | | | | | | | |
|---|---|---|---|---|---|---|---|---|---|---|---|
| rs174575 | miR-1908-! | cis | 5.45E-05 | Nightingale | XL_VLDL_P | NA | G | C | 0.03803 | 0.004446 | 1.20E-18 |
| rs174575 | miR-1908-! | cis | 5.45E-05 | Nightingale | PUFA_pct | NA | G | C | -0.09182 | 0.004449 | 3.00E-96 |
| rs174575 | miR-1908-! | cis | 5.45E-05 | Nightingale | XL_VLDL_C | NA | G | C | -0.04361 | 0.004435 | 7.50E-24 |
| rs174575 | miR-1908-! | cis | 5.45E-05 | Nightingale | M_HDL_TG | NA | G | C | 0.038857 | 0.004568 | 2.40E-18 |
| rs174575 | miR-1908-! | cis | 5.45E-05 | Nightingale | XXL_VLDL_ | NA | G | C | 0.05335 | 0.004439 | 1.70E-35 |
| rs174575 | miR-1908-! | cis | 5.45E-05 | Nightingale | MUFA | NA | G | C | 0.049048 | 0.004578 | 9.40E-28 |
| rs174575 | miR-1908-! | cis | 5.45E-05 | Nightingale | L_VLDL_C_ | NA | G | C | -0.02839 | 0.004518 | 5.60E-11 |
| rs174575 | miR-1908-! | cis | 5.45E-05 | Nightingale | XL_VLDL_F | NA | G | C | -0.05196 | 0.004482 | 8.50E-33 |
| rs174575 | miR-1908-! | cis | 5.45E-05 | Nightingale | XL_VLDL_P | NA | G | C | 0.034698 | 0.004464 | 1.10E-15 |
| rs174575 | miR-1908-! | cis | 5.45E-05 | Nightingale | XL_HDL_FC | NA | G | C | 0.044249 | 0.004101 | 8.60E-31 |
| rs174575 | miR-1908-! | cis | 5.45E-05 | Nightingale | IDL_CE_pct | NA | G | C | -0.02613 | 0.004539 | 4.90E-09 |
| rs174575 | miR-1908-! | cis | 5.45E-05 | Nightingale | L_LDL_TG_ | NA | G | C | 0.059121 | 0.004535 | 1.40E-40 |
| rs174575 | miR-1908-! | cis | 5.45E-05 | Nightingale | Total_L | NA | G | C | -0.02686 | 0.00455 | 1.20E-09 |
| rs174575 | miR-1908-! | cis | 5.45E-05 | Nightingale | VLDL_size | NA | G | C | 0.051664 | 0.004311 | 2.20E-35 |
| rs174575 | miR-1908-! | cis | 5.45E-05 | Nightingale | XS_VLDL_F | NA | G | C | -0.08911 | 0.004591 | 1.20E-86 |
| rs174575 | miR-1908-! | cis | 5.45E-05 | Nightingale | Remnant_( | NA | G | C | -0.03259 | 0.004596 | 1.60E-13 |
| rs174575 | miR-1908-! | cis | 5.45E-05 | Nightingale | XL_HDL_CE | NA | G | C | -0.07182 | 0.004142 | 9.20E-76 |
| rs174575 | miR-1908-! | cis | 5.45E-05 | Nightingale | LA | NA | G | C | 0.069265 | 0.004536 | 9.90E-53 |
| rs174575 | miR-1908-! | cis | 5.45E-05 | Nightingale | HDL_TG | NA | G | C | 0.035301 | 0.004561 | 2.50E-15 |
| rs174575 | miR-1908-! | cis | 5.45E-05 | Nightingale | M_LDL_C | NA | G | C | -0.02582 | 0.004634 | 1.00E-08 |
| rs174575 | miR-1908-! | cis | 5.45E-05 | Nightingale | HDL_C | NA | G | C | -0.04448 | 0.00413 | 2.60E-30 |
| rs174575 | miR-1908-! | cis | 5.45E-05 | Nightingale | HDL_FC | NA | G | C | -0.05136 | 0.004098 | 2.20E-40 |
| rs174575 | miR-1908-! | cis | 5.45E-05 | Nightingale | XL_HDL_C | NA | G | C | -0.07379 | 0.00417 | 1.40E-78 |
| rs174575 | miR-1908-! | cis | 5.45E-05 | Nightingale | L_LDL_FC_ | NA | G | C | -0.06044 | 0.004409 | 1.80E-45 |
| rs174575 | miR-1908-! | cis | 5.45E-05 | Nightingale | XL_HDL_L | NA | G | C | -0.06906 | 0.004156 | 1.60E-69 |
| rs174575 | miR-1908-! | cis | 5.45E-05 | Nightingale | XL_VLDL_T | NA | G | C | 0.044005 | 0.004481 | 1.00E-23 |
| rs174575 | miR-1908-! | cis | 5.45E-05 | Nightingale | XXL_VLDL_ | NA | G | C | 0.055522 | 0.004467 | 1.60E-37 |
| rs174575 | miR-1908-! | cis | 5.45E-05 | Nightingale | L_HDL_FC | NA | G | C | -0.06052 | 0.004054 | 1.30E-56 |
| rs174575 | miR-1908-! | cis | 5.45E-05 | Nightingale | XS_VLDL_L | NA | G | C | -0.03083 | 0.004546 | 7.90E-13 |
| rs174575 | miR-1908-! | cis | 5.45E-05 | Nightingale | XL_VLDL_L | NA | G | C | 0.037187 | 0.00445 | 7.90E-18 |
| rs174575 | miR-1908-! | cis | 5.45E-05 | Nightingale | L_HDL_PL | NA | G | C | -0.0537 | 0.004071 | 2.00E-44 |
| rs174575 | miR-1908-! | cis | 5.45E-05 | Nightingale | S_LDL_CE_ | NA | G | C | 0.042001 | 0.004617 | 1.10E-21 |
| rs174575 | miR-1908-! | cis | 5.45E-05 | Nightingale | L_LDL_C_p | NA | G | C | -0.05922 | 0.004506 | 6.20E-42 |
| rs174575 | miR-1908-! | cis | 5.45E-05 | Nightingale | M_VLDL_F( | NA | G | C | -0.06696 | 0.004398 | 5.30E-56 |
| rs174575 | miR-1908-! | cis | 5.45E-05 | Nightingale | XL_VLDL_C | NA | G | C | 0.026629 | 0.004518 | 9.90E-10 |
| rs174575 | miR-1908-! | cis | 5.45E-05 | Nightingale | L_VLDL_FC | NA | G | C | 0.028064 | 0.004474 | 7.40E-11 |
| rs174575 | miR-1908-! | cis | 5.45E-05 | Nightingale | M_LDL_TG | NA | G | C | 0.02808 | 0.004557 | 3.40E-10 |
| rs174575 | miR-1908-! | cis | 5.45E-05 | Nightingale | Total_PL | NA | G | C | -0.03602 | 0.004404 | 3.40E-17 |
| rs174575 | miR-1908-! | cis | 5.45E-05 | Nightingale | XXL_VLDL_ | NA | G | C | 0.050039 | 0.004455 | 5.20E-31 |
| rs174575 | miR-1908-! | cis | 5.45E-05 | Nightingale | S_HDL_C_p | NA | G | C | -0.03089 | 0.004572 | 1.00E-11 |
| rs174575 | miR-1908-! | cis | 5.45E-05 | Nightingale | L_VLDL_L | NA | G | C | 0.026189 | 0.00447 | 1.40E-09 |
| rs174575 | miR-1908-! | cis | 5.45E-05 | Nightingale | DHA_pct | NA | G | C | -0.21085 | 0.00449 | 0 |
| rs174575 | miR-1908-! | cis | 5.45E-05 | Nightingale | M_HDL_FC | NA | G | C | -0.07402 | 0.004099 | 1.80E-82 |
| rs174575 | miR-1908-! | cis | 5.45E-05 | Nightingale | L_HDL_L | NA | G | C | -0.05486 | 0.004058 | 1.00E-46 |
| rs174575 | miR-1908-! | cis | 5.45E-05 | Nightingale | LDL_C | NA | G | C | -0.03701 | 0.004606 | 8.50E-17 |
| rs174575 | miR-1908-! | cis | 5.45E-05 | Nightingale | Total_C | NA | G | C | -0.04746 | 0.004478 | 4.50E-28 |
| rs174575 | miR-1908-! | cis | 5.45E-05 | Nightingale | Citrate | NA | G | C | -0.02834 | 0.004596 | 2.60E-10 |
| rs174575 | miR-1908-! | cis | 5.45E-05 | Nightingale | non_HDL_( | NA | G | C | -0.03537 | 0.004601 | 1.80E-15 |
| rs174575 | miR-1908-! | cis | 5.45E-05 | Nightingale | L_HDL_CE | NA | G | C | -0.05657 | 0.004084 | 5.90E-49 |
| rs174575 | miR-1908-! | cis | 5.45E-05 | Nightingale | IDL_FC_pct | NA | G | C | -0.07794 | 0.004588 | 3.60E-66 |
| rs174575 | miR-1908-! | cis | 5.45E-05 | Nightingale | M_LDL_TG | NA | G | C | 0.058103 | 0.004567 | 1.60E-38 |
| rs174575 | miR-1908-! | cis | 5.45E-05 | Nightingale | Clinical_LD | NA | G | C | -0.04078 | 0.004599 | 4.60E-20 |
| rs174575 | miR-1908-! | cis | 5.45E-05 | Nightingale | XS_VLDL_P | NA | G | C | 0.03952 | 0.004513 | 2.00E-19 |
| rs174575 | miR-1908-! | cis | 5.45E-05 | Nightingale | S_VLDL_FC | NA | G | C | -0.06039 | 0.004417 | 2.80E-45 |
| rs174575 | miR-1908-! | cis | 5.45E-05 | Nightingale | PUFA_by_I | NA | G | C | -0.12959 | 0.00443 | 2.30E-193 |
| rs174575 | miR-1908-! | cis | 5.45E-05 | Nightingale | TG_by_PG | NA | G | C | 0.048502 | 0.004292 | 9.30E-32 |
| rs174575 | miR-1908-! | cis | 5.45E-05 | Nightingale | XL_HDL_TC | NA | G | C | 0.064858 | 0.004357 | 2.50E-54 |
| rs174575 | miR-1908-! | cis | 5.45E-05 | Nightingale | L_LDL_P | NA | G | C | -0.02798 | 0.00461 | 2.50E-10 |
| rs174575 | miR-1908-! | cis | 5.45E-05 | Nightingale | L_VLDL_CE | NA | G | C | -0.04403 | 0.004426 | 6.60E-25 |
| rs174575 | miR-1908-! | cis | 5.45E-05 | Nightingale | M_VLDL_C | NA | G | C | -0.03392 | 0.004613 | 2.60E-14 |
| rs174575 | miR-1908-! | cis | 5.45E-05 | Nightingale | Total_P | NA | G | C | -0.0266 | 0.004353 | 1.50E-10 |
| rs174575 | miR-1908-! | cis | 5.45E-05 | Nightingale | S_LDL_TG_ | NA | G | C | 0.064698 | 0.004482 | 4.70E-50 |
| rs174575 | miR-1908-! | cis | 5.45E-05 | Nightingale | XS_VLDL_C | NA | G | C | -0.05044 | 0.004489 | 8.50E-32 |

| | | | | | | | | | | | |
|---|---|---|---|---|---|---|---|---|---|---|---|
| rs174575 | miR-1908-5 | cis | 5.45E-05 | Nightingale | S_VLDL_TG | NA | G | C | 0.046024 | 0.004475 | 6.00E-26 |
| rs174575 | miR-1908-5 | cis | 5.45E-05 | Nightingale | L_HDL_C_p | NA | G | C | -0.05116 | 0.00436 | 9.90E-36 |
| rs174575 | miR-1908-5 | cis | 5.45E-05 | Nightingale | XL_HDL_PL | NA | G | C | -0.05053 | 0.004249 | 1.60E-35 |
| rs174575 | miR-1908-5 | cis | 5.45E-05 | Nightingale | XXL_VLDL_ | NA | G | C | 0.048988 | 0.004449 | 4.50E-30 |
| rs174575 | miR-1908-5 | cis | 5.45E-05 | Nightingale | M_HDL_FC | NA | G | C | -0.03403 | 0.004209 | 2.20E-17 |
| rs174575 | miR-1908-5 | cis | 5.45E-05 | Nightingale | M_HDL_PL | NA | G | C | 0.042281 | 0.004387 | 1.20E-23 |
| rs174575 | miR-1908-5 | cis | 5.45E-05 | Nightingale | L_LDL_FC | NA | G | C | -0.05035 | 0.004542 | 5.40E-31 |
| rs174575 | miR-1908-5 | cis | 5.45E-05 | Nightingale | M_LDL_PL_ | NA | G | C | 0.03961 | 0.004623 | 5.40E-18 |
| rs174575 | miR-1908-5 | cis | 5.45E-05 | Nightingale | DHA | NA | G | C | -0.22143 | 0.004457 | 0 |
| rs174575 | miR-1908-5 | cis | 5.45E-05 | Nightingale | L_HDL_C | NA | G | C | -0.0576 | 0.004073 | 4.60E-51 |
| rs174575 | miR-1908-5 | cis | 5.45E-05 | Nightingale | XL_VLDL_F | NA | G | C | 0.03096 | 0.00448 | 9.20E-13 |
| rs174575 | miR-1908-5 | cis | 5.45E-05 | Nightingale | M_LDL_FC_ | NA | G | C | -0.05097 | 0.004431 | 1.40E-32 |
| rs174575 | miR-1908-5 | cis | 5.45E-05 | Nightingale | M_LDL_C_p | NA | G | C | -0.07949 | 0.004629 | 8.60E-67 |
| rs174575 | miR-1908-5 | cis | 5.45E-05 | Nightingale | M_VLDL_C_ | NA | G | C | -0.05802 | 0.004354 | 8.90E-43 |
| rs174575 | miR-1908-5 | cis | 5.45E-05 | Nightingale | Cholines | NA | G | C | -0.02421 | 0.00435 | 4.70E-09 |
| rs174575 | miR-1908-5 | cis | 5.45E-05 | Nightingale | XXL_VLDL_ | NA | G | C | 0.050568 | 0.004456 | 5.80E-32 |
| rs174575 | miR-1908-5 | cis | 5.45E-05 | Nightingale | XXL_VLDL_ | NA | G | C | 0.046188 | 0.004463 | 1.40E-26 |
| rs174575 | miR-1908-5 | cis | 5.45E-05 | Nightingale | SFA_pct | NA | G | C | -0.0325 | 0.004563 | 1.10E-12 |
| rs174575 | miR-1908-5 | cis | 5.45E-05 | Nightingale | LA_pct | NA | G | C | 0.148592 | 0.00452 | 1.70E-238 |
| rs174575 | miR-1908-5 | cis | 5.45E-05 | Nightingale | LDL_FC | NA | G | C | -0.04752 | 0.004576 | 3.00E-27 |
| rs174575 | miR-1908-5 | cis | 5.45E-05 | Nightingale | S_VLDL_TG | NA | G | C | 0.026882 | 0.004488 | 4.40E-10 |
| rs174575 | miR-1908-5 | cis | 5.45E-05 | Nightingale | ApoA1 | NA | G | C | -0.02654 | 0.004233 | 4.20E-11 |
| rs174575 | miR-1908-5 | cis | 5.45E-05 | Nightingale | Phosphogly | NA | G | C | -0.02741 | 0.004381 | 9.40E-11 |
| rs174575 | miR-1908-5 | cis | 5.45E-05 | Nightingale | IDL_P | NA | G | C | -0.03186 | 0.004577 | 4.60E-13 |
| rs174575 | miR-1908-5 | cis | 5.45E-05 | Nightingale | HDL_P | NA | G | C | -0.0227 | 0.004343 | 4.90E-08 |
| rs174575 | miR-1908-5 | cis | 5.45E-05 | Nightingale | M_VLDL_P | NA | G | C | -0.04378 | 0.004438 | 3.50E-25 |
| rs174575 | miR-1908-5 | cis | 5.45E-05 | Nightingale | IDL_CE | NA | G | C | -0.05205 | 0.004465 | 4.60E-34 |
| rs174575 | miR-1908-5 | cis | 5.45E-05 | Nightingale | L_LDL_C | NA | G | C | -0.04278 | 0.004573 | 2.10E-22 |
| rs174575 | miR-1908-5 | cis | 5.45E-05 | Nightingale | ApoB | NA | G | C | -0.02882 | 0.00462 | 1.00E-10 |
| rs174575 | miR-1908-5 | cis | 5.45E-05 | Nightingale | M_HDL_C_ | NA | G | C | -0.04737 | 0.004403 | 3.90E-29 |
| rs174575 | miR-1908-5 | cis | 5.45E-05 | Nightingale | L_VLDL_PL | NA | G | C | 0.034641 | 0.004463 | 9.80E-16 |
| rs174575 | miR-1908-5 | cis | 5.45E-05 | Nightingale | S_LDL_P | NA | G | C | -0.02479 | 0.00463 | 3.70E-08 |
| rs174575 | miR-1908-5 | cis | 5.45E-05 | Nightingale | M_VLDL_C | NA | G | C | -0.04003 | 0.004583 | 1.60E-19 |
| rs174575 | miR-1908-5 | cis | 5.45E-05 | Nightingale | VLDL_TG | NA | G | C | 0.033994 | 0.004451 | 3.90E-15 |
| rs174575 | miR-1908-5 | cis | 5.45E-05 | Nightingale | HDL_L | NA | G | C | -0.03561 | 0.004154 | 1.60E-19 |
| rs174575 | miR-1908-5 | cis | 5.45E-05 | Nightingale | XL_VLDL_C | NA | G | C | -0.04646 | 0.004432 | 8.50E-27 |
| rs174575 | miR-1908-5 | cis | 5.45E-05 | Nightingale | S_LDL_PL | NA | G | C | -0.03858 | 0.004614 | 2.30E-18 |
| rs174575 | miR-1908-5 | cis | 5.45E-05 | Nightingale | M_HDL_C | NA | G | C | -0.02597 | 0.004256 | 1.50E-10 |
| rs174575 | miR-1908-5 | cis | 5.45E-05 | Nightingale | MUFA_pct | NA | G | C | 0.150075 | 0.004426 | 1.10E-260 |
| rs174575 | miR-1908-5 | cis | 5.45E-05 | Nightingale | XL_HDL_FC | NA | G | C | -0.07736 | 0.004309 | 8.60E-80 |
| rs174575 | miR-1908-5 | cis | 5.45E-05 | Nightingale | IDL_PL | NA | G | C | -0.05378 | 0.004463 | 1.50E-36 |
| rs174575 | miR-1908-5 | cis | 5.45E-05 | Nightingale | Omega_3_ | NA | G | C | -0.30318 | 0.004519 | 0 |
| rs174575 | miR-1908-5 | cis | 5.45E-05 | Nightingale | HDL_CE | NA | G | C | -0.04214 | 0.004151 | 4.40E-27 |
| rs174575 | miR-1908-5 | cis | 5.45E-05 | Nightingale | PUFA | NA | G | C | -0.0723 | 0.004511 | 7.00E-61 |
| rs174575 | miR-1908-5 | cis | 5.45E-05 | Nightingale | HDL_PL | NA | G | C | -0.03194 | 0.004182 | 1.10E-15 |
| rs174575 | miR-1908-5 | cis | 5.45E-05 | Nightingale | S_HDL_TG_ | NA | G | C | 0.049999 | 0.004399 | 4.00E-32 |
| rs174575 | miR-1908-5 | cis | 5.45E-05 | Nightingale | L_HDL_TG_ | NA | G | C | 0.057046 | 0.004365 | 7.50E-43 |
| rs174575 | miR-1908-5 | cis | 5.45E-05 | Nightingale | LDL_P | NA | G | C | -0.03055 | 0.004622 | 8.60E-12 |
| rs174575 | miR-1908-5 | cis | 5.45E-05 | Nightingale | XS_VLDL_P | NA | G | C | -0.02595 | 0.004561 | 1.70E-09 |
| rs174575 | miR-1908-5 | cis | 5.45E-05 | Nightingale | IDL_C | NA | G | C | -0.05593 | 0.004466 | 5.70E-39 |
| rs174575 | miR-1908-5 | cis | 5.45E-05 | Nightingale | IDL_FC | NA | G | C | -0.06566 | 0.004477 | 1.90E-52 |
| rs174575 | miR-1908-5 | cis | 5.45E-05 | Nightingale | L_LDL_CE | NA | G | C | -0.03951 | 0.004584 | 3.30E-19 |
| rs174575 | miR-1908-5 | cis | 5.45E-05 | Nightingale | Omega_6_ | NA | G | C | 0.288117 | 0.004558 | 0 |
| rs174575 | miR-1908-5 | cis | 5.45E-05 | Nightingale | L_LDL_L | NA | G | C | -0.03982 | 0.00458 | 1.70E-19 |
| rs174575 | miR-1908-5 | cis | 5.45E-05 | Nightingale | XS_VLDL_C | NA | G | C | -0.05293 | 0.004463 | 3.40E-35 |
| rs174575 | miR-1908-5 | cis | 5.45E-05 | Nightingale | Omega_3 | NA | G | C | -0.26001 | 0.004533 | 0 |
| rs174575 | miR-1908-5 | cis | 5.45E-05 | Nightingale | Total_TG | NA | G | C | 0.033587 | 0.004477 | 9.80E-15 |
| rs174575 | miR-1908-5 | cis | 5.45E-05 | Nightingale | Total_CE | NA | G | C | -0.0486 | 0.004461 | 1.10E-29 |
| rs174575 | miR-1908-5 | cis | 5.45E-05 | Nightingale | IDL_C_pct | NA | G | C | -0.05391 | 0.004516 | 1.50E-34 |
| rs174575 | miR-1908-5 | cis | 5.45E-05 | Nightingale | S_LDL_FC | NA | G | C | -0.04714 | 0.004618 | 2.00E-26 |
| rs174575 | miR-1908-5 | cis | 5.45E-05 | Nightingale | L_VLDL_P | NA | G | C | 0.029176 | 0.004466 | 1.40E-11 |
| rs174575 | miR-1908-5 | cis | 5.45E-05 | Nightingale | Unsaturati | NA | G | C | -0.26419 | 0.00438 | 0 |
| rs174575 | miR-1908-5 | cis | 5.45E-05 | Nightingale | XXL_VLDL_ | NA | G | C | 0.053604 | 0.004458 | 2.10E-35 |

| | | | | | | | | | | | |
|---|---|---|---|---|---|---|---|---|---|---|---|
| rs174575 | miR-1908-5 | cis | 5.45E-05 | Nightingale | S_HDL_FC_ | NA | G | C | -0.07603 | 0.004387 | 2.20E-77 |
| rs174575 | miR-1908-5 | cis | 5.45E-05 | Nightingale | XS_VLDL_F | NA | G | C | -0.04224 | 0.004545 | 3.00E-22 |
| rs174575 | miR-1908-5 | cis | 5.45E-05 | Nightingale | LDL_L | NA | G | C | -0.03353 | 0.00461 | 4.80E-14 |
| rs174575 | miR-1908-5 | cis | 5.45E-05 | Nightingale | XS_VLDL_P | NA | G | C | -0.03228 | 0.004564 | 1.20E-13 |
| rs174576 | miR-1908-5 | cis | 3.13E-08 | Nightingale | SFA | NA | A | C | -0.03124 | 0.004283 | 1.20E-13 |
| rs174576 | miR-1908-5 | cis | 3.13E-08 | Nightingale | Omega_6_ | NA | A | C | 0.366739 | 0.00426 | 0 |
| rs174576 | miR-1908-5 | cis | 3.13E-08 | Nightingale | LDL_CE | NA | A | C | -0.04329 | 0.004311 | 7.60E-25 |
| rs174576 | miR-1908-5 | cis | 3.13E-08 | Nightingale | LDL_PL | NA | A | C | -0.04581 | 0.004312 | 1.70E-27 |
| rs174576 | miR-1908-5 | cis | 3.13E-08 | Nightingale | XL_VLDL_C | NA | A | C | -0.06339 | 0.004141 | 2.00E-54 |
| rs174576 | miR-1908-5 | cis | 3.13E-08 | Nightingale | Omega_3 | NA | A | C | -0.33291 | 0.004236 | 0 |
| rs174576 | miR-1908-5 | cis | 3.13E-08 | Nightingale | XL_VLDL_F | NA | A | C | -0.06721 | 0.004188 | 6.70E-61 |
| rs174576 | miR-1908-5 | cis | 3.13E-08 | Nightingale | Unsaturati | NA | A | C | -0.33678 | 0.004093 | 0 |
| rs174576 | miR-1908-5 | cis | 3.13E-08 | Nightingale | XL_HDL_C | NA | A | C | -0.09869 | 0.003897 | 7.60E-154 |
| rs174576 | miR-1908-5 | cis | 3.13E-08 | Nightingale | HDL_C | NA | A | C | -0.06512 | 0.003859 | 4.00E-69 |
| rs174576 | miR-1908-5 | cis | 3.13E-08 | Nightingale | Cholines | NA | A | C | -0.03724 | 0.004065 | 9.70E-22 |
| rs174576 | miR-1908-5 | cis | 3.13E-08 | Nightingale | L_HDL_CE_ | NA | A | C | -0.05017 | 0.004135 | 4.30E-36 |
| rs174576 | miR-1908-5 | cis | 3.13E-08 | Nightingale | S_VLDL_PL | NA | A | C | -0.0783 | 0.004136 | 9.30E-84 |
| rs174576 | miR-1908-5 | cis | 3.13E-08 | Nightingale | HDL_PL | NA | A | C | -0.04887 | 0.003907 | 1.80E-38 |
| rs174576 | miR-1908-5 | cis | 3.13E-08 | Nightingale | Remnant_( | NA | A | C | -0.04364 | 0.004294 | 1.10E-25 |
| rs174576 | miR-1908-5 | cis | 3.13E-08 | Nightingale | XS_VLDL_F | NA | A | C | -0.11194 | 0.00429 | 3.90E-153 |
| rs174576 | miR-1908-5 | cis | 3.13E-08 | Nightingale | L_VLDL_FC | NA | A | C | 0.040327 | 0.004181 | 5.00E-23 |
| rs174576 | miR-1908-5 | cis | 3.13E-08 | Nightingale | HDL_size | NA | A | C | -0.07265 | 0.00383 | 4.30E-87 |
| rs174576 | miR-1908-5 | cis | 3.13E-08 | Nightingale | XS_VLDL_C | NA | A | C | -0.0979 | 0.004095 | 6.90E-132 |
| rs174576 | miR-1908-5 | cis | 3.13E-08 | Nightingale | Total_FA | NA | A | C | -0.02346 | 0.004288 | 1.60E-08 |
| rs174576 | miR-1908-5 | cis | 3.13E-08 | Nightingale | M_VLDL_T | NA | A | C | 0.031533 | 0.004199 | 1.90E-14 |
| rs174576 | miR-1908-5 | cis | 3.13E-08 | Nightingale | S_LDL_PL_ | NA | A | C | -0.06124 | 0.004313 | 3.40E-48 |
| rs174576 | miR-1908-5 | cis | 3.13E-08 | Nightingale | L_LDL_C_p | NA | A | C | -0.07739 | 0.004211 | 1.30E-77 |
| rs174576 | miR-1908-5 | cis | 3.13E-08 | Nightingale | M_HDL_PL | NA | A | C | 0.058428 | 0.0041 | 5.10E-48 |
| rs174576 | miR-1908-5 | cis | 3.13E-08 | Nightingale | M_HDL_FC | NA | A | C | -0.05197 | 0.003933 | 1.10E-42 |
| rs174576 | miR-1908-5 | cis | 3.13E-08 | Nightingale | ApoB | NA | A | C | -0.0379 | 0.004317 | 1.80E-19 |
| rs174576 | miR-1908-5 | cis | 3.13E-08 | Nightingale | XL_VLDL_C | NA | A | C | 0.037684 | 0.004221 | 7.80E-20 |
| rs174576 | miR-1908-5 | cis | 3.13E-08 | Nightingale | L_HDL_PL | NA | A | C | -0.07629 | 0.003804 | 1.30E-97 |
| rs174576 | miR-1908-5 | cis | 3.13E-08 | Nightingale | M_VLDL_C | NA | A | C | -0.0755 | 0.004058 | 1.60E-80 |
| rs174576 | miR-1908-5 | cis | 3.13E-08 | Nightingale | M_LDL_C_ | NA | A | C | -0.09989 | 0.004325 | 1.30E-118 |
| rs174576 | miR-1908-5 | cis | 3.13E-08 | Nightingale | L_HDL_CE | NA | A | C | -0.07884 | 0.003816 | 3.30E-103 |
| rs174576 | miR-1908-5 | cis | 3.13E-08 | Nightingale | XL_VLDL_C | NA | A | C | 0.030622 | 0.004254 | 2.20E-13 |
| rs174576 | miR-1908-5 | cis | 3.13E-08 | Nightingale | XXL_VLDL_ | NA | A | C | 0.06648 | 0.004163 | 5.60E-60 |
| rs174576 | miR-1908-5 | cis | 3.13E-08 | Nightingale | IDL_CE_pct | NA | A | C | -0.03864 | 0.004241 | 7.00E-20 |
| rs174576 | miR-1908-5 | cis | 3.13E-08 | Nightingale | XS_VLDL_T | NA | A | C | 0.032183 | 0.004256 | 1.30E-13 |
| rs174576 | miR-1908-5 | cis | 3.13E-08 | Nightingale | XL_HDL_PL | NA | A | C | -0.08858 | 0.003885 | 2.20E-125 |
| rs174576 | miR-1908-5 | cis | 3.13E-08 | Nightingale | SFA_pct | NA | A | C | -0.04265 | 0.004264 | 1.50E-23 |
| rs174576 | miR-1908-5 | cis | 3.13E-08 | Nightingale | Tyr | NA | A | C | 0.031862 | 0.004317 | 1.60E-13 |
| rs174576 | miR-1908-5 | cis | 3.13E-08 | Nightingale | S_HDL_C_p | NA | A | C | -0.04244 | 0.004272 | 4.50E-23 |
| rs174576 | miR-1908-5 | cis | 3.13E-08 | Nightingale | LA | NA | A | C | 0.083254 | 0.004239 | 2.40E-86 |
| rs174576 | miR-1908-5 | cis | 3.13E-08 | Nightingale | L_LDL_CE | NA | A | C | -0.05306 | 0.004284 | 3.90E-37 |
| rs174576 | miR-1908-5 | cis | 3.13E-08 | Nightingale | Clinical_LD | NA | A | C | -0.05486 | 0.004297 | 4.70E-39 |
| rs174576 | miR-1908-5 | cis | 3.13E-08 | Nightingale | M_LDL_P | NA | A | C | -0.04599 | 0.004335 | 4.40E-27 |
| rs174576 | miR-1908-5 | cis | 3.13E-08 | Nightingale | M_VLDL_C | NA | A | C | -0.08013 | 0.004068 | 3.70E-90 |
| rs174576 | miR-1908-5 | cis | 3.13E-08 | Nightingale | S_HDL_TG | NA | A | C | 0.06892 | 0.004191 | 1.50E-63 |
| rs174576 | miR-1908-5 | cis | 3.13E-08 | Nightingale | IDL_L | NA | A | C | -0.07202 | 0.004178 | 5.80E-71 |
| rs174576 | miR-1908-5 | cis | 3.13E-08 | Nightingale | M_HDL_P | NA | A | C | -0.03491 | 0.003982 | 7.60E-20 |
| rs174576 | miR-1908-5 | cis | 3.13E-08 | Nightingale | VLDL_size | NA | A | C | 0.072044 | 0.004029 | 5.70E-76 |
| rs174576 | miR-1908-5 | cis | 3.13E-08 | Nightingale | XS_VLDL_C | NA | A | C | -0.09209 | 0.004074 | 2.50E-118 |
| rs174576 | miR-1908-5 | cis | 3.13E-08 | Nightingale | M_HDL_CE | NA | A | C | -0.0473 | 0.004191 | 2.20E-30 |
| rs174576 | miR-1908-5 | cis | 3.13E-08 | Nightingale | M_HDL_TG | NA | A | C | 0.067146 | 0.004145 | 8.70E-62 |
| rs174576 | miR-1908-5 | cis | 3.13E-08 | Nightingale | Total_FC | NA | A | C | -0.05896 | 0.004222 | 7.40E-47 |
| rs174576 | miR-1908-5 | cis | 3.13E-08 | Nightingale | L_LDL_PL | NA | A | C | -0.05464 | 0.00429 | 7.30E-39 |
| rs174576 | miR-1908-5 | cis | 3.13E-08 | Nightingale | XL_HDL_FC | NA | A | C | 0.061086 | 0.003832 | 2.30E-63 |
| rs174576 | miR-1908-5 | cis | 3.13E-08 | Nightingale | XL_HDL_TG | NA | A | C | 0.087258 | 0.004072 | 8.30E-108 |
| rs174576 | miR-1908-5 | cis | 3.13E-08 | Nightingale | M_VLDL_C | NA | A | C | -0.04337 | 0.00431 | 8.20E-25 |
| rs174576 | miR-1908-5 | cis | 3.13E-08 | Nightingale | Total_C | NA | A | C | -0.06494 | 0.004184 | 3.40E-57 |
| rs174576 | miR-1908-5 | cis | 3.13E-08 | Nightingale | XL_VLDL_C | NA | A | C | -0.06048 | 0.004144 | 9.40E-50 |
| rs174576 | miR-1908-5 | cis | 3.13E-08 | Nightingale | XL_HDL_C_ | NA | A | C | 0.02352 | 0.004129 | 6.10E-10 |

| | | | | | | | | | | | |
|---|---|---|---|---|---|---|---|---|---|---|---|
| rs174576 | miR-1908-5 | cis | 3.13E-08 | Nightingale | L_HDL_FC_ | NA | A | C | -0.11153 | 0.004015 | 9.20E-185 |
| rs174576 | miR-1908-5 | cis | 3.13E-08 | Nightingale | L_LDL_FC_ | NA | A | C | -0.08282 | 0.00412 | 2.50E-93 |
| rs174576 | miR-1908-5 | cis | 3.13E-08 | Nightingale | XS_VLDL_T | NA | A | C | 0.097103 | 0.004105 | 1.50E-129 |
| rs174576 | miR-1908-5 | cis | 3.13E-08 | Nightingale | L_HDL_PL_ | NA | A | C | 0.042376 | 0.004183 | 1.10E-25 |
| rs174576 | miR-1908-5 | cis | 3.13E-08 | Nightingale | Total_P | NA | A | C | -0.0408 | 0.004068 | 2.80E-25 |
| rs174576 | miR-1908-5 | cis | 3.13E-08 | Nightingale | L_VLDL_TG | NA | A | C | 0.037204 | 0.004159 | 5.80E-20 |
| rs174576 | miR-1908-5 | cis | 3.13E-08 | Nightingale | L_LDL_FC | NA | A | C | -0.0681 | 0.004244 | 3.00E-61 |
| rs174576 | miR-1908-5 | cis | 3.13E-08 | Nightingale | XXL_VLDL_ | NA | A | C | 0.07166 | 0.004165 | 3.40E-69 |
| rs174576 | miR-1908-5 | cis | 3.13E-08 | Nightingale | XS_VLDL_P | NA | A | C | -0.04281 | 0.004265 | 1.30E-25 |
| rs174576 | miR-1908-5 | cis | 3.13E-08 | Nightingale | S_LDL_FC_ | NA | A | C | -0.08002 | 0.004188 | 1.80E-84 |
| rs174576 | miR-1908-5 | cis | 3.13E-08 | Nightingale | L_HDL_C_p | NA | A | C | -0.06832 | 0.004074 | 1.10E-67 |
| rs174576 | miR-1908-5 | cis | 3.13E-08 | Nightingale | Sphingomy | NA | A | C | -0.1147 | 0.004039 | 2.10E-186 |
| rs174576 | miR-1908-5 | cis | 3.13E-08 | Nightingale | M_LDL_FC | NA | A | C | -0.05216 | 0.004317 | 2.70E-35 |
| rs174576 | miR-1908-5 | cis | 3.13E-08 | Nightingale | M_VLDL_T | NA | A | C | 0.077311 | 0.004082 | 6.50E-84 |
| rs174576 | miR-1908-5 | cis | 3.13E-08 | Nightingale | M_HDL_TG | NA | A | C | 0.050333 | 0.004269 | 6.70E-33 |
| rs174576 | miR-1908-5 | cis | 3.13E-08 | Nightingale | S_VLDL_C_ | NA | A | C | -0.05623 | 0.004205 | 6.00E-42 |
| rs174576 | miR-1908-5 | cis | 3.13E-08 | Nightingale | XL_HDL_P | NA | A | C | -0.089 | 0.00389 | 2.60E-127 |
| rs174576 | miR-1908-5 | cis | 3.13E-08 | Nightingale | MUFA | NA | A | C | 0.063737 | 0.004278 | 1.70E-51 |
| rs174576 | miR-1908-5 | cis | 3.13E-08 | Nightingale | M_LDL_TG | NA | A | C | 0.036941 | 0.004258 | 3.50E-18 |
| rs174576 | miR-1908-5 | cis | 3.13E-08 | Nightingale | XXL_VLDL_ | NA | A | C | 0.073541 | 0.004174 | 3.40E-72 |
| rs174576 | miR-1908-5 | cis | 3.13E-08 | Nightingale | S_VLDL_FC | NA | A | C | -0.08274 | 0.004127 | 2.20E-93 |
| rs174576 | miR-1908-5 | cis | 3.13E-08 | Nightingale | LDL_L | NA | A | C | -0.04513 | 0.004308 | 6.10E-27 |
| rs174576 | miR-1908-5 | cis | 3.13E-08 | Nightingale | XXL_VLDL_ | NA | A | C | -0.02624 | 0.004301 | 2.00E-10 |
| rs174576 | miR-1908-5 | cis | 3.13E-08 | Nightingale | PUFA_by_M | NA | A | C | -0.1698 | 0.00414 | 0 |
| rs174576 | miR-1908-5 | cis | 3.13E-08 | Nightingale | XL_VLDL_T | NA | A | C | 0.056703 | 0.004134 | 1.30E-44 |
| rs174576 | miR-1908-5 | cis | 3.13E-08 | Nightingale | S_LDL_TG_ | NA | A | C | 0.08619 | 0.004188 | 2.80E-97 |
| rs174576 | miR-1908-5 | cis | 3.13E-08 | Nightingale | XL_HDL_L | NA | A | C | -0.0925 | 0.003884 | 9.40E-137 |
| rs174576 | miR-1908-5 | cis | 3.13E-08 | Nightingale | XXL_VLDL_ | NA | A | C | 0.044795 | 0.004346 | 7.50E-26 |
| rs174576 | miR-1908-5 | cis | 3.13E-08 | Nightingale | TG_by_PG | NA | A | C | 0.06901 | 0.004011 | 6.70E-70 |
| rs174576 | miR-1908-5 | cis | 3.13E-08 | Nightingale | M_HDL_CE | NA | A | C | -0.03853 | 0.003992 | 2.10E-23 |
| rs174576 | miR-1908-5 | cis | 3.13E-08 | Nightingale | XS_VLDL_L | NA | A | C | -0.04079 | 0.004248 | 8.40E-24 |
| rs174576 | miR-1908-5 | cis | 3.13E-08 | Nightingale | IDL_PL | NA | A | C | -0.07229 | 0.00417 | 3.60E-72 |
| rs174576 | miR-1908-5 | cis | 3.13E-08 | Nightingale | L_HDL_L | NA | A | C | -0.07737 | 0.003792 | 4.20E-101 |
| rs174576 | miR-1908-5 | cis | 3.13E-08 | Nightingale | XL_VLDL_T | NA | A | C | 0.058825 | 0.004187 | 2.10E-46 |
| rs174576 | miR-1908-5 | cis | 3.13E-08 | Nightingale | Phosphogly | NA | A | C | -0.04109 | 0.004094 | 2.10E-25 |
| rs174576 | miR-1908-5 | cis | 3.13E-08 | Nightingale | XS_VLDL_P | NA | A | C | 0.058667 | 0.004217 | 5.60E-45 |
| rs174576 | miR-1908-5 | cis | 3.13E-08 | Nightingale | M_HDL_FC | NA | A | C | -0.10322 | 0.00383 | 1.20E-174 |
| rs174576 | miR-1908-5 | cis | 3.13E-08 | Nightingale | XS_VLDL_C | NA | A | C | -0.06747 | 0.004195 | 3.80E-62 |
| rs174576 | miR-1908-5 | cis | 3.13E-08 | Nightingale | M_LDL_TG | NA | A | C | 0.075833 | 0.004268 | 5.00E-72 |
| rs174576 | miR-1908-5 | cis | 3.13E-08 | Nightingale | Total_L | NA | A | C | -0.03712 | 0.004252 | 2.00E-19 |
| rs174576 | miR-1908-5 | cis | 3.13E-08 | Nightingale | L_VLDL_PL | NA | A | C | 0.088013 | 0.004195 | 2.60E-101 |
| rs174576 | miR-1908-5 | cis | 3.13E-08 | Nightingale | L_LDL_TG_ | NA | A | C | 0.077771 | 0.004238 | 7.60E-77 |
| rs174576 | miR-1908-5 | cis | 3.13E-08 | Nightingale | XL_VLDL_P | NA | A | C | 0.052837 | 0.004154 | 1.20E-38 |
| rs174576 | miR-1908-5 | cis | 3.13E-08 | Nightingale | L_LDL_L | NA | A | C | -0.05389 | 0.00428 | 2.40E-38 |
| rs174576 | miR-1908-5 | cis | 3.13E-08 | Nightingale | M_LDL_CE | NA | A | C | -0.02601 | 0.004329 | 9.20E-10 |
| rs174576 | miR-1908-5 | cis | 3.13E-08 | Nightingale | L_VLDL_L | NA | A | C | 0.038273 | 0.004177 | 7.30E-21 |
| rs174576 | miR-1908-5 | cis | 3.13E-08 | Nightingale | L_HDL_P | NA | A | C | -0.07793 | 0.003785 | 5.90E-103 |
| rs174576 | miR-1908-5 | cis | 3.13E-08 | Nightingale | S_LDL_PL | NA | A | C | -0.05079 | 0.004312 | 8.50E-34 |
| rs174576 | miR-1908-5 | cis | 3.13E-08 | Nightingale | Total_CE | NA | A | C | -0.06682 | 0.004169 | 6.60E-61 |
| rs174576 | miR-1908-5 | cis | 3.13E-08 | Nightingale | IDL_P | NA | A | C | -0.04486 | 0.004277 | 3.40E-27 |
| rs174576 | miR-1908-5 | cis | 3.13E-08 | Nightingale | IDL_TG_pct | NA | A | C | 0.088203 | 0.004175 | 7.19E-103 |
| rs174576 | miR-1908-5 | cis | 3.13E-08 | Nightingale | XL_HDL_PL | NA | A | C | -0.06676 | 0.003971 | 1.80E-68 |
| rs174576 | miR-1908-5 | cis | 3.13E-08 | Nightingale | PUFA | NA | A | C | -0.09566 | 0.004215 | 1.60E-120 |
| rs174576 | miR-1908-5 | cis | 3.13E-08 | Nightingale | S_VLDL_CE | NA | A | C | -0.02846 | 0.004264 | 1.20E-11 |
| rs174576 | miR-1908-5 | cis | 3.13E-08 | Nightingale | M_VLDL_F | NA | A | C | -0.02884 | 0.004321 | 8.20E-12 |
| rs174576 | miR-1908-5 | cis | 3.13E-08 | Nightingale | S_HDL_TG_ | NA | A | C | 0.069473 | 0.004111 | 3.90E-67 |
| rs174576 | miR-1908-5 | cis | 3.13E-08 | Nightingale | S_LDL_TG | NA | A | C | 0.050738 | 0.004216 | 5.00E-34 |
| rs174576 | miR-1908-5 | cis | 3.13E-08 | Nightingale | Omega_6_ | NA | A | C | 0.02392 | 0.004207 | 8.70E-09 |
| rs174576 | miR-1908-5 | cis | 3.13E-08 | Nightingale | XL_VLDL_L | NA | A | C | 0.051413 | 0.004159 | 1.40E-36 |
| rs174576 | miR-1908-5 | cis | 3.13E-08 | Nightingale | L_HDL_FC | NA | A | C | -0.08459 | 0.003788 | 1.80E-120 |
| rs174576 | miR-1908-5 | cis | 3.13E-08 | Nightingale | IDL_FC | NA | A | C | -0.0893 | 0.004183 | 1.00E-106 |
| rs174576 | miR-1908-5 | cis | 3.13E-08 | Nightingale | LA_pct | NA | A | C | 0.184051 | 0.004224 | 0 |
| rs174576 | miR-1908-5 | cis | 3.13E-08 | Nightingale | S_VLDL_FC | NA | A | C | -0.02763 | 0.004323 | 5.80E-11 |

| | | | | | | | | | | | |
|---|---|---|---|---|---|---|---|---|---|---|---|
| rs174576 | miR-1908-5 | cis | 3.13E-08 | Nightingale | S_LDL_FC | NA | A | C | -0.06247 | 0.004315 | 3.60E-50 |
| rs174576 | miR-1908-5 | cis | 3.13E-08 | Nightingale | S_VLDL_TG | NA | A | C | 0.064162 | 0.004182 | 8.50E-55 |
| rs174576 | miR-1908-5 | cis | 3.13E-08 | Nightingale | Total_PL | NA | A | C | -0.051 | 0.004116 | 2.20E-37 |
| rs174576 | miR-1908-5 | cis | 3.13E-08 | Nightingale | M_LDL_PL_ | NA | A | C | 0.045368 | 0.00432 | 6.30E-27 |
| rs174576 | miR-1908-5 | cis | 3.13E-08 | Nightingale | HDL_P | NA | A | C | -0.03617 | 0.004058 | 3.40E-20 |
| rs174576 | miR-1908-5 | cis | 3.13E-08 | Nightingale | M_LDL_C | NA | A | C | -0.03373 | 0.004331 | 1.90E-15 |
| rs174576 | miR-1908-5 | cis | 3.13E-08 | Nightingale | LDL_TG | NA | A | C | 0.030068 | 0.004274 | 3.30E-12 |
| rs174576 | miR-1908-5 | cis | 3.13E-08 | Nightingale | L_VLDL_PL | NA | A | C | 0.048876 | 0.00417 | 3.20E-33 |
| rs174576 | miR-1908-5 | cis | 3.13E-08 | Nightingale | ApoA1 | NA | A | C | -0.04169 | 0.003955 | 6.10E-28 |
| rs174576 | miR-1908-5 | cis | 3.13E-08 | Nightingale | L_HDL_C | NA | A | C | -0.08038 | 0.003806 | 6.19E-108 |
| rs174576 | miR-1908-5 | cis | 3.13E-08 | Nightingale | L_VLDL_P | NA | A | C | 0.041996 | 0.004173 | 7.60E-25 |
| rs174576 | miR-1908-5 | cis | 3.13E-08 | Nightingale | L_VLDL_C | NA | A | C | 0.029189 | 0.004227 | 1.80E-12 |
| rs174576 | miR-1908-5 | cis | 3.13E-08 | Nightingale | LDL_P | NA | A | C | -0.03963 | 0.004319 | 5.00E-21 |
| rs174576 | miR-1908-5 | cis | 3.13E-08 | Nightingale | MUFA_pct | NA | A | C | 0.196484 | 0.004136 | 0 |
| rs174576 | miR-1908-5 | cis | 3.13E-08 | Nightingale | XXL_VLDL_ | NA | A | C | 0.066057 | 0.004157 | 1.30E-59 |
| rs174576 | miR-1908-5 | cis | 3.13E-08 | Nightingale | XL_HDL_CE | NA | A | C | -0.09666 | 0.00387 | 6.90E-150 |
| rs174576 | miR-1908-5 | cis | 3.13E-08 | Nightingale | HDL_FC | NA | A | C | -0.07407 | 0.003829 | 2.40E-90 |
| rs174576 | miR-1908-5 | cis | 3.13E-08 | Nightingale | L_VLDL_CE | NA | A | C | -0.05819 | 0.004136 | 6.00E-48 |
| rs174576 | miR-1908-5 | cis | 3.13E-08 | Nightingale | DHA_pct | NA | A | C | -0.27181 | 0.004196 | 0 |
| rs174576 | miR-1908-5 | cis | 3.13E-08 | Nightingale | HDL_CE | NA | A | C | -0.06189 | 0.003879 | 7.80E-62 |
| rs174576 | miR-1908-5 | cis | 3.13E-08 | Nightingale | XXL_VLDL_ | NA | A | C | 0.061227 | 0.004171 | 6.30E-51 |
| rs174576 | miR-1908-5 | cis | 3.13E-08 | Nightingale | M_LDL_PL | NA | A | C | -0.02325 | 0.004329 | 3.90E-08 |
| rs174576 | miR-1908-5 | cis | 3.13E-08 | Nightingale | XS_VLDL_C | NA | A | C | -0.07131 | 0.00417 | 6.20E-70 |
| rs174576 | miR-1908-5 | cis | 3.13E-08 | Nightingale | PUFA_pct | NA | A | C | -0.12066 | 0.004157 | 4.00E-187 |
| rs174576 | miR-1908-5 | cis | 3.13E-08 | Nightingale | S_HDL_FC_ | NA | A | C | -0.10169 | 0.0041 | 8.71E-151 |
| rs174576 | miR-1908-5 | cis | 3.13E-08 | Nightingale | S_LDL_CE_ | NA | A | C | 0.056742 | 0.004314 | 1.20E-41 |
| rs174576 | miR-1908-5 | cis | 3.13E-08 | Nightingale | M_HDL_PL | NA | A | C | -0.02218 | 0.004042 | 1.10E-08 |
| rs174576 | miR-1908-5 | cis | 3.13E-08 | Nightingale | Omega_3_ | NA | A | C | -0.38665 | 0.004223 | 0 |
| rs174576 | miR-1908-5 | cis | 3.13E-08 | Nightingale | L_VLDL_C_ | NA | A | C | -0.03771 | 0.004222 | 1.70E-20 |
| rs174576 | miR-1908-5 | cis | 3.13E-08 | Nightingale | S_LDL_C | NA | A | C | -0.03177 | 0.004327 | 5.00E-14 |
| rs174576 | miR-1908-5 | cis | 3.13E-08 | Nightingale | XL_HDL_CE | NA | A | C | -0.0264 | 0.00432 | 2.50E-09 |
| rs174576 | miR-1908-5 | cis | 3.13E-08 | Nightingale | S_LDL_P | NA | A | C | -0.03187 | 0.004326 | 3.90E-14 |
| rs174576 | miR-1908-5 | cis | 3.13E-08 | Nightingale | non_HDL_( | NA | A | C | -0.04741 | 0.004299 | 1.10E-29 |
| rs174576 | miR-1908-5 | cis | 3.13E-08 | Nightingale | XS_VLDL_F | NA | A | C | -0.05524 | 0.004247 | 1.80E-41 |
| rs174576 | miR-1908-5 | cis | 3.13E-08 | Nightingale | IDL_C | NA | A | C | -0.07698 | 0.004173 | 1.80E-80 |
| rs174576 | miR-1908-5 | cis | 3.13E-08 | Nightingale | M_LDL_FC_ | NA | A | C | -0.07172 | 0.004141 | 6.00E-70 |
| rs174576 | miR-1908-5 | cis | 3.13E-08 | Nightingale | L_HDL_TG_ | NA | A | C | 0.07925 | 0.004079 | 3.30E-89 |
| rs174576 | miR-1908-5 | cis | 3.13E-08 | Nightingale | L_LDL_C | NA | A | C | -0.05757 | 0.004274 | 1.30E-43 |
| rs174576 | miR-1908-5 | cis | 3.13E-08 | Nightingale | XXL_VLDL_ | NA | A | C | 0.072131 | 0.004148 | 6.10E-71 |
| rs174576 | miR-1908-5 | cis | 3.13E-08 | Nightingale | M_VLDL_F | NA | A | C | -0.09044 | 0.00411 | 3.30E-113 |
| rs174576 | miR-1908-5 | cis | 3.13E-08 | Nightingale | Total_TG | NA | A | C | 0.046954 | 0.004184 | 2.40E-30 |
| rs174576 | miR-1908-5 | cis | 3.13E-08 | Nightingale | LDL_C | NA | A | C | -0.04948 | 0.004304 | 4.60E-32 |
| rs174576 | miR-1908-5 | cis | 3.13E-08 | Nightingale | M_VLDL_C | NA | A | C | -0.05204 | 0.004283 | 1.60E-35 |
| rs174576 | miR-1908-5 | cis | 3.13E-08 | Nightingale | XS_VLDL_P | NA | A | C | -0.03352 | 0.004262 | 1.20E-16 |
| rs174576 | miR-1908-5 | cis | 3.13E-08 | Nightingale | M_HDL_L | NA | A | C | -0.02833 | 0.004012 | 2.00E-13 |
| rs174576 | miR-1908-5 | cis | 3.13E-08 | Nightingale | XL_VLDL_P | NA | A | C | 0.048372 | 0.004171 | 2.20E-32 |
| rs174576 | miR-1908-5 | cis | 3.13E-08 | Nightingale | S_VLDL_TG | NA | A | C | 0.039252 | 0.004194 | 1.50E-21 |
| rs174576 | miR-1908-5 | cis | 3.13E-08 | Nightingale | L_VLDL_FC | NA | A | C | 0.029231 | 0.004281 | 5.90E-12 |
| rs174576 | miR-1908-5 | cis | 3.13E-08 | Nightingale | XL_VLDL_F | NA | A | C | 0.043581 | 0.004186 | 2.30E-26 |
| rs174576 | miR-1908-5 | cis | 3.13E-08 | Nightingale | XL_HDL_FC | NA | A | C | -0.10184 | 0.004026 | 1.30E-152 |
| rs174576 | miR-1908-5 | cis | 3.13E-08 | Nightingale | L_LDL_P | NA | A | C | -0.03684 | 0.004307 | 1.20E-18 |
| rs174576 | miR-1908-5 | cis | 3.13E-08 | Nightingale | IDL_FC_pct | NA | A | C | -0.10365 | 0.004287 | 1.20E-130 |
| rs174576 | miR-1908-5 | cis | 3.13E-08 | Nightingale | XXL_VLDL_ | NA | A | C | 0.067959 | 0.004164 | 7.30E-63 |
| rs174576 | miR-1908-5 | cis | 3.13E-08 | Nightingale | M_HDL_C | NA | A | C | -0.04145 | 0.003977 | 5.00E-27 |
| rs174576 | miR-1908-5 | cis | 3.13E-08 | Nightingale | IDL_CE | NA | A | C | -0.07202 | 0.004172 | 9.80E-71 |
| rs174576 | miR-1908-5 | cis | 3.13E-08 | Nightingale | M_VLDL_P | NA | A | C | -0.06163 | 0.004147 | 1.70E-53 |
| rs174576 | miR-1908-5 | cis | 3.13E-08 | Nightingale | IDL_C_pct | NA | A | C | -0.0751 | 0.004219 | 1.10E-72 |
| rs174576 | miR-1908-5 | cis | 3.13E-08 | Nightingale | Citrate | NA | A | C | -0.03332 | 0.004295 | 8.00E-16 |
| rs174576 | miR-1908-5 | cis | 3.13E-08 | Nightingale | HDL_L | NA | A | C | -0.05374 | 0.003881 | 7.00E-47 |
| rs174576 | miR-1908-5 | cis | 3.13E-08 | Nightingale | M_HDL_C_ | NA | A | C | -0.06614 | 0.004114 | 3.30E-61 |
| rs174576 | miR-1908-5 | cis | 3.13E-08 | Nightingale | HDL_TG | NA | A | C | 0.04637 | 0.004262 | 4.60E-28 |
| rs174576 | miR-1908-5 | cis | 3.13E-08 | Nightingale | M_LDL_L | NA | A | C | -0.02786 | 0.004329 | 5.40E-11 |
| rs174576 | miR-1908-5 | cis | 3.13E-08 | Nightingale | LDL_FC | NA | A | C | -0.06405 | 0.004276 | 2.10E-53 |

| | | | | | | | | | | | | |
|---|---|---|---|---|---|---|---|---|---|---|---|---|
| rs174576 | miR-1908-5 | cis | | 3.13E-08 | Nightingale | VLDL_TG | NA | A | C | 0.047801 | 0.004159 | 9.50E-32 |
| rs174576 | miR-1908-5 | cis | | 3.13E-08 | Nightingale | S_LDL_L | NA | A | C | -0.03195 | 0.004321 | 3.00E-14 |
| rs174576 | miR-1908-5 | cis | | 3.13E-08 | Nightingale | DHA | NA | A | C | -0.28628 | 0.004165 | 0 |
| rs174576 | miR-1908-5 | cis | | 3.13E-08 | Nightingale | VLDL_L | NA | A | C | 0.02764 | 0.004238 | 3.30E-11 |
| rs174577 | miR-1908-5 | cis | | 1.89E-08 | Nightingale | L_HDL_C | NA | A | C | -0.08014 | 0.003804 | 3.30E-107 |
| rs174577 | miR-1908-5 | cis | | 1.89E-08 | Nightingale | M_VLDL_C | NA | A | C | -0.07998 | 0.004066 | 7.60E-90 |
| rs174577 | miR-1908-5 | cis | | 1.89E-08 | Nightingale | Tyr | NA | A | C | 0.032399 | 0.004314 | 5.90E-14 |
| rs174577 | miR-1908-5 | cis | | 1.89E-08 | Nightingale | S_HDL_C_p | NA | A | C | -0.04254 | 0.00427 | 3.30E-23 |
| rs174577 | miR-1908-5 | cis | | 1.89E-08 | Nightingale | HDL_C | NA | A | C | -0.06513 | 0.003857 | 4.10E-69 |
| rs174577 | miR-1908-5 | cis | | 1.89E-08 | Nightingale | IDL_TG_pct | NA | A | C | 0.088057 | 0.004173 | 1.20E-102 |
| rs174577 | miR-1908-5 | cis | | 1.89E-08 | Nightingale | XS_VLDL_P | NA | A | C | 0.058725 | 0.004215 | 3.60E-45 |
| rs174577 | miR-1908-5 | cis | | 1.89E-08 | Nightingale | L_HDL_FC_ | NA | A | C | -0.11089 | 0.004013 | 1.00E-182 |
| rs174577 | miR-1908-5 | cis | | 1.89E-08 | Nightingale | XS_VLDL_F | NA | A | C | -0.11172 | 0.004287 | 1.10E-152 |
| rs174577 | miR-1908-5 | cis | | 1.89E-08 | Nightingale | L_LDL_TG_ | NA | A | C | 0.077681 | 0.004236 | 8.90E-77 |
| rs174577 | miR-1908-5 | cis | | 1.89E-08 | Nightingale | IDL_PL | NA | A | C | -0.07215 | 0.004168 | 6.20E-72 |
| rs174577 | miR-1908-5 | cis | | 1.89E-08 | Nightingale | M_LDL_P | NA | A | C | -0.04617 | 0.004332 | 2.60E-27 |
| rs174577 | miR-1908-5 | cis | | 1.89E-08 | Nightingale | S_LDL_PL | NA | A | C | -0.05076 | 0.004309 | 7.60E-34 |
| rs174577 | miR-1908-5 | cis | | 1.89E-08 | Nightingale | Sphingomy | NA | A | C | -0.11463 | 0.004037 | 2.40E-186 |
| rs174577 | miR-1908-5 | cis | | 1.89E-08 | Nightingale | IDL_L | NA | A | C | -0.07193 | 0.004176 | 7.30E-71 |
| rs174577 | miR-1908-5 | cis | | 1.89E-08 | Nightingale | M_VLDL_T | NA | A | C | 0.03133 | 0.004197 | 2.60E-14 |
| rs174577 | miR-1908-5 | cis | | 1.89E-08 | Nightingale | PUFA_by_M | NA | A | C | -0.16923 | 0.004138 | 0 |
| rs174577 | miR-1908-5 | cis | | 1.89E-08 | Nightingale | XL_HDL_C | NA | A | C | -0.09815 | 0.003895 | 3.60E-152 |
| rs174577 | miR-1908-5 | cis | | 1.89E-08 | Nightingale | XL_HDL_FC | NA | A | C | -0.10116 | 0.004024 | 1.40E-150 |
| rs174577 | miR-1908-5 | cis | | 1.89E-08 | Nightingale | XL_HDL_L | NA | A | C | -0.09196 | 0.003882 | 3.60E-135 |
| rs174577 | miR-1908-5 | cis | | 1.89E-08 | Nightingale | M_LDL_PL | NA | A | C | -0.02344 | 0.004326 | 2.90E-08 |
| rs174577 | miR-1908-5 | cis | | 1.89E-08 | Nightingale | XL_VLDL_P | NA | A | C | 0.052569 | 0.004152 | 2.80E-38 |
| rs174577 | miR-1908-5 | cis | | 1.89E-08 | Nightingale | L_HDL_FC | NA | A | C | -0.0843 | 0.003786 | 1.40E-119 |
| rs174577 | miR-1908-5 | cis | | 1.89E-08 | Nightingale | S_VLDL_TG | NA | A | C | 0.06418 | 0.00418 | 8.00E-55 |
| rs174577 | miR-1908-5 | cis | | 1.89E-08 | Nightingale | PUFA | NA | A | C | -0.09561 | 0.004213 | 1.80E-120 |
| rs174577 | miR-1908-5 | cis | | 1.89E-08 | Nightingale | VLDL_L | NA | A | C | 0.027436 | 0.004235 | 4.50E-11 |
| rs174577 | miR-1908-5 | cis | | 1.89E-08 | Nightingale | XL_VLDL_T | NA | A | C | 0.058862 | 0.004185 | 1.80E-46 |
| rs174577 | miR-1908-5 | cis | | 1.89E-08 | Nightingale | XL_HDL_PL | NA | A | C | -0.06623 | 0.003968 | 2.20E-67 |
| rs174577 | miR-1908-5 | cis | | 1.89E-08 | Nightingale | M_VLDL_C | NA | A | C | -0.07533 | 0.004055 | 3.50E-80 |
| rs174577 | miR-1908-5 | cis | | 1.89E-08 | Nightingale | HDL_L | NA | A | C | -0.05379 | 0.003879 | 6.00E-47 |
| rs174577 | miR-1908-5 | cis | | 1.89E-08 | Nightingale | LDL_PL | NA | A | C | -0.04591 | 0.00431 | 1.20E-27 |
| rs174577 | miR-1908-5 | cis | | 1.89E-08 | Nightingale | M_HDL_CE | NA | A | C | -0.03876 | 0.003989 | 1.20E-23 |
| rs174577 | miR-1908-5 | cis | | 1.89E-08 | Nightingale | L_HDL_TG_ | NA | A | C | 0.079179 | 0.004076 | 4.80E-89 |
| rs174577 | miR-1908-5 | cis | | 1.89E-08 | Nightingale | XL_VLDL_F | NA | A | C | -0.06728 | 0.004185 | 4.70E-61 |
| rs174577 | miR-1908-5 | cis | | 1.89E-08 | Nightingale | XL_VLDL_C | NA | A | C | -0.0604 | 0.004141 | 1.20E-49 |
| rs174577 | miR-1908-5 | cis | | 1.89E-08 | Nightingale | ApoA1 | NA | A | C | -0.04185 | 0.003953 | 3.90E-28 |
| rs174577 | miR-1908-5 | cis | | 1.89E-08 | Nightingale | XS_VLDL_C | NA | A | C | -0.07112 | 0.004168 | 1.30E-69 |
| rs174577 | miR-1908-5 | cis | | 1.89E-08 | Nightingale | M_LDL_CE | NA | A | C | -0.02625 | 0.004326 | 6.30E-10 |
| rs174577 | miR-1908-5 | cis | | 1.89E-08 | Nightingale | M_HDL_FC | NA | A | C | -0.05209 | 0.003931 | 7.50E-43 |
| rs174577 | miR-1908-5 | cis | | 1.89E-08 | Nightingale | L_HDL_CE | NA | A | C | -0.07861 | 0.003814 | 1.60E-102 |
| rs174577 | miR-1908-5 | cis | | 1.89E-08 | Nightingale | HDL_size | NA | A | C | -0.07235 | 0.003828 | 2.60E-86 |
| rs174577 | miR-1908-5 | cis | | 1.89E-08 | Nightingale | XL_HDL_CE | NA | A | C | -0.09617 | 0.003868 | 2.40E-148 |
| rs174577 | miR-1908-5 | cis | | 1.89E-08 | Nightingale | L_LDL_FC_ | NA | A | C | -0.08252 | 0.004117 | 8.20E-93 |
| rs174577 | miR-1908-5 | cis | | 1.89E-08 | Nightingale | M_VLDL_C | NA | A | C | -0.04341 | 0.004308 | 7.10E-25 |
| rs174577 | miR-1908-5 | cis | | 1.89E-08 | Nightingale | Omega_6_ | NA | A | C | 0.365843 | 0.004257 | 0 |
| rs174577 | miR-1908-5 | cis | | 1.89E-08 | Nightingale | XS_VLDL_P | NA | A | C | -0.03342 | 0.004259 | 1.50E-16 |
| rs174577 | miR-1908-5 | cis | | 1.89E-08 | Nightingale | non_HDL_C | NA | A | C | -0.04747 | 0.004296 | 8.90E-30 |
| rs174577 | miR-1908-5 | cis | | 1.89E-08 | Nightingale | M_HDL_C | NA | A | C | -0.04166 | 0.003975 | 2.80E-27 |
| rs174577 | miR-1908-5 | cis | | 1.89E-08 | Nightingale | XXL_VLDL_ | NA | A | C | 0.071865 | 0.004145 | 1.60E-70 |
| rs174577 | miR-1908-5 | cis | | 1.89E-08 | Nightingale | HDL_P | NA | A | C | -0.03647 | 0.004056 | 1.60E-20 |
| rs174577 | miR-1908-5 | cis | | 1.89E-08 | Nightingale | M_HDL_PL | NA | A | C | -0.02244 | 0.004039 | 7.40E-09 |
| rs174577 | miR-1908-5 | cis | | 1.89E-08 | Nightingale | M_VLDL_T | NA | A | C | 0.077194 | 0.00408 | 1.10E-83 |
| rs174577 | miR-1908-5 | cis | | 1.89E-08 | Nightingale | M_VLDL_C | NA | A | C | -0.05202 | 0.00428 | 1.60E-35 |
| rs174577 | miR-1908-5 | cis | | 1.89E-08 | Nightingale | M_VLDL_F | NA | A | C | -0.02894 | 0.004319 | 6.80E-12 |
| rs174577 | miR-1908-5 | cis | | 1.89E-08 | Nightingale | L_LDL_C_p | NA | A | C | -0.07712 | 0.004209 | 3.50E-77 |
| rs174577 | miR-1908-5 | cis | | 1.89E-08 | Nightingale | Cholines | NA | A | C | -0.03738 | 0.004063 | 7.30E-22 |
| rs174577 | miR-1908-5 | cis | | 1.89E-08 | Nightingale | L_VLDL_FC | NA | A | C | 0.040043 | 0.004178 | 9.60E-23 |
| rs174577 | miR-1908-5 | cis | | 1.89E-08 | Nightingale | S_LDL_L | NA | A | C | -0.03206 | 0.004319 | 2.30E-14 |
| rs174577 | miR-1908-5 | cis | | 1.89E-08 | Nightingale | S_LDL_PL_ | NA | A | C | -0.06071 | 0.004311 | 2.10E-47 |

| | | | | | | | | | | | |
|---|---|---|---|---|---|---|---|---|---|---|---|
| rs174577 | miR-1908-5 | cis | 1.89E-08 | Nightingale | Total_C | NA | A | C | -0.06496 | 0.004182 | 2.80E-57 |
| rs174577 | miR-1908-5 | cis | 1.89E-08 | Nightingale | XL_HDL_C_ | NA | A | C | 0.023146 | 0.004127 | 1.10E-09 |
| rs174577 | miR-1908-5 | cis | 1.89E-08 | Nightingale | Total_L | NA | A | C | -0.03721 | 0.00425 | 1.60E-19 |
| rs174577 | miR-1908-5 | cis | 1.89E-08 | Nightingale | S_VLDL_TG | NA | A | C | 0.039101 | 0.004192 | 2.00E-21 |
| rs174577 | miR-1908-5 | cis | 1.89E-08 | Nightingale | Total_FA | NA | A | C | -0.02361 | 0.004286 | 1.40E-08 |
| rs174577 | miR-1908-5 | cis | 1.89E-08 | Nightingale | XS_VLDL_T | NA | A | C | 0.096829 | 0.004103 | 6.40E-129 |
| rs174577 | miR-1908-5 | cis | 1.89E-08 | Nightingale | Omega_3_ | NA | A | C | -0.38568 | 0.004221 | 0 |
| rs174577 | miR-1908-5 | cis | 1.89E-08 | Nightingale | LA_pct | NA | A | C | 0.183741 | 0.004221 | 0 |
| rs174577 | miR-1908-5 | cis | 1.89E-08 | Nightingale | L_HDL_PL_ | NA | A | C | 0.041925 | 0.004181 | 3.50E-25 |
| rs174577 | miR-1908-5 | cis | 1.89E-08 | Nightingale | S_VLDL_FC | NA | A | C | -0.02775 | 0.004321 | 4.70E-11 |
| rs174577 | miR-1908-5 | cis | 1.89E-08 | Nightingale | L_HDL_L | NA | A | C | -0.07717 | 0.00379 | 1.60E-100 |
| rs174577 | miR-1908-5 | cis | 1.89E-08 | Nightingale | LDL_CE | NA | A | C | -0.0434 | 0.004309 | 5.60E-25 |
| rs174577 | miR-1908-5 | cis | 1.89E-08 | Nightingale | TG_by_PG | NA | A | C | 0.06886 | 0.004009 | 1.20E-69 |
| rs174577 | miR-1908-5 | cis | 1.89E-08 | Nightingale | L_LDL_P | NA | A | C | -0.03684 | 0.004305 | 1.10E-18 |
| rs174577 | miR-1908-5 | cis | 1.89E-08 | Nightingale | Clinical_LD | NA | A | C | -0.05494 | 0.004295 | 3.40E-39 |
| rs174577 | miR-1908-5 | cis | 1.89E-08 | Nightingale | XXL_VLDL_ | NA | A | C | 0.061099 | 0.004168 | 8.40E-51 |
| rs174577 | miR-1908-5 | cis | 1.89E-08 | Nightingale | M_HDL_CE | NA | A | C | -0.0473 | 0.004189 | 2.10E-30 |
| rs174577 | miR-1908-5 | cis | 1.89E-08 | Nightingale | VLDL_TG | NA | A | C | 0.047589 | 0.004157 | 1.60E-31 |
| rs174577 | miR-1908-5 | cis | 1.89E-08 | Nightingale | Omega_6_ | NA | A | C | 0.023968 | 0.004204 | 7.80E-09 |
| rs174577 | miR-1908-5 | cis | 1.89E-08 | Nightingale | M_HDL_PL | NA | A | C | 0.058413 | 0.004097 | 5.30E-48 |
| rs174577 | miR-1908-5 | cis | 1.89E-08 | Nightingale | L_HDL_CE_ | NA | A | C | -0.04997 | 0.004133 | 8.40E-36 |
| rs174577 | miR-1908-5 | cis | 1.89E-08 | Nightingale | XS_VLDL_C | NA | A | C | -0.06731 | 0.004193 | 7.10E-62 |
| rs174577 | miR-1908-5 | cis | 1.89E-08 | Nightingale | Omega_3 | NA | A | C | -0.33215 | 0.004234 | 0 |
| rs174577 | miR-1908-5 | cis | 1.89E-08 | Nightingale | S_VLDL_PL | NA | A | C | -0.07828 | 0.004133 | 1.00E-83 |
| rs174577 | miR-1908-5 | cis | 1.89E-08 | Nightingale | Unsaturati | NA | A | C | -0.33597 | 0.004091 | 0 |
| rs174577 | miR-1908-5 | cis | 1.89E-08 | Nightingale | M_LDL_FC_ | NA | A | C | -0.0714 | 0.004138 | 2.00E-69 |
| rs174577 | miR-1908-5 | cis | 1.89E-08 | Nightingale | Total_P | NA | A | C | -0.04108 | 0.004065 | 1.20E-25 |
| rs174577 | miR-1908-5 | cis | 1.89E-08 | Nightingale | XL_HDL_PL | NA | A | C | -0.08805 | 0.003883 | 6.81E-124 |
| rs174577 | miR-1908-5 | cis | 1.89E-08 | Nightingale | S_VLDL_C_ | NA | A | C | -0.05626 | 0.004203 | 5.50E-42 |
| rs174577 | miR-1908-5 | cis | 1.89E-08 | Nightingale | S_VLDL_CE | NA | A | C | -0.02858 | 0.004262 | 1.00E-11 |
| rs174577 | miR-1908-5 | cis | 1.89E-08 | Nightingale | XS_VLDL_C | NA | A | C | -0.09769 | 0.004093 | 1.80E-131 |
| rs174577 | miR-1908-5 | cis | 1.89E-08 | Nightingale | M_VLDL_P | NA | A | C | -0.06162 | 0.004145 | 1.60E-53 |
| rs174577 | miR-1908-5 | cis | 1.89E-08 | Nightingale | S_LDL_C | NA | A | C | -0.03192 | 0.004324 | 3.70E-14 |
| rs174577 | miR-1908-5 | cis | 1.89E-08 | Nightingale | L_VLDL_L | NA | A | C | 0.038057 | 0.004175 | 1.20E-20 |
| rs174577 | miR-1908-5 | cis | 1.89E-08 | Nightingale | XS_VLDL_L | NA | A | C | -0.0407 | 0.004246 | 1.00E-23 |
| rs174577 | miR-1908-5 | cis | 1.89E-08 | Nightingale | ApoB | NA | A | C | -0.03795 | 0.004314 | 1.60E-19 |
| rs174577 | miR-1908-5 | cis | 1.89E-08 | Nightingale | S_HDL_TG | NA | A | C | 0.068645 | 0.004189 | 4.50E-63 |
| rs174577 | miR-1908-5 | cis | 1.89E-08 | Nightingale | M_LDL_FC | NA | A | C | -0.05227 | 0.004315 | 1.70E-35 |
| rs174577 | miR-1908-5 | cis | 1.89E-08 | Nightingale | DHA | NA | A | C | -0.28569 | 0.004163 | 0 |
| rs174577 | miR-1908-5 | cis | 1.89E-08 | Nightingale | L_LDL_PL | NA | A | C | -0.05473 | 0.004287 | 5.00E-39 |
| rs174577 | miR-1908-5 | cis | 1.89E-08 | Nightingale | L_LDL_C | NA | A | C | -0.05759 | 0.004271 | 1.10E-43 |
| rs174577 | miR-1908-5 | cis | 1.89E-08 | Nightingale | LDL_C | NA | A | C | -0.04956 | 0.004301 | 3.40E-32 |
| rs174577 | miR-1908-5 | cis | 1.89E-08 | Nightingale | XL_VLDL_F | NA | A | C | 0.043326 | 0.004184 | 4.10E-26 |
| rs174577 | miR-1908-5 | cis | 1.89E-08 | Nightingale | Citrate | NA | A | C | -0.03338 | 0.004292 | 8.00E-16 |
| rs174577 | miR-1908-5 | cis | 1.89E-08 | Nightingale | XXL_VLDL_ | NA | A | C | 0.073225 | 0.004172 | 1.20E-71 |
| rs174577 | miR-1908-5 | cis | 1.89E-08 | Nightingale | XS_VLDL_F | NA | A | C | -0.05513 | 0.004244 | 2.50E-41 |
| rs174577 | miR-1908-5 | cis | 1.89E-08 | Nightingale | L_VLDL_PL | NA | A | C | 0.048639 | 0.004168 | 6.10E-33 |
| rs174577 | miR-1908-5 | cis | 1.89E-08 | Nightingale | M_HDL_TG | NA | A | C | 0.050119 | 0.004266 | 1.10E-32 |
| rs174577 | miR-1908-5 | cis | 1.89E-08 | Nightingale | IDL_CE_pct | NA | A | C | -0.03854 | 0.004239 | 7.90E-20 |
| rs174577 | miR-1908-5 | cis | 1.89E-08 | Nightingale | Total_CE | NA | A | C | -0.06685 | 0.004167 | 5.30E-61 |
| rs174577 | miR-1908-5 | cis | 1.89E-08 | Nightingale | S_VLDL_FC | NA | A | C | -0.08263 | 0.004125 | 3.50E-93 |
| rs174577 | miR-1908-5 | cis | 1.89E-08 | Nightingale | M_LDL_C | NA | A | C | -0.03394 | 0.004328 | 1.20E-15 |
| rs174577 | miR-1908-5 | cis | 1.89E-08 | Nightingale | L_HDL_PL | NA | A | C | -0.07613 | 0.003802 | 3.90E-97 |
| rs174577 | miR-1908-5 | cis | 1.89E-08 | Nightingale | Total_FC | NA | A | C | -0.05897 | 0.00422 | 6.40E-47 |
| rs174577 | miR-1908-5 | cis | 1.89E-08 | Nightingale | IDL_C_pct | NA | A | C | -0.07507 | 0.004217 | 9.90E-73 |
| rs174577 | miR-1908-5 | cis | 1.89E-08 | Nightingale | LA | NA | A | C | 0.082961 | 0.004236 | 6.30E-86 |
| rs174577 | miR-1908-5 | cis | 1.89E-08 | Nightingale | XL_VLDL_L | NA | A | C | 0.051193 | 0.004156 | 2.60E-36 |
| rs174577 | miR-1908-5 | cis | 1.89E-08 | Nightingale | XS_VLDL_T | NA | A | C | 0.032063 | 0.004253 | 1.50E-13 |
| rs174577 | miR-1908-5 | cis | 1.89E-08 | Nightingale | VLDL_size | NA | A | C | 0.071635 | 0.004026 | 3.50E-75 |
| rs174577 | miR-1908-5 | cis | 1.89E-08 | Nightingale | HDL_CE | NA | A | C | -0.06193 | 0.003877 | 7.10E-62 |
| rs174577 | miR-1908-5 | cis | 1.89E-08 | Nightingale | L_VLDL_PL | NA | A | C | 0.087798 | 0.004192 | 8.00E-101 |
| rs174577 | miR-1908-5 | cis | 1.89E-08 | Nightingale | HDL_TG | NA | A | C | 0.046227 | 0.004259 | 6.10E-28 |
| rs174577 | miR-1908-5 | cis | 1.89E-08 | Nightingale | XXL_VLDL_ | NA | A | C | 0.071361 | 0.004163 | 1.00E-68 |

| | | | | | | | | | | | | |
|---|---|---|---|---|---|---|---|---|---|---|---|---|
| rs174577 | miR-1908-5 | cis | | 1.89E-08 | Nightingale | XL_HDL_FC | NA | A | C | 0.060844 | 0.00383 | 7.60E-63 |
| rs174577 | miR-1908-5 | cis | | 1.89E-08 | Nightingale | HDL_PL | NA | A | C | -0.04894 | 0.003905 | 1.40E-38 |
| rs174577 | miR-1908-5 | cis | | 1.89E-08 | Nightingale | XXL_VLDL_ | NA | A | C | 0.065811 | 0.004155 | 3.10E-59 |
| rs174577 | miR-1908-5 | cis | | 1.89E-08 | Nightingale | L_HDL_C_p | NA | A | C | -0.068 | 0.004072 | 4.50E-67 |
| rs174577 | miR-1908-5 | cis | | 1.89E-08 | Nightingale | L_VLDL_FC | NA | A | C | 0.028741 | 0.004279 | 1.40E-11 |
| rs174577 | miR-1908-5 | cis | | 1.89E-08 | Nightingale | M_LDL_TG | NA | A | C | 0.075839 | 0.004265 | 3.80E-72 |
| rs174577 | miR-1908-5 | cis | | 1.89E-08 | Nightingale | XL_VLDL_P | NA | A | C | 0.048141 | 0.004169 | 4.00E-32 |
| rs174577 | miR-1908-5 | cis | | 1.89E-08 | Nightingale | XL_VLDL_C | NA | A | C | 0.037446 | 0.004219 | 1.30E-19 |
| rs174577 | miR-1908-5 | cis | | 1.89E-08 | Nightingale | S_HDL_TG_ | NA | A | C | 0.069409 | 0.004108 | 4.70E-67 |
| rs174577 | miR-1908-5 | cis | | 1.89E-08 | Nightingale | L_VLDL_C_ | NA | A | C | -0.038 | 0.00422 | 8.50E-21 |
| rs174577 | miR-1908-5 | cis | | 1.89E-08 | Nightingale | IDL_C | NA | A | C | -0.07691 | 0.004171 | 2.00E-80 |
| rs174577 | miR-1908-5 | cis | | 1.89E-08 | Nightingale | L_VLDL_C | NA | A | C | 0.028961 | 0.004225 | 2.70E-12 |
| rs174577 | miR-1908-5 | cis | | 1.89E-08 | Nightingale | MUFA | NA | A | C | 0.063378 | 0.004276 | 5.00E-51 |
| rs174577 | miR-1908-5 | cis | | 1.89E-08 | Nightingale | LDL_TG | NA | A | C | 0.029903 | 0.004272 | 4.00E-12 |
| rs174577 | miR-1908-5 | cis | | 1.89E-08 | Nightingale | M_HDL_L | NA | A | C | -0.02858 | 0.00401 | 1.30E-13 |
| rs174577 | miR-1908-5 | cis | | 1.89E-08 | Nightingale | XL_HDL_CE | NA | A | C | -0.02664 | 0.004317 | 1.70E-09 |
| rs174577 | miR-1908-5 | cis | | 1.89E-08 | Nightingale | L_LDL_L | NA | A | C | -0.05393 | 0.004278 | 2.00E-38 |
| rs174577 | miR-1908-5 | cis | | 1.89E-08 | Nightingale | M_HDL_TG | NA | A | C | 0.067092 | 0.004142 | 1.10E-61 |
| rs174577 | miR-1908-5 | cis | | 1.89E-08 | Nightingale | M_HDL_P | NA | A | C | -0.03513 | 0.003979 | 4.40E-20 |
| rs174577 | miR-1908-5 | cis | | 1.89E-08 | Nightingale | L_LDL_FC | NA | A | C | -0.06809 | 0.004242 | 2.70E-61 |
| rs174577 | miR-1908-5 | cis | | 1.89E-08 | Nightingale | S_LDL_TG_ | NA | A | C | 0.086081 | 0.004186 | 3.80E-97 |
| rs174577 | miR-1908-5 | cis | | 1.89E-08 | Nightingale | HDL_FC | NA | A | C | -0.07397 | 0.003827 | 4.20E-90 |
| rs174577 | miR-1908-5 | cis | | 1.89E-08 | Nightingale | IDL_FC_pct | NA | A | C | -0.10365 | 0.004285 | 8.71E-131 |
| rs174577 | miR-1908-5 | cis | | 1.89E-08 | Nightingale | MUFA_pct | NA | A | C | 0.195876 | 0.004134 | 0 |
| rs174577 | miR-1908-5 | cis | | 1.89E-08 | Nightingale | LDL_P | NA | A | C | -0.03968 | 0.004317 | 4.20E-21 |
| rs174577 | miR-1908-5 | cis | | 1.89E-08 | Nightingale | M_VLDL_F | NA | A | C | -0.09034 | 0.004107 | 5.20E-113 |
| rs174577 | miR-1908-5 | cis | | 1.89E-08 | Nightingale | LDL_L | NA | A | C | -0.04522 | 0.004305 | 4.50E-27 |
| rs174577 | miR-1908-5 | cis | | 1.89E-08 | Nightingale | Phosphogly | NA | A | C | -0.04122 | 0.004092 | 1.60E-25 |
| rs174577 | miR-1908-5 | cis | | 1.89E-08 | Nightingale | IDL_FC | NA | A | C | -0.08923 | 0.004181 | 1.10E-106 |
| rs174577 | miR-1908-5 | cis | | 1.89E-08 | Nightingale | M_LDL_C_ | NA | A | C | -0.09996 | 0.004323 | 5.40E-119 |
| rs174577 | miR-1908-5 | cis | | 1.89E-08 | Nightingale | M_LDL_TG | NA | A | C | 0.036737 | 0.004256 | 4.70E-18 |
| rs174577 | miR-1908-5 | cis | | 1.89E-08 | Nightingale | Total_PL | NA | A | C | -0.05106 | 0.004113 | 1.80E-37 |
| rs174577 | miR-1908-5 | cis | | 1.89E-08 | Nightingale | XL_VLDL_C | NA | A | C | 0.030395 | 0.004252 | 3.30E-13 |
| rs174577 | miR-1908-5 | cis | | 1.89E-08 | Nightingale | S_LDL_TG | NA | A | C | 0.050548 | 0.004214 | 7.40E-34 |
| rs174577 | miR-1908-5 | cis | | 1.89E-08 | Nightingale | XS_VLDL_C | NA | A | C | -0.09188 | 0.004072 | 6.30E-118 |
| rs174577 | miR-1908-5 | cis | | 1.89E-08 | Nightingale | L_HDL_P | NA | A | C | -0.07771 | 0.003783 | 2.50E-102 |
| rs174577 | miR-1908-5 | cis | | 1.89E-08 | Nightingale | XL_HDL_TG | NA | A | C | 0.086941 | 0.004069 | 4.50E-107 |
| rs174577 | miR-1908-5 | cis | | 1.89E-08 | Nightingale | DHA_pct | NA | A | C | -0.27112 | 0.004193 | 0 |
| rs174577 | miR-1908-5 | cis | | 1.89E-08 | Nightingale | S_LDL_FC_ | NA | A | C | -0.07988 | 0.004186 | 2.90E-84 |
| rs174577 | miR-1908-5 | cis | | 1.89E-08 | Nightingale | M_LDL_PL_ | NA | A | C | 0.045567 | 0.004317 | 3.70E-27 |
| rs174577 | miR-1908-5 | cis | | 1.89E-08 | Nightingale | Remnant_( | NA | A | C | -0.04367 | 0.004292 | 9.40E-26 |
| rs174577 | miR-1908-5 | cis | | 1.89E-08 | Nightingale | IDL_P | NA | A | C | -0.04486 | 0.004274 | 3.20E-27 |
| rs174577 | miR-1908-5 | cis | | 1.89E-08 | Nightingale | XXL_VLDL_ | NA | A | C | 0.067679 | 0.004162 | 2.00E-62 |
| rs174577 | miR-1908-5 | cis | | 1.89E-08 | Nightingale | L_VLDL_P | NA | A | C | 0.041761 | 0.004171 | 1.30E-24 |
| rs174577 | miR-1908-5 | cis | | 1.89E-08 | Nightingale | IDL_CE | NA | A | C | -0.07195 | 0.00417 | 1.10E-70 |
| rs174577 | miR-1908-5 | cis | | 1.89E-08 | Nightingale | PUFA_pct | NA | A | C | -0.1202 | 0.004155 | 6.40E-186 |
| rs174577 | miR-1908-5 | cis | | 1.89E-08 | Nightingale | L_VLDL_TG | NA | A | C | 0.03703 | 0.004156 | 8.40E-20 |
| rs174577 | miR-1908-5 | cis | | 1.89E-08 | Nightingale | XXL_VLDL_ | NA | A | C | 0.044469 | 0.004344 | 1.70E-25 |
| rs174577 | miR-1908-5 | cis | | 1.89E-08 | Nightingale | XL_HDL_P | NA | A | C | -0.08847 | 0.003887 | 8.49E-126 |
| rs174577 | miR-1908-5 | cis | | 1.89E-08 | Nightingale | Total_TG | NA | A | C | 0.046751 | 0.004181 | 3.80E-30 |
| rs174577 | miR-1908-5 | cis | | 1.89E-08 | Nightingale | M_HDL_C_ | NA | A | C | -0.06611 | 0.004112 | 3.90E-61 |
| rs174577 | miR-1908-5 | cis | | 1.89E-08 | Nightingale | M_LDL_L | NA | A | C | -0.02806 | 0.004326 | 3.80E-11 |
| rs174577 | miR-1908-5 | cis | | 1.89E-08 | Nightingale | XXL_VLDL_ | NA | A | C | 0.066277 | 0.004161 | 1.10E-59 |
| rs174577 | miR-1908-5 | cis | | 1.89E-08 | Nightingale | XL_VLDL_T | NA | A | C | 0.05651 | 0.004132 | 2.40E-44 |
| rs174577 | miR-1908-5 | cis | | 1.89E-08 | Nightingale | M_HDL_FC | NA | A | C | -0.10303 | 0.003828 | 4.40E-174 |
| rs174577 | miR-1908-5 | cis | | 1.89E-08 | Nightingale | S_LDL_CE_ | NA | A | C | 0.056281 | 0.004312 | 5.00E-41 |
| rs174577 | miR-1908-5 | cis | | 1.89E-08 | Nightingale | SFA | NA | A | C | -0.03138 | 0.004281 | 1.00E-13 |
| rs174577 | miR-1908-5 | cis | | 1.89E-08 | Nightingale | S_LDL_P | NA | A | C | -0.03195 | 0.004324 | 3.30E-14 |
| rs174577 | miR-1908-5 | cis | | 1.89E-08 | Nightingale | S_HDL_FC_ | NA | A | C | -0.10109 | 0.004097 | 4.60E-149 |
| rs174577 | miR-1908-5 | cis | | 1.89E-08 | Nightingale | XS_VLDL_P | NA | A | C | -0.04273 | 0.004262 | 1.50E-25 |
| rs174577 | miR-1908-5 | cis | | 1.89E-08 | Nightingale | XL_VLDL_C | NA | A | C | -0.06332 | 0.004139 | 2.60E-54 |
| rs174577 | miR-1908-5 | cis | | 1.89E-08 | Nightingale | LDL_FC | NA | A | C | -0.06408 | 0.004274 | 1.70E-53 |
| rs174577 | miR-1908-5 | cis | | 1.89E-08 | Nightingale | XXL_VLDL_ | NA | A | C | -0.02667 | 0.004298 | 9.90E-11 |

| | | | | | | | | | | | | |
|---|---|---|---|---|---|---|---|---|---|---|---|---|
| rs174577 | miR-1908-5 | cis | | 1.89E-08 | Nightingale | L_LDL_CE | NA | A | C | -0.05309 | 0.004281 | 3.30E-37 |
| rs174577 | miR-1908-5 | cis | | 1.89E-08 | Nightingale | S_LDL_FC | NA | A | C | -0.06253 | 0.004313 | 2.50E-50 |
| rs174577 | miR-1908-5 | cis | | 1.89E-08 | Nightingale | L_VLDL_CE | NA | A | C | -0.05829 | 0.004134 | 4.20E-48 |
| rs174577 | miR-1908-5 | cis | | 1.89E-08 | Nightingale | SFA_pct | NA | A | C | -0.04267 | 0.004262 | 1.40E-23 |
| rs174578 | miR-1908-5 | cis | | 2.55E-08 | Nightingale | VLDL_L | NA | A | T | 0.027433 | 0.004235 | 4.80E-11 |
| rs174578 | miR-1908-5 | cis | | 2.55E-08 | Nightingale | L_HDL_FC | NA | A | T | -0.08425 | 0.003785 | 1.80E-119 |
| rs174578 | miR-1908-5 | cis | | 2.55E-08 | Nightingale | M_LDL_CE | NA | A | T | -0.02639 | 0.004326 | 5.20E-10 |
| rs174578 | miR-1908-5 | cis | | 2.55E-08 | Nightingale | XS_VLDL_P | NA | A | T | -0.0334 | 0.004259 | 1.40E-16 |
| rs174578 | miR-1908-5 | cis | | 2.55E-08 | Nightingale | L_LDL_P | NA | A | T | -0.03687 | 0.004304 | 1.10E-18 |
| rs174578 | miR-1908-5 | cis | | 2.55E-08 | Nightingale | Cholines | NA | A | T | -0.03741 | 0.004062 | 6.40E-22 |
| rs174578 | miR-1908-5 | cis | | 2.55E-08 | Nightingale | M_VLDL_C | NA | A | T | -0.08014 | 0.004065 | 3.20E-90 |
| rs174578 | miR-1908-5 | cis | | 2.55E-08 | Nightingale | Omega_6_ | NA | A | T | 0.023821 | 0.004204 | 9.30E-09 |
| rs174578 | miR-1908-5 | cis | | 2.55E-08 | Nightingale | M_LDL_TG | NA | A | T | 0.036787 | 0.004256 | 4.60E-18 |
| rs174578 | miR-1908-5 | cis | | 2.55E-08 | Nightingale | S_LDL_PL | NA | A | T | -0.05083 | 0.004309 | 6.90E-34 |
| rs174578 | miR-1908-5 | cis | | 2.55E-08 | Nightingale | LDL_CE | NA | A | T | -0.04352 | 0.004308 | 4.30E-25 |
| rs174578 | miR-1908-5 | cis | | 2.55E-08 | Nightingale | L_HDL_TG_ | NA | A | T | 0.07925 | 0.004076 | 4.00E-89 |
| rs174578 | miR-1908-5 | cis | | 2.55E-08 | Nightingale | M_HDL_P | NA | A | T | -0.03514 | 0.003979 | 4.20E-20 |
| rs174578 | miR-1908-5 | cis | | 2.55E-08 | Nightingale | IDL_FC_pct | NA | A | T | -0.10377 | 0.004285 | 4.40E-131 |
| rs174578 | miR-1908-5 | cis | | 2.55E-08 | Nightingale | M_VLDL_T | NA | A | T | 0.077364 | 0.004079 | 4.60E-84 |
| rs174578 | miR-1908-5 | cis | | 2.55E-08 | Nightingale | M_LDL_PL | NA | A | T | -0.02356 | 0.004326 | 2.60E-08 |
| rs174578 | miR-1908-5 | cis | | 2.55E-08 | Nightingale | S_VLDL_FC | NA | A | T | -0.08281 | 0.004124 | 1.60E-93 |
| rs174578 | miR-1908-5 | cis | | 2.55E-08 | Nightingale | M_LDL_TG | NA | A | T | 0.076039 | 0.004265 | 1.90E-72 |
| rs174578 | miR-1908-5 | cis | | 2.55E-08 | Nightingale | S_LDL_PL_ | NA | A | T | -0.06059 | 0.00431 | 3.60E-47 |
| rs174578 | miR-1908-5 | cis | | 2.55E-08 | Nightingale | S_VLDL_TG | NA | A | T | 0.039182 | 0.004191 | 1.80E-21 |
| rs174578 | miR-1908-5 | cis | | 2.55E-08 | Nightingale | XL_VLDL_T | NA | A | T | 0.059049 | 0.004184 | 8.90E-47 |
| rs174578 | miR-1908-5 | cis | | 2.55E-08 | Nightingale | HDL_C | NA | A | T | -0.06516 | 0.003857 | 3.50E-69 |
| rs174578 | miR-1908-5 | cis | | 2.55E-08 | Nightingale | LDL_PL | NA | A | T | -0.04604 | 0.004309 | 9.40E-28 |
| rs174578 | miR-1908-5 | cis | | 2.55E-08 | Nightingale | XS_VLDL_L | NA | A | T | -0.04069 | 0.004245 | 9.10E-24 |
| rs174578 | miR-1908-5 | cis | | 2.55E-08 | Nightingale | XL_HDL_C | NA | A | T | -0.09808 | 0.003894 | 5.11E-152 |
| rs174578 | miR-1908-5 | cis | | 2.55E-08 | Nightingale | L_HDL_CE_ | NA | A | T | -0.05012 | 0.004133 | 5.60E-36 |
| rs174578 | miR-1908-5 | cis | | 2.55E-08 | Nightingale | HDL_size | NA | A | T | -0.07224 | 0.003827 | 4.20E-86 |
| rs174578 | miR-1908-5 | cis | | 2.55E-08 | Nightingale | XL_HDL_PL | NA | A | T | -0.06607 | 0.003968 | 4.00E-67 |
| rs174578 | miR-1908-5 | cis | | 2.55E-08 | Nightingale | VLDL_TG | NA | A | T | 0.047632 | 0.004156 | 1.60E-31 |
| rs174578 | miR-1908-5 | cis | | 2.55E-08 | Nightingale | PUFA_by_M | NA | A | T | -0.16925 | 0.004137 | 0 |
| rs174578 | miR-1908-5 | cis | | 2.55E-08 | Nightingale | L_LDL_FC_ | NA | A | T | -0.08262 | 0.004117 | 5.50E-93 |
| rs174578 | miR-1908-5 | cis | | 2.55E-08 | Nightingale | M_HDL_C | NA | A | T | -0.0417 | 0.003974 | 2.40E-27 |
| rs174578 | miR-1908-5 | cis | | 2.55E-08 | Nightingale | L_VLDL_FC | NA | A | T | 0.040039 | 0.004178 | 1.10E-22 |
| rs174578 | miR-1908-5 | cis | | 2.55E-08 | Nightingale | XL_VLDL_F | NA | A | T | 0.043335 | 0.004184 | 4.40E-26 |
| rs174578 | miR-1908-5 | cis | | 2.55E-08 | Nightingale | XS_VLDL_C | NA | A | T | -0.07117 | 0.004168 | 9.10E-70 |
| rs174578 | miR-1908-5 | cis | | 2.55E-08 | Nightingale | S_HDL_C_p | NA | A | T | -0.04275 | 0.004269 | 2.20E-23 |
| rs174578 | miR-1908-5 | cis | | 2.55E-08 | Nightingale | HDL_L | NA | A | T | -0.05377 | 0.003879 | 6.30E-47 |
| rs174578 | miR-1908-5 | cis | | 2.55E-08 | Nightingale | S_HDL_TG | NA | A | T | 0.068695 | 0.004188 | 4.20E-63 |
| rs174578 | miR-1908-5 | cis | | 2.55E-08 | Nightingale | PUFA | NA | A | T | -0.09557 | 0.004212 | 1.90E-120 |
| rs174578 | miR-1908-5 | cis | | 2.55E-08 | Nightingale | L_HDL_PL_ | NA | A | T | 0.042087 | 0.00418 | 2.20E-25 |
| rs174578 | miR-1908-5 | cis | | 2.55E-08 | Nightingale | LDL_P | NA | A | T | -0.03976 | 0.004316 | 3.60E-21 |
| rs174578 | miR-1908-5 | cis | | 2.55E-08 | Nightingale | L_HDL_C_p | NA | A | T | -0.06812 | 0.004071 | 2.60E-67 |
| rs174578 | miR-1908-5 | cis | | 2.55E-08 | Nightingale | HDL_FC | NA | A | T | -0.07393 | 0.003826 | 4.80E-90 |
| rs174578 | miR-1908-5 | cis | | 2.55E-08 | Nightingale | L_LDL_C_p | NA | A | T | -0.07723 | 0.004208 | 2.40E-77 |
| rs174578 | miR-1908-5 | cis | | 2.55E-08 | Nightingale | ApoB | NA | A | T | -0.03803 | 0.004314 | 1.30E-19 |
| rs174578 | miR-1908-5 | cis | | 2.55E-08 | Nightingale | S_LDL_CE_ | NA | A | T | 0.056147 | 0.004311 | 8.60E-41 |
| rs174578 | miR-1908-5 | cis | | 2.55E-08 | Nightingale | XL_VLDL_P | NA | A | T | 0.052585 | 0.004151 | 3.00E-38 |
| rs174578 | miR-1908-5 | cis | | 2.55E-08 | Nightingale | Total_P | NA | A | T | -0.04117 | 0.004065 | 9.70E-26 |
| rs174578 | miR-1908-5 | cis | | 2.55E-08 | Nightingale | Total_FC | NA | A | T | -0.05905 | 0.004219 | 4.70E-47 |
| rs174578 | miR-1908-5 | cis | | 2.55E-08 | Nightingale | M_HDL_FC | NA | A | T | -0.05208 | 0.00393 | 7.40E-43 |
| rs174578 | miR-1908-5 | cis | | 2.55E-08 | Nightingale | M_HDL_PL | NA | A | T | -0.02243 | 0.004039 | 7.40E-09 |
| rs174578 | miR-1908-5 | cis | | 2.55E-08 | Nightingale | XS_VLDL_C | NA | A | T | -0.092 | 0.004071 | 3.50E-118 |
| rs174578 | miR-1908-5 | cis | | 2.55E-08 | Nightingale | HDL_P | NA | A | T | -0.03656 | 0.004056 | 1.30E-20 |
| rs174578 | miR-1908-5 | cis | | 2.55E-08 | Nightingale | DHA_pct | NA | A | T | -0.2709 | 0.004193 | 0 |
| rs174578 | miR-1908-5 | cis | | 2.55E-08 | Nightingale | MUFA_pct | NA | A | T | 0.195888 | 0.004133 | 0 |
| rs174578 | miR-1908-5 | cis | | 2.55E-08 | Nightingale | Sphingomy | NA | A | T | -0.11468 | 0.004036 | 1.30E-186 |
| rs174578 | miR-1908-5 | cis | | 2.55E-08 | Nightingale | M_VLDL_T | NA | A | T | 0.031347 | 0.004196 | 2.70E-14 |
| rs174578 | miR-1908-5 | cis | | 2.55E-08 | Nightingale | S_VLDL_TG | NA | A | T | 0.064373 | 0.004179 | 4.00E-55 |
| rs174578 | miR-1908-5 | cis | | 2.55E-08 | Nightingale | XL_HDL_FC | NA | A | T | -0.10106 | 0.004024 | 2.70E-150 |

| | | | | | | | | | | | |
|---|---|---|---|---|---|---|---|---|---|---|---|
| rs174578 | miR-1908-! | cis | 2.55E-08 | Nightingale | S_LDL_TG_ | NA | A | T | 0.086226 | 0.004185 | 2.10E-97 |
| rs174578 | miR-1908-! | cis | 2.55E-08 | Nightingale | L_LDL_CE | NA | A | T | -0.05321 | 0.004281 | 2.40E-37 |
| rs174578 | miR-1908-! | cis | 2.55E-08 | Nightingale | XS_VLDL_P | NA | A | T | 0.05884 | 0.004214 | 2.70E-45 |
| rs174578 | miR-1908-! | cis | 2.55E-08 | Nightingale | M_LDL_C_ | NA | A | T | -0.1002 | 0.004322 | 1.50E-119 |
| rs174578 | miR-1908-! | cis | 2.55E-08 | Nightingale | M_VLDL_F | NA | A | T | -0.09051 | 0.004107 | 1.80E-113 |
| rs174578 | miR-1908-! | cis | 2.55E-08 | Nightingale | L_VLDL_C_ | NA | A | T | -0.03819 | 0.004219 | 5.40E-21 |
| rs174578 | miR-1908-! | cis | 2.55E-08 | Nightingale | LDL_FC | NA | A | T | -0.06422 | 0.004273 | 1.10E-53 |
| rs174578 | miR-1908-! | cis | 2.55E-08 | Nightingale | Omega_3_ | NA | A | T | -0.38527 | 0.00422 | 0 |
| rs174578 | miR-1908-! | cis | 2.55E-08 | Nightingale | XL_HDL_CE | NA | A | T | -0.09612 | 0.003868 | 3.00E-148 |
| rs174578 | miR-1908-! | cis | 2.55E-08 | Nightingale | S_HDL_TG_ | NA | A | T | 0.069512 | 0.004108 | 3.60E-67 |
| rs174578 | miR-1908-! | cis | 2.55E-08 | Nightingale | IDL_PL | NA | A | T | -0.07222 | 0.004167 | 4.30E-72 |
| rs174578 | miR-1908-! | cis | 2.55E-08 | Nightingale | L_VLDL_CE | NA | A | T | -0.05844 | 0.004133 | 2.20E-48 |
| rs174578 | miR-1908-! | cis | 2.55E-08 | Nightingale | S_LDL_TG | NA | A | T | 0.050601 | 0.004213 | 7.10E-34 |
| rs174578 | miR-1908-! | cis | 2.55E-08 | Nightingale | IDL_FC | NA | A | T | -0.08933 | 0.004181 | 7.00E-107 |
| rs174578 | miR-1908-! | cis | 2.55E-08 | Nightingale | L_HDL_FC_ | NA | A | T | -0.11079 | 0.004012 | 2.20E-182 |
| rs174578 | miR-1908-! | cis | 2.55E-08 | Nightingale | XXL_VLDL_ | NA | A | T | 0.073236 | 0.004171 | 1.30E-71 |
| rs174578 | miR-1908-! | cis | 2.55E-08 | Nightingale | L_LDL_PL | NA | A | T | -0.05487 | 0.004287 | 3.50E-39 |
| rs174578 | miR-1908-! | cis | 2.55E-08 | Nightingale | L_VLDL_PL | NA | A | T | 0.048649 | 0.004167 | 6.60E-33 |
| rs174578 | miR-1908-! | cis | 2.55E-08 | Nightingale | XL_VLDL_C | NA | A | T | -0.06053 | 0.004141 | 7.60E-50 |
| rs174578 | miR-1908-! | cis | 2.55E-08 | Nightingale | XS_VLDL_F | NA | A | T | -0.11184 | 0.004287 | 4.70E-153 |
| rs174578 | miR-1908-! | cis | 2.55E-08 | Nightingale | Total_CE | NA | A | T | -0.06695 | 0.004166 | 3.50E-61 |
| rs174578 | miR-1908-! | cis | 2.55E-08 | Nightingale | IDL_P | NA | A | T | -0.04496 | 0.004274 | 2.50E-27 |
| rs174578 | miR-1908-! | cis | 2.55E-08 | Nightingale | M_VLDL_C | NA | A | T | -0.05216 | 0.00428 | 1.10E-35 |
| rs174578 | miR-1908-! | cis | 2.55E-08 | Nightingale | IDL_L | NA | A | T | -0.07201 | 0.004176 | 5.20E-71 |
| rs174578 | miR-1908-! | cis | 2.55E-08 | Nightingale | M_LDL_PL_ | NA | A | T | 0.045714 | 0.004317 | 2.20E-27 |
| rs174578 | miR-1908-! | cis | 2.55E-08 | Nightingale | XXL_VLDL_ | NA | A | T | 0.066319 | 0.00416 | 1.00E-59 |
| rs174578 | miR-1908-! | cis | 2.55E-08 | Nightingale | VLDL_size | NA | A | T | 0.071619 | 0.004026 | 4.00E-75 |
| rs174578 | miR-1908-! | cis | 2.55E-08 | Nightingale | L_VLDL_FC | NA | A | T | 0.028547 | 0.004278 | 1.90E-11 |
| rs174578 | miR-1908-! | cis | 2.55E-08 | Nightingale | LA_pct | NA | A | T | 0.183393 | 0.004221 | 0 |
| rs174578 | miR-1908-! | cis | 2.55E-08 | Nightingale | XS_VLDL_C | NA | A | T | -0.0978 | 0.004093 | 9.51E-132 |
| rs174578 | miR-1908-! | cis | 2.55E-08 | Nightingale | L_VLDL_TG | NA | A | T | 0.037067 | 0.004156 | 8.10E-20 |
| rs174578 | miR-1908-! | cis | 2.55E-08 | Nightingale | XL_VLDL_T | NA | A | T | 0.05656 | 0.004131 | 2.20E-44 |
| rs174578 | miR-1908-! | cis | 2.55E-08 | Nightingale | L_VLDL_PL | NA | A | T | 0.087758 | 0.004192 | 1.20E-100 |
| rs174578 | miR-1908-! | cis | 2.55E-08 | Nightingale | M_HDL_TG | NA | A | T | 0.050199 | 0.004266 | 1.00E-32 |
| rs174578 | miR-1908-! | cis | 2.55E-08 | Nightingale | XL_HDL_P | NA | A | T | -0.08838 | 0.003887 | 1.40E-125 |
| rs174578 | miR-1908-! | cis | 2.55E-08 | Nightingale | XXL_VLDL_ | NA | A | T | 0.071394 | 0.004162 | 1.10E-68 |
| rs174578 | miR-1908-! | cis | 2.55E-08 | Nightingale | L_VLDL_L | NA | A | T | 0.038069 | 0.004174 | 1.20E-20 |
| rs174578 | miR-1908-! | cis | 2.55E-08 | Nightingale | S_LDL_C | NA | A | T | -0.03203 | 0.004324 | 3.10E-14 |
| rs174578 | miR-1908-! | cis | 2.55E-08 | Nightingale | IDL_TG_pc | NA | A | T | 0.088186 | 0.004173 | 7.19E-103 |
| rs174578 | miR-1908-! | cis | 2.55E-08 | Nightingale | S_LDL_L | NA | A | T | -0.03215 | 0.004318 | 2.10E-14 |
| rs174578 | miR-1908-! | cis | 2.55E-08 | Nightingale | M_HDL_L | NA | A | T | -0.02858 | 0.00401 | 1.20E-13 |
| rs174578 | miR-1908-! | cis | 2.55E-08 | Nightingale | Unsaturati | NA | A | T | -0.33575 | 0.004091 | 0 |
| rs174578 | miR-1908-! | cis | 2.55E-08 | Nightingale | Citrate | NA | A | T | -0.03349 | 0.004292 | 6.70E-16 |
| rs174578 | miR-1908-! | cis | 2.55E-08 | Nightingale | S_LDL_P | NA | A | T | -0.03204 | 0.004323 | 2.80E-14 |
| rs174578 | miR-1908-! | cis | 2.55E-08 | Nightingale | L_HDL_CE | NA | A | T | -0.07862 | 0.003814 | 1.40E-102 |
| rs174578 | miR-1908-! | cis | 2.55E-08 | Nightingale | M_LDL_P | NA | A | T | -0.04634 | 0.004332 | 1.70E-27 |
| rs174578 | miR-1908-! | cis | 2.55E-08 | Nightingale | M_VLDL_C | NA | A | T | -0.07548 | 0.004055 | 1.60E-80 |
| rs174578 | miR-1908-! | cis | 2.55E-08 | Nightingale | L_VLDL_P | NA | A | T | 0.041773 | 0.00417 | 1.40E-24 |
| rs174578 | miR-1908-! | cis | 2.55E-08 | Nightingale | M_HDL_C_ | NA | A | T | -0.06621 | 0.004112 | 2.90E-61 |
| rs174578 | miR-1908-! | cis | 2.55E-08 | Nightingale | M_HDL_CE | NA | A | T | -0.03881 | 0.003989 | 1.00E-23 |
| rs174578 | miR-1908-! | cis | 2.55E-08 | Nightingale | XXL_VLDL_ | NA | A | T | 0.07191 | 0.004145 | 1.50E-70 |
| rs174578 | miR-1908-! | cis | 2.55E-08 | Nightingale | ApoA1 | NA | A | T | -0.04186 | 0.003953 | 3.60E-28 |
| rs174578 | miR-1908-! | cis | 2.55E-08 | Nightingale | M_HDL_TG | NA | A | T | 0.067185 | 0.004142 | 8.90E-62 |
| rs174578 | miR-1908-! | cis | 2.55E-08 | Nightingale | XL_HDL_C_ | NA | A | T | 0.022917 | 0.004126 | 1.50E-09 |
| rs174578 | miR-1908-! | cis | 2.55E-08 | Nightingale | non_HDL_( | NA | A | T | -0.04758 | 0.004296 | 6.80E-30 |
| rs174578 | miR-1908-! | cis | 2.55E-08 | Nightingale | L_HDL_P | NA | A | T | -0.07768 | 0.003783 | 2.70E-102 |
| rs174578 | miR-1908-! | cis | 2.55E-08 | Nightingale | XS_VLDL_P | NA | A | T | -0.04274 | 0.004262 | 1.30E-25 |
| rs174578 | miR-1908-! | cis | 2.55E-08 | Nightingale | Total_PL | NA | A | T | -0.05109 | 0.004113 | 1.60E-37 |
| rs174578 | miR-1908-! | cis | 2.55E-08 | Nightingale | IDL_CE_pct | NA | A | T | -0.03865 | 0.004238 | 6.90E-20 |
| rs174578 | miR-1908-! | cis | 2.55E-08 | Nightingale | XS_VLDL_T | NA | A | T | 0.096949 | 0.004103 | 3.50E-129 |
| rs174578 | miR-1908-! | cis | 2.55E-08 | Nightingale | M_HDL_CE | NA | A | T | -0.04742 | 0.004188 | 1.70E-30 |
| rs174578 | miR-1908-! | cis | 2.55E-08 | Nightingale | XL_HDL_L | NA | A | T | -0.09186 | 0.003881 | 6.10E-135 |
| rs174578 | miR-1908-! | cis | 2.55E-08 | Nightingale | Tyr | NA | A | T | 0.032538 | 0.004314 | 4.60E-14 |

| | | | | | | | | | | | |
|---|---|---|---|---|---|---|---|---|---|---|---|
| rs174578 | miR-1908-5 | cis | 2.55E-08 | Nightingale | M_VLDL_F | NA | A | T | -0.02903 | 0.004319 | 5.70E-12 |
| rs174578 | miR-1908-5 | cis | 2.55E-08 | Nightingale | Total_TG | NA | A | T | 0.04679 | 0.004181 | 3.80E-30 |
| rs174578 | miR-1908-5 | cis | 2.55E-08 | Nightingale | Omega_6_ | NA | A | T | 0.365408 | 0.004257 | 0 |
| rs174578 | miR-1908-5 | cis | 2.55E-08 | Nightingale | XS_VLDL_C | NA | A | T | -0.06734 | 0.004192 | 5.20E-62 |
| rs174578 | miR-1908-5 | cis | 2.55E-08 | Nightingale | Omega_3 | NA | A | T | -0.33179 | 0.004233 | 0 |
| rs174578 | miR-1908-5 | cis | 2.55E-08 | Nightingale | PUFA_pct | NA | A | T | -0.12021 | 0.004155 | 5.60E-186 |
| rs174578 | miR-1908-5 | cis | 2.55E-08 | Nightingale | L_LDL_TG_ | NA | A | T | 0.077829 | 0.004235 | 5.30E-77 |
| rs174578 | miR-1908-5 | cis | 2.55E-08 | Nightingale | XL_VLDL_C | NA | A | T | -0.06344 | 0.004139 | 1.60E-54 |
| rs174578 | miR-1908-5 | cis | 2.55E-08 | Nightingale | XL_VLDL_F | NA | A | T | -0.06736 | 0.004185 | 3.20E-61 |
| rs174578 | miR-1908-5 | cis | 2.55E-08 | Nightingale | XXL_VLDL_ | NA | A | T | -0.0267 | 0.004298 | 9.20E-11 |
| rs174578 | miR-1908-5 | cis | 2.55E-08 | Nightingale | S_LDL_FC | NA | A | T | -0.06266 | 0.004312 | 1.70E-50 |
| rs174578 | miR-1908-5 | cis | 2.55E-08 | Nightingale | M_VLDL_P | NA | A | T | -0.0618 | 0.004144 | 8.10E-54 |
| rs174578 | miR-1908-5 | cis | 2.55E-08 | Nightingale | L_LDL_C | NA | A | T | -0.05771 | 0.004271 | 7.60E-44 |
| rs174578 | miR-1908-5 | cis | 2.55E-08 | Nightingale | L_VLDL_C | NA | A | T | 0.028944 | 0.004224 | 3.00E-12 |
| rs174578 | miR-1908-5 | cis | 2.55E-08 | Nightingale | L_HDL_L | NA | A | T | -0.07713 | 0.00379 | 1.90E-100 |
| rs174578 | miR-1908-5 | cis | 2.55E-08 | Nightingale | XXL_VLDL_ | NA | A | T | 0.065855 | 0.004154 | 2.90E-59 |
| rs174578 | miR-1908-5 | cis | 2.55E-08 | Nightingale | XL_VLDL_L | NA | A | T | 0.051217 | 0.004156 | 2.70E-36 |
| rs174578 | miR-1908-5 | cis | 2.55E-08 | Nightingale | L_HDL_C | NA | A | T | -0.08013 | 0.003803 | 3.30E-107 |
| rs174578 | miR-1908-5 | cis | 2.55E-08 | Nightingale | L_HDL_PL | NA | A | T | -0.07606 | 0.003801 | 5.60E-97 |
| rs174578 | miR-1908-5 | cis | 2.55E-08 | Nightingale | XL_VLDL_P | NA | A | T | 0.048152 | 0.004168 | 4.30E-32 |
| rs174578 | miR-1908-5 | cis | 2.55E-08 | Nightingale | S_VLDL_FC | NA | A | T | -0.02785 | 0.00432 | 4.00E-11 |
| rs174578 | miR-1908-5 | cis | 2.55E-08 | Nightingale | M_LDL_L | NA | A | T | -0.02819 | 0.004326 | 3.20E-11 |
| rs174578 | miR-1908-5 | cis | 2.55E-08 | Nightingale | MUFA | NA | A | T | 0.063414 | 0.004275 | 4.90E-51 |
| rs174578 | miR-1908-5 | cis | 2.55E-08 | Nightingale | S_VLDL_PL | NA | A | T | -0.07845 | 0.004133 | 5.00E-84 |
| rs174578 | miR-1908-5 | cis | 2.55E-08 | Nightingale | XXL_VLDL_ | NA | A | T | 0.044341 | 0.004343 | 2.50E-25 |
| rs174578 | miR-1908-5 | cis | 2.55E-08 | Nightingale | Remnant_( | NA | A | T | -0.04375 | 0.004291 | 7.30E-26 |
| rs174578 | miR-1908-5 | cis | 2.55E-08 | Nightingale | LDL_C | NA | A | T | -0.04969 | 0.004301 | 2.50E-32 |
| rs174578 | miR-1908-5 | cis | 2.55E-08 | Nightingale | M_HDL_FC | NA | A | T | -0.103 | 0.003827 | 5.11E-174 |
| rs174578 | miR-1908-5 | cis | 2.55E-08 | Nightingale | XS_VLDL_T | NA | A | T | 0.032151 | 0.004253 | 1.40E-13 |
| rs174578 | miR-1908-5 | cis | 2.55E-08 | Nightingale | HDL_PL | NA | A | T | -0.0489 | 0.003905 | 1.50E-38 |
| rs174578 | miR-1908-5 | cis | 2.55E-08 | Nightingale | Phosphogly | NA | A | T | -0.04122 | 0.004091 | 1.50E-25 |
| rs174578 | miR-1908-5 | cis | 2.55E-08 | Nightingale | Clinical_LD | NA | A | T | -0.05508 | 0.004294 | 2.30E-39 |
| rs174578 | miR-1908-5 | cis | 2.55E-08 | Nightingale | Total_L | NA | A | T | -0.03725 | 0.004249 | 1.40E-19 |
| rs174578 | miR-1908-5 | cis | 2.55E-08 | Nightingale | S_VLDL_CE | NA | A | T | -0.02878 | 0.004261 | 7.70E-12 |
| rs174578 | miR-1908-5 | cis | 2.55E-08 | Nightingale | Total_FA | NA | A | T | -0.02356 | 0.004285 | 1.40E-08 |
| rs174578 | miR-1908-5 | cis | 2.55E-08 | Nightingale | SFA | NA | A | T | -0.03132 | 0.00428 | 1.00E-13 |
| rs174578 | miR-1908-5 | cis | 2.55E-08 | Nightingale | IDL_CE | NA | A | T | -0.07204 | 0.004169 | 8.00E-71 |
| rs174578 | miR-1908-5 | cis | 2.55E-08 | Nightingale | HDL_TG | NA | A | T | 0.046316 | 0.004259 | 5.60E-28 |
| rs174578 | miR-1908-5 | cis | 2.55E-08 | Nightingale | XL_VLDL_C | NA | A | T | 0.037434 | 0.004218 | 1.40E-19 |
| rs174578 | miR-1908-5 | cis | 2.55E-08 | Nightingale | XL_HDL_TG | NA | A | T | 0.08694 | 0.004069 | 5.70E-107 |
| rs174578 | miR-1908-5 | cis | 2.55E-08 | Nightingale | L_LDL_L | NA | A | T | -0.05405 | 0.004277 | 1.40E-38 |
| rs174578 | miR-1908-5 | cis | 2.55E-08 | Nightingale | S_HDL_FC_ | NA | A | T | -0.10099 | 0.004097 | 8.00E-149 |
| rs174578 | miR-1908-5 | cis | 2.55E-08 | Nightingale | M_LDL_FC | NA | A | T | -0.05242 | 0.004314 | 1.20E-35 |
| rs174578 | miR-1908-5 | cis | 2.55E-08 | Nightingale | M_LDL_C | NA | A | T | -0.03408 | 0.004328 | 9.60E-16 |
| rs174578 | miR-1908-5 | cis | 2.55E-08 | Nightingale | LDL_L | NA | A | T | -0.04534 | 0.004305 | 3.50E-27 |
| rs174578 | miR-1908-5 | cis | 2.55E-08 | Nightingale | XXL_VLDL_ | NA | A | T | 0.067711 | 0.004161 | 2.00E-62 |
| rs174578 | miR-1908-5 | cis | 2.55E-08 | Nightingale | LA | NA | A | T | 0.082794 | 0.004236 | 1.40E-85 |
| rs174578 | miR-1908-5 | cis | 2.55E-08 | Nightingale | TG_by_PG | NA | A | T | 0.06893 | 0.004008 | 1.00E-69 |
| rs174578 | miR-1908-5 | cis | 2.55E-08 | Nightingale | S_VLDL_C_ | NA | A | T | -0.05646 | 0.004202 | 3.00E-42 |
| rs174578 | miR-1908-5 | cis | 2.55E-08 | Nightingale | M_HDL_PL | NA | A | T | 0.058515 | 0.004097 | 4.00E-48 |
| rs174578 | miR-1908-5 | cis | 2.55E-08 | Nightingale | XL_VLDL_C | NA | A | T | 0.030364 | 0.004251 | 3.70E-13 |
| rs174578 | miR-1908-5 | cis | 2.55E-08 | Nightingale | XL_HDL_FC | NA | A | T | 0.06076 | 0.003829 | 1.00E-62 |
| rs174578 | miR-1908-5 | cis | 2.55E-08 | Nightingale | XL_HDL_PL | NA | A | T | -0.08794 | 0.003882 | 1.20E-123 |
| rs174578 | miR-1908-5 | cis | 2.55E-08 | Nightingale | HDL_CE | NA | A | T | -0.06198 | 0.003876 | 5.50E-62 |
| rs174578 | miR-1908-5 | cis | 2.55E-08 | Nightingale | L_LDL_FC | NA | A | T | -0.06823 | 0.004241 | 1.70E-61 |
| rs174578 | miR-1908-5 | cis | 2.55E-08 | Nightingale | DHA | NA | A | T | -0.28546 | 0.004162 | 0 |
| rs174578 | miR-1908-5 | cis | 2.55E-08 | Nightingale | IDL_C_pct | NA | A | T | -0.07518 | 0.004217 | 7.50E-73 |
| rs174578 | miR-1908-5 | cis | 2.55E-08 | Nightingale | Total_C | NA | A | T | -0.06507 | 0.004181 | 2.00E-57 |
| rs174578 | miR-1908-5 | cis | 2.55E-08 | Nightingale | M_LDL_FC_ | NA | A | T | -0.07143 | 0.004138 | 2.00E-69 |
| rs174578 | miR-1908-5 | cis | 2.55E-08 | Nightingale | XL_HDL_CE | NA | A | T | -0.02685 | 0.004317 | 1.30E-09 |
| rs174578 | miR-1908-5 | cis | 2.55E-08 | Nightingale | XS_VLDL_F | NA | A | T | -0.05514 | 0.004244 | 2.10E-41 |
| rs174578 | miR-1908-5 | cis | 2.55E-08 | Nightingale | XXL_VLDL_ | NA | A | T | 0.061126 | 0.004168 | 8.10E-51 |
| rs174578 | miR-1908-5 | cis | 2.55E-08 | Nightingale | SFA_pct | NA | A | T | -0.04263 | 0.004261 | 1.50E-23 |

| | | | | | | | | | | | |
|---|---|---|---|---|---|---|---|---|---|---|---|
| rs174578 | miR-1908-5 | cis | 2.55E-08 | Nightingale | S_LDL_FC_ | NA | A | T | -0.07996 | 0.004186 | 2.40E-84 |
| rs174578 | miR-1908-5 | cis | 2.55E-08 | Nightingale | IDL_C | NA | A | T | -0.07701 | 0.004171 | 1.40E-80 |
| rs174578 | miR-1908-5 | cis | 2.55E-08 | Nightingale | LDL_TG | NA | A | T | 0.029943 | 0.004271 | 4.00E-12 |
| rs174578 | miR-1908-5 | cis | 2.55E-08 | Nightingale | M_VLDL_C | NA | A | T | -0.04353 | 0.004307 | 5.10E-25 |
| rs174580 | miR-1908-5 | cis | 1.81E-08 | Nightingale | LDL_TG | NA | G | A | 0.030243 | 0.004268 | 2.60E-12 |
| rs174580 | miR-1908-5 | cis | 1.81E-08 | Nightingale | XS_VLDL_P | NA | G | A | -0.04255 | 0.004259 | 1.60E-25 |
| rs174580 | miR-1908-5 | cis | 1.81E-08 | Nightingale | Total_CE | NA | G | A | -0.06679 | 0.004163 | 3.90E-61 |
| rs174580 | miR-1908-5 | cis | 1.81E-08 | Nightingale | XXL_VLDL_ | NA | G | A | 0.0722 | 0.004142 | 3.30E-71 |
| rs174580 | miR-1908-5 | cis | 1.81E-08 | Nightingale | XL_VLDL_C | NA | G | A | 0.030518 | 0.004248 | 2.70E-13 |
| rs174580 | miR-1908-5 | cis | 1.81E-08 | Nightingale | L_HDL_FC | NA | G | A | -0.08408 | 0.003783 | 2.10E-119 |
| rs174580 | miR-1908-5 | cis | 1.81E-08 | Nightingale | XL_VLDL_P | NA | G | A | 0.048431 | 0.004165 | 1.70E-32 |
| rs174580 | miR-1908-5 | cis | 1.81E-08 | Nightingale | M_HDL_C_ | NA | G | A | -0.06636 | 0.004109 | 1.20E-61 |
| rs174580 | miR-1908-5 | cis | 1.81E-08 | Nightingale | M_LDL_L | NA | G | A | -0.0281 | 0.004323 | 3.30E-11 |
| rs174580 | miR-1908-5 | cis | 1.81E-08 | Nightingale | XL_HDL_CE | NA | G | A | -0.0271 | 0.004314 | 8.70E-10 |
| rs174580 | miR-1908-5 | cis | 1.81E-08 | Nightingale | S_VLDL_CE | NA | G | A | -0.02894 | 0.004258 | 6.20E-12 |
| rs174580 | miR-1908-5 | cis | 1.81E-08 | Nightingale | XXL_VLDL_ | NA | G | A | -0.02691 | 0.004295 | 6.80E-11 |
| rs174580 | miR-1908-5 | cis | 1.81E-08 | Nightingale | L_LDL_L | NA | G | A | -0.05394 | 0.004274 | 1.40E-38 |
| rs174580 | miR-1908-5 | cis | 1.81E-08 | Nightingale | XL_HDL_P | NA | G | A | -0.0881 | 0.003884 | 3.40E-125 |
| rs174580 | miR-1908-5 | cis | 1.81E-08 | Nightingale | LDL_FC | NA | G | A | -0.06417 | 0.00427 | 8.90E-54 |
| rs174580 | miR-1908-5 | cis | 1.81E-08 | Nightingale | L_LDL_FC_ | NA | G | A | -0.0826 | 0.004114 | 3.40E-93 |
| rs174580 | miR-1908-5 | cis | 1.81E-08 | Nightingale | M_LDL_C | NA | G | A | -0.03398 | 0.004325 | 1.00E-15 |
| rs174580 | miR-1908-5 | cis | 1.81E-08 | Nightingale | LDL_CE | NA | G | A | -0.04341 | 0.004305 | 4.60E-25 |
| rs174580 | miR-1908-5 | cis | 1.81E-08 | Nightingale | IDL_CE | NA | G | A | -0.07199 | 0.004167 | 5.30E-71 |
| rs174580 | miR-1908-5 | cis | 1.81E-08 | Nightingale | L_HDL_P | NA | G | A | -0.07751 | 0.00378 | 3.90E-102 |
| rs174580 | miR-1908-5 | cis | 1.81E-08 | Nightingale | Phosphogly | NA | G | A | -0.04089 | 0.004089 | 2.70E-25 |
| rs174580 | miR-1908-5 | cis | 1.81E-08 | Nightingale | L_LDL_FC | NA | G | A | -0.06814 | 0.004238 | 1.60E-61 |
| rs174580 | miR-1908-5 | cis | 1.81E-08 | Nightingale | L_HDL_PL | NA | G | A | -0.07591 | 0.003799 | 7.10E-97 |
| rs174580 | miR-1908-5 | cis | 1.81E-08 | Nightingale | S_LDL_PL_ | NA | G | A | -0.06093 | 0.004308 | 1.10E-47 |
| rs174580 | miR-1908-5 | cis | 1.81E-08 | Nightingale | L_VLDL_TG | NA | G | A | 0.037343 | 0.004153 | 4.10E-20 |
| rs174580 | miR-1908-5 | cis | 1.81E-08 | Nightingale | M_HDL_L | NA | G | A | -0.02835 | 0.004007 | 1.70E-13 |
| rs174580 | miR-1908-5 | cis | 1.81E-08 | Nightingale | LDL_L | NA | G | A | -0.04524 | 0.004302 | 3.60E-27 |
| rs174580 | miR-1908-5 | cis | 1.81E-08 | Nightingale | IDL_FC | NA | G | A | -0.08916 | 0.004178 | 8.09E-107 |
| rs174580 | miR-1908-5 | cis | 1.81E-08 | Nightingale | M_VLDL_C | NA | G | A | -0.0435 | 0.004305 | 4.50E-25 |
| rs174580 | miR-1908-5 | cis | 1.81E-08 | Nightingale | Unsaturati | NA | G | A | -0.33497 | 0.004088 | 0 |
| rs174580 | miR-1908-5 | cis | 1.81E-08 | Nightingale | XL_VLDL_C | NA | G | A | -0.0636 | 0.004136 | 8.10E-55 |
| rs174580 | miR-1908-5 | cis | 1.81E-08 | Nightingale | L_VLDL_P | NA | G | A | 0.042043 | 0.004168 | 6.50E-25 |
| rs174580 | miR-1908-5 | cis | 1.81E-08 | Nightingale | SFA | NA | G | A | -0.03082 | 0.004277 | 2.20E-13 |
| rs174580 | miR-1908-5 | cis | 1.81E-08 | Nightingale | Total_TG | NA | G | A | 0.047075 | 0.004178 | 1.50E-30 |
| rs174580 | miR-1908-5 | cis | 1.81E-08 | Nightingale | HDL_PL | NA | G | A | -0.04865 | 0.003902 | 2.70E-38 |
| rs174580 | miR-1908-5 | cis | 1.81E-08 | Nightingale | L_VLDL_C_ | NA | G | A | -0.03828 | 0.004216 | 4.40E-21 |
| rs174580 | miR-1908-5 | cis | 1.81E-08 | Nightingale | M_HDL_FC | NA | G | A | -0.05185 | 0.003928 | 1.30E-42 |
| rs174580 | miR-1908-5 | cis | 1.81E-08 | Nightingale | XS_VLDL_L | NA | G | A | -0.04047 | 0.004243 | 1.20E-23 |
| rs174580 | miR-1908-5 | cis | 1.81E-08 | Nightingale | XL_VLDL_P | NA | G | A | 0.052854 | 0.004149 | 1.10E-38 |
| rs174580 | miR-1908-5 | cis | 1.81E-08 | Nightingale | M_LDL_FC_ | NA | G | A | -0.07172 | 0.004135 | 4.00E-70 |
| rs174580 | miR-1908-5 | cis | 1.81E-08 | Nightingale | L_VLDL_C | NA | G | A | 0.029174 | 0.004222 | 1.90E-12 |
| rs174580 | miR-1908-5 | cis | 1.81E-08 | Nightingale | XXL_VLDL_ | NA | G | A | 0.071649 | 0.00416 | 2.80E-69 |
| rs174580 | miR-1908-5 | cis | 1.81E-08 | Nightingale | XXL_VLDL_ | NA | G | A | 0.061419 | 0.004165 | 2.50E-51 |
| rs174580 | miR-1908-5 | cis | 1.81E-08 | Nightingale | HDL_CE | NA | G | A | -0.06185 | 0.003874 | 7.00E-62 |
| rs174580 | miR-1908-5 | cis | 1.81E-08 | Nightingale | L_VLDL_PL | NA | G | A | 0.048878 | 0.004165 | 3.00E-33 |
| rs174580 | miR-1908-5 | cis | 1.81E-08 | Nightingale | M_LDL_PL_ | NA | G | A | 0.045017 | 0.004314 | 1.10E-26 |
| rs174580 | miR-1908-5 | cis | 1.81E-08 | Nightingale | Total_L | NA | G | A | -0.03696 | 0.004246 | 2.00E-19 |
| rs174580 | miR-1908-5 | cis | 1.81E-08 | Nightingale | XXL_VLDL_ | NA | G | A | 0.044547 | 0.00434 | 1.30E-25 |
| rs174580 | miR-1908-5 | cis | 1.81E-08 | Nightingale | IDL_P | NA | G | A | -0.04495 | 0.004271 | 2.00E-27 |
| rs174580 | miR-1908-5 | cis | 1.81E-08 | Nightingale | MUFA_pct | NA | G | A | 0.195664 | 0.004131 | 0 |
| rs174580 | miR-1908-5 | cis | 1.81E-08 | Nightingale | M_HDL_CE | NA | G | A | -0.03866 | 0.003986 | 1.30E-23 |
| rs174580 | miR-1908-5 | cis | 1.81E-08 | Nightingale | LA_pct | NA | G | A | 0.182322 | 0.004218 | 0 |
| rs174580 | miR-1908-5 | cis | 1.81E-08 | Nightingale | ApoB | NA | G | A | -0.03795 | 0.004311 | 1.30E-19 |
| rs174580 | miR-1908-5 | cis | 1.81E-08 | Nightingale | L_HDL_PL_ | NA | G | A | 0.042201 | 0.004178 | 1.50E-25 |
| rs174580 | miR-1908-5 | cis | 1.81E-08 | Nightingale | IDL_CE_pct | NA | G | A | -0.0389 | 0.004235 | 3.30E-20 |
| rs174580 | miR-1908-5 | cis | 1.81E-08 | Nightingale | Omega_6_ | NA | G | A | 0.364104 | 0.004254 | 0 |
| rs174580 | miR-1908-5 | cis | 1.81E-08 | Nightingale | L_LDL_C_p | NA | G | A | -0.07724 | 0.004205 | 1.50E-77 |
| rs174580 | miR-1908-5 | cis | 1.81E-08 | Nightingale | Total_C | NA | G | A | -0.0649 | 0.004178 | 2.30E-57 |
| rs174580 | miR-1908-5 | cis | 1.81E-08 | Nightingale | SFA_pct | NA | G | A | -0.04209 | 0.004258 | 4.80E-23 |

| | | | | | | | | | | | |
|---|---|---|---|---|---|---|---|---|---|---|---|
| rs174580 | miR-1908-5 | cis | 1.81E-08 | Nightingale | XS_VLDL_C | NA | G | A | -0.0921 | 0.004069 | 8.71E-119 |
| rs174580 | miR-1908-5 | cis | 1.81E-08 | Nightingale | XL_HDL_C | NA | G | A | -0.09786 | 0.003892 | 8.49E-152 |
| rs174580 | miR-1908-5 | cis | 1.81E-08 | Nightingale | M_VLDL_T | NA | G | A | 0.077446 | 0.004077 | 2.00E-84 |
| rs174580 | miR-1908-5 | cis | 1.81E-08 | Nightingale | XL_HDL_TG | NA | G | A | 0.087023 | 0.004066 | 2.40E-107 |
| rs174580 | miR-1908-5 | cis | 1.81E-08 | Nightingale | Sphingomy | NA | G | A | -0.11437 | 0.004033 | 3.50E-186 |
| rs174580 | miR-1908-5 | cis | 1.81E-08 | Nightingale | S_HDL_TG | NA | G | A | 0.068942 | 0.004185 | 1.20E-63 |
| rs174580 | miR-1908-5 | cis | 1.81E-08 | Nightingale | Omega_3_ | NA | G | A | -0.38404 | 0.004218 | 0 |
| rs174580 | miR-1908-5 | cis | 1.81E-08 | Nightingale | L_VLDL_CE | NA | G | A | -0.05851 | 0.004131 | 1.60E-48 |
| rs174580 | miR-1908-5 | cis | 1.81E-08 | Nightingale | M_VLDL_P | NA | G | A | -0.06176 | 0.004142 | 6.60E-54 |
| rs174580 | miR-1908-5 | cis | 1.81E-08 | Nightingale | DHA | NA | G | A | -0.28455 | 0.004159 | 0 |
| rs174580 | miR-1908-5 | cis | 1.81E-08 | Nightingale | XL_VLDL_L | NA | G | A | 0.051467 | 0.004153 | 1.10E-36 |
| rs174580 | miR-1908-5 | cis | 1.81E-08 | Nightingale | LDL_PL | NA | G | A | -0.046 | 0.004307 | 8.00E-28 |
| rs174580 | miR-1908-5 | cis | 1.81E-08 | Nightingale | IDL_C_pct | NA | G | A | -0.07526 | 0.004214 | 3.40E-73 |
| rs174580 | miR-1908-5 | cis | 1.81E-08 | Nightingale | S_LDL_FC | NA | G | A | -0.06279 | 0.004309 | 8.20E-51 |
| rs174580 | miR-1908-5 | cis | 1.81E-08 | Nightingale | XL_HDL_C_ | NA | G | A | 0.02271 | 0.004123 | 1.90E-09 |
| rs174580 | miR-1908-5 | cis | 1.81E-08 | Nightingale | L_LDL_TG_ | NA | G | A | 0.07798 | 0.004232 | 2.10E-77 |
| rs174580 | miR-1908-5 | cis | 1.81E-08 | Nightingale | HDL_TG | NA | G | A | 0.046699 | 0.004256 | 2.20E-28 |
| rs174580 | miR-1908-5 | cis | 1.81E-08 | Nightingale | S_HDL_FC_ | NA | G | A | -0.10079 | 0.004094 | 1.50E-148 |
| rs174580 | miR-1908-5 | cis | 1.81E-08 | Nightingale | Total_FC | NA | G | A | -0.05886 | 0.004217 | 5.70E-47 |
| rs174580 | miR-1908-5 | cis | 1.81E-08 | Nightingale | ApoA1 | NA | G | A | -0.04162 | 0.00395 | 5.80E-28 |
| rs174580 | miR-1908-5 | cis | 1.81E-08 | Nightingale | M_HDL_TG | NA | G | A | 0.067401 | 0.004139 | 2.90E-62 |
| rs174580 | miR-1908-5 | cis | 1.81E-08 | Nightingale | M_HDL_P | NA | G | A | -0.03492 | 0.003976 | 6.00E-20 |
| rs174580 | miR-1908-5 | cis | 1.81E-08 | Nightingale | TG_by_PG | NA | G | A | 0.069116 | 0.004006 | 3.10E-70 |
| rs174580 | miR-1908-5 | cis | 1.81E-08 | Nightingale | L_VLDL_L | NA | G | A | 0.038334 | 0.004171 | 6.10E-21 |
| rs174580 | miR-1908-5 | cis | 1.81E-08 | Nightingale | M_VLDL_F | NA | G | A | -0.09049 | 0.004104 | 1.20E-113 |
| rs174580 | miR-1908-5 | cis | 1.81E-08 | Nightingale | Omega_3 | NA | G | A | -0.33058 | 0.004231 | 0 |
| rs174580 | miR-1908-5 | cis | 1.81E-08 | Nightingale | L_LDL_CE | NA | G | A | -0.0531 | 0.004278 | 2.50E-37 |
| rs174580 | miR-1908-5 | cis | 1.81E-08 | Nightingale | XXL_VLDL_ | NA | G | A | 0.066546 | 0.004157 | 3.40E-60 |
| rs174580 | miR-1908-5 | cis | 1.81E-08 | Nightingale | L_LDL_C | NA | G | A | -0.05761 | 0.004268 | 7.50E-44 |
| rs174580 | miR-1908-5 | cis | 1.81E-08 | Nightingale | XL_VLDL_C | NA | G | A | 0.037656 | 0.004216 | 8.10E-20 |
| rs174580 | miR-1908-5 | cis | 1.81E-08 | Nightingale | Tyr | NA | G | A | 0.032715 | 0.004311 | 3.20E-14 |
| rs174580 | miR-1908-5 | cis | 1.81E-08 | Nightingale | L_HDL_L | NA | G | A | -0.07699 | 0.003787 | 2.20E-100 |
| rs174580 | miR-1908-5 | cis | 1.81E-08 | Nightingale | M_HDL_CE | NA | G | A | -0.04765 | 0.004186 | 8.20E-31 |
| rs174580 | miR-1908-5 | cis | 1.81E-08 | Nightingale | S_LDL_TG | NA | G | A | 0.050879 | 0.004211 | 2.90E-34 |
| rs174580 | miR-1908-5 | cis | 1.81E-08 | Nightingale | XL_HDL_FC | NA | G | A | 0.060613 | 0.003827 | 1.30E-62 |
| rs174580 | miR-1908-5 | cis | 1.81E-08 | Nightingale | XL_HDL_PL | NA | G | A | -0.06587 | 0.003965 | 7.20E-67 |
| rs174580 | miR-1908-5 | cis | 1.81E-08 | Nightingale | S_HDL_C_p | NA | G | A | -0.04306 | 0.004266 | 1.00E-23 |
| rs174580 | miR-1908-5 | cis | 1.81E-08 | Nightingale | IDL_L | NA | G | A | -0.07191 | 0.004173 | 4.30E-71 |
| rs174580 | miR-1908-5 | cis | 1.81E-08 | Nightingale | XL_HDL_CE | NA | G | A | -0.09591 | 0.003865 | 4.60E-148 |
| rs174580 | miR-1908-5 | cis | 1.81E-08 | Nightingale | M_VLDL_C | NA | G | A | -0.05223 | 0.004277 | 7.10E-36 |
| rs174580 | miR-1908-5 | cis | 1.81E-08 | Nightingale | XL_VLDL_F | NA | G | A | -0.06725 | 0.004182 | 4.00E-61 |
| rs174580 | miR-1908-5 | cis | 1.81E-08 | Nightingale | XL_VLDL_T | NA | G | A | 0.056774 | 0.004129 | 9.10E-45 |
| rs174580 | miR-1908-5 | cis | 1.81E-08 | Nightingale | M_LDL_TG | NA | G | A | 0.037094 | 0.004253 | 2.60E-18 |
| rs174580 | miR-1908-5 | cis | 1.81E-08 | Nightingale | VLDL_size | NA | G | A | 0.071865 | 0.004023 | 9.40E-76 |
| rs174580 | miR-1908-5 | cis | 1.81E-08 | Nightingale | HDL_size | NA | G | A | -0.07213 | 0.003825 | 4.10E-86 |
| rs174580 | miR-1908-5 | cis | 1.81E-08 | Nightingale | LDL_P | NA | G | A | -0.03969 | 0.004313 | 3.60E-21 |
| rs174580 | miR-1908-5 | cis | 1.81E-08 | Nightingale | XS_VLDL_F | NA | G | A | -0.0549 | 0.004241 | 3.00E-41 |
| rs174580 | miR-1908-5 | cis | 1.81E-08 | Nightingale | HDL_P | NA | G | A | -0.03636 | 0.004053 | 1.80E-20 |
| rs174580 | miR-1908-5 | cis | 1.81E-08 | Nightingale | S_LDL_CE_ | NA | G | A | 0.05656 | 0.004308 | 2.20E-41 |
| rs174580 | miR-1908-5 | cis | 1.81E-08 | Nightingale | XS_VLDL_C | NA | G | A | -0.06717 | 0.004189 | 6.20E-62 |
| rs174580 | miR-1908-5 | cis | 1.81E-08 | Nightingale | S_VLDL_FC | NA | G | A | -0.08297 | 0.004122 | 4.40E-94 |
| rs174580 | miR-1908-5 | cis | 1.81E-08 | Nightingale | XS_VLDL_T | NA | G | A | 0.032428 | 0.00425 | 8.70E-14 |
| rs174580 | miR-1908-5 | cis | 1.81E-08 | Nightingale | XXL_VLDL_ | NA | G | A | 0.068007 | 0.004158 | 4.90E-63 |
| rs174580 | miR-1908-5 | cis | 1.81E-08 | Nightingale | L_VLDL_PL | NA | G | A | 0.087648 | 0.004189 | 1.40E-100 |
| rs174580 | miR-1908-5 | cis | 1.81E-08 | Nightingale | XS_VLDL_C | NA | G | A | -0.09788 | 0.00409 | 2.60E-132 |
| rs174580 | miR-1908-5 | cis | 1.81E-08 | Nightingale | S_HDL_TG_ | NA | G | A | 0.069699 | 0.004105 | 1.20E-67 |
| rs174580 | miR-1908-5 | cis | 1.81E-08 | Nightingale | XXL_VLDL_ | NA | G | A | 0.073452 | 0.004168 | 4.00E-72 |
| rs174580 | miR-1908-5 | cis | 1.81E-08 | Nightingale | LDL_C | NA | G | A | -0.04959 | 0.004298 | 2.50E-32 |
| rs174580 | miR-1908-5 | cis | 1.81E-08 | Nightingale | Cholines | NA | G | A | -0.03715 | 0.00406 | 9.10E-22 |
| rs174580 | miR-1908-5 | cis | 1.81E-08 | Nightingale | L_HDL_CE | NA | G | A | -0.07851 | 0.003811 | 1.40E-102 |
| rs174580 | miR-1908-5 | cis | 1.81E-08 | Nightingale | HDL_L | NA | G | A | -0.05354 | 0.003876 | 1.10E-46 |
| rs174580 | miR-1908-5 | cis | 1.81E-08 | Nightingale | XS_VLDL_F | NA | G | A | -0.11163 | 0.004284 | 9.71E-153 |
| rs174580 | miR-1908-5 | cis | 1.81E-08 | Nightingale | L_HDL_TG_ | NA | G | A | 0.079462 | 0.004073 | 9.20E-90 |

| | | | | | | | | | | | |
|---|---|---|---|---|---|---|---|---|---|---|---|
| rs174580 | miR-1908-! | cis | 1.81E-08 | Nightingale | M_VLDL_F | NA | G | A | -0.02888 | 0.004316 | 6.60E-12 |
| rs174580 | miR-1908-! | cis | 1.81E-08 | Nightingale | DHA_pct | NA | G | A | -0.27026 | 0.00419 | 0 |
| rs174580 | miR-1908-! | cis | 1.81E-08 | Nightingale | S_LDL_C | NA | G | A | -0.03199 | 0.004321 | 2.90E-14 |
| rs174580 | miR-1908-! | cis | 1.81E-08 | Nightingale | IDL_FC_pct | NA | G | A | -0.1034 | 0.004282 | 2.50E-130 |
| rs174580 | miR-1908-! | cis | 1.81E-08 | Nightingale | Total_P | NA | G | A | -0.04097 | 0.004062 | 1.30E-25 |
| rs174580 | miR-1908-! | cis | 1.81E-08 | Nightingale | M_VLDL_C | NA | G | A | -0.08025 | 0.004063 | 1.10E-90 |
| rs174580 | miR-1908-! | cis | 1.81E-08 | Nightingale | M_HDL_PL | NA | G | A | 0.058563 | 0.004094 | 2.90E-48 |
| rs174580 | miR-1908-! | cis | 1.81E-08 | Nightingale | XL_VLDL_F | NA | G | A | 0.043609 | 0.004181 | 2.00E-26 |
| rs174580 | miR-1908-! | cis | 1.81E-08 | Nightingale | M_HDL_FC | NA | G | A | -0.10282 | 0.003825 | 8.39E-174 |
| rs174580 | miR-1908-! | cis | 1.81E-08 | Nightingale | S_LDL_FC_ | NA | G | A | -0.08032 | 0.004183 | 3.40E-85 |
| rs174580 | miR-1908-! | cis | 1.81E-08 | Nightingale | M_VLDL_C | NA | G | A | -0.07564 | 0.004052 | 4.90E-81 |
| rs174580 | miR-1908-! | cis | 1.81E-08 | Nightingale | L_HDL_C | NA | G | A | -0.08001 | 0.003801 | 3.30E-107 |
| rs174580 | miR-1908-! | cis | 1.81E-08 | Nightingale | Total_FA | NA | G | A | -0.02312 | 0.004282 | 2.30E-08 |
| rs174580 | miR-1908-! | cis | 1.81E-08 | Nightingale | XS_VLDL_C | NA | G | A | -0.07102 | 0.004165 | 9.60E-70 |
| rs174580 | miR-1908-! | cis | 1.81E-08 | Nightingale | IDL_PL | NA | G | A | -0.07212 | 0.004165 | 3.70E-72 |
| rs174580 | miR-1908-! | cis | 1.81E-08 | Nightingale | non_HDL_( | NA | G | A | -0.04746 | 0.004293 | 7.30E-30 |
| rs174580 | miR-1908-! | cis | 1.81E-08 | Nightingale | LA | NA | G | A | 0.082595 | 0.004233 | 3.90E-85 |
| rs174580 | miR-1908-! | cis | 1.81E-08 | Nightingale | S_VLDL_TG | NA | G | A | 0.039439 | 0.004189 | 9.20E-22 |
| rs174580 | miR-1908-! | cis | 1.81E-08 | Nightingale | S_VLDL_FC | NA | G | A | -0.02776 | 0.004317 | 4.10E-11 |
| rs174580 | miR-1908-! | cis | 1.81E-08 | Nightingale | Total_PL | NA | G | A | -0.0508 | 0.00411 | 2.60E-37 |
| rs174580 | miR-1908-! | cis | 1.81E-08 | Nightingale | M_LDL_C_ | NA | G | A | -0.09991 | 0.00432 | 4.00E-119 |
| rs174580 | miR-1908-! | cis | 1.81E-08 | Nightingale | XL_HDL_L | NA | G | A | -0.09163 | 0.003879 | 1.20E-134 |
| rs174580 | miR-1908-! | cis | 1.81E-08 | Nightingale | XS_VLDL_P | NA | G | A | 0.058807 | 0.004211 | 2.10E-45 |
| rs174580 | miR-1908-! | cis | 1.81E-08 | Nightingale | XS_VLDL_T | NA | G | A | 0.097059 | 0.0041 | 8.89E-130 |
| rs174580 | miR-1908-! | cis | 1.81E-08 | Nightingale | S_VLDL_C_ | NA | G | A | -0.05662 | 0.0042 | 1.60E-42 |
| rs174580 | miR-1908-! | cis | 1.81E-08 | Nightingale | M_HDL_PL | NA | G | A | -0.02219 | 0.004036 | 9.60E-09 |
| rs174580 | miR-1908-! | cis | 1.81E-08 | Nightingale | L_HDL_CE_ | NA | G | A | -0.05029 | 0.00413 | 3.00E-36 |
| rs174580 | miR-1908-! | cis | 1.81E-08 | Nightingale | M_HDL_TG | NA | G | A | 0.050562 | 0.004263 | 3.90E-33 |
| rs174580 | miR-1908-! | cis | 1.81E-08 | Nightingale | VLDL_TG | NA | G | A | 0.047881 | 0.004154 | 6.80E-32 |
| rs174580 | miR-1908-! | cis | 1.81E-08 | Nightingale | VLDL_L | NA | G | A | 0.027698 | 0.004232 | 3.00E-11 |
| rs174580 | miR-1908-! | cis | 1.81E-08 | Nightingale | S_VLDL_PL | NA | G | A | -0.07865 | 0.00413 | 1.30E-84 |
| rs174580 | miR-1908-! | cis | 1.81E-08 | Nightingale | XXL_VLDL_ | NA | G | A | 0.066156 | 0.004152 | 7.10E-60 |
| rs174580 | miR-1908-! | cis | 1.81E-08 | Nightingale | Omega_6_ | NA | G | A | 0.023186 | 0.004201 | 2.20E-08 |
| rs174580 | miR-1908-! | cis | 1.81E-08 | Nightingale | XL_HDL_PL | NA | G | A | -0.08771 | 0.00388 | 2.40E-123 |
| rs174580 | miR-1908-! | cis | 1.81E-08 | Nightingale | PUFA_pct | NA | G | A | -0.12036 | 0.004152 | 1.30E-186 |
| rs174580 | miR-1908-! | cis | 1.81E-08 | Nightingale | Clinical_LD | NA | G | A | -0.05504 | 0.004292 | 1.90E-39 |
| rs174580 | miR-1908-! | cis | 1.81E-08 | Nightingale | L_LDL_PL | NA | G | A | -0.05482 | 0.004284 | 2.90E-39 |
| rs174580 | miR-1908-! | cis | 1.81E-08 | Nightingale | S_LDL_P | NA | G | A | -0.03189 | 0.00432 | 3.30E-14 |
| rs174580 | miR-1908-! | cis | 1.81E-08 | Nightingale | L_VLDL_FC | NA | G | A | 0.040281 | 0.004175 | 5.50E-23 |
| rs174580 | miR-1908-! | cis | 1.81E-08 | Nightingale | M_HDL_C | NA | G | A | -0.04153 | 0.003971 | 3.20E-27 |
| rs174580 | miR-1908-! | cis | 1.81E-08 | Nightingale | IDL_TG_pct | NA | G | A | 0.088322 | 0.00417 | 2.20E-103 |
| rs174580 | miR-1908-! | cis | 1.81E-08 | Nightingale | L_HDL_C_p | NA | G | A | -0.06827 | 0.004068 | 1.10E-67 |
| rs174580 | miR-1908-! | cis | 1.81E-08 | Nightingale | M_LDL_CE | NA | G | A | -0.02624 | 0.004323 | 5.80E-10 |
| rs174580 | miR-1908-! | cis | 1.81E-08 | Nightingale | M_LDL_PL | NA | G | A | -0.02355 | 0.004323 | 2.40E-08 |
| rs174580 | miR-1908-! | cis | 1.81E-08 | Nightingale | M_LDL_P | NA | G | A | -0.04616 | 0.004329 | 2.20E-27 |
| rs174580 | miR-1908-! | cis | 1.81E-08 | Nightingale | Remnant_( | NA | G | A | -0.04361 | 0.004289 | 7.80E-26 |
| rs174580 | miR-1908-! | cis | 1.81E-08 | Nightingale | MUFA | NA | G | A | 0.063642 | 0.004272 | 2.10E-51 |
| rs174580 | miR-1908-! | cis | 1.81E-08 | Nightingale | HDL_C | NA | G | A | -0.06501 | 0.003854 | 4.70E-69 |
| rs174580 | miR-1908-! | cis | 1.81E-08 | Nightingale | Citrate | NA | G | A | -0.03366 | 0.004289 | 5.20E-16 |
| rs174580 | miR-1908-! | cis | 1.81E-08 | Nightingale | XL_HDL_FC | NA | G | A | -0.1008 | 0.004021 | 6.81E-150 |
| rs174580 | miR-1908-! | cis | 1.81E-08 | Nightingale | M_LDL_TG | NA | G | A | 0.076209 | 0.004262 | 7.60E-73 |
| rs174580 | miR-1908-! | cis | 1.81E-08 | Nightingale | IDL_C | NA | G | A | -0.07692 | 0.004168 | 1.00E-80 |
| rs174580 | miR-1908-! | cis | 1.81E-08 | Nightingale | L_VLDL_FC | NA | G | A | 0.028539 | 0.004275 | 1.90E-11 |
| rs174580 | miR-1908-! | cis | 1.81E-08 | Nightingale | HDL_FC | NA | G | A | -0.0737 | 0.003824 | 9.70E-90 |
| rs174580 | miR-1908-! | cis | 1.81E-08 | Nightingale | S_LDL_L | NA | G | A | -0.03211 | 0.004316 | 2.00E-14 |
| rs174580 | miR-1908-! | cis | 1.81E-08 | Nightingale | S_LDL_TG_ | NA | G | A | 0.086473 | 0.004182 | 4.30E-98 |
| rs174580 | miR-1908-! | cis | 1.81E-08 | Nightingale | S_LDL_PL | NA | G | A | -0.05087 | 0.004306 | 5.20E-34 |
| rs174580 | miR-1908-! | cis | 1.81E-08 | Nightingale | PUFA_by_I | NA | G | A | -0.16917 | 0.004134 | 0 |
| rs174580 | miR-1908-! | cis | 1.81E-08 | Nightingale | XL_VLDL_T | NA | G | A | 0.059121 | 0.004181 | 6.60E-47 |
| rs174580 | miR-1908-! | cis | 1.81E-08 | Nightingale | L_LDL_P | NA | G | A | -0.03686 | 0.004302 | 9.70E-19 |
| rs174580 | miR-1908-! | cis | 1.81E-08 | Nightingale | XS_VLDL_P | NA | G | A | -0.03319 | 0.004256 | 1.70E-16 |
| rs174580 | miR-1908-! | cis | 1.81E-08 | Nightingale | PUFA | NA | G | A | -0.0951 | 0.004209 | 1.10E-119 |
| rs174580 | miR-1908-! | cis | 1.81E-08 | Nightingale | M_VLDL_T | NA | G | A | 0.031583 | 0.004194 | 1.70E-14 |

| | | | | | | | | | | | | |
|---|---|---|---|---|---|---|---|---|---|---|---|---|
| rs174580 | miR-1908-5 | cis | | 1.81E-08 | Nightingale | S_VLDL_TG | NA | G | A | 0.064546 | 0.004176 | 1.70E-55 |
| rs174580 | miR-1908-5 | cis | | 1.81E-08 | Nightingale | M_LDL_FC | NA | G | A | -0.05243 | 0.004311 | 8.90E-36 |
| rs174580 | miR-1908-5 | cis | | 1.81E-08 | Nightingale | XL_VLDL_C | NA | G | A | -0.06074 | 0.004138 | 3.30E-50 |
| rs174580 | miR-1908-5 | cis | | 1.81E-08 | Nightingale | L_HDL_FC_ | NA | G | A | -0.11071 | 0.00401 | 1.60E-182 |
| rs174581 | miR-1908-5 | cis | | 1.68E-08 | Nightingale | M_LDL_C | NA | A | G | -0.03403 | 0.004325 | 9.20E-16 |
| rs174581 | miR-1908-5 | cis | | 1.68E-08 | Nightingale | IDL_CE | NA | A | G | -0.07209 | 0.004167 | 3.50E-71 |
| rs174581 | miR-1908-5 | cis | | 1.68E-08 | Nightingale | XL_VLDL_P | NA | A | G | 0.05282 | 0.004149 | 1.20E-38 |
| rs174581 | miR-1908-5 | cis | | 1.68E-08 | Nightingale | M_HDL_P | NA | A | G | -0.03494 | 0.003977 | 5.80E-20 |
| rs174581 | miR-1908-5 | cis | | 1.68E-08 | Nightingale | S_LDL_FC | NA | A | G | -0.06284 | 0.00431 | 7.10E-51 |
| rs174581 | miR-1908-5 | cis | | 1.68E-08 | Nightingale | L_VLDL_P | NA | A | G | 0.042007 | 0.004168 | 7.40E-25 |
| rs174581 | miR-1908-5 | cis | | 1.68E-08 | Nightingale | HDL_PL | NA | A | G | -0.0487 | 0.003902 | 2.40E-38 |
| rs174581 | miR-1908-5 | cis | | 1.68E-08 | Nightingale | LDL_CE | NA | A | G | -0.04347 | 0.004306 | 4.00E-25 |
| rs174581 | miR-1908-5 | cis | | 1.68E-08 | Nightingale | M_HDL_TG | NA | A | G | 0.067344 | 0.004139 | 4.10E-62 |
| rs174581 | miR-1908-5 | cis | | 1.68E-08 | Nightingale | LA_pct | NA | A | G | 0.182297 | 0.004219 | 0 |
| rs174581 | miR-1908-5 | cis | | 1.68E-08 | Nightingale | M_HDL_C_ | NA | A | G | -0.06632 | 0.004109 | 1.60E-61 |
| rs174581 | miR-1908-5 | cis | | 1.68E-08 | Nightingale | XL_VLDL_C | NA | A | G | 0.030462 | 0.004249 | 3.10E-13 |
| rs174581 | miR-1908-5 | cis | | 1.68E-08 | Nightingale | XXL_VLDL_ | NA | A | G | -0.02687 | 0.004296 | 7.20E-11 |
| rs174581 | miR-1908-5 | cis | | 1.68E-08 | Nightingale | L_LDL_FC | NA | A | G | -0.06821 | 0.004239 | 1.20E-61 |
| rs174581 | miR-1908-5 | cis | | 1.68E-08 | Nightingale | XXL_VLDL_ | NA | A | G | 0.04455 | 0.004341 | 1.40E-25 |
| rs174581 | miR-1908-5 | cis | | 1.68E-08 | Nightingale | XS_VLDL_C | NA | A | G | -0.09214 | 0.004069 | 7.80E-119 |
| rs174581 | miR-1908-5 | cis | | 1.68E-08 | Nightingale | S_HDL_TG | NA | A | G | 0.0689 | 0.004186 | 1.60E-63 |
| rs174581 | miR-1908-5 | cis | | 1.68E-08 | Nightingale | LDL_FC | NA | A | G | -0.06424 | 0.004271 | 7.30E-54 |
| rs174581 | miR-1908-5 | cis | | 1.68E-08 | Nightingale | S_LDL_PL_ | NA | A | G | -0.06092 | 0.004308 | 1.20E-47 |
| rs174581 | miR-1908-5 | cis | | 1.68E-08 | Nightingale | Total_CE | NA | A | G | -0.06687 | 0.004164 | 2.90E-61 |
| rs174581 | miR-1908-5 | cis | | 1.68E-08 | Nightingale | XXL_VLDL_ | NA | A | G | 0.071617 | 0.00416 | 3.40E-69 |
| rs174581 | miR-1908-5 | cis | | 1.68E-08 | Nightingale | XL_HDL_P | NA | A | G | -0.08817 | 0.003885 | 2.50E-125 |
| rs174581 | miR-1908-5 | cis | | 1.68E-08 | Nightingale | LDL_TG | NA | A | G | 0.030162 | 0.004269 | 3.00E-12 |
| rs174581 | miR-1908-5 | cis | | 1.68E-08 | Nightingale | S_VLDL_CE | NA | A | G | -0.02897 | 0.004259 | 5.80E-12 |
| rs174581 | miR-1908-5 | cis | | 1.68E-08 | Nightingale | XL_HDL_TG | NA | A | G | 0.087015 | 0.004067 | 2.80E-107 |
| rs174581 | miR-1908-5 | cis | | 1.68E-08 | Nightingale | M_LDL_L | NA | A | G | -0.02815 | 0.004323 | 3.00E-11 |
| rs174581 | miR-1908-5 | cis | | 1.68E-08 | Nightingale | LDL_P | NA | A | G | -0.03976 | 0.004314 | 3.10E-21 |
| rs174581 | miR-1908-5 | cis | | 1.68E-08 | Nightingale | XL_VLDL_C | NA | A | G | 0.037607 | 0.004216 | 9.30E-20 |
| rs174581 | miR-1908-5 | cis | | 1.68E-08 | Nightingale | ApoB | NA | A | G | -0.03803 | 0.004311 | 1.10E-19 |
| rs174581 | miR-1908-5 | cis | | 1.68E-08 | Nightingale | L_LDL_FC_ | NA | A | G | -0.08262 | 0.004114 | 3.40E-93 |
| rs174581 | miR-1908-5 | cis | | 1.68E-08 | Nightingale | XL_VLDL_P | NA | A | G | 0.048387 | 0.004166 | 2.00E-32 |
| rs174581 | miR-1908-5 | cis | | 1.68E-08 | Nightingale | L_HDL_C | NA | A | G | -0.08005 | 0.003801 | 3.00E-107 |
| rs174581 | miR-1908-5 | cis | | 1.68E-08 | Nightingale | M_LDL_FC | NA | A | G | -0.05248 | 0.004312 | 7.90E-36 |
| rs174581 | miR-1908-5 | cis | | 1.68E-08 | Nightingale | L_LDL_C_p | NA | A | G | -0.07722 | 0.004206 | 1.80E-77 |
| rs174581 | miR-1908-5 | cis | | 1.68E-08 | Nightingale | XL_HDL_L | NA | A | G | -0.09169 | 0.003879 | 8.59E-135 |
| rs174581 | miR-1908-5 | cis | | 1.68E-08 | Nightingale | L_LDL_CE | NA | A | G | -0.05317 | 0.004278 | 2.00E-37 |
| rs174581 | miR-1908-5 | cis | | 1.68E-08 | Nightingale | L_HDL_PL_ | NA | A | G | 0.042248 | 0.004178 | 1.40E-25 |
| rs174581 | miR-1908-5 | cis | | 1.68E-08 | Nightingale | Total_L | NA | A | G | -0.03705 | 0.004247 | 1.70E-19 |
| rs174581 | miR-1908-5 | cis | | 1.68E-08 | Nightingale | XL_HDL_FC | NA | A | G | 0.060647 | 0.003827 | 1.20E-62 |
| rs174581 | miR-1908-5 | cis | | 1.68E-08 | Nightingale | Total_FC | NA | A | G | -0.05896 | 0.004217 | 4.20E-47 |
| rs174581 | miR-1908-5 | cis | | 1.68E-08 | Nightingale | IDL_CE_pct | NA | A | G | -0.03893 | 0.004236 | 3.20E-20 |
| rs174581 | miR-1908-5 | cis | | 1.68E-08 | Nightingale | M_LDL_PL_ | NA | A | G | 0.045024 | 0.004314 | 1.10E-26 |
| rs174581 | miR-1908-5 | cis | | 1.68E-08 | Nightingale | L_VLDL_CE | NA | A | G | -0.0585 | 0.004131 | 1.70E-48 |
| rs174581 | miR-1908-5 | cis | | 1.68E-08 | Nightingale | XXL_VLDL_ | NA | A | G | 0.06138 | 0.004165 | 3.00E-51 |
| rs174581 | miR-1908-5 | cis | | 1.68E-08 | Nightingale | IDL_C_pct | NA | A | G | -0.07527 | 0.004214 | 3.50E-73 |
| rs174581 | miR-1908-5 | cis | | 1.68E-08 | Nightingale | M_HDL_PL | NA | A | G | 0.058529 | 0.004095 | 3.50E-48 |
| rs174581 | miR-1908-5 | cis | | 1.68E-08 | Nightingale | M_HDL_L | NA | A | G | -0.02838 | 0.004007 | 1.60E-13 |
| rs174581 | miR-1908-5 | cis | | 1.68E-08 | Nightingale | L_LDL_L | NA | A | G | -0.05402 | 0.004275 | 1.10E-38 |
| rs174581 | miR-1908-5 | cis | | 1.68E-08 | Nightingale | Total_TG | NA | A | G | 0.047029 | 0.004178 | 1.80E-30 |
| rs174581 | miR-1908-5 | cis | | 1.68E-08 | Nightingale | L_HDL_L | NA | A | G | -0.07703 | 0.003788 | 1.90E-100 |
| rs174581 | miR-1908-5 | cis | | 1.68E-08 | Nightingale | Omega_3 | NA | A | G | -0.33058 | 0.004231 | 0 |
| rs174581 | miR-1908-5 | cis | | 1.68E-08 | Nightingale | MUFA_pct | NA | A | G | 0.195634 | 0.004131 | 0 |
| rs174581 | miR-1908-5 | cis | | 1.68E-08 | Nightingale | S_VLDL_FC | NA | A | G | -0.083 | 0.004122 | 4.30E-94 |
| rs174581 | miR-1908-5 | cis | | 1.68E-08 | Nightingale | L_HDL_P | NA | A | G | -0.07756 | 0.003781 | 3.30E-102 |
| rs174581 | miR-1908-5 | cis | | 1.68E-08 | Nightingale | S_HDL_C_p | NA | A | G | -0.043 | 0.004267 | 1.30E-23 |
| rs174581 | miR-1908-5 | cis | | 1.68E-08 | Nightingale | L_VLDL_C | NA | A | G | 0.029125 | 0.004222 | 2.10E-12 |
| rs174581 | miR-1908-5 | cis | | 1.68E-08 | Nightingale | Unsaturati | NA | A | G | -0.33495 | 0.004088 | 0 |
| rs174581 | miR-1908-5 | cis | | 1.68E-08 | Nightingale | non_HDL_C | NA | A | G | -0.04754 | 0.004294 | 5.80E-30 |
| rs174581 | miR-1908-5 | cis | | 1.68E-08 | Nightingale | HDL_CE | NA | A | G | -0.06187 | 0.003874 | 7.00E-62 |

| | | | | | | | | | | | |
|---|---|---|---|---|---|---|---|---|---|---|---|
| rs174581 | miR-1908-5 | cis | 1.68E-08 | Nightingale | SFA | NA | A | G | -0.03089 | 0.004278 | 1.90E-13 |
| rs174581 | miR-1908-5 | cis | 1.68E-08 | Nightingale | LDL_C | NA | A | G | -0.04966 | 0.004298 | 2.10E-32 |
| rs174581 | miR-1908-5 | cis | 1.68E-08 | Nightingale | IDL_P | NA | A | G | -0.04505 | 0.004271 | 1.50E-27 |
| rs174581 | miR-1908-5 | cis | 1.68E-08 | Nightingale | L_HDL_FC | NA | A | G | -0.08414 | 0.003783 | 1.70E-119 |
| rs174581 | miR-1908-5 | cis | 1.68E-08 | Nightingale | Omega_3_ | NA | A | G | -0.384 | 0.004218 | 0 |
| rs174581 | miR-1908-5 | cis | 1.68E-08 | Nightingale | XS_VLDL_F | NA | A | G | -0.05502 | 0.004241 | 2.00E-41 |
| rs174581 | miR-1908-5 | cis | 1.68E-08 | Nightingale | IDL_PL | NA | A | G | -0.07223 | 0.004165 | 2.40E-72 |
| rs174581 | miR-1908-5 | cis | 1.68E-08 | Nightingale | TG_by_PG | NA | A | G | 0.069108 | 0.004006 | 3.40E-70 |
| rs174581 | miR-1908-5 | cis | 1.68E-08 | Nightingale | M_HDL_PL | NA | A | G | -0.02222 | 0.004037 | 9.30E-09 |
| rs174581 | miR-1908-5 | cis | 1.68E-08 | Nightingale | XL_HDL_CE | NA | A | G | -0.09596 | 0.003865 | 3.70E-148 |
| rs174581 | miR-1908-5 | cis | 1.68E-08 | Nightingale | DHA | NA | A | G | -0.28454 | 0.00416 | 0 |
| rs174581 | miR-1908-5 | cis | 1.68E-08 | Nightingale | L_VLDL_L | NA | A | G | 0.0383 | 0.004172 | 6.90E-21 |
| rs174581 | miR-1908-5 | cis | 1.68E-08 | Nightingale | S_VLDL_FC | NA | A | G | -0.02784 | 0.004318 | 3.70E-11 |
| rs174581 | miR-1908-5 | cis | 1.68E-08 | Nightingale | Cholines | NA | A | G | -0.03724 | 0.00406 | 7.40E-22 |
| rs174581 | miR-1908-5 | cis | 1.68E-08 | Nightingale | L_HDL_PL | NA | A | G | -0.07595 | 0.003799 | 6.30E-97 |
| rs174581 | miR-1908-5 | cis | 1.68E-08 | Nightingale | DHA_pct | NA | A | G | -0.27021 | 0.00419 | 0 |
| rs174581 | miR-1908-5 | cis | 1.68E-08 | Nightingale | LDL_L | NA | A | G | -0.04531 | 0.004303 | 3.00E-27 |
| rs174581 | miR-1908-5 | cis | 1.68E-08 | Nightingale | XXL_VLDL_ | NA | A | G | 0.072166 | 0.004143 | 4.00E-71 |
| rs174581 | miR-1908-5 | cis | 1.68E-08 | Nightingale | XS_VLDL_L | NA | A | G | -0.04059 | 0.004243 | 8.60E-24 |
| rs174581 | miR-1908-5 | cis | 1.68E-08 | Nightingale | XL_VLDL_L | NA | A | G | 0.051435 | 0.004153 | 1.20E-36 |
| rs174581 | miR-1908-5 | cis | 1.68E-08 | Nightingale | M_HDL_CE | NA | A | G | -0.03866 | 0.003987 | 1.30E-23 |
| rs174581 | miR-1908-5 | cis | 1.68E-08 | Nightingale | HDL_P | NA | A | G | -0.03639 | 0.004053 | 1.80E-20 |
| rs174581 | miR-1908-5 | cis | 1.68E-08 | Nightingale | M_LDL_FC_ | NA | A | G | -0.0717 | 0.004135 | 4.70E-70 |
| rs174581 | miR-1908-5 | cis | 1.68E-08 | Nightingale | L_VLDL_PL | NA | A | G | 0.048834 | 0.004165 | 3.60E-33 |
| rs174581 | miR-1908-5 | cis | 1.68E-08 | Nightingale | XL_HDL_CE | NA | A | G | -0.02705 | 0.004314 | 9.60E-10 |
| rs174581 | miR-1908-5 | cis | 1.68E-08 | Nightingale | XS_VLDL_P | NA | A | G | -0.04267 | 0.004259 | 1.10E-25 |
| rs174581 | miR-1908-5 | cis | 1.68E-08 | Nightingale | SFA_pct | NA | A | G | -0.04212 | 0.004259 | 4.60E-23 |
| rs174581 | miR-1908-5 | cis | 1.68E-08 | Nightingale | PUFA_pct | NA | A | G | -0.12032 | 0.004152 | 1.80E-186 |
| rs174581 | miR-1908-5 | cis | 1.68E-08 | Nightingale | L_LDL_TG_ | NA | A | G | 0.077965 | 0.004233 | 2.30E-77 |
| rs174581 | miR-1908-5 | cis | 1.68E-08 | Nightingale | Phosphogly | NA | A | G | -0.04097 | 0.004089 | 2.10E-25 |
| rs174581 | miR-1908-5 | cis | 1.68E-08 | Nightingale | M_HDL_FC | NA | A | G | -0.05188 | 0.003928 | 1.20E-42 |
| rs174581 | miR-1908-5 | cis | 1.68E-08 | Nightingale | S_LDL_CE_ | NA | A | G | 0.05653 | 0.004309 | 2.50E-41 |
| rs174581 | miR-1908-5 | cis | 1.68E-08 | Nightingale | XL_HDL_C_ | NA | A | G | 0.022774 | 0.004124 | 1.70E-09 |
| rs174581 | miR-1908-5 | cis | 1.68E-08 | Nightingale | Total_C | NA | A | G | -0.06498 | 0.004179 | 1.70E-57 |
| rs174581 | miR-1908-5 | cis | 1.68E-08 | Nightingale | L_HDL_TG_ | NA | A | G | 0.079438 | 0.004073 | 1.20E-89 |
| rs174581 | miR-1908-5 | cis | 1.68E-08 | Nightingale | M_VLDL_F | NA | A | G | -0.02895 | 0.004316 | 5.80E-12 |
| rs174581 | miR-1908-5 | cis | 1.68E-08 | Nightingale | L_VLDL_C_ | NA | A | G | -0.03828 | 0.004217 | 4.30E-21 |
| rs174581 | miR-1908-5 | cis | 1.68E-08 | Nightingale | M_HDL_FC | NA | A | G | -0.10287 | 0.003825 | 6.81E-174 |
| rs174581 | miR-1908-5 | cis | 1.68E-08 | Nightingale | XL_VLDL_C | NA | A | G | -0.06359 | 0.004136 | 8.70E-55 |
| rs174581 | miR-1908-5 | cis | 1.68E-08 | Nightingale | M_HDL_CE | NA | A | G | -0.04758 | 0.004186 | 1.10E-30 |
| rs174581 | miR-1908-5 | cis | 1.68E-08 | Nightingale | XL_VLDL_T | NA | A | G | 0.056744 | 0.004129 | 1.00E-44 |
| rs174581 | miR-1908-5 | cis | 1.68E-08 | Nightingale | XL_VLDL_T | NA | A | G | 0.059129 | 0.004182 | 6.50E-47 |
| rs174581 | miR-1908-5 | cis | 1.68E-08 | Nightingale | XL_HDL_PL | NA | A | G | -0.08777 | 0.00388 | 1.80E-123 |
| rs174581 | miR-1908-5 | cis | 1.68E-08 | Nightingale | S_HDL_FC_ | NA | A | G | -0.10086 | 0.004094 | 1.10E-148 |
| rs174581 | miR-1908-5 | cis | 1.68E-08 | Nightingale | L_VLDL_TG | NA | A | G | 0.037318 | 0.004153 | 4.50E-20 |
| rs174581 | miR-1908-5 | cis | 1.68E-08 | Nightingale | Sphingomy | NA | A | G | -0.11443 | 0.004034 | 2.50E-186 |
| rs174581 | miR-1908-5 | cis | 1.68E-08 | Nightingale | ApoA1 | NA | A | G | -0.04166 | 0.00395 | 5.40E-28 |
| rs174581 | miR-1908-5 | cis | 1.68E-08 | Nightingale | HDL_size | NA | A | G | -0.07218 | 0.003825 | 3.50E-86 |
| rs174581 | miR-1908-5 | cis | 1.68E-08 | Nightingale | XXL_VLDL_ | NA | A | G | 0.067973 | 0.004159 | 5.90E-63 |
| rs174581 | miR-1908-5 | cis | 1.68E-08 | Nightingale | M_VLDL_F | NA | A | G | -0.09053 | 0.004104 | 1.00E-113 |
| rs174581 | miR-1908-5 | cis | 1.68E-08 | Nightingale | HDL_L | NA | A | G | -0.05359 | 0.003876 | 9.80E-47 |
| rs174581 | miR-1908-5 | cis | 1.68E-08 | Nightingale | S_VLDL_C_ | NA | A | G | -0.05665 | 0.0042 | 1.40E-42 |
| rs174581 | miR-1908-5 | cis | 1.68E-08 | Nightingale | Omega_6_ | NA | A | G | 0.023209 | 0.004201 | 2.10E-08 |
| rs174581 | miR-1908-5 | cis | 1.68E-08 | Nightingale | M_HDL_C | NA | A | G | -0.04154 | 0.003972 | 3.20E-27 |
| rs174581 | miR-1908-5 | cis | 1.68E-08 | Nightingale | L_VLDL_FC | NA | A | G | 0.040239 | 0.004175 | 6.30E-23 |
| rs174581 | miR-1908-5 | cis | 1.68E-08 | Nightingale | M_VLDL_C | NA | A | G | -0.04358 | 0.004305 | 3.70E-25 |
| rs174581 | miR-1908-5 | cis | 1.68E-08 | Nightingale | Omega_6_ | NA | A | G | 0.364077 | 0.004254 | 0 |
| rs174581 | miR-1908-5 | cis | 1.68E-08 | Nightingale | S_LDL_C | NA | A | G | -0.03205 | 0.004321 | 2.60E-14 |
| rs174581 | miR-1908-5 | cis | 1.68E-08 | Nightingale | IDL_FC | NA | A | G | -0.08925 | 0.004178 | 5.40E-107 |
| rs174581 | miR-1908-5 | cis | 1.68E-08 | Nightingale | XS_VLDL_T | NA | A | G | 0.032352 | 0.00425 | 1.00E-13 |
| rs174581 | miR-1908-5 | cis | 1.68E-08 | Nightingale | L_LDL_C | NA | A | G | -0.05768 | 0.004268 | 6.10E-44 |
| rs174581 | miR-1908-5 | cis | 1.68E-08 | Nightingale | XS_VLDL_F | NA | A | G | -0.11164 | 0.004284 | 9.79E-153 |
| rs174581 | miR-1908-5 | cis | 1.68E-08 | Nightingale | L_HDL_CE_ | NA | A | G | -0.05026 | 0.00413 | 3.40E-36 |

| | | | | | | | | | | | |
|---|---|---|---|---|---|---|---|---|---|---|---|
| rs174581 | miR-1908-5 | cis | 1.68E-08 | Nightingale | M_VLDL_C | NA | A | G | -0.07566 | 0.004052 | 5.00E-81 |
| rs174581 | miR-1908-5 | cis | 1.68E-08 | Nightingale | IDL_FC_pct | NA | A | G | -0.10336 | 0.004282 | 3.30E-130 |
| rs174581 | miR-1908-5 | cis | 1.68E-08 | Nightingale | Total_P | NA | A | G | -0.04101 | 0.004062 | 1.20E-25 |
| rs174581 | miR-1908-5 | cis | 1.68E-08 | Nightingale | M_VLDL_C | NA | A | G | -0.08027 | 0.004063 | 1.10E-90 |
| rs174581 | miR-1908-5 | cis | 1.68E-08 | Nightingale | PUFA | NA | A | G | -0.09518 | 0.00421 | 7.19E-120 |
| rs174581 | miR-1908-5 | cis | 1.68E-08 | Nightingale | Tyr | NA | A | G | 0.032728 | 0.004311 | 3.20E-14 |
| rs174581 | miR-1908-5 | cis | 1.68E-08 | Nightingale | Remnant_( | NA | A | G | -0.04371 | 0.004289 | 6.00E-26 |
| rs174581 | miR-1908-5 | cis | 1.68E-08 | Nightingale | HDL_TG | NA | A | G | 0.046631 | 0.004256 | 2.70E-28 |
| rs174581 | miR-1908-5 | cis | 1.68E-08 | Nightingale | XS_VLDL_P | NA | A | G | -0.0333 | 0.004256 | 1.40E-16 |
| rs174581 | miR-1908-5 | cis | 1.68E-08 | Nightingale | M_VLDL_T | NA | A | G | 0.077471 | 0.004077 | 1.90E-84 |
| rs174581 | miR-1908-5 | cis | 1.68E-08 | Nightingale | VLDL_TG | NA | A | G | 0.047848 | 0.004154 | 7.70E-32 |
| rs174581 | miR-1908-5 | cis | 1.68E-08 | Nightingale | S_LDL_FC_ | NA | A | G | -0.08027 | 0.004183 | 4.60E-85 |
| rs174581 | miR-1908-5 | cis | 1.68E-08 | Nightingale | L_LDL_P | NA | A | G | -0.03694 | 0.004302 | 8.20E-19 |
| rs174581 | miR-1908-5 | cis | 1.68E-08 | Nightingale | M_LDL_C_ | NA | A | G | -0.09991 | 0.00432 | 4.30E-119 |
| rs174581 | miR-1908-5 | cis | 1.68E-08 | Nightingale | LA | NA | A | G | 0.082505 | 0.004233 | 6.30E-85 |
| rs174581 | miR-1908-5 | cis | 1.68E-08 | Nightingale | MUFA | NA | A | G | 0.063573 | 0.004273 | 2.80E-51 |
| rs174581 | miR-1908-5 | cis | 1.68E-08 | Nightingale | L_HDL_CE | NA | A | G | -0.07854 | 0.003812 | 1.30E-102 |
| rs174581 | miR-1908-5 | cis | 1.68E-08 | Nightingale | XL_VLDL_F | NA | A | G | -0.06726 | 0.004182 | 3.80E-61 |
| rs174581 | miR-1908-5 | cis | 1.68E-08 | Nightingale | L_VLDL_PL | NA | A | G | 0.087593 | 0.004189 | 2.20E-100 |
| rs174581 | miR-1908-5 | cis | 1.68E-08 | Nightingale | L_LDL_PL | NA | A | G | -0.05489 | 0.004284 | 2.40E-39 |
| rs174581 | miR-1908-5 | cis | 1.68E-08 | Nightingale | M_LDL_TG | NA | A | G | 0.037026 | 0.004253 | 3.00E-18 |
| rs174581 | miR-1908-5 | cis | 1.68E-08 | Nightingale | Citrate | NA | A | G | -0.03367 | 0.004289 | 5.10E-16 |
| rs174581 | miR-1908-5 | cis | 1.68E-08 | Nightingale | S_LDL_TG_ | NA | A | G | 0.086459 | 0.004183 | 5.10E-98 |
| rs174581 | miR-1908-5 | cis | 1.68E-08 | Nightingale | HDL_FC | NA | A | G | -0.07375 | 0.003824 | 7.90E-90 |
| rs174581 | miR-1908-5 | cis | 1.68E-08 | Nightingale | Clinical_LD | NA | A | G | -0.0551 | 0.004292 | 1.60E-39 |
| rs174581 | miR-1908-5 | cis | 1.68E-08 | Nightingale | IDL_L | NA | A | G | -0.07202 | 0.004173 | 2.80E-71 |
| rs174581 | miR-1908-5 | cis | 1.68E-08 | Nightingale | XL_HDL_FC | NA | A | G | -0.10087 | 0.004021 | 4.60E-150 |
| rs174581 | miR-1908-5 | cis | 1.68E-08 | Nightingale | S_LDL_L | NA | A | G | -0.03217 | 0.004316 | 1.80E-14 |
| rs174581 | miR-1908-5 | cis | 1.68E-08 | Nightingale | VLDL_L | NA | A | G | 0.027643 | 0.004232 | 3.40E-11 |
| rs174581 | miR-1908-5 | cis | 1.68E-08 | Nightingale | IDL_C | NA | A | G | -0.07702 | 0.004168 | 6.80E-81 |
| rs174581 | miR-1908-5 | cis | 1.68E-08 | Nightingale | PUFA_by_N | NA | A | G | -0.16913 | 0.004135 | 0 |
| rs174581 | miR-1908-5 | cis | 1.68E-08 | Nightingale | L_HDL_FC_ | NA | A | G | -0.11082 | 0.00401 | 8.20E-183 |
| rs174581 | miR-1908-5 | cis | 1.68E-08 | Nightingale | XXL_VLDL_ | NA | A | G | 0.073417 | 0.004169 | 5.00E-72 |
| rs174581 | miR-1908-5 | cis | 1.68E-08 | Nightingale | XS_VLDL_P | NA | A | G | 0.058817 | 0.004212 | 2.10E-45 |
| rs174581 | miR-1908-5 | cis | 1.68E-08 | Nightingale | L_VLDL_FC | NA | A | G | 0.028517 | 0.004276 | 2.00E-11 |
| rs174581 | miR-1908-5 | cis | 1.68E-08 | Nightingale | IDL_TG_pc | NA | A | G | 0.088328 | 0.00417 | 2.40E-103 |
| rs174581 | miR-1908-5 | cis | 1.68E-08 | Nightingale | M_VLDL_C | NA | A | G | -0.05231 | 0.004278 | 5.50E-36 |
| rs174581 | miR-1908-5 | cis | 1.68E-08 | Nightingale | S_VLDL_TG | NA | A | G | 0.039401 | 0.004189 | 1.00E-21 |
| rs174581 | miR-1908-5 | cis | 1.68E-08 | Nightingale | M_HDL_TG | NA | A | G | 0.050495 | 0.004263 | 5.00E-33 |
| rs174581 | miR-1908-5 | cis | 1.68E-08 | Nightingale | XL_HDL_PL | NA | A | G | -0.06592 | 0.003966 | 6.00E-67 |
| rs174581 | miR-1908-5 | cis | 1.68E-08 | Nightingale | M_LDL_CE | NA | A | G | -0.0263 | 0.004323 | 5.30E-10 |
| rs174581 | miR-1908-5 | cis | 1.68E-08 | Nightingale | LDL_PL | NA | A | G | -0.04607 | 0.004307 | 6.70E-28 |
| rs174581 | miR-1908-5 | cis | 1.68E-08 | Nightingale | XS_VLDL_C | NA | A | G | -0.06729 | 0.00419 | 3.70E-62 |
| rs174581 | miR-1908-5 | cis | 1.68E-08 | Nightingale | L_HDL_C_p | NA | A | G | -0.06826 | 0.004069 | 1.20E-67 |
| rs174581 | miR-1908-5 | cis | 1.68E-08 | Nightingale | XXL_VLDL_ | NA | A | G | 0.066509 | 0.004158 | 4.10E-60 |
| rs174581 | miR-1908-5 | cis | 1.68E-08 | Nightingale | XS_VLDL_C | NA | A | G | -0.07115 | 0.004165 | 5.60E-70 |
| rs174581 | miR-1908-5 | cis | 1.68E-08 | Nightingale | M_LDL_P | NA | A | G | -0.04622 | 0.004329 | 1.90E-27 |
| rs174581 | miR-1908-5 | cis | 1.68E-08 | Nightingale | S_VLDL_PL | NA | A | G | -0.07867 | 0.00413 | 1.30E-84 |
| rs174581 | miR-1908-5 | cis | 1.68E-08 | Nightingale | Total_PL | NA | A | G | -0.05089 | 0.00411 | 1.90E-37 |
| rs174581 | miR-1908-5 | cis | 1.68E-08 | Nightingale | XXL_VLDL_ | NA | A | G | 0.066122 | 0.004152 | 8.50E-60 |
| rs174581 | miR-1908-5 | cis | 1.68E-08 | Nightingale | S_LDL_P | NA | A | G | -0.03195 | 0.004321 | 2.90E-14 |
| rs174581 | miR-1908-5 | cis | 1.68E-08 | Nightingale | XL_VLDL_F | NA | A | G | 0.043564 | 0.004181 | 2.30E-26 |
| rs174581 | miR-1908-5 | cis | 1.68E-08 | Nightingale | M_LDL_PL | NA | A | G | -0.02361 | 0.004323 | 2.20E-08 |
| rs174581 | miR-1908-5 | cis | 1.68E-08 | Nightingale | S_HDL_TG_ | NA | A | G | 0.069662 | 0.004106 | 1.60E-67 |
| rs174581 | miR-1908-5 | cis | 1.68E-08 | Nightingale | M_VLDL_T | NA | A | G | 0.031547 | 0.004194 | 1.80E-14 |
| rs174581 | miR-1908-5 | cis | 1.68E-08 | Nightingale | M_VLDL_P | NA | A | G | -0.06181 | 0.004142 | 5.60E-54 |
| rs174581 | miR-1908-5 | cis | 1.68E-08 | Nightingale | XS_VLDL_C | NA | A | G | -0.09792 | 0.00409 | 2.40E-132 |
| rs174581 | miR-1908-5 | cis | 1.68E-08 | Nightingale | VLDL_size | NA | A | G | 0.071881 | 0.004024 | 9.30E-76 |
| rs174581 | miR-1908-5 | cis | 1.68E-08 | Nightingale | S_VLDL_TG | NA | A | G | 0.064572 | 0.004177 | 1.60E-55 |
| rs174581 | miR-1908-5 | cis | 1.68E-08 | Nightingale | S_LDL_PL | NA | A | G | -0.05094 | 0.004306 | 4.20E-34 |
| rs174581 | miR-1908-5 | cis | 1.68E-08 | Nightingale | XS_VLDL_T | NA | A | G | 0.097094 | 0.0041 | 7.91E-130 |
| rs174581 | miR-1908-5 | cis | 1.68E-08 | Nightingale | HDL_C | NA | A | G | -0.06503 | 0.003855 | 4.60E-69 |
| rs174581 | miR-1908-5 | cis | 1.68E-08 | Nightingale | XL_HDL_C | NA | A | G | -0.09792 | 0.003892 | 6.50E-152 |

| | | | | | | | | | | | |
|---|---|---|---|---|---|---|---|---|---|---|---|
| rs174581 | miR-1908-! | cis | 1.68E-08 | Nightingale | Total_FA | NA | A | G | -0.0232 | 0.004283 | 2.10E-08 |
| rs174581 | miR-1908-! | cis | 1.68E-08 | Nightingale | M_LDL_TG | NA | A | G | 0.076197 | 0.004262 | 8.40E-73 |
| rs174581 | miR-1908-! | cis | 1.68E-08 | Nightingale | S_LDL_TG | NA | A | G | 0.050825 | 0.004211 | 3.50E-34 |
| rs174581 | miR-1908-! | cis | 1.68E-08 | Nightingale | XL_VLDL_C | NA | A | G | -0.06072 | 0.004138 | 3.70E-50 |
| rs174584 | miR-1908-! | cis | 1.39E-08 | Nightingale | L_HDL_FC | NA | A | G | -0.08365 | 0.003781 | 2.30E-118 |
| rs174584 | miR-1908-! | cis | 1.39E-08 | Nightingale | M_HDL_FC | NA | A | G | -0.10205 | 0.003823 | 8.71E-172 |
| rs174584 | miR-1908-! | cis | 1.39E-08 | Nightingale | VLDL_TG | NA | A | G | 0.047585 | 0.004151 | 1.70E-31 |
| rs174584 | miR-1908-! | cis | 1.39E-08 | Nightingale | S_HDL_TG_ | NA | A | G | 0.069151 | 0.004103 | 1.20E-66 |
| rs174584 | miR-1908-! | cis | 1.39E-08 | Nightingale | XL_HDL_CE | NA | A | G | -0.02667 | 0.004311 | 1.50E-09 |
| rs174584 | miR-1908-! | cis | 1.39E-08 | Nightingale | XS_VLDL_P | NA | A | G | 0.058084 | 0.004209 | 2.50E-44 |
| rs174584 | miR-1908-! | cis | 1.39E-08 | Nightingale | XL_HDL_FC | NA | A | G | -0.1007 | 0.004019 | 1.30E-149 |
| rs174584 | miR-1908-! | cis | 1.39E-08 | Nightingale | S_LDL_PL | NA | A | G | -0.05068 | 0.004304 | 8.60E-34 |
| rs174584 | miR-1908-! | cis | 1.39E-08 | Nightingale | XS_VLDL_C | NA | A | G | -0.0708 | 0.004163 | 3.10E-69 |
| rs174584 | miR-1908-! | cis | 1.39E-08 | Nightingale | XXL_VLDL_ | NA | A | G | 0.071726 | 0.00414 | 2.50E-70 |
| rs174584 | miR-1908-! | cis | 1.39E-08 | Nightingale | M_LDL_PL | NA | A | G | -0.02333 | 0.004321 | 3.10E-08 |
| rs174584 | miR-1908-! | cis | 1.39E-08 | Nightingale | M_HDL_CE | NA | A | G | -0.03794 | 0.003984 | 5.80E-23 |
| rs174584 | miR-1908-! | cis | 1.39E-08 | Nightingale | L_VLDL_FC | NA | A | G | 0.028107 | 0.004273 | 3.10E-11 |
| rs174584 | miR-1908-! | cis | 1.39E-08 | Nightingale | S_VLDL_TG | NA | A | G | 0.064355 | 0.004174 | 3.80E-55 |
| rs174584 | miR-1908-! | cis | 1.39E-08 | Nightingale | XS_VLDL_T | NA | A | G | 0.032291 | 0.004248 | 1.00E-13 |
| rs174584 | miR-1908-! | cis | 1.39E-08 | Nightingale | ApoA1 | NA | A | G | -0.04096 | 0.003948 | 2.50E-27 |
| rs174584 | miR-1908-! | cis | 1.39E-08 | Nightingale | Total_PL | NA | A | G | -0.05033 | 0.004108 | 9.40E-37 |
| rs174584 | miR-1908-! | cis | 1.39E-08 | Nightingale | M_VLDL_T | NA | A | G | 0.077282 | 0.004074 | 5.00E-84 |
| rs174584 | miR-1908-! | cis | 1.39E-08 | Nightingale | L_HDL_PL_ | NA | A | G | 0.042673 | 0.004175 | 4.30E-26 |
| rs174584 | miR-1908-! | cis | 1.39E-08 | Nightingale | IDL_TG_pc | NA | A | G | 0.087915 | 0.004168 | 2.00E-102 |
| rs174584 | miR-1908-! | cis | 1.39E-08 | Nightingale | L_VLDL_L | NA | A | G | 0.038044 | 0.004169 | 1.30E-20 |
| rs174584 | miR-1908-! | cis | 1.39E-08 | Nightingale | IDL_FC_pct | NA | A | G | -0.1029 | 0.004279 | 2.60E-129 |
| rs174584 | miR-1908-! | cis | 1.39E-08 | Nightingale | M_HDL_TG | NA | A | G | 0.066832 | 0.004137 | 2.60E-61 |
| rs174584 | miR-1908-! | cis | 1.39E-08 | Nightingale | L_HDL_C_p | NA | A | G | -0.06827 | 0.004066 | 8.20E-68 |
| rs174584 | miR-1908-! | cis | 1.39E-08 | Nightingale | IDL_CE_pct | NA | A | G | -0.03859 | 0.004233 | 6.20E-20 |
| rs174584 | miR-1908-! | cis | 1.39E-08 | Nightingale | XS_VLDL_L | NA | A | G | -0.04039 | 0.00424 | 1.90E-23 |
| rs174584 | miR-1908-! | cis | 1.39E-08 | Nightingale | Clinical_LD | NA | A | G | -0.05473 | 0.004289 | 4.80E-39 |
| rs174584 | miR-1908-! | cis | 1.39E-08 | Nightingale | Total_FC | NA | A | G | -0.05857 | 0.004214 | 1.40E-46 |
| rs174584 | miR-1908-! | cis | 1.39E-08 | Nightingale | DHA | NA | A | G | -0.28199 | 0.004157 | 0 |
| rs174584 | miR-1908-! | cis | 1.39E-08 | Nightingale | S_LDL_FC_ | NA | A | G | -0.0796 | 0.004181 | 1.00E-83 |
| rs174584 | miR-1908-! | cis | 1.39E-08 | Nightingale | Omega_6_ | NA | A | G | 0.023059 | 0.004199 | 2.40E-08 |
| rs174584 | miR-1908-! | cis | 1.39E-08 | Nightingale | L_HDL_TG_ | NA | A | G | 0.078897 | 0.004071 | 1.60E-88 |
| rs174584 | miR-1908-! | cis | 1.39E-08 | Nightingale | Total_P | NA | A | G | -0.04026 | 0.00406 | 6.50E-25 |
| rs174584 | miR-1908-! | cis | 1.39E-08 | Nightingale | L_VLDL_PL | NA | A | G | 0.048522 | 0.004162 | 9.10E-33 |
| rs174584 | miR-1908-! | cis | 1.39E-08 | Nightingale | MUFA_pct | NA | A | G | 0.194123 | 0.004128 | 0 |
| rs174584 | miR-1908-! | cis | 1.39E-08 | Nightingale | S_HDL_TG | NA | A | G | 0.068739 | 0.004183 | 2.70E-63 |
| rs174584 | miR-1908-! | cis | 1.39E-08 | Nightingale | S_VLDL_PL | NA | A | G | -0.07831 | 0.004128 | 7.70E-84 |
| rs174584 | miR-1908-! | cis | 1.39E-08 | Nightingale | VLDL_L | NA | A | G | 0.027437 | 0.00423 | 4.90E-11 |
| rs174584 | miR-1908-! | cis | 1.39E-08 | Nightingale | Omega_3 | NA | A | G | -0.32789 | 0.004228 | 0 |
| rs174584 | miR-1908-! | cis | 1.39E-08 | Nightingale | M_VLDL_C | NA | A | G | -0.05218 | 0.004275 | 8.80E-36 |
| rs174584 | miR-1908-! | cis | 1.39E-08 | Nightingale | XXL_VLDL_ | NA | A | G | 0.07124 | 0.004157 | 1.50E-68 |
| rs174584 | miR-1908-! | cis | 1.39E-08 | Nightingale | M_HDL_FC | NA | A | G | -0.05111 | 0.003926 | 1.10E-41 |
| rs174584 | miR-1908-! | cis | 1.39E-08 | Nightingale | LDL_P | NA | A | G | -0.03962 | 0.004311 | 4.50E-21 |
| rs174584 | miR-1908-! | cis | 1.39E-08 | Nightingale | PUFA | NA | A | G | -0.0944 | 0.004207 | 2.80E-118 |
| rs174584 | miR-1908-! | cis | 1.39E-08 | Nightingale | IDL_CE | NA | A | G | -0.0716 | 0.004164 | 2.10E-70 |
| rs174584 | miR-1908-! | cis | 1.39E-08 | Nightingale | XL_HDL_PL | NA | A | G | -0.06579 | 0.003963 | 1.10E-66 |
| rs174584 | miR-1908-! | cis | 1.39E-08 | Nightingale | XL_HDL_C | NA | A | G | -0.09764 | 0.00389 | 2.90E-151 |
| rs174584 | miR-1908-! | cis | 1.39E-08 | Nightingale | M_VLDL_P | NA | A | G | -0.06172 | 0.00414 | 8.20E-54 |
| rs174584 | miR-1908-! | cis | 1.39E-08 | Nightingale | S_LDL_L | NA | A | G | -0.03199 | 0.004313 | 2.40E-14 |
| rs174584 | miR-1908-! | cis | 1.39E-08 | Nightingale | S_LDL_CE_ | NA | A | G | 0.056073 | 0.004306 | 9.00E-41 |
| rs174584 | miR-1908-! | cis | 1.39E-08 | Nightingale | XS_VLDL_C | NA | A | G | -0.09745 | 0.004088 | 5.30E-131 |
| rs174584 | miR-1908-! | cis | 1.39E-08 | Nightingale | XL_HDL_CE | NA | A | G | -0.09566 | 0.003863 | 1.80E-147 |
| rs174584 | miR-1908-! | cis | 1.39E-08 | Nightingale | IDL_FC | NA | A | G | -0.0887 | 0.004176 | 7.60E-106 |
| rs174584 | miR-1908-! | cis | 1.39E-08 | Nightingale | Omega_6_ | NA | A | G | 0.36112 | 0.004251 | 0 |
| rs174584 | miR-1908-! | cis | 1.39E-08 | Nightingale | XL_HDL_PL | NA | A | G | -0.08751 | 0.003878 | 8.49E-123 |
| rs174584 | miR-1908-! | cis | 1.39E-08 | Nightingale | S_VLDL_TG | NA | A | G | 0.039291 | 0.004186 | 1.50E-21 |
| rs174584 | miR-1908-! | cis | 1.39E-08 | Nightingale | XS_VLDL_P | NA | A | G | -0.04242 | 0.004257 | 2.90E-25 |
| rs174584 | miR-1908-! | cis | 1.39E-08 | Nightingale | Citrate | NA | A | G | -0.03362 | 0.004287 | 6.20E-16 |
| rs174584 | miR-1908-! | cis | 1.39E-08 | Nightingale | L_HDL_C | NA | A | G | -0.07959 | 0.003799 | 3.20E-106 |

| | | | | | | | | | | | |
|---|---|---|---|---|---|---|---|---|---|---|---|
| rs174584 | miR-1908-5 | cis | 1.39E-08 | Nightingale | M_HDL_PL | NA | A | G | -0.02154 | 0.004034 | 2.30E-08 |
| rs174584 | miR-1908-5 | cis | 1.39E-08 | Nightingale | XXL_VLDL_ | NA | A | G | 0.061094 | 0.004163 | 7.20E-51 |
| rs174584 | miR-1908-5 | cis | 1.39E-08 | Nightingale | L_LDL_L | NA | A | G | -0.05358 | 0.004272 | 3.70E-38 |
| rs174584 | miR-1908-5 | cis | 1.39E-08 | Nightingale | L_LDL_C | NA | A | G | -0.05724 | 0.004266 | 2.10E-43 |
| rs174584 | miR-1908-5 | cis | 1.39E-08 | Nightingale | DHA_pct | NA | A | G | -0.26787 | 0.004188 | 0 |
| rs174584 | miR-1908-5 | cis | 1.39E-08 | Nightingale | XL_VLDL_C | NA | A | G | 0.030203 | 0.004246 | 4.60E-13 |
| rs174584 | miR-1908-5 | cis | 1.39E-08 | Nightingale | LDL_TG | NA | A | G | 0.030157 | 0.004266 | 3.20E-12 |
| rs174584 | miR-1908-5 | cis | 1.39E-08 | Nightingale | M_HDL_P | NA | A | G | -0.03424 | 0.003974 | 2.20E-19 |
| rs174584 | miR-1908-5 | cis | 1.39E-08 | Nightingale | S_HDL_C_p | NA | A | G | -0.04281 | 0.004264 | 1.60E-23 |
| rs174584 | miR-1908-5 | cis | 1.39E-08 | Nightingale | XL_VLDL_P | NA | A | G | 0.052487 | 0.004146 | 3.60E-38 |
| rs174584 | miR-1908-5 | cis | 1.39E-08 | Nightingale | HDL_L | NA | A | G | -0.05289 | 0.003874 | 7.60E-46 |
| rs174584 | miR-1908-5 | cis | 1.39E-08 | Nightingale | L_VLDL_C | NA | A | G | 0.028896 | 0.004219 | 3.10E-12 |
| rs174584 | miR-1908-5 | cis | 1.39E-08 | Nightingale | M_HDL_C_ | NA | A | G | -0.06586 | 0.004107 | 6.80E-61 |
| rs174584 | miR-1908-5 | cis | 1.39E-08 | Nightingale | HDL_PL | NA | A | G | -0.04799 | 0.0039 | 1.60E-37 |
| rs174584 | miR-1908-5 | cis | 1.39E-08 | Nightingale | L_HDL_L | NA | A | G | -0.07651 | 0.003785 | 2.60E-99 |
| rs174584 | miR-1908-5 | cis | 1.39E-08 | Nightingale | S_HDL_FC_ | NA | A | G | -0.10044 | 0.004092 | 7.80E-148 |
| rs174584 | miR-1908-5 | cis | 1.39E-08 | Nightingale | S_VLDL_CE | NA | A | G | -0.02902 | 0.004256 | 6.20E-12 |
| rs174584 | miR-1908-5 | cis | 1.39E-08 | Nightingale | L_LDL_CE | NA | A | G | -0.05274 | 0.004276 | 6.30E-37 |
| rs174584 | miR-1908-5 | cis | 1.39E-08 | Nightingale | Omega_3_ | NA | A | G | -0.38095 | 0.004215 | 0 |
| rs174584 | miR-1908-5 | cis | 1.39E-08 | Nightingale | XL_VLDL_C | NA | A | G | 0.037302 | 0.004213 | 1.80E-19 |
| rs174584 | miR-1908-5 | cis | 1.39E-08 | Nightingale | M_VLDL_C | NA | A | G | -0.04348 | 0.004302 | 5.40E-25 |
| rs174584 | miR-1908-5 | cis | 1.39E-08 | Nightingale | S_LDL_C | NA | A | G | -0.03186 | 0.004319 | 3.80E-14 |
| rs174584 | miR-1908-5 | cis | 1.39E-08 | Nightingale | S_VLDL_FC | NA | A | G | -0.08259 | 0.004119 | 3.60E-93 |
| rs174584 | miR-1908-5 | cis | 1.39E-08 | Nightingale | XL_VLDL_C | NA | A | G | -0.06319 | 0.004134 | 3.20E-54 |
| rs174584 | miR-1908-5 | cis | 1.39E-08 | Nightingale | IDL_P | NA | A | G | -0.04474 | 0.004269 | 3.60E-27 |
| rs174584 | miR-1908-5 | cis | 1.39E-08 | Nightingale | M_VLDL_T | NA | A | G | 0.031397 | 0.004191 | 2.80E-14 |
| rs174584 | miR-1908-5 | cis | 1.39E-08 | Nightingale | M_VLDL_C | NA | A | G | -0.07545 | 0.00405 | 1.40E-80 |
| rs174584 | miR-1908-5 | cis | 1.39E-08 | Nightingale | XS_VLDL_F | NA | A | G | -0.11118 | 0.004282 | 1.50E-151 |
| rs174584 | miR-1908-5 | cis | 1.39E-08 | Nightingale | HDL_P | NA | A | G | -0.03564 | 0.004051 | 7.30E-20 |
| rs174584 | miR-1908-5 | cis | 1.39E-08 | Nightingale | XS_VLDL_C | NA | A | G | -0.06696 | 0.004187 | 1.80E-61 |
| rs174584 | miR-1908-5 | cis | 1.39E-08 | Nightingale | L_HDL_P | NA | A | G | -0.07709 | 0.003778 | 3.50E-101 |
| rs174584 | miR-1908-5 | cis | 1.39E-08 | Nightingale | M_LDL_CE | NA | A | G | -0.02609 | 0.00432 | 7.30E-10 |
| rs174584 | miR-1908-5 | cis | 1.39E-08 | Nightingale | XS_VLDL_P | NA | A | G | -0.03321 | 0.004253 | 2.00E-16 |
| rs174584 | miR-1908-5 | cis | 1.39E-08 | Nightingale | XL_VLDL_P | NA | A | G | 0.048033 | 0.004163 | 5.50E-32 |
| rs174584 | miR-1908-5 | cis | 1.39E-08 | Nightingale | M_HDL_CE | NA | A | G | -0.04726 | 0.004183 | 2.00E-30 |
| rs174584 | miR-1908-5 | cis | 1.39E-08 | Nightingale | L_HDL_PL | NA | A | G | -0.07536 | 0.003797 | 1.10E-95 |
| rs174584 | miR-1908-5 | cis | 1.39E-08 | Nightingale | XL_HDL_FC | NA | A | G | 0.060306 | 0.003825 | 3.40E-62 |
| rs174584 | miR-1908-5 | cis | 1.39E-08 | Nightingale | XL_HDL_TC | NA | A | G | 0.086604 | 0.004064 | 2.30E-106 |
| rs174584 | miR-1908-5 | cis | 1.39E-08 | Nightingale | M_VLDL_F | NA | A | G | -0.09027 | 0.004102 | 4.30E-113 |
| rs174584 | miR-1908-5 | cis | 1.39E-08 | Nightingale | PUFA_by_I | NA | A | G | -0.16791 | 0.004132 | 0 |
| rs174584 | miR-1908-5 | cis | 1.39E-08 | Nightingale | IDL_PL | NA | A | G | -0.07183 | 0.004162 | 1.10E-71 |
| rs174584 | miR-1908-5 | cis | 1.39E-08 | Nightingale | XXL_VLDL_ | NA | A | G | 0.065745 | 0.004149 | 3.40E-59 |
| rs174584 | miR-1908-5 | cis | 1.39E-08 | Nightingale | XS_VLDL_F | NA | A | G | -0.05474 | 0.004239 | 7.30E-41 |
| rs174584 | miR-1908-5 | cis | 1.39E-08 | Nightingale | SFA_pct | NA | A | G | -0.04147 | 0.004256 | 2.00E-22 |
| rs174584 | miR-1908-5 | cis | 1.39E-08 | Nightingale | M_LDL_L | NA | A | G | -0.02788 | 0.004321 | 4.50E-11 |
| rs174584 | miR-1908-5 | cis | 1.39E-08 | Nightingale | XL_VLDL_C | NA | A | G | -0.06031 | 0.004136 | 1.40E-49 |
| rs174584 | miR-1908-5 | cis | 1.39E-08 | Nightingale | L_VLDL_CE | NA | A | G | -0.05845 | 0.004128 | 2.10E-48 |
| rs174584 | miR-1908-5 | cis | 1.39E-08 | Nightingale | ApoB | NA | A | G | -0.03786 | 0.004309 | 1.70E-19 |
| rs174584 | miR-1908-5 | cis | 1.39E-08 | Nightingale | S_LDL_PL_ | NA | A | G | -0.0607 | 0.004305 | 2.30E-47 |
| rs174584 | miR-1908-5 | cis | 1.39E-08 | Nightingale | M_LDL_FC_ | NA | A | G | -0.07123 | 0.004133 | 3.20E-69 |
| rs174584 | miR-1908-5 | cis | 1.39E-08 | Nightingale | M_HDL_C | NA | A | G | -0.0408 | 0.003969 | 1.70E-26 |
| rs174584 | miR-1908-5 | cis | 1.39E-08 | Nightingale | LA_pct | NA | A | G | 0.18065 | 0.004216 | 0 |
| rs174584 | miR-1908-5 | cis | 1.39E-08 | Nightingale | M_LDL_PL_ | NA | A | G | 0.045076 | 0.004311 | 1.10E-26 |
| rs174584 | miR-1908-5 | cis | 1.39E-08 | Nightingale | S_LDL_FC | NA | A | G | -0.06241 | 0.004307 | 2.80E-50 |
| rs174584 | miR-1908-5 | cis | 1.39E-08 | Nightingale | Total_CE | NA | A | G | -0.06633 | 0.004161 | 1.90E-60 |
| rs174584 | miR-1908-5 | cis | 1.39E-08 | Nightingale | Sphingomy | NA | A | G | -0.11353 | 0.004031 | 6.40E-184 |
| rs174584 | miR-1908-5 | cis | 1.39E-08 | Nightingale | XL_HDL_C_ | NA | A | G | 0.022876 | 0.004121 | 1.50E-09 |
| rs174584 | miR-1908-5 | cis | 1.39E-08 | Nightingale | LDL_C | NA | A | G | -0.04929 | 0.004296 | 5.40E-32 |
| rs174584 | miR-1908-5 | cis | 1.39E-08 | Nightingale | HDL_TG | NA | A | G | 0.046518 | 0.004254 | 3.30E-28 |
| rs174584 | miR-1908-5 | cis | 1.39E-08 | Nightingale | M_VLDL_F | NA | A | G | -0.0289 | 0.004313 | 6.70E-12 |
| rs174584 | miR-1908-5 | cis | 1.39E-08 | Nightingale | IDL_C_pct | NA | A | G | -0.0747 | 0.004212 | 3.60E-72 |
| rs174584 | miR-1908-5 | cis | 1.39E-08 | Nightingale | L_LDL_TG_ | NA | A | G | 0.077624 | 0.00423 | 8.80E-77 |
| rs174584 | miR-1908-5 | cis | 1.39E-08 | Nightingale | Total_L | NA | A | G | -0.0367 | 0.004244 | 3.50E-19 |

| | | | | | | | | | | | |
|---|---|---|---|---|---|---|---|---|---|---|---|
| rs174584 | miR-1908-! | cis | 1.39E-08 | Nightingale | LDL_FC | NA | A | G | -0.06379 | 0.004268 | 2.90E-53 |
| rs174584 | miR-1908-! | cis | 1.39E-08 | Nightingale | M_VLDL_C | NA | A | G | -0.08005 | 0.004061 | 3.60E-90 |
| rs174584 | miR-1908-! | cis | 1.39E-08 | Nightingale | XL_HDL_L | NA | A | G | -0.09142 | 0.003877 | 4.00E-134 |
| rs174584 | miR-1908-! | cis | 1.39E-08 | Nightingale | HDL_size | NA | A | G | -0.07182 | 0.003823 | 1.90E-85 |
| rs174584 | miR-1908-! | cis | 1.39E-08 | Nightingale | L_VLDL_C_ | NA | A | G | -0.03837 | 0.004214 | 3.70E-21 |
| rs174584 | miR-1908-! | cis | 1.39E-08 | Nightingale | IDL_C | NA | A | G | -0.07651 | 0.004166 | 5.20E-80 |
| rs174584 | miR-1908-! | cis | 1.39E-08 | Nightingale | LDL_CE | NA | A | G | -0.04313 | 0.004303 | 8.40E-25 |
| rs174584 | miR-1908-! | cis | 1.39E-08 | Nightingale | XL_HDL_P | NA | A | G | -0.08791 | 0.003882 | 8.79E-125 |
| rs174584 | miR-1908-! | cis | 1.39E-08 | Nightingale | Remnant_( | NA | A | G | -0.04348 | 0.004286 | 1.20E-25 |
| rs174584 | miR-1908-! | cis | 1.39E-08 | Nightingale | L_VLDL_PL | NA | A | G | 0.087144 | 0.004187 | 1.50E-99 |
| rs174584 | miR-1908-! | cis | 1.39E-08 | Nightingale | M_HDL_PL | NA | A | G | 0.05818 | 0.004092 | 6.60E-48 |
| rs174584 | miR-1908-! | cis | 1.39E-08 | Nightingale | XXL_VLDL_ | NA | A | G | 0.066213 | 0.004155 | 1.20E-59 |
| rs174584 | miR-1908-! | cis | 1.39E-08 | Nightingale | L_LDL_P | NA | A | G | -0.03681 | 0.004299 | 1.10E-18 |
| rs174584 | miR-1908-! | cis | 1.39E-08 | Nightingale | L_LDL_C_p | NA | A | G | -0.07688 | 0.004203 | 5.60E-77 |
| rs174584 | miR-1908-! | cis | 1.39E-08 | Nightingale | MUFA | NA | A | G | 0.063152 | 0.00427 | 1.50E-50 |
| rs174584 | miR-1908-! | cis | 1.39E-08 | Nightingale | S_LDL_TG_ | NA | A | G | 0.086002 | 0.00418 | 5.00E-97 |
| rs174584 | miR-1908-! | cis | 1.39E-08 | Nightingale | XL_VLDL_F | NA | A | G | 0.043227 | 0.004179 | 5.30E-26 |
| rs174584 | miR-1908-! | cis | 1.39E-08 | Nightingale | non_HDL_( | NA | A | G | -0.04724 | 0.004291 | 1.30E-29 |
| rs174584 | miR-1908-! | cis | 1.39E-08 | Nightingale | Total_TG | NA | A | G | 0.046774 | 0.004176 | 4.00E-30 |
| rs174584 | miR-1908-! | cis | 1.39E-08 | Nightingale | M_LDL_C_ | NA | A | G | -0.09964 | 0.004317 | 1.30E-118 |
| rs174584 | miR-1908-! | cis | 1.39E-08 | Nightingale | XL_VLDL_L | NA | A | G | 0.051106 | 0.004151 | 3.20E-36 |
| rs174584 | miR-1908-! | cis | 1.39E-08 | Nightingale | XL_VLDL_T | NA | A | G | 0.05646 | 0.004126 | 2.60E-44 |
| rs174584 | miR-1908-! | cis | 1.39E-08 | Nightingale | M_LDL_FC | NA | A | G | -0.05205 | 0.004309 | 2.40E-35 |
| rs174584 | miR-1908-! | cis | 1.39E-08 | Nightingale | Unsaturati | NA | A | G | -0.33247 | 0.004086 | 0 |
| rs174584 | miR-1908-! | cis | 1.39E-08 | Nightingale | TG_by_PG | NA | A | G | 0.068587 | 0.004003 | 3.30E-69 |
| rs174584 | miR-1908-! | cis | 1.39E-08 | Nightingale | VLDL_size | NA | A | G | 0.07137 | 0.004021 | 1.30E-74 |
| rs174584 | miR-1908-! | cis | 1.39E-08 | Nightingale | Phosphogl | NA | A | G | -0.0404 | 0.004086 | 7.50E-25 |
| rs174584 | miR-1908-! | cis | 1.39E-08 | Nightingale | XL_VLDL_T | NA | A | G | 0.058916 | 0.004179 | 1.20E-46 |
| rs174584 | miR-1908-! | cis | 1.39E-08 | Nightingale | HDL_FC | NA | A | G | -0.07305 | 0.003822 | 1.60E-88 |
| rs174584 | miR-1908-! | cis | 1.39E-08 | Nightingale | L_VLDL_P | NA | A | G | 0.041741 | 0.004165 | 1.50E-24 |
| rs174584 | miR-1908-! | cis | 1.39E-08 | Nightingale | IDL_L | NA | A | G | -0.07153 | 0.004171 | 1.80E-70 |
| rs174584 | miR-1908-! | cis | 1.39E-08 | Nightingale | LDL_PL | NA | A | G | -0.0457 | 0.004304 | 1.60E-27 |
| rs174584 | miR-1908-! | cis | 1.39E-08 | Nightingale | L_LDL_FC_ | NA | A | G | -0.08243 | 0.004112 | 1.00E-92 |
| rs174584 | miR-1908-! | cis | 1.39E-08 | Nightingale | XXL_VLDL_ | NA | A | G | 0.072989 | 0.004166 | 2.80E-71 |
| rs174584 | miR-1908-! | cis | 1.39E-08 | Nightingale | LDL_L | NA | A | G | -0.04494 | 0.0043 | 7.10E-27 |
| rs174584 | miR-1908-! | cis | 1.39E-08 | Nightingale | L_HDL_FC_ | NA | A | G | -0.11064 | 0.004008 | 1.90E-182 |
| rs174584 | miR-1908-! | cis | 1.39E-08 | Nightingale | L_VLDL_TG | NA | A | G | 0.037093 | 0.004151 | 7.80E-20 |
| rs174584 | miR-1908-! | cis | 1.39E-08 | Nightingale | LA | NA | A | G | 0.081787 | 0.00423 | 1.70E-83 |
| rs174584 | miR-1908-! | cis | 1.39E-08 | Nightingale | M_LDL_C | NA | A | G | -0.03376 | 0.004322 | 1.60E-15 |
| rs174584 | miR-1908-! | cis | 1.39E-08 | Nightingale | HDL_CE | NA | A | G | -0.0612 | 0.003872 | 7.60E-61 |
| rs174584 | miR-1908-! | cis | 1.39E-08 | Nightingale | PUFA_pct | NA | A | G | -0.1195 | 0.004149 | 4.70E-184 |
| rs174584 | miR-1908-! | cis | 1.39E-08 | Nightingale | Cholines | NA | A | G | -0.03675 | 0.004057 | 2.10E-21 |
| rs174584 | miR-1908-! | cis | 1.39E-08 | Nightingale | L_HDL_CE_ | NA | A | G | -0.0503 | 0.004128 | 2.50E-36 |
| rs174584 | miR-1908-! | cis | 1.39E-08 | Nightingale | L_VLDL_FC | NA | A | G | 0.039976 | 0.004173 | 1.20E-22 |
| rs174584 | miR-1908-! | cis | 1.39E-08 | Nightingale | M_LDL_TG | NA | A | G | 0.075822 | 0.00426 | 3.90E-72 |
| rs174584 | miR-1908-! | cis | 1.39E-08 | Nightingale | M_HDL_L | NA | A | G | -0.02768 | 0.004005 | 4.90E-13 |
| rs174584 | miR-1908-! | cis | 1.39E-08 | Nightingale | Total_C | NA | A | G | -0.06448 | 0.004176 | 8.90E-57 |
| rs174584 | miR-1908-! | cis | 1.39E-08 | Nightingale | L_LDL_FC | NA | A | G | -0.06776 | 0.004236 | 5.40E-61 |
| rs174584 | miR-1908-! | cis | 1.39E-08 | Nightingale | SFA | NA | A | G | -0.03046 | 0.004275 | 3.60E-13 |
| rs174584 | miR-1908-! | cis | 1.39E-08 | Nightingale | XS_VLDL_C | NA | A | G | -0.09169 | 0.004067 | 1.30E-117 |
| rs174584 | miR-1908-! | cis | 1.39E-08 | Nightingale | XXL_VLDL_ | NA | A | G | 0.067641 | 0.004156 | 2.10E-62 |
| rs174584 | miR-1908-! | cis | 1.39E-08 | Nightingale | M_LDL_P | NA | A | G | -0.04602 | 0.004327 | 3.50E-27 |
| rs174584 | miR-1908-! | cis | 1.39E-08 | Nightingale | XS_VLDL_T | NA | A | G | 0.096759 | 0.004098 | 9.79E-129 |
| rs174584 | miR-1908-! | cis | 1.39E-08 | Nightingale | M_HDL_TG | NA | A | G | 0.050433 | 0.004261 | 4.80E-33 |
| rs174584 | miR-1908-! | cis | 1.39E-08 | Nightingale | S_VLDL_FC | NA | A | G | -0.0277 | 0.004315 | 4.70E-11 |
| rs174584 | miR-1908-! | cis | 1.39E-08 | Nightingale | HDL_C | NA | A | G | -0.06434 | 0.003852 | 5.80E-68 |
| rs174584 | miR-1908-! | cis | 1.39E-08 | Nightingale | S_LDL_P | NA | A | G | -0.03187 | 0.004318 | 3.60E-14 |
| rs174584 | miR-1908-! | cis | 1.39E-08 | Nightingale | Total_FA | NA | A | G | -0.02289 | 0.00428 | 2.90E-08 |
| rs174584 | miR-1908-! | cis | 1.39E-08 | Nightingale | S_LDL_TG | NA | A | G | 0.050511 | 0.004208 | 1.00E-33 |
| rs174584 | miR-1908-! | cis | 1.39E-08 | Nightingale | XXL_VLDL_ | NA | A | G | 0.044191 | 0.004338 | 3.00E-25 |
| rs174584 | miR-1908-! | cis | 1.39E-08 | Nightingale | XL_VLDL_F | NA | A | G | -0.06701 | 0.00418 | 1.00E-60 |
| rs174584 | miR-1908-! | cis | 1.39E-08 | Nightingale | L_LDL_PL | NA | A | G | -0.05446 | 0.004282 | 8.10E-39 |
| rs174584 | miR-1908-! | cis | 1.39E-08 | Nightingale | XXL_VLDL_ | NA | A | G | -0.02657 | 0.004293 | 1.00E-10 |

| | | | | | | | | | | | |
|---|---|---|---|---|---|---|---|---|---|---|---|
| rs174584 | miR-1908-5 | cis | 1.39E-08 | Nightingale | S_VLDL_C_ | NA | A | G | -0.0565 | 0.004197 | 2.50E-42 |
| rs174584 | miR-1908-5 | cis | 1.39E-08 | Nightingale | M_LDL_TG | NA | A | G | 0.036968 | 0.00425 | 3.80E-18 |
| rs174584 | miR-1908-5 | cis | 1.39E-08 | Nightingale | L_HDL_CE | NA | A | G | -0.07809 | 0.003809 | 1.20E-101 |
| rs174584 | miR-1908-5 | cis | 1.39E-08 | Nightingale | Tyr | NA | A | G | 0.032771 | 0.004309 | 2.80E-14 |
| rs174592 | miR-1908-5 | cis | 3.81E-09 | Nightingale | M_HDL_FC | NA | G | A | -0.09447 | 0.00379 | 2.40E-149 |
| rs174592 | miR-1908-5 | cis | 3.81E-09 | Nightingale | LDL_C | NA | G | A | -0.04651 | 0.004259 | 6.20E-29 |
| rs174592 | miR-1908-5 | cis | 3.81E-09 | Nightingale | Omega_3 | NA | G | A | -0.31025 | 0.004192 | 0 |
| rs174592 | miR-1908-5 | cis | 3.81E-09 | Nightingale | L_LDL_C_p | NA | G | A | -0.07156 | 0.004168 | 4.00E-68 |
| rs174592 | miR-1908-5 | cis | 3.81E-09 | Nightingale | Omega_6_ | NA | G | A | 0.342031 | 0.004216 | 0 |
| rs174592 | miR-1908-5 | cis | 3.81E-09 | Nightingale | XL_VLDL_C | NA | G | A | -0.05882 | 0.004099 | 3.70E-48 |
| rs174592 | miR-1908-5 | cis | 3.81E-09 | Nightingale | Unsaturati | NA | G | A | -0.31395 | 0.004051 | 0 |
| rs174592 | miR-1908-5 | cis | 3.81E-09 | Nightingale | IDL_CE_pct | NA | G | A | -0.03443 | 0.004197 | 1.10E-16 |
| rs174592 | miR-1908-5 | cis | 3.81E-09 | Nightingale | XS_VLDL_F | NA | G | A | -0.10459 | 0.004246 | 2.80E-137 |
| rs174592 | miR-1908-5 | cis | 3.81E-09 | Nightingale | HDL_P | NA | G | A | -0.03205 | 0.004017 | 1.70E-16 |
| rs174592 | miR-1908-5 | cis | 3.81E-09 | Nightingale | Total_P | NA | G | A | -0.03651 | 0.004026 | 6.40E-21 |
| rs174592 | miR-1908-5 | cis | 3.81E-09 | Nightingale | XL_HDL_L | NA | G | A | -0.08495 | 0.003844 | 1.20E-117 |
| rs174592 | miR-1908-5 | cis | 3.81E-09 | Nightingale | IDL_C | NA | G | A | -0.07134 | 0.00413 | 1.30E-70 |
| rs174592 | miR-1908-5 | cis | 3.81E-09 | Nightingale | M_VLDL_F | NA | G | A | -0.02795 | 0.004277 | 2.50E-11 |
| rs174592 | miR-1908-5 | cis | 3.81E-09 | Nightingale | XS_VLDL_T | NA | G | A | 0.089683 | 0.004063 | 2.20E-113 |
| rs174592 | miR-1908-5 | cis | 3.81E-09 | Nightingale | S_LDL_C | NA | G | A | -0.03034 | 0.004282 | 4.70E-13 |
| rs174592 | miR-1908-5 | cis | 3.81E-09 | Nightingale | XL_VLDL_T | NA | G | A | 0.054893 | 0.004144 | 1.80E-41 |
| rs174592 | miR-1908-5 | cis | 3.81E-09 | Nightingale | M_HDL_L | NA | G | A | -0.02451 | 0.003971 | 1.50E-10 |
| rs174592 | miR-1908-5 | cis | 3.81E-09 | Nightingale | S_HDL_TG_ | NA | G | A | 0.063116 | 0.004068 | 4.30E-57 |
| rs174592 | miR-1908-5 | cis | 3.81E-09 | Nightingale | XS_VLDL_F | NA | G | A | -0.05205 | 0.004203 | 4.60E-37 |
| rs174592 | miR-1908-5 | cis | 3.81E-09 | Nightingale | S_VLDL_TG | NA | G | A | 0.059682 | 0.004139 | 1.30E-48 |
| rs174592 | miR-1908-5 | cis | 3.81E-09 | Nightingale | XXL_VLDL_ | NA | G | A | 0.056948 | 0.004128 | 4.40E-45 |
| rs174592 | miR-1908-5 | cis | 3.81E-09 | Nightingale | TG_by_PG | NA | G | A | 0.062685 | 0.00397 | 3.10E-59 |
| rs174592 | miR-1908-5 | cis | 3.81E-09 | Nightingale | S_LDL_FC_ | NA | G | A | -0.0743 | 0.004145 | 1.70E-74 |
| rs174592 | miR-1908-5 | cis | 3.81E-09 | Nightingale | L_HDL_CE_ | NA | G | A | -0.04544 | 0.004093 | 8.40E-31 |
| rs174592 | miR-1908-5 | cis | 3.81E-09 | Nightingale | M_HDL_PL | NA | G | A | 0.053231 | 0.004058 | 5.50E-41 |
| rs174592 | miR-1908-5 | cis | 3.81E-09 | Nightingale | VLDL_TG | NA | G | A | 0.04337 | 0.004116 | 8.20E-27 |
| rs174592 | miR-1908-5 | cis | 3.81E-09 | Nightingale | L_VLDL_P | NA | G | A | 0.037813 | 0.00413 | 7.90E-21 |
| rs174592 | miR-1908-5 | cis | 3.81E-09 | Nightingale | M_LDL_PL_ | NA | G | A | 0.044926 | 0.004275 | 6.50E-27 |
| rs174592 | miR-1908-5 | cis | 3.81E-09 | Nightingale | M_VLDL_C | NA | G | A | -0.04128 | 0.004266 | 5.40E-23 |
| rs174592 | miR-1908-5 | cis | 3.81E-09 | Nightingale | M_LDL_CE | NA | G | A | -0.02542 | 0.004284 | 1.80E-09 |
| rs174592 | miR-1908-5 | cis | 3.81E-09 | Nightingale | IDL_P | NA | G | A | -0.04162 | 0.004233 | 5.20E-24 |
| rs174592 | miR-1908-5 | cis | 3.81E-09 | Nightingale | M_VLDL_C | NA | G | A | -0.04916 | 0.004239 | 2.20E-32 |
| rs174592 | miR-1908-5 | cis | 3.81E-09 | Nightingale | HDL_size | NA | G | A | -0.06629 | 0.00379 | 7.60E-74 |
| rs174592 | miR-1908-5 | cis | 3.81E-09 | Nightingale | L_VLDL_PL | NA | G | A | 0.044267 | 0.004127 | 4.00E-28 |
| rs174592 | miR-1908-5 | cis | 3.81E-09 | Nightingale | XL_VLDL_F | NA | G | A | -0.06185 | 0.004145 | 6.20E-53 |
| rs174592 | miR-1908-5 | cis | 3.81E-09 | Nightingale | S_HDL_C_p | NA | G | A | -0.03965 | 0.004228 | 7.40E-21 |
| rs174592 | miR-1908-5 | cis | 3.81E-09 | Nightingale | ApoB | NA | G | A | -0.036 | 0.004272 | 6.10E-18 |
| rs174592 | miR-1908-5 | cis | 3.81E-09 | Nightingale | M_LDL_TG | NA | G | A | 0.071083 | 0.004224 | 3.10E-65 |
| rs174592 | miR-1908-5 | cis | 3.81E-09 | Nightingale | SFA_pct | NA | G | A | -0.03984 | 0.00422 | 3.70E-21 |
| rs174592 | miR-1908-5 | cis | 3.81E-09 | Nightingale | XS_VLDL_C | NA | G | A | -0.06665 | 0.004127 | 6.50E-62 |
| rs174592 | miR-1908-5 | cis | 3.81E-09 | Nightingale | M_LDL_FC_ | NA | G | A | -0.06546 | 0.004098 | 1.00E-59 |
| rs174592 | miR-1908-5 | cis | 3.81E-09 | Nightingale | XL_VLDL_C | NA | G | A | 0.033907 | 0.004178 | 1.10E-16 |
| rs174592 | miR-1908-5 | cis | 3.81E-09 | Nightingale | Total_CE | NA | G | A | -0.06171 | 0.004126 | 7.50E-53 |
| rs174592 | miR-1908-5 | cis | 3.81E-09 | Nightingale | S_HDL_FC_ | NA | G | A | -0.09434 | 0.004057 | 4.80E-132 |
| rs174592 | miR-1908-5 | cis | 3.81E-09 | Nightingale | M_HDL_C_ | NA | G | A | -0.06013 | 0.004072 | 5.40E-52 |
| rs174592 | miR-1908-5 | cis | 3.81E-09 | Nightingale | XL_HDL_P | NA | G | A | -0.08154 | 0.003849 | 8.00E-109 |
| rs174592 | miR-1908-5 | cis | 3.81E-09 | Nightingale | HDL_L | NA | G | A | -0.04807 | 0.003841 | 1.80E-38 |
| rs174592 | miR-1908-5 | cis | 3.81E-09 | Nightingale | XL_HDL_TG | NA | G | A | 0.080224 | 0.00403 | 2.00E-93 |
| rs174592 | miR-1908-5 | cis | 3.81E-09 | Nightingale | HDL_PL | NA | G | A | -0.04352 | 0.003867 | 2.20E-31 |
| rs174592 | miR-1908-5 | cis | 3.81E-09 | Nightingale | S_VLDL_FC | NA | G | A | -0.07648 | 0.004085 | 4.60E-82 |
| rs174592 | miR-1908-5 | cis | 3.81E-09 | Nightingale | L_HDL_C_p | NA | G | A | -0.06257 | 0.004032 | 7.70E-59 |
| rs174592 | miR-1908-5 | cis | 3.81E-09 | Nightingale | L_HDL_FC | NA | G | A | -0.0774 | 0.003749 | 8.09E-103 |
| rs174592 | miR-1908-5 | cis | 3.81E-09 | Nightingale | L_HDL_PL_ | NA | G | A | 0.039146 | 0.00414 | 1.60E-22 |
| rs174592 | miR-1908-5 | cis | 3.81E-09 | Nightingale | S_LDL_PL | NA | G | A | -0.04788 | 0.004267 | 9.90E-31 |
| rs174592 | miR-1908-5 | cis | 3.81E-09 | Nightingale | M_VLDL_P | NA | G | A | -0.057 | 0.004105 | 5.20E-47 |
| rs174592 | miR-1908-5 | cis | 3.81E-09 | Nightingale | ApoA1 | NA | G | A | -0.03689 | 0.003915 | 1.30E-22 |
| rs174592 | miR-1908-5 | cis | 3.81E-09 | Nightingale | XL_VLDL_C | NA | G | A | -0.05635 | 0.004101 | 3.00E-44 |
| rs174592 | miR-1908-5 | cis | 3.81E-09 | Nightingale | XS_VLDL_C | NA | G | A | -0.09028 | 0.004053 | 2.40E-115 |

| | | | | | | | | | | | |
|---|---|---|---|---|---|---|---|---|---|---|---|
| rs174592 | miR-1908-5 | cis | 3.81E-09 | Nightingale | Clinical_LD | NA | G | A | -0.05146 | 0.004253 | 4.10E-35 |
| rs174592 | miR-1908-5 | cis | 3.81E-09 | Nightingale | L_VLDL_CE | NA | G | A | -0.05435 | 0.004093 | 1.10E-42 |
| rs174592 | miR-1908-5 | cis | 3.81E-09 | Nightingale | XXL_VLDL_ | NA | G | A | 0.039478 | 0.004301 | 6.90E-21 |
| rs174592 | miR-1908-5 | cis | 3.81E-09 | Nightingale | Total_PL | NA | G | A | -0.04649 | 0.004073 | 8.40E-32 |
| rs174592 | miR-1908-5 | cis | 3.81E-09 | Nightingale | L_VLDL_C_ | NA | G | A | -0.03597 | 0.004179 | 7.20E-19 |
| rs174592 | miR-1908-5 | cis | 3.81E-09 | Nightingale | L_VLDL_L | NA | G | A | 0.034299 | 0.004134 | 2.30E-17 |
| rs174592 | miR-1908-5 | cis | 3.81E-09 | Nightingale | Total_L | NA | G | A | -0.0342 | 0.004208 | 6.40E-17 |
| rs174592 | miR-1908-5 | cis | 3.81E-09 | Nightingale | L_HDL_C | NA | G | A | -0.07351 | 0.003767 | 3.20E-92 |
| rs174592 | miR-1908-5 | cis | 3.81E-09 | Nightingale | L_HDL_FC_ | NA | G | A | -0.10371 | 0.003974 | 1.20E-162 |
| rs174592 | miR-1908-5 | cis | 3.81E-09 | Nightingale | M_HDL_P | NA | G | A | -0.03057 | 0.003941 | 7.90E-16 |
| rs174592 | miR-1908-5 | cis | 3.81E-09 | Nightingale | S_VLDL_PL | NA | G | A | -0.07242 | 0.004093 | 1.00E-73 |
| rs174592 | miR-1908-5 | cis | 3.81E-09 | Nightingale | Omega_6_ | NA | G | A | 0.022972 | 0.004163 | 2.10E-08 |
| rs174592 | miR-1908-5 | cis | 3.81E-09 | Nightingale | S_LDL_TG_ | NA | G | A | 0.08011 | 0.004145 | 3.10E-86 |
| rs174592 | miR-1908-5 | cis | 3.81E-09 | Nightingale | XL_HDL_CE | NA | G | A | -0.02395 | 0.004275 | 3.60E-08 |
| rs174592 | miR-1908-5 | cis | 3.81E-09 | Nightingale | LDL_TG | NA | G | A | 0.027833 | 0.00423 | 4.40E-11 |
| rs174592 | miR-1908-5 | cis | 3.81E-09 | Nightingale | XL_HDL_FC | NA | G | A | -0.09358 | 0.003985 | 1.10E-131 |
| rs174592 | miR-1908-5 | cis | 3.81E-09 | Nightingale | M_LDL_C_ | NA | G | A | -0.09517 | 0.004281 | 2.00E-110 |
| rs174592 | miR-1908-5 | cis | 3.81E-09 | Nightingale | non_HDL_( | NA | G | A | -0.04456 | 0.004255 | 9.40E-27 |
| rs174592 | miR-1908-5 | cis | 3.81E-09 | Nightingale | XXL_VLDL_ | NA | G | A | 0.066663 | 0.004105 | 5.40E-62 |
| rs174592 | miR-1908-5 | cis | 3.81E-09 | Nightingale | XL_HDL_CE | NA | G | A | -0.0888 | 0.00383 | 2.50E-129 |
| rs174592 | miR-1908-5 | cis | 3.81E-09 | Nightingale | VLDL_size | NA | G | A | 0.065901 | 0.003987 | 8.80E-65 |
| rs174592 | miR-1908-5 | cis | 3.81E-09 | Nightingale | S_LDL_PL_ | NA | G | A | -0.05643 | 0.004269 | 4.10E-42 |
| rs174592 | miR-1908-5 | cis | 3.81E-09 | Nightingale | L_VLDL_C | NA | G | A | 0.025826 | 0.004184 | 2.70E-10 |
| rs174592 | miR-1908-5 | cis | 3.81E-09 | Nightingale | XS_VLDL_P | NA | G | A | 0.053821 | 0.004174 | 7.70E-39 |
| rs174592 | miR-1908-5 | cis | 3.81E-09 | Nightingale | XS_VLDL_C | NA | G | A | -0.06318 | 0.004152 | 4.20E-55 |
| rs174592 | miR-1908-5 | cis | 3.81E-09 | Nightingale | LDL_L | NA | G | A | -0.04245 | 0.004264 | 2.30E-24 |
| rs174592 | miR-1908-5 | cis | 3.81E-09 | Nightingale | L_HDL_P | NA | G | A | -0.07119 | 0.003746 | 1.40E-87 |
| rs174592 | miR-1908-5 | cis | 3.81E-09 | Nightingale | L_HDL_PL | NA | G | A | -0.06957 | 0.003765 | 9.50E-83 |
| rs174592 | miR-1908-5 | cis | 3.81E-09 | Nightingale | XL_VLDL_L | NA | G | A | 0.046742 | 0.004116 | 4.20E-31 |
| rs174592 | miR-1908-5 | cis | 3.81E-09 | Nightingale | HDL_FC | NA | G | A | -0.06715 | 0.003789 | 7.90E-76 |
| rs174592 | miR-1908-5 | cis | 3.81E-09 | Nightingale | IDL_C_pct | NA | G | A | -0.06861 | 0.004176 | 9.90E-63 |
| rs174592 | miR-1908-5 | cis | 3.81E-09 | Nightingale | XL_VLDL_P | NA | G | A | 0.043948 | 0.004128 | 1.40E-27 |
| rs174592 | miR-1908-5 | cis | 3.81E-09 | Nightingale | XS_VLDL_L | NA | G | A | -0.03856 | 0.004205 | 2.70E-21 |
| rs174592 | miR-1908-5 | cis | 3.81E-09 | Nightingale | Total_C | NA | G | A | -0.06006 | 0.004141 | 9.20E-50 |
| rs174592 | miR-1908-5 | cis | 3.81E-09 | Nightingale | XL_HDL_PL | NA | G | A | -0.08137 | 0.003845 | 8.89E-108 |
| rs174592 | miR-1908-5 | cis | 3.81E-09 | Nightingale | IDL_CE | NA | G | A | -0.06661 | 0.004129 | 7.10E-62 |
| rs174592 | miR-1908-5 | cis | 3.81E-09 | Nightingale | L_VLDL_FC | NA | G | A | 0.036183 | 0.004138 | 3.10E-19 |
| rs174592 | miR-1908-5 | cis | 3.81E-09 | Nightingale | LDL_FC | NA | G | A | -0.05977 | 0.004232 | 1.60E-47 |
| rs174592 | miR-1908-5 | cis | 3.81E-09 | Nightingale | HDL_TG | NA | G | A | 0.042812 | 0.004218 | 8.00E-25 |
| rs174592 | miR-1908-5 | cis | 3.81E-09 | Nightingale | XL_HDL_FC | NA | G | A | 0.055786 | 0.003793 | 4.70E-54 |
| rs174592 | miR-1908-5 | cis | 3.81E-09 | Nightingale | L_LDL_PL | NA | G | A | -0.05124 | 0.004246 | 6.90E-35 |
| rs174592 | miR-1908-5 | cis | 3.81E-09 | Nightingale | L_LDL_C | NA | G | A | -0.05376 | 0.00423 | 8.20E-39 |
| rs174592 | miR-1908-5 | cis | 3.81E-09 | Nightingale | XL_VLDL_C | NA | G | A | 0.027277 | 0.00421 | 3.70E-11 |
| rs174592 | miR-1908-5 | cis | 3.81E-09 | Nightingale | M_LDL_P | NA | G | A | -0.04477 | 0.00429 | 3.60E-26 |
| rs174592 | miR-1908-5 | cis | 3.81E-09 | Nightingale | VLDL_L | NA | G | A | 0.02465 | 0.004194 | 2.20E-09 |
| rs174592 | miR-1908-5 | cis | 3.81E-09 | Nightingale | SFA | NA | G | A | -0.0288 | 0.004239 | 5.20E-12 |
| rs174592 | miR-1908-5 | cis | 3.81E-09 | Nightingale | S_VLDL_TG | NA | G | A | 0.035297 | 0.004151 | 5.30E-18 |
| rs174592 | miR-1908-5 | cis | 3.81E-09 | Nightingale | XL_HDL_PL | NA | G | A | -0.06076 | 0.00393 | 9.40E-58 |
| rs174592 | miR-1908-5 | cis | 3.81E-09 | Nightingale | XS_VLDL_T | NA | G | A | 0.02907 | 0.004212 | 9.50E-12 |
| rs174592 | miR-1908-5 | cis | 3.81E-09 | Nightingale | L_LDL_FC | NA | G | A | -0.06331 | 0.0042 | 4.20E-54 |
| rs174592 | miR-1908-5 | cis | 3.81E-09 | Nightingale | XXL_VLDL_ | NA | G | A | 0.061014 | 0.004114 | 4.50E-52 |
| rs174592 | miR-1908-5 | cis | 3.81E-09 | Nightingale | S_LDL_TG | NA | G | A | 0.046602 | 0.004173 | 1.30E-29 |
| rs174592 | miR-1908-5 | cis | 3.81E-09 | Nightingale | S_LDL_CE_ | NA | G | A | 0.052094 | 0.00427 | 3.70E-36 |
| rs174592 | miR-1908-5 | cis | 3.81E-09 | Nightingale | XXL_VLDL_ | NA | G | A | 0.062703 | 0.004121 | 9.70E-55 |
| rs174592 | miR-1908-5 | cis | 3.81E-09 | Nightingale | Remnant_( | NA | G | A | -0.04098 | 0.00425 | 3.70E-23 |
| rs174592 | miR-1908-5 | cis | 3.81E-09 | Nightingale | M_LDL_FC | NA | G | A | -0.04911 | 0.004273 | 5.70E-32 |
| rs174592 | miR-1908-5 | cis | 3.81E-09 | Nightingale | L_LDL_TG_ | NA | G | A | 0.072358 | 0.004194 | 1.30E-68 |
| rs174592 | miR-1908-5 | cis | 3.81E-09 | Nightingale | HDL_C | NA | G | A | -0.05874 | 0.00382 | 1.20E-57 |
| rs174592 | miR-1908-5 | cis | 3.81E-09 | Nightingale | S_LDL_L | NA | G | A | -0.03046 | 0.004277 | 3.20E-13 |
| rs174592 | miR-1908-5 | cis | 3.81E-09 | Nightingale | MUFA_pct | NA | G | A | 0.182382 | 0.004093 | 0 |
| rs174592 | miR-1908-5 | cis | 3.81E-09 | Nightingale | IDL_L | NA | G | A | -0.06683 | 0.004135 | 3.00E-62 |
| rs174592 | miR-1908-5 | cis | 3.81E-09 | Nightingale | XL_HDL_C_ | NA | G | A | 0.021331 | 0.004086 | 2.20E-08 |
| rs174592 | miR-1908-5 | cis | 3.81E-09 | Nightingale | L_HDL_TG_ | NA | G | A | 0.072846 | 0.004037 | 2.00E-77 |

| | | | | | | | | | | | |
|---|---|---|---|---|---|---|---|---|---|---|---|
| rs174592 | miR-1908-5 | cis | 3.81E-09 | Nightingale | L_VLDL_PL | NA | G | A | 0.081278 | 0.004152 | 5.00E-89 |
| rs174592 | miR-1908-5 | cis | 3.81E-09 | Nightingale | PUFA_pct | NA | G | A | -0.11185 | 0.004114 | 5.20E-164 |
| rs174592 | miR-1908-5 | cis | 3.81E-09 | Nightingale | M_HDL_C | NA | G | A | -0.03667 | 0.003936 | 7.50E-22 |
| rs174592 | miR-1908-5 | cis | 3.81E-09 | Nightingale | L_LDL_L | NA | G | A | -0.05036 | 0.004236 | 3.30E-34 |
| rs174592 | miR-1908-5 | cis | 3.81E-09 | Nightingale | XL_VLDL_T | NA | G | A | 0.05174 | 0.004091 | 4.70E-38 |
| rs174592 | miR-1908-5 | cis | 3.81E-09 | Nightingale | IDL_FC | NA | G | A | -0.08314 | 0.00414 | 2.30E-94 |
| rs174592 | miR-1908-5 | cis | 3.81E-09 | Nightingale | M_HDL_TG | NA | G | A | 0.046517 | 0.004225 | 4.90E-29 |
| rs174592 | miR-1908-5 | cis | 3.81E-09 | Nightingale | L_VLDL_TG | NA | G | A | 0.03338 | 0.004116 | 1.40E-16 |
| rs174592 | miR-1908-5 | cis | 3.81E-09 | Nightingale | S_VLDL_CE | NA | G | A | -0.0272 | 0.00422 | 1.10E-10 |
| rs174592 | miR-1908-5 | cis | 3.81E-09 | Nightingale | M_LDL_TG | NA | G | A | 0.034096 | 0.004215 | 3.00E-16 |
| rs174592 | miR-1908-5 | cis | 3.81E-09 | Nightingale | L_LDL_P | NA | G | A | -0.03466 | 0.004263 | 6.30E-17 |
| rs174592 | miR-1908-5 | cis | 3.81E-09 | Nightingale | L_HDL_L | NA | G | A | -0.07064 | 0.003753 | 5.90E-86 |
| rs174592 | miR-1908-5 | cis | 3.81E-09 | Nightingale | M_VLDL_C | NA | G | A | -0.074 | 0.004026 | 5.20E-79 |
| rs174592 | miR-1908-5 | cis | 3.81E-09 | Nightingale | XXL_VLDL_ | NA | G | A | -0.02519 | 0.004257 | 9.30E-10 |
| rs174592 | miR-1908-5 | cis | 3.81E-09 | Nightingale | LA_pct | NA | G | A | 0.172569 | 0.00418 | 0 |
| rs174592 | miR-1908-5 | cis | 3.81E-09 | Nightingale | IDL_PL | NA | G | A | -0.06738 | 0.004127 | 1.10E-63 |
| rs174592 | miR-1908-5 | cis | 3.81E-09 | Nightingale | DHA | NA | G | A | -0.26604 | 0.004122 | 0 |
| rs174592 | miR-1908-5 | cis | 3.81E-09 | Nightingale | IDL_FC_pct | NA | G | A | -0.09715 | 0.004243 | 6.30E-118 |
| rs174592 | miR-1908-5 | cis | 3.81E-09 | Nightingale | HDL_CE | NA | G | A | -0.05574 | 0.003839 | 1.90E-51 |
| rs174592 | miR-1908-5 | cis | 3.81E-09 | Nightingale | Omega_3_ | NA | G | A | -0.36059 | 0.00418 | 0 |
| rs174592 | miR-1908-5 | cis | 3.81E-09 | Nightingale | L_HDL_CE | NA | G | A | -0.07209 | 0.003777 | 2.90E-88 |
| rs174592 | miR-1908-5 | cis | 3.81E-09 | Nightingale | XXL_VLDL_ | NA | G | A | 0.061656 | 0.00412 | 9.30E-53 |
| rs174592 | miR-1908-5 | cis | 3.81E-09 | Nightingale | M_LDL_C | NA | G | A | -0.03245 | 0.004286 | 1.50E-14 |
| rs174592 | miR-1908-5 | cis | 3.81E-09 | Nightingale | M_HDL_CE | NA | G | A | -0.03396 | 0.00395 | 8.10E-19 |
| rs174592 | miR-1908-5 | cis | 3.81E-09 | Nightingale | Sphingomy | NA | G | A | -0.10617 | 0.003997 | 8.49E-163 |
| rs174592 | miR-1908-5 | cis | 3.81E-09 | Nightingale | M_VLDL_T | NA | G | A | 0.071424 | 0.00404 | 1.50E-73 |
| rs174592 | miR-1908-5 | cis | 3.81E-09 | Nightingale | XS_VLDL_P | NA | G | A | -0.04054 | 0.004221 | 4.70E-23 |
| rs174592 | miR-1908-5 | cis | 3.81E-09 | Nightingale | LDL_P | NA | G | A | -0.03778 | 0.004275 | 1.60E-19 |
| rs174592 | miR-1908-5 | cis | 3.81E-09 | Nightingale | Citrate | NA | G | A | -0.03097 | 0.00425 | 3.50E-14 |
| rs174592 | miR-1908-5 | cis | 3.81E-09 | Nightingale | Total_TG | NA | G | A | 0.042715 | 0.004141 | 8.50E-26 |
| rs174592 | miR-1908-5 | cis | 3.81E-09 | Nightingale | XS_VLDL_C | NA | G | A | -0.08473 | 0.004032 | 4.50E-103 |
| rs174592 | miR-1908-5 | cis | 3.81E-09 | Nightingale | S_VLDL_FC | NA | G | A | -0.02693 | 0.004278 | 1.50E-10 |
| rs174592 | miR-1908-5 | cis | 3.81E-09 | Nightingale | XL_VLDL_P | NA | G | A | 0.048052 | 0.004111 | 7.10E-33 |
| rs174592 | miR-1908-5 | cis | 3.81E-09 | Nightingale | M_HDL_TG | NA | G | A | 0.061074 | 0.004102 | 1.30E-52 |
| rs174592 | miR-1908-5 | cis | 3.81E-09 | Nightingale | M_LDL_L | NA | G | A | -0.02687 | 0.004284 | 2.10E-10 |
| rs174592 | miR-1908-5 | cis | 3.81E-09 | Nightingale | LDL_CE | NA | G | A | -0.04086 | 0.004267 | 1.40E-22 |
| rs174592 | miR-1908-5 | cis | 3.81E-09 | Nightingale | XL_VLDL_F | NA | G | A | 0.039487 | 0.004143 | 2.10E-22 |
| rs174592 | miR-1908-5 | cis | 3.81E-09 | Nightingale | MUFA | NA | G | A | 0.059325 | 0.004234 | 9.20E-46 |
| rs174592 | miR-1908-5 | cis | 3.81E-09 | Nightingale | XL_HDL_C | NA | G | A | -0.09064 | 0.003857 | 9.51E-133 |
| rs174592 | miR-1908-5 | cis | 3.81E-09 | Nightingale | M_HDL_CE | NA | G | A | -0.04275 | 0.004148 | 1.10E-25 |
| rs174592 | miR-1908-5 | cis | 3.81E-09 | Nightingale | PUFA_by_M | NA | G | A | -0.15755 | 0.004097 | 0 |
| rs174592 | miR-1908-5 | cis | 3.81E-09 | Nightingale | M_HDL_FC | NA | G | A | -0.04651 | 0.003893 | 4.50E-35 |
| rs174592 | miR-1908-5 | cis | 3.81E-09 | Nightingale | M_VLDL_C | NA | G | A | -0.06956 | 0.004016 | 2.70E-70 |
| rs174592 | miR-1908-5 | cis | 3.81E-09 | Nightingale | S_HDL_TG | NA | G | A | 0.063145 | 0.004148 | 9.70E-55 |
| rs174592 | miR-1908-5 | cis | 3.81E-09 | Nightingale | XS_VLDL_P | NA | G | A | -0.03187 | 0.004218 | 5.80E-15 |
| rs174592 | miR-1908-5 | cis | 3.81E-09 | Nightingale | XXL_VLDL_ | NA | G | A | 0.067806 | 0.004131 | 8.00E-63 |
| rs174592 | miR-1908-5 | cis | 3.81E-09 | Nightingale | PUFA | NA | G | A | -0.0886 | 0.004172 | 1.50E-105 |
| rs174592 | miR-1908-5 | cis | 3.81E-09 | Nightingale | S_LDL_FC | NA | G | A | -0.05874 | 0.004271 | 2.30E-45 |
| rs174592 | miR-1908-5 | cis | 3.81E-09 | Nightingale | L_LDL_FC_ | NA | G | A | -0.07548 | 0.004077 | 6.30E-80 |
| rs174592 | miR-1908-5 | cis | 3.81E-09 | Nightingale | Phosphogly | NA | G | A | -0.03688 | 0.004052 | 4.80E-21 |
| rs174592 | miR-1908-5 | cis | 3.81E-09 | Nightingale | LDL_PL | NA | G | A | -0.04314 | 0.004268 | 6.00E-25 |
| rs174592 | miR-1908-5 | cis | 3.81E-09 | Nightingale | LA | NA | G | A | 0.078408 | 0.004195 | 7.90E-79 |
| rs174592 | miR-1908-5 | cis | 3.81E-09 | Nightingale | Cholines | NA | G | A | -0.03326 | 0.004023 | 7.70E-18 |
| rs174592 | miR-1908-5 | cis | 3.81E-09 | Nightingale | L_LDL_CE | NA | G | A | -0.04964 | 0.00424 | 3.50E-33 |
| rs174592 | miR-1908-5 | cis | 3.81E-09 | Nightingale | Total_FC | NA | G | A | -0.05473 | 0.004179 | 4.30E-41 |
| rs174592 | miR-1908-5 | cis | 3.81E-09 | Nightingale | XXL_VLDL_ | NA | G | A | 0.06621 | 0.004122 | 1.90E-60 |
| rs174592 | miR-1908-5 | cis | 3.81E-09 | Nightingale | L_VLDL_FC | NA | G | A | 0.026001 | 0.004237 | 4.00E-10 |
| rs174592 | miR-1908-5 | cis | 3.81E-09 | Nightingale | Tyr | NA | G | A | 0.029117 | 0.004272 | 9.40E-12 |
| rs174592 | miR-1908-5 | cis | 3.81E-09 | Nightingale | S_VLDL_C_ | NA | G | A | -0.05249 | 0.004162 | 2.00E-37 |
| rs174592 | miR-1908-5 | cis | 3.81E-09 | Nightingale | S_LDL_P | NA | G | A | -0.0307 | 0.004282 | 2.30E-13 |
| rs174592 | miR-1908-5 | cis | 3.81E-09 | Nightingale | DHA_pct | NA | G | A | -0.25265 | 0.004152 | 0 |
| rs174592 | miR-1908-5 | cis | 3.81E-09 | Nightingale | M_VLDL_F | NA | G | A | -0.08401 | 0.004067 | 5.20E-100 |
| rs174592 | miR-1908-5 | cis | 3.81E-09 | Nightingale | M_VLDL_T | NA | G | A | 0.028036 | 0.004156 | 7.10E-12 |

| | | | | | | | | | | | |
|---|---|---|---|---|---|---|---|---|---|---|---|
| rs174592 | miR-1908-5 | cis | 3.81E-09 | Nightingale | IDL_TG_pct | NA | G | A | 0.081375 | 0.004132 | 2.90E-90 |
| rs174601 | miR-1908-5 | cis | 5.98E-09 | Nightingale | M_LDL_L | NA | T | C | -0.02718 | 0.004271 | 8.50E-11 |
| rs174601 | miR-1908-5 | cis | 5.98E-09 | Nightingale | L_LDL_C | NA | T | C | -0.05378 | 0.004216 | 2.40E-39 |
| rs174601 | miR-1908-5 | cis | 5.98E-09 | Nightingale | DHA | NA | T | C | -0.26115 | 0.004109 | 0 |
| rs174601 | miR-1908-5 | cis | 5.98E-09 | Nightingale | S_HDL_TG | NA | T | C | 0.062725 | 0.004135 | 2.20E-54 |
| rs174601 | miR-1908-5 | cis | 5.98E-09 | Nightingale | XXL_VLDL_ | NA | T | C | 0.040581 | 0.004288 | 4.40E-22 |
| rs174601 | miR-1908-5 | cis | 5.98E-09 | Nightingale | M_HDL_TG | NA | T | C | 0.060927 | 0.004089 | 1.20E-52 |
| rs174601 | miR-1908-5 | cis | 5.98E-09 | Nightingale | S_VLDL_FC | NA | T | C | -0.07605 | 0.004072 | 1.00E-81 |
| rs174601 | miR-1908-5 | cis | 5.98E-09 | Nightingale | XL_VLDL_C | NA | T | C | 0.026586 | 0.004197 | 1.00E-10 |
| rs174601 | miR-1908-5 | cis | 5.98E-09 | Nightingale | XL_HDL_C | NA | T | C | -0.0895 | 0.003845 | 5.20E-130 |
| rs174601 | miR-1908-5 | cis | 5.98E-09 | Nightingale | Cholines | NA | T | C | -0.03334 | 0.00401 | 5.30E-18 |
| rs174601 | miR-1908-5 | cis | 5.98E-09 | Nightingale | Total_PL | NA | T | C | -0.04633 | 0.004061 | 8.40E-32 |
| rs174601 | miR-1908-5 | cis | 5.98E-09 | Nightingale | L_HDL_C | NA | T | C | -0.07259 | 0.003755 | 4.70E-90 |
| rs174601 | miR-1908-5 | cis | 5.98E-09 | Nightingale | L_VLDL_FC | NA | T | C | 0.025724 | 0.004224 | 5.40E-10 |
| rs174601 | miR-1908-5 | cis | 5.98E-09 | Nightingale | L_HDL_FC | NA | T | C | -0.07637 | 0.003737 | 3.30E-100 |
| rs174601 | miR-1908-5 | cis | 5.98E-09 | Nightingale | L_LDL_FC_ | NA | T | C | -0.07475 | 0.004064 | 3.80E-79 |
| rs174601 | miR-1908-5 | cis | 5.98E-09 | Nightingale | S_HDL_TG_ | NA | T | C | 0.062882 | 0.004056 | 6.70E-57 |
| rs174601 | miR-1908-5 | cis | 5.98E-09 | Nightingale | HDL_size | NA | T | C | -0.06517 | 0.003778 | 2.50E-71 |
| rs174601 | miR-1908-5 | cis | 5.98E-09 | Nightingale | XL_VLDL_F | NA | T | C | 0.038765 | 0.00413 | 8.40E-22 |
| rs174601 | miR-1908-5 | cis | 5.98E-09 | Nightingale | M_LDL_C | NA | T | C | -0.03264 | 0.004273 | 6.30E-15 |
| rs174601 | miR-1908-5 | cis | 5.98E-09 | Nightingale | VLDL_TG | NA | T | C | 0.042539 | 0.004104 | 5.10E-26 |
| rs174601 | miR-1908-5 | cis | 5.98E-09 | Nightingale | XL_HDL_CE | NA | T | C | -0.02478 | 0.004262 | 8.70E-09 |
| rs174601 | miR-1908-5 | cis | 5.98E-09 | Nightingale | XL_VLDL_L | NA | T | C | 0.045868 | 0.004103 | 3.30E-30 |
| rs174601 | miR-1908-5 | cis | 5.98E-09 | Nightingale | S_LDL_CE_ | NA | T | C | 0.051435 | 0.004256 | 1.40E-35 |
| rs174601 | miR-1908-5 | cis | 5.98E-09 | Nightingale | L_LDL_L | NA | T | C | -0.05039 | 0.004223 | 1.10E-34 |
| rs174601 | miR-1908-5 | cis | 5.98E-09 | Nightingale | HDL_L | NA | T | C | -0.04756 | 0.003829 | 1.60E-37 |
| rs174601 | miR-1908-5 | cis | 5.98E-09 | Nightingale | M_HDL_L | NA | T | C | -0.02426 | 0.003959 | 2.70E-10 |
| rs174601 | miR-1908-5 | cis | 5.98E-09 | Nightingale | LDL_TG | NA | T | C | 0.027217 | 0.004217 | 1.00E-10 |
| rs174601 | miR-1908-5 | cis | 5.98E-09 | Nightingale | M_VLDL_P | NA | T | C | -0.05637 | 0.004092 | 2.30E-46 |
| rs174601 | miR-1908-5 | cis | 5.98E-09 | Nightingale | L_HDL_PL | NA | T | C | -0.06851 | 0.003753 | 3.10E-80 |
| rs174601 | miR-1908-5 | cis | 5.98E-09 | Nightingale | L_LDL_P | NA | T | C | -0.03501 | 0.00425 | 1.70E-17 |
| rs174601 | miR-1908-5 | cis | 5.98E-09 | Nightingale | L_HDL_C_p | NA | T | C | -0.06224 | 0.004019 | 2.10E-58 |
| rs174601 | miR-1908-5 | cis | 5.98E-09 | Nightingale | L_VLDL_C | NA | T | C | 0.025271 | 0.004171 | 5.90E-10 |
| rs174601 | miR-1908-5 | cis | 5.98E-09 | Nightingale | DHA_pct | NA | T | C | -0.24779 | 0.004139 | 0 |
| rs174601 | miR-1908-5 | cis | 5.98E-09 | Nightingale | Citrate | NA | T | C | -0.03126 | 0.004237 | 2.10E-14 |
| rs174601 | miR-1908-5 | cis | 5.98E-09 | Nightingale | M_VLDL_T | NA | T | C | 0.070677 | 0.004027 | 1.30E-72 |
| rs174601 | miR-1908-5 | cis | 5.98E-09 | Nightingale | SFA | NA | T | C | -0.02853 | 0.004226 | 7.50E-12 |
| rs174601 | miR-1908-5 | cis | 5.98E-09 | Nightingale | XXL_VLDL_ | NA | T | C | 0.065747 | 0.004092 | 7.20E-61 |
| rs174601 | miR-1908-5 | cis | 5.98E-09 | Nightingale | Remnant_( | NA | T | C | -0.04077 | 0.004237 | 3.40E-23 |
| rs174601 | miR-1908-5 | cis | 5.98E-09 | Nightingale | S_VLDL_TG | NA | T | C | 0.034917 | 0.004138 | 9.30E-18 |
| rs174601 | miR-1908-5 | cis | 5.98E-09 | Nightingale | XL_HDL_FC | NA | T | C | 0.054935 | 0.003781 | 1.90E-52 |
| rs174601 | miR-1908-5 | cis | 5.98E-09 | Nightingale | Sphingomy | NA | T | C | -0.10515 | 0.003985 | 8.09E-161 |
| rs174601 | miR-1908-5 | cis | 5.98E-09 | Nightingale | IDL_C_pct | NA | T | C | -0.06832 | 0.004163 | 7.10E-63 |
| rs174601 | miR-1908-5 | cis | 5.98E-09 | Nightingale | XS_VLDL_P | NA | T | C | -0.03122 | 0.004205 | 1.70E-14 |
| rs174601 | miR-1908-5 | cis | 5.98E-09 | Nightingale | HDL_C | NA | T | C | -0.05821 | 0.003808 | 1.60E-56 |
| rs174601 | miR-1908-5 | cis | 5.98E-09 | Nightingale | LDL_C | NA | T | C | -0.04664 | 0.004246 | 1.70E-29 |
| rs174601 | miR-1908-5 | cis | 5.98E-09 | Nightingale | XS_VLDL_F | NA | T | C | -0.10331 | 0.004232 | 6.00E-135 |
| rs174601 | miR-1908-5 | cis | 5.98E-09 | Nightingale | S_LDL_P | NA | T | C | -0.03075 | 0.004268 | 1.50E-13 |
| rs174601 | miR-1908-5 | cis | 5.98E-09 | Nightingale | M_HDL_FC | NA | T | C | -0.04602 | 0.00388 | 2.70E-34 |
| rs174601 | miR-1908-5 | cis | 5.98E-09 | Nightingale | HDL_P | NA | T | C | -0.03202 | 0.004004 | 2.00E-16 |
| rs174601 | miR-1908-5 | cis | 5.98E-09 | Nightingale | non_HDL_( | NA | T | C | -0.04451 | 0.004241 | 4.90E-27 |
| rs174601 | miR-1908-5 | cis | 5.98E-09 | Nightingale | L_HDL_L | NA | T | C | -0.06964 | 0.003742 | 1.50E-83 |
| rs174601 | miR-1908-5 | cis | 5.98E-09 | Nightingale | L_VLDL_CE | NA | T | C | -0.05334 | 0.004081 | 1.20E-41 |
| rs174601 | miR-1908-5 | cis | 5.98E-09 | Nightingale | S_LDL_PL | NA | T | C | -0.04826 | 0.004254 | 1.20E-31 |
| rs174601 | miR-1908-5 | cis | 5.98E-09 | Nightingale | L_HDL_CE | NA | T | C | -0.07121 | 0.003765 | 3.00E-86 |
| rs174601 | miR-1908-5 | cis | 5.98E-09 | Nightingale | S_LDL_TG | NA | T | C | 0.045562 | 0.00416 | 1.30E-28 |
| rs174601 | miR-1908-5 | cis | 5.98E-09 | Nightingale | M_LDL_FC | NA | T | C | -0.04923 | 0.004259 | 1.40E-32 |
| rs174601 | miR-1908-5 | cis | 5.98E-09 | Nightingale | XXL_VLDL_ | NA | T | C | -0.0239 | 0.004244 | 5.60E-09 |
| rs174601 | miR-1908-5 | cis | 5.98E-09 | Nightingale | XL_VLDL_C | NA | T | C | -0.05815 | 0.004087 | 1.60E-47 |
| rs174601 | miR-1908-5 | cis | 5.98E-09 | Nightingale | Total_P | NA | T | C | -0.03651 | 0.004013 | 6.40E-21 |
| rs174601 | miR-1908-5 | cis | 5.98E-09 | Nightingale | LDL_CE | NA | T | C | -0.04105 | 0.004253 | 4.30E-23 |
| rs174601 | miR-1908-5 | cis | 5.98E-09 | Nightingale | HDL_TG | NA | T | C | 0.042878 | 0.004205 | 4.00E-25 |
| rs174601 | miR-1908-5 | cis | 5.98E-09 | Nightingale | Total_FC | NA | T | C | -0.05456 | 0.004166 | 3.10E-41 |

| | | | | | | | | | | | |
|---|---|---|---|---|---|---|---|---|---|---|---|
| rs174601 | miR-1908-! | cis | 5.98E-09 | Nightingale | S_VLDL_FC | NA | T | C | -0.02686 | 0.004265 | 1.20E-10 |
| rs174601 | miR-1908-! | cis | 5.98E-09 | Nightingale | L_VLDL_TG | NA | T | C | 0.032565 | 0.004103 | 6.60E-16 |
| rs174601 | miR-1908-! | cis | 5.98E-09 | Nightingale | MUFA_pct | NA | T | C | 0.178541 | 0.004081 | 0 |
| rs174601 | miR-1908-! | cis | 5.98E-09 | Nightingale | M_LDL_TG | NA | T | C | 0.070423 | 0.004211 | 1.10E-64 |
| rs174601 | miR-1908-! | cis | 5.98E-09 | Nightingale | S_HDL_FC_ | NA | T | C | -0.09324 | 0.004045 | 1.20E-129 |
| rs174601 | miR-1908-! | cis | 5.98E-09 | Nightingale | M_HDL_TG | NA | T | C | 0.046639 | 0.004212 | 2.30E-29 |
| rs174601 | miR-1908-! | cis | 5.98E-09 | Nightingale | XXL_VLDL_ | NA | T | C | 0.066739 | 0.004118 | 1.90E-61 |
| rs174601 | miR-1908-! | cis | 5.98E-09 | Nightingale | IDL_FC | NA | T | C | -0.08237 | 0.004127 | 1.70E-93 |
| rs174601 | miR-1908-! | cis | 5.98E-09 | Nightingale | S_VLDL_C_ | NA | T | C | -0.05205 | 0.004149 | 4.00E-37 |
| rs174601 | miR-1908-! | cis | 5.98E-09 | Nightingale | PUFA | NA | T | C | -0.08762 | 0.004159 | 3.90E-104 |
| rs174601 | miR-1908-! | cis | 5.98E-09 | Nightingale | S_LDL_FC | NA | T | C | -0.05882 | 0.004257 | 4.20E-46 |
| rs174601 | miR-1908-! | cis | 5.98E-09 | Nightingale | IDL_P | NA | T | C | -0.04142 | 0.00422 | 4.40E-24 |
| rs174601 | miR-1908-! | cis | 5.98E-09 | Nightingale | XXL_VLDL_ | NA | T | C | 0.065194 | 0.00411 | 3.70E-59 |
| rs174601 | miR-1908-! | cis | 5.98E-09 | Nightingale | L_VLDL_FC | NA | T | C | 0.03544 | 0.004125 | 1.30E-18 |
| rs174601 | miR-1908-! | cis | 5.98E-09 | Nightingale | VLDL_L | NA | T | C | 0.024041 | 0.004181 | 5.10E-09 |
| rs174601 | miR-1908-! | cis | 5.98E-09 | Nightingale | Tyr | NA | T | C | 0.028631 | 0.004259 | 1.80E-11 |
| rs174601 | miR-1908-! | cis | 5.98E-09 | Nightingale | SFA_pct | NA | T | C | -0.03841 | 0.004207 | 6.80E-20 |
| rs174601 | miR-1908-! | cis | 5.98E-09 | Nightingale | L_LDL_C_p | NA | T | C | -0.07145 | 0.004155 | 2.50E-68 |
| rs174601 | miR-1908-! | cis | 5.98E-09 | Nightingale | XL_VLDL_P | NA | T | C | 0.04315 | 0.004115 | 8.40E-27 |
| rs174601 | miR-1908-! | cis | 5.98E-09 | Nightingale | XXL_VLDL_ | NA | T | C | 0.060086 | 0.004102 | 5.30E-51 |
| rs174601 | miR-1908-! | cis | 5.98E-09 | Nightingale | S_VLDL_CE | NA | T | C | -0.02677 | 0.004207 | 1.70E-10 |
| rs174601 | miR-1908-! | cis | 5.98E-09 | Nightingale | LA_pct | NA | T | C | 0.168299 | 0.004167 | 0 |
| rs174601 | miR-1908-! | cis | 5.98E-09 | Nightingale | M_LDL_PL_ | NA | T | C | 0.04298 | 0.004262 | 9.60E-25 |
| rs174601 | miR-1908-! | cis | 5.98E-09 | Nightingale | XS_VLDL_P | NA | T | C | 0.052676 | 0.004161 | 1.60E-37 |
| rs174601 | miR-1908-! | cis | 5.98E-09 | Nightingale | S_LDL_PL_ | NA | T | C | -0.05576 | 0.004256 | 1.60E-41 |
| rs174601 | miR-1908-! | cis | 5.98E-09 | Nightingale | S_LDL_L | NA | T | C | -0.03095 | 0.004264 | 8.20E-14 |
| rs174601 | miR-1908-! | cis | 5.98E-09 | Nightingale | XL_VLDL_P | NA | T | C | 0.047208 | 0.004099 | 5.60E-32 |
| rs174601 | miR-1908-! | cis | 5.98E-09 | Nightingale | XL_VLDL_T | NA | T | C | 0.050896 | 0.004079 | 4.10E-37 |
| rs174601 | miR-1908-! | cis | 5.98E-09 | Nightingale | M_HDL_FC | NA | T | C | -0.09342 | 0.003779 | 2.40E-146 |
| rs174601 | miR-1908-! | cis | 5.98E-09 | Nightingale | L_LDL_CE | NA | T | C | -0.04975 | 0.004227 | 9.60E-34 |
| rs174601 | miR-1908-! | cis | 5.98E-09 | Nightingale | XS_VLDL_P | NA | T | C | -0.03975 | 0.004207 | 2.20E-22 |
| rs174601 | miR-1908-! | cis | 5.98E-09 | Nightingale | XS_VLDL_C | NA | T | C | -0.08376 | 0.00402 | 1.40E-101 |
| rs174601 | miR-1908-! | cis | 5.98E-09 | Nightingale | XL_HDL_PL | NA | T | C | -0.08013 | 0.003833 | 8.61E-105 |
| rs174601 | miR-1908-! | cis | 5.98E-09 | Nightingale | TG_by_PG | NA | T | C | 0.06182 | 0.003957 | 5.30E-58 |
| rs174601 | miR-1908-! | cis | 5.98E-09 | Nightingale | HDL_CE | NA | T | C | -0.05527 | 0.003827 | 1.60E-50 |
| rs174601 | miR-1908-! | cis | 5.98E-09 | Nightingale | L_LDL_PL | NA | T | C | -0.05116 | 0.004232 | 3.10E-35 |
| rs174601 | miR-1908-! | cis | 5.98E-09 | Nightingale | ApoB | NA | T | C | -0.03606 | 0.004259 | 3.00E-18 |
| rs174601 | miR-1908-! | cis | 5.98E-09 | Nightingale | HDL_PL | NA | T | C | -0.043 | 0.003855 | 1.50E-30 |
| rs174601 | miR-1908-! | cis | 5.98E-09 | Nightingale | MUFA | NA | T | C | 0.057671 | 0.004221 | 1.40E-43 |
| rs174601 | miR-1908-! | cis | 5.98E-09 | Nightingale | IDL_PL | NA | T | C | -0.06678 | 0.004114 | 4.20E-63 |
| rs174601 | miR-1908-! | cis | 5.98E-09 | Nightingale | L_VLDL_C_ | NA | T | C | -0.03523 | 0.004166 | 2.30E-18 |
| rs174601 | miR-1908-! | cis | 5.98E-09 | Nightingale | XL_HDL_CE | NA | T | C | -0.08771 | 0.003819 | 9.71E-127 |
| rs174601 | miR-1908-! | cis | 5.98E-09 | Nightingale | L_HDL_CE_ | NA | T | C | -0.0454 | 0.00408 | 7.80E-31 |
| rs174601 | miR-1908-! | cis | 5.98E-09 | Nightingale | M_VLDL_F | NA | T | C | -0.08292 | 0.004055 | 3.40E-98 |
| rs174601 | miR-1908-! | cis | 5.98E-09 | Nightingale | IDL_TG_pc | NA | T | C | 0.080822 | 0.00412 | 7.70E-90 |
| rs174601 | miR-1908-! | cis | 5.98E-09 | Nightingale | M_VLDL_C | NA | T | C | -0.06889 | 0.004003 | 1.80E-69 |
| rs174601 | miR-1908-! | cis | 5.98E-09 | Nightingale | S_LDL_TG_ | NA | T | C | 0.079309 | 0.004132 | 2.30E-85 |
| rs174601 | miR-1908-! | cis | 5.98E-09 | Nightingale | Phosphogl | NA | T | C | -0.03674 | 0.004039 | 5.10E-21 |
| rs174601 | miR-1908-! | cis | 5.98E-09 | Nightingale | M_VLDL_C | NA | T | C | -0.07323 | 0.004014 | 5.60E-78 |
| rs174601 | miR-1908-! | cis | 5.98E-09 | Nightingale | S_HDL_C_p | NA | T | C | -0.03998 | 0.004215 | 2.40E-21 |
| rs174601 | miR-1908-! | cis | 5.98E-09 | Nightingale | M_VLDL_C | NA | T | C | -0.04893 | 0.004226 | 1.90E-32 |
| rs174601 | miR-1908-! | cis | 5.98E-09 | Nightingale | M_LDL_TG | NA | T | C | 0.033275 | 0.004202 | 1.20E-15 |
| rs174601 | miR-1908-! | cis | 5.98E-09 | Nightingale | M_LDL_PL | NA | T | C | -0.02288 | 0.004271 | 3.90E-08 |
| rs174601 | miR-1908-! | cis | 5.98E-09 | Nightingale | S_VLDL_PL | NA | T | C | -0.07215 | 0.00408 | 1.20E-73 |
| rs174601 | miR-1908-! | cis | 5.98E-09 | Nightingale | LDL_PL | NA | T | C | -0.04331 | 0.004255 | 1.70E-25 |
| rs174601 | miR-1908-! | cis | 5.98E-09 | Nightingale | Total_C | NA | T | C | -0.05989 | 0.004128 | 5.70E-50 |
| rs174601 | miR-1908-! | cis | 5.98E-09 | Nightingale | M_LDL_FC_ | NA | T | C | -0.06491 | 0.004085 | 2.30E-59 |
| rs174601 | miR-1908-! | cis | 5.98E-09 | Nightingale | LDL_P | NA | T | C | -0.03792 | 0.004261 | 6.30E-20 |
| rs174601 | miR-1908-! | cis | 5.98E-09 | Nightingale | XXL_VLDL_ | NA | T | C | 0.061826 | 0.004108 | 9.50E-54 |
| rs174601 | miR-1908-! | cis | 5.98E-09 | Nightingale | PUFA_by_I | NA | T | C | -0.15437 | 0.004084 | 1.69825184 |
| rs174601 | miR-1908-! | cis | 5.98E-09 | Nightingale | LDL_L | NA | T | C | -0.04262 | 0.00425 | 7.00E-25 |
| rs174601 | miR-1908-! | cis | 5.98E-09 | Nightingale | M_HDL_PL | NA | T | C | 0.053029 | 0.004045 | 8.00E-41 |
| rs174601 | miR-1908-! | cis | 5.98E-09 | Nightingale | Omega_6_ | NA | T | C | 0.334882 | 0.004202 | 0 |

| | | | | | | | | | | | |
|---|---|---|---|---|---|---|---|---|---|---|---|
| rs174601 | miR-1908-! | cis | 5.98E-09 | Nightingale | XS_VLDL_L | NA | T | C | -0.03777 | 0.004192 | 1.20E-20 |
| rs174601 | miR-1908-! | cis | 5.98E-09 | Nightingale | XS_VLDL_C | NA | T | C | -0.06562 | 0.004115 | 1.60E-60 |
| rs174601 | miR-1908-! | cis | 5.98E-09 | Nightingale | L_VLDL_L | NA | T | C | 0.033544 | 0.004121 | 9.90E-17 |
| rs174601 | miR-1908-! | cis | 5.98E-09 | Nightingale | XL_VLDL_F | NA | T | C | -0.06085 | 0.004132 | 1.40E-51 |
| rs174601 | miR-1908-! | cis | 5.98E-09 | Nightingale | Total_CE | NA | T | C | -0.06154 | 0.004113 | 4.60E-53 |
| rs174601 | miR-1908-! | cis | 5.98E-09 | Nightingale | S_LDL_C | NA | T | C | -0.03079 | 0.004269 | 1.30E-13 |
| rs174601 | miR-1908-! | cis | 5.98E-09 | Nightingale | XS_VLDL_T | NA | T | C | 0.029093 | 0.004199 | 7.20E-12 |
| rs174601 | miR-1908-! | cis | 5.98E-09 | Nightingale | XL_HDL_FC | NA | T | C | -0.09236 | 0.003972 | 4.40E-129 |
| rs174601 | miR-1908-! | cis | 5.98E-09 | Nightingale | XL_VLDL_T | NA | T | C | 0.054261 | 0.004132 | 6.10E-41 |
| rs174601 | miR-1908-! | cis | 5.98E-09 | Nightingale | M_VLDL_T | NA | T | C | 0.027322 | 0.004143 | 2.30E-11 |
| rs174601 | miR-1908-! | cis | 5.98E-09 | Nightingale | M_LDL_P | NA | T | C | -0.04448 | 0.004277 | 4.20E-26 |
| rs174601 | miR-1908-! | cis | 5.98E-09 | Nightingale | Omega_3_ | NA | T | C | -0.35311 | 0.004167 | 0 |
| rs174601 | miR-1908-! | cis | 5.98E-09 | Nightingale | M_VLDL_C | NA | T | C | -0.04114 | 0.004253 | 3.80E-23 |
| rs174601 | miR-1908-! | cis | 5.98E-09 | Nightingale | XXL_VLDL_ | NA | T | C | 0.060727 | 0.004107 | 9.90E-52 |
| rs174601 | miR-1908-! | cis | 5.98E-09 | Nightingale | XS_VLDL_C | NA | T | C | -0.08923 | 0.004041 | 1.20E-113 |
| rs174601 | miR-1908-! | cis | 5.98E-09 | Nightingale | M_HDL_CE | NA | T | C | -0.03371 | 0.003938 | 1.90E-18 |
| rs174601 | miR-1908-! | cis | 5.98E-09 | Nightingale | Total_L | NA | T | C | -0.0343 | 0.004195 | 3.70E-17 |
| rs174601 | miR-1908-! | cis | 5.98E-09 | Nightingale | IDL_C | NA | T | C | -0.07079 | 0.004118 | 3.10E-70 |
| rs174601 | miR-1908-! | cis | 5.98E-09 | Nightingale | XL_HDL_P | NA | T | C | -0.08048 | 0.003838 | 2.20E-106 |
| rs174601 | miR-1908-! | cis | 5.98E-09 | Nightingale | XS_VLDL_F | NA | T | C | -0.0511 | 0.00419 | 4.50E-36 |
| rs174601 | miR-1908-! | cis | 5.98E-09 | Nightingale | XXL_VLDL_ | NA | T | C | 0.055932 | 0.004115 | 5.50E-44 |
| rs174601 | miR-1908-! | cis | 5.98E-09 | Nightingale | XL_HDL_PL | NA | T | C | -0.0595 | 0.003917 | 1.40E-55 |
| rs174601 | miR-1908-! | cis | 5.98E-09 | Nightingale | L_VLDL_P | NA | T | C | 0.03703 | 0.004117 | 4.00E-20 |
| rs174601 | miR-1908-! | cis | 5.98E-09 | Nightingale | M_HDL_CE | NA | T | C | -0.04289 | 0.004135 | 6.00E-26 |
| rs174601 | miR-1908-! | cis | 5.98E-09 | Nightingale | XS_VLDL_T | NA | T | C | 0.088734 | 0.004051 | 6.30E-112 |
| rs174601 | miR-1908-! | cis | 5.98E-09 | Nightingale | M_LDL_C_ | NA | T | C | -0.09331 | 0.004268 | 6.70E-107 |
| rs174601 | miR-1908-! | cis | 5.98E-09 | Nightingale | Unsaturati | NA | T | C | -0.30756 | 0.004038 | 0 |
| rs174601 | miR-1908-! | cis | 5.98E-09 | Nightingale | PUFA_pct | NA | T | C | -0.10979 | 0.004102 | 2.90E-159 |
| rs174601 | miR-1908-! | cis | 5.98E-09 | Nightingale | M_HDL_C | NA | T | C | -0.03638 | 0.003924 | 2.10E-21 |
| rs174601 | miR-1908-! | cis | 5.98E-09 | Nightingale | Omega_3 | NA | T | C | -0.30411 | 0.004179 | 0 |
| rs174601 | miR-1908-! | cis | 5.98E-09 | Nightingale | HDL_FC | NA | T | C | -0.06644 | 0.003778 | 3.70E-74 |
| rs174601 | miR-1908-! | cis | 5.98E-09 | Nightingale | Clinical_LD | NA | T | C | -0.05145 | 0.00424 | 1.40E-35 |
| rs174601 | miR-1908-! | cis | 5.98E-09 | Nightingale | L_HDL_PL_ | NA | T | C | 0.039073 | 0.004127 | 1.70E-22 |
| rs174601 | miR-1908-! | cis | 5.98E-09 | Nightingale | IDL_FC_pct | NA | T | C | -0.09597 | 0.00423 | 6.19E-116 |
| rs174601 | miR-1908-! | cis | 5.98E-09 | Nightingale | LA | NA | T | C | 0.075795 | 0.004182 | 3.60E-74 |
| rs174601 | miR-1908-! | cis | 5.98E-09 | Nightingale | LDL_FC | NA | T | C | -0.0597 | 0.004219 | 5.10E-48 |
| rs174601 | miR-1908-! | cis | 5.98E-09 | Nightingale | M_LDL_CE | NA | T | C | -0.02563 | 0.004271 | 9.20E-10 |
| rs174601 | miR-1908-! | cis | 5.98E-09 | Nightingale | M_HDL_C_ | NA | T | C | -0.05998 | 0.004059 | 5.60E-52 |
| rs174601 | miR-1908-! | cis | 5.98E-09 | Nightingale | L_HDL_FC_ | NA | T | C | -0.10263 | 0.003962 | 1.40E-160 |
| rs174601 | miR-1908-! | cis | 5.98E-09 | Nightingale | L_LDL_TG_ | NA | T | C | 0.071919 | 0.004181 | 2.00E-68 |
| rs174601 | miR-1908-! | cis | 5.98E-09 | Nightingale | M_HDL_P | NA | T | C | -0.03024 | 0.003928 | 2.00E-15 |
| rs174601 | miR-1908-! | cis | 5.98E-09 | Nightingale | XL_HDL_TC | NA | T | C | 0.079147 | 0.004017 | 1.80E-91 |
| rs174601 | miR-1908-! | cis | 5.98E-09 | Nightingale | S_VLDL_TG | NA | T | C | 0.059289 | 0.004126 | 2.40E-48 |
| rs174601 | miR-1908-! | cis | 5.98E-09 | Nightingale | Total_TG | NA | T | C | 0.041941 | 0.004128 | 4.20E-25 |
| rs174601 | miR-1908-! | cis | 5.98E-09 | Nightingale | S_LDL_FC_ | NA | T | C | -0.07348 | 0.004132 | 1.10E-73 |
| rs174601 | miR-1908-! | cis | 5.98E-09 | Nightingale | IDL_CE | NA | T | C | -0.06614 | 0.004116 | 1.20E-61 |
| rs174601 | miR-1908-! | cis | 5.98E-09 | Nightingale | IDL_L | NA | T | C | -0.0663 | 0.004122 | 6.80E-62 |
| rs174601 | miR-1908-! | cis | 5.98E-09 | Nightingale | L_HDL_P | NA | T | C | -0.07024 | 0.003735 | 2.80E-85 |
| rs174601 | miR-1908-! | cis | 5.98E-09 | Nightingale | XL_VLDL_C | NA | T | C | -0.05581 | 0.004089 | 7.80E-44 |
| rs174601 | miR-1908-! | cis | 5.98E-09 | Nightingale | XL_VLDL_C | NA | T | C | 0.033171 | 0.004165 | 3.80E-16 |
| rs174601 | miR-1908-! | cis | 5.98E-09 | Nightingale | XL_HDL_L | NA | T | C | -0.08374 | 0.003832 | 1.00E-114 |
| rs174601 | miR-1908-! | cis | 5.98E-09 | Nightingale | VLDL_size | NA | T | C | 0.064467 | 0.003975 | 2.00E-62 |
| rs174601 | miR-1908-! | cis | 5.98E-09 | Nightingale | IDL_CE_pct | NA | T | C | -0.03468 | 0.004184 | 3.80E-17 |
| rs174601 | miR-1908-! | cis | 5.98E-09 | Nightingale | L_LDL_FC | NA | T | C | -0.06312 | 0.004187 | 1.90E-54 |
| rs174601 | miR-1908-! | cis | 5.98E-09 | Nightingale | ApoA1 | NA | T | C | -0.03655 | 0.003902 | 3.90E-22 |
| rs174601 | miR-1908-! | cis | 5.98E-09 | Nightingale | L_VLDL_PL | NA | T | C | 0.080547 | 0.004139 | 7.70E-88 |
| rs174601 | miR-1908-! | cis | 5.98E-09 | Nightingale | L_HDL_TG_ | NA | T | C | 0.072366 | 0.004024 | 7.10E-77 |
| rs174601 | miR-1908-! | cis | 5.98E-09 | Nightingale | M_VLDL_F | NA | T | C | -0.02793 | 0.004264 | 1.90E-11 |
| rs174601 | miR-1908-! | cis | 5.98E-09 | Nightingale | L_VLDL_PL | NA | T | C | 0.043499 | 0.004114 | 2.40E-27 |
| rs174601 | miR-1908-! | cis | 5.98E-09 | Nightingale | XS_VLDL_C | NA | T | C | -0.06216 | 0.004139 | 8.30E-54 |
| rs1881116 | miR-339-5[ | cis | 0.000162 | Nightingale | XS_VLDL_C | NA | T | C | -0.02393 | 0.004313 | 1.70E-08 |
| rs1881116 | miR-339-5[ | cis | 0.000162 | Nightingale | XS_VLDL_C | NA | T | C | -0.02363 | 0.004339 | 3.20E-08 |
| rs2072113 | miR-1908-! | cis | 3.35E-05 | Nightingale | VLDL_size | NA | T | C | 0.083694 | 0.006009 | 9.40E-46 |

| | | | | | | | | | | | |
|---|---|---|---|---|---|---|---|---|---|---|---|
| rs2072113 | miR-1908-5 | cis | 3.35E-05 | Nightingale | M_VLDL_C | NA | T | C | -0.08911 | 0.006068 | 7.50E-50 |
| rs2072113 | miR-1908-5 | cis | 3.35E-05 | Nightingale | L_HDL_L | NA | T | C | -0.09169 | 0.005656 | 1.50E-61 |
| rs2072113 | miR-1908-5 | cis | 3.35E-05 | Nightingale | L_LDL_FC | NA | T | C | -0.07533 | 0.00633 | 7.50E-34 |
| rs2072113 | miR-1908-5 | cis | 3.35E-05 | Nightingale | XL_VLDL_F | NA | T | C | -0.06958 | 0.006245 | 8.10E-30 |
| rs2072113 | miR-1908-5 | cis | 3.35E-05 | Nightingale | S_LDL_PL_ | NA | T | C | -0.062 | 0.006433 | 3.80E-21 |
| rs2072113 | miR-1908-5 | cis | 3.35E-05 | Nightingale | L_LDL_FC_ | NA | T | C | -0.09436 | 0.006144 | 9.40E-54 |
| rs2072113 | miR-1908-5 | cis | 3.35E-05 | Nightingale | PUFA_pct | NA | T | C | -0.12736 | 0.006201 | 5.70E-94 |
| rs2072113 | miR-1908-5 | cis | 3.35E-05 | Nightingale | M_HDL_TG | NA | T | C | 0.054462 | 0.006367 | 2.90E-17 |
| rs2072113 | miR-1908-5 | cis | 3.35E-05 | Nightingale | S_HDL_TG | NA | T | C | 0.076213 | 0.006251 | 3.30E-34 |
| rs2072113 | miR-1908-5 | cis | 3.35E-05 | Nightingale | Total_TG | NA | T | C | 0.05696 | 0.00624 | 9.40E-20 |
| rs2072113 | miR-1908-5 | cis | 3.35E-05 | Nightingale | Unsaturati | NA | T | C | -0.33096 | 0.006106 | 0 |
| rs2072113 | miR-1908-5 | cis | 3.35E-05 | Nightingale | M_HDL_CE | NA | T | C | -0.06105 | 0.006251 | 2.40E-22 |
| rs2072113 | miR-1908-5 | cis | 3.35E-05 | Nightingale | XS_VLDL_C | NA | T | C | -0.06825 | 0.006257 | 4.60E-29 |
| rs2072113 | miR-1908-5 | cis | 3.35E-05 | Nightingale | M_HDL_CE | NA | T | C | -0.05689 | 0.005953 | 2.00E-22 |
| rs2072113 | miR-1908-5 | cis | 3.35E-05 | Nightingale | L_LDL_CE | NA | T | C | -0.05777 | 0.006389 | 2.20E-20 |
| rs2072113 | miR-1908-5 | cis | 3.35E-05 | Nightingale | L_HDL_CE | NA | T | C | -0.09264 | 0.005692 | 2.20E-61 |
| rs2072113 | miR-1908-5 | cis | 3.35E-05 | Nightingale | S_VLDL_FC | NA | T | C | -0.09224 | 0.006155 | 2.30E-51 |
| rs2072113 | miR-1908-5 | cis | 3.35E-05 | Nightingale | IDL_P | NA | T | C | -0.05039 | 0.006379 | 6.50E-16 |
| rs2072113 | miR-1908-5 | cis | 3.35E-05 | Nightingale | L_VLDL_FC | NA | T | C | 0.05075 | 0.006235 | 6.50E-16 |
| rs2072113 | miR-1908-5 | cis | 3.35E-05 | Nightingale | XXL_VLDL_ | NA | T | C | 0.080432 | 0.006186 | 5.10E-39 |
| rs2072113 | miR-1908-5 | cis | 3.35E-05 | Nightingale | HDL_PL | NA | T | C | -0.06466 | 0.005828 | 4.40E-30 |
| rs2072113 | miR-1908-5 | cis | 3.35E-05 | Nightingale | LDL_L | NA | T | C | -0.04943 | 0.006425 | 3.30E-15 |
| rs2072113 | miR-1908-5 | cis | 3.35E-05 | Nightingale | HDL_size | NA | T | C | -0.08443 | 0.005712 | 9.10E-52 |
| rs2072113 | miR-1908-5 | cis | 3.35E-05 | Nightingale | LDL_P | NA | T | C | -0.03949 | 0.006442 | 3.50E-10 |
| rs2072113 | miR-1908-5 | cis | 3.35E-05 | Nightingale | L_VLDL_TG | NA | T | C | 0.047832 | 0.006203 | 1.10E-14 |
| rs2072113 | miR-1908-5 | cis | 3.35E-05 | Nightingale | HDL_CE | NA | T | C | -0.07992 | 0.005785 | 1.80E-44 |
| rs2072113 | miR-1908-5 | cis | 3.35E-05 | Nightingale | HDL_C | NA | T | C | -0.08302 | 0.005756 | 1.30E-48 |
| rs2072113 | miR-1908-5 | cis | 3.35E-05 | Nightingale | MUFA | NA | T | C | 0.066889 | 0.006381 | 1.70E-25 |
| rs2072113 | miR-1908-5 | cis | 3.35E-05 | Nightingale | ApoA1 | NA | T | C | -0.05799 | 0.005899 | 5.50E-24 |
| rs2072113 | miR-1908-5 | cis | 3.35E-05 | Nightingale | Total_CE | NA | T | C | -0.07567 | 0.006218 | 3.00E-35 |
| rs2072113 | miR-1908-5 | cis | 3.35E-05 | Nightingale | L_VLDL_PL | NA | T | C | 0.059922 | 0.00622 | 7.60E-22 |
| rs2072113 | miR-1908-5 | cis | 3.35E-05 | Nightingale | XL_VLDL_C | NA | T | C | -0.07092 | 0.006176 | 5.00E-31 |
| rs2072113 | miR-1908-5 | cis | 3.35E-05 | Nightingale | M_LDL_P | NA | T | C | -0.0453 | 0.006465 | 5.80E-13 |
| rs2072113 | miR-1908-5 | cis | 3.35E-05 | Nightingale | PUFA_by_M | NA | T | C | -0.17726 | 0.006175 | 5.70E-184 |
| rs2072113 | miR-1908-5 | cis | 3.35E-05 | Nightingale | L_HDL_CE_ | NA | T | C | -0.05933 | 0.006168 | 1.80E-21 |
| rs2072113 | miR-1908-5 | cis | 3.35E-05 | Nightingale | XL_HDL_C | NA | T | C | -0.10583 | 0.005812 | 1.60E-76 |
| rs2072113 | miR-1908-5 | cis | 3.35E-05 | Nightingale | IDL_TG_pct | NA | T | C | 0.096624 | 0.006228 | 8.60E-55 |
| rs2072113 | miR-1908-5 | cis | 3.35E-05 | Nightingale | XL_VLDL_L | NA | T | C | 0.061225 | 0.006203 | 5.60E-23 |
| rs2072113 | miR-1908-5 | cis | 3.35E-05 | Nightingale | ApoB | NA | T | C | -0.03816 | 0.006438 | 1.20E-09 |
| rs2072113 | miR-1908-5 | cis | 3.35E-05 | Nightingale | XS_VLDL_T | NA | T | C | 0.038768 | 0.006347 | 2.50E-09 |
| rs2072113 | miR-1908-5 | cis | 3.35E-05 | Nightingale | S_HDL_FC_ | NA | T | C | -0.10692 | 0.006114 | 1.30E-71 |
| rs2072113 | miR-1908-5 | cis | 3.35E-05 | Nightingale | LDL_FC | NA | T | C | -0.07087 | 0.006378 | 1.10E-29 |
| rs2072113 | miR-1908-5 | cis | 3.35E-05 | Nightingale | XXL_VLDL_ | NA | T | C | 0.07311 | 0.006209 | 2.10E-32 |
| rs2072113 | miR-1908-5 | cis | 3.35E-05 | Nightingale | S_LDL_FC | NA | T | C | -0.06797 | 0.006436 | 4.30E-27 |
| rs2072113 | miR-1908-5 | cis | 3.35E-05 | Nightingale | Phosphogly | NA | T | C | -0.05121 | 0.006107 | 2.00E-18 |
| rs2072113 | miR-1908-5 | cis | 3.35E-05 | Nightingale | LDL_PL | NA | T | C | -0.05079 | 0.006432 | 7.40E-16 |
| rs2072113 | miR-1908-5 | cis | 3.35E-05 | Nightingale | LA | NA | T | C | 0.071811 | 0.006322 | 5.70E-30 |
| rs2072113 | miR-1908-5 | cis | 3.35E-05 | Nightingale | S_VLDL_C_ | NA | T | C | -0.0635 | 0.006272 | 2.30E-24 |
| rs2072113 | miR-1908-5 | cis | 3.35E-05 | Nightingale | L_HDL_P | NA | T | C | -0.0917 | 0.005646 | 1.10E-61 |
| rs2072113 | miR-1908-5 | cis | 3.35E-05 | Nightingale | M_HDL_P | NA | T | C | -0.05184 | 0.005939 | 2.20E-19 |
| rs2072113 | miR-1908-5 | cis | 3.35E-05 | Nightingale | IDL_FC | NA | T | C | -0.09813 | 0.006239 | 1.20E-57 |
| rs2072113 | miR-1908-5 | cis | 3.35E-05 | Nightingale | M_HDL_FC | NA | T | C | -0.06903 | 0.005866 | 2.10E-33 |
| rs2072113 | miR-1908-5 | cis | 3.35E-05 | Nightingale | XL_HDL_CE | NA | T | C | -0.04081 | 0.006443 | 4.10E-10 |
| rs2072113 | miR-1908-5 | cis | 3.35E-05 | Nightingale | DHA | NA | T | C | -0.28437 | 0.006213 | 0 |
| rs2072113 | miR-1908-5 | cis | 3.35E-05 | Nightingale | SFA_pct | NA | T | C | -0.04153 | 0.00636 | 6.60E-11 |
| rs2072113 | miR-1908-5 | cis | 3.35E-05 | Nightingale | XS_VLDL_P | NA | T | C | -0.04127 | 0.00636 | 1.40E-11 |
| rs2072113 | miR-1908-5 | cis | 3.35E-05 | Nightingale | M_VLDL_T | NA | T | C | 0.041864 | 0.006263 | 4.00E-11 |
| rs2072113 | miR-1908-5 | cis | 3.35E-05 | Nightingale | Total_FC | NA | T | C | -0.06392 | 0.006298 | 2.40E-25 |
| rs2072113 | miR-1908-5 | cis | 3.35E-05 | Nightingale | Sphingomy | NA | T | C | -0.12156 | 0.006024 | 5.10E-93 |
| rs2072113 | miR-1908-5 | cis | 3.35E-05 | Nightingale | M_VLDL_C | NA | T | C | -0.0854 | 0.006052 | 4.30E-46 |
| rs2072113 | miR-1908-5 | cis | 3.35E-05 | Nightingale | L_LDL_PL | NA | T | C | -0.06084 | 0.006398 | 2.20E-22 |
| rs2072113 | miR-1908-5 | cis | 3.35E-05 | Nightingale | L_HDL_FC | NA | T | C | -0.09798 | 0.005649 | 7.00E-70 |
| rs2072113 | miR-1908-5 | cis | 3.35E-05 | Nightingale | IDL_PL | NA | T | C | -0.07629 | 0.00622 | 3.40E-36 |

| | | | | | | | | | | | | |
|---|---|---|---|---|---|---|---|---|---|---|---|---|
| rs2072113 | miR-1908-5 | cis | | 3.35E-05 | Nightingale | PUFA | NA | T | C | -0.09922 | 0.006287 | 5.50E-60 |
| rs2072113 | miR-1908-5 | cis | | 3.35E-05 | Nightingale | DHA_pct | NA | T | C | -0.2698 | 0.006258 | 0 |
| rs2072113 | miR-1908-5 | cis | | 3.35E-05 | Nightingale | M_HDL_PL | NA | T | C | 0.071743 | 0.006115 | 9.70E-32 |
| rs2072113 | miR-1908-5 | cis | | 3.35E-05 | Nightingale | Tyr | NA | T | C | 0.035596 | 0.00644 | 3.20E-08 |
| rs2072113 | miR-1908-5 | cis | | 3.35E-05 | Nightingale | M_LDL_FC | NA | T | C | -0.05817 | 0.006439 | 2.50E-20 |
| rs2072113 | miR-1908-5 | cis | | 3.35E-05 | Nightingale | M_HDL_TG | NA | T | C | 0.082368 | 0.006182 | 2.00E-40 |
| rs2072113 | miR-1908-5 | cis | | 3.35E-05 | Nightingale | XL_HDL_PL | NA | T | C | -0.09379 | 0.005795 | 1.10E-61 |
| rs2072113 | miR-1908-5 | cis | | 3.35E-05 | Nightingale | S_VLDL_PL | NA | T | C | -0.08904 | 0.006168 | 6.80E-48 |
| rs2072113 | miR-1908-5 | cis | | 3.35E-05 | Nightingale | TG_by_PG | NA | T | C | 0.084339 | 0.005983 | 6.40E-46 |
| rs2072113 | miR-1908-5 | cis | | 3.35E-05 | Nightingale | XL_VLDL_C | NA | T | C | 0.039156 | 0.006345 | 1.90E-09 |
| rs2072113 | miR-1908-5 | cis | | 3.35E-05 | Nightingale | IDL_L | NA | T | C | -0.07886 | 0.006232 | 2.20E-38 |
| rs2072113 | miR-1908-5 | cis | | 3.35E-05 | Nightingale | Cholines | NA | T | C | -0.0485 | 0.006063 | 8.50E-17 |
| rs2072113 | miR-1908-5 | cis | | 3.35E-05 | Nightingale | S_HDL_TG_ | NA | T | C | 0.08355 | 0.006131 | 1.20E-42 |
| rs2072113 | miR-1908-5 | cis | | 3.35E-05 | Nightingale | M_LDL_C | NA | T | C | -0.03594 | 0.006459 | 9.90E-09 |
| rs2072113 | miR-1908-5 | cis | | 3.35E-05 | Nightingale | XS_VLDL_L | NA | T | C | -0.03831 | 0.006336 | 3.00E-10 |
| rs2072113 | miR-1908-5 | cis | | 3.35E-05 | Nightingale | L_LDL_L | NA | T | C | -0.05934 | 0.006384 | 1.80E-21 |
| rs2072113 | miR-1908-5 | cis | | 3.35E-05 | Nightingale | XXL_VLDL_ | NA | T | C | 0.079061 | 0.006213 | 4.20E-37 |
| rs2072113 | miR-1908-5 | cis | | 3.35E-05 | Nightingale | XS_VLDL_P | NA | T | C | 0.075283 | 0.00629 | 2.00E-33 |
| rs2072113 | miR-1908-5 | cis | | 3.35E-05 | Nightingale | L_HDL_FC_ | NA | T | C | -0.11703 | 0.005989 | 3.80E-89 |
| rs2072113 | miR-1908-5 | cis | | 3.35E-05 | Nightingale | MUFA_pct | NA | T | C | 0.204011 | 0.00617 | 8.00E-245 |
| rs2072113 | miR-1908-5 | cis | | 3.35E-05 | Nightingale | XL_HDL_L | NA | T | C | -0.09866 | 0.005793 | 9.60E-68 |
| rs2072113 | miR-1908-5 | cis | | 3.35E-05 | Nightingale | XXL_VLDL_ | NA | T | C | 0.080473 | 0.006225 | 4.80E-38 |
| rs2072113 | miR-1908-5 | cis | | 3.35E-05 | Nightingale | LDL_CE | NA | T | C | -0.04666 | 0.00643 | 9.90E-14 |
| rs2072113 | miR-1908-5 | cis | | 3.35E-05 | Nightingale | L_VLDL_PL | NA | T | C | 0.096048 | 0.006256 | 1.90E-53 |
| rs2072113 | miR-1908-5 | cis | | 3.35E-05 | Nightingale | M_VLDL_P | NA | T | C | -0.06762 | 0.006186 | 1.20E-28 |
| rs2072113 | miR-1908-5 | cis | | 3.35E-05 | Nightingale | Omega_3_ | NA | T | C | -0.37525 | 0.006299 | 0 |
| rs2072113 | miR-1908-5 | cis | | 3.35E-05 | Nightingale | IDL_CE_pct | NA | T | C | -0.04744 | 0.006326 | 5.40E-14 |
| rs2072113 | miR-1908-5 | cis | | 3.35E-05 | Nightingale | XS_VLDL_T | NA | T | C | 0.103364 | 0.006123 | 7.60E-65 |
| rs2072113 | miR-1908-5 | cis | | 3.35E-05 | Nightingale | XL_VLDL_P | NA | T | C | 0.063192 | 0.006196 | 2.20E-24 |
| rs2072113 | miR-1908-5 | cis | | 3.35E-05 | Nightingale | LA_pct | NA | T | C | 0.167109 | 0.0063 | 8.30E-156 |
| rs2072113 | miR-1908-5 | cis | | 3.35E-05 | Nightingale | L_HDL_PL_ | NA | T | C | 0.041382 | 0.006239 | 6.00E-11 |
| rs2072113 | miR-1908-5 | cis | | 3.35E-05 | Nightingale | L_HDL_C_p | NA | T | C | -0.07723 | 0.006076 | 7.70E-37 |
| rs2072113 | miR-1908-5 | cis | | 3.35E-05 | Nightingale | XS_VLDL_C | NA | T | C | -0.10716 | 0.006108 | 7.70E-70 |
| rs2072113 | miR-1908-5 | cis | | 3.35E-05 | Nightingale | L_LDL_TG_ | NA | T | C | 0.083793 | 0.006321 | 2.10E-40 |
| rs2072113 | miR-1908-5 | cis | | 3.35E-05 | Nightingale | S_LDL_PL | NA | T | C | -0.05291 | 0.006431 | 7.00E-17 |
| rs2072113 | miR-1908-5 | cis | | 3.35E-05 | Nightingale | XL_VLDL_T | NA | T | C | 0.062265 | 0.006244 | 4.80E-24 |
| rs2072113 | miR-1908-5 | cis | | 3.35E-05 | Nightingale | XS_VLDL_F | NA | T | C | -0.10958 | 0.006398 | 3.30E-66 |
| rs2072113 | miR-1908-5 | cis | | 3.35E-05 | Nightingale | M_HDL_FC | NA | T | C | -0.11925 | 0.005712 | 1.20E-100 |
| rs2072113 | miR-1908-5 | cis | | 3.35E-05 | Nightingale | XL_HDL_FC | NA | T | C | 0.070626 | 0.005716 | 6.00E-37 |
| rs2072113 | miR-1908-5 | cis | | 3.35E-05 | Nightingale | XL_HDL_CE | NA | T | C | -0.10527 | 0.005773 | 8.70E-77 |
| rs2072113 | miR-1908-5 | cis | | 3.35E-05 | Nightingale | S_VLDL_TG | NA | T | C | 0.072623 | 0.006237 | 1.20E-31 |
| rs2072113 | miR-1908-5 | cis | | 3.35E-05 | Nightingale | L_VLDL_C_ | NA | T | C | -0.03881 | 0.006297 | 1.60E-10 |
| rs2072113 | miR-1908-5 | cis | | 3.35E-05 | Nightingale | HDL_L | NA | T | C | -0.07038 | 0.005789 | 1.60E-35 |
| rs2072113 | miR-1908-5 | cis | | 3.35E-05 | Nightingale | S_LDL_CE_ | NA | T | C | 0.060972 | 0.006435 | 1.30E-20 |
| rs2072113 | miR-1908-5 | cis | | 3.35E-05 | Nightingale | M_HDL_C_ | NA | T | C | -0.08135 | 0.006137 | 1.40E-40 |
| rs2072113 | miR-1908-5 | cis | | 3.35E-05 | Nightingale | S_HDL_C_p | NA | T | C | -0.05303 | 0.006372 | 2.60E-16 |
| rs2072113 | miR-1908-5 | cis | | 3.35E-05 | Nightingale | Total_L | NA | T | C | -0.04106 | 0.006342 | 1.80E-11 |
| rs2072113 | miR-1908-5 | cis | | 3.35E-05 | Nightingale | Total_C | NA | T | C | -0.07272 | 0.00624 | 1.30E-32 |
| rs2072113 | miR-1908-5 | cis | | 3.35E-05 | Nightingale | XXL_VLDL_ | NA | T | C | 0.07421 | 0.0062 | 2.10E-33 |
| rs2072113 | miR-1908-5 | cis | | 3.35E-05 | Nightingale | HDL_TG | NA | T | C | 0.051771 | 0.006357 | 7.00E-16 |
| rs2072113 | miR-1908-5 | cis | | 3.35E-05 | Nightingale | M_HDL_L | NA | T | C | -0.045 | 0.005985 | 7.60E-15 |
| rs2072113 | miR-1908-5 | cis | | 3.35E-05 | Nightingale | M_VLDL_C | NA | T | C | -0.05324 | 0.006388 | 2.70E-17 |
| rs2072113 | miR-1908-5 | cis | | 3.35E-05 | Nightingale | S_LDL_TG | NA | T | C | 0.05752 | 0.006289 | 1.10E-19 |
| rs2072113 | miR-1908-5 | cis | | 3.35E-05 | Nightingale | L_VLDL_C | NA | T | C | 0.0399 | 0.006305 | 5.60E-10 |
| rs2072113 | miR-1908-5 | cis | | 3.35E-05 | Nightingale | L_VLDL_CE | NA | T | C | -0.05952 | 0.006169 | 2.80E-23 |
| rs2072113 | miR-1908-5 | cis | | 3.35E-05 | Nightingale | M_LDL_PL_ | NA | T | C | 0.034266 | 0.006443 | 1.70E-08 |
| rs2072113 | miR-1908-5 | cis | | 3.35E-05 | Nightingale | XS_VLDL_C | NA | T | C | -0.1024 | 0.006077 | 1.30E-64 |
| rs2072113 | miR-1908-5 | cis | | 3.35E-05 | Nightingale | Remnant_( | NA | T | C | -0.04471 | 0.006405 | 7.10E-13 |
| rs2072113 | miR-1908-5 | cis | | 3.35E-05 | Nightingale | L_VLDL_L | NA | T | C | 0.049412 | 0.00623 | 3.30E-15 |
| rs2072113 | miR-1908-5 | cis | | 3.35E-05 | Nightingale | L_VLDL_P | NA | T | C | 0.052739 | 0.006224 | 3.80E-17 |
| rs2072113 | miR-1908-5 | cis | | 3.35E-05 | Nightingale | XL_HDL_TG | NA | T | C | 0.097273 | 0.006073 | 2.10E-58 |
| rs2072113 | miR-1908-5 | cis | | 3.35E-05 | Nightingale | M_VLDL_T | NA | T | C | 0.085779 | 0.006088 | 3.40E-46 |
| rs2072113 | miR-1908-5 | cis | | 3.35E-05 | Nightingale | M_LDL_C_ | NA | T | C | -0.09922 | 0.006451 | 2.30E-54 |

| | | | | | | | | | | | |
|---|---|---|---|---|---|---|---|---|---|---|---|
| rs2072113 | miR-1908-5 | cis | 3.35E-05 | Nightingale | XL_HDL_PL | NA | T | C | -0.06828 | 0.005922 | 7.10E-33 |
| rs2072113 | miR-1908-5 | cis | 3.35E-05 | Nightingale | M_HDL_PL | NA | T | C | -0.03796 | 0.006028 | 5.90E-11 |
| rs2072113 | miR-1908-5 | cis | 3.35E-05 | Nightingale | XS_VLDL_C | NA | T | C | -0.07339 | 0.00622 | 9.30E-34 |
| rs2072113 | miR-1908-5 | cis | 3.35E-05 | Nightingale | L_LDL_C_p | NA | T | C | -0.08209 | 0.006281 | 6.50E-39 |
| rs2072113 | miR-1908-5 | cis | 3.35E-05 | Nightingale | LDL_C | NA | T | C | -0.05385 | 0.006419 | 9.30E-18 |
| rs2072113 | miR-1908-5 | cis | 3.35E-05 | Nightingale | Omega_3 | NA | T | C | -0.3243 | 0.006319 | 0 |
| rs2072113 | miR-1908-5 | cis | 3.35E-05 | Nightingale | XL_VLDL_C | NA | T | C | 0.047199 | 0.006296 | 1.50E-13 |
| rs2072113 | miR-1908-5 | cis | 3.35E-05 | Nightingale | L_HDL_TG_ | NA | T | C | 0.094914 | 0.006083 | 4.00E-55 |
| rs2072113 | miR-1908-5 | cis | 3.35E-05 | Nightingale | M_VLDL_F | NA | T | C | -0.09648 | 0.006129 | 1.40E-57 |
| rs2072113 | miR-1908-5 | cis | 3.35E-05 | Nightingale | XXL_VLDL_ | NA | T | C | 0.075118 | 0.00621 | 7.40E-34 |
| rs2072113 | miR-1908-5 | cis | 3.35E-05 | Nightingale | XS_VLDL_F | NA | T | C | -0.05288 | 0.006334 | 7.30E-18 |
| rs2072113 | miR-1908-5 | cis | 3.35E-05 | Nightingale | L_LDL_P | NA | T | C | -0.03724 | 0.006424 | 3.20E-09 |
| rs2072113 | miR-1908-5 | cis | 3.35E-05 | Nightingale | XL_HDL_P | NA | T | C | -0.0957 | 0.005801 | 1.30E-63 |
| rs2072113 | miR-1908-5 | cis | 3.35E-05 | Nightingale | L_HDL_PL | NA | T | C | -0.091 | 0.005673 | 1.30E-60 |
| rs2072113 | miR-1908-5 | cis | 3.35E-05 | Nightingale | IDL_FC_pct | NA | T | C | -0.11593 | 0.006395 | 1.60E-73 |
| rs2072113 | miR-1908-5 | cis | 3.35E-05 | Nightingale | Omega_6_ | NA | T | C | 0.353301 | 0.006353 | 0 |
| rs2072113 | miR-1908-5 | cis | 3.35E-05 | Nightingale | HDL_P | NA | T | C | -0.05267 | 0.006053 | 5.40E-19 |
| rs2072113 | miR-1908-5 | cis | 3.35E-05 | Nightingale | S_LDL_FC_ | NA | T | C | -0.09191 | 0.006247 | 1.70E-49 |
| rs2072113 | miR-1908-5 | cis | 3.35E-05 | Nightingale | Clinical_LD | NA | T | C | -0.0602 | 0.00641 | 9.10E-22 |
| rs2072113 | miR-1908-5 | cis | 3.35E-05 | Nightingale | L_LDL_C | NA | T | C | -0.06301 | 0.006374 | 5.20E-24 |
| rs2072113 | miR-1908-5 | cis | 3.35E-05 | Nightingale | M_VLDL_C | NA | T | C | -0.04217 | 0.006429 | 2.80E-11 |
| rs2072113 | miR-1908-5 | cis | 3.35E-05 | Nightingale | VLDL_TG | NA | T | C | 0.058123 | 0.006203 | 7.70E-21 |
| rs2072113 | miR-1908-5 | cis | 3.35E-05 | Nightingale | Total_P | NA | T | C | -0.05624 | 0.006067 | 2.00E-21 |
| rs2072113 | miR-1908-5 | cis | 3.35E-05 | Nightingale | XL_VLDL_F | NA | T | C | 0.054036 | 0.006244 | 9.50E-18 |
| rs2072113 | miR-1908-5 | cis | 3.35E-05 | Nightingale | VLDL_L | NA | T | C | 0.036854 | 0.006321 | 1.30E-08 |
| rs2072113 | miR-1908-5 | cis | 3.35E-05 | Nightingale | S_VLDL_TG | NA | T | C | 0.049779 | 0.006256 | 4.00E-15 |
| rs2072113 | miR-1908-5 | cis | 3.35E-05 | Nightingale | IDL_C | NA | T | C | -0.08527 | 0.006225 | 2.60E-44 |
| rs2072113 | miR-1908-5 | cis | 3.35E-05 | Nightingale | IDL_CE | NA | T | C | -0.08002 | 0.006223 | 3.00E-39 |
| rs2072113 | miR-1908-5 | cis | 3.35E-05 | Nightingale | L_HDL_C | NA | T | C | -0.09424 | 0.005677 | 7.60E-64 |
| rs2072113 | miR-1908-5 | cis | 3.35E-05 | Nightingale | non_HDL_( | NA | T | C | -0.05037 | 0.006412 | 7.90E-16 |
| rs2072113 | miR-1908-5 | cis | 3.35E-05 | Nightingale | M_LDL_TG | NA | T | C | 0.040576 | 0.006351 | 2.30E-10 |
| rs2072113 | miR-1908-5 | cis | 3.35E-05 | Nightingale | M_LDL_FC_ | NA | T | C | -0.0854 | 0.006175 | 5.60E-44 |
| rs2072113 | miR-1908-5 | cis | 3.35E-05 | Nightingale | XL_VLDL_T | NA | T | C | 0.066289 | 0.006166 | 2.90E-27 |
| rs2072113 | miR-1908-5 | cis | 3.35E-05 | Nightingale | Total_PL | NA | T | C | -0.05915 | 0.006138 | 2.30E-23 |
| rs2072113 | miR-1908-5 | cis | 3.35E-05 | Nightingale | XL_VLDL_P | NA | T | C | 0.058758 | 0.006221 | 5.60E-21 |
| rs2072113 | miR-1908-5 | cis | 3.35E-05 | Nightingale | XXL_VLDL_ | NA | T | C | 0.067126 | 0.006221 | 1.70E-27 |
| rs2072113 | miR-1908-5 | cis | 3.35E-05 | Nightingale | S_LDL_TG_ | NA | T | C | 0.09463 | 0.006246 | 5.10E-52 |
| rs2072113 | miR-1908-5 | cis | 3.35E-05 | Nightingale | XXL_VLDL_ | NA | T | C | 0.055537 | 0.006478 | 9.60E-18 |
| rs2072113 | miR-1908-5 | cis | 3.35E-05 | Nightingale | M_HDL_C | NA | T | C | -0.05969 | 0.005931 | 9.80E-25 |
| rs2072113 | miR-1908-5 | cis | 3.35E-05 | Nightingale | M_LDL_TG | NA | T | C | 0.081081 | 0.006365 | 1.40E-37 |
| rs2072113 | miR-1908-5 | cis | 3.35E-05 | Nightingale | IDL_C_pct | NA | T | C | -0.08772 | 0.006293 | 1.50E-44 |
| rs2072113 | miR-1908-5 | cis | 3.35E-05 | Nightingale | HDL_FC | NA | T | C | -0.09011 | 0.005711 | 4.00E-58 |
| rs2072113 | miR-1908-5 | cis | 3.35E-05 | Nightingale | XL_HDL_FC | NA | T | C | -0.10404 | 0.006005 | 1.00E-69 |
| rs2072113 | miR-1908-5 | cis | 3.35E-05 | Nightingale | XL_VLDL_C | NA | T | C | -0.06924 | 0.006179 | 1.00E-29 |
| rs2072114 | miR-1908-5 | cis | 1.34E-05 | Nightingale | ApoB | NA | G | A | -0.03633 | 0.006275 | 1.80E-09 |
| rs2072114 | miR-1908-5 | cis | 1.34E-05 | Nightingale | IDL_FC | NA | G | A | -0.09515 | 0.006082 | 1.80E-57 |
| rs2072114 | miR-1908-5 | cis | 1.34E-05 | Nightingale | LDL_P | NA | G | A | -0.03784 | 0.006279 | 4.50E-10 |
| rs2072114 | miR-1908-5 | cis | 1.34E-05 | Nightingale | S_VLDL_FC | NA | G | A | -0.0905 | 0.006 | 4.80E-52 |
| rs2072114 | miR-1908-5 | cis | 1.34E-05 | Nightingale | L_VLDL_C | NA | G | A | 0.039819 | 0.006146 | 2.90E-10 |
| rs2072114 | miR-1908-5 | cis | 1.34E-05 | Nightingale | M_VLDL_T | NA | G | A | 0.084337 | 0.005934 | 4.50E-47 |
| rs2072114 | miR-1908-5 | cis | 1.34E-05 | Nightingale | LDL_FC | NA | G | A | -0.06866 | 0.006216 | 1.80E-29 |
| rs2072114 | miR-1908-5 | cis | 1.34E-05 | Nightingale | MUFA | NA | G | A | 0.067396 | 0.00622 | 8.50E-27 |
| rs2072114 | miR-1908-5 | cis | 1.34E-05 | Nightingale | XL_VLDL_C | NA | G | A | -0.07029 | 0.006019 | 6.10E-32 |
| rs2072114 | miR-1908-5 | cis | 1.34E-05 | Nightingale | ApoA1 | NA | G | A | -0.05361 | 0.00575 | 4.80E-22 |
| rs2072114 | miR-1908-5 | cis | 1.34E-05 | Nightingale | XXL_VLDL_ | NA | G | A | 0.080155 | 0.00603 | 1.70E-40 |
| rs2072114 | miR-1908-5 | cis | 1.34E-05 | Nightingale | M_HDL_C_ | NA | G | A | -0.07912 | 0.005981 | 2.30E-40 |
| rs2072114 | miR-1908-5 | cis | 1.34E-05 | Nightingale | M_HDL_P | NA | G | A | -0.0477 | 0.005788 | 9.90E-18 |
| rs2072114 | miR-1908-5 | cis | 1.34E-05 | Nightingale | Unsaturati | NA | G | A | -0.3266 | 0.005951 | 0 |
| rs2072114 | miR-1908-5 | cis | 1.34E-05 | Nightingale | LDL_L | NA | G | A | -0.04731 | 0.006263 | 6.50E-15 |
| rs2072114 | miR-1908-5 | cis | 1.34E-05 | Nightingale | Total_TG | NA | G | A | 0.057171 | 0.006082 | 1.30E-20 |
| rs2072114 | miR-1908-5 | cis | 1.34E-05 | Nightingale | HDL_CE | NA | G | A | -0.07533 | 0.005639 | 6.50E-42 |
| rs2072114 | miR-1908-5 | cis | 1.34E-05 | Nightingale | L_HDL_L | NA | G | A | -0.08689 | 0.005513 | 9.90E-59 |
| rs2072114 | miR-1908-5 | cis | 1.34E-05 | Nightingale | L_LDL_L | NA | G | A | -0.05698 | 0.006222 | 3.60E-21 |

| | | | | | | | | | | | |
|---|---|---|---|---|---|---|---|---|---|---|---|
| rs2072114 | miR-1908-5 | cis | 1.34E-05 | Nightingale | XXL_VLDL_ | NA | G | A | 0.078993 | 0.006055 | 1.00E-38 |
| rs2072114 | miR-1908-5 | cis | 1.34E-05 | Nightingale | S_LDL_FC | NA | G | A | -0.06617 | 0.006273 | 5.30E-27 |
| rs2072114 | miR-1908-5 | cis | 1.34E-05 | Nightingale | S_HDL_FC_ | NA | G | A | -0.10392 | 0.00596 | 8.40E-72 |
| rs2072114 | miR-1908-5 | cis | 1.34E-05 | Nightingale | IDL_P | NA | G | A | -0.04784 | 0.006218 | 2.10E-15 |
| rs2072114 | miR-1908-5 | cis | 1.34E-05 | Nightingale | Omega_3_ | NA | G | A | -0.37131 | 0.00614 | 0 |
| rs2072114 | miR-1908-5 | cis | 1.34E-05 | Nightingale | XS_VLDL_L | NA | G | A | -0.03594 | 0.006176 | 5.40E-10 |
| rs2072114 | miR-1908-5 | cis | 1.34E-05 | Nightingale | L_HDL_CE | NA | G | A | -0.08806 | 0.005548 | 7.30E-59 |
| rs2072114 | miR-1908-5 | cis | 1.34E-05 | Nightingale | LDL_PL | NA | G | A | -0.04875 | 0.006269 | 1.40E-15 |
| rs2072114 | miR-1908-5 | cis | 1.34E-05 | Nightingale | XL_HDL_C | NA | G | A | -0.10223 | 0.005665 | 1.30E-75 |
| rs2072114 | miR-1908-5 | cis | 1.34E-05 | Nightingale | M_HDL_TG | NA | G | A | 0.080288 | 0.006025 | 2.60E-40 |
| rs2072114 | miR-1908-5 | cis | 1.34E-05 | Nightingale | XS_VLDL_T | NA | G | A | 0.039309 | 0.006187 | 1.10E-09 |
| rs2072114 | miR-1908-5 | cis | 1.34E-05 | Nightingale | IDL_PL | NA | G | A | -0.07342 | 0.006062 | 7.20E-36 |
| rs2072114 | miR-1908-5 | cis | 1.34E-05 | Nightingale | IDL_CE_pct | NA | G | A | -0.04494 | 0.006166 | 4.80E-13 |
| rs2072114 | miR-1908-5 | cis | 1.34E-05 | Nightingale | L_VLDL_PL | NA | G | A | 0.059401 | 0.006062 | 2.80E-22 |
| rs2072114 | miR-1908-5 | cis | 1.34E-05 | Nightingale | LA | NA | G | A | 0.074032 | 0.006162 | 7.40E-33 |
| rs2072114 | miR-1908-5 | cis | 1.34E-05 | Nightingale | M_HDL_FC | NA | G | A | -0.06449 | 0.005718 | 4.00E-31 |
| rs2072114 | miR-1908-5 | cis | 1.34E-05 | Nightingale | L_HDL_PL_ | NA | G | A | 0.040374 | 0.006081 | 4.50E-11 |
| rs2072114 | miR-1908-5 | cis | 1.34E-05 | Nightingale | Total_PL | NA | G | A | -0.055 | 0.005983 | 7.90E-22 |
| rs2072114 | miR-1908-5 | cis | 1.34E-05 | Nightingale | S_LDL_CE_ | NA | G | A | 0.06034 | 0.006272 | 4.80E-21 |
| rs2072114 | miR-1908-5 | cis | 1.34E-05 | Nightingale | L_LDL_FC | NA | G | A | -0.07295 | 0.00617 | 1.20E-33 |
| rs2072114 | miR-1908-5 | cis | 1.34E-05 | Nightingale | Cholines | NA | G | A | -0.04441 | 0.00591 | 2.50E-15 |
| rs2072114 | miR-1908-5 | cis | 1.34E-05 | Nightingale | HDL_PL | NA | G | A | -0.05999 | 0.00568 | 1.00E-27 |
| rs2072114 | miR-1908-5 | cis | 1.34E-05 | Nightingale | DHA | NA | G | A | -0.2803 | 0.006055 | 0 |
| rs2072114 | miR-1908-5 | cis | 1.34E-05 | Nightingale | L_HDL_TG_ | NA | G | A | 0.092072 | 0.005929 | 1.30E-54 |
| rs2072114 | miR-1908-5 | cis | 1.34E-05 | Nightingale | XL_VLDL_L | NA | G | A | 0.061044 | 0.006046 | 9.40E-24 |
| rs2072114 | miR-1908-5 | cis | 1.34E-05 | Nightingale | XS_VLDL_C | NA | G | A | -0.07059 | 0.006063 | 1.70E-33 |
| rs2072114 | miR-1908-5 | cis | 1.34E-05 | Nightingale | S_VLDL_C_ | NA | G | A | -0.06281 | 0.006113 | 4.00E-25 |
| rs2072114 | miR-1908-5 | cis | 1.34E-05 | Nightingale | M_HDL_CE | NA | G | A | -0.06006 | 0.006093 | 1.40E-22 |
| rs2072114 | miR-1908-5 | cis | 1.34E-05 | Nightingale | SFA_pct | NA | G | A | -0.03965 | 0.006199 | 1.60E-10 |
| rs2072114 | miR-1908-5 | cis | 1.34E-05 | Nightingale | MUFA_pct | NA | G | A | 0.20069 | 0.006013 | 6.61E-249 |
| rs2072114 | miR-1908-5 | cis | 1.34E-05 | Nightingale | Total_FC | NA | G | A | -0.06059 | 0.006138 | 1.50E-24 |
| rs2072114 | miR-1908-5 | cis | 1.34E-05 | Nightingale | Total_CE | NA | G | A | -0.07215 | 0.006061 | 3.40E-34 |
| rs2072114 | miR-1908-5 | cis | 1.34E-05 | Nightingale | Phosphogly | NA | G | A | -0.04708 | 0.005952 | 7.00E-17 |
| rs2072114 | miR-1908-5 | cis | 1.34E-05 | Nightingale | IDL_TG_pct | NA | G | A | 0.094638 | 0.00607 | 5.30E-55 |
| rs2072114 | miR-1908-5 | cis | 1.34E-05 | Nightingale | M_VLDL_T | NA | G | A | 0.042287 | 0.006105 | 1.10E-11 |
| rs2072114 | miR-1908-5 | cis | 1.34E-05 | Nightingale | S_HDL_TG_ | NA | G | A | 0.081318 | 0.005976 | 2.60E-42 |
| rs2072114 | miR-1908-5 | cis | 1.34E-05 | Nightingale | XL_HDL_CE | NA | G | A | -0.04031 | 0.00628 | 3.70E-10 |
| rs2072114 | miR-1908-5 | cis | 1.34E-05 | Nightingale | XS_VLDL_F | NA | G | A | -0.05038 | 0.006174 | 1.20E-17 |
| rs2072114 | miR-1908-5 | cis | 1.34E-05 | Nightingale | XL_HDL_P | NA | G | A | -0.09192 | 0.005654 | 3.00E-62 |
| rs2072114 | miR-1908-5 | cis | 1.34E-05 | Nightingale | L_HDL_C_p | NA | G | A | -0.07495 | 0.005922 | 9.70E-37 |
| rs2072114 | miR-1908-5 | cis | 1.34E-05 | Nightingale | L_HDL_CE_ | NA | G | A | -0.05775 | 0.006012 | 1.60E-21 |
| rs2072114 | miR-1908-5 | cis | 1.34E-05 | Nightingale | S_LDL_TG | NA | G | A | 0.057853 | 0.00613 | 1.70E-20 |
| rs2072114 | miR-1908-5 | cis | 1.34E-05 | Nightingale | M_LDL_FC | NA | G | A | -0.05632 | 0.006276 | 3.70E-20 |
| rs2072114 | miR-1908-5 | cis | 1.34E-05 | Nightingale | Remnant_( | NA | G | A | -0.04229 | 0.006243 | 1.60E-12 |
| rs2072114 | miR-1908-5 | cis | 1.34E-05 | Nightingale | S_LDL_PL_ | NA | G | A | -0.06117 | 0.006271 | 1.40E-21 |
| rs2072114 | miR-1908-5 | cis | 1.34E-05 | Nightingale | PUFA_by_M | NA | G | A | -0.17462 | 0.006019 | 2.30E-187 |
| rs2072114 | miR-1908-5 | cis | 1.34E-05 | Nightingale | M_VLDL_C | NA | G | A | -0.08385 | 0.005899 | 9.70E-47 |
| rs2072114 | miR-1908-5 | cis | 1.34E-05 | Nightingale | HDL_P | NA | G | A | -0.04869 | 0.0059 | 1.60E-17 |
| rs2072114 | miR-1908-5 | cis | 1.34E-05 | Nightingale | XL_HDL_CE | NA | G | A | -0.10142 | 0.005627 | 1.70E-75 |
| rs2072114 | miR-1908-5 | cis | 1.34E-05 | Nightingale | M_HDL_CE | NA | G | A | -0.0529 | 0.005803 | 9.20E-21 |
| rs2072114 | miR-1908-5 | cis | 1.34E-05 | Nightingale | S_LDL_PL | NA | G | A | -0.05087 | 0.006268 | 1.50E-16 |
| rs2072114 | miR-1908-5 | cis | 1.34E-05 | Nightingale | HDL_L | NA | G | A | -0.06565 | 0.005642 | 5.80E-33 |
| rs2072114 | miR-1908-5 | cis | 1.34E-05 | Nightingale | XS_VLDL_P | NA | G | A | 0.072745 | 0.006131 | 1.00E-32 |
| rs2072114 | miR-1908-5 | cis | 1.34E-05 | Nightingale | XS_VLDL_T | NA | G | A | 0.101372 | 0.005968 | 9.60E-66 |
| rs2072114 | miR-1908-5 | cis | 1.34E-05 | Nightingale | LA_pct | NA | G | A | 0.166439 | 0.006141 | 1.10E-162 |
| rs2072114 | miR-1908-5 | cis | 1.34E-05 | Nightingale | S_HDL_C_p | NA | G | A | -0.05342 | 0.00621 | 4.90E-17 |
| rs2072114 | miR-1908-5 | cis | 1.34E-05 | Nightingale | XS_VLDL_P | NA | G | A | -0.039 | 0.0062 | 2.20E-11 |
| rs2072114 | miR-1908-5 | cis | 1.34E-05 | Nightingale | L_LDL_CE | NA | G | A | -0.05539 | 0.006228 | 5.00E-20 |
| rs2072114 | miR-1908-5 | cis | 1.34E-05 | Nightingale | XS_VLDL_C | NA | G | A | -0.10477 | 0.005954 | 3.30E-70 |
| rs2072114 | miR-1908-5 | cis | 1.34E-05 | Nightingale | LDL_CE | NA | G | A | -0.04454 | 0.006267 | 2.10E-13 |
| rs2072114 | miR-1908-5 | cis | 1.34E-05 | Nightingale | HDL_size | NA | G | A | -0.07997 | 0.005568 | 2.50E-49 |
| rs2072114 | miR-1908-5 | cis | 1.34E-05 | Nightingale | XL_VLDL_F | NA | G | A | -0.06801 | 0.006087 | 3.90E-30 |
| rs2072114 | miR-1908-5 | cis | 1.34E-05 | Nightingale | HDL_C | NA | G | A | -0.07827 | 0.005611 | 7.60E-46 |

| | | | | | | | | | | | |
|---|---|---|---|---|---|---|---|---|---|---|---|
| rs2072114 | miR-1908-5 | cis | 1.34E-05 | Nightingale | M_LDL_C | NA | G | A | -0.03426 | 0.006296 | 1.60E-08 |
| rs2072114 | miR-1908-5 | cis | 1.34E-05 | Nightingale | XS_VLDL_C | NA | G | A | -0.09995 | 0.005923 | 1.10E-64 |
| rs2072114 | miR-1908-5 | cis | 1.34E-05 | Nightingale | L_HDL_FC | NA | G | A | -0.09327 | 0.005507 | 3.30E-67 |
| rs2072114 | miR-1908-5 | cis | 1.34E-05 | Nightingale | XL_VLDL_P | NA | G | A | 0.058508 | 0.006064 | 1.30E-21 |
| rs2072114 | miR-1908-5 | cis | 1.34E-05 | Nightingale | L_VLDL_L | NA | G | A | 0.049487 | 0.006072 | 8.00E-16 |
| rs2072114 | miR-1908-5 | cis | 1.34E-05 | Nightingale | XL_HDL_L | NA | G | A | -0.09499 | 0.005646 | 1.70E-66 |
| rs2072114 | miR-1908-5 | cis | 1.34E-05 | Nightingale | L_HDL_PL | NA | G | A | -0.08616 | 0.00553 | 9.10E-58 |
| rs2072114 | miR-1908-5 | cis | 1.34E-05 | Nightingale | Sphingomy | NA | G | A | -0.11803 | 0.005872 | 6.70E-93 |
| rs2072114 | miR-1908-5 | cis | 1.34E-05 | Nightingale | Tyr | NA | G | A | 0.03534 | 0.006277 | 1.80E-08 |
| rs2072114 | miR-1908-5 | cis | 1.34E-05 | Nightingale | S_LDL_TG_ | NA | G | A | 0.093773 | 0.006088 | 2.60E-53 |
| rs2072114 | miR-1908-5 | cis | 1.34E-05 | Nightingale | XS_VLDL_C | NA | G | A | -0.0655 | 0.006098 | 8.40E-29 |
| rs2072114 | miR-1908-5 | cis | 1.34E-05 | Nightingale | M_VLDL_C | NA | G | A | -0.0513 | 0.006226 | 4.00E-17 |
| rs2072114 | miR-1908-5 | cis | 1.34E-05 | Nightingale | DHA_pct | NA | G | A | -0.26728 | 0.0061 | 0 |
| rs2072114 | miR-1908-5 | cis | 1.34E-05 | Nightingale | L_LDL_FC_ | NA | G | A | -0.09267 | 0.005989 | 2.30E-54 |
| rs2072114 | miR-1908-5 | cis | 1.34E-05 | Nightingale | XS_VLDL_F | NA | G | A | -0.10824 | 0.006236 | 4.70E-68 |
| rs2072114 | miR-1908-5 | cis | 1.34E-05 | Nightingale | M_HDL_FC | NA | G | A | -0.1141 | 0.005568 | 1.70E-97 |
| rs2072114 | miR-1908-5 | cis | 1.34E-05 | Nightingale | L_HDL_FC_ | NA | G | A | -0.11325 | 0.005837 | 1.20E-88 |
| rs2072114 | miR-1908-5 | cis | 1.34E-05 | Nightingale | PUFA_pct | NA | G | A | -0.12587 | 0.006044 | 2.80E-96 |
| rs2072114 | miR-1908-5 | cis | 1.34E-05 | Nightingale | L_VLDL_FC | NA | G | A | 0.050401 | 0.006078 | 2.90E-16 |
| rs2072114 | miR-1908-5 | cis | 1.34E-05 | Nightingale | XL_HDL_FC | NA | G | A | 0.066205 | 0.005571 | 1.30E-34 |
| rs2072114 | miR-1908-5 | cis | 1.34E-05 | Nightingale | XL_HDL_PL | NA | G | A | -0.09021 | 0.005648 | 1.60E-60 |
| rs2072114 | miR-1908-5 | cis | 1.34E-05 | Nightingale | XL_HDL_PL | NA | G | A | -0.06556 | 0.005772 | 3.50E-32 |
| rs2072114 | miR-1908-5 | cis | 1.34E-05 | Nightingale | XL_VLDL_T | NA | G | A | 0.066156 | 0.00601 | 2.70E-28 |
| rs2072114 | miR-1908-5 | cis | 1.34E-05 | Nightingale | M_HDL_PL | NA | G | A | 0.069642 | 0.00596 | 1.60E-31 |
| rs2072114 | miR-1908-5 | cis | 1.34E-05 | Nightingale | IDL_L | NA | G | A | -0.07573 | 0.006074 | 9.70E-38 |
| rs2072114 | miR-1908-5 | cis | 1.34E-05 | Nightingale | L_LDL_PL | NA | G | A | -0.05857 | 0.006236 | 4.60E-22 |
| rs2072114 | miR-1908-5 | cis | 1.34E-05 | Nightingale | M_VLDL_P | NA | G | A | -0.0665 | 0.006029 | 2.60E-29 |
| rs2072114 | miR-1908-5 | cis | 1.34E-05 | Nightingale | XXL_VLDL_ | NA | G | A | 0.067706 | 0.006063 | 3.20E-29 |
| rs2072114 | miR-1908-5 | cis | 1.34E-05 | Nightingale | L_VLDL_CE | NA | G | A | -0.0596 | 0.006013 | 1.50E-24 |
| rs2072114 | miR-1908-5 | cis | 1.34E-05 | Nightingale | M_HDL_PL | NA | G | A | -0.03408 | 0.005876 | 1.00E-09 |
| rs2072114 | miR-1908-5 | cis | 1.34E-05 | Nightingale | IDL_CE | NA | G | A | -0.07673 | 0.006065 | 2.50E-38 |
| rs2072114 | miR-1908-5 | cis | 1.34E-05 | Nightingale | S_VLDL_CE | NA | G | A | -0.03349 | 0.006199 | 5.00E-08 |
| rs2072114 | miR-1908-5 | cis | 1.34E-05 | Nightingale | XXL_VLDL_ | NA | G | A | 0.073613 | 0.006052 | 2.50E-34 |
| rs2072114 | miR-1908-5 | cis | 1.34E-05 | Nightingale | L_LDL_TG_ | NA | G | A | 0.082319 | 0.006161 | 9.90E-41 |
| rs2072114 | miR-1908-5 | cis | 1.34E-05 | Nightingale | M_LDL_PL | NA | G | A | 0.032736 | 0.00628 | 2.40E-08 |
| rs2072114 | miR-1908-5 | cis | 1.34E-05 | Nightingale | Omega_6_ | NA | G | A | 0.349718 | 0.006193 | 0 |
| rs2072114 | miR-1908-5 | cis | 1.34E-05 | Nightingale | VLDL_size | NA | G | A | 0.082293 | 0.005857 | 2.30E-46 |
| rs2072114 | miR-1908-5 | cis | 1.34E-05 | Nightingale | XL_VLDL_C | NA | G | A | 0.047149 | 0.006137 | 5.60E-14 |
| rs2072114 | miR-1908-5 | cis | 1.34E-05 | Nightingale | L_VLDL_C_ | NA | G | A | -0.03992 | 0.006138 | 1.40E-11 |
| rs2072114 | miR-1908-5 | cis | 1.34E-05 | Nightingale | XL_HDL_TG | NA | G | A | 0.095448 | 0.005919 | 4.60E-59 |
| rs2072114 | miR-1908-5 | cis | 1.34E-05 | Nightingale | S_HDL_TG | NA | G | A | 0.075221 | 0.006093 | 8.80E-35 |
| rs2072114 | miR-1908-5 | cis | 1.34E-05 | Nightingale | L_LDL_C_p | NA | G | A | -0.07998 | 0.006122 | 8.30E-39 |
| rs2072114 | miR-1908-5 | cis | 1.34E-05 | Nightingale | Omega_3 | NA | G | A | -0.31989 | 0.006159 | 0 |
| rs2072114 | miR-1908-5 | cis | 1.34E-05 | Nightingale | XL_VLDL_P | NA | G | A | 0.062765 | 0.006039 | 5.00E-25 |
| rs2072114 | miR-1908-5 | cis | 1.34E-05 | Nightingale | Total_P | NA | G | A | -0.05221 | 0.005913 | 6.80E-20 |
| rs2072114 | miR-1908-5 | cis | 1.34E-05 | Nightingale | Total_L | NA | G | A | -0.03774 | 0.006182 | 1.10E-10 |
| rs2072114 | miR-1908-5 | cis | 1.34E-05 | Nightingale | L_VLDL_PL | NA | G | A | 0.093411 | 0.006098 | 5.90E-53 |
| rs2072114 | miR-1908-5 | cis | 1.34E-05 | Nightingale | M_HDL_L | NA | G | A | -0.04102 | 0.005833 | 2.00E-13 |
| rs2072114 | miR-1908-5 | cis | 1.34E-05 | Nightingale | S_VLDL_PL | NA | G | A | -0.08725 | 0.006012 | 2.00E-48 |
| rs2072114 | miR-1908-5 | cis | 1.34E-05 | Nightingale | PUFA | NA | G | A | -0.09524 | 0.006128 | 5.50E-59 |
| rs2072114 | miR-1908-5 | cis | 1.34E-05 | Nightingale | M_HDL_C | NA | G | A | -0.05556 | 0.005781 | 6.10E-23 |
| rs2072114 | miR-1908-5 | cis | 1.34E-05 | Nightingale | M_LDL_FC | NA | G | A | -0.08414 | 0.006019 | 1.80E-44 |
| rs2072114 | miR-1908-5 | cis | 1.34E-05 | Nightingale | IDL_C | NA | G | A | -0.08204 | 0.006067 | 1.50E-43 |
| rs2072114 | miR-1908-5 | cis | 1.34E-05 | Nightingale | S_LDL_FC_ | NA | G | A | -0.09125 | 0.006089 | 8.90E-51 |
| rs2072114 | miR-1908-5 | cis | 1.34E-05 | Nightingale | M_VLDL_F | NA | G | A | -0.09496 | 0.005974 | 7.30E-59 |
| rs2072114 | miR-1908-5 | cis | 1.34E-05 | Nightingale | TG_by_PG | NA | G | A | 0.082454 | 0.005831 | 5.50E-46 |
| rs2072114 | miR-1908-5 | cis | 1.34E-05 | Nightingale | S_VLDL_TG | NA | G | A | 0.071581 | 0.00608 | 2.00E-32 |
| rs2072114 | miR-1908-5 | cis | 1.34E-05 | Nightingale | IDL_FC_pct | NA | G | A | -0.1152 | 0.006233 | 4.10E-76 |
| rs2072114 | miR-1908-5 | cis | 1.34E-05 | Nightingale | XL_VLDL_C | NA | G | A | -0.06878 | 0.006022 | 1.00E-30 |
| rs2072114 | miR-1908-5 | cis | 1.34E-05 | Nightingale | HDL_TG | NA | G | A | 0.052351 | 0.006196 | 1.10E-16 |
| rs2072114 | miR-1908-5 | cis | 1.34E-05 | Nightingale | L_LDL_C | NA | G | A | -0.06062 | 0.006213 | 1.10E-23 |
| rs2072114 | miR-1908-5 | cis | 1.34E-05 | Nightingale | XXL_VLDL_ | NA | G | A | 0.075172 | 0.006053 | 2.20E-35 |
| rs2072114 | miR-1908-5 | cis | 1.34E-05 | Nightingale | IDL_C_pct | NA | G | A | -0.08529 | 0.006134 | 6.10E-44 |

| | | | | | | | | | | | |
|---|---|---|---|---|---|---|---|---|---|---|---|
| rs2072114 | miR-1908-5 | cis | 1.34E-05 | Nightingale | L_HDL_P | NA | G | A | -0.08691 | 0.005503 | 8.50E-59 |
| rs2072114 | miR-1908-5 | cis | 1.34E-05 | Nightingale | M_VLDL_C | NA | G | A | -0.04026 | 0.006266 | 4.40E-11 |
| rs2072114 | miR-1908-5 | cis | 1.34E-05 | Nightingale | L_HDL_C | NA | G | A | -0.08961 | 0.005533 | 2.90E-61 |
| rs2072114 | miR-1908-5 | cis | 1.34E-05 | Nightingale | LDL_C | NA | G | A | -0.05167 | 0.006257 | 2.00E-17 |
| rs2072114 | miR-1908-5 | cis | 1.34E-05 | Nightingale | Total_C | NA | G | A | -0.06924 | 0.006083 | 1.40E-31 |
| rs2072114 | miR-1908-5 | cis | 1.34E-05 | Nightingale | VLDL_L | NA | G | A | 0.037401 | 0.006161 | 4.20E-09 |
| rs2072114 | miR-1908-5 | cis | 1.34E-05 | Nightingale | L_VLDL_TG | NA | G | A | 0.048247 | 0.006046 | 1.80E-15 |
| rs2072114 | miR-1908-5 | cis | 1.34E-05 | Nightingale | XL_VLDL_F | NA | G | A | 0.054007 | 0.006086 | 2.10E-18 |
| rs2072114 | miR-1908-5 | cis | 1.34E-05 | Nightingale | non_HDL_C | NA | G | A | -0.04802 | 0.00625 | 1.80E-15 |
| rs2072114 | miR-1908-5 | cis | 1.34E-05 | Nightingale | VLDL_TG | NA | G | A | 0.05827 | 0.006047 | 9.40E-22 |
| rs2072114 | miR-1908-5 | cis | 1.34E-05 | Nightingale | XL_VLDL_C | NA | G | A | 0.039127 | 0.006184 | 9.90E-10 |
| rs2072114 | miR-1908-5 | cis | 1.34E-05 | Nightingale | Clinical_LD | NA | G | A | -0.05805 | 0.006247 | 1.70E-21 |
| rs2072114 | miR-1908-5 | cis | 1.34E-05 | Nightingale | XL_HDL_FC | NA | G | A | -0.10147 | 0.005853 | 4.30E-70 |
| rs2072114 | miR-1908-5 | cis | 1.34E-05 | Nightingale | S_VLDL_TG | NA | G | A | 0.049709 | 0.006097 | 1.40E-15 |
| rs2072114 | miR-1908-5 | cis | 1.34E-05 | Nightingale | XXL_VLDL_ | NA | G | A | 0.053098 | 0.006312 | 5.00E-17 |
| rs2072114 | miR-1908-5 | cis | 1.34E-05 | Nightingale | XXL_VLDL_ | NA | G | A | 0.080319 | 0.006068 | 1.30E-39 |
| rs2072114 | miR-1908-5 | cis | 1.34E-05 | Nightingale | HDL_FC | NA | G | A | -0.08507 | 0.005566 | 5.30E-55 |
| rs2072114 | miR-1908-5 | cis | 1.34E-05 | Nightingale | M_LDL_C_p | NA | G | A | -0.09707 | 0.006288 | 1.40E-54 |
| rs2072114 | miR-1908-5 | cis | 1.34E-05 | Nightingale | M_LDL_P | NA | G | A | -0.04343 | 0.006302 | 9.30E-13 |
| rs2072114 | miR-1908-5 | cis | 1.34E-05 | Nightingale | L_VLDL_P | NA | G | A | 0.052538 | 0.006067 | 1.20E-17 |
| rs2072114 | miR-1908-5 | cis | 1.34E-05 | Nightingale | M_LDL_TG | NA | G | A | 0.08009 | 0.006204 | 2.90E-38 |
| rs2072114 | miR-1908-5 | cis | 1.34E-05 | Nightingale | M_HDL_TG | NA | G | A | 0.055063 | 0.006206 | 3.20E-18 |
| rs2072114 | miR-1908-5 | cis | 1.34E-05 | Nightingale | XL_VLDL_T | NA | G | A | 0.062127 | 0.006086 | 4.00E-25 |
| rs2072114 | miR-1908-5 | cis | 1.34E-05 | Nightingale | M_VLDL_C | NA | G | A | -0.08758 | 0.005914 | 1.00E-50 |
| rs2072114 | miR-1908-5 | cis | 1.34E-05 | Nightingale | L_LDL_P | NA | G | A | -0.03576 | 0.006262 | 3.70E-09 |
| rs2072114 | miR-1908-5 | cis | 1.34E-05 | Nightingale | M_LDL_TG | NA | G | A | 0.041056 | 0.006191 | 9.40E-11 |
| rs2072114 | miR-1908-5 | cis | 1.34E-05 | Nightingale | XXL_VLDL_ | NA | G | A | 0.074358 | 0.006043 | 5.30E-35 |
| rs2140578 | miR-339-5p | cis | 0.000599 | Nightingale | L_VLDL_C_ | NA | C | T | -0.02151 | 0.004195 | 3.60E-08 |
| rs2140578 | miR-339-5p | cis | 0.000599 | Nightingale | L_VLDL_CE | NA | C | T | -0.02216 | 0.00411 | 1.80E-08 |
| rs2280725 | miR-339-5p | cis | 1.20E-08 | Nightingale | M_VLDL_T | NA | T | G | 0.022309 | 0.003928 | 1.70E-08 |
| rs2280725 | miR-339-5p | cis | 1.20E-08 | Nightingale | XL_VLDL_T | NA | T | G | 0.022941 | 0.00403 | 6.40E-09 |
| rs2280725 | miR-339-5p | cis | 1.20E-08 | Nightingale | XL_VLDL_F | NA | T | G | -0.0221 | 0.004031 | 4.20E-08 |
| rs2280725 | miR-339-5p | cis | 1.20E-08 | Nightingale | M_VLDL_C | NA | T | G | -0.02156 | 0.003915 | 4.30E-08 |
| rs2280725 | miR-339-5p | cis | 1.20E-08 | Nightingale | L_VLDL_CE | NA | T | G | -0.02455 | 0.003979 | 5.30E-10 |
| rs2280725 | miR-339-5p | cis | 1.20E-08 | Nightingale | M_VLDL_F | NA | T | G | -0.02288 | 0.003955 | 9.90E-09 |
| rs2280725 | miR-339-5p | cis | 1.20E-08 | Nightingale | M_VLDL_P | NA | T | G | -0.02362 | 0.003991 | 3.60E-09 |
| rs2280725 | miR-339-5p | cis | 1.20E-08 | Nightingale | L_VLDL_C_ | NA | T | G | -0.02489 | 0.004062 | 3.00E-10 |
| rs2363279 | miR-339-5p | cis | 4.17E-07 | Nightingale | L_VLDL_CE | NA | G | A | -0.02368 | 0.004 | 3.60E-09 |
| rs2363279 | miR-339-5p | cis | 4.17E-07 | Nightingale | L_VLDL_C_ | NA | G | A | -0.02329 | 0.004084 | 5.30E-09 |
| rs2363285 | miR-339-5p | cis | 1.14E-08 | Nightingale | M_VLDL_T | NA | T | C | 0.022574 | 0.003931 | 1.20E-08 |
| rs2363285 | miR-339-5p | cis | 1.14E-08 | Nightingale | M_VLDL_C | NA | T | C | -0.02184 | 0.003918 | 2.90E-08 |
| rs2363285 | miR-339-5p | cis | 1.14E-08 | Nightingale | XL_VLDL_C | NA | T | C | -0.02175 | 0.003989 | 4.10E-08 |
| rs2363285 | miR-339-5p | cis | 1.14E-08 | Nightingale | M_VLDL_F | NA | T | C | -0.02313 | 0.003957 | 7.40E-09 |
| rs2363285 | miR-339-5p | cis | 1.14E-08 | Nightingale | XL_VLDL_T | NA | T | C | 0.023269 | 0.004033 | 4.00E-09 |
| rs2363285 | miR-339-5p | cis | 1.14E-08 | Nightingale | M_VLDL_P | NA | T | C | -0.02378 | 0.003993 | 3.10E-09 |
| rs2363285 | miR-339-5p | cis | 1.14E-08 | Nightingale | XL_VLDL_F | NA | T | C | -0.0224 | 0.004033 | 2.70E-08 |
| rs2363285 | miR-339-5p | cis | 1.14E-08 | Nightingale | L_VLDL_CE | NA | T | C | -0.02468 | 0.003981 | 4.40E-10 |
| rs2363285 | miR-339-5p | cis | 1.14E-08 | Nightingale | L_VLDL_C_ | NA | T | C | -0.02488 | 0.004064 | 3.00E-10 |
| rs2391174 | miR-148a-3 | cis | 0.000164 | Nightingale | M_LDL_FC_ | NA | G | A | 0.031028 | 0.004526 | 6.50E-13 |
| rs2391174 | miR-148a-3 | cis | 0.000164 | Nightingale | XL_VLDL_C | NA | G | A | 0.027017 | 0.004526 | 1.60E-10 |
| rs2391174 | miR-148a-3 | cis | 0.000164 | Nightingale | XL_VLDL_P | NA | G | A | -0.0261 | 0.004559 | 3.90E-09 |
| rs2391174 | miR-148a-3 | cis | 0.000164 | Nightingale | L_HDL_CE_ | NA | G | A | 0.026741 | 0.00452 | 1.40E-09 |
| rs2391174 | miR-148a-3 | cis | 0.000164 | Nightingale | S_HDL_C_p | NA | G | A | 0.028162 | 0.004669 | 3.00E-10 |
| rs2391174 | miR-148a-3 | cis | 0.000164 | Nightingale | XXL_VLDL_ | NA | G | A | -0.02954 | 0.00455 | 1.50E-11 |
| rs2391174 | miR-148a-3 | cis | 0.000164 | Nightingale | L_VLDL_L | NA | G | A | -0.02635 | 0.004565 | 4.20E-09 |
| rs2391174 | miR-148a-3 | cis | 0.000164 | Nightingale | M_HDL_TG | NA | G | A | -0.0291 | 0.004666 | 1.10E-10 |
| rs2391174 | miR-148a-3 | cis | 0.000164 | Nightingale | L_HDL_TG_ | NA | G | A | -0.02541 | 0.004461 | 8.60E-09 |
| rs2391174 | miR-148a-3 | cis | 0.000164 | Nightingale | S_VLDL_C_ | NA | G | A | 0.027048 | 0.004595 | 7.70E-10 |
| rs2391174 | miR-148a-3 | cis | 0.000164 | Nightingale | M_HDL_C_ | NA | G | A | 0.025045 | 0.004499 | 8.30E-09 |
| rs2391174 | miR-148a-3 | cis | 0.000164 | Nightingale | S_HDL_TG | NA | G | A | -0.02779 | 0.004582 | 4.70E-10 |
| rs2391174 | miR-148a-3 | cis | 0.000164 | Nightingale | XL_VLDL_F | NA | G | A | -0.02555 | 0.004576 | 9.70E-09 |
| rs2391174 | miR-148a-3 | cis | 0.000164 | Nightingale | M_VLDL_T | NA | G | A | -0.02656 | 0.004589 | 3.80E-09 |
| rs2391174 | miR-148a-3 | cis | 0.000164 | Nightingale | L_HDL_C_p | NA | G | A | 0.026837 | 0.004454 | 4.80E-10 |

| | | | | | | | | | | | | |
|---|---|---|---|---|---|---|---|---|---|---|---|---|
| rs2391174 | miR-148a-3 | cis | | 0.000164 | Nightingale | M_HDL_TG | NA | G | A | -0.02538 | 0.004533 | 1.00E-08 |
| rs2391174 | miR-148a-3 | cis | | 0.000164 | Nightingale | L_LDL_FC_ | NA | G | A | 0.027834 | 0.004504 | 2.60E-10 |
| rs2391174 | miR-148a-3 | cis | | 0.000164 | Nightingale | XXL_VLDL_ | NA | G | A | -0.02828 | 0.004554 | 1.10E-10 |
| rs2391174 | miR-148a-3 | cis | | 0.000164 | Nightingale | M_VLDL_T | NA | G | A | -0.02947 | 0.004461 | 2.10E-12 |
| rs2391174 | miR-148a-3 | cis | | 0.000164 | Nightingale | XXL_VLDL_ | NA | G | A | -0.02925 | 0.004558 | 1.90E-11 |
| rs2391174 | miR-148a-3 | cis | | 0.000164 | Nightingale | XS_VLDL_C | NA | G | A | 0.023928 | 0.004477 | 4.10E-08 |
| rs2391174 | miR-148a-3 | cis | | 0.000164 | Nightingale | XXL_VLDL_ | NA | G | A | -0.0292 | 0.004544 | 2.80E-11 |
| rs2391174 | miR-148a-3 | cis | | 0.000164 | Nightingale | XL_HDL_TG | NA | G | A | -0.02705 | 0.004452 | 3.50E-10 |
| rs2391174 | miR-148a-3 | cis | | 0.000164 | Nightingale | M_VLDL_C | NA | G | A | 0.031741 | 0.004435 | 3.80E-14 |
| rs2391174 | miR-148a-3 | cis | | 0.000164 | Nightingale | M_VLDL_C | NA | G | A | 0.030719 | 0.004446 | 2.30E-13 |
| rs2391174 | miR-148a-3 | cis | | 0.000164 | Nightingale | TG_by_PG | NA | G | A | -0.02625 | 0.004385 | 1.10E-09 |
| rs2391174 | miR-148a-3 | cis | | 0.000164 | Nightingale | XL_VLDL_L | NA | G | A | -0.0274 | 0.004545 | 6.00E-10 |
| rs2391174 | miR-148a-3 | cis | | 0.000164 | Nightingale | HDL_TG | NA | G | A | -0.0282 | 0.004659 | 4.70E-10 |
| rs2391174 | miR-148a-3 | cis | | 0.000164 | Nightingale | Omega_6_ | NA | G | A | 0.028647 | 0.004596 | 4.00E-10 |
| rs2391174 | miR-148a-3 | cis | | 0.000164 | Nightingale | XXL_VLDL_ | NA | G | A | -0.02934 | 0.004534 | 1.70E-11 |
| rs2391174 | miR-148a-3 | cis | | 0.000164 | Nightingale | LA_pct | NA | G | A | 0.025122 | 0.00462 | 3.50E-08 |
| rs2391174 | miR-148a-3 | cis | | 0.000164 | Nightingale | S_LDL_FC_ | NA | G | A | 0.031334 | 0.004579 | 1.10E-12 |
| rs2391174 | miR-148a-3 | cis | | 0.000164 | Nightingale | XS_VLDL_T | NA | G | A | -0.02536 | 0.004488 | 4.50E-09 |
| rs2391174 | miR-148a-3 | cis | | 0.000164 | Nightingale | XXL_VLDL_ | NA | G | A | -0.02926 | 0.004552 | 2.30E-11 |
| rs2391174 | miR-148a-3 | cis | | 0.000164 | Nightingale | XL_VLDL_P | NA | G | A | -0.02735 | 0.004541 | 5.50E-10 |
| rs2391174 | miR-148a-3 | cis | | 0.000164 | Nightingale | S_HDL_TG_ | NA | G | A | -0.02444 | 0.004495 | 3.60E-08 |
| rs2391174 | miR-148a-3 | cis | | 0.000164 | Nightingale | VLDL_TG | NA | G | A | -0.02907 | 0.004546 | 5.40E-11 |
| rs2391174 | miR-148a-3 | cis | | 0.000164 | Nightingale | XXL_VLDL_ | NA | G | A | -0.02665 | 0.004563 | 1.20E-09 |
| rs2391174 | miR-148a-3 | cis | | 0.000164 | Nightingale | XL_VLDL_C | NA | G | A | 0.027824 | 0.004528 | 4.30E-11 |
| rs2391174 | miR-148a-3 | cis | | 0.000164 | Nightingale | M_VLDL_F | NA | G | A | 0.026263 | 0.004491 | 3.00E-10 |
| rs2391174 | miR-148a-3 | cis | | 0.000164 | Nightingale | S_VLDL_FC | NA | G | A | 0.027496 | 0.004511 | 1.40E-10 |
| rs2391174 | miR-148a-3 | cis | | 0.000164 | Nightingale | S_VLDL_TG | NA | G | A | -0.02781 | 0.00457 | 2.20E-10 |
| rs2391174 | miR-148a-3 | cis | | 0.000164 | Nightingale | L_VLDL_PL | NA | G | A | -0.02584 | 0.004558 | 6.40E-09 |
| rs2391174 | miR-148a-3 | cis | | 0.000164 | Nightingale | M_HDL_CE | NA | G | A | 0.026572 | 0.004582 | 3.00E-09 |
| rs2391174 | miR-148a-3 | cis | | 0.000164 | Nightingale | S_VLDL_TG | NA | G | A | -0.02805 | 0.004585 | 4.90E-10 |
| rs2391174 | miR-148a-3 | cis | | 0.000164 | Nightingale | L_VLDL_P | NA | G | A | -0.02631 | 0.004561 | 3.90E-09 |
| rs2391174 | miR-148a-3 | cis | | 0.000164 | Nightingale | XL_VLDL_T | NA | G | A | -0.02903 | 0.004518 | 3.10E-11 |
| rs2391174 | miR-148a-3 | cis | | 0.000164 | Nightingale | S_VLDL_PL | NA | G | A | 0.028289 | 0.004521 | 6.30E-11 |
| rs2391174 | miR-148a-3 | cis | | 0.000164 | Nightingale | L_VLDL_FC | NA | G | A | -0.02445 | 0.00457 | 4.70E-08 |
| rs2391174 | miR-148a-3 | cis | | 0.000164 | Nightingale | L_VLDL_TG | NA | G | A | -0.02791 | 0.004545 | 3.40E-10 |
| rs2391174 | miR-148a-3 | cis | | 0.000164 | Nightingale | XL_VLDL_T | NA | G | A | -0.02533 | 0.004576 | 3.10E-09 |
| rs2391174 | miR-148a-3 | cis | | 0.000164 | Nightingale | S_LDL_TG_ | NA | G | A | -0.02966 | 0.004579 | 2.20E-11 |
| rs2391174 | miR-148a-3 | cis | | 0.000164 | Nightingale | VLDL_size | NA | G | A | -0.02755 | 0.004403 | 3.90E-11 |
| rs2391174 | miR-148a-3 | cis | | 0.000164 | Nightingale | Total_TG | NA | G | A | -0.02876 | 0.004573 | 1.10E-10 |
| rs2524299 | miR-1908-5 | cis | | 4.82E-05 | Nightingale | L_HDL_FC_ | NA | T | A | -0.11519 | 0.005907 | 2.20E-88 |
| rs2524299 | miR-1908-5 | cis | | 4.82E-05 | Nightingale | L_LDL_FC_ | NA | T | A | -0.0928 | 0.006061 | 1.30E-53 |
| rs2524299 | miR-1908-5 | cis | | 4.82E-05 | Nightingale | XS_VLDL_L | NA | T | A | -0.03712 | 0.00625 | 5.50E-10 |
| rs2524299 | miR-1908-5 | cis | | 4.82E-05 | Nightingale | S_HDL_TG | NA | T | A | 0.075609 | 0.006166 | 1.40E-34 |
| rs2524299 | miR-1908-5 | cis | | 4.82E-05 | Nightingale | M_HDL_P | NA | T | A | -0.05075 | 0.005858 | 7.40E-19 |
| rs2524299 | miR-1908-5 | cis | | 4.82E-05 | Nightingale | L_LDL_FC | NA | T | A | -0.07358 | 0.006244 | 2.30E-33 |
| rs2524299 | miR-1908-5 | cis | | 4.82E-05 | Nightingale | S_LDL_TG | NA | T | A | 0.057127 | 0.006203 | 4.20E-20 |
| rs2524299 | miR-1908-5 | cis | | 4.82E-05 | Nightingale | L_VLDL_TG | NA | T | A | 0.046831 | 0.006119 | 1.70E-14 |
| rs2524299 | miR-1908-5 | cis | | 4.82E-05 | Nightingale | XXL_VLDL_ | NA | T | A | 0.079569 | 0.006103 | 2.00E-39 |
| rs2524299 | miR-1908-5 | cis | | 4.82E-05 | Nightingale | L_LDL_PL | NA | T | A | -0.05925 | 0.006311 | 5.30E-22 |
| rs2524299 | miR-1908-5 | cis | | 4.82E-05 | Nightingale | XL_VLDL_L | NA | T | A | 0.060267 | 0.006118 | 5.80E-23 |
| rs2524299 | miR-1908-5 | cis | | 4.82E-05 | Nightingale | MUFA | NA | T | A | 0.066964 | 0.006295 | 2.20E-26 |
| rs2524299 | miR-1908-5 | cis | | 4.82E-05 | Nightingale | Remnant_( | NA | T | A | -0.04363 | 0.006318 | 9.70E-13 |
| rs2524299 | miR-1908-5 | cis | | 4.82E-05 | Nightingale | XL_HDL_CE | NA | T | A | -0.03989 | 0.006355 | 4.90E-10 |
| rs2524299 | miR-1908-5 | cis | | 4.82E-05 | Nightingale | L_HDL_P | NA | T | A | -0.09038 | 0.005569 | 2.60E-61 |
| rs2524299 | miR-1908-5 | cis | | 4.82E-05 | Nightingale | S_VLDL_FC | NA | T | A | -0.09051 | 0.006072 | 4.20E-51 |
| rs2524299 | miR-1908-5 | cis | | 4.82E-05 | Nightingale | XL_VLDL_F | NA | T | A | -0.06748 | 0.00616 | 4.80E-29 |
| rs2524299 | miR-1908-5 | cis | | 4.82E-05 | Nightingale | M_HDL_TG | NA | T | A | 0.053926 | 0.00628 | 1.50E-17 |
| rs2524299 | miR-1908-5 | cis | | 4.82E-05 | Nightingale | XL_VLDL_P | NA | T | A | 0.062301 | 0.006112 | 2.00E-24 |
| rs2524299 | miR-1908-5 | cis | | 4.82E-05 | Nightingale | M_LDL_C | NA | T | A | -0.0348 | 0.006371 | 1.50E-08 |
| rs2524299 | miR-1908-5 | cis | | 4.82E-05 | Nightingale | LA_pct | NA | T | A | 0.165286 | 0.006215 | 1.60E-156 |
| rs2524299 | miR-1908-5 | cis | | 4.82E-05 | Nightingale | LA | NA | T | A | 0.071555 | 0.006236 | 1.30E-30 |
| rs2524299 | miR-1908-5 | cis | | 4.82E-05 | Nightingale | IDL_FC | NA | T | A | -0.09617 | 0.006155 | 3.30E-57 |
| rs2524299 | miR-1908-5 | cis | | 4.82E-05 | Nightingale | L_HDL_L | NA | T | A | -0.09039 | 0.00558 | 3.80E-61 |

| | | | | | | | | | | | | |
|---|---|---|---|---|---|---|---|---|---|---|---|---|
| rs2524299 | miR-1908-5 | cis | | 4.82E-05 | Nightingale | M_HDL_CE | NA | T | A | -0.05569 | 0.005873 | 6.30E-22 |
| rs2524299 | miR-1908-5 | cis | | 4.82E-05 | Nightingale | IDL_CE_pct | NA | T | A | -0.04733 | 0.00624 | 1.80E-14 |
| rs2524299 | miR-1908-5 | cis | | 4.82E-05 | Nightingale | L_HDL_TG_ | NA | T | A | 0.093651 | 0.006001 | 3.90E-55 |
| rs2524299 | miR-1908-5 | cis | | 4.82E-05 | Nightingale | L_VLDL_PL | NA | T | A | 0.059109 | 0.006135 | 7.00E-22 |
| rs2524299 | miR-1908-5 | cis | | 4.82E-05 | Nightingale | XXL_VLDL_ | NA | T | A | 0.074052 | 0.006126 | 4.90E-34 |
| rs2524299 | miR-1908-5 | cis | | 4.82E-05 | Nightingale | XS_VLDL_T | NA | T | A | 0.101821 | 0.00604 | 7.00E-65 |
| rs2524299 | miR-1908-5 | cis | | 4.82E-05 | Nightingale | XXL_VLDL_ | NA | T | A | 0.077964 | 0.006128 | 2.50E-37 |
| rs2524299 | miR-1908-5 | cis | | 4.82E-05 | Nightingale | M_LDL_FC | NA | T | A | -0.05675 | 0.006352 | 4.80E-20 |
| rs2524299 | miR-1908-5 | cis | | 4.82E-05 | Nightingale | PUFA_pct | NA | T | A | -0.12715 | 0.006117 | 1.80E-96 |
| rs2524299 | miR-1908-5 | cis | | 4.82E-05 | Nightingale | M_LDL_P | NA | T | A | -0.04412 | 0.006378 | 9.60E-13 |
| rs2524299 | miR-1908-5 | cis | | 4.82E-05 | Nightingale | HDL_L | NA | T | A | -0.06905 | 0.00571 | 7.70E-35 |
| rs2524299 | miR-1908-5 | cis | | 4.82E-05 | Nightingale | S_LDL_CE_ | NA | T | A | 0.060165 | 0.006347 | 7.70E-21 |
| rs2524299 | miR-1908-5 | cis | | 4.82E-05 | Nightingale | Omega_6_ | NA | T | A | 0.350946 | 0.006267 | 0 |
| rs2524299 | miR-1908-5 | cis | | 4.82E-05 | Nightingale | L_HDL_PL_ | NA | T | A | 0.040494 | 0.006155 | 1.10E-10 |
| rs2524299 | miR-1908-5 | cis | | 4.82E-05 | Nightingale | LDL_FC | NA | T | A | -0.06922 | 0.006291 | 2.60E-29 |
| rs2524299 | miR-1908-5 | cis | | 4.82E-05 | Nightingale | Total_C | NA | T | A | -0.07104 | 0.006156 | 5.00E-32 |
| rs2524299 | miR-1908-5 | cis | | 4.82E-05 | Nightingale | XL_VLDL_C | NA | T | A | 0.046661 | 0.006211 | 1.20E-13 |
| rs2524299 | miR-1908-5 | cis | | 4.82E-05 | Nightingale | L_VLDL_CE | NA | T | A | -0.05797 | 0.006085 | 6.40E-23 |
| rs2524299 | miR-1908-5 | cis | | 4.82E-05 | Nightingale | S_HDL_C_p | NA | T | A | -0.05175 | 0.006285 | 3.50E-16 |
| rs2524299 | miR-1908-5 | cis | | 4.82E-05 | Nightingale | Omega_3 | NA | T | A | -0.32186 | 0.006233 | 0 |
| rs2524299 | miR-1908-5 | cis | | 4.82E-05 | Nightingale | ApoB | NA | T | A | -0.037 | 0.006351 | 1.90E-09 |
| rs2524299 | miR-1908-5 | cis | | 4.82E-05 | Nightingale | LDL_PL | NA | T | A | -0.04927 | 0.006345 | 1.60E-15 |
| rs2524299 | miR-1908-5 | cis | | 4.82E-05 | Nightingale | M_HDL_PL | NA | T | A | -0.03711 | 0.005946 | 1.50E-10 |
| rs2524299 | miR-1908-5 | cis | | 4.82E-05 | Nightingale | L_LDL_TG_ | NA | T | A | 0.082764 | 0.006235 | 9.80E-41 |
| rs2524299 | miR-1908-5 | cis | | 4.82E-05 | Nightingale | M_HDL_C | NA | T | A | -0.05845 | 0.005851 | 3.50E-24 |
| rs2524299 | miR-1908-5 | cis | | 4.82E-05 | Nightingale | XS_VLDL_C | NA | T | A | -0.07201 | 0.006136 | 1.20E-33 |
| rs2524299 | miR-1908-5 | cis | | 4.82E-05 | Nightingale | Omega_3_ | NA | T | A | -0.37287 | 0.006214 | 0 |
| rs2524299 | miR-1908-5 | cis | | 4.82E-05 | Nightingale | M_HDL_CE | NA | T | A | -0.05981 | 0.006167 | 3.60E-22 |
| rs2524299 | miR-1908-5 | cis | | 4.82E-05 | Nightingale | Total_L | NA | T | A | -0.03986 | 0.006256 | 4.10E-11 |
| rs2524299 | miR-1908-5 | cis | | 4.82E-05 | Nightingale | HDL_CE | NA | T | A | -0.07849 | 0.005707 | 7.50E-44 |
| rs2524299 | miR-1908-5 | cis | | 4.82E-05 | Nightingale | XS_VLDL_T | NA | T | A | 0.038707 | 0.006261 | 1.30E-09 |
| rs2524299 | miR-1908-5 | cis | | 4.82E-05 | Nightingale | IDL_P | NA | T | A | -0.04949 | 0.006292 | 6.00E-16 |
| rs2524299 | miR-1908-5 | cis | | 4.82E-05 | Nightingale | S_LDL_PL_ | NA | T | A | -0.06082 | 0.006346 | 3.70E-21 |
| rs2524299 | miR-1908-5 | cis | | 4.82E-05 | Nightingale | S_VLDL_PL | NA | T | A | -0.08734 | 0.006085 | 1.20E-47 |
| rs2524299 | miR-1908-5 | cis | | 4.82E-05 | Nightingale | L_HDL_CE_ | NA | T | A | -0.0582 | 0.006084 | 3.70E-21 |
| rs2524299 | miR-1908-5 | cis | | 4.82E-05 | Nightingale | XS_VLDL_C | NA | T | A | -0.1012 | 0.005994 | 5.20E-65 |
| rs2524299 | miR-1908-5 | cis | | 4.82E-05 | Nightingale | L_VLDL_C | NA | T | A | 0.039464 | 0.00622 | 4.80E-10 |
| rs2524299 | miR-1908-5 | cis | | 4.82E-05 | Nightingale | TG_by_PG | NA | T | A | 0.082974 | 0.005902 | 6.60E-46 |
| rs2524299 | miR-1908-5 | cis | | 4.82E-05 | Nightingale | XL_HDL_PL | NA | T | A | -0.09261 | 0.005716 | 2.10E-61 |
| rs2524299 | miR-1908-5 | cis | | 4.82E-05 | Nightingale | M_HDL_FC | NA | T | A | -0.06769 | 0.005787 | 1.10E-32 |
| rs2524299 | miR-1908-5 | cis | | 4.82E-05 | Nightingale | Phosphogly | NA | T | A | -0.04995 | 0.006024 | 7.80E-18 |
| rs2524299 | miR-1908-5 | cis | | 4.82E-05 | Nightingale | LDL_L | NA | T | A | -0.04791 | 0.006338 | 7.60E-15 |
| rs2524299 | miR-1908-5 | cis | | 4.82E-05 | Nightingale | XS_VLDL_C | NA | T | A | -0.10582 | 0.006025 | 3.70E-70 |
| rs2524299 | miR-1908-5 | cis | | 4.82E-05 | Nightingale | L_LDL_L | NA | T | A | -0.05766 | 0.006297 | 5.30E-21 |
| rs2524299 | miR-1908-5 | cis | | 4.82E-05 | Nightingale | Sphingomy | NA | T | A | -0.11941 | 0.005943 | 2.20E-92 |
| rs2524299 | miR-1908-5 | cis | | 4.82E-05 | Nightingale | L_HDL_FC | NA | T | A | -0.09656 | 0.005573 | 1.80E-69 |
| rs2524299 | miR-1908-5 | cis | | 4.82E-05 | Nightingale | M_HDL_PL | NA | T | A | 0.07017 | 0.006032 | 2.60E-31 |
| rs2524299 | miR-1908-5 | cis | | 4.82E-05 | Nightingale | S_VLDL_C_ | NA | T | A | -0.06161 | 0.006187 | 8.30E-24 |
| rs2524299 | miR-1908-5 | cis | | 4.82E-05 | Nightingale | IDL_PL_pct | NA | T | A | 0.034578 | 0.006383 | 5.00E-08 |
| rs2524299 | miR-1908-5 | cis | | 4.82E-05 | Nightingale | M_LDL_PL_ | NA | T | A | 0.03444 | 0.006355 | 1.30E-08 |
| rs2524299 | miR-1908-5 | cis | | 4.82E-05 | Nightingale | L_VLDL_FC | NA | T | A | 0.050156 | 0.006151 | 5.30E-16 |
| rs2524299 | miR-1908-5 | cis | | 4.82E-05 | Nightingale | M_VLDL_F | NA | T | A | -0.09426 | 0.006046 | 8.40E-57 |
| rs2524299 | miR-1908-5 | cis | | 4.82E-05 | Nightingale | IDL_C | NA | T | A | -0.08354 | 0.00614 | 5.80E-44 |
| rs2524299 | miR-1908-5 | cis | | 4.82E-05 | Nightingale | XL_HDL_L | NA | T | A | -0.09735 | 0.005714 | 1.70E-67 |
| rs2524299 | miR-1908-5 | cis | | 4.82E-05 | Nightingale | HDL_TG | NA | T | A | 0.051317 | 0.00627 | 3.70E-16 |
| rs2524299 | miR-1908-5 | cis | | 4.82E-05 | Nightingale | S_HDL_FC_ | NA | T | A | -0.10493 | 0.006031 | 6.90E-71 |
| rs2524299 | miR-1908-5 | cis | | 4.82E-05 | Nightingale | LDL_C | NA | T | A | -0.05235 | 0.006332 | 2.10E-17 |
| rs2524299 | miR-1908-5 | cis | | 4.82E-05 | Nightingale | Unsaturati | NA | T | A | -0.3287 | 0.006023 | 0 |
| rs2524299 | miR-1908-5 | cis | | 4.82E-05 | Nightingale | VLDL_size | NA | T | A | 0.082145 | 0.005927 | 2.60E-45 |
| rs2524299 | miR-1908-5 | cis | | 4.82E-05 | Nightingale | LDL_CE | NA | T | A | -0.04524 | 0.006343 | 2.10E-13 |
| rs2524299 | miR-1908-5 | cis | | 4.82E-05 | Nightingale | L_VLDL_L | NA | T | A | 0.048593 | 0.006145 | 3.80E-15 |
| rs2524299 | miR-1908-5 | cis | | 4.82E-05 | Nightingale | Cholines | NA | T | A | -0.04716 | 0.005981 | 3.40E-16 |
| rs2524299 | miR-1908-5 | cis | | 4.82E-05 | Nightingale | M_VLDL_T | NA | T | A | 0.041113 | 0.006178 | 4.50E-11 |

| | | | | | | | | | | | |
|---|---|---|---|---|---|---|---|---|---|---|---|
| rs2524299 | miR-1908-! | cis | 4.82E-05 | Nightingale | ApoA1 | NA | T | A | -0.05674 | 0.005819 | 2.40E-23 |
| rs2524299 | miR-1908-! | cis | 4.82E-05 | Nightingale | M_VLDL_C | NA | T | A | -0.04127 | 0.006342 | 3.10E-11 |
| rs2524299 | miR-1908-! | cis | 4.82E-05 | Nightingale | XS_VLDL_C | NA | T | A | -0.06688 | 0.006172 | 7.00E-29 |
| rs2524299 | miR-1908-! | cis | 4.82E-05 | Nightingale | L_LDL_C | NA | T | A | -0.06135 | 0.006288 | 1.50E-23 |
| rs2524299 | miR-1908-! | cis | 4.82E-05 | Nightingale | Total_CE | NA | T | A | -0.07394 | 0.006134 | 1.20E-34 |
| rs2524299 | miR-1908-! | cis | 4.82E-05 | Nightingale | IDL_TG_pct | NA | T | A | 0.095365 | 0.006143 | 3.60E-55 |
| rs2524299 | miR-1908-! | cis | 4.82E-05 | Nightingale | Total_TG | NA | T | A | 0.056193 | 0.006155 | 7.20E-20 |
| rs2524299 | miR-1908-! | cis | 4.82E-05 | Nightingale | PUFA | NA | T | A | -0.09805 | 0.006202 | 5.90E-60 |
| rs2524299 | miR-1908-! | cis | 4.82E-05 | Nightingale | HDL_PL | NA | T | A | -0.06346 | 0.005749 | 1.90E-29 |
| rs2524299 | miR-1908-! | cis | 4.82E-05 | Nightingale | Total_P | NA | T | A | -0.05484 | 0.005984 | 8.60E-21 |
| rs2524299 | miR-1908-! | cis | 4.82E-05 | Nightingale | XL_HDL_FC | NA | T | A | 0.06923 | 0.005638 | 4.60E-36 |
| rs2524299 | miR-1908-! | cis | 4.82E-05 | Nightingale | S_HDL_TG_ | NA | T | A | 0.082567 | 0.006048 | 5.40E-43 |
| rs2524299 | miR-1908-! | cis | 4.82E-05 | Nightingale | M_VLDL_P | NA | T | A | -0.06569 | 0.006102 | 4.80E-28 |
| rs2524299 | miR-1908-! | cis | 4.82E-05 | Nightingale | L_LDL_C_p | NA | T | A | -0.08105 | 0.006196 | 3.90E-39 |
| rs2524299 | miR-1908-! | cis | 4.82E-05 | Nightingale | M_VLDL_C | NA | T | A | -0.08374 | 0.00597 | 8.10E-46 |
| rs2524299 | miR-1908-! | cis | 4.82E-05 | Nightingale | VLDL_TG | NA | T | A | 0.057169 | 0.006119 | 8.00E-21 |
| rs2524299 | miR-1908-! | cis | 4.82E-05 | Nightingale | XL_HDL_P | NA | T | A | -0.09427 | 0.005723 | 2.40E-63 |
| rs2524299 | miR-1908-! | cis | 4.82E-05 | Nightingale | L_HDL_PL | NA | T | A | -0.08967 | 0.005597 | 4.10E-60 |
| rs2524299 | miR-1908-! | cis | 4.82E-05 | Nightingale | LDL_P | NA | T | A | -0.0382 | 0.006355 | 6.30E-10 |
| rs2524299 | miR-1908-! | cis | 4.82E-05 | Nightingale | XXL_VLDL_ | NA | T | A | 0.073113 | 0.006116 | 1.90E-33 |
| rs2524299 | miR-1908-! | cis | 4.82E-05 | Nightingale | L_VLDL_P | NA | T | A | 0.051964 | 0.00614 | 3.70E-17 |
| rs2524299 | miR-1908-! | cis | 4.82E-05 | Nightingale | PUFA_by_I | NA | T | A | -0.17656 | 0.006091 | 6.00E-188 |
| rs2524299 | miR-1908-! | cis | 4.82E-05 | Nightingale | HDL_P | NA | T | A | -0.0514 | 0.005971 | 1.80E-18 |
| rs2524299 | miR-1908-! | cis | 4.82E-05 | Nightingale | XL_HDL_TG | NA | T | A | 0.096196 | 0.005991 | 1.10E-58 |
| rs2524299 | miR-1908-! | cis | 4.82E-05 | Nightingale | L_HDL_CE | NA | T | A | -0.09135 | 0.005615 | 4.50E-61 |
| rs2524299 | miR-1908-! | cis | 4.82E-05 | Nightingale | M_HDL_TG | NA | T | A | 0.081173 | 0.006098 | 1.80E-40 |
| rs2524299 | miR-1908-! | cis | 4.82E-05 | Nightingale | S_LDL_PL | NA | T | A | -0.0513 | 0.006344 | 1.60E-16 |
| rs2524299 | miR-1908-! | cis | 4.82E-05 | Nightingale | XS_VLDL_P | NA | T | A | 0.07541 | 0.006204 | 1.60E-34 |
| rs2524299 | miR-1908-! | cis | 4.82E-05 | Nightingale | HDL_size | NA | T | A | -0.08344 | 0.005635 | 1.60E-51 |
| rs2524299 | miR-1908-! | cis | 4.82E-05 | Nightingale | DHA | NA | T | A | -0.28229 | 0.006128 | 0 |
| rs2524299 | miR-1908-! | cis | 4.82E-05 | Nightingale | M_LDL_TG | NA | T | A | 0.040854 | 0.006265 | 8.30E-11 |
| rs2524299 | miR-1908-! | cis | 4.82E-05 | Nightingale | SFA_pct | NA | T | A | -0.04052 | 0.006274 | 1.10E-10 |
| rs2524299 | miR-1908-! | cis | 4.82E-05 | Nightingale | XL_VLDL_C | NA | T | A | 0.038761 | 0.006259 | 1.60E-09 |
| rs2524299 | miR-1908-! | cis | 4.82E-05 | Nightingale | L_VLDL_C_ | NA | T | A | -0.03731 | 0.006212 | 3.00E-10 |
| rs2524299 | miR-1908-! | cis | 4.82E-05 | Nightingale | XL_VLDL_T | NA | T | A | 0.065043 | 0.006082 | 5.30E-27 |
| rs2524299 | miR-1908-! | cis | 4.82E-05 | Nightingale | HDL_C | NA | T | A | -0.08152 | 0.005678 | 6.80E-48 |
| rs2524299 | miR-1908-! | cis | 4.82E-05 | Nightingale | Total_FC | NA | T | A | -0.0624 | 0.006212 | 6.50E-25 |
| rs2524299 | miR-1908-! | cis | 4.82E-05 | Nightingale | L_HDL_C | NA | T | A | -0.0929 | 0.0056 | 1.60E-63 |
| rs2524299 | miR-1908-! | cis | 4.82E-05 | Nightingale | IDL_C_pct | NA | T | A | -0.08682 | 0.006208 | 3.40E-45 |
| rs2524299 | miR-1908-! | cis | 4.82E-05 | Nightingale | L_LDL_CE | NA | T | A | -0.05615 | 0.006303 | 6.10E-20 |
| rs2524299 | miR-1908-! | cis | 4.82E-05 | Nightingale | M_VLDL_T | NA | T | A | 0.083858 | 0.006006 | 1.10E-45 |
| rs2524299 | miR-1908-! | cis | 4.82E-05 | Nightingale | M_HDL_L | NA | T | A | -0.04398 | 0.005903 | 2.30E-14 |
| rs2524299 | miR-1908-! | cis | 4.82E-05 | Nightingale | M_VLDL_C | NA | T | A | -0.05219 | 0.006301 | 3.20E-17 |
| rs2524299 | miR-1908-! | cis | 4.82E-05 | Nightingale | XL_HDL_FC | NA | T | A | -0.10264 | 0.005924 | 1.70E-69 |
| rs2524299 | miR-1908-! | cis | 4.82E-05 | Nightingale | M_HDL_FC | NA | T | A | -0.11742 | 0.005635 | 2.80E-100 |
| rs2524299 | miR-1908-! | cis | 4.82E-05 | Nightingale | Total_PL | NA | T | A | -0.05766 | 0.006055 | 1.20E-22 |
| rs2524299 | miR-1908-! | cis | 4.82E-05 | Nightingale | XL_HDL_C | NA | T | A | -0.10445 | 0.005734 | 2.20E-76 |
| rs2524299 | miR-1908-! | cis | 4.82E-05 | Nightingale | M_LDL_FC | NA | T | A | -0.08417 | 0.006092 | 2.70E-44 |
| rs2524299 | miR-1908-! | cis | 4.82E-05 | Nightingale | S_VLDL_TG | NA | T | A | 0.070751 | 0.006153 | 4.10E-31 |
| rs2524299 | miR-1908-! | cis | 4.82E-05 | Nightingale | XXL_VLDL_ | NA | T | A | 0.055852 | 0.006392 | 2.00E-18 |
| rs2524299 | miR-1908-! | cis | 4.82E-05 | Nightingale | M_HDL_C_ | NA | T | A | -0.07995 | 0.006053 | 2.20E-40 |
| rs2524299 | miR-1908-! | cis | 4.82E-05 | Nightingale | L_VLDL_PL | NA | T | A | 0.095379 | 0.006172 | 3.30E-54 |
| rs2524299 | miR-1908-! | cis | 4.82E-05 | Nightingale | XL_VLDL_C | NA | T | A | -0.06914 | 0.006092 | 1.40E-30 |
| rs2524299 | miR-1908-! | cis | 4.82E-05 | Nightingale | non_HDL_( | NA | T | A | -0.04903 | 0.006325 | 1.40E-15 |
| rs2524299 | miR-1908-! | cis | 4.82E-05 | Nightingale | XXL_VLDL_ | NA | T | A | 0.079366 | 0.006141 | 2.60E-38 |
| rs2524299 | miR-1908-! | cis | 4.82E-05 | Nightingale | XS_VLDL_F | NA | T | A | -0.10767 | 0.006311 | 6.80E-66 |
| rs2524299 | miR-1908-! | cis | 4.82E-05 | Nightingale | XL_VLDL_F | NA | T | A | 0.053238 | 0.006159 | 8.60E-18 |
| rs2524299 | miR-1908-! | cis | 4.82E-05 | Nightingale | DHA_pct | NA | T | A | -0.26834 | 0.006173 | 0 |
| rs2524299 | miR-1908-! | cis | 4.82E-05 | Nightingale | XXL_VLDL_ | NA | T | A | 0.071937 | 0.006125 | 2.20E-32 |
| rs2524299 | miR-1908-! | cis | 4.82E-05 | Nightingale | Clinical_LD | NA | T | A | -0.05871 | 0.006323 | 1.80E-21 |
| rs2524299 | miR-1908-! | cis | 4.82E-05 | Nightingale | XL_HDL_CE | NA | T | A | -0.10389 | 0.005694 | 1.20E-76 |
| rs2524299 | miR-1908-! | cis | 4.82E-05 | Nightingale | XS_VLDL_P | NA | T | A | -0.04004 | 0.006274 | 2.60E-11 |
| rs2524299 | miR-1908-! | cis | 4.82E-05 | Nightingale | XL_VLDL_C | NA | T | A | -0.06746 | 0.006095 | 2.80E-29 |

| | | | | | | | | | | | |
|---|---|---|---|---|---|---|---|---|---|---|---|
| rs2524299 | miR-1908-5 | cis | 4.82E-05 | Nightingale | L_LDL_P | NA | T | A | -0.03587 | 0.006337 | 6.20E-09 |
| rs2524299 | miR-1908-5 | cis | 4.82E-05 | Nightingale | M_LDL_C_ | NA | T | A | -0.09843 | 0.006364 | 4.00E-55 |
| rs2524299 | miR-1908-5 | cis | 4.82E-05 | Nightingale | L_HDL_C_p | NA | T | A | -0.07599 | 0.005994 | 1.70E-36 |
| rs2524299 | miR-1908-5 | cis | 4.82E-05 | Nightingale | M_VLDL_C | NA | T | A | -0.08725 | 0.005985 | 2.00E-49 |
| rs2524299 | miR-1908-5 | cis | 4.82E-05 | Nightingale | XXL_VLDL_ | NA | T | A | 0.066147 | 0.006136 | 1.40E-27 |
| rs2524299 | miR-1908-5 | cis | 4.82E-05 | Nightingale | IDL_L | NA | T | A | -0.07711 | 0.006147 | 6.00E-38 |
| rs2524299 | miR-1908-5 | cis | 4.82E-05 | Nightingale | IDL_PL | NA | T | A | -0.07452 | 0.006135 | 1.00E-35 |
| rs2524299 | miR-1908-5 | cis | 4.82E-05 | Nightingale | S_LDL_TG_ | NA | T | A | 0.093068 | 0.006162 | 3.80E-52 |
| rs2524299 | miR-1908-5 | cis | 4.82E-05 | Nightingale | XS_VLDL_F | NA | T | A | -0.05157 | 0.006248 | 1.40E-17 |
| rs2524299 | miR-1908-5 | cis | 4.82E-05 | Nightingale | S_LDL_FC_ | NA | T | A | -0.09096 | 0.006162 | 2.60E-50 |
| rs2524299 | miR-1908-5 | cis | 4.82E-05 | Nightingale | M_LDL_TG | NA | T | A | 0.079919 | 0.006279 | 9.30E-38 |
| rs2524299 | miR-1908-5 | cis | 4.82E-05 | Nightingale | IDL_CE | NA | T | A | -0.07839 | 0.006138 | 5.90E-39 |
| rs2524299 | miR-1908-5 | cis | 4.82E-05 | Nightingale | XL_HDL_PL | NA | T | A | -0.06766 | 0.005842 | 1.00E-32 |
| rs2524299 | miR-1908-5 | cis | 4.82E-05 | Nightingale | IDL_FC_pct | NA | T | A | -0.11401 | 0.006308 | 3.20E-73 |
| rs2524299 | miR-1908-5 | cis | 4.82E-05 | Nightingale | XL_VLDL_T | NA | T | A | 0.060582 | 0.006159 | 1.20E-23 |
| rs2524299 | miR-1908-5 | cis | 4.82E-05 | Nightingale | VLDL_L | NA | T | A | 0.036428 | 0.006235 | 1.10E-08 |
| rs2524299 | miR-1908-5 | cis | 4.82E-05 | Nightingale | S_LDL_FC | NA | T | A | -0.06667 | 0.006349 | 5.40E-27 |
| rs2524299 | miR-1908-5 | cis | 4.82E-05 | Nightingale | S_VLDL_TG | NA | T | A | 0.049086 | 0.006171 | 3.10E-15 |
| rs2524299 | miR-1908-5 | cis | 4.82E-05 | Nightingale | HDL_FC | NA | T | A | -0.0885 | 0.005633 | 2.90E-57 |
| rs2524299 | miR-1908-5 | cis | 4.82E-05 | Nightingale | MUFA_pct | NA | T | A | 0.203037 | 0.006086 | 1.10E-249 |
| rs2524299 | miR-1908-5 | cis | 4.82E-05 | Nightingale | XL_VLDL_P | NA | T | A | 0.058001 | 0.006137 | 4.30E-21 |
| rs2649650 | miR-130a-3 | cis | 0.000449 | Nightingale | Phe | NA | T | C | 0.050788 | 0.00489 | 2.90E-25 |
| rs2649662 | miR-130a-3 | cis | 0.000704 | Nightingale | Phe | NA | C | A | 0.048905 | 0.004923 | 3.00E-23 |
| rs2649663 | miR-130a-3 | cis | 0.000808 | Nightingale | Phe | NA | A | G | 0.049248 | 0.004946 | 2.40E-23 |
| rs2727270 | miR-1908-5 | cis | 4.45E-05 | Nightingale | M_HDL_PL | NA | T | C | 0.071901 | 0.006124 | 8.00E-32 |
| rs2727270 | miR-1908-5 | cis | 4.45E-05 | Nightingale | L_HDL_FC_ | NA | T | C | -0.11754 | 0.005998 | 1.10E-89 |
| rs2727270 | miR-1908-5 | cis | 4.45E-05 | Nightingale | LA_pct | NA | T | C | 0.16778 | 0.00631 | 1.40E-156 |
| rs2727270 | miR-1908-5 | cis | 4.45E-05 | Nightingale | S_VLDL_C_ | NA | T | C | -0.06366 | 0.006282 | 1.60E-24 |
| rs2727270 | miR-1908-5 | cis | 4.45E-05 | Nightingale | XL_VLDL_P | NA | T | C | 0.063442 | 0.006206 | 1.70E-24 |
| rs2727270 | miR-1908-5 | cis | 4.45E-05 | Nightingale | M_HDL_P | NA | T | C | -0.05146 | 0.005948 | 4.60E-19 |
| rs2727270 | miR-1908-5 | cis | 4.45E-05 | Nightingale | XS_VLDL_T | NA | T | C | 0.038731 | 0.006358 | 2.80E-09 |
| rs2727270 | miR-1908-5 | cis | 4.45E-05 | Nightingale | LDL_FC | NA | T | C | -0.07078 | 0.006388 | 1.30E-29 |
| rs2727270 | miR-1908-5 | cis | 4.45E-05 | Nightingale | HDL_CE | NA | T | C | -0.07976 | 0.005794 | 4.00E-44 |
| rs2727270 | miR-1908-5 | cis | 4.45E-05 | Nightingale | L_VLDL_PL | NA | T | C | 0.060152 | 0.00623 | 6.10E-22 |
| rs2727270 | miR-1908-5 | cis | 4.45E-05 | Nightingale | L_VLDL_CE | NA | T | C | -0.06008 | 0.006179 | 1.10E-23 |
| rs2727270 | miR-1908-5 | cis | 4.45E-05 | Nightingale | Sphingomy | NA | T | C | -0.12156 | 0.006034 | 7.00E-93 |
| rs2727270 | miR-1908-5 | cis | 4.45E-05 | Nightingale | M_LDL_P | NA | T | C | -0.04533 | 0.006476 | 5.40E-13 |
| rs2727270 | miR-1908-5 | cis | 4.45E-05 | Nightingale | Omega_3_ | NA | T | C | -0.37745 | 0.006309 | 0 |
| rs2727270 | miR-1908-5 | cis | 4.45E-05 | Nightingale | M_LDL_PL_ | NA | T | C | 0.034452 | 0.006453 | 1.60E-08 |
| rs2727270 | miR-1908-5 | cis | 4.45E-05 | Nightingale | M_HDL_TG | NA | T | C | 0.054864 | 0.006377 | 1.80E-17 |
| rs2727270 | miR-1908-5 | cis | 4.45E-05 | Nightingale | S_VLDL_FC | NA | T | C | -0.09221 | 0.006165 | 2.70E-51 |
| rs2727270 | miR-1908-5 | cis | 4.45E-05 | Nightingale | L_HDL_CE_ | NA | T | C | -0.05965 | 0.006178 | 1.30E-21 |
| rs2727270 | miR-1908-5 | cis | 4.45E-05 | Nightingale | Tyr | NA | T | C | 0.035353 | 0.00645 | 4.20E-08 |
| rs2727270 | miR-1908-5 | cis | 4.45E-05 | Nightingale | XL_HDL_P | NA | T | C | -0.09616 | 0.00581 | 6.00E-64 |
| rs2727270 | miR-1908-5 | cis | 4.45E-05 | Nightingale | S_HDL_C_p | NA | T | C | -0.05334 | 0.006382 | 1.70E-16 |
| rs2727270 | miR-1908-5 | cis | 4.45E-05 | Nightingale | XL_VLDL_F | NA | T | C | -0.07017 | 0.006255 | 3.30E-30 |
| rs2727270 | miR-1908-5 | cis | 4.45E-05 | Nightingale | XXL_VLDL_ | NA | T | C | 0.080636 | 0.006196 | 4.10E-39 |
| rs2727270 | miR-1908-5 | cis | 4.45E-05 | Nightingale | VLDL_size | NA | T | C | 0.083976 | 0.006018 | 5.90E-46 |
| rs2727270 | miR-1908-5 | cis | 4.45E-05 | Nightingale | L_VLDL_PL | NA | T | C | 0.096039 | 0.006266 | 2.70E-53 |
| rs2727270 | miR-1908-5 | cis | 4.45E-05 | Nightingale | XL_VLDL_C | NA | T | C | 0.047383 | 0.006306 | 1.40E-13 |
| rs2727270 | miR-1908-5 | cis | 4.45E-05 | Nightingale | XXL_VLDL_ | NA | T | C | 0.075335 | 0.00622 | 5.60E-34 |
| rs2727270 | miR-1908-5 | cis | 4.45E-05 | Nightingale | M_LDL_TG | NA | T | C | 0.081189 | 0.006375 | 1.20E-37 |
| rs2727270 | miR-1908-5 | cis | 4.45E-05 | Nightingale | LDL_PL | NA | T | C | -0.05064 | 0.006442 | 8.10E-16 |
| rs2727270 | miR-1908-5 | cis | 4.45E-05 | Nightingale | L_LDL_FC | NA | T | C | -0.07522 | 0.00634 | 9.60E-34 |
| rs2727270 | miR-1908-5 | cis | 4.45E-05 | Nightingale | M_HDL_PL | NA | T | C | -0.03751 | 0.006038 | 1.10E-10 |
| rs2727270 | miR-1908-5 | cis | 4.45E-05 | Nightingale | Remnant_( | NA | T | C | -0.04457 | 0.006415 | 7.50E-13 |
| rs2727270 | miR-1908-5 | cis | 4.45E-05 | Nightingale | XL_HDL_PL | NA | T | C | -0.06887 | 0.005932 | 3.30E-33 |
| rs2727270 | miR-1908-5 | cis | 4.45E-05 | Nightingale | L_HDL_C_p | NA | T | C | -0.07769 | 0.006086 | 3.80E-37 |
| rs2727270 | miR-1908-5 | cis | 4.45E-05 | Nightingale | S_LDL_PL | NA | T | C | -0.05284 | 0.006441 | 7.00E-17 |
| rs2727270 | miR-1908-5 | cis | 4.45E-05 | Nightingale | S_VLDL_PL | NA | T | C | -0.08905 | 0.006178 | 6.90E-48 |
| rs2727270 | miR-1908-5 | cis | 4.45E-05 | Nightingale | XL_HDL_TG | NA | T | C | 0.097748 | 0.006083 | 9.10E-59 |
| rs2727270 | miR-1908-5 | cis | 4.45E-05 | Nightingale | IDL_CE_pct | NA | T | C | -0.04684 | 0.006336 | 1.20E-13 |
| rs2727270 | miR-1908-5 | cis | 4.45E-05 | Nightingale | HDL_L | NA | T | C | -0.07011 | 0.005798 | 3.80E-35 |

| | | | | | | | | | | | |
|---|---|---|---|---|---|---|---|---|---|---|---|
| rs2727270 | miR-1908-5 | cis | 4.45E-05 | Nightingale | Total_L | NA | T | C | -0.04078 | 0.006352 | 2.30E-11 |
| rs2727270 | miR-1908-5 | cis | 4.45E-05 | Nightingale | TG_by_PG | NA | T | C | 0.084504 | 0.005992 | 5.30E-46 |
| rs2727270 | miR-1908-5 | cis | 4.45E-05 | Nightingale | IDL_FC_pct | NA | T | C | -0.11621 | 0.006405 | 1.10E-73 |
| rs2727270 | miR-1908-5 | cis | 4.45E-05 | Nightingale | L_LDL_C_p | NA | T | C | -0.08215 | 0.006291 | 6.20E-39 |
| rs2727270 | miR-1908-5 | cis | 4.45E-05 | Nightingale | XXL_VLDL_ | NA | T | C | 0.055002 | 0.006488 | 2.30E-17 |
| rs2727270 | miR-1908-5 | cis | 4.45E-05 | Nightingale | XS_VLDL_C | NA | T | C | -0.10246 | 0.006086 | 1.50E-64 |
| rs2727270 | miR-1908-5 | cis | 4.45E-05 | Nightingale | PUFA_pct | NA | T | C | -0.12833 | 0.006211 | 4.00E-95 |
| rs2727270 | miR-1908-5 | cis | 4.45E-05 | Nightingale | L_HDL_PL_ | NA | T | C | 0.041691 | 0.006249 | 4.70E-11 |
| rs2727270 | miR-1908-5 | cis | 4.45E-05 | Nightingale | XS_VLDL_C | NA | T | C | -0.1072 | 0.006118 | 9.00E-70 |
| rs2727270 | miR-1908-5 | cis | 4.45E-05 | Nightingale | M_VLDL_T | NA | T | C | 0.04214 | 0.006273 | 3.10E-11 |
| rs2727270 | miR-1908-5 | cis | 4.45E-05 | Nightingale | XXL_VLDL_ | NA | T | C | 0.073525 | 0.006219 | 1.10E-32 |
| rs2727270 | miR-1908-5 | cis | 4.45E-05 | Nightingale | IDL_FC | NA | T | C | -0.09813 | 0.006249 | 1.30E-57 |
| rs2727270 | miR-1908-5 | cis | 4.45E-05 | Nightingale | DHA_pct | NA | T | C | -0.27143 | 0.006268 | 0 |
| rs2727270 | miR-1908-5 | cis | 4.45E-05 | Nightingale | HDL_FC | NA | T | C | -0.08996 | 0.00572 | 1.00E-57 |
| rs2727270 | miR-1908-5 | cis | 4.45E-05 | Nightingale | HDL_TG | NA | T | C | 0.052114 | 0.006367 | 4.70E-16 |
| rs2727270 | miR-1908-5 | cis | 4.45E-05 | Nightingale | XS_VLDL_C | NA | T | C | -0.07351 | 0.00623 | 7.30E-34 |
| rs2727270 | miR-1908-5 | cis | 4.45E-05 | Nightingale | S_LDL_PL_ | NA | T | C | -0.06219 | 0.006444 | 3.50E-21 |
| rs2727270 | miR-1908-5 | cis | 4.45E-05 | Nightingale | Omega_6_ | NA | T | C | 0.355335 | 0.006363 | 0 |
| rs2727270 | miR-1908-5 | cis | 4.45E-05 | Nightingale | M_LDL_C | NA | T | C | -0.03585 | 0.006469 | 9.70E-09 |
| rs2727270 | miR-1908-5 | cis | 4.45E-05 | Nightingale | S_VLDL_TG | NA | T | C | 0.072737 | 0.006247 | 8.90E-32 |
| rs2727270 | miR-1908-5 | cis | 4.45E-05 | Nightingale | L_VLDL_L | NA | T | C | 0.04967 | 0.00624 | 2.60E-15 |
| rs2727270 | miR-1908-5 | cis | 4.45E-05 | Nightingale | PUFA | NA | T | C | -0.09956 | 0.006297 | 2.40E-60 |
| rs2727270 | miR-1908-5 | cis | 4.45E-05 | Nightingale | L_VLDL_TG | NA | T | C | 0.048166 | 0.006212 | 7.90E-15 |
| rs2727270 | miR-1908-5 | cis | 4.45E-05 | Nightingale | XS_VLDL_T | NA | T | C | 0.103418 | 0.006133 | 8.30E-65 |
| rs2727270 | miR-1908-5 | cis | 4.45E-05 | Nightingale | M_HDL_CE | NA | T | C | -0.05658 | 0.005963 | 4.20E-22 |
| rs2727270 | miR-1908-5 | cis | 4.45E-05 | Nightingale | M_HDL_C | NA | T | C | -0.05937 | 0.005941 | 2.20E-24 |
| rs2727270 | miR-1908-5 | cis | 4.45E-05 | Nightingale | L_LDL_C | NA | T | C | -0.0629 | 0.006384 | 6.00E-24 |
| rs2727270 | miR-1908-5 | cis | 4.45E-05 | Nightingale | ApoB | NA | T | C | -0.03808 | 0.006448 | 1.20E-09 |
| rs2727270 | miR-1908-5 | cis | 4.45E-05 | Nightingale | XL_HDL_L | NA | T | C | -0.09921 | 0.005802 | 3.50E-68 |
| rs2727270 | miR-1908-5 | cis | 4.45E-05 | Nightingale | M_LDL_C_ | NA | T | C | -0.0997 | 0.006462 | 1.00E-54 |
| rs2727270 | miR-1908-5 | cis | 4.45E-05 | Nightingale | XL_VLDL_T | NA | T | C | 0.066581 | 0.006176 | 2.00E-27 |
| rs2727270 | miR-1908-5 | cis | 4.45E-05 | Nightingale | XL_VLDL_C | NA | T | C | 0.03934 | 0.006355 | 1.80E-09 |
| rs2727270 | miR-1908-5 | cis | 4.45E-05 | Nightingale | XS_VLDL_L | NA | T | C | -0.0384 | 0.006346 | 2.50E-10 |
| rs2727270 | miR-1908-5 | cis | 4.45E-05 | Nightingale | S_LDL_TG | NA | T | C | 0.057731 | 0.006299 | 9.40E-20 |
| rs2727270 | miR-1908-5 | cis | 4.45E-05 | Nightingale | M_HDL_CE | NA | T | C | -0.0613 | 0.006261 | 1.70E-22 |
| rs2727270 | miR-1908-5 | cis | 4.45E-05 | Nightingale | XL_HDL_FC | NA | T | C | -0.10449 | 0.006015 | 4.40E-70 |
| rs2727270 | miR-1908-5 | cis | 4.45E-05 | Nightingale | L_LDL_FC_ | NA | T | C | -0.09411 | 0.006154 | 2.60E-53 |
| rs2727270 | miR-1908-5 | cis | 4.45E-05 | Nightingale | L_VLDL_C | NA | T | C | 0.040067 | 0.006315 | 5.30E-10 |
| rs2727270 | miR-1908-5 | cis | 4.45E-05 | Nightingale | XL_HDL_PL | NA | T | C | -0.09436 | 0.005804 | 3.90E-62 |
| rs2727270 | miR-1908-5 | cis | 4.45E-05 | Nightingale | M_HDL_FC | NA | T | C | -0.0687 | 0.005875 | 5.20E-33 |
| rs2727270 | miR-1908-5 | cis | 4.45E-05 | Nightingale | HDL_C | NA | T | C | -0.08285 | 0.005765 | 3.00E-48 |
| rs2727270 | miR-1908-5 | cis | 4.45E-05 | Nightingale | XXL_VLDL_ | NA | T | C | 0.074502 | 0.00621 | 1.40E-33 |
| rs2727270 | miR-1908-5 | cis | 4.45E-05 | Nightingale | Omega_3 | NA | T | C | -0.3261 | 0.006328 | 0 |
| rs2727270 | miR-1908-5 | cis | 4.45E-05 | Nightingale | VLDL_L | NA | T | C | 0.037047 | 0.006331 | 1.20E-08 |
| rs2727270 | miR-1908-5 | cis | 4.45E-05 | Nightingale | L_VLDL_C_ | NA | T | C | -0.03944 | 0.006307 | 7.80E-11 |
| rs2727270 | miR-1908-5 | cis | 4.45E-05 | Nightingale | M_VLDL_P | NA | T | C | -0.06778 | 0.006195 | 8.70E-29 |
| rs2727270 | miR-1908-5 | cis | 4.45E-05 | Nightingale | S_HDL_TG_ | NA | T | C | 0.083582 | 0.006141 | 1.50E-42 |
| rs2727270 | miR-1908-5 | cis | 4.45E-05 | Nightingale | IDL_L | NA | T | C | -0.07876 | 0.006242 | 2.60E-38 |
| rs2727270 | miR-1908-5 | cis | 4.45E-05 | Nightingale | S_LDL_CE_ | NA | T | C | 0.061213 | 0.006445 | 1.10E-20 |
| rs2727270 | miR-1908-5 | cis | 4.45E-05 | Nightingale | IDL_CE | NA | T | C | -0.07984 | 0.006233 | 4.60E-39 |
| rs2727270 | miR-1908-5 | cis | 4.45E-05 | Nightingale | LDL_C | NA | T | C | -0.05374 | 0.006429 | 1.00E-17 |
| rs2727270 | miR-1908-5 | cis | 4.45E-05 | Nightingale | IDL_TG_pct | NA | T | C | 0.096573 | 0.006237 | 1.20E-54 |
| rs2727270 | miR-1908-5 | cis | 4.45E-05 | Nightingale | L_VLDL_FC | NA | T | C | 0.050948 | 0.006245 | 5.80E-16 |
| rs2727270 | miR-1908-5 | cis | 4.45E-05 | Nightingale | ApoA1 | NA | T | C | -0.0576 | 0.005909 | 1.30E-23 |
| rs2727270 | miR-1908-5 | cis | 4.45E-05 | Nightingale | XXL_VLDL_ | NA | T | C | 0.07937 | 0.006222 | 2.70E-37 |
| rs2727270 | miR-1908-5 | cis | 4.45E-05 | Nightingale | PUFA_by_N | NA | T | C | -0.17803 | 0.006185 | 4.80E-185 |
| rs2727270 | miR-1908-5 | cis | 4.45E-05 | Nightingale | M_VLDL_C | NA | T | C | -0.0894 | 0.006077 | 4.20E-50 |
| rs2727270 | miR-1908-5 | cis | 4.45E-05 | Nightingale | Cholines | NA | T | C | -0.04819 | 0.006073 | 1.30E-16 |
| rs2727270 | miR-1908-5 | cis | 4.45E-05 | Nightingale | L_HDL_FC | NA | T | C | -0.09824 | 0.005658 | 6.50E-70 |
| rs2727270 | miR-1908-5 | cis | 4.45E-05 | Nightingale | L_HDL_L | NA | T | C | -0.09185 | 0.005665 | 1.70E-61 |
| rs2727270 | miR-1908-5 | cis | 4.45E-05 | Nightingale | non_HDL_( | NA | T | C | -0.05023 | 0.006422 | 8.70E-16 |
| rs2727270 | miR-1908-5 | cis | 4.45E-05 | Nightingale | XS_VLDL_P | NA | T | C | 0.075399 | 0.0063 | 2.00E-33 |
| rs2727270 | miR-1908-5 | cis | 4.45E-05 | Nightingale | LA | NA | T | C | 0.072285 | 0.006332 | 3.50E-30 |

| | | | | | | | | | | | |
|---|---|---|---|---|---|---|---|---|---|---|---|
| rs2727270 | miR-1908-5 | cis | 4.45E-05 | Nightingale | L_LDL_TG_ | NA | T | C | 0.083863 | 0.006331 | 2.00E-40 |
| rs2727270 | miR-1908-5 | cis | 4.45E-05 | Nightingale | M_HDL_L | NA | T | C | -0.04459 | 0.005994 | 1.50E-14 |
| rs2727270 | miR-1908-5 | cis | 4.45E-05 | Nightingale | XXL_VLDL_ | NA | T | C | 0.067508 | 0.00623 | 9.30E-28 |
| rs2727270 | miR-1908-5 | cis | 4.45E-05 | Nightingale | M_HDL_C_ | NA | T | C | -0.08152 | 0.006146 | 1.10E-40 |
| rs2727270 | miR-1908-5 | cis | 4.45E-05 | Nightingale | XL_HDL_CE | NA | T | C | -0.04068 | 0.006453 | 4.50E-10 |
| rs2727270 | miR-1908-5 | cis | 4.45E-05 | Nightingale | Total_C | NA | T | C | -0.07255 | 0.00625 | 1.80E-32 |
| rs2727270 | miR-1908-5 | cis | 4.45E-05 | Nightingale | IDL_PL | NA | T | C | -0.07632 | 0.00623 | 3.10E-36 |
| rs2727270 | miR-1908-5 | cis | 4.45E-05 | Nightingale | XL_VLDL_T | NA | T | C | 0.062721 | 0.006254 | 2.30E-24 |
| rs2727270 | miR-1908-5 | cis | 4.45E-05 | Nightingale | XL_HDL_CE | NA | T | C | -0.10575 | 0.005782 | 3.60E-77 |
| rs2727270 | miR-1908-5 | cis | 4.45E-05 | Nightingale | LDL_CE | NA | T | C | -0.04655 | 0.00644 | 1.00E-13 |
| rs2727270 | miR-1908-5 | cis | 4.45E-05 | Nightingale | MUFA | NA | T | C | 0.067253 | 0.006391 | 1.10E-25 |
| rs2727270 | miR-1908-5 | cis | 4.45E-05 | Nightingale | S_HDL_TG | NA | T | C | 0.076476 | 0.006261 | 2.50E-34 |
| rs2727270 | miR-1908-5 | cis | 4.45E-05 | Nightingale | DHA | NA | T | C | -0.28587 | 0.006222 | 0 |
| rs2727270 | miR-1908-5 | cis | 4.45E-05 | Nightingale | S_LDL_TG_ | NA | T | C | 0.094803 | 0.006256 | 3.70E-52 |
| rs2727270 | miR-1908-5 | cis | 4.45E-05 | Nightingale | IDL_C_pct | NA | T | C | -0.08734 | 0.006303 | 4.40E-44 |
| rs2727270 | miR-1908-5 | cis | 4.45E-05 | Nightingale | L_LDL_L | NA | T | C | -0.0592 | 0.006394 | 2.10E-21 |
| rs2727270 | miR-1908-5 | cis | 4.45E-05 | Nightingale | S_VLDL_TG | NA | T | C | 0.049849 | 0.006266 | 4.00E-15 |
| rs2727270 | miR-1908-5 | cis | 4.45E-05 | Nightingale | XL_VLDL_P | NA | T | C | 0.058962 | 0.006231 | 4.70E-21 |
| rs2727270 | miR-1908-5 | cis | 4.45E-05 | Nightingale | L_HDL_TG_ | NA | T | C | 0.09524 | 0.006093 | 2.50E-55 |
| rs2727270 | miR-1908-5 | cis | 4.45E-05 | Nightingale | L_HDL_C | NA | T | C | -0.09447 | 0.005686 | 6.60E-64 |
| rs2727270 | miR-1908-5 | cis | 4.45E-05 | Nightingale | VLDL_TG | NA | T | C | 0.058397 | 0.006213 | 5.50E-21 |
| rs2727270 | miR-1908-5 | cis | 4.45E-05 | Nightingale | Total_FC | NA | T | C | -0.06379 | 0.006308 | 2.90E-25 |
| rs2727270 | miR-1908-5 | cis | 4.45E-05 | Nightingale | L_HDL_P | NA | T | C | -0.09191 | 0.005655 | 1.10E-61 |
| rs2727270 | miR-1908-5 | cis | 4.45E-05 | Nightingale | M_VLDL_C | NA | T | C | -0.05336 | 0.006398 | 2.30E-17 |
| rs2727270 | miR-1908-5 | cis | 4.45E-05 | Nightingale | HDL_P | NA | T | C | -0.05223 | 0.006063 | 1.20E-18 |
| rs2727270 | miR-1908-5 | cis | 4.45E-05 | Nightingale | L_LDL_PL | NA | T | C | -0.06071 | 0.006408 | 2.60E-22 |
| rs2727270 | miR-1908-5 | cis | 4.45E-05 | Nightingale | XS_VLDL_F | NA | T | C | -0.05294 | 0.006344 | 6.10E-18 |
| rs2727270 | miR-1908-5 | cis | 4.45E-05 | Nightingale | Total_P | NA | T | C | -0.05582 | 0.006076 | 4.40E-21 |
| rs2727270 | miR-1908-5 | cis | 4.45E-05 | Nightingale | IDL_P | NA | T | C | -0.05017 | 0.006389 | 8.50E-16 |
| rs2727270 | miR-1908-5 | cis | 4.45E-05 | Nightingale | M_VLDL_F | NA | T | C | -0.09672 | 0.006139 | 9.00E-58 |
| rs2727270 | miR-1908-5 | cis | 4.45E-05 | Nightingale | M_LDL_FC_ | NA | T | C | -0.08543 | 0.006185 | 6.20E-44 |
| rs2727270 | miR-1908-5 | cis | 4.45E-05 | Nightingale | Unsaturati | NA | T | C | -0.33273 | 0.006115 | 0 |
| rs2727270 | miR-1908-5 | cis | 4.45E-05 | Nightingale | L_LDL_CE | NA | T | C | -0.05766 | 0.006399 | 2.50E-20 |
| rs2727270 | miR-1908-5 | cis | 4.45E-05 | Nightingale | XL_VLDL_F | NA | T | C | 0.054215 | 0.006254 | 8.40E-18 |
| rs2727270 | miR-1908-5 | cis | 4.45E-05 | Nightingale | L_HDL_CE | NA | T | C | -0.09287 | 0.005701 | 1.90E-61 |
| rs2727270 | miR-1908-5 | cis | 4.45E-05 | Nightingale | M_VLDL_C | NA | T | C | -0.08572 | 0.006061 | 2.30E-46 |
| rs2727270 | miR-1908-5 | cis | 4.45E-05 | Nightingale | M_VLDL_C | NA | T | C | -0.04218 | 0.006439 | 2.60E-11 |
| rs2727270 | miR-1908-5 | cis | 4.45E-05 | Nightingale | S_LDL_FC_ | NA | T | C | -0.09213 | 0.006257 | 1.10E-49 |
| rs2727270 | miR-1908-5 | cis | 4.45E-05 | Nightingale | L_LDL_P | NA | T | C | -0.0372 | 0.006435 | 3.00E-09 |
| rs2727270 | miR-1908-5 | cis | 4.45E-05 | Nightingale | XL_HDL_FC | NA | T | C | 0.071062 | 0.005725 | 3.30E-37 |
| rs2727270 | miR-1908-5 | cis | 4.45E-05 | Nightingale | S_LDL_FC | NA | T | C | -0.06795 | 0.006446 | 4.10E-27 |
| rs2727270 | miR-1908-5 | cis | 4.45E-05 | Nightingale | S_HDL_FC_ | NA | T | C | -0.10718 | 0.006124 | 9.40E-72 |
| rs2727270 | miR-1908-5 | cis | 4.45E-05 | Nightingale | Total_CE | NA | T | C | -0.07549 | 0.006228 | 4.40E-35 |
| rs2727270 | miR-1908-5 | cis | 4.45E-05 | Nightingale | IDL_C | NA | T | C | -0.08513 | 0.006235 | 3.80E-44 |
| rs2727270 | miR-1908-5 | cis | 4.45E-05 | Nightingale | LDL_L | NA | T | C | -0.04929 | 0.006435 | 3.60E-15 |
| rs2727270 | miR-1908-5 | cis | 4.45E-05 | Nightingale | XL_HDL_C | NA | T | C | -0.10632 | 0.005822 | 6.20E-77 |
| rs2727270 | miR-1908-5 | cis | 4.45E-05 | Nightingale | XS_VLDL_F | NA | T | C | -0.10954 | 0.006408 | 4.50E-66 |
| rs2727270 | miR-1908-5 | cis | 4.45E-05 | Nightingale | L_VLDL_P | NA | T | C | 0.052988 | 0.006234 | 3.00E-17 |
| rs2727270 | miR-1908-5 | cis | 4.45E-05 | Nightingale | M_HDL_FC | NA | T | C | -0.11913 | 0.005721 | 4.00E-100 |
| rs2727270 | miR-1908-5 | cis | 4.45E-05 | Nightingale | M_LDL_TG | NA | T | C | 0.040786 | 0.006362 | 2.10E-10 |
| rs2727270 | miR-1908-5 | cis | 4.45E-05 | Nightingale | SFA_pct | NA | T | C | -0.04074 | 0.00637 | 1.60E-10 |
| rs2727270 | miR-1908-5 | cis | 4.45E-05 | Nightingale | XL_VLDL_L | NA | T | C | 0.061469 | 0.006212 | 4.30E-23 |
| rs2727270 | miR-1908-5 | cis | 4.45E-05 | Nightingale | Total_TG | NA | T | C | 0.057273 | 0.00625 | 6.50E-20 |
| rs2727270 | miR-1908-5 | cis | 4.45E-05 | Nightingale | Total_PL | NA | T | C | -0.05887 | 0.006148 | 3.70E-23 |
| rs2727270 | miR-1908-5 | cis | 4.45E-05 | Nightingale | M_LDL_FC | NA | T | C | -0.0581 | 0.006449 | 2.60E-20 |
| rs2727270 | miR-1908-5 | cis | 4.45E-05 | Nightingale | HDL_PL | NA | T | C | -0.06435 | 0.005837 | 1.00E-29 |
| rs2727270 | miR-1908-5 | cis | 4.45E-05 | Nightingale | XS_VLDL_P | NA | T | C | -0.04133 | 0.006371 | 1.20E-11 |
| rs2727270 | miR-1908-5 | cis | 4.45E-05 | Nightingale | L_HDL_PL | NA | T | C | -0.09111 | 0.005682 | 1.80E-60 |
| rs2727270 | miR-1908-5 | cis | 4.45E-05 | Nightingale | XS_VLDL_C | NA | T | C | -0.06835 | 0.006267 | 3.60E-29 |
| rs2727270 | miR-1908-5 | cis | 4.45E-05 | Nightingale | Clinical_LD | NA | T | C | -0.06007 | 0.00642 | 1.00E-21 |
| rs2727270 | miR-1908-5 | cis | 4.45E-05 | Nightingale | LDL_P | NA | T | C | -0.03947 | 0.006452 | 3.30E-10 |
| rs2727270 | miR-1908-5 | cis | 4.45E-05 | Nightingale | HDL_size | NA | T | C | -0.08464 | 0.005721 | 9.80E-52 |
| rs2727270 | miR-1908-5 | cis | 4.45E-05 | Nightingale | M_VLDL_T | NA | T | C | 0.086052 | 0.006098 | 1.90E-46 |

| | | | | | | | | | | | |
|---|---|---|---|---|---|---|---|---|---|---|---|
| rs2727270 | miR-1908-5 | cis | 4.45E-05 | Nightingale | XXL_VLDL_ | NA | T | C | 0.080819 | 0.006235 | 2.90E-38 |
| rs2727270 | miR-1908-5 | cis | 4.45E-05 | Nightingale | M_HDL_TG | NA | T | C | 0.082535 | 0.006191 | 1.70E-40 |
| rs2727270 | miR-1908-5 | cis | 4.45E-05 | Nightingale | Phosphogly | NA | T | C | -0.05093 | 0.006116 | 3.00E-18 |
| rs2727270 | miR-1908-5 | cis | 4.45E-05 | Nightingale | XL_VLDL_C | NA | T | C | -0.06952 | 0.006189 | 6.10E-30 |
| rs2727270 | miR-1908-5 | cis | 4.45E-05 | Nightingale | XL_VLDL_C | NA | T | C | -0.07129 | 0.006186 | 2.60E-31 |
| rs2727270 | miR-1908-5 | cis | 4.45E-05 | Nightingale | MUFA_pct | NA | T | C | 0.204636 | 0.006179 | 1.20E-245 |
| rs2727271 | miR-1908-5 | cis | 4.23E-05 | Nightingale | XS_VLDL_C | NA | T | A | -0.10251 | 0.006085 | 1.30E-64 |
| rs2727271 | miR-1908-5 | cis | 4.23E-05 | Nightingale | IDL_FC | NA | T | A | -0.09821 | 0.006248 | 1.10E-57 |
| rs2727271 | miR-1908-5 | cis | 4.23E-05 | Nightingale | S_VLDL_C_ | NA | T | A | -0.0637 | 0.006281 | 1.50E-24 |
| rs2727271 | miR-1908-5 | cis | 4.23E-05 | Nightingale | M_HDL_TG | NA | T | A | 0.054817 | 0.006376 | 1.90E-17 |
| rs2727271 | miR-1908-5 | cis | 4.23E-05 | Nightingale | M_HDL_FC | NA | T | A | -0.06878 | 0.005875 | 4.30E-33 |
| rs2727271 | miR-1908-5 | cis | 4.23E-05 | Nightingale | HDL_L | NA | T | A | -0.07019 | 0.005797 | 3.10E-35 |
| rs2727271 | miR-1908-5 | cis | 4.23E-05 | Nightingale | XS_VLDL_L | NA | T | A | -0.03847 | 0.006345 | 2.30E-10 |
| rs2727271 | miR-1908-5 | cis | 4.23E-05 | Nightingale | Omega_3_ | NA | T | A | -0.37744 | 0.006308 | 0 |
| rs2727271 | miR-1908-5 | cis | 4.23E-05 | Nightingale | M_HDL_C | NA | T | A | -0.05944 | 0.00594 | 1.90E-24 |
| rs2727271 | miR-1908-5 | cis | 4.23E-05 | Nightingale | M_HDL_PL | NA | T | A | -0.03758 | 0.006037 | 9.70E-11 |
| rs2727271 | miR-1908-5 | cis | 4.23E-05 | Nightingale | S_LDL_PL_ | NA | T | A | -0.06214 | 0.006443 | 3.70E-21 |
| rs2727271 | miR-1908-5 | cis | 4.23E-05 | Nightingale | XS_VLDL_T | NA | T | A | 0.103445 | 0.006132 | 7.50E-65 |
| rs2727271 | miR-1908-5 | cis | 4.23E-05 | Nightingale | Sphingomy | NA | T | A | -0.12167 | 0.006033 | 4.60E-93 |
| rs2727271 | miR-1908-5 | cis | 4.23E-05 | Nightingale | L_VLDL_PL | NA | T | A | 0.060151 | 0.006229 | 6.00E-22 |
| rs2727271 | miR-1908-5 | cis | 4.23E-05 | Nightingale | Omega_6_ | NA | T | A | 0.355321 | 0.006362 | 0 |
| rs2727271 | miR-1908-5 | cis | 4.23E-05 | Nightingale | XL_VLDL_C | NA | T | A | 0.039323 | 0.006354 | 1.80E-09 |
| rs2727271 | miR-1908-5 | cis | 4.23E-05 | Nightingale | LA_pct | NA | T | A | 0.167661 | 0.006309 | 2.10E-156 |
| rs2727271 | miR-1908-5 | cis | 4.23E-05 | Nightingale | L_VLDL_TG | NA | T | A | 0.04818 | 0.006212 | 7.80E-15 |
| rs2727271 | miR-1908-5 | cis | 4.23E-05 | Nightingale | DHA_pct | NA | T | A | -0.27145 | 0.006267 | 0 |
| rs2727271 | miR-1908-5 | cis | 4.23E-05 | Nightingale | XL_VLDL_P | NA | T | A | 0.063452 | 0.006205 | 1.60E-24 |
| rs2727271 | miR-1908-5 | cis | 4.23E-05 | Nightingale | L_LDL_C_p | NA | T | A | -0.08221 | 0.00629 | 5.40E-39 |
| rs2727271 | miR-1908-5 | cis | 4.23E-05 | Nightingale | Total_L | NA | T | A | -0.04087 | 0.006351 | 2.10E-11 |
| rs2727271 | miR-1908-5 | cis | 4.23E-05 | Nightingale | M_HDL_PL | NA | T | A | 0.071948 | 0.006124 | 7.20E-32 |
| rs2727271 | miR-1908-5 | cis | 4.23E-05 | Nightingale | Tyr | NA | T | A | 0.035294 | 0.006449 | 4.40E-08 |
| rs2727271 | miR-1908-5 | cis | 4.23E-05 | Nightingale | XS_VLDL_C | NA | T | A | -0.10724 | 0.006117 | 7.80E-70 |
| rs2727271 | miR-1908-5 | cis | 4.23E-05 | Nightingale | XL_HDL_PL | NA | T | A | -0.06886 | 0.005931 | 3.30E-33 |
| rs2727271 | miR-1908-5 | cis | 4.23E-05 | Nightingale | S_VLDL_PL | NA | T | A | -0.08909 | 0.006177 | 6.20E-48 |
| rs2727271 | miR-1908-5 | cis | 4.23E-05 | Nightingale | IDL_CE_pct | NA | T | A | -0.04689 | 0.006335 | 1.10E-13 |
| rs2727271 | miR-1908-5 | cis | 4.23E-05 | Nightingale | Cholines | NA | T | A | -0.0483 | 0.006072 | 1.10E-16 |
| rs2727271 | miR-1908-5 | cis | 4.23E-05 | Nightingale | HDL_TG | NA | T | A | 0.052066 | 0.006366 | 4.90E-16 |
| rs2727271 | miR-1908-5 | cis | 4.23E-05 | Nightingale | L_HDL_FC | NA | T | A | -0.09828 | 0.005658 | 5.30E-70 |
| rs2727271 | miR-1908-5 | cis | 4.23E-05 | Nightingale | Total_C | NA | T | A | -0.07265 | 0.006249 | 1.50E-32 |
| rs2727271 | miR-1908-5 | cis | 4.23E-05 | Nightingale | TG_by_PG | NA | T | A | 0.08455 | 0.005991 | 4.70E-46 |
| rs2727271 | miR-1908-5 | cis | 4.23E-05 | Nightingale | XS_VLDL_T | NA | T | A | 0.038685 | 0.006357 | 2.90E-09 |
| rs2727271 | miR-1908-5 | cis | 4.23E-05 | Nightingale | L_VLDL_PL | NA | T | A | 0.09602 | 0.006265 | 2.70E-53 |
| rs2727271 | miR-1908-5 | cis | 4.23E-05 | Nightingale | MUFA | NA | T | A | 0.06719 | 0.00639 | 1.20E-25 |
| rs2727271 | miR-1908-5 | cis | 4.23E-05 | Nightingale | M_HDL_CE | NA | T | A | -0.05665 | 0.005962 | 3.70E-22 |
| rs2727271 | miR-1908-5 | cis | 4.23E-05 | Nightingale | L_HDL_CE | NA | T | A | -0.09292 | 0.0057 | 1.60E-61 |
| rs2727271 | miR-1908-5 | cis | 4.23E-05 | Nightingale | IDL_PL | NA | T | A | -0.07642 | 0.006229 | 2.50E-36 |
| rs2727271 | miR-1908-5 | cis | 4.23E-05 | Nightingale | S_LDL_PL | NA | T | A | -0.05289 | 0.00644 | 6.50E-17 |
| rs2727271 | miR-1908-5 | cis | 4.23E-05 | Nightingale | XL_VLDL_T | NA | T | A | 0.062759 | 0.006253 | 2.10E-24 |
| rs2727271 | miR-1908-5 | cis | 4.23E-05 | Nightingale | ApoB | NA | T | A | -0.03815 | 0.006447 | 1.20E-09 |
| rs2727271 | miR-1908-5 | cis | 4.23E-05 | Nightingale | S_HDL_TG | NA | T | A | 0.076451 | 0.00626 | 2.60E-34 |
| rs2727271 | miR-1908-5 | cis | 4.23E-05 | Nightingale | M_VLDL_C | NA | T | A | -0.08945 | 0.006076 | 3.70E-50 |
| rs2727271 | miR-1908-5 | cis | 4.23E-05 | Nightingale | L_HDL_TG_ | NA | T | A | 0.095233 | 0.006092 | 2.40E-55 |
| rs2727271 | miR-1908-5 | cis | 4.23E-05 | Nightingale | LDL_C | NA | T | A | -0.05383 | 0.006428 | 8.80E-18 |
| rs2727271 | miR-1908-5 | cis | 4.23E-05 | Nightingale | LDL_FC | NA | T | A | -0.07086 | 0.006387 | 1.10E-29 |
| rs2727271 | miR-1908-5 | cis | 4.23E-05 | Nightingale | LA | NA | T | A | 0.072165 | 0.006331 | 4.40E-30 |
| rs2727271 | miR-1908-5 | cis | 4.23E-05 | Nightingale | L_HDL_FC_ | NA | T | A | -0.11759 | 0.005997 | 9.10E-90 |
| rs2727271 | miR-1908-5 | cis | 4.23E-05 | Nightingale | M_HDL_P | NA | T | A | -0.05153 | 0.005947 | 4.10E-19 |
| rs2727271 | miR-1908-5 | cis | 4.23E-05 | Nightingale | M_HDL_CE | NA | T | A | -0.0613 | 0.00626 | 1.70E-22 |
| rs2727271 | miR-1908-5 | cis | 4.23E-05 | Nightingale | Remnant_( | NA | T | A | -0.04465 | 0.006414 | 6.90E-13 |
| rs2727271 | miR-1908-5 | cis | 4.23E-05 | Nightingale | ApoA1 | NA | T | A | -0.05768 | 0.005908 | 1.10E-23 |
| rs2727271 | miR-1908-5 | cis | 4.23E-05 | Nightingale | XXL_VLDL_ | NA | T | A | 0.080665 | 0.006195 | 3.70E-39 |
| rs2727271 | miR-1908-5 | cis | 4.23E-05 | Nightingale | L_HDL_CE_ | NA | T | A | -0.05965 | 0.006177 | 1.30E-21 |
| rs2727271 | miR-1908-5 | cis | 4.23E-05 | Nightingale | XS_VLDL_C | NA | T | A | -0.06843 | 0.006266 | 3.10E-29 |
| rs2727271 | miR-1908-5 | cis | 4.23E-05 | Nightingale | XL_HDL_L | NA | T | A | -0.09924 | 0.005801 | 3.00E-68 |

| | | | | | | | | | | | | |
|---|---|---|---|---|---|---|---|---|---|---|---|---|
| rs2727271 | miR-1908-5 | cis | | 4.23E-05 | Nightingale | S_VLDL_FC | NA | T | A | -0.09225 | 0.006164 | 2.40E-51 |
| rs2727271 | miR-1908-5 | cis | | 4.23E-05 | Nightingale | XS_VLDL_C | NA | T | A | -0.07359 | 0.006229 | 6.20E-34 |
| rs2727271 | miR-1908-5 | cis | | 4.23E-05 | Nightingale | M_LDL_P | NA | T | A | -0.04539 | 0.006475 | 5.00E-13 |
| rs2727271 | miR-1908-5 | cis | | 4.23E-05 | Nightingale | LDL_P | NA | T | A | -0.03954 | 0.006451 | 3.00E-10 |
| rs2727271 | miR-1908-5 | cis | | 4.23E-05 | Nightingale | PUFA_pct | NA | T | A | -0.12835 | 0.00621 | 3.50E-95 |
| rs2727271 | miR-1908-5 | cis | | 4.23E-05 | Nightingale | VLDL_size | NA | T | A | 0.08403 | 0.006017 | 5.20E-46 |
| rs2727271 | miR-1908-5 | cis | | 4.23E-05 | Nightingale | S_LDL_TG | NA | T | A | 0.057706 | 0.006298 | 9.60E-20 |
| rs2727271 | miR-1908-5 | cis | | 4.23E-05 | Nightingale | S_VLDL_TG | NA | T | A | 0.072778 | 0.006246 | 8.20E-32 |
| rs2727271 | miR-1908-5 | cis | | 4.23E-05 | Nightingale | L_LDL_FC_ | NA | T | A | -0.09413 | 0.006153 | 2.40E-53 |
| rs2727271 | miR-1908-5 | cis | | 4.23E-05 | Nightingale | IDL_TG_pct | NA | T | A | 0.096605 | 0.006236 | 1.10E-54 |
| rs2727271 | miR-1908-5 | cis | | 4.23E-05 | Nightingale | PUFA | NA | T | A | -0.09965 | 0.006296 | 1.80E-60 |
| rs2727271 | miR-1908-5 | cis | | 4.23E-05 | Nightingale | L_VLDL_C | NA | T | A | 0.040052 | 0.006314 | 5.40E-10 |
| rs2727271 | miR-1908-5 | cis | | 4.23E-05 | Nightingale | IDL_CE | NA | T | A | -0.07993 | 0.006232 | 3.70E-39 |
| rs2727271 | miR-1908-5 | cis | | 4.23E-05 | Nightingale | XL_HDL_PL | NA | T | A | -0.09439 | 0.005803 | 3.40E-62 |
| rs2727271 | miR-1908-5 | cis | | 4.23E-05 | Nightingale | L_VLDL_FC | NA | T | A | 0.050946 | 0.006244 | 5.70E-16 |
| rs2727271 | miR-1908-5 | cis | | 4.23E-05 | Nightingale | Phosphogly | NA | T | A | -0.05103 | 0.006115 | 2.50E-18 |
| rs2727271 | miR-1908-5 | cis | | 4.23E-05 | Nightingale | Omega_3 | NA | T | A | -0.32612 | 0.006328 | 0 |
| rs2727271 | miR-1908-5 | cis | | 4.23E-05 | Nightingale | XXL_VLDL_ | NA | T | A | 0.073558 | 0.006218 | 1.00E-32 |
| rs2727271 | miR-1908-5 | cis | | 4.23E-05 | Nightingale | L_LDL_PL | NA | T | A | -0.0608 | 0.006407 | 2.20E-22 |
| rs2727271 | miR-1908-5 | cis | | 4.23E-05 | Nightingale | XL_VLDL_L | NA | T | A | 0.061482 | 0.006211 | 4.10E-23 |
| rs2727271 | miR-1908-5 | cis | | 4.23E-05 | Nightingale | L_LDL_FC | NA | T | A | -0.07531 | 0.006339 | 8.00E-34 |
| rs2727271 | miR-1908-5 | cis | | 4.23E-05 | Nightingale | XL_VLDL_P | NA | T | A | 0.058975 | 0.00623 | 4.50E-21 |
| rs2727271 | miR-1908-5 | cis | | 4.23E-05 | Nightingale | XL_HDL_CE | NA | T | A | -0.04071 | 0.006452 | 4.40E-10 |
| rs2727271 | miR-1908-5 | cis | | 4.23E-05 | Nightingale | IDL_C_pct | NA | T | A | -0.08737 | 0.006302 | 4.00E-44 |
| rs2727271 | miR-1908-5 | cis | | 4.23E-05 | Nightingale | L_HDL_PL_ | NA | T | A | 0.041715 | 0.006248 | 4.50E-11 |
| rs2727271 | miR-1908-5 | cis | | 4.23E-05 | Nightingale | XXL_VLDL_ | NA | T | A | 0.067549 | 0.006229 | 8.60E-28 |
| rs2727271 | miR-1908-5 | cis | | 4.23E-05 | Nightingale | M_HDL_L | NA | T | A | -0.04467 | 0.005993 | 1.30E-14 |
| rs2727271 | miR-1908-5 | cis | | 4.23E-05 | Nightingale | L_HDL_C | NA | T | A | -0.09452 | 0.005685 | 5.40E-64 |
| rs2727271 | miR-1908-5 | cis | | 4.23E-05 | Nightingale | S_VLDL_TG | NA | T | A | 0.049825 | 0.006265 | 4.10E-15 |
| rs2727271 | miR-1908-5 | cis | | 4.23E-05 | Nightingale | XL_VLDL_C | NA | T | A | 0.04738 | 0.006305 | 1.40E-13 |
| rs2727271 | miR-1908-5 | cis | | 4.23E-05 | Nightingale | LDL_L | NA | T | A | -0.04938 | 0.006434 | 3.20E-15 |
| rs2727271 | miR-1908-5 | cis | | 4.23E-05 | Nightingale | LDL_PL | NA | T | A | -0.05073 | 0.006441 | 7.20E-16 |
| rs2727271 | miR-1908-5 | cis | | 4.23E-05 | Nightingale | L_HDL_PL | NA | T | A | -0.09116 | 0.005682 | 1.50E-60 |
| rs2727271 | miR-1908-5 | cis | | 4.23E-05 | Nightingale | L_HDL_L | NA | T | A | -0.0919 | 0.005664 | 1.40E-61 |
| rs2727271 | miR-1908-5 | cis | | 4.23E-05 | Nightingale | L_HDL_P | NA | T | A | -0.09196 | 0.005654 | 9.00E-62 |
| rs2727271 | miR-1908-5 | cis | | 4.23E-05 | Nightingale | S_HDL_C_p | NA | T | A | -0.05335 | 0.006381 | 1.70E-16 |
| rs2727271 | miR-1908-5 | cis | | 4.23E-05 | Nightingale | S_LDL_CE_ | NA | T | A | 0.061179 | 0.006444 | 1.20E-20 |
| rs2727271 | miR-1908-5 | cis | | 4.23E-05 | Nightingale | M_VLDL_T | NA | T | A | 0.042128 | 0.006272 | 3.20E-11 |
| rs2727271 | miR-1908-5 | cis | | 4.23E-05 | Nightingale | L_LDL_TG_ | NA | T | A | 0.083896 | 0.00633 | 1.90E-40 |
| rs2727271 | miR-1908-5 | cis | | 4.23E-05 | Nightingale | S_LDL_FC_ | NA | T | A | -0.09216 | 0.006256 | 1.00E-49 |
| rs2727271 | miR-1908-5 | cis | | 4.23E-05 | Nightingale | XXL_VLDL_ | NA | T | A | 0.079394 | 0.006221 | 2.50E-37 |
| rs2727271 | miR-1908-5 | cis | | 4.23E-05 | Nightingale | L_LDL_L | NA | T | A | -0.0593 | 0.006393 | 1.80E-21 |
| rs2727271 | miR-1908-5 | cis | | 4.23E-05 | Nightingale | L_LDL_CE | NA | T | A | -0.05776 | 0.006398 | 2.10E-20 |
| rs2727271 | miR-1908-5 | cis | | 4.23E-05 | Nightingale | M_LDL_C_p | NA | T | A | -0.09973 | 0.006461 | 9.30E-55 |
| rs2727271 | miR-1908-5 | cis | | 4.23E-05 | Nightingale | VLDL_L | NA | T | A | 0.037033 | 0.00633 | 1.20E-08 |
| rs2727271 | miR-1908-5 | cis | | 4.23E-05 | Nightingale | M_LDL_FC | NA | T | A | -0.05819 | 0.006448 | 2.30E-20 |
| rs2727271 | miR-1908-5 | cis | | 4.23E-05 | Nightingale | M_LDL_C | NA | T | A | -0.03593 | 0.006468 | 9.10E-09 |
| rs2727271 | miR-1908-5 | cis | | 4.23E-05 | Nightingale | L_HDL_C_p | NA | T | A | -0.0777 | 0.006085 | 3.60E-37 |
| rs2727271 | miR-1908-5 | cis | | 4.23E-05 | Nightingale | M_VLDL_C | NA | T | A | -0.05344 | 0.006397 | 2.10E-17 |
| rs2727271 | miR-1908-5 | cis | | 4.23E-05 | Nightingale | S_LDL_TG_ | NA | T | A | 0.094826 | 0.006255 | 3.40E-52 |
| rs2727271 | miR-1908-5 | cis | | 4.23E-05 | Nightingale | M_LDL_FC_ | NA | T | A | -0.08546 | 0.006184 | 5.70E-44 |
| rs2727271 | miR-1908-5 | cis | | 4.23E-05 | Nightingale | IDL_L | NA | T | A | -0.07885 | 0.006241 | 2.20E-38 |
| rs2727271 | miR-1908-5 | cis | | 4.23E-05 | Nightingale | XL_VLDL_F | NA | T | A | 0.054227 | 0.006253 | 8.20E-18 |
| rs2727271 | miR-1908-5 | cis | | 4.23E-05 | Nightingale | XL_HDL_FC | NA | T | A | 0.071115 | 0.005724 | 2.80E-37 |
| rs2727271 | miR-1908-5 | cis | | 4.23E-05 | Nightingale | S_HDL_FC_ | NA | T | A | -0.10726 | 0.006123 | 7.10E-72 |
| rs2727271 | miR-1908-5 | cis | | 4.23E-05 | Nightingale | VLDL_TG | NA | T | A | 0.058404 | 0.006212 | 5.50E-21 |
| rs2727271 | miR-1908-5 | cis | | 4.23E-05 | Nightingale | XS_VLDL_P | NA | T | A | -0.0414 | 0.00637 | 1.10E-11 |
| rs2727271 | miR-1908-5 | cis | | 4.23E-05 | Nightingale | HDL_C | NA | T | A | -0.08292 | 0.005764 | 2.40E-48 |
| rs2727271 | miR-1908-5 | cis | | 4.23E-05 | Nightingale | Clinical_LD | NA | T | A | -0.06016 | 0.006419 | 9.10E-22 |
| rs2727271 | miR-1908-5 | cis | | 4.23E-05 | Nightingale | XL_VLDL_T | NA | T | A | 0.066601 | 0.006175 | 1.90E-27 |
| rs2727271 | miR-1908-5 | cis | | 4.23E-05 | Nightingale | XXL_VLDL_ | NA | T | A | 0.075362 | 0.006219 | 5.20E-34 |
| rs2727271 | miR-1908-5 | cis | | 4.23E-05 | Nightingale | M_VLDL_C | NA | T | A | -0.04226 | 0.006438 | 2.40E-11 |
| rs2727271 | miR-1908-5 | cis | | 4.23E-05 | Nightingale | XL_HDL_C | NA | T | A | -0.10636 | 0.005821 | 5.30E-77 |

| | | | | | | | | | | | |
|---|---|---|---|---|---|---|---|---|---|---|---|
| rs2727271 | miR-1908-! | cis | 4.23E-05 | Nightingale | XL_HDL_CE | NA | T | A | -0.10579 | 0.005781 | 3.00E-77 |
| rs2727271 | miR-1908-! | cis | 4.23E-05 | Nightingale | XS_VLDL_F | NA | T | A | -0.05301 | 0.006343 | 5.50E-18 |
| rs2727271 | miR-1908-! | cis | 4.23E-05 | Nightingale | M_VLDL_P | NA | T | A | -0.06784 | 0.006195 | 7.80E-29 |
| rs2727271 | miR-1908-! | cis | 4.23E-05 | Nightingale | XL_HDL_FC | NA | T | A | -0.10451 | 0.006014 | 4.10E-70 |
| rs2727271 | miR-1908-! | cis | 4.23E-05 | Nightingale | M_LDL_TG | NA | T | A | 0.081221 | 0.006374 | 1.10E-37 |
| rs2727271 | miR-1908-! | cis | 4.23E-05 | Nightingale | HDL_PL | NA | T | A | -0.06442 | 0.005836 | 8.50E-30 |
| rs2727271 | miR-1908-! | cis | 4.23E-05 | Nightingale | XL_VLDL_F | NA | T | A | -0.07017 | 0.006254 | 3.30E-30 |
| rs2727271 | miR-1908-! | cis | 4.23E-05 | Nightingale | PUFA_by_I | NA | T | A | -0.17802 | 0.006184 | 4.60E-185 |
| rs2727271 | miR-1908-! | cis | 4.23E-05 | Nightingale | XXL_VLDL_ | NA | T | A | 0.080841 | 0.006234 | 2.70E-38 |
| rs2727271 | miR-1908-! | cis | 4.23E-05 | Nightingale | XS_VLDL_F | NA | T | A | -0.10953 | 0.006407 | 4.60E-66 |
| rs2727271 | miR-1908-! | cis | 4.23E-05 | Nightingale | XXL_VLDL_ | NA | T | A | 0.054954 | 0.006487 | 2.40E-17 |
| rs2727271 | miR-1908-! | cis | 4.23E-05 | Nightingale | L_VLDL_L | NA | T | A | 0.049673 | 0.006239 | 2.50E-15 |
| rs2727271 | miR-1908-! | cis | 4.23E-05 | Nightingale | L_LDL_C | NA | T | A | -0.063 | 0.006383 | 5.10E-24 |
| rs2727271 | miR-1908-! | cis | 4.23E-05 | Nightingale | XL_HDL_P | NA | T | A | -0.09621 | 0.00581 | 5.10E-64 |
| rs2727271 | miR-1908-! | cis | 4.23E-05 | Nightingale | XL_VLDL_C | NA | T | A | -0.07134 | 0.006185 | 2.40E-31 |
| rs2727271 | miR-1908-! | cis | 4.23E-05 | Nightingale | non_HDL_( | NA | T | A | -0.05031 | 0.006421 | 7.80E-16 |
| rs2727271 | miR-1908-! | cis | 4.23E-05 | Nightingale | Unsaturati | NA | T | A | -0.33274 | 0.006114 | 0 |
| rs2727271 | miR-1908-! | cis | 4.23E-05 | Nightingale | Total_CE | NA | T | A | -0.07559 | 0.006227 | 3.60E-35 |
| rs2727271 | miR-1908-! | cis | 4.23E-05 | Nightingale | HDL_FC | NA | T | A | -0.09005 | 0.005719 | 7.60E-58 |
| rs2727271 | miR-1908-! | cis | 4.23E-05 | Nightingale | M_HDL_TG | NA | T | A | 0.082532 | 0.006191 | 1.70E-40 |
| rs2727271 | miR-1908-! | cis | 4.23E-05 | Nightingale | HDL_CE | NA | T | A | -0.07983 | 0.005794 | 3.20E-44 |
| rs2727271 | miR-1908-! | cis | 4.23E-05 | Nightingale | M_HDL_C_ | NA | T | A | -0.08154 | 0.006145 | 1.00E-40 |
| rs2727271 | miR-1908-! | cis | 4.23E-05 | Nightingale | M_VLDL_F | NA | T | A | -0.09676 | 0.006138 | 7.80E-58 |
| rs2727271 | miR-1908-! | cis | 4.23E-05 | Nightingale | IDL_C | NA | T | A | -0.08522 | 0.006234 | 3.10E-44 |
| rs2727271 | miR-1908-! | cis | 4.23E-05 | Nightingale | M_VLDL_C | NA | T | A | -0.08577 | 0.00606 | 2.00E-46 |
| rs2727271 | miR-1908-! | cis | 4.23E-05 | Nightingale | M_LDL_PL_ | NA | T | A | 0.034447 | 0.006452 | 1.60E-08 |
| rs2727271 | miR-1908-! | cis | 4.23E-05 | Nightingale | S_HDL_TG_ | NA | T | A | 0.083593 | 0.00614 | 1.40E-42 |
| rs2727271 | miR-1908-! | cis | 4.23E-05 | Nightingale | MUFA_pct | NA | T | A | 0.204604 | 0.006178 | 1.20E-245 |
| rs2727271 | miR-1908-! | cis | 4.23E-05 | Nightingale | M_HDL_FC | NA | T | A | -0.11922 | 0.005721 | 2.70E-100 |
| rs2727271 | miR-1908-! | cis | 4.23E-05 | Nightingale | Total_FC | NA | T | A | -0.06388 | 0.006307 | 2.40E-25 |
| rs2727271 | miR-1908-! | cis | 4.23E-05 | Nightingale | L_VLDL_P | NA | T | A | 0.052991 | 0.006233 | 2.90E-17 |
| rs2727271 | miR-1908-! | cis | 4.23E-05 | Nightingale | S_LDL_FC | NA | T | A | -0.06802 | 0.006445 | 3.60E-27 |
| rs2727271 | miR-1908-! | cis | 4.23E-05 | Nightingale | HDL_P | NA | T | A | -0.05232 | 0.006062 | 1.00E-18 |
| rs2727271 | miR-1908-! | cis | 4.23E-05 | Nightingale | L_VLDL_CE | NA | T | A | -0.06015 | 0.006178 | 9.80E-24 |
| rs2727271 | miR-1908-! | cis | 4.23E-05 | Nightingale | L_VLDL_C_ | NA | T | A | -0.03951 | 0.006306 | 7.30E-11 |
| rs2727271 | miR-1908-! | cis | 4.23E-05 | Nightingale | LDL_CE | NA | T | A | -0.04664 | 0.006439 | 9.30E-14 |
| rs2727271 | miR-1908-! | cis | 4.23E-05 | Nightingale | L_LDL_P | NA | T | A | -0.03728 | 0.006434 | 2.80E-09 |
| rs2727271 | miR-1908-! | cis | 4.23E-05 | Nightingale | M_LDL_TG | NA | T | A | 0.040733 | 0.006361 | 2.20E-10 |
| rs2727271 | miR-1908-! | cis | 4.23E-05 | Nightingale | Total_TG | NA | T | A | 0.057268 | 0.006249 | 6.50E-20 |
| rs2727271 | miR-1908-! | cis | 4.23E-05 | Nightingale | M_VLDL_T | NA | T | A | 0.086106 | 0.006097 | 1.70E-46 |
| rs2727271 | miR-1908-! | cis | 4.23E-05 | Nightingale | XXL_VLDL_ | NA | T | A | 0.074531 | 0.006209 | 1.30E-33 |
| rs2727271 | miR-1908-! | cis | 4.23E-05 | Nightingale | SFA_pct | NA | T | A | -0.04066 | 0.006369 | 1.70E-10 |
| rs2727271 | miR-1908-! | cis | 4.23E-05 | Nightingale | XL_VLDL_C | NA | T | A | -0.06958 | 0.006188 | 5.60E-30 |
| rs2727271 | miR-1908-! | cis | 4.23E-05 | Nightingale | HDL_size | NA | T | A | -0.08466 | 0.00572 | 8.90E-52 |
| rs2727271 | miR-1908-! | cis | 4.23E-05 | Nightingale | IDL_P | NA | T | A | -0.05024 | 0.006388 | 7.80E-16 |
| rs2727271 | miR-1908-! | cis | 4.23E-05 | Nightingale | DHA | NA | T | A | -0.28592 | 0.006221 | 0 |
| rs2727271 | miR-1908-! | cis | 4.23E-05 | Nightingale | XL_HDL_TG | NA | T | A | 0.097741 | 0.006082 | 8.90E-59 |
| rs2727271 | miR-1908-! | cis | 4.23E-05 | Nightingale | Total_PL | NA | T | A | -0.05896 | 0.006147 | 3.10E-23 |
| rs2727271 | miR-1908-! | cis | 4.23E-05 | Nightingale | IDL_FC_pct | NA | T | A | -0.1162 | 0.006404 | 1.10E-73 |
| rs2727271 | miR-1908-! | cis | 4.23E-05 | Nightingale | Total_P | NA | T | A | -0.05592 | 0.006075 | 3.70E-21 |
| rs2727271 | miR-1908-! | cis | 4.23E-05 | Nightingale | XS_VLDL_P | NA | T | A | 0.075487 | 0.006299 | 1.70E-33 |
| rs2729354 | miR-130a-3 | cis | 0.000809 | Nightingale | Phe | NA | G | A | 0.049202 | 0.004937 | 2.20E-23 |
| rs2729355 | miR-130a-3 | cis | 0.00032 | Nightingale | Phe | NA | G | T | 0.049031 | 0.00493 | 2.60E-23 |
| rs28456 | miR-1908-! | cis | 6.85E-08 | Nightingale | XL_VLDL_F | NA | G | A | 0.044336 | 0.004308 | 5.00E-26 |
| rs28456 | miR-1908-! | cis | 6.85E-08 | Nightingale | Total_C | NA | G | A | -0.06237 | 0.004305 | 3.80E-50 |
| rs28456 | miR-1908-! | cis | 6.85E-08 | Nightingale | XXL_VLDL_ | NA | G | A | 0.061814 | 0.004292 | 2.70E-49 |
| rs28456 | miR-1908-! | cis | 6.85E-08 | Nightingale | LDL_L | NA | G | A | -0.04343 | 0.004433 | 8.50E-24 |
| rs28456 | miR-1908-! | cis | 6.85E-08 | Nightingale | L_HDL_PL | NA | G | A | -0.07351 | 0.003914 | 2.30E-85 |
| rs28456 | miR-1908-! | cis | 6.85E-08 | Nightingale | XL_VLDL_C | NA | G | A | -0.06371 | 0.004262 | 4.30E-52 |
| rs28456 | miR-1908-! | cis | 6.85E-08 | Nightingale | M_LDL_FC_ | NA | G | A | -0.07164 | 0.00426 | 2.40E-66 |
| rs28456 | miR-1908-! | cis | 6.85E-08 | Nightingale | XL_HDL_FC | NA | G | A | -0.09837 | 0.004143 | 9.20E-134 |
| rs28456 | miR-1908-! | cis | 6.85E-08 | Nightingale | L_VLDL_P | NA | G | A | 0.042449 | 0.004294 | 3.60E-24 |
| rs28456 | miR-1908-! | cis | 6.85E-08 | Nightingale | L_LDL_C | NA | G | A | -0.05548 | 0.004397 | 1.50E-38 |

| | | | | | | | | | | | |
|---|---|---|---|---|---|---|---|---|---|---|---|
| rs28456 | miR-1908-5 | cis | 6.85E-08 | Nightingale | XL_HDL_PL | NA | G | A | -0.06394 | 0.004086 | 1.10E-59 |
| rs28456 | miR-1908-5 | cis | 6.85E-08 | Nightingale | S_VLDL_FC | NA | G | A | -0.02635 | 0.004448 | 1.30E-09 |
| rs28456 | miR-1908-5 | cis | 6.85E-08 | Nightingale | XXL_VLDL_ | NA | G | A | 0.073352 | 0.004295 | 2.30E-68 |
| rs28456 | miR-1908-5 | cis | 6.85E-08 | Nightingale | L_LDL_CE | NA | G | A | -0.05104 | 0.004408 | 1.00E-32 |
| rs28456 | miR-1908-5 | cis | 6.85E-08 | Nightingale | Total_TG | NA | G | A | 0.047704 | 0.004305 | 5.60E-30 |
| rs28456 | miR-1908-5 | cis | 6.85E-08 | Nightingale | IDL_C | NA | G | A | -0.074 | 0.004294 | 9.40E-71 |
| rs28456 | miR-1908-5 | cis | 6.85E-08 | Nightingale | HDL_PL | NA | G | A | -0.04646 | 0.004021 | 6.20E-33 |
| rs28456 | miR-1908-5 | cis | 6.85E-08 | Nightingale | XL_HDL_PL | NA | G | A | -0.08557 | 0.003998 | 1.90E-110 |
| rs28456 | miR-1908-5 | cis | 6.85E-08 | Nightingale | XS_VLDL_F | NA | G | A | -0.05253 | 0.00437 | 1.20E-35 |
| rs28456 | miR-1908-5 | cis | 6.85E-08 | Nightingale | L_LDL_C_p | NA | G | A | -0.0752 | 0.004333 | 2.80E-69 |
| rs28456 | miR-1908-5 | cis | 6.85E-08 | Nightingale | XL_VLDL_C | NA | G | A | 0.031234 | 0.004377 | 3.20E-13 |
| rs28456 | miR-1908-5 | cis | 6.85E-08 | Nightingale | Phosphogly | NA | G | A | -0.03832 | 0.004213 | 4.20E-21 |
| rs28456 | miR-1908-5 | cis | 6.85E-08 | Nightingale | LDL_FC | NA | G | A | -0.06216 | 0.0044 | 9.90E-48 |
| rs28456 | miR-1908-5 | cis | 6.85E-08 | Nightingale | Total_CE | NA | G | A | -0.06426 | 0.00429 | 2.00E-53 |
| rs28456 | miR-1908-5 | cis | 6.85E-08 | Nightingale | IDL_P | NA | G | A | -0.04231 | 0.004401 | 2.80E-23 |
| rs28456 | miR-1908-5 | cis | 6.85E-08 | Nightingale | M_VLDL_P | NA | G | A | -0.06006 | 0.004268 | 2.90E-48 |
| rs28456 | miR-1908-5 | cis | 6.85E-08 | Nightingale | IDL_FC | NA | G | A | -0.08647 | 0.004305 | 5.50E-95 |
| rs28456 | miR-1908-5 | cis | 6.85E-08 | Nightingale | M_HDL_C | NA | G | A | -0.03999 | 0.004092 | 9.10E-24 |
| rs28456 | miR-1908-5 | cis | 6.85E-08 | Nightingale | S_LDL_PL | NA | G | A | -0.04925 | 0.004437 | 3.10E-30 |
| rs28456 | miR-1908-5 | cis | 6.85E-08 | Nightingale | IDL_C_pct | NA | G | A | -0.07383 | 0.004342 | 9.30E-67 |
| rs28456 | miR-1908-5 | cis | 6.85E-08 | Nightingale | XS_VLDL_P | NA | G | A | -0.03109 | 0.004385 | 8.80E-14 |
| rs28456 | miR-1908-5 | cis | 6.85E-08 | Nightingale | HDL_P | NA | G | A | -0.03465 | 0.004176 | 1.60E-17 |
| rs28456 | miR-1908-5 | cis | 6.85E-08 | Nightingale | S_VLDL_TG | NA | G | A | 0.038961 | 0.004316 | 1.60E-20 |
| rs28456 | miR-1908-5 | cis | 6.85E-08 | Nightingale | L_HDL_TG_ | NA | G | A | 0.077959 | 0.004197 | 1.30E-81 |
| rs28456 | miR-1908-5 | cis | 6.85E-08 | Nightingale | LDL_P | NA | G | A | -0.03809 | 0.004444 | 1.50E-18 |
| rs28456 | miR-1908-5 | cis | 6.85E-08 | Nightingale | XXL_VLDL_ | NA | G | A | 0.066408 | 0.004278 | 2.40E-57 |
| rs28456 | miR-1908-5 | cis | 6.85E-08 | Nightingale | Tyr | NA | G | A | 0.031018 | 0.004442 | 2.90E-12 |
| rs28456 | miR-1908-5 | cis | 6.85E-08 | Nightingale | L_HDL_P | NA | G | A | -0.07531 | 0.003895 | 2.60E-90 |
| rs28456 | miR-1908-5 | cis | 6.85E-08 | Nightingale | DHA | NA | G | A | -0.28289 | 0.004286 | 0 |
| rs28456 | miR-1908-5 | cis | 6.85E-08 | Nightingale | XXL_VLDL_ | NA | G | A | 0.06806 | 0.004285 | 5.20E-60 |
| rs28456 | miR-1908-5 | cis | 6.85E-08 | Nightingale | L_VLDL_PL | NA | G | A | 0.087332 | 0.004316 | 2.70E-94 |
| rs28456 | miR-1908-5 | cis | 6.85E-08 | Nightingale | XL_HDL_FC | NA | G | A | 0.059366 | 0.003943 | 2.90E-56 |
| rs28456 | miR-1908-5 | cis | 6.85E-08 | Nightingale | SFA | NA | G | A | -0.02872 | 0.004407 | 5.50E-11 |
| rs28456 | miR-1908-5 | cis | 6.85E-08 | Nightingale | M_LDL_TG | NA | G | A | 0.037554 | 0.004382 | 4.20E-18 |
| rs28456 | miR-1908-5 | cis | 6.85E-08 | Nightingale | L_VLDL_FC | NA | G | A | 0.040778 | 0.004302 | 2.10E-22 |
| rs28456 | miR-1908-5 | cis | 6.85E-08 | Nightingale | non_HDL_( | NA | G | A | -0.0453 | 0.004424 | 6.10E-26 |
| rs28456 | miR-1908-5 | cis | 6.85E-08 | Nightingale | IDL_PL | NA | G | A | -0.06923 | 0.004291 | 6.70E-63 |
| rs28456 | miR-1908-5 | cis | 6.85E-08 | Nightingale | Total_P | NA | G | A | -0.03905 | 0.004185 | 7.50E-22 |
| rs28456 | miR-1908-5 | cis | 6.85E-08 | Nightingale | M_HDL_TG | NA | G | A | 0.066996 | 0.004265 | 3.60E-58 |
| rs28456 | miR-1908-5 | cis | 6.85E-08 | Nightingale | SFA_pct | NA | G | A | -0.04044 | 0.004388 | 3.10E-20 |
| rs28456 | miR-1908-5 | cis | 6.85E-08 | Nightingale | XL_HDL_C_ | NA | G | A | 0.021404 | 0.004248 | 3.40E-08 |
| rs28456 | miR-1908-5 | cis | 6.85E-08 | Nightingale | Clinical_LD | NA | G | A | -0.05299 | 0.004422 | 1.10E-34 |
| rs28456 | miR-1908-5 | cis | 6.85E-08 | Nightingale | PUFA | NA | G | A | -0.09269 | 0.004337 | 3.00E-107 |
| rs28456 | miR-1908-5 | cis | 6.85E-08 | Nightingale | M_LDL_L | NA | G | A | -0.0268 | 0.004454 | 9.50E-10 |
| rs28456 | miR-1908-5 | cis | 6.85E-08 | Nightingale | Total_PL | NA | G | A | -0.04803 | 0.004235 | 1.40E-31 |
| rs28456 | miR-1908-5 | cis | 6.85E-08 | Nightingale | M_HDL_P | NA | G | A | -0.03317 | 0.004097 | 4.80E-17 |
| rs28456 | miR-1908-5 | cis | 6.85E-08 | Nightingale | XS_VLDL_P | NA | G | A | 0.058356 | 0.004339 | 3.20E-42 |
| rs28456 | miR-1908-5 | cis | 6.85E-08 | Nightingale | S_VLDL_TG | NA | G | A | 0.062934 | 0.004303 | 4.50E-50 |
| rs28456 | miR-1908-5 | cis | 6.85E-08 | Nightingale | S_HDL_FC_ | NA | G | A | -0.09934 | 0.004218 | 3.40E-135 |
| rs28456 | miR-1908-5 | cis | 6.85E-08 | Nightingale | M_HDL_FC | NA | G | A | -0.04974 | 0.004047 | 6.00E-37 |
| rs28456 | miR-1908-5 | cis | 6.85E-08 | Nightingale | S_LDL_FC | NA | G | A | -0.06115 | 0.00444 | 1.20E-45 |
| rs28456 | miR-1908-5 | cis | 6.85E-08 | Nightingale | S_VLDL_CE | NA | G | A | -0.02801 | 0.004388 | 5.20E-11 |
| rs28456 | miR-1908-5 | cis | 6.85E-08 | Nightingale | Omega_3_ | NA | G | A | -0.38327 | 0.004346 | 0 |
| rs28456 | miR-1908-5 | cis | 6.85E-08 | Nightingale | IDL_TG_pct | NA | G | A | 0.086307 | 0.004296 | 1.50E-93 |
| rs28456 | miR-1908-5 | cis | 6.85E-08 | Nightingale | M_VLDL_C | NA | G | A | -0.0503 | 0.004407 | 1.30E-31 |
| rs28456 | miR-1908-5 | cis | 6.85E-08 | Nightingale | S_LDL_TG_ | NA | G | A | 0.085454 | 0.004309 | 5.30E-91 |
| rs28456 | miR-1908-5 | cis | 6.85E-08 | Nightingale | PUFA_pct | NA | G | A | -0.12001 | 0.004278 | 7.60E-175 |
| rs28456 | miR-1908-5 | cis | 6.85E-08 | Nightingale | XL_VLDL_L | NA | G | A | 0.051963 | 0.004279 | 1.40E-35 |
| rs28456 | miR-1908-5 | cis | 6.85E-08 | Nightingale | XL_VLDL_F | NA | G | A | -0.06731 | 0.004309 | 1.70E-57 |
| rs28456 | miR-1908-5 | cis | 6.85E-08 | Nightingale | L_HDL_C | NA | G | A | -0.07778 | 0.003916 | 6.10E-95 |
| rs28456 | miR-1908-5 | cis | 6.85E-08 | Nightingale | XS_VLDL_C | NA | G | A | -0.09004 | 0.004192 | 1.40E-107 |
| rs28456 | miR-1908-5 | cis | 6.85E-08 | Nightingale | M_VLDL_F( | NA | G | A | -0.08871 | 0.004229 | 6.00E-103 |
| rs28456 | miR-1908-5 | cis | 6.85E-08 | Nightingale | Sphingomy | NA | G | A | -0.11192 | 0.004156 | 1.60E-167 |

| | | | | | | | | | | | |
|---|---|---|---|---|---|---|---|---|---|---|---|
| rs28456 | miR-1908-5 | cis | 6.85E-08 | Nightingale | IDL_FC_pct | NA | G | A | -0.10392 | 0.004412 | 4.70E-124 |
| rs28456 | miR-1908-5 | cis | 6.85E-08 | Nightingale | S_VLDL_PL | NA | G | A | -0.07697 | 0.004255 | 9.80E-77 |
| rs28456 | miR-1908-5 | cis | 6.85E-08 | Nightingale | Cholines | NA | G | A | -0.03464 | 0.004183 | 5.60E-18 |
| rs28456 | miR-1908-5 | cis | 6.85E-08 | Nightingale | XS_VLDL_P | NA | G | A | -0.04051 | 0.004388 | 7.20E-22 |
| rs28456 | miR-1908-5 | cis | 6.85E-08 | Nightingale | S_LDL_C | NA | G | A | -0.03055 | 0.004452 | 1.90E-12 |
| rs28456 | miR-1908-5 | cis | 6.85E-08 | Nightingale | S_HDL_TG_ | NA | G | A | 0.068727 | 0.00423 | 2.60E-62 |
| rs28456 | miR-1908-5 | cis | 6.85E-08 | Nightingale | L_VLDL_C | NA | G | A | 0.02979 | 0.00435 | 2.20E-12 |
| rs28456 | miR-1908-5 | cis | 6.85E-08 | Nightingale | M_HDL_CE | NA | G | A | -0.0485 | 0.004313 | 3.40E-30 |
| rs28456 | miR-1908-5 | cis | 6.85E-08 | Nightingale | S_LDL_CE_ | NA | G | A | 0.05701 | 0.004439 | 1.40E-39 |
| rs28456 | miR-1908-5 | cis | 6.85E-08 | Nightingale | M_HDL_L | NA | G | A | -0.02663 | 0.004129 | 2.40E-11 |
| rs28456 | miR-1908-5 | cis | 6.85E-08 | Nightingale | L_HDL_C_p | NA | G | A | -0.06735 | 0.004192 | 7.40E-62 |
| rs28456 | miR-1908-5 | cis | 6.85E-08 | Nightingale | M_VLDL_T | NA | G | A | 0.032055 | 0.004321 | 2.90E-14 |
| rs28456 | miR-1908-5 | cis | 6.85E-08 | Nightingale | XS_VLDL_C | NA | G | A | -0.06446 | 0.004316 | 6.00E-54 |
| rs28456 | miR-1908-5 | cis | 6.85E-08 | Nightingale | L_HDL_CE | NA | G | A | -0.07633 | 0.003927 | 8.20E-91 |
| rs28456 | miR-1908-5 | cis | 6.85E-08 | Nightingale | XL_VLDL_C | NA | G | A | 0.038358 | 0.004344 | 1.50E-19 |
| rs28456 | miR-1908-5 | cis | 6.85E-08 | Nightingale | S_LDL_PL_ | NA | G | A | -0.06076 | 0.004438 | 8.10E-45 |
| rs28456 | miR-1908-5 | cis | 6.85E-08 | Nightingale | HDL_FC | NA | G | A | -0.07129 | 0.00394 | 1.10E-78 |
| rs28456 | miR-1908-5 | cis | 6.85E-08 | Nightingale | L_VLDL_C_ | NA | G | A | -0.03827 | 0.004344 | 5.20E-20 |
| rs28456 | miR-1908-5 | cis | 6.85E-08 | Nightingale | XL_VLDL_P | NA | G | A | 0.053259 | 0.004275 | 2.30E-37 |
| rs28456 | miR-1908-5 | cis | 6.85E-08 | Nightingale | IDL_L | NA | G | A | -0.06903 | 0.004299 | 4.20E-62 |
| rs28456 | miR-1908-5 | cis | 6.85E-08 | Nightingale | L_LDL_L | NA | G | A | -0.05187 | 0.004404 | 7.90E-34 |
| rs28456 | miR-1908-5 | cis | 6.85E-08 | Nightingale | XXL_VLDL_ | NA | G | A | 0.07163 | 0.004286 | 8.00E-66 |
| rs28456 | miR-1908-5 | cis | 6.85E-08 | Nightingale | S_HDL_TG | NA | G | A | 0.068346 | 0.004313 | 1.10E-59 |
| rs28456 | miR-1908-5 | cis | 6.85E-08 | Nightingale | L_HDL_FC_ | NA | G | A | -0.10769 | 0.004132 | 1.20E-161 |
| rs28456 | miR-1908-5 | cis | 6.85E-08 | Nightingale | XL_VLDL_T | NA | G | A | 0.059346 | 0.004309 | 8.50E-45 |
| rs28456 | miR-1908-5 | cis | 6.85E-08 | Nightingale | L_VLDL_L | NA | G | A | 0.038896 | 0.004298 | 1.70E-20 |
| rs28456 | miR-1908-5 | cis | 6.85E-08 | Nightingale | XS_VLDL_C | NA | G | A | -0.09572 | 0.004214 | 6.40E-120 |
| rs28456 | miR-1908-5 | cis | 6.85E-08 | Nightingale | LDL_CE | NA | G | A | -0.04158 | 0.004436 | 7.10E-22 |
| rs28456 | miR-1908-5 | cis | 6.85E-08 | Nightingale | S_HDL_C_p | NA | G | A | -0.04402 | 0.004396 | 2.90E-23 |
| rs28456 | miR-1908-5 | cis | 6.85E-08 | Nightingale | XL_HDL_L | NA | G | A | -0.08952 | 0.003996 | 1.70E-120 |
| rs28456 | miR-1908-5 | cis | 6.85E-08 | Nightingale | M_HDL_CE | NA | G | A | -0.03727 | 0.004107 | 1.20E-20 |
| rs28456 | miR-1908-5 | cis | 6.85E-08 | Nightingale | M_HDL_TG | NA | G | A | 0.05119 | 0.004392 | 3.40E-32 |
| rs28456 | miR-1908-5 | cis | 6.85E-08 | Nightingale | S_LDL_TG | NA | G | A | 0.05108 | 0.004338 | 4.00E-33 |
| rs28456 | miR-1908-5 | cis | 6.85E-08 | Nightingale | M_LDL_C | NA | G | A | -0.03261 | 0.004456 | 8.70E-14 |
| rs28456 | miR-1908-5 | cis | 6.85E-08 | Nightingale | L_VLDL_PL | NA | G | A | 0.049215 | 0.004291 | 5.20E-32 |
| rs28456 | miR-1908-5 | cis | 6.85E-08 | Nightingale | Citrate | NA | G | A | -0.03344 | 0.004419 | 3.70E-15 |
| rs28456 | miR-1908-5 | cis | 6.85E-08 | Nightingale | XS_VLDL_T | NA | G | A | 0.094576 | 0.004224 | 3.60E-117 |
| rs28456 | miR-1908-5 | cis | 6.85E-08 | Nightingale | L_LDL_FC | NA | G | A | -0.06586 | 0.004367 | 2.30E-54 |
| rs28456 | miR-1908-5 | cis | 6.85E-08 | Nightingale | S_LDL_L | NA | G | A | -0.03068 | 0.004447 | 1.40E-12 |
| rs28456 | miR-1908-5 | cis | 6.85E-08 | Nightingale | LDL_PL | NA | G | A | -0.04425 | 0.004437 | 1.80E-24 |
| rs28456 | miR-1908-5 | cis | 6.85E-08 | Nightingale | S_VLDL_FC | NA | G | A | -0.08092 | 0.004247 | 1.10E-84 |
| rs28456 | miR-1908-5 | cis | 6.85E-08 | Nightingale | Total_L | NA | G | A | -0.0345 | 0.004375 | 3.80E-16 |
| rs28456 | miR-1908-5 | cis | 6.85E-08 | Nightingale | LDL_TG | NA | G | A | 0.030982 | 0.004398 | 1.80E-12 |
| rs28456 | miR-1908-5 | cis | 6.85E-08 | Nightingale | XL_HDL_CE | NA | G | A | -0.02766 | 0.004445 | 1.50E-09 |
| rs28456 | miR-1908-5 | cis | 6.85E-08 | Nightingale | LDL_C | NA | G | A | -0.04771 | 0.004428 | 2.00E-28 |
| rs28456 | miR-1908-5 | cis | 6.85E-08 | Nightingale | XL_HDL_CE | NA | G | A | -0.09396 | 0.003982 | 6.19E-133 |
| rs28456 | miR-1908-5 | cis | 6.85E-08 | Nightingale | IDL_CE_pct | NA | G | A | -0.037 | 0.004364 | 2.20E-17 |
| rs28456 | miR-1908-5 | cis | 6.85E-08 | Nightingale | M_VLDL_C | NA | G | A | -0.07452 | 0.004175 | 3.10E-74 |
| rs28456 | miR-1908-5 | cis | 6.85E-08 | Nightingale | LA | NA | G | A | 0.084287 | 0.004361 | 1.50E-83 |
| rs28456 | miR-1908-5 | cis | 6.85E-08 | Nightingale | LA_pct | NA | G | A | 0.182324 | 0.004346 | 0 |
| rs28456 | miR-1908-5 | cis | 6.85E-08 | Nightingale | ApoB | NA | G | A | -0.03614 | 0.004442 | 5.80E-17 |
| rs28456 | miR-1908-5 | cis | 6.85E-08 | Nightingale | L_LDL_FC_ | NA | G | A | -0.0816 | 0.004239 | 7.90E-86 |
| rs28456 | miR-1908-5 | cis | 6.85E-08 | Nightingale | XS_VLDL_C | NA | G | A | -0.06821 | 0.004291 | 8.40E-61 |
| rs28456 | miR-1908-5 | cis | 6.85E-08 | Nightingale | M_LDL_C_ | NA | G | A | -0.09808 | 0.004451 | 3.50E-108 |
| rs28456 | miR-1908-5 | cis | 6.85E-08 | Nightingale | Total_FC | NA | G | A | -0.05644 | 0.004345 | 6.10E-41 |
| rs28456 | miR-1908-5 | cis | 6.85E-08 | Nightingale | Omega_3 | NA | G | A | -0.32929 | 0.004359 | 0 |
| rs28456 | miR-1908-5 | cis | 6.85E-08 | Nightingale | XS_VLDL_F | NA | G | A | -0.10972 | 0.004414 | 2.10E-139 |
| rs28456 | miR-1908-5 | cis | 6.85E-08 | Nightingale | M_HDL_C_ | NA | G | A | -0.06622 | 0.004234 | 5.90E-58 |
| rs28456 | miR-1908-5 | cis | 6.85E-08 | Nightingale | HDL_C | NA | G | A | -0.06313 | 0.003971 | 4.50E-61 |
| rs28456 | miR-1908-5 | cis | 6.85E-08 | Nightingale | MUFA | NA | G | A | 0.064257 | 0.004402 | 6.10E-50 |
| rs28456 | miR-1908-5 | cis | 6.85E-08 | Nightingale | XL_HDL_C | NA | G | A | -0.0958 | 0.00401 | 2.70E-136 |
| rs28456 | miR-1908-5 | cis | 6.85E-08 | Nightingale | HDL_CE | NA | G | A | -0.06015 | 0.003991 | 6.40E-55 |
| rs28456 | miR-1908-5 | cis | 6.85E-08 | Nightingale | L_HDL_L | NA | G | A | -0.07465 | 0.003902 | 1.50E-88 |

| | | | | | | | | | | | |
|---|---|---|---|---|---|---|---|---|---|---|---|
| rs28456 | miR-1908-5 | cis | 6.85E-08 | Nightingale | XXL_VLDL_ | NA | G | A | 0.072279 | 0.004268 | 9.00E-68 |
| rs28456 | miR-1908-5 | cis | 6.85E-08 | Nightingale | HDL_L | NA | G | A | -0.05148 | 0.003994 | 1.30E-40 |
| rs28456 | miR-1908-5 | cis | 6.85E-08 | Nightingale | L_HDL_FC | NA | G | A | -0.08166 | 0.003898 | 1.40E-105 |
| rs28456 | miR-1908-5 | cis | 6.85E-08 | Nightingale | M_LDL_CE | NA | G | A | -0.02494 | 0.004454 | 1.30E-08 |
| rs28456 | miR-1908-5 | cis | 6.85E-08 | Nightingale | M_VLDL_C | NA | G | A | -0.07893 | 0.004186 | 9.50E-83 |
| rs28456 | miR-1908-5 | cis | 6.85E-08 | Nightingale | L_LDL_P | NA | G | A | -0.03532 | 0.004432 | 1.90E-16 |
| rs28456 | miR-1908-5 | cis | 6.85E-08 | Nightingale | Remnant_( | NA | G | A | -0.04124 | 0.004419 | 5.10E-22 |
| rs28456 | miR-1908-5 | cis | 6.85E-08 | Nightingale | XL_HDL_P | NA | G | A | -0.08606 | 0.004002 | 1.10E-111 |
| rs28456 | miR-1908-5 | cis | 6.85E-08 | Nightingale | XL_VLDL_T | NA | G | A | 0.057236 | 0.004254 | 3.20E-43 |
| rs28456 | miR-1908-5 | cis | 6.85E-08 | Nightingale | S_LDL_P | NA | G | A | -0.03012 | 0.004452 | 4.10E-12 |
| rs28456 | miR-1908-5 | cis | 6.85E-08 | Nightingale | XS_VLDL_L | NA | G | A | -0.03829 | 0.004371 | 4.90E-20 |
| rs28456 | miR-1908-5 | cis | 6.85E-08 | Nightingale | M_LDL_TG | NA | G | A | 0.074968 | 0.004391 | 5.70E-67 |
| rs28456 | miR-1908-5 | cis | 6.85E-08 | Nightingale | IDL_CE | NA | G | A | -0.06902 | 0.004293 | 7.40E-62 |
| rs28456 | miR-1908-5 | cis | 6.85E-08 | Nightingale | DHA_pct | NA | G | A | -0.26957 | 0.004317 | 0 |
| rs28456 | miR-1908-5 | cis | 6.85E-08 | Nightingale | Omega_6_ | NA | G | A | 0.363434 | 0.004383 | 0 |
| rs28456 | miR-1908-5 | cis | 6.85E-08 | Nightingale | S_LDL_FC_ | NA | G | A | -0.08027 | 0.00431 | 9.50E-81 |
| rs28456 | miR-1908-5 | cis | 6.85E-08 | Nightingale | M_HDL_PL | NA | G | A | 0.058933 | 0.004219 | 7.00E-46 |
| rs28456 | miR-1908-5 | cis | 6.85E-08 | Nightingale | VLDL_L | NA | G | A | 0.028634 | 0.004361 | 1.80E-11 |
| rs28456 | miR-1908-5 | cis | 6.85E-08 | Nightingale | XL_VLDL_C | NA | G | A | -0.06094 | 0.004264 | 8.10E-48 |
| rs28456 | miR-1908-5 | cis | 6.85E-08 | Nightingale | L_VLDL_CE | NA | G | A | -0.05816 | 0.004256 | 3.30E-45 |
| rs28456 | miR-1908-5 | cis | 6.85E-08 | Nightingale | ApoA1 | NA | G | A | -0.03971 | 0.00407 | 4.80E-24 |
| rs28456 | miR-1908-5 | cis | 6.85E-08 | Nightingale | TG_by_PG | NA | G | A | 0.068436 | 0.004127 | 2.70E-65 |
| rs28456 | miR-1908-5 | cis | 6.85E-08 | Nightingale | VLDL_TG | NA | G | A | 0.048333 | 0.00428 | 5.30E-31 |
| rs28456 | miR-1908-5 | cis | 6.85E-08 | Nightingale | PUFA_by_I | NA | G | A | -0.16782 | 0.00426 | 0 |
| rs28456 | miR-1908-5 | cis | 6.85E-08 | Nightingale | XXL_VLDL_ | NA | G | A | 0.043186 | 0.00447 | 7.50E-23 |
| rs28456 | miR-1908-5 | cis | 6.85E-08 | Nightingale | L_LDL_TG_ | NA | G | A | 0.076468 | 0.004361 | 1.40E-70 |
| rs28456 | miR-1908-5 | cis | 6.85E-08 | Nightingale | Unsaturati | NA | G | A | -0.33331 | 0.004212 | 0 |
| rs28456 | miR-1908-5 | cis | 6.85E-08 | Nightingale | HDL_TG | NA | G | A | 0.047203 | 0.004385 | 1.30E-27 |
| rs28456 | miR-1908-5 | cis | 6.85E-08 | Nightingale | M_VLDL_T | NA | G | A | 0.076023 | 0.0042 | 1.00E-76 |
| rs28456 | miR-1908-5 | cis | 6.85E-08 | Nightingale | MUFA_pct | NA | G | A | 0.193898 | 0.004256 | 0 |
| rs28456 | miR-1908-5 | cis | 6.85E-08 | Nightingale | M_LDL_P | NA | G | A | -0.04469 | 0.00446 | 2.50E-24 |
| rs28456 | miR-1908-5 | cis | 6.85E-08 | Nightingale | HDL_size | NA | G | A | -0.06998 | 0.003941 | 3.10E-76 |
| rs28456 | miR-1908-5 | cis | 6.85E-08 | Nightingale | M_HDL_FC | NA | G | A | -0.09992 | 0.003941 | 8.79E-154 |
| rs28456 | miR-1908-5 | cis | 6.85E-08 | Nightingale | M_VLDL_F | NA | G | A | -0.02713 | 0.004447 | 4.20E-10 |
| rs28456 | miR-1908-5 | cis | 6.85E-08 | Nightingale | XL_VLDL_P | NA | G | A | 0.048966 | 0.004292 | 1.40E-31 |
| rs28456 | miR-1908-5 | cis | 6.85E-08 | Nightingale | XL_HDL_TG | NA | G | A | 0.085784 | 0.00419 | 6.60E-99 |
| rs28456 | miR-1908-5 | cis | 6.85E-08 | Nightingale | L_HDL_PL_ | NA | G | A | 0.041955 | 0.004304 | 1.40E-23 |
| rs28456 | miR-1908-5 | cis | 6.85E-08 | Nightingale | XXL_VLDL_ | NA | G | A | 0.066833 | 0.004283 | 1.10E-57 |
| rs28456 | miR-1908-5 | cis | 6.85E-08 | Nightingale | XXL_VLDL_ | NA | G | A | -0.0283 | 0.004424 | 2.80E-11 |
| rs28456 | miR-1908-5 | cis | 6.85E-08 | Nightingale | L_HDL_CE_ | NA | G | A | -0.05004 | 0.004255 | 9.20E-34 |
| rs28456 | miR-1908-5 | cis | 6.85E-08 | Nightingale | VLDL_size | NA | G | A | 0.071752 | 0.004146 | 2.80E-71 |
| rs28456 | miR-1908-5 | cis | 6.85E-08 | Nightingale | M_VLDL_C | NA | G | A | -0.04162 | 0.004435 | 7.20E-22 |
| rs28456 | miR-1908-5 | cis | 6.85E-08 | Nightingale | L_VLDL_FC | NA | G | A | 0.02775 | 0.004405 | 3.50E-10 |
| rs28456 | miR-1908-5 | cis | 6.85E-08 | Nightingale | M_LDL_FC | NA | G | A | -0.05096 | 0.004442 | 5.30E-32 |
| rs28456 | miR-1908-5 | cis | 6.85E-08 | Nightingale | XS_VLDL_T | NA | G | A | 0.032532 | 0.004379 | 1.80E-13 |
| rs28456 | miR-1908-5 | cis | 6.85E-08 | Nightingale | S_VLDL_C_ | NA | G | A | -0.05509 | 0.004327 | 2.10E-38 |
| rs28456 | miR-1908-5 | cis | 6.85E-08 | Nightingale | L_VLDL_TG | NA | G | A | 0.038021 | 0.004279 | 8.40E-20 |
| rs28456 | miR-1908-5 | cis | 6.85E-08 | Nightingale | M_LDL_PL_ | NA | G | A | 0.043624 | 0.004445 | 8.60E-24 |
| rs28456 | miR-1908-5 | cis | 6.85E-08 | Nightingale | L_LDL_PL | NA | G | A | -0.0528 | 0.004414 | 1.30E-34 |
| rs2848630 | miR-130a-3 | cis | 0.000839 | Nightingale | Phe | NA | C | T | 0.049264 | 0.00494 | 2.00E-23 |
| rs2949178 | miR-339-5p | cis | 5.58E-05 | Nightingale | M_VLDL_P | NA | C | T | -0.0244 | 0.004057 | 3.50E-10 |
| rs2949178 | miR-339-5p | cis | 5.58E-05 | Nightingale | XL_VLDL_F | NA | C | T | -0.02458 | 0.004099 | 1.20E-09 |
| rs2949178 | miR-339-5p | cis | 5.58E-05 | Nightingale | M_VLDL_T | NA | C | T | 0.02307 | 0.003994 | 2.40E-09 |
| rs2949178 | miR-339-5p | cis | 5.58E-05 | Nightingale | XL_VLDL_T | NA | C | T | 0.023761 | 0.004098 | 1.40E-09 |
| rs2949178 | miR-339-5p | cis | 5.58E-05 | Nightingale | L_VLDL_CE | NA | C | T | -0.02564 | 0.004045 | 4.80E-11 |
| rs2949178 | miR-339-5p | cis | 5.58E-05 | Nightingale | VLDL_size | NA | C | T | 0.023072 | 0.003942 | 1.40E-08 |
| rs2949178 | miR-339-5p | cis | 5.58E-05 | Nightingale | M_VLDL_C | NA | C | T | -0.0217 | 0.00397 | 2.30E-08 |
| rs2949178 | miR-339-5p | cis | 5.58E-05 | Nightingale | M_VLDL_C | NA | C | T | -0.02234 | 0.003981 | 7.90E-09 |
| rs2949178 | miR-339-5p | cis | 5.58E-05 | Nightingale | XL_VLDL_C | NA | C | T | -0.02199 | 0.004054 | 2.80E-08 |
| rs2949178 | miR-339-5p | cis | 5.58E-05 | Nightingale | L_VLDL_C_ | NA | C | T | -0.02566 | 0.004129 | 4.40E-11 |
| rs2949178 | miR-339-5p | cis | 5.58E-05 | Nightingale | M_VLDL_F | NA | C | T | -0.02325 | 0.004021 | 2.50E-09 |
| rs2949179 | miR-339-5p | cis | 5.73E-05 | Nightingale | M_VLDL_C | NA | G | C | -0.0223 | 0.003981 | 8.30E-09 |
| rs2949179 | miR-339-5p | cis | 5.73E-05 | Nightingale | M_VLDL_T | NA | G | C | 0.023028 | 0.003994 | 2.60E-09 |

| | | | | | | | | | | | |
|---|---|---|---|---|---|---|---|---|---|---|---|
| rs2949179 | miR-339-5p | cis | 5.73E-05 | Nightingale | XL_VLDL_F | NA | G | C | -0.02459 | 0.004099 | 1.20E-09 |
| rs2949179 | miR-339-5p | cis | 5.73E-05 | Nightingale | XL_VLDL_C | NA | G | C | -0.02196 | 0.004054 | 2.90E-08 |
| rs2949179 | miR-339-5p | cis | 5.73E-05 | Nightingale | L_VLDL_CE | NA | G | C | -0.02566 | 0.004045 | 4.70E-11 |
| rs2949179 | miR-339-5p | cis | 5.73E-05 | Nightingale | M_VLDL_C | NA | G | C | -0.02166 | 0.00397 | 2.40E-08 |
| rs2949179 | miR-339-5p | cis | 5.73E-05 | Nightingale | M_VLDL_F | NA | G | C | -0.02321 | 0.004021 | 2.60E-09 |
| rs2949179 | miR-339-5p | cis | 5.73E-05 | Nightingale | L_VLDL_C_ | NA | G | C | -0.02568 | 0.004129 | 4.40E-11 |
| rs2949179 | miR-339-5p | cis | 5.73E-05 | Nightingale | XL_VLDL_T | NA | G | C | 0.023726 | 0.004098 | 1.40E-09 |
| rs2949179 | miR-339-5p | cis | 5.73E-05 | Nightingale | M_VLDL_P | NA | G | C | -0.02435 | 0.004057 | 3.70E-10 |
| rs2949179 | miR-339-5p | cis | 5.73E-05 | Nightingale | VLDL_size | NA | G | C | 0.023144 | 0.003942 | 1.20E-08 |
| rs2949180 | miR-339-5p | cis | 5.81E-05 | Nightingale | XL_VLDL_C | NA | A | T | -0.02191 | 0.004056 | 3.30E-08 |
| rs2949180 | miR-339-5p | cis | 5.81E-05 | Nightingale | L_VLDL_C_ | NA | A | T | -0.02554 | 0.004131 | 5.60E-11 |
| rs2949180 | miR-339-5p | cis | 5.81E-05 | Nightingale | M_VLDL_P | NA | A | T | -0.02429 | 0.004059 | 4.40E-10 |
| rs2949180 | miR-339-5p | cis | 5.81E-05 | Nightingale | L_VLDL_CE | NA | A | T | -0.02552 | 0.004047 | 6.00E-11 |
| rs2949180 | miR-339-5p | cis | 5.81E-05 | Nightingale | XL_VLDL_T | NA | A | T | 0.023659 | 0.0041 | 1.60E-09 |
| rs2949180 | miR-339-5p | cis | 5.81E-05 | Nightingale | M_VLDL_F | NA | A | T | -0.02313 | 0.004023 | 3.20E-09 |
| rs2949180 | miR-339-5p | cis | 5.81E-05 | Nightingale | M_VLDL_C | NA | A | T | -0.0216 | 0.003972 | 2.80E-08 |
| rs2949180 | miR-339-5p | cis | 5.81E-05 | Nightingale | XL_VLDL_F | NA | A | T | -0.02445 | 0.004101 | 1.50E-09 |
| rs2949180 | miR-339-5p | cis | 5.81E-05 | Nightingale | M_VLDL_T | NA | A | T | 0.022959 | 0.003996 | 3.00E-09 |
| rs2949180 | miR-339-5p | cis | 5.81E-05 | Nightingale | M_VLDL_C | NA | A | T | -0.02223 | 0.003983 | 9.80E-09 |
| rs2949180 | miR-339-5p | cis | 5.81E-05 | Nightingale | VLDL_size | NA | A | T | 0.022993 | 0.003944 | 1.60E-08 |
| rs2949181 | miR-339-5p | cis | 6.46E-05 | Nightingale | M_VLDL_P | NA | G | A | -0.02438 | 0.004057 | 3.50E-10 |
| rs2949181 | miR-339-5p | cis | 6.46E-05 | Nightingale | XL_VLDL_C | NA | G | A | -0.02197 | 0.004053 | 2.90E-08 |
| rs2949181 | miR-339-5p | cis | 6.46E-05 | Nightingale | L_VLDL_C_ | NA | G | A | -0.02566 | 0.004129 | 4.40E-11 |
| rs2949181 | miR-339-5p | cis | 6.46E-05 | Nightingale | XL_VLDL_F | NA | G | A | -0.02456 | 0.004098 | 1.20E-09 |
| rs2949181 | miR-339-5p | cis | 6.46E-05 | Nightingale | XL_VLDL_T | NA | G | A | 0.023746 | 0.004098 | 1.40E-09 |
| rs2949181 | miR-339-5p | cis | 6.46E-05 | Nightingale | L_VLDL_CE | NA | G | A | -0.02563 | 0.004045 | 4.80E-11 |
| rs2949181 | miR-339-5p | cis | 6.46E-05 | Nightingale | VLDL_size | NA | G | A | 0.023063 | 0.003942 | 1.40E-08 |
| rs2949181 | miR-339-5p | cis | 6.46E-05 | Nightingale | M_VLDL_F | NA | G | A | -0.02323 | 0.004021 | 2.50E-09 |
| rs2949181 | miR-339-5p | cis | 6.46E-05 | Nightingale | M_VLDL_C | NA | G | A | -0.02168 | 0.00397 | 2.40E-08 |
| rs2949181 | miR-339-5p | cis | 6.46E-05 | Nightingale | M_VLDL_T | NA | G | A | 0.023049 | 0.003994 | 2.50E-09 |
| rs2949181 | miR-339-5p | cis | 6.46E-05 | Nightingale | M_VLDL_C | NA | G | A | -0.02232 | 0.00398 | 8.10E-09 |
| rs2949182 | miR-339-5p | cis | 4.80E-05 | Nightingale | L_VLDL_C_ | NA | A | G | -0.02695 | 0.004164 | 7.50E-12 |
| rs2949182 | miR-339-5p | cis | 4.80E-05 | Nightingale | M_VLDL_C | NA | A | G | -0.02278 | 0.004004 | 5.00E-09 |
| rs2949182 | miR-339-5p | cis | 4.80E-05 | Nightingale | XL_VLDL_C | NA | A | G | -0.02324 | 0.004088 | 4.90E-09 |
| rs2949182 | miR-339-5p | cis | 4.80E-05 | Nightingale | XL_VLDL_C | NA | A | G | -0.02192 | 0.00409 | 2.90E-08 |
| rs2949182 | miR-339-5p | cis | 4.80E-05 | Nightingale | M_VLDL_F | NA | A | G | -0.02411 | 0.004055 | 8.30E-10 |
| rs2949182 | miR-339-5p | cis | 4.80E-05 | Nightingale | M_VLDL_C | NA | A | G | -0.02335 | 0.004015 | 1.90E-09 |
| rs2949182 | miR-339-5p | cis | 4.80E-05 | Nightingale | M_VLDL_T | NA | A | G | 0.024066 | 0.004028 | 5.90E-10 |
| rs2949182 | miR-339-5p | cis | 4.80E-05 | Nightingale | L_VLDL_CE | NA | A | G | -0.02714 | 0.004079 | 5.00E-12 |
| rs2949182 | miR-339-5p | cis | 4.80E-05 | Nightingale | M_VLDL_P | NA | A | G | -0.02524 | 0.004092 | 1.30E-10 |
| rs2949182 | miR-339-5p | cis | 4.80E-05 | Nightingale | XL_VLDL_F | NA | A | G | -0.02607 | 0.004133 | 1.40E-10 |
| rs2949182 | miR-339-5p | cis | 4.80E-05 | Nightingale | XL_VLDL_T | NA | A | G | 0.025014 | 0.004133 | 2.30E-10 |
| rs2949182 | miR-339-5p | cis | 4.80E-05 | Nightingale | VLDL_size | NA | A | G | 0.024532 | 0.003976 | 1.80E-09 |
| rs2949183 | miR-339-5p | cis | 5.07E-05 | Nightingale | XL_VLDL_C | NA | A | T | -0.02314 | 0.004088 | 5.60E-09 |
| rs2949183 | miR-339-5p | cis | 5.07E-05 | Nightingale | L_VLDL_C_ | NA | A | T | -0.02686 | 0.004165 | 8.40E-12 |
| rs2949183 | miR-339-5p | cis | 5.07E-05 | Nightingale | XL_VLDL_F | NA | A | T | -0.02598 | 0.004134 | 1.60E-10 |
| rs2949183 | miR-339-5p | cis | 5.07E-05 | Nightingale | L_VLDL_CE | NA | A | T | -0.02706 | 0.00408 | 5.50E-12 |
| rs2949183 | miR-339-5p | cis | 5.07E-05 | Nightingale | M_VLDL_T | NA | A | T | 0.024047 | 0.004028 | 5.90E-10 |
| rs2949183 | miR-339-5p | cis | 5.07E-05 | Nightingale | M_VLDL_F | NA | A | T | -0.0241 | 0.004056 | 8.10E-10 |
| rs2949183 | miR-339-5p | cis | 5.07E-05 | Nightingale | M_VLDL_P | NA | A | T | -0.02525 | 0.004092 | 1.20E-10 |
| rs2949183 | miR-339-5p | cis | 5.07E-05 | Nightingale | VLDL_size | NA | A | T | 0.024494 | 0.003976 | 1.80E-09 |
| rs2949183 | miR-339-5p | cis | 5.07E-05 | Nightingale | M_VLDL_C | NA | A | T | -0.02333 | 0.004015 | 1.90E-09 |
| rs2949183 | miR-339-5p | cis | 5.07E-05 | Nightingale | XL_VLDL_T | NA | A | T | 0.024914 | 0.004133 | 2.60E-10 |
| rs2949183 | miR-339-5p | cis | 5.07E-05 | Nightingale | XL_VLDL_C | NA | A | T | -0.02183 | 0.00409 | 3.20E-08 |
| rs2949183 | miR-339-5p | cis | 5.07E-05 | Nightingale | M_VLDL_C | NA | A | T | -0.02274 | 0.004004 | 5.10E-09 |
| rs2949184 | miR-339-5p | cis | 5.66E-05 | Nightingale | M_VLDL_P | NA | G | T | -0.02444 | 0.004057 | 3.10E-10 |
| rs2949184 | miR-339-5p | cis | 5.66E-05 | Nightingale | L_VLDL_C_ | NA | G | T | -0.02566 | 0.004129 | 4.40E-11 |
| rs2949184 | miR-339-5p | cis | 5.66E-05 | Nightingale | M_VLDL_F | NA | G | T | -0.02332 | 0.004021 | 2.10E-09 |
| rs2949184 | miR-339-5p | cis | 5.66E-05 | Nightingale | M_VLDL_T | NA | G | T | 0.023127 | 0.003994 | 2.20E-09 |
| rs2949184 | miR-339-5p | cis | 5.66E-05 | Nightingale | L_VLDL_CE | NA | G | T | -0.02564 | 0.004045 | 4.70E-11 |
| rs2949184 | miR-339-5p | cis | 5.66E-05 | Nightingale | M_VLDL_C | NA | G | T | -0.02176 | 0.003971 | 2.10E-08 |
| rs2949184 | miR-339-5p | cis | 5.66E-05 | Nightingale | XL_VLDL_T | NA | G | T | 0.023758 | 0.004098 | 1.30E-09 |
| rs2949184 | miR-339-5p | cis | 5.66E-05 | Nightingale | M_VLDL_C | NA | G | T | -0.0224 | 0.003981 | 7.00E-09 |

| | | | | | | | | | | | |
|---|---|---|---|---|---|---|---|---|---|---|---|
| rs2949184 | miR-339-5p | cis | 5.66E-05 | Nightingale | XL_VLDL_C | NA | G | T | -0.022 | 0.004054 | 2.70E-08 |
| rs2949184 | miR-339-5p | cis | 5.66E-05 | Nightingale | XL_VLDL_F | NA | G | T | -0.02458 | 0.004099 | 1.20E-09 |
| rs2949184 | miR-339-5p | cis | 5.66E-05 | Nightingale | VLDL_size | NA | G | T | 0.02309 | 0.003942 | 1.30E-08 |
| rs2949186 | miR-339-5p | cis | 5.74E-05 | Nightingale | M_VLDL_F | NA | C | T | -0.02325 | 0.00402 | 2.40E-09 |
| rs2949186 | miR-339-5p | cis | 5.74E-05 | Nightingale | XL_VLDL_F | NA | C | T | -0.02455 | 0.004097 | 1.20E-09 |
| rs2949186 | miR-339-5p | cis | 5.74E-05 | Nightingale | XL_VLDL_T | NA | C | T | 0.023755 | 0.004097 | 1.30E-09 |
| rs2949186 | miR-339-5p | cis | 5.74E-05 | Nightingale | L_VLDL_C_ | NA | C | T | -0.02577 | 0.004128 | 3.50E-11 |
| rs2949186 | miR-339-5p | cis | 5.74E-05 | Nightingale | VLDL_size | NA | C | T | 0.023081 | 0.003941 | 1.30E-08 |
| rs2949186 | miR-339-5p | cis | 5.74E-05 | Nightingale | XL_VLDL_C | NA | C | T | -0.02198 | 0.004052 | 2.70E-08 |
| rs2949186 | miR-339-5p | cis | 5.74E-05 | Nightingale | M_VLDL_C | NA | C | T | -0.02233 | 0.003979 | 7.70E-09 |
| rs2949186 | miR-339-5p | cis | 5.74E-05 | Nightingale | M_VLDL_T | NA | C | T | 0.023066 | 0.003992 | 2.30E-09 |
| rs2949186 | miR-339-5p | cis | 5.74E-05 | Nightingale | L_VLDL_CE | NA | C | T | -0.02571 | 0.004043 | 4.10E-11 |
| rs2949186 | miR-339-5p | cis | 5.74E-05 | Nightingale | M_VLDL_P | NA | C | T | -0.02443 | 0.004056 | 3.10E-10 |
| rs2949186 | miR-339-5p | cis | 5.74E-05 | Nightingale | M_VLDL_C | NA | C | T | -0.02169 | 0.003969 | 2.20E-08 |
| rs2949187 | miR-339-5p | cis | 5.61E-05 | Nightingale | M_VLDL_T | NA | T | G | 0.0232 | 0.003993 | 1.90E-09 |
| rs2949187 | miR-339-5p | cis | 5.61E-05 | Nightingale | XL_VLDL_F | NA | T | G | -0.02467 | 0.004098 | 1.00E-09 |
| rs2949187 | miR-339-5p | cis | 5.61E-05 | Nightingale | M_VLDL_C | NA | T | G | -0.02185 | 0.00397 | 1.90E-08 |
| rs2949187 | miR-339-5p | cis | 5.61E-05 | Nightingale | L_VLDL_CE | NA | T | G | -0.02588 | 0.004044 | 3.00E-11 |
| rs2949187 | miR-339-5p | cis | 5.61E-05 | Nightingale | XL_VLDL_C | NA | T | G | -0.02218 | 0.004053 | 2.10E-08 |
| rs2949187 | miR-339-5p | cis | 5.61E-05 | Nightingale | M_VLDL_P | NA | T | G | -0.0245 | 0.004057 | 2.80E-10 |
| rs2949187 | miR-339-5p | cis | 5.61E-05 | Nightingale | XL_VLDL_T | NA | T | G | 0.023957 | 0.004098 | 9.80E-10 |
| rs2949187 | miR-339-5p | cis | 5.61E-05 | Nightingale | M_VLDL_C | NA | T | G | -0.02247 | 0.00398 | 6.40E-09 |
| rs2949187 | miR-339-5p | cis | 5.61E-05 | Nightingale | VLDL_size | NA | T | G | 0.023359 | 0.003941 | 9.10E-09 |
| rs2949187 | miR-339-5p | cis | 5.61E-05 | Nightingale | L_VLDL_C_ | NA | T | G | -0.02594 | 0.004129 | 2.60E-11 |
| rs2949187 | miR-339-5p | cis | 5.61E-05 | Nightingale | M_VLDL_F | NA | T | G | -0.02336 | 0.004021 | 2.00E-09 |
| rs2949188 | miR-339-5p | cis | 5.60E-05 | Nightingale | XL_VLDL_F | NA | G | C | -0.02469 | 0.004098 | 9.90E-10 |
| rs2949188 | miR-339-5p | cis | 5.60E-05 | Nightingale | M_VLDL_F | NA | G | C | -0.02337 | 0.00402 | 2.00E-09 |
| rs2949188 | miR-339-5p | cis | 5.60E-05 | Nightingale | M_VLDL_C | NA | G | C | -0.02248 | 0.00398 | 6.40E-09 |
| rs2949188 | miR-339-5p | cis | 5.60E-05 | Nightingale | L_VLDL_C_ | NA | G | C | -0.02597 | 0.004129 | 2.50E-11 |
| rs2949188 | miR-339-5p | cis | 5.60E-05 | Nightingale | L_VLDL_CE | NA | G | C | -0.0259 | 0.004044 | 2.90E-11 |
| rs2949188 | miR-339-5p | cis | 5.60E-05 | Nightingale | XL_VLDL_C | NA | G | C | -0.0222 | 0.004053 | 2.00E-08 |
| rs2949188 | miR-339-5p | cis | 5.60E-05 | Nightingale | XL_VLDL_T | NA | G | C | 0.023977 | 0.004097 | 9.50E-10 |
| rs2949188 | miR-339-5p | cis | 5.60E-05 | Nightingale | M_VLDL_T | NA | G | C | 0.023204 | 0.003993 | 1.90E-09 |
| rs2949188 | miR-339-5p | cis | 5.60E-05 | Nightingale | M_VLDL_P | NA | G | C | -0.0245 | 0.004056 | 2.80E-10 |
| rs2949188 | miR-339-5p | cis | 5.60E-05 | Nightingale | VLDL_size | NA | G | C | 0.023387 | 0.003941 | 8.80E-09 |
| rs2949188 | miR-339-5p | cis | 5.60E-05 | Nightingale | M_VLDL_C | NA | G | C | -0.02185 | 0.00397 | 1.90E-08 |
| rs2949189 | miR-339-5p | cis | 3.39E-05 | Nightingale | M_VLDL_P | NA | G | A | -0.0246 | 0.004057 | 2.50E-10 |
| rs2949189 | miR-339-5p | cis | 3.39E-05 | Nightingale | M_VLDL_T | NA | G | A | 0.023265 | 0.003993 | 1.80E-09 |
| rs2949189 | miR-339-5p | cis | 3.39E-05 | Nightingale | XL_VLDL_F | NA | G | A | -0.02477 | 0.004098 | 8.80E-10 |
| rs2949189 | miR-339-5p | cis | 3.39E-05 | Nightingale | XL_VLDL_T | NA | G | A | 0.024023 | 0.004098 | 9.10E-10 |
| rs2949189 | miR-339-5p | cis | 3.39E-05 | Nightingale | M_VLDL_F | NA | G | A | -0.02346 | 0.004021 | 1.80E-09 |
| rs2949189 | miR-339-5p | cis | 3.39E-05 | Nightingale | XL_VLDL_C | NA | G | A | -0.02223 | 0.004053 | 2.00E-08 |
| rs2949189 | miR-339-5p | cis | 3.39E-05 | Nightingale | M_VLDL_C | NA | G | A | -0.02253 | 0.00398 | 6.20E-09 |
| rs2949189 | miR-339-5p | cis | 3.39E-05 | Nightingale | M_VLDL_C | NA | G | A | -0.02188 | 0.00397 | 1.80E-08 |
| rs2949189 | miR-339-5p | cis | 3.39E-05 | Nightingale | L_VLDL_C_ | NA | G | A | -0.02598 | 0.004129 | 2.50E-11 |
| rs2949189 | miR-339-5p | cis | 3.39E-05 | Nightingale | VLDL_size | NA | G | A | 0.023342 | 0.003941 | 9.50E-09 |
| rs2949189 | miR-339-5p | cis | 3.39E-05 | Nightingale | L_VLDL_CE | NA | G | A | -0.02594 | 0.004044 | 2.80E-11 |
| rs2949190 | miR-339-5p | cis | 5.57E-05 | Nightingale | L_VLDL_C_ | NA | A | T | -0.02594 | 0.004129 | 2.80E-11 |
| rs2949190 | miR-339-5p | cis | 5.57E-05 | Nightingale | M_VLDL_T | NA | A | T | 0.023231 | 0.003993 | 2.00E-09 |
| rs2949190 | miR-339-5p | cis | 5.57E-05 | Nightingale | M_VLDL_C | NA | A | T | -0.02249 | 0.00398 | 6.60E-09 |
| rs2949190 | miR-339-5p | cis | 5.57E-05 | Nightingale | VLDL_size | NA | A | T | 0.02331 | 0.003941 | 1.00E-08 |
| rs2949190 | miR-339-5p | cis | 5.57E-05 | Nightingale | M_VLDL_F | NA | A | T | -0.02342 | 0.00402 | 1.90E-09 |
| rs2949190 | miR-339-5p | cis | 5.57E-05 | Nightingale | XL_VLDL_C | NA | A | T | -0.02219 | 0.004053 | 2.10E-08 |
| rs2949190 | miR-339-5p | cis | 5.57E-05 | Nightingale | M_VLDL_P | NA | A | T | -0.02457 | 0.004057 | 2.70E-10 |
| rs2949190 | miR-339-5p | cis | 5.57E-05 | Nightingale | XL_VLDL_F | NA | A | T | -0.02472 | 0.004098 | 9.70E-10 |
| rs2949190 | miR-339-5p | cis | 5.57E-05 | Nightingale | L_VLDL_CE | NA | A | T | -0.0259 | 0.004044 | 3.10E-11 |
| rs2949190 | miR-339-5p | cis | 5.57E-05 | Nightingale | M_VLDL_C | NA | A | T | -0.02185 | 0.00397 | 2.00E-08 |
| rs2949190 | miR-339-5p | cis | 5.57E-05 | Nightingale | XL_VLDL_T | NA | A | T | 0.02398 | 0.004097 | 9.90E-10 |
| rs2949191 | miR-339-5p | cis | 5.46E-05 | Nightingale | M_VLDL_C | NA | G | C | -0.02249 | 0.00398 | 6.60E-09 |
| rs2949191 | miR-339-5p | cis | 5.46E-05 | Nightingale | XL_VLDL_C | NA | G | C | -0.02219 | 0.004053 | 2.10E-08 |
| rs2949191 | miR-339-5p | cis | 5.46E-05 | Nightingale | VLDL_size | NA | G | C | 0.02331 | 0.003941 | 1.00E-08 |
| rs2949191 | miR-339-5p | cis | 5.46E-05 | Nightingale | M_VLDL_P | NA | G | C | -0.02457 | 0.004056 | 2.70E-10 |
| rs2949191 | miR-339-5p | cis | 5.46E-05 | Nightingale | M_VLDL_T | NA | G | C | 0.023231 | 0.003993 | 2.00E-09 |

| | | | | | | | | | | | |
|---|---|---|---|---|---|---|---|---|---|---|---|
| rs2949191 | miR-339-5p | cis | 5.46E-05 | Nightingale | L_VLDL_C_ | NA | G | C | -0.02594 | 0.004129 | 2.80E-11 |
| rs2949191 | miR-339-5p | cis | 5.46E-05 | Nightingale | XL_VLDL_F | NA | G | C | -0.02472 | 0.004098 | 9.70E-10 |
| rs2949191 | miR-339-5p | cis | 5.46E-05 | Nightingale | M_VLDL_F | NA | G | C | -0.02342 | 0.00402 | 1.90E-09 |
| rs2949191 | miR-339-5p | cis | 5.46E-05 | Nightingale | L_VLDL_CE | NA | G | C | -0.0259 | 0.004044 | 3.10E-11 |
| rs2949191 | miR-339-5p | cis | 5.46E-05 | Nightingale | M_VLDL_C | NA | G | C | -0.02185 | 0.00397 | 2.00E-08 |
| rs2949191 | miR-339-5p | cis | 5.46E-05 | Nightingale | XL_VLDL_T | NA | G | C | 0.02398 | 0.004097 | 9.90E-10 |
| rs2949193 | miR-339-5p | cis | 6.09E-05 | Nightingale | M_VLDL_C | NA | C | T | -0.02246 | 0.00398 | 6.80E-09 |
| rs2949193 | miR-339-5p | cis | 6.09E-05 | Nightingale | L_VLDL_CE | NA | C | T | -0.02585 | 0.004044 | 3.30E-11 |
| rs2949193 | miR-339-5p | cis | 6.09E-05 | Nightingale | VLDL_size | NA | C | T | 0.023322 | 0.003941 | 9.90E-09 |
| rs2949193 | miR-339-5p | cis | 6.09E-05 | Nightingale | XL_VLDL_C | NA | C | T | -0.02217 | 0.004053 | 2.20E-08 |
| rs2949193 | miR-339-5p | cis | 6.09E-05 | Nightingale | XL_VLDL_T | NA | C | T | 0.023945 | 0.004097 | 1.00E-09 |
| rs2949193 | miR-339-5p | cis | 6.09E-05 | Nightingale | XL_VLDL_F | NA | C | T | -0.02468 | 0.004098 | 1.00E-09 |
| rs2949193 | miR-339-5p | cis | 6.09E-05 | Nightingale | M_VLDL_C | NA | C | T | -0.02183 | 0.003969 | 2.00E-08 |
| rs2949193 | miR-339-5p | cis | 6.09E-05 | Nightingale | M_VLDL_F | NA | C | T | -0.02337 | 0.00402 | 2.10E-09 |
| rs2949193 | miR-339-5p | cis | 6.09E-05 | Nightingale | M_VLDL_P | NA | C | T | -0.02451 | 0.004056 | 2.90E-10 |
| rs2949193 | miR-339-5p | cis | 6.09E-05 | Nightingale | M_VLDL_T | NA | C | T | 0.023196 | 0.003993 | 2.00E-09 |
| rs2949193 | miR-339-5p | cis | 6.09E-05 | Nightingale | L_VLDL_C_ | NA | C | T | -0.0259 | 0.004128 | 2.90E-11 |
| rs2949194 | miR-339-5p | cis | 5.41E-05 | Nightingale | M_VLDL_T | NA | G | C | 0.023272 | 0.003993 | 1.80E-09 |
| rs2949194 | miR-339-5p | cis | 5.41E-05 | Nightingale | M_VLDL_F | NA | G | C | -0.02349 | 0.00402 | 1.70E-09 |
| rs2949194 | miR-339-5p | cis | 5.41E-05 | Nightingale | M_VLDL_P | NA | G | C | -0.02466 | 0.004056 | 2.20E-10 |
| rs2949194 | miR-339-5p | cis | 5.41E-05 | Nightingale | XL_VLDL_F | NA | G | C | -0.02483 | 0.004097 | 7.90E-10 |
| rs2949194 | miR-339-5p | cis | 5.41E-05 | Nightingale | M_VLDL_C | NA | G | C | -0.02186 | 0.003969 | 1.80E-08 |
| rs2949194 | miR-339-5p | cis | 5.41E-05 | Nightingale | XL_VLDL_C | NA | G | C | -0.0222 | 0.004052 | 2.00E-08 |
| rs2949194 | miR-339-5p | cis | 5.41E-05 | Nightingale | M_VLDL_C | NA | G | C | -0.02252 | 0.003979 | 6.00E-09 |
| rs2949194 | miR-339-5p | cis | 5.41E-05 | Nightingale | L_VLDL_CE | NA | G | C | -0.02597 | 0.004044 | 2.60E-11 |
| rs2949194 | miR-339-5p | cis | 5.41E-05 | Nightingale | XL_VLDL_T | NA | G | C | 0.024058 | 0.004097 | 8.40E-10 |
| rs2949194 | miR-339-5p | cis | 5.41E-05 | Nightingale | L_VLDL_C_ | NA | G | C | -0.02608 | 0.004128 | 2.10E-11 |
| rs2949194 | miR-339-5p | cis | 5.41E-05 | Nightingale | VLDL_size | NA | G | C | 0.023258 | 0.003941 | 1.00E-08 |
| rs2949197 | miR-339-5p | cis | 6.61E-05 | Nightingale | XL_VLDL_C | NA | T | C | -0.02226 | 0.004053 | 1.90E-08 |
| rs2949197 | miR-339-5p | cis | 6.61E-05 | Nightingale | M_VLDL_C | NA | T | C | -0.02258 | 0.00398 | 5.50E-09 |
| rs2949197 | miR-339-5p | cis | 6.61E-05 | Nightingale | M_VLDL_P | NA | T | C | -0.02473 | 0.004056 | 2.00E-10 |
| rs2949197 | miR-339-5p | cis | 6.61E-05 | Nightingale | M_VLDL_T | NA | T | C | 0.023337 | 0.003993 | 1.60E-09 |
| rs2949197 | miR-339-5p | cis | 6.61E-05 | Nightingale | L_VLDL_C_ | NA | T | C | -0.02613 | 0.004128 | 1.90E-11 |
| rs2949197 | miR-339-5p | cis | 6.61E-05 | Nightingale | M_VLDL_F | NA | T | C | -0.02356 | 0.00402 | 1.50E-09 |
| rs2949197 | miR-339-5p | cis | 6.61E-05 | Nightingale | XL_VLDL_T | NA | T | C | 0.02411 | 0.004097 | 7.80E-10 |
| rs2949197 | miR-339-5p | cis | 6.61E-05 | Nightingale | L_VLDL_CE | NA | T | C | -0.02603 | 0.004044 | 2.40E-11 |
| rs2949197 | miR-339-5p | cis | 6.61E-05 | Nightingale | VLDL_size | NA | T | C | 0.023302 | 0.003941 | 1.00E-08 |
| rs2949197 | miR-339-5p | cis | 6.61E-05 | Nightingale | XL_VLDL_F | NA | T | C | -0.02489 | 0.004098 | 7.20E-10 |
| rs2949197 | miR-339-5p | cis | 6.61E-05 | Nightingale | M_VLDL_C | NA | T | C | -0.02192 | 0.00397 | 1.70E-08 |
| rs2949198 | miR-339-5p | cis | 3.79E-05 | Nightingale | XL_VLDL_C | NA | C | T | -0.0223 | 0.004054 | 1.80E-08 |
| rs2949198 | miR-339-5p | cis | 3.79E-05 | Nightingale | L_VLDL_CE | NA | C | T | -0.02608 | 0.004045 | 2.20E-11 |
| rs2949198 | miR-339-5p | cis | 3.79E-05 | Nightingale | L_VLDL_C_ | NA | C | T | -0.02618 | 0.004129 | 1.80E-11 |
| rs2949198 | miR-339-5p | cis | 3.79E-05 | Nightingale | XL_VLDL_T | NA | C | T | 0.024141 | 0.004098 | 7.60E-10 |
| rs2949198 | miR-339-5p | cis | 3.79E-05 | Nightingale | M_VLDL_P | NA | C | T | -0.02476 | 0.004057 | 1.90E-10 |
| rs2949198 | miR-339-5p | cis | 3.79E-05 | Nightingale | M_VLDL_C | NA | C | T | -0.02196 | 0.00397 | 1.60E-08 |
| rs2949198 | miR-339-5p | cis | 3.79E-05 | Nightingale | XL_VLDL_F | NA | C | T | -0.02493 | 0.004099 | 6.80E-10 |
| rs2949198 | miR-339-5p | cis | 3.79E-05 | Nightingale | M_VLDL_T | NA | C | T | 0.023378 | 0.003994 | 1.50E-09 |
| rs2949198 | miR-339-5p | cis | 3.79E-05 | Nightingale | M_VLDL_C | NA | C | T | -0.02262 | 0.003981 | 5.20E-09 |
| rs2949198 | miR-339-5p | cis | 3.79E-05 | Nightingale | VLDL_size | NA | C | T | 0.023363 | 0.003942 | 9.20E-09 |
| rs2949198 | miR-339-5p | cis | 3.79E-05 | Nightingale | M_VLDL_F | NA | C | T | -0.0236 | 0.004021 | 1.40E-09 |
| rs2949199 | miR-339-5p | cis | 5.29E-05 | Nightingale | M_VLDL_P | NA | A | G | -0.02467 | 0.004056 | 2.20E-10 |
| rs2949199 | miR-339-5p | cis | 5.29E-05 | Nightingale | M_VLDL_C | NA | A | G | -0.02185 | 0.003969 | 1.80E-08 |
| rs2949199 | miR-339-5p | cis | 5.29E-05 | Nightingale | VLDL_size | NA | A | G | 0.023248 | 0.003941 | 1.00E-08 |
| rs2949199 | miR-339-5p | cis | 5.29E-05 | Nightingale | XL_VLDL_F | NA | A | G | -0.02484 | 0.004097 | 7.70E-10 |
| rs2949199 | miR-339-5p | cis | 5.29E-05 | Nightingale | L_VLDL_C_ | NA | A | G | -0.0261 | 0.004128 | 2.00E-11 |
| rs2949199 | miR-339-5p | cis | 5.29E-05 | Nightingale | XL_VLDL_C | NA | A | G | -0.0222 | 0.004052 | 2.00E-08 |
| rs2949199 | miR-339-5p | cis | 5.29E-05 | Nightingale | L_VLDL_CE | NA | A | G | -0.02598 | 0.004044 | 2.50E-11 |
| rs2949199 | miR-339-5p | cis | 5.29E-05 | Nightingale | XL_VLDL_T | NA | A | G | 0.024058 | 0.004097 | 8.30E-10 |
| rs2949199 | miR-339-5p | cis | 5.29E-05 | Nightingale | M_VLDL_F | NA | A | G | -0.0235 | 0.00402 | 1.60E-09 |
| rs2949199 | miR-339-5p | cis | 5.29E-05 | Nightingale | M_VLDL_C | NA | A | G | -0.02252 | 0.003979 | 6.00E-09 |
| rs2949199 | miR-339-5p | cis | 5.29E-05 | Nightingale | M_VLDL_T | NA | A | G | 0.023269 | 0.003993 | 1.80E-09 |
| rs2949200 | miR-339-5p | cis | 5.13E-05 | Nightingale | M_VLDL_P | NA | C | T | -0.02469 | 0.004056 | 2.10E-10 |
| rs2949200 | miR-339-5p | cis | 5.13E-05 | Nightingale | XL_VLDL_T | NA | C | T | 0.024092 | 0.004097 | 7.90E-10 |

| | | | | | | | | | | | |
|---|---|---|---|---|---|---|---|---|---|---|---|
| rs2949200 | miR-339-5p | cis | 5.13E-05 | Nightingale | VLDL_size | NA | C | T | 0.023259 | 0.003941 | 1.00E-08 |
| rs2949200 | miR-339-5p | cis | 5.13E-05 | Nightingale | L_VLDL_C_ | NA | C | T | -0.02614 | 0.004128 | 1.90E-11 |
| rs2949200 | miR-339-5p | cis | 5.13E-05 | Nightingale | M_VLDL_C | NA | C | T | -0.02253 | 0.003979 | 5.90E-09 |
| rs2949200 | miR-339-5p | cis | 5.13E-05 | Nightingale | M_VLDL_C | NA | C | T | -0.02187 | 0.003969 | 1.80E-08 |
| rs2949200 | miR-339-5p | cis | 5.13E-05 | Nightingale | M_VLDL_F | NA | C | T | -0.02352 | 0.00402 | 1.60E-09 |
| rs2949200 | miR-339-5p | cis | 5.13E-05 | Nightingale | XL_VLDL_C | NA | C | T | -0.02223 | 0.004052 | 1.90E-08 |
| rs2949200 | miR-339-5p | cis | 5.13E-05 | Nightingale | M_VLDL_T | NA | C | T | 0.023289 | 0.003992 | 1.70E-09 |
| rs2949200 | miR-339-5p | cis | 5.13E-05 | Nightingale | L_VLDL_CE | NA | C | T | -0.02601 | 0.004043 | 2.40E-11 |
| rs2949200 | miR-339-5p | cis | 5.13E-05 | Nightingale | XL_VLDL_F | NA | C | T | -0.02486 | 0.004097 | 7.50E-10 |
| rs2949201 | miR-339-5p | cis | 4.98E-05 | Nightingale | M_VLDL_P | NA | C | T | -0.02468 | 0.004056 | 2.10E-10 |
| rs2949201 | miR-339-5p | cis | 4.98E-05 | Nightingale | L_VLDL_CE | NA | C | T | -0.026 | 0.004044 | 2.50E-11 |
| rs2949201 | miR-339-5p | cis | 4.98E-05 | Nightingale | XL_VLDL_T | NA | C | T | 0.024073 | 0.004097 | 8.20E-10 |
| rs2949201 | miR-339-5p | cis | 4.98E-05 | Nightingale | XL_VLDL_C | NA | C | T | -0.02221 | 0.004053 | 2.00E-08 |
| rs2949201 | miR-339-5p | cis | 4.98E-05 | Nightingale | M_VLDL_F | NA | C | T | -0.02351 | 0.00402 | 1.60E-09 |
| rs2949201 | miR-339-5p | cis | 4.98E-05 | Nightingale | M_VLDL_C | NA | C | T | -0.02252 | 0.00398 | 6.00E-09 |
| rs2949201 | miR-339-5p | cis | 4.98E-05 | Nightingale | M_VLDL_T | NA | C | T | 0.023274 | 0.003993 | 1.80E-09 |
| rs2949201 | miR-339-5p | cis | 4.98E-05 | Nightingale | VLDL_size | NA | C | T | 0.023243 | 0.003941 | 1.10E-08 |
| rs2949201 | miR-339-5p | cis | 4.98E-05 | Nightingale | XL_VLDL_F | NA | C | T | -0.02485 | 0.004098 | 7.70E-10 |
| rs2949201 | miR-339-5p | cis | 4.98E-05 | Nightingale | L_VLDL_C_ | NA | C | T | -0.02612 | 0.004128 | 2.00E-11 |
| rs2949201 | miR-339-5p | cis | 4.98E-05 | Nightingale | M_VLDL_C | NA | C | T | -0.02185 | 0.003969 | 1.80E-08 |
| rs2949202 | miR-339-5p | cis | 5.12E-05 | Nightingale | M_VLDL_C | NA | A | G | -0.0225 | 0.00398 | 6.20E-09 |
| rs2949202 | miR-339-5p | cis | 5.12E-05 | Nightingale | M_VLDL_T | NA | A | G | 0.023242 | 0.003993 | 1.90E-09 |
| rs2949202 | miR-339-5p | cis | 5.12E-05 | Nightingale | XL_VLDL_T | NA | A | G | 0.024012 | 0.004097 | 9.00E-10 |
| rs2949202 | miR-339-5p | cis | 5.12E-05 | Nightingale | M_VLDL_F | NA | A | G | -0.02347 | 0.00402 | 1.70E-09 |
| rs2949202 | miR-339-5p | cis | 5.12E-05 | Nightingale | L_VLDL_C_ | NA | A | G | -0.026 | 0.004128 | 2.40E-11 |
| rs2949202 | miR-339-5p | cis | 5.12E-05 | Nightingale | M_VLDL_P | NA | A | G | -0.0246 | 0.004056 | 2.40E-10 |
| rs2949202 | miR-339-5p | cis | 5.12E-05 | Nightingale | VLDL_size | NA | A | G | 0.023197 | 0.003941 | 1.10E-08 |
| rs2949202 | miR-339-5p | cis | 5.12E-05 | Nightingale | XL_VLDL_F | NA | A | G | -0.0247 | 0.004098 | 9.60E-10 |
| rs2949202 | miR-339-5p | cis | 5.12E-05 | Nightingale | M_VLDL_C | NA | A | G | -0.02184 | 0.003969 | 1.90E-08 |
| rs2949202 | miR-339-5p | cis | 5.12E-05 | Nightingale | L_VLDL_CE | NA | A | G | -0.02592 | 0.004044 | 2.80E-11 |
| rs2949202 | miR-339-5p | cis | 5.12E-05 | Nightingale | XL_VLDL_C | NA | A | G | -0.02219 | 0.004053 | 2.00E-08 |
| rs2949203 | miR-339-5p | cis | 5.10E-05 | Nightingale | M_VLDL_C | NA | T | C | -0.02189 | 0.003969 | 1.70E-08 |
| rs2949203 | miR-339-5p | cis | 5.10E-05 | Nightingale | XL_VLDL_F | NA | T | C | -0.02487 | 0.004097 | 7.50E-10 |
| rs2949203 | miR-339-5p | cis | 5.10E-05 | Nightingale | XL_VLDL_C | NA | T | C | -0.02224 | 0.004053 | 1.90E-08 |
| rs2949203 | miR-339-5p | cis | 5.10E-05 | Nightingale | VLDL_size | NA | T | C | 0.023297 | 0.003941 | 9.80E-09 |
| rs2949203 | miR-339-5p | cis | 5.10E-05 | Nightingale | M_VLDL_P | NA | T | C | -0.02468 | 0.004056 | 2.20E-10 |
| rs2949203 | miR-339-5p | cis | 5.10E-05 | Nightingale | M_VLDL_F | NA | T | C | -0.02353 | 0.00402 | 1.60E-09 |
| rs2949203 | miR-339-5p | cis | 5.10E-05 | Nightingale | L_VLDL_C_ | NA | T | C | -0.02612 | 0.004128 | 2.00E-11 |
| rs2949203 | miR-339-5p | cis | 5.10E-05 | Nightingale | L_VLDL_CE | NA | T | C | -0.02601 | 0.004044 | 2.50E-11 |
| rs2949203 | miR-339-5p | cis | 5.10E-05 | Nightingale | XL_VLDL_T | NA | T | C | 0.024092 | 0.004097 | 8.10E-10 |
| rs2949203 | miR-339-5p | cis | 5.10E-05 | Nightingale | M_VLDL_T | NA | T | C | 0.023301 | 0.003993 | 1.70E-09 |
| rs2949203 | miR-339-5p | cis | 5.10E-05 | Nightingale | M_VLDL_C | NA | T | C | -0.02255 | 0.00398 | 5.80E-09 |
| rs2949205 | miR-339-5p | cis | 4.85E-05 | Nightingale | M_VLDL_C | NA | T | C | -0.02257 | 0.00398 | 5.60E-09 |
| rs2949205 | miR-339-5p | cis | 4.85E-05 | Nightingale | M_VLDL_T | NA | T | C | 0.023321 | 0.003993 | 1.60E-09 |
| rs2949205 | miR-339-5p | cis | 4.85E-05 | Nightingale | L_VLDL_CE | NA | T | C | -0.02605 | 0.004044 | 2.30E-11 |
| rs2949205 | miR-339-5p | cis | 4.85E-05 | Nightingale | XL_VLDL_C | NA | T | C | -0.02223 | 0.004053 | 1.90E-08 |
| rs2949205 | miR-339-5p | cis | 4.85E-05 | Nightingale | XL_VLDL_F | NA | T | C | -0.02493 | 0.004098 | 6.70E-10 |
| rs2949205 | miR-339-5p | cis | 4.85E-05 | Nightingale | VLDL_size | NA | T | C | 0.023299 | 0.003941 | 9.80E-09 |
| rs2949205 | miR-339-5p | cis | 4.85E-05 | Nightingale | XL_VLDL_T | NA | T | C | 0.024134 | 0.004098 | 7.40E-10 |
| rs2949205 | miR-339-5p | cis | 4.85E-05 | Nightingale | M_VLDL_F | NA | T | C | -0.02355 | 0.004021 | 1.50E-09 |
| rs2949205 | miR-339-5p | cis | 4.85E-05 | Nightingale | M_VLDL_P | NA | T | C | -0.02472 | 0.004057 | 2.00E-10 |
| rs2949205 | miR-339-5p | cis | 4.85E-05 | Nightingale | L_VLDL_C_ | NA | T | C | -0.02617 | 0.004129 | 1.80E-11 |
| rs2949205 | miR-339-5p | cis | 4.85E-05 | Nightingale | M_VLDL_C | NA | T | C | -0.0219 | 0.00397 | 1.70E-08 |
| rs2960829 | miR-339-5p | cis | 4.44E-05 | Nightingale | L_VLDL_C_ | NA | T | C | -0.02622 | 0.004129 | 1.70E-11 |
| rs2960829 | miR-339-5p | cis | 4.44E-05 | Nightingale | M_VLDL_C | NA | T | C | -0.02263 | 0.00398 | 5.10E-09 |
| rs2960829 | miR-339-5p | cis | 4.44E-05 | Nightingale | M_VLDL_C | NA | T | C | -0.02196 | 0.00397 | 1.60E-08 |
| rs2960829 | miR-339-5p | cis | 4.44E-05 | Nightingale | L_VLDL_CE | NA | T | C | -0.0261 | 0.004045 | 2.10E-11 |
| rs2960829 | miR-339-5p | cis | 4.44E-05 | Nightingale | M_VLDL_F | NA | T | C | -0.02363 | 0.004021 | 1.30E-09 |
| rs2960829 | miR-339-5p | cis | 4.44E-05 | Nightingale | XL_VLDL_F | NA | T | C | -0.02497 | 0.004098 | 6.40E-10 |
| rs2960829 | miR-339-5p | cis | 4.44E-05 | Nightingale | XL_VLDL_C | NA | T | C | -0.02227 | 0.004053 | 1.80E-08 |
| rs2960829 | miR-339-5p | cis | 4.44E-05 | Nightingale | M_VLDL_P | NA | T | C | -0.02479 | 0.004057 | 1.80E-10 |
| rs2960829 | miR-339-5p | cis | 4.44E-05 | Nightingale | M_VLDL_T | NA | T | C | 0.02339 | 0.003994 | 1.50E-09 |
| rs2960829 | miR-339-5p | cis | 4.44E-05 | Nightingale | XL_VLDL_T | NA | T | C | 0.024179 | 0.004098 | 7.00E-10 |

| | | | | | | | | | | | |
|---|---|---|---|---|---|---|---|---|---|---|---|
| rs2960829 | miR-339-5p | cis | 4.44E-05 | Nightingale | VLDL_size | NA | T | C | 0.023336 | 0.003942 | 9.20E-09 |
| rs2960832 | miR-339-5p | cis | 5.11E-05 | Nightingale | M_VLDL_F | NA | G | A | -0.02351 | 0.00402 | 1.60E-09 |
| rs2960832 | miR-339-5p | cis | 5.11E-05 | Nightingale | M_VLDL_C | NA | G | A | -0.02187 | 0.003969 | 1.80E-08 |
| rs2960832 | miR-339-5p | cis | 5.11E-05 | Nightingale | XL_VLDL_C | NA | G | A | -0.02221 | 0.004052 | 2.00E-08 |
| rs2960832 | miR-339-5p | cis | 5.11E-05 | Nightingale | M_VLDL_C | NA | G | A | -0.02253 | 0.003979 | 6.00E-09 |
| rs2960832 | miR-339-5p | cis | 5.11E-05 | Nightingale | M_VLDL_P | NA | G | A | -0.02467 | 0.004056 | 2.20E-10 |
| rs2960832 | miR-339-5p | cis | 5.11E-05 | Nightingale | XL_VLDL_T | NA | G | A | 0.024067 | 0.004097 | 8.30E-10 |
| rs2960832 | miR-339-5p | cis | 5.11E-05 | Nightingale | L_VLDL_CE | NA | G | A | -0.02598 | 0.004044 | 2.60E-11 |
| rs2960832 | miR-339-5p | cis | 5.11E-05 | Nightingale | M_VLDL_T | NA | G | A | 0.02328 | 0.003993 | 1.70E-09 |
| rs2960832 | miR-339-5p | cis | 5.11E-05 | Nightingale | XL_VLDL_F | NA | G | A | -0.02485 | 0.004097 | 7.70E-10 |
| rs2960832 | miR-339-5p | cis | 5.11E-05 | Nightingale | L_VLDL_C_ | NA | G | A | -0.02609 | 0.004128 | 2.10E-11 |
| rs2960832 | miR-339-5p | cis | 5.11E-05 | Nightingale | VLDL_size | NA | G | A | 0.023242 | 0.003941 | 1.10E-08 |
| rs2960833 | miR-339-5p | cis | 5.72E-05 | Nightingale | M_VLDL_C | NA | C | A | -0.02186 | 0.003969 | 1.80E-08 |
| rs2960833 | miR-339-5p | cis | 5.72E-05 | Nightingale | M_VLDL_C | NA | C | A | -0.02251 | 0.003979 | 6.10E-09 |
| rs2960833 | miR-339-5p | cis | 5.72E-05 | Nightingale | VLDL_size | NA | C | A | 0.023241 | 0.003941 | 1.10E-08 |
| rs2960833 | miR-339-5p | cis | 5.72E-05 | Nightingale | L_VLDL_CE | NA | C | A | -0.02595 | 0.004044 | 2.70E-11 |
| rs2960833 | miR-339-5p | cis | 5.72E-05 | Nightingale | L_VLDL_C_ | NA | C | A | -0.02607 | 0.004128 | 2.20E-11 |
| rs2960833 | miR-339-5p | cis | 5.72E-05 | Nightingale | XL_VLDL_F | NA | C | A | -0.02482 | 0.004097 | 8.00E-10 |
| rs2960833 | miR-339-5p | cis | 5.72E-05 | Nightingale | M_VLDL_F | NA | C | A | -0.02348 | 0.00402 | 1.70E-09 |
| rs2960833 | miR-339-5p | cis | 5.72E-05 | Nightingale | M_VLDL_T | NA | C | A | 0.023264 | 0.003993 | 1.80E-09 |
| rs2960833 | miR-339-5p | cis | 5.72E-05 | Nightingale | XL_VLDL_C | NA | C | A | -0.02219 | 0.004052 | 2.00E-08 |
| rs2960833 | miR-339-5p | cis | 5.72E-05 | Nightingale | M_VLDL_P | NA | C | A | -0.02465 | 0.004056 | 2.20E-10 |
| rs2960833 | miR-339-5p | cis | 5.72E-05 | Nightingale | XL_VLDL_T | NA | C | A | 0.024042 | 0.004097 | 8.60E-10 |
| rs2960836 | miR-339-5p | cis | 5.40E-05 | Nightingale | XL_VLDL_F | NA | G | T | -0.02479 | 0.004097 | 8.40E-10 |
| rs2960836 | miR-339-5p | cis | 5.40E-05 | Nightingale | M_VLDL_T | NA | G | T | 0.023237 | 0.003992 | 1.80E-09 |
| rs2960836 | miR-339-5p | cis | 5.40E-05 | Nightingale | VLDL_size | NA | G | T | 0.023266 | 0.003941 | 1.00E-08 |
| rs2960836 | miR-339-5p | cis | 5.40E-05 | Nightingale | L_VLDL_CE | NA | G | T | -0.02595 | 0.004044 | 2.70E-11 |
| rs2960836 | miR-339-5p | cis | 5.40E-05 | Nightingale | L_VLDL_C_ | NA | G | T | -0.02606 | 0.004128 | 2.20E-11 |
| rs2960836 | miR-339-5p | cis | 5.40E-05 | Nightingale | M_VLDL_P | NA | G | T | -0.02463 | 0.004056 | 2.30E-10 |
| rs2960836 | miR-339-5p | cis | 5.40E-05 | Nightingale | M_VLDL_C | NA | G | T | -0.02249 | 0.003979 | 6.30E-09 |
| rs2960836 | miR-339-5p | cis | 5.40E-05 | Nightingale | XL_VLDL_C | NA | G | T | -0.02217 | 0.004052 | 2.10E-08 |
| rs2960836 | miR-339-5p | cis | 5.40E-05 | Nightingale | M_VLDL_F | NA | G | T | -0.02346 | 0.00402 | 1.70E-09 |
| rs2960836 | miR-339-5p | cis | 5.40E-05 | Nightingale | M_VLDL_C | NA | G | T | -0.02183 | 0.003969 | 1.90E-08 |
| rs2960836 | miR-339-5p | cis | 5.40E-05 | Nightingale | XL_VLDL_T | NA | G | T | 0.024007 | 0.004097 | 9.00E-10 |
| rs2960838 | miR-339-5p | cis | 3.43E-05 | Nightingale | M_VLDL_T | NA | T | C | 0.023265 | 0.003993 | 1.80E-09 |
| rs2960838 | miR-339-5p | cis | 3.43E-05 | Nightingale | M_VLDL_C | NA | T | C | -0.02253 | 0.00398 | 6.10E-09 |
| rs2960838 | miR-339-5p | cis | 3.43E-05 | Nightingale | M_VLDL_P | NA | T | C | -0.0246 | 0.004057 | 2.50E-10 |
| rs2960838 | miR-339-5p | cis | 3.43E-05 | Nightingale | M_VLDL_F | NA | T | C | -0.02346 | 0.004021 | 1.80E-09 |
| rs2960838 | miR-339-5p | cis | 3.43E-05 | Nightingale | M_VLDL_C | NA | T | C | -0.02188 | 0.00397 | 1.80E-08 |
| rs2960838 | miR-339-5p | cis | 3.43E-05 | Nightingale | L_VLDL_CE | NA | T | C | -0.02594 | 0.004044 | 2.80E-11 |
| rs2960838 | miR-339-5p | cis | 3.43E-05 | Nightingale | XL_VLDL_T | NA | T | C | 0.024024 | 0.004098 | 9.10E-10 |
| rs2960838 | miR-339-5p | cis | 3.43E-05 | Nightingale | VLDL_size | NA | T | C | 0.023342 | 0.003941 | 9.50E-09 |
| rs2960838 | miR-339-5p | cis | 3.43E-05 | Nightingale | L_VLDL_C_ | NA | T | C | -0.02598 | 0.004129 | 2.50E-11 |
| rs2960838 | miR-339-5p | cis | 3.43E-05 | Nightingale | XL_VLDL_C | NA | T | C | -0.02223 | 0.004053 | 2.00E-08 |
| rs2960838 | miR-339-5p | cis | 3.43E-05 | Nightingale | XL_VLDL_F | NA | T | C | -0.02477 | 0.004098 | 8.80E-10 |
| rs2960839 | miR-339-5p | cis | 5.58E-05 | Nightingale | VLDL_size | NA | G | A | 0.02334 | 0.003941 | 9.50E-09 |
| rs2960839 | miR-339-5p | cis | 5.58E-05 | Nightingale | M_VLDL_C | NA | G | A | -0.02186 | 0.003969 | 1.90E-08 |
| rs2960839 | miR-339-5p | cis | 5.58E-05 | Nightingale | M_VLDL_F | NA | G | A | -0.02342 | 0.00402 | 1.90E-09 |
| rs2960839 | miR-339-5p | cis | 5.58E-05 | Nightingale | M_VLDL_C | NA | G | A | -0.0225 | 0.00398 | 6.30E-09 |
| rs2960839 | miR-339-5p | cis | 5.58E-05 | Nightingale | M_VLDL_P | NA | G | A | -0.02455 | 0.004056 | 2.60E-10 |
| rs2960839 | miR-339-5p | cis | 5.58E-05 | Nightingale | XL_VLDL_F | NA | G | A | -0.02473 | 0.004098 | 9.50E-10 |
| rs2960839 | miR-339-5p | cis | 5.58E-05 | Nightingale | L_VLDL_CE | NA | G | A | -0.0259 | 0.004044 | 3.00E-11 |
| rs2960839 | miR-339-5p | cis | 5.58E-05 | Nightingale | XL_VLDL_T | NA | G | A | 0.023992 | 0.004097 | 9.50E-10 |
| rs2960839 | miR-339-5p | cis | 5.58E-05 | Nightingale | L_VLDL_C_ | NA | G | A | -0.02594 | 0.004128 | 2.70E-11 |
| rs2960839 | miR-339-5p | cis | 5.58E-05 | Nightingale | XL_VLDL_C | NA | G | A | -0.02221 | 0.004053 | 2.00E-08 |
| rs2960839 | miR-339-5p | cis | 5.58E-05 | Nightingale | M_VLDL_T | NA | G | A | 0.023235 | 0.003993 | 1.90E-09 |
| rs2960840 | miR-339-5p | cis | 5.64E-05 | Nightingale | M_VLDL_T | NA | T | C | 0.023231 | 0.003993 | 1.90E-09 |
| rs2960840 | miR-339-5p | cis | 5.64E-05 | Nightingale | M_VLDL_P | NA | T | C | -0.02453 | 0.004056 | 2.70E-10 |
| rs2960840 | miR-339-5p | cis | 5.64E-05 | Nightingale | L_VLDL_CE | NA | T | C | -0.02589 | 0.004044 | 3.00E-11 |
| rs2960840 | miR-339-5p | cis | 5.64E-05 | Nightingale | VLDL_size | NA | T | C | 0.023383 | 0.003941 | 8.90E-09 |
| rs2960840 | miR-339-5p | cis | 5.64E-05 | Nightingale | M_VLDL_F | NA | T | C | -0.02341 | 0.00402 | 1.90E-09 |
| rs2960840 | miR-339-5p | cis | 5.64E-05 | Nightingale | M_VLDL_C | NA | T | C | -0.02251 | 0.00398 | 6.30E-09 |
| rs2960840 | miR-339-5p | cis | 5.64E-05 | Nightingale | L_VLDL_C_ | NA | T | C | -0.02594 | 0.004128 | 2.70E-11 |

| | | | | | | | | | | | |
|---|---|---|---|---|---|---|---|---|---|---|---|
| rs2960840 | miR-339-5p | cis | 5.64E-05 | Nightingale | XL_VLDL_T | NA | T | C | 0.02399 | 0.004097 | 9.50E-10 |
| rs2960840 | miR-339-5p | cis | 5.64E-05 | Nightingale | M_VLDL_C | NA | T | C | -0.02187 | 0.003969 | 1.80E-08 |
| rs2960840 | miR-339-5p | cis | 5.64E-05 | Nightingale | XL_VLDL_F | NA | T | C | -0.02472 | 0.004098 | 9.60E-10 |
| rs2960840 | miR-339-5p | cis | 5.64E-05 | Nightingale | XL_VLDL_C | NA | T | C | -0.02222 | 0.004053 | 2.00E-08 |
| rs2960841 | miR-339-5p | cis | 5.61E-05 | Nightingale | VLDL_size | NA | G | A | 0.023389 | 0.003941 | 8.70E-09 |
| rs2960841 | miR-339-5p | cis | 5.61E-05 | Nightingale | XL_VLDL_T | NA | G | A | 0.023977 | 0.004097 | 9.50E-10 |
| rs2960841 | miR-339-5p | cis | 5.61E-05 | Nightingale | XL_VLDL_F | NA | G | A | -0.02469 | 0.004098 | 1.00E-09 |
| rs2960841 | miR-339-5p | cis | 5.61E-05 | Nightingale | M_VLDL_C | NA | G | A | -0.02185 | 0.00397 | 1.80E-08 |
| rs2960841 | miR-339-5p | cis | 5.61E-05 | Nightingale | M_VLDL_T | NA | G | A | 0.023209 | 0.003993 | 1.90E-09 |
| rs2960841 | miR-339-5p | cis | 5.61E-05 | Nightingale | XL_VLDL_C | NA | G | A | -0.0222 | 0.004053 | 2.00E-08 |
| rs2960841 | miR-339-5p | cis | 5.61E-05 | Nightingale | M_VLDL_P | NA | G | A | -0.02451 | 0.004056 | 2.80E-10 |
| rs2960841 | miR-339-5p | cis | 5.61E-05 | Nightingale | L_VLDL_CE | NA | G | A | -0.0259 | 0.004044 | 2.90E-11 |
| rs2960841 | miR-339-5p | cis | 5.61E-05 | Nightingale | M_VLDL_C | NA | G | A | -0.02248 | 0.00398 | 6.30E-09 |
| rs2960841 | miR-339-5p | cis | 5.61E-05 | Nightingale | M_VLDL_F | NA | G | A | -0.02337 | 0.00402 | 2.00E-09 |
| rs2960841 | miR-339-5p | cis | 5.61E-05 | Nightingale | L_VLDL_C_ | NA | G | A | -0.02596 | 0.004128 | 2.50E-11 |
| rs2960842 | miR-339-5p | cis | 5.85E-05 | Nightingale | M_VLDL_C | NA | T | G | -0.02234 | 0.003979 | 7.60E-09 |
| rs2960842 | miR-339-5p | cis | 5.85E-05 | Nightingale | XL_VLDL_F | NA | T | G | -0.02453 | 0.004097 | 1.20E-09 |
| rs2960842 | miR-339-5p | cis | 5.85E-05 | Nightingale | M_VLDL_F | NA | T | G | -0.02325 | 0.00402 | 2.40E-09 |
| rs2960842 | miR-339-5p | cis | 5.85E-05 | Nightingale | L_VLDL_C_ | NA | T | G | -0.02571 | 0.004128 | 3.90E-11 |
| rs2960842 | miR-339-5p | cis | 5.85E-05 | Nightingale | XL_VLDL_T | NA | T | G | 0.023746 | 0.004097 | 1.30E-09 |
| rs2960842 | miR-339-5p | cis | 5.85E-05 | Nightingale | M_VLDL_C | NA | T | G | -0.0217 | 0.003969 | 2.20E-08 |
| rs2960842 | miR-339-5p | cis | 5.85E-05 | Nightingale | L_VLDL_CE | NA | T | G | -0.02567 | 0.004044 | 4.40E-11 |
| rs2960842 | miR-339-5p | cis | 5.85E-05 | Nightingale | VLDL_size | NA | T | G | 0.023105 | 0.003941 | 1.30E-08 |
| rs2960842 | miR-339-5p | cis | 5.85E-05 | Nightingale | M_VLDL_T | NA | T | G | 0.023075 | 0.003993 | 2.30E-09 |
| rs2960842 | miR-339-5p | cis | 5.85E-05 | Nightingale | XL_VLDL_C | NA | T | G | -0.022 | 0.004052 | 2.70E-08 |
| rs2960842 | miR-339-5p | cis | 5.85E-05 | Nightingale | M_VLDL_P | NA | T | G | -0.02443 | 0.004056 | 3.20E-10 |
| rs2960843 | miR-339-5p | cis | 5.82E-05 | Nightingale | M_VLDL_P | NA | T | A | -0.02447 | 0.004056 | 3.00E-10 |
| rs2960843 | miR-339-5p | cis | 5.82E-05 | Nightingale | M_VLDL_T | NA | T | A | 0.023108 | 0.003992 | 2.20E-09 |
| rs2960843 | miR-339-5p | cis | 5.82E-05 | Nightingale | M_VLDL_F | NA | T | A | -0.02329 | 0.00402 | 2.30E-09 |
| rs2960843 | miR-339-5p | cis | 5.82E-05 | Nightingale | L_VLDL_CE | NA | T | A | -0.02571 | 0.004044 | 4.20E-11 |
| rs2960843 | miR-339-5p | cis | 5.82E-05 | Nightingale | XL_VLDL_F | NA | T | A | -0.02456 | 0.004097 | 1.20E-09 |
| rs2960843 | miR-339-5p | cis | 5.82E-05 | Nightingale | L_VLDL_C_ | NA | T | A | -0.02575 | 0.004128 | 3.70E-11 |
| rs2960843 | miR-339-5p | cis | 5.82E-05 | Nightingale | VLDL_size | NA | T | A | 0.023095 | 0.003941 | 1.30E-08 |
| rs2960843 | miR-339-5p | cis | 5.82E-05 | Nightingale | XL_VLDL_C | NA | T | A | -0.02203 | 0.004052 | 2.60E-08 |
| rs2960843 | miR-339-5p | cis | 5.82E-05 | Nightingale | M_VLDL_C | NA | T | A | -0.02173 | 0.003969 | 2.20E-08 |
| rs2960843 | miR-339-5p | cis | 5.82E-05 | Nightingale | M_VLDL_C | NA | T | A | -0.02237 | 0.003979 | 7.40E-09 |
| rs2960843 | miR-339-5p | cis | 5.82E-05 | Nightingale | XL_VLDL_T | NA | T | A | 0.023781 | 0.004097 | 1.30E-09 |
| rs2960844 | miR-339-5p | cis | 5.89E-05 | Nightingale | M_VLDL_C | NA | G | A | -0.02173 | 0.003969 | 2.20E-08 |
| rs2960844 | miR-339-5p | cis | 5.89E-05 | Nightingale | M_VLDL_F | NA | G | A | -0.02328 | 0.00402 | 2.30E-09 |
| rs2960844 | miR-339-5p | cis | 5.89E-05 | Nightingale | XL_VLDL_T | NA | G | A | 0.023758 | 0.004097 | 1.30E-09 |
| rs2960844 | miR-339-5p | cis | 5.89E-05 | Nightingale | M_VLDL_T | NA | G | A | 0.023099 | 0.003992 | 2.30E-09 |
| rs2960844 | miR-339-5p | cis | 5.89E-05 | Nightingale | M_VLDL_C | NA | G | A | -0.02236 | 0.003979 | 7.50E-09 |
| rs2960844 | miR-339-5p | cis | 5.89E-05 | Nightingale | M_VLDL_P | NA | G | A | -0.02445 | 0.004056 | 3.10E-10 |
| rs2960844 | miR-339-5p | cis | 5.89E-05 | Nightingale | XL_VLDL_C | NA | G | A | -0.022 | 0.004052 | 2.70E-08 |
| rs2960844 | miR-339-5p | cis | 5.89E-05 | Nightingale | L_VLDL_C_ | NA | G | A | -0.02573 | 0.004128 | 3.80E-11 |
| rs2960844 | miR-339-5p | cis | 5.89E-05 | Nightingale | XL_VLDL_F | NA | G | A | -0.02455 | 0.004097 | 1.20E-09 |
| rs2960844 | miR-339-5p | cis | 5.89E-05 | Nightingale | L_VLDL_CE | NA | G | A | -0.02569 | 0.004043 | 4.30E-11 |
| rs2960844 | miR-339-5p | cis | 5.89E-05 | Nightingale | VLDL_size | NA | G | A | 0.023092 | 0.003941 | 1.30E-08 |
| rs2960845 | miR-339-5p | cis | 4.74E-05 | Nightingale | M_VLDL_C | NA | G | A | -0.02172 | 0.003969 | 2.20E-08 |
| rs2960845 | miR-339-5p | cis | 4.74E-05 | Nightingale | L_VLDL_CE | NA | G | A | -0.02569 | 0.004043 | 4.30E-11 |
| rs2960845 | miR-339-5p | cis | 4.74E-05 | Nightingale | XL_VLDL_C | NA | G | A | -0.022 | 0.004052 | 2.70E-08 |
| rs2960845 | miR-339-5p | cis | 4.74E-05 | Nightingale | L_VLDL_C_ | NA | G | A | -0.02573 | 0.004128 | 3.80E-11 |
| rs2960845 | miR-339-5p | cis | 4.74E-05 | Nightingale | M_VLDL_C | NA | G | A | -0.02236 | 0.003979 | 7.50E-09 |
| rs2960845 | miR-339-5p | cis | 4.74E-05 | Nightingale | VLDL_size | NA | G | A | 0.02309 | 0.003941 | 1.30E-08 |
| rs2960845 | miR-339-5p | cis | 4.74E-05 | Nightingale | XL_VLDL_T | NA | G | A | 0.023756 | 0.004097 | 1.30E-09 |
| rs2960845 | miR-339-5p | cis | 4.74E-05 | Nightingale | M_VLDL_P | NA | G | A | -0.02445 | 0.004056 | 3.10E-10 |
| rs2960845 | miR-339-5p | cis | 4.74E-05 | Nightingale | M_VLDL_T | NA | G | A | 0.023095 | 0.003992 | 2.30E-09 |
| rs2960845 | miR-339-5p | cis | 4.74E-05 | Nightingale | XL_VLDL_F | NA | G | A | -0.02455 | 0.004097 | 1.20E-09 |
| rs2960845 | miR-339-5p | cis | 4.74E-05 | Nightingale | M_VLDL_F | NA | G | A | -0.02327 | 0.00402 | 2.40E-09 |
| rs2960849 | miR-339-5p | cis | 5.55E-05 | Nightingale | L_VLDL_CE | NA | G | C | -0.02583 | 0.004051 | 4.00E-11 |
| rs2960849 | miR-339-5p | cis | 5.55E-05 | Nightingale | XL_VLDL_C | NA | G | C | -0.02219 | 0.004059 | 2.30E-08 |
| rs2960849 | miR-339-5p | cis | 5.55E-05 | Nightingale | XL_VLDL_T | NA | G | C | 0.024013 | 0.004104 | 1.00E-09 |
| rs2960849 | miR-339-5p | cis | 5.55E-05 | Nightingale | M_VLDL_C | NA | G | C | -0.02186 | 0.003976 | 2.00E-08 |

| | | | | | | | | | | | |
|---|---|---|---|---|---|---|---|---|---|---|---|
| rs2960849 | miR-339-5p | cis | 5.55E-05 | Nightingale | M_VLDL_T | NA | G | C | 0.023222 | 0.003999 | 2.20E-09 |
| rs2960849 | miR-339-5p | cis | 5.55E-05 | Nightingale | M_VLDL_P | NA | G | C | -0.02453 | 0.004063 | 3.30E-10 |
| rs2960849 | miR-339-5p | cis | 5.55E-05 | Nightingale | VLDL_size | NA | G | C | 0.022978 | 0.003947 | 1.50E-08 |
| rs2960849 | miR-339-5p | cis | 5.55E-05 | Nightingale | M_VLDL_C | NA | G | C | -0.0225 | 0.003986 | 6.90E-09 |
| rs2960849 | miR-339-5p | cis | 5.55E-05 | Nightingale | XL_VLDL_F | NA | G | C | -0.02489 | 0.004104 | 8.40E-10 |
| rs2960849 | miR-339-5p | cis | 5.55E-05 | Nightingale | L_VLDL_C_ | NA | G | C | -0.02587 | 0.004135 | 3.60E-11 |
| rs2960849 | miR-339-5p | cis | 5.55E-05 | Nightingale | M_VLDL_F | NA | G | C | -0.02343 | 0.004027 | 2.10E-09 |
| rs2960852 | miR-339-5p | cis | 6.25E-05 | Nightingale | M_VLDL_F | NA | A | G | -0.02336 | 0.00402 | 2.10E-09 |
| rs2960852 | miR-339-5p | cis | 6.25E-05 | Nightingale | XL_VLDL_T | NA | A | G | 0.02382 | 0.004097 | 1.20E-09 |
| rs2960852 | miR-339-5p | cis | 6.25E-05 | Nightingale | XL_VLDL_F | NA | A | G | -0.02464 | 0.004098 | 1.10E-09 |
| rs2960852 | miR-339-5p | cis | 6.25E-05 | Nightingale | M_VLDL_P | NA | A | G | -0.02453 | 0.004056 | 2.70E-10 |
| rs2960852 | miR-339-5p | cis | 6.25E-05 | Nightingale | M_VLDL_C | NA | A | G | -0.02241 | 0.00398 | 7.00E-09 |
| rs2960852 | miR-339-5p | cis | 6.25E-05 | Nightingale | VLDL_size | NA | A | G | 0.023088 | 0.003941 | 1.30E-08 |
| rs2960852 | miR-339-5p | cis | 6.25E-05 | Nightingale | L_VLDL_CE | NA | A | G | -0.02573 | 0.004044 | 4.00E-11 |
| rs2960852 | miR-339-5p | cis | 6.25E-05 | Nightingale | XL_VLDL_C | NA | A | G | -0.02203 | 0.004053 | 2.60E-08 |
| rs2960852 | miR-339-5p | cis | 6.25E-05 | Nightingale | M_VLDL_T | NA | A | G | 0.023153 | 0.003993 | 2.10E-09 |
| rs2960852 | miR-339-5p | cis | 6.25E-05 | Nightingale | M_VLDL_C | NA | A | G | -0.02176 | 0.00397 | 2.10E-08 |
| rs2960852 | miR-339-5p | cis | 6.25E-05 | Nightingale | L_VLDL_C_ | NA | A | G | -0.02579 | 0.004128 | 3.50E-11 |
| rs2960853 | miR-339-5p | cis | 6.33E-05 | Nightingale | L_VLDL_C_ | NA | A | T | -0.0258 | 0.004128 | 3.50E-11 |
| rs2960853 | miR-339-5p | cis | 6.33E-05 | Nightingale | M_VLDL_T | NA | A | T | 0.023136 | 0.003993 | 2.10E-09 |
| rs2960853 | miR-339-5p | cis | 6.33E-05 | Nightingale | M_VLDL_C | NA | A | T | -0.02239 | 0.00398 | 7.10E-09 |
| rs2960853 | miR-339-5p | cis | 6.33E-05 | Nightingale | M_VLDL_C | NA | A | T | -0.02174 | 0.003969 | 2.10E-08 |
| rs2960853 | miR-339-5p | cis | 6.33E-05 | Nightingale | XL_VLDL_T | NA | A | T | 0.023813 | 0.004097 | 1.20E-09 |
| rs2960853 | miR-339-5p | cis | 6.33E-05 | Nightingale | XL_VLDL_F | NA | A | T | -0.02463 | 0.004098 | 1.10E-09 |
| rs2960853 | miR-339-5p | cis | 6.33E-05 | Nightingale | L_VLDL_CE | NA | A | T | -0.02574 | 0.004044 | 3.90E-11 |
| rs2960853 | miR-339-5p | cis | 6.33E-05 | Nightingale | XL_VLDL_C | NA | A | T | -0.02201 | 0.004053 | 2.70E-08 |
| rs2960853 | miR-339-5p | cis | 6.33E-05 | Nightingale | M_VLDL_F | NA | A | T | -0.02333 | 0.00402 | 2.10E-09 |
| rs2960853 | miR-339-5p | cis | 6.33E-05 | Nightingale | M_VLDL_P | NA | A | T | -0.02453 | 0.004056 | 2.70E-10 |
| rs2960853 | miR-339-5p | cis | 6.33E-05 | Nightingale | VLDL_size | NA | A | T | 0.023017 | 0.003941 | 1.40E-08 |
| rs2960854 | miR-339-5p | cis | 6.32E-05 | Nightingale | L_VLDL_CE | NA | G | A | -0.02573 | 0.004044 | 3.90E-11 |
| rs2960854 | miR-339-5p | cis | 6.32E-05 | Nightingale | M_VLDL_C | NA | G | A | -0.02239 | 0.00398 | 7.10E-09 |
| rs2960854 | miR-339-5p | cis | 6.32E-05 | Nightingale | M_VLDL_C | NA | G | A | -0.02174 | 0.003969 | 2.10E-08 |
| rs2960854 | miR-339-5p | cis | 6.32E-05 | Nightingale | XL_VLDL_C | NA | G | A | -0.02201 | 0.004053 | 2.70E-08 |
| rs2960854 | miR-339-5p | cis | 6.32E-05 | Nightingale | XL_VLDL_T | NA | G | A | 0.023812 | 0.004097 | 1.20E-09 |
| rs2960854 | miR-339-5p | cis | 6.32E-05 | Nightingale | M_VLDL_P | NA | G | A | -0.02453 | 0.004056 | 2.70E-10 |
| rs2960854 | miR-339-5p | cis | 6.32E-05 | Nightingale | XL_VLDL_F | NA | G | A | -0.02463 | 0.004098 | 1.10E-09 |
| rs2960854 | miR-339-5p | cis | 6.32E-05 | Nightingale | M_VLDL_T | NA | G | A | 0.023135 | 0.003993 | 2.10E-09 |
| rs2960854 | miR-339-5p | cis | 6.32E-05 | Nightingale | M_VLDL_F | NA | G | A | -0.02333 | 0.00402 | 2.10E-09 |
| rs2960854 | miR-339-5p | cis | 6.32E-05 | Nightingale | VLDL_size | NA | G | A | 0.023016 | 0.003941 | 1.40E-08 |
| rs2960854 | miR-339-5p | cis | 6.32E-05 | Nightingale | L_VLDL_C_ | NA | G | A | -0.0258 | 0.004128 | 3.50E-11 |
| rs2960856 | miR-339-5p | cis | 5.11E-05 | Nightingale | XL_VLDL_T | NA | T | C | 0.023611 | 0.004097 | 1.70E-09 |
| rs2960856 | miR-339-5p | cis | 5.11E-05 | Nightingale | VLDL_size | NA | T | C | 0.02308 | 0.003941 | 1.40E-08 |
| rs2960856 | miR-339-5p | cis | 5.11E-05 | Nightingale | M_VLDL_P | NA | T | C | -0.02436 | 0.004056 | 3.60E-10 |
| rs2960856 | miR-339-5p | cis | 5.11E-05 | Nightingale | M_VLDL_C | NA | T | C | -0.02225 | 0.00398 | 9.30E-09 |
| rs2960856 | miR-339-5p | cis | 5.11E-05 | Nightingale | L_VLDL_C_ | NA | T | C | -0.02564 | 0.004128 | 4.30E-11 |
| rs2960856 | miR-339-5p | cis | 5.11E-05 | Nightingale | M_VLDL_C | NA | T | C | -0.0216 | 0.003969 | 2.80E-08 |
| rs2960856 | miR-339-5p | cis | 5.11E-05 | Nightingale | M_VLDL_F | NA | T | C | -0.02318 | 0.00402 | 2.70E-09 |
| rs2960856 | miR-339-5p | cis | 5.11E-05 | Nightingale | L_VLDL_CE | NA | T | C | -0.02561 | 0.004044 | 4.80E-11 |
| rs2960856 | miR-339-5p | cis | 5.11E-05 | Nightingale | M_VLDL_T | NA | T | C | 0.022988 | 0.003993 | 2.80E-09 |
| rs2960856 | miR-339-5p | cis | 5.11E-05 | Nightingale | XL_VLDL_F | NA | T | C | -0.02447 | 0.004098 | 1.40E-09 |
| rs2960856 | miR-339-5p | cis | 5.11E-05 | Nightingale | XL_VLDL_C | NA | T | C | -0.02188 | 0.004053 | 3.30E-08 |
| rs3735686 | miR-339-5p | cis | 5.70E-07 | Nightingale | L_VLDL_C_ | NA | A | G | -0.02214 | 0.004094 | 3.40E-08 |
| rs3735686 | miR-339-5p | cis | 5.70E-07 | Nightingale | L_VLDL_CE | NA | A | G | -0.02223 | 0.00401 | 3.40E-08 |
| rs3802141 | miR-339-5p | cis | 5.44E-05 | Nightingale | M_VLDL_P | NA | C | T | -0.02219 | 0.004084 | 2.60E-08 |
| rs3802141 | miR-339-5p | cis | 5.44E-05 | Nightingale | L_HDL_CE | NA | C | T | -0.02133 | 0.00376 | 2.30E-08 |
| rs3802141 | miR-339-5p | cis | 5.44E-05 | Nightingale | L_HDL_P | NA | C | T | -0.02137 | 0.003729 | 1.20E-08 |
| rs3802141 | miR-339-5p | cis | 5.44E-05 | Nightingale | L_VLDL_CE | NA | C | T | -0.02403 | 0.004071 | 1.30E-09 |
| rs3802141 | miR-339-5p | cis | 5.44E-05 | Nightingale | L_HDL_C | NA | C | T | -0.02147 | 0.00375 | 1.60E-08 |
| rs3802141 | miR-339-5p | cis | 5.44E-05 | Nightingale | L_VLDL_C_ | NA | C | T | -0.02339 | 0.004156 | 3.30E-09 |
| rs3802141 | miR-339-5p | cis | 5.44E-05 | Nightingale | VLDL_size | NA | C | T | 0.023726 | 0.003968 | 1.00E-08 |
| rs3802141 | miR-339-5p | cis | 5.44E-05 | Nightingale | L_HDL_FC | NA | C | T | -0.02155 | 0.003732 | 1.10E-08 |
| rs3802141 | miR-339-5p | cis | 5.44E-05 | Nightingale | L_HDL_L | NA | C | T | -0.0211 | 0.003736 | 2.20E-08 |
| rs3802141 | miR-339-5p | cis | 5.44E-05 | Nightingale | HDL_size | NA | C | T | -0.02166 | 0.003772 | 1.80E-08 |

| | | | | | | | | | | | |
|---|---|---|---|---|---|---|---|---|---|---|---|
| rs4075937 | miR-339-5| | cis | 5.93E-05 | Nightingale | L_VLDL_C_ | NA | A | C | -0.02486 | 0.004113 | 1.50E-10 |
| rs4075937 | miR-339-5| | cis | 5.93E-05 | Nightingale | M_VLDL_T | NA | A | C | 0.022153 | 0.003978 | 8.80E-09 |
| rs4075937 | miR-339-5| | cis | 5.93E-05 | Nightingale | XL_VLDL_T | NA | A | C | 0.022638 | 0.004082 | 6.50E-09 |
| rs4075937 | miR-339-5| | cis | 5.93E-05 | Nightingale | M_VLDL_P | NA | A | C | -0.02369 | 0.004041 | 8.90E-10 |
| rs4075937 | miR-339-5| | cis | 5.93E-05 | Nightingale | XL_VLDL_F | NA | A | C | -0.02309 | 0.004083 | 8.90E-09 |
| rs4075937 | miR-339-5| | cis | 5.93E-05 | Nightingale | VLDL_size | NA | A | C | 0.022078 | 0.003926 | 4.60E-08 |
| rs4075937 | miR-339-5| | cis | 5.93E-05 | Nightingale | L_VLDL_CE | NA | A | C | -0.0245 | 0.004029 | 2.70E-10 |
| rs4075937 | miR-339-5| | cis | 5.93E-05 | Nightingale | M_VLDL_C | NA | A | C | -0.02138 | 0.003965 | 2.90E-08 |
| rs4075937 | miR-339-5| | cis | 5.93E-05 | Nightingale | M_VLDL_F | NA | A | C | -0.02238 | 0.004005 | 8.30E-09 |
| rs4285396 | miR-339-5| | cis | 4.27E-05 | Nightingale | L_HDL_P | NA | C | T | -0.02197 | 0.003758 | 9.10E-09 |
| rs4285396 | miR-339-5| | cis | 4.27E-05 | Nightingale | XL_HDL_P | NA | C | T | -0.02231 | 0.003862 | 2.10E-08 |
| rs4285396 | miR-339-5| | cis | 4.27E-05 | Nightingale | XL_HDL_L | NA | C | T | -0.02246 | 0.003856 | 3.30E-08 |
| rs4285396 | miR-339-5| | cis | 4.27E-05 | Nightingale | L_HDL_CE | NA | C | T | -0.02154 | 0.003789 | 3.30E-08 |
| rs4285396 | miR-339-5| | cis | 4.27E-05 | Nightingale | XL_HDL_PL | NA | C | T | -0.02248 | 0.003857 | 4.20E-08 |
| rs4285396 | miR-339-5| | cis | 4.27E-05 | Nightingale | HDL_size | NA | C | T | -0.02238 | 0.003801 | 1.00E-08 |
| rs4285396 | miR-339-5| | cis | 4.27E-05 | Nightingale | L_HDL_FC | NA | C | T | -0.02238 | 0.003761 | 5.40E-09 |
| rs4285396 | miR-339-5| | cis | 4.27E-05 | Nightingale | M_VLDL_F | NA | C | T | -0.02248 | 0.004079 | 3.20E-08 |
| rs4285396 | miR-339-5| | cis | 4.27E-05 | Nightingale | L_VLDL_C_ | NA | C | T | -0.02397 | 0.004188 | 2.10E-09 |
| rs4285396 | miR-339-5| | cis | 4.27E-05 | Nightingale | L_HDL_L | NA | C | T | -0.0217 | 0.003765 | 1.60E-08 |
| rs4285396 | miR-339-5| | cis | 4.27E-05 | Nightingale | VLDL_size | NA | C | T | 0.023222 | 0.003998 | 3.70E-08 |
| rs4285396 | miR-339-5| | cis | 4.27E-05 | Nightingale | XL_HDL_CE | NA | C | T | -0.02223 | 0.003843 | 4.10E-08 |
| rs4285396 | miR-339-5| | cis | 4.27E-05 | Nightingale | M_VLDL_P | NA | C | T | -0.02326 | 0.004115 | 9.20E-09 |
| rs4285396 | miR-339-5| | cis | 4.27E-05 | Nightingale | L_HDL_C | NA | C | T | -0.02183 | 0.003779 | 1.90E-08 |
| rs4285396 | miR-339-5| | cis | 4.27E-05 | Nightingale | S_HDL_FC_ | NA | C | T | -0.02347 | 0.00407 | 1.10E-08 |
| rs4285396 | miR-339-5| | cis | 4.27E-05 | Nightingale | L_VLDL_CE | NA | C | T | -0.02455 | 0.004103 | 1.10E-09 |
| rs4574742 | miR-148a-3 | cis | 7.37E-05 | Nightingale | S_VLDL_FC | NA | G | A | -0.02353 | 0.004233 | 2.10E-09 |
| rs4574742 | miR-148a-3 | cis | 7.37E-05 | Nightingale | M_HDL_TG | NA | G | A | 0.024224 | 0.004378 | 4.60E-09 |
| rs4574742 | miR-148a-3 | cis | 7.37E-05 | Nightingale | M_VLDL_C | NA | G | A | -0.02421 | 0.004172 | 4.10E-10 |
| rs4574742 | miR-148a-3 | cis | 7.37E-05 | Nightingale | S_LDL_TG_ | NA | G | A | 0.025183 | 0.004296 | 4.70E-10 |
| rs4574742 | miR-148a-3 | cis | 7.37E-05 | Nightingale | M_VLDL_T | NA | G | A | 0.02336 | 0.004186 | 1.90E-09 |
| rs4574742 | miR-148a-3 | cis | 7.37E-05 | Nightingale | S_VLDL_C_ | NA | G | A | -0.02242 | 0.004311 | 3.80E-08 |
| rs4574742 | miR-148a-3 | cis | 7.37E-05 | Nightingale | S_VLDL_TG | NA | G | A | 0.023311 | 0.004288 | 9.60E-09 |
| rs4574742 | miR-148a-3 | cis | 7.37E-05 | Nightingale | S_LDL_FC_ | NA | G | A | -0.02644 | 0.004297 | 5.60E-11 |
| rs4574742 | miR-148a-3 | cis | 7.37E-05 | Nightingale | M_LDL_FC | NA | G | A | -0.02442 | 0.004247 | 6.40E-10 |
| rs4574742 | miR-148a-3 | cis | 7.37E-05 | Nightingale | S_HDL_C_| | NA | G | A | -0.02463 | 0.004381 | 1.40E-09 |
| rs4574742 | miR-148a-3 | cis | 7.37E-05 | Nightingale | M_VLDL_C | NA | G | A | -0.02505 | 0.004161 | 9.50E-11 |
| rs4574742 | miR-148a-3 | cis | 7.37E-05 | Nightingale | S_VLDL_PL | NA | G | A | -0.02427 | 0.004242 | 1.00E-09 |
| rs4574742 | miR-148a-3 | cis | 7.37E-05 | Nightingale | HDL_TG | NA | G | A | 0.023128 | 0.004371 | 2.10E-08 |
| rs4585652 | miR-339-5| | cis | 7.60E-05 | Nightingale | L_VLDL_C_ | NA | G | A | -0.02242 | 0.004164 | 1.30E-08 |
| rs4585652 | miR-339-5| | cis | 7.60E-05 | Nightingale | L_VLDL_CE | NA | G | A | -0.02281 | 0.004079 | 8.50E-09 |
| rs4585652 | miR-339-5| | cis | 7.60E-05 | Nightingale | VLDL_size | NA | G | A | 0.023187 | 0.003975 | 2.90E-08 |
| rs4719841 | miR-148a-3 | cis | 0.000239 | Nightingale | L_HDL_CE_ | NA | G | A | -0.02218 | 0.004019 | 2.60E-09 |
| rs4719841 | miR-148a-3 | cis | 0.000239 | Nightingale | M_LDL_FC | NA | G | A | -0.02772 | 0.004025 | 8.90E-14 |
| rs4719841 | miR-148a-3 | cis | 0.000239 | Nightingale | TG_by_PG | NA | G | A | 0.020112 | 0.003898 | 3.30E-08 |
| rs4719841 | miR-148a-3 | cis | 0.000239 | Nightingale | Total_TG | NA | G | A | 0.021231 | 0.004066 | 3.90E-08 |
| rs4719841 | miR-148a-3 | cis | 0.000239 | Nightingale | M_HDL_CE | NA | G | A | -0.02279 | 0.004074 | 2.10E-09 |
| rs4719841 | miR-148a-3 | cis | 0.000239 | Nightingale | S_LDL_FC_ | NA | G | A | -0.03077 | 0.004072 | 7.50E-16 |
| rs4719841 | miR-148a-3 | cis | 0.000239 | Nightingale | M_VLDL_C | NA | G | A | -0.02857 | 0.003954 | 3.00E-15 |
| rs4719841 | miR-148a-3 | cis | 0.000239 | Nightingale | M_VLDL_T | NA | G | A | 0.027915 | 0.003967 | 1.50E-14 |
| rs4719841 | miR-148a-3 | cis | 0.000239 | Nightingale | XXL_VLDL_ | NA | G | A | 0.021626 | 0.004057 | 2.00E-08 |
| rs4719841 | miR-148a-3 | cis | 0.000239 | Nightingale | S_VLDL_PL | NA | G | A | -0.02827 | 0.00402 | 9.40E-15 |
| rs4719841 | miR-148a-3 | cis | 0.000239 | Nightingale | L_VLDL_CE | NA | G | A | -0.0214 | 0.004017 | 8.00E-09 |
| rs4719841 | miR-148a-3 | cis | 0.000239 | Nightingale | M_VLDL_C | NA | G | A | -0.02899 | 0.003943 | 1.10E-15 |
| rs4719841 | miR-148a-3 | cis | 0.000239 | Nightingale | XL_VLDL_T | NA | G | A | 0.02241 | 0.004018 | 3.90E-09 |
| rs4719841 | miR-148a-3 | cis | 0.000239 | Nightingale | M_HDL_C_ | NA | G | A | -0.02187 | 0.004 | 2.20E-09 |
| rs4719841 | miR-148a-3 | cis | 0.000239 | Nightingale | XS_VLDL_C | NA | G | A | -0.02286 | 0.00396 | 3.30E-10 |
| rs4719841 | miR-148a-3 | cis | 0.000239 | Nightingale | M_HDL_PL | NA | G | A | 0.020636 | 0.003985 | 1.60E-08 |
| rs4719841 | miR-148a-3 | cis | 0.000239 | Nightingale | IDL_TG_pct | NA | G | A | 0.023581 | 0.004059 | 6.00E-10 |
| rs4719841 | miR-148a-3 | cis | 0.000239 | Nightingale | Omega_6_ | NA | G | A | -0.02551 | 0.004087 | 1.40E-10 |
| rs4719841 | miR-148a-3 | cis | 0.000239 | Nightingale | XXL_VLDL_ | NA | G | A | 0.025355 | 0.004046 | 4.70E-11 |
| rs4719841 | miR-148a-3 | cis | 0.000239 | Nightingale | S_HDL_CE_ | NA | G | A | -0.0237 | 0.004141 | 1.20E-09 |
| rs4719841 | miR-148a-3 | cis | 0.000239 | Nightingale | S_VLDL_FC | NA | G | A | -0.0281 | 0.004011 | 9.10E-15 |
| rs4719841 | miR-148a-3 | cis | 0.000239 | Nightingale | LA_pct | NA | G | A | -0.0261 | 0.004107 | 4.80E-11 |

| | | | | | | | | | | | |
|---|---|---|---|---|---|---|---|---|---|---|---|
| rs4719841 | miR-148a-3 | cis | 0.000239 | Nightingale | S_LDL_TG_ | NA | G | A | 0.029399 | 0.004071 | 7.80E-15 |
| rs4719841 | miR-148a-3 | cis | 0.000239 | Nightingale | XL_VLDL_T | NA | G | A | 0.02478 | 0.004069 | 5.80E-11 |
| rs4719841 | miR-148a-3 | cis | 0.000239 | Nightingale | L_LDL_TG_ | NA | G | A | 0.021942 | 0.004119 | 1.70E-08 |
| rs4719841 | miR-148a-3 | cis | 0.000239 | Nightingale | S_VLDL_CE | NA | G | A | -0.02261 | 0.004142 | 5.00E-09 |
| rs4719841 | miR-148a-3 | cis | 0.000239 | Nightingale | S_VLDL_TG | NA | G | A | 0.027552 | 0.004063 | 1.60E-13 |
| rs4719841 | miR-148a-3 | cis | 0.000239 | Nightingale | Tyr | NA | G | A | 0.024052 | 0.004194 | 9.80E-09 |
| rs4719841 | miR-148a-3 | cis | 0.000239 | Nightingale | S_HDL_TG_ | NA | G | A | 0.020474 | 0.003997 | 3.60E-08 |
| rs4719841 | miR-148a-3 | cis | 0.000239 | Nightingale | M_VLDL_F | NA | G | A | -0.02621 | 0.003994 | 6.70E-13 |
| rs4719841 | miR-148a-3 | cis | 0.000239 | Nightingale | M_HDL_TG | NA | G | A | 0.025864 | 0.004149 | 2.60E-11 |
| rs4719841 | miR-148a-3 | cis | 0.000239 | Nightingale | XXL_VLDL_ | NA | G | A | 0.023932 | 0.004049 | 5.30E-10 |
| rs4719841 | miR-148a-3 | cis | 0.000239 | Nightingale | VLDL_TG | NA | G | A | 0.021375 | 0.004042 | 2.70E-08 |
| rs4719841 | miR-148a-3 | cis | 0.000239 | Nightingale | XL_VLDL_C | NA | G | A | -0.02566 | 0.004025 | 5.50E-12 |
| rs4719841 | miR-148a-3 | cis | 0.000239 | Nightingale | XXL_VLDL_ | NA | G | A | 0.025722 | 0.004047 | 2.10E-11 |
| rs4719841 | miR-148a-3 | cis | 0.000239 | Nightingale | S_HDL_PL_ | NA | G | A | 0.020116 | 0.00406 | 2.80E-08 |
| rs4719841 | miR-148a-3 | cis | 0.000239 | Nightingale | XXL_VLDL_ | NA | G | A | 0.025575 | 0.004032 | 2.20E-11 |
| rs4719841 | miR-148a-3 | cis | 0.000239 | Nightingale | HDL_TG | NA | G | A | 0.024239 | 0.004142 | 3.80E-10 |
| rs4719841 | miR-148a-3 | cis | 0.000239 | Nightingale | L_HDL_C_p | NA | G | A | -0.02305 | 0.00396 | 3.00E-10 |
| rs4719841 | miR-148a-3 | cis | 0.000239 | Nightingale | XS_VLDL_T | NA | G | A | 0.024568 | 0.003991 | 1.70E-11 |
| rs4719841 | miR-148a-3 | cis | 0.000239 | Nightingale | XXL_VLDL_ | NA | G | A | 0.024584 | 0.00404 | 1.90E-10 |
| rs4719841 | miR-148a-3 | cis | 0.000239 | Nightingale | VLDL_size | NA | G | A | 0.020242 | 0.003915 | 2.10E-08 |
| rs4719841 | miR-148a-3 | cis | 0.000239 | Nightingale | S_HDL_C_p | NA | G | A | -0.02833 | 0.004151 | 2.00E-13 |
| rs4719841 | miR-148a-3 | cis | 0.000239 | Nightingale | M_VLDL_P | NA | G | A | -0.02329 | 0.004029 | 2.50E-10 |
| rs4719841 | miR-148a-3 | cis | 0.000239 | Nightingale | XL_VLDL_C | NA | G | A | -0.02652 | 0.004027 | 1.00E-12 |
| rs4719841 | miR-148a-3 | cis | 0.000239 | Nightingale | L_LDL_FC_ | NA | G | A | -0.02498 | 0.004005 | 2.20E-11 |
| rs4719841 | miR-148a-3 | cis | 0.000239 | Nightingale | M_LDL_TG | NA | G | A | 0.023117 | 0.004148 | 2.90E-09 |
| rs4719841 | miR-148a-3 | cis | 0.000239 | Nightingale | XL_HDL_TG | NA | G | A | 0.020642 | 0.003959 | 2.20E-08 |
| rs4719841 | miR-148a-3 | cis | 0.000239 | Nightingale | IDL_C_pct | NA | G | A | -0.02124 | 0.004102 | 3.00E-08 |
| rs4719841 | miR-148a-3 | cis | 0.000239 | Nightingale | S_HDL_TG | NA | G | A | 0.022882 | 0.004074 | 2.60E-09 |
| rs4719841 | miR-148a-3 | cis | 0.000239 | Nightingale | S_VLDL_C_ | NA | G | A | -0.02659 | 0.004086 | 1.60E-12 |
| rs4719841 | miR-148a-3 | cis | 0.000239 | Nightingale | XXL_VLDL_ | NA | G | A | 0.024615 | 0.004053 | 1.80E-10 |
| rs4719841 | miR-148a-3 | cis | 0.000239 | Nightingale | XS_VLDL_C | NA | G | A | -0.02353 | 0.003981 | 1.30E-10 |
| rs4719841 | miR-148a-3 | cis | 0.000239 | Nightingale | M_HDL_TG | NA | G | A | 0.021113 | 0.00403 | 1.60E-08 |
| rs4724039 | miR-339-5p | cis | 4.47E-07 | Nightingale | L_VLDL_C_ | NA | C | T | -0.02276 | 0.004083 | 1.40E-08 |
| rs4724039 | miR-339-5p | cis | 4.47E-07 | Nightingale | L_VLDL_CE | NA | C | T | -0.02301 | 0.004 | 1.20E-08 |
| rs4724424 | miR-339-5p | cis | 0.000129 | Nightingale | L_VLDL_CE | NA | C | T | -0.02234 | 0.00412 | 3.20E-08 |
| rs4724424 | miR-339-5p | cis | 0.000129 | Nightingale | L_VLDL_C_ | NA | C | T | -0.02194 | 0.004205 | 4.40E-08 |
| rs489858 | miR-130a-3 | cis | 0.000525 | Nightingale | Phe | NA | C | A | 0.049134 | 0.00493 | 2.10E-23 |
| rs6463504 | miR-339-5p | cis | 6.64E-05 | Nightingale | M_VLDL_T | NA | G | A | 0.023083 | 0.003993 | 2.30E-09 |
| rs6463504 | miR-339-5p | cis | 6.64E-05 | Nightingale | M_VLDL_F | NA | G | A | -0.02328 | 0.00402 | 2.30E-09 |
| rs6463504 | miR-339-5p | cis | 6.64E-05 | Nightingale | L_VLDL_C_ | NA | G | A | -0.02574 | 0.004128 | 3.80E-11 |
| rs6463504 | miR-339-5p | cis | 6.64E-05 | Nightingale | VLDL_size | NA | G | A | 0.02309 | 0.003941 | 1.30E-08 |
| rs6463504 | miR-339-5p | cis | 6.64E-05 | Nightingale | XL_VLDL_T | NA | G | A | 0.023788 | 0.004097 | 1.30E-09 |
| rs6463504 | miR-339-5p | cis | 6.64E-05 | Nightingale | M_VLDL_C | NA | G | A | -0.0217 | 0.003969 | 2.30E-08 |
| rs6463504 | miR-339-5p | cis | 6.64E-05 | Nightingale | XL_VLDL_F | NA | G | A | -0.0246 | 0.004098 | 1.10E-09 |
| rs6463504 | miR-339-5p | cis | 6.64E-05 | Nightingale | M_VLDL_P | NA | G | A | -0.02445 | 0.004056 | 3.10E-10 |
| rs6463504 | miR-339-5p | cis | 6.64E-05 | Nightingale | XL_VLDL_C | NA | G | A | -0.02201 | 0.004053 | 2.70E-08 |
| rs6463504 | miR-339-5p | cis | 6.64E-05 | Nightingale | M_VLDL_C | NA | G | A | -0.02234 | 0.00398 | 7.80E-09 |
| rs6463504 | miR-339-5p | cis | 6.64E-05 | Nightingale | L_VLDL_CE | NA | G | A | -0.0257 | 0.004044 | 4.30E-11 |
| rs6463505 | miR-339-5p | cis | 6.49E-05 | Nightingale | XL_VLDL_F | NA | G | C | -0.02459 | 0.004098 | 1.20E-09 |
| rs6463505 | miR-339-5p | cis | 6.49E-05 | Nightingale | M_VLDL_P | NA | G | C | -0.02443 | 0.004056 | 3.20E-10 |
| rs6463505 | miR-339-5p | cis | 6.49E-05 | Nightingale | M_VLDL_F | NA | G | C | -0.02326 | 0.00402 | 2.40E-09 |
| rs6463505 | miR-339-5p | cis | 6.49E-05 | Nightingale | M_VLDL_C | NA | G | C | -0.0217 | 0.00397 | 2.30E-08 |
| rs6463505 | miR-339-5p | cis | 6.49E-05 | Nightingale | M_VLDL_T | NA | G | C | 0.023076 | 0.003993 | 2.40E-09 |
| rs6463505 | miR-339-5p | cis | 6.49E-05 | Nightingale | M_VLDL_C | NA | G | C | -0.02234 | 0.00398 | 7.80E-09 |
| rs6463505 | miR-339-5p | cis | 6.49E-05 | Nightingale | XL_VLDL_C | NA | G | C | -0.022 | 0.004053 | 2.70E-08 |
| rs6463505 | miR-339-5p | cis | 6.49E-05 | Nightingale | XL_VLDL_T | NA | G | C | 0.02377 | 0.004097 | 1.30E-09 |
| rs6463505 | miR-339-5p | cis | 6.49E-05 | Nightingale | VLDL_size | NA | G | C | 0.023094 | 0.003941 | 1.30E-08 |
| rs6463505 | miR-339-5p | cis | 6.49E-05 | Nightingale | L_VLDL_CE | NA | G | C | -0.02568 | 0.004044 | 4.40E-11 |
| rs6463505 | miR-339-5p | cis | 6.49E-05 | Nightingale | L_VLDL_C_ | NA | G | C | -0.02572 | 0.004128 | 3.90E-11 |
| rs6945636 | miR-339-5p | cis | 0.000133 | Nightingale | L_VLDL_CE | NA | A | T | -0.02419 | 0.004026 | 4.50E-10 |
| rs6945636 | miR-339-5p | cis | 0.000133 | Nightingale | M_VLDL_T | NA | A | T | 0.021826 | 0.003975 | 1.40E-08 |
| rs6945636 | miR-339-5p | cis | 0.000133 | Nightingale | XL_VLDL_T | NA | A | T | 0.022325 | 0.004079 | 1.10E-08 |
| rs6945636 | miR-339-5p | cis | 0.000133 | Nightingale | M_VLDL_C | NA | A | T | -0.02107 | 0.003962 | 4.60E-08 |

| | | | | | | | | | | | |
|---|---|---|---|---|---|---|---|---|---|---|---|
| rs6945636 | miR-339-5p | cis | 0.000133 | Nightingale | M_VLDL_F | NA | A | T | -0.02197 | 0.004002 | 1.50E-08 |
| rs6945636 | miR-339-5p | cis | 0.000133 | Nightingale | XL_VLDL_F | NA | A | T | -0.02263 | 0.004079 | 1.70E-08 |
| rs6945636 | miR-339-5p | cis | 0.000133 | Nightingale | L_VLDL_C_ | NA | A | T | -0.02447 | 0.00411 | 2.80E-10 |
| rs6945636 | miR-339-5p | cis | 0.000133 | Nightingale | M_VLDL_P | NA | A | T | -0.02332 | 0.004038 | 1.60E-09 |
| rs6947257 | miR-339-5p | cis | 1.41E-06 | Nightingale | L_HDL_PL | NA | C | T | -0.02153 | 0.003802 | 2.30E-08 |
| rs6947257 | miR-339-5p | cis | 1.41E-06 | Nightingale | HDL_size | NA | C | T | -0.02202 | 0.003827 | 1.40E-08 |
| rs6947257 | miR-339-5p | cis | 1.41E-06 | Nightingale | L_HDL_FC | NA | C | T | -0.02196 | 0.003786 | 9.50E-09 |
| rs6947257 | miR-339-5p | cis | 1.41E-06 | Nightingale | XL_HDL_FC | NA | C | T | 0.021318 | 0.003828 | 1.50E-08 |
| rs6947257 | miR-339-5p | cis | 1.41E-06 | Nightingale | L_HDL_L | NA | C | T | -0.02178 | 0.00379 | 1.30E-08 |
| rs6947257 | miR-339-5p | cis | 1.41E-06 | Nightingale | L_HDL_P | NA | C | T | -0.0219 | 0.003783 | 1.00E-08 |
| rs6947257 | miR-339-5p | cis | 1.41E-06 | Nightingale | L_HDL_C | NA | C | T | -0.02118 | 0.003804 | 4.10E-08 |
| rs6950319 | miR-339-5p | cis | 9.51E-05 | Nightingale | L_VLDL_CE | NA | G | A | -0.0223 | 0.004095 | 2.40E-08 |
| rs6950319 | miR-339-5p | cis | 9.51E-05 | Nightingale | L_VLDL_C_ | NA | G | A | -0.02196 | 0.00418 | 3.00E-08 |
| rs6951827 | miR-148a-3 | cis | 0.000135 | Nightingale | M_VLDL_F | NA | A | G | 0.025902 | 0.00448 | 4.90E-10 |
| rs6951827 | miR-148a-3 | cis | 0.000135 | Nightingale | Total_TG | NA | A | G | -0.02853 | 0.004562 | 1.40E-10 |
| rs6951827 | miR-148a-3 | cis | 0.000135 | Nightingale | L_HDL_TG_ | NA | A | G | -0.02529 | 0.00445 | 9.70E-09 |
| rs6951827 | miR-148a-3 | cis | 0.000135 | Nightingale | M_HDL_TG | NA | A | G | -0.02893 | 0.004654 | 1.40E-10 |
| rs6951827 | miR-148a-3 | cis | 0.000135 | Nightingale | VLDL_size | NA | A | G | -0.02742 | 0.004392 | 4.40E-11 |
| rs6951827 | miR-148a-3 | cis | 0.000135 | Nightingale | XL_VLDL_C | NA | A | G | 0.027921 | 0.004517 | 3.00E-11 |
| rs6951827 | miR-148a-3 | cis | 0.000135 | Nightingale | S_VLDL_PL | NA | A | G | 0.027608 | 0.004509 | 1.50E-10 |
| rs6951827 | miR-148a-3 | cis | 0.000135 | Nightingale | XL_VLDL_L | NA | A | G | -0.02718 | 0.004534 | 7.60E-10 |
| rs6951827 | miR-148a-3 | cis | 0.000135 | Nightingale | XXL_VLDL_ | NA | A | G | -0.0263 | 0.004552 | 1.80E-09 |
| rs6951827 | miR-148a-3 | cis | 0.000135 | Nightingale | S_HDL_C_p | NA | A | G | 0.027836 | 0.004657 | 4.10E-10 |
| rs6951827 | miR-148a-3 | cis | 0.000135 | Nightingale | L_HDL_CE_ | NA | A | G | 0.026394 | 0.004509 | 2.20E-09 |
| rs6951827 | miR-148a-3 | cis | 0.000135 | Nightingale | L_HDL_C_p | NA | A | G | 0.026483 | 0.004443 | 8.00E-10 |
| rs6951827 | miR-148a-3 | cis | 0.000135 | Nightingale | M_VLDL_T | NA | A | G | -0.02909 | 0.00445 | 3.50E-12 |
| rs6951827 | miR-148a-3 | cis | 0.000135 | Nightingale | M_HDL_CE | NA | A | G | 0.026198 | 0.004571 | 4.70E-09 |
| rs6951827 | miR-148a-3 | cis | 0.000135 | Nightingale | L_VLDL_PL | NA | A | G | -0.02562 | 0.004547 | 8.10E-09 |
| rs6951827 | miR-148a-3 | cis | 0.000135 | Nightingale | XXL_VLDL_ | NA | A | G | -0.02899 | 0.004547 | 2.50E-11 |
| rs6951827 | miR-148a-3 | cis | 0.000135 | Nightingale | M_VLDL_C | NA | A | G | 0.030321 | 0.004435 | 3.90E-13 |
| rs6951827 | miR-148a-3 | cis | 0.000135 | Nightingale | M_HDL_C_ | NA | A | G | 0.024598 | 0.004488 | 1.50E-08 |
| rs6951827 | miR-148a-3 | cis | 0.000135 | Nightingale | XL_VLDL_P | NA | A | G | -0.02711 | 0.004529 | 7.30E-10 |
| rs6951827 | miR-148a-3 | cis | 0.000135 | Nightingale | S_LDL_TG_ | NA | A | G | -0.02896 | 0.004567 | 5.20E-11 |
| rs6951827 | miR-148a-3 | cis | 0.000135 | Nightingale | L_VLDL_L | NA | A | G | -0.02625 | 0.004554 | 4.80E-09 |
| rs6951827 | miR-148a-3 | cis | 0.000135 | Nightingale | XS_VLDL_T | NA | A | G | -0.02434 | 0.004477 | 1.60E-08 |
| rs6951827 | miR-148a-3 | cis | 0.000135 | Nightingale | XXL_VLDL_ | NA | A | G | -0.02894 | 0.00454 | 3.40E-11 |
| rs6951827 | miR-148a-3 | cis | 0.000135 | Nightingale | XXL_VLDL_ | NA | A | G | -0.02796 | 0.004542 | 1.60E-10 |
| rs6951827 | miR-148a-3 | cis | 0.000135 | Nightingale | XXL_VLDL_ | NA | A | G | -0.02894 | 0.004523 | 2.70E-11 |
| rs6951827 | miR-148a-3 | cis | 0.000135 | Nightingale | L_VLDL_TG | NA | A | G | -0.02792 | 0.004533 | 3.20E-10 |
| rs6951827 | miR-148a-3 | cis | 0.000135 | Nightingale | XXL_VLDL_ | NA | A | G | -0.0293 | 0.004539 | 1.90E-11 |
| rs6951827 | miR-148a-3 | cis | 0.000135 | Nightingale | XL_HDL_TG | NA | A | G | -0.02682 | 0.004441 | 4.70E-10 |
| rs6951827 | miR-148a-3 | cis | 0.000135 | Nightingale | XL_VLDL_C | NA | A | G | 0.027062 | 0.004515 | 1.30E-10 |
| rs6951827 | miR-148a-3 | cis | 0.000135 | Nightingale | Omega_6_ | NA | A | G | 0.027829 | 0.004585 | 1.10E-09 |
| rs6951827 | miR-148a-3 | cis | 0.000135 | Nightingale | M_LDL_FC_ | NA | A | G | 0.030362 | 0.004515 | 1.60E-12 |
| rs6951827 | miR-148a-3 | cis | 0.000135 | Nightingale | S_VLDL_FC | NA | A | G | 0.026827 | 0.0045 | 3.20E-10 |
| rs6951827 | miR-148a-3 | cis | 0.000135 | Nightingale | S_HDL_TG | NA | A | G | -0.0273 | 0.004571 | 8.20E-10 |
| rs6951827 | miR-148a-3 | cis | 0.000135 | Nightingale | XXL_VLDL_ | NA | A | G | -0.02891 | 0.004533 | 3.80E-11 |
| rs6951827 | miR-148a-3 | cis | 0.000135 | Nightingale | XL_VLDL_P | NA | A | G | -0.02591 | 0.004548 | 4.80E-09 |
| rs6951827 | miR-148a-3 | cis | 0.000135 | Nightingale | M_HDL_TG | NA | A | G | -0.02502 | 0.004521 | 1.60E-08 |
| rs6951827 | miR-148a-3 | cis | 0.000135 | Nightingale | L_LDL_FC_ | NA | A | G | 0.026752 | 0.004493 | 1.10E-09 |
| rs6951827 | miR-148a-3 | cis | 0.000135 | Nightingale | VLDL_TG | NA | A | G | -0.02889 | 0.004535 | 6.60E-11 |
| rs6951827 | miR-148a-3 | cis | 0.000135 | Nightingale | L_VLDL_P | NA | A | G | -0.0261 | 0.00455 | 5.00E-09 |
| rs6951827 | miR-148a-3 | cis | 0.000135 | Nightingale | XL_VLDL_T | NA | A | G | -0.02522 | 0.004564 | 3.20E-09 |
| rs6951827 | miR-148a-3 | cis | 0.000135 | Nightingale | M_VLDL_T | NA | A | G | -0.0265 | 0.004578 | 4.00E-09 |
| rs6951827 | miR-148a-3 | cis | 0.000135 | Nightingale | S_LDL_FC_ | NA | A | G | 0.030897 | 0.004568 | 1.80E-12 |
| rs6951827 | miR-148a-3 | cis | 0.000135 | Nightingale | S_VLDL_TG | NA | A | G | -0.02735 | 0.004559 | 4.00E-10 |
| rs6951827 | miR-148a-3 | cis | 0.000135 | Nightingale | M_VLDL_C | NA | A | G | 0.031329 | 0.004424 | 6.70E-14 |
| rs6951827 | miR-148a-3 | cis | 0.000135 | Nightingale | XL_VLDL_F | NA | A | G | -0.02537 | 0.004564 | 1.20E-08 |
| rs6951827 | miR-148a-3 | cis | 0.000135 | Nightingale | S_VLDL_C_ | NA | A | G | 0.026677 | 0.004583 | 1.20E-09 |
| rs6951827 | miR-148a-3 | cis | 0.000135 | Nightingale | HDL_TG | NA | A | G | -0.02797 | 0.004647 | 6.30E-10 |
| rs6951827 | miR-148a-3 | cis | 0.000135 | Nightingale | S_VLDL_TG | NA | A | G | -0.02763 | 0.004573 | 7.60E-10 |
| rs6951827 | miR-148a-3 | cis | 0.000135 | Nightingale | XL_VLDL_T | NA | A | G | -0.02887 | 0.004507 | 3.60E-11 |
| rs6951827 | miR-148a-3 | cis | 0.000135 | Nightingale | TG_by_PG | NA | A | G | -0.02585 | 0.004374 | 1.60E-09 |

| | | | | | | | | | | | |
|---|---|---|---|---|---|---|---|---|---|---|---|
| rs6956265 | miR-339-5p | cis | 0.000133 | Nightingale | XL_VLDL_F | NA | A | G | -0.02264 | 0.004079 | 1.70E-08 |
| rs6956265 | miR-339-5p | cis | 0.000133 | Nightingale | M_VLDL_C | NA | A | G | -0.02107 | 0.003962 | 4.60E-08 |
| rs6956265 | miR-339-5p | cis | 0.000133 | Nightingale | M_VLDL_T | NA | A | G | 0.021828 | 0.003975 | 1.40E-08 |
| rs6956265 | miR-339-5p | cis | 0.000133 | Nightingale | M_VLDL_F | NA | A | G | -0.02198 | 0.004002 | 1.50E-08 |
| rs6956265 | miR-339-5p | cis | 0.000133 | Nightingale | M_VLDL_P | NA | A | G | -0.02333 | 0.004038 | 1.60E-09 |
| rs6956265 | miR-339-5p | cis | 0.000133 | Nightingale | L_VLDL_C_ | NA | A | G | -0.02447 | 0.00411 | 2.80E-10 |
| rs6956265 | miR-339-5p | cis | 0.000133 | Nightingale | L_VLDL_CE | NA | A | G | -0.0242 | 0.004026 | 4.40E-10 |
| rs6956265 | miR-339-5p | cis | 0.000133 | Nightingale | XL_VLDL_T | NA | A | G | 0.022331 | 0.004079 | 1.00E-08 |
| rs6967431 | miR-339-5p | cis | 6.25E-05 | Nightingale | M_VLDL_T | NA | A | T | 0.023551 | 0.003999 | 1.10E-09 |
| rs6967431 | miR-339-5p | cis | 6.25E-05 | Nightingale | M_VLDL_P | NA | A | T | -0.02485 | 0.004062 | 1.80E-10 |
| rs6967431 | miR-339-5p | cis | 6.25E-05 | Nightingale | L_VLDL_C_ | NA | A | T | -0.02616 | 0.004134 | 2.10E-11 |
| rs6967431 | miR-339-5p | cis | 6.25E-05 | Nightingale | M_VLDL_F | NA | A | T | -0.02374 | 0.004026 | 1.20E-09 |
| rs6967431 | miR-339-5p | cis | 6.25E-05 | Nightingale | XL_VLDL_C | NA | A | T | -0.02249 | 0.004059 | 1.30E-08 |
| rs6967431 | miR-339-5p | cis | 6.25E-05 | Nightingale | L_VLDL_CE | NA | A | T | -0.02618 | 0.00405 | 2.00E-11 |
| rs6967431 | miR-339-5p | cis | 6.25E-05 | Nightingale | M_VLDL_C | NA | A | T | -0.02218 | 0.003975 | 1.10E-08 |
| rs6967431 | miR-339-5p | cis | 6.25E-05 | Nightingale | VLDL_size | NA | A | T | 0.023652 | 0.003947 | 6.00E-09 |
| rs6967431 | miR-339-5p | cis | 6.25E-05 | Nightingale | XL_VLDL_F | NA | A | T | -0.02509 | 0.004104 | 5.60E-10 |
| rs6967431 | miR-339-5p | cis | 6.25E-05 | Nightingale | XL_VLDL_T | NA | A | T | 0.024229 | 0.004103 | 6.70E-10 |
| rs6967431 | miR-339-5p | cis | 6.25E-05 | Nightingale | M_VLDL_C | NA | A | T | -0.02282 | 0.003986 | 3.80E-09 |
| rs6972326 | miR-339-5p | cis | 5.64E-05 | Nightingale | XL_VLDL_T | NA | T | C | 0.024086 | 0.004097 | 8.10E-10 |
| rs6972326 | miR-339-5p | cis | 5.64E-05 | Nightingale | M_VLDL_C | NA | T | C | -0.02187 | 0.003969 | 1.80E-08 |
| rs6972326 | miR-339-5p | cis | 5.64E-05 | Nightingale | M_VLDL_C | NA | T | C | -0.02254 | 0.003979 | 5.80E-09 |
| rs6972326 | miR-339-5p | cis | 5.64E-05 | Nightingale | L_VLDL_CE | NA | T | C | -0.02601 | 0.004044 | 2.40E-11 |
| rs6972326 | miR-339-5p | cis | 5.64E-05 | Nightingale | L_VLDL_C_ | NA | T | C | -0.02613 | 0.004128 | 1.90E-11 |
| rs6972326 | miR-339-5p | cis | 5.64E-05 | Nightingale | M_VLDL_P | NA | T | C | -0.0247 | 0.004056 | 2.10E-10 |
| rs6972326 | miR-339-5p | cis | 5.64E-05 | Nightingale | M_VLDL_T | NA | T | C | 0.023296 | 0.003992 | 1.70E-09 |
| rs6972326 | miR-339-5p | cis | 5.64E-05 | Nightingale | XL_VLDL_F | NA | T | C | -0.02486 | 0.004097 | 7.60E-10 |
| rs6972326 | miR-339-5p | cis | 5.64E-05 | Nightingale | XL_VLDL_C | NA | T | C | -0.02223 | 0.004052 | 1.90E-08 |
| rs6972326 | miR-339-5p | cis | 5.64E-05 | Nightingale | M_VLDL_F | NA | T | C | -0.02353 | 0.00402 | 1.60E-09 |
| rs6972326 | miR-339-5p | cis | 5.64E-05 | Nightingale | VLDL_size | NA | T | C | 0.02325 | 0.003941 | 1.00E-08 |
| rs6973680 | miR-339-5p | cis | 8.70E-05 | Nightingale | XL_VLDL_F | NA | C | T | -0.023 | 0.004081 | 1.00E-08 |
| rs6973680 | miR-339-5p | cis | 8.70E-05 | Nightingale | L_VLDL_CE | NA | C | T | -0.02444 | 0.004028 | 3.10E-10 |
| rs6973680 | miR-339-5p | cis | 8.70E-05 | Nightingale | M_VLDL_C | NA | C | T | -0.02126 | 0.003964 | 3.50E-08 |
| rs6973680 | miR-339-5p | cis | 8.70E-05 | Nightingale | XL_VLDL_T | NA | C | T | 0.022576 | 0.004081 | 7.30E-09 |
| rs6973680 | miR-339-5p | cis | 8.70E-05 | Nightingale | M_VLDL_F | NA | C | T | -0.0223 | 0.004004 | 9.50E-09 |
| rs6973680 | miR-339-5p | cis | 8.70E-05 | Nightingale | M_VLDL_P | NA | C | T | -0.02368 | 0.00404 | 9.30E-10 |
| rs6973680 | miR-339-5p | cis | 8.70E-05 | Nightingale | M_VLDL_T | NA | C | T | 0.022051 | 0.003977 | 1.00E-08 |
| rs6973680 | miR-339-5p | cis | 8.70E-05 | Nightingale | L_VLDL_C_ | NA | C | T | -0.02481 | 0.004112 | 1.70E-10 |
| rs6978842 | miR-339-5p | cis | 0.00012 | Nightingale | XL_VLDL_F | NA | C | A | -0.02249 | 0.004161 | 4.90E-08 |
| rs6978842 | miR-339-5p | cis | 0.00012 | Nightingale | VLDL_size | NA | C | A | 0.023145 | 0.004002 | 2.70E-08 |
| rs6978842 | miR-339-5p | cis | 0.00012 | Nightingale | M_VLDL_P | NA | C | A | -0.02184 | 0.004119 | 2.80E-08 |
| rs6978842 | miR-339-5p | cis | 0.00012 | Nightingale | L_VLDL_C_ | NA | C | A | -0.02403 | 0.004192 | 1.10E-09 |
| rs6978842 | miR-339-5p | cis | 0.00012 | Nightingale | L_VLDL_CE | NA | C | A | -0.02424 | 0.004106 | 8.90E-10 |
| rs6979076 | miR-339-5p | cis | 1.92E-05 | Nightingale | L_HDL_C | NA | C | T | -0.02195 | 0.003758 | 1.80E-08 |
| rs6979076 | miR-339-5p | cis | 1.92E-05 | Nightingale | VLDL_size | NA | C | T | 0.023359 | 0.003976 | 2.50E-08 |
| rs6979076 | miR-339-5p | cis | 1.92E-05 | Nightingale | L_HDL_L | NA | C | T | -0.02141 | 0.003744 | 2.80E-08 |
| rs6979076 | miR-339-5p | cis | 1.92E-05 | Nightingale | L_VLDL_C_ | NA | C | T | -0.02352 | 0.004165 | 4.00E-09 |
| rs6979076 | miR-339-5p | cis | 1.92E-05 | Nightingale | L_HDL_CE | NA | C | T | -0.02184 | 0.003768 | 2.40E-08 |
| rs6979076 | miR-339-5p | cis | 1.92E-05 | Nightingale | L_VLDL_CE | NA | C | T | -0.02416 | 0.00408 | 1.70E-09 |
| rs6979076 | miR-339-5p | cis | 1.92E-05 | Nightingale | XL_HDL_CE | NA | C | T | -0.02197 | 0.003822 | 4.00E-08 |
| rs6979076 | miR-339-5p | cis | 1.92E-05 | Nightingale | L_HDL_FC | NA | C | T | -0.02191 | 0.003739 | 1.10E-08 |
| rs6979076 | miR-339-5p | cis | 1.92E-05 | Nightingale | L_HDL_P | NA | C | T | -0.02178 | 0.003737 | 1.50E-08 |
| rs6979076 | miR-339-5p | cis | 1.92E-05 | Nightingale | HDL_size | NA | C | T | -0.02148 | 0.00378 | 2.90E-08 |
| rs6980357 | miR-148a-3 | cis | 6.07E-05 | Nightingale | S_VLDL_FC | NA | A | G | -0.02384 | 0.00424 | 1.50E-09 |
| rs6980357 | miR-148a-3 | cis | 6.07E-05 | Nightingale | S_VLDL_TG | NA | A | G | 0.023645 | 0.004295 | 6.80E-09 |
| rs6980357 | miR-148a-3 | cis | 6.07E-05 | Nightingale | S_VLDL_PL | NA | A | G | -0.02464 | 0.004249 | 6.60E-10 |
| rs6980357 | miR-148a-3 | cis | 6.07E-05 | Nightingale | S_LDL_TG_ | NA | A | G | 0.025584 | 0.004303 | 2.80E-10 |
| rs6980357 | miR-148a-3 | cis | 6.07E-05 | Nightingale | S_LDL_FC_ | NA | A | G | -0.02676 | 0.004304 | 3.50E-11 |
| rs6980357 | miR-148a-3 | cis | 6.07E-05 | Nightingale | XXL_VLDL_ | NA | A | G | 0.021987 | 0.004278 | 4.10E-08 |
| rs6980357 | miR-148a-3 | cis | 6.07E-05 | Nightingale | M_VLDL_T | NA | A | G | 0.023599 | 0.004193 | 1.50E-09 |
| rs6980357 | miR-148a-3 | cis | 6.07E-05 | Nightingale | M_HDL_TG | NA | A | G | 0.024396 | 0.004385 | 3.50E-09 |
| rs6980357 | miR-148a-3 | cis | 6.07E-05 | Nightingale | M_LDL_FC | NA | A | G | -0.02464 | 0.004254 | 4.80E-10 |
| rs6980357 | miR-148a-3 | cis | 6.07E-05 | Nightingale | S_VLDL_C_ | NA | A | G | -0.02273 | 0.004319 | 2.80E-08 |

| | | | | | | | | | | | |
|---|---|---|---|---|---|---|---|---|---|---|---|
| rs6980357 | miR-148a-3 | cis | 6.07E-05 | Nightingale | M_VLDL_C | NA | A | G | -0.02444 | 0.004179 | 3.20E-10 |
| rs6980357 | miR-148a-3 | cis | 6.07E-05 | Nightingale | HDL_TG | NA | A | G | 0.023344 | 0.004378 | 1.60E-08 |
| rs6980357 | miR-148a-3 | cis | 6.07E-05 | Nightingale | M_VLDL_C | NA | A | G | -0.02527 | 0.004168 | 7.40E-11 |
| rs6980357 | miR-148a-3 | cis | 6.07E-05 | Nightingale | S_HDL_C_p | NA | A | G | -0.02497 | 0.004388 | 8.30E-10 |
| rs7303336 | miR-339-5p | cis | 0.000129 | Nightingale | L_VLDL_C_ | NA | G | T | -0.02447 | 0.00411 | 2.80E-10 |
| rs7303336 | miR-339-5p | cis | 0.000129 | Nightingale | XL_VLDL_F | NA | G | T | -0.02266 | 0.00408 | 1.70E-08 |
| rs7303336 | miR-339-5p | cis | 0.000129 | Nightingale | L_VLDL_CE | NA | G | T | -0.02419 | 0.004026 | 4.50E-10 |
| rs7303336 | miR-339-5p | cis | 0.000129 | Nightingale | M_VLDL_P | NA | G | T | -0.02332 | 0.004038 | 1.60E-09 |
| rs7303336 | miR-339-5p | cis | 0.000129 | Nightingale | M_VLDL_F | NA | G | T | -0.02197 | 0.004002 | 1.60E-08 |
| rs7303336 | miR-339-5p | cis | 0.000129 | Nightingale | M_VLDL_C | NA | G | T | -0.02106 | 0.003962 | 4.80E-08 |
| rs7303336 | miR-339-5p | cis | 0.000129 | Nightingale | XL_VLDL_T | NA | G | T | 0.022344 | 0.004079 | 1.00E-08 |
| rs7303336 | miR-339-5p | cis | 0.000129 | Nightingale | M_VLDL_T | NA | G | T | 0.021815 | 0.003975 | 1.50E-08 |
| rs7497669 | miR-339-5p | cis | 0.000961 | Nightingale | L_HDL_P | NA | T | C | 0.025308 | 0.004853 | 3.80E-08 |
| rs7497669 | miR-339-5p | cis | 0.000961 | Nightingale | XS_VLDL_C | NA | T | C | 0.028851 | 0.005375 | 2.20E-08 |
| rs7497669 | miR-339-5p | cis | 0.000961 | Nightingale | HDL_size | NA | T | C | 0.027153 | 0.004909 | 5.90E-09 |
| rs7497669 | miR-339-5p | cis | 0.000961 | Nightingale | XL_HDL_C_ | NA | T | C | -0.03135 | 0.00529 | 7.90E-09 |
| rs7497669 | miR-339-5p | cis | 0.000961 | Nightingale | XL_HDL_L | NA | T | C | 0.025883 | 0.004979 | 1.90E-08 |
| rs7497669 | miR-339-5p | cis | 0.000961 | Nightingale | L_HDL_FC | NA | T | C | 0.025318 | 0.004856 | 3.50E-08 |
| rs7497669 | miR-339-5p | cis | 0.000961 | Nightingale | L_HDL_FC_ | NA | T | C | 0.025845 | 0.005148 | 4.60E-08 |
| rs7497669 | miR-339-5p | cis | 0.000961 | Nightingale | XS_VLDL_L | NA | T | C | 0.029544 | 0.005444 | 4.00E-08 |
| rs7497669 | miR-339-5p | cis | 0.000961 | Nightingale | XL_HDL_P | NA | T | C | 0.027489 | 0.004986 | 2.00E-09 |
| rs7497669 | miR-339-5p | cis | 0.000961 | Nightingale | XL_HDL_PL | NA | T | C | 0.026651 | 0.00498 | 9.40E-09 |
| rs7497669 | miR-339-5p | cis | 0.000961 | Nightingale | XS_VLDL_C | NA | T | C | 0.028393 | 0.005344 | 2.50E-08 |
| rs7497669 | miR-339-5p | cis | 0.000961 | Nightingale | S_HDL_FC_ | NA | T | C | 0.03215 | 0.005256 | 3.30E-11 |
| rs7497669 | miR-339-5p | cis | 0.000961 | Nightingale | L_HDL_TG | NA | T | C | 0.037144 | 0.005318 | 1.50E-11 |
| rs7782651 | miR-339-5p | cis | 9.58E-05 | Nightingale | L_VLDL_CE | NA | G | T | -0.02232 | 0.004095 | 2.30E-08 |
| rs7782651 | miR-339-5p | cis | 9.58E-05 | Nightingale | L_VLDL_C_ | NA | G | T | -0.02195 | 0.004181 | 3.10E-08 |
| rs7784043 | miR-339-5p | cis | 0.000138 | Nightingale | L_VLDL_C_ | NA | T | G | -0.02429 | 0.004111 | 3.90E-10 |
| rs7784043 | miR-339-5p | cis | 0.000138 | Nightingale | M_VLDL_F | NA | T | G | -0.02183 | 0.004003 | 2.00E-08 |
| rs7784043 | miR-339-5p | cis | 0.000138 | Nightingale | L_VLDL_CE | NA | T | G | -0.024 | 0.004027 | 6.50E-10 |
| rs7784043 | miR-339-5p | cis | 0.000138 | Nightingale | XL_VLDL_T | NA | T | G | 0.022103 | 0.00408 | 1.50E-08 |
| rs7784043 | miR-339-5p | cis | 0.000138 | Nightingale | XL_VLDL_F | NA | T | G | -0.0225 | 0.004081 | 2.20E-08 |
| rs7784043 | miR-339-5p | cis | 0.000138 | Nightingale | M_VLDL_P | NA | T | G | -0.02322 | 0.004039 | 1.90E-09 |
| rs7784043 | miR-339-5p | cis | 0.000138 | Nightingale | M_VLDL_T | NA | T | G | 0.021652 | 0.003976 | 1.90E-08 |
| rs7784059 | miR-339-5p | cis | 0.000138 | Nightingale | L_VLDL_C_ | NA | A | G | -0.02431 | 0.004111 | 4.00E-10 |
| rs7784059 | miR-339-5p | cis | 0.000138 | Nightingale | M_VLDL_P | NA | A | G | -0.02324 | 0.004039 | 1.90E-09 |
| rs7784059 | miR-339-5p | cis | 0.000138 | Nightingale | XL_VLDL_F | NA | A | G | -0.02255 | 0.00408 | 2.00E-08 |
| rs7784059 | miR-339-5p | cis | 0.000138 | Nightingale | L_VLDL_CE | NA | A | G | -0.02405 | 0.004027 | 6.10E-10 |
| rs7784059 | miR-339-5p | cis | 0.000138 | Nightingale | M_VLDL_T | NA | A | G | 0.021689 | 0.003976 | 1.80E-08 |
| rs7784059 | miR-339-5p | cis | 0.000138 | Nightingale | M_VLDL_F | NA | A | G | -0.02188 | 0.004003 | 1.80E-08 |
| rs7784059 | miR-339-5p | cis | 0.000138 | Nightingale | XL_VLDL_T | NA | A | G | 0.022156 | 0.00408 | 1.40E-08 |
| rs7784202 | miR-339-5p | cis | 0.000138 | Nightingale | M_VLDL_T | NA | C | G | 0.021666 | 0.003977 | 1.90E-08 |
| rs7784202 | miR-339-5p | cis | 0.000138 | Nightingale | M_VLDL_P | NA | C | G | -0.02323 | 0.00404 | 1.90E-09 |
| rs7784202 | miR-339-5p | cis | 0.000138 | Nightingale | XL_VLDL_F | NA | C | G | -0.02253 | 0.004081 | 2.10E-08 |
| rs7784202 | miR-339-5p | cis | 0.000138 | Nightingale | L_VLDL_C_ | NA | C | G | -0.02425 | 0.004112 | 4.20E-10 |
| rs7784202 | miR-339-5p | cis | 0.000138 | Nightingale | L_VLDL_CE | NA | C | G | -0.02397 | 0.004028 | 6.70E-10 |
| rs7784202 | miR-339-5p | cis | 0.000138 | Nightingale | XL_VLDL_T | NA | C | G | 0.022116 | 0.004081 | 1.50E-08 |
| rs7784202 | miR-339-5p | cis | 0.000138 | Nightingale | M_VLDL_F | NA | C | G | -0.02184 | 0.004004 | 1.90E-08 |
| rs7784334 | miR-339-5p | cis | 5.02E-05 | Nightingale | M_VLDL_C | NA | A | G | -0.02263 | 0.003986 | 5.10E-09 |
| rs7784334 | miR-339-5p | cis | 5.02E-05 | Nightingale | XL_VLDL_F | NA | A | G | -0.02506 | 0.004104 | 6.00E-10 |
| rs7784334 | miR-339-5p | cis | 5.02E-05 | Nightingale | M_VLDL_F | NA | A | G | -0.02355 | 0.004027 | 1.60E-09 |
| rs7784334 | miR-339-5p | cis | 5.02E-05 | Nightingale | M_VLDL_P | NA | A | G | -0.02468 | 0.004063 | 2.30E-10 |
| rs7784334 | miR-339-5p | cis | 5.02E-05 | Nightingale | XL_VLDL_C | NA | A | G | -0.02241 | 0.004059 | 1.60E-08 |
| rs7784334 | miR-339-5p | cis | 5.02E-05 | Nightingale | VLDL_size | NA | A | G | 0.023183 | 0.003948 | 1.20E-08 |
| rs7784334 | miR-339-5p | cis | 5.02E-05 | Nightingale | M_VLDL_T | NA | A | G | 0.023366 | 0.003999 | 1.50E-09 |
| rs7784334 | miR-339-5p | cis | 5.02E-05 | Nightingale | M_VLDL_C | NA | A | G | -0.02199 | 0.003976 | 1.50E-08 |
| rs7784334 | miR-339-5p | cis | 5.02E-05 | Nightingale | L_VLDL_C_ | NA | A | G | -0.02608 | 0.004135 | 2.30E-11 |
| rs7784334 | miR-339-5p | cis | 5.02E-05 | Nightingale | XL_VLDL_T | NA | A | G | 0.024162 | 0.004104 | 7.60E-10 |
| rs7784334 | miR-339-5p | cis | 5.02E-05 | Nightingale | L_VLDL_CE | NA | A | G | -0.02606 | 0.004051 | 2.40E-11 |
| rs7784502 | miR-339-5p | cis | 0.000136 | Nightingale | XL_VLDL_F | NA | A | G | -0.02247 | 0.004082 | 2.30E-08 |
| rs7784502 | miR-339-5p | cis | 0.000136 | Nightingale | XL_VLDL_T | NA | A | G | 0.02204 | 0.004082 | 1.70E-08 |
| rs7784502 | miR-339-5p | cis | 0.000136 | Nightingale | L_VLDL_CE | NA | A | G | -0.02392 | 0.004029 | 7.40E-10 |
| rs7784502 | miR-339-5p | cis | 0.000136 | Nightingale | M_VLDL_F | NA | A | G | -0.02182 | 0.004005 | 2.00E-08 |

| | | | | | | | | | | | |
|---|---|---|---|---|---|---|---|---|---|---|---|
| rs7784502 | miR-339-5p | cis | 0.000136 | Nightingale | M_VLDL_T | NA | A | G | 0.021623 | 0.003978 | 2.00E-08 |
| rs7784502 | miR-339-5p | cis | 0.000136 | Nightingale | M_VLDL_P | NA | A | G | -0.02319 | 0.004041 | 2.00E-09 |
| rs7784502 | miR-339-5p | cis | 0.000136 | Nightingale | L_VLDL_C_ | NA | A | G | -0.0242 | 0.004113 | 4.50E-10 |
| rs7784647 | miR-339-5p | cis | 5.84E-05 | Nightingale | VLDL_size | NA | A | G | 0.022943 | 0.003944 | 1.70E-08 |
| rs7784647 | miR-339-5p | cis | 5.84E-05 | Nightingale | M_VLDL_T | NA | A | G | 0.022987 | 0.003996 | 2.90E-09 |
| rs7784647 | miR-339-5p | cis | 5.84E-05 | Nightingale | L_VLDL_C_ | NA | A | G | -0.02557 | 0.004131 | 5.20E-11 |
| rs7784647 | miR-339-5p | cis | 5.84E-05 | Nightingale | XL_VLDL_C | NA | A | G | -0.02189 | 0.004056 | 3.40E-08 |
| rs7784647 | miR-339-5p | cis | 5.84E-05 | Nightingale | XL_VLDL_F | NA | A | G | -0.02452 | 0.004101 | 1.40E-09 |
| rs7784647 | miR-339-5p | cis | 5.84E-05 | Nightingale | M_VLDL_C | NA | A | G | -0.02226 | 0.003983 | 9.40E-09 |
| rs7784647 | miR-339-5p | cis | 5.84E-05 | Nightingale | L_VLDL_CE | NA | A | G | -0.02554 | 0.004047 | 5.90E-11 |
| rs7784647 | miR-339-5p | cis | 5.84E-05 | Nightingale | M_VLDL_F | NA | A | G | -0.02319 | 0.004023 | 2.80E-09 |
| rs7784647 | miR-339-5p | cis | 5.84E-05 | Nightingale | M_VLDL_P | NA | A | G | -0.02433 | 0.004059 | 3.90E-10 |
| rs7784647 | miR-339-5p | cis | 5.84E-05 | Nightingale | XL_VLDL_T | NA | A | G | 0.023679 | 0.0041 | 1.60E-09 |
| rs7784647 | miR-339-5p | cis | 5.84E-05 | Nightingale | M_VLDL_C | NA | A | G | -0.02161 | 0.003972 | 2.80E-08 |
| rs7789194 | miR-148a-3 | cis | 4.89E-05 | Nightingale | S_VLDL_C_ | NA | C | T | -0.02583 | 0.004236 | 1.50E-10 |
| rs7789194 | miR-148a-3 | cis | 4.89E-05 | Nightingale | S_HDL_C_p | NA | C | T | -0.02647 | 0.004304 | 8.20E-11 |
| rs7789194 | miR-148a-3 | cis | 4.89E-05 | Nightingale | L_LDL_FC_ | NA | C | T | -0.02284 | 0.004152 | 9.00E-09 |
| rs7789194 | miR-148a-3 | cis | 4.89E-05 | Nightingale | M_VLDL_F | NA | C | T | -0.02387 | 0.004141 | 4.00E-10 |
| rs7789194 | miR-148a-3 | cis | 4.89E-05 | Nightingale | S_VLDL_FC | NA | C | T | -0.02641 | 0.004159 | 1.20E-11 |
| rs7789194 | miR-148a-3 | cis | 4.89E-05 | Nightingale | XL_VLDL_C | NA | C | T | -0.02234 | 0.004173 | 5.10E-09 |
| rs7789194 | miR-148a-3 | cis | 4.89E-05 | Nightingale | M_VLDL_C | NA | C | T | -0.02757 | 0.004088 | 4.40E-13 |
| rs7789194 | miR-148a-3 | cis | 4.89E-05 | Nightingale | M_LDL_FC | NA | C | T | -0.02553 | 0.004173 | 5.50E-11 |
| rs7789194 | miR-148a-3 | cis | 4.89E-05 | Nightingale | M_VLDL_T | NA | C | T | 0.026205 | 0.004113 | 7.10E-12 |
| rs7789194 | miR-148a-3 | cis | 4.89E-05 | Nightingale | S_LDL_FC_ | NA | C | T | -0.02783 | 0.004222 | 3.60E-12 |
| rs7789194 | miR-148a-3 | cis | 4.89E-05 | Nightingale | M_HDL_TG | NA | C | T | 0.024361 | 0.004301 | 2.40E-09 |
| rs7789194 | miR-148a-3 | cis | 4.89E-05 | Nightingale | S_LDL_TG_ | NA | C | T | 0.026894 | 0.004221 | 2.10E-11 |
| rs7789194 | miR-148a-3 | cis | 4.89E-05 | Nightingale | XL_VLDL_C | NA | C | T | -0.02332 | 0.004175 | 1.10E-09 |
| rs7789194 | miR-148a-3 | cis | 4.89E-05 | Nightingale | S_HDL_CE_ | NA | C | T | -0.02312 | 0.004293 | 2.70E-08 |
| rs7789194 | miR-148a-3 | cis | 4.89E-05 | Nightingale | M_VLDL_C | NA | C | T | -0.02692 | 0.004099 | 1.60E-12 |
| rs7789194 | miR-148a-3 | cis | 4.89E-05 | Nightingale | S_VLDL_TG | NA | C | T | 0.026636 | 0.004213 | 3.40E-11 |
| rs7789194 | miR-148a-3 | cis | 4.89E-05 | Nightingale | L_HDL_C_p | NA | C | T | -0.02154 | 0.004106 | 2.40E-08 |
| rs7789194 | miR-148a-3 | cis | 4.89E-05 | Nightingale | M_VLDL_P | NA | C | T | -0.0216 | 0.004178 | 2.50E-08 |
| rs7789194 | miR-148a-3 | cis | 4.89E-05 | Nightingale | XS_VLDL_T | NA | C | T | 0.022845 | 0.004138 | 6.80E-09 |
| rs7789194 | miR-148a-3 | cis | 4.89E-05 | Nightingale | XL_VLDL_T | NA | C | T | 0.021648 | 0.004219 | 3.20E-08 |
| rs7789194 | miR-148a-3 | cis | 4.89E-05 | Nightingale | XXL_VLDL_ | NA | C | T | 0.021488 | 0.00418 | 3.30E-08 |
| rs7789194 | miR-148a-3 | cis | 4.89E-05 | Nightingale | HDL_TG | NA | C | T | 0.023287 | 0.004295 | 1.20E-08 |
| rs7789194 | miR-148a-3 | cis | 4.89E-05 | Nightingale | S_VLDL_CE | NA | C | T | -0.02256 | 0.004295 | 4.20E-08 |
| rs7789194 | miR-148a-3 | cis | 4.89E-05 | Nightingale | XXL_VLDL_ | NA | C | T | 0.021564 | 0.004196 | 4.00E-08 |
| rs7789194 | miR-148a-3 | cis | 4.89E-05 | Nightingale | S_VLDL_PL | NA | C | T | -0.02711 | 0.004168 | 5.70E-12 |
| rs7798572 | miR-339-5p | cis | 4.86E-05 | Nightingale | M_VLDL_F | NA | T | C | -0.02328 | 0.004026 | 2.30E-09 |
| rs7798572 | miR-339-5p | cis | 4.86E-05 | Nightingale | L_VLDL_CE | NA | T | C | -0.02583 | 0.00405 | 3.60E-11 |
| rs7798572 | miR-339-5p | cis | 4.86E-05 | Nightingale | XL_VLDL_T | NA | T | C | 0.023818 | 0.004103 | 1.30E-09 |
| rs7798572 | miR-339-5p | cis | 4.86E-05 | Nightingale | XL_VLDL_C | NA | T | C | -0.02206 | 0.004058 | 2.50E-08 |
| rs7798572 | miR-339-5p | cis | 4.86E-05 | Nightingale | M_VLDL_C | NA | T | C | -0.0217 | 0.003975 | 2.20E-08 |
| rs7798572 | miR-339-5p | cis | 4.86E-05 | Nightingale | L_VLDL_C_ | NA | T | C | -0.02584 | 0.004134 | 3.40E-11 |
| rs7798572 | miR-339-5p | cis | 4.86E-05 | Nightingale | M_VLDL_C | NA | T | C | -0.02235 | 0.003985 | 7.60E-09 |
| rs7798572 | miR-339-5p | cis | 4.86E-05 | Nightingale | XL_VLDL_F | NA | T | C | -0.02467 | 0.004103 | 1.10E-09 |
| rs7798572 | miR-339-5p | cis | 4.86E-05 | Nightingale | VLDL_size | NA | T | C | 0.023118 | 0.003946 | 1.20E-08 |
| rs7798572 | miR-339-5p | cis | 4.86E-05 | Nightingale | M_VLDL_T | NA | T | C | 0.023076 | 0.003998 | 2.30E-09 |
| rs7798572 | miR-339-5p | cis | 4.86E-05 | Nightingale | M_VLDL_P | NA | T | C | -0.02441 | 0.004062 | 3.40E-10 |
| rs7799900 | miR-339-5p | cis | 6.24E-05 | Nightingale | VLDL_size | NA | A | G | 0.02306 | 0.00394 | 1.30E-08 |
| rs7799900 | miR-339-5p | cis | 6.24E-05 | Nightingale | XL_VLDL_F | NA | A | G | -0.02455 | 0.004097 | 1.20E-09 |
| rs7799900 | miR-339-5p | cis | 6.24E-05 | Nightingale | L_VLDL_CE | NA | A | G | -0.02567 | 0.004043 | 4.40E-11 |
| rs7799900 | miR-339-5p | cis | 6.24E-05 | Nightingale | M_VLDL_T | NA | A | G | 0.023058 | 0.003992 | 2.40E-09 |
| rs7799900 | miR-339-5p | cis | 6.24E-05 | Nightingale | XL_VLDL_C | NA | A | G | -0.02197 | 0.004052 | 2.80E-08 |
| rs7799900 | miR-339-5p | cis | 6.24E-05 | Nightingale | M_VLDL_F | NA | A | G | -0.02324 | 0.004019 | 2.40E-09 |
| rs7799900 | miR-339-5p | cis | 6.24E-05 | Nightingale | XL_VLDL_T | NA | A | G | 0.023734 | 0.004096 | 1.40E-09 |
| rs7799900 | miR-339-5p | cis | 6.24E-05 | Nightingale | M_VLDL_C | NA | A | G | -0.02232 | 0.003979 | 7.90E-09 |
| rs7799900 | miR-339-5p | cis | 6.24E-05 | Nightingale | M_VLDL_P | NA | A | G | -0.02443 | 0.004055 | 3.20E-10 |
| rs7799900 | miR-339-5p | cis | 6.24E-05 | Nightingale | L_VLDL_C_ | NA | A | G | -0.02571 | 0.004127 | 3.90E-11 |
| rs7799900 | miR-339-5p | cis | 6.24E-05 | Nightingale | M_VLDL_C | NA | A | G | -0.02168 | 0.003968 | 2.30E-08 |
| rs7800060 | miR-339-5p | cis | 6.21E-05 | Nightingale | L_VLDL_C_ | NA | A | G | -0.02571 | 0.004127 | 3.90E-11 |
| rs7800060 | miR-339-5p | cis | 6.21E-05 | Nightingale | M_VLDL_T | NA | A | G | 0.023059 | 0.003992 | 2.40E-09 |

| | | | | | | | | | | | |
|---|---|---|---|---|---|---|---|---|---|---|---|
| rs7800060 | miR-339-5p | cis | 6.21E-05 | Nightingale | L_VLDL_CE | NA | A | G | -0.02567 | 0.004043 | 4.40E-11 |
| rs7800060 | miR-339-5p | cis | 6.21E-05 | Nightingale | M_VLDL_C | NA | A | G | -0.02168 | 0.003969 | 2.30E-08 |
| rs7800060 | miR-339-5p | cis | 6.21E-05 | Nightingale | M_VLDL_P | NA | A | G | -0.02443 | 0.004055 | 3.20E-10 |
| rs7800060 | miR-339-5p | cis | 6.21E-05 | Nightingale | XL_VLDL_T | NA | A | G | 0.023735 | 0.004096 | 1.40E-09 |
| rs7800060 | miR-339-5p | cis | 6.21E-05 | Nightingale | M_VLDL_FC | NA | A | G | -0.02324 | 0.004019 | 2.40E-09 |
| rs7800060 | miR-339-5p | cis | 6.21E-05 | Nightingale | XL_VLDL_F | NA | A | G | -0.02455 | 0.004097 | 1.20E-09 |
| rs7800060 | miR-339-5p | cis | 6.21E-05 | Nightingale | VLDL_size | NA | A | G | 0.023061 | 0.00394 | 1.30E-08 |
| rs7800060 | miR-339-5p | cis | 6.21E-05 | Nightingale | M_VLDL_C | NA | A | G | -0.02232 | 0.003979 | 7.90E-09 |
| rs7800060 | miR-339-5p | cis | 6.21E-05 | Nightingale | XL_VLDL_C | NA | A | G | -0.02197 | 0.004052 | 2.80E-08 |
| rs7806251 | miR-148a-3 | cis | 0.000107 | Nightingale | Omega_6_ | NA | C | T | 0.028096 | 0.004582 | 7.30E-10 |
| rs7806251 | miR-148a-3 | cis | 0.000107 | Nightingale | TG_by_PG | NA | C | T | -0.02593 | 0.004371 | 1.40E-09 |
| rs7806251 | miR-148a-3 | cis | 0.000107 | Nightingale | L_VLDL_L | NA | C | T | -0.02625 | 0.004551 | 4.50E-09 |
| rs7806251 | miR-148a-3 | cis | 0.000107 | Nightingale | XL_VLDL_P | NA | C | T | -0.02708 | 0.004527 | 7.20E-10 |
| rs7806251 | miR-148a-3 | cis | 0.000107 | Nightingale | M_VLDL_T | NA | C | T | -0.02993 | 0.004447 | 9.90E-13 |
| rs7806251 | miR-148a-3 | cis | 0.000107 | Nightingale | XL_VLDL_C | NA | C | T | 0.027467 | 0.004512 | 7.30E-11 |
| rs7806251 | miR-148a-3 | cis | 0.000107 | Nightingale | XXL_VLDL_ | NA | C | T | -0.02887 | 0.004544 | 2.80E-11 |
| rs7806251 | miR-148a-3 | cis | 0.000107 | Nightingale | L_LDL_FC_ | NA | C | T | 0.027478 | 0.00449 | 4.20E-10 |
| rs7806251 | miR-148a-3 | cis | 0.000107 | Nightingale | XXL_VLDL_ | NA | C | T | -0.02796 | 0.00454 | 1.60E-10 |
| rs7806251 | miR-148a-3 | cis | 0.000107 | Nightingale | L_HDL_CE_ | NA | C | T | 0.026721 | 0.004506 | 1.30E-09 |
| rs7806251 | miR-148a-3 | cis | 0.000107 | Nightingale | S_HDL_TG | NA | C | T | -0.02757 | 0.004568 | 5.10E-10 |
| rs7806251 | miR-148a-3 | cis | 0.000107 | Nightingale | M_HDL_CE | NA | C | T | 0.026276 | 0.004568 | 3.50E-09 |
| rs7806251 | miR-148a-3 | cis | 0.000107 | Nightingale | L_HDL_TG_ | NA | C | T | -0.0255 | 0.004447 | 6.30E-09 |
| rs7806251 | miR-148a-3 | cis | 0.000107 | Nightingale | M_HDL_TG | NA | C | T | -0.02929 | 0.004651 | 8.20E-11 |
| rs7806251 | miR-148a-3 | cis | 0.000107 | Nightingale | XXL_VLDL_ | NA | C | T | -0.02925 | 0.004536 | 1.90E-11 |
| rs7806251 | miR-148a-3 | cis | 0.000107 | Nightingale | XL_HDL_TG | NA | C | T | -0.02715 | 0.004438 | 2.60E-10 |
| rs7806251 | miR-148a-3 | cis | 0.000107 | Nightingale | M_VLDL_FC | NA | C | T | 0.026768 | 0.004478 | 1.60E-10 |
| rs7806251 | miR-148a-3 | cis | 0.000107 | Nightingale | XXL_VLDL_ | NA | C | T | -0.02626 | 0.004549 | 1.80E-09 |
| rs7806251 | miR-148a-3 | cis | 0.000107 | Nightingale | S_LDL_FC_ | NA | C | T | 0.031161 | 0.004565 | 1.20E-12 |
| rs7806251 | miR-148a-3 | cis | 0.000107 | Nightingale | M_LDL_FC_ | NA | C | T | 0.030758 | 0.004512 | 8.30E-13 |
| rs7806251 | miR-148a-3 | cis | 0.000107 | Nightingale | S_VLDL_TG | NA | C | T | -0.02829 | 0.004556 | 1.10E-10 |
| rs7806251 | miR-148a-3 | cis | 0.000107 | Nightingale | M_VLDL_C | NA | C | T | 0.031114 | 0.004433 | 1.10E-13 |
| rs7806251 | miR-148a-3 | cis | 0.000107 | Nightingale | XL_VLDL_T | NA | C | T | -0.02895 | 0.004504 | 3.10E-11 |
| rs7806251 | miR-148a-3 | cis | 0.000107 | Nightingale | XL_VLDL_L | NA | C | T | -0.02714 | 0.004531 | 7.60E-10 |
| rs7806251 | miR-148a-3 | cis | 0.000107 | Nightingale | S_VLDL_PL | NA | C | T | 0.028447 | 0.004507 | 4.60E-11 |
| rs7806251 | miR-148a-3 | cis | 0.000107 | Nightingale | L_VLDL_CE | NA | C | T | 0.023034 | 0.004504 | 3.10E-08 |
| rs7806251 | miR-148a-3 | cis | 0.000107 | Nightingale | M_VLDL_P | NA | C | T | 0.023264 | 0.004518 | 4.30E-08 |
| rs7806251 | miR-148a-3 | cis | 0.000107 | Nightingale | M_VLDL_T | NA | C | T | -0.02664 | 0.004575 | 3.30E-09 |
| rs7806251 | miR-148a-3 | cis | 0.000107 | Nightingale | L_HDL_C_p | NA | C | T | 0.026933 | 0.00444 | 4.10E-10 |
| rs7806251 | miR-148a-3 | cis | 0.000107 | Nightingale | L_VLDL_TG | NA | C | T | -0.02801 | 0.004531 | 2.80E-10 |
| rs7806251 | miR-148a-3 | cis | 0.000107 | Nightingale | S_VLDL_TG | NA | C | T | -0.02796 | 0.004571 | 4.60E-10 |
| rs7806251 | miR-148a-3 | cis | 0.000107 | Nightingale | L_VLDL_P | NA | C | T | -0.02607 | 0.004547 | 4.90E-09 |
| rs7806251 | miR-148a-3 | cis | 0.000107 | Nightingale | XXL_VLDL_ | NA | C | T | -0.02898 | 0.004538 | 3.10E-11 |
| rs7806251 | miR-148a-3 | cis | 0.000107 | Nightingale | XL_VLDL_P | NA | C | T | -0.02579 | 0.004545 | 5.30E-09 |
| rs7806251 | miR-148a-3 | cis | 0.000107 | Nightingale | M_HDL_C_ | NA | C | T | 0.02466 | 0.004485 | 1.10E-08 |
| rs7806251 | miR-148a-3 | cis | 0.000107 | Nightingale | XXL_VLDL_ | NA | C | T | -0.02893 | 0.00453 | 3.60E-11 |
| rs7806251 | miR-148a-3 | cis | 0.000107 | Nightingale | XL_VLDL_C | NA | C | T | 0.028281 | 0.004515 | 1.90E-11 |
| rs7806251 | miR-148a-3 | cis | 0.000107 | Nightingale | L_VLDL_PL | NA | C | T | -0.02557 | 0.004544 | 8.20E-09 |
| rs7806251 | miR-148a-3 | cis | 0.000107 | Nightingale | M_VLDL_C | NA | C | T | 0.032047 | 0.004421 | 2.00E-14 |
| rs7806251 | miR-148a-3 | cis | 0.000107 | Nightingale | XXL_VLDL_ | NA | C | T | -0.02897 | 0.00452 | 2.50E-11 |
| rs7806251 | miR-148a-3 | cis | 0.000107 | Nightingale | S_VLDL_FC | NA | C | T | 0.027636 | 0.004497 | 1.10E-10 |
| rs7806251 | miR-148a-3 | cis | 0.000107 | Nightingale | S_VLDL_CE | NA | C | T | 0.024877 | 0.004644 | 3.10E-08 |
| rs7806251 | miR-148a-3 | cis | 0.000107 | Nightingale | XL_VLDL_T | NA | C | T | -0.02576 | 0.004562 | 1.60E-09 |
| rs7806251 | miR-148a-3 | cis | 0.000107 | Nightingale | VLDL_size | NA | C | T | -0.02756 | 0.004389 | 3.50E-11 |
| rs7806251 | miR-148a-3 | cis | 0.000107 | Nightingale | M_HDL_TG | NA | C | T | -0.02509 | 0.004519 | 1.20E-08 |
| rs7806251 | miR-148a-3 | cis | 0.000107 | Nightingale | S_LDL_TG_ | NA | C | T | -0.02943 | 0.004565 | 2.60E-11 |
| rs7806251 | miR-148a-3 | cis | 0.000107 | Nightingale | XS_VLDL_T | NA | C | T | -0.02518 | 0.004474 | 5.60E-09 |
| rs7806251 | miR-148a-3 | cis | 0.000107 | Nightingale | XL_VLDL_F | NA | C | T | -0.02525 | 0.004562 | 1.30E-08 |
| rs7806251 | miR-148a-3 | cis | 0.000107 | Nightingale | S_HDL_C_p | NA | C | T | 0.028052 | 0.004654 | 2.90E-10 |
| rs7806251 | miR-148a-3 | cis | 0.000107 | Nightingale | S_VLDL_C_ | NA | C | T | 0.027637 | 0.004581 | 3.40E-10 |
| rs7806251 | miR-148a-3 | cis | 0.000107 | Nightingale | HDL_TG | NA | C | T | -0.0283 | 0.004644 | 4.00E-10 |
| rs7806251 | miR-148a-3 | cis | 0.000107 | Nightingale | VLDL_TG | NA | C | T | -0.02898 | 0.004532 | 5.60E-11 |
| rs7806251 | miR-148a-3 | cis | 0.000107 | Nightingale | Total_TG | NA | C | T | -0.02863 | 0.004559 | 1.20E-10 |
| rs7806746 | miR-339-5p | cis | 5.61E-07 | Nightingale | L_HDL_FC | NA | C | A | -0.0205 | 0.003661 | 3.70E-08 |

| | | | | | | | | | | | |
|---|---|---|---|---|---|---|---|---|---|---|---|
| rs7809203 | miR-339-5p | cis | 1.20E-08 | Nightingale | XL_VLDL_F | NA | C | T | -0.02205 | 0.004031 | 4.40E-08 |
| rs7809203 | miR-339-5p | cis | 1.20E-08 | Nightingale | L_VLDL_C_ | NA | C | T | -0.02487 | 0.004062 | 3.10E-10 |
| rs7809203 | miR-339-5p | cis | 1.20E-08 | Nightingale | M_VLDL_C | NA | C | T | -0.0215 | 0.003916 | 4.60E-08 |
| rs7809203 | miR-339-5p | cis | 1.20E-08 | Nightingale | M_VLDL_P | NA | C | T | -0.02354 | 0.003991 | 4.00E-09 |
| rs7809203 | miR-339-5p | cis | 1.20E-08 | Nightingale | M_VLDL_F | NA | C | T | -0.02282 | 0.003955 | 1.10E-08 |
| rs7809203 | miR-339-5p | cis | 1.20E-08 | Nightingale | XL_VLDL_T | NA | C | T | 0.022893 | 0.004031 | 7.00E-09 |
| rs7809203 | miR-339-5p | cis | 1.20E-08 | Nightingale | L_VLDL_CE | NA | C | T | -0.02453 | 0.003979 | 5.50E-10 |
| rs7809203 | miR-339-5p | cis | 1.20E-08 | Nightingale | M_VLDL_T | NA | C | T | 0.022246 | 0.003929 | 1.90E-08 |
| rs7809667 | miR-339-5p | cis | 6.41E-05 | Nightingale | L_VLDL_C_ | NA | C | G | -0.02572 | 0.004128 | 4.00E-11 |
| rs7809667 | miR-339-5p | cis | 6.41E-05 | Nightingale | XL_VLDL_T | NA | C | G | 0.023759 | 0.004097 | 1.30E-09 |
| rs7809667 | miR-339-5p | cis | 6.41E-05 | Nightingale | M_VLDL_P | NA | C | G | -0.02443 | 0.004056 | 3.20E-10 |
| rs7809667 | miR-339-5p | cis | 6.41E-05 | Nightingale | XL_VLDL_C | NA | C | G | -0.02199 | 0.004053 | 2.80E-08 |
| rs7809667 | miR-339-5p | cis | 6.41E-05 | Nightingale | XL_VLDL_F | NA | C | G | -0.02458 | 0.004098 | 1.20E-09 |
| rs7809667 | miR-339-5p | cis | 6.41E-05 | Nightingale | M_VLDL_F | NA | C | G | -0.02326 | 0.00402 | 2.40E-09 |
| rs7809667 | miR-339-5p | cis | 6.41E-05 | Nightingale | M_VLDL_T | NA | C | G | 0.023068 | 0.003993 | 2.40E-09 |
| rs7809667 | miR-339-5p | cis | 6.41E-05 | Nightingale | VLDL_size | NA | C | G | 0.023084 | 0.003941 | 1.30E-08 |
| rs7809667 | miR-339-5p | cis | 6.41E-05 | Nightingale | L_VLDL_CE | NA | C | G | -0.02568 | 0.004044 | 4.40E-11 |
| rs7809667 | miR-339-5p | cis | 6.41E-05 | Nightingale | M_VLDL_C | NA | C | G | -0.02233 | 0.00398 | 7.90E-09 |
| rs7809667 | miR-339-5p | cis | 6.41E-05 | Nightingale | M_VLDL_C | NA | C | G | -0.02169 | 0.003969 | 2.30E-08 |
| rs868833 | miR-339-5p | cis | 0.000102 | Nightingale | XS_VLDL_C | NA | G | C | -0.02345 | 0.004334 | 3.60E-08 |
| rs868833 | miR-339-5p | cis | 0.000102 | Nightingale | XS_VLDL_C | NA | G | C | -0.02375 | 0.004309 | 1.90E-08 |
| rs9639523 | miR-148a-3 | cis | 2.92E-05 | Nightingale | M_VLDL_C | NA | T | G | -0.02808 | 0.004084 | 1.50E-13 |
| rs9639523 | miR-148a-3 | cis | 2.92E-05 | Nightingale | XS_VLDL_C | NA | T | G | -0.02159 | 0.004102 | 3.70E-08 |
| rs9639523 | miR-148a-3 | cis | 2.92E-05 | Nightingale | S_LDL_TG_ | NA | T | G | 0.027805 | 0.004217 | 4.40E-12 |
| rs9639523 | miR-148a-3 | cis | 2.92E-05 | Nightingale | Omega_6_ | NA | T | G | -0.0227 | 0.004233 | 4.50E-08 |
| rs9639523 | miR-148a-3 | cis | 2.92E-05 | Nightingale | M_LDL_FC | NA | T | G | -0.02621 | 0.004169 | 1.80E-11 |
| rs9639523 | miR-148a-3 | cis | 2.92E-05 | Nightingale | XL_VLDL_C | NA | T | G | -0.0241 | 0.004171 | 2.90E-10 |
| rs9639523 | miR-148a-3 | cis | 2.92E-05 | Nightingale | S_VLDL_PL | NA | T | G | -0.02776 | 0.004164 | 1.70E-12 |
| rs9639523 | miR-148a-3 | cis | 2.92E-05 | Nightingale | S_HDL_C_p | NA | T | G | -0.02729 | 0.0043 | 2.20E-11 |
| rs9639523 | miR-148a-3 | cis | 2.92E-05 | Nightingale | S_VLDL_C_ | NA | T | G | -0.02674 | 0.004232 | 3.20E-11 |
| rs9639523 | miR-148a-3 | cis | 2.92E-05 | Nightingale | XXL_VLDL_ | NA | T | G | 0.0217 | 0.004191 | 2.90E-08 |
| rs9639523 | miR-148a-3 | cis | 2.92E-05 | Nightingale | L_HDL_C_p | NA | T | G | -0.02185 | 0.004102 | 1.60E-08 |
| rs9639523 | miR-148a-3 | cis | 2.92E-05 | Nightingale | S_HDL_CE_ | NA | T | G | -0.02413 | 0.004289 | 6.90E-09 |
| rs9639523 | miR-148a-3 | cis | 2.92E-05 | Nightingale | M_VLDL_C | NA | T | G | -0.02747 | 0.004095 | 5.40E-13 |
| rs9639523 | miR-148a-3 | cis | 2.92E-05 | Nightingale | M_HDL_TG | NA | T | G | 0.024967 | 0.004297 | 9.40E-10 |
| rs9639523 | miR-148a-3 | cis | 2.92E-05 | Nightingale | S_LDL_FC_ | NA | T | G | -0.02867 | 0.004218 | 8.30E-13 |
| rs9639523 | miR-148a-3 | cis | 2.92E-05 | Nightingale | XL_VLDL_T | NA | T | G | 0.022527 | 0.004215 | 8.80E-09 |
| rs9639523 | miR-148a-3 | cis | 2.92E-05 | Nightingale | XL_VLDL_C | NA | T | G | -0.02313 | 0.004169 | 1.50E-09 |
| rs9639523 | miR-148a-3 | cis | 2.92E-05 | Nightingale | L_HDL_CE_ | NA | T | G | -0.02188 | 0.004163 | 3.00E-08 |
| rs9639523 | miR-148a-3 | cis | 2.92E-05 | Nightingale | HDL_TG | NA | T | G | 0.024002 | 0.004291 | 4.30E-09 |
| rs9639523 | miR-148a-3 | cis | 2.92E-05 | Nightingale | M_VLDL_P | NA | T | G | -0.02205 | 0.004174 | 1.30E-08 |
| rs9639523 | miR-148a-3 | cis | 2.92E-05 | Nightingale | M_VLDL_T | NA | T | G | 0.026738 | 0.004109 | 2.60E-12 |
| rs9639523 | miR-148a-3 | cis | 2.92E-05 | Nightingale | S_VLDL_TG | NA | T | G | 0.027483 | 0.004209 | 7.40E-12 |
| rs9639523 | miR-148a-3 | cis | 2.92E-05 | Nightingale | XXL_VLDL_ | NA | T | G | 0.021984 | 0.004176 | 1.70E-08 |
| rs9639523 | miR-148a-3 | cis | 2.92E-05 | Nightingale | S_VLDL_FC | NA | T | G | -0.02712 | 0.004155 | 3.20E-12 |
| rs9639523 | miR-148a-3 | cis | 2.92E-05 | Nightingale | XS_VLDL_C | NA | T | G | -0.02225 | 0.004123 | 1.60E-08 |
| rs9639523 | miR-148a-3 | cis | 2.92E-05 | Nightingale | M_VLDL_F | NA | T | G | -0.02445 | 0.004137 | 1.50E-10 |
| rs9639523 | miR-148a-3 | cis | 2.92E-05 | Nightingale | XXL_VLDL_ | NA | T | G | 0.022139 | 0.004192 | 1.80E-08 |
| rs9639523 | miR-148a-3 | cis | 2.92E-05 | Nightingale | S_VLDL_CE | NA | T | G | -0.02352 | 0.004291 | 1.20E-08 |
| rs9639523 | miR-148a-3 | cis | 2.92E-05 | Nightingale | L_LDL_FC_ | NA | T | G | -0.02374 | 0.004148 | 2.20E-09 |
| rs9639523 | miR-148a-3 | cis | 2.92E-05 | Nightingale | XS_VLDL_T | NA | T | G | 0.023668 | 0.004134 | 1.80E-09 |
| rs99780 | miR-1908-5 | cis | 1.74E-08 | Nightingale | XL_VLDL_F | NA | T | C | 0.043018 | 0.004187 | 1.00E-25 |
| rs99780 | miR-1908-5 | cis | 1.74E-08 | Nightingale | L_HDL_P | NA | T | C | -0.07766 | 0.003786 | 1.50E-101 |
| rs99780 | miR-1908-5 | cis | 1.74E-08 | Nightingale | IDL_FC | NA | T | C | -0.0897 | 0.004184 | 7.19E-108 |
| rs99780 | miR-1908-5 | cis | 1.74E-08 | Nightingale | HDL_FC | NA | T | C | -0.07398 | 0.00383 | 1.00E-89 |
| rs99780 | miR-1908-5 | cis | 1.74E-08 | Nightingale | L_LDL_L | NA | T | C | -0.0542 | 0.004281 | 6.60E-39 |
| rs99780 | miR-1908-5 | cis | 1.74E-08 | Nightingale | L_VLDL_L | NA | T | C | 0.037739 | 0.004178 | 3.00E-20 |
| rs99780 | miR-1908-5 | cis | 1.74E-08 | Nightingale | L_HDL_CE | NA | T | C | -0.07842 | 0.003817 | 2.10E-101 |
| rs99780 | miR-1908-5 | cis | 1.74E-08 | Nightingale | IDL_C_pct | NA | T | C | -0.07488 | 0.00422 | 1.60E-72 |
| rs99780 | miR-1908-5 | cis | 1.74E-08 | Nightingale | S_LDL_TG_ | NA | T | C | 0.085897 | 0.004189 | 6.60E-97 |
| rs99780 | miR-1908-5 | cis | 1.74E-08 | Nightingale | M_VLDL_T | NA | T | C | 0.030937 | 0.0042 | 6.50E-14 |
| rs99780 | miR-1908-5 | cis | 1.74E-08 | Nightingale | LA_pct | NA | T | C | 0.185657 | 0.004225 | 0 |
| rs99780 | miR-1908-5 | cis | 1.74E-08 | Nightingale | L_HDL_C_p | NA | T | C | -0.06755 | 0.004075 | 5.50E-66 |

| | | | | | | | | | | | |
|---|---|---|---|---|---|---|---|---|---|---|---|
| rs99780 | miR-1908-5 | cis | 1.74E-08 | Nightingale | PUFA_pct | NA | T | C | -0.12081 | 0.004158 | 1.90E-187 |
| rs99780 | miR-1908-5 | cis | 1.74E-08 | Nightingale | Total_FA | NA | T | C | -0.0242 | 0.004289 | 6.30E-09 |
| rs99780 | miR-1908-5 | cis | 1.74E-08 | Nightingale | L_VLDL_C | NA | T | C | 0.028564 | 0.004228 | 6.00E-12 |
| rs99780 | miR-1908-5 | cis | 1.74E-08 | Nightingale | L_HDL_CE_ | NA | T | C | -0.04927 | 0.004136 | 9.10E-35 |
| rs99780 | miR-1908-5 | cis | 1.74E-08 | Nightingale | HDL_C | NA | T | C | -0.06485 | 0.00386 | 2.80E-68 |
| rs99780 | miR-1908-5 | cis | 1.74E-08 | Nightingale | HDL_size | NA | T | C | -0.0724 | 0.00383 | 7.40E-86 |
| rs99780 | miR-1908-5 | cis | 1.74E-08 | Nightingale | IDL_L | NA | T | C | -0.07232 | 0.004179 | 1.10E-71 |
| rs99780 | miR-1908-5 | cis | 1.74E-08 | Nightingale | L_VLDL_FC | NA | T | C | 0.039714 | 0.004181 | 2.60E-22 |
| rs99780 | miR-1908-5 | cis | 1.74E-08 | Nightingale | ApoB | NA | T | C | -0.03847 | 0.004317 | 4.00E-20 |
| rs99780 | miR-1908-5 | cis | 1.74E-08 | Nightingale | XS_VLDL_C | NA | T | C | -0.09759 | 0.004096 | 2.90E-131 |
| rs99780 | miR-1908-5 | cis | 1.74E-08 | Nightingale | S_LDL_L | NA | T | C | -0.03253 | 0.004322 | 7.70E-15 |
| rs99780 | miR-1908-5 | cis | 1.74E-08 | Nightingale | XXL_VLDL_ | NA | T | C | 0.065618 | 0.004158 | 6.60E-59 |
| rs99780 | miR-1908-5 | cis | 1.74E-08 | Nightingale | L_VLDL_PL | NA | T | C | 0.087806 | 0.004195 | 8.49E-101 |
| rs99780 | miR-1908-5 | cis | 1.74E-08 | Nightingale | S_LDL_P | NA | T | C | -0.03254 | 0.004327 | 9.60E-15 |
| rs99780 | miR-1908-5 | cis | 1.74E-08 | Nightingale | M_LDL_C_ | NA | T | C | -0.10022 | 0.004326 | 1.10E-119 |
| rs99780 | miR-1908-5 | cis | 1.74E-08 | Nightingale | S_HDL_FC_ | NA | T | C | -0.10207 | 0.0041 | 2.20E-151 |
| rs99780 | miR-1908-5 | cis | 1.74E-08 | Nightingale | M_HDL_TG | NA | T | C | 0.066539 | 0.004145 | 1.00E-60 |
| rs99780 | miR-1908-5 | cis | 1.74E-08 | Nightingale | LDL_PL | NA | T | C | -0.04624 | 0.004313 | 3.70E-28 |
| rs99780 | miR-1908-5 | cis | 1.74E-08 | Nightingale | M_HDL_C_ | NA | T | C | -0.06566 | 0.004115 | 2.80E-60 |
| rs99780 | miR-1908-5 | cis | 1.74E-08 | Nightingale | M_LDL_P | NA | T | C | -0.04683 | 0.004336 | 3.60E-28 |
| rs99780 | miR-1908-5 | cis | 1.74E-08 | Nightingale | L_HDL_PL_ | NA | T | C | 0.041766 | 0.004184 | 6.10E-25 |
| rs99780 | miR-1908-5 | cis | 1.74E-08 | Nightingale | LDL_CE | NA | T | C | -0.04369 | 0.004312 | 2.10E-25 |
| rs99780 | miR-1908-5 | cis | 1.74E-08 | Nightingale | XS_VLDL_T | NA | T | C | 0.096744 | 0.004106 | 8.71E-129 |
| rs99780 | miR-1908-5 | cis | 1.74E-08 | Nightingale | M_LDL_TG | NA | T | C | 0.036194 | 0.004259 | 1.30E-17 |
| rs99780 | miR-1908-5 | cis | 1.74E-08 | Nightingale | M_LDL_CE | NA | T | C | -0.02662 | 0.004329 | 3.00E-10 |
| rs99780 | miR-1908-5 | cis | 1.74E-08 | Nightingale | M_VLDL_P | NA | T | C | -0.06201 | 0.004148 | 3.70E-54 |
| rs99780 | miR-1908-5 | cis | 1.74E-08 | Nightingale | S_HDL_TG_ | NA | T | C | 0.068895 | 0.004111 | 4.60E-66 |
| rs99780 | miR-1908-5 | cis | 1.74E-08 | Nightingale | L_VLDL_C_ | NA | T | C | -0.03818 | 0.004223 | 6.40E-21 |
| rs99780 | miR-1908-5 | cis | 1.74E-08 | Nightingale | L_HDL_FC | NA | T | C | -0.08431 | 0.003788 | 7.91E-119 |
| rs99780 | miR-1908-5 | cis | 1.74E-08 | Nightingale | L_LDL_CE | NA | T | C | -0.05332 | 0.004285 | 1.30E-37 |
| rs99780 | miR-1908-5 | cis | 1.74E-08 | Nightingale | XS_VLDL_T | NA | T | C | 0.031296 | 0.004257 | 5.00E-13 |
| rs99780 | miR-1908-5 | cis | 1.74E-08 | Nightingale | XL_VLDL_L | NA | T | C | 0.050973 | 0.004159 | 6.10E-36 |
| rs99780 | miR-1908-5 | cis | 1.74E-08 | Nightingale | L_LDL_PL | NA | T | C | -0.05501 | 0.00429 | 1.50E-39 |
| rs99780 | miR-1908-5 | cis | 1.74E-08 | Nightingale | XL_HDL_P | NA | T | C | -0.0888 | 0.00389 | 6.81E-126 |
| rs99780 | miR-1908-5 | cis | 1.74E-08 | Nightingale | Cholines | NA | T | C | -0.0374 | 0.004066 | 8.60E-22 |
| rs99780 | miR-1908-5 | cis | 1.74E-08 | Nightingale | M_LDL_TG | NA | T | C | 0.07558 | 0.004268 | 5.70E-72 |
| rs99780 | miR-1908-5 | cis | 1.74E-08 | Nightingale | S_HDL_TG | NA | T | C | 0.068399 | 0.004192 | 1.40E-62 |
| rs99780 | miR-1908-5 | cis | 1.74E-08 | Nightingale | HDL_CE | NA | T | C | -0.06157 | 0.003879 | 6.00E-61 |
| rs99780 | miR-1908-5 | cis | 1.74E-08 | Nightingale | Omega_3_ | NA | T | C | -0.38918 | 0.004224 | 0 |
| rs99780 | miR-1908-5 | cis | 1.74E-08 | Nightingale | M_VLDL_C | NA | T | C | -0.05234 | 0.004284 | 4.40E-36 |
| rs99780 | miR-1908-5 | cis | 1.74E-08 | Nightingale | XL_HDL_CE | NA | T | C | -0.09639 | 0.003871 | 4.70E-148 |
| rs99780 | miR-1908-5 | cis | 1.74E-08 | Nightingale | S_VLDL_FC | NA | T | C | -0.02832 | 0.004324 | 1.60E-11 |
| rs99780 | miR-1908-5 | cis | 1.74E-08 | Nightingale | L_LDL_FC_ | NA | T | C | -0.08229 | 0.00412 | 3.70E-92 |
| rs99780 | miR-1908-5 | cis | 1.74E-08 | Nightingale | LDL_FC | NA | T | C | -0.06426 | 0.004277 | 5.30E-54 |
| rs99780 | miR-1908-5 | cis | 1.74E-08 | Nightingale | MUFA | NA | T | C | 0.06339 | 0.004279 | 5.70E-51 |
| rs99780 | miR-1908-5 | cis | 1.74E-08 | Nightingale | Phosphogly | NA | T | C | -0.04137 | 0.004095 | 1.40E-25 |
| rs99780 | miR-1908-5 | cis | 1.74E-08 | Nightingale | S_LDL_CE_ | NA | T | C | 0.0567 | 0.004315 | 1.80E-41 |
| rs99780 | miR-1908-5 | cis | 1.74E-08 | Nightingale | M_LDL_FC | NA | T | C | -0.05247 | 0.004318 | 6.40E-36 |
| rs99780 | miR-1908-5 | cis | 1.74E-08 | Nightingale | IDL_CE_pct | NA | T | C | -0.03803 | 0.004242 | 2.00E-19 |
| rs99780 | miR-1908-5 | cis | 1.74E-08 | Nightingale | XL_HDL_C | NA | T | C | -0.09842 | 0.003898 | 4.80E-152 |
| rs99780 | miR-1908-5 | cis | 1.74E-08 | Nightingale | HDL_L | NA | T | C | -0.05359 | 0.003882 | 2.20E-46 |
| rs99780 | miR-1908-5 | cis | 1.74E-08 | Nightingale | PUFA_by_M | NA | T | C | -0.1701 | 0.004141 | 0 |
| rs99780 | miR-1908-5 | cis | 1.74E-08 | Nightingale | VLDL_TG | NA | T | C | 0.047212 | 0.00416 | 5.70E-31 |
| rs99780 | miR-1908-5 | cis | 1.74E-08 | Nightingale | L_HDL_L | NA | T | C | -0.07707 | 0.003793 | 1.30E-99 |
| rs99780 | miR-1908-5 | cis | 1.74E-08 | Nightingale | S_VLDL_CE | NA | T | C | -0.02834 | 0.004265 | 1.30E-11 |
| rs99780 | miR-1908-5 | cis | 1.74E-08 | Nightingale | M_HDL_PL | NA | T | C | -0.02208 | 0.004042 | 1.50E-08 |
| rs99780 | miR-1908-5 | cis | 1.74E-08 | Nightingale | M_LDL_C | NA | T | C | -0.03428 | 0.004331 | 5.10E-16 |
| rs99780 | miR-1908-5 | cis | 1.74E-08 | Nightingale | XS_VLDL_P | NA | T | C | 0.058386 | 0.004218 | 1.10E-44 |
| rs99780 | miR-1908-5 | cis | 1.74E-08 | Nightingale | XS_VLDL_L | NA | T | C | -0.04153 | 0.004249 | 1.50E-24 |
| rs99780 | miR-1908-5 | cis | 1.74E-08 | Nightingale | VLDL_size | NA | T | C | 0.071726 | 0.004029 | 4.90E-75 |
| rs99780 | miR-1908-5 | cis | 1.74E-08 | Nightingale | M_HDL_CE | NA | T | C | -0.04668 | 0.004192 | 1.10E-29 |
| rs99780 | miR-1908-5 | cis | 1.74E-08 | Nightingale | S_VLDL_PL | NA | T | C | -0.07818 | 0.004136 | 1.10E-83 |
| rs99780 | miR-1908-5 | cis | 1.74E-08 | Nightingale | XXL_VLDL_ | NA | T | C | 0.071671 | 0.004149 | 3.80E-70 |

| | | | | | | | | | | | |
|---|---|---|---|---|---|---|---|---|---|---|---|
| rs99780 | miR-1908-5 | cis | 1.74E-08 | Nightingale | XL_VLDL_P | NA | T | C | 0.04785 | 0.004172 | 1.10E-31 |
| rs99780 | miR-1908-5 | cis | 1.74E-08 | Nightingale | XXL_VLDL_ | NA | T | C | 0.067465 | 0.004165 | 4.70E-62 |
| rs99780 | miR-1908-5 | cis | 1.74E-08 | Nightingale | PUFA | NA | T | C | -0.09659 | 0.004216 | 8.89E-123 |
| rs99780 | miR-1908-5 | cis | 1.74E-08 | Nightingale | HDL_PL | NA | T | C | -0.04874 | 0.003908 | 4.20E-38 |
| rs99780 | miR-1908-5 | cis | 1.74E-08 | Nightingale | L_LDL_FC | NA | T | C | -0.06824 | 0.004245 | 9.70E-62 |
| rs99780 | miR-1908-5 | cis | 1.74E-08 | Nightingale | S_LDL_PL | NA | T | C | -0.0514 | 0.004313 | 7.70E-35 |
| rs99780 | miR-1908-5 | cis | 1.74E-08 | Nightingale | non_HDL_( | NA | T | C | -0.04786 | 0.0043 | 2.40E-30 |
| rs99780 | miR-1908-5 | cis | 1.74E-08 | Nightingale | M_HDL_FC | NA | T | C | -0.05186 | 0.003934 | 2.50E-42 |
| rs99780 | miR-1908-5 | cis | 1.74E-08 | Nightingale | IDL_FC_pct | NA | T | C | -0.10403 | 0.004288 | 8.89E-132 |
| rs99780 | miR-1908-5 | cis | 1.74E-08 | Nightingale | DHA | NA | T | C | -0.28834 | 0.004166 | 0 |
| rs99780 | miR-1908-5 | cis | 1.74E-08 | Nightingale | Citrate | NA | T | C | -0.03299 | 0.004295 | 1.80E-15 |
| rs99780 | miR-1908-5 | cis | 1.74E-08 | Nightingale | S_LDL_FC | NA | T | C | -0.06283 | 0.004316 | 4.60E-51 |
| rs99780 | miR-1908-5 | cis | 1.74E-08 | Nightingale | M_VLDL_T | NA | T | C | 0.077386 | 0.004083 | 4.50E-84 |
| rs99780 | miR-1908-5 | cis | 1.74E-08 | Nightingale | XS_VLDL_P | NA | T | C | -0.03429 | 0.004262 | 2.90E-17 |
| rs99780 | miR-1908-5 | cis | 1.74E-08 | Nightingale | IDL_PL | NA | T | C | -0.0726 | 0.004171 | 8.10E-73 |
| rs99780 | miR-1908-5 | cis | 1.74E-08 | Nightingale | XS_VLDL_C | NA | T | C | -0.07169 | 0.004171 | 1.30E-70 |
| rs99780 | miR-1908-5 | cis | 1.74E-08 | Nightingale | LDL_C | NA | T | C | -0.04983 | 0.004304 | 1.20E-32 |
| rs99780 | miR-1908-5 | cis | 1.74E-08 | Nightingale | M_VLDL_C | NA | T | C | -0.0801 | 0.004069 | 4.10E-90 |
| rs99780 | miR-1908-5 | cis | 1.74E-08 | Nightingale | M_HDL_CE | NA | T | C | -0.03824 | 0.003992 | 4.90E-23 |
| rs99780 | miR-1908-5 | cis | 1.74E-08 | Nightingale | XL_VLDL_P | NA | T | C | 0.052343 | 0.004155 | 6.90E-38 |
| rs99780 | miR-1908-5 | cis | 1.74E-08 | Nightingale | IDL_C | NA | T | C | -0.0772 | 0.004174 | 3.50E-81 |
| rs99780 | miR-1908-5 | cis | 1.74E-08 | Nightingale | L_VLDL_FC | NA | T | C | 0.028675 | 0.004282 | 1.60E-11 |
| rs99780 | miR-1908-5 | cis | 1.74E-08 | Nightingale | M_HDL_TG | NA | T | C | 0.049865 | 0.004269 | 1.80E-32 |
| rs99780 | miR-1908-5 | cis | 1.74E-08 | Nightingale | S_HDL_C_p | NA | T | C | -0.04207 | 0.004273 | 7.40E-23 |
| rs99780 | miR-1908-5 | cis | 1.74E-08 | Nightingale | XL_HDL_CE | NA | T | C | -0.02628 | 0.00432 | 2.40E-09 |
| rs99780 | miR-1908-5 | cis | 1.74E-08 | Nightingale | XS_VLDL_F | NA | T | C | -0.056 | 0.004247 | 1.50E-42 |
| rs99780 | miR-1908-5 | cis | 1.74E-08 | Nightingale | XL_VLDL_C | NA | T | C | -0.06048 | 0.004144 | 9.10E-50 |
| rs99780 | miR-1908-5 | cis | 1.74E-08 | Nightingale | Remnant_( | NA | T | C | -0.04416 | 0.004295 | 2.20E-26 |
| rs99780 | miR-1908-5 | cis | 1.74E-08 | Nightingale | M_HDL_C | NA | T | C | -0.04119 | 0.003978 | 1.10E-26 |
| rs99780 | miR-1908-5 | cis | 1.74E-08 | Nightingale | M_VLDL_F | NA | T | C | -0.02949 | 0.004322 | 2.30E-12 |
| rs99780 | miR-1908-5 | cis | 1.74E-08 | Nightingale | S_VLDL_FC | NA | T | C | -0.08249 | 0.004128 | 5.40E-93 |
| rs99780 | miR-1908-5 | cis | 1.74E-08 | Nightingale | L_HDL_TG_ | NA | T | C | 0.078565 | 0.004079 | 1.30E-87 |
| rs99780 | miR-1908-5 | cis | 1.74E-08 | Nightingale | M_HDL_P | NA | T | C | -0.03472 | 0.003982 | 1.40E-19 |
| rs99780 | miR-1908-5 | cis | 1.74E-08 | Nightingale | IDL_P | NA | T | C | -0.04526 | 0.004278 | 8.20E-28 |
| rs99780 | miR-1908-5 | cis | 1.74E-08 | Nightingale | MUFA_pct | NA | T | C | 0.196937 | 0.004137 | 0 |
| rs99780 | miR-1908-5 | cis | 1.74E-08 | Nightingale | S_LDL_FC_ | NA | T | C | -0.0797 | 0.004189 | 5.00E-84 |
| rs99780 | miR-1908-5 | cis | 1.74E-08 | Nightingale | M_VLDL_C | NA | T | C | -0.04386 | 0.004311 | 1.70E-25 |
| rs99780 | miR-1908-5 | cis | 1.74E-08 | Nightingale | XL_VLDL_C | NA | T | C | -0.06345 | 0.004142 | 1.70E-54 |
| rs99780 | miR-1908-5 | cis | 1.74E-08 | Nightingale | LDL_TG | NA | T | C | 0.029305 | 0.004275 | 9.80E-12 |
| rs99780 | miR-1908-5 | cis | 1.74E-08 | Nightingale | L_LDL_C | NA | T | C | -0.0578 | 0.004274 | 4.00E-44 |
| rs99780 | miR-1908-5 | cis | 1.74E-08 | Nightingale | XL_HDL_FC | NA | T | C | -0.10165 | 0.004027 | 3.50E-151 |
| rs99780 | miR-1908-5 | cis | 1.74E-08 | Nightingale | L_LDL_P | NA | T | C | -0.0372 | 0.004308 | 4.10E-19 |
| rs99780 | miR-1908-5 | cis | 1.74E-08 | Nightingale | XL_HDL_C_ | NA | T | C | 0.023257 | 0.00413 | 1.20E-09 |
| rs99780 | miR-1908-5 | cis | 1.74E-08 | Nightingale | XS_VLDL_C | NA | T | C | -0.06798 | 0.004196 | 5.10E-63 |
| rs99780 | miR-1908-5 | cis | 1.74E-08 | Nightingale | IDL_TG_pct | NA | T | C | 0.08776 | 0.004176 | 3.60E-102 |
| rs99780 | miR-1908-5 | cis | 1.74E-08 | Nightingale | L_VLDL_P | NA | T | C | 0.041418 | 0.004174 | 3.80E-24 |
| rs99780 | miR-1908-5 | cis | 1.74E-08 | Nightingale | Total_FC | NA | T | C | -0.05936 | 0.004223 | 1.40E-47 |
| rs99780 | miR-1908-5 | cis | 1.74E-08 | Nightingale | M_HDL_PL | NA | T | C | 0.058098 | 0.0041 | 2.00E-47 |
| rs99780 | miR-1908-5 | cis | 1.74E-08 | Nightingale | XL_HDL_PL | NA | T | C | -0.06626 | 0.003971 | 4.80E-67 |
| rs99780 | miR-1908-5 | cis | 1.74E-08 | Nightingale | M_VLDL_F | NA | T | C | -0.0907 | 0.00411 | 7.19E-114 |
| rs99780 | miR-1908-5 | cis | 1.74E-08 | Nightingale | LDL_L | NA | T | C | -0.04554 | 0.004309 | 1.50E-27 |
| rs99780 | miR-1908-5 | cis | 1.74E-08 | Nightingale | HDL_TG | NA | T | C | 0.045827 | 0.004263 | 1.30E-27 |
| rs99780 | miR-1908-5 | cis | 1.74E-08 | Nightingale | SFA | NA | T | C | -0.03201 | 0.004284 | 3.70E-14 |
| rs99780 | miR-1908-5 | cis | 1.74E-08 | Nightingale | ApoA1 | NA | T | C | -0.04154 | 0.003956 | 1.20E-27 |
| rs99780 | miR-1908-5 | cis | 1.74E-08 | Nightingale | S_LDL_TG | NA | T | C | 0.050064 | 0.004217 | 2.90E-33 |
| rs99780 | miR-1908-5 | cis | 1.74E-08 | Nightingale | XXL_VLDL_ | NA | T | C | 0.071224 | 0.004166 | 1.90E-68 |
| rs99780 | miR-1908-5 | cis | 1.74E-08 | Nightingale | XL_VLDL_C | NA | T | C | 0.037113 | 0.004222 | 3.10E-19 |
| rs99780 | miR-1908-5 | cis | 1.74E-08 | Nightingale | XL_HDL_TG | NA | T | C | 0.08659 | 0.004072 | 5.79E-106 |
| rs99780 | miR-1908-5 | cis | 1.74E-08 | Nightingale | Omega_6_ | NA | T | C | 0.369359 | 0.00426 | 0 |
| rs99780 | miR-1908-5 | cis | 1.74E-08 | Nightingale | S_LDL_PL_ | NA | T | C | -0.06123 | 0.004314 | 4.90E-48 |
| rs99780 | miR-1908-5 | cis | 1.74E-08 | Nightingale | XXL_VLDL_ | NA | T | C | 0.060936 | 0.004171 | 1.70E-50 |
| rs99780 | miR-1908-5 | cis | 1.74E-08 | Nightingale | Omega_6_ | NA | T | C | 0.024717 | 0.004207 | 2.70E-09 |
| rs99780 | miR-1908-5 | cis | 1.74E-08 | Nightingale | XXL_VLDL_ | NA | T | C | 0.066163 | 0.004164 | 1.80E-59 |

| | | | | | | | | | | | |
|---|---|---|---|---|---|---|---|---|---|---|---|
| rs99780 | miR-1908-! | cis | 1.74E-08 | Nightingale | XS_VLDL_C | NA | T | C | -0.09171 | 0.004075 | 1.70E-117 |
| rs99780 | miR-1908-! | cis | 1.74E-08 | Nightingale | LA | NA | T | C | 0.083574 | 0.004239 | 5.70E-87 |
| rs99780 | miR-1908-! | cis | 1.74E-08 | Nightingale | M_LDL_PL_ | NA | T | C | 0.046368 | 0.00432 | 6.10E-28 |
| rs99780 | miR-1908-! | cis | 1.74E-08 | Nightingale | XL_VLDL_T | NA | T | C | 0.056345 | 0.004135 | 5.30E-44 |
| rs99780 | miR-1908-! | cis | 1.74E-08 | Nightingale | XXL_VLDL_ | NA | T | C | 0.073119 | 0.004175 | 2.00E-71 |
| rs99780 | miR-1908-! | cis | 1.74E-08 | Nightingale | SFA_pct | NA | T | C | -0.04314 | 0.004265 | 4.70E-24 |
| rs99780 | miR-1908-! | cis | 1.74E-08 | Nightingale | Total_PL | NA | T | C | -0.0513 | 0.004116 | 1.10E-37 |
| rs99780 | miR-1908-! | cis | 1.74E-08 | Nightingale | L_VLDL_PL | NA | T | C | 0.048336 | 0.004171 | 1.90E-32 |
| rs99780 | miR-1908-! | cis | 1.74E-08 | Nightingale | XL_HDL_L | NA | T | C | -0.09222 | 0.003885 | 5.30E-135 |
| rs99780 | miR-1908-! | cis | 1.74E-08 | Nightingale | M_VLDL_C | NA | T | C | -0.07536 | 0.004058 | 2.90E-80 |
| rs99780 | miR-1908-! | cis | 1.74E-08 | Nightingale | L_VLDL_CE | NA | T | C | -0.05859 | 0.004137 | 1.70E-48 |
| rs99780 | miR-1908-! | cis | 1.74E-08 | Nightingale | IDL_CE | NA | T | C | -0.07217 | 0.004173 | 2.70E-71 |
| rs99780 | miR-1908-! | cis | 1.74E-08 | Nightingale | M_HDL_FC | NA | T | C | -0.10327 | 0.003831 | 2.80E-174 |
| rs99780 | miR-1908-! | cis | 1.74E-08 | Nightingale | DHA_pct | NA | T | C | -0.27345 | 0.004196 | 0 |
| rs99780 | miR-1908-! | cis | 1.74E-08 | Nightingale | XL_HDL_FC | NA | T | C | 0.060576 | 0.003833 | 8.80E-62 |
| rs99780 | miR-1908-! | cis | 1.74E-08 | Nightingale | M_LDL_FC_ | NA | T | C | -0.07116 | 0.004141 | 5.60E-69 |
| rs99780 | miR-1908-! | cis | 1.74E-08 | Nightingale | XL_HDL_PL | NA | T | C | -0.08827 | 0.003886 | 1.20E-123 |
| rs99780 | miR-1908-! | cis | 1.74E-08 | Nightingale | S_VLDL_TG | NA | T | C | 0.038454 | 0.004195 | 9.60E-21 |
| rs99780 | miR-1908-! | cis | 1.74E-08 | Nightingale | Total_P | NA | T | C | -0.04075 | 0.004068 | 3.60E-25 |
| rs99780 | miR-1908-! | cis | 1.74E-08 | Nightingale | S_VLDL_TG | NA | T | C | 0.064001 | 0.004183 | 1.30E-54 |
| rs99780 | miR-1908-! | cis | 1.74E-08 | Nightingale | Sphingomy | NA | T | C | -0.11517 | 0.00404 | 5.60E-188 |
| rs99780 | miR-1908-! | cis | 1.74E-08 | Nightingale | XL_VLDL_F | NA | T | C | -0.06753 | 0.004188 | 2.40E-61 |
| rs99780 | miR-1908-! | cis | 1.74E-08 | Nightingale | S_VLDL_C_ | NA | T | C | -0.05604 | 0.004206 | 9.10E-42 |
| rs99780 | miR-1908-! | cis | 1.74E-08 | Nightingale | XS_VLDL_P | NA | T | C | -0.04359 | 0.004265 | 1.80E-26 |
| rs99780 | miR-1908-! | cis | 1.74E-08 | Nightingale | M_LDL_PL | NA | T | C | -0.0237 | 0.00433 | 1.70E-08 |
| rs99780 | miR-1908-! | cis | 1.74E-08 | Nightingale | VLDL_L | NA | T | C | 0.026972 | 0.004239 | 1.00E-10 |
| rs99780 | miR-1908-! | cis | 1.74E-08 | Nightingale | L_VLDL_TG | NA | T | C | 0.036767 | 0.004159 | 1.90E-19 |
| rs99780 | miR-1908-! | cis | 1.74E-08 | Nightingale | XL_VLDL_T | NA | T | C | 0.058984 | 0.004188 | 1.30E-46 |
| rs99780 | miR-1908-! | cis | 1.74E-08 | Nightingale | Total_TG | NA | T | C | 0.046306 | 0.004184 | 1.40E-29 |
| rs99780 | miR-1908-! | cis | 1.74E-08 | Nightingale | Clinical_LD | NA | T | C | -0.0552 | 0.004298 | 1.00E-39 |
| rs99780 | miR-1908-! | cis | 1.74E-08 | Nightingale | XS_VLDL_F | NA | T | C | -0.11222 | 0.00429 | 2.90E-154 |
| rs99780 | miR-1908-! | cis | 1.74E-08 | Nightingale | Total_CE | NA | T | C | -0.06703 | 0.00417 | 2.10E-61 |
| rs99780 | miR-1908-! | cis | 1.74E-08 | Nightingale | XXL_VLDL_ | NA | T | C | 0.04402 | 0.004347 | 4.20E-25 |
| rs99780 | miR-1908-! | cis | 1.74E-08 | Nightingale | L_HDL_FC_ | NA | T | C | -0.1116 | 0.004016 | 2.20E-184 |
| rs99780 | miR-1908-! | cis | 1.74E-08 | Nightingale | XL_VLDL_C | NA | T | C | 0.030055 | 0.004255 | 7.30E-13 |
| rs99780 | miR-1908-! | cis | 1.74E-08 | Nightingale | L_HDL_PL | NA | T | C | -0.07604 | 0.003805 | 2.80E-96 |
| rs99780 | miR-1908-! | cis | 1.74E-08 | Nightingale | M_HDL_L | NA | T | C | -0.02819 | 0.004013 | 3.30E-13 |
| rs99780 | miR-1908-! | cis | 1.74E-08 | Nightingale | Total_L | NA | T | C | -0.03758 | 0.004253 | 7.00E-20 |
| rs99780 | miR-1908-! | cis | 1.74E-08 | Nightingale | TG_by_PG | NA | T | C | 0.068426 | 0.004012 | 1.20E-68 |
| rs99780 | miR-1908-! | cis | 1.74E-08 | Nightingale | L_HDL_C | NA | T | C | -0.07999 | 0.003807 | 3.80E-106 |
| rs99780 | miR-1908-! | cis | 1.74E-08 | Nightingale | LDL_P | NA | T | C | -0.04017 | 0.00432 | 1.10E-21 |
| rs99780 | miR-1908-! | cis | 1.74E-08 | Nightingale | M_LDL_L | NA | T | C | -0.0284 | 0.004329 | 1.80E-11 |
| rs99780 | miR-1908-! | cis | 1.74E-08 | Nightingale | L_LDL_TG_ | NA | T | C | 0.077298 | 0.004239 | 2.60E-76 |
| rs99780 | miR-1908-! | cis | 1.74E-08 | Nightingale | Omega_3 | NA | T | C | -0.33534 | 0.004237 | 0 |
| rs99780 | miR-1908-! | cis | 1.74E-08 | Nightingale | Tyr | NA | T | C | 0.032775 | 0.004317 | 3.20E-14 |
| rs99780 | miR-1908-! | cis | 1.74E-08 | Nightingale | HDL_P | NA | T | C | -0.03606 | 0.004059 | 5.10E-20 |
| rs99780 | miR-1908-! | cis | 1.74E-08 | Nightingale | S_LDL_C | NA | T | C | -0.03228 | 0.004327 | 1.50E-14 |
| rs99780 | miR-1908-! | cis | 1.74E-08 | Nightingale | Total_C | NA | T | C | -0.06521 | 0.004185 | 9.10E-58 |
| rs99780 | miR-1908-! | cis | 1.74E-08 | Nightingale | L_LDL_C_p | NA | T | C | -0.07666 | 0.004212 | 2.20E-76 |
| rs99780 | miR-1908-! | cis | 1.74E-08 | Nightingale | Unsaturati | NA | T | C | -0.33855 | 0.004094 | 0 |
| rs99780 | miR-1908-! | cis | 1.74E-08 | Nightingale | XXL_VLDL_ | NA | T | C | -0.02677 | 0.004301 | 9.90E-11 |
| rs1124406 | miR-10b-5 | trans | 5.93E-20 | Nightingale | IDL_P | NA | T | C | 0.06104 | 0.006396 | 3.40E-22 |
| rs1124406 | miR-10b-5 | trans | 5.93E-20 | Nightingale | S_VLDL_CE | NA | T | C | 0.057155 | 0.006456 | 1.40E-19 |
| rs1124406 | miR-10b-5 | trans | 5.93E-20 | Nightingale | S_VLDL_C_ | NA | T | C | 0.064596 | 0.006291 | 1.20E-24 |
| rs1124406 | miR-10b-5 | trans | 5.93E-20 | Nightingale | L_VLDL_C_ | NA | T | C | 0.075758 | 0.006313 | 3.40E-34 |
| rs1124406 | miR-10b-5 | trans | 5.93E-20 | Nightingale | Remnant_( | NA | T | C | 0.054534 | 0.006422 | 2.50E-18 |
| rs1124406 | miR-10b-5 | trans | 5.93E-20 | Nightingale | VLDL_CE | NA | T | C | 0.051536 | 0.006466 | 3.80E-16 |
| rs1124406 | miR-10b-5 | trans | 5.93E-20 | Nightingale | HDL_size | NA | T | C | -0.03651 | 0.005731 | 2.40E-10 |
| rs1124406 | miR-10b-5 | trans | 5.93E-20 | Nightingale | XL_VLDL_T | NA | T | C | -0.06387 | 0.006264 | 1.70E-24 |
| rs1124406 | miR-10b-5 | trans | 5.93E-20 | Nightingale | IDL_C | NA | T | C | 0.053448 | 0.006243 | 3.60E-18 |
| rs1124406 | miR-10b-5 | trans | 5.93E-20 | Nightingale | S_LDL_L | NA | T | C | 0.045394 | 0.006463 | 2.30E-12 |
| rs1124406 | miR-10b-5 | trans | 5.93E-20 | Nightingale | M_VLDL_P | NA | T | C | 0.04293 | 0.006454 | 1.10E-11 |
| rs1124406 | miR-10b-5 | trans | 5.93E-20 | Nightingale | ApoB_by_/ | NA | T | C | 0.037666 | 0.006366 | 4.90E-09 |

| | | | | | | | | | | | |
|---|---|---|---|---|---|---|---|---|---|---|---|
| rs1124406 | miR-10b-5p | trans | 5.93E-20 | Nightingale | LDL_C | NA | T | C | 0.055905 | 0.006436 | 2.60E-18 |
| rs1124406 | miR-10b-5p | trans | 5.93E-20 | Nightingale | M_LDL_FC | NA | T | C | 0.0589 | 0.006456 | 7.50E-20 |
| rs1124406 | miR-10b-5p | trans | 5.93E-20 | Nightingale | S_VLDL_L | NA | T | C | 0.036073 | 0.006401 | 5.10E-09 |
| rs1124406 | miR-10b-5p | trans | 5.93E-20 | Nightingale | ApoB | NA | T | C | 0.052104 | 0.006456 | 2.80E-16 |
| rs1124406 | miR-10b-5p | trans | 5.93E-20 | Nightingale | S_HDL_P | NA | T | C | 0.057723 | 0.006465 | 1.80E-19 |
| rs1124406 | miR-10b-5p | trans | 5.93E-20 | Nightingale | L_VLDL_FC | NA | T | C | 0.077668 | 0.006402 | 1.40E-34 |
| rs1124406 | miR-10b-5p | trans | 5.93E-20 | Nightingale | M_LDL_C_ | NA | T | C | 0.038266 | 0.006468 | 2.90E-09 |
| rs1124406 | miR-10b-5p | trans | 5.93E-20 | Nightingale | S_LDL_P | NA | T | C | 0.044511 | 0.00647 | 4.30E-12 |
| rs1124406 | miR-10b-5p | trans | 5.93E-20 | Nightingale | M_LDL_L | NA | T | C | 0.054884 | 0.006473 | 1.40E-17 |
| rs1124406 | miR-10b-5p | trans | 5.93E-20 | Nightingale | L_LDL_L | NA | T | C | 0.056834 | 0.006401 | 2.60E-19 |
| rs1124406 | miR-10b-5p | trans | 5.93E-20 | Nightingale | XXL_VLDL_ | NA | T | C | 0.042267 | 0.006513 | 5.50E-12 |
| rs1124406 | miR-10b-5p | trans | 5.93E-20 | Nightingale | M_VLDL_P | NA | T | C | 0.041502 | 0.006448 | 5.70E-11 |
| rs1124406 | miR-10b-5p | trans | 5.93E-20 | Nightingale | M_LDL_TG | NA | T | C | -0.045 | 0.006384 | 2.40E-12 |
| rs1124406 | miR-10b-5p | trans | 5.93E-20 | Nightingale | LDL_CE | NA | T | C | 0.054664 | 0.006447 | 1.30E-17 |
| rs1124406 | miR-10b-5p | trans | 5.93E-20 | Nightingale | S_VLDL_PL | NA | T | C | 0.048525 | 0.006452 | 1.30E-14 |
| rs1124406 | miR-10b-5p | trans | 5.93E-20 | Nightingale | IDL_FC | NA | T | C | 0.057238 | 0.006258 | 1.80E-20 |
| rs1124406 | miR-10b-5p | trans | 5.93E-20 | Nightingale | XS_VLDL_C | NA | T | C | 0.034946 | 0.006131 | 2.70E-08 |
| rs1124406 | miR-10b-5p | trans | 5.93E-20 | Nightingale | XS_VLDL_L | NA | T | C | 0.045503 | 0.006354 | 5.20E-14 |
| rs1124406 | miR-10b-5p | trans | 5.93E-20 | Nightingale | XL_VLDL_F | NA | T | C | 0.049854 | 0.006265 | 6.40E-16 |
| rs1124406 | miR-10b-5p | trans | 5.93E-20 | Nightingale | M_VLDL_T | NA | T | C | -0.05577 | 0.006108 | 1.10E-19 |
| rs1124406 | miR-10b-5p | trans | 5.93E-20 | Nightingale | XXL_VLDL_ | NA | T | C | 0.044692 | 0.006443 | 6.10E-13 |
| rs1124406 | miR-10b-5p | trans | 5.93E-20 | Nightingale | S_LDL_CE_ | NA | T | C | 0.04825 | 0.006453 | 3.00E-15 |
| rs1124406 | miR-10b-5p | trans | 5.93E-20 | Nightingale | XXL_VLDL_ | NA | T | C | 0.055793 | 0.006484 | 1.50E-18 |
| rs1124406 | miR-10b-5p | trans | 5.93E-20 | Nightingale | Cholines | NA | T | C | 0.032924 | 0.006079 | 7.10E-09 |
| rs1124406 | miR-10b-5p | trans | 5.93E-20 | Nightingale | XL_HDL_PL | NA | T | C | -0.0361 | 0.005815 | 7.50E-10 |
| rs1124406 | miR-10b-5p | trans | 5.93E-20 | Nightingale | IDL_L | NA | T | C | 0.053744 | 0.00625 | 1.80E-18 |
| rs1124406 | miR-10b-5p | trans | 5.93E-20 | Nightingale | L_VLDL_TG | NA | T | C | -0.07788 | 0.006433 | 2.00E-35 |
| rs1124406 | miR-10b-5p | trans | 5.93E-20 | Nightingale | LDL_L | NA | T | C | 0.055941 | 0.006442 | 2.10E-18 |
| rs1124406 | miR-10b-5p | trans | 5.93E-20 | Nightingale | Total_P | NA | T | C | 0.035466 | 0.006084 | 9.10E-10 |
| rs1124406 | miR-10b-5p | trans | 5.93E-20 | Nightingale | S_VLDL_TG | NA | T | C | -0.05894 | 0.006257 | 7.20E-21 |
| rs1124406 | miR-10b-5p | trans | 5.93E-20 | Nightingale | Pyruvate | NA | T | C | 0.037852 | 0.006474 | 5.00E-09 |
| rs1124406 | miR-10b-5p | trans | 5.93E-20 | Nightingale | S_HDL_FC | NA | T | C | 0.042891 | 0.006345 | 2.30E-12 |
| rs1124406 | miR-10b-5p | trans | 5.93E-20 | Nightingale | M_VLDL_F | NA | T | C | 0.048475 | 0.006463 | 2.30E-14 |
| rs1124406 | miR-10b-5p | trans | 5.93E-20 | Nightingale | L_LDL_PL | NA | T | C | 0.061894 | 0.006415 | 2.20E-22 |
| rs1124406 | miR-10b-5p | trans | 5.93E-20 | Nightingale | S_HDL_L | NA | T | C | 0.047864 | 0.006442 | 2.50E-14 |
| rs1124406 | miR-10b-5p | trans | 5.93E-20 | Nightingale | S_VLDL_CE | NA | T | C | 0.072236 | 0.006377 | 1.20E-29 |
| rs1124406 | miR-10b-5p | trans | 5.93E-20 | Nightingale | Total_PL | NA | T | C | 0.036937 | 0.006155 | 1.90E-10 |
| rs1124406 | miR-10b-5p | trans | 5.93E-20 | Nightingale | S_HDL_PL | NA | T | C | 0.041448 | 0.006408 | 1.70E-11 |
| rs1124406 | miR-10b-5p | trans | 5.93E-20 | Nightingale | M_LDL_CE | NA | T | C | 0.052966 | 0.006473 | 1.50E-16 |
| rs1124406 | miR-10b-5p | trans | 5.93E-20 | Nightingale | IDL_CE | NA | T | C | 0.051692 | 0.006241 | 4.10E-17 |
| rs1124406 | miR-10b-5p | trans | 5.93E-20 | Nightingale | S_VLDL_FC | NA | T | C | 0.056517 | 0.006465 | 6.10E-19 |
| rs1124406 | miR-10b-5p | trans | 5.93E-20 | Nightingale | S_VLDL_P | NA | T | C | 0.033907 | 0.006383 | 3.40E-08 |
| rs1124406 | miR-10b-5p | trans | 5.93E-20 | Nightingale | IDL_PL | NA | T | C | 0.0529 | 0.006238 | 5.00E-18 |
| rs1124406 | miR-10b-5p | trans | 5.93E-20 | Nightingale | M_VLDL_C | NA | T | C | 0.057005 | 0.006446 | 3.00E-19 |
| rs1124406 | miR-10b-5p | trans | 5.93E-20 | Nightingale | XS_VLDL_C | NA | T | C | 0.052182 | 0.006274 | 7.70E-18 |
| rs1124406 | miR-10b-5p | trans | 5.93E-20 | Nightingale | VLDL_P | NA | T | C | 0.037618 | 0.006429 | 1.60E-09 |
| rs1124406 | miR-10b-5p | trans | 5.93E-20 | Nightingale | L_LDL_C | NA | T | C | 0.055396 | 0.006391 | 2.70E-18 |
| rs1124406 | miR-10b-5p | trans | 5.93E-20 | Nightingale | S_VLDL_FC | NA | T | C | 0.045895 | 0.006177 | 1.90E-13 |
| rs1124406 | miR-10b-5p | trans | 5.93E-20 | Nightingale | XL_HDL_PL | NA | T | C | -0.04518 | 0.005939 | 5.80E-14 |
| rs1124406 | miR-10b-5p | trans | 5.93E-20 | Nightingale | M_LDL_PL | NA | T | C | 0.055933 | 0.006473 | 3.60E-18 |
| rs1124406 | miR-10b-5p | trans | 5.93E-20 | Nightingale | M_LDL_C | NA | T | C | 0.055638 | 0.006476 | 5.60E-18 |
| rs1124406 | miR-10b-5p | trans | 5.93E-20 | Nightingale | S_LDL_PL | NA | T | C | 0.036793 | 0.006449 | 1.40E-08 |
| rs1124406 | miR-10b-5p | trans | 5.93E-20 | Nightingale | LDL_PL | NA | T | C | 0.057963 | 0.006449 | 1.80E-19 |
| rs1124406 | miR-10b-5p | trans | 5.93E-20 | Nightingale | PUFA | NA | T | C | 0.034601 | 0.006305 | 6.50E-09 |
| rs1124406 | miR-10b-5p | trans | 5.93E-20 | Nightingale | L_VLDL_CE | NA | T | C | 0.066389 | 0.006185 | 1.80E-27 |
| rs1124406 | miR-10b-5p | trans | 5.93E-20 | Nightingale | XL_HDL_C_ | NA | T | C | 0.039885 | 0.006173 | 1.50E-10 |
| rs1124406 | miR-10b-5p | trans | 5.93E-20 | Nightingale | S_HDL_C | NA | T | C | 0.05454 | 0.006453 | 1.20E-17 |
| rs1124406 | miR-10b-5p | trans | 5.93E-20 | Nightingale | Omega_6 | NA | T | C | 0.039298 | 0.006312 | 4.70E-11 |
| rs1124406 | miR-10b-5p | trans | 5.93E-20 | Nightingale | XS_VLDL_P | NA | T | C | 0.050744 | 0.006378 | 8.80E-17 |
| rs1124406 | miR-10b-5p | trans | 5.93E-20 | Nightingale | M_LDL_P | NA | T | C | 0.052266 | 0.006482 | 4.10E-16 |
| rs1124406 | miR-10b-5p | trans | 5.93E-20 | Nightingale | M_VLDL_C | NA | T | C | 0.060552 | 0.006405 | 2.00E-21 |
| rs1124406 | miR-10b-5p | trans | 5.93E-20 | Nightingale | M_VLDL_C | NA | T | C | 0.049499 | 0.006073 | 5.10E-16 |
| rs1124406 | miR-10b-5p | trans | 5.93E-20 | Nightingale | XS_VLDL_P | NA | T | C | 0.04195 | 0.006374 | 5.80E-12 |

| | | | | | | | | | | | |
|---|---|---|---|---|---|---|---|---|---|---|---|
| rs1124406 | miR-10b-5p | trans | 5.93E-20 | Nightingale | M_VLDL_P | NA | T | C | 0.064576 | 0.006204 | 1.90E-25 |
| rs1124406 | miR-10b-5p | trans | 5.93E-20 | Nightingale | LA | NA | T | C | 0.040211 | 0.006339 | 3.90E-11 |
| rs1124406 | miR-10b-5p | trans | 5.93E-20 | Nightingale | M_VLDL_F | NA | T | C | 0.058665 | 0.00615 | 1.90E-21 |
| rs1124406 | miR-10b-5p | trans | 5.93E-20 | Nightingale | S_HDL_CE | NA | T | C | 0.0557 | 0.006474 | 4.10E-18 |
| rs1124406 | miR-10b-5p | trans | 5.93E-20 | Nightingale | L_LDL_P | NA | T | C | 0.047255 | 0.006442 | 1.90E-13 |
| rs1124406 | miR-10b-5p | trans | 5.93E-20 | Nightingale | XL_HDL_L | NA | T | C | -0.03301 | 0.005813 | 1.30E-08 |
| rs1124406 | miR-10b-5p | trans | 5.93E-20 | Nightingale | Clinical_LD | NA | T | C | 0.059497 | 0.006426 | 1.60E-20 |
| rs1124406 | miR-10b-5p | trans | 5.93E-20 | Nightingale | S_LDL_C | NA | T | C | 0.050552 | 0.00647 | 5.10E-15 |
| rs1124406 | miR-10b-5p | trans | 5.93E-20 | Nightingale | non_HDL_( | NA | T | C | 0.056382 | 0.006429 | 4.50E-19 |
| rs1124406 | miR-10b-5p | trans | 5.93E-20 | Nightingale | L_LDL_CE | NA | T | C | 0.05436 | 0.006406 | 1.20E-17 |
| rs1124406 | miR-10b-5p | trans | 5.93E-20 | Nightingale | Total_CE | NA | T | C | 0.050589 | 0.006235 | 1.10E-16 |
| rs1124406 | miR-10b-5p | trans | 5.93E-20 | Nightingale | XL_VLDL_C | NA | T | C | 0.05912 | 0.0062 | 1.20E-21 |
| rs1124406 | miR-10b-5p | trans | 5.93E-20 | Nightingale | XL_VLDL_P | NA | T | C | 0.048305 | 0.006472 | 1.30E-14 |
| rs1124406 | miR-10b-5p | trans | 5.93E-20 | Nightingale | Total_C | NA | T | C | 0.050807 | 0.006257 | 8.90E-17 |
| rs1124406 | miR-10b-5p | trans | 5.93E-20 | Nightingale | S_VLDL_PL | NA | T | C | 0.043289 | 0.006189 | 4.50E-12 |
| rs1124406 | miR-10b-5p | trans | 5.93E-20 | Nightingale | XXL_VLDL_ | NA | T | C | 0.055537 | 0.00652 | 4.30E-18 |
| rs1124406 | miR-10b-5p | trans | 5.93E-20 | Nightingale | L_VLDL_PL | NA | T | C | 0.035899 | 0.006279 | 1.80E-09 |
| rs1124406 | miR-10b-5p | trans | 5.93E-20 | Nightingale | XS_VLDL_F | NA | T | C | 0.043006 | 0.006418 | 1.10E-10 |
| rs1124406 | miR-10b-5p | trans | 5.93E-20 | Nightingale | Total_FC | NA | T | C | 0.050299 | 0.006315 | 1.90E-16 |
| rs1124406 | miR-10b-5p | trans | 5.93E-20 | Nightingale | S_VLDL_C | NA | T | C | 0.057388 | 0.006463 | 1.50E-19 |
| rs1124406 | miR-10b-5p | trans | 5.93E-20 | Nightingale | LDL_FC | NA | T | C | 0.057058 | 0.006395 | 5.40E-19 |
| rs1124406 | miR-10b-5p | trans | 5.93E-20 | Nightingale | LDL_P | NA | T | C | 0.049361 | 0.006459 | 1.40E-14 |
| rs1124406 | miR-10b-5p | trans | 5.93E-20 | Nightingale | Sphingomy | NA | T | C | 0.039791 | 0.006042 | 3.30E-11 |
| rs1124406 | miR-10b-5p | trans | 5.93E-20 | Nightingale | Total_L | NA | T | C | 0.040305 | 0.006359 | 4.00E-11 |
| rs1124406 | miR-10b-5p | trans | 5.93E-20 | Nightingale | S_LDL_TG_ | NA | T | C | -0.0362 | 0.006268 | 1.50E-08 |
| rs1124406 | miR-10b-5p | trans | 5.93E-20 | Nightingale | VLDL_C | NA | T | C | 0.044521 | 0.006455 | 1.50E-12 |
| rs1124406 | miR-10b-5p | trans | 5.93E-20 | Nightingale | XS_VLDL_C | NA | T | C | 0.05245 | 0.006238 | 4.10E-18 |
| rs1124406 | miR-10b-5p | trans | 5.93E-20 | Nightingale | XXL_VLDL_ | NA | T | C | -0.05885 | 0.006526 | 1.40E-20 |
| rs1124406 | miR-10b-5p | trans | 5.93E-20 | Nightingale | S_LDL_FC | NA | T | C | 0.048287 | 0.006454 | 1.20E-13 |
| rs1124406 | miR-10b-5p | trans | 5.93E-20 | Nightingale | XS_VLDL_F | NA | T | C | 0.049513 | 0.006352 | 3.50E-16 |
| rs1124406 | miR-10b-5p | trans | 5.93E-20 | Nightingale | XL_VLDL_C | NA | T | C | 0.058597 | 0.006197 | 3.20E-21 |
| rs1124406 | miR-10b-5p | trans | 5.93E-20 | Nightingale | M_VLDL_C | NA | T | C | 0.052338 | 0.006089 | 1.20E-17 |
| rs1124406 | miR-10b-5p | trans | 5.93E-20 | Nightingale | S_LDL_PL_ | NA | T | C | -0.04931 | 0.006452 | 1.90E-15 |
| rs1124406 | miR-10b-5p | trans | 5.93E-20 | Nightingale | L_LDL_FC | NA | T | C | 0.056807 | 0.006347 | 2.60E-19 |
| rs1124406 | miR-10b-5p | trans | 5.93E-20 | Nightingale | S_LDL_CE | NA | T | C | 0.049443 | 0.006467 | 1.40E-14 |
| rs1124406 | miR-10b-5p | trans | 5.93E-20 | Nightingale | S_LDL_C_p | NA | T | C | 0.071544 | 0.006489 | 1.40E-28 |
| rs2073826 | miR-10a-5p | trans | 9.81E-13 | Nightingale | M_VLDL_C | NA | T | G | -0.02564 | 0.004125 | 3.00E-10 |
| rs2073826 | miR-10a-5p | trans | 9.81E-13 | Nightingale | M_VLDL_C | NA | T | G | -0.0243 | 0.003921 | 4.50E-09 |
| rs2073826 | miR-10a-5p | trans | 9.81E-13 | Nightingale | XXL_VLDL_ | NA | T | G | -0.02527 | 0.004169 | 1.70E-09 |
| rs2073826 | miR-10a-5p | trans | 9.81E-13 | Nightingale | L_LDL_PL | NA | T | G | -0.0243 | 0.004131 | 3.00E-09 |
| rs2073826 | miR-10a-5p | trans | 9.81E-13 | Nightingale | M_VLDL_P | NA | T | G | -0.02867 | 0.003995 | 1.50E-12 |
| rs2073826 | miR-10a-5p | trans | 9.81E-13 | Nightingale | XXL_VLDL_ | NA | T | G | 0.025994 | 0.004196 | 3.20E-10 |
| rs2073826 | miR-10a-5p | trans | 9.81E-13 | Nightingale | L_VLDL_TG | NA | T | G | 0.033026 | 0.004143 | 4.10E-16 |
| rs2073826 | miR-10a-5p | trans | 9.81E-13 | Nightingale | L_VLDL_FC | NA | T | G | -0.0311 | 0.004123 | 2.00E-14 |
| rs2073826 | miR-10a-5p | trans | 9.81E-13 | Nightingale | IDL_L | NA | T | G | -0.02234 | 0.004025 | 3.20E-08 |
| rs2073826 | miR-10a-5p | trans | 9.81E-13 | Nightingale | S_VLDL_CE | NA | T | G | -0.03199 | 0.004107 | 1.10E-14 |
| rs2073826 | miR-10a-5p | trans | 9.81E-13 | Nightingale | M_VLDL_T | NA | T | G | 0.025651 | 0.003934 | 4.80E-10 |
| rs2073826 | miR-10a-5p | trans | 9.81E-13 | Nightingale | L_VLDL_CE | NA | T | G | -0.03304 | 0.003983 | 3.30E-16 |
| rs2073826 | miR-10a-5p | trans | 9.81E-13 | Nightingale | S_VLDL_TG | NA | T | G | 0.026207 | 0.004029 | 4.00E-10 |
| rs2073826 | miR-10a-5p | trans | 9.81E-13 | Nightingale | S_VLDL_C_ | NA | T | G | -0.02855 | 0.004051 | 8.10E-12 |
| rs2073826 | miR-10a-5p | trans | 9.81E-13 | Nightingale | Remnant_( | NA | T | G | -0.02176 | 0.004135 | 5.00E-08 |
| rs2073826 | miR-10a-5p | trans | 9.81E-13 | Nightingale | XL_VLDL_C | NA | T | G | -0.02645 | 0.003993 | 4.10E-10 |
| rs2073826 | miR-10a-5p | trans | 9.81E-13 | Nightingale | M_LDL_P | NA | T | G | -0.02329 | 0.004174 | 1.20E-08 |
| rs2073826 | miR-10a-5p | trans | 9.81E-13 | Nightingale | M_VLDL_C | NA | T | G | -0.02333 | 0.004151 | 6.90E-09 |
| rs2073826 | miR-10a-5p | trans | 9.81E-13 | Nightingale | XXL_VLDL_ | NA | T | G | -0.02438 | 0.004192 | 8.20E-09 |
| rs2073826 | miR-10a-5p | trans | 9.81E-13 | Nightingale | XS_VLDL_C | NA | T | G | -0.02383 | 0.004017 | 2.00E-09 |
| rs2073826 | miR-10a-5p | trans | 9.81E-13 | Nightingale | IDL_C | NA | T | G | -0.02271 | 0.00402 | 1.90E-08 |
| rs2073826 | miR-10a-5p | trans | 9.81E-13 | Nightingale | M_LDL_C_p | NA | T | G | -0.0251 | 0.004165 | 2.00E-09 |
| rs2073826 | miR-10a-5p | trans | 9.81E-13 | Nightingale | XL_VLDL_F | NA | T | G | -0.02522 | 0.004035 | 9.30E-10 |
| rs2073826 | miR-10a-5p | trans | 9.81E-13 | Nightingale | S_VLDL_C | NA | T | G | -0.02205 | 0.004162 | 3.80E-08 |
| rs2073826 | miR-10a-5p | trans | 9.81E-13 | Nightingale | M_VLDL_F | NA | T | G | -0.02753 | 0.00396 | 1.70E-11 |
| rs2073826 | miR-10a-5p | trans | 9.81E-13 | Nightingale | XS_VLDL_C | NA | T | G | -0.02337 | 0.00404 | 4.30E-09 |
| rs2073826 | miR-10a-5p | trans | 9.81E-13 | Nightingale | XXL_VLDL_ | NA | T | G | -0.02299 | 0.004143 | 3.50E-08 |

| | | | | | | | | | | | |
|---|---|---|---|---|---|---|---|---|---|---|---|
| rs2073826 | miR-10a-5p | trans | 9.81E-13 | Nightingale | IDL_PL | NA | T | G | -0.0221 | 0.004017 | 3.70E-08 |
| rs2073826 | miR-10a-5p | trans | 9.81E-13 | Nightingale | XL_VLDL_C | NA | T | G | -0.02688 | 0.003991 | 1.70E-10 |
| rs2073826 | miR-10a-5p | trans | 9.81E-13 | Nightingale | S_LDL_C_p | NA | T | G | -0.03097 | 0.004179 | 5.80E-14 |
| rs2073826 | miR-10a-5p | trans | 9.81E-13 | Nightingale | Clinical_LD | NA | T | G | -0.02321 | 0.004138 | 1.80E-08 |
| rs2073826 | miR-10a-5p | trans | 9.81E-13 | Nightingale | IDL_P | NA | T | G | -0.02376 | 0.004119 | 5.40E-09 |
| rs2073826 | miR-10a-5p | trans | 9.81E-13 | Nightingale | XL_VLDL_T | NA | T | G | 0.030115 | 0.004034 | 5.00E-13 |
| rs2073826 | miR-10a-5p | trans | 9.81E-13 | Nightingale | S_VLDL_CE | NA | T | G | -0.02184 | 0.004157 | 4.30E-08 |
| rs2073826 | miR-10a-5p | trans | 9.81E-13 | Nightingale | IDL_FC | NA | T | G | -0.02522 | 0.00403 | 4.40E-10 |
| rs2073826 | miR-10a-5p | trans | 9.81E-13 | Nightingale | L_VLDL_C_ | NA | T | G | -0.03589 | 0.004065 | 1.40E-18 |
| rs2073826 | miR-10a-5p | trans | 9.81E-13 | Nightingale | XL_VLDL_P | NA | T | G | -0.02215 | 0.004169 | 3.20E-08 |
| rs2073826 | miR-10a-5p | trans | 9.81E-13 | Nightingale | M_VLDL_C | NA | T | G | -0.02288 | 0.003911 | 3.90E-08 |
| rs2073826 | miR-126-3p | trans | 2.24E-18 | Nightingale | M_VLDL_C | NA | T | G | -0.02564 | 0.004125 | 3.00E-10 |
| rs2073826 | miR-126-3p | trans | 2.24E-18 | Nightingale | M_VLDL_C | NA | T | G | -0.0243 | 0.003921 | 4.50E-09 |
| rs2073826 | miR-126-3p | trans | 2.24E-18 | Nightingale | XXL_VLDL_ | NA | T | G | -0.02527 | 0.004169 | 1.70E-09 |
| rs2073826 | miR-126-3p | trans | 2.24E-18 | Nightingale | L_LDL_PL | NA | T | G | -0.0243 | 0.004131 | 3.00E-09 |
| rs2073826 | miR-126-3p | trans | 2.24E-18 | Nightingale | M_VLDL_P | NA | T | G | -0.02867 | 0.003995 | 1.50E-12 |
| rs2073826 | miR-126-3p | trans | 2.24E-18 | Nightingale | XXL_VLDL_ | NA | T | G | 0.025994 | 0.004196 | 3.20E-10 |
| rs2073826 | miR-126-3p | trans | 2.24E-18 | Nightingale | L_VLDL_TG | NA | T | G | 0.033026 | 0.004143 | 4.10E-16 |
| rs2073826 | miR-126-3p | trans | 2.24E-18 | Nightingale | L_VLDL_FC | NA | T | G | -0.0311 | 0.004123 | 2.00E-14 |
| rs2073826 | miR-126-3p | trans | 2.24E-18 | Nightingale | IDL_L | NA | T | G | -0.02234 | 0.004025 | 3.20E-08 |
| rs2073826 | miR-126-3p | trans | 2.24E-18 | Nightingale | S_VLDL_CE | NA | T | G | -0.03199 | 0.004107 | 1.10E-14 |
| rs2073826 | miR-126-3p | trans | 2.24E-18 | Nightingale | M_VLDL_T | NA | T | G | 0.025651 | 0.003934 | 4.80E-10 |
| rs2073826 | miR-126-3p | trans | 2.24E-18 | Nightingale | L_VLDL_CE | NA | T | G | -0.03304 | 0.003983 | 3.30E-16 |
| rs2073826 | miR-126-3p | trans | 2.24E-18 | Nightingale | S_VLDL_TG | NA | T | G | 0.026207 | 0.004029 | 4.00E-10 |
| rs2073826 | miR-126-3p | trans | 2.24E-18 | Nightingale | S_VLDL_C_ | NA | T | G | -0.02855 | 0.004051 | 8.10E-12 |
| rs2073826 | miR-126-3p | trans | 2.24E-18 | Nightingale | Remnant_( | NA | T | G | -0.02176 | 0.004135 | 5.00E-08 |
| rs2073826 | miR-126-3p | trans | 2.24E-18 | Nightingale | XL_VLDL_C | NA | T | G | -0.02645 | 0.003993 | 4.10E-10 |
| rs2073826 | miR-126-3p | trans | 2.24E-18 | Nightingale | M_LDL_P | NA | T | G | -0.02329 | 0.004174 | 1.20E-08 |
| rs2073826 | miR-126-3p | trans | 2.24E-18 | Nightingale | M_VLDL_C | NA | T | G | -0.02333 | 0.004151 | 6.90E-09 |
| rs2073826 | miR-126-3p | trans | 2.24E-18 | Nightingale | XXL_VLDL_ | NA | T | G | -0.02438 | 0.004192 | 8.20E-09 |
| rs2073826 | miR-126-3p | trans | 2.24E-18 | Nightingale | XS_VLDL_C | NA | T | G | -0.02383 | 0.004017 | 2.00E-09 |
| rs2073826 | miR-126-3p | trans | 2.24E-18 | Nightingale | IDL_C | NA | T | G | -0.02271 | 0.00402 | 1.90E-08 |
| rs2073826 | miR-126-3p | trans | 2.24E-18 | Nightingale | M_LDL_C_ | NA | T | G | -0.0251 | 0.004165 | 2.00E-09 |
| rs2073826 | miR-126-3p | trans | 2.24E-18 | Nightingale | XL_VLDL_F | NA | T | G | -0.02522 | 0.004035 | 9.30E-10 |
| rs2073826 | miR-126-3p | trans | 2.24E-18 | Nightingale | S_VLDL_C | NA | T | G | -0.02205 | 0.004162 | 3.80E-08 |
| rs2073826 | miR-126-3p | trans | 2.24E-18 | Nightingale | M_VLDL_F | NA | T | G | -0.02753 | 0.00396 | 1.70E-11 |
| rs2073826 | miR-126-3p | trans | 2.24E-18 | Nightingale | XS_VLDL_C | NA | T | G | -0.02337 | 0.00404 | 4.30E-09 |
| rs2073826 | miR-126-3p | trans | 2.24E-18 | Nightingale | XXL_VLDL_ | NA | T | G | -0.02299 | 0.004143 | 3.50E-08 |
| rs2073826 | miR-126-3p | trans | 2.24E-18 | Nightingale | IDL_PL | NA | T | G | -0.0221 | 0.004017 | 3.70E-08 |
| rs2073826 | miR-126-3p | trans | 2.24E-18 | Nightingale | XL_VLDL_C | NA | T | G | -0.02688 | 0.003991 | 1.70E-10 |
| rs2073826 | miR-126-3p | trans | 2.24E-18 | Nightingale | S_LDL_C_p | NA | T | G | -0.03097 | 0.004179 | 5.80E-14 |
| rs2073826 | miR-126-3p | trans | 2.24E-18 | Nightingale | Clinical_LD | NA | T | G | -0.02321 | 0.004138 | 1.80E-08 |
| rs2073826 | miR-126-3p | trans | 2.24E-18 | Nightingale | IDL_P | NA | T | G | -0.02376 | 0.004119 | 5.40E-09 |
| rs2073826 | miR-126-3p | trans | 2.24E-18 | Nightingale | XL_VLDL_T | NA | T | G | 0.030115 | 0.004034 | 5.00E-13 |
| rs2073826 | miR-126-3p | trans | 2.24E-18 | Nightingale | S_VLDL_CE | NA | T | G | -0.02184 | 0.004157 | 4.30E-08 |
| rs2073826 | miR-126-3p | trans | 2.24E-18 | Nightingale | IDL_FC | NA | T | G | -0.02522 | 0.00403 | 4.40E-10 |
| rs2073826 | miR-126-3p | trans | 2.24E-18 | Nightingale | L_VLDL_C_ | NA | T | G | -0.03589 | 0.004065 | 1.40E-18 |
| rs2073826 | miR-126-3p | trans | 2.24E-18 | Nightingale | XL_VLDL_P | NA | T | G | -0.02215 | 0.004169 | 3.20E-08 |
| rs2073826 | miR-126-3p | trans | 2.24E-18 | Nightingale | M_VLDL_C | NA | T | G | -0.02288 | 0.003911 | 3.90E-08 |
| rs2073827 | miR-10a-5p | trans | 8.39E-13 | Nightingale | Clinical_LD | NA | C | G | -0.02314 | 0.004137 | 2.00E-08 |
| rs2073827 | miR-10a-5p | trans | 8.39E-13 | Nightingale | S_VLDL_C_ | NA | C | G | -0.02855 | 0.00405 | 7.80E-12 |
| rs2073827 | miR-10a-5p | trans | 8.39E-13 | Nightingale | XL_VLDL_C | NA | C | G | -0.02697 | 0.00399 | 1.40E-10 |
| rs2073827 | miR-10a-5p | trans | 8.39E-13 | Nightingale | M_VLDL_T | NA | C | G | 0.025628 | 0.003932 | 5.00E-10 |
| rs2073827 | miR-10a-5p | trans | 8.39E-13 | Nightingale | XS_VLDL_C | NA | C | G | -0.02367 | 0.004016 | 2.50E-09 |
| rs2073827 | miR-10a-5p | trans | 8.39E-13 | Nightingale | XXL_VLDL_ | NA | C | G | -0.0254 | 0.004168 | 1.30E-09 |
| rs2073827 | miR-10a-5p | trans | 8.39E-13 | Nightingale | M_VLDL_C | NA | C | G | -0.02561 | 0.004124 | 3.20E-10 |
| rs2073827 | miR-10a-5p | trans | 8.39E-13 | Nightingale | IDL_L | NA | C | G | -0.0222 | 0.004024 | 3.90E-08 |
| rs2073827 | miR-10a-5p | trans | 8.39E-13 | Nightingale | S_LDL_C_p | NA | C | G | -0.03105 | 0.004178 | 4.90E-14 |
| rs2073827 | miR-10a-5p | trans | 8.39E-13 | Nightingale | IDL_P | NA | C | G | -0.02362 | 0.004118 | 6.60E-09 |
| rs2073827 | miR-10a-5p | trans | 8.39E-13 | Nightingale | XXL_VLDL_ | NA | C | G | 0.026138 | 0.004195 | 2.40E-10 |
| rs2073827 | miR-10a-5p | trans | 8.39E-13 | Nightingale | S_VLDL_TG | NA | C | G | 0.026226 | 0.004028 | 3.80E-10 |
| rs2073827 | miR-10a-5p | trans | 8.39E-13 | Nightingale | XXL_VLDL_ | NA | C | G | -0.02313 | 0.004142 | 2.70E-08 |
| rs2073827 | miR-10a-5p | trans | 8.39E-13 | Nightingale | L_VLDL_C_ | NA | C | G | -0.03583 | 0.004064 | 1.40E-18 |

| | | | | | | | | | | | |
|---|---|---|---|---|---|---|---|---|---|---|---|
| rs2073827 | miR-10a-5p | trans | 8.39E-13 | Nightingale | M_VLDL_F | NA | C | G | -0.02746 | 0.003959 | 2.00E-11 |
| rs2073827 | miR-10a-5p | trans | 8.39E-13 | Nightingale | XL_VLDL_F | NA | C | G | -0.02522 | 0.004034 | 9.00E-10 |
| rs2073827 | miR-10a-5p | trans | 8.39E-13 | Nightingale | S_VLDL_C | NA | C | G | -0.02194 | 0.004161 | 4.50E-08 |
| rs2073827 | miR-10a-5p | trans | 8.39E-13 | Nightingale | M_VLDL_C | NA | C | G | -0.02291 | 0.00391 | 3.80E-08 |
| rs2073827 | miR-10a-5p | trans | 8.39E-13 | Nightingale | XL_VLDL_P | NA | C | G | -0.02208 | 0.004168 | 3.60E-08 |
| rs2073827 | miR-10a-5p | trans | 8.39E-13 | Nightingale | XL_VLDL_T | NA | C | G | 0.030177 | 0.004033 | 4.30E-13 |
| rs2073827 | miR-10a-5p | trans | 8.39E-13 | Nightingale | XS_VLDL_C | NA | C | G | -0.0232 | 0.004039 | 5.60E-09 |
| rs2073827 | miR-10a-5p | trans | 8.39E-13 | Nightingale | IDL_PL | NA | C | G | -0.02196 | 0.004016 | 4.50E-08 |
| rs2073827 | miR-10a-5p | trans | 8.39E-13 | Nightingale | M_VLDL_C | NA | C | G | -0.02327 | 0.00415 | 7.80E-09 |
| rs2073827 | miR-10a-5p | trans | 8.39E-13 | Nightingale | L_LDL_PL | NA | C | G | -0.02421 | 0.00413 | 3.40E-09 |
| rs2073827 | miR-10a-5p | trans | 8.39E-13 | Nightingale | M_LDL_C_ | NA | C | G | -0.0251 | 0.004164 | 1.90E-09 |
| rs2073827 | miR-10a-5p | trans | 8.39E-13 | Nightingale | IDL_C | NA | C | G | -0.02259 | 0.004019 | 2.30E-08 |
| rs2073827 | miR-10a-5p | trans | 8.39E-13 | Nightingale | L_VLDL_TG | NA | C | G | 0.032936 | 0.004142 | 4.70E-16 |
| rs2073827 | miR-10a-5p | trans | 8.39E-13 | Nightingale | XL_VLDL_C | NA | C | G | -0.02655 | 0.003992 | 3.40E-10 |
| rs2073827 | miR-10a-5p | trans | 8.39E-13 | Nightingale | M_VLDL_P | NA | C | G | -0.02857 | 0.003994 | 1.80E-12 |
| rs2073827 | miR-10a-5p | trans | 8.39E-13 | Nightingale | L_VLDL_CE | NA | C | G | -0.03303 | 0.003982 | 3.20E-16 |
| rs2073827 | miR-10a-5p | trans | 8.39E-13 | Nightingale | M_VLDL_C | NA | C | G | -0.0243 | 0.00392 | 4.60E-09 |
| rs2073827 | miR-10a-5p | trans | 8.39E-13 | Nightingale | S_VLDL_CE | NA | C | G | -0.03195 | 0.004105 | 1.10E-14 |
| rs2073827 | miR-10a-5p | trans | 8.39E-13 | Nightingale | XXL_VLDL_ | NA | C | G | -0.02446 | 0.004191 | 6.80E-09 |
| rs2073827 | miR-10a-5p | trans | 8.39E-13 | Nightingale | IDL_FC | NA | C | G | -0.0251 | 0.004029 | 5.20E-10 |
| rs2073827 | miR-10a-5p | trans | 8.39E-13 | Nightingale | L_VLDL_FC | NA | C | G | -0.031 | 0.004121 | 2.50E-14 |
| rs2073827 | miR-10a-5p | trans | 8.39E-13 | Nightingale | M_LDL_P | NA | C | G | -0.02313 | 0.004173 | 1.50E-08 |
| rs2073827 | miR-126-3p | trans | 2.37E-18 | Nightingale | Clinical_LD | NA | C | G | -0.02314 | 0.004137 | 2.00E-08 |
| rs2073827 | miR-126-3p | trans | 2.37E-18 | Nightingale | S_VLDL_C_ | NA | C | G | -0.02855 | 0.00405 | 7.80E-12 |
| rs2073827 | miR-126-3p | trans | 2.37E-18 | Nightingale | XL_VLDL_C | NA | C | G | -0.02697 | 0.00399 | 1.40E-10 |
| rs2073827 | miR-126-3p | trans | 2.37E-18 | Nightingale | M_VLDL_T | NA | C | G | 0.025628 | 0.003932 | 5.00E-10 |
| rs2073827 | miR-126-3p | trans | 2.37E-18 | Nightingale | XS_VLDL_C | NA | C | G | -0.02367 | 0.004016 | 2.50E-09 |
| rs2073827 | miR-126-3p | trans | 2.37E-18 | Nightingale | XXL_VLDL_ | NA | C | G | -0.0254 | 0.004168 | 1.30E-09 |
| rs2073827 | miR-126-3p | trans | 2.37E-18 | Nightingale | M_VLDL_C | NA | C | G | -0.02561 | 0.004124 | 3.20E-10 |
| rs2073827 | miR-126-3p | trans | 2.37E-18 | Nightingale | IDL_L | NA | C | G | -0.0222 | 0.004024 | 3.90E-08 |
| rs2073827 | miR-126-3p | trans | 2.37E-18 | Nightingale | S_LDL_C_p | NA | C | G | -0.03105 | 0.004178 | 4.90E-14 |
| rs2073827 | miR-126-3p | trans | 2.37E-18 | Nightingale | IDL_P | NA | C | G | -0.02362 | 0.004118 | 6.60E-09 |
| rs2073827 | miR-126-3p | trans | 2.37E-18 | Nightingale | XXL_VLDL_ | NA | C | G | 0.026138 | 0.004195 | 2.40E-10 |
| rs2073827 | miR-126-3p | trans | 2.37E-18 | Nightingale | S_VLDL_TG | NA | C | G | 0.026226 | 0.004028 | 3.80E-10 |
| rs2073827 | miR-126-3p | trans | 2.37E-18 | Nightingale | XXL_VLDL_ | NA | C | G | -0.02313 | 0.004142 | 2.70E-08 |
| rs2073827 | miR-126-3p | trans | 2.37E-18 | Nightingale | L_VLDL_C_ | NA | C | G | -0.03583 | 0.004064 | 1.40E-18 |
| rs2073827 | miR-126-3p | trans | 2.37E-18 | Nightingale | M_VLDL_F | NA | C | G | -0.02746 | 0.003959 | 2.00E-11 |
| rs2073827 | miR-126-3p | trans | 2.37E-18 | Nightingale | XL_VLDL_F | NA | C | G | -0.02522 | 0.004034 | 9.00E-10 |
| rs2073827 | miR-126-3p | trans | 2.37E-18 | Nightingale | S_VLDL_C | NA | C | G | -0.02194 | 0.004161 | 4.50E-08 |
| rs2073827 | miR-126-3p | trans | 2.37E-18 | Nightingale | M_VLDL_C | NA | C | G | -0.02291 | 0.00391 | 3.80E-08 |
| rs2073827 | miR-126-3p | trans | 2.37E-18 | Nightingale | XL_VLDL_P | NA | C | G | -0.02208 | 0.004168 | 3.60E-08 |
| rs2073827 | miR-126-3p | trans | 2.37E-18 | Nightingale | XL_VLDL_T | NA | C | G | 0.030177 | 0.004033 | 4.30E-13 |
| rs2073827 | miR-126-3p | trans | 2.37E-18 | Nightingale | XS_VLDL_C | NA | C | G | -0.0232 | 0.004039 | 5.60E-09 |
| rs2073827 | miR-126-3p | trans | 2.37E-18 | Nightingale | IDL_PL | NA | C | G | -0.02196 | 0.004016 | 4.50E-08 |
| rs2073827 | miR-126-3p | trans | 2.37E-18 | Nightingale | M_VLDL_C | NA | C | G | -0.02327 | 0.00415 | 7.80E-09 |
| rs2073827 | miR-126-3p | trans | 2.37E-18 | Nightingale | L_LDL_PL | NA | C | G | -0.02421 | 0.00413 | 3.40E-09 |
| rs2073827 | miR-126-3p | trans | 2.37E-18 | Nightingale | M_LDL_C_ | NA | C | G | -0.0251 | 0.004164 | 1.90E-09 |
| rs2073827 | miR-126-3p | trans | 2.37E-18 | Nightingale | IDL_C | NA | C | G | -0.02259 | 0.004019 | 2.30E-08 |
| rs2073827 | miR-126-3p | trans | 2.37E-18 | Nightingale | L_VLDL_TG | NA | C | G | 0.032936 | 0.004142 | 4.70E-16 |
| rs2073827 | miR-126-3p | trans | 2.37E-18 | Nightingale | XL_VLDL_C | NA | C | G | -0.02655 | 0.003992 | 3.40E-10 |
| rs2073827 | miR-126-3p | trans | 2.37E-18 | Nightingale | M_VLDL_P | NA | C | G | -0.02857 | 0.003994 | 1.80E-12 |
| rs2073827 | miR-126-3p | trans | 2.37E-18 | Nightingale | L_VLDL_CE | NA | C | G | -0.03303 | 0.003982 | 3.20E-16 |
| rs2073827 | miR-126-3p | trans | 2.37E-18 | Nightingale | M_VLDL_C | NA | C | G | -0.0243 | 0.00392 | 4.60E-09 |
| rs2073827 | miR-126-3p | trans | 2.37E-18 | Nightingale | S_VLDL_CE | NA | C | G | -0.03195 | 0.004105 | 1.10E-14 |
| rs2073827 | miR-126-3p | trans | 2.37E-18 | Nightingale | XXL_VLDL_ | NA | C | G | -0.02446 | 0.004191 | 6.80E-09 |
| rs2073827 | miR-126-3p | trans | 2.37E-18 | Nightingale | IDL_FC | NA | C | G | -0.0251 | 0.004029 | 5.20E-10 |
| rs2073827 | miR-126-3p | trans | 2.37E-18 | Nightingale | L_VLDL_FC | NA | C | G | -0.031 | 0.004121 | 2.50E-14 |
| rs2073827 | miR-126-3p | trans | 2.37E-18 | Nightingale | M_LDL_P | NA | C | G | -0.02313 | 0.004173 | 1.50E-08 |
| rs2073828 | miR-126-3p | trans | 2.37E-18 | Nightingale | M_VLDL_T | NA | A | G | 0.025621 | 0.003932 | 5.10E-10 |
| rs2073828 | miR-126-3p | trans | 2.37E-18 | Nightingale | S_VLDL_C_ | NA | A | G | -0.02855 | 0.00405 | 7.90E-12 |
| rs2073828 | miR-126-3p | trans | 2.37E-18 | Nightingale | XL_VLDL_F | NA | A | G | -0.02519 | 0.004034 | 9.50E-10 |
| rs2073828 | miR-126-3p | trans | 2.37E-18 | Nightingale | IDL_L | NA | A | G | -0.02221 | 0.004024 | 3.90E-08 |
| rs2073828 | miR-126-3p | trans | 2.37E-18 | Nightingale | S_VLDL_CE | NA | A | G | -0.03195 | 0.004105 | 1.10E-14 |

| | | | | | | | | | | | |
|---|---|---|---|---|---|---|---|---|---|---|---|
| rs2073828 | miR-126-3p | trans | 2.37E-18 | Nightingale | S_VLDL_TG | NA | A | G | 0.026226 | 0.004028 | 3.90E-10 |
| rs2073828 | miR-126-3p | trans | 2.37E-18 | Nightingale | XXL_VLDL_ | NA | A | G | -0.02443 | 0.004191 | 7.20E-09 |
| rs2073828 | miR-126-3p | trans | 2.37E-18 | Nightingale | XL_VLDL_P | NA | A | G | -0.02205 | 0.004168 | 3.70E-08 |
| rs2073828 | miR-126-3p | trans | 2.37E-18 | Nightingale | XS_VLDL_C | NA | A | G | -0.02366 | 0.004016 | 2.50E-09 |
| rs2073828 | miR-126-3p | trans | 2.37E-18 | Nightingale | Clinical_LD | NA | A | G | -0.02314 | 0.004137 | 2.00E-08 |
| rs2073828 | miR-126-3p | trans | 2.37E-18 | Nightingale | IDL_FC | NA | A | G | -0.0251 | 0.004029 | 5.30E-10 |
| rs2073828 | miR-126-3p | trans | 2.37E-18 | Nightingale | XXL_VLDL_ | NA | A | G | -0.02536 | 0.004168 | 1.40E-09 |
| rs2073828 | miR-126-3p | trans | 2.37E-18 | Nightingale | S_LDL_C_p | NA | A | G | -0.03102 | 0.004177 | 5.10E-14 |
| rs2073828 | miR-126-3p | trans | 2.37E-18 | Nightingale | L_VLDL_TG | NA | A | G | 0.032911 | 0.004142 | 5.00E-16 |
| rs2073828 | miR-126-3p | trans | 2.37E-18 | Nightingale | L_VLDL_CE | NA | A | G | -0.033 | 0.003982 | 3.40E-16 |
| rs2073828 | miR-126-3p | trans | 2.37E-18 | Nightingale | S_VLDL_C | NA | A | G | -0.02193 | 0.004161 | 4.60E-08 |
| rs2073828 | miR-126-3p | trans | 2.37E-18 | Nightingale | M_LDL_C_p | NA | A | G | -0.02508 | 0.004164 | 2.00E-09 |
| rs2073828 | miR-126-3p | trans | 2.37E-18 | Nightingale | XXL_VLDL_ | NA | A | G | -0.0231 | 0.004142 | 2.90E-08 |
| rs2073828 | miR-126-3p | trans | 2.37E-18 | Nightingale | M_VLDL_F | NA | A | G | -0.02745 | 0.003959 | 2.00E-11 |
| rs2073828 | miR-126-3p | trans | 2.37E-18 | Nightingale | M_VLDL_C | NA | A | G | -0.02429 | 0.00392 | 4.60E-09 |
| rs2073828 | miR-126-3p | trans | 2.37E-18 | Nightingale | IDL_PL | NA | A | G | -0.02195 | 0.004016 | 4.50E-08 |
| rs2073828 | miR-126-3p | trans | 2.37E-18 | Nightingale | L_LDL_PL | NA | A | G | -0.02421 | 0.00413 | 3.40E-09 |
| rs2073828 | miR-126-3p | trans | 2.37E-18 | Nightingale | M_LDL_P | NA | A | G | -0.02313 | 0.004173 | 1.50E-08 |
| rs2073828 | miR-126-3p | trans | 2.37E-18 | Nightingale | IDL_P | NA | A | G | -0.02364 | 0.004118 | 6.50E-09 |
| rs2073828 | miR-126-3p | trans | 2.37E-18 | Nightingale | M_VLDL_C | NA | A | G | -0.02559 | 0.004124 | 3.30E-10 |
| rs2073828 | miR-126-3p | trans | 2.37E-18 | Nightingale | XS_VLDL_C | NA | A | G | -0.02319 | 0.004039 | 5.60E-09 |
| rs2073828 | miR-126-3p | trans | 2.37E-18 | Nightingale | L_VLDL_C_ | NA | A | G | -0.03581 | 0.004064 | 1.50E-18 |
| rs2073828 | miR-126-3p | trans | 2.37E-18 | Nightingale | XL_VLDL_C | NA | A | G | -0.02695 | 0.00399 | 1.50E-10 |
| rs2073828 | miR-126-3p | trans | 2.37E-18 | Nightingale | M_VLDL_P | NA | A | G | -0.02856 | 0.003994 | 1.80E-12 |
| rs2073828 | miR-126-3p | trans | 2.37E-18 | Nightingale | M_VLDL_C | NA | A | G | -0.02326 | 0.00415 | 8.00E-09 |
| rs2073828 | miR-126-3p | trans | 2.37E-18 | Nightingale | XXL_VLDL_ | NA | A | G | 0.026108 | 0.004195 | 2.50E-10 |
| rs2073828 | miR-126-3p | trans | 2.37E-18 | Nightingale | XL_VLDL_T | NA | A | G | 0.030159 | 0.004033 | 4.50E-13 |
| rs2073828 | miR-126-3p | trans | 2.37E-18 | Nightingale | L_VLDL_FC | NA | A | G | -0.03098 | 0.004121 | 2.60E-14 |
| rs2073828 | miR-126-3p | trans | 2.37E-18 | Nightingale | M_VLDL_C | NA | A | G | -0.0229 | 0.003909 | 3.80E-08 |
| rs2073828 | miR-126-3p | trans | 2.37E-18 | Nightingale | XL_VLDL_C | NA | A | G | -0.02654 | 0.003992 | 3.50E-10 |
| rs2073828 | miR-126-3p | trans | 2.37E-18 | Nightingale | IDL_C | NA | A | G | -0.0226 | 0.004019 | 2.30E-08 |
| rs2073828 | miR-10a-5p | trans | 8.39E-13 | Nightingale | M_VLDL_T | NA | A | G | 0.025621 | 0.003932 | 5.10E-10 |
| rs2073828 | miR-10a-5p | trans | 8.39E-13 | Nightingale | S_VLDL_C_ | NA | A | G | -0.02855 | 0.00405 | 7.90E-12 |
| rs2073828 | miR-10a-5p | trans | 8.39E-13 | Nightingale | XL_VLDL_F | NA | A | G | -0.02519 | 0.004034 | 9.50E-10 |
| rs2073828 | miR-10a-5p | trans | 8.39E-13 | Nightingale | IDL_L | NA | A | G | -0.02221 | 0.004024 | 3.90E-08 |
| rs2073828 | miR-10a-5p | trans | 8.39E-13 | Nightingale | S_VLDL_CE | NA | A | G | -0.03195 | 0.004105 | 1.10E-14 |
| rs2073828 | miR-10a-5p | trans | 8.39E-13 | Nightingale | S_VLDL_TG | NA | A | G | 0.026226 | 0.004028 | 3.90E-10 |
| rs2073828 | miR-10a-5p | trans | 8.39E-13 | Nightingale | XXL_VLDL_ | NA | A | G | -0.02443 | 0.004191 | 7.20E-09 |
| rs2073828 | miR-10a-5p | trans | 8.39E-13 | Nightingale | XL_VLDL_P | NA | A | G | -0.02205 | 0.004168 | 3.70E-08 |
| rs2073828 | miR-10a-5p | trans | 8.39E-13 | Nightingale | XS_VLDL_C | NA | A | G | -0.02366 | 0.004016 | 2.50E-09 |
| rs2073828 | miR-10a-5p | trans | 8.39E-13 | Nightingale | Clinical_LD | NA | A | G | -0.02314 | 0.004137 | 2.00E-08 |
| rs2073828 | miR-10a-5p | trans | 8.39E-13 | Nightingale | IDL_FC | NA | A | G | -0.0251 | 0.004029 | 5.30E-10 |
| rs2073828 | miR-10a-5p | trans | 8.39E-13 | Nightingale | XXL_VLDL_ | NA | A | G | -0.02536 | 0.004168 | 1.40E-09 |
| rs2073828 | miR-10a-5p | trans | 8.39E-13 | Nightingale | S_LDL_C_p | NA | A | G | -0.03102 | 0.004177 | 5.10E-14 |
| rs2073828 | miR-10a-5p | trans | 8.39E-13 | Nightingale | L_VLDL_TG | NA | A | G | 0.032911 | 0.004142 | 5.00E-16 |
| rs2073828 | miR-10a-5p | trans | 8.39E-13 | Nightingale | L_VLDL_CE | NA | A | G | -0.033 | 0.003982 | 3.40E-16 |
| rs2073828 | miR-10a-5p | trans | 8.39E-13 | Nightingale | S_VLDL_C | NA | A | G | -0.02193 | 0.004161 | 4.60E-08 |
| rs2073828 | miR-10a-5p | trans | 8.39E-13 | Nightingale | M_LDL_C_p | NA | A | G | -0.02508 | 0.004164 | 2.00E-09 |
| rs2073828 | miR-10a-5p | trans | 8.39E-13 | Nightingale | XXL_VLDL_ | NA | A | G | -0.0231 | 0.004142 | 2.90E-08 |
| rs2073828 | miR-10a-5p | trans | 8.39E-13 | Nightingale | M_VLDL_F | NA | A | G | -0.02745 | 0.003959 | 2.00E-11 |
| rs2073828 | miR-10a-5p | trans | 8.39E-13 | Nightingale | M_VLDL_C | NA | A | G | -0.02429 | 0.00392 | 4.60E-09 |
| rs2073828 | miR-10a-5p | trans | 8.39E-13 | Nightingale | IDL_PL | NA | A | G | -0.02195 | 0.004016 | 4.50E-08 |
| rs2073828 | miR-10a-5p | trans | 8.39E-13 | Nightingale | L_LDL_PL | NA | A | G | -0.02421 | 0.00413 | 3.40E-09 |
| rs2073828 | miR-10a-5p | trans | 8.39E-13 | Nightingale | M_LDL_P | NA | A | G | -0.02313 | 0.004173 | 1.50E-08 |
| rs2073828 | miR-10a-5p | trans | 8.39E-13 | Nightingale | IDL_P | NA | A | G | -0.02364 | 0.004118 | 6.50E-09 |
| rs2073828 | miR-10a-5p | trans | 8.39E-13 | Nightingale | M_VLDL_C | NA | A | G | -0.02559 | 0.004124 | 3.30E-10 |
| rs2073828 | miR-10a-5p | trans | 8.39E-13 | Nightingale | XS_VLDL_C | NA | A | G | -0.02319 | 0.004039 | 5.60E-09 |
| rs2073828 | miR-10a-5p | trans | 8.39E-13 | Nightingale | L_VLDL_C_ | NA | A | G | -0.03581 | 0.004064 | 1.50E-18 |
| rs2073828 | miR-10a-5p | trans | 8.39E-13 | Nightingale | XL_VLDL_C | NA | A | G | -0.02695 | 0.00399 | 1.50E-10 |
| rs2073828 | miR-10a-5p | trans | 8.39E-13 | Nightingale | M_VLDL_P | NA | A | G | -0.02856 | 0.003994 | 1.80E-12 |
| rs2073828 | miR-10a-5p | trans | 8.39E-13 | Nightingale | M_VLDL_C | NA | A | G | -0.02326 | 0.00415 | 8.00E-09 |
| rs2073828 | miR-10a-5p | trans | 8.39E-13 | Nightingale | XXL_VLDL_ | NA | A | G | 0.026108 | 0.004195 | 2.50E-10 |
| rs2073828 | miR-10a-5p | trans | 8.39E-13 | Nightingale | XL_VLDL_T | NA | A | G | 0.030159 | 0.004033 | 4.50E-13 |

| | | | | | | | | | | | |
|---|---|---|---|---|---|---|---|---|---|---|---|
| rs2073828 | miR-10a-5p | trans | 8.39E-13 | Nightingale | L_VLDL_FC | NA | A | G | -0.03098 | 0.004121 | 2.60E-14 |
| rs2073828 | miR-10a-5p | trans | 8.39E-13 | Nightingale | M_VLDL_C | NA | A | G | -0.0229 | 0.003909 | 3.80E-08 |
| rs2073828 | miR-10a-5p | trans | 8.39E-13 | Nightingale | XL_VLDL_C | NA | A | G | -0.02654 | 0.003992 | 3.50E-10 |
| rs2073828 | miR-10a-5p | trans | 8.39E-13 | Nightingale | IDL_C | NA | A | G | -0.0226 | 0.004019 | 2.30E-08 |
| rs2343592 | miR-145-5p | trans | 1.93E-08 | Nightingale | Lactate | NA | G | A | -0.02909 | 0.004655 | 4.10E-10 |
| rs2343592 | miR-145-5p | trans | 1.93E-08 | Nightingale | Pyruvate | NA | G | A | -0.03424 | 0.004656 | 1.90E-13 |
| rs2343592 | miR-143-3p | trans | 5.91E-11 | Nightingale | Lactate | NA | G | A | -0.02909 | 0.004655 | 4.10E-10 |
| rs2343592 | miR-143-3p | trans | 5.91E-11 | Nightingale | Pyruvate | NA | G | A | -0.03424 | 0.004656 | 1.90E-13 |
| rs2519093 | let-7c-5p | trans | 4.94E-22 | Nightingale | S_HDL_CE | NA | T | C | 0.053505 | 0.005345 | 7.30E-24 |
| rs2519093 | let-7c-5p | trans | 4.94E-22 | Nightingale | L_LDL_TG_ | NA | T | C | -0.04178 | 0.005234 | 5.50E-15 |
| rs2519093 | let-7c-5p | trans | 4.94E-22 | Nightingale | M_VLDL_P | NA | T | C | 0.04488 | 0.005328 | 1.70E-18 |
| rs2519093 | let-7c-5p | trans | 4.94E-22 | Nightingale | IDL_C | NA | T | C | 0.060311 | 0.005154 | 6.00E-32 |
| rs2519093 | let-7c-5p | trans | 4.94E-22 | Nightingale | M_VLDL_C | NA | T | C | 0.066274 | 0.005288 | 7.60E-37 |
| rs2519093 | let-7c-5p | trans | 4.94E-22 | Nightingale | Total_FC | NA | T | C | 0.055093 | 0.005213 | 7.90E-28 |
| rs2519093 | let-7c-5p | trans | 4.94E-22 | Nightingale | S_VLDL_C_ | NA | T | C | 0.070806 | 0.005193 | 5.60E-42 |
| rs2519093 | let-7c-5p | trans | 4.94E-22 | Nightingale | LA_pct | NA | T | C | 0.031876 | 0.005218 | 1.10E-09 |
| rs2519093 | let-7c-5p | trans | 4.94E-22 | Nightingale | S_VLDL_L | NA | T | C | 0.035666 | 0.005285 | 4.30E-13 |
| rs2519093 | let-7c-5p | trans | 4.94E-22 | Nightingale | M_VLDL_C | NA | T | C | 0.061815 | 0.005321 | 1.60E-32 |
| rs2519093 | let-7c-5p | trans | 4.94E-22 | Nightingale | XS_VLDL_F | NA | T | C | 0.051132 | 0.005298 | 6.20E-21 |
| rs2519093 | let-7c-5p | trans | 4.94E-22 | Nightingale | IDL_P | NA | T | C | 0.065267 | 0.00528 | 1.10E-35 |
| rs2519093 | let-7c-5p | trans | 4.94E-22 | Nightingale | S_VLDL_CE | NA | T | C | 0.05821 | 0.005329 | 1.10E-29 |
| rs2519093 | let-7c-5p | trans | 4.94E-22 | Nightingale | XS_VLDL_L | NA | T | C | 0.048684 | 0.005245 | 1.40E-22 |
| rs2519093 | let-7c-5p | trans | 4.94E-22 | Nightingale | S_LDL_CE | NA | T | C | 0.051776 | 0.005339 | 4.00E-23 |
| rs2519093 | let-7c-5p | trans | 4.94E-22 | Nightingale | IDL_L | NA | T | C | 0.059774 | 0.00516 | 1.10E-31 |
| rs2519093 | let-7c-5p | trans | 4.94E-22 | Nightingale | Total_CE | NA | T | C | 0.05575 | 0.005147 | 2.40E-28 |
| rs2519093 | let-7c-5p | trans | 4.94E-22 | Nightingale | Sphingomy | NA | T | C | 0.047648 | 0.004989 | 2.20E-21 |
| rs2519093 | let-7c-5p | trans | 4.94E-22 | Nightingale | PUFA | NA | T | C | 0.036949 | 0.005205 | 1.30E-13 |
| rs2519093 | let-7c-5p | trans | 4.94E-22 | Nightingale | XS_VLDL_C | NA | T | C | 0.040361 | 0.005035 | 1.00E-14 |
| rs2519093 | let-7c-5p | trans | 4.94E-22 | Nightingale | IDL_CE | NA | T | C | 0.058439 | 0.005152 | 4.70E-30 |
| rs2519093 | let-7c-5p | trans | 4.94E-22 | Nightingale | VLDL_CE | NA | T | C | 0.053783 | 0.005338 | 2.10E-25 |
| rs2519093 | let-7c-5p | trans | 4.94E-22 | Nightingale | S_HDL_PL | NA | T | C | 0.037688 | 0.00529 | 1.40E-13 |
| rs2519093 | let-7c-5p | trans | 4.94E-22 | Nightingale | Omega_6 | NA | T | C | 0.041735 | 0.005211 | 5.40E-17 |
| rs2519093 | let-7c-5p | trans | 4.94E-22 | Nightingale | S_HDL_C | NA | T | C | 0.052616 | 0.005327 | 2.20E-23 |
| rs2519093 | let-7c-5p | trans | 4.94E-22 | Nightingale | M_VLDL_C | NA | T | C | 0.056108 | 0.005013 | 3.90E-28 |
| rs2519093 | let-7c-5p | trans | 4.94E-22 | Nightingale | GlycA | NA | T | C | 0.031449 | 0.005314 | 4.90E-09 |
| rs2519093 | let-7c-5p | trans | 4.94E-22 | Nightingale | XS_VLDL_C | NA | T | C | 0.043421 | 0.005061 | 7.70E-17 |
| rs2519093 | let-7c-5p | trans | 4.94E-22 | Nightingale | M_VLDL_P | NA | T | C | 0.043347 | 0.005323 | 1.90E-17 |
| rs2519093 | let-7c-5p | trans | 4.94E-22 | Nightingale | XS_VLDL_P | NA | T | C | -0.02949 | 0.005207 | 5.00E-08 |
| rs2519093 | let-7c-5p | trans | 4.94E-22 | Nightingale | S_VLDL_PL | NA | T | C | 0.050234 | 0.00511 | 4.10E-22 |
| rs2519093 | let-7c-5p | trans | 4.94E-22 | Nightingale | L_LDL_C | NA | T | C | 0.060158 | 0.005276 | 1.10E-30 |
| rs2519093 | let-7c-5p | trans | 4.94E-22 | Nightingale | Total_L | NA | T | C | 0.042371 | 0.00525 | 4.30E-17 |
| rs2519093 | let-7c-5p | trans | 4.94E-22 | Nightingale | Phosphogly | NA | T | C | 0.031931 | 0.005054 | 3.40E-11 |
| rs2519093 | let-7c-5p | trans | 4.94E-22 | Nightingale | L_VLDL_C_ | NA | T | C | 0.082094 | 0.005212 | 9.00E-57 |
| rs2519093 | let-7c-5p | trans | 4.94E-22 | Nightingale | L_VLDL_CE | NA | T | C | 0.031525 | 0.005274 | 1.30E-10 |
| rs2519093 | let-7c-5p | trans | 4.94E-22 | Nightingale | VLDL_P | NA | T | C | 0.038356 | 0.005308 | 2.10E-14 |
| rs2519093 | let-7c-5p | trans | 4.94E-22 | Nightingale | M_LDL_P | NA | T | C | 0.057192 | 0.005351 | 1.30E-27 |
| rs2519093 | let-7c-5p | trans | 4.94E-22 | Nightingale | S_VLDL_FC | NA | T | C | 0.052723 | 0.005099 | 2.50E-24 |
| rs2519093 | let-7c-5p | trans | 4.94E-22 | Nightingale | S_HDL_P | NA | T | C | 0.055391 | 0.005337 | 1.10E-25 |
| rs2519093 | let-7c-5p | trans | 4.94E-22 | Nightingale | S_LDL_L | NA | T | C | 0.048584 | 0.005335 | 3.30E-20 |
| rs2519093 | let-7c-5p | trans | 4.94E-22 | Nightingale | VLDL_FC | NA | T | C | 0.033679 | 0.005297 | 1.10E-11 |
| rs2519093 | let-7c-5p | trans | 4.94E-22 | Nightingale | XL_VLDL_C | NA | T | C | 0.029731 | 0.005254 | 9.50E-10 |
| rs2519093 | let-7c-5p | trans | 4.94E-22 | Nightingale | XXL_VLDL_ | NA | T | C | 0.046178 | 0.005317 | 1.50E-18 |
| rs2519093 | let-7c-5p | trans | 4.94E-22 | Nightingale | XXL_VLDL_ | NA | T | C | -0.05831 | 0.005385 | 5.10E-28 |
| rs2519093 | let-7c-5p | trans | 4.94E-22 | Nightingale | LDL_P | NA | T | C | 0.053373 | 0.005333 | 2.30E-24 |
| rs2519093 | let-7c-5p | trans | 4.94E-22 | Nightingale | XS_VLDL_P | NA | T | C | 0.044763 | 0.005262 | 2.50E-19 |
| rs2519093 | let-7c-5p | trans | 4.94E-22 | Nightingale | ApoB | NA | T | C | 0.055887 | 0.005329 | 1.10E-26 |
| rs2519093 | let-7c-5p | trans | 4.94E-22 | Nightingale | L_LDL_CE | NA | T | C | 0.058613 | 0.005289 | 3.70E-29 |
| rs2519093 | let-7c-5p | trans | 4.94E-22 | Nightingale | S_LDL_C | NA | T | C | 0.05408 | 0.005341 | 1.20E-24 |
| rs2519093 | let-7c-5p | trans | 4.94E-22 | Nightingale | M_VLDL_F | NA | T | C | 0.05187 | 0.005335 | 1.30E-23 |
| rs2519093 | let-7c-5p | trans | 4.94E-22 | Nightingale | L_LDL_PL | NA | T | C | 0.066759 | 0.005296 | 3.60E-37 |
| rs2519093 | let-7c-5p | trans | 4.94E-22 | Nightingale | IDL_PL | NA | T | C | 0.058969 | 0.005149 | 2.40E-31 |
| rs2519093 | let-7c-5p | trans | 4.94E-22 | Nightingale | XXL_VLDL_ | NA | T | C | 0.056193 | 0.00538 | 1.00E-25 |
| rs2519093 | let-7c-5p | trans | 4.94E-22 | Nightingale | LDL_CE | NA | T | C | 0.0584 | 0.005322 | 9.20E-29 |

| | | | | | | | | | | | |
|---|---|---|---|---|---|---|---|---|---|---|---|
| rs2519093 | let-7c-5p | trans | 4.94E-22 | Nightingale | S_VLDL_C | NA | T | C | 0.059191 | 0.005336 | 2.80E-30 |
| rs2519093 | let-7c-5p | trans | 4.94E-22 | Nightingale | LDL_L | NA | T | C | 0.05971 | 0.005318 | 6.80E-30 |
| rs2519093 | let-7c-5p | trans | 4.94E-22 | Nightingale | L_LDL_L | NA | T | C | 0.061108 | 0.005284 | 1.20E-31 |
| rs2519093 | let-7c-5p | trans | 4.94E-22 | Nightingale | S_LDL_FC | NA | T | C | 0.054719 | 0.005328 | 1.90E-24 |
| rs2519093 | let-7c-5p | trans | 4.94E-22 | Nightingale | HDL_size | NA | T | C | -0.03268 | 0.004731 | 9.30E-13 |
| rs2519093 | let-7c-5p | trans | 4.94E-22 | Nightingale | XS_VLDL_T | NA | T | C | -0.04276 | 0.005074 | 2.50E-16 |
| rs2519093 | let-7c-5p | trans | 4.94E-22 | Nightingale | XS_VLDL_P | NA | T | C | 0.053945 | 0.005265 | 8.60E-27 |
| rs2519093 | let-7c-5p | trans | 4.94E-22 | Nightingale | Total_C | NA | T | C | 0.055882 | 0.005166 | 2.10E-28 |
| rs2519093 | let-7c-5p | trans | 4.94E-22 | Nightingale | Total_P | NA | T | C | 0.03671 | 0.005022 | 4.80E-14 |
| rs2519093 | let-7c-5p | trans | 4.94E-22 | Nightingale | VLDL_C | NA | T | C | 0.045887 | 0.005329 | 2.40E-19 |
| rs2519093 | let-7c-5p | trans | 4.94E-22 | Nightingale | IDL_FC | NA | T | C | 0.064239 | 0.005166 | 4.20E-36 |
| rs2519093 | let-7c-5p | trans | 4.94E-22 | Nightingale | M_LDL_TG | NA | T | C | -0.05344 | 0.00527 | 6.00E-24 |
| rs2519093 | let-7c-5p | trans | 4.94E-22 | Nightingale | XXL_VLDL_ | NA | T | C | 0.057015 | 0.005351 | 1.20E-26 |
| rs2519093 | let-7c-5p | trans | 4.94E-22 | Nightingale | Clinical_LD | NA | T | C | 0.064699 | 0.005305 | 1.10E-34 |
| rs2519093 | let-7c-5p | trans | 4.94E-22 | Nightingale | S_HDL_FC | NA | T | C | 0.041872 | 0.005238 | 1.90E-16 |
| rs2519093 | let-7c-5p | trans | 4.94E-22 | Nightingale | XL_HDL_L | NA | T | C | -0.02733 | 0.004799 | 1.80E-09 |
| rs2519093 | let-7c-5p | trans | 4.94E-22 | Nightingale | L_LDL_FC | NA | T | C | 0.062862 | 0.00524 | 2.40E-33 |
| rs2519093 | let-7c-5p | trans | 4.94E-22 | Nightingale | M_VLDL_F | NA | T | C | 0.067606 | 0.005077 | 8.00E-40 |
| rs2519093 | let-7c-5p | trans | 4.94E-22 | Nightingale | S_LDL_PL_ | NA | T | C | -0.04499 | 0.005326 | 8.80E-20 |
| rs2519093 | let-7c-5p | trans | 4.94E-22 | Nightingale | Citrate | NA | T | C | 0.031335 | 0.005301 | 1.70E-09 |
| rs2519093 | let-7c-5p | trans | 4.94E-22 | Nightingale | M_LDL_CE | NA | T | C | 0.055668 | 0.005343 | 1.90E-26 |
| rs2519093 | let-7c-5p | trans | 4.94E-22 | Nightingale | Phosphatic | NA | T | C | 0.031111 | 0.005 | 6.70E-11 |
| rs2519093 | let-7c-5p | trans | 4.94E-22 | Nightingale | Cholines | NA | T | C | 0.035715 | 0.005018 | 1.00E-13 |
| rs2519093 | let-7c-5p | trans | 4.94E-22 | Nightingale | IDL_C_pct | NA | T | C | 0.037575 | 0.005213 | 4.50E-12 |
| rs2519093 | let-7c-5p | trans | 4.94E-22 | Nightingale | XL_HDL_PL | NA | T | C | -0.0414 | 0.004903 | 9.60E-18 |
| rs2519093 | let-7c-5p | trans | 4.94E-22 | Nightingale | LA | NA | T | C | 0.042082 | 0.005233 | 4.40E-17 |
| rs2519093 | let-7c-5p | trans | 4.94E-22 | Nightingale | S_LDL_P | NA | T | C | 0.048597 | 0.005341 | 1.60E-20 |
| rs2519093 | let-7c-5p | trans | 4.94E-22 | Nightingale | M_VLDL_L | NA | T | C | 0.032196 | 0.0053 | 1.00E-10 |
| rs2519093 | let-7c-5p | trans | 4.94E-22 | Nightingale | XL_HDL_PL | NA | T | C | -0.03044 | 0.0048 | 4.90E-11 |
| rs2519093 | let-7c-5p | trans | 4.94E-22 | Nightingale | XL_VLDL_P | NA | T | C | 0.056118 | 0.005345 | 2.80E-27 |
| rs2519093 | let-7c-5p | trans | 4.94E-22 | Nightingale | XS_VLDL_F | NA | T | C | 0.053855 | 0.005244 | 8.50E-27 |
| rs2519093 | let-7c-5p | trans | 4.94E-22 | Nightingale | non_HDL_( | NA | T | C | 0.060625 | 0.005307 | 2.30E-31 |
| rs2519093 | let-7c-5p | trans | 4.94E-22 | Nightingale | XL_VLDL_T | NA | T | C | -0.06925 | 0.005173 | 9.80E-41 |
| rs2519093 | let-7c-5p | trans | 4.94E-22 | Nightingale | IDL_TG_pc | NA | T | C | -0.03897 | 0.005159 | 2.10E-13 |
| rs2519093 | let-7c-5p | trans | 4.94E-22 | Nightingale | Total_PL | NA | T | C | 0.039528 | 0.005081 | 4.00E-16 |
| rs2519093 | let-7c-5p | trans | 4.94E-22 | Nightingale | M_VLDL_T | NA | T | C | -0.06313 | 0.005043 | 4.00E-35 |
| rs2519093 | let-7c-5p | trans | 4.94E-22 | Nightingale | L_VLDL_FC | NA | T | C | 0.076264 | 0.005285 | 5.50E-48 |
| rs2519093 | let-7c-5p | trans | 4.94E-22 | Nightingale | S_VLDL_P | NA | T | C | 0.033454 | 0.005269 | 6.70E-12 |
| rs2519093 | let-7c-5p | trans | 4.94E-22 | Nightingale | S_LDL_PL | NA | T | C | 0.041418 | 0.005324 | 6.40E-15 |
| rs2519093 | let-7c-5p | trans | 4.94E-22 | Nightingale | XS_VLDL_C | NA | T | C | 0.058552 | 0.00515 | 2.40E-31 |
| rs2519093 | let-7c-5p | trans | 4.94E-22 | Nightingale | L_VLDL_CE | NA | T | C | 0.074537 | 0.005107 | 7.40E-49 |
| rs2519093 | let-7c-5p | trans | 4.94E-22 | Nightingale | XL_HDL_C_ | NA | T | C | 0.038739 | 0.005096 | 1.00E-13 |
| rs2519093 | let-7c-5p | trans | 4.94E-22 | Nightingale | M_LDL_C | NA | T | C | 0.058986 | 0.005346 | 4.10E-29 |
| rs2519093 | let-7c-5p | trans | 4.94E-22 | Nightingale | S_LDL_TG_ | NA | T | C | -0.0438 | 0.005175 | 2.30E-16 |
| rs2519093 | let-7c-5p | trans | 4.94E-22 | Nightingale | XL_VLDL_C | NA | T | C | 0.062611 | 0.00512 | 1.50E-33 |
| rs2519093 | let-7c-5p | trans | 4.94E-22 | Nightingale | S_VLDL_FC | NA | T | C | 0.059655 | 0.005337 | 2.30E-30 |
| rs2519093 | let-7c-5p | trans | 4.94E-22 | Nightingale | IDL_FC_pct | NA | T | C | 0.041553 | 0.005295 | 1.40E-14 |
| rs2519093 | let-7c-5p | trans | 4.94E-22 | Nightingale | M_LDL_PL | NA | T | C | 0.058342 | 0.005344 | 1.80E-28 |
| rs2519093 | let-7c-5p | trans | 4.94E-22 | Nightingale | L_VLDL_PL | NA | T | C | 0.028334 | 0.005183 | 2.20E-09 |
| rs2519093 | let-7c-5p | trans | 4.94E-22 | Nightingale | L_LDL_P | NA | T | C | 0.050604 | 0.005318 | 4.80E-22 |
| rs2519093 | let-7c-5p | trans | 4.94E-22 | Nightingale | LDL_PL | NA | T | C | 0.06213 | 0.005324 | 5.60E-32 |
| rs2519093 | let-7c-5p | trans | 4.94E-22 | Nightingale | S_VLDL_TG | NA | T | C | -0.06553 | 0.005165 | 2.20E-36 |
| rs2519093 | let-7c-5p | trans | 4.94E-22 | Nightingale | Pyruvate | NA | T | C | 0.034476 | 0.005345 | 1.10E-10 |
| rs2519093 | let-7c-5p | trans | 4.94E-22 | Nightingale | ApoB_by_/ | NA | T | C | 0.040245 | 0.005256 | 9.90E-15 |
| rs2519093 | let-7c-5p | trans | 4.94E-22 | Nightingale | XL_VLDL_F | NA | T | C | 0.056835 | 0.005174 | 2.50E-28 |
| rs2519093 | let-7c-5p | trans | 4.94E-22 | Nightingale | S_VLDL_PL | NA | T | C | 0.050243 | 0.005326 | 1.00E-22 |
| rs2519093 | let-7c-5p | trans | 4.94E-22 | Nightingale | M_VLDL_P | NA | T | C | 0.070901 | 0.005121 | 2.00E-43 |
| rs2519093 | let-7c-5p | trans | 4.94E-22 | Nightingale | S_LDL_CE_ | NA | T | C | 0.04485 | 0.005327 | 8.40E-20 |
| rs2519093 | let-7c-5p | trans | 4.94E-22 | Nightingale | XXL_VLDL_ | NA | T | C | 0.036769 | 0.005374 | 2.00E-13 |
| rs2519093 | let-7c-5p | trans | 4.94E-22 | Nightingale | M_VLDL_C | NA | T | C | 0.059763 | 0.005026 | 1.20E-31 |
| rs2519093 | let-7c-5p | trans | 4.94E-22 | Nightingale | XS_VLDL_C | NA | T | C | 0.057788 | 0.00518 | 1.80E-30 |
| rs2519093 | let-7c-5p | trans | 4.94E-22 | Nightingale | S_HDL_L | NA | T | C | 0.04414 | 0.005318 | 1.60E-17 |
| rs2519093 | let-7c-5p | trans | 4.94E-22 | Nightingale | S_LDL_C_p | NA | T | C | 0.075956 | 0.005357 | 8.10E-47 |

| | | | | | | | | | | | |
|---|---|---|---|---|---|---|---|---|---|---|---|
| rs2519093 | let-7c-5p | trans | 4.94E-22 | Nightingale | M_LDL_C_ | NA | T | C | 0.047547 | 0.00534 | 3.80E-19 |
| rs2519093 | let-7c-5p | trans | 4.94E-22 | Nightingale | XL_VLDL_C | NA | T | C | 0.062776 | 0.005117 | 8.10E-34 |
| rs2519093 | let-7c-5p | trans | 4.94E-22 | Nightingale | LDL_FC | NA | T | C | 0.062925 | 0.005279 | 9.00E-33 |
| rs2519093 | let-7c-5p | trans | 4.94E-22 | Nightingale | L_VLDL_TG | NA | T | C | -0.08007 | 0.005311 | 4.90E-53 |
| rs2519093 | let-7c-5p | trans | 4.94E-22 | Nightingale | S_HDL_PL_ | NA | T | C | -0.03292 | 0.005159 | 7.60E-10 |
| rs2519093 | let-7c-5p | trans | 4.94E-22 | Nightingale | HDL_P | NA | T | C | 0.028786 | 0.005011 | 3.00E-09 |
| rs2519093 | let-7c-5p | trans | 4.94E-22 | Nightingale | LDL_C | NA | T | C | 0.060241 | 0.005313 | 2.20E-30 |
| rs2519093 | let-7c-5p | trans | 4.94E-22 | Nightingale | L_LDL_FC_ | NA | T | C | 0.030917 | 0.005091 | 1.20E-08 |
| rs2519093 | let-7c-5p | trans | 4.94E-22 | Nightingale | M_LDL_L | NA | T | C | 0.057522 | 0.005344 | 6.90E-28 |
| rs2519093 | let-7c-5p | trans | 4.94E-22 | Nightingale | XL_HDL_CE | NA | T | C | 0.030201 | 0.005332 | 3.60E-08 |
| rs2519093 | let-7c-5p | trans | 4.94E-22 | Nightingale | M_LDL_FC | NA | T | C | 0.063872 | 0.00533 | 2.80E-33 |
| rs2519093 | let-7c-5p | trans | 4.94E-22 | Nightingale | S_VLDL_CE | NA | T | C | 0.076957 | 0.005265 | 7.60E-48 |
| rs2519093 | let-7c-5p | trans | 4.94E-22 | Nightingale | Remnant_( | NA | T | C | 0.058623 | 0.005302 | 4.00E-30 |
| rs2519093 | miR-195-5p | trans | 7.49E-18 | Nightingale | S_HDL_CE | NA | T | C | 0.053505 | 0.005345 | 7.30E-24 |
| rs2519093 | miR-195-5p | trans | 7.49E-18 | Nightingale | L_LDL_TG_ | NA | T | C | -0.04178 | 0.005234 | 5.50E-15 |
| rs2519093 | miR-195-5p | trans | 7.49E-18 | Nightingale | M_VLDL_P | NA | T | C | 0.04488 | 0.005328 | 1.70E-18 |
| rs2519093 | miR-195-5p | trans | 7.49E-18 | Nightingale | IDL_C | NA | T | C | 0.060311 | 0.005154 | 6.00E-32 |
| rs2519093 | miR-195-5p | trans | 7.49E-18 | Nightingale | M_VLDL_C | NA | T | C | 0.066274 | 0.005288 | 7.60E-37 |
| rs2519093 | miR-195-5p | trans | 7.49E-18 | Nightingale | Total_FC | NA | T | C | 0.055093 | 0.005213 | 7.90E-28 |
| rs2519093 | miR-195-5p | trans | 7.49E-18 | Nightingale | S_VLDL_C_ | NA | T | C | 0.070806 | 0.005193 | 5.60E-42 |
| rs2519093 | miR-195-5p | trans | 7.49E-18 | Nightingale | LA_pct | NA | T | C | 0.031876 | 0.005218 | 1.10E-09 |
| rs2519093 | miR-195-5p | trans | 7.49E-18 | Nightingale | S_VLDL_L | NA | T | C | 0.035666 | 0.005285 | 4.30E-13 |
| rs2519093 | miR-195-5p | trans | 7.49E-18 | Nightingale | M_VLDL_C | NA | T | C | 0.061815 | 0.005321 | 1.60E-32 |
| rs2519093 | miR-195-5p | trans | 7.49E-18 | Nightingale | XS_VLDL_F | NA | T | C | 0.051132 | 0.005298 | 6.20E-21 |
| rs2519093 | miR-195-5p | trans | 7.49E-18 | Nightingale | IDL_P | NA | T | C | 0.065267 | 0.00528 | 1.10E-35 |
| rs2519093 | miR-195-5p | trans | 7.49E-18 | Nightingale | S_VLDL_CE | NA | T | C | 0.05821 | 0.005329 | 1.10E-29 |
| rs2519093 | miR-195-5p | trans | 7.49E-18 | Nightingale | XS_VLDL_L | NA | T | C | 0.048684 | 0.005245 | 1.40E-22 |
| rs2519093 | miR-195-5p | trans | 7.49E-18 | Nightingale | S_LDL_CE | NA | T | C | 0.051776 | 0.005339 | 4.00E-23 |
| rs2519093 | miR-195-5p | trans | 7.49E-18 | Nightingale | IDL_L | NA | T | C | 0.059774 | 0.00516 | 1.10E-31 |
| rs2519093 | miR-195-5p | trans | 7.49E-18 | Nightingale | Total_CE | NA | T | C | 0.05575 | 0.005147 | 2.40E-28 |
| rs2519093 | miR-195-5p | trans | 7.49E-18 | Nightingale | Sphingomy | NA | T | C | 0.047648 | 0.004989 | 2.20E-21 |
| rs2519093 | miR-195-5p | trans | 7.49E-18 | Nightingale | PUFA | NA | T | C | 0.036949 | 0.005205 | 1.30E-13 |
| rs2519093 | miR-195-5p | trans | 7.49E-18 | Nightingale | XS_VLDL_C | NA | T | C | 0.040361 | 0.005035 | 1.00E-14 |
| rs2519093 | miR-195-5p | trans | 7.49E-18 | Nightingale | IDL_CE | NA | T | C | 0.058439 | 0.005152 | 4.70E-30 |
| rs2519093 | miR-195-5p | trans | 7.49E-18 | Nightingale | VLDL_CE | NA | T | C | 0.053783 | 0.005338 | 2.10E-25 |
| rs2519093 | miR-195-5p | trans | 7.49E-18 | Nightingale | S_HDL_PL | NA | T | C | 0.037688 | 0.00529 | 1.40E-13 |
| rs2519093 | miR-195-5p | trans | 7.49E-18 | Nightingale | Omega_6 | NA | T | C | 0.041735 | 0.005211 | 5.40E-17 |
| rs2519093 | miR-195-5p | trans | 7.49E-18 | Nightingale | S_HDL_C | NA | T | C | 0.052616 | 0.005327 | 2.20E-23 |
| rs2519093 | miR-195-5p | trans | 7.49E-18 | Nightingale | M_VLDL_C | NA | T | C | 0.056108 | 0.005013 | 3.90E-28 |
| rs2519093 | miR-195-5p | trans | 7.49E-18 | Nightingale | GlycA | NA | T | C | 0.031449 | 0.005314 | 4.90E-09 |
| rs2519093 | miR-195-5p | trans | 7.49E-18 | Nightingale | XS_VLDL_C | NA | T | C | 0.043421 | 0.005061 | 7.70E-17 |
| rs2519093 | miR-195-5p | trans | 7.49E-18 | Nightingale | M_VLDL_P | NA | T | C | 0.043347 | 0.005323 | 1.90E-17 |
| rs2519093 | miR-195-5p | trans | 7.49E-18 | Nightingale | XS_VLDL_P | NA | T | C | -0.02949 | 0.005207 | 5.00E-08 |
| rs2519093 | miR-195-5p | trans | 7.49E-18 | Nightingale | S_VLDL_PL | NA | T | C | 0.050234 | 0.00511 | 4.10E-22 |
| rs2519093 | miR-195-5p | trans | 7.49E-18 | Nightingale | L_LDL_C | NA | T | C | 0.060158 | 0.005276 | 1.10E-30 |
| rs2519093 | miR-195-5p | trans | 7.49E-18 | Nightingale | Total_L | NA | T | C | 0.042371 | 0.00525 | 4.30E-17 |
| rs2519093 | miR-195-5p | trans | 7.49E-18 | Nightingale | Phosphogl | NA | T | C | 0.031931 | 0.005054 | 3.40E-11 |
| rs2519093 | miR-195-5p | trans | 7.49E-18 | Nightingale | L_VLDL_C_ | NA | T | C | 0.082094 | 0.005212 | 9.00E-57 |
| rs2519093 | miR-195-5p | trans | 7.49E-18 | Nightingale | L_VLDL_CE | NA | T | C | 0.031525 | 0.005274 | 1.30E-10 |
| rs2519093 | miR-195-5p | trans | 7.49E-18 | Nightingale | VLDL_P | NA | T | C | 0.038356 | 0.005308 | 2.10E-14 |
| rs2519093 | miR-195-5p | trans | 7.49E-18 | Nightingale | M_LDL_P | NA | T | C | 0.057192 | 0.005351 | 1.30E-27 |
| rs2519093 | miR-195-5p | trans | 7.49E-18 | Nightingale | S_VLDL_FC | NA | T | C | 0.052723 | 0.005099 | 2.50E-24 |
| rs2519093 | miR-195-5p | trans | 7.49E-18 | Nightingale | S_HDL_P | NA | T | C | 0.055391 | 0.005337 | 1.10E-25 |
| rs2519093 | miR-195-5p | trans | 7.49E-18 | Nightingale | S_LDL_L | NA | T | C | 0.048584 | 0.005335 | 3.30E-20 |
| rs2519093 | miR-195-5p | trans | 7.49E-18 | Nightingale | VLDL_FC | NA | T | C | 0.033679 | 0.005297 | 1.10E-11 |
| rs2519093 | miR-195-5p | trans | 7.49E-18 | Nightingale | XL_VLDL_C | NA | T | C | 0.029731 | 0.005254 | 9.50E-10 |
| rs2519093 | miR-195-5p | trans | 7.49E-18 | Nightingale | XXL_VLDL_ | NA | T | C | 0.046178 | 0.005317 | 1.50E-18 |
| rs2519093 | miR-195-5p | trans | 7.49E-18 | Nightingale | XXL_VLDL_ | NA | T | C | -0.05831 | 0.005385 | 5.10E-28 |
| rs2519093 | miR-195-5p | trans | 7.49E-18 | Nightingale | LDL_P | NA | T | C | 0.053373 | 0.005333 | 2.30E-24 |
| rs2519093 | miR-195-5p | trans | 7.49E-18 | Nightingale | XS_VLDL_P | NA | T | C | 0.044763 | 0.005262 | 2.50E-19 |
| rs2519093 | miR-195-5p | trans | 7.49E-18 | Nightingale | ApoB | NA | T | C | 0.055887 | 0.005329 | 1.10E-26 |
| rs2519093 | miR-195-5p | trans | 7.49E-18 | Nightingale | L_LDL_CE | NA | T | C | 0.058613 | 0.005289 | 3.70E-29 |
| rs2519093 | miR-195-5p | trans | 7.49E-18 | Nightingale | S_LDL_C | NA | T | C | 0.05408 | 0.005341 | 1.20E-24 |

| | | | | | | | | | | | |
|---|---|---|---|---|---|---|---|---|---|---|---|
| rs2519093 | miR-195-5p | trans | 7.49E-18 | Nightingale | M_VLDL_F | NA | T | C | 0.05187 | 0.005335 | 1.30E-23 |
| rs2519093 | miR-195-5p | trans | 7.49E-18 | Nightingale | L_LDL_PL | NA | T | C | 0.066759 | 0.005296 | 3.60E-37 |
| rs2519093 | miR-195-5p | trans | 7.49E-18 | Nightingale | IDL_PL | NA | T | C | 0.058969 | 0.005149 | 2.40E-31 |
| rs2519093 | miR-195-5p | trans | 7.49E-18 | Nightingale | XXL_VLDL_ | NA | T | C | 0.056193 | 0.00538 | 1.00E-25 |
| rs2519093 | miR-195-5p | trans | 7.49E-18 | Nightingale | LDL_CE | NA | T | C | 0.0584 | 0.005322 | 9.20E-29 |
| rs2519093 | miR-195-5p | trans | 7.49E-18 | Nightingale | S_VLDL_C | NA | T | C | 0.059191 | 0.005336 | 2.80E-30 |
| rs2519093 | miR-195-5p | trans | 7.49E-18 | Nightingale | LDL_L | NA | T | C | 0.05971 | 0.005318 | 6.80E-30 |
| rs2519093 | miR-195-5p | trans | 7.49E-18 | Nightingale | L_LDL_L | NA | T | C | 0.061108 | 0.005284 | 1.20E-31 |
| rs2519093 | miR-195-5p | trans | 7.49E-18 | Nightingale | S_LDL_FC | NA | T | C | 0.054719 | 0.005328 | 1.90E-24 |
| rs2519093 | miR-195-5p | trans | 7.49E-18 | Nightingale | HDL_size | NA | T | C | -0.03268 | 0.004731 | 9.30E-13 |
| rs2519093 | miR-195-5p | trans | 7.49E-18 | Nightingale | XS_VLDL_T | NA | T | C | -0.04276 | 0.005074 | 2.50E-16 |
| rs2519093 | miR-195-5p | trans | 7.49E-18 | Nightingale | XS_VLDL_P | NA | T | C | 0.053945 | 0.005265 | 8.60E-27 |
| rs2519093 | miR-195-5p | trans | 7.49E-18 | Nightingale | Total_C | NA | T | C | 0.055882 | 0.005166 | 2.10E-28 |
| rs2519093 | miR-195-5p | trans | 7.49E-18 | Nightingale | Total_P | NA | T | C | 0.03671 | 0.005022 | 4.80E-14 |
| rs2519093 | miR-195-5p | trans | 7.49E-18 | Nightingale | VLDL_C | NA | T | C | 0.045887 | 0.005329 | 2.40E-19 |
| rs2519093 | miR-195-5p | trans | 7.49E-18 | Nightingale | IDL_FC | NA | T | C | 0.064239 | 0.005166 | 4.20E-36 |
| rs2519093 | miR-195-5p | trans | 7.49E-18 | Nightingale | M_LDL_TG | NA | T | C | -0.05344 | 0.00527 | 6.00E-24 |
| rs2519093 | miR-195-5p | trans | 7.49E-18 | Nightingale | XXL_VLDL_ | NA | T | C | 0.057015 | 0.005351 | 1.20E-26 |
| rs2519093 | miR-195-5p | trans | 7.49E-18 | Nightingale | Clinical_LD | NA | T | C | 0.064699 | 0.005305 | 1.10E-34 |
| rs2519093 | miR-195-5p | trans | 7.49E-18 | Nightingale | S_HDL_FC | NA | T | C | 0.041872 | 0.005238 | 1.90E-16 |
| rs2519093 | miR-195-5p | trans | 7.49E-18 | Nightingale | XL_HDL_L | NA | T | C | -0.02733 | 0.004799 | 1.80E-09 |
| rs2519093 | miR-195-5p | trans | 7.49E-18 | Nightingale | L_LDL_FC | NA | T | C | 0.062862 | 0.00524 | 2.40E-33 |
| rs2519093 | miR-195-5p | trans | 7.49E-18 | Nightingale | M_VLDL_F | NA | T | C | 0.067606 | 0.005077 | 8.00E-40 |
| rs2519093 | miR-195-5p | trans | 7.49E-18 | Nightingale | S_LDL_PL_ | NA | T | C | -0.04499 | 0.005326 | 8.80E-20 |
| rs2519093 | miR-195-5p | trans | 7.49E-18 | Nightingale | Citrate | NA | T | C | 0.031335 | 0.005301 | 1.70E-09 |
| rs2519093 | miR-195-5p | trans | 7.49E-18 | Nightingale | M_LDL_CE | NA | T | C | 0.055668 | 0.005343 | 1.90E-26 |
| rs2519093 | miR-195-5p | trans | 7.49E-18 | Nightingale | Phosphatic | NA | T | C | 0.031111 | 0.005 | 6.70E-11 |
| rs2519093 | miR-195-5p | trans | 7.49E-18 | Nightingale | Cholines | NA | T | C | 0.035715 | 0.005018 | 1.00E-13 |
| rs2519093 | miR-195-5p | trans | 7.49E-18 | Nightingale | IDL_C_pct | NA | T | C | 0.037575 | 0.005213 | 4.50E-12 |
| rs2519093 | miR-195-5p | trans | 7.49E-18 | Nightingale | XL_HDL_PL | NA | T | C | -0.0414 | 0.004903 | 9.60E-18 |
| rs2519093 | miR-195-5p | trans | 7.49E-18 | Nightingale | LA | NA | T | C | 0.042082 | 0.005233 | 4.40E-17 |
| rs2519093 | miR-195-5p | trans | 7.49E-18 | Nightingale | S_LDL_P | NA | T | C | 0.048597 | 0.005341 | 1.60E-20 |
| rs2519093 | miR-195-5p | trans | 7.49E-18 | Nightingale | M_VLDL_L | NA | T | C | 0.032196 | 0.0053 | 1.00E-10 |
| rs2519093 | miR-195-5p | trans | 7.49E-18 | Nightingale | XL_HDL_PL | NA | T | C | -0.03044 | 0.0048 | 4.90E-11 |
| rs2519093 | miR-195-5p | trans | 7.49E-18 | Nightingale | XL_VLDL_P | NA | T | C | 0.056118 | 0.005345 | 2.80E-27 |
| rs2519093 | miR-195-5p | trans | 7.49E-18 | Nightingale | XS_VLDL_F | NA | T | C | 0.053855 | 0.005244 | 8.50E-27 |
| rs2519093 | miR-195-5p | trans | 7.49E-18 | Nightingale | non_HDL_( | NA | T | C | 0.060625 | 0.005307 | 2.30E-31 |
| rs2519093 | miR-195-5p | trans | 7.49E-18 | Nightingale | XL_VLDL_T | NA | T | C | -0.06925 | 0.005173 | 9.80E-41 |
| rs2519093 | miR-195-5p | trans | 7.49E-18 | Nightingale | IDL_TG_pct | NA | T | C | -0.03897 | 0.005159 | 2.10E-13 |
| rs2519093 | miR-195-5p | trans | 7.49E-18 | Nightingale | Total_PL | NA | T | C | 0.039528 | 0.005081 | 4.00E-16 |
| rs2519093 | miR-195-5p | trans | 7.49E-18 | Nightingale | M_VLDL_T | NA | T | C | -0.06313 | 0.005043 | 4.00E-35 |
| rs2519093 | miR-195-5p | trans | 7.49E-18 | Nightingale | L_VLDL_FC | NA | T | C | 0.076264 | 0.005285 | 5.50E-48 |
| rs2519093 | miR-195-5p | trans | 7.49E-18 | Nightingale | S_VLDL_P | NA | T | C | 0.033454 | 0.005269 | 6.70E-12 |
| rs2519093 | miR-195-5p | trans | 7.49E-18 | Nightingale | S_LDL_PL | NA | T | C | 0.041418 | 0.005324 | 6.40E-15 |
| rs2519093 | miR-195-5p | trans | 7.49E-18 | Nightingale | XS_VLDL_C | NA | T | C | 0.058552 | 0.00515 | 2.40E-31 |
| rs2519093 | miR-195-5p | trans | 7.49E-18 | Nightingale | L_VLDL_CE | NA | T | C | 0.074537 | 0.005107 | 7.40E-49 |
| rs2519093 | miR-195-5p | trans | 7.49E-18 | Nightingale | XL_HDL_C_ | NA | T | C | 0.038739 | 0.005096 | 1.00E-13 |
| rs2519093 | miR-195-5p | trans | 7.49E-18 | Nightingale | M_LDL_C | NA | T | C | 0.058986 | 0.005346 | 4.10E-29 |
| rs2519093 | miR-195-5p | trans | 7.49E-18 | Nightingale | S_LDL_TG_ | NA | T | C | -0.0438 | 0.005175 | 2.30E-16 |
| rs2519093 | miR-195-5p | trans | 7.49E-18 | Nightingale | XL_VLDL_C | NA | T | C | 0.062611 | 0.00512 | 1.50E-33 |
| rs2519093 | miR-195-5p | trans | 7.49E-18 | Nightingale | S_VLDL_FC | NA | T | C | 0.059655 | 0.005337 | 2.30E-30 |
| rs2519093 | miR-195-5p | trans | 7.49E-18 | Nightingale | IDL_FC_pct | NA | T | C | 0.041553 | 0.005295 | 1.40E-14 |
| rs2519093 | miR-195-5p | trans | 7.49E-18 | Nightingale | M_LDL_PL | NA | T | C | 0.058342 | 0.005344 | 1.80E-28 |
| rs2519093 | miR-195-5p | trans | 7.49E-18 | Nightingale | L_VLDL_PL | NA | T | C | 0.028334 | 0.005183 | 2.20E-09 |
| rs2519093 | miR-195-5p | trans | 7.49E-18 | Nightingale | L_LDL_P | NA | T | C | 0.050604 | 0.005318 | 4.80E-22 |
| rs2519093 | miR-195-5p | trans | 7.49E-18 | Nightingale | LDL_PL | NA | T | C | 0.06213 | 0.005324 | 5.60E-32 |
| rs2519093 | miR-195-5p | trans | 7.49E-18 | Nightingale | S_VLDL_TG | NA | T | C | -0.06553 | 0.005165 | 2.20E-36 |
| rs2519093 | miR-195-5p | trans | 7.49E-18 | Nightingale | Pyruvate | NA | T | C | 0.034476 | 0.005345 | 1.10E-10 |
| rs2519093 | miR-195-5p | trans | 7.49E-18 | Nightingale | ApoB_by_A | NA | T | C | 0.040245 | 0.005256 | 9.90E-15 |
| rs2519093 | miR-195-5p | trans | 7.49E-18 | Nightingale | XL_VLDL_F | NA | T | C | 0.056835 | 0.005174 | 2.50E-28 |
| rs2519093 | miR-195-5p | trans | 7.49E-18 | Nightingale | S_VLDL_PL | NA | T | C | 0.050243 | 0.005326 | 1.00E-22 |
| rs2519093 | miR-195-5p | trans | 7.49E-18 | Nightingale | M_VLDL_P | NA | T | C | 0.070901 | 0.005121 | 2.00E-43 |
| rs2519093 | miR-195-5p | trans | 7.49E-18 | Nightingale | S_LDL_CE_ | NA | T | C | 0.04485 | 0.005327 | 8.40E-20 |

| | | | | | | | | | | | |
|---|---|---|---|---|---|---|---|---|---|---|---|
| rs2519093 | miR-195-5p | trans | 7.49E-18 | Nightingale | XXL_VLDL_ | NA | T | C | 0.036769 | 0.005374 | 2.00E-13 |
| rs2519093 | miR-195-5p | trans | 7.49E-18 | Nightingale | M_VLDL_C | NA | T | C | 0.059763 | 0.005026 | 1.20E-31 |
| rs2519093 | miR-195-5p | trans | 7.49E-18 | Nightingale | XS_VLDL_C | NA | T | C | 0.057788 | 0.00518 | 1.80E-30 |
| rs2519093 | miR-195-5p | trans | 7.49E-18 | Nightingale | S_HDL_L | NA | T | C | 0.04414 | 0.005318 | 1.60E-17 |
| rs2519093 | miR-195-5p | trans | 7.49E-18 | Nightingale | S_LDL_C_p | NA | T | C | 0.075956 | 0.005357 | 8.10E-47 |
| rs2519093 | miR-195-5p | trans | 7.49E-18 | Nightingale | M_LDL_C_ | NA | T | C | 0.047547 | 0.00534 | 3.80E-19 |
| rs2519093 | miR-195-5p | trans | 7.49E-18 | Nightingale | XL_VLDL_C | NA | T | C | 0.062776 | 0.005117 | 8.10E-34 |
| rs2519093 | miR-195-5p | trans | 7.49E-18 | Nightingale | LDL_FC | NA | T | C | 0.062925 | 0.005279 | 9.00E-33 |
| rs2519093 | miR-195-5p | trans | 7.49E-18 | Nightingale | L_VLDL_TG | NA | T | C | -0.08007 | 0.005311 | 4.90E-53 |
| rs2519093 | miR-195-5p | trans | 7.49E-18 | Nightingale | S_HDL_PL_ | NA | T | C | -0.03292 | 0.005159 | 7.60E-10 |
| rs2519093 | miR-195-5p | trans | 7.49E-18 | Nightingale | HDL_P | NA | T | C | 0.028786 | 0.005011 | 3.00E-09 |
| rs2519093 | miR-195-5p | trans | 7.49E-18 | Nightingale | LDL_C | NA | T | C | 0.060241 | 0.005313 | 2.20E-30 |
| rs2519093 | miR-195-5p | trans | 7.49E-18 | Nightingale | L_LDL_FC_ | NA | T | C | 0.030917 | 0.005091 | 1.20E-08 |
| rs2519093 | miR-195-5p | trans | 7.49E-18 | Nightingale | M_LDL_L | NA | T | C | 0.057522 | 0.005344 | 6.90E-28 |
| rs2519093 | miR-195-5p | trans | 7.49E-18 | Nightingale | XL_HDL_CE | NA | T | C | 0.030201 | 0.005332 | 3.60E-08 |
| rs2519093 | miR-195-5p | trans | 7.49E-18 | Nightingale | M_LDL_FC | NA | T | C | 0.063872 | 0.00533 | 2.80E-33 |
| rs2519093 | miR-195-5p | trans | 7.49E-18 | Nightingale | S_VLDL_CE | NA | T | C | 0.076957 | 0.005265 | 7.60E-48 |
| rs2519093 | miR-195-5p | trans | 7.49E-18 | Nightingale | Remnant_( | NA | T | C | 0.058623 | 0.005302 | 4.00E-30 |
| rs2519093 | let-7b-5p | trans | 1.82E-25 | Nightingale | S_HDL_CE | NA | T | C | 0.053505 | 0.005345 | 7.30E-24 |
| rs2519093 | let-7b-5p | trans | 1.82E-25 | Nightingale | L_LDL_TG_ | NA | T | C | -0.04178 | 0.005234 | 5.50E-15 |
| rs2519093 | let-7b-5p | trans | 1.82E-25 | Nightingale | M_VLDL_P | NA | T | C | 0.04488 | 0.005328 | 1.70E-18 |
| rs2519093 | let-7b-5p | trans | 1.82E-25 | Nightingale | IDL_C | NA | T | C | 0.060311 | 0.005154 | 6.00E-32 |
| rs2519093 | let-7b-5p | trans | 1.82E-25 | Nightingale | M_VLDL_C | NA | T | C | 0.066274 | 0.005288 | 7.60E-37 |
| rs2519093 | let-7b-5p | trans | 1.82E-25 | Nightingale | Total_FC | NA | T | C | 0.055093 | 0.005213 | 7.90E-28 |
| rs2519093 | let-7b-5p | trans | 1.82E-25 | Nightingale | S_VLDL_C_ | NA | T | C | 0.070806 | 0.005193 | 5.60E-42 |
| rs2519093 | let-7b-5p | trans | 1.82E-25 | Nightingale | LA_pct | NA | T | C | 0.031876 | 0.005218 | 1.10E-09 |
| rs2519093 | let-7b-5p | trans | 1.82E-25 | Nightingale | S_VLDL_L | NA | T | C | 0.035666 | 0.005285 | 4.30E-13 |
| rs2519093 | let-7b-5p | trans | 1.82E-25 | Nightingale | M_VLDL_C | NA | T | C | 0.061815 | 0.005321 | 1.60E-32 |
| rs2519093 | let-7b-5p | trans | 1.82E-25 | Nightingale | XS_VLDL_F | NA | T | C | 0.051132 | 0.005298 | 6.20E-21 |
| rs2519093 | let-7b-5p | trans | 1.82E-25 | Nightingale | IDL_P | NA | T | C | 0.065267 | 0.00528 | 1.10E-35 |
| rs2519093 | let-7b-5p | trans | 1.82E-25 | Nightingale | S_VLDL_CE | NA | T | C | 0.05821 | 0.005329 | 1.10E-29 |
| rs2519093 | let-7b-5p | trans | 1.82E-25 | Nightingale | XS_VLDL_L | NA | T | C | 0.048684 | 0.005245 | 1.40E-22 |
| rs2519093 | let-7b-5p | trans | 1.82E-25 | Nightingale | S_LDL_CE | NA | T | C | 0.051776 | 0.005339 | 4.00E-23 |
| rs2519093 | let-7b-5p | trans | 1.82E-25 | Nightingale | IDL_L | NA | T | C | 0.059774 | 0.00516 | 1.10E-31 |
| rs2519093 | let-7b-5p | trans | 1.82E-25 | Nightingale | Total_CE | NA | T | C | 0.05575 | 0.005147 | 2.40E-28 |
| rs2519093 | let-7b-5p | trans | 1.82E-25 | Nightingale | Sphingomy | NA | T | C | 0.047648 | 0.004989 | 2.20E-21 |
| rs2519093 | let-7b-5p | trans | 1.82E-25 | Nightingale | PUFA | NA | T | C | 0.036949 | 0.005205 | 1.30E-13 |
| rs2519093 | let-7b-5p | trans | 1.82E-25 | Nightingale | XS_VLDL_C | NA | T | C | 0.040361 | 0.005035 | 1.00E-14 |
| rs2519093 | let-7b-5p | trans | 1.82E-25 | Nightingale | IDL_CE | NA | T | C | 0.058439 | 0.005152 | 4.70E-30 |
| rs2519093 | let-7b-5p | trans | 1.82E-25 | Nightingale | VLDL_CE | NA | T | C | 0.053783 | 0.005338 | 2.10E-25 |
| rs2519093 | let-7b-5p | trans | 1.82E-25 | Nightingale | S_HDL_PL | NA | T | C | 0.037688 | 0.00529 | 1.40E-13 |
| rs2519093 | let-7b-5p | trans | 1.82E-25 | Nightingale | Omega_6 | NA | T | C | 0.041735 | 0.005211 | 5.40E-17 |
| rs2519093 | let-7b-5p | trans | 1.82E-25 | Nightingale | S_HDL_C | NA | T | C | 0.052616 | 0.005327 | 2.20E-23 |
| rs2519093 | let-7b-5p | trans | 1.82E-25 | Nightingale | M_VLDL_C | NA | T | C | 0.056108 | 0.005013 | 3.90E-28 |
| rs2519093 | let-7b-5p | trans | 1.82E-25 | Nightingale | GlycA | NA | T | C | 0.031449 | 0.005314 | 4.90E-09 |
| rs2519093 | let-7b-5p | trans | 1.82E-25 | Nightingale | XS_VLDL_C | NA | T | C | 0.043421 | 0.005061 | 7.70E-17 |
| rs2519093 | let-7b-5p | trans | 1.82E-25 | Nightingale | M_VLDL_P | NA | T | C | 0.043347 | 0.005323 | 1.90E-17 |
| rs2519093 | let-7b-5p | trans | 1.82E-25 | Nightingale | XS_VLDL_P | NA | T | C | -0.02949 | 0.005207 | 5.00E-08 |
| rs2519093 | let-7b-5p | trans | 1.82E-25 | Nightingale | S_VLDL_PL | NA | T | C | 0.050234 | 0.00511 | 4.10E-22 |
| rs2519093 | let-7b-5p | trans | 1.82E-25 | Nightingale | L_LDL_C | NA | T | C | 0.060158 | 0.005276 | 1.10E-30 |
| rs2519093 | let-7b-5p | trans | 1.82E-25 | Nightingale | Total_L | NA | T | C | 0.042371 | 0.00525 | 4.30E-17 |
| rs2519093 | let-7b-5p | trans | 1.82E-25 | Nightingale | Phosphogly | NA | T | C | 0.031931 | 0.005054 | 3.40E-11 |
| rs2519093 | let-7b-5p | trans | 1.82E-25 | Nightingale | L_VLDL_C_ | NA | T | C | 0.082094 | 0.005212 | 9.00E-57 |
| rs2519093 | let-7b-5p | trans | 1.82E-25 | Nightingale | L_VLDL_CE | NA | T | C | 0.031525 | 0.005274 | 1.30E-10 |
| rs2519093 | let-7b-5p | trans | 1.82E-25 | Nightingale | VLDL_P | NA | T | C | 0.038356 | 0.005308 | 2.10E-14 |
| rs2519093 | let-7b-5p | trans | 1.82E-25 | Nightingale | M_LDL_P | NA | T | C | 0.057192 | 0.005351 | 1.30E-27 |
| rs2519093 | let-7b-5p | trans | 1.82E-25 | Nightingale | S_VLDL_FC | NA | T | C | 0.052723 | 0.005099 | 2.50E-24 |
| rs2519093 | let-7b-5p | trans | 1.82E-25 | Nightingale | S_HDL_P | NA | T | C | 0.055391 | 0.005337 | 1.10E-25 |
| rs2519093 | let-7b-5p | trans | 1.82E-25 | Nightingale | S_LDL_L | NA | T | C | 0.048584 | 0.005335 | 3.30E-20 |
| rs2519093 | let-7b-5p | trans | 1.82E-25 | Nightingale | VLDL_FC | NA | T | C | 0.033679 | 0.005297 | 1.10E-11 |
| rs2519093 | let-7b-5p | trans | 1.82E-25 | Nightingale | XL_VLDL_C | NA | T | C | 0.029731 | 0.005254 | 9.50E-10 |
| rs2519093 | let-7b-5p | trans | 1.82E-25 | Nightingale | XXL_VLDL_ | NA | T | C | 0.046178 | 0.005317 | 1.50E-18 |
| rs2519093 | let-7b-5p | trans | 1.82E-25 | Nightingale | XXL_VLDL_ | NA | T | C | -0.05831 | 0.005385 | 5.10E-28 |

| | | | | | | | | | | | |
|---|---|---|---|---|---|---|---|---|---|---|---|
| rs2519093 | let-7b-5p | trans | 1.82E-25 | Nightingale | LDL_P | NA | T | C | 0.053373 | 0.005333 | 2.30E-24 |
| rs2519093 | let-7b-5p | trans | 1.82E-25 | Nightingale | XS_VLDL_P | NA | T | C | 0.044763 | 0.005262 | 2.50E-19 |
| rs2519093 | let-7b-5p | trans | 1.82E-25 | Nightingale | ApoB | NA | T | C | 0.055887 | 0.005329 | 1.10E-26 |
| rs2519093 | let-7b-5p | trans | 1.82E-25 | Nightingale | L_LDL_CE | NA | T | C | 0.058613 | 0.005289 | 3.70E-29 |
| rs2519093 | let-7b-5p | trans | 1.82E-25 | Nightingale | S_LDL_C | NA | T | C | 0.05408 | 0.005341 | 1.20E-24 |
| rs2519093 | let-7b-5p | trans | 1.82E-25 | Nightingale | M_VLDL_F | NA | T | C | 0.05187 | 0.005335 | 1.30E-23 |
| rs2519093 | let-7b-5p | trans | 1.82E-25 | Nightingale | L_LDL_PL | NA | T | C | 0.066759 | 0.005296 | 3.60E-37 |
| rs2519093 | let-7b-5p | trans | 1.82E-25 | Nightingale | IDL_PL | NA | T | C | 0.058969 | 0.005149 | 2.40E-31 |
| rs2519093 | let-7b-5p | trans | 1.82E-25 | Nightingale | XXL_VLDL_ | NA | T | C | 0.056193 | 0.00538 | 1.00E-25 |
| rs2519093 | let-7b-5p | trans | 1.82E-25 | Nightingale | LDL_CE | NA | T | C | 0.0584 | 0.005322 | 9.20E-29 |
| rs2519093 | let-7b-5p | trans | 1.82E-25 | Nightingale | S_VLDL_C | NA | T | C | 0.059191 | 0.005336 | 2.80E-30 |
| rs2519093 | let-7b-5p | trans | 1.82E-25 | Nightingale | LDL_L | NA | T | C | 0.05971 | 0.005318 | 6.80E-30 |
| rs2519093 | let-7b-5p | trans | 1.82E-25 | Nightingale | L_LDL_L | NA | T | C | 0.061108 | 0.005284 | 1.20E-31 |
| rs2519093 | let-7b-5p | trans | 1.82E-25 | Nightingale | S_LDL_FC | NA | T | C | 0.054719 | 0.005328 | 1.90E-24 |
| rs2519093 | let-7b-5p | trans | 1.82E-25 | Nightingale | HDL_size | NA | T | C | -0.03268 | 0.004731 | 9.30E-13 |
| rs2519093 | let-7b-5p | trans | 1.82E-25 | Nightingale | XS_VLDL_T | NA | T | C | -0.04276 | 0.005074 | 2.50E-16 |
| rs2519093 | let-7b-5p | trans | 1.82E-25 | Nightingale | XS_VLDL_P | NA | T | C | 0.053945 | 0.005265 | 8.60E-27 |
| rs2519093 | let-7b-5p | trans | 1.82E-25 | Nightingale | Total_C | NA | T | C | 0.055882 | 0.005166 | 2.10E-28 |
| rs2519093 | let-7b-5p | trans | 1.82E-25 | Nightingale | Total_P | NA | T | C | 0.03671 | 0.005022 | 4.80E-14 |
| rs2519093 | let-7b-5p | trans | 1.82E-25 | Nightingale | VLDL_C | NA | T | C | 0.045887 | 0.005329 | 2.40E-19 |
| rs2519093 | let-7b-5p | trans | 1.82E-25 | Nightingale | IDL_FC | NA | T | C | 0.064239 | 0.005166 | 4.20E-36 |
| rs2519093 | let-7b-5p | trans | 1.82E-25 | Nightingale | M_LDL_TG | NA | T | C | -0.05344 | 0.00527 | 6.00E-24 |
| rs2519093 | let-7b-5p | trans | 1.82E-25 | Nightingale | XXL_VLDL_ | NA | T | C | 0.057015 | 0.005351 | 1.20E-26 |
| rs2519093 | let-7b-5p | trans | 1.82E-25 | Nightingale | Clinical_LD | NA | T | C | 0.064699 | 0.005305 | 1.10E-34 |
| rs2519093 | let-7b-5p | trans | 1.82E-25 | Nightingale | S_HDL_FC | NA | T | C | 0.041872 | 0.005238 | 1.90E-16 |
| rs2519093 | let-7b-5p | trans | 1.82E-25 | Nightingale | XL_HDL_L | NA | T | C | -0.02733 | 0.004799 | 1.80E-09 |
| rs2519093 | let-7b-5p | trans | 1.82E-25 | Nightingale | L_LDL_FC | NA | T | C | 0.062862 | 0.00524 | 2.40E-33 |
| rs2519093 | let-7b-5p | trans | 1.82E-25 | Nightingale | M_VLDL_F | NA | T | C | 0.067606 | 0.005077 | 8.00E-40 |
| rs2519093 | let-7b-5p | trans | 1.82E-25 | Nightingale | S_LDL_PL_ | NA | T | C | -0.04499 | 0.005326 | 8.80E-20 |
| rs2519093 | let-7b-5p | trans | 1.82E-25 | Nightingale | Citrate | NA | T | C | 0.031335 | 0.005301 | 1.70E-09 |
| rs2519093 | let-7b-5p | trans | 1.82E-25 | Nightingale | M_LDL_CE | NA | T | C | 0.055668 | 0.005343 | 1.90E-26 |
| rs2519093 | let-7b-5p | trans | 1.82E-25 | Nightingale | Phosphatic | NA | T | C | 0.031111 | 0.005 | 6.70E-11 |
| rs2519093 | let-7b-5p | trans | 1.82E-25 | Nightingale | Cholines | NA | T | C | 0.035715 | 0.005018 | 1.00E-13 |
| rs2519093 | let-7b-5p | trans | 1.82E-25 | Nightingale | IDL_C_pct | NA | T | C | 0.037575 | 0.005213 | 4.50E-12 |
| rs2519093 | let-7b-5p | trans | 1.82E-25 | Nightingale | XL_HDL_PL | NA | T | C | -0.0414 | 0.004903 | 9.60E-18 |
| rs2519093 | let-7b-5p | trans | 1.82E-25 | Nightingale | LA | NA | T | C | 0.042082 | 0.005233 | 4.40E-17 |
| rs2519093 | let-7b-5p | trans | 1.82E-25 | Nightingale | S_LDL_P | NA | T | C | 0.048597 | 0.005341 | 1.60E-20 |
| rs2519093 | let-7b-5p | trans | 1.82E-25 | Nightingale | M_VLDL_L | NA | T | C | 0.032196 | 0.0053 | 1.00E-10 |
| rs2519093 | let-7b-5p | trans | 1.82E-25 | Nightingale | XL_HDL_PL | NA | T | C | -0.03044 | 0.0048 | 4.90E-11 |
| rs2519093 | let-7b-5p | trans | 1.82E-25 | Nightingale | XL_VLDL_P | NA | T | C | 0.056118 | 0.005345 | 2.80E-27 |
| rs2519093 | let-7b-5p | trans | 1.82E-25 | Nightingale | XS_VLDL_F | NA | T | C | 0.053855 | 0.005244 | 8.50E-27 |
| rs2519093 | let-7b-5p | trans | 1.82E-25 | Nightingale | non_HDL_( | NA | T | C | 0.060625 | 0.005307 | 2.30E-31 |
| rs2519093 | let-7b-5p | trans | 1.82E-25 | Nightingale | XL_VLDL_T | NA | T | C | -0.06925 | 0.005173 | 9.80E-41 |
| rs2519093 | let-7b-5p | trans | 1.82E-25 | Nightingale | IDL_TG_pc | NA | T | C | -0.03897 | 0.005159 | 2.10E-13 |
| rs2519093 | let-7b-5p | trans | 1.82E-25 | Nightingale | Total_PL | NA | T | C | 0.039528 | 0.005081 | 4.00E-16 |
| rs2519093 | let-7b-5p | trans | 1.82E-25 | Nightingale | M_VLDL_T | NA | T | C | -0.06313 | 0.005043 | 4.00E-35 |
| rs2519093 | let-7b-5p | trans | 1.82E-25 | Nightingale | L_VLDL_FC | NA | T | C | 0.076264 | 0.005285 | 5.50E-48 |
| rs2519093 | let-7b-5p | trans | 1.82E-25 | Nightingale | S_VLDL_P | NA | T | C | 0.033454 | 0.005269 | 6.70E-12 |
| rs2519093 | let-7b-5p | trans | 1.82E-25 | Nightingale | S_LDL_PL | NA | T | C | 0.041418 | 0.005324 | 6.40E-15 |
| rs2519093 | let-7b-5p | trans | 1.82E-25 | Nightingale | XS_VLDL_C | NA | T | C | 0.058552 | 0.00515 | 2.40E-31 |
| rs2519093 | let-7b-5p | trans | 1.82E-25 | Nightingale | L_VLDL_CE | NA | T | C | 0.074537 | 0.005107 | 7.40E-49 |
| rs2519093 | let-7b-5p | trans | 1.82E-25 | Nightingale | XL_HDL_C_ | NA | T | C | 0.038739 | 0.005096 | 1.00E-13 |
| rs2519093 | let-7b-5p | trans | 1.82E-25 | Nightingale | M_LDL_C | NA | T | C | 0.058986 | 0.005346 | 4.10E-29 |
| rs2519093 | let-7b-5p | trans | 1.82E-25 | Nightingale | S_LDL_TG_ | NA | T | C | -0.0438 | 0.005175 | 2.30E-16 |
| rs2519093 | let-7b-5p | trans | 1.82E-25 | Nightingale | XL_VLDL_C | NA | T | C | 0.062611 | 0.00512 | 1.50E-33 |
| rs2519093 | let-7b-5p | trans | 1.82E-25 | Nightingale | S_VLDL_FC | NA | T | C | 0.059655 | 0.005337 | 2.30E-30 |
| rs2519093 | let-7b-5p | trans | 1.82E-25 | Nightingale | IDL_FC_pct | NA | T | C | 0.041553 | 0.005295 | 1.40E-14 |
| rs2519093 | let-7b-5p | trans | 1.82E-25 | Nightingale | M_LDL_PL | NA | T | C | 0.058342 | 0.005344 | 1.80E-28 |
| rs2519093 | let-7b-5p | trans | 1.82E-25 | Nightingale | L_VLDL_PL | NA | T | C | 0.028334 | 0.005183 | 2.20E-09 |
| rs2519093 | let-7b-5p | trans | 1.82E-25 | Nightingale | L_LDL_P | NA | T | C | 0.050604 | 0.005318 | 4.80E-22 |
| rs2519093 | let-7b-5p | trans | 1.82E-25 | Nightingale | LDL_PL | NA | T | C | 0.06213 | 0.005324 | 5.60E-32 |
| rs2519093 | let-7b-5p | trans | 1.82E-25 | Nightingale | S_VLDL_TG | NA | T | C | -0.06553 | 0.005165 | 2.20E-36 |
| rs2519093 | let-7b-5p | trans | 1.82E-25 | Nightingale | Pyruvate | NA | T | C | 0.034476 | 0.005345 | 1.10E-10 |

| | | | | | | | | | | | |
|---|---|---|---|---|---|---|---|---|---|---|---|
| rs2519093 | let-7b-5p | trans | 1.82E-25 | Nightingale | ApoB_by_A | NA | T | C | 0.040245 | 0.005256 | 9.90E-15 |
| rs2519093 | let-7b-5p | trans | 1.82E-25 | Nightingale | XL_VLDL_F | NA | T | C | 0.056835 | 0.005174 | 2.50E-28 |
| rs2519093 | let-7b-5p | trans | 1.82E-25 | Nightingale | S_VLDL_PL | NA | T | C | 0.050243 | 0.005326 | 1.00E-22 |
| rs2519093 | let-7b-5p | trans | 1.82E-25 | Nightingale | M_VLDL_P | NA | T | C | 0.070901 | 0.005121 | 2.00E-43 |
| rs2519093 | let-7b-5p | trans | 1.82E-25 | Nightingale | S_LDL_CE_ | NA | T | C | 0.04485 | 0.005327 | 8.40E-20 |
| rs2519093 | let-7b-5p | trans | 1.82E-25 | Nightingale | XXL_VLDL_ | NA | T | C | 0.036769 | 0.005374 | 2.00E-13 |
| rs2519093 | let-7b-5p | trans | 1.82E-25 | Nightingale | M_VLDL_C | NA | T | C | 0.059763 | 0.005026 | 1.20E-31 |
| rs2519093 | let-7b-5p | trans | 1.82E-25 | Nightingale | XS_VLDL_C | NA | T | C | 0.057788 | 0.00518 | 1.80E-30 |
| rs2519093 | let-7b-5p | trans | 1.82E-25 | Nightingale | S_HDL_L | NA | T | C | 0.04414 | 0.005318 | 1.60E-17 |
| rs2519093 | let-7b-5p | trans | 1.82E-25 | Nightingale | S_LDL_C_p | NA | T | C | 0.075956 | 0.005357 | 8.10E-47 |
| rs2519093 | let-7b-5p | trans | 1.82E-25 | Nightingale | M_LDL_C_ | NA | T | C | 0.047547 | 0.00534 | 3.80E-19 |
| rs2519093 | let-7b-5p | trans | 1.82E-25 | Nightingale | XL_VLDL_C | NA | T | C | 0.062776 | 0.005117 | 8.10E-34 |
| rs2519093 | let-7b-5p | trans | 1.82E-25 | Nightingale | LDL_FC | NA | T | C | 0.062925 | 0.005279 | 9.00E-33 |
| rs2519093 | let-7b-5p | trans | 1.82E-25 | Nightingale | L_VLDL_TG | NA | T | C | -0.08007 | 0.005311 | 4.90E-53 |
| rs2519093 | let-7b-5p | trans | 1.82E-25 | Nightingale | S_HDL_PL_ | NA | T | C | -0.03292 | 0.005159 | 7.60E-10 |
| rs2519093 | let-7b-5p | trans | 1.82E-25 | Nightingale | HDL_P | NA | T | C | 0.028786 | 0.005011 | 3.00E-09 |
| rs2519093 | let-7b-5p | trans | 1.82E-25 | Nightingale | LDL_C | NA | T | C | 0.060241 | 0.005313 | 2.20E-30 |
| rs2519093 | let-7b-5p | trans | 1.82E-25 | Nightingale | L_LDL_FC_ | NA | T | C | 0.030917 | 0.005091 | 1.20E-08 |
| rs2519093 | let-7b-5p | trans | 1.82E-25 | Nightingale | M_LDL_L | NA | T | C | 0.057522 | 0.005344 | 6.90E-28 |
| rs2519093 | let-7b-5p | trans | 1.82E-25 | Nightingale | XL_HDL_CE | NA | T | C | 0.030201 | 0.005332 | 3.60E-08 |
| rs2519093 | let-7b-5p | trans | 1.82E-25 | Nightingale | M_LDL_FC | NA | T | C | 0.063872 | 0.00533 | 2.80E-33 |
| rs2519093 | let-7b-5p | trans | 1.82E-25 | Nightingale | S_VLDL_CE | NA | T | C | 0.076957 | 0.005265 | 7.60E-48 |
| rs2519093 | let-7b-5p | trans | 1.82E-25 | Nightingale | Remnant_( | NA | T | C | 0.058623 | 0.005302 | 4.00E-30 |
| rs2519093 | miR-126-3p | trans | 1.90E-40 | Nightingale | S_HDL_CE | NA | T | C | 0.053505 | 0.005345 | 7.30E-24 |
| rs2519093 | miR-126-3p | trans | 1.90E-40 | Nightingale | L_LDL_TG_ | NA | T | C | -0.04178 | 0.005234 | 5.50E-15 |
| rs2519093 | miR-126-3p | trans | 1.90E-40 | Nightingale | M_VLDL_P | NA | T | C | 0.04488 | 0.005328 | 1.70E-18 |
| rs2519093 | miR-126-3p | trans | 1.90E-40 | Nightingale | IDL_C | NA | T | C | 0.060311 | 0.005154 | 6.00E-32 |
| rs2519093 | miR-126-3p | trans | 1.90E-40 | Nightingale | M_VLDL_C | NA | T | C | 0.066274 | 0.005288 | 7.60E-37 |
| rs2519093 | miR-126-3p | trans | 1.90E-40 | Nightingale | Total_FC | NA | T | C | 0.055093 | 0.005213 | 7.90E-28 |
| rs2519093 | miR-126-3p | trans | 1.90E-40 | Nightingale | S_VLDL_C_ | NA | T | C | 0.070806 | 0.005193 | 5.60E-42 |
| rs2519093 | miR-126-3p | trans | 1.90E-40 | Nightingale | LA_pct | NA | T | C | 0.031876 | 0.005218 | 1.10E-09 |
| rs2519093 | miR-126-3p | trans | 1.90E-40 | Nightingale | S_VLDL_L | NA | T | C | 0.035666 | 0.005285 | 4.30E-13 |
| rs2519093 | miR-126-3p | trans | 1.90E-40 | Nightingale | M_VLDL_C | NA | T | C | 0.061815 | 0.005321 | 1.60E-32 |
| rs2519093 | miR-126-3p | trans | 1.90E-40 | Nightingale | XS_VLDL_F | NA | T | C | 0.051132 | 0.005298 | 6.20E-21 |
| rs2519093 | miR-126-3p | trans | 1.90E-40 | Nightingale | IDL_P | NA | T | C | 0.065267 | 0.00528 | 1.10E-35 |
| rs2519093 | miR-126-3p | trans | 1.90E-40 | Nightingale | S_VLDL_CE | NA | T | C | 0.05821 | 0.005329 | 1.10E-29 |
| rs2519093 | miR-126-3p | trans | 1.90E-40 | Nightingale | XS_VLDL_L | NA | T | C | 0.048684 | 0.005245 | 1.40E-22 |
| rs2519093 | miR-126-3p | trans | 1.90E-40 | Nightingale | S_LDL_CE | NA | T | C | 0.051776 | 0.005339 | 4.00E-23 |
| rs2519093 | miR-126-3p | trans | 1.90E-40 | Nightingale | IDL_L | NA | T | C | 0.059774 | 0.00516 | 1.10E-31 |
| rs2519093 | miR-126-3p | trans | 1.90E-40 | Nightingale | Total_CE | NA | T | C | 0.05575 | 0.005147 | 2.40E-28 |
| rs2519093 | miR-126-3p | trans | 1.90E-40 | Nightingale | Sphingomy | NA | T | C | 0.047648 | 0.004989 | 2.20E-21 |
| rs2519093 | miR-126-3p | trans | 1.90E-40 | Nightingale | PUFA | NA | T | C | 0.036949 | 0.005205 | 1.30E-13 |
| rs2519093 | miR-126-3p | trans | 1.90E-40 | Nightingale | XS_VLDL_C | NA | T | C | 0.040361 | 0.005035 | 1.00E-14 |
| rs2519093 | miR-126-3p | trans | 1.90E-40 | Nightingale | IDL_CE | NA | T | C | 0.058439 | 0.005152 | 4.70E-30 |
| rs2519093 | miR-126-3p | trans | 1.90E-40 | Nightingale | VLDL_CE | NA | T | C | 0.053783 | 0.005338 | 2.10E-25 |
| rs2519093 | miR-126-3p | trans | 1.90E-40 | Nightingale | S_HDL_PL | NA | T | C | 0.037688 | 0.00529 | 1.40E-13 |
| rs2519093 | miR-126-3p | trans | 1.90E-40 | Nightingale | Omega_6 | NA | T | C | 0.041735 | 0.005211 | 5.40E-17 |
| rs2519093 | miR-126-3p | trans | 1.90E-40 | Nightingale | S_HDL_C | NA | T | C | 0.052616 | 0.005327 | 2.20E-23 |
| rs2519093 | miR-126-3p | trans | 1.90E-40 | Nightingale | M_VLDL_C | NA | T | C | 0.056108 | 0.005013 | 3.90E-28 |
| rs2519093 | miR-126-3p | trans | 1.90E-40 | Nightingale | GlycA | NA | T | C | 0.031449 | 0.005314 | 4.90E-09 |
| rs2519093 | miR-126-3p | trans | 1.90E-40 | Nightingale | XS_VLDL_C | NA | T | C | 0.043421 | 0.005061 | 7.70E-17 |
| rs2519093 | miR-126-3p | trans | 1.90E-40 | Nightingale | M_VLDL_P | NA | T | C | 0.043347 | 0.005323 | 1.90E-17 |
| rs2519093 | miR-126-3p | trans | 1.90E-40 | Nightingale | XS_VLDL_P | NA | T | C | -0.02949 | 0.005207 | 5.00E-08 |
| rs2519093 | miR-126-3p | trans | 1.90E-40 | Nightingale | S_VLDL_PL | NA | T | C | 0.050234 | 0.00511 | 4.10E-22 |
| rs2519093 | miR-126-3p | trans | 1.90E-40 | Nightingale | L_LDL_C | NA | T | C | 0.060158 | 0.005276 | 1.10E-30 |
| rs2519093 | miR-126-3p | trans | 1.90E-40 | Nightingale | Total_L | NA | T | C | 0.042371 | 0.00525 | 4.30E-17 |
| rs2519093 | miR-126-3p | trans | 1.90E-40 | Nightingale | Phosphogly | NA | T | C | 0.031931 | 0.005054 | 3.40E-11 |
| rs2519093 | miR-126-3p | trans | 1.90E-40 | Nightingale | L_VLDL_C_ | NA | T | C | 0.082094 | 0.005212 | 9.00E-57 |
| rs2519093 | miR-126-3p | trans | 1.90E-40 | Nightingale | L_VLDL_CE | NA | T | C | 0.031525 | 0.005274 | 1.30E-10 |
| rs2519093 | miR-126-3p | trans | 1.90E-40 | Nightingale | VLDL_P | NA | T | C | 0.038356 | 0.005308 | 2.10E-14 |
| rs2519093 | miR-126-3p | trans | 1.90E-40 | Nightingale | M_LDL_P | NA | T | C | 0.057192 | 0.005351 | 1.30E-27 |
| rs2519093 | miR-126-3p | trans | 1.90E-40 | Nightingale | S_VLDL_FC | NA | T | C | 0.052723 | 0.005099 | 2.50E-24 |
| rs2519093 | miR-126-3p | trans | 1.90E-40 | Nightingale | S_HDL_P | NA | T | C | 0.055391 | 0.005337 | 1.10E-25 |

| | | | | | | | | | | | |
|---|---|---|---|---|---|---|---|---|---|---|---|
| rs2519093 | miR-126-3p | trans | 1.90E-40 | Nightingale | S_LDL_L | NA | T | C | 0.048584 | 0.005335 | 3.30E-20 |
| rs2519093 | miR-126-3p | trans | 1.90E-40 | Nightingale | VLDL_FC | NA | T | C | 0.033679 | 0.005297 | 1.10E-11 |
| rs2519093 | miR-126-3p | trans | 1.90E-40 | Nightingale | XL_VLDL_C | NA | T | C | 0.029731 | 0.005254 | 9.50E-10 |
| rs2519093 | miR-126-3p | trans | 1.90E-40 | Nightingale | XXL_VLDL_ | NA | T | C | 0.046178 | 0.005317 | 1.50E-18 |
| rs2519093 | miR-126-3p | trans | 1.90E-40 | Nightingale | XXL_VLDL_ | NA | T | C | -0.05831 | 0.005385 | 5.10E-28 |
| rs2519093 | miR-126-3p | trans | 1.90E-40 | Nightingale | LDL_P | NA | T | C | 0.053373 | 0.005333 | 2.30E-24 |
| rs2519093 | miR-126-3p | trans | 1.90E-40 | Nightingale | XS_VLDL_P | NA | T | C | 0.044763 | 0.005262 | 2.50E-19 |
| rs2519093 | miR-126-3p | trans | 1.90E-40 | Nightingale | ApoB | NA | T | C | 0.055887 | 0.005329 | 1.10E-26 |
| rs2519093 | miR-126-3p | trans | 1.90E-40 | Nightingale | L_LDL_CE | NA | T | C | 0.058613 | 0.005289 | 3.70E-29 |
| rs2519093 | miR-126-3p | trans | 1.90E-40 | Nightingale | S_LDL_C | NA | T | C | 0.05408 | 0.005341 | 1.20E-24 |
| rs2519093 | miR-126-3p | trans | 1.90E-40 | Nightingale | M_VLDL_F | NA | T | C | 0.05187 | 0.005335 | 1.30E-23 |
| rs2519093 | miR-126-3p | trans | 1.90E-40 | Nightingale | L_LDL_PL | NA | T | C | 0.066759 | 0.005296 | 3.60E-37 |
| rs2519093 | miR-126-3p | trans | 1.90E-40 | Nightingale | IDL_PL | NA | T | C | 0.058969 | 0.005149 | 2.40E-31 |
| rs2519093 | miR-126-3p | trans | 1.90E-40 | Nightingale | XXL_VLDL_ | NA | T | C | 0.056193 | 0.00538 | 1.00E-25 |
| rs2519093 | miR-126-3p | trans | 1.90E-40 | Nightingale | LDL_CE | NA | T | C | 0.0584 | 0.005322 | 9.20E-29 |
| rs2519093 | miR-126-3p | trans | 1.90E-40 | Nightingale | S_VLDL_C | NA | T | C | 0.059191 | 0.005336 | 2.80E-30 |
| rs2519093 | miR-126-3p | trans | 1.90E-40 | Nightingale | LDL_L | NA | T | C | 0.05971 | 0.005318 | 6.80E-30 |
| rs2519093 | miR-126-3p | trans | 1.90E-40 | Nightingale | L_LDL_L | NA | T | C | 0.061108 | 0.005284 | 1.20E-31 |
| rs2519093 | miR-126-3p | trans | 1.90E-40 | Nightingale | S_LDL_FC | NA | T | C | 0.054719 | 0.005328 | 1.90E-24 |
| rs2519093 | miR-126-3p | trans | 1.90E-40 | Nightingale | HDL_size | NA | T | C | -0.03268 | 0.004731 | 9.30E-13 |
| rs2519093 | miR-126-3p | trans | 1.90E-40 | Nightingale | XS_VLDL_T | NA | T | C | -0.04276 | 0.005074 | 2.50E-16 |
| rs2519093 | miR-126-3p | trans | 1.90E-40 | Nightingale | XS_VLDL_P | NA | T | C | 0.053945 | 0.005265 | 8.60E-27 |
| rs2519093 | miR-126-3p | trans | 1.90E-40 | Nightingale | Total_C | NA | T | C | 0.055882 | 0.005166 | 2.10E-28 |
| rs2519093 | miR-126-3p | trans | 1.90E-40 | Nightingale | Total_P | NA | T | C | 0.03671 | 0.005022 | 4.80E-14 |
| rs2519093 | miR-126-3p | trans | 1.90E-40 | Nightingale | VLDL_C | NA | T | C | 0.045887 | 0.005329 | 2.40E-19 |
| rs2519093 | miR-126-3p | trans | 1.90E-40 | Nightingale | IDL_FC | NA | T | C | 0.064239 | 0.005166 | 4.20E-36 |
| rs2519093 | miR-126-3p | trans | 1.90E-40 | Nightingale | M_LDL_TG | NA | T | C | -0.05344 | 0.00527 | 6.00E-24 |
| rs2519093 | miR-126-3p | trans | 1.90E-40 | Nightingale | XXL_VLDL_ | NA | T | C | 0.057015 | 0.005351 | 1.20E-26 |
| rs2519093 | miR-126-3p | trans | 1.90E-40 | Nightingale | Clinical_LD | NA | T | C | 0.064699 | 0.005305 | 1.10E-34 |
| rs2519093 | miR-126-3p | trans | 1.90E-40 | Nightingale | S_HDL_FC | NA | T | C | 0.041872 | 0.005238 | 1.90E-16 |
| rs2519093 | miR-126-3p | trans | 1.90E-40 | Nightingale | XL_HDL_L | NA | T | C | -0.02733 | 0.004799 | 1.80E-09 |
| rs2519093 | miR-126-3p | trans | 1.90E-40 | Nightingale | L_LDL_FC | NA | T | C | 0.062862 | 0.00524 | 2.40E-33 |
| rs2519093 | miR-126-3p | trans | 1.90E-40 | Nightingale | M_VLDL_F | NA | T | C | 0.067606 | 0.005077 | 8.00E-40 |
| rs2519093 | miR-126-3p | trans | 1.90E-40 | Nightingale | S_LDL_PL_ | NA | T | C | -0.04499 | 0.005326 | 8.80E-20 |
| rs2519093 | miR-126-3p | trans | 1.90E-40 | Nightingale | Citrate | NA | T | C | 0.031335 | 0.005301 | 1.70E-09 |
| rs2519093 | miR-126-3p | trans | 1.90E-40 | Nightingale | M_LDL_CE | NA | T | C | 0.055668 | 0.005343 | 1.90E-26 |
| rs2519093 | miR-126-3p | trans | 1.90E-40 | Nightingale | Phosphatic | NA | T | C | 0.031111 | 0.005 | 6.70E-11 |
| rs2519093 | miR-126-3p | trans | 1.90E-40 | Nightingale | Cholines | NA | T | C | 0.035715 | 0.005018 | 1.00E-13 |
| rs2519093 | miR-126-3p | trans | 1.90E-40 | Nightingale | IDL_C_pct | NA | T | C | 0.037575 | 0.005213 | 4.50E-12 |
| rs2519093 | miR-126-3p | trans | 1.90E-40 | Nightingale | XL_HDL_PL | NA | T | C | -0.0414 | 0.004903 | 9.60E-18 |
| rs2519093 | miR-126-3p | trans | 1.90E-40 | Nightingale | LA | NA | T | C | 0.042082 | 0.005233 | 4.40E-17 |
| rs2519093 | miR-126-3p | trans | 1.90E-40 | Nightingale | S_LDL_P | NA | T | C | 0.048597 | 0.005341 | 1.60E-20 |
| rs2519093 | miR-126-3p | trans | 1.90E-40 | Nightingale | M_VLDL_L | NA | T | C | 0.032196 | 0.0053 | 1.00E-10 |
| rs2519093 | miR-126-3p | trans | 1.90E-40 | Nightingale | XL_HDL_PL | NA | T | C | -0.03044 | 0.0048 | 4.90E-11 |
| rs2519093 | miR-126-3p | trans | 1.90E-40 | Nightingale | XL_VLDL_P | NA | T | C | 0.056118 | 0.005345 | 2.80E-27 |
| rs2519093 | miR-126-3p | trans | 1.90E-40 | Nightingale | XS_VLDL_F | NA | T | C | 0.053855 | 0.005244 | 8.50E-27 |
| rs2519093 | miR-126-3p | trans | 1.90E-40 | Nightingale | non_HDL_( | NA | T | C | 0.060625 | 0.005307 | 2.30E-31 |
| rs2519093 | miR-126-3p | trans | 1.90E-40 | Nightingale | XL_VLDL_T | NA | T | C | -0.06925 | 0.005173 | 9.80E-41 |
| rs2519093 | miR-126-3p | trans | 1.90E-40 | Nightingale | IDL_TG_pc | NA | T | C | -0.03897 | 0.005159 | 2.10E-13 |
| rs2519093 | miR-126-3p | trans | 1.90E-40 | Nightingale | Total_PL | NA | T | C | 0.039528 | 0.005081 | 4.00E-16 |
| rs2519093 | miR-126-3p | trans | 1.90E-40 | Nightingale | M_VLDL_T | NA | T | C | -0.06313 | 0.005043 | 4.00E-35 |
| rs2519093 | miR-126-3p | trans | 1.90E-40 | Nightingale | L_VLDL_FC | NA | T | C | 0.076264 | 0.005285 | 5.50E-48 |
| rs2519093 | miR-126-3p | trans | 1.90E-40 | Nightingale | S_VLDL_P | NA | T | C | 0.033454 | 0.005269 | 6.70E-12 |
| rs2519093 | miR-126-3p | trans | 1.90E-40 | Nightingale | S_LDL_PL | NA | T | C | 0.041418 | 0.005324 | 6.40E-15 |
| rs2519093 | miR-126-3p | trans | 1.90E-40 | Nightingale | XS_VLDL_C | NA | T | C | 0.058552 | 0.00515 | 2.40E-31 |
| rs2519093 | miR-126-3p | trans | 1.90E-40 | Nightingale | L_VLDL_CE | NA | T | C | 0.074537 | 0.005107 | 7.40E-49 |
| rs2519093 | miR-126-3p | trans | 1.90E-40 | Nightingale | XL_HDL_C_ | NA | T | C | 0.038739 | 0.005096 | 1.00E-13 |
| rs2519093 | miR-126-3p | trans | 1.90E-40 | Nightingale | M_LDL_C | NA | T | C | 0.058986 | 0.005346 | 4.10E-29 |
| rs2519093 | miR-126-3p | trans | 1.90E-40 | Nightingale | S_LDL_TG_ | NA | T | C | -0.0438 | 0.005175 | 2.30E-16 |
| rs2519093 | miR-126-3p | trans | 1.90E-40 | Nightingale | XL_VLDL_C | NA | T | C | 0.062611 | 0.00512 | 1.50E-33 |
| rs2519093 | miR-126-3p | trans | 1.90E-40 | Nightingale | S_VLDL_FC | NA | T | C | 0.059655 | 0.005337 | 2.30E-30 |
| rs2519093 | miR-126-3p | trans | 1.90E-40 | Nightingale | IDL_FC_pct | NA | T | C | 0.041553 | 0.005295 | 1.40E-14 |
| rs2519093 | miR-126-3p | trans | 1.90E-40 | Nightingale | M_LDL_PL | NA | T | C | 0.058342 | 0.005344 | 1.80E-28 |

| | | | | | | | | | | | |
|---|---|---|---|---|---|---|---|---|---|---|---|
| rs2519093 | miR-126-3p | trans | 1.90E-40 | Nightingale | L_VLDL_PL | NA | T | C | 0.028334 | 0.005183 | 2.20E-09 |
| rs2519093 | miR-126-3p | trans | 1.90E-40 | Nightingale | L_LDL_P | NA | T | C | 0.050604 | 0.005318 | 4.80E-22 |
| rs2519093 | miR-126-3p | trans | 1.90E-40 | Nightingale | LDL_PL | NA | T | C | 0.06213 | 0.005324 | 5.60E-32 |
| rs2519093 | miR-126-3p | trans | 1.90E-40 | Nightingale | S_VLDL_TG | NA | T | C | -0.06553 | 0.005165 | 2.20E-36 |
| rs2519093 | miR-126-3p | trans | 1.90E-40 | Nightingale | Pyruvate | NA | T | C | 0.034476 | 0.005345 | 1.10E-10 |
| rs2519093 | miR-126-3p | trans | 1.90E-40 | Nightingale | ApoB_by_A | NA | T | C | 0.040245 | 0.005256 | 9.90E-15 |
| rs2519093 | miR-126-3p | trans | 1.90E-40 | Nightingale | XL_VLDL_F | NA | T | C | 0.056835 | 0.005174 | 2.50E-28 |
| rs2519093 | miR-126-3p | trans | 1.90E-40 | Nightingale | S_VLDL_PL | NA | T | C | 0.050243 | 0.005326 | 1.00E-22 |
| rs2519093 | miR-126-3p | trans | 1.90E-40 | Nightingale | M_VLDL_P | NA | T | C | 0.070901 | 0.005121 | 2.00E-43 |
| rs2519093 | miR-126-3p | trans | 1.90E-40 | Nightingale | S_LDL_CE_ | NA | T | C | 0.04485 | 0.005327 | 8.40E-20 |
| rs2519093 | miR-126-3p | trans | 1.90E-40 | Nightingale | XXL_VLDL_ | NA | T | C | 0.036769 | 0.005374 | 2.00E-13 |
| rs2519093 | miR-126-3p | trans | 1.90E-40 | Nightingale | M_VLDL_C | NA | T | C | 0.059763 | 0.005026 | 1.20E-31 |
| rs2519093 | miR-126-3p | trans | 1.90E-40 | Nightingale | XS_VLDL_C | NA | T | C | 0.057788 | 0.00518 | 1.80E-30 |
| rs2519093 | miR-126-3p | trans | 1.90E-40 | Nightingale | S_HDL_L | NA | T | C | 0.04414 | 0.005318 | 1.60E-17 |
| rs2519093 | miR-126-3p | trans | 1.90E-40 | Nightingale | S_LDL_C_p | NA | T | C | 0.075956 | 0.005357 | 8.10E-47 |
| rs2519093 | miR-126-3p | trans | 1.90E-40 | Nightingale | M_LDL_C_ | NA | T | C | 0.047547 | 0.00534 | 3.80E-19 |
| rs2519093 | miR-126-3p | trans | 1.90E-40 | Nightingale | XL_VLDL_C | NA | T | C | 0.062776 | 0.005117 | 8.10E-34 |
| rs2519093 | miR-126-3p | trans | 1.90E-40 | Nightingale | LDL_FC | NA | T | C | 0.062925 | 0.005279 | 9.00E-33 |
| rs2519093 | miR-126-3p | trans | 1.90E-40 | Nightingale | L_VLDL_TG | NA | T | C | -0.08007 | 0.005311 | 4.90E-53 |
| rs2519093 | miR-126-3p | trans | 1.90E-40 | Nightingale | S_HDL_PL_ | NA | T | C | -0.03292 | 0.005159 | 7.60E-10 |
| rs2519093 | miR-126-3p | trans | 1.90E-40 | Nightingale | HDL_P | NA | T | C | 0.028786 | 0.005011 | 3.00E-09 |
| rs2519093 | miR-126-3p | trans | 1.90E-40 | Nightingale | LDL_C | NA | T | C | 0.060241 | 0.005313 | 2.20E-30 |
| rs2519093 | miR-126-3p | trans | 1.90E-40 | Nightingale | L_LDL_FC_ | NA | T | C | 0.030917 | 0.005091 | 1.20E-08 |
| rs2519093 | miR-126-3p | trans | 1.90E-40 | Nightingale | M_LDL_L | NA | T | C | 0.057522 | 0.005344 | 6.90E-28 |
| rs2519093 | miR-126-3p | trans | 1.90E-40 | Nightingale | XL_HDL_CE | NA | T | C | 0.030201 | 0.005332 | 3.60E-08 |
| rs2519093 | miR-126-3p | trans | 1.90E-40 | Nightingale | M_LDL_FC | NA | T | C | 0.063872 | 0.00533 | 2.80E-33 |
| rs2519093 | miR-126-3p | trans | 1.90E-40 | Nightingale | S_VLDL_CE | NA | T | C | 0.076957 | 0.005265 | 7.60E-48 |
| rs2519093 | miR-126-3p | trans | 1.90E-40 | Nightingale | Remnant_( | NA | T | C | 0.058623 | 0.005302 | 4.00E-30 |
| rs2519093 | let-7a-5p | trans | 1.29E-11 | Nightingale | S_HDL_CE | NA | T | C | 0.053505 | 0.005345 | 7.30E-24 |
| rs2519093 | let-7a-5p | trans | 1.29E-11 | Nightingale | L_LDL_TG_ | NA | T | C | -0.04178 | 0.005234 | 5.50E-15 |
| rs2519093 | let-7a-5p | trans | 1.29E-11 | Nightingale | M_VLDL_P | NA | T | C | 0.04488 | 0.005328 | 1.70E-18 |
| rs2519093 | let-7a-5p | trans | 1.29E-11 | Nightingale | IDL_C | NA | T | C | 0.060311 | 0.005154 | 6.00E-32 |
| rs2519093 | let-7a-5p | trans | 1.29E-11 | Nightingale | M_VLDL_C | NA | T | C | 0.066274 | 0.005288 | 7.60E-37 |
| rs2519093 | let-7a-5p | trans | 1.29E-11 | Nightingale | Total_FC | NA | T | C | 0.055093 | 0.005213 | 7.90E-28 |
| rs2519093 | let-7a-5p | trans | 1.29E-11 | Nightingale | S_VLDL_C_ | NA | T | C | 0.070806 | 0.005193 | 5.60E-42 |
| rs2519093 | let-7a-5p | trans | 1.29E-11 | Nightingale | LA_pct | NA | T | C | 0.031876 | 0.005218 | 1.10E-09 |
| rs2519093 | let-7a-5p | trans | 1.29E-11 | Nightingale | S_VLDL_L | NA | T | C | 0.035666 | 0.005285 | 4.30E-13 |
| rs2519093 | let-7a-5p | trans | 1.29E-11 | Nightingale | M_VLDL_C | NA | T | C | 0.061815 | 0.005321 | 1.60E-32 |
| rs2519093 | let-7a-5p | trans | 1.29E-11 | Nightingale | XS_VLDL_F | NA | T | C | 0.051132 | 0.005298 | 6.20E-21 |
| rs2519093 | let-7a-5p | trans | 1.29E-11 | Nightingale | IDL_P | NA | T | C | 0.065267 | 0.00528 | 1.10E-35 |
| rs2519093 | let-7a-5p | trans | 1.29E-11 | Nightingale | S_VLDL_CE | NA | T | C | 0.05821 | 0.005329 | 1.10E-29 |
| rs2519093 | let-7a-5p | trans | 1.29E-11 | Nightingale | XS_VLDL_L | NA | T | C | 0.048684 | 0.005245 | 1.40E-22 |
| rs2519093 | let-7a-5p | trans | 1.29E-11 | Nightingale | S_LDL_CE | NA | T | C | 0.051776 | 0.005339 | 4.00E-23 |
| rs2519093 | let-7a-5p | trans | 1.29E-11 | Nightingale | IDL_L | NA | T | C | 0.059774 | 0.00516 | 1.10E-31 |
| rs2519093 | let-7a-5p | trans | 1.29E-11 | Nightingale | Total_CE | NA | T | C | 0.05575 | 0.005147 | 2.40E-28 |
| rs2519093 | let-7a-5p | trans | 1.29E-11 | Nightingale | Sphingomy | NA | T | C | 0.047648 | 0.004989 | 2.20E-21 |
| rs2519093 | let-7a-5p | trans | 1.29E-11 | Nightingale | PUFA | NA | T | C | 0.036949 | 0.005205 | 1.30E-13 |
| rs2519093 | let-7a-5p | trans | 1.29E-11 | Nightingale | XS_VLDL_C | NA | T | C | 0.040361 | 0.005035 | 1.00E-14 |
| rs2519093 | let-7a-5p | trans | 1.29E-11 | Nightingale | IDL_CE | NA | T | C | 0.058439 | 0.005152 | 4.70E-30 |
| rs2519093 | let-7a-5p | trans | 1.29E-11 | Nightingale | VLDL_CE | NA | T | C | 0.053783 | 0.005338 | 2.10E-25 |
| rs2519093 | let-7a-5p | trans | 1.29E-11 | Nightingale | S_HDL_PL | NA | T | C | 0.037688 | 0.00529 | 1.40E-13 |
| rs2519093 | let-7a-5p | trans | 1.29E-11 | Nightingale | Omega_6 | NA | T | C | 0.041735 | 0.005211 | 5.40E-17 |
| rs2519093 | let-7a-5p | trans | 1.29E-11 | Nightingale | S_HDL_C | NA | T | C | 0.052616 | 0.005327 | 2.20E-23 |
| rs2519093 | let-7a-5p | trans | 1.29E-11 | Nightingale | M_VLDL_C | NA | T | C | 0.056108 | 0.005013 | 3.90E-28 |
| rs2519093 | let-7a-5p | trans | 1.29E-11 | Nightingale | GlycA | NA | T | C | 0.031449 | 0.005314 | 4.90E-09 |
| rs2519093 | let-7a-5p | trans | 1.29E-11 | Nightingale | XS_VLDL_C | NA | T | C | 0.043421 | 0.005061 | 7.70E-17 |
| rs2519093 | let-7a-5p | trans | 1.29E-11 | Nightingale | M_VLDL_P | NA | T | C | 0.043347 | 0.005323 | 1.90E-17 |
| rs2519093 | let-7a-5p | trans | 1.29E-11 | Nightingale | XS_VLDL_P | NA | T | C | -0.02949 | 0.005207 | 5.00E-08 |
| rs2519093 | let-7a-5p | trans | 1.29E-11 | Nightingale | S_VLDL_PL | NA | T | C | 0.050234 | 0.00511 | 4.10E-22 |
| rs2519093 | let-7a-5p | trans | 1.29E-11 | Nightingale | L_LDL_C | NA | T | C | 0.060158 | 0.005276 | 1.10E-30 |
| rs2519093 | let-7a-5p | trans | 1.29E-11 | Nightingale | Total_L | NA | T | C | 0.042371 | 0.00525 | 4.30E-17 |
| rs2519093 | let-7a-5p | trans | 1.29E-11 | Nightingale | Phosphogly | NA | T | C | 0.031931 | 0.005054 | 3.40E-11 |
| rs2519093 | let-7a-5p | trans | 1.29E-11 | Nightingale | L_VLDL_C_ | NA | T | C | 0.082094 | 0.005212 | 9.00E-57 |

| | | | | | | | | | | | |
|---|---|---|---|---|---|---|---|---|---|---|---|
| rs2519093 | let-7a-5p | trans | 1.29E-11 | Nightingale | L_VLDL_CE | NA | T | C | 0.031525 | 0.005274 | 1.30E-10 |
| rs2519093 | let-7a-5p | trans | 1.29E-11 | Nightingale | VLDL_P | NA | T | C | 0.038356 | 0.005308 | 2.10E-14 |
| rs2519093 | let-7a-5p | trans | 1.29E-11 | Nightingale | M_LDL_P | NA | T | C | 0.057192 | 0.005351 | 1.30E-27 |
| rs2519093 | let-7a-5p | trans | 1.29E-11 | Nightingale | S_VLDL_FC | NA | T | C | 0.052723 | 0.005099 | 2.50E-24 |
| rs2519093 | let-7a-5p | trans | 1.29E-11 | Nightingale | S_HDL_P | NA | T | C | 0.055391 | 0.005337 | 1.10E-25 |
| rs2519093 | let-7a-5p | trans | 1.29E-11 | Nightingale | S_LDL_L | NA | T | C | 0.048584 | 0.005335 | 3.30E-20 |
| rs2519093 | let-7a-5p | trans | 1.29E-11 | Nightingale | VLDL_FC | NA | T | C | 0.033679 | 0.005297 | 1.10E-11 |
| rs2519093 | let-7a-5p | trans | 1.29E-11 | Nightingale | XL_VLDL_C | NA | T | C | 0.029731 | 0.005254 | 9.50E-10 |
| rs2519093 | let-7a-5p | trans | 1.29E-11 | Nightingale | XXL_VLDL_ | NA | T | C | 0.046178 | 0.005317 | 1.50E-18 |
| rs2519093 | let-7a-5p | trans | 1.29E-11 | Nightingale | XXL_VLDL_ | NA | T | C | -0.05831 | 0.005385 | 5.10E-28 |
| rs2519093 | let-7a-5p | trans | 1.29E-11 | Nightingale | LDL_P | NA | T | C | 0.053373 | 0.005333 | 2.30E-24 |
| rs2519093 | let-7a-5p | trans | 1.29E-11 | Nightingale | XS_VLDL_P | NA | T | C | 0.044763 | 0.005262 | 2.50E-19 |
| rs2519093 | let-7a-5p | trans | 1.29E-11 | Nightingale | ApoB | NA | T | C | 0.055887 | 0.005329 | 1.10E-26 |
| rs2519093 | let-7a-5p | trans | 1.29E-11 | Nightingale | L_LDL_CE | NA | T | C | 0.058613 | 0.005289 | 3.70E-29 |
| rs2519093 | let-7a-5p | trans | 1.29E-11 | Nightingale | S_LDL_C | NA | T | C | 0.05408 | 0.005341 | 1.20E-24 |
| rs2519093 | let-7a-5p | trans | 1.29E-11 | Nightingale | M_VLDL_F | NA | T | C | 0.05187 | 0.005335 | 1.30E-23 |
| rs2519093 | let-7a-5p | trans | 1.29E-11 | Nightingale | L_LDL_PL | NA | T | C | 0.066759 | 0.005296 | 3.60E-37 |
| rs2519093 | let-7a-5p | trans | 1.29E-11 | Nightingale | IDL_PL | NA | T | C | 0.058969 | 0.005149 | 2.40E-31 |
| rs2519093 | let-7a-5p | trans | 1.29E-11 | Nightingale | XXL_VLDL_ | NA | T | C | 0.056193 | 0.00538 | 1.00E-25 |
| rs2519093 | let-7a-5p | trans | 1.29E-11 | Nightingale | LDL_CE | NA | T | C | 0.0584 | 0.005322 | 9.20E-29 |
| rs2519093 | let-7a-5p | trans | 1.29E-11 | Nightingale | S_VLDL_C | NA | T | C | 0.059191 | 0.005336 | 2.80E-30 |
| rs2519093 | let-7a-5p | trans | 1.29E-11 | Nightingale | LDL_L | NA | T | C | 0.05971 | 0.005318 | 6.80E-30 |
| rs2519093 | let-7a-5p | trans | 1.29E-11 | Nightingale | L_LDL_L | NA | T | C | 0.061108 | 0.005284 | 1.20E-31 |
| rs2519093 | let-7a-5p | trans | 1.29E-11 | Nightingale | S_LDL_FC | NA | T | C | 0.054719 | 0.005328 | 1.90E-24 |
| rs2519093 | let-7a-5p | trans | 1.29E-11 | Nightingale | HDL_size | NA | T | C | -0.03268 | 0.004731 | 9.30E-13 |
| rs2519093 | let-7a-5p | trans | 1.29E-11 | Nightingale | XS_VLDL_T | NA | T | C | -0.04276 | 0.005074 | 2.50E-16 |
| rs2519093 | let-7a-5p | trans | 1.29E-11 | Nightingale | XS_VLDL_P | NA | T | C | 0.053945 | 0.005265 | 8.60E-27 |
| rs2519093 | let-7a-5p | trans | 1.29E-11 | Nightingale | Total_C | NA | T | C | 0.055882 | 0.005166 | 2.10E-28 |
| rs2519093 | let-7a-5p | trans | 1.29E-11 | Nightingale | Total_P | NA | T | C | 0.03671 | 0.005022 | 4.80E-14 |
| rs2519093 | let-7a-5p | trans | 1.29E-11 | Nightingale | VLDL_C | NA | T | C | 0.045887 | 0.005329 | 2.40E-19 |
| rs2519093 | let-7a-5p | trans | 1.29E-11 | Nightingale | IDL_FC | NA | T | C | 0.064239 | 0.005166 | 4.20E-36 |
| rs2519093 | let-7a-5p | trans | 1.29E-11 | Nightingale | M_LDL_TG | NA | T | C | -0.05344 | 0.00527 | 6.00E-24 |
| rs2519093 | let-7a-5p | trans | 1.29E-11 | Nightingale | XXL_VLDL_ | NA | T | C | 0.057015 | 0.005351 | 1.20E-26 |
| rs2519093 | let-7a-5p | trans | 1.29E-11 | Nightingale | Clinical_LD | NA | T | C | 0.064699 | 0.005305 | 1.10E-34 |
| rs2519093 | let-7a-5p | trans | 1.29E-11 | Nightingale | S_HDL_FC | NA | T | C | 0.041872 | 0.005238 | 1.90E-16 |
| rs2519093 | let-7a-5p | trans | 1.29E-11 | Nightingale | XL_HDL_L | NA | T | C | -0.02733 | 0.004799 | 1.80E-09 |
| rs2519093 | let-7a-5p | trans | 1.29E-11 | Nightingale | L_LDL_FC | NA | T | C | 0.062862 | 0.00524 | 2.40E-33 |
| rs2519093 | let-7a-5p | trans | 1.29E-11 | Nightingale | M_VLDL_F | NA | T | C | 0.067606 | 0.005077 | 8.00E-40 |
| rs2519093 | let-7a-5p | trans | 1.29E-11 | Nightingale | S_LDL_PL_ | NA | T | C | -0.04499 | 0.005326 | 8.80E-20 |
| rs2519093 | let-7a-5p | trans | 1.29E-11 | Nightingale | Citrate | NA | T | C | 0.031335 | 0.005301 | 1.70E-09 |
| rs2519093 | let-7a-5p | trans | 1.29E-11 | Nightingale | M_LDL_CE | NA | T | C | 0.055668 | 0.005343 | 1.90E-26 |
| rs2519093 | let-7a-5p | trans | 1.29E-11 | Nightingale | Phosphatic | NA | T | C | 0.031111 | 0.005 | 6.70E-11 |
| rs2519093 | let-7a-5p | trans | 1.29E-11 | Nightingale | Cholines | NA | T | C | 0.035715 | 0.005018 | 1.00E-13 |
| rs2519093 | let-7a-5p | trans | 1.29E-11 | Nightingale | IDL_C_pct | NA | T | C | 0.037575 | 0.005213 | 4.50E-12 |
| rs2519093 | let-7a-5p | trans | 1.29E-11 | Nightingale | XL_HDL_PL | NA | T | C | -0.0414 | 0.004903 | 9.60E-18 |
| rs2519093 | let-7a-5p | trans | 1.29E-11 | Nightingale | LA | NA | T | C | 0.042082 | 0.005233 | 4.40E-17 |
| rs2519093 | let-7a-5p | trans | 1.29E-11 | Nightingale | S_LDL_P | NA | T | C | 0.048597 | 0.005341 | 1.60E-20 |
| rs2519093 | let-7a-5p | trans | 1.29E-11 | Nightingale | M_VLDL_L | NA | T | C | 0.032196 | 0.0053 | 1.00E-10 |
| rs2519093 | let-7a-5p | trans | 1.29E-11 | Nightingale | XL_HDL_PL | NA | T | C | -0.03044 | 0.0048 | 4.90E-11 |
| rs2519093 | let-7a-5p | trans | 1.29E-11 | Nightingale | XL_VLDL_P | NA | T | C | 0.056118 | 0.005345 | 2.80E-27 |
| rs2519093 | let-7a-5p | trans | 1.29E-11 | Nightingale | XS_VLDL_F | NA | T | C | 0.053855 | 0.005244 | 8.50E-27 |
| rs2519093 | let-7a-5p | trans | 1.29E-11 | Nightingale | non_HDL_( | NA | T | C | 0.060625 | 0.005307 | 2.30E-31 |
| rs2519093 | let-7a-5p | trans | 1.29E-11 | Nightingale | XL_VLDL_T | NA | T | C | -0.06925 | 0.005173 | 9.80E-41 |
| rs2519093 | let-7a-5p | trans | 1.29E-11 | Nightingale | IDL_TG_pc | NA | T | C | -0.03897 | 0.005159 | 2.10E-13 |
| rs2519093 | let-7a-5p | trans | 1.29E-11 | Nightingale | Total_PL | NA | T | C | 0.039528 | 0.005081 | 4.00E-16 |
| rs2519093 | let-7a-5p | trans | 1.29E-11 | Nightingale | M_VLDL_T | NA | T | C | -0.06313 | 0.005043 | 4.00E-35 |
| rs2519093 | let-7a-5p | trans | 1.29E-11 | Nightingale | L_VLDL_FC | NA | T | C | 0.076264 | 0.005285 | 5.50E-48 |
| rs2519093 | let-7a-5p | trans | 1.29E-11 | Nightingale | S_VLDL_P | NA | T | C | 0.033454 | 0.005269 | 6.70E-12 |
| rs2519093 | let-7a-5p | trans | 1.29E-11 | Nightingale | S_LDL_PL | NA | T | C | 0.041418 | 0.005324 | 6.40E-15 |
| rs2519093 | let-7a-5p | trans | 1.29E-11 | Nightingale | XS_VLDL_C | NA | T | C | 0.058552 | 0.00515 | 2.40E-31 |
| rs2519093 | let-7a-5p | trans | 1.29E-11 | Nightingale | L_VLDL_CE | NA | T | C | 0.074537 | 0.005107 | 7.40E-49 |
| rs2519093 | let-7a-5p | trans | 1.29E-11 | Nightingale | XL_HDL_C_ | NA | T | C | 0.038739 | 0.005096 | 1.00E-13 |
| rs2519093 | let-7a-5p | trans | 1.29E-11 | Nightingale | M_LDL_C | NA | T | C | 0.058986 | 0.005346 | 4.10E-29 |

| | | | | | | | | | | | |
|---|---|---|---|---|---|---|---|---|---|---|---|
| rs2519093 | let-7a-5p | trans | 1.29E-11 | Nightingale | S_LDL_TG_ | NA | T | C | -0.0438 | 0.005175 | 2.30E-16 |
| rs2519093 | let-7a-5p | trans | 1.29E-11 | Nightingale | XL_VLDL_C | NA | T | C | 0.062611 | 0.00512 | 1.50E-33 |
| rs2519093 | let-7a-5p | trans | 1.29E-11 | Nightingale | S_VLDL_FC | NA | T | C | 0.059655 | 0.005337 | 2.30E-30 |
| rs2519093 | let-7a-5p | trans | 1.29E-11 | Nightingale | IDL_FC_pct | NA | T | C | 0.041553 | 0.005295 | 1.40E-14 |
| rs2519093 | let-7a-5p | trans | 1.29E-11 | Nightingale | M_LDL_PL | NA | T | C | 0.058342 | 0.005344 | 1.80E-28 |
| rs2519093 | let-7a-5p | trans | 1.29E-11 | Nightingale | L_VLDL_PL | NA | T | C | 0.028334 | 0.005183 | 2.20E-09 |
| rs2519093 | let-7a-5p | trans | 1.29E-11 | Nightingale | L_LDL_P | NA | T | C | 0.050604 | 0.005318 | 4.80E-22 |
| rs2519093 | let-7a-5p | trans | 1.29E-11 | Nightingale | LDL_PL | NA | T | C | 0.06213 | 0.005324 | 5.60E-32 |
| rs2519093 | let-7a-5p | trans | 1.29E-11 | Nightingale | S_VLDL_TG | NA | T | C | -0.06553 | 0.005165 | 2.20E-36 |
| rs2519093 | let-7a-5p | trans | 1.29E-11 | Nightingale | Pyruvate | NA | T | C | 0.034476 | 0.005345 | 1.10E-10 |
| rs2519093 | let-7a-5p | trans | 1.29E-11 | Nightingale | ApoB_by_A | NA | T | C | 0.040245 | 0.005256 | 9.90E-15 |
| rs2519093 | let-7a-5p | trans | 1.29E-11 | Nightingale | XL_VLDL_F | NA | T | C | 0.056835 | 0.005174 | 2.50E-28 |
| rs2519093 | let-7a-5p | trans | 1.29E-11 | Nightingale | S_VLDL_PL | NA | T | C | 0.050243 | 0.005326 | 1.00E-22 |
| rs2519093 | let-7a-5p | trans | 1.29E-11 | Nightingale | M_VLDL_P | NA | T | C | 0.070901 | 0.005121 | 2.00E-43 |
| rs2519093 | let-7a-5p | trans | 1.29E-11 | Nightingale | S_LDL_CE_ | NA | T | C | 0.04485 | 0.005327 | 8.40E-20 |
| rs2519093 | let-7a-5p | trans | 1.29E-11 | Nightingale | XXL_VLDL_ | NA | T | C | 0.036769 | 0.005374 | 2.00E-13 |
| rs2519093 | let-7a-5p | trans | 1.29E-11 | Nightingale | M_VLDL_C | NA | T | C | 0.059763 | 0.005026 | 1.20E-31 |
| rs2519093 | let-7a-5p | trans | 1.29E-11 | Nightingale | XS_VLDL_C | NA | T | C | 0.057788 | 0.00518 | 1.80E-30 |
| rs2519093 | let-7a-5p | trans | 1.29E-11 | Nightingale | S_HDL_L | NA | T | C | 0.04414 | 0.005318 | 1.60E-17 |
| rs2519093 | let-7a-5p | trans | 1.29E-11 | Nightingale | S_LDL_C_p | NA | T | C | 0.075956 | 0.005357 | 8.10E-47 |
| rs2519093 | let-7a-5p | trans | 1.29E-11 | Nightingale | M_LDL_C_ | NA | T | C | 0.047547 | 0.00534 | 3.80E-19 |
| rs2519093 | let-7a-5p | trans | 1.29E-11 | Nightingale | XL_VLDL_C | NA | T | C | 0.062776 | 0.005117 | 8.10E-34 |
| rs2519093 | let-7a-5p | trans | 1.29E-11 | Nightingale | LDL_FC | NA | T | C | 0.062925 | 0.005279 | 9.00E-33 |
| rs2519093 | let-7a-5p | trans | 1.29E-11 | Nightingale | L_VLDL_TG | NA | T | C | -0.08007 | 0.005311 | 4.90E-53 |
| rs2519093 | let-7a-5p | trans | 1.29E-11 | Nightingale | S_HDL_PL_ | NA | T | C | -0.03292 | 0.005159 | 7.60E-10 |
| rs2519093 | let-7a-5p | trans | 1.29E-11 | Nightingale | HDL_P | NA | T | C | 0.028786 | 0.005011 | 3.00E-09 |
| rs2519093 | let-7a-5p | trans | 1.29E-11 | Nightingale | LDL_C | NA | T | C | 0.060241 | 0.005313 | 2.20E-30 |
| rs2519093 | let-7a-5p | trans | 1.29E-11 | Nightingale | L_LDL_FC_ | NA | T | C | 0.030917 | 0.005091 | 1.20E-08 |
| rs2519093 | let-7a-5p | trans | 1.29E-11 | Nightingale | M_LDL_L | NA | T | C | 0.057522 | 0.005344 | 6.90E-28 |
| rs2519093 | let-7a-5p | trans | 1.29E-11 | Nightingale | XL_HDL_CE | NA | T | C | 0.030201 | 0.005332 | 3.60E-08 |
| rs2519093 | let-7a-5p | trans | 1.29E-11 | Nightingale | M_LDL_FC | NA | T | C | 0.063872 | 0.00533 | 2.80E-33 |
| rs2519093 | let-7a-5p | trans | 1.29E-11 | Nightingale | S_VLDL_CE | NA | T | C | 0.076957 | 0.005265 | 7.60E-48 |
| rs2519093 | let-7a-5p | trans | 1.29E-11 | Nightingale | Remnant_( | NA | T | C | 0.058623 | 0.005302 | 4.00E-30 |
| rs2519093 | miR-10b-5p | trans | 3.13E-35 | Nightingale | S_HDL_CE | NA | T | C | 0.053505 | 0.005345 | 7.30E-24 |
| rs2519093 | miR-10b-5p | trans | 3.13E-35 | Nightingale | L_LDL_TG_ | NA | T | C | -0.04178 | 0.005234 | 5.50E-15 |
| rs2519093 | miR-10b-5p | trans | 3.13E-35 | Nightingale | M_VLDL_P | NA | T | C | 0.04488 | 0.005328 | 1.70E-18 |
| rs2519093 | miR-10b-5p | trans | 3.13E-35 | Nightingale | IDL_C | NA | T | C | 0.060311 | 0.005154 | 6.00E-32 |
| rs2519093 | miR-10b-5p | trans | 3.13E-35 | Nightingale | M_VLDL_C | NA | T | C | 0.066274 | 0.005288 | 7.60E-37 |
| rs2519093 | miR-10b-5p | trans | 3.13E-35 | Nightingale | Total_FC | NA | T | C | 0.055093 | 0.005213 | 7.90E-28 |
| rs2519093 | miR-10b-5p | trans | 3.13E-35 | Nightingale | S_VLDL_C_ | NA | T | C | 0.070806 | 0.005193 | 5.60E-42 |
| rs2519093 | miR-10b-5p | trans | 3.13E-35 | Nightingale | LA_pct | NA | T | C | 0.031876 | 0.005218 | 1.10E-09 |
| rs2519093 | miR-10b-5p | trans | 3.13E-35 | Nightingale | S_VLDL_L | NA | T | C | 0.035666 | 0.005285 | 4.30E-13 |
| rs2519093 | miR-10b-5p | trans | 3.13E-35 | Nightingale | M_VLDL_C | NA | T | C | 0.061815 | 0.005321 | 1.60E-32 |
| rs2519093 | miR-10b-5p | trans | 3.13E-35 | Nightingale | XS_VLDL_F | NA | T | C | 0.051132 | 0.005298 | 6.20E-21 |
| rs2519093 | miR-10b-5p | trans | 3.13E-35 | Nightingale | IDL_P | NA | T | C | 0.065267 | 0.00528 | 1.10E-35 |
| rs2519093 | miR-10b-5p | trans | 3.13E-35 | Nightingale | S_VLDL_CE | NA | T | C | 0.05821 | 0.005329 | 1.10E-29 |
| rs2519093 | miR-10b-5p | trans | 3.13E-35 | Nightingale | XS_VLDL_L | NA | T | C | 0.048684 | 0.005245 | 1.40E-22 |
| rs2519093 | miR-10b-5p | trans | 3.13E-35 | Nightingale | S_LDL_CE | NA | T | C | 0.051776 | 0.005339 | 4.00E-23 |
| rs2519093 | miR-10b-5p | trans | 3.13E-35 | Nightingale | IDL_L | NA | T | C | 0.059774 | 0.00516 | 1.10E-31 |
| rs2519093 | miR-10b-5p | trans | 3.13E-35 | Nightingale | Total_CE | NA | T | C | 0.05575 | 0.005147 | 2.40E-28 |
| rs2519093 | miR-10b-5p | trans | 3.13E-35 | Nightingale | Sphingomy | NA | T | C | 0.047648 | 0.004989 | 2.20E-21 |
| rs2519093 | miR-10b-5p | trans | 3.13E-35 | Nightingale | PUFA | NA | T | C | 0.036949 | 0.005205 | 1.30E-13 |
| rs2519093 | miR-10b-5p | trans | 3.13E-35 | Nightingale | XS_VLDL_C | NA | T | C | 0.040361 | 0.005035 | 1.00E-14 |
| rs2519093 | miR-10b-5p | trans | 3.13E-35 | Nightingale | IDL_CE | NA | T | C | 0.058439 | 0.005152 | 4.70E-30 |
| rs2519093 | miR-10b-5p | trans | 3.13E-35 | Nightingale | VLDL_CE | NA | T | C | 0.053783 | 0.005338 | 2.10E-25 |
| rs2519093 | miR-10b-5p | trans | 3.13E-35 | Nightingale | S_HDL_PL | NA | T | C | 0.037688 | 0.00529 | 1.40E-13 |
| rs2519093 | miR-10b-5p | trans | 3.13E-35 | Nightingale | Omega_6 | NA | T | C | 0.041735 | 0.005211 | 5.40E-17 |
| rs2519093 | miR-10b-5p | trans | 3.13E-35 | Nightingale | S_HDL_C | NA | T | C | 0.052616 | 0.005327 | 2.20E-23 |
| rs2519093 | miR-10b-5p | trans | 3.13E-35 | Nightingale | M_VLDL_C | NA | T | C | 0.056108 | 0.005013 | 3.90E-28 |
| rs2519093 | miR-10b-5p | trans | 3.13E-35 | Nightingale | GlycA | NA | T | C | 0.031449 | 0.005314 | 4.90E-09 |
| rs2519093 | miR-10b-5p | trans | 3.13E-35 | Nightingale | XS_VLDL_C | NA | T | C | 0.043421 | 0.005061 | 7.70E-17 |
| rs2519093 | miR-10b-5p | trans | 3.13E-35 | Nightingale | M_VLDL_P | NA | T | C | 0.043347 | 0.005323 | 1.90E-17 |
| rs2519093 | miR-10b-5p | trans | 3.13E-35 | Nightingale | XS_VLDL_P | NA | T | C | -0.02949 | 0.005207 | 5.00E-08 |

| | | | | | | | | | | | | |
|---|---|---|---|---|---|---|---|---|---|---|---|---|
| rs2519093 | miR-10b-5p | trans | | 3.13E-35 | Nightingale | S_VLDL_PL | NA | T | C | 0.050234 | 0.00511 | 4.10E-22 |
| rs2519093 | miR-10b-5p | trans | | 3.13E-35 | Nightingale | L_LDL_C | NA | T | C | 0.060158 | 0.005276 | 1.10E-30 |
| rs2519093 | miR-10b-5p | trans | | 3.13E-35 | Nightingale | Total_L | NA | T | C | 0.042371 | 0.00525 | 4.30E-17 |
| rs2519093 | miR-10b-5p | trans | | 3.13E-35 | Nightingale | Phosphogly | NA | T | C | 0.031931 | 0.005054 | 3.40E-11 |
| rs2519093 | miR-10b-5p | trans | | 3.13E-35 | Nightingale | L_VLDL_C_ | NA | T | C | 0.082094 | 0.005212 | 9.00E-57 |
| rs2519093 | miR-10b-5p | trans | | 3.13E-35 | Nightingale | L_VLDL_CE | NA | T | C | 0.031525 | 0.005274 | 1.30E-10 |
| rs2519093 | miR-10b-5p | trans | | 3.13E-35 | Nightingale | VLDL_P | NA | T | C | 0.038356 | 0.005308 | 2.10E-14 |
| rs2519093 | miR-10b-5p | trans | | 3.13E-35 | Nightingale | M_LDL_P | NA | T | C | 0.057192 | 0.005351 | 1.30E-27 |
| rs2519093 | miR-10b-5p | trans | | 3.13E-35 | Nightingale | S_VLDL_FC | NA | T | C | 0.052723 | 0.005099 | 2.50E-24 |
| rs2519093 | miR-10b-5p | trans | | 3.13E-35 | Nightingale | S_HDL_P | NA | T | C | 0.055391 | 0.005337 | 1.10E-25 |
| rs2519093 | miR-10b-5p | trans | | 3.13E-35 | Nightingale | S_LDL_L | NA | T | C | 0.048584 | 0.005335 | 3.30E-20 |
| rs2519093 | miR-10b-5p | trans | | 3.13E-35 | Nightingale | VLDL_FC | NA | T | C | 0.033679 | 0.005297 | 1.10E-11 |
| rs2519093 | miR-10b-5p | trans | | 3.13E-35 | Nightingale | XL_VLDL_C | NA | T | C | 0.029731 | 0.005254 | 9.50E-10 |
| rs2519093 | miR-10b-5p | trans | | 3.13E-35 | Nightingale | XXL_VLDL_ | NA | T | C | 0.046178 | 0.005317 | 1.50E-18 |
| rs2519093 | miR-10b-5p | trans | | 3.13E-35 | Nightingale | XXL_VLDL_ | NA | T | C | -0.05831 | 0.005385 | 5.10E-28 |
| rs2519093 | miR-10b-5p | trans | | 3.13E-35 | Nightingale | LDL_P | NA | T | C | 0.053373 | 0.005333 | 2.30E-24 |
| rs2519093 | miR-10b-5p | trans | | 3.13E-35 | Nightingale | XS_VLDL_P | NA | T | C | 0.044763 | 0.005262 | 2.50E-19 |
| rs2519093 | miR-10b-5p | trans | | 3.13E-35 | Nightingale | ApoB | NA | T | C | 0.055887 | 0.005329 | 1.10E-26 |
| rs2519093 | miR-10b-5p | trans | | 3.13E-35 | Nightingale | L_LDL_CE | NA | T | C | 0.058613 | 0.005289 | 3.70E-29 |
| rs2519093 | miR-10b-5p | trans | | 3.13E-35 | Nightingale | S_LDL_C | NA | T | C | 0.05408 | 0.005341 | 1.20E-24 |
| rs2519093 | miR-10b-5p | trans | | 3.13E-35 | Nightingale | M_VLDL_F | NA | T | C | 0.05187 | 0.005335 | 1.30E-23 |
| rs2519093 | miR-10b-5p | trans | | 3.13E-35 | Nightingale | L_LDL_PL | NA | T | C | 0.066759 | 0.005296 | 3.60E-37 |
| rs2519093 | miR-10b-5p | trans | | 3.13E-35 | Nightingale | IDL_PL | NA | T | C | 0.058969 | 0.005149 | 2.40E-31 |
| rs2519093 | miR-10b-5p | trans | | 3.13E-35 | Nightingale | XXL_VLDL_ | NA | T | C | 0.056193 | 0.00538 | 1.00E-25 |
| rs2519093 | miR-10b-5p | trans | | 3.13E-35 | Nightingale | LDL_CE | NA | T | C | 0.0584 | 0.005322 | 9.20E-29 |
| rs2519093 | miR-10b-5p | trans | | 3.13E-35 | Nightingale | S_VLDL_C | NA | T | C | 0.059191 | 0.005336 | 2.80E-30 |
| rs2519093 | miR-10b-5p | trans | | 3.13E-35 | Nightingale | LDL_L | NA | T | C | 0.05971 | 0.005318 | 6.80E-30 |
| rs2519093 | miR-10b-5p | trans | | 3.13E-35 | Nightingale | L_LDL_L | NA | T | C | 0.061108 | 0.005284 | 1.20E-31 |
| rs2519093 | miR-10b-5p | trans | | 3.13E-35 | Nightingale | S_LDL_FC | NA | T | C | 0.054719 | 0.005328 | 1.90E-24 |
| rs2519093 | miR-10b-5p | trans | | 3.13E-35 | Nightingale | HDL_size | NA | T | C | -0.03268 | 0.004731 | 9.30E-13 |
| rs2519093 | miR-10b-5p | trans | | 3.13E-35 | Nightingale | XS_VLDL_T | NA | T | C | -0.04276 | 0.005074 | 2.50E-16 |
| rs2519093 | miR-10b-5p | trans | | 3.13E-35 | Nightingale | XS_VLDL_P | NA | T | C | 0.053945 | 0.005265 | 8.60E-27 |
| rs2519093 | miR-10b-5p | trans | | 3.13E-35 | Nightingale | Total_C | NA | T | C | 0.055882 | 0.005166 | 2.10E-28 |
| rs2519093 | miR-10b-5p | trans | | 3.13E-35 | Nightingale | Total_P | NA | T | C | 0.03671 | 0.005022 | 4.80E-14 |
| rs2519093 | miR-10b-5p | trans | | 3.13E-35 | Nightingale | VLDL_C | NA | T | C | 0.045887 | 0.005329 | 2.40E-19 |
| rs2519093 | miR-10b-5p | trans | | 3.13E-35 | Nightingale | IDL_FC | NA | T | C | 0.064239 | 0.005166 | 4.20E-36 |
| rs2519093 | miR-10b-5p | trans | | 3.13E-35 | Nightingale | M_LDL_TG | NA | T | C | -0.05344 | 0.00527 | 6.00E-24 |
| rs2519093 | miR-10b-5p | trans | | 3.13E-35 | Nightingale | XXL_VLDL_ | NA | T | C | 0.057015 | 0.005351 | 1.20E-26 |
| rs2519093 | miR-10b-5p | trans | | 3.13E-35 | Nightingale | Clinical_LD | NA | T | C | 0.064699 | 0.005305 | 1.10E-34 |
| rs2519093 | miR-10b-5p | trans | | 3.13E-35 | Nightingale | S_HDL_FC | NA | T | C | 0.041872 | 0.005238 | 1.90E-16 |
| rs2519093 | miR-10b-5p | trans | | 3.13E-35 | Nightingale | XL_HDL_L | NA | T | C | -0.02733 | 0.004799 | 1.80E-09 |
| rs2519093 | miR-10b-5p | trans | | 3.13E-35 | Nightingale | L_LDL_FC | NA | T | C | 0.062862 | 0.00524 | 2.40E-33 |
| rs2519093 | miR-10b-5p | trans | | 3.13E-35 | Nightingale | M_VLDL_F | NA | T | C | 0.067606 | 0.005077 | 8.00E-40 |
| rs2519093 | miR-10b-5p | trans | | 3.13E-35 | Nightingale | S_LDL_PL_ | NA | T | C | -0.04499 | 0.005326 | 8.80E-20 |
| rs2519093 | miR-10b-5p | trans | | 3.13E-35 | Nightingale | Citrate | NA | T | C | 0.031335 | 0.005301 | 1.70E-09 |
| rs2519093 | miR-10b-5p | trans | | 3.13E-35 | Nightingale | M_LDL_CE | NA | T | C | 0.055668 | 0.005343 | 1.90E-26 |
| rs2519093 | miR-10b-5p | trans | | 3.13E-35 | Nightingale | Phosphatic | NA | T | C | 0.031111 | 0.005 | 6.70E-11 |
| rs2519093 | miR-10b-5p | trans | | 3.13E-35 | Nightingale | Cholines | NA | T | C | 0.035715 | 0.005018 | 1.00E-13 |
| rs2519093 | miR-10b-5p | trans | | 3.13E-35 | Nightingale | IDL_C_pct | NA | T | C | 0.037575 | 0.005213 | 4.50E-12 |
| rs2519093 | miR-10b-5p | trans | | 3.13E-35 | Nightingale | XL_HDL_PL | NA | T | C | -0.0414 | 0.004903 | 9.60E-18 |
| rs2519093 | miR-10b-5p | trans | | 3.13E-35 | Nightingale | LA | NA | T | C | 0.042082 | 0.005233 | 4.40E-17 |
| rs2519093 | miR-10b-5p | trans | | 3.13E-35 | Nightingale | S_LDL_P | NA | T | C | 0.048597 | 0.005341 | 1.60E-20 |
| rs2519093 | miR-10b-5p | trans | | 3.13E-35 | Nightingale | M_VLDL_L | NA | T | C | 0.032196 | 0.0053 | 1.00E-10 |
| rs2519093 | miR-10b-5p | trans | | 3.13E-35 | Nightingale | XL_HDL_PL | NA | T | C | -0.03044 | 0.0048 | 4.90E-11 |
| rs2519093 | miR-10b-5p | trans | | 3.13E-35 | Nightingale | XL_VLDL_P | NA | T | C | 0.056118 | 0.005345 | 2.80E-27 |
| rs2519093 | miR-10b-5p | trans | | 3.13E-35 | Nightingale | XS_VLDL_F | NA | T | C | 0.053855 | 0.005244 | 8.50E-27 |
| rs2519093 | miR-10b-5p | trans | | 3.13E-35 | Nightingale | non_HDL_( | NA | T | C | 0.060625 | 0.005307 | 2.30E-31 |
| rs2519093 | miR-10b-5p | trans | | 3.13E-35 | Nightingale | XL_VLDL_T | NA | T | C | -0.06925 | 0.005173 | 9.80E-41 |
| rs2519093 | miR-10b-5p | trans | | 3.13E-35 | Nightingale | IDL_TG_pc | NA | T | C | -0.03897 | 0.005159 | 2.10E-13 |
| rs2519093 | miR-10b-5p | trans | | 3.13E-35 | Nightingale | Total_PL | NA | T | C | 0.039528 | 0.005081 | 4.00E-16 |
| rs2519093 | miR-10b-5p | trans | | 3.13E-35 | Nightingale | M_VLDL_T | NA | T | C | -0.06313 | 0.005043 | 4.00E-35 |
| rs2519093 | miR-10b-5p | trans | | 3.13E-35 | Nightingale | L_VLDL_FC | NA | T | C | 0.076264 | 0.005285 | 5.50E-48 |
| rs2519093 | miR-10b-5p | trans | | 3.13E-35 | Nightingale | S_VLDL_P | NA | T | C | 0.033454 | 0.005269 | 6.70E-12 |

| | | | | | | | | | | | |
|---|---|---|---|---|---|---|---|---|---|---|---|
| rs2519093 | miR-10b-5p | trans | 3.13E-35 | Nightingale | S_LDL_PL | NA | T | C | 0.041418 | 0.005324 | 6.40E-15 |
| rs2519093 | miR-10b-5p | trans | 3.13E-35 | Nightingale | XS_VLDL_C | NA | T | C | 0.058552 | 0.00515 | 2.40E-31 |
| rs2519093 | miR-10b-5p | trans | 3.13E-35 | Nightingale | L_VLDL_CE | NA | T | C | 0.074537 | 0.005107 | 7.40E-49 |
| rs2519093 | miR-10b-5p | trans | 3.13E-35 | Nightingale | XL_HDL_C_ | NA | T | C | 0.038739 | 0.005096 | 1.00E-13 |
| rs2519093 | miR-10b-5p | trans | 3.13E-35 | Nightingale | M_LDL_C | NA | T | C | 0.058986 | 0.005346 | 4.10E-29 |
| rs2519093 | miR-10b-5p | trans | 3.13E-35 | Nightingale | S_LDL_TG_ | NA | T | C | -0.0438 | 0.005175 | 2.30E-16 |
| rs2519093 | miR-10b-5p | trans | 3.13E-35 | Nightingale | XL_VLDL_C | NA | T | C | 0.062611 | 0.00512 | 1.50E-33 |
| rs2519093 | miR-10b-5p | trans | 3.13E-35 | Nightingale | S_VLDL_FC | NA | T | C | 0.059655 | 0.005337 | 2.30E-30 |
| rs2519093 | miR-10b-5p | trans | 3.13E-35 | Nightingale | IDL_FC_pct | NA | T | C | 0.041553 | 0.005295 | 1.40E-14 |
| rs2519093 | miR-10b-5p | trans | 3.13E-35 | Nightingale | M_LDL_PL | NA | T | C | 0.058342 | 0.005344 | 1.80E-28 |
| rs2519093 | miR-10b-5p | trans | 3.13E-35 | Nightingale | L_VLDL_PL | NA | T | C | 0.028334 | 0.005183 | 2.20E-09 |
| rs2519093 | miR-10b-5p | trans | 3.13E-35 | Nightingale | L_LDL_P | NA | T | C | 0.050604 | 0.005318 | 4.80E-22 |
| rs2519093 | miR-10b-5p | trans | 3.13E-35 | Nightingale | LDL_PL | NA | T | C | 0.06213 | 0.005324 | 5.60E-32 |
| rs2519093 | miR-10b-5p | trans | 3.13E-35 | Nightingale | S_VLDL_TG | NA | T | C | -0.06553 | 0.005165 | 2.20E-36 |
| rs2519093 | miR-10b-5p | trans | 3.13E-35 | Nightingale | Pyruvate | NA | T | C | 0.034476 | 0.005345 | 1.10E-10 |
| rs2519093 | miR-10b-5p | trans | 3.13E-35 | Nightingale | ApoB_by_A | NA | T | C | 0.040245 | 0.005256 | 9.90E-15 |
| rs2519093 | miR-10b-5p | trans | 3.13E-35 | Nightingale | XL_VLDL_F | NA | T | C | 0.056835 | 0.005174 | 2.50E-28 |
| rs2519093 | miR-10b-5p | trans | 3.13E-35 | Nightingale | S_VLDL_PL | NA | T | C | 0.050243 | 0.005326 | 1.00E-22 |
| rs2519093 | miR-10b-5p | trans | 3.13E-35 | Nightingale | M_VLDL_P | NA | T | C | 0.070901 | 0.005121 | 2.00E-43 |
| rs2519093 | miR-10b-5p | trans | 3.13E-35 | Nightingale | S_LDL_CE_ | NA | T | C | 0.04485 | 0.005327 | 8.40E-20 |
| rs2519093 | miR-10b-5p | trans | 3.13E-35 | Nightingale | XXL_VLDL_ | NA | T | C | 0.036769 | 0.005374 | 2.00E-13 |
| rs2519093 | miR-10b-5p | trans | 3.13E-35 | Nightingale | M_VLDL_C | NA | T | C | 0.059763 | 0.005026 | 1.20E-31 |
| rs2519093 | miR-10b-5p | trans | 3.13E-35 | Nightingale | XS_VLDL_C | NA | T | C | 0.057788 | 0.00518 | 1.80E-30 |
| rs2519093 | miR-10b-5p | trans | 3.13E-35 | Nightingale | S_HDL_L | NA | T | C | 0.04414 | 0.005318 | 1.60E-17 |
| rs2519093 | miR-10b-5p | trans | 3.13E-35 | Nightingale | S_LDL_C_p | NA | T | C | 0.075956 | 0.005357 | 8.10E-47 |
| rs2519093 | miR-10b-5p | trans | 3.13E-35 | Nightingale | M_LDL_C_p | NA | T | C | 0.047547 | 0.00534 | 3.80E-19 |
| rs2519093 | miR-10b-5p | trans | 3.13E-35 | Nightingale | XL_VLDL_C | NA | T | C | 0.062776 | 0.005117 | 8.10E-34 |
| rs2519093 | miR-10b-5p | trans | 3.13E-35 | Nightingale | LDL_FC | NA | T | C | 0.062925 | 0.005279 | 9.00E-33 |
| rs2519093 | miR-10b-5p | trans | 3.13E-35 | Nightingale | L_VLDL_TG | NA | T | C | -0.08007 | 0.005311 | 4.90E-53 |
| rs2519093 | miR-10b-5p | trans | 3.13E-35 | Nightingale | S_HDL_PL_ | NA | T | C | -0.03292 | 0.005159 | 7.60E-10 |
| rs2519093 | miR-10b-5p | trans | 3.13E-35 | Nightingale | HDL_P | NA | T | C | 0.028786 | 0.005011 | 3.00E-09 |
| rs2519093 | miR-10b-5p | trans | 3.13E-35 | Nightingale | LDL_C | NA | T | C | 0.060241 | 0.005313 | 2.20E-30 |
| rs2519093 | miR-10b-5p | trans | 3.13E-35 | Nightingale | L_LDL_FC_ | NA | T | C | 0.030917 | 0.005091 | 1.20E-08 |
| rs2519093 | miR-10b-5p | trans | 3.13E-35 | Nightingale | M_LDL_L | NA | T | C | 0.057522 | 0.005344 | 6.90E-28 |
| rs2519093 | miR-10b-5p | trans | 3.13E-35 | Nightingale | XL_HDL_CE | NA | T | C | 0.030201 | 0.005332 | 3.60E-08 |
| rs2519093 | miR-10b-5p | trans | 3.13E-35 | Nightingale | M_LDL_FC | NA | T | C | 0.063872 | 0.00533 | 2.80E-33 |
| rs2519093 | miR-10b-5p | trans | 3.13E-35 | Nightingale | S_VLDL_CE | NA | T | C | 0.076957 | 0.005265 | 7.60E-48 |
| rs2519093 | miR-10b-5p | trans | 3.13E-35 | Nightingale | Remnant_C | NA | T | C | 0.058623 | 0.005302 | 4.00E-30 |
| rs2519093 | miR-139-5p | trans | 1.63E-36 | Nightingale | S_HDL_CE | NA | T | C | 0.053505 | 0.005345 | 7.30E-24 |
| rs2519093 | miR-139-5p | trans | 1.63E-36 | Nightingale | L_LDL_TG_ | NA | T | C | -0.04178 | 0.005234 | 5.50E-15 |
| rs2519093 | miR-139-5p | trans | 1.63E-36 | Nightingale | M_VLDL_P | NA | T | C | 0.04488 | 0.005328 | 1.70E-18 |
| rs2519093 | miR-139-5p | trans | 1.63E-36 | Nightingale | IDL_C | NA | T | C | 0.060311 | 0.005154 | 6.00E-32 |
| rs2519093 | miR-139-5p | trans | 1.63E-36 | Nightingale | M_VLDL_C | NA | T | C | 0.066274 | 0.005288 | 7.60E-37 |
| rs2519093 | miR-139-5p | trans | 1.63E-36 | Nightingale | Total_FC | NA | T | C | 0.055093 | 0.005213 | 7.90E-28 |
| rs2519093 | miR-139-5p | trans | 1.63E-36 | Nightingale | S_VLDL_C_ | NA | T | C | 0.070806 | 0.005193 | 5.60E-42 |
| rs2519093 | miR-139-5p | trans | 1.63E-36 | Nightingale | LA_pct | NA | T | C | 0.031876 | 0.005218 | 1.10E-09 |
| rs2519093 | miR-139-5p | trans | 1.63E-36 | Nightingale | S_VLDL_L | NA | T | C | 0.035666 | 0.005285 | 4.30E-13 |
| rs2519093 | miR-139-5p | trans | 1.63E-36 | Nightingale | M_VLDL_C | NA | T | C | 0.061815 | 0.005321 | 1.60E-32 |
| rs2519093 | miR-139-5p | trans | 1.63E-36 | Nightingale | XS_VLDL_F | NA | T | C | 0.051132 | 0.005298 | 6.20E-21 |
| rs2519093 | miR-139-5p | trans | 1.63E-36 | Nightingale | IDL_P | NA | T | C | 0.065267 | 0.00528 | 1.10E-35 |
| rs2519093 | miR-139-5p | trans | 1.63E-36 | Nightingale | S_VLDL_CE | NA | T | C | 0.05821 | 0.005329 | 1.10E-29 |
| rs2519093 | miR-139-5p | trans | 1.63E-36 | Nightingale | XS_VLDL_L | NA | T | C | 0.048684 | 0.005245 | 1.40E-22 |
| rs2519093 | miR-139-5p | trans | 1.63E-36 | Nightingale | S_LDL_CE | NA | T | C | 0.051776 | 0.005339 | 4.00E-23 |
| rs2519093 | miR-139-5p | trans | 1.63E-36 | Nightingale | IDL_L | NA | T | C | 0.059774 | 0.00516 | 1.10E-31 |
| rs2519093 | miR-139-5p | trans | 1.63E-36 | Nightingale | Total_CE | NA | T | C | 0.05575 | 0.005147 | 2.40E-28 |
| rs2519093 | miR-139-5p | trans | 1.63E-36 | Nightingale | Sphingomy | NA | T | C | 0.047648 | 0.004989 | 2.20E-21 |
| rs2519093 | miR-139-5p | trans | 1.63E-36 | Nightingale | PUFA | NA | T | C | 0.036949 | 0.005205 | 1.30E-13 |
| rs2519093 | miR-139-5p | trans | 1.63E-36 | Nightingale | XS_VLDL_C | NA | T | C | 0.040361 | 0.005035 | 1.00E-14 |
| rs2519093 | miR-139-5p | trans | 1.63E-36 | Nightingale | IDL_CE | NA | T | C | 0.058439 | 0.005152 | 4.70E-30 |
| rs2519093 | miR-139-5p | trans | 1.63E-36 | Nightingale | VLDL_CE | NA | T | C | 0.053783 | 0.005338 | 2.10E-25 |
| rs2519093 | miR-139-5p | trans | 1.63E-36 | Nightingale | S_HDL_PL | NA | T | C | 0.037688 | 0.00529 | 1.40E-13 |
| rs2519093 | miR-139-5p | trans | 1.63E-36 | Nightingale | Omega_6 | NA | T | C | 0.041735 | 0.005211 | 5.40E-17 |
| rs2519093 | miR-139-5p | trans | 1.63E-36 | Nightingale | S_HDL_C | NA | T | C | 0.052616 | 0.005327 | 2.20E-23 |

| | | | | | | | | | | | |
|---|---|---|---|---|---|---|---|---|---|---|---|
| rs2519093 | miR-139-5p | trans | 1.63E-36 | Nightingale | M_VLDL_C | NA | T | C | 0.056108 | 0.005013 | 3.90E-28 |
| rs2519093 | miR-139-5p | trans | 1.63E-36 | Nightingale | GlycA | NA | T | C | 0.031449 | 0.005314 | 4.90E-09 |
| rs2519093 | miR-139-5p | trans | 1.63E-36 | Nightingale | XS_VLDL_C | NA | T | C | 0.043421 | 0.005061 | 7.70E-17 |
| rs2519093 | miR-139-5p | trans | 1.63E-36 | Nightingale | M_VLDL_P | NA | T | C | 0.043347 | 0.005323 | 1.90E-17 |
| rs2519093 | miR-139-5p | trans | 1.63E-36 | Nightingale | XS_VLDL_P | NA | T | C | -0.02949 | 0.005207 | 5.00E-08 |
| rs2519093 | miR-139-5p | trans | 1.63E-36 | Nightingale | S_VLDL_PL | NA | T | C | 0.050234 | 0.00511 | 4.10E-22 |
| rs2519093 | miR-139-5p | trans | 1.63E-36 | Nightingale | L_LDL_C | NA | T | C | 0.060158 | 0.005276 | 1.10E-30 |
| rs2519093 | miR-139-5p | trans | 1.63E-36 | Nightingale | Total_L | NA | T | C | 0.042371 | 0.00525 | 4.30E-17 |
| rs2519093 | miR-139-5p | trans | 1.63E-36 | Nightingale | Phosphogly | NA | T | C | 0.031931 | 0.005054 | 3.40E-11 |
| rs2519093 | miR-139-5p | trans | 1.63E-36 | Nightingale | L_VLDL_C_ | NA | T | C | 0.082094 | 0.005212 | 9.00E-57 |
| rs2519093 | miR-139-5p | trans | 1.63E-36 | Nightingale | L_VLDL_CE | NA | T | C | 0.031525 | 0.005274 | 1.30E-10 |
| rs2519093 | miR-139-5p | trans | 1.63E-36 | Nightingale | VLDL_P | NA | T | C | 0.038356 | 0.005308 | 2.10E-14 |
| rs2519093 | miR-139-5p | trans | 1.63E-36 | Nightingale | M_LDL_P | NA | T | C | 0.057192 | 0.005351 | 1.30E-27 |
| rs2519093 | miR-139-5p | trans | 1.63E-36 | Nightingale | S_VLDL_FC | NA | T | C | 0.052723 | 0.005099 | 2.50E-24 |
| rs2519093 | miR-139-5p | trans | 1.63E-36 | Nightingale | S_HDL_P | NA | T | C | 0.055391 | 0.005337 | 1.10E-25 |
| rs2519093 | miR-139-5p | trans | 1.63E-36 | Nightingale | S_LDL_L | NA | T | C | 0.048584 | 0.005335 | 3.30E-20 |
| rs2519093 | miR-139-5p | trans | 1.63E-36 | Nightingale | VLDL_FC | NA | T | C | 0.033679 | 0.005297 | 1.10E-11 |
| rs2519093 | miR-139-5p | trans | 1.63E-36 | Nightingale | XL_VLDL_C | NA | T | C | 0.029731 | 0.005254 | 9.50E-10 |
| rs2519093 | miR-139-5p | trans | 1.63E-36 | Nightingale | XXL_VLDL_ | NA | T | C | 0.046178 | 0.005317 | 1.50E-18 |
| rs2519093 | miR-139-5p | trans | 1.63E-36 | Nightingale | XXL_VLDL_ | NA | T | C | -0.05831 | 0.005385 | 5.10E-28 |
| rs2519093 | miR-139-5p | trans | 1.63E-36 | Nightingale | LDL_P | NA | T | C | 0.053373 | 0.005333 | 2.30E-24 |
| rs2519093 | miR-139-5p | trans | 1.63E-36 | Nightingale | XS_VLDL_P | NA | T | C | 0.044763 | 0.005262 | 2.50E-19 |
| rs2519093 | miR-139-5p | trans | 1.63E-36 | Nightingale | ApoB | NA | T | C | 0.055887 | 0.005329 | 1.10E-26 |
| rs2519093 | miR-139-5p | trans | 1.63E-36 | Nightingale | L_LDL_CE | NA | T | C | 0.058613 | 0.005289 | 3.70E-29 |
| rs2519093 | miR-139-5p | trans | 1.63E-36 | Nightingale | S_LDL_C | NA | T | C | 0.05408 | 0.005341 | 1.20E-24 |
| rs2519093 | miR-139-5p | trans | 1.63E-36 | Nightingale | M_VLDL_F( | NA | T | C | 0.05187 | 0.005335 | 1.30E-23 |
| rs2519093 | miR-139-5p | trans | 1.63E-36 | Nightingale | L_LDL_PL | NA | T | C | 0.066759 | 0.005296 | 3.60E-37 |
| rs2519093 | miR-139-5p | trans | 1.63E-36 | Nightingale | IDL_PL | NA | T | C | 0.058969 | 0.005149 | 2.40E-31 |
| rs2519093 | miR-139-5p | trans | 1.63E-36 | Nightingale | XXL_VLDL_ | NA | T | C | 0.056193 | 0.00538 | 1.00E-25 |
| rs2519093 | miR-139-5p | trans | 1.63E-36 | Nightingale | LDL_CE | NA | T | C | 0.0584 | 0.005322 | 9.20E-29 |
| rs2519093 | miR-139-5p | trans | 1.63E-36 | Nightingale | S_VLDL_C | NA | T | C | 0.059191 | 0.005336 | 2.80E-30 |
| rs2519093 | miR-139-5p | trans | 1.63E-36 | Nightingale | LDL_L | NA | T | C | 0.05971 | 0.005318 | 6.80E-30 |
| rs2519093 | miR-139-5p | trans | 1.63E-36 | Nightingale | L_LDL_L | NA | T | C | 0.061108 | 0.005284 | 1.20E-31 |
| rs2519093 | miR-139-5p | trans | 1.63E-36 | Nightingale | S_LDL_FC | NA | T | C | 0.054719 | 0.005328 | 1.90E-24 |
| rs2519093 | miR-139-5p | trans | 1.63E-36 | Nightingale | HDL_size | NA | T | C | -0.03268 | 0.004731 | 9.30E-13 |
| rs2519093 | miR-139-5p | trans | 1.63E-36 | Nightingale | XS_VLDL_T | NA | T | C | -0.04276 | 0.005074 | 2.50E-16 |
| rs2519093 | miR-139-5p | trans | 1.63E-36 | Nightingale | XS_VLDL_P | NA | T | C | 0.053945 | 0.005265 | 8.60E-27 |
| rs2519093 | miR-139-5p | trans | 1.63E-36 | Nightingale | Total_C | NA | T | C | 0.055882 | 0.005166 | 2.10E-28 |
| rs2519093 | miR-139-5p | trans | 1.63E-36 | Nightingale | Total_P | NA | T | C | 0.03671 | 0.005022 | 4.80E-14 |
| rs2519093 | miR-139-5p | trans | 1.63E-36 | Nightingale | VLDL_C | NA | T | C | 0.045887 | 0.005329 | 2.40E-19 |
| rs2519093 | miR-139-5p | trans | 1.63E-36 | Nightingale | IDL_FC | NA | T | C | 0.064239 | 0.005166 | 4.20E-36 |
| rs2519093 | miR-139-5p | trans | 1.63E-36 | Nightingale | M_LDL_TG | NA | T | C | -0.05344 | 0.00527 | 6.00E-24 |
| rs2519093 | miR-139-5p | trans | 1.63E-36 | Nightingale | XXL_VLDL_ | NA | T | C | 0.057015 | 0.005351 | 1.20E-26 |
| rs2519093 | miR-139-5p | trans | 1.63E-36 | Nightingale | Clinical_LD | NA | T | C | 0.064699 | 0.005305 | 1.10E-34 |
| rs2519093 | miR-139-5p | trans | 1.63E-36 | Nightingale | S_HDL_FC | NA | T | C | 0.041872 | 0.005238 | 1.90E-16 |
| rs2519093 | miR-139-5p | trans | 1.63E-36 | Nightingale | XL_HDL_L | NA | T | C | -0.02733 | 0.004799 | 1.80E-09 |
| rs2519093 | miR-139-5p | trans | 1.63E-36 | Nightingale | L_LDL_FC | NA | T | C | 0.062862 | 0.00524 | 2.40E-33 |
| rs2519093 | miR-139-5p | trans | 1.63E-36 | Nightingale | M_VLDL_F( | NA | T | C | 0.067606 | 0.005077 | 8.00E-40 |
| rs2519093 | miR-139-5p | trans | 1.63E-36 | Nightingale | S_LDL_PL_ | NA | T | C | -0.04499 | 0.005326 | 8.80E-20 |
| rs2519093 | miR-139-5p | trans | 1.63E-36 | Nightingale | Citrate | NA | T | C | 0.031335 | 0.005301 | 1.70E-09 |
| rs2519093 | miR-139-5p | trans | 1.63E-36 | Nightingale | M_LDL_CE | NA | T | C | 0.055668 | 0.005343 | 1.90E-26 |
| rs2519093 | miR-139-5p | trans | 1.63E-36 | Nightingale | Phosphatic | NA | T | C | 0.031111 | 0.005 | 6.70E-11 |
| rs2519093 | miR-139-5p | trans | 1.63E-36 | Nightingale | Cholines | NA | T | C | 0.035715 | 0.005018 | 1.00E-13 |
| rs2519093 | miR-139-5p | trans | 1.63E-36 | Nightingale | IDL_C_pct | NA | T | C | 0.037575 | 0.005213 | 4.50E-12 |
| rs2519093 | miR-139-5p | trans | 1.63E-36 | Nightingale | XL_HDL_PL | NA | T | C | -0.0414 | 0.004903 | 9.60E-18 |
| rs2519093 | miR-139-5p | trans | 1.63E-36 | Nightingale | LA | NA | T | C | 0.042082 | 0.005233 | 4.40E-17 |
| rs2519093 | miR-139-5p | trans | 1.63E-36 | Nightingale | S_LDL_P | NA | T | C | 0.048597 | 0.005341 | 1.60E-20 |
| rs2519093 | miR-139-5p | trans | 1.63E-36 | Nightingale | M_VLDL_L | NA | T | C | 0.032196 | 0.0053 | 1.00E-10 |
| rs2519093 | miR-139-5p | trans | 1.63E-36 | Nightingale | XL_HDL_PL | NA | T | C | -0.03044 | 0.0048 | 4.90E-11 |
| rs2519093 | miR-139-5p | trans | 1.63E-36 | Nightingale | XL_VLDL_P | NA | T | C | 0.056118 | 0.005345 | 2.80E-27 |
| rs2519093 | miR-139-5p | trans | 1.63E-36 | Nightingale | XS_VLDL_F | NA | T | C | 0.053855 | 0.005244 | 8.50E-27 |
| rs2519093 | miR-139-5p | trans | 1.63E-36 | Nightingale | non_HDL_( | NA | T | C | 0.060625 | 0.005307 | 2.30E-31 |
| rs2519093 | miR-139-5p | trans | 1.63E-36 | Nightingale | XL_VLDL_T | NA | T | C | -0.06925 | 0.005173 | 9.80E-41 |

| | | | | | | | | | | | |
|---|---|---|---|---|---|---|---|---|---|---|---|
| rs2519093 | miR-139-5p | trans | 1.63E-36 | Nightingale | IDL_TG_pct | NA | T | C | -0.03897 | 0.005159 | 2.10E-13 |
| rs2519093 | miR-139-5p | trans | 1.63E-36 | Nightingale | Total_PL | NA | T | C | 0.039528 | 0.005081 | 4.00E-16 |
| rs2519093 | miR-139-5p | trans | 1.63E-36 | Nightingale | M_VLDL_T | NA | T | C | -0.06313 | 0.005043 | 4.00E-35 |
| rs2519093 | miR-139-5p | trans | 1.63E-36 | Nightingale | L_VLDL_FC | NA | T | C | 0.076264 | 0.005285 | 5.50E-48 |
| rs2519093 | miR-139-5p | trans | 1.63E-36 | Nightingale | S_VLDL_P | NA | T | C | 0.033454 | 0.005269 | 6.70E-12 |
| rs2519093 | miR-139-5p | trans | 1.63E-36 | Nightingale | S_LDL_PL | NA | T | C | 0.041418 | 0.005324 | 6.40E-15 |
| rs2519093 | miR-139-5p | trans | 1.63E-36 | Nightingale | XS_VLDL_C | NA | T | C | 0.058552 | 0.00515 | 2.40E-31 |
| rs2519093 | miR-139-5p | trans | 1.63E-36 | Nightingale | L_VLDL_CE | NA | T | C | 0.074537 | 0.005107 | 7.40E-49 |
| rs2519093 | miR-139-5p | trans | 1.63E-36 | Nightingale | XL_HDL_C_ | NA | T | C | 0.038739 | 0.005096 | 1.00E-13 |
| rs2519093 | miR-139-5p | trans | 1.63E-36 | Nightingale | M_LDL_C | NA | T | C | 0.058986 | 0.005346 | 4.10E-29 |
| rs2519093 | miR-139-5p | trans | 1.63E-36 | Nightingale | S_LDL_TG_ | NA | T | C | -0.0438 | 0.005175 | 2.30E-16 |
| rs2519093 | miR-139-5p | trans | 1.63E-36 | Nightingale | XL_VLDL_C | NA | T | C | 0.062611 | 0.00512 | 1.50E-33 |
| rs2519093 | miR-139-5p | trans | 1.63E-36 | Nightingale | S_VLDL_FC | NA | T | C | 0.059655 | 0.005337 | 2.30E-30 |
| rs2519093 | miR-139-5p | trans | 1.63E-36 | Nightingale | IDL_FC_pct | NA | T | C | 0.041553 | 0.005295 | 1.40E-14 |
| rs2519093 | miR-139-5p | trans | 1.63E-36 | Nightingale | M_LDL_PL | NA | T | C | 0.058342 | 0.005344 | 1.80E-28 |
| rs2519093 | miR-139-5p | trans | 1.63E-36 | Nightingale | L_VLDL_PL | NA | T | C | 0.028334 | 0.005183 | 2.20E-09 |
| rs2519093 | miR-139-5p | trans | 1.63E-36 | Nightingale | L_LDL_P | NA | T | C | 0.050604 | 0.005318 | 4.80E-22 |
| rs2519093 | miR-139-5p | trans | 1.63E-36 | Nightingale | LDL_PL | NA | T | C | 0.06213 | 0.005324 | 5.60E-32 |
| rs2519093 | miR-139-5p | trans | 1.63E-36 | Nightingale | S_VLDL_TG | NA | T | C | -0.06553 | 0.005165 | 2.20E-36 |
| rs2519093 | miR-139-5p | trans | 1.63E-36 | Nightingale | Pyruvate | NA | T | C | 0.034476 | 0.005345 | 1.10E-10 |
| rs2519093 | miR-139-5p | trans | 1.63E-36 | Nightingale | ApoB_by_A | NA | T | C | 0.040245 | 0.005256 | 9.90E-15 |
| rs2519093 | miR-139-5p | trans | 1.63E-36 | Nightingale | XL_VLDL_F | NA | T | C | 0.056835 | 0.005174 | 2.50E-28 |
| rs2519093 | miR-139-5p | trans | 1.63E-36 | Nightingale | S_VLDL_PL | NA | T | C | 0.050243 | 0.005326 | 1.00E-22 |
| rs2519093 | miR-139-5p | trans | 1.63E-36 | Nightingale | M_VLDL_P | NA | T | C | 0.070901 | 0.005121 | 2.00E-43 |
| rs2519093 | miR-139-5p | trans | 1.63E-36 | Nightingale | S_LDL_CE_ | NA | T | C | 0.04485 | 0.005327 | 8.40E-20 |
| rs2519093 | miR-139-5p | trans | 1.63E-36 | Nightingale | XXL_VLDL_ | NA | T | C | 0.036769 | 0.005374 | 2.00E-13 |
| rs2519093 | miR-139-5p | trans | 1.63E-36 | Nightingale | M_VLDL_C | NA | T | C | 0.059763 | 0.005026 | 1.20E-31 |
| rs2519093 | miR-139-5p | trans | 1.63E-36 | Nightingale | XS_VLDL_C | NA | T | C | 0.057788 | 0.00518 | 1.80E-30 |
| rs2519093 | miR-139-5p | trans | 1.63E-36 | Nightingale | S_HDL_L | NA | T | C | 0.04414 | 0.005318 | 1.60E-17 |
| rs2519093 | miR-139-5p | trans | 1.63E-36 | Nightingale | S_LDL_C_p | NA | T | C | 0.075956 | 0.005357 | 8.10E-47 |
| rs2519093 | miR-139-5p | trans | 1.63E-36 | Nightingale | M_LDL_C_ | NA | T | C | 0.047547 | 0.00534 | 3.80E-19 |
| rs2519093 | miR-139-5p | trans | 1.63E-36 | Nightingale | XL_VLDL_C | NA | T | C | 0.062776 | 0.005117 | 8.10E-34 |
| rs2519093 | miR-139-5p | trans | 1.63E-36 | Nightingale | LDL_FC | NA | T | C | 0.062925 | 0.005279 | 9.00E-33 |
| rs2519093 | miR-139-5p | trans | 1.63E-36 | Nightingale | L_VLDL_TG | NA | T | C | -0.08007 | 0.005311 | 4.90E-53 |
| rs2519093 | miR-139-5p | trans | 1.63E-36 | Nightingale | S_HDL_PL_ | NA | T | C | -0.03292 | 0.005159 | 7.60E-10 |
| rs2519093 | miR-139-5p | trans | 1.63E-36 | Nightingale | HDL_P | NA | T | C | 0.028786 | 0.005011 | 3.00E-09 |
| rs2519093 | miR-139-5p | trans | 1.63E-36 | Nightingale | LDL_C | NA | T | C | 0.060241 | 0.005313 | 2.20E-30 |
| rs2519093 | miR-139-5p | trans | 1.63E-36 | Nightingale | L_LDL_FC_ | NA | T | C | 0.030917 | 0.005091 | 1.20E-08 |
| rs2519093 | miR-139-5p | trans | 1.63E-36 | Nightingale | M_LDL_L | NA | T | C | 0.057522 | 0.005344 | 6.90E-28 |
| rs2519093 | miR-139-5p | trans | 1.63E-36 | Nightingale | XL_HDL_CE | NA | T | C | 0.030201 | 0.005332 | 3.60E-08 |
| rs2519093 | miR-139-5p | trans | 1.63E-36 | Nightingale | M_LDL_FC | NA | T | C | 0.063872 | 0.00533 | 2.80E-33 |
| rs2519093 | miR-139-5p | trans | 1.63E-36 | Nightingale | S_VLDL_CE | NA | T | C | 0.076957 | 0.005265 | 7.60E-48 |
| rs2519093 | miR-139-5p | trans | 1.63E-36 | Nightingale | Remnant_( | NA | T | C | 0.058623 | 0.005302 | 4.00E-30 |
| rs2769071 | let-7c-5p | trans | 3.76E-23 | Nightingale | S_LDL_C_p | NA | G | A | 0.045281 | 0.004444 | 7.30E-25 |
| rs2769071 | let-7c-5p | trans | 3.76E-23 | Nightingale | S_LDL_P | NA | G | A | 0.029553 | 0.004431 | 3.40E-11 |
| rs2769071 | let-7c-5p | trans | 3.76E-23 | Nightingale | L_VLDL_C_ | NA | G | A | 0.051993 | 0.004324 | 7.60E-34 |
| rs2769071 | let-7c-5p | trans | 3.76E-23 | Nightingale | GlycA | NA | G | A | 0.028001 | 0.004408 | 1.50E-10 |
| rs2769071 | let-7c-5p | trans | 3.76E-23 | Nightingale | Total_C | NA | G | A | 0.032411 | 0.004286 | 3.20E-14 |
| rs2769071 | let-7c-5p | trans | 3.76E-23 | Nightingale | M_VLDL_C | NA | G | A | 0.033528 | 0.004159 | 2.40E-15 |
| rs2769071 | let-7c-5p | trans | 3.76E-23 | Nightingale | M_LDL_L | NA | G | A | 0.033001 | 0.004433 | 1.10E-13 |
| rs2769071 | let-7c-5p | trans | 3.76E-23 | Nightingale | M_VLDL_P | NA | G | A | 0.043923 | 0.004249 | 8.10E-25 |
| rs2769071 | let-7c-5p | trans | 3.76E-23 | Nightingale | IDL_FC | NA | G | A | 0.038256 | 0.004286 | 1.50E-18 |
| rs2769071 | let-7c-5p | trans | 3.76E-23 | Nightingale | VLDL_P | NA | G | A | 0.026645 | 0.004403 | 4.40E-10 |
| rs2769071 | let-7c-5p | trans | 3.76E-23 | Nightingale | M_LDL_P | NA | G | A | 0.035884 | 0.004439 | 7.10E-16 |
| rs2769071 | let-7c-5p | trans | 3.76E-23 | Nightingale | XL_VLDL_T | NA | G | A | -0.04368 | 0.004291 | 3.40E-24 |
| rs2769071 | let-7c-5p | trans | 3.76E-23 | Nightingale | S_VLDL_C_ | NA | G | A | 0.041808 | 0.004308 | 6.50E-22 |
| rs2769071 | let-7c-5p | trans | 3.76E-23 | Nightingale | M_LDL_C | NA | G | A | 0.033958 | 0.004435 | 2.30E-14 |
| rs2769071 | let-7c-5p | trans | 3.76E-23 | Nightingale | L_LDL_C | NA | G | A | 0.032304 | 0.004377 | 2.70E-13 |
| rs2769071 | let-7c-5p | trans | 3.76E-23 | Nightingale | IDL_PL | NA | G | A | 0.035362 | 0.004272 | 3.30E-16 |
| rs2769071 | let-7c-5p | trans | 3.76E-23 | Nightingale | Sphingomy | NA | G | A | 0.029058 | 0.004139 | 6.70E-12 |
| rs2769071 | let-7c-5p | trans | 3.76E-23 | Nightingale | S_HDL_L | NA | G | A | 0.026342 | 0.004412 | 1.50E-09 |
| rs2769071 | let-7c-5p | trans | 3.76E-23 | Nightingale | Clinical_LD | NA | G | A | 0.036447 | 0.004401 | 2.90E-16 |
| rs2769071 | let-7c-5p | trans | 3.76E-23 | Nightingale | M_VLDL_P | NA | G | A | 0.029124 | 0.00442 | 2.10E-11 |

| | | | | | | | | | | | |
|---|---|---|---|---|---|---|---|---|---|---|---|
| rs2769071 | let-7c-5p | trans | 3.76E-23 | Nightingale | ApoB_by_A | NA | G | A | 0.024951 | 0.00436 | 1.40E-08 |
| rs2769071 | let-7c-5p | trans | 3.76E-23 | Nightingale | XS_VLDL_C | NA | G | A | 0.037835 | 0.004272 | 2.00E-18 |
| rs2769071 | let-7c-5p | trans | 3.76E-23 | Nightingale | XS_VLDL_P | NA | G | A | 0.036739 | 0.004368 | 1.30E-17 |
| rs2769071 | let-7c-5p | trans | 3.76E-23 | Nightingale | IDL_L | NA | G | A | 0.035297 | 0.00428 | 4.70E-16 |
| rs2769071 | let-7c-5p | trans | 3.76E-23 | Nightingale | S_VLDL_PL | NA | G | A | 0.032773 | 0.004419 | 4.20E-14 |
| rs2769071 | let-7c-5p | trans | 3.76E-23 | Nightingale | VLDL_FC | NA | G | A | 0.024155 | 0.004395 | 1.10E-08 |
| rs2769071 | let-7c-5p | trans | 3.76E-23 | Nightingale | Omega_6 | NA | G | A | 0.025917 | 0.004323 | 5.60E-10 |
| rs2769071 | let-7c-5p | trans | 3.76E-23 | Nightingale | VLDL_C | NA | G | A | 0.031294 | 0.004421 | 5.00E-13 |
| rs2769071 | let-7c-5p | trans | 3.76E-23 | Nightingale | XXL_VLDL_ | NA | G | A | -0.03561 | 0.004465 | 2.70E-16 |
| rs2769071 | let-7c-5p | trans | 3.76E-23 | Nightingale | LDL_L | NA | G | A | 0.033285 | 0.004412 | 7.70E-14 |
| rs2769071 | let-7c-5p | trans | 3.76E-23 | Nightingale | XXL_VLDL_ | NA | G | A | 0.024963 | 0.004456 | 1.60E-09 |
| rs2769071 | let-7c-5p | trans | 3.76E-23 | Nightingale | XXL_VLDL_ | NA | G | A | 0.034743 | 0.004437 | 3.40E-15 |
| rs2769071 | let-7c-5p | trans | 3.76E-23 | Nightingale | S_LDL_CE | NA | G | A | 0.030512 | 0.004429 | 6.20E-12 |
| rs2769071 | let-7c-5p | trans | 3.76E-23 | Nightingale | ApoB | NA | G | A | 0.033916 | 0.004421 | 2.20E-14 |
| rs2769071 | let-7c-5p | trans | 3.76E-23 | Nightingale | XL_VLDL_C | NA | G | A | 0.038066 | 0.004245 | 6.90E-19 |
| rs2769071 | let-7c-5p | trans | 3.76E-23 | Nightingale | M_VLDL_C | NA | G | A | 0.040819 | 0.004387 | 2.50E-20 |
| rs2769071 | let-7c-5p | trans | 3.76E-23 | Nightingale | S_VLDL_FC | NA | G | A | 0.029307 | 0.00423 | 1.20E-11 |
| rs2769071 | let-7c-5p | trans | 3.76E-23 | Nightingale | XXL_VLDL_ | NA | G | A | 0.030722 | 0.004408 | 1.30E-12 |
| rs2769071 | let-7c-5p | trans | 3.76E-23 | Nightingale | XL_VLDL_P | NA | G | A | 0.038703 | 0.004434 | 7.10E-19 |
| rs2769071 | let-7c-5p | trans | 3.76E-23 | Nightingale | Total_CE | NA | G | A | 0.031783 | 0.00427 | 9.40E-14 |
| rs2769071 | let-7c-5p | trans | 3.76E-23 | Nightingale | S_LDL_PL_ | NA | G | A | -0.03002 | 0.004419 | 2.90E-13 |
| rs2769071 | let-7c-5p | trans | 3.76E-23 | Nightingale | Total_FC | NA | G | A | 0.033293 | 0.004325 | 6.50E-15 |
| rs2769071 | let-7c-5p | trans | 3.76E-23 | Nightingale | M_LDL_C_ | NA | G | A | 0.031139 | 0.00443 | 3.20E-12 |
| rs2769071 | let-7c-5p | trans | 3.76E-23 | Nightingale | IDL_CE | NA | G | A | 0.033536 | 0.004274 | 1.30E-14 |
| rs2769071 | let-7c-5p | trans | 3.76E-23 | Nightingale | S_LDL_CE_ | NA | G | A | 0.030898 | 0.004419 | 5.50E-14 |
| rs2769071 | let-7c-5p | trans | 3.76E-23 | Nightingale | LDL_FC | NA | G | A | 0.033607 | 0.00438 | 5.60E-14 |
| rs2769071 | let-7c-5p | trans | 3.76E-23 | Nightingale | L_LDL_P | NA | G | A | 0.029139 | 0.004412 | 6.50E-11 |
| rs2769071 | let-7c-5p | trans | 3.76E-23 | Nightingale | S_HDL_CE | NA | G | A | 0.02862 | 0.004434 | 1.40E-10 |
| rs2769071 | let-7c-5p | trans | 3.76E-23 | Nightingale | XS_VLDL_L | NA | G | A | 0.033869 | 0.004352 | 1.90E-15 |
| rs2769071 | let-7c-5p | trans | 3.76E-23 | Nightingale | L_VLDL_CE | NA | G | A | 0.047208 | 0.004236 | 3.80E-29 |
| rs2769071 | let-7c-5p | trans | 3.76E-23 | Nightingale | Remnant_( | NA | G | A | 0.036871 | 0.004398 | 3.70E-17 |
| rs2769071 | let-7c-5p | trans | 3.76E-23 | Nightingale | S_LDL_L | NA | G | A | 0.027711 | 0.004426 | 8.50E-10 |
| rs2769071 | let-7c-5p | trans | 3.76E-23 | Nightingale | XS_VLDL_C | NA | G | A | 0.038033 | 0.004297 | 1.30E-18 |
| rs2769071 | let-7c-5p | trans | 3.76E-23 | Nightingale | S_VLDL_FC | NA | G | A | 0.03789 | 0.004428 | 7.60E-18 |
| rs2769071 | let-7c-5p | trans | 3.76E-23 | Nightingale | IDL_C | NA | G | A | 0.034977 | 0.004276 | 9.40E-16 |
| rs2769071 | let-7c-5p | trans | 3.76E-23 | Nightingale | S_HDL_FC | NA | G | A | 0.023537 | 0.004346 | 4.30E-08 |
| rs2769071 | let-7c-5p | trans | 3.76E-23 | Nightingale | LDL_C | NA | G | A | 0.033173 | 0.004408 | 9.30E-14 |
| rs2769071 | let-7c-5p | trans | 3.76E-23 | Nightingale | M_LDL_CE | NA | G | A | 0.032948 | 0.004433 | 1.00E-13 |
| rs2769071 | let-7c-5p | trans | 3.76E-23 | Nightingale | IDL_P | NA | G | A | 0.039976 | 0.004381 | 1.40E-19 |
| rs2769071 | let-7c-5p | trans | 3.76E-23 | Nightingale | LDL_P | NA | G | A | 0.03177 | 0.004424 | 1.10E-12 |
| rs2769071 | let-7c-5p | trans | 3.76E-23 | Nightingale | S_HDL_C | NA | G | A | 0.028424 | 0.00442 | 1.70E-10 |
| rs2769071 | let-7c-5p | trans | 3.76E-23 | Nightingale | LDL_PL | NA | G | A | 0.034963 | 0.004417 | 5.00E-15 |
| rs2769071 | let-7c-5p | trans | 3.76E-23 | Nightingale | M_VLDL_P | NA | G | A | 0.028288 | 0.004416 | 6.40E-11 |
| rs2769071 | let-7c-5p | trans | 3.76E-23 | Nightingale | L_LDL_FC | NA | G | A | 0.033608 | 0.004347 | 3.00E-14 |
| rs2769071 | let-7c-5p | trans | 3.76E-23 | Nightingale | Total_PL | NA | G | A | 0.025022 | 0.004216 | 9.60E-10 |
| rs2769071 | let-7c-5p | trans | 3.76E-23 | Nightingale | Total_L | NA | G | A | 0.026409 | 0.004355 | 4.30E-10 |
| rs2769071 | let-7c-5p | trans | 3.76E-23 | Nightingale | S_HDL_P | NA | G | A | 0.030619 | 0.004428 | 5.20E-12 |
| rs2769071 | let-7c-5p | trans | 3.76E-23 | Nightingale | L_LDL_PL | NA | G | A | 0.037972 | 0.004393 | 9.60E-18 |
| rs2769071 | let-7c-5p | trans | 3.76E-23 | Nightingale | XS_VLDL_F | NA | G | A | 0.03143 | 0.004395 | 2.50E-12 |
| rs2769071 | let-7c-5p | trans | 3.76E-23 | Nightingale | VLDL_CE | NA | G | A | 0.035775 | 0.004428 | 3.10E-16 |
| rs2769071 | let-7c-5p | trans | 3.76E-23 | Nightingale | L_LDL_CE | NA | G | A | 0.031552 | 0.004388 | 1.00E-12 |
| rs2769071 | let-7c-5p | trans | 3.76E-23 | Nightingale | S_VLDL_TG | NA | G | A | -0.03806 | 0.004285 | 1.70E-18 |
| rs2769071 | let-7c-5p | trans | 3.76E-23 | Nightingale | S_VLDL_PL | NA | G | A | 0.027684 | 0.004239 | 2.00E-10 |
| rs2769071 | let-7c-5p | trans | 3.76E-23 | Nightingale | S_VLDL_CE | NA | G | A | 0.046931 | 0.004368 | 1.60E-26 |
| rs2769071 | let-7c-5p | trans | 3.76E-23 | Nightingale | M_LDL_FC | NA | G | A | 0.034307 | 0.004421 | 2.10E-14 |
| rs2769071 | let-7c-5p | trans | 3.76E-23 | Nightingale | M_VLDL_C | NA | G | A | 0.036101 | 0.00417 | 1.50E-17 |
| rs2769071 | let-7c-5p | trans | 3.76E-23 | Nightingale | L_VLDL_TG | NA | G | A | -0.05094 | 0.004406 | 9.70E-32 |
| rs2769071 | let-7c-5p | trans | 3.76E-23 | Nightingale | M_VLDL_F | NA | G | A | 0.041951 | 0.004212 | 4.70E-23 |
| rs2769071 | let-7c-5p | trans | 3.76E-23 | Nightingale | M_VLDL_F | NA | G | A | 0.033361 | 0.004426 | 3.20E-14 |
| rs2769071 | let-7c-5p | trans | 3.76E-23 | Nightingale | XL_VLDL_F | NA | G | A | 0.036999 | 0.004292 | 3.20E-18 |
| rs2769071 | let-7c-5p | trans | 3.76E-23 | Nightingale | S_VLDL_CE | NA | G | A | 0.038158 | 0.004421 | 1.70E-18 |
| rs2769071 | let-7c-5p | trans | 3.76E-23 | Nightingale | S_LDL_C | NA | G | A | 0.030907 | 0.004431 | 5.80E-12 |
| rs2769071 | let-7c-5p | trans | 3.76E-23 | Nightingale | XL_VLDL_C | NA | G | A | 0.037208 | 0.004247 | 4.50E-18 |

| | | | | | | | | | | | |
|---|---|---|---|---|---|---|---|---|---|---|---|
| rs2769071 | let-7c-5p | trans | 3.76E-23 | Nightingale | Citrate | NA | G | A | 0.026789 | 0.004398 | 7.90E-10 |
| rs2769071 | let-7c-5p | trans | 3.76E-23 | Nightingale | XXL_VLDL_ | NA | G | A | 0.033072 | 0.004461 | 9.80E-14 |
| rs2769071 | let-7c-5p | trans | 3.76E-23 | Nightingale | S_VLDL_L | NA | G | A | 0.024677 | 0.004384 | 3.70E-09 |
| rs2769071 | let-7c-5p | trans | 3.76E-23 | Nightingale | S_HDL_PL | NA | G | A | 0.023848 | 0.004389 | 3.00E-08 |
| rs2769071 | let-7c-5p | trans | 3.76E-23 | Nightingale | M_LDL_TG | NA | G | A | -0.02754 | 0.004372 | 3.80E-10 |
| rs2769071 | let-7c-5p | trans | 3.76E-23 | Nightingale | non_HDL_( | NA | G | A | 0.035686 | 0.004403 | 5.30E-16 |
| rs2769071 | let-7c-5p | trans | 3.76E-23 | Nightingale | M_VLDL_C | NA | G | A | 0.038687 | 0.004415 | 2.00E-18 |
| rs2769071 | let-7c-5p | trans | 3.76E-23 | Nightingale | XS_VLDL_F | NA | G | A | 0.037113 | 0.00435 | 6.00E-18 |
| rs2769071 | let-7c-5p | trans | 3.76E-23 | Nightingale | L_VLDL_FC | NA | G | A | 0.048347 | 0.004385 | 8.70E-29 |
| rs2769071 | let-7c-5p | trans | 3.76E-23 | Nightingale | Cholines | NA | G | A | 0.022376 | 0.004163 | 4.50E-08 |
| rs2769071 | let-7c-5p | trans | 3.76E-23 | Nightingale | LA | NA | G | A | 0.024942 | 0.004341 | 3.50E-09 |
| rs2769071 | let-7c-5p | trans | 3.76E-23 | Nightingale | S_LDL_FC | NA | G | A | 0.028535 | 0.00442 | 4.40E-10 |
| rs2769071 | let-7c-5p | trans | 3.76E-23 | Nightingale | PUFA | NA | G | A | 0.023736 | 0.004318 | 1.40E-08 |
| rs2769071 | let-7c-5p | trans | 3.76E-23 | Nightingale | XL_HDL_PL | NA | G | A | -0.02261 | 0.004067 | 1.40E-08 |
| rs2769071 | let-7c-5p | trans | 3.76E-23 | Nightingale | LDL_CE | NA | G | A | 0.032533 | 0.004415 | 2.30E-13 |
| rs2769071 | let-7c-5p | trans | 3.76E-23 | Nightingale | M_VLDL_T | NA | G | A | -0.03845 | 0.004184 | 9.20E-20 |
| rs2769071 | let-7c-5p | trans | 3.76E-23 | Nightingale | L_LDL_L | NA | G | A | 0.033538 | 0.004384 | 3.10E-14 |
| rs2769071 | let-7c-5p | trans | 3.76E-23 | Nightingale | XS_VLDL_P | NA | G | A | 0.032167 | 0.004365 | 4.90E-14 |
| rs2769071 | let-7c-5p | trans | 3.76E-23 | Nightingale | IDL_FC_pct | NA | G | A | 0.025676 | 0.004393 | 1.20E-08 |
| rs2769071 | let-7c-5p | trans | 3.76E-23 | Nightingale | S_VLDL_C | NA | G | A | 0.038352 | 0.004427 | 1.90E-18 |
| rs2769071 | let-7c-5p | trans | 3.76E-23 | Nightingale | S_VLDL_P | NA | G | A | 0.023474 | 0.004372 | 1.60E-08 |
| rs2769071 | let-7c-5p | trans | 3.76E-23 | Nightingale | M_LDL_PL | NA | G | A | 0.032427 | 0.004433 | 2.80E-13 |
| rs2769071 | miR-10b-5p | trans | 3.48E-35 | Nightingale | S_LDL_C_p | NA | G | A | 0.045281 | 0.004444 | 7.30E-25 |
| rs2769071 | miR-10b-5p | trans | 3.48E-35 | Nightingale | S_LDL_P | NA | G | A | 0.029553 | 0.004431 | 3.40E-11 |
| rs2769071 | miR-10b-5p | trans | 3.48E-35 | Nightingale | L_VLDL_C_ | NA | G | A | 0.051993 | 0.004324 | 7.60E-34 |
| rs2769071 | miR-10b-5p | trans | 3.48E-35 | Nightingale | GlycA | NA | G | A | 0.028001 | 0.004408 | 1.50E-10 |
| rs2769071 | miR-10b-5p | trans | 3.48E-35 | Nightingale | Total_C | NA | G | A | 0.032411 | 0.004286 | 3.20E-14 |
| rs2769071 | miR-10b-5p | trans | 3.48E-35 | Nightingale | M_VLDL_C | NA | G | A | 0.033528 | 0.004159 | 2.40E-15 |
| rs2769071 | miR-10b-5p | trans | 3.48E-35 | Nightingale | M_LDL_L | NA | G | A | 0.033001 | 0.004433 | 1.10E-13 |
| rs2769071 | miR-10b-5p | trans | 3.48E-35 | Nightingale | M_VLDL_P | NA | G | A | 0.043923 | 0.004249 | 8.10E-25 |
| rs2769071 | miR-10b-5p | trans | 3.48E-35 | Nightingale | IDL_FC | NA | G | A | 0.038256 | 0.004286 | 1.50E-18 |
| rs2769071 | miR-10b-5p | trans | 3.48E-35 | Nightingale | VLDL_P | NA | G | A | 0.026645 | 0.004403 | 4.40E-10 |
| rs2769071 | miR-10b-5p | trans | 3.48E-35 | Nightingale | M_LDL_P | NA | G | A | 0.035884 | 0.004439 | 7.10E-16 |
| rs2769071 | miR-10b-5p | trans | 3.48E-35 | Nightingale | XL_VLDL_T | NA | G | A | -0.04368 | 0.004291 | 3.40E-24 |
| rs2769071 | miR-10b-5p | trans | 3.48E-35 | Nightingale | S_VLDL_C_ | NA | G | A | 0.041808 | 0.004308 | 6.50E-22 |
| rs2769071 | miR-10b-5p | trans | 3.48E-35 | Nightingale | M_LDL_C | NA | G | A | 0.033958 | 0.004435 | 2.30E-14 |
| rs2769071 | miR-10b-5p | trans | 3.48E-35 | Nightingale | L_LDL_C | NA | G | A | 0.032304 | 0.004377 | 2.70E-13 |
| rs2769071 | miR-10b-5p | trans | 3.48E-35 | Nightingale | IDL_PL | NA | G | A | 0.035362 | 0.004272 | 3.30E-16 |
| rs2769071 | miR-10b-5p | trans | 3.48E-35 | Nightingale | Sphingomy | NA | G | A | 0.029058 | 0.004139 | 6.70E-12 |
| rs2769071 | miR-10b-5p | trans | 3.48E-35 | Nightingale | S_HDL_L | NA | G | A | 0.026342 | 0.004412 | 1.50E-09 |
| rs2769071 | miR-10b-5p | trans | 3.48E-35 | Nightingale | Clinical_LD | NA | G | A | 0.036447 | 0.004401 | 2.90E-16 |
| rs2769071 | miR-10b-5p | trans | 3.48E-35 | Nightingale | M_VLDL_P | NA | G | A | 0.029124 | 0.00442 | 2.10E-11 |
| rs2769071 | miR-10b-5p | trans | 3.48E-35 | Nightingale | ApoB_by_/ | NA | G | A | 0.024951 | 0.00436 | 1.40E-08 |
| rs2769071 | miR-10b-5p | trans | 3.48E-35 | Nightingale | XS_VLDL_C | NA | G | A | 0.037835 | 0.004272 | 2.00E-18 |
| rs2769071 | miR-10b-5p | trans | 3.48E-35 | Nightingale | XS_VLDL_P | NA | G | A | 0.036739 | 0.004368 | 1.30E-17 |
| rs2769071 | miR-10b-5p | trans | 3.48E-35 | Nightingale | IDL_L | NA | G | A | 0.035297 | 0.00428 | 4.70E-16 |
| rs2769071 | miR-10b-5p | trans | 3.48E-35 | Nightingale | S_VLDL_PL | NA | G | A | 0.032773 | 0.004419 | 4.20E-14 |
| rs2769071 | miR-10b-5p | trans | 3.48E-35 | Nightingale | VLDL_FC | NA | G | A | 0.024155 | 0.004395 | 1.10E-08 |
| rs2769071 | miR-10b-5p | trans | 3.48E-35 | Nightingale | Omega_6 | NA | G | A | 0.025917 | 0.004323 | 5.60E-10 |
| rs2769071 | miR-10b-5p | trans | 3.48E-35 | Nightingale | VLDL_C | NA | G | A | 0.031294 | 0.004421 | 5.00E-13 |
| rs2769071 | miR-10b-5p | trans | 3.48E-35 | Nightingale | XXL_VLDL_ | NA | G | A | -0.03561 | 0.004465 | 2.70E-16 |
| rs2769071 | miR-10b-5p | trans | 3.48E-35 | Nightingale | LDL_L | NA | G | A | 0.033285 | 0.004412 | 7.70E-14 |
| rs2769071 | miR-10b-5p | trans | 3.48E-35 | Nightingale | XXL_VLDL_ | NA | G | A | 0.024963 | 0.004456 | 1.60E-09 |
| rs2769071 | miR-10b-5p | trans | 3.48E-35 | Nightingale | XXL_VLDL_ | NA | G | A | 0.034743 | 0.004437 | 3.40E-15 |
| rs2769071 | miR-10b-5p | trans | 3.48E-35 | Nightingale | S_LDL_CE | NA | G | A | 0.030512 | 0.004429 | 6.20E-12 |
| rs2769071 | miR-10b-5p | trans | 3.48E-35 | Nightingale | ApoB | NA | G | A | 0.033916 | 0.004421 | 2.20E-14 |
| rs2769071 | miR-10b-5p | trans | 3.48E-35 | Nightingale | XL_VLDL_C | NA | G | A | 0.038066 | 0.004245 | 6.90E-19 |
| rs2769071 | miR-10b-5p | trans | 3.48E-35 | Nightingale | M_VLDL_C | NA | G | A | 0.040819 | 0.004387 | 2.50E-20 |
| rs2769071 | miR-10b-5p | trans | 3.48E-35 | Nightingale | S_VLDL_FC | NA | G | A | 0.029307 | 0.00423 | 1.20E-11 |
| rs2769071 | miR-10b-5p | trans | 3.48E-35 | Nightingale | XXL_VLDL_ | NA | G | A | 0.030722 | 0.004408 | 1.30E-12 |
| rs2769071 | miR-10b-5p | trans | 3.48E-35 | Nightingale | XL_VLDL_P | NA | G | A | 0.038703 | 0.004434 | 7.10E-19 |
| rs2769071 | miR-10b-5p | trans | 3.48E-35 | Nightingale | Total_CE | NA | G | A | 0.031783 | 0.00427 | 9.40E-14 |
| rs2769071 | miR-10b-5p | trans | 3.48E-35 | Nightingale | S_LDL_PL_ | NA | G | A | -0.03002 | 0.004419 | 2.90E-13 |

| | | | | | | | | | | | |
|---|---|---|---|---|---|---|---|---|---|---|---|
| rs2769071 | miR-10b-5p | trans | 3.48E-35 | Nightingale | Total_FC | NA | G | A | 0.033293 | 0.004325 | 6.50E-15 |
| rs2769071 | miR-10b-5p | trans | 3.48E-35 | Nightingale | M_LDL_C_ | NA | G | A | 0.031139 | 0.00443 | 3.20E-12 |
| rs2769071 | miR-10b-5p | trans | 3.48E-35 | Nightingale | IDL_CE | NA | G | A | 0.033536 | 0.004274 | 1.30E-14 |
| rs2769071 | miR-10b-5p | trans | 3.48E-35 | Nightingale | S_LDL_CE_ | NA | G | A | 0.030898 | 0.004419 | 5.50E-14 |
| rs2769071 | miR-10b-5p | trans | 3.48E-35 | Nightingale | LDL_FC | NA | G | A | 0.033607 | 0.00438 | 5.60E-14 |
| rs2769071 | miR-10b-5p | trans | 3.48E-35 | Nightingale | L_LDL_P | NA | G | A | 0.029139 | 0.004412 | 6.50E-11 |
| rs2769071 | miR-10b-5p | trans | 3.48E-35 | Nightingale | S_HDL_CE | NA | G | A | 0.02862 | 0.004434 | 1.40E-10 |
| rs2769071 | miR-10b-5p | trans | 3.48E-35 | Nightingale | XS_VLDL_L | NA | G | A | 0.033869 | 0.004352 | 1.90E-15 |
| rs2769071 | miR-10b-5p | trans | 3.48E-35 | Nightingale | L_VLDL_CE | NA | G | A | 0.047208 | 0.004236 | 3.80E-29 |
| rs2769071 | miR-10b-5p | trans | 3.48E-35 | Nightingale | Remnant_( | NA | G | A | 0.036871 | 0.004398 | 3.70E-17 |
| rs2769071 | miR-10b-5p | trans | 3.48E-35 | Nightingale | S_LDL_L | NA | G | A | 0.027711 | 0.004426 | 8.50E-10 |
| rs2769071 | miR-10b-5p | trans | 3.48E-35 | Nightingale | XS_VLDL_C | NA | G | A | 0.038033 | 0.004297 | 1.30E-18 |
| rs2769071 | miR-10b-5p | trans | 3.48E-35 | Nightingale | S_VLDL_FC | NA | G | A | 0.03789 | 0.004428 | 7.60E-18 |
| rs2769071 | miR-10b-5p | trans | 3.48E-35 | Nightingale | IDL_C | NA | G | A | 0.034977 | 0.004276 | 9.40E-16 |
| rs2769071 | miR-10b-5p | trans | 3.48E-35 | Nightingale | S_HDL_FC | NA | G | A | 0.023537 | 0.004346 | 4.30E-08 |
| rs2769071 | miR-10b-5p | trans | 3.48E-35 | Nightingale | LDL_C | NA | G | A | 0.033173 | 0.004408 | 9.30E-14 |
| rs2769071 | miR-10b-5p | trans | 3.48E-35 | Nightingale | M_LDL_CE | NA | G | A | 0.032948 | 0.004433 | 1.00E-13 |
| rs2769071 | miR-10b-5p | trans | 3.48E-35 | Nightingale | IDL_P | NA | G | A | 0.039976 | 0.004381 | 1.40E-19 |
| rs2769071 | miR-10b-5p | trans | 3.48E-35 | Nightingale | LDL_P | NA | G | A | 0.03177 | 0.004424 | 1.10E-12 |
| rs2769071 | miR-10b-5p | trans | 3.48E-35 | Nightingale | S_HDL_C | NA | G | A | 0.028424 | 0.00442 | 1.70E-10 |
| rs2769071 | miR-10b-5p | trans | 3.48E-35 | Nightingale | LDL_PL | NA | G | A | 0.034963 | 0.004417 | 5.00E-15 |
| rs2769071 | miR-10b-5p | trans | 3.48E-35 | Nightingale | M_VLDL_P | NA | G | A | 0.028288 | 0.004416 | 6.40E-11 |
| rs2769071 | miR-10b-5p | trans | 3.48E-35 | Nightingale | L_LDL_FC | NA | G | A | 0.033608 | 0.004347 | 3.00E-14 |
| rs2769071 | miR-10b-5p | trans | 3.48E-35 | Nightingale | Total_PL | NA | G | A | 0.025022 | 0.004216 | 9.60E-10 |
| rs2769071 | miR-10b-5p | trans | 3.48E-35 | Nightingale | Total_L | NA | G | A | 0.026409 | 0.004355 | 4.30E-10 |
| rs2769071 | miR-10b-5p | trans | 3.48E-35 | Nightingale | S_HDL_P | NA | G | A | 0.030619 | 0.004428 | 5.20E-12 |
| rs2769071 | miR-10b-5p | trans | 3.48E-35 | Nightingale | L_LDL_PL | NA | G | A | 0.037972 | 0.004393 | 9.60E-18 |
| rs2769071 | miR-10b-5p | trans | 3.48E-35 | Nightingale | XS_VLDL_F | NA | G | A | 0.03143 | 0.004395 | 2.50E-12 |
| rs2769071 | miR-10b-5p | trans | 3.48E-35 | Nightingale | VLDL_CE | NA | G | A | 0.035775 | 0.004428 | 3.10E-16 |
| rs2769071 | miR-10b-5p | trans | 3.48E-35 | Nightingale | L_LDL_CE | NA | G | A | 0.031552 | 0.004388 | 1.00E-12 |
| rs2769071 | miR-10b-5p | trans | 3.48E-35 | Nightingale | S_VLDL_TG | NA | G | A | -0.03806 | 0.004285 | 1.70E-18 |
| rs2769071 | miR-10b-5p | trans | 3.48E-35 | Nightingale | S_VLDL_PL | NA | G | A | 0.027684 | 0.004239 | 2.00E-10 |
| rs2769071 | miR-10b-5p | trans | 3.48E-35 | Nightingale | S_VLDL_CE | NA | G | A | 0.046931 | 0.004368 | 1.60E-26 |
| rs2769071 | miR-10b-5p | trans | 3.48E-35 | Nightingale | M_LDL_FC | NA | G | A | 0.034307 | 0.004421 | 2.10E-14 |
| rs2769071 | miR-10b-5p | trans | 3.48E-35 | Nightingale | M_VLDL_C | NA | G | A | 0.036101 | 0.00417 | 1.50E-17 |
| rs2769071 | miR-10b-5p | trans | 3.48E-35 | Nightingale | L_VLDL_TG | NA | G | A | -0.05094 | 0.004406 | 9.70E-32 |
| rs2769071 | miR-10b-5p | trans | 3.48E-35 | Nightingale | M_VLDL_F | NA | G | A | 0.041951 | 0.004212 | 4.70E-23 |
| rs2769071 | miR-10b-5p | trans | 3.48E-35 | Nightingale | M_VLDL_F | NA | G | A | 0.033361 | 0.004426 | 3.20E-14 |
| rs2769071 | miR-10b-5p | trans | 3.48E-35 | Nightingale | XL_VLDL_F | NA | G | A | 0.036999 | 0.004292 | 3.20E-18 |
| rs2769071 | miR-10b-5p | trans | 3.48E-35 | Nightingale | S_VLDL_CE | NA | G | A | 0.038158 | 0.004421 | 1.70E-18 |
| rs2769071 | miR-10b-5p | trans | 3.48E-35 | Nightingale | S_LDL_C | NA | G | A | 0.030907 | 0.004431 | 5.80E-12 |
| rs2769071 | miR-10b-5p | trans | 3.48E-35 | Nightingale | XL_VLDL_C | NA | G | A | 0.037208 | 0.004247 | 4.50E-18 |
| rs2769071 | miR-10b-5p | trans | 3.48E-35 | Nightingale | Citrate | NA | G | A | 0.026789 | 0.004398 | 7.90E-10 |
| rs2769071 | miR-10b-5p | trans | 3.48E-35 | Nightingale | XXL_VLDL_ | NA | G | A | 0.033072 | 0.004461 | 9.80E-14 |
| rs2769071 | miR-10b-5p | trans | 3.48E-35 | Nightingale | S_VLDL_L | NA | G | A | 0.024677 | 0.004384 | 3.70E-09 |
| rs2769071 | miR-10b-5p | trans | 3.48E-35 | Nightingale | S_HDL_PL | NA | G | A | 0.023848 | 0.004389 | 3.00E-08 |
| rs2769071 | miR-10b-5p | trans | 3.48E-35 | Nightingale | M_LDL_TG | NA | G | A | -0.02754 | 0.004372 | 3.80E-10 |
| rs2769071 | miR-10b-5p | trans | 3.48E-35 | Nightingale | non_HDL_( | NA | G | A | 0.035686 | 0.004403 | 5.30E-16 |
| rs2769071 | miR-10b-5p | trans | 3.48E-35 | Nightingale | M_VLDL_C | NA | G | A | 0.038687 | 0.004415 | 2.00E-18 |
| rs2769071 | miR-10b-5p | trans | 3.48E-35 | Nightingale | XS_VLDL_F | NA | G | A | 0.037113 | 0.00435 | 6.00E-18 |
| rs2769071 | miR-10b-5p | trans | 3.48E-35 | Nightingale | L_VLDL_FC | NA | G | A | 0.048347 | 0.004385 | 8.70E-29 |
| rs2769071 | miR-10b-5p | trans | 3.48E-35 | Nightingale | Cholines | NA | G | A | 0.022376 | 0.004163 | 4.50E-08 |
| rs2769071 | miR-10b-5p | trans | 3.48E-35 | Nightingale | LA | NA | G | A | 0.024942 | 0.004341 | 3.50E-09 |
| rs2769071 | miR-10b-5p | trans | 3.48E-35 | Nightingale | S_LDL_FC | NA | G | A | 0.028535 | 0.00442 | 4.40E-10 |
| rs2769071 | miR-10b-5p | trans | 3.48E-35 | Nightingale | PUFA | NA | G | A | 0.023736 | 0.004318 | 1.40E-08 |
| rs2769071 | miR-10b-5p | trans | 3.48E-35 | Nightingale | XL_HDL_PL | NA | G | A | -0.02261 | 0.004067 | 1.40E-08 |
| rs2769071 | miR-10b-5p | trans | 3.48E-35 | Nightingale | LDL_CE | NA | G | A | 0.032533 | 0.004415 | 2.30E-13 |
| rs2769071 | miR-10b-5p | trans | 3.48E-35 | Nightingale | M_VLDL_T | NA | G | A | -0.03845 | 0.004184 | 9.20E-20 |
| rs2769071 | miR-10b-5p | trans | 3.48E-35 | Nightingale | L_LDL_L | NA | G | A | 0.033538 | 0.004384 | 3.10E-14 |
| rs2769071 | miR-10b-5p | trans | 3.48E-35 | Nightingale | XS_VLDL_P | NA | G | A | 0.032167 | 0.004365 | 4.90E-14 |
| rs2769071 | miR-10b-5p | trans | 3.48E-35 | Nightingale | IDL_FC_pct | NA | G | A | 0.025676 | 0.004393 | 1.20E-08 |
| rs2769071 | miR-10b-5p | trans | 3.48E-35 | Nightingale | S_VLDL_C | NA | G | A | 0.038352 | 0.004427 | 1.90E-18 |
| rs2769071 | miR-10b-5p | trans | 3.48E-35 | Nightingale | S_VLDL_P | NA | G | A | 0.023474 | 0.004372 | 1.60E-08 |

| | | | | | | | | | | | |
|---|---|---|---|---|---|---|---|---|---|---|---|
| rs2769071 | miR-10b-5p | trans | 3.48E-35 | Nightingale | M_LDL_PL | NA | G | A | 0.032427 | 0.004433 | 2.80E-13 |
| rs2769071 | miR-126-3p | trans | 4.06E-39 | Nightingale | S_LDL_C_p | NA | G | A | 0.045281 | 0.004444 | 7.30E-25 |
| rs2769071 | miR-126-3p | trans | 4.06E-39 | Nightingale | S_LDL_P | NA | G | A | 0.029553 | 0.004431 | 3.40E-11 |
| rs2769071 | miR-126-3p | trans | 4.06E-39 | Nightingale | L_VLDL_C_ | NA | G | A | 0.051993 | 0.004324 | 7.60E-34 |
| rs2769071 | miR-126-3p | trans | 4.06E-39 | Nightingale | GlycA | NA | G | A | 0.028001 | 0.004408 | 1.50E-10 |
| rs2769071 | miR-126-3p | trans | 4.06E-39 | Nightingale | Total_C | NA | G | A | 0.032411 | 0.004286 | 3.20E-14 |
| rs2769071 | miR-126-3p | trans | 4.06E-39 | Nightingale | M_VLDL_C | NA | G | A | 0.033528 | 0.004159 | 2.40E-15 |
| rs2769071 | miR-126-3p | trans | 4.06E-39 | Nightingale | M_LDL_L | NA | G | A | 0.033001 | 0.004433 | 1.10E-13 |
| rs2769071 | miR-126-3p | trans | 4.06E-39 | Nightingale | M_VLDL_P | NA | G | A | 0.043923 | 0.004249 | 8.10E-25 |
| rs2769071 | miR-126-3p | trans | 4.06E-39 | Nightingale | IDL_FC | NA | G | A | 0.038256 | 0.004286 | 1.50E-18 |
| rs2769071 | miR-126-3p | trans | 4.06E-39 | Nightingale | VLDL_P | NA | G | A | 0.026645 | 0.004403 | 4.40E-10 |
| rs2769071 | miR-126-3p | trans | 4.06E-39 | Nightingale | M_LDL_P | NA | G | A | 0.035884 | 0.004439 | 7.10E-16 |
| rs2769071 | miR-126-3p | trans | 4.06E-39 | Nightingale | XL_VLDL_T | NA | G | A | -0.04368 | 0.004291 | 3.40E-24 |
| rs2769071 | miR-126-3p | trans | 4.06E-39 | Nightingale | S_VLDL_C_ | NA | G | A | 0.041808 | 0.004308 | 6.50E-22 |
| rs2769071 | miR-126-3p | trans | 4.06E-39 | Nightingale | M_LDL_C | NA | G | A | 0.033958 | 0.004435 | 2.30E-14 |
| rs2769071 | miR-126-3p | trans | 4.06E-39 | Nightingale | L_LDL_C | NA | G | A | 0.032304 | 0.004377 | 2.70E-13 |
| rs2769071 | miR-126-3p | trans | 4.06E-39 | Nightingale | IDL_PL | NA | G | A | 0.035362 | 0.004272 | 3.30E-16 |
| rs2769071 | miR-126-3p | trans | 4.06E-39 | Nightingale | Sphingomy | NA | G | A | 0.029058 | 0.004139 | 6.70E-12 |
| rs2769071 | miR-126-3p | trans | 4.06E-39 | Nightingale | S_HDL_L | NA | G | A | 0.026342 | 0.004412 | 1.50E-09 |
| rs2769071 | miR-126-3p | trans | 4.06E-39 | Nightingale | Clinical_LD | NA | G | A | 0.036447 | 0.004401 | 2.90E-16 |
| rs2769071 | miR-126-3p | trans | 4.06E-39 | Nightingale | M_VLDL_P | NA | G | A | 0.029124 | 0.00442 | 2.10E-11 |
| rs2769071 | miR-126-3p | trans | 4.06E-39 | Nightingale | ApoB_by_A | NA | G | A | 0.024951 | 0.00436 | 1.40E-08 |
| rs2769071 | miR-126-3p | trans | 4.06E-39 | Nightingale | XS_VLDL_C | NA | G | A | 0.037835 | 0.004272 | 2.00E-18 |
| rs2769071 | miR-126-3p | trans | 4.06E-39 | Nightingale | XS_VLDL_P | NA | G | A | 0.036739 | 0.004368 | 1.30E-17 |
| rs2769071 | miR-126-3p | trans | 4.06E-39 | Nightingale | IDL_L | NA | G | A | 0.035297 | 0.00428 | 4.70E-16 |
| rs2769071 | miR-126-3p | trans | 4.06E-39 | Nightingale | S_VLDL_PL | NA | G | A | 0.032773 | 0.004419 | 4.20E-14 |
| rs2769071 | miR-126-3p | trans | 4.06E-39 | Nightingale | VLDL_FC | NA | G | A | 0.024155 | 0.004395 | 1.10E-08 |
| rs2769071 | miR-126-3p | trans | 4.06E-39 | Nightingale | Omega_6 | NA | G | A | 0.025917 | 0.004323 | 5.60E-10 |
| rs2769071 | miR-126-3p | trans | 4.06E-39 | Nightingale | VLDL_C | NA | G | A | 0.031294 | 0.004421 | 5.00E-13 |
| rs2769071 | miR-126-3p | trans | 4.06E-39 | Nightingale | XXL_VLDL_ | NA | G | A | -0.03561 | 0.004465 | 2.70E-16 |
| rs2769071 | miR-126-3p | trans | 4.06E-39 | Nightingale | LDL_L | NA | G | A | 0.033285 | 0.004412 | 7.70E-14 |
| rs2769071 | miR-126-3p | trans | 4.06E-39 | Nightingale | XXL_VLDL_ | NA | G | A | 0.024963 | 0.004456 | 1.60E-09 |
| rs2769071 | miR-126-3p | trans | 4.06E-39 | Nightingale | XXL_VLDL_ | NA | G | A | 0.034743 | 0.004437 | 3.40E-15 |
| rs2769071 | miR-126-3p | trans | 4.06E-39 | Nightingale | S_LDL_CE | NA | G | A | 0.030512 | 0.004429 | 6.20E-12 |
| rs2769071 | miR-126-3p | trans | 4.06E-39 | Nightingale | ApoB | NA | G | A | 0.033916 | 0.004421 | 2.20E-14 |
| rs2769071 | miR-126-3p | trans | 4.06E-39 | Nightingale | XL_VLDL_C | NA | G | A | 0.038066 | 0.004245 | 6.90E-19 |
| rs2769071 | miR-126-3p | trans | 4.06E-39 | Nightingale | M_VLDL_C | NA | G | A | 0.040819 | 0.004387 | 2.50E-20 |
| rs2769071 | miR-126-3p | trans | 4.06E-39 | Nightingale | S_VLDL_FC | NA | G | A | 0.029307 | 0.00423 | 1.20E-11 |
| rs2769071 | miR-126-3p | trans | 4.06E-39 | Nightingale | XXL_VLDL_ | NA | G | A | 0.030722 | 0.004408 | 1.30E-12 |
| rs2769071 | miR-126-3p | trans | 4.06E-39 | Nightingale | XL_VLDL_P | NA | G | A | 0.038703 | 0.004434 | 7.10E-19 |
| rs2769071 | miR-126-3p | trans | 4.06E-39 | Nightingale | Total_CE | NA | G | A | 0.031783 | 0.00427 | 9.40E-14 |
| rs2769071 | miR-126-3p | trans | 4.06E-39 | Nightingale | S_LDL_PL_ | NA | G | A | -0.03002 | 0.004419 | 2.90E-13 |
| rs2769071 | miR-126-3p | trans | 4.06E-39 | Nightingale | Total_FC | NA | G | A | 0.033293 | 0.004325 | 6.50E-15 |
| rs2769071 | miR-126-3p | trans | 4.06E-39 | Nightingale | M_LDL_C_p | NA | G | A | 0.031139 | 0.00443 | 3.20E-12 |
| rs2769071 | miR-126-3p | trans | 4.06E-39 | Nightingale | IDL_CE | NA | G | A | 0.033536 | 0.004274 | 1.30E-14 |
| rs2769071 | miR-126-3p | trans | 4.06E-39 | Nightingale | S_LDL_CE_ | NA | G | A | 0.030898 | 0.004419 | 5.50E-14 |
| rs2769071 | miR-126-3p | trans | 4.06E-39 | Nightingale | LDL_FC | NA | G | A | 0.033607 | 0.00438 | 5.60E-14 |
| rs2769071 | miR-126-3p | trans | 4.06E-39 | Nightingale | L_LDL_P | NA | G | A | 0.029139 | 0.004412 | 6.50E-11 |
| rs2769071 | miR-126-3p | trans | 4.06E-39 | Nightingale | S_HDL_CE | NA | G | A | 0.02862 | 0.004434 | 1.40E-10 |
| rs2769071 | miR-126-3p | trans | 4.06E-39 | Nightingale | XS_VLDL_L | NA | G | A | 0.033869 | 0.004352 | 1.90E-15 |
| rs2769071 | miR-126-3p | trans | 4.06E-39 | Nightingale | L_VLDL_CE | NA | G | A | 0.047208 | 0.004236 | 3.80E-29 |
| rs2769071 | miR-126-3p | trans | 4.06E-39 | Nightingale | Remnant_C | NA | G | A | 0.036871 | 0.004398 | 3.70E-17 |
| rs2769071 | miR-126-3p | trans | 4.06E-39 | Nightingale | S_LDL_L | NA | G | A | 0.027711 | 0.004426 | 8.50E-10 |
| rs2769071 | miR-126-3p | trans | 4.06E-39 | Nightingale | XS_VLDL_C | NA | G | A | 0.038033 | 0.004297 | 1.30E-18 |
| rs2769071 | miR-126-3p | trans | 4.06E-39 | Nightingale | S_VLDL_FC | NA | G | A | 0.03789 | 0.004428 | 7.60E-18 |
| rs2769071 | miR-126-3p | trans | 4.06E-39 | Nightingale | IDL_C | NA | G | A | 0.034977 | 0.004276 | 9.40E-16 |
| rs2769071 | miR-126-3p | trans | 4.06E-39 | Nightingale | S_HDL_FC | NA | G | A | 0.023537 | 0.004346 | 4.30E-08 |
| rs2769071 | miR-126-3p | trans | 4.06E-39 | Nightingale | LDL_C | NA | G | A | 0.033173 | 0.004408 | 9.30E-14 |
| rs2769071 | miR-126-3p | trans | 4.06E-39 | Nightingale | M_LDL_CE | NA | G | A | 0.032948 | 0.004433 | 1.00E-13 |
| rs2769071 | miR-126-3p | trans | 4.06E-39 | Nightingale | IDL_P | NA | G | A | 0.039976 | 0.004381 | 1.40E-19 |
| rs2769071 | miR-126-3p | trans | 4.06E-39 | Nightingale | LDL_P | NA | G | A | 0.03177 | 0.004424 | 1.10E-12 |
| rs2769071 | miR-126-3p | trans | 4.06E-39 | Nightingale | S_HDL_C | NA | G | A | 0.028424 | 0.00442 | 1.70E-10 |
| rs2769071 | miR-126-3p | trans | 4.06E-39 | Nightingale | LDL_PL | NA | G | A | 0.034963 | 0.004417 | 5.00E-15 |

| | | | | | | | | | | | |
|---|---|---|---|---|---|---|---|---|---|---|---|
| rs2769071 | miR-126-3p | trans | 4.06E-39 | Nightingale | M_VLDL_P | NA | G | A | 0.028288 | 0.004416 | 6.40E-11 |
| rs2769071 | miR-126-3p | trans | 4.06E-39 | Nightingale | L_LDL_FC | NA | G | A | 0.033608 | 0.004347 | 3.00E-14 |
| rs2769071 | miR-126-3p | trans | 4.06E-39 | Nightingale | Total_PL | NA | G | A | 0.025022 | 0.004216 | 9.60E-10 |
| rs2769071 | miR-126-3p | trans | 4.06E-39 | Nightingale | Total_L | NA | G | A | 0.026409 | 0.004355 | 4.30E-10 |
| rs2769071 | miR-126-3p | trans | 4.06E-39 | Nightingale | S_HDL_P | NA | G | A | 0.030619 | 0.004428 | 5.20E-12 |
| rs2769071 | miR-126-3p | trans | 4.06E-39 | Nightingale | L_LDL_PL | NA | G | A | 0.037972 | 0.004393 | 9.60E-18 |
| rs2769071 | miR-126-3p | trans | 4.06E-39 | Nightingale | XS_VLDL_F | NA | G | A | 0.03143 | 0.004395 | 2.50E-12 |
| rs2769071 | miR-126-3p | trans | 4.06E-39 | Nightingale | VLDL_CE | NA | G | A | 0.035775 | 0.004428 | 3.10E-16 |
| rs2769071 | miR-126-3p | trans | 4.06E-39 | Nightingale | L_LDL_CE | NA | G | A | 0.031552 | 0.004388 | 1.00E-12 |
| rs2769071 | miR-126-3p | trans | 4.06E-39 | Nightingale | S_VLDL_TG | NA | G | A | -0.03806 | 0.004285 | 1.70E-18 |
| rs2769071 | miR-126-3p | trans | 4.06E-39 | Nightingale | S_VLDL_PL | NA | G | A | 0.027684 | 0.004239 | 2.00E-10 |
| rs2769071 | miR-126-3p | trans | 4.06E-39 | Nightingale | S_VLDL_CE | NA | G | A | 0.046931 | 0.004368 | 1.60E-26 |
| rs2769071 | miR-126-3p | trans | 4.06E-39 | Nightingale | M_LDL_FC | NA | G | A | 0.034307 | 0.004421 | 2.10E-14 |
| rs2769071 | miR-126-3p | trans | 4.06E-39 | Nightingale | M_VLDL_C | NA | G | A | 0.036101 | 0.00417 | 1.50E-17 |
| rs2769071 | miR-126-3p | trans | 4.06E-39 | Nightingale | L_VLDL_TG | NA | G | A | -0.05094 | 0.004406 | 9.70E-32 |
| rs2769071 | miR-126-3p | trans | 4.06E-39 | Nightingale | M_VLDL_F | NA | G | A | 0.041951 | 0.004212 | 4.70E-23 |
| rs2769071 | miR-126-3p | trans | 4.06E-39 | Nightingale | M_VLDL_F | NA | G | A | 0.033361 | 0.004426 | 3.20E-14 |
| rs2769071 | miR-126-3p | trans | 4.06E-39 | Nightingale | XL_VLDL_F | NA | G | A | 0.036999 | 0.004292 | 3.20E-18 |
| rs2769071 | miR-126-3p | trans | 4.06E-39 | Nightingale | S_VLDL_CE | NA | G | A | 0.038158 | 0.004421 | 1.70E-18 |
| rs2769071 | miR-126-3p | trans | 4.06E-39 | Nightingale | S_LDL_C | NA | G | A | 0.030907 | 0.004431 | 5.80E-12 |
| rs2769071 | miR-126-3p | trans | 4.06E-39 | Nightingale | XL_VLDL_C | NA | G | A | 0.037208 | 0.004247 | 4.50E-18 |
| rs2769071 | miR-126-3p | trans | 4.06E-39 | Nightingale | Citrate | NA | G | A | 0.026789 | 0.004398 | 7.90E-10 |
| rs2769071 | miR-126-3p | trans | 4.06E-39 | Nightingale | XXL_VLDL_ | NA | G | A | 0.033072 | 0.004461 | 9.80E-14 |
| rs2769071 | miR-126-3p | trans | 4.06E-39 | Nightingale | S_VLDL_L | NA | G | A | 0.024677 | 0.004384 | 3.70E-09 |
| rs2769071 | miR-126-3p | trans | 4.06E-39 | Nightingale | S_HDL_PL | NA | G | A | 0.023848 | 0.004389 | 3.00E-08 |
| rs2769071 | miR-126-3p | trans | 4.06E-39 | Nightingale | M_LDL_TG | NA | G | A | -0.02754 | 0.004372 | 3.80E-10 |
| rs2769071 | miR-126-3p | trans | 4.06E-39 | Nightingale | non_HDL_( | NA | G | A | 0.035686 | 0.004403 | 5.30E-16 |
| rs2769071 | miR-126-3p | trans | 4.06E-39 | Nightingale | M_VLDL_C | NA | G | A | 0.038687 | 0.004415 | 2.00E-18 |
| rs2769071 | miR-126-3p | trans | 4.06E-39 | Nightingale | XS_VLDL_F | NA | G | A | 0.037113 | 0.00435 | 6.00E-18 |
| rs2769071 | miR-126-3p | trans | 4.06E-39 | Nightingale | L_VLDL_FC | NA | G | A | 0.048347 | 0.004385 | 8.70E-29 |
| rs2769071 | miR-126-3p | trans | 4.06E-39 | Nightingale | Cholines | NA | G | A | 0.022376 | 0.004163 | 4.50E-08 |
| rs2769071 | miR-126-3p | trans | 4.06E-39 | Nightingale | LA | NA | G | A | 0.024942 | 0.004341 | 3.50E-09 |
| rs2769071 | miR-126-3p | trans | 4.06E-39 | Nightingale | S_LDL_FC | NA | G | A | 0.028535 | 0.00442 | 4.40E-10 |
| rs2769071 | miR-126-3p | trans | 4.06E-39 | Nightingale | PUFA | NA | G | A | 0.023736 | 0.004318 | 1.40E-08 |
| rs2769071 | miR-126-3p | trans | 4.06E-39 | Nightingale | XL_HDL_PL | NA | G | A | -0.02261 | 0.004067 | 1.40E-08 |
| rs2769071 | miR-126-3p | trans | 4.06E-39 | Nightingale | LDL_CE | NA | G | A | 0.032533 | 0.004415 | 2.30E-13 |
| rs2769071 | miR-126-3p | trans | 4.06E-39 | Nightingale | M_VLDL_T | NA | G | A | -0.03845 | 0.004184 | 9.20E-20 |
| rs2769071 | miR-126-3p | trans | 4.06E-39 | Nightingale | L_LDL_L | NA | G | A | 0.033538 | 0.004384 | 3.10E-14 |
| rs2769071 | miR-126-3p | trans | 4.06E-39 | Nightingale | XS_VLDL_P | NA | G | A | 0.032167 | 0.004365 | 4.90E-14 |
| rs2769071 | miR-126-3p | trans | 4.06E-39 | Nightingale | IDL_FC_pct | NA | G | A | 0.025676 | 0.004393 | 1.20E-08 |
| rs2769071 | miR-126-3p | trans | 4.06E-39 | Nightingale | S_VLDL_C | NA | G | A | 0.038352 | 0.004427 | 1.90E-18 |
| rs2769071 | miR-126-3p | trans | 4.06E-39 | Nightingale | S_VLDL_P | NA | G | A | 0.023474 | 0.004372 | 1.60E-08 |
| rs2769071 | miR-126-3p | trans | 4.06E-39 | Nightingale | M_LDL_PL | NA | G | A | 0.032427 | 0.004433 | 2.80E-13 |
| rs2769071 | let-7b-5p | trans | 9.93E-27 | Nightingale | S_LDL_C_p | NA | G | A | 0.045281 | 0.004444 | 7.30E-25 |
| rs2769071 | let-7b-5p | trans | 9.93E-27 | Nightingale | S_LDL_P | NA | G | A | 0.029553 | 0.004431 | 3.40E-11 |
| rs2769071 | let-7b-5p | trans | 9.93E-27 | Nightingale | L_VLDL_C_ | NA | G | A | 0.051993 | 0.004324 | 7.60E-34 |
| rs2769071 | let-7b-5p | trans | 9.93E-27 | Nightingale | GlycA | NA | G | A | 0.028001 | 0.004408 | 1.50E-10 |
| rs2769071 | let-7b-5p | trans | 9.93E-27 | Nightingale | Total_C | NA | G | A | 0.032411 | 0.004286 | 3.20E-14 |
| rs2769071 | let-7b-5p | trans | 9.93E-27 | Nightingale | M_VLDL_C | NA | G | A | 0.033528 | 0.004159 | 2.40E-15 |
| rs2769071 | let-7b-5p | trans | 9.93E-27 | Nightingale | M_LDL_L | NA | G | A | 0.033001 | 0.004433 | 1.10E-13 |
| rs2769071 | let-7b-5p | trans | 9.93E-27 | Nightingale | M_VLDL_P | NA | G | A | 0.043923 | 0.004249 | 8.10E-25 |
| rs2769071 | let-7b-5p | trans | 9.93E-27 | Nightingale | IDL_FC | NA | G | A | 0.038256 | 0.004286 | 1.50E-18 |
| rs2769071 | let-7b-5p | trans | 9.93E-27 | Nightingale | VLDL_P | NA | G | A | 0.026645 | 0.004403 | 4.40E-10 |
| rs2769071 | let-7b-5p | trans | 9.93E-27 | Nightingale | M_LDL_P | NA | G | A | 0.035884 | 0.004439 | 7.10E-16 |
| rs2769071 | let-7b-5p | trans | 9.93E-27 | Nightingale | XL_VLDL_T | NA | G | A | -0.04368 | 0.004291 | 3.40E-24 |
| rs2769071 | let-7b-5p | trans | 9.93E-27 | Nightingale | S_VLDL_C_ | NA | G | A | 0.041808 | 0.004308 | 6.50E-22 |
| rs2769071 | let-7b-5p | trans | 9.93E-27 | Nightingale | M_LDL_C | NA | G | A | 0.033958 | 0.004435 | 2.30E-14 |
| rs2769071 | let-7b-5p | trans | 9.93E-27 | Nightingale | L_LDL_C | NA | G | A | 0.032304 | 0.004377 | 2.70E-13 |
| rs2769071 | let-7b-5p | trans | 9.93E-27 | Nightingale | IDL_PL | NA | G | A | 0.035362 | 0.004272 | 3.30E-16 |
| rs2769071 | let-7b-5p | trans | 9.93E-27 | Nightingale | Sphingomy | NA | G | A | 0.029058 | 0.004139 | 6.70E-12 |
| rs2769071 | let-7b-5p | trans | 9.93E-27 | Nightingale | S_HDL_L | NA | G | A | 0.026342 | 0.004412 | 1.50E-09 |
| rs2769071 | let-7b-5p | trans | 9.93E-27 | Nightingale | Clinical_LD | NA | G | A | 0.036447 | 0.004401 | 2.90E-16 |
| rs2769071 | let-7b-5p | trans | 9.93E-27 | Nightingale | M_VLDL_P | NA | G | A | 0.029124 | 0.00442 | 2.10E-11 |

| | | | | | | | | | | | |
|---|---|---|---|---|---|---|---|---|---|---|---|
| rs2769071 | let-7b-5p | trans | 9.93E-27 | Nightingale | ApoB_by_A | NA | G | A | 0.024951 | 0.00436 | 1.40E-08 |
| rs2769071 | let-7b-5p | trans | 9.93E-27 | Nightingale | XS_VLDL_C | NA | G | A | 0.037835 | 0.004272 | 2.00E-18 |
| rs2769071 | let-7b-5p | trans | 9.93E-27 | Nightingale | XS_VLDL_P | NA | G | A | 0.036739 | 0.004368 | 1.30E-17 |
| rs2769071 | let-7b-5p | trans | 9.93E-27 | Nightingale | IDL_L | NA | G | A | 0.035297 | 0.00428 | 4.70E-16 |
| rs2769071 | let-7b-5p | trans | 9.93E-27 | Nightingale | S_VLDL_PL | NA | G | A | 0.032773 | 0.004419 | 4.20E-14 |
| rs2769071 | let-7b-5p | trans | 9.93E-27 | Nightingale | VLDL_FC | NA | G | A | 0.024155 | 0.004395 | 1.10E-08 |
| rs2769071 | let-7b-5p | trans | 9.93E-27 | Nightingale | Omega_6 | NA | G | A | 0.025917 | 0.004323 | 5.60E-10 |
| rs2769071 | let-7b-5p | trans | 9.93E-27 | Nightingale | VLDL_C | NA | G | A | 0.031294 | 0.004421 | 5.00E-13 |
| rs2769071 | let-7b-5p | trans | 9.93E-27 | Nightingale | XXL_VLDL_ | NA | G | A | -0.03561 | 0.004465 | 2.70E-16 |
| rs2769071 | let-7b-5p | trans | 9.93E-27 | Nightingale | LDL_L | NA | G | A | 0.033285 | 0.004412 | 7.70E-14 |
| rs2769071 | let-7b-5p | trans | 9.93E-27 | Nightingale | XXL_VLDL_ | NA | G | A | 0.024963 | 0.004456 | 1.60E-09 |
| rs2769071 | let-7b-5p | trans | 9.93E-27 | Nightingale | XXL_VLDL_ | NA | G | A | 0.034743 | 0.004437 | 3.40E-15 |
| rs2769071 | let-7b-5p | trans | 9.93E-27 | Nightingale | S_LDL_CE | NA | G | A | 0.030512 | 0.004429 | 6.20E-12 |
| rs2769071 | let-7b-5p | trans | 9.93E-27 | Nightingale | ApoB | NA | G | A | 0.033916 | 0.004421 | 2.20E-14 |
| rs2769071 | let-7b-5p | trans | 9.93E-27 | Nightingale | XL_VLDL_C | NA | G | A | 0.038066 | 0.004245 | 6.90E-19 |
| rs2769071 | let-7b-5p | trans | 9.93E-27 | Nightingale | M_VLDL_C | NA | G | A | 0.040819 | 0.004387 | 2.50E-20 |
| rs2769071 | let-7b-5p | trans | 9.93E-27 | Nightingale | S_VLDL_FC | NA | G | A | 0.029307 | 0.00423 | 1.20E-11 |
| rs2769071 | let-7b-5p | trans | 9.93E-27 | Nightingale | XXL_VLDL_ | NA | G | A | 0.030722 | 0.004408 | 1.30E-12 |
| rs2769071 | let-7b-5p | trans | 9.93E-27 | Nightingale | XL_VLDL_P | NA | G | A | 0.038703 | 0.004434 | 7.10E-19 |
| rs2769071 | let-7b-5p | trans | 9.93E-27 | Nightingale | Total_CE | NA | G | A | 0.031783 | 0.00427 | 9.40E-14 |
| rs2769071 | let-7b-5p | trans | 9.93E-27 | Nightingale | S_LDL_PL_ | NA | G | A | -0.03002 | 0.004419 | 2.90E-13 |
| rs2769071 | let-7b-5p | trans | 9.93E-27 | Nightingale | Total_FC | NA | G | A | 0.033293 | 0.004325 | 6.50E-15 |
| rs2769071 | let-7b-5p | trans | 9.93E-27 | Nightingale | M_LDL_C_ | NA | G | A | 0.031139 | 0.00443 | 3.20E-12 |
| rs2769071 | let-7b-5p | trans | 9.93E-27 | Nightingale | IDL_CE | NA | G | A | 0.033536 | 0.004274 | 1.30E-14 |
| rs2769071 | let-7b-5p | trans | 9.93E-27 | Nightingale | S_LDL_CE_ | NA | G | A | 0.030898 | 0.004419 | 5.50E-14 |
| rs2769071 | let-7b-5p | trans | 9.93E-27 | Nightingale | LDL_FC | NA | G | A | 0.033607 | 0.00438 | 5.60E-14 |
| rs2769071 | let-7b-5p | trans | 9.93E-27 | Nightingale | L_LDL_P | NA | G | A | 0.029139 | 0.004412 | 6.50E-11 |
| rs2769071 | let-7b-5p | trans | 9.93E-27 | Nightingale | S_HDL_CE | NA | G | A | 0.02862 | 0.004434 | 1.40E-10 |
| rs2769071 | let-7b-5p | trans | 9.93E-27 | Nightingale | XS_VLDL_L | NA | G | A | 0.033869 | 0.004352 | 1.90E-15 |
| rs2769071 | let-7b-5p | trans | 9.93E-27 | Nightingale | L_VLDL_CE | NA | G | A | 0.047208 | 0.004236 | 3.80E-29 |
| rs2769071 | let-7b-5p | trans | 9.93E-27 | Nightingale | Remnant_( | NA | G | A | 0.036871 | 0.004398 | 3.70E-17 |
| rs2769071 | let-7b-5p | trans | 9.93E-27 | Nightingale | S_LDL_L | NA | G | A | 0.027711 | 0.004426 | 8.50E-10 |
| rs2769071 | let-7b-5p | trans | 9.93E-27 | Nightingale | XS_VLDL_C | NA | G | A | 0.038033 | 0.004297 | 1.30E-18 |
| rs2769071 | let-7b-5p | trans | 9.93E-27 | Nightingale | S_VLDL_FC | NA | G | A | 0.03789 | 0.004428 | 7.60E-18 |
| rs2769071 | let-7b-5p | trans | 9.93E-27 | Nightingale | IDL_C | NA | G | A | 0.034977 | 0.004276 | 9.40E-16 |
| rs2769071 | let-7b-5p | trans | 9.93E-27 | Nightingale | S_HDL_FC | NA | G | A | 0.023537 | 0.004346 | 4.30E-08 |
| rs2769071 | let-7b-5p | trans | 9.93E-27 | Nightingale | LDL_C | NA | G | A | 0.033173 | 0.004408 | 9.30E-14 |
| rs2769071 | let-7b-5p | trans | 9.93E-27 | Nightingale | M_LDL_CE | NA | G | A | 0.032948 | 0.004433 | 1.00E-13 |
| rs2769071 | let-7b-5p | trans | 9.93E-27 | Nightingale | IDL_P | NA | G | A | 0.039976 | 0.004381 | 1.40E-19 |
| rs2769071 | let-7b-5p | trans | 9.93E-27 | Nightingale | LDL_P | NA | G | A | 0.03177 | 0.004424 | 1.10E-12 |
| rs2769071 | let-7b-5p | trans | 9.93E-27 | Nightingale | S_HDL_C | NA | G | A | 0.028424 | 0.00442 | 1.70E-10 |
| rs2769071 | let-7b-5p | trans | 9.93E-27 | Nightingale | LDL_PL | NA | G | A | 0.034963 | 0.004417 | 5.00E-15 |
| rs2769071 | let-7b-5p | trans | 9.93E-27 | Nightingale | M_VLDL_P | NA | G | A | 0.028288 | 0.004416 | 6.40E-11 |
| rs2769071 | let-7b-5p | trans | 9.93E-27 | Nightingale | L_LDL_FC | NA | G | A | 0.033608 | 0.004347 | 3.00E-14 |
| rs2769071 | let-7b-5p | trans | 9.93E-27 | Nightingale | Total_PL | NA | G | A | 0.025022 | 0.004216 | 9.60E-10 |
| rs2769071 | let-7b-5p | trans | 9.93E-27 | Nightingale | Total_L | NA | G | A | 0.026409 | 0.004355 | 4.30E-10 |
| rs2769071 | let-7b-5p | trans | 9.93E-27 | Nightingale | S_HDL_P | NA | G | A | 0.030619 | 0.004428 | 5.20E-12 |
| rs2769071 | let-7b-5p | trans | 9.93E-27 | Nightingale | L_LDL_PL | NA | G | A | 0.037972 | 0.004393 | 9.60E-18 |
| rs2769071 | let-7b-5p | trans | 9.93E-27 | Nightingale | XS_VLDL_F | NA | G | A | 0.03143 | 0.004395 | 2.50E-12 |
| rs2769071 | let-7b-5p | trans | 9.93E-27 | Nightingale | VLDL_CE | NA | G | A | 0.035775 | 0.004428 | 3.10E-16 |
| rs2769071 | let-7b-5p | trans | 9.93E-27 | Nightingale | L_LDL_CE | NA | G | A | 0.031552 | 0.004388 | 1.00E-12 |
| rs2769071 | let-7b-5p | trans | 9.93E-27 | Nightingale | S_VLDL_TG | NA | G | A | -0.03806 | 0.004285 | 1.70E-18 |
| rs2769071 | let-7b-5p | trans | 9.93E-27 | Nightingale | S_VLDL_PL | NA | G | A | 0.027684 | 0.004239 | 2.00E-10 |
| rs2769071 | let-7b-5p | trans | 9.93E-27 | Nightingale | S_VLDL_CE | NA | G | A | 0.046931 | 0.004368 | 1.60E-26 |
| rs2769071 | let-7b-5p | trans | 9.93E-27 | Nightingale | M_LDL_FC | NA | G | A | 0.034307 | 0.004421 | 2.10E-14 |
| rs2769071 | let-7b-5p | trans | 9.93E-27 | Nightingale | M_VLDL_C_ | NA | G | A | 0.036101 | 0.00417 | 1.50E-17 |
| rs2769071 | let-7b-5p | trans | 9.93E-27 | Nightingale | L_VLDL_TG | NA | G | A | -0.05094 | 0.004406 | 9.70E-32 |
| rs2769071 | let-7b-5p | trans | 9.93E-27 | Nightingale | M_VLDL_F | NA | G | A | 0.041951 | 0.004212 | 4.70E-23 |
| rs2769071 | let-7b-5p | trans | 9.93E-27 | Nightingale | M_VLDL_F | NA | G | A | 0.033361 | 0.004426 | 3.20E-14 |
| rs2769071 | let-7b-5p | trans | 9.93E-27 | Nightingale | XL_VLDL_F | NA | G | A | 0.036999 | 0.004292 | 3.20E-18 |
| rs2769071 | let-7b-5p | trans | 9.93E-27 | Nightingale | S_VLDL_CE | NA | G | A | 0.038158 | 0.004421 | 1.70E-18 |
| rs2769071 | let-7b-5p | trans | 9.93E-27 | Nightingale | S_LDL_C | NA | G | A | 0.030907 | 0.004431 | 5.80E-12 |
| rs2769071 | let-7b-5p | trans | 9.93E-27 | Nightingale | XL_VLDL_C | NA | G | A | 0.037208 | 0.004247 | 4.50E-18 |

| | | | | | | | | | | | |
|---|---|---|---|---|---|---|---|---|---|---|---|
| rs2769071 | let-7b-5p | trans | 9.93E-27 | Nightingale | Citrate | NA | G | A | 0.026789 | 0.004398 | 7.90E-10 |
| rs2769071 | let-7b-5p | trans | 9.93E-27 | Nightingale | XXL_VLDL_ | NA | G | A | 0.033072 | 0.004461 | 9.80E-14 |
| rs2769071 | let-7b-5p | trans | 9.93E-27 | Nightingale | S_VLDL_L | NA | G | A | 0.024677 | 0.004384 | 3.70E-09 |
| rs2769071 | let-7b-5p | trans | 9.93E-27 | Nightingale | S_HDL_PL | NA | G | A | 0.023848 | 0.004389 | 3.00E-08 |
| rs2769071 | let-7b-5p | trans | 9.93E-27 | Nightingale | M_LDL_TG | NA | G | A | -0.02754 | 0.004372 | 3.80E-10 |
| rs2769071 | let-7b-5p | trans | 9.93E-27 | Nightingale | non_HDL_( | NA | G | A | 0.035686 | 0.004403 | 5.30E-16 |
| rs2769071 | let-7b-5p | trans | 9.93E-27 | Nightingale | M_VLDL_C | NA | G | A | 0.038687 | 0.004415 | 2.00E-18 |
| rs2769071 | let-7b-5p | trans | 9.93E-27 | Nightingale | XS_VLDL_F | NA | G | A | 0.037113 | 0.00435 | 6.00E-18 |
| rs2769071 | let-7b-5p | trans | 9.93E-27 | Nightingale | L_VLDL_FC | NA | G | A | 0.048347 | 0.004385 | 8.70E-29 |
| rs2769071 | let-7b-5p | trans | 9.93E-27 | Nightingale | Cholines | NA | G | A | 0.022376 | 0.004163 | 4.50E-08 |
| rs2769071 | let-7b-5p | trans | 9.93E-27 | Nightingale | LA | NA | G | A | 0.024942 | 0.004341 | 3.50E-09 |
| rs2769071 | let-7b-5p | trans | 9.93E-27 | Nightingale | S_LDL_FC | NA | G | A | 0.028535 | 0.00442 | 4.40E-10 |
| rs2769071 | let-7b-5p | trans | 9.93E-27 | Nightingale | PUFA | NA | G | A | 0.023736 | 0.004318 | 1.40E-08 |
| rs2769071 | let-7b-5p | trans | 9.93E-27 | Nightingale | XL_HDL_PL | NA | G | A | -0.02261 | 0.004067 | 1.40E-08 |
| rs2769071 | let-7b-5p | trans | 9.93E-27 | Nightingale | LDL_CE | NA | G | A | 0.032533 | 0.004415 | 2.30E-13 |
| rs2769071 | let-7b-5p | trans | 9.93E-27 | Nightingale | M_VLDL_T | NA | G | A | -0.03845 | 0.004184 | 9.20E-20 |
| rs2769071 | let-7b-5p | trans | 9.93E-27 | Nightingale | L_LDL_L | NA | G | A | 0.033538 | 0.004384 | 3.10E-14 |
| rs2769071 | let-7b-5p | trans | 9.93E-27 | Nightingale | XS_VLDL_P | NA | G | A | 0.032167 | 0.004365 | 4.90E-14 |
| rs2769071 | let-7b-5p | trans | 9.93E-27 | Nightingale | IDL_FC_pct | NA | G | A | 0.025676 | 0.004393 | 1.20E-08 |
| rs2769071 | let-7b-5p | trans | 9.93E-27 | Nightingale | S_VLDL_C | NA | G | A | 0.038352 | 0.004427 | 1.90E-18 |
| rs2769071 | let-7b-5p | trans | 9.93E-27 | Nightingale | S_VLDL_P | NA | G | A | 0.023474 | 0.004372 | 1.60E-08 |
| rs2769071 | let-7b-5p | trans | 9.93E-27 | Nightingale | M_LDL_PL | NA | G | A | 0.032427 | 0.004433 | 2.80E-13 |
| rs2769071 | miR-139-5p | trans | 1.14E-41 | Nightingale | S_LDL_C_p | NA | G | A | 0.045281 | 0.004444 | 7.30E-25 |
| rs2769071 | miR-139-5p | trans | 1.14E-41 | Nightingale | S_LDL_P | NA | G | A | 0.029553 | 0.004431 | 3.40E-11 |
| rs2769071 | miR-139-5p | trans | 1.14E-41 | Nightingale | L_VLDL_C_ | NA | G | A | 0.051993 | 0.004324 | 7.60E-34 |
| rs2769071 | miR-139-5p | trans | 1.14E-41 | Nightingale | GlycA | NA | G | A | 0.028001 | 0.004408 | 1.50E-10 |
| rs2769071 | miR-139-5p | trans | 1.14E-41 | Nightingale | Total_C | NA | G | A | 0.032411 | 0.004286 | 3.20E-14 |
| rs2769071 | miR-139-5p | trans | 1.14E-41 | Nightingale | M_VLDL_C | NA | G | A | 0.033528 | 0.004159 | 2.40E-15 |
| rs2769071 | miR-139-5p | trans | 1.14E-41 | Nightingale | M_LDL_L | NA | G | A | 0.033001 | 0.004433 | 1.10E-13 |
| rs2769071 | miR-139-5p | trans | 1.14E-41 | Nightingale | M_VLDL_P | NA | G | A | 0.043923 | 0.004249 | 8.10E-25 |
| rs2769071 | miR-139-5p | trans | 1.14E-41 | Nightingale | IDL_FC | NA | G | A | 0.038256 | 0.004286 | 1.50E-18 |
| rs2769071 | miR-139-5p | trans | 1.14E-41 | Nightingale | VLDL_P | NA | G | A | 0.026645 | 0.004403 | 4.40E-10 |
| rs2769071 | miR-139-5p | trans | 1.14E-41 | Nightingale | M_LDL_P | NA | G | A | 0.035884 | 0.004439 | 7.10E-16 |
| rs2769071 | miR-139-5p | trans | 1.14E-41 | Nightingale | XL_VLDL_T | NA | G | A | -0.04368 | 0.004291 | 3.40E-24 |
| rs2769071 | miR-139-5p | trans | 1.14E-41 | Nightingale | S_VLDL_C_ | NA | G | A | 0.041808 | 0.004308 | 6.50E-22 |
| rs2769071 | miR-139-5p | trans | 1.14E-41 | Nightingale | M_LDL_C | NA | G | A | 0.033958 | 0.004435 | 2.30E-14 |
| rs2769071 | miR-139-5p | trans | 1.14E-41 | Nightingale | L_LDL_C | NA | G | A | 0.032304 | 0.004377 | 2.70E-13 |
| rs2769071 | miR-139-5p | trans | 1.14E-41 | Nightingale | IDL_PL | NA | G | A | 0.035362 | 0.004272 | 3.30E-16 |
| rs2769071 | miR-139-5p | trans | 1.14E-41 | Nightingale | Sphingomy | NA | G | A | 0.029058 | 0.004139 | 6.70E-12 |
| rs2769071 | miR-139-5p | trans | 1.14E-41 | Nightingale | S_HDL_L | NA | G | A | 0.026342 | 0.004412 | 1.50E-09 |
| rs2769071 | miR-139-5p | trans | 1.14E-41 | Nightingale | Clinical_LD | NA | G | A | 0.036447 | 0.004401 | 2.90E-16 |
| rs2769071 | miR-139-5p | trans | 1.14E-41 | Nightingale | M_VLDL_P | NA | G | A | 0.029124 | 0.00442 | 2.10E-11 |
| rs2769071 | miR-139-5p | trans | 1.14E-41 | Nightingale | ApoB_by_/ | NA | G | A | 0.024951 | 0.00436 | 1.40E-08 |
| rs2769071 | miR-139-5p | trans | 1.14E-41 | Nightingale | XS_VLDL_C | NA | G | A | 0.037835 | 0.004272 | 2.00E-18 |
| rs2769071 | miR-139-5p | trans | 1.14E-41 | Nightingale | XS_VLDL_P | NA | G | A | 0.036739 | 0.004368 | 1.30E-17 |
| rs2769071 | miR-139-5p | trans | 1.14E-41 | Nightingale | IDL_L | NA | G | A | 0.035297 | 0.00428 | 4.70E-16 |
| rs2769071 | miR-139-5p | trans | 1.14E-41 | Nightingale | S_VLDL_PL | NA | G | A | 0.032773 | 0.004419 | 4.20E-14 |
| rs2769071 | miR-139-5p | trans | 1.14E-41 | Nightingale | VLDL_FC | NA | G | A | 0.024155 | 0.004395 | 1.10E-08 |
| rs2769071 | miR-139-5p | trans | 1.14E-41 | Nightingale | Omega_6 | NA | G | A | 0.025917 | 0.004323 | 5.60E-10 |
| rs2769071 | miR-139-5p | trans | 1.14E-41 | Nightingale | VLDL_C | NA | G | A | 0.031294 | 0.004421 | 5.00E-13 |
| rs2769071 | miR-139-5p | trans | 1.14E-41 | Nightingale | XXL_VLDL_ | NA | G | A | -0.03561 | 0.004465 | 2.70E-16 |
| rs2769071 | miR-139-5p | trans | 1.14E-41 | Nightingale | LDL_L | NA | G | A | 0.033285 | 0.004412 | 7.70E-14 |
| rs2769071 | miR-139-5p | trans | 1.14E-41 | Nightingale | XXL_VLDL_ | NA | G | A | 0.024963 | 0.004456 | 1.60E-09 |
| rs2769071 | miR-139-5p | trans | 1.14E-41 | Nightingale | XXL_VLDL_ | NA | G | A | 0.034743 | 0.004437 | 3.40E-15 |
| rs2769071 | miR-139-5p | trans | 1.14E-41 | Nightingale | S_LDL_CE | NA | G | A | 0.030512 | 0.004429 | 6.20E-12 |
| rs2769071 | miR-139-5p | trans | 1.14E-41 | Nightingale | ApoB | NA | G | A | 0.033916 | 0.004421 | 2.20E-14 |
| rs2769071 | miR-139-5p | trans | 1.14E-41 | Nightingale | XL_VLDL_C | NA | G | A | 0.038066 | 0.004245 | 6.90E-19 |
| rs2769071 | miR-139-5p | trans | 1.14E-41 | Nightingale | M_VLDL_C | NA | G | A | 0.040819 | 0.004387 | 2.50E-20 |
| rs2769071 | miR-139-5p | trans | 1.14E-41 | Nightingale | S_VLDL_FC | NA | G | A | 0.029307 | 0.00423 | 1.20E-11 |
| rs2769071 | miR-139-5p | trans | 1.14E-41 | Nightingale | XXL_VLDL_ | NA | G | A | 0.030722 | 0.004408 | 1.30E-12 |
| rs2769071 | miR-139-5p | trans | 1.14E-41 | Nightingale | XL_VLDL_P | NA | G | A | 0.038703 | 0.004434 | 7.10E-19 |
| rs2769071 | miR-139-5p | trans | 1.14E-41 | Nightingale | Total_CE | NA | G | A | 0.031783 | 0.00427 | 9.40E-14 |
| rs2769071 | miR-139-5p | trans | 1.14E-41 | Nightingale | S_LDL_PL_ | NA | G | A | -0.03002 | 0.004419 | 2.90E-13 |

| | | | | | | | | | | | |
|---|---|---|---|---|---|---|---|---|---|---|---|
| rs2769071 | miR-139-5p | trans | 1.14E-41 | Nightingale | Total_FC | NA | G | A | 0.033293 | 0.004325 | 6.50E-15 |
| rs2769071 | miR-139-5p | trans | 1.14E-41 | Nightingale | M_LDL_C_p | NA | G | A | 0.031139 | 0.00443 | 3.20E-12 |
| rs2769071 | miR-139-5p | trans | 1.14E-41 | Nightingale | IDL_CE | NA | G | A | 0.033536 | 0.004274 | 1.30E-14 |
| rs2769071 | miR-139-5p | trans | 1.14E-41 | Nightingale | S_LDL_CE_ | NA | G | A | 0.030898 | 0.004419 | 5.50E-14 |
| rs2769071 | miR-139-5p | trans | 1.14E-41 | Nightingale | LDL_FC | NA | G | A | 0.033607 | 0.00438 | 5.60E-14 |
| rs2769071 | miR-139-5p | trans | 1.14E-41 | Nightingale | L_LDL_P | NA | G | A | 0.029139 | 0.004412 | 6.50E-11 |
| rs2769071 | miR-139-5p | trans | 1.14E-41 | Nightingale | S_HDL_CE | NA | G | A | 0.02862 | 0.004434 | 1.40E-10 |
| rs2769071 | miR-139-5p | trans | 1.14E-41 | Nightingale | XS_VLDL_L | NA | G | A | 0.033869 | 0.004352 | 1.90E-15 |
| rs2769071 | miR-139-5p | trans | 1.14E-41 | Nightingale | L_VLDL_CE | NA | G | A | 0.047208 | 0.004236 | 3.80E-29 |
| rs2769071 | miR-139-5p | trans | 1.14E-41 | Nightingale | Remnant_( | NA | G | A | 0.036871 | 0.004398 | 3.70E-17 |
| rs2769071 | miR-139-5p | trans | 1.14E-41 | Nightingale | S_LDL_L | NA | G | A | 0.027711 | 0.004426 | 8.50E-10 |
| rs2769071 | miR-139-5p | trans | 1.14E-41 | Nightingale | XS_VLDL_C | NA | G | A | 0.038033 | 0.004297 | 1.30E-18 |
| rs2769071 | miR-139-5p | trans | 1.14E-41 | Nightingale | S_VLDL_FC | NA | G | A | 0.03789 | 0.004428 | 7.60E-18 |
| rs2769071 | miR-139-5p | trans | 1.14E-41 | Nightingale | IDL_C | NA | G | A | 0.034977 | 0.004276 | 9.40E-16 |
| rs2769071 | miR-139-5p | trans | 1.14E-41 | Nightingale | S_HDL_FC | NA | G | A | 0.023537 | 0.004346 | 4.30E-08 |
| rs2769071 | miR-139-5p | trans | 1.14E-41 | Nightingale | LDL_C | NA | G | A | 0.033173 | 0.004408 | 9.30E-14 |
| rs2769071 | miR-139-5p | trans | 1.14E-41 | Nightingale | M_LDL_CE | NA | G | A | 0.032948 | 0.004433 | 1.00E-13 |
| rs2769071 | miR-139-5p | trans | 1.14E-41 | Nightingale | IDL_P | NA | G | A | 0.039976 | 0.004381 | 1.40E-19 |
| rs2769071 | miR-139-5p | trans | 1.14E-41 | Nightingale | LDL_P | NA | G | A | 0.03177 | 0.004424 | 1.10E-12 |
| rs2769071 | miR-139-5p | trans | 1.14E-41 | Nightingale | S_HDL_C | NA | G | A | 0.028424 | 0.00442 | 1.70E-10 |
| rs2769071 | miR-139-5p | trans | 1.14E-41 | Nightingale | LDL_PL | NA | G | A | 0.034963 | 0.004417 | 5.00E-15 |
| rs2769071 | miR-139-5p | trans | 1.14E-41 | Nightingale | M_VLDL_P | NA | G | A | 0.028288 | 0.004416 | 6.40E-11 |
| rs2769071 | miR-139-5p | trans | 1.14E-41 | Nightingale | L_LDL_FC | NA | G | A | 0.033608 | 0.004347 | 3.00E-14 |
| rs2769071 | miR-139-5p | trans | 1.14E-41 | Nightingale | Total_PL | NA | G | A | 0.025022 | 0.004216 | 9.60E-10 |
| rs2769071 | miR-139-5p | trans | 1.14E-41 | Nightingale | Total_L | NA | G | A | 0.026409 | 0.004355 | 4.30E-10 |
| rs2769071 | miR-139-5p | trans | 1.14E-41 | Nightingale | S_HDL_P | NA | G | A | 0.030619 | 0.004428 | 5.20E-12 |
| rs2769071 | miR-139-5p | trans | 1.14E-41 | Nightingale | L_LDL_PL | NA | G | A | 0.037972 | 0.004393 | 9.60E-18 |
| rs2769071 | miR-139-5p | trans | 1.14E-41 | Nightingale | XS_VLDL_F | NA | G | A | 0.03143 | 0.004395 | 2.50E-12 |
| rs2769071 | miR-139-5p | trans | 1.14E-41 | Nightingale | VLDL_CE | NA | G | A | 0.035775 | 0.004428 | 3.10E-16 |
| rs2769071 | miR-139-5p | trans | 1.14E-41 | Nightingale | L_LDL_CE | NA | G | A | 0.031552 | 0.004388 | 1.00E-12 |
| rs2769071 | miR-139-5p | trans | 1.14E-41 | Nightingale | S_VLDL_TG | NA | G | A | -0.03806 | 0.004285 | 1.70E-18 |
| rs2769071 | miR-139-5p | trans | 1.14E-41 | Nightingale | S_VLDL_PL | NA | G | A | 0.027684 | 0.004239 | 2.00E-10 |
| rs2769071 | miR-139-5p | trans | 1.14E-41 | Nightingale | S_VLDL_CE | NA | G | A | 0.046931 | 0.004368 | 1.60E-26 |
| rs2769071 | miR-139-5p | trans | 1.14E-41 | Nightingale | M_LDL_FC | NA | G | A | 0.034307 | 0.004421 | 2.10E-14 |
| rs2769071 | miR-139-5p | trans | 1.14E-41 | Nightingale | M_VLDL_C | NA | G | A | 0.036101 | 0.00417 | 1.50E-17 |
| rs2769071 | miR-139-5p | trans | 1.14E-41 | Nightingale | L_VLDL_TG | NA | G | A | -0.05094 | 0.004406 | 9.70E-32 |
| rs2769071 | miR-139-5p | trans | 1.14E-41 | Nightingale | M_VLDL_F | NA | G | A | 0.041951 | 0.004212 | 4.70E-23 |
| rs2769071 | miR-139-5p | trans | 1.14E-41 | Nightingale | M_VLDL_F | NA | G | A | 0.033361 | 0.004426 | 3.20E-14 |
| rs2769071 | miR-139-5p | trans | 1.14E-41 | Nightingale | XL_VLDL_F | NA | G | A | 0.036999 | 0.004292 | 3.20E-18 |
| rs2769071 | miR-139-5p | trans | 1.14E-41 | Nightingale | S_VLDL_CE | NA | G | A | 0.038158 | 0.004421 | 1.70E-18 |
| rs2769071 | miR-139-5p | trans | 1.14E-41 | Nightingale | S_LDL_C | NA | G | A | 0.030907 | 0.004431 | 5.80E-12 |
| rs2769071 | miR-139-5p | trans | 1.14E-41 | Nightingale | XL_VLDL_C | NA | G | A | 0.037208 | 0.004247 | 4.50E-18 |
| rs2769071 | miR-139-5p | trans | 1.14E-41 | Nightingale | Citrate | NA | G | A | 0.026789 | 0.004398 | 7.90E-10 |
| rs2769071 | miR-139-5p | trans | 1.14E-41 | Nightingale | XXL_VLDL_ | NA | G | A | 0.033072 | 0.004461 | 9.80E-14 |
| rs2769071 | miR-139-5p | trans | 1.14E-41 | Nightingale | S_VLDL_L | NA | G | A | 0.024677 | 0.004384 | 3.70E-09 |
| rs2769071 | miR-139-5p | trans | 1.14E-41 | Nightingale | S_HDL_PL | NA | G | A | 0.023848 | 0.004389 | 3.00E-08 |
| rs2769071 | miR-139-5p | trans | 1.14E-41 | Nightingale | M_LDL_TG | NA | G | A | -0.02754 | 0.004372 | 3.80E-10 |
| rs2769071 | miR-139-5p | trans | 1.14E-41 | Nightingale | non_HDL_( | NA | G | A | 0.035686 | 0.004403 | 5.30E-16 |
| rs2769071 | miR-139-5p | trans | 1.14E-41 | Nightingale | M_VLDL_C | NA | G | A | 0.038687 | 0.004415 | 2.00E-18 |
| rs2769071 | miR-139-5p | trans | 1.14E-41 | Nightingale | XS_VLDL_F | NA | G | A | 0.037113 | 0.00435 | 6.00E-18 |
| rs2769071 | miR-139-5p | trans | 1.14E-41 | Nightingale | L_VLDL_FC | NA | G | A | 0.048347 | 0.004385 | 8.70E-29 |
| rs2769071 | miR-139-5p | trans | 1.14E-41 | Nightingale | Cholines | NA | G | A | 0.022376 | 0.004163 | 4.50E-08 |
| rs2769071 | miR-139-5p | trans | 1.14E-41 | Nightingale | LA | NA | G | A | 0.024942 | 0.004341 | 3.50E-09 |
| rs2769071 | miR-139-5p | trans | 1.14E-41 | Nightingale | S_LDL_FC | NA | G | A | 0.028535 | 0.00442 | 4.40E-10 |
| rs2769071 | miR-139-5p | trans | 1.14E-41 | Nightingale | PUFA | NA | G | A | 0.023736 | 0.004318 | 1.40E-08 |
| rs2769071 | miR-139-5p | trans | 1.14E-41 | Nightingale | XL_HDL_PL | NA | G | A | -0.02261 | 0.004067 | 1.40E-08 |
| rs2769071 | miR-139-5p | trans | 1.14E-41 | Nightingale | LDL_CE | NA | G | A | 0.032533 | 0.004415 | 2.30E-13 |
| rs2769071 | miR-139-5p | trans | 1.14E-41 | Nightingale | M_VLDL_T | NA | G | A | -0.03845 | 0.004184 | 9.20E-20 |
| rs2769071 | miR-139-5p | trans | 1.14E-41 | Nightingale | L_LDL_L | NA | G | A | 0.033538 | 0.004384 | 3.10E-14 |
| rs2769071 | miR-139-5p | trans | 1.14E-41 | Nightingale | XS_VLDL_P | NA | G | A | 0.032167 | 0.004365 | 4.90E-14 |
| rs2769071 | miR-139-5p | trans | 1.14E-41 | Nightingale | IDL_FC_pct | NA | G | A | 0.025676 | 0.004393 | 1.20E-08 |
| rs2769071 | miR-139-5p | trans | 1.14E-41 | Nightingale | S_VLDL_C | NA | G | A | 0.038352 | 0.004427 | 1.90E-18 |
| rs2769071 | miR-139-5p | trans | 1.14E-41 | Nightingale | S_VLDL_P | NA | G | A | 0.023474 | 0.004372 | 1.60E-08 |

| | | | | | | | | | | | |
|---|---|---|---|---|---|---|---|---|---|---|---|
| rs2769071 | miR-139-5p | trans | 1.14E-41 | Nightingale | M_LDL_PL | NA | G | A | 0.032427 | 0.004433 | 2.80E-13 |
| rs2769071 | miR-195-5p | trans | 3.25E-17 | Nightingale | S_LDL_C_p | NA | G | A | 0.045281 | 0.004444 | 7.30E-25 |
| rs2769071 | miR-195-5p | trans | 3.25E-17 | Nightingale | S_LDL_P | NA | G | A | 0.029553 | 0.004431 | 3.40E-11 |
| rs2769071 | miR-195-5p | trans | 3.25E-17 | Nightingale | L_VLDL_C_ | NA | G | A | 0.051993 | 0.004324 | 7.60E-34 |
| rs2769071 | miR-195-5p | trans | 3.25E-17 | Nightingale | GlycA | NA | G | A | 0.028001 | 0.004408 | 1.50E-10 |
| rs2769071 | miR-195-5p | trans | 3.25E-17 | Nightingale | Total_C | NA | G | A | 0.032411 | 0.004286 | 3.20E-14 |
| rs2769071 | miR-195-5p | trans | 3.25E-17 | Nightingale | M_VLDL_C | NA | G | A | 0.033528 | 0.004159 | 2.40E-15 |
| rs2769071 | miR-195-5p | trans | 3.25E-17 | Nightingale | M_LDL_L | NA | G | A | 0.033001 | 0.004433 | 1.10E-13 |
| rs2769071 | miR-195-5p | trans | 3.25E-17 | Nightingale | M_VLDL_P | NA | G | A | 0.043923 | 0.004249 | 8.10E-25 |
| rs2769071 | miR-195-5p | trans | 3.25E-17 | Nightingale | IDL_FC | NA | G | A | 0.038256 | 0.004286 | 1.50E-18 |
| rs2769071 | miR-195-5p | trans | 3.25E-17 | Nightingale | VLDL_P | NA | G | A | 0.026645 | 0.004403 | 4.40E-10 |
| rs2769071 | miR-195-5p | trans | 3.25E-17 | Nightingale | M_LDL_P | NA | G | A | 0.035884 | 0.004439 | 7.10E-16 |
| rs2769071 | miR-195-5p | trans | 3.25E-17 | Nightingale | XL_VLDL_T | NA | G | A | -0.04368 | 0.004291 | 3.40E-24 |
| rs2769071 | miR-195-5p | trans | 3.25E-17 | Nightingale | S_VLDL_C_ | NA | G | A | 0.041808 | 0.004308 | 6.50E-22 |
| rs2769071 | miR-195-5p | trans | 3.25E-17 | Nightingale | M_LDL_C | NA | G | A | 0.033958 | 0.004435 | 2.30E-14 |
| rs2769071 | miR-195-5p | trans | 3.25E-17 | Nightingale | L_LDL_C | NA | G | A | 0.032304 | 0.004377 | 2.70E-13 |
| rs2769071 | miR-195-5p | trans | 3.25E-17 | Nightingale | IDL_PL | NA | G | A | 0.035362 | 0.004272 | 3.30E-16 |
| rs2769071 | miR-195-5p | trans | 3.25E-17 | Nightingale | Sphingomy | NA | G | A | 0.029058 | 0.004139 | 6.70E-12 |
| rs2769071 | miR-195-5p | trans | 3.25E-17 | Nightingale | S_HDL_L | NA | G | A | 0.026342 | 0.004412 | 1.50E-09 |
| rs2769071 | miR-195-5p | trans | 3.25E-17 | Nightingale | Clinical_LD | NA | G | A | 0.036447 | 0.004401 | 2.90E-16 |
| rs2769071 | miR-195-5p | trans | 3.25E-17 | Nightingale | M_VLDL_P | NA | G | A | 0.029124 | 0.00442 | 2.10E-11 |
| rs2769071 | miR-195-5p | trans | 3.25E-17 | Nightingale | ApoB_by_A | NA | G | A | 0.024951 | 0.00436 | 1.40E-08 |
| rs2769071 | miR-195-5p | trans | 3.25E-17 | Nightingale | XS_VLDL_C | NA | G | A | 0.037835 | 0.004272 | 2.00E-18 |
| rs2769071 | miR-195-5p | trans | 3.25E-17 | Nightingale | XS_VLDL_P | NA | G | A | 0.036739 | 0.004368 | 1.30E-17 |
| rs2769071 | miR-195-5p | trans | 3.25E-17 | Nightingale | IDL_L | NA | G | A | 0.035297 | 0.00428 | 4.70E-16 |
| rs2769071 | miR-195-5p | trans | 3.25E-17 | Nightingale | S_VLDL_PL | NA | G | A | 0.032773 | 0.004419 | 4.20E-14 |
| rs2769071 | miR-195-5p | trans | 3.25E-17 | Nightingale | VLDL_FC | NA | G | A | 0.024155 | 0.004395 | 1.10E-08 |
| rs2769071 | miR-195-5p | trans | 3.25E-17 | Nightingale | Omega_6 | NA | G | A | 0.025917 | 0.004323 | 5.60E-10 |
| rs2769071 | miR-195-5p | trans | 3.25E-17 | Nightingale | VLDL_C | NA | G | A | 0.031294 | 0.004421 | 5.00E-13 |
| rs2769071 | miR-195-5p | trans | 3.25E-17 | Nightingale | XXL_VLDL_ | NA | G | A | -0.03561 | 0.004465 | 2.70E-16 |
| rs2769071 | miR-195-5p | trans | 3.25E-17 | Nightingale | LDL_L | NA | G | A | 0.033285 | 0.004412 | 7.70E-14 |
| rs2769071 | miR-195-5p | trans | 3.25E-17 | Nightingale | XXL_VLDL_ | NA | G | A | 0.024963 | 0.004456 | 1.60E-09 |
| rs2769071 | miR-195-5p | trans | 3.25E-17 | Nightingale | XXL_VLDL_ | NA | G | A | 0.034743 | 0.004437 | 3.40E-15 |
| rs2769071 | miR-195-5p | trans | 3.25E-17 | Nightingale | S_LDL_CE | NA | G | A | 0.030512 | 0.004429 | 6.20E-12 |
| rs2769071 | miR-195-5p | trans | 3.25E-17 | Nightingale | ApoB | NA | G | A | 0.033916 | 0.004421 | 2.20E-14 |
| rs2769071 | miR-195-5p | trans | 3.25E-17 | Nightingale | XL_VLDL_C | NA | G | A | 0.038066 | 0.004245 | 6.90E-19 |
| rs2769071 | miR-195-5p | trans | 3.25E-17 | Nightingale | M_VLDL_C | NA | G | A | 0.040819 | 0.004387 | 2.50E-20 |
| rs2769071 | miR-195-5p | trans | 3.25E-17 | Nightingale | S_VLDL_FC | NA | G | A | 0.029307 | 0.00423 | 1.20E-11 |
| rs2769071 | miR-195-5p | trans | 3.25E-17 | Nightingale | XXL_VLDL_ | NA | G | A | 0.030722 | 0.004408 | 1.30E-12 |
| rs2769071 | miR-195-5p | trans | 3.25E-17 | Nightingale | XL_VLDL_P | NA | G | A | 0.038703 | 0.004434 | 7.10E-19 |
| rs2769071 | miR-195-5p | trans | 3.25E-17 | Nightingale | Total_CE | NA | G | A | 0.031783 | 0.00427 | 9.40E-14 |
| rs2769071 | miR-195-5p | trans | 3.25E-17 | Nightingale | S_LDL_PL_ | NA | G | A | -0.03002 | 0.004419 | 2.90E-13 |
| rs2769071 | miR-195-5p | trans | 3.25E-17 | Nightingale | Total_FC | NA | G | A | 0.033293 | 0.004325 | 6.50E-15 |
| rs2769071 | miR-195-5p | trans | 3.25E-17 | Nightingale | M_LDL_C_p | NA | G | A | 0.031139 | 0.00443 | 3.20E-12 |
| rs2769071 | miR-195-5p | trans | 3.25E-17 | Nightingale | IDL_CE | NA | G | A | 0.033536 | 0.004274 | 1.30E-14 |
| rs2769071 | miR-195-5p | trans | 3.25E-17 | Nightingale | S_LDL_CE_ | NA | G | A | 0.030898 | 0.004419 | 5.50E-14 |
| rs2769071 | miR-195-5p | trans | 3.25E-17 | Nightingale | LDL_FC | NA | G | A | 0.033607 | 0.00438 | 5.60E-14 |
| rs2769071 | miR-195-5p | trans | 3.25E-17 | Nightingale | L_LDL_P | NA | G | A | 0.029139 | 0.004412 | 6.50E-11 |
| rs2769071 | miR-195-5p | trans | 3.25E-17 | Nightingale | S_HDL_CE | NA | G | A | 0.02862 | 0.004434 | 1.40E-10 |
| rs2769071 | miR-195-5p | trans | 3.25E-17 | Nightingale | XS_VLDL_L | NA | G | A | 0.033869 | 0.004352 | 1.90E-15 |
| rs2769071 | miR-195-5p | trans | 3.25E-17 | Nightingale | L_VLDL_CE | NA | G | A | 0.047208 | 0.004236 | 3.80E-29 |
| rs2769071 | miR-195-5p | trans | 3.25E-17 | Nightingale | Remnant_C | NA | G | A | 0.036871 | 0.004398 | 3.70E-17 |
| rs2769071 | miR-195-5p | trans | 3.25E-17 | Nightingale | S_LDL_L | NA | G | A | 0.027711 | 0.004426 | 8.50E-10 |
| rs2769071 | miR-195-5p | trans | 3.25E-17 | Nightingale | XS_VLDL_C | NA | G | A | 0.038033 | 0.004297 | 1.30E-18 |
| rs2769071 | miR-195-5p | trans | 3.25E-17 | Nightingale | S_VLDL_FC | NA | G | A | 0.03789 | 0.004428 | 7.60E-18 |
| rs2769071 | miR-195-5p | trans | 3.25E-17 | Nightingale | IDL_C | NA | G | A | 0.034977 | 0.004276 | 9.40E-16 |
| rs2769071 | miR-195-5p | trans | 3.25E-17 | Nightingale | S_HDL_FC | NA | G | A | 0.023537 | 0.004346 | 4.30E-08 |
| rs2769071 | miR-195-5p | trans | 3.25E-17 | Nightingale | LDL_C | NA | G | A | 0.033173 | 0.004408 | 9.30E-14 |
| rs2769071 | miR-195-5p | trans | 3.25E-17 | Nightingale | M_LDL_CE | NA | G | A | 0.032948 | 0.004433 | 1.00E-13 |
| rs2769071 | miR-195-5p | trans | 3.25E-17 | Nightingale | IDL_P | NA | G | A | 0.039976 | 0.004381 | 1.40E-19 |
| rs2769071 | miR-195-5p | trans | 3.25E-17 | Nightingale | LDL_P | NA | G | A | 0.03177 | 0.004424 | 1.10E-12 |
| rs2769071 | miR-195-5p | trans | 3.25E-17 | Nightingale | S_HDL_C | NA | G | A | 0.028424 | 0.00442 | 1.70E-10 |
| rs2769071 | miR-195-5p | trans | 3.25E-17 | Nightingale | LDL_PL | NA | G | A | 0.034963 | 0.004417 | 5.00E-15 |

| | | | | | | | | | | | |
|---|---|---|---|---|---|---|---|---|---|---|---|
| rs2769071 | miR-195-5p | trans | 3.25E-17 | Nightingale | M_VLDL_P | NA | G | A | 0.028288 | 0.004416 | 6.40E-11 |
| rs2769071 | miR-195-5p | trans | 3.25E-17 | Nightingale | L_LDL_FC | NA | G | A | 0.033608 | 0.004347 | 3.00E-14 |
| rs2769071 | miR-195-5p | trans | 3.25E-17 | Nightingale | Total_PL | NA | G | A | 0.025022 | 0.004216 | 9.60E-10 |
| rs2769071 | miR-195-5p | trans | 3.25E-17 | Nightingale | Total_L | NA | G | A | 0.026409 | 0.004355 | 4.30E-10 |
| rs2769071 | miR-195-5p | trans | 3.25E-17 | Nightingale | S_HDL_P | NA | G | A | 0.030619 | 0.004428 | 5.20E-12 |
| rs2769071 | miR-195-5p | trans | 3.25E-17 | Nightingale | L_LDL_PL | NA | G | A | 0.037972 | 0.004393 | 9.60E-18 |
| rs2769071 | miR-195-5p | trans | 3.25E-17 | Nightingale | XS_VLDL_F | NA | G | A | 0.03143 | 0.004395 | 2.50E-12 |
| rs2769071 | miR-195-5p | trans | 3.25E-17 | Nightingale | VLDL_CE | NA | G | A | 0.035775 | 0.004428 | 3.10E-16 |
| rs2769071 | miR-195-5p | trans | 3.25E-17 | Nightingale | L_LDL_CE | NA | G | A | 0.031552 | 0.004388 | 1.00E-12 |
| rs2769071 | miR-195-5p | trans | 3.25E-17 | Nightingale | S_VLDL_TG | NA | G | A | -0.03806 | 0.004285 | 1.70E-18 |
| rs2769071 | miR-195-5p | trans | 3.25E-17 | Nightingale | S_VLDL_PL | NA | G | A | 0.027684 | 0.004239 | 2.00E-10 |
| rs2769071 | miR-195-5p | trans | 3.25E-17 | Nightingale | S_VLDL_CE | NA | G | A | 0.046931 | 0.004368 | 1.60E-26 |
| rs2769071 | miR-195-5p | trans | 3.25E-17 | Nightingale | M_LDL_FC | NA | G | A | 0.034307 | 0.004421 | 2.10E-14 |
| rs2769071 | miR-195-5p | trans | 3.25E-17 | Nightingale | M_VLDL_C | NA | G | A | 0.036101 | 0.00417 | 1.50E-17 |
| rs2769071 | miR-195-5p | trans | 3.25E-17 | Nightingale | L_VLDL_TG | NA | G | A | -0.05094 | 0.004406 | 9.70E-32 |
| rs2769071 | miR-195-5p | trans | 3.25E-17 | Nightingale | M_VLDL_F | NA | G | A | 0.041951 | 0.004212 | 4.70E-23 |
| rs2769071 | miR-195-5p | trans | 3.25E-17 | Nightingale | M_VLDL_F | NA | G | A | 0.033361 | 0.004426 | 3.20E-14 |
| rs2769071 | miR-195-5p | trans | 3.25E-17 | Nightingale | XL_VLDL_F | NA | G | A | 0.036999 | 0.004292 | 3.20E-18 |
| rs2769071 | miR-195-5p | trans | 3.25E-17 | Nightingale | S_VLDL_CE | NA | G | A | 0.038158 | 0.004421 | 1.70E-18 |
| rs2769071 | miR-195-5p | trans | 3.25E-17 | Nightingale | S_LDL_C | NA | G | A | 0.030907 | 0.004431 | 5.80E-12 |
| rs2769071 | miR-195-5p | trans | 3.25E-17 | Nightingale | XL_VLDL_C | NA | G | A | 0.037208 | 0.004247 | 4.50E-18 |
| rs2769071 | miR-195-5p | trans | 3.25E-17 | Nightingale | Citrate | NA | G | A | 0.026789 | 0.004398 | 7.90E-10 |
| rs2769071 | miR-195-5p | trans | 3.25E-17 | Nightingale | XXL_VLDL_ | NA | G | A | 0.033072 | 0.004461 | 9.80E-14 |
| rs2769071 | miR-195-5p | trans | 3.25E-17 | Nightingale | S_VLDL_L | NA | G | A | 0.024677 | 0.004384 | 3.70E-09 |
| rs2769071 | miR-195-5p | trans | 3.25E-17 | Nightingale | S_HDL_PL | NA | G | A | 0.023848 | 0.004389 | 3.00E-08 |
| rs2769071 | miR-195-5p | trans | 3.25E-17 | Nightingale | M_LDL_TG | NA | G | A | -0.02754 | 0.004372 | 3.80E-10 |
| rs2769071 | miR-195-5p | trans | 3.25E-17 | Nightingale | non_HDL_( | NA | G | A | 0.035686 | 0.004403 | 5.30E-16 |
| rs2769071 | miR-195-5p | trans | 3.25E-17 | Nightingale | M_VLDL_C | NA | G | A | 0.038687 | 0.004415 | 2.00E-18 |
| rs2769071 | miR-195-5p | trans | 3.25E-17 | Nightingale | XS_VLDL_F | NA | G | A | 0.037113 | 0.00435 | 6.00E-18 |
| rs2769071 | miR-195-5p | trans | 3.25E-17 | Nightingale | L_VLDL_FC | NA | G | A | 0.048347 | 0.004385 | 8.70E-29 |
| rs2769071 | miR-195-5p | trans | 3.25E-17 | Nightingale | Cholines | NA | G | A | 0.022376 | 0.004163 | 4.50E-08 |
| rs2769071 | miR-195-5p | trans | 3.25E-17 | Nightingale | LA | NA | G | A | 0.024942 | 0.004341 | 3.50E-09 |
| rs2769071 | miR-195-5p | trans | 3.25E-17 | Nightingale | S_LDL_FC | NA | G | A | 0.028535 | 0.00442 | 4.40E-10 |
| rs2769071 | miR-195-5p | trans | 3.25E-17 | Nightingale | PUFA | NA | G | A | 0.023736 | 0.004318 | 1.40E-08 |
| rs2769071 | miR-195-5p | trans | 3.25E-17 | Nightingale | XL_HDL_PL | NA | G | A | -0.02261 | 0.004067 | 1.40E-08 |
| rs2769071 | miR-195-5p | trans | 3.25E-17 | Nightingale | LDL_CE | NA | G | A | 0.032533 | 0.004415 | 2.30E-13 |
| rs2769071 | miR-195-5p | trans | 3.25E-17 | Nightingale | M_VLDL_T | NA | G | A | -0.03845 | 0.004184 | 9.20E-20 |
| rs2769071 | miR-195-5p | trans | 3.25E-17 | Nightingale | L_LDL_L | NA | G | A | 0.033538 | 0.004384 | 3.10E-14 |
| rs2769071 | miR-195-5p | trans | 3.25E-17 | Nightingale | XS_VLDL_P | NA | G | A | 0.032167 | 0.004365 | 4.90E-14 |
| rs2769071 | miR-195-5p | trans | 3.25E-17 | Nightingale | IDL_FC_pct | NA | G | A | 0.025676 | 0.004393 | 1.20E-08 |
| rs2769071 | miR-195-5p | trans | 3.25E-17 | Nightingale | S_VLDL_C | NA | G | A | 0.038352 | 0.004427 | 1.90E-18 |
| rs2769071 | miR-195-5p | trans | 3.25E-17 | Nightingale | S_VLDL_P | NA | G | A | 0.023474 | 0.004372 | 1.60E-08 |
| rs2769071 | miR-195-5p | trans | 3.25E-17 | Nightingale | M_LDL_PL | NA | G | A | 0.032427 | 0.004433 | 2.80E-13 |
| rs3482677! | miR-145-5p | trans | 3.62E-08 | Nightingale | Lactate | NA | T | G | -0.02913 | 0.004663 | 4.20E-10 |
| rs3482677! | miR-145-5p | trans | 3.62E-08 | Nightingale | Pyruvate | NA | T | G | -0.03396 | 0.004664 | 3.30E-13 |
| rs3482677! | miR-143-3p | trans | 9.26E-11 | Nightingale | Lactate | NA | T | G | -0.02913 | 0.004663 | 4.20E-10 |
| rs3482677! | miR-143-3p | trans | 9.26E-11 | Nightingale | Pyruvate | NA | T | G | -0.03396 | 0.004664 | 3.30E-13 |
| rs4734879 | miR-143-3p | trans | 8.35E-11 | Nightingale | Lactate | NA | G | A | -0.02915 | 0.004612 | 2.60E-10 |
| rs4734879 | miR-143-3p | trans | 8.35E-11 | Nightingale | Pyruvate | NA | G | A | -0.03392 | 0.004613 | 1.90E-13 |
| rs4734879 | miR-145-5p | trans | 2.62E-08 | Nightingale | Lactate | NA | G | A | -0.02915 | 0.004612 | 2.60E-10 |
| rs4734879 | miR-145-5p | trans | 2.62E-08 | Nightingale | Pyruvate | NA | G | A | -0.03392 | 0.004613 | 1.90E-13 |
| rs473533 | miR-10b-5p | trans | 1.17E-19 | Nightingale | M_VLDL_C | NA | C | T | 0.021435 | 0.003907 | 1.10E-08 |
| rs473533 | miR-10b-5p | trans | 1.17E-19 | Nightingale | XL_VLDL_C | NA | C | T | 0.022528 | 0.003977 | 5.00E-09 |
| rs473533 | miR-10b-5p | trans | 1.17E-19 | Nightingale | L_VLDL_TG | NA | C | T | -0.02538 | 0.004128 | 6.60E-10 |
| rs473533 | miR-10b-5p | trans | 1.17E-19 | Nightingale | XL_VLDL_C | NA | C | T | 0.022065 | 0.003979 | 9.30E-09 |
| rs473533 | miR-10b-5p | trans | 1.17E-19 | Nightingale | S_VLDL_C_ | NA | C | T | 0.023309 | 0.004037 | 3.60E-09 |
| rs473533 | miR-10b-5p | trans | 1.17E-19 | Nightingale | S_VLDL_CE | NA | C | T | 0.02534 | 0.004092 | 3.70E-10 |
| rs473533 | miR-10b-5p | trans | 1.17E-19 | Nightingale | S_VLDL_TG | NA | C | T | -0.0215 | 0.004015 | 4.00E-08 |
| rs473533 | miR-10b-5p | trans | 1.17E-19 | Nightingale | IDL_P | NA | C | T | 0.023145 | 0.004105 | 2.00E-08 |
| rs473533 | miR-10b-5p | trans | 1.17E-19 | Nightingale | IDL_FC | NA | C | T | 0.022145 | 0.004016 | 3.80E-08 |
| rs473533 | miR-10b-5p | trans | 1.17E-19 | Nightingale | XL_VLDL_F | NA | C | T | 0.021873 | 0.00402 | 1.50E-08 |
| rs473533 | miR-10b-5p | trans | 1.17E-19 | Nightingale | M_VLDL_P | NA | C | T | 0.024495 | 0.003981 | 4.30E-10 |
| rs473533 | miR-10b-5p | trans | 1.17E-19 | Nightingale | XL_VLDL_T | NA | C | T | -0.02534 | 0.00402 | 9.10E-11 |

| | | | | | | | | | | | |
|---|---|---|---|---|---|---|---|---|---|---|---|
| rs473533 | miR-10b-5p | trans | 1.17E-19 | Nightingale | M_VLDL_T | NA | C | T | -0.02243 | 0.00392 | 3.10E-09 |
| rs473533 | miR-10b-5p | trans | 1.17E-19 | Nightingale | M_VLDL_F | NA | C | T | 0.024736 | 0.003946 | 1.50E-10 |
| rs473533 | miR-10b-5p | trans | 1.17E-19 | Nightingale | L_VLDL_CE | NA | C | T | 0.026348 | 0.003969 | 1.20E-11 |
| rs473533 | miR-10b-5p | trans | 1.17E-19 | Nightingale | L_VLDL_FC | NA | C | T | 0.023753 | 0.004108 | 6.30E-09 |
| rs473533 | miR-10b-5p | trans | 1.17E-19 | Nightingale | S_LDL_C_p | NA | C | T | 0.026871 | 0.004164 | 1.10E-10 |
| rs473533 | miR-10b-5p | trans | 1.17E-19 | Nightingale | L_VLDL_C_ | NA | C | T | 0.02817 | 0.004051 | 1.80E-12 |
| rs476410 | miR-10b-5p | trans | 1.29E-19 | Nightingale | XL_VLDL_C | NA | C | G | 0.022034 | 0.003979 | 9.70E-09 |
| rs476410 | miR-10b-5p | trans | 1.29E-19 | Nightingale | L_VLDL_CE | NA | C | G | 0.02631 | 0.00397 | 1.30E-11 |
| rs476410 | miR-10b-5p | trans | 1.29E-19 | Nightingale | S_VLDL_CE | NA | C | G | 0.02536 | 0.004093 | 3.40E-10 |
| rs476410 | miR-10b-5p | trans | 1.29E-19 | Nightingale | M_VLDL_F | NA | C | G | 0.024681 | 0.003947 | 1.60E-10 |
| rs476410 | miR-10b-5p | trans | 1.29E-19 | Nightingale | IDL_P | NA | C | G | 0.023086 | 0.004105 | 2.10E-08 |
| rs476410 | miR-10b-5p | trans | 1.29E-19 | Nightingale | S_VLDL_C_ | NA | C | G | 0.0233 | 0.004037 | 3.60E-09 |
| rs476410 | miR-10b-5p | trans | 1.29E-19 | Nightingale | S_LDL_C_p | NA | C | G | 0.026895 | 0.004164 | 1.10E-10 |
| rs476410 | miR-10b-5p | trans | 1.29E-19 | Nightingale | M_VLDL_T | NA | C | G | -0.02238 | 0.00392 | 3.30E-09 |
| rs476410 | miR-10b-5p | trans | 1.29E-19 | Nightingale | L_VLDL_C_ | NA | C | G | 0.028156 | 0.004052 | 1.80E-12 |
| rs476410 | miR-10b-5p | trans | 1.29E-19 | Nightingale | XL_VLDL_T | NA | C | G | -0.02528 | 0.00402 | 1.00E-10 |
| rs476410 | miR-10b-5p | trans | 1.29E-19 | Nightingale | L_VLDL_TG | NA | C | G | -0.02543 | 0.004129 | 5.90E-10 |
| rs476410 | miR-10b-5p | trans | 1.29E-19 | Nightingale | XL_VLDL_C | NA | C | G | 0.022479 | 0.003977 | 5.40E-09 |
| rs476410 | miR-10b-5p | trans | 1.29E-19 | Nightingale | L_VLDL_FC | NA | C | G | 0.02378 | 0.004108 | 5.90E-09 |
| rs476410 | miR-10b-5p | trans | 1.29E-19 | Nightingale | S_VLDL_TG | NA | C | G | -0.02148 | 0.004015 | 4.00E-08 |
| rs476410 | miR-10b-5p | trans | 1.29E-19 | Nightingale | M_VLDL_P | NA | C | G | 0.024446 | 0.003981 | 4.60E-10 |
| rs476410 | miR-10b-5p | trans | 1.29E-19 | Nightingale | IDL_FC | NA | C | G | 0.022099 | 0.004016 | 4.00E-08 |
| rs476410 | miR-10b-5p | trans | 1.29E-19 | Nightingale | M_VLDL_C | NA | C | G | 0.021383 | 0.003907 | 1.20E-08 |
| rs476410 | miR-10b-5p | trans | 1.29E-19 | Nightingale | XL_VLDL_F | NA | C | G | 0.021757 | 0.004021 | 1.80E-08 |
| rs491626 | let-7b-5p | trans | 8.39E-27 | Nightingale | VLDL_CE | NA | T | C | 0.035794 | 0.004426 | 3.00E-16 |
| rs491626 | let-7b-5p | trans | 8.39E-27 | Nightingale | S_LDL_FC | NA | T | C | 0.028572 | 0.004418 | 4.20E-10 |
| rs491626 | let-7b-5p | trans | 8.39E-27 | Nightingale | IDL_FC | NA | T | C | 0.038266 | 0.004284 | 1.40E-18 |
| rs491626 | let-7b-5p | trans | 8.39E-27 | Nightingale | S_HDL_L | NA | T | C | 0.026306 | 0.00441 | 1.50E-09 |
| rs491626 | let-7b-5p | trans | 8.39E-27 | Nightingale | S_VLDL_TG | NA | T | C | -0.03813 | 0.004283 | 1.40E-18 |
| rs491626 | let-7b-5p | trans | 8.39E-27 | Nightingale | VLDL_P | NA | T | C | 0.026641 | 0.004401 | 4.40E-10 |
| rs491626 | let-7b-5p | trans | 8.39E-27 | Nightingale | L_LDL_CE | NA | T | C | 0.031579 | 0.004385 | 9.70E-13 |
| rs491626 | let-7b-5p | trans | 8.39E-27 | Nightingale | M_VLDL_P | NA | T | C | 0.044066 | 0.004247 | 5.50E-25 |
| rs491626 | let-7b-5p | trans | 8.39E-27 | Nightingale | L_LDL_P | NA | T | C | 0.029186 | 0.00441 | 6.10E-11 |
| rs491626 | let-7b-5p | trans | 8.39E-27 | Nightingale | M_LDL_L | NA | T | C | 0.033042 | 0.004431 | 1.00E-13 |
| rs491626 | let-7b-5p | trans | 8.39E-27 | Nightingale | M_VLDL_P | NA | T | C | 0.029134 | 0.004418 | 2.00E-11 |
| rs491626 | let-7b-5p | trans | 8.39E-27 | Nightingale | XS_VLDL_P | NA | T | C | 0.036714 | 0.004366 | 1.40E-17 |
| rs491626 | let-7b-5p | trans | 8.39E-27 | Nightingale | M_LDL_C | NA | T | C | 0.033997 | 0.004433 | 2.10E-14 |
| rs491626 | let-7b-5p | trans | 8.39E-27 | Nightingale | Total_FC | NA | T | C | 0.033268 | 0.004323 | 6.70E-15 |
| rs491626 | let-7b-5p | trans | 8.39E-27 | Nightingale | IDL_P | NA | T | C | 0.039995 | 0.004378 | 1.30E-19 |
| rs491626 | let-7b-5p | trans | 8.39E-27 | Nightingale | S_HDL_CE | NA | T | C | 0.028644 | 0.004432 | 1.30E-10 |
| rs491626 | let-7b-5p | trans | 8.39E-27 | Nightingale | IDL_C | NA | T | C | 0.034966 | 0.004274 | 9.30E-16 |
| rs491626 | let-7b-5p | trans | 8.39E-27 | Nightingale | Clinical_LD | NA | T | C | 0.036485 | 0.004399 | 2.70E-16 |
| rs491626 | let-7b-5p | trans | 8.39E-27 | Nightingale | Total_CE | NA | T | C | 0.03176 | 0.004268 | 9.50E-14 |
| rs491626 | let-7b-5p | trans | 8.39E-27 | Nightingale | M_LDL_C_ | NA | T | C | 0.031163 | 0.004428 | 2.90E-12 |
| rs491626 | let-7b-5p | trans | 8.39E-27 | Nightingale | S_VLDL_PL | NA | T | C | 0.032785 | 0.004417 | 4.20E-14 |
| rs491626 | let-7b-5p | trans | 8.39E-27 | Nightingale | XS_VLDL_F | NA | T | C | 0.03143 | 0.004393 | 2.40E-12 |
| rs491626 | let-7b-5p | trans | 8.39E-27 | Nightingale | XS_VLDL_P | NA | T | C | 0.032127 | 0.004363 | 5.30E-14 |
| rs491626 | let-7b-5p | trans | 8.39E-27 | Nightingale | XXL_VLDL_ | NA | T | C | 0.025011 | 0.004454 | 1.50E-09 |
| rs491626 | let-7b-5p | trans | 8.39E-27 | Nightingale | S_HDL_PL | NA | T | C | 0.023794 | 0.004387 | 3.20E-08 |
| rs491626 | let-7b-5p | trans | 8.39E-27 | Nightingale | LDL_C | NA | T | C | 0.033205 | 0.004406 | 8.70E-14 |
| rs491626 | let-7b-5p | trans | 8.39E-27 | Nightingale | XL_VLDL_P | NA | T | C | 0.038844 | 0.004432 | 5.20E-19 |
| rs491626 | let-7b-5p | trans | 8.39E-27 | Nightingale | VLDL_FC | NA | T | C | 0.024164 | 0.004392 | 1.10E-08 |
| rs491626 | let-7b-5p | trans | 8.39E-27 | Nightingale | S_HDL_FC | NA | T | C | 0.023468 | 0.004344 | 4.50E-08 |
| rs491626 | let-7b-5p | trans | 8.39E-27 | Nightingale | S_VLDL_C_ | NA | T | C | 0.041891 | 0.004306 | 5.20E-22 |
| rs491626 | let-7b-5p | trans | 8.39E-27 | Nightingale | M_VLDL_C | NA | T | C | 0.033636 | 0.004157 | 1.90E-15 |
| rs491626 | let-7b-5p | trans | 8.39E-27 | Nightingale | M_LDL_PL | NA | T | C | 0.032477 | 0.004431 | 2.60E-13 |
| rs491626 | let-7b-5p | trans | 8.39E-27 | Nightingale | Total_C | NA | T | C | 0.032387 | 0.004284 | 3.30E-14 |
| rs491626 | let-7b-5p | trans | 8.39E-27 | Nightingale | L_VLDL_C_ | NA | T | C | 0.052205 | 0.004322 | 3.90E-34 |
| rs491626 | let-7b-5p | trans | 8.39E-27 | Nightingale | L_VLDL_TG | NA | T | C | -0.05115 | 0.004404 | 5.30E-32 |
| rs491626 | let-7b-5p | trans | 8.39E-27 | Nightingale | S_LDL_P | NA | T | C | 0.029597 | 0.004429 | 3.20E-11 |
| rs491626 | let-7b-5p | trans | 8.39E-27 | Nightingale | M_VLDL_C | NA | T | C | 0.036214 | 0.004168 | 1.20E-17 |
| rs491626 | let-7b-5p | trans | 8.39E-27 | Nightingale | VLDL_C | NA | T | C | 0.031308 | 0.004419 | 4.80E-13 |
| rs491626 | let-7b-5p | trans | 8.39E-27 | Nightingale | XXL_VLDL_ | NA | T | C | 0.030859 | 0.004406 | 1.10E-12 |

| | | | | | | | | | | | |
|---|---|---|---|---|---|---|---|---|---|---|---|
| rs491626 | let-7b-5p | trans | 8.39E-27 | Nightingale | L_LDL_PL | NA | T | C | 0.038001 | 0.004391 | 8.90E-18 |
| rs491626 | let-7b-5p | trans | 8.39E-27 | Nightingale | S_VLDL_P | NA | T | C | 0.023472 | 0.004369 | 1.60E-08 |
| rs491626 | let-7b-5p | trans | 8.39E-27 | Nightingale | Total_PL | NA | T | C | 0.024943 | 0.004214 | 1.00E-09 |
| rs491626 | let-7b-5p | trans | 8.39E-27 | Nightingale | IDL_FC_pct | NA | T | C | 0.025773 | 0.004391 | 1.10E-08 |
| rs491626 | let-7b-5p | trans | 8.39E-27 | Nightingale | IDL_CE | NA | T | C | 0.033518 | 0.004272 | 1.30E-14 |
| rs491626 | let-7b-5p | trans | 8.39E-27 | Nightingale | L_LDL_L | NA | T | C | 0.033562 | 0.004382 | 2.90E-14 |
| rs491626 | let-7b-5p | trans | 8.39E-27 | Nightingale | XL_VLDL_C | NA | T | C | 0.037359 | 0.004245 | 3.10E-18 |
| rs491626 | let-7b-5p | trans | 8.39E-27 | Nightingale | XS_VLDL_C | NA | T | C | 0.038004 | 0.004295 | 1.30E-18 |
| rs491626 | let-7b-5p | trans | 8.39E-27 | Nightingale | S_VLDL_PL | NA | T | C | 0.027741 | 0.004237 | 1.80E-10 |
| rs491626 | let-7b-5p | trans | 8.39E-27 | Nightingale | XXL_VLDL_ | NA | T | C | 0.034842 | 0.004434 | 2.90E-15 |
| rs491626 | let-7b-5p | trans | 8.39E-27 | Nightingale | XS_VLDL_F | NA | T | C | 0.037079 | 0.004348 | 6.40E-18 |
| rs491626 | let-7b-5p | trans | 8.39E-27 | Nightingale | GlycA | NA | T | C | 0.028021 | 0.004406 | 1.40E-10 |
| rs491626 | let-7b-5p | trans | 8.39E-27 | Nightingale | S_LDL_CE | NA | T | C | 0.030557 | 0.004427 | 5.80E-12 |
| rs491626 | let-7b-5p | trans | 8.39E-27 | Nightingale | LDL_FC | NA | T | C | 0.033643 | 0.004378 | 5.20E-14 |
| rs491626 | let-7b-5p | trans | 8.39E-27 | Nightingale | Total_L | NA | T | C | 0.026362 | 0.004353 | 4.50E-10 |
| rs491626 | let-7b-5p | trans | 8.39E-27 | Nightingale | L_LDL_FC | NA | T | C | 0.033636 | 0.004345 | 2.80E-14 |
| rs491626 | let-7b-5p | trans | 8.39E-27 | Nightingale | LA | NA | T | C | 0.024924 | 0.004339 | 3.60E-09 |
| rs491626 | let-7b-5p | trans | 8.39E-27 | Nightingale | PUFA | NA | T | C | 0.023737 | 0.004316 | 1.40E-08 |
| rs491626 | let-7b-5p | trans | 8.39E-27 | Nightingale | XS_VLDL_C | NA | T | C | 0.037809 | 0.00427 | 2.10E-18 |
| rs491626 | let-7b-5p | trans | 8.39E-27 | Nightingale | S_VLDL_C | NA | T | C | 0.03838 | 0.004424 | 1.80E-18 |
| rs491626 | let-7b-5p | trans | 8.39E-27 | Nightingale | S_VLDL_L | NA | T | C | 0.024675 | 0.004382 | 3.70E-09 |
| rs491626 | let-7b-5p | trans | 8.39E-27 | Nightingale | Sphingomy | NA | T | C | 0.02903 | 0.004137 | 6.60E-12 |
| rs491626 | let-7b-5p | trans | 8.39E-27 | Nightingale | Omega_6 | NA | T | C | 0.025911 | 0.004321 | 5.60E-10 |
| rs491626 | let-7b-5p | trans | 8.39E-27 | Nightingale | M_VLDL_F | NA | T | C | 0.042073 | 0.00421 | 3.40E-23 |
| rs491626 | let-7b-5p | trans | 8.39E-27 | Nightingale | M_LDL_FC | NA | T | C | 0.034358 | 0.004419 | 1.90E-14 |
| rs491626 | let-7b-5p | trans | 8.39E-27 | Nightingale | M_VLDL_P | NA | T | C | 0.028301 | 0.004414 | 6.30E-11 |
| rs491626 | let-7b-5p | trans | 8.39E-27 | Nightingale | M_VLDL_T | NA | T | C | -0.03857 | 0.004181 | 6.70E-20 |
| rs491626 | let-7b-5p | trans | 8.39E-27 | Nightingale | S_VLDL_CE | NA | T | C | 0.047036 | 0.004365 | 1.20E-26 |
| rs491626 | let-7b-5p | trans | 8.39E-27 | Nightingale | Remnant_( | NA | T | C | 0.036878 | 0.004396 | 3.60E-17 |
| rs491626 | let-7b-5p | trans | 8.39E-27 | Nightingale | L_VLDL_FC | NA | T | C | 0.048596 | 0.004382 | 4.50E-29 |
| rs491626 | let-7b-5p | trans | 8.39E-27 | Nightingale | Citrate | NA | T | C | 0.026826 | 0.004396 | 7.40E-10 |
| rs491626 | let-7b-5p | trans | 8.39E-27 | Nightingale | M_VLDL_C | NA | T | C | 0.040852 | 0.004385 | 2.30E-20 |
| rs491626 | let-7b-5p | trans | 8.39E-27 | Nightingale | ApoB_by_A | NA | T | C | 0.025049 | 0.004358 | 1.30E-08 |
| rs491626 | let-7b-5p | trans | 8.39E-27 | Nightingale | IDL_L | NA | T | C | 0.035282 | 0.004278 | 4.70E-16 |
| rs491626 | let-7b-5p | trans | 8.39E-27 | Nightingale | XL_VLDL_T | NA | T | C | -0.04393 | 0.004289 | 1.70E-24 |
| rs491626 | let-7b-5p | trans | 8.39E-27 | Nightingale | XL_VLDL_F | NA | T | C | 0.037203 | 0.00429 | 2.00E-18 |
| rs491626 | let-7b-5p | trans | 8.39E-27 | Nightingale | IDL_PL | NA | T | C | 0.035356 | 0.00427 | 3.30E-16 |
| rs491626 | let-7b-5p | trans | 8.39E-27 | Nightingale | M_VLDL_F | NA | T | C | 0.033378 | 0.004424 | 3.10E-14 |
| rs491626 | let-7b-5p | trans | 8.39E-27 | Nightingale | LDL_PL | NA | T | C | 0.034999 | 0.004415 | 4.70E-15 |
| rs491626 | let-7b-5p | trans | 8.39E-27 | Nightingale | S_VLDL_CE | NA | T | C | 0.038189 | 0.004419 | 1.60E-18 |
| rs491626 | let-7b-5p | trans | 8.39E-27 | Nightingale | XL_VLDL_C | NA | T | C | 0.03823 | 0.004243 | 4.60E-19 |
| rs491626 | let-7b-5p | trans | 8.39E-27 | Nightingale | LDL_L | NA | T | C | 0.033316 | 0.00441 | 7.30E-14 |
| rs491626 | let-7b-5p | trans | 8.39E-27 | Nightingale | M_LDL_P | NA | T | C | 0.0359 | 0.004437 | 6.80E-16 |
| rs491626 | let-7b-5p | trans | 8.39E-27 | Nightingale | Cholines | NA | T | C | 0.022312 | 0.004161 | 4.70E-08 |
| rs491626 | let-7b-5p | trans | 8.39E-27 | Nightingale | S_VLDL_FC | NA | T | C | 0.029359 | 0.004228 | 1.10E-11 |
| rs491626 | let-7b-5p | trans | 8.39E-27 | Nightingale | S_LDL_L | NA | T | C | 0.02775 | 0.004424 | 8.20E-10 |
| rs491626 | let-7b-5p | trans | 8.39E-27 | Nightingale | S_HDL_P | NA | T | C | 0.030621 | 0.004425 | 5.00E-12 |
| rs491626 | let-7b-5p | trans | 8.39E-27 | Nightingale | XXL_VLDL_ | NA | T | C | -0.03574 | 0.004463 | 2.10E-16 |
| rs491626 | let-7b-5p | trans | 8.39E-27 | Nightingale | L_VLDL_CE | NA | T | C | 0.047349 | 0.004234 | 2.40E-29 |
| rs491626 | let-7b-5p | trans | 8.39E-27 | Nightingale | LDL_CE | NA | T | C | 0.032565 | 0.004413 | 2.20E-13 |
| rs491626 | let-7b-5p | trans | 8.39E-27 | Nightingale | XXL_VLDL_ | NA | T | C | 0.033133 | 0.004459 | 8.80E-14 |
| rs491626 | let-7b-5p | trans | 8.39E-27 | Nightingale | S_LDL_CE_ | NA | T | C | 0.030951 | 0.004417 | 4.80E-14 |
| rs491626 | let-7b-5p | trans | 8.39E-27 | Nightingale | S_LDL_C | NA | T | C | 0.030952 | 0.004429 | 5.50E-12 |
| rs491626 | let-7b-5p | trans | 8.39E-27 | Nightingale | non_HDL_( | NA | T | C | 0.035708 | 0.004401 | 5.00E-16 |
| rs491626 | let-7b-5p | trans | 8.39E-27 | Nightingale | M_LDL_CE | NA | T | C | 0.032982 | 0.004431 | 9.50E-14 |
| rs491626 | let-7b-5p | trans | 8.39E-27 | Nightingale | S_HDL_C | NA | T | C | 0.028422 | 0.004417 | 1.70E-10 |
| rs491626 | let-7b-5p | trans | 8.39E-27 | Nightingale | S_LDL_PL_ | NA | T | C | -0.03007 | 0.004417 | 2.60E-13 |
| rs491626 | let-7b-5p | trans | 8.39E-27 | Nightingale | LDL_P | NA | T | C | 0.031809 | 0.004422 | 1.00E-12 |
| rs491626 | let-7b-5p | trans | 8.39E-27 | Nightingale | ApoB | NA | T | C | 0.033947 | 0.004419 | 2.10E-14 |
| rs491626 | let-7b-5p | trans | 8.39E-27 | Nightingale | S_VLDL_FC | NA | T | C | 0.037912 | 0.004426 | 7.30E-18 |
| rs491626 | let-7b-5p | trans | 8.39E-27 | Nightingale | M_LDL_TG | NA | T | C | -0.02762 | 0.00437 | 3.20E-10 |
| rs491626 | let-7b-5p | trans | 8.39E-27 | Nightingale | XL_HDL_PL | NA | T | C | -0.02274 | 0.004065 | 1.20E-08 |
| rs491626 | let-7b-5p | trans | 8.39E-27 | Nightingale | M_VLDL_C | NA | T | C | 0.038718 | 0.004413 | 1.90E-18 |

| | | | | | | | | | | | |
|---|---|---|---|---|---|---|---|---|---|---|---|
| rs491626 | let-7b-5p | trans | 8.39E-27 | Nightingale | S_LDL_C_p | NA | T | C | 0.045367 | 0.004442 | 5.40E-25 |
| rs491626 | let-7b-5p | trans | 8.39E-27 | Nightingale | XS_VLDL_L | NA | T | C | 0.03383 | 0.00435 | 2.00E-15 |
| rs491626 | let-7b-5p | trans | 8.39E-27 | Nightingale | L_LDL_C | NA | T | C | 0.032332 | 0.004375 | 2.60E-13 |
| rs491626 | let-7c-5p | trans | 2.01E-23 | Nightingale | VLDL_CE | NA | T | C | 0.035794 | 0.004426 | 3.00E-16 |
| rs491626 | let-7c-5p | trans | 2.01E-23 | Nightingale | S_LDL_FC | NA | T | C | 0.028572 | 0.004418 | 4.20E-10 |
| rs491626 | let-7c-5p | trans | 2.01E-23 | Nightingale | IDL_FC | NA | T | C | 0.038266 | 0.004284 | 1.40E-18 |
| rs491626 | let-7c-5p | trans | 2.01E-23 | Nightingale | S_HDL_L | NA | T | C | 0.026306 | 0.00441 | 1.50E-09 |
| rs491626 | let-7c-5p | trans | 2.01E-23 | Nightingale | S_VLDL_TG | NA | T | C | -0.03813 | 0.004283 | 1.40E-18 |
| rs491626 | let-7c-5p | trans | 2.01E-23 | Nightingale | VLDL_P | NA | T | C | 0.026641 | 0.004401 | 4.40E-10 |
| rs491626 | let-7c-5p | trans | 2.01E-23 | Nightingale | L_LDL_CE | NA | T | C | 0.031579 | 0.004385 | 9.70E-13 |
| rs491626 | let-7c-5p | trans | 2.01E-23 | Nightingale | M_VLDL_P | NA | T | C | 0.044066 | 0.004247 | 5.50E-25 |
| rs491626 | let-7c-5p | trans | 2.01E-23 | Nightingale | L_LDL_P | NA | T | C | 0.029186 | 0.00441 | 6.10E-11 |
| rs491626 | let-7c-5p | trans | 2.01E-23 | Nightingale | M_LDL_L | NA | T | C | 0.033042 | 0.004431 | 1.00E-13 |
| rs491626 | let-7c-5p | trans | 2.01E-23 | Nightingale | M_VLDL_P | NA | T | C | 0.029134 | 0.004418 | 2.00E-11 |
| rs491626 | let-7c-5p | trans | 2.01E-23 | Nightingale | XS_VLDL_P | NA | T | C | 0.036714 | 0.004366 | 1.40E-17 |
| rs491626 | let-7c-5p | trans | 2.01E-23 | Nightingale | M_LDL_C | NA | T | C | 0.033997 | 0.004433 | 2.10E-14 |
| rs491626 | let-7c-5p | trans | 2.01E-23 | Nightingale | Total_FC | NA | T | C | 0.033268 | 0.004323 | 6.70E-15 |
| rs491626 | let-7c-5p | trans | 2.01E-23 | Nightingale | IDL_P | NA | T | C | 0.039995 | 0.004378 | 1.30E-19 |
| rs491626 | let-7c-5p | trans | 2.01E-23 | Nightingale | S_HDL_CE | NA | T | C | 0.028644 | 0.004432 | 1.30E-10 |
| rs491626 | let-7c-5p | trans | 2.01E-23 | Nightingale | IDL_C | NA | T | C | 0.034966 | 0.004274 | 9.30E-16 |
| rs491626 | let-7c-5p | trans | 2.01E-23 | Nightingale | Clinical_LD | NA | T | C | 0.036485 | 0.004399 | 2.70E-16 |
| rs491626 | let-7c-5p | trans | 2.01E-23 | Nightingale | Total_CE | NA | T | C | 0.03176 | 0.004268 | 9.50E-14 |
| rs491626 | let-7c-5p | trans | 2.01E-23 | Nightingale | M_LDL_C_ | NA | T | C | 0.031163 | 0.004428 | 2.90E-12 |
| rs491626 | let-7c-5p | trans | 2.01E-23 | Nightingale | S_VLDL_PL | NA | T | C | 0.032785 | 0.004417 | 4.20E-14 |
| rs491626 | let-7c-5p | trans | 2.01E-23 | Nightingale | XS_VLDL_F | NA | T | C | 0.03143 | 0.004393 | 2.40E-12 |
| rs491626 | let-7c-5p | trans | 2.01E-23 | Nightingale | XS_VLDL_P | NA | T | C | 0.032127 | 0.004363 | 5.30E-14 |
| rs491626 | let-7c-5p | trans | 2.01E-23 | Nightingale | XXL_VLDL_ | NA | T | C | 0.025011 | 0.004454 | 1.50E-09 |
| rs491626 | let-7c-5p | trans | 2.01E-23 | Nightingale | S_HDL_PL | NA | T | C | 0.023794 | 0.004387 | 3.20E-08 |
| rs491626 | let-7c-5p | trans | 2.01E-23 | Nightingale | LDL_C | NA | T | C | 0.033205 | 0.004406 | 8.70E-14 |
| rs491626 | let-7c-5p | trans | 2.01E-23 | Nightingale | XL_VLDL_P | NA | T | C | 0.038844 | 0.004432 | 5.20E-19 |
| rs491626 | let-7c-5p | trans | 2.01E-23 | Nightingale | VLDL_FC | NA | T | C | 0.024164 | 0.004392 | 1.10E-08 |
| rs491626 | let-7c-5p | trans | 2.01E-23 | Nightingale | S_HDL_FC | NA | T | C | 0.023468 | 0.004344 | 4.50E-08 |
| rs491626 | let-7c-5p | trans | 2.01E-23 | Nightingale | S_VLDL_C_ | NA | T | C | 0.041891 | 0.004306 | 5.20E-22 |
| rs491626 | let-7c-5p | trans | 2.01E-23 | Nightingale | M_VLDL_C | NA | T | C | 0.033636 | 0.004157 | 1.90E-15 |
| rs491626 | let-7c-5p | trans | 2.01E-23 | Nightingale | M_LDL_PL | NA | T | C | 0.032477 | 0.004431 | 2.60E-13 |
| rs491626 | let-7c-5p | trans | 2.01E-23 | Nightingale | Total_C | NA | T | C | 0.032387 | 0.004284 | 3.30E-14 |
| rs491626 | let-7c-5p | trans | 2.01E-23 | Nightingale | L_VLDL_C_ | NA | T | C | 0.052205 | 0.004322 | 3.90E-34 |
| rs491626 | let-7c-5p | trans | 2.01E-23 | Nightingale | L_VLDL_TG | NA | T | C | -0.05115 | 0.004404 | 5.30E-32 |
| rs491626 | let-7c-5p | trans | 2.01E-23 | Nightingale | S_LDL_P | NA | T | C | 0.029597 | 0.004429 | 3.20E-11 |
| rs491626 | let-7c-5p | trans | 2.01E-23 | Nightingale | M_VLDL_C | NA | T | C | 0.036214 | 0.004168 | 1.20E-17 |
| rs491626 | let-7c-5p | trans | 2.01E-23 | Nightingale | VLDL_C | NA | T | C | 0.031308 | 0.004419 | 4.80E-13 |
| rs491626 | let-7c-5p | trans | 2.01E-23 | Nightingale | XXL_VLDL_ | NA | T | C | 0.030859 | 0.004406 | 1.10E-12 |
| rs491626 | let-7c-5p | trans | 2.01E-23 | Nightingale | L_LDL_PL | NA | T | C | 0.038001 | 0.004391 | 8.90E-18 |
| rs491626 | let-7c-5p | trans | 2.01E-23 | Nightingale | S_VLDL_P | NA | T | C | 0.023472 | 0.004369 | 1.60E-08 |
| rs491626 | let-7c-5p | trans | 2.01E-23 | Nightingale | Total_PL | NA | T | C | 0.024943 | 0.004214 | 1.00E-09 |
| rs491626 | let-7c-5p | trans | 2.01E-23 | Nightingale | IDL_FC_pct | NA | T | C | 0.025773 | 0.004391 | 1.10E-08 |
| rs491626 | let-7c-5p | trans | 2.01E-23 | Nightingale | IDL_CE | NA | T | C | 0.033518 | 0.004272 | 1.30E-14 |
| rs491626 | let-7c-5p | trans | 2.01E-23 | Nightingale | L_LDL_L | NA | T | C | 0.033562 | 0.004382 | 2.90E-14 |
| rs491626 | let-7c-5p | trans | 2.01E-23 | Nightingale | XL_VLDL_C | NA | T | C | 0.037359 | 0.004245 | 3.10E-18 |
| rs491626 | let-7c-5p | trans | 2.01E-23 | Nightingale | XS_VLDL_C | NA | T | C | 0.038004 | 0.004295 | 1.30E-18 |
| rs491626 | let-7c-5p | trans | 2.01E-23 | Nightingale | S_VLDL_PL | NA | T | C | 0.027741 | 0.004237 | 1.80E-10 |
| rs491626 | let-7c-5p | trans | 2.01E-23 | Nightingale | XXL_VLDL_ | NA | T | C | 0.034842 | 0.004434 | 2.90E-15 |
| rs491626 | let-7c-5p | trans | 2.01E-23 | Nightingale | XS_VLDL_F | NA | T | C | 0.037079 | 0.004348 | 6.40E-18 |
| rs491626 | let-7c-5p | trans | 2.01E-23 | Nightingale | GlycA | NA | T | C | 0.028021 | 0.004406 | 1.40E-10 |
| rs491626 | let-7c-5p | trans | 2.01E-23 | Nightingale | S_LDL_CE | NA | T | C | 0.030557 | 0.004427 | 5.80E-12 |
| rs491626 | let-7c-5p | trans | 2.01E-23 | Nightingale | LDL_FC | NA | T | C | 0.033643 | 0.004378 | 5.20E-14 |
| rs491626 | let-7c-5p | trans | 2.01E-23 | Nightingale | Total_L | NA | T | C | 0.026362 | 0.004353 | 4.50E-10 |
| rs491626 | let-7c-5p | trans | 2.01E-23 | Nightingale | L_LDL_FC | NA | T | C | 0.033636 | 0.004345 | 2.80E-14 |
| rs491626 | let-7c-5p | trans | 2.01E-23 | Nightingale | LA | NA | T | C | 0.024924 | 0.004339 | 3.60E-09 |
| rs491626 | let-7c-5p | trans | 2.01E-23 | Nightingale | PUFA | NA | T | C | 0.023737 | 0.004316 | 1.40E-08 |
| rs491626 | let-7c-5p | trans | 2.01E-23 | Nightingale | XS_VLDL_C | NA | T | C | 0.037809 | 0.00427 | 2.10E-18 |
| rs491626 | let-7c-5p | trans | 2.01E-23 | Nightingale | S_VLDL_C | NA | T | C | 0.03838 | 0.004424 | 1.80E-18 |
| rs491626 | let-7c-5p | trans | 2.01E-23 | Nightingale | S_VLDL_L | NA | T | C | 0.024675 | 0.004382 | 3.70E-09 |

| | | | | | | | | | | | |
|---|---|---|---|---|---|---|---|---|---|---|---|
| rs491626 | let-7c-5p | trans | 2.01E-23 | Nightingale | Sphingomy | NA | T | C | 0.02903 | 0.004137 | 6.60E-12 |
| rs491626 | let-7c-5p | trans | 2.01E-23 | Nightingale | Omega_6 | NA | T | C | 0.025911 | 0.004321 | 5.60E-10 |
| rs491626 | let-7c-5p | trans | 2.01E-23 | Nightingale | M_VLDL_F | NA | T | C | 0.042073 | 0.00421 | 3.40E-23 |
| rs491626 | let-7c-5p | trans | 2.01E-23 | Nightingale | M_LDL_FC | NA | T | C | 0.034358 | 0.004419 | 1.90E-14 |
| rs491626 | let-7c-5p | trans | 2.01E-23 | Nightingale | M_VLDL_P | NA | T | C | 0.028301 | 0.004414 | 6.30E-11 |
| rs491626 | let-7c-5p | trans | 2.01E-23 | Nightingale | M_VLDL_T | NA | T | C | -0.03857 | 0.004181 | 6.70E-20 |
| rs491626 | let-7c-5p | trans | 2.01E-23 | Nightingale | S_VLDL_CE | NA | T | C | 0.047036 | 0.004365 | 1.20E-26 |
| rs491626 | let-7c-5p | trans | 2.01E-23 | Nightingale | Remnant_( | NA | T | C | 0.036878 | 0.004396 | 3.60E-17 |
| rs491626 | let-7c-5p | trans | 2.01E-23 | Nightingale | L_VLDL_FC | NA | T | C | 0.048596 | 0.004382 | 4.50E-29 |
| rs491626 | let-7c-5p | trans | 2.01E-23 | Nightingale | Citrate | NA | T | C | 0.026826 | 0.004396 | 7.40E-10 |
| rs491626 | let-7c-5p | trans | 2.01E-23 | Nightingale | M_VLDL_C | NA | T | C | 0.040852 | 0.004385 | 2.30E-20 |
| rs491626 | let-7c-5p | trans | 2.01E-23 | Nightingale | ApoB_by_/ | NA | T | C | 0.025049 | 0.004358 | 1.30E-08 |
| rs491626 | let-7c-5p | trans | 2.01E-23 | Nightingale | IDL_L | NA | T | C | 0.035282 | 0.004278 | 4.70E-16 |
| rs491626 | let-7c-5p | trans | 2.01E-23 | Nightingale | XL_VLDL_T | NA | T | C | -0.04393 | 0.004289 | 1.70E-24 |
| rs491626 | let-7c-5p | trans | 2.01E-23 | Nightingale | XL_VLDL_F | NA | T | C | 0.037203 | 0.00429 | 2.00E-18 |
| rs491626 | let-7c-5p | trans | 2.01E-23 | Nightingale | IDL_PL | NA | T | C | 0.035356 | 0.00427 | 3.30E-16 |
| rs491626 | let-7c-5p | trans | 2.01E-23 | Nightingale | M_VLDL_F | NA | T | C | 0.033378 | 0.004424 | 3.10E-14 |
| rs491626 | let-7c-5p | trans | 2.01E-23 | Nightingale | LDL_PL | NA | T | C | 0.034999 | 0.004415 | 4.70E-15 |
| rs491626 | let-7c-5p | trans | 2.01E-23 | Nightingale | S_VLDL_CE | NA | T | C | 0.038189 | 0.004419 | 1.60E-18 |
| rs491626 | let-7c-5p | trans | 2.01E-23 | Nightingale | XL_VLDL_C | NA | T | C | 0.03823 | 0.004243 | 4.60E-19 |
| rs491626 | let-7c-5p | trans | 2.01E-23 | Nightingale | LDL_L | NA | T | C | 0.033316 | 0.00441 | 7.30E-14 |
| rs491626 | let-7c-5p | trans | 2.01E-23 | Nightingale | M_LDL_P | NA | T | C | 0.0359 | 0.004437 | 6.80E-16 |
| rs491626 | let-7c-5p | trans | 2.01E-23 | Nightingale | Cholines | NA | T | C | 0.022312 | 0.004161 | 4.70E-08 |
| rs491626 | let-7c-5p | trans | 2.01E-23 | Nightingale | S_VLDL_FC | NA | T | C | 0.029359 | 0.004228 | 1.10E-11 |
| rs491626 | let-7c-5p | trans | 2.01E-23 | Nightingale | S_LDL_L | NA | T | C | 0.02775 | 0.004424 | 8.20E-10 |
| rs491626 | let-7c-5p | trans | 2.01E-23 | Nightingale | S_HDL_P | NA | T | C | 0.030621 | 0.004425 | 5.00E-12 |
| rs491626 | let-7c-5p | trans | 2.01E-23 | Nightingale | XXL_VLDL_ | NA | T | C | -0.03574 | 0.004463 | 2.10E-16 |
| rs491626 | let-7c-5p | trans | 2.01E-23 | Nightingale | L_VLDL_CE | NA | T | C | 0.047349 | 0.004234 | 2.40E-29 |
| rs491626 | let-7c-5p | trans | 2.01E-23 | Nightingale | LDL_CE | NA | T | C | 0.032565 | 0.004413 | 2.20E-13 |
| rs491626 | let-7c-5p | trans | 2.01E-23 | Nightingale | XXL_VLDL_ | NA | T | C | 0.033133 | 0.004459 | 8.80E-14 |
| rs491626 | let-7c-5p | trans | 2.01E-23 | Nightingale | S_LDL_CE_ | NA | T | C | 0.030951 | 0.004417 | 4.80E-14 |
| rs491626 | let-7c-5p | trans | 2.01E-23 | Nightingale | S_LDL_C | NA | T | C | 0.030952 | 0.004429 | 5.50E-12 |
| rs491626 | let-7c-5p | trans | 2.01E-23 | Nightingale | non_HDL_( | NA | T | C | 0.035708 | 0.004401 | 5.00E-16 |
| rs491626 | let-7c-5p | trans | 2.01E-23 | Nightingale | M_LDL_CE | NA | T | C | 0.032982 | 0.004431 | 9.50E-14 |
| rs491626 | let-7c-5p | trans | 2.01E-23 | Nightingale | S_HDL_C | NA | T | C | 0.028422 | 0.004417 | 1.70E-10 |
| rs491626 | let-7c-5p | trans | 2.01E-23 | Nightingale | S_LDL_PL_ | NA | T | C | -0.03007 | 0.004417 | 2.60E-13 |
| rs491626 | let-7c-5p | trans | 2.01E-23 | Nightingale | LDL_P | NA | T | C | 0.031809 | 0.004422 | 1.00E-12 |
| rs491626 | let-7c-5p | trans | 2.01E-23 | Nightingale | ApoB | NA | T | C | 0.033947 | 0.004419 | 2.10E-14 |
| rs491626 | let-7c-5p | trans | 2.01E-23 | Nightingale | S_VLDL_FC | NA | T | C | 0.037912 | 0.004426 | 7.30E-18 |
| rs491626 | let-7c-5p | trans | 2.01E-23 | Nightingale | M_LDL_TG | NA | T | C | -0.02762 | 0.00437 | 3.20E-10 |
| rs491626 | let-7c-5p | trans | 2.01E-23 | Nightingale | XL_HDL_PL | NA | T | C | -0.02274 | 0.004065 | 1.20E-08 |
| rs491626 | let-7c-5p | trans | 2.01E-23 | Nightingale | M_VLDL_C | NA | T | C | 0.038718 | 0.004413 | 1.90E-18 |
| rs491626 | let-7c-5p | trans | 2.01E-23 | Nightingale | S_LDL_C_p | NA | T | C | 0.045367 | 0.004442 | 5.40E-25 |
| rs491626 | let-7c-5p | trans | 2.01E-23 | Nightingale | XS_VLDL_L | NA | T | C | 0.03383 | 0.00435 | 2.00E-15 |
| rs491626 | let-7c-5p | trans | 2.01E-23 | Nightingale | L_LDL_C | NA | T | C | 0.032332 | 0.004375 | 2.60E-13 |
| rs491626 | miR-195-5p | trans | 2.32E-17 | Nightingale | VLDL_CE | NA | T | C | 0.035794 | 0.004426 | 3.00E-16 |
| rs491626 | miR-195-5p | trans | 2.32E-17 | Nightingale | S_LDL_FC | NA | T | C | 0.028572 | 0.004418 | 4.20E-10 |
| rs491626 | miR-195-5p | trans | 2.32E-17 | Nightingale | IDL_FC | NA | T | C | 0.038266 | 0.004284 | 1.40E-18 |
| rs491626 | miR-195-5p | trans | 2.32E-17 | Nightingale | S_HDL_L | NA | T | C | 0.026306 | 0.00441 | 1.50E-09 |
| rs491626 | miR-195-5p | trans | 2.32E-17 | Nightingale | S_VLDL_TG | NA | T | C | -0.03813 | 0.004283 | 1.40E-18 |
| rs491626 | miR-195-5p | trans | 2.32E-17 | Nightingale | VLDL_P | NA | T | C | 0.026641 | 0.004401 | 4.40E-10 |
| rs491626 | miR-195-5p | trans | 2.32E-17 | Nightingale | L_LDL_CE | NA | T | C | 0.031579 | 0.004385 | 9.70E-13 |
| rs491626 | miR-195-5p | trans | 2.32E-17 | Nightingale | M_VLDL_P | NA | T | C | 0.044066 | 0.004247 | 5.50E-25 |
| rs491626 | miR-195-5p | trans | 2.32E-17 | Nightingale | L_LDL_P | NA | T | C | 0.029186 | 0.00441 | 6.10E-11 |
| rs491626 | miR-195-5p | trans | 2.32E-17 | Nightingale | M_LDL_L | NA | T | C | 0.033042 | 0.004431 | 1.00E-13 |
| rs491626 | miR-195-5p | trans | 2.32E-17 | Nightingale | M_VLDL_P | NA | T | C | 0.029134 | 0.004418 | 2.00E-11 |
| rs491626 | miR-195-5p | trans | 2.32E-17 | Nightingale | XS_VLDL_P | NA | T | C | 0.036714 | 0.004366 | 1.40E-17 |
| rs491626 | miR-195-5p | trans | 2.32E-17 | Nightingale | M_LDL_C | NA | T | C | 0.033997 | 0.004433 | 2.10E-14 |
| rs491626 | miR-195-5p | trans | 2.32E-17 | Nightingale | Total_FC | NA | T | C | 0.033268 | 0.004323 | 6.70E-15 |
| rs491626 | miR-195-5p | trans | 2.32E-17 | Nightingale | IDL_P | NA | T | C | 0.039995 | 0.004378 | 1.30E-19 |
| rs491626 | miR-195-5p | trans | 2.32E-17 | Nightingale | S_HDL_CE | NA | T | C | 0.028644 | 0.004432 | 1.30E-10 |
| rs491626 | miR-195-5p | trans | 2.32E-17 | Nightingale | IDL_C | NA | T | C | 0.034966 | 0.004274 | 9.30E-16 |
| rs491626 | miR-195-5p | trans | 2.32E-17 | Nightingale | Clinical_LD | NA | T | C | 0.036485 | 0.004399 | 2.70E-16 |

| | | | | | | | | | | | |
|---|---|---|---|---|---|---|---|---|---|---|---|
| rs491626 | miR-195-5p | trans | 2.32E-17 | Nightingale | Total_CE | NA | T | C | 0.03176 | 0.004268 | 9.50E-14 |
| rs491626 | miR-195-5p | trans | 2.32E-17 | Nightingale | M_LDL_C_ | NA | T | C | 0.031163 | 0.004428 | 2.90E-12 |
| rs491626 | miR-195-5p | trans | 2.32E-17 | Nightingale | S_VLDL_PL | NA | T | C | 0.032785 | 0.004417 | 4.20E-14 |
| rs491626 | miR-195-5p | trans | 2.32E-17 | Nightingale | XS_VLDL_F | NA | T | C | 0.03143 | 0.004393 | 2.40E-12 |
| rs491626 | miR-195-5p | trans | 2.32E-17 | Nightingale | XS_VLDL_P | NA | T | C | 0.032127 | 0.004363 | 5.30E-14 |
| rs491626 | miR-195-5p | trans | 2.32E-17 | Nightingale | XXL_VLDL_ | NA | T | C | 0.025011 | 0.004454 | 1.50E-09 |
| rs491626 | miR-195-5p | trans | 2.32E-17 | Nightingale | S_HDL_PL | NA | T | C | 0.023794 | 0.004387 | 3.20E-08 |
| rs491626 | miR-195-5p | trans | 2.32E-17 | Nightingale | LDL_C | NA | T | C | 0.033205 | 0.004406 | 8.70E-14 |
| rs491626 | miR-195-5p | trans | 2.32E-17 | Nightingale | XL_VLDL_P | NA | T | C | 0.038844 | 0.004432 | 5.20E-19 |
| rs491626 | miR-195-5p | trans | 2.32E-17 | Nightingale | VLDL_FC | NA | T | C | 0.024164 | 0.004392 | 1.10E-08 |
| rs491626 | miR-195-5p | trans | 2.32E-17 | Nightingale | S_HDL_FC | NA | T | C | 0.023468 | 0.004344 | 4.50E-08 |
| rs491626 | miR-195-5p | trans | 2.32E-17 | Nightingale | S_VLDL_C_ | NA | T | C | 0.041891 | 0.004306 | 5.20E-22 |
| rs491626 | miR-195-5p | trans | 2.32E-17 | Nightingale | M_VLDL_C | NA | T | C | 0.033636 | 0.004157 | 1.90E-15 |
| rs491626 | miR-195-5p | trans | 2.32E-17 | Nightingale | M_LDL_PL | NA | T | C | 0.032477 | 0.004431 | 2.60E-13 |
| rs491626 | miR-195-5p | trans | 2.32E-17 | Nightingale | Total_C | NA | T | C | 0.032387 | 0.004284 | 3.30E-14 |
| rs491626 | miR-195-5p | trans | 2.32E-17 | Nightingale | L_VLDL_C_ | NA | T | C | 0.052205 | 0.004322 | 3.90E-34 |
| rs491626 | miR-195-5p | trans | 2.32E-17 | Nightingale | L_VLDL_TG | NA | T | C | -0.05115 | 0.004404 | 5.30E-32 |
| rs491626 | miR-195-5p | trans | 2.32E-17 | Nightingale | S_LDL_P | NA | T | C | 0.029597 | 0.004429 | 3.20E-11 |
| rs491626 | miR-195-5p | trans | 2.32E-17 | Nightingale | M_VLDL_C | NA | T | C | 0.036214 | 0.004168 | 1.20E-17 |
| rs491626 | miR-195-5p | trans | 2.32E-17 | Nightingale | VLDL_C | NA | T | C | 0.031308 | 0.004419 | 4.80E-13 |
| rs491626 | miR-195-5p | trans | 2.32E-17 | Nightingale | XXL_VLDL_ | NA | T | C | 0.030859 | 0.004406 | 1.10E-12 |
| rs491626 | miR-195-5p | trans | 2.32E-17 | Nightingale | L_LDL_PL | NA | T | C | 0.038001 | 0.004391 | 8.90E-18 |
| rs491626 | miR-195-5p | trans | 2.32E-17 | Nightingale | S_VLDL_P | NA | T | C | 0.023472 | 0.004369 | 1.60E-08 |
| rs491626 | miR-195-5p | trans | 2.32E-17 | Nightingale | Total_PL | NA | T | C | 0.024943 | 0.004214 | 1.00E-09 |
| rs491626 | miR-195-5p | trans | 2.32E-17 | Nightingale | IDL_FC_pct | NA | T | C | 0.025773 | 0.004391 | 1.10E-08 |
| rs491626 | miR-195-5p | trans | 2.32E-17 | Nightingale | IDL_CE | NA | T | C | 0.033518 | 0.004272 | 1.30E-14 |
| rs491626 | miR-195-5p | trans | 2.32E-17 | Nightingale | L_LDL_L | NA | T | C | 0.033562 | 0.004382 | 2.90E-14 |
| rs491626 | miR-195-5p | trans | 2.32E-17 | Nightingale | XL_VLDL_C | NA | T | C | 0.037359 | 0.004245 | 3.10E-18 |
| rs491626 | miR-195-5p | trans | 2.32E-17 | Nightingale | XS_VLDL_C | NA | T | C | 0.038004 | 0.004295 | 1.30E-18 |
| rs491626 | miR-195-5p | trans | 2.32E-17 | Nightingale | S_VLDL_PL | NA | T | C | 0.027741 | 0.004237 | 1.80E-10 |
| rs491626 | miR-195-5p | trans | 2.32E-17 | Nightingale | XXL_VLDL_ | NA | T | C | 0.034842 | 0.004434 | 2.90E-15 |
| rs491626 | miR-195-5p | trans | 2.32E-17 | Nightingale | XS_VLDL_F | NA | T | C | 0.037079 | 0.004348 | 6.40E-18 |
| rs491626 | miR-195-5p | trans | 2.32E-17 | Nightingale | GlycA | NA | T | C | 0.028021 | 0.004406 | 1.40E-10 |
| rs491626 | miR-195-5p | trans | 2.32E-17 | Nightingale | S_LDL_CE | NA | T | C | 0.030557 | 0.004427 | 5.80E-12 |
| rs491626 | miR-195-5p | trans | 2.32E-17 | Nightingale | LDL_FC | NA | T | C | 0.033643 | 0.004378 | 5.20E-14 |
| rs491626 | miR-195-5p | trans | 2.32E-17 | Nightingale | Total_L | NA | T | C | 0.026362 | 0.004353 | 4.50E-10 |
| rs491626 | miR-195-5p | trans | 2.32E-17 | Nightingale | L_LDL_FC | NA | T | C | 0.033636 | 0.004345 | 2.80E-14 |
| rs491626 | miR-195-5p | trans | 2.32E-17 | Nightingale | LA | NA | T | C | 0.024924 | 0.004339 | 3.60E-09 |
| rs491626 | miR-195-5p | trans | 2.32E-17 | Nightingale | PUFA | NA | T | C | 0.023737 | 0.004316 | 1.40E-08 |
| rs491626 | miR-195-5p | trans | 2.32E-17 | Nightingale | XS_VLDL_C | NA | T | C | 0.037809 | 0.00427 | 2.10E-18 |
| rs491626 | miR-195-5p | trans | 2.32E-17 | Nightingale | S_VLDL_C | NA | T | C | 0.03838 | 0.004424 | 1.80E-18 |
| rs491626 | miR-195-5p | trans | 2.32E-17 | Nightingale | S_VLDL_L | NA | T | C | 0.024675 | 0.004382 | 3.70E-09 |
| rs491626 | miR-195-5p | trans | 2.32E-17 | Nightingale | Sphingomy | NA | T | C | 0.02903 | 0.004137 | 6.60E-12 |
| rs491626 | miR-195-5p | trans | 2.32E-17 | Nightingale | Omega_6 | NA | T | C | 0.025911 | 0.004321 | 5.60E-10 |
| rs491626 | miR-195-5p | trans | 2.32E-17 | Nightingale | M_VLDL_F | NA | T | C | 0.042073 | 0.00421 | 3.40E-23 |
| rs491626 | miR-195-5p | trans | 2.32E-17 | Nightingale | M_LDL_FC | NA | T | C | 0.034358 | 0.004419 | 1.90E-14 |
| rs491626 | miR-195-5p | trans | 2.32E-17 | Nightingale | M_VLDL_P | NA | T | C | 0.028301 | 0.004414 | 6.30E-11 |
| rs491626 | miR-195-5p | trans | 2.32E-17 | Nightingale | M_VLDL_T | NA | T | C | -0.03857 | 0.004181 | 6.70E-20 |
| rs491626 | miR-195-5p | trans | 2.32E-17 | Nightingale | S_VLDL_CE | NA | T | C | 0.047036 | 0.004365 | 1.20E-26 |
| rs491626 | miR-195-5p | trans | 2.32E-17 | Nightingale | Remnant_( | NA | T | C | 0.036878 | 0.004396 | 3.60E-17 |
| rs491626 | miR-195-5p | trans | 2.32E-17 | Nightingale | L_VLDL_FC | NA | T | C | 0.048596 | 0.004382 | 4.50E-29 |
| rs491626 | miR-195-5p | trans | 2.32E-17 | Nightingale | Citrate | NA | T | C | 0.026826 | 0.004396 | 7.40E-10 |
| rs491626 | miR-195-5p | trans | 2.32E-17 | Nightingale | M_VLDL_C | NA | T | C | 0.040852 | 0.004385 | 2.30E-20 |
| rs491626 | miR-195-5p | trans | 2.32E-17 | Nightingale | ApoB_by_A | NA | T | C | 0.025049 | 0.004358 | 1.30E-08 |
| rs491626 | miR-195-5p | trans | 2.32E-17 | Nightingale | IDL_L | NA | T | C | 0.035282 | 0.004278 | 4.70E-16 |
| rs491626 | miR-195-5p | trans | 2.32E-17 | Nightingale | XL_VLDL_T | NA | T | C | -0.04393 | 0.004289 | 1.70E-24 |
| rs491626 | miR-195-5p | trans | 2.32E-17 | Nightingale | XL_VLDL_F | NA | T | C | 0.037203 | 0.00429 | 2.00E-18 |
| rs491626 | miR-195-5p | trans | 2.32E-17 | Nightingale | IDL_PL | NA | T | C | 0.035356 | 0.00427 | 3.30E-16 |
| rs491626 | miR-195-5p | trans | 2.32E-17 | Nightingale | M_VLDL_F | NA | T | C | 0.033378 | 0.004424 | 3.10E-14 |
| rs491626 | miR-195-5p | trans | 2.32E-17 | Nightingale | LDL_PL | NA | T | C | 0.034999 | 0.004415 | 4.70E-15 |
| rs491626 | miR-195-5p | trans | 2.32E-17 | Nightingale | S_VLDL_CE | NA | T | C | 0.038189 | 0.004419 | 1.60E-18 |
| rs491626 | miR-195-5p | trans | 2.32E-17 | Nightingale | XL_VLDL_C | NA | T | C | 0.03823 | 0.004243 | 4.60E-19 |
| rs491626 | miR-195-5p | trans | 2.32E-17 | Nightingale | LDL_L | NA | T | C | 0.033316 | 0.00441 | 7.30E-14 |

| | | | | | | | | | | | |
|---|---|---|---|---|---|---|---|---|---|---|---|
| rs491626 | miR-195-5p | trans | 2.32E-17 | Nightingale | M_LDL_P | NA | T | C | 0.0359 | 0.004437 | 6.80E-16 |
| rs491626 | miR-195-5p | trans | 2.32E-17 | Nightingale | Cholines | NA | T | C | 0.022312 | 0.004161 | 4.70E-08 |
| rs491626 | miR-195-5p | trans | 2.32E-17 | Nightingale | S_VLDL_FC | NA | T | C | 0.029359 | 0.004228 | 1.10E-11 |
| rs491626 | miR-195-5p | trans | 2.32E-17 | Nightingale | S_LDL_L | NA | T | C | 0.02775 | 0.004424 | 8.20E-10 |
| rs491626 | miR-195-5p | trans | 2.32E-17 | Nightingale | S_HDL_P | NA | T | C | 0.030621 | 0.004425 | 5.00E-12 |
| rs491626 | miR-195-5p | trans | 2.32E-17 | Nightingale | XXL_VLDL_ | NA | T | C | -0.03574 | 0.004463 | 2.10E-16 |
| rs491626 | miR-195-5p | trans | 2.32E-17 | Nightingale | L_VLDL_CE | NA | T | C | 0.047349 | 0.004234 | 2.40E-29 |
| rs491626 | miR-195-5p | trans | 2.32E-17 | Nightingale | LDL_CE | NA | T | C | 0.032565 | 0.004413 | 2.20E-13 |
| rs491626 | miR-195-5p | trans | 2.32E-17 | Nightingale | XXL_VLDL_ | NA | T | C | 0.033133 | 0.004459 | 8.80E-14 |
| rs491626 | miR-195-5p | trans | 2.32E-17 | Nightingale | S_LDL_CE_ | NA | T | C | 0.030951 | 0.004417 | 4.80E-14 |
| rs491626 | miR-195-5p | trans | 2.32E-17 | Nightingale | S_LDL_C | NA | T | C | 0.030952 | 0.004429 | 5.50E-12 |
| rs491626 | miR-195-5p | trans | 2.32E-17 | Nightingale | non_HDL_( | NA | T | C | 0.035708 | 0.004401 | 5.00E-16 |
| rs491626 | miR-195-5p | trans | 2.32E-17 | Nightingale | M_LDL_CE | NA | T | C | 0.032982 | 0.004431 | 9.50E-14 |
| rs491626 | miR-195-5p | trans | 2.32E-17 | Nightingale | S_HDL_C | NA | T | C | 0.028422 | 0.004417 | 1.70E-10 |
| rs491626 | miR-195-5p | trans | 2.32E-17 | Nightingale | S_LDL_PL_ | NA | T | C | -0.03007 | 0.004417 | 2.60E-13 |
| rs491626 | miR-195-5p | trans | 2.32E-17 | Nightingale | LDL_P | NA | T | C | 0.031809 | 0.004422 | 1.00E-12 |
| rs491626 | miR-195-5p | trans | 2.32E-17 | Nightingale | ApoB | NA | T | C | 0.033947 | 0.004419 | 2.10E-14 |
| rs491626 | miR-195-5p | trans | 2.32E-17 | Nightingale | S_VLDL_FC | NA | T | C | 0.037912 | 0.004426 | 7.30E-18 |
| rs491626 | miR-195-5p | trans | 2.32E-17 | Nightingale | M_LDL_TG | NA | T | C | -0.02762 | 0.00437 | 3.20E-10 |
| rs491626 | miR-195-5p | trans | 2.32E-17 | Nightingale | XL_HDL_PL | NA | T | C | -0.02274 | 0.004065 | 1.20E-08 |
| rs491626 | miR-195-5p | trans | 2.32E-17 | Nightingale | M_VLDL_C | NA | T | C | 0.038718 | 0.004413 | 1.90E-18 |
| rs491626 | miR-195-5p | trans | 2.32E-17 | Nightingale | S_LDL_C_p | NA | T | C | 0.045367 | 0.004442 | 5.40E-25 |
| rs491626 | miR-195-5p | trans | 2.32E-17 | Nightingale | XS_VLDL_L | NA | T | C | 0.03383 | 0.00435 | 2.00E-15 |
| rs491626 | miR-195-5p | trans | 2.32E-17 | Nightingale | L_LDL_C | NA | T | C | 0.032332 | 0.004375 | 2.60E-13 |
| rs491626 | miR-139-5p | trans | 8.32E-42 | Nightingale | VLDL_CE | NA | T | C | 0.035794 | 0.004426 | 3.00E-16 |
| rs491626 | miR-139-5p | trans | 8.32E-42 | Nightingale | S_LDL_FC | NA | T | C | 0.028572 | 0.004418 | 4.20E-10 |
| rs491626 | miR-139-5p | trans | 8.32E-42 | Nightingale | IDL_FC | NA | T | C | 0.038266 | 0.004284 | 1.40E-18 |
| rs491626 | miR-139-5p | trans | 8.32E-42 | Nightingale | S_HDL_L | NA | T | C | 0.026306 | 0.00441 | 1.50E-09 |
| rs491626 | miR-139-5p | trans | 8.32E-42 | Nightingale | S_VLDL_TG | NA | T | C | -0.03813 | 0.004283 | 1.40E-18 |
| rs491626 | miR-139-5p | trans | 8.32E-42 | Nightingale | VLDL_P | NA | T | C | 0.026641 | 0.004401 | 4.40E-10 |
| rs491626 | miR-139-5p | trans | 8.32E-42 | Nightingale | L_LDL_CE | NA | T | C | 0.031579 | 0.004385 | 9.70E-13 |
| rs491626 | miR-139-5p | trans | 8.32E-42 | Nightingale | M_VLDL_P | NA | T | C | 0.044066 | 0.004247 | 5.50E-25 |
| rs491626 | miR-139-5p | trans | 8.32E-42 | Nightingale | L_LDL_P | NA | T | C | 0.029186 | 0.00441 | 6.10E-11 |
| rs491626 | miR-139-5p | trans | 8.32E-42 | Nightingale | M_LDL_L | NA | T | C | 0.033042 | 0.004431 | 1.00E-13 |
| rs491626 | miR-139-5p | trans | 8.32E-42 | Nightingale | M_VLDL_P | NA | T | C | 0.029134 | 0.004418 | 2.00E-11 |
| rs491626 | miR-139-5p | trans | 8.32E-42 | Nightingale | XS_VLDL_P | NA | T | C | 0.036714 | 0.004366 | 1.40E-17 |
| rs491626 | miR-139-5p | trans | 8.32E-42 | Nightingale | M_LDL_C | NA | T | C | 0.033997 | 0.004433 | 2.10E-14 |
| rs491626 | miR-139-5p | trans | 8.32E-42 | Nightingale | Total_FC | NA | T | C | 0.033268 | 0.004323 | 6.70E-15 |
| rs491626 | miR-139-5p | trans | 8.32E-42 | Nightingale | IDL_P | NA | T | C | 0.039995 | 0.004378 | 1.30E-19 |
| rs491626 | miR-139-5p | trans | 8.32E-42 | Nightingale | S_HDL_CE | NA | T | C | 0.028644 | 0.004432 | 1.30E-10 |
| rs491626 | miR-139-5p | trans | 8.32E-42 | Nightingale | IDL_C | NA | T | C | 0.034966 | 0.004274 | 9.30E-16 |
| rs491626 | miR-139-5p | trans | 8.32E-42 | Nightingale | Clinical_LD | NA | T | C | 0.036485 | 0.004399 | 2.70E-16 |
| rs491626 | miR-139-5p | trans | 8.32E-42 | Nightingale | Total_CE | NA | T | C | 0.03176 | 0.004268 | 9.50E-14 |
| rs491626 | miR-139-5p | trans | 8.32E-42 | Nightingale | M_LDL_C_ | NA | T | C | 0.031163 | 0.004428 | 2.90E-12 |
| rs491626 | miR-139-5p | trans | 8.32E-42 | Nightingale | S_VLDL_PL | NA | T | C | 0.032785 | 0.004417 | 4.20E-14 |
| rs491626 | miR-139-5p | trans | 8.32E-42 | Nightingale | XS_VLDL_F | NA | T | C | 0.03143 | 0.004393 | 2.40E-12 |
| rs491626 | miR-139-5p | trans | 8.32E-42 | Nightingale | XS_VLDL_P | NA | T | C | 0.032127 | 0.004363 | 5.30E-14 |
| rs491626 | miR-139-5p | trans | 8.32E-42 | Nightingale | XXL_VLDL_ | NA | T | C | 0.025011 | 0.004454 | 1.50E-09 |
| rs491626 | miR-139-5p | trans | 8.32E-42 | Nightingale | S_HDL_PL | NA | T | C | 0.023794 | 0.004387 | 3.20E-08 |
| rs491626 | miR-139-5p | trans | 8.32E-42 | Nightingale | LDL_C | NA | T | C | 0.033205 | 0.004406 | 8.70E-14 |
| rs491626 | miR-139-5p | trans | 8.32E-42 | Nightingale | XL_VLDL_P | NA | T | C | 0.038844 | 0.004432 | 5.20E-19 |
| rs491626 | miR-139-5p | trans | 8.32E-42 | Nightingale | VLDL_FC | NA | T | C | 0.024164 | 0.004392 | 1.10E-08 |
| rs491626 | miR-139-5p | trans | 8.32E-42 | Nightingale | S_HDL_FC | NA | T | C | 0.023468 | 0.004344 | 4.50E-08 |
| rs491626 | miR-139-5p | trans | 8.32E-42 | Nightingale | S_VLDL_C_ | NA | T | C | 0.041891 | 0.004306 | 5.20E-22 |
| rs491626 | miR-139-5p | trans | 8.32E-42 | Nightingale | M_VLDL_C | NA | T | C | 0.033636 | 0.004157 | 1.90E-15 |
| rs491626 | miR-139-5p | trans | 8.32E-42 | Nightingale | M_LDL_PL | NA | T | C | 0.032477 | 0.004431 | 2.60E-13 |
| rs491626 | miR-139-5p | trans | 8.32E-42 | Nightingale | Total_C | NA | T | C | 0.032387 | 0.004284 | 3.30E-14 |
| rs491626 | miR-139-5p | trans | 8.32E-42 | Nightingale | L_VLDL_C_ | NA | T | C | 0.052205 | 0.004322 | 3.90E-34 |
| rs491626 | miR-139-5p | trans | 8.32E-42 | Nightingale | L_VLDL_TG | NA | T | C | -0.05115 | 0.004404 | 5.30E-32 |
| rs491626 | miR-139-5p | trans | 8.32E-42 | Nightingale | S_LDL_P | NA | T | C | 0.029597 | 0.004429 | 3.20E-11 |
| rs491626 | miR-139-5p | trans | 8.32E-42 | Nightingale | M_VLDL_C | NA | T | C | 0.036214 | 0.004168 | 1.20E-17 |
| rs491626 | miR-139-5p | trans | 8.32E-42 | Nightingale | VLDL_C | NA | T | C | 0.031308 | 0.004419 | 4.80E-13 |
| rs491626 | miR-139-5p | trans | 8.32E-42 | Nightingale | XXL_VLDL_ | NA | T | C | 0.030859 | 0.004406 | 1.10E-12 |

| | | | | | | | | | | | |
|---|---|---|---|---|---|---|---|---|---|---|---|
| rs491626 | miR-139-5p | trans | 8.32E-42 | Nightingale | L_LDL_PL | NA | T | C | 0.038001 | 0.004391 | 8.90E-18 |
| rs491626 | miR-139-5p | trans | 8.32E-42 | Nightingale | S_VLDL_P | NA | T | C | 0.023472 | 0.004369 | 1.60E-08 |
| rs491626 | miR-139-5p | trans | 8.32E-42 | Nightingale | Total_PL | NA | T | C | 0.024943 | 0.004214 | 1.00E-09 |
| rs491626 | miR-139-5p | trans | 8.32E-42 | Nightingale | IDL_FC_pct | NA | T | C | 0.025773 | 0.004391 | 1.10E-08 |
| rs491626 | miR-139-5p | trans | 8.32E-42 | Nightingale | IDL_CE | NA | T | C | 0.033518 | 0.004272 | 1.30E-14 |
| rs491626 | miR-139-5p | trans | 8.32E-42 | Nightingale | L_LDL_L | NA | T | C | 0.033562 | 0.004382 | 2.90E-14 |
| rs491626 | miR-139-5p | trans | 8.32E-42 | Nightingale | XL_VLDL_C | NA | T | C | 0.037359 | 0.004245 | 3.10E-18 |
| rs491626 | miR-139-5p | trans | 8.32E-42 | Nightingale | XS_VLDL_C | NA | T | C | 0.038004 | 0.004295 | 1.30E-18 |
| rs491626 | miR-139-5p | trans | 8.32E-42 | Nightingale | S_VLDL_PL | NA | T | C | 0.027741 | 0.004237 | 1.80E-10 |
| rs491626 | miR-139-5p | trans | 8.32E-42 | Nightingale | XXL_VLDL_ | NA | T | C | 0.034842 | 0.004434 | 2.90E-15 |
| rs491626 | miR-139-5p | trans | 8.32E-42 | Nightingale | XS_VLDL_F | NA | T | C | 0.037079 | 0.004348 | 6.40E-18 |
| rs491626 | miR-139-5p | trans | 8.32E-42 | Nightingale | GlycA | NA | T | C | 0.028021 | 0.004406 | 1.40E-10 |
| rs491626 | miR-139-5p | trans | 8.32E-42 | Nightingale | S_LDL_CE | NA | T | C | 0.030557 | 0.004427 | 5.80E-12 |
| rs491626 | miR-139-5p | trans | 8.32E-42 | Nightingale | LDL_FC | NA | T | C | 0.033643 | 0.004378 | 5.20E-14 |
| rs491626 | miR-139-5p | trans | 8.32E-42 | Nightingale | Total_L | NA | T | C | 0.026362 | 0.004353 | 4.50E-10 |
| rs491626 | miR-139-5p | trans | 8.32E-42 | Nightingale | L_LDL_FC | NA | T | C | 0.033636 | 0.004345 | 2.80E-14 |
| rs491626 | miR-139-5p | trans | 8.32E-42 | Nightingale | LA | NA | T | C | 0.024924 | 0.004339 | 3.60E-09 |
| rs491626 | miR-139-5p | trans | 8.32E-42 | Nightingale | PUFA | NA | T | C | 0.023737 | 0.004316 | 1.40E-08 |
| rs491626 | miR-139-5p | trans | 8.32E-42 | Nightingale | XS_VLDL_C | NA | T | C | 0.037809 | 0.00427 | 2.10E-18 |
| rs491626 | miR-139-5p | trans | 8.32E-42 | Nightingale | S_VLDL_C | NA | T | C | 0.03838 | 0.004424 | 1.80E-18 |
| rs491626 | miR-139-5p | trans | 8.32E-42 | Nightingale | S_VLDL_L | NA | T | C | 0.024675 | 0.004382 | 3.70E-09 |
| rs491626 | miR-139-5p | trans | 8.32E-42 | Nightingale | Sphingomy | NA | T | C | 0.02903 | 0.004137 | 6.60E-12 |
| rs491626 | miR-139-5p | trans | 8.32E-42 | Nightingale | Omega_6 | NA | T | C | 0.025911 | 0.004321 | 5.60E-10 |
| rs491626 | miR-139-5p | trans | 8.32E-42 | Nightingale | M_VLDL_F | NA | T | C | 0.042073 | 0.00421 | 3.40E-23 |
| rs491626 | miR-139-5p | trans | 8.32E-42 | Nightingale | M_LDL_FC | NA | T | C | 0.034358 | 0.004419 | 1.90E-14 |
| rs491626 | miR-139-5p | trans | 8.32E-42 | Nightingale | M_VLDL_P | NA | T | C | 0.028301 | 0.004414 | 6.30E-11 |
| rs491626 | miR-139-5p | trans | 8.32E-42 | Nightingale | M_VLDL_T | NA | T | C | -0.03857 | 0.004181 | 6.70E-20 |
| rs491626 | miR-139-5p | trans | 8.32E-42 | Nightingale | S_VLDL_CE | NA | T | C | 0.047036 | 0.004365 | 1.20E-26 |
| rs491626 | miR-139-5p | trans | 8.32E-42 | Nightingale | Remnant_( | NA | T | C | 0.036878 | 0.004396 | 3.60E-17 |
| rs491626 | miR-139-5p | trans | 8.32E-42 | Nightingale | L_VLDL_FC | NA | T | C | 0.048596 | 0.004382 | 4.50E-29 |
| rs491626 | miR-139-5p | trans | 8.32E-42 | Nightingale | Citrate | NA | T | C | 0.026826 | 0.004396 | 7.40E-10 |
| rs491626 | miR-139-5p | trans | 8.32E-42 | Nightingale | M_VLDL_C | NA | T | C | 0.040852 | 0.004385 | 2.30E-20 |
| rs491626 | miR-139-5p | trans | 8.32E-42 | Nightingale | ApoB_by_A | NA | T | C | 0.025049 | 0.004358 | 1.30E-08 |
| rs491626 | miR-139-5p | trans | 8.32E-42 | Nightingale | IDL_L | NA | T | C | 0.035282 | 0.004278 | 4.70E-16 |
| rs491626 | miR-139-5p | trans | 8.32E-42 | Nightingale | XL_VLDL_T | NA | T | C | -0.04393 | 0.004289 | 1.70E-24 |
| rs491626 | miR-139-5p | trans | 8.32E-42 | Nightingale | XL_VLDL_F | NA | T | C | 0.037203 | 0.00429 | 2.00E-18 |
| rs491626 | miR-139-5p | trans | 8.32E-42 | Nightingale | IDL_PL | NA | T | C | 0.035356 | 0.00427 | 3.30E-16 |
| rs491626 | miR-139-5p | trans | 8.32E-42 | Nightingale | M_VLDL_F | NA | T | C | 0.033378 | 0.004424 | 3.10E-14 |
| rs491626 | miR-139-5p | trans | 8.32E-42 | Nightingale | LDL_PL | NA | T | C | 0.034999 | 0.004415 | 4.70E-15 |
| rs491626 | miR-139-5p | trans | 8.32E-42 | Nightingale | S_VLDL_CE | NA | T | C | 0.038189 | 0.004419 | 1.60E-18 |
| rs491626 | miR-139-5p | trans | 8.32E-42 | Nightingale | XL_VLDL_C | NA | T | C | 0.03823 | 0.004243 | 4.60E-19 |
| rs491626 | miR-139-5p | trans | 8.32E-42 | Nightingale | LDL_L | NA | T | C | 0.033316 | 0.00441 | 7.30E-14 |
| rs491626 | miR-139-5p | trans | 8.32E-42 | Nightingale | M_LDL_P | NA | T | C | 0.0359 | 0.004437 | 6.80E-16 |
| rs491626 | miR-139-5p | trans | 8.32E-42 | Nightingale | Cholines | NA | T | C | 0.022312 | 0.004161 | 4.70E-08 |
| rs491626 | miR-139-5p | trans | 8.32E-42 | Nightingale | S_VLDL_FC | NA | T | C | 0.029359 | 0.004228 | 1.10E-11 |
| rs491626 | miR-139-5p | trans | 8.32E-42 | Nightingale | S_LDL_L | NA | T | C | 0.02775 | 0.004424 | 8.20E-10 |
| rs491626 | miR-139-5p | trans | 8.32E-42 | Nightingale | S_HDL_P | NA | T | C | 0.030621 | 0.004425 | 5.00E-12 |
| rs491626 | miR-139-5p | trans | 8.32E-42 | Nightingale | XXL_VLDL_ | NA | T | C | -0.03574 | 0.004463 | 2.10E-16 |
| rs491626 | miR-139-5p | trans | 8.32E-42 | Nightingale | L_VLDL_CE | NA | T | C | 0.047349 | 0.004234 | 2.40E-29 |
| rs491626 | miR-139-5p | trans | 8.32E-42 | Nightingale | LDL_CE | NA | T | C | 0.032565 | 0.004413 | 2.20E-13 |
| rs491626 | miR-139-5p | trans | 8.32E-42 | Nightingale | XXL_VLDL_ | NA | T | C | 0.033133 | 0.004459 | 8.80E-14 |
| rs491626 | miR-139-5p | trans | 8.32E-42 | Nightingale | S_LDL_CE_ | NA | T | C | 0.030951 | 0.004417 | 4.80E-14 |
| rs491626 | miR-139-5p | trans | 8.32E-42 | Nightingale | S_LDL_C | NA | T | C | 0.030952 | 0.004429 | 5.50E-12 |
| rs491626 | miR-139-5p | trans | 8.32E-42 | Nightingale | non_HDL_( | NA | T | C | 0.035708 | 0.004401 | 5.00E-16 |
| rs491626 | miR-139-5p | trans | 8.32E-42 | Nightingale | M_LDL_CE | NA | T | C | 0.032982 | 0.004431 | 9.50E-14 |
| rs491626 | miR-139-5p | trans | 8.32E-42 | Nightingale | S_HDL_C | NA | T | C | 0.028422 | 0.004417 | 1.70E-10 |
| rs491626 | miR-139-5p | trans | 8.32E-42 | Nightingale | S_LDL_PL_ | NA | T | C | -0.03007 | 0.004417 | 2.60E-13 |
| rs491626 | miR-139-5p | trans | 8.32E-42 | Nightingale | LDL_P | NA | T | C | 0.031809 | 0.004422 | 1.00E-12 |
| rs491626 | miR-139-5p | trans | 8.32E-42 | Nightingale | ApoB | NA | T | C | 0.033947 | 0.004419 | 2.10E-14 |
| rs491626 | miR-139-5p | trans | 8.32E-42 | Nightingale | S_VLDL_FC | NA | T | C | 0.037912 | 0.004426 | 7.30E-18 |
| rs491626 | miR-139-5p | trans | 8.32E-42 | Nightingale | M_LDL_TG | NA | T | C | -0.02762 | 0.00437 | 3.20E-10 |
| rs491626 | miR-139-5p | trans | 8.32E-42 | Nightingale | XL_HDL_PL | NA | T | C | -0.02274 | 0.004065 | 1.20E-08 |
| rs491626 | miR-139-5p | trans | 8.32E-42 | Nightingale | M_VLDL_C | NA | T | C | 0.038718 | 0.004413 | 1.90E-18 |

| | | | | | | | | | | | |
|---|---|---|---|---|---|---|---|---|---|---|---|
| rs491626 | miR-139-5| trans | 8.32E-42 | Nightingale | S_LDL_C_p | NA | T | C | 0.045367 | 0.004442 | 5.40E-25 |
| rs491626 | miR-139-5| trans | 8.32E-42 | Nightingale | XS_VLDL_L | NA | T | C | 0.03383 | 0.00435 | 2.00E-15 |
| rs491626 | miR-139-5| trans | 8.32E-42 | Nightingale | L_LDL_C | NA | T | C | 0.032332 | 0.004375 | 2.60E-13 |
| rs491626 | miR-10b-5| trans | 3.14E-35 | Nightingale | VLDL_CE | NA | T | C | 0.035794 | 0.004426 | 3.00E-16 |
| rs491626 | miR-10b-5| trans | 3.14E-35 | Nightingale | S_LDL_FC | NA | T | C | 0.028572 | 0.004418 | 4.20E-10 |
| rs491626 | miR-10b-5| trans | 3.14E-35 | Nightingale | IDL_FC | NA | T | C | 0.038266 | 0.004284 | 1.40E-18 |
| rs491626 | miR-10b-5| trans | 3.14E-35 | Nightingale | S_HDL_L | NA | T | C | 0.026306 | 0.00441 | 1.50E-09 |
| rs491626 | miR-10b-5| trans | 3.14E-35 | Nightingale | S_VLDL_TG | NA | T | C | -0.03813 | 0.004283 | 1.40E-18 |
| rs491626 | miR-10b-5| trans | 3.14E-35 | Nightingale | VLDL_P | NA | T | C | 0.026641 | 0.004401 | 4.40E-10 |
| rs491626 | miR-10b-5| trans | 3.14E-35 | Nightingale | L_LDL_CE | NA | T | C | 0.031579 | 0.004385 | 9.70E-13 |
| rs491626 | miR-10b-5| trans | 3.14E-35 | Nightingale | M_VLDL_P | NA | T | C | 0.044066 | 0.004247 | 5.50E-25 |
| rs491626 | miR-10b-5| trans | 3.14E-35 | Nightingale | L_LDL_P | NA | T | C | 0.029186 | 0.00441 | 6.10E-11 |
| rs491626 | miR-10b-5| trans | 3.14E-35 | Nightingale | M_LDL_L | NA | T | C | 0.033042 | 0.004431 | 1.00E-13 |
| rs491626 | miR-10b-5| trans | 3.14E-35 | Nightingale | M_VLDL_P | NA | T | C | 0.029134 | 0.004418 | 2.00E-11 |
| rs491626 | miR-10b-5| trans | 3.14E-35 | Nightingale | XS_VLDL_P | NA | T | C | 0.036714 | 0.004366 | 1.40E-17 |
| rs491626 | miR-10b-5| trans | 3.14E-35 | Nightingale | M_LDL_C | NA | T | C | 0.033997 | 0.004433 | 2.10E-14 |
| rs491626 | miR-10b-5| trans | 3.14E-35 | Nightingale | Total_FC | NA | T | C | 0.033268 | 0.004323 | 6.70E-15 |
| rs491626 | miR-10b-5| trans | 3.14E-35 | Nightingale | IDL_P | NA | T | C | 0.039995 | 0.004378 | 1.30E-19 |
| rs491626 | miR-10b-5| trans | 3.14E-35 | Nightingale | S_HDL_CE | NA | T | C | 0.028644 | 0.004432 | 1.30E-10 |
| rs491626 | miR-10b-5| trans | 3.14E-35 | Nightingale | IDL_C | NA | T | C | 0.034966 | 0.004274 | 9.30E-16 |
| rs491626 | miR-10b-5| trans | 3.14E-35 | Nightingale | Clinical_LD | NA | T | C | 0.036485 | 0.004399 | 2.70E-16 |
| rs491626 | miR-10b-5| trans | 3.14E-35 | Nightingale | Total_CE | NA | T | C | 0.03176 | 0.004268 | 9.50E-14 |
| rs491626 | miR-10b-5| trans | 3.14E-35 | Nightingale | M_LDL_C_| | NA | T | C | 0.031163 | 0.004428 | 2.90E-12 |
| rs491626 | miR-10b-5| trans | 3.14E-35 | Nightingale | S_VLDL_PL | NA | T | C | 0.032785 | 0.004417 | 4.20E-14 |
| rs491626 | miR-10b-5| trans | 3.14E-35 | Nightingale | XS_VLDL_F | NA | T | C | 0.03143 | 0.004393 | 2.40E-12 |
| rs491626 | miR-10b-5| trans | 3.14E-35 | Nightingale | XS_VLDL_P | NA | T | C | 0.032127 | 0.004363 | 5.30E-14 |
| rs491626 | miR-10b-5| trans | 3.14E-35 | Nightingale | XXL_VLDL_ | NA | T | C | 0.025011 | 0.004454 | 1.50E-09 |
| rs491626 | miR-10b-5| trans | 3.14E-35 | Nightingale | S_HDL_PL | NA | T | C | 0.023794 | 0.004387 | 3.20E-08 |
| rs491626 | miR-10b-5| trans | 3.14E-35 | Nightingale | LDL_C | NA | T | C | 0.033205 | 0.004406 | 8.70E-14 |
| rs491626 | miR-10b-5| trans | 3.14E-35 | Nightingale | XL_VLDL_P | NA | T | C | 0.038844 | 0.004432 | 5.20E-19 |
| rs491626 | miR-10b-5| trans | 3.14E-35 | Nightingale | VLDL_FC | NA | T | C | 0.024164 | 0.004392 | 1.10E-08 |
| rs491626 | miR-10b-5| trans | 3.14E-35 | Nightingale | S_HDL_FC | NA | T | C | 0.023468 | 0.004344 | 4.50E-08 |
| rs491626 | miR-10b-5| trans | 3.14E-35 | Nightingale | S_VLDL_C_ | NA | T | C | 0.041891 | 0.004306 | 5.20E-22 |
| rs491626 | miR-10b-5| trans | 3.14E-35 | Nightingale | M_VLDL_C | NA | T | C | 0.033636 | 0.004157 | 1.90E-15 |
| rs491626 | miR-10b-5| trans | 3.14E-35 | Nightingale | M_LDL_PL | NA | T | C | 0.032477 | 0.004431 | 2.60E-13 |
| rs491626 | miR-10b-5| trans | 3.14E-35 | Nightingale | Total_C | NA | T | C | 0.032387 | 0.004284 | 3.30E-14 |
| rs491626 | miR-10b-5| trans | 3.14E-35 | Nightingale | L_VLDL_C_ | NA | T | C | 0.052205 | 0.004322 | 3.90E-34 |
| rs491626 | miR-10b-5| trans | 3.14E-35 | Nightingale | L_VLDL_TG | NA | T | C | -0.05115 | 0.004404 | 5.30E-32 |
| rs491626 | miR-10b-5| trans | 3.14E-35 | Nightingale | S_LDL_P | NA | T | C | 0.029597 | 0.004429 | 3.20E-11 |
| rs491626 | miR-10b-5| trans | 3.14E-35 | Nightingale | M_VLDL_C | NA | T | C | 0.036214 | 0.004168 | 1.20E-17 |
| rs491626 | miR-10b-5| trans | 3.14E-35 | Nightingale | VLDL_C | NA | T | C | 0.031308 | 0.004419 | 4.80E-13 |
| rs491626 | miR-10b-5| trans | 3.14E-35 | Nightingale | XXL_VLDL_ | NA | T | C | 0.030859 | 0.004406 | 1.10E-12 |
| rs491626 | miR-10b-5| trans | 3.14E-35 | Nightingale | L_LDL_PL | NA | T | C | 0.038001 | 0.004391 | 8.90E-18 |
| rs491626 | miR-10b-5| trans | 3.14E-35 | Nightingale | S_VLDL_P | NA | T | C | 0.023472 | 0.004369 | 1.60E-08 |
| rs491626 | miR-10b-5| trans | 3.14E-35 | Nightingale | Total_PL | NA | T | C | 0.024943 | 0.004214 | 1.00E-09 |
| rs491626 | miR-10b-5| trans | 3.14E-35 | Nightingale | IDL_FC_pct | NA | T | C | 0.025773 | 0.004391 | 1.10E-08 |
| rs491626 | miR-10b-5| trans | 3.14E-35 | Nightingale | IDL_CE | NA | T | C | 0.033518 | 0.004272 | 1.30E-14 |
| rs491626 | miR-10b-5| trans | 3.14E-35 | Nightingale | L_LDL_L | NA | T | C | 0.033562 | 0.004382 | 2.90E-14 |
| rs491626 | miR-10b-5| trans | 3.14E-35 | Nightingale | XL_VLDL_C | NA | T | C | 0.037359 | 0.004245 | 3.10E-18 |
| rs491626 | miR-10b-5| trans | 3.14E-35 | Nightingale | XS_VLDL_C | NA | T | C | 0.038004 | 0.004295 | 1.30E-18 |
| rs491626 | miR-10b-5| trans | 3.14E-35 | Nightingale | S_VLDL_PL | NA | T | C | 0.027741 | 0.004237 | 1.80E-10 |
| rs491626 | miR-10b-5| trans | 3.14E-35 | Nightingale | XXL_VLDL_ | NA | T | C | 0.034842 | 0.004434 | 2.90E-15 |
| rs491626 | miR-10b-5| trans | 3.14E-35 | Nightingale | XS_VLDL_F | NA | T | C | 0.037079 | 0.004348 | 6.40E-18 |
| rs491626 | miR-10b-5| trans | 3.14E-35 | Nightingale | GlycA | NA | T | C | 0.028021 | 0.004406 | 1.40E-10 |
| rs491626 | miR-10b-5| trans | 3.14E-35 | Nightingale | S_LDL_CE | NA | T | C | 0.030557 | 0.004427 | 5.80E-12 |
| rs491626 | miR-10b-5| trans | 3.14E-35 | Nightingale | LDL_FC | NA | T | C | 0.033643 | 0.004378 | 5.20E-14 |
| rs491626 | miR-10b-5| trans | 3.14E-35 | Nightingale | Total_L | NA | T | C | 0.026362 | 0.004353 | 4.50E-10 |
| rs491626 | miR-10b-5| trans | 3.14E-35 | Nightingale | L_LDL_FC | NA | T | C | 0.033636 | 0.004345 | 2.80E-14 |
| rs491626 | miR-10b-5| trans | 3.14E-35 | Nightingale | LA | NA | T | C | 0.024924 | 0.004339 | 3.60E-09 |
| rs491626 | miR-10b-5| trans | 3.14E-35 | Nightingale | PUFA | NA | T | C | 0.023737 | 0.004316 | 1.40E-08 |
| rs491626 | miR-10b-5| trans | 3.14E-35 | Nightingale | XS_VLDL_C | NA | T | C | 0.037809 | 0.00427 | 2.10E-18 |
| rs491626 | miR-10b-5| trans | 3.14E-35 | Nightingale | S_VLDL_C | NA | T | C | 0.03838 | 0.004424 | 1.80E-18 |
| rs491626 | miR-10b-5| trans | 3.14E-35 | Nightingale | S_VLDL_L | NA | T | C | 0.024675 | 0.004382 | 3.70E-09 |

| | | | | | | | | | | | |
|---|---|---|---|---|---|---|---|---|---|---|---|
| rs491626 | miR-10b-5p | trans | 3.14E-35 | Nightingale | Sphingomy | NA | T | C | 0.02903 | 0.004137 | 6.60E-12 |
| rs491626 | miR-10b-5p | trans | 3.14E-35 | Nightingale | Omega_6 | NA | T | C | 0.025911 | 0.004321 | 5.60E-10 |
| rs491626 | miR-10b-5p | trans | 3.14E-35 | Nightingale | M_VLDL_F | NA | T | C | 0.042073 | 0.00421 | 3.40E-23 |
| rs491626 | miR-10b-5p | trans | 3.14E-35 | Nightingale | M_LDL_FC | NA | T | C | 0.034358 | 0.004419 | 1.90E-14 |
| rs491626 | miR-10b-5p | trans | 3.14E-35 | Nightingale | M_VLDL_P | NA | T | C | 0.028301 | 0.004414 | 6.30E-11 |
| rs491626 | miR-10b-5p | trans | 3.14E-35 | Nightingale | M_VLDL_T | NA | T | C | -0.03857 | 0.004181 | 6.70E-20 |
| rs491626 | miR-10b-5p | trans | 3.14E-35 | Nightingale | S_VLDL_CE | NA | T | C | 0.047036 | 0.004365 | 1.20E-26 |
| rs491626 | miR-10b-5p | trans | 3.14E-35 | Nightingale | Remnant_( | NA | T | C | 0.036878 | 0.004396 | 3.60E-17 |
| rs491626 | miR-10b-5p | trans | 3.14E-35 | Nightingale | L_VLDL_FC | NA | T | C | 0.048596 | 0.004382 | 4.50E-29 |
| rs491626 | miR-10b-5p | trans | 3.14E-35 | Nightingale | Citrate | NA | T | C | 0.026826 | 0.004396 | 7.40E-10 |
| rs491626 | miR-10b-5p | trans | 3.14E-35 | Nightingale | M_VLDL_C | NA | T | C | 0.040852 | 0.004385 | 2.30E-20 |
| rs491626 | miR-10b-5p | trans | 3.14E-35 | Nightingale | ApoB_by_A | NA | T | C | 0.025049 | 0.004358 | 1.30E-08 |
| rs491626 | miR-10b-5p | trans | 3.14E-35 | Nightingale | IDL_L | NA | T | C | 0.035282 | 0.004278 | 4.70E-16 |
| rs491626 | miR-10b-5p | trans | 3.14E-35 | Nightingale | XL_VLDL_T | NA | T | C | -0.04393 | 0.004289 | 1.70E-24 |
| rs491626 | miR-10b-5p | trans | 3.14E-35 | Nightingale | XL_VLDL_F | NA | T | C | 0.037203 | 0.00429 | 2.00E-18 |
| rs491626 | miR-10b-5p | trans | 3.14E-35 | Nightingale | IDL_PL | NA | T | C | 0.035356 | 0.00427 | 3.30E-16 |
| rs491626 | miR-10b-5p | trans | 3.14E-35 | Nightingale | M_VLDL_F | NA | T | C | 0.033378 | 0.004424 | 3.10E-14 |
| rs491626 | miR-10b-5p | trans | 3.14E-35 | Nightingale | LDL_PL | NA | T | C | 0.034999 | 0.004415 | 4.70E-15 |
| rs491626 | miR-10b-5p | trans | 3.14E-35 | Nightingale | S_VLDL_CE | NA | T | C | 0.038189 | 0.004419 | 1.60E-18 |
| rs491626 | miR-10b-5p | trans | 3.14E-35 | Nightingale | XL_VLDL_C | NA | T | C | 0.03823 | 0.004243 | 4.60E-19 |
| rs491626 | miR-10b-5p | trans | 3.14E-35 | Nightingale | LDL_L | NA | T | C | 0.033316 | 0.00441 | 7.30E-14 |
| rs491626 | miR-10b-5p | trans | 3.14E-35 | Nightingale | M_LDL_P | NA | T | C | 0.0359 | 0.004437 | 6.80E-16 |
| rs491626 | miR-10b-5p | trans | 3.14E-35 | Nightingale | Cholines | NA | T | C | 0.022312 | 0.004161 | 4.70E-08 |
| rs491626 | miR-10b-5p | trans | 3.14E-35 | Nightingale | S_VLDL_FC | NA | T | C | 0.029359 | 0.004228 | 1.10E-11 |
| rs491626 | miR-10b-5p | trans | 3.14E-35 | Nightingale | S_LDL_L | NA | T | C | 0.02775 | 0.004424 | 8.20E-10 |
| rs491626 | miR-10b-5p | trans | 3.14E-35 | Nightingale | S_HDL_P | NA | T | C | 0.030621 | 0.004425 | 5.00E-12 |
| rs491626 | miR-10b-5p | trans | 3.14E-35 | Nightingale | XXL_VLDL_ | NA | T | C | -0.03574 | 0.004463 | 2.10E-16 |
| rs491626 | miR-10b-5p | trans | 3.14E-35 | Nightingale | L_VLDL_CE | NA | T | C | 0.047349 | 0.004234 | 2.40E-29 |
| rs491626 | miR-10b-5p | trans | 3.14E-35 | Nightingale | LDL_CE | NA | T | C | 0.032565 | 0.004413 | 2.20E-13 |
| rs491626 | miR-10b-5p | trans | 3.14E-35 | Nightingale | XXL_VLDL_ | NA | T | C | 0.033133 | 0.004459 | 8.80E-14 |
| rs491626 | miR-10b-5p | trans | 3.14E-35 | Nightingale | S_LDL_CE_ | NA | T | C | 0.030951 | 0.004417 | 4.80E-14 |
| rs491626 | miR-10b-5p | trans | 3.14E-35 | Nightingale | S_LDL_C | NA | T | C | 0.030952 | 0.004429 | 5.50E-12 |
| rs491626 | miR-10b-5p | trans | 3.14E-35 | Nightingale | non_HDL_( | NA | T | C | 0.035708 | 0.004401 | 5.00E-16 |
| rs491626 | miR-10b-5p | trans | 3.14E-35 | Nightingale | M_LDL_CE | NA | T | C | 0.032982 | 0.004431 | 9.50E-14 |
| rs491626 | miR-10b-5p | trans | 3.14E-35 | Nightingale | S_HDL_C | NA | T | C | 0.028422 | 0.004417 | 1.70E-10 |
| rs491626 | miR-10b-5p | trans | 3.14E-35 | Nightingale | S_LDL_PL_ | NA | T | C | -0.03007 | 0.004417 | 2.60E-13 |
| rs491626 | miR-10b-5p | trans | 3.14E-35 | Nightingale | LDL_P | NA | T | C | 0.031809 | 0.004422 | 1.00E-12 |
| rs491626 | miR-10b-5p | trans | 3.14E-35 | Nightingale | ApoB | NA | T | C | 0.033947 | 0.004419 | 2.10E-14 |
| rs491626 | miR-10b-5p | trans | 3.14E-35 | Nightingale | S_VLDL_FC | NA | T | C | 0.037912 | 0.004426 | 7.30E-18 |
| rs491626 | miR-10b-5p | trans | 3.14E-35 | Nightingale | M_LDL_TG | NA | T | C | -0.02762 | 0.00437 | 3.20E-10 |
| rs491626 | miR-10b-5p | trans | 3.14E-35 | Nightingale | XL_HDL_PL | NA | T | C | -0.02274 | 0.004065 | 1.20E-08 |
| rs491626 | miR-10b-5p | trans | 3.14E-35 | Nightingale | M_VLDL_C | NA | T | C | 0.038718 | 0.004413 | 1.90E-18 |
| rs491626 | miR-10b-5p | trans | 3.14E-35 | Nightingale | S_LDL_C_p | NA | T | C | 0.045367 | 0.004442 | 5.40E-25 |
| rs491626 | miR-10b-5p | trans | 3.14E-35 | Nightingale | XS_VLDL_L | NA | T | C | 0.03383 | 0.00435 | 2.00E-15 |
| rs491626 | miR-10b-5p | trans | 3.14E-35 | Nightingale | L_LDL_C | NA | T | C | 0.032332 | 0.004375 | 2.60E-13 |
| rs491626 | miR-126-3p | trans | 2.10E-39 | Nightingale | VLDL_CE | NA | T | C | 0.035794 | 0.004426 | 3.00E-16 |
| rs491626 | miR-126-3p | trans | 2.10E-39 | Nightingale | S_LDL_FC | NA | T | C | 0.028572 | 0.004418 | 4.20E-10 |
| rs491626 | miR-126-3p | trans | 2.10E-39 | Nightingale | IDL_FC | NA | T | C | 0.038266 | 0.004284 | 1.40E-18 |
| rs491626 | miR-126-3p | trans | 2.10E-39 | Nightingale | S_HDL_L | NA | T | C | 0.026306 | 0.00441 | 1.50E-09 |
| rs491626 | miR-126-3p | trans | 2.10E-39 | Nightingale | S_VLDL_TG | NA | T | C | -0.03813 | 0.004283 | 1.40E-18 |
| rs491626 | miR-126-3p | trans | 2.10E-39 | Nightingale | VLDL_P | NA | T | C | 0.026641 | 0.004401 | 4.40E-10 |
| rs491626 | miR-126-3p | trans | 2.10E-39 | Nightingale | L_LDL_CE | NA | T | C | 0.031579 | 0.004385 | 9.70E-13 |
| rs491626 | miR-126-3p | trans | 2.10E-39 | Nightingale | M_VLDL_P | NA | T | C | 0.044066 | 0.004247 | 5.50E-25 |
| rs491626 | miR-126-3p | trans | 2.10E-39 | Nightingale | L_LDL_P | NA | T | C | 0.029186 | 0.00441 | 6.10E-11 |
| rs491626 | miR-126-3p | trans | 2.10E-39 | Nightingale | M_LDL_L | NA | T | C | 0.033042 | 0.004431 | 1.00E-13 |
| rs491626 | miR-126-3p | trans | 2.10E-39 | Nightingale | M_VLDL_P | NA | T | C | 0.029134 | 0.004418 | 2.00E-11 |
| rs491626 | miR-126-3p | trans | 2.10E-39 | Nightingale | XS_VLDL_P | NA | T | C | 0.036714 | 0.004366 | 1.40E-17 |
| rs491626 | miR-126-3p | trans | 2.10E-39 | Nightingale | M_LDL_C | NA | T | C | 0.033997 | 0.004433 | 2.10E-14 |
| rs491626 | miR-126-3p | trans | 2.10E-39 | Nightingale | Total_FC | NA | T | C | 0.033268 | 0.004323 | 6.70E-15 |
| rs491626 | miR-126-3p | trans | 2.10E-39 | Nightingale | IDL_P | NA | T | C | 0.039995 | 0.004378 | 1.30E-19 |
| rs491626 | miR-126-3p | trans | 2.10E-39 | Nightingale | S_HDL_CE | NA | T | C | 0.028644 | 0.004432 | 1.30E-10 |
| rs491626 | miR-126-3p | trans | 2.10E-39 | Nightingale | IDL_C | NA | T | C | 0.034966 | 0.004274 | 9.30E-16 |
| rs491626 | miR-126-3p | trans | 2.10E-39 | Nightingale | Clinical_LD | NA | T | C | 0.036485 | 0.004399 | 2.70E-16 |

| | | | | | | | | | | | |
|---|---|---|---|---|---|---|---|---|---|---|---|
| rs491626 | miR-126-3p | trans | 2.10E-39 | Nightingale | Total_CE | NA | T | C | 0.03176 | 0.004268 | 9.50E-14 |
| rs491626 | miR-126-3p | trans | 2.10E-39 | Nightingale | M_LDL_C_ | NA | T | C | 0.031163 | 0.004428 | 2.90E-12 |
| rs491626 | miR-126-3p | trans | 2.10E-39 | Nightingale | S_VLDL_PL | NA | T | C | 0.032785 | 0.004417 | 4.20E-14 |
| rs491626 | miR-126-3p | trans | 2.10E-39 | Nightingale | XS_VLDL_F | NA | T | C | 0.03143 | 0.004393 | 2.40E-12 |
| rs491626 | miR-126-3p | trans | 2.10E-39 | Nightingale | XS_VLDL_P | NA | T | C | 0.032127 | 0.004363 | 5.30E-14 |
| rs491626 | miR-126-3p | trans | 2.10E-39 | Nightingale | XXL_VLDL_ | NA | T | C | 0.025011 | 0.004454 | 1.50E-09 |
| rs491626 | miR-126-3p | trans | 2.10E-39 | Nightingale | S_HDL_PL | NA | T | C | 0.023794 | 0.004387 | 3.20E-08 |
| rs491626 | miR-126-3p | trans | 2.10E-39 | Nightingale | LDL_C | NA | T | C | 0.033205 | 0.004406 | 8.70E-14 |
| rs491626 | miR-126-3p | trans | 2.10E-39 | Nightingale | XL_VLDL_P | NA | T | C | 0.038844 | 0.004432 | 5.20E-19 |
| rs491626 | miR-126-3p | trans | 2.10E-39 | Nightingale | VLDL_FC | NA | T | C | 0.024164 | 0.004392 | 1.10E-08 |
| rs491626 | miR-126-3p | trans | 2.10E-39 | Nightingale | S_HDL_FC | NA | T | C | 0.023468 | 0.004344 | 4.50E-08 |
| rs491626 | miR-126-3p | trans | 2.10E-39 | Nightingale | S_VLDL_C_ | NA | T | C | 0.041891 | 0.004306 | 5.20E-22 |
| rs491626 | miR-126-3p | trans | 2.10E-39 | Nightingale | M_VLDL_C | NA | T | C | 0.033636 | 0.004157 | 1.90E-15 |
| rs491626 | miR-126-3p | trans | 2.10E-39 | Nightingale | M_LDL_PL | NA | T | C | 0.032477 | 0.004431 | 2.60E-13 |
| rs491626 | miR-126-3p | trans | 2.10E-39 | Nightingale | Total_C | NA | T | C | 0.032387 | 0.004284 | 3.30E-14 |
| rs491626 | miR-126-3p | trans | 2.10E-39 | Nightingale | L_VLDL_C_ | NA | T | C | 0.052205 | 0.004322 | 3.90E-34 |
| rs491626 | miR-126-3p | trans | 2.10E-39 | Nightingale | L_VLDL_TG | NA | T | C | -0.05115 | 0.004404 | 5.30E-32 |
| rs491626 | miR-126-3p | trans | 2.10E-39 | Nightingale | S_LDL_P | NA | T | C | 0.029597 | 0.004429 | 3.20E-11 |
| rs491626 | miR-126-3p | trans | 2.10E-39 | Nightingale | M_VLDL_C | NA | T | C | 0.036214 | 0.004168 | 1.20E-17 |
| rs491626 | miR-126-3p | trans | 2.10E-39 | Nightingale | VLDL_C | NA | T | C | 0.031308 | 0.004419 | 4.80E-13 |
| rs491626 | miR-126-3p | trans | 2.10E-39 | Nightingale | XXL_VLDL_ | NA | T | C | 0.030859 | 0.004406 | 1.10E-12 |
| rs491626 | miR-126-3p | trans | 2.10E-39 | Nightingale | L_LDL_PL | NA | T | C | 0.038001 | 0.004391 | 8.90E-18 |
| rs491626 | miR-126-3p | trans | 2.10E-39 | Nightingale | S_VLDL_P | NA | T | C | 0.023472 | 0.004369 | 1.60E-08 |
| rs491626 | miR-126-3p | trans | 2.10E-39 | Nightingale | Total_PL | NA | T | C | 0.024943 | 0.004214 | 1.00E-09 |
| rs491626 | miR-126-3p | trans | 2.10E-39 | Nightingale | IDL_FC_pct | NA | T | C | 0.025773 | 0.004391 | 1.10E-08 |
| rs491626 | miR-126-3p | trans | 2.10E-39 | Nightingale | IDL_CE | NA | T | C | 0.033518 | 0.004272 | 1.30E-14 |
| rs491626 | miR-126-3p | trans | 2.10E-39 | Nightingale | L_LDL_L | NA | T | C | 0.033562 | 0.004382 | 2.90E-14 |
| rs491626 | miR-126-3p | trans | 2.10E-39 | Nightingale | XL_VLDL_C | NA | T | C | 0.037359 | 0.004245 | 3.10E-18 |
| rs491626 | miR-126-3p | trans | 2.10E-39 | Nightingale | XS_VLDL_C | NA | T | C | 0.038004 | 0.004295 | 1.30E-18 |
| rs491626 | miR-126-3p | trans | 2.10E-39 | Nightingale | S_VLDL_PL | NA | T | C | 0.027741 | 0.004237 | 1.80E-10 |
| rs491626 | miR-126-3p | trans | 2.10E-39 | Nightingale | XXL_VLDL_ | NA | T | C | 0.034842 | 0.004434 | 2.90E-15 |
| rs491626 | miR-126-3p | trans | 2.10E-39 | Nightingale | XS_VLDL_F | NA | T | C | 0.037079 | 0.004348 | 6.40E-18 |
| rs491626 | miR-126-3p | trans | 2.10E-39 | Nightingale | GlycA | NA | T | C | 0.028021 | 0.004406 | 1.40E-10 |
| rs491626 | miR-126-3p | trans | 2.10E-39 | Nightingale | S_LDL_CE | NA | T | C | 0.030557 | 0.004427 | 5.80E-12 |
| rs491626 | miR-126-3p | trans | 2.10E-39 | Nightingale | LDL_FC | NA | T | C | 0.033643 | 0.004378 | 5.20E-14 |
| rs491626 | miR-126-3p | trans | 2.10E-39 | Nightingale | Total_L | NA | T | C | 0.026362 | 0.004353 | 4.50E-10 |
| rs491626 | miR-126-3p | trans | 2.10E-39 | Nightingale | L_LDL_FC | NA | T | C | 0.033636 | 0.004345 | 2.80E-14 |
| rs491626 | miR-126-3p | trans | 2.10E-39 | Nightingale | LA | NA | T | C | 0.024924 | 0.004339 | 3.60E-09 |
| rs491626 | miR-126-3p | trans | 2.10E-39 | Nightingale | PUFA | NA | T | C | 0.023737 | 0.004316 | 1.40E-08 |
| rs491626 | miR-126-3p | trans | 2.10E-39 | Nightingale | XS_VLDL_C | NA | T | C | 0.037809 | 0.00427 | 2.10E-18 |
| rs491626 | miR-126-3p | trans | 2.10E-39 | Nightingale | S_VLDL_C | NA | T | C | 0.03838 | 0.004424 | 1.80E-18 |
| rs491626 | miR-126-3p | trans | 2.10E-39 | Nightingale | S_VLDL_L | NA | T | C | 0.024675 | 0.004382 | 3.70E-09 |
| rs491626 | miR-126-3p | trans | 2.10E-39 | Nightingale | Sphingomy | NA | T | C | 0.02903 | 0.004137 | 6.60E-12 |
| rs491626 | miR-126-3p | trans | 2.10E-39 | Nightingale | Omega_6 | NA | T | C | 0.025911 | 0.004321 | 5.60E-10 |
| rs491626 | miR-126-3p | trans | 2.10E-39 | Nightingale | M_VLDL_F | NA | T | C | 0.042073 | 0.00421 | 3.40E-23 |
| rs491626 | miR-126-3p | trans | 2.10E-39 | Nightingale | M_LDL_FC | NA | T | C | 0.034358 | 0.004419 | 1.90E-14 |
| rs491626 | miR-126-3p | trans | 2.10E-39 | Nightingale | M_VLDL_P | NA | T | C | 0.028301 | 0.004414 | 6.30E-11 |
| rs491626 | miR-126-3p | trans | 2.10E-39 | Nightingale | M_VLDL_T | NA | T | C | -0.03857 | 0.004181 | 6.70E-20 |
| rs491626 | miR-126-3p | trans | 2.10E-39 | Nightingale | S_VLDL_CE | NA | T | C | 0.047036 | 0.004365 | 1.20E-26 |
| rs491626 | miR-126-3p | trans | 2.10E-39 | Nightingale | Remnant_( | NA | T | C | 0.036878 | 0.004396 | 3.60E-17 |
| rs491626 | miR-126-3p | trans | 2.10E-39 | Nightingale | L_VLDL_FC | NA | T | C | 0.048596 | 0.004382 | 4.50E-29 |
| rs491626 | miR-126-3p | trans | 2.10E-39 | Nightingale | Citrate | NA | T | C | 0.026826 | 0.004396 | 7.40E-10 |
| rs491626 | miR-126-3p | trans | 2.10E-39 | Nightingale | M_VLDL_C | NA | T | C | 0.040852 | 0.004385 | 2.30E-20 |
| rs491626 | miR-126-3p | trans | 2.10E-39 | Nightingale | ApoB_by_/ | NA | T | C | 0.025049 | 0.004358 | 1.30E-08 |
| rs491626 | miR-126-3p | trans | 2.10E-39 | Nightingale | IDL_L | NA | T | C | 0.035282 | 0.004278 | 4.70E-16 |
| rs491626 | miR-126-3p | trans | 2.10E-39 | Nightingale | XL_VLDL_T | NA | T | C | -0.04393 | 0.004289 | 1.70E-24 |
| rs491626 | miR-126-3p | trans | 2.10E-39 | Nightingale | XL_VLDL_F | NA | T | C | 0.037203 | 0.00429 | 2.00E-18 |
| rs491626 | miR-126-3p | trans | 2.10E-39 | Nightingale | IDL_PL | NA | T | C | 0.035356 | 0.00427 | 3.30E-16 |
| rs491626 | miR-126-3p | trans | 2.10E-39 | Nightingale | M_VLDL_F | NA | T | C | 0.033378 | 0.004424 | 3.10E-14 |
| rs491626 | miR-126-3p | trans | 2.10E-39 | Nightingale | LDL_PL | NA | T | C | 0.034999 | 0.004415 | 4.70E-15 |
| rs491626 | miR-126-3p | trans | 2.10E-39 | Nightingale | S_VLDL_CE | NA | T | C | 0.038189 | 0.004419 | 1.60E-18 |
| rs491626 | miR-126-3p | trans | 2.10E-39 | Nightingale | XL_VLDL_C | NA | T | C | 0.03823 | 0.004243 | 4.60E-19 |
| rs491626 | miR-126-3p | trans | 2.10E-39 | Nightingale | LDL_L | NA | T | C | 0.033316 | 0.00441 | 7.30E-14 |

| | | | | | | | | | | | |
|---|---|---|---|---|---|---|---|---|---|---|---|
| rs491626 | miR-126-3p | trans | 2.10E-39 | Nightingale | M_LDL_P | NA | T | C | 0.0359 | 0.004437 | 6.80E-16 |
| rs491626 | miR-126-3p | trans | 2.10E-39 | Nightingale | Cholines | NA | T | C | 0.022312 | 0.004161 | 4.70E-08 |
| rs491626 | miR-126-3p | trans | 2.10E-39 | Nightingale | S_VLDL_FC | NA | T | C | 0.029359 | 0.004228 | 1.10E-11 |
| rs491626 | miR-126-3p | trans | 2.10E-39 | Nightingale | S_LDL_L | NA | T | C | 0.02775 | 0.004424 | 8.20E-10 |
| rs491626 | miR-126-3p | trans | 2.10E-39 | Nightingale | S_HDL_P | NA | T | C | 0.030621 | 0.004425 | 5.00E-12 |
| rs491626 | miR-126-3p | trans | 2.10E-39 | Nightingale | XXL_VLDL_ | NA | T | C | -0.03574 | 0.004463 | 2.10E-16 |
| rs491626 | miR-126-3p | trans | 2.10E-39 | Nightingale | L_VLDL_CE | NA | T | C | 0.047349 | 0.004234 | 2.40E-29 |
| rs491626 | miR-126-3p | trans | 2.10E-39 | Nightingale | LDL_CE | NA | T | C | 0.032565 | 0.004413 | 2.20E-13 |
| rs491626 | miR-126-3p | trans | 2.10E-39 | Nightingale | XXL_VLDL_ | NA | T | C | 0.033133 | 0.004459 | 8.80E-14 |
| rs491626 | miR-126-3p | trans | 2.10E-39 | Nightingale | S_LDL_CE_ | NA | T | C | 0.030951 | 0.004417 | 4.80E-14 |
| rs491626 | miR-126-3p | trans | 2.10E-39 | Nightingale | S_LDL_C | NA | T | C | 0.030952 | 0.004429 | 5.50E-12 |
| rs491626 | miR-126-3p | trans | 2.10E-39 | Nightingale | non_HDL_( | NA | T | C | 0.035708 | 0.004401 | 5.00E-16 |
| rs491626 | miR-126-3p | trans | 2.10E-39 | Nightingale | M_LDL_CE | NA | T | C | 0.032982 | 0.004431 | 9.50E-14 |
| rs491626 | miR-126-3p | trans | 2.10E-39 | Nightingale | S_HDL_C | NA | T | C | 0.028422 | 0.004417 | 1.70E-10 |
| rs491626 | miR-126-3p | trans | 2.10E-39 | Nightingale | S_LDL_PL_ | NA | T | C | -0.03007 | 0.004417 | 2.60E-13 |
| rs491626 | miR-126-3p | trans | 2.10E-39 | Nightingale | LDL_P | NA | T | C | 0.031809 | 0.004422 | 1.00E-12 |
| rs491626 | miR-126-3p | trans | 2.10E-39 | Nightingale | ApoB | NA | T | C | 0.033947 | 0.004419 | 2.10E-14 |
| rs491626 | miR-126-3p | trans | 2.10E-39 | Nightingale | S_VLDL_FC | NA | T | C | 0.037912 | 0.004426 | 7.30E-18 |
| rs491626 | miR-126-3p | trans | 2.10E-39 | Nightingale | M_LDL_TG | NA | T | C | -0.02762 | 0.00437 | 3.20E-10 |
| rs491626 | miR-126-3p | trans | 2.10E-39 | Nightingale | XL_HDL_PL | NA | T | C | -0.02274 | 0.004065 | 1.20E-08 |
| rs491626 | miR-126-3p | trans | 2.10E-39 | Nightingale | M_VLDL_C | NA | T | C | 0.038718 | 0.004413 | 1.90E-18 |
| rs491626 | miR-126-3p | trans | 2.10E-39 | Nightingale | S_LDL_C_p | NA | T | C | 0.045367 | 0.004442 | 5.40E-25 |
| rs491626 | miR-126-3p | trans | 2.10E-39 | Nightingale | XS_VLDL_L | NA | T | C | 0.03383 | 0.00435 | 2.00E-15 |
| rs491626 | miR-126-3p | trans | 2.10E-39 | Nightingale | L_LDL_C | NA | T | C | 0.032332 | 0.004375 | 2.60E-13 |
| rs492488 | let-7c-5p | trans | 2.67E-23 | Nightingale | VLDL_CE | NA | A | G | 0.035795 | 0.004426 | 3.00E-16 |
| rs492488 | let-7c-5p | trans | 2.67E-23 | Nightingale | S_VLDL_CE | NA | A | G | 0.047037 | 0.004365 | 1.20E-26 |
| rs492488 | let-7c-5p | trans | 2.67E-23 | Nightingale | VLDL_FC | NA | A | G | 0.024164 | 0.004392 | 1.10E-08 |
| rs492488 | let-7c-5p | trans | 2.67E-23 | Nightingale | Sphingomy | NA | A | G | 0.029031 | 0.004137 | 6.60E-12 |
| rs492488 | let-7c-5p | trans | 2.67E-23 | Nightingale | IDL_FC | NA | A | G | 0.038268 | 0.004284 | 1.40E-18 |
| rs492488 | let-7c-5p | trans | 2.67E-23 | Nightingale | VLDL_P | NA | A | G | 0.026641 | 0.004401 | 4.40E-10 |
| rs492488 | let-7c-5p | trans | 2.67E-23 | Nightingale | Total_L | NA | A | G | 0.026363 | 0.004353 | 4.50E-10 |
| rs492488 | let-7c-5p | trans | 2.67E-23 | Nightingale | XXL_VLDL_ | NA | A | G | 0.034841 | 0.004434 | 2.90E-15 |
| rs492488 | let-7c-5p | trans | 2.67E-23 | Nightingale | XS_VLDL_P | NA | A | G | 0.032128 | 0.004363 | 5.30E-14 |
| rs492488 | let-7c-5p | trans | 2.67E-23 | Nightingale | M_LDL_L | NA | A | G | 0.033043 | 0.004431 | 1.00E-13 |
| rs492488 | let-7c-5p | trans | 2.67E-23 | Nightingale | S_VLDL_P | NA | A | G | 0.023473 | 0.004369 | 1.60E-08 |
| rs492488 | let-7c-5p | trans | 2.67E-23 | Nightingale | LDL_C | NA | A | G | 0.033206 | 0.004406 | 8.70E-14 |
| rs492488 | let-7c-5p | trans | 2.67E-23 | Nightingale | Remnant_( | NA | A | G | 0.036879 | 0.004396 | 3.60E-17 |
| rs492488 | let-7c-5p | trans | 2.67E-23 | Nightingale | L_VLDL_TG | NA | A | G | -0.05115 | 0.004404 | 5.30E-32 |
| rs492488 | let-7c-5p | trans | 2.67E-23 | Nightingale | GlycA | NA | A | G | 0.028022 | 0.004406 | 1.40E-10 |
| rs492488 | let-7c-5p | trans | 2.67E-23 | Nightingale | S_HDL_C | NA | A | G | 0.028422 | 0.004417 | 1.70E-10 |
| rs492488 | let-7c-5p | trans | 2.67E-23 | Nightingale | S_HDL_L | NA | A | G | 0.026306 | 0.00441 | 1.50E-09 |
| rs492488 | let-7c-5p | trans | 2.67E-23 | Nightingale | Total_CE | NA | A | G | 0.031761 | 0.004268 | 9.50E-14 |
| rs492488 | let-7c-5p | trans | 2.67E-23 | Nightingale | S_VLDL_TG | NA | A | G | -0.03813 | 0.004283 | 1.40E-18 |
| rs492488 | let-7c-5p | trans | 2.67E-23 | Nightingale | S_HDL_FC | NA | A | G | 0.023468 | 0.004344 | 4.50E-08 |
| rs492488 | let-7c-5p | trans | 2.67E-23 | Nightingale | IDL_FC_pct | NA | A | G | 0.025772 | 0.004391 | 1.10E-08 |
| rs492488 | let-7c-5p | trans | 2.67E-23 | Nightingale | M_VLDL_F( | NA | A | G | 0.033379 | 0.004424 | 3.10E-14 |
| rs492488 | let-7c-5p | trans | 2.67E-23 | Nightingale | S_VLDL_C_ | NA | A | G | 0.041892 | 0.004306 | 5.20E-22 |
| rs492488 | let-7c-5p | trans | 2.67E-23 | Nightingale | IDL_C | NA | A | G | 0.034968 | 0.004274 | 9.20E-16 |
| rs492488 | let-7c-5p | trans | 2.67E-23 | Nightingale | XS_VLDL_C | NA | A | G | 0.03781 | 0.00427 | 2.10E-18 |
| rs492488 | let-7c-5p | trans | 2.67E-23 | Nightingale | S_VLDL_FC | NA | A | G | 0.02936 | 0.004228 | 1.10E-11 |
| rs492488 | let-7c-5p | trans | 2.67E-23 | Nightingale | M_VLDL_C | NA | A | G | 0.03872 | 0.004413 | 1.90E-18 |
| rs492488 | let-7c-5p | trans | 2.67E-23 | Nightingale | M_VLDL_F( | NA | A | G | 0.042074 | 0.00421 | 3.40E-23 |
| rs492488 | let-7c-5p | trans | 2.67E-23 | Nightingale | XS_VLDL_F | NA | A | G | 0.03143 | 0.004393 | 2.40E-12 |
| rs492488 | let-7c-5p | trans | 2.67E-23 | Nightingale | L_LDL_FC | NA | A | G | 0.033638 | 0.004345 | 2.80E-14 |
| rs492488 | let-7c-5p | trans | 2.67E-23 | Nightingale | L_VLDL_FC | NA | A | G | 0.048596 | 0.004382 | 4.50E-29 |
| rs492488 | let-7c-5p | trans | 2.67E-23 | Nightingale | Total_PL | NA | A | G | 0.024943 | 0.004214 | 1.00E-09 |
| rs492488 | let-7c-5p | trans | 2.67E-23 | Nightingale | S_LDL_P | NA | A | G | 0.029597 | 0.004429 | 3.20E-11 |
| rs492488 | let-7c-5p | trans | 2.67E-23 | Nightingale | XS_VLDL_F | NA | A | G | 0.03708 | 0.004348 | 6.40E-18 |
| rs492488 | let-7c-5p | trans | 2.67E-23 | Nightingale | IDL_PL | NA | A | G | 0.035357 | 0.00427 | 3.30E-16 |
| rs492488 | let-7c-5p | trans | 2.67E-23 | Nightingale | XL_HDL_PL | NA | A | G | -0.02274 | 0.004065 | 1.20E-08 |
| rs492488 | let-7c-5p | trans | 2.67E-23 | Nightingale | M_VLDL_C_ | NA | A | G | 0.036215 | 0.004168 | 1.20E-17 |
| rs492488 | let-7c-5p | trans | 2.67E-23 | Nightingale | XXL_VLDL_ | NA | A | G | 0.025012 | 0.004454 | 1.50E-09 |
| rs492488 | let-7c-5p | trans | 2.67E-23 | Nightingale | S_LDL_PL_ | NA | A | G | -0.03007 | 0.004417 | 2.60E-13 |

| | | | | | | | | | | | |
|---|---|---|---|---|---|---|---|---|---|---|---|
| rs492488 | let-7c-5p | trans | 2.67E-23 | Nightingale | S_VLDL_C | NA | A | G | 0.038381 | 0.004424 | 1.80E-18 |
| rs492488 | let-7c-5p | trans | 2.67E-23 | Nightingale | XL_VLDL_C | NA | A | G | 0.037359 | 0.004245 | 3.10E-18 |
| rs492488 | let-7c-5p | trans | 2.67E-23 | Nightingale | M_LDL_C_ | NA | A | G | 0.031163 | 0.004428 | 2.90E-12 |
| rs492488 | let-7c-5p | trans | 2.67E-23 | Nightingale | S_LDL_C_p | NA | A | G | 0.045367 | 0.004442 | 5.40E-25 |
| rs492488 | let-7c-5p | trans | 2.67E-23 | Nightingale | XXL_VLDL_ | NA | A | G | -0.03574 | 0.004463 | 2.10E-16 |
| rs492488 | let-7c-5p | trans | 2.67E-23 | Nightingale | LA | NA | A | G | 0.024925 | 0.004339 | 3.60E-09 |
| rs492488 | let-7c-5p | trans | 2.67E-23 | Nightingale | L_LDL_P | NA | A | G | 0.029187 | 0.00441 | 6.10E-11 |
| rs492488 | let-7c-5p | trans | 2.67E-23 | Nightingale | M_LDL_P | NA | A | G | 0.035901 | 0.004437 | 6.80E-16 |
| rs492488 | let-7c-5p | trans | 2.67E-23 | Nightingale | M_LDL_TG | NA | A | G | -0.02762 | 0.00437 | 3.20E-10 |
| rs492488 | let-7c-5p | trans | 2.67E-23 | Nightingale | L_VLDL_C_ | NA | A | G | 0.052206 | 0.004322 | 3.90E-34 |
| rs492488 | let-7c-5p | trans | 2.67E-23 | Nightingale | LDL_FC | NA | A | G | 0.033644 | 0.004378 | 5.20E-14 |
| rs492488 | let-7c-5p | trans | 2.67E-23 | Nightingale | LDL_L | NA | A | G | 0.033317 | 0.00441 | 7.30E-14 |
| rs492488 | let-7c-5p | trans | 2.67E-23 | Nightingale | PUFA | NA | A | G | 0.023739 | 0.004316 | 1.40E-08 |
| rs492488 | let-7c-5p | trans | 2.67E-23 | Nightingale | VLDL_C | NA | A | G | 0.031309 | 0.004419 | 4.80E-13 |
| rs492488 | let-7c-5p | trans | 2.67E-23 | Nightingale | XL_VLDL_F | NA | A | G | 0.037204 | 0.00429 | 2.00E-18 |
| rs492488 | let-7c-5p | trans | 2.67E-23 | Nightingale | M_VLDL_P | NA | A | G | 0.044067 | 0.004247 | 5.40E-25 |
| rs492488 | let-7c-5p | trans | 2.67E-23 | Nightingale | L_VLDL_CE | NA | A | G | 0.04735 | 0.004234 | 2.40E-29 |
| rs492488 | let-7c-5p | trans | 2.67E-23 | Nightingale | XS_VLDL_P | NA | A | G | 0.036714 | 0.004366 | 1.40E-17 |
| rs492488 | let-7c-5p | trans | 2.67E-23 | Nightingale | non_HDL_( | NA | A | G | 0.035709 | 0.004401 | 5.00E-16 |
| rs492488 | let-7c-5p | trans | 2.67E-23 | Nightingale | M_LDL_PL | NA | A | G | 0.032478 | 0.004431 | 2.60E-13 |
| rs492488 | let-7c-5p | trans | 2.67E-23 | Nightingale | M_VLDL_C | NA | A | G | 0.033637 | 0.004157 | 1.90E-15 |
| rs492488 | let-7c-5p | trans | 2.67E-23 | Nightingale | IDL_CE | NA | A | G | 0.03352 | 0.004272 | 1.30E-14 |
| rs492488 | let-7c-5p | trans | 2.67E-23 | Nightingale | Total_FC | NA | A | G | 0.03327 | 0.004323 | 6.70E-15 |
| rs492488 | let-7c-5p | trans | 2.67E-23 | Nightingale | XL_VLDL_C | NA | A | G | 0.03823 | 0.004243 | 4.60E-19 |
| rs492488 | let-7c-5p | trans | 2.67E-23 | Nightingale | L_LDL_PL | NA | A | G | 0.038002 | 0.004391 | 8.90E-18 |
| rs492488 | let-7c-5p | trans | 2.67E-23 | Nightingale | IDL_P | NA | A | G | 0.039997 | 0.004378 | 1.30E-19 |
| rs492488 | let-7c-5p | trans | 2.67E-23 | Nightingale | S_LDL_FC | NA | A | G | 0.028573 | 0.004418 | 4.20E-10 |
| rs492488 | let-7c-5p | trans | 2.67E-23 | Nightingale | M_LDL_C | NA | A | G | 0.033998 | 0.004433 | 2.10E-14 |
| rs492488 | let-7c-5p | trans | 2.67E-23 | Nightingale | S_HDL_PL | NA | A | G | 0.023794 | 0.004387 | 3.20E-08 |
| rs492488 | let-7c-5p | trans | 2.67E-23 | Nightingale | XS_VLDL_L | NA | A | G | 0.033831 | 0.00435 | 2.00E-15 |
| rs492488 | let-7c-5p | trans | 2.67E-23 | Nightingale | XL_VLDL_P | NA | A | G | 0.038845 | 0.004432 | 5.20E-19 |
| rs492488 | let-7c-5p | trans | 2.67E-23 | Nightingale | S_LDL_L | NA | A | G | 0.027751 | 0.004424 | 8.20E-10 |
| rs492488 | let-7c-5p | trans | 2.67E-23 | Nightingale | ApoB_by_/ | NA | A | G | 0.02505 | 0.004358 | 1.30E-08 |
| rs492488 | let-7c-5p | trans | 2.67E-23 | Nightingale | XL_VLDL_T | NA | A | G | -0.04393 | 0.004289 | 1.70E-24 |
| rs492488 | let-7c-5p | trans | 2.67E-23 | Nightingale | S_VLDL_L | NA | A | G | 0.024676 | 0.004382 | 3.70E-09 |
| rs492488 | let-7c-5p | trans | 2.67E-23 | Nightingale | XXL_VLDL_ | NA | A | G | 0.033132 | 0.004459 | 8.80E-14 |
| rs492488 | let-7c-5p | trans | 2.67E-23 | Nightingale | S_VLDL_PL | NA | A | G | 0.032786 | 0.004417 | 4.20E-14 |
| rs492488 | let-7c-5p | trans | 2.67E-23 | Nightingale | LDL_P | NA | A | G | 0.03181 | 0.004422 | 1.00E-12 |
| rs492488 | let-7c-5p | trans | 2.67E-23 | Nightingale | S_VLDL_PL | NA | A | G | 0.027742 | 0.004237 | 1.80E-10 |
| rs492488 | let-7c-5p | trans | 2.67E-23 | Nightingale | LDL_PL | NA | A | G | 0.035001 | 0.004415 | 4.70E-15 |
| rs492488 | let-7c-5p | trans | 2.67E-23 | Nightingale | XXL_VLDL_ | NA | A | G | 0.030859 | 0.004406 | 1.10E-12 |
| rs492488 | let-7c-5p | trans | 2.67E-23 | Nightingale | L_LDL_C | NA | A | G | 0.032333 | 0.004375 | 2.50E-13 |
| rs492488 | let-7c-5p | trans | 2.67E-23 | Nightingale | Total_C | NA | A | G | 0.032388 | 0.004284 | 3.20E-14 |
| rs492488 | let-7c-5p | trans | 2.67E-23 | Nightingale | M_VLDL_P | NA | A | G | 0.028302 | 0.004414 | 6.30E-11 |
| rs492488 | let-7c-5p | trans | 2.67E-23 | Nightingale | S_LDL_CE | NA | A | G | 0.030558 | 0.004427 | 5.80E-12 |
| rs492488 | let-7c-5p | trans | 2.67E-23 | Nightingale | LDL_CE | NA | A | G | 0.032566 | 0.004413 | 2.20E-13 |
| rs492488 | let-7c-5p | trans | 2.67E-23 | Nightingale | Clinical_LD | NA | A | G | 0.036486 | 0.004399 | 2.70E-16 |
| rs492488 | let-7c-5p | trans | 2.67E-23 | Nightingale | S_VLDL_CE | NA | A | G | 0.038189 | 0.004419 | 1.60E-18 |
| rs492488 | let-7c-5p | trans | 2.67E-23 | Nightingale | Cholines | NA | A | G | 0.022312 | 0.004161 | 4.70E-08 |
| rs492488 | let-7c-5p | trans | 2.67E-23 | Nightingale | M_VLDL_C | NA | A | G | 0.040853 | 0.004385 | 2.30E-20 |
| rs492488 | let-7c-5p | trans | 2.67E-23 | Nightingale | S_VLDL_FC | NA | A | G | 0.037913 | 0.004426 | 7.30E-18 |
| rs492488 | let-7c-5p | trans | 2.67E-23 | Nightingale | L_LDL_L | NA | A | G | 0.033564 | 0.004382 | 2.90E-14 |
| rs492488 | let-7c-5p | trans | 2.67E-23 | Nightingale | M_VLDL_P | NA | A | G | 0.029135 | 0.004418 | 2.00E-11 |
| rs492488 | let-7c-5p | trans | 2.67E-23 | Nightingale | S_HDL_CE | NA | A | G | 0.028644 | 0.004432 | 1.30E-10 |
| rs492488 | let-7c-5p | trans | 2.67E-23 | Nightingale | M_LDL_CE | NA | A | G | 0.032983 | 0.004431 | 9.50E-14 |
| rs492488 | let-7c-5p | trans | 2.67E-23 | Nightingale | S_LDL_C | NA | A | G | 0.030953 | 0.004429 | 5.50E-12 |
| rs492488 | let-7c-5p | trans | 2.67E-23 | Nightingale | S_HDL_P | NA | A | G | 0.030622 | 0.004425 | 5.00E-12 |
| rs492488 | let-7c-5p | trans | 2.67E-23 | Nightingale | ApoB | NA | A | G | 0.033948 | 0.004419 | 2.10E-14 |
| rs492488 | let-7c-5p | trans | 2.67E-23 | Nightingale | Citrate | NA | A | G | 0.026826 | 0.004396 | 7.40E-10 |
| rs492488 | let-7c-5p | trans | 2.67E-23 | Nightingale | M_VLDL_T | NA | A | G | -0.03857 | 0.004181 | 6.70E-20 |
| rs492488 | let-7c-5p | trans | 2.67E-23 | Nightingale | XS_VLDL_C | NA | A | G | 0.038005 | 0.004295 | 1.30E-18 |
| rs492488 | let-7c-5p | trans | 2.67E-23 | Nightingale | S_LDL_CE_ | NA | A | G | 0.03095 | 0.004417 | 4.80E-14 |
| rs492488 | let-7c-5p | trans | 2.67E-23 | Nightingale | M_LDL_FC | NA | A | G | 0.034359 | 0.004419 | 1.90E-14 |

| | | | | | | | | | | | |
|---|---|---|---|---|---|---|---|---|---|---|---|
| rs492488 | let-7c-5p | trans | 2.67E-23 | Nightingale | IDL_L | NA | A | G | 0.035284 | 0.004278 | 4.70E-16 |
| rs492488 | let-7c-5p | trans | 2.67E-23 | Nightingale | L_LDL_CE | NA | A | G | 0.031581 | 0.004385 | 9.70E-13 |
| rs492488 | let-7c-5p | trans | 2.67E-23 | Nightingale | Omega_6 | NA | A | G | 0.025912 | 0.004321 | 5.60E-10 |
| rs492488 | miR-195-5p | trans | 2.54E-17 | Nightingale | VLDL_CE | NA | A | G | 0.035795 | 0.004426 | 3.00E-16 |
| rs492488 | miR-195-5p | trans | 2.54E-17 | Nightingale | S_VLDL_CE | NA | A | G | 0.047037 | 0.004365 | 1.20E-26 |
| rs492488 | miR-195-5p | trans | 2.54E-17 | Nightingale | VLDL_FC | NA | A | G | 0.024164 | 0.004392 | 1.10E-08 |
| rs492488 | miR-195-5p | trans | 2.54E-17 | Nightingale | Sphingomy | NA | A | G | 0.029031 | 0.004137 | 6.60E-12 |
| rs492488 | miR-195-5p | trans | 2.54E-17 | Nightingale | IDL_FC | NA | A | G | 0.038268 | 0.004284 | 1.40E-18 |
| rs492488 | miR-195-5p | trans | 2.54E-17 | Nightingale | VLDL_P | NA | A | G | 0.026641 | 0.004401 | 4.40E-10 |
| rs492488 | miR-195-5p | trans | 2.54E-17 | Nightingale | Total_L | NA | A | G | 0.026363 | 0.004353 | 4.50E-10 |
| rs492488 | miR-195-5p | trans | 2.54E-17 | Nightingale | XXL_VLDL_ | NA | A | G | 0.034841 | 0.004434 | 2.90E-15 |
| rs492488 | miR-195-5p | trans | 2.54E-17 | Nightingale | XS_VLDL_P | NA | A | G | 0.032128 | 0.004363 | 5.30E-14 |
| rs492488 | miR-195-5p | trans | 2.54E-17 | Nightingale | M_LDL_L | NA | A | G | 0.033043 | 0.004431 | 1.00E-13 |
| rs492488 | miR-195-5p | trans | 2.54E-17 | Nightingale | S_VLDL_P | NA | A | G | 0.023473 | 0.004369 | 1.60E-08 |
| rs492488 | miR-195-5p | trans | 2.54E-17 | Nightingale | LDL_C | NA | A | G | 0.033206 | 0.004406 | 8.70E-14 |
| rs492488 | miR-195-5p | trans | 2.54E-17 | Nightingale | Remnant_( | NA | A | G | 0.036879 | 0.004396 | 3.60E-17 |
| rs492488 | miR-195-5p | trans | 2.54E-17 | Nightingale | L_VLDL_TG | NA | A | G | -0.05115 | 0.004404 | 5.30E-32 |
| rs492488 | miR-195-5p | trans | 2.54E-17 | Nightingale | GlycA | NA | A | G | 0.028022 | 0.004406 | 1.40E-10 |
| rs492488 | miR-195-5p | trans | 2.54E-17 | Nightingale | S_HDL_C | NA | A | G | 0.028422 | 0.004417 | 1.70E-10 |
| rs492488 | miR-195-5p | trans | 2.54E-17 | Nightingale | S_HDL_L | NA | A | G | 0.026306 | 0.00441 | 1.50E-09 |
| rs492488 | miR-195-5p | trans | 2.54E-17 | Nightingale | Total_CE | NA | A | G | 0.031761 | 0.004268 | 9.50E-14 |
| rs492488 | miR-195-5p | trans | 2.54E-17 | Nightingale | S_VLDL_TG | NA | A | G | -0.03813 | 0.004283 | 1.40E-18 |
| rs492488 | miR-195-5p | trans | 2.54E-17 | Nightingale | S_HDL_FC | NA | A | G | 0.023468 | 0.004344 | 4.50E-08 |
| rs492488 | miR-195-5p | trans | 2.54E-17 | Nightingale | IDL_FC_pct | NA | A | G | 0.025772 | 0.004391 | 1.10E-08 |
| rs492488 | miR-195-5p | trans | 2.54E-17 | Nightingale | M_VLDL_F( | NA | A | G | 0.033379 | 0.004424 | 3.10E-14 |
| rs492488 | miR-195-5p | trans | 2.54E-17 | Nightingale | S_VLDL_C_ | NA | A | G | 0.041892 | 0.004306 | 5.20E-22 |
| rs492488 | miR-195-5p | trans | 2.54E-17 | Nightingale | IDL_C | NA | A | G | 0.034968 | 0.004274 | 9.20E-16 |
| rs492488 | miR-195-5p | trans | 2.54E-17 | Nightingale | XS_VLDL_C | NA | A | G | 0.03781 | 0.00427 | 2.10E-18 |
| rs492488 | miR-195-5p | trans | 2.54E-17 | Nightingale | S_VLDL_FC | NA | A | G | 0.02936 | 0.004228 | 1.10E-11 |
| rs492488 | miR-195-5p | trans | 2.54E-17 | Nightingale | M_VLDL_C | NA | A | G | 0.03872 | 0.004413 | 1.90E-18 |
| rs492488 | miR-195-5p | trans | 2.54E-17 | Nightingale | M_VLDL_F( | NA | A | G | 0.042074 | 0.00421 | 3.40E-23 |
| rs492488 | miR-195-5p | trans | 2.54E-17 | Nightingale | XS_VLDL_F | NA | A | G | 0.03143 | 0.004393 | 2.40E-12 |
| rs492488 | miR-195-5p | trans | 2.54E-17 | Nightingale | L_LDL_FC | NA | A | G | 0.033638 | 0.004345 | 2.80E-14 |
| rs492488 | miR-195-5p | trans | 2.54E-17 | Nightingale | L_VLDL_FC | NA | A | G | 0.048596 | 0.004382 | 4.50E-29 |
| rs492488 | miR-195-5p | trans | 2.54E-17 | Nightingale | Total_PL | NA | A | G | 0.024943 | 0.004214 | 1.00E-09 |
| rs492488 | miR-195-5p | trans | 2.54E-17 | Nightingale | S_LDL_P | NA | A | G | 0.029597 | 0.004429 | 3.20E-11 |
| rs492488 | miR-195-5p | trans | 2.54E-17 | Nightingale | XS_VLDL_F | NA | A | G | 0.03708 | 0.004348 | 6.40E-18 |
| rs492488 | miR-195-5p | trans | 2.54E-17 | Nightingale | IDL_PL | NA | A | G | 0.035357 | 0.00427 | 3.30E-16 |
| rs492488 | miR-195-5p | trans | 2.54E-17 | Nightingale | XL_HDL_PL | NA | A | G | -0.02274 | 0.004065 | 1.20E-08 |
| rs492488 | miR-195-5p | trans | 2.54E-17 | Nightingale | M_VLDL_C_ | NA | A | G | 0.036215 | 0.004168 | 1.20E-17 |
| rs492488 | miR-195-5p | trans | 2.54E-17 | Nightingale | XXL_VLDL_ | NA | A | G | 0.025012 | 0.004454 | 1.50E-09 |
| rs492488 | miR-195-5p | trans | 2.54E-17 | Nightingale | S_LDL_PL_ | NA | A | G | -0.03007 | 0.004417 | 2.60E-13 |
| rs492488 | miR-195-5p | trans | 2.54E-17 | Nightingale | S_VLDL_C | NA | A | G | 0.038381 | 0.004424 | 1.80E-18 |
| rs492488 | miR-195-5p | trans | 2.54E-17 | Nightingale | XL_VLDL_C | NA | A | G | 0.037359 | 0.004245 | 3.10E-18 |
| rs492488 | miR-195-5p | trans | 2.54E-17 | Nightingale | M_LDL_C_ | NA | A | G | 0.031163 | 0.004428 | 2.90E-12 |
| rs492488 | miR-195-5p | trans | 2.54E-17 | Nightingale | S_LDL_C_p | NA | A | G | 0.045367 | 0.004442 | 5.40E-25 |
| rs492488 | miR-195-5p | trans | 2.54E-17 | Nightingale | XXL_VLDL_ | NA | A | G | -0.03574 | 0.004463 | 2.10E-16 |
| rs492488 | miR-195-5p | trans | 2.54E-17 | Nightingale | LA | NA | A | G | 0.024925 | 0.004339 | 3.60E-09 |
| rs492488 | miR-195-5p | trans | 2.54E-17 | Nightingale | L_LDL_P | NA | A | G | 0.029187 | 0.00441 | 6.10E-11 |
| rs492488 | miR-195-5p | trans | 2.54E-17 | Nightingale | M_LDL_P | NA | A | G | 0.035901 | 0.004437 | 6.80E-16 |
| rs492488 | miR-195-5p | trans | 2.54E-17 | Nightingale | M_LDL_TG | NA | A | G | -0.02762 | 0.00437 | 3.20E-10 |
| rs492488 | miR-195-5p | trans | 2.54E-17 | Nightingale | L_VLDL_C_ | NA | A | G | 0.052206 | 0.004322 | 3.90E-34 |
| rs492488 | miR-195-5p | trans | 2.54E-17 | Nightingale | LDL_FC | NA | A | G | 0.033644 | 0.004378 | 5.20E-14 |
| rs492488 | miR-195-5p | trans | 2.54E-17 | Nightingale | LDL_L | NA | A | G | 0.033317 | 0.00441 | 7.30E-14 |
| rs492488 | miR-195-5p | trans | 2.54E-17 | Nightingale | PUFA | NA | A | G | 0.023739 | 0.004316 | 1.40E-08 |
| rs492488 | miR-195-5p | trans | 2.54E-17 | Nightingale | VLDL_C | NA | A | G | 0.031309 | 0.004419 | 4.80E-13 |
| rs492488 | miR-195-5p | trans | 2.54E-17 | Nightingale | XL_VLDL_F | NA | A | G | 0.037204 | 0.00429 | 2.00E-18 |
| rs492488 | miR-195-5p | trans | 2.54E-17 | Nightingale | M_VLDL_P | NA | A | G | 0.044067 | 0.004247 | 5.40E-25 |
| rs492488 | miR-195-5p | trans | 2.54E-17 | Nightingale | L_VLDL_CE | NA | A | G | 0.04735 | 0.004234 | 2.40E-29 |
| rs492488 | miR-195-5p | trans | 2.54E-17 | Nightingale | XS_VLDL_P | NA | A | G | 0.036714 | 0.004366 | 1.40E-17 |
| rs492488 | miR-195-5p | trans | 2.54E-17 | Nightingale | non_HDL_( | NA | A | G | 0.035709 | 0.004401 | 5.00E-16 |
| rs492488 | miR-195-5p | trans | 2.54E-17 | Nightingale | M_LDL_PL | NA | A | G | 0.032478 | 0.004431 | 2.60E-13 |
| rs492488 | miR-195-5p | trans | 2.54E-17 | Nightingale | M_VLDL_C | NA | A | G | 0.033637 | 0.004157 | 1.90E-15 |

| | | | | | | | | | | | |
|---|---|---|---|---|---|---|---|---|---|---|---|
| rs492488 | miR-195-5p | trans | 2.54E-17 | Nightingale | IDL_CE | NA | A | G | 0.03352 | 0.004272 | 1.30E-14 |
| rs492488 | miR-195-5p | trans | 2.54E-17 | Nightingale | Total_FC | NA | A | G | 0.03327 | 0.004323 | 6.70E-15 |
| rs492488 | miR-195-5p | trans | 2.54E-17 | Nightingale | XL_VLDL_C | NA | A | G | 0.03823 | 0.004243 | 4.60E-19 |
| rs492488 | miR-195-5p | trans | 2.54E-17 | Nightingale | L_LDL_PL | NA | A | G | 0.038002 | 0.004391 | 8.90E-18 |
| rs492488 | miR-195-5p | trans | 2.54E-17 | Nightingale | IDL_P | NA | A | G | 0.039997 | 0.004378 | 1.30E-19 |
| rs492488 | miR-195-5p | trans | 2.54E-17 | Nightingale | S_LDL_FC | NA | A | G | 0.028573 | 0.004418 | 4.20E-10 |
| rs492488 | miR-195-5p | trans | 2.54E-17 | Nightingale | M_LDL_C | NA | A | G | 0.033998 | 0.004433 | 2.10E-14 |
| rs492488 | miR-195-5p | trans | 2.54E-17 | Nightingale | S_HDL_PL | NA | A | G | 0.023794 | 0.004387 | 3.20E-08 |
| rs492488 | miR-195-5p | trans | 2.54E-17 | Nightingale | XS_VLDL_L | NA | A | G | 0.033831 | 0.00435 | 2.00E-15 |
| rs492488 | miR-195-5p | trans | 2.54E-17 | Nightingale | XL_VLDL_P | NA | A | G | 0.038845 | 0.004432 | 5.20E-19 |
| rs492488 | miR-195-5p | trans | 2.54E-17 | Nightingale | S_LDL_L | NA | A | G | 0.027751 | 0.004424 | 8.20E-10 |
| rs492488 | miR-195-5p | trans | 2.54E-17 | Nightingale | ApoB_by_A | NA | A | G | 0.02505 | 0.004358 | 1.30E-08 |
| rs492488 | miR-195-5p | trans | 2.54E-17 | Nightingale | XL_VLDL_T | NA | A | G | -0.04393 | 0.004289 | 1.70E-24 |
| rs492488 | miR-195-5p | trans | 2.54E-17 | Nightingale | S_VLDL_L | NA | A | G | 0.024676 | 0.004382 | 3.70E-09 |
| rs492488 | miR-195-5p | trans | 2.54E-17 | Nightingale | XXL_VLDL_ | NA | A | G | 0.033132 | 0.004459 | 8.80E-14 |
| rs492488 | miR-195-5p | trans | 2.54E-17 | Nightingale | S_VLDL_PL | NA | A | G | 0.032786 | 0.004417 | 4.20E-14 |
| rs492488 | miR-195-5p | trans | 2.54E-17 | Nightingale | LDL_P | NA | A | G | 0.03181 | 0.004422 | 1.00E-12 |
| rs492488 | miR-195-5p | trans | 2.54E-17 | Nightingale | S_VLDL_PL | NA | A | G | 0.027742 | 0.004237 | 1.80E-10 |
| rs492488 | miR-195-5p | trans | 2.54E-17 | Nightingale | LDL_PL | NA | A | G | 0.035001 | 0.004415 | 4.70E-15 |
| rs492488 | miR-195-5p | trans | 2.54E-17 | Nightingale | XXL_VLDL_ | NA | A | G | 0.030859 | 0.004406 | 1.10E-12 |
| rs492488 | miR-195-5p | trans | 2.54E-17 | Nightingale | L_LDL_C | NA | A | G | 0.032333 | 0.004375 | 2.50E-13 |
| rs492488 | miR-195-5p | trans | 2.54E-17 | Nightingale | Total_C | NA | A | G | 0.032388 | 0.004284 | 3.20E-14 |
| rs492488 | miR-195-5p | trans | 2.54E-17 | Nightingale | M_VLDL_P | NA | A | G | 0.028302 | 0.004414 | 6.30E-11 |
| rs492488 | miR-195-5p | trans | 2.54E-17 | Nightingale | S_LDL_CE | NA | A | G | 0.030558 | 0.004427 | 5.80E-12 |
| rs492488 | miR-195-5p | trans | 2.54E-17 | Nightingale | LDL_CE | NA | A | G | 0.032566 | 0.004413 | 2.20E-13 |
| rs492488 | miR-195-5p | trans | 2.54E-17 | Nightingale | Clinical_LD | NA | A | G | 0.036486 | 0.004399 | 2.70E-16 |
| rs492488 | miR-195-5p | trans | 2.54E-17 | Nightingale | S_VLDL_CE | NA | A | G | 0.038189 | 0.004419 | 1.60E-18 |
| rs492488 | miR-195-5p | trans | 2.54E-17 | Nightingale | Cholines | NA | A | G | 0.022312 | 0.004161 | 4.70E-08 |
| rs492488 | miR-195-5p | trans | 2.54E-17 | Nightingale | M_VLDL_C | NA | A | G | 0.040853 | 0.004385 | 2.30E-20 |
| rs492488 | miR-195-5p | trans | 2.54E-17 | Nightingale | S_VLDL_FC | NA | A | G | 0.037913 | 0.004426 | 7.30E-18 |
| rs492488 | miR-195-5p | trans | 2.54E-17 | Nightingale | L_LDL_L | NA | A | G | 0.033564 | 0.004382 | 2.90E-14 |
| rs492488 | miR-195-5p | trans | 2.54E-17 | Nightingale | M_VLDL_P | NA | A | G | 0.029135 | 0.004418 | 2.00E-11 |
| rs492488 | miR-195-5p | trans | 2.54E-17 | Nightingale | S_HDL_CE | NA | A | G | 0.028644 | 0.004432 | 1.30E-10 |
| rs492488 | miR-195-5p | trans | 2.54E-17 | Nightingale | M_LDL_CE | NA | A | G | 0.032983 | 0.004431 | 9.50E-14 |
| rs492488 | miR-195-5p | trans | 2.54E-17 | Nightingale | S_LDL_C | NA | A | G | 0.030953 | 0.004429 | 5.50E-12 |
| rs492488 | miR-195-5p | trans | 2.54E-17 | Nightingale | S_HDL_P | NA | A | G | 0.030622 | 0.004425 | 5.00E-12 |
| rs492488 | miR-195-5p | trans | 2.54E-17 | Nightingale | ApoB | NA | A | G | 0.033948 | 0.004419 | 2.10E-14 |
| rs492488 | miR-195-5p | trans | 2.54E-17 | Nightingale | Citrate | NA | A | G | 0.026826 | 0.004396 | 7.40E-10 |
| rs492488 | miR-195-5p | trans | 2.54E-17 | Nightingale | M_VLDL_T | NA | A | G | -0.03857 | 0.004181 | 6.70E-20 |
| rs492488 | miR-195-5p | trans | 2.54E-17 | Nightingale | XS_VLDL_C | NA | A | G | 0.038005 | 0.004295 | 1.30E-18 |
| rs492488 | miR-195-5p | trans | 2.54E-17 | Nightingale | S_LDL_CE_ | NA | A | G | 0.03095 | 0.004417 | 4.80E-14 |
| rs492488 | miR-195-5p | trans | 2.54E-17 | Nightingale | M_LDL_FC | NA | A | G | 0.034359 | 0.004419 | 1.90E-14 |
| rs492488 | miR-195-5p | trans | 2.54E-17 | Nightingale | IDL_L | NA | A | G | 0.035284 | 0.004278 | 4.70E-16 |
| rs492488 | miR-195-5p | trans | 2.54E-17 | Nightingale | L_LDL_CE | NA | A | G | 0.031581 | 0.004385 | 9.70E-13 |
| rs492488 | miR-195-5p | trans | 2.54E-17 | Nightingale | Omega_6 | NA | A | G | 0.025912 | 0.004321 | 5.60E-10 |
| rs492488 | miR-126-3p | trans | 2.58E-39 | Nightingale | VLDL_CE | NA | A | G | 0.035795 | 0.004426 | 3.00E-16 |
| rs492488 | miR-126-3p | trans | 2.58E-39 | Nightingale | S_VLDL_CE | NA | A | G | 0.047037 | 0.004365 | 1.20E-26 |
| rs492488 | miR-126-3p | trans | 2.58E-39 | Nightingale | VLDL_FC | NA | A | G | 0.024164 | 0.004392 | 1.10E-08 |
| rs492488 | miR-126-3p | trans | 2.58E-39 | Nightingale | Sphingomy | NA | A | G | 0.029031 | 0.004137 | 6.60E-12 |
| rs492488 | miR-126-3p | trans | 2.58E-39 | Nightingale | IDL_FC | NA | A | G | 0.038268 | 0.004284 | 1.40E-18 |
| rs492488 | miR-126-3p | trans | 2.58E-39 | Nightingale | VLDL_P | NA | A | G | 0.026641 | 0.004401 | 4.40E-10 |
| rs492488 | miR-126-3p | trans | 2.58E-39 | Nightingale | Total_L | NA | A | G | 0.026363 | 0.004353 | 4.50E-10 |
| rs492488 | miR-126-3p | trans | 2.58E-39 | Nightingale | XXL_VLDL_ | NA | A | G | 0.034841 | 0.004434 | 2.90E-15 |
| rs492488 | miR-126-3p | trans | 2.58E-39 | Nightingale | XS_VLDL_P | NA | A | G | 0.032128 | 0.004363 | 5.30E-14 |
| rs492488 | miR-126-3p | trans | 2.58E-39 | Nightingale | M_LDL_L | NA | A | G | 0.033043 | 0.004431 | 1.00E-13 |
| rs492488 | miR-126-3p | trans | 2.58E-39 | Nightingale | S_VLDL_P | NA | A | G | 0.023473 | 0.004369 | 1.60E-08 |
| rs492488 | miR-126-3p | trans | 2.58E-39 | Nightingale | LDL_C | NA | A | G | 0.033206 | 0.004406 | 8.70E-14 |
| rs492488 | miR-126-3p | trans | 2.58E-39 | Nightingale | Remnant_( | NA | A | G | 0.036879 | 0.004396 | 3.60E-17 |
| rs492488 | miR-126-3p | trans | 2.58E-39 | Nightingale | L_VLDL_TG | NA | A | G | -0.05115 | 0.004404 | 5.30E-32 |
| rs492488 | miR-126-3p | trans | 2.58E-39 | Nightingale | GlycA | NA | A | G | 0.028022 | 0.004406 | 1.40E-10 |
| rs492488 | miR-126-3p | trans | 2.58E-39 | Nightingale | S_HDL_C | NA | A | G | 0.028422 | 0.004417 | 1.70E-10 |
| rs492488 | miR-126-3p | trans | 2.58E-39 | Nightingale | S_HDL_L | NA | A | G | 0.026306 | 0.00441 | 1.50E-09 |
| rs492488 | miR-126-3p | trans | 2.58E-39 | Nightingale | Total_CE | NA | A | G | 0.031761 | 0.004268 | 9.50E-14 |

| | | | | | | | | | | | |
|---|---|---|---|---|---|---|---|---|---|---|---|
| rs492488 | miR-126-3p | trans | 2.58E-39 | Nightingale | S_VLDL_TG | NA | A | G | -0.03813 | 0.004283 | 1.40E-18 |
| rs492488 | miR-126-3p | trans | 2.58E-39 | Nightingale | S_HDL_FC | NA | A | G | 0.023468 | 0.004344 | 4.50E-08 |
| rs492488 | miR-126-3p | trans | 2.58E-39 | Nightingale | IDL_FC_pct | NA | A | G | 0.025772 | 0.004391 | 1.10E-08 |
| rs492488 | miR-126-3p | trans | 2.58E-39 | Nightingale | M_VLDL_F | NA | A | G | 0.033379 | 0.004424 | 3.10E-14 |
| rs492488 | miR-126-3p | trans | 2.58E-39 | Nightingale | S_VLDL_C_ | NA | A | G | 0.041892 | 0.004306 | 5.20E-22 |
| rs492488 | miR-126-3p | trans | 2.58E-39 | Nightingale | IDL_C | NA | A | G | 0.034968 | 0.004274 | 9.20E-16 |
| rs492488 | miR-126-3p | trans | 2.58E-39 | Nightingale | XS_VLDL_C | NA | A | G | 0.03781 | 0.00427 | 2.10E-18 |
| rs492488 | miR-126-3p | trans | 2.58E-39 | Nightingale | S_VLDL_FC | NA | A | G | 0.02936 | 0.004228 | 1.10E-11 |
| rs492488 | miR-126-3p | trans | 2.58E-39 | Nightingale | M_VLDL_C | NA | A | G | 0.03872 | 0.004413 | 1.90E-18 |
| rs492488 | miR-126-3p | trans | 2.58E-39 | Nightingale | M_VLDL_F | NA | A | G | 0.042074 | 0.00421 | 3.40E-23 |
| rs492488 | miR-126-3p | trans | 2.58E-39 | Nightingale | XS_VLDL_F | NA | A | G | 0.03143 | 0.004393 | 2.40E-12 |
| rs492488 | miR-126-3p | trans | 2.58E-39 | Nightingale | L_LDL_FC | NA | A | G | 0.033638 | 0.004345 | 2.80E-14 |
| rs492488 | miR-126-3p | trans | 2.58E-39 | Nightingale | L_VLDL_FC | NA | A | G | 0.048596 | 0.004382 | 4.50E-29 |
| rs492488 | miR-126-3p | trans | 2.58E-39 | Nightingale | Total_PL | NA | A | G | 0.024943 | 0.004214 | 1.00E-09 |
| rs492488 | miR-126-3p | trans | 2.58E-39 | Nightingale | S_LDL_P | NA | A | G | 0.029597 | 0.004429 | 3.20E-11 |
| rs492488 | miR-126-3p | trans | 2.58E-39 | Nightingale | XS_VLDL_F | NA | A | G | 0.03708 | 0.004348 | 6.40E-18 |
| rs492488 | miR-126-3p | trans | 2.58E-39 | Nightingale | IDL_PL | NA | A | G | 0.035357 | 0.00427 | 3.30E-16 |
| rs492488 | miR-126-3p | trans | 2.58E-39 | Nightingale | XL_HDL_PL | NA | A | G | -0.02274 | 0.004065 | 1.20E-08 |
| rs492488 | miR-126-3p | trans | 2.58E-39 | Nightingale | M_VLDL_C | NA | A | G | 0.036215 | 0.004168 | 1.20E-17 |
| rs492488 | miR-126-3p | trans | 2.58E-39 | Nightingale | XXL_VLDL_ | NA | A | G | 0.025012 | 0.004454 | 1.50E-09 |
| rs492488 | miR-126-3p | trans | 2.58E-39 | Nightingale | S_LDL_PL_ | NA | A | G | -0.03007 | 0.004417 | 2.60E-13 |
| rs492488 | miR-126-3p | trans | 2.58E-39 | Nightingale | S_VLDL_C | NA | A | G | 0.038381 | 0.004424 | 1.80E-18 |
| rs492488 | miR-126-3p | trans | 2.58E-39 | Nightingale | XL_VLDL_C | NA | A | G | 0.037359 | 0.004245 | 3.10E-18 |
| rs492488 | miR-126-3p | trans | 2.58E-39 | Nightingale | M_LDL_C_ | NA | A | G | 0.031163 | 0.004428 | 2.90E-12 |
| rs492488 | miR-126-3p | trans | 2.58E-39 | Nightingale | S_LDL_C_p | NA | A | G | 0.045367 | 0.004442 | 5.40E-25 |
| rs492488 | miR-126-3p | trans | 2.58E-39 | Nightingale | XXL_VLDL_ | NA | A | G | -0.03574 | 0.004463 | 2.10E-16 |
| rs492488 | miR-126-3p | trans | 2.58E-39 | Nightingale | LA | NA | A | G | 0.024925 | 0.004339 | 3.60E-09 |
| rs492488 | miR-126-3p | trans | 2.58E-39 | Nightingale | L_LDL_P | NA | A | G | 0.029187 | 0.00441 | 6.10E-11 |
| rs492488 | miR-126-3p | trans | 2.58E-39 | Nightingale | M_LDL_P | NA | A | G | 0.035901 | 0.004437 | 6.80E-16 |
| rs492488 | miR-126-3p | trans | 2.58E-39 | Nightingale | M_LDL_TG | NA | A | G | -0.02762 | 0.00437 | 3.20E-10 |
| rs492488 | miR-126-3p | trans | 2.58E-39 | Nightingale | L_VLDL_C_ | NA | A | G | 0.052206 | 0.004322 | 3.90E-34 |
| rs492488 | miR-126-3p | trans | 2.58E-39 | Nightingale | LDL_FC | NA | A | G | 0.033644 | 0.004378 | 5.20E-14 |
| rs492488 | miR-126-3p | trans | 2.58E-39 | Nightingale | LDL_L | NA | A | G | 0.033317 | 0.00441 | 7.30E-14 |
| rs492488 | miR-126-3p | trans | 2.58E-39 | Nightingale | PUFA | NA | A | G | 0.023739 | 0.004316 | 1.40E-08 |
| rs492488 | miR-126-3p | trans | 2.58E-39 | Nightingale | VLDL_C | NA | A | G | 0.031309 | 0.004419 | 4.80E-13 |
| rs492488 | miR-126-3p | trans | 2.58E-39 | Nightingale | XL_VLDL_F | NA | A | G | 0.037204 | 0.00429 | 2.00E-18 |
| rs492488 | miR-126-3p | trans | 2.58E-39 | Nightingale | M_VLDL_P | NA | A | G | 0.044067 | 0.004247 | 5.40E-25 |
| rs492488 | miR-126-3p | trans | 2.58E-39 | Nightingale | L_VLDL_CE | NA | A | G | 0.04735 | 0.004234 | 2.40E-29 |
| rs492488 | miR-126-3p | trans | 2.58E-39 | Nightingale | XS_VLDL_P | NA | A | G | 0.036714 | 0.004366 | 1.40E-17 |
| rs492488 | miR-126-3p | trans | 2.58E-39 | Nightingale | non_HDL_( | NA | A | G | 0.035709 | 0.004401 | 5.00E-16 |
| rs492488 | miR-126-3p | trans | 2.58E-39 | Nightingale | M_LDL_PL | NA | A | G | 0.032478 | 0.004431 | 2.60E-13 |
| rs492488 | miR-126-3p | trans | 2.58E-39 | Nightingale | M_VLDL_C | NA | A | G | 0.033637 | 0.004157 | 1.90E-15 |
| rs492488 | miR-126-3p | trans | 2.58E-39 | Nightingale | IDL_CE | NA | A | G | 0.03352 | 0.004272 | 1.30E-14 |
| rs492488 | miR-126-3p | trans | 2.58E-39 | Nightingale | Total_FC | NA | A | G | 0.03327 | 0.004323 | 6.70E-15 |
| rs492488 | miR-126-3p | trans | 2.58E-39 | Nightingale | XL_VLDL_C | NA | A | G | 0.03823 | 0.004243 | 4.60E-19 |
| rs492488 | miR-126-3p | trans | 2.58E-39 | Nightingale | L_LDL_PL | NA | A | G | 0.038002 | 0.004391 | 8.90E-18 |
| rs492488 | miR-126-3p | trans | 2.58E-39 | Nightingale | IDL_P | NA | A | G | 0.039997 | 0.004378 | 1.30E-19 |
| rs492488 | miR-126-3p | trans | 2.58E-39 | Nightingale | S_LDL_FC | NA | A | G | 0.028573 | 0.004418 | 4.20E-10 |
| rs492488 | miR-126-3p | trans | 2.58E-39 | Nightingale | M_LDL_C | NA | A | G | 0.033998 | 0.004433 | 2.10E-14 |
| rs492488 | miR-126-3p | trans | 2.58E-39 | Nightingale | S_HDL_PL | NA | A | G | 0.023794 | 0.004387 | 3.20E-08 |
| rs492488 | miR-126-3p | trans | 2.58E-39 | Nightingale | XS_VLDL_L | NA | A | G | 0.033831 | 0.00435 | 2.00E-15 |
| rs492488 | miR-126-3p | trans | 2.58E-39 | Nightingale | XL_VLDL_P | NA | A | G | 0.038845 | 0.004432 | 5.20E-19 |
| rs492488 | miR-126-3p | trans | 2.58E-39 | Nightingale | S_LDL_L | NA | A | G | 0.027751 | 0.004424 | 8.20E-10 |
| rs492488 | miR-126-3p | trans | 2.58E-39 | Nightingale | ApoB_by_A | NA | A | G | 0.02505 | 0.004358 | 1.30E-08 |
| rs492488 | miR-126-3p | trans | 2.58E-39 | Nightingale | XL_VLDL_T | NA | A | G | -0.04393 | 0.004289 | 1.70E-24 |
| rs492488 | miR-126-3p | trans | 2.58E-39 | Nightingale | S_VLDL_L | NA | A | G | 0.024676 | 0.004382 | 3.70E-09 |
| rs492488 | miR-126-3p | trans | 2.58E-39 | Nightingale | XXL_VLDL_ | NA | A | G | 0.033132 | 0.004459 | 8.80E-14 |
| rs492488 | miR-126-3p | trans | 2.58E-39 | Nightingale | S_VLDL_PL | NA | A | G | 0.032786 | 0.004417 | 4.20E-14 |
| rs492488 | miR-126-3p | trans | 2.58E-39 | Nightingale | LDL_P | NA | A | G | 0.03181 | 0.004422 | 1.00E-12 |
| rs492488 | miR-126-3p | trans | 2.58E-39 | Nightingale | S_VLDL_PL | NA | A | G | 0.027742 | 0.004237 | 1.80E-10 |
| rs492488 | miR-126-3p | trans | 2.58E-39 | Nightingale | LDL_PL | NA | A | G | 0.035001 | 0.004415 | 4.70E-15 |
| rs492488 | miR-126-3p | trans | 2.58E-39 | Nightingale | XXL_VLDL_ | NA | A | G | 0.030859 | 0.004406 | 1.10E-12 |
| rs492488 | miR-126-3p | trans | 2.58E-39 | Nightingale | L_LDL_C | NA | A | G | 0.032333 | 0.004375 | 2.50E-13 |

| | | | | | | | | | | | |
|---|---|---|---|---|---|---|---|---|---|---|---|
| rs492488 | miR-126-3p | trans | 2.58E-39 | Nightingale | Total_C | NA | A | G | 0.032388 | 0.004284 | 3.20E-14 |
| rs492488 | miR-126-3p | trans | 2.58E-39 | Nightingale | M_VLDL_P | NA | A | G | 0.028302 | 0.004414 | 6.30E-11 |
| rs492488 | miR-126-3p | trans | 2.58E-39 | Nightingale | S_LDL_CE | NA | A | G | 0.030558 | 0.004427 | 5.80E-12 |
| rs492488 | miR-126-3p | trans | 2.58E-39 | Nightingale | LDL_CE | NA | A | G | 0.032566 | 0.004413 | 2.20E-13 |
| rs492488 | miR-126-3p | trans | 2.58E-39 | Nightingale | Clinical_LD | NA | A | G | 0.036486 | 0.004399 | 2.70E-16 |
| rs492488 | miR-126-3p | trans | 2.58E-39 | Nightingale | S_VLDL_CE | NA | A | G | 0.038189 | 0.004419 | 1.60E-18 |
| rs492488 | miR-126-3p | trans | 2.58E-39 | Nightingale | Cholines | NA | A | G | 0.022312 | 0.004161 | 4.70E-08 |
| rs492488 | miR-126-3p | trans | 2.58E-39 | Nightingale | M_VLDL_C | NA | A | G | 0.040853 | 0.004385 | 2.30E-20 |
| rs492488 | miR-126-3p | trans | 2.58E-39 | Nightingale | S_VLDL_FC | NA | A | G | 0.037913 | 0.004426 | 7.30E-18 |
| rs492488 | miR-126-3p | trans | 2.58E-39 | Nightingale | L_LDL_L | NA | A | G | 0.033564 | 0.004382 | 2.90E-14 |
| rs492488 | miR-126-3p | trans | 2.58E-39 | Nightingale | M_VLDL_P | NA | A | G | 0.029135 | 0.004418 | 2.00E-11 |
| rs492488 | miR-126-3p | trans | 2.58E-39 | Nightingale | S_HDL_CE | NA | A | G | 0.028644 | 0.004432 | 1.30E-10 |
| rs492488 | miR-126-3p | trans | 2.58E-39 | Nightingale | M_LDL_CE | NA | A | G | 0.032983 | 0.004431 | 9.50E-14 |
| rs492488 | miR-126-3p | trans | 2.58E-39 | Nightingale | S_LDL_C | NA | A | G | 0.030953 | 0.004429 | 5.50E-12 |
| rs492488 | miR-126-3p | trans | 2.58E-39 | Nightingale | S_HDL_P | NA | A | G | 0.030622 | 0.004425 | 5.00E-12 |
| rs492488 | miR-126-3p | trans | 2.58E-39 | Nightingale | ApoB | NA | A | G | 0.033948 | 0.004419 | 2.10E-14 |
| rs492488 | miR-126-3p | trans | 2.58E-39 | Nightingale | Citrate | NA | A | G | 0.026826 | 0.004396 | 7.40E-10 |
| rs492488 | miR-126-3p | trans | 2.58E-39 | Nightingale | M_VLDL_T | NA | A | G | -0.03857 | 0.004181 | 6.70E-20 |
| rs492488 | miR-126-3p | trans | 2.58E-39 | Nightingale | XS_VLDL_C | NA | A | G | 0.038005 | 0.004295 | 1.30E-18 |
| rs492488 | miR-126-3p | trans | 2.58E-39 | Nightingale | S_LDL_CE_ | NA | A | G | 0.03095 | 0.004417 | 4.80E-14 |
| rs492488 | miR-126-3p | trans | 2.58E-39 | Nightingale | M_LDL_FC | NA | A | G | 0.034359 | 0.004419 | 1.90E-14 |
| rs492488 | miR-126-3p | trans | 2.58E-39 | Nightingale | IDL_L | NA | A | G | 0.035284 | 0.004278 | 4.70E-16 |
| rs492488 | miR-126-3p | trans | 2.58E-39 | Nightingale | L_LDL_CE | NA | A | G | 0.031581 | 0.004385 | 9.70E-13 |
| rs492488 | miR-126-3p | trans | 2.58E-39 | Nightingale | Omega_6 | NA | A | G | 0.025912 | 0.004321 | 5.60E-10 |
| rs492488 | miR-139-5p | trans | 8.23E-42 | Nightingale | VLDL_CE | NA | A | G | 0.035795 | 0.004426 | 3.00E-16 |
| rs492488 | miR-139-5p | trans | 8.23E-42 | Nightingale | S_VLDL_CE | NA | A | G | 0.047037 | 0.004365 | 1.20E-26 |
| rs492488 | miR-139-5p | trans | 8.23E-42 | Nightingale | VLDL_FC | NA | A | G | 0.024164 | 0.004392 | 1.10E-08 |
| rs492488 | miR-139-5p | trans | 8.23E-42 | Nightingale | Sphingomy | NA | A | G | 0.029031 | 0.004137 | 6.60E-12 |
| rs492488 | miR-139-5p | trans | 8.23E-42 | Nightingale | IDL_FC | NA | A | G | 0.038268 | 0.004284 | 1.40E-18 |
| rs492488 | miR-139-5p | trans | 8.23E-42 | Nightingale | VLDL_P | NA | A | G | 0.026641 | 0.004401 | 4.40E-10 |
| rs492488 | miR-139-5p | trans | 8.23E-42 | Nightingale | Total_L | NA | A | G | 0.026363 | 0.004353 | 4.50E-10 |
| rs492488 | miR-139-5p | trans | 8.23E-42 | Nightingale | XXL_VLDL_ | NA | A | G | 0.034841 | 0.004434 | 2.90E-15 |
| rs492488 | miR-139-5p | trans | 8.23E-42 | Nightingale | XS_VLDL_P | NA | A | G | 0.032128 | 0.004363 | 5.30E-14 |
| rs492488 | miR-139-5p | trans | 8.23E-42 | Nightingale | M_LDL_L | NA | A | G | 0.033043 | 0.004431 | 1.00E-13 |
| rs492488 | miR-139-5p | trans | 8.23E-42 | Nightingale | S_VLDL_P | NA | A | G | 0.023473 | 0.004369 | 1.60E-08 |
| rs492488 | miR-139-5p | trans | 8.23E-42 | Nightingale | LDL_C | NA | A | G | 0.033206 | 0.004406 | 8.70E-14 |
| rs492488 | miR-139-5p | trans | 8.23E-42 | Nightingale | Remnant_( | NA | A | G | 0.036879 | 0.004396 | 3.60E-17 |
| rs492488 | miR-139-5p | trans | 8.23E-42 | Nightingale | L_VLDL_TG | NA | A | G | -0.05115 | 0.004404 | 5.30E-32 |
| rs492488 | miR-139-5p | trans | 8.23E-42 | Nightingale | GlycA | NA | A | G | 0.028022 | 0.004406 | 1.40E-10 |
| rs492488 | miR-139-5p | trans | 8.23E-42 | Nightingale | S_HDL_C | NA | A | G | 0.028422 | 0.004417 | 1.70E-10 |
| rs492488 | miR-139-5p | trans | 8.23E-42 | Nightingale | S_HDL_L | NA | A | G | 0.026306 | 0.00441 | 1.50E-09 |
| rs492488 | miR-139-5p | trans | 8.23E-42 | Nightingale | Total_CE | NA | A | G | 0.031761 | 0.004268 | 9.50E-14 |
| rs492488 | miR-139-5p | trans | 8.23E-42 | Nightingale | S_VLDL_TG | NA | A | G | -0.03813 | 0.004283 | 1.40E-18 |
| rs492488 | miR-139-5p | trans | 8.23E-42 | Nightingale | S_HDL_FC | NA | A | G | 0.023468 | 0.004344 | 4.50E-08 |
| rs492488 | miR-139-5p | trans | 8.23E-42 | Nightingale | IDL_FC_pct | NA | A | G | 0.025772 | 0.004391 | 1.10E-08 |
| rs492488 | miR-139-5p | trans | 8.23E-42 | Nightingale | M_VLDL_F | NA | A | G | 0.033379 | 0.004424 | 3.10E-14 |
| rs492488 | miR-139-5p | trans | 8.23E-42 | Nightingale | S_VLDL_C_ | NA | A | G | 0.041892 | 0.004306 | 5.20E-22 |
| rs492488 | miR-139-5p | trans | 8.23E-42 | Nightingale | IDL_C | NA | A | G | 0.034968 | 0.004274 | 9.20E-16 |
| rs492488 | miR-139-5p | trans | 8.23E-42 | Nightingale | XS_VLDL_C | NA | A | G | 0.03781 | 0.00427 | 2.10E-18 |
| rs492488 | miR-139-5p | trans | 8.23E-42 | Nightingale | S_VLDL_FC | NA | A | G | 0.02936 | 0.004228 | 1.10E-11 |
| rs492488 | miR-139-5p | trans | 8.23E-42 | Nightingale | M_VLDL_C | NA | A | G | 0.03872 | 0.004413 | 1.90E-18 |
| rs492488 | miR-139-5p | trans | 8.23E-42 | Nightingale | M_VLDL_F | NA | A | G | 0.042074 | 0.00421 | 3.40E-23 |
| rs492488 | miR-139-5p | trans | 8.23E-42 | Nightingale | XS_VLDL_F | NA | A | G | 0.03143 | 0.004393 | 2.40E-12 |
| rs492488 | miR-139-5p | trans | 8.23E-42 | Nightingale | L_LDL_FC | NA | A | G | 0.033638 | 0.004345 | 2.80E-14 |
| rs492488 | miR-139-5p | trans | 8.23E-42 | Nightingale | L_VLDL_FC | NA | A | G | 0.048596 | 0.004382 | 4.50E-29 |
| rs492488 | miR-139-5p | trans | 8.23E-42 | Nightingale | Total_PL | NA | A | G | 0.024943 | 0.004214 | 1.00E-09 |
| rs492488 | miR-139-5p | trans | 8.23E-42 | Nightingale | S_LDL_P | NA | A | G | 0.029597 | 0.004429 | 3.20E-11 |
| rs492488 | miR-139-5p | trans | 8.23E-42 | Nightingale | XS_VLDL_F | NA | A | G | 0.03708 | 0.004348 | 6.40E-18 |
| rs492488 | miR-139-5p | trans | 8.23E-42 | Nightingale | IDL_PL | NA | A | G | 0.035357 | 0.00427 | 3.30E-16 |
| rs492488 | miR-139-5p | trans | 8.23E-42 | Nightingale | XL_HDL_PL | NA | A | G | -0.02274 | 0.004065 | 1.20E-08 |
| rs492488 | miR-139-5p | trans | 8.23E-42 | Nightingale | M_VLDL_C_ | NA | A | G | 0.036215 | 0.004168 | 1.20E-17 |
| rs492488 | miR-139-5p | trans | 8.23E-42 | Nightingale | XXL_VLDL_ | NA | A | G | 0.025012 | 0.004454 | 1.50E-09 |
| rs492488 | miR-139-5p | trans | 8.23E-42 | Nightingale | S_LDL_PL_ | NA | A | G | -0.03007 | 0.004417 | 2.60E-13 |

| | | | | | | | | | | | |
|---|---|---|---|---|---|---|---|---|---|---|---|
| rs492488 | miR-139-5| trans | 8.23E-42 | Nightingale | S_VLDL_C | NA | A | G | 0.038381 | 0.004424 | 1.80E-18 |
| rs492488 | miR-139-5| trans | 8.23E-42 | Nightingale | XL_VLDL_C | NA | A | G | 0.037359 | 0.004245 | 3.10E-18 |
| rs492488 | miR-139-5| trans | 8.23E-42 | Nightingale | M_LDL_C_| | NA | A | G | 0.031163 | 0.004428 | 2.90E-12 |
| rs492488 | miR-139-5| trans | 8.23E-42 | Nightingale | S_LDL_C_p | NA | A | G | 0.045367 | 0.004442 | 5.40E-25 |
| rs492488 | miR-139-5| trans | 8.23E-42 | Nightingale | XXL_VLDL_ | NA | A | G | -0.03574 | 0.004463 | 2.10E-16 |
| rs492488 | miR-139-5| trans | 8.23E-42 | Nightingale | LA | NA | A | G | 0.024925 | 0.004339 | 3.60E-09 |
| rs492488 | miR-139-5| trans | 8.23E-42 | Nightingale | L_LDL_P | NA | A | G | 0.029187 | 0.00441 | 6.10E-11 |
| rs492488 | miR-139-5| trans | 8.23E-42 | Nightingale | M_LDL_P | NA | A | G | 0.035901 | 0.004437 | 6.80E-16 |
| rs492488 | miR-139-5| trans | 8.23E-42 | Nightingale | M_LDL_TG | NA | A | G | -0.02762 | 0.00437 | 3.20E-10 |
| rs492488 | miR-139-5| trans | 8.23E-42 | Nightingale | L_VLDL_C_ | NA | A | G | 0.052206 | 0.004322 | 3.90E-34 |
| rs492488 | miR-139-5| trans | 8.23E-42 | Nightingale | LDL_FC | NA | A | G | 0.033644 | 0.004378 | 5.20E-14 |
| rs492488 | miR-139-5| trans | 8.23E-42 | Nightingale | LDL_L | NA | A | G | 0.033317 | 0.00441 | 7.30E-14 |
| rs492488 | miR-139-5| trans | 8.23E-42 | Nightingale | PUFA | NA | A | G | 0.023739 | 0.004316 | 1.40E-08 |
| rs492488 | miR-139-5| trans | 8.23E-42 | Nightingale | VLDL_C | NA | A | G | 0.031309 | 0.004419 | 4.80E-13 |
| rs492488 | miR-139-5| trans | 8.23E-42 | Nightingale | XL_VLDL_F | NA | A | G | 0.037204 | 0.00429 | 2.00E-18 |
| rs492488 | miR-139-5| trans | 8.23E-42 | Nightingale | M_VLDL_P | NA | A | G | 0.044067 | 0.004247 | 5.40E-25 |
| rs492488 | miR-139-5| trans | 8.23E-42 | Nightingale | L_VLDL_CE | NA | A | G | 0.04735 | 0.004234 | 2.40E-29 |
| rs492488 | miR-139-5| trans | 8.23E-42 | Nightingale | XS_VLDL_P | NA | A | G | 0.036714 | 0.004366 | 1.40E-17 |
| rs492488 | miR-139-5| trans | 8.23E-42 | Nightingale | non_HDL_( | NA | A | G | 0.035709 | 0.004401 | 5.00E-16 |
| rs492488 | miR-139-5| trans | 8.23E-42 | Nightingale | M_LDL_PL | NA | A | G | 0.032478 | 0.004431 | 2.60E-13 |
| rs492488 | miR-139-5| trans | 8.23E-42 | Nightingale | M_VLDL_C | NA | A | G | 0.033637 | 0.004157 | 1.90E-15 |
| rs492488 | miR-139-5| trans | 8.23E-42 | Nightingale | IDL_CE | NA | A | G | 0.03352 | 0.004272 | 1.30E-14 |
| rs492488 | miR-139-5| trans | 8.23E-42 | Nightingale | Total_FC | NA | A | G | 0.03327 | 0.004323 | 6.70E-15 |
| rs492488 | miR-139-5| trans | 8.23E-42 | Nightingale | XL_VLDL_C | NA | A | G | 0.03823 | 0.004243 | 4.60E-19 |
| rs492488 | miR-139-5| trans | 8.23E-42 | Nightingale | L_LDL_PL | NA | A | G | 0.038002 | 0.004391 | 8.90E-18 |
| rs492488 | miR-139-5| trans | 8.23E-42 | Nightingale | IDL_P | NA | A | G | 0.039997 | 0.004378 | 1.30E-19 |
| rs492488 | miR-139-5| trans | 8.23E-42 | Nightingale | S_LDL_FC | NA | A | G | 0.028573 | 0.004418 | 4.20E-10 |
| rs492488 | miR-139-5| trans | 8.23E-42 | Nightingale | M_LDL_C | NA | A | G | 0.033998 | 0.004433 | 2.10E-14 |
| rs492488 | miR-139-5| trans | 8.23E-42 | Nightingale | S_HDL_PL | NA | A | G | 0.023794 | 0.004387 | 3.20E-08 |
| rs492488 | miR-139-5| trans | 8.23E-42 | Nightingale | XS_VLDL_L | NA | A | G | 0.033831 | 0.00435 | 2.00E-15 |
| rs492488 | miR-139-5| trans | 8.23E-42 | Nightingale | XL_VLDL_P | NA | A | G | 0.038845 | 0.004432 | 5.20E-19 |
| rs492488 | miR-139-5| trans | 8.23E-42 | Nightingale | S_LDL_L | NA | A | G | 0.027751 | 0.004424 | 8.20E-10 |
| rs492488 | miR-139-5| trans | 8.23E-42 | Nightingale | ApoB_by_/ | NA | A | G | 0.02505 | 0.004358 | 1.30E-08 |
| rs492488 | miR-139-5| trans | 8.23E-42 | Nightingale | XL_VLDL_T | NA | A | G | -0.04393 | 0.004289 | 1.70E-24 |
| rs492488 | miR-139-5| trans | 8.23E-42 | Nightingale | S_VLDL_L | NA | A | G | 0.024676 | 0.004382 | 3.70E-09 |
| rs492488 | miR-139-5| trans | 8.23E-42 | Nightingale | XXL_VLDL_ | NA | A | G | 0.033132 | 0.004459 | 8.80E-14 |
| rs492488 | miR-139-5| trans | 8.23E-42 | Nightingale | S_VLDL_PL | NA | A | G | 0.032786 | 0.004417 | 4.20E-14 |
| rs492488 | miR-139-5| trans | 8.23E-42 | Nightingale | LDL_P | NA | A | G | 0.03181 | 0.004422 | 1.00E-12 |
| rs492488 | miR-139-5| trans | 8.23E-42 | Nightingale | S_VLDL_PL | NA | A | G | 0.027742 | 0.004237 | 1.80E-10 |
| rs492488 | miR-139-5| trans | 8.23E-42 | Nightingale | LDL_PL | NA | A | G | 0.035001 | 0.004415 | 4.70E-15 |
| rs492488 | miR-139-5| trans | 8.23E-42 | Nightingale | XXL_VLDL_ | NA | A | G | 0.030859 | 0.004406 | 1.10E-12 |
| rs492488 | miR-139-5| trans | 8.23E-42 | Nightingale | L_LDL_C | NA | A | G | 0.032333 | 0.004375 | 2.50E-13 |
| rs492488 | miR-139-5| trans | 8.23E-42 | Nightingale | Total_C | NA | A | G | 0.032388 | 0.004284 | 3.20E-14 |
| rs492488 | miR-139-5| trans | 8.23E-42 | Nightingale | M_VLDL_P | NA | A | G | 0.028302 | 0.004414 | 6.30E-11 |
| rs492488 | miR-139-5| trans | 8.23E-42 | Nightingale | S_LDL_CE | NA | A | G | 0.030558 | 0.004427 | 5.80E-12 |
| rs492488 | miR-139-5| trans | 8.23E-42 | Nightingale | LDL_CE | NA | A | G | 0.032566 | 0.004413 | 2.20E-13 |
| rs492488 | miR-139-5| trans | 8.23E-42 | Nightingale | Clinical_LD | NA | A | G | 0.036486 | 0.004399 | 2.70E-16 |
| rs492488 | miR-139-5| trans | 8.23E-42 | Nightingale | S_VLDL_CE | NA | A | G | 0.038189 | 0.004419 | 1.60E-18 |
| rs492488 | miR-139-5| trans | 8.23E-42 | Nightingale | Cholines | NA | A | G | 0.022312 | 0.004161 | 4.70E-08 |
| rs492488 | miR-139-5| trans | 8.23E-42 | Nightingale | M_VLDL_C | NA | A | G | 0.040853 | 0.004385 | 2.30E-20 |
| rs492488 | miR-139-5| trans | 8.23E-42 | Nightingale | S_VLDL_FC | NA | A | G | 0.037913 | 0.004426 | 7.30E-18 |
| rs492488 | miR-139-5| trans | 8.23E-42 | Nightingale | L_LDL_L | NA | A | G | 0.033564 | 0.004382 | 2.90E-14 |
| rs492488 | miR-139-5| trans | 8.23E-42 | Nightingale | M_VLDL_P | NA | A | G | 0.029135 | 0.004418 | 2.00E-11 |
| rs492488 | miR-139-5| trans | 8.23E-42 | Nightingale | S_HDL_CE | NA | A | G | 0.028644 | 0.004432 | 1.30E-10 |
| rs492488 | miR-139-5| trans | 8.23E-42 | Nightingale | M_LDL_CE | NA | A | G | 0.032983 | 0.004431 | 9.50E-14 |
| rs492488 | miR-139-5| trans | 8.23E-42 | Nightingale | S_LDL_C | NA | A | G | 0.030953 | 0.004429 | 5.50E-12 |
| rs492488 | miR-139-5| trans | 8.23E-42 | Nightingale | S_HDL_P | NA | A | G | 0.030622 | 0.004425 | 5.00E-12 |
| rs492488 | miR-139-5| trans | 8.23E-42 | Nightingale | ApoB | NA | A | G | 0.033948 | 0.004419 | 2.10E-14 |
| rs492488 | miR-139-5| trans | 8.23E-42 | Nightingale | Citrate | NA | A | G | 0.026826 | 0.004396 | 7.40E-10 |
| rs492488 | miR-139-5| trans | 8.23E-42 | Nightingale | M_VLDL_T | NA | A | G | -0.03857 | 0.004181 | 6.70E-20 |
| rs492488 | miR-139-5| trans | 8.23E-42 | Nightingale | XS_VLDL_C | NA | A | G | 0.038005 | 0.004295 | 1.30E-18 |
| rs492488 | miR-139-5| trans | 8.23E-42 | Nightingale | S_LDL_CE_ | NA | A | G | 0.03095 | 0.004417 | 4.80E-14 |
| rs492488 | miR-139-5| trans | 8.23E-42 | Nightingale | M_LDL_FC | NA | A | G | 0.034359 | 0.004419 | 1.90E-14 |

| | | | | | | | | | | | |
|---|---|---|---|---|---|---|---|---|---|---|---|
| rs492488 | miR-139-5ʅ | trans | 8.23E-42 | Nightingale | IDL_L | NA | A | G | 0.035284 | 0.004278 | 4.70E-16 |
| rs492488 | miR-139-5ʅ | trans | 8.23E-42 | Nightingale | L_LDL_CE | NA | A | G | 0.031581 | 0.004385 | 9.70E-13 |
| rs492488 | miR-139-5ʅ | trans | 8.23E-42 | Nightingale | Omega_6 | NA | A | G | 0.025912 | 0.004321 | 5.60E-10 |
| rs492488 | let-7b-5p | trans | 1.07E-26 | Nightingale | VLDL_CE | NA | A | G | 0.035795 | 0.004426 | 3.00E-16 |
| rs492488 | let-7b-5p | trans | 1.07E-26 | Nightingale | S_VLDL_CE | NA | A | G | 0.047037 | 0.004365 | 1.20E-26 |
| rs492488 | let-7b-5p | trans | 1.07E-26 | Nightingale | VLDL_FC | NA | A | G | 0.024164 | 0.004392 | 1.10E-08 |
| rs492488 | let-7b-5p | trans | 1.07E-26 | Nightingale | Sphingomy | NA | A | G | 0.029031 | 0.004137 | 6.60E-12 |
| rs492488 | let-7b-5p | trans | 1.07E-26 | Nightingale | IDL_FC | NA | A | G | 0.038268 | 0.004284 | 1.40E-18 |
| rs492488 | let-7b-5p | trans | 1.07E-26 | Nightingale | VLDL_P | NA | A | G | 0.026641 | 0.004401 | 4.40E-10 |
| rs492488 | let-7b-5p | trans | 1.07E-26 | Nightingale | Total_L | NA | A | G | 0.026363 | 0.004353 | 4.50E-10 |
| rs492488 | let-7b-5p | trans | 1.07E-26 | Nightingale | XXL_VLDL_ | NA | A | G | 0.034841 | 0.004434 | 2.90E-15 |
| rs492488 | let-7b-5p | trans | 1.07E-26 | Nightingale | XS_VLDL_P | NA | A | G | 0.032128 | 0.004363 | 5.30E-14 |
| rs492488 | let-7b-5p | trans | 1.07E-26 | Nightingale | M_LDL_L | NA | A | G | 0.033043 | 0.004431 | 1.00E-13 |
| rs492488 | let-7b-5p | trans | 1.07E-26 | Nightingale | S_VLDL_P | NA | A | G | 0.023473 | 0.004369 | 1.60E-08 |
| rs492488 | let-7b-5p | trans | 1.07E-26 | Nightingale | LDL_C | NA | A | G | 0.033206 | 0.004406 | 8.70E-14 |
| rs492488 | let-7b-5p | trans | 1.07E-26 | Nightingale | Remnant_( | NA | A | G | 0.036879 | 0.004396 | 3.60E-17 |
| rs492488 | let-7b-5p | trans | 1.07E-26 | Nightingale | L_VLDL_TG | NA | A | G | -0.05115 | 0.004404 | 5.30E-32 |
| rs492488 | let-7b-5p | trans | 1.07E-26 | Nightingale | GlycA | NA | A | G | 0.028022 | 0.004406 | 1.40E-10 |
| rs492488 | let-7b-5p | trans | 1.07E-26 | Nightingale | S_HDL_C | NA | A | G | 0.028422 | 0.004417 | 1.70E-10 |
| rs492488 | let-7b-5p | trans | 1.07E-26 | Nightingale | S_HDL_L | NA | A | G | 0.026306 | 0.00441 | 1.50E-09 |
| rs492488 | let-7b-5p | trans | 1.07E-26 | Nightingale | Total_CE | NA | A | G | 0.031761 | 0.004268 | 9.50E-14 |
| rs492488 | let-7b-5p | trans | 1.07E-26 | Nightingale | S_VLDL_TG | NA | A | G | -0.03813 | 0.004283 | 1.40E-18 |
| rs492488 | let-7b-5p | trans | 1.07E-26 | Nightingale | S_HDL_FC | NA | A | G | 0.023468 | 0.004344 | 4.50E-08 |
| rs492488 | let-7b-5p | trans | 1.07E-26 | Nightingale | IDL_FC_pct | NA | A | G | 0.025772 | 0.004391 | 1.10E-08 |
| rs492488 | let-7b-5p | trans | 1.07E-26 | Nightingale | M_VLDL_F | NA | A | G | 0.033379 | 0.004424 | 3.10E-14 |
| rs492488 | let-7b-5p | trans | 1.07E-26 | Nightingale | S_VLDL_C_ | NA | A | G | 0.041892 | 0.004306 | 5.20E-22 |
| rs492488 | let-7b-5p | trans | 1.07E-26 | Nightingale | IDL_C | NA | A | G | 0.034968 | 0.004274 | 9.20E-16 |
| rs492488 | let-7b-5p | trans | 1.07E-26 | Nightingale | XS_VLDL_C | NA | A | G | 0.03781 | 0.00427 | 2.10E-18 |
| rs492488 | let-7b-5p | trans | 1.07E-26 | Nightingale | S_VLDL_FC | NA | A | G | 0.02936 | 0.004228 | 1.10E-11 |
| rs492488 | let-7b-5p | trans | 1.07E-26 | Nightingale | M_VLDL_C | NA | A | G | 0.03872 | 0.004413 | 1.90E-18 |
| rs492488 | let-7b-5p | trans | 1.07E-26 | Nightingale | M_VLDL_F | NA | A | G | 0.042074 | 0.00421 | 3.40E-23 |
| rs492488 | let-7b-5p | trans | 1.07E-26 | Nightingale | XS_VLDL_F | NA | A | G | 0.03143 | 0.004393 | 2.40E-12 |
| rs492488 | let-7b-5p | trans | 1.07E-26 | Nightingale | L_LDL_FC | NA | A | G | 0.033638 | 0.004345 | 2.80E-14 |
| rs492488 | let-7b-5p | trans | 1.07E-26 | Nightingale | L_VLDL_FC | NA | A | G | 0.048596 | 0.004382 | 4.50E-29 |
| rs492488 | let-7b-5p | trans | 1.07E-26 | Nightingale | Total_PL | NA | A | G | 0.024943 | 0.004214 | 1.00E-09 |
| rs492488 | let-7b-5p | trans | 1.07E-26 | Nightingale | S_LDL_P | NA | A | G | 0.029597 | 0.004429 | 3.20E-11 |
| rs492488 | let-7b-5p | trans | 1.07E-26 | Nightingale | XS_VLDL_F | NA | A | G | 0.03708 | 0.004348 | 6.40E-18 |
| rs492488 | let-7b-5p | trans | 1.07E-26 | Nightingale | IDL_PL | NA | A | G | 0.035357 | 0.00427 | 3.30E-16 |
| rs492488 | let-7b-5p | trans | 1.07E-26 | Nightingale | XL_HDL_PL | NA | A | G | -0.02274 | 0.004065 | 1.20E-08 |
| rs492488 | let-7b-5p | trans | 1.07E-26 | Nightingale | M_VLDL_C | NA | A | G | 0.036215 | 0.004168 | 1.20E-17 |
| rs492488 | let-7b-5p | trans | 1.07E-26 | Nightingale | XXL_VLDL_ | NA | A | G | 0.025012 | 0.004454 | 1.50E-09 |
| rs492488 | let-7b-5p | trans | 1.07E-26 | Nightingale | S_LDL_PL_ | NA | A | G | -0.03007 | 0.004417 | 2.60E-13 |
| rs492488 | let-7b-5p | trans | 1.07E-26 | Nightingale | S_VLDL_C | NA | A | G | 0.038381 | 0.004424 | 1.80E-18 |
| rs492488 | let-7b-5p | trans | 1.07E-26 | Nightingale | XL_VLDL_C | NA | A | G | 0.037359 | 0.004245 | 3.10E-18 |
| rs492488 | let-7b-5p | trans | 1.07E-26 | Nightingale | M_LDL_C_ | NA | A | G | 0.031163 | 0.004428 | 2.90E-12 |
| rs492488 | let-7b-5p | trans | 1.07E-26 | Nightingale | S_LDL_C_p | NA | A | G | 0.045367 | 0.004442 | 5.40E-25 |
| rs492488 | let-7b-5p | trans | 1.07E-26 | Nightingale | XXL_VLDL_ | NA | A | G | -0.03574 | 0.004463 | 2.10E-16 |
| rs492488 | let-7b-5p | trans | 1.07E-26 | Nightingale | LA | NA | A | G | 0.024925 | 0.004339 | 3.60E-09 |
| rs492488 | let-7b-5p | trans | 1.07E-26 | Nightingale | L_LDL_P | NA | A | G | 0.029187 | 0.00441 | 6.10E-11 |
| rs492488 | let-7b-5p | trans | 1.07E-26 | Nightingale | M_LDL_P | NA | A | G | 0.035901 | 0.004437 | 6.80E-16 |
| rs492488 | let-7b-5p | trans | 1.07E-26 | Nightingale | M_LDL_TG | NA | A | G | -0.02762 | 0.00437 | 3.20E-10 |
| rs492488 | let-7b-5p | trans | 1.07E-26 | Nightingale | L_VLDL_C_ | NA | A | G | 0.052206 | 0.004322 | 3.90E-34 |
| rs492488 | let-7b-5p | trans | 1.07E-26 | Nightingale | LDL_FC | NA | A | G | 0.033644 | 0.004378 | 5.20E-14 |
| rs492488 | let-7b-5p | trans | 1.07E-26 | Nightingale | LDL_L | NA | A | G | 0.033317 | 0.00441 | 7.30E-14 |
| rs492488 | let-7b-5p | trans | 1.07E-26 | Nightingale | PUFA | NA | A | G | 0.023739 | 0.004316 | 1.40E-08 |
| rs492488 | let-7b-5p | trans | 1.07E-26 | Nightingale | VLDL_C | NA | A | G | 0.031309 | 0.004419 | 4.80E-13 |
| rs492488 | let-7b-5p | trans | 1.07E-26 | Nightingale | XL_VLDL_F | NA | A | G | 0.037204 | 0.00429 | 2.00E-18 |
| rs492488 | let-7b-5p | trans | 1.07E-26 | Nightingale | M_VLDL_P | NA | A | G | 0.044067 | 0.004247 | 5.40E-25 |
| rs492488 | let-7b-5p | trans | 1.07E-26 | Nightingale | L_VLDL_CE | NA | A | G | 0.04735 | 0.004234 | 2.40E-29 |
| rs492488 | let-7b-5p | trans | 1.07E-26 | Nightingale | XS_VLDL_P | NA | A | G | 0.036714 | 0.004366 | 1.40E-17 |
| rs492488 | let-7b-5p | trans | 1.07E-26 | Nightingale | non_HDL_( | NA | A | G | 0.035709 | 0.004401 | 5.00E-16 |
| rs492488 | let-7b-5p | trans | 1.07E-26 | Nightingale | M_LDL_PL | NA | A | G | 0.032478 | 0.004431 | 2.60E-13 |
| rs492488 | let-7b-5p | trans | 1.07E-26 | Nightingale | M_VLDL_C | NA | A | G | 0.033637 | 0.004157 | 1.90E-15 |

| | | | | | | | | | | | |
|---|---|---|---|---|---|---|---|---|---|---|---|
| rs492488 | let-7b-5p | trans | 1.07E-26 | Nightingale | IDL_CE | NA | A | G | 0.03352 | 0.004272 | 1.30E-14 |
| rs492488 | let-7b-5p | trans | 1.07E-26 | Nightingale | Total_FC | NA | A | G | 0.03327 | 0.004323 | 6.70E-15 |
| rs492488 | let-7b-5p | trans | 1.07E-26 | Nightingale | XL_VLDL_C | NA | A | G | 0.03823 | 0.004243 | 4.60E-19 |
| rs492488 | let-7b-5p | trans | 1.07E-26 | Nightingale | L_LDL_PL | NA | A | G | 0.038002 | 0.004391 | 8.90E-18 |
| rs492488 | let-7b-5p | trans | 1.07E-26 | Nightingale | IDL_P | NA | A | G | 0.039997 | 0.004378 | 1.30E-19 |
| rs492488 | let-7b-5p | trans | 1.07E-26 | Nightingale | S_LDL_FC | NA | A | G | 0.028573 | 0.004418 | 4.20E-10 |
| rs492488 | let-7b-5p | trans | 1.07E-26 | Nightingale | M_LDL_C | NA | A | G | 0.033998 | 0.004433 | 2.10E-14 |
| rs492488 | let-7b-5p | trans | 1.07E-26 | Nightingale | S_HDL_PL | NA | A | G | 0.023794 | 0.004387 | 3.20E-08 |
| rs492488 | let-7b-5p | trans | 1.07E-26 | Nightingale | XS_VLDL_L | NA | A | G | 0.033831 | 0.00435 | 2.00E-15 |
| rs492488 | let-7b-5p | trans | 1.07E-26 | Nightingale | XL_VLDL_P | NA | A | G | 0.038845 | 0.004432 | 5.20E-19 |
| rs492488 | let-7b-5p | trans | 1.07E-26 | Nightingale | S_LDL_L | NA | A | G | 0.027751 | 0.004424 | 8.20E-10 |
| rs492488 | let-7b-5p | trans | 1.07E-26 | Nightingale | ApoB_by_A | NA | A | G | 0.02505 | 0.004358 | 1.30E-08 |
| rs492488 | let-7b-5p | trans | 1.07E-26 | Nightingale | XL_VLDL_T | NA | A | G | -0.04393 | 0.004289 | 1.70E-24 |
| rs492488 | let-7b-5p | trans | 1.07E-26 | Nightingale | S_VLDL_L | NA | A | G | 0.024676 | 0.004382 | 3.70E-09 |
| rs492488 | let-7b-5p | trans | 1.07E-26 | Nightingale | XXL_VLDL_ | NA | A | G | 0.033132 | 0.004459 | 8.80E-14 |
| rs492488 | let-7b-5p | trans | 1.07E-26 | Nightingale | S_VLDL_PL | NA | A | G | 0.032786 | 0.004417 | 4.20E-14 |
| rs492488 | let-7b-5p | trans | 1.07E-26 | Nightingale | LDL_P | NA | A | G | 0.03181 | 0.004422 | 1.00E-12 |
| rs492488 | let-7b-5p | trans | 1.07E-26 | Nightingale | S_VLDL_PL | NA | A | G | 0.027742 | 0.004237 | 1.80E-10 |
| rs492488 | let-7b-5p | trans | 1.07E-26 | Nightingale | LDL_PL | NA | A | G | 0.035001 | 0.004415 | 4.70E-15 |
| rs492488 | let-7b-5p | trans | 1.07E-26 | Nightingale | XXL_VLDL_ | NA | A | G | 0.030859 | 0.004406 | 1.10E-12 |
| rs492488 | let-7b-5p | trans | 1.07E-26 | Nightingale | L_LDL_C | NA | A | G | 0.032333 | 0.004375 | 2.50E-13 |
| rs492488 | let-7b-5p | trans | 1.07E-26 | Nightingale | Total_C | NA | A | G | 0.032388 | 0.004284 | 3.20E-14 |
| rs492488 | let-7b-5p | trans | 1.07E-26 | Nightingale | M_VLDL_P | NA | A | G | 0.028302 | 0.004414 | 6.30E-11 |
| rs492488 | let-7b-5p | trans | 1.07E-26 | Nightingale | S_LDL_CE | NA | A | G | 0.030558 | 0.004427 | 5.80E-12 |
| rs492488 | let-7b-5p | trans | 1.07E-26 | Nightingale | LDL_CE | NA | A | G | 0.032566 | 0.004413 | 2.20E-13 |
| rs492488 | let-7b-5p | trans | 1.07E-26 | Nightingale | Clinical_LD | NA | A | G | 0.036486 | 0.004399 | 2.70E-16 |
| rs492488 | let-7b-5p | trans | 1.07E-26 | Nightingale | S_VLDL_CE | NA | A | G | 0.038189 | 0.004419 | 1.60E-18 |
| rs492488 | let-7b-5p | trans | 1.07E-26 | Nightingale | Cholines | NA | A | G | 0.022312 | 0.004161 | 4.70E-08 |
| rs492488 | let-7b-5p | trans | 1.07E-26 | Nightingale | M_VLDL_C | NA | A | G | 0.040853 | 0.004385 | 2.30E-20 |
| rs492488 | let-7b-5p | trans | 1.07E-26 | Nightingale | S_VLDL_FC | NA | A | G | 0.037913 | 0.004426 | 7.30E-18 |
| rs492488 | let-7b-5p | trans | 1.07E-26 | Nightingale | L_LDL_L | NA | A | G | 0.033564 | 0.004382 | 2.90E-14 |
| rs492488 | let-7b-5p | trans | 1.07E-26 | Nightingale | M_VLDL_P | NA | A | G | 0.029135 | 0.004418 | 2.00E-11 |
| rs492488 | let-7b-5p | trans | 1.07E-26 | Nightingale | S_HDL_CE | NA | A | G | 0.028644 | 0.004432 | 1.30E-10 |
| rs492488 | let-7b-5p | trans | 1.07E-26 | Nightingale | M_LDL_CE | NA | A | G | 0.032983 | 0.004431 | 9.50E-14 |
| rs492488 | let-7b-5p | trans | 1.07E-26 | Nightingale | S_LDL_C | NA | A | G | 0.030953 | 0.004429 | 5.50E-12 |
| rs492488 | let-7b-5p | trans | 1.07E-26 | Nightingale | S_HDL_P | NA | A | G | 0.030622 | 0.004425 | 5.00E-12 |
| rs492488 | let-7b-5p | trans | 1.07E-26 | Nightingale | ApoB | NA | A | G | 0.033948 | 0.004419 | 2.10E-14 |
| rs492488 | let-7b-5p | trans | 1.07E-26 | Nightingale | Citrate | NA | A | G | 0.026826 | 0.004396 | 7.40E-10 |
| rs492488 | let-7b-5p | trans | 1.07E-26 | Nightingale | M_VLDL_T | NA | A | G | -0.03857 | 0.004181 | 6.70E-20 |
| rs492488 | let-7b-5p | trans | 1.07E-26 | Nightingale | XS_VLDL_C | NA | A | G | 0.038005 | 0.004295 | 1.30E-18 |
| rs492488 | let-7b-5p | trans | 1.07E-26 | Nightingale | S_LDL_CE_ | NA | A | G | 0.03095 | 0.004417 | 4.80E-14 |
| rs492488 | let-7b-5p | trans | 1.07E-26 | Nightingale | M_LDL_FC | NA | A | G | 0.034359 | 0.004419 | 1.90E-14 |
| rs492488 | let-7b-5p | trans | 1.07E-26 | Nightingale | IDL_L | NA | A | G | 0.035284 | 0.004278 | 4.70E-16 |
| rs492488 | let-7b-5p | trans | 1.07E-26 | Nightingale | L_LDL_CE | NA | A | G | 0.031581 | 0.004385 | 9.70E-13 |
| rs492488 | let-7b-5p | trans | 1.07E-26 | Nightingale | Omega_6 | NA | A | G | 0.025912 | 0.004321 | 5.60E-10 |
| rs492488 | miR-10b-5p | trans | 3.26E-35 | Nightingale | VLDL_CE | NA | A | G | 0.035795 | 0.004426 | 3.00E-16 |
| rs492488 | miR-10b-5p | trans | 3.26E-35 | Nightingale | S_VLDL_CE | NA | A | G | 0.047037 | 0.004365 | 1.20E-26 |
| rs492488 | miR-10b-5p | trans | 3.26E-35 | Nightingale | VLDL_FC | NA | A | G | 0.024164 | 0.004392 | 1.10E-08 |
| rs492488 | miR-10b-5p | trans | 3.26E-35 | Nightingale | Sphingomy | NA | A | G | 0.029031 | 0.004137 | 6.60E-12 |
| rs492488 | miR-10b-5p | trans | 3.26E-35 | Nightingale | IDL_FC | NA | A | G | 0.038268 | 0.004284 | 1.40E-18 |
| rs492488 | miR-10b-5p | trans | 3.26E-35 | Nightingale | VLDL_P | NA | A | G | 0.026641 | 0.004401 | 4.40E-10 |
| rs492488 | miR-10b-5p | trans | 3.26E-35 | Nightingale | Total_L | NA | A | G | 0.026363 | 0.004353 | 4.50E-10 |
| rs492488 | miR-10b-5p | trans | 3.26E-35 | Nightingale | XXL_VLDL_ | NA | A | G | 0.034841 | 0.004434 | 2.90E-15 |
| rs492488 | miR-10b-5p | trans | 3.26E-35 | Nightingale | XS_VLDL_P | NA | A | G | 0.032128 | 0.004363 | 5.30E-14 |
| rs492488 | miR-10b-5p | trans | 3.26E-35 | Nightingale | M_LDL_L | NA | A | G | 0.033043 | 0.004431 | 1.00E-13 |
| rs492488 | miR-10b-5p | trans | 3.26E-35 | Nightingale | S_VLDL_P | NA | A | G | 0.023473 | 0.004369 | 1.60E-08 |
| rs492488 | miR-10b-5p | trans | 3.26E-35 | Nightingale | LDL_C | NA | A | G | 0.033206 | 0.004406 | 8.70E-14 |
| rs492488 | miR-10b-5p | trans | 3.26E-35 | Nightingale | Remnant_( | NA | A | G | 0.036879 | 0.004396 | 3.60E-17 |
| rs492488 | miR-10b-5p | trans | 3.26E-35 | Nightingale | L_VLDL_TG | NA | A | G | -0.05115 | 0.004404 | 5.30E-32 |
| rs492488 | miR-10b-5p | trans | 3.26E-35 | Nightingale | GlycA | NA | A | G | 0.028022 | 0.004406 | 1.40E-10 |
| rs492488 | miR-10b-5p | trans | 3.26E-35 | Nightingale | S_HDL_C | NA | A | G | 0.028422 | 0.004417 | 1.70E-10 |
| rs492488 | miR-10b-5p | trans | 3.26E-35 | Nightingale | S_HDL_L | NA | A | G | 0.026306 | 0.00441 | 1.50E-09 |
| rs492488 | miR-10b-5p | trans | 3.26E-35 | Nightingale | Total_CE | NA | A | G | 0.031761 | 0.004268 | 9.50E-14 |

| | | | | | | | | | | | |
|---|---|---|---|---|---|---|---|---|---|---|---|
| rs492488 | miR-10b-5p | trans | 3.26E-35 | Nightingale | S_VLDL_TG | NA | A | G | -0.03813 | 0.004283 | 1.40E-18 |
| rs492488 | miR-10b-5p | trans | 3.26E-35 | Nightingale | S_HDL_FC | NA | A | G | 0.023468 | 0.004344 | 4.50E-08 |
| rs492488 | miR-10b-5p | trans | 3.26E-35 | Nightingale | IDL_FC_pct | NA | A | G | 0.025772 | 0.004391 | 1.10E-08 |
| rs492488 | miR-10b-5p | trans | 3.26E-35 | Nightingale | M_VLDL_F | NA | A | G | 0.033379 | 0.004424 | 3.10E-14 |
| rs492488 | miR-10b-5p | trans | 3.26E-35 | Nightingale | S_VLDL_C_ | NA | A | G | 0.041892 | 0.004306 | 5.20E-22 |
| rs492488 | miR-10b-5p | trans | 3.26E-35 | Nightingale | IDL_C | NA | A | G | 0.034968 | 0.004274 | 9.20E-16 |
| rs492488 | miR-10b-5p | trans | 3.26E-35 | Nightingale | XS_VLDL_C | NA | A | G | 0.03781 | 0.00427 | 2.10E-18 |
| rs492488 | miR-10b-5p | trans | 3.26E-35 | Nightingale | S_VLDL_FC | NA | A | G | 0.02936 | 0.004228 | 1.10E-11 |
| rs492488 | miR-10b-5p | trans | 3.26E-35 | Nightingale | M_VLDL_C | NA | A | G | 0.03872 | 0.004413 | 1.90E-18 |
| rs492488 | miR-10b-5p | trans | 3.26E-35 | Nightingale | M_VLDL_F | NA | A | G | 0.042074 | 0.00421 | 3.40E-23 |
| rs492488 | miR-10b-5p | trans | 3.26E-35 | Nightingale | XS_VLDL_F | NA | A | G | 0.03143 | 0.004393 | 2.40E-12 |
| rs492488 | miR-10b-5p | trans | 3.26E-35 | Nightingale | L_LDL_FC | NA | A | G | 0.033638 | 0.004345 | 2.80E-14 |
| rs492488 | miR-10b-5p | trans | 3.26E-35 | Nightingale | L_VLDL_FC | NA | A | G | 0.048596 | 0.004382 | 4.50E-29 |
| rs492488 | miR-10b-5p | trans | 3.26E-35 | Nightingale | Total_PL | NA | A | G | 0.024943 | 0.004214 | 1.00E-09 |
| rs492488 | miR-10b-5p | trans | 3.26E-35 | Nightingale | S_LDL_P | NA | A | G | 0.029597 | 0.004429 | 3.20E-11 |
| rs492488 | miR-10b-5p | trans | 3.26E-35 | Nightingale | XS_VLDL_F | NA | A | G | 0.03708 | 0.004348 | 6.40E-18 |
| rs492488 | miR-10b-5p | trans | 3.26E-35 | Nightingale | IDL_PL | NA | A | G | 0.035357 | 0.00427 | 3.30E-16 |
| rs492488 | miR-10b-5p | trans | 3.26E-35 | Nightingale | XL_HDL_PL | NA | A | G | -0.02274 | 0.004065 | 1.20E-08 |
| rs492488 | miR-10b-5p | trans | 3.26E-35 | Nightingale | M_VLDL_C | NA | A | G | 0.036215 | 0.004168 | 1.20E-17 |
| rs492488 | miR-10b-5p | trans | 3.26E-35 | Nightingale | XXL_VLDL_ | NA | A | G | 0.025012 | 0.004454 | 1.50E-09 |
| rs492488 | miR-10b-5p | trans | 3.26E-35 | Nightingale | S_LDL_PL_ | NA | A | G | -0.03007 | 0.004417 | 2.60E-13 |
| rs492488 | miR-10b-5p | trans | 3.26E-35 | Nightingale | S_VLDL_C | NA | A | G | 0.038381 | 0.004424 | 1.80E-18 |
| rs492488 | miR-10b-5p | trans | 3.26E-35 | Nightingale | XL_VLDL_C | NA | A | G | 0.037359 | 0.004245 | 3.10E-18 |
| rs492488 | miR-10b-5p | trans | 3.26E-35 | Nightingale | M_LDL_C_ | NA | A | G | 0.031163 | 0.004428 | 2.90E-12 |
| rs492488 | miR-10b-5p | trans | 3.26E-35 | Nightingale | S_LDL_C_p | NA | A | G | 0.045367 | 0.004442 | 5.40E-25 |
| rs492488 | miR-10b-5p | trans | 3.26E-35 | Nightingale | XXL_VLDL_ | NA | A | G | -0.03574 | 0.004463 | 2.10E-16 |
| rs492488 | miR-10b-5p | trans | 3.26E-35 | Nightingale | LA | NA | A | G | 0.024925 | 0.004339 | 3.60E-09 |
| rs492488 | miR-10b-5p | trans | 3.26E-35 | Nightingale | L_LDL_P | NA | A | G | 0.029187 | 0.00441 | 6.10E-11 |
| rs492488 | miR-10b-5p | trans | 3.26E-35 | Nightingale | M_LDL_P | NA | A | G | 0.035901 | 0.004437 | 6.80E-16 |
| rs492488 | miR-10b-5p | trans | 3.26E-35 | Nightingale | M_LDL_TG | NA | A | G | -0.02762 | 0.00437 | 3.20E-10 |
| rs492488 | miR-10b-5p | trans | 3.26E-35 | Nightingale | L_VLDL_C_ | NA | A | G | 0.052206 | 0.004322 | 3.90E-34 |
| rs492488 | miR-10b-5p | trans | 3.26E-35 | Nightingale | LDL_FC | NA | A | G | 0.033644 | 0.004378 | 5.20E-14 |
| rs492488 | miR-10b-5p | trans | 3.26E-35 | Nightingale | LDL_L | NA | A | G | 0.033317 | 0.00441 | 7.30E-14 |
| rs492488 | miR-10b-5p | trans | 3.26E-35 | Nightingale | PUFA | NA | A | G | 0.023739 | 0.004316 | 1.40E-08 |
| rs492488 | miR-10b-5p | trans | 3.26E-35 | Nightingale | VLDL_C | NA | A | G | 0.031309 | 0.004419 | 4.80E-13 |
| rs492488 | miR-10b-5p | trans | 3.26E-35 | Nightingale | XL_VLDL_F | NA | A | G | 0.037204 | 0.00429 | 2.00E-18 |
| rs492488 | miR-10b-5p | trans | 3.26E-35 | Nightingale | M_VLDL_P | NA | A | G | 0.044067 | 0.004247 | 5.40E-25 |
| rs492488 | miR-10b-5p | trans | 3.26E-35 | Nightingale | L_VLDL_CE | NA | A | G | 0.04735 | 0.004234 | 2.40E-29 |
| rs492488 | miR-10b-5p | trans | 3.26E-35 | Nightingale | XS_VLDL_P | NA | A | G | 0.036714 | 0.004366 | 1.40E-17 |
| rs492488 | miR-10b-5p | trans | 3.26E-35 | Nightingale | non_HDL_( | NA | A | G | 0.035709 | 0.004401 | 5.00E-16 |
| rs492488 | miR-10b-5p | trans | 3.26E-35 | Nightingale | M_LDL_PL | NA | A | G | 0.032478 | 0.004431 | 2.60E-13 |
| rs492488 | miR-10b-5p | trans | 3.26E-35 | Nightingale | M_VLDL_C | NA | A | G | 0.033637 | 0.004157 | 1.90E-15 |
| rs492488 | miR-10b-5p | trans | 3.26E-35 | Nightingale | IDL_CE | NA | A | G | 0.03352 | 0.004272 | 1.30E-14 |
| rs492488 | miR-10b-5p | trans | 3.26E-35 | Nightingale | Total_FC | NA | A | G | 0.03327 | 0.004323 | 6.70E-15 |
| rs492488 | miR-10b-5p | trans | 3.26E-35 | Nightingale | XL_VLDL_C | NA | A | G | 0.03823 | 0.004243 | 4.60E-19 |
| rs492488 | miR-10b-5p | trans | 3.26E-35 | Nightingale | L_LDL_PL | NA | A | G | 0.038002 | 0.004391 | 8.90E-18 |
| rs492488 | miR-10b-5p | trans | 3.26E-35 | Nightingale | IDL_P | NA | A | G | 0.039997 | 0.004378 | 1.30E-19 |
| rs492488 | miR-10b-5p | trans | 3.26E-35 | Nightingale | S_LDL_FC | NA | A | G | 0.028573 | 0.004418 | 4.20E-10 |
| rs492488 | miR-10b-5p | trans | 3.26E-35 | Nightingale | M_LDL_C | NA | A | G | 0.033998 | 0.004433 | 2.10E-14 |
| rs492488 | miR-10b-5p | trans | 3.26E-35 | Nightingale | S_HDL_PL | NA | A | G | 0.023794 | 0.004387 | 3.20E-08 |
| rs492488 | miR-10b-5p | trans | 3.26E-35 | Nightingale | XS_VLDL_L | NA | A | G | 0.033831 | 0.00435 | 2.00E-15 |
| rs492488 | miR-10b-5p | trans | 3.26E-35 | Nightingale | XL_VLDL_P | NA | A | G | 0.038845 | 0.004432 | 5.20E-19 |
| rs492488 | miR-10b-5p | trans | 3.26E-35 | Nightingale | S_LDL_L | NA | A | G | 0.027751 | 0.004424 | 8.20E-10 |
| rs492488 | miR-10b-5p | trans | 3.26E-35 | Nightingale | ApoB_by_/ | NA | A | G | 0.02505 | 0.004358 | 1.30E-08 |
| rs492488 | miR-10b-5p | trans | 3.26E-35 | Nightingale | XL_VLDL_T | NA | A | G | -0.04393 | 0.004289 | 1.70E-24 |
| rs492488 | miR-10b-5p | trans | 3.26E-35 | Nightingale | S_VLDL_L | NA | A | G | 0.024676 | 0.004382 | 3.70E-09 |
| rs492488 | miR-10b-5p | trans | 3.26E-35 | Nightingale | XXL_VLDL_ | NA | A | G | 0.033132 | 0.004459 | 8.80E-14 |
| rs492488 | miR-10b-5p | trans | 3.26E-35 | Nightingale | S_VLDL_PL | NA | A | G | 0.032786 | 0.004417 | 4.20E-14 |
| rs492488 | miR-10b-5p | trans | 3.26E-35 | Nightingale | LDL_P | NA | A | G | 0.03181 | 0.004422 | 1.00E-12 |
| rs492488 | miR-10b-5p | trans | 3.26E-35 | Nightingale | S_VLDL_PL | NA | A | G | 0.027742 | 0.004237 | 1.80E-10 |
| rs492488 | miR-10b-5p | trans | 3.26E-35 | Nightingale | LDL_PL | NA | A | G | 0.035001 | 0.004415 | 4.70E-15 |
| rs492488 | miR-10b-5p | trans | 3.26E-35 | Nightingale | XXL_VLDL_ | NA | A | G | 0.030859 | 0.004406 | 1.10E-12 |
| rs492488 | miR-10b-5p | trans | 3.26E-35 | Nightingale | L_LDL_C | NA | A | G | 0.032333 | 0.004375 | 2.50E-13 |

| | | | | | | | | | | | |
|---|---|---|---|---|---|---|---|---|---|---|---|
| rs492488 | miR-10b-5p | trans | 3.26E-35 | Nightingale | Total_C | NA | A | G | 0.032388 | 0.004284 | 3.20E-14 |
| rs492488 | miR-10b-5p | trans | 3.26E-35 | Nightingale | M_VLDL_P | NA | A | G | 0.028302 | 0.004414 | 6.30E-11 |
| rs492488 | miR-10b-5p | trans | 3.26E-35 | Nightingale | S_LDL_CE | NA | A | G | 0.030558 | 0.004427 | 5.80E-12 |
| rs492488 | miR-10b-5p | trans | 3.26E-35 | Nightingale | LDL_CE | NA | A | G | 0.032566 | 0.004413 | 2.20E-13 |
| rs492488 | miR-10b-5p | trans | 3.26E-35 | Nightingale | Clinical_LD | NA | A | G | 0.036486 | 0.004399 | 2.70E-16 |
| rs492488 | miR-10b-5p | trans | 3.26E-35 | Nightingale | S_VLDL_CE | NA | A | G | 0.038189 | 0.004419 | 1.60E-18 |
| rs492488 | miR-10b-5p | trans | 3.26E-35 | Nightingale | Cholines | NA | A | G | 0.022312 | 0.004161 | 4.70E-08 |
| rs492488 | miR-10b-5p | trans | 3.26E-35 | Nightingale | M_VLDL_C | NA | A | G | 0.040853 | 0.004385 | 2.30E-20 |
| rs492488 | miR-10b-5p | trans | 3.26E-35 | Nightingale | S_VLDL_FC | NA | A | G | 0.037913 | 0.004426 | 7.30E-18 |
| rs492488 | miR-10b-5p | trans | 3.26E-35 | Nightingale | L_LDL_L | NA | A | G | 0.033564 | 0.004382 | 2.90E-14 |
| rs492488 | miR-10b-5p | trans | 3.26E-35 | Nightingale | M_VLDL_P | NA | A | G | 0.029135 | 0.004418 | 2.00E-11 |
| rs492488 | miR-10b-5p | trans | 3.26E-35 | Nightingale | S_HDL_CE | NA | A | G | 0.028644 | 0.004432 | 1.30E-10 |
| rs492488 | miR-10b-5p | trans | 3.26E-35 | Nightingale | M_LDL_CE | NA | A | G | 0.032983 | 0.004431 | 9.50E-14 |
| rs492488 | miR-10b-5p | trans | 3.26E-35 | Nightingale | S_LDL_C | NA | A | G | 0.030953 | 0.004429 | 5.50E-12 |
| rs492488 | miR-10b-5p | trans | 3.26E-35 | Nightingale | S_HDL_P | NA | A | G | 0.030622 | 0.004425 | 5.00E-12 |
| rs492488 | miR-10b-5p | trans | 3.26E-35 | Nightingale | ApoB | NA | A | G | 0.033948 | 0.004419 | 2.10E-14 |
| rs492488 | miR-10b-5p | trans | 3.26E-35 | Nightingale | Citrate | NA | A | G | 0.026826 | 0.004396 | 7.40E-10 |
| rs492488 | miR-10b-5p | trans | 3.26E-35 | Nightingale | M_VLDL_T | NA | A | G | -0.03857 | 0.004181 | 6.70E-20 |
| rs492488 | miR-10b-5p | trans | 3.26E-35 | Nightingale | XS_VLDL_C | NA | A | G | 0.038005 | 0.004295 | 1.30E-18 |
| rs492488 | miR-10b-5p | trans | 3.26E-35 | Nightingale | S_LDL_CE_ | NA | A | G | 0.03095 | 0.004417 | 4.80E-14 |
| rs492488 | miR-10b-5p | trans | 3.26E-35 | Nightingale | M_LDL_FC | NA | A | G | 0.034359 | 0.004419 | 1.90E-14 |
| rs492488 | miR-10b-5p | trans | 3.26E-35 | Nightingale | IDL_L | NA | A | G | 0.035284 | 0.004278 | 4.70E-16 |
| rs492488 | miR-10b-5p | trans | 3.26E-35 | Nightingale | L_LDL_CE | NA | A | G | 0.031581 | 0.004385 | 9.70E-13 |
| rs492488 | miR-10b-5p | trans | 3.26E-35 | Nightingale | Omega_6 | NA | A | G | 0.025912 | 0.004321 | 5.60E-10 |
| rs493246 | miR-10b-5p | trans | 3.23E-35 | Nightingale | IDL_FC | NA | A | G | 0.038267 | 0.004284 | 1.40E-18 |
| rs493246 | miR-10b-5p | trans | 3.23E-35 | Nightingale | LDL_CE | NA | A | G | 0.032566 | 0.004413 | 2.20E-13 |
| rs493246 | miR-10b-5p | trans | 3.23E-35 | Nightingale | L_VLDL_CE | NA | A | G | 0.04735 | 0.004234 | 2.40E-29 |
| rs493246 | miR-10b-5p | trans | 3.23E-35 | Nightingale | M_LDL_C_ | NA | A | G | 0.031162 | 0.004428 | 2.90E-12 |
| rs493246 | miR-10b-5p | trans | 3.23E-35 | Nightingale | XS_VLDL_F | NA | A | G | 0.03143 | 0.004393 | 2.40E-12 |
| rs493246 | miR-10b-5p | trans | 3.23E-35 | Nightingale | S_HDL_FC | NA | A | G | 0.023468 | 0.004344 | 4.50E-08 |
| rs493246 | miR-10b-5p | trans | 3.23E-35 | Nightingale | S_LDL_L | NA | A | G | 0.027751 | 0.004424 | 8.20E-10 |
| rs493246 | miR-10b-5p | trans | 3.23E-35 | Nightingale | LDL_L | NA | A | G | 0.033316 | 0.00441 | 7.30E-14 |
| rs493246 | miR-10b-5p | trans | 3.23E-35 | Nightingale | M_LDL_L | NA | A | G | 0.033043 | 0.004431 | 1.00E-13 |
| rs493246 | miR-10b-5p | trans | 3.23E-35 | Nightingale | M_LDL_C | NA | A | G | 0.033998 | 0.004433 | 2.10E-14 |
| rs493246 | miR-10b-5p | trans | 3.23E-35 | Nightingale | Clinical_LD | NA | A | G | 0.036486 | 0.004399 | 2.70E-16 |
| rs493246 | miR-10b-5p | trans | 3.23E-35 | Nightingale | S_VLDL_FC | NA | A | G | 0.02936 | 0.004228 | 1.10E-11 |
| rs493246 | miR-10b-5p | trans | 3.23E-35 | Nightingale | IDL_FC_pct | NA | A | G | 0.025772 | 0.004391 | 1.10E-08 |
| rs493246 | miR-10b-5p | trans | 3.23E-35 | Nightingale | M_VLDL_P | NA | A | G | 0.044066 | 0.004247 | 5.50E-25 |
| rs493246 | miR-10b-5p | trans | 3.23E-35 | Nightingale | Total_CE | NA | A | G | 0.031761 | 0.004268 | 9.50E-14 |
| rs493246 | miR-10b-5p | trans | 3.23E-35 | Nightingale | M_VLDL_F | NA | A | G | 0.033378 | 0.004424 | 3.10E-14 |
| rs493246 | miR-10b-5p | trans | 3.23E-35 | Nightingale | IDL_C | NA | A | G | 0.034968 | 0.004274 | 9.20E-16 |
| rs493246 | miR-10b-5p | trans | 3.23E-35 | Nightingale | LDL_PL | NA | A | G | 0.035 | 0.004415 | 4.70E-15 |
| rs493246 | miR-10b-5p | trans | 3.23E-35 | Nightingale | M_VLDL_P | NA | A | G | 0.029134 | 0.004418 | 2.00E-11 |
| rs493246 | miR-10b-5p | trans | 3.23E-35 | Nightingale | XL_VLDL_P | NA | A | G | 0.038845 | 0.004432 | 5.20E-19 |
| rs493246 | miR-10b-5p | trans | 3.23E-35 | Nightingale | XXL_VLDL_ | NA | A | G | 0.034841 | 0.004434 | 2.90E-15 |
| rs493246 | miR-10b-5p | trans | 3.23E-35 | Nightingale | XS_VLDL_F | NA | A | G | 0.03708 | 0.004348 | 6.40E-18 |
| rs493246 | miR-10b-5p | trans | 3.23E-35 | Nightingale | GlycA | NA | A | G | 0.028022 | 0.004406 | 1.40E-10 |
| rs493246 | miR-10b-5p | trans | 3.23E-35 | Nightingale | XL_VLDL_C | NA | A | G | 0.03823 | 0.004243 | 4.60E-19 |
| rs493246 | miR-10b-5p | trans | 3.23E-35 | Nightingale | M_VLDL_C | NA | A | G | 0.040853 | 0.004385 | 2.30E-20 |
| rs493246 | miR-10b-5p | trans | 3.23E-35 | Nightingale | Cholines | NA | A | G | 0.022312 | 0.004161 | 4.70E-08 |
| rs493246 | miR-10b-5p | trans | 3.23E-35 | Nightingale | S_HDL_CE | NA | A | G | 0.028644 | 0.004432 | 1.30E-10 |
| rs493246 | miR-10b-5p | trans | 3.23E-35 | Nightingale | M_VLDL_C | NA | A | G | 0.038719 | 0.004413 | 1.90E-18 |
| rs493246 | miR-10b-5p | trans | 3.23E-35 | Nightingale | XS_VLDL_C | NA | A | G | 0.03781 | 0.00427 | 2.10E-18 |
| rs493246 | miR-10b-5p | trans | 3.23E-35 | Nightingale | VLDL_CE | NA | A | G | 0.035794 | 0.004426 | 3.00E-16 |
| rs493246 | miR-10b-5p | trans | 3.23E-35 | Nightingale | S_HDL_L | NA | A | G | 0.026306 | 0.00441 | 1.50E-09 |
| rs493246 | miR-10b-5p | trans | 3.23E-35 | Nightingale | Total_FC | NA | A | G | 0.033269 | 0.004323 | 6.70E-15 |
| rs493246 | miR-10b-5p | trans | 3.23E-35 | Nightingale | M_VLDL_F | NA | A | G | 0.042074 | 0.00421 | 3.40E-23 |
| rs493246 | miR-10b-5p | trans | 3.23E-35 | Nightingale | S_VLDL_L | NA | A | G | 0.024676 | 0.004382 | 3.70E-09 |
| rs493246 | miR-10b-5p | trans | 3.23E-35 | Nightingale | S_LDL_PL_ | NA | A | G | -0.03007 | 0.004417 | 2.60E-13 |
| rs493246 | miR-10b-5p | trans | 3.23E-35 | Nightingale | XS_VLDL_P | NA | A | G | 0.032128 | 0.004363 | 5.30E-14 |
| rs493246 | miR-10b-5p | trans | 3.23E-35 | Nightingale | Sphingomy | NA | A | G | 0.029031 | 0.004137 | 6.60E-12 |
| rs493246 | miR-10b-5p | trans | 3.23E-35 | Nightingale | S_LDL_C_p | NA | A | G | 0.045367 | 0.004442 | 5.40E-25 |
| rs493246 | miR-10b-5p | trans | 3.23E-35 | Nightingale | VLDL_FC | NA | A | G | 0.024164 | 0.004392 | 1.10E-08 |

| | | | | | | | | | | | |
|---|---|---|---|---|---|---|---|---|---|---|---|
| rs493246 | miR-10b-5| trans | 3.23E-35 | Nightingale | XL_VLDL_C | NA | A | G | 0.037359 | 0.004245 | 3.10E-18 |
| rs493246 | miR-10b-5| trans | 3.23E-35 | Nightingale | L_VLDL_C_ | NA | A | G | 0.052205 | 0.004322 | 3.90E-34 |
| rs493246 | miR-10b-5| trans | 3.23E-35 | Nightingale | Total_C | NA | A | G | 0.032388 | 0.004284 | 3.20E-14 |
| rs493246 | miR-10b-5| trans | 3.23E-35 | Nightingale | non_HDL_( | NA | A | G | 0.035709 | 0.004401 | 5.00E-16 |
| rs493246 | miR-10b-5| trans | 3.23E-35 | Nightingale | IDL_P | NA | A | G | 0.039997 | 0.004378 | 1.30E-19 |
| rs493246 | miR-10b-5| trans | 3.23E-35 | Nightingale | S_LDL_FC | NA | A | G | 0.028573 | 0.004418 | 4.20E-10 |
| rs493246 | miR-10b-5| trans | 3.23E-35 | Nightingale | XXL_VLDL_ | NA | A | G | 0.025012 | 0.004454 | 1.50E-09 |
| rs493246 | miR-10b-5| trans | 3.23E-35 | Nightingale | L_LDL_PL | NA | A | G | 0.038002 | 0.004391 | 8.90E-18 |
| rs493246 | miR-10b-5| trans | 3.23E-35 | Nightingale | L_VLDL_TG | NA | A | G | -0.05115 | 0.004404 | 5.30E-32 |
| rs493246 | miR-10b-5| trans | 3.23E-35 | Nightingale | S_VLDL_PL | NA | A | G | 0.032786 | 0.004417 | 4.20E-14 |
| rs493246 | miR-10b-5| trans | 3.23E-35 | Nightingale | IDL_PL | NA | A | G | 0.035357 | 0.00427 | 3.30E-16 |
| rs493246 | miR-10b-5| trans | 3.23E-35 | Nightingale | M_VLDL_C | NA | A | G | 0.036214 | 0.004168 | 1.20E-17 |
| rs493246 | miR-10b-5| trans | 3.23E-35 | Nightingale | XL_HDL_PL | NA | A | G | -0.02274 | 0.004065 | 1.20E-08 |
| rs493246 | miR-10b-5| trans | 3.23E-35 | Nightingale | XXL_VLDL_ | NA | A | G | 0.030859 | 0.004406 | 1.10E-12 |
| rs493246 | miR-10b-5| trans | 3.23E-35 | Nightingale | XS_VLDL_C | NA | A | G | 0.038005 | 0.004295 | 1.30E-18 |
| rs493246 | miR-10b-5| trans | 3.23E-35 | Nightingale | M_LDL_TG | NA | A | G | -0.02762 | 0.00437 | 3.20E-10 |
| rs493246 | miR-10b-5| trans | 3.23E-35 | Nightingale | S_VLDL_CE | NA | A | G | 0.047037 | 0.004365 | 1.20E-26 |
| rs493246 | miR-10b-5| trans | 3.23E-35 | Nightingale | S_HDL_C | NA | A | G | 0.028422 | 0.004417 | 1.70E-10 |
| rs493246 | miR-10b-5| trans | 3.23E-35 | Nightingale | LDL_P | NA | A | G | 0.031809 | 0.004422 | 1.00E-12 |
| rs493246 | miR-10b-5| trans | 3.23E-35 | Nightingale | PUFA | NA | A | G | 0.023738 | 0.004316 | 1.40E-08 |
| rs493246 | miR-10b-5| trans | 3.23E-35 | Nightingale | M_VLDL_T( | NA | A | G | -0.03857 | 0.004181 | 6.70E-20 |
| rs493246 | miR-10b-5| trans | 3.23E-35 | Nightingale | VLDL_C | NA | A | G | 0.031309 | 0.004419 | 4.80E-13 |
| rs493246 | miR-10b-5| trans | 3.23E-35 | Nightingale | VLDL_P | NA | A | G | 0.026641 | 0.004401 | 4.40E-10 |
| rs493246 | miR-10b-5| trans | 3.23E-35 | Nightingale | S_HDL_PL | NA | A | G | 0.023794 | 0.004387 | 3.20E-08 |
| rs493246 | miR-10b-5| trans | 3.23E-35 | Nightingale | XL_VLDL_T | NA | A | G | -0.04393 | 0.004289 | 1.70E-24 |
| rs493246 | miR-10b-5| trans | 3.23E-35 | Nightingale | LDL_FC | NA | A | G | 0.033644 | 0.004378 | 5.20E-14 |
| rs493246 | miR-10b-5| trans | 3.23E-35 | Nightingale | M_LDL_FC | NA | A | G | 0.034359 | 0.004419 | 1.90E-14 |
| rs493246 | miR-10b-5| trans | 3.23E-35 | Nightingale | M_LDL_PL | NA | A | G | 0.032478 | 0.004431 | 2.60E-13 |
| rs493246 | miR-10b-5| trans | 3.23E-35 | Nightingale | Omega_6 | NA | A | G | 0.025912 | 0.004321 | 5.60E-10 |
| rs493246 | miR-10b-5| trans | 3.23E-35 | Nightingale | Total_PL | NA | A | G | 0.024943 | 0.004214 | 1.00E-09 |
| rs493246 | miR-10b-5| trans | 3.23E-35 | Nightingale | S_VLDL_C_ | NA | A | G | 0.041892 | 0.004306 | 5.20E-22 |
| rs493246 | miR-10b-5| trans | 3.23E-35 | Nightingale | S_LDL_CE | NA | A | G | 0.030558 | 0.004427 | 5.80E-12 |
| rs493246 | miR-10b-5| trans | 3.23E-35 | Nightingale | L_LDL_P | NA | A | G | 0.029187 | 0.00441 | 6.10E-11 |
| rs493246 | miR-10b-5| trans | 3.23E-35 | Nightingale | XS_VLDL_P | NA | A | G | 0.036714 | 0.004366 | 1.40E-17 |
| rs493246 | miR-10b-5| trans | 3.23E-35 | Nightingale | LDL_C | NA | A | G | 0.033206 | 0.004406 | 8.70E-14 |
| rs493246 | miR-10b-5| trans | 3.23E-35 | Nightingale | S_VLDL_PL | NA | A | G | 0.027742 | 0.004237 | 1.80E-10 |
| rs493246 | miR-10b-5| trans | 3.23E-35 | Nightingale | M_LDL_P | NA | A | G | 0.0359 | 0.004437 | 6.80E-16 |
| rs493246 | miR-10b-5| trans | 3.23E-35 | Nightingale | S_LDL_C | NA | A | G | 0.030953 | 0.004429 | 5.50E-12 |
| rs493246 | miR-10b-5| trans | 3.23E-35 | Nightingale | XXL_VLDL_ | NA | A | G | 0.033132 | 0.004459 | 8.80E-14 |
| rs493246 | miR-10b-5| trans | 3.23E-35 | Nightingale | S_LDL_CE_ | NA | A | G | 0.03095 | 0.004417 | 4.80E-14 |
| rs493246 | miR-10b-5| trans | 3.23E-35 | Nightingale | LA | NA | A | G | 0.024925 | 0.004339 | 3.60E-09 |
| rs493246 | miR-10b-5| trans | 3.23E-35 | Nightingale | Remnant_( | NA | A | G | 0.036878 | 0.004396 | 3.60E-17 |
| rs493246 | miR-10b-5| trans | 3.23E-35 | Nightingale | L_VLDL_FC | NA | A | G | 0.048596 | 0.004382 | 4.50E-29 |
| rs493246 | miR-10b-5| trans | 3.23E-35 | Nightingale | S_VLDL_FC | NA | A | G | 0.037913 | 0.004426 | 7.30E-18 |
| rs493246 | miR-10b-5| trans | 3.23E-35 | Nightingale | S_LDL_P | NA | A | G | 0.029597 | 0.004429 | 3.20E-11 |
| rs493246 | miR-10b-5| trans | 3.23E-35 | Nightingale | M_LDL_CE | NA | A | G | 0.032983 | 0.004431 | 9.50E-14 |
| rs493246 | miR-10b-5| trans | 3.23E-35 | Nightingale | Total_L | NA | A | G | 0.026363 | 0.004353 | 4.50E-10 |
| rs493246 | miR-10b-5| trans | 3.23E-35 | Nightingale | L_LDL_L | NA | A | G | 0.033563 | 0.004382 | 2.90E-14 |
| rs493246 | miR-10b-5| trans | 3.23E-35 | Nightingale | M_VLDL_C | NA | A | G | 0.033637 | 0.004157 | 1.90E-15 |
| rs493246 | miR-10b-5| trans | 3.23E-35 | Nightingale | ApoB_by_/ | NA | A | G | 0.02505 | 0.004358 | 1.30E-08 |
| rs493246 | miR-10b-5| trans | 3.23E-35 | Nightingale | S_VLDL_C | NA | A | G | 0.038381 | 0.004424 | 1.80E-18 |
| rs493246 | miR-10b-5| trans | 3.23E-35 | Nightingale | S_VLDL_TG | NA | A | G | -0.03813 | 0.004283 | 1.40E-18 |
| rs493246 | miR-10b-5| trans | 3.23E-35 | Nightingale | ApoB | NA | A | G | 0.033948 | 0.004419 | 2.10E-14 |
| rs493246 | miR-10b-5| trans | 3.23E-35 | Nightingale | IDL_L | NA | A | G | 0.035283 | 0.004278 | 4.70E-16 |
| rs493246 | miR-10b-5| trans | 3.23E-35 | Nightingale | S_VLDL_P | NA | A | G | 0.023473 | 0.004369 | 1.60E-08 |
| rs493246 | miR-10b-5| trans | 3.23E-35 | Nightingale | L_LDL_C | NA | A | G | 0.032333 | 0.004375 | 2.50E-13 |
| rs493246 | miR-10b-5| trans | 3.23E-35 | Nightingale | IDL_CE | NA | A | G | 0.03352 | 0.004272 | 1.30E-14 |
| rs493246 | miR-10b-5| trans | 3.23E-35 | Nightingale | XL_VLDL_F | NA | A | G | 0.037204 | 0.00429 | 2.00E-18 |
| rs493246 | miR-10b-5| trans | 3.23E-35 | Nightingale | XXL_VLDL_ | NA | A | G | -0.03574 | 0.004463 | 2.10E-16 |
| rs493246 | miR-10b-5| trans | 3.23E-35 | Nightingale | XS_VLDL_L | NA | A | G | 0.033831 | 0.00435 | 2.00E-15 |
| rs493246 | miR-10b-5| trans | 3.23E-35 | Nightingale | M_VLDL_P | NA | A | G | 0.028301 | 0.004414 | 6.30E-11 |
| rs493246 | miR-10b-5| trans | 3.23E-35 | Nightingale | S_HDL_P | NA | A | G | 0.030621 | 0.004425 | 5.00E-12 |
| rs493246 | miR-10b-5| trans | 3.23E-35 | Nightingale | S_VLDL_CE | NA | A | G | 0.038189 | 0.004419 | 1.60E-18 |

| | | | | | | | | | | | |
|---|---|---|---|---|---|---|---|---|---|---|---|
| rs493246 | miR-10b-5p | trans | 3.23E-35 | Nightingale | L_LDL_FC | NA | A | G | 0.033637 | 0.004345 | 2.80E-14 |
| rs493246 | miR-10b-5p | trans | 3.23E-35 | Nightingale | Citrate | NA | A | G | 0.026826 | 0.004396 | 7.40E-10 |
| rs493246 | miR-10b-5p | trans | 3.23E-35 | Nightingale | L_LDL_CE | NA | A | G | 0.03158 | 0.004385 | 9.70E-13 |
| rs493246 | miR-195-5p | trans | 2.42E-17 | Nightingale | IDL_FC | NA | A | G | 0.038267 | 0.004284 | 1.40E-18 |
| rs493246 | miR-195-5p | trans | 2.42E-17 | Nightingale | LDL_CE | NA | A | G | 0.032566 | 0.004413 | 2.20E-13 |
| rs493246 | miR-195-5p | trans | 2.42E-17 | Nightingale | L_VLDL_CE | NA | A | G | 0.04735 | 0.004234 | 2.40E-29 |
| rs493246 | miR-195-5p | trans | 2.42E-17 | Nightingale | M_LDL_C_ | NA | A | G | 0.031162 | 0.004428 | 2.90E-12 |
| rs493246 | miR-195-5p | trans | 2.42E-17 | Nightingale | XS_VLDL_F | NA | A | G | 0.03143 | 0.004393 | 2.40E-12 |
| rs493246 | miR-195-5p | trans | 2.42E-17 | Nightingale | S_HDL_FC | NA | A | G | 0.023468 | 0.004344 | 4.50E-08 |
| rs493246 | miR-195-5p | trans | 2.42E-17 | Nightingale | S_LDL_L | NA | A | G | 0.027751 | 0.004424 | 8.20E-10 |
| rs493246 | miR-195-5p | trans | 2.42E-17 | Nightingale | LDL_L | NA | A | G | 0.033316 | 0.00441 | 7.30E-14 |
| rs493246 | miR-195-5p | trans | 2.42E-17 | Nightingale | M_LDL_L | NA | A | G | 0.033043 | 0.004431 | 1.00E-13 |
| rs493246 | miR-195-5p | trans | 2.42E-17 | Nightingale | M_LDL_C | NA | A | G | 0.033998 | 0.004433 | 2.10E-14 |
| rs493246 | miR-195-5p | trans | 2.42E-17 | Nightingale | Clinical_LD | NA | A | G | 0.036486 | 0.004399 | 2.70E-16 |
| rs493246 | miR-195-5p | trans | 2.42E-17 | Nightingale | S_VLDL_FC | NA | A | G | 0.02936 | 0.004228 | 1.10E-11 |
| rs493246 | miR-195-5p | trans | 2.42E-17 | Nightingale | IDL_FC_pct | NA | A | G | 0.025772 | 0.004391 | 1.10E-08 |
| rs493246 | miR-195-5p | trans | 2.42E-17 | Nightingale | M_VLDL_P | NA | A | G | 0.044066 | 0.004247 | 5.50E-25 |
| rs493246 | miR-195-5p | trans | 2.42E-17 | Nightingale | Total_CE | NA | A | G | 0.031761 | 0.004268 | 9.50E-14 |
| rs493246 | miR-195-5p | trans | 2.42E-17 | Nightingale | M_VLDL_F | NA | A | G | 0.033378 | 0.004424 | 3.10E-14 |
| rs493246 | miR-195-5p | trans | 2.42E-17 | Nightingale | IDL_C | NA | A | G | 0.034968 | 0.004274 | 9.20E-16 |
| rs493246 | miR-195-5p | trans | 2.42E-17 | Nightingale | LDL_PL | NA | A | G | 0.035 | 0.004415 | 4.70E-15 |
| rs493246 | miR-195-5p | trans | 2.42E-17 | Nightingale | M_VLDL_P | NA | A | G | 0.029134 | 0.004418 | 2.00E-11 |
| rs493246 | miR-195-5p | trans | 2.42E-17 | Nightingale | XL_VLDL_P | NA | A | G | 0.038845 | 0.004432 | 5.20E-19 |
| rs493246 | miR-195-5p | trans | 2.42E-17 | Nightingale | XXL_VLDL_ | NA | A | G | 0.034841 | 0.004434 | 2.90E-15 |
| rs493246 | miR-195-5p | trans | 2.42E-17 | Nightingale | XS_VLDL_F | NA | A | G | 0.03708 | 0.004348 | 6.40E-18 |
| rs493246 | miR-195-5p | trans | 2.42E-17 | Nightingale | GlycA | NA | A | G | 0.028022 | 0.004406 | 1.40E-10 |
| rs493246 | miR-195-5p | trans | 2.42E-17 | Nightingale | XL_VLDL_C | NA | A | G | 0.03823 | 0.004243 | 4.60E-19 |
| rs493246 | miR-195-5p | trans | 2.42E-17 | Nightingale | M_VLDL_C | NA | A | G | 0.040853 | 0.004385 | 2.30E-20 |
| rs493246 | miR-195-5p | trans | 2.42E-17 | Nightingale | Cholines | NA | A | G | 0.022312 | 0.004161 | 4.70E-08 |
| rs493246 | miR-195-5p | trans | 2.42E-17 | Nightingale | S_HDL_CE | NA | A | G | 0.028644 | 0.004432 | 1.30E-10 |
| rs493246 | miR-195-5p | trans | 2.42E-17 | Nightingale | M_VLDL_C | NA | A | G | 0.038719 | 0.004413 | 1.90E-18 |
| rs493246 | miR-195-5p | trans | 2.42E-17 | Nightingale | XS_VLDL_C | NA | A | G | 0.03781 | 0.00427 | 2.10E-18 |
| rs493246 | miR-195-5p | trans | 2.42E-17 | Nightingale | VLDL_CE | NA | A | G | 0.035794 | 0.004426 | 3.00E-16 |
| rs493246 | miR-195-5p | trans | 2.42E-17 | Nightingale | S_HDL_L | NA | A | G | 0.026306 | 0.00441 | 1.50E-09 |
| rs493246 | miR-195-5p | trans | 2.42E-17 | Nightingale | Total_FC | NA | A | G | 0.033269 | 0.004323 | 6.70E-15 |
| rs493246 | miR-195-5p | trans | 2.42E-17 | Nightingale | M_VLDL_F | NA | A | G | 0.042074 | 0.00421 | 3.40E-23 |
| rs493246 | miR-195-5p | trans | 2.42E-17 | Nightingale | S_VLDL_L | NA | A | G | 0.024676 | 0.004382 | 3.70E-09 |
| rs493246 | miR-195-5p | trans | 2.42E-17 | Nightingale | S_LDL_PL_ | NA | A | G | -0.03007 | 0.004417 | 2.60E-13 |
| rs493246 | miR-195-5p | trans | 2.42E-17 | Nightingale | XS_VLDL_P | NA | A | G | 0.032128 | 0.004363 | 5.30E-14 |
| rs493246 | miR-195-5p | trans | 2.42E-17 | Nightingale | Sphingomy | NA | A | G | 0.029031 | 0.004137 | 6.60E-12 |
| rs493246 | miR-195-5p | trans | 2.42E-17 | Nightingale | S_LDL_C_p | NA | A | G | 0.045367 | 0.004442 | 5.40E-25 |
| rs493246 | miR-195-5p | trans | 2.42E-17 | Nightingale | VLDL_FC | NA | A | G | 0.024164 | 0.004392 | 1.10E-08 |
| rs493246 | miR-195-5p | trans | 2.42E-17 | Nightingale | XL_VLDL_C | NA | A | G | 0.037359 | 0.004245 | 3.10E-18 |
| rs493246 | miR-195-5p | trans | 2.42E-17 | Nightingale | L_VLDL_C_ | NA | A | G | 0.052205 | 0.004322 | 3.90E-34 |
| rs493246 | miR-195-5p | trans | 2.42E-17 | Nightingale | Total_C | NA | A | G | 0.032388 | 0.004284 | 3.20E-14 |
| rs493246 | miR-195-5p | trans | 2.42E-17 | Nightingale | non_HDL_( | NA | A | G | 0.035709 | 0.004401 | 5.00E-16 |
| rs493246 | miR-195-5p | trans | 2.42E-17 | Nightingale | IDL_P | NA | A | G | 0.039997 | 0.004378 | 1.30E-19 |
| rs493246 | miR-195-5p | trans | 2.42E-17 | Nightingale | S_LDL_FC | NA | A | G | 0.028573 | 0.004418 | 4.20E-10 |
| rs493246 | miR-195-5p | trans | 2.42E-17 | Nightingale | XXL_VLDL_ | NA | A | G | 0.025012 | 0.004454 | 1.50E-09 |
| rs493246 | miR-195-5p | trans | 2.42E-17 | Nightingale | L_LDL_PL | NA | A | G | 0.038002 | 0.004391 | 8.90E-18 |
| rs493246 | miR-195-5p | trans | 2.42E-17 | Nightingale | L_VLDL_TG | NA | A | G | -0.05115 | 0.004404 | 5.30E-32 |
| rs493246 | miR-195-5p | trans | 2.42E-17 | Nightingale | S_VLDL_PL | NA | A | G | 0.032786 | 0.004417 | 4.20E-14 |
| rs493246 | miR-195-5p | trans | 2.42E-17 | Nightingale | IDL_PL | NA | A | G | 0.035357 | 0.00427 | 3.30E-16 |
| rs493246 | miR-195-5p | trans | 2.42E-17 | Nightingale | M_VLDL_C | NA | A | G | 0.036214 | 0.004168 | 1.20E-17 |
| rs493246 | miR-195-5p | trans | 2.42E-17 | Nightingale | XL_HDL_PL | NA | A | G | -0.02274 | 0.004065 | 1.20E-08 |
| rs493246 | miR-195-5p | trans | 2.42E-17 | Nightingale | XXL_VLDL_ | NA | A | G | 0.030859 | 0.004406 | 1.10E-12 |
| rs493246 | miR-195-5p | trans | 2.42E-17 | Nightingale | XS_VLDL_C | NA | A | G | 0.038005 | 0.004295 | 1.30E-18 |
| rs493246 | miR-195-5p | trans | 2.42E-17 | Nightingale | M_LDL_TG | NA | A | G | -0.02762 | 0.00437 | 3.20E-10 |
| rs493246 | miR-195-5p | trans | 2.42E-17 | Nightingale | S_VLDL_CE | NA | A | G | 0.047037 | 0.004365 | 1.20E-26 |
| rs493246 | miR-195-5p | trans | 2.42E-17 | Nightingale | S_HDL_C | NA | A | G | 0.028422 | 0.004417 | 1.70E-10 |
| rs493246 | miR-195-5p | trans | 2.42E-17 | Nightingale | LDL_P | NA | A | G | 0.031809 | 0.004422 | 1.00E-12 |
| rs493246 | miR-195-5p | trans | 2.42E-17 | Nightingale | PUFA | NA | A | G | 0.023738 | 0.004316 | 1.40E-08 |
| rs493246 | miR-195-5p | trans | 2.42E-17 | Nightingale | M_VLDL_T | NA | A | G | -0.03857 | 0.004181 | 6.70E-20 |

| | | | | | | | | | | | |
|---|---|---|---|---|---|---|---|---|---|---|---|
| rs493246 | miR-195-5p | trans | 2.42E-17 | Nightingale | VLDL_C | NA | A | G | 0.031309 | 0.004419 | 4.80E-13 |
| rs493246 | miR-195-5p | trans | 2.42E-17 | Nightingale | VLDL_P | NA | A | G | 0.026641 | 0.004401 | 4.40E-10 |
| rs493246 | miR-195-5p | trans | 2.42E-17 | Nightingale | S_HDL_PL | NA | A | G | 0.023794 | 0.004387 | 3.20E-08 |
| rs493246 | miR-195-5p | trans | 2.42E-17 | Nightingale | XL_VLDL_T | NA | A | G | -0.04393 | 0.004289 | 1.70E-24 |
| rs493246 | miR-195-5p | trans | 2.42E-17 | Nightingale | LDL_FC | NA | A | G | 0.033644 | 0.004378 | 5.20E-14 |
| rs493246 | miR-195-5p | trans | 2.42E-17 | Nightingale | M_LDL_FC | NA | A | G | 0.034359 | 0.004419 | 1.90E-14 |
| rs493246 | miR-195-5p | trans | 2.42E-17 | Nightingale | M_LDL_PL | NA | A | G | 0.032478 | 0.004431 | 2.60E-13 |
| rs493246 | miR-195-5p | trans | 2.42E-17 | Nightingale | Omega_6 | NA | A | G | 0.025912 | 0.004321 | 5.60E-10 |
| rs493246 | miR-195-5p | trans | 2.42E-17 | Nightingale | Total_PL | NA | A | G | 0.024943 | 0.004214 | 1.00E-09 |
| rs493246 | miR-195-5p | trans | 2.42E-17 | Nightingale | S_VLDL_C_ | NA | A | G | 0.041892 | 0.004306 | 5.20E-22 |
| rs493246 | miR-195-5p | trans | 2.42E-17 | Nightingale | S_LDL_CE | NA | A | G | 0.030558 | 0.004427 | 5.80E-12 |
| rs493246 | miR-195-5p | trans | 2.42E-17 | Nightingale | L_LDL_P | NA | A | G | 0.029187 | 0.00441 | 6.10E-11 |
| rs493246 | miR-195-5p | trans | 2.42E-17 | Nightingale | XS_VLDL_P | NA | A | G | 0.036714 | 0.004366 | 1.40E-17 |
| rs493246 | miR-195-5p | trans | 2.42E-17 | Nightingale | LDL_C | NA | A | G | 0.033206 | 0.004406 | 8.70E-14 |
| rs493246 | miR-195-5p | trans | 2.42E-17 | Nightingale | S_VLDL_PL | NA | A | G | 0.027742 | 0.004237 | 1.80E-10 |
| rs493246 | miR-195-5p | trans | 2.42E-17 | Nightingale | M_LDL_P | NA | A | G | 0.0359 | 0.004437 | 6.80E-16 |
| rs493246 | miR-195-5p | trans | 2.42E-17 | Nightingale | S_LDL_C | NA | A | G | 0.030953 | 0.004429 | 5.50E-12 |
| rs493246 | miR-195-5p | trans | 2.42E-17 | Nightingale | XXL_VLDL_ | NA | A | G | 0.033132 | 0.004459 | 8.80E-14 |
| rs493246 | miR-195-5p | trans | 2.42E-17 | Nightingale | S_LDL_CE_ | NA | A | G | 0.03095 | 0.004417 | 4.80E-14 |
| rs493246 | miR-195-5p | trans | 2.42E-17 | Nightingale | LA | NA | A | G | 0.024925 | 0.004339 | 3.60E-09 |
| rs493246 | miR-195-5p | trans | 2.42E-17 | Nightingale | Remnant_( | NA | A | G | 0.036878 | 0.004396 | 3.60E-17 |
| rs493246 | miR-195-5p | trans | 2.42E-17 | Nightingale | L_VLDL_FC | NA | A | G | 0.048596 | 0.004382 | 4.50E-29 |
| rs493246 | miR-195-5p | trans | 2.42E-17 | Nightingale | S_VLDL_FC | NA | A | G | 0.037913 | 0.004426 | 7.30E-18 |
| rs493246 | miR-195-5p | trans | 2.42E-17 | Nightingale | S_LDL_P | NA | A | G | 0.029597 | 0.004429 | 3.20E-11 |
| rs493246 | miR-195-5p | trans | 2.42E-17 | Nightingale | M_LDL_CE | NA | A | G | 0.032983 | 0.004431 | 9.50E-14 |
| rs493246 | miR-195-5p | trans | 2.42E-17 | Nightingale | Total_L | NA | A | G | 0.026363 | 0.004353 | 4.50E-10 |
| rs493246 | miR-195-5p | trans | 2.42E-17 | Nightingale | L_LDL_L | NA | A | G | 0.033563 | 0.004382 | 2.90E-14 |
| rs493246 | miR-195-5p | trans | 2.42E-17 | Nightingale | M_VLDL_C | NA | A | G | 0.033637 | 0.004157 | 1.90E-15 |
| rs493246 | miR-195-5p | trans | 2.42E-17 | Nightingale | ApoB_by_/ | NA | A | G | 0.02505 | 0.004358 | 1.30E-08 |
| rs493246 | miR-195-5p | trans | 2.42E-17 | Nightingale | S_VLDL_C | NA | A | G | 0.038381 | 0.004424 | 1.80E-18 |
| rs493246 | miR-195-5p | trans | 2.42E-17 | Nightingale | S_VLDL_TG | NA | A | G | -0.03813 | 0.004283 | 1.40E-18 |
| rs493246 | miR-195-5p | trans | 2.42E-17 | Nightingale | ApoB | NA | A | G | 0.033948 | 0.004419 | 2.10E-14 |
| rs493246 | miR-195-5p | trans | 2.42E-17 | Nightingale | IDL_L | NA | A | G | 0.035283 | 0.004278 | 4.70E-16 |
| rs493246 | miR-195-5p | trans | 2.42E-17 | Nightingale | S_VLDL_P | NA | A | G | 0.023473 | 0.004369 | 1.60E-08 |
| rs493246 | miR-195-5p | trans | 2.42E-17 | Nightingale | L_LDL_C | NA | A | G | 0.032333 | 0.004375 | 2.50E-13 |
| rs493246 | miR-195-5p | trans | 2.42E-17 | Nightingale | IDL_CE | NA | A | G | 0.03352 | 0.004272 | 1.30E-14 |
| rs493246 | miR-195-5p | trans | 2.42E-17 | Nightingale | XL_VLDL_F | NA | A | G | 0.037204 | 0.00429 | 2.00E-18 |
| rs493246 | miR-195-5p | trans | 2.42E-17 | Nightingale | XXL_VLDL_ | NA | A | G | -0.03574 | 0.004463 | 2.10E-16 |
| rs493246 | miR-195-5p | trans | 2.42E-17 | Nightingale | XS_VLDL_L | NA | A | G | 0.033831 | 0.00435 | 2.00E-15 |
| rs493246 | miR-195-5p | trans | 2.42E-17 | Nightingale | M_VLDL_P | NA | A | G | 0.028301 | 0.004414 | 6.30E-11 |
| rs493246 | miR-195-5p | trans | 2.42E-17 | Nightingale | S_HDL_P | NA | A | G | 0.030621 | 0.004425 | 5.00E-12 |
| rs493246 | miR-195-5p | trans | 2.42E-17 | Nightingale | S_VLDL_CE | NA | A | G | 0.038189 | 0.004419 | 1.60E-18 |
| rs493246 | miR-195-5p | trans | 2.42E-17 | Nightingale | L_LDL_FC | NA | A | G | 0.033637 | 0.004345 | 2.80E-14 |
| rs493246 | miR-195-5p | trans | 2.42E-17 | Nightingale | Citrate | NA | A | G | 0.026826 | 0.004396 | 7.40E-10 |
| rs493246 | miR-195-5p | trans | 2.42E-17 | Nightingale | L_LDL_CE | NA | A | G | 0.03158 | 0.004385 | 9.70E-13 |
| rs493246 | miR-126-3p | trans | 2.55E-39 | Nightingale | IDL_FC | NA | A | G | 0.038267 | 0.004284 | 1.40E-18 |
| rs493246 | miR-126-3p | trans | 2.55E-39 | Nightingale | LDL_CE | NA | A | G | 0.032566 | 0.004413 | 2.20E-13 |
| rs493246 | miR-126-3p | trans | 2.55E-39 | Nightingale | L_VLDL_CE | NA | A | G | 0.04735 | 0.004234 | 2.40E-29 |
| rs493246 | miR-126-3p | trans | 2.55E-39 | Nightingale | M_LDL_C_ | NA | A | G | 0.031162 | 0.004428 | 2.90E-12 |
| rs493246 | miR-126-3p | trans | 2.55E-39 | Nightingale | XS_VLDL_F | NA | A | G | 0.03143 | 0.004393 | 2.40E-12 |
| rs493246 | miR-126-3p | trans | 2.55E-39 | Nightingale | S_HDL_FC | NA | A | G | 0.023468 | 0.004344 | 4.50E-08 |
| rs493246 | miR-126-3p | trans | 2.55E-39 | Nightingale | S_LDL_L | NA | A | G | 0.027751 | 0.004424 | 8.20E-10 |
| rs493246 | miR-126-3p | trans | 2.55E-39 | Nightingale | LDL_L | NA | A | G | 0.033316 | 0.00441 | 7.30E-14 |
| rs493246 | miR-126-3p | trans | 2.55E-39 | Nightingale | M_LDL_L | NA | A | G | 0.033043 | 0.004431 | 1.00E-13 |
| rs493246 | miR-126-3p | trans | 2.55E-39 | Nightingale | M_LDL_C | NA | A | G | 0.033998 | 0.004433 | 2.10E-14 |
| rs493246 | miR-126-3p | trans | 2.55E-39 | Nightingale | Clinical_LD | NA | A | G | 0.036486 | 0.004399 | 2.70E-16 |
| rs493246 | miR-126-3p | trans | 2.55E-39 | Nightingale | S_VLDL_FC | NA | A | G | 0.02936 | 0.004228 | 1.10E-11 |
| rs493246 | miR-126-3p | trans | 2.55E-39 | Nightingale | IDL_FC_pct | NA | A | G | 0.025772 | 0.004391 | 1.10E-08 |
| rs493246 | miR-126-3p | trans | 2.55E-39 | Nightingale | M_VLDL_P | NA | A | G | 0.044066 | 0.004247 | 5.50E-25 |
| rs493246 | miR-126-3p | trans | 2.55E-39 | Nightingale | Total_CE | NA | A | G | 0.031761 | 0.004268 | 9.50E-14 |
| rs493246 | miR-126-3p | trans | 2.55E-39 | Nightingale | M_VLDL_F | NA | A | G | 0.033378 | 0.004424 | 3.10E-14 |
| rs493246 | miR-126-3p | trans | 2.55E-39 | Nightingale | IDL_C | NA | A | G | 0.034968 | 0.004274 | 9.20E-16 |
| rs493246 | miR-126-3p | trans | 2.55E-39 | Nightingale | LDL_PL | NA | A | G | 0.035 | 0.004415 | 4.70E-15 |

| | | | | | | | | | | | |
|---|---|---|---|---|---|---|---|---|---|---|---|
| rs493246 | miR-126-3p | trans | 2.55E-39 | Nightingale | M_VLDL_P | NA | A | G | 0.029134 | 0.004418 | 2.00E-11 |
| rs493246 | miR-126-3p | trans | 2.55E-39 | Nightingale | XL_VLDL_P | NA | A | G | 0.038845 | 0.004432 | 5.20E-19 |
| rs493246 | miR-126-3p | trans | 2.55E-39 | Nightingale | XXL_VLDL_ | NA | A | G | 0.034841 | 0.004434 | 2.90E-15 |
| rs493246 | miR-126-3p | trans | 2.55E-39 | Nightingale | XS_VLDL_F | NA | A | G | 0.03708 | 0.004348 | 6.40E-18 |
| rs493246 | miR-126-3p | trans | 2.55E-39 | Nightingale | GlycA | NA | A | G | 0.028022 | 0.004406 | 1.40E-10 |
| rs493246 | miR-126-3p | trans | 2.55E-39 | Nightingale | XL_VLDL_C | NA | A | G | 0.03823 | 0.004243 | 4.60E-19 |
| rs493246 | miR-126-3p | trans | 2.55E-39 | Nightingale | M_VLDL_C | NA | A | G | 0.040853 | 0.004385 | 2.30E-20 |
| rs493246 | miR-126-3p | trans | 2.55E-39 | Nightingale | Cholines | NA | A | G | 0.022312 | 0.004161 | 4.70E-08 |
| rs493246 | miR-126-3p | trans | 2.55E-39 | Nightingale | S_HDL_CE | NA | A | G | 0.028644 | 0.004432 | 1.30E-10 |
| rs493246 | miR-126-3p | trans | 2.55E-39 | Nightingale | M_VLDL_C | NA | A | G | 0.038719 | 0.004413 | 1.90E-18 |
| rs493246 | miR-126-3p | trans | 2.55E-39 | Nightingale | XS_VLDL_C | NA | A | G | 0.03781 | 0.00427 | 2.10E-18 |
| rs493246 | miR-126-3p | trans | 2.55E-39 | Nightingale | VLDL_CE | NA | A | G | 0.035794 | 0.004426 | 3.00E-16 |
| rs493246 | miR-126-3p | trans | 2.55E-39 | Nightingale | S_HDL_L | NA | A | G | 0.026306 | 0.00441 | 1.50E-09 |
| rs493246 | miR-126-3p | trans | 2.55E-39 | Nightingale | Total_FC | NA | A | G | 0.033269 | 0.004323 | 6.70E-15 |
| rs493246 | miR-126-3p | trans | 2.55E-39 | Nightingale | M_VLDL_F | NA | A | G | 0.042074 | 0.00421 | 3.40E-23 |
| rs493246 | miR-126-3p | trans | 2.55E-39 | Nightingale | S_VLDL_L | NA | A | G | 0.024676 | 0.004382 | 3.70E-09 |
| rs493246 | miR-126-3p | trans | 2.55E-39 | Nightingale | S_LDL_PL_ | NA | A | G | -0.03007 | 0.004417 | 2.60E-13 |
| rs493246 | miR-126-3p | trans | 2.55E-39 | Nightingale | XS_VLDL_P | NA | A | G | 0.032128 | 0.004363 | 5.30E-14 |
| rs493246 | miR-126-3p | trans | 2.55E-39 | Nightingale | Sphingomy | NA | A | G | 0.029031 | 0.004137 | 6.60E-12 |
| rs493246 | miR-126-3p | trans | 2.55E-39 | Nightingale | S_LDL_C_p | NA | A | G | 0.045367 | 0.004442 | 5.40E-25 |
| rs493246 | miR-126-3p | trans | 2.55E-39 | Nightingale | VLDL_FC | NA | A | G | 0.024164 | 0.004392 | 1.10E-08 |
| rs493246 | miR-126-3p | trans | 2.55E-39 | Nightingale | XL_VLDL_C | NA | A | G | 0.037359 | 0.004245 | 3.10E-18 |
| rs493246 | miR-126-3p | trans | 2.55E-39 | Nightingale | L_VLDL_C_ | NA | A | G | 0.052205 | 0.004322 | 3.90E-34 |
| rs493246 | miR-126-3p | trans | 2.55E-39 | Nightingale | Total_C | NA | A | G | 0.032388 | 0.004284 | 3.20E-14 |
| rs493246 | miR-126-3p | trans | 2.55E-39 | Nightingale | non_HDL_C | NA | A | G | 0.035709 | 0.004401 | 5.00E-16 |
| rs493246 | miR-126-3p | trans | 2.55E-39 | Nightingale | IDL_P | NA | A | G | 0.039997 | 0.004378 | 1.30E-19 |
| rs493246 | miR-126-3p | trans | 2.55E-39 | Nightingale | S_LDL_FC | NA | A | G | 0.028573 | 0.004418 | 4.20E-10 |
| rs493246 | miR-126-3p | trans | 2.55E-39 | Nightingale | XXL_VLDL_ | NA | A | G | 0.025012 | 0.004454 | 1.50E-09 |
| rs493246 | miR-126-3p | trans | 2.55E-39 | Nightingale | L_LDL_PL | NA | A | G | 0.038002 | 0.004391 | 8.90E-18 |
| rs493246 | miR-126-3p | trans | 2.55E-39 | Nightingale | L_VLDL_TG | NA | A | G | -0.05115 | 0.004404 | 5.30E-32 |
| rs493246 | miR-126-3p | trans | 2.55E-39 | Nightingale | S_VLDL_PL | NA | A | G | 0.032786 | 0.004417 | 4.20E-14 |
| rs493246 | miR-126-3p | trans | 2.55E-39 | Nightingale | IDL_PL | NA | A | G | 0.035357 | 0.00427 | 3.30E-16 |
| rs493246 | miR-126-3p | trans | 2.55E-39 | Nightingale | M_VLDL_C | NA | A | G | 0.036214 | 0.004168 | 1.20E-17 |
| rs493246 | miR-126-3p | trans | 2.55E-39 | Nightingale | XL_HDL_PL | NA | A | G | -0.02274 | 0.004065 | 1.20E-08 |
| rs493246 | miR-126-3p | trans | 2.55E-39 | Nightingale | XXL_VLDL_ | NA | A | G | 0.030859 | 0.004406 | 1.10E-12 |
| rs493246 | miR-126-3p | trans | 2.55E-39 | Nightingale | XS_VLDL_C | NA | A | G | 0.038005 | 0.004295 | 1.30E-18 |
| rs493246 | miR-126-3p | trans | 2.55E-39 | Nightingale | M_LDL_TG | NA | A | G | -0.02762 | 0.00437 | 3.20E-10 |
| rs493246 | miR-126-3p | trans | 2.55E-39 | Nightingale | S_VLDL_CE | NA | A | G | 0.047037 | 0.004365 | 1.20E-26 |
| rs493246 | miR-126-3p | trans | 2.55E-39 | Nightingale | S_HDL_C | NA | A | G | 0.028422 | 0.004417 | 1.70E-10 |
| rs493246 | miR-126-3p | trans | 2.55E-39 | Nightingale | LDL_P | NA | A | G | 0.031809 | 0.004422 | 1.00E-12 |
| rs493246 | miR-126-3p | trans | 2.55E-39 | Nightingale | PUFA | NA | A | G | 0.023738 | 0.004316 | 1.40E-08 |
| rs493246 | miR-126-3p | trans | 2.55E-39 | Nightingale | M_VLDL_T | NA | A | G | -0.03857 | 0.004181 | 6.70E-20 |
| rs493246 | miR-126-3p | trans | 2.55E-39 | Nightingale | VLDL_C | NA | A | G | 0.031309 | 0.004419 | 4.80E-13 |
| rs493246 | miR-126-3p | trans | 2.55E-39 | Nightingale | VLDL_P | NA | A | G | 0.026641 | 0.004401 | 4.40E-10 |
| rs493246 | miR-126-3p | trans | 2.55E-39 | Nightingale | S_HDL_PL | NA | A | G | 0.023794 | 0.004387 | 3.20E-08 |
| rs493246 | miR-126-3p | trans | 2.55E-39 | Nightingale | XL_VLDL_T | NA | A | G | -0.04393 | 0.004289 | 1.70E-24 |
| rs493246 | miR-126-3p | trans | 2.55E-39 | Nightingale | LDL_FC | NA | A | G | 0.033644 | 0.004378 | 5.20E-14 |
| rs493246 | miR-126-3p | trans | 2.55E-39 | Nightingale | M_LDL_FC | NA | A | G | 0.034359 | 0.004419 | 1.90E-14 |
| rs493246 | miR-126-3p | trans | 2.55E-39 | Nightingale | M_LDL_PL | NA | A | G | 0.032478 | 0.004431 | 2.60E-13 |
| rs493246 | miR-126-3p | trans | 2.55E-39 | Nightingale | Omega_6 | NA | A | G | 0.025912 | 0.004321 | 5.60E-10 |
| rs493246 | miR-126-3p | trans | 2.55E-39 | Nightingale | Total_PL | NA | A | G | 0.024943 | 0.004214 | 1.00E-09 |
| rs493246 | miR-126-3p | trans | 2.55E-39 | Nightingale | S_VLDL_C_ | NA | A | G | 0.041892 | 0.004306 | 5.20E-22 |
| rs493246 | miR-126-3p | trans | 2.55E-39 | Nightingale | S_LDL_CE | NA | A | G | 0.030558 | 0.004427 | 5.80E-12 |
| rs493246 | miR-126-3p | trans | 2.55E-39 | Nightingale | L_LDL_P | NA | A | G | 0.029187 | 0.00441 | 6.10E-11 |
| rs493246 | miR-126-3p | trans | 2.55E-39 | Nightingale | XS_VLDL_P | NA | A | G | 0.036714 | 0.004366 | 1.40E-17 |
| rs493246 | miR-126-3p | trans | 2.55E-39 | Nightingale | LDL_C | NA | A | G | 0.033206 | 0.004406 | 8.70E-14 |
| rs493246 | miR-126-3p | trans | 2.55E-39 | Nightingale | S_VLDL_PL | NA | A | G | 0.027742 | 0.004237 | 1.80E-10 |
| rs493246 | miR-126-3p | trans | 2.55E-39 | Nightingale | M_LDL_P | NA | A | G | 0.0359 | 0.004437 | 6.80E-16 |
| rs493246 | miR-126-3p | trans | 2.55E-39 | Nightingale | S_LDL_C | NA | A | G | 0.030953 | 0.004429 | 5.50E-12 |
| rs493246 | miR-126-3p | trans | 2.55E-39 | Nightingale | XXL_VLDL_ | NA | A | G | 0.033132 | 0.004459 | 8.80E-14 |
| rs493246 | miR-126-3p | trans | 2.55E-39 | Nightingale | S_LDL_CE_ | NA | A | G | 0.03095 | 0.004417 | 4.80E-14 |
| rs493246 | miR-126-3p | trans | 2.55E-39 | Nightingale | LA | NA | A | G | 0.024925 | 0.004339 | 3.60E-09 |
| rs493246 | miR-126-3p | trans | 2.55E-39 | Nightingale | Remnant_C | NA | A | G | 0.036878 | 0.004396 | 3.60E-17 |

| | | | | | | | | | | | |
|---|---|---|---|---|---|---|---|---|---|---|---|
| rs493246 | miR-126-3p | trans | 2.55E-39 | Nightingale | L_VLDL_FC | NA | A | G | 0.048596 | 0.004382 | 4.50E-29 |
| rs493246 | miR-126-3p | trans | 2.55E-39 | Nightingale | S_VLDL_FC | NA | A | G | 0.037913 | 0.004426 | 7.30E-18 |
| rs493246 | miR-126-3p | trans | 2.55E-39 | Nightingale | S_LDL_P | NA | A | G | 0.029597 | 0.004429 | 3.20E-11 |
| rs493246 | miR-126-3p | trans | 2.55E-39 | Nightingale | M_LDL_CE | NA | A | G | 0.032983 | 0.004431 | 9.50E-14 |
| rs493246 | miR-126-3p | trans | 2.55E-39 | Nightingale | Total_L | NA | A | G | 0.026363 | 0.004353 | 4.50E-10 |
| rs493246 | miR-126-3p | trans | 2.55E-39 | Nightingale | L_LDL_L | NA | A | G | 0.033563 | 0.004382 | 2.90E-14 |
| rs493246 | miR-126-3p | trans | 2.55E-39 | Nightingale | M_VLDL_C | NA | A | G | 0.033637 | 0.004157 | 1.90E-15 |
| rs493246 | miR-126-3p | trans | 2.55E-39 | Nightingale | ApoB_by_A | NA | A | G | 0.02505 | 0.004358 | 1.30E-08 |
| rs493246 | miR-126-3p | trans | 2.55E-39 | Nightingale | S_VLDL_C | NA | A | G | 0.038381 | 0.004424 | 1.80E-18 |
| rs493246 | miR-126-3p | trans | 2.55E-39 | Nightingale | S_VLDL_TG | NA | A | G | -0.03813 | 0.004283 | 1.40E-18 |
| rs493246 | miR-126-3p | trans | 2.55E-39 | Nightingale | ApoB | NA | A | G | 0.033948 | 0.004419 | 2.10E-14 |
| rs493246 | miR-126-3p | trans | 2.55E-39 | Nightingale | IDL_L | NA | A | G | 0.035283 | 0.004278 | 4.70E-16 |
| rs493246 | miR-126-3p | trans | 2.55E-39 | Nightingale | S_VLDL_P | NA | A | G | 0.023473 | 0.004369 | 1.60E-08 |
| rs493246 | miR-126-3p | trans | 2.55E-39 | Nightingale | L_LDL_C | NA | A | G | 0.032333 | 0.004375 | 2.50E-13 |
| rs493246 | miR-126-3p | trans | 2.55E-39 | Nightingale | IDL_CE | NA | A | G | 0.03352 | 0.004272 | 1.30E-14 |
| rs493246 | miR-126-3p | trans | 2.55E-39 | Nightingale | XL_VLDL_P | NA | A | G | 0.037204 | 0.00429 | 2.00E-18 |
| rs493246 | miR-126-3p | trans | 2.55E-39 | Nightingale | XXL_VLDL_ | NA | A | G | -0.03574 | 0.004463 | 2.10E-16 |
| rs493246 | miR-126-3p | trans | 2.55E-39 | Nightingale | XS_VLDL_L | NA | A | G | 0.033831 | 0.00435 | 2.00E-15 |
| rs493246 | miR-126-3p | trans | 2.55E-39 | Nightingale | M_VLDL_P | NA | A | G | 0.028301 | 0.004414 | 6.30E-11 |
| rs493246 | miR-126-3p | trans | 2.55E-39 | Nightingale | S_HDL_P | NA | A | G | 0.030621 | 0.004425 | 5.00E-12 |
| rs493246 | miR-126-3p | trans | 2.55E-39 | Nightingale | S_VLDL_CE | NA | A | G | 0.038189 | 0.004419 | 1.60E-18 |
| rs493246 | miR-126-3p | trans | 2.55E-39 | Nightingale | L_LDL_FC | NA | A | G | 0.033637 | 0.004345 | 2.80E-14 |
| rs493246 | miR-126-3p | trans | 2.55E-39 | Nightingale | Citrate | NA | A | G | 0.026826 | 0.004396 | 7.40E-10 |
| rs493246 | miR-126-3p | trans | 2.55E-39 | Nightingale | L_LDL_CE | NA | A | G | 0.03158 | 0.004385 | 9.70E-13 |
| rs493246 | miR-139-5p | trans | 8.06E-42 | Nightingale | IDL_FC | NA | A | G | 0.038267 | 0.004284 | 1.40E-18 |
| rs493246 | miR-139-5p | trans | 8.06E-42 | Nightingale | LDL_CE | NA | A | G | 0.032566 | 0.004413 | 2.20E-13 |
| rs493246 | miR-139-5p | trans | 8.06E-42 | Nightingale | L_VLDL_CE | NA | A | G | 0.04735 | 0.004234 | 2.40E-29 |
| rs493246 | miR-139-5p | trans | 8.06E-42 | Nightingale | M_LDL_C_p | NA | A | G | 0.031162 | 0.004428 | 2.90E-12 |
| rs493246 | miR-139-5p | trans | 8.06E-42 | Nightingale | XS_VLDL_F | NA | A | G | 0.03143 | 0.004393 | 2.40E-12 |
| rs493246 | miR-139-5p | trans | 8.06E-42 | Nightingale | S_HDL_FC | NA | A | G | 0.023468 | 0.004344 | 4.50E-08 |
| rs493246 | miR-139-5p | trans | 8.06E-42 | Nightingale | S_LDL_L | NA | A | G | 0.027751 | 0.004424 | 8.20E-10 |
| rs493246 | miR-139-5p | trans | 8.06E-42 | Nightingale | LDL_L | NA | A | G | 0.033316 | 0.00441 | 7.30E-14 |
| rs493246 | miR-139-5p | trans | 8.06E-42 | Nightingale | M_LDL_L | NA | A | G | 0.033043 | 0.004431 | 1.00E-13 |
| rs493246 | miR-139-5p | trans | 8.06E-42 | Nightingale | M_LDL_C | NA | A | G | 0.033998 | 0.004433 | 2.10E-14 |
| rs493246 | miR-139-5p | trans | 8.06E-42 | Nightingale | Clinical_LD | NA | A | G | 0.036486 | 0.004399 | 2.70E-16 |
| rs493246 | miR-139-5p | trans | 8.06E-42 | Nightingale | S_VLDL_FC | NA | A | G | 0.02936 | 0.004228 | 1.10E-11 |
| rs493246 | miR-139-5p | trans | 8.06E-42 | Nightingale | IDL_FC_pct | NA | A | G | 0.025772 | 0.004391 | 1.10E-08 |
| rs493246 | miR-139-5p | trans | 8.06E-42 | Nightingale | M_VLDL_P | NA | A | G | 0.044066 | 0.004247 | 5.50E-25 |
| rs493246 | miR-139-5p | trans | 8.06E-42 | Nightingale | Total_CE | NA | A | G | 0.031761 | 0.004268 | 9.50E-14 |
| rs493246 | miR-139-5p | trans | 8.06E-42 | Nightingale | M_VLDL_F | NA | A | G | 0.033378 | 0.004424 | 3.10E-14 |
| rs493246 | miR-139-5p | trans | 8.06E-42 | Nightingale | IDL_C | NA | A | G | 0.034968 | 0.004274 | 9.20E-16 |
| rs493246 | miR-139-5p | trans | 8.06E-42 | Nightingale | LDL_PL | NA | A | G | 0.035 | 0.004415 | 4.70E-15 |
| rs493246 | miR-139-5p | trans | 8.06E-42 | Nightingale | M_VLDL_P | NA | A | G | 0.029134 | 0.004418 | 2.00E-11 |
| rs493246 | miR-139-5p | trans | 8.06E-42 | Nightingale | XL_VLDL_P | NA | A | G | 0.038845 | 0.004432 | 5.20E-19 |
| rs493246 | miR-139-5p | trans | 8.06E-42 | Nightingale | XXL_VLDL_ | NA | A | G | 0.034841 | 0.004434 | 2.90E-15 |
| rs493246 | miR-139-5p | trans | 8.06E-42 | Nightingale | XS_VLDL_F | NA | A | G | 0.03708 | 0.004348 | 6.40E-18 |
| rs493246 | miR-139-5p | trans | 8.06E-42 | Nightingale | GlycA | NA | A | G | 0.028022 | 0.004406 | 1.40E-10 |
| rs493246 | miR-139-5p | trans | 8.06E-42 | Nightingale | XL_VLDL_C | NA | A | G | 0.03823 | 0.004243 | 4.60E-19 |
| rs493246 | miR-139-5p | trans | 8.06E-42 | Nightingale | M_VLDL_C | NA | A | G | 0.040853 | 0.004385 | 2.30E-20 |
| rs493246 | miR-139-5p | trans | 8.06E-42 | Nightingale | Cholines | NA | A | G | 0.022312 | 0.004161 | 4.70E-08 |
| rs493246 | miR-139-5p | trans | 8.06E-42 | Nightingale | S_HDL_CE | NA | A | G | 0.028644 | 0.004432 | 1.30E-10 |
| rs493246 | miR-139-5p | trans | 8.06E-42 | Nightingale | M_VLDL_C | NA | A | G | 0.038719 | 0.004413 | 1.90E-18 |
| rs493246 | miR-139-5p | trans | 8.06E-42 | Nightingale | XS_VLDL_C | NA | A | G | 0.03781 | 0.00427 | 2.10E-18 |
| rs493246 | miR-139-5p | trans | 8.06E-42 | Nightingale | VLDL_CE | NA | A | G | 0.035794 | 0.004426 | 3.00E-16 |
| rs493246 | miR-139-5p | trans | 8.06E-42 | Nightingale | S_HDL_L | NA | A | G | 0.026306 | 0.00441 | 1.50E-09 |
| rs493246 | miR-139-5p | trans | 8.06E-42 | Nightingale | Total_FC | NA | A | G | 0.033269 | 0.004323 | 6.70E-15 |
| rs493246 | miR-139-5p | trans | 8.06E-42 | Nightingale | M_VLDL_F | NA | A | G | 0.042074 | 0.00421 | 3.40E-23 |
| rs493246 | miR-139-5p | trans | 8.06E-42 | Nightingale | S_VLDL_L | NA | A | G | 0.024676 | 0.004382 | 3.70E-09 |
| rs493246 | miR-139-5p | trans | 8.06E-42 | Nightingale | S_LDL_PL_ | NA | A | G | -0.03007 | 0.004417 | 2.60E-13 |
| rs493246 | miR-139-5p | trans | 8.06E-42 | Nightingale | XS_VLDL_P | NA | A | G | 0.032128 | 0.004363 | 5.30E-14 |
| rs493246 | miR-139-5p | trans | 8.06E-42 | Nightingale | Sphingomy | NA | A | G | 0.029031 | 0.004137 | 6.60E-12 |
| rs493246 | miR-139-5p | trans | 8.06E-42 | Nightingale | S_LDL_C_p | NA | A | G | 0.045367 | 0.004442 | 5.40E-25 |
| rs493246 | miR-139-5p | trans | 8.06E-42 | Nightingale | VLDL_FC | NA | A | G | 0.024164 | 0.004392 | 1.10E-08 |

| | | | | | | | | | | | |
|---|---|---|---|---|---|---|---|---|---|---|---|
| rs493246 | miR-139-5p | trans | 8.06E-42 | Nightingale | XL_VLDL_C | NA | A | G | 0.037359 | 0.004245 | 3.10E-18 |
| rs493246 | miR-139-5p | trans | 8.06E-42 | Nightingale | L_VLDL_C_ | NA | A | G | 0.052205 | 0.004322 | 3.90E-34 |
| rs493246 | miR-139-5p | trans | 8.06E-42 | Nightingale | Total_C | NA | A | G | 0.032388 | 0.004284 | 3.20E-14 |
| rs493246 | miR-139-5p | trans | 8.06E-42 | Nightingale | non_HDL_( | NA | A | G | 0.035709 | 0.004401 | 5.00E-16 |
| rs493246 | miR-139-5p | trans | 8.06E-42 | Nightingale | IDL_P | NA | A | G | 0.039997 | 0.004378 | 1.30E-19 |
| rs493246 | miR-139-5p | trans | 8.06E-42 | Nightingale | S_LDL_FC | NA | A | G | 0.028573 | 0.004418 | 4.20E-10 |
| rs493246 | miR-139-5p | trans | 8.06E-42 | Nightingale | XXL_VLDL_ | NA | A | G | 0.025012 | 0.004454 | 1.50E-09 |
| rs493246 | miR-139-5p | trans | 8.06E-42 | Nightingale | L_LDL_PL | NA | A | G | 0.038002 | 0.004391 | 8.90E-18 |
| rs493246 | miR-139-5p | trans | 8.06E-42 | Nightingale | L_VLDL_TG | NA | A | G | -0.05115 | 0.004404 | 5.30E-32 |
| rs493246 | miR-139-5p | trans | 8.06E-42 | Nightingale | S_VLDL_PL | NA | A | G | 0.032786 | 0.004417 | 4.20E-14 |
| rs493246 | miR-139-5p | trans | 8.06E-42 | Nightingale | IDL_PL | NA | A | G | 0.035357 | 0.00427 | 3.30E-16 |
| rs493246 | miR-139-5p | trans | 8.06E-42 | Nightingale | M_VLDL_C | NA | A | G | 0.036214 | 0.004168 | 1.20E-17 |
| rs493246 | miR-139-5p | trans | 8.06E-42 | Nightingale | XL_HDL_PL | NA | A | G | -0.02274 | 0.004065 | 1.20E-08 |
| rs493246 | miR-139-5p | trans | 8.06E-42 | Nightingale | XXL_VLDL_ | NA | A | G | 0.030859 | 0.004406 | 1.10E-12 |
| rs493246 | miR-139-5p | trans | 8.06E-42 | Nightingale | XS_VLDL_C | NA | A | G | 0.038005 | 0.004295 | 1.30E-18 |
| rs493246 | miR-139-5p | trans | 8.06E-42 | Nightingale | M_LDL_TG | NA | A | G | -0.02762 | 0.00437 | 3.20E-10 |
| rs493246 | miR-139-5p | trans | 8.06E-42 | Nightingale | S_VLDL_CE | NA | A | G | 0.047037 | 0.004365 | 1.20E-26 |
| rs493246 | miR-139-5p | trans | 8.06E-42 | Nightingale | S_HDL_C | NA | A | G | 0.028422 | 0.004417 | 1.70E-10 |
| rs493246 | miR-139-5p | trans | 8.06E-42 | Nightingale | LDL_P | NA | A | G | 0.031809 | 0.004422 | 1.00E-12 |
| rs493246 | miR-139-5p | trans | 8.06E-42 | Nightingale | PUFA | NA | A | G | 0.023738 | 0.004316 | 1.40E-08 |
| rs493246 | miR-139-5p | trans | 8.06E-42 | Nightingale | M_VLDL_T | NA | A | G | -0.03857 | 0.004181 | 6.70E-20 |
| rs493246 | miR-139-5p | trans | 8.06E-42 | Nightingale | VLDL_C | NA | A | G | 0.031309 | 0.004419 | 4.80E-13 |
| rs493246 | miR-139-5p | trans | 8.06E-42 | Nightingale | VLDL_P | NA | A | G | 0.026641 | 0.004401 | 4.40E-10 |
| rs493246 | miR-139-5p | trans | 8.06E-42 | Nightingale | S_HDL_PL | NA | A | G | 0.023794 | 0.004387 | 3.20E-08 |
| rs493246 | miR-139-5p | trans | 8.06E-42 | Nightingale | XL_VLDL_T | NA | A | G | -0.04393 | 0.004289 | 1.70E-24 |
| rs493246 | miR-139-5p | trans | 8.06E-42 | Nightingale | LDL_FC | NA | A | G | 0.033644 | 0.004378 | 5.20E-14 |
| rs493246 | miR-139-5p | trans | 8.06E-42 | Nightingale | M_LDL_FC | NA | A | G | 0.034359 | 0.004419 | 1.90E-14 |
| rs493246 | miR-139-5p | trans | 8.06E-42 | Nightingale | M_LDL_PL | NA | A | G | 0.032478 | 0.004431 | 2.60E-13 |
| rs493246 | miR-139-5p | trans | 8.06E-42 | Nightingale | Omega_6 | NA | A | G | 0.025912 | 0.004321 | 5.60E-10 |
| rs493246 | miR-139-5p | trans | 8.06E-42 | Nightingale | Total_PL | NA | A | G | 0.024943 | 0.004214 | 1.00E-09 |
| rs493246 | miR-139-5p | trans | 8.06E-42 | Nightingale | S_VLDL_C_ | NA | A | G | 0.041892 | 0.004306 | 5.20E-22 |
| rs493246 | miR-139-5p | trans | 8.06E-42 | Nightingale | S_LDL_CE | NA | A | G | 0.030558 | 0.004427 | 5.80E-12 |
| rs493246 | miR-139-5p | trans | 8.06E-42 | Nightingale | L_LDL_P | NA | A | G | 0.029187 | 0.00441 | 6.10E-11 |
| rs493246 | miR-139-5p | trans | 8.06E-42 | Nightingale | XS_VLDL_P | NA | A | G | 0.036714 | 0.004366 | 1.40E-17 |
| rs493246 | miR-139-5p | trans | 8.06E-42 | Nightingale | LDL_C | NA | A | G | 0.033206 | 0.004406 | 8.70E-14 |
| rs493246 | miR-139-5p | trans | 8.06E-42 | Nightingale | S_VLDL_PL | NA | A | G | 0.027742 | 0.004237 | 1.80E-10 |
| rs493246 | miR-139-5p | trans | 8.06E-42 | Nightingale | M_LDL_P | NA | A | G | 0.0359 | 0.004437 | 6.80E-16 |
| rs493246 | miR-139-5p | trans | 8.06E-42 | Nightingale | S_LDL_C | NA | A | G | 0.030953 | 0.004429 | 5.50E-12 |
| rs493246 | miR-139-5p | trans | 8.06E-42 | Nightingale | XXL_VLDL_ | NA | A | G | 0.033132 | 0.004459 | 8.80E-14 |
| rs493246 | miR-139-5p | trans | 8.06E-42 | Nightingale | S_LDL_CE_ | NA | A | G | 0.03095 | 0.004417 | 4.80E-14 |
| rs493246 | miR-139-5p | trans | 8.06E-42 | Nightingale | LA | NA | A | G | 0.024925 | 0.004339 | 3.60E-09 |
| rs493246 | miR-139-5p | trans | 8.06E-42 | Nightingale | Remnant_( | NA | A | G | 0.036878 | 0.004396 | 3.60E-17 |
| rs493246 | miR-139-5p | trans | 8.06E-42 | Nightingale | L_VLDL_FC | NA | A | G | 0.048596 | 0.004382 | 4.50E-29 |
| rs493246 | miR-139-5p | trans | 8.06E-42 | Nightingale | S_VLDL_FC | NA | A | G | 0.037913 | 0.004426 | 7.30E-18 |
| rs493246 | miR-139-5p | trans | 8.06E-42 | Nightingale | S_LDL_P | NA | A | G | 0.029597 | 0.004429 | 3.20E-11 |
| rs493246 | miR-139-5p | trans | 8.06E-42 | Nightingale | M_LDL_CE | NA | A | G | 0.032983 | 0.004431 | 9.50E-14 |
| rs493246 | miR-139-5p | trans | 8.06E-42 | Nightingale | Total_L | NA | A | G | 0.026363 | 0.004353 | 4.50E-10 |
| rs493246 | miR-139-5p | trans | 8.06E-42 | Nightingale | L_LDL_L | NA | A | G | 0.033563 | 0.004382 | 2.90E-14 |
| rs493246 | miR-139-5p | trans | 8.06E-42 | Nightingale | M_VLDL_C | NA | A | G | 0.033637 | 0.004157 | 1.90E-15 |
| rs493246 | miR-139-5p | trans | 8.06E-42 | Nightingale | ApoB_by_/ | NA | A | G | 0.02505 | 0.004358 | 1.30E-08 |
| rs493246 | miR-139-5p | trans | 8.06E-42 | Nightingale | S_VLDL_C | NA | A | G | 0.038381 | 0.004424 | 1.80E-18 |
| rs493246 | miR-139-5p | trans | 8.06E-42 | Nightingale | S_VLDL_TG | NA | A | G | -0.03813 | 0.004283 | 1.40E-18 |
| rs493246 | miR-139-5p | trans | 8.06E-42 | Nightingale | ApoB | NA | A | G | 0.033948 | 0.004419 | 2.10E-14 |
| rs493246 | miR-139-5p | trans | 8.06E-42 | Nightingale | IDL_L | NA | A | G | 0.035283 | 0.004278 | 4.70E-16 |
| rs493246 | miR-139-5p | trans | 8.06E-42 | Nightingale | S_VLDL_P | NA | A | G | 0.023473 | 0.004369 | 1.60E-08 |
| rs493246 | miR-139-5p | trans | 8.06E-42 | Nightingale | L_LDL_C | NA | A | G | 0.032333 | 0.004375 | 2.50E-13 |
| rs493246 | miR-139-5p | trans | 8.06E-42 | Nightingale | IDL_CE | NA | A | G | 0.03352 | 0.004272 | 1.30E-14 |
| rs493246 | miR-139-5p | trans | 8.06E-42 | Nightingale | XL_VLDL_F | NA | A | G | 0.037204 | 0.00429 | 2.00E-18 |
| rs493246 | miR-139-5p | trans | 8.06E-42 | Nightingale | XXL_VLDL_ | NA | A | G | -0.03574 | 0.004463 | 2.10E-16 |
| rs493246 | miR-139-5p | trans | 8.06E-42 | Nightingale | XS_VLDL_L | NA | A | G | 0.033831 | 0.00435 | 2.00E-15 |
| rs493246 | miR-139-5p | trans | 8.06E-42 | Nightingale | M_VLDL_P | NA | A | G | 0.028301 | 0.004414 | 6.30E-11 |
| rs493246 | miR-139-5p | trans | 8.06E-42 | Nightingale | S_HDL_P | NA | A | G | 0.030621 | 0.004425 | 5.00E-12 |
| rs493246 | miR-139-5p | trans | 8.06E-42 | Nightingale | S_VLDL_CE | NA | A | G | 0.038189 | 0.004419 | 1.60E-18 |

| | | | | | | | | | | | |
|---|---|---|---|---|---|---|---|---|---|---|---|
| rs493246 | miR-139-5p | trans | 8.06E-42 | Nightingale | L_LDL_FC | NA | A | G | 0.033637 | 0.004345 | 2.80E-14 |
| rs493246 | miR-139-5p | trans | 8.06E-42 | Nightingale | Citrate | NA | A | G | 0.026826 | 0.004396 | 7.40E-10 |
| rs493246 | miR-139-5p | trans | 8.06E-42 | Nightingale | L_LDL_CE | NA | A | G | 0.03158 | 0.004385 | 9.70E-13 |
| rs493246 | let-7b-5p | trans | 1.09E-26 | Nightingale | IDL_FC | NA | A | G | 0.038267 | 0.004284 | 1.40E-18 |
| rs493246 | let-7b-5p | trans | 1.09E-26 | Nightingale | LDL_CE | NA | A | G | 0.032566 | 0.004413 | 2.20E-13 |
| rs493246 | let-7b-5p | trans | 1.09E-26 | Nightingale | L_VLDL_CE | NA | A | G | 0.04735 | 0.004234 | 2.40E-29 |
| rs493246 | let-7b-5p | trans | 1.09E-26 | Nightingale | M_LDL_C_ | NA | A | G | 0.031162 | 0.004428 | 2.90E-12 |
| rs493246 | let-7b-5p | trans | 1.09E-26 | Nightingale | XS_VLDL_F | NA | A | G | 0.03143 | 0.004393 | 2.40E-12 |
| rs493246 | let-7b-5p | trans | 1.09E-26 | Nightingale | S_HDL_FC | NA | A | G | 0.023468 | 0.004344 | 4.50E-08 |
| rs493246 | let-7b-5p | trans | 1.09E-26 | Nightingale | S_LDL_L | NA | A | G | 0.027751 | 0.004424 | 8.20E-10 |
| rs493246 | let-7b-5p | trans | 1.09E-26 | Nightingale | LDL_L | NA | A | G | 0.033316 | 0.00441 | 7.30E-14 |
| rs493246 | let-7b-5p | trans | 1.09E-26 | Nightingale | M_LDL_L | NA | A | G | 0.033043 | 0.004431 | 1.00E-13 |
| rs493246 | let-7b-5p | trans | 1.09E-26 | Nightingale | M_LDL_C | NA | A | G | 0.033998 | 0.004433 | 2.10E-14 |
| rs493246 | let-7b-5p | trans | 1.09E-26 | Nightingale | Clinical_LD | NA | A | G | 0.036486 | 0.004399 | 2.70E-16 |
| rs493246 | let-7b-5p | trans | 1.09E-26 | Nightingale | S_VLDL_FC | NA | A | G | 0.02936 | 0.004228 | 1.10E-11 |
| rs493246 | let-7b-5p | trans | 1.09E-26 | Nightingale | IDL_FC_pct | NA | A | G | 0.025772 | 0.004391 | 1.10E-08 |
| rs493246 | let-7b-5p | trans | 1.09E-26 | Nightingale | M_VLDL_P | NA | A | G | 0.044066 | 0.004247 | 5.50E-25 |
| rs493246 | let-7b-5p | trans | 1.09E-26 | Nightingale | Total_CE | NA | A | G | 0.031761 | 0.004268 | 9.50E-14 |
| rs493246 | let-7b-5p | trans | 1.09E-26 | Nightingale | M_VLDL_F | NA | A | G | 0.033378 | 0.004424 | 3.10E-14 |
| rs493246 | let-7b-5p | trans | 1.09E-26 | Nightingale | IDL_C | NA | A | G | 0.034968 | 0.004274 | 9.20E-16 |
| rs493246 | let-7b-5p | trans | 1.09E-26 | Nightingale | LDL_PL | NA | A | G | 0.035 | 0.004415 | 4.70E-15 |
| rs493246 | let-7b-5p | trans | 1.09E-26 | Nightingale | M_VLDL_P | NA | A | G | 0.029134 | 0.004418 | 2.00E-11 |
| rs493246 | let-7b-5p | trans | 1.09E-26 | Nightingale | XL_VLDL_P | NA | A | G | 0.038845 | 0.004432 | 5.20E-19 |
| rs493246 | let-7b-5p | trans | 1.09E-26 | Nightingale | XXL_VLDL_ | NA | A | G | 0.034841 | 0.004434 | 2.90E-15 |
| rs493246 | let-7b-5p | trans | 1.09E-26 | Nightingale | XS_VLDL_F | NA | A | G | 0.03708 | 0.004348 | 6.40E-18 |
| rs493246 | let-7b-5p | trans | 1.09E-26 | Nightingale | GlycA | NA | A | G | 0.028022 | 0.004406 | 1.40E-10 |
| rs493246 | let-7b-5p | trans | 1.09E-26 | Nightingale | XL_VLDL_C | NA | A | G | 0.03823 | 0.004243 | 4.60E-19 |
| rs493246 | let-7b-5p | trans | 1.09E-26 | Nightingale | M_VLDL_C | NA | A | G | 0.040853 | 0.004385 | 2.30E-20 |
| rs493246 | let-7b-5p | trans | 1.09E-26 | Nightingale | Cholines | NA | A | G | 0.022312 | 0.004161 | 4.70E-08 |
| rs493246 | let-7b-5p | trans | 1.09E-26 | Nightingale | S_HDL_CE | NA | A | G | 0.028644 | 0.004432 | 1.30E-10 |
| rs493246 | let-7b-5p | trans | 1.09E-26 | Nightingale | M_VLDL_C | NA | A | G | 0.038719 | 0.004413 | 1.90E-18 |
| rs493246 | let-7b-5p | trans | 1.09E-26 | Nightingale | XS_VLDL_C | NA | A | G | 0.03781 | 0.00427 | 2.10E-18 |
| rs493246 | let-7b-5p | trans | 1.09E-26 | Nightingale | VLDL_CE | NA | A | G | 0.035794 | 0.004426 | 3.00E-16 |
| rs493246 | let-7b-5p | trans | 1.09E-26 | Nightingale | S_HDL_L | NA | A | G | 0.026306 | 0.00441 | 1.50E-09 |
| rs493246 | let-7b-5p | trans | 1.09E-26 | Nightingale | Total_FC | NA | A | G | 0.033269 | 0.004323 | 6.70E-15 |
| rs493246 | let-7b-5p | trans | 1.09E-26 | Nightingale | M_VLDL_F | NA | A | G | 0.042074 | 0.00421 | 3.40E-23 |
| rs493246 | let-7b-5p | trans | 1.09E-26 | Nightingale | S_VLDL_L | NA | A | G | 0.024676 | 0.004382 | 3.70E-09 |
| rs493246 | let-7b-5p | trans | 1.09E-26 | Nightingale | S_LDL_PL_ | NA | A | G | -0.03007 | 0.004417 | 2.60E-13 |
| rs493246 | let-7b-5p | trans | 1.09E-26 | Nightingale | XS_VLDL_P | NA | A | G | 0.032128 | 0.004363 | 5.30E-14 |
| rs493246 | let-7b-5p | trans | 1.09E-26 | Nightingale | Sphingomy | NA | A | G | 0.029031 | 0.004137 | 6.60E-12 |
| rs493246 | let-7b-5p | trans | 1.09E-26 | Nightingale | S_LDL_C_p | NA | A | G | 0.045367 | 0.004442 | 5.40E-25 |
| rs493246 | let-7b-5p | trans | 1.09E-26 | Nightingale | VLDL_FC | NA | A | G | 0.024164 | 0.004392 | 1.10E-08 |
| rs493246 | let-7b-5p | trans | 1.09E-26 | Nightingale | XL_VLDL_C | NA | A | G | 0.037359 | 0.004245 | 3.10E-18 |
| rs493246 | let-7b-5p | trans | 1.09E-26 | Nightingale | L_VLDL_C_ | NA | A | G | 0.052205 | 0.004322 | 3.90E-34 |
| rs493246 | let-7b-5p | trans | 1.09E-26 | Nightingale | Total_C | NA | A | G | 0.032388 | 0.004284 | 3.20E-14 |
| rs493246 | let-7b-5p | trans | 1.09E-26 | Nightingale | non_HDL_( | NA | A | G | 0.035709 | 0.004401 | 5.00E-16 |
| rs493246 | let-7b-5p | trans | 1.09E-26 | Nightingale | IDL_P | NA | A | G | 0.039997 | 0.004378 | 1.30E-19 |
| rs493246 | let-7b-5p | trans | 1.09E-26 | Nightingale | S_LDL_FC | NA | A | G | 0.028573 | 0.004418 | 4.20E-10 |
| rs493246 | let-7b-5p | trans | 1.09E-26 | Nightingale | XXL_VLDL_ | NA | A | G | 0.025012 | 0.004454 | 1.50E-09 |
| rs493246 | let-7b-5p | trans | 1.09E-26 | Nightingale | L_LDL_PL | NA | A | G | 0.038002 | 0.004391 | 8.90E-18 |
| rs493246 | let-7b-5p | trans | 1.09E-26 | Nightingale | L_VLDL_TG | NA | A | G | -0.05115 | 0.004404 | 5.30E-32 |
| rs493246 | let-7b-5p | trans | 1.09E-26 | Nightingale | S_VLDL_PL | NA | A | G | 0.032786 | 0.004417 | 4.20E-14 |
| rs493246 | let-7b-5p | trans | 1.09E-26 | Nightingale | IDL_PL | NA | A | G | 0.035357 | 0.00427 | 3.30E-16 |
| rs493246 | let-7b-5p | trans | 1.09E-26 | Nightingale | M_VLDL_C | NA | A | G | 0.036214 | 0.004168 | 1.20E-17 |
| rs493246 | let-7b-5p | trans | 1.09E-26 | Nightingale | XL_HDL_PL | NA | A | G | -0.02274 | 0.004065 | 1.20E-08 |
| rs493246 | let-7b-5p | trans | 1.09E-26 | Nightingale | XXL_VLDL_ | NA | A | G | 0.030859 | 0.004406 | 1.10E-12 |
| rs493246 | let-7b-5p | trans | 1.09E-26 | Nightingale | XS_VLDL_C | NA | A | G | 0.038005 | 0.004295 | 1.30E-18 |
| rs493246 | let-7b-5p | trans | 1.09E-26 | Nightingale | M_LDL_TG | NA | A | G | -0.02762 | 0.00437 | 3.20E-10 |
| rs493246 | let-7b-5p | trans | 1.09E-26 | Nightingale | S_VLDL_CE | NA | A | G | 0.047037 | 0.004365 | 1.20E-26 |
| rs493246 | let-7b-5p | trans | 1.09E-26 | Nightingale | S_HDL_C | NA | A | G | 0.028422 | 0.004417 | 1.70E-10 |
| rs493246 | let-7b-5p | trans | 1.09E-26 | Nightingale | LDL_P | NA | A | G | 0.031809 | 0.004422 | 1.00E-12 |
| rs493246 | let-7b-5p | trans | 1.09E-26 | Nightingale | PUFA | NA | A | G | 0.023738 | 0.004316 | 1.40E-08 |
| rs493246 | let-7b-5p | trans | 1.09E-26 | Nightingale | M_VLDL_T | NA | A | G | -0.03857 | 0.004181 | 6.70E-20 |

| | | | | | | | | | | | |
|---|---|---|---|---|---|---|---|---|---|---|---|
| rs493246 | let-7b-5p | trans | 1.09E-26 | Nightingale | VLDL_C | NA | A | G | 0.031309 | 0.004419 | 4.80E-13 |
| rs493246 | let-7b-5p | trans | 1.09E-26 | Nightingale | VLDL_P | NA | A | G | 0.026641 | 0.004401 | 4.40E-10 |
| rs493246 | let-7b-5p | trans | 1.09E-26 | Nightingale | S_HDL_PL | NA | A | G | 0.023794 | 0.004387 | 3.20E-08 |
| rs493246 | let-7b-5p | trans | 1.09E-26 | Nightingale | XL_VLDL_T | NA | A | G | -0.04393 | 0.004289 | 1.70E-24 |
| rs493246 | let-7b-5p | trans | 1.09E-26 | Nightingale | LDL_FC | NA | A | G | 0.033644 | 0.004378 | 5.20E-14 |
| rs493246 | let-7b-5p | trans | 1.09E-26 | Nightingale | M_LDL_FC | NA | A | G | 0.034359 | 0.004419 | 1.90E-14 |
| rs493246 | let-7b-5p | trans | 1.09E-26 | Nightingale | M_LDL_PL | NA | A | G | 0.032478 | 0.004431 | 2.60E-13 |
| rs493246 | let-7b-5p | trans | 1.09E-26 | Nightingale | Omega_6 | NA | A | G | 0.025912 | 0.004321 | 5.60E-10 |
| rs493246 | let-7b-5p | trans | 1.09E-26 | Nightingale | Total_PL | NA | A | G | 0.024943 | 0.004214 | 1.00E-09 |
| rs493246 | let-7b-5p | trans | 1.09E-26 | Nightingale | S_VLDL_C_ | NA | A | G | 0.041892 | 0.004306 | 5.20E-22 |
| rs493246 | let-7b-5p | trans | 1.09E-26 | Nightingale | S_LDL_CE | NA | A | G | 0.030558 | 0.004427 | 5.80E-12 |
| rs493246 | let-7b-5p | trans | 1.09E-26 | Nightingale | L_LDL_P | NA | A | G | 0.029187 | 0.00441 | 6.10E-11 |
| rs493246 | let-7b-5p | trans | 1.09E-26 | Nightingale | XS_VLDL_P | NA | A | G | 0.036714 | 0.004366 | 1.40E-17 |
| rs493246 | let-7b-5p | trans | 1.09E-26 | Nightingale | LDL_C | NA | A | G | 0.033206 | 0.004406 | 8.70E-14 |
| rs493246 | let-7b-5p | trans | 1.09E-26 | Nightingale | S_VLDL_PL | NA | A | G | 0.027742 | 0.004237 | 1.80E-10 |
| rs493246 | let-7b-5p | trans | 1.09E-26 | Nightingale | M_LDL_P | NA | A | G | 0.0359 | 0.004437 | 6.80E-16 |
| rs493246 | let-7b-5p | trans | 1.09E-26 | Nightingale | S_LDL_C | NA | A | G | 0.030953 | 0.004429 | 5.50E-12 |
| rs493246 | let-7b-5p | trans | 1.09E-26 | Nightingale | XXL_VLDL_ | NA | A | G | 0.033132 | 0.004459 | 8.80E-14 |
| rs493246 | let-7b-5p | trans | 1.09E-26 | Nightingale | S_LDL_CE_ | NA | A | G | 0.03095 | 0.004417 | 4.80E-14 |
| rs493246 | let-7b-5p | trans | 1.09E-26 | Nightingale | LA | NA | A | G | 0.024925 | 0.004339 | 3.60E-09 |
| rs493246 | let-7b-5p | trans | 1.09E-26 | Nightingale | Remnant_( | NA | A | G | 0.036878 | 0.004396 | 3.60E-17 |
| rs493246 | let-7b-5p | trans | 1.09E-26 | Nightingale | L_VLDL_FC | NA | A | G | 0.048596 | 0.004382 | 4.50E-29 |
| rs493246 | let-7b-5p | trans | 1.09E-26 | Nightingale | S_VLDL_FC | NA | A | G | 0.037913 | 0.004426 | 7.30E-18 |
| rs493246 | let-7b-5p | trans | 1.09E-26 | Nightingale | S_LDL_P | NA | A | G | 0.029597 | 0.004429 | 3.20E-11 |
| rs493246 | let-7b-5p | trans | 1.09E-26 | Nightingale | M_LDL_CE | NA | A | G | 0.032983 | 0.004431 | 9.50E-14 |
| rs493246 | let-7b-5p | trans | 1.09E-26 | Nightingale | Total_L | NA | A | G | 0.026363 | 0.004353 | 4.50E-10 |
| rs493246 | let-7b-5p | trans | 1.09E-26 | Nightingale | L_LDL_L | NA | A | G | 0.033563 | 0.004382 | 2.90E-14 |
| rs493246 | let-7b-5p | trans | 1.09E-26 | Nightingale | M_VLDL_C | NA | A | G | 0.033637 | 0.004157 | 1.90E-15 |
| rs493246 | let-7b-5p | trans | 1.09E-26 | Nightingale | ApoB_by_/ | NA | A | G | 0.02505 | 0.004358 | 1.30E-08 |
| rs493246 | let-7b-5p | trans | 1.09E-26 | Nightingale | S_VLDL_C | NA | A | G | 0.038381 | 0.004424 | 1.80E-18 |
| rs493246 | let-7b-5p | trans | 1.09E-26 | Nightingale | S_VLDL_TG | NA | A | G | -0.03813 | 0.004283 | 1.40E-18 |
| rs493246 | let-7b-5p | trans | 1.09E-26 | Nightingale | ApoB | NA | A | G | 0.033948 | 0.004419 | 2.10E-14 |
| rs493246 | let-7b-5p | trans | 1.09E-26 | Nightingale | IDL_L | NA | A | G | 0.035283 | 0.004278 | 4.70E-16 |
| rs493246 | let-7b-5p | trans | 1.09E-26 | Nightingale | S_VLDL_P | NA | A | G | 0.023473 | 0.004369 | 1.60E-08 |
| rs493246 | let-7b-5p | trans | 1.09E-26 | Nightingale | L_LDL_C | NA | A | G | 0.032333 | 0.004375 | 2.50E-13 |
| rs493246 | let-7b-5p | trans | 1.09E-26 | Nightingale | IDL_CE | NA | A | G | 0.03352 | 0.004272 | 1.30E-14 |
| rs493246 | let-7b-5p | trans | 1.09E-26 | Nightingale | XL_VLDL_F | NA | A | G | 0.037204 | 0.00429 | 2.00E-18 |
| rs493246 | let-7b-5p | trans | 1.09E-26 | Nightingale | XXL_VLDL_ | NA | A | G | -0.03574 | 0.004463 | 2.10E-16 |
| rs493246 | let-7b-5p | trans | 1.09E-26 | Nightingale | XS_VLDL_L | NA | A | G | 0.033831 | 0.00435 | 2.00E-15 |
| rs493246 | let-7b-5p | trans | 1.09E-26 | Nightingale | M_VLDL_P | NA | A | G | 0.028301 | 0.004414 | 6.30E-11 |
| rs493246 | let-7b-5p | trans | 1.09E-26 | Nightingale | S_HDL_P | NA | A | G | 0.030621 | 0.004425 | 5.00E-12 |
| rs493246 | let-7b-5p | trans | 1.09E-26 | Nightingale | S_VLDL_CE | NA | A | G | 0.038189 | 0.004419 | 1.60E-18 |
| rs493246 | let-7b-5p | trans | 1.09E-26 | Nightingale | L_LDL_FC | NA | A | G | 0.033637 | 0.004345 | 2.80E-14 |
| rs493246 | let-7b-5p | trans | 1.09E-26 | Nightingale | Citrate | NA | A | G | 0.026826 | 0.004396 | 7.40E-10 |
| rs493246 | let-7b-5p | trans | 1.09E-26 | Nightingale | L_LDL_CE | NA | A | G | 0.03158 | 0.004385 | 9.70E-13 |
| rs493246 | let-7c-5p | trans | 2.70E-23 | Nightingale | IDL_FC | NA | A | G | 0.038267 | 0.004284 | 1.40E-18 |
| rs493246 | let-7c-5p | trans | 2.70E-23 | Nightingale | LDL_CE | NA | A | G | 0.032566 | 0.004413 | 2.20E-13 |
| rs493246 | let-7c-5p | trans | 2.70E-23 | Nightingale | L_VLDL_CE | NA | A | G | 0.04735 | 0.004234 | 2.40E-29 |
| rs493246 | let-7c-5p | trans | 2.70E-23 | Nightingale | M_LDL_C_\| | NA | A | G | 0.031162 | 0.004428 | 2.90E-12 |
| rs493246 | let-7c-5p | trans | 2.70E-23 | Nightingale | XS_VLDL_F | NA | A | G | 0.03143 | 0.004393 | 2.40E-12 |
| rs493246 | let-7c-5p | trans | 2.70E-23 | Nightingale | S_HDL_FC | NA | A | G | 0.023468 | 0.004344 | 4.50E-08 |
| rs493246 | let-7c-5p | trans | 2.70E-23 | Nightingale | S_LDL_L | NA | A | G | 0.027751 | 0.004424 | 8.20E-10 |
| rs493246 | let-7c-5p | trans | 2.70E-23 | Nightingale | LDL_L | NA | A | G | 0.033316 | 0.00441 | 7.30E-14 |
| rs493246 | let-7c-5p | trans | 2.70E-23 | Nightingale | M_LDL_L | NA | A | G | 0.033043 | 0.004431 | 1.00E-13 |
| rs493246 | let-7c-5p | trans | 2.70E-23 | Nightingale | M_LDL_C | NA | A | G | 0.033998 | 0.004433 | 2.10E-14 |
| rs493246 | let-7c-5p | trans | 2.70E-23 | Nightingale | Clinical_LD | NA | A | G | 0.036486 | 0.004399 | 2.70E-16 |
| rs493246 | let-7c-5p | trans | 2.70E-23 | Nightingale | S_VLDL_FC | NA | A | G | 0.02936 | 0.004228 | 1.10E-11 |
| rs493246 | let-7c-5p | trans | 2.70E-23 | Nightingale | IDL_FC_pct | NA | A | G | 0.025772 | 0.004391 | 1.10E-08 |
| rs493246 | let-7c-5p | trans | 2.70E-23 | Nightingale | M_VLDL_P | NA | A | G | 0.044066 | 0.004247 | 5.50E-25 |
| rs493246 | let-7c-5p | trans | 2.70E-23 | Nightingale | Total_CE | NA | A | G | 0.031761 | 0.004268 | 9.50E-14 |
| rs493246 | let-7c-5p | trans | 2.70E-23 | Nightingale | M_VLDL_F( | NA | A | G | 0.033378 | 0.004424 | 3.10E-14 |
| rs493246 | let-7c-5p | trans | 2.70E-23 | Nightingale | IDL_C | NA | A | G | 0.034968 | 0.004274 | 9.20E-16 |
| rs493246 | let-7c-5p | trans | 2.70E-23 | Nightingale | LDL_PL | NA | A | G | 0.035 | 0.004415 | 4.70E-15 |

| | | | | | | | | | | | |
|---|---|---|---|---|---|---|---|---|---|---|---|
| rs493246 | let-7c-5p | trans | 2.70E-23 | Nightingale | M_VLDL_P | NA | A | G | 0.029134 | 0.004418 | 2.00E-11 |
| rs493246 | let-7c-5p | trans | 2.70E-23 | Nightingale | XL_VLDL_P | NA | A | G | 0.038845 | 0.004432 | 5.20E-19 |
| rs493246 | let-7c-5p | trans | 2.70E-23 | Nightingale | XXL_VLDL_ | NA | A | G | 0.034841 | 0.004434 | 2.90E-15 |
| rs493246 | let-7c-5p | trans | 2.70E-23 | Nightingale | XS_VLDL_F | NA | A | G | 0.03708 | 0.004348 | 6.40E-18 |
| rs493246 | let-7c-5p | trans | 2.70E-23 | Nightingale | GlycA | NA | A | G | 0.028022 | 0.004406 | 1.40E-10 |
| rs493246 | let-7c-5p | trans | 2.70E-23 | Nightingale | XL_VLDL_C | NA | A | G | 0.03823 | 0.004243 | 4.60E-19 |
| rs493246 | let-7c-5p | trans | 2.70E-23 | Nightingale | M_VLDL_C | NA | A | G | 0.040853 | 0.004385 | 2.30E-20 |
| rs493246 | let-7c-5p | trans | 2.70E-23 | Nightingale | Cholines | NA | A | G | 0.022312 | 0.004161 | 4.70E-08 |
| rs493246 | let-7c-5p | trans | 2.70E-23 | Nightingale | S_HDL_CE | NA | A | G | 0.028644 | 0.004432 | 1.30E-10 |
| rs493246 | let-7c-5p | trans | 2.70E-23 | Nightingale | M_VLDL_C | NA | A | G | 0.038719 | 0.004413 | 1.90E-18 |
| rs493246 | let-7c-5p | trans | 2.70E-23 | Nightingale | XS_VLDL_C | NA | A | G | 0.03781 | 0.00427 | 2.10E-18 |
| rs493246 | let-7c-5p | trans | 2.70E-23 | Nightingale | VLDL_CE | NA | A | G | 0.035794 | 0.004426 | 3.00E-16 |
| rs493246 | let-7c-5p | trans | 2.70E-23 | Nightingale | S_HDL_L | NA | A | G | 0.026306 | 0.00441 | 1.50E-09 |
| rs493246 | let-7c-5p | trans | 2.70E-23 | Nightingale | Total_FC | NA | A | G | 0.033269 | 0.004323 | 6.70E-15 |
| rs493246 | let-7c-5p | trans | 2.70E-23 | Nightingale | M_VLDL_F | NA | A | G | 0.042074 | 0.00421 | 3.40E-23 |
| rs493246 | let-7c-5p | trans | 2.70E-23 | Nightingale | S_VLDL_L | NA | A | G | 0.024676 | 0.004382 | 3.70E-09 |
| rs493246 | let-7c-5p | trans | 2.70E-23 | Nightingale | S_LDL_PL_ | NA | A | G | -0.03007 | 0.004417 | 2.60E-13 |
| rs493246 | let-7c-5p | trans | 2.70E-23 | Nightingale | XS_VLDL_P | NA | A | G | 0.032128 | 0.004363 | 5.30E-14 |
| rs493246 | let-7c-5p | trans | 2.70E-23 | Nightingale | Sphingomy | NA | A | G | 0.029031 | 0.004137 | 6.60E-12 |
| rs493246 | let-7c-5p | trans | 2.70E-23 | Nightingale | S_LDL_C_p | NA | A | G | 0.045367 | 0.004442 | 5.40E-25 |
| rs493246 | let-7c-5p | trans | 2.70E-23 | Nightingale | VLDL_FC | NA | A | G | 0.024164 | 0.004392 | 1.10E-08 |
| rs493246 | let-7c-5p | trans | 2.70E-23 | Nightingale | XL_VLDL_C | NA | A | G | 0.037359 | 0.004245 | 3.10E-18 |
| rs493246 | let-7c-5p | trans | 2.70E-23 | Nightingale | L_VLDL_C_ | NA | A | G | 0.052205 | 0.004322 | 3.90E-34 |
| rs493246 | let-7c-5p | trans | 2.70E-23 | Nightingale | Total_C | NA | A | G | 0.032388 | 0.004284 | 3.20E-14 |
| rs493246 | let-7c-5p | trans | 2.70E-23 | Nightingale | non_HDL_( | NA | A | G | 0.035709 | 0.004401 | 5.00E-16 |
| rs493246 | let-7c-5p | trans | 2.70E-23 | Nightingale | IDL_P | NA | A | G | 0.039997 | 0.004378 | 1.30E-19 |
| rs493246 | let-7c-5p | trans | 2.70E-23 | Nightingale | S_LDL_FC | NA | A | G | 0.028573 | 0.004418 | 4.20E-10 |
| rs493246 | let-7c-5p | trans | 2.70E-23 | Nightingale | XXL_VLDL_ | NA | A | G | 0.025012 | 0.004454 | 1.50E-09 |
| rs493246 | let-7c-5p | trans | 2.70E-23 | Nightingale | L_LDL_PL | NA | A | G | 0.038002 | 0.004391 | 8.90E-18 |
| rs493246 | let-7c-5p | trans | 2.70E-23 | Nightingale | L_VLDL_TG | NA | A | G | -0.05115 | 0.004404 | 5.30E-32 |
| rs493246 | let-7c-5p | trans | 2.70E-23 | Nightingale | S_VLDL_PL | NA | A | G | 0.032786 | 0.004417 | 4.20E-14 |
| rs493246 | let-7c-5p | trans | 2.70E-23 | Nightingale | IDL_PL | NA | A | G | 0.035357 | 0.00427 | 3.30E-16 |
| rs493246 | let-7c-5p | trans | 2.70E-23 | Nightingale | M_VLDL_C | NA | A | G | 0.036214 | 0.004168 | 1.20E-17 |
| rs493246 | let-7c-5p | trans | 2.70E-23 | Nightingale | XL_HDL_PL | NA | A | G | -0.02274 | 0.004065 | 1.20E-08 |
| rs493246 | let-7c-5p | trans | 2.70E-23 | Nightingale | XXL_VLDL_ | NA | A | G | 0.030859 | 0.004406 | 1.10E-12 |
| rs493246 | let-7c-5p | trans | 2.70E-23 | Nightingale | XS_VLDL_C | NA | A | G | 0.038005 | 0.004295 | 1.30E-18 |
| rs493246 | let-7c-5p | trans | 2.70E-23 | Nightingale | M_LDL_TG | NA | A | G | -0.02762 | 0.00437 | 3.20E-10 |
| rs493246 | let-7c-5p | trans | 2.70E-23 | Nightingale | S_VLDL_CE | NA | A | G | 0.047037 | 0.004365 | 1.20E-26 |
| rs493246 | let-7c-5p | trans | 2.70E-23 | Nightingale | S_HDL_C | NA | A | G | 0.028422 | 0.004417 | 1.70E-10 |
| rs493246 | let-7c-5p | trans | 2.70E-23 | Nightingale | LDL_P | NA | A | G | 0.031809 | 0.004422 | 1.00E-12 |
| rs493246 | let-7c-5p | trans | 2.70E-23 | Nightingale | PUFA | NA | A | G | 0.023738 | 0.004316 | 1.40E-08 |
| rs493246 | let-7c-5p | trans | 2.70E-23 | Nightingale | M_VLDL_T | NA | A | G | -0.03857 | 0.004181 | 6.70E-20 |
| rs493246 | let-7c-5p | trans | 2.70E-23 | Nightingale | VLDL_C | NA | A | G | 0.031309 | 0.004419 | 4.80E-13 |
| rs493246 | let-7c-5p | trans | 2.70E-23 | Nightingale | VLDL_P | NA | A | G | 0.026641 | 0.004401 | 4.40E-10 |
| rs493246 | let-7c-5p | trans | 2.70E-23 | Nightingale | S_HDL_PL | NA | A | G | 0.023794 | 0.004387 | 3.20E-08 |
| rs493246 | let-7c-5p | trans | 2.70E-23 | Nightingale | XL_VLDL_T | NA | A | G | -0.04393 | 0.004289 | 1.70E-24 |
| rs493246 | let-7c-5p | trans | 2.70E-23 | Nightingale | LDL_FC | NA | A | G | 0.033644 | 0.004378 | 5.20E-14 |
| rs493246 | let-7c-5p | trans | 2.70E-23 | Nightingale | M_LDL_FC | NA | A | G | 0.034359 | 0.004419 | 1.90E-14 |
| rs493246 | let-7c-5p | trans | 2.70E-23 | Nightingale | M_LDL_PL | NA | A | G | 0.032478 | 0.004431 | 2.60E-13 |
| rs493246 | let-7c-5p | trans | 2.70E-23 | Nightingale | Omega_6 | NA | A | G | 0.025912 | 0.004321 | 5.60E-10 |
| rs493246 | let-7c-5p | trans | 2.70E-23 | Nightingale | Total_PL | NA | A | G | 0.024943 | 0.004214 | 1.00E-09 |
| rs493246 | let-7c-5p | trans | 2.70E-23 | Nightingale | S_VLDL_C_ | NA | A | G | 0.041892 | 0.004306 | 5.20E-22 |
| rs493246 | let-7c-5p | trans | 2.70E-23 | Nightingale | S_LDL_CE | NA | A | G | 0.030558 | 0.004427 | 5.80E-12 |
| rs493246 | let-7c-5p | trans | 2.70E-23 | Nightingale | L_LDL_P | NA | A | G | 0.029187 | 0.00441 | 6.10E-11 |
| rs493246 | let-7c-5p | trans | 2.70E-23 | Nightingale | XS_VLDL_P | NA | A | G | 0.036714 | 0.004366 | 1.40E-17 |
| rs493246 | let-7c-5p | trans | 2.70E-23 | Nightingale | LDL_C | NA | A | G | 0.033206 | 0.004406 | 8.70E-14 |
| rs493246 | let-7c-5p | trans | 2.70E-23 | Nightingale | S_VLDL_PL | NA | A | G | 0.027742 | 0.004237 | 1.80E-10 |
| rs493246 | let-7c-5p | trans | 2.70E-23 | Nightingale | M_LDL_P | NA | A | G | 0.0359 | 0.004437 | 6.80E-16 |
| rs493246 | let-7c-5p | trans | 2.70E-23 | Nightingale | S_LDL_C | NA | A | G | 0.030953 | 0.004429 | 5.50E-12 |
| rs493246 | let-7c-5p | trans | 2.70E-23 | Nightingale | XXL_VLDL_ | NA | A | G | 0.033132 | 0.004459 | 8.80E-14 |
| rs493246 | let-7c-5p | trans | 2.70E-23 | Nightingale | S_LDL_CE_ | NA | A | G | 0.03095 | 0.004417 | 4.80E-14 |
| rs493246 | let-7c-5p | trans | 2.70E-23 | Nightingale | LA | NA | A | G | 0.024925 | 0.004339 | 3.60E-09 |
| rs493246 | let-7c-5p | trans | 2.70E-23 | Nightingale | Remnant_( | NA | A | G | 0.036878 | 0.004396 | 3.60E-17 |

| | | | | | | | | | | | |
|---|---|---|---|---|---|---|---|---|---|---|---|
| rs493246 | let-7c-5p | trans | 2.70E-23 | Nightingale | L_VLDL_FC | NA | A | G | 0.048596 | 0.004382 | 4.50E-29 |
| rs493246 | let-7c-5p | trans | 2.70E-23 | Nightingale | S_VLDL_FC | NA | A | G | 0.037913 | 0.004426 | 7.30E-18 |
| rs493246 | let-7c-5p | trans | 2.70E-23 | Nightingale | S_LDL_P | NA | A | G | 0.029597 | 0.004429 | 3.20E-11 |
| rs493246 | let-7c-5p | trans | 2.70E-23 | Nightingale | M_LDL_CE | NA | A | G | 0.032983 | 0.004431 | 9.50E-14 |
| rs493246 | let-7c-5p | trans | 2.70E-23 | Nightingale | Total_L | NA | A | G | 0.026363 | 0.004353 | 4.50E-10 |
| rs493246 | let-7c-5p | trans | 2.70E-23 | Nightingale | L_LDL_L | NA | A | G | 0.033563 | 0.004382 | 2.90E-14 |
| rs493246 | let-7c-5p | trans | 2.70E-23 | Nightingale | M_VLDL_C | NA | A | G | 0.033637 | 0.004157 | 1.90E-15 |
| rs493246 | let-7c-5p | trans | 2.70E-23 | Nightingale | ApoB_by_A | NA | A | G | 0.02505 | 0.004358 | 1.30E-08 |
| rs493246 | let-7c-5p | trans | 2.70E-23 | Nightingale | S_VLDL_C | NA | A | G | 0.038381 | 0.004424 | 1.80E-18 |
| rs493246 | let-7c-5p | trans | 2.70E-23 | Nightingale | S_VLDL_TG | NA | A | G | -0.03813 | 0.004283 | 1.40E-18 |
| rs493246 | let-7c-5p | trans | 2.70E-23 | Nightingale | ApoB | NA | A | G | 0.033948 | 0.004419 | 2.10E-14 |
| rs493246 | let-7c-5p | trans | 2.70E-23 | Nightingale | IDL_L | NA | A | G | 0.035283 | 0.004278 | 4.70E-16 |
| rs493246 | let-7c-5p | trans | 2.70E-23 | Nightingale | S_VLDL_P | NA | A | G | 0.023473 | 0.004369 | 1.60E-08 |
| rs493246 | let-7c-5p | trans | 2.70E-23 | Nightingale | L_LDL_C | NA | A | G | 0.032333 | 0.004375 | 2.50E-13 |
| rs493246 | let-7c-5p | trans | 2.70E-23 | Nightingale | IDL_CE | NA | A | G | 0.03352 | 0.004272 | 1.30E-14 |
| rs493246 | let-7c-5p | trans | 2.70E-23 | Nightingale | XL_VLDL_F | NA | A | G | 0.037204 | 0.00429 | 2.00E-18 |
| rs493246 | let-7c-5p | trans | 2.70E-23 | Nightingale | XXL_VLDL_ | NA | A | G | -0.03574 | 0.004463 | 2.10E-16 |
| rs493246 | let-7c-5p | trans | 2.70E-23 | Nightingale | XS_VLDL_L | NA | A | G | 0.033831 | 0.00435 | 2.00E-15 |
| rs493246 | let-7c-5p | trans | 2.70E-23 | Nightingale | M_VLDL_P | NA | A | G | 0.028301 | 0.004414 | 6.30E-11 |
| rs493246 | let-7c-5p | trans | 2.70E-23 | Nightingale | S_HDL_P | NA | A | G | 0.030621 | 0.004425 | 5.00E-12 |
| rs493246 | let-7c-5p | trans | 2.70E-23 | Nightingale | S_VLDL_CE | NA | A | G | 0.038189 | 0.004419 | 1.60E-18 |
| rs493246 | let-7c-5p | trans | 2.70E-23 | Nightingale | L_LDL_FC | NA | A | G | 0.033637 | 0.004345 | 2.80E-14 |
| rs493246 | let-7c-5p | trans | 2.70E-23 | Nightingale | Citrate | NA | A | G | 0.026826 | 0.004396 | 7.40E-10 |
| rs493246 | let-7c-5p | trans | 2.70E-23 | Nightingale | L_LDL_CE | NA | A | G | 0.03158 | 0.004385 | 9.70E-13 |
| rs494242 | miR-10b-5p | trans | 1.47E-34 | Nightingale | VLDL_P | NA | T | C | 0.026586 | 0.004324 | 1.90E-10 |
| rs494242 | miR-10b-5p | trans | 1.47E-34 | Nightingale | M_VLDL_C | NA | T | C | 0.033835 | 0.004094 | 2.70E-16 |
| rs494242 | miR-10b-5p | trans | 1.47E-34 | Nightingale | M_LDL_C_ | NA | T | C | 0.026574 | 0.00435 | 1.60E-09 |
| rs494242 | miR-10b-5p | trans | 1.47E-34 | Nightingale | M_LDL_FC | NA | T | C | 0.032299 | 0.004341 | 2.30E-13 |
| rs494242 | miR-10b-5p | trans | 1.47E-34 | Nightingale | XS_VLDL_P | NA | T | C | 0.035812 | 0.004289 | 9.60E-18 |
| rs494242 | miR-10b-5p | trans | 1.47E-34 | Nightingale | Total_CE | NA | T | C | 0.029414 | 0.004193 | 1.80E-12 |
| rs494242 | miR-10b-5p | trans | 1.47E-34 | Nightingale | GlycA | NA | T | C | 0.025353 | 0.004328 | 4.70E-09 |
| rs494242 | miR-10b-5p | trans | 1.47E-34 | Nightingale | M_VLDL_P | NA | T | C | 0.027954 | 0.004336 | 4.50E-11 |
| rs494242 | miR-10b-5p | trans | 1.47E-34 | Nightingale | XS_VLDL_F | NA | T | C | 0.029107 | 0.004316 | 3.20E-11 |
| rs494242 | miR-10b-5p | trans | 1.47E-34 | Nightingale | IDL_PL | NA | T | C | 0.033595 | 0.004195 | 1.90E-15 |
| rs494242 | miR-10b-5p | trans | 1.47E-34 | Nightingale | XS_VLDL_P | NA | T | C | 0.031685 | 0.004286 | 1.90E-14 |
| rs494242 | miR-10b-5p | trans | 1.47E-34 | Nightingale | M_LDL_C | NA | T | C | 0.032117 | 0.004355 | 2.20E-13 |
| rs494242 | miR-10b-5p | trans | 1.47E-34 | Nightingale | XS_VLDL_C | NA | T | C | 0.036408 | 0.004195 | 4.00E-18 |
| rs494242 | miR-10b-5p | trans | 1.47E-34 | Nightingale | IDL_C | NA | T | C | 0.03332 | 0.004198 | 4.70E-15 |
| rs494242 | miR-10b-5p | trans | 1.47E-34 | Nightingale | Total_C | NA | T | C | 0.030169 | 0.004208 | 4.70E-13 |
| rs494242 | miR-10b-5p | trans | 1.47E-34 | Nightingale | S_LDL_P | NA | T | C | 0.028983 | 0.004351 | 2.90E-11 |
| rs494242 | miR-10b-5p | trans | 1.47E-34 | Nightingale | M_VLDL_F | NA | T | C | 0.032542 | 0.004346 | 3.80E-14 |
| rs494242 | miR-10b-5p | trans | 1.47E-34 | Nightingale | XXL_VLDL_ | NA | T | C | 0.033442 | 0.004356 | 8.80E-15 |
| rs494242 | miR-10b-5p | trans | 1.47E-34 | Nightingale | S_HDL_L | NA | T | C | 0.024003 | 0.004332 | 2.60E-08 |
| rs494242 | miR-10b-5p | trans | 1.47E-34 | Nightingale | M_LDL_P | NA | T | C | 0.034022 | 0.004359 | 7.20E-15 |
| rs494242 | miR-10b-5p | trans | 1.47E-34 | Nightingale | XL_HDL_PL | NA | T | C | -0.0218 | 0.003994 | 4.60E-08 |
| rs494242 | miR-10b-5p | trans | 1.47E-34 | Nightingale | XS_VLDL_F | NA | T | C | 0.035906 | 0.004271 | 7.30E-18 |
| rs494242 | miR-10b-5p | trans | 1.47E-34 | Nightingale | VLDL_FC | NA | T | C | 0.024475 | 0.004315 | 3.40E-09 |
| rs494242 | miR-10b-5p | trans | 1.47E-34 | Nightingale | S_VLDL_FC | NA | T | C | 0.036668 | 0.004348 | 1.70E-17 |
| rs494242 | miR-10b-5p | trans | 1.47E-34 | Nightingale | ApoB | NA | T | C | 0.032952 | 0.004341 | 3.30E-14 |
| rs494242 | miR-10b-5p | trans | 1.47E-34 | Nightingale | M_VLDL_C | NA | T | C | 0.031337 | 0.004084 | 3.30E-14 |
| rs494242 | miR-10b-5p | trans | 1.47E-34 | Nightingale | S_LDL_C_p | NA | T | C | 0.041866 | 0.004364 | 5.60E-22 |
| rs494242 | miR-10b-5p | trans | 1.47E-34 | Nightingale | L_VLDL_TG | NA | T | C | -0.04923 | 0.004326 | 6.60E-31 |
| rs494242 | miR-10b-5p | trans | 1.47E-34 | Nightingale | LDL_CE | NA | T | C | 0.030645 | 0.004335 | 2.00E-12 |
| rs494242 | miR-10b-5p | trans | 1.47E-34 | Nightingale | Remnant_( | NA | T | C | 0.035863 | 0.004319 | 5.10E-17 |
| rs494242 | miR-10b-5p | trans | 1.47E-34 | Nightingale | S_LDL_L | NA | T | C | 0.026918 | 0.004346 | 1.20E-09 |
| rs494242 | miR-10b-5p | trans | 1.47E-34 | Nightingale | S_VLDL_TG | NA | T | C | -0.03597 | 0.004208 | 1.90E-17 |
| rs494242 | miR-10b-5p | trans | 1.47E-34 | Nightingale | L_VLDL_CE | NA | T | C | 0.044066 | 0.00416 | 8.10E-27 |
| rs494242 | miR-10b-5p | trans | 1.47E-34 | Nightingale | M_LDL_L | NA | T | C | 0.031415 | 0.004353 | 6.70E-13 |
| rs494242 | miR-10b-5p | trans | 1.47E-34 | Nightingale | LDL_P | NA | T | C | 0.030684 | 0.004344 | 2.10E-12 |
| rs494242 | miR-10b-5p | trans | 1.47E-34 | Nightingale | LDL_C | NA | T | C | 0.031211 | 0.004328 | 9.20E-13 |
| rs494242 | miR-10b-5p | trans | 1.47E-34 | Nightingale | M_VLDL_P | NA | T | C | 0.042089 | 0.004172 | 6.40E-24 |
| rs494242 | miR-10b-5p | trans | 1.47E-34 | Nightingale | LA | NA | T | C | 0.02331 | 0.004263 | 1.50E-08 |
| rs494242 | miR-10b-5p | trans | 1.47E-34 | Nightingale | L_VLDL_CE | NA | T | C | 0.022783 | 0.004296 | 2.80E-08 |

| | | | | | | | | | | | |
|---|---|---|---|---|---|---|---|---|---|---|---|
| rs494242 | miR-10b-5p | trans | 1.47E-34 | Nightingale | L_LDL_C | NA | T | C | 0.030249 | 0.004298 | 2.90E-12 |
| rs494242 | miR-10b-5p | trans | 1.47E-34 | Nightingale | XXL_VLDL_ | NA | T | C | 0.023509 | 0.004375 | 5.40E-09 |
| rs494242 | miR-10b-5p | trans | 1.47E-34 | Nightingale | S_LDL_PL_ | NA | T | C | -0.02809 | 0.004339 | 4.70E-12 |
| rs494242 | miR-10b-5p | trans | 1.47E-34 | Nightingale | VLDL_C | NA | T | C | 0.031089 | 0.004341 | 2.10E-13 |
| rs494242 | miR-10b-5p | trans | 1.47E-34 | Nightingale | XL_VLDL_P | NA | T | C | 0.039093 | 0.004353 | 7.70E-20 |
| rs494242 | miR-10b-5p | trans | 1.47E-34 | Nightingale | non_HDL_( | NA | T | C | 0.034165 | 0.004323 | 2.30E-15 |
| rs494242 | miR-10b-5p | trans | 1.47E-34 | Nightingale | S_LDL_FC | NA | T | C | 0.027211 | 0.00434 | 1.30E-09 |
| rs494242 | miR-10b-5p | trans | 1.47E-34 | Nightingale | LDL_L | NA | T | C | 0.031492 | 0.004332 | 5.60E-13 |
| rs494242 | miR-10b-5p | trans | 1.47E-34 | Nightingale | XL_VLDL_C | NA | T | C | 0.034568 | 0.00417 | 1.60E-16 |
| rs494242 | miR-10b-5p | trans | 1.47E-34 | Nightingale | IDL_FC | NA | T | C | 0.035991 | 0.004208 | 3.10E-17 |
| rs494242 | miR-10b-5p | trans | 1.47E-34 | Nightingale | XL_VLDL_C | NA | T | C | 0.035582 | 0.004168 | 1.80E-17 |
| rs494242 | miR-10b-5p | trans | 1.47E-34 | Nightingale | S_VLDL_C | NA | T | C | 0.03738 | 0.004346 | 2.50E-18 |
| rs494242 | miR-10b-5p | trans | 1.47E-34 | Nightingale | L_LDL_L | NA | T | C | 0.031549 | 0.004304 | 3.10E-13 |
| rs494242 | miR-10b-5p | trans | 1.47E-34 | Nightingale | XXL_VLDL_ | NA | T | C | 0.031802 | 0.00438 | 2.10E-13 |
| rs494242 | miR-10b-5p | trans | 1.47E-34 | Nightingale | M_LDL_TG | NA | T | C | -0.0251 | 0.004293 | 8.80E-09 |
| rs494242 | miR-10b-5p | trans | 1.47E-34 | Nightingale | S_VLDL_CE | NA | T | C | 0.037335 | 0.004341 | 1.70E-18 |
| rs494242 | miR-10b-5p | trans | 1.47E-34 | Nightingale | S_VLDL_P | NA | T | C | 0.023424 | 0.004292 | 9.40E-09 |
| rs494242 | miR-10b-5p | trans | 1.47E-34 | Nightingale | S_VLDL_PL | NA | T | C | 0.032066 | 0.004339 | 4.60E-14 |
| rs494242 | miR-10b-5p | trans | 1.47E-34 | Nightingale | M_VLDL_F | NA | T | C | 0.039367 | 0.004136 | 2.20E-21 |
| rs494242 | miR-10b-5p | trans | 1.47E-34 | Nightingale | S_VLDL_CE | NA | T | C | 0.045132 | 0.004288 | 1.50E-25 |
| rs494242 | miR-10b-5p | trans | 1.47E-34 | Nightingale | XXL_VLDL_ | NA | T | C | -0.03443 | 0.004384 | 6.30E-16 |
| rs494242 | miR-10b-5p | trans | 1.47E-34 | Nightingale | L_VLDL_C_ | NA | T | C | 0.049215 | 0.004246 | 9.30E-32 |
| rs494242 | miR-10b-5p | trans | 1.47E-34 | Nightingale | S_VLDL_PL | NA | T | C | 0.025717 | 0.004162 | 1.50E-09 |
| rs494242 | miR-10b-5p | trans | 1.47E-34 | Nightingale | IDL_CE | NA | T | C | 0.032104 | 0.004197 | 4.20E-14 |
| rs494242 | miR-10b-5p | trans | 1.47E-34 | Nightingale | XL_VLDL_T | NA | T | C | -0.04154 | 0.004213 | 5.00E-23 |
| rs494242 | miR-10b-5p | trans | 1.47E-34 | Nightingale | VLDL_CE | NA | T | C | 0.03517 | 0.004348 | 2.00E-16 |
| rs494242 | miR-10b-5p | trans | 1.47E-34 | Nightingale | Sphingomy | NA | T | C | 0.025942 | 0.004064 | 3.10E-10 |
| rs494242 | miR-10b-5p | trans | 1.47E-34 | Nightingale | L_LDL_P | NA | T | C | 0.028344 | 0.004332 | 7.50E-11 |
| rs494242 | miR-10b-5p | trans | 1.47E-34 | Nightingale | S_VLDL_L | NA | T | C | 0.02453 | 0.004305 | 2.40E-09 |
| rs494242 | miR-10b-5p | trans | 1.47E-34 | Nightingale | XS_VLDL_L | NA | T | C | 0.033036 | 0.004273 | 1.30E-15 |
| rs494242 | miR-10b-5p | trans | 1.47E-34 | Nightingale | L_VLDL_FC | NA | T | C | 0.046929 | 0.004305 | 4.10E-28 |
| rs494242 | miR-10b-5p | trans | 1.47E-34 | Nightingale | L_LDL_FC | NA | T | C | 0.031491 | 0.004268 | 4.00E-13 |
| rs494242 | miR-10b-5p | trans | 1.47E-34 | Nightingale | Total_PL | NA | T | C | 0.023039 | 0.004139 | 7.30E-09 |
| rs494242 | miR-10b-5p | trans | 1.47E-34 | Nightingale | L_VLDL_PL | NA | T | C | 0.021646 | 0.004222 | 4.50E-08 |
| rs494242 | miR-10b-5p | trans | 1.47E-34 | Nightingale | M_LDL_PL | NA | T | C | 0.031199 | 0.004353 | 9.60E-13 |
| rs494242 | miR-10b-5p | trans | 1.47E-34 | Nightingale | Clinical_LD | NA | T | C | 0.034576 | 0.004322 | 2.70E-15 |
| rs494242 | miR-10b-5p | trans | 1.47E-34 | Nightingale | S_LDL_CE_ | NA | T | C | 0.028871 | 0.004339 | 1.00E-12 |
| rs494242 | miR-10b-5p | trans | 1.47E-34 | Nightingale | S_VLDL_FC | NA | T | C | 0.027143 | 0.004154 | 1.20E-10 |
| rs494242 | miR-10b-5p | trans | 1.47E-34 | Nightingale | S_HDL_CE | NA | T | C | 0.02571 | 0.004354 | 6.80E-09 |
| rs494242 | miR-10b-5p | trans | 1.47E-34 | Nightingale | M_VLDL_P | NA | T | C | 0.028782 | 0.00434 | 1.30E-11 |
| rs494242 | miR-10b-5p | trans | 1.47E-34 | Nightingale | M_VLDL_C | NA | T | C | 0.039214 | 0.004307 | 1.30E-19 |
| rs494242 | miR-10b-5p | trans | 1.47E-34 | Nightingale | Total_L | NA | T | C | 0.025049 | 0.004276 | 1.40E-09 |
| rs494242 | miR-10b-5p | trans | 1.47E-34 | Nightingale | IDL_L | NA | T | C | 0.03368 | 0.004203 | 2.20E-15 |
| rs494242 | miR-10b-5p | trans | 1.47E-34 | Nightingale | S_VLDL_C_ | NA | T | C | 0.03972 | 0.00423 | 8.60E-21 |
| rs494242 | miR-10b-5p | trans | 1.47E-34 | Nightingale | ApoB_by_/ | NA | T | C | 0.025679 | 0.004281 | 2.10E-09 |
| rs494242 | miR-10b-5p | trans | 1.47E-34 | Nightingale | LDL_PL | NA | T | C | 0.03334 | 0.004337 | 2.90E-14 |
| rs494242 | miR-10b-5p | trans | 1.47E-34 | Nightingale | M_VLDL_T | NA | T | C | -0.03622 | 0.004108 | 1.60E-18 |
| rs494242 | miR-10b-5p | trans | 1.47E-34 | Nightingale | S_HDL_P | NA | T | C | 0.027834 | 0.004347 | 2.60E-10 |
| rs494242 | miR-10b-5p | trans | 1.47E-34 | Nightingale | XL_VLDL_F | NA | T | C | 0.035197 | 0.004214 | 1.80E-17 |
| rs494242 | miR-10b-5p | trans | 1.47E-34 | Nightingale | XXL_VLDL_ | NA | T | C | 0.029266 | 0.004328 | 5.20E-12 |
| rs494242 | miR-10b-5p | trans | 1.47E-34 | Nightingale | Total_FC | NA | T | C | 0.031409 | 0.004246 | 4.40E-14 |
| rs494242 | miR-10b-5p | trans | 1.47E-34 | Nightingale | XS_VLDL_C | NA | T | C | 0.036659 | 0.004219 | 2.30E-18 |
| rs494242 | miR-10b-5p | trans | 1.47E-34 | Nightingale | M_VLDL_C | NA | T | C | 0.037361 | 0.004335 | 5.20E-18 |
| rs494242 | miR-10b-5p | trans | 1.47E-34 | Nightingale | Citrate | NA | T | C | 0.024682 | 0.004318 | 6.10E-09 |
| rs494242 | miR-10b-5p | trans | 1.47E-34 | Nightingale | LDL_FC | NA | T | C | 0.031566 | 0.004301 | 6.40E-13 |
| rs494242 | miR-10b-5p | trans | 1.47E-34 | Nightingale | L_LDL_CE | NA | T | C | 0.029548 | 0.004308 | 9.50E-12 |
| rs494242 | miR-10b-5p | trans | 1.47E-34 | Nightingale | L_LDL_PL | NA | T | C | 0.03594 | 0.004314 | 1.30E-16 |
| rs494242 | miR-10b-5p | trans | 1.47E-34 | Nightingale | S_HDL_C | NA | T | C | 0.025407 | 0.004339 | 9.00E-09 |
| rs494242 | miR-10b-5p | trans | 1.47E-34 | Nightingale | S_LDL_CE | NA | T | C | 0.029469 | 0.004349 | 1.40E-11 |
| rs494242 | miR-10b-5p | trans | 1.47E-34 | Nightingale | Omega_6 | NA | T | C | 0.024204 | 0.004245 | 2.90E-09 |
| rs494242 | miR-10b-5p | trans | 1.47E-34 | Nightingale | IDL_P | NA | T | C | 0.03904 | 0.004301 | 1.80E-19 |
| rs494242 | miR-10b-5p | trans | 1.47E-34 | Nightingale | M_LDL_CE | NA | T | C | 0.031239 | 0.004353 | 8.20E-13 |
| rs494242 | miR-10b-5p | trans | 1.47E-34 | Nightingale | S_LDL_C | NA | T | C | 0.029721 | 0.004351 | 1.50E-11 |

| | | | | | | | | | | | |
|---|---|---|---|---|---|---|---|---|---|---|---|
| rs494242 | miR-126-3p | trans | 9.85E-38 | Nightingale | VLDL_P | NA | T | C | 0.026586 | 0.004324 | 1.90E-10 |
| rs494242 | miR-126-3p | trans | 9.85E-38 | Nightingale | M_VLDL_C | NA | T | C | 0.033835 | 0.004094 | 2.70E-16 |
| rs494242 | miR-126-3p | trans | 9.85E-38 | Nightingale | M_LDL_C_ | NA | T | C | 0.026574 | 0.00435 | 1.60E-09 |
| rs494242 | miR-126-3p | trans | 9.85E-38 | Nightingale | M_LDL_FC | NA | T | C | 0.032299 | 0.004341 | 2.30E-13 |
| rs494242 | miR-126-3p | trans | 9.85E-38 | Nightingale | XS_VLDL_P | NA | T | C | 0.035812 | 0.004289 | 9.60E-18 |
| rs494242 | miR-126-3p | trans | 9.85E-38 | Nightingale | Total_CE | NA | T | C | 0.029414 | 0.004193 | 1.80E-12 |
| rs494242 | miR-126-3p | trans | 9.85E-38 | Nightingale | GlycA | NA | T | C | 0.025353 | 0.004328 | 4.70E-09 |
| rs494242 | miR-126-3p | trans | 9.85E-38 | Nightingale | M_VLDL_P | NA | T | C | 0.027954 | 0.004336 | 4.50E-11 |
| rs494242 | miR-126-3p | trans | 9.85E-38 | Nightingale | XS_VLDL_F | NA | T | C | 0.029107 | 0.004316 | 3.20E-11 |
| rs494242 | miR-126-3p | trans | 9.85E-38 | Nightingale | IDL_PL | NA | T | C | 0.033595 | 0.004195 | 1.90E-15 |
| rs494242 | miR-126-3p | trans | 9.85E-38 | Nightingale | XS_VLDL_P | NA | T | C | 0.031685 | 0.004286 | 1.90E-14 |
| rs494242 | miR-126-3p | trans | 9.85E-38 | Nightingale | M_LDL_C | NA | T | C | 0.032117 | 0.004355 | 2.20E-13 |
| rs494242 | miR-126-3p | trans | 9.85E-38 | Nightingale | XS_VLDL_C | NA | T | C | 0.036408 | 0.004195 | 4.00E-18 |
| rs494242 | miR-126-3p | trans | 9.85E-38 | Nightingale | IDL_C | NA | T | C | 0.03332 | 0.004198 | 4.70E-15 |
| rs494242 | miR-126-3p | trans | 9.85E-38 | Nightingale | Total_C | NA | T | C | 0.030169 | 0.004208 | 4.70E-13 |
| rs494242 | miR-126-3p | trans | 9.85E-38 | Nightingale | S_LDL_P | NA | T | C | 0.028983 | 0.004351 | 2.90E-11 |
| rs494242 | miR-126-3p | trans | 9.85E-38 | Nightingale | M_VLDL_F | NA | T | C | 0.032542 | 0.004346 | 3.80E-14 |
| rs494242 | miR-126-3p | trans | 9.85E-38 | Nightingale | XXL_VLDL_ | NA | T | C | 0.033442 | 0.004356 | 8.80E-15 |
| rs494242 | miR-126-3p | trans | 9.85E-38 | Nightingale | S_HDL_L | NA | T | C | 0.024003 | 0.004332 | 2.60E-08 |
| rs494242 | miR-126-3p | trans | 9.85E-38 | Nightingale | M_LDL_P | NA | T | C | 0.034022 | 0.004359 | 7.20E-15 |
| rs494242 | miR-126-3p | trans | 9.85E-38 | Nightingale | XL_HDL_PL | NA | T | C | -0.0218 | 0.003994 | 4.60E-08 |
| rs494242 | miR-126-3p | trans | 9.85E-38 | Nightingale | XS_VLDL_F | NA | T | C | 0.035906 | 0.004271 | 7.30E-18 |
| rs494242 | miR-126-3p | trans | 9.85E-38 | Nightingale | VLDL_FC | NA | T | C | 0.024475 | 0.004315 | 3.40E-09 |
| rs494242 | miR-126-3p | trans | 9.85E-38 | Nightingale | S_VLDL_FC | NA | T | C | 0.036668 | 0.004348 | 1.70E-17 |
| rs494242 | miR-126-3p | trans | 9.85E-38 | Nightingale | ApoB | NA | T | C | 0.032952 | 0.004341 | 3.30E-14 |
| rs494242 | miR-126-3p | trans | 9.85E-38 | Nightingale | M_VLDL_C | NA | T | C | 0.031337 | 0.004084 | 3.30E-14 |
| rs494242 | miR-126-3p | trans | 9.85E-38 | Nightingale | S_LDL_C_p | NA | T | C | 0.041866 | 0.004364 | 5.60E-22 |
| rs494242 | miR-126-3p | trans | 9.85E-38 | Nightingale | L_VLDL_TG | NA | T | C | -0.04923 | 0.004326 | 6.60E-31 |
| rs494242 | miR-126-3p | trans | 9.85E-38 | Nightingale | LDL_CE | NA | T | C | 0.030645 | 0.004335 | 2.00E-12 |
| rs494242 | miR-126-3p | trans | 9.85E-38 | Nightingale | Remnant_( | NA | T | C | 0.035863 | 0.004319 | 5.10E-17 |
| rs494242 | miR-126-3p | trans | 9.85E-38 | Nightingale | S_LDL_L | NA | T | C | 0.026918 | 0.004346 | 1.20E-09 |
| rs494242 | miR-126-3p | trans | 9.85E-38 | Nightingale | S_VLDL_TG | NA | T | C | -0.03597 | 0.004208 | 1.90E-17 |
| rs494242 | miR-126-3p | trans | 9.85E-38 | Nightingale | L_VLDL_CE | NA | T | C | 0.044066 | 0.00416 | 8.10E-27 |
| rs494242 | miR-126-3p | trans | 9.85E-38 | Nightingale | M_LDL_L | NA | T | C | 0.031415 | 0.004353 | 6.70E-13 |
| rs494242 | miR-126-3p | trans | 9.85E-38 | Nightingale | LDL_P | NA | T | C | 0.030684 | 0.004344 | 2.10E-12 |
| rs494242 | miR-126-3p | trans | 9.85E-38 | Nightingale | LDL_C | NA | T | C | 0.031211 | 0.004328 | 9.20E-13 |
| rs494242 | miR-126-3p | trans | 9.85E-38 | Nightingale | M_VLDL_P | NA | T | C | 0.042089 | 0.004172 | 6.40E-24 |
| rs494242 | miR-126-3p | trans | 9.85E-38 | Nightingale | LA | NA | T | C | 0.02331 | 0.004263 | 1.50E-08 |
| rs494242 | miR-126-3p | trans | 9.85E-38 | Nightingale | L_VLDL_CE | NA | T | C | 0.022783 | 0.004296 | 2.80E-08 |
| rs494242 | miR-126-3p | trans | 9.85E-38 | Nightingale | L_LDL_C | NA | T | C | 0.030249 | 0.004298 | 2.90E-12 |
| rs494242 | miR-126-3p | trans | 9.85E-38 | Nightingale | XXL_VLDL_ | NA | T | C | 0.023509 | 0.004375 | 5.40E-09 |
| rs494242 | miR-126-3p | trans | 9.85E-38 | Nightingale | S_LDL_PL_ | NA | T | C | -0.02809 | 0.004339 | 4.70E-12 |
| rs494242 | miR-126-3p | trans | 9.85E-38 | Nightingale | VLDL_C | NA | T | C | 0.031089 | 0.004341 | 2.10E-13 |
| rs494242 | miR-126-3p | trans | 9.85E-38 | Nightingale | XL_VLDL_P | NA | T | C | 0.039093 | 0.004353 | 7.70E-20 |
| rs494242 | miR-126-3p | trans | 9.85E-38 | Nightingale | non_HDL_( | NA | T | C | 0.034165 | 0.004323 | 2.30E-15 |
| rs494242 | miR-126-3p | trans | 9.85E-38 | Nightingale | S_LDL_FC | NA | T | C | 0.027211 | 0.00434 | 1.30E-09 |
| rs494242 | miR-126-3p | trans | 9.85E-38 | Nightingale | LDL_L | NA | T | C | 0.031492 | 0.004332 | 5.60E-13 |
| rs494242 | miR-126-3p | trans | 9.85E-38 | Nightingale | XL_VLDL_C | NA | T | C | 0.034568 | 0.00417 | 1.60E-16 |
| rs494242 | miR-126-3p | trans | 9.85E-38 | Nightingale | IDL_FC | NA | T | C | 0.035991 | 0.004208 | 3.10E-17 |
| rs494242 | miR-126-3p | trans | 9.85E-38 | Nightingale | XL_VLDL_C | NA | T | C | 0.035582 | 0.004168 | 1.80E-17 |
| rs494242 | miR-126-3p | trans | 9.85E-38 | Nightingale | S_VLDL_C | NA | T | C | 0.03738 | 0.004346 | 2.50E-18 |
| rs494242 | miR-126-3p | trans | 9.85E-38 | Nightingale | L_LDL_L | NA | T | C | 0.031549 | 0.004304 | 3.10E-13 |
| rs494242 | miR-126-3p | trans | 9.85E-38 | Nightingale | XXL_VLDL_ | NA | T | C | 0.031802 | 0.00438 | 2.10E-13 |
| rs494242 | miR-126-3p | trans | 9.85E-38 | Nightingale | M_LDL_TG | NA | T | C | -0.0251 | 0.004293 | 8.80E-09 |
| rs494242 | miR-126-3p | trans | 9.85E-38 | Nightingale | S_VLDL_CE | NA | T | C | 0.037335 | 0.004341 | 1.70E-18 |
| rs494242 | miR-126-3p | trans | 9.85E-38 | Nightingale | S_VLDL_P | NA | T | C | 0.023424 | 0.004292 | 9.40E-09 |
| rs494242 | miR-126-3p | trans | 9.85E-38 | Nightingale | S_VLDL_PL | NA | T | C | 0.032066 | 0.004339 | 4.60E-14 |
| rs494242 | miR-126-3p | trans | 9.85E-38 | Nightingale | M_VLDL_F | NA | T | C | 0.039367 | 0.004136 | 2.20E-21 |
| rs494242 | miR-126-3p | trans | 9.85E-38 | Nightingale | S_VLDL_CE | NA | T | C | 0.045132 | 0.004288 | 1.50E-25 |
| rs494242 | miR-126-3p | trans | 9.85E-38 | Nightingale | XXL_VLDL_ | NA | T | C | -0.03443 | 0.004384 | 6.30E-16 |
| rs494242 | miR-126-3p | trans | 9.85E-38 | Nightingale | L_VLDL_C_ | NA | T | C | 0.049215 | 0.004246 | 9.30E-32 |
| rs494242 | miR-126-3p | trans | 9.85E-38 | Nightingale | S_VLDL_PL | NA | T | C | 0.025717 | 0.004162 | 1.50E-09 |
| rs494242 | miR-126-3p | trans | 9.85E-38 | Nightingale | IDL_CE | NA | T | C | 0.032104 | 0.004197 | 4.20E-14 |

| | | | | | | | | | | | |
|---|---|---|---|---|---|---|---|---|---|---|---|
| rs494242 | miR-126-3| | trans | 9.85E-38 | Nightingale | XL_VLDL_T | NA | T | C | -0.04154 | 0.004213 | 5.00E-23 |
| rs494242 | miR-126-3| | trans | 9.85E-38 | Nightingale | VLDL_CE | NA | T | C | 0.03517 | 0.004348 | 2.00E-16 |
| rs494242 | miR-126-3| | trans | 9.85E-38 | Nightingale | Sphingomy | NA | T | C | 0.025942 | 0.004064 | 3.10E-10 |
| rs494242 | miR-126-3| | trans | 9.85E-38 | Nightingale | L_LDL_P | NA | T | C | 0.028344 | 0.004332 | 7.50E-11 |
| rs494242 | miR-126-3| | trans | 9.85E-38 | Nightingale | S_VLDL_L | NA | T | C | 0.02453 | 0.004305 | 2.40E-09 |
| rs494242 | miR-126-3| | trans | 9.85E-38 | Nightingale | XS_VLDL_L | NA | T | C | 0.033036 | 0.004273 | 1.30E-15 |
| rs494242 | miR-126-3| | trans | 9.85E-38 | Nightingale | L_VLDL_FC | NA | T | C | 0.046929 | 0.004305 | 4.10E-28 |
| rs494242 | miR-126-3| | trans | 9.85E-38 | Nightingale | L_LDL_FC | NA | T | C | 0.031491 | 0.004268 | 4.00E-13 |
| rs494242 | miR-126-3| | trans | 9.85E-38 | Nightingale | Total_PL | NA | T | C | 0.023039 | 0.004139 | 7.30E-09 |
| rs494242 | miR-126-3| | trans | 9.85E-38 | Nightingale | L_VLDL_PL | NA | T | C | 0.021646 | 0.004222 | 4.50E-08 |
| rs494242 | miR-126-3| | trans | 9.85E-38 | Nightingale | M_LDL_PL | NA | T | C | 0.031199 | 0.004353 | 9.60E-13 |
| rs494242 | miR-126-3| | trans | 9.85E-38 | Nightingale | Clinical_LD | NA | T | C | 0.034576 | 0.004322 | 2.70E-15 |
| rs494242 | miR-126-3| | trans | 9.85E-38 | Nightingale | S_LDL_CE_ | NA | T | C | 0.028871 | 0.004339 | 1.00E-12 |
| rs494242 | miR-126-3| | trans | 9.85E-38 | Nightingale | S_VLDL_FC | NA | T | C | 0.027143 | 0.004154 | 1.20E-10 |
| rs494242 | miR-126-3| | trans | 9.85E-38 | Nightingale | S_HDL_CE | NA | T | C | 0.02571 | 0.004354 | 6.80E-09 |
| rs494242 | miR-126-3| | trans | 9.85E-38 | Nightingale | M_VLDL_P | NA | T | C | 0.028782 | 0.00434 | 1.30E-11 |
| rs494242 | miR-126-3| | trans | 9.85E-38 | Nightingale | M_VLDL_C | NA | T | C | 0.039214 | 0.004307 | 1.30E-19 |
| rs494242 | miR-126-3| | trans | 9.85E-38 | Nightingale | Total_L | NA | T | C | 0.025049 | 0.004276 | 1.40E-09 |
| rs494242 | miR-126-3| | trans | 9.85E-38 | Nightingale | IDL_L | NA | T | C | 0.03368 | 0.004203 | 2.20E-15 |
| rs494242 | miR-126-3| | trans | 9.85E-38 | Nightingale | S_VLDL_C_ | NA | T | C | 0.03972 | 0.00423 | 8.60E-21 |
| rs494242 | miR-126-3| | trans | 9.85E-38 | Nightingale | ApoB_by_/ | NA | T | C | 0.025679 | 0.004281 | 2.10E-09 |
| rs494242 | miR-126-3| | trans | 9.85E-38 | Nightingale | LDL_PL | NA | T | C | 0.03334 | 0.004337 | 2.90E-14 |
| rs494242 | miR-126-3| | trans | 9.85E-38 | Nightingale | M_VLDL_T | NA | T | C | -0.03622 | 0.004108 | 1.60E-18 |
| rs494242 | miR-126-3| | trans | 9.85E-38 | Nightingale | S_HDL_P | NA | T | C | 0.027834 | 0.004347 | 2.60E-10 |
| rs494242 | miR-126-3| | trans | 9.85E-38 | Nightingale | XL_VLDL_F | NA | T | C | 0.035197 | 0.004214 | 1.80E-17 |
| rs494242 | miR-126-3| | trans | 9.85E-38 | Nightingale | XXL_VLDL_ | NA | T | C | 0.029266 | 0.004328 | 5.20E-12 |
| rs494242 | miR-126-3| | trans | 9.85E-38 | Nightingale | Total_FC | NA | T | C | 0.031409 | 0.004246 | 4.40E-14 |
| rs494242 | miR-126-3| | trans | 9.85E-38 | Nightingale | XS_VLDL_C | NA | T | C | 0.036659 | 0.004219 | 2.30E-18 |
| rs494242 | miR-126-3| | trans | 9.85E-38 | Nightingale | M_VLDL_C | NA | T | C | 0.037361 | 0.004335 | 5.20E-18 |
| rs494242 | miR-126-3| | trans | 9.85E-38 | Nightingale | Citrate | NA | T | C | 0.024682 | 0.004318 | 6.10E-09 |
| rs494242 | miR-126-3| | trans | 9.85E-38 | Nightingale | LDL_FC | NA | T | C | 0.031566 | 0.004301 | 6.40E-13 |
| rs494242 | miR-126-3| | trans | 9.85E-38 | Nightingale | L_LDL_CE | NA | T | C | 0.029548 | 0.004308 | 9.50E-12 |
| rs494242 | miR-126-3| | trans | 9.85E-38 | Nightingale | L_LDL_PL | NA | T | C | 0.03594 | 0.004314 | 1.30E-16 |
| rs494242 | miR-126-3| | trans | 9.85E-38 | Nightingale | S_HDL_C | NA | T | C | 0.025407 | 0.004339 | 9.00E-09 |
| rs494242 | miR-126-3| | trans | 9.85E-38 | Nightingale | S_LDL_CE | NA | T | C | 0.029469 | 0.004349 | 1.40E-11 |
| rs494242 | miR-126-3| | trans | 9.85E-38 | Nightingale | Omega_6 | NA | T | C | 0.024204 | 0.004245 | 2.90E-09 |
| rs494242 | miR-126-3| | trans | 9.85E-38 | Nightingale | IDL_P | NA | T | C | 0.03904 | 0.004301 | 1.80E-19 |
| rs494242 | miR-126-3| | trans | 9.85E-38 | Nightingale | M_LDL_CE | NA | T | C | 0.031239 | 0.004353 | 8.20E-13 |
| rs494242 | miR-126-3| | trans | 9.85E-38 | Nightingale | S_LDL_C | NA | T | C | 0.029721 | 0.004351 | 1.50E-11 |
| rs494242 | let-7b-5p | trans | 1.74E-26 | Nightingale | VLDL_P | NA | T | C | 0.026586 | 0.004324 | 1.90E-10 |
| rs494242 | let-7b-5p | trans | 1.74E-26 | Nightingale | M_VLDL_C | NA | T | C | 0.033835 | 0.004094 | 2.70E-16 |
| rs494242 | let-7b-5p | trans | 1.74E-26 | Nightingale | M_LDL_C_| | NA | T | C | 0.026574 | 0.00435 | 1.60E-09 |
| rs494242 | let-7b-5p | trans | 1.74E-26 | Nightingale | M_LDL_FC | NA | T | C | 0.032299 | 0.004341 | 2.30E-13 |
| rs494242 | let-7b-5p | trans | 1.74E-26 | Nightingale | XS_VLDL_P | NA | T | C | 0.035812 | 0.004289 | 9.60E-18 |
| rs494242 | let-7b-5p | trans | 1.74E-26 | Nightingale | Total_CE | NA | T | C | 0.029414 | 0.004193 | 1.80E-12 |
| rs494242 | let-7b-5p | trans | 1.74E-26 | Nightingale | GlycA | NA | T | C | 0.025353 | 0.004328 | 4.70E-09 |
| rs494242 | let-7b-5p | trans | 1.74E-26 | Nightingale | M_VLDL_P | NA | T | C | 0.027954 | 0.004336 | 4.50E-11 |
| rs494242 | let-7b-5p | trans | 1.74E-26 | Nightingale | XS_VLDL_F | NA | T | C | 0.029107 | 0.004316 | 3.20E-11 |
| rs494242 | let-7b-5p | trans | 1.74E-26 | Nightingale | IDL_PL | NA | T | C | 0.033595 | 0.004195 | 1.90E-15 |
| rs494242 | let-7b-5p | trans | 1.74E-26 | Nightingale | XS_VLDL_P | NA | T | C | 0.031685 | 0.004286 | 1.90E-14 |
| rs494242 | let-7b-5p | trans | 1.74E-26 | Nightingale | M_LDL_C | NA | T | C | 0.032117 | 0.004355 | 2.20E-13 |
| rs494242 | let-7b-5p | trans | 1.74E-26 | Nightingale | XS_VLDL_C | NA | T | C | 0.036408 | 0.004195 | 4.00E-18 |
| rs494242 | let-7b-5p | trans | 1.74E-26 | Nightingale | IDL_C | NA | T | C | 0.03332 | 0.004198 | 4.70E-15 |
| rs494242 | let-7b-5p | trans | 1.74E-26 | Nightingale | Total_C | NA | T | C | 0.030169 | 0.004208 | 4.70E-13 |
| rs494242 | let-7b-5p | trans | 1.74E-26 | Nightingale | S_LDL_P | NA | T | C | 0.028983 | 0.004351 | 2.90E-11 |
| rs494242 | let-7b-5p | trans | 1.74E-26 | Nightingale | M_VLDL_F | NA | T | C | 0.032542 | 0.004346 | 3.80E-14 |
| rs494242 | let-7b-5p | trans | 1.74E-26 | Nightingale | XXL_VLDL_ | NA | T | C | 0.033442 | 0.004356 | 8.80E-15 |
| rs494242 | let-7b-5p | trans | 1.74E-26 | Nightingale | S_HDL_L | NA | T | C | 0.024003 | 0.004332 | 2.60E-08 |
| rs494242 | let-7b-5p | trans | 1.74E-26 | Nightingale | M_LDL_P | NA | T | C | 0.034022 | 0.004359 | 7.20E-15 |
| rs494242 | let-7b-5p | trans | 1.74E-26 | Nightingale | XL_HDL_PL | NA | T | C | -0.0218 | 0.003994 | 4.60E-08 |
| rs494242 | let-7b-5p | trans | 1.74E-26 | Nightingale | XS_VLDL_F | NA | T | C | 0.035906 | 0.004271 | 7.30E-18 |
| rs494242 | let-7b-5p | trans | 1.74E-26 | Nightingale | VLDL_FC | NA | T | C | 0.024475 | 0.004315 | 3.40E-09 |
| rs494242 | let-7b-5p | trans | 1.74E-26 | Nightingale | S_VLDL_FC | NA | T | C | 0.036668 | 0.004348 | 1.70E-17 |

| | | | | | | | | | | | |
|---|---|---|---|---|---|---|---|---|---|---|---|
| rs494242 | let-7b-5p | trans | 1.74E-26 | Nightingale | ApoB | NA | T | C | 0.032952 | 0.004341 | 3.30E-14 |
| rs494242 | let-7b-5p | trans | 1.74E-26 | Nightingale | M_VLDL_C | NA | T | C | 0.031337 | 0.004084 | 3.30E-14 |
| rs494242 | let-7b-5p | trans | 1.74E-26 | Nightingale | S_LDL_C_p | NA | T | C | 0.041866 | 0.004364 | 5.60E-22 |
| rs494242 | let-7b-5p | trans | 1.74E-26 | Nightingale | L_VLDL_TG | NA | T | C | -0.04923 | 0.004326 | 6.60E-31 |
| rs494242 | let-7b-5p | trans | 1.74E-26 | Nightingale | LDL_CE | NA | T | C | 0.030645 | 0.004335 | 2.00E-12 |
| rs494242 | let-7b-5p | trans | 1.74E-26 | Nightingale | Remnant_( | NA | T | C | 0.035863 | 0.004319 | 5.10E-17 |
| rs494242 | let-7b-5p | trans | 1.74E-26 | Nightingale | S_LDL_L | NA | T | C | 0.026918 | 0.004346 | 1.20E-09 |
| rs494242 | let-7b-5p | trans | 1.74E-26 | Nightingale | S_VLDL_TG | NA | T | C | -0.03597 | 0.004208 | 1.90E-17 |
| rs494242 | let-7b-5p | trans | 1.74E-26 | Nightingale | L_VLDL_CE | NA | T | C | 0.044066 | 0.00416 | 8.10E-27 |
| rs494242 | let-7b-5p | trans | 1.74E-26 | Nightingale | M_LDL_L | NA | T | C | 0.031415 | 0.004353 | 6.70E-13 |
| rs494242 | let-7b-5p | trans | 1.74E-26 | Nightingale | LDL_P | NA | T | C | 0.030684 | 0.004344 | 2.10E-12 |
| rs494242 | let-7b-5p | trans | 1.74E-26 | Nightingale | LDL_C | NA | T | C | 0.031211 | 0.004328 | 9.20E-13 |
| rs494242 | let-7b-5p | trans | 1.74E-26 | Nightingale | M_VLDL_P | NA | T | C | 0.042089 | 0.004172 | 6.40E-24 |
| rs494242 | let-7b-5p | trans | 1.74E-26 | Nightingale | LA | NA | T | C | 0.02331 | 0.004263 | 1.50E-08 |
| rs494242 | let-7b-5p | trans | 1.74E-26 | Nightingale | L_VLDL_CE | NA | T | C | 0.022783 | 0.004296 | 2.80E-08 |
| rs494242 | let-7b-5p | trans | 1.74E-26 | Nightingale | L_LDL_C | NA | T | C | 0.030249 | 0.004298 | 2.90E-12 |
| rs494242 | let-7b-5p | trans | 1.74E-26 | Nightingale | XXL_VLDL_ | NA | T | C | 0.023509 | 0.004375 | 5.40E-09 |
| rs494242 | let-7b-5p | trans | 1.74E-26 | Nightingale | S_LDL_PL_ | NA | T | C | -0.02809 | 0.004339 | 4.70E-12 |
| rs494242 | let-7b-5p | trans | 1.74E-26 | Nightingale | VLDL_C | NA | T | C | 0.031089 | 0.004341 | 2.10E-13 |
| rs494242 | let-7b-5p | trans | 1.74E-26 | Nightingale | XL_VLDL_P | NA | T | C | 0.039093 | 0.004353 | 7.70E-20 |
| rs494242 | let-7b-5p | trans | 1.74E-26 | Nightingale | non_HDL_( | NA | T | C | 0.034165 | 0.004323 | 2.30E-15 |
| rs494242 | let-7b-5p | trans | 1.74E-26 | Nightingale | S_LDL_FC | NA | T | C | 0.027211 | 0.00434 | 1.30E-09 |
| rs494242 | let-7b-5p | trans | 1.74E-26 | Nightingale | LDL_L | NA | T | C | 0.031492 | 0.004332 | 5.60E-13 |
| rs494242 | let-7b-5p | trans | 1.74E-26 | Nightingale | XL_VLDL_C | NA | T | C | 0.034568 | 0.00417 | 1.60E-16 |
| rs494242 | let-7b-5p | trans | 1.74E-26 | Nightingale | IDL_FC | NA | T | C | 0.035991 | 0.004208 | 3.10E-17 |
| rs494242 | let-7b-5p | trans | 1.74E-26 | Nightingale | XL_VLDL_C | NA | T | C | 0.035582 | 0.004168 | 1.80E-17 |
| rs494242 | let-7b-5p | trans | 1.74E-26 | Nightingale | S_VLDL_C | NA | T | C | 0.03738 | 0.004346 | 2.50E-18 |
| rs494242 | let-7b-5p | trans | 1.74E-26 | Nightingale | L_LDL_L | NA | T | C | 0.031549 | 0.004304 | 3.10E-13 |
| rs494242 | let-7b-5p | trans | 1.74E-26 | Nightingale | XXL_VLDL_ | NA | T | C | 0.031802 | 0.00438 | 2.10E-13 |
| rs494242 | let-7b-5p | trans | 1.74E-26 | Nightingale | M_LDL_TG | NA | T | C | -0.0251 | 0.004293 | 8.80E-09 |
| rs494242 | let-7b-5p | trans | 1.74E-26 | Nightingale | S_VLDL_CE | NA | T | C | 0.037335 | 0.004341 | 1.70E-18 |
| rs494242 | let-7b-5p | trans | 1.74E-26 | Nightingale | S_VLDL_P | NA | T | C | 0.023424 | 0.004292 | 9.40E-09 |
| rs494242 | let-7b-5p | trans | 1.74E-26 | Nightingale | S_VLDL_PL | NA | T | C | 0.032066 | 0.004339 | 4.60E-14 |
| rs494242 | let-7b-5p | trans | 1.74E-26 | Nightingale | M_VLDL_F( | NA | T | C | 0.039367 | 0.004136 | 2.20E-21 |
| rs494242 | let-7b-5p | trans | 1.74E-26 | Nightingale | S_VLDL_CE | NA | T | C | 0.045132 | 0.004288 | 1.50E-25 |
| rs494242 | let-7b-5p | trans | 1.74E-26 | Nightingale | XXL_VLDL_ | NA | T | C | -0.03443 | 0.004384 | 6.30E-16 |
| rs494242 | let-7b-5p | trans | 1.74E-26 | Nightingale | L_VLDL_C_ | NA | T | C | 0.049215 | 0.004246 | 9.30E-32 |
| rs494242 | let-7b-5p | trans | 1.74E-26 | Nightingale | S_VLDL_PL | NA | T | C | 0.025717 | 0.004162 | 1.50E-09 |
| rs494242 | let-7b-5p | trans | 1.74E-26 | Nightingale | IDL_CE | NA | T | C | 0.032104 | 0.004197 | 4.20E-14 |
| rs494242 | let-7b-5p | trans | 1.74E-26 | Nightingale | XL_VLDL_T | NA | T | C | -0.04154 | 0.004213 | 5.00E-23 |
| rs494242 | let-7b-5p | trans | 1.74E-26 | Nightingale | VLDL_CE | NA | T | C | 0.03517 | 0.004348 | 2.00E-16 |
| rs494242 | let-7b-5p | trans | 1.74E-26 | Nightingale | Sphingomy | NA | T | C | 0.025942 | 0.004064 | 3.10E-10 |
| rs494242 | let-7b-5p | trans | 1.74E-26 | Nightingale | L_LDL_P | NA | T | C | 0.028344 | 0.004332 | 7.50E-11 |
| rs494242 | let-7b-5p | trans | 1.74E-26 | Nightingale | S_VLDL_L | NA | T | C | 0.02453 | 0.004305 | 2.40E-09 |
| rs494242 | let-7b-5p | trans | 1.74E-26 | Nightingale | XS_VLDL_L | NA | T | C | 0.033036 | 0.004273 | 1.30E-15 |
| rs494242 | let-7b-5p | trans | 1.74E-26 | Nightingale | L_VLDL_FC | NA | T | C | 0.046929 | 0.004305 | 4.10E-28 |
| rs494242 | let-7b-5p | trans | 1.74E-26 | Nightingale | L_LDL_FC | NA | T | C | 0.031491 | 0.004268 | 4.00E-13 |
| rs494242 | let-7b-5p | trans | 1.74E-26 | Nightingale | Total_PL | NA | T | C | 0.023039 | 0.004139 | 7.30E-09 |
| rs494242 | let-7b-5p | trans | 1.74E-26 | Nightingale | L_VLDL_PL | NA | T | C | 0.021646 | 0.004222 | 4.50E-08 |
| rs494242 | let-7b-5p | trans | 1.74E-26 | Nightingale | M_LDL_PL | NA | T | C | 0.031199 | 0.004353 | 9.60E-13 |
| rs494242 | let-7b-5p | trans | 1.74E-26 | Nightingale | Clinical_LD | NA | T | C | 0.034576 | 0.004322 | 2.70E-15 |
| rs494242 | let-7b-5p | trans | 1.74E-26 | Nightingale | S_LDL_CE_ | NA | T | C | 0.028871 | 0.004339 | 1.00E-12 |
| rs494242 | let-7b-5p | trans | 1.74E-26 | Nightingale | S_VLDL_FC | NA | T | C | 0.027143 | 0.004154 | 1.20E-10 |
| rs494242 | let-7b-5p | trans | 1.74E-26 | Nightingale | S_HDL_CE | NA | T | C | 0.02571 | 0.004354 | 6.80E-09 |
| rs494242 | let-7b-5p | trans | 1.74E-26 | Nightingale | M_VLDL_P | NA | T | C | 0.028782 | 0.00434 | 1.30E-11 |
| rs494242 | let-7b-5p | trans | 1.74E-26 | Nightingale | M_VLDL_C | NA | T | C | 0.039214 | 0.004307 | 1.30E-19 |
| rs494242 | let-7b-5p | trans | 1.74E-26 | Nightingale | Total_L | NA | T | C | 0.025049 | 0.004276 | 1.40E-09 |
| rs494242 | let-7b-5p | trans | 1.74E-26 | Nightingale | IDL_L | NA | T | C | 0.03368 | 0.004203 | 2.20E-15 |
| rs494242 | let-7b-5p | trans | 1.74E-26 | Nightingale | S_VLDL_C_ | NA | T | C | 0.03972 | 0.00423 | 8.60E-21 |
| rs494242 | let-7b-5p | trans | 1.74E-26 | Nightingale | ApoB_by_/ | NA | T | C | 0.025679 | 0.004281 | 2.10E-09 |
| rs494242 | let-7b-5p | trans | 1.74E-26 | Nightingale | LDL_PL | NA | T | C | 0.03334 | 0.004337 | 2.90E-14 |
| rs494242 | let-7b-5p | trans | 1.74E-26 | Nightingale | M_VLDL_T( | NA | T | C | -0.03622 | 0.004108 | 1.60E-18 |
| rs494242 | let-7b-5p | trans | 1.74E-26 | Nightingale | S_HDL_P | NA | T | C | 0.027834 | 0.004347 | 2.60E-10 |

| | | | | | | | | | | | |
|---|---|---|---|---|---|---|---|---|---|---|---|
| rs494242 | let-7b-5p | trans | 1.74E-26 | Nightingale | XL_VLDL_F | NA | T | C | 0.035197 | 0.004214 | 1.80E-17 |
| rs494242 | let-7b-5p | trans | 1.74E-26 | Nightingale | XXL_VLDL_ | NA | T | C | 0.029266 | 0.004328 | 5.20E-12 |
| rs494242 | let-7b-5p | trans | 1.74E-26 | Nightingale | Total_FC | NA | T | C | 0.031409 | 0.004246 | 4.40E-14 |
| rs494242 | let-7b-5p | trans | 1.74E-26 | Nightingale | XS_VLDL_C | NA | T | C | 0.036659 | 0.004219 | 2.30E-18 |
| rs494242 | let-7b-5p | trans | 1.74E-26 | Nightingale | M_VLDL_C | NA | T | C | 0.037361 | 0.004335 | 5.20E-18 |
| rs494242 | let-7b-5p | trans | 1.74E-26 | Nightingale | Citrate | NA | T | C | 0.024682 | 0.004318 | 6.10E-09 |
| rs494242 | let-7b-5p | trans | 1.74E-26 | Nightingale | LDL_FC | NA | T | C | 0.031566 | 0.004301 | 6.40E-13 |
| rs494242 | let-7b-5p | trans | 1.74E-26 | Nightingale | L_LDL_CE | NA | T | C | 0.029548 | 0.004308 | 9.50E-12 |
| rs494242 | let-7b-5p | trans | 1.74E-26 | Nightingale | L_LDL_PL | NA | T | C | 0.03594 | 0.004314 | 1.30E-16 |
| rs494242 | let-7b-5p | trans | 1.74E-26 | Nightingale | S_HDL_C | NA | T | C | 0.025407 | 0.004339 | 9.00E-09 |
| rs494242 | let-7b-5p | trans | 1.74E-26 | Nightingale | S_LDL_CE | NA | T | C | 0.029469 | 0.004349 | 1.40E-11 |
| rs494242 | let-7b-5p | trans | 1.74E-26 | Nightingale | Omega_6 | NA | T | C | 0.024204 | 0.004245 | 2.90E-09 |
| rs494242 | let-7b-5p | trans | 1.74E-26 | Nightingale | IDL_P | NA | T | C | 0.03904 | 0.004301 | 1.80E-19 |
| rs494242 | let-7b-5p | trans | 1.74E-26 | Nightingale | M_LDL_CE | NA | T | C | 0.031239 | 0.004353 | 8.20E-13 |
| rs494242 | let-7b-5p | trans | 1.74E-26 | Nightingale | S_LDL_C | NA | T | C | 0.029721 | 0.004351 | 1.50E-11 |
| rs494242 | let-7c-5p | trans | 2.75E-23 | Nightingale | VLDL_P | NA | T | C | 0.026586 | 0.004324 | 1.90E-10 |
| rs494242 | let-7c-5p | trans | 2.75E-23 | Nightingale | M_VLDL_C | NA | T | C | 0.033835 | 0.004094 | 2.70E-16 |
| rs494242 | let-7c-5p | trans | 2.75E-23 | Nightingale | M_LDL_C_ | NA | T | C | 0.026574 | 0.00435 | 1.60E-09 |
| rs494242 | let-7c-5p | trans | 2.75E-23 | Nightingale | M_LDL_FC | NA | T | C | 0.032299 | 0.004341 | 2.30E-13 |
| rs494242 | let-7c-5p | trans | 2.75E-23 | Nightingale | XS_VLDL_P | NA | T | C | 0.035812 | 0.004289 | 9.60E-18 |
| rs494242 | let-7c-5p | trans | 2.75E-23 | Nightingale | Total_CE | NA | T | C | 0.029414 | 0.004193 | 1.80E-12 |
| rs494242 | let-7c-5p | trans | 2.75E-23 | Nightingale | GlycA | NA | T | C | 0.025353 | 0.004328 | 4.70E-09 |
| rs494242 | let-7c-5p | trans | 2.75E-23 | Nightingale | M_VLDL_P | NA | T | C | 0.027954 | 0.004336 | 4.50E-11 |
| rs494242 | let-7c-5p | trans | 2.75E-23 | Nightingale | XS_VLDL_F | NA | T | C | 0.029107 | 0.004316 | 3.20E-11 |
| rs494242 | let-7c-5p | trans | 2.75E-23 | Nightingale | IDL_PL | NA | T | C | 0.033595 | 0.004195 | 1.90E-15 |
| rs494242 | let-7c-5p | trans | 2.75E-23 | Nightingale | XS_VLDL_P | NA | T | C | 0.031685 | 0.004286 | 1.90E-14 |
| rs494242 | let-7c-5p | trans | 2.75E-23 | Nightingale | M_LDL_C | NA | T | C | 0.032117 | 0.004355 | 2.20E-13 |
| rs494242 | let-7c-5p | trans | 2.75E-23 | Nightingale | XS_VLDL_C | NA | T | C | 0.036408 | 0.004195 | 4.00E-18 |
| rs494242 | let-7c-5p | trans | 2.75E-23 | Nightingale | IDL_C | NA | T | C | 0.03332 | 0.004198 | 4.70E-15 |
| rs494242 | let-7c-5p | trans | 2.75E-23 | Nightingale | Total_C | NA | T | C | 0.030169 | 0.004208 | 4.70E-13 |
| rs494242 | let-7c-5p | trans | 2.75E-23 | Nightingale | S_LDL_P | NA | T | C | 0.028983 | 0.004351 | 2.90E-11 |
| rs494242 | let-7c-5p | trans | 2.75E-23 | Nightingale | M_VLDL_F | NA | T | C | 0.032542 | 0.004346 | 3.80E-14 |
| rs494242 | let-7c-5p | trans | 2.75E-23 | Nightingale | XXL_VLDL_ | NA | T | C | 0.033442 | 0.004356 | 8.80E-15 |
| rs494242 | let-7c-5p | trans | 2.75E-23 | Nightingale | S_HDL_L | NA | T | C | 0.024003 | 0.004332 | 2.60E-08 |
| rs494242 | let-7c-5p | trans | 2.75E-23 | Nightingale | M_LDL_P | NA | T | C | 0.034022 | 0.004359 | 7.20E-15 |
| rs494242 | let-7c-5p | trans | 2.75E-23 | Nightingale | XL_HDL_PL | NA | T | C | -0.0218 | 0.003994 | 4.60E-08 |
| rs494242 | let-7c-5p | trans | 2.75E-23 | Nightingale | XS_VLDL_F | NA | T | C | 0.035906 | 0.004271 | 7.30E-18 |
| rs494242 | let-7c-5p | trans | 2.75E-23 | Nightingale | VLDL_FC | NA | T | C | 0.024475 | 0.004315 | 3.40E-09 |
| rs494242 | let-7c-5p | trans | 2.75E-23 | Nightingale | S_VLDL_FC | NA | T | C | 0.036668 | 0.004348 | 1.70E-17 |
| rs494242 | let-7c-5p | trans | 2.75E-23 | Nightingale | ApoB | NA | T | C | 0.032952 | 0.004341 | 3.30E-14 |
| rs494242 | let-7c-5p | trans | 2.75E-23 | Nightingale | M_VLDL_C | NA | T | C | 0.031337 | 0.004084 | 3.30E-14 |
| rs494242 | let-7c-5p | trans | 2.75E-23 | Nightingale | S_LDL_C_p | NA | T | C | 0.041866 | 0.004364 | 5.60E-22 |
| rs494242 | let-7c-5p | trans | 2.75E-23 | Nightingale | L_VLDL_TG | NA | T | C | -0.04923 | 0.004326 | 6.60E-31 |
| rs494242 | let-7c-5p | trans | 2.75E-23 | Nightingale | LDL_CE | NA | T | C | 0.030645 | 0.004335 | 2.00E-12 |
| rs494242 | let-7c-5p | trans | 2.75E-23 | Nightingale | Remnant_( | NA | T | C | 0.035863 | 0.004319 | 5.10E-17 |
| rs494242 | let-7c-5p | trans | 2.75E-23 | Nightingale | S_LDL_L | NA | T | C | 0.026918 | 0.004346 | 1.20E-09 |
| rs494242 | let-7c-5p | trans | 2.75E-23 | Nightingale | S_VLDL_TG | NA | T | C | -0.03597 | 0.004208 | 1.90E-17 |
| rs494242 | let-7c-5p | trans | 2.75E-23 | Nightingale | L_VLDL_CE | NA | T | C | 0.044066 | 0.00416 | 8.10E-27 |
| rs494242 | let-7c-5p | trans | 2.75E-23 | Nightingale | M_LDL_L | NA | T | C | 0.031415 | 0.004353 | 6.70E-13 |
| rs494242 | let-7c-5p | trans | 2.75E-23 | Nightingale | LDL_P | NA | T | C | 0.030684 | 0.004344 | 2.10E-12 |
| rs494242 | let-7c-5p | trans | 2.75E-23 | Nightingale | LDL_C | NA | T | C | 0.031211 | 0.004328 | 9.20E-13 |
| rs494242 | let-7c-5p | trans | 2.75E-23 | Nightingale | M_VLDL_P | NA | T | C | 0.042089 | 0.004172 | 6.40E-24 |
| rs494242 | let-7c-5p | trans | 2.75E-23 | Nightingale | LA | NA | T | C | 0.02331 | 0.004263 | 1.50E-08 |
| rs494242 | let-7c-5p | trans | 2.75E-23 | Nightingale | L_VLDL_CE | NA | T | C | 0.022783 | 0.004296 | 2.80E-08 |
| rs494242 | let-7c-5p | trans | 2.75E-23 | Nightingale | L_LDL_C | NA | T | C | 0.030249 | 0.004298 | 2.90E-12 |
| rs494242 | let-7c-5p | trans | 2.75E-23 | Nightingale | XXL_VLDL_ | NA | T | C | 0.023509 | 0.004375 | 5.40E-09 |
| rs494242 | let-7c-5p | trans | 2.75E-23 | Nightingale | S_LDL_PL_ | NA | T | C | -0.02809 | 0.004339 | 4.70E-12 |
| rs494242 | let-7c-5p | trans | 2.75E-23 | Nightingale | VLDL_C | NA | T | C | 0.031089 | 0.004341 | 2.10E-13 |
| rs494242 | let-7c-5p | trans | 2.75E-23 | Nightingale | XL_VLDL_P | NA | T | C | 0.039093 | 0.004353 | 7.70E-20 |
| rs494242 | let-7c-5p | trans | 2.75E-23 | Nightingale | non_HDL_( | NA | T | C | 0.034165 | 0.004323 | 2.30E-15 |
| rs494242 | let-7c-5p | trans | 2.75E-23 | Nightingale | S_LDL_FC | NA | T | C | 0.027211 | 0.00434 | 1.30E-09 |
| rs494242 | let-7c-5p | trans | 2.75E-23 | Nightingale | LDL_L | NA | T | C | 0.031492 | 0.004332 | 5.60E-13 |
| rs494242 | let-7c-5p | trans | 2.75E-23 | Nightingale | XL_VLDL_C | NA | T | C | 0.034568 | 0.00417 | 1.60E-16 |

| | | | | | | | | | | | |
|---|---|---|---|---|---|---|---|---|---|---|---|
| rs494242 | let-7c-5p | trans | 2.75E-23 | Nightingale | IDL_FC | NA | T | C | 0.035991 | 0.004208 | 3.10E-17 |
| rs494242 | let-7c-5p | trans | 2.75E-23 | Nightingale | XL_VLDL_C | NA | T | C | 0.035582 | 0.004168 | 1.80E-17 |
| rs494242 | let-7c-5p | trans | 2.75E-23 | Nightingale | S_VLDL_C | NA | T | C | 0.03738 | 0.004346 | 2.50E-18 |
| rs494242 | let-7c-5p | trans | 2.75E-23 | Nightingale | L_LDL_L | NA | T | C | 0.031549 | 0.004304 | 3.10E-13 |
| rs494242 | let-7c-5p | trans | 2.75E-23 | Nightingale | XXL_VLDL_ | NA | T | C | 0.031802 | 0.00438 | 2.10E-13 |
| rs494242 | let-7c-5p | trans | 2.75E-23 | Nightingale | M_LDL_TG | NA | T | C | -0.0251 | 0.004293 | 8.80E-09 |
| rs494242 | let-7c-5p | trans | 2.75E-23 | Nightingale | S_VLDL_CE | NA | T | C | 0.037335 | 0.004341 | 1.70E-18 |
| rs494242 | let-7c-5p | trans | 2.75E-23 | Nightingale | S_VLDL_P | NA | T | C | 0.023424 | 0.004292 | 9.40E-09 |
| rs494242 | let-7c-5p | trans | 2.75E-23 | Nightingale | S_VLDL_PL | NA | T | C | 0.032066 | 0.004339 | 4.60E-14 |
| rs494242 | let-7c-5p | trans | 2.75E-23 | Nightingale | M_VLDL_F | NA | T | C | 0.039367 | 0.004136 | 2.20E-21 |
| rs494242 | let-7c-5p | trans | 2.75E-23 | Nightingale | S_VLDL_CE | NA | T | C | 0.045132 | 0.004288 | 1.50E-25 |
| rs494242 | let-7c-5p | trans | 2.75E-23 | Nightingale | XXL_VLDL_ | NA | T | C | -0.03443 | 0.004384 | 6.30E-16 |
| rs494242 | let-7c-5p | trans | 2.75E-23 | Nightingale | L_VLDL_C_ | NA | T | C | 0.049215 | 0.004246 | 9.30E-32 |
| rs494242 | let-7c-5p | trans | 2.75E-23 | Nightingale | S_VLDL_PL | NA | T | C | 0.025717 | 0.004162 | 1.50E-09 |
| rs494242 | let-7c-5p | trans | 2.75E-23 | Nightingale | IDL_CE | NA | T | C | 0.032104 | 0.004197 | 4.20E-14 |
| rs494242 | let-7c-5p | trans | 2.75E-23 | Nightingale | XL_VLDL_T | NA | T | C | -0.04154 | 0.004213 | 5.00E-23 |
| rs494242 | let-7c-5p | trans | 2.75E-23 | Nightingale | VLDL_CE | NA | T | C | 0.03517 | 0.004348 | 2.00E-16 |
| rs494242 | let-7c-5p | trans | 2.75E-23 | Nightingale | Sphingomy | NA | T | C | 0.025942 | 0.004064 | 3.10E-10 |
| rs494242 | let-7c-5p | trans | 2.75E-23 | Nightingale | L_LDL_P | NA | T | C | 0.028344 | 0.004332 | 7.50E-11 |
| rs494242 | let-7c-5p | trans | 2.75E-23 | Nightingale | S_VLDL_L | NA | T | C | 0.02453 | 0.004305 | 2.40E-09 |
| rs494242 | let-7c-5p | trans | 2.75E-23 | Nightingale | XS_VLDL_L | NA | T | C | 0.033036 | 0.004273 | 1.30E-15 |
| rs494242 | let-7c-5p | trans | 2.75E-23 | Nightingale | L_VLDL_FC | NA | T | C | 0.046929 | 0.004305 | 4.10E-28 |
| rs494242 | let-7c-5p | trans | 2.75E-23 | Nightingale | L_LDL_FC | NA | T | C | 0.031491 | 0.004268 | 4.00E-13 |
| rs494242 | let-7c-5p | trans | 2.75E-23 | Nightingale | Total_PL | NA | T | C | 0.023039 | 0.004139 | 7.30E-09 |
| rs494242 | let-7c-5p | trans | 2.75E-23 | Nightingale | L_VLDL_PL | NA | T | C | 0.021646 | 0.004222 | 4.50E-08 |
| rs494242 | let-7c-5p | trans | 2.75E-23 | Nightingale | M_LDL_PL | NA | T | C | 0.031199 | 0.004353 | 9.60E-13 |
| rs494242 | let-7c-5p | trans | 2.75E-23 | Nightingale | Clinical_LD | NA | T | C | 0.034576 | 0.004322 | 2.70E-15 |
| rs494242 | let-7c-5p | trans | 2.75E-23 | Nightingale | S_LDL_CE_ | NA | T | C | 0.028871 | 0.004339 | 1.00E-12 |
| rs494242 | let-7c-5p | trans | 2.75E-23 | Nightingale | S_VLDL_FC | NA | T | C | 0.027143 | 0.004154 | 1.20E-10 |
| rs494242 | let-7c-5p | trans | 2.75E-23 | Nightingale | S_HDL_CE | NA | T | C | 0.02571 | 0.004354 | 6.80E-09 |
| rs494242 | let-7c-5p | trans | 2.75E-23 | Nightingale | M_VLDL_P | NA | T | C | 0.028782 | 0.00434 | 1.30E-11 |
| rs494242 | let-7c-5p | trans | 2.75E-23 | Nightingale | M_VLDL_C | NA | T | C | 0.039214 | 0.004307 | 1.30E-19 |
| rs494242 | let-7c-5p | trans | 2.75E-23 | Nightingale | Total_L | NA | T | C | 0.025049 | 0.004276 | 1.40E-09 |
| rs494242 | let-7c-5p | trans | 2.75E-23 | Nightingale | IDL_L | NA | T | C | 0.03368 | 0.004203 | 2.20E-15 |
| rs494242 | let-7c-5p | trans | 2.75E-23 | Nightingale | S_VLDL_C_ | NA | T | C | 0.03972 | 0.00423 | 8.60E-21 |
| rs494242 | let-7c-5p | trans | 2.75E-23 | Nightingale | ApoB_by_A | NA | T | C | 0.025679 | 0.004281 | 2.10E-09 |
| rs494242 | let-7c-5p | trans | 2.75E-23 | Nightingale | LDL_PL | NA | T | C | 0.03334 | 0.004337 | 2.90E-14 |
| rs494242 | let-7c-5p | trans | 2.75E-23 | Nightingale | M_VLDL_T | NA | T | C | -0.03622 | 0.004108 | 1.60E-18 |
| rs494242 | let-7c-5p | trans | 2.75E-23 | Nightingale | S_HDL_P | NA | T | C | 0.027834 | 0.004347 | 2.60E-10 |
| rs494242 | let-7c-5p | trans | 2.75E-23 | Nightingale | XL_VLDL_F | NA | T | C | 0.035197 | 0.004214 | 1.80E-17 |
| rs494242 | let-7c-5p | trans | 2.75E-23 | Nightingale | XXL_VLDL_ | NA | T | C | 0.029266 | 0.004328 | 5.20E-12 |
| rs494242 | let-7c-5p | trans | 2.75E-23 | Nightingale | Total_FC | NA | T | C | 0.031409 | 0.004246 | 4.40E-14 |
| rs494242 | let-7c-5p | trans | 2.75E-23 | Nightingale | XS_VLDL_C | NA | T | C | 0.036659 | 0.004219 | 2.30E-18 |
| rs494242 | let-7c-5p | trans | 2.75E-23 | Nightingale | M_VLDL_C | NA | T | C | 0.037361 | 0.004335 | 5.20E-18 |
| rs494242 | let-7c-5p | trans | 2.75E-23 | Nightingale | Citrate | NA | T | C | 0.024682 | 0.004318 | 6.10E-09 |
| rs494242 | let-7c-5p | trans | 2.75E-23 | Nightingale | LDL_FC | NA | T | C | 0.031566 | 0.004301 | 6.40E-13 |
| rs494242 | let-7c-5p | trans | 2.75E-23 | Nightingale | L_LDL_CE | NA | T | C | 0.029548 | 0.004308 | 9.50E-12 |
| rs494242 | let-7c-5p | trans | 2.75E-23 | Nightingale | L_LDL_PL | NA | T | C | 0.03594 | 0.004314 | 1.30E-16 |
| rs494242 | let-7c-5p | trans | 2.75E-23 | Nightingale | S_HDL_C | NA | T | C | 0.025407 | 0.004339 | 9.00E-09 |
| rs494242 | let-7c-5p | trans | 2.75E-23 | Nightingale | S_LDL_CE | NA | T | C | 0.029469 | 0.004349 | 1.40E-11 |
| rs494242 | let-7c-5p | trans | 2.75E-23 | Nightingale | Omega_6 | NA | T | C | 0.024204 | 0.004245 | 2.90E-09 |
| rs494242 | let-7c-5p | trans | 2.75E-23 | Nightingale | IDL_P | NA | T | C | 0.03904 | 0.004301 | 1.80E-19 |
| rs494242 | let-7c-5p | trans | 2.75E-23 | Nightingale | M_LDL_CE | NA | T | C | 0.031239 | 0.004353 | 8.20E-13 |
| rs494242 | let-7c-5p | trans | 2.75E-23 | Nightingale | S_LDL_C | NA | T | C | 0.029721 | 0.004351 | 1.50E-11 |
| rs494242 | let-7a-5p | trans | 7.78E-13 | Nightingale | VLDL_P | NA | T | C | 0.026586 | 0.004324 | 1.90E-10 |
| rs494242 | let-7a-5p | trans | 7.78E-13 | Nightingale | M_VLDL_C | NA | T | C | 0.033835 | 0.004094 | 2.70E-16 |
| rs494242 | let-7a-5p | trans | 7.78E-13 | Nightingale | M_LDL_C_ | NA | T | C | 0.026574 | 0.00435 | 1.60E-09 |
| rs494242 | let-7a-5p | trans | 7.78E-13 | Nightingale | M_LDL_FC | NA | T | C | 0.032299 | 0.004341 | 2.30E-13 |
| rs494242 | let-7a-5p | trans | 7.78E-13 | Nightingale | XS_VLDL_P | NA | T | C | 0.035812 | 0.004289 | 9.60E-18 |
| rs494242 | let-7a-5p | trans | 7.78E-13 | Nightingale | Total_CE | NA | T | C | 0.029414 | 0.004193 | 1.80E-12 |
| rs494242 | let-7a-5p | trans | 7.78E-13 | Nightingale | GlycA | NA | T | C | 0.025353 | 0.004328 | 4.70E-09 |
| rs494242 | let-7a-5p | trans | 7.78E-13 | Nightingale | M_VLDL_P | NA | T | C | 0.027954 | 0.004336 | 4.50E-11 |
| rs494242 | let-7a-5p | trans | 7.78E-13 | Nightingale | XS_VLDL_F | NA | T | C | 0.029107 | 0.004316 | 3.20E-11 |

| | | | | | | | | | | | |
|---|---|---|---|---|---|---|---|---|---|---|---|
| rs494242 | let-7a-5p | trans | 7.78E-13 | Nightingale | IDL_PL | NA | T | C | 0.033595 | 0.004195 | 1.90E-15 |
| rs494242 | let-7a-5p | trans | 7.78E-13 | Nightingale | XS_VLDL_P | NA | T | C | 0.031685 | 0.004286 | 1.90E-14 |
| rs494242 | let-7a-5p | trans | 7.78E-13 | Nightingale | M_LDL_C | NA | T | C | 0.032117 | 0.004355 | 2.20E-13 |
| rs494242 | let-7a-5p | trans | 7.78E-13 | Nightingale | XS_VLDL_C | NA | T | C | 0.036408 | 0.004195 | 4.00E-18 |
| rs494242 | let-7a-5p | trans | 7.78E-13 | Nightingale | IDL_C | NA | T | C | 0.03332 | 0.004198 | 4.70E-15 |
| rs494242 | let-7a-5p | trans | 7.78E-13 | Nightingale | Total_C | NA | T | C | 0.030169 | 0.004208 | 4.70E-13 |
| rs494242 | let-7a-5p | trans | 7.78E-13 | Nightingale | S_LDL_P | NA | T | C | 0.028983 | 0.004351 | 2.90E-11 |
| rs494242 | let-7a-5p | trans | 7.78E-13 | Nightingale | M_VLDL_F | NA | T | C | 0.032542 | 0.004346 | 3.80E-14 |
| rs494242 | let-7a-5p | trans | 7.78E-13 | Nightingale | XXL_VLDL_ | NA | T | C | 0.033442 | 0.004356 | 8.80E-15 |
| rs494242 | let-7a-5p | trans | 7.78E-13 | Nightingale | S_HDL_L | NA | T | C | 0.024003 | 0.004332 | 2.60E-08 |
| rs494242 | let-7a-5p | trans | 7.78E-13 | Nightingale | M_LDL_P | NA | T | C | 0.034022 | 0.004359 | 7.20E-15 |
| rs494242 | let-7a-5p | trans | 7.78E-13 | Nightingale | XL_HDL_PL | NA | T | C | -0.0218 | 0.003994 | 4.60E-08 |
| rs494242 | let-7a-5p | trans | 7.78E-13 | Nightingale | XS_VLDL_F | NA | T | C | 0.035906 | 0.004271 | 7.30E-18 |
| rs494242 | let-7a-5p | trans | 7.78E-13 | Nightingale | VLDL_FC | NA | T | C | 0.024475 | 0.004315 | 3.40E-09 |
| rs494242 | let-7a-5p | trans | 7.78E-13 | Nightingale | S_VLDL_FC | NA | T | C | 0.036668 | 0.004348 | 1.70E-17 |
| rs494242 | let-7a-5p | trans | 7.78E-13 | Nightingale | ApoB | NA | T | C | 0.032952 | 0.004341 | 3.30E-14 |
| rs494242 | let-7a-5p | trans | 7.78E-13 | Nightingale | M_VLDL_C | NA | T | C | 0.031337 | 0.004084 | 3.30E-14 |
| rs494242 | let-7a-5p | trans | 7.78E-13 | Nightingale | S_LDL_C_p | NA | T | C | 0.041866 | 0.004364 | 5.60E-22 |
| rs494242 | let-7a-5p | trans | 7.78E-13 | Nightingale | L_VLDL_TG | NA | T | C | -0.04923 | 0.004326 | 6.60E-31 |
| rs494242 | let-7a-5p | trans | 7.78E-13 | Nightingale | LDL_CE | NA | T | C | 0.030645 | 0.004335 | 2.00E-12 |
| rs494242 | let-7a-5p | trans | 7.78E-13 | Nightingale | Remnant_( | NA | T | C | 0.035863 | 0.004319 | 5.10E-17 |
| rs494242 | let-7a-5p | trans | 7.78E-13 | Nightingale | S_LDL_L | NA | T | C | 0.026918 | 0.004346 | 1.20E-09 |
| rs494242 | let-7a-5p | trans | 7.78E-13 | Nightingale | S_VLDL_TG | NA | T | C | -0.03597 | 0.004208 | 1.90E-17 |
| rs494242 | let-7a-5p | trans | 7.78E-13 | Nightingale | L_VLDL_CE | NA | T | C | 0.044066 | 0.00416 | 8.10E-27 |
| rs494242 | let-7a-5p | trans | 7.78E-13 | Nightingale | M_LDL_L | NA | T | C | 0.031415 | 0.004353 | 6.70E-13 |
| rs494242 | let-7a-5p | trans | 7.78E-13 | Nightingale | LDL_P | NA | T | C | 0.030684 | 0.004344 | 2.10E-12 |
| rs494242 | let-7a-5p | trans | 7.78E-13 | Nightingale | LDL_C | NA | T | C | 0.031211 | 0.004328 | 9.20E-13 |
| rs494242 | let-7a-5p | trans | 7.78E-13 | Nightingale | M_VLDL_P | NA | T | C | 0.042089 | 0.004172 | 6.40E-24 |
| rs494242 | let-7a-5p | trans | 7.78E-13 | Nightingale | LA | NA | T | C | 0.02331 | 0.004263 | 1.50E-08 |
| rs494242 | let-7a-5p | trans | 7.78E-13 | Nightingale | L_VLDL_CE | NA | T | C | 0.022783 | 0.004296 | 2.80E-08 |
| rs494242 | let-7a-5p | trans | 7.78E-13 | Nightingale | L_LDL_C | NA | T | C | 0.030249 | 0.004298 | 2.90E-12 |
| rs494242 | let-7a-5p | trans | 7.78E-13 | Nightingale | XXL_VLDL_ | NA | T | C | 0.023509 | 0.004375 | 5.40E-09 |
| rs494242 | let-7a-5p | trans | 7.78E-13 | Nightingale | S_LDL_PL_ | NA | T | C | -0.02809 | 0.004339 | 4.70E-12 |
| rs494242 | let-7a-5p | trans | 7.78E-13 | Nightingale | VLDL_C | NA | T | C | 0.031089 | 0.004341 | 2.10E-13 |
| rs494242 | let-7a-5p | trans | 7.78E-13 | Nightingale | XL_VLDL_P | NA | T | C | 0.039093 | 0.004353 | 7.70E-20 |
| rs494242 | let-7a-5p | trans | 7.78E-13 | Nightingale | non_HDL_( | NA | T | C | 0.034165 | 0.004323 | 2.30E-15 |
| rs494242 | let-7a-5p | trans | 7.78E-13 | Nightingale | S_LDL_FC | NA | T | C | 0.027211 | 0.00434 | 1.30E-09 |
| rs494242 | let-7a-5p | trans | 7.78E-13 | Nightingale | LDL_L | NA | T | C | 0.031492 | 0.004332 | 5.60E-13 |
| rs494242 | let-7a-5p | trans | 7.78E-13 | Nightingale | XL_VLDL_C | NA | T | C | 0.034568 | 0.00417 | 1.60E-16 |
| rs494242 | let-7a-5p | trans | 7.78E-13 | Nightingale | IDL_FC | NA | T | C | 0.035991 | 0.004208 | 3.10E-17 |
| rs494242 | let-7a-5p | trans | 7.78E-13 | Nightingale | XL_VLDL_C | NA | T | C | 0.035582 | 0.004168 | 1.80E-17 |
| rs494242 | let-7a-5p | trans | 7.78E-13 | Nightingale | S_VLDL_C | NA | T | C | 0.03738 | 0.004346 | 2.50E-18 |
| rs494242 | let-7a-5p | trans | 7.78E-13 | Nightingale | L_LDL_L | NA | T | C | 0.031549 | 0.004304 | 3.10E-13 |
| rs494242 | let-7a-5p | trans | 7.78E-13 | Nightingale | XXL_VLDL_ | NA | T | C | 0.031802 | 0.00438 | 2.10E-13 |
| rs494242 | let-7a-5p | trans | 7.78E-13 | Nightingale | M_LDL_TG | NA | T | C | -0.0251 | 0.004293 | 8.80E-09 |
| rs494242 | let-7a-5p | trans | 7.78E-13 | Nightingale | S_VLDL_CE | NA | T | C | 0.037335 | 0.004341 | 1.70E-18 |
| rs494242 | let-7a-5p | trans | 7.78E-13 | Nightingale | S_VLDL_P | NA | T | C | 0.023424 | 0.004292 | 9.40E-09 |
| rs494242 | let-7a-5p | trans | 7.78E-13 | Nightingale | S_VLDL_PL | NA | T | C | 0.032066 | 0.004339 | 4.60E-14 |
| rs494242 | let-7a-5p | trans | 7.78E-13 | Nightingale | M_VLDL_F | NA | T | C | 0.039367 | 0.004136 | 2.20E-21 |
| rs494242 | let-7a-5p | trans | 7.78E-13 | Nightingale | S_VLDL_CE | NA | T | C | 0.045132 | 0.004288 | 1.50E-25 |
| rs494242 | let-7a-5p | trans | 7.78E-13 | Nightingale | XXL_VLDL_ | NA | T | C | -0.03443 | 0.004384 | 6.30E-16 |
| rs494242 | let-7a-5p | trans | 7.78E-13 | Nightingale | L_VLDL_C_ | NA | T | C | 0.049215 | 0.004246 | 9.30E-32 |
| rs494242 | let-7a-5p | trans | 7.78E-13 | Nightingale | S_VLDL_PL | NA | T | C | 0.025717 | 0.004162 | 1.50E-09 |
| rs494242 | let-7a-5p | trans | 7.78E-13 | Nightingale | IDL_CE | NA | T | C | 0.032104 | 0.004197 | 4.20E-14 |
| rs494242 | let-7a-5p | trans | 7.78E-13 | Nightingale | XL_VLDL_T | NA | T | C | -0.04154 | 0.004213 | 5.00E-23 |
| rs494242 | let-7a-5p | trans | 7.78E-13 | Nightingale | VLDL_CE | NA | T | C | 0.03517 | 0.004348 | 2.00E-16 |
| rs494242 | let-7a-5p | trans | 7.78E-13 | Nightingale | Sphingomy | NA | T | C | 0.025942 | 0.004064 | 3.10E-10 |
| rs494242 | let-7a-5p | trans | 7.78E-13 | Nightingale | L_LDL_P | NA | T | C | 0.028344 | 0.004332 | 7.50E-11 |
| rs494242 | let-7a-5p | trans | 7.78E-13 | Nightingale | S_VLDL_L | NA | T | C | 0.02453 | 0.004305 | 2.40E-09 |
| rs494242 | let-7a-5p | trans | 7.78E-13 | Nightingale | XS_VLDL_L | NA | T | C | 0.033036 | 0.004273 | 1.30E-15 |
| rs494242 | let-7a-5p | trans | 7.78E-13 | Nightingale | L_VLDL_FC | NA | T | C | 0.046929 | 0.004305 | 4.10E-28 |
| rs494242 | let-7a-5p | trans | 7.78E-13 | Nightingale | L_LDL_FC | NA | T | C | 0.031491 | 0.004268 | 4.00E-13 |
| rs494242 | let-7a-5p | trans | 7.78E-13 | Nightingale | Total_PL | NA | T | C | 0.023039 | 0.004139 | 7.30E-09 |

| | | | | | | | | | | | |
|---|---|---|---|---|---|---|---|---|---|---|---|
| rs494242 | let-7a-5p | trans | 7.78E-13 | Nightingale | L_VLDL_PL | NA | T | C | 0.021646 | 0.004222 | 4.50E-08 |
| rs494242 | let-7a-5p | trans | 7.78E-13 | Nightingale | M_LDL_PL | NA | T | C | 0.031199 | 0.004353 | 9.60E-13 |
| rs494242 | let-7a-5p | trans | 7.78E-13 | Nightingale | Clinical_LD | NA | T | C | 0.034576 | 0.004322 | 2.70E-15 |
| rs494242 | let-7a-5p | trans | 7.78E-13 | Nightingale | S_LDL_CE_ | NA | T | C | 0.028871 | 0.004339 | 1.00E-12 |
| rs494242 | let-7a-5p | trans | 7.78E-13 | Nightingale | S_VLDL_FC | NA | T | C | 0.027143 | 0.004154 | 1.20E-10 |
| rs494242 | let-7a-5p | trans | 7.78E-13 | Nightingale | S_HDL_CE | NA | T | C | 0.02571 | 0.004354 | 6.80E-09 |
| rs494242 | let-7a-5p | trans | 7.78E-13 | Nightingale | M_VLDL_P | NA | T | C | 0.028782 | 0.00434 | 1.30E-11 |
| rs494242 | let-7a-5p | trans | 7.78E-13 | Nightingale | M_VLDL_C | NA | T | C | 0.039214 | 0.004307 | 1.30E-19 |
| rs494242 | let-7a-5p | trans | 7.78E-13 | Nightingale | Total_L | NA | T | C | 0.025049 | 0.004276 | 1.40E-09 |
| rs494242 | let-7a-5p | trans | 7.78E-13 | Nightingale | IDL_L | NA | T | C | 0.03368 | 0.004203 | 2.20E-15 |
| rs494242 | let-7a-5p | trans | 7.78E-13 | Nightingale | S_VLDL_C_ | NA | T | C | 0.03972 | 0.00423 | 8.60E-21 |
| rs494242 | let-7a-5p | trans | 7.78E-13 | Nightingale | ApoB_by_A | NA | T | C | 0.025679 | 0.004281 | 2.10E-09 |
| rs494242 | let-7a-5p | trans | 7.78E-13 | Nightingale | LDL_PL | NA | T | C | 0.03334 | 0.004337 | 2.90E-14 |
| rs494242 | let-7a-5p | trans | 7.78E-13 | Nightingale | M_VLDL_T | NA | T | C | -0.03622 | 0.004108 | 1.60E-18 |
| rs494242 | let-7a-5p | trans | 7.78E-13 | Nightingale | S_HDL_P | NA | T | C | 0.027834 | 0.004347 | 2.60E-10 |
| rs494242 | let-7a-5p | trans | 7.78E-13 | Nightingale | XL_VLDL_F | NA | T | C | 0.035197 | 0.004214 | 1.80E-17 |
| rs494242 | let-7a-5p | trans | 7.78E-13 | Nightingale | XXL_VLDL_ | NA | T | C | 0.029266 | 0.004328 | 5.20E-12 |
| rs494242 | let-7a-5p | trans | 7.78E-13 | Nightingale | Total_FC | NA | T | C | 0.031409 | 0.004246 | 4.40E-14 |
| rs494242 | let-7a-5p | trans | 7.78E-13 | Nightingale | XS_VLDL_C | NA | T | C | 0.036659 | 0.004219 | 2.30E-18 |
| rs494242 | let-7a-5p | trans | 7.78E-13 | Nightingale | M_VLDL_C | NA | T | C | 0.037361 | 0.004335 | 5.20E-18 |
| rs494242 | let-7a-5p | trans | 7.78E-13 | Nightingale | Citrate | NA | T | C | 0.024682 | 0.004318 | 6.10E-09 |
| rs494242 | let-7a-5p | trans | 7.78E-13 | Nightingale | LDL_FC | NA | T | C | 0.031566 | 0.004301 | 6.40E-13 |
| rs494242 | let-7a-5p | trans | 7.78E-13 | Nightingale | L_LDL_CE | NA | T | C | 0.029548 | 0.004308 | 9.50E-12 |
| rs494242 | let-7a-5p | trans | 7.78E-13 | Nightingale | L_LDL_PL | NA | T | C | 0.03594 | 0.004314 | 1.30E-16 |
| rs494242 | let-7a-5p | trans | 7.78E-13 | Nightingale | S_HDL_C | NA | T | C | 0.025407 | 0.004339 | 9.00E-09 |
| rs494242 | let-7a-5p | trans | 7.78E-13 | Nightingale | S_LDL_CE | NA | T | C | 0.029469 | 0.004349 | 1.40E-11 |
| rs494242 | let-7a-5p | trans | 7.78E-13 | Nightingale | Omega_6 | NA | T | C | 0.024204 | 0.004245 | 2.90E-09 |
| rs494242 | let-7a-5p | trans | 7.78E-13 | Nightingale | IDL_P | NA | T | C | 0.03904 | 0.004301 | 1.80E-19 |
| rs494242 | let-7a-5p | trans | 7.78E-13 | Nightingale | M_LDL_CE | NA | T | C | 0.031239 | 0.004353 | 8.20E-13 |
| rs494242 | let-7a-5p | trans | 7.78E-13 | Nightingale | S_LDL_C | NA | T | C | 0.029721 | 0.004351 | 1.50E-11 |
| rs494242 | miR-139-5p | trans | 1.11E-40 | Nightingale | VLDL_P | NA | T | C | 0.026586 | 0.004324 | 1.90E-10 |
| rs494242 | miR-139-5p | trans | 1.11E-40 | Nightingale | M_VLDL_C | NA | T | C | 0.033835 | 0.004094 | 2.70E-16 |
| rs494242 | miR-139-5p | trans | 1.11E-40 | Nightingale | M_LDL_C_ | NA | T | C | 0.026574 | 0.00435 | 1.60E-09 |
| rs494242 | miR-139-5p | trans | 1.11E-40 | Nightingale | M_LDL_FC | NA | T | C | 0.032299 | 0.004341 | 2.30E-13 |
| rs494242 | miR-139-5p | trans | 1.11E-40 | Nightingale | XS_VLDL_P | NA | T | C | 0.035812 | 0.004289 | 9.60E-18 |
| rs494242 | miR-139-5p | trans | 1.11E-40 | Nightingale | Total_CE | NA | T | C | 0.029414 | 0.004193 | 1.80E-12 |
| rs494242 | miR-139-5p | trans | 1.11E-40 | Nightingale | GlycA | NA | T | C | 0.025353 | 0.004328 | 4.70E-09 |
| rs494242 | miR-139-5p | trans | 1.11E-40 | Nightingale | M_VLDL_P | NA | T | C | 0.027954 | 0.004336 | 4.50E-11 |
| rs494242 | miR-139-5p | trans | 1.11E-40 | Nightingale | XS_VLDL_F | NA | T | C | 0.029107 | 0.004316 | 3.20E-11 |
| rs494242 | miR-139-5p | trans | 1.11E-40 | Nightingale | IDL_PL | NA | T | C | 0.033595 | 0.004195 | 1.90E-15 |
| rs494242 | miR-139-5p | trans | 1.11E-40 | Nightingale | XS_VLDL_P | NA | T | C | 0.031685 | 0.004286 | 1.90E-14 |
| rs494242 | miR-139-5p | trans | 1.11E-40 | Nightingale | M_LDL_C | NA | T | C | 0.032117 | 0.004355 | 2.20E-13 |
| rs494242 | miR-139-5p | trans | 1.11E-40 | Nightingale | XS_VLDL_C | NA | T | C | 0.036408 | 0.004195 | 4.00E-18 |
| rs494242 | miR-139-5p | trans | 1.11E-40 | Nightingale | IDL_C | NA | T | C | 0.03332 | 0.004198 | 4.70E-15 |
| rs494242 | miR-139-5p | trans | 1.11E-40 | Nightingale | Total_C | NA | T | C | 0.030169 | 0.004208 | 4.70E-13 |
| rs494242 | miR-139-5p | trans | 1.11E-40 | Nightingale | S_LDL_P | NA | T | C | 0.028983 | 0.004351 | 2.90E-11 |
| rs494242 | miR-139-5p | trans | 1.11E-40 | Nightingale | M_VLDL_F | NA | T | C | 0.032542 | 0.004346 | 3.80E-14 |
| rs494242 | miR-139-5p | trans | 1.11E-40 | Nightingale | XXL_VLDL_ | NA | T | C | 0.033442 | 0.004356 | 8.80E-15 |
| rs494242 | miR-139-5p | trans | 1.11E-40 | Nightingale | S_HDL_L | NA | T | C | 0.024003 | 0.004332 | 2.60E-08 |
| rs494242 | miR-139-5p | trans | 1.11E-40 | Nightingale | M_LDL_P | NA | T | C | 0.034022 | 0.004359 | 7.20E-15 |
| rs494242 | miR-139-5p | trans | 1.11E-40 | Nightingale | XL_HDL_PL | NA | T | C | -0.0218 | 0.003994 | 4.60E-08 |
| rs494242 | miR-139-5p | trans | 1.11E-40 | Nightingale | XS_VLDL_F | NA | T | C | 0.035906 | 0.004271 | 7.30E-18 |
| rs494242 | miR-139-5p | trans | 1.11E-40 | Nightingale | VLDL_FC | NA | T | C | 0.024475 | 0.004315 | 3.40E-09 |
| rs494242 | miR-139-5p | trans | 1.11E-40 | Nightingale | S_VLDL_FC | NA | T | C | 0.036668 | 0.004348 | 1.70E-17 |
| rs494242 | miR-139-5p | trans | 1.11E-40 | Nightingale | ApoB | NA | T | C | 0.032952 | 0.004341 | 3.30E-14 |
| rs494242 | miR-139-5p | trans | 1.11E-40 | Nightingale | M_VLDL_C | NA | T | C | 0.031337 | 0.004084 | 3.30E-14 |
| rs494242 | miR-139-5p | trans | 1.11E-40 | Nightingale | S_LDL_C_p | NA | T | C | 0.041866 | 0.004364 | 5.60E-22 |
| rs494242 | miR-139-5p | trans | 1.11E-40 | Nightingale | L_VLDL_TG | NA | T | C | -0.04923 | 0.004326 | 6.60E-31 |
| rs494242 | miR-139-5p | trans | 1.11E-40 | Nightingale | LDL_CE | NA | T | C | 0.030645 | 0.004335 | 2.00E-12 |
| rs494242 | miR-139-5p | trans | 1.11E-40 | Nightingale | Remnant_( | NA | T | C | 0.035863 | 0.004319 | 5.10E-17 |
| rs494242 | miR-139-5p | trans | 1.11E-40 | Nightingale | S_LDL_L | NA | T | C | 0.026918 | 0.004346 | 1.20E-09 |
| rs494242 | miR-139-5p | trans | 1.11E-40 | Nightingale | S_VLDL_TG | NA | T | C | -0.03597 | 0.004208 | 1.90E-17 |
| rs494242 | miR-139-5p | trans | 1.11E-40 | Nightingale | L_VLDL_CE | NA | T | C | 0.044066 | 0.00416 | 8.10E-27 |

| | | | | | | | | | | | |
|---|---|---|---|---|---|---|---|---|---|---|---|
| rs494242 | miR-139-5p | trans | 1.11E-40 | Nightingale | M_LDL_L | NA | T | C | 0.031415 | 0.004353 | 6.70E-13 |
| rs494242 | miR-139-5p | trans | 1.11E-40 | Nightingale | LDL_P | NA | T | C | 0.030684 | 0.004344 | 2.10E-12 |
| rs494242 | miR-139-5p | trans | 1.11E-40 | Nightingale | LDL_C | NA | T | C | 0.031211 | 0.004328 | 9.20E-13 |
| rs494242 | miR-139-5p | trans | 1.11E-40 | Nightingale | M_VLDL_P | NA | T | C | 0.042089 | 0.004172 | 6.40E-24 |
| rs494242 | miR-139-5p | trans | 1.11E-40 | Nightingale | LA | NA | T | C | 0.02331 | 0.004263 | 1.50E-08 |
| rs494242 | miR-139-5p | trans | 1.11E-40 | Nightingale | L_VLDL_CE | NA | T | C | 0.022783 | 0.004296 | 2.80E-08 |
| rs494242 | miR-139-5p | trans | 1.11E-40 | Nightingale | L_LDL_C | NA | T | C | 0.030249 | 0.004298 | 2.90E-12 |
| rs494242 | miR-139-5p | trans | 1.11E-40 | Nightingale | XXL_VLDL_ | NA | T | C | 0.023509 | 0.004375 | 5.40E-09 |
| rs494242 | miR-139-5p | trans | 1.11E-40 | Nightingale | S_LDL_PL_ | NA | T | C | -0.02809 | 0.004339 | 4.70E-12 |
| rs494242 | miR-139-5p | trans | 1.11E-40 | Nightingale | VLDL_C | NA | T | C | 0.031089 | 0.004341 | 2.10E-13 |
| rs494242 | miR-139-5p | trans | 1.11E-40 | Nightingale | XL_VLDL_P | NA | T | C | 0.039093 | 0.004353 | 7.70E-20 |
| rs494242 | miR-139-5p | trans | 1.11E-40 | Nightingale | non_HDL_( | NA | T | C | 0.034165 | 0.004323 | 2.30E-15 |
| rs494242 | miR-139-5p | trans | 1.11E-40 | Nightingale | S_LDL_FC | NA | T | C | 0.027211 | 0.00434 | 1.30E-09 |
| rs494242 | miR-139-5p | trans | 1.11E-40 | Nightingale | LDL_L | NA | T | C | 0.031492 | 0.004332 | 5.60E-13 |
| rs494242 | miR-139-5p | trans | 1.11E-40 | Nightingale | XL_VLDL_C | NA | T | C | 0.034568 | 0.00417 | 1.60E-16 |
| rs494242 | miR-139-5p | trans | 1.11E-40 | Nightingale | IDL_FC | NA | T | C | 0.035991 | 0.004208 | 3.10E-17 |
| rs494242 | miR-139-5p | trans | 1.11E-40 | Nightingale | XL_VLDL_C | NA | T | C | 0.035582 | 0.004168 | 1.80E-17 |
| rs494242 | miR-139-5p | trans | 1.11E-40 | Nightingale | S_VLDL_C | NA | T | C | 0.03738 | 0.004346 | 2.50E-18 |
| rs494242 | miR-139-5p | trans | 1.11E-40 | Nightingale | L_LDL_L | NA | T | C | 0.031549 | 0.004304 | 3.10E-13 |
| rs494242 | miR-139-5p | trans | 1.11E-40 | Nightingale | XXL_VLDL_ | NA | T | C | 0.031802 | 0.00438 | 2.10E-13 |
| rs494242 | miR-139-5p | trans | 1.11E-40 | Nightingale | M_LDL_TG | NA | T | C | -0.0251 | 0.004293 | 8.80E-09 |
| rs494242 | miR-139-5p | trans | 1.11E-40 | Nightingale | S_VLDL_CE | NA | T | C | 0.037335 | 0.004341 | 1.70E-18 |
| rs494242 | miR-139-5p | trans | 1.11E-40 | Nightingale | S_VLDL_P | NA | T | C | 0.023424 | 0.004292 | 9.40E-09 |
| rs494242 | miR-139-5p | trans | 1.11E-40 | Nightingale | S_VLDL_PL | NA | T | C | 0.032066 | 0.004339 | 4.60E-14 |
| rs494242 | miR-139-5p | trans | 1.11E-40 | Nightingale | M_VLDL_F | NA | T | C | 0.039367 | 0.004136 | 2.20E-21 |
| rs494242 | miR-139-5p | trans | 1.11E-40 | Nightingale | S_VLDL_CE | NA | T | C | 0.045132 | 0.004288 | 1.50E-25 |
| rs494242 | miR-139-5p | trans | 1.11E-40 | Nightingale | XXL_VLDL_ | NA | T | C | -0.03443 | 0.004384 | 6.30E-16 |
| rs494242 | miR-139-5p | trans | 1.11E-40 | Nightingale | L_VLDL_C_ | NA | T | C | 0.049215 | 0.004246 | 9.30E-32 |
| rs494242 | miR-139-5p | trans | 1.11E-40 | Nightingale | S_VLDL_PL | NA | T | C | 0.025717 | 0.004162 | 1.50E-09 |
| rs494242 | miR-139-5p | trans | 1.11E-40 | Nightingale | IDL_CE | NA | T | C | 0.032104 | 0.004197 | 4.20E-14 |
| rs494242 | miR-139-5p | trans | 1.11E-40 | Nightingale | XL_VLDL_T | NA | T | C | -0.04154 | 0.004213 | 5.00E-23 |
| rs494242 | miR-139-5p | trans | 1.11E-40 | Nightingale | VLDL_CE | NA | T | C | 0.03517 | 0.004348 | 2.00E-16 |
| rs494242 | miR-139-5p | trans | 1.11E-40 | Nightingale | Sphingomy | NA | T | C | 0.025942 | 0.004064 | 3.10E-10 |
| rs494242 | miR-139-5p | trans | 1.11E-40 | Nightingale | L_LDL_P | NA | T | C | 0.028344 | 0.004332 | 7.50E-11 |
| rs494242 | miR-139-5p | trans | 1.11E-40 | Nightingale | S_VLDL_L | NA | T | C | 0.02453 | 0.004305 | 2.40E-09 |
| rs494242 | miR-139-5p | trans | 1.11E-40 | Nightingale | XS_VLDL_L | NA | T | C | 0.033036 | 0.004273 | 1.30E-15 |
| rs494242 | miR-139-5p | trans | 1.11E-40 | Nightingale | L_VLDL_FC | NA | T | C | 0.046929 | 0.004305 | 4.10E-28 |
| rs494242 | miR-139-5p | trans | 1.11E-40 | Nightingale | L_LDL_FC | NA | T | C | 0.031491 | 0.004268 | 4.00E-13 |
| rs494242 | miR-139-5p | trans | 1.11E-40 | Nightingale | Total_PL | NA | T | C | 0.023039 | 0.004139 | 7.30E-09 |
| rs494242 | miR-139-5p | trans | 1.11E-40 | Nightingale | L_VLDL_PL | NA | T | C | 0.021646 | 0.004222 | 4.50E-08 |
| rs494242 | miR-139-5p | trans | 1.11E-40 | Nightingale | M_LDL_PL | NA | T | C | 0.031199 | 0.004353 | 9.60E-13 |
| rs494242 | miR-139-5p | trans | 1.11E-40 | Nightingale | Clinical_LD | NA | T | C | 0.034576 | 0.004322 | 2.70E-15 |
| rs494242 | miR-139-5p | trans | 1.11E-40 | Nightingale | S_LDL_CE_ | NA | T | C | 0.028871 | 0.004339 | 1.00E-12 |
| rs494242 | miR-139-5p | trans | 1.11E-40 | Nightingale | S_VLDL_FC | NA | T | C | 0.027143 | 0.004154 | 1.20E-10 |
| rs494242 | miR-139-5p | trans | 1.11E-40 | Nightingale | S_HDL_CE | NA | T | C | 0.02571 | 0.004354 | 6.80E-09 |
| rs494242 | miR-139-5p | trans | 1.11E-40 | Nightingale | M_VLDL_P | NA | T | C | 0.028782 | 0.00434 | 1.30E-11 |
| rs494242 | miR-139-5p | trans | 1.11E-40 | Nightingale | M_VLDL_C | NA | T | C | 0.039214 | 0.004307 | 1.30E-19 |
| rs494242 | miR-139-5p | trans | 1.11E-40 | Nightingale | Total_L | NA | T | C | 0.025049 | 0.004276 | 1.40E-09 |
| rs494242 | miR-139-5p | trans | 1.11E-40 | Nightingale | IDL_L | NA | T | C | 0.03368 | 0.004203 | 2.20E-15 |
| rs494242 | miR-139-5p | trans | 1.11E-40 | Nightingale | S_VLDL_C_ | NA | T | C | 0.03972 | 0.00423 | 8.60E-21 |
| rs494242 | miR-139-5p | trans | 1.11E-40 | Nightingale | ApoB_by_A | NA | T | C | 0.025679 | 0.004281 | 2.10E-09 |
| rs494242 | miR-139-5p | trans | 1.11E-40 | Nightingale | LDL_PL | NA | T | C | 0.03334 | 0.004337 | 2.90E-14 |
| rs494242 | miR-139-5p | trans | 1.11E-40 | Nightingale | M_VLDL_T | NA | T | C | -0.03622 | 0.004108 | 1.60E-18 |
| rs494242 | miR-139-5p | trans | 1.11E-40 | Nightingale | S_HDL_P | NA | T | C | 0.027834 | 0.004347 | 2.60E-10 |
| rs494242 | miR-139-5p | trans | 1.11E-40 | Nightingale | XL_VLDL_F | NA | T | C | 0.035197 | 0.004214 | 1.80E-17 |
| rs494242 | miR-139-5p | trans | 1.11E-40 | Nightingale | XXL_VLDL_ | NA | T | C | 0.029266 | 0.004328 | 5.20E-12 |
| rs494242 | miR-139-5p | trans | 1.11E-40 | Nightingale | Total_FC | NA | T | C | 0.031409 | 0.004246 | 4.40E-14 |
| rs494242 | miR-139-5p | trans | 1.11E-40 | Nightingale | XS_VLDL_C | NA | T | C | 0.036659 | 0.004219 | 2.30E-18 |
| rs494242 | miR-139-5p | trans | 1.11E-40 | Nightingale | M_VLDL_C | NA | T | C | 0.037361 | 0.004335 | 5.20E-18 |
| rs494242 | miR-139-5p | trans | 1.11E-40 | Nightingale | Citrate | NA | T | C | 0.024682 | 0.004318 | 6.10E-09 |
| rs494242 | miR-139-5p | trans | 1.11E-40 | Nightingale | LDL_FC | NA | T | C | 0.031566 | 0.004301 | 6.40E-13 |
| rs494242 | miR-139-5p | trans | 1.11E-40 | Nightingale | L_LDL_CE | NA | T | C | 0.029548 | 0.004308 | 9.50E-12 |
| rs494242 | miR-139-5p | trans | 1.11E-40 | Nightingale | L_LDL_PL | NA | T | C | 0.03594 | 0.004314 | 1.30E-16 |

| | | | | | | | | | | | |
|---|---|---|---|---|---|---|---|---|---|---|---|
| rs494242 | miR-139-5p | trans | 1.11E-40 | Nightingale | S_HDL_C | NA | T | C | 0.025407 | 0.004339 | 9.00E-09 |
| rs494242 | miR-139-5p | trans | 1.11E-40 | Nightingale | S_LDL_CE | NA | T | C | 0.029469 | 0.004349 | 1.40E-11 |
| rs494242 | miR-139-5p | trans | 1.11E-40 | Nightingale | Omega_6 | NA | T | C | 0.024204 | 0.004245 | 2.90E-09 |
| rs494242 | miR-139-5p | trans | 1.11E-40 | Nightingale | IDL_P | NA | T | C | 0.03904 | 0.004301 | 1.80E-19 |
| rs494242 | miR-139-5p | trans | 1.11E-40 | Nightingale | M_LDL_CE | NA | T | C | 0.031239 | 0.004353 | 8.20E-13 |
| rs494242 | miR-139-5p | trans | 1.11E-40 | Nightingale | S_LDL_C | NA | T | C | 0.029721 | 0.004351 | 1.50E-11 |
| rs495203 | miR-10b-5p | trans | 3.15E-35 | Nightingale | L_VLDL_CE | NA | T | C | 0.047347 | 0.004234 | 2.40E-29 |
| rs495203 | miR-10b-5p | trans | 3.15E-35 | Nightingale | L_VLDL_FC | NA | T | C | 0.048592 | 0.004383 | 4.60E-29 |
| rs495203 | miR-10b-5p | trans | 3.15E-35 | Nightingale | XXL_VLDL_ | NA | T | C | -0.03574 | 0.004463 | 2.10E-16 |
| rs495203 | miR-10b-5p | trans | 3.15E-35 | Nightingale | IDL_FC | NA | T | C | 0.03826 | 0.004284 | 1.40E-18 |
| rs495203 | miR-10b-5p | trans | 3.15E-35 | Nightingale | Clinical_LD | NA | T | C | 0.036481 | 0.004399 | 2.70E-16 |
| rs495203 | miR-10b-5p | trans | 3.15E-35 | Nightingale | S_HDL_L | NA | T | C | 0.026311 | 0.00441 | 1.50E-09 |
| rs495203 | miR-10b-5p | trans | 3.15E-35 | Nightingale | PUFA | NA | T | C | 0.023739 | 0.004316 | 1.40E-08 |
| rs495203 | miR-10b-5p | trans | 3.15E-35 | Nightingale | XL_VLDL_F | NA | T | C | 0.0372 | 0.00429 | 2.00E-18 |
| rs495203 | miR-10b-5p | trans | 3.15E-35 | Nightingale | XS_VLDL_L | NA | T | C | 0.033828 | 0.00435 | 2.00E-15 |
| rs495203 | miR-10b-5p | trans | 3.15E-35 | Nightingale | S_HDL_C | NA | T | C | 0.028426 | 0.004417 | 1.70E-10 |
| rs495203 | miR-10b-5p | trans | 3.15E-35 | Nightingale | S_LDL_C | NA | T | C | 0.03095 | 0.004429 | 5.50E-12 |
| rs495203 | miR-10b-5p | trans | 3.15E-35 | Nightingale | VLDL_C | NA | T | C | 0.031307 | 0.004419 | 4.90E-13 |
| rs495203 | miR-10b-5p | trans | 3.15E-35 | Nightingale | XS_VLDL_P | NA | T | C | 0.036711 | 0.004366 | 1.40E-17 |
| rs495203 | miR-10b-5p | trans | 3.15E-35 | Nightingale | IDL_P | NA | T | C | 0.039988 | 0.004378 | 1.30E-19 |
| rs495203 | miR-10b-5p | trans | 3.15E-35 | Nightingale | XL_VLDL_T | NA | T | C | -0.04392 | 0.004289 | 1.70E-24 |
| rs495203 | miR-10b-5p | trans | 3.15E-35 | Nightingale | L_LDL_PL | NA | T | C | 0.037997 | 0.004391 | 9.00E-18 |
| rs495203 | miR-10b-5p | trans | 3.15E-35 | Nightingale | M_LDL_C | NA | T | C | 0.033996 | 0.004433 | 2.10E-14 |
| rs495203 | miR-10b-5p | trans | 3.15E-35 | Nightingale | S_VLDL_CE | NA | T | C | 0.047029 | 0.004365 | 1.20E-26 |
| rs495203 | miR-10b-5p | trans | 3.15E-35 | Nightingale | XS_VLDL_F | NA | T | C | 0.037076 | 0.004348 | 6.50E-18 |
| rs495203 | miR-10b-5p | trans | 3.15E-35 | Nightingale | IDL_C | NA | T | C | 0.03496 | 0.004274 | 9.40E-16 |
| rs495203 | miR-10b-5p | trans | 3.15E-35 | Nightingale | M_LDL_FC | NA | T | C | 0.034355 | 0.004419 | 1.90E-14 |
| rs495203 | miR-10b-5p | trans | 3.15E-35 | Nightingale | ApoB | NA | T | C | 0.033945 | 0.004419 | 2.10E-14 |
| rs495203 | miR-10b-5p | trans | 3.15E-35 | Nightingale | VLDL_CE | NA | T | C | 0.035791 | 0.004426 | 3.00E-16 |
| rs495203 | miR-10b-5p | trans | 3.15E-35 | Nightingale | M_VLDL_F | NA | T | C | 0.033377 | 0.004424 | 3.10E-14 |
| rs495203 | miR-10b-5p | trans | 3.15E-35 | Nightingale | LDL_L | NA | T | C | 0.033314 | 0.00441 | 7.30E-14 |
| rs495203 | miR-10b-5p | trans | 3.15E-35 | Nightingale | S_HDL_CE | NA | T | C | 0.028647 | 0.004432 | 1.30E-10 |
| rs495203 | miR-10b-5p | trans | 3.15E-35 | Nightingale | S_HDL_P | NA | T | C | 0.030626 | 0.004425 | 5.00E-12 |
| rs495203 | miR-10b-5p | trans | 3.15E-35 | Nightingale | M_LDL_TG | NA | T | C | -0.02762 | 0.00437 | 3.20E-10 |
| rs495203 | miR-10b-5p | trans | 3.15E-35 | Nightingale | M_LDL_C_ | NA | T | C | 0.031158 | 0.004428 | 2.90E-12 |
| rs495203 | miR-10b-5p | trans | 3.15E-35 | Nightingale | S_VLDL_C_ | NA | T | C | 0.041885 | 0.004306 | 5.20E-22 |
| rs495203 | miR-10b-5p | trans | 3.15E-35 | Nightingale | XL_VLDL_P | NA | T | C | 0.038836 | 0.004432 | 5.30E-19 |
| rs495203 | miR-10b-5p | trans | 3.15E-35 | Nightingale | M_LDL_L | NA | T | C | 0.033041 | 0.004431 | 1.00E-13 |
| rs495203 | miR-10b-5p | trans | 3.15E-35 | Nightingale | LDL_P | NA | T | C | 0.031807 | 0.004422 | 1.00E-12 |
| rs495203 | miR-10b-5p | trans | 3.15E-35 | Nightingale | LDL_C | NA | T | C | 0.033203 | 0.004406 | 8.80E-14 |
| rs495203 | miR-10b-5p | trans | 3.15E-35 | Nightingale | Total_PL | NA | T | C | 0.024944 | 0.004214 | 1.00E-09 |
| rs495203 | miR-10b-5p | trans | 3.15E-35 | Nightingale | Total_FC | NA | T | C | 0.033266 | 0.004323 | 6.70E-15 |
| rs495203 | miR-10b-5p | trans | 3.15E-35 | Nightingale | Cholines | NA | T | C | 0.022313 | 0.004161 | 4.70E-08 |
| rs495203 | miR-10b-5p | trans | 3.15E-35 | Nightingale | S_HDL_FC | NA | T | C | 0.023473 | 0.004344 | 4.50E-08 |
| rs495203 | miR-10b-5p | trans | 3.15E-35 | Nightingale | XS_VLDL_F | NA | T | C | 0.031421 | 0.004393 | 2.50E-12 |
| rs495203 | miR-10b-5p | trans | 3.15E-35 | Nightingale | S_VLDL_PL | NA | T | C | 0.027738 | 0.004237 | 1.80E-10 |
| rs495203 | miR-10b-5p | trans | 3.15E-35 | Nightingale | XXL_VLDL_ | NA | T | C | 0.033132 | 0.004459 | 8.80E-14 |
| rs495203 | miR-10b-5p | trans | 3.15E-35 | Nightingale | XS_VLDL_P | NA | T | C | 0.032126 | 0.004363 | 5.30E-14 |
| rs495203 | miR-10b-5p | trans | 3.15E-35 | Nightingale | S_VLDL_PL | NA | T | C | 0.032786 | 0.004417 | 4.20E-14 |
| rs495203 | miR-10b-5p | trans | 3.15E-35 | Nightingale | XS_VLDL_C | NA | T | C | 0.037999 | 0.004295 | 1.30E-18 |
| rs495203 | miR-10b-5p | trans | 3.15E-35 | Nightingale | M_LDL_CE | NA | T | C | 0.032982 | 0.004431 | 9.60E-14 |
| rs495203 | miR-10b-5p | trans | 3.15E-35 | Nightingale | LDL_FC | NA | T | C | 0.033639 | 0.004378 | 5.20E-14 |
| rs495203 | miR-10b-5p | trans | 3.15E-35 | Nightingale | LA | NA | T | C | 0.024923 | 0.004339 | 3.60E-09 |
| rs495203 | miR-10b-5p | trans | 3.15E-35 | Nightingale | Sphingomy | NA | T | C | 0.029029 | 0.004137 | 6.60E-12 |
| rs495203 | miR-10b-5p | trans | 3.15E-35 | Nightingale | M_VLDL_C | NA | T | C | 0.040848 | 0.004385 | 2.30E-20 |
| rs495203 | miR-10b-5p | trans | 3.15E-35 | Nightingale | S_LDL_PL_ | NA | T | C | -0.03007 | 0.004417 | 2.60E-13 |
| rs495203 | miR-10b-5p | trans | 3.15E-35 | Nightingale | S_VLDL_L | NA | T | C | 0.024678 | 0.004382 | 3.70E-09 |
| rs495203 | miR-10b-5p | trans | 3.15E-35 | Nightingale | S_HDL_PL | NA | T | C | 0.023799 | 0.004387 | 3.20E-08 |
| rs495203 | miR-10b-5p | trans | 3.15E-35 | Nightingale | S_VLDL_P | NA | T | C | 0.023475 | 0.004369 | 1.60E-08 |
| rs495203 | miR-10b-5p | trans | 3.15E-35 | Nightingale | XS_VLDL_C | NA | T | C | 0.037803 | 0.00427 | 2.10E-18 |
| rs495203 | miR-10b-5p | trans | 3.15E-35 | Nightingale | S_VLDL_C | NA | T | C | 0.03838 | 0.004424 | 1.80E-18 |
| rs495203 | miR-10b-5p | trans | 3.15E-35 | Nightingale | non_HDL_( | NA | T | C | 0.035705 | 0.004401 | 5.00E-16 |
| rs495203 | miR-10b-5p | trans | 3.15E-35 | Nightingale | Total_CE | NA | T | C | 0.031758 | 0.004268 | 9.50E-14 |

| | | | | | | | | | | | |
|---|---|---|---|---|---|---|---|---|---|---|---|
| rs495203 | miR-10b-5p | trans | 3.15E-35 | Nightingale | M_VLDL_T | NA | T | C | -0.03856 | 0.004182 | 6.80E-20 |
| rs495203 | miR-10b-5p | trans | 3.15E-35 | Nightingale | M_VLDL_C | NA | T | C | 0.038716 | 0.004413 | 1.90E-18 |
| rs495203 | miR-10b-5p | trans | 3.15E-35 | Nightingale | M_VLDL_P | NA | T | C | 0.029134 | 0.004418 | 2.00E-11 |
| rs495203 | miR-10b-5p | trans | 3.15E-35 | Nightingale | XL_VLDL_C | NA | T | C | 0.037358 | 0.004245 | 3.10E-18 |
| rs495203 | miR-10b-5p | trans | 3.15E-35 | Nightingale | L_LDL_L | NA | T | C | 0.03356 | 0.004382 | 3.00E-14 |
| rs495203 | miR-10b-5p | trans | 3.15E-35 | Nightingale | XXL_VLDL_ | NA | T | C | 0.025016 | 0.004454 | 1.50E-09 |
| rs495203 | miR-10b-5p | trans | 3.15E-35 | Nightingale | M_VLDL_P | NA | T | C | 0.028301 | 0.004414 | 6.30E-11 |
| rs495203 | miR-10b-5p | trans | 3.15E-35 | Nightingale | M_LDL_P | NA | T | C | 0.035898 | 0.004437 | 6.80E-16 |
| rs495203 | miR-10b-5p | trans | 3.15E-35 | Nightingale | IDL_CE | NA | T | C | 0.033513 | 0.004272 | 1.30E-14 |
| rs495203 | miR-10b-5p | trans | 3.15E-35 | Nightingale | S_VLDL_FC | NA | T | C | 0.037912 | 0.004426 | 7.30E-18 |
| rs495203 | miR-10b-5p | trans | 3.15E-35 | Nightingale | IDL_FC_pct | NA | T | C | 0.025765 | 0.004391 | 1.10E-08 |
| rs495203 | miR-10b-5p | trans | 3.15E-35 | Nightingale | S_VLDL_FC | NA | T | C | 0.029355 | 0.004228 | 1.10E-11 |
| rs495203 | miR-10b-5p | trans | 3.15E-35 | Nightingale | GlycA | NA | T | C | 0.028026 | 0.004406 | 1.40E-10 |
| rs495203 | miR-10b-5p | trans | 3.15E-35 | Nightingale | M_VLDL_P | NA | T | C | 0.04406 | 0.004247 | 5.50E-25 |
| rs495203 | miR-10b-5p | trans | 3.15E-35 | Nightingale | L_LDL_CE | NA | T | C | 0.031578 | 0.004385 | 9.80E-13 |
| rs495203 | miR-10b-5p | trans | 3.15E-35 | Nightingale | VLDL_P | NA | T | C | 0.026641 | 0.004401 | 4.40E-10 |
| rs495203 | miR-10b-5p | trans | 3.15E-35 | Nightingale | L_LDL_P | NA | T | C | 0.029185 | 0.00441 | 6.10E-11 |
| rs495203 | miR-10b-5p | trans | 3.15E-35 | Nightingale | LDL_PL | NA | T | C | 0.034997 | 0.004415 | 4.70E-15 |
| rs495203 | miR-10b-5p | trans | 3.15E-35 | Nightingale | S_LDL_CE | NA | T | C | 0.030556 | 0.004427 | 5.80E-12 |
| rs495203 | miR-10b-5p | trans | 3.15E-35 | Nightingale | Total_L | NA | T | C | 0.026363 | 0.004353 | 4.50E-10 |
| rs495203 | miR-10b-5p | trans | 3.15E-35 | Nightingale | Omega_6 | NA | T | C | 0.02591 | 0.004321 | 5.60E-10 |
| rs495203 | miR-10b-5p | trans | 3.15E-35 | Nightingale | M_VLDL_F | NA | T | C | 0.042068 | 0.00421 | 3.50E-23 |
| rs495203 | miR-10b-5p | trans | 3.15E-35 | Nightingale | Total_C | NA | T | C | 0.032385 | 0.004284 | 3.30E-14 |
| rs495203 | miR-10b-5p | trans | 3.15E-35 | Nightingale | XXL_VLDL_ | NA | T | C | 0.034842 | 0.004434 | 2.90E-15 |
| rs495203 | miR-10b-5p | trans | 3.15E-35 | Nightingale | L_LDL_C | NA | T | C | 0.03233 | 0.004375 | 2.60E-13 |
| rs495203 | miR-10b-5p | trans | 3.15E-35 | Nightingale | Remnant_( | NA | T | C | 0.036874 | 0.004396 | 3.70E-17 |
| rs495203 | miR-10b-5p | trans | 3.15E-35 | Nightingale | S_LDL_FC | NA | T | C | 0.028569 | 0.004418 | 4.30E-10 |
| rs495203 | miR-10b-5p | trans | 3.15E-35 | Nightingale | VLDL_FC | NA | T | C | 0.024164 | 0.004392 | 1.10E-08 |
| rs495203 | miR-10b-5p | trans | 3.15E-35 | Nightingale | M_VLDL_C | NA | T | C | 0.033632 | 0.004157 | 1.90E-15 |
| rs495203 | miR-10b-5p | trans | 3.15E-35 | Nightingale | ApoB_by_A | NA | T | C | 0.025045 | 0.004358 | 1.30E-08 |
| rs495203 | miR-10b-5p | trans | 3.15E-35 | Nightingale | S_LDL_P | NA | T | C | 0.029594 | 0.004429 | 3.20E-11 |
| rs495203 | miR-10b-5p | trans | 3.15E-35 | Nightingale | S_LDL_C_p | NA | T | C | 0.045365 | 0.004442 | 5.50E-25 |
| rs495203 | miR-10b-5p | trans | 3.15E-35 | Nightingale | LDL_CE | NA | T | C | 0.032564 | 0.004413 | 2.20E-13 |
| rs495203 | miR-10b-5p | trans | 3.15E-35 | Nightingale | IDL_L | NA | T | C | 0.035277 | 0.004278 | 4.80E-16 |
| rs495203 | miR-10b-5p | trans | 3.15E-35 | Nightingale | M_VLDL_C | NA | T | C | 0.036209 | 0.004168 | 1.20E-17 |
| rs495203 | miR-10b-5p | trans | 3.15E-35 | Nightingale | L_VLDL_TG | NA | T | C | -0.05115 | 0.004404 | 5.30E-32 |
| rs495203 | miR-10b-5p | trans | 3.15E-35 | Nightingale | Citrate | NA | T | C | 0.026828 | 0.004396 | 7.30E-10 |
| rs495203 | miR-10b-5p | trans | 3.15E-35 | Nightingale | S_LDL_CE_ | NA | T | C | 0.03095 | 0.004417 | 4.90E-14 |
| rs495203 | miR-10b-5p | trans | 3.15E-35 | Nightingale | IDL_PL | NA | T | C | 0.035351 | 0.00427 | 3.30E-16 |
| rs495203 | miR-10b-5p | trans | 3.15E-35 | Nightingale | L_VLDL_C_ | NA | T | C | 0.052202 | 0.004322 | 4.00E-34 |
| rs495203 | miR-10b-5p | trans | 3.15E-35 | Nightingale | M_LDL_PL | NA | T | C | 0.032476 | 0.004431 | 2.60E-13 |
| rs495203 | miR-10b-5p | trans | 3.15E-35 | Nightingale | XL_VLDL_C | NA | T | C | 0.038228 | 0.004243 | 4.60E-19 |
| rs495203 | miR-10b-5p | trans | 3.15E-35 | Nightingale | S_LDL_L | NA | T | C | 0.027749 | 0.004424 | 8.20E-10 |
| rs495203 | miR-10b-5p | trans | 3.15E-35 | Nightingale | S_VLDL_CE | NA | T | C | 0.038188 | 0.004419 | 1.60E-18 |
| rs495203 | miR-10b-5p | trans | 3.15E-35 | Nightingale | XL_HDL_PL | NA | T | C | -0.02275 | 0.004065 | 1.20E-08 |
| rs495203 | miR-10b-5p | trans | 3.15E-35 | Nightingale | XXL_VLDL_ | NA | T | C | 0.030861 | 0.004406 | 1.10E-12 |
| rs495203 | miR-10b-5p | trans | 3.15E-35 | Nightingale | L_LDL_FC | NA | T | C | 0.033632 | 0.004345 | 2.80E-14 |
| rs495203 | miR-10b-5p | trans | 3.15E-35 | Nightingale | S_VLDL_TG | NA | T | C | -0.03813 | 0.004283 | 1.40E-18 |
| rs495203 | miR-195-5p | trans | 2.34E-17 | Nightingale | L_VLDL_CE | NA | T | C | 0.047347 | 0.004234 | 2.40E-29 |
| rs495203 | miR-195-5p | trans | 2.34E-17 | Nightingale | L_VLDL_FC | NA | T | C | 0.048592 | 0.004383 | 4.60E-29 |
| rs495203 | miR-195-5p | trans | 2.34E-17 | Nightingale | XXL_VLDL_ | NA | T | C | -0.03574 | 0.004463 | 2.10E-16 |
| rs495203 | miR-195-5p | trans | 2.34E-17 | Nightingale | IDL_FC | NA | T | C | 0.03826 | 0.004284 | 1.40E-18 |
| rs495203 | miR-195-5p | trans | 2.34E-17 | Nightingale | Clinical_LD | NA | T | C | 0.036481 | 0.004399 | 2.70E-16 |
| rs495203 | miR-195-5p | trans | 2.34E-17 | Nightingale | S_HDL_L | NA | T | C | 0.026311 | 0.00441 | 1.50E-09 |
| rs495203 | miR-195-5p | trans | 2.34E-17 | Nightingale | PUFA | NA | T | C | 0.023739 | 0.004316 | 1.40E-08 |
| rs495203 | miR-195-5p | trans | 2.34E-17 | Nightingale | XL_VLDL_F | NA | T | C | 0.0372 | 0.00429 | 2.00E-18 |
| rs495203 | miR-195-5p | trans | 2.34E-17 | Nightingale | XS_VLDL_L | NA | T | C | 0.033828 | 0.00435 | 2.00E-15 |
| rs495203 | miR-195-5p | trans | 2.34E-17 | Nightingale | S_HDL_C | NA | T | C | 0.028426 | 0.004417 | 1.70E-10 |
| rs495203 | miR-195-5p | trans | 2.34E-17 | Nightingale | S_LDL_C | NA | T | C | 0.03095 | 0.004429 | 5.50E-12 |
| rs495203 | miR-195-5p | trans | 2.34E-17 | Nightingale | VLDL_C | NA | T | C | 0.031307 | 0.004419 | 4.90E-13 |
| rs495203 | miR-195-5p | trans | 2.34E-17 | Nightingale | XS_VLDL_P | NA | T | C | 0.036711 | 0.004366 | 1.40E-17 |
| rs495203 | miR-195-5p | trans | 2.34E-17 | Nightingale | IDL_P | NA | T | C | 0.039988 | 0.004378 | 1.30E-19 |
| rs495203 | miR-195-5p | trans | 2.34E-17 | Nightingale | XL_VLDL_T | NA | T | C | -0.04392 | 0.004289 | 1.70E-24 |

| | | | | | | | | | | | |
|---|---|---|---|---|---|---|---|---|---|---|---|
| rs495203 | miR-195-5p | trans | 2.34E-17 | Nightingale | L_LDL_PL | NA | T | C | 0.037997 | 0.004391 | 9.00E-18 |
| rs495203 | miR-195-5p | trans | 2.34E-17 | Nightingale | M_LDL_C | NA | T | C | 0.033996 | 0.004433 | 2.10E-14 |
| rs495203 | miR-195-5p | trans | 2.34E-17 | Nightingale | S_VLDL_CE | NA | T | C | 0.047029 | 0.004365 | 1.20E-26 |
| rs495203 | miR-195-5p | trans | 2.34E-17 | Nightingale | XS_VLDL_F | NA | T | C | 0.037076 | 0.004348 | 6.50E-18 |
| rs495203 | miR-195-5p | trans | 2.34E-17 | Nightingale | IDL_C | NA | T | C | 0.03496 | 0.004274 | 9.40E-16 |
| rs495203 | miR-195-5p | trans | 2.34E-17 | Nightingale | M_LDL_FC | NA | T | C | 0.034355 | 0.004419 | 1.90E-14 |
| rs495203 | miR-195-5p | trans | 2.34E-17 | Nightingale | ApoB | NA | T | C | 0.033945 | 0.004419 | 2.10E-14 |
| rs495203 | miR-195-5p | trans | 2.34E-17 | Nightingale | VLDL_CE | NA | T | C | 0.035791 | 0.004426 | 3.00E-16 |
| rs495203 | miR-195-5p | trans | 2.34E-17 | Nightingale | M_VLDL_F | NA | T | C | 0.033377 | 0.004424 | 3.10E-14 |
| rs495203 | miR-195-5p | trans | 2.34E-17 | Nightingale | LDL_L | NA | T | C | 0.033314 | 0.00441 | 7.30E-14 |
| rs495203 | miR-195-5p | trans | 2.34E-17 | Nightingale | S_HDL_CE | NA | T | C | 0.028647 | 0.004432 | 1.30E-10 |
| rs495203 | miR-195-5p | trans | 2.34E-17 | Nightingale | S_HDL_P | NA | T | C | 0.030626 | 0.004425 | 5.00E-12 |
| rs495203 | miR-195-5p | trans | 2.34E-17 | Nightingale | M_LDL_TG | NA | T | C | -0.02762 | 0.00437 | 3.20E-10 |
| rs495203 | miR-195-5p | trans | 2.34E-17 | Nightingale | M_LDL_C_ | NA | T | C | 0.031158 | 0.004428 | 2.90E-12 |
| rs495203 | miR-195-5p | trans | 2.34E-17 | Nightingale | S_VLDL_C_ | NA | T | C | 0.041885 | 0.004306 | 5.20E-22 |
| rs495203 | miR-195-5p | trans | 2.34E-17 | Nightingale | XL_VLDL_P | NA | T | C | 0.038836 | 0.004432 | 5.30E-19 |
| rs495203 | miR-195-5p | trans | 2.34E-17 | Nightingale | M_LDL_L | NA | T | C | 0.033041 | 0.004431 | 1.00E-13 |
| rs495203 | miR-195-5p | trans | 2.34E-17 | Nightingale | LDL_P | NA | T | C | 0.031807 | 0.004422 | 1.00E-12 |
| rs495203 | miR-195-5p | trans | 2.34E-17 | Nightingale | LDL_C | NA | T | C | 0.033203 | 0.004406 | 8.80E-14 |
| rs495203 | miR-195-5p | trans | 2.34E-17 | Nightingale | Total_PL | NA | T | C | 0.024944 | 0.004214 | 1.00E-09 |
| rs495203 | miR-195-5p | trans | 2.34E-17 | Nightingale | Total_FC | NA | T | C | 0.033266 | 0.004323 | 6.70E-15 |
| rs495203 | miR-195-5p | trans | 2.34E-17 | Nightingale | Cholines | NA | T | C | 0.022313 | 0.004161 | 4.70E-08 |
| rs495203 | miR-195-5p | trans | 2.34E-17 | Nightingale | S_HDL_FC | NA | T | C | 0.023473 | 0.004344 | 4.50E-08 |
| rs495203 | miR-195-5p | trans | 2.34E-17 | Nightingale | XS_VLDL_F | NA | T | C | 0.031421 | 0.004393 | 2.50E-12 |
| rs495203 | miR-195-5p | trans | 2.34E-17 | Nightingale | S_VLDL_PL | NA | T | C | 0.027738 | 0.004237 | 1.80E-10 |
| rs495203 | miR-195-5p | trans | 2.34E-17 | Nightingale | XXL_VLDL_ | NA | T | C | 0.033132 | 0.004459 | 8.80E-14 |
| rs495203 | miR-195-5p | trans | 2.34E-17 | Nightingale | XS_VLDL_P | NA | T | C | 0.032126 | 0.004363 | 5.30E-14 |
| rs495203 | miR-195-5p | trans | 2.34E-17 | Nightingale | S_VLDL_PL | NA | T | C | 0.032786 | 0.004417 | 4.20E-14 |
| rs495203 | miR-195-5p | trans | 2.34E-17 | Nightingale | XS_VLDL_C | NA | T | C | 0.037999 | 0.004295 | 1.30E-18 |
| rs495203 | miR-195-5p | trans | 2.34E-17 | Nightingale | M_LDL_CE | NA | T | C | 0.032982 | 0.004431 | 9.60E-14 |
| rs495203 | miR-195-5p | trans | 2.34E-17 | Nightingale | LDL_FC | NA | T | C | 0.033639 | 0.004378 | 5.20E-14 |
| rs495203 | miR-195-5p | trans | 2.34E-17 | Nightingale | LA | NA | T | C | 0.024923 | 0.004339 | 3.60E-09 |
| rs495203 | miR-195-5p | trans | 2.34E-17 | Nightingale | Sphingomy | NA | T | C | 0.029029 | 0.004137 | 6.60E-12 |
| rs495203 | miR-195-5p | trans | 2.34E-17 | Nightingale | M_VLDL_C | NA | T | C | 0.040848 | 0.004385 | 2.30E-20 |
| rs495203 | miR-195-5p | trans | 2.34E-17 | Nightingale | S_LDL_PL_ | NA | T | C | -0.03007 | 0.004417 | 2.60E-13 |
| rs495203 | miR-195-5p | trans | 2.34E-17 | Nightingale | S_VLDL_L | NA | T | C | 0.024678 | 0.004382 | 3.70E-09 |
| rs495203 | miR-195-5p | trans | 2.34E-17 | Nightingale | S_HDL_PL | NA | T | C | 0.023799 | 0.004387 | 3.20E-08 |
| rs495203 | miR-195-5p | trans | 2.34E-17 | Nightingale | S_VLDL_P | NA | T | C | 0.023475 | 0.004369 | 1.60E-08 |
| rs495203 | miR-195-5p | trans | 2.34E-17 | Nightingale | XS_VLDL_C | NA | T | C | 0.037803 | 0.00427 | 2.10E-18 |
| rs495203 | miR-195-5p | trans | 2.34E-17 | Nightingale | S_VLDL_C | NA | T | C | 0.03838 | 0.004424 | 1.80E-18 |
| rs495203 | miR-195-5p | trans | 2.34E-17 | Nightingale | non_HDL_( | NA | T | C | 0.035705 | 0.004401 | 5.00E-16 |
| rs495203 | miR-195-5p | trans | 2.34E-17 | Nightingale | Total_CE | NA | T | C | 0.031758 | 0.004268 | 9.50E-14 |
| rs495203 | miR-195-5p | trans | 2.34E-17 | Nightingale | M_VLDL_T | NA | T | C | -0.03856 | 0.004182 | 6.80E-20 |
| rs495203 | miR-195-5p | trans | 2.34E-17 | Nightingale | M_VLDL_C | NA | T | C | 0.038716 | 0.004413 | 1.90E-18 |
| rs495203 | miR-195-5p | trans | 2.34E-17 | Nightingale | M_VLDL_P | NA | T | C | 0.029134 | 0.004418 | 2.00E-11 |
| rs495203 | miR-195-5p | trans | 2.34E-17 | Nightingale | XL_VLDL_C | NA | T | C | 0.037358 | 0.004245 | 3.10E-18 |
| rs495203 | miR-195-5p | trans | 2.34E-17 | Nightingale | L_LDL_L | NA | T | C | 0.03356 | 0.004382 | 3.00E-14 |
| rs495203 | miR-195-5p | trans | 2.34E-17 | Nightingale | XXL_VLDL_ | NA | T | C | 0.025016 | 0.004454 | 1.50E-09 |
| rs495203 | miR-195-5p | trans | 2.34E-17 | Nightingale | M_VLDL_P | NA | T | C | 0.028301 | 0.004414 | 6.30E-11 |
| rs495203 | miR-195-5p | trans | 2.34E-17 | Nightingale | M_LDL_P | NA | T | C | 0.035898 | 0.004437 | 6.80E-16 |
| rs495203 | miR-195-5p | trans | 2.34E-17 | Nightingale | IDL_CE | NA | T | C | 0.033513 | 0.004272 | 1.30E-14 |
| rs495203 | miR-195-5p | trans | 2.34E-17 | Nightingale | S_VLDL_FC | NA | T | C | 0.037912 | 0.004426 | 7.30E-18 |
| rs495203 | miR-195-5p | trans | 2.34E-17 | Nightingale | IDL_FC_pct | NA | T | C | 0.025765 | 0.004391 | 1.10E-08 |
| rs495203 | miR-195-5p | trans | 2.34E-17 | Nightingale | S_VLDL_FC | NA | T | C | 0.029355 | 0.004228 | 1.10E-11 |
| rs495203 | miR-195-5p | trans | 2.34E-17 | Nightingale | GlycA | NA | T | C | 0.028026 | 0.004406 | 1.40E-10 |
| rs495203 | miR-195-5p | trans | 2.34E-17 | Nightingale | M_VLDL_P | NA | T | C | 0.04406 | 0.004247 | 5.50E-25 |
| rs495203 | miR-195-5p | trans | 2.34E-17 | Nightingale | L_LDL_CE | NA | T | C | 0.031578 | 0.004385 | 9.80E-13 |
| rs495203 | miR-195-5p | trans | 2.34E-17 | Nightingale | VLDL_P | NA | T | C | 0.026641 | 0.004401 | 4.40E-10 |
| rs495203 | miR-195-5p | trans | 2.34E-17 | Nightingale | L_LDL_P | NA | T | C | 0.029185 | 0.00441 | 6.10E-11 |
| rs495203 | miR-195-5p | trans | 2.34E-17 | Nightingale | LDL_PL | NA | T | C | 0.034997 | 0.004415 | 4.70E-15 |
| rs495203 | miR-195-5p | trans | 2.34E-17 | Nightingale | S_LDL_CE | NA | T | C | 0.030556 | 0.004427 | 5.80E-12 |
| rs495203 | miR-195-5p | trans | 2.34E-17 | Nightingale | Total_L | NA | T | C | 0.026363 | 0.004353 | 4.50E-10 |
| rs495203 | miR-195-5p | trans | 2.34E-17 | Nightingale | Omega_6 | NA | T | C | 0.02591 | 0.004321 | 5.60E-10 |

| | | | | | | | | | | | |
|---|---|---|---|---|---|---|---|---|---|---|---|
| rs495203 | miR-195-5p | trans | 2.34E-17 | Nightingale | M_VLDL_F | NA | T | C | 0.042068 | 0.00421 | 3.50E-23 |
| rs495203 | miR-195-5p | trans | 2.34E-17 | Nightingale | Total_C | NA | T | C | 0.032385 | 0.004284 | 3.30E-14 |
| rs495203 | miR-195-5p | trans | 2.34E-17 | Nightingale | XXL_VLDL_ | NA | T | C | 0.034842 | 0.004434 | 2.90E-15 |
| rs495203 | miR-195-5p | trans | 2.34E-17 | Nightingale | L_LDL_C | NA | T | C | 0.03233 | 0.004375 | 2.60E-13 |
| rs495203 | miR-195-5p | trans | 2.34E-17 | Nightingale | Remnant_( | NA | T | C | 0.036874 | 0.004396 | 3.70E-17 |
| rs495203 | miR-195-5p | trans | 2.34E-17 | Nightingale | S_LDL_FC | NA | T | C | 0.028569 | 0.004418 | 4.30E-10 |
| rs495203 | miR-195-5p | trans | 2.34E-17 | Nightingale | VLDL_FC | NA | T | C | 0.024164 | 0.004392 | 1.10E-08 |
| rs495203 | miR-195-5p | trans | 2.34E-17 | Nightingale | M_VLDL_C | NA | T | C | 0.033632 | 0.004157 | 1.90E-15 |
| rs495203 | miR-195-5p | trans | 2.34E-17 | Nightingale | ApoB_by_A | NA | T | C | 0.025045 | 0.004358 | 1.30E-08 |
| rs495203 | miR-195-5p | trans | 2.34E-17 | Nightingale | S_LDL_P | NA | T | C | 0.029594 | 0.004429 | 3.20E-11 |
| rs495203 | miR-195-5p | trans | 2.34E-17 | Nightingale | S_LDL_C_p | NA | T | C | 0.045365 | 0.004442 | 5.50E-25 |
| rs495203 | miR-195-5p | trans | 2.34E-17 | Nightingale | LDL_CE | NA | T | C | 0.032564 | 0.004413 | 2.20E-13 |
| rs495203 | miR-195-5p | trans | 2.34E-17 | Nightingale | IDL_L | NA | T | C | 0.035277 | 0.004278 | 4.80E-16 |
| rs495203 | miR-195-5p | trans | 2.34E-17 | Nightingale | M_VLDL_C | NA | T | C | 0.036209 | 0.004168 | 1.20E-17 |
| rs495203 | miR-195-5p | trans | 2.34E-17 | Nightingale | L_VLDL_TG | NA | T | C | -0.05115 | 0.004404 | 5.30E-32 |
| rs495203 | miR-195-5p | trans | 2.34E-17 | Nightingale | Citrate | NA | T | C | 0.026828 | 0.004396 | 7.30E-10 |
| rs495203 | miR-195-5p | trans | 2.34E-17 | Nightingale | S_LDL_CE_ | NA | T | C | 0.03095 | 0.004417 | 4.90E-14 |
| rs495203 | miR-195-5p | trans | 2.34E-17 | Nightingale | IDL_PL | NA | T | C | 0.035351 | 0.00427 | 3.30E-16 |
| rs495203 | miR-195-5p | trans | 2.34E-17 | Nightingale | L_VLDL_C_ | NA | T | C | 0.052202 | 0.004322 | 4.00E-34 |
| rs495203 | miR-195-5p | trans | 2.34E-17 | Nightingale | M_LDL_PL | NA | T | C | 0.032476 | 0.004431 | 2.60E-13 |
| rs495203 | miR-195-5p | trans | 2.34E-17 | Nightingale | XL_VLDL_C | NA | T | C | 0.038228 | 0.004243 | 4.60E-19 |
| rs495203 | miR-195-5p | trans | 2.34E-17 | Nightingale | S_LDL_L | NA | T | C | 0.027749 | 0.004424 | 8.20E-10 |
| rs495203 | miR-195-5p | trans | 2.34E-17 | Nightingale | S_VLDL_CE | NA | T | C | 0.038188 | 0.004419 | 1.60E-18 |
| rs495203 | miR-195-5p | trans | 2.34E-17 | Nightingale | XL_HDL_PL | NA | T | C | -0.02275 | 0.004065 | 1.20E-08 |
| rs495203 | miR-195-5p | trans | 2.34E-17 | Nightingale | XXL_VLDL_ | NA | T | C | 0.030861 | 0.004406 | 1.10E-12 |
| rs495203 | miR-195-5p | trans | 2.34E-17 | Nightingale | L_LDL_FC | NA | T | C | 0.033632 | 0.004345 | 2.80E-14 |
| rs495203 | miR-195-5p | trans | 2.34E-17 | Nightingale | S_VLDL_TG | NA | T | C | -0.03813 | 0.004283 | 1.40E-18 |
| rs495203 | miR-126-3p | trans | 2.48E-39 | Nightingale | L_VLDL_CE | NA | T | C | 0.047347 | 0.004234 | 2.40E-29 |
| rs495203 | miR-126-3p | trans | 2.48E-39 | Nightingale | L_VLDL_FC | NA | T | C | 0.048592 | 0.004383 | 4.60E-29 |
| rs495203 | miR-126-3p | trans | 2.48E-39 | Nightingale | XXL_VLDL_ | NA | T | C | -0.03574 | 0.004463 | 2.10E-16 |
| rs495203 | miR-126-3p | trans | 2.48E-39 | Nightingale | IDL_FC | NA | T | C | 0.03826 | 0.004284 | 1.40E-18 |
| rs495203 | miR-126-3p | trans | 2.48E-39 | Nightingale | Clinical_LD | NA | T | C | 0.036481 | 0.004399 | 2.70E-16 |
| rs495203 | miR-126-3p | trans | 2.48E-39 | Nightingale | S_HDL_L | NA | T | C | 0.026311 | 0.00441 | 1.50E-09 |
| rs495203 | miR-126-3p | trans | 2.48E-39 | Nightingale | PUFA | NA | T | C | 0.023739 | 0.004316 | 1.40E-08 |
| rs495203 | miR-126-3p | trans | 2.48E-39 | Nightingale | XL_VLDL_F | NA | T | C | 0.0372 | 0.00429 | 2.00E-18 |
| rs495203 | miR-126-3p | trans | 2.48E-39 | Nightingale | XS_VLDL_L | NA | T | C | 0.033828 | 0.00435 | 2.00E-15 |
| rs495203 | miR-126-3p | trans | 2.48E-39 | Nightingale | S_HDL_C | NA | T | C | 0.028426 | 0.004417 | 1.70E-10 |
| rs495203 | miR-126-3p | trans | 2.48E-39 | Nightingale | S_LDL_C | NA | T | C | 0.03095 | 0.004429 | 5.50E-12 |
| rs495203 | miR-126-3p | trans | 2.48E-39 | Nightingale | VLDL_C | NA | T | C | 0.031307 | 0.004419 | 4.90E-13 |
| rs495203 | miR-126-3p | trans | 2.48E-39 | Nightingale | XS_VLDL_P | NA | T | C | 0.036711 | 0.004366 | 1.40E-17 |
| rs495203 | miR-126-3p | trans | 2.48E-39 | Nightingale | IDL_P | NA | T | C | 0.039988 | 0.004378 | 1.30E-19 |
| rs495203 | miR-126-3p | trans | 2.48E-39 | Nightingale | XL_VLDL_T | NA | T | C | -0.04392 | 0.004289 | 1.70E-24 |
| rs495203 | miR-126-3p | trans | 2.48E-39 | Nightingale | L_LDL_PL | NA | T | C | 0.037997 | 0.004391 | 9.00E-18 |
| rs495203 | miR-126-3p | trans | 2.48E-39 | Nightingale | M_LDL_C | NA | T | C | 0.033996 | 0.004433 | 2.10E-14 |
| rs495203 | miR-126-3p | trans | 2.48E-39 | Nightingale | S_VLDL_CE | NA | T | C | 0.047029 | 0.004365 | 1.20E-26 |
| rs495203 | miR-126-3p | trans | 2.48E-39 | Nightingale | XS_VLDL_F | NA | T | C | 0.037076 | 0.004348 | 6.50E-18 |
| rs495203 | miR-126-3p | trans | 2.48E-39 | Nightingale | IDL_C | NA | T | C | 0.03496 | 0.004274 | 9.40E-16 |
| rs495203 | miR-126-3p | trans | 2.48E-39 | Nightingale | M_LDL_FC | NA | T | C | 0.034355 | 0.004419 | 1.90E-14 |
| rs495203 | miR-126-3p | trans | 2.48E-39 | Nightingale | ApoB | NA | T | C | 0.033945 | 0.004419 | 2.10E-14 |
| rs495203 | miR-126-3p | trans | 2.48E-39 | Nightingale | VLDL_CE | NA | T | C | 0.035791 | 0.004426 | 3.00E-16 |
| rs495203 | miR-126-3p | trans | 2.48E-39 | Nightingale | M_VLDL_F | NA | T | C | 0.033377 | 0.004424 | 3.10E-14 |
| rs495203 | miR-126-3p | trans | 2.48E-39 | Nightingale | LDL_L | NA | T | C | 0.033314 | 0.00441 | 7.30E-14 |
| rs495203 | miR-126-3p | trans | 2.48E-39 | Nightingale | S_HDL_CE | NA | T | C | 0.028647 | 0.004432 | 1.30E-10 |
| rs495203 | miR-126-3p | trans | 2.48E-39 | Nightingale | S_HDL_P | NA | T | C | 0.030626 | 0.004425 | 5.00E-12 |
| rs495203 | miR-126-3p | trans | 2.48E-39 | Nightingale | M_LDL_TG | NA | T | C | -0.02762 | 0.00437 | 3.20E-10 |
| rs495203 | miR-126-3p | trans | 2.48E-39 | Nightingale | M_LDL_C_ | NA | T | C | 0.031158 | 0.004428 | 2.90E-12 |
| rs495203 | miR-126-3p | trans | 2.48E-39 | Nightingale | S_VLDL_C_ | NA | T | C | 0.041885 | 0.004306 | 5.20E-22 |
| rs495203 | miR-126-3p | trans | 2.48E-39 | Nightingale | XL_VLDL_P | NA | T | C | 0.038836 | 0.004432 | 5.30E-19 |
| rs495203 | miR-126-3p | trans | 2.48E-39 | Nightingale | M_LDL_L | NA | T | C | 0.033041 | 0.004431 | 1.00E-13 |
| rs495203 | miR-126-3p | trans | 2.48E-39 | Nightingale | LDL_P | NA | T | C | 0.031807 | 0.004422 | 1.00E-12 |
| rs495203 | miR-126-3p | trans | 2.48E-39 | Nightingale | LDL_C | NA | T | C | 0.033203 | 0.004406 | 8.80E-14 |
| rs495203 | miR-126-3p | trans | 2.48E-39 | Nightingale | Total_PL | NA | T | C | 0.024944 | 0.004214 | 1.00E-09 |
| rs495203 | miR-126-3p | trans | 2.48E-39 | Nightingale | Total_FC | NA | T | C | 0.033266 | 0.004323 | 6.70E-15 |

| | | | | | | | | | | | |
|---|---|---|---|---|---|---|---|---|---|---|---|
| rs495203 | miR-126-3p | trans | 2.48E-39 | Nightingale | Cholines | NA | T | C | 0.022313 | 0.004161 | 4.70E-08 |
| rs495203 | miR-126-3p | trans | 2.48E-39 | Nightingale | S_HDL_FC | NA | T | C | 0.023473 | 0.004344 | 4.50E-08 |
| rs495203 | miR-126-3p | trans | 2.48E-39 | Nightingale | XS_VLDL_F | NA | T | C | 0.031421 | 0.004393 | 2.50E-12 |
| rs495203 | miR-126-3p | trans | 2.48E-39 | Nightingale | S_VLDL_PL | NA | T | C | 0.027738 | 0.004237 | 1.80E-10 |
| rs495203 | miR-126-3p | trans | 2.48E-39 | Nightingale | XXL_VLDL_ | NA | T | C | 0.033132 | 0.004459 | 8.80E-14 |
| rs495203 | miR-126-3p | trans | 2.48E-39 | Nightingale | XS_VLDL_P | NA | T | C | 0.032126 | 0.004363 | 5.30E-14 |
| rs495203 | miR-126-3p | trans | 2.48E-39 | Nightingale | S_VLDL_PL | NA | T | C | 0.032786 | 0.004417 | 4.20E-14 |
| rs495203 | miR-126-3p | trans | 2.48E-39 | Nightingale | XS_VLDL_C | NA | T | C | 0.037999 | 0.004295 | 1.30E-18 |
| rs495203 | miR-126-3p | trans | 2.48E-39 | Nightingale | M_LDL_CE | NA | T | C | 0.032982 | 0.004431 | 9.60E-14 |
| rs495203 | miR-126-3p | trans | 2.48E-39 | Nightingale | LDL_FC | NA | T | C | 0.033639 | 0.004378 | 5.20E-14 |
| rs495203 | miR-126-3p | trans | 2.48E-39 | Nightingale | LA | NA | T | C | 0.024923 | 0.004339 | 3.60E-09 |
| rs495203 | miR-126-3p | trans | 2.48E-39 | Nightingale | Sphingomy | NA | T | C | 0.029029 | 0.004137 | 6.60E-12 |
| rs495203 | miR-126-3p | trans | 2.48E-39 | Nightingale | M_VLDL_C | NA | T | C | 0.040848 | 0.004385 | 2.30E-20 |
| rs495203 | miR-126-3p | trans | 2.48E-39 | Nightingale | S_LDL_PL_ | NA | T | C | -0.03007 | 0.004417 | 2.60E-13 |
| rs495203 | miR-126-3p | trans | 2.48E-39 | Nightingale | S_VLDL_L | NA | T | C | 0.024678 | 0.004382 | 3.70E-09 |
| rs495203 | miR-126-3p | trans | 2.48E-39 | Nightingale | S_HDL_PL | NA | T | C | 0.023799 | 0.004387 | 3.20E-08 |
| rs495203 | miR-126-3p | trans | 2.48E-39 | Nightingale | S_VLDL_P | NA | T | C | 0.023475 | 0.004369 | 1.60E-08 |
| rs495203 | miR-126-3p | trans | 2.48E-39 | Nightingale | XS_VLDL_C | NA | T | C | 0.037803 | 0.00427 | 2.10E-18 |
| rs495203 | miR-126-3p | trans | 2.48E-39 | Nightingale | S_VLDL_C | NA | T | C | 0.03838 | 0.004424 | 1.80E-18 |
| rs495203 | miR-126-3p | trans | 2.48E-39 | Nightingale | non_HDL_( | NA | T | C | 0.035705 | 0.004401 | 5.00E-16 |
| rs495203 | miR-126-3p | trans | 2.48E-39 | Nightingale | Total_CE | NA | T | C | 0.031758 | 0.004268 | 9.50E-14 |
| rs495203 | miR-126-3p | trans | 2.48E-39 | Nightingale | M_VLDL_T | NA | T | C | -0.03856 | 0.004182 | 6.80E-20 |
| rs495203 | miR-126-3p | trans | 2.48E-39 | Nightingale | M_VLDL_C | NA | T | C | 0.038716 | 0.004413 | 1.90E-18 |
| rs495203 | miR-126-3p | trans | 2.48E-39 | Nightingale | M_VLDL_P | NA | T | C | 0.029134 | 0.004418 | 2.00E-11 |
| rs495203 | miR-126-3p | trans | 2.48E-39 | Nightingale | XL_VLDL_C | NA | T | C | 0.037358 | 0.004245 | 3.10E-18 |
| rs495203 | miR-126-3p | trans | 2.48E-39 | Nightingale | L_LDL_L | NA | T | C | 0.03356 | 0.004382 | 3.00E-14 |
| rs495203 | miR-126-3p | trans | 2.48E-39 | Nightingale | XXL_VLDL_ | NA | T | C | 0.025016 | 0.004454 | 1.50E-09 |
| rs495203 | miR-126-3p | trans | 2.48E-39 | Nightingale | M_VLDL_P | NA | T | C | 0.028301 | 0.004414 | 6.30E-11 |
| rs495203 | miR-126-3p | trans | 2.48E-39 | Nightingale | M_LDL_P | NA | T | C | 0.035898 | 0.004437 | 6.80E-16 |
| rs495203 | miR-126-3p | trans | 2.48E-39 | Nightingale | IDL_CE | NA | T | C | 0.033513 | 0.004272 | 1.30E-14 |
| rs495203 | miR-126-3p | trans | 2.48E-39 | Nightingale | S_VLDL_FC | NA | T | C | 0.037912 | 0.004426 | 7.30E-18 |
| rs495203 | miR-126-3p | trans | 2.48E-39 | Nightingale | IDL_FC_pct | NA | T | C | 0.025765 | 0.004391 | 1.10E-08 |
| rs495203 | miR-126-3p | trans | 2.48E-39 | Nightingale | S_VLDL_FC | NA | T | C | 0.029355 | 0.004228 | 1.10E-11 |
| rs495203 | miR-126-3p | trans | 2.48E-39 | Nightingale | GlycA | NA | T | C | 0.028026 | 0.004406 | 1.40E-10 |
| rs495203 | miR-126-3p | trans | 2.48E-39 | Nightingale | M_VLDL_P | NA | T | C | 0.04406 | 0.004247 | 5.50E-25 |
| rs495203 | miR-126-3p | trans | 2.48E-39 | Nightingale | L_LDL_CE | NA | T | C | 0.031578 | 0.004385 | 9.80E-13 |
| rs495203 | miR-126-3p | trans | 2.48E-39 | Nightingale | VLDL_P | NA | T | C | 0.026641 | 0.004401 | 4.40E-10 |
| rs495203 | miR-126-3p | trans | 2.48E-39 | Nightingale | L_LDL_P | NA | T | C | 0.029185 | 0.00441 | 6.10E-11 |
| rs495203 | miR-126-3p | trans | 2.48E-39 | Nightingale | LDL_PL | NA | T | C | 0.034997 | 0.004415 | 4.70E-15 |
| rs495203 | miR-126-3p | trans | 2.48E-39 | Nightingale | S_LDL_CE | NA | T | C | 0.030556 | 0.004427 | 5.80E-12 |
| rs495203 | miR-126-3p | trans | 2.48E-39 | Nightingale | Total_L | NA | T | C | 0.026363 | 0.004353 | 4.50E-10 |
| rs495203 | miR-126-3p | trans | 2.48E-39 | Nightingale | Omega_6 | NA | T | C | 0.02591 | 0.004321 | 5.60E-10 |
| rs495203 | miR-126-3p | trans | 2.48E-39 | Nightingale | M_VLDL_F | NA | T | C | 0.042068 | 0.00421 | 3.50E-23 |
| rs495203 | miR-126-3p | trans | 2.48E-39 | Nightingale | Total_C | NA | T | C | 0.032385 | 0.004284 | 3.30E-14 |
| rs495203 | miR-126-3p | trans | 2.48E-39 | Nightingale | XXL_VLDL_ | NA | T | C | 0.034842 | 0.004434 | 2.90E-15 |
| rs495203 | miR-126-3p | trans | 2.48E-39 | Nightingale | L_LDL_C | NA | T | C | 0.03233 | 0.004375 | 2.60E-13 |
| rs495203 | miR-126-3p | trans | 2.48E-39 | Nightingale | Remnant_( | NA | T | C | 0.036874 | 0.004396 | 3.70E-17 |
| rs495203 | miR-126-3p | trans | 2.48E-39 | Nightingale | S_LDL_FC | NA | T | C | 0.028569 | 0.004418 | 4.30E-10 |
| rs495203 | miR-126-3p | trans | 2.48E-39 | Nightingale | VLDL_FC | NA | T | C | 0.024164 | 0.004392 | 1.10E-08 |
| rs495203 | miR-126-3p | trans | 2.48E-39 | Nightingale | M_VLDL_C | NA | T | C | 0.033632 | 0.004157 | 1.90E-15 |
| rs495203 | miR-126-3p | trans | 2.48E-39 | Nightingale | ApoB_by_A | NA | T | C | 0.025045 | 0.004358 | 1.30E-08 |
| rs495203 | miR-126-3p | trans | 2.48E-39 | Nightingale | S_LDL_P | NA | T | C | 0.029594 | 0.004429 | 3.20E-11 |
| rs495203 | miR-126-3p | trans | 2.48E-39 | Nightingale | S_LDL_C_p | NA | T | C | 0.045365 | 0.004442 | 5.50E-25 |
| rs495203 | miR-126-3p | trans | 2.48E-39 | Nightingale | LDL_CE | NA | T | C | 0.032564 | 0.004413 | 2.20E-13 |
| rs495203 | miR-126-3p | trans | 2.48E-39 | Nightingale | IDL_L | NA | T | C | 0.035277 | 0.004278 | 4.80E-16 |
| rs495203 | miR-126-3p | trans | 2.48E-39 | Nightingale | M_VLDL_C | NA | T | C | 0.036209 | 0.004168 | 1.20E-17 |
| rs495203 | miR-126-3p | trans | 2.48E-39 | Nightingale | L_VLDL_TG | NA | T | C | -0.05115 | 0.004404 | 5.30E-32 |
| rs495203 | miR-126-3p | trans | 2.48E-39 | Nightingale | Citrate | NA | T | C | 0.026828 | 0.004396 | 7.30E-10 |
| rs495203 | miR-126-3p | trans | 2.48E-39 | Nightingale | S_LDL_CE_ | NA | T | C | 0.03095 | 0.004417 | 4.90E-14 |
| rs495203 | miR-126-3p | trans | 2.48E-39 | Nightingale | IDL_PL | NA | T | C | 0.035351 | 0.00427 | 3.30E-16 |
| rs495203 | miR-126-3p | trans | 2.48E-39 | Nightingale | L_VLDL_C_ | NA | T | C | 0.052202 | 0.004322 | 4.00E-34 |
| rs495203 | miR-126-3p | trans | 2.48E-39 | Nightingale | M_LDL_PL | NA | T | C | 0.032476 | 0.004431 | 2.60E-13 |
| rs495203 | miR-126-3p | trans | 2.48E-39 | Nightingale | XL_VLDL_C | NA | T | C | 0.038228 | 0.004243 | 4.60E-19 |

| | | | | | | | | | | | |
|---|---|---|---|---|---|---|---|---|---|---|---|
| rs495203 | miR-126-3p | trans | 2.48E-39 | Nightingale | S_LDL_L | NA | T | C | 0.027749 | 0.004424 | 8.20E-10 |
| rs495203 | miR-126-3p | trans | 2.48E-39 | Nightingale | S_VLDL_CE | NA | T | C | 0.038188 | 0.004419 | 1.60E-18 |
| rs495203 | miR-126-3p | trans | 2.48E-39 | Nightingale | XL_HDL_PL | NA | T | C | -0.02275 | 0.004065 | 1.20E-08 |
| rs495203 | miR-126-3p | trans | 2.48E-39 | Nightingale | XXL_VLDL_ | NA | T | C | 0.030861 | 0.004406 | 1.10E-12 |
| rs495203 | miR-126-3p | trans | 2.48E-39 | Nightingale | L_LDL_FC | NA | T | C | 0.033632 | 0.004345 | 2.80E-14 |
| rs495203 | miR-126-3p | trans | 2.48E-39 | Nightingale | S_VLDL_TG | NA | T | C | -0.03813 | 0.004283 | 1.40E-18 |
| rs495203 | miR-139-5p | trans | 8.37E-42 | Nightingale | L_VLDL_CE | NA | T | C | 0.047347 | 0.004234 | 2.40E-29 |
| rs495203 | miR-139-5p | trans | 8.37E-42 | Nightingale | L_VLDL_FC | NA | T | C | 0.048592 | 0.004383 | 4.60E-29 |
| rs495203 | miR-139-5p | trans | 8.37E-42 | Nightingale | XXL_VLDL_ | NA | T | C | -0.03574 | 0.004463 | 2.10E-16 |
| rs495203 | miR-139-5p | trans | 8.37E-42 | Nightingale | IDL_FC | NA | T | C | 0.03826 | 0.004284 | 1.40E-18 |
| rs495203 | miR-139-5p | trans | 8.37E-42 | Nightingale | Clinical_LD | NA | T | C | 0.036481 | 0.004399 | 2.70E-16 |
| rs495203 | miR-139-5p | trans | 8.37E-42 | Nightingale | S_HDL_L | NA | T | C | 0.026311 | 0.00441 | 1.50E-09 |
| rs495203 | miR-139-5p | trans | 8.37E-42 | Nightingale | PUFA | NA | T | C | 0.023739 | 0.004316 | 1.40E-08 |
| rs495203 | miR-139-5p | trans | 8.37E-42 | Nightingale | XL_VLDL_F | NA | T | C | 0.0372 | 0.00429 | 2.00E-18 |
| rs495203 | miR-139-5p | trans | 8.37E-42 | Nightingale | XS_VLDL_L | NA | T | C | 0.033828 | 0.00435 | 2.00E-15 |
| rs495203 | miR-139-5p | trans | 8.37E-42 | Nightingale | S_HDL_C | NA | T | C | 0.028426 | 0.004417 | 1.70E-10 |
| rs495203 | miR-139-5p | trans | 8.37E-42 | Nightingale | S_LDL_C | NA | T | C | 0.03095 | 0.004429 | 5.50E-12 |
| rs495203 | miR-139-5p | trans | 8.37E-42 | Nightingale | VLDL_C | NA | T | C | 0.031307 | 0.004419 | 4.90E-13 |
| rs495203 | miR-139-5p | trans | 8.37E-42 | Nightingale | XS_VLDL_P | NA | T | C | 0.036711 | 0.004366 | 1.40E-17 |
| rs495203 | miR-139-5p | trans | 8.37E-42 | Nightingale | IDL_P | NA | T | C | 0.039988 | 0.004378 | 1.30E-19 |
| rs495203 | miR-139-5p | trans | 8.37E-42 | Nightingale | XL_VLDL_T | NA | T | C | -0.04392 | 0.004289 | 1.70E-24 |
| rs495203 | miR-139-5p | trans | 8.37E-42 | Nightingale | L_LDL_PL | NA | T | C | 0.037997 | 0.004391 | 9.00E-18 |
| rs495203 | miR-139-5p | trans | 8.37E-42 | Nightingale | M_LDL_C | NA | T | C | 0.033996 | 0.004433 | 2.10E-14 |
| rs495203 | miR-139-5p | trans | 8.37E-42 | Nightingale | S_VLDL_CE | NA | T | C | 0.047029 | 0.004365 | 1.20E-26 |
| rs495203 | miR-139-5p | trans | 8.37E-42 | Nightingale | XS_VLDL_F | NA | T | C | 0.037076 | 0.004348 | 6.50E-18 |
| rs495203 | miR-139-5p | trans | 8.37E-42 | Nightingale | IDL_C | NA | T | C | 0.03496 | 0.004274 | 9.40E-16 |
| rs495203 | miR-139-5p | trans | 8.37E-42 | Nightingale | M_LDL_FC | NA | T | C | 0.034355 | 0.004419 | 1.90E-14 |
| rs495203 | miR-139-5p | trans | 8.37E-42 | Nightingale | ApoB | NA | T | C | 0.033945 | 0.004419 | 2.10E-14 |
| rs495203 | miR-139-5p | trans | 8.37E-42 | Nightingale | VLDL_CE | NA | T | C | 0.035791 | 0.004426 | 3.00E-16 |
| rs495203 | miR-139-5p | trans | 8.37E-42 | Nightingale | M_VLDL_F | NA | T | C | 0.033377 | 0.004424 | 3.10E-14 |
| rs495203 | miR-139-5p | trans | 8.37E-42 | Nightingale | LDL_L | NA | T | C | 0.033314 | 0.00441 | 7.30E-14 |
| rs495203 | miR-139-5p | trans | 8.37E-42 | Nightingale | S_HDL_CE | NA | T | C | 0.028647 | 0.004432 | 1.30E-10 |
| rs495203 | miR-139-5p | trans | 8.37E-42 | Nightingale | S_HDL_P | NA | T | C | 0.030626 | 0.004425 | 5.00E-12 |
| rs495203 | miR-139-5p | trans | 8.37E-42 | Nightingale | M_LDL_TG | NA | T | C | -0.02762 | 0.00437 | 3.20E-10 |
| rs495203 | miR-139-5p | trans | 8.37E-42 | Nightingale | M_LDL_C_ | NA | T | C | 0.031158 | 0.004428 | 2.90E-12 |
| rs495203 | miR-139-5p | trans | 8.37E-42 | Nightingale | S_VLDL_C_ | NA | T | C | 0.041885 | 0.004306 | 5.20E-22 |
| rs495203 | miR-139-5p | trans | 8.37E-42 | Nightingale | XL_VLDL_P | NA | T | C | 0.038836 | 0.004432 | 5.30E-19 |
| rs495203 | miR-139-5p | trans | 8.37E-42 | Nightingale | M_LDL_L | NA | T | C | 0.033041 | 0.004431 | 1.00E-13 |
| rs495203 | miR-139-5p | trans | 8.37E-42 | Nightingale | LDL_P | NA | T | C | 0.031807 | 0.004422 | 1.00E-12 |
| rs495203 | miR-139-5p | trans | 8.37E-42 | Nightingale | LDL_C | NA | T | C | 0.033203 | 0.004406 | 8.80E-14 |
| rs495203 | miR-139-5p | trans | 8.37E-42 | Nightingale | Total_PL | NA | T | C | 0.024944 | 0.004214 | 1.00E-09 |
| rs495203 | miR-139-5p | trans | 8.37E-42 | Nightingale | Total_FC | NA | T | C | 0.033266 | 0.004323 | 6.70E-15 |
| rs495203 | miR-139-5p | trans | 8.37E-42 | Nightingale | Cholines | NA | T | C | 0.022313 | 0.004161 | 4.70E-08 |
| rs495203 | miR-139-5p | trans | 8.37E-42 | Nightingale | S_HDL_FC | NA | T | C | 0.023473 | 0.004344 | 4.50E-08 |
| rs495203 | miR-139-5p | trans | 8.37E-42 | Nightingale | XS_VLDL_F | NA | T | C | 0.031421 | 0.004393 | 2.50E-12 |
| rs495203 | miR-139-5p | trans | 8.37E-42 | Nightingale | S_VLDL_PL | NA | T | C | 0.027738 | 0.004237 | 1.80E-10 |
| rs495203 | miR-139-5p | trans | 8.37E-42 | Nightingale | XXL_VLDL_ | NA | T | C | 0.033132 | 0.004459 | 8.80E-14 |
| rs495203 | miR-139-5p | trans | 8.37E-42 | Nightingale | XS_VLDL_P | NA | T | C | 0.032126 | 0.004363 | 5.30E-14 |
| rs495203 | miR-139-5p | trans | 8.37E-42 | Nightingale | S_VLDL_PL | NA | T | C | 0.032786 | 0.004417 | 4.20E-14 |
| rs495203 | miR-139-5p | trans | 8.37E-42 | Nightingale | XS_VLDL_C | NA | T | C | 0.037999 | 0.004295 | 1.30E-18 |
| rs495203 | miR-139-5p | trans | 8.37E-42 | Nightingale | M_LDL_CE | NA | T | C | 0.032982 | 0.004431 | 9.60E-14 |
| rs495203 | miR-139-5p | trans | 8.37E-42 | Nightingale | LDL_FC | NA | T | C | 0.033639 | 0.004378 | 5.20E-14 |
| rs495203 | miR-139-5p | trans | 8.37E-42 | Nightingale | LA | NA | T | C | 0.024923 | 0.004339 | 3.60E-09 |
| rs495203 | miR-139-5p | trans | 8.37E-42 | Nightingale | Sphingomy | NA | T | C | 0.029029 | 0.004137 | 6.60E-12 |
| rs495203 | miR-139-5p | trans | 8.37E-42 | Nightingale | M_VLDL_C | NA | T | C | 0.040848 | 0.004385 | 2.30E-20 |
| rs495203 | miR-139-5p | trans | 8.37E-42 | Nightingale | S_LDL_PL_ | NA | T | C | -0.03007 | 0.004417 | 2.60E-13 |
| rs495203 | miR-139-5p | trans | 8.37E-42 | Nightingale | S_VLDL_L | NA | T | C | 0.024678 | 0.004382 | 3.70E-09 |
| rs495203 | miR-139-5p | trans | 8.37E-42 | Nightingale | S_HDL_PL | NA | T | C | 0.023799 | 0.004387 | 3.20E-08 |
| rs495203 | miR-139-5p | trans | 8.37E-42 | Nightingale | S_VLDL_P | NA | T | C | 0.023475 | 0.004369 | 1.60E-08 |
| rs495203 | miR-139-5p | trans | 8.37E-42 | Nightingale | XS_VLDL_C | NA | T | C | 0.037803 | 0.00427 | 2.10E-18 |
| rs495203 | miR-139-5p | trans | 8.37E-42 | Nightingale | S_VLDL_C | NA | T | C | 0.03838 | 0.004424 | 1.80E-18 |
| rs495203 | miR-139-5p | trans | 8.37E-42 | Nightingale | non_HDL_( | NA | T | C | 0.035705 | 0.004401 | 5.00E-16 |
| rs495203 | miR-139-5p | trans | 8.37E-42 | Nightingale | Total_CE | NA | T | C | 0.031758 | 0.004268 | 9.50E-14 |

| | | | | | | | | | | | |
|---|---|---|---|---|---|---|---|---|---|---|---|
| rs495203 | miR-139-5p | trans | 8.37E-42 | Nightingale | M_VLDL_T | NA | T | C | -0.03856 | 0.004182 | 6.80E-20 |
| rs495203 | miR-139-5p | trans | 8.37E-42 | Nightingale | M_VLDL_C | NA | T | C | 0.038716 | 0.004413 | 1.90E-18 |
| rs495203 | miR-139-5p | trans | 8.37E-42 | Nightingale | M_VLDL_P | NA | T | C | 0.029134 | 0.004418 | 2.00E-11 |
| rs495203 | miR-139-5p | trans | 8.37E-42 | Nightingale | XL_VLDL_C | NA | T | C | 0.037358 | 0.004245 | 3.10E-18 |
| rs495203 | miR-139-5p | trans | 8.37E-42 | Nightingale | L_LDL_L | NA | T | C | 0.03356 | 0.004382 | 3.00E-14 |
| rs495203 | miR-139-5p | trans | 8.37E-42 | Nightingale | XXL_VLDL_ | NA | T | C | 0.025016 | 0.004454 | 1.50E-09 |
| rs495203 | miR-139-5p | trans | 8.37E-42 | Nightingale | M_VLDL_P | NA | T | C | 0.028301 | 0.004414 | 6.30E-11 |
| rs495203 | miR-139-5p | trans | 8.37E-42 | Nightingale | M_LDL_P | NA | T | C | 0.035898 | 0.004437 | 6.80E-16 |
| rs495203 | miR-139-5p | trans | 8.37E-42 | Nightingale | IDL_CE | NA | T | C | 0.033513 | 0.004272 | 1.30E-14 |
| rs495203 | miR-139-5p | trans | 8.37E-42 | Nightingale | S_VLDL_FC | NA | T | C | 0.037912 | 0.004426 | 7.30E-18 |
| rs495203 | miR-139-5p | trans | 8.37E-42 | Nightingale | IDL_FC_pct | NA | T | C | 0.025765 | 0.004391 | 1.10E-08 |
| rs495203 | miR-139-5p | trans | 8.37E-42 | Nightingale | S_VLDL_FC | NA | T | C | 0.029355 | 0.004228 | 1.10E-11 |
| rs495203 | miR-139-5p | trans | 8.37E-42 | Nightingale | GlycA | NA | T | C | 0.028026 | 0.004406 | 1.40E-10 |
| rs495203 | miR-139-5p | trans | 8.37E-42 | Nightingale | M_VLDL_P | NA | T | C | 0.04406 | 0.004247 | 5.50E-25 |
| rs495203 | miR-139-5p | trans | 8.37E-42 | Nightingale | L_LDL_CE | NA | T | C | 0.031578 | 0.004385 | 9.80E-13 |
| rs495203 | miR-139-5p | trans | 8.37E-42 | Nightingale | VLDL_P | NA | T | C | 0.026641 | 0.004401 | 4.40E-10 |
| rs495203 | miR-139-5p | trans | 8.37E-42 | Nightingale | L_LDL_P | NA | T | C | 0.029185 | 0.00441 | 6.10E-11 |
| rs495203 | miR-139-5p | trans | 8.37E-42 | Nightingale | LDL_PL | NA | T | C | 0.034997 | 0.004415 | 4.70E-15 |
| rs495203 | miR-139-5p | trans | 8.37E-42 | Nightingale | S_LDL_CE | NA | T | C | 0.030556 | 0.004427 | 5.80E-12 |
| rs495203 | miR-139-5p | trans | 8.37E-42 | Nightingale | Total_L | NA | T | C | 0.026363 | 0.004353 | 4.50E-10 |
| rs495203 | miR-139-5p | trans | 8.37E-42 | Nightingale | Omega_6 | NA | T | C | 0.02591 | 0.004321 | 5.60E-10 |
| rs495203 | miR-139-5p | trans | 8.37E-42 | Nightingale | M_VLDL_F | NA | T | C | 0.042068 | 0.00421 | 3.50E-23 |
| rs495203 | miR-139-5p | trans | 8.37E-42 | Nightingale | Total_C | NA | T | C | 0.032385 | 0.004284 | 3.30E-14 |
| rs495203 | miR-139-5p | trans | 8.37E-42 | Nightingale | XXL_VLDL_ | NA | T | C | 0.034842 | 0.004434 | 2.90E-15 |
| rs495203 | miR-139-5p | trans | 8.37E-42 | Nightingale | L_LDL_C | NA | T | C | 0.03233 | 0.004375 | 2.60E-13 |
| rs495203 | miR-139-5p | trans | 8.37E-42 | Nightingale | Remnant_( | NA | T | C | 0.036874 | 0.004396 | 3.70E-17 |
| rs495203 | miR-139-5p | trans | 8.37E-42 | Nightingale | S_LDL_FC | NA | T | C | 0.028569 | 0.004418 | 4.30E-10 |
| rs495203 | miR-139-5p | trans | 8.37E-42 | Nightingale | VLDL_FC | NA | T | C | 0.024164 | 0.004392 | 1.10E-08 |
| rs495203 | miR-139-5p | trans | 8.37E-42 | Nightingale | M_VLDL_C | NA | T | C | 0.033632 | 0.004157 | 1.90E-15 |
| rs495203 | miR-139-5p | trans | 8.37E-42 | Nightingale | ApoB_by_A | NA | T | C | 0.025045 | 0.004358 | 1.30E-08 |
| rs495203 | miR-139-5p | trans | 8.37E-42 | Nightingale | S_LDL_P | NA | T | C | 0.029594 | 0.004429 | 3.20E-11 |
| rs495203 | miR-139-5p | trans | 8.37E-42 | Nightingale | S_LDL_C_p | NA | T | C | 0.045365 | 0.004442 | 5.50E-25 |
| rs495203 | miR-139-5p | trans | 8.37E-42 | Nightingale | LDL_CE | NA | T | C | 0.032564 | 0.004413 | 2.20E-13 |
| rs495203 | miR-139-5p | trans | 8.37E-42 | Nightingale | IDL_L | NA | T | C | 0.035277 | 0.004278 | 4.80E-16 |
| rs495203 | miR-139-5p | trans | 8.37E-42 | Nightingale | M_VLDL_C | NA | T | C | 0.036209 | 0.004168 | 1.20E-17 |
| rs495203 | miR-139-5p | trans | 8.37E-42 | Nightingale | L_VLDL_TG | NA | T | C | -0.05115 | 0.004404 | 5.30E-32 |
| rs495203 | miR-139-5p | trans | 8.37E-42 | Nightingale | Citrate | NA | T | C | 0.026828 | 0.004396 | 7.30E-10 |
| rs495203 | miR-139-5p | trans | 8.37E-42 | Nightingale | S_LDL_CE_ | NA | T | C | 0.03095 | 0.004417 | 4.90E-14 |
| rs495203 | miR-139-5p | trans | 8.37E-42 | Nightingale | IDL_PL | NA | T | C | 0.035351 | 0.00427 | 3.30E-16 |
| rs495203 | miR-139-5p | trans | 8.37E-42 | Nightingale | L_VLDL_C_ | NA | T | C | 0.052202 | 0.004322 | 4.00E-34 |
| rs495203 | miR-139-5p | trans | 8.37E-42 | Nightingale | M_LDL_PL | NA | T | C | 0.032476 | 0.004431 | 2.60E-13 |
| rs495203 | miR-139-5p | trans | 8.37E-42 | Nightingale | XL_VLDL_C | NA | T | C | 0.038228 | 0.004243 | 4.60E-19 |
| rs495203 | miR-139-5p | trans | 8.37E-42 | Nightingale | S_LDL_L | NA | T | C | 0.027749 | 0.004424 | 8.20E-10 |
| rs495203 | miR-139-5p | trans | 8.37E-42 | Nightingale | S_VLDL_CE | NA | T | C | 0.038188 | 0.004419 | 1.60E-18 |
| rs495203 | miR-139-5p | trans | 8.37E-42 | Nightingale | XL_HDL_PL | NA | T | C | -0.02275 | 0.004065 | 1.20E-08 |
| rs495203 | miR-139-5p | trans | 8.37E-42 | Nightingale | XXL_VLDL_ | NA | T | C | 0.030861 | 0.004406 | 1.10E-12 |
| rs495203 | miR-139-5p | trans | 8.37E-42 | Nightingale | L_LDL_FC | NA | T | C | 0.033632 | 0.004345 | 2.80E-14 |
| rs495203 | miR-139-5p | trans | 8.37E-42 | Nightingale | S_VLDL_TG | NA | T | C | -0.03813 | 0.004283 | 1.40E-18 |
| rs495203 | let-7b-5p | trans | 1.06E-26 | Nightingale | L_VLDL_CE | NA | T | C | 0.047347 | 0.004234 | 2.40E-29 |
| rs495203 | let-7b-5p | trans | 1.06E-26 | Nightingale | L_VLDL_FC | NA | T | C | 0.048592 | 0.004383 | 4.60E-29 |
| rs495203 | let-7b-5p | trans | 1.06E-26 | Nightingale | XXL_VLDL_ | NA | T | C | -0.03574 | 0.004463 | 2.10E-16 |
| rs495203 | let-7b-5p | trans | 1.06E-26 | Nightingale | IDL_FC | NA | T | C | 0.03826 | 0.004284 | 1.40E-18 |
| rs495203 | let-7b-5p | trans | 1.06E-26 | Nightingale | Clinical_LD | NA | T | C | 0.036481 | 0.004399 | 2.70E-16 |
| rs495203 | let-7b-5p | trans | 1.06E-26 | Nightingale | S_HDL_L | NA | T | C | 0.026311 | 0.00441 | 1.50E-09 |
| rs495203 | let-7b-5p | trans | 1.06E-26 | Nightingale | PUFA | NA | T | C | 0.023739 | 0.004316 | 1.40E-08 |
| rs495203 | let-7b-5p | trans | 1.06E-26 | Nightingale | XL_VLDL_F | NA | T | C | 0.0372 | 0.00429 | 2.00E-18 |
| rs495203 | let-7b-5p | trans | 1.06E-26 | Nightingale | XS_VLDL_L | NA | T | C | 0.033828 | 0.00435 | 2.00E-15 |
| rs495203 | let-7b-5p | trans | 1.06E-26 | Nightingale | S_HDL_C | NA | T | C | 0.028426 | 0.004417 | 1.70E-10 |
| rs495203 | let-7b-5p | trans | 1.06E-26 | Nightingale | S_LDL_C | NA | T | C | 0.03095 | 0.004429 | 5.50E-12 |
| rs495203 | let-7b-5p | trans | 1.06E-26 | Nightingale | VLDL_C | NA | T | C | 0.031307 | 0.004419 | 4.90E-13 |
| rs495203 | let-7b-5p | trans | 1.06E-26 | Nightingale | XS_VLDL_P | NA | T | C | 0.036711 | 0.004366 | 1.40E-17 |
| rs495203 | let-7b-5p | trans | 1.06E-26 | Nightingale | IDL_P | NA | T | C | 0.039988 | 0.004378 | 1.30E-19 |
| rs495203 | let-7b-5p | trans | 1.06E-26 | Nightingale | XL_VLDL_T | NA | T | C | -0.04392 | 0.004289 | 1.70E-24 |

| | | | | | | | | | | | |
|---|---|---|---|---|---|---|---|---|---|---|---|
| rs495203 | let-7b-5p | trans | 1.06E-26 | Nightingale | L_LDL_PL | NA | T | C | 0.037997 | 0.004391 | 9.00E-18 |
| rs495203 | let-7b-5p | trans | 1.06E-26 | Nightingale | M_LDL_C | NA | T | C | 0.033996 | 0.004433 | 2.10E-14 |
| rs495203 | let-7b-5p | trans | 1.06E-26 | Nightingale | S_VLDL_CE | NA | T | C | 0.047029 | 0.004365 | 1.20E-26 |
| rs495203 | let-7b-5p | trans | 1.06E-26 | Nightingale | XS_VLDL_F | NA | T | C | 0.037076 | 0.004348 | 6.50E-18 |
| rs495203 | let-7b-5p | trans | 1.06E-26 | Nightingale | IDL_C | NA | T | C | 0.03496 | 0.004274 | 9.40E-16 |
| rs495203 | let-7b-5p | trans | 1.06E-26 | Nightingale | M_LDL_FC | NA | T | C | 0.034355 | 0.004419 | 1.90E-14 |
| rs495203 | let-7b-5p | trans | 1.06E-26 | Nightingale | ApoB | NA | T | C | 0.033945 | 0.004419 | 2.10E-14 |
| rs495203 | let-7b-5p | trans | 1.06E-26 | Nightingale | VLDL_CE | NA | T | C | 0.035791 | 0.004426 | 3.00E-16 |
| rs495203 | let-7b-5p | trans | 1.06E-26 | Nightingale | M_VLDL_F | NA | T | C | 0.033377 | 0.004424 | 3.10E-14 |
| rs495203 | let-7b-5p | trans | 1.06E-26 | Nightingale | LDL_L | NA | T | C | 0.033314 | 0.00441 | 7.30E-14 |
| rs495203 | let-7b-5p | trans | 1.06E-26 | Nightingale | S_HDL_CE | NA | T | C | 0.028647 | 0.004432 | 1.30E-10 |
| rs495203 | let-7b-5p | trans | 1.06E-26 | Nightingale | S_HDL_P | NA | T | C | 0.030626 | 0.004425 | 5.00E-12 |
| rs495203 | let-7b-5p | trans | 1.06E-26 | Nightingale | M_LDL_TG | NA | T | C | -0.02762 | 0.00437 | 3.20E-10 |
| rs495203 | let-7b-5p | trans | 1.06E-26 | Nightingale | M_LDL_C_ | NA | T | C | 0.031158 | 0.004428 | 2.90E-12 |
| rs495203 | let-7b-5p | trans | 1.06E-26 | Nightingale | S_VLDL_C_ | NA | T | C | 0.041885 | 0.004306 | 5.20E-22 |
| rs495203 | let-7b-5p | trans | 1.06E-26 | Nightingale | XL_VLDL_P | NA | T | C | 0.038836 | 0.004432 | 5.30E-19 |
| rs495203 | let-7b-5p | trans | 1.06E-26 | Nightingale | M_LDL_L | NA | T | C | 0.033041 | 0.004431 | 1.00E-13 |
| rs495203 | let-7b-5p | trans | 1.06E-26 | Nightingale | LDL_P | NA | T | C | 0.031807 | 0.004422 | 1.00E-12 |
| rs495203 | let-7b-5p | trans | 1.06E-26 | Nightingale | LDL_C | NA | T | C | 0.033203 | 0.004406 | 8.80E-14 |
| rs495203 | let-7b-5p | trans | 1.06E-26 | Nightingale | Total_PL | NA | T | C | 0.024944 | 0.004214 | 1.00E-09 |
| rs495203 | let-7b-5p | trans | 1.06E-26 | Nightingale | Total_FC | NA | T | C | 0.033266 | 0.004323 | 6.70E-15 |
| rs495203 | let-7b-5p | trans | 1.06E-26 | Nightingale | Cholines | NA | T | C | 0.022313 | 0.004161 | 4.70E-08 |
| rs495203 | let-7b-5p | trans | 1.06E-26 | Nightingale | S_HDL_FC | NA | T | C | 0.023473 | 0.004344 | 4.50E-08 |
| rs495203 | let-7b-5p | trans | 1.06E-26 | Nightingale | XS_VLDL_F | NA | T | C | 0.031421 | 0.004393 | 2.50E-12 |
| rs495203 | let-7b-5p | trans | 1.06E-26 | Nightingale | S_VLDL_PL | NA | T | C | 0.027738 | 0.004237 | 1.80E-10 |
| rs495203 | let-7b-5p | trans | 1.06E-26 | Nightingale | XXL_VLDL_ | NA | T | C | 0.033132 | 0.004459 | 8.80E-14 |
| rs495203 | let-7b-5p | trans | 1.06E-26 | Nightingale | XS_VLDL_P | NA | T | C | 0.032126 | 0.004363 | 5.30E-14 |
| rs495203 | let-7b-5p | trans | 1.06E-26 | Nightingale | S_VLDL_PL | NA | T | C | 0.032786 | 0.004417 | 4.20E-14 |
| rs495203 | let-7b-5p | trans | 1.06E-26 | Nightingale | XS_VLDL_C | NA | T | C | 0.037999 | 0.004295 | 1.30E-18 |
| rs495203 | let-7b-5p | trans | 1.06E-26 | Nightingale | M_LDL_CE | NA | T | C | 0.032982 | 0.004431 | 9.60E-14 |
| rs495203 | let-7b-5p | trans | 1.06E-26 | Nightingale | LDL_FC | NA | T | C | 0.033639 | 0.004378 | 5.20E-14 |
| rs495203 | let-7b-5p | trans | 1.06E-26 | Nightingale | LA | NA | T | C | 0.024923 | 0.004339 | 3.60E-09 |
| rs495203 | let-7b-5p | trans | 1.06E-26 | Nightingale | Sphingomy | NA | T | C | 0.029029 | 0.004137 | 6.60E-12 |
| rs495203 | let-7b-5p | trans | 1.06E-26 | Nightingale | M_VLDL_C | NA | T | C | 0.040848 | 0.004385 | 2.30E-20 |
| rs495203 | let-7b-5p | trans | 1.06E-26 | Nightingale | S_LDL_PL_ | NA | T | C | -0.03007 | 0.004417 | 2.60E-13 |
| rs495203 | let-7b-5p | trans | 1.06E-26 | Nightingale | S_VLDL_L | NA | T | C | 0.024678 | 0.004382 | 3.70E-09 |
| rs495203 | let-7b-5p | trans | 1.06E-26 | Nightingale | S_HDL_PL | NA | T | C | 0.023799 | 0.004387 | 3.20E-08 |
| rs495203 | let-7b-5p | trans | 1.06E-26 | Nightingale | S_VLDL_P | NA | T | C | 0.023475 | 0.004369 | 1.60E-08 |
| rs495203 | let-7b-5p | trans | 1.06E-26 | Nightingale | XS_VLDL_C | NA | T | C | 0.037803 | 0.00427 | 2.10E-18 |
| rs495203 | let-7b-5p | trans | 1.06E-26 | Nightingale | S_VLDL_C | NA | T | C | 0.03838 | 0.004424 | 1.80E-18 |
| rs495203 | let-7b-5p | trans | 1.06E-26 | Nightingale | non_HDL_( | NA | T | C | 0.035705 | 0.004401 | 5.00E-16 |
| rs495203 | let-7b-5p | trans | 1.06E-26 | Nightingale | Total_CE | NA | T | C | 0.031758 | 0.004268 | 9.50E-14 |
| rs495203 | let-7b-5p | trans | 1.06E-26 | Nightingale | M_VLDL_T | NA | T | C | -0.03856 | 0.004182 | 6.80E-20 |
| rs495203 | let-7b-5p | trans | 1.06E-26 | Nightingale | M_VLDL_C | NA | T | C | 0.038716 | 0.004413 | 1.90E-18 |
| rs495203 | let-7b-5p | trans | 1.06E-26 | Nightingale | M_VLDL_P | NA | T | C | 0.029134 | 0.004418 | 2.00E-11 |
| rs495203 | let-7b-5p | trans | 1.06E-26 | Nightingale | XL_VLDL_C | NA | T | C | 0.037358 | 0.004245 | 3.10E-18 |
| rs495203 | let-7b-5p | trans | 1.06E-26 | Nightingale | L_LDL_L | NA | T | C | 0.03356 | 0.004382 | 3.00E-14 |
| rs495203 | let-7b-5p | trans | 1.06E-26 | Nightingale | XXL_VLDL_ | NA | T | C | 0.025016 | 0.004454 | 1.50E-09 |
| rs495203 | let-7b-5p | trans | 1.06E-26 | Nightingale | M_VLDL_P | NA | T | C | 0.028301 | 0.004414 | 6.30E-11 |
| rs495203 | let-7b-5p | trans | 1.06E-26 | Nightingale | M_LDL_P | NA | T | C | 0.035898 | 0.004437 | 6.80E-16 |
| rs495203 | let-7b-5p | trans | 1.06E-26 | Nightingale | IDL_CE | NA | T | C | 0.033513 | 0.004272 | 1.30E-14 |
| rs495203 | let-7b-5p | trans | 1.06E-26 | Nightingale | S_VLDL_FC | NA | T | C | 0.037912 | 0.004426 | 7.30E-18 |
| rs495203 | let-7b-5p | trans | 1.06E-26 | Nightingale | IDL_FC_pct | NA | T | C | 0.025765 | 0.004391 | 1.10E-08 |
| rs495203 | let-7b-5p | trans | 1.06E-26 | Nightingale | S_VLDL_FC | NA | T | C | 0.029355 | 0.004228 | 1.10E-11 |
| rs495203 | let-7b-5p | trans | 1.06E-26 | Nightingale | GlycA | NA | T | C | 0.028026 | 0.004406 | 1.40E-10 |
| rs495203 | let-7b-5p | trans | 1.06E-26 | Nightingale | M_VLDL_P | NA | T | C | 0.04406 | 0.004247 | 5.50E-25 |
| rs495203 | let-7b-5p | trans | 1.06E-26 | Nightingale | L_LDL_CE | NA | T | C | 0.031578 | 0.004385 | 9.80E-13 |
| rs495203 | let-7b-5p | trans | 1.06E-26 | Nightingale | VLDL_P | NA | T | C | 0.026641 | 0.004401 | 4.40E-10 |
| rs495203 | let-7b-5p | trans | 1.06E-26 | Nightingale | L_LDL_P | NA | T | C | 0.029185 | 0.00441 | 6.10E-11 |
| rs495203 | let-7b-5p | trans | 1.06E-26 | Nightingale | LDL_PL | NA | T | C | 0.034997 | 0.004415 | 4.70E-15 |
| rs495203 | let-7b-5p | trans | 1.06E-26 | Nightingale | S_LDL_CE | NA | T | C | 0.030556 | 0.004427 | 5.80E-12 |
| rs495203 | let-7b-5p | trans | 1.06E-26 | Nightingale | Total_L | NA | T | C | 0.026363 | 0.004353 | 4.50E-10 |
| rs495203 | let-7b-5p | trans | 1.06E-26 | Nightingale | Omega_6 | NA | T | C | 0.02591 | 0.004321 | 5.60E-10 |

| | | | | | | | | | | | |
|---|---|---|---|---|---|---|---|---|---|---|---|
| rs495203 | let-7b-5p | trans | 1.06E-26 | Nightingale | M_VLDL_F | NA | T | C | 0.042068 | 0.00421 | 3.50E-23 |
| rs495203 | let-7b-5p | trans | 1.06E-26 | Nightingale | Total_C | NA | T | C | 0.032385 | 0.004284 | 3.30E-14 |
| rs495203 | let-7b-5p | trans | 1.06E-26 | Nightingale | XXL_VLDL_ | NA | T | C | 0.034842 | 0.004434 | 2.90E-15 |
| rs495203 | let-7b-5p | trans | 1.06E-26 | Nightingale | L_LDL_C | NA | T | C | 0.03233 | 0.004375 | 2.60E-13 |
| rs495203 | let-7b-5p | trans | 1.06E-26 | Nightingale | Remnant_( | NA | T | C | 0.036874 | 0.004396 | 3.70E-17 |
| rs495203 | let-7b-5p | trans | 1.06E-26 | Nightingale | S_LDL_FC | NA | T | C | 0.028569 | 0.004418 | 4.30E-10 |
| rs495203 | let-7b-5p | trans | 1.06E-26 | Nightingale | VLDL_FC | NA | T | C | 0.024164 | 0.004392 | 1.10E-08 |
| rs495203 | let-7b-5p | trans | 1.06E-26 | Nightingale | M_VLDL_C | NA | T | C | 0.033632 | 0.004157 | 1.90E-15 |
| rs495203 | let-7b-5p | trans | 1.06E-26 | Nightingale | ApoB_by_/ | NA | T | C | 0.025045 | 0.004358 | 1.30E-08 |
| rs495203 | let-7b-5p | trans | 1.06E-26 | Nightingale | S_LDL_P | NA | T | C | 0.029594 | 0.004429 | 3.20E-11 |
| rs495203 | let-7b-5p | trans | 1.06E-26 | Nightingale | S_LDL_C_p | NA | T | C | 0.045365 | 0.004442 | 5.50E-25 |
| rs495203 | let-7b-5p | trans | 1.06E-26 | Nightingale | LDL_CE | NA | T | C | 0.032564 | 0.004413 | 2.20E-13 |
| rs495203 | let-7b-5p | trans | 1.06E-26 | Nightingale | IDL_L | NA | T | C | 0.035277 | 0.004278 | 4.80E-16 |
| rs495203 | let-7b-5p | trans | 1.06E-26 | Nightingale | M_VLDL_C | NA | T | C | 0.036209 | 0.004168 | 1.20E-17 |
| rs495203 | let-7b-5p | trans | 1.06E-26 | Nightingale | L_VLDL_TG | NA | T | C | -0.05115 | 0.004404 | 5.30E-32 |
| rs495203 | let-7b-5p | trans | 1.06E-26 | Nightingale | Citrate | NA | T | C | 0.026828 | 0.004396 | 7.30E-10 |
| rs495203 | let-7b-5p | trans | 1.06E-26 | Nightingale | S_LDL_CE_ | NA | T | C | 0.03095 | 0.004417 | 4.90E-14 |
| rs495203 | let-7b-5p | trans | 1.06E-26 | Nightingale | IDL_PL | NA | T | C | 0.035351 | 0.00427 | 3.30E-16 |
| rs495203 | let-7b-5p | trans | 1.06E-26 | Nightingale | L_VLDL_C_ | NA | T | C | 0.052202 | 0.004322 | 4.00E-34 |
| rs495203 | let-7b-5p | trans | 1.06E-26 | Nightingale | M_LDL_PL | NA | T | C | 0.032476 | 0.004431 | 2.60E-13 |
| rs495203 | let-7b-5p | trans | 1.06E-26 | Nightingale | XL_VLDL_C | NA | T | C | 0.038228 | 0.004243 | 4.60E-19 |
| rs495203 | let-7b-5p | trans | 1.06E-26 | Nightingale | S_LDL_L | NA | T | C | 0.027749 | 0.004424 | 8.20E-10 |
| rs495203 | let-7b-5p | trans | 1.06E-26 | Nightingale | S_VLDL_CE | NA | T | C | 0.038188 | 0.004419 | 1.60E-18 |
| rs495203 | let-7b-5p | trans | 1.06E-26 | Nightingale | XL_HDL_PL | NA | T | C | -0.02275 | 0.004065 | 1.20E-08 |
| rs495203 | let-7b-5p | trans | 1.06E-26 | Nightingale | XXL_VLDL_ | NA | T | C | 0.030861 | 0.004406 | 1.10E-12 |
| rs495203 | let-7b-5p | trans | 1.06E-26 | Nightingale | L_LDL_FC | NA | T | C | 0.033632 | 0.004345 | 2.80E-14 |
| rs495203 | let-7b-5p | trans | 1.06E-26 | Nightingale | S_VLDL_TG | NA | T | C | -0.03813 | 0.004283 | 1.40E-18 |
| rs495203 | let-7c-5p | trans | 2.73E-23 | Nightingale | L_VLDL_CE | NA | T | C | 0.047347 | 0.004234 | 2.40E-29 |
| rs495203 | let-7c-5p | trans | 2.73E-23 | Nightingale | L_VLDL_FC | NA | T | C | 0.048592 | 0.004383 | 4.60E-29 |
| rs495203 | let-7c-5p | trans | 2.73E-23 | Nightingale | XXL_VLDL_ | NA | T | C | -0.03574 | 0.004463 | 2.10E-16 |
| rs495203 | let-7c-5p | trans | 2.73E-23 | Nightingale | IDL_FC | NA | T | C | 0.03826 | 0.004284 | 1.40E-18 |
| rs495203 | let-7c-5p | trans | 2.73E-23 | Nightingale | Clinical_LD | NA | T | C | 0.036481 | 0.004399 | 2.70E-16 |
| rs495203 | let-7c-5p | trans | 2.73E-23 | Nightingale | S_HDL_L | NA | T | C | 0.026311 | 0.00441 | 1.50E-09 |
| rs495203 | let-7c-5p | trans | 2.73E-23 | Nightingale | PUFA | NA | T | C | 0.023739 | 0.004316 | 1.40E-08 |
| rs495203 | let-7c-5p | trans | 2.73E-23 | Nightingale | XL_VLDL_F | NA | T | C | 0.0372 | 0.00429 | 2.00E-18 |
| rs495203 | let-7c-5p | trans | 2.73E-23 | Nightingale | XS_VLDL_L | NA | T | C | 0.033828 | 0.00435 | 2.00E-15 |
| rs495203 | let-7c-5p | trans | 2.73E-23 | Nightingale | S_HDL_C | NA | T | C | 0.028426 | 0.004417 | 1.70E-10 |
| rs495203 | let-7c-5p | trans | 2.73E-23 | Nightingale | S_LDL_C | NA | T | C | 0.03095 | 0.004429 | 5.50E-12 |
| rs495203 | let-7c-5p | trans | 2.73E-23 | Nightingale | VLDL_C | NA | T | C | 0.031307 | 0.004419 | 4.90E-13 |
| rs495203 | let-7c-5p | trans | 2.73E-23 | Nightingale | XS_VLDL_P | NA | T | C | 0.036711 | 0.004366 | 1.40E-17 |
| rs495203 | let-7c-5p | trans | 2.73E-23 | Nightingale | IDL_P | NA | T | C | 0.039988 | 0.004378 | 1.30E-19 |
| rs495203 | let-7c-5p | trans | 2.73E-23 | Nightingale | XL_VLDL_T | NA | T | C | -0.04392 | 0.004289 | 1.70E-24 |
| rs495203 | let-7c-5p | trans | 2.73E-23 | Nightingale | L_LDL_PL | NA | T | C | 0.037997 | 0.004391 | 9.00E-18 |
| rs495203 | let-7c-5p | trans | 2.73E-23 | Nightingale | M_LDL_C | NA | T | C | 0.033996 | 0.004433 | 2.10E-14 |
| rs495203 | let-7c-5p | trans | 2.73E-23 | Nightingale | S_VLDL_CE | NA | T | C | 0.047029 | 0.004365 | 1.20E-26 |
| rs495203 | let-7c-5p | trans | 2.73E-23 | Nightingale | XS_VLDL_F | NA | T | C | 0.037076 | 0.004348 | 6.50E-18 |
| rs495203 | let-7c-5p | trans | 2.73E-23 | Nightingale | IDL_C | NA | T | C | 0.03496 | 0.004274 | 9.40E-16 |
| rs495203 | let-7c-5p | trans | 2.73E-23 | Nightingale | M_LDL_FC | NA | T | C | 0.034355 | 0.004419 | 1.90E-14 |
| rs495203 | let-7c-5p | trans | 2.73E-23 | Nightingale | ApoB | NA | T | C | 0.033945 | 0.004419 | 2.10E-14 |
| rs495203 | let-7c-5p | trans | 2.73E-23 | Nightingale | VLDL_CE | NA | T | C | 0.035791 | 0.004426 | 3.00E-16 |
| rs495203 | let-7c-5p | trans | 2.73E-23 | Nightingale | M_VLDL_F | NA | T | C | 0.033377 | 0.004424 | 3.10E-14 |
| rs495203 | let-7c-5p | trans | 2.73E-23 | Nightingale | LDL_L | NA | T | C | 0.033314 | 0.00441 | 7.30E-14 |
| rs495203 | let-7c-5p | trans | 2.73E-23 | Nightingale | S_HDL_CE | NA | T | C | 0.028647 | 0.004432 | 1.30E-10 |
| rs495203 | let-7c-5p | trans | 2.73E-23 | Nightingale | S_HDL_P | NA | T | C | 0.030626 | 0.004425 | 5.00E-12 |
| rs495203 | let-7c-5p | trans | 2.73E-23 | Nightingale | M_LDL_TG | NA | T | C | -0.02762 | 0.00437 | 3.20E-10 |
| rs495203 | let-7c-5p | trans | 2.73E-23 | Nightingale | M_LDL_C_ | NA | T | C | 0.031158 | 0.004428 | 2.90E-12 |
| rs495203 | let-7c-5p | trans | 2.73E-23 | Nightingale | S_VLDL_C_ | NA | T | C | 0.041885 | 0.004306 | 5.20E-22 |
| rs495203 | let-7c-5p | trans | 2.73E-23 | Nightingale | XL_VLDL_P | NA | T | C | 0.038836 | 0.004432 | 5.30E-19 |
| rs495203 | let-7c-5p | trans | 2.73E-23 | Nightingale | M_LDL_L | NA | T | C | 0.033041 | 0.004431 | 1.00E-13 |
| rs495203 | let-7c-5p | trans | 2.73E-23 | Nightingale | LDL_P | NA | T | C | 0.031807 | 0.004422 | 1.00E-12 |
| rs495203 | let-7c-5p | trans | 2.73E-23 | Nightingale | LDL_C | NA | T | C | 0.033203 | 0.004406 | 8.80E-14 |
| rs495203 | let-7c-5p | trans | 2.73E-23 | Nightingale | Total_PL | NA | T | C | 0.024944 | 0.004214 | 1.00E-09 |
| rs495203 | let-7c-5p | trans | 2.73E-23 | Nightingale | Total_FC | NA | T | C | 0.033266 | 0.004323 | 6.70E-15 |

| | | | | | | | | | | | |
|---|---|---|---|---|---|---|---|---|---|---|---|
| rs495203 | let-7c-5p | trans | 2.73E-23 | Nightingale | Cholines | NA | T | C | 0.022313 | 0.004161 | 4.70E-08 |
| rs495203 | let-7c-5p | trans | 2.73E-23 | Nightingale | S_HDL_FC | NA | T | C | 0.023473 | 0.004344 | 4.50E-08 |
| rs495203 | let-7c-5p | trans | 2.73E-23 | Nightingale | XS_VLDL_F | NA | T | C | 0.031421 | 0.004393 | 2.50E-12 |
| rs495203 | let-7c-5p | trans | 2.73E-23 | Nightingale | S_VLDL_PL | NA | T | C | 0.027738 | 0.004237 | 1.80E-10 |
| rs495203 | let-7c-5p | trans | 2.73E-23 | Nightingale | XXL_VLDL_ | NA | T | C | 0.033132 | 0.004459 | 8.80E-14 |
| rs495203 | let-7c-5p | trans | 2.73E-23 | Nightingale | XS_VLDL_P | NA | T | C | 0.032126 | 0.004363 | 5.30E-14 |
| rs495203 | let-7c-5p | trans | 2.73E-23 | Nightingale | S_VLDL_PL | NA | T | C | 0.032786 | 0.004417 | 4.20E-14 |
| rs495203 | let-7c-5p | trans | 2.73E-23 | Nightingale | XS_VLDL_C | NA | T | C | 0.037999 | 0.004295 | 1.30E-18 |
| rs495203 | let-7c-5p | trans | 2.73E-23 | Nightingale | M_LDL_CE | NA | T | C | 0.032982 | 0.004431 | 9.60E-14 |
| rs495203 | let-7c-5p | trans | 2.73E-23 | Nightingale | LDL_FC | NA | T | C | 0.033639 | 0.004378 | 5.20E-14 |
| rs495203 | let-7c-5p | trans | 2.73E-23 | Nightingale | LA | NA | T | C | 0.024923 | 0.004339 | 3.60E-09 |
| rs495203 | let-7c-5p | trans | 2.73E-23 | Nightingale | Sphingomy | NA | T | C | 0.029029 | 0.004137 | 6.60E-12 |
| rs495203 | let-7c-5p | trans | 2.73E-23 | Nightingale | M_VLDL_C | NA | T | C | 0.040848 | 0.004385 | 2.30E-20 |
| rs495203 | let-7c-5p | trans | 2.73E-23 | Nightingale | S_LDL_PL_ | NA | T | C | -0.03007 | 0.004417 | 2.60E-13 |
| rs495203 | let-7c-5p | trans | 2.73E-23 | Nightingale | S_VLDL_L | NA | T | C | 0.024678 | 0.004382 | 3.70E-09 |
| rs495203 | let-7c-5p | trans | 2.73E-23 | Nightingale | S_HDL_PL | NA | T | C | 0.023799 | 0.004387 | 3.20E-08 |
| rs495203 | let-7c-5p | trans | 2.73E-23 | Nightingale | S_VLDL_P | NA | T | C | 0.023475 | 0.004369 | 1.60E-08 |
| rs495203 | let-7c-5p | trans | 2.73E-23 | Nightingale | XS_VLDL_C | NA | T | C | 0.037803 | 0.00427 | 2.10E-18 |
| rs495203 | let-7c-5p | trans | 2.73E-23 | Nightingale | S_VLDL_C | NA | T | C | 0.03838 | 0.004424 | 1.80E-18 |
| rs495203 | let-7c-5p | trans | 2.73E-23 | Nightingale | non_HDL_( | NA | T | C | 0.035705 | 0.004401 | 5.00E-16 |
| rs495203 | let-7c-5p | trans | 2.73E-23 | Nightingale | Total_CE | NA | T | C | 0.031758 | 0.004268 | 9.50E-14 |
| rs495203 | let-7c-5p | trans | 2.73E-23 | Nightingale | M_VLDL_T | NA | T | C | -0.03856 | 0.004182 | 6.80E-20 |
| rs495203 | let-7c-5p | trans | 2.73E-23 | Nightingale | M_VLDL_C | NA | T | C | 0.038716 | 0.004413 | 1.90E-18 |
| rs495203 | let-7c-5p | trans | 2.73E-23 | Nightingale | M_VLDL_P | NA | T | C | 0.029134 | 0.004418 | 2.00E-11 |
| rs495203 | let-7c-5p | trans | 2.73E-23 | Nightingale | XL_VLDL_C | NA | T | C | 0.037358 | 0.004245 | 3.10E-18 |
| rs495203 | let-7c-5p | trans | 2.73E-23 | Nightingale | L_LDL_L | NA | T | C | 0.03356 | 0.004382 | 3.00E-14 |
| rs495203 | let-7c-5p | trans | 2.73E-23 | Nightingale | XXL_VLDL_ | NA | T | C | 0.025016 | 0.004454 | 1.50E-09 |
| rs495203 | let-7c-5p | trans | 2.73E-23 | Nightingale | M_VLDL_P | NA | T | C | 0.028301 | 0.004414 | 6.30E-11 |
| rs495203 | let-7c-5p | trans | 2.73E-23 | Nightingale | M_LDL_P | NA | T | C | 0.035898 | 0.004437 | 6.80E-16 |
| rs495203 | let-7c-5p | trans | 2.73E-23 | Nightingale | IDL_CE | NA | T | C | 0.033513 | 0.004272 | 1.30E-14 |
| rs495203 | let-7c-5p | trans | 2.73E-23 | Nightingale | S_VLDL_FC | NA | T | C | 0.037912 | 0.004426 | 7.30E-18 |
| rs495203 | let-7c-5p | trans | 2.73E-23 | Nightingale | IDL_FC_pct | NA | T | C | 0.025765 | 0.004391 | 1.10E-08 |
| rs495203 | let-7c-5p | trans | 2.73E-23 | Nightingale | S_VLDL_FC | NA | T | C | 0.029355 | 0.004228 | 1.10E-11 |
| rs495203 | let-7c-5p | trans | 2.73E-23 | Nightingale | GlycA | NA | T | C | 0.028026 | 0.004406 | 1.40E-10 |
| rs495203 | let-7c-5p | trans | 2.73E-23 | Nightingale | M_VLDL_P | NA | T | C | 0.04406 | 0.004247 | 5.50E-25 |
| rs495203 | let-7c-5p | trans | 2.73E-23 | Nightingale | L_LDL_CE | NA | T | C | 0.031578 | 0.004385 | 9.80E-13 |
| rs495203 | let-7c-5p | trans | 2.73E-23 | Nightingale | VLDL_P | NA | T | C | 0.026641 | 0.004401 | 4.40E-10 |
| rs495203 | let-7c-5p | trans | 2.73E-23 | Nightingale | L_LDL_P | NA | T | C | 0.029185 | 0.00441 | 6.10E-11 |
| rs495203 | let-7c-5p | trans | 2.73E-23 | Nightingale | LDL_PL | NA | T | C | 0.034997 | 0.004415 | 4.70E-15 |
| rs495203 | let-7c-5p | trans | 2.73E-23 | Nightingale | S_LDL_CE | NA | T | C | 0.030556 | 0.004427 | 5.80E-12 |
| rs495203 | let-7c-5p | trans | 2.73E-23 | Nightingale | Total_L | NA | T | C | 0.026363 | 0.004353 | 4.50E-10 |
| rs495203 | let-7c-5p | trans | 2.73E-23 | Nightingale | Omega_6 | NA | T | C | 0.02591 | 0.004321 | 5.60E-10 |
| rs495203 | let-7c-5p | trans | 2.73E-23 | Nightingale | M_VLDL_F | NA | T | C | 0.042068 | 0.00421 | 3.50E-23 |
| rs495203 | let-7c-5p | trans | 2.73E-23 | Nightingale | Total_C | NA | T | C | 0.032385 | 0.004284 | 3.30E-14 |
| rs495203 | let-7c-5p | trans | 2.73E-23 | Nightingale | XXL_VLDL_ | NA | T | C | 0.034842 | 0.004434 | 2.90E-15 |
| rs495203 | let-7c-5p | trans | 2.73E-23 | Nightingale | L_LDL_C | NA | T | C | 0.03233 | 0.004375 | 2.60E-13 |
| rs495203 | let-7c-5p | trans | 2.73E-23 | Nightingale | Remnant_( | NA | T | C | 0.036874 | 0.004396 | 3.70E-17 |
| rs495203 | let-7c-5p | trans | 2.73E-23 | Nightingale | S_LDL_FC | NA | T | C | 0.028569 | 0.004418 | 4.30E-10 |
| rs495203 | let-7c-5p | trans | 2.73E-23 | Nightingale | VLDL_FC | NA | T | C | 0.024164 | 0.004392 | 1.10E-08 |
| rs495203 | let-7c-5p | trans | 2.73E-23 | Nightingale | M_VLDL_C | NA | T | C | 0.033632 | 0.004157 | 1.90E-15 |
| rs495203 | let-7c-5p | trans | 2.73E-23 | Nightingale | ApoB_by_/ | NA | T | C | 0.025045 | 0.004358 | 1.30E-08 |
| rs495203 | let-7c-5p | trans | 2.73E-23 | Nightingale | S_LDL_P | NA | T | C | 0.029594 | 0.004429 | 3.20E-11 |
| rs495203 | let-7c-5p | trans | 2.73E-23 | Nightingale | S_LDL_C_p | NA | T | C | 0.045365 | 0.004442 | 5.50E-25 |
| rs495203 | let-7c-5p | trans | 2.73E-23 | Nightingale | LDL_CE | NA | T | C | 0.032564 | 0.004413 | 2.20E-13 |
| rs495203 | let-7c-5p | trans | 2.73E-23 | Nightingale | IDL_L | NA | T | C | 0.035277 | 0.004278 | 4.80E-16 |
| rs495203 | let-7c-5p | trans | 2.73E-23 | Nightingale | M_VLDL_C | NA | T | C | 0.036209 | 0.004168 | 1.20E-17 |
| rs495203 | let-7c-5p | trans | 2.73E-23 | Nightingale | L_VLDL_TG | NA | T | C | -0.05115 | 0.004404 | 5.30E-32 |
| rs495203 | let-7c-5p | trans | 2.73E-23 | Nightingale | Citrate | NA | T | C | 0.026828 | 0.004396 | 7.30E-10 |
| rs495203 | let-7c-5p | trans | 2.73E-23 | Nightingale | S_LDL_CE_ | NA | T | C | 0.03095 | 0.004417 | 4.90E-14 |
| rs495203 | let-7c-5p | trans | 2.73E-23 | Nightingale | IDL_PL | NA | T | C | 0.035351 | 0.00427 | 3.30E-16 |
| rs495203 | let-7c-5p | trans | 2.73E-23 | Nightingale | L_VLDL_C_ | NA | T | C | 0.052202 | 0.004322 | 4.00E-34 |
| rs495203 | let-7c-5p | trans | 2.73E-23 | Nightingale | M_LDL_PL | NA | T | C | 0.032476 | 0.004431 | 2.60E-13 |
| rs495203 | let-7c-5p | trans | 2.73E-23 | Nightingale | XL_VLDL_C | NA | T | C | 0.038228 | 0.004243 | 4.60E-19 |

| | | | | | | | | | | | |
|---|---|---|---|---|---|---|---|---|---|---|---|
| rs495203 | let-7c-5p | trans | 2.73E-23 | Nightingale | S_LDL_L | NA | T | C | 0.027749 | 0.004424 | 8.20E-10 |
| rs495203 | let-7c-5p | trans | 2.73E-23 | Nightingale | S_VLDL_CE | NA | T | C | 0.038188 | 0.004419 | 1.60E-18 |
| rs495203 | let-7c-5p | trans | 2.73E-23 | Nightingale | XL_HDL_PL | NA | T | C | -0.02275 | 0.004065 | 1.20E-08 |
| rs495203 | let-7c-5p | trans | 2.73E-23 | Nightingale | XXL_VLDL_ | NA | T | C | 0.030861 | 0.004406 | 1.10E-12 |
| rs495203 | let-7c-5p | trans | 2.73E-23 | Nightingale | L_LDL_FC | NA | T | C | 0.033632 | 0.004345 | 2.80E-14 |
| rs495203 | let-7c-5p | trans | 2.73E-23 | Nightingale | S_VLDL_TG | NA | T | C | -0.03813 | 0.004283 | 1.40E-18 |
| rs495828 | miR-139-5p | trans | 1.46E-34 | Nightingale | Total_PL | NA | T | G | 0.034823 | 0.004833 | 2.70E-14 |
| rs495828 | miR-139-5p | trans | 1.46E-34 | Nightingale | M_LDL_TG | NA | T | G | -0.0451 | 0.005013 | 5.60E-19 |
| rs495828 | miR-139-5p | trans | 1.46E-34 | Nightingale | LDL_C | NA | T | G | 0.053495 | 0.005054 | 7.00E-27 |
| rs495828 | miR-139-5p | trans | 1.46E-34 | Nightingale | S_VLDL_TG | NA | T | G | -0.05749 | 0.004913 | 1.10E-31 |
| rs495828 | miR-139-5p | trans | 1.46E-34 | Nightingale | XXL_VLDL_ | NA | T | G | 0.050865 | 0.005089 | 7.70E-24 |
| rs495828 | miR-139-5p | trans | 1.46E-34 | Nightingale | S_LDL_CE_ | NA | T | G | 0.039219 | 0.005067 | 7.30E-17 |
| rs495828 | miR-139-5p | trans | 1.46E-34 | Nightingale | Remnant_( | NA | T | G | 0.053715 | 0.005043 | 1.70E-28 |
| rs495828 | miR-139-5p | trans | 1.46E-34 | Nightingale | S_VLDL_CE | NA | T | G | 0.053551 | 0.005069 | 3.30E-28 |
| rs495828 | miR-139-5p | trans | 1.46E-34 | Nightingale | S_HDL_PL | NA | T | G | 0.03225 | 0.005032 | 3.90E-11 |
| rs495828 | miR-139-5p | trans | 1.46E-34 | Nightingale | ApoB_by_A | NA | T | G | 0.038647 | 0.004999 | 2.60E-15 |
| rs495828 | miR-139-5p | trans | 1.46E-34 | Nightingale | Total_P | NA | T | G | 0.029852 | 0.004777 | 9.80E-11 |
| rs495828 | miR-139-5p | trans | 1.46E-34 | Nightingale | XL_HDL_PL | NA | T | G | -0.0273 | 0.004566 | 8.00E-10 |
| rs495828 | miR-139-5p | trans | 1.46E-34 | Nightingale | S_VLDL_PL | NA | T | G | 0.046443 | 0.005066 | 9.30E-22 |
| rs495828 | miR-139-5p | trans | 1.46E-34 | Nightingale | S_LDL_P | NA | T | G | 0.045039 | 0.00508 | 7.80E-20 |
| rs495828 | miR-139-5p | trans | 1.46E-34 | Nightingale | M_VLDL_C | NA | T | G | 0.059565 | 0.005029 | 2.00E-33 |
| rs495828 | miR-139-5p | trans | 1.46E-34 | Nightingale | IDL_C | NA | T | G | 0.054084 | 0.004902 | 6.00E-29 |
| rs495828 | miR-139-5p | trans | 1.46E-34 | Nightingale | LDL_CE | NA | T | G | 0.05194 | 0.005062 | 1.60E-25 |
| rs495828 | miR-139-5p | trans | 1.46E-34 | Nightingale | M_LDL_FC | NA | T | G | 0.056635 | 0.005069 | 2.70E-29 |
| rs495828 | miR-139-5p | trans | 1.46E-34 | Nightingale | L_VLDL_C_ | NA | T | G | 0.072298 | 0.004957 | 3.00E-49 |
| rs495828 | miR-139-5p | trans | 1.46E-34 | Nightingale | Total_C | NA | T | G | 0.04931 | 0.004913 | 5.30E-25 |
| rs495828 | miR-139-5p | trans | 1.46E-34 | Nightingale | VLDL_CE | NA | T | G | 0.049812 | 0.005077 | 1.60E-24 |
| rs495828 | miR-139-5p | trans | 1.46E-34 | Nightingale | M_LDL_C_ | NA | T | G | 0.037099 | 0.005079 | 2.30E-13 |
| rs495828 | miR-139-5p | trans | 1.46E-34 | Nightingale | XXL_VLDL_ | NA | T | G | 0.032415 | 0.005111 | 7.40E-12 |
| rs495828 | miR-139-5p | trans | 1.46E-34 | Nightingale | XL_HDL_L | NA | T | G | -0.02457 | 0.004565 | 1.70E-08 |
| rs495828 | miR-139-5p | trans | 1.46E-34 | Nightingale | XL_HDL_PL | NA | T | G | -0.03691 | 0.004663 | 2.10E-15 |
| rs495828 | miR-139-5p | trans | 1.46E-34 | Nightingale | S_LDL_PL | NA | T | G | 0.038404 | 0.005064 | 1.60E-14 |
| rs495828 | miR-139-5p | trans | 1.46E-34 | Nightingale | S_HDL_PL_ | NA | T | G | -0.02857 | 0.004907 | 1.60E-08 |
| rs495828 | miR-139-5p | trans | 1.46E-34 | Nightingale | LDL_P | NA | T | G | 0.04853 | 0.005072 | 1.00E-22 |
| rs495828 | miR-139-5p | trans | 1.46E-34 | Nightingale | M_VLDL_P | NA | T | G | 0.040531 | 0.005063 | 4.70E-17 |
| rs495828 | miR-139-5p | trans | 1.46E-34 | Nightingale | L_VLDL_FC | NA | T | G | 0.068377 | 0.005027 | 7.00E-43 |
| rs495828 | miR-139-5p | trans | 1.46E-34 | Nightingale | VLDL_PL | NA | T | G | 0.026027 | 0.005015 | 1.40E-08 |
| rs495828 | miR-139-5p | trans | 1.46E-34 | Nightingale | XS_VLDL_P | NA | T | G | 0.041918 | 0.005005 | 2.50E-19 |
| rs495828 | miR-139-5p | trans | 1.46E-34 | Nightingale | XXL_VLDL_ | NA | T | G | -0.05232 | 0.005121 | 3.00E-25 |
| rs495828 | miR-139-5p | trans | 1.46E-34 | Nightingale | L_LDL_C | NA | T | G | 0.053211 | 0.005018 | 5.20E-27 |
| rs495828 | miR-139-5p | trans | 1.46E-34 | Nightingale | M_VLDL_C | NA | T | G | 0.048831 | 0.004768 | 4.00E-24 |
| rs495828 | miR-139-5p | trans | 1.46E-34 | Nightingale | L_VLDL_TG | NA | T | G | -0.07156 | 0.005052 | 2.50E-47 |
| rs495828 | miR-139-5p | trans | 1.46E-34 | Nightingale | IDL_L | NA | T | G | 0.053765 | 0.004907 | 7.10E-29 |
| rs495828 | miR-139-5p | trans | 1.46E-34 | Nightingale | M_LDL_CE | NA | T | G | 0.049782 | 0.005082 | 1.40E-23 |
| rs495828 | miR-139-5p | trans | 1.46E-34 | Nightingale | Clinical_LD | NA | T | G | 0.057708 | 0.005046 | 6.80E-31 |
| rs495828 | miR-139-5p | trans | 1.46E-34 | Nightingale | ApoB | NA | T | G | 0.051118 | 0.005069 | 3.80E-25 |
| rs495828 | miR-139-5p | trans | 1.46E-34 | Nightingale | M_LDL_L | NA | T | G | 0.051656 | 0.005083 | 4.40E-25 |
| rs495828 | miR-139-5p | trans | 1.46E-34 | Nightingale | IDL_FC_pct | NA | T | G | 0.032918 | 0.005036 | 1.40E-10 |
| rs495828 | miR-139-5p | trans | 1.46E-34 | Nightingale | Cholines | NA | T | G | 0.030838 | 0.004773 | 1.00E-11 |
| rs495828 | miR-139-5p | trans | 1.46E-34 | Nightingale | XS_VLDL_T | NA | T | G | -0.0369 | 0.004826 | 6.80E-14 |
| rs495828 | miR-139-5p | trans | 1.46E-34 | Nightingale | M_VLDL_T | NA | T | G | -0.05541 | 0.004796 | 1.50E-30 |
| rs495828 | miR-139-5p | trans | 1.46E-34 | Nightingale | XS_VLDL_C | NA | T | G | 0.052409 | 0.004927 | 1.50E-28 |
| rs495828 | miR-139-5p | trans | 1.46E-34 | Nightingale | S_HDL_L | NA | T | G | 0.03789 | 0.005058 | 2.30E-14 |
| rs495828 | miR-139-5p | trans | 1.46E-34 | Nightingale | S_LDL_C | NA | T | G | 0.04899 | 0.005081 | 1.10E-22 |
| rs495828 | miR-139-5p | trans | 1.46E-34 | Nightingale | S_LDL_PL_ | NA | T | G | -0.03948 | 0.005066 | 7.80E-17 |
| rs495828 | miR-139-5p | trans | 1.46E-34 | Nightingale | IDL_CE | NA | T | G | 0.052671 | 0.0049 | 1.60E-27 |
| rs495828 | miR-139-5p | trans | 1.46E-34 | Nightingale | XL_HDL_C_ | NA | T | G | 0.033312 | 0.004847 | 3.10E-11 |
| rs495828 | miR-139-5p | trans | 1.46E-34 | Nightingale | L_VLDL_CE | NA | T | G | 0.065015 | 0.004857 | 7.50E-42 |
| rs495828 | miR-139-5p | trans | 1.46E-34 | Nightingale | XL_VLDL_T | NA | T | G | -0.061 | 0.00492 | 1.60E-35 |
| rs495828 | miR-139-5p | trans | 1.46E-34 | Nightingale | LDL_PL | NA | T | G | 0.055772 | 0.005064 | 7.10E-29 |
| rs495828 | miR-139-5p | trans | 1.46E-34 | Nightingale | XS_VLDL_P | NA | T | G | 0.04977 | 0.005008 | 5.90E-26 |
| rs495828 | miR-139-5p | trans | 1.46E-34 | Nightingale | M_VLDL_C_ | NA | T | G | 0.052185 | 0.004781 | 2.80E-27 |
| rs495828 | miR-139-5p | trans | 1.46E-34 | Nightingale | L_LDL_FC | NA | T | G | 0.055548 | 0.004984 | 3.50E-29 |

| | | | | | | | | | | | |
|---|---|---|---|---|---|---|---|---|---|---|---|
| rs495828 | miR-139-5ƥ | trans | 1.46E-34 | Nightingale | VLDL_C | NA | T | G | 0.043128 | 0.005069 | 3.50E-19 |
| rs495828 | miR-139-5ƥ | trans | 1.46E-34 | Nightingale | Sphingomy | NA | T | G | 0.040453 | 0.004745 | 1.10E-17 |
| rs495828 | miR-139-5ƥ | trans | 1.46E-34 | Nightingale | XL_VLDL_C | NA | T | G | 0.0539 | 0.00487 | 4.50E-28 |
| rs495828 | miR-139-5ƥ | trans | 1.46E-34 | Nightingale | M_VLDL_P | NA | T | G | 0.063345 | 0.004871 | 6.40E-39 |
| rs495828 | miR-139-5ƥ | trans | 1.46E-34 | Nightingale | IDL_C_pct | NA | T | G | 0.03253 | 0.004958 | 3.20E-10 |
| rs495828 | miR-139-5ƥ | trans | 1.46E-34 | Nightingale | PUFA | NA | T | G | 0.032704 | 0.004951 | 3.80E-12 |
| rs495828 | miR-139-5ƥ | trans | 1.46E-34 | Nightingale | S_VLDL_CE | NA | T | G | 0.0685 | 0.005007 | 2.10E-42 |
| rs495828 | miR-139-5ƥ | trans | 1.46E-34 | Nightingale | L_LDL_P | NA | T | G | 0.046207 | 0.005058 | 8.20E-21 |
| rs495828 | miR-139-5ƥ | trans | 1.46E-34 | Nightingale | L_LDL_L | NA | T | G | 0.054281 | 0.005026 | 4.40E-28 |
| rs495828 | miR-139-5ƥ | trans | 1.46E-34 | Nightingale | IDL_PL | NA | T | G | 0.052753 | 0.004898 | 2.50E-28 |
| rs495828 | miR-139-5ƥ | trans | 1.46E-34 | Nightingale | LDL_L | NA | T | G | 0.053322 | 0.005058 | 9.10E-27 |
| rs495828 | miR-139-5ƥ | trans | 1.46E-34 | Nightingale | M_VLDL_C | NA | T | G | 0.056 | 0.005061 | 4.20E-30 |
| rs495828 | miR-139-5ƥ | trans | 1.46E-34 | Nightingale | S_HDL_FC | NA | T | G | 0.035359 | 0.004983 | 3.00E-13 |
| rs495828 | miR-139-5ƥ | trans | 1.46E-34 | Nightingale | L_LDL_CE | NA | T | G | 0.051875 | 0.00503 | 9.70E-26 |
| rs495828 | miR-139-5ƥ | trans | 1.46E-34 | Nightingale | S_HDL_CE | NA | T | G | 0.045125 | 0.005083 | 8.90E-19 |
| rs495828 | miR-139-5ƥ | trans | 1.46E-34 | Nightingale | L_VLDL_CE | NA | T | G | 0.030432 | 0.005016 | 6.30E-11 |
| rs495828 | miR-139-5ƥ | trans | 1.46E-34 | Nightingale | L_LDL_PL | NA | T | G | 0.059441 | 0.005037 | 5.50E-33 |
| rs495828 | miR-139-5ƥ | trans | 1.46E-34 | Nightingale | S_VLDL_FC | NA | T | G | 0.045509 | 0.00485 | 1.30E-20 |
| rs495828 | miR-139-5ƥ | trans | 1.46E-34 | Nightingale | XL_VLDL_F | NA | T | G | 0.05038 | 0.004921 | 3.60E-25 |
| rs495828 | miR-139-5ƥ | trans | 1.46E-34 | Nightingale | M_VLDL_F | NA | T | G | 0.059176 | 0.004829 | 2.10E-34 |
| rs495828 | miR-139-5ƥ | trans | 1.46E-34 | Nightingale | XXL_VLDL_ | NA | T | G | 0.040705 | 0.005056 | 3.40E-16 |
| rs495828 | miR-139-5ƥ | trans | 1.46E-34 | Nightingale | S_VLDL_P | NA | T | G | 0.031959 | 0.005012 | 4.90E-12 |
| rs495828 | miR-139-5ƥ | trans | 1.46E-34 | Nightingale | S_VLDL_FC | NA | T | G | 0.054306 | 0.005076 | 2.60E-28 |
| rs495828 | miR-139-5ƥ | trans | 1.46E-34 | Nightingale | XS_VLDL_C | NA | T | G | 0.037019 | 0.004814 | 5.50E-14 |
| rs495828 | miR-139-5ƥ | trans | 1.46E-34 | Nightingale | Pyruvate | NA | T | G | 0.029634 | 0.005084 | 5.60E-09 |
| rs495828 | miR-139-5ƥ | trans | 1.46E-34 | Nightingale | M_VLDL_P | NA | T | G | 0.041928 | 0.005068 | 4.10E-18 |
| rs495828 | miR-139-5ƥ | trans | 1.46E-34 | Nightingale | Total_FC | NA | T | G | 0.049237 | 0.004958 | 3.30E-25 |
| rs495828 | miR-139-5ƥ | trans | 1.46E-34 | Nightingale | VLDL_FC | NA | T | G | 0.032606 | 0.005038 | 3.90E-12 |
| rs495828 | miR-139-5ƥ | trans | 1.46E-34 | Nightingale | XS_VLDL_F | NA | T | G | 0.049238 | 0.004987 | 1.30E-25 |
| rs495828 | miR-139-5ƥ | trans | 1.46E-34 | Nightingale | XXL_VLDL_ | NA | T | G | 0.050116 | 0.005116 | 4.00E-23 |
| rs495828 | miR-139-5ƥ | trans | 1.46E-34 | Nightingale | S_VLDL_C | NA | T | G | 0.054248 | 0.005075 | 1.40E-28 |
| rs495828 | miR-139-5ƥ | trans | 1.46E-34 | Nightingale | LA | NA | T | G | 0.037741 | 0.004977 | 1.40E-15 |
| rs495828 | miR-139-5ƥ | trans | 1.46E-34 | Nightingale | M_VLDL_F | NA | T | G | 0.047692 | 0.005075 | 1.90E-22 |
| rs495828 | miR-139-5ƥ | trans | 1.46E-34 | Nightingale | M_LDL_C | NA | T | G | 0.052602 | 0.005085 | 7.60E-26 |
| rs495828 | miR-139-5ƥ | trans | 1.46E-34 | Nightingale | L_HDL_TG_ | NA | T | G | 0.024075 | 0.004795 | 1.30E-08 |
| rs495828 | miR-139-5ƥ | trans | 1.46E-34 | Nightingale | XS_VLDL_L | NA | T | G | 0.045042 | 0.004989 | 4.90E-22 |
| rs495828 | miR-139-5ƥ | trans | 1.46E-34 | Nightingale | IDL_FC | NA | T | G | 0.056848 | 0.004914 | 9.10E-32 |
| rs495828 | miR-139-5ƥ | trans | 1.46E-34 | Nightingale | S_LDL_L | NA | T | G | 0.044532 | 0.005075 | 4.30E-19 |
| rs495828 | miR-139-5ƥ | trans | 1.46E-34 | Nightingale | IDL_TG_pc | NA | T | G | -0.03284 | 0.004907 | 7.40E-11 |
| rs495828 | miR-139-5ƥ | trans | 1.46E-34 | Nightingale | S_LDL_CE | NA | T | G | 0.047169 | 0.005078 | 1.80E-21 |
| rs495828 | miR-139-5ƥ | trans | 1.46E-34 | Nightingale | Total_CE | NA | T | G | 0.048939 | 0.004896 | 1.10E-24 |
| rs495828 | miR-139-5ƥ | trans | 1.46E-34 | Nightingale | Phosphatic | NA | T | G | 0.027047 | 0.004756 | 1.70E-09 |
| rs495828 | miR-139-5ƥ | trans | 1.46E-34 | Nightingale | LDL_FC | NA | T | G | 0.055702 | 0.005021 | 9.40E-29 |
| rs495828 | miR-139-5ƥ | trans | 1.46E-34 | Nightingale | XL_VLDL_P | NA | T | G | 0.052707 | 0.005084 | 1.40E-26 |
| rs495828 | miR-139-5ƥ | trans | 1.46E-34 | Nightingale | M_LDL_PL | NA | T | G | 0.052849 | 0.005083 | 4.60E-26 |
| rs495828 | miR-139-5ƥ | trans | 1.46E-34 | Nightingale | S_VLDL_PL | NA | T | G | 0.043474 | 0.00486 | 7.60E-19 |
| rs495828 | miR-139-5ƥ | trans | 1.46E-34 | Nightingale | S_HDL_P | NA | T | G | 0.047417 | 0.005076 | 8.50E-21 |
| rs495828 | miR-139-5ƥ | trans | 1.46E-34 | Nightingale | L_VLDL_PL | NA | T | G | 0.027644 | 0.00493 | 6.40E-10 |
| rs495828 | miR-139-5ƥ | trans | 1.46E-34 | Nightingale | XS_VLDL_F | NA | T | G | 0.043854 | 0.005039 | 1.70E-17 |
| rs495828 | miR-139-5ƥ | trans | 1.46E-34 | Nightingale | Phosphogly | NA | T | G | 0.027772 | 0.004807 | 9.60E-10 |
| rs495828 | miR-139-5ƥ | trans | 1.46E-34 | Nightingale | HDL_size | NA | T | G | -0.03057 | 0.0045 | 3.10E-12 |
| rs495828 | miR-139-5ƥ | trans | 1.46E-34 | Nightingale | S_VLDL_L | NA | T | G | 0.033815 | 0.005026 | 4.60E-13 |
| rs495828 | miR-139-5ƥ | trans | 1.46E-34 | Nightingale | S_LDL_FC | NA | T | G | 0.048985 | 0.005068 | 4.70E-22 |
| rs495828 | miR-139-5ƥ | trans | 1.46E-34 | Nightingale | Omega_6 | NA | T | G | 0.037091 | 0.004956 | 2.90E-15 |
| rs495828 | miR-139-5ƥ | trans | 1.46E-34 | Nightingale | Total_L | NA | T | G | 0.03839 | 0.004993 | 7.90E-16 |
| rs495828 | miR-139-5ƥ | trans | 1.46E-34 | Nightingale | S_LDL_C_p | NA | T | G | 0.065319 | 0.005095 | 3.20E-38 |
| rs495828 | miR-139-5ƥ | trans | 1.46E-34 | Nightingale | XL_VLDL_C | NA | T | G | 0.054421 | 0.004867 | 1.00E-28 |
| rs495828 | miR-139-5ƥ | trans | 1.46E-34 | Nightingale | non_HDL_( | NA | T | G | 0.05466 | 0.005048 | 1.20E-28 |
| rs495828 | miR-139-5ƥ | trans | 1.46E-34 | Nightingale | XS_VLDL_C | NA | T | G | 0.034393 | 0.004789 | 3.10E-12 |
| rs495828 | miR-139-5ƥ | trans | 1.46E-34 | Nightingale | IDL_P | NA | T | G | 0.059807 | 0.005022 | 1.60E-33 |
| rs495828 | miR-139-5ƥ | trans | 1.46E-34 | Nightingale | VLDL_P | NA | T | G | 0.0365 | 0.005048 | 1.40E-14 |
| rs495828 | miR-139-5ƥ | trans | 1.46E-34 | Nightingale | S_VLDL_C_ | NA | T | G | 0.062388 | 0.00494 | 1.10E-36 |
| rs495828 | miR-139-5ƥ | trans | 1.46E-34 | Nightingale | XL_VLDL_C | NA | T | G | 0.029159 | 0.004997 | 2.80E-10 |

| | | | | | | | | | | | |
|---|---|---|---|---|---|---|---|---|---|---|---|
| rs495828 | miR-139-5p | trans | 1.46E-34 | Nightingale | M_LDL_P | NA | T | G | 0.051176 | 0.00509 | 1.00E-24 |
| rs495828 | miR-139-5p | trans | 1.46E-34 | Nightingale | L_LDL_TG_ | NA | T | G | -0.0346 | 0.004978 | 1.30E-11 |
| rs495828 | miR-139-5p | trans | 1.46E-34 | Nightingale | S_HDL_C | NA | T | G | 0.044417 | 0.005067 | 1.80E-18 |
| rs495828 | miR-139-5p | trans | 1.46E-34 | Nightingale | Citrate | NA | T | G | 0.027426 | 0.005043 | 2.00E-08 |
| rs495828 | miR-139-5p | trans | 1.46E-34 | Nightingale | S_LDL_TG_ | NA | T | G | -0.0367 | 0.004922 | 5.00E-13 |
| rs495828 | miR-139-5p | trans | 1.46E-34 | Nightingale | M_VLDL_L | NA | T | G | 0.030732 | 0.005041 | 7.30E-11 |
| rs495828 | miR-139-5p | trans | 1.46E-34 | Nightingale | XS_VLDL_C | NA | T | G | 0.052929 | 0.004898 | 3.40E-29 |
| rs495828 | miR-126-3p | trans | 2.00E-37 | Nightingale | Total_PL | NA | T | G | 0.034823 | 0.004833 | 2.70E-14 |
| rs495828 | miR-126-3p | trans | 2.00E-37 | Nightingale | M_LDL_TG | NA | T | G | -0.0451 | 0.005013 | 5.60E-19 |
| rs495828 | miR-126-3p | trans | 2.00E-37 | Nightingale | LDL_C | NA | T | G | 0.053495 | 0.005054 | 7.00E-27 |
| rs495828 | miR-126-3p | trans | 2.00E-37 | Nightingale | S_VLDL_TG | NA | T | G | -0.05749 | 0.004913 | 1.10E-31 |
| rs495828 | miR-126-3p | trans | 2.00E-37 | Nightingale | XXL_VLDL_ | NA | T | G | 0.050865 | 0.005089 | 7.70E-24 |
| rs495828 | miR-126-3p | trans | 2.00E-37 | Nightingale | S_LDL_CE_ | NA | T | G | 0.039219 | 0.005067 | 7.30E-17 |
| rs495828 | miR-126-3p | trans | 2.00E-37 | Nightingale | Remnant_( | NA | T | G | 0.053715 | 0.005043 | 1.70E-28 |
| rs495828 | miR-126-3p | trans | 2.00E-37 | Nightingale | S_VLDL_CE | NA | T | G | 0.053551 | 0.005069 | 3.30E-28 |
| rs495828 | miR-126-3p | trans | 2.00E-37 | Nightingale | S_HDL_PL | NA | T | G | 0.03225 | 0.005032 | 3.90E-11 |
| rs495828 | miR-126-3p | trans | 2.00E-37 | Nightingale | ApoB_by_/ | NA | T | G | 0.038647 | 0.004999 | 2.60E-15 |
| rs495828 | miR-126-3p | trans | 2.00E-37 | Nightingale | Total_P | NA | T | G | 0.029852 | 0.004777 | 9.80E-11 |
| rs495828 | miR-126-3p | trans | 2.00E-37 | Nightingale | XL_HDL_PL | NA | T | G | -0.0273 | 0.004566 | 8.00E-10 |
| rs495828 | miR-126-3p | trans | 2.00E-37 | Nightingale | S_VLDL_PL | NA | T | G | 0.046443 | 0.005066 | 9.30E-22 |
| rs495828 | miR-126-3p | trans | 2.00E-37 | Nightingale | S_LDL_P | NA | T | G | 0.045039 | 0.00508 | 7.80E-20 |
| rs495828 | miR-126-3p | trans | 2.00E-37 | Nightingale | M_VLDL_C | NA | T | G | 0.059565 | 0.005029 | 2.00E-33 |
| rs495828 | miR-126-3p | trans | 2.00E-37 | Nightingale | IDL_C | NA | T | G | 0.054084 | 0.004902 | 6.00E-29 |
| rs495828 | miR-126-3p | trans | 2.00E-37 | Nightingale | LDL_CE | NA | T | G | 0.05194 | 0.005062 | 1.60E-25 |
| rs495828 | miR-126-3p | trans | 2.00E-37 | Nightingale | M_LDL_FC | NA | T | G | 0.056635 | 0.005069 | 2.70E-29 |
| rs495828 | miR-126-3p | trans | 2.00E-37 | Nightingale | L_VLDL_C_ | NA | T | G | 0.072298 | 0.004957 | 3.00E-49 |
| rs495828 | miR-126-3p | trans | 2.00E-37 | Nightingale | Total_C | NA | T | G | 0.04931 | 0.004913 | 5.30E-25 |
| rs495828 | miR-126-3p | trans | 2.00E-37 | Nightingale | VLDL_CE | NA | T | G | 0.049812 | 0.005077 | 1.60E-24 |
| rs495828 | miR-126-3p | trans | 2.00E-37 | Nightingale | M_LDL_C_ | NA | T | G | 0.037099 | 0.005079 | 2.30E-13 |
| rs495828 | miR-126-3p | trans | 2.00E-37 | Nightingale | XXL_VLDL_ | NA | T | G | 0.032415 | 0.005111 | 7.40E-12 |
| rs495828 | miR-126-3p | trans | 2.00E-37 | Nightingale | XL_HDL_L | NA | T | G | -0.02457 | 0.004565 | 1.70E-08 |
| rs495828 | miR-126-3p | trans | 2.00E-37 | Nightingale | XL_HDL_PL | NA | T | G | -0.03691 | 0.004663 | 2.10E-15 |
| rs495828 | miR-126-3p | trans | 2.00E-37 | Nightingale | S_LDL_PL | NA | T | G | 0.038404 | 0.005064 | 1.60E-14 |
| rs495828 | miR-126-3p | trans | 2.00E-37 | Nightingale | S_HDL_PL_ | NA | T | G | -0.02857 | 0.004907 | 1.60E-08 |
| rs495828 | miR-126-3p | trans | 2.00E-37 | Nightingale | LDL_P | NA | T | G | 0.04853 | 0.005072 | 1.00E-22 |
| rs495828 | miR-126-3p | trans | 2.00E-37 | Nightingale | M_VLDL_P | NA | T | G | 0.040531 | 0.005063 | 4.70E-17 |
| rs495828 | miR-126-3p | trans | 2.00E-37 | Nightingale | L_VLDL_FC | NA | T | G | 0.068377 | 0.005027 | 7.00E-43 |
| rs495828 | miR-126-3p | trans | 2.00E-37 | Nightingale | VLDL_PL | NA | T | G | 0.026027 | 0.005015 | 1.40E-08 |
| rs495828 | miR-126-3p | trans | 2.00E-37 | Nightingale | XS_VLDL_P | NA | T | G | 0.041918 | 0.005005 | 2.50E-19 |
| rs495828 | miR-126-3p | trans | 2.00E-37 | Nightingale | XXL_VLDL_ | NA | T | G | -0.05232 | 0.005121 | 3.00E-25 |
| rs495828 | miR-126-3p | trans | 2.00E-37 | Nightingale | L_LDL_C | NA | T | G | 0.053211 | 0.005018 | 5.20E-27 |
| rs495828 | miR-126-3p | trans | 2.00E-37 | Nightingale | M_VLDL_C | NA | T | G | 0.048831 | 0.004768 | 4.00E-24 |
| rs495828 | miR-126-3p | trans | 2.00E-37 | Nightingale | L_VLDL_TG | NA | T | G | -0.07156 | 0.005052 | 2.50E-47 |
| rs495828 | miR-126-3p | trans | 2.00E-37 | Nightingale | IDL_L | NA | T | G | 0.053765 | 0.004907 | 7.10E-29 |
| rs495828 | miR-126-3p | trans | 2.00E-37 | Nightingale | M_LDL_CE | NA | T | G | 0.049782 | 0.005082 | 1.40E-23 |
| rs495828 | miR-126-3p | trans | 2.00E-37 | Nightingale | Clinical_LD | NA | T | G | 0.057708 | 0.005046 | 6.80E-31 |
| rs495828 | miR-126-3p | trans | 2.00E-37 | Nightingale | ApoB | NA | T | G | 0.051118 | 0.005069 | 3.80E-25 |
| rs495828 | miR-126-3p | trans | 2.00E-37 | Nightingale | M_LDL_L | NA | T | G | 0.051656 | 0.005083 | 4.40E-25 |
| rs495828 | miR-126-3p | trans | 2.00E-37 | Nightingale | IDL_FC_pct | NA | T | G | 0.032918 | 0.005036 | 1.40E-10 |
| rs495828 | miR-126-3p | trans | 2.00E-37 | Nightingale | Cholines | NA | T | G | 0.030838 | 0.004773 | 1.00E-11 |
| rs495828 | miR-126-3p | trans | 2.00E-37 | Nightingale | XS_VLDL_T | NA | T | G | -0.0369 | 0.004826 | 6.80E-14 |
| rs495828 | miR-126-3p | trans | 2.00E-37 | Nightingale | M_VLDL_T | NA | T | G | -0.05541 | 0.004796 | 1.50E-30 |
| rs495828 | miR-126-3p | trans | 2.00E-37 | Nightingale | XS_VLDL_C | NA | T | G | 0.052409 | 0.004927 | 1.50E-28 |
| rs495828 | miR-126-3p | trans | 2.00E-37 | Nightingale | S_HDL_L | NA | T | G | 0.03789 | 0.005058 | 2.30E-14 |
| rs495828 | miR-126-3p | trans | 2.00E-37 | Nightingale | S_LDL_C | NA | T | G | 0.04899 | 0.005081 | 1.10E-22 |
| rs495828 | miR-126-3p | trans | 2.00E-37 | Nightingale | S_LDL_PL_ | NA | T | G | -0.03948 | 0.005066 | 7.80E-17 |
| rs495828 | miR-126-3p | trans | 2.00E-37 | Nightingale | IDL_CE | NA | T | G | 0.052671 | 0.0049 | 1.60E-27 |
| rs495828 | miR-126-3p | trans | 2.00E-37 | Nightingale | XL_HDL_C_ | NA | T | G | 0.033312 | 0.004847 | 3.10E-11 |
| rs495828 | miR-126-3p | trans | 2.00E-37 | Nightingale | L_VLDL_CE | NA | T | G | 0.065015 | 0.004857 | 7.50E-42 |
| rs495828 | miR-126-3p | trans | 2.00E-37 | Nightingale | XL_VLDL_T | NA | T | G | -0.061 | 0.00492 | 1.60E-35 |
| rs495828 | miR-126-3p | trans | 2.00E-37 | Nightingale | LDL_PL | NA | T | G | 0.055772 | 0.005064 | 7.10E-29 |
| rs495828 | miR-126-3p | trans | 2.00E-37 | Nightingale | XS_VLDL_P | NA | T | G | 0.04977 | 0.005008 | 5.90E-26 |
| rs495828 | miR-126-3p | trans | 2.00E-37 | Nightingale | M_VLDL_C | NA | T | G | 0.052185 | 0.004781 | 2.80E-27 |

| | | | | | | | | | | | |
|---|---|---|---|---|---|---|---|---|---|---|---|
| rs495828 | miR-126-3p | trans | 2.00E-37 | Nightingale | L_LDL_FC | NA | T | G | 0.055548 | 0.004984 | 3.50E-29 |
| rs495828 | miR-126-3p | trans | 2.00E-37 | Nightingale | VLDL_C | NA | T | G | 0.043128 | 0.005069 | 3.50E-19 |
| rs495828 | miR-126-3p | trans | 2.00E-37 | Nightingale | Sphingomy | NA | T | G | 0.040453 | 0.004745 | 1.10E-17 |
| rs495828 | miR-126-3p | trans | 2.00E-37 | Nightingale | XL_VLDL_C | NA | T | G | 0.0539 | 0.00487 | 4.50E-28 |
| rs495828 | miR-126-3p | trans | 2.00E-37 | Nightingale | M_VLDL_P | NA | T | G | 0.063345 | 0.004871 | 6.40E-39 |
| rs495828 | miR-126-3p | trans | 2.00E-37 | Nightingale | IDL_C_pct | NA | T | G | 0.03253 | 0.004958 | 3.20E-10 |
| rs495828 | miR-126-3p | trans | 2.00E-37 | Nightingale | PUFA | NA | T | G | 0.032704 | 0.004951 | 3.80E-12 |
| rs495828 | miR-126-3p | trans | 2.00E-37 | Nightingale | S_VLDL_CE | NA | T | G | 0.0685 | 0.005007 | 2.10E-42 |
| rs495828 | miR-126-3p | trans | 2.00E-37 | Nightingale | L_LDL_P | NA | T | G | 0.046207 | 0.005058 | 8.20E-21 |
| rs495828 | miR-126-3p | trans | 2.00E-37 | Nightingale | L_LDL_L | NA | T | G | 0.054281 | 0.005026 | 4.40E-28 |
| rs495828 | miR-126-3p | trans | 2.00E-37 | Nightingale | IDL_PL | NA | T | G | 0.052753 | 0.004898 | 2.50E-28 |
| rs495828 | miR-126-3p | trans | 2.00E-37 | Nightingale | LDL_L | NA | T | G | 0.053322 | 0.005058 | 9.10E-27 |
| rs495828 | miR-126-3p | trans | 2.00E-37 | Nightingale | M_VLDL_C | NA | T | G | 0.056 | 0.005061 | 4.20E-30 |
| rs495828 | miR-126-3p | trans | 2.00E-37 | Nightingale | S_HDL_FC | NA | T | G | 0.035359 | 0.004983 | 3.00E-13 |
| rs495828 | miR-126-3p | trans | 2.00E-37 | Nightingale | L_LDL_CE | NA | T | G | 0.051875 | 0.00503 | 9.70E-26 |
| rs495828 | miR-126-3p | trans | 2.00E-37 | Nightingale | S_HDL_CE | NA | T | G | 0.045125 | 0.005083 | 8.90E-19 |
| rs495828 | miR-126-3p | trans | 2.00E-37 | Nightingale | L_VLDL_CE | NA | T | G | 0.030432 | 0.005016 | 6.30E-11 |
| rs495828 | miR-126-3p | trans | 2.00E-37 | Nightingale | L_LDL_PL | NA | T | G | 0.059441 | 0.005037 | 5.50E-33 |
| rs495828 | miR-126-3p | trans | 2.00E-37 | Nightingale | S_VLDL_FC | NA | T | G | 0.045509 | 0.00485 | 1.30E-20 |
| rs495828 | miR-126-3p | trans | 2.00E-37 | Nightingale | XL_VLDL_F | NA | T | G | 0.05038 | 0.004921 | 3.60E-25 |
| rs495828 | miR-126-3p | trans | 2.00E-37 | Nightingale | M_VLDL_F | NA | T | G | 0.059176 | 0.004829 | 2.10E-34 |
| rs495828 | miR-126-3p | trans | 2.00E-37 | Nightingale | XXL_VLDL_ | NA | T | G | 0.040705 | 0.005056 | 3.40E-16 |
| rs495828 | miR-126-3p | trans | 2.00E-37 | Nightingale | S_VLDL_P | NA | T | G | 0.031959 | 0.005012 | 4.90E-12 |
| rs495828 | miR-126-3p | trans | 2.00E-37 | Nightingale | S_VLDL_FC | NA | T | G | 0.054306 | 0.005076 | 2.60E-28 |
| rs495828 | miR-126-3p | trans | 2.00E-37 | Nightingale | XS_VLDL_C | NA | T | G | 0.037019 | 0.004814 | 5.50E-14 |
| rs495828 | miR-126-3p | trans | 2.00E-37 | Nightingale | Pyruvate | NA | T | G | 0.029634 | 0.005084 | 5.60E-09 |
| rs495828 | miR-126-3p | trans | 2.00E-37 | Nightingale | M_VLDL_P | NA | T | G | 0.041928 | 0.005068 | 4.10E-18 |
| rs495828 | miR-126-3p | trans | 2.00E-37 | Nightingale | Total_FC | NA | T | G | 0.049237 | 0.004958 | 3.30E-25 |
| rs495828 | miR-126-3p | trans | 2.00E-37 | Nightingale | VLDL_FC | NA | T | G | 0.032606 | 0.005038 | 3.90E-12 |
| rs495828 | miR-126-3p | trans | 2.00E-37 | Nightingale | XS_VLDL_F | NA | T | G | 0.049238 | 0.004987 | 1.30E-25 |
| rs495828 | miR-126-3p | trans | 2.00E-37 | Nightingale | XXL_VLDL_ | NA | T | G | 0.050116 | 0.005116 | 4.00E-23 |
| rs495828 | miR-126-3p | trans | 2.00E-37 | Nightingale | S_VLDL_C | NA | T | G | 0.054248 | 0.005075 | 1.40E-28 |
| rs495828 | miR-126-3p | trans | 2.00E-37 | Nightingale | LA | NA | T | G | 0.037741 | 0.004977 | 1.40E-15 |
| rs495828 | miR-126-3p | trans | 2.00E-37 | Nightingale | M_VLDL_F | NA | T | G | 0.047692 | 0.005075 | 1.90E-22 |
| rs495828 | miR-126-3p | trans | 2.00E-37 | Nightingale | M_LDL_C | NA | T | G | 0.052602 | 0.005085 | 7.60E-26 |
| rs495828 | miR-126-3p | trans | 2.00E-37 | Nightingale | L_HDL_TG_ | NA | T | G | 0.024075 | 0.004795 | 1.30E-08 |
| rs495828 | miR-126-3p | trans | 2.00E-37 | Nightingale | XS_VLDL_L | NA | T | G | 0.045042 | 0.004989 | 4.90E-22 |
| rs495828 | miR-126-3p | trans | 2.00E-37 | Nightingale | IDL_FC | NA | T | G | 0.056848 | 0.004914 | 9.10E-32 |
| rs495828 | miR-126-3p | trans | 2.00E-37 | Nightingale | S_LDL_L | NA | T | G | 0.044532 | 0.005075 | 4.30E-19 |
| rs495828 | miR-126-3p | trans | 2.00E-37 | Nightingale | IDL_TG_pc | NA | T | G | -0.03284 | 0.004907 | 7.40E-11 |
| rs495828 | miR-126-3p | trans | 2.00E-37 | Nightingale | S_LDL_CE | NA | T | G | 0.047169 | 0.005078 | 1.80E-21 |
| rs495828 | miR-126-3p | trans | 2.00E-37 | Nightingale | Total_CE | NA | T | G | 0.048939 | 0.004896 | 1.10E-24 |
| rs495828 | miR-126-3p | trans | 2.00E-37 | Nightingale | Phosphatic | NA | T | G | 0.027047 | 0.004756 | 1.70E-09 |
| rs495828 | miR-126-3p | trans | 2.00E-37 | Nightingale | LDL_FC | NA | T | G | 0.055702 | 0.005021 | 9.40E-29 |
| rs495828 | miR-126-3p | trans | 2.00E-37 | Nightingale | XL_VLDL_P | NA | T | G | 0.052707 | 0.005084 | 1.40E-26 |
| rs495828 | miR-126-3p | trans | 2.00E-37 | Nightingale | M_LDL_PL | NA | T | G | 0.052849 | 0.005083 | 4.60E-26 |
| rs495828 | miR-126-3p | trans | 2.00E-37 | Nightingale | S_VLDL_PL | NA | T | G | 0.043474 | 0.00486 | 7.60E-19 |
| rs495828 | miR-126-3p | trans | 2.00E-37 | Nightingale | S_HDL_P | NA | T | G | 0.047417 | 0.005076 | 8.50E-21 |
| rs495828 | miR-126-3p | trans | 2.00E-37 | Nightingale | L_VLDL_PL | NA | T | G | 0.027644 | 0.00493 | 6.40E-10 |
| rs495828 | miR-126-3p | trans | 2.00E-37 | Nightingale | XS_VLDL_F | NA | T | G | 0.043854 | 0.005039 | 1.70E-17 |
| rs495828 | miR-126-3p | trans | 2.00E-37 | Nightingale | Phosphogly | NA | T | G | 0.027772 | 0.004807 | 9.60E-10 |
| rs495828 | miR-126-3p | trans | 2.00E-37 | Nightingale | HDL_size | NA | T | G | -0.03057 | 0.0045 | 3.10E-12 |
| rs495828 | miR-126-3p | trans | 2.00E-37 | Nightingale | S_VLDL_L | NA | T | G | 0.033815 | 0.005026 | 4.60E-13 |
| rs495828 | miR-126-3p | trans | 2.00E-37 | Nightingale | S_LDL_FC | NA | T | G | 0.048985 | 0.005068 | 4.70E-22 |
| rs495828 | miR-126-3p | trans | 2.00E-37 | Nightingale | Omega_6 | NA | T | G | 0.037091 | 0.004956 | 2.90E-15 |
| rs495828 | miR-126-3p | trans | 2.00E-37 | Nightingale | Total_L | NA | T | G | 0.03839 | 0.004993 | 7.90E-16 |
| rs495828 | miR-126-3p | trans | 2.00E-37 | Nightingale | S_LDL_C_p | NA | T | G | 0.065319 | 0.005095 | 3.20E-38 |
| rs495828 | miR-126-3p | trans | 2.00E-37 | Nightingale | XL_VLDL_C | NA | T | G | 0.054421 | 0.004867 | 1.00E-28 |
| rs495828 | miR-126-3p | trans | 2.00E-37 | Nightingale | non_HDL_( | NA | T | G | 0.05466 | 0.005048 | 1.20E-28 |
| rs495828 | miR-126-3p | trans | 2.00E-37 | Nightingale | XS_VLDL_C | NA | T | G | 0.034393 | 0.004789 | 3.10E-12 |
| rs495828 | miR-126-3p | trans | 2.00E-37 | Nightingale | IDL_P | NA | T | G | 0.059807 | 0.005022 | 1.60E-33 |
| rs495828 | miR-126-3p | trans | 2.00E-37 | Nightingale | VLDL_P | NA | T | G | 0.0365 | 0.005048 | 1.40E-14 |
| rs495828 | miR-126-3p | trans | 2.00E-37 | Nightingale | S_VLDL_C_ | NA | T | G | 0.062388 | 0.00494 | 1.10E-36 |

| | | | | | | | | | | | |
|---|---|---|---|---|---|---|---|---|---|---|---|
| rs495828 | miR-126-3| trans | 2.00E-37 | Nightingale | XL_VLDL_C | NA | T | G | 0.029159 | 0.004997 | 2.80E-10 |
| rs495828 | miR-126-3| trans | 2.00E-37 | Nightingale | M_LDL_P | NA | T | G | 0.051176 | 0.00509 | 1.00E-24 |
| rs495828 | miR-126-3| trans | 2.00E-37 | Nightingale | L_LDL_TG_ | NA | T | G | -0.0346 | 0.004978 | 1.30E-11 |
| rs495828 | miR-126-3| trans | 2.00E-37 | Nightingale | S_HDL_C | NA | T | G | 0.044417 | 0.005067 | 1.80E-18 |
| rs495828 | miR-126-3| trans | 2.00E-37 | Nightingale | Citrate | NA | T | G | 0.027426 | 0.005043 | 2.00E-08 |
| rs495828 | miR-126-3| trans | 2.00E-37 | Nightingale | S_LDL_TG_ | NA | T | G | -0.0367 | 0.004922 | 5.00E-13 |
| rs495828 | miR-126-3| trans | 2.00E-37 | Nightingale | M_VLDL_L | NA | T | G | 0.030732 | 0.005041 | 7.30E-11 |
| rs495828 | miR-126-3| trans | 2.00E-37 | Nightingale | XS_VLDL_C | NA | T | G | 0.052929 | 0.004898 | 3.40E-29 |
| rs495828 | miR-10b-5| trans | 2.56E-34 | Nightingale | Total_PL | NA | T | G | 0.034823 | 0.004833 | 2.70E-14 |
| rs495828 | miR-10b-5| trans | 2.56E-34 | Nightingale | M_LDL_TG | NA | T | G | -0.0451 | 0.005013 | 5.60E-19 |
| rs495828 | miR-10b-5| trans | 2.56E-34 | Nightingale | LDL_C | NA | T | G | 0.053495 | 0.005054 | 7.00E-27 |
| rs495828 | miR-10b-5| trans | 2.56E-34 | Nightingale | S_VLDL_TG | NA | T | G | -0.05749 | 0.004913 | 1.10E-31 |
| rs495828 | miR-10b-5| trans | 2.56E-34 | Nightingale | XXL_VLDL_ | NA | T | G | 0.050865 | 0.005089 | 7.70E-24 |
| rs495828 | miR-10b-5| trans | 2.56E-34 | Nightingale | S_LDL_CE_ | NA | T | G | 0.039219 | 0.005067 | 7.30E-17 |
| rs495828 | miR-10b-5| trans | 2.56E-34 | Nightingale | Remnant_( | NA | T | G | 0.053715 | 0.005043 | 1.70E-28 |
| rs495828 | miR-10b-5| trans | 2.56E-34 | Nightingale | S_VLDL_CE | NA | T | G | 0.053551 | 0.005069 | 3.30E-28 |
| rs495828 | miR-10b-5| trans | 2.56E-34 | Nightingale | S_HDL_PL | NA | T | G | 0.03225 | 0.005032 | 3.90E-11 |
| rs495828 | miR-10b-5| trans | 2.56E-34 | Nightingale | ApoB_by_/ | NA | T | G | 0.038647 | 0.004999 | 2.60E-15 |
| rs495828 | miR-10b-5| trans | 2.56E-34 | Nightingale | Total_P | NA | T | G | 0.029852 | 0.004777 | 9.80E-11 |
| rs495828 | miR-10b-5| trans | 2.56E-34 | Nightingale | XL_HDL_PL | NA | T | G | -0.0273 | 0.004566 | 8.00E-10 |
| rs495828 | miR-10b-5| trans | 2.56E-34 | Nightingale | S_VLDL_PL | NA | T | G | 0.046443 | 0.005066 | 9.30E-22 |
| rs495828 | miR-10b-5| trans | 2.56E-34 | Nightingale | S_LDL_P | NA | T | G | 0.045039 | 0.00508 | 7.80E-20 |
| rs495828 | miR-10b-5| trans | 2.56E-34 | Nightingale | M_VLDL_C | NA | T | G | 0.059565 | 0.005029 | 2.00E-33 |
| rs495828 | miR-10b-5| trans | 2.56E-34 | Nightingale | IDL_C | NA | T | G | 0.054084 | 0.004902 | 6.00E-29 |
| rs495828 | miR-10b-5| trans | 2.56E-34 | Nightingale | LDL_CE | NA | T | G | 0.05194 | 0.005062 | 1.60E-25 |
| rs495828 | miR-10b-5| trans | 2.56E-34 | Nightingale | M_LDL_FC | NA | T | G | 0.056635 | 0.005069 | 2.70E-29 |
| rs495828 | miR-10b-5| trans | 2.56E-34 | Nightingale | L_VLDL_C_ | NA | T | G | 0.072298 | 0.004957 | 3.00E-49 |
| rs495828 | miR-10b-5| trans | 2.56E-34 | Nightingale | Total_C | NA | T | G | 0.04931 | 0.004913 | 5.30E-25 |
| rs495828 | miR-10b-5| trans | 2.56E-34 | Nightingale | VLDL_CE | NA | T | G | 0.049812 | 0.005077 | 1.60E-24 |
| rs495828 | miR-10b-5| trans | 2.56E-34 | Nightingale | M_LDL_C_| | NA | T | G | 0.037099 | 0.005079 | 2.30E-13 |
| rs495828 | miR-10b-5| trans | 2.56E-34 | Nightingale | XXL_VLDL_ | NA | T | G | 0.032415 | 0.005111 | 7.40E-12 |
| rs495828 | miR-10b-5| trans | 2.56E-34 | Nightingale | XL_HDL_L | NA | T | G | -0.02457 | 0.004565 | 1.70E-08 |
| rs495828 | miR-10b-5| trans | 2.56E-34 | Nightingale | XL_HDL_PL | NA | T | G | -0.03691 | 0.004663 | 2.10E-15 |
| rs495828 | miR-10b-5| trans | 2.56E-34 | Nightingale | S_LDL_PL | NA | T | G | 0.038404 | 0.005064 | 1.60E-14 |
| rs495828 | miR-10b-5| trans | 2.56E-34 | Nightingale | S_HDL_PL_ | NA | T | G | -0.02857 | 0.004907 | 1.60E-08 |
| rs495828 | miR-10b-5| trans | 2.56E-34 | Nightingale | LDL_P | NA | T | G | 0.04853 | 0.005072 | 1.00E-22 |
| rs495828 | miR-10b-5| trans | 2.56E-34 | Nightingale | M_VLDL_P | NA | T | G | 0.040531 | 0.005063 | 4.70E-17 |
| rs495828 | miR-10b-5| trans | 2.56E-34 | Nightingale | L_VLDL_FC | NA | T | G | 0.068377 | 0.005027 | 7.00E-43 |
| rs495828 | miR-10b-5| trans | 2.56E-34 | Nightingale | VLDL_PL | NA | T | G | 0.026027 | 0.005015 | 1.40E-08 |
| rs495828 | miR-10b-5| trans | 2.56E-34 | Nightingale | XS_VLDL_P | NA | T | G | 0.041918 | 0.005005 | 2.50E-19 |
| rs495828 | miR-10b-5| trans | 2.56E-34 | Nightingale | XXL_VLDL_ | NA | T | G | -0.05232 | 0.005121 | 3.00E-25 |
| rs495828 | miR-10b-5| trans | 2.56E-34 | Nightingale | L_LDL_C | NA | T | G | 0.053211 | 0.005018 | 5.20E-27 |
| rs495828 | miR-10b-5| trans | 2.56E-34 | Nightingale | M_VLDL_C | NA | T | G | 0.048831 | 0.004768 | 4.00E-24 |
| rs495828 | miR-10b-5| trans | 2.56E-34 | Nightingale | L_VLDL_TG | NA | T | G | -0.07156 | 0.005052 | 2.50E-47 |
| rs495828 | miR-10b-5| trans | 2.56E-34 | Nightingale | IDL_L | NA | T | G | 0.053765 | 0.004907 | 7.10E-29 |
| rs495828 | miR-10b-5| trans | 2.56E-34 | Nightingale | M_LDL_CE | NA | T | G | 0.049782 | 0.005082 | 1.40E-23 |
| rs495828 | miR-10b-5| trans | 2.56E-34 | Nightingale | Clinical_LD | NA | T | G | 0.057708 | 0.005046 | 6.80E-31 |
| rs495828 | miR-10b-5| trans | 2.56E-34 | Nightingale | ApoB | NA | T | G | 0.051118 | 0.005069 | 3.80E-25 |
| rs495828 | miR-10b-5| trans | 2.56E-34 | Nightingale | M_LDL_L | NA | T | G | 0.051656 | 0.005083 | 4.40E-25 |
| rs495828 | miR-10b-5| trans | 2.56E-34 | Nightingale | IDL_FC_pct | NA | T | G | 0.032918 | 0.005036 | 1.40E-10 |
| rs495828 | miR-10b-5| trans | 2.56E-34 | Nightingale | Cholines | NA | T | G | 0.030838 | 0.004773 | 1.00E-11 |
| rs495828 | miR-10b-5| trans | 2.56E-34 | Nightingale | XS_VLDL_T | NA | T | G | -0.0369 | 0.004826 | 6.80E-14 |
| rs495828 | miR-10b-5| trans | 2.56E-34 | Nightingale | M_VLDL_T | NA | T | G | -0.05541 | 0.004796 | 1.50E-30 |
| rs495828 | miR-10b-5| trans | 2.56E-34 | Nightingale | XS_VLDL_C | NA | T | G | 0.052409 | 0.004927 | 1.50E-28 |
| rs495828 | miR-10b-5| trans | 2.56E-34 | Nightingale | S_HDL_L | NA | T | G | 0.03789 | 0.005058 | 2.30E-14 |
| rs495828 | miR-10b-5| trans | 2.56E-34 | Nightingale | S_LDL_C | NA | T | G | 0.04899 | 0.005081 | 1.10E-22 |
| rs495828 | miR-10b-5| trans | 2.56E-34 | Nightingale | S_LDL_PL_ | NA | T | G | -0.03948 | 0.005066 | 7.80E-17 |
| rs495828 | miR-10b-5| trans | 2.56E-34 | Nightingale | IDL_CE | NA | T | G | 0.052671 | 0.0049 | 1.60E-27 |
| rs495828 | miR-10b-5| trans | 2.56E-34 | Nightingale | XL_HDL_C_ | NA | T | G | 0.033312 | 0.004847 | 3.10E-11 |
| rs495828 | miR-10b-5| trans | 2.56E-34 | Nightingale | L_VLDL_CE | NA | T | G | 0.065015 | 0.004857 | 7.50E-42 |
| rs495828 | miR-10b-5| trans | 2.56E-34 | Nightingale | XL_VLDL_T | NA | T | G | -0.061 | 0.00492 | 1.60E-35 |
| rs495828 | miR-10b-5| trans | 2.56E-34 | Nightingale | LDL_PL | NA | T | G | 0.055772 | 0.005064 | 7.10E-29 |
| rs495828 | miR-10b-5| trans | 2.56E-34 | Nightingale | XS_VLDL_P | NA | T | G | 0.04977 | 0.005008 | 5.90E-26 |

| | | | | | | | | | | | |
|---|---|---|---|---|---|---|---|---|---|---|---|
| rs495828 | miR-10b-5p | trans | 2.56E-34 | Nightingale | M_VLDL_C | NA | T | G | 0.052185 | 0.004781 | 2.80E-27 |
| rs495828 | miR-10b-5p | trans | 2.56E-34 | Nightingale | L_LDL_FC | NA | T | G | 0.055548 | 0.004984 | 3.50E-29 |
| rs495828 | miR-10b-5p | trans | 2.56E-34 | Nightingale | VLDL_C | NA | T | G | 0.043128 | 0.005069 | 3.50E-19 |
| rs495828 | miR-10b-5p | trans | 2.56E-34 | Nightingale | Sphingomy | NA | T | G | 0.040453 | 0.004745 | 1.10E-17 |
| rs495828 | miR-10b-5p | trans | 2.56E-34 | Nightingale | XL_VLDL_C | NA | T | G | 0.0539 | 0.00487 | 4.50E-28 |
| rs495828 | miR-10b-5p | trans | 2.56E-34 | Nightingale | M_VLDL_P | NA | T | G | 0.063345 | 0.004871 | 6.40E-39 |
| rs495828 | miR-10b-5p | trans | 2.56E-34 | Nightingale | IDL_C_pct | NA | T | G | 0.03253 | 0.004958 | 3.20E-10 |
| rs495828 | miR-10b-5p | trans | 2.56E-34 | Nightingale | PUFA | NA | T | G | 0.032704 | 0.004951 | 3.80E-12 |
| rs495828 | miR-10b-5p | trans | 2.56E-34 | Nightingale | S_VLDL_CE | NA | T | G | 0.0685 | 0.005007 | 2.10E-42 |
| rs495828 | miR-10b-5p | trans | 2.56E-34 | Nightingale | L_LDL_P | NA | T | G | 0.046207 | 0.005058 | 8.20E-21 |
| rs495828 | miR-10b-5p | trans | 2.56E-34 | Nightingale | L_LDL_L | NA | T | G | 0.054281 | 0.005026 | 4.40E-28 |
| rs495828 | miR-10b-5p | trans | 2.56E-34 | Nightingale | IDL_PL | NA | T | G | 0.052753 | 0.004898 | 2.50E-28 |
| rs495828 | miR-10b-5p | trans | 2.56E-34 | Nightingale | LDL_L | NA | T | G | 0.053322 | 0.005058 | 9.10E-27 |
| rs495828 | miR-10b-5p | trans | 2.56E-34 | Nightingale | M_VLDL_C | NA | T | G | 0.056 | 0.005061 | 4.20E-30 |
| rs495828 | miR-10b-5p | trans | 2.56E-34 | Nightingale | S_HDL_FC | NA | T | G | 0.035359 | 0.004983 | 3.00E-13 |
| rs495828 | miR-10b-5p | trans | 2.56E-34 | Nightingale | L_LDL_CE | NA | T | G | 0.051875 | 0.00503 | 9.70E-26 |
| rs495828 | miR-10b-5p | trans | 2.56E-34 | Nightingale | S_HDL_CE | NA | T | G | 0.045125 | 0.005083 | 8.90E-19 |
| rs495828 | miR-10b-5p | trans | 2.56E-34 | Nightingale | L_VLDL_CE | NA | T | G | 0.030432 | 0.005016 | 6.30E-11 |
| rs495828 | miR-10b-5p | trans | 2.56E-34 | Nightingale | L_LDL_PL | NA | T | G | 0.059441 | 0.005037 | 5.50E-33 |
| rs495828 | miR-10b-5p | trans | 2.56E-34 | Nightingale | S_VLDL_FC | NA | T | G | 0.045509 | 0.00485 | 1.30E-20 |
| rs495828 | miR-10b-5p | trans | 2.56E-34 | Nightingale | XL_VLDL_F | NA | T | G | 0.05038 | 0.004921 | 3.60E-25 |
| rs495828 | miR-10b-5p | trans | 2.56E-34 | Nightingale | M_VLDL_F | NA | T | G | 0.059176 | 0.004829 | 2.10E-34 |
| rs495828 | miR-10b-5p | trans | 2.56E-34 | Nightingale | XXL_VLDL_ | NA | T | G | 0.040705 | 0.005056 | 3.40E-16 |
| rs495828 | miR-10b-5p | trans | 2.56E-34 | Nightingale | S_VLDL_P | NA | T | G | 0.031959 | 0.005012 | 4.90E-12 |
| rs495828 | miR-10b-5p | trans | 2.56E-34 | Nightingale | S_VLDL_FC | NA | T | G | 0.054306 | 0.005076 | 2.60E-28 |
| rs495828 | miR-10b-5p | trans | 2.56E-34 | Nightingale | XS_VLDL_C | NA | T | G | 0.037019 | 0.004814 | 5.50E-14 |
| rs495828 | miR-10b-5p | trans | 2.56E-34 | Nightingale | Pyruvate | NA | T | G | 0.029634 | 0.005084 | 5.60E-09 |
| rs495828 | miR-10b-5p | trans | 2.56E-34 | Nightingale | M_VLDL_P | NA | T | G | 0.041928 | 0.005068 | 4.10E-18 |
| rs495828 | miR-10b-5p | trans | 2.56E-34 | Nightingale | Total_FC | NA | T | G | 0.049237 | 0.004958 | 3.30E-25 |
| rs495828 | miR-10b-5p | trans | 2.56E-34 | Nightingale | VLDL_FC | NA | T | G | 0.032606 | 0.005038 | 3.90E-12 |
| rs495828 | miR-10b-5p | trans | 2.56E-34 | Nightingale | XS_VLDL_F | NA | T | G | 0.049238 | 0.004987 | 1.30E-25 |
| rs495828 | miR-10b-5p | trans | 2.56E-34 | Nightingale | XXL_VLDL_ | NA | T | G | 0.050116 | 0.005116 | 4.00E-23 |
| rs495828 | miR-10b-5p | trans | 2.56E-34 | Nightingale | S_VLDL_C | NA | T | G | 0.054248 | 0.005075 | 1.40E-28 |
| rs495828 | miR-10b-5p | trans | 2.56E-34 | Nightingale | LA | NA | T | G | 0.037741 | 0.004977 | 1.40E-15 |
| rs495828 | miR-10b-5p | trans | 2.56E-34 | Nightingale | M_VLDL_F | NA | T | G | 0.047692 | 0.005075 | 1.90E-22 |
| rs495828 | miR-10b-5p | trans | 2.56E-34 | Nightingale | M_LDL_C | NA | T | G | 0.052602 | 0.005085 | 7.60E-26 |
| rs495828 | miR-10b-5p | trans | 2.56E-34 | Nightingale | L_HDL_TG_ | NA | T | G | 0.024075 | 0.004795 | 1.30E-08 |
| rs495828 | miR-10b-5p | trans | 2.56E-34 | Nightingale | XS_VLDL_L | NA | T | G | 0.045042 | 0.004989 | 4.90E-22 |
| rs495828 | miR-10b-5p | trans | 2.56E-34 | Nightingale | IDL_FC | NA | T | G | 0.056848 | 0.004914 | 9.10E-32 |
| rs495828 | miR-10b-5p | trans | 2.56E-34 | Nightingale | S_LDL_L | NA | T | G | 0.044532 | 0.005075 | 4.30E-19 |
| rs495828 | miR-10b-5p | trans | 2.56E-34 | Nightingale | IDL_TG_pct | NA | T | G | -0.03284 | 0.004907 | 7.40E-11 |
| rs495828 | miR-10b-5p | trans | 2.56E-34 | Nightingale | S_LDL_CE | NA | T | G | 0.047169 | 0.005078 | 1.80E-21 |
| rs495828 | miR-10b-5p | trans | 2.56E-34 | Nightingale | Total_CE | NA | T | G | 0.048939 | 0.004896 | 1.10E-24 |
| rs495828 | miR-10b-5p | trans | 2.56E-34 | Nightingale | Phosphatic | NA | T | G | 0.027047 | 0.004756 | 1.70E-09 |
| rs495828 | miR-10b-5p | trans | 2.56E-34 | Nightingale | LDL_FC | NA | T | G | 0.055702 | 0.005021 | 9.40E-29 |
| rs495828 | miR-10b-5p | trans | 2.56E-34 | Nightingale | XL_VLDL_P | NA | T | G | 0.052707 | 0.005084 | 1.40E-26 |
| rs495828 | miR-10b-5p | trans | 2.56E-34 | Nightingale | M_LDL_PL | NA | T | G | 0.052849 | 0.005083 | 4.60E-26 |
| rs495828 | miR-10b-5p | trans | 2.56E-34 | Nightingale | S_VLDL_PL | NA | T | G | 0.043474 | 0.00486 | 7.60E-19 |
| rs495828 | miR-10b-5p | trans | 2.56E-34 | Nightingale | S_HDL_P | NA | T | G | 0.047417 | 0.005076 | 8.50E-21 |
| rs495828 | miR-10b-5p | trans | 2.56E-34 | Nightingale | L_VLDL_PL | NA | T | G | 0.027644 | 0.00493 | 6.40E-10 |
| rs495828 | miR-10b-5p | trans | 2.56E-34 | Nightingale | XS_VLDL_F | NA | T | G | 0.043854 | 0.005039 | 1.70E-17 |
| rs495828 | miR-10b-5p | trans | 2.56E-34 | Nightingale | Phosphogly | NA | T | G | 0.027772 | 0.004807 | 9.60E-10 |
| rs495828 | miR-10b-5p | trans | 2.56E-34 | Nightingale | HDL_size | NA | T | G | -0.03057 | 0.0045 | 3.10E-12 |
| rs495828 | miR-10b-5p | trans | 2.56E-34 | Nightingale | S_VLDL_L | NA | T | G | 0.033815 | 0.005026 | 4.60E-13 |
| rs495828 | miR-10b-5p | trans | 2.56E-34 | Nightingale | S_LDL_FC | NA | T | G | 0.048985 | 0.005068 | 4.70E-22 |
| rs495828 | miR-10b-5p | trans | 2.56E-34 | Nightingale | Omega_6 | NA | T | G | 0.037091 | 0.004956 | 2.90E-15 |
| rs495828 | miR-10b-5p | trans | 2.56E-34 | Nightingale | Total_L | NA | T | G | 0.03839 | 0.004993 | 7.90E-16 |
| rs495828 | miR-10b-5p | trans | 2.56E-34 | Nightingale | S_LDL_C_p | NA | T | G | 0.065319 | 0.005095 | 3.20E-38 |
| rs495828 | miR-10b-5p | trans | 2.56E-34 | Nightingale | XL_VLDL_C | NA | T | G | 0.054421 | 0.004867 | 1.00E-28 |
| rs495828 | miR-10b-5p | trans | 2.56E-34 | Nightingale | non_HDL_( | NA | T | G | 0.05466 | 0.005048 | 1.20E-28 |
| rs495828 | miR-10b-5p | trans | 2.56E-34 | Nightingale | XS_VLDL_C | NA | T | G | 0.034393 | 0.004789 | 3.10E-12 |
| rs495828 | miR-10b-5p | trans | 2.56E-34 | Nightingale | IDL_P | NA | T | G | 0.059807 | 0.005022 | 1.60E-33 |
| rs495828 | miR-10b-5p | trans | 2.56E-34 | Nightingale | VLDL_P | NA | T | G | 0.0365 | 0.005048 | 1.40E-14 |

| | | | | | | | | | | | |
|---|---|---|---|---|---|---|---|---|---|---|---|
| rs495828 | miR-10b-5p | trans | 2.56E-34 | Nightingale | S_VLDL_C_ | NA | T | G | 0.062388 | 0.00494 | 1.10E-36 |
| rs495828 | miR-10b-5p | trans | 2.56E-34 | Nightingale | XL_VLDL_C | NA | T | G | 0.029159 | 0.004997 | 2.80E-10 |
| rs495828 | miR-10b-5p | trans | 2.56E-34 | Nightingale | M_LDL_P | NA | T | G | 0.051176 | 0.00509 | 1.00E-24 |
| rs495828 | miR-10b-5p | trans | 2.56E-34 | Nightingale | L_LDL_TG_ | NA | T | G | -0.0346 | 0.004978 | 1.30E-11 |
| rs495828 | miR-10b-5p | trans | 2.56E-34 | Nightingale | S_HDL_C | NA | T | G | 0.044417 | 0.005067 | 1.80E-18 |
| rs495828 | miR-10b-5p | trans | 2.56E-34 | Nightingale | Citrate | NA | T | G | 0.027426 | 0.005043 | 2.00E-08 |
| rs495828 | miR-10b-5p | trans | 2.56E-34 | Nightingale | S_LDL_TG_ | NA | T | G | -0.0367 | 0.004922 | 5.00E-13 |
| rs495828 | miR-10b-5p | trans | 2.56E-34 | Nightingale | M_VLDL_L | NA | T | G | 0.030732 | 0.005041 | 7.30E-11 |
| rs495828 | miR-10b-5p | trans | 2.56E-34 | Nightingale | XS_VLDL_C | NA | T | G | 0.052929 | 0.004898 | 3.40E-29 |
| rs495828 | let-7a-5p | trans | 1.53E-12 | Nightingale | Total_PL | NA | T | G | 0.034823 | 0.004833 | 2.70E-14 |
| rs495828 | let-7a-5p | trans | 1.53E-12 | Nightingale | M_LDL_TG | NA | T | G | -0.0451 | 0.005013 | 5.60E-19 |
| rs495828 | let-7a-5p | trans | 1.53E-12 | Nightingale | LDL_C | NA | T | G | 0.053495 | 0.005054 | 7.00E-27 |
| rs495828 | let-7a-5p | trans | 1.53E-12 | Nightingale | S_VLDL_TG | NA | T | G | -0.05749 | 0.004913 | 1.10E-31 |
| rs495828 | let-7a-5p | trans | 1.53E-12 | Nightingale | XXL_VLDL_ | NA | T | G | 0.050865 | 0.005089 | 7.70E-24 |
| rs495828 | let-7a-5p | trans | 1.53E-12 | Nightingale | S_LDL_CE_ | NA | T | G | 0.039219 | 0.005067 | 7.30E-17 |
| rs495828 | let-7a-5p | trans | 1.53E-12 | Nightingale | Remnant_( | NA | T | G | 0.053715 | 0.005043 | 1.70E-28 |
| rs495828 | let-7a-5p | trans | 1.53E-12 | Nightingale | S_VLDL_CE | NA | T | G | 0.053551 | 0.005069 | 3.30E-28 |
| rs495828 | let-7a-5p | trans | 1.53E-12 | Nightingale | S_HDL_PL | NA | T | G | 0.03225 | 0.005032 | 3.90E-11 |
| rs495828 | let-7a-5p | trans | 1.53E-12 | Nightingale | ApoB_by_A | NA | T | G | 0.038647 | 0.004999 | 2.60E-15 |
| rs495828 | let-7a-5p | trans | 1.53E-12 | Nightingale | Total_P | NA | T | G | 0.029852 | 0.004777 | 9.80E-11 |
| rs495828 | let-7a-5p | trans | 1.53E-12 | Nightingale | XL_HDL_PL | NA | T | G | -0.0273 | 0.004566 | 8.00E-10 |
| rs495828 | let-7a-5p | trans | 1.53E-12 | Nightingale | S_VLDL_PL | NA | T | G | 0.046443 | 0.005066 | 9.30E-22 |
| rs495828 | let-7a-5p | trans | 1.53E-12 | Nightingale | S_LDL_P | NA | T | G | 0.045039 | 0.00508 | 7.80E-20 |
| rs495828 | let-7a-5p | trans | 1.53E-12 | Nightingale | M_VLDL_C | NA | T | G | 0.059565 | 0.005029 | 2.00E-33 |
| rs495828 | let-7a-5p | trans | 1.53E-12 | Nightingale | IDL_C | NA | T | G | 0.054084 | 0.004902 | 6.00E-29 |
| rs495828 | let-7a-5p | trans | 1.53E-12 | Nightingale | LDL_CE | NA | T | G | 0.05194 | 0.005062 | 1.60E-25 |
| rs495828 | let-7a-5p | trans | 1.53E-12 | Nightingale | M_LDL_FC | NA | T | G | 0.056635 | 0.005069 | 2.70E-29 |
| rs495828 | let-7a-5p | trans | 1.53E-12 | Nightingale | L_VLDL_C_ | NA | T | G | 0.072298 | 0.004957 | 3.00E-49 |
| rs495828 | let-7a-5p | trans | 1.53E-12 | Nightingale | Total_C | NA | T | G | 0.04931 | 0.004913 | 5.30E-25 |
| rs495828 | let-7a-5p | trans | 1.53E-12 | Nightingale | VLDL_CE | NA | T | G | 0.049812 | 0.005077 | 1.60E-24 |
| rs495828 | let-7a-5p | trans | 1.53E-12 | Nightingale | M_LDL_C_ | NA | T | G | 0.037099 | 0.005079 | 2.30E-13 |
| rs495828 | let-7a-5p | trans | 1.53E-12 | Nightingale | XXL_VLDL_ | NA | T | G | 0.032415 | 0.005111 | 7.40E-12 |
| rs495828 | let-7a-5p | trans | 1.53E-12 | Nightingale | XL_HDL_L | NA | T | G | -0.02457 | 0.004565 | 1.70E-08 |
| rs495828 | let-7a-5p | trans | 1.53E-12 | Nightingale | XL_HDL_PL | NA | T | G | -0.03691 | 0.004663 | 2.10E-15 |
| rs495828 | let-7a-5p | trans | 1.53E-12 | Nightingale | S_LDL_PL | NA | T | G | 0.038404 | 0.005064 | 1.60E-14 |
| rs495828 | let-7a-5p | trans | 1.53E-12 | Nightingale | S_HDL_PL_ | NA | T | G | -0.02857 | 0.004907 | 1.60E-08 |
| rs495828 | let-7a-5p | trans | 1.53E-12 | Nightingale | LDL_P | NA | T | G | 0.04853 | 0.005072 | 1.00E-22 |
| rs495828 | let-7a-5p | trans | 1.53E-12 | Nightingale | M_VLDL_P | NA | T | G | 0.040531 | 0.005063 | 4.70E-17 |
| rs495828 | let-7a-5p | trans | 1.53E-12 | Nightingale | L_VLDL_FC | NA | T | G | 0.068377 | 0.005027 | 7.00E-43 |
| rs495828 | let-7a-5p | trans | 1.53E-12 | Nightingale | VLDL_PL | NA | T | G | 0.026027 | 0.005015 | 1.40E-08 |
| rs495828 | let-7a-5p | trans | 1.53E-12 | Nightingale | XS_VLDL_P | NA | T | G | 0.041918 | 0.005005 | 2.50E-19 |
| rs495828 | let-7a-5p | trans | 1.53E-12 | Nightingale | XXL_VLDL_ | NA | T | G | -0.05232 | 0.005121 | 3.00E-25 |
| rs495828 | let-7a-5p | trans | 1.53E-12 | Nightingale | L_LDL_C | NA | T | G | 0.053211 | 0.005018 | 5.20E-27 |
| rs495828 | let-7a-5p | trans | 1.53E-12 | Nightingale | M_VLDL_C | NA | T | G | 0.048831 | 0.004768 | 4.00E-24 |
| rs495828 | let-7a-5p | trans | 1.53E-12 | Nightingale | L_VLDL_TG | NA | T | G | -0.07156 | 0.005052 | 2.50E-47 |
| rs495828 | let-7a-5p | trans | 1.53E-12 | Nightingale | IDL_L | NA | T | G | 0.053765 | 0.004907 | 7.10E-29 |
| rs495828 | let-7a-5p | trans | 1.53E-12 | Nightingale | M_LDL_CE | NA | T | G | 0.049782 | 0.005082 | 1.40E-23 |
| rs495828 | let-7a-5p | trans | 1.53E-12 | Nightingale | Clinical_LD | NA | T | G | 0.057708 | 0.005046 | 6.80E-31 |
| rs495828 | let-7a-5p | trans | 1.53E-12 | Nightingale | ApoB | NA | T | G | 0.051118 | 0.005069 | 3.80E-25 |
| rs495828 | let-7a-5p | trans | 1.53E-12 | Nightingale | M_LDL_L | NA | T | G | 0.051656 | 0.005083 | 4.40E-25 |
| rs495828 | let-7a-5p | trans | 1.53E-12 | Nightingale | IDL_FC_pct | NA | T | G | 0.032918 | 0.005036 | 1.40E-10 |
| rs495828 | let-7a-5p | trans | 1.53E-12 | Nightingale | Cholines | NA | T | G | 0.030838 | 0.004773 | 1.00E-11 |
| rs495828 | let-7a-5p | trans | 1.53E-12 | Nightingale | XS_VLDL_T | NA | T | G | -0.0369 | 0.004826 | 6.80E-14 |
| rs495828 | let-7a-5p | trans | 1.53E-12 | Nightingale | M_VLDL_T | NA | T | G | -0.05541 | 0.004796 | 1.50E-30 |
| rs495828 | let-7a-5p | trans | 1.53E-12 | Nightingale | XS_VLDL_C | NA | T | G | 0.052409 | 0.004927 | 1.50E-28 |
| rs495828 | let-7a-5p | trans | 1.53E-12 | Nightingale | S_HDL_L | NA | T | G | 0.03789 | 0.005058 | 2.30E-14 |
| rs495828 | let-7a-5p | trans | 1.53E-12 | Nightingale | S_LDL_C | NA | T | G | 0.04899 | 0.005081 | 1.10E-22 |
| rs495828 | let-7a-5p | trans | 1.53E-12 | Nightingale | S_LDL_PL_ | NA | T | G | -0.03948 | 0.005066 | 7.80E-17 |
| rs495828 | let-7a-5p | trans | 1.53E-12 | Nightingale | IDL_CE | NA | T | G | 0.052671 | 0.0049 | 1.60E-27 |
| rs495828 | let-7a-5p | trans | 1.53E-12 | Nightingale | XL_HDL_C_ | NA | T | G | 0.033312 | 0.004847 | 3.10E-11 |
| rs495828 | let-7a-5p | trans | 1.53E-12 | Nightingale | L_VLDL_CE | NA | T | G | 0.065015 | 0.004857 | 7.50E-42 |
| rs495828 | let-7a-5p | trans | 1.53E-12 | Nightingale | XL_VLDL_T | NA | T | G | -0.061 | 0.00492 | 1.60E-35 |
| rs495828 | let-7a-5p | trans | 1.53E-12 | Nightingale | LDL_PL | NA | T | G | 0.055772 | 0.005064 | 7.10E-29 |

| | | | | | | | | | | | |
|---|---|---|---|---|---|---|---|---|---|---|---|
| rs495828 | let-7a-5p | trans | 1.53E-12 | Nightingale | XS_VLDL_P | NA | T | G | 0.04977 | 0.005008 | 5.90E-26 |
| rs495828 | let-7a-5p | trans | 1.53E-12 | Nightingale | M_VLDL_C | NA | T | G | 0.052185 | 0.004781 | 2.80E-27 |
| rs495828 | let-7a-5p | trans | 1.53E-12 | Nightingale | L_LDL_FC | NA | T | G | 0.055548 | 0.004984 | 3.50E-29 |
| rs495828 | let-7a-5p | trans | 1.53E-12 | Nightingale | VLDL_C | NA | T | G | 0.043128 | 0.005069 | 3.50E-19 |
| rs495828 | let-7a-5p | trans | 1.53E-12 | Nightingale | Sphingomy | NA | T | G | 0.040453 | 0.004745 | 1.10E-17 |
| rs495828 | let-7a-5p | trans | 1.53E-12 | Nightingale | XL_VLDL_C | NA | T | G | 0.0539 | 0.00487 | 4.50E-28 |
| rs495828 | let-7a-5p | trans | 1.53E-12 | Nightingale | M_VLDL_P | NA | T | G | 0.063345 | 0.004871 | 6.40E-39 |
| rs495828 | let-7a-5p | trans | 1.53E-12 | Nightingale | IDL_C_pct | NA | T | G | 0.03253 | 0.004958 | 3.20E-10 |
| rs495828 | let-7a-5p | trans | 1.53E-12 | Nightingale | PUFA | NA | T | G | 0.032704 | 0.004951 | 3.80E-12 |
| rs495828 | let-7a-5p | trans | 1.53E-12 | Nightingale | S_VLDL_CE | NA | T | G | 0.0685 | 0.005007 | 2.10E-42 |
| rs495828 | let-7a-5p | trans | 1.53E-12 | Nightingale | L_LDL_P | NA | T | G | 0.046207 | 0.005058 | 8.20E-21 |
| rs495828 | let-7a-5p | trans | 1.53E-12 | Nightingale | L_LDL_L | NA | T | G | 0.054281 | 0.005026 | 4.40E-28 |
| rs495828 | let-7a-5p | trans | 1.53E-12 | Nightingale | IDL_PL | NA | T | G | 0.052753 | 0.004898 | 2.50E-28 |
| rs495828 | let-7a-5p | trans | 1.53E-12 | Nightingale | LDL_L | NA | T | G | 0.053322 | 0.005058 | 9.10E-27 |
| rs495828 | let-7a-5p | trans | 1.53E-12 | Nightingale | M_VLDL_C | NA | T | G | 0.056 | 0.005061 | 4.20E-30 |
| rs495828 | let-7a-5p | trans | 1.53E-12 | Nightingale | S_HDL_FC | NA | T | G | 0.035359 | 0.004983 | 3.00E-13 |
| rs495828 | let-7a-5p | trans | 1.53E-12 | Nightingale | L_LDL_CE | NA | T | G | 0.051875 | 0.00503 | 9.70E-26 |
| rs495828 | let-7a-5p | trans | 1.53E-12 | Nightingale | S_HDL_CE | NA | T | G | 0.045125 | 0.005083 | 8.90E-19 |
| rs495828 | let-7a-5p | trans | 1.53E-12 | Nightingale | L_VLDL_CE | NA | T | G | 0.030432 | 0.005016 | 6.30E-11 |
| rs495828 | let-7a-5p | trans | 1.53E-12 | Nightingale | L_LDL_PL | NA | T | G | 0.059441 | 0.005037 | 5.50E-33 |
| rs495828 | let-7a-5p | trans | 1.53E-12 | Nightingale | S_VLDL_FC | NA | T | G | 0.045509 | 0.00485 | 1.30E-20 |
| rs495828 | let-7a-5p | trans | 1.53E-12 | Nightingale | XL_VLDL_F | NA | T | G | 0.05038 | 0.004921 | 3.60E-25 |
| rs495828 | let-7a-5p | trans | 1.53E-12 | Nightingale | M_VLDL_F | NA | T | G | 0.059176 | 0.004829 | 2.10E-34 |
| rs495828 | let-7a-5p | trans | 1.53E-12 | Nightingale | XXL_VLDL_ | NA | T | G | 0.040705 | 0.005056 | 3.40E-16 |
| rs495828 | let-7a-5p | trans | 1.53E-12 | Nightingale | S_VLDL_P | NA | T | G | 0.031959 | 0.005012 | 4.90E-12 |
| rs495828 | let-7a-5p | trans | 1.53E-12 | Nightingale | S_VLDL_FC | NA | T | G | 0.054306 | 0.005076 | 2.60E-28 |
| rs495828 | let-7a-5p | trans | 1.53E-12 | Nightingale | XS_VLDL_C | NA | T | G | 0.037019 | 0.004814 | 5.50E-14 |
| rs495828 | let-7a-5p | trans | 1.53E-12 | Nightingale | Pyruvate | NA | T | G | 0.029634 | 0.005084 | 5.60E-09 |
| rs495828 | let-7a-5p | trans | 1.53E-12 | Nightingale | M_VLDL_P | NA | T | G | 0.041928 | 0.005068 | 4.10E-18 |
| rs495828 | let-7a-5p | trans | 1.53E-12 | Nightingale | Total_FC | NA | T | G | 0.049237 | 0.004958 | 3.30E-25 |
| rs495828 | let-7a-5p | trans | 1.53E-12 | Nightingale | VLDL_FC | NA | T | G | 0.032606 | 0.005038 | 3.90E-12 |
| rs495828 | let-7a-5p | trans | 1.53E-12 | Nightingale | XS_VLDL_F | NA | T | G | 0.049238 | 0.004987 | 1.30E-25 |
| rs495828 | let-7a-5p | trans | 1.53E-12 | Nightingale | XXL_VLDL_ | NA | T | G | 0.050116 | 0.005116 | 4.00E-23 |
| rs495828 | let-7a-5p | trans | 1.53E-12 | Nightingale | S_VLDL_C | NA | T | G | 0.054248 | 0.005075 | 1.40E-28 |
| rs495828 | let-7a-5p | trans | 1.53E-12 | Nightingale | LA | NA | T | G | 0.037741 | 0.004977 | 1.40E-15 |
| rs495828 | let-7a-5p | trans | 1.53E-12 | Nightingale | M_VLDL_F | NA | T | G | 0.047692 | 0.005075 | 1.90E-22 |
| rs495828 | let-7a-5p | trans | 1.53E-12 | Nightingale | M_LDL_C | NA | T | G | 0.052602 | 0.005085 | 7.60E-26 |
| rs495828 | let-7a-5p | trans | 1.53E-12 | Nightingale | L_HDL_TG_ | NA | T | G | 0.024075 | 0.004795 | 1.30E-08 |
| rs495828 | let-7a-5p | trans | 1.53E-12 | Nightingale | XS_VLDL_L | NA | T | G | 0.045042 | 0.004989 | 4.90E-22 |
| rs495828 | let-7a-5p | trans | 1.53E-12 | Nightingale | IDL_FC | NA | T | G | 0.056848 | 0.004914 | 9.10E-32 |
| rs495828 | let-7a-5p | trans | 1.53E-12 | Nightingale | S_LDL_L | NA | T | G | 0.044532 | 0.005075 | 4.30E-19 |
| rs495828 | let-7a-5p | trans | 1.53E-12 | Nightingale | IDL_TG_pc | NA | T | G | -0.03284 | 0.004907 | 7.40E-11 |
| rs495828 | let-7a-5p | trans | 1.53E-12 | Nightingale | S_LDL_CE | NA | T | G | 0.047169 | 0.005078 | 1.80E-21 |
| rs495828 | let-7a-5p | trans | 1.53E-12 | Nightingale | Total_CE | NA | T | G | 0.048939 | 0.004896 | 1.10E-24 |
| rs495828 | let-7a-5p | trans | 1.53E-12 | Nightingale | Phosphatic | NA | T | G | 0.027047 | 0.004756 | 1.70E-09 |
| rs495828 | let-7a-5p | trans | 1.53E-12 | Nightingale | LDL_FC | NA | T | G | 0.055702 | 0.005021 | 9.40E-29 |
| rs495828 | let-7a-5p | trans | 1.53E-12 | Nightingale | XL_VLDL_P | NA | T | G | 0.052707 | 0.005084 | 1.40E-26 |
| rs495828 | let-7a-5p | trans | 1.53E-12 | Nightingale | M_LDL_PL | NA | T | G | 0.052849 | 0.005083 | 4.60E-26 |
| rs495828 | let-7a-5p | trans | 1.53E-12 | Nightingale | S_VLDL_PL | NA | T | G | 0.043474 | 0.00486 | 7.60E-19 |
| rs495828 | let-7a-5p | trans | 1.53E-12 | Nightingale | S_HDL_P | NA | T | G | 0.047417 | 0.005076 | 8.50E-21 |
| rs495828 | let-7a-5p | trans | 1.53E-12 | Nightingale | L_VLDL_PL | NA | T | G | 0.027644 | 0.00493 | 6.40E-10 |
| rs495828 | let-7a-5p | trans | 1.53E-12 | Nightingale | XS_VLDL_F | NA | T | G | 0.043854 | 0.005039 | 1.70E-17 |
| rs495828 | let-7a-5p | trans | 1.53E-12 | Nightingale | Phosphogly | NA | T | G | 0.027772 | 0.004807 | 9.60E-10 |
| rs495828 | let-7a-5p | trans | 1.53E-12 | Nightingale | HDL_size | NA | T | G | -0.03057 | 0.0045 | 3.10E-12 |
| rs495828 | let-7a-5p | trans | 1.53E-12 | Nightingale | S_VLDL_L | NA | T | G | 0.033815 | 0.005026 | 4.60E-13 |
| rs495828 | let-7a-5p | trans | 1.53E-12 | Nightingale | S_LDL_FC | NA | T | G | 0.048985 | 0.005068 | 4.70E-22 |
| rs495828 | let-7a-5p | trans | 1.53E-12 | Nightingale | Omega_6 | NA | T | G | 0.037091 | 0.004956 | 2.90E-15 |
| rs495828 | let-7a-5p | trans | 1.53E-12 | Nightingale | Total_L | NA | T | G | 0.03839 | 0.004993 | 7.90E-16 |
| rs495828 | let-7a-5p | trans | 1.53E-12 | Nightingale | S_LDL_C_p | NA | T | G | 0.065319 | 0.005095 | 3.20E-38 |
| rs495828 | let-7a-5p | trans | 1.53E-12 | Nightingale | XL_VLDL_C | NA | T | G | 0.054421 | 0.004867 | 1.00E-28 |
| rs495828 | let-7a-5p | trans | 1.53E-12 | Nightingale | non_HDL_( | NA | T | G | 0.05466 | 0.005048 | 1.20E-28 |
| rs495828 | let-7a-5p | trans | 1.53E-12 | Nightingale | XS_VLDL_C | NA | T | G | 0.034393 | 0.004789 | 3.10E-12 |
| rs495828 | let-7a-5p | trans | 1.53E-12 | Nightingale | IDL_P | NA | T | G | 0.059807 | 0.005022 | 1.60E-33 |

| | | | | | | | | | | | |
|---|---|---|---|---|---|---|---|---|---|---|---|
| rs495828 | let-7a-5p | trans | 1.53E-12 | Nightingale | VLDL_P | NA | T | G | 0.0365 | 0.005048 | 1.40E-14 |
| rs495828 | let-7a-5p | trans | 1.53E-12 | Nightingale | S_VLDL_C_ | NA | T | G | 0.062388 | 0.00494 | 1.10E-36 |
| rs495828 | let-7a-5p | trans | 1.53E-12 | Nightingale | XL_VLDL_C | NA | T | G | 0.029159 | 0.004997 | 2.80E-10 |
| rs495828 | let-7a-5p | trans | 1.53E-12 | Nightingale | M_LDL_P | NA | T | G | 0.051176 | 0.00509 | 1.00E-24 |
| rs495828 | let-7a-5p | trans | 1.53E-12 | Nightingale | L_LDL_TG_ | NA | T | G | -0.0346 | 0.004978 | 1.30E-11 |
| rs495828 | let-7a-5p | trans | 1.53E-12 | Nightingale | S_HDL_C | NA | T | G | 0.044417 | 0.005067 | 1.80E-18 |
| rs495828 | let-7a-5p | trans | 1.53E-12 | Nightingale | Citrate | NA | T | G | 0.027426 | 0.005043 | 2.00E-08 |
| rs495828 | let-7a-5p | trans | 1.53E-12 | Nightingale | S_LDL_TG_ | NA | T | G | -0.0367 | 0.004922 | 5.00E-13 |
| rs495828 | let-7a-5p | trans | 1.53E-12 | Nightingale | M_VLDL_L | NA | T | G | 0.030732 | 0.005041 | 7.30E-11 |
| rs495828 | let-7a-5p | trans | 1.53E-12 | Nightingale | XS_VLDL_C | NA | T | G | 0.052929 | 0.004898 | 3.40E-29 |
| rs4962040 | miR-10a-5p | trans | 5.47E-13 | Nightingale | L_VLDL_CE | NA | G | A | -0.03218 | 0.004009 | 1.70E-15 |
| rs4962040 | miR-10a-5p | trans | 5.47E-13 | Nightingale | M_VLDL_C | NA | G | A | -0.02344 | 0.004178 | 5.50E-09 |
| rs4962040 | miR-10a-5p | trans | 5.47E-13 | Nightingale | S_VLDL_C_ | NA | G | A | -0.0284 | 0.004078 | 1.00E-11 |
| rs4962040 | miR-10a-5p | trans | 5.47E-13 | Nightingale | XL_VLDL_C | NA | G | A | -0.02571 | 0.004019 | 1.30E-09 |
| rs4962040 | miR-10a-5p | trans | 5.47E-13 | Nightingale | XS_VLDL_F | NA | G | A | -0.02146 | 0.004117 | 3.10E-08 |
| rs4962040 | miR-10a-5p | trans | 5.47E-13 | Nightingale | M_VLDL_C | NA | G | A | -0.02389 | 0.003947 | 7.60E-09 |
| rs4962040 | miR-10a-5p | trans | 5.47E-13 | Nightingale | L_VLDL_C_ | NA | G | A | -0.03539 | 0.004092 | 4.50E-18 |
| rs4962040 | miR-10a-5p | trans | 5.47E-13 | Nightingale | XXL_VLDL_ | NA | G | A | -0.02321 | 0.00417 | 2.80E-08 |
| rs4962040 | miR-10a-5p | trans | 5.47E-13 | Nightingale | M_VLDL_C | NA | G | A | -0.02564 | 0.004152 | 3.20E-10 |
| rs4962040 | miR-10a-5p | trans | 5.47E-13 | Nightingale | XS_VLDL_C | NA | G | A | -0.02339 | 0.004067 | 2.60E-09 |
| rs4962040 | miR-10a-5p | trans | 5.47E-13 | Nightingale | IDL_PL | NA | G | A | -0.02176 | 0.004043 | 4.90E-08 |
| rs4962040 | miR-10a-5p | trans | 5.47E-13 | Nightingale | L_VLDL_TG | NA | G | A | 0.033079 | 0.00417 | 4.10E-16 |
| rs4962040 | miR-10a-5p | trans | 5.47E-13 | Nightingale | M_VLDL_P | NA | G | A | -0.02877 | 0.004021 | 1.10E-12 |
| rs4962040 | miR-10a-5p | trans | 5.47E-13 | Nightingale | S_LDL_C_p | NA | G | A | -0.02967 | 0.004206 | 1.30E-12 |
| rs4962040 | miR-10a-5p | trans | 5.47E-13 | Nightingale | S_VLDL_C | NA | G | A | -0.02227 | 0.004189 | 2.50E-08 |
| rs4962040 | miR-10a-5p | trans | 5.47E-13 | Nightingale | XL_VLDL_C | NA | G | A | -0.02629 | 0.004017 | 4.00E-10 |
| rs4962040 | miR-10a-5p | trans | 5.47E-13 | Nightingale | IDL_L | NA | G | A | -0.02222 | 0.004051 | 3.50E-08 |
| rs4962040 | miR-10a-5p | trans | 5.47E-13 | Nightingale | S_VLDL_CE | NA | G | A | -0.03211 | 0.004134 | 1.10E-14 |
| rs4962040 | miR-10a-5p | trans | 5.47E-13 | Nightingale | XL_VLDL_T | NA | G | A | 0.029913 | 0.004061 | 6.90E-13 |
| rs4962040 | miR-10a-5p | trans | 5.47E-13 | Nightingale | S_VLDL_CE | NA | G | A | -0.02212 | 0.004184 | 2.50E-08 |
| rs4962040 | miR-10a-5p | trans | 5.47E-13 | Nightingale | S_VLDL_FC | NA | G | A | -0.02203 | 0.004191 | 4.70E-08 |
| rs4962040 | miR-10a-5p | trans | 5.47E-13 | Nightingale | Remnant_( | NA | G | A | -0.02207 | 0.004163 | 2.90E-08 |
| rs4962040 | miR-10a-5p | trans | 5.47E-13 | Nightingale | M_LDL_P | NA | G | A | -0.02283 | 0.004202 | 2.70E-08 |
| rs4962040 | miR-10a-5p | trans | 5.47E-13 | Nightingale | IDL_P | NA | G | A | -0.02436 | 0.004146 | 2.30E-09 |
| rs4962040 | miR-10a-5p | trans | 5.47E-13 | Nightingale | L_LDL_PL | NA | G | A | -0.02385 | 0.004158 | 6.50E-09 |
| rs4962040 | miR-10a-5p | trans | 5.47E-13 | Nightingale | XL_VLDL_F | NA | G | A | -0.02514 | 0.004061 | 8.50E-10 |
| rs4962040 | miR-10a-5p | trans | 5.47E-13 | Nightingale | IDL_FC | NA | G | A | -0.02463 | 0.004056 | 1.10E-09 |
| rs4962040 | miR-10a-5p | trans | 5.47E-13 | Nightingale | XS_VLDL_P | NA | G | A | -0.0212 | 0.004134 | 4.00E-08 |
| rs4962040 | miR-10a-5p | trans | 5.47E-13 | Nightingale | S_VLDL_TG | NA | G | A | 0.025983 | 0.004056 | 5.20E-10 |
| rs4962040 | miR-10a-5p | trans | 5.47E-13 | Nightingale | XXL_VLDL_ | NA | G | A | 0.026512 | 0.004224 | 1.50E-10 |
| rs4962040 | miR-10a-5p | trans | 5.47E-13 | Nightingale | XXL_VLDL_ | NA | G | A | -0.02587 | 0.004197 | 6.90E-10 |
| rs4962040 | miR-10a-5p | trans | 5.47E-13 | Nightingale | M_VLDL_T | NA | G | A | 0.025336 | 0.003959 | 6.90E-10 |
| rs4962040 | miR-10a-5p | trans | 5.47E-13 | Nightingale | Clinical_LD | NA | G | A | -0.02286 | 0.004166 | 3.40E-08 |
| rs4962040 | miR-10a-5p | trans | 5.47E-13 | Nightingale | XL_VLDL_P | NA | G | A | -0.0236 | 0.004196 | 4.90E-09 |
| rs4962040 | miR-10a-5p | trans | 5.47E-13 | Nightingale | L_VLDL_FC | NA | G | A | -0.03125 | 0.00415 | 2.20E-14 |
| rs4962040 | miR-10a-5p | trans | 5.47E-13 | Nightingale | XXL_VLDL_ | NA | G | A | -0.02489 | 0.00422 | 3.50E-09 |
| rs4962040 | miR-10a-5p | trans | 5.47E-13 | Nightingale | XS_VLDL_C | NA | G | A | -0.02383 | 0.004043 | 1.20E-09 |
| rs4962040 | miR-10a-5p | trans | 5.47E-13 | Nightingale | IDL_C | NA | G | A | -0.02259 | 0.004047 | 2.10E-08 |
| rs4962040 | miR-10a-5p | trans | 5.47E-13 | Nightingale | M_VLDL_F | NA | G | A | -0.02699 | 0.003986 | 4.00E-11 |
| rs4962040 | miR-126-3p | trans | 2.08E-18 | Nightingale | L_VLDL_CE | NA | G | A | -0.03218 | 0.004009 | 1.70E-15 |
| rs4962040 | miR-126-3p | trans | 2.08E-18 | Nightingale | M_VLDL_C | NA | G | A | -0.02344 | 0.004178 | 5.50E-09 |
| rs4962040 | miR-126-3p | trans | 2.08E-18 | Nightingale | S_VLDL_C_ | NA | G | A | -0.0284 | 0.004078 | 1.00E-11 |
| rs4962040 | miR-126-3p | trans | 2.08E-18 | Nightingale | XL_VLDL_C | NA | G | A | -0.02571 | 0.004019 | 1.30E-09 |
| rs4962040 | miR-126-3p | trans | 2.08E-18 | Nightingale | XS_VLDL_F | NA | G | A | -0.02146 | 0.004117 | 3.10E-08 |
| rs4962040 | miR-126-3p | trans | 2.08E-18 | Nightingale | M_VLDL_C | NA | G | A | -0.02389 | 0.003947 | 7.60E-09 |
| rs4962040 | miR-126-3p | trans | 2.08E-18 | Nightingale | L_VLDL_C_ | NA | G | A | -0.03539 | 0.004092 | 4.50E-18 |
| rs4962040 | miR-126-3p | trans | 2.08E-18 | Nightingale | XXL_VLDL_ | NA | G | A | -0.02321 | 0.00417 | 2.80E-08 |
| rs4962040 | miR-126-3p | trans | 2.08E-18 | Nightingale | M_VLDL_C | NA | G | A | -0.02564 | 0.004152 | 3.20E-10 |
| rs4962040 | miR-126-3p | trans | 2.08E-18 | Nightingale | XS_VLDL_C | NA | G | A | -0.02339 | 0.004067 | 2.60E-09 |
| rs4962040 | miR-126-3p | trans | 2.08E-18 | Nightingale | IDL_PL | NA | G | A | -0.02176 | 0.004043 | 4.90E-08 |
| rs4962040 | miR-126-3p | trans | 2.08E-18 | Nightingale | L_VLDL_TG | NA | G | A | 0.033079 | 0.00417 | 4.10E-16 |
| rs4962040 | miR-126-3p | trans | 2.08E-18 | Nightingale | M_VLDL_P | NA | G | A | -0.02877 | 0.004021 | 1.10E-12 |
| rs4962040 | miR-126-3p | trans | 2.08E-18 | Nightingale | S_LDL_C_p | NA | G | A | -0.02967 | 0.004206 | 1.30E-12 |

| | | | | | | | | | | | |
|---|---|---|---|---|---|---|---|---|---|---|---|
| rs4962040 | miR-126-3p | trans | 2.08E-18 | Nightingale | S_VLDL_C | NA | G | A | -0.02227 | 0.004189 | 2.50E-08 |
| rs4962040 | miR-126-3p | trans | 2.08E-18 | Nightingale | XL_VLDL_C | NA | G | A | -0.02629 | 0.004017 | 4.00E-10 |
| rs4962040 | miR-126-3p | trans | 2.08E-18 | Nightingale | IDL_L | NA | G | A | -0.02222 | 0.004051 | 3.50E-08 |
| rs4962040 | miR-126-3p | trans | 2.08E-18 | Nightingale | S_VLDL_CE | NA | G | A | -0.03211 | 0.004134 | 1.10E-14 |
| rs4962040 | miR-126-3p | trans | 2.08E-18 | Nightingale | XL_VLDL_T | NA | G | A | 0.029913 | 0.004061 | 6.90E-13 |
| rs4962040 | miR-126-3p | trans | 2.08E-18 | Nightingale | S_VLDL_CE | NA | G | A | -0.02212 | 0.004184 | 2.50E-08 |
| rs4962040 | miR-126-3p | trans | 2.08E-18 | Nightingale | S_VLDL_FC | NA | G | A | -0.02203 | 0.004191 | 4.70E-08 |
| rs4962040 | miR-126-3p | trans | 2.08E-18 | Nightingale | Remnant_( | NA | G | A | -0.02207 | 0.004163 | 2.90E-08 |
| rs4962040 | miR-126-3p | trans | 2.08E-18 | Nightingale | M_LDL_P | NA | G | A | -0.02283 | 0.004202 | 2.70E-08 |
| rs4962040 | miR-126-3p | trans | 2.08E-18 | Nightingale | IDL_P | NA | G | A | -0.02436 | 0.004146 | 2.30E-09 |
| rs4962040 | miR-126-3p | trans | 2.08E-18 | Nightingale | L_LDL_PL | NA | G | A | -0.02385 | 0.004158 | 6.50E-09 |
| rs4962040 | miR-126-3p | trans | 2.08E-18 | Nightingale | XL_VLDL_F | NA | G | A | -0.02514 | 0.004061 | 8.50E-10 |
| rs4962040 | miR-126-3p | trans | 2.08E-18 | Nightingale | IDL_FC | NA | G | A | -0.02463 | 0.004056 | 1.10E-09 |
| rs4962040 | miR-126-3p | trans | 2.08E-18 | Nightingale | XS_VLDL_P | NA | G | A | -0.0212 | 0.004134 | 4.00E-08 |
| rs4962040 | miR-126-3p | trans | 2.08E-18 | Nightingale | S_VLDL_TG | NA | G | A | 0.025983 | 0.004056 | 5.20E-10 |
| rs4962040 | miR-126-3p | trans | 2.08E-18 | Nightingale | XXL_VLDL_ | NA | G | A | 0.026512 | 0.004224 | 1.50E-10 |
| rs4962040 | miR-126-3p | trans | 2.08E-18 | Nightingale | XXL_VLDL_ | NA | G | A | -0.02587 | 0.004197 | 6.90E-10 |
| rs4962040 | miR-126-3p | trans | 2.08E-18 | Nightingale | M_VLDL_T | NA | G | A | 0.025336 | 0.003959 | 6.90E-10 |
| rs4962040 | miR-126-3p | trans | 2.08E-18 | Nightingale | Clinical_LD | NA | G | A | -0.02286 | 0.004166 | 3.40E-08 |
| rs4962040 | miR-126-3p | trans | 2.08E-18 | Nightingale | XL_VLDL_P | NA | G | A | -0.0236 | 0.004196 | 4.90E-09 |
| rs4962040 | miR-126-3p | trans | 2.08E-18 | Nightingale | L_VLDL_FC | NA | G | A | -0.03125 | 0.00415 | 2.20E-14 |
| rs4962040 | miR-126-3p | trans | 2.08E-18 | Nightingale | XXL_VLDL_ | NA | G | A | -0.02489 | 0.00422 | 3.50E-09 |
| rs4962040 | miR-126-3p | trans | 2.08E-18 | Nightingale | XS_VLDL_C | NA | G | A | -0.02383 | 0.004043 | 1.20E-09 |
| rs4962040 | miR-126-3p | trans | 2.08E-18 | Nightingale | IDL_C | NA | G | A | -0.02259 | 0.004047 | 2.10E-08 |
| rs4962040 | miR-126-3p | trans | 2.08E-18 | Nightingale | M_VLDL_F | NA | G | A | -0.02699 | 0.003986 | 4.00E-11 |
| rs529565 | miR-195-5p | trans | 3.57E-18 | Nightingale | M_LDL_PL | NA | C | T | 0.03206 | 0.004437 | 4.30E-13 |
| rs529565 | miR-195-5p | trans | 3.57E-18 | Nightingale | S_LDL_L | NA | C | T | 0.027285 | 0.00443 | 1.30E-09 |
| rs529565 | miR-195-5p | trans | 3.57E-18 | Nightingale | XXL_VLDL_ | NA | C | T | 0.031056 | 0.004412 | 9.30E-13 |
| rs529565 | miR-195-5p | trans | 3.57E-18 | Nightingale | VLDL_C | NA | C | T | 0.030948 | 0.004425 | 6.40E-13 |
| rs529565 | miR-195-5p | trans | 3.57E-18 | Nightingale | S_LDL_CE_ | NA | C | T | 0.031515 | 0.004423 | 1.80E-14 |
| rs529565 | miR-195-5p | trans | 3.57E-18 | Nightingale | M_VLDL_P | NA | C | T | 0.043673 | 0.004253 | 1.80E-24 |
| rs529565 | miR-195-5p | trans | 3.57E-18 | Nightingale | LDL_P | NA | C | T | 0.031312 | 0.004428 | 1.80E-12 |
| rs529565 | miR-195-5p | trans | 3.57E-18 | Nightingale | S_HDL_P | NA | C | T | 0.030973 | 0.004432 | 3.30E-12 |
| rs529565 | miR-195-5p | trans | 3.57E-18 | Nightingale | XL_VLDL_P | NA | C | T | 0.038314 | 0.004438 | 1.50E-18 |
| rs529565 | miR-195-5p | trans | 3.57E-18 | Nightingale | S_VLDL_L | NA | C | T | 0.02438 | 0.004388 | 4.50E-09 |
| rs529565 | miR-195-5p | trans | 3.57E-18 | Nightingale | VLDL_FC | NA | C | T | 0.02385 | 0.004399 | 1.30E-08 |
| rs529565 | miR-195-5p | trans | 3.57E-18 | Nightingale | Citrate | NA | C | T | 0.027701 | 0.004402 | 2.10E-10 |
| rs529565 | miR-195-5p | trans | 3.57E-18 | Nightingale | M_VLDL_C | NA | C | T | 0.036055 | 0.004174 | 2.10E-17 |
| rs529565 | miR-195-5p | trans | 3.57E-18 | Nightingale | XL_VLDL_F | NA | C | T | 0.036987 | 0.004296 | 3.90E-18 |
| rs529565 | miR-195-5p | trans | 3.57E-18 | Nightingale | M_LDL_C | NA | C | T | 0.033658 | 0.004439 | 3.10E-14 |
| rs529565 | miR-195-5p | trans | 3.57E-18 | Nightingale | S_VLDL_CE | NA | C | T | 0.046913 | 0.004372 | 2.40E-26 |
| rs529565 | miR-195-5p | trans | 3.57E-18 | Nightingale | M_VLDL_C | NA | C | T | 0.038197 | 0.004419 | 4.00E-18 |
| rs529565 | miR-195-5p | trans | 3.57E-18 | Nightingale | S_LDL_P | NA | C | T | 0.029187 | 0.004435 | 4.60E-11 |
| rs529565 | miR-195-5p | trans | 3.57E-18 | Nightingale | IDL_FC_pct | NA | C | T | 0.025499 | 0.004397 | 1.50E-08 |
| rs529565 | miR-195-5p | trans | 3.57E-18 | Nightingale | Sphingomy | NA | C | T | 0.02867 | 0.004142 | 1.30E-11 |
| rs529565 | miR-195-5p | trans | 3.57E-18 | Nightingale | Total_FC | NA | C | T | 0.032841 | 0.004329 | 1.20E-14 |
| rs529565 | miR-195-5p | trans | 3.57E-18 | Nightingale | Clinical_LD | NA | C | T | 0.035972 | 0.004405 | 5.80E-16 |
| rs529565 | miR-195-5p | trans | 3.57E-18 | Nightingale | L_LDL_L | NA | C | T | 0.033109 | 0.004388 | 5.60E-14 |
| rs529565 | miR-195-5p | trans | 3.57E-18 | Nightingale | LDL_CE | NA | C | T | 0.032164 | 0.004419 | 3.50E-13 |
| rs529565 | miR-195-5p | trans | 3.57E-18 | Nightingale | S_VLDL_TG | NA | C | T | -0.03791 | 0.004289 | 2.80E-18 |
| rs529565 | miR-195-5p | trans | 3.57E-18 | Nightingale | S_LDL_PL_ | NA | C | T | -0.03064 | 0.004423 | 9.40E-14 |
| rs529565 | miR-195-5p | trans | 3.57E-18 | Nightingale | VLDL_CE | NA | C | T | 0.035403 | 0.004432 | 4.50E-16 |
| rs529565 | miR-195-5p | trans | 3.57E-18 | Nightingale | L_LDL_CE | NA | C | T | 0.03113 | 0.004392 | 1.70E-12 |
| rs529565 | miR-195-5p | trans | 3.57E-18 | Nightingale | L_VLDL_TG | NA | C | T | -0.05092 | 0.00441 | 1.60E-31 |
| rs529565 | miR-195-5p | trans | 3.57E-18 | Nightingale | S_VLDL_C_ | NA | C | T | 0.041702 | 0.004312 | 1.10E-21 |
| rs529565 | miR-195-5p | trans | 3.57E-18 | Nightingale | LDL_FC | NA | C | T | 0.033085 | 0.004384 | 1.10E-13 |
| rs529565 | miR-195-5p | trans | 3.57E-18 | Nightingale | XXL_VLDL_ | NA | C | T | -0.03595 | 0.004469 | 1.80E-16 |
| rs529565 | miR-195-5p | trans | 3.57E-18 | Nightingale | S_VLDL_C | NA | C | T | 0.038015 | 0.004431 | 2.60E-18 |
| rs529565 | miR-195-5p | trans | 3.57E-18 | Nightingale | IDL_L | NA | C | T | 0.034696 | 0.004284 | 1.30E-15 |
| rs529565 | miR-195-5p | trans | 3.57E-18 | Nightingale | IDL_PL | NA | C | T | 0.034834 | 0.004276 | 7.90E-16 |
| rs529565 | miR-195-5p | trans | 3.57E-18 | Nightingale | non_HDL_( | NA | C | T | 0.035225 | 0.004407 | 1.00E-15 |
| rs529565 | miR-195-5p | trans | 3.57E-18 | Nightingale | S_VLDL_FC | NA | C | T | 0.02911 | 0.004234 | 1.70E-11 |
| rs529565 | miR-195-5p | trans | 3.57E-18 | Nightingale | S_LDL_CE | NA | C | T | 0.030231 | 0.004433 | 7.50E-12 |

| | | | | | | | | | | | |
|---|---|---|---|---|---|---|---|---|---|---|---|
| rs529565 | miR-195-5p | trans | 3.57E-18 | Nightingale | XS_VLDL_C | NA | C | T | 0.037263 | 0.004276 | 5.70E-18 |
| rs529565 | miR-195-5p | trans | 3.57E-18 | Nightingale | Total_PL | NA | C | T | 0.024805 | 0.004219 | 1.20E-09 |
| rs529565 | miR-195-5p | trans | 3.57E-18 | Nightingale | M_LDL_TG | NA | C | T | -0.02724 | 0.004376 | 5.60E-10 |
| rs529565 | miR-195-5p | trans | 3.57E-18 | Nightingale | S_LDL_C | NA | C | T | 0.030526 | 0.004435 | 8.70E-12 |
| rs529565 | miR-195-5p | trans | 3.57E-18 | Nightingale | M_VLDL_F | NA | C | T | 0.041684 | 0.004216 | 1.10E-22 |
| rs529565 | miR-195-5p | trans | 3.57E-18 | Nightingale | M_VLDL_C | NA | C | T | 0.040303 | 0.004391 | 5.90E-20 |
| rs529565 | miR-195-5p | trans | 3.57E-18 | Nightingale | XL_HDL_PL | NA | C | T | -0.02275 | 0.004071 | 1.10E-08 |
| rs529565 | miR-195-5p | trans | 3.57E-18 | Nightingale | LDL_L | NA | C | T | 0.032885 | 0.004416 | 1.30E-13 |
| rs529565 | miR-195-5p | trans | 3.57E-18 | Nightingale | Remnant_( | NA | C | T | 0.036369 | 0.004402 | 7.40E-17 |
| rs529565 | miR-195-5p | trans | 3.57E-18 | Nightingale | IDL_FC | NA | C | T | 0.037662 | 0.00429 | 4.70E-18 |
| rs529565 | miR-195-5p | trans | 3.57E-18 | Nightingale | LDL_PL | NA | C | T | 0.034514 | 0.004421 | 9.20E-15 |
| rs529565 | miR-195-5p | trans | 3.57E-18 | Nightingale | M_LDL_CE | NA | C | T | 0.032712 | 0.004437 | 1.20E-13 |
| rs529565 | miR-195-5p | trans | 3.57E-18 | Nightingale | S_VLDL_P | NA | C | T | 0.023183 | 0.004376 | 1.90E-08 |
| rs529565 | miR-195-5p | trans | 3.57E-18 | Nightingale | L_VLDL_CE | NA | C | T | 0.047309 | 0.00424 | 5.10E-29 |
| rs529565 | miR-195-5p | trans | 3.57E-18 | Nightingale | L_LDL_PL | NA | C | T | 0.03752 | 0.004398 | 1.90E-17 |
| rs529565 | miR-195-5p | trans | 3.57E-18 | Nightingale | S_HDL_C | NA | C | T | 0.02876 | 0.004424 | 1.20E-10 |
| rs529565 | miR-195-5p | trans | 3.57E-18 | Nightingale | XS_VLDL_F | NA | C | T | 0.036631 | 0.004354 | 1.20E-17 |
| rs529565 | miR-195-5p | trans | 3.57E-18 | Nightingale | XL_VLDL_C | NA | C | T | 0.037516 | 0.004251 | 3.10E-18 |
| rs529565 | miR-195-5p | trans | 3.57E-18 | Nightingale | S_VLDL_PL | NA | C | T | 0.027457 | 0.004243 | 2.90E-10 |
| rs529565 | miR-195-5p | trans | 3.57E-18 | Nightingale | M_VLDL_P | NA | C | T | 0.027887 | 0.00442 | 8.70E-11 |
| rs529565 | miR-195-5p | trans | 3.57E-18 | Nightingale | LA | NA | C | T | 0.024778 | 0.004345 | 3.90E-09 |
| rs529565 | miR-195-5p | trans | 3.57E-18 | Nightingale | IDL_CE | NA | C | T | 0.03286 | 0.004278 | 4.10E-14 |
| rs529565 | miR-195-5p | trans | 3.57E-18 | Nightingale | XS_VLDL_C | NA | C | T | 0.037489 | 0.004301 | 3.30E-18 |
| rs529565 | miR-195-5p | trans | 3.57E-18 | Nightingale | VLDL_P | NA | C | T | 0.026295 | 0.004407 | 5.50E-10 |
| rs529565 | miR-195-5p | trans | 3.57E-18 | Nightingale | M_VLDL_T | NA | C | T | -0.03834 | 0.004187 | 1.50E-19 |
| rs529565 | miR-195-5p | trans | 3.57E-18 | Nightingale | L_LDL_P | NA | C | T | 0.028589 | 0.004416 | 1.20E-10 |
| rs529565 | miR-195-5p | trans | 3.57E-18 | Nightingale | S_HDL_PL | NA | C | T | 0.024302 | 0.004393 | 1.90E-08 |
| rs529565 | miR-195-5p | trans | 3.57E-18 | Nightingale | Total_CE | NA | C | T | 0.031352 | 0.004274 | 1.80E-13 |
| rs529565 | miR-195-5p | trans | 3.57E-18 | Nightingale | M_LDL_C_ | NA | C | T | 0.030966 | 0.004434 | 4.00E-12 |
| rs529565 | miR-195-5p | trans | 3.57E-18 | Nightingale | XXL_VLDL_ | NA | C | T | 0.034955 | 0.00444 | 2.80E-15 |
| rs529565 | miR-195-5p | trans | 3.57E-18 | Nightingale | XS_VLDL_P | NA | C | T | 0.036324 | 0.004372 | 2.20E-17 |
| rs529565 | miR-195-5p | trans | 3.57E-18 | Nightingale | S_LDL_FC | NA | C | T | 0.027889 | 0.004424 | 9.30E-10 |
| rs529565 | miR-195-5p | trans | 3.57E-18 | Nightingale | L_VLDL_FC | NA | C | T | 0.048532 | 0.004389 | 7.40E-29 |
| rs529565 | miR-195-5p | trans | 3.57E-18 | Nightingale | Omega_6 | NA | C | T | 0.025773 | 0.004327 | 6.10E-10 |
| rs529565 | miR-195-5p | trans | 3.57E-18 | Nightingale | L_LDL_FC | NA | C | T | 0.033087 | 0.004351 | 6.50E-14 |
| rs529565 | miR-195-5p | trans | 3.57E-18 | Nightingale | XS_VLDL_P | NA | C | T | 0.031742 | 0.004369 | 8.00E-14 |
| rs529565 | miR-195-5p | trans | 3.57E-18 | Nightingale | S_HDL_CE | NA | C | T | 0.028978 | 0.004438 | 8.90E-11 |
| rs529565 | miR-195-5p | trans | 3.57E-18 | Nightingale | GlycA | NA | C | T | 0.027732 | 0.004412 | 2.40E-10 |
| rs529565 | miR-195-5p | trans | 3.57E-18 | Nightingale | Total_L | NA | C | T | 0.026125 | 0.004359 | 5.60E-10 |
| rs529565 | miR-195-5p | trans | 3.57E-18 | Nightingale | S_HDL_FC | NA | C | T | 0.023726 | 0.00435 | 3.60E-08 |
| rs529565 | miR-195-5p | trans | 3.57E-18 | Nightingale | L_LDL_C | NA | C | T | 0.031856 | 0.004381 | 4.80E-13 |
| rs529565 | miR-195-5p | trans | 3.57E-18 | Nightingale | LDL_C | NA | C | T | 0.032762 | 0.004412 | 1.60E-13 |
| rs529565 | miR-195-5p | trans | 3.57E-18 | Nightingale | M_VLDL_P | NA | C | T | 0.02872 | 0.004424 | 2.80E-11 |
| rs529565 | miR-195-5p | trans | 3.57E-18 | Nightingale | S_VLDL_CE | NA | C | T | 0.037878 | 0.004425 | 2.20E-18 |
| rs529565 | miR-195-5p | trans | 3.57E-18 | Nightingale | XL_VLDL_T | NA | C | T | -0.04378 | 0.004295 | 3.70E-24 |
| rs529565 | miR-195-5p | trans | 3.57E-18 | Nightingale | S_VLDL_FC | NA | C | T | 0.03746 | 0.004432 | 1.30E-17 |
| rs529565 | miR-195-5p | trans | 3.57E-18 | Nightingale | XXL_VLDL_ | NA | C | T | 0.025677 | 0.004459 | 7.00E-10 |
| rs529565 | miR-195-5p | trans | 3.57E-18 | Nightingale | L_VLDL_C_ | NA | C | T | 0.05212 | 0.004328 | 9.90E-34 |
| rs529565 | miR-195-5p | trans | 3.57E-18 | Nightingale | IDL_P | NA | C | T | 0.039431 | 0.004385 | 3.50E-19 |
| rs529565 | miR-195-5p | trans | 3.57E-18 | Nightingale | XS_VLDL_L | NA | C | T | 0.033423 | 0.004356 | 3.30E-15 |
| rs529565 | miR-195-5p | trans | 3.57E-18 | Nightingale | M_VLDL_C | NA | C | T | 0.033552 | 0.004163 | 2.80E-15 |
| rs529565 | miR-195-5p | trans | 3.57E-18 | Nightingale | XL_VLDL_C | NA | C | T | 0.038352 | 0.004249 | 5.20E-19 |
| rs529565 | miR-195-5p | trans | 3.57E-18 | Nightingale | Total_C | NA | C | T | 0.031972 | 0.00429 | 6.30E-14 |
| rs529565 | miR-195-5p | trans | 3.57E-18 | Nightingale | M_LDL_FC | NA | C | T | 0.033834 | 0.004426 | 4.00E-14 |
| rs529565 | miR-195-5p | trans | 3.57E-18 | Nightingale | M_VLDL_F | NA | C | T | 0.032904 | 0.00443 | 5.40E-14 |
| rs529565 | miR-195-5p | trans | 3.57E-18 | Nightingale | ApoB | NA | C | T | 0.033444 | 0.004425 | 3.90E-14 |
| rs529565 | miR-195-5p | trans | 3.57E-18 | Nightingale | ApoB_by_/ | NA | C | T | 0.02449 | 0.004364 | 2.40E-08 |
| rs529565 | miR-195-5p | trans | 3.57E-18 | Nightingale | S_LDL_C_p | NA | C | T | 0.045563 | 0.004448 | 4.00E-25 |
| rs529565 | miR-195-5p | trans | 3.57E-18 | Nightingale | PUFA | NA | C | T | 0.02347 | 0.004322 | 1.80E-08 |
| rs529565 | miR-195-5p | trans | 3.57E-18 | Nightingale | S_HDL_L | NA | C | T | 0.02679 | 0.004416 | 8.80E-10 |
| rs529565 | miR-195-5p | trans | 3.57E-18 | Nightingale | S_VLDL_PL | NA | C | T | 0.03238 | 0.004423 | 6.00E-14 |
| rs529565 | miR-195-5p | trans | 3.57E-18 | Nightingale | M_LDL_L | NA | C | T | 0.032702 | 0.004437 | 1.40E-13 |
| rs529565 | miR-195-5p | trans | 3.57E-18 | Nightingale | M_LDL_P | NA | C | T | 0.035668 | 0.004443 | 8.20E-16 |

| | | | | | | | | | | | |
|---|---|---|---|---|---|---|---|---|---|---|---|
| rs529565 | miR-195-5p | trans | 3.57E-18 | Nightingale | XS_VLDL_F | NA | C | T | 0.030891 | 0.004399 | 5.80E-12 |
| rs529565 | miR-195-5p | trans | 3.57E-18 | Nightingale | XXL_VLDL_ | NA | C | T | 0.033086 | 0.004464 | 1.10E-13 |
| rs529565 | miR-195-5p | trans | 3.57E-18 | Nightingale | IDL_C | NA | C | T | 0.034316 | 0.00428 | 3.00E-15 |
| rs529565 | miR-139-5p | trans | 2.82E-43 | Nightingale | M_LDL_PL | NA | C | T | 0.03206 | 0.004437 | 4.30E-13 |
| rs529565 | miR-139-5p | trans | 2.82E-43 | Nightingale | S_LDL_L | NA | C | T | 0.027285 | 0.00443 | 1.30E-09 |
| rs529565 | miR-139-5p | trans | 2.82E-43 | Nightingale | XXL_VLDL_ | NA | C | T | 0.031056 | 0.004412 | 9.30E-13 |
| rs529565 | miR-139-5p | trans | 2.82E-43 | Nightingale | VLDL_C | NA | C | T | 0.030948 | 0.004425 | 6.40E-13 |
| rs529565 | miR-139-5p | trans | 2.82E-43 | Nightingale | S_LDL_CE_ | NA | C | T | 0.031515 | 0.004423 | 1.80E-14 |
| rs529565 | miR-139-5p | trans | 2.82E-43 | Nightingale | M_VLDL_P | NA | C | T | 0.043673 | 0.004253 | 1.80E-24 |
| rs529565 | miR-139-5p | trans | 2.82E-43 | Nightingale | LDL_P | NA | C | T | 0.031312 | 0.004428 | 1.80E-12 |
| rs529565 | miR-139-5p | trans | 2.82E-43 | Nightingale | S_HDL_P | NA | C | T | 0.030973 | 0.004432 | 3.30E-12 |
| rs529565 | miR-139-5p | trans | 2.82E-43 | Nightingale | XL_VLDL_P | NA | C | T | 0.038314 | 0.004438 | 1.50E-18 |
| rs529565 | miR-139-5p | trans | 2.82E-43 | Nightingale | S_VLDL_L | NA | C | T | 0.02438 | 0.004388 | 4.50E-09 |
| rs529565 | miR-139-5p | trans | 2.82E-43 | Nightingale | VLDL_FC | NA | C | T | 0.02385 | 0.004399 | 1.30E-08 |
| rs529565 | miR-139-5p | trans | 2.82E-43 | Nightingale | Citrate | NA | C | T | 0.027701 | 0.004402 | 2.10E-10 |
| rs529565 | miR-139-5p | trans | 2.82E-43 | Nightingale | M_VLDL_C_ | NA | C | T | 0.036055 | 0.004174 | 2.10E-17 |
| rs529565 | miR-139-5p | trans | 2.82E-43 | Nightingale | XL_VLDL_F | NA | C | T | 0.036987 | 0.004296 | 3.90E-18 |
| rs529565 | miR-139-5p | trans | 2.82E-43 | Nightingale | M_LDL_C | NA | C | T | 0.033658 | 0.004439 | 3.10E-14 |
| rs529565 | miR-139-5p | trans | 2.82E-43 | Nightingale | S_VLDL_CE | NA | C | T | 0.046913 | 0.004372 | 2.40E-26 |
| rs529565 | miR-139-5p | trans | 2.82E-43 | Nightingale | M_VLDL_C | NA | C | T | 0.038197 | 0.004419 | 4.00E-18 |
| rs529565 | miR-139-5p | trans | 2.82E-43 | Nightingale | S_LDL_P | NA | C | T | 0.029187 | 0.004435 | 4.60E-11 |
| rs529565 | miR-139-5p | trans | 2.82E-43 | Nightingale | IDL_FC_pct | NA | C | T | 0.025499 | 0.004397 | 1.50E-08 |
| rs529565 | miR-139-5p | trans | 2.82E-43 | Nightingale | Sphingomy | NA | C | T | 0.02867 | 0.004142 | 1.30E-11 |
| rs529565 | miR-139-5p | trans | 2.82E-43 | Nightingale | Total_FC | NA | C | T | 0.032841 | 0.004329 | 1.20E-14 |
| rs529565 | miR-139-5p | trans | 2.82E-43 | Nightingale | Clinical_LD | NA | C | T | 0.035972 | 0.004405 | 5.80E-16 |
| rs529565 | miR-139-5p | trans | 2.82E-43 | Nightingale | L_LDL_L | NA | C | T | 0.033109 | 0.004388 | 5.60E-14 |
| rs529565 | miR-139-5p | trans | 2.82E-43 | Nightingale | LDL_CE | NA | C | T | 0.032164 | 0.004419 | 3.50E-13 |
| rs529565 | miR-139-5p | trans | 2.82E-43 | Nightingale | S_VLDL_TG | NA | C | T | -0.03791 | 0.004289 | 2.80E-18 |
| rs529565 | miR-139-5p | trans | 2.82E-43 | Nightingale | S_LDL_PL_ | NA | C | T | -0.03064 | 0.004423 | 9.40E-14 |
| rs529565 | miR-139-5p | trans | 2.82E-43 | Nightingale | VLDL_CE | NA | C | T | 0.035403 | 0.004432 | 4.50E-16 |
| rs529565 | miR-139-5p | trans | 2.82E-43 | Nightingale | L_LDL_CE | NA | C | T | 0.03113 | 0.004392 | 1.70E-12 |
| rs529565 | miR-139-5p | trans | 2.82E-43 | Nightingale | L_VLDL_TG | NA | C | T | -0.05092 | 0.00441 | 1.60E-31 |
| rs529565 | miR-139-5p | trans | 2.82E-43 | Nightingale | S_VLDL_C_ | NA | C | T | 0.041702 | 0.004312 | 1.10E-21 |
| rs529565 | miR-139-5p | trans | 2.82E-43 | Nightingale | LDL_FC | NA | C | T | 0.033085 | 0.004384 | 1.10E-13 |
| rs529565 | miR-139-5p | trans | 2.82E-43 | Nightingale | XXL_VLDL_ | NA | C | T | -0.03595 | 0.004469 | 1.80E-16 |
| rs529565 | miR-139-5p | trans | 2.82E-43 | Nightingale | S_VLDL_C | NA | C | T | 0.038015 | 0.004431 | 2.60E-18 |
| rs529565 | miR-139-5p | trans | 2.82E-43 | Nightingale | IDL_L | NA | C | T | 0.034696 | 0.004284 | 1.30E-15 |
| rs529565 | miR-139-5p | trans | 2.82E-43 | Nightingale | IDL_PL | NA | C | T | 0.034834 | 0.004276 | 7.90E-16 |
| rs529565 | miR-139-5p | trans | 2.82E-43 | Nightingale | non_HDL_( | NA | C | T | 0.035225 | 0.004407 | 1.00E-15 |
| rs529565 | miR-139-5p | trans | 2.82E-43 | Nightingale | S_VLDL_FC | NA | C | T | 0.02911 | 0.004234 | 1.70E-11 |
| rs529565 | miR-139-5p | trans | 2.82E-43 | Nightingale | S_LDL_CE | NA | C | T | 0.030231 | 0.004433 | 7.50E-12 |
| rs529565 | miR-139-5p | trans | 2.82E-43 | Nightingale | XS_VLDL_C | NA | C | T | 0.037263 | 0.004276 | 5.70E-18 |
| rs529565 | miR-139-5p | trans | 2.82E-43 | Nightingale | Total_PL | NA | C | T | 0.024805 | 0.004219 | 1.20E-09 |
| rs529565 | miR-139-5p | trans | 2.82E-43 | Nightingale | M_LDL_TG | NA | C | T | -0.02724 | 0.004376 | 5.60E-10 |
| rs529565 | miR-139-5p | trans | 2.82E-43 | Nightingale | S_LDL_C | NA | C | T | 0.030526 | 0.004435 | 8.70E-12 |
| rs529565 | miR-139-5p | trans | 2.82E-43 | Nightingale | M_VLDL_F | NA | C | T | 0.041684 | 0.004216 | 1.10E-22 |
| rs529565 | miR-139-5p | trans | 2.82E-43 | Nightingale | M_VLDL_C | NA | C | T | 0.040303 | 0.004391 | 5.90E-20 |
| rs529565 | miR-139-5p | trans | 2.82E-43 | Nightingale | XL_HDL_PL | NA | C | T | -0.02275 | 0.004071 | 1.10E-08 |
| rs529565 | miR-139-5p | trans | 2.82E-43 | Nightingale | LDL_L | NA | C | T | 0.032885 | 0.004416 | 1.30E-13 |
| rs529565 | miR-139-5p | trans | 2.82E-43 | Nightingale | Remnant_( | NA | C | T | 0.036369 | 0.004402 | 7.40E-17 |
| rs529565 | miR-139-5p | trans | 2.82E-43 | Nightingale | IDL_FC | NA | C | T | 0.037662 | 0.00429 | 4.70E-18 |
| rs529565 | miR-139-5p | trans | 2.82E-43 | Nightingale | LDL_PL | NA | C | T | 0.034514 | 0.004421 | 9.20E-15 |
| rs529565 | miR-139-5p | trans | 2.82E-43 | Nightingale | M_LDL_CE | NA | C | T | 0.032712 | 0.004437 | 1.20E-13 |
| rs529565 | miR-139-5p | trans | 2.82E-43 | Nightingale | S_VLDL_P | NA | C | T | 0.023183 | 0.004376 | 1.90E-08 |
| rs529565 | miR-139-5p | trans | 2.82E-43 | Nightingale | L_VLDL_CE | NA | C | T | 0.047309 | 0.00424 | 5.10E-29 |
| rs529565 | miR-139-5p | trans | 2.82E-43 | Nightingale | L_LDL_PL | NA | C | T | 0.03752 | 0.004398 | 1.90E-17 |
| rs529565 | miR-139-5p | trans | 2.82E-43 | Nightingale | S_HDL_C | NA | C | T | 0.02876 | 0.004424 | 1.20E-10 |
| rs529565 | miR-139-5p | trans | 2.82E-43 | Nightingale | XS_VLDL_F | NA | C | T | 0.036631 | 0.004354 | 1.20E-17 |
| rs529565 | miR-139-5p | trans | 2.82E-43 | Nightingale | XL_VLDL_C | NA | C | T | 0.037516 | 0.004251 | 3.10E-18 |
| rs529565 | miR-139-5p | trans | 2.82E-43 | Nightingale | S_VLDL_PL | NA | C | T | 0.027457 | 0.004243 | 2.90E-10 |
| rs529565 | miR-139-5p | trans | 2.82E-43 | Nightingale | M_VLDL_P | NA | C | T | 0.027887 | 0.00442 | 8.70E-11 |
| rs529565 | miR-139-5p | trans | 2.82E-43 | Nightingale | LA | NA | C | T | 0.024778 | 0.004345 | 3.90E-09 |
| rs529565 | miR-139-5p | trans | 2.82E-43 | Nightingale | IDL_CE | NA | C | T | 0.03286 | 0.004278 | 4.10E-14 |

| | | | | | | | | | | | |
|---|---|---|---|---|---|---|---|---|---|---|---|
| rs529565 | miR-139-5p | trans | 2.82E-43 | Nightingale | XS_VLDL_C | NA | C | T | 0.037489 | 0.004301 | 3.30E-18 |
| rs529565 | miR-139-5p | trans | 2.82E-43 | Nightingale | VLDL_P | NA | C | T | 0.026295 | 0.004407 | 5.50E-10 |
| rs529565 | miR-139-5p | trans | 2.82E-43 | Nightingale | M_VLDL_T | NA | C | T | -0.03834 | 0.004187 | 1.50E-19 |
| rs529565 | miR-139-5p | trans | 2.82E-43 | Nightingale | L_LDL_P | NA | C | T | 0.028589 | 0.004416 | 1.20E-10 |
| rs529565 | miR-139-5p | trans | 2.82E-43 | Nightingale | S_HDL_PL | NA | C | T | 0.024302 | 0.004393 | 1.90E-08 |
| rs529565 | miR-139-5p | trans | 2.82E-43 | Nightingale | Total_CE | NA | C | T | 0.031352 | 0.004274 | 1.80E-13 |
| rs529565 | miR-139-5p | trans | 2.82E-43 | Nightingale | M_LDL_C_ | NA | C | T | 0.030966 | 0.004434 | 4.00E-12 |
| rs529565 | miR-139-5p | trans | 2.82E-43 | Nightingale | XXL_VLDL_ | NA | C | T | 0.034955 | 0.00444 | 2.80E-15 |
| rs529565 | miR-139-5p | trans | 2.82E-43 | Nightingale | XS_VLDL_P | NA | C | T | 0.036324 | 0.004372 | 2.20E-17 |
| rs529565 | miR-139-5p | trans | 2.82E-43 | Nightingale | S_LDL_FC | NA | C | T | 0.027889 | 0.004424 | 9.30E-10 |
| rs529565 | miR-139-5p | trans | 2.82E-43 | Nightingale | L_VLDL_FC | NA | C | T | 0.048532 | 0.004389 | 7.40E-29 |
| rs529565 | miR-139-5p | trans | 2.82E-43 | Nightingale | Omega_6 | NA | C | T | 0.025773 | 0.004327 | 6.10E-10 |
| rs529565 | miR-139-5p | trans | 2.82E-43 | Nightingale | L_LDL_FC | NA | C | T | 0.033087 | 0.004351 | 6.50E-14 |
| rs529565 | miR-139-5p | trans | 2.82E-43 | Nightingale | XS_VLDL_P | NA | C | T | 0.031742 | 0.004369 | 8.00E-14 |
| rs529565 | miR-139-5p | trans | 2.82E-43 | Nightingale | S_HDL_CE | NA | C | T | 0.028978 | 0.004438 | 8.90E-11 |
| rs529565 | miR-139-5p | trans | 2.82E-43 | Nightingale | GlycA | NA | C | T | 0.027732 | 0.004412 | 2.40E-10 |
| rs529565 | miR-139-5p | trans | 2.82E-43 | Nightingale | Total_L | NA | C | T | 0.026125 | 0.004359 | 5.60E-10 |
| rs529565 | miR-139-5p | trans | 2.82E-43 | Nightingale | S_HDL_FC | NA | C | T | 0.023726 | 0.00435 | 3.60E-08 |
| rs529565 | miR-139-5p | trans | 2.82E-43 | Nightingale | L_LDL_C | NA | C | T | 0.031856 | 0.004381 | 4.80E-13 |
| rs529565 | miR-139-5p | trans | 2.82E-43 | Nightingale | LDL_C | NA | C | T | 0.032762 | 0.004412 | 1.60E-13 |
| rs529565 | miR-139-5p | trans | 2.82E-43 | Nightingale | M_VLDL_P | NA | C | T | 0.02872 | 0.004424 | 2.80E-11 |
| rs529565 | miR-139-5p | trans | 2.82E-43 | Nightingale | S_VLDL_CE | NA | C | T | 0.037878 | 0.004425 | 2.20E-18 |
| rs529565 | miR-139-5p | trans | 2.82E-43 | Nightingale | XL_VLDL_T | NA | C | T | -0.04378 | 0.004295 | 3.70E-24 |
| rs529565 | miR-139-5p | trans | 2.82E-43 | Nightingale | S_VLDL_FC | NA | C | T | 0.03746 | 0.004432 | 1.30E-17 |
| rs529565 | miR-139-5p | trans | 2.82E-43 | Nightingale | XXL_VLDL_ | NA | C | T | 0.025677 | 0.004459 | 7.00E-10 |
| rs529565 | miR-139-5p | trans | 2.82E-43 | Nightingale | L_VLDL_C_ | NA | C | T | 0.05212 | 0.004328 | 9.90E-34 |
| rs529565 | miR-139-5p | trans | 2.82E-43 | Nightingale | IDL_P | NA | C | T | 0.039431 | 0.004385 | 3.50E-19 |
| rs529565 | miR-139-5p | trans | 2.82E-43 | Nightingale | XS_VLDL_L | NA | C | T | 0.033423 | 0.004356 | 3.30E-15 |
| rs529565 | miR-139-5p | trans | 2.82E-43 | Nightingale | M_VLDL_C | NA | C | T | 0.033552 | 0.004163 | 2.80E-15 |
| rs529565 | miR-139-5p | trans | 2.82E-43 | Nightingale | XL_VLDL_C | NA | C | T | 0.038352 | 0.004249 | 5.20E-19 |
| rs529565 | miR-139-5p | trans | 2.82E-43 | Nightingale | Total_C | NA | C | T | 0.031972 | 0.00429 | 6.30E-14 |
| rs529565 | miR-139-5p | trans | 2.82E-43 | Nightingale | M_LDL_FC | NA | C | T | 0.033834 | 0.004426 | 4.00E-14 |
| rs529565 | miR-139-5p | trans | 2.82E-43 | Nightingale | M_VLDL_F | NA | C | T | 0.032904 | 0.00443 | 5.40E-14 |
| rs529565 | miR-139-5p | trans | 2.82E-43 | Nightingale | ApoB | NA | C | T | 0.033444 | 0.004425 | 3.90E-14 |
| rs529565 | miR-139-5p | trans | 2.82E-43 | Nightingale | ApoB_by_A | NA | C | T | 0.02449 | 0.004364 | 2.40E-08 |
| rs529565 | miR-139-5p | trans | 2.82E-43 | Nightingale | S_LDL_C_p | NA | C | T | 0.045563 | 0.004448 | 4.00E-25 |
| rs529565 | miR-139-5p | trans | 2.82E-43 | Nightingale | PUFA | NA | C | T | 0.02347 | 0.004322 | 1.80E-08 |
| rs529565 | miR-139-5p | trans | 2.82E-43 | Nightingale | S_HDL_L | NA | C | T | 0.02679 | 0.004416 | 8.80E-10 |
| rs529565 | miR-139-5p | trans | 2.82E-43 | Nightingale | S_VLDL_PL | NA | C | T | 0.03238 | 0.004423 | 6.00E-14 |
| rs529565 | miR-139-5p | trans | 2.82E-43 | Nightingale | M_LDL_L | NA | C | T | 0.032702 | 0.004437 | 1.40E-13 |
| rs529565 | miR-139-5p | trans | 2.82E-43 | Nightingale | M_LDL_P | NA | C | T | 0.035668 | 0.004443 | 8.20E-16 |
| rs529565 | miR-139-5p | trans | 2.82E-43 | Nightingale | XS_VLDL_F | NA | C | T | 0.030891 | 0.004399 | 5.80E-12 |
| rs529565 | miR-139-5p | trans | 2.82E-43 | Nightingale | XXL_VLDL_ | NA | C | T | 0.033086 | 0.004464 | 1.10E-13 |
| rs529565 | miR-139-5p | trans | 2.82E-43 | Nightingale | IDL_C | NA | C | T | 0.034316 | 0.00428 | 3.00E-15 |
| rs529565 | let-7c-5p | trans | 3.42E-24 | Nightingale | M_LDL_PL | NA | C | T | 0.03206 | 0.004437 | 4.30E-13 |
| rs529565 | let-7c-5p | trans | 3.42E-24 | Nightingale | S_LDL_L | NA | C | T | 0.027285 | 0.00443 | 1.30E-09 |
| rs529565 | let-7c-5p | trans | 3.42E-24 | Nightingale | XXL_VLDL_ | NA | C | T | 0.031056 | 0.004412 | 9.30E-13 |
| rs529565 | let-7c-5p | trans | 3.42E-24 | Nightingale | VLDL_C | NA | C | T | 0.030948 | 0.004425 | 6.40E-13 |
| rs529565 | let-7c-5p | trans | 3.42E-24 | Nightingale | S_LDL_CE_ | NA | C | T | 0.031515 | 0.004423 | 1.80E-14 |
| rs529565 | let-7c-5p | trans | 3.42E-24 | Nightingale | M_VLDL_P | NA | C | T | 0.043673 | 0.004253 | 1.80E-24 |
| rs529565 | let-7c-5p | trans | 3.42E-24 | Nightingale | LDL_P | NA | C | T | 0.031312 | 0.004428 | 1.80E-12 |
| rs529565 | let-7c-5p | trans | 3.42E-24 | Nightingale | S_HDL_P | NA | C | T | 0.030973 | 0.004432 | 3.30E-12 |
| rs529565 | let-7c-5p | trans | 3.42E-24 | Nightingale | XL_VLDL_P | NA | C | T | 0.038314 | 0.004438 | 1.50E-18 |
| rs529565 | let-7c-5p | trans | 3.42E-24 | Nightingale | S_VLDL_L | NA | C | T | 0.02438 | 0.004388 | 4.50E-09 |
| rs529565 | let-7c-5p | trans | 3.42E-24 | Nightingale | VLDL_FC | NA | C | T | 0.02385 | 0.004399 | 1.30E-08 |
| rs529565 | let-7c-5p | trans | 3.42E-24 | Nightingale | Citrate | NA | C | T | 0.027701 | 0.004402 | 2.10E-10 |
| rs529565 | let-7c-5p | trans | 3.42E-24 | Nightingale | M_VLDL_C | NA | C | T | 0.036055 | 0.004174 | 2.10E-17 |
| rs529565 | let-7c-5p | trans | 3.42E-24 | Nightingale | XL_VLDL_F | NA | C | T | 0.036987 | 0.004296 | 3.90E-18 |
| rs529565 | let-7c-5p | trans | 3.42E-24 | Nightingale | M_LDL_C | NA | C | T | 0.033658 | 0.004439 | 3.10E-14 |
| rs529565 | let-7c-5p | trans | 3.42E-24 | Nightingale | S_VLDL_CE | NA | C | T | 0.046913 | 0.004372 | 2.40E-26 |
| rs529565 | let-7c-5p | trans | 3.42E-24 | Nightingale | M_VLDL_C | NA | C | T | 0.038197 | 0.004419 | 4.00E-18 |
| rs529565 | let-7c-5p | trans | 3.42E-24 | Nightingale | S_LDL_P | NA | C | T | 0.029187 | 0.004435 | 4.60E-11 |
| rs529565 | let-7c-5p | trans | 3.42E-24 | Nightingale | IDL_FC_pct | NA | C | T | 0.025499 | 0.004397 | 1.50E-08 |

| | | | | | | | | | | | |
|---|---|---|---|---|---|---|---|---|---|---|---|
| rs529565 | let-7c-5p | trans | 3.42E-24 | Nightingale | Sphingomy | NA | C | T | 0.02867 | 0.004142 | 1.30E-11 |
| rs529565 | let-7c-5p | trans | 3.42E-24 | Nightingale | Total_FC | NA | C | T | 0.032841 | 0.004329 | 1.20E-14 |
| rs529565 | let-7c-5p | trans | 3.42E-24 | Nightingale | Clinical_LD | NA | C | T | 0.035972 | 0.004405 | 5.80E-16 |
| rs529565 | let-7c-5p | trans | 3.42E-24 | Nightingale | L_LDL_L | NA | C | T | 0.033109 | 0.004388 | 5.60E-14 |
| rs529565 | let-7c-5p | trans | 3.42E-24 | Nightingale | LDL_CE | NA | C | T | 0.032164 | 0.004419 | 3.50E-13 |
| rs529565 | let-7c-5p | trans | 3.42E-24 | Nightingale | S_VLDL_TG | NA | C | T | -0.03791 | 0.004289 | 2.80E-18 |
| rs529565 | let-7c-5p | trans | 3.42E-24 | Nightingale | S_LDL_PL_ | NA | C | T | -0.03064 | 0.004423 | 9.40E-14 |
| rs529565 | let-7c-5p | trans | 3.42E-24 | Nightingale | VLDL_CE | NA | C | T | 0.035403 | 0.004432 | 4.50E-16 |
| rs529565 | let-7c-5p | trans | 3.42E-24 | Nightingale | L_LDL_CE | NA | C | T | 0.03113 | 0.004392 | 1.70E-12 |
| rs529565 | let-7c-5p | trans | 3.42E-24 | Nightingale | L_VLDL_TG | NA | C | T | -0.05092 | 0.00441 | 1.60E-31 |
| rs529565 | let-7c-5p | trans | 3.42E-24 | Nightingale | S_VLDL_C_ | NA | C | T | 0.041702 | 0.004312 | 1.10E-21 |
| rs529565 | let-7c-5p | trans | 3.42E-24 | Nightingale | LDL_FC | NA | C | T | 0.033085 | 0.004384 | 1.10E-13 |
| rs529565 | let-7c-5p | trans | 3.42E-24 | Nightingale | XXL_VLDL_ | NA | C | T | -0.03595 | 0.004469 | 1.80E-16 |
| rs529565 | let-7c-5p | trans | 3.42E-24 | Nightingale | S_VLDL_C | NA | C | T | 0.038015 | 0.004431 | 2.60E-18 |
| rs529565 | let-7c-5p | trans | 3.42E-24 | Nightingale | IDL_L | NA | C | T | 0.034696 | 0.004284 | 1.30E-15 |
| rs529565 | let-7c-5p | trans | 3.42E-24 | Nightingale | IDL_PL | NA | C | T | 0.034834 | 0.004276 | 7.90E-16 |
| rs529565 | let-7c-5p | trans | 3.42E-24 | Nightingale | non_HDL_( | NA | C | T | 0.035225 | 0.004407 | 1.00E-15 |
| rs529565 | let-7c-5p | trans | 3.42E-24 | Nightingale | S_VLDL_FC | NA | C | T | 0.02911 | 0.004234 | 1.70E-11 |
| rs529565 | let-7c-5p | trans | 3.42E-24 | Nightingale | S_LDL_CE | NA | C | T | 0.030231 | 0.004433 | 7.50E-12 |
| rs529565 | let-7c-5p | trans | 3.42E-24 | Nightingale | XS_VLDL_C | NA | C | T | 0.037263 | 0.004276 | 5.70E-18 |
| rs529565 | let-7c-5p | trans | 3.42E-24 | Nightingale | Total_PL | NA | C | T | 0.024805 | 0.004219 | 1.20E-09 |
| rs529565 | let-7c-5p | trans | 3.42E-24 | Nightingale | M_LDL_TG | NA | C | T | -0.02724 | 0.004376 | 5.60E-10 |
| rs529565 | let-7c-5p | trans | 3.42E-24 | Nightingale | S_LDL_C | NA | C | T | 0.030526 | 0.004435 | 8.70E-12 |
| rs529565 | let-7c-5p | trans | 3.42E-24 | Nightingale | M_VLDL_F | NA | C | T | 0.041684 | 0.004216 | 1.10E-22 |
| rs529565 | let-7c-5p | trans | 3.42E-24 | Nightingale | M_VLDL_C | NA | C | T | 0.040303 | 0.004391 | 5.90E-20 |
| rs529565 | let-7c-5p | trans | 3.42E-24 | Nightingale | XL_HDL_PL | NA | C | T | -0.02275 | 0.004071 | 1.10E-08 |
| rs529565 | let-7c-5p | trans | 3.42E-24 | Nightingale | LDL_L | NA | C | T | 0.032885 | 0.004416 | 1.30E-13 |
| rs529565 | let-7c-5p | trans | 3.42E-24 | Nightingale | Remnant_( | NA | C | T | 0.036369 | 0.004402 | 7.40E-17 |
| rs529565 | let-7c-5p | trans | 3.42E-24 | Nightingale | IDL_FC | NA | C | T | 0.037662 | 0.00429 | 4.70E-18 |
| rs529565 | let-7c-5p | trans | 3.42E-24 | Nightingale | LDL_PL | NA | C | T | 0.034514 | 0.004421 | 9.20E-15 |
| rs529565 | let-7c-5p | trans | 3.42E-24 | Nightingale | M_LDL_CE | NA | C | T | 0.032712 | 0.004437 | 1.20E-13 |
| rs529565 | let-7c-5p | trans | 3.42E-24 | Nightingale | S_VLDL_P | NA | C | T | 0.023183 | 0.004376 | 1.90E-08 |
| rs529565 | let-7c-5p | trans | 3.42E-24 | Nightingale | L_VLDL_CE | NA | C | T | 0.047309 | 0.00424 | 5.10E-29 |
| rs529565 | let-7c-5p | trans | 3.42E-24 | Nightingale | L_LDL_PL | NA | C | T | 0.03752 | 0.004398 | 1.90E-17 |
| rs529565 | let-7c-5p | trans | 3.42E-24 | Nightingale | S_HDL_C | NA | C | T | 0.02876 | 0.004424 | 1.20E-10 |
| rs529565 | let-7c-5p | trans | 3.42E-24 | Nightingale | XS_VLDL_F | NA | C | T | 0.036631 | 0.004354 | 1.20E-17 |
| rs529565 | let-7c-5p | trans | 3.42E-24 | Nightingale | XL_VLDL_C | NA | C | T | 0.037516 | 0.004251 | 3.10E-18 |
| rs529565 | let-7c-5p | trans | 3.42E-24 | Nightingale | S_VLDL_PL | NA | C | T | 0.027457 | 0.004243 | 2.90E-10 |
| rs529565 | let-7c-5p | trans | 3.42E-24 | Nightingale | M_VLDL_P | NA | C | T | 0.027887 | 0.00442 | 8.70E-11 |
| rs529565 | let-7c-5p | trans | 3.42E-24 | Nightingale | LA | NA | C | T | 0.024778 | 0.004345 | 3.90E-09 |
| rs529565 | let-7c-5p | trans | 3.42E-24 | Nightingale | IDL_CE | NA | C | T | 0.03286 | 0.004278 | 4.10E-14 |
| rs529565 | let-7c-5p | trans | 3.42E-24 | Nightingale | XS_VLDL_C | NA | C | T | 0.037489 | 0.004301 | 3.30E-18 |
| rs529565 | let-7c-5p | trans | 3.42E-24 | Nightingale | VLDL_P | NA | C | T | 0.026295 | 0.004407 | 5.50E-10 |
| rs529565 | let-7c-5p | trans | 3.42E-24 | Nightingale | M_VLDL_T | NA | C | T | -0.03834 | 0.004187 | 1.50E-19 |
| rs529565 | let-7c-5p | trans | 3.42E-24 | Nightingale | L_LDL_P | NA | C | T | 0.028589 | 0.004416 | 1.20E-10 |
| rs529565 | let-7c-5p | trans | 3.42E-24 | Nightingale | S_HDL_PL | NA | C | T | 0.024302 | 0.004393 | 1.90E-08 |
| rs529565 | let-7c-5p | trans | 3.42E-24 | Nightingale | Total_CE | NA | C | T | 0.031352 | 0.004274 | 1.80E-13 |
| rs529565 | let-7c-5p | trans | 3.42E-24 | Nightingale | M_LDL_C_ | NA | C | T | 0.030966 | 0.004434 | 4.00E-12 |
| rs529565 | let-7c-5p | trans | 3.42E-24 | Nightingale | XXL_VLDL_ | NA | C | T | 0.034955 | 0.00444 | 2.80E-15 |
| rs529565 | let-7c-5p | trans | 3.42E-24 | Nightingale | XS_VLDL_P | NA | C | T | 0.036324 | 0.004372 | 2.20E-17 |
| rs529565 | let-7c-5p | trans | 3.42E-24 | Nightingale | S_LDL_FC | NA | C | T | 0.027889 | 0.004424 | 9.30E-10 |
| rs529565 | let-7c-5p | trans | 3.42E-24 | Nightingale | L_VLDL_FC | NA | C | T | 0.048532 | 0.004389 | 7.40E-29 |
| rs529565 | let-7c-5p | trans | 3.42E-24 | Nightingale | Omega_6 | NA | C | T | 0.025773 | 0.004327 | 6.10E-10 |
| rs529565 | let-7c-5p | trans | 3.42E-24 | Nightingale | L_LDL_FC | NA | C | T | 0.033087 | 0.004351 | 6.50E-14 |
| rs529565 | let-7c-5p | trans | 3.42E-24 | Nightingale | XS_VLDL_P | NA | C | T | 0.031742 | 0.004369 | 8.00E-14 |
| rs529565 | let-7c-5p | trans | 3.42E-24 | Nightingale | S_HDL_CE | NA | C | T | 0.028978 | 0.004438 | 8.90E-11 |
| rs529565 | let-7c-5p | trans | 3.42E-24 | Nightingale | GlycA | NA | C | T | 0.027732 | 0.004412 | 2.40E-10 |
| rs529565 | let-7c-5p | trans | 3.42E-24 | Nightingale | Total_L | NA | C | T | 0.026125 | 0.004359 | 5.60E-10 |
| rs529565 | let-7c-5p | trans | 3.42E-24 | Nightingale | S_HDL_FC | NA | C | T | 0.023726 | 0.00435 | 3.60E-08 |
| rs529565 | let-7c-5p | trans | 3.42E-24 | Nightingale | L_LDL_C | NA | C | T | 0.031856 | 0.004381 | 4.80E-13 |
| rs529565 | let-7c-5p | trans | 3.42E-24 | Nightingale | LDL_C | NA | C | T | 0.032762 | 0.004412 | 1.60E-13 |
| rs529565 | let-7c-5p | trans | 3.42E-24 | Nightingale | M_VLDL_P | NA | C | T | 0.02872 | 0.004424 | 2.80E-11 |
| rs529565 | let-7c-5p | trans | 3.42E-24 | Nightingale | S_VLDL_CE | NA | C | T | 0.037878 | 0.004425 | 2.20E-18 |

| | | | | | | | | | | | |
|---|---|---|---|---|---|---|---|---|---|---|---|
| rs529565 | let-7c-5p | trans | 3.42E-24 | Nightingale | XL_VLDL_T | NA | C | T | -0.04378 | 0.004295 | 3.70E-24 |
| rs529565 | let-7c-5p | trans | 3.42E-24 | Nightingale | S_VLDL_FC | NA | C | T | 0.03746 | 0.004432 | 1.30E-17 |
| rs529565 | let-7c-5p | trans | 3.42E-24 | Nightingale | XXL_VLDL_ | NA | C | T | 0.025677 | 0.004459 | 7.00E-10 |
| rs529565 | let-7c-5p | trans | 3.42E-24 | Nightingale | L_VLDL_C_ | NA | C | T | 0.05212 | 0.004328 | 9.90E-34 |
| rs529565 | let-7c-5p | trans | 3.42E-24 | Nightingale | IDL_P | NA | C | T | 0.039431 | 0.004385 | 3.50E-19 |
| rs529565 | let-7c-5p | trans | 3.42E-24 | Nightingale | XS_VLDL_L | NA | C | T | 0.033423 | 0.004356 | 3.30E-15 |
| rs529565 | let-7c-5p | trans | 3.42E-24 | Nightingale | M_VLDL_C | NA | C | T | 0.033552 | 0.004163 | 2.80E-15 |
| rs529565 | let-7c-5p | trans | 3.42E-24 | Nightingale | XL_VLDL_C | NA | C | T | 0.038352 | 0.004249 | 5.20E-19 |
| rs529565 | let-7c-5p | trans | 3.42E-24 | Nightingale | Total_C | NA | C | T | 0.031972 | 0.00429 | 6.30E-14 |
| rs529565 | let-7c-5p | trans | 3.42E-24 | Nightingale | M_LDL_FC | NA | C | T | 0.033834 | 0.004426 | 4.00E-14 |
| rs529565 | let-7c-5p | trans | 3.42E-24 | Nightingale | M_VLDL_F | NA | C | T | 0.032904 | 0.00443 | 5.40E-14 |
| rs529565 | let-7c-5p | trans | 3.42E-24 | Nightingale | ApoB | NA | C | T | 0.033444 | 0.004425 | 3.90E-14 |
| rs529565 | let-7c-5p | trans | 3.42E-24 | Nightingale | ApoB_by_ | NA | C | T | 0.02449 | 0.004364 | 2.40E-08 |
| rs529565 | let-7c-5p | trans | 3.42E-24 | Nightingale | S_LDL_C_p | NA | C | T | 0.045563 | 0.004448 | 4.00E-25 |
| rs529565 | let-7c-5p | trans | 3.42E-24 | Nightingale | PUFA | NA | C | T | 0.02347 | 0.004322 | 1.80E-08 |
| rs529565 | let-7c-5p | trans | 3.42E-24 | Nightingale | S_HDL_L | NA | C | T | 0.02679 | 0.004416 | 8.80E-10 |
| rs529565 | let-7c-5p | trans | 3.42E-24 | Nightingale | S_VLDL_PL | NA | C | T | 0.03238 | 0.004423 | 6.00E-14 |
| rs529565 | let-7c-5p | trans | 3.42E-24 | Nightingale | M_LDL_L | NA | C | T | 0.032702 | 0.004437 | 1.40E-13 |
| rs529565 | let-7c-5p | trans | 3.42E-24 | Nightingale | M_LDL_P | NA | C | T | 0.035668 | 0.004443 | 8.20E-16 |
| rs529565 | let-7c-5p | trans | 3.42E-24 | Nightingale | XS_VLDL_F | NA | C | T | 0.030891 | 0.004399 | 5.80E-12 |
| rs529565 | let-7c-5p | trans | 3.42E-24 | Nightingale | XXL_VLDL_ | NA | C | T | 0.033086 | 0.004464 | 1.10E-13 |
| rs529565 | let-7c-5p | trans | 3.42E-24 | Nightingale | IDL_C | NA | C | T | 0.034316 | 0.00428 | 3.00E-15 |
| rs529565 | miR-10b-5p | trans | 4.06E-36 | Nightingale | M_LDL_PL | NA | C | T | 0.03206 | 0.004437 | 4.30E-13 |
| rs529565 | miR-10b-5p | trans | 4.06E-36 | Nightingale | S_LDL_L | NA | C | T | 0.027285 | 0.00443 | 1.30E-09 |
| rs529565 | miR-10b-5p | trans | 4.06E-36 | Nightingale | XXL_VLDL_ | NA | C | T | 0.031056 | 0.004412 | 9.30E-13 |
| rs529565 | miR-10b-5p | trans | 4.06E-36 | Nightingale | VLDL_C | NA | C | T | 0.030948 | 0.004425 | 6.40E-13 |
| rs529565 | miR-10b-5p | trans | 4.06E-36 | Nightingale | S_LDL_CE_ | NA | C | T | 0.031515 | 0.004423 | 1.80E-14 |
| rs529565 | miR-10b-5p | trans | 4.06E-36 | Nightingale | M_VLDL_P | NA | C | T | 0.043673 | 0.004253 | 1.80E-24 |
| rs529565 | miR-10b-5p | trans | 4.06E-36 | Nightingale | LDL_P | NA | C | T | 0.031312 | 0.004428 | 1.80E-12 |
| rs529565 | miR-10b-5p | trans | 4.06E-36 | Nightingale | S_HDL_P | NA | C | T | 0.030973 | 0.004432 | 3.30E-12 |
| rs529565 | miR-10b-5p | trans | 4.06E-36 | Nightingale | XL_VLDL_P | NA | C | T | 0.038314 | 0.004438 | 1.50E-18 |
| rs529565 | miR-10b-5p | trans | 4.06E-36 | Nightingale | S_VLDL_L | NA | C | T | 0.02438 | 0.004388 | 4.50E-09 |
| rs529565 | miR-10b-5p | trans | 4.06E-36 | Nightingale | VLDL_FC | NA | C | T | 0.02385 | 0.004399 | 1.30E-08 |
| rs529565 | miR-10b-5p | trans | 4.06E-36 | Nightingale | Citrate | NA | C | T | 0.027701 | 0.004402 | 2.10E-10 |
| rs529565 | miR-10b-5p | trans | 4.06E-36 | Nightingale | M_VLDL_C | NA | C | T | 0.036055 | 0.004174 | 2.10E-17 |
| rs529565 | miR-10b-5p | trans | 4.06E-36 | Nightingale | XL_VLDL_F | NA | C | T | 0.036987 | 0.004296 | 3.90E-18 |
| rs529565 | miR-10b-5p | trans | 4.06E-36 | Nightingale | M_LDL_C | NA | C | T | 0.033658 | 0.004439 | 3.10E-14 |
| rs529565 | miR-10b-5p | trans | 4.06E-36 | Nightingale | S_VLDL_CE | NA | C | T | 0.046913 | 0.004372 | 2.40E-26 |
| rs529565 | miR-10b-5p | trans | 4.06E-36 | Nightingale | M_VLDL_C | NA | C | T | 0.038197 | 0.004419 | 4.00E-18 |
| rs529565 | miR-10b-5p | trans | 4.06E-36 | Nightingale | S_LDL_P | NA | C | T | 0.029187 | 0.004435 | 4.60E-11 |
| rs529565 | miR-10b-5p | trans | 4.06E-36 | Nightingale | IDL_FC_pct | NA | C | T | 0.025499 | 0.004397 | 1.50E-08 |
| rs529565 | miR-10b-5p | trans | 4.06E-36 | Nightingale | Sphingomy | NA | C | T | 0.02867 | 0.004142 | 1.30E-11 |
| rs529565 | miR-10b-5p | trans | 4.06E-36 | Nightingale | Total_FC | NA | C | T | 0.032841 | 0.004329 | 1.20E-14 |
| rs529565 | miR-10b-5p | trans | 4.06E-36 | Nightingale | Clinical_LD | NA | C | T | 0.035972 | 0.004405 | 5.80E-16 |
| rs529565 | miR-10b-5p | trans | 4.06E-36 | Nightingale | L_LDL_L | NA | C | T | 0.033109 | 0.004388 | 5.60E-14 |
| rs529565 | miR-10b-5p | trans | 4.06E-36 | Nightingale | LDL_CE | NA | C | T | 0.032164 | 0.004419 | 3.50E-13 |
| rs529565 | miR-10b-5p | trans | 4.06E-36 | Nightingale | S_VLDL_TG | NA | C | T | -0.03791 | 0.004289 | 2.80E-18 |
| rs529565 | miR-10b-5p | trans | 4.06E-36 | Nightingale | S_LDL_PL_ | NA | C | T | -0.03064 | 0.004423 | 9.40E-14 |
| rs529565 | miR-10b-5p | trans | 4.06E-36 | Nightingale | VLDL_CE | NA | C | T | 0.035403 | 0.004432 | 4.50E-16 |
| rs529565 | miR-10b-5p | trans | 4.06E-36 | Nightingale | L_LDL_CE | NA | C | T | 0.03113 | 0.004392 | 1.70E-12 |
| rs529565 | miR-10b-5p | trans | 4.06E-36 | Nightingale | L_VLDL_TG | NA | C | T | -0.05092 | 0.00441 | 1.60E-31 |
| rs529565 | miR-10b-5p | trans | 4.06E-36 | Nightingale | S_VLDL_C_ | NA | C | T | 0.041702 | 0.004312 | 1.10E-21 |
| rs529565 | miR-10b-5p | trans | 4.06E-36 | Nightingale | LDL_FC | NA | C | T | 0.033085 | 0.004384 | 1.10E-13 |
| rs529565 | miR-10b-5p | trans | 4.06E-36 | Nightingale | XXL_VLDL_ | NA | C | T | -0.03595 | 0.004469 | 1.80E-16 |
| rs529565 | miR-10b-5p | trans | 4.06E-36 | Nightingale | S_VLDL_C | NA | C | T | 0.038015 | 0.004431 | 2.60E-18 |
| rs529565 | miR-10b-5p | trans | 4.06E-36 | Nightingale | IDL_L | NA | C | T | 0.034696 | 0.004284 | 1.30E-15 |
| rs529565 | miR-10b-5p | trans | 4.06E-36 | Nightingale | IDL_PL | NA | C | T | 0.034834 | 0.004276 | 7.90E-16 |
| rs529565 | miR-10b-5p | trans | 4.06E-36 | Nightingale | non_HDL_ | NA | C | T | 0.035225 | 0.004407 | 1.00E-15 |
| rs529565 | miR-10b-5p | trans | 4.06E-36 | Nightingale | S_VLDL_FC | NA | C | T | 0.02911 | 0.004234 | 1.70E-11 |
| rs529565 | miR-10b-5p | trans | 4.06E-36 | Nightingale | S_LDL_CE | NA | C | T | 0.030231 | 0.004433 | 7.50E-12 |
| rs529565 | miR-10b-5p | trans | 4.06E-36 | Nightingale | XS_VLDL_C | NA | C | T | 0.037263 | 0.004276 | 5.70E-18 |
| rs529565 | miR-10b-5p | trans | 4.06E-36 | Nightingale | Total_PL | NA | C | T | 0.024805 | 0.004219 | 1.20E-09 |
| rs529565 | miR-10b-5p | trans | 4.06E-36 | Nightingale | M_LDL_TG | NA | C | T | -0.02724 | 0.004376 | 5.60E-10 |

| | | | | | | | | | | | |
|---|---|---|---|---|---|---|---|---|---|---|---|
| rs529565 | miR-10b-5p | trans | 4.06E-36 | Nightingale | S_LDL_C | NA | C | T | 0.030526 | 0.004435 | 8.70E-12 |
| rs529565 | miR-10b-5p | trans | 4.06E-36 | Nightingale | M_VLDL_F | NA | C | T | 0.041684 | 0.004216 | 1.10E-22 |
| rs529565 | miR-10b-5p | trans | 4.06E-36 | Nightingale | M_VLDL_C | NA | C | T | 0.040303 | 0.004391 | 5.90E-20 |
| rs529565 | miR-10b-5p | trans | 4.06E-36 | Nightingale | XL_HDL_PL | NA | C | T | -0.02275 | 0.004071 | 1.10E-08 |
| rs529565 | miR-10b-5p | trans | 4.06E-36 | Nightingale | LDL_L | NA | C | T | 0.032885 | 0.004416 | 1.30E-13 |
| rs529565 | miR-10b-5p | trans | 4.06E-36 | Nightingale | Remnant_( | NA | C | T | 0.036369 | 0.004402 | 7.40E-17 |
| rs529565 | miR-10b-5p | trans | 4.06E-36 | Nightingale | IDL_FC | NA | C | T | 0.037662 | 0.00429 | 4.70E-18 |
| rs529565 | miR-10b-5p | trans | 4.06E-36 | Nightingale | LDL_PL | NA | C | T | 0.034514 | 0.004421 | 9.20E-15 |
| rs529565 | miR-10b-5p | trans | 4.06E-36 | Nightingale | M_LDL_CE | NA | C | T | 0.032712 | 0.004437 | 1.20E-13 |
| rs529565 | miR-10b-5p | trans | 4.06E-36 | Nightingale | S_VLDL_P | NA | C | T | 0.023183 | 0.004376 | 1.90E-08 |
| rs529565 | miR-10b-5p | trans | 4.06E-36 | Nightingale | L_VLDL_CE | NA | C | T | 0.047309 | 0.00424 | 5.10E-29 |
| rs529565 | miR-10b-5p | trans | 4.06E-36 | Nightingale | L_LDL_PL | NA | C | T | 0.03752 | 0.004398 | 1.90E-17 |
| rs529565 | miR-10b-5p | trans | 4.06E-36 | Nightingale | S_HDL_C | NA | C | T | 0.02876 | 0.004424 | 1.20E-10 |
| rs529565 | miR-10b-5p | trans | 4.06E-36 | Nightingale | XS_VLDL_F | NA | C | T | 0.036631 | 0.004354 | 1.20E-17 |
| rs529565 | miR-10b-5p | trans | 4.06E-36 | Nightingale | XL_VLDL_C | NA | C | T | 0.037516 | 0.004251 | 3.10E-18 |
| rs529565 | miR-10b-5p | trans | 4.06E-36 | Nightingale | S_VLDL_PL | NA | C | T | 0.027457 | 0.004243 | 2.90E-10 |
| rs529565 | miR-10b-5p | trans | 4.06E-36 | Nightingale | M_VLDL_P | NA | C | T | 0.027887 | 0.00442 | 8.70E-11 |
| rs529565 | miR-10b-5p | trans | 4.06E-36 | Nightingale | LA | NA | C | T | 0.024778 | 0.004345 | 3.90E-09 |
| rs529565 | miR-10b-5p | trans | 4.06E-36 | Nightingale | IDL_CE | NA | C | T | 0.03286 | 0.004278 | 4.10E-14 |
| rs529565 | miR-10b-5p | trans | 4.06E-36 | Nightingale | XS_VLDL_C | NA | C | T | 0.037489 | 0.004301 | 3.30E-18 |
| rs529565 | miR-10b-5p | trans | 4.06E-36 | Nightingale | VLDL_P | NA | C | T | 0.026295 | 0.004407 | 5.50E-10 |
| rs529565 | miR-10b-5p | trans | 4.06E-36 | Nightingale | M_VLDL_T | NA | C | T | -0.03834 | 0.004187 | 1.50E-19 |
| rs529565 | miR-10b-5p | trans | 4.06E-36 | Nightingale | L_LDL_P | NA | C | T | 0.028589 | 0.004416 | 1.20E-10 |
| rs529565 | miR-10b-5p | trans | 4.06E-36 | Nightingale | S_HDL_PL | NA | C | T | 0.024302 | 0.004393 | 1.90E-08 |
| rs529565 | miR-10b-5p | trans | 4.06E-36 | Nightingale | Total_CE | NA | C | T | 0.031352 | 0.004274 | 1.80E-13 |
| rs529565 | miR-10b-5p | trans | 4.06E-36 | Nightingale | M_LDL_C_ | NA | C | T | 0.030966 | 0.004434 | 4.00E-12 |
| rs529565 | miR-10b-5p | trans | 4.06E-36 | Nightingale | XXL_VLDL_ | NA | C | T | 0.034955 | 0.00444 | 2.80E-15 |
| rs529565 | miR-10b-5p | trans | 4.06E-36 | Nightingale | XS_VLDL_P | NA | C | T | 0.036324 | 0.004372 | 2.20E-17 |
| rs529565 | miR-10b-5p | trans | 4.06E-36 | Nightingale | S_LDL_FC | NA | C | T | 0.027889 | 0.004424 | 9.30E-10 |
| rs529565 | miR-10b-5p | trans | 4.06E-36 | Nightingale | L_VLDL_FC | NA | C | T | 0.048532 | 0.004389 | 7.40E-29 |
| rs529565 | miR-10b-5p | trans | 4.06E-36 | Nightingale | Omega_6 | NA | C | T | 0.025773 | 0.004327 | 6.10E-10 |
| rs529565 | miR-10b-5p | trans | 4.06E-36 | Nightingale | L_LDL_FC | NA | C | T | 0.033087 | 0.004351 | 6.50E-14 |
| rs529565 | miR-10b-5p | trans | 4.06E-36 | Nightingale | XS_VLDL_P | NA | C | T | 0.031742 | 0.004369 | 8.00E-14 |
| rs529565 | miR-10b-5p | trans | 4.06E-36 | Nightingale | S_HDL_CE | NA | C | T | 0.028978 | 0.004438 | 8.90E-11 |
| rs529565 | miR-10b-5p | trans | 4.06E-36 | Nightingale | GlycA | NA | C | T | 0.027732 | 0.004412 | 2.40E-10 |
| rs529565 | miR-10b-5p | trans | 4.06E-36 | Nightingale | Total_L | NA | C | T | 0.026125 | 0.004359 | 5.60E-10 |
| rs529565 | miR-10b-5p | trans | 4.06E-36 | Nightingale | S_HDL_FC | NA | C | T | 0.023726 | 0.00435 | 3.60E-08 |
| rs529565 | miR-10b-5p | trans | 4.06E-36 | Nightingale | L_LDL_C | NA | C | T | 0.031856 | 0.004381 | 4.80E-13 |
| rs529565 | miR-10b-5p | trans | 4.06E-36 | Nightingale | LDL_C | NA | C | T | 0.032762 | 0.004412 | 1.60E-13 |
| rs529565 | miR-10b-5p | trans | 4.06E-36 | Nightingale | M_VLDL_P | NA | C | T | 0.02872 | 0.004424 | 2.80E-11 |
| rs529565 | miR-10b-5p | trans | 4.06E-36 | Nightingale | S_VLDL_CE | NA | C | T | 0.037878 | 0.004425 | 2.20E-18 |
| rs529565 | miR-10b-5p | trans | 4.06E-36 | Nightingale | XL_VLDL_T | NA | C | T | -0.04378 | 0.004295 | 3.70E-24 |
| rs529565 | miR-10b-5p | trans | 4.06E-36 | Nightingale | S_VLDL_FC | NA | C | T | 0.03746 | 0.004432 | 1.30E-17 |
| rs529565 | miR-10b-5p | trans | 4.06E-36 | Nightingale | XXL_VLDL_ | NA | C | T | 0.025677 | 0.004459 | 7.00E-10 |
| rs529565 | miR-10b-5p | trans | 4.06E-36 | Nightingale | L_VLDL_C_ | NA | C | T | 0.05212 | 0.004328 | 9.90E-34 |
| rs529565 | miR-10b-5p | trans | 4.06E-36 | Nightingale | IDL_P | NA | C | T | 0.039431 | 0.004385 | 3.50E-19 |
| rs529565 | miR-10b-5p | trans | 4.06E-36 | Nightingale | XS_VLDL_L | NA | C | T | 0.033423 | 0.004356 | 3.30E-15 |
| rs529565 | miR-10b-5p | trans | 4.06E-36 | Nightingale | M_VLDL_C | NA | C | T | 0.033552 | 0.004163 | 2.80E-15 |
| rs529565 | miR-10b-5p | trans | 4.06E-36 | Nightingale | XL_VLDL_C | NA | C | T | 0.038352 | 0.004249 | 5.20E-19 |
| rs529565 | miR-10b-5p | trans | 4.06E-36 | Nightingale | Total_C | NA | C | T | 0.031972 | 0.00429 | 6.30E-14 |
| rs529565 | miR-10b-5p | trans | 4.06E-36 | Nightingale | M_LDL_FC | NA | C | T | 0.033834 | 0.004426 | 4.00E-14 |
| rs529565 | miR-10b-5p | trans | 4.06E-36 | Nightingale | M_VLDL_F | NA | C | T | 0.032904 | 0.00443 | 5.40E-14 |
| rs529565 | miR-10b-5p | trans | 4.06E-36 | Nightingale | ApoB | NA | C | T | 0.033444 | 0.004425 | 3.90E-14 |
| rs529565 | miR-10b-5p | trans | 4.06E-36 | Nightingale | ApoB_by_A | NA | C | T | 0.02449 | 0.004364 | 2.40E-08 |
| rs529565 | miR-10b-5p | trans | 4.06E-36 | Nightingale | S_LDL_C_p | NA | C | T | 0.045563 | 0.004448 | 4.00E-25 |
| rs529565 | miR-10b-5p | trans | 4.06E-36 | Nightingale | PUFA | NA | C | T | 0.02347 | 0.004322 | 1.80E-08 |
| rs529565 | miR-10b-5p | trans | 4.06E-36 | Nightingale | S_HDL_L | NA | C | T | 0.02679 | 0.004416 | 8.80E-10 |
| rs529565 | miR-10b-5p | trans | 4.06E-36 | Nightingale | S_VLDL_PL | NA | C | T | 0.03238 | 0.004423 | 6.00E-14 |
| rs529565 | miR-10b-5p | trans | 4.06E-36 | Nightingale | M_LDL_L | NA | C | T | 0.032702 | 0.004437 | 1.40E-13 |
| rs529565 | miR-10b-5p | trans | 4.06E-36 | Nightingale | M_LDL_P | NA | C | T | 0.035668 | 0.004443 | 8.20E-16 |
| rs529565 | miR-10b-5p | trans | 4.06E-36 | Nightingale | XS_VLDL_F | NA | C | T | 0.030891 | 0.004399 | 5.80E-12 |
| rs529565 | miR-10b-5p | trans | 4.06E-36 | Nightingale | XXL_VLDL_ | NA | C | T | 0.033086 | 0.004464 | 1.10E-13 |
| rs529565 | miR-10b-5p | trans | 4.06E-36 | Nightingale | IDL_C | NA | C | T | 0.034316 | 0.00428 | 3.00E-15 |

| | | | | | | | | | | | |
|---|---|---|---|---|---|---|---|---|---|---|---|
| rs529565 | miR-126-3p | trans | 8.91E-41 | Nightingale | M_LDL_PL | NA | C | T | 0.03206 | 0.004437 | 4.30E-13 |
| rs529565 | miR-126-3p | trans | 8.91E-41 | Nightingale | S_LDL_L | NA | C | T | 0.027285 | 0.00443 | 1.30E-09 |
| rs529565 | miR-126-3p | trans | 8.91E-41 | Nightingale | XXL_VLDL_ | NA | C | T | 0.031056 | 0.004412 | 9.30E-13 |
| rs529565 | miR-126-3p | trans | 8.91E-41 | Nightingale | VLDL_C | NA | C | T | 0.030948 | 0.004425 | 6.40E-13 |
| rs529565 | miR-126-3p | trans | 8.91E-41 | Nightingale | S_LDL_CE_ | NA | C | T | 0.031515 | 0.004423 | 1.80E-14 |
| rs529565 | miR-126-3p | trans | 8.91E-41 | Nightingale | M_VLDL_P | NA | C | T | 0.043673 | 0.004253 | 1.80E-24 |
| rs529565 | miR-126-3p | trans | 8.91E-41 | Nightingale | LDL_P | NA | C | T | 0.031312 | 0.004428 | 1.80E-12 |
| rs529565 | miR-126-3p | trans | 8.91E-41 | Nightingale | S_HDL_P | NA | C | T | 0.030973 | 0.004432 | 3.30E-12 |
| rs529565 | miR-126-3p | trans | 8.91E-41 | Nightingale | XL_VLDL_P | NA | C | T | 0.038314 | 0.004438 | 1.50E-18 |
| rs529565 | miR-126-3p | trans | 8.91E-41 | Nightingale | S_VLDL_L | NA | C | T | 0.02438 | 0.004388 | 4.50E-09 |
| rs529565 | miR-126-3p | trans | 8.91E-41 | Nightingale | VLDL_FC | NA | C | T | 0.02385 | 0.004399 | 1.30E-08 |
| rs529565 | miR-126-3p | trans | 8.91E-41 | Nightingale | Citrate | NA | C | T | 0.027701 | 0.004402 | 2.10E-10 |
| rs529565 | miR-126-3p | trans | 8.91E-41 | Nightingale | M_VLDL_C | NA | C | T | 0.036055 | 0.004174 | 2.10E-17 |
| rs529565 | miR-126-3p | trans | 8.91E-41 | Nightingale | XL_VLDL_F | NA | C | T | 0.036987 | 0.004296 | 3.90E-18 |
| rs529565 | miR-126-3p | trans | 8.91E-41 | Nightingale | M_LDL_C | NA | C | T | 0.033658 | 0.004439 | 3.10E-14 |
| rs529565 | miR-126-3p | trans | 8.91E-41 | Nightingale | S_VLDL_CE | NA | C | T | 0.046913 | 0.004372 | 2.40E-26 |
| rs529565 | miR-126-3p | trans | 8.91E-41 | Nightingale | M_VLDL_C | NA | C | T | 0.038197 | 0.004419 | 4.00E-18 |
| rs529565 | miR-126-3p | trans | 8.91E-41 | Nightingale | S_LDL_P | NA | C | T | 0.029187 | 0.004435 | 4.60E-11 |
| rs529565 | miR-126-3p | trans | 8.91E-41 | Nightingale | IDL_FC_pct | NA | C | T | 0.025499 | 0.004397 | 1.50E-08 |
| rs529565 | miR-126-3p | trans | 8.91E-41 | Nightingale | Sphingomy | NA | C | T | 0.02867 | 0.004142 | 1.30E-11 |
| rs529565 | miR-126-3p | trans | 8.91E-41 | Nightingale | Total_FC | NA | C | T | 0.032841 | 0.004329 | 1.20E-14 |
| rs529565 | miR-126-3p | trans | 8.91E-41 | Nightingale | Clinical_LD | NA | C | T | 0.035972 | 0.004405 | 5.80E-16 |
| rs529565 | miR-126-3p | trans | 8.91E-41 | Nightingale | L_LDL_L | NA | C | T | 0.033109 | 0.004388 | 5.60E-14 |
| rs529565 | miR-126-3p | trans | 8.91E-41 | Nightingale | LDL_CE | NA | C | T | 0.032164 | 0.004419 | 3.50E-13 |
| rs529565 | miR-126-3p | trans | 8.91E-41 | Nightingale | S_VLDL_TG | NA | C | T | -0.03791 | 0.004289 | 2.80E-18 |
| rs529565 | miR-126-3p | trans | 8.91E-41 | Nightingale | S_LDL_PL_ | NA | C | T | -0.03064 | 0.004423 | 9.40E-14 |
| rs529565 | miR-126-3p | trans | 8.91E-41 | Nightingale | VLDL_CE | NA | C | T | 0.035403 | 0.004432 | 4.50E-16 |
| rs529565 | miR-126-3p | trans | 8.91E-41 | Nightingale | L_LDL_CE | NA | C | T | 0.03113 | 0.004392 | 1.70E-12 |
| rs529565 | miR-126-3p | trans | 8.91E-41 | Nightingale | L_VLDL_TG | NA | C | T | -0.05092 | 0.00441 | 1.60E-31 |
| rs529565 | miR-126-3p | trans | 8.91E-41 | Nightingale | S_VLDL_C_ | NA | C | T | 0.041702 | 0.004312 | 1.10E-21 |
| rs529565 | miR-126-3p | trans | 8.91E-41 | Nightingale | LDL_FC | NA | C | T | 0.033085 | 0.004384 | 1.10E-13 |
| rs529565 | miR-126-3p | trans | 8.91E-41 | Nightingale | XXL_VLDL_ | NA | C | T | -0.03595 | 0.004469 | 1.80E-16 |
| rs529565 | miR-126-3p | trans | 8.91E-41 | Nightingale | S_VLDL_C | NA | C | T | 0.038015 | 0.004431 | 2.60E-18 |
| rs529565 | miR-126-3p | trans | 8.91E-41 | Nightingale | IDL_L | NA | C | T | 0.034696 | 0.004284 | 1.30E-15 |
| rs529565 | miR-126-3p | trans | 8.91E-41 | Nightingale | IDL_PL | NA | C | T | 0.034834 | 0.004276 | 7.90E-16 |
| rs529565 | miR-126-3p | trans | 8.91E-41 | Nightingale | non_HDL_( | NA | C | T | 0.035225 | 0.004407 | 1.00E-15 |
| rs529565 | miR-126-3p | trans | 8.91E-41 | Nightingale | S_VLDL_FC | NA | C | T | 0.02911 | 0.004234 | 1.70E-11 |
| rs529565 | miR-126-3p | trans | 8.91E-41 | Nightingale | S_LDL_CE | NA | C | T | 0.030231 | 0.004433 | 7.50E-12 |
| rs529565 | miR-126-3p | trans | 8.91E-41 | Nightingale | XS_VLDL_C | NA | C | T | 0.037263 | 0.004276 | 5.70E-18 |
| rs529565 | miR-126-3p | trans | 8.91E-41 | Nightingale | Total_PL | NA | C | T | 0.024805 | 0.004219 | 1.20E-09 |
| rs529565 | miR-126-3p | trans | 8.91E-41 | Nightingale | M_LDL_TG | NA | C | T | -0.02724 | 0.004376 | 5.60E-10 |
| rs529565 | miR-126-3p | trans | 8.91E-41 | Nightingale | S_LDL_C | NA | C | T | 0.030526 | 0.004435 | 8.70E-12 |
| rs529565 | miR-126-3p | trans | 8.91E-41 | Nightingale | M_VLDL_F | NA | C | T | 0.041684 | 0.004216 | 1.10E-22 |
| rs529565 | miR-126-3p | trans | 8.91E-41 | Nightingale | M_VLDL_C | NA | C | T | 0.040303 | 0.004391 | 5.90E-20 |
| rs529565 | miR-126-3p | trans | 8.91E-41 | Nightingale | XL_HDL_PL | NA | C | T | -0.02275 | 0.004071 | 1.10E-08 |
| rs529565 | miR-126-3p | trans | 8.91E-41 | Nightingale | LDL_L | NA | C | T | 0.032885 | 0.004416 | 1.30E-13 |
| rs529565 | miR-126-3p | trans | 8.91E-41 | Nightingale | Remnant_( | NA | C | T | 0.036369 | 0.004402 | 7.40E-17 |
| rs529565 | miR-126-3p | trans | 8.91E-41 | Nightingale | IDL_FC | NA | C | T | 0.037662 | 0.00429 | 4.70E-18 |
| rs529565 | miR-126-3p | trans | 8.91E-41 | Nightingale | LDL_PL | NA | C | T | 0.034514 | 0.004421 | 9.20E-15 |
| rs529565 | miR-126-3p | trans | 8.91E-41 | Nightingale | M_LDL_CE | NA | C | T | 0.032712 | 0.004437 | 1.20E-13 |
| rs529565 | miR-126-3p | trans | 8.91E-41 | Nightingale | S_VLDL_P | NA | C | T | 0.023183 | 0.004376 | 1.90E-08 |
| rs529565 | miR-126-3p | trans | 8.91E-41 | Nightingale | L_VLDL_CE | NA | C | T | 0.047309 | 0.00424 | 5.10E-29 |
| rs529565 | miR-126-3p | trans | 8.91E-41 | Nightingale | L_LDL_PL | NA | C | T | 0.03752 | 0.004398 | 1.90E-17 |
| rs529565 | miR-126-3p | trans | 8.91E-41 | Nightingale | S_HDL_C | NA | C | T | 0.02876 | 0.004424 | 1.20E-10 |
| rs529565 | miR-126-3p | trans | 8.91E-41 | Nightingale | XS_VLDL_F | NA | C | T | 0.036631 | 0.004354 | 1.20E-17 |
| rs529565 | miR-126-3p | trans | 8.91E-41 | Nightingale | XL_VLDL_C | NA | C | T | 0.037516 | 0.004251 | 3.10E-18 |
| rs529565 | miR-126-3p | trans | 8.91E-41 | Nightingale | S_VLDL_PL | NA | C | T | 0.027457 | 0.004243 | 2.90E-10 |
| rs529565 | miR-126-3p | trans | 8.91E-41 | Nightingale | M_VLDL_P | NA | C | T | 0.027887 | 0.00442 | 8.70E-11 |
| rs529565 | miR-126-3p | trans | 8.91E-41 | Nightingale | LA | NA | C | T | 0.024778 | 0.004345 | 3.90E-09 |
| rs529565 | miR-126-3p | trans | 8.91E-41 | Nightingale | IDL_CE | NA | C | T | 0.03286 | 0.004278 | 4.10E-14 |
| rs529565 | miR-126-3p | trans | 8.91E-41 | Nightingale | XS_VLDL_C | NA | C | T | 0.037489 | 0.004301 | 3.30E-18 |
| rs529565 | miR-126-3p | trans | 8.91E-41 | Nightingale | VLDL_P | NA | C | T | 0.026295 | 0.004407 | 5.50E-10 |
| rs529565 | miR-126-3p | trans | 8.91E-41 | Nightingale | M_VLDL_T | NA | C | T | -0.03834 | 0.004187 | 1.50E-19 |

| | | | | | | | | | | | |
|---|---|---|---|---|---|---|---|---|---|---|---|
| rs529565 | miR-126-3p | trans | 8.91E-41 | Nightingale | L_LDL_P | NA | C | T | 0.028589 | 0.004416 | 1.20E-10 |
| rs529565 | miR-126-3p | trans | 8.91E-41 | Nightingale | S_HDL_PL | NA | C | T | 0.024302 | 0.004393 | 1.90E-08 |
| rs529565 | miR-126-3p | trans | 8.91E-41 | Nightingale | Total_CE | NA | C | T | 0.031352 | 0.004274 | 1.80E-13 |
| rs529565 | miR-126-3p | trans | 8.91E-41 | Nightingale | M_LDL_C_ | NA | C | T | 0.030966 | 0.004434 | 4.00E-12 |
| rs529565 | miR-126-3p | trans | 8.91E-41 | Nightingale | XXL_VLDL_ | NA | C | T | 0.034955 | 0.00444 | 2.80E-15 |
| rs529565 | miR-126-3p | trans | 8.91E-41 | Nightingale | XS_VLDL_P | NA | C | T | 0.036324 | 0.004372 | 2.20E-17 |
| rs529565 | miR-126-3p | trans | 8.91E-41 | Nightingale | S_LDL_FC | NA | C | T | 0.027889 | 0.004424 | 9.30E-10 |
| rs529565 | miR-126-3p | trans | 8.91E-41 | Nightingale | L_VLDL_FC | NA | C | T | 0.048532 | 0.004389 | 7.40E-29 |
| rs529565 | miR-126-3p | trans | 8.91E-41 | Nightingale | Omega_6 | NA | C | T | 0.025773 | 0.004327 | 6.10E-10 |
| rs529565 | miR-126-3p | trans | 8.91E-41 | Nightingale | L_LDL_FC | NA | C | T | 0.033087 | 0.004351 | 6.50E-14 |
| rs529565 | miR-126-3p | trans | 8.91E-41 | Nightingale | XS_VLDL_P | NA | C | T | 0.031742 | 0.004369 | 8.00E-14 |
| rs529565 | miR-126-3p | trans | 8.91E-41 | Nightingale | S_HDL_CE | NA | C | T | 0.028978 | 0.004438 | 8.90E-11 |
| rs529565 | miR-126-3p | trans | 8.91E-41 | Nightingale | GlycA | NA | C | T | 0.027732 | 0.004412 | 2.40E-10 |
| rs529565 | miR-126-3p | trans | 8.91E-41 | Nightingale | Total_L | NA | C | T | 0.026125 | 0.004359 | 5.60E-10 |
| rs529565 | miR-126-3p | trans | 8.91E-41 | Nightingale | S_HDL_FC | NA | C | T | 0.023726 | 0.00435 | 3.60E-08 |
| rs529565 | miR-126-3p | trans | 8.91E-41 | Nightingale | L_LDL_C | NA | C | T | 0.031856 | 0.004381 | 4.80E-13 |
| rs529565 | miR-126-3p | trans | 8.91E-41 | Nightingale | LDL_C | NA | C | T | 0.032762 | 0.004412 | 1.60E-13 |
| rs529565 | miR-126-3p | trans | 8.91E-41 | Nightingale | M_VLDL_P | NA | C | T | 0.02872 | 0.004424 | 2.80E-11 |
| rs529565 | miR-126-3p | trans | 8.91E-41 | Nightingale | S_VLDL_CE | NA | C | T | 0.037878 | 0.004425 | 2.20E-18 |
| rs529565 | miR-126-3p | trans | 8.91E-41 | Nightingale | XL_VLDL_T | NA | C | T | -0.04378 | 0.004295 | 3.70E-24 |
| rs529565 | miR-126-3p | trans | 8.91E-41 | Nightingale | S_VLDL_FC | NA | C | T | 0.03746 | 0.004432 | 1.30E-17 |
| rs529565 | miR-126-3p | trans | 8.91E-41 | Nightingale | XXL_VLDL_ | NA | C | T | 0.025677 | 0.004459 | 7.00E-10 |
| rs529565 | miR-126-3p | trans | 8.91E-41 | Nightingale | L_VLDL_C_ | NA | C | T | 0.05212 | 0.004328 | 9.90E-34 |
| rs529565 | miR-126-3p | trans | 8.91E-41 | Nightingale | IDL_P | NA | C | T | 0.039431 | 0.004385 | 3.50E-19 |
| rs529565 | miR-126-3p | trans | 8.91E-41 | Nightingale | XS_VLDL_L | NA | C | T | 0.033423 | 0.004356 | 3.30E-15 |
| rs529565 | miR-126-3p | trans | 8.91E-41 | Nightingale | M_VLDL_C | NA | C | T | 0.033552 | 0.004163 | 2.80E-15 |
| rs529565 | miR-126-3p | trans | 8.91E-41 | Nightingale | XL_VLDL_C | NA | C | T | 0.038352 | 0.004249 | 5.20E-19 |
| rs529565 | miR-126-3p | trans | 8.91E-41 | Nightingale | Total_C | NA | C | T | 0.031972 | 0.00429 | 6.30E-14 |
| rs529565 | miR-126-3p | trans | 8.91E-41 | Nightingale | M_LDL_FC | NA | C | T | 0.033834 | 0.004426 | 4.00E-14 |
| rs529565 | miR-126-3p | trans | 8.91E-41 | Nightingale | M_VLDL_F | NA | C | T | 0.032904 | 0.00443 | 5.40E-14 |
| rs529565 | miR-126-3p | trans | 8.91E-41 | Nightingale | ApoB | NA | C | T | 0.033444 | 0.004425 | 3.90E-14 |
| rs529565 | miR-126-3p | trans | 8.91E-41 | Nightingale | ApoB_by_A | NA | C | T | 0.02449 | 0.004364 | 2.40E-08 |
| rs529565 | miR-126-3p | trans | 8.91E-41 | Nightingale | S_LDL_C_p | NA | C | T | 0.045563 | 0.004448 | 4.00E-25 |
| rs529565 | miR-126-3p | trans | 8.91E-41 | Nightingale | PUFA | NA | C | T | 0.02347 | 0.004322 | 1.80E-08 |
| rs529565 | miR-126-3p | trans | 8.91E-41 | Nightingale | S_HDL_L | NA | C | T | 0.02679 | 0.004416 | 8.80E-10 |
| rs529565 | miR-126-3p | trans | 8.91E-41 | Nightingale | S_VLDL_PL | NA | C | T | 0.03238 | 0.004423 | 6.00E-14 |
| rs529565 | miR-126-3p | trans | 8.91E-41 | Nightingale | M_LDL_L | NA | C | T | 0.032702 | 0.004437 | 1.40E-13 |
| rs529565 | miR-126-3p | trans | 8.91E-41 | Nightingale | M_LDL_P | NA | C | T | 0.035668 | 0.004443 | 8.20E-16 |
| rs529565 | miR-126-3p | trans | 8.91E-41 | Nightingale | XS_VLDL_F | NA | C | T | 0.030891 | 0.004399 | 5.80E-12 |
| rs529565 | miR-126-3p | trans | 8.91E-41 | Nightingale | XXL_VLDL_ | NA | C | T | 0.033086 | 0.004464 | 1.10E-13 |
| rs529565 | miR-126-3p | trans | 8.91E-41 | Nightingale | IDL_C | NA | C | T | 0.034316 | 0.00428 | 3.00E-15 |
| rs529565 | let-7b-5p | trans | 4.77E-28 | Nightingale | M_LDL_PL | NA | C | T | 0.03206 | 0.004437 | 4.30E-13 |
| rs529565 | let-7b-5p | trans | 4.77E-28 | Nightingale | S_LDL_L | NA | C | T | 0.027285 | 0.00443 | 1.30E-09 |
| rs529565 | let-7b-5p | trans | 4.77E-28 | Nightingale | XXL_VLDL_ | NA | C | T | 0.031056 | 0.004412 | 9.30E-13 |
| rs529565 | let-7b-5p | trans | 4.77E-28 | Nightingale | VLDL_C | NA | C | T | 0.030948 | 0.004425 | 6.40E-13 |
| rs529565 | let-7b-5p | trans | 4.77E-28 | Nightingale | S_LDL_CE_ | NA | C | T | 0.031515 | 0.004423 | 1.80E-14 |
| rs529565 | let-7b-5p | trans | 4.77E-28 | Nightingale | M_VLDL_P | NA | C | T | 0.043673 | 0.004253 | 1.80E-24 |
| rs529565 | let-7b-5p | trans | 4.77E-28 | Nightingale | LDL_P | NA | C | T | 0.031312 | 0.004428 | 1.80E-12 |
| rs529565 | let-7b-5p | trans | 4.77E-28 | Nightingale | S_HDL_P | NA | C | T | 0.030973 | 0.004432 | 3.30E-12 |
| rs529565 | let-7b-5p | trans | 4.77E-28 | Nightingale | XL_VLDL_P | NA | C | T | 0.038314 | 0.004438 | 1.50E-18 |
| rs529565 | let-7b-5p | trans | 4.77E-28 | Nightingale | S_VLDL_L | NA | C | T | 0.02438 | 0.004388 | 4.50E-09 |
| rs529565 | let-7b-5p | trans | 4.77E-28 | Nightingale | VLDL_FC | NA | C | T | 0.02385 | 0.004399 | 1.30E-08 |
| rs529565 | let-7b-5p | trans | 4.77E-28 | Nightingale | Citrate | NA | C | T | 0.027701 | 0.004402 | 2.10E-10 |
| rs529565 | let-7b-5p | trans | 4.77E-28 | Nightingale | M_VLDL_C | NA | C | T | 0.036055 | 0.004174 | 2.10E-17 |
| rs529565 | let-7b-5p | trans | 4.77E-28 | Nightingale | XL_VLDL_F | NA | C | T | 0.036987 | 0.004296 | 3.90E-18 |
| rs529565 | let-7b-5p | trans | 4.77E-28 | Nightingale | M_LDL_C | NA | C | T | 0.033658 | 0.004439 | 3.10E-14 |
| rs529565 | let-7b-5p | trans | 4.77E-28 | Nightingale | S_VLDL_CE | NA | C | T | 0.046913 | 0.004372 | 2.40E-26 |
| rs529565 | let-7b-5p | trans | 4.77E-28 | Nightingale | M_VLDL_C | NA | C | T | 0.038197 | 0.004419 | 4.00E-18 |
| rs529565 | let-7b-5p | trans | 4.77E-28 | Nightingale | S_LDL_P | NA | C | T | 0.029187 | 0.004435 | 4.60E-11 |
| rs529565 | let-7b-5p | trans | 4.77E-28 | Nightingale | IDL_FC_pct | NA | C | T | 0.025499 | 0.004397 | 1.50E-08 |
| rs529565 | let-7b-5p | trans | 4.77E-28 | Nightingale | Sphingomy | NA | C | T | 0.02867 | 0.004142 | 1.30E-11 |
| rs529565 | let-7b-5p | trans | 4.77E-28 | Nightingale | Total_FC | NA | C | T | 0.032841 | 0.004329 | 1.20E-14 |
| rs529565 | let-7b-5p | trans | 4.77E-28 | Nightingale | Clinical_LD | NA | C | T | 0.035972 | 0.004405 | 5.80E-16 |

| | | | | | | | | | | | |
|---|---|---|---|---|---|---|---|---|---|---|---|
| rs529565 | let-7b-5p | trans | 4.77E-28 | Nightingale | L_LDL_L | NA | C | T | 0.033109 | 0.004388 | 5.60E-14 |
| rs529565 | let-7b-5p | trans | 4.77E-28 | Nightingale | LDL_CE | NA | C | T | 0.032164 | 0.004419 | 3.50E-13 |
| rs529565 | let-7b-5p | trans | 4.77E-28 | Nightingale | S_VLDL_TG | NA | C | T | -0.03791 | 0.004289 | 2.80E-18 |
| rs529565 | let-7b-5p | trans | 4.77E-28 | Nightingale | S_LDL_PL_ | NA | C | T | -0.03064 | 0.004423 | 9.40E-14 |
| rs529565 | let-7b-5p | trans | 4.77E-28 | Nightingale | VLDL_CE | NA | C | T | 0.035403 | 0.004432 | 4.50E-16 |
| rs529565 | let-7b-5p | trans | 4.77E-28 | Nightingale | L_LDL_CE | NA | C | T | 0.03113 | 0.004392 | 1.70E-12 |
| rs529565 | let-7b-5p | trans | 4.77E-28 | Nightingale | L_VLDL_TG | NA | C | T | -0.05092 | 0.00441 | 1.60E-31 |
| rs529565 | let-7b-5p | trans | 4.77E-28 | Nightingale | S_VLDL_C_ | NA | C | T | 0.041702 | 0.004312 | 1.10E-21 |
| rs529565 | let-7b-5p | trans | 4.77E-28 | Nightingale | LDL_FC | NA | C | T | 0.033085 | 0.004384 | 1.10E-13 |
| rs529565 | let-7b-5p | trans | 4.77E-28 | Nightingale | XXL_VLDL_ | NA | C | T | -0.03595 | 0.004469 | 1.80E-16 |
| rs529565 | let-7b-5p | trans | 4.77E-28 | Nightingale | S_VLDL_C | NA | C | T | 0.038015 | 0.004431 | 2.60E-18 |
| rs529565 | let-7b-5p | trans | 4.77E-28 | Nightingale | IDL_L | NA | C | T | 0.034696 | 0.004284 | 1.30E-15 |
| rs529565 | let-7b-5p | trans | 4.77E-28 | Nightingale | IDL_PL | NA | C | T | 0.034834 | 0.004276 | 7.90E-16 |
| rs529565 | let-7b-5p | trans | 4.77E-28 | Nightingale | non_HDL_( | NA | C | T | 0.035225 | 0.004407 | 1.00E-15 |
| rs529565 | let-7b-5p | trans | 4.77E-28 | Nightingale | S_VLDL_FC | NA | C | T | 0.02911 | 0.004234 | 1.70E-11 |
| rs529565 | let-7b-5p | trans | 4.77E-28 | Nightingale | S_LDL_CE | NA | C | T | 0.030231 | 0.004433 | 7.50E-12 |
| rs529565 | let-7b-5p | trans | 4.77E-28 | Nightingale | XS_VLDL_C | NA | C | T | 0.037263 | 0.004276 | 5.70E-18 |
| rs529565 | let-7b-5p | trans | 4.77E-28 | Nightingale | Total_PL | NA | C | T | 0.024805 | 0.004219 | 1.20E-09 |
| rs529565 | let-7b-5p | trans | 4.77E-28 | Nightingale | M_LDL_TG | NA | C | T | -0.02724 | 0.004376 | 5.60E-10 |
| rs529565 | let-7b-5p | trans | 4.77E-28 | Nightingale | S_LDL_C | NA | C | T | 0.030526 | 0.004435 | 8.70E-12 |
| rs529565 | let-7b-5p | trans | 4.77E-28 | Nightingale | M_VLDL_F | NA | C | T | 0.041684 | 0.004216 | 1.10E-22 |
| rs529565 | let-7b-5p | trans | 4.77E-28 | Nightingale | M_VLDL_C | NA | C | T | 0.040303 | 0.004391 | 5.90E-20 |
| rs529565 | let-7b-5p | trans | 4.77E-28 | Nightingale | XL_HDL_PL | NA | C | T | -0.02275 | 0.004071 | 1.10E-08 |
| rs529565 | let-7b-5p | trans | 4.77E-28 | Nightingale | LDL_L | NA | C | T | 0.032885 | 0.004416 | 1.30E-13 |
| rs529565 | let-7b-5p | trans | 4.77E-28 | Nightingale | Remnant_( | NA | C | T | 0.036369 | 0.004402 | 7.40E-17 |
| rs529565 | let-7b-5p | trans | 4.77E-28 | Nightingale | IDL_FC | NA | C | T | 0.037662 | 0.00429 | 4.70E-18 |
| rs529565 | let-7b-5p | trans | 4.77E-28 | Nightingale | LDL_PL | NA | C | T | 0.034514 | 0.004421 | 9.20E-15 |
| rs529565 | let-7b-5p | trans | 4.77E-28 | Nightingale | M_LDL_CE | NA | C | T | 0.032712 | 0.004437 | 1.20E-13 |
| rs529565 | let-7b-5p | trans | 4.77E-28 | Nightingale | S_VLDL_P | NA | C | T | 0.023183 | 0.004376 | 1.90E-08 |
| rs529565 | let-7b-5p | trans | 4.77E-28 | Nightingale | L_VLDL_CE | NA | C | T | 0.047309 | 0.00424 | 5.10E-29 |
| rs529565 | let-7b-5p | trans | 4.77E-28 | Nightingale | L_LDL_PL | NA | C | T | 0.03752 | 0.004398 | 1.90E-17 |
| rs529565 | let-7b-5p | trans | 4.77E-28 | Nightingale | S_HDL_C | NA | C | T | 0.02876 | 0.004424 | 1.20E-10 |
| rs529565 | let-7b-5p | trans | 4.77E-28 | Nightingale | XS_VLDL_F | NA | C | T | 0.036631 | 0.004354 | 1.20E-17 |
| rs529565 | let-7b-5p | trans | 4.77E-28 | Nightingale | XL_VLDL_C | NA | C | T | 0.037516 | 0.004251 | 3.10E-18 |
| rs529565 | let-7b-5p | trans | 4.77E-28 | Nightingale | S_VLDL_PL | NA | C | T | 0.027457 | 0.004243 | 2.90E-10 |
| rs529565 | let-7b-5p | trans | 4.77E-28 | Nightingale | M_VLDL_P | NA | C | T | 0.027887 | 0.00442 | 8.70E-11 |
| rs529565 | let-7b-5p | trans | 4.77E-28 | Nightingale | LA | NA | C | T | 0.024778 | 0.004345 | 3.90E-09 |
| rs529565 | let-7b-5p | trans | 4.77E-28 | Nightingale | IDL_CE | NA | C | T | 0.03286 | 0.004278 | 4.10E-14 |
| rs529565 | let-7b-5p | trans | 4.77E-28 | Nightingale | XS_VLDL_C | NA | C | T | 0.037489 | 0.004301 | 3.30E-18 |
| rs529565 | let-7b-5p | trans | 4.77E-28 | Nightingale | VLDL_P | NA | C | T | 0.026295 | 0.004407 | 5.50E-10 |
| rs529565 | let-7b-5p | trans | 4.77E-28 | Nightingale | M_VLDL_T | NA | C | T | -0.03834 | 0.004187 | 1.50E-19 |
| rs529565 | let-7b-5p | trans | 4.77E-28 | Nightingale | L_LDL_P | NA | C | T | 0.028589 | 0.004416 | 1.20E-10 |
| rs529565 | let-7b-5p | trans | 4.77E-28 | Nightingale | S_HDL_PL | NA | C | T | 0.024302 | 0.004393 | 1.90E-08 |
| rs529565 | let-7b-5p | trans | 4.77E-28 | Nightingale | Total_CE | NA | C | T | 0.031352 | 0.004274 | 1.80E-13 |
| rs529565 | let-7b-5p | trans | 4.77E-28 | Nightingale | M_LDL_C_ | NA | C | T | 0.030966 | 0.004434 | 4.00E-12 |
| rs529565 | let-7b-5p | trans | 4.77E-28 | Nightingale | XXL_VLDL_ | NA | C | T | 0.034955 | 0.00444 | 2.80E-15 |
| rs529565 | let-7b-5p | trans | 4.77E-28 | Nightingale | XS_VLDL_P | NA | C | T | 0.036324 | 0.004372 | 2.20E-17 |
| rs529565 | let-7b-5p | trans | 4.77E-28 | Nightingale | S_LDL_FC | NA | C | T | 0.027889 | 0.004424 | 9.30E-10 |
| rs529565 | let-7b-5p | trans | 4.77E-28 | Nightingale | L_VLDL_FC | NA | C | T | 0.048532 | 0.004389 | 7.40E-29 |
| rs529565 | let-7b-5p | trans | 4.77E-28 | Nightingale | Omega_6 | NA | C | T | 0.025773 | 0.004327 | 6.10E-10 |
| rs529565 | let-7b-5p | trans | 4.77E-28 | Nightingale | L_LDL_FC | NA | C | T | 0.033087 | 0.004351 | 6.50E-14 |
| rs529565 | let-7b-5p | trans | 4.77E-28 | Nightingale | XS_VLDL_P | NA | C | T | 0.031742 | 0.004369 | 8.00E-14 |
| rs529565 | let-7b-5p | trans | 4.77E-28 | Nightingale | S_HDL_CE | NA | C | T | 0.028978 | 0.004438 | 8.90E-11 |
| rs529565 | let-7b-5p | trans | 4.77E-28 | Nightingale | GlycA | NA | C | T | 0.027732 | 0.004412 | 2.40E-10 |
| rs529565 | let-7b-5p | trans | 4.77E-28 | Nightingale | Total_L | NA | C | T | 0.026125 | 0.004359 | 5.60E-10 |
| rs529565 | let-7b-5p | trans | 4.77E-28 | Nightingale | S_HDL_FC | NA | C | T | 0.023726 | 0.00435 | 3.60E-08 |
| rs529565 | let-7b-5p | trans | 4.77E-28 | Nightingale | L_LDL_C | NA | C | T | 0.031856 | 0.004381 | 4.80E-13 |
| rs529565 | let-7b-5p | trans | 4.77E-28 | Nightingale | LDL_C | NA | C | T | 0.032762 | 0.004412 | 1.60E-13 |
| rs529565 | let-7b-5p | trans | 4.77E-28 | Nightingale | M_VLDL_P | NA | C | T | 0.02872 | 0.004424 | 2.80E-11 |
| rs529565 | let-7b-5p | trans | 4.77E-28 | Nightingale | S_VLDL_CE | NA | C | T | 0.037878 | 0.004425 | 2.20E-18 |
| rs529565 | let-7b-5p | trans | 4.77E-28 | Nightingale | XL_VLDL_T | NA | C | T | -0.04378 | 0.004295 | 3.70E-24 |
| rs529565 | let-7b-5p | trans | 4.77E-28 | Nightingale | S_VLDL_FC | NA | C | T | 0.03746 | 0.004432 | 1.30E-17 |
| rs529565 | let-7b-5p | trans | 4.77E-28 | Nightingale | XXL_VLDL_ | NA | C | T | 0.025677 | 0.004459 | 7.00E-10 |

| | | | | | | | | | | | |
|---|---|---|---|---|---|---|---|---|---|---|---|
| rs529565 | let-7b-5p | trans | 4.77E-28 | Nightingale | L_VLDL_C_ | NA | C | T | 0.05212 | 0.004328 | 9.90E-34 |
| rs529565 | let-7b-5p | trans | 4.77E-28 | Nightingale | IDL_P | NA | C | T | 0.039431 | 0.004385 | 3.50E-19 |
| rs529565 | let-7b-5p | trans | 4.77E-28 | Nightingale | XS_VLDL_L | NA | C | T | 0.033423 | 0.004356 | 3.30E-15 |
| rs529565 | let-7b-5p | trans | 4.77E-28 | Nightingale | M_VLDL_C | NA | C | T | 0.033552 | 0.004163 | 2.80E-15 |
| rs529565 | let-7b-5p | trans | 4.77E-28 | Nightingale | XL_VLDL_C | NA | C | T | 0.038352 | 0.004249 | 5.20E-19 |
| rs529565 | let-7b-5p | trans | 4.77E-28 | Nightingale | Total_C | NA | C | T | 0.031972 | 0.00429 | 6.30E-14 |
| rs529565 | let-7b-5p | trans | 4.77E-28 | Nightingale | M_LDL_FC | NA | C | T | 0.033834 | 0.004426 | 4.00E-14 |
| rs529565 | let-7b-5p | trans | 4.77E-28 | Nightingale | M_VLDL_F | NA | C | T | 0.032904 | 0.00443 | 5.40E-14 |
| rs529565 | let-7b-5p | trans | 4.77E-28 | Nightingale | ApoB | NA | C | T | 0.033444 | 0.004425 | 3.90E-14 |
| rs529565 | let-7b-5p | trans | 4.77E-28 | Nightingale | ApoB_by_/ | NA | C | T | 0.02449 | 0.004364 | 2.40E-08 |
| rs529565 | let-7b-5p | trans | 4.77E-28 | Nightingale | S_LDL_C_p | NA | C | T | 0.045563 | 0.004448 | 4.00E-25 |
| rs529565 | let-7b-5p | trans | 4.77E-28 | Nightingale | PUFA | NA | C | T | 0.02347 | 0.004322 | 1.80E-08 |
| rs529565 | let-7b-5p | trans | 4.77E-28 | Nightingale | S_HDL_L | NA | C | T | 0.02679 | 0.004416 | 8.80E-10 |
| rs529565 | let-7b-5p | trans | 4.77E-28 | Nightingale | S_VLDL_PL | NA | C | T | 0.03238 | 0.004423 | 6.00E-14 |
| rs529565 | let-7b-5p | trans | 4.77E-28 | Nightingale | M_LDL_L | NA | C | T | 0.032702 | 0.004437 | 1.40E-13 |
| rs529565 | let-7b-5p | trans | 4.77E-28 | Nightingale | M_LDL_P | NA | C | T | 0.035668 | 0.004443 | 8.20E-16 |
| rs529565 | let-7b-5p | trans | 4.77E-28 | Nightingale | XS_VLDL_F | NA | C | T | 0.030891 | 0.004399 | 5.80E-12 |
| rs529565 | let-7b-5p | trans | 4.77E-28 | Nightingale | XXL_VLDL_ | NA | C | T | 0.033086 | 0.004464 | 1.10E-13 |
| rs529565 | let-7b-5p | trans | 4.77E-28 | Nightingale | IDL_C | NA | C | T | 0.034316 | 0.00428 | 3.00E-15 |
| rs532436 | miR-126-3p | trans | 2.49E-40 | Nightingale | XS_VLDL_T | NA | A | G | -0.0425 | 0.005069 | 3.40E-16 |
| rs532436 | miR-126-3p | trans | 2.49E-40 | Nightingale | M_VLDL_P | NA | A | G | 0.070479 | 0.005117 | 5.70E-43 |
| rs532436 | miR-126-3p | trans | 2.49E-40 | Nightingale | L_LDL_C | NA | A | G | 0.059903 | 0.005272 | 1.70E-30 |
| rs532436 | miR-126-3p | trans | 2.49E-40 | Nightingale | LA_pct | NA | A | G | 0.032246 | 0.005214 | 6.90E-10 |
| rs532436 | miR-126-3p | trans | 2.49E-40 | Nightingale | S_VLDL_C | NA | A | G | 0.058922 | 0.005331 | 3.90E-30 |
| rs532436 | miR-126-3p | trans | 2.49E-40 | Nightingale | S_LDL_L | NA | A | G | 0.0484 | 0.005331 | 3.80E-20 |
| rs532436 | miR-126-3p | trans | 2.49E-40 | Nightingale | L_VLDL_FC | NA | A | G | 0.075634 | 0.005281 | 2.30E-47 |
| rs532436 | miR-126-3p | trans | 2.49E-40 | Nightingale | M_LDL_PL | NA | A | G | 0.058078 | 0.005339 | 2.50E-28 |
| rs532436 | miR-126-3p | trans | 2.49E-40 | Nightingale | L_LDL_L | NA | A | G | 0.060847 | 0.00528 | 1.80E-31 |
| rs532436 | miR-126-3p | trans | 2.49E-40 | Nightingale | XL_VLDL_C | NA | A | G | 0.062307 | 0.005115 | 2.90E-33 |
| rs532436 | miR-126-3p | trans | 2.49E-40 | Nightingale | S_VLDL_L | NA | A | G | 0.035519 | 0.00528 | 4.90E-13 |
| rs532436 | miR-126-3p | trans | 2.49E-40 | Nightingale | S_LDL_FC | NA | A | G | 0.054532 | 0.005323 | 2.40E-24 |
| rs532436 | miR-126-3p | trans | 2.49E-40 | Nightingale | M_LDL_P | NA | A | G | 0.057013 | 0.005347 | 1.50E-27 |
| rs532436 | miR-126-3p | trans | 2.49E-40 | Nightingale | M_VLDL_C | NA | A | G | 0.0595 | 0.005022 | 1.90E-31 |
| rs532436 | miR-126-3p | trans | 2.49E-40 | Nightingale | S_LDL_P | NA | A | G | 0.048388 | 0.005337 | 1.90E-20 |
| rs532436 | miR-126-3p | trans | 2.49E-40 | Nightingale | XL_HDL_L | NA | A | G | -0.02706 | 0.004795 | 2.40E-09 |
| rs532436 | miR-126-3p | trans | 2.49E-40 | Nightingale | S_LDL_CE_ | NA | A | G | 0.044561 | 0.005323 | 1.40E-19 |
| rs532436 | miR-126-3p | trans | 2.49E-40 | Nightingale | S_HDL_C | NA | A | G | 0.052394 | 0.005323 | 3.50E-23 |
| rs532436 | miR-126-3p | trans | 2.49E-40 | Nightingale | S_LDL_C_p | NA | A | G | 0.075576 | 0.005352 | 2.10E-46 |
| rs532436 | miR-126-3p | trans | 2.49E-40 | Nightingale | LDL_FC | NA | A | G | 0.062671 | 0.005275 | 1.40E-32 |
| rs532436 | miR-126-3p | trans | 2.49E-40 | Nightingale | XS_VLDL_C | NA | A | G | 0.040138 | 0.005031 | 1.40E-14 |
| rs532436 | miR-126-3p | trans | 2.49E-40 | Nightingale | XS_VLDL_P | NA | A | G | 0.044567 | 0.005257 | 3.00E-19 |
| rs532436 | miR-126-3p | trans | 2.49E-40 | Nightingale | LDL_L | NA | A | G | 0.059463 | 0.005314 | 9.50E-30 |
| rs532436 | miR-126-3p | trans | 2.49E-40 | Nightingale | Clinical_LD | NA | A | G | 0.064426 | 0.005301 | 1.80E-34 |
| rs532436 | miR-126-3p | trans | 2.49E-40 | Nightingale | Citrate | NA | A | G | 0.031277 | 0.005297 | 1.60E-09 |
| rs532436 | miR-126-3p | trans | 2.49E-40 | Nightingale | XS_VLDL_P | NA | A | G | 0.05371 | 0.005261 | 1.20E-26 |
| rs532436 | miR-126-3p | trans | 2.49E-40 | Nightingale | LDL_CE | NA | A | G | 0.058166 | 0.005318 | 1.20E-28 |
| rs532436 | miR-126-3p | trans | 2.49E-40 | Nightingale | Remnant_( | NA | A | G | 0.058377 | 0.005297 | 5.50E-30 |
| rs532436 | miR-126-3p | trans | 2.49E-40 | Nightingale | Cholines | NA | A | G | 0.035678 | 0.005014 | 1.10E-13 |
| rs532436 | miR-126-3p | trans | 2.49E-40 | Nightingale | HDL_size | NA | A | G | -0.03226 | 0.004727 | 1.70E-12 |
| rs532436 | miR-126-3p | trans | 2.49E-40 | Nightingale | S_HDL_L | NA | A | G | 0.043965 | 0.005313 | 2.20E-17 |
| rs532436 | miR-126-3p | trans | 2.49E-40 | Nightingale | S_VLDL_FC | NA | A | G | 0.059408 | 0.005333 | 3.00E-30 |
| rs532436 | miR-126-3p | trans | 2.49E-40 | Nightingale | XXL_VLDL_ | NA | A | G | 0.046201 | 0.005312 | 1.30E-18 |
| rs532436 | miR-126-3p | trans | 2.49E-40 | Nightingale | XS_VLDL_C | NA | A | G | 0.057562 | 0.005175 | 2.50E-30 |
| rs532436 | miR-126-3p | trans | 2.49E-40 | Nightingale | M_VLDL_F | NA | A | G | 0.067258 | 0.005073 | 1.70E-39 |
| rs532436 | miR-126-3p | trans | 2.49E-40 | Nightingale | IDL_TG_pc | NA | A | G | -0.03868 | 0.005155 | 3.10E-13 |
| rs532436 | miR-126-3p | trans | 2.49E-40 | Nightingale | XS_VLDL_P | NA | A | G | -0.02951 | 0.005203 | 5.00E-08 |
| rs532436 | miR-126-3p | trans | 2.49E-40 | Nightingale | M_VLDL_L | NA | A | G | 0.032097 | 0.005296 | 1.10E-10 |
| rs532436 | miR-126-3p | trans | 2.49E-40 | Nightingale | LA | NA | A | G | 0.042219 | 0.005229 | 3.30E-17 |
| rs532436 | miR-126-3p | trans | 2.49E-40 | Nightingale | M_VLDL_C | NA | A | G | 0.06605 | 0.005283 | 1.00E-36 |
| rs532436 | miR-126-3p | trans | 2.49E-40 | Nightingale | Total_P | NA | A | G | 0.036654 | 0.005018 | 5.50E-14 |
| rs532436 | miR-126-3p | trans | 2.49E-40 | Nightingale | Total_L | NA | A | G | 0.042255 | 0.005245 | 4.90E-17 |
| rs532436 | miR-126-3p | trans | 2.49E-40 | Nightingale | M_LDL_TG | NA | A | G | -0.0531 | 0.005266 | 1.10E-23 |
| rs532436 | miR-126-3p | trans | 2.49E-40 | Nightingale | L_LDL_P | NA | A | G | 0.050357 | 0.005314 | 6.40E-22 |

| | | | | | | | | | | | |
|---|---|---|---|---|---|---|---|---|---|---|---|
| rs532436 | miR-126-3p | trans | 2.49E-40 | Nightingale | S_LDL_CE | NA | A | G | 0.051567 | 0.005334 | 4.80E-23 |
| rs532436 | miR-126-3p | trans | 2.49E-40 | Nightingale | S_LDL_TG_ | NA | A | G | -0.04355 | 0.00517 | 3.00E-16 |
| rs532436 | miR-126-3p | trans | 2.49E-40 | Nightingale | M_LDL_L | NA | A | G | 0.057299 | 0.005339 | 8.90E-28 |
| rs532436 | miR-126-3p | trans | 2.49E-40 | Nightingale | IDL_FC_pct | NA | A | G | 0.041213 | 0.00529 | 2.10E-14 |
| rs532436 | miR-126-3p | trans | 2.49E-40 | Nightingale | Phosphogly | NA | A | G | 0.031891 | 0.00505 | 3.60E-11 |
| rs532436 | miR-126-3p | trans | 2.49E-40 | Nightingale | XL_VLDL_F | NA | A | G | 0.056773 | 0.005169 | 2.60E-28 |
| rs532436 | miR-126-3p | trans | 2.49E-40 | Nightingale | S_HDL_FC | NA | A | G | 0.041762 | 0.005234 | 2.40E-16 |
| rs532436 | miR-126-3p | trans | 2.49E-40 | Nightingale | IDL_FC | NA | A | G | 0.063966 | 0.005162 | 7.70E-36 |
| rs532436 | miR-126-3p | trans | 2.49E-40 | Nightingale | S_VLDL_PL | NA | A | G | 0.050032 | 0.005322 | 1.30E-22 |
| rs532436 | miR-126-3p | trans | 2.49E-40 | Nightingale | IDL_C_pct | NA | A | G | 0.037343 | 0.005209 | 6.30E-12 |
| rs532436 | miR-126-3p | trans | 2.49E-40 | Nightingale | L_VLDL_CE | NA | A | G | 0.074326 | 0.005102 | 1.30E-48 |
| rs532436 | miR-126-3p | trans | 2.49E-40 | Nightingale | L_LDL_CE | NA | A | G | 0.058358 | 0.005284 | 5.40E-29 |
| rs532436 | miR-126-3p | trans | 2.49E-40 | Nightingale | S_VLDL_CE | NA | A | G | 0.076497 | 0.00526 | 2.10E-47 |
| rs532436 | miR-126-3p | trans | 2.49E-40 | Nightingale | XL_HDL_CE | NA | A | G | 0.030066 | 0.005327 | 4.20E-08 |
| rs532436 | miR-126-3p | trans | 2.49E-40 | Nightingale | XS_VLDL_F | NA | A | G | 0.053623 | 0.005239 | 1.10E-26 |
| rs532436 | miR-126-3p | trans | 2.49E-40 | Nightingale | M_LDL_C_ | NA | A | G | 0.047449 | 0.005336 | 4.30E-19 |
| rs532436 | miR-126-3p | trans | 2.49E-40 | Nightingale | S_VLDL_P | NA | A | G | 0.033337 | 0.005265 | 7.40E-12 |
| rs532436 | miR-126-3p | trans | 2.49E-40 | Nightingale | S_VLDL_FC | NA | A | G | 0.05247 | 0.005095 | 3.60E-24 |
| rs532436 | miR-126-3p | trans | 2.49E-40 | Nightingale | L_VLDL_TG | NA | A | G | -0.0795 | 0.005307 | 2.00E-52 |
| rs532436 | miR-126-3p | trans | 2.49E-40 | Nightingale | GlycA | NA | A | G | 0.031177 | 0.005309 | 6.60E-09 |
| rs532436 | miR-126-3p | trans | 2.49E-40 | Nightingale | L_VLDL_CE | NA | A | G | 0.031425 | 0.005269 | 1.40E-10 |
| rs532436 | miR-126-3p | trans | 2.49E-40 | Nightingale | M_VLDL_C | NA | A | G | 0.061586 | 0.005317 | 2.10E-32 |
| rs532436 | miR-126-3p | trans | 2.49E-40 | Nightingale | IDL_L | NA | A | G | 0.059551 | 0.005155 | 1.70E-31 |
| rs532436 | miR-126-3p | trans | 2.49E-40 | Nightingale | IDL_CE | NA | A | G | 0.058228 | 0.005148 | 7.40E-30 |
| rs532436 | miR-126-3p | trans | 2.49E-40 | Nightingale | S_VLDL_PL | NA | A | G | 0.049979 | 0.005105 | 5.70E-22 |
| rs532436 | miR-126-3p | trans | 2.49E-40 | Nightingale | M_LDL_CE | NA | A | G | 0.055488 | 0.005339 | 2.20E-26 |
| rs532436 | miR-126-3p | trans | 2.49E-40 | Nightingale | XL_HDL_C_ | NA | A | G | 0.038356 | 0.005092 | 1.70E-13 |
| rs532436 | miR-126-3p | trans | 2.49E-40 | Nightingale | VLDL_C | NA | A | G | 0.045684 | 0.005324 | 2.90E-19 |
| rs532436 | miR-126-3p | trans | 2.49E-40 | Nightingale | IDL_C | NA | A | G | 0.060082 | 0.005149 | 9.90E-32 |
| rs532436 | miR-126-3p | trans | 2.49E-40 | Nightingale | S_VLDL_TG | NA | A | G | -0.06518 | 0.005161 | 4.40E-36 |
| rs532436 | miR-126-3p | trans | 2.49E-40 | Nightingale | ApoB_by_A | NA | A | G | 0.039971 | 0.005251 | 1.30E-14 |
| rs532436 | miR-126-3p | trans | 2.49E-40 | Nightingale | VLDL_P | NA | A | G | 0.038195 | 0.005303 | 2.40E-14 |
| rs532436 | miR-126-3p | trans | 2.49E-40 | Nightingale | Total_CE | NA | A | G | 0.05558 | 0.005143 | 3.40E-28 |
| rs532436 | miR-126-3p | trans | 2.49E-40 | Nightingale | L_LDL_FC_ | NA | A | G | 0.030877 | 0.005087 | 1.20E-08 |
| rs532436 | miR-126-3p | trans | 2.49E-40 | Nightingale | VLDL_CE | NA | A | G | 0.053545 | 0.005333 | 2.70E-25 |
| rs532436 | miR-126-3p | trans | 2.49E-40 | Nightingale | M_VLDL_C | NA | A | G | 0.055885 | 0.005009 | 5.70E-28 |
| rs532436 | miR-126-3p | trans | 2.49E-40 | Nightingale | XXL_VLDL_ | NA | A | G | -0.05817 | 0.005381 | 5.80E-28 |
| rs532436 | miR-126-3p | trans | 2.49E-40 | Nightingale | IDL_PL | NA | A | G | 0.05875 | 0.005145 | 3.70E-31 |
| rs532436 | miR-126-3p | trans | 2.49E-40 | Nightingale | S_HDL_PL_ | NA | A | G | -0.03259 | 0.005155 | 1.00E-09 |
| rs532436 | miR-126-3p | trans | 2.49E-40 | Nightingale | S_LDL_PL | NA | A | G | 0.041297 | 0.00532 | 6.80E-15 |
| rs532436 | miR-126-3p | trans | 2.49E-40 | Nightingale | non_HDL_( | NA | A | G | 0.060373 | 0.005303 | 3.40E-31 |
| rs532436 | miR-126-3p | trans | 2.49E-40 | Nightingale | M_VLDL_P | NA | A | G | 0.044715 | 0.005324 | 1.90E-18 |
| rs532436 | miR-126-3p | trans | 2.49E-40 | Nightingale | Sphingomy | NA | A | G | 0.047389 | 0.004984 | 3.70E-21 |
| rs532436 | miR-126-3p | trans | 2.49E-40 | Nightingale | LDL_PL | NA | A | G | 0.06187 | 0.005319 | 7.90E-32 |
| rs532436 | miR-126-3p | trans | 2.49E-40 | Nightingale | L_VLDL_PL | NA | A | G | 0.027951 | 0.005179 | 3.30E-09 |
| rs532436 | miR-126-3p | trans | 2.49E-40 | Nightingale | S_HDL_PL | NA | A | G | 0.037551 | 0.005286 | 1.80E-13 |
| rs532436 | miR-126-3p | trans | 2.49E-40 | Nightingale | XS_VLDL_C | NA | A | G | 0.043168 | 0.005057 | 1.10E-16 |
| rs532436 | miR-126-3p | trans | 2.49E-40 | Nightingale | IDL_P | NA | A | G | 0.06498 | 0.005276 | 1.90E-35 |
| rs532436 | miR-126-3p | trans | 2.49E-40 | Nightingale | S_HDL_CE | NA | A | G | 0.053255 | 0.00534 | 1.20E-23 |
| rs532436 | miR-126-3p | trans | 2.49E-40 | Nightingale | L_LDL_FC | NA | A | G | 0.062609 | 0.005236 | 3.80E-33 |
| rs532436 | miR-126-3p | trans | 2.49E-40 | Nightingale | PUFA | NA | A | G | 0.036816 | 0.005201 | 1.50E-13 |
| rs532436 | miR-126-3p | trans | 2.49E-40 | Nightingale | XS_VLDL_C | NA | A | G | 0.058331 | 0.005145 | 3.40E-31 |
| rs532436 | miR-126-3p | trans | 2.49E-40 | Nightingale | ApoB | NA | A | G | 0.055646 | 0.005325 | 1.40E-26 |
| rs532436 | miR-126-3p | trans | 2.49E-40 | Nightingale | M_VLDL_F | NA | A | G | 0.051674 | 0.005331 | 1.50E-23 |
| rs532436 | miR-126-3p | trans | 2.49E-40 | Nightingale | S_LDL_C | NA | A | G | 0.053873 | 0.005337 | 1.50E-24 |
| rs532436 | miR-126-3p | trans | 2.49E-40 | Nightingale | XL_VLDL_C | NA | A | G | 0.062531 | 0.005113 | 1.30E-33 |
| rs532436 | miR-126-3p | trans | 2.49E-40 | Nightingale | XXL_VLDL_ | NA | A | G | 0.055922 | 0.005375 | 1.40E-25 |
| rs532436 | miR-126-3p | trans | 2.49E-40 | Nightingale | Omega_6 | NA | A | G | 0.0417 | 0.005207 | 5.30E-17 |
| rs532436 | miR-126-3p | trans | 2.49E-40 | Nightingale | LDL_P | NA | A | G | 0.053144 | 0.005328 | 2.90E-24 |
| rs532436 | miR-126-3p | trans | 2.49E-40 | Nightingale | XXL_VLDL_ | NA | A | G | 0.056832 | 0.005346 | 1.50E-26 |
| rs532436 | miR-126-3p | trans | 2.49E-40 | Nightingale | Pyruvate | NA | A | G | 0.034156 | 0.00534 | 1.60E-10 |
| rs532436 | miR-126-3p | trans | 2.49E-40 | Nightingale | Total_PL | NA | A | G | 0.03947 | 0.005077 | 4.30E-16 |
| rs532436 | miR-126-3p | trans | 2.49E-40 | Nightingale | L_LDL_TG_ | NA | A | G | -0.04149 | 0.00523 | 8.10E-15 |

| | | | | | | | | | | | |
|---|---|---|---|---|---|---|---|---|---|---|---|
| rs532436 | miR-126-3p | trans | 2.49E-40 | Nightingale | XL_VLDL_T | NA | A | G | -0.06894 | 0.005168 | 1.90E-40 |
| rs532436 | miR-126-3p | trans | 2.49E-40 | Nightingale | XS_VLDL_L | NA | A | G | 0.048489 | 0.005241 | 1.70E-22 |
| rs532436 | miR-126-3p | trans | 2.49E-40 | Nightingale | HDL_P | NA | A | G | 0.028757 | 0.005007 | 3.30E-09 |
| rs532436 | miR-126-3p | trans | 2.49E-40 | Nightingale | S_VLDL_C_ | NA | A | G | 0.070418 | 0.005189 | 1.30E-41 |
| rs532436 | miR-126-3p | trans | 2.49E-40 | Nightingale | Phosphatic | NA | A | G | 0.031132 | 0.004996 | 6.60E-11 |
| rs532436 | miR-126-3p | trans | 2.49E-40 | Nightingale | XXL_VLDL_ | NA | A | G | 0.036801 | 0.00537 | 1.70E-13 |
| rs532436 | miR-126-3p | trans | 2.49E-40 | Nightingale | Total_FC | NA | A | G | 0.054936 | 0.005209 | 1.00E-27 |
| rs532436 | miR-126-3p | trans | 2.49E-40 | Nightingale | S_LDL_PL_ | NA | A | G | -0.04468 | 0.005322 | 1.60E-19 |
| rs532436 | miR-126-3p | trans | 2.49E-40 | Nightingale | M_VLDL_T | NA | A | G | -0.06283 | 0.005038 | 7.40E-35 |
| rs532436 | miR-126-3p | trans | 2.49E-40 | Nightingale | LDL_C | NA | A | G | 0.060001 | 0.005309 | 3.10E-30 |
| rs532436 | miR-126-3p | trans | 2.49E-40 | Nightingale | Total_C | NA | A | G | 0.055716 | 0.005162 | 2.80E-28 |
| rs532436 | miR-126-3p | trans | 2.49E-40 | Nightingale | S_VLDL_CE | NA | A | G | 0.057936 | 0.005325 | 1.60E-29 |
| rs532436 | miR-126-3p | trans | 2.49E-40 | Nightingale | S_HDL_P | NA | A | G | 0.055157 | 0.005332 | 1.70E-25 |
| rs532436 | miR-126-3p | trans | 2.49E-40 | Nightingale | L_LDL_PL | NA | A | G | 0.066479 | 0.005291 | 5.80E-37 |
| rs532436 | miR-126-3p | trans | 2.49E-40 | Nightingale | M_LDL_FC | NA | A | G | 0.063598 | 0.005325 | 4.40E-33 |
| rs532436 | miR-126-3p | trans | 2.49E-40 | Nightingale | XL_HDL_PL | NA | A | G | -0.04102 | 0.004899 | 1.70E-17 |
| rs532436 | miR-126-3p | trans | 2.49E-40 | Nightingale | M_LDL_C | NA | A | G | 0.05878 | 0.005341 | 5.20E-29 |
| rs532436 | miR-126-3p | trans | 2.49E-40 | Nightingale | M_VLDL_P | NA | A | G | 0.043187 | 0.005319 | 2.10E-17 |
| rs532436 | miR-126-3p | trans | 2.49E-40 | Nightingale | XL_VLDL_P | NA | A | G | 0.055667 | 0.00534 | 6.50E-27 |
| rs532436 | miR-126-3p | trans | 2.49E-40 | Nightingale | VLDL_FC | NA | A | G | 0.033528 | 0.005293 | 1.30E-11 |
| rs532436 | miR-126-3p | trans | 2.49E-40 | Nightingale | L_VLDL_C_ | NA | A | G | 0.08172 | 0.005208 | 2.50E-56 |
| rs532436 | miR-126-3p | trans | 2.49E-40 | Nightingale | XL_VLDL_C | NA | A | G | 0.029577 | 0.005249 | 1.10E-09 |
| rs532436 | miR-126-3p | trans | 2.49E-40 | Nightingale | XS_VLDL_F | NA | A | G | 0.050786 | 0.005294 | 9.70E-21 |
| rs532436 | miR-126-3p | trans | 2.49E-40 | Nightingale | XL_HDL_PL | NA | A | G | -0.03014 | 0.004796 | 6.90E-11 |
| rs532436 | miR-10b-5p | trans | 5.20E-36 | Nightingale | XS_VLDL_T | NA | A | G | -0.0425 | 0.005069 | 3.40E-16 |
| rs532436 | miR-10b-5p | trans | 5.20E-36 | Nightingale | M_VLDL_P | NA | A | G | 0.070479 | 0.005117 | 5.70E-43 |
| rs532436 | miR-10b-5p | trans | 5.20E-36 | Nightingale | L_LDL_C | NA | A | G | 0.059903 | 0.005272 | 1.70E-30 |
| rs532436 | miR-10b-5p | trans | 5.20E-36 | Nightingale | LA_pct | NA | A | G | 0.032246 | 0.005214 | 6.90E-10 |
| rs532436 | miR-10b-5p | trans | 5.20E-36 | Nightingale | S_VLDL_C | NA | A | G | 0.058922 | 0.005331 | 3.90E-30 |
| rs532436 | miR-10b-5p | trans | 5.20E-36 | Nightingale | S_LDL_L | NA | A | G | 0.0484 | 0.005331 | 3.80E-20 |
| rs532436 | miR-10b-5p | trans | 5.20E-36 | Nightingale | L_VLDL_FC | NA | A | G | 0.075634 | 0.005281 | 2.30E-47 |
| rs532436 | miR-10b-5p | trans | 5.20E-36 | Nightingale | M_LDL_PL | NA | A | G | 0.058078 | 0.005339 | 2.50E-28 |
| rs532436 | miR-10b-5p | trans | 5.20E-36 | Nightingale | L_LDL_L | NA | A | G | 0.060847 | 0.00528 | 1.80E-31 |
| rs532436 | miR-10b-5p | trans | 5.20E-36 | Nightingale | XL_VLDL_C | NA | A | G | 0.062307 | 0.005115 | 2.90E-33 |
| rs532436 | miR-10b-5p | trans | 5.20E-36 | Nightingale | S_VLDL_L | NA | A | G | 0.035519 | 0.00528 | 4.90E-13 |
| rs532436 | miR-10b-5p | trans | 5.20E-36 | Nightingale | S_LDL_FC | NA | A | G | 0.054532 | 0.005323 | 2.40E-24 |
| rs532436 | miR-10b-5p | trans | 5.20E-36 | Nightingale | M_LDL_P | NA | A | G | 0.057013 | 0.005347 | 1.50E-27 |
| rs532436 | miR-10b-5p | trans | 5.20E-36 | Nightingale | M_VLDL_C | NA | A | G | 0.0595 | 0.005022 | 1.90E-31 |
| rs532436 | miR-10b-5p | trans | 5.20E-36 | Nightingale | S_LDL_P | NA | A | G | 0.048388 | 0.005337 | 1.90E-20 |
| rs532436 | miR-10b-5p | trans | 5.20E-36 | Nightingale | XL_HDL_L | NA | A | G | -0.02706 | 0.004795 | 2.40E-09 |
| rs532436 | miR-10b-5p | trans | 5.20E-36 | Nightingale | S_LDL_CE_ | NA | A | G | 0.044561 | 0.005323 | 1.40E-19 |
| rs532436 | miR-10b-5p | trans | 5.20E-36 | Nightingale | S_HDL_C | NA | A | G | 0.052394 | 0.005323 | 3.50E-23 |
| rs532436 | miR-10b-5p | trans | 5.20E-36 | Nightingale | S_LDL_C_p | NA | A | G | 0.075576 | 0.005352 | 2.10E-46 |
| rs532436 | miR-10b-5p | trans | 5.20E-36 | Nightingale | LDL_FC | NA | A | G | 0.062671 | 0.005275 | 1.40E-32 |
| rs532436 | miR-10b-5p | trans | 5.20E-36 | Nightingale | XS_VLDL_C | NA | A | G | 0.040138 | 0.005031 | 1.40E-14 |
| rs532436 | miR-10b-5p | trans | 5.20E-36 | Nightingale | XS_VLDL_P | NA | A | G | 0.044567 | 0.005257 | 3.00E-19 |
| rs532436 | miR-10b-5p | trans | 5.20E-36 | Nightingale | LDL_L | NA | A | G | 0.059463 | 0.005314 | 9.50E-30 |
| rs532436 | miR-10b-5p | trans | 5.20E-36 | Nightingale | Clinical_LD | NA | A | G | 0.064426 | 0.005301 | 1.80E-34 |
| rs532436 | miR-10b-5p | trans | 5.20E-36 | Nightingale | Citrate | NA | A | G | 0.031277 | 0.005297 | 1.60E-09 |
| rs532436 | miR-10b-5p | trans | 5.20E-36 | Nightingale | XS_VLDL_P | NA | A | G | 0.05371 | 0.005261 | 1.20E-26 |
| rs532436 | miR-10b-5p | trans | 5.20E-36 | Nightingale | LDL_CE | NA | A | G | 0.058166 | 0.005318 | 1.20E-28 |
| rs532436 | miR-10b-5p | trans | 5.20E-36 | Nightingale | Remnant_( | NA | A | G | 0.058377 | 0.005297 | 5.50E-30 |
| rs532436 | miR-10b-5p | trans | 5.20E-36 | Nightingale | Cholines | NA | A | G | 0.035678 | 0.005014 | 1.10E-13 |
| rs532436 | miR-10b-5p | trans | 5.20E-36 | Nightingale | HDL_size | NA | A | G | -0.03226 | 0.004727 | 1.70E-12 |
| rs532436 | miR-10b-5p | trans | 5.20E-36 | Nightingale | S_HDL_L | NA | A | G | 0.043965 | 0.005313 | 2.20E-17 |
| rs532436 | miR-10b-5p | trans | 5.20E-36 | Nightingale | S_VLDL_FC | NA | A | G | 0.059408 | 0.005333 | 3.00E-30 |
| rs532436 | miR-10b-5p | trans | 5.20E-36 | Nightingale | XXL_VLDL_ | NA | A | G | 0.046201 | 0.005312 | 1.30E-18 |
| rs532436 | miR-10b-5p | trans | 5.20E-36 | Nightingale | XS_VLDL_C | NA | A | G | 0.057562 | 0.005175 | 2.50E-30 |
| rs532436 | miR-10b-5p | trans | 5.20E-36 | Nightingale | M_VLDL_F | NA | A | G | 0.067258 | 0.005073 | 1.70E-39 |
| rs532436 | miR-10b-5p | trans | 5.20E-36 | Nightingale | IDL_TG_pct | NA | A | G | -0.03868 | 0.005155 | 3.10E-13 |
| rs532436 | miR-10b-5p | trans | 5.20E-36 | Nightingale | XS_VLDL_P | NA | A | G | -0.02951 | 0.005203 | 5.00E-08 |
| rs532436 | miR-10b-5p | trans | 5.20E-36 | Nightingale | M_VLDL_L | NA | A | G | 0.032097 | 0.005296 | 1.10E-10 |
| rs532436 | miR-10b-5p | trans | 5.20E-36 | Nightingale | LA | NA | A | G | 0.042219 | 0.005229 | 3.30E-17 |

| | | | | | | | | | | | |
|---|---|---|---|---|---|---|---|---|---|---|---|
| rs532436 | miR-10b-5p | trans | 5.20E-36 | Nightingale | M_VLDL_C | NA | A | G | 0.06605 | 0.005283 | 1.00E-36 |
| rs532436 | miR-10b-5p | trans | 5.20E-36 | Nightingale | Total_P | NA | A | G | 0.036654 | 0.005018 | 5.50E-14 |
| rs532436 | miR-10b-5p | trans | 5.20E-36 | Nightingale | Total_L | NA | A | G | 0.042255 | 0.005245 | 4.90E-17 |
| rs532436 | miR-10b-5p | trans | 5.20E-36 | Nightingale | M_LDL_TG | NA | A | G | -0.0531 | 0.005266 | 1.10E-23 |
| rs532436 | miR-10b-5p | trans | 5.20E-36 | Nightingale | L_LDL_P | NA | A | G | 0.050357 | 0.005314 | 6.40E-22 |
| rs532436 | miR-10b-5p | trans | 5.20E-36 | Nightingale | S_LDL_CE | NA | A | G | 0.051567 | 0.005334 | 4.80E-23 |
| rs532436 | miR-10b-5p | trans | 5.20E-36 | Nightingale | S_LDL_TG_ | NA | A | G | -0.04355 | 0.00517 | 3.00E-16 |
| rs532436 | miR-10b-5p | trans | 5.20E-36 | Nightingale | M_LDL_L | NA | A | G | 0.057299 | 0.005339 | 8.90E-28 |
| rs532436 | miR-10b-5p | trans | 5.20E-36 | Nightingale | IDL_FC_pct | NA | A | G | 0.041213 | 0.00529 | 2.10E-14 |
| rs532436 | miR-10b-5p | trans | 5.20E-36 | Nightingale | Phosphogly | NA | A | G | 0.031891 | 0.00505 | 3.60E-11 |
| rs532436 | miR-10b-5p | trans | 5.20E-36 | Nightingale | XL_VLDL_P | NA | A | G | 0.056773 | 0.005169 | 2.60E-28 |
| rs532436 | miR-10b-5p | trans | 5.20E-36 | Nightingale | S_HDL_FC | NA | A | G | 0.041762 | 0.005234 | 2.40E-16 |
| rs532436 | miR-10b-5p | trans | 5.20E-36 | Nightingale | IDL_FC | NA | A | G | 0.063966 | 0.005162 | 7.70E-36 |
| rs532436 | miR-10b-5p | trans | 5.20E-36 | Nightingale | S_VLDL_PL | NA | A | G | 0.050032 | 0.005322 | 1.30E-22 |
| rs532436 | miR-10b-5p | trans | 5.20E-36 | Nightingale | IDL_C_pct | NA | A | G | 0.037343 | 0.005209 | 6.30E-12 |
| rs532436 | miR-10b-5p | trans | 5.20E-36 | Nightingale | L_VLDL_CE | NA | A | G | 0.074326 | 0.005102 | 1.30E-48 |
| rs532436 | miR-10b-5p | trans | 5.20E-36 | Nightingale | L_LDL_CE | NA | A | G | 0.058358 | 0.005284 | 5.40E-29 |
| rs532436 | miR-10b-5p | trans | 5.20E-36 | Nightingale | S_VLDL_CE | NA | A | G | 0.076497 | 0.00526 | 2.10E-47 |
| rs532436 | miR-10b-5p | trans | 5.20E-36 | Nightingale | XL_HDL_CE | NA | A | G | 0.030066 | 0.005327 | 4.20E-08 |
| rs532436 | miR-10b-5p | trans | 5.20E-36 | Nightingale | XS_VLDL_P | NA | A | G | 0.053623 | 0.005239 | 1.10E-26 |
| rs532436 | miR-10b-5p | trans | 5.20E-36 | Nightingale | M_LDL_C_p | NA | A | G | 0.047449 | 0.005336 | 4.30E-19 |
| rs532436 | miR-10b-5p | trans | 5.20E-36 | Nightingale | S_VLDL_P | NA | A | G | 0.033337 | 0.005265 | 7.40E-12 |
| rs532436 | miR-10b-5p | trans | 5.20E-36 | Nightingale | S_VLDL_FC | NA | A | G | 0.05247 | 0.005095 | 3.60E-24 |
| rs532436 | miR-10b-5p | trans | 5.20E-36 | Nightingale | L_VLDL_TG | NA | A | G | -0.0795 | 0.005307 | 2.00E-52 |
| rs532436 | miR-10b-5p | trans | 5.20E-36 | Nightingale | GlycA | NA | A | G | 0.031177 | 0.005309 | 6.60E-09 |
| rs532436 | miR-10b-5p | trans | 5.20E-36 | Nightingale | L_VLDL_CE | NA | A | G | 0.031425 | 0.005269 | 1.40E-10 |
| rs532436 | miR-10b-5p | trans | 5.20E-36 | Nightingale | M_VLDL_C | NA | A | G | 0.061586 | 0.005317 | 2.10E-32 |
| rs532436 | miR-10b-5p | trans | 5.20E-36 | Nightingale | IDL_L | NA | A | G | 0.059551 | 0.005155 | 1.70E-31 |
| rs532436 | miR-10b-5p | trans | 5.20E-36 | Nightingale | IDL_CE | NA | A | G | 0.058228 | 0.005148 | 7.40E-30 |
| rs532436 | miR-10b-5p | trans | 5.20E-36 | Nightingale | S_VLDL_PL | NA | A | G | 0.049979 | 0.005105 | 5.70E-22 |
| rs532436 | miR-10b-5p | trans | 5.20E-36 | Nightingale | M_LDL_CE | NA | A | G | 0.055488 | 0.005339 | 2.20E-26 |
| rs532436 | miR-10b-5p | trans | 5.20E-36 | Nightingale | XL_HDL_C_ | NA | A | G | 0.038356 | 0.005092 | 1.70E-13 |
| rs532436 | miR-10b-5p | trans | 5.20E-36 | Nightingale | VLDL_C | NA | A | G | 0.045684 | 0.005324 | 2.90E-19 |
| rs532436 | miR-10b-5p | trans | 5.20E-36 | Nightingale | IDL_C | NA | A | G | 0.060082 | 0.005149 | 9.90E-32 |
| rs532436 | miR-10b-5p | trans | 5.20E-36 | Nightingale | S_VLDL_TG | NA | A | G | -0.06518 | 0.005161 | 4.40E-36 |
| rs532436 | miR-10b-5p | trans | 5.20E-36 | Nightingale | ApoB_by_A | NA | A | G | 0.039971 | 0.005251 | 1.30E-14 |
| rs532436 | miR-10b-5p | trans | 5.20E-36 | Nightingale | VLDL_P | NA | A | G | 0.038195 | 0.005303 | 2.40E-14 |
| rs532436 | miR-10b-5p | trans | 5.20E-36 | Nightingale | Total_CE | NA | A | G | 0.05558 | 0.005143 | 3.40E-28 |
| rs532436 | miR-10b-5p | trans | 5.20E-36 | Nightingale | L_LDL_FC_ | NA | A | G | 0.030877 | 0.005087 | 1.20E-08 |
| rs532436 | miR-10b-5p | trans | 5.20E-36 | Nightingale | VLDL_CE | NA | A | G | 0.053545 | 0.005333 | 2.70E-25 |
| rs532436 | miR-10b-5p | trans | 5.20E-36 | Nightingale | M_VLDL_C | NA | A | G | 0.055885 | 0.005009 | 5.70E-28 |
| rs532436 | miR-10b-5p | trans | 5.20E-36 | Nightingale | XXL_VLDL_ | NA | A | G | -0.05817 | 0.005381 | 5.80E-28 |
| rs532436 | miR-10b-5p | trans | 5.20E-36 | Nightingale | IDL_PL | NA | A | G | 0.05875 | 0.005145 | 3.70E-31 |
| rs532436 | miR-10b-5p | trans | 5.20E-36 | Nightingale | S_HDL_PL_ | NA | A | G | -0.03259 | 0.005155 | 1.00E-09 |
| rs532436 | miR-10b-5p | trans | 5.20E-36 | Nightingale | S_LDL_PL | NA | A | G | 0.041297 | 0.00532 | 6.80E-15 |
| rs532436 | miR-10b-5p | trans | 5.20E-36 | Nightingale | non_HDL_( | NA | A | G | 0.060373 | 0.005303 | 3.40E-31 |
| rs532436 | miR-10b-5p | trans | 5.20E-36 | Nightingale | M_VLDL_P | NA | A | G | 0.044715 | 0.005324 | 1.90E-18 |
| rs532436 | miR-10b-5p | trans | 5.20E-36 | Nightingale | Sphingomy | NA | A | G | 0.047389 | 0.004984 | 3.70E-21 |
| rs532436 | miR-10b-5p | trans | 5.20E-36 | Nightingale | LDL_PL | NA | A | G | 0.06187 | 0.005319 | 7.90E-32 |
| rs532436 | miR-10b-5p | trans | 5.20E-36 | Nightingale | L_VLDL_PL | NA | A | G | 0.027951 | 0.005179 | 3.30E-09 |
| rs532436 | miR-10b-5p | trans | 5.20E-36 | Nightingale | S_HDL_PL | NA | A | G | 0.037551 | 0.005286 | 1.80E-13 |
| rs532436 | miR-10b-5p | trans | 5.20E-36 | Nightingale | XS_VLDL_C | NA | A | G | 0.043168 | 0.005057 | 1.10E-16 |
| rs532436 | miR-10b-5p | trans | 5.20E-36 | Nightingale | IDL_P | NA | A | G | 0.06498 | 0.005276 | 1.90E-35 |
| rs532436 | miR-10b-5p | trans | 5.20E-36 | Nightingale | S_HDL_CE | NA | A | G | 0.053255 | 0.00534 | 1.20E-23 |
| rs532436 | miR-10b-5p | trans | 5.20E-36 | Nightingale | L_LDL_FC | NA | A | G | 0.062609 | 0.005236 | 3.80E-33 |
| rs532436 | miR-10b-5p | trans | 5.20E-36 | Nightingale | PUFA | NA | A | G | 0.036816 | 0.005201 | 1.50E-13 |
| rs532436 | miR-10b-5p | trans | 5.20E-36 | Nightingale | XS_VLDL_C | NA | A | G | 0.058331 | 0.005145 | 3.40E-31 |
| rs532436 | miR-10b-5p | trans | 5.20E-36 | Nightingale | ApoB | NA | A | G | 0.055646 | 0.005325 | 1.40E-26 |
| rs532436 | miR-10b-5p | trans | 5.20E-36 | Nightingale | M_VLDL_F( | NA | A | G | 0.051674 | 0.005331 | 1.50E-23 |
| rs532436 | miR-10b-5p | trans | 5.20E-36 | Nightingale | S_LDL_C | NA | A | G | 0.053873 | 0.005337 | 1.50E-24 |
| rs532436 | miR-10b-5p | trans | 5.20E-36 | Nightingale | XL_VLDL_C | NA | A | G | 0.062531 | 0.005113 | 1.30E-33 |
| rs532436 | miR-10b-5p | trans | 5.20E-36 | Nightingale | XXL_VLDL_ | NA | A | G | 0.055922 | 0.005375 | 1.40E-25 |
| rs532436 | miR-10b-5p | trans | 5.20E-36 | Nightingale | Omega_6 | NA | A | G | 0.0417 | 0.005207 | 5.30E-17 |

| | | | | | | | | | | | |
|---|---|---|---|---|---|---|---|---|---|---|---|
| rs532436 | miR-10b-5p | trans | 5.20E-36 | Nightingale | LDL_P | NA | A | G | 0.053144 | 0.005328 | 2.90E-24 |
| rs532436 | miR-10b-5p | trans | 5.20E-36 | Nightingale | XXL_VLDL_ | NA | A | G | 0.056832 | 0.005346 | 1.50E-26 |
| rs532436 | miR-10b-5p | trans | 5.20E-36 | Nightingale | Pyruvate | NA | A | G | 0.034156 | 0.00534 | 1.60E-10 |
| rs532436 | miR-10b-5p | trans | 5.20E-36 | Nightingale | Total_PL | NA | A | G | 0.03947 | 0.005077 | 4.30E-16 |
| rs532436 | miR-10b-5p | trans | 5.20E-36 | Nightingale | L_LDL_TG_ | NA | A | G | -0.04149 | 0.00523 | 8.10E-15 |
| rs532436 | miR-10b-5p | trans | 5.20E-36 | Nightingale | XL_VLDL_T | NA | A | G | -0.06894 | 0.005168 | 1.90E-40 |
| rs532436 | miR-10b-5p | trans | 5.20E-36 | Nightingale | XS_VLDL_L | NA | A | G | 0.048489 | 0.005241 | 1.70E-22 |
| rs532436 | miR-10b-5p | trans | 5.20E-36 | Nightingale | HDL_P | NA | A | G | 0.028757 | 0.005007 | 3.30E-09 |
| rs532436 | miR-10b-5p | trans | 5.20E-36 | Nightingale | S_VLDL_C_ | NA | A | G | 0.070418 | 0.005189 | 1.30E-41 |
| rs532436 | miR-10b-5p | trans | 5.20E-36 | Nightingale | Phosphatic | NA | A | G | 0.031132 | 0.004996 | 6.60E-11 |
| rs532436 | miR-10b-5p | trans | 5.20E-36 | Nightingale | XXL_VLDL_ | NA | A | G | 0.036801 | 0.00537 | 1.70E-13 |
| rs532436 | miR-10b-5p | trans | 5.20E-36 | Nightingale | Total_FC | NA | A | G | 0.054936 | 0.005209 | 1.00E-27 |
| rs532436 | miR-10b-5p | trans | 5.20E-36 | Nightingale | S_LDL_PL_ | NA | A | G | -0.04468 | 0.005322 | 1.60E-19 |
| rs532436 | miR-10b-5p | trans | 5.20E-36 | Nightingale | M_VLDL_T | NA | A | G | -0.06283 | 0.005038 | 7.40E-35 |
| rs532436 | miR-10b-5p | trans | 5.20E-36 | Nightingale | LDL_C | NA | A | G | 0.060001 | 0.005309 | 3.10E-30 |
| rs532436 | miR-10b-5p | trans | 5.20E-36 | Nightingale | Total_C | NA | A | G | 0.055716 | 0.005162 | 2.80E-28 |
| rs532436 | miR-10b-5p | trans | 5.20E-36 | Nightingale | S_VLDL_CE | NA | A | G | 0.057936 | 0.005325 | 1.60E-29 |
| rs532436 | miR-10b-5p | trans | 5.20E-36 | Nightingale | S_HDL_P | NA | A | G | 0.055157 | 0.005332 | 1.70E-25 |
| rs532436 | miR-10b-5p | trans | 5.20E-36 | Nightingale | L_LDL_PL | NA | A | G | 0.066479 | 0.005291 | 5.80E-37 |
| rs532436 | miR-10b-5p | trans | 5.20E-36 | Nightingale | M_LDL_FC | NA | A | G | 0.063598 | 0.005325 | 4.40E-33 |
| rs532436 | miR-10b-5p | trans | 5.20E-36 | Nightingale | XL_HDL_PL | NA | A | G | -0.04102 | 0.004899 | 1.70E-17 |
| rs532436 | miR-10b-5p | trans | 5.20E-36 | Nightingale | M_LDL_C | NA | A | G | 0.05878 | 0.005341 | 5.20E-29 |
| rs532436 | miR-10b-5p | trans | 5.20E-36 | Nightingale | M_VLDL_P | NA | A | G | 0.043187 | 0.005319 | 2.10E-17 |
| rs532436 | miR-10b-5p | trans | 5.20E-36 | Nightingale | XL_VLDL_P | NA | A | G | 0.055667 | 0.00534 | 6.50E-27 |
| rs532436 | miR-10b-5p | trans | 5.20E-36 | Nightingale | VLDL_FC | NA | A | G | 0.033528 | 0.005293 | 1.30E-11 |
| rs532436 | miR-10b-5p | trans | 5.20E-36 | Nightingale | L_VLDL_C_ | NA | A | G | 0.08172 | 0.005208 | 2.50E-56 |
| rs532436 | miR-10b-5p | trans | 5.20E-36 | Nightingale | XL_VLDL_C | NA | A | G | 0.029577 | 0.005249 | 1.10E-09 |
| rs532436 | miR-10b-5p | trans | 5.20E-36 | Nightingale | XS_VLDL_F | NA | A | G | 0.050786 | 0.005294 | 9.70E-21 |
| rs532436 | miR-10b-5p | trans | 5.20E-36 | Nightingale | XL_HDL_PL | NA | A | G | -0.03014 | 0.004796 | 6.90E-11 |
| rs532436 | let-7c-5p | trans | 2.13E-22 | Nightingale | XS_VLDL_T | NA | A | G | -0.0425 | 0.005069 | 3.40E-16 |
| rs532436 | let-7c-5p | trans | 2.13E-22 | Nightingale | M_VLDL_P | NA | A | G | 0.070479 | 0.005117 | 5.70E-43 |
| rs532436 | let-7c-5p | trans | 2.13E-22 | Nightingale | L_LDL_C | NA | A | G | 0.059903 | 0.005272 | 1.70E-30 |
| rs532436 | let-7c-5p | trans | 2.13E-22 | Nightingale | LA_pct | NA | A | G | 0.032246 | 0.005214 | 6.90E-10 |
| rs532436 | let-7c-5p | trans | 2.13E-22 | Nightingale | S_VLDL_C | NA | A | G | 0.058922 | 0.005331 | 3.90E-30 |
| rs532436 | let-7c-5p | trans | 2.13E-22 | Nightingale | S_LDL_L | NA | A | G | 0.0484 | 0.005331 | 3.80E-20 |
| rs532436 | let-7c-5p | trans | 2.13E-22 | Nightingale | L_VLDL_FC | NA | A | G | 0.075634 | 0.005281 | 2.30E-47 |
| rs532436 | let-7c-5p | trans | 2.13E-22 | Nightingale | M_LDL_PL | NA | A | G | 0.058078 | 0.005339 | 2.50E-28 |
| rs532436 | let-7c-5p | trans | 2.13E-22 | Nightingale | L_LDL_L | NA | A | G | 0.060847 | 0.00528 | 1.80E-31 |
| rs532436 | let-7c-5p | trans | 2.13E-22 | Nightingale | XL_VLDL_C | NA | A | G | 0.062307 | 0.005115 | 2.90E-33 |
| rs532436 | let-7c-5p | trans | 2.13E-22 | Nightingale | S_VLDL_L | NA | A | G | 0.035519 | 0.00528 | 4.90E-13 |
| rs532436 | let-7c-5p | trans | 2.13E-22 | Nightingale | S_LDL_FC | NA | A | G | 0.054532 | 0.005323 | 2.40E-24 |
| rs532436 | let-7c-5p | trans | 2.13E-22 | Nightingale | M_LDL_P | NA | A | G | 0.057013 | 0.005347 | 1.50E-27 |
| rs532436 | let-7c-5p | trans | 2.13E-22 | Nightingale | M_VLDL_C | NA | A | G | 0.0595 | 0.005022 | 1.90E-31 |
| rs532436 | let-7c-5p | trans | 2.13E-22 | Nightingale | S_LDL_P | NA | A | G | 0.048388 | 0.005337 | 1.90E-20 |
| rs532436 | let-7c-5p | trans | 2.13E-22 | Nightingale | XL_HDL_L | NA | A | G | -0.02706 | 0.004795 | 2.40E-09 |
| rs532436 | let-7c-5p | trans | 2.13E-22 | Nightingale | S_LDL_CE_ | NA | A | G | 0.044561 | 0.005323 | 1.40E-19 |
| rs532436 | let-7c-5p | trans | 2.13E-22 | Nightingale | S_HDL_C | NA | A | G | 0.052394 | 0.005323 | 3.50E-23 |
| rs532436 | let-7c-5p | trans | 2.13E-22 | Nightingale | S_LDL_C_p | NA | A | G | 0.075576 | 0.005352 | 2.10E-46 |
| rs532436 | let-7c-5p | trans | 2.13E-22 | Nightingale | LDL_FC | NA | A | G | 0.062671 | 0.005275 | 1.40E-32 |
| rs532436 | let-7c-5p | trans | 2.13E-22 | Nightingale | XS_VLDL_C | NA | A | G | 0.040138 | 0.005031 | 1.40E-14 |
| rs532436 | let-7c-5p | trans | 2.13E-22 | Nightingale | XS_VLDL_P | NA | A | G | 0.044567 | 0.005257 | 3.00E-19 |
| rs532436 | let-7c-5p | trans | 2.13E-22 | Nightingale | LDL_L | NA | A | G | 0.059463 | 0.005314 | 9.50E-30 |
| rs532436 | let-7c-5p | trans | 2.13E-22 | Nightingale | Clinical_LD | NA | A | G | 0.064426 | 0.005301 | 1.80E-34 |
| rs532436 | let-7c-5p | trans | 2.13E-22 | Nightingale | Citrate | NA | A | G | 0.031277 | 0.005297 | 1.60E-09 |
| rs532436 | let-7c-5p | trans | 2.13E-22 | Nightingale | XS_VLDL_P | NA | A | G | 0.05371 | 0.005261 | 1.20E-26 |
| rs532436 | let-7c-5p | trans | 2.13E-22 | Nightingale | LDL_CE | NA | A | G | 0.058166 | 0.005318 | 1.20E-28 |
| rs532436 | let-7c-5p | trans | 2.13E-22 | Nightingale | Remnant_( | NA | A | G | 0.058377 | 0.005297 | 5.50E-30 |
| rs532436 | let-7c-5p | trans | 2.13E-22 | Nightingale | Cholines | NA | A | G | 0.035678 | 0.005014 | 1.10E-13 |
| rs532436 | let-7c-5p | trans | 2.13E-22 | Nightingale | HDL_size | NA | A | G | -0.03226 | 0.004727 | 1.70E-12 |
| rs532436 | let-7c-5p | trans | 2.13E-22 | Nightingale | S_HDL_L | NA | A | G | 0.043965 | 0.005313 | 2.20E-17 |
| rs532436 | let-7c-5p | trans | 2.13E-22 | Nightingale | S_VLDL_FC | NA | A | G | 0.059408 | 0.005333 | 3.00E-30 |
| rs532436 | let-7c-5p | trans | 2.13E-22 | Nightingale | XXL_VLDL_ | NA | A | G | 0.046201 | 0.005312 | 1.30E-18 |
| rs532436 | let-7c-5p | trans | 2.13E-22 | Nightingale | XS_VLDL_C | NA | A | G | 0.057562 | 0.005175 | 2.50E-30 |

| | | | | | | | | | | | |
|---|---|---|---|---|---|---|---|---|---|---|---|
| rs532436 | let-7c-5p | trans | 2.13E-22 | Nightingale | M_VLDL_F | NA | A | G | 0.067258 | 0.005073 | 1.70E-39 |
| rs532436 | let-7c-5p | trans | 2.13E-22 | Nightingale | IDL_TG_pc | NA | A | G | -0.03868 | 0.005155 | 3.10E-13 |
| rs532436 | let-7c-5p | trans | 2.13E-22 | Nightingale | XS_VLDL_P | NA | A | G | -0.02951 | 0.005203 | 5.00E-08 |
| rs532436 | let-7c-5p | trans | 2.13E-22 | Nightingale | M_VLDL_L | NA | A | G | 0.032097 | 0.005296 | 1.10E-10 |
| rs532436 | let-7c-5p | trans | 2.13E-22 | Nightingale | LA | NA | A | G | 0.042219 | 0.005229 | 3.30E-17 |
| rs532436 | let-7c-5p | trans | 2.13E-22 | Nightingale | M_VLDL_C | NA | A | G | 0.06605 | 0.005283 | 1.00E-36 |
| rs532436 | let-7c-5p | trans | 2.13E-22 | Nightingale | Total_P | NA | A | G | 0.036654 | 0.005018 | 5.50E-14 |
| rs532436 | let-7c-5p | trans | 2.13E-22 | Nightingale | Total_L | NA | A | G | 0.042255 | 0.005245 | 4.90E-17 |
| rs532436 | let-7c-5p | trans | 2.13E-22 | Nightingale | M_LDL_TG | NA | A | G | -0.0531 | 0.005266 | 1.10E-23 |
| rs532436 | let-7c-5p | trans | 2.13E-22 | Nightingale | L_LDL_P | NA | A | G | 0.050357 | 0.005314 | 6.40E-22 |
| rs532436 | let-7c-5p | trans | 2.13E-22 | Nightingale | S_LDL_CE | NA | A | G | 0.051567 | 0.005334 | 4.80E-23 |
| rs532436 | let-7c-5p | trans | 2.13E-22 | Nightingale | S_LDL_TG_ | NA | A | G | -0.04355 | 0.00517 | 3.00E-16 |
| rs532436 | let-7c-5p | trans | 2.13E-22 | Nightingale | M_LDL_L | NA | A | G | 0.057299 | 0.005339 | 8.90E-28 |
| rs532436 | let-7c-5p | trans | 2.13E-22 | Nightingale | IDL_FC_pct | NA | A | G | 0.041213 | 0.00529 | 2.10E-14 |
| rs532436 | let-7c-5p | trans | 2.13E-22 | Nightingale | Phosphogl | NA | A | G | 0.031891 | 0.00505 | 3.60E-11 |
| rs532436 | let-7c-5p | trans | 2.13E-22 | Nightingale | XL_VLDL_F | NA | A | G | 0.056773 | 0.005169 | 2.60E-28 |
| rs532436 | let-7c-5p | trans | 2.13E-22 | Nightingale | S_HDL_FC | NA | A | G | 0.041762 | 0.005234 | 2.40E-16 |
| rs532436 | let-7c-5p | trans | 2.13E-22 | Nightingale | IDL_FC | NA | A | G | 0.063966 | 0.005162 | 7.70E-36 |
| rs532436 | let-7c-5p | trans | 2.13E-22 | Nightingale | S_VLDL_PL | NA | A | G | 0.050032 | 0.005322 | 1.30E-22 |
| rs532436 | let-7c-5p | trans | 2.13E-22 | Nightingale | IDL_C_pct | NA | A | G | 0.037343 | 0.005209 | 6.30E-12 |
| rs532436 | let-7c-5p | trans | 2.13E-22 | Nightingale | L_VLDL_CE | NA | A | G | 0.074326 | 0.005102 | 1.30E-48 |
| rs532436 | let-7c-5p | trans | 2.13E-22 | Nightingale | L_LDL_CE | NA | A | G | 0.058358 | 0.005284 | 5.40E-29 |
| rs532436 | let-7c-5p | trans | 2.13E-22 | Nightingale | S_VLDL_CE | NA | A | G | 0.076497 | 0.00526 | 2.10E-47 |
| rs532436 | let-7c-5p | trans | 2.13E-22 | Nightingale | XL_HDL_CE | NA | A | G | 0.030066 | 0.005327 | 4.20E-08 |
| rs532436 | let-7c-5p | trans | 2.13E-22 | Nightingale | XS_VLDL_F | NA | A | G | 0.053623 | 0.005239 | 1.10E-26 |
| rs532436 | let-7c-5p | trans | 2.13E-22 | Nightingale | M_LDL_C_ | NA | A | G | 0.047449 | 0.005336 | 4.30E-19 |
| rs532436 | let-7c-5p | trans | 2.13E-22 | Nightingale | S_VLDL_P | NA | A | G | 0.033337 | 0.005265 | 7.40E-12 |
| rs532436 | let-7c-5p | trans | 2.13E-22 | Nightingale | S_VLDL_FC | NA | A | G | 0.05247 | 0.005095 | 3.60E-24 |
| rs532436 | let-7c-5p | trans | 2.13E-22 | Nightingale | L_VLDL_TG | NA | A | G | -0.0795 | 0.005307 | 2.00E-52 |
| rs532436 | let-7c-5p | trans | 2.13E-22 | Nightingale | GlycA | NA | A | G | 0.031177 | 0.005309 | 6.60E-09 |
| rs532436 | let-7c-5p | trans | 2.13E-22 | Nightingale | L_VLDL_CE | NA | A | G | 0.031425 | 0.005269 | 1.40E-10 |
| rs532436 | let-7c-5p | trans | 2.13E-22 | Nightingale | M_VLDL_C | NA | A | G | 0.061586 | 0.005317 | 2.10E-32 |
| rs532436 | let-7c-5p | trans | 2.13E-22 | Nightingale | IDL_L | NA | A | G | 0.059551 | 0.005155 | 1.70E-31 |
| rs532436 | let-7c-5p | trans | 2.13E-22 | Nightingale | IDL_CE | NA | A | G | 0.058228 | 0.005148 | 7.40E-30 |
| rs532436 | let-7c-5p | trans | 2.13E-22 | Nightingale | S_VLDL_PL | NA | A | G | 0.049979 | 0.005105 | 5.70E-22 |
| rs532436 | let-7c-5p | trans | 2.13E-22 | Nightingale | M_LDL_CE | NA | A | G | 0.055488 | 0.005339 | 2.20E-26 |
| rs532436 | let-7c-5p | trans | 2.13E-22 | Nightingale | XL_HDL_C_ | NA | A | G | 0.038356 | 0.005092 | 1.70E-13 |
| rs532436 | let-7c-5p | trans | 2.13E-22 | Nightingale | VLDL_C | NA | A | G | 0.045684 | 0.005324 | 2.90E-19 |
| rs532436 | let-7c-5p | trans | 2.13E-22 | Nightingale | IDL_C | NA | A | G | 0.060082 | 0.005149 | 9.90E-32 |
| rs532436 | let-7c-5p | trans | 2.13E-22 | Nightingale | S_VLDL_TG | NA | A | G | -0.06518 | 0.005161 | 4.40E-36 |
| rs532436 | let-7c-5p | trans | 2.13E-22 | Nightingale | ApoB_by_A | NA | A | G | 0.039971 | 0.005251 | 1.30E-14 |
| rs532436 | let-7c-5p | trans | 2.13E-22 | Nightingale | VLDL_P | NA | A | G | 0.038195 | 0.005303 | 2.40E-14 |
| rs532436 | let-7c-5p | trans | 2.13E-22 | Nightingale | Total_CE | NA | A | G | 0.05558 | 0.005143 | 3.40E-28 |
| rs532436 | let-7c-5p | trans | 2.13E-22 | Nightingale | L_LDL_FC_ | NA | A | G | 0.030877 | 0.005087 | 1.20E-08 |
| rs532436 | let-7c-5p | trans | 2.13E-22 | Nightingale | VLDL_CE | NA | A | G | 0.053545 | 0.005333 | 2.70E-25 |
| rs532436 | let-7c-5p | trans | 2.13E-22 | Nightingale | M_VLDL_C | NA | A | G | 0.055885 | 0.005009 | 5.70E-28 |
| rs532436 | let-7c-5p | trans | 2.13E-22 | Nightingale | XXL_VLDL_ | NA | A | G | -0.05817 | 0.005381 | 5.80E-28 |
| rs532436 | let-7c-5p | trans | 2.13E-22 | Nightingale | IDL_PL | NA | A | G | 0.05875 | 0.005145 | 3.70E-31 |
| rs532436 | let-7c-5p | trans | 2.13E-22 | Nightingale | S_HDL_PL_ | NA | A | G | -0.03259 | 0.005155 | 1.00E-09 |
| rs532436 | let-7c-5p | trans | 2.13E-22 | Nightingale | S_LDL_PL | NA | A | G | 0.041297 | 0.00532 | 6.80E-15 |
| rs532436 | let-7c-5p | trans | 2.13E-22 | Nightingale | non_HDL_( | NA | A | G | 0.060373 | 0.005303 | 3.40E-31 |
| rs532436 | let-7c-5p | trans | 2.13E-22 | Nightingale | M_VLDL_P | NA | A | G | 0.044715 | 0.005324 | 1.90E-18 |
| rs532436 | let-7c-5p | trans | 2.13E-22 | Nightingale | Sphingomy | NA | A | G | 0.047389 | 0.004984 | 3.70E-21 |
| rs532436 | let-7c-5p | trans | 2.13E-22 | Nightingale | LDL_PL | NA | A | G | 0.06187 | 0.005319 | 7.90E-32 |
| rs532436 | let-7c-5p | trans | 2.13E-22 | Nightingale | L_VLDL_PL | NA | A | G | 0.027951 | 0.005179 | 3.30E-09 |
| rs532436 | let-7c-5p | trans | 2.13E-22 | Nightingale | S_HDL_PL | NA | A | G | 0.037551 | 0.005286 | 1.80E-13 |
| rs532436 | let-7c-5p | trans | 2.13E-22 | Nightingale | XS_VLDL_C | NA | A | G | 0.043168 | 0.005057 | 1.10E-16 |
| rs532436 | let-7c-5p | trans | 2.13E-22 | Nightingale | IDL_P | NA | A | G | 0.06498 | 0.005276 | 1.90E-35 |
| rs532436 | let-7c-5p | trans | 2.13E-22 | Nightingale | S_HDL_CE | NA | A | G | 0.053255 | 0.00534 | 1.20E-23 |
| rs532436 | let-7c-5p | trans | 2.13E-22 | Nightingale | L_LDL_FC | NA | A | G | 0.062609 | 0.005236 | 3.80E-33 |
| rs532436 | let-7c-5p | trans | 2.13E-22 | Nightingale | PUFA | NA | A | G | 0.036816 | 0.005201 | 1.50E-13 |
| rs532436 | let-7c-5p | trans | 2.13E-22 | Nightingale | XS_VLDL_C | NA | A | G | 0.058331 | 0.005145 | 3.40E-31 |
| rs532436 | let-7c-5p | trans | 2.13E-22 | Nightingale | ApoB | NA | A | G | 0.055646 | 0.005325 | 1.40E-26 |

| | | | | | | | | | | | |
|---|---|---|---|---|---|---|---|---|---|---|---|
| rs532436 | let-7c-5p | trans | 2.13E-22 | Nightingale | M_VLDL_F | NA | A | G | 0.051674 | 0.005331 | 1.50E-23 |
| rs532436 | let-7c-5p | trans | 2.13E-22 | Nightingale | S_LDL_C | NA | A | G | 0.053873 | 0.005337 | 1.50E-24 |
| rs532436 | let-7c-5p | trans | 2.13E-22 | Nightingale | XL_VLDL_C | NA | A | G | 0.062531 | 0.005113 | 1.30E-33 |
| rs532436 | let-7c-5p | trans | 2.13E-22 | Nightingale | XXL_VLDL_ | NA | A | G | 0.055922 | 0.005375 | 1.40E-25 |
| rs532436 | let-7c-5p | trans | 2.13E-22 | Nightingale | Omega_6 | NA | A | G | 0.0417 | 0.005207 | 5.30E-17 |
| rs532436 | let-7c-5p | trans | 2.13E-22 | Nightingale | LDL_P | NA | A | G | 0.053144 | 0.005328 | 2.90E-24 |
| rs532436 | let-7c-5p | trans | 2.13E-22 | Nightingale | XXL_VLDL_ | NA | A | G | 0.056832 | 0.005346 | 1.50E-26 |
| rs532436 | let-7c-5p | trans | 2.13E-22 | Nightingale | Pyruvate | NA | A | G | 0.034156 | 0.00534 | 1.60E-10 |
| rs532436 | let-7c-5p | trans | 2.13E-22 | Nightingale | Total_PL | NA | A | G | 0.03947 | 0.005077 | 4.30E-16 |
| rs532436 | let-7c-5p | trans | 2.13E-22 | Nightingale | L_LDL_TG_ | NA | A | G | -0.04149 | 0.00523 | 8.10E-15 |
| rs532436 | let-7c-5p | trans | 2.13E-22 | Nightingale | XL_VLDL_T | NA | A | G | -0.06894 | 0.005168 | 1.90E-40 |
| rs532436 | let-7c-5p | trans | 2.13E-22 | Nightingale | XS_VLDL_L | NA | A | G | 0.048489 | 0.005241 | 1.70E-22 |
| rs532436 | let-7c-5p | trans | 2.13E-22 | Nightingale | HDL_P | NA | A | G | 0.028757 | 0.005007 | 3.30E-09 |
| rs532436 | let-7c-5p | trans | 2.13E-22 | Nightingale | S_VLDL_C_ | NA | A | G | 0.070418 | 0.005189 | 1.30E-41 |
| rs532436 | let-7c-5p | trans | 2.13E-22 | Nightingale | Phosphatic | NA | A | G | 0.031132 | 0.004996 | 6.60E-11 |
| rs532436 | let-7c-5p | trans | 2.13E-22 | Nightingale | XXL_VLDL_ | NA | A | G | 0.036801 | 0.00537 | 1.70E-13 |
| rs532436 | let-7c-5p | trans | 2.13E-22 | Nightingale | Total_FC | NA | A | G | 0.054936 | 0.005209 | 1.00E-27 |
| rs532436 | let-7c-5p | trans | 2.13E-22 | Nightingale | S_LDL_PL_ | NA | A | G | -0.04468 | 0.005322 | 1.60E-19 |
| rs532436 | let-7c-5p | trans | 2.13E-22 | Nightingale | M_VLDL_T | NA | A | G | -0.06283 | 0.005038 | 7.40E-35 |
| rs532436 | let-7c-5p | trans | 2.13E-22 | Nightingale | LDL_C | NA | A | G | 0.060001 | 0.005309 | 3.10E-30 |
| rs532436 | let-7c-5p | trans | 2.13E-22 | Nightingale | Total_C | NA | A | G | 0.055716 | 0.005162 | 2.80E-28 |
| rs532436 | let-7c-5p | trans | 2.13E-22 | Nightingale | S_VLDL_CE | NA | A | G | 0.057936 | 0.005325 | 1.60E-29 |
| rs532436 | let-7c-5p | trans | 2.13E-22 | Nightingale | S_HDL_P | NA | A | G | 0.055157 | 0.005332 | 1.70E-25 |
| rs532436 | let-7c-5p | trans | 2.13E-22 | Nightingale | L_LDL_PL | NA | A | G | 0.066479 | 0.005291 | 5.80E-37 |
| rs532436 | let-7c-5p | trans | 2.13E-22 | Nightingale | M_LDL_FC | NA | A | G | 0.063598 | 0.005325 | 4.40E-33 |
| rs532436 | let-7c-5p | trans | 2.13E-22 | Nightingale | XL_HDL_PL | NA | A | G | -0.04102 | 0.004899 | 1.70E-17 |
| rs532436 | let-7c-5p | trans | 2.13E-22 | Nightingale | M_LDL_C | NA | A | G | 0.05878 | 0.005341 | 5.20E-29 |
| rs532436 | let-7c-5p | trans | 2.13E-22 | Nightingale | M_VLDL_P | NA | A | G | 0.043187 | 0.005319 | 2.10E-17 |
| rs532436 | let-7c-5p | trans | 2.13E-22 | Nightingale | XL_VLDL_P | NA | A | G | 0.055667 | 0.00534 | 6.50E-27 |
| rs532436 | let-7c-5p | trans | 2.13E-22 | Nightingale | VLDL_FC | NA | A | G | 0.033528 | 0.005293 | 1.30E-11 |
| rs532436 | let-7c-5p | trans | 2.13E-22 | Nightingale | L_VLDL_C_ | NA | A | G | 0.08172 | 0.005208 | 2.50E-56 |
| rs532436 | let-7c-5p | trans | 2.13E-22 | Nightingale | XL_VLDL_C | NA | A | G | 0.029577 | 0.005249 | 1.10E-09 |
| rs532436 | let-7c-5p | trans | 2.13E-22 | Nightingale | XS_VLDL_F | NA | A | G | 0.050786 | 0.005294 | 9.70E-21 |
| rs532436 | let-7c-5p | trans | 2.13E-22 | Nightingale | XL_HDL_PL | NA | A | G | -0.03014 | 0.004796 | 6.90E-11 |
| rs532436 | let-7b-5p | trans | 1.04E-25 | Nightingale | XS_VLDL_T | NA | A | G | -0.0425 | 0.005069 | 3.40E-16 |
| rs532436 | let-7b-5p | trans | 1.04E-25 | Nightingale | M_VLDL_P | NA | A | G | 0.070479 | 0.005117 | 5.70E-43 |
| rs532436 | let-7b-5p | trans | 1.04E-25 | Nightingale | L_LDL_C | NA | A | G | 0.059903 | 0.005272 | 1.70E-30 |
| rs532436 | let-7b-5p | trans | 1.04E-25 | Nightingale | LA_pct | NA | A | G | 0.032246 | 0.005214 | 6.90E-10 |
| rs532436 | let-7b-5p | trans | 1.04E-25 | Nightingale | S_VLDL_C | NA | A | G | 0.058922 | 0.005331 | 3.90E-30 |
| rs532436 | let-7b-5p | trans | 1.04E-25 | Nightingale | S_LDL_L | NA | A | G | 0.0484 | 0.005331 | 3.80E-20 |
| rs532436 | let-7b-5p | trans | 1.04E-25 | Nightingale | L_VLDL_FC | NA | A | G | 0.075634 | 0.005281 | 2.30E-47 |
| rs532436 | let-7b-5p | trans | 1.04E-25 | Nightingale | M_LDL_PL | NA | A | G | 0.058078 | 0.005339 | 2.50E-28 |
| rs532436 | let-7b-5p | trans | 1.04E-25 | Nightingale | L_LDL_L | NA | A | G | 0.060847 | 0.00528 | 1.80E-31 |
| rs532436 | let-7b-5p | trans | 1.04E-25 | Nightingale | XL_VLDL_C | NA | A | G | 0.062307 | 0.005115 | 2.90E-33 |
| rs532436 | let-7b-5p | trans | 1.04E-25 | Nightingale | S_VLDL_L | NA | A | G | 0.035519 | 0.00528 | 4.90E-13 |
| rs532436 | let-7b-5p | trans | 1.04E-25 | Nightingale | S_LDL_FC | NA | A | G | 0.054532 | 0.005323 | 2.40E-24 |
| rs532436 | let-7b-5p | trans | 1.04E-25 | Nightingale | M_LDL_P | NA | A | G | 0.057013 | 0.005347 | 1.50E-27 |
| rs532436 | let-7b-5p | trans | 1.04E-25 | Nightingale | M_VLDL_C | NA | A | G | 0.0595 | 0.005022 | 1.90E-31 |
| rs532436 | let-7b-5p | trans | 1.04E-25 | Nightingale | S_LDL_P | NA | A | G | 0.048388 | 0.005337 | 1.90E-20 |
| rs532436 | let-7b-5p | trans | 1.04E-25 | Nightingale | XL_HDL_L | NA | A | G | -0.02706 | 0.004795 | 2.40E-09 |
| rs532436 | let-7b-5p | trans | 1.04E-25 | Nightingale | S_LDL_CE_ | NA | A | G | 0.044561 | 0.005323 | 1.40E-19 |
| rs532436 | let-7b-5p | trans | 1.04E-25 | Nightingale | S_HDL_C | NA | A | G | 0.052394 | 0.005323 | 3.50E-23 |
| rs532436 | let-7b-5p | trans | 1.04E-25 | Nightingale | S_LDL_C_p | NA | A | G | 0.075576 | 0.005352 | 2.10E-46 |
| rs532436 | let-7b-5p | trans | 1.04E-25 | Nightingale | LDL_FC | NA | A | G | 0.062671 | 0.005275 | 1.40E-32 |
| rs532436 | let-7b-5p | trans | 1.04E-25 | Nightingale | XS_VLDL_C | NA | A | G | 0.040138 | 0.005031 | 1.40E-14 |
| rs532436 | let-7b-5p | trans | 1.04E-25 | Nightingale | XS_VLDL_P | NA | A | G | 0.044567 | 0.005257 | 3.00E-19 |
| rs532436 | let-7b-5p | trans | 1.04E-25 | Nightingale | LDL_L | NA | A | G | 0.059463 | 0.005314 | 9.50E-30 |
| rs532436 | let-7b-5p | trans | 1.04E-25 | Nightingale | Clinical_LD | NA | A | G | 0.064426 | 0.005301 | 1.80E-34 |
| rs532436 | let-7b-5p | trans | 1.04E-25 | Nightingale | Citrate | NA | A | G | 0.031277 | 0.005297 | 1.60E-09 |
| rs532436 | let-7b-5p | trans | 1.04E-25 | Nightingale | XS_VLDL_P | NA | A | G | 0.05371 | 0.005261 | 1.20E-26 |
| rs532436 | let-7b-5p | trans | 1.04E-25 | Nightingale | LDL_CE | NA | A | G | 0.058166 | 0.005318 | 1.20E-28 |
| rs532436 | let-7b-5p | trans | 1.04E-25 | Nightingale | Remnant_( | NA | A | G | 0.058377 | 0.005297 | 5.50E-30 |
| rs532436 | let-7b-5p | trans | 1.04E-25 | Nightingale | Cholines | NA | A | G | 0.035678 | 0.005014 | 1.10E-13 |

| | | | | | | | | | | | |
|---|---|---|---|---|---|---|---|---|---|---|---|
| rs532436 | let-7b-5p | trans | 1.04E-25 | Nightingale | HDL_size | NA | A | G | -0.03226 | 0.004727 | 1.70E-12 |
| rs532436 | let-7b-5p | trans | 1.04E-25 | Nightingale | S_HDL_L | NA | A | G | 0.043965 | 0.005313 | 2.20E-17 |
| rs532436 | let-7b-5p | trans | 1.04E-25 | Nightingale | S_VLDL_FC | NA | A | G | 0.059408 | 0.005333 | 3.00E-30 |
| rs532436 | let-7b-5p | trans | 1.04E-25 | Nightingale | XXL_VLDL_ | NA | A | G | 0.046201 | 0.005312 | 1.30E-18 |
| rs532436 | let-7b-5p | trans | 1.04E-25 | Nightingale | XS_VLDL_C | NA | A | G | 0.057562 | 0.005175 | 2.50E-30 |
| rs532436 | let-7b-5p | trans | 1.04E-25 | Nightingale | M_VLDL_F | NA | A | G | 0.067258 | 0.005073 | 1.70E-39 |
| rs532436 | let-7b-5p | trans | 1.04E-25 | Nightingale | IDL_TG_pc | NA | A | G | -0.03868 | 0.005155 | 3.10E-13 |
| rs532436 | let-7b-5p | trans | 1.04E-25 | Nightingale | XS_VLDL_P | NA | A | G | -0.02951 | 0.005203 | 5.00E-08 |
| rs532436 | let-7b-5p | trans | 1.04E-25 | Nightingale | M_VLDL_L | NA | A | G | 0.032097 | 0.005296 | 1.10E-10 |
| rs532436 | let-7b-5p | trans | 1.04E-25 | Nightingale | LA | NA | A | G | 0.042219 | 0.005229 | 3.30E-17 |
| rs532436 | let-7b-5p | trans | 1.04E-25 | Nightingale | M_VLDL_C | NA | A | G | 0.06605 | 0.005283 | 1.00E-36 |
| rs532436 | let-7b-5p | trans | 1.04E-25 | Nightingale | Total_P | NA | A | G | 0.036654 | 0.005018 | 5.50E-14 |
| rs532436 | let-7b-5p | trans | 1.04E-25 | Nightingale | Total_L | NA | A | G | 0.042255 | 0.005245 | 4.90E-17 |
| rs532436 | let-7b-5p | trans | 1.04E-25 | Nightingale | M_LDL_TG | NA | A | G | -0.0531 | 0.005266 | 1.10E-23 |
| rs532436 | let-7b-5p | trans | 1.04E-25 | Nightingale | L_LDL_P | NA | A | G | 0.050357 | 0.005314 | 6.40E-22 |
| rs532436 | let-7b-5p | trans | 1.04E-25 | Nightingale | S_LDL_CE | NA | A | G | 0.051567 | 0.005334 | 4.80E-23 |
| rs532436 | let-7b-5p | trans | 1.04E-25 | Nightingale | S_LDL_TG_ | NA | A | G | -0.04355 | 0.00517 | 3.00E-16 |
| rs532436 | let-7b-5p | trans | 1.04E-25 | Nightingale | M_LDL_L | NA | A | G | 0.057299 | 0.005339 | 8.90E-28 |
| rs532436 | let-7b-5p | trans | 1.04E-25 | Nightingale | IDL_FC_pct | NA | A | G | 0.041213 | 0.00529 | 2.10E-14 |
| rs532436 | let-7b-5p | trans | 1.04E-25 | Nightingale | Phosphogly | NA | A | G | 0.031891 | 0.00505 | 3.60E-11 |
| rs532436 | let-7b-5p | trans | 1.04E-25 | Nightingale | XL_VLDL_F | NA | A | G | 0.056773 | 0.005169 | 2.60E-28 |
| rs532436 | let-7b-5p | trans | 1.04E-25 | Nightingale | S_HDL_FC | NA | A | G | 0.041762 | 0.005234 | 2.40E-16 |
| rs532436 | let-7b-5p | trans | 1.04E-25 | Nightingale | IDL_FC | NA | A | G | 0.063966 | 0.005162 | 7.70E-36 |
| rs532436 | let-7b-5p | trans | 1.04E-25 | Nightingale | S_VLDL_PL | NA | A | G | 0.050032 | 0.005322 | 1.30E-22 |
| rs532436 | let-7b-5p | trans | 1.04E-25 | Nightingale | IDL_C_pct | NA | A | G | 0.037343 | 0.005209 | 6.30E-12 |
| rs532436 | let-7b-5p | trans | 1.04E-25 | Nightingale | L_VLDL_CE | NA | A | G | 0.074326 | 0.005102 | 1.30E-48 |
| rs532436 | let-7b-5p | trans | 1.04E-25 | Nightingale | L_LDL_CE | NA | A | G | 0.058358 | 0.005284 | 5.40E-29 |
| rs532436 | let-7b-5p | trans | 1.04E-25 | Nightingale | S_VLDL_CE | NA | A | G | 0.076497 | 0.00526 | 2.10E-47 |
| rs532436 | let-7b-5p | trans | 1.04E-25 | Nightingale | XL_HDL_CE | NA | A | G | 0.030066 | 0.005327 | 4.20E-08 |
| rs532436 | let-7b-5p | trans | 1.04E-25 | Nightingale | XS_VLDL_F | NA | A | G | 0.053623 | 0.005239 | 1.10E-26 |
| rs532436 | let-7b-5p | trans | 1.04E-25 | Nightingale | M_LDL_C_ | NA | A | G | 0.047449 | 0.005336 | 4.30E-19 |
| rs532436 | let-7b-5p | trans | 1.04E-25 | Nightingale | S_VLDL_P | NA | A | G | 0.033337 | 0.005265 | 7.40E-12 |
| rs532436 | let-7b-5p | trans | 1.04E-25 | Nightingale | S_VLDL_FC | NA | A | G | 0.05247 | 0.005095 | 3.60E-24 |
| rs532436 | let-7b-5p | trans | 1.04E-25 | Nightingale | L_VLDL_TG | NA | A | G | -0.0795 | 0.005307 | 2.00E-52 |
| rs532436 | let-7b-5p | trans | 1.04E-25 | Nightingale | GlycA | NA | A | G | 0.031177 | 0.005309 | 6.60E-09 |
| rs532436 | let-7b-5p | trans | 1.04E-25 | Nightingale | L_VLDL_CE | NA | A | G | 0.031425 | 0.005269 | 1.40E-10 |
| rs532436 | let-7b-5p | trans | 1.04E-25 | Nightingale | M_VLDL_C | NA | A | G | 0.061586 | 0.005317 | 2.10E-32 |
| rs532436 | let-7b-5p | trans | 1.04E-25 | Nightingale | IDL_L | NA | A | G | 0.059551 | 0.005155 | 1.70E-31 |
| rs532436 | let-7b-5p | trans | 1.04E-25 | Nightingale | IDL_CE | NA | A | G | 0.058228 | 0.005148 | 7.40E-30 |
| rs532436 | let-7b-5p | trans | 1.04E-25 | Nightingale | S_VLDL_PL | NA | A | G | 0.049979 | 0.005105 | 5.70E-22 |
| rs532436 | let-7b-5p | trans | 1.04E-25 | Nightingale | M_LDL_CE | NA | A | G | 0.055488 | 0.005339 | 2.20E-26 |
| rs532436 | let-7b-5p | trans | 1.04E-25 | Nightingale | XL_HDL_C_ | NA | A | G | 0.038356 | 0.005092 | 1.70E-13 |
| rs532436 | let-7b-5p | trans | 1.04E-25 | Nightingale | VLDL_C | NA | A | G | 0.045684 | 0.005324 | 2.90E-19 |
| rs532436 | let-7b-5p | trans | 1.04E-25 | Nightingale | IDL_C | NA | A | G | 0.060082 | 0.005149 | 9.90E-32 |
| rs532436 | let-7b-5p | trans | 1.04E-25 | Nightingale | S_VLDL_TG | NA | A | G | -0.06518 | 0.005161 | 4.40E-36 |
| rs532436 | let-7b-5p | trans | 1.04E-25 | Nightingale | ApoB_by_A | NA | A | G | 0.039971 | 0.005251 | 1.30E-14 |
| rs532436 | let-7b-5p | trans | 1.04E-25 | Nightingale | VLDL_P | NA | A | G | 0.038195 | 0.005303 | 2.40E-14 |
| rs532436 | let-7b-5p | trans | 1.04E-25 | Nightingale | Total_CE | NA | A | G | 0.05558 | 0.005143 | 3.40E-28 |
| rs532436 | let-7b-5p | trans | 1.04E-25 | Nightingale | L_LDL_FC_ | NA | A | G | 0.030877 | 0.005087 | 1.20E-08 |
| rs532436 | let-7b-5p | trans | 1.04E-25 | Nightingale | VLDL_CE | NA | A | G | 0.053545 | 0.005333 | 2.70E-25 |
| rs532436 | let-7b-5p | trans | 1.04E-25 | Nightingale | M_VLDL_C | NA | A | G | 0.055885 | 0.005009 | 5.70E-28 |
| rs532436 | let-7b-5p | trans | 1.04E-25 | Nightingale | XXL_VLDL_ | NA | A | G | -0.05817 | 0.005381 | 5.80E-28 |
| rs532436 | let-7b-5p | trans | 1.04E-25 | Nightingale | IDL_PL | NA | A | G | 0.05875 | 0.005145 | 3.70E-31 |
| rs532436 | let-7b-5p | trans | 1.04E-25 | Nightingale | S_HDL_PL_ | NA | A | G | -0.03259 | 0.005155 | 1.00E-09 |
| rs532436 | let-7b-5p | trans | 1.04E-25 | Nightingale | S_LDL_PL | NA | A | G | 0.041297 | 0.00532 | 6.80E-15 |
| rs532436 | let-7b-5p | trans | 1.04E-25 | Nightingale | non_HDL_( | NA | A | G | 0.060373 | 0.005303 | 3.40E-31 |
| rs532436 | let-7b-5p | trans | 1.04E-25 | Nightingale | M_VLDL_P | NA | A | G | 0.044715 | 0.005324 | 1.90E-18 |
| rs532436 | let-7b-5p | trans | 1.04E-25 | Nightingale | Sphingomy | NA | A | G | 0.047389 | 0.004984 | 3.70E-21 |
| rs532436 | let-7b-5p | trans | 1.04E-25 | Nightingale | LDL_PL | NA | A | G | 0.06187 | 0.005319 | 7.90E-32 |
| rs532436 | let-7b-5p | trans | 1.04E-25 | Nightingale | L_VLDL_PL | NA | A | G | 0.027951 | 0.005179 | 3.30E-09 |
| rs532436 | let-7b-5p | trans | 1.04E-25 | Nightingale | S_HDL_PL | NA | A | G | 0.037551 | 0.005286 | 1.80E-13 |
| rs532436 | let-7b-5p | trans | 1.04E-25 | Nightingale | XS_VLDL_C | NA | A | G | 0.043168 | 0.005057 | 1.10E-16 |
| rs532436 | let-7b-5p | trans | 1.04E-25 | Nightingale | IDL_P | NA | A | G | 0.06498 | 0.005276 | 1.90E-35 |

| | | | | | | | | | | | |
|---|---|---|---|---|---|---|---|---|---|---|---|
| rs532436 | let-7b-5p | trans | 1.04E-25 | Nightingale | S_HDL_CE | NA | A | G | 0.053255 | 0.00534 | 1.20E-23 |
| rs532436 | let-7b-5p | trans | 1.04E-25 | Nightingale | L_LDL_FC | NA | A | G | 0.062609 | 0.005236 | 3.80E-33 |
| rs532436 | let-7b-5p | trans | 1.04E-25 | Nightingale | PUFA | NA | A | G | 0.036816 | 0.005201 | 1.50E-13 |
| rs532436 | let-7b-5p | trans | 1.04E-25 | Nightingale | XS_VLDL_C | NA | A | G | 0.058331 | 0.005145 | 3.40E-31 |
| rs532436 | let-7b-5p | trans | 1.04E-25 | Nightingale | ApoB | NA | A | G | 0.055646 | 0.005325 | 1.40E-26 |
| rs532436 | let-7b-5p | trans | 1.04E-25 | Nightingale | M_VLDL_F | NA | A | G | 0.051674 | 0.005331 | 1.50E-23 |
| rs532436 | let-7b-5p | trans | 1.04E-25 | Nightingale | S_LDL_C | NA | A | G | 0.053873 | 0.005337 | 1.50E-24 |
| rs532436 | let-7b-5p | trans | 1.04E-25 | Nightingale | XL_VLDL_C | NA | A | G | 0.062531 | 0.005113 | 1.30E-33 |
| rs532436 | let-7b-5p | trans | 1.04E-25 | Nightingale | XXL_VLDL_ | NA | A | G | 0.055922 | 0.005375 | 1.40E-25 |
| rs532436 | let-7b-5p | trans | 1.04E-25 | Nightingale | Omega_6 | NA | A | G | 0.0417 | 0.005207 | 5.30E-17 |
| rs532436 | let-7b-5p | trans | 1.04E-25 | Nightingale | LDL_P | NA | A | G | 0.053144 | 0.005328 | 2.90E-24 |
| rs532436 | let-7b-5p | trans | 1.04E-25 | Nightingale | XXL_VLDL_ | NA | A | G | 0.056832 | 0.005346 | 1.50E-26 |
| rs532436 | let-7b-5p | trans | 1.04E-25 | Nightingale | Pyruvate | NA | A | G | 0.034156 | 0.00534 | 1.60E-10 |
| rs532436 | let-7b-5p | trans | 1.04E-25 | Nightingale | Total_PL | NA | A | G | 0.03947 | 0.005077 | 4.30E-16 |
| rs532436 | let-7b-5p | trans | 1.04E-25 | Nightingale | L_LDL_TG_ | NA | A | G | -0.04149 | 0.00523 | 8.10E-15 |
| rs532436 | let-7b-5p | trans | 1.04E-25 | Nightingale | XL_VLDL_T | NA | A | G | -0.06894 | 0.005168 | 1.90E-40 |
| rs532436 | let-7b-5p | trans | 1.04E-25 | Nightingale | XS_VLDL_L | NA | A | G | 0.048489 | 0.005241 | 1.70E-22 |
| rs532436 | let-7b-5p | trans | 1.04E-25 | Nightingale | HDL_P | NA | A | G | 0.028757 | 0.005007 | 3.30E-09 |
| rs532436 | let-7b-5p | trans | 1.04E-25 | Nightingale | S_VLDL_C_ | NA | A | G | 0.070418 | 0.005189 | 1.30E-41 |
| rs532436 | let-7b-5p | trans | 1.04E-25 | Nightingale | Phosphatic | NA | A | G | 0.031132 | 0.004996 | 6.60E-11 |
| rs532436 | let-7b-5p | trans | 1.04E-25 | Nightingale | XXL_VLDL_ | NA | A | G | 0.036801 | 0.00537 | 1.70E-13 |
| rs532436 | let-7b-5p | trans | 1.04E-25 | Nightingale | Total_FC | NA | A | G | 0.054936 | 0.005209 | 1.00E-27 |
| rs532436 | let-7b-5p | trans | 1.04E-25 | Nightingale | S_LDL_PL_ | NA | A | G | -0.04468 | 0.005322 | 1.60E-19 |
| rs532436 | let-7b-5p | trans | 1.04E-25 | Nightingale | M_VLDL_T | NA | A | G | -0.06283 | 0.005038 | 7.40E-35 |
| rs532436 | let-7b-5p | trans | 1.04E-25 | Nightingale | LDL_C | NA | A | G | 0.060001 | 0.005309 | 3.10E-30 |
| rs532436 | let-7b-5p | trans | 1.04E-25 | Nightingale | Total_C | NA | A | G | 0.055716 | 0.005162 | 2.80E-28 |
| rs532436 | let-7b-5p | trans | 1.04E-25 | Nightingale | S_VLDL_CE | NA | A | G | 0.057936 | 0.005325 | 1.60E-29 |
| rs532436 | let-7b-5p | trans | 1.04E-25 | Nightingale | S_HDL_P | NA | A | G | 0.055157 | 0.005332 | 1.70E-25 |
| rs532436 | let-7b-5p | trans | 1.04E-25 | Nightingale | L_LDL_PL | NA | A | G | 0.066479 | 0.005291 | 5.80E-37 |
| rs532436 | let-7b-5p | trans | 1.04E-25 | Nightingale | M_LDL_FC | NA | A | G | 0.063598 | 0.005325 | 4.40E-33 |
| rs532436 | let-7b-5p | trans | 1.04E-25 | Nightingale | XL_HDL_PL | NA | A | G | -0.04102 | 0.004899 | 1.70E-17 |
| rs532436 | let-7b-5p | trans | 1.04E-25 | Nightingale | M_LDL_C | NA | A | G | 0.05878 | 0.005341 | 5.20E-29 |
| rs532436 | let-7b-5p | trans | 1.04E-25 | Nightingale | M_VLDL_P | NA | A | G | 0.043187 | 0.005319 | 2.10E-17 |
| rs532436 | let-7b-5p | trans | 1.04E-25 | Nightingale | XL_VLDL_P | NA | A | G | 0.055667 | 0.00534 | 6.50E-27 |
| rs532436 | let-7b-5p | trans | 1.04E-25 | Nightingale | VLDL_FC | NA | A | G | 0.033528 | 0.005293 | 1.30E-11 |
| rs532436 | let-7b-5p | trans | 1.04E-25 | Nightingale | L_VLDL_C_ | NA | A | G | 0.08172 | 0.005208 | 2.50E-56 |
| rs532436 | let-7b-5p | trans | 1.04E-25 | Nightingale | XL_VLDL_C | NA | A | G | 0.029577 | 0.005249 | 1.10E-09 |
| rs532436 | let-7b-5p | trans | 1.04E-25 | Nightingale | XS_VLDL_F | NA | A | G | 0.050786 | 0.005294 | 9.70E-21 |
| rs532436 | let-7b-5p | trans | 1.04E-25 | Nightingale | XL_HDL_PL | NA | A | G | -0.03014 | 0.004796 | 6.90E-11 |
| rs532436 | miR-139-5p | trans | 6.95E-37 | Nightingale | XS_VLDL_T | NA | A | G | -0.0425 | 0.005069 | 3.40E-16 |
| rs532436 | miR-139-5p | trans | 6.95E-37 | Nightingale | M_VLDL_P | NA | A | G | 0.070479 | 0.005117 | 5.70E-43 |
| rs532436 | miR-139-5p | trans | 6.95E-37 | Nightingale | L_LDL_C | NA | A | G | 0.059903 | 0.005272 | 1.70E-30 |
| rs532436 | miR-139-5p | trans | 6.95E-37 | Nightingale | LA_pct | NA | A | G | 0.032246 | 0.005214 | 6.90E-10 |
| rs532436 | miR-139-5p | trans | 6.95E-37 | Nightingale | S_VLDL_C | NA | A | G | 0.058922 | 0.005331 | 3.90E-30 |
| rs532436 | miR-139-5p | trans | 6.95E-37 | Nightingale | S_LDL_L | NA | A | G | 0.0484 | 0.005331 | 3.80E-20 |
| rs532436 | miR-139-5p | trans | 6.95E-37 | Nightingale | L_VLDL_FC | NA | A | G | 0.075634 | 0.005281 | 2.30E-47 |
| rs532436 | miR-139-5p | trans | 6.95E-37 | Nightingale | M_LDL_PL | NA | A | G | 0.058078 | 0.005339 | 2.50E-28 |
| rs532436 | miR-139-5p | trans | 6.95E-37 | Nightingale | L_LDL_L | NA | A | G | 0.060847 | 0.00528 | 1.80E-31 |
| rs532436 | miR-139-5p | trans | 6.95E-37 | Nightingale | XL_VLDL_C | NA | A | G | 0.062307 | 0.005115 | 2.90E-33 |
| rs532436 | miR-139-5p | trans | 6.95E-37 | Nightingale | S_VLDL_L | NA | A | G | 0.035519 | 0.00528 | 4.90E-13 |
| rs532436 | miR-139-5p | trans | 6.95E-37 | Nightingale | S_LDL_FC | NA | A | G | 0.054532 | 0.005323 | 2.40E-24 |
| rs532436 | miR-139-5p | trans | 6.95E-37 | Nightingale | M_LDL_P | NA | A | G | 0.057013 | 0.005347 | 1.50E-27 |
| rs532436 | miR-139-5p | trans | 6.95E-37 | Nightingale | M_VLDL_C | NA | A | G | 0.0595 | 0.005022 | 1.90E-31 |
| rs532436 | miR-139-5p | trans | 6.95E-37 | Nightingale | S_LDL_P | NA | A | G | 0.048388 | 0.005337 | 1.90E-20 |
| rs532436 | miR-139-5p | trans | 6.95E-37 | Nightingale | XL_HDL_L | NA | A | G | -0.02706 | 0.004795 | 2.40E-09 |
| rs532436 | miR-139-5p | trans | 6.95E-37 | Nightingale | S_LDL_CE_ | NA | A | G | 0.044561 | 0.005323 | 1.40E-19 |
| rs532436 | miR-139-5p | trans | 6.95E-37 | Nightingale | S_HDL_C | NA | A | G | 0.052394 | 0.005323 | 3.50E-23 |
| rs532436 | miR-139-5p | trans | 6.95E-37 | Nightingale | S_LDL_C_p | NA | A | G | 0.075576 | 0.005352 | 2.10E-46 |
| rs532436 | miR-139-5p | trans | 6.95E-37 | Nightingale | LDL_FC | NA | A | G | 0.062671 | 0.005275 | 1.40E-32 |
| rs532436 | miR-139-5p | trans | 6.95E-37 | Nightingale | XS_VLDL_C | NA | A | G | 0.040138 | 0.005031 | 1.40E-14 |
| rs532436 | miR-139-5p | trans | 6.95E-37 | Nightingale | XS_VLDL_P | NA | A | G | 0.044567 | 0.005257 | 3.00E-19 |
| rs532436 | miR-139-5p | trans | 6.95E-37 | Nightingale | LDL_L | NA | A | G | 0.059463 | 0.005314 | 9.50E-30 |
| rs532436 | miR-139-5p | trans | 6.95E-37 | Nightingale | Clinical_LD | NA | A | G | 0.064426 | 0.005301 | 1.80E-34 |

| | | | | | | | | | | | |
|---|---|---|---|---|---|---|---|---|---|---|---|
| rs532436 | miR-139-5p | trans | 6.95E-37 | Nightingale | Citrate | NA | A | G | 0.031277 | 0.005297 | 1.60E-09 |
| rs532436 | miR-139-5p | trans | 6.95E-37 | Nightingale | XS_VLDL_P | NA | A | G | 0.05371 | 0.005261 | 1.20E-26 |
| rs532436 | miR-139-5p | trans | 6.95E-37 | Nightingale | LDL_CE | NA | A | G | 0.058166 | 0.005318 | 1.20E-28 |
| rs532436 | miR-139-5p | trans | 6.95E-37 | Nightingale | Remnant_( | NA | A | G | 0.058377 | 0.005297 | 5.50E-30 |
| rs532436 | miR-139-5p | trans | 6.95E-37 | Nightingale | Cholines | NA | A | G | 0.035678 | 0.005014 | 1.10E-13 |
| rs532436 | miR-139-5p | trans | 6.95E-37 | Nightingale | HDL_size | NA | A | G | -0.03226 | 0.004727 | 1.70E-12 |
| rs532436 | miR-139-5p | trans | 6.95E-37 | Nightingale | S_HDL_L | NA | A | G | 0.043965 | 0.005313 | 2.20E-17 |
| rs532436 | miR-139-5p | trans | 6.95E-37 | Nightingale | S_VLDL_FC | NA | A | G | 0.059408 | 0.005333 | 3.00E-30 |
| rs532436 | miR-139-5p | trans | 6.95E-37 | Nightingale | XXL_VLDL_ | NA | A | G | 0.046201 | 0.005312 | 1.30E-18 |
| rs532436 | miR-139-5p | trans | 6.95E-37 | Nightingale | XS_VLDL_C | NA | A | G | 0.057562 | 0.005175 | 2.50E-30 |
| rs532436 | miR-139-5p | trans | 6.95E-37 | Nightingale | M_VLDL_F | NA | A | G | 0.067258 | 0.005073 | 1.70E-39 |
| rs532436 | miR-139-5p | trans | 6.95E-37 | Nightingale | IDL_TG_pc | NA | A | G | -0.03868 | 0.005155 | 3.10E-13 |
| rs532436 | miR-139-5p | trans | 6.95E-37 | Nightingale | XS_VLDL_P | NA | A | G | -0.02951 | 0.005203 | 5.00E-08 |
| rs532436 | miR-139-5p | trans | 6.95E-37 | Nightingale | M_VLDL_L | NA | A | G | 0.032097 | 0.005296 | 1.10E-10 |
| rs532436 | miR-139-5p | trans | 6.95E-37 | Nightingale | LA | NA | A | G | 0.042219 | 0.005229 | 3.30E-17 |
| rs532436 | miR-139-5p | trans | 6.95E-37 | Nightingale | M_VLDL_C | NA | A | G | 0.06605 | 0.005283 | 1.00E-36 |
| rs532436 | miR-139-5p | trans | 6.95E-37 | Nightingale | Total_P | NA | A | G | 0.036654 | 0.005018 | 5.50E-14 |
| rs532436 | miR-139-5p | trans | 6.95E-37 | Nightingale | Total_L | NA | A | G | 0.042255 | 0.005245 | 4.90E-17 |
| rs532436 | miR-139-5p | trans | 6.95E-37 | Nightingale | M_LDL_TG | NA | A | G | -0.0531 | 0.005266 | 1.10E-23 |
| rs532436 | miR-139-5p | trans | 6.95E-37 | Nightingale | L_LDL_P | NA | A | G | 0.050357 | 0.005314 | 6.40E-22 |
| rs532436 | miR-139-5p | trans | 6.95E-37 | Nightingale | S_LDL_CE | NA | A | G | 0.051567 | 0.005334 | 4.80E-23 |
| rs532436 | miR-139-5p | trans | 6.95E-37 | Nightingale | S_LDL_TG_ | NA | A | G | -0.04355 | 0.00517 | 3.00E-16 |
| rs532436 | miR-139-5p | trans | 6.95E-37 | Nightingale | M_LDL_L | NA | A | G | 0.057299 | 0.005339 | 8.90E-28 |
| rs532436 | miR-139-5p | trans | 6.95E-37 | Nightingale | IDL_FC_pct | NA | A | G | 0.041213 | 0.00529 | 2.10E-14 |
| rs532436 | miR-139-5p | trans | 6.95E-37 | Nightingale | Phosphogly | NA | A | G | 0.031891 | 0.00505 | 3.60E-11 |
| rs532436 | miR-139-5p | trans | 6.95E-37 | Nightingale | XL_VLDL_F | NA | A | G | 0.056773 | 0.005169 | 2.60E-28 |
| rs532436 | miR-139-5p | trans | 6.95E-37 | Nightingale | S_HDL_FC | NA | A | G | 0.041762 | 0.005234 | 2.40E-16 |
| rs532436 | miR-139-5p | trans | 6.95E-37 | Nightingale | IDL_FC | NA | A | G | 0.063966 | 0.005162 | 7.70E-36 |
| rs532436 | miR-139-5p | trans | 6.95E-37 | Nightingale | S_VLDL_PL | NA | A | G | 0.050032 | 0.005322 | 1.30E-22 |
| rs532436 | miR-139-5p | trans | 6.95E-37 | Nightingale | IDL_C_pct | NA | A | G | 0.037343 | 0.005209 | 6.30E-12 |
| rs532436 | miR-139-5p | trans | 6.95E-37 | Nightingale | L_VLDL_CE | NA | A | G | 0.074326 | 0.005102 | 1.30E-48 |
| rs532436 | miR-139-5p | trans | 6.95E-37 | Nightingale | L_LDL_CE | NA | A | G | 0.058358 | 0.005284 | 5.40E-29 |
| rs532436 | miR-139-5p | trans | 6.95E-37 | Nightingale | S_VLDL_CE | NA | A | G | 0.076497 | 0.00526 | 2.10E-47 |
| rs532436 | miR-139-5p | trans | 6.95E-37 | Nightingale | XL_HDL_CE | NA | A | G | 0.030066 | 0.005327 | 4.20E-08 |
| rs532436 | miR-139-5p | trans | 6.95E-37 | Nightingale | XS_VLDL_F | NA | A | G | 0.053623 | 0.005239 | 1.10E-26 |
| rs532436 | miR-139-5p | trans | 6.95E-37 | Nightingale | M_LDL_C_ | NA | A | G | 0.047449 | 0.005336 | 4.30E-19 |
| rs532436 | miR-139-5p | trans | 6.95E-37 | Nightingale | S_VLDL_P | NA | A | G | 0.033337 | 0.005265 | 7.40E-12 |
| rs532436 | miR-139-5p | trans | 6.95E-37 | Nightingale | S_VLDL_FC | NA | A | G | 0.05247 | 0.005095 | 3.60E-24 |
| rs532436 | miR-139-5p | trans | 6.95E-37 | Nightingale | L_VLDL_TG | NA | A | G | -0.0795 | 0.005307 | 2.00E-52 |
| rs532436 | miR-139-5p | trans | 6.95E-37 | Nightingale | GlycA | NA | A | G | 0.031177 | 0.005309 | 6.60E-09 |
| rs532436 | miR-139-5p | trans | 6.95E-37 | Nightingale | L_VLDL_CE | NA | A | G | 0.031425 | 0.005269 | 1.40E-10 |
| rs532436 | miR-139-5p | trans | 6.95E-37 | Nightingale | M_VLDL_C | NA | A | G | 0.061586 | 0.005317 | 2.10E-32 |
| rs532436 | miR-139-5p | trans | 6.95E-37 | Nightingale | IDL_L | NA | A | G | 0.059551 | 0.005155 | 1.70E-31 |
| rs532436 | miR-139-5p | trans | 6.95E-37 | Nightingale | IDL_CE | NA | A | G | 0.058228 | 0.005148 | 7.40E-30 |
| rs532436 | miR-139-5p | trans | 6.95E-37 | Nightingale | S_VLDL_PL | NA | A | G | 0.049979 | 0.005105 | 5.70E-22 |
| rs532436 | miR-139-5p | trans | 6.95E-37 | Nightingale | M_LDL_CE | NA | A | G | 0.055488 | 0.005339 | 2.20E-26 |
| rs532436 | miR-139-5p | trans | 6.95E-37 | Nightingale | XL_HDL_C_ | NA | A | G | 0.038356 | 0.005092 | 1.70E-13 |
| rs532436 | miR-139-5p | trans | 6.95E-37 | Nightingale | VLDL_C | NA | A | G | 0.045684 | 0.005324 | 2.90E-19 |
| rs532436 | miR-139-5p | trans | 6.95E-37 | Nightingale | IDL_C | NA | A | G | 0.060082 | 0.005149 | 9.90E-32 |
| rs532436 | miR-139-5p | trans | 6.95E-37 | Nightingale | S_VLDL_TG | NA | A | G | -0.06518 | 0.005161 | 4.40E-36 |
| rs532436 | miR-139-5p | trans | 6.95E-37 | Nightingale | ApoB_by_A | NA | A | G | 0.039971 | 0.005251 | 1.30E-14 |
| rs532436 | miR-139-5p | trans | 6.95E-37 | Nightingale | VLDL_P | NA | A | G | 0.038195 | 0.005303 | 2.40E-14 |
| rs532436 | miR-139-5p | trans | 6.95E-37 | Nightingale | Total_CE | NA | A | G | 0.05558 | 0.005143 | 3.40E-28 |
| rs532436 | miR-139-5p | trans | 6.95E-37 | Nightingale | L_LDL_FC_ | NA | A | G | 0.030877 | 0.005087 | 1.20E-08 |
| rs532436 | miR-139-5p | trans | 6.95E-37 | Nightingale | VLDL_CE | NA | A | G | 0.053545 | 0.005333 | 2.70E-25 |
| rs532436 | miR-139-5p | trans | 6.95E-37 | Nightingale | M_VLDL_C | NA | A | G | 0.055885 | 0.005009 | 5.70E-28 |
| rs532436 | miR-139-5p | trans | 6.95E-37 | Nightingale | XXL_VLDL_ | NA | A | G | -0.05817 | 0.005381 | 5.80E-28 |
| rs532436 | miR-139-5p | trans | 6.95E-37 | Nightingale | IDL_PL | NA | A | G | 0.05875 | 0.005145 | 3.70E-31 |
| rs532436 | miR-139-5p | trans | 6.95E-37 | Nightingale | S_HDL_PL_ | NA | A | G | -0.03259 | 0.005155 | 1.00E-09 |
| rs532436 | miR-139-5p | trans | 6.95E-37 | Nightingale | S_LDL_PL | NA | A | G | 0.041297 | 0.00532 | 6.80E-15 |
| rs532436 | miR-139-5p | trans | 6.95E-37 | Nightingale | non_HDL_( | NA | A | G | 0.060373 | 0.005303 | 3.40E-31 |
| rs532436 | miR-139-5p | trans | 6.95E-37 | Nightingale | M_VLDL_P | NA | A | G | 0.044715 | 0.005324 | 1.90E-18 |
| rs532436 | miR-139-5p | trans | 6.95E-37 | Nightingale | Sphingomy | NA | A | G | 0.047389 | 0.004984 | 3.70E-21 |

| | | | | | | | | | | | |
|---|---|---|---|---|---|---|---|---|---|---|---|
| rs532436 | miR-139-5p | trans | 6.95E-37 | Nightingale | LDL_PL | NA | A | G | 0.06187 | 0.005319 | 7.90E-32 |
| rs532436 | miR-139-5p | trans | 6.95E-37 | Nightingale | L_VLDL_PL | NA | A | G | 0.027951 | 0.005179 | 3.30E-09 |
| rs532436 | miR-139-5p | trans | 6.95E-37 | Nightingale | S_HDL_PL | NA | A | G | 0.037551 | 0.005286 | 1.80E-13 |
| rs532436 | miR-139-5p | trans | 6.95E-37 | Nightingale | XS_VLDL_C | NA | A | G | 0.043168 | 0.005057 | 1.10E-16 |
| rs532436 | miR-139-5p | trans | 6.95E-37 | Nightingale | IDL_P | NA | A | G | 0.06498 | 0.005276 | 1.90E-35 |
| rs532436 | miR-139-5p | trans | 6.95E-37 | Nightingale | S_HDL_CE | NA | A | G | 0.053255 | 0.00534 | 1.20E-23 |
| rs532436 | miR-139-5p | trans | 6.95E-37 | Nightingale | L_LDL_FC | NA | A | G | 0.062609 | 0.005236 | 3.80E-33 |
| rs532436 | miR-139-5p | trans | 6.95E-37 | Nightingale | PUFA | NA | A | G | 0.036816 | 0.005201 | 1.50E-13 |
| rs532436 | miR-139-5p | trans | 6.95E-37 | Nightingale | XS_VLDL_C | NA | A | G | 0.058331 | 0.005145 | 3.40E-31 |
| rs532436 | miR-139-5p | trans | 6.95E-37 | Nightingale | ApoB | NA | A | G | 0.055646 | 0.005325 | 1.40E-26 |
| rs532436 | miR-139-5p | trans | 6.95E-37 | Nightingale | M_VLDL_F | NA | A | G | 0.051674 | 0.005331 | 1.50E-23 |
| rs532436 | miR-139-5p | trans | 6.95E-37 | Nightingale | S_LDL_C | NA | A | G | 0.053873 | 0.005337 | 1.50E-24 |
| rs532436 | miR-139-5p | trans | 6.95E-37 | Nightingale | XL_VLDL_C | NA | A | G | 0.062531 | 0.005113 | 1.30E-33 |
| rs532436 | miR-139-5p | trans | 6.95E-37 | Nightingale | XXL_VLDL_ | NA | A | G | 0.055922 | 0.005375 | 1.40E-25 |
| rs532436 | miR-139-5p | trans | 6.95E-37 | Nightingale | Omega_6 | NA | A | G | 0.0417 | 0.005207 | 5.30E-17 |
| rs532436 | miR-139-5p | trans | 6.95E-37 | Nightingale | LDL_P | NA | A | G | 0.053144 | 0.005328 | 2.90E-24 |
| rs532436 | miR-139-5p | trans | 6.95E-37 | Nightingale | XXL_VLDL_ | NA | A | G | 0.056832 | 0.005346 | 1.50E-26 |
| rs532436 | miR-139-5p | trans | 6.95E-37 | Nightingale | Pyruvate | NA | A | G | 0.034156 | 0.00534 | 1.60E-10 |
| rs532436 | miR-139-5p | trans | 6.95E-37 | Nightingale | Total_PL | NA | A | G | 0.03947 | 0.005077 | 4.30E-16 |
| rs532436 | miR-139-5p | trans | 6.95E-37 | Nightingale | L_LDL_TG_ | NA | A | G | -0.04149 | 0.00523 | 8.10E-15 |
| rs532436 | miR-139-5p | trans | 6.95E-37 | Nightingale | XL_VLDL_T | NA | A | G | -0.06894 | 0.005168 | 1.90E-40 |
| rs532436 | miR-139-5p | trans | 6.95E-37 | Nightingale | XS_VLDL_L | NA | A | G | 0.048489 | 0.005241 | 1.70E-22 |
| rs532436 | miR-139-5p | trans | 6.95E-37 | Nightingale | HDL_P | NA | A | G | 0.028757 | 0.005007 | 3.30E-09 |
| rs532436 | miR-139-5p | trans | 6.95E-37 | Nightingale | S_VLDL_C_ | NA | A | G | 0.070418 | 0.005189 | 1.30E-41 |
| rs532436 | miR-139-5p | trans | 6.95E-37 | Nightingale | Phosphatic | NA | A | G | 0.031132 | 0.004996 | 6.60E-11 |
| rs532436 | miR-139-5p | trans | 6.95E-37 | Nightingale | XXL_VLDL_ | NA | A | G | 0.036801 | 0.00537 | 1.70E-13 |
| rs532436 | miR-139-5p | trans | 6.95E-37 | Nightingale | Total_FC | NA | A | G | 0.054936 | 0.005209 | 1.00E-27 |
| rs532436 | miR-139-5p | trans | 6.95E-37 | Nightingale | S_LDL_PL_ | NA | A | G | -0.04468 | 0.005322 | 1.60E-19 |
| rs532436 | miR-139-5p | trans | 6.95E-37 | Nightingale | M_VLDL_T | NA | A | G | -0.06283 | 0.005038 | 7.40E-35 |
| rs532436 | miR-139-5p | trans | 6.95E-37 | Nightingale | LDL_C | NA | A | G | 0.060001 | 0.005309 | 3.10E-30 |
| rs532436 | miR-139-5p | trans | 6.95E-37 | Nightingale | Total_C | NA | A | G | 0.055716 | 0.005162 | 2.80E-28 |
| rs532436 | miR-139-5p | trans | 6.95E-37 | Nightingale | S_VLDL_CE | NA | A | G | 0.057936 | 0.005325 | 1.60E-29 |
| rs532436 | miR-139-5p | trans | 6.95E-37 | Nightingale | S_HDL_P | NA | A | G | 0.055157 | 0.005332 | 1.70E-25 |
| rs532436 | miR-139-5p | trans | 6.95E-37 | Nightingale | L_LDL_PL | NA | A | G | 0.066479 | 0.005291 | 5.80E-37 |
| rs532436 | miR-139-5p | trans | 6.95E-37 | Nightingale | M_LDL_FC | NA | A | G | 0.063598 | 0.005325 | 4.40E-33 |
| rs532436 | miR-139-5p | trans | 6.95E-37 | Nightingale | XL_HDL_PL | NA | A | G | -0.04102 | 0.004899 | 1.70E-17 |
| rs532436 | miR-139-5p | trans | 6.95E-37 | Nightingale | M_LDL_C | NA | A | G | 0.05878 | 0.005341 | 5.20E-29 |
| rs532436 | miR-139-5p | trans | 6.95E-37 | Nightingale | M_VLDL_P | NA | A | G | 0.043187 | 0.005319 | 2.10E-17 |
| rs532436 | miR-139-5p | trans | 6.95E-37 | Nightingale | XL_VLDL_P | NA | A | G | 0.055667 | 0.00534 | 6.50E-27 |
| rs532436 | miR-139-5p | trans | 6.95E-37 | Nightingale | VLDL_FC | NA | A | G | 0.033528 | 0.005293 | 1.30E-11 |
| rs532436 | miR-139-5p | trans | 6.95E-37 | Nightingale | L_VLDL_C_ | NA | A | G | 0.08172 | 0.005208 | 2.50E-56 |
| rs532436 | miR-139-5p | trans | 6.95E-37 | Nightingale | XL_VLDL_C | NA | A | G | 0.029577 | 0.005249 | 1.10E-09 |
| rs532436 | miR-139-5p | trans | 6.95E-37 | Nightingale | XS_VLDL_F | NA | A | G | 0.050786 | 0.005294 | 9.70E-21 |
| rs532436 | miR-139-5p | trans | 6.95E-37 | Nightingale | XL_HDL_PL | NA | A | G | -0.03014 | 0.004796 | 6.90E-11 |
| rs532436 | miR-195-5p | trans | 3.89E-18 | Nightingale | XS_VLDL_T | NA | A | G | -0.0425 | 0.005069 | 3.40E-16 |
| rs532436 | miR-195-5p | trans | 3.89E-18 | Nightingale | M_VLDL_P | NA | A | G | 0.070479 | 0.005117 | 5.70E-43 |
| rs532436 | miR-195-5p | trans | 3.89E-18 | Nightingale | L_LDL_C | NA | A | G | 0.059903 | 0.005272 | 1.70E-30 |
| rs532436 | miR-195-5p | trans | 3.89E-18 | Nightingale | LA_pct | NA | A | G | 0.032246 | 0.005214 | 6.90E-10 |
| rs532436 | miR-195-5p | trans | 3.89E-18 | Nightingale | S_VLDL_C | NA | A | G | 0.058922 | 0.005331 | 3.90E-30 |
| rs532436 | miR-195-5p | trans | 3.89E-18 | Nightingale | S_LDL_L | NA | A | G | 0.0484 | 0.005331 | 3.80E-20 |
| rs532436 | miR-195-5p | trans | 3.89E-18 | Nightingale | L_VLDL_FC | NA | A | G | 0.075634 | 0.005281 | 2.30E-47 |
| rs532436 | miR-195-5p | trans | 3.89E-18 | Nightingale | M_LDL_PL | NA | A | G | 0.058078 | 0.005339 | 2.50E-28 |
| rs532436 | miR-195-5p | trans | 3.89E-18 | Nightingale | L_LDL_L | NA | A | G | 0.060847 | 0.00528 | 1.80E-31 |
| rs532436 | miR-195-5p | trans | 3.89E-18 | Nightingale | XL_VLDL_C | NA | A | G | 0.062307 | 0.005115 | 2.90E-33 |
| rs532436 | miR-195-5p | trans | 3.89E-18 | Nightingale | S_VLDL_L | NA | A | G | 0.035519 | 0.00528 | 4.90E-13 |
| rs532436 | miR-195-5p | trans | 3.89E-18 | Nightingale | S_LDL_FC | NA | A | G | 0.054532 | 0.005323 | 2.40E-24 |
| rs532436 | miR-195-5p | trans | 3.89E-18 | Nightingale | M_LDL_P | NA | A | G | 0.057013 | 0.005347 | 1.50E-27 |
| rs532436 | miR-195-5p | trans | 3.89E-18 | Nightingale | M_VLDL_C | NA | A | G | 0.0595 | 0.005022 | 1.90E-31 |
| rs532436 | miR-195-5p | trans | 3.89E-18 | Nightingale | S_LDL_P | NA | A | G | 0.048388 | 0.005337 | 1.90E-20 |
| rs532436 | miR-195-5p | trans | 3.89E-18 | Nightingale | XL_HDL_L | NA | A | G | -0.02706 | 0.004795 | 2.40E-09 |
| rs532436 | miR-195-5p | trans | 3.89E-18 | Nightingale | S_LDL_CE_ | NA | A | G | 0.044561 | 0.005323 | 1.40E-19 |
| rs532436 | miR-195-5p | trans | 3.89E-18 | Nightingale | S_HDL_C | NA | A | G | 0.052394 | 0.005323 | 3.50E-23 |
| rs532436 | miR-195-5p | trans | 3.89E-18 | Nightingale | S_LDL_C_p | NA | A | G | 0.075576 | 0.005352 | 2.10E-46 |

| | | | | | | | | | | | |
|---|---|---|---|---|---|---|---|---|---|---|---|
| rs532436 | miR-195-5p | trans | 3.89E-18 | Nightingale | LDL_FC | NA | A | G | 0.062671 | 0.005275 | 1.40E-32 |
| rs532436 | miR-195-5p | trans | 3.89E-18 | Nightingale | XS_VLDL_C | NA | A | G | 0.040138 | 0.005031 | 1.40E-14 |
| rs532436 | miR-195-5p | trans | 3.89E-18 | Nightingale | XS_VLDL_P | NA | A | G | 0.044567 | 0.005257 | 3.00E-19 |
| rs532436 | miR-195-5p | trans | 3.89E-18 | Nightingale | LDL_L | NA | A | G | 0.059463 | 0.005314 | 9.50E-30 |
| rs532436 | miR-195-5p | trans | 3.89E-18 | Nightingale | Clinical_LD | NA | A | G | 0.064426 | 0.005301 | 1.80E-34 |
| rs532436 | miR-195-5p | trans | 3.89E-18 | Nightingale | Citrate | NA | A | G | 0.031277 | 0.005297 | 1.60E-09 |
| rs532436 | miR-195-5p | trans | 3.89E-18 | Nightingale | XS_VLDL_P | NA | A | G | 0.05371 | 0.005261 | 1.20E-26 |
| rs532436 | miR-195-5p | trans | 3.89E-18 | Nightingale | LDL_CE | NA | A | G | 0.058166 | 0.005318 | 1.20E-28 |
| rs532436 | miR-195-5p | trans | 3.89E-18 | Nightingale | Remnant_( | NA | A | G | 0.058377 | 0.005297 | 5.50E-30 |
| rs532436 | miR-195-5p | trans | 3.89E-18 | Nightingale | Cholines | NA | A | G | 0.035678 | 0.005014 | 1.10E-13 |
| rs532436 | miR-195-5p | trans | 3.89E-18 | Nightingale | HDL_size | NA | A | G | -0.03226 | 0.004727 | 1.70E-12 |
| rs532436 | miR-195-5p | trans | 3.89E-18 | Nightingale | S_HDL_L | NA | A | G | 0.043965 | 0.005313 | 2.20E-17 |
| rs532436 | miR-195-5p | trans | 3.89E-18 | Nightingale | S_VLDL_FC | NA | A | G | 0.059408 | 0.005333 | 3.00E-30 |
| rs532436 | miR-195-5p | trans | 3.89E-18 | Nightingale | XXL_VLDL_ | NA | A | G | 0.046201 | 0.005312 | 1.30E-18 |
| rs532436 | miR-195-5p | trans | 3.89E-18 | Nightingale | XS_VLDL_C | NA | A | G | 0.057562 | 0.005175 | 2.50E-30 |
| rs532436 | miR-195-5p | trans | 3.89E-18 | Nightingale | M_VLDL_F | NA | A | G | 0.067258 | 0.005073 | 1.70E-39 |
| rs532436 | miR-195-5p | trans | 3.89E-18 | Nightingale | IDL_TG_pc | NA | A | G | -0.03868 | 0.005155 | 3.10E-13 |
| rs532436 | miR-195-5p | trans | 3.89E-18 | Nightingale | XS_VLDL_P | NA | A | G | -0.02951 | 0.005203 | 5.00E-08 |
| rs532436 | miR-195-5p | trans | 3.89E-18 | Nightingale | M_VLDL_L | NA | A | G | 0.032097 | 0.005296 | 1.10E-10 |
| rs532436 | miR-195-5p | trans | 3.89E-18 | Nightingale | LA | NA | A | G | 0.042219 | 0.005229 | 3.30E-17 |
| rs532436 | miR-195-5p | trans | 3.89E-18 | Nightingale | M_VLDL_C | NA | A | G | 0.06605 | 0.005283 | 1.00E-36 |
| rs532436 | miR-195-5p | trans | 3.89E-18 | Nightingale | Total_P | NA | A | G | 0.036654 | 0.005018 | 5.50E-14 |
| rs532436 | miR-195-5p | trans | 3.89E-18 | Nightingale | Total_L | NA | A | G | 0.042255 | 0.005245 | 4.90E-17 |
| rs532436 | miR-195-5p | trans | 3.89E-18 | Nightingale | M_LDL_TG | NA | A | G | -0.0531 | 0.005266 | 1.10E-23 |
| rs532436 | miR-195-5p | trans | 3.89E-18 | Nightingale | L_LDL_P | NA | A | G | 0.050357 | 0.005314 | 6.40E-22 |
| rs532436 | miR-195-5p | trans | 3.89E-18 | Nightingale | S_LDL_CE | NA | A | G | 0.051567 | 0.005334 | 4.80E-23 |
| rs532436 | miR-195-5p | trans | 3.89E-18 | Nightingale | S_LDL_TG_ | NA | A | G | -0.04355 | 0.00517 | 3.00E-16 |
| rs532436 | miR-195-5p | trans | 3.89E-18 | Nightingale | M_LDL_L | NA | A | G | 0.057299 | 0.005339 | 8.90E-28 |
| rs532436 | miR-195-5p | trans | 3.89E-18 | Nightingale | IDL_FC_pct | NA | A | G | 0.041213 | 0.00529 | 2.10E-14 |
| rs532436 | miR-195-5p | trans | 3.89E-18 | Nightingale | Phosphogly | NA | A | G | 0.031891 | 0.00505 | 3.60E-11 |
| rs532436 | miR-195-5p | trans | 3.89E-18 | Nightingale | XL_VLDL_F | NA | A | G | 0.056773 | 0.005169 | 2.60E-28 |
| rs532436 | miR-195-5p | trans | 3.89E-18 | Nightingale | S_HDL_FC | NA | A | G | 0.041762 | 0.005234 | 2.40E-16 |
| rs532436 | miR-195-5p | trans | 3.89E-18 | Nightingale | IDL_FC | NA | A | G | 0.063966 | 0.005162 | 7.70E-36 |
| rs532436 | miR-195-5p | trans | 3.89E-18 | Nightingale | S_VLDL_PL | NA | A | G | 0.050032 | 0.005322 | 1.30E-22 |
| rs532436 | miR-195-5p | trans | 3.89E-18 | Nightingale | IDL_C_pct | NA | A | G | 0.037343 | 0.005209 | 6.30E-12 |
| rs532436 | miR-195-5p | trans | 3.89E-18 | Nightingale | L_VLDL_CE | NA | A | G | 0.074326 | 0.005102 | 1.30E-48 |
| rs532436 | miR-195-5p | trans | 3.89E-18 | Nightingale | L_LDL_CE | NA | A | G | 0.058358 | 0.005284 | 5.40E-29 |
| rs532436 | miR-195-5p | trans | 3.89E-18 | Nightingale | S_VLDL_CE | NA | A | G | 0.076497 | 0.00526 | 2.10E-47 |
| rs532436 | miR-195-5p | trans | 3.89E-18 | Nightingale | XL_HDL_CE | NA | A | G | 0.030066 | 0.005327 | 4.20E-08 |
| rs532436 | miR-195-5p | trans | 3.89E-18 | Nightingale | XS_VLDL_F | NA | A | G | 0.053623 | 0.005239 | 1.10E-26 |
| rs532436 | miR-195-5p | trans | 3.89E-18 | Nightingale | M_LDL_C_ | NA | A | G | 0.047449 | 0.005336 | 4.30E-19 |
| rs532436 | miR-195-5p | trans | 3.89E-18 | Nightingale | S_VLDL_P | NA | A | G | 0.033337 | 0.005265 | 7.40E-12 |
| rs532436 | miR-195-5p | trans | 3.89E-18 | Nightingale | S_VLDL_FC | NA | A | G | 0.05247 | 0.005095 | 3.60E-24 |
| rs532436 | miR-195-5p | trans | 3.89E-18 | Nightingale | L_VLDL_TG | NA | A | G | -0.0795 | 0.005307 | 2.00E-52 |
| rs532436 | miR-195-5p | trans | 3.89E-18 | Nightingale | GlycA | NA | A | G | 0.031177 | 0.005309 | 6.60E-09 |
| rs532436 | miR-195-5p | trans | 3.89E-18 | Nightingale | L_VLDL_CE | NA | A | G | 0.031425 | 0.005269 | 1.40E-10 |
| rs532436 | miR-195-5p | trans | 3.89E-18 | Nightingale | M_VLDL_C | NA | A | G | 0.061586 | 0.005317 | 2.10E-32 |
| rs532436 | miR-195-5p | trans | 3.89E-18 | Nightingale | IDL_L | NA | A | G | 0.059551 | 0.005155 | 1.70E-31 |
| rs532436 | miR-195-5p | trans | 3.89E-18 | Nightingale | IDL_CE | NA | A | G | 0.058228 | 0.005148 | 7.40E-30 |
| rs532436 | miR-195-5p | trans | 3.89E-18 | Nightingale | S_VLDL_PL | NA | A | G | 0.049979 | 0.005105 | 5.70E-22 |
| rs532436 | miR-195-5p | trans | 3.89E-18 | Nightingale | M_LDL_CE | NA | A | G | 0.055488 | 0.005339 | 2.20E-26 |
| rs532436 | miR-195-5p | trans | 3.89E-18 | Nightingale | XL_HDL_C_ | NA | A | G | 0.038356 | 0.005092 | 1.70E-13 |
| rs532436 | miR-195-5p | trans | 3.89E-18 | Nightingale | VLDL_C | NA | A | G | 0.045684 | 0.005324 | 2.90E-19 |
| rs532436 | miR-195-5p | trans | 3.89E-18 | Nightingale | IDL_C | NA | A | G | 0.060082 | 0.005149 | 9.90E-32 |
| rs532436 | miR-195-5p | trans | 3.89E-18 | Nightingale | S_VLDL_TG | NA | A | G | -0.06518 | 0.005161 | 4.40E-36 |
| rs532436 | miR-195-5p | trans | 3.89E-18 | Nightingale | ApoB_by_A | NA | A | G | 0.039971 | 0.005251 | 1.30E-14 |
| rs532436 | miR-195-5p | trans | 3.89E-18 | Nightingale | VLDL_P | NA | A | G | 0.038195 | 0.005303 | 2.40E-14 |
| rs532436 | miR-195-5p | trans | 3.89E-18 | Nightingale | Total_CE | NA | A | G | 0.05558 | 0.005143 | 3.40E-28 |
| rs532436 | miR-195-5p | trans | 3.89E-18 | Nightingale | L_LDL_FC_ | NA | A | G | 0.030877 | 0.005087 | 1.20E-08 |
| rs532436 | miR-195-5p | trans | 3.89E-18 | Nightingale | VLDL_CE | NA | A | G | 0.053545 | 0.005333 | 2.70E-25 |
| rs532436 | miR-195-5p | trans | 3.89E-18 | Nightingale | M_VLDL_C | NA | A | G | 0.055885 | 0.005009 | 5.70E-28 |
| rs532436 | miR-195-5p | trans | 3.89E-18 | Nightingale | XXL_VLDL_ | NA | A | G | -0.05817 | 0.005381 | 5.80E-28 |
| rs532436 | miR-195-5p | trans | 3.89E-18 | Nightingale | IDL_PL | NA | A | G | 0.05875 | 0.005145 | 3.70E-31 |

| | | | | | | | | | | | |
|---|---|---|---|---|---|---|---|---|---|---|---|
| rs532436 | miR-195-5p | trans | 3.89E-18 | Nightingale | S_HDL_PL_ | NA | A | G | -0.03259 | 0.005155 | 1.00E-09 |
| rs532436 | miR-195-5p | trans | 3.89E-18 | Nightingale | S_LDL_PL | NA | A | G | 0.041297 | 0.00532 | 6.80E-15 |
| rs532436 | miR-195-5p | trans | 3.89E-18 | Nightingale | non_HDL_( | NA | A | G | 0.060373 | 0.005303 | 3.40E-31 |
| rs532436 | miR-195-5p | trans | 3.89E-18 | Nightingale | M_VLDL_P | NA | A | G | 0.044715 | 0.005324 | 1.90E-18 |
| rs532436 | miR-195-5p | trans | 3.89E-18 | Nightingale | Sphingomy | NA | A | G | 0.047389 | 0.004984 | 3.70E-21 |
| rs532436 | miR-195-5p | trans | 3.89E-18 | Nightingale | LDL_PL | NA | A | G | 0.06187 | 0.005319 | 7.90E-32 |
| rs532436 | miR-195-5p | trans | 3.89E-18 | Nightingale | L_VLDL_PL | NA | A | G | 0.027951 | 0.005179 | 3.30E-09 |
| rs532436 | miR-195-5p | trans | 3.89E-18 | Nightingale | S_HDL_PL | NA | A | G | 0.037551 | 0.005286 | 1.80E-13 |
| rs532436 | miR-195-5p | trans | 3.89E-18 | Nightingale | XS_VLDL_C | NA | A | G | 0.043168 | 0.005057 | 1.10E-16 |
| rs532436 | miR-195-5p | trans | 3.89E-18 | Nightingale | IDL_P | NA | A | G | 0.06498 | 0.005276 | 1.90E-35 |
| rs532436 | miR-195-5p | trans | 3.89E-18 | Nightingale | S_HDL_CE | NA | A | G | 0.053255 | 0.00534 | 1.20E-23 |
| rs532436 | miR-195-5p | trans | 3.89E-18 | Nightingale | L_LDL_FC | NA | A | G | 0.062609 | 0.005236 | 3.80E-33 |
| rs532436 | miR-195-5p | trans | 3.89E-18 | Nightingale | PUFA | NA | A | G | 0.036816 | 0.005201 | 1.50E-13 |
| rs532436 | miR-195-5p | trans | 3.89E-18 | Nightingale | XS_VLDL_C | NA | A | G | 0.058331 | 0.005145 | 3.40E-31 |
| rs532436 | miR-195-5p | trans | 3.89E-18 | Nightingale | ApoB | NA | A | G | 0.055646 | 0.005325 | 1.40E-26 |
| rs532436 | miR-195-5p | trans | 3.89E-18 | Nightingale | M_VLDL_F | NA | A | G | 0.051674 | 0.005331 | 1.50E-23 |
| rs532436 | miR-195-5p | trans | 3.89E-18 | Nightingale | S_LDL_C | NA | A | G | 0.053873 | 0.005337 | 1.50E-24 |
| rs532436 | miR-195-5p | trans | 3.89E-18 | Nightingale | XL_VLDL_C | NA | A | G | 0.062531 | 0.005113 | 1.30E-33 |
| rs532436 | miR-195-5p | trans | 3.89E-18 | Nightingale | XXL_VLDL_ | NA | A | G | 0.055922 | 0.005375 | 1.40E-25 |
| rs532436 | miR-195-5p | trans | 3.89E-18 | Nightingale | Omega_6 | NA | A | G | 0.0417 | 0.005207 | 5.30E-17 |
| rs532436 | miR-195-5p | trans | 3.89E-18 | Nightingale | LDL_P | NA | A | G | 0.053144 | 0.005328 | 2.90E-24 |
| rs532436 | miR-195-5p | trans | 3.89E-18 | Nightingale | XXL_VLDL_ | NA | A | G | 0.056832 | 0.005346 | 1.50E-26 |
| rs532436 | miR-195-5p | trans | 3.89E-18 | Nightingale | Pyruvate | NA | A | G | 0.034156 | 0.00534 | 1.60E-10 |
| rs532436 | miR-195-5p | trans | 3.89E-18 | Nightingale | Total_PL | NA | A | G | 0.03947 | 0.005077 | 4.30E-16 |
| rs532436 | miR-195-5p | trans | 3.89E-18 | Nightingale | L_LDL_TG_ | NA | A | G | -0.04149 | 0.00523 | 8.10E-15 |
| rs532436 | miR-195-5p | trans | 3.89E-18 | Nightingale | XL_VLDL_T | NA | A | G | -0.06894 | 0.005168 | 1.90E-40 |
| rs532436 | miR-195-5p | trans | 3.89E-18 | Nightingale | XS_VLDL_L | NA | A | G | 0.048489 | 0.005241 | 1.70E-22 |
| rs532436 | miR-195-5p | trans | 3.89E-18 | Nightingale | HDL_P | NA | A | G | 0.028757 | 0.005007 | 3.30E-09 |
| rs532436 | miR-195-5p | trans | 3.89E-18 | Nightingale | S_VLDL_C_ | NA | A | G | 0.070418 | 0.005189 | 1.30E-41 |
| rs532436 | miR-195-5p | trans | 3.89E-18 | Nightingale | Phosphatic | NA | A | G | 0.031132 | 0.004996 | 6.60E-11 |
| rs532436 | miR-195-5p | trans | 3.89E-18 | Nightingale | XXL_VLDL_ | NA | A | G | 0.036801 | 0.00537 | 1.70E-13 |
| rs532436 | miR-195-5p | trans | 3.89E-18 | Nightingale | Total_FC | NA | A | G | 0.054936 | 0.005209 | 1.00E-27 |
| rs532436 | miR-195-5p | trans | 3.89E-18 | Nightingale | S_LDL_PL_ | NA | A | G | -0.04468 | 0.005322 | 1.60E-19 |
| rs532436 | miR-195-5p | trans | 3.89E-18 | Nightingale | M_VLDL_T | NA | A | G | -0.06283 | 0.005038 | 7.40E-35 |
| rs532436 | miR-195-5p | trans | 3.89E-18 | Nightingale | LDL_C | NA | A | G | 0.060001 | 0.005309 | 3.10E-30 |
| rs532436 | miR-195-5p | trans | 3.89E-18 | Nightingale | Total_C | NA | A | G | 0.055716 | 0.005162 | 2.80E-28 |
| rs532436 | miR-195-5p | trans | 3.89E-18 | Nightingale | S_VLDL_CE | NA | A | G | 0.057936 | 0.005325 | 1.60E-29 |
| rs532436 | miR-195-5p | trans | 3.89E-18 | Nightingale | S_HDL_P | NA | A | G | 0.055157 | 0.005332 | 1.70E-25 |
| rs532436 | miR-195-5p | trans | 3.89E-18 | Nightingale | L_LDL_PL | NA | A | G | 0.066479 | 0.005291 | 5.80E-37 |
| rs532436 | miR-195-5p | trans | 3.89E-18 | Nightingale | M_LDL_FC | NA | A | G | 0.063598 | 0.005325 | 4.40E-33 |
| rs532436 | miR-195-5p | trans | 3.89E-18 | Nightingale | XL_HDL_PL | NA | A | G | -0.04102 | 0.004899 | 1.70E-17 |
| rs532436 | miR-195-5p | trans | 3.89E-18 | Nightingale | M_LDL_C | NA | A | G | 0.05878 | 0.005341 | 5.20E-29 |
| rs532436 | miR-195-5p | trans | 3.89E-18 | Nightingale | M_VLDL_P | NA | A | G | 0.043187 | 0.005319 | 2.10E-17 |
| rs532436 | miR-195-5p | trans | 3.89E-18 | Nightingale | XL_VLDL_P | NA | A | G | 0.055667 | 0.00534 | 6.50E-27 |
| rs532436 | miR-195-5p | trans | 3.89E-18 | Nightingale | VLDL_FC | NA | A | G | 0.033528 | 0.005293 | 1.30E-11 |
| rs532436 | miR-195-5p | trans | 3.89E-18 | Nightingale | L_VLDL_C_ | NA | A | G | 0.08172 | 0.005208 | 2.50E-56 |
| rs532436 | miR-195-5p | trans | 3.89E-18 | Nightingale | XL_VLDL_C | NA | A | G | 0.029577 | 0.005249 | 1.10E-09 |
| rs532436 | miR-195-5p | trans | 3.89E-18 | Nightingale | XS_VLDL_F | NA | A | G | 0.050786 | 0.005294 | 9.70E-21 |
| rs532436 | miR-195-5p | trans | 3.89E-18 | Nightingale | XL_HDL_PL | NA | A | G | -0.03014 | 0.004796 | 6.90E-11 |
| rs543040 | miR-195-5p | trans | 2.56E-17 | Nightingale | IDL_CE | NA | T | A | 0.03344 | 0.00428 | 1.60E-14 |
| rs543040 | miR-195-5p | trans | 2.56E-17 | Nightingale | M_VLDL_T | NA | T | A | -0.03835 | 0.004189 | 1.20E-19 |
| rs543040 | miR-195-5p | trans | 2.56E-17 | Nightingale | M_LDL_C | NA | T | A | 0.034069 | 0.004441 | 2.10E-14 |
| rs543040 | miR-195-5p | trans | 2.56E-17 | Nightingale | S_VLDL_L | NA | T | A | 0.024881 | 0.00439 | 3.10E-09 |
| rs543040 | miR-195-5p | trans | 2.56E-17 | Nightingale | GlycA | NA | T | A | 0.028404 | 0.004414 | 8.90E-11 |
| rs543040 | miR-195-5p | trans | 2.56E-17 | Nightingale | S_LDL_L | NA | T | A | 0.027837 | 0.004432 | 7.80E-10 |
| rs543040 | miR-195-5p | trans | 2.56E-17 | Nightingale | S_VLDL_TG | NA | T | A | -0.03792 | 0.004291 | 2.60E-18 |
| rs543040 | miR-195-5p | trans | 2.56E-17 | Nightingale | S_VLDL_CE | NA | T | A | 0.038284 | 0.004427 | 1.60E-18 |
| rs543040 | miR-195-5p | trans | 2.56E-17 | Nightingale | Total_FC | NA | T | A | 0.033359 | 0.004331 | 6.10E-15 |
| rs543040 | miR-195-5p | trans | 2.56E-17 | Nightingale | L_LDL_FC | NA | T | A | 0.033578 | 0.004353 | 3.40E-14 |
| rs543040 | miR-195-5p | trans | 2.56E-17 | Nightingale | L_LDL_PL | NA | T | A | 0.03798 | 0.004399 | 1.10E-17 |
| rs543040 | miR-195-5p | trans | 2.56E-17 | Nightingale | M_LDL_CE | NA | T | A | 0.033102 | 0.004439 | 8.90E-14 |
| rs543040 | miR-195-5p | trans | 2.56E-17 | Nightingale | LA | NA | T | A | 0.025008 | 0.004347 | 3.40E-09 |
| rs543040 | miR-195-5p | trans | 2.56E-17 | Nightingale | XS_VLDL_P | NA | T | A | 0.032244 | 0.004371 | 4.80E-14 |

| | | | | | | | | | | | |
|---|---|---|---|---|---|---|---|---|---|---|---|
| rs543040 | miR-195-5p | trans | 2.56E-17 | Nightingale | M_VLDL_F( | NA | T | A | 0.033543 | 0.004432 | 2.70E-14 |
| rs543040 | miR-195-5p | trans | 2.56E-17 | Nightingale | S_HDL_CE | NA | T | A | 0.028568 | 0.00444 | 1.60E-10 |
| rs543040 | miR-195-5p | trans | 2.56E-17 | Nightingale | S_LDL_CE_ | NA | T | A | 0.031287 | 0.004425 | 3.30E-14 |
| rs543040 | miR-195-5p | trans | 2.56E-17 | Nightingale | S_VLDL_P | NA | T | A | 0.023677 | 0.004377 | 1.40E-08 |
| rs543040 | miR-195-5p | trans | 2.56E-17 | Nightingale | IDL_P | NA | T | A | 0.039934 | 0.004386 | 1.70E-19 |
| rs543040 | miR-195-5p | trans | 2.56E-17 | Nightingale | XXL_VLDL_ | NA | T | A | 0.030507 | 0.004414 | 2.00E-12 |
| rs543040 | miR-195-5p | trans | 2.56E-17 | Nightingale | M_VLDL_C | NA | T | A | 0.040846 | 0.004393 | 2.70E-20 |
| rs543040 | miR-195-5p | trans | 2.56E-17 | Nightingale | S_LDL_CE | NA | T | A | 0.030692 | 0.004435 | 5.20E-12 |
| rs543040 | miR-195-5p | trans | 2.56E-17 | Nightingale | VLDL_P | NA | T | A | 0.02687 | 0.004409 | 3.40E-10 |
| rs543040 | miR-195-5p | trans | 2.56E-17 | Nightingale | S_VLDL_C_ | NA | T | A | 0.041674 | 0.004314 | 1.10E-21 |
| rs543040 | miR-195-5p | trans | 2.56E-17 | Nightingale | M_LDL_TG | NA | T | A | -0.02749 | 0.004378 | 4.30E-10 |
| rs543040 | miR-195-5p | trans | 2.56E-17 | Nightingale | Omega_6 | NA | T | A | 0.025996 | 0.004329 | 5.10E-10 |
| rs543040 | miR-195-5p | trans | 2.56E-17 | Nightingale | L_VLDL_FC | NA | T | A | 0.048505 | 0.004391 | 8.10E-29 |
| rs543040 | miR-195-5p | trans | 2.56E-17 | Nightingale | Cholines | NA | T | A | 0.022428 | 0.004169 | 4.00E-08 |
| rs543040 | miR-195-5p | trans | 2.56E-17 | Nightingale | XL_VLDL_C | NA | T | A | 0.037056 | 0.004253 | 6.40E-18 |
| rs543040 | miR-195-5p | trans | 2.56E-17 | Nightingale | M_VLDL_P | NA | T | A | 0.029334 | 0.004426 | 1.70E-11 |
| rs543040 | miR-195-5p | trans | 2.56E-17 | Nightingale | XL_VLDL_P | NA | T | A | 0.03891 | 0.00444 | 5.80E-19 |
| rs543040 | miR-195-5p | trans | 2.56E-17 | Nightingale | Total_CE | NA | T | A | 0.031773 | 0.004276 | 9.70E-14 |
| rs543040 | miR-195-5p | trans | 2.56E-17 | Nightingale | LDL_PL | NA | T | A | 0.035004 | 0.004423 | 5.10E-15 |
| rs543040 | miR-195-5p | trans | 2.56E-17 | Nightingale | ApoB_by_A | NA | T | A | 0.025114 | 0.004366 | 1.30E-08 |
| rs543040 | miR-195-5p | trans | 2.56E-17 | Nightingale | XL_HDL_PL | NA | T | A | -0.02279 | 0.004073 | 1.20E-08 |
| rs543040 | miR-195-5p | trans | 2.56E-17 | Nightingale | PUFA | NA | T | A | 0.023843 | 0.004324 | 1.30E-08 |
| rs543040 | miR-195-5p | trans | 2.56E-17 | Nightingale | L_LDL_P | NA | T | A | 0.02926 | 0.004418 | 6.00E-11 |
| rs543040 | miR-195-5p | trans | 2.56E-17 | Nightingale | XS_VLDL_C | NA | T | A | 0.037825 | 0.004278 | 2.30E-18 |
| rs543040 | miR-195-5p | trans | 2.56E-17 | Nightingale | L_VLDL_CE | NA | T | A | 0.047001 | 0.004242 | 7.50E-29 |
| rs543040 | miR-195-5p | trans | 2.56E-17 | Nightingale | XXL_VLDL_ | NA | T | A | 0.024559 | 0.004462 | 3.20E-09 |
| rs543040 | miR-195-5p | trans | 2.56E-17 | Nightingale | L_VLDL_C_ | NA | T | A | 0.051898 | 0.00433 | 1.30E-33 |
| rs543040 | miR-195-5p | trans | 2.56E-17 | Nightingale | S_HDL_P | NA | T | A | 0.030617 | 0.004434 | 5.60E-12 |
| rs543040 | miR-195-5p | trans | 2.56E-17 | Nightingale | M_VLDL_C | NA | T | A | 0.033408 | 0.004165 | 3.20E-15 |
| rs543040 | miR-195-5p | trans | 2.56E-17 | Nightingale | LDL_L | NA | T | A | 0.033354 | 0.004418 | 7.50E-14 |
| rs543040 | miR-195-5p | trans | 2.56E-17 | Nightingale | XXL_VLDL_ | NA | T | A | 0.034686 | 0.004443 | 4.10E-15 |
| rs543040 | miR-195-5p | trans | 2.56E-17 | Nightingale | M_LDL_FC | NA | T | A | 0.034292 | 0.004427 | 2.40E-14 |
| rs543040 | miR-195-5p | trans | 2.56E-17 | Nightingale | S_VLDL_C | NA | T | A | 0.038477 | 0.004432 | 1.80E-18 |
| rs543040 | miR-195-5p | trans | 2.56E-17 | Nightingale | Citrate | NA | T | A | 0.027118 | 0.004404 | 5.00E-10 |
| rs543040 | miR-195-5p | trans | 2.56E-17 | Nightingale | IDL_PL | NA | T | A | 0.035357 | 0.004278 | 3.60E-16 |
| rs543040 | miR-195-5p | trans | 2.56E-17 | Nightingale | S_HDL_C | NA | T | A | 0.028373 | 0.004426 | 2.00E-10 |
| rs543040 | miR-195-5p | trans | 2.56E-17 | Nightingale | Sphingomy | NA | T | A | 0.029085 | 0.004144 | 6.00E-12 |
| rs543040 | miR-195-5p | trans | 2.56E-17 | Nightingale | non_HDL_( | NA | T | A | 0.035777 | 0.004409 | 5.00E-16 |
| rs543040 | miR-195-5p | trans | 2.56E-17 | Nightingale | M_LDL_L | NA | T | A | 0.033119 | 0.004439 | 9.90E-14 |
| rs543040 | miR-195-5p | trans | 2.56E-17 | Nightingale | Total_C | NA | T | A | 0.032423 | 0.004291 | 3.20E-14 |
| rs543040 | miR-195-5p | trans | 2.56E-17 | Nightingale | L_LDL_CE | NA | T | A | 0.03161 | 0.004393 | 1.00E-12 |
| rs543040 | miR-195-5p | trans | 2.56E-17 | Nightingale | M_VLDL_C | NA | T | A | 0.038795 | 0.004421 | 1.90E-18 |
| rs543040 | miR-195-5p | trans | 2.56E-17 | Nightingale | S_VLDL_FC | NA | T | A | 0.029186 | 0.004236 | 1.50E-11 |
| rs543040 | miR-195-5p | trans | 2.56E-17 | Nightingale | S_LDL_C_p | NA | T | A | 0.045363 | 0.00445 | 7.00E-25 |
| rs543040 | miR-195-5p | trans | 2.56E-17 | Nightingale | LDL_CE | NA | T | A | 0.032631 | 0.004421 | 2.10E-13 |
| rs543040 | miR-195-5p | trans | 2.56E-17 | Nightingale | XXL_VLDL_ | NA | T | A | 0.033103 | 0.004467 | 1.00E-13 |
| rs543040 | miR-195-5p | trans | 2.56E-17 | Nightingale | S_LDL_P | NA | T | A | 0.029725 | 0.004437 | 2.90E-11 |
| rs543040 | miR-195-5p | trans | 2.56E-17 | Nightingale | IDL_L | NA | T | A | 0.035244 | 0.004286 | 5.60E-16 |
| rs543040 | miR-195-5p | trans | 2.56E-17 | Nightingale | S_HDL_FC | NA | T | A | 0.023503 | 0.004352 | 4.50E-08 |
| rs543040 | miR-195-5p | trans | 2.56E-17 | Nightingale | S_HDL_PL | NA | T | A | 0.02389 | 0.004395 | 3.00E-08 |
| rs543040 | miR-195-5p | trans | 2.56E-17 | Nightingale | L_LDL_L | NA | T | A | 0.033574 | 0.00439 | 3.20E-14 |
| rs543040 | miR-195-5p | trans | 2.56E-17 | Nightingale | S_VLDL_CE | NA | T | A | 0.046815 | 0.004373 | 2.90E-26 |
| rs543040 | miR-195-5p | trans | 2.56E-17 | Nightingale | LDL_C | NA | T | A | 0.033236 | 0.004414 | 9.10E-14 |
| rs543040 | miR-195-5p | trans | 2.56E-17 | Nightingale | S_LDL_C | NA | T | A | 0.03102 | 0.004437 | 5.40E-12 |
| rs543040 | miR-195-5p | trans | 2.56E-17 | Nightingale | M_VLDL_P | NA | T | A | 0.02852 | 0.004422 | 5.10E-11 |
| rs543040 | miR-195-5p | trans | 2.56E-17 | Nightingale | L_LDL_C | NA | T | A | 0.032339 | 0.004383 | 2.70E-13 |
| rs543040 | miR-195-5p | trans | 2.56E-17 | Nightingale | M_LDL_PL | NA | T | A | 0.032515 | 0.004439 | 2.70E-13 |
| rs543040 | miR-195-5p | trans | 2.56E-17 | Nightingale | IDL_C | NA | T | A | 0.034894 | 0.004281 | 1.20E-15 |
| rs543040 | miR-195-5p | trans | 2.56E-17 | Nightingale | M_VLDL_C | NA | T | A | 0.035992 | 0.004176 | 2.10E-17 |
| rs543040 | miR-195-5p | trans | 2.56E-17 | Nightingale | LDL_FC | NA | T | A | 0.033577 | 0.004386 | 6.30E-14 |
| rs543040 | miR-195-5p | trans | 2.56E-17 | Nightingale | XXL_VLDL_ | NA | T | A | -0.0356 | 0.004471 | 3.10E-16 |
| rs543040 | miR-195-5p | trans | 2.56E-17 | Nightingale | M_LDL_C_ | NA | T | A | 0.031281 | 0.004436 | 2.60E-12 |
| rs543040 | miR-195-5p | trans | 2.56E-17 | Nightingale | L_VLDL_TG | NA | T | A | -0.05098 | 0.004412 | 1.20E-31 |

| | | | | | | | | | | | |
|---|---|---|---|---|---|---|---|---|---|---|---|
| rs543040 | miR-195-5p | trans | 2.56E-17 | Nightingale | XS_VLDL_F | NA | T | A | 0.031254 | 0.004401 | 3.50E-12 |
| rs543040 | miR-195-5p | trans | 2.56E-17 | Nightingale | S_LDL_FC | NA | T | A | 0.028463 | 0.004426 | 5.20E-10 |
| rs543040 | miR-195-5p | trans | 2.56E-17 | Nightingale | M_VLDL_P | NA | T | A | 0.043865 | 0.004254 | 1.10E-24 |
| rs543040 | miR-195-5p | trans | 2.56E-17 | Nightingale | S_LDL_PL_ | NA | T | A | -0.03037 | 0.004425 | 1.90E-13 |
| rs543040 | miR-195-5p | trans | 2.56E-17 | Nightingale | XS_VLDL_L | NA | T | A | 0.033935 | 0.004358 | 1.80E-15 |
| rs543040 | miR-195-5p | trans | 2.56E-17 | Nightingale | VLDL_CE | NA | T | A | 0.035975 | 0.004434 | 2.50E-16 |
| rs543040 | miR-195-5p | trans | 2.56E-17 | Nightingale | VLDL_FC | NA | T | A | 0.024444 | 0.004401 | 8.20E-09 |
| rs543040 | miR-195-5p | trans | 2.56E-17 | Nightingale | S_VLDL_PL | NA | T | A | 0.032933 | 0.004425 | 3.70E-14 |
| rs543040 | miR-195-5p | trans | 2.56E-17 | Nightingale | M_VLDL_F | NA | T | A | 0.041872 | 0.004217 | 6.40E-23 |
| rs543040 | miR-195-5p | trans | 2.56E-17 | Nightingale | VLDL_C | NA | T | A | 0.031535 | 0.004427 | 3.80E-13 |
| rs543040 | miR-195-5p | trans | 2.56E-17 | Nightingale | S_HDL_L | NA | T | A | 0.026372 | 0.004418 | 1.50E-09 |
| rs543040 | miR-195-5p | trans | 2.56E-17 | Nightingale | Clinical_LD | NA | T | A | 0.036464 | 0.004407 | 3.10E-16 |
| rs543040 | miR-195-5p | trans | 2.56E-17 | Nightingale | XL_VLDL_C | NA | T | A | 0.037947 | 0.004251 | 9.30E-19 |
| rs543040 | miR-195-5p | trans | 2.56E-17 | Nightingale | M_LDL_P | NA | T | A | 0.036025 | 0.004445 | 6.20E-16 |
| rs543040 | miR-195-5p | trans | 2.56E-17 | Nightingale | IDL_FC | NA | T | A | 0.038215 | 0.004292 | 1.80E-18 |
| rs543040 | miR-195-5p | trans | 2.56E-17 | Nightingale | Total_PL | NA | T | A | 0.025082 | 0.004221 | 8.60E-10 |
| rs543040 | miR-195-5p | trans | 2.56E-17 | Nightingale | S_VLDL_FC | NA | T | A | 0.038005 | 0.004434 | 7.30E-18 |
| rs543040 | miR-195-5p | trans | 2.56E-17 | Nightingale | IDL_FC_pct | NA | T | A | 0.025728 | 0.004399 | 1.20E-08 |
| rs543040 | miR-195-5p | trans | 2.56E-17 | Nightingale | XS_VLDL_F | NA | T | A | 0.037149 | 0.004356 | 6.40E-18 |
| rs543040 | miR-195-5p | trans | 2.56E-17 | Nightingale | XL_VLDL_F | NA | T | A | 0.036996 | 0.004298 | 3.40E-18 |
| rs543040 | miR-195-5p | trans | 2.56E-17 | Nightingale | Remnant_( | NA | T | A | 0.036984 | 0.004404 | 3.40E-17 |
| rs543040 | miR-195-5p | trans | 2.56E-17 | Nightingale | XL_VLDL_T | NA | T | A | -0.04371 | 0.004297 | 3.50E-24 |
| rs543040 | miR-195-5p | trans | 2.56E-17 | Nightingale | Total_L | NA | T | A | 0.02656 | 0.004361 | 3.50E-10 |
| rs543040 | miR-195-5p | trans | 2.56E-17 | Nightingale | XS_VLDL_C | NA | T | A | 0.038037 | 0.004303 | 1.40E-18 |
| rs543040 | miR-195-5p | trans | 2.56E-17 | Nightingale | XS_VLDL_P | NA | T | A | 0.036804 | 0.004374 | 1.30E-17 |
| rs543040 | miR-195-5p | trans | 2.56E-17 | Nightingale | S_VLDL_PL | NA | T | A | 0.02757 | 0.004245 | 2.50E-10 |
| rs543040 | miR-195-5p | trans | 2.56E-17 | Nightingale | LDL_P | NA | T | A | 0.031905 | 0.00443 | 9.70E-13 |
| rs543040 | miR-195-5p | trans | 2.56E-17 | Nightingale | ApoB | NA | T | A | 0.034032 | 0.004427 | 2.10E-14 |
| rs543040 | miR-139-5p | trans | 2.94E-41 | Nightingale | IDL_CE | NA | T | A | 0.03344 | 0.00428 | 1.60E-14 |
| rs543040 | miR-139-5p | trans | 2.94E-41 | Nightingale | M_VLDL_T | NA | T | A | -0.03835 | 0.004189 | 1.20E-19 |
| rs543040 | miR-139-5p | trans | 2.94E-41 | Nightingale | M_LDL_C | NA | T | A | 0.034069 | 0.004441 | 2.10E-14 |
| rs543040 | miR-139-5p | trans | 2.94E-41 | Nightingale | S_VLDL_L | NA | T | A | 0.024881 | 0.00439 | 3.10E-09 |
| rs543040 | miR-139-5p | trans | 2.94E-41 | Nightingale | GlycA | NA | T | A | 0.028404 | 0.004414 | 8.90E-11 |
| rs543040 | miR-139-5p | trans | 2.94E-41 | Nightingale | S_LDL_L | NA | T | A | 0.027837 | 0.004432 | 7.80E-10 |
| rs543040 | miR-139-5p | trans | 2.94E-41 | Nightingale | S_VLDL_TG | NA | T | A | -0.03792 | 0.004291 | 2.60E-18 |
| rs543040 | miR-139-5p | trans | 2.94E-41 | Nightingale | S_VLDL_CE | NA | T | A | 0.038284 | 0.004427 | 1.60E-18 |
| rs543040 | miR-139-5p | trans | 2.94E-41 | Nightingale | Total_FC | NA | T | A | 0.033359 | 0.004331 | 6.10E-15 |
| rs543040 | miR-139-5p | trans | 2.94E-41 | Nightingale | L_LDL_FC | NA | T | A | 0.033578 | 0.004353 | 3.40E-14 |
| rs543040 | miR-139-5p | trans | 2.94E-41 | Nightingale | L_LDL_PL | NA | T | A | 0.03798 | 0.004399 | 1.10E-17 |
| rs543040 | miR-139-5p | trans | 2.94E-41 | Nightingale | M_LDL_CE | NA | T | A | 0.033102 | 0.004439 | 8.90E-14 |
| rs543040 | miR-139-5p | trans | 2.94E-41 | Nightingale | LA | NA | T | A | 0.025008 | 0.004347 | 3.40E-09 |
| rs543040 | miR-139-5p | trans | 2.94E-41 | Nightingale | XS_VLDL_P | NA | T | A | 0.032244 | 0.004371 | 4.80E-14 |
| rs543040 | miR-139-5p | trans | 2.94E-41 | Nightingale | M_VLDL_F | NA | T | A | 0.033543 | 0.004432 | 2.70E-14 |
| rs543040 | miR-139-5p | trans | 2.94E-41 | Nightingale | S_HDL_CE | NA | T | A | 0.028568 | 0.00444 | 1.60E-10 |
| rs543040 | miR-139-5p | trans | 2.94E-41 | Nightingale | S_LDL_CE_ | NA | T | A | 0.031287 | 0.004425 | 3.30E-14 |
| rs543040 | miR-139-5p | trans | 2.94E-41 | Nightingale | S_VLDL_P | NA | T | A | 0.023677 | 0.004377 | 1.40E-08 |
| rs543040 | miR-139-5p | trans | 2.94E-41 | Nightingale | IDL_P | NA | T | A | 0.039934 | 0.004386 | 1.70E-19 |
| rs543040 | miR-139-5p | trans | 2.94E-41 | Nightingale | XXL_VLDL_ | NA | T | A | 0.030507 | 0.004414 | 2.00E-12 |
| rs543040 | miR-139-5p | trans | 2.94E-41 | Nightingale | M_VLDL_C | NA | T | A | 0.040846 | 0.004393 | 2.70E-20 |
| rs543040 | miR-139-5p | trans | 2.94E-41 | Nightingale | S_LDL_CE | NA | T | A | 0.030692 | 0.004435 | 5.20E-12 |
| rs543040 | miR-139-5p | trans | 2.94E-41 | Nightingale | VLDL_P | NA | T | A | 0.02687 | 0.004409 | 3.40E-10 |
| rs543040 | miR-139-5p | trans | 2.94E-41 | Nightingale | S_VLDL_C_ | NA | T | A | 0.041674 | 0.004314 | 1.10E-21 |
| rs543040 | miR-139-5p | trans | 2.94E-41 | Nightingale | M_LDL_TG | NA | T | A | -0.02749 | 0.004378 | 4.30E-10 |
| rs543040 | miR-139-5p | trans | 2.94E-41 | Nightingale | Omega_6 | NA | T | A | 0.025996 | 0.004329 | 5.10E-10 |
| rs543040 | miR-139-5p | trans | 2.94E-41 | Nightingale | L_VLDL_FC | NA | T | A | 0.048505 | 0.004391 | 8.10E-29 |
| rs543040 | miR-139-5p | trans | 2.94E-41 | Nightingale | Cholines | NA | T | A | 0.022428 | 0.004169 | 4.00E-08 |
| rs543040 | miR-139-5p | trans | 2.94E-41 | Nightingale | XL_VLDL_C | NA | T | A | 0.037056 | 0.004253 | 6.40E-18 |
| rs543040 | miR-139-5p | trans | 2.94E-41 | Nightingale | M_VLDL_P | NA | T | A | 0.029334 | 0.004426 | 1.70E-11 |
| rs543040 | miR-139-5p | trans | 2.94E-41 | Nightingale | XL_VLDL_P | NA | T | A | 0.03891 | 0.00444 | 5.80E-19 |
| rs543040 | miR-139-5p | trans | 2.94E-41 | Nightingale | Total_CE | NA | T | A | 0.031773 | 0.004276 | 9.70E-14 |
| rs543040 | miR-139-5p | trans | 2.94E-41 | Nightingale | LDL_PL | NA | T | A | 0.035004 | 0.004423 | 5.10E-15 |
| rs543040 | miR-139-5p | trans | 2.94E-41 | Nightingale | ApoB_by_A | NA | T | A | 0.025114 | 0.004366 | 1.30E-08 |
| rs543040 | miR-139-5p | trans | 2.94E-41 | Nightingale | XL_HDL_PL | NA | T | A | -0.02279 | 0.004073 | 1.20E-08 |

| | | | | | | | | | | | |
|---|---|---|---|---|---|---|---|---|---|---|---|
| rs543040 | miR-139-5p | trans | 2.94E-41 | Nightingale | PUFA | NA | T | A | 0.023843 | 0.004324 | 1.30E-08 |
| rs543040 | miR-139-5p | trans | 2.94E-41 | Nightingale | L_LDL_P | NA | T | A | 0.02926 | 0.004418 | 6.00E-11 |
| rs543040 | miR-139-5p | trans | 2.94E-41 | Nightingale | XS_VLDL_C | NA | T | A | 0.037825 | 0.004278 | 2.30E-18 |
| rs543040 | miR-139-5p | trans | 2.94E-41 | Nightingale | L_VLDL_CE | NA | T | A | 0.047001 | 0.004242 | 7.50E-29 |
| rs543040 | miR-139-5p | trans | 2.94E-41 | Nightingale | XXL_VLDL_ | NA | T | A | 0.024559 | 0.004462 | 3.20E-09 |
| rs543040 | miR-139-5p | trans | 2.94E-41 | Nightingale | L_VLDL_C_ | NA | T | A | 0.051898 | 0.00433 | 1.30E-33 |
| rs543040 | miR-139-5p | trans | 2.94E-41 | Nightingale | S_HDL_P | NA | T | A | 0.030617 | 0.004434 | 5.60E-12 |
| rs543040 | miR-139-5p | trans | 2.94E-41 | Nightingale | M_VLDL_C | NA | T | A | 0.033408 | 0.004165 | 3.20E-15 |
| rs543040 | miR-139-5p | trans | 2.94E-41 | Nightingale | LDL_L | NA | T | A | 0.033354 | 0.004418 | 7.50E-14 |
| rs543040 | miR-139-5p | trans | 2.94E-41 | Nightingale | XXL_VLDL_ | NA | T | A | 0.034686 | 0.004443 | 4.10E-15 |
| rs543040 | miR-139-5p | trans | 2.94E-41 | Nightingale | M_LDL_FC | NA | T | A | 0.034292 | 0.004427 | 2.40E-14 |
| rs543040 | miR-139-5p | trans | 2.94E-41 | Nightingale | S_VLDL_C | NA | T | A | 0.038477 | 0.004432 | 1.80E-18 |
| rs543040 | miR-139-5p | trans | 2.94E-41 | Nightingale | Citrate | NA | T | A | 0.027118 | 0.004404 | 5.00E-10 |
| rs543040 | miR-139-5p | trans | 2.94E-41 | Nightingale | IDL_PL | NA | T | A | 0.035357 | 0.004278 | 3.60E-16 |
| rs543040 | miR-139-5p | trans | 2.94E-41 | Nightingale | S_HDL_C | NA | T | A | 0.028373 | 0.004426 | 2.00E-10 |
| rs543040 | miR-139-5p | trans | 2.94E-41 | Nightingale | Sphingomy | NA | T | A | 0.029085 | 0.004144 | 6.00E-12 |
| rs543040 | miR-139-5p | trans | 2.94E-41 | Nightingale | non_HDL_( | NA | T | A | 0.035777 | 0.004409 | 5.00E-16 |
| rs543040 | miR-139-5p | trans | 2.94E-41 | Nightingale | M_LDL_L | NA | T | A | 0.033119 | 0.004439 | 9.90E-14 |
| rs543040 | miR-139-5p | trans | 2.94E-41 | Nightingale | Total_C | NA | T | A | 0.032423 | 0.004291 | 3.20E-14 |
| rs543040 | miR-139-5p | trans | 2.94E-41 | Nightingale | L_LDL_CE | NA | T | A | 0.03161 | 0.004393 | 1.00E-12 |
| rs543040 | miR-139-5p | trans | 2.94E-41 | Nightingale | M_VLDL_C | NA | T | A | 0.038795 | 0.004421 | 1.90E-18 |
| rs543040 | miR-139-5p | trans | 2.94E-41 | Nightingale | S_VLDL_FC | NA | T | A | 0.029186 | 0.004236 | 1.50E-11 |
| rs543040 | miR-139-5p | trans | 2.94E-41 | Nightingale | S_LDL_C_p | NA | T | A | 0.045363 | 0.00445 | 7.00E-25 |
| rs543040 | miR-139-5p | trans | 2.94E-41 | Nightingale | LDL_CE | NA | T | A | 0.032631 | 0.004421 | 2.10E-13 |
| rs543040 | miR-139-5p | trans | 2.94E-41 | Nightingale | XXL_VLDL_ | NA | T | A | 0.033103 | 0.004467 | 1.00E-13 |
| rs543040 | miR-139-5p | trans | 2.94E-41 | Nightingale | S_LDL_P | NA | T | A | 0.029725 | 0.004437 | 2.90E-11 |
| rs543040 | miR-139-5p | trans | 2.94E-41 | Nightingale | IDL_L | NA | T | A | 0.035244 | 0.004286 | 5.60E-16 |
| rs543040 | miR-139-5p | trans | 2.94E-41 | Nightingale | S_HDL_FC | NA | T | A | 0.023503 | 0.004352 | 4.50E-08 |
| rs543040 | miR-139-5p | trans | 2.94E-41 | Nightingale | S_HDL_PL | NA | T | A | 0.02389 | 0.004395 | 3.00E-08 |
| rs543040 | miR-139-5p | trans | 2.94E-41 | Nightingale | L_LDL_L | NA | T | A | 0.033574 | 0.00439 | 3.20E-14 |
| rs543040 | miR-139-5p | trans | 2.94E-41 | Nightingale | S_VLDL_CE | NA | T | A | 0.046815 | 0.004373 | 2.90E-26 |
| rs543040 | miR-139-5p | trans | 2.94E-41 | Nightingale | LDL_C | NA | T | A | 0.033236 | 0.004414 | 9.10E-14 |
| rs543040 | miR-139-5p | trans | 2.94E-41 | Nightingale | S_LDL_C | NA | T | A | 0.03102 | 0.004437 | 5.40E-12 |
| rs543040 | miR-139-5p | trans | 2.94E-41 | Nightingale | M_VLDL_P | NA | T | A | 0.02852 | 0.004422 | 5.10E-11 |
| rs543040 | miR-139-5p | trans | 2.94E-41 | Nightingale | L_LDL_C | NA | T | A | 0.032339 | 0.004383 | 2.70E-13 |
| rs543040 | miR-139-5p | trans | 2.94E-41 | Nightingale | M_LDL_PL | NA | T | A | 0.032515 | 0.004439 | 2.70E-13 |
| rs543040 | miR-139-5p | trans | 2.94E-41 | Nightingale | IDL_C | NA | T | A | 0.034894 | 0.004281 | 1.20E-15 |
| rs543040 | miR-139-5p | trans | 2.94E-41 | Nightingale | M_VLDL_C | NA | T | A | 0.035992 | 0.004176 | 2.10E-17 |
| rs543040 | miR-139-5p | trans | 2.94E-41 | Nightingale | LDL_FC | NA | T | A | 0.033577 | 0.004386 | 6.30E-14 |
| rs543040 | miR-139-5p | trans | 2.94E-41 | Nightingale | XXL_VLDL_ | NA | T | A | -0.0356 | 0.004471 | 3.10E-16 |
| rs543040 | miR-139-5p | trans | 2.94E-41 | Nightingale | M_LDL_C_ | NA | T | A | 0.031281 | 0.004436 | 2.60E-12 |
| rs543040 | miR-139-5p | trans | 2.94E-41 | Nightingale | L_VLDL_TG | NA | T | A | -0.05098 | 0.004412 | 1.20E-31 |
| rs543040 | miR-139-5p | trans | 2.94E-41 | Nightingale | XS_VLDL_F | NA | T | A | 0.031254 | 0.004401 | 3.50E-12 |
| rs543040 | miR-139-5p | trans | 2.94E-41 | Nightingale | S_LDL_FC | NA | T | A | 0.028463 | 0.004426 | 5.20E-10 |
| rs543040 | miR-139-5p | trans | 2.94E-41 | Nightingale | M_VLDL_P | NA | T | A | 0.043865 | 0.004254 | 1.10E-24 |
| rs543040 | miR-139-5p | trans | 2.94E-41 | Nightingale | S_LDL_PL_ | NA | T | A | -0.03037 | 0.004425 | 1.90E-13 |
| rs543040 | miR-139-5p | trans | 2.94E-41 | Nightingale | XS_VLDL_L | NA | T | A | 0.033935 | 0.004358 | 1.80E-15 |
| rs543040 | miR-139-5p | trans | 2.94E-41 | Nightingale | VLDL_CE | NA | T | A | 0.035975 | 0.004434 | 2.50E-16 |
| rs543040 | miR-139-5p | trans | 2.94E-41 | Nightingale | VLDL_FC | NA | T | A | 0.024444 | 0.004401 | 8.20E-09 |
| rs543040 | miR-139-5p | trans | 2.94E-41 | Nightingale | S_VLDL_PL | NA | T | A | 0.032933 | 0.004425 | 3.70E-14 |
| rs543040 | miR-139-5p | trans | 2.94E-41 | Nightingale | M_VLDL_F | NA | T | A | 0.041872 | 0.004217 | 6.40E-23 |
| rs543040 | miR-139-5p | trans | 2.94E-41 | Nightingale | VLDL_C | NA | T | A | 0.031535 | 0.004427 | 3.80E-13 |
| rs543040 | miR-139-5p | trans | 2.94E-41 | Nightingale | S_HDL_L | NA | T | A | 0.026372 | 0.004418 | 1.50E-09 |
| rs543040 | miR-139-5p | trans | 2.94E-41 | Nightingale | Clinical_LD | NA | T | A | 0.036464 | 0.004407 | 3.10E-16 |
| rs543040 | miR-139-5p | trans | 2.94E-41 | Nightingale | XL_VLDL_C | NA | T | A | 0.037947 | 0.004251 | 9.30E-19 |
| rs543040 | miR-139-5p | trans | 2.94E-41 | Nightingale | M_LDL_P | NA | T | A | 0.036025 | 0.004445 | 6.20E-16 |
| rs543040 | miR-139-5p | trans | 2.94E-41 | Nightingale | IDL_FC | NA | T | A | 0.038215 | 0.004292 | 1.80E-18 |
| rs543040 | miR-139-5p | trans | 2.94E-41 | Nightingale | Total_PL | NA | T | A | 0.025082 | 0.004221 | 8.60E-10 |
| rs543040 | miR-139-5p | trans | 2.94E-41 | Nightingale | S_VLDL_FC | NA | T | A | 0.038005 | 0.004434 | 7.30E-18 |
| rs543040 | miR-139-5p | trans | 2.94E-41 | Nightingale | IDL_FC_pct | NA | T | A | 0.025728 | 0.004399 | 1.20E-08 |
| rs543040 | miR-139-5p | trans | 2.94E-41 | Nightingale | XS_VLDL_F | NA | T | A | 0.037149 | 0.004356 | 6.40E-18 |
| rs543040 | miR-139-5p | trans | 2.94E-41 | Nightingale | XL_VLDL_F | NA | T | A | 0.036996 | 0.004298 | 3.40E-18 |
| rs543040 | miR-139-5p | trans | 2.94E-41 | Nightingale | Remnant_( | NA | T | A | 0.036984 | 0.004404 | 3.40E-17 |

| | | | | | | | | | | | |
|---|---|---|---|---|---|---|---|---|---|---|---|
| rs543040 | miR-139-5p | trans | 2.94E-41 | Nightingale | XL_VLDL_T | NA | T | A | -0.04371 | 0.004297 | 3.50E-24 |
| rs543040 | miR-139-5p | trans | 2.94E-41 | Nightingale | Total_L | NA | T | A | 0.02656 | 0.004361 | 3.50E-10 |
| rs543040 | miR-139-5p | trans | 2.94E-41 | Nightingale | XS_VLDL_C | NA | T | A | 0.038037 | 0.004303 | 1.40E-18 |
| rs543040 | miR-139-5p | trans | 2.94E-41 | Nightingale | XS_VLDL_P | NA | T | A | 0.036804 | 0.004374 | 1.30E-17 |
| rs543040 | miR-139-5p | trans | 2.94E-41 | Nightingale | S_VLDL_PL | NA | T | A | 0.02757 | 0.004245 | 2.50E-10 |
| rs543040 | miR-139-5p | trans | 2.94E-41 | Nightingale | LDL_P | NA | T | A | 0.031905 | 0.00443 | 9.70E-13 |
| rs543040 | miR-139-5p | trans | 2.94E-41 | Nightingale | ApoB | NA | T | A | 0.034032 | 0.004427 | 2.10E-14 |
| rs543040 | miR-10b-5p | trans | 7.68E-35 | Nightingale | IDL_CE | NA | T | A | 0.03344 | 0.00428 | 1.60E-14 |
| rs543040 | miR-10b-5p | trans | 7.68E-35 | Nightingale | M_VLDL_T | NA | T | A | -0.03835 | 0.004189 | 1.20E-19 |
| rs543040 | miR-10b-5p | trans | 7.68E-35 | Nightingale | M_LDL_C | NA | T | A | 0.034069 | 0.004441 | 2.10E-14 |
| rs543040 | miR-10b-5p | trans | 7.68E-35 | Nightingale | S_VLDL_L | NA | T | A | 0.024881 | 0.00439 | 3.10E-09 |
| rs543040 | miR-10b-5p | trans | 7.68E-35 | Nightingale | GlycA | NA | T | A | 0.028404 | 0.004414 | 8.90E-11 |
| rs543040 | miR-10b-5p | trans | 7.68E-35 | Nightingale | S_LDL_L | NA | T | A | 0.027837 | 0.004432 | 7.80E-10 |
| rs543040 | miR-10b-5p | trans | 7.68E-35 | Nightingale | S_VLDL_TG | NA | T | A | -0.03792 | 0.004291 | 2.60E-18 |
| rs543040 | miR-10b-5p | trans | 7.68E-35 | Nightingale | S_VLDL_CE | NA | T | A | 0.038284 | 0.004427 | 1.60E-18 |
| rs543040 | miR-10b-5p | trans | 7.68E-35 | Nightingale | Total_FC | NA | T | A | 0.033359 | 0.004331 | 6.10E-15 |
| rs543040 | miR-10b-5p | trans | 7.68E-35 | Nightingale | L_LDL_FC | NA | T | A | 0.033578 | 0.004353 | 3.40E-14 |
| rs543040 | miR-10b-5p | trans | 7.68E-35 | Nightingale | L_LDL_PL | NA | T | A | 0.03798 | 0.004399 | 1.10E-17 |
| rs543040 | miR-10b-5p | trans | 7.68E-35 | Nightingale | M_LDL_CE | NA | T | A | 0.033102 | 0.004439 | 8.90E-14 |
| rs543040 | miR-10b-5p | trans | 7.68E-35 | Nightingale | LA | NA | T | A | 0.025008 | 0.004347 | 3.40E-09 |
| rs543040 | miR-10b-5p | trans | 7.68E-35 | Nightingale | XS_VLDL_P | NA | T | A | 0.032244 | 0.004371 | 4.80E-14 |
| rs543040 | miR-10b-5p | trans | 7.68E-35 | Nightingale | M_VLDL_F | NA | T | A | 0.033543 | 0.004432 | 2.70E-14 |
| rs543040 | miR-10b-5p | trans | 7.68E-35 | Nightingale | S_HDL_CE | NA | T | A | 0.028568 | 0.00444 | 1.60E-10 |
| rs543040 | miR-10b-5p | trans | 7.68E-35 | Nightingale | S_LDL_CE_ | NA | T | A | 0.031287 | 0.004425 | 3.30E-14 |
| rs543040 | miR-10b-5p | trans | 7.68E-35 | Nightingale | S_VLDL_P | NA | T | A | 0.023677 | 0.004377 | 1.40E-08 |
| rs543040 | miR-10b-5p | trans | 7.68E-35 | Nightingale | IDL_P | NA | T | A | 0.039934 | 0.004386 | 1.70E-19 |
| rs543040 | miR-10b-5p | trans | 7.68E-35 | Nightingale | XXL_VLDL_ | NA | T | A | 0.030507 | 0.004414 | 2.00E-12 |
| rs543040 | miR-10b-5p | trans | 7.68E-35 | Nightingale | M_VLDL_C | NA | T | A | 0.040846 | 0.004393 | 2.70E-20 |
| rs543040 | miR-10b-5p | trans | 7.68E-35 | Nightingale | S_LDL_CE | NA | T | A | 0.030692 | 0.004435 | 5.20E-12 |
| rs543040 | miR-10b-5p | trans | 7.68E-35 | Nightingale | VLDL_P | NA | T | A | 0.02687 | 0.004409 | 3.40E-10 |
| rs543040 | miR-10b-5p | trans | 7.68E-35 | Nightingale | S_VLDL_C_ | NA | T | A | 0.041674 | 0.004314 | 1.10E-21 |
| rs543040 | miR-10b-5p | trans | 7.68E-35 | Nightingale | M_LDL_TG | NA | T | A | -0.02749 | 0.004378 | 4.30E-10 |
| rs543040 | miR-10b-5p | trans | 7.68E-35 | Nightingale | Omega_6 | NA | T | A | 0.025996 | 0.004329 | 5.10E-10 |
| rs543040 | miR-10b-5p | trans | 7.68E-35 | Nightingale | L_VLDL_FC | NA | T | A | 0.048505 | 0.004391 | 8.10E-29 |
| rs543040 | miR-10b-5p | trans | 7.68E-35 | Nightingale | Cholines | NA | T | A | 0.022428 | 0.004169 | 4.00E-08 |
| rs543040 | miR-10b-5p | trans | 7.68E-35 | Nightingale | XL_VLDL_C | NA | T | A | 0.037056 | 0.004253 | 6.40E-18 |
| rs543040 | miR-10b-5p | trans | 7.68E-35 | Nightingale | M_VLDL_P | NA | T | A | 0.029334 | 0.004426 | 1.70E-11 |
| rs543040 | miR-10b-5p | trans | 7.68E-35 | Nightingale | XL_VLDL_P | NA | T | A | 0.03891 | 0.00444 | 5.80E-19 |
| rs543040 | miR-10b-5p | trans | 7.68E-35 | Nightingale | Total_CE | NA | T | A | 0.031773 | 0.004276 | 9.70E-14 |
| rs543040 | miR-10b-5p | trans | 7.68E-35 | Nightingale | LDL_PL | NA | T | A | 0.035004 | 0.004423 | 5.10E-15 |
| rs543040 | miR-10b-5p | trans | 7.68E-35 | Nightingale | ApoB_by_A | NA | T | A | 0.025114 | 0.004366 | 1.30E-08 |
| rs543040 | miR-10b-5p | trans | 7.68E-35 | Nightingale | XL_HDL_PL | NA | T | A | -0.02279 | 0.004073 | 1.20E-08 |
| rs543040 | miR-10b-5p | trans | 7.68E-35 | Nightingale | PUFA | NA | T | A | 0.023843 | 0.004324 | 1.30E-08 |
| rs543040 | miR-10b-5p | trans | 7.68E-35 | Nightingale | L_LDL_P | NA | T | A | 0.02926 | 0.004418 | 6.00E-11 |
| rs543040 | miR-10b-5p | trans | 7.68E-35 | Nightingale | XS_VLDL_C | NA | T | A | 0.037825 | 0.004278 | 2.30E-18 |
| rs543040 | miR-10b-5p | trans | 7.68E-35 | Nightingale | L_VLDL_CE | NA | T | A | 0.047001 | 0.004242 | 7.50E-29 |
| rs543040 | miR-10b-5p | trans | 7.68E-35 | Nightingale | XXL_VLDL_ | NA | T | A | 0.024559 | 0.004462 | 3.20E-09 |
| rs543040 | miR-10b-5p | trans | 7.68E-35 | Nightingale | L_VLDL_C_ | NA | T | A | 0.051898 | 0.00433 | 1.30E-33 |
| rs543040 | miR-10b-5p | trans | 7.68E-35 | Nightingale | S_HDL_P | NA | T | A | 0.030617 | 0.004434 | 5.60E-12 |
| rs543040 | miR-10b-5p | trans | 7.68E-35 | Nightingale | M_VLDL_C | NA | T | A | 0.033408 | 0.004165 | 3.20E-15 |
| rs543040 | miR-10b-5p | trans | 7.68E-35 | Nightingale | LDL_L | NA | T | A | 0.033354 | 0.004418 | 7.50E-14 |
| rs543040 | miR-10b-5p | trans | 7.68E-35 | Nightingale | XXL_VLDL_ | NA | T | A | 0.034686 | 0.004443 | 4.10E-15 |
| rs543040 | miR-10b-5p | trans | 7.68E-35 | Nightingale | M_LDL_FC | NA | T | A | 0.034292 | 0.004427 | 2.40E-14 |
| rs543040 | miR-10b-5p | trans | 7.68E-35 | Nightingale | S_VLDL_C | NA | T | A | 0.038477 | 0.004432 | 1.80E-18 |
| rs543040 | miR-10b-5p | trans | 7.68E-35 | Nightingale | Citrate | NA | T | A | 0.027118 | 0.004404 | 5.00E-10 |
| rs543040 | miR-10b-5p | trans | 7.68E-35 | Nightingale | IDL_PL | NA | T | A | 0.035357 | 0.004278 | 3.60E-16 |
| rs543040 | miR-10b-5p | trans | 7.68E-35 | Nightingale | S_HDL_C | NA | T | A | 0.028373 | 0.004426 | 2.00E-10 |
| rs543040 | miR-10b-5p | trans | 7.68E-35 | Nightingale | Sphingomy | NA | T | A | 0.029085 | 0.004144 | 6.00E-12 |
| rs543040 | miR-10b-5p | trans | 7.68E-35 | Nightingale | non_HDL_( | NA | T | A | 0.035777 | 0.004409 | 5.00E-16 |
| rs543040 | miR-10b-5p | trans | 7.68E-35 | Nightingale | M_LDL_L | NA | T | A | 0.033119 | 0.004439 | 9.90E-14 |
| rs543040 | miR-10b-5p | trans | 7.68E-35 | Nightingale | Total_C | NA | T | A | 0.032423 | 0.004291 | 3.20E-14 |
| rs543040 | miR-10b-5p | trans | 7.68E-35 | Nightingale | L_LDL_CE | NA | T | A | 0.03161 | 0.004393 | 1.00E-12 |
| rs543040 | miR-10b-5p | trans | 7.68E-35 | Nightingale | M_VLDL_C | NA | T | A | 0.038795 | 0.004421 | 1.90E-18 |

| | | | | | | | | | | | |
|---|---|---|---|---|---|---|---|---|---|---|---|
| rs543040 | miR-10b-5p | trans | 7.68E-35 | Nightingale | S_VLDL_FC | NA | T | A | 0.029186 | 0.004236 | 1.50E-11 |
| rs543040 | miR-10b-5p | trans | 7.68E-35 | Nightingale | S_LDL_C_p | NA | T | A | 0.045363 | 0.00445 | 7.00E-25 |
| rs543040 | miR-10b-5p | trans | 7.68E-35 | Nightingale | LDL_CE | NA | T | A | 0.032631 | 0.004421 | 2.10E-13 |
| rs543040 | miR-10b-5p | trans | 7.68E-35 | Nightingale | XXL_VLDL_ | NA | T | A | 0.033103 | 0.004467 | 1.00E-13 |
| rs543040 | miR-10b-5p | trans | 7.68E-35 | Nightingale | S_LDL_P | NA | T | A | 0.029725 | 0.004437 | 2.90E-11 |
| rs543040 | miR-10b-5p | trans | 7.68E-35 | Nightingale | IDL_L | NA | T | A | 0.035244 | 0.004286 | 5.60E-16 |
| rs543040 | miR-10b-5p | trans | 7.68E-35 | Nightingale | S_HDL_FC | NA | T | A | 0.023503 | 0.004352 | 4.50E-08 |
| rs543040 | miR-10b-5p | trans | 7.68E-35 | Nightingale | S_HDL_PL | NA | T | A | 0.02389 | 0.004395 | 3.00E-08 |
| rs543040 | miR-10b-5p | trans | 7.68E-35 | Nightingale | L_LDL_L | NA | T | A | 0.033574 | 0.00439 | 3.20E-14 |
| rs543040 | miR-10b-5p | trans | 7.68E-35 | Nightingale | S_VLDL_CE | NA | T | A | 0.046815 | 0.004373 | 2.90E-26 |
| rs543040 | miR-10b-5p | trans | 7.68E-35 | Nightingale | LDL_C | NA | T | A | 0.033236 | 0.004414 | 9.10E-14 |
| rs543040 | miR-10b-5p | trans | 7.68E-35 | Nightingale | S_LDL_C | NA | T | A | 0.03102 | 0.004437 | 5.40E-12 |
| rs543040 | miR-10b-5p | trans | 7.68E-35 | Nightingale | M_VLDL_P | NA | T | A | 0.02852 | 0.004422 | 5.10E-11 |
| rs543040 | miR-10b-5p | trans | 7.68E-35 | Nightingale | L_LDL_C | NA | T | A | 0.032339 | 0.004383 | 2.70E-13 |
| rs543040 | miR-10b-5p | trans | 7.68E-35 | Nightingale | M_LDL_PL | NA | T | A | 0.032515 | 0.004439 | 2.70E-13 |
| rs543040 | miR-10b-5p | trans | 7.68E-35 | Nightingale | IDL_C | NA | T | A | 0.034894 | 0.004281 | 1.20E-15 |
| rs543040 | miR-10b-5p | trans | 7.68E-35 | Nightingale | M_VLDL_C | NA | T | A | 0.035992 | 0.004176 | 2.10E-17 |
| rs543040 | miR-10b-5p | trans | 7.68E-35 | Nightingale | LDL_FC | NA | T | A | 0.033577 | 0.004386 | 6.30E-14 |
| rs543040 | miR-10b-5p | trans | 7.68E-35 | Nightingale | XXL_VLDL_ | NA | T | A | -0.0356 | 0.004471 | 3.10E-16 |
| rs543040 | miR-10b-5p | trans | 7.68E-35 | Nightingale | M_LDL_C_ | NA | T | A | 0.031281 | 0.004436 | 2.60E-12 |
| rs543040 | miR-10b-5p | trans | 7.68E-35 | Nightingale | L_VLDL_TG | NA | T | A | -0.05098 | 0.004412 | 1.20E-31 |
| rs543040 | miR-10b-5p | trans | 7.68E-35 | Nightingale | XS_VLDL_F | NA | T | A | 0.031254 | 0.004401 | 3.50E-12 |
| rs543040 | miR-10b-5p | trans | 7.68E-35 | Nightingale | S_LDL_FC | NA | T | A | 0.028463 | 0.004426 | 5.20E-10 |
| rs543040 | miR-10b-5p | trans | 7.68E-35 | Nightingale | M_VLDL_P | NA | T | A | 0.043865 | 0.004254 | 1.10E-24 |
| rs543040 | miR-10b-5p | trans | 7.68E-35 | Nightingale | S_LDL_PL_ | NA | T | A | -0.03037 | 0.004425 | 1.90E-13 |
| rs543040 | miR-10b-5p | trans | 7.68E-35 | Nightingale | XS_VLDL_L | NA | T | A | 0.033935 | 0.004358 | 1.80E-15 |
| rs543040 | miR-10b-5p | trans | 7.68E-35 | Nightingale | VLDL_CE | NA | T | A | 0.035975 | 0.004434 | 2.50E-16 |
| rs543040 | miR-10b-5p | trans | 7.68E-35 | Nightingale | VLDL_FC | NA | T | A | 0.024444 | 0.004401 | 8.20E-09 |
| rs543040 | miR-10b-5p | trans | 7.68E-35 | Nightingale | S_VLDL_PL | NA | T | A | 0.032933 | 0.004425 | 3.70E-14 |
| rs543040 | miR-10b-5p | trans | 7.68E-35 | Nightingale | M_VLDL_F | NA | T | A | 0.041872 | 0.004217 | 6.40E-23 |
| rs543040 | miR-10b-5p | trans | 7.68E-35 | Nightingale | VLDL_C | NA | T | A | 0.031535 | 0.004427 | 3.80E-13 |
| rs543040 | miR-10b-5p | trans | 7.68E-35 | Nightingale | S_HDL_L | NA | T | A | 0.026372 | 0.004418 | 1.50E-09 |
| rs543040 | miR-10b-5p | trans | 7.68E-35 | Nightingale | Clinical_LD | NA | T | A | 0.036464 | 0.004407 | 3.10E-16 |
| rs543040 | miR-10b-5p | trans | 7.68E-35 | Nightingale | XL_VLDL_C | NA | T | A | 0.037947 | 0.004251 | 9.30E-19 |
| rs543040 | miR-10b-5p | trans | 7.68E-35 | Nightingale | M_LDL_P | NA | T | A | 0.036025 | 0.004445 | 6.20E-16 |
| rs543040 | miR-10b-5p | trans | 7.68E-35 | Nightingale | IDL_FC | NA | T | A | 0.038215 | 0.004292 | 1.80E-18 |
| rs543040 | miR-10b-5p | trans | 7.68E-35 | Nightingale | Total_PL | NA | T | A | 0.025082 | 0.004221 | 8.60E-10 |
| rs543040 | miR-10b-5p | trans | 7.68E-35 | Nightingale | S_VLDL_FC | NA | T | A | 0.038005 | 0.004434 | 7.30E-18 |
| rs543040 | miR-10b-5p | trans | 7.68E-35 | Nightingale | IDL_FC_pct | NA | T | A | 0.025728 | 0.004399 | 1.20E-08 |
| rs543040 | miR-10b-5p | trans | 7.68E-35 | Nightingale | XS_VLDL_F | NA | T | A | 0.037149 | 0.004356 | 6.40E-18 |
| rs543040 | miR-10b-5p | trans | 7.68E-35 | Nightingale | XL_VLDL_F | NA | T | A | 0.036996 | 0.004298 | 3.40E-18 |
| rs543040 | miR-10b-5p | trans | 7.68E-35 | Nightingale | Remnant_( | NA | T | A | 0.036984 | 0.004404 | 3.40E-17 |
| rs543040 | miR-10b-5p | trans | 7.68E-35 | Nightingale | XL_VLDL_T | NA | T | A | -0.04371 | 0.004297 | 3.50E-24 |
| rs543040 | miR-10b-5p | trans | 7.68E-35 | Nightingale | Total_L | NA | T | A | 0.02656 | 0.004361 | 3.50E-10 |
| rs543040 | miR-10b-5p | trans | 7.68E-35 | Nightingale | XS_VLDL_C | NA | T | A | 0.038037 | 0.004303 | 1.40E-18 |
| rs543040 | miR-10b-5p | trans | 7.68E-35 | Nightingale | XS_VLDL_P | NA | T | A | 0.036804 | 0.004374 | 1.30E-17 |
| rs543040 | miR-10b-5p | trans | 7.68E-35 | Nightingale | S_VLDL_PL | NA | T | A | 0.02757 | 0.004245 | 2.50E-10 |
| rs543040 | miR-10b-5p | trans | 7.68E-35 | Nightingale | LDL_P | NA | T | A | 0.031905 | 0.00443 | 9.70E-13 |
| rs543040 | miR-10b-5p | trans | 7.68E-35 | Nightingale | ApoB | NA | T | A | 0.034032 | 0.004427 | 2.10E-14 |
| rs543040 | let-7b-5p | trans | 1.13E-26 | Nightingale | IDL_CE | NA | T | A | 0.03344 | 0.00428 | 1.60E-14 |
| rs543040 | let-7b-5p | trans | 1.13E-26 | Nightingale | M_VLDL_T | NA | T | A | -0.03835 | 0.004189 | 1.20E-19 |
| rs543040 | let-7b-5p | trans | 1.13E-26 | Nightingale | M_LDL_C | NA | T | A | 0.034069 | 0.004441 | 2.10E-14 |
| rs543040 | let-7b-5p | trans | 1.13E-26 | Nightingale | S_VLDL_L | NA | T | A | 0.024881 | 0.00439 | 3.10E-09 |
| rs543040 | let-7b-5p | trans | 1.13E-26 | Nightingale | GlycA | NA | T | A | 0.028404 | 0.004414 | 8.90E-11 |
| rs543040 | let-7b-5p | trans | 1.13E-26 | Nightingale | S_LDL_L | NA | T | A | 0.027837 | 0.004432 | 7.80E-10 |
| rs543040 | let-7b-5p | trans | 1.13E-26 | Nightingale | S_VLDL_TG | NA | T | A | -0.03792 | 0.004291 | 2.60E-18 |
| rs543040 | let-7b-5p | trans | 1.13E-26 | Nightingale | S_VLDL_CE | NA | T | A | 0.038284 | 0.004427 | 1.60E-18 |
| rs543040 | let-7b-5p | trans | 1.13E-26 | Nightingale | Total_FC | NA | T | A | 0.033359 | 0.004331 | 6.10E-15 |
| rs543040 | let-7b-5p | trans | 1.13E-26 | Nightingale | L_LDL_FC | NA | T | A | 0.033578 | 0.004353 | 3.40E-14 |
| rs543040 | let-7b-5p | trans | 1.13E-26 | Nightingale | L_LDL_PL | NA | T | A | 0.03798 | 0.004399 | 1.10E-17 |
| rs543040 | let-7b-5p | trans | 1.13E-26 | Nightingale | M_LDL_CE | NA | T | A | 0.033102 | 0.004439 | 8.90E-14 |
| rs543040 | let-7b-5p | trans | 1.13E-26 | Nightingale | LA | NA | T | A | 0.025008 | 0.004347 | 3.40E-09 |
| rs543040 | let-7b-5p | trans | 1.13E-26 | Nightingale | XS_VLDL_P | NA | T | A | 0.032244 | 0.004371 | 4.80E-14 |

| | | | | | | | | | | | |
|---|---|---|---|---|---|---|---|---|---|---|---|
| rs543040 | let-7b-5p | trans | 1.13E-26 | Nightingale | M_VLDL_F | NA | T | A | 0.033543 | 0.004432 | 2.70E-14 |
| rs543040 | let-7b-5p | trans | 1.13E-26 | Nightingale | S_HDL_CE | NA | T | A | 0.028568 | 0.00444 | 1.60E-10 |
| rs543040 | let-7b-5p | trans | 1.13E-26 | Nightingale | S_LDL_CE_ | NA | T | A | 0.031287 | 0.004425 | 3.30E-14 |
| rs543040 | let-7b-5p | trans | 1.13E-26 | Nightingale | S_VLDL_P | NA | T | A | 0.023677 | 0.004377 | 1.40E-08 |
| rs543040 | let-7b-5p | trans | 1.13E-26 | Nightingale | IDL_P | NA | T | A | 0.039934 | 0.004386 | 1.70E-19 |
| rs543040 | let-7b-5p | trans | 1.13E-26 | Nightingale | XXL_VLDL_ | NA | T | A | 0.030507 | 0.004414 | 2.00E-12 |
| rs543040 | let-7b-5p | trans | 1.13E-26 | Nightingale | M_VLDL_C | NA | T | A | 0.040846 | 0.004393 | 2.70E-20 |
| rs543040 | let-7b-5p | trans | 1.13E-26 | Nightingale | S_LDL_CE | NA | T | A | 0.030692 | 0.004435 | 5.20E-12 |
| rs543040 | let-7b-5p | trans | 1.13E-26 | Nightingale | VLDL_P | NA | T | A | 0.02687 | 0.004409 | 3.40E-10 |
| rs543040 | let-7b-5p | trans | 1.13E-26 | Nightingale | S_VLDL_C_ | NA | T | A | 0.041674 | 0.004314 | 1.10E-21 |
| rs543040 | let-7b-5p | trans | 1.13E-26 | Nightingale | M_LDL_TG | NA | T | A | -0.02749 | 0.004378 | 4.30E-10 |
| rs543040 | let-7b-5p | trans | 1.13E-26 | Nightingale | Omega_6 | NA | T | A | 0.025996 | 0.004329 | 5.10E-10 |
| rs543040 | let-7b-5p | trans | 1.13E-26 | Nightingale | L_VLDL_FC | NA | T | A | 0.048505 | 0.004391 | 8.10E-29 |
| rs543040 | let-7b-5p | trans | 1.13E-26 | Nightingale | Cholines | NA | T | A | 0.022428 | 0.004169 | 4.00E-08 |
| rs543040 | let-7b-5p | trans | 1.13E-26 | Nightingale | XL_VLDL_C | NA | T | A | 0.037056 | 0.004253 | 6.40E-18 |
| rs543040 | let-7b-5p | trans | 1.13E-26 | Nightingale | M_VLDL_P | NA | T | A | 0.029334 | 0.004426 | 1.70E-11 |
| rs543040 | let-7b-5p | trans | 1.13E-26 | Nightingale | XL_VLDL_P | NA | T | A | 0.03891 | 0.00444 | 5.80E-19 |
| rs543040 | let-7b-5p | trans | 1.13E-26 | Nightingale | Total_CE | NA | T | A | 0.031773 | 0.004276 | 9.70E-14 |
| rs543040 | let-7b-5p | trans | 1.13E-26 | Nightingale | LDL_PL | NA | T | A | 0.035004 | 0.004423 | 5.10E-15 |
| rs543040 | let-7b-5p | trans | 1.13E-26 | Nightingale | ApoB_by_A | NA | T | A | 0.025114 | 0.004366 | 1.30E-08 |
| rs543040 | let-7b-5p | trans | 1.13E-26 | Nightingale | XL_HDL_PL | NA | T | A | -0.02279 | 0.004073 | 1.20E-08 |
| rs543040 | let-7b-5p | trans | 1.13E-26 | Nightingale | PUFA | NA | T | A | 0.023843 | 0.004324 | 1.30E-08 |
| rs543040 | let-7b-5p | trans | 1.13E-26 | Nightingale | L_LDL_P | NA | T | A | 0.02926 | 0.004418 | 6.00E-11 |
| rs543040 | let-7b-5p | trans | 1.13E-26 | Nightingale | XS_VLDL_C | NA | T | A | 0.037825 | 0.004278 | 2.30E-18 |
| rs543040 | let-7b-5p | trans | 1.13E-26 | Nightingale | L_VLDL_CE | NA | T | A | 0.047001 | 0.004242 | 7.50E-29 |
| rs543040 | let-7b-5p | trans | 1.13E-26 | Nightingale | XXL_VLDL_ | NA | T | A | 0.024559 | 0.004462 | 3.20E-09 |
| rs543040 | let-7b-5p | trans | 1.13E-26 | Nightingale | L_VLDL_C_ | NA | T | A | 0.051898 | 0.00433 | 1.30E-33 |
| rs543040 | let-7b-5p | trans | 1.13E-26 | Nightingale | S_HDL_P | NA | T | A | 0.030617 | 0.004434 | 5.60E-12 |
| rs543040 | let-7b-5p | trans | 1.13E-26 | Nightingale | M_VLDL_C | NA | T | A | 0.033408 | 0.004165 | 3.20E-15 |
| rs543040 | let-7b-5p | trans | 1.13E-26 | Nightingale | LDL_L | NA | T | A | 0.033354 | 0.004418 | 7.50E-14 |
| rs543040 | let-7b-5p | trans | 1.13E-26 | Nightingale | XXL_VLDL_ | NA | T | A | 0.034686 | 0.004443 | 4.10E-15 |
| rs543040 | let-7b-5p | trans | 1.13E-26 | Nightingale | M_LDL_FC | NA | T | A | 0.034292 | 0.004427 | 2.40E-14 |
| rs543040 | let-7b-5p | trans | 1.13E-26 | Nightingale | S_VLDL_C | NA | T | A | 0.038477 | 0.004432 | 1.80E-18 |
| rs543040 | let-7b-5p | trans | 1.13E-26 | Nightingale | Citrate | NA | T | A | 0.027118 | 0.004404 | 5.00E-10 |
| rs543040 | let-7b-5p | trans | 1.13E-26 | Nightingale | IDL_PL | NA | T | A | 0.035357 | 0.004278 | 3.60E-16 |
| rs543040 | let-7b-5p | trans | 1.13E-26 | Nightingale | S_HDL_C | NA | T | A | 0.028373 | 0.004426 | 2.00E-10 |
| rs543040 | let-7b-5p | trans | 1.13E-26 | Nightingale | Sphingomy | NA | T | A | 0.029085 | 0.004144 | 6.00E-12 |
| rs543040 | let-7b-5p | trans | 1.13E-26 | Nightingale | non_HDL_( | NA | T | A | 0.035777 | 0.004409 | 5.00E-16 |
| rs543040 | let-7b-5p | trans | 1.13E-26 | Nightingale | M_LDL_L | NA | T | A | 0.033119 | 0.004439 | 9.90E-14 |
| rs543040 | let-7b-5p | trans | 1.13E-26 | Nightingale | Total_C | NA | T | A | 0.032423 | 0.004291 | 3.20E-14 |
| rs543040 | let-7b-5p | trans | 1.13E-26 | Nightingale | L_LDL_CE | NA | T | A | 0.03161 | 0.004393 | 1.00E-12 |
| rs543040 | let-7b-5p | trans | 1.13E-26 | Nightingale | M_VLDL_C | NA | T | A | 0.038795 | 0.004421 | 1.90E-18 |
| rs543040 | let-7b-5p | trans | 1.13E-26 | Nightingale | S_VLDL_FC | NA | T | A | 0.029186 | 0.004236 | 1.50E-11 |
| rs543040 | let-7b-5p | trans | 1.13E-26 | Nightingale | S_LDL_C_p | NA | T | A | 0.045363 | 0.00445 | 7.00E-25 |
| rs543040 | let-7b-5p | trans | 1.13E-26 | Nightingale | LDL_CE | NA | T | A | 0.032631 | 0.004421 | 2.10E-13 |
| rs543040 | let-7b-5p | trans | 1.13E-26 | Nightingale | XXL_VLDL_ | NA | T | A | 0.033103 | 0.004467 | 1.00E-13 |
| rs543040 | let-7b-5p | trans | 1.13E-26 | Nightingale | S_LDL_P | NA | T | A | 0.029725 | 0.004437 | 2.90E-11 |
| rs543040 | let-7b-5p | trans | 1.13E-26 | Nightingale | IDL_L | NA | T | A | 0.035244 | 0.004286 | 5.60E-16 |
| rs543040 | let-7b-5p | trans | 1.13E-26 | Nightingale | S_HDL_FC | NA | T | A | 0.023503 | 0.004352 | 4.50E-08 |
| rs543040 | let-7b-5p | trans | 1.13E-26 | Nightingale | S_HDL_PL | NA | T | A | 0.02389 | 0.004395 | 3.00E-08 |
| rs543040 | let-7b-5p | trans | 1.13E-26 | Nightingale | L_LDL_L | NA | T | A | 0.033574 | 0.00439 | 3.20E-14 |
| rs543040 | let-7b-5p | trans | 1.13E-26 | Nightingale | S_VLDL_CE | NA | T | A | 0.046815 | 0.004373 | 2.90E-26 |
| rs543040 | let-7b-5p | trans | 1.13E-26 | Nightingale | LDL_C | NA | T | A | 0.033236 | 0.004414 | 9.10E-14 |
| rs543040 | let-7b-5p | trans | 1.13E-26 | Nightingale | S_LDL_C | NA | T | A | 0.03102 | 0.004437 | 5.40E-12 |
| rs543040 | let-7b-5p | trans | 1.13E-26 | Nightingale | M_VLDL_P | NA | T | A | 0.02852 | 0.004422 | 5.10E-11 |
| rs543040 | let-7b-5p | trans | 1.13E-26 | Nightingale | L_LDL_C | NA | T | A | 0.032339 | 0.004383 | 2.70E-13 |
| rs543040 | let-7b-5p | trans | 1.13E-26 | Nightingale | M_LDL_PL | NA | T | A | 0.032515 | 0.004439 | 2.70E-13 |
| rs543040 | let-7b-5p | trans | 1.13E-26 | Nightingale | IDL_C | NA | T | A | 0.034894 | 0.004281 | 1.20E-15 |
| rs543040 | let-7b-5p | trans | 1.13E-26 | Nightingale | M_VLDL_C | NA | T | A | 0.035992 | 0.004176 | 2.10E-17 |
| rs543040 | let-7b-5p | trans | 1.13E-26 | Nightingale | LDL_FC | NA | T | A | 0.033577 | 0.004386 | 6.30E-14 |
| rs543040 | let-7b-5p | trans | 1.13E-26 | Nightingale | XXL_VLDL_ | NA | T | A | -0.0356 | 0.004471 | 3.10E-16 |
| rs543040 | let-7b-5p | trans | 1.13E-26 | Nightingale | M_LDL_C_ | NA | T | A | 0.031281 | 0.004436 | 2.60E-12 |
| rs543040 | let-7b-5p | trans | 1.13E-26 | Nightingale | L_VLDL_TG | NA | T | A | -0.05098 | 0.004412 | 1.20E-31 |

| | | | | | | | | | | | |
|---|---|---|---|---|---|---|---|---|---|---|---|
| rs543040 | let-7b-5p | trans | 1.13E-26 | Nightingale | XS_VLDL_F | NA | T | A | 0.031254 | 0.004401 | 3.50E-12 |
| rs543040 | let-7b-5p | trans | 1.13E-26 | Nightingale | S_LDL_FC | NA | T | A | 0.028463 | 0.004426 | 5.20E-10 |
| rs543040 | let-7b-5p | trans | 1.13E-26 | Nightingale | M_VLDL_P | NA | T | A | 0.043865 | 0.004254 | 1.10E-24 |
| rs543040 | let-7b-5p | trans | 1.13E-26 | Nightingale | S_LDL_PL_ | NA | T | A | -0.03037 | 0.004425 | 1.90E-13 |
| rs543040 | let-7b-5p | trans | 1.13E-26 | Nightingale | XS_VLDL_L | NA | T | A | 0.033935 | 0.004358 | 1.80E-15 |
| rs543040 | let-7b-5p | trans | 1.13E-26 | Nightingale | VLDL_CE | NA | T | A | 0.035975 | 0.004434 | 2.50E-16 |
| rs543040 | let-7b-5p | trans | 1.13E-26 | Nightingale | VLDL_FC | NA | T | A | 0.024444 | 0.004401 | 8.20E-09 |
| rs543040 | let-7b-5p | trans | 1.13E-26 | Nightingale | S_VLDL_PL | NA | T | A | 0.032933 | 0.004425 | 3.70E-14 |
| rs543040 | let-7b-5p | trans | 1.13E-26 | Nightingale | M_VLDL_F | NA | T | A | 0.041872 | 0.004217 | 6.40E-23 |
| rs543040 | let-7b-5p | trans | 1.13E-26 | Nightingale | VLDL_C | NA | T | A | 0.031535 | 0.004427 | 3.80E-13 |
| rs543040 | let-7b-5p | trans | 1.13E-26 | Nightingale | S_HDL_L | NA | T | A | 0.026372 | 0.004418 | 1.50E-09 |
| rs543040 | let-7b-5p | trans | 1.13E-26 | Nightingale | Clinical_LD | NA | T | A | 0.036464 | 0.004407 | 3.10E-16 |
| rs543040 | let-7b-5p | trans | 1.13E-26 | Nightingale | XL_VLDL_C | NA | T | A | 0.037947 | 0.004251 | 9.30E-19 |
| rs543040 | let-7b-5p | trans | 1.13E-26 | Nightingale | M_LDL_P | NA | T | A | 0.036025 | 0.004445 | 6.20E-16 |
| rs543040 | let-7b-5p | trans | 1.13E-26 | Nightingale | IDL_FC | NA | T | A | 0.038215 | 0.004292 | 1.80E-18 |
| rs543040 | let-7b-5p | trans | 1.13E-26 | Nightingale | Total_PL | NA | T | A | 0.025082 | 0.004221 | 8.60E-10 |
| rs543040 | let-7b-5p | trans | 1.13E-26 | Nightingale | S_VLDL_FC | NA | T | A | 0.038005 | 0.004434 | 7.30E-18 |
| rs543040 | let-7b-5p | trans | 1.13E-26 | Nightingale | IDL_FC_pct | NA | T | A | 0.025728 | 0.004399 | 1.20E-08 |
| rs543040 | let-7b-5p | trans | 1.13E-26 | Nightingale | XS_VLDL_F | NA | T | A | 0.037149 | 0.004356 | 6.40E-18 |
| rs543040 | let-7b-5p | trans | 1.13E-26 | Nightingale | XL_VLDL_F | NA | T | A | 0.036996 | 0.004298 | 3.40E-18 |
| rs543040 | let-7b-5p | trans | 1.13E-26 | Nightingale | Remnant_( | NA | T | A | 0.036984 | 0.004404 | 3.40E-17 |
| rs543040 | let-7b-5p | trans | 1.13E-26 | Nightingale | XL_VLDL_T | NA | T | A | -0.04371 | 0.004297 | 3.50E-24 |
| rs543040 | let-7b-5p | trans | 1.13E-26 | Nightingale | Total_L | NA | T | A | 0.02656 | 0.004361 | 3.50E-10 |
| rs543040 | let-7b-5p | trans | 1.13E-26 | Nightingale | XS_VLDL_C | NA | T | A | 0.038037 | 0.004303 | 1.40E-18 |
| rs543040 | let-7b-5p | trans | 1.13E-26 | Nightingale | XS_VLDL_P | NA | T | A | 0.036804 | 0.004374 | 1.30E-17 |
| rs543040 | let-7b-5p | trans | 1.13E-26 | Nightingale | S_VLDL_PL | NA | T | A | 0.02757 | 0.004245 | 2.50E-10 |
| rs543040 | let-7b-5p | trans | 1.13E-26 | Nightingale | LDL_P | NA | T | A | 0.031905 | 0.00443 | 9.70E-13 |
| rs543040 | let-7b-5p | trans | 1.13E-26 | Nightingale | ApoB | NA | T | A | 0.034032 | 0.004427 | 2.10E-14 |
| rs543040 | miR-126-3p | trans | 6.96E-39 | Nightingale | IDL_CE | NA | T | A | 0.03344 | 0.00428 | 1.60E-14 |
| rs543040 | miR-126-3p | trans | 6.96E-39 | Nightingale | M_VLDL_T | NA | T | A | -0.03835 | 0.004189 | 1.20E-19 |
| rs543040 | miR-126-3p | trans | 6.96E-39 | Nightingale | M_LDL_C | NA | T | A | 0.034069 | 0.004441 | 2.10E-14 |
| rs543040 | miR-126-3p | trans | 6.96E-39 | Nightingale | S_VLDL_L | NA | T | A | 0.024881 | 0.00439 | 3.10E-09 |
| rs543040 | miR-126-3p | trans | 6.96E-39 | Nightingale | GlycA | NA | T | A | 0.028404 | 0.004414 | 8.90E-11 |
| rs543040 | miR-126-3p | trans | 6.96E-39 | Nightingale | S_LDL_L | NA | T | A | 0.027837 | 0.004432 | 7.80E-10 |
| rs543040 | miR-126-3p | trans | 6.96E-39 | Nightingale | S_VLDL_TG | NA | T | A | -0.03792 | 0.004291 | 2.60E-18 |
| rs543040 | miR-126-3p | trans | 6.96E-39 | Nightingale | S_VLDL_CE | NA | T | A | 0.038284 | 0.004427 | 1.60E-18 |
| rs543040 | miR-126-3p | trans | 6.96E-39 | Nightingale | Total_FC | NA | T | A | 0.033359 | 0.004331 | 6.10E-15 |
| rs543040 | miR-126-3p | trans | 6.96E-39 | Nightingale | L_LDL_FC | NA | T | A | 0.033578 | 0.004353 | 3.40E-14 |
| rs543040 | miR-126-3p | trans | 6.96E-39 | Nightingale | L_LDL_PL | NA | T | A | 0.03798 | 0.004399 | 1.10E-17 |
| rs543040 | miR-126-3p | trans | 6.96E-39 | Nightingale | M_LDL_CE | NA | T | A | 0.033102 | 0.004439 | 8.90E-14 |
| rs543040 | miR-126-3p | trans | 6.96E-39 | Nightingale | LA | NA | T | A | 0.025008 | 0.004347 | 3.40E-09 |
| rs543040 | miR-126-3p | trans | 6.96E-39 | Nightingale | XS_VLDL_P | NA | T | A | 0.032244 | 0.004371 | 4.80E-14 |
| rs543040 | miR-126-3p | trans | 6.96E-39 | Nightingale | M_VLDL_F | NA | T | A | 0.033543 | 0.004432 | 2.70E-14 |
| rs543040 | miR-126-3p | trans | 6.96E-39 | Nightingale | S_HDL_CE | NA | T | A | 0.028568 | 0.00444 | 1.60E-10 |
| rs543040 | miR-126-3p | trans | 6.96E-39 | Nightingale | S_LDL_CE_ | NA | T | A | 0.031287 | 0.004425 | 3.30E-14 |
| rs543040 | miR-126-3p | trans | 6.96E-39 | Nightingale | S_VLDL_P | NA | T | A | 0.023677 | 0.004377 | 1.40E-08 |
| rs543040 | miR-126-3p | trans | 6.96E-39 | Nightingale | IDL_P | NA | T | A | 0.039934 | 0.004386 | 1.70E-19 |
| rs543040 | miR-126-3p | trans | 6.96E-39 | Nightingale | XXL_VLDL_ | NA | T | A | 0.030507 | 0.004414 | 2.00E-12 |
| rs543040 | miR-126-3p | trans | 6.96E-39 | Nightingale | M_VLDL_C | NA | T | A | 0.040846 | 0.004393 | 2.70E-20 |
| rs543040 | miR-126-3p | trans | 6.96E-39 | Nightingale | S_LDL_CE | NA | T | A | 0.030692 | 0.004435 | 5.20E-12 |
| rs543040 | miR-126-3p | trans | 6.96E-39 | Nightingale | VLDL_P | NA | T | A | 0.02687 | 0.004409 | 3.40E-10 |
| rs543040 | miR-126-3p | trans | 6.96E-39 | Nightingale | S_VLDL_C_ | NA | T | A | 0.041674 | 0.004314 | 1.10E-21 |
| rs543040 | miR-126-3p | trans | 6.96E-39 | Nightingale | M_LDL_TG | NA | T | A | -0.02749 | 0.004378 | 4.30E-10 |
| rs543040 | miR-126-3p | trans | 6.96E-39 | Nightingale | Omega_6 | NA | T | A | 0.025996 | 0.004329 | 5.10E-10 |
| rs543040 | miR-126-3p | trans | 6.96E-39 | Nightingale | L_VLDL_FC | NA | T | A | 0.048505 | 0.004391 | 8.10E-29 |
| rs543040 | miR-126-3p | trans | 6.96E-39 | Nightingale | Cholines | NA | T | A | 0.022428 | 0.004169 | 4.00E-08 |
| rs543040 | miR-126-3p | trans | 6.96E-39 | Nightingale | XL_VLDL_C | NA | T | A | 0.037056 | 0.004253 | 6.40E-18 |
| rs543040 | miR-126-3p | trans | 6.96E-39 | Nightingale | M_VLDL_P | NA | T | A | 0.029334 | 0.004426 | 1.70E-11 |
| rs543040 | miR-126-3p | trans | 6.96E-39 | Nightingale | XL_VLDL_P | NA | T | A | 0.03891 | 0.00444 | 5.80E-19 |
| rs543040 | miR-126-3p | trans | 6.96E-39 | Nightingale | Total_CE | NA | T | A | 0.031773 | 0.004276 | 9.70E-14 |
| rs543040 | miR-126-3p | trans | 6.96E-39 | Nightingale | LDL_PL | NA | T | A | 0.035004 | 0.004423 | 5.10E-15 |
| rs543040 | miR-126-3p | trans | 6.96E-39 | Nightingale | ApoB_by_A | NA | T | A | 0.025114 | 0.004366 | 1.30E-08 |
| rs543040 | miR-126-3p | trans | 6.96E-39 | Nightingale | XL_HDL_PL | NA | T | A | -0.02279 | 0.004073 | 1.20E-08 |

| | | | | | | | | | | | |
|---|---|---|---|---|---|---|---|---|---|---|---|
| rs543040 | miR-126-3p | trans | 6.96E-39 | Nightingale | PUFA | NA | T | A | 0.023843 | 0.004324 | 1.30E-08 |
| rs543040 | miR-126-3p | trans | 6.96E-39 | Nightingale | L_LDL_P | NA | T | A | 0.02926 | 0.004418 | 6.00E-11 |
| rs543040 | miR-126-3p | trans | 6.96E-39 | Nightingale | XS_VLDL_C | NA | T | A | 0.037825 | 0.004278 | 2.30E-18 |
| rs543040 | miR-126-3p | trans | 6.96E-39 | Nightingale | L_VLDL_CE | NA | T | A | 0.047001 | 0.004242 | 7.50E-29 |
| rs543040 | miR-126-3p | trans | 6.96E-39 | Nightingale | XXL_VLDL_ | NA | T | A | 0.024559 | 0.004462 | 3.20E-09 |
| rs543040 | miR-126-3p | trans | 6.96E-39 | Nightingale | L_VLDL_C_ | NA | T | A | 0.051898 | 0.00433 | 1.30E-33 |
| rs543040 | miR-126-3p | trans | 6.96E-39 | Nightingale | S_HDL_P | NA | T | A | 0.030617 | 0.004434 | 5.60E-12 |
| rs543040 | miR-126-3p | trans | 6.96E-39 | Nightingale | M_VLDL_C | NA | T | A | 0.033408 | 0.004165 | 3.20E-15 |
| rs543040 | miR-126-3p | trans | 6.96E-39 | Nightingale | LDL_L | NA | T | A | 0.033354 | 0.004418 | 7.50E-14 |
| rs543040 | miR-126-3p | trans | 6.96E-39 | Nightingale | XXL_VLDL_ | NA | T | A | 0.034686 | 0.004443 | 4.10E-15 |
| rs543040 | miR-126-3p | trans | 6.96E-39 | Nightingale | M_LDL_FC | NA | T | A | 0.034292 | 0.004427 | 2.40E-14 |
| rs543040 | miR-126-3p | trans | 6.96E-39 | Nightingale | S_VLDL_C | NA | T | A | 0.038477 | 0.004432 | 1.80E-18 |
| rs543040 | miR-126-3p | trans | 6.96E-39 | Nightingale | Citrate | NA | T | A | 0.027118 | 0.004404 | 5.00E-10 |
| rs543040 | miR-126-3p | trans | 6.96E-39 | Nightingale | IDL_PL | NA | T | A | 0.035357 | 0.004278 | 3.60E-16 |
| rs543040 | miR-126-3p | trans | 6.96E-39 | Nightingale | S_HDL_C | NA | T | A | 0.028373 | 0.004426 | 2.00E-10 |
| rs543040 | miR-126-3p | trans | 6.96E-39 | Nightingale | Sphingomy | NA | T | A | 0.029085 | 0.004144 | 6.00E-12 |
| rs543040 | miR-126-3p | trans | 6.96E-39 | Nightingale | non_HDL_( | NA | T | A | 0.035777 | 0.004409 | 5.00E-16 |
| rs543040 | miR-126-3p | trans | 6.96E-39 | Nightingale | M_LDL_L | NA | T | A | 0.033119 | 0.004439 | 9.90E-14 |
| rs543040 | miR-126-3p | trans | 6.96E-39 | Nightingale | Total_C | NA | T | A | 0.032423 | 0.004291 | 3.20E-14 |
| rs543040 | miR-126-3p | trans | 6.96E-39 | Nightingale | L_LDL_CE | NA | T | A | 0.03161 | 0.004393 | 1.00E-12 |
| rs543040 | miR-126-3p | trans | 6.96E-39 | Nightingale | M_VLDL_C | NA | T | A | 0.038795 | 0.004421 | 1.90E-18 |
| rs543040 | miR-126-3p | trans | 6.96E-39 | Nightingale | S_VLDL_FC | NA | T | A | 0.029186 | 0.004236 | 1.50E-11 |
| rs543040 | miR-126-3p | trans | 6.96E-39 | Nightingale | S_LDL_C_p | NA | T | A | 0.045363 | 0.00445 | 7.00E-25 |
| rs543040 | miR-126-3p | trans | 6.96E-39 | Nightingale | LDL_CE | NA | T | A | 0.032631 | 0.004421 | 2.10E-13 |
| rs543040 | miR-126-3p | trans | 6.96E-39 | Nightingale | XXL_VLDL_ | NA | T | A | 0.033103 | 0.004467 | 1.00E-13 |
| rs543040 | miR-126-3p | trans | 6.96E-39 | Nightingale | S_LDL_P | NA | T | A | 0.029725 | 0.004437 | 2.90E-11 |
| rs543040 | miR-126-3p | trans | 6.96E-39 | Nightingale | IDL_L | NA | T | A | 0.035244 | 0.004286 | 5.60E-16 |
| rs543040 | miR-126-3p | trans | 6.96E-39 | Nightingale | S_HDL_FC | NA | T | A | 0.023503 | 0.004352 | 4.50E-08 |
| rs543040 | miR-126-3p | trans | 6.96E-39 | Nightingale | S_HDL_PL | NA | T | A | 0.02389 | 0.004395 | 3.00E-08 |
| rs543040 | miR-126-3p | trans | 6.96E-39 | Nightingale | L_LDL_L | NA | T | A | 0.033574 | 0.00439 | 3.20E-14 |
| rs543040 | miR-126-3p | trans | 6.96E-39 | Nightingale | S_VLDL_CE | NA | T | A | 0.046815 | 0.004373 | 2.90E-26 |
| rs543040 | miR-126-3p | trans | 6.96E-39 | Nightingale | LDL_C | NA | T | A | 0.033236 | 0.004414 | 9.10E-14 |
| rs543040 | miR-126-3p | trans | 6.96E-39 | Nightingale | S_LDL_C | NA | T | A | 0.03102 | 0.004437 | 5.40E-12 |
| rs543040 | miR-126-3p | trans | 6.96E-39 | Nightingale | M_VLDL_P | NA | T | A | 0.02852 | 0.004422 | 5.10E-11 |
| rs543040 | miR-126-3p | trans | 6.96E-39 | Nightingale | L_LDL_C | NA | T | A | 0.032339 | 0.004383 | 2.70E-13 |
| rs543040 | miR-126-3p | trans | 6.96E-39 | Nightingale | M_LDL_PL | NA | T | A | 0.032515 | 0.004439 | 2.70E-13 |
| rs543040 | miR-126-3p | trans | 6.96E-39 | Nightingale | IDL_C | NA | T | A | 0.034894 | 0.004281 | 1.20E-15 |
| rs543040 | miR-126-3p | trans | 6.96E-39 | Nightingale | M_VLDL_C | NA | T | A | 0.035992 | 0.004176 | 2.10E-17 |
| rs543040 | miR-126-3p | trans | 6.96E-39 | Nightingale | LDL_FC | NA | T | A | 0.033577 | 0.004386 | 6.30E-14 |
| rs543040 | miR-126-3p | trans | 6.96E-39 | Nightingale | XXL_VLDL_ | NA | T | A | -0.0356 | 0.004471 | 3.10E-16 |
| rs543040 | miR-126-3p | trans | 6.96E-39 | Nightingale | M_LDL_C_ | NA | T | A | 0.031281 | 0.004436 | 2.60E-12 |
| rs543040 | miR-126-3p | trans | 6.96E-39 | Nightingale | L_VLDL_TG | NA | T | A | -0.05098 | 0.004412 | 1.20E-31 |
| rs543040 | miR-126-3p | trans | 6.96E-39 | Nightingale | XS_VLDL_F | NA | T | A | 0.031254 | 0.004401 | 3.50E-12 |
| rs543040 | miR-126-3p | trans | 6.96E-39 | Nightingale | S_LDL_FC | NA | T | A | 0.028463 | 0.004426 | 5.20E-10 |
| rs543040 | miR-126-3p | trans | 6.96E-39 | Nightingale | M_VLDL_P | NA | T | A | 0.043865 | 0.004254 | 1.10E-24 |
| rs543040 | miR-126-3p | trans | 6.96E-39 | Nightingale | S_LDL_PL_ | NA | T | A | -0.03037 | 0.004425 | 1.90E-13 |
| rs543040 | miR-126-3p | trans | 6.96E-39 | Nightingale | XS_VLDL_L | NA | T | A | 0.033935 | 0.004358 | 1.80E-15 |
| rs543040 | miR-126-3p | trans | 6.96E-39 | Nightingale | VLDL_CE | NA | T | A | 0.035975 | 0.004434 | 2.50E-16 |
| rs543040 | miR-126-3p | trans | 6.96E-39 | Nightingale | VLDL_FC | NA | T | A | 0.024444 | 0.004401 | 8.20E-09 |
| rs543040 | miR-126-3p | trans | 6.96E-39 | Nightingale | S_VLDL_PL | NA | T | A | 0.032933 | 0.004425 | 3.70E-14 |
| rs543040 | miR-126-3p | trans | 6.96E-39 | Nightingale | M_VLDL_F | NA | T | A | 0.041872 | 0.004217 | 6.40E-23 |
| rs543040 | miR-126-3p | trans | 6.96E-39 | Nightingale | VLDL_C | NA | T | A | 0.031535 | 0.004427 | 3.80E-13 |
| rs543040 | miR-126-3p | trans | 6.96E-39 | Nightingale | S_HDL_L | NA | T | A | 0.026372 | 0.004418 | 1.50E-09 |
| rs543040 | miR-126-3p | trans | 6.96E-39 | Nightingale | Clinical_LD | NA | T | A | 0.036464 | 0.004407 | 3.10E-16 |
| rs543040 | miR-126-3p | trans | 6.96E-39 | Nightingale | XL_VLDL_C | NA | T | A | 0.037947 | 0.004251 | 9.30E-19 |
| rs543040 | miR-126-3p | trans | 6.96E-39 | Nightingale | M_LDL_P | NA | T | A | 0.036025 | 0.004445 | 6.20E-16 |
| rs543040 | miR-126-3p | trans | 6.96E-39 | Nightingale | IDL_FC | NA | T | A | 0.038215 | 0.004292 | 1.80E-18 |
| rs543040 | miR-126-3p | trans | 6.96E-39 | Nightingale | Total_PL | NA | T | A | 0.025082 | 0.004221 | 8.60E-10 |
| rs543040 | miR-126-3p | trans | 6.96E-39 | Nightingale | S_VLDL_FC | NA | T | A | 0.038005 | 0.004434 | 7.30E-18 |
| rs543040 | miR-126-3p | trans | 6.96E-39 | Nightingale | IDL_FC_pct | NA | T | A | 0.025728 | 0.004399 | 1.20E-08 |
| rs543040 | miR-126-3p | trans | 6.96E-39 | Nightingale | XS_VLDL_F | NA | T | A | 0.037149 | 0.004356 | 6.40E-18 |
| rs543040 | miR-126-3p | trans | 6.96E-39 | Nightingale | XL_VLDL_F | NA | T | A | 0.036996 | 0.004298 | 3.40E-18 |
| rs543040 | miR-126-3p | trans | 6.96E-39 | Nightingale | Remnant_( | NA | T | A | 0.036984 | 0.004404 | 3.40E-17 |

| | | | | | | | | | | | |
|---|---|---|---|---|---|---|---|---|---|---|---|
| rs543040 | miR-126-3p | trans | 6.96E-39 | Nightingale | XL_VLDL_T | NA | T | A | -0.04371 | 0.004297 | 3.50E-24 |
| rs543040 | miR-126-3p | trans | 6.96E-39 | Nightingale | Total_L | NA | T | A | 0.02656 | 0.004361 | 3.50E-10 |
| rs543040 | miR-126-3p | trans | 6.96E-39 | Nightingale | XS_VLDL_C | NA | T | A | 0.038037 | 0.004303 | 1.40E-18 |
| rs543040 | miR-126-3p | trans | 6.96E-39 | Nightingale | XS_VLDL_P | NA | T | A | 0.036804 | 0.004374 | 1.30E-17 |
| rs543040 | miR-126-3p | trans | 6.96E-39 | Nightingale | S_VLDL_PL | NA | T | A | 0.02757 | 0.004245 | 2.50E-10 |
| rs543040 | miR-126-3p | trans | 6.96E-39 | Nightingale | LDL_P | NA | T | A | 0.031905 | 0.00443 | 9.70E-13 |
| rs543040 | miR-126-3p | trans | 6.96E-39 | Nightingale | ApoB | NA | T | A | 0.034032 | 0.004427 | 2.10E-14 |
| rs543040 | let-7c-5p | trans | 6.26E-23 | Nightingale | IDL_CE | NA | T | A | 0.03344 | 0.00428 | 1.60E-14 |
| rs543040 | let-7c-5p | trans | 6.26E-23 | Nightingale | M_VLDL_T | NA | T | A | -0.03835 | 0.004189 | 1.20E-19 |
| rs543040 | let-7c-5p | trans | 6.26E-23 | Nightingale | M_LDL_C | NA | T | A | 0.034069 | 0.004441 | 2.10E-14 |
| rs543040 | let-7c-5p | trans | 6.26E-23 | Nightingale | S_VLDL_L | NA | T | A | 0.024881 | 0.00439 | 3.10E-09 |
| rs543040 | let-7c-5p | trans | 6.26E-23 | Nightingale | GlycA | NA | T | A | 0.028404 | 0.004414 | 8.90E-11 |
| rs543040 | let-7c-5p | trans | 6.26E-23 | Nightingale | S_LDL_L | NA | T | A | 0.027837 | 0.004432 | 7.80E-10 |
| rs543040 | let-7c-5p | trans | 6.26E-23 | Nightingale | S_VLDL_TG | NA | T | A | -0.03792 | 0.004291 | 2.60E-18 |
| rs543040 | let-7c-5p | trans | 6.26E-23 | Nightingale | S_VLDL_CE | NA | T | A | 0.038284 | 0.004427 | 1.60E-18 |
| rs543040 | let-7c-5p | trans | 6.26E-23 | Nightingale | Total_FC | NA | T | A | 0.033359 | 0.004331 | 6.10E-15 |
| rs543040 | let-7c-5p | trans | 6.26E-23 | Nightingale | L_LDL_FC | NA | T | A | 0.033578 | 0.004353 | 3.40E-14 |
| rs543040 | let-7c-5p | trans | 6.26E-23 | Nightingale | L_LDL_PL | NA | T | A | 0.03798 | 0.004399 | 1.10E-17 |
| rs543040 | let-7c-5p | trans | 6.26E-23 | Nightingale | M_LDL_CE | NA | T | A | 0.033102 | 0.004439 | 8.90E-14 |
| rs543040 | let-7c-5p | trans | 6.26E-23 | Nightingale | LA | NA | T | A | 0.025008 | 0.004347 | 3.40E-09 |
| rs543040 | let-7c-5p | trans | 6.26E-23 | Nightingale | XS_VLDL_P | NA | T | A | 0.032244 | 0.004371 | 4.80E-14 |
| rs543040 | let-7c-5p | trans | 6.26E-23 | Nightingale | M_VLDL_F | NA | T | A | 0.033543 | 0.004432 | 2.70E-14 |
| rs543040 | let-7c-5p | trans | 6.26E-23 | Nightingale | S_HDL_CE | NA | T | A | 0.028568 | 0.00444 | 1.60E-10 |
| rs543040 | let-7c-5p | trans | 6.26E-23 | Nightingale | S_LDL_CE_ | NA | T | A | 0.031287 | 0.004425 | 3.30E-14 |
| rs543040 | let-7c-5p | trans | 6.26E-23 | Nightingale | S_VLDL_P | NA | T | A | 0.023677 | 0.004377 | 1.40E-08 |
| rs543040 | let-7c-5p | trans | 6.26E-23 | Nightingale | IDL_P | NA | T | A | 0.039934 | 0.004386 | 1.70E-19 |
| rs543040 | let-7c-5p | trans | 6.26E-23 | Nightingale | XXL_VLDL_ | NA | T | A | 0.030507 | 0.004414 | 2.00E-12 |
| rs543040 | let-7c-5p | trans | 6.26E-23 | Nightingale | M_VLDL_C | NA | T | A | 0.040846 | 0.004393 | 2.70E-20 |
| rs543040 | let-7c-5p | trans | 6.26E-23 | Nightingale | S_LDL_CE | NA | T | A | 0.030692 | 0.004435 | 5.20E-12 |
| rs543040 | let-7c-5p | trans | 6.26E-23 | Nightingale | VLDL_P | NA | T | A | 0.02687 | 0.004409 | 3.40E-10 |
| rs543040 | let-7c-5p | trans | 6.26E-23 | Nightingale | S_VLDL_C_ | NA | T | A | 0.041674 | 0.004314 | 1.10E-21 |
| rs543040 | let-7c-5p | trans | 6.26E-23 | Nightingale | M_LDL_TG | NA | T | A | -0.02749 | 0.004378 | 4.30E-10 |
| rs543040 | let-7c-5p | trans | 6.26E-23 | Nightingale | Omega_6 | NA | T | A | 0.025996 | 0.004329 | 5.10E-10 |
| rs543040 | let-7c-5p | trans | 6.26E-23 | Nightingale | L_VLDL_FC | NA | T | A | 0.048505 | 0.004391 | 8.10E-29 |
| rs543040 | let-7c-5p | trans | 6.26E-23 | Nightingale | Cholines | NA | T | A | 0.022428 | 0.004169 | 4.00E-08 |
| rs543040 | let-7c-5p | trans | 6.26E-23 | Nightingale | XL_VLDL_C | NA | T | A | 0.037056 | 0.004253 | 6.40E-18 |
| rs543040 | let-7c-5p | trans | 6.26E-23 | Nightingale | M_VLDL_P | NA | T | A | 0.029334 | 0.004426 | 1.70E-11 |
| rs543040 | let-7c-5p | trans | 6.26E-23 | Nightingale | XL_VLDL_P | NA | T | A | 0.03891 | 0.00444 | 5.80E-19 |
| rs543040 | let-7c-5p | trans | 6.26E-23 | Nightingale | Total_CE | NA | T | A | 0.031773 | 0.004276 | 9.70E-14 |
| rs543040 | let-7c-5p | trans | 6.26E-23 | Nightingale | LDL_PL | NA | T | A | 0.035004 | 0.004423 | 5.10E-15 |
| rs543040 | let-7c-5p | trans | 6.26E-23 | Nightingale | ApoB_by_A | NA | T | A | 0.025114 | 0.004366 | 1.30E-08 |
| rs543040 | let-7c-5p | trans | 6.26E-23 | Nightingale | XL_HDL_PL | NA | T | A | -0.02279 | 0.004073 | 1.20E-08 |
| rs543040 | let-7c-5p | trans | 6.26E-23 | Nightingale | PUFA | NA | T | A | 0.023843 | 0.004324 | 1.30E-08 |
| rs543040 | let-7c-5p | trans | 6.26E-23 | Nightingale | L_LDL_P | NA | T | A | 0.02926 | 0.004418 | 6.00E-11 |
| rs543040 | let-7c-5p | trans | 6.26E-23 | Nightingale | XS_VLDL_C | NA | T | A | 0.037825 | 0.004278 | 2.30E-18 |
| rs543040 | let-7c-5p | trans | 6.26E-23 | Nightingale | L_VLDL_CE | NA | T | A | 0.047001 | 0.004242 | 7.50E-29 |
| rs543040 | let-7c-5p | trans | 6.26E-23 | Nightingale | XXL_VLDL_ | NA | T | A | 0.024559 | 0.004462 | 3.20E-09 |
| rs543040 | let-7c-5p | trans | 6.26E-23 | Nightingale | L_VLDL_C_ | NA | T | A | 0.051898 | 0.00433 | 1.30E-33 |
| rs543040 | let-7c-5p | trans | 6.26E-23 | Nightingale | S_HDL_P | NA | T | A | 0.030617 | 0.004434 | 5.60E-12 |
| rs543040 | let-7c-5p | trans | 6.26E-23 | Nightingale | M_VLDL_C | NA | T | A | 0.033408 | 0.004165 | 3.20E-15 |
| rs543040 | let-7c-5p | trans | 6.26E-23 | Nightingale | LDL_L | NA | T | A | 0.033354 | 0.004418 | 7.50E-14 |
| rs543040 | let-7c-5p | trans | 6.26E-23 | Nightingale | XXL_VLDL_ | NA | T | A | 0.034686 | 0.004443 | 4.10E-15 |
| rs543040 | let-7c-5p | trans | 6.26E-23 | Nightingale | M_LDL_FC | NA | T | A | 0.034292 | 0.004427 | 2.40E-14 |
| rs543040 | let-7c-5p | trans | 6.26E-23 | Nightingale | S_VLDL_C | NA | T | A | 0.038477 | 0.004432 | 1.80E-18 |
| rs543040 | let-7c-5p | trans | 6.26E-23 | Nightingale | Citrate | NA | T | A | 0.027118 | 0.004404 | 5.00E-10 |
| rs543040 | let-7c-5p | trans | 6.26E-23 | Nightingale | IDL_PL | NA | T | A | 0.035357 | 0.004278 | 3.60E-16 |
| rs543040 | let-7c-5p | trans | 6.26E-23 | Nightingale | S_HDL_C | NA | T | A | 0.028373 | 0.004426 | 2.00E-10 |
| rs543040 | let-7c-5p | trans | 6.26E-23 | Nightingale | Sphingomy | NA | T | A | 0.029085 | 0.004144 | 6.00E-12 |
| rs543040 | let-7c-5p | trans | 6.26E-23 | Nightingale | non_HDL_( | NA | T | A | 0.035777 | 0.004409 | 5.00E-16 |
| rs543040 | let-7c-5p | trans | 6.26E-23 | Nightingale | M_LDL_L | NA | T | A | 0.033119 | 0.004439 | 9.90E-14 |
| rs543040 | let-7c-5p | trans | 6.26E-23 | Nightingale | Total_C | NA | T | A | 0.032423 | 0.004291 | 3.20E-14 |
| rs543040 | let-7c-5p | trans | 6.26E-23 | Nightingale | L_LDL_CE | NA | T | A | 0.03161 | 0.004393 | 1.00E-12 |
| rs543040 | let-7c-5p | trans | 6.26E-23 | Nightingale | M_VLDL_C | NA | T | A | 0.038795 | 0.004421 | 1.90E-18 |

| | | | | | | | | | | | |
|---|---|---|---|---|---|---|---|---|---|---|---|
| rs543040 | let-7c-5p | trans | 6.26E-23 | Nightingale | S_VLDL_FC | NA | T | A | 0.029186 | 0.004236 | 1.50E-11 |
| rs543040 | let-7c-5p | trans | 6.26E-23 | Nightingale | S_LDL_C_p | NA | T | A | 0.045363 | 0.00445 | 7.00E-25 |
| rs543040 | let-7c-5p | trans | 6.26E-23 | Nightingale | LDL_CE | NA | T | A | 0.032631 | 0.004421 | 2.10E-13 |
| rs543040 | let-7c-5p | trans | 6.26E-23 | Nightingale | XXL_VLDL_ | NA | T | A | 0.033103 | 0.004467 | 1.00E-13 |
| rs543040 | let-7c-5p | trans | 6.26E-23 | Nightingale | S_LDL_P | NA | T | A | 0.029725 | 0.004437 | 2.90E-11 |
| rs543040 | let-7c-5p | trans | 6.26E-23 | Nightingale | IDL_L | NA | T | A | 0.035244 | 0.004286 | 5.60E-16 |
| rs543040 | let-7c-5p | trans | 6.26E-23 | Nightingale | S_HDL_FC | NA | T | A | 0.023503 | 0.004352 | 4.50E-08 |
| rs543040 | let-7c-5p | trans | 6.26E-23 | Nightingale | S_HDL_PL | NA | T | A | 0.02389 | 0.004395 | 3.00E-08 |
| rs543040 | let-7c-5p | trans | 6.26E-23 | Nightingale | L_LDL_L | NA | T | A | 0.033574 | 0.00439 | 3.20E-14 |
| rs543040 | let-7c-5p | trans | 6.26E-23 | Nightingale | S_VLDL_CE | NA | T | A | 0.046815 | 0.004373 | 2.90E-26 |
| rs543040 | let-7c-5p | trans | 6.26E-23 | Nightingale | LDL_C | NA | T | A | 0.033236 | 0.004414 | 9.10E-14 |
| rs543040 | let-7c-5p | trans | 6.26E-23 | Nightingale | S_LDL_C | NA | T | A | 0.03102 | 0.004437 | 5.40E-12 |
| rs543040 | let-7c-5p | trans | 6.26E-23 | Nightingale | M_VLDL_P | NA | T | A | 0.02852 | 0.004422 | 5.10E-11 |
| rs543040 | let-7c-5p | trans | 6.26E-23 | Nightingale | L_LDL_C | NA | T | A | 0.032339 | 0.004383 | 2.70E-13 |
| rs543040 | let-7c-5p | trans | 6.26E-23 | Nightingale | M_LDL_PL | NA | T | A | 0.032515 | 0.004439 | 2.70E-13 |
| rs543040 | let-7c-5p | trans | 6.26E-23 | Nightingale | IDL_C | NA | T | A | 0.034894 | 0.004281 | 1.20E-15 |
| rs543040 | let-7c-5p | trans | 6.26E-23 | Nightingale | M_VLDL_C | NA | T | A | 0.035992 | 0.004176 | 2.10E-17 |
| rs543040 | let-7c-5p | trans | 6.26E-23 | Nightingale | LDL_FC | NA | T | A | 0.033577 | 0.004386 | 6.30E-14 |
| rs543040 | let-7c-5p | trans | 6.26E-23 | Nightingale | XXL_VLDL_ | NA | T | A | -0.0356 | 0.004471 | 3.10E-16 |
| rs543040 | let-7c-5p | trans | 6.26E-23 | Nightingale | M_LDL_C_ | NA | T | A | 0.031281 | 0.004436 | 2.60E-12 |
| rs543040 | let-7c-5p | trans | 6.26E-23 | Nightingale | L_VLDL_TG | NA | T | A | -0.05098 | 0.004412 | 1.20E-31 |
| rs543040 | let-7c-5p | trans | 6.26E-23 | Nightingale | XS_VLDL_F | NA | T | A | 0.031254 | 0.004401 | 3.50E-12 |
| rs543040 | let-7c-5p | trans | 6.26E-23 | Nightingale | S_LDL_FC | NA | T | A | 0.028463 | 0.004426 | 5.20E-10 |
| rs543040 | let-7c-5p | trans | 6.26E-23 | Nightingale | M_VLDL_P | NA | T | A | 0.043865 | 0.004254 | 1.10E-24 |
| rs543040 | let-7c-5p | trans | 6.26E-23 | Nightingale | S_LDL_PL_ | NA | T | A | -0.03037 | 0.004425 | 1.90E-13 |
| rs543040 | let-7c-5p | trans | 6.26E-23 | Nightingale | XS_VLDL_L | NA | T | A | 0.033935 | 0.004358 | 1.80E-15 |
| rs543040 | let-7c-5p | trans | 6.26E-23 | Nightingale | VLDL_CE | NA | T | A | 0.035975 | 0.004434 | 2.50E-16 |
| rs543040 | let-7c-5p | trans | 6.26E-23 | Nightingale | VLDL_FC | NA | T | A | 0.024444 | 0.004401 | 8.20E-09 |
| rs543040 | let-7c-5p | trans | 6.26E-23 | Nightingale | S_VLDL_PL | NA | T | A | 0.032933 | 0.004425 | 3.70E-14 |
| rs543040 | let-7c-5p | trans | 6.26E-23 | Nightingale | M_VLDL_F | NA | T | A | 0.041872 | 0.004217 | 6.40E-23 |
| rs543040 | let-7c-5p | trans | 6.26E-23 | Nightingale | VLDL_C | NA | T | A | 0.031535 | 0.004427 | 3.80E-13 |
| rs543040 | let-7c-5p | trans | 6.26E-23 | Nightingale | S_HDL_L | NA | T | A | 0.026372 | 0.004418 | 1.50E-09 |
| rs543040 | let-7c-5p | trans | 6.26E-23 | Nightingale | Clinical_LD | NA | T | A | 0.036464 | 0.004407 | 3.10E-16 |
| rs543040 | let-7c-5p | trans | 6.26E-23 | Nightingale | XL_VLDL_C | NA | T | A | 0.037947 | 0.004251 | 9.30E-19 |
| rs543040 | let-7c-5p | trans | 6.26E-23 | Nightingale | M_LDL_P | NA | T | A | 0.036025 | 0.004445 | 6.20E-16 |
| rs543040 | let-7c-5p | trans | 6.26E-23 | Nightingale | IDL_FC | NA | T | A | 0.038215 | 0.004292 | 1.80E-18 |
| rs543040 | let-7c-5p | trans | 6.26E-23 | Nightingale | Total_PL | NA | T | A | 0.025082 | 0.004221 | 8.60E-10 |
| rs543040 | let-7c-5p | trans | 6.26E-23 | Nightingale | S_VLDL_FC | NA | T | A | 0.038005 | 0.004434 | 7.30E-18 |
| rs543040 | let-7c-5p | trans | 6.26E-23 | Nightingale | IDL_FC_pct | NA | T | A | 0.025728 | 0.004399 | 1.20E-08 |
| rs543040 | let-7c-5p | trans | 6.26E-23 | Nightingale | XS_VLDL_F | NA | T | A | 0.037149 | 0.004356 | 6.40E-18 |
| rs543040 | let-7c-5p | trans | 6.26E-23 | Nightingale | XL_VLDL_F | NA | T | A | 0.036996 | 0.004298 | 3.40E-18 |
| rs543040 | let-7c-5p | trans | 6.26E-23 | Nightingale | Remnant_( | NA | T | A | 0.036984 | 0.004404 | 3.40E-17 |
| rs543040 | let-7c-5p | trans | 6.26E-23 | Nightingale | XL_VLDL_T | NA | T | A | -0.04371 | 0.004297 | 3.50E-24 |
| rs543040 | let-7c-5p | trans | 6.26E-23 | Nightingale | Total_L | NA | T | A | 0.02656 | 0.004361 | 3.50E-10 |
| rs543040 | let-7c-5p | trans | 6.26E-23 | Nightingale | XS_VLDL_C | NA | T | A | 0.038037 | 0.004303 | 1.40E-18 |
| rs543040 | let-7c-5p | trans | 6.26E-23 | Nightingale | XS_VLDL_P | NA | T | A | 0.036804 | 0.004374 | 1.30E-17 |
| rs543040 | let-7c-5p | trans | 6.26E-23 | Nightingale | S_VLDL_PL | NA | T | A | 0.02757 | 0.004245 | 2.50E-10 |
| rs543040 | let-7c-5p | trans | 6.26E-23 | Nightingale | LDL_P | NA | T | A | 0.031905 | 0.00443 | 9.70E-13 |
| rs543040 | let-7c-5p | trans | 6.26E-23 | Nightingale | ApoB | NA | T | A | 0.034032 | 0.004427 | 2.10E-14 |
| rs543968 | let-7b-5p | trans | 5.69E-26 | Nightingale | IDL_L | NA | C | T | 0.03366 | 0.004203 | 2.30E-15 |
| rs543968 | let-7b-5p | trans | 5.69E-26 | Nightingale | S_VLDL_C_ | NA | C | T | 0.039736 | 0.00423 | 8.30E-21 |
| rs543968 | let-7b-5p | trans | 5.69E-26 | Nightingale | S_HDL_C | NA | C | T | 0.02538 | 0.004339 | 9.40E-09 |
| rs543968 | let-7b-5p | trans | 5.69E-26 | Nightingale | M_VLDL_C | NA | C | T | 0.037341 | 0.004335 | 5.40E-18 |
| rs543968 | let-7b-5p | trans | 5.69E-26 | Nightingale | LDL_L | NA | C | T | 0.031465 | 0.004332 | 5.80E-13 |
| rs543968 | let-7b-5p | trans | 5.69E-26 | Nightingale | XS_VLDL_P | NA | C | T | 0.031657 | 0.004286 | 2.00E-14 |
| rs543968 | let-7b-5p | trans | 5.69E-26 | Nightingale | L_VLDL_C_ | NA | C | T | 0.049226 | 0.004245 | 8.90E-32 |
| rs543968 | let-7b-5p | trans | 5.69E-26 | Nightingale | M_LDL_TG | NA | C | T | -0.02512 | 0.004293 | 8.60E-09 |
| rs543968 | let-7b-5p | trans | 5.69E-26 | Nightingale | L_LDL_P | NA | C | T | 0.028317 | 0.004332 | 7.80E-11 |
| rs543968 | let-7b-5p | trans | 5.69E-26 | Nightingale | M_LDL_CE | NA | C | T | 0.031212 | 0.004353 | 8.50E-13 |
| rs543968 | let-7b-5p | trans | 5.69E-26 | Nightingale | XS_VLDL_P | NA | C | T | 0.035785 | 0.004289 | 1.00E-17 |
| rs543968 | let-7b-5p | trans | 5.69E-26 | Nightingale | M_VLDL_F | NA | C | T | 0.032515 | 0.004346 | 3.90E-14 |
| rs543968 | let-7b-5p | trans | 5.69E-26 | Nightingale | M_VLDL_C | NA | C | T | 0.031348 | 0.004084 | 3.30E-14 |
| rs543968 | let-7b-5p | trans | 5.69E-26 | Nightingale | LDL_PL | NA | C | T | 0.033318 | 0.004337 | 3.00E-14 |

| | | | | | | | | | | | |
|---|---|---|---|---|---|---|---|---|---|---|---|
| rs543968 | let-7b-5p | trans | 5.69E-26 | Nightingale | LDL_CE | NA | C | T | 0.030617 | 0.004335 | 2.10E-12 |
| rs543968 | let-7b-5p | trans | 5.69E-26 | Nightingale | M_VLDL_T | NA | C | T | -0.03623 | 0.004108 | 1.50E-18 |
| rs543968 | let-7b-5p | trans | 5.69E-26 | Nightingale | S_LDL_L | NA | C | T | 0.026893 | 0.004346 | 1.20E-09 |
| rs543968 | let-7b-5p | trans | 5.69E-26 | Nightingale | IDL_PL | NA | C | T | 0.033573 | 0.004195 | 2.00E-15 |
| rs543968 | let-7b-5p | trans | 5.69E-26 | Nightingale | LDL_C | NA | C | T | 0.031186 | 0.004328 | 9.60E-13 |
| rs543968 | let-7b-5p | trans | 5.69E-26 | Nightingale | XS_VLDL_F | NA | C | T | 0.029134 | 0.004316 | 3.10E-11 |
| rs543968 | let-7b-5p | trans | 5.69E-26 | Nightingale | Clinical_LD | NA | C | T | 0.034557 | 0.004322 | 2.70E-15 |
| rs543968 | let-7b-5p | trans | 5.69E-26 | Nightingale | non_HDL_( | NA | C | T | 0.03414 | 0.004323 | 2.40E-15 |
| rs543968 | let-7b-5p | trans | 5.69E-26 | Nightingale | M_VLDL_F | NA | C | T | 0.039372 | 0.004135 | 2.20E-21 |
| rs543968 | let-7b-5p | trans | 5.69E-26 | Nightingale | XXL_VLDL_ | NA | C | T | 0.029268 | 0.004328 | 5.10E-12 |
| rs543968 | let-7b-5p | trans | 5.69E-26 | Nightingale | M_LDL_PL | NA | C | T | 0.031174 | 0.004353 | 9.90E-13 |
| rs543968 | let-7b-5p | trans | 5.69E-26 | Nightingale | L_VLDL_FC | NA | C | T | 0.046941 | 0.004305 | 3.90E-28 |
| rs543968 | let-7b-5p | trans | 5.69E-26 | Nightingale | ApoB | NA | C | T | 0.032928 | 0.004341 | 3.40E-14 |
| rs543968 | let-7b-5p | trans | 5.69E-26 | Nightingale | IDL_C | NA | C | T | 0.033305 | 0.004198 | 4.90E-15 |
| rs543968 | let-7b-5p | trans | 5.69E-26 | Nightingale | XL_VLDL_P | NA | C | T | 0.039106 | 0.004353 | 7.40E-20 |
| rs543968 | let-7b-5p | trans | 5.69E-26 | Nightingale | S_LDL_CE | NA | C | T | 0.029442 | 0.004349 | 1.50E-11 |
| rs543968 | let-7b-5p | trans | 5.69E-26 | Nightingale | L_LDL_C | NA | C | T | 0.030224 | 0.004298 | 3.00E-12 |
| rs543968 | let-7b-5p | trans | 5.69E-26 | Nightingale | Total_FC | NA | C | T | 0.031378 | 0.004246 | 4.70E-14 |
| rs543968 | let-7b-5p | trans | 5.69E-26 | Nightingale | S_HDL_P | NA | C | T | 0.027805 | 0.004347 | 2.70E-10 |
| rs543968 | let-7b-5p | trans | 5.69E-26 | Nightingale | XXL_VLDL_ | NA | C | T | 0.023502 | 0.004375 | 5.40E-09 |
| rs543968 | let-7b-5p | trans | 5.69E-26 | Nightingale | LDL_FC | NA | C | T | 0.031548 | 0.0043 | 6.60E-13 |
| rs543968 | let-7b-5p | trans | 5.69E-26 | Nightingale | S_VLDL_TG | NA | C | T | -0.03598 | 0.004208 | 1.90E-17 |
| rs543968 | let-7b-5p | trans | 5.69E-26 | Nightingale | L_VLDL_PL | NA | C | T | 0.021645 | 0.004222 | 4.40E-08 |
| rs543968 | let-7b-5p | trans | 5.69E-26 | Nightingale | IDL_FC | NA | C | T | 0.035978 | 0.004208 | 3.20E-17 |
| rs543968 | let-7b-5p | trans | 5.69E-26 | Nightingale | S_VLDL_P | NA | C | T | 0.02339 | 0.004292 | 9.70E-09 |
| rs543968 | let-7b-5p | trans | 5.69E-26 | Nightingale | XXL_VLDL_ | NA | C | T | 0.033437 | 0.004356 | 8.80E-15 |
| rs543968 | let-7b-5p | trans | 5.69E-26 | Nightingale | IDL_P | NA | C | T | 0.039032 | 0.004301 | 1.80E-19 |
| rs543968 | let-7b-5p | trans | 5.69E-26 | Nightingale | S_VLDL_FC | NA | C | T | 0.027154 | 0.004154 | 1.20E-10 |
| rs543968 | let-7b-5p | trans | 5.69E-26 | Nightingale | L_VLDL_CE | NA | C | T | 0.022756 | 0.004296 | 2.90E-08 |
| rs543968 | let-7b-5p | trans | 5.69E-26 | Nightingale | S_VLDL_L | NA | C | T | 0.024496 | 0.004305 | 2.50E-09 |
| rs543968 | let-7b-5p | trans | 5.69E-26 | Nightingale | XS_VLDL_L | NA | C | T | 0.033006 | 0.004273 | 1.40E-15 |
| rs543968 | let-7b-5p | trans | 5.69E-26 | Nightingale | L_LDL_FC | NA | C | T | 0.031473 | 0.004268 | 4.10E-13 |
| rs543968 | let-7b-5p | trans | 5.69E-26 | Nightingale | XXL_VLDL_ | NA | C | T | 0.03179 | 0.00438 | 2.10E-13 |
| rs543968 | let-7b-5p | trans | 5.69E-26 | Nightingale | GlycA | NA | C | T | 0.025329 | 0.004328 | 4.80E-09 |
| rs543968 | let-7b-5p | trans | 5.69E-26 | Nightingale | LA | NA | C | T | 0.023263 | 0.004263 | 1.60E-08 |
| rs543968 | let-7b-5p | trans | 5.69E-26 | Nightingale | L_VLDL_TG | NA | C | T | -0.04923 | 0.004326 | 6.40E-31 |
| rs543968 | let-7b-5p | trans | 5.69E-26 | Nightingale | XXL_VLDL_ | NA | C | T | -0.03443 | 0.004384 | 6.30E-16 |
| rs543968 | let-7b-5p | trans | 5.69E-26 | Nightingale | L_LDL_L | NA | C | T | 0.031523 | 0.004304 | 3.20E-13 |
| rs543968 | let-7b-5p | trans | 5.69E-26 | Nightingale | S_VLDL_PL | NA | C | T | 0.025728 | 0.004162 | 1.50E-09 |
| rs543968 | let-7b-5p | trans | 5.69E-26 | Nightingale | XL_VLDL_C | NA | C | T | 0.034575 | 0.00417 | 1.50E-16 |
| rs543968 | let-7b-5p | trans | 5.69E-26 | Nightingale | XS_VLDL_C | NA | C | T | 0.036391 | 0.004195 | 4.10E-18 |
| rs543968 | let-7b-5p | trans | 5.69E-26 | Nightingale | Total_PL | NA | C | T | 0.022997 | 0.004139 | 7.80E-09 |
| rs543968 | let-7b-5p | trans | 5.69E-26 | Nightingale | Omega_6 | NA | C | T | 0.024156 | 0.004245 | 3.10E-09 |
| rs543968 | let-7b-5p | trans | 5.69E-26 | Nightingale | XS_VLDL_F | NA | C | T | 0.035882 | 0.004271 | 7.50E-18 |
| rs543968 | let-7b-5p | trans | 5.69E-26 | Nightingale | Total_CE | NA | C | T | 0.029384 | 0.004193 | 1.80E-12 |
| rs543968 | let-7b-5p | trans | 5.69E-26 | Nightingale | S_VLDL_PL | NA | C | T | 0.032035 | 0.004339 | 4.90E-14 |
| rs543968 | let-7b-5p | trans | 5.69E-26 | Nightingale | XL_HDL_PL | NA | C | T | -0.02181 | 0.003994 | 4.60E-08 |
| rs543968 | let-7b-5p | trans | 5.69E-26 | Nightingale | L_LDL_PL | NA | C | T | 0.035919 | 0.004314 | 1.30E-16 |
| rs543968 | let-7b-5p | trans | 5.69E-26 | Nightingale | S_LDL_CE_ | NA | C | T | 0.028844 | 0.004339 | 1.10E-12 |
| rs543968 | let-7b-5p | trans | 5.69E-26 | Nightingale | M_LDL_C | NA | C | T | 0.032091 | 0.004355 | 2.30E-13 |
| rs543968 | let-7b-5p | trans | 5.69E-26 | Nightingale | M_LDL_L | NA | C | T | 0.031388 | 0.004353 | 7.00E-13 |
| rs543968 | let-7b-5p | trans | 5.69E-26 | Nightingale | XL_VLDL_C | NA | C | T | 0.03559 | 0.004168 | 1.70E-17 |
| rs543968 | let-7b-5p | trans | 5.69E-26 | Nightingale | S_HDL_CE | NA | C | T | 0.02569 | 0.004354 | 7.00E-09 |
| rs543968 | let-7b-5p | trans | 5.69E-26 | Nightingale | S_VLDL_CE | NA | C | T | 0.037311 | 0.004341 | 1.70E-18 |
| rs543968 | let-7b-5p | trans | 5.69E-26 | Nightingale | IDL_CE | NA | C | T | 0.032088 | 0.004197 | 4.30E-14 |
| rs543968 | let-7b-5p | trans | 5.69E-26 | Nightingale | Citrate | NA | C | T | 0.024678 | 0.004318 | 6.10E-09 |
| rs543968 | let-7b-5p | trans | 5.69E-26 | Nightingale | VLDL_C | NA | C | T | 0.031062 | 0.004341 | 2.10E-13 |
| rs543968 | let-7b-5p | trans | 5.69E-26 | Nightingale | M_LDL_C_ | NA | C | T | 0.026582 | 0.00435 | 1.60E-09 |
| rs543968 | let-7b-5p | trans | 5.69E-26 | Nightingale | XL_VLDL_F | NA | C | T | 0.035204 | 0.004214 | 1.80E-17 |
| rs543968 | let-7b-5p | trans | 5.69E-26 | Nightingale | VLDL_P | NA | C | T | 0.026555 | 0.004324 | 2.00E-10 |
| rs543968 | let-7b-5p | trans | 5.69E-26 | Nightingale | XL_VLDL_T | NA | C | T | -0.04155 | 0.004213 | 4.90E-23 |
| rs543968 | let-7b-5p | trans | 5.69E-26 | Nightingale | M_VLDL_P | NA | C | T | 0.027926 | 0.004336 | 4.70E-11 |
| rs543968 | let-7b-5p | trans | 5.69E-26 | Nightingale | M_LDL_P | NA | C | T | 0.034001 | 0.004359 | 7.50E-15 |

| | | | | | | | | | | | |
|---|---|---|---|---|---|---|---|---|---|---|---|
| rs543968 | let-7b-5p | trans | 5.69E-26 | Nightingale | M_VLDL_P | NA | C | T | 0.042093 | 0.004172 | 6.30E-24 |
| rs543968 | let-7b-5p | trans | 5.69E-26 | Nightingale | S_VLDL_FC | NA | C | T | 0.036642 | 0.004347 | 1.70E-17 |
| rs543968 | let-7b-5p | trans | 5.69E-26 | Nightingale | Total_L | NA | C | T | 0.025008 | 0.004276 | 1.50E-09 |
| rs543968 | let-7b-5p | trans | 5.69E-26 | Nightingale | S_VLDL_CE | NA | C | T | 0.045151 | 0.004288 | 1.40E-25 |
| rs543968 | let-7b-5p | trans | 5.69E-26 | Nightingale | S_LDL_PL_ | NA | C | T | -0.02806 | 0.004339 | 4.90E-12 |
| rs543968 | let-7b-5p | trans | 5.69E-26 | Nightingale | S_LDL_FC | NA | C | T | 0.0272 | 0.00434 | 1.30E-09 |
| rs543968 | let-7b-5p | trans | 5.69E-26 | Nightingale | VLDL_CE | NA | C | T | 0.035144 | 0.004348 | 2.10E-16 |
| rs543968 | let-7b-5p | trans | 5.69E-26 | Nightingale | Sphingomy | NA | C | T | 0.025917 | 0.004063 | 3.30E-10 |
| rs543968 | let-7b-5p | trans | 5.69E-26 | Nightingale | L_LDL_CE | NA | C | T | 0.02952 | 0.004308 | 9.90E-12 |
| rs543968 | let-7b-5p | trans | 5.69E-26 | Nightingale | Remnant_( | NA | C | T | 0.035838 | 0.004319 | 5.30E-17 |
| rs543968 | let-7b-5p | trans | 5.69E-26 | Nightingale | Total_C | NA | C | T | 0.030138 | 0.004208 | 5.00E-13 |
| rs543968 | let-7b-5p | trans | 5.69E-26 | Nightingale | S_LDL_C_p | NA | C | T | 0.041864 | 0.004364 | 5.60E-22 |
| rs543968 | let-7b-5p | trans | 5.69E-26 | Nightingale | VLDL_FC | NA | C | T | 0.024446 | 0.004315 | 3.60E-09 |
| rs543968 | let-7b-5p | trans | 5.69E-26 | Nightingale | ApoB_by_/ | NA | C | T | 0.025679 | 0.004281 | 2.00E-09 |
| rs543968 | let-7b-5p | trans | 5.69E-26 | Nightingale | S_LDL_P | NA | C | T | 0.028959 | 0.004351 | 3.00E-11 |
| rs543968 | let-7b-5p | trans | 5.69E-26 | Nightingale | S_LDL_C | NA | C | T | 0.029698 | 0.004351 | 1.60E-11 |
| rs543968 | let-7b-5p | trans | 5.69E-26 | Nightingale | L_VLDL_CE | NA | C | T | 0.04407 | 0.00416 | 7.90E-27 |
| rs543968 | let-7b-5p | trans | 5.69E-26 | Nightingale | LDL_P | NA | C | T | 0.030659 | 0.004344 | 2.20E-12 |
| rs543968 | let-7b-5p | trans | 5.69E-26 | Nightingale | M_VLDL_C | NA | C | T | 0.0392 | 0.004307 | 1.30E-19 |
| rs543968 | let-7b-5p | trans | 5.69E-26 | Nightingale | M_LDL_FC | NA | C | T | 0.032281 | 0.004341 | 2.40E-13 |
| rs543968 | let-7b-5p | trans | 5.69E-26 | Nightingale | M_VLDL_C_ | NA | C | T | 0.033845 | 0.004094 | 2.70E-16 |
| rs543968 | let-7b-5p | trans | 5.69E-26 | Nightingale | XS_VLDL_C | NA | C | T | 0.036639 | 0.004219 | 2.40E-18 |
| rs543968 | let-7b-5p | trans | 5.69E-26 | Nightingale | M_VLDL_P | NA | C | T | 0.028753 | 0.00434 | 1.40E-11 |
| rs543968 | let-7b-5p | trans | 5.69E-26 | Nightingale | S_VLDL_C | NA | C | T | 0.037355 | 0.004346 | 2.60E-18 |
| rs543968 | let-7b-5p | trans | 5.69E-26 | Nightingale | S_HDL_L | NA | C | T | 0.023969 | 0.004332 | 2.70E-08 |
| rs543968 | let-7c-5p | trans | 4.68E-23 | Nightingale | IDL_L | NA | C | T | 0.03366 | 0.004203 | 2.30E-15 |
| rs543968 | let-7c-5p | trans | 4.68E-23 | Nightingale | S_VLDL_C_ | NA | C | T | 0.039736 | 0.00423 | 8.30E-21 |
| rs543968 | let-7c-5p | trans | 4.68E-23 | Nightingale | S_HDL_C | NA | C | T | 0.02538 | 0.004339 | 9.40E-09 |
| rs543968 | let-7c-5p | trans | 4.68E-23 | Nightingale | M_VLDL_C | NA | C | T | 0.037341 | 0.004335 | 5.40E-18 |
| rs543968 | let-7c-5p | trans | 4.68E-23 | Nightingale | LDL_L | NA | C | T | 0.031465 | 0.004332 | 5.80E-13 |
| rs543968 | let-7c-5p | trans | 4.68E-23 | Nightingale | XS_VLDL_P | NA | C | T | 0.031657 | 0.004286 | 2.00E-14 |
| rs543968 | let-7c-5p | trans | 4.68E-23 | Nightingale | L_VLDL_C_ | NA | C | T | 0.049226 | 0.004245 | 8.90E-32 |
| rs543968 | let-7c-5p | trans | 4.68E-23 | Nightingale | M_LDL_TG | NA | C | T | -0.02512 | 0.004293 | 8.60E-09 |
| rs543968 | let-7c-5p | trans | 4.68E-23 | Nightingale | L_LDL_P | NA | C | T | 0.028317 | 0.004332 | 7.80E-11 |
| rs543968 | let-7c-5p | trans | 4.68E-23 | Nightingale | M_LDL_CE | NA | C | T | 0.031212 | 0.004353 | 8.50E-13 |
| rs543968 | let-7c-5p | trans | 4.68E-23 | Nightingale | XS_VLDL_P | NA | C | T | 0.035785 | 0.004289 | 1.00E-17 |
| rs543968 | let-7c-5p | trans | 4.68E-23 | Nightingale | M_VLDL_F( | NA | C | T | 0.032515 | 0.004346 | 3.90E-14 |
| rs543968 | let-7c-5p | trans | 4.68E-23 | Nightingale | M_VLDL_C | NA | C | T | 0.031348 | 0.004084 | 3.30E-14 |
| rs543968 | let-7c-5p | trans | 4.68E-23 | Nightingale | LDL_PL | NA | C | T | 0.033318 | 0.004337 | 3.00E-14 |
| rs543968 | let-7c-5p | trans | 4.68E-23 | Nightingale | LDL_CE | NA | C | T | 0.030617 | 0.004335 | 2.10E-12 |
| rs543968 | let-7c-5p | trans | 4.68E-23 | Nightingale | M_VLDL_T( | NA | C | T | -0.03623 | 0.004108 | 1.50E-18 |
| rs543968 | let-7c-5p | trans | 4.68E-23 | Nightingale | S_LDL_L | NA | C | T | 0.026893 | 0.004346 | 1.20E-09 |
| rs543968 | let-7c-5p | trans | 4.68E-23 | Nightingale | IDL_PL | NA | C | T | 0.033573 | 0.004195 | 2.00E-15 |
| rs543968 | let-7c-5p | trans | 4.68E-23 | Nightingale | LDL_C | NA | C | T | 0.031186 | 0.004328 | 9.60E-13 |
| rs543968 | let-7c-5p | trans | 4.68E-23 | Nightingale | XS_VLDL_F | NA | C | T | 0.029134 | 0.004316 | 3.10E-11 |
| rs543968 | let-7c-5p | trans | 4.68E-23 | Nightingale | Clinical_LD | NA | C | T | 0.034557 | 0.004322 | 2.70E-15 |
| rs543968 | let-7c-5p | trans | 4.68E-23 | Nightingale | non_HDL_( | NA | C | T | 0.03414 | 0.004323 | 2.40E-15 |
| rs543968 | let-7c-5p | trans | 4.68E-23 | Nightingale | M_VLDL_F( | NA | C | T | 0.039372 | 0.004135 | 2.20E-21 |
| rs543968 | let-7c-5p | trans | 4.68E-23 | Nightingale | XXL_VLDL_ | NA | C | T | 0.029268 | 0.004328 | 5.10E-12 |
| rs543968 | let-7c-5p | trans | 4.68E-23 | Nightingale | M_LDL_PL | NA | C | T | 0.031174 | 0.004353 | 9.90E-13 |
| rs543968 | let-7c-5p | trans | 4.68E-23 | Nightingale | L_VLDL_FC | NA | C | T | 0.046941 | 0.004305 | 3.90E-28 |
| rs543968 | let-7c-5p | trans | 4.68E-23 | Nightingale | ApoB | NA | C | T | 0.032928 | 0.004341 | 3.40E-14 |
| rs543968 | let-7c-5p | trans | 4.68E-23 | Nightingale | IDL_C | NA | C | T | 0.033305 | 0.004198 | 4.90E-15 |
| rs543968 | let-7c-5p | trans | 4.68E-23 | Nightingale | XL_VLDL_P | NA | C | T | 0.039106 | 0.004353 | 7.40E-20 |
| rs543968 | let-7c-5p | trans | 4.68E-23 | Nightingale | S_LDL_CE | NA | C | T | 0.029442 | 0.004349 | 1.50E-11 |
| rs543968 | let-7c-5p | trans | 4.68E-23 | Nightingale | L_LDL_C | NA | C | T | 0.030224 | 0.004298 | 3.00E-12 |
| rs543968 | let-7c-5p | trans | 4.68E-23 | Nightingale | Total_FC | NA | C | T | 0.031378 | 0.004246 | 4.70E-14 |
| rs543968 | let-7c-5p | trans | 4.68E-23 | Nightingale | S_HDL_P | NA | C | T | 0.027805 | 0.004347 | 2.70E-10 |
| rs543968 | let-7c-5p | trans | 4.68E-23 | Nightingale | XXL_VLDL_ | NA | C | T | 0.023502 | 0.004375 | 5.40E-09 |
| rs543968 | let-7c-5p | trans | 4.68E-23 | Nightingale | LDL_FC | NA | C | T | 0.031548 | 0.0043 | 6.60E-13 |
| rs543968 | let-7c-5p | trans | 4.68E-23 | Nightingale | S_VLDL_TG | NA | C | T | -0.03598 | 0.004208 | 1.90E-17 |
| rs543968 | let-7c-5p | trans | 4.68E-23 | Nightingale | L_VLDL_PL | NA | C | T | 0.021645 | 0.004222 | 4.40E-08 |
| rs543968 | let-7c-5p | trans | 4.68E-23 | Nightingale | IDL_FC | NA | C | T | 0.035978 | 0.004208 | 3.20E-17 |

| | | | | | | | | | | | |
|---|---|---|---|---|---|---|---|---|---|---|---|
| rs543968 | let-7c-5p | trans | 4.68E-23 | Nightingale | S_VLDL_P | NA | C | T | 0.02339 | 0.004292 | 9.70E-09 |
| rs543968 | let-7c-5p | trans | 4.68E-23 | Nightingale | XXL_VLDL_ | NA | C | T | 0.033437 | 0.004356 | 8.80E-15 |
| rs543968 | let-7c-5p | trans | 4.68E-23 | Nightingale | IDL_P | NA | C | T | 0.039032 | 0.004301 | 1.80E-19 |
| rs543968 | let-7c-5p | trans | 4.68E-23 | Nightingale | S_VLDL_FC | NA | C | T | 0.027154 | 0.004154 | 1.20E-10 |
| rs543968 | let-7c-5p | trans | 4.68E-23 | Nightingale | L_VLDL_CE | NA | C | T | 0.022756 | 0.004296 | 2.90E-08 |
| rs543968 | let-7c-5p | trans | 4.68E-23 | Nightingale | S_VLDL_L | NA | C | T | 0.024496 | 0.004305 | 2.50E-09 |
| rs543968 | let-7c-5p | trans | 4.68E-23 | Nightingale | XS_VLDL_L | NA | C | T | 0.033006 | 0.004273 | 1.40E-15 |
| rs543968 | let-7c-5p | trans | 4.68E-23 | Nightingale | L_LDL_FC | NA | C | T | 0.031473 | 0.004268 | 4.10E-13 |
| rs543968 | let-7c-5p | trans | 4.68E-23 | Nightingale | XXL_VLDL_ | NA | C | T | 0.03179 | 0.00438 | 2.10E-13 |
| rs543968 | let-7c-5p | trans | 4.68E-23 | Nightingale | GlycA | NA | C | T | 0.025329 | 0.004328 | 4.80E-09 |
| rs543968 | let-7c-5p | trans | 4.68E-23 | Nightingale | LA | NA | C | T | 0.023263 | 0.004263 | 1.60E-08 |
| rs543968 | let-7c-5p | trans | 4.68E-23 | Nightingale | L_VLDL_TG | NA | C | T | -0.04923 | 0.004326 | 6.40E-31 |
| rs543968 | let-7c-5p | trans | 4.68E-23 | Nightingale | XXL_VLDL_ | NA | C | T | -0.03443 | 0.004384 | 6.30E-16 |
| rs543968 | let-7c-5p | trans | 4.68E-23 | Nightingale | L_LDL_L | NA | C | T | 0.031523 | 0.004304 | 3.20E-13 |
| rs543968 | let-7c-5p | trans | 4.68E-23 | Nightingale | S_VLDL_PL | NA | C | T | 0.025728 | 0.004162 | 1.50E-09 |
| rs543968 | let-7c-5p | trans | 4.68E-23 | Nightingale | XL_VLDL_C | NA | C | T | 0.034575 | 0.00417 | 1.50E-16 |
| rs543968 | let-7c-5p | trans | 4.68E-23 | Nightingale | XS_VLDL_C | NA | C | T | 0.036391 | 0.004195 | 4.10E-18 |
| rs543968 | let-7c-5p | trans | 4.68E-23 | Nightingale | Total_PL | NA | C | T | 0.022997 | 0.004139 | 7.80E-09 |
| rs543968 | let-7c-5p | trans | 4.68E-23 | Nightingale | Omega_6 | NA | C | T | 0.024156 | 0.004245 | 3.10E-09 |
| rs543968 | let-7c-5p | trans | 4.68E-23 | Nightingale | XS_VLDL_F | NA | C | T | 0.035882 | 0.004271 | 7.50E-18 |
| rs543968 | let-7c-5p | trans | 4.68E-23 | Nightingale | Total_CE | NA | C | T | 0.029384 | 0.004193 | 1.80E-12 |
| rs543968 | let-7c-5p | trans | 4.68E-23 | Nightingale | S_VLDL_PL | NA | C | T | 0.032035 | 0.004339 | 4.90E-14 |
| rs543968 | let-7c-5p | trans | 4.68E-23 | Nightingale | XL_HDL_PL | NA | C | T | -0.02181 | 0.003994 | 4.60E-08 |
| rs543968 | let-7c-5p | trans | 4.68E-23 | Nightingale | L_LDL_PL | NA | C | T | 0.035919 | 0.004314 | 1.30E-16 |
| rs543968 | let-7c-5p | trans | 4.68E-23 | Nightingale | S_LDL_CE_ | NA | C | T | 0.028844 | 0.004339 | 1.10E-12 |
| rs543968 | let-7c-5p | trans | 4.68E-23 | Nightingale | M_LDL_C | NA | C | T | 0.032091 | 0.004355 | 2.30E-13 |
| rs543968 | let-7c-5p | trans | 4.68E-23 | Nightingale | M_LDL_L | NA | C | T | 0.031388 | 0.004353 | 7.00E-13 |
| rs543968 | let-7c-5p | trans | 4.68E-23 | Nightingale | XL_VLDL_C | NA | C | T | 0.03559 | 0.004168 | 1.70E-17 |
| rs543968 | let-7c-5p | trans | 4.68E-23 | Nightingale | S_HDL_CE | NA | C | T | 0.02569 | 0.004354 | 7.00E-09 |
| rs543968 | let-7c-5p | trans | 4.68E-23 | Nightingale | S_VLDL_CE | NA | C | T | 0.037311 | 0.004341 | 1.70E-18 |
| rs543968 | let-7c-5p | trans | 4.68E-23 | Nightingale | IDL_CE | NA | C | T | 0.032088 | 0.004197 | 4.30E-14 |
| rs543968 | let-7c-5p | trans | 4.68E-23 | Nightingale | Citrate | NA | C | T | 0.024678 | 0.004318 | 6.10E-09 |
| rs543968 | let-7c-5p | trans | 4.68E-23 | Nightingale | VLDL_C | NA | C | T | 0.031062 | 0.004341 | 2.10E-13 |
| rs543968 | let-7c-5p | trans | 4.68E-23 | Nightingale | M_LDL_C_ | NA | C | T | 0.026582 | 0.00435 | 1.60E-09 |
| rs543968 | let-7c-5p | trans | 4.68E-23 | Nightingale | XL_VLDL_F | NA | C | T | 0.035204 | 0.004214 | 1.80E-17 |
| rs543968 | let-7c-5p | trans | 4.68E-23 | Nightingale | VLDL_P | NA | C | T | 0.026555 | 0.004324 | 2.00E-10 |
| rs543968 | let-7c-5p | trans | 4.68E-23 | Nightingale | XL_VLDL_T | NA | C | T | -0.04155 | 0.004213 | 4.90E-23 |
| rs543968 | let-7c-5p | trans | 4.68E-23 | Nightingale | M_VLDL_P | NA | C | T | 0.027926 | 0.004336 | 4.70E-11 |
| rs543968 | let-7c-5p | trans | 4.68E-23 | Nightingale | M_LDL_P | NA | C | T | 0.034001 | 0.004359 | 7.50E-15 |
| rs543968 | let-7c-5p | trans | 4.68E-23 | Nightingale | M_VLDL_P | NA | C | T | 0.042093 | 0.004172 | 6.30E-24 |
| rs543968 | let-7c-5p | trans | 4.68E-23 | Nightingale | S_VLDL_FC | NA | C | T | 0.036642 | 0.004347 | 1.70E-17 |
| rs543968 | let-7c-5p | trans | 4.68E-23 | Nightingale | Total_L | NA | C | T | 0.025008 | 0.004276 | 1.50E-09 |
| rs543968 | let-7c-5p | trans | 4.68E-23 | Nightingale | S_VLDL_CE | NA | C | T | 0.045151 | 0.004288 | 1.40E-25 |
| rs543968 | let-7c-5p | trans | 4.68E-23 | Nightingale | S_LDL_PL_ | NA | C | T | -0.02806 | 0.004339 | 4.90E-12 |
| rs543968 | let-7c-5p | trans | 4.68E-23 | Nightingale | S_LDL_FC | NA | C | T | 0.0272 | 0.00434 | 1.30E-09 |
| rs543968 | let-7c-5p | trans | 4.68E-23 | Nightingale | VLDL_CE | NA | C | T | 0.035144 | 0.004348 | 2.10E-16 |
| rs543968 | let-7c-5p | trans | 4.68E-23 | Nightingale | Sphingomy | NA | C | T | 0.025917 | 0.004063 | 3.30E-10 |
| rs543968 | let-7c-5p | trans | 4.68E-23 | Nightingale | L_LDL_CE | NA | C | T | 0.02952 | 0.004308 | 9.90E-12 |
| rs543968 | let-7c-5p | trans | 4.68E-23 | Nightingale | Remnant_( | NA | C | T | 0.035838 | 0.004319 | 5.30E-17 |
| rs543968 | let-7c-5p | trans | 4.68E-23 | Nightingale | Total_C | NA | C | T | 0.030138 | 0.004208 | 5.00E-13 |
| rs543968 | let-7c-5p | trans | 4.68E-23 | Nightingale | S_LDL_C_p | NA | C | T | 0.041864 | 0.004364 | 5.60E-22 |
| rs543968 | let-7c-5p | trans | 4.68E-23 | Nightingale | VLDL_FC | NA | C | T | 0.024446 | 0.004315 | 3.60E-09 |
| rs543968 | let-7c-5p | trans | 4.68E-23 | Nightingale | ApoB_by_/ | NA | C | T | 0.025679 | 0.004281 | 2.00E-09 |
| rs543968 | let-7c-5p | trans | 4.68E-23 | Nightingale | S_LDL_P | NA | C | T | 0.028959 | 0.004351 | 3.00E-11 |
| rs543968 | let-7c-5p | trans | 4.68E-23 | Nightingale | S_LDL_C | NA | C | T | 0.029698 | 0.004351 | 1.60E-11 |
| rs543968 | let-7c-5p | trans | 4.68E-23 | Nightingale | L_VLDL_CE | NA | C | T | 0.04407 | 0.00416 | 7.90E-27 |
| rs543968 | let-7c-5p | trans | 4.68E-23 | Nightingale | LDL_P | NA | C | T | 0.030659 | 0.004344 | 2.20E-12 |
| rs543968 | let-7c-5p | trans | 4.68E-23 | Nightingale | M_VLDL_C | NA | C | T | 0.0392 | 0.004307 | 1.30E-19 |
| rs543968 | let-7c-5p | trans | 4.68E-23 | Nightingale | M_LDL_FC | NA | C | T | 0.032281 | 0.004341 | 2.40E-13 |
| rs543968 | let-7c-5p | trans | 4.68E-23 | Nightingale | M_VLDL_C | NA | C | T | 0.033845 | 0.004094 | 2.70E-16 |
| rs543968 | let-7c-5p | trans | 4.68E-23 | Nightingale | XS_VLDL_C | NA | C | T | 0.036639 | 0.004219 | 2.40E-18 |
| rs543968 | let-7c-5p | trans | 4.68E-23 | Nightingale | M_VLDL_P | NA | C | T | 0.028753 | 0.00434 | 1.40E-11 |
| rs543968 | let-7c-5p | trans | 4.68E-23 | Nightingale | S_VLDL_C | NA | C | T | 0.037355 | 0.004346 | 2.60E-18 |

| | | | | | | | | | | | |
|---|---|---|---|---|---|---|---|---|---|---|---|
| rs543968 | let-7c-5p | trans | 4.68E-23 | Nightingale | S_HDL_L | NA | C | T | 0.023969 | 0.004332 | 2.70E-08 |
| rs543968 | miR-139-5p | trans | 4.23E-40 | Nightingale | IDL_L | NA | C | T | 0.03366 | 0.004203 | 2.30E-15 |
| rs543968 | miR-139-5p | trans | 4.23E-40 | Nightingale | S_VLDL_C_ | NA | C | T | 0.039736 | 0.00423 | 8.30E-21 |
| rs543968 | miR-139-5p | trans | 4.23E-40 | Nightingale | S_HDL_C | NA | C | T | 0.02538 | 0.004339 | 9.40E-09 |
| rs543968 | miR-139-5p | trans | 4.23E-40 | Nightingale | M_VLDL_C | NA | C | T | 0.037341 | 0.004335 | 5.40E-18 |
| rs543968 | miR-139-5p | trans | 4.23E-40 | Nightingale | LDL_L | NA | C | T | 0.031465 | 0.004332 | 5.80E-13 |
| rs543968 | miR-139-5p | trans | 4.23E-40 | Nightingale | XS_VLDL_P | NA | C | T | 0.031657 | 0.004286 | 2.00E-14 |
| rs543968 | miR-139-5p | trans | 4.23E-40 | Nightingale | L_VLDL_C_ | NA | C | T | 0.049226 | 0.004245 | 8.90E-32 |
| rs543968 | miR-139-5p | trans | 4.23E-40 | Nightingale | M_LDL_TG | NA | C | T | -0.02512 | 0.004293 | 8.60E-09 |
| rs543968 | miR-139-5p | trans | 4.23E-40 | Nightingale | L_LDL_P | NA | C | T | 0.028317 | 0.004332 | 7.80E-11 |
| rs543968 | miR-139-5p | trans | 4.23E-40 | Nightingale | M_LDL_CE | NA | C | T | 0.031212 | 0.004353 | 8.50E-13 |
| rs543968 | miR-139-5p | trans | 4.23E-40 | Nightingale | XS_VLDL_P | NA | C | T | 0.035785 | 0.004289 | 1.00E-17 |
| rs543968 | miR-139-5p | trans | 4.23E-40 | Nightingale | M_VLDL_F | NA | C | T | 0.032515 | 0.004346 | 3.90E-14 |
| rs543968 | miR-139-5p | trans | 4.23E-40 | Nightingale | M_VLDL_C | NA | C | T | 0.031348 | 0.004084 | 3.30E-14 |
| rs543968 | miR-139-5p | trans | 4.23E-40 | Nightingale | LDL_PL | NA | C | T | 0.033318 | 0.004337 | 3.00E-14 |
| rs543968 | miR-139-5p | trans | 4.23E-40 | Nightingale | LDL_CE | NA | C | T | 0.030617 | 0.004335 | 2.10E-12 |
| rs543968 | miR-139-5p | trans | 4.23E-40 | Nightingale | M_VLDL_T | NA | C | T | -0.03623 | 0.004108 | 1.50E-18 |
| rs543968 | miR-139-5p | trans | 4.23E-40 | Nightingale | S_LDL_L | NA | C | T | 0.026893 | 0.004346 | 1.20E-09 |
| rs543968 | miR-139-5p | trans | 4.23E-40 | Nightingale | IDL_PL | NA | C | T | 0.033573 | 0.004195 | 2.00E-15 |
| rs543968 | miR-139-5p | trans | 4.23E-40 | Nightingale | LDL_C | NA | C | T | 0.031186 | 0.004328 | 9.60E-13 |
| rs543968 | miR-139-5p | trans | 4.23E-40 | Nightingale | XS_VLDL_F | NA | C | T | 0.029134 | 0.004316 | 3.10E-11 |
| rs543968 | miR-139-5p | trans | 4.23E-40 | Nightingale | Clinical_LD | NA | C | T | 0.034557 | 0.004322 | 2.70E-15 |
| rs543968 | miR-139-5p | trans | 4.23E-40 | Nightingale | non_HDL_( | NA | C | T | 0.03414 | 0.004323 | 2.40E-15 |
| rs543968 | miR-139-5p | trans | 4.23E-40 | Nightingale | M_VLDL_F | NA | C | T | 0.039372 | 0.004135 | 2.20E-21 |
| rs543968 | miR-139-5p | trans | 4.23E-40 | Nightingale | XXL_VLDL_ | NA | C | T | 0.029268 | 0.004328 | 5.10E-12 |
| rs543968 | miR-139-5p | trans | 4.23E-40 | Nightingale | M_LDL_PL | NA | C | T | 0.031174 | 0.004353 | 9.90E-13 |
| rs543968 | miR-139-5p | trans | 4.23E-40 | Nightingale | L_VLDL_FC | NA | C | T | 0.046941 | 0.004305 | 3.90E-28 |
| rs543968 | miR-139-5p | trans | 4.23E-40 | Nightingale | ApoB | NA | C | T | 0.032928 | 0.004341 | 3.40E-14 |
| rs543968 | miR-139-5p | trans | 4.23E-40 | Nightingale | IDL_C | NA | C | T | 0.033305 | 0.004198 | 4.90E-15 |
| rs543968 | miR-139-5p | trans | 4.23E-40 | Nightingale | XL_VLDL_P | NA | C | T | 0.039106 | 0.004353 | 7.40E-20 |
| rs543968 | miR-139-5p | trans | 4.23E-40 | Nightingale | S_LDL_CE | NA | C | T | 0.029442 | 0.004349 | 1.50E-11 |
| rs543968 | miR-139-5p | trans | 4.23E-40 | Nightingale | L_LDL_C | NA | C | T | 0.030224 | 0.004298 | 3.00E-12 |
| rs543968 | miR-139-5p | trans | 4.23E-40 | Nightingale | Total_FC | NA | C | T | 0.031378 | 0.004246 | 4.70E-14 |
| rs543968 | miR-139-5p | trans | 4.23E-40 | Nightingale | S_HDL_P | NA | C | T | 0.027805 | 0.004347 | 2.70E-10 |
| rs543968 | miR-139-5p | trans | 4.23E-40 | Nightingale | XXL_VLDL_ | NA | C | T | 0.023502 | 0.004375 | 5.40E-09 |
| rs543968 | miR-139-5p | trans | 4.23E-40 | Nightingale | LDL_FC | NA | C | T | 0.031548 | 0.0043 | 6.60E-13 |
| rs543968 | miR-139-5p | trans | 4.23E-40 | Nightingale | S_VLDL_TG | NA | C | T | -0.03598 | 0.004208 | 1.90E-17 |
| rs543968 | miR-139-5p | trans | 4.23E-40 | Nightingale | L_VLDL_PL | NA | C | T | 0.021645 | 0.004222 | 4.40E-08 |
| rs543968 | miR-139-5p | trans | 4.23E-40 | Nightingale | IDL_FC | NA | C | T | 0.035978 | 0.004208 | 3.20E-17 |
| rs543968 | miR-139-5p | trans | 4.23E-40 | Nightingale | S_VLDL_P | NA | C | T | 0.02339 | 0.004292 | 9.70E-09 |
| rs543968 | miR-139-5p | trans | 4.23E-40 | Nightingale | XXL_VLDL_ | NA | C | T | 0.033437 | 0.004356 | 8.80E-15 |
| rs543968 | miR-139-5p | trans | 4.23E-40 | Nightingale | IDL_P | NA | C | T | 0.039032 | 0.004301 | 1.80E-19 |
| rs543968 | miR-139-5p | trans | 4.23E-40 | Nightingale | S_VLDL_FC | NA | C | T | 0.027154 | 0.004154 | 1.20E-10 |
| rs543968 | miR-139-5p | trans | 4.23E-40 | Nightingale | L_VLDL_CE | NA | C | T | 0.022756 | 0.004296 | 2.90E-08 |
| rs543968 | miR-139-5p | trans | 4.23E-40 | Nightingale | S_VLDL_L | NA | C | T | 0.024496 | 0.004305 | 2.50E-09 |
| rs543968 | miR-139-5p | trans | 4.23E-40 | Nightingale | XS_VLDL_L | NA | C | T | 0.033006 | 0.004273 | 1.40E-15 |
| rs543968 | miR-139-5p | trans | 4.23E-40 | Nightingale | L_LDL_FC | NA | C | T | 0.031473 | 0.004268 | 4.10E-13 |
| rs543968 | miR-139-5p | trans | 4.23E-40 | Nightingale | XXL_VLDL_ | NA | C | T | 0.03179 | 0.00438 | 2.10E-13 |
| rs543968 | miR-139-5p | trans | 4.23E-40 | Nightingale | GlycA | NA | C | T | 0.025329 | 0.004328 | 4.80E-09 |
| rs543968 | miR-139-5p | trans | 4.23E-40 | Nightingale | LA | NA | C | T | 0.023263 | 0.004263 | 1.60E-08 |
| rs543968 | miR-139-5p | trans | 4.23E-40 | Nightingale | L_VLDL_TG | NA | C | T | -0.04923 | 0.004326 | 6.40E-31 |
| rs543968 | miR-139-5p | trans | 4.23E-40 | Nightingale | XXL_VLDL_ | NA | C | T | -0.03443 | 0.004384 | 6.30E-16 |
| rs543968 | miR-139-5p | trans | 4.23E-40 | Nightingale | L_LDL_L | NA | C | T | 0.031523 | 0.004304 | 3.20E-13 |
| rs543968 | miR-139-5p | trans | 4.23E-40 | Nightingale | S_VLDL_PL | NA | C | T | 0.025728 | 0.004162 | 1.50E-09 |
| rs543968 | miR-139-5p | trans | 4.23E-40 | Nightingale | XL_VLDL_C | NA | C | T | 0.034575 | 0.00417 | 1.50E-16 |
| rs543968 | miR-139-5p | trans | 4.23E-40 | Nightingale | XS_VLDL_C | NA | C | T | 0.036391 | 0.004195 | 4.10E-18 |
| rs543968 | miR-139-5p | trans | 4.23E-40 | Nightingale | Total_PL | NA | C | T | 0.022997 | 0.004139 | 7.80E-09 |
| rs543968 | miR-139-5p | trans | 4.23E-40 | Nightingale | Omega_6 | NA | C | T | 0.024156 | 0.004245 | 3.10E-09 |
| rs543968 | miR-139-5p | trans | 4.23E-40 | Nightingale | XS_VLDL_F | NA | C | T | 0.035882 | 0.004271 | 7.50E-18 |
| rs543968 | miR-139-5p | trans | 4.23E-40 | Nightingale | Total_CE | NA | C | T | 0.029384 | 0.004193 | 1.80E-12 |
| rs543968 | miR-139-5p | trans | 4.23E-40 | Nightingale | S_VLDL_PL | NA | C | T | 0.032035 | 0.004339 | 4.90E-14 |
| rs543968 | miR-139-5p | trans | 4.23E-40 | Nightingale | XL_HDL_PL | NA | C | T | -0.02181 | 0.003994 | 4.60E-08 |
| rs543968 | miR-139-5p | trans | 4.23E-40 | Nightingale | L_LDL_PL | NA | C | T | 0.035919 | 0.004314 | 1.30E-16 |

| | | | | | | | | | | | |
|---|---|---|---|---|---|---|---|---|---|---|---|
| rs543968 | miR-139-5p | trans | 4.23E-40 | Nightingale | S_LDL_CE_ | NA | C | T | 0.028844 | 0.004339 | 1.10E-12 |
| rs543968 | miR-139-5p | trans | 4.23E-40 | Nightingale | M_LDL_C | NA | C | T | 0.032091 | 0.004355 | 2.30E-13 |
| rs543968 | miR-139-5p | trans | 4.23E-40 | Nightingale | M_LDL_L | NA | C | T | 0.031388 | 0.004353 | 7.00E-13 |
| rs543968 | miR-139-5p | trans | 4.23E-40 | Nightingale | XL_VLDL_C | NA | C | T | 0.03559 | 0.004168 | 1.70E-17 |
| rs543968 | miR-139-5p | trans | 4.23E-40 | Nightingale | S_HDL_CE | NA | C | T | 0.02569 | 0.004354 | 7.00E-09 |
| rs543968 | miR-139-5p | trans | 4.23E-40 | Nightingale | S_VLDL_CE | NA | C | T | 0.037311 | 0.004341 | 1.70E-18 |
| rs543968 | miR-139-5p | trans | 4.23E-40 | Nightingale | IDL_CE | NA | C | T | 0.032088 | 0.004197 | 4.30E-14 |
| rs543968 | miR-139-5p | trans | 4.23E-40 | Nightingale | Citrate | NA | C | T | 0.024678 | 0.004318 | 6.10E-09 |
| rs543968 | miR-139-5p | trans | 4.23E-40 | Nightingale | VLDL_C | NA | C | T | 0.031062 | 0.004341 | 2.10E-13 |
| rs543968 | miR-139-5p | trans | 4.23E-40 | Nightingale | M_LDL_C_ | NA | C | T | 0.026582 | 0.00435 | 1.60E-09 |
| rs543968 | miR-139-5p | trans | 4.23E-40 | Nightingale | XL_VLDL_F | NA | C | T | 0.035204 | 0.004214 | 1.80E-17 |
| rs543968 | miR-139-5p | trans | 4.23E-40 | Nightingale | VLDL_P | NA | C | T | 0.026555 | 0.004324 | 2.00E-10 |
| rs543968 | miR-139-5p | trans | 4.23E-40 | Nightingale | XL_VLDL_T | NA | C | T | -0.04155 | 0.004213 | 4.90E-23 |
| rs543968 | miR-139-5p | trans | 4.23E-40 | Nightingale | M_VLDL_P | NA | C | T | 0.027926 | 0.004336 | 4.70E-11 |
| rs543968 | miR-139-5p | trans | 4.23E-40 | Nightingale | M_LDL_P | NA | C | T | 0.034001 | 0.004359 | 7.50E-15 |
| rs543968 | miR-139-5p | trans | 4.23E-40 | Nightingale | M_VLDL_P | NA | C | T | 0.042093 | 0.004172 | 6.30E-24 |
| rs543968 | miR-139-5p | trans | 4.23E-40 | Nightingale | S_VLDL_FC | NA | C | T | 0.036642 | 0.004347 | 1.70E-17 |
| rs543968 | miR-139-5p | trans | 4.23E-40 | Nightingale | Total_L | NA | C | T | 0.025008 | 0.004276 | 1.50E-09 |
| rs543968 | miR-139-5p | trans | 4.23E-40 | Nightingale | S_VLDL_CE | NA | C | T | 0.045151 | 0.004288 | 1.40E-25 |
| rs543968 | miR-139-5p | trans | 4.23E-40 | Nightingale | S_LDL_PL_ | NA | C | T | -0.02806 | 0.004339 | 4.90E-12 |
| rs543968 | miR-139-5p | trans | 4.23E-40 | Nightingale | S_LDL_FC | NA | C | T | 0.0272 | 0.00434 | 1.30E-09 |
| rs543968 | miR-139-5p | trans | 4.23E-40 | Nightingale | VLDL_CE | NA | C | T | 0.035144 | 0.004348 | 2.10E-16 |
| rs543968 | miR-139-5p | trans | 4.23E-40 | Nightingale | Sphingomy | NA | C | T | 0.025917 | 0.004063 | 3.30E-10 |
| rs543968 | miR-139-5p | trans | 4.23E-40 | Nightingale | L_LDL_CE | NA | C | T | 0.02952 | 0.004308 | 9.90E-12 |
| rs543968 | miR-139-5p | trans | 4.23E-40 | Nightingale | Remnant_( | NA | C | T | 0.035838 | 0.004319 | 5.30E-17 |
| rs543968 | miR-139-5p | trans | 4.23E-40 | Nightingale | Total_C | NA | C | T | 0.030138 | 0.004208 | 5.00E-13 |
| rs543968 | miR-139-5p | trans | 4.23E-40 | Nightingale | S_LDL_C_p | NA | C | T | 0.041864 | 0.004364 | 5.60E-22 |
| rs543968 | miR-139-5p | trans | 4.23E-40 | Nightingale | VLDL_FC | NA | C | T | 0.024446 | 0.004315 | 3.60E-09 |
| rs543968 | miR-139-5p | trans | 4.23E-40 | Nightingale | ApoB_by_/ | NA | C | T | 0.025679 | 0.004281 | 2.00E-09 |
| rs543968 | miR-139-5p | trans | 4.23E-40 | Nightingale | S_LDL_P | NA | C | T | 0.028959 | 0.004351 | 3.00E-11 |
| rs543968 | miR-139-5p | trans | 4.23E-40 | Nightingale | S_LDL_C | NA | C | T | 0.029698 | 0.004351 | 1.60E-11 |
| rs543968 | miR-139-5p | trans | 4.23E-40 | Nightingale | L_VLDL_CE | NA | C | T | 0.04407 | 0.00416 | 7.90E-27 |
| rs543968 | miR-139-5p | trans | 4.23E-40 | Nightingale | LDL_P | NA | C | T | 0.030659 | 0.004344 | 2.20E-12 |
| rs543968 | miR-139-5p | trans | 4.23E-40 | Nightingale | M_VLDL_C | NA | C | T | 0.0392 | 0.004307 | 1.30E-19 |
| rs543968 | miR-139-5p | trans | 4.23E-40 | Nightingale | M_LDL_FC | NA | C | T | 0.032281 | 0.004341 | 2.40E-13 |
| rs543968 | miR-139-5p | trans | 4.23E-40 | Nightingale | M_VLDL_C | NA | C | T | 0.033845 | 0.004094 | 2.70E-16 |
| rs543968 | miR-139-5p | trans | 4.23E-40 | Nightingale | XS_VLDL_C | NA | C | T | 0.036639 | 0.004219 | 2.40E-18 |
| rs543968 | miR-139-5p | trans | 4.23E-40 | Nightingale | M_VLDL_P | NA | C | T | 0.028753 | 0.00434 | 1.40E-11 |
| rs543968 | miR-139-5p | trans | 4.23E-40 | Nightingale | S_VLDL_C | NA | C | T | 0.037355 | 0.004346 | 2.60E-18 |
| rs543968 | miR-139-5p | trans | 4.23E-40 | Nightingale | S_HDL_L | NA | C | T | 0.023969 | 0.004332 | 2.70E-08 |
| rs543968 | miR-126-3p | trans | 9.52E-37 | Nightingale | IDL_L | NA | C | T | 0.03366 | 0.004203 | 2.30E-15 |
| rs543968 | miR-126-3p | trans | 9.52E-37 | Nightingale | S_VLDL_C_ | NA | C | T | 0.039736 | 0.00423 | 8.30E-21 |
| rs543968 | miR-126-3p | trans | 9.52E-37 | Nightingale | S_HDL_C | NA | C | T | 0.02538 | 0.004339 | 9.40E-09 |
| rs543968 | miR-126-3p | trans | 9.52E-37 | Nightingale | M_VLDL_C | NA | C | T | 0.037341 | 0.004335 | 5.40E-18 |
| rs543968 | miR-126-3p | trans | 9.52E-37 | Nightingale | LDL_L | NA | C | T | 0.031465 | 0.004332 | 5.80E-13 |
| rs543968 | miR-126-3p | trans | 9.52E-37 | Nightingale | XS_VLDL_P | NA | C | T | 0.031657 | 0.004286 | 2.00E-14 |
| rs543968 | miR-126-3p | trans | 9.52E-37 | Nightingale | L_VLDL_C_ | NA | C | T | 0.049226 | 0.004245 | 8.90E-32 |
| rs543968 | miR-126-3p | trans | 9.52E-37 | Nightingale | M_LDL_TG | NA | C | T | -0.02512 | 0.004293 | 8.60E-09 |
| rs543968 | miR-126-3p | trans | 9.52E-37 | Nightingale | L_LDL_P | NA | C | T | 0.028317 | 0.004332 | 7.80E-11 |
| rs543968 | miR-126-3p | trans | 9.52E-37 | Nightingale | M_LDL_CE | NA | C | T | 0.031212 | 0.004353 | 8.50E-13 |
| rs543968 | miR-126-3p | trans | 9.52E-37 | Nightingale | XS_VLDL_P | NA | C | T | 0.035785 | 0.004289 | 1.00E-17 |
| rs543968 | miR-126-3p | trans | 9.52E-37 | Nightingale | M_VLDL_F | NA | C | T | 0.032515 | 0.004346 | 3.90E-14 |
| rs543968 | miR-126-3p | trans | 9.52E-37 | Nightingale | M_VLDL_C | NA | C | T | 0.031348 | 0.004084 | 3.30E-14 |
| rs543968 | miR-126-3p | trans | 9.52E-37 | Nightingale | LDL_PL | NA | C | T | 0.033318 | 0.004337 | 3.00E-14 |
| rs543968 | miR-126-3p | trans | 9.52E-37 | Nightingale | LDL_CE | NA | C | T | 0.030617 | 0.004335 | 2.10E-12 |
| rs543968 | miR-126-3p | trans | 9.52E-37 | Nightingale | M_VLDL_T | NA | C | T | -0.03623 | 0.004108 | 1.50E-18 |
| rs543968 | miR-126-3p | trans | 9.52E-37 | Nightingale | S_LDL_L | NA | C | T | 0.026893 | 0.004346 | 1.20E-09 |
| rs543968 | miR-126-3p | trans | 9.52E-37 | Nightingale | IDL_PL | NA | C | T | 0.033573 | 0.004195 | 2.00E-15 |
| rs543968 | miR-126-3p | trans | 9.52E-37 | Nightingale | LDL_C | NA | C | T | 0.031186 | 0.004328 | 9.60E-13 |
| rs543968 | miR-126-3p | trans | 9.52E-37 | Nightingale | XS_VLDL_F | NA | C | T | 0.029134 | 0.004316 | 3.10E-11 |
| rs543968 | miR-126-3p | trans | 9.52E-37 | Nightingale | Clinical_LD | NA | C | T | 0.034557 | 0.004322 | 2.70E-15 |
| rs543968 | miR-126-3p | trans | 9.52E-37 | Nightingale | non_HDL_( | NA | C | T | 0.03414 | 0.004323 | 2.40E-15 |
| rs543968 | miR-126-3p | trans | 9.52E-37 | Nightingale | M_VLDL_F | NA | C | T | 0.039372 | 0.004135 | 2.20E-21 |

| | | | | | | | | | | | |
|---|---|---|---|---|---|---|---|---|---|---|---|
| rs543968 | miR-126-3p | trans | 9.52E-37 | Nightingale | XXL_VLDL_ | NA | C | T | 0.029268 | 0.004328 | 5.10E-12 |
| rs543968 | miR-126-3p | trans | 9.52E-37 | Nightingale | M_LDL_PL | NA | C | T | 0.031174 | 0.004353 | 9.90E-13 |
| rs543968 | miR-126-3p | trans | 9.52E-37 | Nightingale | L_VLDL_FC | NA | C | T | 0.046941 | 0.004305 | 3.90E-28 |
| rs543968 | miR-126-3p | trans | 9.52E-37 | Nightingale | ApoB | NA | C | T | 0.032928 | 0.004341 | 3.40E-14 |
| rs543968 | miR-126-3p | trans | 9.52E-37 | Nightingale | IDL_C | NA | C | T | 0.033305 | 0.004198 | 4.90E-15 |
| rs543968 | miR-126-3p | trans | 9.52E-37 | Nightingale | XL_VLDL_P | NA | C | T | 0.039106 | 0.004353 | 7.40E-20 |
| rs543968 | miR-126-3p | trans | 9.52E-37 | Nightingale | S_LDL_CE | NA | C | T | 0.029442 | 0.004349 | 1.50E-11 |
| rs543968 | miR-126-3p | trans | 9.52E-37 | Nightingale | L_LDL_C | NA | C | T | 0.030224 | 0.004298 | 3.00E-12 |
| rs543968 | miR-126-3p | trans | 9.52E-37 | Nightingale | Total_FC | NA | C | T | 0.031378 | 0.004246 | 4.70E-14 |
| rs543968 | miR-126-3p | trans | 9.52E-37 | Nightingale | S_HDL_P | NA | C | T | 0.027805 | 0.004347 | 2.70E-10 |
| rs543968 | miR-126-3p | trans | 9.52E-37 | Nightingale | XXL_VLDL_ | NA | C | T | 0.023502 | 0.004375 | 5.40E-09 |
| rs543968 | miR-126-3p | trans | 9.52E-37 | Nightingale | LDL_FC | NA | C | T | 0.031548 | 0.0043 | 6.60E-13 |
| rs543968 | miR-126-3p | trans | 9.52E-37 | Nightingale | S_VLDL_TG | NA | C | T | -0.03598 | 0.004208 | 1.90E-17 |
| rs543968 | miR-126-3p | trans | 9.52E-37 | Nightingale | L_VLDL_PL | NA | C | T | 0.021645 | 0.004222 | 4.40E-08 |
| rs543968 | miR-126-3p | trans | 9.52E-37 | Nightingale | IDL_FC | NA | C | T | 0.035978 | 0.004208 | 3.20E-17 |
| rs543968 | miR-126-3p | trans | 9.52E-37 | Nightingale | S_VLDL_P | NA | C | T | 0.02339 | 0.004292 | 9.70E-09 |
| rs543968 | miR-126-3p | trans | 9.52E-37 | Nightingale | XXL_VLDL_ | NA | C | T | 0.033437 | 0.004356 | 8.80E-15 |
| rs543968 | miR-126-3p | trans | 9.52E-37 | Nightingale | IDL_P | NA | C | T | 0.039032 | 0.004301 | 1.80E-19 |
| rs543968 | miR-126-3p | trans | 9.52E-37 | Nightingale | S_VLDL_FC | NA | C | T | 0.027154 | 0.004154 | 1.20E-10 |
| rs543968 | miR-126-3p | trans | 9.52E-37 | Nightingale | L_VLDL_CE | NA | C | T | 0.022756 | 0.004296 | 2.90E-08 |
| rs543968 | miR-126-3p | trans | 9.52E-37 | Nightingale | S_VLDL_L | NA | C | T | 0.024496 | 0.004305 | 2.50E-09 |
| rs543968 | miR-126-3p | trans | 9.52E-37 | Nightingale | XS_VLDL_L | NA | C | T | 0.033006 | 0.004273 | 1.40E-15 |
| rs543968 | miR-126-3p | trans | 9.52E-37 | Nightingale | L_LDL_FC | NA | C | T | 0.031473 | 0.004268 | 4.10E-13 |
| rs543968 | miR-126-3p | trans | 9.52E-37 | Nightingale | XXL_VLDL_ | NA | C | T | 0.03179 | 0.00438 | 2.10E-13 |
| rs543968 | miR-126-3p | trans | 9.52E-37 | Nightingale | GlycA | NA | C | T | 0.025329 | 0.004328 | 4.80E-09 |
| rs543968 | miR-126-3p | trans | 9.52E-37 | Nightingale | LA | NA | C | T | 0.023263 | 0.004263 | 1.60E-08 |
| rs543968 | miR-126-3p | trans | 9.52E-37 | Nightingale | L_VLDL_TG | NA | C | T | -0.04923 | 0.004326 | 6.40E-31 |
| rs543968 | miR-126-3p | trans | 9.52E-37 | Nightingale | XXL_VLDL_ | NA | C | T | -0.03443 | 0.004384 | 6.30E-16 |
| rs543968 | miR-126-3p | trans | 9.52E-37 | Nightingale | L_LDL_L | NA | C | T | 0.031523 | 0.004304 | 3.20E-13 |
| rs543968 | miR-126-3p | trans | 9.52E-37 | Nightingale | S_VLDL_PL | NA | C | T | 0.025728 | 0.004162 | 1.50E-09 |
| rs543968 | miR-126-3p | trans | 9.52E-37 | Nightingale | XL_VLDL_C | NA | C | T | 0.034575 | 0.00417 | 1.50E-16 |
| rs543968 | miR-126-3p | trans | 9.52E-37 | Nightingale | XS_VLDL_C | NA | C | T | 0.036391 | 0.004195 | 4.10E-18 |
| rs543968 | miR-126-3p | trans | 9.52E-37 | Nightingale | Total_PL | NA | C | T | 0.022997 | 0.004139 | 7.80E-09 |
| rs543968 | miR-126-3p | trans | 9.52E-37 | Nightingale | Omega_6 | NA | C | T | 0.024156 | 0.004245 | 3.10E-09 |
| rs543968 | miR-126-3p | trans | 9.52E-37 | Nightingale | XS_VLDL_F | NA | C | T | 0.035882 | 0.004271 | 7.50E-18 |
| rs543968 | miR-126-3p | trans | 9.52E-37 | Nightingale | Total_CE | NA | C | T | 0.029384 | 0.004193 | 1.80E-12 |
| rs543968 | miR-126-3p | trans | 9.52E-37 | Nightingale | S_VLDL_PL | NA | C | T | 0.032035 | 0.004339 | 4.90E-14 |
| rs543968 | miR-126-3p | trans | 9.52E-37 | Nightingale | XL_HDL_PL | NA | C | T | -0.02181 | 0.003994 | 4.60E-08 |
| rs543968 | miR-126-3p | trans | 9.52E-37 | Nightingale | L_LDL_PL | NA | C | T | 0.035919 | 0.004314 | 1.30E-16 |
| rs543968 | miR-126-3p | trans | 9.52E-37 | Nightingale | S_LDL_CE_ | NA | C | T | 0.028844 | 0.004339 | 1.10E-12 |
| rs543968 | miR-126-3p | trans | 9.52E-37 | Nightingale | M_LDL_C | NA | C | T | 0.032091 | 0.004355 | 2.30E-13 |
| rs543968 | miR-126-3p | trans | 9.52E-37 | Nightingale | M_LDL_L | NA | C | T | 0.031388 | 0.004353 | 7.00E-13 |
| rs543968 | miR-126-3p | trans | 9.52E-37 | Nightingale | XL_VLDL_C | NA | C | T | 0.03559 | 0.004168 | 1.70E-17 |
| rs543968 | miR-126-3p | trans | 9.52E-37 | Nightingale | S_HDL_CE | NA | C | T | 0.02569 | 0.004354 | 7.00E-09 |
| rs543968 | miR-126-3p | trans | 9.52E-37 | Nightingale | S_VLDL_CE | NA | C | T | 0.037311 | 0.004341 | 1.70E-18 |
| rs543968 | miR-126-3p | trans | 9.52E-37 | Nightingale | IDL_CE | NA | C | T | 0.032088 | 0.004197 | 4.30E-14 |
| rs543968 | miR-126-3p | trans | 9.52E-37 | Nightingale | Citrate | NA | C | T | 0.024678 | 0.004318 | 6.10E-09 |
| rs543968 | miR-126-3p | trans | 9.52E-37 | Nightingale | VLDL_C | NA | C | T | 0.031062 | 0.004341 | 2.10E-13 |
| rs543968 | miR-126-3p | trans | 9.52E-37 | Nightingale | M_LDL_C_ | NA | C | T | 0.026582 | 0.00435 | 1.60E-09 |
| rs543968 | miR-126-3p | trans | 9.52E-37 | Nightingale | XL_VLDL_F | NA | C | T | 0.035204 | 0.004214 | 1.80E-17 |
| rs543968 | miR-126-3p | trans | 9.52E-37 | Nightingale | VLDL_P | NA | C | T | 0.026555 | 0.004324 | 2.00E-10 |
| rs543968 | miR-126-3p | trans | 9.52E-37 | Nightingale | XL_VLDL_T | NA | C | T | -0.04155 | 0.004213 | 4.90E-23 |
| rs543968 | miR-126-3p | trans | 9.52E-37 | Nightingale | M_VLDL_P | NA | C | T | 0.027926 | 0.004336 | 4.70E-11 |
| rs543968 | miR-126-3p | trans | 9.52E-37 | Nightingale | M_LDL_P | NA | C | T | 0.034001 | 0.004359 | 7.50E-15 |
| rs543968 | miR-126-3p | trans | 9.52E-37 | Nightingale | M_VLDL_P | NA | C | T | 0.042093 | 0.004172 | 6.30E-24 |
| rs543968 | miR-126-3p | trans | 9.52E-37 | Nightingale | S_VLDL_FC | NA | C | T | 0.036642 | 0.004347 | 1.70E-17 |
| rs543968 | miR-126-3p | trans | 9.52E-37 | Nightingale | Total_L | NA | C | T | 0.025008 | 0.004276 | 1.50E-09 |
| rs543968 | miR-126-3p | trans | 9.52E-37 | Nightingale | S_VLDL_CE | NA | C | T | 0.045151 | 0.004288 | 1.40E-25 |
| rs543968 | miR-126-3p | trans | 9.52E-37 | Nightingale | S_LDL_PL_ | NA | C | T | -0.02806 | 0.004339 | 4.90E-12 |
| rs543968 | miR-126-3p | trans | 9.52E-37 | Nightingale | S_LDL_FC | NA | C | T | 0.0272 | 0.00434 | 1.30E-09 |
| rs543968 | miR-126-3p | trans | 9.52E-37 | Nightingale | VLDL_CE | NA | C | T | 0.035144 | 0.004348 | 2.10E-16 |
| rs543968 | miR-126-3p | trans | 9.52E-37 | Nightingale | Sphingomy | NA | C | T | 0.025917 | 0.004063 | 3.30E-10 |
| rs543968 | miR-126-3p | trans | 9.52E-37 | Nightingale | L_LDL_CE | NA | C | T | 0.02952 | 0.004308 | 9.90E-12 |

| | | | | | | | | | | | | |
|---|---|---|---|---|---|---|---|---|---|---|---|---|
| rs543968 | miR-126-3p | trans | | 9.52E-37 | Nightingale | Remnant_( | NA | C | T | 0.035838 | 0.004319 | 5.30E-17 |
| rs543968 | miR-126-3p | trans | | 9.52E-37 | Nightingale | Total_C | NA | C | T | 0.030138 | 0.004208 | 5.00E-13 |
| rs543968 | miR-126-3p | trans | | 9.52E-37 | Nightingale | S_LDL_C_p | NA | C | T | 0.041864 | 0.004364 | 5.60E-22 |
| rs543968 | miR-126-3p | trans | | 9.52E-37 | Nightingale | VLDL_FC | NA | C | T | 0.024446 | 0.004315 | 3.60E-09 |
| rs543968 | miR-126-3p | trans | | 9.52E-37 | Nightingale | ApoB_by_A | NA | C | T | 0.025679 | 0.004281 | 2.00E-09 |
| rs543968 | miR-126-3p | trans | | 9.52E-37 | Nightingale | S_LDL_P | NA | C | T | 0.028959 | 0.004351 | 3.00E-11 |
| rs543968 | miR-126-3p | trans | | 9.52E-37 | Nightingale | S_LDL_C | NA | C | T | 0.029698 | 0.004351 | 1.60E-11 |
| rs543968 | miR-126-3p | trans | | 9.52E-37 | Nightingale | L_VLDL_CE | NA | C | T | 0.04407 | 0.00416 | 7.90E-27 |
| rs543968 | miR-126-3p | trans | | 9.52E-37 | Nightingale | LDL_P | NA | C | T | 0.030659 | 0.004344 | 2.20E-12 |
| rs543968 | miR-126-3p | trans | | 9.52E-37 | Nightingale | M_VLDL_C | NA | C | T | 0.0392 | 0.004307 | 1.30E-19 |
| rs543968 | miR-126-3p | trans | | 9.52E-37 | Nightingale | M_LDL_FC | NA | C | T | 0.032281 | 0.004341 | 2.40E-13 |
| rs543968 | miR-126-3p | trans | | 9.52E-37 | Nightingale | M_VLDL_C | NA | C | T | 0.033845 | 0.004094 | 2.70E-16 |
| rs543968 | miR-126-3p | trans | | 9.52E-37 | Nightingale | XS_VLDL_C | NA | C | T | 0.036639 | 0.004219 | 2.40E-18 |
| rs543968 | miR-126-3p | trans | | 9.52E-37 | Nightingale | M_VLDL_P | NA | C | T | 0.028753 | 0.00434 | 1.40E-11 |
| rs543968 | miR-126-3p | trans | | 9.52E-37 | Nightingale | S_VLDL_C | NA | C | T | 0.037355 | 0.004346 | 2.60E-18 |
| rs543968 | miR-126-3p | trans | | 9.52E-37 | Nightingale | S_HDL_L | NA | C | T | 0.023969 | 0.004332 | 2.70E-08 |
| rs543968 | miR-10b-5p | trans | | 3.05E-34 | Nightingale | IDL_L | NA | C | T | 0.03366 | 0.004203 | 2.30E-15 |
| rs543968 | miR-10b-5p | trans | | 3.05E-34 | Nightingale | S_VLDL_C_ | NA | C | T | 0.039736 | 0.00423 | 8.30E-21 |
| rs543968 | miR-10b-5p | trans | | 3.05E-34 | Nightingale | S_HDL_C | NA | C | T | 0.02538 | 0.004339 | 9.40E-09 |
| rs543968 | miR-10b-5p | trans | | 3.05E-34 | Nightingale | M_VLDL_C | NA | C | T | 0.037341 | 0.004335 | 5.40E-18 |
| rs543968 | miR-10b-5p | trans | | 3.05E-34 | Nightingale | LDL_L | NA | C | T | 0.031465 | 0.004332 | 5.80E-13 |
| rs543968 | miR-10b-5p | trans | | 3.05E-34 | Nightingale | XS_VLDL_P | NA | C | T | 0.031657 | 0.004286 | 2.00E-14 |
| rs543968 | miR-10b-5p | trans | | 3.05E-34 | Nightingale | L_VLDL_C_ | NA | C | T | 0.049226 | 0.004245 | 8.90E-32 |
| rs543968 | miR-10b-5p | trans | | 3.05E-34 | Nightingale | M_LDL_TG | NA | C | T | -0.02512 | 0.004293 | 8.60E-09 |
| rs543968 | miR-10b-5p | trans | | 3.05E-34 | Nightingale | L_LDL_P | NA | C | T | 0.028317 | 0.004332 | 7.80E-11 |
| rs543968 | miR-10b-5p | trans | | 3.05E-34 | Nightingale | M_LDL_CE | NA | C | T | 0.031212 | 0.004353 | 8.50E-13 |
| rs543968 | miR-10b-5p | trans | | 3.05E-34 | Nightingale | XS_VLDL_P | NA | C | T | 0.035785 | 0.004289 | 1.00E-17 |
| rs543968 | miR-10b-5p | trans | | 3.05E-34 | Nightingale | M_VLDL_F | NA | C | T | 0.032515 | 0.004346 | 3.90E-14 |
| rs543968 | miR-10b-5p | trans | | 3.05E-34 | Nightingale | M_VLDL_C | NA | C | T | 0.031348 | 0.004084 | 3.30E-14 |
| rs543968 | miR-10b-5p | trans | | 3.05E-34 | Nightingale | LDL_PL | NA | C | T | 0.033318 | 0.004337 | 3.00E-14 |
| rs543968 | miR-10b-5p | trans | | 3.05E-34 | Nightingale | LDL_CE | NA | C | T | 0.030617 | 0.004335 | 2.10E-12 |
| rs543968 | miR-10b-5p | trans | | 3.05E-34 | Nightingale | M_VLDL_T | NA | C | T | -0.03623 | 0.004108 | 1.50E-18 |
| rs543968 | miR-10b-5p | trans | | 3.05E-34 | Nightingale | S_LDL_L | NA | C | T | 0.026893 | 0.004346 | 1.20E-09 |
| rs543968 | miR-10b-5p | trans | | 3.05E-34 | Nightingale | IDL_PL | NA | C | T | 0.033573 | 0.004195 | 2.00E-15 |
| rs543968 | miR-10b-5p | trans | | 3.05E-34 | Nightingale | LDL_C | NA | C | T | 0.031186 | 0.004328 | 9.60E-13 |
| rs543968 | miR-10b-5p | trans | | 3.05E-34 | Nightingale | XS_VLDL_F | NA | C | T | 0.029134 | 0.004316 | 3.10E-11 |
| rs543968 | miR-10b-5p | trans | | 3.05E-34 | Nightingale | Clinical_LD | NA | C | T | 0.034557 | 0.004322 | 2.70E-15 |
| rs543968 | miR-10b-5p | trans | | 3.05E-34 | Nightingale | non_HDL_( | NA | C | T | 0.03414 | 0.004323 | 2.40E-15 |
| rs543968 | miR-10b-5p | trans | | 3.05E-34 | Nightingale | M_VLDL_F | NA | C | T | 0.039372 | 0.004135 | 2.20E-21 |
| rs543968 | miR-10b-5p | trans | | 3.05E-34 | Nightingale | XXL_VLDL_ | NA | C | T | 0.029268 | 0.004328 | 5.10E-12 |
| rs543968 | miR-10b-5p | trans | | 3.05E-34 | Nightingale | M_LDL_PL | NA | C | T | 0.031174 | 0.004353 | 9.90E-13 |
| rs543968 | miR-10b-5p | trans | | 3.05E-34 | Nightingale | L_VLDL_FC | NA | C | T | 0.046941 | 0.004305 | 3.90E-28 |
| rs543968 | miR-10b-5p | trans | | 3.05E-34 | Nightingale | ApoB | NA | C | T | 0.032928 | 0.004341 | 3.40E-14 |
| rs543968 | miR-10b-5p | trans | | 3.05E-34 | Nightingale | IDL_C | NA | C | T | 0.033305 | 0.004198 | 4.90E-15 |
| rs543968 | miR-10b-5p | trans | | 3.05E-34 | Nightingale | XL_VLDL_P | NA | C | T | 0.039106 | 0.004353 | 7.40E-20 |
| rs543968 | miR-10b-5p | trans | | 3.05E-34 | Nightingale | S_LDL_CE | NA | C | T | 0.029442 | 0.004349 | 1.50E-11 |
| rs543968 | miR-10b-5p | trans | | 3.05E-34 | Nightingale | L_LDL_C | NA | C | T | 0.030224 | 0.004298 | 3.00E-12 |
| rs543968 | miR-10b-5p | trans | | 3.05E-34 | Nightingale | Total_FC | NA | C | T | 0.031378 | 0.004246 | 4.70E-14 |
| rs543968 | miR-10b-5p | trans | | 3.05E-34 | Nightingale | S_HDL_P | NA | C | T | 0.027805 | 0.004347 | 2.70E-10 |
| rs543968 | miR-10b-5p | trans | | 3.05E-34 | Nightingale | XXL_VLDL_ | NA | C | T | 0.023502 | 0.004375 | 5.40E-09 |
| rs543968 | miR-10b-5p | trans | | 3.05E-34 | Nightingale | LDL_FC | NA | C | T | 0.031548 | 0.0043 | 6.60E-13 |
| rs543968 | miR-10b-5p | trans | | 3.05E-34 | Nightingale | S_VLDL_TG | NA | C | T | -0.03598 | 0.004208 | 1.90E-17 |
| rs543968 | miR-10b-5p | trans | | 3.05E-34 | Nightingale | L_VLDL_PL | NA | C | T | 0.021645 | 0.004222 | 4.40E-08 |
| rs543968 | miR-10b-5p | trans | | 3.05E-34 | Nightingale | IDL_FC | NA | C | T | 0.035978 | 0.004208 | 3.20E-17 |
| rs543968 | miR-10b-5p | trans | | 3.05E-34 | Nightingale | S_VLDL_P | NA | C | T | 0.02339 | 0.004292 | 9.70E-09 |
| rs543968 | miR-10b-5p | trans | | 3.05E-34 | Nightingale | XXL_VLDL_ | NA | C | T | 0.033437 | 0.004356 | 8.80E-15 |
| rs543968 | miR-10b-5p | trans | | 3.05E-34 | Nightingale | IDL_P | NA | C | T | 0.039032 | 0.004301 | 1.80E-19 |
| rs543968 | miR-10b-5p | trans | | 3.05E-34 | Nightingale | S_VLDL_FC | NA | C | T | 0.027154 | 0.004154 | 1.20E-10 |
| rs543968 | miR-10b-5p | trans | | 3.05E-34 | Nightingale | L_VLDL_CE | NA | C | T | 0.022756 | 0.004296 | 2.90E-08 |
| rs543968 | miR-10b-5p | trans | | 3.05E-34 | Nightingale | S_VLDL_L | NA | C | T | 0.024496 | 0.004305 | 2.50E-09 |
| rs543968 | miR-10b-5p | trans | | 3.05E-34 | Nightingale | XS_VLDL_L | NA | C | T | 0.033006 | 0.004273 | 1.40E-15 |
| rs543968 | miR-10b-5p | trans | | 3.05E-34 | Nightingale | L_LDL_FC | NA | C | T | 0.031473 | 0.004268 | 4.10E-13 |
| rs543968 | miR-10b-5p | trans | | 3.05E-34 | Nightingale | XXL_VLDL_ | NA | C | T | 0.03179 | 0.00438 | 2.10E-13 |

| | | | | | | | | | | | |
|---|---|---|---|---|---|---|---|---|---|---|---|
| rs543968 | miR-10b-5p | trans | 3.05E-34 | Nightingale | GlycA | NA | C | T | 0.025329 | 0.004328 | 4.80E-09 |
| rs543968 | miR-10b-5p | trans | 3.05E-34 | Nightingale | LA | NA | C | T | 0.023263 | 0.004263 | 1.60E-08 |
| rs543968 | miR-10b-5p | trans | 3.05E-34 | Nightingale | L_VLDL_TG | NA | C | T | -0.04923 | 0.004326 | 6.40E-31 |
| rs543968 | miR-10b-5p | trans | 3.05E-34 | Nightingale | XXL_VLDL_ | NA | C | T | -0.03443 | 0.004384 | 6.30E-16 |
| rs543968 | miR-10b-5p | trans | 3.05E-34 | Nightingale | L_LDL_L | NA | C | T | 0.031523 | 0.004304 | 3.20E-13 |
| rs543968 | miR-10b-5p | trans | 3.05E-34 | Nightingale | S_VLDL_PL | NA | C | T | 0.025728 | 0.004162 | 1.50E-09 |
| rs543968 | miR-10b-5p | trans | 3.05E-34 | Nightingale | XL_VLDL_C | NA | C | T | 0.034575 | 0.00417 | 1.50E-16 |
| rs543968 | miR-10b-5p | trans | 3.05E-34 | Nightingale | XS_VLDL_C | NA | C | T | 0.036391 | 0.004195 | 4.10E-18 |
| rs543968 | miR-10b-5p | trans | 3.05E-34 | Nightingale | Total_PL | NA | C | T | 0.022997 | 0.004139 | 7.80E-09 |
| rs543968 | miR-10b-5p | trans | 3.05E-34 | Nightingale | Omega_6 | NA | C | T | 0.024156 | 0.004245 | 3.10E-09 |
| rs543968 | miR-10b-5p | trans | 3.05E-34 | Nightingale | XS_VLDL_F | NA | C | T | 0.035882 | 0.004271 | 7.50E-18 |
| rs543968 | miR-10b-5p | trans | 3.05E-34 | Nightingale | Total_CE | NA | C | T | 0.029384 | 0.004193 | 1.80E-12 |
| rs543968 | miR-10b-5p | trans | 3.05E-34 | Nightingale | S_VLDL_PL | NA | C | T | 0.032035 | 0.004339 | 4.90E-14 |
| rs543968 | miR-10b-5p | trans | 3.05E-34 | Nightingale | XL_HDL_PL | NA | C | T | -0.02181 | 0.003994 | 4.60E-08 |
| rs543968 | miR-10b-5p | trans | 3.05E-34 | Nightingale | L_LDL_PL | NA | C | T | 0.035919 | 0.004314 | 1.30E-16 |
| rs543968 | miR-10b-5p | trans | 3.05E-34 | Nightingale | S_LDL_CE_ | NA | C | T | 0.028844 | 0.004339 | 1.10E-12 |
| rs543968 | miR-10b-5p | trans | 3.05E-34 | Nightingale | M_LDL_C | NA | C | T | 0.032091 | 0.004355 | 2.30E-13 |
| rs543968 | miR-10b-5p | trans | 3.05E-34 | Nightingale | M_LDL_L | NA | C | T | 0.031388 | 0.004353 | 7.00E-13 |
| rs543968 | miR-10b-5p | trans | 3.05E-34 | Nightingale | XL_VLDL_C | NA | C | T | 0.03559 | 0.004168 | 1.70E-17 |
| rs543968 | miR-10b-5p | trans | 3.05E-34 | Nightingale | S_HDL_CE | NA | C | T | 0.02569 | 0.004354 | 7.00E-09 |
| rs543968 | miR-10b-5p | trans | 3.05E-34 | Nightingale | S_VLDL_CE | NA | C | T | 0.037311 | 0.004341 | 1.70E-18 |
| rs543968 | miR-10b-5p | trans | 3.05E-34 | Nightingale | IDL_CE | NA | C | T | 0.032088 | 0.004197 | 4.30E-14 |
| rs543968 | miR-10b-5p | trans | 3.05E-34 | Nightingale | Citrate | NA | C | T | 0.024678 | 0.004318 | 6.10E-09 |
| rs543968 | miR-10b-5p | trans | 3.05E-34 | Nightingale | VLDL_C | NA | C | T | 0.031062 | 0.004341 | 2.10E-13 |
| rs543968 | miR-10b-5p | trans | 3.05E-34 | Nightingale | M_LDL_C_ | NA | C | T | 0.026582 | 0.00435 | 1.60E-09 |
| rs543968 | miR-10b-5p | trans | 3.05E-34 | Nightingale | XL_VLDL_F | NA | C | T | 0.035204 | 0.004214 | 1.80E-17 |
| rs543968 | miR-10b-5p | trans | 3.05E-34 | Nightingale | VLDL_P | NA | C | T | 0.026555 | 0.004324 | 2.00E-10 |
| rs543968 | miR-10b-5p | trans | 3.05E-34 | Nightingale | XL_VLDL_T | NA | C | T | -0.04155 | 0.004213 | 4.90E-23 |
| rs543968 | miR-10b-5p | trans | 3.05E-34 | Nightingale | M_VLDL_P | NA | C | T | 0.027926 | 0.004336 | 4.70E-11 |
| rs543968 | miR-10b-5p | trans | 3.05E-34 | Nightingale | M_LDL_P | NA | C | T | 0.034001 | 0.004359 | 7.50E-15 |
| rs543968 | miR-10b-5p | trans | 3.05E-34 | Nightingale | M_VLDL_P | NA | C | T | 0.042093 | 0.004172 | 6.30E-24 |
| rs543968 | miR-10b-5p | trans | 3.05E-34 | Nightingale | S_VLDL_FC | NA | C | T | 0.036642 | 0.004347 | 1.70E-17 |
| rs543968 | miR-10b-5p | trans | 3.05E-34 | Nightingale | Total_L | NA | C | T | 0.025008 | 0.004276 | 1.50E-09 |
| rs543968 | miR-10b-5p | trans | 3.05E-34 | Nightingale | S_VLDL_CE | NA | C | T | 0.045151 | 0.004288 | 1.40E-25 |
| rs543968 | miR-10b-5p | trans | 3.05E-34 | Nightingale | S_LDL_PL_ | NA | C | T | -0.02806 | 0.004339 | 4.90E-12 |
| rs543968 | miR-10b-5p | trans | 3.05E-34 | Nightingale | S_LDL_FC | NA | C | T | 0.0272 | 0.00434 | 1.30E-09 |
| rs543968 | miR-10b-5p | trans | 3.05E-34 | Nightingale | VLDL_CE | NA | C | T | 0.035144 | 0.004348 | 2.10E-16 |
| rs543968 | miR-10b-5p | trans | 3.05E-34 | Nightingale | Sphingomy | NA | C | T | 0.025917 | 0.004063 | 3.30E-10 |
| rs543968 | miR-10b-5p | trans | 3.05E-34 | Nightingale | L_LDL_CE | NA | C | T | 0.02952 | 0.004308 | 9.90E-12 |
| rs543968 | miR-10b-5p | trans | 3.05E-34 | Nightingale | Remnant_( | NA | C | T | 0.035838 | 0.004319 | 5.30E-17 |
| rs543968 | miR-10b-5p | trans | 3.05E-34 | Nightingale | Total_C | NA | C | T | 0.030138 | 0.004208 | 5.00E-13 |
| rs543968 | miR-10b-5p | trans | 3.05E-34 | Nightingale | S_LDL_C_p | NA | C | T | 0.041864 | 0.004364 | 5.60E-22 |
| rs543968 | miR-10b-5p | trans | 3.05E-34 | Nightingale | VLDL_FC | NA | C | T | 0.024446 | 0.004315 | 3.60E-09 |
| rs543968 | miR-10b-5p | trans | 3.05E-34 | Nightingale | ApoB_by_A | NA | C | T | 0.025679 | 0.004281 | 2.00E-09 |
| rs543968 | miR-10b-5p | trans | 3.05E-34 | Nightingale | S_LDL_P | NA | C | T | 0.028959 | 0.004351 | 3.00E-11 |
| rs543968 | miR-10b-5p | trans | 3.05E-34 | Nightingale | S_LDL_C | NA | C | T | 0.029698 | 0.004351 | 1.60E-11 |
| rs543968 | miR-10b-5p | trans | 3.05E-34 | Nightingale | L_VLDL_CE | NA | C | T | 0.04407 | 0.00416 | 7.90E-27 |
| rs543968 | miR-10b-5p | trans | 3.05E-34 | Nightingale | LDL_P | NA | C | T | 0.030659 | 0.004344 | 2.20E-12 |
| rs543968 | miR-10b-5p | trans | 3.05E-34 | Nightingale | M_VLDL_C | NA | C | T | 0.0392 | 0.004307 | 1.30E-19 |
| rs543968 | miR-10b-5p | trans | 3.05E-34 | Nightingale | M_LDL_FC | NA | C | T | 0.032281 | 0.004341 | 2.40E-13 |
| rs543968 | miR-10b-5p | trans | 3.05E-34 | Nightingale | M_VLDL_C | NA | C | T | 0.033845 | 0.004094 | 2.70E-16 |
| rs543968 | miR-10b-5p | trans | 3.05E-34 | Nightingale | XS_VLDL_C | NA | C | T | 0.036639 | 0.004219 | 2.40E-18 |
| rs543968 | miR-10b-5p | trans | 3.05E-34 | Nightingale | M_VLDL_P | NA | C | T | 0.028753 | 0.00434 | 1.40E-11 |
| rs543968 | miR-10b-5p | trans | 3.05E-34 | Nightingale | S_VLDL_C | NA | C | T | 0.037355 | 0.004346 | 2.60E-18 |
| rs543968 | miR-10b-5p | trans | 3.05E-34 | Nightingale | S_HDL_L | NA | C | T | 0.023969 | 0.004332 | 2.70E-08 |
| rs544873 | let-7b-5p | trans | 1.88E-26 | Nightingale | IDL_L | NA | A | G | 0.03366 | 0.004203 | 2.30E-15 |
| rs544873 | let-7b-5p | trans | 1.88E-26 | Nightingale | S_LDL_L | NA | A | G | 0.026893 | 0.004346 | 1.20E-09 |
| rs544873 | let-7b-5p | trans | 1.88E-26 | Nightingale | M_VLDL_C | NA | A | G | 0.0392 | 0.004307 | 1.30E-19 |
| rs544873 | let-7b-5p | trans | 1.88E-26 | Nightingale | IDL_CE | NA | A | G | 0.032088 | 0.004197 | 4.30E-14 |
| rs544873 | let-7b-5p | trans | 1.88E-26 | Nightingale | XXL_VLDL_ | NA | A | G | -0.03443 | 0.004384 | 6.30E-16 |
| rs544873 | let-7b-5p | trans | 1.88E-26 | Nightingale | Clinical_LD | NA | A | G | 0.034557 | 0.004322 | 2.70E-15 |
| rs544873 | let-7b-5p | trans | 1.88E-26 | Nightingale | L_LDL_C | NA | A | G | 0.030224 | 0.004298 | 3.00E-12 |
| rs544873 | let-7b-5p | trans | 1.88E-26 | Nightingale | M_VLDL_C | NA | A | G | 0.033845 | 0.004094 | 2.70E-16 |

| | | | | | | | | | | | |
|---|---|---|---|---|---|---|---|---|---|---|---|
| rs544873 | let-7b-5p | trans | 1.88E-26 | Nightingale | M_LDL_PL | NA | A | G | 0.031174 | 0.004353 | 9.90E-13 |
| rs544873 | let-7b-5p | trans | 1.88E-26 | Nightingale | S_HDL_P | NA | A | G | 0.027805 | 0.004347 | 2.70E-10 |
| rs544873 | let-7b-5p | trans | 1.88E-26 | Nightingale | Sphingomy | NA | A | G | 0.025917 | 0.004063 | 3.30E-10 |
| rs544873 | let-7b-5p | trans | 1.88E-26 | Nightingale | VLDL_P | NA | A | G | 0.026555 | 0.004324 | 2.00E-10 |
| rs544873 | let-7b-5p | trans | 1.88E-26 | Nightingale | IDL_P | NA | A | G | 0.039032 | 0.004301 | 1.80E-19 |
| rs544873 | let-7b-5p | trans | 1.88E-26 | Nightingale | L_VLDL_FC | NA | A | G | 0.046941 | 0.004305 | 3.90E-28 |
| rs544873 | let-7b-5p | trans | 1.88E-26 | Nightingale | M_VLDL_C | NA | A | G | 0.031348 | 0.004084 | 3.30E-14 |
| rs544873 | let-7b-5p | trans | 1.88E-26 | Nightingale | S_VLDL_CE | NA | A | G | 0.037311 | 0.004341 | 1.70E-18 |
| rs544873 | let-7b-5p | trans | 1.88E-26 | Nightingale | IDL_PL | NA | A | G | 0.033573 | 0.004195 | 2.00E-15 |
| rs544873 | let-7b-5p | trans | 1.88E-26 | Nightingale | XS_VLDL_F | NA | A | G | 0.029134 | 0.004316 | 3.10E-11 |
| rs544873 | let-7b-5p | trans | 1.88E-26 | Nightingale | Total_L | NA | A | G | 0.025008 | 0.004276 | 1.50E-09 |
| rs544873 | let-7b-5p | trans | 1.88E-26 | Nightingale | XL_HDL_PL | NA | A | G | -0.02181 | 0.003994 | 4.60E-08 |
| rs544873 | let-7b-5p | trans | 1.88E-26 | Nightingale | GlycA | NA | A | G | 0.025329 | 0.004328 | 4.80E-09 |
| rs544873 | let-7b-5p | trans | 1.88E-26 | Nightingale | L_VLDL_PL | NA | A | G | 0.021645 | 0.004222 | 4.40E-08 |
| rs544873 | let-7b-5p | trans | 1.88E-26 | Nightingale | L_LDL_P | NA | A | G | 0.028317 | 0.004332 | 7.80E-11 |
| rs544873 | let-7b-5p | trans | 1.88E-26 | Nightingale | IDL_FC | NA | A | G | 0.035978 | 0.004208 | 3.20E-17 |
| rs544873 | let-7b-5p | trans | 1.88E-26 | Nightingale | VLDL_FC | NA | A | G | 0.024446 | 0.004315 | 3.60E-09 |
| rs544873 | let-7b-5p | trans | 1.88E-26 | Nightingale | IDL_C | NA | A | G | 0.033305 | 0.004198 | 4.90E-15 |
| rs544873 | let-7b-5p | trans | 1.88E-26 | Nightingale | M_LDL_FC | NA | A | G | 0.032281 | 0.004341 | 2.40E-13 |
| rs544873 | let-7b-5p | trans | 1.88E-26 | Nightingale | S_VLDL_CE | NA | A | G | 0.045151 | 0.004288 | 1.40E-25 |
| rs544873 | let-7b-5p | trans | 1.88E-26 | Nightingale | M_LDL_C | NA | A | G | 0.032091 | 0.004355 | 2.30E-13 |
| rs544873 | let-7b-5p | trans | 1.88E-26 | Nightingale | XS_VLDL_P | NA | A | G | 0.031657 | 0.004286 | 2.00E-14 |
| rs544873 | let-7b-5p | trans | 1.88E-26 | Nightingale | M_VLDL_F | NA | A | G | 0.032515 | 0.004346 | 3.90E-14 |
| rs544873 | let-7b-5p | trans | 1.88E-26 | Nightingale | L_VLDL_CE | NA | A | G | 0.04407 | 0.00416 | 7.90E-27 |
| rs544873 | let-7b-5p | trans | 1.88E-26 | Nightingale | Omega_6 | NA | A | G | 0.024156 | 0.004245 | 3.10E-09 |
| rs544873 | let-7b-5p | trans | 1.88E-26 | Nightingale | XL_VLDL_P | NA | A | G | 0.039106 | 0.004353 | 7.40E-20 |
| rs544873 | let-7b-5p | trans | 1.88E-26 | Nightingale | S_LDL_C_p | NA | A | G | 0.041864 | 0.004364 | 5.60E-22 |
| rs544873 | let-7b-5p | trans | 1.88E-26 | Nightingale | VLDL_CE | NA | A | G | 0.035144 | 0.004348 | 2.10E-16 |
| rs544873 | let-7b-5p | trans | 1.88E-26 | Nightingale | L_LDL_CE | NA | A | G | 0.02952 | 0.004308 | 9.90E-12 |
| rs544873 | let-7b-5p | trans | 1.88E-26 | Nightingale | LDL_L | NA | A | G | 0.031465 | 0.004332 | 5.80E-13 |
| rs544873 | let-7b-5p | trans | 1.88E-26 | Nightingale | XS_VLDL_L | NA | A | G | 0.033006 | 0.004273 | 1.40E-15 |
| rs544873 | let-7b-5p | trans | 1.88E-26 | Nightingale | S_VLDL_C | NA | A | G | 0.037355 | 0.004346 | 2.60E-18 |
| rs544873 | let-7b-5p | trans | 1.88E-26 | Nightingale | M_LDL_CE | NA | A | G | 0.031212 | 0.004353 | 8.50E-13 |
| rs544873 | let-7b-5p | trans | 1.88E-26 | Nightingale | S_VLDL_FC | NA | A | G | 0.027154 | 0.004154 | 1.20E-10 |
| rs544873 | let-7b-5p | trans | 1.88E-26 | Nightingale | XS_VLDL_C | NA | A | G | 0.036391 | 0.004195 | 4.10E-18 |
| rs544873 | let-7b-5p | trans | 1.88E-26 | Nightingale | L_VLDL_CE | NA | A | G | 0.022756 | 0.004296 | 2.90E-08 |
| rs544873 | let-7b-5p | trans | 1.88E-26 | Nightingale | S_VLDL_PL | NA | A | G | 0.032035 | 0.004339 | 4.90E-14 |
| rs544873 | let-7b-5p | trans | 1.88E-26 | Nightingale | XXL_VLDL_ | NA | A | G | 0.023502 | 0.004375 | 5.40E-09 |
| rs544873 | let-7b-5p | trans | 1.88E-26 | Nightingale | Remnant_( | NA | A | G | 0.035838 | 0.004319 | 5.30E-17 |
| rs544873 | let-7b-5p | trans | 1.88E-26 | Nightingale | S_VLDL_P | NA | A | G | 0.02339 | 0.004292 | 9.70E-09 |
| rs544873 | let-7b-5p | trans | 1.88E-26 | Nightingale | Total_C | NA | A | G | 0.030138 | 0.004208 | 5.00E-13 |
| rs544873 | let-7b-5p | trans | 1.88E-26 | Nightingale | VLDL_C | NA | A | G | 0.031062 | 0.004341 | 2.10E-13 |
| rs544873 | let-7b-5p | trans | 1.88E-26 | Nightingale | Total_CE | NA | A | G | 0.029384 | 0.004193 | 1.80E-12 |
| rs544873 | let-7b-5p | trans | 1.88E-26 | Nightingale | M_LDL_C_ | NA | A | G | 0.026582 | 0.00435 | 1.60E-09 |
| rs544873 | let-7b-5p | trans | 1.88E-26 | Nightingale | S_VLDL_TG | NA | A | G | -0.03598 | 0.004208 | 1.90E-17 |
| rs544873 | let-7b-5p | trans | 1.88E-26 | Nightingale | S_LDL_CE_ | NA | A | G | 0.028844 | 0.004339 | 1.10E-12 |
| rs544873 | let-7b-5p | trans | 1.88E-26 | Nightingale | S_HDL_L | NA | A | G | 0.023969 | 0.004332 | 2.70E-08 |
| rs544873 | let-7b-5p | trans | 1.88E-26 | Nightingale | LDL_FC | NA | A | G | 0.031548 | 0.0043 | 6.60E-13 |
| rs544873 | let-7b-5p | trans | 1.88E-26 | Nightingale | LDL_CE | NA | A | G | 0.030617 | 0.004335 | 2.10E-12 |
| rs544873 | let-7b-5p | trans | 1.88E-26 | Nightingale | XS_VLDL_F | NA | A | G | 0.035882 | 0.004271 | 7.50E-18 |
| rs544873 | let-7b-5p | trans | 1.88E-26 | Nightingale | M_LDL_TG | NA | A | G | -0.02512 | 0.004293 | 8.60E-09 |
| rs544873 | let-7b-5p | trans | 1.88E-26 | Nightingale | LDL_PL | NA | A | G | 0.033318 | 0.004337 | 3.00E-14 |
| rs544873 | let-7b-5p | trans | 1.88E-26 | Nightingale | M_LDL_P | NA | A | G | 0.034001 | 0.004359 | 7.50E-15 |
| rs544873 | let-7b-5p | trans | 1.88E-26 | Nightingale | Citrate | NA | A | G | 0.024678 | 0.004318 | 6.10E-09 |
| rs544873 | let-7b-5p | trans | 1.88E-26 | Nightingale | XXL_VLDL_ | NA | A | G | 0.029268 | 0.004328 | 5.10E-12 |
| rs544873 | let-7b-5p | trans | 1.88E-26 | Nightingale | S_LDL_FC | NA | A | G | 0.0272 | 0.00434 | 1.30E-09 |
| rs544873 | let-7b-5p | trans | 1.88E-26 | Nightingale | LDL_C | NA | A | G | 0.031186 | 0.004328 | 9.60E-13 |
| rs544873 | let-7b-5p | trans | 1.88E-26 | Nightingale | ApoB_by_/ | NA | A | G | 0.025679 | 0.004281 | 2.00E-09 |
| rs544873 | let-7b-5p | trans | 1.88E-26 | Nightingale | S_VLDL_FC | NA | A | G | 0.036642 | 0.004347 | 1.70E-17 |
| rs544873 | let-7b-5p | trans | 1.88E-26 | Nightingale | LA | NA | A | G | 0.023263 | 0.004263 | 1.60E-08 |
| rs544873 | let-7b-5p | trans | 1.88E-26 | Nightingale | non_HDL_( | NA | A | G | 0.03414 | 0.004323 | 2.40E-15 |
| rs544873 | let-7b-5p | trans | 1.88E-26 | Nightingale | LDL_P | NA | A | G | 0.030659 | 0.004344 | 2.20E-12 |
| rs544873 | let-7b-5p | trans | 1.88E-26 | Nightingale | M_VLDL_C | NA | A | G | 0.037341 | 0.004335 | 5.40E-18 |

| | | | | | | | | | | | |
|---|---|---|---|---|---|---|---|---|---|---|---|
| rs544873 | let-7b-5p | trans | 1.88E-26 | Nightingale | Total_PL | NA | A | G | 0.022997 | 0.004139 | 7.80E-09 |
| rs544873 | let-7b-5p | trans | 1.88E-26 | Nightingale | M_VLDL_T | NA | A | G | -0.03623 | 0.004108 | 1.50E-18 |
| rs544873 | let-7b-5p | trans | 1.88E-26 | Nightingale | S_HDL_C | NA | A | G | 0.02538 | 0.004339 | 9.40E-09 |
| rs544873 | let-7b-5p | trans | 1.88E-26 | Nightingale | L_LDL_FC | NA | A | G | 0.031473 | 0.004268 | 4.10E-13 |
| rs544873 | let-7b-5p | trans | 1.88E-26 | Nightingale | S_LDL_P | NA | A | G | 0.028959 | 0.004351 | 3.00E-11 |
| rs544873 | let-7b-5p | trans | 1.88E-26 | Nightingale | S_VLDL_L | NA | A | G | 0.024496 | 0.004305 | 2.50E-09 |
| rs544873 | let-7b-5p | trans | 1.88E-26 | Nightingale | XL_VLDL_F | NA | A | G | 0.035204 | 0.004214 | 1.80E-17 |
| rs544873 | let-7b-5p | trans | 1.88E-26 | Nightingale | L_VLDL_TG | NA | A | G | -0.04923 | 0.004326 | 6.40E-31 |
| rs544873 | let-7b-5p | trans | 1.88E-26 | Nightingale | L_LDL_PL | NA | A | G | 0.035919 | 0.004314 | 1.30E-16 |
| rs544873 | let-7b-5p | trans | 1.88E-26 | Nightingale | S_LDL_PL_ | NA | A | G | -0.02806 | 0.004339 | 4.90E-12 |
| rs544873 | let-7b-5p | trans | 1.88E-26 | Nightingale | S_VLDL_C_ | NA | A | G | 0.039736 | 0.00423 | 8.30E-21 |
| rs544873 | let-7b-5p | trans | 1.88E-26 | Nightingale | L_VLDL_C_ | NA | A | G | 0.049226 | 0.004245 | 8.90E-32 |
| rs544873 | let-7b-5p | trans | 1.88E-26 | Nightingale | XXL_VLDL_ | NA | A | G | 0.033437 | 0.004356 | 8.80E-15 |
| rs544873 | let-7b-5p | trans | 1.88E-26 | Nightingale | XL_VLDL_C | NA | A | G | 0.03559 | 0.004168 | 1.70E-17 |
| rs544873 | let-7b-5p | trans | 1.88E-26 | Nightingale | XL_VLDL_C | NA | A | G | 0.034575 | 0.00417 | 1.50E-16 |
| rs544873 | let-7b-5p | trans | 1.88E-26 | Nightingale | M_VLDL_P | NA | A | G | 0.028753 | 0.00434 | 1.40E-11 |
| rs544873 | let-7b-5p | trans | 1.88E-26 | Nightingale | XS_VLDL_C | NA | A | G | 0.036639 | 0.004219 | 2.40E-18 |
| rs544873 | let-7b-5p | trans | 1.88E-26 | Nightingale | M_LDL_L | NA | A | G | 0.031388 | 0.004353 | 7.00E-13 |
| rs544873 | let-7b-5p | trans | 1.88E-26 | Nightingale | M_VLDL_P | NA | A | G | 0.042093 | 0.004172 | 6.30E-24 |
| rs544873 | let-7b-5p | trans | 1.88E-26 | Nightingale | M_VLDL_P | NA | A | G | 0.027926 | 0.004336 | 4.70E-11 |
| rs544873 | let-7b-5p | trans | 1.88E-26 | Nightingale | M_VLDL_F | NA | A | G | 0.039372 | 0.004135 | 2.20E-21 |
| rs544873 | let-7b-5p | trans | 1.88E-26 | Nightingale | L_LDL_L | NA | A | G | 0.031523 | 0.004304 | 3.20E-13 |
| rs544873 | let-7b-5p | trans | 1.88E-26 | Nightingale | S_LDL_C | NA | A | G | 0.029698 | 0.004351 | 1.60E-11 |
| rs544873 | let-7b-5p | trans | 1.88E-26 | Nightingale | XL_VLDL_T | NA | A | G | -0.04155 | 0.004213 | 4.90E-23 |
| rs544873 | let-7b-5p | trans | 1.88E-26 | Nightingale | Total_FC | NA | A | G | 0.031378 | 0.004246 | 4.70E-14 |
| rs544873 | let-7b-5p | trans | 1.88E-26 | Nightingale | XS_VLDL_P | NA | A | G | 0.035785 | 0.004289 | 1.00E-17 |
| rs544873 | let-7b-5p | trans | 1.88E-26 | Nightingale | XXL_VLDL_ | NA | A | G | 0.03179 | 0.00438 | 2.10E-13 |
| rs544873 | let-7b-5p | trans | 1.88E-26 | Nightingale | S_VLDL_PL | NA | A | G | 0.025728 | 0.004162 | 1.50E-09 |
| rs544873 | let-7b-5p | trans | 1.88E-26 | Nightingale | S_LDL_CE | NA | A | G | 0.029442 | 0.004349 | 1.50E-11 |
| rs544873 | let-7b-5p | trans | 1.88E-26 | Nightingale | S_HDL_CE | NA | A | G | 0.02569 | 0.004354 | 7.00E-09 |
| rs544873 | let-7b-5p | trans | 1.88E-26 | Nightingale | ApoB | NA | A | G | 0.032928 | 0.004341 | 3.40E-14 |
| rs544873 | miR-10b-5p | trans | 1.70E-34 | Nightingale | IDL_L | NA | A | G | 0.03366 | 0.004203 | 2.30E-15 |
| rs544873 | miR-10b-5p | trans | 1.70E-34 | Nightingale | S_LDL_L | NA | A | G | 0.026893 | 0.004346 | 1.20E-09 |
| rs544873 | miR-10b-5p | trans | 1.70E-34 | Nightingale | M_VLDL_C | NA | A | G | 0.0392 | 0.004307 | 1.30E-19 |
| rs544873 | miR-10b-5p | trans | 1.70E-34 | Nightingale | IDL_CE | NA | A | G | 0.032088 | 0.004197 | 4.30E-14 |
| rs544873 | miR-10b-5p | trans | 1.70E-34 | Nightingale | XXL_VLDL_ | NA | A | G | -0.03443 | 0.004384 | 6.30E-16 |
| rs544873 | miR-10b-5p | trans | 1.70E-34 | Nightingale | Clinical_LD | NA | A | G | 0.034557 | 0.004322 | 2.70E-15 |
| rs544873 | miR-10b-5p | trans | 1.70E-34 | Nightingale | L_LDL_C | NA | A | G | 0.030224 | 0.004298 | 3.00E-12 |
| rs544873 | miR-10b-5p | trans | 1.70E-34 | Nightingale | M_VLDL_C | NA | A | G | 0.033845 | 0.004094 | 2.70E-16 |
| rs544873 | miR-10b-5p | trans | 1.70E-34 | Nightingale | M_LDL_PL | NA | A | G | 0.031174 | 0.004353 | 9.90E-13 |
| rs544873 | miR-10b-5p | trans | 1.70E-34 | Nightingale | S_HDL_P | NA | A | G | 0.027805 | 0.004347 | 2.70E-10 |
| rs544873 | miR-10b-5p | trans | 1.70E-34 | Nightingale | Sphingomy | NA | A | G | 0.025917 | 0.004063 | 3.30E-10 |
| rs544873 | miR-10b-5p | trans | 1.70E-34 | Nightingale | VLDL_P | NA | A | G | 0.026555 | 0.004324 | 2.00E-10 |
| rs544873 | miR-10b-5p | trans | 1.70E-34 | Nightingale | IDL_P | NA | A | G | 0.039032 | 0.004301 | 1.80E-19 |
| rs544873 | miR-10b-5p | trans | 1.70E-34 | Nightingale | L_VLDL_FC | NA | A | G | 0.046941 | 0.004305 | 3.90E-28 |
| rs544873 | miR-10b-5p | trans | 1.70E-34 | Nightingale | M_VLDL_C | NA | A | G | 0.031348 | 0.004084 | 3.30E-14 |
| rs544873 | miR-10b-5p | trans | 1.70E-34 | Nightingale | S_VLDL_CE | NA | A | G | 0.037311 | 0.004341 | 1.70E-18 |
| rs544873 | miR-10b-5p | trans | 1.70E-34 | Nightingale | IDL_PL | NA | A | G | 0.033573 | 0.004195 | 2.00E-15 |
| rs544873 | miR-10b-5p | trans | 1.70E-34 | Nightingale | XS_VLDL_F | NA | A | G | 0.029134 | 0.004316 | 3.10E-11 |
| rs544873 | miR-10b-5p | trans | 1.70E-34 | Nightingale | Total_L | NA | A | G | 0.025008 | 0.004276 | 1.50E-09 |
| rs544873 | miR-10b-5p | trans | 1.70E-34 | Nightingale | XL_HDL_PL | NA | A | G | -0.02181 | 0.003994 | 4.60E-08 |
| rs544873 | miR-10b-5p | trans | 1.70E-34 | Nightingale | GlycA | NA | A | G | 0.025329 | 0.004328 | 4.80E-09 |
| rs544873 | miR-10b-5p | trans | 1.70E-34 | Nightingale | L_VLDL_PL | NA | A | G | 0.021645 | 0.004222 | 4.40E-08 |
| rs544873 | miR-10b-5p | trans | 1.70E-34 | Nightingale | L_LDL_P | NA | A | G | 0.028317 | 0.004332 | 7.80E-11 |
| rs544873 | miR-10b-5p | trans | 1.70E-34 | Nightingale | IDL_FC | NA | A | G | 0.035978 | 0.004208 | 3.20E-17 |
| rs544873 | miR-10b-5p | trans | 1.70E-34 | Nightingale | VLDL_FC | NA | A | G | 0.024446 | 0.004315 | 3.60E-09 |
| rs544873 | miR-10b-5p | trans | 1.70E-34 | Nightingale | IDL_C | NA | A | G | 0.033305 | 0.004198 | 4.90E-15 |
| rs544873 | miR-10b-5p | trans | 1.70E-34 | Nightingale | M_LDL_FC | NA | A | G | 0.032281 | 0.004341 | 2.40E-13 |
| rs544873 | miR-10b-5p | trans | 1.70E-34 | Nightingale | S_VLDL_CE | NA | A | G | 0.045151 | 0.004288 | 1.40E-25 |
| rs544873 | miR-10b-5p | trans | 1.70E-34 | Nightingale | M_LDL_C | NA | A | G | 0.032091 | 0.004355 | 2.30E-13 |
| rs544873 | miR-10b-5p | trans | 1.70E-34 | Nightingale | XS_VLDL_P | NA | A | G | 0.031657 | 0.004286 | 2.00E-14 |
| rs544873 | miR-10b-5p | trans | 1.70E-34 | Nightingale | M_VLDL_F | NA | A | G | 0.032515 | 0.004346 | 3.90E-14 |
| rs544873 | miR-10b-5p | trans | 1.70E-34 | Nightingale | L_VLDL_CE | NA | A | G | 0.04407 | 0.00416 | 7.90E-27 |

| | | | | | | | | | | | |
|---|---|---|---|---|---|---|---|---|---|---|---|
| rs544873 | miR-10b-5p | trans | 1.70E-34 | Nightingale | Omega_6 | NA | A | G | 0.024156 | 0.004245 | 3.10E-09 |
| rs544873 | miR-10b-5p | trans | 1.70E-34 | Nightingale | XL_VLDL_P | NA | A | G | 0.039106 | 0.004353 | 7.40E-20 |
| rs544873 | miR-10b-5p | trans | 1.70E-34 | Nightingale | S_LDL_C_p | NA | A | G | 0.041864 | 0.004364 | 5.60E-22 |
| rs544873 | miR-10b-5p | trans | 1.70E-34 | Nightingale | VLDL_CE | NA | A | G | 0.035144 | 0.004348 | 2.10E-16 |
| rs544873 | miR-10b-5p | trans | 1.70E-34 | Nightingale | L_LDL_CE | NA | A | G | 0.02952 | 0.004308 | 9.90E-12 |
| rs544873 | miR-10b-5p | trans | 1.70E-34 | Nightingale | LDL_L | NA | A | G | 0.031465 | 0.004332 | 5.80E-13 |
| rs544873 | miR-10b-5p | trans | 1.70E-34 | Nightingale | XS_VLDL_L | NA | A | G | 0.033006 | 0.004273 | 1.40E-15 |
| rs544873 | miR-10b-5p | trans | 1.70E-34 | Nightingale | S_VLDL_C | NA | A | G | 0.037355 | 0.004346 | 2.60E-18 |
| rs544873 | miR-10b-5p | trans | 1.70E-34 | Nightingale | M_LDL_CE | NA | A | G | 0.031212 | 0.004353 | 8.50E-13 |
| rs544873 | miR-10b-5p | trans | 1.70E-34 | Nightingale | S_VLDL_FC | NA | A | G | 0.027154 | 0.004154 | 1.20E-10 |
| rs544873 | miR-10b-5p | trans | 1.70E-34 | Nightingale | XS_VLDL_C | NA | A | G | 0.036391 | 0.004195 | 4.10E-18 |
| rs544873 | miR-10b-5p | trans | 1.70E-34 | Nightingale | L_VLDL_CE | NA | A | G | 0.022756 | 0.004296 | 2.90E-08 |
| rs544873 | miR-10b-5p | trans | 1.70E-34 | Nightingale | S_VLDL_PL | NA | A | G | 0.032035 | 0.004339 | 4.90E-14 |
| rs544873 | miR-10b-5p | trans | 1.70E-34 | Nightingale | XXL_VLDL_ | NA | A | G | 0.023502 | 0.004375 | 5.40E-09 |
| rs544873 | miR-10b-5p | trans | 1.70E-34 | Nightingale | Remnant_( | NA | A | G | 0.035838 | 0.004319 | 5.30E-17 |
| rs544873 | miR-10b-5p | trans | 1.70E-34 | Nightingale | S_VLDL_P | NA | A | G | 0.02339 | 0.004292 | 9.70E-09 |
| rs544873 | miR-10b-5p | trans | 1.70E-34 | Nightingale | Total_C | NA | A | G | 0.030138 | 0.004208 | 5.00E-13 |
| rs544873 | miR-10b-5p | trans | 1.70E-34 | Nightingale | VLDL_C | NA | A | G | 0.031062 | 0.004341 | 2.10E-13 |
| rs544873 | miR-10b-5p | trans | 1.70E-34 | Nightingale | Total_CE | NA | A | G | 0.029384 | 0.004193 | 1.80E-12 |
| rs544873 | miR-10b-5p | trans | 1.70E-34 | Nightingale | M_LDL_C_ | NA | A | G | 0.026582 | 0.00435 | 1.60E-09 |
| rs544873 | miR-10b-5p | trans | 1.70E-34 | Nightingale | S_VLDL_TG | NA | A | G | -0.03598 | 0.004208 | 1.90E-17 |
| rs544873 | miR-10b-5p | trans | 1.70E-34 | Nightingale | S_LDL_CE_ | NA | A | G | 0.028844 | 0.004339 | 1.10E-12 |
| rs544873 | miR-10b-5p | trans | 1.70E-34 | Nightingale | S_HDL_L | NA | A | G | 0.023969 | 0.004332 | 2.70E-08 |
| rs544873 | miR-10b-5p | trans | 1.70E-34 | Nightingale | LDL_FC | NA | A | G | 0.031548 | 0.0043 | 6.60E-13 |
| rs544873 | miR-10b-5p | trans | 1.70E-34 | Nightingale | LDL_CE | NA | A | G | 0.030617 | 0.004335 | 2.10E-12 |
| rs544873 | miR-10b-5p | trans | 1.70E-34 | Nightingale | XS_VLDL_F | NA | A | G | 0.035882 | 0.004271 | 7.50E-18 |
| rs544873 | miR-10b-5p | trans | 1.70E-34 | Nightingale | M_LDL_TG | NA | A | G | -0.02512 | 0.004293 | 8.60E-09 |
| rs544873 | miR-10b-5p | trans | 1.70E-34 | Nightingale | LDL_PL | NA | A | G | 0.033318 | 0.004337 | 3.00E-14 |
| rs544873 | miR-10b-5p | trans | 1.70E-34 | Nightingale | M_LDL_P | NA | A | G | 0.034001 | 0.004359 | 7.50E-15 |
| rs544873 | miR-10b-5p | trans | 1.70E-34 | Nightingale | Citrate | NA | A | G | 0.024678 | 0.004318 | 6.10E-09 |
| rs544873 | miR-10b-5p | trans | 1.70E-34 | Nightingale | XXL_VLDL_ | NA | A | G | 0.029268 | 0.004328 | 5.10E-12 |
| rs544873 | miR-10b-5p | trans | 1.70E-34 | Nightingale | S_LDL_FC | NA | A | G | 0.0272 | 0.00434 | 1.30E-09 |
| rs544873 | miR-10b-5p | trans | 1.70E-34 | Nightingale | LDL_C | NA | A | G | 0.031186 | 0.004328 | 9.60E-13 |
| rs544873 | miR-10b-5p | trans | 1.70E-34 | Nightingale | ApoB_by_A | NA | A | G | 0.025679 | 0.004281 | 2.00E-09 |
| rs544873 | miR-10b-5p | trans | 1.70E-34 | Nightingale | S_VLDL_FC | NA | A | G | 0.036642 | 0.004347 | 1.70E-17 |
| rs544873 | miR-10b-5p | trans | 1.70E-34 | Nightingale | LA | NA | A | G | 0.023263 | 0.004263 | 1.60E-08 |
| rs544873 | miR-10b-5p | trans | 1.70E-34 | Nightingale | non_HDL_( | NA | A | G | 0.03414 | 0.004323 | 2.40E-15 |
| rs544873 | miR-10b-5p | trans | 1.70E-34 | Nightingale | LDL_P | NA | A | G | 0.030659 | 0.004344 | 2.20E-12 |
| rs544873 | miR-10b-5p | trans | 1.70E-34 | Nightingale | M_VLDL_C | NA | A | G | 0.037341 | 0.004335 | 5.40E-18 |
| rs544873 | miR-10b-5p | trans | 1.70E-34 | Nightingale | Total_PL | NA | A | G | 0.022997 | 0.004139 | 7.80E-09 |
| rs544873 | miR-10b-5p | trans | 1.70E-34 | Nightingale | M_VLDL_T | NA | A | G | -0.03623 | 0.004108 | 1.50E-18 |
| rs544873 | miR-10b-5p | trans | 1.70E-34 | Nightingale | S_HDL_C | NA | A | G | 0.02538 | 0.004339 | 9.40E-09 |
| rs544873 | miR-10b-5p | trans | 1.70E-34 | Nightingale | L_LDL_FC | NA | A | G | 0.031473 | 0.004268 | 4.10E-13 |
| rs544873 | miR-10b-5p | trans | 1.70E-34 | Nightingale | S_LDL_P | NA | A | G | 0.028959 | 0.004351 | 3.00E-11 |
| rs544873 | miR-10b-5p | trans | 1.70E-34 | Nightingale | S_VLDL_L | NA | A | G | 0.024496 | 0.004305 | 2.50E-09 |
| rs544873 | miR-10b-5p | trans | 1.70E-34 | Nightingale | XL_VLDL_F | NA | A | G | 0.035204 | 0.004214 | 1.80E-17 |
| rs544873 | miR-10b-5p | trans | 1.70E-34 | Nightingale | L_VLDL_TG | NA | A | G | -0.04923 | 0.004326 | 6.40E-31 |
| rs544873 | miR-10b-5p | trans | 1.70E-34 | Nightingale | L_LDL_PL | NA | A | G | 0.035919 | 0.004314 | 1.30E-16 |
| rs544873 | miR-10b-5p | trans | 1.70E-34 | Nightingale | S_LDL_PL_ | NA | A | G | -0.02806 | 0.004339 | 4.90E-12 |
| rs544873 | miR-10b-5p | trans | 1.70E-34 | Nightingale | S_VLDL_C_ | NA | A | G | 0.039736 | 0.00423 | 8.30E-21 |
| rs544873 | miR-10b-5p | trans | 1.70E-34 | Nightingale | L_VLDL_C_ | NA | A | G | 0.049226 | 0.004245 | 8.90E-32 |
| rs544873 | miR-10b-5p | trans | 1.70E-34 | Nightingale | XXL_VLDL_ | NA | A | G | 0.033437 | 0.004356 | 8.80E-15 |
| rs544873 | miR-10b-5p | trans | 1.70E-34 | Nightingale | XL_VLDL_C | NA | A | G | 0.03559 | 0.004168 | 1.70E-17 |
| rs544873 | miR-10b-5p | trans | 1.70E-34 | Nightingale | XL_VLDL_C | NA | A | G | 0.034575 | 0.00417 | 1.50E-16 |
| rs544873 | miR-10b-5p | trans | 1.70E-34 | Nightingale | M_VLDL_P | NA | A | G | 0.028753 | 0.00434 | 1.40E-11 |
| rs544873 | miR-10b-5p | trans | 1.70E-34 | Nightingale | XS_VLDL_C | NA | A | G | 0.036639 | 0.004219 | 2.40E-18 |
| rs544873 | miR-10b-5p | trans | 1.70E-34 | Nightingale | M_LDL_L | NA | A | G | 0.031388 | 0.004353 | 7.00E-13 |
| rs544873 | miR-10b-5p | trans | 1.70E-34 | Nightingale | M_VLDL_P | NA | A | G | 0.042093 | 0.004172 | 6.30E-24 |
| rs544873 | miR-10b-5p | trans | 1.70E-34 | Nightingale | M_VLDL_P | NA | A | G | 0.027926 | 0.004336 | 4.70E-11 |
| rs544873 | miR-10b-5p | trans | 1.70E-34 | Nightingale | M_VLDL_F | NA | A | G | 0.039372 | 0.004135 | 2.20E-21 |
| rs544873 | miR-10b-5p | trans | 1.70E-34 | Nightingale | L_LDL_L | NA | A | G | 0.031523 | 0.004304 | 3.20E-13 |
| rs544873 | miR-10b-5p | trans | 1.70E-34 | Nightingale | S_LDL_C | NA | A | G | 0.029698 | 0.004351 | 1.60E-11 |
| rs544873 | miR-10b-5p | trans | 1.70E-34 | Nightingale | XL_VLDL_T | NA | A | G | -0.04155 | 0.004213 | 4.90E-23 |

| | | | | | | | | | | | |
|---|---|---|---|---|---|---|---|---|---|---|---|
| rs544873 | miR-10b-5p | trans | 1.70E-34 | Nightingale | Total_FC | NA | A | G | 0.031378 | 0.004246 | 4.70E-14 |
| rs544873 | miR-10b-5p | trans | 1.70E-34 | Nightingale | XS_VLDL_P | NA | A | G | 0.035785 | 0.004289 | 1.00E-17 |
| rs544873 | miR-10b-5p | trans | 1.70E-34 | Nightingale | XXL_VLDL_ | NA | A | G | 0.03179 | 0.00438 | 2.10E-13 |
| rs544873 | miR-10b-5p | trans | 1.70E-34 | Nightingale | S_VLDL_PL | NA | A | G | 0.025728 | 0.004162 | 1.50E-09 |
| rs544873 | miR-10b-5p | trans | 1.70E-34 | Nightingale | S_LDL_CE | NA | A | G | 0.029442 | 0.004349 | 1.50E-11 |
| rs544873 | miR-10b-5p | trans | 1.70E-34 | Nightingale | S_HDL_CE | NA | A | G | 0.02569 | 0.004354 | 7.00E-09 |
| rs544873 | miR-10b-5p | trans | 1.70E-34 | Nightingale | ApoB | NA | A | G | 0.032928 | 0.004341 | 3.40E-14 |
| rs544873 | let-7a-5p | trans | 8.38E-13 | Nightingale | IDL_L | NA | A | G | 0.03366 | 0.004203 | 2.30E-15 |
| rs544873 | let-7a-5p | trans | 8.38E-13 | Nightingale | S_LDL_L | NA | A | G | 0.026893 | 0.004346 | 1.20E-09 |
| rs544873 | let-7a-5p | trans | 8.38E-13 | Nightingale | M_VLDL_C | NA | A | G | 0.0392 | 0.004307 | 1.30E-19 |
| rs544873 | let-7a-5p | trans | 8.38E-13 | Nightingale | IDL_CE | NA | A | G | 0.032088 | 0.004197 | 4.30E-14 |
| rs544873 | let-7a-5p | trans | 8.38E-13 | Nightingale | XXL_VLDL_ | NA | A | G | -0.03443 | 0.004384 | 6.30E-16 |
| rs544873 | let-7a-5p | trans | 8.38E-13 | Nightingale | Clinical_LD | NA | A | G | 0.034557 | 0.004322 | 2.70E-15 |
| rs544873 | let-7a-5p | trans | 8.38E-13 | Nightingale | L_LDL_C | NA | A | G | 0.030224 | 0.004298 | 3.00E-12 |
| rs544873 | let-7a-5p | trans | 8.38E-13 | Nightingale | M_VLDL_C | NA | A | G | 0.033845 | 0.004094 | 2.70E-16 |
| rs544873 | let-7a-5p | trans | 8.38E-13 | Nightingale | M_LDL_PL | NA | A | G | 0.031174 | 0.004353 | 9.90E-13 |
| rs544873 | let-7a-5p | trans | 8.38E-13 | Nightingale | S_HDL_P | NA | A | G | 0.027805 | 0.004347 | 2.70E-10 |
| rs544873 | let-7a-5p | trans | 8.38E-13 | Nightingale | Sphingomy | NA | A | G | 0.025917 | 0.004063 | 3.30E-10 |
| rs544873 | let-7a-5p | trans | 8.38E-13 | Nightingale | VLDL_P | NA | A | G | 0.026555 | 0.004324 | 2.00E-10 |
| rs544873 | let-7a-5p | trans | 8.38E-13 | Nightingale | IDL_P | NA | A | G | 0.039032 | 0.004301 | 1.80E-19 |
| rs544873 | let-7a-5p | trans | 8.38E-13 | Nightingale | L_VLDL_FC | NA | A | G | 0.046941 | 0.004305 | 3.90E-28 |
| rs544873 | let-7a-5p | trans | 8.38E-13 | Nightingale | M_VLDL_C | NA | A | G | 0.031348 | 0.004084 | 3.30E-14 |
| rs544873 | let-7a-5p | trans | 8.38E-13 | Nightingale | S_VLDL_CE | NA | A | G | 0.037311 | 0.004341 | 1.70E-18 |
| rs544873 | let-7a-5p | trans | 8.38E-13 | Nightingale | IDL_PL | NA | A | G | 0.033573 | 0.004195 | 2.00E-15 |
| rs544873 | let-7a-5p | trans | 8.38E-13 | Nightingale | XS_VLDL_F | NA | A | G | 0.029134 | 0.004316 | 3.10E-11 |
| rs544873 | let-7a-5p | trans | 8.38E-13 | Nightingale | Total_L | NA | A | G | 0.025008 | 0.004276 | 1.50E-09 |
| rs544873 | let-7a-5p | trans | 8.38E-13 | Nightingale | XL_HDL_PL | NA | A | G | -0.02181 | 0.003994 | 4.60E-08 |
| rs544873 | let-7a-5p | trans | 8.38E-13 | Nightingale | GlycA | NA | A | G | 0.025329 | 0.004328 | 4.80E-09 |
| rs544873 | let-7a-5p | trans | 8.38E-13 | Nightingale | L_VLDL_PL | NA | A | G | 0.021645 | 0.004222 | 4.40E-08 |
| rs544873 | let-7a-5p | trans | 8.38E-13 | Nightingale | L_LDL_P | NA | A | G | 0.028317 | 0.004332 | 7.80E-11 |
| rs544873 | let-7a-5p | trans | 8.38E-13 | Nightingale | IDL_FC | NA | A | G | 0.035978 | 0.004208 | 3.20E-17 |
| rs544873 | let-7a-5p | trans | 8.38E-13 | Nightingale | VLDL_FC | NA | A | G | 0.024446 | 0.004315 | 3.60E-09 |
| rs544873 | let-7a-5p | trans | 8.38E-13 | Nightingale | IDL_C | NA | A | G | 0.033305 | 0.004198 | 4.90E-15 |
| rs544873 | let-7a-5p | trans | 8.38E-13 | Nightingale | M_LDL_FC | NA | A | G | 0.032281 | 0.004341 | 2.40E-13 |
| rs544873 | let-7a-5p | trans | 8.38E-13 | Nightingale | S_VLDL_CE | NA | A | G | 0.045151 | 0.004288 | 1.40E-25 |
| rs544873 | let-7a-5p | trans | 8.38E-13 | Nightingale | M_LDL_C | NA | A | G | 0.032091 | 0.004355 | 2.30E-13 |
| rs544873 | let-7a-5p | trans | 8.38E-13 | Nightingale | XS_VLDL_P | NA | A | G | 0.031657 | 0.004286 | 2.00E-14 |
| rs544873 | let-7a-5p | trans | 8.38E-13 | Nightingale | M_VLDL_F | NA | A | G | 0.032515 | 0.004346 | 3.90E-14 |
| rs544873 | let-7a-5p | trans | 8.38E-13 | Nightingale | L_VLDL_CE | NA | A | G | 0.04407 | 0.00416 | 7.90E-27 |
| rs544873 | let-7a-5p | trans | 8.38E-13 | Nightingale | Omega_6 | NA | A | G | 0.024156 | 0.004245 | 3.10E-09 |
| rs544873 | let-7a-5p | trans | 8.38E-13 | Nightingale | XL_VLDL_P | NA | A | G | 0.039106 | 0.004353 | 7.40E-20 |
| rs544873 | let-7a-5p | trans | 8.38E-13 | Nightingale | S_LDL_C_p | NA | A | G | 0.041864 | 0.004364 | 5.60E-22 |
| rs544873 | let-7a-5p | trans | 8.38E-13 | Nightingale | VLDL_CE | NA | A | G | 0.035144 | 0.004348 | 2.10E-16 |
| rs544873 | let-7a-5p | trans | 8.38E-13 | Nightingale | L_LDL_CE | NA | A | G | 0.02952 | 0.004308 | 9.90E-12 |
| rs544873 | let-7a-5p | trans | 8.38E-13 | Nightingale | LDL_L | NA | A | G | 0.031465 | 0.004332 | 5.80E-13 |
| rs544873 | let-7a-5p | trans | 8.38E-13 | Nightingale | XS_VLDL_L | NA | A | G | 0.033006 | 0.004273 | 1.40E-15 |
| rs544873 | let-7a-5p | trans | 8.38E-13 | Nightingale | S_VLDL_C | NA | A | G | 0.037355 | 0.004346 | 2.60E-18 |
| rs544873 | let-7a-5p | trans | 8.38E-13 | Nightingale | M_LDL_CE | NA | A | G | 0.031212 | 0.004353 | 8.50E-13 |
| rs544873 | let-7a-5p | trans | 8.38E-13 | Nightingale | S_VLDL_FC | NA | A | G | 0.027154 | 0.004154 | 1.20E-10 |
| rs544873 | let-7a-5p | trans | 8.38E-13 | Nightingale | XS_VLDL_C | NA | A | G | 0.036391 | 0.004195 | 4.10E-18 |
| rs544873 | let-7a-5p | trans | 8.38E-13 | Nightingale | L_VLDL_CE | NA | A | G | 0.022756 | 0.004296 | 2.90E-08 |
| rs544873 | let-7a-5p | trans | 8.38E-13 | Nightingale | S_VLDL_PL | NA | A | G | 0.032035 | 0.004339 | 4.90E-14 |
| rs544873 | let-7a-5p | trans | 8.38E-13 | Nightingale | XXL_VLDL_ | NA | A | G | 0.023502 | 0.004375 | 5.40E-09 |
| rs544873 | let-7a-5p | trans | 8.38E-13 | Nightingale | Remnant_( | NA | A | G | 0.035838 | 0.004319 | 5.30E-17 |
| rs544873 | let-7a-5p | trans | 8.38E-13 | Nightingale | S_VLDL_P | NA | A | G | 0.02339 | 0.004292 | 9.70E-09 |
| rs544873 | let-7a-5p | trans | 8.38E-13 | Nightingale | Total_C | NA | A | G | 0.030138 | 0.004208 | 5.00E-13 |
| rs544873 | let-7a-5p | trans | 8.38E-13 | Nightingale | VLDL_C | NA | A | G | 0.031062 | 0.004341 | 2.10E-13 |
| rs544873 | let-7a-5p | trans | 8.38E-13 | Nightingale | Total_CE | NA | A | G | 0.029384 | 0.004193 | 1.80E-12 |
| rs544873 | let-7a-5p | trans | 8.38E-13 | Nightingale | M_LDL_C_ | NA | A | G | 0.026582 | 0.00435 | 1.60E-09 |
| rs544873 | let-7a-5p | trans | 8.38E-13 | Nightingale | S_VLDL_TG | NA | A | G | -0.03598 | 0.004208 | 1.90E-17 |
| rs544873 | let-7a-5p | trans | 8.38E-13 | Nightingale | S_LDL_CE_ | NA | A | G | 0.028844 | 0.004339 | 1.10E-12 |
| rs544873 | let-7a-5p | trans | 8.38E-13 | Nightingale | S_HDL_L | NA | A | G | 0.023969 | 0.004332 | 2.70E-08 |
| rs544873 | let-7a-5p | trans | 8.38E-13 | Nightingale | LDL_FC | NA | A | G | 0.031548 | 0.0043 | 6.60E-13 |

| | | | | | | | | | | | |
|---|---|---|---|---|---|---|---|---|---|---|---|
| rs544873 | let-7a-5p | trans | 8.38E-13 | Nightingale | LDL_CE | NA | A | G | 0.030617 | 0.004335 | 2.10E-12 |
| rs544873 | let-7a-5p | trans | 8.38E-13 | Nightingale | XS_VLDL_F | NA | A | G | 0.035882 | 0.004271 | 7.50E-18 |
| rs544873 | let-7a-5p | trans | 8.38E-13 | Nightingale | M_LDL_TG | NA | A | G | -0.02512 | 0.004293 | 8.60E-09 |
| rs544873 | let-7a-5p | trans | 8.38E-13 | Nightingale | LDL_PL | NA | A | G | 0.033318 | 0.004337 | 3.00E-14 |
| rs544873 | let-7a-5p | trans | 8.38E-13 | Nightingale | M_LDL_P | NA | A | G | 0.034001 | 0.004359 | 7.50E-15 |
| rs544873 | let-7a-5p | trans | 8.38E-13 | Nightingale | Citrate | NA | A | G | 0.024678 | 0.004318 | 6.10E-09 |
| rs544873 | let-7a-5p | trans | 8.38E-13 | Nightingale | XXL_VLDL_ | NA | A | G | 0.029268 | 0.004328 | 5.10E-12 |
| rs544873 | let-7a-5p | trans | 8.38E-13 | Nightingale | S_LDL_FC | NA | A | G | 0.0272 | 0.00434 | 1.30E-09 |
| rs544873 | let-7a-5p | trans | 8.38E-13 | Nightingale | LDL_C | NA | A | G | 0.031186 | 0.004328 | 9.60E-13 |
| rs544873 | let-7a-5p | trans | 8.38E-13 | Nightingale | ApoB_by_A | NA | A | G | 0.025679 | 0.004281 | 2.00E-09 |
| rs544873 | let-7a-5p | trans | 8.38E-13 | Nightingale | S_VLDL_FC | NA | A | G | 0.036642 | 0.004347 | 1.70E-17 |
| rs544873 | let-7a-5p | trans | 8.38E-13 | Nightingale | LA | NA | A | G | 0.023263 | 0.004263 | 1.60E-08 |
| rs544873 | let-7a-5p | trans | 8.38E-13 | Nightingale | non_HDL_( | NA | A | G | 0.03414 | 0.004323 | 2.40E-15 |
| rs544873 | let-7a-5p | trans | 8.38E-13 | Nightingale | LDL_P | NA | A | G | 0.030659 | 0.004344 | 2.20E-12 |
| rs544873 | let-7a-5p | trans | 8.38E-13 | Nightingale | M_VLDL_C | NA | A | G | 0.037341 | 0.004335 | 5.40E-18 |
| rs544873 | let-7a-5p | trans | 8.38E-13 | Nightingale | Total_PL | NA | A | G | 0.022997 | 0.004139 | 7.80E-09 |
| rs544873 | let-7a-5p | trans | 8.38E-13 | Nightingale | M_VLDL_T | NA | A | G | -0.03623 | 0.004108 | 1.50E-18 |
| rs544873 | let-7a-5p | trans | 8.38E-13 | Nightingale | S_HDL_C | NA | A | G | 0.02538 | 0.004339 | 9.40E-09 |
| rs544873 | let-7a-5p | trans | 8.38E-13 | Nightingale | L_LDL_FC | NA | A | G | 0.031473 | 0.004268 | 4.10E-13 |
| rs544873 | let-7a-5p | trans | 8.38E-13 | Nightingale | S_LDL_P | NA | A | G | 0.028959 | 0.004351 | 3.00E-11 |
| rs544873 | let-7a-5p | trans | 8.38E-13 | Nightingale | S_VLDL_L | NA | A | G | 0.024496 | 0.004305 | 2.50E-09 |
| rs544873 | let-7a-5p | trans | 8.38E-13 | Nightingale | XL_VLDL_F | NA | A | G | 0.035204 | 0.004214 | 1.80E-17 |
| rs544873 | let-7a-5p | trans | 8.38E-13 | Nightingale | L_VLDL_TG | NA | A | G | -0.04923 | 0.004326 | 6.40E-31 |
| rs544873 | let-7a-5p | trans | 8.38E-13 | Nightingale | L_LDL_PL | NA | A | G | 0.035919 | 0.004314 | 1.30E-16 |
| rs544873 | let-7a-5p | trans | 8.38E-13 | Nightingale | S_LDL_PL_ | NA | A | G | -0.02806 | 0.004339 | 4.90E-12 |
| rs544873 | let-7a-5p | trans | 8.38E-13 | Nightingale | S_VLDL_C_ | NA | A | G | 0.039736 | 0.00423 | 8.30E-21 |
| rs544873 | let-7a-5p | trans | 8.38E-13 | Nightingale | L_VLDL_C_ | NA | A | G | 0.049226 | 0.004245 | 8.90E-32 |
| rs544873 | let-7a-5p | trans | 8.38E-13 | Nightingale | XXL_VLDL_ | NA | A | G | 0.033437 | 0.004356 | 8.80E-15 |
| rs544873 | let-7a-5p | trans | 8.38E-13 | Nightingale | XL_VLDL_C | NA | A | G | 0.03559 | 0.004168 | 1.70E-17 |
| rs544873 | let-7a-5p | trans | 8.38E-13 | Nightingale | XL_VLDL_C | NA | A | G | 0.034575 | 0.00417 | 1.50E-16 |
| rs544873 | let-7a-5p | trans | 8.38E-13 | Nightingale | M_VLDL_P | NA | A | G | 0.028753 | 0.00434 | 1.40E-11 |
| rs544873 | let-7a-5p | trans | 8.38E-13 | Nightingale | XS_VLDL_C | NA | A | G | 0.036639 | 0.004219 | 2.40E-18 |
| rs544873 | let-7a-5p | trans | 8.38E-13 | Nightingale | M_LDL_L | NA | A | G | 0.031388 | 0.004353 | 7.00E-13 |
| rs544873 | let-7a-5p | trans | 8.38E-13 | Nightingale | M_VLDL_P | NA | A | G | 0.042093 | 0.004172 | 6.30E-24 |
| rs544873 | let-7a-5p | trans | 8.38E-13 | Nightingale | M_VLDL_P | NA | A | G | 0.027926 | 0.004336 | 4.70E-11 |
| rs544873 | let-7a-5p | trans | 8.38E-13 | Nightingale | M_VLDL_F | NA | A | G | 0.039372 | 0.004135 | 2.20E-21 |
| rs544873 | let-7a-5p | trans | 8.38E-13 | Nightingale | L_LDL_L | NA | A | G | 0.031523 | 0.004304 | 3.20E-13 |
| rs544873 | let-7a-5p | trans | 8.38E-13 | Nightingale | S_LDL_C | NA | A | G | 0.029698 | 0.004351 | 1.60E-11 |
| rs544873 | let-7a-5p | trans | 8.38E-13 | Nightingale | XL_VLDL_T | NA | A | G | -0.04155 | 0.004213 | 4.90E-23 |
| rs544873 | let-7a-5p | trans | 8.38E-13 | Nightingale | Total_FC | NA | A | G | 0.031378 | 0.004246 | 4.70E-14 |
| rs544873 | let-7a-5p | trans | 8.38E-13 | Nightingale | XS_VLDL_P | NA | A | G | 0.035785 | 0.004289 | 1.00E-17 |
| rs544873 | let-7a-5p | trans | 8.38E-13 | Nightingale | XXL_VLDL_ | NA | A | G | 0.03179 | 0.00438 | 2.10E-13 |
| rs544873 | let-7a-5p | trans | 8.38E-13 | Nightingale | S_VLDL_PL | NA | A | G | 0.025728 | 0.004162 | 1.50E-09 |
| rs544873 | let-7a-5p | trans | 8.38E-13 | Nightingale | S_LDL_CE | NA | A | G | 0.029442 | 0.004349 | 1.50E-11 |
| rs544873 | let-7a-5p | trans | 8.38E-13 | Nightingale | S_HDL_CE | NA | A | G | 0.02569 | 0.004354 | 7.00E-09 |
| rs544873 | let-7a-5p | trans | 8.38E-13 | Nightingale | ApoB | NA | A | G | 0.032928 | 0.004341 | 3.40E-14 |
| rs544873 | miR-139-5p | trans | 1.34E-40 | Nightingale | IDL_L | NA | A | G | 0.03366 | 0.004203 | 2.30E-15 |
| rs544873 | miR-139-5p | trans | 1.34E-40 | Nightingale | S_LDL_L | NA | A | G | 0.026893 | 0.004346 | 1.20E-09 |
| rs544873 | miR-139-5p | trans | 1.34E-40 | Nightingale | M_VLDL_C | NA | A | G | 0.0392 | 0.004307 | 1.30E-19 |
| rs544873 | miR-139-5p | trans | 1.34E-40 | Nightingale | IDL_CE | NA | A | G | 0.032088 | 0.004197 | 4.30E-14 |
| rs544873 | miR-139-5p | trans | 1.34E-40 | Nightingale | XXL_VLDL_ | NA | A | G | -0.03443 | 0.004384 | 6.30E-16 |
| rs544873 | miR-139-5p | trans | 1.34E-40 | Nightingale | Clinical_LD | NA | A | G | 0.034557 | 0.004322 | 2.70E-15 |
| rs544873 | miR-139-5p | trans | 1.34E-40 | Nightingale | L_LDL_C | NA | A | G | 0.030224 | 0.004298 | 3.00E-12 |
| rs544873 | miR-139-5p | trans | 1.34E-40 | Nightingale | M_VLDL_C | NA | A | G | 0.033845 | 0.004094 | 2.70E-16 |
| rs544873 | miR-139-5p | trans | 1.34E-40 | Nightingale | M_LDL_PL | NA | A | G | 0.031174 | 0.004353 | 9.90E-13 |
| rs544873 | miR-139-5p | trans | 1.34E-40 | Nightingale | S_HDL_P | NA | A | G | 0.027805 | 0.004347 | 2.70E-10 |
| rs544873 | miR-139-5p | trans | 1.34E-40 | Nightingale | Sphingomy | NA | A | G | 0.025917 | 0.004063 | 3.30E-10 |
| rs544873 | miR-139-5p | trans | 1.34E-40 | Nightingale | VLDL_P | NA | A | G | 0.026555 | 0.004324 | 2.00E-10 |
| rs544873 | miR-139-5p | trans | 1.34E-40 | Nightingale | IDL_P | NA | A | G | 0.039032 | 0.004301 | 1.80E-19 |
| rs544873 | miR-139-5p | trans | 1.34E-40 | Nightingale | L_VLDL_FC | NA | A | G | 0.046941 | 0.004305 | 3.90E-28 |
| rs544873 | miR-139-5p | trans | 1.34E-40 | Nightingale | M_VLDL_C | NA | A | G | 0.031348 | 0.004084 | 3.30E-14 |
| rs544873 | miR-139-5p | trans | 1.34E-40 | Nightingale | S_VLDL_CE | NA | A | G | 0.037311 | 0.004341 | 1.70E-18 |
| rs544873 | miR-139-5p | trans | 1.34E-40 | Nightingale | IDL_PL | NA | A | G | 0.033573 | 0.004195 | 2.00E-15 |

| | | | | | | | | | | | |
|---|---|---|---|---|---|---|---|---|---|---|---|
| rs544873 | miR-139-5p | trans | 1.34E-40 | Nightingale | XS_VLDL_F | NA | A | G | 0.029134 | 0.004316 | 3.10E-11 |
| rs544873 | miR-139-5p | trans | 1.34E-40 | Nightingale | Total_L | NA | A | G | 0.025008 | 0.004276 | 1.50E-09 |
| rs544873 | miR-139-5p | trans | 1.34E-40 | Nightingale | XL_HDL_PL | NA | A | G | -0.02181 | 0.003994 | 4.60E-08 |
| rs544873 | miR-139-5p | trans | 1.34E-40 | Nightingale | GlycA | NA | A | G | 0.025329 | 0.004328 | 4.80E-09 |
| rs544873 | miR-139-5p | trans | 1.34E-40 | Nightingale | L_VLDL_PL | NA | A | G | 0.021645 | 0.004222 | 4.40E-08 |
| rs544873 | miR-139-5p | trans | 1.34E-40 | Nightingale | L_LDL_P | NA | A | G | 0.028317 | 0.004332 | 7.80E-11 |
| rs544873 | miR-139-5p | trans | 1.34E-40 | Nightingale | IDL_FC | NA | A | G | 0.035978 | 0.004208 | 3.20E-17 |
| rs544873 | miR-139-5p | trans | 1.34E-40 | Nightingale | VLDL_FC | NA | A | G | 0.024446 | 0.004315 | 3.60E-09 |
| rs544873 | miR-139-5p | trans | 1.34E-40 | Nightingale | IDL_C | NA | A | G | 0.033305 | 0.004198 | 4.90E-15 |
| rs544873 | miR-139-5p | trans | 1.34E-40 | Nightingale | M_LDL_FC | NA | A | G | 0.032281 | 0.004341 | 2.40E-13 |
| rs544873 | miR-139-5p | trans | 1.34E-40 | Nightingale | S_VLDL_CE | NA | A | G | 0.045151 | 0.004288 | 1.40E-25 |
| rs544873 | miR-139-5p | trans | 1.34E-40 | Nightingale | M_LDL_C | NA | A | G | 0.032091 | 0.004355 | 2.30E-13 |
| rs544873 | miR-139-5p | trans | 1.34E-40 | Nightingale | XS_VLDL_P | NA | A | G | 0.031657 | 0.004286 | 2.00E-14 |
| rs544873 | miR-139-5p | trans | 1.34E-40 | Nightingale | M_VLDL_F | NA | A | G | 0.032515 | 0.004346 | 3.90E-14 |
| rs544873 | miR-139-5p | trans | 1.34E-40 | Nightingale | L_VLDL_CE | NA | A | G | 0.04407 | 0.00416 | 7.90E-27 |
| rs544873 | miR-139-5p | trans | 1.34E-40 | Nightingale | Omega_6 | NA | A | G | 0.024156 | 0.004245 | 3.10E-09 |
| rs544873 | miR-139-5p | trans | 1.34E-40 | Nightingale | XL_VLDL_P | NA | A | G | 0.039106 | 0.004353 | 7.40E-20 |
| rs544873 | miR-139-5p | trans | 1.34E-40 | Nightingale | S_LDL_C_p | NA | A | G | 0.041864 | 0.004364 | 5.60E-22 |
| rs544873 | miR-139-5p | trans | 1.34E-40 | Nightingale | VLDL_CE | NA | A | G | 0.035144 | 0.004348 | 2.10E-16 |
| rs544873 | miR-139-5p | trans | 1.34E-40 | Nightingale | L_LDL_CE | NA | A | G | 0.02952 | 0.004308 | 9.90E-12 |
| rs544873 | miR-139-5p | trans | 1.34E-40 | Nightingale | LDL_L | NA | A | G | 0.031465 | 0.004332 | 5.80E-13 |
| rs544873 | miR-139-5p | trans | 1.34E-40 | Nightingale | XS_VLDL_L | NA | A | G | 0.033006 | 0.004273 | 1.40E-15 |
| rs544873 | miR-139-5p | trans | 1.34E-40 | Nightingale | S_VLDL_C | NA | A | G | 0.037355 | 0.004346 | 2.60E-18 |
| rs544873 | miR-139-5p | trans | 1.34E-40 | Nightingale | M_LDL_CE | NA | A | G | 0.031212 | 0.004353 | 8.50E-13 |
| rs544873 | miR-139-5p | trans | 1.34E-40 | Nightingale | S_VLDL_FC | NA | A | G | 0.027154 | 0.004154 | 1.20E-10 |
| rs544873 | miR-139-5p | trans | 1.34E-40 | Nightingale | XS_VLDL_C | NA | A | G | 0.036391 | 0.004195 | 4.10E-18 |
| rs544873 | miR-139-5p | trans | 1.34E-40 | Nightingale | L_VLDL_CE | NA | A | G | 0.022756 | 0.004296 | 2.90E-08 |
| rs544873 | miR-139-5p | trans | 1.34E-40 | Nightingale | S_VLDL_PL | NA | A | G | 0.032035 | 0.004339 | 4.90E-14 |
| rs544873 | miR-139-5p | trans | 1.34E-40 | Nightingale | XXL_VLDL_ | NA | A | G | 0.023502 | 0.004375 | 5.40E-09 |
| rs544873 | miR-139-5p | trans | 1.34E-40 | Nightingale | Remnant_( | NA | A | G | 0.035838 | 0.004319 | 5.30E-17 |
| rs544873 | miR-139-5p | trans | 1.34E-40 | Nightingale | S_VLDL_P | NA | A | G | 0.02339 | 0.004292 | 9.70E-09 |
| rs544873 | miR-139-5p | trans | 1.34E-40 | Nightingale | Total_C | NA | A | G | 0.030138 | 0.004208 | 5.00E-13 |
| rs544873 | miR-139-5p | trans | 1.34E-40 | Nightingale | VLDL_C | NA | A | G | 0.031062 | 0.004341 | 2.10E-13 |
| rs544873 | miR-139-5p | trans | 1.34E-40 | Nightingale | Total_CE | NA | A | G | 0.029384 | 0.004193 | 1.80E-12 |
| rs544873 | miR-139-5p | trans | 1.34E-40 | Nightingale | M_LDL_C_p | NA | A | G | 0.026582 | 0.00435 | 1.60E-09 |
| rs544873 | miR-139-5p | trans | 1.34E-40 | Nightingale | S_VLDL_TG | NA | A | G | -0.03598 | 0.004208 | 1.90E-17 |
| rs544873 | miR-139-5p | trans | 1.34E-40 | Nightingale | S_LDL_CE_ | NA | A | G | 0.028844 | 0.004339 | 1.10E-12 |
| rs544873 | miR-139-5p | trans | 1.34E-40 | Nightingale | S_HDL_L | NA | A | G | 0.023969 | 0.004332 | 2.70E-08 |
| rs544873 | miR-139-5p | trans | 1.34E-40 | Nightingale | LDL_FC | NA | A | G | 0.031548 | 0.0043 | 6.60E-13 |
| rs544873 | miR-139-5p | trans | 1.34E-40 | Nightingale | LDL_CE | NA | A | G | 0.030617 | 0.004335 | 2.10E-12 |
| rs544873 | miR-139-5p | trans | 1.34E-40 | Nightingale | XS_VLDL_F | NA | A | G | 0.035882 | 0.004271 | 7.50E-18 |
| rs544873 | miR-139-5p | trans | 1.34E-40 | Nightingale | M_LDL_TG | NA | A | G | -0.02512 | 0.004293 | 8.60E-09 |
| rs544873 | miR-139-5p | trans | 1.34E-40 | Nightingale | LDL_PL | NA | A | G | 0.033318 | 0.004337 | 3.00E-14 |
| rs544873 | miR-139-5p | trans | 1.34E-40 | Nightingale | M_LDL_P | NA | A | G | 0.034001 | 0.004359 | 7.50E-15 |
| rs544873 | miR-139-5p | trans | 1.34E-40 | Nightingale | Citrate | NA | A | G | 0.024678 | 0.004318 | 6.10E-09 |
| rs544873 | miR-139-5p | trans | 1.34E-40 | Nightingale | XXL_VLDL_ | NA | A | G | 0.029268 | 0.004328 | 5.10E-12 |
| rs544873 | miR-139-5p | trans | 1.34E-40 | Nightingale | S_LDL_FC | NA | A | G | 0.0272 | 0.00434 | 1.30E-09 |
| rs544873 | miR-139-5p | trans | 1.34E-40 | Nightingale | LDL_C | NA | A | G | 0.031186 | 0.004328 | 9.60E-13 |
| rs544873 | miR-139-5p | trans | 1.34E-40 | Nightingale | ApoB_by_A | NA | A | G | 0.025679 | 0.004281 | 2.00E-09 |
| rs544873 | miR-139-5p | trans | 1.34E-40 | Nightingale | S_VLDL_FC | NA | A | G | 0.036642 | 0.004347 | 1.70E-17 |
| rs544873 | miR-139-5p | trans | 1.34E-40 | Nightingale | LA | NA | A | G | 0.023263 | 0.004263 | 1.60E-08 |
| rs544873 | miR-139-5p | trans | 1.34E-40 | Nightingale | non_HDL_( | NA | A | G | 0.03414 | 0.004323 | 2.40E-15 |
| rs544873 | miR-139-5p | trans | 1.34E-40 | Nightingale | LDL_P | NA | A | G | 0.030659 | 0.004344 | 2.20E-12 |
| rs544873 | miR-139-5p | trans | 1.34E-40 | Nightingale | M_VLDL_C | NA | A | G | 0.037341 | 0.004335 | 5.40E-18 |
| rs544873 | miR-139-5p | trans | 1.34E-40 | Nightingale | Total_PL | NA | A | G | 0.022997 | 0.004139 | 7.80E-09 |
| rs544873 | miR-139-5p | trans | 1.34E-40 | Nightingale | M_VLDL_T | NA | A | G | -0.03623 | 0.004108 | 1.50E-18 |
| rs544873 | miR-139-5p | trans | 1.34E-40 | Nightingale | S_HDL_C | NA | A | G | 0.02538 | 0.004339 | 9.40E-09 |
| rs544873 | miR-139-5p | trans | 1.34E-40 | Nightingale | L_LDL_FC | NA | A | G | 0.031473 | 0.004268 | 4.10E-13 |
| rs544873 | miR-139-5p | trans | 1.34E-40 | Nightingale | S_LDL_P | NA | A | G | 0.028959 | 0.004351 | 3.00E-11 |
| rs544873 | miR-139-5p | trans | 1.34E-40 | Nightingale | S_VLDL_L | NA | A | G | 0.024496 | 0.004305 | 2.50E-09 |
| rs544873 | miR-139-5p | trans | 1.34E-40 | Nightingale | XL_VLDL_F | NA | A | G | 0.035204 | 0.004214 | 1.80E-17 |
| rs544873 | miR-139-5p | trans | 1.34E-40 | Nightingale | L_VLDL_TG | NA | A | G | -0.04923 | 0.004326 | 6.40E-31 |
| rs544873 | miR-139-5p | trans | 1.34E-40 | Nightingale | L_LDL_PL | NA | A | G | 0.035919 | 0.004314 | 1.30E-16 |

| | | | | | | | | | | | |
|---|---|---|---|---|---|---|---|---|---|---|---|
| rs544873 | miR-139-5p | trans | 1.34E-40 | Nightingale | S_LDL_PL_ | NA | A | G | -0.02806 | 0.004339 | 4.90E-12 |
| rs544873 | miR-139-5p | trans | 1.34E-40 | Nightingale | S_VLDL_C_ | NA | A | G | 0.039736 | 0.00423 | 8.30E-21 |
| rs544873 | miR-139-5p | trans | 1.34E-40 | Nightingale | L_VLDL_C_ | NA | A | G | 0.049226 | 0.004245 | 8.90E-32 |
| rs544873 | miR-139-5p | trans | 1.34E-40 | Nightingale | XXL_VLDL_ | NA | A | G | 0.033437 | 0.004356 | 8.80E-15 |
| rs544873 | miR-139-5p | trans | 1.34E-40 | Nightingale | XL_VLDL_C | NA | A | G | 0.03559 | 0.004168 | 1.70E-17 |
| rs544873 | miR-139-5p | trans | 1.34E-40 | Nightingale | XL_VLDL_C | NA | A | G | 0.034575 | 0.00417 | 1.50E-16 |
| rs544873 | miR-139-5p | trans | 1.34E-40 | Nightingale | M_VLDL_P | NA | A | G | 0.028753 | 0.00434 | 1.40E-11 |
| rs544873 | miR-139-5p | trans | 1.34E-40 | Nightingale | XS_VLDL_C | NA | A | G | 0.036639 | 0.004219 | 2.40E-18 |
| rs544873 | miR-139-5p | trans | 1.34E-40 | Nightingale | M_LDL_L | NA | A | G | 0.031388 | 0.004353 | 7.00E-13 |
| rs544873 | miR-139-5p | trans | 1.34E-40 | Nightingale | M_VLDL_P | NA | A | G | 0.042093 | 0.004172 | 6.30E-24 |
| rs544873 | miR-139-5p | trans | 1.34E-40 | Nightingale | M_VLDL_P | NA | A | G | 0.027926 | 0.004336 | 4.70E-11 |
| rs544873 | miR-139-5p | trans | 1.34E-40 | Nightingale | M_VLDL_F | NA | A | G | 0.039372 | 0.004135 | 2.20E-21 |
| rs544873 | miR-139-5p | trans | 1.34E-40 | Nightingale | L_LDL_L | NA | A | G | 0.031523 | 0.004304 | 3.20E-13 |
| rs544873 | miR-139-5p | trans | 1.34E-40 | Nightingale | S_LDL_C | NA | A | G | 0.029698 | 0.004351 | 1.60E-11 |
| rs544873 | miR-139-5p | trans | 1.34E-40 | Nightingale | XL_VLDL_T | NA | A | G | -0.04155 | 0.004213 | 4.90E-23 |
| rs544873 | miR-139-5p | trans | 1.34E-40 | Nightingale | Total_FC | NA | A | G | 0.031378 | 0.004246 | 4.70E-14 |
| rs544873 | miR-139-5p | trans | 1.34E-40 | Nightingale | XS_VLDL_P | NA | A | G | 0.035785 | 0.004289 | 1.00E-17 |
| rs544873 | miR-139-5p | trans | 1.34E-40 | Nightingale | XXL_VLDL_ | NA | A | G | 0.03179 | 0.00438 | 2.10E-13 |
| rs544873 | miR-139-5p | trans | 1.34E-40 | Nightingale | S_VLDL_PL | NA | A | G | 0.025728 | 0.004162 | 1.50E-09 |
| rs544873 | miR-139-5p | trans | 1.34E-40 | Nightingale | S_LDL_CE | NA | A | G | 0.029442 | 0.004349 | 1.50E-11 |
| rs544873 | miR-139-5p | trans | 1.34E-40 | Nightingale | S_HDL_CE | NA | A | G | 0.02569 | 0.004354 | 7.00E-09 |
| rs544873 | miR-139-5p | trans | 1.34E-40 | Nightingale | ApoB | NA | A | G | 0.032928 | 0.004341 | 3.40E-14 |
| rs544873 | miR-126-3p | trans | 1.31E-37 | Nightingale | IDL_L | NA | A | G | 0.03366 | 0.004203 | 2.30E-15 |
| rs544873 | miR-126-3p | trans | 1.31E-37 | Nightingale | S_LDL_L | NA | A | G | 0.026893 | 0.004346 | 1.20E-09 |
| rs544873 | miR-126-3p | trans | 1.31E-37 | Nightingale | M_VLDL_C | NA | A | G | 0.0392 | 0.004307 | 1.30E-19 |
| rs544873 | miR-126-3p | trans | 1.31E-37 | Nightingale | IDL_CE | NA | A | G | 0.032088 | 0.004197 | 4.30E-14 |
| rs544873 | miR-126-3p | trans | 1.31E-37 | Nightingale | XXL_VLDL_ | NA | A | G | -0.03443 | 0.004384 | 6.30E-16 |
| rs544873 | miR-126-3p | trans | 1.31E-37 | Nightingale | Clinical_LD | NA | A | G | 0.034557 | 0.004322 | 2.70E-15 |
| rs544873 | miR-126-3p | trans | 1.31E-37 | Nightingale | L_LDL_C | NA | A | G | 0.030224 | 0.004298 | 3.00E-12 |
| rs544873 | miR-126-3p | trans | 1.31E-37 | Nightingale | M_VLDL_C | NA | A | G | 0.033845 | 0.004094 | 2.70E-16 |
| rs544873 | miR-126-3p | trans | 1.31E-37 | Nightingale | M_LDL_PL | NA | A | G | 0.031174 | 0.004353 | 9.90E-13 |
| rs544873 | miR-126-3p | trans | 1.31E-37 | Nightingale | S_HDL_P | NA | A | G | 0.027805 | 0.004347 | 2.70E-10 |
| rs544873 | miR-126-3p | trans | 1.31E-37 | Nightingale | Sphingomy | NA | A | G | 0.025917 | 0.004063 | 3.30E-10 |
| rs544873 | miR-126-3p | trans | 1.31E-37 | Nightingale | VLDL_P | NA | A | G | 0.026555 | 0.004324 | 2.00E-10 |
| rs544873 | miR-126-3p | trans | 1.31E-37 | Nightingale | IDL_P | NA | A | G | 0.039032 | 0.004301 | 1.80E-19 |
| rs544873 | miR-126-3p | trans | 1.31E-37 | Nightingale | L_VLDL_FC | NA | A | G | 0.046941 | 0.004305 | 3.90E-28 |
| rs544873 | miR-126-3p | trans | 1.31E-37 | Nightingale | M_VLDL_C | NA | A | G | 0.031348 | 0.004084 | 3.30E-14 |
| rs544873 | miR-126-3p | trans | 1.31E-37 | Nightingale | S_VLDL_CE | NA | A | G | 0.037311 | 0.004341 | 1.70E-18 |
| rs544873 | miR-126-3p | trans | 1.31E-37 | Nightingale | IDL_PL | NA | A | G | 0.033573 | 0.004195 | 2.00E-15 |
| rs544873 | miR-126-3p | trans | 1.31E-37 | Nightingale | XS_VLDL_F | NA | A | G | 0.029134 | 0.004316 | 3.10E-11 |
| rs544873 | miR-126-3p | trans | 1.31E-37 | Nightingale | Total_L | NA | A | G | 0.025008 | 0.004276 | 1.50E-09 |
| rs544873 | miR-126-3p | trans | 1.31E-37 | Nightingale | XL_HDL_PL | NA | A | G | -0.02181 | 0.003994 | 4.60E-08 |
| rs544873 | miR-126-3p | trans | 1.31E-37 | Nightingale | GlycA | NA | A | G | 0.025329 | 0.004328 | 4.80E-09 |
| rs544873 | miR-126-3p | trans | 1.31E-37 | Nightingale | L_VLDL_PL | NA | A | G | 0.021645 | 0.004222 | 4.40E-08 |
| rs544873 | miR-126-3p | trans | 1.31E-37 | Nightingale | L_LDL_P | NA | A | G | 0.028317 | 0.004332 | 7.80E-11 |
| rs544873 | miR-126-3p | trans | 1.31E-37 | Nightingale | IDL_FC | NA | A | G | 0.035978 | 0.004208 | 3.20E-17 |
| rs544873 | miR-126-3p | trans | 1.31E-37 | Nightingale | VLDL_FC | NA | A | G | 0.024446 | 0.004315 | 3.60E-09 |
| rs544873 | miR-126-3p | trans | 1.31E-37 | Nightingale | IDL_C | NA | A | G | 0.033305 | 0.004198 | 4.90E-15 |
| rs544873 | miR-126-3p | trans | 1.31E-37 | Nightingale | M_LDL_FC | NA | A | G | 0.032281 | 0.004341 | 2.40E-13 |
| rs544873 | miR-126-3p | trans | 1.31E-37 | Nightingale | S_VLDL_CE | NA | A | G | 0.045151 | 0.004288 | 1.40E-25 |
| rs544873 | miR-126-3p | trans | 1.31E-37 | Nightingale | M_LDL_C | NA | A | G | 0.032091 | 0.004355 | 2.30E-13 |
| rs544873 | miR-126-3p | trans | 1.31E-37 | Nightingale | XS_VLDL_P | NA | A | G | 0.031657 | 0.004286 | 2.00E-14 |
| rs544873 | miR-126-3p | trans | 1.31E-37 | Nightingale | M_VLDL_F | NA | A | G | 0.032515 | 0.004346 | 3.90E-14 |
| rs544873 | miR-126-3p | trans | 1.31E-37 | Nightingale | L_VLDL_CE | NA | A | G | 0.04407 | 0.00416 | 7.90E-27 |
| rs544873 | miR-126-3p | trans | 1.31E-37 | Nightingale | Omega_6 | NA | A | G | 0.024156 | 0.004245 | 3.10E-09 |
| rs544873 | miR-126-3p | trans | 1.31E-37 | Nightingale | XL_VLDL_P | NA | A | G | 0.039106 | 0.004353 | 7.40E-20 |
| rs544873 | miR-126-3p | trans | 1.31E-37 | Nightingale | S_LDL_C_p | NA | A | G | 0.041864 | 0.004364 | 5.60E-22 |
| rs544873 | miR-126-3p | trans | 1.31E-37 | Nightingale | VLDL_CE | NA | A | G | 0.035144 | 0.004348 | 2.10E-16 |
| rs544873 | miR-126-3p | trans | 1.31E-37 | Nightingale | L_LDL_CE | NA | A | G | 0.02952 | 0.004308 | 9.90E-12 |
| rs544873 | miR-126-3p | trans | 1.31E-37 | Nightingale | LDL_L | NA | A | G | 0.031465 | 0.004332 | 5.80E-13 |
| rs544873 | miR-126-3p | trans | 1.31E-37 | Nightingale | XS_VLDL_L | NA | A | G | 0.033006 | 0.004273 | 1.40E-15 |
| rs544873 | miR-126-3p | trans | 1.31E-37 | Nightingale | S_VLDL_C | NA | A | G | 0.037355 | 0.004346 | 2.60E-18 |
| rs544873 | miR-126-3p | trans | 1.31E-37 | Nightingale | M_LDL_CE | NA | A | G | 0.031212 | 0.004353 | 8.50E-13 |

| | | | | | | | | | | | |
|---|---|---|---|---|---|---|---|---|---|---|---|
| rs544873 | miR-126-3p | trans | 1.31E-37 | Nightingale | S_VLDL_FC | NA | A | G | 0.027154 | 0.004154 | 1.20E-10 |
| rs544873 | miR-126-3p | trans | 1.31E-37 | Nightingale | XS_VLDL_C | NA | A | G | 0.036391 | 0.004195 | 4.10E-18 |
| rs544873 | miR-126-3p | trans | 1.31E-37 | Nightingale | L_VLDL_CE | NA | A | G | 0.022756 | 0.004296 | 2.90E-08 |
| rs544873 | miR-126-3p | trans | 1.31E-37 | Nightingale | S_VLDL_PL | NA | A | G | 0.032035 | 0.004339 | 4.90E-14 |
| rs544873 | miR-126-3p | trans | 1.31E-37 | Nightingale | XXL_VLDL_ | NA | A | G | 0.023502 | 0.004375 | 5.40E-09 |
| rs544873 | miR-126-3p | trans | 1.31E-37 | Nightingale | Remnant_( | NA | A | G | 0.035838 | 0.004319 | 5.30E-17 |
| rs544873 | miR-126-3p | trans | 1.31E-37 | Nightingale | S_VLDL_P | NA | A | G | 0.02339 | 0.004292 | 9.70E-09 |
| rs544873 | miR-126-3p | trans | 1.31E-37 | Nightingale | Total_C | NA | A | G | 0.030138 | 0.004208 | 5.00E-13 |
| rs544873 | miR-126-3p | trans | 1.31E-37 | Nightingale | VLDL_C | NA | A | G | 0.031062 | 0.004341 | 2.10E-13 |
| rs544873 | miR-126-3p | trans | 1.31E-37 | Nightingale | Total_CE | NA | A | G | 0.029384 | 0.004193 | 1.80E-12 |
| rs544873 | miR-126-3p | trans | 1.31E-37 | Nightingale | M_LDL_C_ | NA | A | G | 0.026582 | 0.00435 | 1.60E-09 |
| rs544873 | miR-126-3p | trans | 1.31E-37 | Nightingale | S_VLDL_TG | NA | A | G | -0.03598 | 0.004208 | 1.90E-17 |
| rs544873 | miR-126-3p | trans | 1.31E-37 | Nightingale | S_LDL_CE_ | NA | A | G | 0.028844 | 0.004339 | 1.10E-12 |
| rs544873 | miR-126-3p | trans | 1.31E-37 | Nightingale | S_HDL_L | NA | A | G | 0.023969 | 0.004332 | 2.70E-08 |
| rs544873 | miR-126-3p | trans | 1.31E-37 | Nightingale | LDL_FC | NA | A | G | 0.031548 | 0.0043 | 6.60E-13 |
| rs544873 | miR-126-3p | trans | 1.31E-37 | Nightingale | LDL_CE | NA | A | G | 0.030617 | 0.004335 | 2.10E-12 |
| rs544873 | miR-126-3p | trans | 1.31E-37 | Nightingale | XS_VLDL_F | NA | A | G | 0.035882 | 0.004271 | 7.50E-18 |
| rs544873 | miR-126-3p | trans | 1.31E-37 | Nightingale | M_LDL_TG | NA | A | G | -0.02512 | 0.004293 | 8.60E-09 |
| rs544873 | miR-126-3p | trans | 1.31E-37 | Nightingale | LDL_PL | NA | A | G | 0.033318 | 0.004337 | 3.00E-14 |
| rs544873 | miR-126-3p | trans | 1.31E-37 | Nightingale | M_LDL_P | NA | A | G | 0.034001 | 0.004359 | 7.50E-15 |
| rs544873 | miR-126-3p | trans | 1.31E-37 | Nightingale | Citrate | NA | A | G | 0.024678 | 0.004318 | 6.10E-09 |
| rs544873 | miR-126-3p | trans | 1.31E-37 | Nightingale | XXL_VLDL_ | NA | A | G | 0.029268 | 0.004328 | 5.10E-12 |
| rs544873 | miR-126-3p | trans | 1.31E-37 | Nightingale | S_LDL_FC | NA | A | G | 0.0272 | 0.00434 | 1.30E-09 |
| rs544873 | miR-126-3p | trans | 1.31E-37 | Nightingale | LDL_C | NA | A | G | 0.031186 | 0.004328 | 9.60E-13 |
| rs544873 | miR-126-3p | trans | 1.31E-37 | Nightingale | ApoB_by_A | NA | A | G | 0.025679 | 0.004281 | 2.00E-09 |
| rs544873 | miR-126-3p | trans | 1.31E-37 | Nightingale | S_VLDL_FC | NA | A | G | 0.036642 | 0.004347 | 1.70E-17 |
| rs544873 | miR-126-3p | trans | 1.31E-37 | Nightingale | LA | NA | A | G | 0.023263 | 0.004263 | 1.60E-08 |
| rs544873 | miR-126-3p | trans | 1.31E-37 | Nightingale | non_HDL_( | NA | A | G | 0.03414 | 0.004323 | 2.40E-15 |
| rs544873 | miR-126-3p | trans | 1.31E-37 | Nightingale | LDL_P | NA | A | G | 0.030659 | 0.004344 | 2.20E-12 |
| rs544873 | miR-126-3p | trans | 1.31E-37 | Nightingale | M_VLDL_C | NA | A | G | 0.037341 | 0.004335 | 5.40E-18 |
| rs544873 | miR-126-3p | trans | 1.31E-37 | Nightingale | Total_PL | NA | A | G | 0.022997 | 0.004139 | 7.80E-09 |
| rs544873 | miR-126-3p | trans | 1.31E-37 | Nightingale | M_VLDL_T | NA | A | G | -0.03623 | 0.004108 | 1.50E-18 |
| rs544873 | miR-126-3p | trans | 1.31E-37 | Nightingale | S_HDL_C | NA | A | G | 0.02538 | 0.004339 | 9.40E-09 |
| rs544873 | miR-126-3p | trans | 1.31E-37 | Nightingale | L_LDL_FC | NA | A | G | 0.031473 | 0.004268 | 4.10E-13 |
| rs544873 | miR-126-3p | trans | 1.31E-37 | Nightingale | S_LDL_P | NA | A | G | 0.028959 | 0.004351 | 3.00E-11 |
| rs544873 | miR-126-3p | trans | 1.31E-37 | Nightingale | S_VLDL_L | NA | A | G | 0.024496 | 0.004305 | 2.50E-09 |
| rs544873 | miR-126-3p | trans | 1.31E-37 | Nightingale | XL_VLDL_F | NA | A | G | 0.035204 | 0.004214 | 1.80E-17 |
| rs544873 | miR-126-3p | trans | 1.31E-37 | Nightingale | L_VLDL_TG | NA | A | G | -0.04923 | 0.004326 | 6.40E-31 |
| rs544873 | miR-126-3p | trans | 1.31E-37 | Nightingale | L_LDL_PL | NA | A | G | 0.035919 | 0.004314 | 1.30E-16 |
| rs544873 | miR-126-3p | trans | 1.31E-37 | Nightingale | S_LDL_PL_ | NA | A | G | -0.02806 | 0.004339 | 4.90E-12 |
| rs544873 | miR-126-3p | trans | 1.31E-37 | Nightingale | S_VLDL_C_ | NA | A | G | 0.039736 | 0.00423 | 8.30E-21 |
| rs544873 | miR-126-3p | trans | 1.31E-37 | Nightingale | L_VLDL_C_ | NA | A | G | 0.049226 | 0.004245 | 8.90E-32 |
| rs544873 | miR-126-3p | trans | 1.31E-37 | Nightingale | XXL_VLDL_ | NA | A | G | 0.033437 | 0.004356 | 8.80E-15 |
| rs544873 | miR-126-3p | trans | 1.31E-37 | Nightingale | XL_VLDL_C | NA | A | G | 0.03559 | 0.004168 | 1.70E-17 |
| rs544873 | miR-126-3p | trans | 1.31E-37 | Nightingale | XL_VLDL_C | NA | A | G | 0.034575 | 0.00417 | 1.50E-16 |
| rs544873 | miR-126-3p | trans | 1.31E-37 | Nightingale | M_VLDL_P | NA | A | G | 0.028753 | 0.00434 | 1.40E-11 |
| rs544873 | miR-126-3p | trans | 1.31E-37 | Nightingale | XS_VLDL_C | NA | A | G | 0.036639 | 0.004219 | 2.40E-18 |
| rs544873 | miR-126-3p | trans | 1.31E-37 | Nightingale | M_LDL_L | NA | A | G | 0.031388 | 0.004353 | 7.00E-13 |
| rs544873 | miR-126-3p | trans | 1.31E-37 | Nightingale | M_VLDL_P | NA | A | G | 0.042093 | 0.004172 | 6.30E-24 |
| rs544873 | miR-126-3p | trans | 1.31E-37 | Nightingale | M_VLDL_P | NA | A | G | 0.027926 | 0.004336 | 4.70E-11 |
| rs544873 | miR-126-3p | trans | 1.31E-37 | Nightingale | M_VLDL_F | NA | A | G | 0.039372 | 0.004135 | 2.20E-21 |
| rs544873 | miR-126-3p | trans | 1.31E-37 | Nightingale | L_LDL_L | NA | A | G | 0.031523 | 0.004304 | 3.20E-13 |
| rs544873 | miR-126-3p | trans | 1.31E-37 | Nightingale | S_LDL_C | NA | A | G | 0.029698 | 0.004351 | 1.60E-11 |
| rs544873 | miR-126-3p | trans | 1.31E-37 | Nightingale | XL_VLDL_T | NA | A | G | -0.04155 | 0.004213 | 4.90E-23 |
| rs544873 | miR-126-3p | trans | 1.31E-37 | Nightingale | Total_FC | NA | A | G | 0.031378 | 0.004246 | 4.70E-14 |
| rs544873 | miR-126-3p | trans | 1.31E-37 | Nightingale | XS_VLDL_P | NA | A | G | 0.035785 | 0.004289 | 1.00E-17 |
| rs544873 | miR-126-3p | trans | 1.31E-37 | Nightingale | XXL_VLDL_ | NA | A | G | 0.03179 | 0.00438 | 2.10E-13 |
| rs544873 | miR-126-3p | trans | 1.31E-37 | Nightingale | S_VLDL_PL | NA | A | G | 0.025728 | 0.004162 | 1.50E-09 |
| rs544873 | miR-126-3p | trans | 1.31E-37 | Nightingale | S_LDL_CE | NA | A | G | 0.029442 | 0.004349 | 1.50E-11 |
| rs544873 | miR-126-3p | trans | 1.31E-37 | Nightingale | S_HDL_CE | NA | A | G | 0.02569 | 0.004354 | 7.00E-09 |
| rs544873 | miR-126-3p | trans | 1.31E-37 | Nightingale | ApoB | NA | A | G | 0.032928 | 0.004341 | 3.40E-14 |
| rs544873 | let-7c-5p | trans | 3.11E-23 | Nightingale | IDL_L | NA | A | G | 0.03366 | 0.004203 | 2.30E-15 |
| rs544873 | let-7c-5p | trans | 3.11E-23 | Nightingale | S_LDL_L | NA | A | G | 0.026893 | 0.004346 | 1.20E-09 |

| | | | | | | | | | | | |
|---|---|---|---|---|---|---|---|---|---|---|---|
| rs544873 | let-7c-5p | trans | 3.11E-23 | Nightingale | M_VLDL_C | NA | A | G | 0.0392 | 0.004307 | 1.30E-19 |
| rs544873 | let-7c-5p | trans | 3.11E-23 | Nightingale | IDL_CE | NA | A | G | 0.032088 | 0.004197 | 4.30E-14 |
| rs544873 | let-7c-5p | trans | 3.11E-23 | Nightingale | XXL_VLDL_ | NA | A | G | -0.03443 | 0.004384 | 6.30E-16 |
| rs544873 | let-7c-5p | trans | 3.11E-23 | Nightingale | Clinical_LD | NA | A | G | 0.034557 | 0.004322 | 2.70E-15 |
| rs544873 | let-7c-5p | trans | 3.11E-23 | Nightingale | L_LDL_C | NA | A | G | 0.030224 | 0.004298 | 3.00E-12 |
| rs544873 | let-7c-5p | trans | 3.11E-23 | Nightingale | M_VLDL_C | NA | A | G | 0.033845 | 0.004094 | 2.70E-16 |
| rs544873 | let-7c-5p | trans | 3.11E-23 | Nightingale | M_LDL_PL | NA | A | G | 0.031174 | 0.004353 | 9.90E-13 |
| rs544873 | let-7c-5p | trans | 3.11E-23 | Nightingale | S_HDL_P | NA | A | G | 0.027805 | 0.004347 | 2.70E-10 |
| rs544873 | let-7c-5p | trans | 3.11E-23 | Nightingale | Sphingomy | NA | A | G | 0.025917 | 0.004063 | 3.30E-10 |
| rs544873 | let-7c-5p | trans | 3.11E-23 | Nightingale | VLDL_P | NA | A | G | 0.026555 | 0.004324 | 2.00E-10 |
| rs544873 | let-7c-5p | trans | 3.11E-23 | Nightingale | IDL_P | NA | A | G | 0.039032 | 0.004301 | 1.80E-19 |
| rs544873 | let-7c-5p | trans | 3.11E-23 | Nightingale | L_VLDL_FC | NA | A | G | 0.046941 | 0.004305 | 3.90E-28 |
| rs544873 | let-7c-5p | trans | 3.11E-23 | Nightingale | M_VLDL_C | NA | A | G | 0.031348 | 0.004084 | 3.30E-14 |
| rs544873 | let-7c-5p | trans | 3.11E-23 | Nightingale | S_VLDL_CE | NA | A | G | 0.037311 | 0.004341 | 1.70E-18 |
| rs544873 | let-7c-5p | trans | 3.11E-23 | Nightingale | IDL_PL | NA | A | G | 0.033573 | 0.004195 | 2.00E-15 |
| rs544873 | let-7c-5p | trans | 3.11E-23 | Nightingale | XS_VLDL_F | NA | A | G | 0.029134 | 0.004316 | 3.10E-11 |
| rs544873 | let-7c-5p | trans | 3.11E-23 | Nightingale | Total_L | NA | A | G | 0.025008 | 0.004276 | 1.50E-09 |
| rs544873 | let-7c-5p | trans | 3.11E-23 | Nightingale | XL_HDL_PL | NA | A | G | -0.02181 | 0.003994 | 4.60E-08 |
| rs544873 | let-7c-5p | trans | 3.11E-23 | Nightingale | GlycA | NA | A | G | 0.025329 | 0.004328 | 4.80E-09 |
| rs544873 | let-7c-5p | trans | 3.11E-23 | Nightingale | L_VLDL_PL | NA | A | G | 0.021645 | 0.004222 | 4.40E-08 |
| rs544873 | let-7c-5p | trans | 3.11E-23 | Nightingale | L_LDL_P | NA | A | G | 0.028317 | 0.004332 | 7.80E-11 |
| rs544873 | let-7c-5p | trans | 3.11E-23 | Nightingale | IDL_FC | NA | A | G | 0.035978 | 0.004208 | 3.20E-17 |
| rs544873 | let-7c-5p | trans | 3.11E-23 | Nightingale | VLDL_FC | NA | A | G | 0.024446 | 0.004315 | 3.60E-09 |
| rs544873 | let-7c-5p | trans | 3.11E-23 | Nightingale | IDL_C | NA | A | G | 0.033305 | 0.004198 | 4.90E-15 |
| rs544873 | let-7c-5p | trans | 3.11E-23 | Nightingale | M_LDL_FC | NA | A | G | 0.032281 | 0.004341 | 2.40E-13 |
| rs544873 | let-7c-5p | trans | 3.11E-23 | Nightingale | S_VLDL_CE | NA | A | G | 0.045151 | 0.004288 | 1.40E-25 |
| rs544873 | let-7c-5p | trans | 3.11E-23 | Nightingale | M_LDL_C | NA | A | G | 0.032091 | 0.004355 | 2.30E-13 |
| rs544873 | let-7c-5p | trans | 3.11E-23 | Nightingale | XS_VLDL_P | NA | A | G | 0.031657 | 0.004286 | 2.00E-14 |
| rs544873 | let-7c-5p | trans | 3.11E-23 | Nightingale | M_VLDL_F | NA | A | G | 0.032515 | 0.004346 | 3.90E-14 |
| rs544873 | let-7c-5p | trans | 3.11E-23 | Nightingale | L_VLDL_CE | NA | A | G | 0.04407 | 0.00416 | 7.90E-27 |
| rs544873 | let-7c-5p | trans | 3.11E-23 | Nightingale | Omega_6 | NA | A | G | 0.024156 | 0.004245 | 3.10E-09 |
| rs544873 | let-7c-5p | trans | 3.11E-23 | Nightingale | XL_VLDL_P | NA | A | G | 0.039106 | 0.004353 | 7.40E-20 |
| rs544873 | let-7c-5p | trans | 3.11E-23 | Nightingale | S_LDL_C_p | NA | A | G | 0.041864 | 0.004364 | 5.60E-22 |
| rs544873 | let-7c-5p | trans | 3.11E-23 | Nightingale | VLDL_CE | NA | A | G | 0.035144 | 0.004348 | 2.10E-16 |
| rs544873 | let-7c-5p | trans | 3.11E-23 | Nightingale | L_LDL_CE | NA | A | G | 0.02952 | 0.004308 | 9.90E-12 |
| rs544873 | let-7c-5p | trans | 3.11E-23 | Nightingale | LDL_L | NA | A | G | 0.031465 | 0.004332 | 5.80E-13 |
| rs544873 | let-7c-5p | trans | 3.11E-23 | Nightingale | XS_VLDL_L | NA | A | G | 0.033006 | 0.004273 | 1.40E-15 |
| rs544873 | let-7c-5p | trans | 3.11E-23 | Nightingale | S_VLDL_C | NA | A | G | 0.037355 | 0.004346 | 2.60E-18 |
| rs544873 | let-7c-5p | trans | 3.11E-23 | Nightingale | M_LDL_CE | NA | A | G | 0.031212 | 0.004353 | 8.50E-13 |
| rs544873 | let-7c-5p | trans | 3.11E-23 | Nightingale | S_VLDL_FC | NA | A | G | 0.027154 | 0.004154 | 1.20E-10 |
| rs544873 | let-7c-5p | trans | 3.11E-23 | Nightingale | XS_VLDL_C | NA | A | G | 0.036391 | 0.004195 | 4.10E-18 |
| rs544873 | let-7c-5p | trans | 3.11E-23 | Nightingale | L_VLDL_CE | NA | A | G | 0.022756 | 0.004296 | 2.90E-08 |
| rs544873 | let-7c-5p | trans | 3.11E-23 | Nightingale | S_VLDL_PL | NA | A | G | 0.032035 | 0.004339 | 4.90E-14 |
| rs544873 | let-7c-5p | trans | 3.11E-23 | Nightingale | XXL_VLDL_ | NA | A | G | 0.023502 | 0.004375 | 5.40E-09 |
| rs544873 | let-7c-5p | trans | 3.11E-23 | Nightingale | Remnant_( | NA | A | G | 0.035838 | 0.004319 | 5.30E-17 |
| rs544873 | let-7c-5p | trans | 3.11E-23 | Nightingale | S_VLDL_P | NA | A | G | 0.02339 | 0.004292 | 9.70E-09 |
| rs544873 | let-7c-5p | trans | 3.11E-23 | Nightingale | Total_C | NA | A | G | 0.030138 | 0.004208 | 5.00E-13 |
| rs544873 | let-7c-5p | trans | 3.11E-23 | Nightingale | VLDL_C | NA | A | G | 0.031062 | 0.004341 | 2.10E-13 |
| rs544873 | let-7c-5p | trans | 3.11E-23 | Nightingale | Total_CE | NA | A | G | 0.029384 | 0.004193 | 1.80E-12 |
| rs544873 | let-7c-5p | trans | 3.11E-23 | Nightingale | M_LDL_C_ | NA | A | G | 0.026582 | 0.00435 | 1.60E-09 |
| rs544873 | let-7c-5p | trans | 3.11E-23 | Nightingale | S_VLDL_TG | NA | A | G | -0.03598 | 0.004208 | 1.90E-17 |
| rs544873 | let-7c-5p | trans | 3.11E-23 | Nightingale | S_LDL_CE_ | NA | A | G | 0.028844 | 0.004339 | 1.10E-12 |
| rs544873 | let-7c-5p | trans | 3.11E-23 | Nightingale | S_HDL_L | NA | A | G | 0.023969 | 0.004332 | 2.70E-08 |
| rs544873 | let-7c-5p | trans | 3.11E-23 | Nightingale | LDL_FC | NA | A | G | 0.031548 | 0.0043 | 6.60E-13 |
| rs544873 | let-7c-5p | trans | 3.11E-23 | Nightingale | LDL_CE | NA | A | G | 0.030617 | 0.004335 | 2.10E-12 |
| rs544873 | let-7c-5p | trans | 3.11E-23 | Nightingale | XS_VLDL_F | NA | A | G | 0.035882 | 0.004271 | 7.50E-18 |
| rs544873 | let-7c-5p | trans | 3.11E-23 | Nightingale | M_LDL_TG | NA | A | G | -0.02512 | 0.004293 | 8.60E-09 |
| rs544873 | let-7c-5p | trans | 3.11E-23 | Nightingale | LDL_PL | NA | A | G | 0.033318 | 0.004337 | 3.00E-14 |
| rs544873 | let-7c-5p | trans | 3.11E-23 | Nightingale | M_LDL_P | NA | A | G | 0.034001 | 0.004359 | 7.50E-15 |
| rs544873 | let-7c-5p | trans | 3.11E-23 | Nightingale | Citrate | NA | A | G | 0.024678 | 0.004318 | 6.10E-09 |
| rs544873 | let-7c-5p | trans | 3.11E-23 | Nightingale | XXL_VLDL_ | NA | A | G | 0.029268 | 0.004328 | 5.10E-12 |
| rs544873 | let-7c-5p | trans | 3.11E-23 | Nightingale | S_LDL_FC | NA | A | G | 0.0272 | 0.00434 | 1.30E-09 |
| rs544873 | let-7c-5p | trans | 3.11E-23 | Nightingale | LDL_C | NA | A | G | 0.031186 | 0.004328 | 9.60E-13 |

| | | | | | | | | | | | |
|---|---|---|---|---|---|---|---|---|---|---|---|
| rs544873 | let-7c-5p | trans | 3.11E-23 | Nightingale | ApoB_by_A | NA | A | G | 0.025679 | 0.004281 | 2.00E-09 |
| rs544873 | let-7c-5p | trans | 3.11E-23 | Nightingale | S_VLDL_FC | NA | A | G | 0.036642 | 0.004347 | 1.70E-17 |
| rs544873 | let-7c-5p | trans | 3.11E-23 | Nightingale | LA | NA | A | G | 0.023263 | 0.004263 | 1.60E-08 |
| rs544873 | let-7c-5p | trans | 3.11E-23 | Nightingale | non_HDL_( | NA | A | G | 0.03414 | 0.004323 | 2.40E-15 |
| rs544873 | let-7c-5p | trans | 3.11E-23 | Nightingale | LDL_P | NA | A | G | 0.030659 | 0.004344 | 2.20E-12 |
| rs544873 | let-7c-5p | trans | 3.11E-23 | Nightingale | M_VLDL_C | NA | A | G | 0.037341 | 0.004335 | 5.40E-18 |
| rs544873 | let-7c-5p | trans | 3.11E-23 | Nightingale | Total_PL | NA | A | G | 0.022997 | 0.004139 | 7.80E-09 |
| rs544873 | let-7c-5p | trans | 3.11E-23 | Nightingale | M_VLDL_T | NA | A | G | -0.03623 | 0.004108 | 1.50E-18 |
| rs544873 | let-7c-5p | trans | 3.11E-23 | Nightingale | S_HDL_C | NA | A | G | 0.02538 | 0.004339 | 9.40E-09 |
| rs544873 | let-7c-5p | trans | 3.11E-23 | Nightingale | L_LDL_FC | NA | A | G | 0.031473 | 0.004268 | 4.10E-13 |
| rs544873 | let-7c-5p | trans | 3.11E-23 | Nightingale | S_LDL_P | NA | A | G | 0.028959 | 0.004351 | 3.00E-11 |
| rs544873 | let-7c-5p | trans | 3.11E-23 | Nightingale | S_VLDL_L | NA | A | G | 0.024496 | 0.004305 | 2.50E-09 |
| rs544873 | let-7c-5p | trans | 3.11E-23 | Nightingale | XL_VLDL_F | NA | A | G | 0.035204 | 0.004214 | 1.80E-17 |
| rs544873 | let-7c-5p | trans | 3.11E-23 | Nightingale | L_VLDL_TG | NA | A | G | -0.04923 | 0.004326 | 6.40E-31 |
| rs544873 | let-7c-5p | trans | 3.11E-23 | Nightingale | L_LDL_PL | NA | A | G | 0.035919 | 0.004314 | 1.30E-16 |
| rs544873 | let-7c-5p | trans | 3.11E-23 | Nightingale | S_LDL_PL_ | NA | A | G | -0.02806 | 0.004339 | 4.90E-12 |
| rs544873 | let-7c-5p | trans | 3.11E-23 | Nightingale | S_VLDL_C_ | NA | A | G | 0.039736 | 0.00423 | 8.30E-21 |
| rs544873 | let-7c-5p | trans | 3.11E-23 | Nightingale | L_VLDL_C_ | NA | A | G | 0.049226 | 0.004245 | 8.90E-32 |
| rs544873 | let-7c-5p | trans | 3.11E-23 | Nightingale | XXL_VLDL_ | NA | A | G | 0.033437 | 0.004356 | 8.80E-15 |
| rs544873 | let-7c-5p | trans | 3.11E-23 | Nightingale | XL_VLDL_C | NA | A | G | 0.03559 | 0.004168 | 1.70E-17 |
| rs544873 | let-7c-5p | trans | 3.11E-23 | Nightingale | XL_VLDL_C | NA | A | G | 0.034575 | 0.00417 | 1.50E-16 |
| rs544873 | let-7c-5p | trans | 3.11E-23 | Nightingale | M_VLDL_P | NA | A | G | 0.028753 | 0.00434 | 1.40E-11 |
| rs544873 | let-7c-5p | trans | 3.11E-23 | Nightingale | XS_VLDL_C | NA | A | G | 0.036639 | 0.004219 | 2.40E-18 |
| rs544873 | let-7c-5p | trans | 3.11E-23 | Nightingale | M_LDL_L | NA | A | G | 0.031388 | 0.004353 | 7.00E-13 |
| rs544873 | let-7c-5p | trans | 3.11E-23 | Nightingale | M_VLDL_P | NA | A | G | 0.042093 | 0.004172 | 6.30E-24 |
| rs544873 | let-7c-5p | trans | 3.11E-23 | Nightingale | M_VLDL_P | NA | A | G | 0.027926 | 0.004336 | 4.70E-11 |
| rs544873 | let-7c-5p | trans | 3.11E-23 | Nightingale | M_VLDL_F | NA | A | G | 0.039372 | 0.004135 | 2.20E-21 |
| rs544873 | let-7c-5p | trans | 3.11E-23 | Nightingale | L_LDL_L | NA | A | G | 0.031523 | 0.004304 | 3.20E-13 |
| rs544873 | let-7c-5p | trans | 3.11E-23 | Nightingale | S_LDL_C | NA | A | G | 0.029698 | 0.004351 | 1.60E-11 |
| rs544873 | let-7c-5p | trans | 3.11E-23 | Nightingale | XL_VLDL_T | NA | A | G | -0.04155 | 0.004213 | 4.90E-23 |
| rs544873 | let-7c-5p | trans | 3.11E-23 | Nightingale | Total_FC | NA | A | G | 0.031378 | 0.004246 | 4.70E-14 |
| rs544873 | let-7c-5p | trans | 3.11E-23 | Nightingale | XS_VLDL_P | NA | A | G | 0.035785 | 0.004289 | 1.00E-17 |
| rs544873 | let-7c-5p | trans | 3.11E-23 | Nightingale | XXL_VLDL_ | NA | A | G | 0.03179 | 0.00438 | 2.10E-13 |
| rs544873 | let-7c-5p | trans | 3.11E-23 | Nightingale | S_VLDL_PL | NA | A | G | 0.025728 | 0.004162 | 1.50E-09 |
| rs544873 | let-7c-5p | trans | 3.11E-23 | Nightingale | S_LDL_CE | NA | A | G | 0.029442 | 0.004349 | 1.50E-11 |
| rs544873 | let-7c-5p | trans | 3.11E-23 | Nightingale | S_HDL_CE | NA | A | G | 0.02569 | 0.004354 | 7.00E-09 |
| rs544873 | let-7c-5p | trans | 3.11E-23 | Nightingale | ApoB | NA | A | G | 0.032928 | 0.004341 | 3.40E-14 |
| rs545971 | miR-139-5p | trans | 8.40E-42 | Nightingale | M_LDL_C_ | NA | T | C | 0.031165 | 0.004428 | 2.90E-12 |
| rs545971 | miR-139-5p | trans | 8.40E-42 | Nightingale | LDL_FC | NA | T | C | 0.033635 | 0.004378 | 5.20E-14 |
| rs545971 | miR-139-5p | trans | 8.40E-42 | Nightingale | GlycA | NA | T | C | 0.028013 | 0.004406 | 1.40E-10 |
| rs545971 | miR-139-5p | trans | 8.40E-42 | Nightingale | XS_VLDL_P | NA | T | C | 0.032112 | 0.004363 | 5.40E-14 |
| rs545971 | miR-139-5p | trans | 8.40E-42 | Nightingale | IDL_PL | NA | T | C | 0.035341 | 0.00427 | 3.30E-16 |
| rs545971 | miR-139-5p | trans | 8.40E-42 | Nightingale | VLDL_P | NA | T | C | 0.026633 | 0.004401 | 4.40E-10 |
| rs545971 | miR-139-5p | trans | 8.40E-42 | Nightingale | M_VLDL_C | NA | T | C | 0.036205 | 0.004168 | 1.20E-17 |
| rs545971 | miR-139-5p | trans | 8.40E-42 | Nightingale | XS_VLDL_L | NA | T | C | 0.033813 | 0.00435 | 2.10E-15 |
| rs545971 | miR-139-5p | trans | 8.40E-42 | Nightingale | M_VLDL_F | NA | T | C | 0.033372 | 0.004424 | 3.10E-14 |
| rs545971 | miR-139-5p | trans | 8.40E-42 | Nightingale | LDL_L | NA | T | C | 0.033306 | 0.00441 | 7.40E-14 |
| rs545971 | miR-139-5p | trans | 8.40E-42 | Nightingale | LDL_CE | NA | T | C | 0.032556 | 0.004413 | 2.20E-13 |
| rs545971 | miR-139-5p | trans | 8.40E-42 | Nightingale | IDL_L | NA | T | C | 0.035268 | 0.004278 | 4.80E-16 |
| rs545971 | miR-139-5p | trans | 8.40E-42 | Nightingale | S_VLDL_CE | NA | T | C | 0.047034 | 0.004365 | 1.20E-26 |
| rs545971 | miR-139-5p | trans | 8.40E-42 | Nightingale | XL_VLDL_F | NA | T | C | 0.037195 | 0.00429 | 2.00E-18 |
| rs545971 | miR-139-5p | trans | 8.40E-42 | Nightingale | IDL_FC | NA | T | C | 0.038254 | 0.004284 | 1.50E-18 |
| rs545971 | miR-139-5p | trans | 8.40E-42 | Nightingale | XXL_VLDL_ | NA | T | C | 0.030847 | 0.004406 | 1.10E-12 |
| rs545971 | miR-139-5p | trans | 8.40E-42 | Nightingale | S_VLDL_CE | NA | T | C | 0.038182 | 0.004419 | 1.60E-18 |
| rs545971 | miR-139-5p | trans | 8.40E-42 | Nightingale | S_HDL_L | NA | T | C | 0.026297 | 0.00441 | 1.50E-09 |
| rs545971 | miR-139-5p | trans | 8.40E-42 | Nightingale | S_LDL_FC | NA | T | C | 0.028568 | 0.004418 | 4.20E-10 |
| rs545971 | miR-139-5p | trans | 8.40E-42 | Nightingale | L_VLDL_CE | NA | T | C | 0.047336 | 0.004234 | 2.40E-29 |
| rs545971 | miR-139-5p | trans | 8.40E-42 | Nightingale | S_VLDL_C_ | NA | T | C | 0.041889 | 0.004306 | 5.20E-22 |
| rs545971 | miR-139-5p | trans | 8.40E-42 | Nightingale | Citrate | NA | T | C | 0.026829 | 0.004396 | 7.30E-10 |
| rs545971 | miR-139-5p | trans | 8.40E-42 | Nightingale | L_LDL_C | NA | T | C | 0.032322 | 0.004375 | 2.60E-13 |
| rs545971 | miR-139-5p | trans | 8.40E-42 | Nightingale | XS_VLDL_F | NA | T | C | 0.031441 | 0.004393 | 2.40E-12 |
| rs545971 | miR-139-5p | trans | 8.40E-42 | Nightingale | LDL_C | NA | T | C | 0.033196 | 0.004406 | 8.80E-14 |
| rs545971 | miR-139-5p | trans | 8.40E-42 | Nightingale | IDL_P | NA | T | C | 0.039988 | 0.004378 | 1.30E-19 |

| | | | | | | | | | | | |
|---|---|---|---|---|---|---|---|---|---|---|---|
| rs545971 | miR-139-5p | trans | 8.40E-42 | Nightingale | Clinical_LD | NA | T | C | 0.036477 | 0.004399 | 2.70E-16 |
| rs545971 | miR-139-5p | trans | 8.40E-42 | Nightingale | S_VLDL_FC | NA | T | C | 0.037905 | 0.004426 | 7.30E-18 |
| rs545971 | miR-139-5p | trans | 8.40E-42 | Nightingale | S_HDL_PL | NA | T | C | 0.023784 | 0.004386 | 3.20E-08 |
| rs545971 | miR-139-5p | trans | 8.40E-42 | Nightingale | M_LDL_FC | NA | T | C | 0.034352 | 0.004419 | 1.90E-14 |
| rs545971 | miR-139-5p | trans | 8.40E-42 | Nightingale | M_VLDL_P | NA | T | C | 0.028296 | 0.004414 | 6.30E-11 |
| rs545971 | miR-139-5p | trans | 8.40E-42 | Nightingale | XXL_VLDL_ | NA | T | C | -0.03573 | 0.004463 | 2.20E-16 |
| rs545971 | miR-139-5p | trans | 8.40E-42 | Nightingale | M_VLDL_F | NA | T | C | 0.042064 | 0.00421 | 3.50E-23 |
| rs545971 | miR-139-5p | trans | 8.40E-42 | Nightingale | LDL_PL | NA | T | C | 0.034991 | 0.004415 | 4.70E-15 |
| rs545971 | miR-139-5p | trans | 8.40E-42 | Nightingale | L_LDL_P | NA | T | C | 0.029177 | 0.00441 | 6.20E-11 |
| rs545971 | miR-139-5p | trans | 8.40E-42 | Nightingale | Sphingomy | NA | T | C | 0.029016 | 0.004136 | 6.80E-12 |
| rs545971 | miR-139-5p | trans | 8.40E-42 | Nightingale | PUFA | NA | T | C | 0.023713 | 0.004316 | 1.40E-08 |
| rs545971 | miR-139-5p | trans | 8.40E-42 | Nightingale | S_HDL_P | NA | T | C | 0.030615 | 0.004425 | 5.10E-12 |
| rs545971 | miR-139-5p | trans | 8.40E-42 | Nightingale | XL_VLDL_C | NA | T | C | 0.037351 | 0.004245 | 3.10E-18 |
| rs545971 | miR-139-5p | trans | 8.40E-42 | Nightingale | M_LDL_P | NA | T | C | 0.035895 | 0.004437 | 6.80E-16 |
| rs545971 | miR-139-5p | trans | 8.40E-42 | Nightingale | L_LDL_PL | NA | T | C | 0.037992 | 0.004391 | 9.00E-18 |
| rs545971 | miR-139-5p | trans | 8.40E-42 | Nightingale | Total_L | NA | T | C | 0.026346 | 0.004353 | 4.60E-10 |
| rs545971 | miR-139-5p | trans | 8.40E-42 | Nightingale | M_LDL_PL | NA | T | C | 0.032471 | 0.004431 | 2.60E-13 |
| rs545971 | miR-139-5p | trans | 8.40E-42 | Nightingale | M_LDL_TG | NA | T | C | -0.02763 | 0.00437 | 3.20E-10 |
| rs545971 | miR-139-5p | trans | 8.40E-42 | Nightingale | XL_HDL_PL | NA | T | C | -0.02277 | 0.004065 | 1.10E-08 |
| rs545971 | miR-139-5p | trans | 8.40E-42 | Nightingale | M_VLDL_T | NA | T | C | -0.03856 | 0.004181 | 6.80E-20 |
| rs545971 | miR-139-5p | trans | 8.40E-42 | Nightingale | S_VLDL_PL | NA | T | C | 0.032777 | 0.004417 | 4.20E-14 |
| rs545971 | miR-139-5p | trans | 8.40E-42 | Nightingale | Total_C | NA | T | C | 0.032372 | 0.004284 | 3.30E-14 |
| rs545971 | miR-139-5p | trans | 8.40E-42 | Nightingale | L_VLDL_TG | NA | T | C | -0.05114 | 0.004404 | 5.30E-32 |
| rs545971 | miR-139-5p | trans | 8.40E-42 | Nightingale | S_VLDL_TG | NA | T | C | -0.03813 | 0.004283 | 1.40E-18 |
| rs545971 | miR-139-5p | trans | 8.40E-42 | Nightingale | Cholines | NA | T | C | 0.022288 | 0.004161 | 4.90E-08 |
| rs545971 | miR-139-5p | trans | 8.40E-42 | Nightingale | L_LDL_L | NA | T | C | 0.033551 | 0.004382 | 3.00E-14 |
| rs545971 | miR-139-5p | trans | 8.40E-42 | Nightingale | XL_VLDL_P | NA | T | C | 0.038847 | 0.004432 | 5.10E-19 |
| rs545971 | miR-139-5p | trans | 8.40E-42 | Nightingale | VLDL_FC | NA | T | C | 0.024159 | 0.004392 | 1.10E-08 |
| rs545971 | miR-139-5p | trans | 8.40E-42 | Nightingale | S_HDL_FC | NA | T | C | 0.02345 | 0.004344 | 4.60E-08 |
| rs545971 | miR-139-5p | trans | 8.40E-42 | Nightingale | S_HDL_C | NA | T | C | 0.028415 | 0.004417 | 1.70E-10 |
| rs545971 | miR-139-5p | trans | 8.40E-42 | Nightingale | S_LDL_L | NA | T | C | 0.027745 | 0.004424 | 8.20E-10 |
| rs545971 | miR-139-5p | trans | 8.40E-42 | Nightingale | S_LDL_C | NA | T | C | 0.030946 | 0.004429 | 5.50E-12 |
| rs545971 | miR-139-5p | trans | 8.40E-42 | Nightingale | S_VLDL_PL | NA | T | C | 0.02774 | 0.004237 | 1.80E-10 |
| rs545971 | miR-139-5p | trans | 8.40E-42 | Nightingale | M_VLDL_C | NA | T | C | 0.033628 | 0.004157 | 1.90E-15 |
| rs545971 | miR-139-5p | trans | 8.40E-42 | Nightingale | S_VLDL_C | NA | T | C | 0.038373 | 0.004424 | 1.80E-18 |
| rs545971 | miR-139-5p | trans | 8.40E-42 | Nightingale | non_HDL_( | NA | T | C | 0.035698 | 0.004401 | 5.10E-16 |
| rs545971 | miR-139-5p | trans | 8.40E-42 | Nightingale | XL_VLDL_T | NA | T | C | -0.04392 | 0.004289 | 1.70E-24 |
| rs545971 | miR-139-5p | trans | 8.40E-42 | Nightingale | XS_VLDL_F | NA | T | C | 0.037065 | 0.004348 | 6.50E-18 |
| rs545971 | miR-139-5p | trans | 8.40E-42 | Nightingale | XXL_VLDL_ | NA | T | C | 0.025004 | 0.004454 | 1.50E-09 |
| rs545971 | miR-139-5p | trans | 8.40E-42 | Nightingale | S_LDL_CE_ | NA | T | C | 0.030946 | 0.004417 | 4.90E-14 |
| rs545971 | miR-139-5p | trans | 8.40E-42 | Nightingale | XXL_VLDL_ | NA | T | C | 0.034827 | 0.004434 | 2.90E-15 |
| rs545971 | miR-139-5p | trans | 8.40E-42 | Nightingale | XL_VLDL_C | NA | T | C | 0.038222 | 0.004243 | 4.70E-19 |
| rs545971 | miR-139-5p | trans | 8.40E-42 | Nightingale | M_VLDL_C | NA | T | C | 0.038711 | 0.004413 | 1.90E-18 |
| rs545971 | miR-139-5p | trans | 8.40E-42 | Nightingale | S_VLDL_FC | NA | T | C | 0.029358 | 0.004228 | 1.10E-11 |
| rs545971 | miR-139-5p | trans | 8.40E-42 | Nightingale | IDL_C | NA | T | C | 0.034955 | 0.004273 | 9.40E-16 |
| rs545971 | miR-139-5p | trans | 8.40E-42 | Nightingale | Total_CE | NA | T | C | 0.031745 | 0.004268 | 9.70E-14 |
| rs545971 | miR-139-5p | trans | 8.40E-42 | Nightingale | L_VLDL_FC | NA | T | C | 0.048588 | 0.004382 | 4.50E-29 |
| rs545971 | miR-139-5p | trans | 8.40E-42 | Nightingale | Remnant_( | NA | T | C | 0.036867 | 0.004396 | 3.70E-17 |
| rs545971 | miR-139-5p | trans | 8.40E-42 | Nightingale | VLDL_CE | NA | T | C | 0.035786 | 0.004426 | 3.00E-16 |
| rs545971 | miR-139-5p | trans | 8.40E-42 | Nightingale | XS_VLDL_P | NA | T | C | 0.036698 | 0.004366 | 1.40E-17 |
| rs545971 | miR-139-5p | trans | 8.40E-42 | Nightingale | M_VLDL_P | NA | T | C | 0.029128 | 0.004418 | 2.00E-11 |
| rs545971 | miR-139-5p | trans | 8.40E-42 | Nightingale | IDL_CE | NA | T | C | 0.033507 | 0.004272 | 1.30E-14 |
| rs545971 | miR-139-5p | trans | 8.40E-42 | Nightingale | M_LDL_L | NA | T | C | 0.033035 | 0.004431 | 1.00E-13 |
| rs545971 | miR-139-5p | trans | 8.40E-42 | Nightingale | XS_VLDL_C | NA | T | C | 0.037794 | 0.00427 | 2.10E-18 |
| rs545971 | miR-139-5p | trans | 8.40E-42 | Nightingale | S_LDL_PL_ | NA | T | C | -0.03006 | 0.004417 | 2.60E-13 |
| rs545971 | miR-139-5p | trans | 8.40E-42 | Nightingale | L_LDL_FC | NA | T | C | 0.033627 | 0.004345 | 2.90E-14 |
| rs545971 | miR-139-5p | trans | 8.40E-42 | Nightingale | S_LDL_C_p | NA | T | C | 0.045366 | 0.004442 | 5.40E-25 |
| rs545971 | miR-139-5p | trans | 8.40E-42 | Nightingale | M_LDL_CE | NA | T | C | 0.032977 | 0.004431 | 9.60E-14 |
| rs545971 | miR-139-5p | trans | 8.40E-42 | Nightingale | S_LDL_P | NA | T | C | 0.029589 | 0.004429 | 3.20E-11 |
| rs545971 | miR-139-5p | trans | 8.40E-42 | Nightingale | L_VLDL_C_ | NA | T | C | 0.052192 | 0.004322 | 4.00E-34 |
| rs545971 | miR-139-5p | trans | 8.40E-42 | Nightingale | M_VLDL_P | NA | T | C | 0.044056 | 0.004247 | 5.50E-25 |
| rs545971 | miR-139-5p | trans | 8.40E-42 | Nightingale | M_LDL_C | NA | T | C | 0.033991 | 0.004433 | 2.10E-14 |
| rs545971 | miR-139-5p | trans | 8.40E-42 | Nightingale | S_HDL_CE | NA | T | C | 0.02864 | 0.004432 | 1.30E-10 |

| | | | | | | | | | | | |
|---|---|---|---|---|---|---|---|---|---|---|---|
| rs545971 | miR-139-5p | trans | 8.40E-42 | Nightingale | S_VLDL_P | NA | T | C | 0.023465 | 0.004369 | 1.70E-08 |
| rs545971 | miR-139-5p | trans | 8.40E-42 | Nightingale | ApoB_by_A | NA | T | C | 0.025055 | 0.004358 | 1.30E-08 |
| rs545971 | miR-139-5p | trans | 8.40E-42 | Nightingale | LA | NA | T | C | 0.024903 | 0.004339 | 3.70E-09 |
| rs545971 | miR-139-5p | trans | 8.40E-42 | Nightingale | L_LDL_CE | NA | T | C | 0.031569 | 0.004385 | 9.80E-13 |
| rs545971 | miR-139-5p | trans | 8.40E-42 | Nightingale | XXL_VLDL_ | NA | T | C | 0.033119 | 0.004459 | 9.00E-14 |
| rs545971 | miR-139-5p | trans | 8.40E-42 | Nightingale | ApoB | NA | T | C | 0.033939 | 0.004419 | 2.10E-14 |
| rs545971 | miR-139-5p | trans | 8.40E-42 | Nightingale | IDL_FC_pct | NA | T | C | 0.025773 | 0.004391 | 1.10E-08 |
| rs545971 | miR-139-5p | trans | 8.40E-42 | Nightingale | VLDL_C | NA | T | C | 0.031302 | 0.004419 | 4.90E-13 |
| rs545971 | miR-139-5p | trans | 8.40E-42 | Nightingale | XS_VLDL_C | NA | T | C | 0.037989 | 0.004295 | 1.30E-18 |
| rs545971 | miR-139-5p | trans | 8.40E-42 | Nightingale | S_VLDL_L | NA | T | C | 0.024668 | 0.004382 | 3.70E-09 |
| rs545971 | miR-139-5p | trans | 8.40E-42 | Nightingale | LDL_P | NA | T | C | 0.031801 | 0.004422 | 1.00E-12 |
| rs545971 | miR-139-5p | trans | 8.40E-42 | Nightingale | S_LDL_CE | NA | T | C | 0.030551 | 0.004427 | 5.80E-12 |
| rs545971 | miR-139-5p | trans | 8.40E-42 | Nightingale | M_VLDL_C | NA | T | C | 0.040844 | 0.004385 | 2.30E-20 |
| rs545971 | miR-139-5p | trans | 8.40E-42 | Nightingale | Total_PL | NA | T | C | 0.024922 | 0.004213 | 1.10E-09 |
| rs545971 | miR-139-5p | trans | 8.40E-42 | Nightingale | Omega_6 | NA | T | C | 0.025889 | 0.004321 | 5.80E-10 |
| rs545971 | miR-139-5p | trans | 8.40E-42 | Nightingale | Total_FC | NA | T | C | 0.033253 | 0.004323 | 6.80E-15 |
| rs545971 | let-7c-5p | trans | 2.87E-23 | Nightingale | M_LDL_C_p | NA | T | C | 0.031165 | 0.004428 | 2.90E-12 |
| rs545971 | let-7c-5p | trans | 2.87E-23 | Nightingale | LDL_FC | NA | T | C | 0.033635 | 0.004378 | 5.20E-14 |
| rs545971 | let-7c-5p | trans | 2.87E-23 | Nightingale | GlycA | NA | T | C | 0.028013 | 0.004406 | 1.40E-10 |
| rs545971 | let-7c-5p | trans | 2.87E-23 | Nightingale | XS_VLDL_P | NA | T | C | 0.032112 | 0.004363 | 5.40E-14 |
| rs545971 | let-7c-5p | trans | 2.87E-23 | Nightingale | IDL_PL | NA | T | C | 0.035341 | 0.00427 | 3.30E-16 |
| rs545971 | let-7c-5p | trans | 2.87E-23 | Nightingale | VLDL_P | NA | T | C | 0.026633 | 0.004401 | 4.40E-10 |
| rs545971 | let-7c-5p | trans | 2.87E-23 | Nightingale | M_VLDL_C | NA | T | C | 0.036205 | 0.004168 | 1.20E-17 |
| rs545971 | let-7c-5p | trans | 2.87E-23 | Nightingale | XS_VLDL_L | NA | T | C | 0.033813 | 0.00435 | 2.10E-15 |
| rs545971 | let-7c-5p | trans | 2.87E-23 | Nightingale | M_VLDL_F | NA | T | C | 0.033372 | 0.004424 | 3.10E-14 |
| rs545971 | let-7c-5p | trans | 2.87E-23 | Nightingale | LDL_L | NA | T | C | 0.033306 | 0.00441 | 7.40E-14 |
| rs545971 | let-7c-5p | trans | 2.87E-23 | Nightingale | LDL_CE | NA | T | C | 0.032556 | 0.004413 | 2.20E-13 |
| rs545971 | let-7c-5p | trans | 2.87E-23 | Nightingale | IDL_L | NA | T | C | 0.035268 | 0.004278 | 4.80E-16 |
| rs545971 | let-7c-5p | trans | 2.87E-23 | Nightingale | S_VLDL_CE | NA | T | C | 0.047034 | 0.004365 | 1.20E-26 |
| rs545971 | let-7c-5p | trans | 2.87E-23 | Nightingale | XL_VLDL_F | NA | T | C | 0.037195 | 0.00429 | 2.00E-18 |
| rs545971 | let-7c-5p | trans | 2.87E-23 | Nightingale | IDL_FC | NA | T | C | 0.038254 | 0.004284 | 1.50E-18 |
| rs545971 | let-7c-5p | trans | 2.87E-23 | Nightingale | XXL_VLDL_ | NA | T | C | 0.030847 | 0.004406 | 1.10E-12 |
| rs545971 | let-7c-5p | trans | 2.87E-23 | Nightingale | S_VLDL_CE | NA | T | C | 0.038182 | 0.004419 | 1.60E-18 |
| rs545971 | let-7c-5p | trans | 2.87E-23 | Nightingale | S_HDL_L | NA | T | C | 0.026297 | 0.00441 | 1.50E-09 |
| rs545971 | let-7c-5p | trans | 2.87E-23 | Nightingale | S_LDL_FC | NA | T | C | 0.028568 | 0.004418 | 4.20E-10 |
| rs545971 | let-7c-5p | trans | 2.87E-23 | Nightingale | L_VLDL_CE | NA | T | C | 0.047336 | 0.004234 | 2.40E-29 |
| rs545971 | let-7c-5p | trans | 2.87E-23 | Nightingale | S_VLDL_C_ | NA | T | C | 0.041889 | 0.004306 | 5.20E-22 |
| rs545971 | let-7c-5p | trans | 2.87E-23 | Nightingale | Citrate | NA | T | C | 0.026829 | 0.004396 | 7.30E-10 |
| rs545971 | let-7c-5p | trans | 2.87E-23 | Nightingale | L_LDL_C | NA | T | C | 0.032322 | 0.004375 | 2.60E-13 |
| rs545971 | let-7c-5p | trans | 2.87E-23 | Nightingale | XS_VLDL_F | NA | T | C | 0.031441 | 0.004393 | 2.40E-12 |
| rs545971 | let-7c-5p | trans | 2.87E-23 | Nightingale | LDL_C | NA | T | C | 0.033196 | 0.004406 | 8.80E-14 |
| rs545971 | let-7c-5p | trans | 2.87E-23 | Nightingale | IDL_P | NA | T | C | 0.039988 | 0.004378 | 1.30E-19 |
| rs545971 | let-7c-5p | trans | 2.87E-23 | Nightingale | Clinical_LD | NA | T | C | 0.036477 | 0.004399 | 2.70E-16 |
| rs545971 | let-7c-5p | trans | 2.87E-23 | Nightingale | S_VLDL_FC | NA | T | C | 0.037905 | 0.004426 | 7.30E-18 |
| rs545971 | let-7c-5p | trans | 2.87E-23 | Nightingale | S_HDL_PL | NA | T | C | 0.023784 | 0.004386 | 3.20E-08 |
| rs545971 | let-7c-5p | trans | 2.87E-23 | Nightingale | M_LDL_FC | NA | T | C | 0.034352 | 0.004419 | 1.90E-14 |
| rs545971 | let-7c-5p | trans | 2.87E-23 | Nightingale | M_VLDL_P | NA | T | C | 0.028296 | 0.004414 | 6.30E-11 |
| rs545971 | let-7c-5p | trans | 2.87E-23 | Nightingale | XXL_VLDL_ | NA | T | C | -0.03573 | 0.004463 | 2.20E-16 |
| rs545971 | let-7c-5p | trans | 2.87E-23 | Nightingale | M_VLDL_F | NA | T | C | 0.042064 | 0.00421 | 3.50E-23 |
| rs545971 | let-7c-5p | trans | 2.87E-23 | Nightingale | LDL_PL | NA | T | C | 0.034991 | 0.004415 | 4.70E-15 |
| rs545971 | let-7c-5p | trans | 2.87E-23 | Nightingale | L_LDL_P | NA | T | C | 0.029177 | 0.00441 | 6.20E-11 |
| rs545971 | let-7c-5p | trans | 2.87E-23 | Nightingale | Sphingomy | NA | T | C | 0.029016 | 0.004136 | 6.80E-12 |
| rs545971 | let-7c-5p | trans | 2.87E-23 | Nightingale | PUFA | NA | T | C | 0.023713 | 0.004316 | 1.40E-08 |
| rs545971 | let-7c-5p | trans | 2.87E-23 | Nightingale | S_HDL_P | NA | T | C | 0.030615 | 0.004425 | 5.10E-12 |
| rs545971 | let-7c-5p | trans | 2.87E-23 | Nightingale | XL_VLDL_C | NA | T | C | 0.037351 | 0.004245 | 3.10E-18 |
| rs545971 | let-7c-5p | trans | 2.87E-23 | Nightingale | M_LDL_P | NA | T | C | 0.035895 | 0.004437 | 6.80E-16 |
| rs545971 | let-7c-5p | trans | 2.87E-23 | Nightingale | L_LDL_PL | NA | T | C | 0.037992 | 0.004391 | 9.00E-18 |
| rs545971 | let-7c-5p | trans | 2.87E-23 | Nightingale | Total_L | NA | T | C | 0.026346 | 0.004353 | 4.60E-10 |
| rs545971 | let-7c-5p | trans | 2.87E-23 | Nightingale | M_LDL_PL | NA | T | C | 0.032471 | 0.004431 | 2.60E-13 |
| rs545971 | let-7c-5p | trans | 2.87E-23 | Nightingale | M_LDL_TG | NA | T | C | -0.02763 | 0.00437 | 3.20E-10 |
| rs545971 | let-7c-5p | trans | 2.87E-23 | Nightingale | XL_HDL_PL | NA | T | C | -0.02277 | 0.004065 | 1.10E-08 |
| rs545971 | let-7c-5p | trans | 2.87E-23 | Nightingale | M_VLDL_T | NA | T | C | -0.03856 | 0.004181 | 6.80E-20 |
| rs545971 | let-7c-5p | trans | 2.87E-23 | Nightingale | S_VLDL_PL | NA | T | C | 0.032777 | 0.004417 | 4.20E-14 |

| | | | | | | | | | | | |
|---|---|---|---|---|---|---|---|---|---|---|---|
| rs545971 | let-7c-5p | trans | 2.87E-23 | Nightingale | Total_C | NA | T | C | 0.032372 | 0.004284 | 3.30E-14 |
| rs545971 | let-7c-5p | trans | 2.87E-23 | Nightingale | L_VLDL_TG | NA | T | C | -0.05114 | 0.004404 | 5.30E-32 |
| rs545971 | let-7c-5p | trans | 2.87E-23 | Nightingale | S_VLDL_TG | NA | T | C | -0.03813 | 0.004283 | 1.40E-18 |
| rs545971 | let-7c-5p | trans | 2.87E-23 | Nightingale | Cholines | NA | T | C | 0.022288 | 0.004161 | 4.90E-08 |
| rs545971 | let-7c-5p | trans | 2.87E-23 | Nightingale | L_LDL_L | NA | T | C | 0.033551 | 0.004382 | 3.00E-14 |
| rs545971 | let-7c-5p | trans | 2.87E-23 | Nightingale | XL_VLDL_P | NA | T | C | 0.038847 | 0.004432 | 5.10E-19 |
| rs545971 | let-7c-5p | trans | 2.87E-23 | Nightingale | VLDL_FC | NA | T | C | 0.024159 | 0.004392 | 1.10E-08 |
| rs545971 | let-7c-5p | trans | 2.87E-23 | Nightingale | S_HDL_FC | NA | T | C | 0.02345 | 0.004344 | 4.60E-08 |
| rs545971 | let-7c-5p | trans | 2.87E-23 | Nightingale | S_HDL_C | NA | T | C | 0.028415 | 0.004417 | 1.70E-10 |
| rs545971 | let-7c-5p | trans | 2.87E-23 | Nightingale | S_LDL_L | NA | T | C | 0.027745 | 0.004424 | 8.20E-10 |
| rs545971 | let-7c-5p | trans | 2.87E-23 | Nightingale | S_LDL_C | NA | T | C | 0.030946 | 0.004429 | 5.50E-12 |
| rs545971 | let-7c-5p | trans | 2.87E-23 | Nightingale | S_VLDL_PL | NA | T | C | 0.02774 | 0.004237 | 1.80E-10 |
| rs545971 | let-7c-5p | trans | 2.87E-23 | Nightingale | M_VLDL_C | NA | T | C | 0.033628 | 0.004157 | 1.90E-15 |
| rs545971 | let-7c-5p | trans | 2.87E-23 | Nightingale | S_VLDL_C | NA | T | C | 0.038373 | 0.004424 | 1.80E-18 |
| rs545971 | let-7c-5p | trans | 2.87E-23 | Nightingale | non_HDL_( | NA | T | C | 0.035698 | 0.004401 | 5.10E-16 |
| rs545971 | let-7c-5p | trans | 2.87E-23 | Nightingale | XL_VLDL_T | NA | T | C | -0.04392 | 0.004289 | 1.70E-24 |
| rs545971 | let-7c-5p | trans | 2.87E-23 | Nightingale | XS_VLDL_F | NA | T | C | 0.037065 | 0.004348 | 6.50E-18 |
| rs545971 | let-7c-5p | trans | 2.87E-23 | Nightingale | XXL_VLDL_ | NA | T | C | 0.025004 | 0.004454 | 1.50E-09 |
| rs545971 | let-7c-5p | trans | 2.87E-23 | Nightingale | S_LDL_CE_ | NA | T | C | 0.030946 | 0.004417 | 4.90E-14 |
| rs545971 | let-7c-5p | trans | 2.87E-23 | Nightingale | XXL_VLDL_ | NA | T | C | 0.034827 | 0.004434 | 2.90E-15 |
| rs545971 | let-7c-5p | trans | 2.87E-23 | Nightingale | XL_VLDL_C | NA | T | C | 0.038222 | 0.004243 | 4.70E-19 |
| rs545971 | let-7c-5p | trans | 2.87E-23 | Nightingale | M_VLDL_C | NA | T | C | 0.038711 | 0.004413 | 1.90E-18 |
| rs545971 | let-7c-5p | trans | 2.87E-23 | Nightingale | S_VLDL_FC | NA | T | C | 0.029358 | 0.004228 | 1.10E-11 |
| rs545971 | let-7c-5p | trans | 2.87E-23 | Nightingale | IDL_C | NA | T | C | 0.034955 | 0.004273 | 9.40E-16 |
| rs545971 | let-7c-5p | trans | 2.87E-23 | Nightingale | Total_CE | NA | T | C | 0.031745 | 0.004268 | 9.70E-14 |
| rs545971 | let-7c-5p | trans | 2.87E-23 | Nightingale | L_VLDL_FC | NA | T | C | 0.048588 | 0.004382 | 4.50E-29 |
| rs545971 | let-7c-5p | trans | 2.87E-23 | Nightingale | Remnant_( | NA | T | C | 0.036867 | 0.004396 | 3.70E-17 |
| rs545971 | let-7c-5p | trans | 2.87E-23 | Nightingale | VLDL_CE | NA | T | C | 0.035786 | 0.004426 | 3.00E-16 |
| rs545971 | let-7c-5p | trans | 2.87E-23 | Nightingale | XS_VLDL_P | NA | T | C | 0.036698 | 0.004366 | 1.40E-17 |
| rs545971 | let-7c-5p | trans | 2.87E-23 | Nightingale | M_VLDL_P | NA | T | C | 0.029128 | 0.004418 | 2.00E-11 |
| rs545971 | let-7c-5p | trans | 2.87E-23 | Nightingale | IDL_CE | NA | T | C | 0.033507 | 0.004272 | 1.30E-14 |
| rs545971 | let-7c-5p | trans | 2.87E-23 | Nightingale | M_LDL_L | NA | T | C | 0.033035 | 0.004431 | 1.00E-13 |
| rs545971 | let-7c-5p | trans | 2.87E-23 | Nightingale | XS_VLDL_C | NA | T | C | 0.037794 | 0.00427 | 2.10E-18 |
| rs545971 | let-7c-5p | trans | 2.87E-23 | Nightingale | S_LDL_PL_ | NA | T | C | -0.03006 | 0.004417 | 2.60E-13 |
| rs545971 | let-7c-5p | trans | 2.87E-23 | Nightingale | L_LDL_FC | NA | T | C | 0.033627 | 0.004345 | 2.90E-14 |
| rs545971 | let-7c-5p | trans | 2.87E-23 | Nightingale | S_LDL_C_p | NA | T | C | 0.045366 | 0.004442 | 5.40E-25 |
| rs545971 | let-7c-5p | trans | 2.87E-23 | Nightingale | M_LDL_CE | NA | T | C | 0.032977 | 0.004431 | 9.60E-14 |
| rs545971 | let-7c-5p | trans | 2.87E-23 | Nightingale | S_LDL_P | NA | T | C | 0.029589 | 0.004429 | 3.20E-11 |
| rs545971 | let-7c-5p | trans | 2.87E-23 | Nightingale | L_VLDL_C_ | NA | T | C | 0.052192 | 0.004322 | 4.00E-34 |
| rs545971 | let-7c-5p | trans | 2.87E-23 | Nightingale | M_VLDL_P | NA | T | C | 0.044056 | 0.004247 | 5.50E-25 |
| rs545971 | let-7c-5p | trans | 2.87E-23 | Nightingale | M_LDL_C | NA | T | C | 0.033991 | 0.004433 | 2.10E-14 |
| rs545971 | let-7c-5p | trans | 2.87E-23 | Nightingale | S_HDL_CE | NA | T | C | 0.02864 | 0.004432 | 1.30E-10 |
| rs545971 | let-7c-5p | trans | 2.87E-23 | Nightingale | S_VLDL_P | NA | T | C | 0.023465 | 0.004369 | 1.70E-08 |
| rs545971 | let-7c-5p | trans | 2.87E-23 | Nightingale | ApoB_by_/ | NA | T | C | 0.025055 | 0.004358 | 1.30E-08 |
| rs545971 | let-7c-5p | trans | 2.87E-23 | Nightingale | LA | NA | T | C | 0.024903 | 0.004339 | 3.70E-09 |
| rs545971 | let-7c-5p | trans | 2.87E-23 | Nightingale | L_LDL_CE | NA | T | C | 0.031569 | 0.004385 | 9.80E-13 |
| rs545971 | let-7c-5p | trans | 2.87E-23 | Nightingale | XXL_VLDL_ | NA | T | C | 0.033119 | 0.004459 | 9.00E-14 |
| rs545971 | let-7c-5p | trans | 2.87E-23 | Nightingale | ApoB | NA | T | C | 0.033939 | 0.004419 | 2.10E-14 |
| rs545971 | let-7c-5p | trans | 2.87E-23 | Nightingale | IDL_FC_pct | NA | T | C | 0.025773 | 0.004391 | 1.10E-08 |
| rs545971 | let-7c-5p | trans | 2.87E-23 | Nightingale | VLDL_C | NA | T | C | 0.031302 | 0.004419 | 4.90E-13 |
| rs545971 | let-7c-5p | trans | 2.87E-23 | Nightingale | XS_VLDL_C | NA | T | C | 0.037989 | 0.004295 | 1.30E-18 |
| rs545971 | let-7c-5p | trans | 2.87E-23 | Nightingale | S_VLDL_L | NA | T | C | 0.024668 | 0.004382 | 3.70E-09 |
| rs545971 | let-7c-5p | trans | 2.87E-23 | Nightingale | LDL_P | NA | T | C | 0.031801 | 0.004422 | 1.00E-12 |
| rs545971 | let-7c-5p | trans | 2.87E-23 | Nightingale | S_LDL_CE | NA | T | C | 0.030551 | 0.004427 | 5.80E-12 |
| rs545971 | let-7c-5p | trans | 2.87E-23 | Nightingale | M_VLDL_C | NA | T | C | 0.040844 | 0.004385 | 2.30E-20 |
| rs545971 | let-7c-5p | trans | 2.87E-23 | Nightingale | Total_PL | NA | T | C | 0.024922 | 0.004213 | 1.10E-09 |
| rs545971 | let-7c-5p | trans | 2.87E-23 | Nightingale | Omega_6 | NA | T | C | 0.025889 | 0.004321 | 5.80E-10 |
| rs545971 | let-7c-5p | trans | 2.87E-23 | Nightingale | Total_FC | NA | T | C | 0.033253 | 0.004323 | 6.80E-15 |
| rs545971 | let-7b-5p | trans | 1.11E-26 | Nightingale | M_LDL_C_ | NA | T | C | 0.031165 | 0.004428 | 2.90E-12 |
| rs545971 | let-7b-5p | trans | 1.11E-26 | Nightingale | LDL_FC | NA | T | C | 0.033635 | 0.004378 | 5.20E-14 |
| rs545971 | let-7b-5p | trans | 1.11E-26 | Nightingale | GlycA | NA | T | C | 0.028013 | 0.004406 | 1.40E-10 |
| rs545971 | let-7b-5p | trans | 1.11E-26 | Nightingale | XS_VLDL_P | NA | T | C | 0.032112 | 0.004363 | 5.40E-14 |
| rs545971 | let-7b-5p | trans | 1.11E-26 | Nightingale | IDL_PL | NA | T | C | 0.035341 | 0.00427 | 3.30E-16 |

| | | | | | | | | | | | |
|---|---|---|---|---|---|---|---|---|---|---|---|
| rs545971 | let-7b-5p | trans | 1.11E-26 | Nightingale | VLDL_P | NA | T | C | 0.026633 | 0.004401 | 4.40E-10 |
| rs545971 | let-7b-5p | trans | 1.11E-26 | Nightingale | M_VLDL_C | NA | T | C | 0.036205 | 0.004168 | 1.20E-17 |
| rs545971 | let-7b-5p | trans | 1.11E-26 | Nightingale | XS_VLDL_L | NA | T | C | 0.033813 | 0.00435 | 2.10E-15 |
| rs545971 | let-7b-5p | trans | 1.11E-26 | Nightingale | M_VLDL_F | NA | T | C | 0.033372 | 0.004424 | 3.10E-14 |
| rs545971 | let-7b-5p | trans | 1.11E-26 | Nightingale | LDL_L | NA | T | C | 0.033306 | 0.00441 | 7.40E-14 |
| rs545971 | let-7b-5p | trans | 1.11E-26 | Nightingale | LDL_CE | NA | T | C | 0.032556 | 0.004413 | 2.20E-13 |
| rs545971 | let-7b-5p | trans | 1.11E-26 | Nightingale | IDL_L | NA | T | C | 0.035268 | 0.004278 | 4.80E-16 |
| rs545971 | let-7b-5p | trans | 1.11E-26 | Nightingale | S_VLDL_CE | NA | T | C | 0.047034 | 0.004365 | 1.20E-26 |
| rs545971 | let-7b-5p | trans | 1.11E-26 | Nightingale | XL_VLDL_F | NA | T | C | 0.037195 | 0.00429 | 2.00E-18 |
| rs545971 | let-7b-5p | trans | 1.11E-26 | Nightingale | IDL_FC | NA | T | C | 0.038254 | 0.004284 | 1.50E-18 |
| rs545971 | let-7b-5p | trans | 1.11E-26 | Nightingale | XXL_VLDL_ | NA | T | C | 0.030847 | 0.004406 | 1.10E-12 |
| rs545971 | let-7b-5p | trans | 1.11E-26 | Nightingale | S_VLDL_CE | NA | T | C | 0.038182 | 0.004419 | 1.60E-18 |
| rs545971 | let-7b-5p | trans | 1.11E-26 | Nightingale | S_HDL_L | NA | T | C | 0.026297 | 0.00441 | 1.50E-09 |
| rs545971 | let-7b-5p | trans | 1.11E-26 | Nightingale | S_LDL_FC | NA | T | C | 0.028568 | 0.004418 | 4.20E-10 |
| rs545971 | let-7b-5p | trans | 1.11E-26 | Nightingale | L_VLDL_CE | NA | T | C | 0.047336 | 0.004234 | 2.40E-29 |
| rs545971 | let-7b-5p | trans | 1.11E-26 | Nightingale | S_VLDL_C_ | NA | T | C | 0.041889 | 0.004306 | 5.20E-22 |
| rs545971 | let-7b-5p | trans | 1.11E-26 | Nightingale | Citrate | NA | T | C | 0.026829 | 0.004396 | 7.30E-10 |
| rs545971 | let-7b-5p | trans | 1.11E-26 | Nightingale | L_LDL_C | NA | T | C | 0.032322 | 0.004375 | 2.60E-13 |
| rs545971 | let-7b-5p | trans | 1.11E-26 | Nightingale | XS_VLDL_F | NA | T | C | 0.031441 | 0.004393 | 2.40E-12 |
| rs545971 | let-7b-5p | trans | 1.11E-26 | Nightingale | LDL_C | NA | T | C | 0.033196 | 0.004406 | 8.80E-14 |
| rs545971 | let-7b-5p | trans | 1.11E-26 | Nightingale | IDL_P | NA | T | C | 0.039988 | 0.004378 | 1.30E-19 |
| rs545971 | let-7b-5p | trans | 1.11E-26 | Nightingale | Clinical_LD | NA | T | C | 0.036477 | 0.004399 | 2.70E-16 |
| rs545971 | let-7b-5p | trans | 1.11E-26 | Nightingale | S_VLDL_FC | NA | T | C | 0.037905 | 0.004426 | 7.30E-18 |
| rs545971 | let-7b-5p | trans | 1.11E-26 | Nightingale | S_HDL_PL | NA | T | C | 0.023784 | 0.004386 | 3.20E-08 |
| rs545971 | let-7b-5p | trans | 1.11E-26 | Nightingale | M_LDL_FC | NA | T | C | 0.034352 | 0.004419 | 1.90E-14 |
| rs545971 | let-7b-5p | trans | 1.11E-26 | Nightingale | M_VLDL_P | NA | T | C | 0.028296 | 0.004414 | 6.30E-11 |
| rs545971 | let-7b-5p | trans | 1.11E-26 | Nightingale | XXL_VLDL_ | NA | T | C | -0.03573 | 0.004463 | 2.20E-16 |
| rs545971 | let-7b-5p | trans | 1.11E-26 | Nightingale | M_VLDL_F | NA | T | C | 0.042064 | 0.00421 | 3.50E-23 |
| rs545971 | let-7b-5p | trans | 1.11E-26 | Nightingale | LDL_PL | NA | T | C | 0.034991 | 0.004415 | 4.70E-15 |
| rs545971 | let-7b-5p | trans | 1.11E-26 | Nightingale | L_LDL_P | NA | T | C | 0.029177 | 0.00441 | 6.20E-11 |
| rs545971 | let-7b-5p | trans | 1.11E-26 | Nightingale | Sphingomy | NA | T | C | 0.029016 | 0.004136 | 6.80E-12 |
| rs545971 | let-7b-5p | trans | 1.11E-26 | Nightingale | PUFA | NA | T | C | 0.023713 | 0.004316 | 1.40E-08 |
| rs545971 | let-7b-5p | trans | 1.11E-26 | Nightingale | S_HDL_P | NA | T | C | 0.030615 | 0.004425 | 5.10E-12 |
| rs545971 | let-7b-5p | trans | 1.11E-26 | Nightingale | XL_VLDL_C | NA | T | C | 0.037351 | 0.004245 | 3.10E-18 |
| rs545971 | let-7b-5p | trans | 1.11E-26 | Nightingale | M_LDL_P | NA | T | C | 0.035895 | 0.004437 | 6.80E-16 |
| rs545971 | let-7b-5p | trans | 1.11E-26 | Nightingale | L_LDL_PL | NA | T | C | 0.037992 | 0.004391 | 9.00E-18 |
| rs545971 | let-7b-5p | trans | 1.11E-26 | Nightingale | Total_L | NA | T | C | 0.026346 | 0.004353 | 4.60E-10 |
| rs545971 | let-7b-5p | trans | 1.11E-26 | Nightingale | M_LDL_PL | NA | T | C | 0.032471 | 0.004431 | 2.60E-13 |
| rs545971 | let-7b-5p | trans | 1.11E-26 | Nightingale | M_LDL_TG | NA | T | C | -0.02763 | 0.00437 | 3.20E-10 |
| rs545971 | let-7b-5p | trans | 1.11E-26 | Nightingale | XL_HDL_PL | NA | T | C | -0.02277 | 0.004065 | 1.10E-08 |
| rs545971 | let-7b-5p | trans | 1.11E-26 | Nightingale | M_VLDL_T | NA | T | C | -0.03856 | 0.004181 | 6.80E-20 |
| rs545971 | let-7b-5p | trans | 1.11E-26 | Nightingale | S_VLDL_PL | NA | T | C | 0.032777 | 0.004417 | 4.20E-14 |
| rs545971 | let-7b-5p | trans | 1.11E-26 | Nightingale | Total_C | NA | T | C | 0.032372 | 0.004284 | 3.30E-14 |
| rs545971 | let-7b-5p | trans | 1.11E-26 | Nightingale | L_VLDL_TG | NA | T | C | -0.05114 | 0.004404 | 5.30E-32 |
| rs545971 | let-7b-5p | trans | 1.11E-26 | Nightingale | S_VLDL_TG | NA | T | C | -0.03813 | 0.004283 | 1.40E-18 |
| rs545971 | let-7b-5p | trans | 1.11E-26 | Nightingale | Cholines | NA | T | C | 0.022288 | 0.004161 | 4.90E-08 |
| rs545971 | let-7b-5p | trans | 1.11E-26 | Nightingale | L_LDL_L | NA | T | C | 0.033551 | 0.004382 | 3.00E-14 |
| rs545971 | let-7b-5p | trans | 1.11E-26 | Nightingale | XL_VLDL_P | NA | T | C | 0.038847 | 0.004432 | 5.10E-19 |
| rs545971 | let-7b-5p | trans | 1.11E-26 | Nightingale | VLDL_FC | NA | T | C | 0.024159 | 0.004392 | 1.10E-08 |
| rs545971 | let-7b-5p | trans | 1.11E-26 | Nightingale | S_HDL_FC | NA | T | C | 0.02345 | 0.004344 | 4.60E-08 |
| rs545971 | let-7b-5p | trans | 1.11E-26 | Nightingale | S_HDL_C | NA | T | C | 0.028415 | 0.004417 | 1.70E-10 |
| rs545971 | let-7b-5p | trans | 1.11E-26 | Nightingale | S_LDL_L | NA | T | C | 0.027745 | 0.004424 | 8.20E-10 |
| rs545971 | let-7b-5p | trans | 1.11E-26 | Nightingale | S_LDL_C | NA | T | C | 0.030946 | 0.004429 | 5.50E-12 |
| rs545971 | let-7b-5p | trans | 1.11E-26 | Nightingale | S_VLDL_PL | NA | T | C | 0.02774 | 0.004237 | 1.80E-10 |
| rs545971 | let-7b-5p | trans | 1.11E-26 | Nightingale | M_VLDL_C | NA | T | C | 0.033628 | 0.004157 | 1.90E-15 |
| rs545971 | let-7b-5p | trans | 1.11E-26 | Nightingale | S_VLDL_C | NA | T | C | 0.038373 | 0.004424 | 1.80E-18 |
| rs545971 | let-7b-5p | trans | 1.11E-26 | Nightingale | non_HDL_( | NA | T | C | 0.035698 | 0.004401 | 5.10E-16 |
| rs545971 | let-7b-5p | trans | 1.11E-26 | Nightingale | XL_VLDL_T | NA | T | C | -0.04392 | 0.004289 | 1.70E-24 |
| rs545971 | let-7b-5p | trans | 1.11E-26 | Nightingale | XS_VLDL_F | NA | T | C | 0.037065 | 0.004348 | 6.50E-18 |
| rs545971 | let-7b-5p | trans | 1.11E-26 | Nightingale | XXL_VLDL_ | NA | T | C | 0.025004 | 0.004454 | 1.50E-09 |
| rs545971 | let-7b-5p | trans | 1.11E-26 | Nightingale | S_LDL_CE_ | NA | T | C | 0.030946 | 0.004417 | 4.90E-14 |
| rs545971 | let-7b-5p | trans | 1.11E-26 | Nightingale | XXL_VLDL_ | NA | T | C | 0.034827 | 0.004434 | 2.90E-15 |
| rs545971 | let-7b-5p | trans | 1.11E-26 | Nightingale | XL_VLDL_C | NA | T | C | 0.038222 | 0.004243 | 4.70E-19 |

| | | | | | | | | | | | |
|---|---|---|---|---|---|---|---|---|---|---|---|
| rs545971 | let-7b-5p | trans | 1.11E-26 | Nightingale | M_VLDL_C | NA | T | C | 0.038711 | 0.004413 | 1.90E-18 |
| rs545971 | let-7b-5p | trans | 1.11E-26 | Nightingale | S_VLDL_FC | NA | T | C | 0.029358 | 0.004228 | 1.10E-11 |
| rs545971 | let-7b-5p | trans | 1.11E-26 | Nightingale | IDL_C | NA | T | C | 0.034955 | 0.004273 | 9.40E-16 |
| rs545971 | let-7b-5p | trans | 1.11E-26 | Nightingale | Total_CE | NA | T | C | 0.031745 | 0.004268 | 9.70E-14 |
| rs545971 | let-7b-5p | trans | 1.11E-26 | Nightingale | L_VLDL_FC | NA | T | C | 0.048588 | 0.004382 | 4.50E-29 |
| rs545971 | let-7b-5p | trans | 1.11E-26 | Nightingale | Remnant_( | NA | T | C | 0.036867 | 0.004396 | 3.70E-17 |
| rs545971 | let-7b-5p | trans | 1.11E-26 | Nightingale | VLDL_CE | NA | T | C | 0.035786 | 0.004426 | 3.00E-16 |
| rs545971 | let-7b-5p | trans | 1.11E-26 | Nightingale | XS_VLDL_P | NA | T | C | 0.036698 | 0.004366 | 1.40E-17 |
| rs545971 | let-7b-5p | trans | 1.11E-26 | Nightingale | M_VLDL_P | NA | T | C | 0.029128 | 0.004418 | 2.00E-11 |
| rs545971 | let-7b-5p | trans | 1.11E-26 | Nightingale | IDL_CE | NA | T | C | 0.033507 | 0.004272 | 1.30E-14 |
| rs545971 | let-7b-5p | trans | 1.11E-26 | Nightingale | M_LDL_L | NA | T | C | 0.033035 | 0.004431 | 1.00E-13 |
| rs545971 | let-7b-5p | trans | 1.11E-26 | Nightingale | XS_VLDL_C | NA | T | C | 0.037794 | 0.00427 | 2.10E-18 |
| rs545971 | let-7b-5p | trans | 1.11E-26 | Nightingale | S_LDL_PL_ | NA | T | C | -0.03006 | 0.004417 | 2.60E-13 |
| rs545971 | let-7b-5p | trans | 1.11E-26 | Nightingale | L_LDL_FC | NA | T | C | 0.033627 | 0.004345 | 2.90E-14 |
| rs545971 | let-7b-5p | trans | 1.11E-26 | Nightingale | S_LDL_C_p | NA | T | C | 0.045366 | 0.004442 | 5.40E-25 |
| rs545971 | let-7b-5p | trans | 1.11E-26 | Nightingale | M_LDL_CE | NA | T | C | 0.032977 | 0.004431 | 9.60E-14 |
| rs545971 | let-7b-5p | trans | 1.11E-26 | Nightingale | S_LDL_P | NA | T | C | 0.029589 | 0.004429 | 3.20E-11 |
| rs545971 | let-7b-5p | trans | 1.11E-26 | Nightingale | L_VLDL_C_ | NA | T | C | 0.052192 | 0.004322 | 4.00E-34 |
| rs545971 | let-7b-5p | trans | 1.11E-26 | Nightingale | M_VLDL_P | NA | T | C | 0.044056 | 0.004247 | 5.50E-25 |
| rs545971 | let-7b-5p | trans | 1.11E-26 | Nightingale | M_LDL_C | NA | T | C | 0.033991 | 0.004433 | 2.10E-14 |
| rs545971 | let-7b-5p | trans | 1.11E-26 | Nightingale | S_HDL_CE | NA | T | C | 0.02864 | 0.004432 | 1.30E-10 |
| rs545971 | let-7b-5p | trans | 1.11E-26 | Nightingale | S_VLDL_P | NA | T | C | 0.023465 | 0.004369 | 1.70E-08 |
| rs545971 | let-7b-5p | trans | 1.11E-26 | Nightingale | ApoB_by_A | NA | T | C | 0.025055 | 0.004358 | 1.30E-08 |
| rs545971 | let-7b-5p | trans | 1.11E-26 | Nightingale | LA | NA | T | C | 0.024903 | 0.004339 | 3.70E-09 |
| rs545971 | let-7b-5p | trans | 1.11E-26 | Nightingale | L_LDL_CE | NA | T | C | 0.031569 | 0.004385 | 9.80E-13 |
| rs545971 | let-7b-5p | trans | 1.11E-26 | Nightingale | XXL_VLDL_ | NA | T | C | 0.033119 | 0.004459 | 9.00E-14 |
| rs545971 | let-7b-5p | trans | 1.11E-26 | Nightingale | ApoB | NA | T | C | 0.033939 | 0.004419 | 2.10E-14 |
| rs545971 | let-7b-5p | trans | 1.11E-26 | Nightingale | IDL_FC_pct | NA | T | C | 0.025773 | 0.004391 | 1.10E-08 |
| rs545971 | let-7b-5p | trans | 1.11E-26 | Nightingale | VLDL_C | NA | T | C | 0.031302 | 0.004419 | 4.90E-13 |
| rs545971 | let-7b-5p | trans | 1.11E-26 | Nightingale | XS_VLDL_C | NA | T | C | 0.037989 | 0.004295 | 1.30E-18 |
| rs545971 | let-7b-5p | trans | 1.11E-26 | Nightingale | S_VLDL_L | NA | T | C | 0.024668 | 0.004382 | 3.70E-09 |
| rs545971 | let-7b-5p | trans | 1.11E-26 | Nightingale | LDL_P | NA | T | C | 0.031801 | 0.004422 | 1.00E-12 |
| rs545971 | let-7b-5p | trans | 1.11E-26 | Nightingale | S_LDL_CE | NA | T | C | 0.030551 | 0.004427 | 5.80E-12 |
| rs545971 | let-7b-5p | trans | 1.11E-26 | Nightingale | M_VLDL_C | NA | T | C | 0.040844 | 0.004385 | 2.30E-20 |
| rs545971 | let-7b-5p | trans | 1.11E-26 | Nightingale | Total_PL | NA | T | C | 0.024922 | 0.004213 | 1.10E-09 |
| rs545971 | let-7b-5p | trans | 1.11E-26 | Nightingale | Omega_6 | NA | T | C | 0.025889 | 0.004321 | 5.80E-10 |
| rs545971 | let-7b-5p | trans | 1.11E-26 | Nightingale | Total_FC | NA | T | C | 0.033253 | 0.004323 | 6.80E-15 |
| rs545971 | miR-195-5p | trans | 2.54E-17 | Nightingale | M_LDL_C_ | NA | T | C | 0.031165 | 0.004428 | 2.90E-12 |
| rs545971 | miR-195-5p | trans | 2.54E-17 | Nightingale | LDL_FC | NA | T | C | 0.033635 | 0.004378 | 5.20E-14 |
| rs545971 | miR-195-5p | trans | 2.54E-17 | Nightingale | GlycA | NA | T | C | 0.028013 | 0.004406 | 1.40E-10 |
| rs545971 | miR-195-5p | trans | 2.54E-17 | Nightingale | XS_VLDL_P | NA | T | C | 0.032112 | 0.004363 | 5.40E-14 |
| rs545971 | miR-195-5p | trans | 2.54E-17 | Nightingale | IDL_PL | NA | T | C | 0.035341 | 0.00427 | 3.30E-16 |
| rs545971 | miR-195-5p | trans | 2.54E-17 | Nightingale | VLDL_P | NA | T | C | 0.026633 | 0.004401 | 4.40E-10 |
| rs545971 | miR-195-5p | trans | 2.54E-17 | Nightingale | M_VLDL_C | NA | T | C | 0.036205 | 0.004168 | 1.20E-17 |
| rs545971 | miR-195-5p | trans | 2.54E-17 | Nightingale | XS_VLDL_L | NA | T | C | 0.033813 | 0.00435 | 2.10E-15 |
| rs545971 | miR-195-5p | trans | 2.54E-17 | Nightingale | M_VLDL_F | NA | T | C | 0.033372 | 0.004424 | 3.10E-14 |
| rs545971 | miR-195-5p | trans | 2.54E-17 | Nightingale | LDL_L | NA | T | C | 0.033306 | 0.00441 | 7.40E-14 |
| rs545971 | miR-195-5p | trans | 2.54E-17 | Nightingale | LDL_CE | NA | T | C | 0.032556 | 0.004413 | 2.20E-13 |
| rs545971 | miR-195-5p | trans | 2.54E-17 | Nightingale | IDL_L | NA | T | C | 0.035268 | 0.004278 | 4.80E-16 |
| rs545971 | miR-195-5p | trans | 2.54E-17 | Nightingale | S_VLDL_CE | NA | T | C | 0.047034 | 0.004365 | 1.20E-26 |
| rs545971 | miR-195-5p | trans | 2.54E-17 | Nightingale | XL_VLDL_F | NA | T | C | 0.037195 | 0.00429 | 2.00E-18 |
| rs545971 | miR-195-5p | trans | 2.54E-17 | Nightingale | IDL_FC | NA | T | C | 0.038254 | 0.004284 | 1.50E-18 |
| rs545971 | miR-195-5p | trans | 2.54E-17 | Nightingale | XXL_VLDL_ | NA | T | C | 0.030847 | 0.004406 | 1.10E-12 |
| rs545971 | miR-195-5p | trans | 2.54E-17 | Nightingale | S_VLDL_CE | NA | T | C | 0.038182 | 0.004419 | 1.60E-18 |
| rs545971 | miR-195-5p | trans | 2.54E-17 | Nightingale | S_HDL_L | NA | T | C | 0.026297 | 0.00441 | 1.50E-09 |
| rs545971 | miR-195-5p | trans | 2.54E-17 | Nightingale | S_LDL_FC | NA | T | C | 0.028568 | 0.004418 | 4.20E-10 |
| rs545971 | miR-195-5p | trans | 2.54E-17 | Nightingale | L_VLDL_CE | NA | T | C | 0.047336 | 0.004234 | 2.40E-29 |
| rs545971 | miR-195-5p | trans | 2.54E-17 | Nightingale | S_VLDL_C_ | NA | T | C | 0.041889 | 0.004306 | 5.20E-22 |
| rs545971 | miR-195-5p | trans | 2.54E-17 | Nightingale | Citrate | NA | T | C | 0.026829 | 0.004396 | 7.30E-10 |
| rs545971 | miR-195-5p | trans | 2.54E-17 | Nightingale | L_LDL_C | NA | T | C | 0.032322 | 0.004375 | 2.60E-13 |
| rs545971 | miR-195-5p | trans | 2.54E-17 | Nightingale | XS_VLDL_F | NA | T | C | 0.031441 | 0.004393 | 2.40E-12 |
| rs545971 | miR-195-5p | trans | 2.54E-17 | Nightingale | LDL_C | NA | T | C | 0.033196 | 0.004406 | 8.80E-14 |
| rs545971 | miR-195-5p | trans | 2.54E-17 | Nightingale | IDL_P | NA | T | C | 0.039988 | 0.004378 | 1.30E-19 |

| | | | | | | | | | | | |
|---|---|---|---|---|---|---|---|---|---|---|---|
| rs545971 | miR-195-5p | trans | 2.54E-17 | Nightingale | Clinical_LD | NA | T | C | 0.036477 | 0.004399 | 2.70E-16 |
| rs545971 | miR-195-5p | trans | 2.54E-17 | Nightingale | S_VLDL_FC | NA | T | C | 0.037905 | 0.004426 | 7.30E-18 |
| rs545971 | miR-195-5p | trans | 2.54E-17 | Nightingale | S_HDL_PL | NA | T | C | 0.023784 | 0.004386 | 3.20E-08 |
| rs545971 | miR-195-5p | trans | 2.54E-17 | Nightingale | M_LDL_FC | NA | T | C | 0.034352 | 0.004419 | 1.90E-14 |
| rs545971 | miR-195-5p | trans | 2.54E-17 | Nightingale | M_VLDL_P | NA | T | C | 0.028296 | 0.004414 | 6.30E-11 |
| rs545971 | miR-195-5p | trans | 2.54E-17 | Nightingale | XXL_VLDL_ | NA | T | C | -0.03573 | 0.004463 | 2.20E-16 |
| rs545971 | miR-195-5p | trans | 2.54E-17 | Nightingale | M_VLDL_F | NA | T | C | 0.042064 | 0.00421 | 3.50E-23 |
| rs545971 | miR-195-5p | trans | 2.54E-17 | Nightingale | LDL_PL | NA | T | C | 0.034991 | 0.004415 | 4.70E-15 |
| rs545971 | miR-195-5p | trans | 2.54E-17 | Nightingale | L_LDL_P | NA | T | C | 0.029177 | 0.00441 | 6.20E-11 |
| rs545971 | miR-195-5p | trans | 2.54E-17 | Nightingale | Sphingomy | NA | T | C | 0.029016 | 0.004136 | 6.80E-12 |
| rs545971 | miR-195-5p | trans | 2.54E-17 | Nightingale | PUFA | NA | T | C | 0.023713 | 0.004316 | 1.40E-08 |
| rs545971 | miR-195-5p | trans | 2.54E-17 | Nightingale | S_HDL_P | NA | T | C | 0.030615 | 0.004425 | 5.10E-12 |
| rs545971 | miR-195-5p | trans | 2.54E-17 | Nightingale | XL_VLDL_C | NA | T | C | 0.037351 | 0.004245 | 3.10E-18 |
| rs545971 | miR-195-5p | trans | 2.54E-17 | Nightingale | M_LDL_P | NA | T | C | 0.035895 | 0.004437 | 6.80E-16 |
| rs545971 | miR-195-5p | trans | 2.54E-17 | Nightingale | L_LDL_PL | NA | T | C | 0.037992 | 0.004391 | 9.00E-18 |
| rs545971 | miR-195-5p | trans | 2.54E-17 | Nightingale | Total_L | NA | T | C | 0.026346 | 0.004353 | 4.60E-10 |
| rs545971 | miR-195-5p | trans | 2.54E-17 | Nightingale | M_LDL_PL | NA | T | C | 0.032471 | 0.004431 | 2.60E-13 |
| rs545971 | miR-195-5p | trans | 2.54E-17 | Nightingale | M_LDL_TG | NA | T | C | -0.02763 | 0.00437 | 3.20E-10 |
| rs545971 | miR-195-5p | trans | 2.54E-17 | Nightingale | XL_HDL_PL | NA | T | C | -0.02277 | 0.004065 | 1.10E-08 |
| rs545971 | miR-195-5p | trans | 2.54E-17 | Nightingale | M_VLDL_T | NA | T | C | -0.03856 | 0.004181 | 6.80E-20 |
| rs545971 | miR-195-5p | trans | 2.54E-17 | Nightingale | S_VLDL_PL | NA | T | C | 0.032777 | 0.004417 | 4.20E-14 |
| rs545971 | miR-195-5p | trans | 2.54E-17 | Nightingale | Total_C | NA | T | C | 0.032372 | 0.004284 | 3.30E-14 |
| rs545971 | miR-195-5p | trans | 2.54E-17 | Nightingale | L_VLDL_TG | NA | T | C | -0.05114 | 0.004404 | 5.30E-32 |
| rs545971 | miR-195-5p | trans | 2.54E-17 | Nightingale | S_VLDL_TG | NA | T | C | -0.03813 | 0.004283 | 1.40E-18 |
| rs545971 | miR-195-5p | trans | 2.54E-17 | Nightingale | Cholines | NA | T | C | 0.022288 | 0.004161 | 4.90E-08 |
| rs545971 | miR-195-5p | trans | 2.54E-17 | Nightingale | L_LDL_L | NA | T | C | 0.033551 | 0.004382 | 3.00E-14 |
| rs545971 | miR-195-5p | trans | 2.54E-17 | Nightingale | XL_VLDL_P | NA | T | C | 0.038847 | 0.004432 | 5.10E-19 |
| rs545971 | miR-195-5p | trans | 2.54E-17 | Nightingale | VLDL_FC | NA | T | C | 0.024159 | 0.004392 | 1.10E-08 |
| rs545971 | miR-195-5p | trans | 2.54E-17 | Nightingale | S_HDL_FC | NA | T | C | 0.02345 | 0.004344 | 4.60E-08 |
| rs545971 | miR-195-5p | trans | 2.54E-17 | Nightingale | S_HDL_C | NA | T | C | 0.028415 | 0.004417 | 1.70E-10 |
| rs545971 | miR-195-5p | trans | 2.54E-17 | Nightingale | S_LDL_L | NA | T | C | 0.027745 | 0.004424 | 8.20E-10 |
| rs545971 | miR-195-5p | trans | 2.54E-17 | Nightingale | S_LDL_C | NA | T | C | 0.030946 | 0.004429 | 5.50E-12 |
| rs545971 | miR-195-5p | trans | 2.54E-17 | Nightingale | S_VLDL_PL | NA | T | C | 0.02774 | 0.004237 | 1.80E-10 |
| rs545971 | miR-195-5p | trans | 2.54E-17 | Nightingale | M_VLDL_C | NA | T | C | 0.033628 | 0.004157 | 1.90E-15 |
| rs545971 | miR-195-5p | trans | 2.54E-17 | Nightingale | S_VLDL_C | NA | T | C | 0.038373 | 0.004424 | 1.80E-18 |
| rs545971 | miR-195-5p | trans | 2.54E-17 | Nightingale | non_HDL_( | NA | T | C | 0.035698 | 0.004401 | 5.10E-16 |
| rs545971 | miR-195-5p | trans | 2.54E-17 | Nightingale | XL_VLDL_T | NA | T | C | -0.04392 | 0.004289 | 1.70E-24 |
| rs545971 | miR-195-5p | trans | 2.54E-17 | Nightingale | XS_VLDL_F | NA | T | C | 0.037065 | 0.004348 | 6.50E-18 |
| rs545971 | miR-195-5p | trans | 2.54E-17 | Nightingale | XXL_VLDL_ | NA | T | C | 0.025004 | 0.004454 | 1.50E-09 |
| rs545971 | miR-195-5p | trans | 2.54E-17 | Nightingale | S_LDL_CE_ | NA | T | C | 0.030946 | 0.004417 | 4.90E-14 |
| rs545971 | miR-195-5p | trans | 2.54E-17 | Nightingale | XXL_VLDL_ | NA | T | C | 0.034827 | 0.004434 | 2.90E-15 |
| rs545971 | miR-195-5p | trans | 2.54E-17 | Nightingale | XL_VLDL_C | NA | T | C | 0.038222 | 0.004243 | 4.70E-19 |
| rs545971 | miR-195-5p | trans | 2.54E-17 | Nightingale | M_VLDL_C | NA | T | C | 0.038711 | 0.004413 | 1.90E-18 |
| rs545971 | miR-195-5p | trans | 2.54E-17 | Nightingale | S_VLDL_FC | NA | T | C | 0.029358 | 0.004228 | 1.10E-11 |
| rs545971 | miR-195-5p | trans | 2.54E-17 | Nightingale | IDL_C | NA | T | C | 0.034955 | 0.004273 | 9.40E-16 |
| rs545971 | miR-195-5p | trans | 2.54E-17 | Nightingale | Total_CE | NA | T | C | 0.031745 | 0.004268 | 9.70E-14 |
| rs545971 | miR-195-5p | trans | 2.54E-17 | Nightingale | L_VLDL_FC | NA | T | C | 0.048588 | 0.004382 | 4.50E-29 |
| rs545971 | miR-195-5p | trans | 2.54E-17 | Nightingale | Remnant_( | NA | T | C | 0.036867 | 0.004396 | 3.70E-17 |
| rs545971 | miR-195-5p | trans | 2.54E-17 | Nightingale | VLDL_CE | NA | T | C | 0.035786 | 0.004426 | 3.00E-16 |
| rs545971 | miR-195-5p | trans | 2.54E-17 | Nightingale | XS_VLDL_P | NA | T | C | 0.036698 | 0.004366 | 1.40E-17 |
| rs545971 | miR-195-5p | trans | 2.54E-17 | Nightingale | M_VLDL_P | NA | T | C | 0.029128 | 0.004418 | 2.00E-11 |
| rs545971 | miR-195-5p | trans | 2.54E-17 | Nightingale | IDL_CE | NA | T | C | 0.033507 | 0.004272 | 1.30E-14 |
| rs545971 | miR-195-5p | trans | 2.54E-17 | Nightingale | M_LDL_L | NA | T | C | 0.033035 | 0.004431 | 1.00E-13 |
| rs545971 | miR-195-5p | trans | 2.54E-17 | Nightingale | XS_VLDL_C | NA | T | C | 0.037794 | 0.00427 | 2.10E-18 |
| rs545971 | miR-195-5p | trans | 2.54E-17 | Nightingale | S_LDL_PL_ | NA | T | C | -0.03006 | 0.004417 | 2.60E-13 |
| rs545971 | miR-195-5p | trans | 2.54E-17 | Nightingale | L_LDL_FC | NA | T | C | 0.033627 | 0.004345 | 2.90E-14 |
| rs545971 | miR-195-5p | trans | 2.54E-17 | Nightingale | S_LDL_C_p | NA | T | C | 0.045366 | 0.004442 | 5.40E-25 |
| rs545971 | miR-195-5p | trans | 2.54E-17 | Nightingale | M_LDL_CE | NA | T | C | 0.032977 | 0.004431 | 9.60E-14 |
| rs545971 | miR-195-5p | trans | 2.54E-17 | Nightingale | S_LDL_P | NA | T | C | 0.029589 | 0.004429 | 3.20E-11 |
| rs545971 | miR-195-5p | trans | 2.54E-17 | Nightingale | L_VLDL_C_ | NA | T | C | 0.052192 | 0.004322 | 4.00E-34 |
| rs545971 | miR-195-5p | trans | 2.54E-17 | Nightingale | M_VLDL_P | NA | T | C | 0.044056 | 0.004247 | 5.50E-25 |
| rs545971 | miR-195-5p | trans | 2.54E-17 | Nightingale | M_LDL_C | NA | T | C | 0.033991 | 0.004433 | 2.10E-14 |
| rs545971 | miR-195-5p | trans | 2.54E-17 | Nightingale | S_HDL_CE | NA | T | C | 0.02864 | 0.004432 | 1.30E-10 |

| | | | | | | | | | | | |
|---|---|---|---|---|---|---|---|---|---|---|---|
| rs545971 | miR-195-5p | trans | 2.54E-17 | Nightingale | S_VLDL_P | NA | T | C | 0.023465 | 0.004369 | 1.70E-08 |
| rs545971 | miR-195-5p | trans | 2.54E-17 | Nightingale | ApoB_by_A | NA | T | C | 0.025055 | 0.004358 | 1.30E-08 |
| rs545971 | miR-195-5p | trans | 2.54E-17 | Nightingale | LA | NA | T | C | 0.024903 | 0.004339 | 3.70E-09 |
| rs545971 | miR-195-5p | trans | 2.54E-17 | Nightingale | L_LDL_CE | NA | T | C | 0.031569 | 0.004385 | 9.80E-13 |
| rs545971 | miR-195-5p | trans | 2.54E-17 | Nightingale | XXL_VLDL_ | NA | T | C | 0.033119 | 0.004459 | 9.00E-14 |
| rs545971 | miR-195-5p | trans | 2.54E-17 | Nightingale | ApoB | NA | T | C | 0.033939 | 0.004419 | 2.10E-14 |
| rs545971 | miR-195-5p | trans | 2.54E-17 | Nightingale | IDL_FC_pct | NA | T | C | 0.025773 | 0.004391 | 1.10E-08 |
| rs545971 | miR-195-5p | trans | 2.54E-17 | Nightingale | VLDL_C | NA | T | C | 0.031302 | 0.004419 | 4.90E-13 |
| rs545971 | miR-195-5p | trans | 2.54E-17 | Nightingale | XS_VLDL_C | NA | T | C | 0.037989 | 0.004295 | 1.30E-18 |
| rs545971 | miR-195-5p | trans | 2.54E-17 | Nightingale | S_VLDL_L | NA | T | C | 0.024668 | 0.004382 | 3.70E-09 |
| rs545971 | miR-195-5p | trans | 2.54E-17 | Nightingale | LDL_P | NA | T | C | 0.031801 | 0.004422 | 1.00E-12 |
| rs545971 | miR-195-5p | trans | 2.54E-17 | Nightingale | S_LDL_CE | NA | T | C | 0.030551 | 0.004427 | 5.80E-12 |
| rs545971 | miR-195-5p | trans | 2.54E-17 | Nightingale | M_VLDL_C | NA | T | C | 0.040844 | 0.004385 | 2.30E-20 |
| rs545971 | miR-195-5p | trans | 2.54E-17 | Nightingale | Total_PL | NA | T | C | 0.024922 | 0.004213 | 1.10E-09 |
| rs545971 | miR-195-5p | trans | 2.54E-17 | Nightingale | Omega_6 | NA | T | C | 0.025889 | 0.004321 | 5.80E-10 |
| rs545971 | miR-195-5p | trans | 2.54E-17 | Nightingale | Total_FC | NA | T | C | 0.033253 | 0.004323 | 6.80E-15 |
| rs545971 | miR-10b-5p | trans | 3.43E-35 | Nightingale | M_LDL_C_ | NA | T | C | 0.031165 | 0.004428 | 2.90E-12 |
| rs545971 | miR-10b-5p | trans | 3.43E-35 | Nightingale | LDL_FC | NA | T | C | 0.033635 | 0.004378 | 5.20E-14 |
| rs545971 | miR-10b-5p | trans | 3.43E-35 | Nightingale | GlycA | NA | T | C | 0.028013 | 0.004406 | 1.40E-10 |
| rs545971 | miR-10b-5p | trans | 3.43E-35 | Nightingale | XS_VLDL_P | NA | T | C | 0.032112 | 0.004363 | 5.40E-14 |
| rs545971 | miR-10b-5p | trans | 3.43E-35 | Nightingale | IDL_PL | NA | T | C | 0.035341 | 0.00427 | 3.30E-16 |
| rs545971 | miR-10b-5p | trans | 3.43E-35 | Nightingale | VLDL_P | NA | T | C | 0.026633 | 0.004401 | 4.40E-10 |
| rs545971 | miR-10b-5p | trans | 3.43E-35 | Nightingale | M_VLDL_C | NA | T | C | 0.036205 | 0.004168 | 1.20E-17 |
| rs545971 | miR-10b-5p | trans | 3.43E-35 | Nightingale | XS_VLDL_L | NA | T | C | 0.033813 | 0.00435 | 2.10E-15 |
| rs545971 | miR-10b-5p | trans | 3.43E-35 | Nightingale | M_VLDL_F | NA | T | C | 0.033372 | 0.004424 | 3.10E-14 |
| rs545971 | miR-10b-5p | trans | 3.43E-35 | Nightingale | LDL_L | NA | T | C | 0.033306 | 0.00441 | 7.40E-14 |
| rs545971 | miR-10b-5p | trans | 3.43E-35 | Nightingale | LDL_CE | NA | T | C | 0.032556 | 0.004413 | 2.20E-13 |
| rs545971 | miR-10b-5p | trans | 3.43E-35 | Nightingale | IDL_L | NA | T | C | 0.035268 | 0.004278 | 4.80E-16 |
| rs545971 | miR-10b-5p | trans | 3.43E-35 | Nightingale | S_VLDL_CE | NA | T | C | 0.047034 | 0.004365 | 1.20E-26 |
| rs545971 | miR-10b-5p | trans | 3.43E-35 | Nightingale | XL_VLDL_F | NA | T | C | 0.037195 | 0.00429 | 2.00E-18 |
| rs545971 | miR-10b-5p | trans | 3.43E-35 | Nightingale | IDL_FC | NA | T | C | 0.038254 | 0.004284 | 1.50E-18 |
| rs545971 | miR-10b-5p | trans | 3.43E-35 | Nightingale | XXL_VLDL_ | NA | T | C | 0.030847 | 0.004406 | 1.10E-12 |
| rs545971 | miR-10b-5p | trans | 3.43E-35 | Nightingale | S_VLDL_CE | NA | T | C | 0.038182 | 0.004419 | 1.60E-18 |
| rs545971 | miR-10b-5p | trans | 3.43E-35 | Nightingale | S_HDL_L | NA | T | C | 0.026297 | 0.00441 | 1.50E-09 |
| rs545971 | miR-10b-5p | trans | 3.43E-35 | Nightingale | S_LDL_FC | NA | T | C | 0.028568 | 0.004418 | 4.20E-10 |
| rs545971 | miR-10b-5p | trans | 3.43E-35 | Nightingale | L_VLDL_CE | NA | T | C | 0.047336 | 0.004234 | 2.40E-29 |
| rs545971 | miR-10b-5p | trans | 3.43E-35 | Nightingale | S_VLDL_C_ | NA | T | C | 0.041889 | 0.004306 | 5.20E-22 |
| rs545971 | miR-10b-5p | trans | 3.43E-35 | Nightingale | Citrate | NA | T | C | 0.026829 | 0.004396 | 7.30E-10 |
| rs545971 | miR-10b-5p | trans | 3.43E-35 | Nightingale | L_LDL_C | NA | T | C | 0.032322 | 0.004375 | 2.60E-13 |
| rs545971 | miR-10b-5p | trans | 3.43E-35 | Nightingale | XS_VLDL_F | NA | T | C | 0.031441 | 0.004393 | 2.40E-12 |
| rs545971 | miR-10b-5p | trans | 3.43E-35 | Nightingale | LDL_C | NA | T | C | 0.033196 | 0.004406 | 8.80E-14 |
| rs545971 | miR-10b-5p | trans | 3.43E-35 | Nightingale | IDL_P | NA | T | C | 0.039988 | 0.004378 | 1.30E-19 |
| rs545971 | miR-10b-5p | trans | 3.43E-35 | Nightingale | Clinical_LD | NA | T | C | 0.036477 | 0.004399 | 2.70E-16 |
| rs545971 | miR-10b-5p | trans | 3.43E-35 | Nightingale | S_VLDL_FC | NA | T | C | 0.037905 | 0.004426 | 7.30E-18 |
| rs545971 | miR-10b-5p | trans | 3.43E-35 | Nightingale | S_HDL_PL | NA | T | C | 0.023784 | 0.004386 | 3.20E-08 |
| rs545971 | miR-10b-5p | trans | 3.43E-35 | Nightingale | M_LDL_FC | NA | T | C | 0.034352 | 0.004419 | 1.90E-14 |
| rs545971 | miR-10b-5p | trans | 3.43E-35 | Nightingale | M_VLDL_P | NA | T | C | 0.028296 | 0.004414 | 6.30E-11 |
| rs545971 | miR-10b-5p | trans | 3.43E-35 | Nightingale | XXL_VLDL_ | NA | T | C | -0.03573 | 0.004463 | 2.20E-16 |
| rs545971 | miR-10b-5p | trans | 3.43E-35 | Nightingale | M_VLDL_F | NA | T | C | 0.042064 | 0.00421 | 3.50E-23 |
| rs545971 | miR-10b-5p | trans | 3.43E-35 | Nightingale | LDL_PL | NA | T | C | 0.034991 | 0.004415 | 4.70E-15 |
| rs545971 | miR-10b-5p | trans | 3.43E-35 | Nightingale | L_LDL_P | NA | T | C | 0.029177 | 0.00441 | 6.20E-11 |
| rs545971 | miR-10b-5p | trans | 3.43E-35 | Nightingale | Sphingomy | NA | T | C | 0.029016 | 0.004136 | 6.80E-12 |
| rs545971 | miR-10b-5p | trans | 3.43E-35 | Nightingale | PUFA | NA | T | C | 0.023713 | 0.004316 | 1.40E-08 |
| rs545971 | miR-10b-5p | trans | 3.43E-35 | Nightingale | S_HDL_P | NA | T | C | 0.030615 | 0.004425 | 5.10E-12 |
| rs545971 | miR-10b-5p | trans | 3.43E-35 | Nightingale | XL_VLDL_C | NA | T | C | 0.037351 | 0.004245 | 3.10E-18 |
| rs545971 | miR-10b-5p | trans | 3.43E-35 | Nightingale | M_LDL_P | NA | T | C | 0.035895 | 0.004437 | 6.80E-16 |
| rs545971 | miR-10b-5p | trans | 3.43E-35 | Nightingale | L_LDL_PL | NA | T | C | 0.037992 | 0.004391 | 9.00E-18 |
| rs545971 | miR-10b-5p | trans | 3.43E-35 | Nightingale | Total_L | NA | T | C | 0.026346 | 0.004353 | 4.60E-10 |
| rs545971 | miR-10b-5p | trans | 3.43E-35 | Nightingale | M_LDL_PL | NA | T | C | 0.032471 | 0.004431 | 2.60E-13 |
| rs545971 | miR-10b-5p | trans | 3.43E-35 | Nightingale | M_LDL_TG | NA | T | C | -0.02763 | 0.00437 | 3.20E-10 |
| rs545971 | miR-10b-5p | trans | 3.43E-35 | Nightingale | XL_HDL_PL | NA | T | C | -0.02277 | 0.004065 | 1.10E-08 |
| rs545971 | miR-10b-5p | trans | 3.43E-35 | Nightingale | M_VLDL_T | NA | T | C | -0.03856 | 0.004181 | 6.80E-20 |
| rs545971 | miR-10b-5p | trans | 3.43E-35 | Nightingale | S_VLDL_PL | NA | T | C | 0.032777 | 0.004417 | 4.20E-14 |

| | | | | | | | | | | | |
|---|---|---|---|---|---|---|---|---|---|---|---|
| rs545971 | miR-10b-5p | trans | 3.43E-35 | Nightingale | Total_C | NA | T | C | 0.032372 | 0.004284 | 3.30E-14 |
| rs545971 | miR-10b-5p | trans | 3.43E-35 | Nightingale | L_VLDL_TG | NA | T | C | -0.05114 | 0.004404 | 5.30E-32 |
| rs545971 | miR-10b-5p | trans | 3.43E-35 | Nightingale | S_VLDL_TG | NA | T | C | -0.03813 | 0.004283 | 1.40E-18 |
| rs545971 | miR-10b-5p | trans | 3.43E-35 | Nightingale | Cholines | NA | T | C | 0.022288 | 0.004161 | 4.90E-08 |
| rs545971 | miR-10b-5p | trans | 3.43E-35 | Nightingale | L_LDL_L | NA | T | C | 0.033551 | 0.004382 | 3.00E-14 |
| rs545971 | miR-10b-5p | trans | 3.43E-35 | Nightingale | XL_VLDL_P | NA | T | C | 0.038847 | 0.004432 | 5.10E-19 |
| rs545971 | miR-10b-5p | trans | 3.43E-35 | Nightingale | VLDL_FC | NA | T | C | 0.024159 | 0.004392 | 1.10E-08 |
| rs545971 | miR-10b-5p | trans | 3.43E-35 | Nightingale | S_HDL_FC | NA | T | C | 0.02345 | 0.004344 | 4.60E-08 |
| rs545971 | miR-10b-5p | trans | 3.43E-35 | Nightingale | S_HDL_C | NA | T | C | 0.028415 | 0.004417 | 1.70E-10 |
| rs545971 | miR-10b-5p | trans | 3.43E-35 | Nightingale | S_LDL_L | NA | T | C | 0.027745 | 0.004424 | 8.20E-10 |
| rs545971 | miR-10b-5p | trans | 3.43E-35 | Nightingale | S_LDL_C | NA | T | C | 0.030946 | 0.004429 | 5.50E-12 |
| rs545971 | miR-10b-5p | trans | 3.43E-35 | Nightingale | S_VLDL_PL | NA | T | C | 0.02774 | 0.004237 | 1.80E-10 |
| rs545971 | miR-10b-5p | trans | 3.43E-35 | Nightingale | M_VLDL_C | NA | T | C | 0.033628 | 0.004157 | 1.90E-15 |
| rs545971 | miR-10b-5p | trans | 3.43E-35 | Nightingale | S_VLDL_C | NA | T | C | 0.038373 | 0.004424 | 1.80E-18 |
| rs545971 | miR-10b-5p | trans | 3.43E-35 | Nightingale | non_HDL_( | NA | T | C | 0.035698 | 0.004401 | 5.10E-16 |
| rs545971 | miR-10b-5p | trans | 3.43E-35 | Nightingale | XL_VLDL_T | NA | T | C | -0.04392 | 0.004289 | 1.70E-24 |
| rs545971 | miR-10b-5p | trans | 3.43E-35 | Nightingale | XS_VLDL_F | NA | T | C | 0.037065 | 0.004348 | 6.50E-18 |
| rs545971 | miR-10b-5p | trans | 3.43E-35 | Nightingale | XXL_VLDL_ | NA | T | C | 0.025004 | 0.004454 | 1.50E-09 |
| rs545971 | miR-10b-5p | trans | 3.43E-35 | Nightingale | S_LDL_CE_ | NA | T | C | 0.030946 | 0.004417 | 4.90E-14 |
| rs545971 | miR-10b-5p | trans | 3.43E-35 | Nightingale | XXL_VLDL_ | NA | T | C | 0.034827 | 0.004434 | 2.90E-15 |
| rs545971 | miR-10b-5p | trans | 3.43E-35 | Nightingale | XL_VLDL_C | NA | T | C | 0.038222 | 0.004243 | 4.70E-19 |
| rs545971 | miR-10b-5p | trans | 3.43E-35 | Nightingale | M_VLDL_C | NA | T | C | 0.038711 | 0.004413 | 1.90E-18 |
| rs545971 | miR-10b-5p | trans | 3.43E-35 | Nightingale | S_VLDL_FC | NA | T | C | 0.029358 | 0.004228 | 1.10E-11 |
| rs545971 | miR-10b-5p | trans | 3.43E-35 | Nightingale | IDL_C | NA | T | C | 0.034955 | 0.004273 | 9.40E-16 |
| rs545971 | miR-10b-5p | trans | 3.43E-35 | Nightingale | Total_CE | NA | T | C | 0.031745 | 0.004268 | 9.70E-14 |
| rs545971 | miR-10b-5p | trans | 3.43E-35 | Nightingale | L_VLDL_FC | NA | T | C | 0.048588 | 0.004382 | 4.50E-29 |
| rs545971 | miR-10b-5p | trans | 3.43E-35 | Nightingale | Remnant_( | NA | T | C | 0.036867 | 0.004396 | 3.70E-17 |
| rs545971 | miR-10b-5p | trans | 3.43E-35 | Nightingale | VLDL_CE | NA | T | C | 0.035786 | 0.004426 | 3.00E-16 |
| rs545971 | miR-10b-5p | trans | 3.43E-35 | Nightingale | XS_VLDL_P | NA | T | C | 0.036698 | 0.004366 | 1.40E-17 |
| rs545971 | miR-10b-5p | trans | 3.43E-35 | Nightingale | M_VLDL_P | NA | T | C | 0.029128 | 0.004418 | 2.00E-11 |
| rs545971 | miR-10b-5p | trans | 3.43E-35 | Nightingale | IDL_CE | NA | T | C | 0.033507 | 0.004272 | 1.30E-14 |
| rs545971 | miR-10b-5p | trans | 3.43E-35 | Nightingale | M_LDL_L | NA | T | C | 0.033035 | 0.004431 | 1.00E-13 |
| rs545971 | miR-10b-5p | trans | 3.43E-35 | Nightingale | XS_VLDL_C | NA | T | C | 0.037794 | 0.00427 | 2.10E-18 |
| rs545971 | miR-10b-5p | trans | 3.43E-35 | Nightingale | S_LDL_PL_ | NA | T | C | -0.03006 | 0.004417 | 2.60E-13 |
| rs545971 | miR-10b-5p | trans | 3.43E-35 | Nightingale | L_LDL_FC | NA | T | C | 0.033627 | 0.004345 | 2.90E-14 |
| rs545971 | miR-10b-5p | trans | 3.43E-35 | Nightingale | S_LDL_C_p | NA | T | C | 0.045366 | 0.004442 | 5.40E-25 |
| rs545971 | miR-10b-5p | trans | 3.43E-35 | Nightingale | M_LDL_CE | NA | T | C | 0.032977 | 0.004431 | 9.60E-14 |
| rs545971 | miR-10b-5p | trans | 3.43E-35 | Nightingale | S_LDL_P | NA | T | C | 0.029589 | 0.004429 | 3.20E-11 |
| rs545971 | miR-10b-5p | trans | 3.43E-35 | Nightingale | L_VLDL_C_ | NA | T | C | 0.052192 | 0.004322 | 4.00E-34 |
| rs545971 | miR-10b-5p | trans | 3.43E-35 | Nightingale | M_VLDL_P | NA | T | C | 0.044056 | 0.004247 | 5.50E-25 |
| rs545971 | miR-10b-5p | trans | 3.43E-35 | Nightingale | M_LDL_C | NA | T | C | 0.033991 | 0.004433 | 2.10E-14 |
| rs545971 | miR-10b-5p | trans | 3.43E-35 | Nightingale | S_HDL_CE | NA | T | C | 0.02864 | 0.004432 | 1.30E-10 |
| rs545971 | miR-10b-5p | trans | 3.43E-35 | Nightingale | S_VLDL_P | NA | T | C | 0.023465 | 0.004369 | 1.70E-08 |
| rs545971 | miR-10b-5p | trans | 3.43E-35 | Nightingale | ApoB_by_A | NA | T | C | 0.025055 | 0.004358 | 1.30E-08 |
| rs545971 | miR-10b-5p | trans | 3.43E-35 | Nightingale | LA | NA | T | C | 0.024903 | 0.004339 | 3.70E-09 |
| rs545971 | miR-10b-5p | trans | 3.43E-35 | Nightingale | L_LDL_CE | NA | T | C | 0.031569 | 0.004385 | 9.80E-13 |
| rs545971 | miR-10b-5p | trans | 3.43E-35 | Nightingale | XXL_VLDL_ | NA | T | C | 0.033119 | 0.004459 | 9.00E-14 |
| rs545971 | miR-10b-5p | trans | 3.43E-35 | Nightingale | ApoB | NA | T | C | 0.033939 | 0.004419 | 2.10E-14 |
| rs545971 | miR-10b-5p | trans | 3.43E-35 | Nightingale | IDL_FC_pct | NA | T | C | 0.025773 | 0.004391 | 1.10E-08 |
| rs545971 | miR-10b-5p | trans | 3.43E-35 | Nightingale | VLDL_C | NA | T | C | 0.031302 | 0.004419 | 4.90E-13 |
| rs545971 | miR-10b-5p | trans | 3.43E-35 | Nightingale | XS_VLDL_C | NA | T | C | 0.037989 | 0.004295 | 1.30E-18 |
| rs545971 | miR-10b-5p | trans | 3.43E-35 | Nightingale | S_VLDL_L | NA | T | C | 0.024668 | 0.004382 | 3.70E-09 |
| rs545971 | miR-10b-5p | trans | 3.43E-35 | Nightingale | LDL_P | NA | T | C | 0.031801 | 0.004422 | 1.00E-12 |
| rs545971 | miR-10b-5p | trans | 3.43E-35 | Nightingale | S_LDL_CE | NA | T | C | 0.030551 | 0.004427 | 5.80E-12 |
| rs545971 | miR-10b-5p | trans | 3.43E-35 | Nightingale | M_VLDL_C | NA | T | C | 0.040844 | 0.004385 | 2.30E-20 |
| rs545971 | miR-10b-5p | trans | 3.43E-35 | Nightingale | Total_PL | NA | T | C | 0.024922 | 0.004213 | 1.10E-09 |
| rs545971 | miR-10b-5p | trans | 3.43E-35 | Nightingale | Omega_6 | NA | T | C | 0.025889 | 0.004321 | 5.80E-10 |
| rs545971 | miR-10b-5p | trans | 3.43E-35 | Nightingale | Total_FC | NA | T | C | 0.033253 | 0.004323 | 6.80E-15 |
| rs545971 | miR-126-3p | trans | 2.76E-39 | Nightingale | M_LDL_C_p | NA | T | C | 0.031165 | 0.004428 | 2.90E-12 |
| rs545971 | miR-126-3p | trans | 2.76E-39 | Nightingale | LDL_FC | NA | T | C | 0.033635 | 0.004378 | 5.20E-14 |
| rs545971 | miR-126-3p | trans | 2.76E-39 | Nightingale | GlycA | NA | T | C | 0.028013 | 0.004406 | 1.40E-10 |
| rs545971 | miR-126-3p | trans | 2.76E-39 | Nightingale | XS_VLDL_P | NA | T | C | 0.032112 | 0.004363 | 5.40E-14 |
| rs545971 | miR-126-3p | trans | 2.76E-39 | Nightingale | IDL_PL | NA | T | C | 0.035341 | 0.00427 | 3.30E-16 |

| | | | | | | | | | | | | |
|---|---|---|---|---|---|---|---|---|---|---|---|---|
| rs545971 | miR-126-3p | trans | | 2.76E-39 | Nightingale | VLDL_P | NA | T | C | 0.026633 | 0.004401 | 4.40E-10 |
| rs545971 | miR-126-3p | trans | | 2.76E-39 | Nightingale | M_VLDL_C | NA | T | C | 0.036205 | 0.004168 | 1.20E-17 |
| rs545971 | miR-126-3p | trans | | 2.76E-39 | Nightingale | XS_VLDL_L | NA | T | C | 0.033813 | 0.00435 | 2.10E-15 |
| rs545971 | miR-126-3p | trans | | 2.76E-39 | Nightingale | M_VLDL_F | NA | T | C | 0.033372 | 0.004424 | 3.10E-14 |
| rs545971 | miR-126-3p | trans | | 2.76E-39 | Nightingale | LDL_L | NA | T | C | 0.033306 | 0.00441 | 7.40E-14 |
| rs545971 | miR-126-3p | trans | | 2.76E-39 | Nightingale | LDL_CE | NA | T | C | 0.032556 | 0.004413 | 2.20E-13 |
| rs545971 | miR-126-3p | trans | | 2.76E-39 | Nightingale | IDL_L | NA | T | C | 0.035268 | 0.004278 | 4.80E-16 |
| rs545971 | miR-126-3p | trans | | 2.76E-39 | Nightingale | S_VLDL_CE | NA | T | C | 0.047034 | 0.004365 | 1.20E-26 |
| rs545971 | miR-126-3p | trans | | 2.76E-39 | Nightingale | XL_VLDL_F | NA | T | C | 0.037195 | 0.00429 | 2.00E-18 |
| rs545971 | miR-126-3p | trans | | 2.76E-39 | Nightingale | IDL_FC | NA | T | C | 0.038254 | 0.004284 | 1.50E-18 |
| rs545971 | miR-126-3p | trans | | 2.76E-39 | Nightingale | XXL_VLDL_ | NA | T | C | 0.030847 | 0.004406 | 1.10E-12 |
| rs545971 | miR-126-3p | trans | | 2.76E-39 | Nightingale | S_VLDL_CE | NA | T | C | 0.038182 | 0.004419 | 1.60E-18 |
| rs545971 | miR-126-3p | trans | | 2.76E-39 | Nightingale | S_HDL_L | NA | T | C | 0.026297 | 0.00441 | 1.50E-09 |
| rs545971 | miR-126-3p | trans | | 2.76E-39 | Nightingale | S_LDL_FC | NA | T | C | 0.028568 | 0.004418 | 4.20E-10 |
| rs545971 | miR-126-3p | trans | | 2.76E-39 | Nightingale | L_VLDL_CE | NA | T | C | 0.047336 | 0.004234 | 2.40E-29 |
| rs545971 | miR-126-3p | trans | | 2.76E-39 | Nightingale | S_VLDL_C_ | NA | T | C | 0.041889 | 0.004306 | 5.20E-22 |
| rs545971 | miR-126-3p | trans | | 2.76E-39 | Nightingale | Citrate | NA | T | C | 0.026829 | 0.004396 | 7.30E-10 |
| rs545971 | miR-126-3p | trans | | 2.76E-39 | Nightingale | L_LDL_C | NA | T | C | 0.032322 | 0.004375 | 2.60E-13 |
| rs545971 | miR-126-3p | trans | | 2.76E-39 | Nightingale | XS_VLDL_F | NA | T | C | 0.031441 | 0.004393 | 2.40E-12 |
| rs545971 | miR-126-3p | trans | | 2.76E-39 | Nightingale | LDL_C | NA | T | C | 0.033196 | 0.004406 | 8.80E-14 |
| rs545971 | miR-126-3p | trans | | 2.76E-39 | Nightingale | IDL_P | NA | T | C | 0.039988 | 0.004378 | 1.30E-19 |
| rs545971 | miR-126-3p | trans | | 2.76E-39 | Nightingale | Clinical_LD | NA | T | C | 0.036477 | 0.004399 | 2.70E-16 |
| rs545971 | miR-126-3p | trans | | 2.76E-39 | Nightingale | S_VLDL_FC | NA | T | C | 0.037905 | 0.004426 | 7.30E-18 |
| rs545971 | miR-126-3p | trans | | 2.76E-39 | Nightingale | S_HDL_PL | NA | T | C | 0.023784 | 0.004386 | 3.20E-08 |
| rs545971 | miR-126-3p | trans | | 2.76E-39 | Nightingale | M_LDL_FC | NA | T | C | 0.034352 | 0.004419 | 1.90E-14 |
| rs545971 | miR-126-3p | trans | | 2.76E-39 | Nightingale | M_VLDL_P | NA | T | C | 0.028296 | 0.004414 | 6.30E-11 |
| rs545971 | miR-126-3p | trans | | 2.76E-39 | Nightingale | XXL_VLDL_ | NA | T | C | -0.03573 | 0.004463 | 2.20E-16 |
| rs545971 | miR-126-3p | trans | | 2.76E-39 | Nightingale | M_VLDL_F | NA | T | C | 0.042064 | 0.00421 | 3.50E-23 |
| rs545971 | miR-126-3p | trans | | 2.76E-39 | Nightingale | LDL_PL | NA | T | C | 0.034991 | 0.004415 | 4.70E-15 |
| rs545971 | miR-126-3p | trans | | 2.76E-39 | Nightingale | L_LDL_P | NA | T | C | 0.029177 | 0.00441 | 6.20E-11 |
| rs545971 | miR-126-3p | trans | | 2.76E-39 | Nightingale | Sphingomy | NA | T | C | 0.029016 | 0.004136 | 6.80E-12 |
| rs545971 | miR-126-3p | trans | | 2.76E-39 | Nightingale | PUFA | NA | T | C | 0.023713 | 0.004316 | 1.40E-08 |
| rs545971 | miR-126-3p | trans | | 2.76E-39 | Nightingale | S_HDL_P | NA | T | C | 0.030615 | 0.004425 | 5.10E-12 |
| rs545971 | miR-126-3p | trans | | 2.76E-39 | Nightingale | XL_VLDL_C | NA | T | C | 0.037351 | 0.004245 | 3.10E-18 |
| rs545971 | miR-126-3p | trans | | 2.76E-39 | Nightingale | M_LDL_P | NA | T | C | 0.035895 | 0.004437 | 6.80E-16 |
| rs545971 | miR-126-3p | trans | | 2.76E-39 | Nightingale | L_LDL_PL | NA | T | C | 0.037992 | 0.004391 | 9.00E-18 |
| rs545971 | miR-126-3p | trans | | 2.76E-39 | Nightingale | Total_L | NA | T | C | 0.026346 | 0.004353 | 4.60E-10 |
| rs545971 | miR-126-3p | trans | | 2.76E-39 | Nightingale | M_LDL_PL | NA | T | C | 0.032471 | 0.004431 | 2.60E-13 |
| rs545971 | miR-126-3p | trans | | 2.76E-39 | Nightingale | M_LDL_TG | NA | T | C | -0.02763 | 0.00437 | 3.20E-10 |
| rs545971 | miR-126-3p | trans | | 2.76E-39 | Nightingale | XL_HDL_PL | NA | T | C | -0.02277 | 0.004065 | 1.10E-08 |
| rs545971 | miR-126-3p | trans | | 2.76E-39 | Nightingale | M_VLDL_T | NA | T | C | -0.03856 | 0.004181 | 6.80E-20 |
| rs545971 | miR-126-3p | trans | | 2.76E-39 | Nightingale | S_VLDL_PL | NA | T | C | 0.032777 | 0.004417 | 4.20E-14 |
| rs545971 | miR-126-3p | trans | | 2.76E-39 | Nightingale | Total_C | NA | T | C | 0.032372 | 0.004284 | 3.30E-14 |
| rs545971 | miR-126-3p | trans | | 2.76E-39 | Nightingale | L_VLDL_TG | NA | T | C | -0.05114 | 0.004404 | 5.30E-32 |
| rs545971 | miR-126-3p | trans | | 2.76E-39 | Nightingale | S_VLDL_TG | NA | T | C | -0.03813 | 0.004283 | 1.40E-18 |
| rs545971 | miR-126-3p | trans | | 2.76E-39 | Nightingale | Cholines | NA | T | C | 0.022288 | 0.004161 | 4.90E-08 |
| rs545971 | miR-126-3p | trans | | 2.76E-39 | Nightingale | L_LDL_L | NA | T | C | 0.033551 | 0.004382 | 3.00E-14 |
| rs545971 | miR-126-3p | trans | | 2.76E-39 | Nightingale | XL_VLDL_P | NA | T | C | 0.038847 | 0.004432 | 5.10E-19 |
| rs545971 | miR-126-3p | trans | | 2.76E-39 | Nightingale | VLDL_FC | NA | T | C | 0.024159 | 0.004392 | 1.10E-08 |
| rs545971 | miR-126-3p | trans | | 2.76E-39 | Nightingale | S_HDL_FC | NA | T | C | 0.02345 | 0.004344 | 4.60E-08 |
| rs545971 | miR-126-3p | trans | | 2.76E-39 | Nightingale | S_HDL_C | NA | T | C | 0.028415 | 0.004417 | 1.70E-10 |
| rs545971 | miR-126-3p | trans | | 2.76E-39 | Nightingale | S_LDL_L | NA | T | C | 0.027745 | 0.004424 | 8.20E-10 |
| rs545971 | miR-126-3p | trans | | 2.76E-39 | Nightingale | S_LDL_C | NA | T | C | 0.030946 | 0.004429 | 5.50E-12 |
| rs545971 | miR-126-3p | trans | | 2.76E-39 | Nightingale | S_VLDL_PL | NA | T | C | 0.02774 | 0.004237 | 1.80E-10 |
| rs545971 | miR-126-3p | trans | | 2.76E-39 | Nightingale | M_VLDL_C | NA | T | C | 0.033628 | 0.004157 | 1.90E-15 |
| rs545971 | miR-126-3p | trans | | 2.76E-39 | Nightingale | S_VLDL_C | NA | T | C | 0.038373 | 0.004424 | 1.80E-18 |
| rs545971 | miR-126-3p | trans | | 2.76E-39 | Nightingale | non_HDL_( | NA | T | C | 0.035698 | 0.004401 | 5.10E-16 |
| rs545971 | miR-126-3p | trans | | 2.76E-39 | Nightingale | XL_VLDL_T | NA | T | C | -0.04392 | 0.004289 | 1.70E-24 |
| rs545971 | miR-126-3p | trans | | 2.76E-39 | Nightingale | XS_VLDL_F | NA | T | C | 0.037065 | 0.004348 | 6.50E-18 |
| rs545971 | miR-126-3p | trans | | 2.76E-39 | Nightingale | XXL_VLDL_ | NA | T | C | 0.025004 | 0.004454 | 1.50E-09 |
| rs545971 | miR-126-3p | trans | | 2.76E-39 | Nightingale | S_LDL_CE_ | NA | T | C | 0.030946 | 0.004417 | 4.90E-14 |
| rs545971 | miR-126-3p | trans | | 2.76E-39 | Nightingale | XXL_VLDL_ | NA | T | C | 0.034827 | 0.004434 | 2.90E-15 |
| rs545971 | miR-126-3p | trans | | 2.76E-39 | Nightingale | XL_VLDL_C | NA | T | C | 0.038222 | 0.004243 | 4.70E-19 |

| | | | | | | | | | | | |
|---|---|---|---|---|---|---|---|---|---|---|---|
| rs545971 | miR-126-3p | trans | 2.76E-39 | Nightingale | M_VLDL_C | NA | T | C | 0.038711 | 0.004413 | 1.90E-18 |
| rs545971 | miR-126-3p | trans | 2.76E-39 | Nightingale | S_VLDL_FC | NA | T | C | 0.029358 | 0.004228 | 1.10E-11 |
| rs545971 | miR-126-3p | trans | 2.76E-39 | Nightingale | IDL_C | NA | T | C | 0.034955 | 0.004273 | 9.40E-16 |
| rs545971 | miR-126-3p | trans | 2.76E-39 | Nightingale | Total_CE | NA | T | C | 0.031745 | 0.004268 | 9.70E-14 |
| rs545971 | miR-126-3p | trans | 2.76E-39 | Nightingale | L_VLDL_FC | NA | T | C | 0.048588 | 0.004382 | 4.50E-29 |
| rs545971 | miR-126-3p | trans | 2.76E-39 | Nightingale | Remnant_( | NA | T | C | 0.036867 | 0.004396 | 3.70E-17 |
| rs545971 | miR-126-3p | trans | 2.76E-39 | Nightingale | VLDL_CE | NA | T | C | 0.035786 | 0.004426 | 3.00E-16 |
| rs545971 | miR-126-3p | trans | 2.76E-39 | Nightingale | XS_VLDL_P | NA | T | C | 0.036698 | 0.004366 | 1.40E-17 |
| rs545971 | miR-126-3p | trans | 2.76E-39 | Nightingale | M_VLDL_P | NA | T | C | 0.029128 | 0.004418 | 2.00E-11 |
| rs545971 | miR-126-3p | trans | 2.76E-39 | Nightingale | IDL_CE | NA | T | C | 0.033507 | 0.004272 | 1.30E-14 |
| rs545971 | miR-126-3p | trans | 2.76E-39 | Nightingale | M_LDL_L | NA | T | C | 0.033035 | 0.004431 | 1.00E-13 |
| rs545971 | miR-126-3p | trans | 2.76E-39 | Nightingale | XS_VLDL_C | NA | T | C | 0.037794 | 0.00427 | 2.10E-18 |
| rs545971 | miR-126-3p | trans | 2.76E-39 | Nightingale | S_LDL_PL_ | NA | T | C | -0.03006 | 0.004417 | 2.60E-13 |
| rs545971 | miR-126-3p | trans | 2.76E-39 | Nightingale | L_LDL_FC | NA | T | C | 0.033627 | 0.004345 | 2.90E-14 |
| rs545971 | miR-126-3p | trans | 2.76E-39 | Nightingale | S_LDL_C_p | NA | T | C | 0.045366 | 0.004442 | 5.40E-25 |
| rs545971 | miR-126-3p | trans | 2.76E-39 | Nightingale | M_LDL_CE | NA | T | C | 0.032977 | 0.004431 | 9.60E-14 |
| rs545971 | miR-126-3p | trans | 2.76E-39 | Nightingale | S_LDL_P | NA | T | C | 0.029589 | 0.004429 | 3.20E-11 |
| rs545971 | miR-126-3p | trans | 2.76E-39 | Nightingale | L_VLDL_C_ | NA | T | C | 0.052192 | 0.004322 | 4.00E-34 |
| rs545971 | miR-126-3p | trans | 2.76E-39 | Nightingale | M_VLDL_P | NA | T | C | 0.044056 | 0.004247 | 5.50E-25 |
| rs545971 | miR-126-3p | trans | 2.76E-39 | Nightingale | M_LDL_C | NA | T | C | 0.033991 | 0.004433 | 2.10E-14 |
| rs545971 | miR-126-3p | trans | 2.76E-39 | Nightingale | S_HDL_CE | NA | T | C | 0.02864 | 0.004432 | 1.30E-10 |
| rs545971 | miR-126-3p | trans | 2.76E-39 | Nightingale | S_VLDL_P | NA | T | C | 0.023465 | 0.004369 | 1.70E-08 |
| rs545971 | miR-126-3p | trans | 2.76E-39 | Nightingale | ApoB_by_A | NA | T | C | 0.025055 | 0.004358 | 1.30E-08 |
| rs545971 | miR-126-3p | trans | 2.76E-39 | Nightingale | LA | NA | T | C | 0.024903 | 0.004339 | 3.70E-09 |
| rs545971 | miR-126-3p | trans | 2.76E-39 | Nightingale | L_LDL_CE | NA | T | C | 0.031569 | 0.004385 | 9.80E-13 |
| rs545971 | miR-126-3p | trans | 2.76E-39 | Nightingale | XXL_VLDL_ | NA | T | C | 0.033119 | 0.004459 | 9.00E-14 |
| rs545971 | miR-126-3p | trans | 2.76E-39 | Nightingale | ApoB | NA | T | C | 0.033939 | 0.004419 | 2.10E-14 |
| rs545971 | miR-126-3p | trans | 2.76E-39 | Nightingale | IDL_FC_pct | NA | T | C | 0.025773 | 0.004391 | 1.10E-08 |
| rs545971 | miR-126-3p | trans | 2.76E-39 | Nightingale | VLDL_C | NA | T | C | 0.031302 | 0.004419 | 4.90E-13 |
| rs545971 | miR-126-3p | trans | 2.76E-39 | Nightingale | XS_VLDL_C | NA | T | C | 0.037989 | 0.004295 | 1.30E-18 |
| rs545971 | miR-126-3p | trans | 2.76E-39 | Nightingale | S_VLDL_L | NA | T | C | 0.024668 | 0.004382 | 3.70E-09 |
| rs545971 | miR-126-3p | trans | 2.76E-39 | Nightingale | LDL_P | NA | T | C | 0.031801 | 0.004422 | 1.00E-12 |
| rs545971 | miR-126-3p | trans | 2.76E-39 | Nightingale | S_LDL_CE | NA | T | C | 0.030551 | 0.004427 | 5.80E-12 |
| rs545971 | miR-126-3p | trans | 2.76E-39 | Nightingale | M_VLDL_C | NA | T | C | 0.040844 | 0.004385 | 2.30E-20 |
| rs545971 | miR-126-3p | trans | 2.76E-39 | Nightingale | Total_PL | NA | T | C | 0.024922 | 0.004213 | 1.10E-09 |
| rs545971 | miR-126-3p | trans | 2.76E-39 | Nightingale | Omega_6 | NA | T | C | 0.025889 | 0.004321 | 5.80E-10 |
| rs545971 | miR-126-3p | trans | 2.76E-39 | Nightingale | Total_FC | NA | T | C | 0.033253 | 0.004323 | 6.80E-15 |
| rs550057 | miR-139-5p | trans | 5.52E-32 | Nightingale | S_LDL_P | NA | T | C | 0.040014 | 0.004743 | 7.70E-18 |
| rs550057 | miR-139-5p | trans | 5.52E-32 | Nightingale | S_LDL_C_p | NA | T | C | 0.059583 | 0.004757 | 3.20E-37 |
| rs550057 | miR-139-5p | trans | 5.52E-32 | Nightingale | XXL_VLDL_ | NA | T | C | 0.040046 | 0.004752 | 1.60E-17 |
| rs550057 | miR-139-5p | trans | 5.52E-32 | Nightingale | L_LDL_PL | NA | T | C | 0.052492 | 0.004703 | 1.00E-29 |
| rs550057 | miR-139-5p | trans | 5.52E-32 | Nightingale | IDL_CE | NA | T | C | 0.046199 | 0.004575 | 1.80E-24 |
| rs550057 | miR-139-5p | trans | 5.52E-32 | Nightingale | S_LDL_C | NA | T | C | 0.043673 | 0.004743 | 1.20E-20 |
| rs550057 | miR-139-5p | trans | 5.52E-32 | Nightingale | XL_VLDL_C | NA | T | C | 0.025867 | 0.004666 | 2.80E-09 |
| rs550057 | miR-139-5p | trans | 5.52E-32 | Nightingale | M_LDL_CE | NA | T | C | 0.044363 | 0.004745 | 9.80E-22 |
| rs550057 | miR-139-5p | trans | 5.52E-32 | Nightingale | XS_VLDL_T | NA | T | C | -0.03521 | 0.004505 | 1.60E-14 |
| rs550057 | miR-139-5p | trans | 5.52E-32 | Nightingale | L_LDL_CE | NA | T | C | 0.045326 | 0.004697 | 1.00E-22 |
| rs550057 | miR-139-5p | trans | 5.52E-32 | Nightingale | Omega_6 | NA | T | C | 0.03221 | 0.004628 | 1.30E-13 |
| rs550057 | miR-139-5p | trans | 5.52E-32 | Nightingale | S_LDL_PL_ | NA | T | C | -0.03504 | 0.00473 | 8.30E-16 |
| rs550057 | miR-139-5p | trans | 5.52E-32 | Nightingale | LDL_L | NA | T | C | 0.046916 | 0.004723 | 6.60E-24 |
| rs550057 | miR-139-5p | trans | 5.52E-32 | Nightingale | S_HDL_C | NA | T | C | 0.037754 | 0.004731 | 1.40E-15 |
| rs550057 | miR-139-5p | trans | 5.52E-32 | Nightingale | IDL_C | NA | T | C | 0.047774 | 0.004577 | 5.10E-26 |
| rs550057 | miR-139-5p | trans | 5.52E-32 | Nightingale | S_HDL_CE | NA | T | C | 0.038488 | 0.004746 | 6.00E-16 |
| rs550057 | miR-139-5p | trans | 5.52E-32 | Nightingale | S_LDL_CE | NA | T | C | 0.04208 | 0.004741 | 1.20E-19 |
| rs550057 | miR-139-5p | trans | 5.52E-32 | Nightingale | XS_VLDL_C | NA | T | C | 0.032181 | 0.004471 | 2.20E-12 |
| rs550057 | miR-139-5p | trans | 5.52E-32 | Nightingale | L_LDL_P | NA | T | C | 0.040754 | 0.004723 | 2.20E-18 |
| rs550057 | miR-139-5p | trans | 5.52E-32 | Nightingale | IDL_L | NA | T | C | 0.047298 | 0.004582 | 1.10E-25 |
| rs550057 | miR-139-5p | trans | 5.52E-32 | Nightingale | L_LDL_TG_ | NA | T | C | -0.03317 | 0.004648 | 1.40E-12 |
| rs550057 | miR-139-5p | trans | 5.52E-32 | Nightingale | S_HDL_P | NA | T | C | 0.040281 | 0.004739 | 1.30E-17 |
| rs550057 | miR-139-5p | trans | 5.52E-32 | Nightingale | M_LDL_P | NA | T | C | 0.046745 | 0.004752 | 9.50E-24 |
| rs550057 | miR-139-5p | trans | 5.52E-32 | Nightingale | XL_HDL_PL | NA | T | C | -0.03225 | 0.004354 | 1.10E-13 |
| rs550057 | miR-139-5p | trans | 5.52E-32 | Nightingale | M_VLDL_F | NA | T | C | 0.054228 | 0.004508 | 3.10E-33 |
| rs550057 | miR-139-5p | trans | 5.52E-32 | Nightingale | M_VLDL_P | NA | T | C | 0.036129 | 0.004727 | 1.70E-15 |

| | | | | | | | | | | | |
|---|---|---|---|---|---|---|---|---|---|---|---|
| rs550057 | miR-139-5p | trans | 5.52E-32 | Nightingale | L_LDL_L | NA | T | C | 0.047551 | 0.004693 | 6.80E-25 |
| rs550057 | miR-139-5p | trans | 5.52E-32 | Nightingale | XXL_VLDL_ | NA | T | C | -0.04063 | 0.004782 | 2.80E-18 |
| rs550057 | miR-139-5p | trans | 5.52E-32 | Nightingale | XXL_VLDL_ | NA | T | C | 0.025317 | 0.004772 | 7.10E-09 |
| rs550057 | miR-139-5p | trans | 5.52E-32 | Nightingale | S_VLDL_L | NA | T | C | 0.029085 | 0.004693 | 3.50E-11 |
| rs550057 | miR-139-5p | trans | 5.52E-32 | Nightingale | XS_VLDL_C | NA | T | C | 0.046509 | 0.0046 | 3.90E-25 |
| rs550057 | miR-139-5p | trans | 5.52E-32 | Nightingale | XS_VLDL_P | NA | T | C | 0.043776 | 0.004676 | 1.80E-22 |
| rs550057 | miR-139-5p | trans | 5.52E-32 | Nightingale | M_LDL_C_ | NA | T | C | 0.038134 | 0.004742 | 4.30E-16 |
| rs550057 | miR-139-5p | trans | 5.52E-32 | Nightingale | LDL_FC | NA | T | C | 0.049134 | 0.004688 | 7.00E-26 |
| rs550057 | miR-139-5p | trans | 5.52E-32 | Nightingale | Cholines | NA | T | C | 0.02627 | 0.004457 | 3.90E-10 |
| rs550057 | miR-139-5p | trans | 5.52E-32 | Nightingale | LDL_PL | NA | T | C | 0.049193 | 0.004728 | 6.40E-26 |
| rs550057 | miR-139-5p | trans | 5.52E-32 | Nightingale | LDL_CE | NA | T | C | 0.04573 | 0.004726 | 6.60E-23 |
| rs550057 | miR-139-5p | trans | 5.52E-32 | Nightingale | S_VLDL_CE | NA | T | C | 0.047282 | 0.004733 | 6.20E-25 |
| rs550057 | miR-139-5p | trans | 5.52E-32 | Nightingale | ApoB_by_A | NA | T | C | 0.034854 | 0.004667 | 5.50E-14 |
| rs550057 | miR-139-5p | trans | 5.52E-32 | Nightingale | XS_VLDL_F | NA | T | C | 0.043814 | 0.004657 | 1.60E-22 |
| rs550057 | miR-139-5p | trans | 5.52E-32 | Nightingale | S_LDL_FC | NA | T | C | 0.043269 | 0.004731 | 8.00E-20 |
| rs550057 | miR-139-5p | trans | 5.52E-32 | Nightingale | S_LDL_L | NA | T | C | 0.039564 | 0.004738 | 3.50E-17 |
| rs550057 | miR-139-5p | trans | 5.52E-32 | Nightingale | M_LDL_FC | NA | T | C | 0.050019 | 0.004733 | 1.70E-26 |
| rs550057 | miR-139-5p | trans | 5.52E-32 | Nightingale | M_LDL_TG | NA | T | C | -0.04255 | 0.00468 | 8.30E-20 |
| rs550057 | miR-139-5p | trans | 5.52E-32 | Nightingale | L_LDL_C | NA | T | C | 0.046611 | 0.004685 | 5.60E-24 |
| rs550057 | miR-139-5p | trans | 5.52E-32 | Nightingale | L_VLDL_C_ | NA | T | C | 0.064893 | 0.004628 | 1.20E-45 |
| rs550057 | miR-139-5p | trans | 5.52E-32 | Nightingale | Remnant_C | NA | T | C | 0.047674 | 0.004708 | 1.40E-25 |
| rs550057 | miR-139-5p | trans | 5.52E-32 | Nightingale | M_VLDL_T | NA | T | C | -0.05036 | 0.004478 | 4.10E-29 |
| rs550057 | miR-139-5p | trans | 5.52E-32 | Nightingale | XS_VLDL_L | NA | T | C | 0.039312 | 0.004658 | 8.60E-19 |
| rs550057 | miR-139-5p | trans | 5.52E-32 | Nightingale | XXL_VLDL_ | NA | T | C | 0.033136 | 0.004721 | 8.10E-13 |
| rs550057 | miR-139-5p | trans | 5.52E-32 | Nightingale | M_VLDL_C | NA | T | C | 0.0536 | 0.004696 | 7.40E-31 |
| rs550057 | miR-139-5p | trans | 5.52E-32 | Nightingale | IDL_FC | NA | T | C | 0.051144 | 0.004588 | 1.70E-29 |
| rs550057 | miR-139-5p | trans | 5.52E-32 | Nightingale | S_LDL_CE_ | NA | T | C | 0.03581 | 0.004731 | 2.00E-16 |
| rs550057 | miR-139-5p | trans | 5.52E-32 | Nightingale | M_VLDL_C | NA | T | C | 0.044675 | 0.004452 | 2.60E-23 |
| rs550057 | miR-139-5p | trans | 5.52E-32 | Nightingale | IDL_FC_pct | NA | T | C | 0.034868 | 0.004702 | 4.70E-13 |
| rs550057 | miR-139-5p | trans | 5.52E-32 | Nightingale | M_VLDL_P | NA | T | C | 0.03714 | 0.004732 | 3.20E-16 |
| rs550057 | miR-139-5p | trans | 5.52E-32 | Nightingale | LDL_P | NA | T | C | 0.043296 | 0.004735 | 1.40E-20 |
| rs550057 | miR-139-5p | trans | 5.52E-32 | Nightingale | VLDL_FC | NA | T | C | 0.02878 | 0.004704 | 9.50E-11 |
| rs550057 | miR-139-5p | trans | 5.52E-32 | Nightingale | S_VLDL_C_ | NA | T | C | 0.056984 | 0.004612 | 6.60E-35 |
| rs550057 | miR-139-5p | trans | 5.52E-32 | Nightingale | LDL_C | NA | T | C | 0.047148 | 0.004718 | 3.90E-24 |
| rs550057 | miR-139-5p | trans | 5.52E-32 | Nightingale | S_HDL_L | NA | T | C | 0.0316 | 0.004722 | 5.20E-12 |
| rs550057 | miR-139-5p | trans | 5.52E-32 | Nightingale | S_VLDL_PL | NA | T | C | 0.040569 | 0.004538 | 1.10E-18 |
| rs550057 | miR-139-5p | trans | 5.52E-32 | Nightingale | HDL_size | NA | T | C | -0.02598 | 0.004202 | 4.40E-10 |
| rs550057 | miR-139-5p | trans | 5.52E-32 | Nightingale | Phosphogly | NA | T | C | 0.023356 | 0.004489 | 3.00E-08 |
| rs550057 | miR-139-5p | trans | 5.52E-32 | Nightingale | XL_VLDL_C | NA | T | C | 0.048624 | 0.004544 | 3.40E-26 |
| rs550057 | miR-139-5p | trans | 5.52E-32 | Nightingale | S_HDL_PL | NA | T | C | 0.026717 | 0.004698 | 2.80E-09 |
| rs550057 | miR-139-5p | trans | 5.52E-32 | Nightingale | Total_C | NA | T | C | 0.043514 | 0.004588 | 1.70E-22 |
| rs550057 | miR-139-5p | trans | 5.52E-32 | Nightingale | XL_VLDL_C | NA | T | C | 0.048184 | 0.004547 | 1.10E-25 |
| rs550057 | miR-139-5p | trans | 5.52E-32 | Nightingale | Citrate | NA | T | C | 0.026542 | 0.004708 | 8.90E-09 |
| rs550057 | miR-139-5p | trans | 5.52E-32 | Nightingale | M_LDL_L | NA | T | C | 0.045701 | 0.004745 | 8.50E-23 |
| rs550057 | miR-139-5p | trans | 5.52E-32 | Nightingale | S_HDL_FC | NA | T | C | 0.029798 | 0.004652 | 2.70E-11 |
| rs550057 | miR-139-5p | trans | 5.52E-32 | Nightingale | IDL_PL | NA | T | C | 0.046477 | 0.004573 | 4.80E-25 |
| rs550057 | miR-139-5p | trans | 5.52E-32 | Nightingale | Total_PL | NA | T | C | 0.030079 | 0.004512 | 1.20E-12 |
| rs550057 | miR-139-5p | trans | 5.52E-32 | Nightingale | S_LDL_PL | NA | T | C | 0.034085 | 0.004728 | 4.70E-13 |
| rs550057 | miR-139-5p | trans | 5.52E-32 | Nightingale | IDL_P | NA | T | C | 0.053052 | 0.004689 | 2.00E-30 |
| rs550057 | miR-139-5p | trans | 5.52E-32 | Nightingale | Total_P | NA | T | C | 0.025587 | 0.00446 | 1.70E-09 |
| rs550057 | miR-139-5p | trans | 5.52E-32 | Nightingale | M_VLDL_C | NA | T | C | 0.050328 | 0.004726 | 1.60E-27 |
| rs550057 | miR-139-5p | trans | 5.52E-32 | Nightingale | L_VLDL_CE | NA | T | C | 0.027055 | 0.004683 | 7.80E-10 |
| rs550057 | miR-139-5p | trans | 5.52E-32 | Nightingale | S_VLDL_FC | NA | T | C | 0.042888 | 0.004528 | 6.60E-21 |
| rs550057 | miR-139-5p | trans | 5.52E-32 | Nightingale | VLDL_CE | NA | T | C | 0.044291 | 0.00474 | 5.90E-22 |
| rs550057 | miR-139-5p | trans | 5.52E-32 | Nightingale | XS_VLDL_C | NA | T | C | 0.046905 | 0.004573 | 1.30E-25 |
| rs550057 | miR-139-5p | trans | 5.52E-32 | Nightingale | L_VLDL_TG | NA | T | C | -0.06228 | 0.004716 | 4.90E-41 |
| rs550057 | miR-139-5p | trans | 5.52E-32 | Nightingale | M_LDL_PL | NA | T | C | 0.046345 | 0.004746 | 2.60E-23 |
| rs550057 | miR-139-5p | trans | 5.52E-32 | Nightingale | L_VLDL_FC | NA | T | C | 0.06018 | 0.004693 | 3.20E-38 |
| rs550057 | miR-139-5p | trans | 5.52E-32 | Nightingale | Total_L | NA | T | C | 0.033368 | 0.004662 | 4.10E-14 |
| rs550057 | miR-139-5p | trans | 5.52E-32 | Nightingale | Sphingomy | NA | T | C | 0.03676 | 0.00443 | 6.30E-17 |
| rs550057 | miR-139-5p | trans | 5.52E-32 | Nightingale | M_VLDL_F | NA | T | C | 0.042701 | 0.004738 | 2.00E-20 |
| rs550057 | miR-139-5p | trans | 5.52E-32 | Nightingale | XS_VLDL_C | NA | T | C | 0.035005 | 0.004494 | 2.30E-14 |
| rs550057 | miR-139-5p | trans | 5.52E-32 | Nightingale | IDL_C_pct | NA | T | C | 0.031128 | 0.004629 | 3.40E-11 |

| | | | | | | | | | | | |
|---|---|---|---|---|---|---|---|---|---|---|---|
| rs550057 | miR-139-5p | trans | 5.52E-32 | Nightingale | XL_VLDL_T | NA | T | C | -0.05437 | 0.004594 | 3.30E-32 |
| rs550057 | miR-139-5p | trans | 5.52E-32 | Nightingale | M_LDL_C | NA | T | C | 0.046775 | 0.004747 | 1.00E-23 |
| rs550057 | miR-139-5p | trans | 5.52E-32 | Nightingale | L_VLDL_CE | NA | T | C | 0.058972 | 0.004535 | 2.60E-39 |
| rs550057 | miR-139-5p | trans | 5.52E-32 | Nightingale | S_VLDL_C | NA | T | C | 0.048078 | 0.004738 | 1.80E-25 |
| rs550057 | miR-139-5p | trans | 5.52E-32 | Nightingale | M_VLDL_P | NA | T | C | 0.05684 | 0.004548 | 4.30E-36 |
| rs550057 | miR-139-5p | trans | 5.52E-32 | Nightingale | PUFA | NA | T | C | 0.029052 | 0.004622 | 2.60E-11 |
| rs550057 | miR-139-5p | trans | 5.52E-32 | Nightingale | M_VLDL_C | NA | T | C | 0.047633 | 0.004464 | 3.30E-26 |
| rs550057 | miR-139-5p | trans | 5.52E-32 | Nightingale | XL_VLDL_P | NA | T | C | 0.046629 | 0.004747 | 7.70E-24 |
| rs550057 | miR-139-5p | trans | 5.52E-32 | Nightingale | Total_FC | NA | T | C | 0.043696 | 0.004629 | 8.00E-23 |
| rs550057 | miR-139-5p | trans | 5.52E-32 | Nightingale | S_VLDL_CE | NA | T | C | 0.061711 | 0.004675 | 2.00E-39 |
| rs550057 | miR-139-5p | trans | 5.52E-32 | Nightingale | non_HDL_( | NA | T | C | 0.048348 | 0.004713 | 9.80E-26 |
| rs550057 | miR-139-5p | trans | 5.52E-32 | Nightingale | S_VLDL_TG | NA | T | C | -0.05282 | 0.004587 | 2.10E-30 |
| rs550057 | miR-139-5p | trans | 5.52E-32 | Nightingale | XXL_VLDL_ | NA | T | C | 0.040178 | 0.004778 | 1.80E-17 |
| rs550057 | miR-139-5p | trans | 5.52E-32 | Nightingale | L_LDL_FC | NA | T | C | 0.048951 | 0.004653 | 3.20E-26 |
| rs550057 | miR-139-5p | trans | 5.52E-32 | Nightingale | IDL_TG_pc | NA | T | C | -0.03163 | 0.004581 | 7.70E-12 |
| rs550057 | miR-139-5p | trans | 5.52E-32 | Nightingale | XS_VLDL_F | NA | T | C | 0.043373 | 0.004705 | 1.40E-19 |
| rs550057 | miR-139-5p | trans | 5.52E-32 | Nightingale | Glucose | NA | T | C | 0.026619 | 0.00475 | 2.10E-08 |
| rs550057 | miR-139-5p | trans | 5.52E-32 | Nightingale | M_VLDL_L | NA | T | C | 0.026949 | 0.004707 | 1.30E-09 |
| rs550057 | miR-139-5p | trans | 5.52E-32 | Nightingale | Total_CE | NA | T | C | 0.043093 | 0.004571 | 3.60E-22 |
| rs550057 | miR-139-5p | trans | 5.52E-32 | Nightingale | GlycA | NA | T | C | 0.026618 | 0.004719 | 1.00E-08 |
| rs550057 | miR-139-5p | trans | 5.52E-32 | Nightingale | S_VLDL_PL | NA | T | C | 0.040787 | 0.00473 | 3.40E-19 |
| rs550057 | miR-139-5p | trans | 5.52E-32 | Nightingale | Clinical_LD | NA | T | C | 0.051215 | 0.004711 | 4.80E-28 |
| rs550057 | miR-139-5p | trans | 5.52E-32 | Nightingale | XL_VLDL_F | NA | T | C | 0.04531 | 0.004594 | 2.40E-23 |
| rs550057 | miR-139-5p | trans | 5.52E-32 | Nightingale | S_VLDL_P | NA | T | C | 0.027467 | 0.004679 | 2.70E-10 |
| rs550057 | miR-139-5p | trans | 5.52E-32 | Nightingale | LA | NA | T | C | 0.032534 | 0.004647 | 1.40E-13 |
| rs550057 | miR-139-5p | trans | 5.52E-32 | Nightingale | VLDL_P | NA | T | C | 0.031898 | 0.004713 | 1.00E-12 |
| rs550057 | miR-139-5p | trans | 5.52E-32 | Nightingale | XL_HDL_C_ | NA | T | C | 0.03137 | 0.004525 | 1.90E-11 |
| rs550057 | miR-139-5p | trans | 5.52E-32 | Nightingale | S_LDL_TG_ | NA | T | C | -0.03461 | 0.004595 | 2.30E-13 |
| rs550057 | miR-139-5p | trans | 5.52E-32 | Nightingale | XS_VLDL_P | NA | T | C | 0.03659 | 0.004673 | 2.30E-16 |
| rs550057 | miR-139-5p | trans | 5.52E-32 | Nightingale | VLDL_C | NA | T | C | 0.038243 | 0.004732 | 4.20E-17 |
| rs550057 | miR-139-5p | trans | 5.52E-32 | Nightingale | ApoB | NA | T | C | 0.045452 | 0.004733 | 1.20E-22 |
| rs550057 | miR-139-5p | trans | 5.52E-32 | Nightingale | Pyruvate | NA | T | C | 0.027561 | 0.004747 | 6.40E-09 |
| rs550057 | miR-139-5p | trans | 5.52E-32 | Nightingale | S_VLDL_FC | NA | T | C | 0.048419 | 0.00474 | 1.40E-25 |
| rs550057 | miR-139-5p | trans | 5.52E-32 | Nightingale | XL_HDL_PL | NA | T | C | -0.02299 | 0.004263 | 5.00E-08 |
| rs550057 | miR-10b-5p | trans | 1.61E-27 | Nightingale | S_LDL_P | NA | T | C | 0.040014 | 0.004743 | 7.70E-18 |
| rs550057 | miR-10b-5p | trans | 1.61E-27 | Nightingale | S_LDL_C_p | NA | T | C | 0.059583 | 0.004757 | 3.20E-37 |
| rs550057 | miR-10b-5p | trans | 1.61E-27 | Nightingale | XXL_VLDL_ | NA | T | C | 0.040046 | 0.004752 | 1.60E-17 |
| rs550057 | miR-10b-5p | trans | 1.61E-27 | Nightingale | L_LDL_PL | NA | T | C | 0.052492 | 0.004703 | 1.00E-29 |
| rs550057 | miR-10b-5p | trans | 1.61E-27 | Nightingale | IDL_CE | NA | T | C | 0.046199 | 0.004575 | 1.80E-24 |
| rs550057 | miR-10b-5p | trans | 1.61E-27 | Nightingale | S_LDL_C | NA | T | C | 0.043673 | 0.004743 | 1.20E-20 |
| rs550057 | miR-10b-5p | trans | 1.61E-27 | Nightingale | XL_VLDL_C | NA | T | C | 0.025867 | 0.004666 | 2.80E-09 |
| rs550057 | miR-10b-5p | trans | 1.61E-27 | Nightingale | M_LDL_CE | NA | T | C | 0.044363 | 0.004745 | 9.80E-22 |
| rs550057 | miR-10b-5p | trans | 1.61E-27 | Nightingale | XS_VLDL_T | NA | T | C | -0.03521 | 0.004505 | 1.60E-14 |
| rs550057 | miR-10b-5p | trans | 1.61E-27 | Nightingale | L_LDL_CE | NA | T | C | 0.045326 | 0.004697 | 1.00E-22 |
| rs550057 | miR-10b-5p | trans | 1.61E-27 | Nightingale | Omega_6 | NA | T | C | 0.03221 | 0.004628 | 1.30E-13 |
| rs550057 | miR-10b-5p | trans | 1.61E-27 | Nightingale | S_LDL_PL_ | NA | T | C | -0.03504 | 0.00473 | 8.30E-16 |
| rs550057 | miR-10b-5p | trans | 1.61E-27 | Nightingale | LDL_L | NA | T | C | 0.046916 | 0.004723 | 6.60E-24 |
| rs550057 | miR-10b-5p | trans | 1.61E-27 | Nightingale | S_HDL_C | NA | T | C | 0.037754 | 0.004731 | 1.40E-15 |
| rs550057 | miR-10b-5p | trans | 1.61E-27 | Nightingale | IDL_C | NA | T | C | 0.047774 | 0.004577 | 5.10E-26 |
| rs550057 | miR-10b-5p | trans | 1.61E-27 | Nightingale | S_HDL_CE | NA | T | C | 0.038488 | 0.004746 | 6.00E-16 |
| rs550057 | miR-10b-5p | trans | 1.61E-27 | Nightingale | S_LDL_CE | NA | T | C | 0.04208 | 0.004741 | 1.20E-19 |
| rs550057 | miR-10b-5p | trans | 1.61E-27 | Nightingale | XS_VLDL_C | NA | T | C | 0.032181 | 0.004471 | 2.20E-12 |
| rs550057 | miR-10b-5p | trans | 1.61E-27 | Nightingale | L_LDL_P | NA | T | C | 0.040754 | 0.004723 | 2.20E-18 |
| rs550057 | miR-10b-5p | trans | 1.61E-27 | Nightingale | IDL_L | NA | T | C | 0.047298 | 0.004582 | 1.10E-25 |
| rs550057 | miR-10b-5p | trans | 1.61E-27 | Nightingale | L_LDL_TG_ | NA | T | C | -0.03317 | 0.004648 | 1.40E-12 |
| rs550057 | miR-10b-5p | trans | 1.61E-27 | Nightingale | S_HDL_P | NA | T | C | 0.040281 | 0.004739 | 1.30E-17 |
| rs550057 | miR-10b-5p | trans | 1.61E-27 | Nightingale | M_LDL_P | NA | T | C | 0.046745 | 0.004752 | 9.50E-24 |
| rs550057 | miR-10b-5p | trans | 1.61E-27 | Nightingale | XL_HDL_PL | NA | T | C | -0.03225 | 0.004354 | 1.10E-13 |
| rs550057 | miR-10b-5p | trans | 1.61E-27 | Nightingale | M_VLDL_F | NA | T | C | 0.054228 | 0.004508 | 3.10E-33 |
| rs550057 | miR-10b-5p | trans | 1.61E-27 | Nightingale | M_VLDL_P | NA | T | C | 0.036129 | 0.004727 | 1.70E-15 |
| rs550057 | miR-10b-5p | trans | 1.61E-27 | Nightingale | L_LDL_L | NA | T | C | 0.047551 | 0.004693 | 6.80E-25 |
| rs550057 | miR-10b-5p | trans | 1.61E-27 | Nightingale | XXL_VLDL_ | NA | T | C | -0.04063 | 0.004782 | 2.80E-18 |
| rs550057 | miR-10b-5p | trans | 1.61E-27 | Nightingale | XXL_VLDL_ | NA | T | C | 0.025317 | 0.004772 | 7.10E-09 |

| | | | | | | | | | | | |
|---|---|---|---|---|---|---|---|---|---|---|---|
| rs550057 | miR-10b-5p | trans | 1.61E-27 | Nightingale | S_VLDL_L | NA | T | C | 0.029085 | 0.004693 | 3.50E-11 |
| rs550057 | miR-10b-5p | trans | 1.61E-27 | Nightingale | XS_VLDL_C | NA | T | C | 0.046509 | 0.0046 | 3.90E-25 |
| rs550057 | miR-10b-5p | trans | 1.61E-27 | Nightingale | XS_VLDL_P | NA | T | C | 0.043776 | 0.004676 | 1.80E-22 |
| rs550057 | miR-10b-5p | trans | 1.61E-27 | Nightingale | M_LDL_C_ | NA | T | C | 0.038134 | 0.004742 | 4.30E-16 |
| rs550057 | miR-10b-5p | trans | 1.61E-27 | Nightingale | LDL_FC | NA | T | C | 0.049134 | 0.004688 | 7.00E-26 |
| rs550057 | miR-10b-5p | trans | 1.61E-27 | Nightingale | Cholines | NA | T | C | 0.02627 | 0.004457 | 3.90E-10 |
| rs550057 | miR-10b-5p | trans | 1.61E-27 | Nightingale | LDL_PL | NA | T | C | 0.049193 | 0.004728 | 6.40E-26 |
| rs550057 | miR-10b-5p | trans | 1.61E-27 | Nightingale | LDL_CE | NA | T | C | 0.04573 | 0.004726 | 6.60E-23 |
| rs550057 | miR-10b-5p | trans | 1.61E-27 | Nightingale | S_VLDL_CE | NA | T | C | 0.047282 | 0.004733 | 6.20E-25 |
| rs550057 | miR-10b-5p | trans | 1.61E-27 | Nightingale | ApoB_by_A | NA | T | C | 0.034854 | 0.004667 | 5.50E-14 |
| rs550057 | miR-10b-5p | trans | 1.61E-27 | Nightingale | XS_VLDL_F | NA | T | C | 0.043814 | 0.004657 | 1.60E-22 |
| rs550057 | miR-10b-5p | trans | 1.61E-27 | Nightingale | S_LDL_FC | NA | T | C | 0.043269 | 0.004731 | 8.00E-20 |
| rs550057 | miR-10b-5p | trans | 1.61E-27 | Nightingale | S_LDL_L | NA | T | C | 0.039564 | 0.004738 | 3.50E-17 |
| rs550057 | miR-10b-5p | trans | 1.61E-27 | Nightingale | M_LDL_FC | NA | T | C | 0.050019 | 0.004733 | 1.70E-26 |
| rs550057 | miR-10b-5p | trans | 1.61E-27 | Nightingale | M_LDL_TG | NA | T | C | -0.04255 | 0.00468 | 8.30E-20 |
| rs550057 | miR-10b-5p | trans | 1.61E-27 | Nightingale | L_LDL_C | NA | T | C | 0.046611 | 0.004685 | 5.60E-24 |
| rs550057 | miR-10b-5p | trans | 1.61E-27 | Nightingale | L_VLDL_C_ | NA | T | C | 0.064893 | 0.004628 | 1.20E-45 |
| rs550057 | miR-10b-5p | trans | 1.61E-27 | Nightingale | Remnant_( | NA | T | C | 0.047674 | 0.004708 | 1.40E-25 |
| rs550057 | miR-10b-5p | trans | 1.61E-27 | Nightingale | M_VLDL_T | NA | T | C | -0.05036 | 0.004478 | 4.10E-29 |
| rs550057 | miR-10b-5p | trans | 1.61E-27 | Nightingale | XS_VLDL_L | NA | T | C | 0.039312 | 0.004658 | 8.60E-19 |
| rs550057 | miR-10b-5p | trans | 1.61E-27 | Nightingale | XXL_VLDL_ | NA | T | C | 0.033136 | 0.004721 | 8.10E-13 |
| rs550057 | miR-10b-5p | trans | 1.61E-27 | Nightingale | M_VLDL_C | NA | T | C | 0.0536 | 0.004696 | 7.40E-31 |
| rs550057 | miR-10b-5p | trans | 1.61E-27 | Nightingale | IDL_FC | NA | T | C | 0.051144 | 0.004588 | 1.70E-29 |
| rs550057 | miR-10b-5p | trans | 1.61E-27 | Nightingale | S_LDL_CE_ | NA | T | C | 0.03581 | 0.004731 | 2.00E-16 |
| rs550057 | miR-10b-5p | trans | 1.61E-27 | Nightingale | M_VLDL_C | NA | T | C | 0.044675 | 0.004452 | 2.60E-23 |
| rs550057 | miR-10b-5p | trans | 1.61E-27 | Nightingale | IDL_FC_pct | NA | T | C | 0.034868 | 0.004702 | 4.70E-13 |
| rs550057 | miR-10b-5p | trans | 1.61E-27 | Nightingale | M_VLDL_P | NA | T | C | 0.03714 | 0.004732 | 3.20E-16 |
| rs550057 | miR-10b-5p | trans | 1.61E-27 | Nightingale | LDL_P | NA | T | C | 0.043296 | 0.004735 | 1.40E-20 |
| rs550057 | miR-10b-5p | trans | 1.61E-27 | Nightingale | VLDL_FC | NA | T | C | 0.02878 | 0.004704 | 9.50E-11 |
| rs550057 | miR-10b-5p | trans | 1.61E-27 | Nightingale | S_VLDL_C_ | NA | T | C | 0.056984 | 0.004612 | 6.60E-35 |
| rs550057 | miR-10b-5p | trans | 1.61E-27 | Nightingale | LDL_C | NA | T | C | 0.047148 | 0.004718 | 3.90E-24 |
| rs550057 | miR-10b-5p | trans | 1.61E-27 | Nightingale | S_HDL_L | NA | T | C | 0.0316 | 0.004722 | 5.20E-12 |
| rs550057 | miR-10b-5p | trans | 1.61E-27 | Nightingale | S_VLDL_PL | NA | T | C | 0.040569 | 0.004538 | 1.10E-18 |
| rs550057 | miR-10b-5p | trans | 1.61E-27 | Nightingale | HDL_size | NA | T | C | -0.02598 | 0.004202 | 4.40E-10 |
| rs550057 | miR-10b-5p | trans | 1.61E-27 | Nightingale | Phosphogly | NA | T | C | 0.023356 | 0.004489 | 3.00E-08 |
| rs550057 | miR-10b-5p | trans | 1.61E-27 | Nightingale | XL_VLDL_C | NA | T | C | 0.048624 | 0.004544 | 3.40E-26 |
| rs550057 | miR-10b-5p | trans | 1.61E-27 | Nightingale | S_HDL_PL | NA | T | C | 0.026717 | 0.004698 | 2.80E-09 |
| rs550057 | miR-10b-5p | trans | 1.61E-27 | Nightingale | Total_C | NA | T | C | 0.043514 | 0.004588 | 1.70E-22 |
| rs550057 | miR-10b-5p | trans | 1.61E-27 | Nightingale | XL_VLDL_C | NA | T | C | 0.048184 | 0.004547 | 1.10E-25 |
| rs550057 | miR-10b-5p | trans | 1.61E-27 | Nightingale | Citrate | NA | T | C | 0.026542 | 0.004708 | 8.90E-09 |
| rs550057 | miR-10b-5p | trans | 1.61E-27 | Nightingale | M_LDL_L | NA | T | C | 0.045701 | 0.004745 | 8.50E-23 |
| rs550057 | miR-10b-5p | trans | 1.61E-27 | Nightingale | S_HDL_FC | NA | T | C | 0.029798 | 0.004652 | 2.70E-11 |
| rs550057 | miR-10b-5p | trans | 1.61E-27 | Nightingale | IDL_PL | NA | T | C | 0.046477 | 0.004573 | 4.80E-25 |
| rs550057 | miR-10b-5p | trans | 1.61E-27 | Nightingale | Total_PL | NA | T | C | 0.030079 | 0.004512 | 1.20E-12 |
| rs550057 | miR-10b-5p | trans | 1.61E-27 | Nightingale | S_LDL_PL | NA | T | C | 0.034085 | 0.004728 | 4.70E-13 |
| rs550057 | miR-10b-5p | trans | 1.61E-27 | Nightingale | IDL_P | NA | T | C | 0.053052 | 0.004689 | 2.00E-30 |
| rs550057 | miR-10b-5p | trans | 1.61E-27 | Nightingale | Total_P | NA | T | C | 0.025587 | 0.00446 | 1.70E-09 |
| rs550057 | miR-10b-5p | trans | 1.61E-27 | Nightingale | M_VLDL_C | NA | T | C | 0.050328 | 0.004726 | 1.60E-27 |
| rs550057 | miR-10b-5p | trans | 1.61E-27 | Nightingale | L_VLDL_CE | NA | T | C | 0.027055 | 0.004683 | 7.80E-10 |
| rs550057 | miR-10b-5p | trans | 1.61E-27 | Nightingale | S_VLDL_FC | NA | T | C | 0.042888 | 0.004528 | 6.60E-21 |
| rs550057 | miR-10b-5p | trans | 1.61E-27 | Nightingale | VLDL_CE | NA | T | C | 0.044291 | 0.00474 | 5.90E-22 |
| rs550057 | miR-10b-5p | trans | 1.61E-27 | Nightingale | XS_VLDL_C | NA | T | C | 0.046905 | 0.004573 | 1.30E-25 |
| rs550057 | miR-10b-5p | trans | 1.61E-27 | Nightingale | L_VLDL_TG | NA | T | C | -0.06228 | 0.004716 | 4.90E-41 |
| rs550057 | miR-10b-5p | trans | 1.61E-27 | Nightingale | M_LDL_PL | NA | T | C | 0.046345 | 0.004746 | 2.60E-23 |
| rs550057 | miR-10b-5p | trans | 1.61E-27 | Nightingale | L_VLDL_FC | NA | T | C | 0.06018 | 0.004693 | 3.20E-38 |
| rs550057 | miR-10b-5p | trans | 1.61E-27 | Nightingale | Total_L | NA | T | C | 0.033368 | 0.004662 | 4.10E-14 |
| rs550057 | miR-10b-5p | trans | 1.61E-27 | Nightingale | Sphingomy | NA | T | C | 0.03676 | 0.00443 | 6.30E-17 |
| rs550057 | miR-10b-5p | trans | 1.61E-27 | Nightingale | M_VLDL_F | NA | T | C | 0.042701 | 0.004738 | 2.00E-20 |
| rs550057 | miR-10b-5p | trans | 1.61E-27 | Nightingale | XS_VLDL_C | NA | T | C | 0.035005 | 0.004494 | 2.30E-14 |
| rs550057 | miR-10b-5p | trans | 1.61E-27 | Nightingale | IDL_C_pct | NA | T | C | 0.031128 | 0.004629 | 3.40E-11 |
| rs550057 | miR-10b-5p | trans | 1.61E-27 | Nightingale | XL_VLDL_T | NA | T | C | -0.05437 | 0.004594 | 3.30E-32 |
| rs550057 | miR-10b-5p | trans | 1.61E-27 | Nightingale | M_LDL_C | NA | T | C | 0.046775 | 0.004747 | 1.00E-23 |
| rs550057 | miR-10b-5p | trans | 1.61E-27 | Nightingale | L_VLDL_CE | NA | T | C | 0.058972 | 0.004535 | 2.60E-39 |

| | | | | | | | | | | | |
|---|---|---|---|---|---|---|---|---|---|---|---|
| rs550057 | miR-10b-5p | trans | 1.61E-27 | Nightingale | S_VLDL_C | NA | T | C | 0.048078 | 0.004738 | 1.80E-25 |
| rs550057 | miR-10b-5p | trans | 1.61E-27 | Nightingale | M_VLDL_P | NA | T | C | 0.05684 | 0.004548 | 4.30E-36 |
| rs550057 | miR-10b-5p | trans | 1.61E-27 | Nightingale | PUFA | NA | T | C | 0.029052 | 0.004622 | 2.60E-11 |
| rs550057 | miR-10b-5p | trans | 1.61E-27 | Nightingale | M_VLDL_C | NA | T | C | 0.047633 | 0.004464 | 3.30E-26 |
| rs550057 | miR-10b-5p | trans | 1.61E-27 | Nightingale | XL_VLDL_P | NA | T | C | 0.046629 | 0.004747 | 7.70E-24 |
| rs550057 | miR-10b-5p | trans | 1.61E-27 | Nightingale | Total_FC | NA | T | C | 0.043696 | 0.004629 | 8.00E-23 |
| rs550057 | miR-10b-5p | trans | 1.61E-27 | Nightingale | S_VLDL_CE | NA | T | C | 0.061711 | 0.004675 | 2.00E-39 |
| rs550057 | miR-10b-5p | trans | 1.61E-27 | Nightingale | non_HDL_( | NA | T | C | 0.048348 | 0.004713 | 9.80E-26 |
| rs550057 | miR-10b-5p | trans | 1.61E-27 | Nightingale | S_VLDL_TG | NA | T | C | -0.05282 | 0.004587 | 2.10E-30 |
| rs550057 | miR-10b-5p | trans | 1.61E-27 | Nightingale | XXL_VLDL_ | NA | T | C | 0.040178 | 0.004778 | 1.80E-17 |
| rs550057 | miR-10b-5p | trans | 1.61E-27 | Nightingale | L_LDL_FC | NA | T | C | 0.048951 | 0.004653 | 3.20E-26 |
| rs550057 | miR-10b-5p | trans | 1.61E-27 | Nightingale | IDL_TG_pc | NA | T | C | -0.03163 | 0.004581 | 7.70E-12 |
| rs550057 | miR-10b-5p | trans | 1.61E-27 | Nightingale | XS_VLDL_F | NA | T | C | 0.043373 | 0.004705 | 1.40E-19 |
| rs550057 | miR-10b-5p | trans | 1.61E-27 | Nightingale | Glucose | NA | T | C | 0.026619 | 0.00475 | 2.10E-08 |
| rs550057 | miR-10b-5p | trans | 1.61E-27 | Nightingale | M_VLDL_L | NA | T | C | 0.026949 | 0.004707 | 1.30E-09 |
| rs550057 | miR-10b-5p | trans | 1.61E-27 | Nightingale | Total_CE | NA | T | C | 0.043093 | 0.004571 | 3.60E-22 |
| rs550057 | miR-10b-5p | trans | 1.61E-27 | Nightingale | GlycA | NA | T | C | 0.026618 | 0.004719 | 1.00E-08 |
| rs550057 | miR-10b-5p | trans | 1.61E-27 | Nightingale | S_VLDL_PL | NA | T | C | 0.040787 | 0.00473 | 3.40E-19 |
| rs550057 | miR-10b-5p | trans | 1.61E-27 | Nightingale | Clinical_LD | NA | T | C | 0.051215 | 0.004711 | 4.80E-28 |
| rs550057 | miR-10b-5p | trans | 1.61E-27 | Nightingale | XL_VLDL_F | NA | T | C | 0.04531 | 0.004594 | 2.40E-23 |
| rs550057 | miR-10b-5p | trans | 1.61E-27 | Nightingale | S_VLDL_P | NA | T | C | 0.027467 | 0.004679 | 2.70E-10 |
| rs550057 | miR-10b-5p | trans | 1.61E-27 | Nightingale | LA | NA | T | C | 0.032534 | 0.004647 | 1.40E-13 |
| rs550057 | miR-10b-5p | trans | 1.61E-27 | Nightingale | VLDL_P | NA | T | C | 0.031898 | 0.004713 | 1.00E-12 |
| rs550057 | miR-10b-5p | trans | 1.61E-27 | Nightingale | XL_HDL_C_ | NA | T | C | 0.03137 | 0.004525 | 1.90E-11 |
| rs550057 | miR-10b-5p | trans | 1.61E-27 | Nightingale | S_LDL_TG_ | NA | T | C | -0.03461 | 0.004595 | 2.30E-13 |
| rs550057 | miR-10b-5p | trans | 1.61E-27 | Nightingale | XS_VLDL_P | NA | T | C | 0.03659 | 0.004673 | 2.30E-16 |
| rs550057 | miR-10b-5p | trans | 1.61E-27 | Nightingale | VLDL_C | NA | T | C | 0.038243 | 0.004732 | 4.20E-17 |
| rs550057 | miR-10b-5p | trans | 1.61E-27 | Nightingale | ApoB | NA | T | C | 0.045452 | 0.004733 | 1.20E-22 |
| rs550057 | miR-10b-5p | trans | 1.61E-27 | Nightingale | Pyruvate | NA | T | C | 0.027561 | 0.004747 | 6.40E-09 |
| rs550057 | miR-10b-5p | trans | 1.61E-27 | Nightingale | S_VLDL_FC | NA | T | C | 0.048419 | 0.00474 | 1.40E-25 |
| rs550057 | miR-10b-5p | trans | 1.61E-27 | Nightingale | XL_HDL_PL | NA | T | C | -0.02299 | 0.004263 | 5.00E-08 |
| rs550057 | let-7b-5p | trans | 2.70E-19 | Nightingale | S_LDL_P | NA | T | C | 0.040014 | 0.004743 | 7.70E-18 |
| rs550057 | let-7b-5p | trans | 2.70E-19 | Nightingale | S_LDL_C_p | NA | T | C | 0.059583 | 0.004757 | 3.20E-37 |
| rs550057 | let-7b-5p | trans | 2.70E-19 | Nightingale | XXL_VLDL_ | NA | T | C | 0.040046 | 0.004752 | 1.60E-17 |
| rs550057 | let-7b-5p | trans | 2.70E-19 | Nightingale | L_LDL_PL | NA | T | C | 0.052492 | 0.004703 | 1.00E-29 |
| rs550057 | let-7b-5p | trans | 2.70E-19 | Nightingale | IDL_CE | NA | T | C | 0.046199 | 0.004575 | 1.80E-24 |
| rs550057 | let-7b-5p | trans | 2.70E-19 | Nightingale | S_LDL_C | NA | T | C | 0.043673 | 0.004743 | 1.20E-20 |
| rs550057 | let-7b-5p | trans | 2.70E-19 | Nightingale | XL_VLDL_C | NA | T | C | 0.025867 | 0.004666 | 2.80E-09 |
| rs550057 | let-7b-5p | trans | 2.70E-19 | Nightingale | M_LDL_CE | NA | T | C | 0.044363 | 0.004745 | 9.80E-22 |
| rs550057 | let-7b-5p | trans | 2.70E-19 | Nightingale | XS_VLDL_T | NA | T | C | -0.03521 | 0.004505 | 1.60E-14 |
| rs550057 | let-7b-5p | trans | 2.70E-19 | Nightingale | L_LDL_CE | NA | T | C | 0.045326 | 0.004697 | 1.00E-22 |
| rs550057 | let-7b-5p | trans | 2.70E-19 | Nightingale | Omega_6 | NA | T | C | 0.03221 | 0.004628 | 1.30E-13 |
| rs550057 | let-7b-5p | trans | 2.70E-19 | Nightingale | S_LDL_PL_ | NA | T | C | -0.03504 | 0.00473 | 8.30E-16 |
| rs550057 | let-7b-5p | trans | 2.70E-19 | Nightingale | LDL_L | NA | T | C | 0.046916 | 0.004723 | 6.60E-24 |
| rs550057 | let-7b-5p | trans | 2.70E-19 | Nightingale | S_HDL_C | NA | T | C | 0.037754 | 0.004731 | 1.40E-15 |
| rs550057 | let-7b-5p | trans | 2.70E-19 | Nightingale | IDL_C | NA | T | C | 0.047774 | 0.004577 | 5.10E-26 |
| rs550057 | let-7b-5p | trans | 2.70E-19 | Nightingale | S_HDL_CE | NA | T | C | 0.038488 | 0.004746 | 6.00E-16 |
| rs550057 | let-7b-5p | trans | 2.70E-19 | Nightingale | S_LDL_CE | NA | T | C | 0.04208 | 0.004741 | 1.20E-19 |
| rs550057 | let-7b-5p | trans | 2.70E-19 | Nightingale | XS_VLDL_C | NA | T | C | 0.032181 | 0.004471 | 2.20E-12 |
| rs550057 | let-7b-5p | trans | 2.70E-19 | Nightingale | L_LDL_P | NA | T | C | 0.040754 | 0.004723 | 2.20E-18 |
| rs550057 | let-7b-5p | trans | 2.70E-19 | Nightingale | IDL_L | NA | T | C | 0.047298 | 0.004582 | 1.10E-25 |
| rs550057 | let-7b-5p | trans | 2.70E-19 | Nightingale | L_LDL_TG_ | NA | T | C | -0.03317 | 0.004648 | 1.40E-12 |
| rs550057 | let-7b-5p | trans | 2.70E-19 | Nightingale | S_HDL_P | NA | T | C | 0.040281 | 0.004739 | 1.30E-17 |
| rs550057 | let-7b-5p | trans | 2.70E-19 | Nightingale | M_LDL_P | NA | T | C | 0.046745 | 0.004752 | 9.50E-24 |
| rs550057 | let-7b-5p | trans | 2.70E-19 | Nightingale | XL_HDL_PL | NA | T | C | -0.03225 | 0.004354 | 1.10E-13 |
| rs550057 | let-7b-5p | trans | 2.70E-19 | Nightingale | M_VLDL_F | NA | T | C | 0.054228 | 0.004508 | 3.10E-33 |
| rs550057 | let-7b-5p | trans | 2.70E-19 | Nightingale | M_VLDL_P | NA | T | C | 0.036129 | 0.004727 | 1.70E-15 |
| rs550057 | let-7b-5p | trans | 2.70E-19 | Nightingale | L_LDL_L | NA | T | C | 0.047551 | 0.004693 | 6.80E-25 |
| rs550057 | let-7b-5p | trans | 2.70E-19 | Nightingale | XXL_VLDL_ | NA | T | C | -0.04063 | 0.004782 | 2.80E-18 |
| rs550057 | let-7b-5p | trans | 2.70E-19 | Nightingale | XXL_VLDL_ | NA | T | C | 0.025317 | 0.004772 | 7.10E-09 |
| rs550057 | let-7b-5p | trans | 2.70E-19 | Nightingale | S_VLDL_L | NA | T | C | 0.029085 | 0.004693 | 3.50E-11 |
| rs550057 | let-7b-5p | trans | 2.70E-19 | Nightingale | XS_VLDL_C | NA | T | C | 0.046509 | 0.0046 | 3.90E-25 |
| rs550057 | let-7b-5p | trans | 2.70E-19 | Nightingale | XS_VLDL_P | NA | T | C | 0.043776 | 0.004676 | 1.80E-22 |

| | | | | | | | | | | | |
|---|---|---|---|---|---|---|---|---|---|---|---|
| rs550057 | let-7b-5p | trans | 2.70E-19 | Nightingale | M_LDL_C_ | NA | T | C | 0.038134 | 0.004742 | 4.30E-16 |
| rs550057 | let-7b-5p | trans | 2.70E-19 | Nightingale | LDL_FC | NA | T | C | 0.049134 | 0.004688 | 7.00E-26 |
| rs550057 | let-7b-5p | trans | 2.70E-19 | Nightingale | Cholines | NA | T | C | 0.02627 | 0.004457 | 3.90E-10 |
| rs550057 | let-7b-5p | trans | 2.70E-19 | Nightingale | LDL_PL | NA | T | C | 0.049193 | 0.004728 | 6.40E-26 |
| rs550057 | let-7b-5p | trans | 2.70E-19 | Nightingale | LDL_CE | NA | T | C | 0.04573 | 0.004726 | 6.60E-23 |
| rs550057 | let-7b-5p | trans | 2.70E-19 | Nightingale | S_VLDL_CE | NA | T | C | 0.047282 | 0.004733 | 6.20E-25 |
| rs550057 | let-7b-5p | trans | 2.70E-19 | Nightingale | ApoB_by_/ | NA | T | C | 0.034854 | 0.004667 | 5.50E-14 |
| rs550057 | let-7b-5p | trans | 2.70E-19 | Nightingale | XS_VLDL_F | NA | T | C | 0.043814 | 0.004657 | 1.60E-22 |
| rs550057 | let-7b-5p | trans | 2.70E-19 | Nightingale | S_LDL_FC | NA | T | C | 0.043269 | 0.004731 | 8.00E-20 |
| rs550057 | let-7b-5p | trans | 2.70E-19 | Nightingale | S_LDL_L | NA | T | C | 0.039564 | 0.004738 | 3.50E-17 |
| rs550057 | let-7b-5p | trans | 2.70E-19 | Nightingale | M_LDL_FC | NA | T | C | 0.050019 | 0.004733 | 1.70E-26 |
| rs550057 | let-7b-5p | trans | 2.70E-19 | Nightingale | M_LDL_TG | NA | T | C | -0.04255 | 0.00468 | 8.30E-20 |
| rs550057 | let-7b-5p | trans | 2.70E-19 | Nightingale | L_LDL_C | NA | T | C | 0.046611 | 0.004685 | 5.60E-24 |
| rs550057 | let-7b-5p | trans | 2.70E-19 | Nightingale | L_VLDL_C_ | NA | T | C | 0.064893 | 0.004628 | 1.20E-45 |
| rs550057 | let-7b-5p | trans | 2.70E-19 | Nightingale | Remnant_( | NA | T | C | 0.047674 | 0.004708 | 1.40E-25 |
| rs550057 | let-7b-5p | trans | 2.70E-19 | Nightingale | M_VLDL_T | NA | T | C | -0.05036 | 0.004478 | 4.10E-29 |
| rs550057 | let-7b-5p | trans | 2.70E-19 | Nightingale | XS_VLDL_L | NA | T | C | 0.039312 | 0.004658 | 8.60E-19 |
| rs550057 | let-7b-5p | trans | 2.70E-19 | Nightingale | XXL_VLDL_ | NA | T | C | 0.033136 | 0.004721 | 8.10E-13 |
| rs550057 | let-7b-5p | trans | 2.70E-19 | Nightingale | M_VLDL_C | NA | T | C | 0.0536 | 0.004696 | 7.40E-31 |
| rs550057 | let-7b-5p | trans | 2.70E-19 | Nightingale | IDL_FC | NA | T | C | 0.051144 | 0.004588 | 1.70E-29 |
| rs550057 | let-7b-5p | trans | 2.70E-19 | Nightingale | S_LDL_CE_ | NA | T | C | 0.03581 | 0.004731 | 2.00E-16 |
| rs550057 | let-7b-5p | trans | 2.70E-19 | Nightingale | M_VLDL_C | NA | T | C | 0.044675 | 0.004452 | 2.60E-23 |
| rs550057 | let-7b-5p | trans | 2.70E-19 | Nightingale | IDL_FC_pct | NA | T | C | 0.034868 | 0.004702 | 4.70E-13 |
| rs550057 | let-7b-5p | trans | 2.70E-19 | Nightingale | M_VLDL_P | NA | T | C | 0.03714 | 0.004732 | 3.20E-16 |
| rs550057 | let-7b-5p | trans | 2.70E-19 | Nightingale | LDL_P | NA | T | C | 0.043296 | 0.004735 | 1.40E-20 |
| rs550057 | let-7b-5p | trans | 2.70E-19 | Nightingale | VLDL_FC | NA | T | C | 0.02878 | 0.004704 | 9.50E-11 |
| rs550057 | let-7b-5p | trans | 2.70E-19 | Nightingale | S_VLDL_C_ | NA | T | C | 0.056984 | 0.004612 | 6.60E-35 |
| rs550057 | let-7b-5p | trans | 2.70E-19 | Nightingale | LDL_C | NA | T | C | 0.047148 | 0.004718 | 3.90E-24 |
| rs550057 | let-7b-5p | trans | 2.70E-19 | Nightingale | S_HDL_L | NA | T | C | 0.0316 | 0.004722 | 5.20E-12 |
| rs550057 | let-7b-5p | trans | 2.70E-19 | Nightingale | S_VLDL_PL | NA | T | C | 0.040569 | 0.004538 | 1.10E-18 |
| rs550057 | let-7b-5p | trans | 2.70E-19 | Nightingale | HDL_size | NA | T | C | -0.02598 | 0.004202 | 4.40E-10 |
| rs550057 | let-7b-5p | trans | 2.70E-19 | Nightingale | Phosphogly | NA | T | C | 0.023356 | 0.004489 | 3.00E-08 |
| rs550057 | let-7b-5p | trans | 2.70E-19 | Nightingale | XL_VLDL_C | NA | T | C | 0.048624 | 0.004544 | 3.40E-26 |
| rs550057 | let-7b-5p | trans | 2.70E-19 | Nightingale | S_HDL_PL | NA | T | C | 0.026717 | 0.004698 | 2.80E-09 |
| rs550057 | let-7b-5p | trans | 2.70E-19 | Nightingale | Total_C | NA | T | C | 0.043514 | 0.004588 | 1.70E-22 |
| rs550057 | let-7b-5p | trans | 2.70E-19 | Nightingale | XL_VLDL_C | NA | T | C | 0.048184 | 0.004547 | 1.10E-25 |
| rs550057 | let-7b-5p | trans | 2.70E-19 | Nightingale | Citrate | NA | T | C | 0.026542 | 0.004708 | 8.90E-09 |
| rs550057 | let-7b-5p | trans | 2.70E-19 | Nightingale | M_LDL_L | NA | T | C | 0.045701 | 0.004745 | 8.50E-23 |
| rs550057 | let-7b-5p | trans | 2.70E-19 | Nightingale | S_HDL_FC | NA | T | C | 0.029798 | 0.004652 | 2.70E-11 |
| rs550057 | let-7b-5p | trans | 2.70E-19 | Nightingale | IDL_PL | NA | T | C | 0.046477 | 0.004573 | 4.80E-25 |
| rs550057 | let-7b-5p | trans | 2.70E-19 | Nightingale | Total_PL | NA | T | C | 0.030079 | 0.004512 | 1.20E-12 |
| rs550057 | let-7b-5p | trans | 2.70E-19 | Nightingale | S_LDL_PL | NA | T | C | 0.034085 | 0.004728 | 4.70E-13 |
| rs550057 | let-7b-5p | trans | 2.70E-19 | Nightingale | IDL_P | NA | T | C | 0.053052 | 0.004689 | 2.00E-30 |
| rs550057 | let-7b-5p | trans | 2.70E-19 | Nightingale | Total_P | NA | T | C | 0.025587 | 0.00446 | 1.70E-09 |
| rs550057 | let-7b-5p | trans | 2.70E-19 | Nightingale | M_VLDL_C | NA | T | C | 0.050328 | 0.004726 | 1.60E-27 |
| rs550057 | let-7b-5p | trans | 2.70E-19 | Nightingale | L_VLDL_CE | NA | T | C | 0.027055 | 0.004683 | 7.80E-10 |
| rs550057 | let-7b-5p | trans | 2.70E-19 | Nightingale | S_VLDL_FC | NA | T | C | 0.042888 | 0.004528 | 6.60E-21 |
| rs550057 | let-7b-5p | trans | 2.70E-19 | Nightingale | VLDL_CE | NA | T | C | 0.044291 | 0.00474 | 5.90E-22 |
| rs550057 | let-7b-5p | trans | 2.70E-19 | Nightingale | XS_VLDL_C | NA | T | C | 0.046905 | 0.004573 | 1.30E-25 |
| rs550057 | let-7b-5p | trans | 2.70E-19 | Nightingale | L_VLDL_TG | NA | T | C | -0.06228 | 0.004716 | 4.90E-41 |
| rs550057 | let-7b-5p | trans | 2.70E-19 | Nightingale | M_LDL_PL | NA | T | C | 0.046345 | 0.004746 | 2.60E-23 |
| rs550057 | let-7b-5p | trans | 2.70E-19 | Nightingale | L_VLDL_FC | NA | T | C | 0.06018 | 0.004693 | 3.20E-38 |
| rs550057 | let-7b-5p | trans | 2.70E-19 | Nightingale | Total_L | NA | T | C | 0.033368 | 0.004662 | 4.10E-14 |
| rs550057 | let-7b-5p | trans | 2.70E-19 | Nightingale | Sphingomy | NA | T | C | 0.03676 | 0.00443 | 6.30E-17 |
| rs550057 | let-7b-5p | trans | 2.70E-19 | Nightingale | M_VLDL_F | NA | T | C | 0.042701 | 0.004738 | 2.00E-20 |
| rs550057 | let-7b-5p | trans | 2.70E-19 | Nightingale | XS_VLDL_C | NA | T | C | 0.035005 | 0.004494 | 2.30E-14 |
| rs550057 | let-7b-5p | trans | 2.70E-19 | Nightingale | IDL_C_pct | NA | T | C | 0.031128 | 0.004629 | 3.40E-11 |
| rs550057 | let-7b-5p | trans | 2.70E-19 | Nightingale | XL_VLDL_T | NA | T | C | -0.05437 | 0.004594 | 3.30E-32 |
| rs550057 | let-7b-5p | trans | 2.70E-19 | Nightingale | M_LDL_C | NA | T | C | 0.046775 | 0.004747 | 1.00E-23 |
| rs550057 | let-7b-5p | trans | 2.70E-19 | Nightingale | L_VLDL_CE | NA | T | C | 0.058972 | 0.004535 | 2.60E-39 |
| rs550057 | let-7b-5p | trans | 2.70E-19 | Nightingale | S_VLDL_C | NA | T | C | 0.048078 | 0.004738 | 1.80E-25 |
| rs550057 | let-7b-5p | trans | 2.70E-19 | Nightingale | M_VLDL_P | NA | T | C | 0.05684 | 0.004548 | 4.30E-36 |
| rs550057 | let-7b-5p | trans | 2.70E-19 | Nightingale | PUFA | NA | T | C | 0.029052 | 0.004622 | 2.60E-11 |

| | | | | | | | | | | | |
|---|---|---|---|---|---|---|---|---|---|---|---|
| rs550057 | let-7b-5p | trans | 2.70E-19 | Nightingale | M_VLDL_C | NA | T | C | 0.047633 | 0.004464 | 3.30E-26 |
| rs550057 | let-7b-5p | trans | 2.70E-19 | Nightingale | XL_VLDL_P | NA | T | C | 0.046629 | 0.004747 | 7.70E-24 |
| rs550057 | let-7b-5p | trans | 2.70E-19 | Nightingale | Total_FC | NA | T | C | 0.043696 | 0.004629 | 8.00E-23 |
| rs550057 | let-7b-5p | trans | 2.70E-19 | Nightingale | S_VLDL_CE | NA | T | C | 0.061711 | 0.004675 | 2.00E-39 |
| rs550057 | let-7b-5p | trans | 2.70E-19 | Nightingale | non_HDL_( | NA | T | C | 0.048348 | 0.004713 | 9.80E-26 |
| rs550057 | let-7b-5p | trans | 2.70E-19 | Nightingale | S_VLDL_TG | NA | T | C | -0.05282 | 0.004587 | 2.10E-30 |
| rs550057 | let-7b-5p | trans | 2.70E-19 | Nightingale | XXL_VLDL_ | NA | T | C | 0.040178 | 0.004778 | 1.80E-17 |
| rs550057 | let-7b-5p | trans | 2.70E-19 | Nightingale | L_LDL_FC | NA | T | C | 0.048951 | 0.004653 | 3.20E-26 |
| rs550057 | let-7b-5p | trans | 2.70E-19 | Nightingale | IDL_TG_pc | NA | T | C | -0.03163 | 0.004581 | 7.70E-12 |
| rs550057 | let-7b-5p | trans | 2.70E-19 | Nightingale | XS_VLDL_F | NA | T | C | 0.043373 | 0.004705 | 1.40E-19 |
| rs550057 | let-7b-5p | trans | 2.70E-19 | Nightingale | Glucose | NA | T | C | 0.026619 | 0.00475 | 2.10E-08 |
| rs550057 | let-7b-5p | trans | 2.70E-19 | Nightingale | M_VLDL_L | NA | T | C | 0.026949 | 0.004707 | 1.30E-09 |
| rs550057 | let-7b-5p | trans | 2.70E-19 | Nightingale | Total_CE | NA | T | C | 0.043093 | 0.004571 | 3.60E-22 |
| rs550057 | let-7b-5p | trans | 2.70E-19 | Nightingale | GlycA | NA | T | C | 0.026618 | 0.004719 | 1.00E-08 |
| rs550057 | let-7b-5p | trans | 2.70E-19 | Nightingale | S_VLDL_PL | NA | T | C | 0.040787 | 0.00473 | 3.40E-19 |
| rs550057 | let-7b-5p | trans | 2.70E-19 | Nightingale | Clinical_LD | NA | T | C | 0.051215 | 0.004711 | 4.80E-28 |
| rs550057 | let-7b-5p | trans | 2.70E-19 | Nightingale | XL_VLDL_F | NA | T | C | 0.04531 | 0.004594 | 2.40E-23 |
| rs550057 | let-7b-5p | trans | 2.70E-19 | Nightingale | S_VLDL_P | NA | T | C | 0.027467 | 0.004679 | 2.70E-10 |
| rs550057 | let-7b-5p | trans | 2.70E-19 | Nightingale | LA | NA | T | C | 0.032534 | 0.004647 | 1.40E-13 |
| rs550057 | let-7b-5p | trans | 2.70E-19 | Nightingale | VLDL_P | NA | T | C | 0.031898 | 0.004713 | 1.00E-12 |
| rs550057 | let-7b-5p | trans | 2.70E-19 | Nightingale | XL_HDL_C_ | NA | T | C | 0.03137 | 0.004525 | 1.90E-11 |
| rs550057 | let-7b-5p | trans | 2.70E-19 | Nightingale | S_LDL_TG_ | NA | T | C | -0.03461 | 0.004595 | 2.30E-13 |
| rs550057 | let-7b-5p | trans | 2.70E-19 | Nightingale | XS_VLDL_P | NA | T | C | 0.03659 | 0.004673 | 2.30E-16 |
| rs550057 | let-7b-5p | trans | 2.70E-19 | Nightingale | VLDL_C | NA | T | C | 0.038243 | 0.004732 | 4.20E-17 |
| rs550057 | let-7b-5p | trans | 2.70E-19 | Nightingale | ApoB | NA | T | C | 0.045452 | 0.004733 | 1.20E-22 |
| rs550057 | let-7b-5p | trans | 2.70E-19 | Nightingale | Pyruvate | NA | T | C | 0.027561 | 0.004747 | 6.40E-09 |
| rs550057 | let-7b-5p | trans | 2.70E-19 | Nightingale | S_VLDL_FC | NA | T | C | 0.048419 | 0.00474 | 1.40E-25 |
| rs550057 | let-7b-5p | trans | 2.70E-19 | Nightingale | XL_HDL_PL | NA | T | C | -0.02299 | 0.004263 | 5.00E-08 |
| rs550057 | miR-126-3p | trans | 5.01E-31 | Nightingale | S_LDL_P | NA | T | C | 0.040014 | 0.004743 | 7.70E-18 |
| rs550057 | miR-126-3p | trans | 5.01E-31 | Nightingale | S_LDL_C_p | NA | T | C | 0.059583 | 0.004757 | 3.20E-37 |
| rs550057 | miR-126-3p | trans | 5.01E-31 | Nightingale | XXL_VLDL_ | NA | T | C | 0.040046 | 0.004752 | 1.60E-17 |
| rs550057 | miR-126-3p | trans | 5.01E-31 | Nightingale | L_LDL_PL | NA | T | C | 0.052492 | 0.004703 | 1.00E-29 |
| rs550057 | miR-126-3p | trans | 5.01E-31 | Nightingale | IDL_CE | NA | T | C | 0.046199 | 0.004575 | 1.80E-24 |
| rs550057 | miR-126-3p | trans | 5.01E-31 | Nightingale | S_LDL_C | NA | T | C | 0.043673 | 0.004743 | 1.20E-20 |
| rs550057 | miR-126-3p | trans | 5.01E-31 | Nightingale | XL_VLDL_C | NA | T | C | 0.025867 | 0.004666 | 2.80E-09 |
| rs550057 | miR-126-3p | trans | 5.01E-31 | Nightingale | M_LDL_CE | NA | T | C | 0.044363 | 0.004745 | 9.80E-22 |
| rs550057 | miR-126-3p | trans | 5.01E-31 | Nightingale | XS_VLDL_T | NA | T | C | -0.03521 | 0.004505 | 1.60E-14 |
| rs550057 | miR-126-3p | trans | 5.01E-31 | Nightingale | L_LDL_CE | NA | T | C | 0.045326 | 0.004697 | 1.00E-22 |
| rs550057 | miR-126-3p | trans | 5.01E-31 | Nightingale | Omega_6 | NA | T | C | 0.03221 | 0.004628 | 1.30E-13 |
| rs550057 | miR-126-3p | trans | 5.01E-31 | Nightingale | S_LDL_PL_ | NA | T | C | -0.03504 | 0.00473 | 8.30E-16 |
| rs550057 | miR-126-3p | trans | 5.01E-31 | Nightingale | LDL_L | NA | T | C | 0.046916 | 0.004723 | 6.60E-24 |
| rs550057 | miR-126-3p | trans | 5.01E-31 | Nightingale | S_HDL_C | NA | T | C | 0.037754 | 0.004731 | 1.40E-15 |
| rs550057 | miR-126-3p | trans | 5.01E-31 | Nightingale | IDL_C | NA | T | C | 0.047774 | 0.004577 | 5.10E-26 |
| rs550057 | miR-126-3p | trans | 5.01E-31 | Nightingale | S_HDL_CE | NA | T | C | 0.038488 | 0.004746 | 6.00E-16 |
| rs550057 | miR-126-3p | trans | 5.01E-31 | Nightingale | S_LDL_CE | NA | T | C | 0.04208 | 0.004741 | 1.20E-19 |
| rs550057 | miR-126-3p | trans | 5.01E-31 | Nightingale | XS_VLDL_C | NA | T | C | 0.032181 | 0.004471 | 2.20E-12 |
| rs550057 | miR-126-3p | trans | 5.01E-31 | Nightingale | L_LDL_P | NA | T | C | 0.040754 | 0.004723 | 2.20E-18 |
| rs550057 | miR-126-3p | trans | 5.01E-31 | Nightingale | IDL_L | NA | T | C | 0.047298 | 0.004582 | 1.10E-25 |
| rs550057 | miR-126-3p | trans | 5.01E-31 | Nightingale | L_LDL_TG_ | NA | T | C | -0.03317 | 0.004648 | 1.40E-12 |
| rs550057 | miR-126-3p | trans | 5.01E-31 | Nightingale | S_HDL_P | NA | T | C | 0.040281 | 0.004739 | 1.30E-17 |
| rs550057 | miR-126-3p | trans | 5.01E-31 | Nightingale | M_LDL_P | NA | T | C | 0.046745 | 0.004752 | 9.50E-24 |
| rs550057 | miR-126-3p | trans | 5.01E-31 | Nightingale | XL_HDL_PL | NA | T | C | -0.03225 | 0.004354 | 1.10E-13 |
| rs550057 | miR-126-3p | trans | 5.01E-31 | Nightingale | M_VLDL_F | NA | T | C | 0.054228 | 0.004508 | 3.10E-33 |
| rs550057 | miR-126-3p | trans | 5.01E-31 | Nightingale | M_VLDL_P | NA | T | C | 0.036129 | 0.004727 | 1.70E-15 |
| rs550057 | miR-126-3p | trans | 5.01E-31 | Nightingale | L_LDL_L | NA | T | C | 0.047551 | 0.004693 | 6.80E-25 |
| rs550057 | miR-126-3p | trans | 5.01E-31 | Nightingale | XXL_VLDL_ | NA | T | C | -0.04063 | 0.004782 | 2.80E-18 |
| rs550057 | miR-126-3p | trans | 5.01E-31 | Nightingale | XXL_VLDL_ | NA | T | C | 0.025317 | 0.004772 | 7.10E-09 |
| rs550057 | miR-126-3p | trans | 5.01E-31 | Nightingale | S_VLDL_L | NA | T | C | 0.029085 | 0.004693 | 3.50E-11 |
| rs550057 | miR-126-3p | trans | 5.01E-31 | Nightingale | XS_VLDL_C | NA | T | C | 0.046509 | 0.0046 | 3.90E-25 |
| rs550057 | miR-126-3p | trans | 5.01E-31 | Nightingale | XS_VLDL_P | NA | T | C | 0.043776 | 0.004676 | 1.80E-22 |
| rs550057 | miR-126-3p | trans | 5.01E-31 | Nightingale | M_LDL_C_ | NA | T | C | 0.038134 | 0.004742 | 4.30E-16 |
| rs550057 | miR-126-3p | trans | 5.01E-31 | Nightingale | LDL_FC | NA | T | C | 0.049134 | 0.004688 | 7.00E-26 |
| rs550057 | miR-126-3p | trans | 5.01E-31 | Nightingale | Cholines | NA | T | C | 0.02627 | 0.004457 | 3.90E-10 |

| | | | | | | | | | | | |
|---|---|---|---|---|---|---|---|---|---|---|---|
| rs550057 | miR-126-3p | trans | 5.01E-31 | Nightingale | LDL_PL | NA | T | C | 0.049193 | 0.004728 | 6.40E-26 |
| rs550057 | miR-126-3p | trans | 5.01E-31 | Nightingale | LDL_CE | NA | T | C | 0.04573 | 0.004726 | 6.60E-23 |
| rs550057 | miR-126-3p | trans | 5.01E-31 | Nightingale | S_VLDL_CE | NA | T | C | 0.047282 | 0.004733 | 6.20E-25 |
| rs550057 | miR-126-3p | trans | 5.01E-31 | Nightingale | ApoB_by_A | NA | T | C | 0.034854 | 0.004667 | 5.50E-14 |
| rs550057 | miR-126-3p | trans | 5.01E-31 | Nightingale | XS_VLDL_F | NA | T | C | 0.043814 | 0.004657 | 1.60E-22 |
| rs550057 | miR-126-3p | trans | 5.01E-31 | Nightingale | S_LDL_FC | NA | T | C | 0.043269 | 0.004731 | 8.00E-20 |
| rs550057 | miR-126-3p | trans | 5.01E-31 | Nightingale | S_LDL_L | NA | T | C | 0.039564 | 0.004738 | 3.50E-17 |
| rs550057 | miR-126-3p | trans | 5.01E-31 | Nightingale | M_LDL_FC | NA | T | C | 0.050019 | 0.004733 | 1.70E-26 |
| rs550057 | miR-126-3p | trans | 5.01E-31 | Nightingale | M_LDL_TG | NA | T | C | -0.04255 | 0.00468 | 8.30E-20 |
| rs550057 | miR-126-3p | trans | 5.01E-31 | Nightingale | L_LDL_C | NA | T | C | 0.046611 | 0.004685 | 5.60E-24 |
| rs550057 | miR-126-3p | trans | 5.01E-31 | Nightingale | L_VLDL_C_ | NA | T | C | 0.064893 | 0.004628 | 1.20E-45 |
| rs550057 | miR-126-3p | trans | 5.01E-31 | Nightingale | Remnant_( | NA | T | C | 0.047674 | 0.004708 | 1.40E-25 |
| rs550057 | miR-126-3p | trans | 5.01E-31 | Nightingale | M_VLDL_T | NA | T | C | -0.05036 | 0.004478 | 4.10E-29 |
| rs550057 | miR-126-3p | trans | 5.01E-31 | Nightingale | XS_VLDL_L | NA | T | C | 0.039312 | 0.004658 | 8.60E-19 |
| rs550057 | miR-126-3p | trans | 5.01E-31 | Nightingale | XXL_VLDL_ | NA | T | C | 0.033136 | 0.004721 | 8.10E-13 |
| rs550057 | miR-126-3p | trans | 5.01E-31 | Nightingale | M_VLDL_C | NA | T | C | 0.0536 | 0.004696 | 7.40E-31 |
| rs550057 | miR-126-3p | trans | 5.01E-31 | Nightingale | IDL_FC | NA | T | C | 0.051144 | 0.004588 | 1.70E-29 |
| rs550057 | miR-126-3p | trans | 5.01E-31 | Nightingale | S_LDL_CE_ | NA | T | C | 0.03581 | 0.004731 | 2.00E-16 |
| rs550057 | miR-126-3p | trans | 5.01E-31 | Nightingale | M_VLDL_C | NA | T | C | 0.044675 | 0.004452 | 2.60E-23 |
| rs550057 | miR-126-3p | trans | 5.01E-31 | Nightingale | IDL_FC_pct | NA | T | C | 0.034868 | 0.004702 | 4.70E-13 |
| rs550057 | miR-126-3p | trans | 5.01E-31 | Nightingale | M_VLDL_P | NA | T | C | 0.03714 | 0.004732 | 3.20E-16 |
| rs550057 | miR-126-3p | trans | 5.01E-31 | Nightingale | LDL_P | NA | T | C | 0.043296 | 0.004735 | 1.40E-20 |
| rs550057 | miR-126-3p | trans | 5.01E-31 | Nightingale | VLDL_FC | NA | T | C | 0.02878 | 0.004704 | 9.50E-11 |
| rs550057 | miR-126-3p | trans | 5.01E-31 | Nightingale | S_VLDL_C_ | NA | T | C | 0.056984 | 0.004612 | 6.60E-35 |
| rs550057 | miR-126-3p | trans | 5.01E-31 | Nightingale | LDL_C | NA | T | C | 0.047148 | 0.004718 | 3.90E-24 |
| rs550057 | miR-126-3p | trans | 5.01E-31 | Nightingale | S_HDL_L | NA | T | C | 0.0316 | 0.004722 | 5.20E-12 |
| rs550057 | miR-126-3p | trans | 5.01E-31 | Nightingale | S_VLDL_PL | NA | T | C | 0.040569 | 0.004538 | 1.10E-18 |
| rs550057 | miR-126-3p | trans | 5.01E-31 | Nightingale | HDL_size | NA | T | C | -0.02598 | 0.004202 | 4.40E-10 |
| rs550057 | miR-126-3p | trans | 5.01E-31 | Nightingale | Phosphogly | NA | T | C | 0.023356 | 0.004489 | 3.00E-08 |
| rs550057 | miR-126-3p | trans | 5.01E-31 | Nightingale | XL_VLDL_C | NA | T | C | 0.048624 | 0.004544 | 3.40E-26 |
| rs550057 | miR-126-3p | trans | 5.01E-31 | Nightingale | S_HDL_PL | NA | T | C | 0.026717 | 0.004698 | 2.80E-09 |
| rs550057 | miR-126-3p | trans | 5.01E-31 | Nightingale | Total_C | NA | T | C | 0.043514 | 0.004588 | 1.70E-22 |
| rs550057 | miR-126-3p | trans | 5.01E-31 | Nightingale | XL_VLDL_C | NA | T | C | 0.048184 | 0.004547 | 1.10E-25 |
| rs550057 | miR-126-3p | trans | 5.01E-31 | Nightingale | Citrate | NA | T | C | 0.026542 | 0.004708 | 8.90E-09 |
| rs550057 | miR-126-3p | trans | 5.01E-31 | Nightingale | M_LDL_L | NA | T | C | 0.045701 | 0.004745 | 8.50E-23 |
| rs550057 | miR-126-3p | trans | 5.01E-31 | Nightingale | S_HDL_FC | NA | T | C | 0.029798 | 0.004652 | 2.70E-11 |
| rs550057 | miR-126-3p | trans | 5.01E-31 | Nightingale | IDL_PL | NA | T | C | 0.046477 | 0.004573 | 4.80E-25 |
| rs550057 | miR-126-3p | trans | 5.01E-31 | Nightingale | Total_PL | NA | T | C | 0.030079 | 0.004512 | 1.20E-12 |
| rs550057 | miR-126-3p | trans | 5.01E-31 | Nightingale | S_LDL_PL | NA | T | C | 0.034085 | 0.004728 | 4.70E-13 |
| rs550057 | miR-126-3p | trans | 5.01E-31 | Nightingale | IDL_P | NA | T | C | 0.053052 | 0.004689 | 2.00E-30 |
| rs550057 | miR-126-3p | trans | 5.01E-31 | Nightingale | Total_P | NA | T | C | 0.025587 | 0.00446 | 1.70E-09 |
| rs550057 | miR-126-3p | trans | 5.01E-31 | Nightingale | M_VLDL_C | NA | T | C | 0.050328 | 0.004726 | 1.60E-27 |
| rs550057 | miR-126-3p | trans | 5.01E-31 | Nightingale | L_VLDL_CE | NA | T | C | 0.027055 | 0.004683 | 7.80E-10 |
| rs550057 | miR-126-3p | trans | 5.01E-31 | Nightingale | S_VLDL_FC | NA | T | C | 0.042888 | 0.004528 | 6.60E-21 |
| rs550057 | miR-126-3p | trans | 5.01E-31 | Nightingale | VLDL_CE | NA | T | C | 0.044291 | 0.00474 | 5.90E-22 |
| rs550057 | miR-126-3p | trans | 5.01E-31 | Nightingale | XS_VLDL_C | NA | T | C | 0.046905 | 0.004573 | 1.30E-25 |
| rs550057 | miR-126-3p | trans | 5.01E-31 | Nightingale | L_VLDL_TG | NA | T | C | -0.06228 | 0.004716 | 4.90E-41 |
| rs550057 | miR-126-3p | trans | 5.01E-31 | Nightingale | M_LDL_PL | NA | T | C | 0.046345 | 0.004746 | 2.60E-23 |
| rs550057 | miR-126-3p | trans | 5.01E-31 | Nightingale | L_VLDL_FC | NA | T | C | 0.06018 | 0.004693 | 3.20E-38 |
| rs550057 | miR-126-3p | trans | 5.01E-31 | Nightingale | Total_L | NA | T | C | 0.033368 | 0.004662 | 4.10E-14 |
| rs550057 | miR-126-3p | trans | 5.01E-31 | Nightingale | Sphingomy | NA | T | C | 0.03676 | 0.00443 | 6.30E-17 |
| rs550057 | miR-126-3p | trans | 5.01E-31 | Nightingale | M_VLDL_F | NA | T | C | 0.042701 | 0.004738 | 2.00E-20 |
| rs550057 | miR-126-3p | trans | 5.01E-31 | Nightingale | XS_VLDL_C | NA | T | C | 0.035005 | 0.004494 | 2.30E-14 |
| rs550057 | miR-126-3p | trans | 5.01E-31 | Nightingale | IDL_C_pct | NA | T | C | 0.031128 | 0.004629 | 3.40E-11 |
| rs550057 | miR-126-3p | trans | 5.01E-31 | Nightingale | XL_VLDL_T | NA | T | C | -0.05437 | 0.004594 | 3.30E-32 |
| rs550057 | miR-126-3p | trans | 5.01E-31 | Nightingale | M_LDL_C | NA | T | C | 0.046775 | 0.004747 | 1.00E-23 |
| rs550057 | miR-126-3p | trans | 5.01E-31 | Nightingale | L_VLDL_CE | NA | T | C | 0.058972 | 0.004535 | 2.60E-39 |
| rs550057 | miR-126-3p | trans | 5.01E-31 | Nightingale | S_VLDL_C | NA | T | C | 0.048078 | 0.004738 | 1.80E-25 |
| rs550057 | miR-126-3p | trans | 5.01E-31 | Nightingale | M_VLDL_P | NA | T | C | 0.05684 | 0.004548 | 4.30E-36 |
| rs550057 | miR-126-3p | trans | 5.01E-31 | Nightingale | PUFA | NA | T | C | 0.029052 | 0.004622 | 2.60E-11 |
| rs550057 | miR-126-3p | trans | 5.01E-31 | Nightingale | M_VLDL_C_ | NA | T | C | 0.047633 | 0.004464 | 3.30E-26 |
| rs550057 | miR-126-3p | trans | 5.01E-31 | Nightingale | XL_VLDL_P | NA | T | C | 0.046629 | 0.004747 | 7.70E-24 |
| rs550057 | miR-126-3p | trans | 5.01E-31 | Nightingale | Total_FC | NA | T | C | 0.043696 | 0.004629 | 8.00E-23 |

| | | | | | | | | | | | |
|---|---|---|---|---|---|---|---|---|---|---|---|
| rs550057 | miR-126-3p | trans | 5.01E-31 | Nightingale | S_VLDL_CE | NA | T | C | 0.061711 | 0.004675 | 2.00E-39 |
| rs550057 | miR-126-3p | trans | 5.01E-31 | Nightingale | non_HDL_( | NA | T | C | 0.048348 | 0.004713 | 9.80E-26 |
| rs550057 | miR-126-3p | trans | 5.01E-31 | Nightingale | S_VLDL_TG | NA | T | C | -0.05282 | 0.004587 | 2.10E-30 |
| rs550057 | miR-126-3p | trans | 5.01E-31 | Nightingale | XXL_VLDL_ | NA | T | C | 0.040178 | 0.004778 | 1.80E-17 |
| rs550057 | miR-126-3p | trans | 5.01E-31 | Nightingale | L_LDL_FC | NA | T | C | 0.048951 | 0.004653 | 3.20E-26 |
| rs550057 | miR-126-3p | trans | 5.01E-31 | Nightingale | IDL_TG_pc | NA | T | C | -0.03163 | 0.004581 | 7.70E-12 |
| rs550057 | miR-126-3p | trans | 5.01E-31 | Nightingale | XS_VLDL_F | NA | T | C | 0.043373 | 0.004705 | 1.40E-19 |
| rs550057 | miR-126-3p | trans | 5.01E-31 | Nightingale | Glucose | NA | T | C | 0.026619 | 0.00475 | 2.10E-08 |
| rs550057 | miR-126-3p | trans | 5.01E-31 | Nightingale | M_VLDL_L | NA | T | C | 0.026949 | 0.004707 | 1.30E-09 |
| rs550057 | miR-126-3p | trans | 5.01E-31 | Nightingale | Total_CE | NA | T | C | 0.043093 | 0.004571 | 3.60E-22 |
| rs550057 | miR-126-3p | trans | 5.01E-31 | Nightingale | GlycA | NA | T | C | 0.026618 | 0.004719 | 1.00E-08 |
| rs550057 | miR-126-3p | trans | 5.01E-31 | Nightingale | S_VLDL_PL | NA | T | C | 0.040787 | 0.00473 | 3.40E-19 |
| rs550057 | miR-126-3p | trans | 5.01E-31 | Nightingale | Clinical_LD | NA | T | C | 0.051215 | 0.004711 | 4.80E-28 |
| rs550057 | miR-126-3p | trans | 5.01E-31 | Nightingale | XL_VLDL_F | NA | T | C | 0.04531 | 0.004594 | 2.40E-23 |
| rs550057 | miR-126-3p | trans | 5.01E-31 | Nightingale | S_VLDL_P | NA | T | C | 0.027467 | 0.004679 | 2.70E-10 |
| rs550057 | miR-126-3p | trans | 5.01E-31 | Nightingale | LA | NA | T | C | 0.032534 | 0.004647 | 1.40E-13 |
| rs550057 | miR-126-3p | trans | 5.01E-31 | Nightingale | VLDL_P | NA | T | C | 0.031898 | 0.004713 | 1.00E-12 |
| rs550057 | miR-126-3p | trans | 5.01E-31 | Nightingale | XL_HDL_C_ | NA | T | C | 0.03137 | 0.004525 | 1.90E-11 |
| rs550057 | miR-126-3p | trans | 5.01E-31 | Nightingale | S_LDL_TG_ | NA | T | C | -0.03461 | 0.004595 | 2.30E-13 |
| rs550057 | miR-126-3p | trans | 5.01E-31 | Nightingale | XS_VLDL_P | NA | T | C | 0.03659 | 0.004673 | 2.30E-16 |
| rs550057 | miR-126-3p | trans | 5.01E-31 | Nightingale | VLDL_C | NA | T | C | 0.038243 | 0.004732 | 4.20E-17 |
| rs550057 | miR-126-3p | trans | 5.01E-31 | Nightingale | ApoB | NA | T | C | 0.045452 | 0.004733 | 1.20E-22 |
| rs550057 | miR-126-3p | trans | 5.01E-31 | Nightingale | Pyruvate | NA | T | C | 0.027561 | 0.004747 | 6.40E-09 |
| rs550057 | miR-126-3p | trans | 5.01E-31 | Nightingale | S_VLDL_FC | NA | T | C | 0.048419 | 0.00474 | 1.40E-25 |
| rs550057 | miR-126-3p | trans | 5.01E-31 | Nightingale | XL_HDL_PL | NA | T | C | -0.02299 | 0.004263 | 5.00E-08 |
| rs554833 | miR-139-5p | trans | 6.24E-42 | Nightingale | S_LDL_C | NA | T | C | 0.03085 | 0.004429 | 6.20E-12 |
| rs554833 | miR-139-5p | trans | 6.24E-42 | Nightingale | Remnant_( | NA | T | C | 0.036771 | 0.004396 | 4.30E-17 |
| rs554833 | miR-139-5p | trans | 6.24E-42 | Nightingale | S_VLDL_CE | NA | T | C | 0.038101 | 0.004419 | 1.80E-18 |
| rs554833 | miR-139-5p | trans | 6.24E-42 | Nightingale | LDL_P | NA | T | C | 0.031693 | 0.004422 | 1.20E-12 |
| rs554833 | miR-139-5p | trans | 6.24E-42 | Nightingale | M_VLDL_P | NA | T | C | 0.029057 | 0.004418 | 2.20E-11 |
| rs554833 | miR-139-5p | trans | 6.24E-42 | Nightingale | L_LDL_PL | NA | T | C | 0.037892 | 0.004391 | 1.10E-17 |
| rs554833 | miR-139-5p | trans | 6.24E-42 | Nightingale | XXL_VLDL_ | NA | T | C | -0.03558 | 0.004463 | 2.80E-16 |
| rs554833 | miR-139-5p | trans | 6.24E-42 | Nightingale | M_VLDL_P | NA | T | C | 0.043907 | 0.004247 | 8.30E-25 |
| rs554833 | miR-139-5p | trans | 6.24E-42 | Nightingale | LDL_CE | NA | T | C | 0.032457 | 0.004413 | 2.50E-13 |
| rs554833 | miR-139-5p | trans | 6.24E-42 | Nightingale | M_VLDL_C | NA | T | C | 0.038627 | 0.004413 | 2.20E-18 |
| rs554833 | miR-139-5p | trans | 6.24E-42 | Nightingale | Sphingomy | NA | T | C | 0.028916 | 0.004136 | 8.10E-12 |
| rs554833 | miR-139-5p | trans | 6.24E-42 | Nightingale | S_HDL_C | NA | T | C | 0.028389 | 0.004417 | 1.80E-10 |
| rs554833 | miR-139-5p | trans | 6.24E-42 | Nightingale | M_LDL_TG | NA | T | C | -0.02757 | 0.00437 | 3.50E-10 |
| rs554833 | miR-139-5p | trans | 6.24E-42 | Nightingale | L_LDL_CE | NA | T | C | 0.031473 | 0.004385 | 1.10E-12 |
| rs554833 | miR-139-5p | trans | 6.24E-42 | Nightingale | XS_VLDL_L | NA | T | C | 0.033736 | 0.00435 | 2.30E-15 |
| rs554833 | miR-139-5p | trans | 6.24E-42 | Nightingale | XS_VLDL_P | NA | T | C | 0.032029 | 0.004363 | 6.00E-14 |
| rs554833 | miR-139-5p | trans | 6.24E-42 | Nightingale | Omega_6 | NA | T | C | 0.025802 | 0.004321 | 6.50E-10 |
| rs554833 | miR-139-5p | trans | 6.24E-42 | Nightingale | ApoB_by_/ | NA | T | C | 0.024942 | 0.004358 | 1.40E-08 |
| rs554833 | miR-139-5p | trans | 6.24E-42 | Nightingale | Citrate | NA | T | C | 0.026896 | 0.004396 | 6.50E-10 |
| rs554833 | miR-139-5p | trans | 6.24E-42 | Nightingale | M_VLDL_F | NA | T | C | 0.041958 | 0.00421 | 4.60E-23 |
| rs554833 | miR-139-5p | trans | 6.24E-42 | Nightingale | IDL_L | NA | T | C | 0.035176 | 0.004278 | 5.80E-16 |
| rs554833 | miR-139-5p | trans | 6.24E-42 | Nightingale | VLDL_FC | NA | T | C | 0.024084 | 0.004392 | 1.20E-08 |
| rs554833 | miR-139-5p | trans | 6.24E-42 | Nightingale | PUFA | NA | T | C | 0.023632 | 0.004316 | 1.60E-08 |
| rs554833 | miR-139-5p | trans | 6.24E-42 | Nightingale | Total_FC | NA | T | C | 0.033181 | 0.004323 | 7.70E-15 |
| rs554833 | miR-139-5p | trans | 6.24E-42 | Nightingale | LA | NA | T | C | 0.024875 | 0.004339 | 3.80E-09 |
| rs554833 | miR-139-5p | trans | 6.24E-42 | Nightingale | S_LDL_FC | NA | T | C | 0.028501 | 0.004418 | 4.50E-10 |
| rs554833 | miR-139-5p | trans | 6.24E-42 | Nightingale | M_LDL_CE | NA | T | C | 0.032879 | 0.004431 | 1.10E-13 |
| rs554833 | miR-139-5p | trans | 6.24E-42 | Nightingale | L_VLDL_CE | NA | T | C | 0.047245 | 0.004234 | 3.20E-29 |
| rs554833 | miR-139-5p | trans | 6.24E-42 | Nightingale | XL_VLDL_C | NA | T | C | 0.037257 | 0.004245 | 3.80E-18 |
| rs554833 | miR-139-5p | trans | 6.24E-42 | Nightingale | S_HDL_P | NA | T | C | 0.030576 | 0.004425 | 5.40E-12 |
| rs554833 | miR-139-5p | trans | 6.24E-42 | Nightingale | LDL_L | NA | T | C | 0.033207 | 0.00441 | 8.50E-14 |
| rs554833 | miR-139-5p | trans | 6.24E-42 | Nightingale | IDL_P | NA | T | C | 0.039913 | 0.004378 | 1.50E-19 |
| rs554833 | miR-139-5p | trans | 6.24E-42 | Nightingale | Clinical_LD | NA | T | C | 0.036378 | 0.004399 | 3.20E-16 |
| rs554833 | miR-139-5p | trans | 6.24E-42 | Nightingale | S_LDL_PL_ | NA | T | C | -0.03004 | 0.004417 | 2.80E-13 |
| rs554833 | miR-139-5p | trans | 6.24E-42 | Nightingale | XL_VLDL_F | NA | T | C | 0.037098 | 0.00429 | 2.50E-18 |
| rs554833 | miR-139-5p | trans | 6.24E-42 | Nightingale | S_HDL_CE | NA | T | C | 0.028614 | 0.004432 | 1.40E-10 |
| rs554833 | miR-139-5p | trans | 6.24E-42 | Nightingale | S_VLDL_C_ | NA | T | C | 0.041797 | 0.004306 | 6.50E-22 |
| rs554833 | miR-139-5p | trans | 6.24E-42 | Nightingale | M_LDL_FC | NA | T | C | 0.03427 | 0.004419 | 2.20E-14 |

| | | | | | | | | | | | |
|---|---|---|---|---|---|---|---|---|---|---|---|
| rs554833 | miR-139-5p | trans | 6.24E-42 | Nightingale | Total_PL | NA | T | C | 0.024871 | 0.004213 | 1.20E-09 |
| rs554833 | miR-139-5p | trans | 6.24E-42 | Nightingale | M_VLDL_T | NA | T | C | -0.03846 | 0.004181 | 8.80E-20 |
| rs554833 | miR-139-5p | trans | 6.24E-42 | Nightingale | IDL_CE | NA | T | C | 0.033425 | 0.004272 | 1.50E-14 |
| rs554833 | miR-139-5p | trans | 6.24E-42 | Nightingale | non_HDL_( | NA | T | C | 0.035598 | 0.004401 | 6.00E-16 |
| rs554833 | miR-139-5p | trans | 6.24E-42 | Nightingale | L_VLDL_FC | NA | T | C | 0.048348 | 0.004382 | 8.40E-29 |
| rs554833 | miR-139-5p | trans | 6.24E-42 | Nightingale | M_VLDL_C | NA | T | C | 0.040762 | 0.004385 | 2.60E-20 |
| rs554833 | miR-139-5p | trans | 6.24E-42 | Nightingale | M_VLDL_C | NA | T | C | 0.033552 | 0.004157 | 2.20E-15 |
| rs554833 | miR-139-5p | trans | 6.24E-42 | Nightingale | VLDL_P | NA | T | C | 0.026565 | 0.004401 | 4.70E-10 |
| rs554833 | miR-139-5p | trans | 6.24E-42 | Nightingale | IDL_C | NA | T | C | 0.03487 | 0.004273 | 1.10E-15 |
| rs554833 | miR-139-5p | trans | 6.24E-42 | Nightingale | Total_L | NA | T | C | 0.026282 | 0.004353 | 5.00E-10 |
| rs554833 | miR-139-5p | trans | 6.24E-42 | Nightingale | L_LDL_P | NA | T | C | 0.029065 | 0.00441 | 7.10E-11 |
| rs554833 | miR-139-5p | trans | 6.24E-42 | Nightingale | IDL_FC | NA | T | C | 0.038165 | 0.004284 | 1.80E-18 |
| rs554833 | miR-139-5p | trans | 6.24E-42 | Nightingale | IDL_FC_pct | NA | T | C | 0.025732 | 0.004391 | 1.10E-08 |
| rs554833 | miR-139-5p | trans | 6.24E-42 | Nightingale | XXL_VLDL_ | NA | T | C | 0.0307 | 0.004406 | 1.40E-12 |
| rs554833 | miR-139-5p | trans | 6.24E-42 | Nightingale | LDL_PL | NA | T | C | 0.034891 | 0.004415 | 5.50E-15 |
| rs554833 | miR-139-5p | trans | 6.24E-42 | Nightingale | Total_CE | NA | T | C | 0.031671 | 0.004268 | 1.10E-13 |
| rs554833 | miR-139-5p | trans | 6.24E-42 | Nightingale | S_VLDL_TG | NA | T | C | -0.03805 | 0.004283 | 1.70E-18 |
| rs554833 | miR-139-5p | trans | 6.24E-42 | Nightingale | Total_C | NA | T | C | 0.032298 | 0.004284 | 3.80E-14 |
| rs554833 | miR-139-5p | trans | 6.24E-42 | Nightingale | L_LDL_L | NA | T | C | 0.033453 | 0.004382 | 3.50E-14 |
| rs554833 | miR-139-5p | trans | 6.24E-42 | Nightingale | L_LDL_FC | NA | T | C | 0.033535 | 0.004345 | 3.30E-14 |
| rs554833 | miR-139-5p | trans | 6.24E-42 | Nightingale | VLDL_CE | NA | T | C | 0.035694 | 0.004426 | 3.40E-16 |
| rs554833 | miR-139-5p | trans | 6.24E-42 | Nightingale | M_VLDL_P | NA | T | C | 0.028221 | 0.004414 | 6.80E-11 |
| rs554833 | miR-139-5p | trans | 6.24E-42 | Nightingale | S_HDL_FC | NA | T | C | 0.023434 | 0.004344 | 4.80E-08 |
| rs554833 | miR-139-5p | trans | 6.24E-42 | Nightingale | S_HDL_L | NA | T | C | 0.026261 | 0.00441 | 1.60E-09 |
| rs554833 | miR-139-5p | trans | 6.24E-42 | Nightingale | XS_VLDL_F | NA | T | C | 0.031395 | 0.004393 | 2.50E-12 |
| rs554833 | miR-139-5p | trans | 6.24E-42 | Nightingale | M_LDL_C | NA | T | C | 0.033897 | 0.004433 | 2.40E-14 |
| rs554833 | miR-139-5p | trans | 6.24E-42 | Nightingale | M_LDL_P | NA | T | C | 0.035797 | 0.004437 | 7.90E-16 |
| rs554833 | miR-139-5p | trans | 6.24E-42 | Nightingale | XS_VLDL_C | NA | T | C | 0.037904 | 0.004295 | 1.60E-18 |
| rs554833 | miR-139-5p | trans | 6.24E-42 | Nightingale | L_LDL_C | NA | T | C | 0.032226 | 0.004375 | 3.00E-13 |
| rs554833 | miR-139-5p | trans | 6.24E-42 | Nightingale | M_VLDL_F | NA | T | C | 0.033298 | 0.004424 | 3.40E-14 |
| rs554833 | miR-139-5p | trans | 6.24E-42 | Nightingale | XL_VLDL_T | NA | T | C | -0.04375 | 0.004289 | 2.60E-24 |
| rs554833 | miR-139-5p | trans | 6.24E-42 | Nightingale | L_VLDL_TG | NA | T | C | -0.05094 | 0.004404 | 9.30E-32 |
| rs554833 | miR-139-5p | trans | 6.24E-42 | Nightingale | M_VLDL_C_ | NA | T | C | 0.03612 | 0.004168 | 1.40E-17 |
| rs554833 | miR-139-5p | trans | 6.24E-42 | Nightingale | M_LDL_L | NA | T | C | 0.03294 | 0.004431 | 1.10E-13 |
| rs554833 | miR-139-5p | trans | 6.24E-42 | Nightingale | XXL_VLDL_ | NA | T | C | 0.033029 | 0.004459 | 1.00E-13 |
| rs554833 | miR-139-5p | trans | 6.24E-42 | Nightingale | S_VLDL_PL | NA | T | C | 0.027679 | 0.004237 | 2.00E-10 |
| rs554833 | miR-139-5p | trans | 6.24E-42 | Nightingale | VLDL_C | NA | T | C | 0.031216 | 0.004419 | 5.50E-13 |
| rs554833 | miR-139-5p | trans | 6.24E-42 | Nightingale | S_VLDL_PL | NA | T | C | 0.032705 | 0.004417 | 4.50E-14 |
| rs554833 | miR-139-5p | trans | 6.24E-42 | Nightingale | S_VLDL_CE | NA | T | C | 0.046918 | 0.004365 | 1.60E-26 |
| rs554833 | miR-139-5p | trans | 6.24E-42 | Nightingale | M_LDL_C_ | NA | T | C | 0.03113 | 0.004428 | 3.10E-12 |
| rs554833 | miR-139-5p | trans | 6.24E-42 | Nightingale | XXL_VLDL_ | NA | T | C | 0.034703 | 0.004434 | 3.60E-15 |
| rs554833 | miR-139-5p | trans | 6.24E-42 | Nightingale | S_LDL_CE | NA | T | C | 0.030448 | 0.004427 | 6.60E-12 |
| rs554833 | miR-139-5p | trans | 6.24E-42 | Nightingale | XXL_VLDL_ | NA | T | C | 0.024904 | 0.004454 | 1.70E-09 |
| rs554833 | miR-139-5p | trans | 6.24E-42 | Nightingale | XL_VLDL_C | NA | T | C | 0.03813 | 0.004243 | 5.70E-19 |
| rs554833 | miR-139-5p | trans | 6.24E-42 | Nightingale | S_HDL_PL | NA | T | C | 0.023749 | 0.004386 | 3.40E-08 |
| rs554833 | miR-139-5p | trans | 6.24E-42 | Nightingale | LDL_C | NA | T | C | 0.0331 | 0.004406 | 1.00E-13 |
| rs554833 | miR-139-5p | trans | 6.24E-42 | Nightingale | XL_VLDL_P | NA | T | C | 0.038618 | 0.004432 | 8.20E-19 |
| rs554833 | miR-139-5p | trans | 6.24E-42 | Nightingale | XS_VLDL_C | NA | T | C | 0.037708 | 0.00427 | 2.50E-18 |
| rs554833 | miR-139-5p | trans | 6.24E-42 | Nightingale | LDL_FC | NA | T | C | 0.033547 | 0.004378 | 6.00E-14 |
| rs554833 | miR-139-5p | trans | 6.24E-42 | Nightingale | XS_VLDL_P | NA | T | C | 0.036622 | 0.004366 | 1.60E-17 |
| rs554833 | miR-139-5p | trans | 6.24E-42 | Nightingale | IDL_PL | NA | T | C | 0.035232 | 0.00427 | 4.10E-16 |
| rs554833 | miR-139-5p | trans | 6.24E-42 | Nightingale | S_LDL_C_p | NA | T | C | 0.045327 | 0.004442 | 6.00E-25 |
| rs554833 | miR-139-5p | trans | 6.24E-42 | Nightingale | ApoB | NA | T | C | 0.033838 | 0.004419 | 2.40E-14 |
| rs554833 | miR-139-5p | trans | 6.24E-42 | Nightingale | XL_HDL_PL | NA | T | C | -0.02272 | 0.004065 | 1.20E-08 |
| rs554833 | miR-139-5p | trans | 6.24E-42 | Nightingale | S_LDL_CE_ | NA | T | C | 0.030859 | 0.004417 | 5.70E-14 |
| rs554833 | miR-139-5p | trans | 6.24E-42 | Nightingale | XS_VLDL_F | NA | T | C | 0.036985 | 0.004348 | 7.40E-18 |
| rs554833 | miR-139-5p | trans | 6.24E-42 | Nightingale | S_VLDL_FC | NA | T | C | 0.037825 | 0.004426 | 8.10E-18 |
| rs554833 | miR-139-5p | trans | 6.24E-42 | Nightingale | S_VLDL_L | NA | T | C | 0.024612 | 0.004382 | 4.00E-09 |
| rs554833 | miR-139-5p | trans | 6.24E-42 | Nightingale | S_VLDL_P | NA | T | C | 0.023421 | 0.004369 | 1.70E-08 |
| rs554833 | miR-139-5p | trans | 6.24E-42 | Nightingale | L_VLDL_C_ | NA | T | C | 0.05203 | 0.004322 | 6.60E-34 |
| rs554833 | miR-139-5p | trans | 6.24E-42 | Nightingale | M_LDL_PL | NA | T | C | 0.032377 | 0.004431 | 3.00E-13 |
| rs554833 | miR-139-5p | trans | 6.24E-42 | Nightingale | S_LDL_L | NA | T | C | 0.027648 | 0.004424 | 9.10E-10 |
| rs554833 | miR-139-5p | trans | 6.24E-42 | Nightingale | S_VLDL_C | NA | T | C | 0.038292 | 0.004424 | 2.00E-18 |

| | | | | | | | | | | | | |
|---|---|---|---|---|---|---|---|---|---|---|---|---|
| rs554833 | miR-139-5p | trans | | 6.24E-42 | Nightingale | S_VLDL_FC | NA | T | C | 0.029304 | 0.004228 | 1.10E-11 |
| rs554833 | miR-139-5p | trans | | 6.24E-42 | Nightingale | S_LDL_P | NA | T | C | 0.029501 | 0.004429 | 3.50E-11 |
| rs554833 | miR-139-5p | trans | | 6.24E-42 | Nightingale | GlycA | NA | T | C | 0.027847 | 0.004406 | 1.80E-10 |
| rs554833 | miR-10b-5p | trans | | 3.22E-35 | Nightingale | S_LDL_C | NA | T | C | 0.03085 | 0.004429 | 6.20E-12 |
| rs554833 | miR-10b-5p | trans | | 3.22E-35 | Nightingale | Remnant_( | NA | T | C | 0.036771 | 0.004396 | 4.30E-17 |
| rs554833 | miR-10b-5p | trans | | 3.22E-35 | Nightingale | S_VLDL_CE | NA | T | C | 0.038101 | 0.004419 | 1.80E-18 |
| rs554833 | miR-10b-5p | trans | | 3.22E-35 | Nightingale | LDL_P | NA | T | C | 0.031693 | 0.004422 | 1.20E-12 |
| rs554833 | miR-10b-5p | trans | | 3.22E-35 | Nightingale | M_VLDL_P | NA | T | C | 0.029057 | 0.004418 | 2.20E-11 |
| rs554833 | miR-10b-5p | trans | | 3.22E-35 | Nightingale | L_LDL_PL | NA | T | C | 0.037892 | 0.004391 | 1.10E-17 |
| rs554833 | miR-10b-5p | trans | | 3.22E-35 | Nightingale | XXL_VLDL_ | NA | T | C | -0.03558 | 0.004463 | 2.80E-16 |
| rs554833 | miR-10b-5p | trans | | 3.22E-35 | Nightingale | M_VLDL_P | NA | T | C | 0.043907 | 0.004247 | 8.30E-25 |
| rs554833 | miR-10b-5p | trans | | 3.22E-35 | Nightingale | LDL_CE | NA | T | C | 0.032457 | 0.004413 | 2.50E-13 |
| rs554833 | miR-10b-5p | trans | | 3.22E-35 | Nightingale | M_VLDL_C | NA | T | C | 0.038627 | 0.004413 | 2.20E-18 |
| rs554833 | miR-10b-5p | trans | | 3.22E-35 | Nightingale | Sphingomy | NA | T | C | 0.028916 | 0.004136 | 8.10E-12 |
| rs554833 | miR-10b-5p | trans | | 3.22E-35 | Nightingale | S_HDL_C | NA | T | C | 0.028389 | 0.004417 | 1.80E-10 |
| rs554833 | miR-10b-5p | trans | | 3.22E-35 | Nightingale | M_LDL_TG | NA | T | C | -0.02757 | 0.00437 | 3.50E-10 |
| rs554833 | miR-10b-5p | trans | | 3.22E-35 | Nightingale | L_LDL_CE | NA | T | C | 0.031473 | 0.004385 | 1.10E-12 |
| rs554833 | miR-10b-5p | trans | | 3.22E-35 | Nightingale | XS_VLDL_L | NA | T | C | 0.033736 | 0.00435 | 2.30E-15 |
| rs554833 | miR-10b-5p | trans | | 3.22E-35 | Nightingale | XS_VLDL_P | NA | T | C | 0.032029 | 0.004363 | 6.00E-14 |
| rs554833 | miR-10b-5p | trans | | 3.22E-35 | Nightingale | Omega_6 | NA | T | C | 0.025802 | 0.004321 | 6.50E-10 |
| rs554833 | miR-10b-5p | trans | | 3.22E-35 | Nightingale | ApoB_by_/ | NA | T | C | 0.024942 | 0.004358 | 1.40E-08 |
| rs554833 | miR-10b-5p | trans | | 3.22E-35 | Nightingale | Citrate | NA | T | C | 0.026896 | 0.004396 | 6.50E-10 |
| rs554833 | miR-10b-5p | trans | | 3.22E-35 | Nightingale | M_VLDL_F | NA | T | C | 0.041958 | 0.00421 | 4.60E-23 |
| rs554833 | miR-10b-5p | trans | | 3.22E-35 | Nightingale | IDL_L | NA | T | C | 0.035176 | 0.004278 | 5.80E-16 |
| rs554833 | miR-10b-5p | trans | | 3.22E-35 | Nightingale | VLDL_FC | NA | T | C | 0.024084 | 0.004392 | 1.20E-08 |
| rs554833 | miR-10b-5p | trans | | 3.22E-35 | Nightingale | PUFA | NA | T | C | 0.023632 | 0.004316 | 1.60E-08 |
| rs554833 | miR-10b-5p | trans | | 3.22E-35 | Nightingale | Total_FC | NA | T | C | 0.033181 | 0.004323 | 7.70E-15 |
| rs554833 | miR-10b-5p | trans | | 3.22E-35 | Nightingale | LA | NA | T | C | 0.024875 | 0.004339 | 3.80E-09 |
| rs554833 | miR-10b-5p | trans | | 3.22E-35 | Nightingale | S_LDL_FC | NA | T | C | 0.028501 | 0.004418 | 4.50E-10 |
| rs554833 | miR-10b-5p | trans | | 3.22E-35 | Nightingale | M_LDL_CE | NA | T | C | 0.032879 | 0.004431 | 1.10E-13 |
| rs554833 | miR-10b-5p | trans | | 3.22E-35 | Nightingale | L_VLDL_CE | NA | T | C | 0.047245 | 0.004234 | 3.20E-29 |
| rs554833 | miR-10b-5p | trans | | 3.22E-35 | Nightingale | XL_VLDL_C | NA | T | C | 0.037257 | 0.004245 | 3.80E-18 |
| rs554833 | miR-10b-5p | trans | | 3.22E-35 | Nightingale | S_HDL_P | NA | T | C | 0.030576 | 0.004425 | 5.40E-12 |
| rs554833 | miR-10b-5p | trans | | 3.22E-35 | Nightingale | LDL_L | NA | T | C | 0.033207 | 0.00441 | 8.50E-14 |
| rs554833 | miR-10b-5p | trans | | 3.22E-35 | Nightingale | IDL_P | NA | T | C | 0.039913 | 0.004378 | 1.50E-19 |
| rs554833 | miR-10b-5p | trans | | 3.22E-35 | Nightingale | Clinical_LD | NA | T | C | 0.036378 | 0.004399 | 3.20E-16 |
| rs554833 | miR-10b-5p | trans | | 3.22E-35 | Nightingale | S_LDL_PL_ | NA | T | C | -0.03004 | 0.004417 | 2.80E-13 |
| rs554833 | miR-10b-5p | trans | | 3.22E-35 | Nightingale | XL_VLDL_F | NA | T | C | 0.037098 | 0.00429 | 2.50E-18 |
| rs554833 | miR-10b-5p | trans | | 3.22E-35 | Nightingale | S_HDL_CE | NA | T | C | 0.028614 | 0.004432 | 1.40E-10 |
| rs554833 | miR-10b-5p | trans | | 3.22E-35 | Nightingale | S_VLDL_C_ | NA | T | C | 0.041797 | 0.004306 | 6.50E-22 |
| rs554833 | miR-10b-5p | trans | | 3.22E-35 | Nightingale | M_LDL_FC | NA | T | C | 0.03427 | 0.004419 | 2.20E-14 |
| rs554833 | miR-10b-5p | trans | | 3.22E-35 | Nightingale | Total_PL | NA | T | C | 0.024871 | 0.004213 | 1.20E-09 |
| rs554833 | miR-10b-5p | trans | | 3.22E-35 | Nightingale | M_VLDL_T | NA | T | C | -0.03846 | 0.004181 | 8.80E-20 |
| rs554833 | miR-10b-5p | trans | | 3.22E-35 | Nightingale | IDL_CE | NA | T | C | 0.033425 | 0.004272 | 1.50E-14 |
| rs554833 | miR-10b-5p | trans | | 3.22E-35 | Nightingale | non_HDL_( | NA | T | C | 0.035598 | 0.004401 | 6.00E-16 |
| rs554833 | miR-10b-5p | trans | | 3.22E-35 | Nightingale | L_VLDL_FC | NA | T | C | 0.048348 | 0.004382 | 8.40E-29 |
| rs554833 | miR-10b-5p | trans | | 3.22E-35 | Nightingale | M_VLDL_C | NA | T | C | 0.040762 | 0.004385 | 2.60E-20 |
| rs554833 | miR-10b-5p | trans | | 3.22E-35 | Nightingale | M_VLDL_C | NA | T | C | 0.033552 | 0.004157 | 2.20E-15 |
| rs554833 | miR-10b-5p | trans | | 3.22E-35 | Nightingale | VLDL_P | NA | T | C | 0.026565 | 0.004401 | 4.70E-10 |
| rs554833 | miR-10b-5p | trans | | 3.22E-35 | Nightingale | IDL_C | NA | T | C | 0.03487 | 0.004273 | 1.10E-15 |
| rs554833 | miR-10b-5p | trans | | 3.22E-35 | Nightingale | Total_L | NA | T | C | 0.026282 | 0.004353 | 5.00E-10 |
| rs554833 | miR-10b-5p | trans | | 3.22E-35 | Nightingale | L_LDL_P | NA | T | C | 0.029065 | 0.00441 | 7.10E-11 |
| rs554833 | miR-10b-5p | trans | | 3.22E-35 | Nightingale | IDL_FC | NA | T | C | 0.038165 | 0.004284 | 1.80E-18 |
| rs554833 | miR-10b-5p | trans | | 3.22E-35 | Nightingale | IDL_FC_pct | NA | T | C | 0.025732 | 0.004391 | 1.10E-08 |
| rs554833 | miR-10b-5p | trans | | 3.22E-35 | Nightingale | XXL_VLDL_ | NA | T | C | 0.0307 | 0.004406 | 1.40E-12 |
| rs554833 | miR-10b-5p | trans | | 3.22E-35 | Nightingale | LDL_PL | NA | T | C | 0.034891 | 0.004415 | 5.50E-15 |
| rs554833 | miR-10b-5p | trans | | 3.22E-35 | Nightingale | Total_CE | NA | T | C | 0.031671 | 0.004268 | 1.10E-13 |
| rs554833 | miR-10b-5p | trans | | 3.22E-35 | Nightingale | S_VLDL_TG | NA | T | C | -0.03805 | 0.004283 | 1.70E-18 |
| rs554833 | miR-10b-5p | trans | | 3.22E-35 | Nightingale | Total_C | NA | T | C | 0.032298 | 0.004284 | 3.80E-14 |
| rs554833 | miR-10b-5p | trans | | 3.22E-35 | Nightingale | L_LDL_L | NA | T | C | 0.033453 | 0.004382 | 3.50E-14 |
| rs554833 | miR-10b-5p | trans | | 3.22E-35 | Nightingale | L_LDL_FC | NA | T | C | 0.033535 | 0.004345 | 3.30E-14 |
| rs554833 | miR-10b-5p | trans | | 3.22E-35 | Nightingale | VLDL_CE | NA | T | C | 0.035694 | 0.004426 | 3.40E-16 |
| rs554833 | miR-10b-5p | trans | | 3.22E-35 | Nightingale | M_VLDL_P | NA | T | C | 0.028221 | 0.004414 | 6.80E-11 |

| | | | | | | | | | | | |
|---|---|---|---|---|---|---|---|---|---|---|---|
| rs554833 | miR-10b-5p | trans | 3.22E-35 | Nightingale | S_HDL_FC | NA | T | C | 0.023434 | 0.004344 | 4.80E-08 |
| rs554833 | miR-10b-5p | trans | 3.22E-35 | Nightingale | S_HDL_L | NA | T | C | 0.026261 | 0.00441 | 1.60E-09 |
| rs554833 | miR-10b-5p | trans | 3.22E-35 | Nightingale | XS_VLDL_F | NA | T | C | 0.031395 | 0.004393 | 2.50E-12 |
| rs554833 | miR-10b-5p | trans | 3.22E-35 | Nightingale | M_LDL_C | NA | T | C | 0.033897 | 0.004433 | 2.40E-14 |
| rs554833 | miR-10b-5p | trans | 3.22E-35 | Nightingale | M_LDL_P | NA | T | C | 0.035797 | 0.004437 | 7.90E-16 |
| rs554833 | miR-10b-5p | trans | 3.22E-35 | Nightingale | XS_VLDL_C | NA | T | C | 0.037904 | 0.004295 | 1.60E-18 |
| rs554833 | miR-10b-5p | trans | 3.22E-35 | Nightingale | L_LDL_C | NA | T | C | 0.032226 | 0.004375 | 3.00E-13 |
| rs554833 | miR-10b-5p | trans | 3.22E-35 | Nightingale | M_VLDL_F | NA | T | C | 0.033298 | 0.004424 | 3.40E-14 |
| rs554833 | miR-10b-5p | trans | 3.22E-35 | Nightingale | XL_VLDL_T | NA | T | C | -0.04375 | 0.004289 | 2.60E-24 |
| rs554833 | miR-10b-5p | trans | 3.22E-35 | Nightingale | L_VLDL_TG | NA | T | C | -0.05094 | 0.004404 | 9.30E-32 |
| rs554833 | miR-10b-5p | trans | 3.22E-35 | Nightingale | M_VLDL_C | NA | T | C | 0.03612 | 0.004168 | 1.40E-17 |
| rs554833 | miR-10b-5p | trans | 3.22E-35 | Nightingale | M_LDL_L | NA | T | C | 0.03294 | 0.004431 | 1.10E-13 |
| rs554833 | miR-10b-5p | trans | 3.22E-35 | Nightingale | XXL_VLDL_ | NA | T | C | 0.033029 | 0.004459 | 1.00E-13 |
| rs554833 | miR-10b-5p | trans | 3.22E-35 | Nightingale | S_VLDL_PL | NA | T | C | 0.027679 | 0.004237 | 2.00E-10 |
| rs554833 | miR-10b-5p | trans | 3.22E-35 | Nightingale | VLDL_C | NA | T | C | 0.031216 | 0.004419 | 5.50E-13 |
| rs554833 | miR-10b-5p | trans | 3.22E-35 | Nightingale | S_VLDL_PL | NA | T | C | 0.032705 | 0.004417 | 4.50E-14 |
| rs554833 | miR-10b-5p | trans | 3.22E-35 | Nightingale | S_VLDL_CE | NA | T | C | 0.046918 | 0.004365 | 1.60E-26 |
| rs554833 | miR-10b-5p | trans | 3.22E-35 | Nightingale | M_LDL_C_ | NA | T | C | 0.03113 | 0.004428 | 3.10E-12 |
| rs554833 | miR-10b-5p | trans | 3.22E-35 | Nightingale | XXL_VLDL_ | NA | T | C | 0.034703 | 0.004434 | 3.60E-15 |
| rs554833 | miR-10b-5p | trans | 3.22E-35 | Nightingale | S_LDL_CE | NA | T | C | 0.030448 | 0.004427 | 6.60E-12 |
| rs554833 | miR-10b-5p | trans | 3.22E-35 | Nightingale | XXL_VLDL_ | NA | T | C | 0.024904 | 0.004454 | 1.70E-09 |
| rs554833 | miR-10b-5p | trans | 3.22E-35 | Nightingale | XL_VLDL_C | NA | T | C | 0.03813 | 0.004243 | 5.70E-19 |
| rs554833 | miR-10b-5p | trans | 3.22E-35 | Nightingale | S_HDL_PL | NA | T | C | 0.023749 | 0.004386 | 3.40E-08 |
| rs554833 | miR-10b-5p | trans | 3.22E-35 | Nightingale | LDL_C | NA | T | C | 0.0331 | 0.004406 | 1.00E-13 |
| rs554833 | miR-10b-5p | trans | 3.22E-35 | Nightingale | XL_VLDL_P | NA | T | C | 0.038618 | 0.004432 | 8.20E-19 |
| rs554833 | miR-10b-5p | trans | 3.22E-35 | Nightingale | XS_VLDL_C | NA | T | C | 0.037708 | 0.00427 | 2.50E-18 |
| rs554833 | miR-10b-5p | trans | 3.22E-35 | Nightingale | LDL_FC | NA | T | C | 0.033547 | 0.004378 | 6.00E-14 |
| rs554833 | miR-10b-5p | trans | 3.22E-35 | Nightingale | XS_VLDL_P | NA | T | C | 0.036622 | 0.004366 | 1.60E-17 |
| rs554833 | miR-10b-5p | trans | 3.22E-35 | Nightingale | IDL_PL | NA | T | C | 0.035232 | 0.00427 | 4.10E-16 |
| rs554833 | miR-10b-5p | trans | 3.22E-35 | Nightingale | S_LDL_C_p | NA | T | C | 0.045327 | 0.004442 | 6.00E-25 |
| rs554833 | miR-10b-5p | trans | 3.22E-35 | Nightingale | ApoB | NA | T | C | 0.033838 | 0.004419 | 2.40E-14 |
| rs554833 | miR-10b-5p | trans | 3.22E-35 | Nightingale | XL_HDL_PL | NA | T | C | -0.02272 | 0.004065 | 1.20E-08 |
| rs554833 | miR-10b-5p | trans | 3.22E-35 | Nightingale | S_LDL_CE_ | NA | T | C | 0.030859 | 0.004417 | 5.70E-14 |
| rs554833 | miR-10b-5p | trans | 3.22E-35 | Nightingale | XS_VLDL_F | NA | T | C | 0.036985 | 0.004348 | 7.40E-18 |
| rs554833 | miR-10b-5p | trans | 3.22E-35 | Nightingale | S_VLDL_FC | NA | T | C | 0.037825 | 0.004426 | 8.10E-18 |
| rs554833 | miR-10b-5p | trans | 3.22E-35 | Nightingale | S_VLDL_L | NA | T | C | 0.024612 | 0.004382 | 4.00E-09 |
| rs554833 | miR-10b-5p | trans | 3.22E-35 | Nightingale | S_VLDL_P | NA | T | C | 0.023421 | 0.004369 | 1.70E-08 |
| rs554833 | miR-10b-5p | trans | 3.22E-35 | Nightingale | L_VLDL_C_ | NA | T | C | 0.05203 | 0.004322 | 6.60E-34 |
| rs554833 | miR-10b-5p | trans | 3.22E-35 | Nightingale | M_LDL_PL | NA | T | C | 0.032377 | 0.004431 | 3.00E-13 |
| rs554833 | miR-10b-5p | trans | 3.22E-35 | Nightingale | S_LDL_L | NA | T | C | 0.027648 | 0.004424 | 9.10E-10 |
| rs554833 | miR-10b-5p | trans | 3.22E-35 | Nightingale | S_VLDL_C | NA | T | C | 0.038292 | 0.004424 | 2.00E-18 |
| rs554833 | miR-10b-5p | trans | 3.22E-35 | Nightingale | S_VLDL_FC | NA | T | C | 0.029304 | 0.004228 | 1.10E-11 |
| rs554833 | miR-10b-5p | trans | 3.22E-35 | Nightingale | S_LDL_P | NA | T | C | 0.029501 | 0.004429 | 3.50E-11 |
| rs554833 | miR-10b-5p | trans | 3.22E-35 | Nightingale | GlycA | NA | T | C | 0.027847 | 0.004406 | 1.80E-10 |
| rs554833 | let-7b-5p | trans | 8.42E-27 | Nightingale | S_LDL_C | NA | T | C | 0.03085 | 0.004429 | 6.20E-12 |
| rs554833 | let-7b-5p | trans | 8.42E-27 | Nightingale | Remnant_( | NA | T | C | 0.036771 | 0.004396 | 4.30E-17 |
| rs554833 | let-7b-5p | trans | 8.42E-27 | Nightingale | S_VLDL_CE | NA | T | C | 0.038101 | 0.004419 | 1.80E-18 |
| rs554833 | let-7b-5p | trans | 8.42E-27 | Nightingale | LDL_P | NA | T | C | 0.031693 | 0.004422 | 1.20E-12 |
| rs554833 | let-7b-5p | trans | 8.42E-27 | Nightingale | M_VLDL_P | NA | T | C | 0.029057 | 0.004418 | 2.20E-11 |
| rs554833 | let-7b-5p | trans | 8.42E-27 | Nightingale | L_LDL_PL | NA | T | C | 0.037892 | 0.004391 | 1.10E-17 |
| rs554833 | let-7b-5p | trans | 8.42E-27 | Nightingale | XXL_VLDL_ | NA | T | C | -0.03558 | 0.004463 | 2.80E-16 |
| rs554833 | let-7b-5p | trans | 8.42E-27 | Nightingale | M_VLDL_P | NA | T | C | 0.043907 | 0.004247 | 8.30E-25 |
| rs554833 | let-7b-5p | trans | 8.42E-27 | Nightingale | LDL_CE | NA | T | C | 0.032457 | 0.004413 | 2.50E-13 |
| rs554833 | let-7b-5p | trans | 8.42E-27 | Nightingale | M_VLDL_C | NA | T | C | 0.038627 | 0.004413 | 2.20E-18 |
| rs554833 | let-7b-5p | trans | 8.42E-27 | Nightingale | Sphingomy | NA | T | C | 0.028916 | 0.004136 | 8.10E-12 |
| rs554833 | let-7b-5p | trans | 8.42E-27 | Nightingale | S_HDL_C | NA | T | C | 0.028389 | 0.004417 | 1.80E-10 |
| rs554833 | let-7b-5p | trans | 8.42E-27 | Nightingale | M_LDL_TG | NA | T | C | -0.02757 | 0.00437 | 3.50E-10 |
| rs554833 | let-7b-5p | trans | 8.42E-27 | Nightingale | L_LDL_CE | NA | T | C | 0.031473 | 0.004385 | 1.10E-12 |
| rs554833 | let-7b-5p | trans | 8.42E-27 | Nightingale | XS_VLDL_L | NA | T | C | 0.033736 | 0.00435 | 2.30E-15 |
| rs554833 | let-7b-5p | trans | 8.42E-27 | Nightingale | XS_VLDL_P | NA | T | C | 0.032029 | 0.004363 | 6.00E-14 |
| rs554833 | let-7b-5p | trans | 8.42E-27 | Nightingale | Omega_6 | NA | T | C | 0.025802 | 0.004321 | 6.50E-10 |
| rs554833 | let-7b-5p | trans | 8.42E-27 | Nightingale | ApoB_by_A | NA | T | C | 0.024942 | 0.004358 | 1.40E-08 |
| rs554833 | let-7b-5p | trans | 8.42E-27 | Nightingale | Citrate | NA | T | C | 0.026896 | 0.004396 | 6.50E-10 |

| | | | | | | | | | | | |
|---|---|---|---|---|---|---|---|---|---|---|---|
| rs554833 | let-7b-5p | trans | 8.42E-27 | Nightingale | M_VLDL_F | NA | T | C | 0.041958 | 0.00421 | 4.60E-23 |
| rs554833 | let-7b-5p | trans | 8.42E-27 | Nightingale | IDL_L | NA | T | C | 0.035176 | 0.004278 | 5.80E-16 |
| rs554833 | let-7b-5p | trans | 8.42E-27 | Nightingale | VLDL_FC | NA | T | C | 0.024084 | 0.004392 | 1.20E-08 |
| rs554833 | let-7b-5p | trans | 8.42E-27 | Nightingale | PUFA | NA | T | C | 0.023632 | 0.004316 | 1.60E-08 |
| rs554833 | let-7b-5p | trans | 8.42E-27 | Nightingale | Total_FC | NA | T | C | 0.033181 | 0.004323 | 7.70E-15 |
| rs554833 | let-7b-5p | trans | 8.42E-27 | Nightingale | LA | NA | T | C | 0.024875 | 0.004339 | 3.80E-09 |
| rs554833 | let-7b-5p | trans | 8.42E-27 | Nightingale | S_LDL_FC | NA | T | C | 0.028501 | 0.004418 | 4.50E-10 |
| rs554833 | let-7b-5p | trans | 8.42E-27 | Nightingale | M_LDL_CE | NA | T | C | 0.032879 | 0.004431 | 1.10E-13 |
| rs554833 | let-7b-5p | trans | 8.42E-27 | Nightingale | L_VLDL_CE | NA | T | C | 0.047245 | 0.004234 | 3.20E-29 |
| rs554833 | let-7b-5p | trans | 8.42E-27 | Nightingale | XL_VLDL_C | NA | T | C | 0.037257 | 0.004245 | 3.80E-18 |
| rs554833 | let-7b-5p | trans | 8.42E-27 | Nightingale | S_HDL_P | NA | T | C | 0.030576 | 0.004425 | 5.40E-12 |
| rs554833 | let-7b-5p | trans | 8.42E-27 | Nightingale | LDL_L | NA | T | C | 0.033207 | 0.00441 | 8.50E-14 |
| rs554833 | let-7b-5p | trans | 8.42E-27 | Nightingale | IDL_P | NA | T | C | 0.039913 | 0.004378 | 1.50E-19 |
| rs554833 | let-7b-5p | trans | 8.42E-27 | Nightingale | Clinical_LD | NA | T | C | 0.036378 | 0.004399 | 3.20E-16 |
| rs554833 | let-7b-5p | trans | 8.42E-27 | Nightingale | S_LDL_PL_ | NA | T | C | -0.03004 | 0.004417 | 2.80E-13 |
| rs554833 | let-7b-5p | trans | 8.42E-27 | Nightingale | XL_VLDL_F | NA | T | C | 0.037098 | 0.00429 | 2.50E-18 |
| rs554833 | let-7b-5p | trans | 8.42E-27 | Nightingale | S_HDL_CE | NA | T | C | 0.028614 | 0.004432 | 1.40E-10 |
| rs554833 | let-7b-5p | trans | 8.42E-27 | Nightingale | S_VLDL_C_ | NA | T | C | 0.041797 | 0.004306 | 6.50E-22 |
| rs554833 | let-7b-5p | trans | 8.42E-27 | Nightingale | M_LDL_FC | NA | T | C | 0.03427 | 0.004419 | 2.20E-14 |
| rs554833 | let-7b-5p | trans | 8.42E-27 | Nightingale | Total_PL | NA | T | C | 0.024871 | 0.004213 | 1.20E-09 |
| rs554833 | let-7b-5p | trans | 8.42E-27 | Nightingale | M_VLDL_T | NA | T | C | -0.03846 | 0.004181 | 8.80E-20 |
| rs554833 | let-7b-5p | trans | 8.42E-27 | Nightingale | IDL_CE | NA | T | C | 0.033425 | 0.004272 | 1.50E-14 |
| rs554833 | let-7b-5p | trans | 8.42E-27 | Nightingale | non_HDL_( | NA | T | C | 0.035598 | 0.004401 | 6.00E-16 |
| rs554833 | let-7b-5p | trans | 8.42E-27 | Nightingale | L_VLDL_FC | NA | T | C | 0.048348 | 0.004382 | 8.40E-29 |
| rs554833 | let-7b-5p | trans | 8.42E-27 | Nightingale | M_VLDL_C | NA | T | C | 0.040762 | 0.004385 | 2.60E-20 |
| rs554833 | let-7b-5p | trans | 8.42E-27 | Nightingale | M_VLDL_C | NA | T | C | 0.033552 | 0.004157 | 2.20E-15 |
| rs554833 | let-7b-5p | trans | 8.42E-27 | Nightingale | VLDL_P | NA | T | C | 0.026565 | 0.004401 | 4.70E-10 |
| rs554833 | let-7b-5p | trans | 8.42E-27 | Nightingale | IDL_C | NA | T | C | 0.03487 | 0.004273 | 1.10E-15 |
| rs554833 | let-7b-5p | trans | 8.42E-27 | Nightingale | Total_L | NA | T | C | 0.026282 | 0.004353 | 5.00E-10 |
| rs554833 | let-7b-5p | trans | 8.42E-27 | Nightingale | L_LDL_P | NA | T | C | 0.029065 | 0.00441 | 7.10E-11 |
| rs554833 | let-7b-5p | trans | 8.42E-27 | Nightingale | IDL_FC | NA | T | C | 0.038165 | 0.004284 | 1.80E-18 |
| rs554833 | let-7b-5p | trans | 8.42E-27 | Nightingale | IDL_FC_pct | NA | T | C | 0.025732 | 0.004391 | 1.10E-08 |
| rs554833 | let-7b-5p | trans | 8.42E-27 | Nightingale | XXL_VLDL_ | NA | T | C | 0.0307 | 0.004406 | 1.40E-12 |
| rs554833 | let-7b-5p | trans | 8.42E-27 | Nightingale | LDL_PL | NA | T | C | 0.034891 | 0.004415 | 5.50E-15 |
| rs554833 | let-7b-5p | trans | 8.42E-27 | Nightingale | Total_CE | NA | T | C | 0.031671 | 0.004268 | 1.10E-13 |
| rs554833 | let-7b-5p | trans | 8.42E-27 | Nightingale | S_VLDL_TG | NA | T | C | -0.03805 | 0.004283 | 1.70E-18 |
| rs554833 | let-7b-5p | trans | 8.42E-27 | Nightingale | Total_C | NA | T | C | 0.032298 | 0.004284 | 3.80E-14 |
| rs554833 | let-7b-5p | trans | 8.42E-27 | Nightingale | L_LDL_L | NA | T | C | 0.033453 | 0.004382 | 3.50E-14 |
| rs554833 | let-7b-5p | trans | 8.42E-27 | Nightingale | L_LDL_FC | NA | T | C | 0.033535 | 0.004345 | 3.30E-14 |
| rs554833 | let-7b-5p | trans | 8.42E-27 | Nightingale | VLDL_CE | NA | T | C | 0.035694 | 0.004426 | 3.40E-16 |
| rs554833 | let-7b-5p | trans | 8.42E-27 | Nightingale | M_VLDL_P | NA | T | C | 0.028221 | 0.004414 | 6.80E-11 |
| rs554833 | let-7b-5p | trans | 8.42E-27 | Nightingale | S_HDL_FC | NA | T | C | 0.023434 | 0.004344 | 4.80E-08 |
| rs554833 | let-7b-5p | trans | 8.42E-27 | Nightingale | S_HDL_L | NA | T | C | 0.026261 | 0.00441 | 1.60E-09 |
| rs554833 | let-7b-5p | trans | 8.42E-27 | Nightingale | XS_VLDL_F | NA | T | C | 0.031395 | 0.004393 | 2.50E-12 |
| rs554833 | let-7b-5p | trans | 8.42E-27 | Nightingale | M_LDL_C | NA | T | C | 0.033897 | 0.004433 | 2.40E-14 |
| rs554833 | let-7b-5p | trans | 8.42E-27 | Nightingale | M_LDL_P | NA | T | C | 0.035797 | 0.004437 | 7.90E-16 |
| rs554833 | let-7b-5p | trans | 8.42E-27 | Nightingale | XS_VLDL_C | NA | T | C | 0.037904 | 0.004295 | 1.60E-18 |
| rs554833 | let-7b-5p | trans | 8.42E-27 | Nightingale | L_LDL_C | NA | T | C | 0.032226 | 0.004375 | 3.00E-13 |
| rs554833 | let-7b-5p | trans | 8.42E-27 | Nightingale | M_VLDL_F | NA | T | C | 0.033298 | 0.004424 | 3.40E-14 |
| rs554833 | let-7b-5p | trans | 8.42E-27 | Nightingale | XL_VLDL_T | NA | T | C | -0.04375 | 0.004289 | 2.60E-24 |
| rs554833 | let-7b-5p | trans | 8.42E-27 | Nightingale | L_VLDL_TG | NA | T | C | -0.05094 | 0.004404 | 9.30E-32 |
| rs554833 | let-7b-5p | trans | 8.42E-27 | Nightingale | M_VLDL_C | NA | T | C | 0.03612 | 0.004168 | 1.40E-17 |
| rs554833 | let-7b-5p | trans | 8.42E-27 | Nightingale | M_LDL_L | NA | T | C | 0.03294 | 0.004431 | 1.10E-13 |
| rs554833 | let-7b-5p | trans | 8.42E-27 | Nightingale | XXL_VLDL_ | NA | T | C | 0.033029 | 0.004459 | 1.00E-13 |
| rs554833 | let-7b-5p | trans | 8.42E-27 | Nightingale | S_VLDL_PL | NA | T | C | 0.027679 | 0.004237 | 2.00E-10 |
| rs554833 | let-7b-5p | trans | 8.42E-27 | Nightingale | VLDL_C | NA | T | C | 0.031216 | 0.004419 | 5.50E-13 |
| rs554833 | let-7b-5p | trans | 8.42E-27 | Nightingale | S_VLDL_PL | NA | T | C | 0.032705 | 0.004417 | 4.50E-14 |
| rs554833 | let-7b-5p | trans | 8.42E-27 | Nightingale | S_VLDL_CE | NA | T | C | 0.046918 | 0.004365 | 1.60E-26 |
| rs554833 | let-7b-5p | trans | 8.42E-27 | Nightingale | M_LDL_C_ | NA | T | C | 0.03113 | 0.004428 | 3.10E-12 |
| rs554833 | let-7b-5p | trans | 8.42E-27 | Nightingale | XXL_VLDL_ | NA | T | C | 0.034703 | 0.004434 | 3.60E-15 |
| rs554833 | let-7b-5p | trans | 8.42E-27 | Nightingale | S_LDL_CE | NA | T | C | 0.030448 | 0.004427 | 6.60E-12 |
| rs554833 | let-7b-5p | trans | 8.42E-27 | Nightingale | XXL_VLDL_ | NA | T | C | 0.024904 | 0.004454 | 1.70E-09 |
| rs554833 | let-7b-5p | trans | 8.42E-27 | Nightingale | XL_VLDL_C | NA | T | C | 0.03813 | 0.004243 | 5.70E-19 |

| | | | | | | | | | | | |
|---|---|---|---|---|---|---|---|---|---|---|---|
| rs554833 | let-7b-5p | trans | 8.42E-27 | Nightingale | S_HDL_PL | NA | T | C | 0.023749 | 0.004386 | 3.40E-08 |
| rs554833 | let-7b-5p | trans | 8.42E-27 | Nightingale | LDL_C | NA | T | C | 0.0331 | 0.004406 | 1.00E-13 |
| rs554833 | let-7b-5p | trans | 8.42E-27 | Nightingale | XL_VLDL_P | NA | T | C | 0.038618 | 0.004432 | 8.20E-19 |
| rs554833 | let-7b-5p | trans | 8.42E-27 | Nightingale | XS_VLDL_C | NA | T | C | 0.037708 | 0.00427 | 2.50E-18 |
| rs554833 | let-7b-5p | trans | 8.42E-27 | Nightingale | LDL_FC | NA | T | C | 0.033547 | 0.004378 | 6.00E-14 |
| rs554833 | let-7b-5p | trans | 8.42E-27 | Nightingale | XS_VLDL_P | NA | T | C | 0.036622 | 0.004366 | 1.60E-17 |
| rs554833 | let-7b-5p | trans | 8.42E-27 | Nightingale | IDL_PL | NA | T | C | 0.035232 | 0.00427 | 4.10E-16 |
| rs554833 | let-7b-5p | trans | 8.42E-27 | Nightingale | S_LDL_C_p | NA | T | C | 0.045327 | 0.004442 | 6.00E-25 |
| rs554833 | let-7b-5p | trans | 8.42E-27 | Nightingale | ApoB | NA | T | C | 0.033838 | 0.004419 | 2.40E-14 |
| rs554833 | let-7b-5p | trans | 8.42E-27 | Nightingale | XL_HDL_PL | NA | T | C | -0.02272 | 0.004065 | 1.20E-08 |
| rs554833 | let-7b-5p | trans | 8.42E-27 | Nightingale | S_LDL_CE_ | NA | T | C | 0.030859 | 0.004417 | 5.70E-14 |
| rs554833 | let-7b-5p | trans | 8.42E-27 | Nightingale | XS_VLDL_F | NA | T | C | 0.036985 | 0.004348 | 7.40E-18 |
| rs554833 | let-7b-5p | trans | 8.42E-27 | Nightingale | S_VLDL_FC | NA | T | C | 0.037825 | 0.004426 | 8.10E-18 |
| rs554833 | let-7b-5p | trans | 8.42E-27 | Nightingale | S_VLDL_L | NA | T | C | 0.024612 | 0.004382 | 4.00E-09 |
| rs554833 | let-7b-5p | trans | 8.42E-27 | Nightingale | S_VLDL_P | NA | T | C | 0.023421 | 0.004369 | 1.70E-08 |
| rs554833 | let-7b-5p | trans | 8.42E-27 | Nightingale | L_VLDL_C_ | NA | T | C | 0.05203 | 0.004322 | 6.60E-34 |
| rs554833 | let-7b-5p | trans | 8.42E-27 | Nightingale | M_LDL_PL | NA | T | C | 0.032377 | 0.004431 | 3.00E-13 |
| rs554833 | let-7b-5p | trans | 8.42E-27 | Nightingale | S_LDL_L | NA | T | C | 0.027648 | 0.004424 | 9.10E-10 |
| rs554833 | let-7b-5p | trans | 8.42E-27 | Nightingale | S_VLDL_C | NA | T | C | 0.038292 | 0.004424 | 2.00E-18 |
| rs554833 | let-7b-5p | trans | 8.42E-27 | Nightingale | S_VLDL_FC | NA | T | C | 0.029304 | 0.004228 | 1.10E-11 |
| rs554833 | let-7b-5p | trans | 8.42E-27 | Nightingale | S_LDL_P | NA | T | C | 0.029501 | 0.004429 | 3.50E-11 |
| rs554833 | let-7b-5p | trans | 8.42E-27 | Nightingale | GlycA | NA | T | C | 0.027847 | 0.004406 | 1.80E-10 |
| rs554833 | miR-126-3p | trans | 2.40E-39 | Nightingale | S_LDL_C | NA | T | C | 0.03085 | 0.004429 | 6.20E-12 |
| rs554833 | miR-126-3p | trans | 2.40E-39 | Nightingale | Remnant_( | NA | T | C | 0.036771 | 0.004396 | 4.30E-17 |
| rs554833 | miR-126-3p | trans | 2.40E-39 | Nightingale | S_VLDL_CE | NA | T | C | 0.038101 | 0.004419 | 1.80E-18 |
| rs554833 | miR-126-3p | trans | 2.40E-39 | Nightingale | LDL_P | NA | T | C | 0.031693 | 0.004422 | 1.20E-12 |
| rs554833 | miR-126-3p | trans | 2.40E-39 | Nightingale | M_VLDL_P | NA | T | C | 0.029057 | 0.004418 | 2.20E-11 |
| rs554833 | miR-126-3p | trans | 2.40E-39 | Nightingale | L_LDL_PL | NA | T | C | 0.037892 | 0.004391 | 1.10E-17 |
| rs554833 | miR-126-3p | trans | 2.40E-39 | Nightingale | XXL_VLDL_ | NA | T | C | -0.03558 | 0.004463 | 2.80E-16 |
| rs554833 | miR-126-3p | trans | 2.40E-39 | Nightingale | M_VLDL_P | NA | T | C | 0.043907 | 0.004247 | 8.30E-25 |
| rs554833 | miR-126-3p | trans | 2.40E-39 | Nightingale | LDL_CE | NA | T | C | 0.032457 | 0.004413 | 2.50E-13 |
| rs554833 | miR-126-3p | trans | 2.40E-39 | Nightingale | M_VLDL_C | NA | T | C | 0.038627 | 0.004413 | 2.20E-18 |
| rs554833 | miR-126-3p | trans | 2.40E-39 | Nightingale | Sphingomy | NA | T | C | 0.028916 | 0.004136 | 8.10E-12 |
| rs554833 | miR-126-3p | trans | 2.40E-39 | Nightingale | S_HDL_C | NA | T | C | 0.028389 | 0.004417 | 1.80E-10 |
| rs554833 | miR-126-3p | trans | 2.40E-39 | Nightingale | M_LDL_TG | NA | T | C | -0.02757 | 0.00437 | 3.50E-10 |
| rs554833 | miR-126-3p | trans | 2.40E-39 | Nightingale | L_LDL_CE | NA | T | C | 0.031473 | 0.004385 | 1.10E-12 |
| rs554833 | miR-126-3p | trans | 2.40E-39 | Nightingale | XS_VLDL_L | NA | T | C | 0.033736 | 0.00435 | 2.30E-15 |
| rs554833 | miR-126-3p | trans | 2.40E-39 | Nightingale | XS_VLDL_P | NA | T | C | 0.032029 | 0.004363 | 6.00E-14 |
| rs554833 | miR-126-3p | trans | 2.40E-39 | Nightingale | Omega_6 | NA | T | C | 0.025802 | 0.004321 | 6.50E-10 |
| rs554833 | miR-126-3p | trans | 2.40E-39 | Nightingale | ApoB_by_A | NA | T | C | 0.024942 | 0.004358 | 1.40E-08 |
| rs554833 | miR-126-3p | trans | 2.40E-39 | Nightingale | Citrate | NA | T | C | 0.026896 | 0.004396 | 6.50E-10 |
| rs554833 | miR-126-3p | trans | 2.40E-39 | Nightingale | M_VLDL_F | NA | T | C | 0.041958 | 0.00421 | 4.60E-23 |
| rs554833 | miR-126-3p | trans | 2.40E-39 | Nightingale | IDL_L | NA | T | C | 0.035176 | 0.004278 | 5.80E-16 |
| rs554833 | miR-126-3p | trans | 2.40E-39 | Nightingale | VLDL_FC | NA | T | C | 0.024084 | 0.004392 | 1.20E-08 |
| rs554833 | miR-126-3p | trans | 2.40E-39 | Nightingale | PUFA | NA | T | C | 0.023632 | 0.004316 | 1.60E-08 |
| rs554833 | miR-126-3p | trans | 2.40E-39 | Nightingale | Total_FC | NA | T | C | 0.033181 | 0.004323 | 7.70E-15 |
| rs554833 | miR-126-3p | trans | 2.40E-39 | Nightingale | LA | NA | T | C | 0.024875 | 0.004339 | 3.80E-09 |
| rs554833 | miR-126-3p | trans | 2.40E-39 | Nightingale | S_LDL_FC | NA | T | C | 0.028501 | 0.004418 | 4.50E-10 |
| rs554833 | miR-126-3p | trans | 2.40E-39 | Nightingale | M_LDL_CE | NA | T | C | 0.032879 | 0.004431 | 1.10E-13 |
| rs554833 | miR-126-3p | trans | 2.40E-39 | Nightingale | L_VLDL_CE | NA | T | C | 0.047245 | 0.004234 | 3.20E-29 |
| rs554833 | miR-126-3p | trans | 2.40E-39 | Nightingale | XL_VLDL_C | NA | T | C | 0.037257 | 0.004245 | 3.80E-18 |
| rs554833 | miR-126-3p | trans | 2.40E-39 | Nightingale | S_HDL_P | NA | T | C | 0.030576 | 0.004425 | 5.40E-12 |
| rs554833 | miR-126-3p | trans | 2.40E-39 | Nightingale | LDL_L | NA | T | C | 0.033207 | 0.00441 | 8.50E-14 |
| rs554833 | miR-126-3p | trans | 2.40E-39 | Nightingale | IDL_P | NA | T | C | 0.039913 | 0.004378 | 1.50E-19 |
| rs554833 | miR-126-3p | trans | 2.40E-39 | Nightingale | Clinical_LD | NA | T | C | 0.036378 | 0.004399 | 3.20E-16 |
| rs554833 | miR-126-3p | trans | 2.40E-39 | Nightingale | S_LDL_PL_ | NA | T | C | -0.03004 | 0.004417 | 2.80E-13 |
| rs554833 | miR-126-3p | trans | 2.40E-39 | Nightingale | XL_VLDL_F | NA | T | C | 0.037098 | 0.00429 | 2.50E-18 |
| rs554833 | miR-126-3p | trans | 2.40E-39 | Nightingale | S_HDL_CE | NA | T | C | 0.028614 | 0.004432 | 1.40E-10 |
| rs554833 | miR-126-3p | trans | 2.40E-39 | Nightingale | S_VLDL_C_ | NA | T | C | 0.041797 | 0.004306 | 6.50E-22 |
| rs554833 | miR-126-3p | trans | 2.40E-39 | Nightingale | M_LDL_FC | NA | T | C | 0.03427 | 0.004419 | 2.20E-14 |
| rs554833 | miR-126-3p | trans | 2.40E-39 | Nightingale | Total_PL | NA | T | C | 0.024871 | 0.004213 | 1.20E-09 |
| rs554833 | miR-126-3p | trans | 2.40E-39 | Nightingale | M_VLDL_T | NA | T | C | -0.03846 | 0.004181 | 8.80E-20 |
| rs554833 | miR-126-3p | trans | 2.40E-39 | Nightingale | IDL_CE | NA | T | C | 0.033425 | 0.004272 | 1.50E-14 |

| | | | | | | | | | | | |
|---|---|---|---|---|---|---|---|---|---|---|---|
| rs554833 | miR-126-3p | trans | 2.40E-39 | Nightingale | non_HDL_( | NA | T | C | 0.035598 | 0.004401 | 6.00E-16 |
| rs554833 | miR-126-3p | trans | 2.40E-39 | Nightingale | L_VLDL_FC | NA | T | C | 0.048348 | 0.004382 | 8.40E-29 |
| rs554833 | miR-126-3p | trans | 2.40E-39 | Nightingale | M_VLDL_C | NA | T | C | 0.040762 | 0.004385 | 2.60E-20 |
| rs554833 | miR-126-3p | trans | 2.40E-39 | Nightingale | M_VLDL_C | NA | T | C | 0.033552 | 0.004157 | 2.20E-15 |
| rs554833 | miR-126-3p | trans | 2.40E-39 | Nightingale | VLDL_P | NA | T | C | 0.026565 | 0.004401 | 4.70E-10 |
| rs554833 | miR-126-3p | trans | 2.40E-39 | Nightingale | IDL_C | NA | T | C | 0.03487 | 0.004273 | 1.10E-15 |
| rs554833 | miR-126-3p | trans | 2.40E-39 | Nightingale | Total_L | NA | T | C | 0.026282 | 0.004353 | 5.00E-10 |
| rs554833 | miR-126-3p | trans | 2.40E-39 | Nightingale | L_LDL_P | NA | T | C | 0.029065 | 0.00441 | 7.10E-11 |
| rs554833 | miR-126-3p | trans | 2.40E-39 | Nightingale | IDL_FC | NA | T | C | 0.038165 | 0.004284 | 1.80E-18 |
| rs554833 | miR-126-3p | trans | 2.40E-39 | Nightingale | IDL_FC_pct | NA | T | C | 0.025732 | 0.004391 | 1.10E-08 |
| rs554833 | miR-126-3p | trans | 2.40E-39 | Nightingale | XXL_VLDL_ | NA | T | C | 0.0307 | 0.004406 | 1.40E-12 |
| rs554833 | miR-126-3p | trans | 2.40E-39 | Nightingale | LDL_PL | NA | T | C | 0.034891 | 0.004415 | 5.50E-15 |
| rs554833 | miR-126-3p | trans | 2.40E-39 | Nightingale | Total_CE | NA | T | C | 0.031671 | 0.004268 | 1.10E-13 |
| rs554833 | miR-126-3p | trans | 2.40E-39 | Nightingale | S_VLDL_TG | NA | T | C | -0.03805 | 0.004283 | 1.70E-18 |
| rs554833 | miR-126-3p | trans | 2.40E-39 | Nightingale | Total_C | NA | T | C | 0.032298 | 0.004284 | 3.80E-14 |
| rs554833 | miR-126-3p | trans | 2.40E-39 | Nightingale | L_LDL_L | NA | T | C | 0.033453 | 0.004382 | 3.50E-14 |
| rs554833 | miR-126-3p | trans | 2.40E-39 | Nightingale | L_LDL_FC | NA | T | C | 0.033535 | 0.004345 | 3.30E-14 |
| rs554833 | miR-126-3p | trans | 2.40E-39 | Nightingale | VLDL_CE | NA | T | C | 0.035694 | 0.004426 | 3.40E-16 |
| rs554833 | miR-126-3p | trans | 2.40E-39 | Nightingale | M_VLDL_P | NA | T | C | 0.028221 | 0.004414 | 6.80E-11 |
| rs554833 | miR-126-3p | trans | 2.40E-39 | Nightingale | S_HDL_FC | NA | T | C | 0.023434 | 0.004344 | 4.80E-08 |
| rs554833 | miR-126-3p | trans | 2.40E-39 | Nightingale | S_HDL_L | NA | T | C | 0.026261 | 0.00441 | 1.60E-09 |
| rs554833 | miR-126-3p | trans | 2.40E-39 | Nightingale | XS_VLDL_F | NA | T | C | 0.031395 | 0.004393 | 2.50E-12 |
| rs554833 | miR-126-3p | trans | 2.40E-39 | Nightingale | M_LDL_C | NA | T | C | 0.033897 | 0.004433 | 2.40E-14 |
| rs554833 | miR-126-3p | trans | 2.40E-39 | Nightingale | M_LDL_P | NA | T | C | 0.035797 | 0.004437 | 7.90E-16 |
| rs554833 | miR-126-3p | trans | 2.40E-39 | Nightingale | XS_VLDL_C | NA | T | C | 0.037904 | 0.004295 | 1.60E-18 |
| rs554833 | miR-126-3p | trans | 2.40E-39 | Nightingale | L_LDL_C | NA | T | C | 0.032226 | 0.004375 | 3.00E-13 |
| rs554833 | miR-126-3p | trans | 2.40E-39 | Nightingale | M_VLDL_F | NA | T | C | 0.033298 | 0.004424 | 3.40E-14 |
| rs554833 | miR-126-3p | trans | 2.40E-39 | Nightingale | XL_VLDL_T | NA | T | C | -0.04375 | 0.004289 | 2.60E-24 |
| rs554833 | miR-126-3p | trans | 2.40E-39 | Nightingale | L_VLDL_TG | NA | T | C | -0.05094 | 0.004404 | 9.30E-32 |
| rs554833 | miR-126-3p | trans | 2.40E-39 | Nightingale | M_VLDL_C | NA | T | C | 0.03612 | 0.004168 | 1.40E-17 |
| rs554833 | miR-126-3p | trans | 2.40E-39 | Nightingale | M_LDL_L | NA | T | C | 0.03294 | 0.004431 | 1.10E-13 |
| rs554833 | miR-126-3p | trans | 2.40E-39 | Nightingale | XXL_VLDL_ | NA | T | C | 0.033029 | 0.004459 | 1.00E-13 |
| rs554833 | miR-126-3p | trans | 2.40E-39 | Nightingale | S_VLDL_PL | NA | T | C | 0.027679 | 0.004237 | 2.00E-10 |
| rs554833 | miR-126-3p | trans | 2.40E-39 | Nightingale | VLDL_C | NA | T | C | 0.031216 | 0.004419 | 5.50E-13 |
| rs554833 | miR-126-3p | trans | 2.40E-39 | Nightingale | S_VLDL_PL | NA | T | C | 0.032705 | 0.004417 | 4.50E-14 |
| rs554833 | miR-126-3p | trans | 2.40E-39 | Nightingale | S_VLDL_CE | NA | T | C | 0.046918 | 0.004365 | 1.60E-26 |
| rs554833 | miR-126-3p | trans | 2.40E-39 | Nightingale | M_LDL_C_ | NA | T | C | 0.03113 | 0.004428 | 3.10E-12 |
| rs554833 | miR-126-3p | trans | 2.40E-39 | Nightingale | XXL_VLDL_ | NA | T | C | 0.034703 | 0.004434 | 3.60E-15 |
| rs554833 | miR-126-3p | trans | 2.40E-39 | Nightingale | S_LDL_CE | NA | T | C | 0.030448 | 0.004427 | 6.60E-12 |
| rs554833 | miR-126-3p | trans | 2.40E-39 | Nightingale | XXL_VLDL_ | NA | T | C | 0.024904 | 0.004454 | 1.70E-09 |
| rs554833 | miR-126-3p | trans | 2.40E-39 | Nightingale | XL_VLDL_C | NA | T | C | 0.03813 | 0.004243 | 5.70E-19 |
| rs554833 | miR-126-3p | trans | 2.40E-39 | Nightingale | S_HDL_PL | NA | T | C | 0.023749 | 0.004386 | 3.40E-08 |
| rs554833 | miR-126-3p | trans | 2.40E-39 | Nightingale | LDL_C | NA | T | C | 0.0331 | 0.004406 | 1.00E-13 |
| rs554833 | miR-126-3p | trans | 2.40E-39 | Nightingale | XL_VLDL_P | NA | T | C | 0.038618 | 0.004432 | 8.20E-19 |
| rs554833 | miR-126-3p | trans | 2.40E-39 | Nightingale | XS_VLDL_C | NA | T | C | 0.037708 | 0.00427 | 2.50E-18 |
| rs554833 | miR-126-3p | trans | 2.40E-39 | Nightingale | LDL_FC | NA | T | C | 0.033547 | 0.004378 | 6.00E-14 |
| rs554833 | miR-126-3p | trans | 2.40E-39 | Nightingale | XS_VLDL_P | NA | T | C | 0.036622 | 0.004366 | 1.60E-17 |
| rs554833 | miR-126-3p | trans | 2.40E-39 | Nightingale | IDL_PL | NA | T | C | 0.035232 | 0.00427 | 4.10E-16 |
| rs554833 | miR-126-3p | trans | 2.40E-39 | Nightingale | S_LDL_C_p | NA | T | C | 0.045327 | 0.004442 | 6.00E-25 |
| rs554833 | miR-126-3p | trans | 2.40E-39 | Nightingale | ApoB | NA | T | C | 0.033838 | 0.004419 | 2.40E-14 |
| rs554833 | miR-126-3p | trans | 2.40E-39 | Nightingale | XL_HDL_PL | NA | T | C | -0.02272 | 0.004065 | 1.20E-08 |
| rs554833 | miR-126-3p | trans | 2.40E-39 | Nightingale | S_LDL_CE_ | NA | T | C | 0.030859 | 0.004417 | 5.70E-14 |
| rs554833 | miR-126-3p | trans | 2.40E-39 | Nightingale | XS_VLDL_F | NA | T | C | 0.036985 | 0.004348 | 7.40E-18 |
| rs554833 | miR-126-3p | trans | 2.40E-39 | Nightingale | S_VLDL_FC | NA | T | C | 0.037825 | 0.004426 | 8.10E-18 |
| rs554833 | miR-126-3p | trans | 2.40E-39 | Nightingale | S_VLDL_L | NA | T | C | 0.024612 | 0.004382 | 4.00E-09 |
| rs554833 | miR-126-3p | trans | 2.40E-39 | Nightingale | S_VLDL_P | NA | T | C | 0.023421 | 0.004369 | 1.70E-08 |
| rs554833 | miR-126-3p | trans | 2.40E-39 | Nightingale | L_VLDL_C_ | NA | T | C | 0.05203 | 0.004322 | 6.60E-34 |
| rs554833 | miR-126-3p | trans | 2.40E-39 | Nightingale | M_LDL_PL | NA | T | C | 0.032377 | 0.004431 | 3.00E-13 |
| rs554833 | miR-126-3p | trans | 2.40E-39 | Nightingale | S_LDL_L | NA | T | C | 0.027648 | 0.004424 | 9.10E-10 |
| rs554833 | miR-126-3p | trans | 2.40E-39 | Nightingale | S_VLDL_C | NA | T | C | 0.038292 | 0.004424 | 2.00E-18 |
| rs554833 | miR-126-3p | trans | 2.40E-39 | Nightingale | S_VLDL_FC | NA | T | C | 0.029304 | 0.004228 | 1.10E-11 |
| rs554833 | miR-126-3p | trans | 2.40E-39 | Nightingale | S_LDL_P | NA | T | C | 0.029501 | 0.004429 | 3.50E-11 |
| rs554833 | miR-126-3p | trans | 2.40E-39 | Nightingale | GlycA | NA | T | C | 0.027847 | 0.004406 | 1.80E-10 |

| | | | | | | | | | | | |
|---|---|---|---|---|---|---|---|---|---|---|---|
| rs554833 | let-7c-5p | trans | 2.46E-23 | Nightingale | S_LDL_C | NA | T | C | 0.03085 | 0.004429 | 6.20E-12 |
| rs554833 | let-7c-5p | trans | 2.46E-23 | Nightingale | Remnant_( | NA | T | C | 0.036771 | 0.004396 | 4.30E-17 |
| rs554833 | let-7c-5p | trans | 2.46E-23 | Nightingale | S_VLDL_CE | NA | T | C | 0.038101 | 0.004419 | 1.80E-18 |
| rs554833 | let-7c-5p | trans | 2.46E-23 | Nightingale | LDL_P | NA | T | C | 0.031693 | 0.004422 | 1.20E-12 |
| rs554833 | let-7c-5p | trans | 2.46E-23 | Nightingale | M_VLDL_P | NA | T | C | 0.029057 | 0.004418 | 2.20E-11 |
| rs554833 | let-7c-5p | trans | 2.46E-23 | Nightingale | L_LDL_PL | NA | T | C | 0.037892 | 0.004391 | 1.10E-17 |
| rs554833 | let-7c-5p | trans | 2.46E-23 | Nightingale | XXL_VLDL_ | NA | T | C | -0.03558 | 0.004463 | 2.80E-16 |
| rs554833 | let-7c-5p | trans | 2.46E-23 | Nightingale | M_VLDL_P | NA | T | C | 0.043907 | 0.004247 | 8.30E-25 |
| rs554833 | let-7c-5p | trans | 2.46E-23 | Nightingale | LDL_CE | NA | T | C | 0.032457 | 0.004413 | 2.50E-13 |
| rs554833 | let-7c-5p | trans | 2.46E-23 | Nightingale | M_VLDL_C | NA | T | C | 0.038627 | 0.004413 | 2.20E-18 |
| rs554833 | let-7c-5p | trans | 2.46E-23 | Nightingale | Sphingomy | NA | T | C | 0.028916 | 0.004136 | 8.10E-12 |
| rs554833 | let-7c-5p | trans | 2.46E-23 | Nightingale | S_HDL_C | NA | T | C | 0.028389 | 0.004417 | 1.80E-10 |
| rs554833 | let-7c-5p | trans | 2.46E-23 | Nightingale | M_LDL_TG | NA | T | C | -0.02757 | 0.00437 | 3.50E-10 |
| rs554833 | let-7c-5p | trans | 2.46E-23 | Nightingale | L_LDL_CE | NA | T | C | 0.031473 | 0.004385 | 1.10E-12 |
| rs554833 | let-7c-5p | trans | 2.46E-23 | Nightingale | XS_VLDL_L | NA | T | C | 0.033736 | 0.00435 | 2.30E-15 |
| rs554833 | let-7c-5p | trans | 2.46E-23 | Nightingale | XS_VLDL_P | NA | T | C | 0.032029 | 0.004363 | 6.00E-14 |
| rs554833 | let-7c-5p | trans | 2.46E-23 | Nightingale | Omega_6 | NA | T | C | 0.025802 | 0.004321 | 6.50E-10 |
| rs554833 | let-7c-5p | trans | 2.46E-23 | Nightingale | ApoB_by_/ | NA | T | C | 0.024942 | 0.004358 | 1.40E-08 |
| rs554833 | let-7c-5p | trans | 2.46E-23 | Nightingale | Citrate | NA | T | C | 0.026896 | 0.004396 | 6.50E-10 |
| rs554833 | let-7c-5p | trans | 2.46E-23 | Nightingale | M_VLDL_F | NA | T | C | 0.041958 | 0.00421 | 4.60E-23 |
| rs554833 | let-7c-5p | trans | 2.46E-23 | Nightingale | IDL_L | NA | T | C | 0.035176 | 0.004278 | 5.80E-16 |
| rs554833 | let-7c-5p | trans | 2.46E-23 | Nightingale | VLDL_FC | NA | T | C | 0.024084 | 0.004392 | 1.20E-08 |
| rs554833 | let-7c-5p | trans | 2.46E-23 | Nightingale | PUFA | NA | T | C | 0.023632 | 0.004316 | 1.60E-08 |
| rs554833 | let-7c-5p | trans | 2.46E-23 | Nightingale | Total_FC | NA | T | C | 0.033181 | 0.004323 | 7.70E-15 |
| rs554833 | let-7c-5p | trans | 2.46E-23 | Nightingale | LA | NA | T | C | 0.024875 | 0.004339 | 3.80E-09 |
| rs554833 | let-7c-5p | trans | 2.46E-23 | Nightingale | S_LDL_FC | NA | T | C | 0.028501 | 0.004418 | 4.50E-10 |
| rs554833 | let-7c-5p | trans | 2.46E-23 | Nightingale | M_LDL_CE | NA | T | C | 0.032879 | 0.004431 | 1.10E-13 |
| rs554833 | let-7c-5p | trans | 2.46E-23 | Nightingale | L_VLDL_CE | NA | T | C | 0.047245 | 0.004234 | 3.20E-29 |
| rs554833 | let-7c-5p | trans | 2.46E-23 | Nightingale | XL_VLDL_C | NA | T | C | 0.037257 | 0.004245 | 3.80E-18 |
| rs554833 | let-7c-5p | trans | 2.46E-23 | Nightingale | S_HDL_P | NA | T | C | 0.030576 | 0.004425 | 5.40E-12 |
| rs554833 | let-7c-5p | trans | 2.46E-23 | Nightingale | LDL_L | NA | T | C | 0.033207 | 0.00441 | 8.50E-14 |
| rs554833 | let-7c-5p | trans | 2.46E-23 | Nightingale | IDL_P | NA | T | C | 0.039913 | 0.004378 | 1.50E-19 |
| rs554833 | let-7c-5p | trans | 2.46E-23 | Nightingale | Clinical_LD | NA | T | C | 0.036378 | 0.004399 | 3.20E-16 |
| rs554833 | let-7c-5p | trans | 2.46E-23 | Nightingale | S_LDL_PL_ | NA | T | C | -0.03004 | 0.004417 | 2.80E-13 |
| rs554833 | let-7c-5p | trans | 2.46E-23 | Nightingale | XL_VLDL_F | NA | T | C | 0.037098 | 0.00429 | 2.50E-18 |
| rs554833 | let-7c-5p | trans | 2.46E-23 | Nightingale | S_HDL_CE | NA | T | C | 0.028614 | 0.004432 | 1.40E-10 |
| rs554833 | let-7c-5p | trans | 2.46E-23 | Nightingale | S_VLDL_C_ | NA | T | C | 0.041797 | 0.004306 | 6.50E-22 |
| rs554833 | let-7c-5p | trans | 2.46E-23 | Nightingale | M_LDL_FC | NA | T | C | 0.03427 | 0.004419 | 2.20E-14 |
| rs554833 | let-7c-5p | trans | 2.46E-23 | Nightingale | Total_PL | NA | T | C | 0.024871 | 0.004213 | 1.20E-09 |
| rs554833 | let-7c-5p | trans | 2.46E-23 | Nightingale | M_VLDL_T | NA | T | C | -0.03846 | 0.004181 | 8.80E-20 |
| rs554833 | let-7c-5p | trans | 2.46E-23 | Nightingale | IDL_CE | NA | T | C | 0.033425 | 0.004272 | 1.50E-14 |
| rs554833 | let-7c-5p | trans | 2.46E-23 | Nightingale | non_HDL_( | NA | T | C | 0.035598 | 0.004401 | 6.00E-16 |
| rs554833 | let-7c-5p | trans | 2.46E-23 | Nightingale | L_VLDL_FC | NA | T | C | 0.048348 | 0.004382 | 8.40E-29 |
| rs554833 | let-7c-5p | trans | 2.46E-23 | Nightingale | M_VLDL_C | NA | T | C | 0.040762 | 0.004385 | 2.60E-20 |
| rs554833 | let-7c-5p | trans | 2.46E-23 | Nightingale | M_VLDL_C | NA | T | C | 0.033552 | 0.004157 | 2.20E-15 |
| rs554833 | let-7c-5p | trans | 2.46E-23 | Nightingale | VLDL_P | NA | T | C | 0.026565 | 0.004401 | 4.70E-10 |
| rs554833 | let-7c-5p | trans | 2.46E-23 | Nightingale | IDL_C | NA | T | C | 0.03487 | 0.004273 | 1.10E-15 |
| rs554833 | let-7c-5p | trans | 2.46E-23 | Nightingale | Total_L | NA | T | C | 0.026282 | 0.004353 | 5.00E-10 |
| rs554833 | let-7c-5p | trans | 2.46E-23 | Nightingale | L_LDL_P | NA | T | C | 0.029065 | 0.00441 | 7.10E-11 |
| rs554833 | let-7c-5p | trans | 2.46E-23 | Nightingale | IDL_FC | NA | T | C | 0.038165 | 0.004284 | 1.80E-18 |
| rs554833 | let-7c-5p | trans | 2.46E-23 | Nightingale | IDL_FC_pct | NA | T | C | 0.025732 | 0.004391 | 1.10E-08 |
| rs554833 | let-7c-5p | trans | 2.46E-23 | Nightingale | XXL_VLDL_ | NA | T | C | 0.0307 | 0.004406 | 1.40E-12 |
| rs554833 | let-7c-5p | trans | 2.46E-23 | Nightingale | LDL_PL | NA | T | C | 0.034891 | 0.004415 | 5.50E-15 |
| rs554833 | let-7c-5p | trans | 2.46E-23 | Nightingale | Total_CE | NA | T | C | 0.031671 | 0.004268 | 1.10E-13 |
| rs554833 | let-7c-5p | trans | 2.46E-23 | Nightingale | S_VLDL_TG | NA | T | C | -0.03805 | 0.004283 | 1.70E-18 |
| rs554833 | let-7c-5p | trans | 2.46E-23 | Nightingale | Total_C | NA | T | C | 0.032298 | 0.004284 | 3.80E-14 |
| rs554833 | let-7c-5p | trans | 2.46E-23 | Nightingale | L_LDL_L | NA | T | C | 0.033453 | 0.004382 | 3.50E-14 |
| rs554833 | let-7c-5p | trans | 2.46E-23 | Nightingale | L_LDL_FC | NA | T | C | 0.033535 | 0.004345 | 3.30E-14 |
| rs554833 | let-7c-5p | trans | 2.46E-23 | Nightingale | VLDL_CE | NA | T | C | 0.035694 | 0.004426 | 3.40E-16 |
| rs554833 | let-7c-5p | trans | 2.46E-23 | Nightingale | M_VLDL_P | NA | T | C | 0.028221 | 0.004414 | 6.80E-11 |
| rs554833 | let-7c-5p | trans | 2.46E-23 | Nightingale | S_HDL_FC | NA | T | C | 0.023434 | 0.004344 | 4.80E-08 |
| rs554833 | let-7c-5p | trans | 2.46E-23 | Nightingale | S_HDL_L | NA | T | C | 0.026261 | 0.00441 | 1.60E-09 |
| rs554833 | let-7c-5p | trans | 2.46E-23 | Nightingale | XS_VLDL_F | NA | T | C | 0.031395 | 0.004393 | 2.50E-12 |

| | | | | | | | | | | | |
|---|---|---|---|---|---|---|---|---|---|---|---|
| rs554833 | let-7c-5p | trans | 2.46E-23 | Nightingale | M_LDL_C | NA | T | C | 0.033897 | 0.004433 | 2.40E-14 |
| rs554833 | let-7c-5p | trans | 2.46E-23 | Nightingale | M_LDL_P | NA | T | C | 0.035797 | 0.004437 | 7.90E-16 |
| rs554833 | let-7c-5p | trans | 2.46E-23 | Nightingale | XS_VLDL_C | NA | T | C | 0.037904 | 0.004295 | 1.60E-18 |
| rs554833 | let-7c-5p | trans | 2.46E-23 | Nightingale | L_LDL_C | NA | T | C | 0.032226 | 0.004375 | 3.00E-13 |
| rs554833 | let-7c-5p | trans | 2.46E-23 | Nightingale | M_VLDL_F | NA | T | C | 0.033298 | 0.004424 | 3.40E-14 |
| rs554833 | let-7c-5p | trans | 2.46E-23 | Nightingale | XL_VLDL_T | NA | T | C | -0.04375 | 0.004289 | 2.60E-24 |
| rs554833 | let-7c-5p | trans | 2.46E-23 | Nightingale | L_VLDL_TG | NA | T | C | -0.05094 | 0.004404 | 9.30E-32 |
| rs554833 | let-7c-5p | trans | 2.46E-23 | Nightingale | M_VLDL_C | NA | T | C | 0.03612 | 0.004168 | 1.40E-17 |
| rs554833 | let-7c-5p | trans | 2.46E-23 | Nightingale | M_LDL_L | NA | T | C | 0.03294 | 0.004431 | 1.10E-13 |
| rs554833 | let-7c-5p | trans | 2.46E-23 | Nightingale | XXL_VLDL_ | NA | T | C | 0.033029 | 0.004459 | 1.00E-13 |
| rs554833 | let-7c-5p | trans | 2.46E-23 | Nightingale | S_VLDL_PL | NA | T | C | 0.027679 | 0.004237 | 2.00E-10 |
| rs554833 | let-7c-5p | trans | 2.46E-23 | Nightingale | VLDL_C | NA | T | C | 0.031216 | 0.004419 | 5.50E-13 |
| rs554833 | let-7c-5p | trans | 2.46E-23 | Nightingale | S_VLDL_PL | NA | T | C | 0.032705 | 0.004417 | 4.50E-14 |
| rs554833 | let-7c-5p | trans | 2.46E-23 | Nightingale | S_VLDL_CE | NA | T | C | 0.046918 | 0.004365 | 1.60E-26 |
| rs554833 | let-7c-5p | trans | 2.46E-23 | Nightingale | M_LDL_C_ | NA | T | C | 0.03113 | 0.004428 | 3.10E-12 |
| rs554833 | let-7c-5p | trans | 2.46E-23 | Nightingale | XXL_VLDL_ | NA | T | C | 0.034703 | 0.004434 | 3.60E-15 |
| rs554833 | let-7c-5p | trans | 2.46E-23 | Nightingale | S_LDL_CE | NA | T | C | 0.030448 | 0.004427 | 6.60E-12 |
| rs554833 | let-7c-5p | trans | 2.46E-23 | Nightingale | XXL_VLDL_ | NA | T | C | 0.024904 | 0.004454 | 1.70E-09 |
| rs554833 | let-7c-5p | trans | 2.46E-23 | Nightingale | XL_VLDL_C | NA | T | C | 0.03813 | 0.004243 | 5.70E-19 |
| rs554833 | let-7c-5p | trans | 2.46E-23 | Nightingale | S_HDL_PL | NA | T | C | 0.023749 | 0.004386 | 3.40E-08 |
| rs554833 | let-7c-5p | trans | 2.46E-23 | Nightingale | LDL_C | NA | T | C | 0.0331 | 0.004406 | 1.00E-13 |
| rs554833 | let-7c-5p | trans | 2.46E-23 | Nightingale | XL_VLDL_P | NA | T | C | 0.038618 | 0.004432 | 8.20E-19 |
| rs554833 | let-7c-5p | trans | 2.46E-23 | Nightingale | XS_VLDL_C | NA | T | C | 0.037708 | 0.00427 | 2.50E-18 |
| rs554833 | let-7c-5p | trans | 2.46E-23 | Nightingale | LDL_FC | NA | T | C | 0.033547 | 0.004378 | 6.00E-14 |
| rs554833 | let-7c-5p | trans | 2.46E-23 | Nightingale | XS_VLDL_P | NA | T | C | 0.036622 | 0.004366 | 1.60E-17 |
| rs554833 | let-7c-5p | trans | 2.46E-23 | Nightingale | IDL_PL | NA | T | C | 0.035232 | 0.00427 | 4.10E-16 |
| rs554833 | let-7c-5p | trans | 2.46E-23 | Nightingale | S_LDL_C_p | NA | T | C | 0.045327 | 0.004442 | 6.00E-25 |
| rs554833 | let-7c-5p | trans | 2.46E-23 | Nightingale | ApoB | NA | T | C | 0.033838 | 0.004419 | 2.40E-14 |
| rs554833 | let-7c-5p | trans | 2.46E-23 | Nightingale | XL_HDL_PL | NA | T | C | -0.02272 | 0.004065 | 1.20E-08 |
| rs554833 | let-7c-5p | trans | 2.46E-23 | Nightingale | S_LDL_CE_ | NA | T | C | 0.030859 | 0.004417 | 5.70E-14 |
| rs554833 | let-7c-5p | trans | 2.46E-23 | Nightingale | XS_VLDL_F | NA | T | C | 0.036985 | 0.004348 | 7.40E-18 |
| rs554833 | let-7c-5p | trans | 2.46E-23 | Nightingale | S_VLDL_FC | NA | T | C | 0.037825 | 0.004426 | 8.10E-18 |
| rs554833 | let-7c-5p | trans | 2.46E-23 | Nightingale | S_VLDL_L | NA | T | C | 0.024612 | 0.004382 | 4.00E-09 |
| rs554833 | let-7c-5p | trans | 2.46E-23 | Nightingale | S_VLDL_P | NA | T | C | 0.023421 | 0.004369 | 1.70E-08 |
| rs554833 | let-7c-5p | trans | 2.46E-23 | Nightingale | L_VLDL_C_ | NA | T | C | 0.05203 | 0.004322 | 6.60E-34 |
| rs554833 | let-7c-5p | trans | 2.46E-23 | Nightingale | M_LDL_PL | NA | T | C | 0.032377 | 0.004431 | 3.00E-13 |
| rs554833 | let-7c-5p | trans | 2.46E-23 | Nightingale | S_LDL_L | NA | T | C | 0.027648 | 0.004424 | 9.10E-10 |
| rs554833 | let-7c-5p | trans | 2.46E-23 | Nightingale | S_VLDL_C | NA | T | C | 0.038292 | 0.004424 | 2.00E-18 |
| rs554833 | let-7c-5p | trans | 2.46E-23 | Nightingale | S_VLDL_FC | NA | T | C | 0.029304 | 0.004228 | 1.10E-11 |
| rs554833 | let-7c-5p | trans | 2.46E-23 | Nightingale | S_LDL_P | NA | T | C | 0.029501 | 0.004429 | 3.50E-11 |
| rs554833 | let-7c-5p | trans | 2.46E-23 | Nightingale | GlycA | NA | T | C | 0.027847 | 0.004406 | 1.80E-10 |
| rs554833 | miR-195-5p | trans | 2.11E-17 | Nightingale | S_LDL_C | NA | T | C | 0.03085 | 0.004429 | 6.20E-12 |
| rs554833 | miR-195-5p | trans | 2.11E-17 | Nightingale | Remnant_( | NA | T | C | 0.036771 | 0.004396 | 4.30E-17 |
| rs554833 | miR-195-5p | trans | 2.11E-17 | Nightingale | S_VLDL_CE | NA | T | C | 0.038101 | 0.004419 | 1.80E-18 |
| rs554833 | miR-195-5p | trans | 2.11E-17 | Nightingale | LDL_P | NA | T | C | 0.031693 | 0.004422 | 1.20E-12 |
| rs554833 | miR-195-5p | trans | 2.11E-17 | Nightingale | M_VLDL_P | NA | T | C | 0.029057 | 0.004418 | 2.20E-11 |
| rs554833 | miR-195-5p | trans | 2.11E-17 | Nightingale | L_LDL_PL | NA | T | C | 0.037892 | 0.004391 | 1.10E-17 |
| rs554833 | miR-195-5p | trans | 2.11E-17 | Nightingale | XXL_VLDL_ | NA | T | C | -0.03558 | 0.004463 | 2.80E-16 |
| rs554833 | miR-195-5p | trans | 2.11E-17 | Nightingale | M_VLDL_P | NA | T | C | 0.043907 | 0.004247 | 8.30E-25 |
| rs554833 | miR-195-5p | trans | 2.11E-17 | Nightingale | LDL_CE | NA | T | C | 0.032457 | 0.004413 | 2.50E-13 |
| rs554833 | miR-195-5p | trans | 2.11E-17 | Nightingale | M_VLDL_C | NA | T | C | 0.038627 | 0.004413 | 2.20E-18 |
| rs554833 | miR-195-5p | trans | 2.11E-17 | Nightingale | Sphingomy | NA | T | C | 0.028916 | 0.004136 | 8.10E-12 |
| rs554833 | miR-195-5p | trans | 2.11E-17 | Nightingale | S_HDL_C | NA | T | C | 0.028389 | 0.004417 | 1.80E-10 |
| rs554833 | miR-195-5p | trans | 2.11E-17 | Nightingale | M_LDL_TG | NA | T | C | -0.02757 | 0.00437 | 3.50E-10 |
| rs554833 | miR-195-5p | trans | 2.11E-17 | Nightingale | L_LDL_CE | NA | T | C | 0.031473 | 0.004385 | 1.10E-12 |
| rs554833 | miR-195-5p | trans | 2.11E-17 | Nightingale | XS_VLDL_L | NA | T | C | 0.033736 | 0.00435 | 2.30E-15 |
| rs554833 | miR-195-5p | trans | 2.11E-17 | Nightingale | XS_VLDL_P | NA | T | C | 0.032029 | 0.004363 | 6.00E-14 |
| rs554833 | miR-195-5p | trans | 2.11E-17 | Nightingale | Omega_6 | NA | T | C | 0.025802 | 0.004321 | 6.50E-10 |
| rs554833 | miR-195-5p | trans | 2.11E-17 | Nightingale | ApoB_by_/ | NA | T | C | 0.024942 | 0.004358 | 1.40E-08 |
| rs554833 | miR-195-5p | trans | 2.11E-17 | Nightingale | Citrate | NA | T | C | 0.026896 | 0.004396 | 6.50E-10 |
| rs554833 | miR-195-5p | trans | 2.11E-17 | Nightingale | M_VLDL_F | NA | T | C | 0.041958 | 0.00421 | 4.60E-23 |
| rs554833 | miR-195-5p | trans | 2.11E-17 | Nightingale | IDL_L | NA | T | C | 0.035176 | 0.004278 | 5.80E-16 |
| rs554833 | miR-195-5p | trans | 2.11E-17 | Nightingale | VLDL_FC | NA | T | C | 0.024084 | 0.004392 | 1.20E-08 |

| | | | | | | | | | | | |
|---|---|---|---|---|---|---|---|---|---|---|---|
| rs554833 | miR-195-5p | trans | 2.11E-17 | Nightingale | PUFA | NA | T | C | 0.023632 | 0.004316 | 1.60E-08 |
| rs554833 | miR-195-5p | trans | 2.11E-17 | Nightingale | Total_FC | NA | T | C | 0.033181 | 0.004323 | 7.70E-15 |
| rs554833 | miR-195-5p | trans | 2.11E-17 | Nightingale | LA | NA | T | C | 0.024875 | 0.004339 | 3.80E-09 |
| rs554833 | miR-195-5p | trans | 2.11E-17 | Nightingale | S_LDL_FC | NA | T | C | 0.028501 | 0.004418 | 4.50E-10 |
| rs554833 | miR-195-5p | trans | 2.11E-17 | Nightingale | M_LDL_CE | NA | T | C | 0.032879 | 0.004431 | 1.10E-13 |
| rs554833 | miR-195-5p | trans | 2.11E-17 | Nightingale | L_VLDL_CE | NA | T | C | 0.047245 | 0.004234 | 3.20E-29 |
| rs554833 | miR-195-5p | trans | 2.11E-17 | Nightingale | XL_VLDL_C | NA | T | C | 0.037257 | 0.004245 | 3.80E-18 |
| rs554833 | miR-195-5p | trans | 2.11E-17 | Nightingale | S_HDL_P | NA | T | C | 0.030576 | 0.004425 | 5.40E-12 |
| rs554833 | miR-195-5p | trans | 2.11E-17 | Nightingale | LDL_L | NA | T | C | 0.033207 | 0.00441 | 8.50E-14 |
| rs554833 | miR-195-5p | trans | 2.11E-17 | Nightingale | IDL_P | NA | T | C | 0.039913 | 0.004378 | 1.50E-19 |
| rs554833 | miR-195-5p | trans | 2.11E-17 | Nightingale | Clinical_LD | NA | T | C | 0.036378 | 0.004399 | 3.20E-16 |
| rs554833 | miR-195-5p | trans | 2.11E-17 | Nightingale | S_LDL_PL_ | NA | T | C | -0.03004 | 0.004417 | 2.80E-13 |
| rs554833 | miR-195-5p | trans | 2.11E-17 | Nightingale | XL_VLDL_F | NA | T | C | 0.037098 | 0.00429 | 2.50E-18 |
| rs554833 | miR-195-5p | trans | 2.11E-17 | Nightingale | S_HDL_CE | NA | T | C | 0.028614 | 0.004432 | 1.40E-10 |
| rs554833 | miR-195-5p | trans | 2.11E-17 | Nightingale | S_VLDL_C_ | NA | T | C | 0.041797 | 0.004306 | 6.50E-22 |
| rs554833 | miR-195-5p | trans | 2.11E-17 | Nightingale | M_LDL_FC | NA | T | C | 0.03427 | 0.004419 | 2.20E-14 |
| rs554833 | miR-195-5p | trans | 2.11E-17 | Nightingale | Total_PL | NA | T | C | 0.024871 | 0.004213 | 1.20E-09 |
| rs554833 | miR-195-5p | trans | 2.11E-17 | Nightingale | M_VLDL_T | NA | T | C | -0.03846 | 0.004181 | 8.80E-20 |
| rs554833 | miR-195-5p | trans | 2.11E-17 | Nightingale | IDL_CE | NA | T | C | 0.033425 | 0.004272 | 1.50E-14 |
| rs554833 | miR-195-5p | trans | 2.11E-17 | Nightingale | non_HDL_( | NA | T | C | 0.035598 | 0.004401 | 6.00E-16 |
| rs554833 | miR-195-5p | trans | 2.11E-17 | Nightingale | L_VLDL_FC | NA | T | C | 0.048348 | 0.004382 | 8.40E-29 |
| rs554833 | miR-195-5p | trans | 2.11E-17 | Nightingale | M_VLDL_C | NA | T | C | 0.040762 | 0.004385 | 2.60E-20 |
| rs554833 | miR-195-5p | trans | 2.11E-17 | Nightingale | M_VLDL_C | NA | T | C | 0.033552 | 0.004157 | 2.20E-15 |
| rs554833 | miR-195-5p | trans | 2.11E-17 | Nightingale | VLDL_P | NA | T | C | 0.026565 | 0.004401 | 4.70E-10 |
| rs554833 | miR-195-5p | trans | 2.11E-17 | Nightingale | IDL_C | NA | T | C | 0.03487 | 0.004273 | 1.10E-15 |
| rs554833 | miR-195-5p | trans | 2.11E-17 | Nightingale | Total_L | NA | T | C | 0.026282 | 0.004353 | 5.00E-10 |
| rs554833 | miR-195-5p | trans | 2.11E-17 | Nightingale | L_LDL_P | NA | T | C | 0.029065 | 0.00441 | 7.10E-11 |
| rs554833 | miR-195-5p | trans | 2.11E-17 | Nightingale | IDL_FC | NA | T | C | 0.038165 | 0.004284 | 1.80E-18 |
| rs554833 | miR-195-5p | trans | 2.11E-17 | Nightingale | IDL_FC_pct | NA | T | C | 0.025732 | 0.004391 | 1.10E-08 |
| rs554833 | miR-195-5p | trans | 2.11E-17 | Nightingale | XXL_VLDL_ | NA | T | C | 0.0307 | 0.004406 | 1.40E-12 |
| rs554833 | miR-195-5p | trans | 2.11E-17 | Nightingale | LDL_PL | NA | T | C | 0.034891 | 0.004415 | 5.50E-15 |
| rs554833 | miR-195-5p | trans | 2.11E-17 | Nightingale | Total_CE | NA | T | C | 0.031671 | 0.004268 | 1.10E-13 |
| rs554833 | miR-195-5p | trans | 2.11E-17 | Nightingale | S_VLDL_TG | NA | T | C | -0.03805 | 0.004283 | 1.70E-18 |
| rs554833 | miR-195-5p | trans | 2.11E-17 | Nightingale | Total_C | NA | T | C | 0.032298 | 0.004284 | 3.80E-14 |
| rs554833 | miR-195-5p | trans | 2.11E-17 | Nightingale | L_LDL_L | NA | T | C | 0.033453 | 0.004382 | 3.50E-14 |
| rs554833 | miR-195-5p | trans | 2.11E-17 | Nightingale | L_LDL_FC | NA | T | C | 0.033535 | 0.004345 | 3.30E-14 |
| rs554833 | miR-195-5p | trans | 2.11E-17 | Nightingale | VLDL_CE | NA | T | C | 0.035694 | 0.004426 | 3.40E-16 |
| rs554833 | miR-195-5p | trans | 2.11E-17 | Nightingale | M_VLDL_P | NA | T | C | 0.028221 | 0.004414 | 6.80E-11 |
| rs554833 | miR-195-5p | trans | 2.11E-17 | Nightingale | S_HDL_FC | NA | T | C | 0.023434 | 0.004344 | 4.80E-08 |
| rs554833 | miR-195-5p | trans | 2.11E-17 | Nightingale | S_HDL_L | NA | T | C | 0.026261 | 0.00441 | 1.60E-09 |
| rs554833 | miR-195-5p | trans | 2.11E-17 | Nightingale | XS_VLDL_F | NA | T | C | 0.031395 | 0.004393 | 2.50E-12 |
| rs554833 | miR-195-5p | trans | 2.11E-17 | Nightingale | M_LDL_C | NA | T | C | 0.033897 | 0.004433 | 2.40E-14 |
| rs554833 | miR-195-5p | trans | 2.11E-17 | Nightingale | M_LDL_P | NA | T | C | 0.035797 | 0.004437 | 7.90E-16 |
| rs554833 | miR-195-5p | trans | 2.11E-17 | Nightingale | XS_VLDL_C | NA | T | C | 0.037904 | 0.004295 | 1.60E-18 |
| rs554833 | miR-195-5p | trans | 2.11E-17 | Nightingale | L_LDL_C | NA | T | C | 0.032226 | 0.004375 | 3.00E-13 |
| rs554833 | miR-195-5p | trans | 2.11E-17 | Nightingale | M_VLDL_F | NA | T | C | 0.033298 | 0.004424 | 3.40E-14 |
| rs554833 | miR-195-5p | trans | 2.11E-17 | Nightingale | XL_VLDL_T | NA | T | C | -0.04375 | 0.004289 | 2.60E-24 |
| rs554833 | miR-195-5p | trans | 2.11E-17 | Nightingale | L_VLDL_TG | NA | T | C | -0.05094 | 0.004404 | 9.30E-32 |
| rs554833 | miR-195-5p | trans | 2.11E-17 | Nightingale | M_VLDL_C | NA | T | C | 0.03612 | 0.004168 | 1.40E-17 |
| rs554833 | miR-195-5p | trans | 2.11E-17 | Nightingale | M_LDL_L | NA | T | C | 0.03294 | 0.004431 | 1.10E-13 |
| rs554833 | miR-195-5p | trans | 2.11E-17 | Nightingale | XXL_VLDL_ | NA | T | C | 0.033029 | 0.004459 | 1.00E-13 |
| rs554833 | miR-195-5p | trans | 2.11E-17 | Nightingale | S_VLDL_PL | NA | T | C | 0.027679 | 0.004237 | 2.00E-10 |
| rs554833 | miR-195-5p | trans | 2.11E-17 | Nightingale | VLDL_C | NA | T | C | 0.031216 | 0.004419 | 5.50E-13 |
| rs554833 | miR-195-5p | trans | 2.11E-17 | Nightingale | S_VLDL_PL | NA | T | C | 0.032705 | 0.004417 | 4.50E-14 |
| rs554833 | miR-195-5p | trans | 2.11E-17 | Nightingale | S_VLDL_CE | NA | T | C | 0.046918 | 0.004365 | 1.60E-26 |
| rs554833 | miR-195-5p | trans | 2.11E-17 | Nightingale | M_LDL_C_ | NA | T | C | 0.03113 | 0.004428 | 3.10E-12 |
| rs554833 | miR-195-5p | trans | 2.11E-17 | Nightingale | XXL_VLDL_ | NA | T | C | 0.034703 | 0.004434 | 3.60E-15 |
| rs554833 | miR-195-5p | trans | 2.11E-17 | Nightingale | S_LDL_CE | NA | T | C | 0.030448 | 0.004427 | 6.60E-12 |
| rs554833 | miR-195-5p | trans | 2.11E-17 | Nightingale | XXL_VLDL_ | NA | T | C | 0.024904 | 0.004454 | 1.70E-09 |
| rs554833 | miR-195-5p | trans | 2.11E-17 | Nightingale | XL_VLDL_C | NA | T | C | 0.03813 | 0.004243 | 5.70E-19 |
| rs554833 | miR-195-5p | trans | 2.11E-17 | Nightingale | S_HDL_PL | NA | T | C | 0.023749 | 0.004386 | 3.40E-08 |
| rs554833 | miR-195-5p | trans | 2.11E-17 | Nightingale | LDL_C | NA | T | C | 0.0331 | 0.004406 | 1.00E-13 |
| rs554833 | miR-195-5p | trans | 2.11E-17 | Nightingale | XL_VLDL_P | NA | T | C | 0.038618 | 0.004432 | 8.20E-19 |

| | | | | | | | | | | | |
|---|---|---|---|---|---|---|---|---|---|---|---|
| rs554833 | miR-195-5p | trans | 2.11E-17 | Nightingale | XS_VLDL_C | NA | T | C | 0.037708 | 0.00427 | 2.50E-18 |
| rs554833 | miR-195-5p | trans | 2.11E-17 | Nightingale | LDL_FC | NA | T | C | 0.033547 | 0.004378 | 6.00E-14 |
| rs554833 | miR-195-5p | trans | 2.11E-17 | Nightingale | XS_VLDL_P | NA | T | C | 0.036622 | 0.004366 | 1.60E-17 |
| rs554833 | miR-195-5p | trans | 2.11E-17 | Nightingale | IDL_PL | NA | T | C | 0.035232 | 0.00427 | 4.10E-16 |
| rs554833 | miR-195-5p | trans | 2.11E-17 | Nightingale | S_LDL_C_p | NA | T | C | 0.045327 | 0.004442 | 6.00E-25 |
| rs554833 | miR-195-5p | trans | 2.11E-17 | Nightingale | ApoB | NA | T | C | 0.033838 | 0.004419 | 2.40E-14 |
| rs554833 | miR-195-5p | trans | 2.11E-17 | Nightingale | XL_HDL_PL | NA | T | C | -0.02272 | 0.004065 | 1.20E-08 |
| rs554833 | miR-195-5p | trans | 2.11E-17 | Nightingale | S_LDL_CE_ | NA | T | C | 0.030859 | 0.004417 | 5.70E-14 |
| rs554833 | miR-195-5p | trans | 2.11E-17 | Nightingale | XS_VLDL_F | NA | T | C | 0.036985 | 0.004348 | 7.40E-18 |
| rs554833 | miR-195-5p | trans | 2.11E-17 | Nightingale | S_VLDL_FC | NA | T | C | 0.037825 | 0.004426 | 8.10E-18 |
| rs554833 | miR-195-5p | trans | 2.11E-17 | Nightingale | S_VLDL_L | NA | T | C | 0.024612 | 0.004382 | 4.00E-09 |
| rs554833 | miR-195-5p | trans | 2.11E-17 | Nightingale | S_VLDL_P | NA | T | C | 0.023421 | 0.004369 | 1.70E-08 |
| rs554833 | miR-195-5p | trans | 2.11E-17 | Nightingale | L_VLDL_C_ | NA | T | C | 0.05203 | 0.004322 | 6.60E-34 |
| rs554833 | miR-195-5p | trans | 2.11E-17 | Nightingale | M_LDL_PL | NA | T | C | 0.032377 | 0.004431 | 3.00E-13 |
| rs554833 | miR-195-5p | trans | 2.11E-17 | Nightingale | S_LDL_L | NA | T | C | 0.027648 | 0.004424 | 9.10E-10 |
| rs554833 | miR-195-5p | trans | 2.11E-17 | Nightingale | S_VLDL_C | NA | T | C | 0.038292 | 0.004424 | 2.00E-18 |
| rs554833 | miR-195-5p | trans | 2.11E-17 | Nightingale | S_VLDL_FC | NA | T | C | 0.029304 | 0.004228 | 1.10E-11 |
| rs554833 | miR-195-5p | trans | 2.11E-17 | Nightingale | S_LDL_P | NA | T | C | 0.029501 | 0.004429 | 3.50E-11 |
| rs554833 | miR-195-5p | trans | 2.11E-17 | Nightingale | GlycA | NA | T | C | 0.027847 | 0.004406 | 1.80E-10 |
| rs581107 | miR-10b-5p | trans | 1.11E-19 | Nightingale | S_VLDL_CE | NA | T | C | 0.025458 | 0.004092 | 3.00E-10 |
| rs581107 | miR-10b-5p | trans | 1.11E-19 | Nightingale | L_VLDL_CE | NA | T | C | 0.02637 | 0.003969 | 1.20E-11 |
| rs581107 | miR-10b-5p | trans | 1.11E-19 | Nightingale | XL_VLDL_C | NA | T | C | 0.022055 | 0.003978 | 9.20E-09 |
| rs581107 | miR-10b-5p | trans | 1.11E-19 | Nightingale | M_VLDL_T | NA | T | C | -0.02242 | 0.00392 | 3.10E-09 |
| rs581107 | miR-10b-5p | trans | 1.11E-19 | Nightingale | XL_VLDL_F | NA | T | C | 0.021915 | 0.00402 | 1.40E-08 |
| rs581107 | miR-10b-5p | trans | 1.11E-19 | Nightingale | M_VLDL_C | NA | T | C | 0.02141 | 0.003907 | 1.10E-08 |
| rs581107 | miR-10b-5p | trans | 1.11E-19 | Nightingale | XL_VLDL_C | NA | T | C | 0.022527 | 0.003976 | 4.90E-09 |
| rs581107 | miR-10b-5p | trans | 1.11E-19 | Nightingale | M_VLDL_F | NA | T | C | 0.024737 | 0.003946 | 1.40E-10 |
| rs581107 | miR-10b-5p | trans | 1.11E-19 | Nightingale | IDL_FC | NA | T | C | 0.022228 | 0.004016 | 3.40E-08 |
| rs581107 | miR-10b-5p | trans | 1.11E-19 | Nightingale | L_VLDL_TG | NA | T | C | -0.0255 | 0.004128 | 5.80E-10 |
| rs581107 | miR-10b-5p | trans | 1.11E-19 | Nightingale | S_VLDL_C_ | NA | T | C | 0.023366 | 0.004037 | 3.20E-09 |
| rs581107 | miR-10b-5p | trans | 1.11E-19 | Nightingale | L_VLDL_FC | NA | T | C | 0.023867 | 0.004108 | 5.70E-09 |
| rs581107 | miR-10b-5p | trans | 1.11E-19 | Nightingale | IDL_P | NA | T | C | 0.023312 | 0.004104 | 1.60E-08 |
| rs581107 | miR-10b-5p | trans | 1.11E-19 | Nightingale | XL_VLDL_T | NA | T | C | -0.02536 | 0.004019 | 8.80E-11 |
| rs581107 | miR-10b-5p | trans | 1.11E-19 | Nightingale | M_VLDL_P | NA | T | C | 0.024521 | 0.003981 | 4.10E-10 |
| rs581107 | miR-10b-5p | trans | 1.11E-19 | Nightingale | S_LDL_C_p | NA | T | C | 0.027026 | 0.004164 | 8.80E-11 |
| rs581107 | miR-10b-5p | trans | 1.11E-19 | Nightingale | L_VLDL_C_ | NA | T | C | 0.028219 | 0.004051 | 1.60E-12 |
| rs581107 | miR-10b-5p | trans | 1.11E-19 | Nightingale | S_VLDL_TG | NA | T | C | -0.02153 | 0.004015 | 3.70E-08 |
| rs582094 | let-7c-5p | trans | 2.74E-23 | Nightingale | XS_VLDL_P | NA | T | A | 0.036606 | 0.004372 | 1.90E-17 |
| rs582094 | let-7c-5p | trans | 2.74E-23 | Nightingale | S_LDL_C | NA | T | A | 0.030792 | 0.004435 | 7.70E-12 |
| rs582094 | let-7c-5p | trans | 2.74E-23 | Nightingale | ApoB_by_A | NA | T | A | 0.024975 | 0.004364 | 1.60E-08 |
| rs582094 | let-7c-5p | trans | 2.74E-23 | Nightingale | M_VLDL_T | NA | T | A | -0.03854 | 0.004187 | 8.80E-20 |
| rs582094 | let-7c-5p | trans | 2.74E-23 | Nightingale | L_LDL_FC | NA | T | A | 0.033451 | 0.004351 | 4.40E-14 |
| rs582094 | let-7c-5p | trans | 2.74E-23 | Nightingale | Total_C | NA | T | A | 0.032199 | 0.00429 | 4.90E-14 |
| rs582094 | let-7c-5p | trans | 2.74E-23 | Nightingale | S_LDL_C_p | NA | T | A | 0.045184 | 0.004448 | 9.90E-25 |
| rs582094 | let-7c-5p | trans | 2.74E-23 | Nightingale | IDL_CE | NA | T | A | 0.033389 | 0.004278 | 1.80E-14 |
| rs582094 | let-7c-5p | trans | 2.74E-23 | Nightingale | XS_VLDL_F | NA | T | A | 0.036947 | 0.004354 | 9.50E-18 |
| rs582094 | let-7c-5p | trans | 2.74E-23 | Nightingale | Total_FC | NA | T | A | 0.03308 | 0.004329 | 1.00E-14 |
| rs582094 | let-7c-5p | trans | 2.74E-23 | Nightingale | S_VLDL_CE | NA | T | A | 0.046942 | 0.004372 | 1.80E-26 |
| rs582094 | let-7c-5p | trans | 2.74E-23 | Nightingale | M_LDL_P | NA | T | A | 0.035786 | 0.004444 | 9.30E-16 |
| rs582094 | let-7c-5p | trans | 2.74E-23 | Nightingale | M_VLDL_C | NA | T | A | 0.033603 | 0.004163 | 2.40E-15 |
| rs582094 | let-7c-5p | trans | 2.74E-23 | Nightingale | IDL_L | NA | T | A | 0.03514 | 0.004284 | 7.00E-16 |
| rs582094 | let-7c-5p | trans | 2.74E-23 | Nightingale | IDL_PL | NA | T | A | 0.035198 | 0.004276 | 4.90E-16 |
| rs582094 | let-7c-5p | trans | 2.74E-23 | Nightingale | XXL_VLDL_ | NA | T | A | 0.032961 | 0.004465 | 1.30E-13 |
| rs582094 | let-7c-5p | trans | 2.74E-23 | Nightingale | IDL_FC_pct | NA | T | A | 0.025717 | 0.004397 | 1.20E-08 |
| rs582094 | let-7c-5p | trans | 2.74E-23 | Nightingale | VLDL_C | NA | T | A | 0.031173 | 0.004425 | 6.70E-13 |
| rs582094 | let-7c-5p | trans | 2.74E-23 | Nightingale | M_VLDL_C | NA | T | A | 0.036181 | 0.004174 | 1.50E-17 |
| rs582094 | let-7c-5p | trans | 2.74E-23 | Nightingale | S_LDL_P | NA | T | A | 0.029487 | 0.004435 | 4.10E-11 |
| rs582094 | let-7c-5p | trans | 2.74E-23 | Nightingale | LDL_L | NA | T | A | 0.033121 | 0.004416 | 1.10E-13 |
| rs582094 | let-7c-5p | trans | 2.74E-23 | Nightingale | Total_L | NA | T | A | 0.026203 | 0.004359 | 5.90E-10 |
| rs582094 | let-7c-5p | trans | 2.74E-23 | Nightingale | LA | NA | T | A | 0.024685 | 0.004345 | 5.20E-09 |
| rs582094 | let-7c-5p | trans | 2.74E-23 | Nightingale | M_VLDL_P | NA | T | A | 0.028969 | 0.004424 | 2.80E-11 |
| rs582094 | let-7c-5p | trans | 2.74E-23 | Nightingale | XS_VLDL_C | NA | T | A | 0.037867 | 0.004301 | 2.00E-18 |
| rs582094 | let-7c-5p | trans | 2.74E-23 | Nightingale | ApoB | NA | T | A | 0.033786 | 0.004425 | 3.10E-14 |

| | | | | | | | | | | | |
|---|---|---|---|---|---|---|---|---|---|---|---|
| rs582094 | let-7c-5p | trans | 2.74E-23 | Nightingale | GlycA | NA | T | A | 0.027998 | 0.004412 | 1.60E-10 |
| rs582094 | let-7c-5p | trans | 2.74E-23 | Nightingale | S_VLDL_FC | NA | T | A | 0.029218 | 0.004234 | 1.60E-11 |
| rs582094 | let-7c-5p | trans | 2.74E-23 | Nightingale | L_VLDL_FC | NA | T | A | 0.048617 | 0.004389 | 5.10E-29 |
| rs582094 | let-7c-5p | trans | 2.74E-23 | Nightingale | VLDL_FC | NA | T | A | 0.024055 | 0.004399 | 1.30E-08 |
| rs582094 | let-7c-5p | trans | 2.74E-23 | Nightingale | M_LDL_FC | NA | T | A | 0.034163 | 0.004426 | 3.00E-14 |
| rs582094 | let-7c-5p | trans | 2.74E-23 | Nightingale | L_LDL_L | NA | T | A | 0.033355 | 0.004388 | 4.70E-14 |
| rs582094 | let-7c-5p | trans | 2.74E-23 | Nightingale | M_VLDL_P | NA | T | A | 0.028139 | 0.00442 | 8.60E-11 |
| rs582094 | let-7c-5p | trans | 2.74E-23 | Nightingale | Remnant_( | NA | T | A | 0.036725 | 0.004402 | 5.60E-17 |
| rs582094 | let-7c-5p | trans | 2.74E-23 | Nightingale | S_VLDL_CE | NA | T | A | 0.038034 | 0.004425 | 2.50E-18 |
| rs582094 | let-7c-5p | trans | 2.74E-23 | Nightingale | XXL_VLDL_ | NA | T | A | 0.03468 | 0.004441 | 4.10E-15 |
| rs582094 | let-7c-5p | trans | 2.74E-23 | Nightingale | S_VLDL_L | NA | T | A | 0.024549 | 0.004388 | 4.70E-09 |
| rs582094 | let-7c-5p | trans | 2.74E-23 | Nightingale | M_LDL_C_ | NA | T | A | 0.031036 | 0.004434 | 3.90E-12 |
| rs582094 | let-7c-5p | trans | 2.74E-23 | Nightingale | L_LDL_C | NA | T | A | 0.032117 | 0.004381 | 4.00E-13 |
| rs582094 | let-7c-5p | trans | 2.74E-23 | Nightingale | M_VLDL_C | NA | T | A | 0.040673 | 0.004391 | 3.90E-20 |
| rs582094 | let-7c-5p | trans | 2.74E-23 | Nightingale | L_VLDL_C_ | NA | T | A | 0.052204 | 0.004328 | 4.90E-34 |
| rs582094 | let-7c-5p | trans | 2.74E-23 | Nightingale | XS_VLDL_C | NA | T | A | 0.037674 | 0.004276 | 3.10E-18 |
| rs582094 | let-7c-5p | trans | 2.74E-23 | Nightingale | M_LDL_CE | NA | T | A | 0.032814 | 0.004437 | 1.40E-13 |
| rs582094 | let-7c-5p | trans | 2.74E-23 | Nightingale | S_HDL_L | NA | T | A | 0.02611 | 0.004416 | 2.10E-09 |
| rs582094 | let-7c-5p | trans | 2.74E-23 | Nightingale | M_VLDL_F | NA | T | A | 0.042037 | 0.004216 | 4.50E-23 |
| rs582094 | let-7c-5p | trans | 2.74E-23 | Nightingale | S_VLDL_C_ | NA | T | A | 0.041764 | 0.004312 | 8.50E-22 |
| rs582094 | let-7c-5p | trans | 2.74E-23 | Nightingale | VLDL_CE | NA | T | A | 0.035642 | 0.004432 | 4.50E-16 |
| rs582094 | let-7c-5p | trans | 2.74E-23 | Nightingale | IDL_C | NA | T | A | 0.034832 | 0.00428 | 1.40E-15 |
| rs582094 | let-7c-5p | trans | 2.74E-23 | Nightingale | XL_VLDL_P | NA | T | A | 0.038942 | 0.004438 | 4.80E-19 |
| rs582094 | let-7c-5p | trans | 2.74E-23 | Nightingale | L_LDL_P | NA | T | A | 0.028971 | 0.004416 | 9.20E-11 |
| rs582094 | let-7c-5p | trans | 2.74E-23 | Nightingale | XL_VLDL_F | NA | T | A | 0.037159 | 0.004296 | 2.40E-18 |
| rs582094 | let-7c-5p | trans | 2.74E-23 | Nightingale | XL_VLDL_T | NA | T | A | -0.04392 | 0.004295 | 2.20E-24 |
| rs582094 | let-7c-5p | trans | 2.74E-23 | Nightingale | L_LDL_PL | NA | T | A | 0.037813 | 0.004398 | 1.50E-17 |
| rs582094 | let-7c-5p | trans | 2.74E-23 | Nightingale | L_VLDL_CE | NA | T | A | 0.047319 | 0.00424 | 3.20E-29 |
| rs582094 | let-7c-5p | trans | 2.74E-23 | Nightingale | M_LDL_L | NA | T | A | 0.032867 | 0.004437 | 1.50E-13 |
| rs582094 | let-7c-5p | trans | 2.74E-23 | Nightingale | S_VLDL_FC | NA | T | A | 0.037726 | 0.004432 | 1.20E-17 |
| rs582094 | let-7c-5p | trans | 2.74E-23 | Nightingale | S_VLDL_PL | NA | T | A | 0.032628 | 0.004423 | 6.00E-14 |
| rs582094 | let-7c-5p | trans | 2.74E-23 | Nightingale | XS_VLDL_F | NA | T | A | 0.03125 | 0.0044 | 3.70E-12 |
| rs582094 | let-7c-5p | trans | 2.74E-23 | Nightingale | S_VLDL_C | NA | T | A | 0.038213 | 0.004431 | 2.80E-18 |
| rs582094 | let-7c-5p | trans | 2.74E-23 | Nightingale | XL_VLDL_C | NA | T | A | 0.038128 | 0.004249 | 6.90E-19 |
| rs582094 | let-7c-5p | trans | 2.74E-23 | Nightingale | S_LDL_PL_ | NA | T | A | -0.02995 | 0.004423 | 3.30E-13 |
| rs582094 | let-7c-5p | trans | 2.74E-23 | Nightingale | LDL_PL | NA | T | A | 0.03482 | 0.004421 | 7.30E-15 |
| rs582094 | let-7c-5p | trans | 2.74E-23 | Nightingale | non_HDL_( | NA | T | A | 0.035527 | 0.004407 | 7.90E-16 |
| rs582094 | let-7c-5p | trans | 2.74E-23 | Nightingale | XXL_VLDL_ | NA | T | A | 0.025093 | 0.00446 | 1.40E-09 |
| rs582094 | let-7c-5p | trans | 2.74E-23 | Nightingale | M_LDL_PL | NA | T | A | 0.032301 | 0.004437 | 3.90E-13 |
| rs582094 | let-7c-5p | trans | 2.74E-23 | Nightingale | XL_VLDL_C | NA | T | A | 0.037247 | 0.004251 | 4.70E-18 |
| rs582094 | let-7c-5p | trans | 2.74E-23 | Nightingale | XS_VLDL_P | NA | T | A | 0.032031 | 0.004369 | 6.90E-14 |
| rs582094 | let-7c-5p | trans | 2.74E-23 | Nightingale | M_VLDL_F | NA | T | A | 0.033201 | 0.00443 | 4.70E-14 |
| rs582094 | let-7c-5p | trans | 2.74E-23 | Nightingale | S_HDL_PL | NA | T | A | 0.023614 | 0.004393 | 4.10E-08 |
| rs582094 | let-7c-5p | trans | 2.74E-23 | Nightingale | XXL_VLDL_ | NA | T | A | 0.030797 | 0.004412 | 1.20E-12 |
| rs582094 | let-7c-5p | trans | 2.74E-23 | Nightingale | S_LDL_FC | NA | T | A | 0.028398 | 0.004424 | 6.00E-10 |
| rs582094 | let-7c-5p | trans | 2.74E-23 | Nightingale | PUFA | NA | T | A | 0.023572 | 0.004322 | 1.80E-08 |
| rs582094 | let-7c-5p | trans | 2.74E-23 | Nightingale | LDL_FC | NA | T | A | 0.033455 | 0.004384 | 8.10E-14 |
| rs582094 | let-7c-5p | trans | 2.74E-23 | Nightingale | LDL_P | NA | T | A | 0.03163 | 0.004428 | 1.50E-12 |
| rs582094 | let-7c-5p | trans | 2.74E-23 | Nightingale | L_VLDL_TG | NA | T | A | -0.05112 | 0.00441 | 7.10E-32 |
| rs582094 | let-7c-5p | trans | 2.74E-23 | Nightingale | IDL_FC | NA | T | A | 0.038121 | 0.00429 | 2.20E-18 |
| rs582094 | let-7c-5p | trans | 2.74E-23 | Nightingale | LDL_C | NA | T | A | 0.033002 | 0.004412 | 1.40E-13 |
| rs582094 | let-7c-5p | trans | 2.74E-23 | Nightingale | Total_CE | NA | T | A | 0.031574 | 0.004274 | 1.40E-13 |
| rs582094 | let-7c-5p | trans | 2.74E-23 | Nightingale | M_LDL_TG | NA | T | A | -0.02744 | 0.004376 | 4.70E-10 |
| rs582094 | let-7c-5p | trans | 2.74E-23 | Nightingale | S_LDL_CE_ | NA | T | A | 0.03085 | 0.004424 | 6.10E-14 |
| rs582094 | let-7c-5p | trans | 2.74E-23 | Nightingale | VLDL_P | NA | T | A | 0.026524 | 0.004407 | 5.50E-10 |
| rs582094 | let-7c-5p | trans | 2.74E-23 | Nightingale | S_HDL_CE | NA | T | A | 0.028434 | 0.004438 | 1.90E-10 |
| rs582094 | let-7c-5p | trans | 2.74E-23 | Nightingale | S_VLDL_PL | NA | T | A | 0.027637 | 0.004243 | 2.50E-10 |
| rs582094 | let-7c-5p | trans | 2.74E-23 | Nightingale | M_VLDL_C | NA | T | A | 0.038533 | 0.004419 | 3.20E-18 |
| rs582094 | let-7c-5p | trans | 2.74E-23 | Nightingale | Sphingomy | NA | T | A | 0.028892 | 0.004142 | 8.90E-12 |
| rs582094 | let-7c-5p | trans | 2.74E-23 | Nightingale | M_VLDL_P | NA | T | A | 0.044033 | 0.004253 | 7.10E-25 |
| rs582094 | let-7c-5p | trans | 2.74E-23 | Nightingale | LDL_CE | NA | T | A | 0.032359 | 0.004419 | 3.30E-13 |
| rs582094 | let-7c-5p | trans | 2.74E-23 | Nightingale | XXL_VLDL_ | NA | T | A | -0.03566 | 0.004469 | 2.70E-16 |
| rs582094 | let-7c-5p | trans | 2.74E-23 | Nightingale | IDL_P | NA | T | A | 0.039908 | 0.004385 | 1.90E-19 |

| | | | | | | | | | | | |
|---|---|---|---|---|---|---|---|---|---|---|---|
| rs582094 | let-7c-5p | trans | 2.74E-23 | Nightingale | Total_PL | NA | T | A | 0.024785 | 0.00422 | 1.40E-09 |
| rs582094 | let-7c-5p | trans | 2.74E-23 | Nightingale | S_LDL_L | NA | T | A | 0.027606 | 0.00443 | 1.10E-09 |
| rs582094 | let-7c-5p | trans | 2.74E-23 | Nightingale | S_VLDL_TG | NA | T | A | -0.03801 | 0.004289 | 2.20E-18 |
| rs582094 | let-7c-5p | trans | 2.74E-23 | Nightingale | L_LDL_CE | NA | T | A | 0.031356 | 0.004392 | 1.50E-12 |
| rs582094 | let-7c-5p | trans | 2.74E-23 | Nightingale | S_VLDL_P | NA | T | A | 0.023349 | 0.004376 | 2.00E-08 |
| rs582094 | let-7c-5p | trans | 2.74E-23 | Nightingale | S_LDL_CE | NA | T | A | 0.030409 | 0.004433 | 7.90E-12 |
| rs582094 | let-7c-5p | trans | 2.74E-23 | Nightingale | XL_HDL_PL | NA | T | A | -0.02265 | 0.004071 | 1.40E-08 |
| rs582094 | let-7c-5p | trans | 2.74E-23 | Nightingale | XS_VLDL_L | NA | T | A | 0.033719 | 0.004356 | 2.70E-15 |
| rs582094 | let-7c-5p | trans | 2.74E-23 | Nightingale | Citrate | NA | T | A | 0.02682 | 0.004402 | 7.90E-10 |
| rs582094 | let-7c-5p | trans | 2.74E-23 | Nightingale | Clinical_LD | NA | T | A | 0.0363 | 0.004406 | 4.30E-16 |
| rs582094 | let-7c-5p | trans | 2.74E-23 | Nightingale | Omega_6 | NA | T | A | 0.025694 | 0.004327 | 8.00E-10 |
| rs582094 | let-7c-5p | trans | 2.74E-23 | Nightingale | S_HDL_C | NA | T | A | 0.028202 | 0.004424 | 2.50E-10 |
| rs582094 | let-7c-5p | trans | 2.74E-23 | Nightingale | M_LDL_C | NA | T | A | 0.033818 | 0.004439 | 3.20E-14 |
| rs582094 | let-7c-5p | trans | 2.74E-23 | Nightingale | S_HDL_P | NA | T | A | 0.030405 | 0.004432 | 7.50E-12 |
| rs582094 | miR-126-3p | trans | 2.46E-39 | Nightingale | XS_VLDL_P | NA | T | A | 0.036606 | 0.004372 | 1.90E-17 |
| rs582094 | miR-126-3p | trans | 2.46E-39 | Nightingale | S_LDL_C | NA | T | A | 0.030792 | 0.004435 | 7.70E-12 |
| rs582094 | miR-126-3p | trans | 2.46E-39 | Nightingale | ApoB_by_A | NA | T | A | 0.024975 | 0.004364 | 1.60E-08 |
| rs582094 | miR-126-3p | trans | 2.46E-39 | Nightingale | M_VLDL_T | NA | T | A | -0.03854 | 0.004187 | 8.80E-20 |
| rs582094 | miR-126-3p | trans | 2.46E-39 | Nightingale | L_LDL_FC | NA | T | A | 0.033451 | 0.004351 | 4.40E-14 |
| rs582094 | miR-126-3p | trans | 2.46E-39 | Nightingale | Total_C | NA | T | A | 0.032199 | 0.00429 | 4.90E-14 |
| rs582094 | miR-126-3p | trans | 2.46E-39 | Nightingale | S_LDL_C_p | NA | T | A | 0.045184 | 0.004448 | 9.90E-25 |
| rs582094 | miR-126-3p | trans | 2.46E-39 | Nightingale | IDL_CE | NA | T | A | 0.033389 | 0.004278 | 1.80E-14 |
| rs582094 | miR-126-3p | trans | 2.46E-39 | Nightingale | XS_VLDL_F | NA | T | A | 0.036947 | 0.004354 | 9.50E-18 |
| rs582094 | miR-126-3p | trans | 2.46E-39 | Nightingale | Total_FC | NA | T | A | 0.03308 | 0.004329 | 1.00E-14 |
| rs582094 | miR-126-3p | trans | 2.46E-39 | Nightingale | S_VLDL_CE | NA | T | A | 0.046942 | 0.004372 | 1.80E-26 |
| rs582094 | miR-126-3p | trans | 2.46E-39 | Nightingale | M_LDL_P | NA | T | A | 0.035786 | 0.004444 | 9.30E-16 |
| rs582094 | miR-126-3p | trans | 2.46E-39 | Nightingale | M_VLDL_C | NA | T | A | 0.033603 | 0.004163 | 2.40E-15 |
| rs582094 | miR-126-3p | trans | 2.46E-39 | Nightingale | IDL_L | NA | T | A | 0.03514 | 0.004284 | 7.00E-16 |
| rs582094 | miR-126-3p | trans | 2.46E-39 | Nightingale | IDL_PL | NA | T | A | 0.035198 | 0.004276 | 4.90E-16 |
| rs582094 | miR-126-3p | trans | 2.46E-39 | Nightingale | XXL_VLDL_ | NA | T | A | 0.032961 | 0.004465 | 1.30E-13 |
| rs582094 | miR-126-3p | trans | 2.46E-39 | Nightingale | IDL_FC_pct | NA | T | A | 0.025717 | 0.004397 | 1.20E-08 |
| rs582094 | miR-126-3p | trans | 2.46E-39 | Nightingale | VLDL_C | NA | T | A | 0.031173 | 0.004425 | 6.70E-13 |
| rs582094 | miR-126-3p | trans | 2.46E-39 | Nightingale | M_VLDL_C | NA | T | A | 0.036181 | 0.004174 | 1.50E-17 |
| rs582094 | miR-126-3p | trans | 2.46E-39 | Nightingale | S_LDL_P | NA | T | A | 0.029487 | 0.004435 | 4.10E-11 |
| rs582094 | miR-126-3p | trans | 2.46E-39 | Nightingale | LDL_L | NA | T | A | 0.033121 | 0.004416 | 1.10E-13 |
| rs582094 | miR-126-3p | trans | 2.46E-39 | Nightingale | Total_L | NA | T | A | 0.026203 | 0.004359 | 5.90E-10 |
| rs582094 | miR-126-3p | trans | 2.46E-39 | Nightingale | LA | NA | T | A | 0.024685 | 0.004345 | 5.20E-09 |
| rs582094 | miR-126-3p | trans | 2.46E-39 | Nightingale | M_VLDL_P | NA | T | A | 0.028969 | 0.004424 | 2.80E-11 |
| rs582094 | miR-126-3p | trans | 2.46E-39 | Nightingale | XS_VLDL_C | NA | T | A | 0.037867 | 0.004301 | 2.00E-18 |
| rs582094 | miR-126-3p | trans | 2.46E-39 | Nightingale | ApoB | NA | T | A | 0.033786 | 0.004425 | 3.10E-14 |
| rs582094 | miR-126-3p | trans | 2.46E-39 | Nightingale | GlycA | NA | T | A | 0.027998 | 0.004412 | 1.60E-10 |
| rs582094 | miR-126-3p | trans | 2.46E-39 | Nightingale | S_VLDL_FC | NA | T | A | 0.029218 | 0.004234 | 1.60E-11 |
| rs582094 | miR-126-3p | trans | 2.46E-39 | Nightingale | L_VLDL_FC | NA | T | A | 0.048617 | 0.004389 | 5.10E-29 |
| rs582094 | miR-126-3p | trans | 2.46E-39 | Nightingale | VLDL_FC | NA | T | A | 0.024055 | 0.004399 | 1.30E-08 |
| rs582094 | miR-126-3p | trans | 2.46E-39 | Nightingale | M_LDL_FC | NA | T | A | 0.034163 | 0.004426 | 3.00E-14 |
| rs582094 | miR-126-3p | trans | 2.46E-39 | Nightingale | L_LDL_L | NA | T | A | 0.033355 | 0.004388 | 4.70E-14 |
| rs582094 | miR-126-3p | trans | 2.46E-39 | Nightingale | M_VLDL_P | NA | T | A | 0.028139 | 0.00442 | 8.60E-11 |
| rs582094 | miR-126-3p | trans | 2.46E-39 | Nightingale | Remnant_( | NA | T | A | 0.036725 | 0.004402 | 5.60E-17 |
| rs582094 | miR-126-3p | trans | 2.46E-39 | Nightingale | S_VLDL_CE | NA | T | A | 0.038034 | 0.004425 | 2.50E-18 |
| rs582094 | miR-126-3p | trans | 2.46E-39 | Nightingale | XXL_VLDL_ | NA | T | A | 0.03468 | 0.004441 | 4.10E-15 |
| rs582094 | miR-126-3p | trans | 2.46E-39 | Nightingale | S_VLDL_L | NA | T | A | 0.024549 | 0.004388 | 4.70E-09 |
| rs582094 | miR-126-3p | trans | 2.46E-39 | Nightingale | M_LDL_C_ | NA | T | A | 0.031036 | 0.004434 | 3.90E-12 |
| rs582094 | miR-126-3p | trans | 2.46E-39 | Nightingale | L_LDL_C | NA | T | A | 0.032117 | 0.004381 | 4.00E-13 |
| rs582094 | miR-126-3p | trans | 2.46E-39 | Nightingale | M_VLDL_C | NA | T | A | 0.040673 | 0.004391 | 3.90E-20 |
| rs582094 | miR-126-3p | trans | 2.46E-39 | Nightingale | L_VLDL_C_ | NA | T | A | 0.052204 | 0.004328 | 4.90E-34 |
| rs582094 | miR-126-3p | trans | 2.46E-39 | Nightingale | XS_VLDL_C | NA | T | A | 0.037674 | 0.004276 | 3.10E-18 |
| rs582094 | miR-126-3p | trans | 2.46E-39 | Nightingale | M_LDL_CE | NA | T | A | 0.032814 | 0.004437 | 1.40E-13 |
| rs582094 | miR-126-3p | trans | 2.46E-39 | Nightingale | S_HDL_L | NA | T | A | 0.02611 | 0.004416 | 2.10E-09 |
| rs582094 | miR-126-3p | trans | 2.46E-39 | Nightingale | M_VLDL_F | NA | T | A | 0.042037 | 0.004216 | 4.50E-23 |
| rs582094 | miR-126-3p | trans | 2.46E-39 | Nightingale | S_VLDL_C_ | NA | T | A | 0.041764 | 0.004312 | 8.50E-22 |
| rs582094 | miR-126-3p | trans | 2.46E-39 | Nightingale | VLDL_CE | NA | T | A | 0.035642 | 0.004432 | 4.50E-16 |
| rs582094 | miR-126-3p | trans | 2.46E-39 | Nightingale | IDL_C | NA | T | A | 0.034832 | 0.00428 | 1.40E-15 |
| rs582094 | miR-126-3p | trans | 2.46E-39 | Nightingale | XL_VLDL_P | NA | T | A | 0.038942 | 0.004438 | 4.80E-19 |

| | | | | | | | | | | | |
|---|---|---|---|---|---|---|---|---|---|---|---|
| rs582094 | miR-126-3p | trans | 2.46E-39 | Nightingale | L_LDL_P | NA | T | A | 0.028971 | 0.004416 | 9.20E-11 |
| rs582094 | miR-126-3p | trans | 2.46E-39 | Nightingale | XL_VLDL_F | NA | T | A | 0.037159 | 0.004296 | 2.40E-18 |
| rs582094 | miR-126-3p | trans | 2.46E-39 | Nightingale | XL_VLDL_T | NA | T | A | -0.04392 | 0.004295 | 2.20E-24 |
| rs582094 | miR-126-3p | trans | 2.46E-39 | Nightingale | L_LDL_PL | NA | T | A | 0.037813 | 0.004398 | 1.50E-17 |
| rs582094 | miR-126-3p | trans | 2.46E-39 | Nightingale | L_VLDL_CE | NA | T | A | 0.047319 | 0.00424 | 3.20E-29 |
| rs582094 | miR-126-3p | trans | 2.46E-39 | Nightingale | M_LDL_L | NA | T | A | 0.032867 | 0.004437 | 1.50E-13 |
| rs582094 | miR-126-3p | trans | 2.46E-39 | Nightingale | S_VLDL_FC | NA | T | A | 0.037726 | 0.004432 | 1.20E-17 |
| rs582094 | miR-126-3p | trans | 2.46E-39 | Nightingale | S_VLDL_PL | NA | T | A | 0.032628 | 0.004423 | 6.00E-14 |
| rs582094 | miR-126-3p | trans | 2.46E-39 | Nightingale | XS_VLDL_F | NA | T | A | 0.03125 | 0.0044 | 3.70E-12 |
| rs582094 | miR-126-3p | trans | 2.46E-39 | Nightingale | S_VLDL_C | NA | T | A | 0.038213 | 0.004431 | 2.80E-18 |
| rs582094 | miR-126-3p | trans | 2.46E-39 | Nightingale | XL_VLDL_C | NA | T | A | 0.038128 | 0.004249 | 6.90E-19 |
| rs582094 | miR-126-3p | trans | 2.46E-39 | Nightingale | S_LDL_PL_ | NA | T | A | -0.02995 | 0.004423 | 3.30E-13 |
| rs582094 | miR-126-3p | trans | 2.46E-39 | Nightingale | LDL_PL | NA | T | A | 0.03482 | 0.004421 | 7.30E-15 |
| rs582094 | miR-126-3p | trans | 2.46E-39 | Nightingale | non_HDL_( | NA | T | A | 0.035527 | 0.004407 | 7.90E-16 |
| rs582094 | miR-126-3p | trans | 2.46E-39 | Nightingale | XXL_VLDL_ | NA | T | A | 0.025093 | 0.00446 | 1.40E-09 |
| rs582094 | miR-126-3p | trans | 2.46E-39 | Nightingale | M_LDL_PL | NA | T | A | 0.032301 | 0.004437 | 3.90E-13 |
| rs582094 | miR-126-3p | trans | 2.46E-39 | Nightingale | XL_VLDL_C | NA | T | A | 0.037247 | 0.004251 | 4.70E-18 |
| rs582094 | miR-126-3p | trans | 2.46E-39 | Nightingale | XS_VLDL_P | NA | T | A | 0.032031 | 0.004369 | 6.90E-14 |
| rs582094 | miR-126-3p | trans | 2.46E-39 | Nightingale | M_VLDL_F | NA | T | A | 0.033201 | 0.00443 | 4.70E-14 |
| rs582094 | miR-126-3p | trans | 2.46E-39 | Nightingale | S_HDL_PL | NA | T | A | 0.023614 | 0.004393 | 4.10E-08 |
| rs582094 | miR-126-3p | trans | 2.46E-39 | Nightingale | XXL_VLDL_ | NA | T | A | 0.030797 | 0.004412 | 1.20E-12 |
| rs582094 | miR-126-3p | trans | 2.46E-39 | Nightingale | S_LDL_FC | NA | T | A | 0.028398 | 0.004424 | 6.00E-10 |
| rs582094 | miR-126-3p | trans | 2.46E-39 | Nightingale | PUFA | NA | T | A | 0.023572 | 0.004322 | 1.80E-08 |
| rs582094 | miR-126-3p | trans | 2.46E-39 | Nightingale | LDL_FC | NA | T | A | 0.033455 | 0.004384 | 8.10E-14 |
| rs582094 | miR-126-3p | trans | 2.46E-39 | Nightingale | LDL_P | NA | T | A | 0.03163 | 0.004428 | 1.50E-12 |
| rs582094 | miR-126-3p | trans | 2.46E-39 | Nightingale | L_VLDL_TG | NA | T | A | -0.05112 | 0.00441 | 7.10E-32 |
| rs582094 | miR-126-3p | trans | 2.46E-39 | Nightingale | IDL_FC | NA | T | A | 0.038121 | 0.00429 | 2.20E-18 |
| rs582094 | miR-126-3p | trans | 2.46E-39 | Nightingale | LDL_C | NA | T | A | 0.033002 | 0.004412 | 1.40E-13 |
| rs582094 | miR-126-3p | trans | 2.46E-39 | Nightingale | Total_CE | NA | T | A | 0.031574 | 0.004274 | 1.40E-13 |
| rs582094 | miR-126-3p | trans | 2.46E-39 | Nightingale | M_LDL_TG | NA | T | A | -0.02744 | 0.004376 | 4.70E-10 |
| rs582094 | miR-126-3p | trans | 2.46E-39 | Nightingale | S_LDL_CE_ | NA | T | A | 0.03085 | 0.004424 | 6.10E-14 |
| rs582094 | miR-126-3p | trans | 2.46E-39 | Nightingale | VLDL_P | NA | T | A | 0.026524 | 0.004407 | 5.50E-10 |
| rs582094 | miR-126-3p | trans | 2.46E-39 | Nightingale | S_HDL_CE | NA | T | A | 0.028434 | 0.004438 | 1.90E-10 |
| rs582094 | miR-126-3p | trans | 2.46E-39 | Nightingale | S_VLDL_PL | NA | T | A | 0.027637 | 0.004243 | 2.50E-10 |
| rs582094 | miR-126-3p | trans | 2.46E-39 | Nightingale | M_VLDL_C | NA | T | A | 0.038533 | 0.004419 | 3.20E-18 |
| rs582094 | miR-126-3p | trans | 2.46E-39 | Nightingale | Sphingomy | NA | T | A | 0.028892 | 0.004142 | 8.90E-12 |
| rs582094 | miR-126-3p | trans | 2.46E-39 | Nightingale | M_VLDL_P | NA | T | A | 0.044033 | 0.004253 | 7.10E-25 |
| rs582094 | miR-126-3p | trans | 2.46E-39 | Nightingale | LDL_CE | NA | T | A | 0.032359 | 0.004419 | 3.30E-13 |
| rs582094 | miR-126-3p | trans | 2.46E-39 | Nightingale | XXL_VLDL_ | NA | T | A | -0.03566 | 0.004469 | 2.70E-16 |
| rs582094 | miR-126-3p | trans | 2.46E-39 | Nightingale | IDL_P | NA | T | A | 0.039908 | 0.004385 | 1.90E-19 |
| rs582094 | miR-126-3p | trans | 2.46E-39 | Nightingale | Total_PL | NA | T | A | 0.024785 | 0.00422 | 1.40E-09 |
| rs582094 | miR-126-3p | trans | 2.46E-39 | Nightingale | S_LDL_L | NA | T | A | 0.027606 | 0.00443 | 1.10E-09 |
| rs582094 | miR-126-3p | trans | 2.46E-39 | Nightingale | S_VLDL_TG | NA | T | A | -0.03801 | 0.004289 | 2.20E-18 |
| rs582094 | miR-126-3p | trans | 2.46E-39 | Nightingale | L_LDL_CE | NA | T | A | 0.031356 | 0.004392 | 1.50E-12 |
| rs582094 | miR-126-3p | trans | 2.46E-39 | Nightingale | S_VLDL_P | NA | T | A | 0.023349 | 0.004376 | 2.00E-08 |
| rs582094 | miR-126-3p | trans | 2.46E-39 | Nightingale | S_LDL_CE | NA | T | A | 0.030409 | 0.004433 | 7.90E-12 |
| rs582094 | miR-126-3p | trans | 2.46E-39 | Nightingale | XL_HDL_PL | NA | T | A | -0.02265 | 0.004071 | 1.40E-08 |
| rs582094 | miR-126-3p | trans | 2.46E-39 | Nightingale | XS_VLDL_L | NA | T | A | 0.033719 | 0.004356 | 2.70E-15 |
| rs582094 | miR-126-3p | trans | 2.46E-39 | Nightingale | Citrate | NA | T | A | 0.02682 | 0.004402 | 7.90E-10 |
| rs582094 | miR-126-3p | trans | 2.46E-39 | Nightingale | Clinical_LD | NA | T | A | 0.0363 | 0.004406 | 4.30E-16 |
| rs582094 | miR-126-3p | trans | 2.46E-39 | Nightingale | Omega_6 | NA | T | A | 0.025694 | 0.004327 | 8.00E-10 |
| rs582094 | miR-126-3p | trans | 2.46E-39 | Nightingale | S_HDL_C | NA | T | A | 0.028202 | 0.004424 | 2.50E-10 |
| rs582094 | miR-126-3p | trans | 2.46E-39 | Nightingale | M_LDL_C | NA | T | A | 0.033818 | 0.004439 | 3.20E-14 |
| rs582094 | miR-126-3p | trans | 2.46E-39 | Nightingale | S_HDL_P | NA | T | A | 0.030405 | 0.004432 | 7.50E-12 |
| rs582094 | miR-195-5p | trans | 2.17E-17 | Nightingale | XS_VLDL_P | NA | T | A | 0.036606 | 0.004372 | 1.90E-17 |
| rs582094 | miR-195-5p | trans | 2.17E-17 | Nightingale | S_LDL_C | NA | T | A | 0.030792 | 0.004435 | 7.70E-12 |
| rs582094 | miR-195-5p | trans | 2.17E-17 | Nightingale | ApoB_by_/ | NA | T | A | 0.024975 | 0.004364 | 1.60E-08 |
| rs582094 | miR-195-5p | trans | 2.17E-17 | Nightingale | M_VLDL_T | NA | T | A | -0.03854 | 0.004187 | 8.80E-20 |
| rs582094 | miR-195-5p | trans | 2.17E-17 | Nightingale | L_LDL_FC | NA | T | A | 0.033451 | 0.004351 | 4.40E-14 |
| rs582094 | miR-195-5p | trans | 2.17E-17 | Nightingale | Total_C | NA | T | A | 0.032199 | 0.00429 | 4.90E-14 |
| rs582094 | miR-195-5p | trans | 2.17E-17 | Nightingale | S_LDL_C_p | NA | T | A | 0.045184 | 0.004448 | 9.90E-25 |
| rs582094 | miR-195-5p | trans | 2.17E-17 | Nightingale | IDL_CE | NA | T | A | 0.033389 | 0.004278 | 1.80E-14 |
| rs582094 | miR-195-5p | trans | 2.17E-17 | Nightingale | XS_VLDL_F | NA | T | A | 0.036947 | 0.004354 | 9.50E-18 |

| | | | | | | | | | | | |
|---|---|---|---|---|---|---|---|---|---|---|---|
| rs582094 | miR-195-5p | trans | 2.17E-17 | Nightingale | Total_FC | NA | T | A | 0.03308 | 0.004329 | 1.00E-14 |
| rs582094 | miR-195-5p | trans | 2.17E-17 | Nightingale | S_VLDL_CE | NA | T | A | 0.046942 | 0.004372 | 1.80E-26 |
| rs582094 | miR-195-5p | trans | 2.17E-17 | Nightingale | M_LDL_P | NA | T | A | 0.035786 | 0.004444 | 9.30E-16 |
| rs582094 | miR-195-5p | trans | 2.17E-17 | Nightingale | M_VLDL_C | NA | T | A | 0.033603 | 0.004163 | 2.40E-15 |
| rs582094 | miR-195-5p | trans | 2.17E-17 | Nightingale | IDL_L | NA | T | A | 0.03514 | 0.004284 | 7.00E-16 |
| rs582094 | miR-195-5p | trans | 2.17E-17 | Nightingale | IDL_PL | NA | T | A | 0.035198 | 0.004276 | 4.90E-16 |
| rs582094 | miR-195-5p | trans | 2.17E-17 | Nightingale | XXL_VLDL_ | NA | T | A | 0.032961 | 0.004465 | 1.30E-13 |
| rs582094 | miR-195-5p | trans | 2.17E-17 | Nightingale | IDL_FC_pct | NA | T | A | 0.025717 | 0.004397 | 1.20E-08 |
| rs582094 | miR-195-5p | trans | 2.17E-17 | Nightingale | VLDL_C | NA | T | A | 0.031173 | 0.004425 | 6.70E-13 |
| rs582094 | miR-195-5p | trans | 2.17E-17 | Nightingale | M_VLDL_C_ | NA | T | A | 0.036181 | 0.004174 | 1.50E-17 |
| rs582094 | miR-195-5p | trans | 2.17E-17 | Nightingale | S_LDL_P | NA | T | A | 0.029487 | 0.004435 | 4.10E-11 |
| rs582094 | miR-195-5p | trans | 2.17E-17 | Nightingale | LDL_L | NA | T | A | 0.033121 | 0.004416 | 1.10E-13 |
| rs582094 | miR-195-5p | trans | 2.17E-17 | Nightingale | Total_L | NA | T | A | 0.026203 | 0.004359 | 5.90E-10 |
| rs582094 | miR-195-5p | trans | 2.17E-17 | Nightingale | LA | NA | T | A | 0.024685 | 0.004345 | 5.20E-09 |
| rs582094 | miR-195-5p | trans | 2.17E-17 | Nightingale | M_VLDL_P | NA | T | A | 0.028969 | 0.004424 | 2.80E-11 |
| rs582094 | miR-195-5p | trans | 2.17E-17 | Nightingale | XS_VLDL_C | NA | T | A | 0.037867 | 0.004301 | 2.00E-18 |
| rs582094 | miR-195-5p | trans | 2.17E-17 | Nightingale | ApoB | NA | T | A | 0.033786 | 0.004425 | 3.10E-14 |
| rs582094 | miR-195-5p | trans | 2.17E-17 | Nightingale | GlycA | NA | T | A | 0.027998 | 0.004412 | 1.60E-10 |
| rs582094 | miR-195-5p | trans | 2.17E-17 | Nightingale | S_VLDL_FC | NA | T | A | 0.029218 | 0.004234 | 1.60E-11 |
| rs582094 | miR-195-5p | trans | 2.17E-17 | Nightingale | L_VLDL_FC | NA | T | A | 0.048617 | 0.004389 | 5.10E-29 |
| rs582094 | miR-195-5p | trans | 2.17E-17 | Nightingale | VLDL_FC | NA | T | A | 0.024055 | 0.004399 | 1.30E-08 |
| rs582094 | miR-195-5p | trans | 2.17E-17 | Nightingale | M_LDL_FC | NA | T | A | 0.034163 | 0.004426 | 3.00E-14 |
| rs582094 | miR-195-5p | trans | 2.17E-17 | Nightingale | L_LDL_L | NA | T | A | 0.033355 | 0.004388 | 4.70E-14 |
| rs582094 | miR-195-5p | trans | 2.17E-17 | Nightingale | M_VLDL_P | NA | T | A | 0.028139 | 0.00442 | 8.60E-11 |
| rs582094 | miR-195-5p | trans | 2.17E-17 | Nightingale | Remnant_C | NA | T | A | 0.036725 | 0.004402 | 5.60E-17 |
| rs582094 | miR-195-5p | trans | 2.17E-17 | Nightingale | S_VLDL_CE | NA | T | A | 0.038034 | 0.004425 | 2.50E-18 |
| rs582094 | miR-195-5p | trans | 2.17E-17 | Nightingale | XXL_VLDL_ | NA | T | A | 0.03468 | 0.004441 | 4.10E-15 |
| rs582094 | miR-195-5p | trans | 2.17E-17 | Nightingale | S_VLDL_L | NA | T | A | 0.024549 | 0.004388 | 4.70E-09 |
| rs582094 | miR-195-5p | trans | 2.17E-17 | Nightingale | M_LDL_C_p | NA | T | A | 0.031036 | 0.004434 | 3.90E-12 |
| rs582094 | miR-195-5p | trans | 2.17E-17 | Nightingale | L_LDL_C | NA | T | A | 0.032117 | 0.004381 | 4.00E-13 |
| rs582094 | miR-195-5p | trans | 2.17E-17 | Nightingale | M_VLDL_C | NA | T | A | 0.040673 | 0.004391 | 3.90E-20 |
| rs582094 | miR-195-5p | trans | 2.17E-17 | Nightingale | L_VLDL_C_ | NA | T | A | 0.052204 | 0.004328 | 4.90E-34 |
| rs582094 | miR-195-5p | trans | 2.17E-17 | Nightingale | XS_VLDL_C | NA | T | A | 0.037674 | 0.004276 | 3.10E-18 |
| rs582094 | miR-195-5p | trans | 2.17E-17 | Nightingale | M_LDL_CE | NA | T | A | 0.032814 | 0.004437 | 1.40E-13 |
| rs582094 | miR-195-5p | trans | 2.17E-17 | Nightingale | S_HDL_L | NA | T | A | 0.02611 | 0.004416 | 2.10E-09 |
| rs582094 | miR-195-5p | trans | 2.17E-17 | Nightingale | M_VLDL_F | NA | T | A | 0.042037 | 0.004216 | 4.50E-23 |
| rs582094 | miR-195-5p | trans | 2.17E-17 | Nightingale | S_VLDL_C_ | NA | T | A | 0.041764 | 0.004312 | 8.50E-22 |
| rs582094 | miR-195-5p | trans | 2.17E-17 | Nightingale | VLDL_CE | NA | T | A | 0.035642 | 0.004432 | 4.50E-16 |
| rs582094 | miR-195-5p | trans | 2.17E-17 | Nightingale | IDL_C | NA | T | A | 0.034832 | 0.00428 | 1.40E-15 |
| rs582094 | miR-195-5p | trans | 2.17E-17 | Nightingale | XL_VLDL_P | NA | T | A | 0.038942 | 0.004438 | 4.80E-19 |
| rs582094 | miR-195-5p | trans | 2.17E-17 | Nightingale | L_LDL_P | NA | T | A | 0.028971 | 0.004416 | 9.20E-11 |
| rs582094 | miR-195-5p | trans | 2.17E-17 | Nightingale | XL_VLDL_F | NA | T | A | 0.037159 | 0.004296 | 2.40E-18 |
| rs582094 | miR-195-5p | trans | 2.17E-17 | Nightingale | XL_VLDL_T | NA | T | A | -0.04392 | 0.004295 | 2.20E-24 |
| rs582094 | miR-195-5p | trans | 2.17E-17 | Nightingale | L_LDL_PL | NA | T | A | 0.037813 | 0.004398 | 1.50E-17 |
| rs582094 | miR-195-5p | trans | 2.17E-17 | Nightingale | L_VLDL_CE | NA | T | A | 0.047319 | 0.00424 | 3.20E-29 |
| rs582094 | miR-195-5p | trans | 2.17E-17 | Nightingale | M_LDL_L | NA | T | A | 0.032867 | 0.004437 | 1.50E-13 |
| rs582094 | miR-195-5p | trans | 2.17E-17 | Nightingale | S_VLDL_FC | NA | T | A | 0.037726 | 0.004432 | 1.20E-17 |
| rs582094 | miR-195-5p | trans | 2.17E-17 | Nightingale | S_VLDL_PL | NA | T | A | 0.032628 | 0.004423 | 6.00E-14 |
| rs582094 | miR-195-5p | trans | 2.17E-17 | Nightingale | XS_VLDL_F | NA | T | A | 0.03125 | 0.0044 | 3.70E-12 |
| rs582094 | miR-195-5p | trans | 2.17E-17 | Nightingale | S_VLDL_C | NA | T | A | 0.038213 | 0.004431 | 2.80E-18 |
| rs582094 | miR-195-5p | trans | 2.17E-17 | Nightingale | XL_VLDL_C | NA | T | A | 0.038128 | 0.004249 | 6.90E-19 |
| rs582094 | miR-195-5p | trans | 2.17E-17 | Nightingale | S_LDL_PL_ | NA | T | A | -0.02995 | 0.004423 | 3.30E-13 |
| rs582094 | miR-195-5p | trans | 2.17E-17 | Nightingale | LDL_PL | NA | T | A | 0.03482 | 0.004421 | 7.30E-15 |
| rs582094 | miR-195-5p | trans | 2.17E-17 | Nightingale | non_HDL_C | NA | T | A | 0.035527 | 0.004407 | 7.90E-16 |
| rs582094 | miR-195-5p | trans | 2.17E-17 | Nightingale | XXL_VLDL_ | NA | T | A | 0.025093 | 0.00446 | 1.40E-09 |
| rs582094 | miR-195-5p | trans | 2.17E-17 | Nightingale | M_LDL_PL | NA | T | A | 0.032301 | 0.004437 | 3.90E-13 |
| rs582094 | miR-195-5p | trans | 2.17E-17 | Nightingale | XL_VLDL_C | NA | T | A | 0.037247 | 0.004251 | 4.70E-18 |
| rs582094 | miR-195-5p | trans | 2.17E-17 | Nightingale | XS_VLDL_P | NA | T | A | 0.032031 | 0.004369 | 6.90E-14 |
| rs582094 | miR-195-5p | trans | 2.17E-17 | Nightingale | M_VLDL_F | NA | T | A | 0.033201 | 0.00443 | 4.70E-14 |
| rs582094 | miR-195-5p | trans | 2.17E-17 | Nightingale | S_HDL_PL | NA | T | A | 0.023614 | 0.004393 | 4.10E-08 |
| rs582094 | miR-195-5p | trans | 2.17E-17 | Nightingale | XXL_VLDL_ | NA | T | A | 0.030797 | 0.004412 | 1.20E-12 |
| rs582094 | miR-195-5p | trans | 2.17E-17 | Nightingale | S_LDL_FC | NA | T | A | 0.028398 | 0.004424 | 6.00E-10 |
| rs582094 | miR-195-5p | trans | 2.17E-17 | Nightingale | PUFA | NA | T | A | 0.023572 | 0.004322 | 1.80E-08 |

| | | | | | | | | | | | |
|---|---|---|---|---|---|---|---|---|---|---|---|
| rs582094 | miR-195-5ɾ | trans | 2.17E-17 | Nightingale | LDL_FC | NA | T | A | 0.033455 | 0.004384 | 8.10E-14 |
| rs582094 | miR-195-5ɾ | trans | 2.17E-17 | Nightingale | LDL_P | NA | T | A | 0.03163 | 0.004428 | 1.50E-12 |
| rs582094 | miR-195-5ɾ | trans | 2.17E-17 | Nightingale | L_VLDL_TG | NA | T | A | -0.05112 | 0.00441 | 7.10E-32 |
| rs582094 | miR-195-5ɾ | trans | 2.17E-17 | Nightingale | IDL_FC | NA | T | A | 0.038121 | 0.00429 | 2.20E-18 |
| rs582094 | miR-195-5ɾ | trans | 2.17E-17 | Nightingale | LDL_C | NA | T | A | 0.033002 | 0.004412 | 1.40E-13 |
| rs582094 | miR-195-5ɾ | trans | 2.17E-17 | Nightingale | Total_CE | NA | T | A | 0.031574 | 0.004274 | 1.40E-13 |
| rs582094 | miR-195-5ɾ | trans | 2.17E-17 | Nightingale | M_LDL_TG | NA | T | A | -0.02744 | 0.004376 | 4.70E-10 |
| rs582094 | miR-195-5ɾ | trans | 2.17E-17 | Nightingale | S_LDL_CE_ | NA | T | A | 0.03085 | 0.004424 | 6.10E-14 |
| rs582094 | miR-195-5ɾ | trans | 2.17E-17 | Nightingale | VLDL_P | NA | T | A | 0.026524 | 0.004407 | 5.50E-10 |
| rs582094 | miR-195-5ɾ | trans | 2.17E-17 | Nightingale | S_HDL_CE | NA | T | A | 0.028434 | 0.004438 | 1.90E-10 |
| rs582094 | miR-195-5ɾ | trans | 2.17E-17 | Nightingale | S_VLDL_PL | NA | T | A | 0.027637 | 0.004243 | 2.50E-10 |
| rs582094 | miR-195-5ɾ | trans | 2.17E-17 | Nightingale | M_VLDL_C | NA | T | A | 0.038533 | 0.004419 | 3.20E-18 |
| rs582094 | miR-195-5ɾ | trans | 2.17E-17 | Nightingale | Sphingomy | NA | T | A | 0.028892 | 0.004142 | 8.90E-12 |
| rs582094 | miR-195-5ɾ | trans | 2.17E-17 | Nightingale | M_VLDL_P | NA | T | A | 0.044033 | 0.004253 | 7.10E-25 |
| rs582094 | miR-195-5ɾ | trans | 2.17E-17 | Nightingale | LDL_CE | NA | T | A | 0.032359 | 0.004419 | 3.30E-13 |
| rs582094 | miR-195-5ɾ | trans | 2.17E-17 | Nightingale | XXL_VLDL_ | NA | T | A | -0.03566 | 0.004469 | 2.70E-16 |
| rs582094 | miR-195-5ɾ | trans | 2.17E-17 | Nightingale | IDL_P | NA | T | A | 0.039908 | 0.004385 | 1.90E-19 |
| rs582094 | miR-195-5ɾ | trans | 2.17E-17 | Nightingale | Total_PL | NA | T | A | 0.024785 | 0.00422 | 1.40E-09 |
| rs582094 | miR-195-5ɾ | trans | 2.17E-17 | Nightingale | S_LDL_L | NA | T | A | 0.027606 | 0.00443 | 1.10E-09 |
| rs582094 | miR-195-5ɾ | trans | 2.17E-17 | Nightingale | S_VLDL_TG | NA | T | A | -0.03801 | 0.004289 | 2.20E-18 |
| rs582094 | miR-195-5ɾ | trans | 2.17E-17 | Nightingale | L_LDL_CE | NA | T | A | 0.031356 | 0.004392 | 1.50E-12 |
| rs582094 | miR-195-5ɾ | trans | 2.17E-17 | Nightingale | S_VLDL_P | NA | T | A | 0.023349 | 0.004376 | 2.00E-08 |
| rs582094 | miR-195-5ɾ | trans | 2.17E-17 | Nightingale | S_LDL_CE | NA | T | A | 0.030409 | 0.004433 | 7.90E-12 |
| rs582094 | miR-195-5ɾ | trans | 2.17E-17 | Nightingale | XL_HDL_PL | NA | T | A | -0.02265 | 0.004071 | 1.40E-08 |
| rs582094 | miR-195-5ɾ | trans | 2.17E-17 | Nightingale | XS_VLDL_L | NA | T | A | 0.033719 | 0.004356 | 2.70E-15 |
| rs582094 | miR-195-5ɾ | trans | 2.17E-17 | Nightingale | Citrate | NA | T | A | 0.02682 | 0.004402 | 7.90E-10 |
| rs582094 | miR-195-5ɾ | trans | 2.17E-17 | Nightingale | Clinical_LD | NA | T | A | 0.0363 | 0.004406 | 4.30E-16 |
| rs582094 | miR-195-5ɾ | trans | 2.17E-17 | Nightingale | Omega_6 | NA | T | A | 0.025694 | 0.004327 | 8.00E-10 |
| rs582094 | miR-195-5ɾ | trans | 2.17E-17 | Nightingale | S_HDL_C | NA | T | A | 0.028202 | 0.004424 | 2.50E-10 |
| rs582094 | miR-195-5ɾ | trans | 2.17E-17 | Nightingale | M_LDL_C | NA | T | A | 0.033818 | 0.004439 | 3.20E-14 |
| rs582094 | miR-195-5ɾ | trans | 2.17E-17 | Nightingale | S_HDL_P | NA | T | A | 0.030405 | 0.004432 | 7.50E-12 |
| rs582094 | let-7b-5p | trans | 9.59E-27 | Nightingale | XS_VLDL_P | NA | T | A | 0.036606 | 0.004372 | 1.90E-17 |
| rs582094 | let-7b-5p | trans | 9.59E-27 | Nightingale | S_LDL_C | NA | T | A | 0.030792 | 0.004435 | 7.70E-12 |
| rs582094 | let-7b-5p | trans | 9.59E-27 | Nightingale | ApoB_by_A | NA | T | A | 0.024975 | 0.004364 | 1.60E-08 |
| rs582094 | let-7b-5p | trans | 9.59E-27 | Nightingale | M_VLDL_T | NA | T | A | -0.03854 | 0.004187 | 8.80E-20 |
| rs582094 | let-7b-5p | trans | 9.59E-27 | Nightingale | L_LDL_FC | NA | T | A | 0.033451 | 0.004351 | 4.40E-14 |
| rs582094 | let-7b-5p | trans | 9.59E-27 | Nightingale | Total_C | NA | T | A | 0.032199 | 0.00429 | 4.90E-14 |
| rs582094 | let-7b-5p | trans | 9.59E-27 | Nightingale | S_LDL_C_p | NA | T | A | 0.045184 | 0.004448 | 9.90E-25 |
| rs582094 | let-7b-5p | trans | 9.59E-27 | Nightingale | IDL_CE | NA | T | A | 0.033389 | 0.004278 | 1.80E-14 |
| rs582094 | let-7b-5p | trans | 9.59E-27 | Nightingale | XS_VLDL_F | NA | T | A | 0.036947 | 0.004354 | 9.50E-18 |
| rs582094 | let-7b-5p | trans | 9.59E-27 | Nightingale | Total_FC | NA | T | A | 0.03308 | 0.004329 | 1.00E-14 |
| rs582094 | let-7b-5p | trans | 9.59E-27 | Nightingale | S_VLDL_CE | NA | T | A | 0.046942 | 0.004372 | 1.80E-26 |
| rs582094 | let-7b-5p | trans | 9.59E-27 | Nightingale | M_LDL_P | NA | T | A | 0.035786 | 0.004444 | 9.30E-16 |
| rs582094 | let-7b-5p | trans | 9.59E-27 | Nightingale | M_VLDL_C | NA | T | A | 0.033603 | 0.004163 | 2.40E-15 |
| rs582094 | let-7b-5p | trans | 9.59E-27 | Nightingale | IDL_L | NA | T | A | 0.03514 | 0.004284 | 7.00E-16 |
| rs582094 | let-7b-5p | trans | 9.59E-27 | Nightingale | IDL_PL | NA | T | A | 0.035198 | 0.004276 | 4.90E-16 |
| rs582094 | let-7b-5p | trans | 9.59E-27 | Nightingale | XXL_VLDL_ | NA | T | A | 0.032961 | 0.004465 | 1.30E-13 |
| rs582094 | let-7b-5p | trans | 9.59E-27 | Nightingale | IDL_FC_pct | NA | T | A | 0.025717 | 0.004397 | 1.20E-08 |
| rs582094 | let-7b-5p | trans | 9.59E-27 | Nightingale | VLDL_C | NA | T | A | 0.031173 | 0.004425 | 6.70E-13 |
| rs582094 | let-7b-5p | trans | 9.59E-27 | Nightingale | M_VLDL_C | NA | T | A | 0.036181 | 0.004174 | 1.50E-17 |
| rs582094 | let-7b-5p | trans | 9.59E-27 | Nightingale | S_LDL_P | NA | T | A | 0.029487 | 0.004435 | 4.10E-11 |
| rs582094 | let-7b-5p | trans | 9.59E-27 | Nightingale | LDL_L | NA | T | A | 0.033121 | 0.004416 | 1.10E-13 |
| rs582094 | let-7b-5p | trans | 9.59E-27 | Nightingale | Total_L | NA | T | A | 0.026203 | 0.004359 | 5.90E-10 |
| rs582094 | let-7b-5p | trans | 9.59E-27 | Nightingale | LA | NA | T | A | 0.024685 | 0.004345 | 5.20E-09 |
| rs582094 | let-7b-5p | trans | 9.59E-27 | Nightingale | M_VLDL_P | NA | T | A | 0.028969 | 0.004424 | 2.80E-11 |
| rs582094 | let-7b-5p | trans | 9.59E-27 | Nightingale | XS_VLDL_C | NA | T | A | 0.037867 | 0.004301 | 2.00E-18 |
| rs582094 | let-7b-5p | trans | 9.59E-27 | Nightingale | ApoB | NA | T | A | 0.033786 | 0.004425 | 3.10E-14 |
| rs582094 | let-7b-5p | trans | 9.59E-27 | Nightingale | GlycA | NA | T | A | 0.027998 | 0.004412 | 1.60E-10 |
| rs582094 | let-7b-5p | trans | 9.59E-27 | Nightingale | S_VLDL_FC | NA | T | A | 0.029218 | 0.004234 | 1.60E-11 |
| rs582094 | let-7b-5p | trans | 9.59E-27 | Nightingale | L_VLDL_FC | NA | T | A | 0.048617 | 0.004389 | 5.10E-29 |
| rs582094 | let-7b-5p | trans | 9.59E-27 | Nightingale | VLDL_FC | NA | T | A | 0.024055 | 0.004399 | 1.30E-08 |
| rs582094 | let-7b-5p | trans | 9.59E-27 | Nightingale | M_LDL_FC | NA | T | A | 0.034163 | 0.004426 | 3.00E-14 |
| rs582094 | let-7b-5p | trans | 9.59E-27 | Nightingale | L_LDL_L | NA | T | A | 0.033355 | 0.004388 | 4.70E-14 |

| | | | | | | | | | | | |
|---|---|---|---|---|---|---|---|---|---|---|---|
| rs582094 | let-7b-5p | trans | 9.59E-27 | Nightingale | M_VLDL_P | NA | T | A | 0.028139 | 0.00442 | 8.60E-11 |
| rs582094 | let-7b-5p | trans | 9.59E-27 | Nightingale | Remnant_( | NA | T | A | 0.036725 | 0.004402 | 5.60E-17 |
| rs582094 | let-7b-5p | trans | 9.59E-27 | Nightingale | S_VLDL_CE | NA | T | A | 0.038034 | 0.004425 | 2.50E-18 |
| rs582094 | let-7b-5p | trans | 9.59E-27 | Nightingale | XXL_VLDL_ | NA | T | A | 0.03468 | 0.004441 | 4.10E-15 |
| rs582094 | let-7b-5p | trans | 9.59E-27 | Nightingale | S_VLDL_L | NA | T | A | 0.024549 | 0.004388 | 4.70E-09 |
| rs582094 | let-7b-5p | trans | 9.59E-27 | Nightingale | M_LDL_C_ | NA | T | A | 0.031036 | 0.004434 | 3.90E-12 |
| rs582094 | let-7b-5p | trans | 9.59E-27 | Nightingale | L_LDL_C | NA | T | A | 0.032117 | 0.004381 | 4.00E-13 |
| rs582094 | let-7b-5p | trans | 9.59E-27 | Nightingale | M_VLDL_C | NA | T | A | 0.040673 | 0.004391 | 3.90E-20 |
| rs582094 | let-7b-5p | trans | 9.59E-27 | Nightingale | L_VLDL_C_ | NA | T | A | 0.052204 | 0.004328 | 4.90E-34 |
| rs582094 | let-7b-5p | trans | 9.59E-27 | Nightingale | XS_VLDL_C | NA | T | A | 0.037674 | 0.004276 | 3.10E-18 |
| rs582094 | let-7b-5p | trans | 9.59E-27 | Nightingale | M_LDL_CE | NA | T | A | 0.032814 | 0.004437 | 1.40E-13 |
| rs582094 | let-7b-5p | trans | 9.59E-27 | Nightingale | S_HDL_L | NA | T | A | 0.02611 | 0.004416 | 2.10E-09 |
| rs582094 | let-7b-5p | trans | 9.59E-27 | Nightingale | M_VLDL_F | NA | T | A | 0.042037 | 0.004216 | 4.50E-23 |
| rs582094 | let-7b-5p | trans | 9.59E-27 | Nightingale | S_VLDL_C_ | NA | T | A | 0.041764 | 0.004312 | 8.50E-22 |
| rs582094 | let-7b-5p | trans | 9.59E-27 | Nightingale | VLDL_CE | NA | T | A | 0.035642 | 0.004432 | 4.50E-16 |
| rs582094 | let-7b-5p | trans | 9.59E-27 | Nightingale | IDL_C | NA | T | A | 0.034832 | 0.00428 | 1.40E-15 |
| rs582094 | let-7b-5p | trans | 9.59E-27 | Nightingale | XL_VLDL_P | NA | T | A | 0.038942 | 0.004438 | 4.80E-19 |
| rs582094 | let-7b-5p | trans | 9.59E-27 | Nightingale | L_LDL_P | NA | T | A | 0.028971 | 0.004416 | 9.20E-11 |
| rs582094 | let-7b-5p | trans | 9.59E-27 | Nightingale | XL_VLDL_F | NA | T | A | 0.037159 | 0.004296 | 2.40E-18 |
| rs582094 | let-7b-5p | trans | 9.59E-27 | Nightingale | XL_VLDL_T | NA | T | A | -0.04392 | 0.004295 | 2.20E-24 |
| rs582094 | let-7b-5p | trans | 9.59E-27 | Nightingale | L_LDL_PL | NA | T | A | 0.037813 | 0.004398 | 1.50E-17 |
| rs582094 | let-7b-5p | trans | 9.59E-27 | Nightingale | L_VLDL_CE | NA | T | A | 0.047319 | 0.00424 | 3.20E-29 |
| rs582094 | let-7b-5p | trans | 9.59E-27 | Nightingale | M_LDL_L | NA | T | A | 0.032867 | 0.004437 | 1.50E-13 |
| rs582094 | let-7b-5p | trans | 9.59E-27 | Nightingale | S_VLDL_FC | NA | T | A | 0.037726 | 0.004432 | 1.20E-17 |
| rs582094 | let-7b-5p | trans | 9.59E-27 | Nightingale | S_VLDL_PL | NA | T | A | 0.032628 | 0.004423 | 6.00E-14 |
| rs582094 | let-7b-5p | trans | 9.59E-27 | Nightingale | XS_VLDL_F | NA | T | A | 0.03125 | 0.0044 | 3.70E-12 |
| rs582094 | let-7b-5p | trans | 9.59E-27 | Nightingale | S_VLDL_C | NA | T | A | 0.038213 | 0.004431 | 2.80E-18 |
| rs582094 | let-7b-5p | trans | 9.59E-27 | Nightingale | XL_VLDL_C | NA | T | A | 0.038128 | 0.004249 | 6.90E-19 |
| rs582094 | let-7b-5p | trans | 9.59E-27 | Nightingale | S_LDL_PL_ | NA | T | A | -0.02995 | 0.004423 | 3.30E-13 |
| rs582094 | let-7b-5p | trans | 9.59E-27 | Nightingale | LDL_PL | NA | T | A | 0.03482 | 0.004421 | 7.30E-15 |
| rs582094 | let-7b-5p | trans | 9.59E-27 | Nightingale | non_HDL_( | NA | T | A | 0.035527 | 0.004407 | 7.90E-16 |
| rs582094 | let-7b-5p | trans | 9.59E-27 | Nightingale | XXL_VLDL_ | NA | T | A | 0.025093 | 0.00446 | 1.40E-09 |
| rs582094 | let-7b-5p | trans | 9.59E-27 | Nightingale | M_LDL_PL | NA | T | A | 0.032301 | 0.004437 | 3.90E-13 |
| rs582094 | let-7b-5p | trans | 9.59E-27 | Nightingale | XL_VLDL_C | NA | T | A | 0.037247 | 0.004251 | 4.70E-18 |
| rs582094 | let-7b-5p | trans | 9.59E-27 | Nightingale | XS_VLDL_P | NA | T | A | 0.032031 | 0.004369 | 6.90E-14 |
| rs582094 | let-7b-5p | trans | 9.59E-27 | Nightingale | M_VLDL_F | NA | T | A | 0.033201 | 0.00443 | 4.70E-14 |
| rs582094 | let-7b-5p | trans | 9.59E-27 | Nightingale | S_HDL_PL | NA | T | A | 0.023614 | 0.004393 | 4.10E-08 |
| rs582094 | let-7b-5p | trans | 9.59E-27 | Nightingale | XXL_VLDL_ | NA | T | A | 0.030797 | 0.004412 | 1.20E-12 |
| rs582094 | let-7b-5p | trans | 9.59E-27 | Nightingale | S_LDL_FC | NA | T | A | 0.028398 | 0.004424 | 6.00E-10 |
| rs582094 | let-7b-5p | trans | 9.59E-27 | Nightingale | PUFA | NA | T | A | 0.023572 | 0.004322 | 1.80E-08 |
| rs582094 | let-7b-5p | trans | 9.59E-27 | Nightingale | LDL_FC | NA | T | A | 0.033455 | 0.004384 | 8.10E-14 |
| rs582094 | let-7b-5p | trans | 9.59E-27 | Nightingale | LDL_P | NA | T | A | 0.03163 | 0.004428 | 1.50E-12 |
| rs582094 | let-7b-5p | trans | 9.59E-27 | Nightingale | L_VLDL_TG | NA | T | A | -0.05112 | 0.00441 | 7.10E-32 |
| rs582094 | let-7b-5p | trans | 9.59E-27 | Nightingale | IDL_FC | NA | T | A | 0.038121 | 0.00429 | 2.20E-18 |
| rs582094 | let-7b-5p | trans | 9.59E-27 | Nightingale | LDL_C | NA | T | A | 0.033002 | 0.004412 | 1.40E-13 |
| rs582094 | let-7b-5p | trans | 9.59E-27 | Nightingale | Total_CE | NA | T | A | 0.031574 | 0.004274 | 1.40E-13 |
| rs582094 | let-7b-5p | trans | 9.59E-27 | Nightingale | M_LDL_TG | NA | T | A | -0.02744 | 0.004376 | 4.70E-10 |
| rs582094 | let-7b-5p | trans | 9.59E-27 | Nightingale | S_LDL_CE_ | NA | T | A | 0.03085 | 0.004424 | 6.10E-14 |
| rs582094 | let-7b-5p | trans | 9.59E-27 | Nightingale | VLDL_P | NA | T | A | 0.026524 | 0.004407 | 5.50E-10 |
| rs582094 | let-7b-5p | trans | 9.59E-27 | Nightingale | S_HDL_CE | NA | T | A | 0.028434 | 0.004438 | 1.90E-10 |
| rs582094 | let-7b-5p | trans | 9.59E-27 | Nightingale | S_VLDL_PL | NA | T | A | 0.027637 | 0.004243 | 2.50E-10 |
| rs582094 | let-7b-5p | trans | 9.59E-27 | Nightingale | M_VLDL_C | NA | T | A | 0.038533 | 0.004419 | 3.20E-18 |
| rs582094 | let-7b-5p | trans | 9.59E-27 | Nightingale | Sphingomy | NA | T | A | 0.028892 | 0.004142 | 8.90E-12 |
| rs582094 | let-7b-5p | trans | 9.59E-27 | Nightingale | M_VLDL_P | NA | T | A | 0.044033 | 0.004253 | 7.10E-25 |
| rs582094 | let-7b-5p | trans | 9.59E-27 | Nightingale | LDL_CE | NA | T | A | 0.032359 | 0.004419 | 3.30E-13 |
| rs582094 | let-7b-5p | trans | 9.59E-27 | Nightingale | XXL_VLDL_ | NA | T | A | -0.03566 | 0.004469 | 2.70E-16 |
| rs582094 | let-7b-5p | trans | 9.59E-27 | Nightingale | IDL_P | NA | T | A | 0.039908 | 0.004385 | 1.90E-19 |
| rs582094 | let-7b-5p | trans | 9.59E-27 | Nightingale | Total_PL | NA | T | A | 0.024785 | 0.00422 | 1.40E-09 |
| rs582094 | let-7b-5p | trans | 9.59E-27 | Nightingale | S_LDL_L | NA | T | A | 0.027606 | 0.00443 | 1.10E-09 |
| rs582094 | let-7b-5p | trans | 9.59E-27 | Nightingale | S_VLDL_TG | NA | T | A | -0.03801 | 0.004289 | 2.20E-18 |
| rs582094 | let-7b-5p | trans | 9.59E-27 | Nightingale | L_LDL_CE | NA | T | A | 0.031356 | 0.004392 | 1.50E-12 |
| rs582094 | let-7b-5p | trans | 9.59E-27 | Nightingale | S_VLDL_P | NA | T | A | 0.023349 | 0.004376 | 2.00E-08 |
| rs582094 | let-7b-5p | trans | 9.59E-27 | Nightingale | S_LDL_CE | NA | T | A | 0.030409 | 0.004433 | 7.90E-12 |

| | | | | | | | | | | | |
|---|---|---|---|---|---|---|---|---|---|---|---|
| rs582094 | let-7b-5p | trans | 9.59E-27 | Nightingale | XL_HDL_PL | NA | T | A | -0.02265 | 0.004071 | 1.40E-08 |
| rs582094 | let-7b-5p | trans | 9.59E-27 | Nightingale | XS_VLDL_L | NA | T | A | 0.033719 | 0.004356 | 2.70E-15 |
| rs582094 | let-7b-5p | trans | 9.59E-27 | Nightingale | Citrate | NA | T | A | 0.02682 | 0.004402 | 7.90E-10 |
| rs582094 | let-7b-5p | trans | 9.59E-27 | Nightingale | Clinical_LD | NA | T | A | 0.0363 | 0.004406 | 4.30E-16 |
| rs582094 | let-7b-5p | trans | 9.59E-27 | Nightingale | Omega_6 | NA | T | A | 0.025694 | 0.004327 | 8.00E-10 |
| rs582094 | let-7b-5p | trans | 9.59E-27 | Nightingale | S_HDL_C | NA | T | A | 0.028202 | 0.004424 | 2.50E-10 |
| rs582094 | let-7b-5p | trans | 9.59E-27 | Nightingale | M_LDL_C | NA | T | A | 0.033818 | 0.004439 | 3.20E-14 |
| rs582094 | let-7b-5p | trans | 9.59E-27 | Nightingale | S_HDL_P | NA | T | A | 0.030405 | 0.004432 | 7.50E-12 |
| rs582094 | miR-10b-5p | trans | 3.25E-35 | Nightingale | XS_VLDL_P | NA | T | A | 0.036606 | 0.004372 | 1.90E-17 |
| rs582094 | miR-10b-5p | trans | 3.25E-35 | Nightingale | S_LDL_C | NA | T | A | 0.030792 | 0.004435 | 7.70E-12 |
| rs582094 | miR-10b-5p | trans | 3.25E-35 | Nightingale | ApoB_by_A | NA | T | A | 0.024975 | 0.004364 | 1.60E-08 |
| rs582094 | miR-10b-5p | trans | 3.25E-35 | Nightingale | M_VLDL_T | NA | T | A | -0.03854 | 0.004187 | 8.80E-20 |
| rs582094 | miR-10b-5p | trans | 3.25E-35 | Nightingale | L_LDL_FC | NA | T | A | 0.033451 | 0.004351 | 4.40E-14 |
| rs582094 | miR-10b-5p | trans | 3.25E-35 | Nightingale | Total_C | NA | T | A | 0.032199 | 0.00429 | 4.90E-14 |
| rs582094 | miR-10b-5p | trans | 3.25E-35 | Nightingale | S_LDL_C_p | NA | T | A | 0.045184 | 0.004448 | 9.90E-25 |
| rs582094 | miR-10b-5p | trans | 3.25E-35 | Nightingale | IDL_CE | NA | T | A | 0.033389 | 0.004278 | 1.80E-14 |
| rs582094 | miR-10b-5p | trans | 3.25E-35 | Nightingale | XS_VLDL_F | NA | T | A | 0.036947 | 0.004354 | 9.50E-18 |
| rs582094 | miR-10b-5p | trans | 3.25E-35 | Nightingale | Total_FC | NA | T | A | 0.03308 | 0.004329 | 1.00E-14 |
| rs582094 | miR-10b-5p | trans | 3.25E-35 | Nightingale | S_VLDL_CE | NA | T | A | 0.046942 | 0.004372 | 1.80E-26 |
| rs582094 | miR-10b-5p | trans | 3.25E-35 | Nightingale | M_LDL_P | NA | T | A | 0.035786 | 0.004444 | 9.30E-16 |
| rs582094 | miR-10b-5p | trans | 3.25E-35 | Nightingale | M_VLDL_C | NA | T | A | 0.033603 | 0.004163 | 2.40E-15 |
| rs582094 | miR-10b-5p | trans | 3.25E-35 | Nightingale | IDL_L | NA | T | A | 0.03514 | 0.004284 | 7.00E-16 |
| rs582094 | miR-10b-5p | trans | 3.25E-35 | Nightingale | IDL_PL | NA | T | A | 0.035198 | 0.004276 | 4.90E-16 |
| rs582094 | miR-10b-5p | trans | 3.25E-35 | Nightingale | XXL_VLDL_ | NA | T | A | 0.032961 | 0.004465 | 1.30E-13 |
| rs582094 | miR-10b-5p | trans | 3.25E-35 | Nightingale | IDL_FC_pct | NA | T | A | 0.025717 | 0.004397 | 1.20E-08 |
| rs582094 | miR-10b-5p | trans | 3.25E-35 | Nightingale | VLDL_C | NA | T | A | 0.031173 | 0.004425 | 6.70E-13 |
| rs582094 | miR-10b-5p | trans | 3.25E-35 | Nightingale | M_VLDL_C | NA | T | A | 0.036181 | 0.004174 | 1.50E-17 |
| rs582094 | miR-10b-5p | trans | 3.25E-35 | Nightingale | S_LDL_P | NA | T | A | 0.029487 | 0.004435 | 4.10E-11 |
| rs582094 | miR-10b-5p | trans | 3.25E-35 | Nightingale | LDL_L | NA | T | A | 0.033121 | 0.004416 | 1.10E-13 |
| rs582094 | miR-10b-5p | trans | 3.25E-35 | Nightingale | Total_L | NA | T | A | 0.026203 | 0.004359 | 5.90E-10 |
| rs582094 | miR-10b-5p | trans | 3.25E-35 | Nightingale | LA | NA | T | A | 0.024685 | 0.004345 | 5.20E-09 |
| rs582094 | miR-10b-5p | trans | 3.25E-35 | Nightingale | M_VLDL_P | NA | T | A | 0.028969 | 0.004424 | 2.80E-11 |
| rs582094 | miR-10b-5p | trans | 3.25E-35 | Nightingale | XS_VLDL_C | NA | T | A | 0.037867 | 0.004301 | 2.00E-18 |
| rs582094 | miR-10b-5p | trans | 3.25E-35 | Nightingale | ApoB | NA | T | A | 0.033786 | 0.004425 | 3.10E-14 |
| rs582094 | miR-10b-5p | trans | 3.25E-35 | Nightingale | GlycA | NA | T | A | 0.027998 | 0.004412 | 1.60E-10 |
| rs582094 | miR-10b-5p | trans | 3.25E-35 | Nightingale | S_VLDL_FC | NA | T | A | 0.029218 | 0.004234 | 1.60E-11 |
| rs582094 | miR-10b-5p | trans | 3.25E-35 | Nightingale | L_VLDL_FC | NA | T | A | 0.048617 | 0.004389 | 5.10E-29 |
| rs582094 | miR-10b-5p | trans | 3.25E-35 | Nightingale | VLDL_FC | NA | T | A | 0.024055 | 0.004399 | 1.30E-08 |
| rs582094 | miR-10b-5p | trans | 3.25E-35 | Nightingale | M_LDL_FC | NA | T | A | 0.034163 | 0.004426 | 3.00E-14 |
| rs582094 | miR-10b-5p | trans | 3.25E-35 | Nightingale | L_LDL_L | NA | T | A | 0.033355 | 0.004388 | 4.70E-14 |
| rs582094 | miR-10b-5p | trans | 3.25E-35 | Nightingale | M_VLDL_P | NA | T | A | 0.028139 | 0.00442 | 8.60E-11 |
| rs582094 | miR-10b-5p | trans | 3.25E-35 | Nightingale | Remnant_( | NA | T | A | 0.036725 | 0.004402 | 5.60E-17 |
| rs582094 | miR-10b-5p | trans | 3.25E-35 | Nightingale | S_VLDL_CE | NA | T | A | 0.038034 | 0.004425 | 2.50E-18 |
| rs582094 | miR-10b-5p | trans | 3.25E-35 | Nightingale | XXL_VLDL_ | NA | T | A | 0.03468 | 0.004441 | 4.10E-15 |
| rs582094 | miR-10b-5p | trans | 3.25E-35 | Nightingale | S_VLDL_L | NA | T | A | 0.024549 | 0.004388 | 4.70E-09 |
| rs582094 | miR-10b-5p | trans | 3.25E-35 | Nightingale | M_LDL_C_p | NA | T | A | 0.031036 | 0.004434 | 3.90E-12 |
| rs582094 | miR-10b-5p | trans | 3.25E-35 | Nightingale | L_LDL_C | NA | T | A | 0.032117 | 0.004381 | 4.00E-13 |
| rs582094 | miR-10b-5p | trans | 3.25E-35 | Nightingale | M_VLDL_C | NA | T | A | 0.040673 | 0.004391 | 3.90E-20 |
| rs582094 | miR-10b-5p | trans | 3.25E-35 | Nightingale | L_VLDL_C_ | NA | T | A | 0.052204 | 0.004328 | 4.90E-34 |
| rs582094 | miR-10b-5p | trans | 3.25E-35 | Nightingale | XS_VLDL_C | NA | T | A | 0.037674 | 0.004276 | 3.10E-18 |
| rs582094 | miR-10b-5p | trans | 3.25E-35 | Nightingale | M_LDL_CE | NA | T | A | 0.032814 | 0.004437 | 1.40E-13 |
| rs582094 | miR-10b-5p | trans | 3.25E-35 | Nightingale | S_HDL_L | NA | T | A | 0.02611 | 0.004416 | 2.10E-09 |
| rs582094 | miR-10b-5p | trans | 3.25E-35 | Nightingale | M_VLDL_F | NA | T | A | 0.042037 | 0.004216 | 4.50E-23 |
| rs582094 | miR-10b-5p | trans | 3.25E-35 | Nightingale | S_VLDL_C_ | NA | T | A | 0.041764 | 0.004312 | 8.50E-22 |
| rs582094 | miR-10b-5p | trans | 3.25E-35 | Nightingale | VLDL_CE | NA | T | A | 0.035642 | 0.004432 | 4.50E-16 |
| rs582094 | miR-10b-5p | trans | 3.25E-35 | Nightingale | IDL_C | NA | T | A | 0.034832 | 0.00428 | 1.40E-15 |
| rs582094 | miR-10b-5p | trans | 3.25E-35 | Nightingale | XL_VLDL_P | NA | T | A | 0.038942 | 0.004438 | 4.80E-19 |
| rs582094 | miR-10b-5p | trans | 3.25E-35 | Nightingale | L_LDL_P | NA | T | A | 0.028971 | 0.004416 | 9.20E-11 |
| rs582094 | miR-10b-5p | trans | 3.25E-35 | Nightingale | XL_VLDL_F | NA | T | A | 0.037159 | 0.004296 | 2.40E-18 |
| rs582094 | miR-10b-5p | trans | 3.25E-35 | Nightingale | XL_VLDL_T | NA | T | A | -0.04392 | 0.004295 | 2.20E-24 |
| rs582094 | miR-10b-5p | trans | 3.25E-35 | Nightingale | L_LDL_PL | NA | T | A | 0.037813 | 0.004398 | 1.50E-17 |
| rs582094 | miR-10b-5p | trans | 3.25E-35 | Nightingale | L_VLDL_CE | NA | T | A | 0.047319 | 0.00424 | 3.20E-29 |
| rs582094 | miR-10b-5p | trans | 3.25E-35 | Nightingale | M_LDL_L | NA | T | A | 0.032867 | 0.004437 | 1.50E-13 |

| | | | | | | | | | | | |
|---|---|---|---|---|---|---|---|---|---|---|---|
| rs582094 | miR-10b-5¦ | trans | 3.25E-35 | Nightingale | S_VLDL_FC | NA | T | A | 0.037726 | 0.004432 | 1.20E-17 |
| rs582094 | miR-10b-5¦ | trans | 3.25E-35 | Nightingale | S_VLDL_PL | NA | T | A | 0.032628 | 0.004423 | 6.00E-14 |
| rs582094 | miR-10b-5¦ | trans | 3.25E-35 | Nightingale | XS_VLDL_F | NA | T | A | 0.03125 | 0.0044 | 3.70E-12 |
| rs582094 | miR-10b-5¦ | trans | 3.25E-35 | Nightingale | S_VLDL_C | NA | T | A | 0.038213 | 0.004431 | 2.80E-18 |
| rs582094 | miR-10b-5¦ | trans | 3.25E-35 | Nightingale | XL_VLDL_C | NA | T | A | 0.038128 | 0.004249 | 6.90E-19 |
| rs582094 | miR-10b-5¦ | trans | 3.25E-35 | Nightingale | S_LDL_PL_ | NA | T | A | -0.02995 | 0.004423 | 3.30E-13 |
| rs582094 | miR-10b-5¦ | trans | 3.25E-35 | Nightingale | LDL_PL | NA | T | A | 0.03482 | 0.004421 | 7.30E-15 |
| rs582094 | miR-10b-5¦ | trans | 3.25E-35 | Nightingale | non_HDL_( | NA | T | A | 0.035527 | 0.004407 | 7.90E-16 |
| rs582094 | miR-10b-5¦ | trans | 3.25E-35 | Nightingale | XXL_VLDL_ | NA | T | A | 0.025093 | 0.00446 | 1.40E-09 |
| rs582094 | miR-10b-5¦ | trans | 3.25E-35 | Nightingale | M_LDL_PL | NA | T | A | 0.032301 | 0.004437 | 3.90E-13 |
| rs582094 | miR-10b-5¦ | trans | 3.25E-35 | Nightingale | XL_VLDL_C | NA | T | A | 0.037247 | 0.004251 | 4.70E-18 |
| rs582094 | miR-10b-5¦ | trans | 3.25E-35 | Nightingale | XS_VLDL_P | NA | T | A | 0.032031 | 0.004369 | 6.90E-14 |
| rs582094 | miR-10b-5¦ | trans | 3.25E-35 | Nightingale | M_VLDL_F | NA | T | A | 0.033201 | 0.00443 | 4.70E-14 |
| rs582094 | miR-10b-5¦ | trans | 3.25E-35 | Nightingale | S_HDL_PL | NA | T | A | 0.023614 | 0.004393 | 4.10E-08 |
| rs582094 | miR-10b-5¦ | trans | 3.25E-35 | Nightingale | XXL_VLDL_ | NA | T | A | 0.030797 | 0.004412 | 1.20E-12 |
| rs582094 | miR-10b-5¦ | trans | 3.25E-35 | Nightingale | S_LDL_FC | NA | T | A | 0.028398 | 0.004424 | 6.00E-10 |
| rs582094 | miR-10b-5¦ | trans | 3.25E-35 | Nightingale | PUFA | NA | T | A | 0.023572 | 0.004322 | 1.80E-08 |
| rs582094 | miR-10b-5¦ | trans | 3.25E-35 | Nightingale | LDL_FC | NA | T | A | 0.033455 | 0.004384 | 8.10E-14 |
| rs582094 | miR-10b-5¦ | trans | 3.25E-35 | Nightingale | LDL_P | NA | T | A | 0.03163 | 0.004428 | 1.50E-12 |
| rs582094 | miR-10b-5¦ | trans | 3.25E-35 | Nightingale | L_VLDL_TG | NA | T | A | -0.05112 | 0.00441 | 7.10E-32 |
| rs582094 | miR-10b-5¦ | trans | 3.25E-35 | Nightingale | IDL_FC | NA | T | A | 0.038121 | 0.00429 | 2.20E-18 |
| rs582094 | miR-10b-5¦ | trans | 3.25E-35 | Nightingale | LDL_C | NA | T | A | 0.033002 | 0.004412 | 1.40E-13 |
| rs582094 | miR-10b-5¦ | trans | 3.25E-35 | Nightingale | Total_CE | NA | T | A | 0.031574 | 0.004274 | 1.40E-13 |
| rs582094 | miR-10b-5¦ | trans | 3.25E-35 | Nightingale | M_LDL_TG | NA | T | A | -0.02744 | 0.004376 | 4.70E-10 |
| rs582094 | miR-10b-5¦ | trans | 3.25E-35 | Nightingale | S_LDL_CE_ | NA | T | A | 0.03085 | 0.004424 | 6.10E-14 |
| rs582094 | miR-10b-5¦ | trans | 3.25E-35 | Nightingale | VLDL_P | NA | T | A | 0.026524 | 0.004407 | 5.50E-10 |
| rs582094 | miR-10b-5¦ | trans | 3.25E-35 | Nightingale | S_HDL_CE | NA | T | A | 0.028434 | 0.004438 | 1.90E-10 |
| rs582094 | miR-10b-5¦ | trans | 3.25E-35 | Nightingale | S_VLDL_PL | NA | T | A | 0.027637 | 0.004243 | 2.50E-10 |
| rs582094 | miR-10b-5¦ | trans | 3.25E-35 | Nightingale | M_VLDL_C | NA | T | A | 0.038533 | 0.004419 | 3.20E-18 |
| rs582094 | miR-10b-5¦ | trans | 3.25E-35 | Nightingale | Sphingomy | NA | T | A | 0.028892 | 0.004142 | 8.90E-12 |
| rs582094 | miR-10b-5¦ | trans | 3.25E-35 | Nightingale | M_VLDL_P | NA | T | A | 0.044033 | 0.004253 | 7.10E-25 |
| rs582094 | miR-10b-5¦ | trans | 3.25E-35 | Nightingale | LDL_CE | NA | T | A | 0.032359 | 0.004419 | 3.30E-13 |
| rs582094 | miR-10b-5¦ | trans | 3.25E-35 | Nightingale | XXL_VLDL_ | NA | T | A | -0.03566 | 0.004469 | 2.70E-16 |
| rs582094 | miR-10b-5¦ | trans | 3.25E-35 | Nightingale | IDL_P | NA | T | A | 0.039908 | 0.004385 | 1.90E-19 |
| rs582094 | miR-10b-5¦ | trans | 3.25E-35 | Nightingale | Total_PL | NA | T | A | 0.024785 | 0.00422 | 1.40E-09 |
| rs582094 | miR-10b-5¦ | trans | 3.25E-35 | Nightingale | S_LDL_L | NA | T | A | 0.027606 | 0.00443 | 1.10E-09 |
| rs582094 | miR-10b-5¦ | trans | 3.25E-35 | Nightingale | S_VLDL_TG | NA | T | A | -0.03801 | 0.004289 | 2.20E-18 |
| rs582094 | miR-10b-5¦ | trans | 3.25E-35 | Nightingale | L_LDL_CE | NA | T | A | 0.031356 | 0.004392 | 1.50E-12 |
| rs582094 | miR-10b-5¦ | trans | 3.25E-35 | Nightingale | S_VLDL_P | NA | T | A | 0.023349 | 0.004376 | 2.00E-08 |
| rs582094 | miR-10b-5¦ | trans | 3.25E-35 | Nightingale | S_LDL_CE | NA | T | A | 0.030409 | 0.004433 | 7.90E-12 |
| rs582094 | miR-10b-5¦ | trans | 3.25E-35 | Nightingale | XL_HDL_PL | NA | T | A | -0.02265 | 0.004071 | 1.40E-08 |
| rs582094 | miR-10b-5¦ | trans | 3.25E-35 | Nightingale | XS_VLDL_L | NA | T | A | 0.033719 | 0.004356 | 2.70E-15 |
| rs582094 | miR-10b-5¦ | trans | 3.25E-35 | Nightingale | Citrate | NA | T | A | 0.02682 | 0.004402 | 7.90E-10 |
| rs582094 | miR-10b-5¦ | trans | 3.25E-35 | Nightingale | Clinical_LD | NA | T | A | 0.0363 | 0.004406 | 4.30E-16 |
| rs582094 | miR-10b-5¦ | trans | 3.25E-35 | Nightingale | Omega_6 | NA | T | A | 0.025694 | 0.004327 | 8.00E-10 |
| rs582094 | miR-10b-5¦ | trans | 3.25E-35 | Nightingale | S_HDL_C | NA | T | A | 0.028202 | 0.004424 | 2.50E-10 |
| rs582094 | miR-10b-5¦ | trans | 3.25E-35 | Nightingale | M_LDL_C | NA | T | A | 0.033818 | 0.004439 | 3.20E-14 |
| rs582094 | miR-10b-5¦ | trans | 3.25E-35 | Nightingale | S_HDL_P | NA | T | A | 0.030405 | 0.004432 | 7.50E-12 |
| rs582094 | miR-139-5¦ | trans | 7.69E-42 | Nightingale | XS_VLDL_P | NA | T | A | 0.036606 | 0.004372 | 1.90E-17 |
| rs582094 | miR-139-5¦ | trans | 7.69E-42 | Nightingale | S_LDL_C | NA | T | A | 0.030792 | 0.004435 | 7.70E-12 |
| rs582094 | miR-139-5¦ | trans | 7.69E-42 | Nightingale | ApoB_by_A | NA | T | A | 0.024975 | 0.004364 | 1.60E-08 |
| rs582094 | miR-139-5¦ | trans | 7.69E-42 | Nightingale | M_VLDL_T | NA | T | A | -0.03854 | 0.004187 | 8.80E-20 |
| rs582094 | miR-139-5¦ | trans | 7.69E-42 | Nightingale | L_LDL_FC | NA | T | A | 0.033451 | 0.004351 | 4.40E-14 |
| rs582094 | miR-139-5¦ | trans | 7.69E-42 | Nightingale | Total_C | NA | T | A | 0.032199 | 0.00429 | 4.90E-14 |
| rs582094 | miR-139-5¦ | trans | 7.69E-42 | Nightingale | S_LDL_C_p | NA | T | A | 0.045184 | 0.004448 | 9.90E-25 |
| rs582094 | miR-139-5¦ | trans | 7.69E-42 | Nightingale | IDL_CE | NA | T | A | 0.033389 | 0.004278 | 1.80E-14 |
| rs582094 | miR-139-5¦ | trans | 7.69E-42 | Nightingale | XS_VLDL_F | NA | T | A | 0.036947 | 0.004354 | 9.50E-18 |
| rs582094 | miR-139-5¦ | trans | 7.69E-42 | Nightingale | Total_FC | NA | T | A | 0.03308 | 0.004329 | 1.00E-14 |
| rs582094 | miR-139-5¦ | trans | 7.69E-42 | Nightingale | S_VLDL_CE | NA | T | A | 0.046942 | 0.004372 | 1.80E-26 |
| rs582094 | miR-139-5¦ | trans | 7.69E-42 | Nightingale | M_LDL_P | NA | T | A | 0.035786 | 0.004444 | 9.30E-16 |
| rs582094 | miR-139-5¦ | trans | 7.69E-42 | Nightingale | M_VLDL_C | NA | T | A | 0.033603 | 0.004163 | 2.40E-15 |
| rs582094 | miR-139-5¦ | trans | 7.69E-42 | Nightingale | IDL_L | NA | T | A | 0.03514 | 0.004284 | 7.00E-16 |
| rs582094 | miR-139-5¦ | trans | 7.69E-42 | Nightingale | IDL_PL | NA | T | A | 0.035198 | 0.004276 | 4.90E-16 |

| | | | | | | | | | | | |
|---|---|---|---|---|---|---|---|---|---|---|---|
| rs582094 | miR-139-5p | trans | 7.69E-42 | Nightingale | XXL_VLDL_ | NA | T | A | 0.032961 | 0.004465 | 1.30E-13 |
| rs582094 | miR-139-5p | trans | 7.69E-42 | Nightingale | IDL_FC_pct | NA | T | A | 0.025717 | 0.004397 | 1.20E-08 |
| rs582094 | miR-139-5p | trans | 7.69E-42 | Nightingale | VLDL_C | NA | T | A | 0.031173 | 0.004425 | 6.70E-13 |
| rs582094 | miR-139-5p | trans | 7.69E-42 | Nightingale | M_VLDL_C_ | NA | T | A | 0.036181 | 0.004174 | 1.50E-17 |
| rs582094 | miR-139-5p | trans | 7.69E-42 | Nightingale | S_LDL_P | NA | T | A | 0.029487 | 0.004435 | 4.10E-11 |
| rs582094 | miR-139-5p | trans | 7.69E-42 | Nightingale | LDL_L | NA | T | A | 0.033121 | 0.004416 | 1.10E-13 |
| rs582094 | miR-139-5p | trans | 7.69E-42 | Nightingale | Total_L | NA | T | A | 0.026203 | 0.004359 | 5.90E-10 |
| rs582094 | miR-139-5p | trans | 7.69E-42 | Nightingale | LA | NA | T | A | 0.024685 | 0.004345 | 5.20E-09 |
| rs582094 | miR-139-5p | trans | 7.69E-42 | Nightingale | M_VLDL_P | NA | T | A | 0.028969 | 0.004424 | 2.80E-11 |
| rs582094 | miR-139-5p | trans | 7.69E-42 | Nightingale | XS_VLDL_C | NA | T | A | 0.037867 | 0.004301 | 2.00E-18 |
| rs582094 | miR-139-5p | trans | 7.69E-42 | Nightingale | ApoB | NA | T | A | 0.033786 | 0.004425 | 3.10E-14 |
| rs582094 | miR-139-5p | trans | 7.69E-42 | Nightingale | GlycA | NA | T | A | 0.027998 | 0.004412 | 1.60E-10 |
| rs582094 | miR-139-5p | trans | 7.69E-42 | Nightingale | S_VLDL_FC | NA | T | A | 0.029218 | 0.004234 | 1.60E-11 |
| rs582094 | miR-139-5p | trans | 7.69E-42 | Nightingale | L_VLDL_FC | NA | T | A | 0.048617 | 0.004389 | 5.10E-29 |
| rs582094 | miR-139-5p | trans | 7.69E-42 | Nightingale | VLDL_FC | NA | T | A | 0.024055 | 0.004399 | 1.30E-08 |
| rs582094 | miR-139-5p | trans | 7.69E-42 | Nightingale | M_LDL_FC | NA | T | A | 0.034163 | 0.004426 | 3.00E-14 |
| rs582094 | miR-139-5p | trans | 7.69E-42 | Nightingale | L_LDL_L | NA | T | A | 0.033355 | 0.004388 | 4.70E-14 |
| rs582094 | miR-139-5p | trans | 7.69E-42 | Nightingale | M_VLDL_P | NA | T | A | 0.028139 | 0.00442 | 8.60E-11 |
| rs582094 | miR-139-5p | trans | 7.69E-42 | Nightingale | Remnant_( | NA | T | A | 0.036725 | 0.004402 | 5.60E-17 |
| rs582094 | miR-139-5p | trans | 7.69E-42 | Nightingale | S_VLDL_CE | NA | T | A | 0.038034 | 0.004425 | 2.50E-18 |
| rs582094 | miR-139-5p | trans | 7.69E-42 | Nightingale | XXL_VLDL_ | NA | T | A | 0.03468 | 0.004441 | 4.10E-15 |
| rs582094 | miR-139-5p | trans | 7.69E-42 | Nightingale | S_VLDL_L | NA | T | A | 0.024549 | 0.004388 | 4.70E-09 |
| rs582094 | miR-139-5p | trans | 7.69E-42 | Nightingale | M_LDL_C_ | NA | T | A | 0.031036 | 0.004434 | 3.90E-12 |
| rs582094 | miR-139-5p | trans | 7.69E-42 | Nightingale | L_LDL_C | NA | T | A | 0.032117 | 0.004381 | 4.00E-13 |
| rs582094 | miR-139-5p | trans | 7.69E-42 | Nightingale | M_VLDL_C | NA | T | A | 0.040673 | 0.004391 | 3.90E-20 |
| rs582094 | miR-139-5p | trans | 7.69E-42 | Nightingale | L_VLDL_C_ | NA | T | A | 0.052204 | 0.004328 | 4.90E-34 |
| rs582094 | miR-139-5p | trans | 7.69E-42 | Nightingale | XS_VLDL_C | NA | T | A | 0.037674 | 0.004276 | 3.10E-18 |
| rs582094 | miR-139-5p | trans | 7.69E-42 | Nightingale | M_LDL_CE | NA | T | A | 0.032814 | 0.004437 | 1.40E-13 |
| rs582094 | miR-139-5p | trans | 7.69E-42 | Nightingale | S_HDL_L | NA | T | A | 0.02611 | 0.004416 | 2.10E-09 |
| rs582094 | miR-139-5p | trans | 7.69E-42 | Nightingale | M_VLDL_F | NA | T | A | 0.042037 | 0.004216 | 4.50E-23 |
| rs582094 | miR-139-5p | trans | 7.69E-42 | Nightingale | S_VLDL_C_ | NA | T | A | 0.041764 | 0.004312 | 8.50E-22 |
| rs582094 | miR-139-5p | trans | 7.69E-42 | Nightingale | VLDL_CE | NA | T | A | 0.035642 | 0.004432 | 4.50E-16 |
| rs582094 | miR-139-5p | trans | 7.69E-42 | Nightingale | IDL_C | NA | T | A | 0.034832 | 0.00428 | 1.40E-15 |
| rs582094 | miR-139-5p | trans | 7.69E-42 | Nightingale | XL_VLDL_P | NA | T | A | 0.038942 | 0.004438 | 4.80E-19 |
| rs582094 | miR-139-5p | trans | 7.69E-42 | Nightingale | L_LDL_P | NA | T | A | 0.028971 | 0.004416 | 9.20E-11 |
| rs582094 | miR-139-5p | trans | 7.69E-42 | Nightingale | XL_VLDL_F | NA | T | A | 0.037159 | 0.004296 | 2.40E-18 |
| rs582094 | miR-139-5p | trans | 7.69E-42 | Nightingale | XL_VLDL_T | NA | T | A | -0.04392 | 0.004295 | 2.20E-24 |
| rs582094 | miR-139-5p | trans | 7.69E-42 | Nightingale | L_LDL_PL | NA | T | A | 0.037813 | 0.004398 | 1.50E-17 |
| rs582094 | miR-139-5p | trans | 7.69E-42 | Nightingale | L_VLDL_CE | NA | T | A | 0.047319 | 0.00424 | 3.20E-29 |
| rs582094 | miR-139-5p | trans | 7.69E-42 | Nightingale | M_LDL_L | NA | T | A | 0.032867 | 0.004437 | 1.50E-13 |
| rs582094 | miR-139-5p | trans | 7.69E-42 | Nightingale | S_VLDL_FC | NA | T | A | 0.037726 | 0.004432 | 1.20E-17 |
| rs582094 | miR-139-5p | trans | 7.69E-42 | Nightingale | S_VLDL_PL | NA | T | A | 0.032628 | 0.004423 | 6.00E-14 |
| rs582094 | miR-139-5p | trans | 7.69E-42 | Nightingale | XS_VLDL_F | NA | T | A | 0.03125 | 0.0044 | 3.70E-12 |
| rs582094 | miR-139-5p | trans | 7.69E-42 | Nightingale | S_VLDL_C | NA | T | A | 0.038213 | 0.004431 | 2.80E-18 |
| rs582094 | miR-139-5p | trans | 7.69E-42 | Nightingale | XL_VLDL_C | NA | T | A | 0.038128 | 0.004249 | 6.90E-19 |
| rs582094 | miR-139-5p | trans | 7.69E-42 | Nightingale | S_LDL_PL_ | NA | T | A | -0.02995 | 0.004423 | 3.30E-13 |
| rs582094 | miR-139-5p | trans | 7.69E-42 | Nightingale | LDL_PL | NA | T | A | 0.03482 | 0.004421 | 7.30E-15 |
| rs582094 | miR-139-5p | trans | 7.69E-42 | Nightingale | non_HDL_( | NA | T | A | 0.035527 | 0.004407 | 7.90E-16 |
| rs582094 | miR-139-5p | trans | 7.69E-42 | Nightingale | XXL_VLDL_ | NA | T | A | 0.025093 | 0.00446 | 1.40E-09 |
| rs582094 | miR-139-5p | trans | 7.69E-42 | Nightingale | M_LDL_PL | NA | T | A | 0.032301 | 0.004437 | 3.90E-13 |
| rs582094 | miR-139-5p | trans | 7.69E-42 | Nightingale | XL_VLDL_C | NA | T | A | 0.037247 | 0.004251 | 4.70E-18 |
| rs582094 | miR-139-5p | trans | 7.69E-42 | Nightingale | XS_VLDL_P | NA | T | A | 0.032031 | 0.004369 | 6.90E-14 |
| rs582094 | miR-139-5p | trans | 7.69E-42 | Nightingale | M_VLDL_F | NA | T | A | 0.033201 | 0.00443 | 4.70E-14 |
| rs582094 | miR-139-5p | trans | 7.69E-42 | Nightingale | S_HDL_PL | NA | T | A | 0.023614 | 0.004393 | 4.10E-08 |
| rs582094 | miR-139-5p | trans | 7.69E-42 | Nightingale | XXL_VLDL_ | NA | T | A | 0.030797 | 0.004412 | 1.20E-12 |
| rs582094 | miR-139-5p | trans | 7.69E-42 | Nightingale | S_LDL_FC | NA | T | A | 0.028398 | 0.004424 | 6.00E-10 |
| rs582094 | miR-139-5p | trans | 7.69E-42 | Nightingale | PUFA | NA | T | A | 0.023572 | 0.004322 | 1.80E-08 |
| rs582094 | miR-139-5p | trans | 7.69E-42 | Nightingale | LDL_FC | NA | T | A | 0.033455 | 0.004384 | 8.10E-14 |
| rs582094 | miR-139-5p | trans | 7.69E-42 | Nightingale | LDL_P | NA | T | A | 0.03163 | 0.004428 | 1.50E-12 |
| rs582094 | miR-139-5p | trans | 7.69E-42 | Nightingale | L_VLDL_TG | NA | T | A | -0.05112 | 0.00441 | 7.10E-32 |
| rs582094 | miR-139-5p | trans | 7.69E-42 | Nightingale | IDL_FC | NA | T | A | 0.038121 | 0.00429 | 2.20E-18 |
| rs582094 | miR-139-5p | trans | 7.69E-42 | Nightingale | LDL_C | NA | T | A | 0.033002 | 0.004412 | 1.40E-13 |
| rs582094 | miR-139-5p | trans | 7.69E-42 | Nightingale | Total_CE | NA | T | A | 0.031574 | 0.004274 | 1.40E-13 |

| | | | | | | | | | | | |
|---|---|---|---|---|---|---|---|---|---|---|---|
| rs582094 | miR-139-5p | trans | 7.69E-42 | Nightingale | M_LDL_TG | NA | T | A | -0.02744 | 0.004376 | 4.70E-10 |
| rs582094 | miR-139-5p | trans | 7.69E-42 | Nightingale | S_LDL_CE_ | NA | T | A | 0.03085 | 0.004424 | 6.10E-14 |
| rs582094 | miR-139-5p | trans | 7.69E-42 | Nightingale | VLDL_P | NA | T | A | 0.026524 | 0.004407 | 5.50E-10 |
| rs582094 | miR-139-5p | trans | 7.69E-42 | Nightingale | S_HDL_CE | NA | T | A | 0.028434 | 0.004438 | 1.90E-10 |
| rs582094 | miR-139-5p | trans | 7.69E-42 | Nightingale | S_VLDL_PL | NA | T | A | 0.027637 | 0.004243 | 2.50E-10 |
| rs582094 | miR-139-5p | trans | 7.69E-42 | Nightingale | M_VLDL_C | NA | T | A | 0.038533 | 0.004419 | 3.20E-18 |
| rs582094 | miR-139-5p | trans | 7.69E-42 | Nightingale | Sphingomy | NA | T | A | 0.028892 | 0.004142 | 8.90E-12 |
| rs582094 | miR-139-5p | trans | 7.69E-42 | Nightingale | M_VLDL_P | NA | T | A | 0.044033 | 0.004253 | 7.10E-25 |
| rs582094 | miR-139-5p | trans | 7.69E-42 | Nightingale | LDL_CE | NA | T | A | 0.032359 | 0.004419 | 3.30E-13 |
| rs582094 | miR-139-5p | trans | 7.69E-42 | Nightingale | XXL_VLDL_ | NA | T | A | -0.03566 | 0.004469 | 2.70E-16 |
| rs582094 | miR-139-5p | trans | 7.69E-42 | Nightingale | IDL_P | NA | T | A | 0.039908 | 0.004385 | 1.90E-19 |
| rs582094 | miR-139-5p | trans | 7.69E-42 | Nightingale | Total_PL | NA | T | A | 0.024785 | 0.00422 | 1.40E-09 |
| rs582094 | miR-139-5p | trans | 7.69E-42 | Nightingale | S_LDL_L | NA | T | A | 0.027606 | 0.00443 | 1.10E-09 |
| rs582094 | miR-139-5p | trans | 7.69E-42 | Nightingale | S_VLDL_TG | NA | T | A | -0.03801 | 0.004289 | 2.20E-18 |
| rs582094 | miR-139-5p | trans | 7.69E-42 | Nightingale | L_LDL_CE | NA | T | A | 0.031356 | 0.004392 | 1.50E-12 |
| rs582094 | miR-139-5p | trans | 7.69E-42 | Nightingale | S_VLDL_P | NA | T | A | 0.023349 | 0.004376 | 2.00E-08 |
| rs582094 | miR-139-5p | trans | 7.69E-42 | Nightingale | S_LDL_CE | NA | T | A | 0.030409 | 0.004433 | 7.90E-12 |
| rs582094 | miR-139-5p | trans | 7.69E-42 | Nightingale | XL_HDL_PL | NA | T | A | -0.02265 | 0.004071 | 1.40E-08 |
| rs582094 | miR-139-5p | trans | 7.69E-42 | Nightingale | XS_VLDL_L | NA | T | A | 0.033719 | 0.004356 | 2.70E-15 |
| rs582094 | miR-139-5p | trans | 7.69E-42 | Nightingale | Citrate | NA | T | A | 0.02682 | 0.004402 | 7.90E-10 |
| rs582094 | miR-139-5p | trans | 7.69E-42 | Nightingale | Clinical_LD | NA | T | A | 0.0363 | 0.004406 | 4.30E-16 |
| rs582094 | miR-139-5p | trans | 7.69E-42 | Nightingale | Omega_6 | NA | T | A | 0.025694 | 0.004327 | 8.00E-10 |
| rs582094 | miR-139-5p | trans | 7.69E-42 | Nightingale | S_HDL_C | NA | T | A | 0.028202 | 0.004424 | 2.50E-10 |
| rs582094 | miR-139-5p | trans | 7.69E-42 | Nightingale | M_LDL_C | NA | T | A | 0.033818 | 0.004439 | 3.20E-14 |
| rs582094 | miR-139-5p | trans | 7.69E-42 | Nightingale | S_HDL_P | NA | T | A | 0.030405 | 0.004432 | 7.50E-12 |
| rs582118 | miR-10b-5p | trans | 3.28E-35 | Nightingale | S_VLDL_CE | NA | G | A | 0.04702 | 0.004366 | 1.20E-26 |
| rs582118 | miR-10b-5p | trans | 3.28E-35 | Nightingale | S_LDL_P | NA | G | A | 0.029577 | 0.004429 | 3.30E-11 |
| rs582118 | miR-10b-5p | trans | 3.28E-35 | Nightingale | M_LDL_C | NA | G | A | 0.033974 | 0.004433 | 2.20E-14 |
| rs582118 | miR-10b-5p | trans | 3.28E-35 | Nightingale | S_VLDL_FC | NA | G | A | 0.03789 | 0.004426 | 7.60E-18 |
| rs582118 | miR-10b-5p | trans | 3.28E-35 | Nightingale | L_VLDL_C_ | NA | G | A | 0.052199 | 0.004322 | 4.00E-34 |
| rs582118 | miR-10b-5p | trans | 3.28E-35 | Nightingale | S_LDL_C_p | NA | G | A | 0.045354 | 0.004442 | 5.70E-25 |
| rs582118 | miR-10b-5p | trans | 3.28E-35 | Nightingale | M_LDL_P | NA | G | A | 0.035885 | 0.004438 | 6.90E-16 |
| rs582118 | miR-10b-5p | trans | 3.28E-35 | Nightingale | S_LDL_C | NA | G | A | 0.030929 | 0.00443 | 5.60E-12 |
| rs582118 | miR-10b-5p | trans | 3.28E-35 | Nightingale | XS_VLDL_F | NA | G | A | 0.037062 | 0.004348 | 6.70E-18 |
| rs582118 | miR-10b-5p | trans | 3.28E-35 | Nightingale | XS_VLDL_P | NA | G | A | 0.036694 | 0.004367 | 1.40E-17 |
| rs582118 | miR-10b-5p | trans | 3.28E-35 | Nightingale | IDL_C | NA | G | A | 0.034941 | 0.004274 | 9.70E-16 |
| rs582118 | miR-10b-5p | trans | 3.28E-35 | Nightingale | VLDL_C | NA | G | A | 0.031288 | 0.004419 | 5.00E-13 |
| rs582118 | miR-10b-5p | trans | 3.28E-35 | Nightingale | XXL_VLDL_ | NA | G | A | 0.033117 | 0.004459 | 9.10E-14 |
| rs582118 | miR-10b-5p | trans | 3.28E-35 | Nightingale | XL_VLDL_C | NA | G | A | 0.037363 | 0.004245 | 3.10E-18 |
| rs582118 | miR-10b-5p | trans | 3.28E-35 | Nightingale | VLDL_P | NA | G | A | 0.026621 | 0.004402 | 4.50E-10 |
| rs582118 | miR-10b-5p | trans | 3.28E-35 | Nightingale | S_VLDL_FC | NA | G | A | 0.029356 | 0.004229 | 1.10E-11 |
| rs582118 | miR-10b-5p | trans | 3.28E-35 | Nightingale | XXL_VLDL_ | NA | G | A | -0.03572 | 0.004463 | 2.20E-16 |
| rs582118 | miR-10b-5p | trans | 3.28E-35 | Nightingale | L_VLDL_FC | NA | G | A | 0.04857 | 0.004383 | 4.80E-29 |
| rs582118 | miR-10b-5p | trans | 3.28E-35 | Nightingale | IDL_FC_pct | NA | G | A | 0.025779 | 0.004391 | 1.10E-08 |
| rs582118 | miR-10b-5p | trans | 3.28E-35 | Nightingale | M_VLDL_P | NA | G | A | 0.029113 | 0.004418 | 2.10E-11 |
| rs582118 | miR-10b-5p | trans | 3.28E-35 | Nightingale | XL_VLDL_P | NA | G | A | 0.03883 | 0.004432 | 5.30E-19 |
| rs582118 | miR-10b-5p | trans | 3.28E-35 | Nightingale | M_LDL_CE | NA | G | A | 0.03296 | 0.004431 | 9.90E-14 |
| rs582118 | miR-10b-5p | trans | 3.28E-35 | Nightingale | XS_VLDL_C | NA | G | A | 0.03779 | 0.004271 | 2.10E-18 |
| rs582118 | miR-10b-5p | trans | 3.28E-35 | Nightingale | IDL_P | NA | G | A | 0.039958 | 0.004379 | 1.40E-19 |
| rs582118 | miR-10b-5p | trans | 3.28E-35 | Nightingale | ApoB_by_A | NA | G | A | 0.025026 | 0.004358 | 1.30E-08 |
| rs582118 | miR-10b-5p | trans | 3.28E-35 | Nightingale | XS_VLDL_C | NA | G | A | 0.037986 | 0.004295 | 1.40E-18 |
| rs582118 | miR-10b-5p | trans | 3.28E-35 | Nightingale | S_HDL_FC | NA | G | A | 0.023456 | 0.004344 | 4.60E-08 |
| rs582118 | miR-10b-5p | trans | 3.28E-35 | Nightingale | M_VLDL_T | NA | G | A | -0.03857 | 0.004182 | 6.70E-20 |
| rs582118 | miR-10b-5p | trans | 3.28E-35 | Nightingale | S_VLDL_C_ | NA | G | A | 0.041881 | 0.004307 | 5.30E-22 |
| rs582118 | miR-10b-5p | trans | 3.28E-35 | Nightingale | IDL_PL | NA | G | A | 0.035338 | 0.00427 | 3.40E-16 |
| rs582118 | miR-10b-5p | trans | 3.28E-35 | Nightingale | M_VLDL_P | NA | G | A | 0.04406 | 0.004247 | 5.60E-25 |
| rs582118 | miR-10b-5p | trans | 3.28E-35 | Nightingale | XL_VLDL_C | NA | G | A | 0.038233 | 0.004243 | 4.60E-19 |
| rs582118 | miR-10b-5p | trans | 3.28E-35 | Nightingale | L_VLDL_CE | NA | G | A | 0.047351 | 0.004235 | 2.40E-29 |
| rs582118 | miR-10b-5p | trans | 3.28E-35 | Nightingale | XS_VLDL_F | NA | G | A | 0.031428 | 0.004394 | 2.50E-12 |
| rs582118 | miR-10b-5p | trans | 3.28E-35 | Nightingale | S_LDL_PL_ | NA | G | A | -0.03006 | 0.004417 | 2.70E-13 |
| rs582118 | miR-10b-5p | trans | 3.28E-35 | Nightingale | M_VLDL_P | NA | G | A | 0.02828 | 0.004414 | 6.50E-11 |
| rs582118 | miR-10b-5p | trans | 3.28E-35 | Nightingale | S_LDL_CE | NA | G | A | 0.030534 | 0.004428 | 6.00E-12 |
| rs582118 | miR-10b-5p | trans | 3.28E-35 | Nightingale | S_HDL_C | NA | G | A | 0.028404 | 0.004418 | 1.70E-10 |

| | | | | | | | | | | | |
|---|---|---|---|---|---|---|---|---|---|---|---|
| rs582118 | miR-10b-5| trans | 3.28E-35 | Nightingale | M_VLDL_C | NA | G | A | 0.036218 | 0.004168 | 1.20E-17 |
| rs582118 | miR-10b-5| trans | 3.28E-35 | Nightingale | XS_VLDL_L | NA | G | A | 0.033813 | 0.00435 | 2.10E-15 |
| rs582118 | miR-10b-5| trans | 3.28E-35 | Nightingale | M_VLDL_C | NA | G | A | 0.040833 | 0.004385 | 2.40E-20 |
| rs582118 | miR-10b-5| trans | 3.28E-35 | Nightingale | Total_FC | NA | G | A | 0.033252 | 0.004323 | 6.90E-15 |
| rs582118 | miR-10b-5| trans | 3.28E-35 | Nightingale | XS_VLDL_P | NA | G | A | 0.032112 | 0.004364 | 5.50E-14 |
| rs582118 | miR-10b-5| trans | 3.28E-35 | Nightingale | LDL_C | NA | G | A | 0.033183 | 0.004406 | 9.00E-14 |
| rs582118 | miR-10b-5| trans | 3.28E-35 | Nightingale | L_LDL_C | NA | G | A | 0.032311 | 0.004375 | 2.60E-13 |
| rs582118 | miR-10b-5| trans | 3.28E-35 | Nightingale | S_VLDL_PL | NA | G | A | 0.032763 | 0.004417 | 4.30E-14 |
| rs582118 | miR-10b-5| trans | 3.28E-35 | Nightingale | Total_CE | NA | G | A | 0.031745 | 0.004269 | 9.80E-14 |
| rs582118 | miR-10b-5| trans | 3.28E-35 | Nightingale | L_VLDL_TG | NA | G | A | -0.05114 | 0.004404 | 5.60E-32 |
| rs582118 | miR-10b-5| trans | 3.28E-35 | Nightingale | IDL_L | NA | G | A | 0.035258 | 0.004279 | 5.00E-16 |
| rs582118 | miR-10b-5| trans | 3.28E-35 | Nightingale | S_VLDL_CE | NA | G | A | 0.038165 | 0.00442 | 1.70E-18 |
| rs582118 | miR-10b-5| trans | 3.28E-35 | Nightingale | Cholines | NA | G | A | 0.022307 | 0.004162 | 4.80E-08 |
| rs582118 | miR-10b-5| trans | 3.28E-35 | Nightingale | Remnant_( | NA | G | A | 0.036853 | 0.004397 | 3.80E-17 |
| rs582118 | miR-10b-5| trans | 3.28E-35 | Nightingale | Total_L | NA | G | A | 0.026347 | 0.004353 | 4.60E-10 |
| rs582118 | miR-10b-5| trans | 3.28E-35 | Nightingale | M_LDL_C_| | NA | G | A | 0.031166 | 0.004428 | 2.90E-12 |
| rs582118 | miR-10b-5| trans | 3.28E-35 | Nightingale | S_VLDL_PL | NA | G | A | 0.027738 | 0.004237 | 1.80E-10 |
| rs582118 | miR-10b-5| trans | 3.28E-35 | Nightingale | XXL_VLDL_ | NA | G | A | 0.034828 | 0.004435 | 2.90E-15 |
| rs582118 | miR-10b-5| trans | 3.28E-35 | Nightingale | Omega_6 | NA | G | A | 0.025897 | 0.004321 | 5.80E-10 |
| rs582118 | miR-10b-5| trans | 3.28E-35 | Nightingale | ApoB | NA | G | A | 0.033924 | 0.00442 | 2.20E-14 |
| rs582118 | miR-10b-5| trans | 3.28E-35 | Nightingale | L_LDL_FC | NA | G | A | 0.033616 | 0.004345 | 2.90E-14 |
| rs582118 | miR-10b-5| trans | 3.28E-35 | Nightingale | XXL_VLDL_ | NA | G | A | 0.024988 | 0.004454 | 1.50E-09 |
| rs582118 | miR-10b-5| trans | 3.28E-35 | Nightingale | XL_HDL_PL | NA | G | A | -0.02272 | 0.004066 | 1.20E-08 |
| rs582118 | miR-10b-5| trans | 3.28E-35 | Nightingale | S_HDL_L | NA | G | A | 0.026289 | 0.00441 | 1.60E-09 |
| rs582118 | miR-10b-5| trans | 3.28E-35 | Nightingale | LDL_PL | NA | G | A | 0.034973 | 0.004415 | 4.90E-15 |
| rs582118 | miR-10b-5| trans | 3.28E-35 | Nightingale | L_LDL_P | NA | G | A | 0.029166 | 0.00441 | 6.30E-11 |
| rs582118 | miR-10b-5| trans | 3.28E-35 | Nightingale | VLDL_FC | NA | G | A | 0.024145 | 0.004393 | 1.10E-08 |
| rs582118 | miR-10b-5| trans | 3.28E-35 | Nightingale | S_HDL_P | NA | G | A | 0.030602 | 0.004426 | 5.20E-12 |
| rs582118 | miR-10b-5| trans | 3.28E-35 | Nightingale | S_LDL_CE_ | NA | G | A | 0.030939 | 0.004418 | 5.00E-14 |
| rs582118 | miR-10b-5| trans | 3.28E-35 | Nightingale | M_VLDL_F( | NA | G | A | 0.033358 | 0.004424 | 3.20E-14 |
| rs582118 | miR-10b-5| trans | 3.28E-35 | Nightingale | XXL_VLDL_ | NA | G | A | 0.030845 | 0.004407 | 1.10E-12 |
| rs582118 | miR-10b-5| trans | 3.28E-35 | Nightingale | IDL_FC | NA | G | A | 0.038246 | 0.004284 | 1.50E-18 |
| rs582118 | miR-10b-5| trans | 3.28E-35 | Nightingale | LDL_CE | NA | G | A | 0.032543 | 0.004414 | 2.20E-13 |
| rs582118 | miR-10b-5| trans | 3.28E-35 | Nightingale | M_VLDL_F( | NA | G | A | 0.042075 | 0.00421 | 3.40E-23 |
| rs582118 | miR-10b-5| trans | 3.28E-35 | Nightingale | S_VLDL_P | NA | G | A | 0.023453 | 0.00437 | 1.70E-08 |
| rs582118 | miR-10b-5| trans | 3.28E-35 | Nightingale | Citrate | NA | G | A | 0.026837 | 0.004396 | 7.30E-10 |
| rs582118 | miR-10b-5| trans | 3.28E-35 | Nightingale | LDL_L | NA | G | A | 0.033293 | 0.00441 | 7.60E-14 |
| rs582118 | miR-10b-5| trans | 3.28E-35 | Nightingale | Total_C | NA | G | A | 0.032371 | 0.004284 | 3.40E-14 |
| rs582118 | miR-10b-5| trans | 3.28E-35 | Nightingale | S_LDL_L | NA | G | A | 0.027727 | 0.004424 | 8.40E-10 |
| rs582118 | miR-10b-5| trans | 3.28E-35 | Nightingale | L_LDL_L | NA | G | A | 0.033541 | 0.004382 | 3.10E-14 |
| rs582118 | miR-10b-5| trans | 3.28E-35 | Nightingale | S_HDL_PL | NA | G | A | 0.02378 | 0.004387 | 3.30E-08 |
| rs582118 | miR-10b-5| trans | 3.28E-35 | Nightingale | XL_VLDL_T | NA | G | A | -0.04392 | 0.004289 | 1.80E-24 |
| rs582118 | miR-10b-5| trans | 3.28E-35 | Nightingale | L_LDL_PL | NA | G | A | 0.037976 | 0.004392 | 9.40E-18 |
| rs582118 | miR-10b-5| trans | 3.28E-35 | Nightingale | M_VLDL_C | NA | G | A | 0.033641 | 0.004157 | 1.90E-15 |
| rs582118 | miR-10b-5| trans | 3.28E-35 | Nightingale | IDL_CE | NA | G | A | 0.033491 | 0.004273 | 1.40E-14 |
| rs582118 | miR-10b-5| trans | 3.28E-35 | Nightingale | M_LDL_L | NA | G | A | 0.033017 | 0.004431 | 1.00E-13 |
| rs582118 | miR-10b-5| trans | 3.28E-35 | Nightingale | S_LDL_FC | NA | G | A | 0.028549 | 0.004418 | 4.40E-10 |
| rs582118 | miR-10b-5| trans | 3.28E-35 | Nightingale | M_LDL_FC | NA | G | A | 0.034333 | 0.00442 | 2.00E-14 |
| rs582118 | miR-10b-5| trans | 3.28E-35 | Nightingale | Sphingomy | NA | G | A | 0.029028 | 0.004137 | 6.70E-12 |
| rs582118 | miR-10b-5| trans | 3.28E-35 | Nightingale | S_VLDL_TG | NA | G | A | -0.03812 | 0.004283 | 1.50E-18 |
| rs582118 | miR-10b-5| trans | 3.28E-35 | Nightingale | Total_PL | NA | G | A | 0.024934 | 0.004214 | 1.10E-09 |
| rs582118 | miR-10b-5| trans | 3.28E-35 | Nightingale | S_VLDL_L | NA | G | A | 0.024655 | 0.004382 | 3.80E-09 |
| rs582118 | miR-10b-5| trans | 3.28E-35 | Nightingale | GlycA | NA | G | A | 0.028033 | 0.004406 | 1.40E-10 |
| rs582118 | miR-10b-5| trans | 3.28E-35 | Nightingale | M_LDL_PL | NA | G | A | 0.032449 | 0.004432 | 2.70E-13 |
| rs582118 | miR-10b-5| trans | 3.28E-35 | Nightingale | S_HDL_CE | NA | G | A | 0.028625 | 0.004432 | 1.30E-10 |
| rs582118 | miR-10b-5| trans | 3.28E-35 | Nightingale | S_VLDL_C | NA | G | A | 0.038357 | 0.004425 | 1.80E-18 |
| rs582118 | miR-10b-5| trans | 3.28E-35 | Nightingale | XL_VLDL_F | NA | G | A | 0.037203 | 0.00429 | 2.00E-18 |
| rs582118 | miR-10b-5| trans | 3.28E-35 | Nightingale | LDL_P | NA | G | A | 0.03179 | 0.004422 | 1.00E-12 |
| rs582118 | miR-10b-5| trans | 3.28E-35 | Nightingale | LDL_FC | NA | G | A | 0.03362 | 0.004378 | 5.40E-14 |
| rs582118 | miR-10b-5| trans | 3.28E-35 | Nightingale | M_VLDL_C | NA | G | A | 0.038699 | 0.004413 | 2.00E-18 |
| rs582118 | miR-10b-5| trans | 3.28E-35 | Nightingale | L_LDL_CE | NA | G | A | 0.031559 | 0.004386 | 1.00E-12 |
| rs582118 | miR-10b-5| trans | 3.28E-35 | Nightingale | VLDL_CE | NA | G | A | 0.035773 | 0.004426 | 3.10E-16 |
| rs582118 | miR-10b-5| trans | 3.28E-35 | Nightingale | non_HDL_( | NA | G | A | 0.035684 | 0.004401 | 5.20E-16 |

| | | | | | | | | | | | |
|---|---|---|---|---|---|---|---|---|---|---|---|
| rs582118 | miR-10b-5p | trans | 3.28E-35 | Nightingale | M_LDL_TG | NA | G | A | -0.02761 | 0.00437 | 3.20E-10 |
| rs582118 | miR-10b-5p | trans | 3.28E-35 | Nightingale | LA | NA | G | A | 0.024908 | 0.00434 | 3.70E-09 |
| rs582118 | miR-10b-5p | trans | 3.28E-35 | Nightingale | PUFA | NA | G | A | 0.023718 | 0.004316 | 1.40E-08 |
| rs582118 | miR-10b-5p | trans | 3.28E-35 | Nightingale | Clinical_LD | NA | G | A | 0.03646 | 0.0044 | 2.80E-16 |
| rs582118 | miR-139-5p | trans | 7.44E-42 | Nightingale | S_VLDL_CE | NA | G | A | 0.04702 | 0.004366 | 1.20E-26 |
| rs582118 | miR-139-5p | trans | 7.44E-42 | Nightingale | S_LDL_P | NA | G | A | 0.029577 | 0.004429 | 3.30E-11 |
| rs582118 | miR-139-5p | trans | 7.44E-42 | Nightingale | M_LDL_C | NA | G | A | 0.033974 | 0.004433 | 2.20E-14 |
| rs582118 | miR-139-5p | trans | 7.44E-42 | Nightingale | S_VLDL_FC | NA | G | A | 0.03789 | 0.004426 | 7.60E-18 |
| rs582118 | miR-139-5p | trans | 7.44E-42 | Nightingale | L_VLDL_C_ | NA | G | A | 0.052199 | 0.004322 | 4.00E-34 |
| rs582118 | miR-139-5p | trans | 7.44E-42 | Nightingale | S_LDL_C_p | NA | G | A | 0.045354 | 0.004442 | 5.70E-25 |
| rs582118 | miR-139-5p | trans | 7.44E-42 | Nightingale | M_LDL_P | NA | G | A | 0.035885 | 0.004438 | 6.90E-16 |
| rs582118 | miR-139-5p | trans | 7.44E-42 | Nightingale | S_LDL_C | NA | G | A | 0.030929 | 0.00443 | 5.60E-12 |
| rs582118 | miR-139-5p | trans | 7.44E-42 | Nightingale | XS_VLDL_F | NA | G | A | 0.037062 | 0.004348 | 6.70E-18 |
| rs582118 | miR-139-5p | trans | 7.44E-42 | Nightingale | XS_VLDL_P | NA | G | A | 0.036694 | 0.004367 | 1.40E-17 |
| rs582118 | miR-139-5p | trans | 7.44E-42 | Nightingale | IDL_C | NA | G | A | 0.034941 | 0.004274 | 9.70E-16 |
| rs582118 | miR-139-5p | trans | 7.44E-42 | Nightingale | VLDL_C | NA | G | A | 0.031288 | 0.004419 | 5.00E-13 |
| rs582118 | miR-139-5p | trans | 7.44E-42 | Nightingale | XXL_VLDL_ | NA | G | A | 0.033117 | 0.004459 | 9.10E-14 |
| rs582118 | miR-139-5p | trans | 7.44E-42 | Nightingale | XL_VLDL_C | NA | G | A | 0.037363 | 0.004245 | 3.10E-18 |
| rs582118 | miR-139-5p | trans | 7.44E-42 | Nightingale | VLDL_P | NA | G | A | 0.026621 | 0.004402 | 4.50E-10 |
| rs582118 | miR-139-5p | trans | 7.44E-42 | Nightingale | S_VLDL_FC | NA | G | A | 0.029356 | 0.004229 | 1.10E-11 |
| rs582118 | miR-139-5p | trans | 7.44E-42 | Nightingale | XXL_VLDL_ | NA | G | A | -0.03572 | 0.004463 | 2.20E-16 |
| rs582118 | miR-139-5p | trans | 7.44E-42 | Nightingale | L_VLDL_FC | NA | G | A | 0.04857 | 0.004383 | 4.80E-29 |
| rs582118 | miR-139-5p | trans | 7.44E-42 | Nightingale | IDL_FC_pct | NA | G | A | 0.025779 | 0.004391 | 1.10E-08 |
| rs582118 | miR-139-5p | trans | 7.44E-42 | Nightingale | M_VLDL_P | NA | G | A | 0.029113 | 0.004418 | 2.10E-11 |
| rs582118 | miR-139-5p | trans | 7.44E-42 | Nightingale | XL_VLDL_P | NA | G | A | 0.03883 | 0.004432 | 5.30E-19 |
| rs582118 | miR-139-5p | trans | 7.44E-42 | Nightingale | M_LDL_CE | NA | G | A | 0.03296 | 0.004431 | 9.90E-14 |
| rs582118 | miR-139-5p | trans | 7.44E-42 | Nightingale | XS_VLDL_C | NA | G | A | 0.03779 | 0.004271 | 2.10E-18 |
| rs582118 | miR-139-5p | trans | 7.44E-42 | Nightingale | IDL_P | NA | G | A | 0.039958 | 0.004379 | 1.40E-19 |
| rs582118 | miR-139-5p | trans | 7.44E-42 | Nightingale | ApoB_by_A | NA | G | A | 0.025026 | 0.004358 | 1.30E-08 |
| rs582118 | miR-139-5p | trans | 7.44E-42 | Nightingale | XS_VLDL_C | NA | G | A | 0.037986 | 0.004295 | 1.40E-18 |
| rs582118 | miR-139-5p | trans | 7.44E-42 | Nightingale | S_HDL_FC | NA | G | A | 0.023456 | 0.004344 | 4.60E-08 |
| rs582118 | miR-139-5p | trans | 7.44E-42 | Nightingale | M_VLDL_T | NA | G | A | -0.03857 | 0.004182 | 6.70E-20 |
| rs582118 | miR-139-5p | trans | 7.44E-42 | Nightingale | S_VLDL_C_ | NA | G | A | 0.041881 | 0.004307 | 5.30E-22 |
| rs582118 | miR-139-5p | trans | 7.44E-42 | Nightingale | IDL_PL | NA | G | A | 0.035338 | 0.00427 | 3.40E-16 |
| rs582118 | miR-139-5p | trans | 7.44E-42 | Nightingale | M_VLDL_P | NA | G | A | 0.04406 | 0.004247 | 5.60E-25 |
| rs582118 | miR-139-5p | trans | 7.44E-42 | Nightingale | XL_VLDL_C | NA | G | A | 0.038233 | 0.004243 | 4.60E-19 |
| rs582118 | miR-139-5p | trans | 7.44E-42 | Nightingale | L_VLDL_CE | NA | G | A | 0.047351 | 0.004235 | 2.40E-29 |
| rs582118 | miR-139-5p | trans | 7.44E-42 | Nightingale | XS_VLDL_F | NA | G | A | 0.031428 | 0.004394 | 2.50E-12 |
| rs582118 | miR-139-5p | trans | 7.44E-42 | Nightingale | S_LDL_PL_ | NA | G | A | -0.03006 | 0.004417 | 2.70E-13 |
| rs582118 | miR-139-5p | trans | 7.44E-42 | Nightingale | M_VLDL_P | NA | G | A | 0.02828 | 0.004414 | 6.50E-11 |
| rs582118 | miR-139-5p | trans | 7.44E-42 | Nightingale | S_LDL_CE | NA | G | A | 0.030534 | 0.004428 | 6.00E-12 |
| rs582118 | miR-139-5p | trans | 7.44E-42 | Nightingale | S_HDL_C | NA | G | A | 0.028404 | 0.004418 | 1.70E-10 |
| rs582118 | miR-139-5p | trans | 7.44E-42 | Nightingale | M_VLDL_C | NA | G | A | 0.036218 | 0.004168 | 1.20E-17 |
| rs582118 | miR-139-5p | trans | 7.44E-42 | Nightingale | XS_VLDL_L | NA | G | A | 0.033813 | 0.00435 | 2.10E-15 |
| rs582118 | miR-139-5p | trans | 7.44E-42 | Nightingale | M_VLDL_C | NA | G | A | 0.040833 | 0.004385 | 2.40E-20 |
| rs582118 | miR-139-5p | trans | 7.44E-42 | Nightingale | Total_FC | NA | G | A | 0.033252 | 0.004323 | 6.90E-15 |
| rs582118 | miR-139-5p | trans | 7.44E-42 | Nightingale | XS_VLDL_P | NA | G | A | 0.032112 | 0.004364 | 5.50E-14 |
| rs582118 | miR-139-5p | trans | 7.44E-42 | Nightingale | LDL_C | NA | G | A | 0.033183 | 0.004406 | 9.00E-14 |
| rs582118 | miR-139-5p | trans | 7.44E-42 | Nightingale | L_LDL_C | NA | G | A | 0.032311 | 0.004375 | 2.60E-13 |
| rs582118 | miR-139-5p | trans | 7.44E-42 | Nightingale | S_VLDL_PL | NA | G | A | 0.032763 | 0.004417 | 4.30E-14 |
| rs582118 | miR-139-5p | trans | 7.44E-42 | Nightingale | Total_CE | NA | G | A | 0.031745 | 0.004269 | 9.80E-14 |
| rs582118 | miR-139-5p | trans | 7.44E-42 | Nightingale | L_VLDL_TG | NA | G | A | -0.05114 | 0.004404 | 5.60E-32 |
| rs582118 | miR-139-5p | trans | 7.44E-42 | Nightingale | IDL_L | NA | G | A | 0.035258 | 0.004279 | 5.00E-16 |
| rs582118 | miR-139-5p | trans | 7.44E-42 | Nightingale | S_VLDL_CE | NA | G | A | 0.038165 | 0.00442 | 1.70E-18 |
| rs582118 | miR-139-5p | trans | 7.44E-42 | Nightingale | Cholines | NA | G | A | 0.022307 | 0.004162 | 4.80E-08 |
| rs582118 | miR-139-5p | trans | 7.44E-42 | Nightingale | Remnant_( | NA | G | A | 0.036853 | 0.004397 | 3.80E-17 |
| rs582118 | miR-139-5p | trans | 7.44E-42 | Nightingale | Total_L | NA | G | A | 0.026347 | 0.004353 | 4.60E-10 |
| rs582118 | miR-139-5p | trans | 7.44E-42 | Nightingale | M_LDL_C_ | NA | G | A | 0.031166 | 0.004428 | 2.90E-12 |
| rs582118 | miR-139-5p | trans | 7.44E-42 | Nightingale | S_VLDL_PL | NA | G | A | 0.027738 | 0.004237 | 1.80E-10 |
| rs582118 | miR-139-5p | trans | 7.44E-42 | Nightingale | XXL_VLDL_ | NA | G | A | 0.034828 | 0.004435 | 2.90E-15 |
| rs582118 | miR-139-5p | trans | 7.44E-42 | Nightingale | Omega_6 | NA | G | A | 0.025897 | 0.004321 | 5.80E-10 |
| rs582118 | miR-139-5p | trans | 7.44E-42 | Nightingale | ApoB | NA | G | A | 0.033924 | 0.00442 | 2.20E-14 |
| rs582118 | miR-139-5p | trans | 7.44E-42 | Nightingale | L_LDL_FC | NA | G | A | 0.033616 | 0.004345 | 2.90E-14 |

| | | | | | | | | | | | |
|---|---|---|---|---|---|---|---|---|---|---|---|
| rs582118 | miR-139-5p | trans | 7.44E-42 | Nightingale | XXL_VLDL_ | NA | G | A | 0.024988 | 0.004454 | 1.50E-09 |
| rs582118 | miR-139-5p | trans | 7.44E-42 | Nightingale | XL_HDL_PL | NA | G | A | -0.02272 | 0.004066 | 1.20E-08 |
| rs582118 | miR-139-5p | trans | 7.44E-42 | Nightingale | S_HDL_L | NA | G | A | 0.026289 | 0.00441 | 1.60E-09 |
| rs582118 | miR-139-5p | trans | 7.44E-42 | Nightingale | LDL_PL | NA | G | A | 0.034973 | 0.004415 | 4.90E-15 |
| rs582118 | miR-139-5p | trans | 7.44E-42 | Nightingale | L_LDL_P | NA | G | A | 0.029166 | 0.00441 | 6.30E-11 |
| rs582118 | miR-139-5p | trans | 7.44E-42 | Nightingale | VLDL_FC | NA | G | A | 0.024145 | 0.004393 | 1.10E-08 |
| rs582118 | miR-139-5p | trans | 7.44E-42 | Nightingale | S_HDL_P | NA | G | A | 0.030602 | 0.004426 | 5.20E-12 |
| rs582118 | miR-139-5p | trans | 7.44E-42 | Nightingale | S_LDL_CE_ | NA | G | A | 0.030939 | 0.004418 | 5.00E-14 |
| rs582118 | miR-139-5p | trans | 7.44E-42 | Nightingale | M_VLDL_F | NA | G | A | 0.033358 | 0.004424 | 3.20E-14 |
| rs582118 | miR-139-5p | trans | 7.44E-42 | Nightingale | XXL_VLDL_ | NA | G | A | 0.030845 | 0.004407 | 1.10E-12 |
| rs582118 | miR-139-5p | trans | 7.44E-42 | Nightingale | IDL_FC | NA | G | A | 0.038246 | 0.004284 | 1.50E-18 |
| rs582118 | miR-139-5p | trans | 7.44E-42 | Nightingale | LDL_CE | NA | G | A | 0.032543 | 0.004414 | 2.20E-13 |
| rs582118 | miR-139-5p | trans | 7.44E-42 | Nightingale | M_VLDL_F | NA | G | A | 0.042075 | 0.00421 | 3.40E-23 |
| rs582118 | miR-139-5p | trans | 7.44E-42 | Nightingale | S_VLDL_P | NA | G | A | 0.023453 | 0.00437 | 1.70E-08 |
| rs582118 | miR-139-5p | trans | 7.44E-42 | Nightingale | Citrate | NA | G | A | 0.026837 | 0.004396 | 7.30E-10 |
| rs582118 | miR-139-5p | trans | 7.44E-42 | Nightingale | LDL_L | NA | G | A | 0.033293 | 0.00441 | 7.60E-14 |
| rs582118 | miR-139-5p | trans | 7.44E-42 | Nightingale | Total_C | NA | G | A | 0.032371 | 0.004284 | 3.40E-14 |
| rs582118 | miR-139-5p | trans | 7.44E-42 | Nightingale | S_LDL_L | NA | G | A | 0.027727 | 0.004424 | 8.40E-10 |
| rs582118 | miR-139-5p | trans | 7.44E-42 | Nightingale | L_LDL_L | NA | G | A | 0.033541 | 0.004382 | 3.10E-14 |
| rs582118 | miR-139-5p | trans | 7.44E-42 | Nightingale | S_HDL_PL | NA | G | A | 0.02378 | 0.004387 | 3.30E-08 |
| rs582118 | miR-139-5p | trans | 7.44E-42 | Nightingale | XL_VLDL_T | NA | G | A | -0.04392 | 0.004289 | 1.80E-24 |
| rs582118 | miR-139-5p | trans | 7.44E-42 | Nightingale | L_LDL_PL | NA | G | A | 0.037976 | 0.004392 | 9.40E-18 |
| rs582118 | miR-139-5p | trans | 7.44E-42 | Nightingale | M_VLDL_C | NA | G | A | 0.033641 | 0.004157 | 1.90E-15 |
| rs582118 | miR-139-5p | trans | 7.44E-42 | Nightingale | IDL_CE | NA | G | A | 0.033491 | 0.004273 | 1.40E-14 |
| rs582118 | miR-139-5p | trans | 7.44E-42 | Nightingale | M_LDL_L | NA | G | A | 0.033017 | 0.004431 | 1.00E-13 |
| rs582118 | miR-139-5p | trans | 7.44E-42 | Nightingale | S_LDL_FC | NA | G | A | 0.028549 | 0.004418 | 4.40E-10 |
| rs582118 | miR-139-5p | trans | 7.44E-42 | Nightingale | M_LDL_FC | NA | G | A | 0.034333 | 0.00442 | 2.00E-14 |
| rs582118 | miR-139-5p | trans | 7.44E-42 | Nightingale | Sphingomy | NA | G | A | 0.029028 | 0.004137 | 6.70E-12 |
| rs582118 | miR-139-5p | trans | 7.44E-42 | Nightingale | S_VLDL_TG | NA | G | A | -0.03812 | 0.004283 | 1.50E-18 |
| rs582118 | miR-139-5p | trans | 7.44E-42 | Nightingale | Total_PL | NA | G | A | 0.024934 | 0.004214 | 1.10E-09 |
| rs582118 | miR-139-5p | trans | 7.44E-42 | Nightingale | S_VLDL_L | NA | G | A | 0.024655 | 0.004382 | 3.80E-09 |
| rs582118 | miR-139-5p | trans | 7.44E-42 | Nightingale | GlycA | NA | G | A | 0.028033 | 0.004406 | 1.40E-10 |
| rs582118 | miR-139-5p | trans | 7.44E-42 | Nightingale | M_LDL_PL | NA | G | A | 0.032449 | 0.004432 | 2.70E-13 |
| rs582118 | miR-139-5p | trans | 7.44E-42 | Nightingale | S_HDL_CE | NA | G | A | 0.028625 | 0.004432 | 1.30E-10 |
| rs582118 | miR-139-5p | trans | 7.44E-42 | Nightingale | S_VLDL_C | NA | G | A | 0.038357 | 0.004425 | 1.80E-18 |
| rs582118 | miR-139-5p | trans | 7.44E-42 | Nightingale | XL_VLDL_F | NA | G | A | 0.037203 | 0.00429 | 2.00E-18 |
| rs582118 | miR-139-5p | trans | 7.44E-42 | Nightingale | LDL_P | NA | G | A | 0.03179 | 0.004422 | 1.00E-12 |
| rs582118 | miR-139-5p | trans | 7.44E-42 | Nightingale | LDL_FC | NA | G | A | 0.03362 | 0.004378 | 5.40E-14 |
| rs582118 | miR-139-5p | trans | 7.44E-42 | Nightingale | M_VLDL_C | NA | G | A | 0.038699 | 0.004413 | 2.00E-18 |
| rs582118 | miR-139-5p | trans | 7.44E-42 | Nightingale | L_LDL_CE | NA | G | A | 0.031559 | 0.004386 | 1.00E-12 |
| rs582118 | miR-139-5p | trans | 7.44E-42 | Nightingale | VLDL_CE | NA | G | A | 0.035773 | 0.004426 | 3.10E-16 |
| rs582118 | miR-139-5p | trans | 7.44E-42 | Nightingale | non_HDL_( | NA | G | A | 0.035684 | 0.004401 | 5.20E-16 |
| rs582118 | miR-139-5p | trans | 7.44E-42 | Nightingale | M_LDL_TG | NA | G | A | -0.02761 | 0.00437 | 3.20E-10 |
| rs582118 | miR-139-5p | trans | 7.44E-42 | Nightingale | LA | NA | G | A | 0.024908 | 0.00434 | 3.70E-09 |
| rs582118 | miR-139-5p | trans | 7.44E-42 | Nightingale | PUFA | NA | G | A | 0.023718 | 0.004316 | 1.40E-08 |
| rs582118 | miR-139-5p | trans | 7.44E-42 | Nightingale | Clinical_LD | NA | G | A | 0.03646 | 0.0044 | 2.80E-16 |
| rs582118 | miR-126-3p | trans | 2.52E-39 | Nightingale | S_VLDL_CE | NA | G | A | 0.04702 | 0.004366 | 1.20E-26 |
| rs582118 | miR-126-3p | trans | 2.52E-39 | Nightingale | S_LDL_P | NA | G | A | 0.029577 | 0.004429 | 3.30E-11 |
| rs582118 | miR-126-3p | trans | 2.52E-39 | Nightingale | M_LDL_C | NA | G | A | 0.033974 | 0.004433 | 2.20E-14 |
| rs582118 | miR-126-3p | trans | 2.52E-39 | Nightingale | S_VLDL_FC | NA | G | A | 0.03789 | 0.004426 | 7.60E-18 |
| rs582118 | miR-126-3p | trans | 2.52E-39 | Nightingale | L_VLDL_C_ | NA | G | A | 0.052199 | 0.004322 | 4.00E-34 |
| rs582118 | miR-126-3p | trans | 2.52E-39 | Nightingale | S_LDL_C_p | NA | G | A | 0.045354 | 0.004442 | 5.70E-25 |
| rs582118 | miR-126-3p | trans | 2.52E-39 | Nightingale | M_LDL_P | NA | G | A | 0.035885 | 0.004438 | 6.90E-16 |
| rs582118 | miR-126-3p | trans | 2.52E-39 | Nightingale | S_LDL_C | NA | G | A | 0.030929 | 0.00443 | 5.60E-12 |
| rs582118 | miR-126-3p | trans | 2.52E-39 | Nightingale | XS_VLDL_F | NA | G | A | 0.037062 | 0.004348 | 6.70E-18 |
| rs582118 | miR-126-3p | trans | 2.52E-39 | Nightingale | XS_VLDL_P | NA | G | A | 0.036694 | 0.004367 | 1.40E-17 |
| rs582118 | miR-126-3p | trans | 2.52E-39 | Nightingale | IDL_C | NA | G | A | 0.034941 | 0.004274 | 9.70E-16 |
| rs582118 | miR-126-3p | trans | 2.52E-39 | Nightingale | VLDL_C | NA | G | A | 0.031288 | 0.004419 | 5.00E-13 |
| rs582118 | miR-126-3p | trans | 2.52E-39 | Nightingale | XXL_VLDL_ | NA | G | A | 0.033117 | 0.004459 | 9.10E-14 |
| rs582118 | miR-126-3p | trans | 2.52E-39 | Nightingale | XL_VLDL_C | NA | G | A | 0.037363 | 0.004245 | 3.10E-18 |
| rs582118 | miR-126-3p | trans | 2.52E-39 | Nightingale | VLDL_P | NA | G | A | 0.026621 | 0.004402 | 4.50E-10 |
| rs582118 | miR-126-3p | trans | 2.52E-39 | Nightingale | S_VLDL_FC | NA | G | A | 0.029356 | 0.004229 | 1.10E-11 |
| rs582118 | miR-126-3p | trans | 2.52E-39 | Nightingale | XXL_VLDL_ | NA | G | A | -0.03572 | 0.004463 | 2.20E-16 |

| | | | | | | | | | | | |
|---|---|---|---|---|---|---|---|---|---|---|---|
| rs582118 | miR-126-3p | trans | 2.52E-39 | Nightingale | L_VLDL_FC | NA | G | A | 0.04857 | 0.004383 | 4.80E-29 |
| rs582118 | miR-126-3p | trans | 2.52E-39 | Nightingale | IDL_FC_pct | NA | G | A | 0.025779 | 0.004391 | 1.10E-08 |
| rs582118 | miR-126-3p | trans | 2.52E-39 | Nightingale | M_VLDL_P | NA | G | A | 0.029113 | 0.004418 | 2.10E-11 |
| rs582118 | miR-126-3p | trans | 2.52E-39 | Nightingale | XL_VLDL_P | NA | G | A | 0.03883 | 0.004432 | 5.30E-19 |
| rs582118 | miR-126-3p | trans | 2.52E-39 | Nightingale | M_LDL_CE | NA | G | A | 0.03296 | 0.004431 | 9.90E-14 |
| rs582118 | miR-126-3p | trans | 2.52E-39 | Nightingale | XS_VLDL_C | NA | G | A | 0.03779 | 0.004271 | 2.10E-18 |
| rs582118 | miR-126-3p | trans | 2.52E-39 | Nightingale | IDL_P | NA | G | A | 0.039958 | 0.004379 | 1.40E-19 |
| rs582118 | miR-126-3p | trans | 2.52E-39 | Nightingale | ApoB_by_A | NA | G | A | 0.025026 | 0.004358 | 1.30E-08 |
| rs582118 | miR-126-3p | trans | 2.52E-39 | Nightingale | XS_VLDL_C | NA | G | A | 0.037986 | 0.004295 | 1.40E-18 |
| rs582118 | miR-126-3p | trans | 2.52E-39 | Nightingale | S_HDL_FC | NA | G | A | 0.023456 | 0.004344 | 4.60E-08 |
| rs582118 | miR-126-3p | trans | 2.52E-39 | Nightingale | M_VLDL_T | NA | G | A | -0.03857 | 0.004182 | 6.70E-20 |
| rs582118 | miR-126-3p | trans | 2.52E-39 | Nightingale | S_VLDL_C_ | NA | G | A | 0.041881 | 0.004307 | 5.30E-22 |
| rs582118 | miR-126-3p | trans | 2.52E-39 | Nightingale | IDL_PL | NA | G | A | 0.035338 | 0.00427 | 3.40E-16 |
| rs582118 | miR-126-3p | trans | 2.52E-39 | Nightingale | M_VLDL_P | NA | G | A | 0.04406 | 0.004247 | 5.60E-25 |
| rs582118 | miR-126-3p | trans | 2.52E-39 | Nightingale | XL_VLDL_C | NA | G | A | 0.038233 | 0.004243 | 4.60E-19 |
| rs582118 | miR-126-3p | trans | 2.52E-39 | Nightingale | L_VLDL_CE | NA | G | A | 0.047351 | 0.004235 | 2.40E-29 |
| rs582118 | miR-126-3p | trans | 2.52E-39 | Nightingale | XS_VLDL_F | NA | G | A | 0.031428 | 0.004394 | 2.50E-12 |
| rs582118 | miR-126-3p | trans | 2.52E-39 | Nightingale | S_LDL_PL_ | NA | G | A | -0.03006 | 0.004417 | 2.70E-13 |
| rs582118 | miR-126-3p | trans | 2.52E-39 | Nightingale | M_VLDL_P | NA | G | A | 0.02828 | 0.004414 | 6.50E-11 |
| rs582118 | miR-126-3p | trans | 2.52E-39 | Nightingale | S_LDL_CE | NA | G | A | 0.030534 | 0.004428 | 6.00E-12 |
| rs582118 | miR-126-3p | trans | 2.52E-39 | Nightingale | S_HDL_C | NA | G | A | 0.028404 | 0.004418 | 1.70E-10 |
| rs582118 | miR-126-3p | trans | 2.52E-39 | Nightingale | M_VLDL_C | NA | G | A | 0.036218 | 0.004168 | 1.20E-17 |
| rs582118 | miR-126-3p | trans | 2.52E-39 | Nightingale | XS_VLDL_L | NA | G | A | 0.033813 | 0.00435 | 2.10E-15 |
| rs582118 | miR-126-3p | trans | 2.52E-39 | Nightingale | M_VLDL_C | NA | G | A | 0.040833 | 0.004385 | 2.40E-20 |
| rs582118 | miR-126-3p | trans | 2.52E-39 | Nightingale | Total_FC | NA | G | A | 0.033252 | 0.004323 | 6.90E-15 |
| rs582118 | miR-126-3p | trans | 2.52E-39 | Nightingale | XS_VLDL_P | NA | G | A | 0.032112 | 0.004364 | 5.50E-14 |
| rs582118 | miR-126-3p | trans | 2.52E-39 | Nightingale | LDL_C | NA | G | A | 0.033183 | 0.004406 | 9.00E-14 |
| rs582118 | miR-126-3p | trans | 2.52E-39 | Nightingale | L_LDL_C | NA | G | A | 0.032311 | 0.004375 | 2.60E-13 |
| rs582118 | miR-126-3p | trans | 2.52E-39 | Nightingale | S_VLDL_PL | NA | G | A | 0.032763 | 0.004417 | 4.30E-14 |
| rs582118 | miR-126-3p | trans | 2.52E-39 | Nightingale | Total_CE | NA | G | A | 0.031745 | 0.004269 | 9.80E-14 |
| rs582118 | miR-126-3p | trans | 2.52E-39 | Nightingale | L_VLDL_TG | NA | G | A | -0.05114 | 0.004404 | 5.60E-32 |
| rs582118 | miR-126-3p | trans | 2.52E-39 | Nightingale | IDL_L | NA | G | A | 0.035258 | 0.004279 | 5.00E-16 |
| rs582118 | miR-126-3p | trans | 2.52E-39 | Nightingale | S_VLDL_CE | NA | G | A | 0.038165 | 0.00442 | 1.70E-18 |
| rs582118 | miR-126-3p | trans | 2.52E-39 | Nightingale | Cholines | NA | G | A | 0.022307 | 0.004162 | 4.80E-08 |
| rs582118 | miR-126-3p | trans | 2.52E-39 | Nightingale | Remnant_( | NA | G | A | 0.036853 | 0.004397 | 3.80E-17 |
| rs582118 | miR-126-3p | trans | 2.52E-39 | Nightingale | Total_L | NA | G | A | 0.026347 | 0.004353 | 4.60E-10 |
| rs582118 | miR-126-3p | trans | 2.52E-39 | Nightingale | M_LDL_C_ | NA | G | A | 0.031166 | 0.004428 | 2.90E-12 |
| rs582118 | miR-126-3p | trans | 2.52E-39 | Nightingale | S_VLDL_PL | NA | G | A | 0.027738 | 0.004237 | 1.80E-10 |
| rs582118 | miR-126-3p | trans | 2.52E-39 | Nightingale | XXL_VLDL_ | NA | G | A | 0.034828 | 0.004435 | 2.90E-15 |
| rs582118 | miR-126-3p | trans | 2.52E-39 | Nightingale | Omega_6 | NA | G | A | 0.025897 | 0.004321 | 5.80E-10 |
| rs582118 | miR-126-3p | trans | 2.52E-39 | Nightingale | ApoB | NA | G | A | 0.033924 | 0.00442 | 2.20E-14 |
| rs582118 | miR-126-3p | trans | 2.52E-39 | Nightingale | L_LDL_FC | NA | G | A | 0.033616 | 0.004345 | 2.90E-14 |
| rs582118 | miR-126-3p | trans | 2.52E-39 | Nightingale | XXL_VLDL_ | NA | G | A | 0.024988 | 0.004454 | 1.50E-09 |
| rs582118 | miR-126-3p | trans | 2.52E-39 | Nightingale | XL_HDL_PL | NA | G | A | -0.02272 | 0.004066 | 1.20E-08 |
| rs582118 | miR-126-3p | trans | 2.52E-39 | Nightingale | S_HDL_L | NA | G | A | 0.026289 | 0.00441 | 1.60E-09 |
| rs582118 | miR-126-3p | trans | 2.52E-39 | Nightingale | LDL_PL | NA | G | A | 0.034973 | 0.004415 | 4.90E-15 |
| rs582118 | miR-126-3p | trans | 2.52E-39 | Nightingale | L_LDL_P | NA | G | A | 0.029166 | 0.00441 | 6.30E-11 |
| rs582118 | miR-126-3p | trans | 2.52E-39 | Nightingale | VLDL_FC | NA | G | A | 0.024145 | 0.004393 | 1.10E-08 |
| rs582118 | miR-126-3p | trans | 2.52E-39 | Nightingale | S_HDL_P | NA | G | A | 0.030602 | 0.004426 | 5.20E-12 |
| rs582118 | miR-126-3p | trans | 2.52E-39 | Nightingale | S_LDL_CE_ | NA | G | A | 0.030939 | 0.004418 | 5.00E-14 |
| rs582118 | miR-126-3p | trans | 2.52E-39 | Nightingale | M_VLDL_F | NA | G | A | 0.033358 | 0.004424 | 3.20E-14 |
| rs582118 | miR-126-3p | trans | 2.52E-39 | Nightingale | XXL_VLDL_ | NA | G | A | 0.030845 | 0.004407 | 1.10E-12 |
| rs582118 | miR-126-3p | trans | 2.52E-39 | Nightingale | IDL_FC | NA | G | A | 0.038246 | 0.004284 | 1.50E-18 |
| rs582118 | miR-126-3p | trans | 2.52E-39 | Nightingale | LDL_CE | NA | G | A | 0.032543 | 0.004414 | 2.20E-13 |
| rs582118 | miR-126-3p | trans | 2.52E-39 | Nightingale | M_VLDL_F | NA | G | A | 0.042075 | 0.00421 | 3.40E-23 |
| rs582118 | miR-126-3p | trans | 2.52E-39 | Nightingale | S_VLDL_P | NA | G | A | 0.023453 | 0.00437 | 1.70E-08 |
| rs582118 | miR-126-3p | trans | 2.52E-39 | Nightingale | Citrate | NA | G | A | 0.026837 | 0.004396 | 7.30E-10 |
| rs582118 | miR-126-3p | trans | 2.52E-39 | Nightingale | LDL_L | NA | G | A | 0.033293 | 0.00441 | 7.60E-14 |
| rs582118 | miR-126-3p | trans | 2.52E-39 | Nightingale | Total_C | NA | G | A | 0.032371 | 0.004284 | 3.40E-14 |
| rs582118 | miR-126-3p | trans | 2.52E-39 | Nightingale | S_LDL_L | NA | G | A | 0.027727 | 0.004424 | 8.40E-10 |
| rs582118 | miR-126-3p | trans | 2.52E-39 | Nightingale | L_LDL_L | NA | G | A | 0.033541 | 0.004382 | 3.10E-14 |
| rs582118 | miR-126-3p | trans | 2.52E-39 | Nightingale | S_HDL_PL | NA | G | A | 0.02378 | 0.004387 | 3.30E-08 |
| rs582118 | miR-126-3p | trans | 2.52E-39 | Nightingale | XL_VLDL_T | NA | G | A | -0.04392 | 0.004289 | 1.80E-24 |

| | | | | | | | | | | | |
|---|---|---|---|---|---|---|---|---|---|---|---|
| rs582118 | miR-126-3p | trans | 2.52E-39 | Nightingale | L_LDL_PL | NA | G | A | 0.037976 | 0.004392 | 9.40E-18 |
| rs582118 | miR-126-3p | trans | 2.52E-39 | Nightingale | M_VLDL_C | NA | G | A | 0.033641 | 0.004157 | 1.90E-15 |
| rs582118 | miR-126-3p | trans | 2.52E-39 | Nightingale | IDL_CE | NA | G | A | 0.033491 | 0.004273 | 1.40E-14 |
| rs582118 | miR-126-3p | trans | 2.52E-39 | Nightingale | M_LDL_L | NA | G | A | 0.033017 | 0.004431 | 1.00E-13 |
| rs582118 | miR-126-3p | trans | 2.52E-39 | Nightingale | S_LDL_FC | NA | G | A | 0.028549 | 0.004418 | 4.40E-10 |
| rs582118 | miR-126-3p | trans | 2.52E-39 | Nightingale | M_LDL_FC | NA | G | A | 0.034333 | 0.00442 | 2.00E-14 |
| rs582118 | miR-126-3p | trans | 2.52E-39 | Nightingale | Sphingomy | NA | G | A | 0.029028 | 0.004137 | 6.70E-12 |
| rs582118 | miR-126-3p | trans | 2.52E-39 | Nightingale | S_VLDL_TG | NA | G | A | -0.03812 | 0.004283 | 1.50E-18 |
| rs582118 | miR-126-3p | trans | 2.52E-39 | Nightingale | Total_PL | NA | G | A | 0.024934 | 0.004214 | 1.10E-09 |
| rs582118 | miR-126-3p | trans | 2.52E-39 | Nightingale | S_VLDL_L | NA | G | A | 0.024655 | 0.004382 | 3.80E-09 |
| rs582118 | miR-126-3p | trans | 2.52E-39 | Nightingale | GlycA | NA | G | A | 0.028033 | 0.004406 | 1.40E-10 |
| rs582118 | miR-126-3p | trans | 2.52E-39 | Nightingale | M_LDL_PL | NA | G | A | 0.032449 | 0.004432 | 2.70E-13 |
| rs582118 | miR-126-3p | trans | 2.52E-39 | Nightingale | S_HDL_CE | NA | G | A | 0.028625 | 0.004432 | 1.30E-10 |
| rs582118 | miR-126-3p | trans | 2.52E-39 | Nightingale | S_VLDL_C | NA | G | A | 0.038357 | 0.004425 | 1.80E-18 |
| rs582118 | miR-126-3p | trans | 2.52E-39 | Nightingale | XL_VLDL_F | NA | G | A | 0.037203 | 0.00429 | 2.00E-18 |
| rs582118 | miR-126-3p | trans | 2.52E-39 | Nightingale | LDL_P | NA | G | A | 0.03179 | 0.004422 | 1.00E-12 |
| rs582118 | miR-126-3p | trans | 2.52E-39 | Nightingale | LDL_FC | NA | G | A | 0.03362 | 0.004378 | 5.40E-14 |
| rs582118 | miR-126-3p | trans | 2.52E-39 | Nightingale | M_VLDL_C | NA | G | A | 0.038699 | 0.004413 | 2.00E-18 |
| rs582118 | miR-126-3p | trans | 2.52E-39 | Nightingale | L_LDL_CE | NA | G | A | 0.031559 | 0.004386 | 1.00E-12 |
| rs582118 | miR-126-3p | trans | 2.52E-39 | Nightingale | VLDL_CE | NA | G | A | 0.035773 | 0.004426 | 3.10E-16 |
| rs582118 | miR-126-3p | trans | 2.52E-39 | Nightingale | non_HDL_C | NA | G | A | 0.035684 | 0.004401 | 5.20E-16 |
| rs582118 | miR-126-3p | trans | 2.52E-39 | Nightingale | M_LDL_TG | NA | G | A | -0.02761 | 0.00437 | 3.20E-10 |
| rs582118 | miR-126-3p | trans | 2.52E-39 | Nightingale | LA | NA | G | A | 0.024908 | 0.00434 | 3.70E-09 |
| rs582118 | miR-126-3p | trans | 2.52E-39 | Nightingale | PUFA | NA | G | A | 0.023718 | 0.004316 | 1.40E-08 |
| rs582118 | miR-126-3p | trans | 2.52E-39 | Nightingale | Clinical_LD | NA | G | A | 0.03646 | 0.0044 | 2.80E-16 |
| rs582118 | let-7b-5p | trans | 9.58E-27 | Nightingale | S_VLDL_CE | NA | G | A | 0.04702 | 0.004366 | 1.20E-26 |
| rs582118 | let-7b-5p | trans | 9.58E-27 | Nightingale | S_LDL_P | NA | G | A | 0.029577 | 0.004429 | 3.30E-11 |
| rs582118 | let-7b-5p | trans | 9.58E-27 | Nightingale | M_LDL_C | NA | G | A | 0.033974 | 0.004433 | 2.20E-14 |
| rs582118 | let-7b-5p | trans | 9.58E-27 | Nightingale | S_VLDL_FC | NA | G | A | 0.03789 | 0.004426 | 7.60E-18 |
| rs582118 | let-7b-5p | trans | 9.58E-27 | Nightingale | L_VLDL_C_ | NA | G | A | 0.052199 | 0.004322 | 4.00E-34 |
| rs582118 | let-7b-5p | trans | 9.58E-27 | Nightingale | S_LDL_C_p | NA | G | A | 0.045354 | 0.004442 | 5.70E-25 |
| rs582118 | let-7b-5p | trans | 9.58E-27 | Nightingale | M_LDL_P | NA | G | A | 0.035885 | 0.004438 | 6.90E-16 |
| rs582118 | let-7b-5p | trans | 9.58E-27 | Nightingale | S_LDL_C | NA | G | A | 0.030929 | 0.00443 | 5.60E-12 |
| rs582118 | let-7b-5p | trans | 9.58E-27 | Nightingale | XS_VLDL_F | NA | G | A | 0.037062 | 0.004348 | 6.70E-18 |
| rs582118 | let-7b-5p | trans | 9.58E-27 | Nightingale | XS_VLDL_P | NA | G | A | 0.036694 | 0.004367 | 1.40E-17 |
| rs582118 | let-7b-5p | trans | 9.58E-27 | Nightingale | IDL_C | NA | G | A | 0.034941 | 0.004274 | 9.70E-16 |
| rs582118 | let-7b-5p | trans | 9.58E-27 | Nightingale | VLDL_C | NA | G | A | 0.031288 | 0.004419 | 5.00E-13 |
| rs582118 | let-7b-5p | trans | 9.58E-27 | Nightingale | XXL_VLDL_ | NA | G | A | 0.033117 | 0.004459 | 9.10E-14 |
| rs582118 | let-7b-5p | trans | 9.58E-27 | Nightingale | XL_VLDL_C | NA | G | A | 0.037363 | 0.004245 | 3.10E-18 |
| rs582118 | let-7b-5p | trans | 9.58E-27 | Nightingale | VLDL_P | NA | G | A | 0.026621 | 0.004402 | 4.50E-10 |
| rs582118 | let-7b-5p | trans | 9.58E-27 | Nightingale | S_VLDL_FC | NA | G | A | 0.029356 | 0.004229 | 1.10E-11 |
| rs582118 | let-7b-5p | trans | 9.58E-27 | Nightingale | XXL_VLDL_ | NA | G | A | -0.03572 | 0.004463 | 2.20E-16 |
| rs582118 | let-7b-5p | trans | 9.58E-27 | Nightingale | L_VLDL_FC | NA | G | A | 0.04857 | 0.004383 | 4.80E-29 |
| rs582118 | let-7b-5p | trans | 9.58E-27 | Nightingale | IDL_FC_pct | NA | G | A | 0.025779 | 0.004391 | 1.10E-08 |
| rs582118 | let-7b-5p | trans | 9.58E-27 | Nightingale | M_VLDL_P | NA | G | A | 0.029113 | 0.004418 | 2.10E-11 |
| rs582118 | let-7b-5p | trans | 9.58E-27 | Nightingale | XL_VLDL_P | NA | G | A | 0.03883 | 0.004432 | 5.30E-19 |
| rs582118 | let-7b-5p | trans | 9.58E-27 | Nightingale | M_LDL_CE | NA | G | A | 0.03296 | 0.004431 | 9.90E-14 |
| rs582118 | let-7b-5p | trans | 9.58E-27 | Nightingale | XS_VLDL_C | NA | G | A | 0.03779 | 0.004271 | 2.10E-18 |
| rs582118 | let-7b-5p | trans | 9.58E-27 | Nightingale | IDL_P | NA | G | A | 0.039958 | 0.004379 | 1.40E-19 |
| rs582118 | let-7b-5p | trans | 9.58E-27 | Nightingale | ApoB_by_A | NA | G | A | 0.025026 | 0.004358 | 1.30E-08 |
| rs582118 | let-7b-5p | trans | 9.58E-27 | Nightingale | XS_VLDL_C | NA | G | A | 0.037986 | 0.004295 | 1.40E-18 |
| rs582118 | let-7b-5p | trans | 9.58E-27 | Nightingale | S_HDL_FC | NA | G | A | 0.023456 | 0.004344 | 4.60E-08 |
| rs582118 | let-7b-5p | trans | 9.58E-27 | Nightingale | M_VLDL_T | NA | G | A | -0.03857 | 0.004182 | 6.70E-20 |
| rs582118 | let-7b-5p | trans | 9.58E-27 | Nightingale | S_VLDL_C_ | NA | G | A | 0.041881 | 0.004307 | 5.30E-22 |
| rs582118 | let-7b-5p | trans | 9.58E-27 | Nightingale | IDL_PL | NA | G | A | 0.035338 | 0.00427 | 3.40E-16 |
| rs582118 | let-7b-5p | trans | 9.58E-27 | Nightingale | M_VLDL_P | NA | G | A | 0.04406 | 0.004247 | 5.60E-25 |
| rs582118 | let-7b-5p | trans | 9.58E-27 | Nightingale | XL_VLDL_C | NA | G | A | 0.038233 | 0.004243 | 4.60E-19 |
| rs582118 | let-7b-5p | trans | 9.58E-27 | Nightingale | L_VLDL_CE | NA | G | A | 0.047351 | 0.004235 | 2.40E-29 |
| rs582118 | let-7b-5p | trans | 9.58E-27 | Nightingale | XS_VLDL_F | NA | G | A | 0.031428 | 0.004394 | 2.50E-12 |
| rs582118 | let-7b-5p | trans | 9.58E-27 | Nightingale | S_LDL_PL_ | NA | G | A | -0.03006 | 0.004417 | 2.70E-13 |
| rs582118 | let-7b-5p | trans | 9.58E-27 | Nightingale | M_VLDL_P | NA | G | A | 0.02828 | 0.004414 | 6.50E-11 |
| rs582118 | let-7b-5p | trans | 9.58E-27 | Nightingale | S_LDL_CE | NA | G | A | 0.030534 | 0.004428 | 6.00E-12 |
| rs582118 | let-7b-5p | trans | 9.58E-27 | Nightingale | S_HDL_C | NA | G | A | 0.028404 | 0.004418 | 1.70E-10 |

| | | | | | | | | | | | |
|---|---|---|---|---|---|---|---|---|---|---|---|
| rs582118 | let-7b-5p | trans | 9.58E-27 | Nightingale | M_VLDL_C | NA | G | A | 0.036218 | 0.004168 | 1.20E-17 |
| rs582118 | let-7b-5p | trans | 9.58E-27 | Nightingale | XS_VLDL_L | NA | G | A | 0.033813 | 0.00435 | 2.10E-15 |
| rs582118 | let-7b-5p | trans | 9.58E-27 | Nightingale | M_VLDL_C | NA | G | A | 0.040833 | 0.004385 | 2.40E-20 |
| rs582118 | let-7b-5p | trans | 9.58E-27 | Nightingale | Total_FC | NA | G | A | 0.033252 | 0.004323 | 6.90E-15 |
| rs582118 | let-7b-5p | trans | 9.58E-27 | Nightingale | XS_VLDL_P | NA | G | A | 0.032112 | 0.004364 | 5.50E-14 |
| rs582118 | let-7b-5p | trans | 9.58E-27 | Nightingale | LDL_C | NA | G | A | 0.033183 | 0.004406 | 9.00E-14 |
| rs582118 | let-7b-5p | trans | 9.58E-27 | Nightingale | L_LDL_C | NA | G | A | 0.032311 | 0.004375 | 2.60E-13 |
| rs582118 | let-7b-5p | trans | 9.58E-27 | Nightingale | S_VLDL_PL | NA | G | A | 0.032763 | 0.004417 | 4.30E-14 |
| rs582118 | let-7b-5p | trans | 9.58E-27 | Nightingale | Total_CE | NA | G | A | 0.031745 | 0.004269 | 9.80E-14 |
| rs582118 | let-7b-5p | trans | 9.58E-27 | Nightingale | L_VLDL_TG | NA | G | A | -0.05114 | 0.004404 | 5.60E-32 |
| rs582118 | let-7b-5p | trans | 9.58E-27 | Nightingale | IDL_L | NA | G | A | 0.035258 | 0.004279 | 5.00E-16 |
| rs582118 | let-7b-5p | trans | 9.58E-27 | Nightingale | S_VLDL_CE | NA | G | A | 0.038165 | 0.00442 | 1.70E-18 |
| rs582118 | let-7b-5p | trans | 9.58E-27 | Nightingale | Cholines | NA | G | A | 0.022307 | 0.004162 | 4.80E-08 |
| rs582118 | let-7b-5p | trans | 9.58E-27 | Nightingale | Remnant_( | NA | G | A | 0.036853 | 0.004397 | 3.80E-17 |
| rs582118 | let-7b-5p | trans | 9.58E-27 | Nightingale | Total_L | NA | G | A | 0.026347 | 0.004353 | 4.60E-10 |
| rs582118 | let-7b-5p | trans | 9.58E-27 | Nightingale | M_LDL_C_ | NA | G | A | 0.031166 | 0.004428 | 2.90E-12 |
| rs582118 | let-7b-5p | trans | 9.58E-27 | Nightingale | S_VLDL_PL | NA | G | A | 0.027738 | 0.004237 | 1.80E-10 |
| rs582118 | let-7b-5p | trans | 9.58E-27 | Nightingale | XXL_VLDL_ | NA | G | A | 0.034828 | 0.004435 | 2.90E-15 |
| rs582118 | let-7b-5p | trans | 9.58E-27 | Nightingale | Omega_6 | NA | G | A | 0.025897 | 0.004321 | 5.80E-10 |
| rs582118 | let-7b-5p | trans | 9.58E-27 | Nightingale | ApoB | NA | G | A | 0.033924 | 0.00442 | 2.20E-14 |
| rs582118 | let-7b-5p | trans | 9.58E-27 | Nightingale | L_LDL_FC | NA | G | A | 0.033616 | 0.004345 | 2.90E-14 |
| rs582118 | let-7b-5p | trans | 9.58E-27 | Nightingale | XXL_VLDL_ | NA | G | A | 0.024988 | 0.004454 | 1.50E-09 |
| rs582118 | let-7b-5p | trans | 9.58E-27 | Nightingale | XL_HDL_PL | NA | G | A | -0.02272 | 0.004066 | 1.20E-08 |
| rs582118 | let-7b-5p | trans | 9.58E-27 | Nightingale | S_HDL_L | NA | G | A | 0.026289 | 0.00441 | 1.60E-09 |
| rs582118 | let-7b-5p | trans | 9.58E-27 | Nightingale | LDL_PL | NA | G | A | 0.034973 | 0.004415 | 4.90E-15 |
| rs582118 | let-7b-5p | trans | 9.58E-27 | Nightingale | L_LDL_P | NA | G | A | 0.029166 | 0.00441 | 6.30E-11 |
| rs582118 | let-7b-5p | trans | 9.58E-27 | Nightingale | VLDL_FC | NA | G | A | 0.024145 | 0.004393 | 1.10E-08 |
| rs582118 | let-7b-5p | trans | 9.58E-27 | Nightingale | S_HDL_P | NA | G | A | 0.030602 | 0.004426 | 5.20E-12 |
| rs582118 | let-7b-5p | trans | 9.58E-27 | Nightingale | S_LDL_CE_ | NA | G | A | 0.030939 | 0.004418 | 5.00E-14 |
| rs582118 | let-7b-5p | trans | 9.58E-27 | Nightingale | M_VLDL_F( | NA | G | A | 0.033358 | 0.004424 | 3.20E-14 |
| rs582118 | let-7b-5p | trans | 9.58E-27 | Nightingale | XXL_VLDL_ | NA | G | A | 0.030845 | 0.004407 | 1.10E-12 |
| rs582118 | let-7b-5p | trans | 9.58E-27 | Nightingale | IDL_FC | NA | G | A | 0.038246 | 0.004284 | 1.50E-18 |
| rs582118 | let-7b-5p | trans | 9.58E-27 | Nightingale | LDL_CE | NA | G | A | 0.032543 | 0.004414 | 2.20E-13 |
| rs582118 | let-7b-5p | trans | 9.58E-27 | Nightingale | M_VLDL_F( | NA | G | A | 0.042075 | 0.00421 | 3.40E-23 |
| rs582118 | let-7b-5p | trans | 9.58E-27 | Nightingale | S_VLDL_P | NA | G | A | 0.023453 | 0.00437 | 1.70E-08 |
| rs582118 | let-7b-5p | trans | 9.58E-27 | Nightingale | Citrate | NA | G | A | 0.026837 | 0.004396 | 7.30E-10 |
| rs582118 | let-7b-5p | trans | 9.58E-27 | Nightingale | LDL_L | NA | G | A | 0.033293 | 0.00441 | 7.60E-14 |
| rs582118 | let-7b-5p | trans | 9.58E-27 | Nightingale | Total_C | NA | G | A | 0.032371 | 0.004284 | 3.40E-14 |
| rs582118 | let-7b-5p | trans | 9.58E-27 | Nightingale | S_LDL_L | NA | G | A | 0.027727 | 0.004424 | 8.40E-10 |
| rs582118 | let-7b-5p | trans | 9.58E-27 | Nightingale | L_LDL_L | NA | G | A | 0.033541 | 0.004382 | 3.10E-14 |
| rs582118 | let-7b-5p | trans | 9.58E-27 | Nightingale | S_HDL_PL | NA | G | A | 0.02378 | 0.004387 | 3.30E-08 |
| rs582118 | let-7b-5p | trans | 9.58E-27 | Nightingale | XL_VLDL_T | NA | G | A | -0.04392 | 0.004289 | 1.80E-24 |
| rs582118 | let-7b-5p | trans | 9.58E-27 | Nightingale | L_LDL_PL | NA | G | A | 0.037976 | 0.004392 | 9.40E-18 |
| rs582118 | let-7b-5p | trans | 9.58E-27 | Nightingale | M_VLDL_C | NA | G | A | 0.033641 | 0.004157 | 1.90E-15 |
| rs582118 | let-7b-5p | trans | 9.58E-27 | Nightingale | IDL_CE | NA | G | A | 0.033491 | 0.004273 | 1.40E-14 |
| rs582118 | let-7b-5p | trans | 9.58E-27 | Nightingale | M_LDL_L | NA | G | A | 0.033017 | 0.004431 | 1.00E-13 |
| rs582118 | let-7b-5p | trans | 9.58E-27 | Nightingale | S_LDL_FC | NA | G | A | 0.028549 | 0.004418 | 4.40E-10 |
| rs582118 | let-7b-5p | trans | 9.58E-27 | Nightingale | M_LDL_FC | NA | G | A | 0.034333 | 0.00442 | 2.00E-14 |
| rs582118 | let-7b-5p | trans | 9.58E-27 | Nightingale | Sphingomy | NA | G | A | 0.029028 | 0.004137 | 6.70E-12 |
| rs582118 | let-7b-5p | trans | 9.58E-27 | Nightingale | S_VLDL_TG | NA | G | A | -0.03812 | 0.004283 | 1.50E-18 |
| rs582118 | let-7b-5p | trans | 9.58E-27 | Nightingale | Total_PL | NA | G | A | 0.024934 | 0.004214 | 1.10E-09 |
| rs582118 | let-7b-5p | trans | 9.58E-27 | Nightingale | S_VLDL_L | NA | G | A | 0.024655 | 0.004382 | 3.80E-09 |
| rs582118 | let-7b-5p | trans | 9.58E-27 | Nightingale | GlycA | NA | G | A | 0.028033 | 0.004406 | 1.40E-10 |
| rs582118 | let-7b-5p | trans | 9.58E-27 | Nightingale | M_LDL_PL | NA | G | A | 0.032449 | 0.004432 | 2.70E-13 |
| rs582118 | let-7b-5p | trans | 9.58E-27 | Nightingale | S_HDL_CE | NA | G | A | 0.028625 | 0.004432 | 1.30E-10 |
| rs582118 | let-7b-5p | trans | 9.58E-27 | Nightingale | S_VLDL_C | NA | G | A | 0.038357 | 0.004425 | 1.80E-18 |
| rs582118 | let-7b-5p | trans | 9.58E-27 | Nightingale | XL_VLDL_F | NA | G | A | 0.037203 | 0.00429 | 2.00E-18 |
| rs582118 | let-7b-5p | trans | 9.58E-27 | Nightingale | LDL_P | NA | G | A | 0.03179 | 0.004422 | 1.00E-12 |
| rs582118 | let-7b-5p | trans | 9.58E-27 | Nightingale | LDL_FC | NA | G | A | 0.03362 | 0.004378 | 5.40E-14 |
| rs582118 | let-7b-5p | trans | 9.58E-27 | Nightingale | M_VLDL_C | NA | G | A | 0.038699 | 0.004413 | 2.00E-18 |
| rs582118 | let-7b-5p | trans | 9.58E-27 | Nightingale | L_LDL_CE | NA | G | A | 0.031559 | 0.004386 | 1.00E-12 |
| rs582118 | let-7b-5p | trans | 9.58E-27 | Nightingale | VLDL_CE | NA | G | A | 0.035773 | 0.004426 | 3.10E-16 |
| rs582118 | let-7b-5p | trans | 9.58E-27 | Nightingale | non_HDL_( | NA | G | A | 0.035684 | 0.004401 | 5.20E-16 |

| | | | | | | | | | | | |
|---|---|---|---|---|---|---|---|---|---|---|---|
| rs582118 | let-7b-5p | trans | 9.58E-27 | Nightingale | M_LDL_TG | NA | G | A | -0.02761 | 0.00437 | 3.20E-10 |
| rs582118 | let-7b-5p | trans | 9.58E-27 | Nightingale | LA | NA | G | A | 0.024908 | 0.00434 | 3.70E-09 |
| rs582118 | let-7b-5p | trans | 9.58E-27 | Nightingale | PUFA | NA | G | A | 0.023718 | 0.004316 | 1.40E-08 |
| rs582118 | let-7b-5p | trans | 9.58E-27 | Nightingale | Clinical_LD | NA | G | A | 0.03646 | 0.0044 | 2.80E-16 |
| rs582118 | miR-195-5p | trans | 2.17E-17 | Nightingale | S_VLDL_CE | NA | G | A | 0.04702 | 0.004366 | 1.20E-26 |
| rs582118 | miR-195-5p | trans | 2.17E-17 | Nightingale | S_LDL_P | NA | G | A | 0.029577 | 0.004429 | 3.30E-11 |
| rs582118 | miR-195-5p | trans | 2.17E-17 | Nightingale | M_LDL_C | NA | G | A | 0.033974 | 0.004433 | 2.20E-14 |
| rs582118 | miR-195-5p | trans | 2.17E-17 | Nightingale | S_VLDL_FC | NA | G | A | 0.03789 | 0.004426 | 7.60E-18 |
| rs582118 | miR-195-5p | trans | 2.17E-17 | Nightingale | L_VLDL_C_ | NA | G | A | 0.052199 | 0.004322 | 4.00E-34 |
| rs582118 | miR-195-5p | trans | 2.17E-17 | Nightingale | S_LDL_C_p | NA | G | A | 0.045354 | 0.004442 | 5.70E-25 |
| rs582118 | miR-195-5p | trans | 2.17E-17 | Nightingale | M_LDL_P | NA | G | A | 0.035885 | 0.004438 | 6.90E-16 |
| rs582118 | miR-195-5p | trans | 2.17E-17 | Nightingale | S_LDL_C | NA | G | A | 0.030929 | 0.00443 | 5.60E-12 |
| rs582118 | miR-195-5p | trans | 2.17E-17 | Nightingale | XS_VLDL_F | NA | G | A | 0.037062 | 0.004348 | 6.70E-18 |
| rs582118 | miR-195-5p | trans | 2.17E-17 | Nightingale | XS_VLDL_P | NA | G | A | 0.036694 | 0.004367 | 1.40E-17 |
| rs582118 | miR-195-5p | trans | 2.17E-17 | Nightingale | IDL_C | NA | G | A | 0.034941 | 0.004274 | 9.70E-16 |
| rs582118 | miR-195-5p | trans | 2.17E-17 | Nightingale | VLDL_C | NA | G | A | 0.031288 | 0.004419 | 5.00E-13 |
| rs582118 | miR-195-5p | trans | 2.17E-17 | Nightingale | XXL_VLDL_ | NA | G | A | 0.033117 | 0.004459 | 9.10E-14 |
| rs582118 | miR-195-5p | trans | 2.17E-17 | Nightingale | XL_VLDL_C | NA | G | A | 0.037363 | 0.004245 | 3.10E-18 |
| rs582118 | miR-195-5p | trans | 2.17E-17 | Nightingale | VLDL_P | NA | G | A | 0.026621 | 0.004402 | 4.50E-10 |
| rs582118 | miR-195-5p | trans | 2.17E-17 | Nightingale | S_VLDL_FC | NA | G | A | 0.029356 | 0.004229 | 1.10E-11 |
| rs582118 | miR-195-5p | trans | 2.17E-17 | Nightingale | XXL_VLDL_ | NA | G | A | -0.03572 | 0.004463 | 2.20E-16 |
| rs582118 | miR-195-5p | trans | 2.17E-17 | Nightingale | L_VLDL_FC | NA | G | A | 0.04857 | 0.004383 | 4.80E-29 |
| rs582118 | miR-195-5p | trans | 2.17E-17 | Nightingale | IDL_FC_pct | NA | G | A | 0.025779 | 0.004391 | 1.10E-08 |
| rs582118 | miR-195-5p | trans | 2.17E-17 | Nightingale | M_VLDL_P | NA | G | A | 0.029113 | 0.004418 | 2.10E-11 |
| rs582118 | miR-195-5p | trans | 2.17E-17 | Nightingale | XL_VLDL_P | NA | G | A | 0.03883 | 0.004432 | 5.30E-19 |
| rs582118 | miR-195-5p | trans | 2.17E-17 | Nightingale | M_LDL_CE | NA | G | A | 0.03296 | 0.004431 | 9.90E-14 |
| rs582118 | miR-195-5p | trans | 2.17E-17 | Nightingale | XS_VLDL_C | NA | G | A | 0.03779 | 0.004271 | 2.10E-18 |
| rs582118 | miR-195-5p | trans | 2.17E-17 | Nightingale | IDL_P | NA | G | A | 0.039958 | 0.004379 | 1.40E-19 |
| rs582118 | miR-195-5p | trans | 2.17E-17 | Nightingale | ApoB_by_A | NA | G | A | 0.025026 | 0.004358 | 1.30E-08 |
| rs582118 | miR-195-5p | trans | 2.17E-17 | Nightingale | XS_VLDL_C | NA | G | A | 0.037986 | 0.004295 | 1.40E-18 |
| rs582118 | miR-195-5p | trans | 2.17E-17 | Nightingale | S_HDL_FC | NA | G | A | 0.023456 | 0.004344 | 4.60E-08 |
| rs582118 | miR-195-5p | trans | 2.17E-17 | Nightingale | M_VLDL_T | NA | G | A | -0.03857 | 0.004182 | 6.70E-20 |
| rs582118 | miR-195-5p | trans | 2.17E-17 | Nightingale | S_VLDL_C_ | NA | G | A | 0.041881 | 0.004307 | 5.30E-22 |
| rs582118 | miR-195-5p | trans | 2.17E-17 | Nightingale | IDL_PL | NA | G | A | 0.035338 | 0.00427 | 3.40E-16 |
| rs582118 | miR-195-5p | trans | 2.17E-17 | Nightingale | M_VLDL_P | NA | G | A | 0.04406 | 0.004247 | 5.60E-25 |
| rs582118 | miR-195-5p | trans | 2.17E-17 | Nightingale | XL_VLDL_C | NA | G | A | 0.038233 | 0.004243 | 4.60E-19 |
| rs582118 | miR-195-5p | trans | 2.17E-17 | Nightingale | L_VLDL_CE | NA | G | A | 0.047351 | 0.004235 | 2.40E-29 |
| rs582118 | miR-195-5p | trans | 2.17E-17 | Nightingale | XS_VLDL_F | NA | G | A | 0.031428 | 0.004394 | 2.50E-12 |
| rs582118 | miR-195-5p | trans | 2.17E-17 | Nightingale | S_LDL_PL_ | NA | G | A | -0.03006 | 0.004417 | 2.70E-13 |
| rs582118 | miR-195-5p | trans | 2.17E-17 | Nightingale | M_VLDL_P | NA | G | A | 0.02828 | 0.004414 | 6.50E-11 |
| rs582118 | miR-195-5p | trans | 2.17E-17 | Nightingale | S_LDL_CE | NA | G | A | 0.030534 | 0.004428 | 6.00E-12 |
| rs582118 | miR-195-5p | trans | 2.17E-17 | Nightingale | S_HDL_C | NA | G | A | 0.028404 | 0.004418 | 1.70E-10 |
| rs582118 | miR-195-5p | trans | 2.17E-17 | Nightingale | M_VLDL_C | NA | G | A | 0.036218 | 0.004168 | 1.20E-17 |
| rs582118 | miR-195-5p | trans | 2.17E-17 | Nightingale | XS_VLDL_L | NA | G | A | 0.033813 | 0.00435 | 2.10E-15 |
| rs582118 | miR-195-5p | trans | 2.17E-17 | Nightingale | M_VLDL_C | NA | G | A | 0.040833 | 0.004385 | 2.40E-20 |
| rs582118 | miR-195-5p | trans | 2.17E-17 | Nightingale | Total_FC | NA | G | A | 0.033252 | 0.004323 | 6.90E-15 |
| rs582118 | miR-195-5p | trans | 2.17E-17 | Nightingale | XS_VLDL_P | NA | G | A | 0.032112 | 0.004364 | 5.50E-14 |
| rs582118 | miR-195-5p | trans | 2.17E-17 | Nightingale | LDL_C | NA | G | A | 0.033183 | 0.004406 | 9.00E-14 |
| rs582118 | miR-195-5p | trans | 2.17E-17 | Nightingale | L_LDL_C | NA | G | A | 0.032311 | 0.004375 | 2.60E-13 |
| rs582118 | miR-195-5p | trans | 2.17E-17 | Nightingale | S_VLDL_PL | NA | G | A | 0.032763 | 0.004417 | 4.30E-14 |
| rs582118 | miR-195-5p | trans | 2.17E-17 | Nightingale | Total_CE | NA | G | A | 0.031745 | 0.004269 | 9.80E-14 |
| rs582118 | miR-195-5p | trans | 2.17E-17 | Nightingale | L_VLDL_TG | NA | G | A | -0.05114 | 0.004404 | 5.60E-32 |
| rs582118 | miR-195-5p | trans | 2.17E-17 | Nightingale | IDL_L | NA | G | A | 0.035258 | 0.004279 | 5.00E-16 |
| rs582118 | miR-195-5p | trans | 2.17E-17 | Nightingale | S_VLDL_CE | NA | G | A | 0.038165 | 0.00442 | 1.70E-18 |
| rs582118 | miR-195-5p | trans | 2.17E-17 | Nightingale | Cholines | NA | G | A | 0.022307 | 0.004162 | 4.80E-08 |
| rs582118 | miR-195-5p | trans | 2.17E-17 | Nightingale | Remnant_( | NA | G | A | 0.036853 | 0.004397 | 3.80E-17 |
| rs582118 | miR-195-5p | trans | 2.17E-17 | Nightingale | Total_L | NA | G | A | 0.026347 | 0.004353 | 4.60E-10 |
| rs582118 | miR-195-5p | trans | 2.17E-17 | Nightingale | M_LDL_C_ | NA | G | A | 0.031166 | 0.004428 | 2.90E-12 |
| rs582118 | miR-195-5p | trans | 2.17E-17 | Nightingale | S_VLDL_PL | NA | G | A | 0.027738 | 0.004237 | 1.80E-10 |
| rs582118 | miR-195-5p | trans | 2.17E-17 | Nightingale | XXL_VLDL_ | NA | G | A | 0.034828 | 0.004435 | 2.90E-15 |
| rs582118 | miR-195-5p | trans | 2.17E-17 | Nightingale | Omega_6 | NA | G | A | 0.025897 | 0.004321 | 5.80E-10 |
| rs582118 | miR-195-5p | trans | 2.17E-17 | Nightingale | ApoB | NA | G | A | 0.033924 | 0.00442 | 2.20E-14 |
| rs582118 | miR-195-5p | trans | 2.17E-17 | Nightingale | L_LDL_FC | NA | G | A | 0.033616 | 0.004345 | 2.90E-14 |

| | | | | | | | | | | | |
|---|---|---|---|---|---|---|---|---|---|---|---|
| rs582118 | miR-195-5p | trans | 2.17E-17 | Nightingale | XXL_VLDL_ | NA | G | A | 0.024988 | 0.004454 | 1.50E-09 |
| rs582118 | miR-195-5p | trans | 2.17E-17 | Nightingale | XL_HDL_PL | NA | G | A | -0.02272 | 0.004066 | 1.20E-08 |
| rs582118 | miR-195-5p | trans | 2.17E-17 | Nightingale | S_HDL_L | NA | G | A | 0.026289 | 0.00441 | 1.60E-09 |
| rs582118 | miR-195-5p | trans | 2.17E-17 | Nightingale | LDL_PL | NA | G | A | 0.034973 | 0.004415 | 4.90E-15 |
| rs582118 | miR-195-5p | trans | 2.17E-17 | Nightingale | L_LDL_P | NA | G | A | 0.029166 | 0.00441 | 6.30E-11 |
| rs582118 | miR-195-5p | trans | 2.17E-17 | Nightingale | VLDL_FC | NA | G | A | 0.024145 | 0.004393 | 1.10E-08 |
| rs582118 | miR-195-5p | trans | 2.17E-17 | Nightingale | S_HDL_P | NA | G | A | 0.030602 | 0.004426 | 5.20E-12 |
| rs582118 | miR-195-5p | trans | 2.17E-17 | Nightingale | S_LDL_CE_ | NA | G | A | 0.030939 | 0.004418 | 5.00E-14 |
| rs582118 | miR-195-5p | trans | 2.17E-17 | Nightingale | M_VLDL_F | NA | G | A | 0.033358 | 0.004424 | 3.20E-14 |
| rs582118 | miR-195-5p | trans | 2.17E-17 | Nightingale | XXL_VLDL_ | NA | G | A | 0.030845 | 0.004407 | 1.10E-12 |
| rs582118 | miR-195-5p | trans | 2.17E-17 | Nightingale | IDL_FC | NA | G | A | 0.038246 | 0.004284 | 1.50E-18 |
| rs582118 | miR-195-5p | trans | 2.17E-17 | Nightingale | LDL_CE | NA | G | A | 0.032543 | 0.004414 | 2.20E-13 |
| rs582118 | miR-195-5p | trans | 2.17E-17 | Nightingale | M_VLDL_F | NA | G | A | 0.042075 | 0.00421 | 3.40E-23 |
| rs582118 | miR-195-5p | trans | 2.17E-17 | Nightingale | S_VLDL_P | NA | G | A | 0.023453 | 0.00437 | 1.70E-08 |
| rs582118 | miR-195-5p | trans | 2.17E-17 | Nightingale | Citrate | NA | G | A | 0.026837 | 0.004396 | 7.30E-10 |
| rs582118 | miR-195-5p | trans | 2.17E-17 | Nightingale | LDL_L | NA | G | A | 0.033293 | 0.00441 | 7.60E-14 |
| rs582118 | miR-195-5p | trans | 2.17E-17 | Nightingale | Total_C | NA | G | A | 0.032371 | 0.004284 | 3.40E-14 |
| rs582118 | miR-195-5p | trans | 2.17E-17 | Nightingale | S_LDL_L | NA | G | A | 0.027727 | 0.004424 | 8.40E-10 |
| rs582118 | miR-195-5p | trans | 2.17E-17 | Nightingale | L_LDL_L | NA | G | A | 0.033541 | 0.004382 | 3.10E-14 |
| rs582118 | miR-195-5p | trans | 2.17E-17 | Nightingale | S_HDL_PL | NA | G | A | 0.02378 | 0.004387 | 3.30E-08 |
| rs582118 | miR-195-5p | trans | 2.17E-17 | Nightingale | XL_VLDL_T | NA | G | A | -0.04392 | 0.004289 | 1.80E-24 |
| rs582118 | miR-195-5p | trans | 2.17E-17 | Nightingale | L_LDL_PL | NA | G | A | 0.037976 | 0.004392 | 9.40E-18 |
| rs582118 | miR-195-5p | trans | 2.17E-17 | Nightingale | M_VLDL_C | NA | G | A | 0.033641 | 0.004157 | 1.90E-15 |
| rs582118 | miR-195-5p | trans | 2.17E-17 | Nightingale | IDL_CE | NA | G | A | 0.033491 | 0.004273 | 1.40E-14 |
| rs582118 | miR-195-5p | trans | 2.17E-17 | Nightingale | M_LDL_L | NA | G | A | 0.033017 | 0.004431 | 1.00E-13 |
| rs582118 | miR-195-5p | trans | 2.17E-17 | Nightingale | S_LDL_FC | NA | G | A | 0.028549 | 0.004418 | 4.40E-10 |
| rs582118 | miR-195-5p | trans | 2.17E-17 | Nightingale | M_LDL_FC | NA | G | A | 0.034333 | 0.00442 | 2.00E-14 |
| rs582118 | miR-195-5p | trans | 2.17E-17 | Nightingale | Sphingomy | NA | G | A | 0.029028 | 0.004137 | 6.70E-12 |
| rs582118 | miR-195-5p | trans | 2.17E-17 | Nightingale | S_VLDL_TG | NA | G | A | -0.03812 | 0.004283 | 1.50E-18 |
| rs582118 | miR-195-5p | trans | 2.17E-17 | Nightingale | Total_PL | NA | G | A | 0.024934 | 0.004214 | 1.10E-09 |
| rs582118 | miR-195-5p | trans | 2.17E-17 | Nightingale | S_VLDL_L | NA | G | A | 0.024655 | 0.004382 | 3.80E-09 |
| rs582118 | miR-195-5p | trans | 2.17E-17 | Nightingale | GlycA | NA | G | A | 0.028033 | 0.004406 | 1.40E-10 |
| rs582118 | miR-195-5p | trans | 2.17E-17 | Nightingale | M_LDL_PL | NA | G | A | 0.032449 | 0.004432 | 2.70E-13 |
| rs582118 | miR-195-5p | trans | 2.17E-17 | Nightingale | S_HDL_CE | NA | G | A | 0.028625 | 0.004432 | 1.30E-10 |
| rs582118 | miR-195-5p | trans | 2.17E-17 | Nightingale | S_VLDL_C | NA | G | A | 0.038357 | 0.004425 | 1.80E-18 |
| rs582118 | miR-195-5p | trans | 2.17E-17 | Nightingale | XL_VLDL_F | NA | G | A | 0.037203 | 0.00429 | 2.00E-18 |
| rs582118 | miR-195-5p | trans | 2.17E-17 | Nightingale | LDL_P | NA | G | A | 0.03179 | 0.004422 | 1.00E-12 |
| rs582118 | miR-195-5p | trans | 2.17E-17 | Nightingale | LDL_FC | NA | G | A | 0.03362 | 0.004378 | 5.40E-14 |
| rs582118 | miR-195-5p | trans | 2.17E-17 | Nightingale | M_VLDL_C | NA | G | A | 0.038699 | 0.004413 | 2.00E-18 |
| rs582118 | miR-195-5p | trans | 2.17E-17 | Nightingale | L_LDL_CE | NA | G | A | 0.031559 | 0.004386 | 1.00E-12 |
| rs582118 | miR-195-5p | trans | 2.17E-17 | Nightingale | VLDL_CE | NA | G | A | 0.035773 | 0.004426 | 3.10E-16 |
| rs582118 | miR-195-5p | trans | 2.17E-17 | Nightingale | non_HDL_( | NA | G | A | 0.035684 | 0.004401 | 5.20E-16 |
| rs582118 | miR-195-5p | trans | 2.17E-17 | Nightingale | M_LDL_TG | NA | G | A | -0.02761 | 0.00437 | 3.20E-10 |
| rs582118 | miR-195-5p | trans | 2.17E-17 | Nightingale | LA | NA | G | A | 0.024908 | 0.00434 | 3.70E-09 |
| rs582118 | miR-195-5p | trans | 2.17E-17 | Nightingale | PUFA | NA | G | A | 0.023718 | 0.004316 | 1.40E-08 |
| rs582118 | miR-195-5p | trans | 2.17E-17 | Nightingale | Clinical_LD | NA | G | A | 0.03646 | 0.0044 | 2.80E-16 |
| rs582118 | let-7c-5p | trans | 2.66E-23 | Nightingale | S_VLDL_CE | NA | G | A | 0.04702 | 0.004366 | 1.20E-26 |
| rs582118 | let-7c-5p | trans | 2.66E-23 | Nightingale | S_LDL_P | NA | G | A | 0.029577 | 0.004429 | 3.30E-11 |
| rs582118 | let-7c-5p | trans | 2.66E-23 | Nightingale | M_LDL_C | NA | G | A | 0.033974 | 0.004433 | 2.20E-14 |
| rs582118 | let-7c-5p | trans | 2.66E-23 | Nightingale | S_VLDL_FC | NA | G | A | 0.03789 | 0.004426 | 7.60E-18 |
| rs582118 | let-7c-5p | trans | 2.66E-23 | Nightingale | L_VLDL_C_ | NA | G | A | 0.052199 | 0.004322 | 4.00E-34 |
| rs582118 | let-7c-5p | trans | 2.66E-23 | Nightingale | S_LDL_C_p | NA | G | A | 0.045354 | 0.004442 | 5.70E-25 |
| rs582118 | let-7c-5p | trans | 2.66E-23 | Nightingale | M_LDL_P | NA | G | A | 0.035885 | 0.004438 | 6.90E-16 |
| rs582118 | let-7c-5p | trans | 2.66E-23 | Nightingale | S_LDL_C | NA | G | A | 0.030929 | 0.00443 | 5.60E-12 |
| rs582118 | let-7c-5p | trans | 2.66E-23 | Nightingale | XS_VLDL_F | NA | G | A | 0.037062 | 0.004348 | 6.70E-18 |
| rs582118 | let-7c-5p | trans | 2.66E-23 | Nightingale | XS_VLDL_P | NA | G | A | 0.036694 | 0.004367 | 1.40E-17 |
| rs582118 | let-7c-5p | trans | 2.66E-23 | Nightingale | IDL_C | NA | G | A | 0.034941 | 0.004274 | 9.70E-16 |
| rs582118 | let-7c-5p | trans | 2.66E-23 | Nightingale | VLDL_C | NA | G | A | 0.031288 | 0.004419 | 5.00E-13 |
| rs582118 | let-7c-5p | trans | 2.66E-23 | Nightingale | XXL_VLDL_ | NA | G | A | 0.033117 | 0.004459 | 9.10E-14 |
| rs582118 | let-7c-5p | trans | 2.66E-23 | Nightingale | XL_VLDL_C | NA | G | A | 0.037363 | 0.004245 | 3.10E-18 |
| rs582118 | let-7c-5p | trans | 2.66E-23 | Nightingale | VLDL_P | NA | G | A | 0.026621 | 0.004402 | 4.50E-10 |
| rs582118 | let-7c-5p | trans | 2.66E-23 | Nightingale | S_VLDL_FC | NA | G | A | 0.029356 | 0.004229 | 1.10E-11 |
| rs582118 | let-7c-5p | trans | 2.66E-23 | Nightingale | XXL_VLDL_ | NA | G | A | -0.03572 | 0.004463 | 2.20E-16 |

| | | | | | | | | | | | |
|---|---|---|---|---|---|---|---|---|---|---|---|
| rs582118 | let-7c-5p | trans | 2.66E-23 | Nightingale | L_VLDL_FC | NA | G | A | 0.04857 | 0.004383 | 4.80E-29 |
| rs582118 | let-7c-5p | trans | 2.66E-23 | Nightingale | IDL_FC_pct | NA | G | A | 0.025779 | 0.004391 | 1.10E-08 |
| rs582118 | let-7c-5p | trans | 2.66E-23 | Nightingale | M_VLDL_P | NA | G | A | 0.029113 | 0.004418 | 2.10E-11 |
| rs582118 | let-7c-5p | trans | 2.66E-23 | Nightingale | XL_VLDL_P | NA | G | A | 0.03883 | 0.004432 | 5.30E-19 |
| rs582118 | let-7c-5p | trans | 2.66E-23 | Nightingale | M_LDL_CE | NA | G | A | 0.03296 | 0.004431 | 9.90E-14 |
| rs582118 | let-7c-5p | trans | 2.66E-23 | Nightingale | XS_VLDL_C | NA | G | A | 0.03779 | 0.004271 | 2.10E-18 |
| rs582118 | let-7c-5p | trans | 2.66E-23 | Nightingale | IDL_P | NA | G | A | 0.039958 | 0.004379 | 1.40E-19 |
| rs582118 | let-7c-5p | trans | 2.66E-23 | Nightingale | ApoB_by_A | NA | G | A | 0.025026 | 0.004358 | 1.30E-08 |
| rs582118 | let-7c-5p | trans | 2.66E-23 | Nightingale | XS_VLDL_C | NA | G | A | 0.037986 | 0.004295 | 1.40E-18 |
| rs582118 | let-7c-5p | trans | 2.66E-23 | Nightingale | S_HDL_FC | NA | G | A | 0.023456 | 0.004344 | 4.60E-08 |
| rs582118 | let-7c-5p | trans | 2.66E-23 | Nightingale | M_VLDL_T | NA | G | A | -0.03857 | 0.004182 | 6.70E-20 |
| rs582118 | let-7c-5p | trans | 2.66E-23 | Nightingale | S_VLDL_C_ | NA | G | A | 0.041881 | 0.004307 | 5.30E-22 |
| rs582118 | let-7c-5p | trans | 2.66E-23 | Nightingale | IDL_PL | NA | G | A | 0.035338 | 0.00427 | 3.40E-16 |
| rs582118 | let-7c-5p | trans | 2.66E-23 | Nightingale | M_VLDL_P | NA | G | A | 0.04406 | 0.004247 | 5.60E-25 |
| rs582118 | let-7c-5p | trans | 2.66E-23 | Nightingale | XL_VLDL_C | NA | G | A | 0.038233 | 0.004243 | 4.60E-19 |
| rs582118 | let-7c-5p | trans | 2.66E-23 | Nightingale | L_VLDL_CE | NA | G | A | 0.047351 | 0.004235 | 2.40E-29 |
| rs582118 | let-7c-5p | trans | 2.66E-23 | Nightingale | XS_VLDL_F | NA | G | A | 0.031428 | 0.004394 | 2.50E-12 |
| rs582118 | let-7c-5p | trans | 2.66E-23 | Nightingale | S_LDL_PL_ | NA | G | A | -0.03006 | 0.004417 | 2.70E-13 |
| rs582118 | let-7c-5p | trans | 2.66E-23 | Nightingale | M_VLDL_P | NA | G | A | 0.02828 | 0.004414 | 6.50E-11 |
| rs582118 | let-7c-5p | trans | 2.66E-23 | Nightingale | S_LDL_CE | NA | G | A | 0.030534 | 0.004428 | 6.00E-12 |
| rs582118 | let-7c-5p | trans | 2.66E-23 | Nightingale | S_HDL_C | NA | G | A | 0.028404 | 0.004418 | 1.70E-10 |
| rs582118 | let-7c-5p | trans | 2.66E-23 | Nightingale | M_VLDL_C | NA | G | A | 0.036218 | 0.004168 | 1.20E-17 |
| rs582118 | let-7c-5p | trans | 2.66E-23 | Nightingale | XS_VLDL_L | NA | G | A | 0.033813 | 0.00435 | 2.10E-15 |
| rs582118 | let-7c-5p | trans | 2.66E-23 | Nightingale | M_VLDL_C | NA | G | A | 0.040833 | 0.004385 | 2.40E-20 |
| rs582118 | let-7c-5p | trans | 2.66E-23 | Nightingale | Total_FC | NA | G | A | 0.033252 | 0.004323 | 6.90E-15 |
| rs582118 | let-7c-5p | trans | 2.66E-23 | Nightingale | XS_VLDL_P | NA | G | A | 0.032112 | 0.004364 | 5.50E-14 |
| rs582118 | let-7c-5p | trans | 2.66E-23 | Nightingale | LDL_C | NA | G | A | 0.033183 | 0.004406 | 9.00E-14 |
| rs582118 | let-7c-5p | trans | 2.66E-23 | Nightingale | L_LDL_C | NA | G | A | 0.032311 | 0.004375 | 2.60E-13 |
| rs582118 | let-7c-5p | trans | 2.66E-23 | Nightingale | S_VLDL_PL | NA | G | A | 0.032763 | 0.004417 | 4.30E-14 |
| rs582118 | let-7c-5p | trans | 2.66E-23 | Nightingale | Total_CE | NA | G | A | 0.031745 | 0.004269 | 9.80E-14 |
| rs582118 | let-7c-5p | trans | 2.66E-23 | Nightingale | L_VLDL_TG | NA | G | A | -0.05114 | 0.004404 | 5.60E-32 |
| rs582118 | let-7c-5p | trans | 2.66E-23 | Nightingale | IDL_L | NA | G | A | 0.035258 | 0.004279 | 5.00E-16 |
| rs582118 | let-7c-5p | trans | 2.66E-23 | Nightingale | S_VLDL_CE | NA | G | A | 0.038165 | 0.00442 | 1.70E-18 |
| rs582118 | let-7c-5p | trans | 2.66E-23 | Nightingale | Cholines | NA | G | A | 0.022307 | 0.004162 | 4.80E-08 |
| rs582118 | let-7c-5p | trans | 2.66E-23 | Nightingale | Remnant_( | NA | G | A | 0.036853 | 0.004397 | 3.80E-17 |
| rs582118 | let-7c-5p | trans | 2.66E-23 | Nightingale | Total_L | NA | G | A | 0.026347 | 0.004353 | 4.60E-10 |
| rs582118 | let-7c-5p | trans | 2.66E-23 | Nightingale | M_LDL_C_ | NA | G | A | 0.031166 | 0.004428 | 2.90E-12 |
| rs582118 | let-7c-5p | trans | 2.66E-23 | Nightingale | S_VLDL_PL | NA | G | A | 0.027738 | 0.004237 | 1.80E-10 |
| rs582118 | let-7c-5p | trans | 2.66E-23 | Nightingale | XXL_VLDL_ | NA | G | A | 0.034828 | 0.004435 | 2.90E-15 |
| rs582118 | let-7c-5p | trans | 2.66E-23 | Nightingale | Omega_6 | NA | G | A | 0.025897 | 0.004321 | 5.80E-10 |
| rs582118 | let-7c-5p | trans | 2.66E-23 | Nightingale | ApoB | NA | G | A | 0.033924 | 0.00442 | 2.20E-14 |
| rs582118 | let-7c-5p | trans | 2.66E-23 | Nightingale | L_LDL_FC | NA | G | A | 0.033616 | 0.004345 | 2.90E-14 |
| rs582118 | let-7c-5p | trans | 2.66E-23 | Nightingale | XXL_VLDL_ | NA | G | A | 0.024988 | 0.004454 | 1.50E-09 |
| rs582118 | let-7c-5p | trans | 2.66E-23 | Nightingale | XL_HDL_PL | NA | G | A | -0.02272 | 0.004066 | 1.20E-08 |
| rs582118 | let-7c-5p | trans | 2.66E-23 | Nightingale | S_HDL_L | NA | G | A | 0.026289 | 0.00441 | 1.60E-09 |
| rs582118 | let-7c-5p | trans | 2.66E-23 | Nightingale | LDL_PL | NA | G | A | 0.034973 | 0.004415 | 4.90E-15 |
| rs582118 | let-7c-5p | trans | 2.66E-23 | Nightingale | L_LDL_P | NA | G | A | 0.029166 | 0.00441 | 6.30E-11 |
| rs582118 | let-7c-5p | trans | 2.66E-23 | Nightingale | VLDL_FC | NA | G | A | 0.024145 | 0.004393 | 1.10E-08 |
| rs582118 | let-7c-5p | trans | 2.66E-23 | Nightingale | S_HDL_P | NA | G | A | 0.030602 | 0.004426 | 5.20E-12 |
| rs582118 | let-7c-5p | trans | 2.66E-23 | Nightingale | S_LDL_CE_ | NA | G | A | 0.030939 | 0.004418 | 5.00E-14 |
| rs582118 | let-7c-5p | trans | 2.66E-23 | Nightingale | M_VLDL_F | NA | G | A | 0.033358 | 0.004424 | 3.20E-14 |
| rs582118 | let-7c-5p | trans | 2.66E-23 | Nightingale | XXL_VLDL_ | NA | G | A | 0.030845 | 0.004407 | 1.10E-12 |
| rs582118 | let-7c-5p | trans | 2.66E-23 | Nightingale | IDL_FC | NA | G | A | 0.038246 | 0.004284 | 1.50E-18 |
| rs582118 | let-7c-5p | trans | 2.66E-23 | Nightingale | LDL_CE | NA | G | A | 0.032543 | 0.004414 | 2.20E-13 |
| rs582118 | let-7c-5p | trans | 2.66E-23 | Nightingale | M_VLDL_F | NA | G | A | 0.042075 | 0.00421 | 3.40E-23 |
| rs582118 | let-7c-5p | trans | 2.66E-23 | Nightingale | S_VLDL_P | NA | G | A | 0.023453 | 0.00437 | 1.70E-08 |
| rs582118 | let-7c-5p | trans | 2.66E-23 | Nightingale | Citrate | NA | G | A | 0.026837 | 0.004396 | 7.30E-10 |
| rs582118 | let-7c-5p | trans | 2.66E-23 | Nightingale | LDL_L | NA | G | A | 0.033293 | 0.00441 | 7.60E-14 |
| rs582118 | let-7c-5p | trans | 2.66E-23 | Nightingale | Total_C | NA | G | A | 0.032371 | 0.004284 | 3.40E-14 |
| rs582118 | let-7c-5p | trans | 2.66E-23 | Nightingale | S_LDL_L | NA | G | A | 0.027727 | 0.004424 | 8.40E-10 |
| rs582118 | let-7c-5p | trans | 2.66E-23 | Nightingale | L_LDL_L | NA | G | A | 0.033541 | 0.004382 | 3.10E-14 |
| rs582118 | let-7c-5p | trans | 2.66E-23 | Nightingale | S_HDL_PL | NA | G | A | 0.02378 | 0.004387 | 3.30E-08 |
| rs582118 | let-7c-5p | trans | 2.66E-23 | Nightingale | XL_VLDL_T | NA | G | A | -0.04392 | 0.004289 | 1.80E-24 |

| | | | | | | | | | | | |
|---|---|---|---|---|---|---|---|---|---|---|---|
| rs582118 | let-7c-5p | trans | 2.66E-23 | Nightingale | L_LDL_PL | NA | G | A | 0.037976 | 0.004392 | 9.40E-18 |
| rs582118 | let-7c-5p | trans | 2.66E-23 | Nightingale | M_VLDL_C | NA | G | A | 0.033641 | 0.004157 | 1.90E-15 |
| rs582118 | let-7c-5p | trans | 2.66E-23 | Nightingale | IDL_CE | NA | G | A | 0.033491 | 0.004273 | 1.40E-14 |
| rs582118 | let-7c-5p | trans | 2.66E-23 | Nightingale | M_LDL_L | NA | G | A | 0.033017 | 0.004431 | 1.00E-13 |
| rs582118 | let-7c-5p | trans | 2.66E-23 | Nightingale | S_LDL_FC | NA | G | A | 0.028549 | 0.004418 | 4.40E-10 |
| rs582118 | let-7c-5p | trans | 2.66E-23 | Nightingale | M_LDL_FC | NA | G | A | 0.034333 | 0.00442 | 2.00E-14 |
| rs582118 | let-7c-5p | trans | 2.66E-23 | Nightingale | Sphingomy | NA | G | A | 0.029028 | 0.004137 | 6.70E-12 |
| rs582118 | let-7c-5p | trans | 2.66E-23 | Nightingale | S_VLDL_TG | NA | G | A | -0.03812 | 0.004283 | 1.50E-18 |
| rs582118 | let-7c-5p | trans | 2.66E-23 | Nightingale | Total_PL | NA | G | A | 0.024934 | 0.004214 | 1.10E-09 |
| rs582118 | let-7c-5p | trans | 2.66E-23 | Nightingale | S_VLDL_L | NA | G | A | 0.024655 | 0.004382 | 3.80E-09 |
| rs582118 | let-7c-5p | trans | 2.66E-23 | Nightingale | GlycA | NA | G | A | 0.028033 | 0.004406 | 1.40E-10 |
| rs582118 | let-7c-5p | trans | 2.66E-23 | Nightingale | M_LDL_PL | NA | G | A | 0.032449 | 0.004432 | 2.70E-13 |
| rs582118 | let-7c-5p | trans | 2.66E-23 | Nightingale | S_HDL_CE | NA | G | A | 0.028625 | 0.004432 | 1.30E-10 |
| rs582118 | let-7c-5p | trans | 2.66E-23 | Nightingale | S_VLDL_C | NA | G | A | 0.038357 | 0.004425 | 1.80E-18 |
| rs582118 | let-7c-5p | trans | 2.66E-23 | Nightingale | XL_VLDL_F | NA | G | A | 0.037203 | 0.00429 | 2.00E-18 |
| rs582118 | let-7c-5p | trans | 2.66E-23 | Nightingale | LDL_P | NA | G | A | 0.03179 | 0.004422 | 1.00E-12 |
| rs582118 | let-7c-5p | trans | 2.66E-23 | Nightingale | LDL_FC | NA | G | A | 0.03362 | 0.004378 | 5.40E-14 |
| rs582118 | let-7c-5p | trans | 2.66E-23 | Nightingale | M_VLDL_C | NA | G | A | 0.038699 | 0.004413 | 2.00E-18 |
| rs582118 | let-7c-5p | trans | 2.66E-23 | Nightingale | L_LDL_CE | NA | G | A | 0.031559 | 0.004386 | 1.00E-12 |
| rs582118 | let-7c-5p | trans | 2.66E-23 | Nightingale | VLDL_CE | NA | G | A | 0.035773 | 0.004426 | 3.10E-16 |
| rs582118 | let-7c-5p | trans | 2.66E-23 | Nightingale | non_HDL_( | NA | G | A | 0.035684 | 0.004401 | 5.20E-16 |
| rs582118 | let-7c-5p | trans | 2.66E-23 | Nightingale | M_LDL_TG | NA | G | A | -0.02761 | 0.00437 | 3.20E-10 |
| rs582118 | let-7c-5p | trans | 2.66E-23 | Nightingale | LA | NA | G | A | 0.024908 | 0.00434 | 3.70E-09 |
| rs582118 | let-7c-5p | trans | 2.66E-23 | Nightingale | PUFA | NA | G | A | 0.023718 | 0.004316 | 1.40E-08 |
| rs582118 | let-7c-5p | trans | 2.66E-23 | Nightingale | Clinical_LD | NA | G | A | 0.03646 | 0.0044 | 2.80E-16 |
| rs600038 | let-7a-5p | trans | 1.48E-12 | Nightingale | XL_VLDL_C | NA | C | T | 0.054368 | 0.004895 | 2.80E-28 |
| rs600038 | let-7a-5p | trans | 1.48E-12 | Nightingale | M_VLDL_C | NA | C | T | 0.049278 | 0.004794 | 2.70E-24 |
| rs600038 | let-7a-5p | trans | 1.48E-12 | Nightingale | Total_CE | NA | C | T | 0.049009 | 0.004922 | 2.10E-24 |
| rs600038 | let-7a-5p | trans | 1.48E-12 | Nightingale | L_LDL_L | NA | C | T | 0.054273 | 0.005053 | 1.00E-27 |
| rs600038 | let-7a-5p | trans | 1.48E-12 | Nightingale | L_LDL_CE | NA | C | T | 0.051898 | 0.005057 | 2.00E-25 |
| rs600038 | let-7a-5p | trans | 1.48E-12 | Nightingale | L_LDL_C | NA | C | T | 0.053255 | 0.005045 | 1.10E-26 |
| rs600038 | let-7a-5p | trans | 1.48E-12 | Nightingale | Phosphogly | NA | C | T | 0.027754 | 0.004833 | 1.30E-09 |
| rs600038 | let-7a-5p | trans | 1.48E-12 | Nightingale | IDL_FC | NA | C | T | 0.057015 | 0.00494 | 1.50E-31 |
| rs600038 | let-7a-5p | trans | 1.48E-12 | Nightingale | S_VLDL_C | NA | C | T | 0.05403 | 0.005102 | 5.10E-28 |
| rs600038 | let-7a-5p | trans | 1.48E-12 | Nightingale | Total_C | NA | C | T | 0.049326 | 0.00494 | 1.10E-24 |
| rs600038 | let-7a-5p | trans | 1.48E-12 | Nightingale | L_VLDL_PL | NA | C | T | 0.027472 | 0.004957 | 1.00E-09 |
| rs600038 | let-7a-5p | trans | 1.48E-12 | Nightingale | LDL_P | NA | C | T | 0.048287 | 0.005099 | 3.30E-22 |
| rs600038 | let-7a-5p | trans | 1.48E-12 | Nightingale | M_VLDL_C | NA | C | T | 0.055912 | 0.005089 | 1.20E-29 |
| rs600038 | let-7a-5p | trans | 1.48E-12 | Nightingale | M_LDL_PL | NA | C | T | 0.052654 | 0.00511 | 1.50E-25 |
| rs600038 | let-7a-5p | trans | 1.48E-12 | Nightingale | LA | NA | C | T | 0.037469 | 0.005004 | 3.30E-15 |
| rs600038 | let-7a-5p | trans | 1.48E-12 | Nightingale | Citrate | NA | C | T | 0.02733 | 0.00507 | 2.60E-08 |
| rs600038 | let-7a-5p | trans | 1.48E-12 | Nightingale | Pyruvate | NA | C | T | 0.030176 | 0.005111 | 3.60E-09 |
| rs600038 | let-7a-5p | trans | 1.48E-12 | Nightingale | VLDL_P | NA | C | T | 0.03602 | 0.005075 | 4.70E-14 |
| rs600038 | let-7a-5p | trans | 1.48E-12 | Nightingale | Sphingomy | NA | C | T | 0.040531 | 0.00477 | 1.70E-17 |
| rs600038 | let-7a-5p | trans | 1.48E-12 | Nightingale | S_LDL_FC | NA | C | T | 0.049076 | 0.005095 | 7.70E-22 |
| rs600038 | let-7a-5p | trans | 1.48E-12 | Nightingale | LDL_FC | NA | C | T | 0.055801 | 0.005048 | 1.60E-28 |
| rs600038 | let-7a-5p | trans | 1.48E-12 | Nightingale | XS_VLDL_C | NA | C | T | 0.052331 | 0.004953 | 3.80E-28 |
| rs600038 | let-7a-5p | trans | 1.48E-12 | Nightingale | XS_VLDL_P | NA | C | T | 0.041555 | 0.005032 | 8.10E-19 |
| rs600038 | let-7a-5p | trans | 1.48E-12 | Nightingale | L_LDL_FC | NA | C | T | 0.055661 | 0.005011 | 5.90E-29 |
| rs600038 | let-7a-5p | trans | 1.48E-12 | Nightingale | XXL_VLDL_ | NA | C | T | -0.05276 | 0.005149 | 1.90E-25 |
| rs600038 | let-7a-5p | trans | 1.48E-12 | Nightingale | XS_VLDL_C | NA | C | T | 0.052917 | 0.004924 | 7.70E-29 |
| rs600038 | let-7a-5p | trans | 1.48E-12 | Nightingale | S_VLDL_FC | NA | C | T | 0.045985 | 0.004876 | 8.40E-21 |
| rs600038 | let-7a-5p | trans | 1.48E-12 | Nightingale | S_LDL_CE_ | NA | C | T | 0.039177 | 0.005094 | 1.30E-16 |
| rs600038 | let-7a-5p | trans | 1.48E-12 | Nightingale | S_VLDL_C_ | NA | C | T | 0.062934 | 0.004966 | 6.40E-37 |
| rs600038 | let-7a-5p | trans | 1.48E-12 | Nightingale | L_VLDL_CE | NA | C | T | 0.065536 | 0.004883 | 4.90E-42 |
| rs600038 | let-7a-5p | trans | 1.48E-12 | Nightingale | S_VLDL_PL | NA | C | T | 0.043922 | 0.004886 | 5.30E-19 |
| rs600038 | let-7a-5p | trans | 1.48E-12 | Nightingale | XS_VLDL_F | NA | C | T | 0.044491 | 0.005066 | 8.00E-18 |
| rs600038 | let-7a-5p | trans | 1.48E-12 | Nightingale | XL_VLDL_C | NA | C | T | 0.054864 | 0.004893 | 6.90E-29 |
| rs600038 | let-7a-5p | trans | 1.48E-12 | Nightingale | S_LDL_P | NA | C | T | 0.044606 | 0.005107 | 3.00E-19 |
| rs600038 | let-7a-5p | trans | 1.48E-12 | Nightingale | L_LDL_TG_ | NA | C | T | -0.03537 | 0.005005 | 5.10E-12 |
| rs600038 | let-7a-5p | trans | 1.48E-12 | Nightingale | M_VLDL_P | NA | C | T | 0.041549 | 0.005095 | 1.40E-17 |
| rs600038 | let-7a-5p | trans | 1.48E-12 | Nightingale | L_HDL_TG_ | NA | C | T | 0.023502 | 0.004821 | 3.50E-08 |
| rs600038 | let-7a-5p | trans | 1.48E-12 | Nightingale | LDL_C | NA | C | T | 0.053489 | 0.005081 | 1.60E-26 |

| | | | | | | | | | | | |
|---|---|---|---|---|---|---|---|---|---|---|---|
| rs600038 | let-7a-5p | trans | 1.48E-12 | Nightingale | S_VLDL_L | NA | C | T | 0.033361 | 0.005053 | 1.40E-12 |
| rs600038 | let-7a-5p | trans | 1.48E-12 | Nightingale | LDL_L | NA | C | T | 0.053245 | 0.005086 | 2.30E-26 |
| rs600038 | let-7a-5p | trans | 1.48E-12 | Nightingale | M_VLDL_F | NA | C | T | 0.047425 | 0.005102 | 6.00E-22 |
| rs600038 | let-7a-5p | trans | 1.48E-12 | Nightingale | IDL_C | NA | C | T | 0.054083 | 0.004928 | 1.40E-28 |
| rs600038 | let-7a-5p | trans | 1.48E-12 | Nightingale | IDL_L | NA | C | T | 0.053693 | 0.004934 | 1.90E-28 |
| rs600038 | let-7a-5p | trans | 1.48E-12 | Nightingale | Clinical_LD | NA | C | T | 0.057741 | 0.005073 | 1.50E-30 |
| rs600038 | let-7a-5p | trans | 1.48E-12 | Nightingale | L_VLDL_C_ | NA | C | T | 0.072919 | 0.004984 | 1.60E-49 |
| rs600038 | let-7a-5p | trans | 1.48E-12 | Nightingale | Remnant_( | NA | C | T | 0.053469 | 0.00507 | 6.60E-28 |
| rs600038 | let-7a-5p | trans | 1.48E-12 | Nightingale | M_VLDL_L | NA | C | T | 0.030261 | 0.005068 | 2.00E-10 |
| rs600038 | let-7a-5p | trans | 1.48E-12 | Nightingale | XS_VLDL_C | NA | C | T | 0.0349 | 0.004815 | 1.80E-12 |
| rs600038 | let-7a-5p | trans | 1.48E-12 | Nightingale | Cholines | NA | C | T | 0.030838 | 0.004799 | 1.40E-11 |
| rs600038 | let-7a-5p | trans | 1.48E-12 | Nightingale | VLDL_C | NA | C | T | 0.042745 | 0.005096 | 1.20E-18 |
| rs600038 | let-7a-5p | trans | 1.48E-12 | Nightingale | PUFA | NA | C | T | 0.032623 | 0.004977 | 6.00E-12 |
| rs600038 | let-7a-5p | trans | 1.48E-12 | Nightingale | XL_VLDL_F | NA | C | T | 0.050732 | 0.004947 | 2.90E-25 |
| rs600038 | let-7a-5p | trans | 1.48E-12 | Nightingale | VLDL_PL | NA | C | T | 0.025473 | 0.005042 | 3.80E-08 |
| rs600038 | let-7a-5p | trans | 1.48E-12 | Nightingale | ApoB | NA | C | T | 0.050849 | 0.005096 | 1.40E-24 |
| rs600038 | let-7a-5p | trans | 1.48E-12 | Nightingale | Total_P | NA | C | T | 0.03028 | 0.004803 | 7.20E-11 |
| rs600038 | let-7a-5p | trans | 1.48E-12 | Nightingale | S_HDL_CE | NA | C | T | 0.045974 | 0.005111 | 3.30E-19 |
| rs600038 | let-7a-5p | trans | 1.48E-12 | Nightingale | L_LDL_P | NA | C | T | 0.045959 | 0.005086 | 2.50E-20 |
| rs600038 | let-7a-5p | trans | 1.48E-12 | Nightingale | M_LDL_TG | NA | C | T | -0.04594 | 0.00504 | 1.80E-19 |
| rs600038 | let-7a-5p | trans | 1.48E-12 | Nightingale | XL_HDL_L | NA | C | T | -0.02484 | 0.004589 | 1.40E-08 |
| rs600038 | let-7a-5p | trans | 1.48E-12 | Nightingale | L_VLDL_TG | NA | C | T | -0.07208 | 0.005079 | 1.80E-47 |
| rs600038 | let-7a-5p | trans | 1.48E-12 | Nightingale | S_LDL_C_p | NA | C | T | 0.065911 | 0.005123 | 1.80E-38 |
| rs600038 | let-7a-5p | trans | 1.48E-12 | Nightingale | XS_VLDL_T | NA | C | T | -0.03742 | 0.004852 | 4.10E-14 |
| rs600038 | let-7a-5p | trans | 1.48E-12 | Nightingale | Total_FC | NA | C | T | 0.049114 | 0.004985 | 8.70E-25 |
| rs600038 | let-7a-5p | trans | 1.48E-12 | Nightingale | XL_VLDL_T | NA | C | T | -0.06142 | 0.004946 | 1.30E-35 |
| rs600038 | let-7a-5p | trans | 1.48E-12 | Nightingale | S_HDL_L | NA | C | T | 0.038363 | 0.005085 | 1.60E-14 |
| rs600038 | let-7a-5p | trans | 1.48E-12 | Nightingale | IDL_FC_pct | NA | C | T | 0.033912 | 0.005063 | 4.50E-11 |
| rs600038 | let-7a-5p | trans | 1.48E-12 | Nightingale | S_VLDL_CE | NA | C | T | 0.069016 | 0.005034 | 1.40E-42 |
| rs600038 | let-7a-5p | trans | 1.48E-12 | Nightingale | L_VLDL_FC | NA | C | T | 0.068916 | 0.005054 | 4.60E-43 |
| rs600038 | let-7a-5p | trans | 1.48E-12 | Nightingale | XXL_VLDL_ | NA | C | T | 0.041556 | 0.005084 | 1.00E-16 |
| rs600038 | let-7a-5p | trans | 1.48E-12 | Nightingale | Total_L | NA | C | T | 0.038104 | 0.00502 | 2.10E-15 |
| rs600038 | let-7a-5p | trans | 1.48E-12 | Nightingale | M_LDL_P | NA | C | T | 0.051071 | 0.005117 | 2.50E-24 |
| rs600038 | let-7a-5p | trans | 1.48E-12 | Nightingale | M_LDL_L | NA | C | T | 0.051518 | 0.00511 | 1.20E-24 |
| rs600038 | let-7a-5p | trans | 1.48E-12 | Nightingale | ApoB_by_/ | NA | C | T | 0.038216 | 0.005026 | 8.20E-15 |
| rs600038 | let-7a-5p | trans | 1.48E-12 | Nightingale | HDL_size | NA | C | T | -0.03071 | 0.004524 | 3.50E-12 |
| rs600038 | let-7a-5p | trans | 1.48E-12 | Nightingale | IDL_C_pct | NA | C | T | 0.032847 | 0.004985 | 2.60E-10 |
| rs600038 | let-7a-5p | trans | 1.48E-12 | Nightingale | S_LDL_L | NA | C | T | 0.044203 | 0.005102 | 1.30E-18 |
| rs600038 | let-7a-5p | trans | 1.48E-12 | Nightingale | M_LDL_FC | NA | C | T | 0.056713 | 0.005096 | 5.00E-29 |
| rs600038 | let-7a-5p | trans | 1.48E-12 | Nightingale | M_VLDL_F | NA | C | T | 0.059646 | 0.004855 | 1.60E-34 |
| rs600038 | let-7a-5p | trans | 1.48E-12 | Nightingale | IDL_TG_pc | NA | C | T | -0.03341 | 0.004933 | 4.00E-11 |
| rs600038 | let-7a-5p | trans | 1.48E-12 | Nightingale | M_LDL_C_ | NA | C | T | 0.038144 | 0.005106 | 7.20E-14 |
| rs600038 | let-7a-5p | trans | 1.48E-12 | Nightingale | M_VLDL_P | NA | C | T | 0.063603 | 0.004897 | 8.80E-39 |
| rs600038 | let-7a-5p | trans | 1.48E-12 | Nightingale | S_HDL_FC | NA | C | T | 0.035556 | 0.005009 | 3.10E-13 |
| rs600038 | let-7a-5p | trans | 1.48E-12 | Nightingale | IDL_CE | NA | C | T | 0.052609 | 0.004927 | 4.00E-27 |
| rs600038 | let-7a-5p | trans | 1.48E-12 | Nightingale | M_VLDL_C | NA | C | T | 0.052637 | 0.004806 | 2.00E-27 |
| rs600038 | let-7a-5p | trans | 1.48E-12 | Nightingale | M_VLDL_C | NA | C | T | 0.059646 | 0.005056 | 4.30E-33 |
| rs600038 | let-7a-5p | trans | 1.48E-12 | Nightingale | S_VLDL_P | NA | C | T | 0.031474 | 0.005039 | 1.50E-11 |
| rs600038 | let-7a-5p | trans | 1.48E-12 | Nightingale | S_HDL_C | NA | C | T | 0.04513 | 0.005094 | 8.20E-19 |
| rs600038 | let-7a-5p | trans | 1.48E-12 | Nightingale | non_HDL_( | NA | C | T | 0.054532 | 0.005075 | 3.60E-28 |
| rs600038 | let-7a-5p | trans | 1.48E-12 | Nightingale | XL_VLDL_C | NA | C | T | 0.028612 | 0.005024 | 8.00E-10 |
| rs600038 | let-7a-5p | trans | 1.48E-12 | Nightingale | IDL_P | NA | C | T | 0.059653 | 0.005049 | 5.60E-33 |
| rs600038 | let-7a-5p | trans | 1.48E-12 | Nightingale | XXL_VLDL_ | NA | C | T | 0.032858 | 0.005138 | 5.00E-12 |
| rs600038 | let-7a-5p | trans | 1.48E-12 | Nightingale | LDL_PL | NA | C | T | 0.0557 | 0.005091 | 1.90E-28 |
| rs600038 | let-7a-5p | trans | 1.48E-12 | Nightingale | S_VLDL_CE | NA | C | T | 0.053323 | 0.005096 | 1.20E-27 |
| rs600038 | let-7a-5p | trans | 1.48E-12 | Nightingale | S_LDL_C | NA | C | T | 0.04877 | 0.005108 | 3.30E-22 |
| rs600038 | let-7a-5p | trans | 1.48E-12 | Nightingale | IDL_PL | NA | C | T | 0.052744 | 0.004924 | 5.20E-28 |
| rs600038 | let-7a-5p | trans | 1.48E-12 | Nightingale | S_VLDL_PL | NA | C | T | 0.046119 | 0.005093 | 3.30E-21 |
| rs600038 | let-7a-5p | trans | 1.48E-12 | Nightingale | Omega_6 | NA | C | T | 0.037012 | 0.004983 | 5.10E-15 |
| rs600038 | let-7a-5p | trans | 1.48E-12 | Nightingale | XL_HDL_C_ | NA | C | T | 0.033784 | 0.004873 | 2.10E-11 |
| rs600038 | let-7a-5p | trans | 1.48E-12 | Nightingale | M_VLDL_P | NA | C | T | 0.040156 | 0.00509 | 1.50E-16 |
| rs600038 | let-7a-5p | trans | 1.48E-12 | Nightingale | Phosphatic | NA | C | T | 0.027053 | 0.004781 | 2.30E-09 |
| rs600038 | let-7a-5p | trans | 1.48E-12 | Nightingale | Total_PL | NA | C | T | 0.034757 | 0.004859 | 4.60E-14 |

| | | | | | | | | | | | |
|---|---|---|---|---|---|---|---|---|---|---|---|
| rs600038 | let-7a-5p | trans | 1.48E-12 | Nightingale | LDL_CE | NA | C | T | 0.051899 | 0.005089 | 3.60E-25 |
| rs600038 | let-7a-5p | trans | 1.48E-12 | Nightingale | VLDL_CE | NA | C | T | 0.049508 | 0.005104 | 5.70E-24 |
| rs600038 | let-7a-5p | trans | 1.48E-12 | Nightingale | S_VLDL_FC | NA | C | T | 0.054116 | 0.005104 | 8.50E-28 |
| rs600038 | let-7a-5p | trans | 1.48E-12 | Nightingale | XS_VLDL_L | NA | C | T | 0.044732 | 0.005016 | 1.60E-21 |
| rs600038 | let-7a-5p | trans | 1.48E-12 | Nightingale | S_LDL_TG_ | NA | C | T | -0.03742 | 0.004948 | 2.20E-13 |
| rs600038 | let-7a-5p | trans | 1.48E-12 | Nightingale | XL_HDL_PL | NA | C | T | -0.02757 | 0.00459 | 6.70E-10 |
| rs600038 | let-7a-5p | trans | 1.48E-12 | Nightingale | M_LDL_C | NA | C | T | 0.052552 | 0.005112 | 1.70E-25 |
| rs600038 | let-7a-5p | trans | 1.48E-12 | Nightingale | L_LDL_PL | NA | C | T | 0.059497 | 0.005064 | 1.20E-32 |
| rs600038 | let-7a-5p | trans | 1.48E-12 | Nightingale | XS_VLDL_C | NA | C | T | 0.037556 | 0.00484 | 3.10E-14 |
| rs600038 | let-7a-5p | trans | 1.48E-12 | Nightingale | S_LDL_PL | NA | C | T | 0.038067 | 0.005091 | 4.30E-14 |
| rs600038 | let-7a-5p | trans | 1.48E-12 | Nightingale | S_LDL_CE | NA | C | T | 0.046863 | 0.005105 | 5.90E-21 |
| rs600038 | let-7a-5p | trans | 1.48E-12 | Nightingale | XL_VLDL_P | NA | C | T | 0.052743 | 0.005111 | 2.40E-26 |
| rs600038 | let-7a-5p | trans | 1.48E-12 | Nightingale | XXL_VLDL_ | NA | C | T | 0.051211 | 0.005116 | 5.90E-24 |
| rs600038 | let-7a-5p | trans | 1.48E-12 | Nightingale | S_HDL_PL_ | NA | C | T | -0.02877 | 0.004933 | 1.50E-08 |
| rs600038 | let-7a-5p | trans | 1.48E-12 | Nightingale | XS_VLDL_F | NA | C | T | 0.049012 | 0.005014 | 4.20E-25 |
| rs600038 | let-7a-5p | trans | 1.48E-12 | Nightingale | S_HDL_PL | NA | C | T | 0.032674 | 0.005059 | 2.80E-11 |
| rs600038 | let-7a-5p | trans | 1.48E-12 | Nightingale | L_VLDL_CE | NA | C | T | 0.029942 | 0.005043 | 1.80E-10 |
| rs600038 | let-7a-5p | trans | 1.48E-12 | Nightingale | XS_VLDL_P | NA | C | T | 0.04948 | 0.005035 | 2.10E-25 |
| rs600038 | let-7a-5p | trans | 1.48E-12 | Nightingale | VLDL_FC | NA | C | T | 0.032134 | 0.005065 | 1.10E-11 |
| rs600038 | let-7a-5p | trans | 1.48E-12 | Nightingale | S_LDL_PL_ | NA | C | T | -0.0395 | 0.005093 | 1.10E-16 |
| rs600038 | let-7a-5p | trans | 1.48E-12 | Nightingale | XXL_VLDL_ | NA | C | T | 0.050318 | 0.005144 | 4.30E-23 |
| rs600038 | let-7a-5p | trans | 1.48E-12 | Nightingale | XL_HDL_PL | NA | C | T | -0.03707 | 0.004688 | 2.50E-15 |
| rs600038 | let-7a-5p | trans | 1.48E-12 | Nightingale | M_LDL_CE | NA | C | T | 0.049694 | 0.00511 | 3.20E-23 |
| rs600038 | let-7a-5p | trans | 1.48E-12 | Nightingale | S_VLDL_TG | NA | C | T | -0.05802 | 0.004939 | 6.80E-32 |
| rs600038 | let-7a-5p | trans | 1.48E-12 | Nightingale | S_HDL_P | NA | C | T | 0.048052 | 0.005103 | 4.50E-21 |
| rs600038 | let-7a-5p | trans | 1.48E-12 | Nightingale | M_VLDL_T | NA | C | T | -0.05584 | 0.004822 | 1.20E-30 |
| rs600038 | miR-10b-5p | trans | 2.32E-34 | Nightingale | XL_VLDL_C | NA | C | T | 0.054368 | 0.004895 | 2.80E-28 |
| rs600038 | miR-10b-5p | trans | 2.32E-34 | Nightingale | M_VLDL_C | NA | C | T | 0.049278 | 0.004794 | 2.70E-24 |
| rs600038 | miR-10b-5p | trans | 2.32E-34 | Nightingale | Total_CE | NA | C | T | 0.049009 | 0.004922 | 2.10E-24 |
| rs600038 | miR-10b-5p | trans | 2.32E-34 | Nightingale | L_LDL_L | NA | C | T | 0.054273 | 0.005053 | 1.00E-27 |
| rs600038 | miR-10b-5p | trans | 2.32E-34 | Nightingale | L_LDL_CE | NA | C | T | 0.051898 | 0.005057 | 2.00E-25 |
| rs600038 | miR-10b-5p | trans | 2.32E-34 | Nightingale | L_LDL_C | NA | C | T | 0.053255 | 0.005045 | 1.10E-26 |
| rs600038 | miR-10b-5p | trans | 2.32E-34 | Nightingale | Phosphogly | NA | C | T | 0.027754 | 0.004833 | 1.30E-09 |
| rs600038 | miR-10b-5p | trans | 2.32E-34 | Nightingale | IDL_FC | NA | C | T | 0.057015 | 0.00494 | 1.50E-31 |
| rs600038 | miR-10b-5p | trans | 2.32E-34 | Nightingale | S_VLDL_C | NA | C | T | 0.05403 | 0.005102 | 5.10E-28 |
| rs600038 | miR-10b-5p | trans | 2.32E-34 | Nightingale | Total_C | NA | C | T | 0.049326 | 0.00494 | 1.10E-24 |
| rs600038 | miR-10b-5p | trans | 2.32E-34 | Nightingale | L_VLDL_PL | NA | C | T | 0.027472 | 0.004957 | 1.00E-09 |
| rs600038 | miR-10b-5p | trans | 2.32E-34 | Nightingale | LDL_P | NA | C | T | 0.048287 | 0.005099 | 3.30E-22 |
| rs600038 | miR-10b-5p | trans | 2.32E-34 | Nightingale | M_VLDL_C | NA | C | T | 0.055912 | 0.005089 | 1.20E-29 |
| rs600038 | miR-10b-5p | trans | 2.32E-34 | Nightingale | M_LDL_PL | NA | C | T | 0.052654 | 0.00511 | 1.50E-25 |
| rs600038 | miR-10b-5p | trans | 2.32E-34 | Nightingale | LA | NA | C | T | 0.037469 | 0.005004 | 3.30E-15 |
| rs600038 | miR-10b-5p | trans | 2.32E-34 | Nightingale | Citrate | NA | C | T | 0.02733 | 0.00507 | 2.60E-08 |
| rs600038 | miR-10b-5p | trans | 2.32E-34 | Nightingale | Pyruvate | NA | C | T | 0.030176 | 0.005111 | 3.60E-09 |
| rs600038 | miR-10b-5p | trans | 2.32E-34 | Nightingale | VLDL_P | NA | C | T | 0.03602 | 0.005075 | 4.70E-14 |
| rs600038 | miR-10b-5p | trans | 2.32E-34 | Nightingale | Sphingomy | NA | C | T | 0.040531 | 0.00477 | 1.70E-17 |
| rs600038 | miR-10b-5p | trans | 2.32E-34 | Nightingale | S_LDL_FC | NA | C | T | 0.049076 | 0.005095 | 7.70E-22 |
| rs600038 | miR-10b-5p | trans | 2.32E-34 | Nightingale | LDL_FC | NA | C | T | 0.055801 | 0.005048 | 1.60E-28 |
| rs600038 | miR-10b-5p | trans | 2.32E-34 | Nightingale | XS_VLDL_C | NA | C | T | 0.052331 | 0.004953 | 3.80E-28 |
| rs600038 | miR-10b-5p | trans | 2.32E-34 | Nightingale | XS_VLDL_P | NA | C | T | 0.041555 | 0.005032 | 8.10E-19 |
| rs600038 | miR-10b-5p | trans | 2.32E-34 | Nightingale | L_LDL_FC | NA | C | T | 0.055661 | 0.005011 | 5.90E-29 |
| rs600038 | miR-10b-5p | trans | 2.32E-34 | Nightingale | XXL_VLDL_ | NA | C | T | -0.05276 | 0.005149 | 1.90E-25 |
| rs600038 | miR-10b-5p | trans | 2.32E-34 | Nightingale | XS_VLDL_C | NA | C | T | 0.052917 | 0.004924 | 7.70E-29 |
| rs600038 | miR-10b-5p | trans | 2.32E-34 | Nightingale | S_VLDL_FC | NA | C | T | 0.045985 | 0.004876 | 8.40E-21 |
| rs600038 | miR-10b-5p | trans | 2.32E-34 | Nightingale | S_LDL_CE_ | NA | C | T | 0.039177 | 0.005094 | 1.30E-16 |
| rs600038 | miR-10b-5p | trans | 2.32E-34 | Nightingale | S_VLDL_C_ | NA | C | T | 0.062934 | 0.004966 | 6.40E-37 |
| rs600038 | miR-10b-5p | trans | 2.32E-34 | Nightingale | L_VLDL_CE | NA | C | T | 0.065536 | 0.004883 | 4.90E-42 |
| rs600038 | miR-10b-5p | trans | 2.32E-34 | Nightingale | S_VLDL_PL | NA | C | T | 0.043922 | 0.004886 | 5.30E-19 |
| rs600038 | miR-10b-5p | trans | 2.32E-34 | Nightingale | XS_VLDL_F | NA | C | T | 0.044491 | 0.005066 | 8.00E-18 |
| rs600038 | miR-10b-5p | trans | 2.32E-34 | Nightingale | XL_VLDL_C | NA | C | T | 0.054864 | 0.004893 | 6.90E-29 |
| rs600038 | miR-10b-5p | trans | 2.32E-34 | Nightingale | S_LDL_P | NA | C | T | 0.044606 | 0.005107 | 3.00E-19 |
| rs600038 | miR-10b-5p | trans | 2.32E-34 | Nightingale | L_LDL_TG_ | NA | C | T | -0.03537 | 0.005005 | 5.10E-12 |
| rs600038 | miR-10b-5p | trans | 2.32E-34 | Nightingale | M_VLDL_P | NA | C | T | 0.041549 | 0.005095 | 1.40E-17 |
| rs600038 | miR-10b-5p | trans | 2.32E-34 | Nightingale | L_HDL_TG_ | NA | C | T | 0.023502 | 0.004821 | 3.50E-08 |

| | | | | | | | | | | | |
|---|---|---|---|---|---|---|---|---|---|---|---|
| rs600038 | miR-10b-5p | trans | 2.32E-34 | Nightingale | LDL_C | NA | C | T | 0.053489 | 0.005081 | 1.60E-26 |
| rs600038 | miR-10b-5p | trans | 2.32E-34 | Nightingale | S_VLDL_L | NA | C | T | 0.033361 | 0.005053 | 1.40E-12 |
| rs600038 | miR-10b-5p | trans | 2.32E-34 | Nightingale | LDL_L | NA | C | T | 0.053245 | 0.005086 | 2.30E-26 |
| rs600038 | miR-10b-5p | trans | 2.32E-34 | Nightingale | M_VLDL_F | NA | C | T | 0.047425 | 0.005102 | 6.00E-22 |
| rs600038 | miR-10b-5p | trans | 2.32E-34 | Nightingale | IDL_C | NA | C | T | 0.054083 | 0.004928 | 1.40E-28 |
| rs600038 | miR-10b-5p | trans | 2.32E-34 | Nightingale | IDL_L | NA | C | T | 0.053693 | 0.004934 | 1.90E-28 |
| rs600038 | miR-10b-5p | trans | 2.32E-34 | Nightingale | Clinical_LD | NA | C | T | 0.057741 | 0.005073 | 1.50E-30 |
| rs600038 | miR-10b-5p | trans | 2.32E-34 | Nightingale | L_VLDL_C_ | NA | C | T | 0.072919 | 0.004984 | 1.60E-49 |
| rs600038 | miR-10b-5p | trans | 2.32E-34 | Nightingale | Remnant_( | NA | C | T | 0.053469 | 0.00507 | 6.60E-28 |
| rs600038 | miR-10b-5p | trans | 2.32E-34 | Nightingale | M_VLDL_L | NA | C | T | 0.030261 | 0.005068 | 2.00E-10 |
| rs600038 | miR-10b-5p | trans | 2.32E-34 | Nightingale | XS_VLDL_C | NA | C | T | 0.0349 | 0.004815 | 1.80E-12 |
| rs600038 | miR-10b-5p | trans | 2.32E-34 | Nightingale | Cholines | NA | C | T | 0.030838 | 0.004799 | 1.40E-11 |
| rs600038 | miR-10b-5p | trans | 2.32E-34 | Nightingale | VLDL_C | NA | C | T | 0.042745 | 0.005096 | 1.20E-18 |
| rs600038 | miR-10b-5p | trans | 2.32E-34 | Nightingale | PUFA | NA | C | T | 0.032623 | 0.004977 | 6.00E-12 |
| rs600038 | miR-10b-5p | trans | 2.32E-34 | Nightingale | XL_VLDL_F | NA | C | T | 0.050732 | 0.004947 | 2.90E-25 |
| rs600038 | miR-10b-5p | trans | 2.32E-34 | Nightingale | VLDL_PL | NA | C | T | 0.025473 | 0.005042 | 3.80E-08 |
| rs600038 | miR-10b-5p | trans | 2.32E-34 | Nightingale | ApoB | NA | C | T | 0.050849 | 0.005096 | 1.40E-24 |
| rs600038 | miR-10b-5p | trans | 2.32E-34 | Nightingale | Total_P | NA | C | T | 0.03028 | 0.004803 | 7.20E-11 |
| rs600038 | miR-10b-5p | trans | 2.32E-34 | Nightingale | S_HDL_CE | NA | C | T | 0.045974 | 0.005111 | 3.30E-19 |
| rs600038 | miR-10b-5p | trans | 2.32E-34 | Nightingale | L_LDL_P | NA | C | T | 0.045959 | 0.005086 | 2.50E-20 |
| rs600038 | miR-10b-5p | trans | 2.32E-34 | Nightingale | M_LDL_TG | NA | C | T | -0.04594 | 0.00504 | 1.80E-19 |
| rs600038 | miR-10b-5p | trans | 2.32E-34 | Nightingale | XL_HDL_L | NA | C | T | -0.02484 | 0.004589 | 1.40E-08 |
| rs600038 | miR-10b-5p | trans | 2.32E-34 | Nightingale | L_VLDL_TG | NA | C | T | -0.07208 | 0.005079 | 1.80E-47 |
| rs600038 | miR-10b-5p | trans | 2.32E-34 | Nightingale | S_LDL_C_p | NA | C | T | 0.065911 | 0.005123 | 1.80E-38 |
| rs600038 | miR-10b-5p | trans | 2.32E-34 | Nightingale | XS_VLDL_T | NA | C | T | -0.03742 | 0.004852 | 4.10E-14 |
| rs600038 | miR-10b-5p | trans | 2.32E-34 | Nightingale | Total_FC | NA | C | T | 0.049114 | 0.004985 | 8.70E-25 |
| rs600038 | miR-10b-5p | trans | 2.32E-34 | Nightingale | XL_VLDL_T | NA | C | T | -0.06142 | 0.004946 | 1.30E-35 |
| rs600038 | miR-10b-5p | trans | 2.32E-34 | Nightingale | S_HDL_L | NA | C | T | 0.038363 | 0.005085 | 1.60E-14 |
| rs600038 | miR-10b-5p | trans | 2.32E-34 | Nightingale | IDL_FC_pct | NA | C | T | 0.033912 | 0.005063 | 4.50E-11 |
| rs600038 | miR-10b-5p | trans | 2.32E-34 | Nightingale | S_VLDL_CE | NA | C | T | 0.069016 | 0.005034 | 1.40E-42 |
| rs600038 | miR-10b-5p | trans | 2.32E-34 | Nightingale | L_VLDL_FC | NA | C | T | 0.068916 | 0.005054 | 4.60E-43 |
| rs600038 | miR-10b-5p | trans | 2.32E-34 | Nightingale | XXL_VLDL_ | NA | C | T | 0.041556 | 0.005084 | 1.00E-16 |
| rs600038 | miR-10b-5p | trans | 2.32E-34 | Nightingale | Total_L | NA | C | T | 0.038104 | 0.00502 | 2.10E-15 |
| rs600038 | miR-10b-5p | trans | 2.32E-34 | Nightingale | M_LDL_P | NA | C | T | 0.051071 | 0.005117 | 2.50E-24 |
| rs600038 | miR-10b-5p | trans | 2.32E-34 | Nightingale | M_LDL_L | NA | C | T | 0.051518 | 0.00511 | 1.20E-24 |
| rs600038 | miR-10b-5p | trans | 2.32E-34 | Nightingale | ApoB_by_A | NA | C | T | 0.038216 | 0.005026 | 8.20E-15 |
| rs600038 | miR-10b-5p | trans | 2.32E-34 | Nightingale | HDL_size | NA | C | T | -0.03071 | 0.004524 | 3.50E-12 |
| rs600038 | miR-10b-5p | trans | 2.32E-34 | Nightingale | IDL_C_pct | NA | C | T | 0.032847 | 0.004985 | 2.60E-10 |
| rs600038 | miR-10b-5p | trans | 2.32E-34 | Nightingale | S_LDL_L | NA | C | T | 0.044203 | 0.005102 | 1.30E-18 |
| rs600038 | miR-10b-5p | trans | 2.32E-34 | Nightingale | M_LDL_FC | NA | C | T | 0.056713 | 0.005096 | 5.00E-29 |
| rs600038 | miR-10b-5p | trans | 2.32E-34 | Nightingale | M_VLDL_F | NA | C | T | 0.059646 | 0.004855 | 1.60E-34 |
| rs600038 | miR-10b-5p | trans | 2.32E-34 | Nightingale | IDL_TG_pc | NA | C | T | -0.03341 | 0.004933 | 4.00E-11 |
| rs600038 | miR-10b-5p | trans | 2.32E-34 | Nightingale | M_LDL_C_ | NA | C | T | 0.038144 | 0.005106 | 7.20E-14 |
| rs600038 | miR-10b-5p | trans | 2.32E-34 | Nightingale | M_VLDL_P | NA | C | T | 0.063603 | 0.004897 | 8.80E-39 |
| rs600038 | miR-10b-5p | trans | 2.32E-34 | Nightingale | S_HDL_FC | NA | C | T | 0.035556 | 0.005009 | 3.10E-13 |
| rs600038 | miR-10b-5p | trans | 2.32E-34 | Nightingale | IDL_CE | NA | C | T | 0.052609 | 0.004927 | 4.00E-27 |
| rs600038 | miR-10b-5p | trans | 2.32E-34 | Nightingale | M_VLDL_C | NA | C | T | 0.052637 | 0.004806 | 2.00E-27 |
| rs600038 | miR-10b-5p | trans | 2.32E-34 | Nightingale | M_VLDL_C | NA | C | T | 0.059646 | 0.005056 | 4.30E-33 |
| rs600038 | miR-10b-5p | trans | 2.32E-34 | Nightingale | S_VLDL_P | NA | C | T | 0.031474 | 0.005039 | 1.50E-11 |
| rs600038 | miR-10b-5p | trans | 2.32E-34 | Nightingale | S_HDL_C | NA | C | T | 0.04513 | 0.005094 | 8.20E-19 |
| rs600038 | miR-10b-5p | trans | 2.32E-34 | Nightingale | non_HDL_( | NA | C | T | 0.054532 | 0.005075 | 3.60E-28 |
| rs600038 | miR-10b-5p | trans | 2.32E-34 | Nightingale | XL_VLDL_C | NA | C | T | 0.028612 | 0.005024 | 8.00E-10 |
| rs600038 | miR-10b-5p | trans | 2.32E-34 | Nightingale | IDL_P | NA | C | T | 0.059653 | 0.005049 | 5.60E-33 |
| rs600038 | miR-10b-5p | trans | 2.32E-34 | Nightingale | XXL_VLDL_ | NA | C | T | 0.032858 | 0.005138 | 5.00E-12 |
| rs600038 | miR-10b-5p | trans | 2.32E-34 | Nightingale | LDL_PL | NA | C | T | 0.0557 | 0.005091 | 1.90E-28 |
| rs600038 | miR-10b-5p | trans | 2.32E-34 | Nightingale | S_VLDL_CE | NA | C | T | 0.053323 | 0.005096 | 1.20E-27 |
| rs600038 | miR-10b-5p | trans | 2.32E-34 | Nightingale | S_LDL_C | NA | C | T | 0.04877 | 0.005108 | 3.30E-22 |
| rs600038 | miR-10b-5p | trans | 2.32E-34 | Nightingale | IDL_PL | NA | C | T | 0.052744 | 0.004924 | 5.20E-28 |
| rs600038 | miR-10b-5p | trans | 2.32E-34 | Nightingale | S_VLDL_PL | NA | C | T | 0.046119 | 0.005093 | 3.30E-21 |
| rs600038 | miR-10b-5p | trans | 2.32E-34 | Nightingale | Omega_6 | NA | C | T | 0.037012 | 0.004983 | 5.10E-15 |
| rs600038 | miR-10b-5p | trans | 2.32E-34 | Nightingale | XL_HDL_C_ | NA | C | T | 0.033784 | 0.004873 | 2.10E-11 |
| rs600038 | miR-10b-5p | trans | 2.32E-34 | Nightingale | M_VLDL_P | NA | C | T | 0.040156 | 0.00509 | 1.50E-16 |
| rs600038 | miR-10b-5p | trans | 2.32E-34 | Nightingale | Phosphatic | NA | C | T | 0.027053 | 0.004781 | 2.30E-09 |

| | | | | | | | | | | | |
|---|---|---|---|---|---|---|---|---|---|---|---|
| rs600038 | miR-10b-5p | trans | 2.32E-34 | Nightingale | Total_PL | NA | C | T | 0.034757 | 0.004859 | 4.60E-14 |
| rs600038 | miR-10b-5p | trans | 2.32E-34 | Nightingale | LDL_CE | NA | C | T | 0.051899 | 0.005089 | 3.60E-25 |
| rs600038 | miR-10b-5p | trans | 2.32E-34 | Nightingale | VLDL_CE | NA | C | T | 0.049508 | 0.005104 | 5.70E-24 |
| rs600038 | miR-10b-5p | trans | 2.32E-34 | Nightingale | S_VLDL_FC | NA | C | T | 0.054116 | 0.005104 | 8.50E-28 |
| rs600038 | miR-10b-5p | trans | 2.32E-34 | Nightingale | XS_VLDL_L | NA | C | T | 0.044732 | 0.005016 | 1.60E-21 |
| rs600038 | miR-10b-5p | trans | 2.32E-34 | Nightingale | S_LDL_TG_ | NA | C | T | -0.03742 | 0.004948 | 2.20E-13 |
| rs600038 | miR-10b-5p | trans | 2.32E-34 | Nightingale | XL_HDL_PL | NA | C | T | -0.02757 | 0.00459 | 6.70E-10 |
| rs600038 | miR-10b-5p | trans | 2.32E-34 | Nightingale | M_LDL_C | NA | C | T | 0.052552 | 0.005112 | 1.70E-25 |
| rs600038 | miR-10b-5p | trans | 2.32E-34 | Nightingale | L_LDL_PL | NA | C | T | 0.059497 | 0.005064 | 1.20E-32 |
| rs600038 | miR-10b-5p | trans | 2.32E-34 | Nightingale | XS_VLDL_C | NA | C | T | 0.037556 | 0.00484 | 3.10E-14 |
| rs600038 | miR-10b-5p | trans | 2.32E-34 | Nightingale | S_LDL_PL | NA | C | T | 0.038067 | 0.005091 | 4.30E-14 |
| rs600038 | miR-10b-5p | trans | 2.32E-34 | Nightingale | S_LDL_CE | NA | C | T | 0.046863 | 0.005105 | 5.90E-21 |
| rs600038 | miR-10b-5p | trans | 2.32E-34 | Nightingale | XL_VLDL_P | NA | C | T | 0.052743 | 0.005111 | 2.40E-26 |
| rs600038 | miR-10b-5p | trans | 2.32E-34 | Nightingale | XXL_VLDL_ | NA | C | T | 0.051211 | 0.005116 | 5.90E-24 |
| rs600038 | miR-10b-5p | trans | 2.32E-34 | Nightingale | S_HDL_PL_ | NA | C | T | -0.02877 | 0.004933 | 1.50E-08 |
| rs600038 | miR-10b-5p | trans | 2.32E-34 | Nightingale | XS_VLDL_F | NA | C | T | 0.049012 | 0.005014 | 4.20E-25 |
| rs600038 | miR-10b-5p | trans | 2.32E-34 | Nightingale | S_HDL_PL | NA | C | T | 0.032674 | 0.005059 | 2.80E-11 |
| rs600038 | miR-10b-5p | trans | 2.32E-34 | Nightingale | L_VLDL_CE | NA | C | T | 0.029942 | 0.005043 | 1.80E-10 |
| rs600038 | miR-10b-5p | trans | 2.32E-34 | Nightingale | XS_VLDL_P | NA | C | T | 0.04948 | 0.005035 | 2.10E-25 |
| rs600038 | miR-10b-5p | trans | 2.32E-34 | Nightingale | VLDL_FC | NA | C | T | 0.032134 | 0.005065 | 1.10E-11 |
| rs600038 | miR-10b-5p | trans | 2.32E-34 | Nightingale | S_LDL_PL_ | NA | C | T | -0.0395 | 0.005093 | 1.10E-16 |
| rs600038 | miR-10b-5p | trans | 2.32E-34 | Nightingale | XXL_VLDL_ | NA | C | T | 0.050318 | 0.005144 | 4.30E-23 |
| rs600038 | miR-10b-5p | trans | 2.32E-34 | Nightingale | XL_HDL_PL | NA | C | T | -0.03707 | 0.004688 | 2.50E-15 |
| rs600038 | miR-10b-5p | trans | 2.32E-34 | Nightingale | M_LDL_CE | NA | C | T | 0.049694 | 0.00511 | 3.20E-23 |
| rs600038 | miR-10b-5p | trans | 2.32E-34 | Nightingale | S_VLDL_TG | NA | C | T | -0.05802 | 0.004939 | 6.80E-32 |
| rs600038 | miR-10b-5p | trans | 2.32E-34 | Nightingale | S_HDL_P | NA | C | T | 0.048052 | 0.005103 | 4.50E-21 |
| rs600038 | miR-10b-5p | trans | 2.32E-34 | Nightingale | M_VLDL_T | NA | C | T | -0.05584 | 0.004822 | 1.20E-30 |
| rs600038 | miR-126-3p | trans | 1.80E-37 | Nightingale | XL_VLDL_C | NA | C | T | 0.054368 | 0.004895 | 2.80E-28 |
| rs600038 | miR-126-3p | trans | 1.80E-37 | Nightingale | M_VLDL_C | NA | C | T | 0.049278 | 0.004794 | 2.70E-24 |
| rs600038 | miR-126-3p | trans | 1.80E-37 | Nightingale | Total_CE | NA | C | T | 0.049009 | 0.004922 | 2.10E-24 |
| rs600038 | miR-126-3p | trans | 1.80E-37 | Nightingale | L_LDL_L | NA | C | T | 0.054273 | 0.005053 | 1.00E-27 |
| rs600038 | miR-126-3p | trans | 1.80E-37 | Nightingale | L_LDL_CE | NA | C | T | 0.051898 | 0.005057 | 2.00E-25 |
| rs600038 | miR-126-3p | trans | 1.80E-37 | Nightingale | L_LDL_C | NA | C | T | 0.053255 | 0.005045 | 1.10E-26 |
| rs600038 | miR-126-3p | trans | 1.80E-37 | Nightingale | Phosphogly | NA | C | T | 0.027754 | 0.004833 | 1.30E-09 |
| rs600038 | miR-126-3p | trans | 1.80E-37 | Nightingale | IDL_FC | NA | C | T | 0.057015 | 0.00494 | 1.50E-31 |
| rs600038 | miR-126-3p | trans | 1.80E-37 | Nightingale | S_VLDL_C | NA | C | T | 0.05403 | 0.005102 | 5.10E-28 |
| rs600038 | miR-126-3p | trans | 1.80E-37 | Nightingale | Total_C | NA | C | T | 0.049326 | 0.00494 | 1.10E-24 |
| rs600038 | miR-126-3p | trans | 1.80E-37 | Nightingale | L_VLDL_PL | NA | C | T | 0.027472 | 0.004957 | 1.00E-09 |
| rs600038 | miR-126-3p | trans | 1.80E-37 | Nightingale | LDL_P | NA | C | T | 0.048287 | 0.005099 | 3.30E-22 |
| rs600038 | miR-126-3p | trans | 1.80E-37 | Nightingale | M_VLDL_C | NA | C | T | 0.055912 | 0.005089 | 1.20E-29 |
| rs600038 | miR-126-3p | trans | 1.80E-37 | Nightingale | M_LDL_PL | NA | C | T | 0.052654 | 0.00511 | 1.50E-25 |
| rs600038 | miR-126-3p | trans | 1.80E-37 | Nightingale | LA | NA | C | T | 0.037469 | 0.005004 | 3.30E-15 |
| rs600038 | miR-126-3p | trans | 1.80E-37 | Nightingale | Citrate | NA | C | T | 0.02733 | 0.00507 | 2.60E-08 |
| rs600038 | miR-126-3p | trans | 1.80E-37 | Nightingale | Pyruvate | NA | C | T | 0.030176 | 0.005111 | 3.60E-09 |
| rs600038 | miR-126-3p | trans | 1.80E-37 | Nightingale | VLDL_P | NA | C | T | 0.03602 | 0.005075 | 4.70E-14 |
| rs600038 | miR-126-3p | trans | 1.80E-37 | Nightingale | Sphingomy | NA | C | T | 0.040531 | 0.00477 | 1.70E-17 |
| rs600038 | miR-126-3p | trans | 1.80E-37 | Nightingale | S_LDL_FC | NA | C | T | 0.049076 | 0.005095 | 7.70E-22 |
| rs600038 | miR-126-3p | trans | 1.80E-37 | Nightingale | LDL_FC | NA | C | T | 0.055801 | 0.005048 | 1.60E-28 |
| rs600038 | miR-126-3p | trans | 1.80E-37 | Nightingale | XS_VLDL_C | NA | C | T | 0.052331 | 0.004953 | 3.80E-28 |
| rs600038 | miR-126-3p | trans | 1.80E-37 | Nightingale | XS_VLDL_P | NA | C | T | 0.041555 | 0.005032 | 8.10E-19 |
| rs600038 | miR-126-3p | trans | 1.80E-37 | Nightingale | L_LDL_FC | NA | C | T | 0.055661 | 0.005011 | 5.90E-29 |
| rs600038 | miR-126-3p | trans | 1.80E-37 | Nightingale | XXL_VLDL_ | NA | C | T | -0.05276 | 0.005149 | 1.90E-25 |
| rs600038 | miR-126-3p | trans | 1.80E-37 | Nightingale | XS_VLDL_C | NA | C | T | 0.052917 | 0.004924 | 7.70E-29 |
| rs600038 | miR-126-3p | trans | 1.80E-37 | Nightingale | S_VLDL_FC | NA | C | T | 0.045985 | 0.004876 | 8.40E-21 |
| rs600038 | miR-126-3p | trans | 1.80E-37 | Nightingale | S_LDL_CE_ | NA | C | T | 0.039177 | 0.005094 | 1.30E-16 |
| rs600038 | miR-126-3p | trans | 1.80E-37 | Nightingale | S_VLDL_C_ | NA | C | T | 0.062934 | 0.004966 | 6.40E-37 |
| rs600038 | miR-126-3p | trans | 1.80E-37 | Nightingale | L_VLDL_CE | NA | C | T | 0.065536 | 0.004883 | 4.90E-42 |
| rs600038 | miR-126-3p | trans | 1.80E-37 | Nightingale | S_VLDL_PL | NA | C | T | 0.043922 | 0.004886 | 5.30E-19 |
| rs600038 | miR-126-3p | trans | 1.80E-37 | Nightingale | XS_VLDL_F | NA | C | T | 0.044491 | 0.005066 | 8.00E-18 |
| rs600038 | miR-126-3p | trans | 1.80E-37 | Nightingale | XL_VLDL_C | NA | C | T | 0.054864 | 0.004893 | 6.90E-29 |
| rs600038 | miR-126-3p | trans | 1.80E-37 | Nightingale | S_LDL_P | NA | C | T | 0.044606 | 0.005107 | 3.00E-19 |
| rs600038 | miR-126-3p | trans | 1.80E-37 | Nightingale | L_LDL_TG_ | NA | C | T | -0.03537 | 0.005005 | 5.10E-12 |
| rs600038 | miR-126-3p | trans | 1.80E-37 | Nightingale | M_VLDL_P | NA | C | T | 0.041549 | 0.005095 | 1.40E-17 |

| | | | | | | | | | | | |
|---|---|---|---|---|---|---|---|---|---|---|---|
| rs600038 | miR-126-3p | trans | 1.80E-37 | Nightingale | L_HDL_TG_ | NA | C | T | 0.023502 | 0.004821 | 3.50E-08 |
| rs600038 | miR-126-3p | trans | 1.80E-37 | Nightingale | LDL_C | NA | C | T | 0.053489 | 0.005081 | 1.60E-26 |
| rs600038 | miR-126-3p | trans | 1.80E-37 | Nightingale | S_VLDL_L | NA | C | T | 0.033361 | 0.005053 | 1.40E-12 |
| rs600038 | miR-126-3p | trans | 1.80E-37 | Nightingale | LDL_L | NA | C | T | 0.053245 | 0.005086 | 2.30E-26 |
| rs600038 | miR-126-3p | trans | 1.80E-37 | Nightingale | M_VLDL_F | NA | C | T | 0.047425 | 0.005102 | 6.00E-22 |
| rs600038 | miR-126-3p | trans | 1.80E-37 | Nightingale | IDL_C | NA | C | T | 0.054083 | 0.004928 | 1.40E-28 |
| rs600038 | miR-126-3p | trans | 1.80E-37 | Nightingale | IDL_L | NA | C | T | 0.053693 | 0.004934 | 1.90E-28 |
| rs600038 | miR-126-3p | trans | 1.80E-37 | Nightingale | Clinical_LD | NA | C | T | 0.057741 | 0.005073 | 1.50E-30 |
| rs600038 | miR-126-3p | trans | 1.80E-37 | Nightingale | L_VLDL_C_ | NA | C | T | 0.072919 | 0.004984 | 1.60E-49 |
| rs600038 | miR-126-3p | trans | 1.80E-37 | Nightingale | Remnant_( | NA | C | T | 0.053469 | 0.00507 | 6.60E-28 |
| rs600038 | miR-126-3p | trans | 1.80E-37 | Nightingale | M_VLDL_L | NA | C | T | 0.030261 | 0.005068 | 2.00E-10 |
| rs600038 | miR-126-3p | trans | 1.80E-37 | Nightingale | XS_VLDL_C | NA | C | T | 0.0349 | 0.004815 | 1.80E-12 |
| rs600038 | miR-126-3p | trans | 1.80E-37 | Nightingale | Cholines | NA | C | T | 0.030838 | 0.004799 | 1.40E-11 |
| rs600038 | miR-126-3p | trans | 1.80E-37 | Nightingale | VLDL_C | NA | C | T | 0.042745 | 0.005096 | 1.20E-18 |
| rs600038 | miR-126-3p | trans | 1.80E-37 | Nightingale | PUFA | NA | C | T | 0.032623 | 0.004977 | 6.00E-12 |
| rs600038 | miR-126-3p | trans | 1.80E-37 | Nightingale | XL_VLDL_F | NA | C | T | 0.050732 | 0.004947 | 2.90E-25 |
| rs600038 | miR-126-3p | trans | 1.80E-37 | Nightingale | VLDL_PL | NA | C | T | 0.025473 | 0.005042 | 3.80E-08 |
| rs600038 | miR-126-3p | trans | 1.80E-37 | Nightingale | ApoB | NA | C | T | 0.050849 | 0.005096 | 1.40E-24 |
| rs600038 | miR-126-3p | trans | 1.80E-37 | Nightingale | Total_P | NA | C | T | 0.03028 | 0.004803 | 7.20E-11 |
| rs600038 | miR-126-3p | trans | 1.80E-37 | Nightingale | S_HDL_CE | NA | C | T | 0.045974 | 0.005111 | 3.30E-19 |
| rs600038 | miR-126-3p | trans | 1.80E-37 | Nightingale | L_LDL_P | NA | C | T | 0.045959 | 0.005086 | 2.50E-20 |
| rs600038 | miR-126-3p | trans | 1.80E-37 | Nightingale | M_LDL_TG | NA | C | T | -0.04594 | 0.00504 | 1.80E-19 |
| rs600038 | miR-126-3p | trans | 1.80E-37 | Nightingale | XL_HDL_L | NA | C | T | -0.02484 | 0.004589 | 1.40E-08 |
| rs600038 | miR-126-3p | trans | 1.80E-37 | Nightingale | L_VLDL_TG | NA | C | T | -0.07208 | 0.005079 | 1.80E-47 |
| rs600038 | miR-126-3p | trans | 1.80E-37 | Nightingale | S_LDL_C_p | NA | C | T | 0.065911 | 0.005123 | 1.80E-38 |
| rs600038 | miR-126-3p | trans | 1.80E-37 | Nightingale | XS_VLDL_T | NA | C | T | -0.03742 | 0.004852 | 4.10E-14 |
| rs600038 | miR-126-3p | trans | 1.80E-37 | Nightingale | Total_FC | NA | C | T | 0.049114 | 0.004985 | 8.70E-25 |
| rs600038 | miR-126-3p | trans | 1.80E-37 | Nightingale | XL_VLDL_T | NA | C | T | -0.06142 | 0.004946 | 1.30E-35 |
| rs600038 | miR-126-3p | trans | 1.80E-37 | Nightingale | S_HDL_L | NA | C | T | 0.038363 | 0.005085 | 1.60E-14 |
| rs600038 | miR-126-3p | trans | 1.80E-37 | Nightingale | IDL_FC_pct | NA | C | T | 0.033912 | 0.005063 | 4.50E-11 |
| rs600038 | miR-126-3p | trans | 1.80E-37 | Nightingale | S_VLDL_CE | NA | C | T | 0.069016 | 0.005034 | 1.40E-42 |
| rs600038 | miR-126-3p | trans | 1.80E-37 | Nightingale | L_VLDL_FC | NA | C | T | 0.068916 | 0.005054 | 4.60E-43 |
| rs600038 | miR-126-3p | trans | 1.80E-37 | Nightingale | XXL_VLDL_ | NA | C | T | 0.041556 | 0.005084 | 1.00E-16 |
| rs600038 | miR-126-3p | trans | 1.80E-37 | Nightingale | Total_L | NA | C | T | 0.038104 | 0.00502 | 2.10E-15 |
| rs600038 | miR-126-3p | trans | 1.80E-37 | Nightingale | M_LDL_P | NA | C | T | 0.051071 | 0.005117 | 2.50E-24 |
| rs600038 | miR-126-3p | trans | 1.80E-37 | Nightingale | M_LDL_L | NA | C | T | 0.051518 | 0.00511 | 1.20E-24 |
| rs600038 | miR-126-3p | trans | 1.80E-37 | Nightingale | ApoB_by_A | NA | C | T | 0.038216 | 0.005026 | 8.20E-15 |
| rs600038 | miR-126-3p | trans | 1.80E-37 | Nightingale | HDL_size | NA | C | T | -0.03071 | 0.004524 | 3.50E-12 |
| rs600038 | miR-126-3p | trans | 1.80E-37 | Nightingale | IDL_C_pct | NA | C | T | 0.032847 | 0.004985 | 2.60E-10 |
| rs600038 | miR-126-3p | trans | 1.80E-37 | Nightingale | S_LDL_L | NA | C | T | 0.044203 | 0.005102 | 1.30E-18 |
| rs600038 | miR-126-3p | trans | 1.80E-37 | Nightingale | M_LDL_FC | NA | C | T | 0.056713 | 0.005096 | 5.00E-29 |
| rs600038 | miR-126-3p | trans | 1.80E-37 | Nightingale | M_VLDL_F | NA | C | T | 0.059646 | 0.004855 | 1.60E-34 |
| rs600038 | miR-126-3p | trans | 1.80E-37 | Nightingale | IDL_TG_pct | NA | C | T | -0.03341 | 0.004933 | 4.00E-11 |
| rs600038 | miR-126-3p | trans | 1.80E-37 | Nightingale | M_LDL_C_ | NA | C | T | 0.038144 | 0.005106 | 7.20E-14 |
| rs600038 | miR-126-3p | trans | 1.80E-37 | Nightingale | M_VLDL_P | NA | C | T | 0.063603 | 0.004897 | 8.80E-39 |
| rs600038 | miR-126-3p | trans | 1.80E-37 | Nightingale | S_HDL_FC | NA | C | T | 0.035556 | 0.005009 | 3.10E-13 |
| rs600038 | miR-126-3p | trans | 1.80E-37 | Nightingale | IDL_CE | NA | C | T | 0.052609 | 0.004927 | 4.00E-27 |
| rs600038 | miR-126-3p | trans | 1.80E-37 | Nightingale | M_VLDL_C | NA | C | T | 0.052637 | 0.004806 | 2.00E-27 |
| rs600038 | miR-126-3p | trans | 1.80E-37 | Nightingale | M_VLDL_C | NA | C | T | 0.059646 | 0.005056 | 4.30E-33 |
| rs600038 | miR-126-3p | trans | 1.80E-37 | Nightingale | S_VLDL_P | NA | C | T | 0.031474 | 0.005039 | 1.50E-11 |
| rs600038 | miR-126-3p | trans | 1.80E-37 | Nightingale | S_HDL_C | NA | C | T | 0.04513 | 0.005094 | 8.20E-19 |
| rs600038 | miR-126-3p | trans | 1.80E-37 | Nightingale | non_HDL_( | NA | C | T | 0.054532 | 0.005075 | 3.60E-28 |
| rs600038 | miR-126-3p | trans | 1.80E-37 | Nightingale | XL_VLDL_C | NA | C | T | 0.028612 | 0.005024 | 8.00E-10 |
| rs600038 | miR-126-3p | trans | 1.80E-37 | Nightingale | IDL_P | NA | C | T | 0.059653 | 0.005049 | 5.60E-33 |
| rs600038 | miR-126-3p | trans | 1.80E-37 | Nightingale | XXL_VLDL_ | NA | C | T | 0.032858 | 0.005138 | 5.00E-12 |
| rs600038 | miR-126-3p | trans | 1.80E-37 | Nightingale | LDL_PL | NA | C | T | 0.0557 | 0.005091 | 1.90E-28 |
| rs600038 | miR-126-3p | trans | 1.80E-37 | Nightingale | S_VLDL_CE | NA | C | T | 0.053323 | 0.005096 | 1.20E-27 |
| rs600038 | miR-126-3p | trans | 1.80E-37 | Nightingale | S_LDL_C | NA | C | T | 0.04877 | 0.005108 | 3.30E-22 |
| rs600038 | miR-126-3p | trans | 1.80E-37 | Nightingale | IDL_PL | NA | C | T | 0.052744 | 0.004924 | 5.20E-28 |
| rs600038 | miR-126-3p | trans | 1.80E-37 | Nightingale | S_VLDL_PL | NA | C | T | 0.046119 | 0.005093 | 3.30E-21 |
| rs600038 | miR-126-3p | trans | 1.80E-37 | Nightingale | Omega_6 | NA | C | T | 0.037012 | 0.004983 | 5.10E-15 |
| rs600038 | miR-126-3p | trans | 1.80E-37 | Nightingale | XL_HDL_C_ | NA | C | T | 0.033784 | 0.004873 | 2.10E-11 |
| rs600038 | miR-126-3p | trans | 1.80E-37 | Nightingale | M_VLDL_P | NA | C | T | 0.040156 | 0.00509 | 1.50E-16 |

| | | | | | | | | | | | |
|---|---|---|---|---|---|---|---|---|---|---|---|
| rs600038 | miR-126-3p | trans | 1.80E-37 | Nightingale | Phosphatic | NA | C | T | 0.027053 | 0.004781 | 2.30E-09 |
| rs600038 | miR-126-3p | trans | 1.80E-37 | Nightingale | Total_PL | NA | C | T | 0.034757 | 0.004859 | 4.60E-14 |
| rs600038 | miR-126-3p | trans | 1.80E-37 | Nightingale | LDL_CE | NA | C | T | 0.051899 | 0.005089 | 3.60E-25 |
| rs600038 | miR-126-3p | trans | 1.80E-37 | Nightingale | VLDL_CE | NA | C | T | 0.049508 | 0.005104 | 5.70E-24 |
| rs600038 | miR-126-3p | trans | 1.80E-37 | Nightingale | S_VLDL_FC | NA | C | T | 0.054116 | 0.005104 | 8.50E-28 |
| rs600038 | miR-126-3p | trans | 1.80E-37 | Nightingale | XS_VLDL_L | NA | C | T | 0.044732 | 0.005016 | 1.60E-21 |
| rs600038 | miR-126-3p | trans | 1.80E-37 | Nightingale | S_LDL_TG_ | NA | C | T | -0.03742 | 0.004948 | 2.20E-13 |
| rs600038 | miR-126-3p | trans | 1.80E-37 | Nightingale | XL_HDL_PL | NA | C | T | -0.02757 | 0.00459 | 6.70E-10 |
| rs600038 | miR-126-3p | trans | 1.80E-37 | Nightingale | M_LDL_C | NA | C | T | 0.052552 | 0.005112 | 1.70E-25 |
| rs600038 | miR-126-3p | trans | 1.80E-37 | Nightingale | L_LDL_PL | NA | C | T | 0.059497 | 0.005064 | 1.20E-32 |
| rs600038 | miR-126-3p | trans | 1.80E-37 | Nightingale | XS_VLDL_C | NA | C | T | 0.037556 | 0.00484 | 3.10E-14 |
| rs600038 | miR-126-3p | trans | 1.80E-37 | Nightingale | S_LDL_PL | NA | C | T | 0.038067 | 0.005091 | 4.30E-14 |
| rs600038 | miR-126-3p | trans | 1.80E-37 | Nightingale | S_LDL_CE | NA | C | T | 0.046863 | 0.005105 | 5.90E-21 |
| rs600038 | miR-126-3p | trans | 1.80E-37 | Nightingale | XL_VLDL_P | NA | C | T | 0.052743 | 0.005111 | 2.40E-26 |
| rs600038 | miR-126-3p | trans | 1.80E-37 | Nightingale | XXL_VLDL_ | NA | C | T | 0.051211 | 0.005116 | 5.90E-24 |
| rs600038 | miR-126-3p | trans | 1.80E-37 | Nightingale | S_HDL_PL_ | NA | C | T | -0.02877 | 0.004933 | 1.50E-08 |
| rs600038 | miR-126-3p | trans | 1.80E-37 | Nightingale | XS_VLDL_F | NA | C | T | 0.049012 | 0.005014 | 4.20E-25 |
| rs600038 | miR-126-3p | trans | 1.80E-37 | Nightingale | S_HDL_PL | NA | C | T | 0.032674 | 0.005059 | 2.80E-11 |
| rs600038 | miR-126-3p | trans | 1.80E-37 | Nightingale | L_VLDL_CE | NA | C | T | 0.029942 | 0.005043 | 1.80E-10 |
| rs600038 | miR-126-3p | trans | 1.80E-37 | Nightingale | XS_VLDL_P | NA | C | T | 0.04948 | 0.005035 | 2.10E-25 |
| rs600038 | miR-126-3p | trans | 1.80E-37 | Nightingale | VLDL_FC | NA | C | T | 0.032134 | 0.005065 | 1.10E-11 |
| rs600038 | miR-126-3p | trans | 1.80E-37 | Nightingale | S_LDL_PL_ | NA | C | T | -0.0395 | 0.005093 | 1.10E-16 |
| rs600038 | miR-126-3p | trans | 1.80E-37 | Nightingale | XXL_VLDL_ | NA | C | T | 0.050318 | 0.005144 | 4.30E-23 |
| rs600038 | miR-126-3p | trans | 1.80E-37 | Nightingale | XL_HDL_PL | NA | C | T | -0.03707 | 0.004688 | 2.50E-15 |
| rs600038 | miR-126-3p | trans | 1.80E-37 | Nightingale | M_LDL_CE | NA | C | T | 0.049694 | 0.00511 | 3.20E-23 |
| rs600038 | miR-126-3p | trans | 1.80E-37 | Nightingale | S_VLDL_TG | NA | C | T | -0.05802 | 0.004939 | 6.80E-32 |
| rs600038 | miR-126-3p | trans | 1.80E-37 | Nightingale | S_HDL_P | NA | C | T | 0.048052 | 0.005103 | 4.50E-21 |
| rs600038 | miR-126-3p | trans | 1.80E-37 | Nightingale | M_VLDL_T | NA | C | T | -0.05584 | 0.004822 | 1.20E-30 |
| rs600038 | miR-139-5p | trans | 7.68E-35 | Nightingale | XL_VLDL_C | NA | C | T | 0.054368 | 0.004895 | 2.80E-28 |
| rs600038 | miR-139-5p | trans | 7.68E-35 | Nightingale | M_VLDL_C | NA | C | T | 0.049278 | 0.004794 | 2.70E-24 |
| rs600038 | miR-139-5p | trans | 7.68E-35 | Nightingale | Total_CE | NA | C | T | 0.049009 | 0.004922 | 2.10E-24 |
| rs600038 | miR-139-5p | trans | 7.68E-35 | Nightingale | L_LDL_L | NA | C | T | 0.054273 | 0.005053 | 1.00E-27 |
| rs600038 | miR-139-5p | trans | 7.68E-35 | Nightingale | L_LDL_CE | NA | C | T | 0.051898 | 0.005057 | 2.00E-25 |
| rs600038 | miR-139-5p | trans | 7.68E-35 | Nightingale | L_LDL_C | NA | C | T | 0.053255 | 0.005045 | 1.10E-26 |
| rs600038 | miR-139-5p | trans | 7.68E-35 | Nightingale | Phosphogly | NA | C | T | 0.027754 | 0.004833 | 1.30E-09 |
| rs600038 | miR-139-5p | trans | 7.68E-35 | Nightingale | IDL_FC | NA | C | T | 0.057015 | 0.00494 | 1.50E-31 |
| rs600038 | miR-139-5p | trans | 7.68E-35 | Nightingale | S_VLDL_C | NA | C | T | 0.05403 | 0.005102 | 5.10E-28 |
| rs600038 | miR-139-5p | trans | 7.68E-35 | Nightingale | Total_C | NA | C | T | 0.049326 | 0.00494 | 1.10E-24 |
| rs600038 | miR-139-5p | trans | 7.68E-35 | Nightingale | L_VLDL_PL | NA | C | T | 0.027472 | 0.004957 | 1.00E-09 |
| rs600038 | miR-139-5p | trans | 7.68E-35 | Nightingale | LDL_P | NA | C | T | 0.048287 | 0.005099 | 3.30E-22 |
| rs600038 | miR-139-5p | trans | 7.68E-35 | Nightingale | M_VLDL_C | NA | C | T | 0.055912 | 0.005089 | 1.20E-29 |
| rs600038 | miR-139-5p | trans | 7.68E-35 | Nightingale | M_LDL_PL | NA | C | T | 0.052654 | 0.00511 | 1.50E-25 |
| rs600038 | miR-139-5p | trans | 7.68E-35 | Nightingale | LA | NA | C | T | 0.037469 | 0.005004 | 3.30E-15 |
| rs600038 | miR-139-5p | trans | 7.68E-35 | Nightingale | Citrate | NA | C | T | 0.02733 | 0.00507 | 2.60E-08 |
| rs600038 | miR-139-5p | trans | 7.68E-35 | Nightingale | Pyruvate | NA | C | T | 0.030176 | 0.005111 | 3.60E-09 |
| rs600038 | miR-139-5p | trans | 7.68E-35 | Nightingale | VLDL_P | NA | C | T | 0.03602 | 0.005075 | 4.70E-14 |
| rs600038 | miR-139-5p | trans | 7.68E-35 | Nightingale | Sphingomy | NA | C | T | 0.040531 | 0.00477 | 1.70E-17 |
| rs600038 | miR-139-5p | trans | 7.68E-35 | Nightingale | S_LDL_FC | NA | C | T | 0.049076 | 0.005095 | 7.70E-22 |
| rs600038 | miR-139-5p | trans | 7.68E-35 | Nightingale | LDL_FC | NA | C | T | 0.055801 | 0.005048 | 1.60E-28 |
| rs600038 | miR-139-5p | trans | 7.68E-35 | Nightingale | XS_VLDL_C | NA | C | T | 0.052331 | 0.004953 | 3.80E-28 |
| rs600038 | miR-139-5p | trans | 7.68E-35 | Nightingale | XS_VLDL_P | NA | C | T | 0.041555 | 0.005032 | 8.10E-19 |
| rs600038 | miR-139-5p | trans | 7.68E-35 | Nightingale | L_LDL_FC | NA | C | T | 0.055661 | 0.005011 | 5.90E-29 |
| rs600038 | miR-139-5p | trans | 7.68E-35 | Nightingale | XXL_VLDL_ | NA | C | T | -0.05276 | 0.005149 | 1.90E-25 |
| rs600038 | miR-139-5p | trans | 7.68E-35 | Nightingale | XS_VLDL_C | NA | C | T | 0.052917 | 0.004924 | 7.70E-29 |
| rs600038 | miR-139-5p | trans | 7.68E-35 | Nightingale | S_VLDL_FC | NA | C | T | 0.045985 | 0.004876 | 8.40E-21 |
| rs600038 | miR-139-5p | trans | 7.68E-35 | Nightingale | S_LDL_CE_ | NA | C | T | 0.039177 | 0.005094 | 1.30E-16 |
| rs600038 | miR-139-5p | trans | 7.68E-35 | Nightingale | S_VLDL_C_ | NA | C | T | 0.062934 | 0.004966 | 6.40E-37 |
| rs600038 | miR-139-5p | trans | 7.68E-35 | Nightingale | L_VLDL_CE | NA | C | T | 0.065536 | 0.004883 | 4.90E-42 |
| rs600038 | miR-139-5p | trans | 7.68E-35 | Nightingale | S_VLDL_PL | NA | C | T | 0.043922 | 0.004886 | 5.30E-19 |
| rs600038 | miR-139-5p | trans | 7.68E-35 | Nightingale | XS_VLDL_F | NA | C | T | 0.044491 | 0.005066 | 8.00E-18 |
| rs600038 | miR-139-5p | trans | 7.68E-35 | Nightingale | XL_VLDL_C | NA | C | T | 0.054864 | 0.004893 | 6.90E-29 |
| rs600038 | miR-139-5p | trans | 7.68E-35 | Nightingale | S_LDL_P | NA | C | T | 0.044606 | 0.005107 | 3.00E-19 |
| rs600038 | miR-139-5p | trans | 7.68E-35 | Nightingale | L_LDL_TG_ | NA | C | T | -0.03537 | 0.005005 | 5.10E-12 |

| | | | | | | | | | | | |
|---|---|---|---|---|---|---|---|---|---|---|---|
| rs600038 | miR-139-5p | trans | 7.68E-35 | Nightingale | M_VLDL_P | NA | C | T | 0.041549 | 0.005095 | 1.40E-17 |
| rs600038 | miR-139-5p | trans | 7.68E-35 | Nightingale | L_HDL_TG_ | NA | C | T | 0.023502 | 0.004821 | 3.50E-08 |
| rs600038 | miR-139-5p | trans | 7.68E-35 | Nightingale | LDL_C | NA | C | T | 0.053489 | 0.005081 | 1.60E-26 |
| rs600038 | miR-139-5p | trans | 7.68E-35 | Nightingale | S_VLDL_L | NA | C | T | 0.033361 | 0.005053 | 1.40E-12 |
| rs600038 | miR-139-5p | trans | 7.68E-35 | Nightingale | LDL_L | NA | C | T | 0.053245 | 0.005086 | 2.30E-26 |
| rs600038 | miR-139-5p | trans | 7.68E-35 | Nightingale | M_VLDL_F | NA | C | T | 0.047425 | 0.005102 | 6.00E-22 |
| rs600038 | miR-139-5p | trans | 7.68E-35 | Nightingale | IDL_C | NA | C | T | 0.054083 | 0.004928 | 1.40E-28 |
| rs600038 | miR-139-5p | trans | 7.68E-35 | Nightingale | IDL_L | NA | C | T | 0.053693 | 0.004934 | 1.90E-28 |
| rs600038 | miR-139-5p | trans | 7.68E-35 | Nightingale | Clinical_LD | NA | C | T | 0.057741 | 0.005073 | 1.50E-30 |
| rs600038 | miR-139-5p | trans | 7.68E-35 | Nightingale | L_VLDL_C_ | NA | C | T | 0.072919 | 0.004984 | 1.60E-49 |
| rs600038 | miR-139-5p | trans | 7.68E-35 | Nightingale | Remnant_( | NA | C | T | 0.053469 | 0.00507 | 6.60E-28 |
| rs600038 | miR-139-5p | trans | 7.68E-35 | Nightingale | M_VLDL_L | NA | C | T | 0.030261 | 0.005068 | 2.00E-10 |
| rs600038 | miR-139-5p | trans | 7.68E-35 | Nightingale | XS_VLDL_C | NA | C | T | 0.0349 | 0.004815 | 1.80E-12 |
| rs600038 | miR-139-5p | trans | 7.68E-35 | Nightingale | Cholines | NA | C | T | 0.030838 | 0.004799 | 1.40E-11 |
| rs600038 | miR-139-5p | trans | 7.68E-35 | Nightingale | VLDL_C | NA | C | T | 0.042745 | 0.005096 | 1.20E-18 |
| rs600038 | miR-139-5p | trans | 7.68E-35 | Nightingale | PUFA | NA | C | T | 0.032623 | 0.004977 | 6.00E-12 |
| rs600038 | miR-139-5p | trans | 7.68E-35 | Nightingale | XL_VLDL_F | NA | C | T | 0.050732 | 0.004947 | 2.90E-25 |
| rs600038 | miR-139-5p | trans | 7.68E-35 | Nightingale | VLDL_PL | NA | C | T | 0.025473 | 0.005042 | 3.80E-08 |
| rs600038 | miR-139-5p | trans | 7.68E-35 | Nightingale | ApoB | NA | C | T | 0.050849 | 0.005096 | 1.40E-24 |
| rs600038 | miR-139-5p | trans | 7.68E-35 | Nightingale | Total_P | NA | C | T | 0.03028 | 0.004803 | 7.20E-11 |
| rs600038 | miR-139-5p | trans | 7.68E-35 | Nightingale | S_HDL_CE | NA | C | T | 0.045974 | 0.005111 | 3.30E-19 |
| rs600038 | miR-139-5p | trans | 7.68E-35 | Nightingale | L_LDL_P | NA | C | T | 0.045959 | 0.005086 | 2.50E-20 |
| rs600038 | miR-139-5p | trans | 7.68E-35 | Nightingale | M_LDL_TG | NA | C | T | -0.04594 | 0.00504 | 1.80E-19 |
| rs600038 | miR-139-5p | trans | 7.68E-35 | Nightingale | XL_HDL_L | NA | C | T | -0.02484 | 0.004589 | 1.40E-08 |
| rs600038 | miR-139-5p | trans | 7.68E-35 | Nightingale | L_VLDL_TG | NA | C | T | -0.07208 | 0.005079 | 1.80E-47 |
| rs600038 | miR-139-5p | trans | 7.68E-35 | Nightingale | S_LDL_C_p | NA | C | T | 0.065911 | 0.005123 | 1.80E-38 |
| rs600038 | miR-139-5p | trans | 7.68E-35 | Nightingale | XS_VLDL_T | NA | C | T | -0.03742 | 0.004852 | 4.10E-14 |
| rs600038 | miR-139-5p | trans | 7.68E-35 | Nightingale | Total_FC | NA | C | T | 0.049114 | 0.004985 | 8.70E-25 |
| rs600038 | miR-139-5p | trans | 7.68E-35 | Nightingale | XL_VLDL_T | NA | C | T | -0.06142 | 0.004946 | 1.30E-35 |
| rs600038 | miR-139-5p | trans | 7.68E-35 | Nightingale | S_HDL_L | NA | C | T | 0.038363 | 0.005085 | 1.60E-14 |
| rs600038 | miR-139-5p | trans | 7.68E-35 | Nightingale | IDL_FC_pct | NA | C | T | 0.033912 | 0.005063 | 4.50E-11 |
| rs600038 | miR-139-5p | trans | 7.68E-35 | Nightingale | S_VLDL_CE | NA | C | T | 0.069016 | 0.005034 | 1.40E-42 |
| rs600038 | miR-139-5p | trans | 7.68E-35 | Nightingale | L_VLDL_FC | NA | C | T | 0.068916 | 0.005054 | 4.60E-43 |
| rs600038 | miR-139-5p | trans | 7.68E-35 | Nightingale | XXL_VLDL_ | NA | C | T | 0.041556 | 0.005084 | 1.00E-16 |
| rs600038 | miR-139-5p | trans | 7.68E-35 | Nightingale | Total_L | NA | C | T | 0.038104 | 0.00502 | 2.10E-15 |
| rs600038 | miR-139-5p | trans | 7.68E-35 | Nightingale | M_LDL_P | NA | C | T | 0.051071 | 0.005117 | 2.50E-24 |
| rs600038 | miR-139-5p | trans | 7.68E-35 | Nightingale | M_LDL_L | NA | C | T | 0.051518 | 0.00511 | 1.20E-24 |
| rs600038 | miR-139-5p | trans | 7.68E-35 | Nightingale | ApoB_by_A | NA | C | T | 0.038216 | 0.005026 | 8.20E-15 |
| rs600038 | miR-139-5p | trans | 7.68E-35 | Nightingale | HDL_size | NA | C | T | -0.03071 | 0.004524 | 3.50E-12 |
| rs600038 | miR-139-5p | trans | 7.68E-35 | Nightingale | IDL_C_pct | NA | C | T | 0.032847 | 0.004985 | 2.60E-10 |
| rs600038 | miR-139-5p | trans | 7.68E-35 | Nightingale | S_LDL_L | NA | C | T | 0.044203 | 0.005102 | 1.30E-18 |
| rs600038 | miR-139-5p | trans | 7.68E-35 | Nightingale | M_LDL_FC | NA | C | T | 0.056713 | 0.005096 | 5.00E-29 |
| rs600038 | miR-139-5p | trans | 7.68E-35 | Nightingale | M_VLDL_F | NA | C | T | 0.059646 | 0.004855 | 1.60E-34 |
| rs600038 | miR-139-5p | trans | 7.68E-35 | Nightingale | IDL_TG_pc | NA | C | T | -0.03341 | 0.004933 | 4.00E-11 |
| rs600038 | miR-139-5p | trans | 7.68E-35 | Nightingale | M_LDL_C_ | NA | C | T | 0.038144 | 0.005106 | 7.20E-14 |
| rs600038 | miR-139-5p | trans | 7.68E-35 | Nightingale | M_VLDL_P | NA | C | T | 0.063603 | 0.004897 | 8.80E-39 |
| rs600038 | miR-139-5p | trans | 7.68E-35 | Nightingale | S_HDL_FC | NA | C | T | 0.035556 | 0.005009 | 3.10E-13 |
| rs600038 | miR-139-5p | trans | 7.68E-35 | Nightingale | IDL_CE | NA | C | T | 0.052609 | 0.004927 | 4.00E-27 |
| rs600038 | miR-139-5p | trans | 7.68E-35 | Nightingale | M_VLDL_C | NA | C | T | 0.052637 | 0.004806 | 2.00E-27 |
| rs600038 | miR-139-5p | trans | 7.68E-35 | Nightingale | M_VLDL_C | NA | C | T | 0.059646 | 0.005056 | 4.30E-33 |
| rs600038 | miR-139-5p | trans | 7.68E-35 | Nightingale | S_VLDL_P | NA | C | T | 0.031474 | 0.005039 | 1.50E-11 |
| rs600038 | miR-139-5p | trans | 7.68E-35 | Nightingale | S_HDL_C | NA | C | T | 0.04513 | 0.005094 | 8.20E-19 |
| rs600038 | miR-139-5p | trans | 7.68E-35 | Nightingale | non_HDL_( | NA | C | T | 0.054532 | 0.005075 | 3.60E-28 |
| rs600038 | miR-139-5p | trans | 7.68E-35 | Nightingale | XL_VLDL_C | NA | C | T | 0.028612 | 0.005024 | 8.00E-10 |
| rs600038 | miR-139-5p | trans | 7.68E-35 | Nightingale | IDL_P | NA | C | T | 0.059653 | 0.005049 | 5.60E-33 |
| rs600038 | miR-139-5p | trans | 7.68E-35 | Nightingale | XXL_VLDL_ | NA | C | T | 0.032858 | 0.005138 | 5.00E-12 |
| rs600038 | miR-139-5p | trans | 7.68E-35 | Nightingale | LDL_PL | NA | C | T | 0.0557 | 0.005091 | 1.90E-28 |
| rs600038 | miR-139-5p | trans | 7.68E-35 | Nightingale | S_VLDL_CE | NA | C | T | 0.053323 | 0.005096 | 1.20E-27 |
| rs600038 | miR-139-5p | trans | 7.68E-35 | Nightingale | S_LDL_C | NA | C | T | 0.04877 | 0.005108 | 3.30E-22 |
| rs600038 | miR-139-5p | trans | 7.68E-35 | Nightingale | IDL_PL | NA | C | T | 0.052744 | 0.004924 | 5.20E-28 |
| rs600038 | miR-139-5p | trans | 7.68E-35 | Nightingale | S_VLDL_PL | NA | C | T | 0.046119 | 0.005093 | 3.30E-21 |
| rs600038 | miR-139-5p | trans | 7.68E-35 | Nightingale | Omega_6 | NA | C | T | 0.037012 | 0.004983 | 5.10E-15 |
| rs600038 | miR-139-5p | trans | 7.68E-35 | Nightingale | XL_HDL_C_ | NA | C | T | 0.033784 | 0.004873 | 2.10E-11 |

| | | | | | | | | | | | |
|---|---|---|---|---|---|---|---|---|---|---|---|
| rs600038 | miR-139-5p | trans | 7.68E-35 | Nightingale | M_VLDL_P | NA | C | T | 0.040156 | 0.00509 | 1.50E-16 |
| rs600038 | miR-139-5p | trans | 7.68E-35 | Nightingale | Phosphatic | NA | C | T | 0.027053 | 0.004781 | 2.30E-09 |
| rs600038 | miR-139-5p | trans | 7.68E-35 | Nightingale | Total_PL | NA | C | T | 0.034757 | 0.004859 | 4.60E-14 |
| rs600038 | miR-139-5p | trans | 7.68E-35 | Nightingale | LDL_CE | NA | C | T | 0.051899 | 0.005089 | 3.60E-25 |
| rs600038 | miR-139-5p | trans | 7.68E-35 | Nightingale | VLDL_CE | NA | C | T | 0.049508 | 0.005104 | 5.70E-24 |
| rs600038 | miR-139-5p | trans | 7.68E-35 | Nightingale | S_VLDL_FC | NA | C | T | 0.054116 | 0.005104 | 8.50E-28 |
| rs600038 | miR-139-5p | trans | 7.68E-35 | Nightingale | XS_VLDL_L | NA | C | T | 0.044732 | 0.005016 | 1.60E-21 |
| rs600038 | miR-139-5p | trans | 7.68E-35 | Nightingale | S_LDL_TG_ | NA | C | T | -0.03742 | 0.004948 | 2.20E-13 |
| rs600038 | miR-139-5p | trans | 7.68E-35 | Nightingale | XL_HDL_PL | NA | C | T | -0.02757 | 0.00459 | 6.70E-10 |
| rs600038 | miR-139-5p | trans | 7.68E-35 | Nightingale | M_LDL_C | NA | C | T | 0.052552 | 0.005112 | 1.70E-25 |
| rs600038 | miR-139-5p | trans | 7.68E-35 | Nightingale | L_LDL_PL | NA | C | T | 0.059497 | 0.005064 | 1.20E-32 |
| rs600038 | miR-139-5p | trans | 7.68E-35 | Nightingale | XS_VLDL_C | NA | C | T | 0.037556 | 0.00484 | 3.10E-14 |
| rs600038 | miR-139-5p | trans | 7.68E-35 | Nightingale | S_LDL_PL | NA | C | T | 0.038067 | 0.005091 | 4.30E-14 |
| rs600038 | miR-139-5p | trans | 7.68E-35 | Nightingale | S_LDL_CE | NA | C | T | 0.046863 | 0.005105 | 5.90E-21 |
| rs600038 | miR-139-5p | trans | 7.68E-35 | Nightingale | XL_VLDL_P | NA | C | T | 0.052743 | 0.005111 | 2.40E-26 |
| rs600038 | miR-139-5p | trans | 7.68E-35 | Nightingale | XXL_VLDL_ | NA | C | T | 0.051211 | 0.005116 | 5.90E-24 |
| rs600038 | miR-139-5p | trans | 7.68E-35 | Nightingale | S_HDL_PL_ | NA | C | T | -0.02877 | 0.004933 | 1.50E-08 |
| rs600038 | miR-139-5p | trans | 7.68E-35 | Nightingale | XS_VLDL_F | NA | C | T | 0.049012 | 0.005014 | 4.20E-25 |
| rs600038 | miR-139-5p | trans | 7.68E-35 | Nightingale | S_HDL_PL | NA | C | T | 0.032674 | 0.005059 | 2.80E-11 |
| rs600038 | miR-139-5p | trans | 7.68E-35 | Nightingale | L_VLDL_CE | NA | C | T | 0.029942 | 0.005043 | 1.80E-10 |
| rs600038 | miR-139-5p | trans | 7.68E-35 | Nightingale | XS_VLDL_P | NA | C | T | 0.04948 | 0.005035 | 2.10E-25 |
| rs600038 | miR-139-5p | trans | 7.68E-35 | Nightingale | VLDL_FC | NA | C | T | 0.032134 | 0.005065 | 1.10E-11 |
| rs600038 | miR-139-5p | trans | 7.68E-35 | Nightingale | S_LDL_PL_ | NA | C | T | -0.0395 | 0.005093 | 1.10E-16 |
| rs600038 | miR-139-5p | trans | 7.68E-35 | Nightingale | XXL_VLDL_ | NA | C | T | 0.050318 | 0.005144 | 4.30E-23 |
| rs600038 | miR-139-5p | trans | 7.68E-35 | Nightingale | XL_HDL_PL | NA | C | T | -0.03707 | 0.004688 | 2.50E-15 |
| rs600038 | miR-139-5p | trans | 7.68E-35 | Nightingale | M_LDL_CE | NA | C | T | 0.049694 | 0.00511 | 3.20E-23 |
| rs600038 | miR-139-5p | trans | 7.68E-35 | Nightingale | S_VLDL_TG | NA | C | T | -0.05802 | 0.004939 | 6.80E-32 |
| rs600038 | miR-139-5p | trans | 7.68E-35 | Nightingale | S_HDL_P | NA | C | T | 0.048052 | 0.005103 | 4.50E-21 |
| rs600038 | miR-139-5p | trans | 7.68E-35 | Nightingale | M_VLDL_T | NA | C | T | -0.05584 | 0.004822 | 1.20E-30 |
| rs600038 | let-7b-5p | trans | 1.02E-24 | Nightingale | XL_VLDL_C | NA | C | T | 0.054368 | 0.004895 | 2.80E-28 |
| rs600038 | let-7b-5p | trans | 1.02E-24 | Nightingale | M_VLDL_C | NA | C | T | 0.049278 | 0.004794 | 2.70E-24 |
| rs600038 | let-7b-5p | trans | 1.02E-24 | Nightingale | Total_CE | NA | C | T | 0.049009 | 0.004922 | 2.10E-24 |
| rs600038 | let-7b-5p | trans | 1.02E-24 | Nightingale | L_LDL_L | NA | C | T | 0.054273 | 0.005053 | 1.00E-27 |
| rs600038 | let-7b-5p | trans | 1.02E-24 | Nightingale | L_LDL_CE | NA | C | T | 0.051898 | 0.005057 | 2.00E-25 |
| rs600038 | let-7b-5p | trans | 1.02E-24 | Nightingale | L_LDL_C | NA | C | T | 0.053255 | 0.005045 | 1.10E-26 |
| rs600038 | let-7b-5p | trans | 1.02E-24 | Nightingale | Phosphogly | NA | C | T | 0.027754 | 0.004833 | 1.30E-09 |
| rs600038 | let-7b-5p | trans | 1.02E-24 | Nightingale | IDL_FC | NA | C | T | 0.057015 | 0.00494 | 1.50E-31 |
| rs600038 | let-7b-5p | trans | 1.02E-24 | Nightingale | S_VLDL_C | NA | C | T | 0.05403 | 0.005102 | 5.10E-28 |
| rs600038 | let-7b-5p | trans | 1.02E-24 | Nightingale | Total_C | NA | C | T | 0.049326 | 0.00494 | 1.10E-24 |
| rs600038 | let-7b-5p | trans | 1.02E-24 | Nightingale | L_VLDL_PL | NA | C | T | 0.027472 | 0.004957 | 1.00E-09 |
| rs600038 | let-7b-5p | trans | 1.02E-24 | Nightingale | LDL_P | NA | C | T | 0.048287 | 0.005099 | 3.30E-22 |
| rs600038 | let-7b-5p | trans | 1.02E-24 | Nightingale | M_VLDL_C | NA | C | T | 0.055912 | 0.005089 | 1.20E-29 |
| rs600038 | let-7b-5p | trans | 1.02E-24 | Nightingale | M_LDL_PL | NA | C | T | 0.052654 | 0.00511 | 1.50E-25 |
| rs600038 | let-7b-5p | trans | 1.02E-24 | Nightingale | LA | NA | C | T | 0.037469 | 0.005004 | 3.30E-15 |
| rs600038 | let-7b-5p | trans | 1.02E-24 | Nightingale | Citrate | NA | C | T | 0.02733 | 0.00507 | 2.60E-08 |
| rs600038 | let-7b-5p | trans | 1.02E-24 | Nightingale | Pyruvate | NA | C | T | 0.030176 | 0.005111 | 3.60E-09 |
| rs600038 | let-7b-5p | trans | 1.02E-24 | Nightingale | VLDL_P | NA | C | T | 0.03602 | 0.005075 | 4.70E-14 |
| rs600038 | let-7b-5p | trans | 1.02E-24 | Nightingale | Sphingomy | NA | C | T | 0.040531 | 0.00477 | 1.70E-17 |
| rs600038 | let-7b-5p | trans | 1.02E-24 | Nightingale | S_LDL_FC | NA | C | T | 0.049076 | 0.005095 | 7.70E-22 |
| rs600038 | let-7b-5p | trans | 1.02E-24 | Nightingale | LDL_FC | NA | C | T | 0.055801 | 0.005048 | 1.60E-28 |
| rs600038 | let-7b-5p | trans | 1.02E-24 | Nightingale | XS_VLDL_C | NA | C | T | 0.052331 | 0.004953 | 3.80E-28 |
| rs600038 | let-7b-5p | trans | 1.02E-24 | Nightingale | XS_VLDL_P | NA | C | T | 0.041555 | 0.005032 | 8.10E-19 |
| rs600038 | let-7b-5p | trans | 1.02E-24 | Nightingale | L_LDL_FC | NA | C | T | 0.055661 | 0.005011 | 5.90E-29 |
| rs600038 | let-7b-5p | trans | 1.02E-24 | Nightingale | XXL_VLDL_ | NA | C | T | -0.05276 | 0.005149 | 1.90E-25 |
| rs600038 | let-7b-5p | trans | 1.02E-24 | Nightingale | XS_VLDL_C | NA | C | T | 0.052917 | 0.004924 | 7.70E-29 |
| rs600038 | let-7b-5p | trans | 1.02E-24 | Nightingale | S_VLDL_FC | NA | C | T | 0.045985 | 0.004876 | 8.40E-21 |
| rs600038 | let-7b-5p | trans | 1.02E-24 | Nightingale | S_LDL_CE_ | NA | C | T | 0.039177 | 0.005094 | 1.30E-16 |
| rs600038 | let-7b-5p | trans | 1.02E-24 | Nightingale | S_VLDL_C_ | NA | C | T | 0.062934 | 0.004966 | 6.40E-37 |
| rs600038 | let-7b-5p | trans | 1.02E-24 | Nightingale | L_VLDL_CE | NA | C | T | 0.065536 | 0.004883 | 4.90E-42 |
| rs600038 | let-7b-5p | trans | 1.02E-24 | Nightingale | S_VLDL_PL | NA | C | T | 0.043922 | 0.004886 | 5.30E-19 |
| rs600038 | let-7b-5p | trans | 1.02E-24 | Nightingale | XS_VLDL_F | NA | C | T | 0.044491 | 0.005066 | 8.00E-18 |
| rs600038 | let-7b-5p | trans | 1.02E-24 | Nightingale | XL_VLDL_C | NA | C | T | 0.054864 | 0.004893 | 6.90E-29 |
| rs600038 | let-7b-5p | trans | 1.02E-24 | Nightingale | S_LDL_P | NA | C | T | 0.044606 | 0.005107 | 3.00E-19 |

| | | | | | | | | | | | |
|---|---|---|---|---|---|---|---|---|---|---|---|
| rs600038 | let-7b-5p | trans | 1.02E-24 | Nightingale | L_LDL_TG_ | NA | C | T | -0.03537 | 0.005005 | 5.10E-12 |
| rs600038 | let-7b-5p | trans | 1.02E-24 | Nightingale | M_VLDL_P | NA | C | T | 0.041549 | 0.005095 | 1.40E-17 |
| rs600038 | let-7b-5p | trans | 1.02E-24 | Nightingale | L_HDL_TG_ | NA | C | T | 0.023502 | 0.004821 | 3.50E-08 |
| rs600038 | let-7b-5p | trans | 1.02E-24 | Nightingale | LDL_C | NA | C | T | 0.053489 | 0.005081 | 1.60E-26 |
| rs600038 | let-7b-5p | trans | 1.02E-24 | Nightingale | S_VLDL_L | NA | C | T | 0.033361 | 0.005053 | 1.40E-12 |
| rs600038 | let-7b-5p | trans | 1.02E-24 | Nightingale | LDL_L | NA | C | T | 0.053245 | 0.005086 | 2.30E-26 |
| rs600038 | let-7b-5p | trans | 1.02E-24 | Nightingale | M_VLDL_F | NA | C | T | 0.047425 | 0.005102 | 6.00E-22 |
| rs600038 | let-7b-5p | trans | 1.02E-24 | Nightingale | IDL_C | NA | C | T | 0.054083 | 0.004928 | 1.40E-28 |
| rs600038 | let-7b-5p | trans | 1.02E-24 | Nightingale | IDL_L | NA | C | T | 0.053693 | 0.004934 | 1.90E-28 |
| rs600038 | let-7b-5p | trans | 1.02E-24 | Nightingale | Clinical_LD | NA | C | T | 0.057741 | 0.005073 | 1.50E-30 |
| rs600038 | let-7b-5p | trans | 1.02E-24 | Nightingale | L_VLDL_C_ | NA | C | T | 0.072919 | 0.004984 | 1.60E-49 |
| rs600038 | let-7b-5p | trans | 1.02E-24 | Nightingale | Remnant_( | NA | C | T | 0.053469 | 0.00507 | 6.60E-28 |
| rs600038 | let-7b-5p | trans | 1.02E-24 | Nightingale | M_VLDL_L | NA | C | T | 0.030261 | 0.005068 | 2.00E-10 |
| rs600038 | let-7b-5p | trans | 1.02E-24 | Nightingale | XS_VLDL_C | NA | C | T | 0.0349 | 0.004815 | 1.80E-12 |
| rs600038 | let-7b-5p | trans | 1.02E-24 | Nightingale | Cholines | NA | C | T | 0.030838 | 0.004799 | 1.40E-11 |
| rs600038 | let-7b-5p | trans | 1.02E-24 | Nightingale | VLDL_C | NA | C | T | 0.042745 | 0.005096 | 1.20E-18 |
| rs600038 | let-7b-5p | trans | 1.02E-24 | Nightingale | PUFA | NA | C | T | 0.032623 | 0.004977 | 6.00E-12 |
| rs600038 | let-7b-5p | trans | 1.02E-24 | Nightingale | XL_VLDL_F | NA | C | T | 0.050732 | 0.004947 | 2.90E-25 |
| rs600038 | let-7b-5p | trans | 1.02E-24 | Nightingale | VLDL_PL | NA | C | T | 0.025473 | 0.005042 | 3.80E-08 |
| rs600038 | let-7b-5p | trans | 1.02E-24 | Nightingale | ApoB | NA | C | T | 0.050849 | 0.005096 | 1.40E-24 |
| rs600038 | let-7b-5p | trans | 1.02E-24 | Nightingale | Total_P | NA | C | T | 0.03028 | 0.004803 | 7.20E-11 |
| rs600038 | let-7b-5p | trans | 1.02E-24 | Nightingale | S_HDL_CE | NA | C | T | 0.045974 | 0.005111 | 3.30E-19 |
| rs600038 | let-7b-5p | trans | 1.02E-24 | Nightingale | L_LDL_P | NA | C | T | 0.045959 | 0.005086 | 2.50E-20 |
| rs600038 | let-7b-5p | trans | 1.02E-24 | Nightingale | M_LDL_TG | NA | C | T | -0.04594 | 0.00504 | 1.80E-19 |
| rs600038 | let-7b-5p | trans | 1.02E-24 | Nightingale | XL_HDL_L | NA | C | T | -0.02484 | 0.004589 | 1.40E-08 |
| rs600038 | let-7b-5p | trans | 1.02E-24 | Nightingale | L_VLDL_TG | NA | C | T | -0.07208 | 0.005079 | 1.80E-47 |
| rs600038 | let-7b-5p | trans | 1.02E-24 | Nightingale | S_LDL_C_p | NA | C | T | 0.065911 | 0.005123 | 1.80E-38 |
| rs600038 | let-7b-5p | trans | 1.02E-24 | Nightingale | XS_VLDL_T | NA | C | T | -0.03742 | 0.004852 | 4.10E-14 |
| rs600038 | let-7b-5p | trans | 1.02E-24 | Nightingale | Total_FC | NA | C | T | 0.049114 | 0.004985 | 8.70E-25 |
| rs600038 | let-7b-5p | trans | 1.02E-24 | Nightingale | XL_VLDL_T | NA | C | T | -0.06142 | 0.004946 | 1.30E-35 |
| rs600038 | let-7b-5p | trans | 1.02E-24 | Nightingale | S_HDL_L | NA | C | T | 0.038363 | 0.005085 | 1.60E-14 |
| rs600038 | let-7b-5p | trans | 1.02E-24 | Nightingale | IDL_FC_pct | NA | C | T | 0.033912 | 0.005063 | 4.50E-11 |
| rs600038 | let-7b-5p | trans | 1.02E-24 | Nightingale | S_VLDL_CE | NA | C | T | 0.069016 | 0.005034 | 1.40E-42 |
| rs600038 | let-7b-5p | trans | 1.02E-24 | Nightingale | L_VLDL_FC | NA | C | T | 0.068916 | 0.005054 | 4.60E-43 |
| rs600038 | let-7b-5p | trans | 1.02E-24 | Nightingale | XXL_VLDL_ | NA | C | T | 0.041556 | 0.005084 | 1.00E-16 |
| rs600038 | let-7b-5p | trans | 1.02E-24 | Nightingale | Total_L | NA | C | T | 0.038104 | 0.00502 | 2.10E-15 |
| rs600038 | let-7b-5p | trans | 1.02E-24 | Nightingale | M_LDL_P | NA | C | T | 0.051071 | 0.005117 | 2.50E-24 |
| rs600038 | let-7b-5p | trans | 1.02E-24 | Nightingale | M_LDL_L | NA | C | T | 0.051518 | 0.00511 | 1.20E-24 |
| rs600038 | let-7b-5p | trans | 1.02E-24 | Nightingale | ApoB_by_/ | NA | C | T | 0.038216 | 0.005026 | 8.20E-15 |
| rs600038 | let-7b-5p | trans | 1.02E-24 | Nightingale | HDL_size | NA | C | T | -0.03071 | 0.004524 | 3.50E-12 |
| rs600038 | let-7b-5p | trans | 1.02E-24 | Nightingale | IDL_C_pct | NA | C | T | 0.032847 | 0.004985 | 2.60E-10 |
| rs600038 | let-7b-5p | trans | 1.02E-24 | Nightingale | S_LDL_L | NA | C | T | 0.044203 | 0.005102 | 1.30E-18 |
| rs600038 | let-7b-5p | trans | 1.02E-24 | Nightingale | M_LDL_FC | NA | C | T | 0.056713 | 0.005096 | 5.00E-29 |
| rs600038 | let-7b-5p | trans | 1.02E-24 | Nightingale | M_VLDL_F | NA | C | T | 0.059646 | 0.004855 | 1.60E-34 |
| rs600038 | let-7b-5p | trans | 1.02E-24 | Nightingale | IDL_TG_pc | NA | C | T | -0.03341 | 0.004933 | 4.00E-11 |
| rs600038 | let-7b-5p | trans | 1.02E-24 | Nightingale | M_LDL_C_ | NA | C | T | 0.038144 | 0.005106 | 7.20E-14 |
| rs600038 | let-7b-5p | trans | 1.02E-24 | Nightingale | M_VLDL_P | NA | C | T | 0.063603 | 0.004897 | 8.80E-39 |
| rs600038 | let-7b-5p | trans | 1.02E-24 | Nightingale | S_HDL_FC | NA | C | T | 0.035556 | 0.005009 | 3.10E-13 |
| rs600038 | let-7b-5p | trans | 1.02E-24 | Nightingale | IDL_CE | NA | C | T | 0.052609 | 0.004927 | 4.00E-27 |
| rs600038 | let-7b-5p | trans | 1.02E-24 | Nightingale | M_VLDL_C | NA | C | T | 0.052637 | 0.004806 | 2.00E-27 |
| rs600038 | let-7b-5p | trans | 1.02E-24 | Nightingale | M_VLDL_C | NA | C | T | 0.059646 | 0.005056 | 4.30E-33 |
| rs600038 | let-7b-5p | trans | 1.02E-24 | Nightingale | S_VLDL_P | NA | C | T | 0.031474 | 0.005039 | 1.50E-11 |
| rs600038 | let-7b-5p | trans | 1.02E-24 | Nightingale | S_HDL_C | NA | C | T | 0.04513 | 0.005094 | 8.20E-19 |
| rs600038 | let-7b-5p | trans | 1.02E-24 | Nightingale | non_HDL_( | NA | C | T | 0.054532 | 0.005075 | 3.60E-28 |
| rs600038 | let-7b-5p | trans | 1.02E-24 | Nightingale | XL_VLDL_C | NA | C | T | 0.028612 | 0.005024 | 8.00E-10 |
| rs600038 | let-7b-5p | trans | 1.02E-24 | Nightingale | IDL_P | NA | C | T | 0.059653 | 0.005049 | 5.60E-33 |
| rs600038 | let-7b-5p | trans | 1.02E-24 | Nightingale | XXL_VLDL_ | NA | C | T | 0.032858 | 0.005138 | 5.00E-12 |
| rs600038 | let-7b-5p | trans | 1.02E-24 | Nightingale | LDL_PL | NA | C | T | 0.0557 | 0.005091 | 1.90E-28 |
| rs600038 | let-7b-5p | trans | 1.02E-24 | Nightingale | S_VLDL_CE | NA | C | T | 0.053323 | 0.005096 | 1.20E-27 |
| rs600038 | let-7b-5p | trans | 1.02E-24 | Nightingale | S_LDL_C | NA | C | T | 0.04877 | 0.005108 | 3.30E-22 |
| rs600038 | let-7b-5p | trans | 1.02E-24 | Nightingale | IDL_PL | NA | C | T | 0.052744 | 0.004924 | 5.20E-28 |
| rs600038 | let-7b-5p | trans | 1.02E-24 | Nightingale | S_VLDL_PL | NA | C | T | 0.046119 | 0.005093 | 3.30E-21 |
| rs600038 | let-7b-5p | trans | 1.02E-24 | Nightingale | Omega_6 | NA | C | T | 0.037012 | 0.004983 | 5.10E-15 |

| | | | | | | | | | | | |
|---|---|---|---|---|---|---|---|---|---|---|---|
| rs600038 | let-7b-5p | trans | 1.02E-24 | Nightingale | XL_HDL_C_ | NA | C | T | 0.033784 | 0.004873 | 2.10E-11 |
| rs600038 | let-7b-5p | trans | 1.02E-24 | Nightingale | M_VLDL_P | NA | C | T | 0.040156 | 0.00509 | 1.50E-16 |
| rs600038 | let-7b-5p | trans | 1.02E-24 | Nightingale | Phosphatic | NA | C | T | 0.027053 | 0.004781 | 2.30E-09 |
| rs600038 | let-7b-5p | trans | 1.02E-24 | Nightingale | Total_PL | NA | C | T | 0.034757 | 0.004859 | 4.60E-14 |
| rs600038 | let-7b-5p | trans | 1.02E-24 | Nightingale | LDL_CE | NA | C | T | 0.051899 | 0.005089 | 3.60E-25 |
| rs600038 | let-7b-5p | trans | 1.02E-24 | Nightingale | VLDL_CE | NA | C | T | 0.049508 | 0.005104 | 5.70E-24 |
| rs600038 | let-7b-5p | trans | 1.02E-24 | Nightingale | S_VLDL_FC | NA | C | T | 0.054116 | 0.005104 | 8.50E-28 |
| rs600038 | let-7b-5p | trans | 1.02E-24 | Nightingale | XS_VLDL_L | NA | C | T | 0.044732 | 0.005016 | 1.60E-21 |
| rs600038 | let-7b-5p | trans | 1.02E-24 | Nightingale | S_LDL_TG_ | NA | C | T | -0.03742 | 0.004948 | 2.20E-13 |
| rs600038 | let-7b-5p | trans | 1.02E-24 | Nightingale | XL_HDL_PL | NA | C | T | -0.02757 | 0.00459 | 6.70E-10 |
| rs600038 | let-7b-5p | trans | 1.02E-24 | Nightingale | M_LDL_C | NA | C | T | 0.052552 | 0.005112 | 1.70E-25 |
| rs600038 | let-7b-5p | trans | 1.02E-24 | Nightingale | L_LDL_PL | NA | C | T | 0.059497 | 0.005064 | 1.20E-32 |
| rs600038 | let-7b-5p | trans | 1.02E-24 | Nightingale | XS_VLDL_C | NA | C | T | 0.037556 | 0.00484 | 3.10E-14 |
| rs600038 | let-7b-5p | trans | 1.02E-24 | Nightingale | S_LDL_PL | NA | C | T | 0.038067 | 0.005091 | 4.30E-14 |
| rs600038 | let-7b-5p | trans | 1.02E-24 | Nightingale | S_LDL_CE | NA | C | T | 0.046863 | 0.005105 | 5.90E-21 |
| rs600038 | let-7b-5p | trans | 1.02E-24 | Nightingale | XL_VLDL_P | NA | C | T | 0.052743 | 0.005111 | 2.40E-26 |
| rs600038 | let-7b-5p | trans | 1.02E-24 | Nightingale | XXL_VLDL_ | NA | C | T | 0.051211 | 0.005116 | 5.90E-24 |
| rs600038 | let-7b-5p | trans | 1.02E-24 | Nightingale | S_HDL_PL_ | NA | C | T | -0.02877 | 0.004933 | 1.50E-08 |
| rs600038 | let-7b-5p | trans | 1.02E-24 | Nightingale | XS_VLDL_F | NA | C | T | 0.049012 | 0.005014 | 4.20E-25 |
| rs600038 | let-7b-5p | trans | 1.02E-24 | Nightingale | S_HDL_PL | NA | C | T | 0.032674 | 0.005059 | 2.80E-11 |
| rs600038 | let-7b-5p | trans | 1.02E-24 | Nightingale | L_VLDL_CE | NA | C | T | 0.029942 | 0.005043 | 1.80E-10 |
| rs600038 | let-7b-5p | trans | 1.02E-24 | Nightingale | XS_VLDL_P | NA | C | T | 0.04948 | 0.005035 | 2.10E-25 |
| rs600038 | let-7b-5p | trans | 1.02E-24 | Nightingale | VLDL_FC | NA | C | T | 0.032134 | 0.005065 | 1.10E-11 |
| rs600038 | let-7b-5p | trans | 1.02E-24 | Nightingale | S_LDL_PL_ | NA | C | T | -0.0395 | 0.005093 | 1.10E-16 |
| rs600038 | let-7b-5p | trans | 1.02E-24 | Nightingale | XXL_VLDL_ | NA | C | T | 0.050318 | 0.005144 | 4.30E-23 |
| rs600038 | let-7b-5p | trans | 1.02E-24 | Nightingale | XL_HDL_PL | NA | C | T | -0.03707 | 0.004688 | 2.50E-15 |
| rs600038 | let-7b-5p | trans | 1.02E-24 | Nightingale | M_LDL_CE | NA | C | T | 0.049694 | 0.00511 | 3.20E-23 |
| rs600038 | let-7b-5p | trans | 1.02E-24 | Nightingale | S_VLDL_TG | NA | C | T | -0.05802 | 0.004939 | 6.80E-32 |
| rs600038 | let-7b-5p | trans | 1.02E-24 | Nightingale | S_HDL_P | NA | C | T | 0.048052 | 0.005103 | 4.50E-21 |
| rs600038 | let-7b-5p | trans | 1.02E-24 | Nightingale | M_VLDL_T | NA | C | T | -0.05584 | 0.004822 | 1.20E-30 |
| rs612169 | miR-10b-5p | trans | 3.54E-35 | Nightingale | LDL_PL | NA | G | A | 0.034856 | 0.004416 | 6.10E-15 |
| rs612169 | miR-10b-5p | trans | 3.54E-35 | Nightingale | L_LDL_P | NA | G | A | 0.029066 | 0.004411 | 7.10E-11 |
| rs612169 | miR-10b-5p | trans | 3.54E-35 | Nightingale | LDL_FC | NA | G | A | 0.033519 | 0.004379 | 6.60E-14 |
| rs612169 | miR-10b-5p | trans | 3.54E-35 | Nightingale | Citrate | NA | G | A | 0.026782 | 0.004397 | 7.80E-10 |
| rs612169 | miR-10b-5p | trans | 3.54E-35 | Nightingale | M_LDL_TG | NA | G | A | -0.02765 | 0.004371 | 3.20E-10 |
| rs612169 | miR-10b-5p | trans | 3.54E-35 | Nightingale | M_LDL_PL | NA | G | A | 0.032328 | 0.004432 | 3.40E-13 |
| rs612169 | miR-10b-5p | trans | 3.54E-35 | Nightingale | M_LDL_FC | NA | G | A | 0.034215 | 0.00442 | 2.50E-14 |
| rs612169 | miR-10b-5p | trans | 3.54E-35 | Nightingale | Omega_6 | NA | G | A | 0.025843 | 0.004322 | 6.30E-10 |
| rs612169 | miR-10b-5p | trans | 3.54E-35 | Nightingale | LDL_CE | NA | G | A | 0.03244 | 0.004414 | 2.70E-13 |
| rs612169 | miR-10b-5p | trans | 3.54E-35 | Nightingale | IDL_FC_pct | NA | G | A | 0.02565 | 0.004392 | 1.30E-08 |
| rs612169 | miR-10b-5p | trans | 3.54E-35 | Nightingale | S_LDL_FC | NA | G | A | 0.02845 | 0.004419 | 5.20E-10 |
| rs612169 | miR-10b-5p | trans | 3.54E-35 | Nightingale | ApoB | NA | G | A | 0.033778 | 0.00442 | 2.70E-14 |
| rs612169 | miR-10b-5p | trans | 3.54E-35 | Nightingale | S_LDL_L | NA | G | A | 0.027614 | 0.004425 | 9.90E-10 |
| rs612169 | miR-10b-5p | trans | 3.54E-35 | Nightingale | S_HDL_C | NA | G | A | 0.028392 | 0.004418 | 1.80E-10 |
| rs612169 | miR-10b-5p | trans | 3.54E-35 | Nightingale | M_VLDL_C | NA | G | A | 0.036191 | 0.004169 | 1.30E-17 |
| rs612169 | miR-10b-5p | trans | 3.54E-35 | Nightingale | IDL_FC | NA | G | A | 0.038123 | 0.004285 | 2.00E-18 |
| rs612169 | miR-10b-5p | trans | 3.54E-35 | Nightingale | M_LDL_L | NA | G | A | 0.032889 | 0.004432 | 1.30E-13 |
| rs612169 | miR-10b-5p | trans | 3.54E-35 | Nightingale | M_LDL_P | NA | G | A | 0.035713 | 0.004438 | 9.50E-16 |
| rs612169 | miR-10b-5p | trans | 3.54E-35 | Nightingale | XS_VLDL_C | NA | G | A | 0.037817 | 0.004296 | 1.80E-18 |
| rs612169 | miR-10b-5p | trans | 3.54E-35 | Nightingale | S_VLDL_CE | NA | G | A | 0.037935 | 0.00442 | 2.50E-18 |
| rs612169 | miR-10b-5p | trans | 3.54E-35 | Nightingale | VLDL_FC | NA | G | A | 0.023986 | 0.004393 | 1.30E-08 |
| rs612169 | miR-10b-5p | trans | 3.54E-35 | Nightingale | Total_CE | NA | G | A | 0.031681 | 0.004269 | 1.10E-13 |
| rs612169 | miR-10b-5p | trans | 3.54E-35 | Nightingale | VLDL_CE | NA | G | A | 0.035585 | 0.004427 | 4.20E-16 |
| rs612169 | miR-10b-5p | trans | 3.54E-35 | Nightingale | Sphingomy | NA | G | A | 0.028998 | 0.004137 | 7.10E-12 |
| rs612169 | miR-10b-5p | trans | 3.54E-35 | Nightingale | M_LDL_C_ | NA | G | A | 0.031134 | 0.004429 | 3.10E-12 |
| rs612169 | miR-10b-5p | trans | 3.54E-35 | Nightingale | Total_L | NA | G | A | 0.026277 | 0.004354 | 5.10E-10 |
| rs612169 | miR-10b-5p | trans | 3.54E-35 | Nightingale | XL_VLDL_F | NA | G | A | 0.037185 | 0.004291 | 2.00E-18 |
| rs612169 | miR-10b-5p | trans | 3.54E-35 | Nightingale | XS_VLDL_F | NA | G | A | 0.031389 | 0.004394 | 2.70E-12 |
| rs612169 | miR-10b-5p | trans | 3.54E-35 | Nightingale | IDL_C | NA | G | A | 0.034851 | 0.004274 | 1.20E-15 |
| rs612169 | miR-10b-5p | trans | 3.54E-35 | Nightingale | M_VLDL_F | NA | G | A | 0.033192 | 0.004425 | 4.20E-14 |
| rs612169 | miR-10b-5p | trans | 3.54E-35 | Nightingale | S_VLDL_TG | NA | G | A | -0.0381 | 0.004284 | 1.60E-18 |
| rs612169 | miR-10b-5p | trans | 3.54E-35 | Nightingale | S_VLDL_PL | NA | G | A | 0.032582 | 0.004418 | 5.70E-14 |
| rs612169 | miR-10b-5p | trans | 3.54E-35 | Nightingale | L_VLDL_TG | NA | G | A | -0.05092 | 0.004405 | 9.20E-32 |

| | | | | | | | | | | | |
|---|---|---|---|---|---|---|---|---|---|---|---|
| rs612169 | miR-10b-5p | trans | 3.54E-35 | Nightingale | M_VLDL_T | NA | G | A | -0.03853 | 0.004182 | 7.80E-20 |
| rs612169 | miR-10b-5p | trans | 3.54E-35 | Nightingale | S_VLDL_P | NA | G | A | 0.023264 | 0.00437 | 2.10E-08 |
| rs612169 | miR-10b-5p | trans | 3.54E-35 | Nightingale | L_LDL_L | NA | G | A | 0.033441 | 0.004383 | 3.70E-14 |
| rs612169 | miR-10b-5p | trans | 3.54E-35 | Nightingale | XXL_VLDL_ | NA | G | A | 0.03485 | 0.004435 | 2.70E-15 |
| rs612169 | miR-10b-5p | trans | 3.54E-35 | Nightingale | S_VLDL_FC | NA | G | A | 0.029377 | 0.004229 | 1.10E-11 |
| rs612169 | miR-10b-5p | trans | 3.54E-35 | Nightingale | XS_VLDL_P | NA | G | A | 0.036482 | 0.004367 | 2.00E-17 |
| rs612169 | miR-10b-5p | trans | 3.54E-35 | Nightingale | L_LDL_CE | NA | G | A | 0.031474 | 0.004386 | 1.20E-12 |
| rs612169 | miR-10b-5p | trans | 3.54E-35 | Nightingale | L_LDL_C | NA | G | A | 0.032224 | 0.004376 | 3.10E-13 |
| rs612169 | miR-10b-5p | trans | 3.54E-35 | Nightingale | XXL_VLDL_ | NA | G | A | 0.0248 | 0.004455 | 1.90E-09 |
| rs612169 | miR-10b-5p | trans | 3.54E-35 | Nightingale | M_VLDL_C | NA | G | A | 0.033628 | 0.004158 | 2.00E-15 |
| rs612169 | miR-10b-5p | trans | 3.54E-35 | Nightingale | S_VLDL_C | NA | G | A | 0.038142 | 0.004425 | 2.70E-18 |
| rs612169 | miR-10b-5p | trans | 3.54E-35 | Nightingale | M_VLDL_P | NA | G | A | 0.028126 | 0.004415 | 8.00E-11 |
| rs612169 | miR-10b-5p | trans | 3.54E-35 | Nightingale | XXL_VLDL_ | NA | G | A | 0.030915 | 0.004407 | 9.30E-13 |
| rs612169 | miR-10b-5p | trans | 3.54E-35 | Nightingale | S_HDL_L | NA | G | A | 0.026261 | 0.004411 | 1.60E-09 |
| rs612169 | miR-10b-5p | trans | 3.54E-35 | Nightingale | IDL_P | NA | G | A | 0.039804 | 0.004379 | 1.90E-19 |
| rs612169 | miR-10b-5p | trans | 3.54E-35 | Nightingale | S_VLDL_PL | NA | G | A | 0.027786 | 0.004238 | 1.80E-10 |
| rs612169 | miR-10b-5p | trans | 3.54E-35 | Nightingale | S_HDL_FC | NA | G | A | 0.023434 | 0.004345 | 4.80E-08 |
| rs612169 | miR-10b-5p | trans | 3.54E-35 | Nightingale | IDL_CE | NA | G | A | 0.033414 | 0.004273 | 1.60E-14 |
| rs612169 | miR-10b-5p | trans | 3.54E-35 | Nightingale | GlycA | NA | G | A | 0.027887 | 0.004407 | 1.70E-10 |
| rs612169 | miR-10b-5p | trans | 3.54E-35 | Nightingale | M_VLDL_C | NA | G | A | 0.038535 | 0.004414 | 2.70E-18 |
| rs612169 | miR-10b-5p | trans | 3.54E-35 | Nightingale | Clinical_LD | NA | G | A | 0.036343 | 0.0044 | 3.60E-16 |
| rs612169 | miR-10b-5p | trans | 3.54E-35 | Nightingale | M_VLDL_F | NA | G | A | 0.042 | 0.004211 | 4.30E-23 |
| rs612169 | miR-10b-5p | trans | 3.54E-35 | Nightingale | S_VLDL_L | NA | G | A | 0.024472 | 0.004383 | 4.70E-09 |
| rs612169 | miR-10b-5p | trans | 3.54E-35 | Nightingale | IDL_PL | NA | G | A | 0.035229 | 0.004271 | 4.10E-16 |
| rs612169 | miR-10b-5p | trans | 3.54E-35 | Nightingale | L_LDL_PL | NA | G | A | 0.037855 | 0.004392 | 1.20E-17 |
| rs612169 | miR-10b-5p | trans | 3.54E-35 | Nightingale | XS_VLDL_P | NA | G | A | 0.031933 | 0.004364 | 6.80E-14 |
| rs612169 | miR-10b-5p | trans | 3.54E-35 | Nightingale | Total_C | NA | G | A | 0.032298 | 0.004285 | 3.90E-14 |
| rs612169 | miR-10b-5p | trans | 3.54E-35 | Nightingale | ApoB_by_A | NA | G | A | 0.024859 | 0.004359 | 1.60E-08 |
| rs612169 | miR-10b-5p | trans | 3.54E-35 | Nightingale | VLDL_P | NA | G | A | 0.026438 | 0.004402 | 5.60E-10 |
| rs612169 | miR-10b-5p | trans | 3.54E-35 | Nightingale | S_HDL_CE | NA | G | A | 0.028617 | 0.004433 | 1.40E-10 |
| rs612169 | miR-10b-5p | trans | 3.54E-35 | Nightingale | Total_PL | NA | G | A | 0.024903 | 0.004214 | 1.10E-09 |
| rs612169 | miR-10b-5p | trans | 3.54E-35 | Nightingale | S_LDL_C | NA | G | A | 0.030803 | 0.00443 | 6.90E-12 |
| rs612169 | miR-10b-5p | trans | 3.54E-35 | Nightingale | S_LDL_CE | NA | G | A | 0.030405 | 0.004428 | 7.20E-12 |
| rs612169 | miR-10b-5p | trans | 3.54E-35 | Nightingale | IDL_L | NA | G | A | 0.035155 | 0.004279 | 6.10E-16 |
| rs612169 | miR-10b-5p | trans | 3.54E-35 | Nightingale | XS_VLDL_L | NA | G | A | 0.033628 | 0.004351 | 2.70E-15 |
| rs612169 | miR-10b-5p | trans | 3.54E-35 | Nightingale | S_HDL_P | NA | G | A | 0.030563 | 0.004426 | 5.70E-12 |
| rs612169 | miR-10b-5p | trans | 3.54E-35 | Nightingale | S_LDL_PL_ | NA | G | A | -0.02989 | 0.004418 | 3.40E-13 |
| rs612169 | miR-10b-5p | trans | 3.54E-35 | Nightingale | PUFA | NA | G | A | 0.023654 | 0.004317 | 1.60E-08 |
| rs612169 | miR-10b-5p | trans | 3.54E-35 | Nightingale | S_LDL_CE_ | NA | G | A | 0.030784 | 0.004418 | 6.20E-14 |
| rs612169 | miR-10b-5p | trans | 3.54E-35 | Nightingale | XL_HDL_PL | NA | G | A | -0.0226 | 0.004066 | 1.40E-08 |
| rs612169 | miR-10b-5p | trans | 3.54E-35 | Nightingale | S_VLDL_C_ | NA | G | A | 0.041831 | 0.004307 | 6.30E-22 |
| rs612169 | miR-10b-5p | trans | 3.54E-35 | Nightingale | LA | NA | G | A | 0.024865 | 0.00434 | 4.00E-09 |
| rs612169 | miR-10b-5p | trans | 3.54E-35 | Nightingale | non_HDL_( | NA | G | A | 0.035557 | 0.004402 | 6.60E-16 |
| rs612169 | miR-10b-5p | trans | 3.54E-35 | Nightingale | L_VLDL_FC | NA | G | A | 0.04835 | 0.004383 | 7.90E-29 |
| rs612169 | miR-10b-5p | trans | 3.54E-35 | Nightingale | Total_FC | NA | G | A | 0.033156 | 0.004324 | 8.30E-15 |
| rs612169 | miR-10b-5p | trans | 3.54E-35 | Nightingale | M_LDL_C | NA | G | A | 0.033844 | 0.004434 | 2.80E-14 |
| rs612169 | miR-10b-5p | trans | 3.54E-35 | Nightingale | XS_VLDL_C | NA | G | A | 0.037631 | 0.004271 | 2.80E-18 |
| rs612169 | miR-10b-5p | trans | 3.54E-35 | Nightingale | M_VLDL_C | NA | G | A | 0.04068 | 0.004386 | 3.10E-20 |
| rs612169 | miR-10b-5p | trans | 3.54E-35 | Nightingale | S_VLDL_CE | NA | G | A | 0.046907 | 0.004366 | 1.70E-26 |
| rs612169 | miR-10b-5p | trans | 3.54E-35 | Nightingale | M_VLDL_P | NA | G | A | 0.043968 | 0.004248 | 7.10E-25 |
| rs612169 | miR-10b-5p | trans | 3.54E-35 | Nightingale | M_LDL_CE | NA | G | A | 0.03283 | 0.004432 | 1.20E-13 |
| rs612169 | miR-10b-5p | trans | 3.54E-35 | Nightingale | VLDL_C | NA | G | A | 0.031111 | 0.00442 | 6.50E-13 |
| rs612169 | miR-10b-5p | trans | 3.54E-35 | Nightingale | LDL_L | NA | G | A | 0.033183 | 0.004411 | 9.30E-14 |
| rs612169 | miR-10b-5p | trans | 3.54E-35 | Nightingale | S_LDL_P | NA | G | A | 0.029399 | 0.00443 | 4.20E-11 |
| rs612169 | miR-10b-5p | trans | 3.54E-35 | Nightingale | XL_VLDL_T | NA | G | A | -0.0439 | 0.00429 | 1.80E-24 |
| rs612169 | miR-10b-5p | trans | 3.54E-35 | Nightingale | S_HDL_PL | NA | G | A | 0.023764 | 0.004387 | 3.40E-08 |
| rs612169 | miR-10b-5p | trans | 3.54E-35 | Nightingale | S_LDL_C_p | NA | G | A | 0.045206 | 0.004443 | 8.20E-25 |
| rs612169 | miR-10b-5p | trans | 3.54E-35 | Nightingale | XL_VLDL_C | NA | G | A | 0.037396 | 0.004246 | 2.90E-18 |
| rs612169 | miR-10b-5p | trans | 3.54E-35 | Nightingale | L_VLDL_CE | NA | G | A | 0.047313 | 0.004235 | 2.50E-29 |
| rs612169 | miR-10b-5p | trans | 3.54E-35 | Nightingale | XS_VLDL_F | NA | G | A | 0.036875 | 0.004349 | 8.90E-18 |
| rs612169 | miR-10b-5p | trans | 3.54E-35 | Nightingale | Cholines | NA | G | A | 0.022291 | 0.004162 | 4.90E-08 |
| rs612169 | miR-10b-5p | trans | 3.54E-35 | Nightingale | LDL_C | NA | G | A | 0.03308 | 0.004407 | 1.10E-13 |
| rs612169 | miR-10b-5p | trans | 3.54E-35 | Nightingale | L_VLDL_C_ | NA | G | A | 0.052101 | 0.004323 | 5.00E-34 |

| | | | | | | | | | | | |
|---|---|---|---|---|---|---|---|---|---|---|---|
| rs612169 | miR-10b-5p | trans | 3.54E-35 | Nightingale | LDL_P | NA | G | A | 0.031659 | 0.004423 | 1.30E-12 |
| rs612169 | miR-10b-5p | trans | 3.54E-35 | Nightingale | XXL_VLDL_ | NA | G | A | 0.033073 | 0.00446 | 9.30E-14 |
| rs612169 | miR-10b-5p | trans | 3.54E-35 | Nightingale | XL_VLDL_P | NA | G | A | 0.03861 | 0.004433 | 7.90E-19 |
| rs612169 | miR-10b-5p | trans | 3.54E-35 | Nightingale | Remnant_( | NA | G | A | 0.036703 | 0.004397 | 5.00E-17 |
| rs612169 | miR-10b-5p | trans | 3.54E-35 | Nightingale | XL_VLDL_C | NA | G | A | 0.038268 | 0.004244 | 4.10E-19 |
| rs612169 | miR-10b-5p | trans | 3.54E-35 | Nightingale | XXL_VLDL_ | NA | G | A | -0.03569 | 0.004464 | 2.20E-16 |
| rs612169 | miR-10b-5p | trans | 3.54E-35 | Nightingale | M_VLDL_P | NA | G | A | 0.028958 | 0.004419 | 2.60E-11 |
| rs612169 | miR-10b-5p | trans | 3.54E-35 | Nightingale | L_LDL_FC | NA | G | A | 0.033523 | 0.004346 | 3.50E-14 |
| rs612169 | miR-10b-5p | trans | 3.54E-35 | Nightingale | S_VLDL_FC | NA | G | A | 0.037705 | 0.004427 | 1.10E-17 |
| rs612169 | miR-139-5p | trans | 7.83E-42 | Nightingale | LDL_PL | NA | G | A | 0.034856 | 0.004416 | 6.10E-15 |
| rs612169 | miR-139-5p | trans | 7.83E-42 | Nightingale | L_LDL_P | NA | G | A | 0.029066 | 0.004411 | 7.10E-11 |
| rs612169 | miR-139-5p | trans | 7.83E-42 | Nightingale | LDL_FC | NA | G | A | 0.033519 | 0.004379 | 6.60E-14 |
| rs612169 | miR-139-5p | trans | 7.83E-42 | Nightingale | Citrate | NA | G | A | 0.026782 | 0.004397 | 7.80E-10 |
| rs612169 | miR-139-5p | trans | 7.83E-42 | Nightingale | M_LDL_TG | NA | G | A | -0.02765 | 0.004371 | 3.20E-10 |
| rs612169 | miR-139-5p | trans | 7.83E-42 | Nightingale | M_LDL_PL | NA | G | A | 0.032328 | 0.004432 | 3.40E-13 |
| rs612169 | miR-139-5p | trans | 7.83E-42 | Nightingale | M_LDL_FC | NA | G | A | 0.034215 | 0.00442 | 2.50E-14 |
| rs612169 | miR-139-5p | trans | 7.83E-42 | Nightingale | Omega_6 | NA | G | A | 0.025843 | 0.004322 | 6.30E-10 |
| rs612169 | miR-139-5p | trans | 7.83E-42 | Nightingale | LDL_CE | NA | G | A | 0.03244 | 0.004414 | 2.70E-13 |
| rs612169 | miR-139-5p | trans | 7.83E-42 | Nightingale | IDL_FC_pct | NA | G | A | 0.02565 | 0.004392 | 1.30E-08 |
| rs612169 | miR-139-5p | trans | 7.83E-42 | Nightingale | S_LDL_FC | NA | G | A | 0.02845 | 0.004419 | 5.20E-10 |
| rs612169 | miR-139-5p | trans | 7.83E-42 | Nightingale | ApoB | NA | G | A | 0.033778 | 0.00442 | 2.70E-14 |
| rs612169 | miR-139-5p | trans | 7.83E-42 | Nightingale | S_LDL_L | NA | G | A | 0.027614 | 0.004425 | 9.90E-10 |
| rs612169 | miR-139-5p | trans | 7.83E-42 | Nightingale | S_HDL_C | NA | G | A | 0.028392 | 0.004418 | 1.80E-10 |
| rs612169 | miR-139-5p | trans | 7.83E-42 | Nightingale | M_VLDL_C_ | NA | G | A | 0.036191 | 0.004169 | 1.30E-17 |
| rs612169 | miR-139-5p | trans | 7.83E-42 | Nightingale | IDL_FC | NA | G | A | 0.038123 | 0.004285 | 2.00E-18 |
| rs612169 | miR-139-5p | trans | 7.83E-42 | Nightingale | M_LDL_L | NA | G | A | 0.032889 | 0.004432 | 1.30E-13 |
| rs612169 | miR-139-5p | trans | 7.83E-42 | Nightingale | M_LDL_P | NA | G | A | 0.035713 | 0.004438 | 9.50E-16 |
| rs612169 | miR-139-5p | trans | 7.83E-42 | Nightingale | XS_VLDL_C | NA | G | A | 0.037817 | 0.004296 | 1.80E-18 |
| rs612169 | miR-139-5p | trans | 7.83E-42 | Nightingale | S_VLDL_CE | NA | G | A | 0.037935 | 0.00442 | 2.50E-18 |
| rs612169 | miR-139-5p | trans | 7.83E-42 | Nightingale | VLDL_FC | NA | G | A | 0.023986 | 0.004393 | 1.30E-08 |
| rs612169 | miR-139-5p | trans | 7.83E-42 | Nightingale | Total_CE | NA | G | A | 0.031681 | 0.004269 | 1.10E-13 |
| rs612169 | miR-139-5p | trans | 7.83E-42 | Nightingale | VLDL_CE | NA | G | A | 0.035585 | 0.004427 | 4.20E-16 |
| rs612169 | miR-139-5p | trans | 7.83E-42 | Nightingale | Sphingomy | NA | G | A | 0.028998 | 0.004137 | 7.10E-12 |
| rs612169 | miR-139-5p | trans | 7.83E-42 | Nightingale | M_LDL_C_ | NA | G | A | 0.031134 | 0.004429 | 3.10E-12 |
| rs612169 | miR-139-5p | trans | 7.83E-42 | Nightingale | Total_L | NA | G | A | 0.026277 | 0.004354 | 5.10E-10 |
| rs612169 | miR-139-5p | trans | 7.83E-42 | Nightingale | XL_VLDL_F | NA | G | A | 0.037185 | 0.004291 | 2.00E-18 |
| rs612169 | miR-139-5p | trans | 7.83E-42 | Nightingale | XS_VLDL_F | NA | G | A | 0.031389 | 0.004394 | 2.70E-12 |
| rs612169 | miR-139-5p | trans | 7.83E-42 | Nightingale | IDL_C | NA | G | A | 0.034851 | 0.004274 | 1.20E-15 |
| rs612169 | miR-139-5p | trans | 7.83E-42 | Nightingale | M_VLDL_F | NA | G | A | 0.033192 | 0.004425 | 4.20E-14 |
| rs612169 | miR-139-5p | trans | 7.83E-42 | Nightingale | S_VLDL_TG | NA | G | A | -0.0381 | 0.004284 | 1.60E-18 |
| rs612169 | miR-139-5p | trans | 7.83E-42 | Nightingale | S_VLDL_PL | NA | G | A | 0.032582 | 0.004418 | 5.70E-14 |
| rs612169 | miR-139-5p | trans | 7.83E-42 | Nightingale | L_VLDL_TG | NA | G | A | -0.05092 | 0.004405 | 9.20E-32 |
| rs612169 | miR-139-5p | trans | 7.83E-42 | Nightingale | M_VLDL_T | NA | G | A | -0.03853 | 0.004182 | 7.80E-20 |
| rs612169 | miR-139-5p | trans | 7.83E-42 | Nightingale | S_VLDL_P | NA | G | A | 0.023264 | 0.00437 | 2.10E-08 |
| rs612169 | miR-139-5p | trans | 7.83E-42 | Nightingale | L_LDL_L | NA | G | A | 0.033441 | 0.004383 | 3.70E-14 |
| rs612169 | miR-139-5p | trans | 7.83E-42 | Nightingale | XXL_VLDL_ | NA | G | A | 0.03485 | 0.004435 | 2.70E-15 |
| rs612169 | miR-139-5p | trans | 7.83E-42 | Nightingale | S_VLDL_FC | NA | G | A | 0.029377 | 0.004229 | 1.10E-11 |
| rs612169 | miR-139-5p | trans | 7.83E-42 | Nightingale | XS_VLDL_P | NA | G | A | 0.036482 | 0.004367 | 2.00E-17 |
| rs612169 | miR-139-5p | trans | 7.83E-42 | Nightingale | L_LDL_CE | NA | G | A | 0.031474 | 0.004386 | 1.20E-12 |
| rs612169 | miR-139-5p | trans | 7.83E-42 | Nightingale | L_LDL_C | NA | G | A | 0.032224 | 0.004376 | 3.10E-13 |
| rs612169 | miR-139-5p | trans | 7.83E-42 | Nightingale | XXL_VLDL_ | NA | G | A | 0.0248 | 0.004455 | 1.90E-09 |
| rs612169 | miR-139-5p | trans | 7.83E-42 | Nightingale | M_VLDL_C | NA | G | A | 0.033628 | 0.004158 | 2.00E-15 |
| rs612169 | miR-139-5p | trans | 7.83E-42 | Nightingale | S_VLDL_C | NA | G | A | 0.038142 | 0.004425 | 2.70E-18 |
| rs612169 | miR-139-5p | trans | 7.83E-42 | Nightingale | M_VLDL_P | NA | G | A | 0.028126 | 0.004415 | 8.00E-11 |
| rs612169 | miR-139-5p | trans | 7.83E-42 | Nightingale | XXL_VLDL_ | NA | G | A | 0.030915 | 0.004407 | 9.30E-13 |
| rs612169 | miR-139-5p | trans | 7.83E-42 | Nightingale | S_HDL_L | NA | G | A | 0.026261 | 0.004411 | 1.60E-09 |
| rs612169 | miR-139-5p | trans | 7.83E-42 | Nightingale | IDL_P | NA | G | A | 0.039804 | 0.004379 | 1.90E-19 |
| rs612169 | miR-139-5p | trans | 7.83E-42 | Nightingale | S_VLDL_PL | NA | G | A | 0.027786 | 0.004238 | 1.80E-10 |
| rs612169 | miR-139-5p | trans | 7.83E-42 | Nightingale | S_HDL_FC | NA | G | A | 0.023434 | 0.004345 | 4.80E-08 |
| rs612169 | miR-139-5p | trans | 7.83E-42 | Nightingale | IDL_CE | NA | G | A | 0.033414 | 0.004273 | 1.60E-14 |
| rs612169 | miR-139-5p | trans | 7.83E-42 | Nightingale | GlycA | NA | G | A | 0.027887 | 0.004407 | 1.70E-10 |
| rs612169 | miR-139-5p | trans | 7.83E-42 | Nightingale | M_VLDL_C | NA | G | A | 0.038535 | 0.004414 | 2.70E-18 |
| rs612169 | miR-139-5p | trans | 7.83E-42 | Nightingale | Clinical_LD | NA | G | A | 0.036343 | 0.0044 | 3.60E-16 |

| | | | | | | | | | | | |
|---|---|---|---|---|---|---|---|---|---|---|---|
| rs612169 | miR-139-5p | trans | 7.83E-42 | Nightingale | M_VLDL_F( | NA | G | A | 0.042 | 0.004211 | 4.30E-23 |
| rs612169 | miR-139-5p | trans | 7.83E-42 | Nightingale | S_VLDL_L | NA | G | A | 0.024472 | 0.004383 | 4.70E-09 |
| rs612169 | miR-139-5p | trans | 7.83E-42 | Nightingale | IDL_PL | NA | G | A | 0.035229 | 0.004271 | 4.10E-16 |
| rs612169 | miR-139-5p | trans | 7.83E-42 | Nightingale | L_LDL_PL | NA | G | A | 0.037855 | 0.004392 | 1.20E-17 |
| rs612169 | miR-139-5p | trans | 7.83E-42 | Nightingale | XS_VLDL_P | NA | G | A | 0.031933 | 0.004364 | 6.80E-14 |
| rs612169 | miR-139-5p | trans | 7.83E-42 | Nightingale | Total_C | NA | G | A | 0.032298 | 0.004285 | 3.90E-14 |
| rs612169 | miR-139-5p | trans | 7.83E-42 | Nightingale | ApoB_by_A | NA | G | A | 0.024859 | 0.004359 | 1.60E-08 |
| rs612169 | miR-139-5p | trans | 7.83E-42 | Nightingale | VLDL_P | NA | G | A | 0.026438 | 0.004402 | 5.60E-10 |
| rs612169 | miR-139-5p | trans | 7.83E-42 | Nightingale | S_HDL_CE | NA | G | A | 0.028617 | 0.004433 | 1.40E-10 |
| rs612169 | miR-139-5p | trans | 7.83E-42 | Nightingale | Total_PL | NA | G | A | 0.024903 | 0.004214 | 1.10E-09 |
| rs612169 | miR-139-5p | trans | 7.83E-42 | Nightingale | S_LDL_C | NA | G | A | 0.030803 | 0.00443 | 6.90E-12 |
| rs612169 | miR-139-5p | trans | 7.83E-42 | Nightingale | S_LDL_CE | NA | G | A | 0.030405 | 0.004428 | 7.20E-12 |
| rs612169 | miR-139-5p | trans | 7.83E-42 | Nightingale | IDL_L | NA | G | A | 0.035155 | 0.004279 | 6.10E-16 |
| rs612169 | miR-139-5p | trans | 7.83E-42 | Nightingale | XS_VLDL_L | NA | G | A | 0.033628 | 0.004351 | 2.70E-15 |
| rs612169 | miR-139-5p | trans | 7.83E-42 | Nightingale | S_HDL_P | NA | G | A | 0.030563 | 0.004426 | 5.70E-12 |
| rs612169 | miR-139-5p | trans | 7.83E-42 | Nightingale | S_LDL_PL_ | NA | G | A | -0.02989 | 0.004418 | 3.40E-13 |
| rs612169 | miR-139-5p | trans | 7.83E-42 | Nightingale | PUFA | NA | G | A | 0.023654 | 0.004317 | 1.60E-08 |
| rs612169 | miR-139-5p | trans | 7.83E-42 | Nightingale | S_LDL_CE_ | NA | G | A | 0.030784 | 0.004418 | 6.20E-14 |
| rs612169 | miR-139-5p | trans | 7.83E-42 | Nightingale | XL_HDL_PL | NA | G | A | -0.0226 | 0.004066 | 1.40E-08 |
| rs612169 | miR-139-5p | trans | 7.83E-42 | Nightingale | S_VLDL_C_ | NA | G | A | 0.041831 | 0.004307 | 6.30E-22 |
| rs612169 | miR-139-5p | trans | 7.83E-42 | Nightingale | LA | NA | G | A | 0.024865 | 0.00434 | 4.00E-09 |
| rs612169 | miR-139-5p | trans | 7.83E-42 | Nightingale | non_HDL_( | NA | G | A | 0.035557 | 0.004402 | 6.60E-16 |
| rs612169 | miR-139-5p | trans | 7.83E-42 | Nightingale | L_VLDL_FC | NA | G | A | 0.04835 | 0.004383 | 7.90E-29 |
| rs612169 | miR-139-5p | trans | 7.83E-42 | Nightingale | Total_FC | NA | G | A | 0.033156 | 0.004324 | 8.30E-15 |
| rs612169 | miR-139-5p | trans | 7.83E-42 | Nightingale | M_LDL_C | NA | G | A | 0.033844 | 0.004434 | 2.80E-14 |
| rs612169 | miR-139-5p | trans | 7.83E-42 | Nightingale | XS_VLDL_C | NA | G | A | 0.037631 | 0.004271 | 2.80E-18 |
| rs612169 | miR-139-5p | trans | 7.83E-42 | Nightingale | M_VLDL_C | NA | G | A | 0.04068 | 0.004386 | 3.10E-20 |
| rs612169 | miR-139-5p | trans | 7.83E-42 | Nightingale | S_VLDL_CE | NA | G | A | 0.046907 | 0.004366 | 1.70E-26 |
| rs612169 | miR-139-5p | trans | 7.83E-42 | Nightingale | M_VLDL_P | NA | G | A | 0.043968 | 0.004248 | 7.10E-25 |
| rs612169 | miR-139-5p | trans | 7.83E-42 | Nightingale | M_LDL_CE | NA | G | A | 0.03283 | 0.004432 | 1.20E-13 |
| rs612169 | miR-139-5p | trans | 7.83E-42 | Nightingale | VLDL_C | NA | G | A | 0.031111 | 0.00442 | 6.50E-13 |
| rs612169 | miR-139-5p | trans | 7.83E-42 | Nightingale | LDL_L | NA | G | A | 0.033183 | 0.004411 | 9.30E-14 |
| rs612169 | miR-139-5p | trans | 7.83E-42 | Nightingale | S_LDL_P | NA | G | A | 0.029399 | 0.00443 | 4.20E-11 |
| rs612169 | miR-139-5p | trans | 7.83E-42 | Nightingale | XL_VLDL_T | NA | G | A | -0.0439 | 0.00429 | 1.80E-24 |
| rs612169 | miR-139-5p | trans | 7.83E-42 | Nightingale | S_HDL_PL | NA | G | A | 0.023764 | 0.004387 | 3.40E-08 |
| rs612169 | miR-139-5p | trans | 7.83E-42 | Nightingale | S_LDL_C_p | NA | G | A | 0.045206 | 0.004443 | 8.20E-25 |
| rs612169 | miR-139-5p | trans | 7.83E-42 | Nightingale | XL_VLDL_C | NA | G | A | 0.037396 | 0.004246 | 2.90E-18 |
| rs612169 | miR-139-5p | trans | 7.83E-42 | Nightingale | L_VLDL_CE | NA | G | A | 0.047313 | 0.004235 | 2.50E-29 |
| rs612169 | miR-139-5p | trans | 7.83E-42 | Nightingale | XS_VLDL_F | NA | G | A | 0.036875 | 0.004349 | 8.90E-18 |
| rs612169 | miR-139-5p | trans | 7.83E-42 | Nightingale | Cholines | NA | G | A | 0.022291 | 0.004162 | 4.90E-08 |
| rs612169 | miR-139-5p | trans | 7.83E-42 | Nightingale | LDL_C | NA | G | A | 0.03308 | 0.004407 | 1.10E-13 |
| rs612169 | miR-139-5p | trans | 7.83E-42 | Nightingale | L_VLDL_C_ | NA | G | A | 0.052101 | 0.004323 | 5.00E-34 |
| rs612169 | miR-139-5p | trans | 7.83E-42 | Nightingale | LDL_P | NA | G | A | 0.031659 | 0.004423 | 1.30E-12 |
| rs612169 | miR-139-5p | trans | 7.83E-42 | Nightingale | XXL_VLDL_ | NA | G | A | 0.033073 | 0.00446 | 9.30E-14 |
| rs612169 | miR-139-5p | trans | 7.83E-42 | Nightingale | XL_VLDL_P | NA | G | A | 0.03861 | 0.004433 | 7.90E-19 |
| rs612169 | miR-139-5p | trans | 7.83E-42 | Nightingale | Remnant_( | NA | G | A | 0.036703 | 0.004397 | 5.00E-17 |
| rs612169 | miR-139-5p | trans | 7.83E-42 | Nightingale | XL_VLDL_C | NA | G | A | 0.038268 | 0.004244 | 4.10E-19 |
| rs612169 | miR-139-5p | trans | 7.83E-42 | Nightingale | XXL_VLDL_ | NA | G | A | -0.03569 | 0.004464 | 2.20E-16 |
| rs612169 | miR-139-5p | trans | 7.83E-42 | Nightingale | M_VLDL_P | NA | G | A | 0.028958 | 0.004419 | 2.60E-11 |
| rs612169 | miR-139-5p | trans | 7.83E-42 | Nightingale | L_LDL_FC | NA | G | A | 0.033523 | 0.004346 | 3.50E-14 |
| rs612169 | miR-139-5p | trans | 7.83E-42 | Nightingale | S_VLDL_FC | NA | G | A | 0.037705 | 0.004427 | 1.10E-17 |
| rs612169 | miR-126-3p | trans | 2.89E-39 | Nightingale | LDL_PL | NA | G | A | 0.034856 | 0.004416 | 6.10E-15 |
| rs612169 | miR-126-3p | trans | 2.89E-39 | Nightingale | L_LDL_P | NA | G | A | 0.029066 | 0.004411 | 7.10E-11 |
| rs612169 | miR-126-3p | trans | 2.89E-39 | Nightingale | LDL_FC | NA | G | A | 0.033519 | 0.004379 | 6.60E-14 |
| rs612169 | miR-126-3p | trans | 2.89E-39 | Nightingale | Citrate | NA | G | A | 0.026782 | 0.004397 | 7.80E-10 |
| rs612169 | miR-126-3p | trans | 2.89E-39 | Nightingale | M_LDL_TG | NA | G | A | -0.02765 | 0.004371 | 3.20E-10 |
| rs612169 | miR-126-3p | trans | 2.89E-39 | Nightingale | M_LDL_PL | NA | G | A | 0.032328 | 0.004432 | 3.40E-13 |
| rs612169 | miR-126-3p | trans | 2.89E-39 | Nightingale | M_LDL_FC | NA | G | A | 0.034215 | 0.00442 | 2.50E-14 |
| rs612169 | miR-126-3p | trans | 2.89E-39 | Nightingale | Omega_6 | NA | G | A | 0.025843 | 0.004322 | 6.30E-10 |
| rs612169 | miR-126-3p | trans | 2.89E-39 | Nightingale | LDL_CE | NA | G | A | 0.03244 | 0.004414 | 2.70E-13 |
| rs612169 | miR-126-3p | trans | 2.89E-39 | Nightingale | IDL_FC_pct | NA | G | A | 0.02565 | 0.004392 | 1.30E-08 |
| rs612169 | miR-126-3p | trans | 2.89E-39 | Nightingale | S_LDL_FC | NA | G | A | 0.02845 | 0.004419 | 5.20E-10 |
| rs612169 | miR-126-3p | trans | 2.89E-39 | Nightingale | ApoB | NA | G | A | 0.033778 | 0.00442 | 2.70E-14 |

| | | | | | | | | | | | |
|---|---|---|---|---|---|---|---|---|---|---|---|
| rs612169 | miR-126-3p | trans | 2.89E-39 | Nightingale | S_LDL_L | NA | G | A | 0.027614 | 0.004425 | 9.90E-10 |
| rs612169 | miR-126-3p | trans | 2.89E-39 | Nightingale | S_HDL_C | NA | G | A | 0.028392 | 0.004418 | 1.80E-10 |
| rs612169 | miR-126-3p | trans | 2.89E-39 | Nightingale | M_VLDL_C | NA | G | A | 0.036191 | 0.004169 | 1.30E-17 |
| rs612169 | miR-126-3p | trans | 2.89E-39 | Nightingale | IDL_FC | NA | G | A | 0.038123 | 0.004285 | 2.00E-18 |
| rs612169 | miR-126-3p | trans | 2.89E-39 | Nightingale | M_LDL_L | NA | G | A | 0.032889 | 0.004432 | 1.30E-13 |
| rs612169 | miR-126-3p | trans | 2.89E-39 | Nightingale | M_LDL_P | NA | G | A | 0.035713 | 0.004438 | 9.50E-16 |
| rs612169 | miR-126-3p | trans | 2.89E-39 | Nightingale | XS_VLDL_C | NA | G | A | 0.037817 | 0.004296 | 1.80E-18 |
| rs612169 | miR-126-3p | trans | 2.89E-39 | Nightingale | S_VLDL_CE | NA | G | A | 0.037935 | 0.00442 | 2.50E-18 |
| rs612169 | miR-126-3p | trans | 2.89E-39 | Nightingale | VLDL_FC | NA | G | A | 0.023986 | 0.004393 | 1.30E-08 |
| rs612169 | miR-126-3p | trans | 2.89E-39 | Nightingale | Total_CE | NA | G | A | 0.031681 | 0.004269 | 1.10E-13 |
| rs612169 | miR-126-3p | trans | 2.89E-39 | Nightingale | VLDL_CE | NA | G | A | 0.035585 | 0.004427 | 4.20E-16 |
| rs612169 | miR-126-3p | trans | 2.89E-39 | Nightingale | Sphingomy | NA | G | A | 0.028998 | 0.004137 | 7.10E-12 |
| rs612169 | miR-126-3p | trans | 2.89E-39 | Nightingale | M_LDL_C_ | NA | G | A | 0.031134 | 0.004429 | 3.10E-12 |
| rs612169 | miR-126-3p | trans | 2.89E-39 | Nightingale | Total_L | NA | G | A | 0.026277 | 0.004354 | 5.10E-10 |
| rs612169 | miR-126-3p | trans | 2.89E-39 | Nightingale | XL_VLDL_F | NA | G | A | 0.037185 | 0.004291 | 2.00E-18 |
| rs612169 | miR-126-3p | trans | 2.89E-39 | Nightingale | XS_VLDL_F | NA | G | A | 0.031389 | 0.004394 | 2.70E-12 |
| rs612169 | miR-126-3p | trans | 2.89E-39 | Nightingale | IDL_C | NA | G | A | 0.034851 | 0.004274 | 1.20E-15 |
| rs612169 | miR-126-3p | trans | 2.89E-39 | Nightingale | M_VLDL_F | NA | G | A | 0.033192 | 0.004425 | 4.20E-14 |
| rs612169 | miR-126-3p | trans | 2.89E-39 | Nightingale | S_VLDL_TG | NA | G | A | -0.0381 | 0.004284 | 1.60E-18 |
| rs612169 | miR-126-3p | trans | 2.89E-39 | Nightingale | S_VLDL_PL | NA | G | A | 0.032582 | 0.004418 | 5.70E-14 |
| rs612169 | miR-126-3p | trans | 2.89E-39 | Nightingale | L_VLDL_TG | NA | G | A | -0.05092 | 0.004405 | 9.20E-32 |
| rs612169 | miR-126-3p | trans | 2.89E-39 | Nightingale | M_VLDL_T | NA | G | A | -0.03853 | 0.004182 | 7.80E-20 |
| rs612169 | miR-126-3p | trans | 2.89E-39 | Nightingale | S_VLDL_P | NA | G | A | 0.023264 | 0.00437 | 2.10E-08 |
| rs612169 | miR-126-3p | trans | 2.89E-39 | Nightingale | L_LDL_L | NA | G | A | 0.033441 | 0.004383 | 3.70E-14 |
| rs612169 | miR-126-3p | trans | 2.89E-39 | Nightingale | XXL_VLDL_ | NA | G | A | 0.03485 | 0.004435 | 2.70E-15 |
| rs612169 | miR-126-3p | trans | 2.89E-39 | Nightingale | S_VLDL_FC | NA | G | A | 0.029377 | 0.004229 | 1.10E-11 |
| rs612169 | miR-126-3p | trans | 2.89E-39 | Nightingale | XS_VLDL_P | NA | G | A | 0.036482 | 0.004367 | 2.00E-17 |
| rs612169 | miR-126-3p | trans | 2.89E-39 | Nightingale | L_LDL_CE | NA | G | A | 0.031474 | 0.004386 | 1.20E-12 |
| rs612169 | miR-126-3p | trans | 2.89E-39 | Nightingale | L_LDL_C | NA | G | A | 0.032224 | 0.004376 | 3.10E-13 |
| rs612169 | miR-126-3p | trans | 2.89E-39 | Nightingale | XXL_VLDL_ | NA | G | A | 0.0248 | 0.004455 | 1.90E-09 |
| rs612169 | miR-126-3p | trans | 2.89E-39 | Nightingale | M_VLDL_C | NA | G | A | 0.033628 | 0.004158 | 2.00E-15 |
| rs612169 | miR-126-3p | trans | 2.89E-39 | Nightingale | S_VLDL_C | NA | G | A | 0.038142 | 0.004425 | 2.70E-18 |
| rs612169 | miR-126-3p | trans | 2.89E-39 | Nightingale | M_VLDL_P | NA | G | A | 0.028126 | 0.004415 | 8.00E-11 |
| rs612169 | miR-126-3p | trans | 2.89E-39 | Nightingale | XXL_VLDL_ | NA | G | A | 0.030915 | 0.004407 | 9.30E-13 |
| rs612169 | miR-126-3p | trans | 2.89E-39 | Nightingale | S_HDL_L | NA | G | A | 0.026261 | 0.004411 | 1.60E-09 |
| rs612169 | miR-126-3p | trans | 2.89E-39 | Nightingale | IDL_P | NA | G | A | 0.039804 | 0.004379 | 1.90E-19 |
| rs612169 | miR-126-3p | trans | 2.89E-39 | Nightingale | S_VLDL_PL | NA | G | A | 0.027786 | 0.004238 | 1.80E-10 |
| rs612169 | miR-126-3p | trans | 2.89E-39 | Nightingale | S_HDL_FC | NA | G | A | 0.023434 | 0.004345 | 4.80E-08 |
| rs612169 | miR-126-3p | trans | 2.89E-39 | Nightingale | IDL_CE | NA | G | A | 0.033414 | 0.004273 | 1.60E-14 |
| rs612169 | miR-126-3p | trans | 2.89E-39 | Nightingale | GlycA | NA | G | A | 0.027887 | 0.004407 | 1.70E-10 |
| rs612169 | miR-126-3p | trans | 2.89E-39 | Nightingale | M_VLDL_C | NA | G | A | 0.038535 | 0.004414 | 2.70E-18 |
| rs612169 | miR-126-3p | trans | 2.89E-39 | Nightingale | Clinical_LD | NA | G | A | 0.036343 | 0.0044 | 3.60E-16 |
| rs612169 | miR-126-3p | trans | 2.89E-39 | Nightingale | M_VLDL_F | NA | G | A | 0.042 | 0.004211 | 4.30E-23 |
| rs612169 | miR-126-3p | trans | 2.89E-39 | Nightingale | S_VLDL_L | NA | G | A | 0.024472 | 0.004383 | 4.70E-09 |
| rs612169 | miR-126-3p | trans | 2.89E-39 | Nightingale | IDL_PL | NA | G | A | 0.035229 | 0.004271 | 4.10E-16 |
| rs612169 | miR-126-3p | trans | 2.89E-39 | Nightingale | L_LDL_PL | NA | G | A | 0.037855 | 0.004392 | 1.20E-17 |
| rs612169 | miR-126-3p | trans | 2.89E-39 | Nightingale | XS_VLDL_P | NA | G | A | 0.031933 | 0.004364 | 6.80E-14 |
| rs612169 | miR-126-3p | trans | 2.89E-39 | Nightingale | Total_C | NA | G | A | 0.032298 | 0.004285 | 3.90E-14 |
| rs612169 | miR-126-3p | trans | 2.89E-39 | Nightingale | ApoB_by_A | NA | G | A | 0.024859 | 0.004359 | 1.60E-08 |
| rs612169 | miR-126-3p | trans | 2.89E-39 | Nightingale | VLDL_P | NA | G | A | 0.026438 | 0.004402 | 5.60E-10 |
| rs612169 | miR-126-3p | trans | 2.89E-39 | Nightingale | S_HDL_CE | NA | G | A | 0.028617 | 0.004433 | 1.40E-10 |
| rs612169 | miR-126-3p | trans | 2.89E-39 | Nightingale | Total_PL | NA | G | A | 0.024903 | 0.004214 | 1.10E-09 |
| rs612169 | miR-126-3p | trans | 2.89E-39 | Nightingale | S_LDL_C | NA | G | A | 0.030803 | 0.00443 | 6.90E-12 |
| rs612169 | miR-126-3p | trans | 2.89E-39 | Nightingale | S_LDL_CE | NA | G | A | 0.030405 | 0.004428 | 7.20E-12 |
| rs612169 | miR-126-3p | trans | 2.89E-39 | Nightingale | IDL_L | NA | G | A | 0.035155 | 0.004279 | 6.10E-16 |
| rs612169 | miR-126-3p | trans | 2.89E-39 | Nightingale | XS_VLDL_L | NA | G | A | 0.033628 | 0.004351 | 2.70E-15 |
| rs612169 | miR-126-3p | trans | 2.89E-39 | Nightingale | S_HDL_P | NA | G | A | 0.030563 | 0.004426 | 5.70E-12 |
| rs612169 | miR-126-3p | trans | 2.89E-39 | Nightingale | S_LDL_PL_ | NA | G | A | -0.02989 | 0.004418 | 3.40E-13 |
| rs612169 | miR-126-3p | trans | 2.89E-39 | Nightingale | PUFA | NA | G | A | 0.023654 | 0.004317 | 1.60E-08 |
| rs612169 | miR-126-3p | trans | 2.89E-39 | Nightingale | S_LDL_CE_ | NA | G | A | 0.030784 | 0.004418 | 6.20E-14 |
| rs612169 | miR-126-3p | trans | 2.89E-39 | Nightingale | XL_HDL_PL | NA | G | A | -0.0226 | 0.004066 | 1.40E-08 |
| rs612169 | miR-126-3p | trans | 2.89E-39 | Nightingale | S_VLDL_C_ | NA | G | A | 0.041831 | 0.004307 | 6.30E-22 |
| rs612169 | miR-126-3p | trans | 2.89E-39 | Nightingale | LA | NA | G | A | 0.024865 | 0.00434 | 4.00E-09 |

| | | | | | | | | | | | |
|---|---|---|---|---|---|---|---|---|---|---|---|
| rs612169 | miR-126-3p | trans | 2.89E-39 | Nightingale | non_HDL_( | NA | G | A | 0.035557 | 0.004402 | 6.60E-16 |
| rs612169 | miR-126-3p | trans | 2.89E-39 | Nightingale | L_VLDL_FC | NA | G | A | 0.04835 | 0.004383 | 7.90E-29 |
| rs612169 | miR-126-3p | trans | 2.89E-39 | Nightingale | Total_FC | NA | G | A | 0.033156 | 0.004324 | 8.30E-15 |
| rs612169 | miR-126-3p | trans | 2.89E-39 | Nightingale | M_LDL_C | NA | G | A | 0.033844 | 0.004434 | 2.80E-14 |
| rs612169 | miR-126-3p | trans | 2.89E-39 | Nightingale | XS_VLDL_C | NA | G | A | 0.037631 | 0.004271 | 2.80E-18 |
| rs612169 | miR-126-3p | trans | 2.89E-39 | Nightingale | M_VLDL_C | NA | G | A | 0.04068 | 0.004386 | 3.10E-20 |
| rs612169 | miR-126-3p | trans | 2.89E-39 | Nightingale | S_VLDL_CE | NA | G | A | 0.046907 | 0.004366 | 1.70E-26 |
| rs612169 | miR-126-3p | trans | 2.89E-39 | Nightingale | M_VLDL_P | NA | G | A | 0.043968 | 0.004248 | 7.10E-25 |
| rs612169 | miR-126-3p | trans | 2.89E-39 | Nightingale | M_LDL_CE | NA | G | A | 0.03283 | 0.004432 | 1.20E-13 |
| rs612169 | miR-126-3p | trans | 2.89E-39 | Nightingale | VLDL_C | NA | G | A | 0.031111 | 0.00442 | 6.50E-13 |
| rs612169 | miR-126-3p | trans | 2.89E-39 | Nightingale | LDL_L | NA | G | A | 0.033183 | 0.004411 | 9.30E-14 |
| rs612169 | miR-126-3p | trans | 2.89E-39 | Nightingale | S_LDL_P | NA | G | A | 0.029399 | 0.00443 | 4.20E-11 |
| rs612169 | miR-126-3p | trans | 2.89E-39 | Nightingale | XL_VLDL_T | NA | G | A | -0.0439 | 0.00429 | 1.80E-24 |
| rs612169 | miR-126-3p | trans | 2.89E-39 | Nightingale | S_HDL_PL | NA | G | A | 0.023764 | 0.004387 | 3.40E-08 |
| rs612169 | miR-126-3p | trans | 2.89E-39 | Nightingale | S_LDL_C_p | NA | G | A | 0.045206 | 0.004443 | 8.20E-25 |
| rs612169 | miR-126-3p | trans | 2.89E-39 | Nightingale | XL_VLDL_C | NA | G | A | 0.037396 | 0.004246 | 2.90E-18 |
| rs612169 | miR-126-3p | trans | 2.89E-39 | Nightingale | L_VLDL_CE | NA | G | A | 0.047313 | 0.004235 | 2.50E-29 |
| rs612169 | miR-126-3p | trans | 2.89E-39 | Nightingale | XS_VLDL_F | NA | G | A | 0.036875 | 0.004349 | 8.90E-18 |
| rs612169 | miR-126-3p | trans | 2.89E-39 | Nightingale | Cholines | NA | G | A | 0.022291 | 0.004162 | 4.90E-08 |
| rs612169 | miR-126-3p | trans | 2.89E-39 | Nightingale | LDL_C | NA | G | A | 0.03308 | 0.004407 | 1.10E-13 |
| rs612169 | miR-126-3p | trans | 2.89E-39 | Nightingale | L_VLDL_C_ | NA | G | A | 0.052101 | 0.004323 | 5.00E-34 |
| rs612169 | miR-126-3p | trans | 2.89E-39 | Nightingale | LDL_P | NA | G | A | 0.031659 | 0.004423 | 1.30E-12 |
| rs612169 | miR-126-3p | trans | 2.89E-39 | Nightingale | XXL_VLDL_ | NA | G | A | 0.033073 | 0.00446 | 9.30E-14 |
| rs612169 | miR-126-3p | trans | 2.89E-39 | Nightingale | XL_VLDL_P | NA | G | A | 0.03861 | 0.004433 | 7.90E-19 |
| rs612169 | miR-126-3p | trans | 2.89E-39 | Nightingale | Remnant_( | NA | G | A | 0.036703 | 0.004397 | 5.00E-17 |
| rs612169 | miR-126-3p | trans | 2.89E-39 | Nightingale | XL_VLDL_C | NA | G | A | 0.038268 | 0.004244 | 4.10E-19 |
| rs612169 | miR-126-3p | trans | 2.89E-39 | Nightingale | XXL_VLDL_ | NA | G | A | -0.03569 | 0.004464 | 2.20E-16 |
| rs612169 | miR-126-3p | trans | 2.89E-39 | Nightingale | M_VLDL_P | NA | G | A | 0.028958 | 0.004419 | 2.60E-11 |
| rs612169 | miR-126-3p | trans | 2.89E-39 | Nightingale | L_LDL_FC | NA | G | A | 0.033523 | 0.004346 | 3.50E-14 |
| rs612169 | miR-126-3p | trans | 2.89E-39 | Nightingale | S_VLDL_FC | NA | G | A | 0.037705 | 0.004427 | 1.10E-17 |
| rs612169 | let-7b-5p | trans | 1.17E-26 | Nightingale | LDL_PL | NA | G | A | 0.034856 | 0.004416 | 6.10E-15 |
| rs612169 | let-7b-5p | trans | 1.17E-26 | Nightingale | L_LDL_P | NA | G | A | 0.029066 | 0.004411 | 7.10E-11 |
| rs612169 | let-7b-5p | trans | 1.17E-26 | Nightingale | LDL_FC | NA | G | A | 0.033519 | 0.004379 | 6.60E-14 |
| rs612169 | let-7b-5p | trans | 1.17E-26 | Nightingale | Citrate | NA | G | A | 0.026782 | 0.004397 | 7.80E-10 |
| rs612169 | let-7b-5p | trans | 1.17E-26 | Nightingale | M_LDL_TG | NA | G | A | -0.02765 | 0.004371 | 3.20E-10 |
| rs612169 | let-7b-5p | trans | 1.17E-26 | Nightingale | M_LDL_PL | NA | G | A | 0.032328 | 0.004432 | 3.40E-13 |
| rs612169 | let-7b-5p | trans | 1.17E-26 | Nightingale | M_LDL_FC | NA | G | A | 0.034215 | 0.00442 | 2.50E-14 |
| rs612169 | let-7b-5p | trans | 1.17E-26 | Nightingale | Omega_6 | NA | G | A | 0.025843 | 0.004322 | 6.30E-10 |
| rs612169 | let-7b-5p | trans | 1.17E-26 | Nightingale | LDL_CE | NA | G | A | 0.03244 | 0.004414 | 2.70E-13 |
| rs612169 | let-7b-5p | trans | 1.17E-26 | Nightingale | IDL_FC_pct | NA | G | A | 0.02565 | 0.004392 | 1.30E-08 |
| rs612169 | let-7b-5p | trans | 1.17E-26 | Nightingale | S_LDL_FC | NA | G | A | 0.02845 | 0.004419 | 5.20E-10 |
| rs612169 | let-7b-5p | trans | 1.17E-26 | Nightingale | ApoB | NA | G | A | 0.033778 | 0.00442 | 2.70E-14 |
| rs612169 | let-7b-5p | trans | 1.17E-26 | Nightingale | S_LDL_L | NA | G | A | 0.027614 | 0.004425 | 9.90E-10 |
| rs612169 | let-7b-5p | trans | 1.17E-26 | Nightingale | S_HDL_C | NA | G | A | 0.028392 | 0.004418 | 1.80E-10 |
| rs612169 | let-7b-5p | trans | 1.17E-26 | Nightingale | M_VLDL_C_ | NA | G | A | 0.036191 | 0.004169 | 1.30E-17 |
| rs612169 | let-7b-5p | trans | 1.17E-26 | Nightingale | IDL_FC | NA | G | A | 0.038123 | 0.004285 | 2.00E-18 |
| rs612169 | let-7b-5p | trans | 1.17E-26 | Nightingale | M_LDL_L | NA | G | A | 0.032889 | 0.004432 | 1.30E-13 |
| rs612169 | let-7b-5p | trans | 1.17E-26 | Nightingale | M_LDL_P | NA | G | A | 0.035713 | 0.004438 | 9.50E-16 |
| rs612169 | let-7b-5p | trans | 1.17E-26 | Nightingale | XS_VLDL_C | NA | G | A | 0.037817 | 0.004296 | 1.80E-18 |
| rs612169 | let-7b-5p | trans | 1.17E-26 | Nightingale | S_VLDL_CE | NA | G | A | 0.037935 | 0.00442 | 2.50E-18 |
| rs612169 | let-7b-5p | trans | 1.17E-26 | Nightingale | VLDL_FC | NA | G | A | 0.023986 | 0.004393 | 1.30E-08 |
| rs612169 | let-7b-5p | trans | 1.17E-26 | Nightingale | Total_CE | NA | G | A | 0.031681 | 0.004269 | 1.10E-13 |
| rs612169 | let-7b-5p | trans | 1.17E-26 | Nightingale | VLDL_CE | NA | G | A | 0.035585 | 0.004427 | 4.20E-16 |
| rs612169 | let-7b-5p | trans | 1.17E-26 | Nightingale | Sphingomy | NA | G | A | 0.028998 | 0.004137 | 7.10E-12 |
| rs612169 | let-7b-5p | trans | 1.17E-26 | Nightingale | M_LDL_C_ | NA | G | A | 0.031134 | 0.004429 | 3.10E-12 |
| rs612169 | let-7b-5p | trans | 1.17E-26 | Nightingale | Total_L | NA | G | A | 0.026277 | 0.004354 | 5.10E-10 |
| rs612169 | let-7b-5p | trans | 1.17E-26 | Nightingale | XL_VLDL_F | NA | G | A | 0.037185 | 0.004291 | 2.00E-18 |
| rs612169 | let-7b-5p | trans | 1.17E-26 | Nightingale | XS_VLDL_F | NA | G | A | 0.031389 | 0.004394 | 2.70E-12 |
| rs612169 | let-7b-5p | trans | 1.17E-26 | Nightingale | IDL_C | NA | G | A | 0.034851 | 0.004274 | 1.20E-15 |
| rs612169 | let-7b-5p | trans | 1.17E-26 | Nightingale | M_VLDL_F | NA | G | A | 0.033192 | 0.004425 | 4.20E-14 |
| rs612169 | let-7b-5p | trans | 1.17E-26 | Nightingale | S_VLDL_TG | NA | G | A | -0.0381 | 0.004284 | 1.60E-18 |
| rs612169 | let-7b-5p | trans | 1.17E-26 | Nightingale | S_VLDL_PL | NA | G | A | 0.032582 | 0.004418 | 5.70E-14 |
| rs612169 | let-7b-5p | trans | 1.17E-26 | Nightingale | L_VLDL_TG | NA | G | A | -0.05092 | 0.004405 | 9.20E-32 |

| | | | | | | | | | | | |
|---|---|---|---|---|---|---|---|---|---|---|---|
| rs612169 | let-7b-5p | trans | 1.17E-26 | Nightingale | M_VLDL_T | NA | G | A | -0.03853 | 0.004182 | 7.80E-20 |
| rs612169 | let-7b-5p | trans | 1.17E-26 | Nightingale | S_VLDL_P | NA | G | A | 0.023264 | 0.00437 | 2.10E-08 |
| rs612169 | let-7b-5p | trans | 1.17E-26 | Nightingale | L_LDL_L | NA | G | A | 0.033441 | 0.004383 | 3.70E-14 |
| rs612169 | let-7b-5p | trans | 1.17E-26 | Nightingale | XXL_VLDL_ | NA | G | A | 0.03485 | 0.004435 | 2.70E-15 |
| rs612169 | let-7b-5p | trans | 1.17E-26 | Nightingale | S_VLDL_FC | NA | G | A | 0.029377 | 0.004229 | 1.10E-11 |
| rs612169 | let-7b-5p | trans | 1.17E-26 | Nightingale | XS_VLDL_P | NA | G | A | 0.036482 | 0.004367 | 2.00E-17 |
| rs612169 | let-7b-5p | trans | 1.17E-26 | Nightingale | L_LDL_CE | NA | G | A | 0.031474 | 0.004386 | 1.20E-12 |
| rs612169 | let-7b-5p | trans | 1.17E-26 | Nightingale | L_LDL_C | NA | G | A | 0.032224 | 0.004376 | 3.10E-13 |
| rs612169 | let-7b-5p | trans | 1.17E-26 | Nightingale | XXL_VLDL_ | NA | G | A | 0.0248 | 0.004455 | 1.90E-09 |
| rs612169 | let-7b-5p | trans | 1.17E-26 | Nightingale | M_VLDL_C | NA | G | A | 0.033628 | 0.004158 | 2.00E-15 |
| rs612169 | let-7b-5p | trans | 1.17E-26 | Nightingale | S_VLDL_C | NA | G | A | 0.038142 | 0.004425 | 2.70E-18 |
| rs612169 | let-7b-5p | trans | 1.17E-26 | Nightingale | M_VLDL_P | NA | G | A | 0.028126 | 0.004415 | 8.00E-11 |
| rs612169 | let-7b-5p | trans | 1.17E-26 | Nightingale | XXL_VLDL_ | NA | G | A | 0.030915 | 0.004407 | 9.30E-13 |
| rs612169 | let-7b-5p | trans | 1.17E-26 | Nightingale | S_HDL_L | NA | G | A | 0.026261 | 0.004411 | 1.60E-09 |
| rs612169 | let-7b-5p | trans | 1.17E-26 | Nightingale | IDL_P | NA | G | A | 0.039804 | 0.004379 | 1.90E-19 |
| rs612169 | let-7b-5p | trans | 1.17E-26 | Nightingale | S_VLDL_PL | NA | G | A | 0.027786 | 0.004238 | 1.80E-10 |
| rs612169 | let-7b-5p | trans | 1.17E-26 | Nightingale | S_HDL_FC | NA | G | A | 0.023434 | 0.004345 | 4.80E-08 |
| rs612169 | let-7b-5p | trans | 1.17E-26 | Nightingale | IDL_CE | NA | G | A | 0.033414 | 0.004273 | 1.60E-14 |
| rs612169 | let-7b-5p | trans | 1.17E-26 | Nightingale | GlycA | NA | G | A | 0.027887 | 0.004407 | 1.70E-10 |
| rs612169 | let-7b-5p | trans | 1.17E-26 | Nightingale | M_VLDL_C | NA | G | A | 0.038535 | 0.004414 | 2.70E-18 |
| rs612169 | let-7b-5p | trans | 1.17E-26 | Nightingale | Clinical_LD | NA | G | A | 0.036343 | 0.0044 | 3.60E-16 |
| rs612169 | let-7b-5p | trans | 1.17E-26 | Nightingale | M_VLDL_F | NA | G | A | 0.042 | 0.004211 | 4.30E-23 |
| rs612169 | let-7b-5p | trans | 1.17E-26 | Nightingale | S_VLDL_L | NA | G | A | 0.024472 | 0.004383 | 4.70E-09 |
| rs612169 | let-7b-5p | trans | 1.17E-26 | Nightingale | IDL_PL | NA | G | A | 0.035229 | 0.004271 | 4.10E-16 |
| rs612169 | let-7b-5p | trans | 1.17E-26 | Nightingale | L_LDL_PL | NA | G | A | 0.037855 | 0.004392 | 1.20E-17 |
| rs612169 | let-7b-5p | trans | 1.17E-26 | Nightingale | XS_VLDL_P | NA | G | A | 0.031933 | 0.004364 | 6.80E-14 |
| rs612169 | let-7b-5p | trans | 1.17E-26 | Nightingale | Total_C | NA | G | A | 0.032298 | 0.004285 | 3.90E-14 |
| rs612169 | let-7b-5p | trans | 1.17E-26 | Nightingale | ApoB_by_ | NA | G | A | 0.024859 | 0.004359 | 1.60E-08 |
| rs612169 | let-7b-5p | trans | 1.17E-26 | Nightingale | VLDL_P | NA | G | A | 0.026438 | 0.004402 | 5.60E-10 |
| rs612169 | let-7b-5p | trans | 1.17E-26 | Nightingale | S_HDL_CE | NA | G | A | 0.028617 | 0.004433 | 1.40E-10 |
| rs612169 | let-7b-5p | trans | 1.17E-26 | Nightingale | Total_PL | NA | G | A | 0.024903 | 0.004214 | 1.10E-09 |
| rs612169 | let-7b-5p | trans | 1.17E-26 | Nightingale | S_LDL_C | NA | G | A | 0.030803 | 0.00443 | 6.90E-12 |
| rs612169 | let-7b-5p | trans | 1.17E-26 | Nightingale | S_LDL_CE | NA | G | A | 0.030405 | 0.004428 | 7.20E-12 |
| rs612169 | let-7b-5p | trans | 1.17E-26 | Nightingale | IDL_L | NA | G | A | 0.035155 | 0.004279 | 6.10E-16 |
| rs612169 | let-7b-5p | trans | 1.17E-26 | Nightingale | XS_VLDL_L | NA | G | A | 0.033628 | 0.004351 | 2.70E-15 |
| rs612169 | let-7b-5p | trans | 1.17E-26 | Nightingale | S_HDL_P | NA | G | A | 0.030563 | 0.004426 | 5.70E-12 |
| rs612169 | let-7b-5p | trans | 1.17E-26 | Nightingale | S_LDL_PL_ | NA | G | A | -0.02989 | 0.004418 | 3.40E-13 |
| rs612169 | let-7b-5p | trans | 1.17E-26 | Nightingale | PUFA | NA | G | A | 0.023654 | 0.004317 | 1.60E-08 |
| rs612169 | let-7b-5p | trans | 1.17E-26 | Nightingale | S_LDL_CE_ | NA | G | A | 0.030784 | 0.004418 | 6.20E-14 |
| rs612169 | let-7b-5p | trans | 1.17E-26 | Nightingale | XL_HDL_PL | NA | G | A | -0.0226 | 0.004066 | 1.40E-08 |
| rs612169 | let-7b-5p | trans | 1.17E-26 | Nightingale | S_VLDL_C_ | NA | G | A | 0.041831 | 0.004307 | 6.30E-22 |
| rs612169 | let-7b-5p | trans | 1.17E-26 | Nightingale | LA | NA | G | A | 0.024865 | 0.00434 | 4.00E-09 |
| rs612169 | let-7b-5p | trans | 1.17E-26 | Nightingale | non_HDL_( | NA | G | A | 0.035557 | 0.004402 | 6.60E-16 |
| rs612169 | let-7b-5p | trans | 1.17E-26 | Nightingale | L_VLDL_FC | NA | G | A | 0.04835 | 0.004383 | 7.90E-29 |
| rs612169 | let-7b-5p | trans | 1.17E-26 | Nightingale | Total_FC | NA | G | A | 0.033156 | 0.004324 | 8.30E-15 |
| rs612169 | let-7b-5p | trans | 1.17E-26 | Nightingale | M_LDL_C | NA | G | A | 0.033844 | 0.004434 | 2.80E-14 |
| rs612169 | let-7b-5p | trans | 1.17E-26 | Nightingale | XS_VLDL_C | NA | G | A | 0.037631 | 0.004271 | 2.80E-18 |
| rs612169 | let-7b-5p | trans | 1.17E-26 | Nightingale | M_VLDL_C | NA | G | A | 0.04068 | 0.004386 | 3.10E-20 |
| rs612169 | let-7b-5p | trans | 1.17E-26 | Nightingale | S_VLDL_CE | NA | G | A | 0.046907 | 0.004366 | 1.70E-26 |
| rs612169 | let-7b-5p | trans | 1.17E-26 | Nightingale | M_VLDL_P | NA | G | A | 0.043968 | 0.004248 | 7.10E-25 |
| rs612169 | let-7b-5p | trans | 1.17E-26 | Nightingale | M_LDL_CE | NA | G | A | 0.03283 | 0.004432 | 1.20E-13 |
| rs612169 | let-7b-5p | trans | 1.17E-26 | Nightingale | VLDL_C | NA | G | A | 0.031111 | 0.00442 | 6.50E-13 |
| rs612169 | let-7b-5p | trans | 1.17E-26 | Nightingale | LDL_L | NA | G | A | 0.033183 | 0.004411 | 9.30E-14 |
| rs612169 | let-7b-5p | trans | 1.17E-26 | Nightingale | S_LDL_P | NA | G | A | 0.029399 | 0.00443 | 4.20E-11 |
| rs612169 | let-7b-5p | trans | 1.17E-26 | Nightingale | XL_VLDL_T | NA | G | A | -0.0439 | 0.00429 | 1.80E-24 |
| rs612169 | let-7b-5p | trans | 1.17E-26 | Nightingale | S_HDL_PL | NA | G | A | 0.023764 | 0.004387 | 3.40E-08 |
| rs612169 | let-7b-5p | trans | 1.17E-26 | Nightingale | S_LDL_C_p | NA | G | A | 0.045206 | 0.004443 | 8.20E-25 |
| rs612169 | let-7b-5p | trans | 1.17E-26 | Nightingale | XL_VLDL_C | NA | G | A | 0.037396 | 0.004246 | 2.90E-18 |
| rs612169 | let-7b-5p | trans | 1.17E-26 | Nightingale | L_VLDL_CE | NA | G | A | 0.047313 | 0.004235 | 2.50E-29 |
| rs612169 | let-7b-5p | trans | 1.17E-26 | Nightingale | XS_VLDL_F | NA | G | A | 0.036875 | 0.004349 | 8.90E-18 |
| rs612169 | let-7b-5p | trans | 1.17E-26 | Nightingale | Cholines | NA | G | A | 0.022291 | 0.004162 | 4.90E-08 |
| rs612169 | let-7b-5p | trans | 1.17E-26 | Nightingale | LDL_C | NA | G | A | 0.03308 | 0.004407 | 1.10E-13 |
| rs612169 | let-7b-5p | trans | 1.17E-26 | Nightingale | L_VLDL_C_ | NA | G | A | 0.052101 | 0.004323 | 5.00E-34 |

| | | | | | | | | | | | |
|---|---|---|---|---|---|---|---|---|---|---|---|
| rs612169 | let-7b-5p | trans | 1.17E-26 | Nightingale | LDL_P | NA | G | A | 0.031659 | 0.004423 | 1.30E-12 |
| rs612169 | let-7b-5p | trans | 1.17E-26 | Nightingale | XXL_VLDL_ | NA | G | A | 0.033073 | 0.00446 | 9.30E-14 |
| rs612169 | let-7b-5p | trans | 1.17E-26 | Nightingale | XL_VLDL_P | NA | G | A | 0.03861 | 0.004433 | 7.90E-19 |
| rs612169 | let-7b-5p | trans | 1.17E-26 | Nightingale | Remnant_( | NA | G | A | 0.036703 | 0.004397 | 5.00E-17 |
| rs612169 | let-7b-5p | trans | 1.17E-26 | Nightingale | XL_VLDL_C | NA | G | A | 0.038268 | 0.004244 | 4.10E-19 |
| rs612169 | let-7b-5p | trans | 1.17E-26 | Nightingale | XXL_VLDL_ | NA | G | A | -0.03569 | 0.004464 | 2.20E-16 |
| rs612169 | let-7b-5p | trans | 1.17E-26 | Nightingale | M_VLDL_P | NA | G | A | 0.028958 | 0.004419 | 2.60E-11 |
| rs612169 | let-7b-5p | trans | 1.17E-26 | Nightingale | L_LDL_FC | NA | G | A | 0.033523 | 0.004346 | 3.50E-14 |
| rs612169 | let-7b-5p | trans | 1.17E-26 | Nightingale | S_VLDL_FC | NA | G | A | 0.037705 | 0.004427 | 1.10E-17 |
| rs612169 | let-7c-5p | trans | 2.76E-23 | Nightingale | LDL_PL | NA | G | A | 0.034856 | 0.004416 | 6.10E-15 |
| rs612169 | let-7c-5p | trans | 2.76E-23 | Nightingale | L_LDL_P | NA | G | A | 0.029066 | 0.004411 | 7.10E-11 |
| rs612169 | let-7c-5p | trans | 2.76E-23 | Nightingale | LDL_FC | NA | G | A | 0.033519 | 0.004379 | 6.60E-14 |
| rs612169 | let-7c-5p | trans | 2.76E-23 | Nightingale | Citrate | NA | G | A | 0.026782 | 0.004397 | 7.80E-10 |
| rs612169 | let-7c-5p | trans | 2.76E-23 | Nightingale | M_LDL_TG | NA | G | A | -0.02765 | 0.004371 | 3.20E-10 |
| rs612169 | let-7c-5p | trans | 2.76E-23 | Nightingale | M_LDL_PL | NA | G | A | 0.032328 | 0.004432 | 3.40E-13 |
| rs612169 | let-7c-5p | trans | 2.76E-23 | Nightingale | M_LDL_FC | NA | G | A | 0.034215 | 0.00442 | 2.50E-14 |
| rs612169 | let-7c-5p | trans | 2.76E-23 | Nightingale | Omega_6 | NA | G | A | 0.025843 | 0.004322 | 6.30E-10 |
| rs612169 | let-7c-5p | trans | 2.76E-23 | Nightingale | LDL_CE | NA | G | A | 0.03244 | 0.004414 | 2.70E-13 |
| rs612169 | let-7c-5p | trans | 2.76E-23 | Nightingale | IDL_FC_pct | NA | G | A | 0.02565 | 0.004392 | 1.30E-08 |
| rs612169 | let-7c-5p | trans | 2.76E-23 | Nightingale | S_LDL_FC | NA | G | A | 0.02845 | 0.004419 | 5.20E-10 |
| rs612169 | let-7c-5p | trans | 2.76E-23 | Nightingale | ApoB | NA | G | A | 0.033778 | 0.00442 | 2.70E-14 |
| rs612169 | let-7c-5p | trans | 2.76E-23 | Nightingale | S_LDL_L | NA | G | A | 0.027614 | 0.004425 | 9.90E-10 |
| rs612169 | let-7c-5p | trans | 2.76E-23 | Nightingale | S_HDL_C | NA | G | A | 0.028392 | 0.004418 | 1.80E-10 |
| rs612169 | let-7c-5p | trans | 2.76E-23 | Nightingale | M_VLDL_C | NA | G | A | 0.036191 | 0.004169 | 1.30E-17 |
| rs612169 | let-7c-5p | trans | 2.76E-23 | Nightingale | IDL_FC | NA | G | A | 0.038123 | 0.004285 | 2.00E-18 |
| rs612169 | let-7c-5p | trans | 2.76E-23 | Nightingale | M_LDL_L | NA | G | A | 0.032889 | 0.004432 | 1.30E-13 |
| rs612169 | let-7c-5p | trans | 2.76E-23 | Nightingale | M_LDL_P | NA | G | A | 0.035713 | 0.004438 | 9.50E-16 |
| rs612169 | let-7c-5p | trans | 2.76E-23 | Nightingale | XS_VLDL_C | NA | G | A | 0.037817 | 0.004296 | 1.80E-18 |
| rs612169 | let-7c-5p | trans | 2.76E-23 | Nightingale | S_VLDL_CE | NA | G | A | 0.037935 | 0.00442 | 2.50E-18 |
| rs612169 | let-7c-5p | trans | 2.76E-23 | Nightingale | VLDL_FC | NA | G | A | 0.023986 | 0.004393 | 1.30E-08 |
| rs612169 | let-7c-5p | trans | 2.76E-23 | Nightingale | Total_CE | NA | G | A | 0.031681 | 0.004269 | 1.10E-13 |
| rs612169 | let-7c-5p | trans | 2.76E-23 | Nightingale | VLDL_CE | NA | G | A | 0.035585 | 0.004427 | 4.20E-16 |
| rs612169 | let-7c-5p | trans | 2.76E-23 | Nightingale | Sphingomy | NA | G | A | 0.028998 | 0.004137 | 7.10E-12 |
| rs612169 | let-7c-5p | trans | 2.76E-23 | Nightingale | M_LDL_C_ | NA | G | A | 0.031134 | 0.004429 | 3.10E-12 |
| rs612169 | let-7c-5p | trans | 2.76E-23 | Nightingale | Total_L | NA | G | A | 0.026277 | 0.004354 | 5.10E-10 |
| rs612169 | let-7c-5p | trans | 2.76E-23 | Nightingale | XL_VLDL_F | NA | G | A | 0.037185 | 0.004291 | 2.00E-18 |
| rs612169 | let-7c-5p | trans | 2.76E-23 | Nightingale | XS_VLDL_F | NA | G | A | 0.031389 | 0.004394 | 2.70E-12 |
| rs612169 | let-7c-5p | trans | 2.76E-23 | Nightingale | IDL_C | NA | G | A | 0.034851 | 0.004274 | 1.20E-15 |
| rs612169 | let-7c-5p | trans | 2.76E-23 | Nightingale | M_VLDL_F | NA | G | A | 0.033192 | 0.004425 | 4.20E-14 |
| rs612169 | let-7c-5p | trans | 2.76E-23 | Nightingale | S_VLDL_TG | NA | G | A | -0.0381 | 0.004284 | 1.60E-18 |
| rs612169 | let-7c-5p | trans | 2.76E-23 | Nightingale | S_VLDL_PL | NA | G | A | 0.032582 | 0.004418 | 5.70E-14 |
| rs612169 | let-7c-5p | trans | 2.76E-23 | Nightingale | L_VLDL_TG | NA | G | A | -0.05092 | 0.004405 | 9.20E-32 |
| rs612169 | let-7c-5p | trans | 2.76E-23 | Nightingale | M_VLDL_T | NA | G | A | -0.03853 | 0.004182 | 7.80E-20 |
| rs612169 | let-7c-5p | trans | 2.76E-23 | Nightingale | S_VLDL_P | NA | G | A | 0.023264 | 0.00437 | 2.10E-08 |
| rs612169 | let-7c-5p | trans | 2.76E-23 | Nightingale | L_LDL_L | NA | G | A | 0.033441 | 0.004383 | 3.70E-14 |
| rs612169 | let-7c-5p | trans | 2.76E-23 | Nightingale | XXL_VLDL_ | NA | G | A | 0.03485 | 0.004435 | 2.70E-15 |
| rs612169 | let-7c-5p | trans | 2.76E-23 | Nightingale | S_VLDL_FC | NA | G | A | 0.029377 | 0.004229 | 1.10E-11 |
| rs612169 | let-7c-5p | trans | 2.76E-23 | Nightingale | XS_VLDL_P | NA | G | A | 0.036482 | 0.004367 | 2.00E-17 |
| rs612169 | let-7c-5p | trans | 2.76E-23 | Nightingale | L_LDL_CE | NA | G | A | 0.031474 | 0.004386 | 1.20E-12 |
| rs612169 | let-7c-5p | trans | 2.76E-23 | Nightingale | L_LDL_C | NA | G | A | 0.032224 | 0.004376 | 3.10E-13 |
| rs612169 | let-7c-5p | trans | 2.76E-23 | Nightingale | XXL_VLDL_ | NA | G | A | 0.0248 | 0.004455 | 1.90E-09 |
| rs612169 | let-7c-5p | trans | 2.76E-23 | Nightingale | M_VLDL_C | NA | G | A | 0.033628 | 0.004158 | 2.00E-15 |
| rs612169 | let-7c-5p | trans | 2.76E-23 | Nightingale | S_VLDL_C | NA | G | A | 0.038142 | 0.004425 | 2.70E-18 |
| rs612169 | let-7c-5p | trans | 2.76E-23 | Nightingale | M_VLDL_P | NA | G | A | 0.028126 | 0.004415 | 8.00E-11 |
| rs612169 | let-7c-5p | trans | 2.76E-23 | Nightingale | XXL_VLDL_ | NA | G | A | 0.030915 | 0.004407 | 9.30E-13 |
| rs612169 | let-7c-5p | trans | 2.76E-23 | Nightingale | S_HDL_L | NA | G | A | 0.026261 | 0.004411 | 1.60E-09 |
| rs612169 | let-7c-5p | trans | 2.76E-23 | Nightingale | IDL_P | NA | G | A | 0.039804 | 0.004379 | 1.90E-19 |
| rs612169 | let-7c-5p | trans | 2.76E-23 | Nightingale | S_VLDL_PL | NA | G | A | 0.027786 | 0.004238 | 1.80E-10 |
| rs612169 | let-7c-5p | trans | 2.76E-23 | Nightingale | S_HDL_FC | NA | G | A | 0.023434 | 0.004345 | 4.80E-08 |
| rs612169 | let-7c-5p | trans | 2.76E-23 | Nightingale | IDL_CE | NA | G | A | 0.033414 | 0.004273 | 1.60E-14 |
| rs612169 | let-7c-5p | trans | 2.76E-23 | Nightingale | GlycA | NA | G | A | 0.027887 | 0.004407 | 1.70E-10 |
| rs612169 | let-7c-5p | trans | 2.76E-23 | Nightingale | M_VLDL_C | NA | G | A | 0.038535 | 0.004414 | 2.70E-18 |
| rs612169 | let-7c-5p | trans | 2.76E-23 | Nightingale | Clinical_LD | NA | G | A | 0.036343 | 0.0044 | 3.60E-16 |

| | | | | | | | | | | | |
|---|---|---|---|---|---|---|---|---|---|---|---|
| rs612169 | let-7c-5p | trans | 2.76E-23 | Nightingale | M_VLDL_F | NA | G | A | 0.042 | 0.004211 | 4.30E-23 |
| rs612169 | let-7c-5p | trans | 2.76E-23 | Nightingale | S_VLDL_L | NA | G | A | 0.024472 | 0.004383 | 4.70E-09 |
| rs612169 | let-7c-5p | trans | 2.76E-23 | Nightingale | IDL_PL | NA | G | A | 0.035229 | 0.004271 | 4.10E-16 |
| rs612169 | let-7c-5p | trans | 2.76E-23 | Nightingale | L_LDL_PL | NA | G | A | 0.037855 | 0.004392 | 1.20E-17 |
| rs612169 | let-7c-5p | trans | 2.76E-23 | Nightingale | XS_VLDL_P | NA | G | A | 0.031933 | 0.004364 | 6.80E-14 |
| rs612169 | let-7c-5p | trans | 2.76E-23 | Nightingale | Total_C | NA | G | A | 0.032298 | 0.004285 | 3.90E-14 |
| rs612169 | let-7c-5p | trans | 2.76E-23 | Nightingale | ApoB_by_/ | NA | G | A | 0.024859 | 0.004359 | 1.60E-08 |
| rs612169 | let-7c-5p | trans | 2.76E-23 | Nightingale | VLDL_P | NA | G | A | 0.026438 | 0.004402 | 5.60E-10 |
| rs612169 | let-7c-5p | trans | 2.76E-23 | Nightingale | S_HDL_CE | NA | G | A | 0.028617 | 0.004433 | 1.40E-10 |
| rs612169 | let-7c-5p | trans | 2.76E-23 | Nightingale | Total_PL | NA | G | A | 0.024903 | 0.004214 | 1.10E-09 |
| rs612169 | let-7c-5p | trans | 2.76E-23 | Nightingale | S_LDL_C | NA | G | A | 0.030803 | 0.00443 | 6.90E-12 |
| rs612169 | let-7c-5p | trans | 2.76E-23 | Nightingale | S_LDL_CE | NA | G | A | 0.030405 | 0.004428 | 7.20E-12 |
| rs612169 | let-7c-5p | trans | 2.76E-23 | Nightingale | IDL_L | NA | G | A | 0.035155 | 0.004279 | 6.10E-16 |
| rs612169 | let-7c-5p | trans | 2.76E-23 | Nightingale | XS_VLDL_L | NA | G | A | 0.033628 | 0.004351 | 2.70E-15 |
| rs612169 | let-7c-5p | trans | 2.76E-23 | Nightingale | S_HDL_P | NA | G | A | 0.030563 | 0.004426 | 5.70E-12 |
| rs612169 | let-7c-5p | trans | 2.76E-23 | Nightingale | S_LDL_PL_ | NA | G | A | -0.02989 | 0.004418 | 3.40E-13 |
| rs612169 | let-7c-5p | trans | 2.76E-23 | Nightingale | PUFA | NA | G | A | 0.023654 | 0.004317 | 1.60E-08 |
| rs612169 | let-7c-5p | trans | 2.76E-23 | Nightingale | S_LDL_CE_ | NA | G | A | 0.030784 | 0.004418 | 6.20E-14 |
| rs612169 | let-7c-5p | trans | 2.76E-23 | Nightingale | XL_HDL_PL | NA | G | A | -0.0226 | 0.004066 | 1.40E-08 |
| rs612169 | let-7c-5p | trans | 2.76E-23 | Nightingale | S_VLDL_C_ | NA | G | A | 0.041831 | 0.004307 | 6.30E-22 |
| rs612169 | let-7c-5p | trans | 2.76E-23 | Nightingale | LA | NA | G | A | 0.024865 | 0.00434 | 4.00E-09 |
| rs612169 | let-7c-5p | trans | 2.76E-23 | Nightingale | non_HDL_( | NA | G | A | 0.035557 | 0.004402 | 6.60E-16 |
| rs612169 | let-7c-5p | trans | 2.76E-23 | Nightingale | L_VLDL_FC | NA | G | A | 0.04835 | 0.004383 | 7.90E-29 |
| rs612169 | let-7c-5p | trans | 2.76E-23 | Nightingale | Total_FC | NA | G | A | 0.033156 | 0.004324 | 8.30E-15 |
| rs612169 | let-7c-5p | trans | 2.76E-23 | Nightingale | M_LDL_C | NA | G | A | 0.033844 | 0.004434 | 2.80E-14 |
| rs612169 | let-7c-5p | trans | 2.76E-23 | Nightingale | XS_VLDL_C | NA | G | A | 0.037631 | 0.004271 | 2.80E-18 |
| rs612169 | let-7c-5p | trans | 2.76E-23 | Nightingale | M_VLDL_C | NA | G | A | 0.04068 | 0.004386 | 3.10E-20 |
| rs612169 | let-7c-5p | trans | 2.76E-23 | Nightingale | S_VLDL_CE | NA | G | A | 0.046907 | 0.004366 | 1.70E-26 |
| rs612169 | let-7c-5p | trans | 2.76E-23 | Nightingale | M_VLDL_P | NA | G | A | 0.043968 | 0.004248 | 7.10E-25 |
| rs612169 | let-7c-5p | trans | 2.76E-23 | Nightingale | M_LDL_CE | NA | G | A | 0.03283 | 0.004432 | 1.20E-13 |
| rs612169 | let-7c-5p | trans | 2.76E-23 | Nightingale | VLDL_C | NA | G | A | 0.031111 | 0.00442 | 6.50E-13 |
| rs612169 | let-7c-5p | trans | 2.76E-23 | Nightingale | LDL_L | NA | G | A | 0.033183 | 0.004411 | 9.30E-14 |
| rs612169 | let-7c-5p | trans | 2.76E-23 | Nightingale | S_LDL_P | NA | G | A | 0.029399 | 0.00443 | 4.20E-11 |
| rs612169 | let-7c-5p | trans | 2.76E-23 | Nightingale | XL_VLDL_T | NA | G | A | -0.0439 | 0.00429 | 1.80E-24 |
| rs612169 | let-7c-5p | trans | 2.76E-23 | Nightingale | S_HDL_PL | NA | G | A | 0.023764 | 0.004387 | 3.40E-08 |
| rs612169 | let-7c-5p | trans | 2.76E-23 | Nightingale | S_LDL_C_p | NA | G | A | 0.045206 | 0.004443 | 8.20E-25 |
| rs612169 | let-7c-5p | trans | 2.76E-23 | Nightingale | XL_VLDL_C | NA | G | A | 0.037396 | 0.004246 | 2.90E-18 |
| rs612169 | let-7c-5p | trans | 2.76E-23 | Nightingale | L_VLDL_CE | NA | G | A | 0.047313 | 0.004235 | 2.50E-29 |
| rs612169 | let-7c-5p | trans | 2.76E-23 | Nightingale | XS_VLDL_F | NA | G | A | 0.036875 | 0.004349 | 8.90E-18 |
| rs612169 | let-7c-5p | trans | 2.76E-23 | Nightingale | Cholines | NA | G | A | 0.022291 | 0.004162 | 4.90E-08 |
| rs612169 | let-7c-5p | trans | 2.76E-23 | Nightingale | LDL_C | NA | G | A | 0.03308 | 0.004407 | 1.10E-13 |
| rs612169 | let-7c-5p | trans | 2.76E-23 | Nightingale | L_VLDL_C_ | NA | G | A | 0.052101 | 0.004323 | 5.00E-34 |
| rs612169 | let-7c-5p | trans | 2.76E-23 | Nightingale | LDL_P | NA | G | A | 0.031659 | 0.004423 | 1.30E-12 |
| rs612169 | let-7c-5p | trans | 2.76E-23 | Nightingale | XXL_VLDL_ | NA | G | A | 0.033073 | 0.00446 | 9.30E-14 |
| rs612169 | let-7c-5p | trans | 2.76E-23 | Nightingale | XL_VLDL_P | NA | G | A | 0.03861 | 0.004433 | 7.90E-19 |
| rs612169 | let-7c-5p | trans | 2.76E-23 | Nightingale | Remnant_( | NA | G | A | 0.036703 | 0.004397 | 5.00E-17 |
| rs612169 | let-7c-5p | trans | 2.76E-23 | Nightingale | XL_VLDL_C | NA | G | A | 0.038268 | 0.004244 | 4.10E-19 |
| rs612169 | let-7c-5p | trans | 2.76E-23 | Nightingale | XXL_VLDL_ | NA | G | A | -0.03569 | 0.004464 | 2.20E-16 |
| rs612169 | let-7c-5p | trans | 2.76E-23 | Nightingale | M_VLDL_P | NA | G | A | 0.028958 | 0.004419 | 2.60E-11 |
| rs612169 | let-7c-5p | trans | 2.76E-23 | Nightingale | L_LDL_FC | NA | G | A | 0.033523 | 0.004346 | 3.50E-14 |
| rs612169 | let-7c-5p | trans | 2.76E-23 | Nightingale | S_VLDL_FC | NA | G | A | 0.037705 | 0.004427 | 1.10E-17 |
| rs612169 | miR-195-5p | trans | 2.56E-17 | Nightingale | LDL_PL | NA | G | A | 0.034856 | 0.004416 | 6.10E-15 |
| rs612169 | miR-195-5p | trans | 2.56E-17 | Nightingale | L_LDL_P | NA | G | A | 0.029066 | 0.004411 | 7.10E-11 |
| rs612169 | miR-195-5p | trans | 2.56E-17 | Nightingale | LDL_FC | NA | G | A | 0.033519 | 0.004379 | 6.60E-14 |
| rs612169 | miR-195-5p | trans | 2.56E-17 | Nightingale | Citrate | NA | G | A | 0.026782 | 0.004397 | 7.80E-10 |
| rs612169 | miR-195-5p | trans | 2.56E-17 | Nightingale | M_LDL_TG | NA | G | A | -0.02765 | 0.004371 | 3.20E-10 |
| rs612169 | miR-195-5p | trans | 2.56E-17 | Nightingale | M_LDL_PL | NA | G | A | 0.032328 | 0.004432 | 3.40E-13 |
| rs612169 | miR-195-5p | trans | 2.56E-17 | Nightingale | M_LDL_FC | NA | G | A | 0.034215 | 0.00442 | 2.50E-14 |
| rs612169 | miR-195-5p | trans | 2.56E-17 | Nightingale | Omega_6 | NA | G | A | 0.025843 | 0.004322 | 6.30E-10 |
| rs612169 | miR-195-5p | trans | 2.56E-17 | Nightingale | LDL_CE | NA | G | A | 0.03244 | 0.004414 | 2.70E-13 |
| rs612169 | miR-195-5p | trans | 2.56E-17 | Nightingale | IDL_FC_pct | NA | G | A | 0.02565 | 0.004392 | 1.30E-08 |
| rs612169 | miR-195-5p | trans | 2.56E-17 | Nightingale | S_LDL_FC | NA | G | A | 0.02845 | 0.004419 | 5.20E-10 |
| rs612169 | miR-195-5p | trans | 2.56E-17 | Nightingale | ApoB | NA | G | A | 0.033778 | 0.00442 | 2.70E-14 |

| | | | | | | | | | | | |
|---|---|---|---|---|---|---|---|---|---|---|---|
| rs612169 | miR-195-5| | trans | 2.56E-17 | Nightingale | S_LDL_L | NA | G | A | 0.027614 | 0.004425 | 9.90E-10 |
| rs612169 | miR-195-5| | trans | 2.56E-17 | Nightingale | S_HDL_C | NA | G | A | 0.028392 | 0.004418 | 1.80E-10 |
| rs612169 | miR-195-5| | trans | 2.56E-17 | Nightingale | M_VLDL_C | NA | G | A | 0.036191 | 0.004169 | 1.30E-17 |
| rs612169 | miR-195-5| | trans | 2.56E-17 | Nightingale | IDL_FC | NA | G | A | 0.038123 | 0.004285 | 2.00E-18 |
| rs612169 | miR-195-5| | trans | 2.56E-17 | Nightingale | M_LDL_L | NA | G | A | 0.032889 | 0.004432 | 1.30E-13 |
| rs612169 | miR-195-5| | trans | 2.56E-17 | Nightingale | M_LDL_P | NA | G | A | 0.035713 | 0.004438 | 9.50E-16 |
| rs612169 | miR-195-5| | trans | 2.56E-17 | Nightingale | XS_VLDL_C | NA | G | A | 0.037817 | 0.004296 | 1.80E-18 |
| rs612169 | miR-195-5| | trans | 2.56E-17 | Nightingale | S_VLDL_CE | NA | G | A | 0.037935 | 0.00442 | 2.50E-18 |
| rs612169 | miR-195-5| | trans | 2.56E-17 | Nightingale | VLDL_FC | NA | G | A | 0.023986 | 0.004393 | 1.30E-08 |
| rs612169 | miR-195-5| | trans | 2.56E-17 | Nightingale | Total_CE | NA | G | A | 0.031681 | 0.004269 | 1.10E-13 |
| rs612169 | miR-195-5| | trans | 2.56E-17 | Nightingale | VLDL_CE | NA | G | A | 0.035585 | 0.004427 | 4.20E-16 |
| rs612169 | miR-195-5| | trans | 2.56E-17 | Nightingale | Sphingomy | NA | G | A | 0.028998 | 0.004137 | 7.10E-12 |
| rs612169 | miR-195-5| | trans | 2.56E-17 | Nightingale | M_LDL_C_| | NA | G | A | 0.031134 | 0.004429 | 3.10E-12 |
| rs612169 | miR-195-5| | trans | 2.56E-17 | Nightingale | Total_L | NA | G | A | 0.026277 | 0.004354 | 5.10E-10 |
| rs612169 | miR-195-5| | trans | 2.56E-17 | Nightingale | XL_VLDL_F | NA | G | A | 0.037185 | 0.004291 | 2.00E-18 |
| rs612169 | miR-195-5| | trans | 2.56E-17 | Nightingale | XS_VLDL_F | NA | G | A | 0.031389 | 0.004394 | 2.70E-12 |
| rs612169 | miR-195-5| | trans | 2.56E-17 | Nightingale | IDL_C | NA | G | A | 0.034851 | 0.004274 | 1.20E-15 |
| rs612169 | miR-195-5| | trans | 2.56E-17 | Nightingale | M_VLDL_F | NA | G | A | 0.033192 | 0.004425 | 4.20E-14 |
| rs612169 | miR-195-5| | trans | 2.56E-17 | Nightingale | S_VLDL_TG | NA | G | A | -0.0381 | 0.004284 | 1.60E-18 |
| rs612169 | miR-195-5| | trans | 2.56E-17 | Nightingale | S_VLDL_PL | NA | G | A | 0.032582 | 0.004418 | 5.70E-14 |
| rs612169 | miR-195-5| | trans | 2.56E-17 | Nightingale | L_VLDL_TG | NA | G | A | -0.05092 | 0.004405 | 9.20E-32 |
| rs612169 | miR-195-5| | trans | 2.56E-17 | Nightingale | M_VLDL_T | NA | G | A | -0.03853 | 0.004182 | 7.80E-20 |
| rs612169 | miR-195-5| | trans | 2.56E-17 | Nightingale | S_VLDL_P | NA | G | A | 0.023264 | 0.00437 | 2.10E-08 |
| rs612169 | miR-195-5| | trans | 2.56E-17 | Nightingale | L_LDL_L | NA | G | A | 0.033441 | 0.004383 | 3.70E-14 |
| rs612169 | miR-195-5| | trans | 2.56E-17 | Nightingale | XXL_VLDL_ | NA | G | A | 0.03485 | 0.004435 | 2.70E-15 |
| rs612169 | miR-195-5| | trans | 2.56E-17 | Nightingale | S_VLDL_FC | NA | G | A | 0.029377 | 0.004229 | 1.10E-11 |
| rs612169 | miR-195-5| | trans | 2.56E-17 | Nightingale | XS_VLDL_P | NA | G | A | 0.036482 | 0.004367 | 2.00E-17 |
| rs612169 | miR-195-5| | trans | 2.56E-17 | Nightingale | L_LDL_CE | NA | G | A | 0.031474 | 0.004386 | 1.20E-12 |
| rs612169 | miR-195-5| | trans | 2.56E-17 | Nightingale | L_LDL_C | NA | G | A | 0.032224 | 0.004376 | 3.10E-13 |
| rs612169 | miR-195-5| | trans | 2.56E-17 | Nightingale | XXL_VLDL_ | NA | G | A | 0.0248 | 0.004455 | 1.90E-09 |
| rs612169 | miR-195-5| | trans | 2.56E-17 | Nightingale | M_VLDL_C | NA | G | A | 0.033628 | 0.004158 | 2.00E-15 |
| rs612169 | miR-195-5| | trans | 2.56E-17 | Nightingale | S_VLDL_C | NA | G | A | 0.038142 | 0.004425 | 2.70E-18 |
| rs612169 | miR-195-5| | trans | 2.56E-17 | Nightingale | M_VLDL_P | NA | G | A | 0.028126 | 0.004415 | 8.00E-11 |
| rs612169 | miR-195-5| | trans | 2.56E-17 | Nightingale | XXL_VLDL_ | NA | G | A | 0.030915 | 0.004407 | 9.30E-13 |
| rs612169 | miR-195-5| | trans | 2.56E-17 | Nightingale | S_HDL_L | NA | G | A | 0.026261 | 0.004411 | 1.60E-09 |
| rs612169 | miR-195-5| | trans | 2.56E-17 | Nightingale | IDL_P | NA | G | A | 0.039804 | 0.004379 | 1.90E-19 |
| rs612169 | miR-195-5| | trans | 2.56E-17 | Nightingale | S_VLDL_PL | NA | G | A | 0.027786 | 0.004238 | 1.80E-10 |
| rs612169 | miR-195-5| | trans | 2.56E-17 | Nightingale | S_HDL_FC | NA | G | A | 0.023434 | 0.004345 | 4.80E-08 |
| rs612169 | miR-195-5| | trans | 2.56E-17 | Nightingale | IDL_CE | NA | G | A | 0.033414 | 0.004273 | 1.60E-14 |
| rs612169 | miR-195-5| | trans | 2.56E-17 | Nightingale | GlycA | NA | G | A | 0.027887 | 0.004407 | 1.70E-10 |
| rs612169 | miR-195-5| | trans | 2.56E-17 | Nightingale | M_VLDL_C | NA | G | A | 0.038535 | 0.004414 | 2.70E-18 |
| rs612169 | miR-195-5| | trans | 2.56E-17 | Nightingale | Clinical_LD | NA | G | A | 0.036343 | 0.0044 | 3.60E-16 |
| rs612169 | miR-195-5| | trans | 2.56E-17 | Nightingale | M_VLDL_F | NA | G | A | 0.042 | 0.004211 | 4.30E-23 |
| rs612169 | miR-195-5| | trans | 2.56E-17 | Nightingale | S_VLDL_L | NA | G | A | 0.024472 | 0.004383 | 4.70E-09 |
| rs612169 | miR-195-5| | trans | 2.56E-17 | Nightingale | IDL_PL | NA | G | A | 0.035229 | 0.004271 | 4.10E-16 |
| rs612169 | miR-195-5| | trans | 2.56E-17 | Nightingale | L_LDL_PL | NA | G | A | 0.037855 | 0.004392 | 1.20E-17 |
| rs612169 | miR-195-5| | trans | 2.56E-17 | Nightingale | XS_VLDL_P | NA | G | A | 0.031933 | 0.004364 | 6.80E-14 |
| rs612169 | miR-195-5| | trans | 2.56E-17 | Nightingale | Total_C | NA | G | A | 0.032298 | 0.004285 | 3.90E-14 |
| rs612169 | miR-195-5| | trans | 2.56E-17 | Nightingale | ApoB_by_/ | NA | G | A | 0.024859 | 0.004359 | 1.60E-08 |
| rs612169 | miR-195-5| | trans | 2.56E-17 | Nightingale | VLDL_P | NA | G | A | 0.026438 | 0.004402 | 5.60E-10 |
| rs612169 | miR-195-5| | trans | 2.56E-17 | Nightingale | S_HDL_CE | NA | G | A | 0.028617 | 0.004433 | 1.40E-10 |
| rs612169 | miR-195-5| | trans | 2.56E-17 | Nightingale | Total_PL | NA | G | A | 0.024903 | 0.004214 | 1.10E-09 |
| rs612169 | miR-195-5| | trans | 2.56E-17 | Nightingale | S_LDL_C | NA | G | A | 0.030803 | 0.00443 | 6.90E-12 |
| rs612169 | miR-195-5| | trans | 2.56E-17 | Nightingale | S_LDL_CE | NA | G | A | 0.030405 | 0.004428 | 7.20E-12 |
| rs612169 | miR-195-5| | trans | 2.56E-17 | Nightingale | IDL_L | NA | G | A | 0.035155 | 0.004279 | 6.10E-16 |
| rs612169 | miR-195-5| | trans | 2.56E-17 | Nightingale | XS_VLDL_L | NA | G | A | 0.033628 | 0.004351 | 2.70E-15 |
| rs612169 | miR-195-5| | trans | 2.56E-17 | Nightingale | S_HDL_P | NA | G | A | 0.030563 | 0.004426 | 5.70E-12 |
| rs612169 | miR-195-5| | trans | 2.56E-17 | Nightingale | S_LDL_PL_ | NA | G | A | -0.02989 | 0.004418 | 3.40E-13 |
| rs612169 | miR-195-5| | trans | 2.56E-17 | Nightingale | PUFA | NA | G | A | 0.023654 | 0.004317 | 1.60E-08 |
| rs612169 | miR-195-5| | trans | 2.56E-17 | Nightingale | S_LDL_CE_ | NA | G | A | 0.030784 | 0.004418 | 6.20E-14 |
| rs612169 | miR-195-5| | trans | 2.56E-17 | Nightingale | XL_HDL_PL | NA | G | A | -0.0226 | 0.004066 | 1.40E-08 |
| rs612169 | miR-195-5| | trans | 2.56E-17 | Nightingale | S_VLDL_C_ | NA | G | A | 0.041831 | 0.004307 | 6.30E-22 |
| rs612169 | miR-195-5| | trans | 2.56E-17 | Nightingale | LA | NA | G | A | 0.024865 | 0.00434 | 4.00E-09 |

| | | | | | | | | | | | |
|---|---|---|---|---|---|---|---|---|---|---|---|
| rs612169 | miR-195-5p | trans | 2.56E-17 | Nightingale | non_HDL_( | NA | G | A | 0.035557 | 0.004402 | 6.60E-16 |
| rs612169 | miR-195-5p | trans | 2.56E-17 | Nightingale | L_VLDL_FC | NA | G | A | 0.04835 | 0.004383 | 7.90E-29 |
| rs612169 | miR-195-5p | trans | 2.56E-17 | Nightingale | Total_FC | NA | G | A | 0.033156 | 0.004324 | 8.30E-15 |
| rs612169 | miR-195-5p | trans | 2.56E-17 | Nightingale | M_LDL_C | NA | G | A | 0.033844 | 0.004434 | 2.80E-14 |
| rs612169 | miR-195-5p | trans | 2.56E-17 | Nightingale | XS_VLDL_C | NA | G | A | 0.037631 | 0.004271 | 2.80E-18 |
| rs612169 | miR-195-5p | trans | 2.56E-17 | Nightingale | M_VLDL_C | NA | G | A | 0.04068 | 0.004386 | 3.10E-20 |
| rs612169 | miR-195-5p | trans | 2.56E-17 | Nightingale | S_VLDL_CE | NA | G | A | 0.046907 | 0.004366 | 1.70E-26 |
| rs612169 | miR-195-5p | trans | 2.56E-17 | Nightingale | M_VLDL_P | NA | G | A | 0.043968 | 0.004248 | 7.10E-25 |
| rs612169 | miR-195-5p | trans | 2.56E-17 | Nightingale | M_LDL_CE | NA | G | A | 0.03283 | 0.004432 | 1.20E-13 |
| rs612169 | miR-195-5p | trans | 2.56E-17 | Nightingale | VLDL_C | NA | G | A | 0.031111 | 0.00442 | 6.50E-13 |
| rs612169 | miR-195-5p | trans | 2.56E-17 | Nightingale | LDL_L | NA | G | A | 0.033183 | 0.004411 | 9.30E-14 |
| rs612169 | miR-195-5p | trans | 2.56E-17 | Nightingale | S_LDL_P | NA | G | A | 0.029399 | 0.00443 | 4.20E-11 |
| rs612169 | miR-195-5p | trans | 2.56E-17 | Nightingale | XL_VLDL_T | NA | G | A | -0.0439 | 0.00429 | 1.80E-24 |
| rs612169 | miR-195-5p | trans | 2.56E-17 | Nightingale | S_HDL_PL | NA | G | A | 0.023764 | 0.004387 | 3.40E-08 |
| rs612169 | miR-195-5p | trans | 2.56E-17 | Nightingale | S_LDL_C_p | NA | G | A | 0.045206 | 0.004443 | 8.20E-25 |
| rs612169 | miR-195-5p | trans | 2.56E-17 | Nightingale | XL_VLDL_C | NA | G | A | 0.037396 | 0.004246 | 2.90E-18 |
| rs612169 | miR-195-5p | trans | 2.56E-17 | Nightingale | L_VLDL_CE | NA | G | A | 0.047313 | 0.004235 | 2.50E-29 |
| rs612169 | miR-195-5p | trans | 2.56E-17 | Nightingale | XS_VLDL_F | NA | G | A | 0.036875 | 0.004349 | 8.90E-18 |
| rs612169 | miR-195-5p | trans | 2.56E-17 | Nightingale | Cholines | NA | G | A | 0.022291 | 0.004162 | 4.90E-08 |
| rs612169 | miR-195-5p | trans | 2.56E-17 | Nightingale | LDL_C | NA | G | A | 0.03308 | 0.004407 | 1.10E-13 |
| rs612169 | miR-195-5p | trans | 2.56E-17 | Nightingale | L_VLDL_C_ | NA | G | A | 0.052101 | 0.004323 | 5.00E-34 |
| rs612169 | miR-195-5p | trans | 2.56E-17 | Nightingale | LDL_P | NA | G | A | 0.031659 | 0.004423 | 1.30E-12 |
| rs612169 | miR-195-5p | trans | 2.56E-17 | Nightingale | XXL_VLDL_ | NA | G | A | 0.033073 | 0.00446 | 9.30E-14 |
| rs612169 | miR-195-5p | trans | 2.56E-17 | Nightingale | XL_VLDL_P | NA | G | A | 0.03861 | 0.004433 | 7.90E-19 |
| rs612169 | miR-195-5p | trans | 2.56E-17 | Nightingale | Remnant_( | NA | G | A | 0.036703 | 0.004397 | 5.00E-17 |
| rs612169 | miR-195-5p | trans | 2.56E-17 | Nightingale | XL_VLDL_C | NA | G | A | 0.038268 | 0.004244 | 4.10E-19 |
| rs612169 | miR-195-5p | trans | 2.56E-17 | Nightingale | XXL_VLDL_ | NA | G | A | -0.03569 | 0.004464 | 2.20E-16 |
| rs612169 | miR-195-5p | trans | 2.56E-17 | Nightingale | M_VLDL_P | NA | G | A | 0.028958 | 0.004419 | 2.60E-11 |
| rs612169 | miR-195-5p | trans | 2.56E-17 | Nightingale | L_LDL_FC | NA | G | A | 0.033523 | 0.004346 | 3.50E-14 |
| rs612169 | miR-195-5p | trans | 2.56E-17 | Nightingale | S_VLDL_FC | NA | G | A | 0.037705 | 0.004427 | 1.10E-17 |
| rs613534 | miR-10b-5p | trans | 2.68E-34 | Nightingale | S_VLDL_L | NA | G | A | 0.024496 | 0.004305 | 2.50E-09 |
| rs613534 | miR-10b-5p | trans | 2.68E-34 | Nightingale | IDL_L | NA | G | A | 0.03366 | 0.004203 | 2.30E-15 |
| rs613534 | miR-10b-5p | trans | 2.68E-34 | Nightingale | M_VLDL_C | NA | G | A | 0.037341 | 0.004335 | 5.40E-18 |
| rs613534 | miR-10b-5p | trans | 2.68E-34 | Nightingale | S_HDL_C | NA | G | A | 0.02538 | 0.004339 | 9.40E-09 |
| rs613534 | miR-10b-5p | trans | 2.68E-34 | Nightingale | L_VLDL_C_ | NA | G | A | 0.049226 | 0.004245 | 8.90E-32 |
| rs613534 | miR-10b-5p | trans | 2.68E-34 | Nightingale | VLDL_P | NA | G | A | 0.026555 | 0.004324 | 2.00E-10 |
| rs613534 | miR-10b-5p | trans | 2.68E-34 | Nightingale | M_VLDL_P | NA | G | A | 0.027926 | 0.004336 | 4.70E-11 |
| rs613534 | miR-10b-5p | trans | 2.68E-34 | Nightingale | L_LDL_C | NA | G | A | 0.030224 | 0.004298 | 3.00E-12 |
| rs613534 | miR-10b-5p | trans | 2.68E-34 | Nightingale | M_VLDL_F( | NA | G | A | 0.032515 | 0.004346 | 3.90E-14 |
| rs613534 | miR-10b-5p | trans | 2.68E-34 | Nightingale | S_VLDL_TG | NA | G | A | -0.03598 | 0.004208 | 1.90E-17 |
| rs613534 | miR-10b-5p | trans | 2.68E-34 | Nightingale | S_HDL_P | NA | G | A | 0.027805 | 0.004347 | 2.70E-10 |
| rs613534 | miR-10b-5p | trans | 2.68E-34 | Nightingale | XS_VLDL_F | NA | G | A | 0.035882 | 0.004271 | 7.50E-18 |
| rs613534 | miR-10b-5p | trans | 2.68E-34 | Nightingale | L_LDL_L | NA | G | A | 0.031523 | 0.004304 | 3.20E-13 |
| rs613534 | miR-10b-5p | trans | 2.68E-34 | Nightingale | LDL_PL | NA | G | A | 0.033318 | 0.004337 | 3.00E-14 |
| rs613534 | miR-10b-5p | trans | 2.68E-34 | Nightingale | XL_VLDL_P | NA | G | A | 0.039106 | 0.004353 | 7.40E-20 |
| rs613534 | miR-10b-5p | trans | 2.68E-34 | Nightingale | GlycA | NA | G | A | 0.025329 | 0.004328 | 4.80E-09 |
| rs613534 | miR-10b-5p | trans | 2.68E-34 | Nightingale | L_LDL_P | NA | G | A | 0.028317 | 0.004332 | 7.80E-11 |
| rs613534 | miR-10b-5p | trans | 2.68E-34 | Nightingale | L_VLDL_CE | NA | G | A | 0.022756 | 0.004296 | 2.90E-08 |
| rs613534 | miR-10b-5p | trans | 2.68E-34 | Nightingale | M_VLDL_C | NA | G | A | 0.031348 | 0.004084 | 3.30E-14 |
| rs613534 | miR-10b-5p | trans | 2.68E-34 | Nightingale | XXL_VLDL_ | NA | G | A | 0.023502 | 0.004375 | 5.40E-09 |
| rs613534 | miR-10b-5p | trans | 2.68E-34 | Nightingale | IDL_P | NA | G | A | 0.039032 | 0.004301 | 1.80E-19 |
| rs613534 | miR-10b-5p | trans | 2.68E-34 | Nightingale | LDL_P | NA | G | A | 0.030659 | 0.004344 | 2.20E-12 |
| rs613534 | miR-10b-5p | trans | 2.68E-34 | Nightingale | LA | NA | G | A | 0.023263 | 0.004263 | 1.60E-08 |
| rs613534 | miR-10b-5p | trans | 2.68E-34 | Nightingale | non_HDL_( | NA | G | A | 0.03414 | 0.004323 | 2.40E-15 |
| rs613534 | miR-10b-5p | trans | 2.68E-34 | Nightingale | XXL_VLDL_ | NA | G | A | -0.03443 | 0.004384 | 6.30E-16 |
| rs613534 | miR-10b-5p | trans | 2.68E-34 | Nightingale | S_VLDL_P | NA | G | A | 0.02339 | 0.004292 | 9.70E-09 |
| rs613534 | miR-10b-5p | trans | 2.68E-34 | Nightingale | LDL_L | NA | G | A | 0.031465 | 0.004332 | 5.80E-13 |
| rs613534 | miR-10b-5p | trans | 2.68E-34 | Nightingale | S_VLDL_C_ | NA | G | A | 0.039736 | 0.00423 | 8.30E-21 |
| rs613534 | miR-10b-5p | trans | 2.68E-34 | Nightingale | M_LDL_TG | NA | G | A | -0.02512 | 0.004293 | 8.60E-09 |
| rs613534 | miR-10b-5p | trans | 2.68E-34 | Nightingale | XS_VLDL_C | NA | G | A | 0.036639 | 0.004219 | 2.40E-18 |
| rs613534 | miR-10b-5p | trans | 2.68E-34 | Nightingale | S_LDL_P | NA | G | A | 0.028959 | 0.004351 | 3.00E-11 |
| rs613534 | miR-10b-5p | trans | 2.68E-34 | Nightingale | XS_VLDL_F | NA | G | A | 0.029134 | 0.004316 | 3.10E-11 |
| rs613534 | miR-10b-5p | trans | 2.68E-34 | Nightingale | ApoB | NA | G | A | 0.032928 | 0.004341 | 3.40E-14 |

| | | | | | | | | | | | |
|---|---|---|---|---|---|---|---|---|---|---|---|
| rs613534 | miR-10b-5p | trans | 2.68E-34 | Nightingale | XS_VLDL_P | NA | G | A | 0.031657 | 0.004286 | 2.00E-14 |
| rs613534 | miR-10b-5p | trans | 2.68E-34 | Nightingale | Total_C | NA | G | A | 0.030138 | 0.004208 | 5.00E-13 |
| rs613534 | miR-10b-5p | trans | 2.68E-34 | Nightingale | S_VLDL_CE | NA | G | A | 0.037311 | 0.004341 | 1.70E-18 |
| rs613534 | miR-10b-5p | trans | 2.68E-34 | Nightingale | L_LDL_FC | NA | G | A | 0.031473 | 0.004268 | 4.10E-13 |
| rs613534 | miR-10b-5p | trans | 2.68E-34 | Nightingale | S_LDL_CE_ | NA | G | A | 0.028844 | 0.004339 | 1.10E-12 |
| rs613534 | miR-10b-5p | trans | 2.68E-34 | Nightingale | LDL_CE | NA | G | A | 0.030617 | 0.004335 | 2.10E-12 |
| rs613534 | miR-10b-5p | trans | 2.68E-34 | Nightingale | XS_VLDL_C | NA | G | A | 0.036391 | 0.004195 | 4.10E-18 |
| rs613534 | miR-10b-5p | trans | 2.68E-34 | Nightingale | XXL_VLDL_ | NA | G | A | 0.033437 | 0.004356 | 8.80E-15 |
| rs613534 | miR-10b-5p | trans | 2.68E-34 | Nightingale | M_VLDL_F | NA | G | A | 0.039372 | 0.004135 | 2.20E-21 |
| rs613534 | miR-10b-5p | trans | 2.68E-34 | Nightingale | S_HDL_CE | NA | G | A | 0.02569 | 0.004354 | 7.00E-09 |
| rs613534 | miR-10b-5p | trans | 2.68E-34 | Nightingale | S_VLDL_CE | NA | G | A | 0.045151 | 0.004288 | 1.40E-25 |
| rs613534 | miR-10b-5p | trans | 2.68E-34 | Nightingale | M_VLDL_C | NA | G | A | 0.033845 | 0.004094 | 2.70E-16 |
| rs613534 | miR-10b-5p | trans | 2.68E-34 | Nightingale | LDL_FC | NA | G | A | 0.031548 | 0.0043 | 6.60E-13 |
| rs613534 | miR-10b-5p | trans | 2.68E-34 | Nightingale | Omega_6 | NA | G | A | 0.024156 | 0.004245 | 3.10E-09 |
| rs613534 | miR-10b-5p | trans | 2.68E-34 | Nightingale | Total_CE | NA | G | A | 0.029384 | 0.004193 | 1.80E-12 |
| rs613534 | miR-10b-5p | trans | 2.68E-34 | Nightingale | M_VLDL_T | NA | G | A | -0.03623 | 0.004108 | 1.50E-18 |
| rs613534 | miR-10b-5p | trans | 2.68E-34 | Nightingale | ApoB_by_A | NA | G | A | 0.025679 | 0.004281 | 2.00E-09 |
| rs613534 | miR-10b-5p | trans | 2.68E-34 | Nightingale | IDL_C | NA | G | A | 0.033305 | 0.004198 | 4.90E-15 |
| rs613534 | miR-10b-5p | trans | 2.68E-34 | Nightingale | Citrate | NA | G | A | 0.024678 | 0.004318 | 6.10E-09 |
| rs613534 | miR-10b-5p | trans | 2.68E-34 | Nightingale | S_LDL_PL_ | NA | G | A | -0.02806 | 0.004339 | 4.90E-12 |
| rs613534 | miR-10b-5p | trans | 2.68E-34 | Nightingale | L_LDL_PL | NA | G | A | 0.035919 | 0.004314 | 1.30E-16 |
| rs613534 | miR-10b-5p | trans | 2.68E-34 | Nightingale | S_LDL_C | NA | G | A | 0.029698 | 0.004351 | 1.60E-11 |
| rs613534 | miR-10b-5p | trans | 2.68E-34 | Nightingale | Remnant_( | NA | G | A | 0.035838 | 0.004319 | 5.30E-17 |
| rs613534 | miR-10b-5p | trans | 2.68E-34 | Nightingale | S_VLDL_FC | NA | G | A | 0.027154 | 0.004154 | 1.20E-10 |
| rs613534 | miR-10b-5p | trans | 2.68E-34 | Nightingale | Total_PL | NA | G | A | 0.022997 | 0.004139 | 7.80E-09 |
| rs613534 | miR-10b-5p | trans | 2.68E-34 | Nightingale | IDL_PL | NA | G | A | 0.033573 | 0.004195 | 2.00E-15 |
| rs613534 | miR-10b-5p | trans | 2.68E-34 | Nightingale | S_LDL_C_p | NA | G | A | 0.041864 | 0.004364 | 5.60E-22 |
| rs613534 | miR-10b-5p | trans | 2.68E-34 | Nightingale | M_LDL_C | NA | G | A | 0.032091 | 0.004355 | 2.30E-13 |
| rs613534 | miR-10b-5p | trans | 2.68E-34 | Nightingale | S_VLDL_PL | NA | G | A | 0.025728 | 0.004162 | 1.50E-09 |
| rs613534 | miR-10b-5p | trans | 2.68E-34 | Nightingale | M_VLDL_P | NA | G | A | 0.028753 | 0.00434 | 1.40E-11 |
| rs613534 | miR-10b-5p | trans | 2.68E-34 | Nightingale | XL_VLDL_F | NA | G | A | 0.035204 | 0.004214 | 1.80E-17 |
| rs613534 | miR-10b-5p | trans | 2.68E-34 | Nightingale | M_LDL_PL | NA | G | A | 0.031174 | 0.004353 | 9.90E-13 |
| rs613534 | miR-10b-5p | trans | 2.68E-34 | Nightingale | L_VLDL_PL | NA | G | A | 0.021645 | 0.004222 | 4.40E-08 |
| rs613534 | miR-10b-5p | trans | 2.68E-34 | Nightingale | M_LDL_CE | NA | G | A | 0.031212 | 0.004353 | 8.50E-13 |
| rs613534 | miR-10b-5p | trans | 2.68E-34 | Nightingale | XS_VLDL_P | NA | G | A | 0.035785 | 0.004289 | 1.00E-17 |
| rs613534 | miR-10b-5p | trans | 2.68E-34 | Nightingale | M_VLDL_P | NA | G | A | 0.042093 | 0.004172 | 6.30E-24 |
| rs613534 | miR-10b-5p | trans | 2.68E-34 | Nightingale | M_LDL_L | NA | G | A | 0.031388 | 0.004353 | 7.00E-13 |
| rs613534 | miR-10b-5p | trans | 2.68E-34 | Nightingale | S_LDL_CE | NA | G | A | 0.029442 | 0.004349 | 1.50E-11 |
| rs613534 | miR-10b-5p | trans | 2.68E-34 | Nightingale | LDL_C | NA | G | A | 0.031186 | 0.004328 | 9.60E-13 |
| rs613534 | miR-10b-5p | trans | 2.68E-34 | Nightingale | S_VLDL_C | NA | G | A | 0.037355 | 0.004346 | 2.60E-18 |
| rs613534 | miR-10b-5p | trans | 2.68E-34 | Nightingale | IDL_CE | NA | G | A | 0.032088 | 0.004197 | 4.30E-14 |
| rs613534 | miR-10b-5p | trans | 2.68E-34 | Nightingale | S_LDL_L | NA | G | A | 0.026893 | 0.004346 | 1.20E-09 |
| rs613534 | miR-10b-5p | trans | 2.68E-34 | Nightingale | XXL_VLDL_ | NA | G | A | 0.03179 | 0.00438 | 2.10E-13 |
| rs613534 | miR-10b-5p | trans | 2.68E-34 | Nightingale | L_LDL_CE | NA | G | A | 0.02952 | 0.004308 | 9.90E-12 |
| rs613534 | miR-10b-5p | trans | 2.68E-34 | Nightingale | XL_HDL_PL | NA | G | A | -0.02181 | 0.003994 | 4.60E-08 |
| rs613534 | miR-10b-5p | trans | 2.68E-34 | Nightingale | Sphingomy | NA | G | A | 0.025917 | 0.004063 | 3.30E-10 |
| rs613534 | miR-10b-5p | trans | 2.68E-34 | Nightingale | VLDL_C | NA | G | A | 0.031062 | 0.004341 | 2.10E-13 |
| rs613534 | miR-10b-5p | trans | 2.68E-34 | Nightingale | M_LDL_C_ | NA | G | A | 0.026582 | 0.00435 | 1.60E-09 |
| rs613534 | miR-10b-5p | trans | 2.68E-34 | Nightingale | XL_VLDL_T | NA | G | A | -0.04155 | 0.004213 | 4.90E-23 |
| rs613534 | miR-10b-5p | trans | 2.68E-34 | Nightingale | S_LDL_FC | NA | G | A | 0.0272 | 0.00434 | 1.30E-09 |
| rs613534 | miR-10b-5p | trans | 2.68E-34 | Nightingale | Total_L | NA | G | A | 0.025008 | 0.004276 | 1.50E-09 |
| rs613534 | miR-10b-5p | trans | 2.68E-34 | Nightingale | Clinical_LD | NA | G | A | 0.034557 | 0.004322 | 2.70E-15 |
| rs613534 | miR-10b-5p | trans | 2.68E-34 | Nightingale | XXL_VLDL_ | NA | G | A | 0.029268 | 0.004328 | 5.10E-12 |
| rs613534 | miR-10b-5p | trans | 2.68E-34 | Nightingale | XL_VLDL_C | NA | G | A | 0.03559 | 0.004168 | 1.70E-17 |
| rs613534 | miR-10b-5p | trans | 2.68E-34 | Nightingale | M_LDL_P | NA | G | A | 0.034001 | 0.004359 | 7.50E-15 |
| rs613534 | miR-10b-5p | trans | 2.68E-34 | Nightingale | IDL_FC | NA | G | A | 0.035978 | 0.004208 | 3.20E-17 |
| rs613534 | miR-10b-5p | trans | 2.68E-34 | Nightingale | XS_VLDL_L | NA | G | A | 0.033006 | 0.004273 | 1.40E-15 |
| rs613534 | miR-10b-5p | trans | 2.68E-34 | Nightingale | XL_VLDL_C | NA | G | A | 0.034575 | 0.00417 | 1.50E-16 |
| rs613534 | miR-10b-5p | trans | 2.68E-34 | Nightingale | M_LDL_FC | NA | G | A | 0.032281 | 0.004341 | 2.40E-13 |
| rs613534 | miR-10b-5p | trans | 2.68E-34 | Nightingale | S_VLDL_PL | NA | G | A | 0.032035 | 0.004339 | 4.90E-14 |
| rs613534 | miR-10b-5p | trans | 2.68E-34 | Nightingale | L_VLDL_FC | NA | G | A | 0.046941 | 0.004305 | 3.90E-28 |
| rs613534 | miR-10b-5p | trans | 2.68E-34 | Nightingale | S_HDL_L | NA | G | A | 0.023969 | 0.004332 | 2.70E-08 |
| rs613534 | miR-10b-5p | trans | 2.68E-34 | Nightingale | VLDL_CE | NA | G | A | 0.035144 | 0.004348 | 2.10E-16 |

| | | | | | | | | | | | |
|---|---|---|---|---|---|---|---|---|---|---|---|
| rs613534 | miR-10b-5p | trans | 2.68E-34 | Nightingale | L_VLDL_TG | NA | G | A | -0.04923 | 0.004326 | 6.40E-31 |
| rs613534 | miR-10b-5p | trans | 2.68E-34 | Nightingale | L_VLDL_CE | NA | G | A | 0.04407 | 0.00416 | 7.90E-27 |
| rs613534 | miR-10b-5p | trans | 2.68E-34 | Nightingale | S_VLDL_FC | NA | G | A | 0.036642 | 0.004347 | 1.70E-17 |
| rs613534 | miR-10b-5p | trans | 2.68E-34 | Nightingale | M_VLDL_C | NA | G | A | 0.0392 | 0.004307 | 1.30E-19 |
| rs613534 | miR-10b-5p | trans | 2.68E-34 | Nightingale | Total_FC | NA | G | A | 0.031378 | 0.004246 | 4.70E-14 |
| rs613534 | miR-10b-5p | trans | 2.68E-34 | Nightingale | VLDL_FC | NA | G | A | 0.024446 | 0.004315 | 3.60E-09 |
| rs613534 | miR-139-5p | trans | 3.54E-40 | Nightingale | S_VLDL_L | NA | G | A | 0.024496 | 0.004305 | 2.50E-09 |
| rs613534 | miR-139-5p | trans | 3.54E-40 | Nightingale | IDL_L | NA | G | A | 0.03366 | 0.004203 | 2.30E-15 |
| rs613534 | miR-139-5p | trans | 3.54E-40 | Nightingale | M_VLDL_C | NA | G | A | 0.037341 | 0.004335 | 5.40E-18 |
| rs613534 | miR-139-5p | trans | 3.54E-40 | Nightingale | S_HDL_C | NA | G | A | 0.02538 | 0.004339 | 9.40E-09 |
| rs613534 | miR-139-5p | trans | 3.54E-40 | Nightingale | L_VLDL_C_ | NA | G | A | 0.049226 | 0.004245 | 8.90E-32 |
| rs613534 | miR-139-5p | trans | 3.54E-40 | Nightingale | VLDL_P | NA | G | A | 0.026555 | 0.004324 | 2.00E-10 |
| rs613534 | miR-139-5p | trans | 3.54E-40 | Nightingale | M_VLDL_P | NA | G | A | 0.027926 | 0.004336 | 4.70E-11 |
| rs613534 | miR-139-5p | trans | 3.54E-40 | Nightingale | L_LDL_C | NA | G | A | 0.030224 | 0.004298 | 3.00E-12 |
| rs613534 | miR-139-5p | trans | 3.54E-40 | Nightingale | M_VLDL_F | NA | G | A | 0.032515 | 0.004346 | 3.90E-14 |
| rs613534 | miR-139-5p | trans | 3.54E-40 | Nightingale | S_VLDL_TG | NA | G | A | -0.03598 | 0.004208 | 1.90E-17 |
| rs613534 | miR-139-5p | trans | 3.54E-40 | Nightingale | S_HDL_P | NA | G | A | 0.027805 | 0.004347 | 2.70E-10 |
| rs613534 | miR-139-5p | trans | 3.54E-40 | Nightingale | XS_VLDL_F | NA | G | A | 0.035882 | 0.004271 | 7.50E-18 |
| rs613534 | miR-139-5p | trans | 3.54E-40 | Nightingale | L_LDL_L | NA | G | A | 0.031523 | 0.004304 | 3.20E-13 |
| rs613534 | miR-139-5p | trans | 3.54E-40 | Nightingale | LDL_PL | NA | G | A | 0.033318 | 0.004337 | 3.00E-14 |
| rs613534 | miR-139-5p | trans | 3.54E-40 | Nightingale | XL_VLDL_P | NA | G | A | 0.039106 | 0.004353 | 7.40E-20 |
| rs613534 | miR-139-5p | trans | 3.54E-40 | Nightingale | GlycA | NA | G | A | 0.025329 | 0.004328 | 4.80E-09 |
| rs613534 | miR-139-5p | trans | 3.54E-40 | Nightingale | L_LDL_P | NA | G | A | 0.028317 | 0.004332 | 7.80E-11 |
| rs613534 | miR-139-5p | trans | 3.54E-40 | Nightingale | L_VLDL_CE | NA | G | A | 0.022756 | 0.004296 | 2.90E-08 |
| rs613534 | miR-139-5p | trans | 3.54E-40 | Nightingale | M_VLDL_C | NA | G | A | 0.031348 | 0.004084 | 3.30E-14 |
| rs613534 | miR-139-5p | trans | 3.54E-40 | Nightingale | XXL_VLDL_ | NA | G | A | 0.023502 | 0.004375 | 5.40E-09 |
| rs613534 | miR-139-5p | trans | 3.54E-40 | Nightingale | IDL_P | NA | G | A | 0.039032 | 0.004301 | 1.80E-19 |
| rs613534 | miR-139-5p | trans | 3.54E-40 | Nightingale | LDL_P | NA | G | A | 0.030659 | 0.004344 | 2.20E-12 |
| rs613534 | miR-139-5p | trans | 3.54E-40 | Nightingale | LA | NA | G | A | 0.023263 | 0.004263 | 1.60E-08 |
| rs613534 | miR-139-5p | trans | 3.54E-40 | Nightingale | non_HDL_( | NA | G | A | 0.03414 | 0.004323 | 2.40E-15 |
| rs613534 | miR-139-5p | trans | 3.54E-40 | Nightingale | XXL_VLDL_ | NA | G | A | -0.03443 | 0.004384 | 6.30E-16 |
| rs613534 | miR-139-5p | trans | 3.54E-40 | Nightingale | S_VLDL_P | NA | G | A | 0.02339 | 0.004292 | 9.70E-09 |
| rs613534 | miR-139-5p | trans | 3.54E-40 | Nightingale | LDL_L | NA | G | A | 0.031465 | 0.004332 | 5.80E-13 |
| rs613534 | miR-139-5p | trans | 3.54E-40 | Nightingale | S_VLDL_C_ | NA | G | A | 0.039736 | 0.00423 | 8.30E-21 |
| rs613534 | miR-139-5p | trans | 3.54E-40 | Nightingale | M_LDL_TG | NA | G | A | -0.02512 | 0.004293 | 8.60E-09 |
| rs613534 | miR-139-5p | trans | 3.54E-40 | Nightingale | XS_VLDL_C | NA | G | A | 0.036639 | 0.004219 | 2.40E-18 |
| rs613534 | miR-139-5p | trans | 3.54E-40 | Nightingale | S_LDL_P | NA | G | A | 0.028959 | 0.004351 | 3.00E-11 |
| rs613534 | miR-139-5p | trans | 3.54E-40 | Nightingale | XS_VLDL_F | NA | G | A | 0.029134 | 0.004316 | 3.10E-11 |
| rs613534 | miR-139-5p | trans | 3.54E-40 | Nightingale | ApoB | NA | G | A | 0.032928 | 0.004341 | 3.40E-14 |
| rs613534 | miR-139-5p | trans | 3.54E-40 | Nightingale | XS_VLDL_P | NA | G | A | 0.031657 | 0.004286 | 2.00E-14 |
| rs613534 | miR-139-5p | trans | 3.54E-40 | Nightingale | Total_C | NA | G | A | 0.030138 | 0.004208 | 5.00E-13 |
| rs613534 | miR-139-5p | trans | 3.54E-40 | Nightingale | S_VLDL_CE | NA | G | A | 0.037311 | 0.004341 | 1.70E-18 |
| rs613534 | miR-139-5p | trans | 3.54E-40 | Nightingale | L_LDL_FC | NA | G | A | 0.031473 | 0.004268 | 4.10E-13 |
| rs613534 | miR-139-5p | trans | 3.54E-40 | Nightingale | S_LDL_CE_ | NA | G | A | 0.028844 | 0.004339 | 1.10E-12 |
| rs613534 | miR-139-5p | trans | 3.54E-40 | Nightingale | LDL_CE | NA | G | A | 0.030617 | 0.004335 | 2.10E-12 |
| rs613534 | miR-139-5p | trans | 3.54E-40 | Nightingale | XS_VLDL_C | NA | G | A | 0.036391 | 0.004195 | 4.10E-18 |
| rs613534 | miR-139-5p | trans | 3.54E-40 | Nightingale | XXL_VLDL_ | NA | G | A | 0.033437 | 0.004356 | 8.80E-15 |
| rs613534 | miR-139-5p | trans | 3.54E-40 | Nightingale | M_VLDL_F | NA | G | A | 0.039372 | 0.004135 | 2.20E-21 |
| rs613534 | miR-139-5p | trans | 3.54E-40 | Nightingale | S_HDL_CE | NA | G | A | 0.02569 | 0.004354 | 7.00E-09 |
| rs613534 | miR-139-5p | trans | 3.54E-40 | Nightingale | S_VLDL_CE | NA | G | A | 0.045151 | 0.004288 | 1.40E-25 |
| rs613534 | miR-139-5p | trans | 3.54E-40 | Nightingale | M_VLDL_C | NA | G | A | 0.033845 | 0.004094 | 2.70E-16 |
| rs613534 | miR-139-5p | trans | 3.54E-40 | Nightingale | LDL_FC | NA | G | A | 0.031548 | 0.0043 | 6.60E-13 |
| rs613534 | miR-139-5p | trans | 3.54E-40 | Nightingale | Omega_6 | NA | G | A | 0.024156 | 0.004245 | 3.10E-09 |
| rs613534 | miR-139-5p | trans | 3.54E-40 | Nightingale | Total_CE | NA | G | A | 0.029384 | 0.004193 | 1.80E-12 |
| rs613534 | miR-139-5p | trans | 3.54E-40 | Nightingale | M_VLDL_T | NA | G | A | -0.03623 | 0.004108 | 1.50E-18 |
| rs613534 | miR-139-5p | trans | 3.54E-40 | Nightingale | ApoB_by_A | NA | G | A | 0.025679 | 0.004281 | 2.00E-09 |
| rs613534 | miR-139-5p | trans | 3.54E-40 | Nightingale | IDL_C | NA | G | A | 0.033305 | 0.004198 | 4.90E-15 |
| rs613534 | miR-139-5p | trans | 3.54E-40 | Nightingale | Citrate | NA | G | A | 0.024678 | 0.004318 | 6.10E-09 |
| rs613534 | miR-139-5p | trans | 3.54E-40 | Nightingale | S_LDL_PL_ | NA | G | A | -0.02806 | 0.004339 | 4.90E-12 |
| rs613534 | miR-139-5p | trans | 3.54E-40 | Nightingale | L_LDL_PL | NA | G | A | 0.035919 | 0.004314 | 1.30E-16 |
| rs613534 | miR-139-5p | trans | 3.54E-40 | Nightingale | S_LDL_C | NA | G | A | 0.029698 | 0.004351 | 1.60E-11 |
| rs613534 | miR-139-5p | trans | 3.54E-40 | Nightingale | Remnant_( | NA | G | A | 0.035838 | 0.004319 | 5.30E-17 |
| rs613534 | miR-139-5p | trans | 3.54E-40 | Nightingale | S_VLDL_FC | NA | G | A | 0.027154 | 0.004154 | 1.20E-10 |

| | | | | | | | | | | | |
|---|---|---|---|---|---|---|---|---|---|---|---|
| rs613534 | miR-139-5ı | trans | 3.54E-40 | Nightingale | Total_PL | NA | G | A | 0.022997 | 0.004139 | 7.80E-09 |
| rs613534 | miR-139-5ı | trans | 3.54E-40 | Nightingale | IDL_PL | NA | G | A | 0.033573 | 0.004195 | 2.00E-15 |
| rs613534 | miR-139-5ı | trans | 3.54E-40 | Nightingale | S_LDL_C_p | NA | G | A | 0.041864 | 0.004364 | 5.60E-22 |
| rs613534 | miR-139-5ı | trans | 3.54E-40 | Nightingale | M_LDL_C | NA | G | A | 0.032091 | 0.004355 | 2.30E-13 |
| rs613534 | miR-139-5ı | trans | 3.54E-40 | Nightingale | S_VLDL_PL | NA | G | A | 0.025728 | 0.004162 | 1.50E-09 |
| rs613534 | miR-139-5ı | trans | 3.54E-40 | Nightingale | M_VLDL_P | NA | G | A | 0.028753 | 0.00434 | 1.40E-11 |
| rs613534 | miR-139-5ı | trans | 3.54E-40 | Nightingale | XL_VLDL_F | NA | G | A | 0.035204 | 0.004214 | 1.80E-17 |
| rs613534 | miR-139-5ı | trans | 3.54E-40 | Nightingale | M_LDL_PL | NA | G | A | 0.031174 | 0.004353 | 9.90E-13 |
| rs613534 | miR-139-5ı | trans | 3.54E-40 | Nightingale | L_VLDL_PL | NA | G | A | 0.021645 | 0.004222 | 4.40E-08 |
| rs613534 | miR-139-5ı | trans | 3.54E-40 | Nightingale | M_LDL_CE | NA | G | A | 0.031212 | 0.004353 | 8.50E-13 |
| rs613534 | miR-139-5ı | trans | 3.54E-40 | Nightingale | XS_VLDL_P | NA | G | A | 0.035785 | 0.004289 | 1.00E-17 |
| rs613534 | miR-139-5ı | trans | 3.54E-40 | Nightingale | M_VLDL_P | NA | G | A | 0.042093 | 0.004172 | 6.30E-24 |
| rs613534 | miR-139-5ı | trans | 3.54E-40 | Nightingale | M_LDL_L | NA | G | A | 0.031388 | 0.004353 | 7.00E-13 |
| rs613534 | miR-139-5ı | trans | 3.54E-40 | Nightingale | S_LDL_CE | NA | G | A | 0.029442 | 0.004349 | 1.50E-11 |
| rs613534 | miR-139-5ı | trans | 3.54E-40 | Nightingale | LDL_C | NA | G | A | 0.031186 | 0.004328 | 9.60E-13 |
| rs613534 | miR-139-5ı | trans | 3.54E-40 | Nightingale | S_VLDL_C | NA | G | A | 0.037355 | 0.004346 | 2.60E-18 |
| rs613534 | miR-139-5ı | trans | 3.54E-40 | Nightingale | IDL_CE | NA | G | A | 0.032088 | 0.004197 | 4.30E-14 |
| rs613534 | miR-139-5ı | trans | 3.54E-40 | Nightingale | S_LDL_L | NA | G | A | 0.026893 | 0.004346 | 1.20E-09 |
| rs613534 | miR-139-5ı | trans | 3.54E-40 | Nightingale | XXL_VLDL_ | NA | G | A | 0.03179 | 0.00438 | 2.10E-13 |
| rs613534 | miR-139-5ı | trans | 3.54E-40 | Nightingale | L_LDL_CE | NA | G | A | 0.02952 | 0.004308 | 9.90E-12 |
| rs613534 | miR-139-5ı | trans | 3.54E-40 | Nightingale | XL_HDL_PL | NA | G | A | -0.02181 | 0.003994 | 4.60E-08 |
| rs613534 | miR-139-5ı | trans | 3.54E-40 | Nightingale | Sphingomy | NA | G | A | 0.025917 | 0.004063 | 3.30E-10 |
| rs613534 | miR-139-5ı | trans | 3.54E-40 | Nightingale | VLDL_C | NA | G | A | 0.031062 | 0.004341 | 2.10E-13 |
| rs613534 | miR-139-5ı | trans | 3.54E-40 | Nightingale | M_LDL_C_ | NA | G | A | 0.026582 | 0.00435 | 1.60E-09 |
| rs613534 | miR-139-5ı | trans | 3.54E-40 | Nightingale | XL_VLDL_T | NA | G | A | -0.04155 | 0.004213 | 4.90E-23 |
| rs613534 | miR-139-5ı | trans | 3.54E-40 | Nightingale | S_LDL_FC | NA | G | A | 0.0272 | 0.00434 | 1.30E-09 |
| rs613534 | miR-139-5ı | trans | 3.54E-40 | Nightingale | Total_L | NA | G | A | 0.025008 | 0.004276 | 1.50E-09 |
| rs613534 | miR-139-5ı | trans | 3.54E-40 | Nightingale | Clinical_LD | NA | G | A | 0.034557 | 0.004322 | 2.70E-15 |
| rs613534 | miR-139-5ı | trans | 3.54E-40 | Nightingale | XXL_VLDL_ | NA | G | A | 0.029268 | 0.004328 | 5.10E-12 |
| rs613534 | miR-139-5ı | trans | 3.54E-40 | Nightingale | XL_VLDL_C | NA | G | A | 0.03559 | 0.004168 | 1.70E-17 |
| rs613534 | miR-139-5ı | trans | 3.54E-40 | Nightingale | M_LDL_P | NA | G | A | 0.034001 | 0.004359 | 7.50E-15 |
| rs613534 | miR-139-5ı | trans | 3.54E-40 | Nightingale | IDL_FC | NA | G | A | 0.035978 | 0.004208 | 3.20E-17 |
| rs613534 | miR-139-5ı | trans | 3.54E-40 | Nightingale | XS_VLDL_L | NA | G | A | 0.033006 | 0.004273 | 1.40E-15 |
| rs613534 | miR-139-5ı | trans | 3.54E-40 | Nightingale | XL_VLDL_C | NA | G | A | 0.034575 | 0.00417 | 1.50E-16 |
| rs613534 | miR-139-5ı | trans | 3.54E-40 | Nightingale | M_LDL_FC | NA | G | A | 0.032281 | 0.004341 | 2.40E-13 |
| rs613534 | miR-139-5ı | trans | 3.54E-40 | Nightingale | S_VLDL_PL | NA | G | A | 0.032035 | 0.004339 | 4.90E-14 |
| rs613534 | miR-139-5ı | trans | 3.54E-40 | Nightingale | L_VLDL_FC | NA | G | A | 0.046941 | 0.004305 | 3.90E-28 |
| rs613534 | miR-139-5ı | trans | 3.54E-40 | Nightingale | S_HDL_L | NA | G | A | 0.023969 | 0.004332 | 2.70E-08 |
| rs613534 | miR-139-5ı | trans | 3.54E-40 | Nightingale | VLDL_CE | NA | G | A | 0.035144 | 0.004348 | 2.10E-16 |
| rs613534 | miR-139-5ı | trans | 3.54E-40 | Nightingale | L_VLDL_TG | NA | G | A | -0.04923 | 0.004326 | 6.40E-31 |
| rs613534 | miR-139-5ı | trans | 3.54E-40 | Nightingale | L_VLDL_CE | NA | G | A | 0.04407 | 0.00416 | 7.90E-27 |
| rs613534 | miR-139-5ı | trans | 3.54E-40 | Nightingale | S_VLDL_FC | NA | G | A | 0.036642 | 0.004347 | 1.70E-17 |
| rs613534 | miR-139-5ı | trans | 3.54E-40 | Nightingale | M_VLDL_C | NA | G | A | 0.0392 | 0.004307 | 1.30E-19 |
| rs613534 | miR-139-5ı | trans | 3.54E-40 | Nightingale | Total_FC | NA | G | A | 0.031378 | 0.004246 | 4.70E-14 |
| rs613534 | miR-139-5ı | trans | 3.54E-40 | Nightingale | VLDL_FC | NA | G | A | 0.024446 | 0.004315 | 3.60E-09 |
| rs613534 | let-7b-5p | trans | 5.20E-26 | Nightingale | S_VLDL_L | NA | G | A | 0.024496 | 0.004305 | 2.50E-09 |
| rs613534 | let-7b-5p | trans | 5.20E-26 | Nightingale | IDL_L | NA | G | A | 0.03366 | 0.004203 | 2.30E-15 |
| rs613534 | let-7b-5p | trans | 5.20E-26 | Nightingale | M_VLDL_C | NA | G | A | 0.037341 | 0.004335 | 5.40E-18 |
| rs613534 | let-7b-5p | trans | 5.20E-26 | Nightingale | S_HDL_C | NA | G | A | 0.02538 | 0.004339 | 9.40E-09 |
| rs613534 | let-7b-5p | trans | 5.20E-26 | Nightingale | L_VLDL_C_ | NA | G | A | 0.049226 | 0.004245 | 8.90E-32 |
| rs613534 | let-7b-5p | trans | 5.20E-26 | Nightingale | VLDL_P | NA | G | A | 0.026555 | 0.004324 | 2.00E-10 |
| rs613534 | let-7b-5p | trans | 5.20E-26 | Nightingale | M_VLDL_P | NA | G | A | 0.027926 | 0.004336 | 4.70E-11 |
| rs613534 | let-7b-5p | trans | 5.20E-26 | Nightingale | L_LDL_C | NA | G | A | 0.030224 | 0.004298 | 3.00E-12 |
| rs613534 | let-7b-5p | trans | 5.20E-26 | Nightingale | M_VLDL_F | NA | G | A | 0.032515 | 0.004346 | 3.90E-14 |
| rs613534 | let-7b-5p | trans | 5.20E-26 | Nightingale | S_VLDL_TG | NA | G | A | -0.03598 | 0.004208 | 1.90E-17 |
| rs613534 | let-7b-5p | trans | 5.20E-26 | Nightingale | S_HDL_P | NA | G | A | 0.027805 | 0.004347 | 2.70E-10 |
| rs613534 | let-7b-5p | trans | 5.20E-26 | Nightingale | XS_VLDL_F | NA | G | A | 0.035882 | 0.004271 | 7.50E-18 |
| rs613534 | let-7b-5p | trans | 5.20E-26 | Nightingale | L_LDL_L | NA | G | A | 0.031523 | 0.004304 | 3.20E-13 |
| rs613534 | let-7b-5p | trans | 5.20E-26 | Nightingale | LDL_PL | NA | G | A | 0.033318 | 0.004337 | 3.00E-14 |
| rs613534 | let-7b-5p | trans | 5.20E-26 | Nightingale | XL_VLDL_P | NA | G | A | 0.039106 | 0.004353 | 7.40E-20 |
| rs613534 | let-7b-5p | trans | 5.20E-26 | Nightingale | GlycA | NA | G | A | 0.025329 | 0.004328 | 4.80E-09 |
| rs613534 | let-7b-5p | trans | 5.20E-26 | Nightingale | L_LDL_P | NA | G | A | 0.028317 | 0.004332 | 7.80E-11 |
| rs613534 | let-7b-5p | trans | 5.20E-26 | Nightingale | L_VLDL_CE | NA | G | A | 0.022756 | 0.004296 | 2.90E-08 |

| | | | | | | | | | | | |
|---|---|---|---|---|---|---|---|---|---|---|---|
| rs613534 | let-7b-5p | trans | 5.20E-26 | Nightingale | M_VLDL_C | NA | G | A | 0.031348 | 0.004084 | 3.30E-14 |
| rs613534 | let-7b-5p | trans | 5.20E-26 | Nightingale | XXL_VLDL_ | NA | G | A | 0.023502 | 0.004375 | 5.40E-09 |
| rs613534 | let-7b-5p | trans | 5.20E-26 | Nightingale | IDL_P | NA | G | A | 0.039032 | 0.004301 | 1.80E-19 |
| rs613534 | let-7b-5p | trans | 5.20E-26 | Nightingale | LDL_P | NA | G | A | 0.030659 | 0.004344 | 2.20E-12 |
| rs613534 | let-7b-5p | trans | 5.20E-26 | Nightingale | LA | NA | G | A | 0.023263 | 0.004263 | 1.60E-08 |
| rs613534 | let-7b-5p | trans | 5.20E-26 | Nightingale | non_HDL_( | NA | G | A | 0.03414 | 0.004323 | 2.40E-15 |
| rs613534 | let-7b-5p | trans | 5.20E-26 | Nightingale | XXL_VLDL_ | NA | G | A | -0.03443 | 0.004384 | 6.30E-16 |
| rs613534 | let-7b-5p | trans | 5.20E-26 | Nightingale | S_VLDL_P | NA | G | A | 0.02339 | 0.004292 | 9.70E-09 |
| rs613534 | let-7b-5p | trans | 5.20E-26 | Nightingale | LDL_L | NA | G | A | 0.031465 | 0.004332 | 5.80E-13 |
| rs613534 | let-7b-5p | trans | 5.20E-26 | Nightingale | S_VLDL_C_ | NA | G | A | 0.039736 | 0.00423 | 8.30E-21 |
| rs613534 | let-7b-5p | trans | 5.20E-26 | Nightingale | M_LDL_TG | NA | G | A | -0.02512 | 0.004293 | 8.60E-09 |
| rs613534 | let-7b-5p | trans | 5.20E-26 | Nightingale | XS_VLDL_C | NA | G | A | 0.036639 | 0.004219 | 2.40E-18 |
| rs613534 | let-7b-5p | trans | 5.20E-26 | Nightingale | S_LDL_P | NA | G | A | 0.028959 | 0.004351 | 3.00E-11 |
| rs613534 | let-7b-5p | trans | 5.20E-26 | Nightingale | XS_VLDL_F | NA | G | A | 0.029134 | 0.004316 | 3.10E-11 |
| rs613534 | let-7b-5p | trans | 5.20E-26 | Nightingale | ApoB | NA | G | A | 0.032928 | 0.004341 | 3.40E-14 |
| rs613534 | let-7b-5p | trans | 5.20E-26 | Nightingale | XS_VLDL_P | NA | G | A | 0.031657 | 0.004286 | 2.00E-14 |
| rs613534 | let-7b-5p | trans | 5.20E-26 | Nightingale | Total_C | NA | G | A | 0.030138 | 0.004208 | 5.00E-13 |
| rs613534 | let-7b-5p | trans | 5.20E-26 | Nightingale | S_VLDL_CE | NA | G | A | 0.037311 | 0.004341 | 1.70E-18 |
| rs613534 | let-7b-5p | trans | 5.20E-26 | Nightingale | L_LDL_FC | NA | G | A | 0.031473 | 0.004268 | 4.10E-13 |
| rs613534 | let-7b-5p | trans | 5.20E-26 | Nightingale | S_LDL_CE_ | NA | G | A | 0.028844 | 0.004339 | 1.10E-12 |
| rs613534 | let-7b-5p | trans | 5.20E-26 | Nightingale | LDL_CE | NA | G | A | 0.030617 | 0.004335 | 2.10E-12 |
| rs613534 | let-7b-5p | trans | 5.20E-26 | Nightingale | XS_VLDL_C | NA | G | A | 0.036391 | 0.004195 | 4.10E-18 |
| rs613534 | let-7b-5p | trans | 5.20E-26 | Nightingale | XXL_VLDL_ | NA | G | A | 0.033437 | 0.004356 | 8.80E-15 |
| rs613534 | let-7b-5p | trans | 5.20E-26 | Nightingale | M_VLDL_F | NA | G | A | 0.039372 | 0.004135 | 2.20E-21 |
| rs613534 | let-7b-5p | trans | 5.20E-26 | Nightingale | S_HDL_CE | NA | G | A | 0.02569 | 0.004354 | 7.00E-09 |
| rs613534 | let-7b-5p | trans | 5.20E-26 | Nightingale | S_VLDL_CE | NA | G | A | 0.045151 | 0.004288 | 1.40E-25 |
| rs613534 | let-7b-5p | trans | 5.20E-26 | Nightingale | M_VLDL_C | NA | G | A | 0.033845 | 0.004094 | 2.70E-16 |
| rs613534 | let-7b-5p | trans | 5.20E-26 | Nightingale | LDL_FC | NA | G | A | 0.031548 | 0.0043 | 6.60E-13 |
| rs613534 | let-7b-5p | trans | 5.20E-26 | Nightingale | Omega_6 | NA | G | A | 0.024156 | 0.004245 | 3.10E-09 |
| rs613534 | let-7b-5p | trans | 5.20E-26 | Nightingale | Total_CE | NA | G | A | 0.029384 | 0.004193 | 1.80E-12 |
| rs613534 | let-7b-5p | trans | 5.20E-26 | Nightingale | M_VLDL_T | NA | G | A | -0.03623 | 0.004108 | 1.50E-18 |
| rs613534 | let-7b-5p | trans | 5.20E-26 | Nightingale | ApoB_by_/ | NA | G | A | 0.025679 | 0.004281 | 2.00E-09 |
| rs613534 | let-7b-5p | trans | 5.20E-26 | Nightingale | IDL_C | NA | G | A | 0.033305 | 0.004198 | 4.90E-15 |
| rs613534 | let-7b-5p | trans | 5.20E-26 | Nightingale | Citrate | NA | G | A | 0.024678 | 0.004318 | 6.10E-09 |
| rs613534 | let-7b-5p | trans | 5.20E-26 | Nightingale | S_LDL_PL_ | NA | G | A | -0.02806 | 0.004339 | 4.90E-12 |
| rs613534 | let-7b-5p | trans | 5.20E-26 | Nightingale | L_LDL_PL | NA | G | A | 0.035919 | 0.004314 | 1.30E-16 |
| rs613534 | let-7b-5p | trans | 5.20E-26 | Nightingale | S_LDL_C | NA | G | A | 0.029698 | 0.004351 | 1.60E-11 |
| rs613534 | let-7b-5p | trans | 5.20E-26 | Nightingale | Remnant_( | NA | G | A | 0.035838 | 0.004319 | 5.30E-17 |
| rs613534 | let-7b-5p | trans | 5.20E-26 | Nightingale | S_VLDL_FC | NA | G | A | 0.027154 | 0.004154 | 1.20E-10 |
| rs613534 | let-7b-5p | trans | 5.20E-26 | Nightingale | Total_PL | NA | G | A | 0.022997 | 0.004139 | 7.80E-09 |
| rs613534 | let-7b-5p | trans | 5.20E-26 | Nightingale | IDL_PL | NA | G | A | 0.033573 | 0.004195 | 2.00E-15 |
| rs613534 | let-7b-5p | trans | 5.20E-26 | Nightingale | S_LDL_C_p | NA | G | A | 0.041864 | 0.004364 | 5.60E-22 |
| rs613534 | let-7b-5p | trans | 5.20E-26 | Nightingale | M_LDL_C | NA | G | A | 0.032091 | 0.004355 | 2.30E-13 |
| rs613534 | let-7b-5p | trans | 5.20E-26 | Nightingale | S_VLDL_PL | NA | G | A | 0.025728 | 0.004162 | 1.50E-09 |
| rs613534 | let-7b-5p | trans | 5.20E-26 | Nightingale | M_VLDL_P | NA | G | A | 0.028753 | 0.00434 | 1.40E-11 |
| rs613534 | let-7b-5p | trans | 5.20E-26 | Nightingale | XL_VLDL_F | NA | G | A | 0.035204 | 0.004214 | 1.80E-17 |
| rs613534 | let-7b-5p | trans | 5.20E-26 | Nightingale | M_LDL_PL | NA | G | A | 0.031174 | 0.004353 | 9.90E-13 |
| rs613534 | let-7b-5p | trans | 5.20E-26 | Nightingale | L_VLDL_PL | NA | G | A | 0.021645 | 0.004222 | 4.40E-08 |
| rs613534 | let-7b-5p | trans | 5.20E-26 | Nightingale | M_LDL_CE | NA | G | A | 0.031212 | 0.004353 | 8.50E-13 |
| rs613534 | let-7b-5p | trans | 5.20E-26 | Nightingale | XS_VLDL_P | NA | G | A | 0.035785 | 0.004289 | 1.00E-17 |
| rs613534 | let-7b-5p | trans | 5.20E-26 | Nightingale | M_VLDL_P | NA | G | A | 0.042093 | 0.004172 | 6.30E-24 |
| rs613534 | let-7b-5p | trans | 5.20E-26 | Nightingale | M_LDL_L | NA | G | A | 0.031388 | 0.004353 | 7.00E-13 |
| rs613534 | let-7b-5p | trans | 5.20E-26 | Nightingale | S_LDL_CE | NA | G | A | 0.029442 | 0.004349 | 1.50E-11 |
| rs613534 | let-7b-5p | trans | 5.20E-26 | Nightingale | LDL_C | NA | G | A | 0.031186 | 0.004328 | 9.60E-13 |
| rs613534 | let-7b-5p | trans | 5.20E-26 | Nightingale | S_VLDL_C | NA | G | A | 0.037355 | 0.004346 | 2.60E-18 |
| rs613534 | let-7b-5p | trans | 5.20E-26 | Nightingale | IDL_CE | NA | G | A | 0.032088 | 0.004197 | 4.30E-14 |
| rs613534 | let-7b-5p | trans | 5.20E-26 | Nightingale | S_LDL_L | NA | G | A | 0.026893 | 0.004346 | 1.20E-09 |
| rs613534 | let-7b-5p | trans | 5.20E-26 | Nightingale | XXL_VLDL_ | NA | G | A | 0.03179 | 0.00438 | 2.10E-13 |
| rs613534 | let-7b-5p | trans | 5.20E-26 | Nightingale | L_LDL_CE | NA | G | A | 0.02952 | 0.004308 | 9.90E-12 |
| rs613534 | let-7b-5p | trans | 5.20E-26 | Nightingale | XL_HDL_PL | NA | G | A | -0.02181 | 0.003994 | 4.60E-08 |
| rs613534 | let-7b-5p | trans | 5.20E-26 | Nightingale | Sphingomy | NA | G | A | 0.025917 | 0.004063 | 3.30E-10 |
| rs613534 | let-7b-5p | trans | 5.20E-26 | Nightingale | VLDL_C | NA | G | A | 0.031062 | 0.004341 | 2.10E-13 |
| rs613534 | let-7b-5p | trans | 5.20E-26 | Nightingale | M_LDL_C_ | NA | G | A | 0.026582 | 0.00435 | 1.60E-09 |

| | | | | | | | | | | | |
|---|---|---|---|---|---|---|---|---|---|---|---|
| rs613534 | let-7b-5p | trans | 5.20E-26 | Nightingale | XL_VLDL_T | NA | G | A | -0.04155 | 0.004213 | 4.90E-23 |
| rs613534 | let-7b-5p | trans | 5.20E-26 | Nightingale | S_LDL_FC | NA | G | A | 0.0272 | 0.00434 | 1.30E-09 |
| rs613534 | let-7b-5p | trans | 5.20E-26 | Nightingale | Total_L | NA | G | A | 0.025008 | 0.004276 | 1.50E-09 |
| rs613534 | let-7b-5p | trans | 5.20E-26 | Nightingale | Clinical_LD | NA | G | A | 0.034557 | 0.004322 | 2.70E-15 |
| rs613534 | let-7b-5p | trans | 5.20E-26 | Nightingale | XXL_VLDL_ | NA | G | A | 0.029268 | 0.004328 | 5.10E-12 |
| rs613534 | let-7b-5p | trans | 5.20E-26 | Nightingale | XL_VLDL_C | NA | G | A | 0.03559 | 0.004168 | 1.70E-17 |
| rs613534 | let-7b-5p | trans | 5.20E-26 | Nightingale | M_LDL_P | NA | G | A | 0.034001 | 0.004359 | 7.50E-15 |
| rs613534 | let-7b-5p | trans | 5.20E-26 | Nightingale | IDL_FC | NA | G | A | 0.035978 | 0.004208 | 3.20E-17 |
| rs613534 | let-7b-5p | trans | 5.20E-26 | Nightingale | XS_VLDL_L | NA | G | A | 0.033006 | 0.004273 | 1.40E-15 |
| rs613534 | let-7b-5p | trans | 5.20E-26 | Nightingale | XL_VLDL_C | NA | G | A | 0.034575 | 0.00417 | 1.50E-16 |
| rs613534 | let-7b-5p | trans | 5.20E-26 | Nightingale | M_LDL_FC | NA | G | A | 0.032281 | 0.004341 | 2.40E-13 |
| rs613534 | let-7b-5p | trans | 5.20E-26 | Nightingale | S_VLDL_PL | NA | G | A | 0.032035 | 0.004339 | 4.90E-14 |
| rs613534 | let-7b-5p | trans | 5.20E-26 | Nightingale | L_VLDL_FC | NA | G | A | 0.046941 | 0.004305 | 3.90E-28 |
| rs613534 | let-7b-5p | trans | 5.20E-26 | Nightingale | S_HDL_L | NA | G | A | 0.023969 | 0.004332 | 2.70E-08 |
| rs613534 | let-7b-5p | trans | 5.20E-26 | Nightingale | VLDL_CE | NA | G | A | 0.035144 | 0.004348 | 2.10E-16 |
| rs613534 | let-7b-5p | trans | 5.20E-26 | Nightingale | L_VLDL_TG | NA | G | A | -0.04923 | 0.004326 | 6.40E-31 |
| rs613534 | let-7b-5p | trans | 5.20E-26 | Nightingale | L_VLDL_CE | NA | G | A | 0.04407 | 0.00416 | 7.90E-27 |
| rs613534 | let-7b-5p | trans | 5.20E-26 | Nightingale | S_VLDL_FC | NA | G | A | 0.036642 | 0.004347 | 1.70E-17 |
| rs613534 | let-7b-5p | trans | 5.20E-26 | Nightingale | M_VLDL_C | NA | G | A | 0.0392 | 0.004307 | 1.30E-19 |
| rs613534 | let-7b-5p | trans | 5.20E-26 | Nightingale | Total_FC | NA | G | A | 0.031378 | 0.004246 | 4.70E-14 |
| rs613534 | let-7b-5p | trans | 5.20E-26 | Nightingale | VLDL_FC | NA | G | A | 0.024446 | 0.004315 | 3.60E-09 |
| rs613534 | miR-126-3p | trans | 9.46E-37 | Nightingale | S_VLDL_L | NA | G | A | 0.024496 | 0.004305 | 2.50E-09 |
| rs613534 | miR-126-3p | trans | 9.46E-37 | Nightingale | IDL_L | NA | G | A | 0.03366 | 0.004203 | 2.30E-15 |
| rs613534 | miR-126-3p | trans | 9.46E-37 | Nightingale | M_VLDL_C | NA | G | A | 0.037341 | 0.004335 | 5.40E-18 |
| rs613534 | miR-126-3p | trans | 9.46E-37 | Nightingale | S_HDL_C | NA | G | A | 0.02538 | 0.004339 | 9.40E-09 |
| rs613534 | miR-126-3p | trans | 9.46E-37 | Nightingale | L_VLDL_C_ | NA | G | A | 0.049226 | 0.004245 | 8.90E-32 |
| rs613534 | miR-126-3p | trans | 9.46E-37 | Nightingale | VLDL_P | NA | G | A | 0.026555 | 0.004324 | 2.00E-10 |
| rs613534 | miR-126-3p | trans | 9.46E-37 | Nightingale | M_VLDL_P | NA | G | A | 0.027926 | 0.004336 | 4.70E-11 |
| rs613534 | miR-126-3p | trans | 9.46E-37 | Nightingale | L_LDL_C | NA | G | A | 0.030224 | 0.004298 | 3.00E-12 |
| rs613534 | miR-126-3p | trans | 9.46E-37 | Nightingale | M_VLDL_F | NA | G | A | 0.032515 | 0.004346 | 3.90E-14 |
| rs613534 | miR-126-3p | trans | 9.46E-37 | Nightingale | S_VLDL_TG | NA | G | A | -0.03598 | 0.004208 | 1.90E-17 |
| rs613534 | miR-126-3p | trans | 9.46E-37 | Nightingale | S_HDL_P | NA | G | A | 0.027805 | 0.004347 | 2.70E-10 |
| rs613534 | miR-126-3p | trans | 9.46E-37 | Nightingale | XS_VLDL_F | NA | G | A | 0.035882 | 0.004271 | 7.50E-18 |
| rs613534 | miR-126-3p | trans | 9.46E-37 | Nightingale | L_LDL_L | NA | G | A | 0.031523 | 0.004304 | 3.20E-13 |
| rs613534 | miR-126-3p | trans | 9.46E-37 | Nightingale | LDL_PL | NA | G | A | 0.033318 | 0.004337 | 3.00E-14 |
| rs613534 | miR-126-3p | trans | 9.46E-37 | Nightingale | XL_VLDL_P | NA | G | A | 0.039106 | 0.004353 | 7.40E-20 |
| rs613534 | miR-126-3p | trans | 9.46E-37 | Nightingale | GlycA | NA | G | A | 0.025329 | 0.004328 | 4.80E-09 |
| rs613534 | miR-126-3p | trans | 9.46E-37 | Nightingale | L_LDL_P | NA | G | A | 0.028317 | 0.004332 | 7.80E-11 |
| rs613534 | miR-126-3p | trans | 9.46E-37 | Nightingale | L_VLDL_CE | NA | G | A | 0.022756 | 0.004296 | 2.90E-08 |
| rs613534 | miR-126-3p | trans | 9.46E-37 | Nightingale | M_VLDL_C | NA | G | A | 0.031348 | 0.004084 | 3.30E-14 |
| rs613534 | miR-126-3p | trans | 9.46E-37 | Nightingale | XXL_VLDL_ | NA | G | A | 0.023502 | 0.004375 | 5.40E-09 |
| rs613534 | miR-126-3p | trans | 9.46E-37 | Nightingale | IDL_P | NA | G | A | 0.039032 | 0.004301 | 1.80E-19 |
| rs613534 | miR-126-3p | trans | 9.46E-37 | Nightingale | LDL_P | NA | G | A | 0.030659 | 0.004344 | 2.20E-12 |
| rs613534 | miR-126-3p | trans | 9.46E-37 | Nightingale | LA | NA | G | A | 0.023263 | 0.004263 | 1.60E-08 |
| rs613534 | miR-126-3p | trans | 9.46E-37 | Nightingale | non_HDL_( | NA | G | A | 0.03414 | 0.004323 | 2.40E-15 |
| rs613534 | miR-126-3p | trans | 9.46E-37 | Nightingale | XXL_VLDL_ | NA | G | A | -0.03443 | 0.004384 | 6.30E-16 |
| rs613534 | miR-126-3p | trans | 9.46E-37 | Nightingale | S_VLDL_P | NA | G | A | 0.02339 | 0.004292 | 9.70E-09 |
| rs613534 | miR-126-3p | trans | 9.46E-37 | Nightingale | LDL_L | NA | G | A | 0.031465 | 0.004332 | 5.80E-13 |
| rs613534 | miR-126-3p | trans | 9.46E-37 | Nightingale | S_VLDL_C_ | NA | G | A | 0.039736 | 0.00423 | 8.30E-21 |
| rs613534 | miR-126-3p | trans | 9.46E-37 | Nightingale | M_LDL_TG | NA | G | A | -0.02512 | 0.004293 | 8.60E-09 |
| rs613534 | miR-126-3p | trans | 9.46E-37 | Nightingale | XS_VLDL_C | NA | G | A | 0.036639 | 0.004219 | 2.40E-18 |
| rs613534 | miR-126-3p | trans | 9.46E-37 | Nightingale | S_LDL_P | NA | G | A | 0.028959 | 0.004351 | 3.00E-11 |
| rs613534 | miR-126-3p | trans | 9.46E-37 | Nightingale | XS_VLDL_F | NA | G | A | 0.029134 | 0.004316 | 3.10E-11 |
| rs613534 | miR-126-3p | trans | 9.46E-37 | Nightingale | ApoB | NA | G | A | 0.032928 | 0.004341 | 3.40E-14 |
| rs613534 | miR-126-3p | trans | 9.46E-37 | Nightingale | XS_VLDL_P | NA | G | A | 0.031657 | 0.004286 | 2.00E-14 |
| rs613534 | miR-126-3p | trans | 9.46E-37 | Nightingale | Total_C | NA | G | A | 0.030138 | 0.004208 | 5.00E-13 |
| rs613534 | miR-126-3p | trans | 9.46E-37 | Nightingale | S_VLDL_CE | NA | G | A | 0.037311 | 0.004341 | 1.70E-18 |
| rs613534 | miR-126-3p | trans | 9.46E-37 | Nightingale | L_LDL_FC | NA | G | A | 0.031473 | 0.004268 | 4.10E-13 |
| rs613534 | miR-126-3p | trans | 9.46E-37 | Nightingale | S_LDL_CE_ | NA | G | A | 0.028844 | 0.004339 | 1.10E-12 |
| rs613534 | miR-126-3p | trans | 9.46E-37 | Nightingale | LDL_CE | NA | G | A | 0.030617 | 0.004335 | 2.10E-12 |
| rs613534 | miR-126-3p | trans | 9.46E-37 | Nightingale | XS_VLDL_C | NA | G | A | 0.036391 | 0.004195 | 4.10E-18 |
| rs613534 | miR-126-3p | trans | 9.46E-37 | Nightingale | XXL_VLDL_ | NA | G | A | 0.033437 | 0.004356 | 8.80E-15 |
| rs613534 | miR-126-3p | trans | 9.46E-37 | Nightingale | M_VLDL_F | NA | G | A | 0.039372 | 0.004135 | 2.20E-21 |

| | | | | | | | | | | | |
|---|---|---|---|---|---|---|---|---|---|---|---|
| rs613534 | miR-126-3p | trans | 9.46E-37 | Nightingale | S_HDL_CE | NA | G | A | 0.02569 | 0.004354 | 7.00E-09 |
| rs613534 | miR-126-3p | trans | 9.46E-37 | Nightingale | S_VLDL_CE | NA | G | A | 0.045151 | 0.004288 | 1.40E-25 |
| rs613534 | miR-126-3p | trans | 9.46E-37 | Nightingale | M_VLDL_C | NA | G | A | 0.033845 | 0.004094 | 2.70E-16 |
| rs613534 | miR-126-3p | trans | 9.46E-37 | Nightingale | LDL_FC | NA | G | A | 0.031548 | 0.0043 | 6.60E-13 |
| rs613534 | miR-126-3p | trans | 9.46E-37 | Nightingale | Omega_6 | NA | G | A | 0.024156 | 0.004245 | 3.10E-09 |
| rs613534 | miR-126-3p | trans | 9.46E-37 | Nightingale | Total_CE | NA | G | A | 0.029384 | 0.004193 | 1.80E-12 |
| rs613534 | miR-126-3p | trans | 9.46E-37 | Nightingale | M_VLDL_T | NA | G | A | -0.03623 | 0.004108 | 1.50E-18 |
| rs613534 | miR-126-3p | trans | 9.46E-37 | Nightingale | ApoB_by_A | NA | G | A | 0.025679 | 0.004281 | 2.00E-09 |
| rs613534 | miR-126-3p | trans | 9.46E-37 | Nightingale | IDL_C | NA | G | A | 0.033305 | 0.004198 | 4.90E-15 |
| rs613534 | miR-126-3p | trans | 9.46E-37 | Nightingale | Citrate | NA | G | A | 0.024678 | 0.004318 | 6.10E-09 |
| rs613534 | miR-126-3p | trans | 9.46E-37 | Nightingale | S_LDL_PL_ | NA | G | A | -0.02806 | 0.004339 | 4.90E-12 |
| rs613534 | miR-126-3p | trans | 9.46E-37 | Nightingale | L_LDL_PL | NA | G | A | 0.035919 | 0.004314 | 1.30E-16 |
| rs613534 | miR-126-3p | trans | 9.46E-37 | Nightingale | S_LDL_C | NA | G | A | 0.029698 | 0.004351 | 1.60E-11 |
| rs613534 | miR-126-3p | trans | 9.46E-37 | Nightingale | Remnant_( | NA | G | A | 0.035838 | 0.004319 | 5.30E-17 |
| rs613534 | miR-126-3p | trans | 9.46E-37 | Nightingale | S_VLDL_FC | NA | G | A | 0.027154 | 0.004154 | 1.20E-10 |
| rs613534 | miR-126-3p | trans | 9.46E-37 | Nightingale | Total_PL | NA | G | A | 0.022997 | 0.004139 | 7.80E-09 |
| rs613534 | miR-126-3p | trans | 9.46E-37 | Nightingale | IDL_PL | NA | G | A | 0.033573 | 0.004195 | 2.00E-15 |
| rs613534 | miR-126-3p | trans | 9.46E-37 | Nightingale | S_LDL_C_p | NA | G | A | 0.041864 | 0.004364 | 5.60E-22 |
| rs613534 | miR-126-3p | trans | 9.46E-37 | Nightingale | M_LDL_C | NA | G | A | 0.032091 | 0.004355 | 2.30E-13 |
| rs613534 | miR-126-3p | trans | 9.46E-37 | Nightingale | S_VLDL_PL | NA | G | A | 0.025728 | 0.004162 | 1.50E-09 |
| rs613534 | miR-126-3p | trans | 9.46E-37 | Nightingale | M_VLDL_P | NA | G | A | 0.028753 | 0.00434 | 1.40E-11 |
| rs613534 | miR-126-3p | trans | 9.46E-37 | Nightingale | XL_VLDL_F | NA | G | A | 0.035204 | 0.004214 | 1.80E-17 |
| rs613534 | miR-126-3p | trans | 9.46E-37 | Nightingale | M_LDL_PL | NA | G | A | 0.031174 | 0.004353 | 9.90E-13 |
| rs613534 | miR-126-3p | trans | 9.46E-37 | Nightingale | L_VLDL_PL | NA | G | A | 0.021645 | 0.004222 | 4.40E-08 |
| rs613534 | miR-126-3p | trans | 9.46E-37 | Nightingale | M_LDL_CE | NA | G | A | 0.031212 | 0.004353 | 8.50E-13 |
| rs613534 | miR-126-3p | trans | 9.46E-37 | Nightingale | XS_VLDL_P | NA | G | A | 0.035785 | 0.004289 | 1.00E-17 |
| rs613534 | miR-126-3p | trans | 9.46E-37 | Nightingale | M_VLDL_P | NA | G | A | 0.042093 | 0.004172 | 6.30E-24 |
| rs613534 | miR-126-3p | trans | 9.46E-37 | Nightingale | M_LDL_L | NA | G | A | 0.031388 | 0.004353 | 7.00E-13 |
| rs613534 | miR-126-3p | trans | 9.46E-37 | Nightingale | S_LDL_CE | NA | G | A | 0.029442 | 0.004349 | 1.50E-11 |
| rs613534 | miR-126-3p | trans | 9.46E-37 | Nightingale | LDL_C | NA | G | A | 0.031186 | 0.004328 | 9.60E-13 |
| rs613534 | miR-126-3p | trans | 9.46E-37 | Nightingale | S_VLDL_C | NA | G | A | 0.037355 | 0.004346 | 2.60E-18 |
| rs613534 | miR-126-3p | trans | 9.46E-37 | Nightingale | IDL_CE | NA | G | A | 0.032088 | 0.004197 | 4.30E-14 |
| rs613534 | miR-126-3p | trans | 9.46E-37 | Nightingale | S_LDL_L | NA | G | A | 0.026893 | 0.004346 | 1.20E-09 |
| rs613534 | miR-126-3p | trans | 9.46E-37 | Nightingale | XXL_VLDL_ | NA | G | A | 0.03179 | 0.00438 | 2.10E-13 |
| rs613534 | miR-126-3p | trans | 9.46E-37 | Nightingale | L_LDL_CE | NA | G | A | 0.02952 | 0.004308 | 9.90E-12 |
| rs613534 | miR-126-3p | trans | 9.46E-37 | Nightingale | XL_HDL_PL | NA | G | A | -0.02181 | 0.003994 | 4.60E-08 |
| rs613534 | miR-126-3p | trans | 9.46E-37 | Nightingale | Sphingomy | NA | G | A | 0.025917 | 0.004063 | 3.30E-10 |
| rs613534 | miR-126-3p | trans | 9.46E-37 | Nightingale | VLDL_C | NA | G | A | 0.031062 | 0.004341 | 2.10E-13 |
| rs613534 | miR-126-3p | trans | 9.46E-37 | Nightingale | M_LDL_C_ | NA | G | A | 0.026582 | 0.00435 | 1.60E-09 |
| rs613534 | miR-126-3p | trans | 9.46E-37 | Nightingale | XL_VLDL_T | NA | G | A | -0.04155 | 0.004213 | 4.90E-23 |
| rs613534 | miR-126-3p | trans | 9.46E-37 | Nightingale | S_LDL_FC | NA | G | A | 0.0272 | 0.00434 | 1.30E-09 |
| rs613534 | miR-126-3p | trans | 9.46E-37 | Nightingale | Total_L | NA | G | A | 0.025008 | 0.004276 | 1.50E-09 |
| rs613534 | miR-126-3p | trans | 9.46E-37 | Nightingale | Clinical_LD | NA | G | A | 0.034557 | 0.004322 | 2.70E-15 |
| rs613534 | miR-126-3p | trans | 9.46E-37 | Nightingale | XXL_VLDL_ | NA | G | A | 0.029268 | 0.004328 | 5.10E-12 |
| rs613534 | miR-126-3p | trans | 9.46E-37 | Nightingale | XL_VLDL_C | NA | G | A | 0.03559 | 0.004168 | 1.70E-17 |
| rs613534 | miR-126-3p | trans | 9.46E-37 | Nightingale | M_LDL_P | NA | G | A | 0.034001 | 0.004359 | 7.50E-15 |
| rs613534 | miR-126-3p | trans | 9.46E-37 | Nightingale | IDL_FC | NA | G | A | 0.035978 | 0.004208 | 3.20E-17 |
| rs613534 | miR-126-3p | trans | 9.46E-37 | Nightingale | XS_VLDL_L | NA | G | A | 0.033006 | 0.004273 | 1.40E-15 |
| rs613534 | miR-126-3p | trans | 9.46E-37 | Nightingale | XL_VLDL_C | NA | G | A | 0.034575 | 0.00417 | 1.50E-16 |
| rs613534 | miR-126-3p | trans | 9.46E-37 | Nightingale | M_LDL_FC | NA | G | A | 0.032281 | 0.004341 | 2.40E-13 |
| rs613534 | miR-126-3p | trans | 9.46E-37 | Nightingale | S_VLDL_PL | NA | G | A | 0.032035 | 0.004339 | 4.90E-14 |
| rs613534 | miR-126-3p | trans | 9.46E-37 | Nightingale | L_VLDL_FC | NA | G | A | 0.046941 | 0.004305 | 3.90E-28 |
| rs613534 | miR-126-3p | trans | 9.46E-37 | Nightingale | S_HDL_L | NA | G | A | 0.023969 | 0.004332 | 2.70E-08 |
| rs613534 | miR-126-3p | trans | 9.46E-37 | Nightingale | VLDL_CE | NA | G | A | 0.035144 | 0.004348 | 2.10E-16 |
| rs613534 | miR-126-3p | trans | 9.46E-37 | Nightingale | L_VLDL_TG | NA | G | A | -0.04923 | 0.004326 | 6.40E-31 |
| rs613534 | miR-126-3p | trans | 9.46E-37 | Nightingale | L_VLDL_CE | NA | G | A | 0.04407 | 0.00416 | 7.90E-27 |
| rs613534 | miR-126-3p | trans | 9.46E-37 | Nightingale | S_VLDL_FC | NA | G | A | 0.036642 | 0.004347 | 1.70E-17 |
| rs613534 | miR-126-3p | trans | 9.46E-37 | Nightingale | M_VLDL_C | NA | G | A | 0.0392 | 0.004307 | 1.30E-19 |
| rs613534 | miR-126-3p | trans | 9.46E-37 | Nightingale | Total_FC | NA | G | A | 0.031378 | 0.004246 | 4.70E-14 |
| rs613534 | miR-126-3p | trans | 9.46E-37 | Nightingale | VLDL_FC | NA | G | A | 0.024446 | 0.004315 | 3.60E-09 |
| rs613534 | let-7c-5p | trans | 4.30E-23 | Nightingale | S_VLDL_L | NA | G | A | 0.024496 | 0.004305 | 2.50E-09 |
| rs613534 | let-7c-5p | trans | 4.30E-23 | Nightingale | IDL_L | NA | G | A | 0.03366 | 0.004203 | 2.30E-15 |
| rs613534 | let-7c-5p | trans | 4.30E-23 | Nightingale | M_VLDL_C | NA | G | A | 0.037341 | 0.004335 | 5.40E-18 |

| | | | | | | | | | | | |
|---|---|---|---|---|---|---|---|---|---|---|---|
| rs613534 | let-7c-5p | trans | 4.30E-23 | Nightingale | S_HDL_C | NA | G | A | 0.02538 | 0.004339 | 9.40E-09 |
| rs613534 | let-7c-5p | trans | 4.30E-23 | Nightingale | L_VLDL_C_ | NA | G | A | 0.049226 | 0.004245 | 8.90E-32 |
| rs613534 | let-7c-5p | trans | 4.30E-23 | Nightingale | VLDL_P | NA | G | A | 0.026555 | 0.004324 | 2.00E-10 |
| rs613534 | let-7c-5p | trans | 4.30E-23 | Nightingale | M_VLDL_P | NA | G | A | 0.027926 | 0.004336 | 4.70E-11 |
| rs613534 | let-7c-5p | trans | 4.30E-23 | Nightingale | L_LDL_C | NA | G | A | 0.030224 | 0.004298 | 3.00E-12 |
| rs613534 | let-7c-5p | trans | 4.30E-23 | Nightingale | M_VLDL_F | NA | G | A | 0.032515 | 0.004346 | 3.90E-14 |
| rs613534 | let-7c-5p | trans | 4.30E-23 | Nightingale | S_VLDL_TG | NA | G | A | -0.03598 | 0.004208 | 1.90E-17 |
| rs613534 | let-7c-5p | trans | 4.30E-23 | Nightingale | S_HDL_P | NA | G | A | 0.027805 | 0.004347 | 2.70E-10 |
| rs613534 | let-7c-5p | trans | 4.30E-23 | Nightingale | XS_VLDL_F | NA | G | A | 0.035882 | 0.004271 | 7.50E-18 |
| rs613534 | let-7c-5p | trans | 4.30E-23 | Nightingale | L_LDL_L | NA | G | A | 0.031523 | 0.004304 | 3.20E-13 |
| rs613534 | let-7c-5p | trans | 4.30E-23 | Nightingale | LDL_PL | NA | G | A | 0.033318 | 0.004337 | 3.00E-14 |
| rs613534 | let-7c-5p | trans | 4.30E-23 | Nightingale | XL_VLDL_P | NA | G | A | 0.039106 | 0.004353 | 7.40E-20 |
| rs613534 | let-7c-5p | trans | 4.30E-23 | Nightingale | GlycA | NA | G | A | 0.025329 | 0.004328 | 4.80E-09 |
| rs613534 | let-7c-5p | trans | 4.30E-23 | Nightingale | L_LDL_P | NA | G | A | 0.028317 | 0.004332 | 7.80E-11 |
| rs613534 | let-7c-5p | trans | 4.30E-23 | Nightingale | L_VLDL_CE | NA | G | A | 0.022756 | 0.004296 | 2.90E-08 |
| rs613534 | let-7c-5p | trans | 4.30E-23 | Nightingale | M_VLDL_C | NA | G | A | 0.031348 | 0.004084 | 3.30E-14 |
| rs613534 | let-7c-5p | trans | 4.30E-23 | Nightingale | XXL_VLDL_ | NA | G | A | 0.023502 | 0.004375 | 5.40E-09 |
| rs613534 | let-7c-5p | trans | 4.30E-23 | Nightingale | IDL_P | NA | G | A | 0.039032 | 0.004301 | 1.80E-19 |
| rs613534 | let-7c-5p | trans | 4.30E-23 | Nightingale | LDL_P | NA | G | A | 0.030659 | 0.004344 | 2.20E-12 |
| rs613534 | let-7c-5p | trans | 4.30E-23 | Nightingale | LA | NA | G | A | 0.023263 | 0.004263 | 1.60E-08 |
| rs613534 | let-7c-5p | trans | 4.30E-23 | Nightingale | non_HDL_( | NA | G | A | 0.03414 | 0.004323 | 2.40E-15 |
| rs613534 | let-7c-5p | trans | 4.30E-23 | Nightingale | XXL_VLDL_ | NA | G | A | -0.03443 | 0.004384 | 6.30E-16 |
| rs613534 | let-7c-5p | trans | 4.30E-23 | Nightingale | S_VLDL_P | NA | G | A | 0.02339 | 0.004292 | 9.70E-09 |
| rs613534 | let-7c-5p | trans | 4.30E-23 | Nightingale | LDL_L | NA | G | A | 0.031465 | 0.004332 | 5.80E-13 |
| rs613534 | let-7c-5p | trans | 4.30E-23 | Nightingale | S_VLDL_C_ | NA | G | A | 0.039736 | 0.00423 | 8.30E-21 |
| rs613534 | let-7c-5p | trans | 4.30E-23 | Nightingale | M_LDL_TG | NA | G | A | -0.02512 | 0.004293 | 8.60E-09 |
| rs613534 | let-7c-5p | trans | 4.30E-23 | Nightingale | XS_VLDL_C | NA | G | A | 0.036639 | 0.004219 | 2.40E-18 |
| rs613534 | let-7c-5p | trans | 4.30E-23 | Nightingale | S_LDL_P | NA | G | A | 0.028959 | 0.004351 | 3.00E-11 |
| rs613534 | let-7c-5p | trans | 4.30E-23 | Nightingale | XS_VLDL_F | NA | G | A | 0.029134 | 0.004316 | 3.10E-11 |
| rs613534 | let-7c-5p | trans | 4.30E-23 | Nightingale | ApoB | NA | G | A | 0.032928 | 0.004341 | 3.40E-14 |
| rs613534 | let-7c-5p | trans | 4.30E-23 | Nightingale | XS_VLDL_P | NA | G | A | 0.031657 | 0.004286 | 2.00E-14 |
| rs613534 | let-7c-5p | trans | 4.30E-23 | Nightingale | Total_C | NA | G | A | 0.030138 | 0.004208 | 5.00E-13 |
| rs613534 | let-7c-5p | trans | 4.30E-23 | Nightingale | S_VLDL_CE | NA | G | A | 0.037311 | 0.004341 | 1.70E-18 |
| rs613534 | let-7c-5p | trans | 4.30E-23 | Nightingale | L_LDL_FC | NA | G | A | 0.031473 | 0.004268 | 4.10E-13 |
| rs613534 | let-7c-5p | trans | 4.30E-23 | Nightingale | S_LDL_CE_ | NA | G | A | 0.028844 | 0.004339 | 1.10E-12 |
| rs613534 | let-7c-5p | trans | 4.30E-23 | Nightingale | LDL_CE | NA | G | A | 0.030617 | 0.004335 | 2.10E-12 |
| rs613534 | let-7c-5p | trans | 4.30E-23 | Nightingale | XS_VLDL_C | NA | G | A | 0.036391 | 0.004195 | 4.10E-18 |
| rs613534 | let-7c-5p | trans | 4.30E-23 | Nightingale | XXL_VLDL_ | NA | G | A | 0.033437 | 0.004356 | 8.80E-15 |
| rs613534 | let-7c-5p | trans | 4.30E-23 | Nightingale | M_VLDL_F | NA | G | A | 0.039372 | 0.004135 | 2.20E-21 |
| rs613534 | let-7c-5p | trans | 4.30E-23 | Nightingale | S_HDL_CE | NA | G | A | 0.02569 | 0.004354 | 7.00E-09 |
| rs613534 | let-7c-5p | trans | 4.30E-23 | Nightingale | S_VLDL_CE | NA | G | A | 0.045151 | 0.004288 | 1.40E-25 |
| rs613534 | let-7c-5p | trans | 4.30E-23 | Nightingale | M_VLDL_C | NA | G | A | 0.033845 | 0.004094 | 2.70E-16 |
| rs613534 | let-7c-5p | trans | 4.30E-23 | Nightingale | LDL_FC | NA | G | A | 0.031548 | 0.0043 | 6.60E-13 |
| rs613534 | let-7c-5p | trans | 4.30E-23 | Nightingale | Omega_6 | NA | G | A | 0.024156 | 0.004245 | 3.10E-09 |
| rs613534 | let-7c-5p | trans | 4.30E-23 | Nightingale | Total_CE | NA | G | A | 0.029384 | 0.004193 | 1.80E-12 |
| rs613534 | let-7c-5p | trans | 4.30E-23 | Nightingale | M_VLDL_T | NA | G | A | -0.03623 | 0.004108 | 1.50E-18 |
| rs613534 | let-7c-5p | trans | 4.30E-23 | Nightingale | ApoB_by_/ | NA | G | A | 0.025679 | 0.004281 | 2.00E-09 |
| rs613534 | let-7c-5p | trans | 4.30E-23 | Nightingale | IDL_C | NA | G | A | 0.033305 | 0.004198 | 4.90E-15 |
| rs613534 | let-7c-5p | trans | 4.30E-23 | Nightingale | Citrate | NA | G | A | 0.024678 | 0.004318 | 6.10E-09 |
| rs613534 | let-7c-5p | trans | 4.30E-23 | Nightingale | S_LDL_PL_ | NA | G | A | -0.02806 | 0.004339 | 4.90E-12 |
| rs613534 | let-7c-5p | trans | 4.30E-23 | Nightingale | L_LDL_PL | NA | G | A | 0.035919 | 0.004314 | 1.30E-16 |
| rs613534 | let-7c-5p | trans | 4.30E-23 | Nightingale | S_LDL_C | NA | G | A | 0.029698 | 0.004351 | 1.60E-11 |
| rs613534 | let-7c-5p | trans | 4.30E-23 | Nightingale | Remnant_( | NA | G | A | 0.035838 | 0.004319 | 5.30E-17 |
| rs613534 | let-7c-5p | trans | 4.30E-23 | Nightingale | S_VLDL_FC | NA | G | A | 0.027154 | 0.004154 | 1.20E-10 |
| rs613534 | let-7c-5p | trans | 4.30E-23 | Nightingale | Total_PL | NA | G | A | 0.022997 | 0.004139 | 7.80E-09 |
| rs613534 | let-7c-5p | trans | 4.30E-23 | Nightingale | IDL_PL | NA | G | A | 0.033573 | 0.004195 | 2.00E-15 |
| rs613534 | let-7c-5p | trans | 4.30E-23 | Nightingale | S_LDL_C_p | NA | G | A | 0.041864 | 0.004364 | 5.60E-22 |
| rs613534 | let-7c-5p | trans | 4.30E-23 | Nightingale | M_LDL_C | NA | G | A | 0.032091 | 0.004355 | 2.30E-13 |
| rs613534 | let-7c-5p | trans | 4.30E-23 | Nightingale | S_VLDL_PL | NA | G | A | 0.025728 | 0.004162 | 1.50E-09 |
| rs613534 | let-7c-5p | trans | 4.30E-23 | Nightingale | M_VLDL_P | NA | G | A | 0.028753 | 0.00434 | 1.40E-11 |
| rs613534 | let-7c-5p | trans | 4.30E-23 | Nightingale | XL_VLDL_F | NA | G | A | 0.035204 | 0.004214 | 1.80E-17 |
| rs613534 | let-7c-5p | trans | 4.30E-23 | Nightingale | M_LDL_PL | NA | G | A | 0.031174 | 0.004353 | 9.90E-13 |
| rs613534 | let-7c-5p | trans | 4.30E-23 | Nightingale | L_VLDL_PL | NA | G | A | 0.021645 | 0.004222 | 4.40E-08 |

| | | | | | | | | | | | |
|---|---|---|---|---|---|---|---|---|---|---|---|
| rs613534 | let-7c-5p | trans | 4.30E-23 | Nightingale | M_LDL_CE | NA | G | A | 0.031212 | 0.004353 | 8.50E-13 |
| rs613534 | let-7c-5p | trans | 4.30E-23 | Nightingale | XS_VLDL_P | NA | G | A | 0.035785 | 0.004289 | 1.00E-17 |
| rs613534 | let-7c-5p | trans | 4.30E-23 | Nightingale | M_VLDL_P | NA | G | A | 0.042093 | 0.004172 | 6.30E-24 |
| rs613534 | let-7c-5p | trans | 4.30E-23 | Nightingale | M_LDL_L | NA | G | A | 0.031388 | 0.004353 | 7.00E-13 |
| rs613534 | let-7c-5p | trans | 4.30E-23 | Nightingale | S_LDL_CE | NA | G | A | 0.029442 | 0.004349 | 1.50E-11 |
| rs613534 | let-7c-5p | trans | 4.30E-23 | Nightingale | LDL_C | NA | G | A | 0.031186 | 0.004328 | 9.60E-13 |
| rs613534 | let-7c-5p | trans | 4.30E-23 | Nightingale | S_VLDL_C | NA | G | A | 0.037355 | 0.004346 | 2.60E-18 |
| rs613534 | let-7c-5p | trans | 4.30E-23 | Nightingale | IDL_CE | NA | G | A | 0.032088 | 0.004197 | 4.30E-14 |
| rs613534 | let-7c-5p | trans | 4.30E-23 | Nightingale | S_LDL_L | NA | G | A | 0.026893 | 0.004346 | 1.20E-09 |
| rs613534 | let-7c-5p | trans | 4.30E-23 | Nightingale | XXL_VLDL_ | NA | G | A | 0.03179 | 0.00438 | 2.10E-13 |
| rs613534 | let-7c-5p | trans | 4.30E-23 | Nightingale | L_LDL_CE | NA | G | A | 0.02952 | 0.004308 | 9.90E-12 |
| rs613534 | let-7c-5p | trans | 4.30E-23 | Nightingale | XL_HDL_PL | NA | G | A | -0.02181 | 0.003994 | 4.60E-08 |
| rs613534 | let-7c-5p | trans | 4.30E-23 | Nightingale | Sphingomy | NA | G | A | 0.025917 | 0.004063 | 3.30E-10 |
| rs613534 | let-7c-5p | trans | 4.30E-23 | Nightingale | VLDL_C | NA | G | A | 0.031062 | 0.004341 | 2.10E-13 |
| rs613534 | let-7c-5p | trans | 4.30E-23 | Nightingale | M_LDL_C_ | NA | G | A | 0.026582 | 0.00435 | 1.60E-09 |
| rs613534 | let-7c-5p | trans | 4.30E-23 | Nightingale | XL_VLDL_T | NA | G | A | -0.04155 | 0.004213 | 4.90E-23 |
| rs613534 | let-7c-5p | trans | 4.30E-23 | Nightingale | S_LDL_FC | NA | G | A | 0.0272 | 0.00434 | 1.30E-09 |
| rs613534 | let-7c-5p | trans | 4.30E-23 | Nightingale | Total_L | NA | G | A | 0.025008 | 0.004276 | 1.50E-09 |
| rs613534 | let-7c-5p | trans | 4.30E-23 | Nightingale | Clinical_LD | NA | G | A | 0.034557 | 0.004322 | 2.70E-15 |
| rs613534 | let-7c-5p | trans | 4.30E-23 | Nightingale | XXL_VLDL_ | NA | G | A | 0.029268 | 0.004328 | 5.10E-12 |
| rs613534 | let-7c-5p | trans | 4.30E-23 | Nightingale | XL_VLDL_C | NA | G | A | 0.03559 | 0.004168 | 1.70E-17 |
| rs613534 | let-7c-5p | trans | 4.30E-23 | Nightingale | M_LDL_P | NA | G | A | 0.034001 | 0.004359 | 7.50E-15 |
| rs613534 | let-7c-5p | trans | 4.30E-23 | Nightingale | IDL_FC | NA | G | A | 0.035978 | 0.004208 | 3.20E-17 |
| rs613534 | let-7c-5p | trans | 4.30E-23 | Nightingale | XS_VLDL_L | NA | G | A | 0.033006 | 0.004273 | 1.40E-15 |
| rs613534 | let-7c-5p | trans | 4.30E-23 | Nightingale | XL_VLDL_C | NA | G | A | 0.034575 | 0.00417 | 1.50E-16 |
| rs613534 | let-7c-5p | trans | 4.30E-23 | Nightingale | M_LDL_FC | NA | G | A | 0.032281 | 0.004341 | 2.40E-13 |
| rs613534 | let-7c-5p | trans | 4.30E-23 | Nightingale | S_VLDL_PL | NA | G | A | 0.032035 | 0.004339 | 4.90E-14 |
| rs613534 | let-7c-5p | trans | 4.30E-23 | Nightingale | L_VLDL_FC | NA | G | A | 0.046941 | 0.004305 | 3.90E-28 |
| rs613534 | let-7c-5p | trans | 4.30E-23 | Nightingale | S_HDL_L | NA | G | A | 0.023969 | 0.004332 | 2.70E-08 |
| rs613534 | let-7c-5p | trans | 4.30E-23 | Nightingale | VLDL_CE | NA | G | A | 0.035144 | 0.004348 | 2.10E-16 |
| rs613534 | let-7c-5p | trans | 4.30E-23 | Nightingale | L_VLDL_TG | NA | G | A | -0.04923 | 0.004326 | 6.40E-31 |
| rs613534 | let-7c-5p | trans | 4.30E-23 | Nightingale | L_VLDL_CE | NA | G | A | 0.04407 | 0.00416 | 7.90E-27 |
| rs613534 | let-7c-5p | trans | 4.30E-23 | Nightingale | S_VLDL_FC | NA | G | A | 0.036642 | 0.004347 | 1.70E-17 |
| rs613534 | let-7c-5p | trans | 4.30E-23 | Nightingale | M_VLDL_C | NA | G | A | 0.0392 | 0.004307 | 1.30E-19 |
| rs613534 | let-7c-5p | trans | 4.30E-23 | Nightingale | Total_FC | NA | G | A | 0.031378 | 0.004246 | 4.70E-14 |
| rs613534 | let-7c-5p | trans | 4.30E-23 | Nightingale | VLDL_FC | NA | G | A | 0.024446 | 0.004315 | 3.60E-09 |
| rs630014 | miR-126-3p | trans | 3.15E-19 | Nightingale | XL_VLDL_T | NA | G | A | -0.02212 | 0.004004 | 4.20E-08 |
| rs630014 | miR-126-3p | trans | 3.15E-19 | Nightingale | S_VLDL_CE | NA | G | A | 0.023822 | 0.004127 | 7.60E-09 |
| rs630014 | miR-126-3p | trans | 3.15E-19 | Nightingale | S_VLDL_FC | NA | G | A | 0.023102 | 0.004133 | 4.30E-08 |
| rs630014 | miR-126-3p | trans | 3.15E-19 | Nightingale | L_VLDL_CE | NA | G | A | 0.024915 | 0.003954 | 2.20E-10 |
| rs630014 | miR-126-3p | trans | 3.15E-19 | Nightingale | XL_VLDL_P | NA | G | A | 0.023649 | 0.004138 | 1.20E-08 |
| rs630014 | miR-126-3p | trans | 3.15E-19 | Nightingale | M_VLDL_F | NA | G | A | 0.022041 | 0.003932 | 3.60E-08 |
| rs630014 | miR-126-3p | trans | 3.15E-19 | Nightingale | L_VLDL_FC | NA | G | A | 0.029769 | 0.004093 | 1.90E-13 |
| rs630014 | miR-126-3p | trans | 3.15E-19 | Nightingale | S_VLDL_C | NA | G | A | 0.023768 | 0.004132 | 1.10E-08 |
| rs630014 | miR-126-3p | trans | 3.15E-19 | Nightingale | S_LDL_C_p | NA | G | A | 0.023635 | 0.004148 | 4.70E-09 |
| rs630014 | miR-126-3p | trans | 3.15E-19 | Nightingale | L_VLDL_C_ | NA | G | A | 0.028245 | 0.004036 | 1.50E-12 |
| rs630014 | miR-126-3p | trans | 3.15E-19 | Nightingale | L_VLDL_TG | NA | G | A | -0.03063 | 0.004113 | 4.80E-14 |
| rs630014 | miR-126-3p | trans | 3.15E-19 | Nightingale | S_VLDL_CE | NA | G | A | 0.02462 | 0.004077 | 5.10E-09 |
| rs630014 | miR-126-3p | trans | 3.15E-19 | Nightingale | M_VLDL_P | NA | G | A | 0.023825 | 0.003966 | 4.30E-09 |
| rs630014 | miR-10b-5p | trans | 2.46E-16 | Nightingale | XL_VLDL_T | NA | G | A | -0.02212 | 0.004004 | 4.20E-08 |
| rs630014 | miR-10b-5p | trans | 2.46E-16 | Nightingale | S_VLDL_CE | NA | G | A | 0.023822 | 0.004127 | 7.60E-09 |
| rs630014 | miR-10b-5p | trans | 2.46E-16 | Nightingale | S_VLDL_FC | NA | G | A | 0.023102 | 0.004133 | 4.30E-08 |
| rs630014 | miR-10b-5p | trans | 2.46E-16 | Nightingale | L_VLDL_CE | NA | G | A | 0.024915 | 0.003954 | 2.20E-10 |
| rs630014 | miR-10b-5p | trans | 2.46E-16 | Nightingale | XL_VLDL_P | NA | G | A | 0.023649 | 0.004138 | 1.20E-08 |
| rs630014 | miR-10b-5p | trans | 2.46E-16 | Nightingale | M_VLDL_F | NA | G | A | 0.022041 | 0.003932 | 3.60E-08 |
| rs630014 | miR-10b-5p | trans | 2.46E-16 | Nightingale | L_VLDL_FC | NA | G | A | 0.029769 | 0.004093 | 1.90E-13 |
| rs630014 | miR-10b-5p | trans | 2.46E-16 | Nightingale | S_VLDL_C | NA | G | A | 0.023768 | 0.004132 | 1.10E-08 |
| rs630014 | miR-10b-5p | trans | 2.46E-16 | Nightingale | S_LDL_C_p | NA | G | A | 0.023635 | 0.004148 | 4.70E-09 |
| rs630014 | miR-10b-5p | trans | 2.46E-16 | Nightingale | L_VLDL_C_ | NA | G | A | 0.028245 | 0.004036 | 1.50E-12 |
| rs630014 | miR-10b-5p | trans | 2.46E-16 | Nightingale | L_VLDL_TG | NA | G | A | -0.03063 | 0.004113 | 4.80E-14 |
| rs630014 | miR-10b-5p | trans | 2.46E-16 | Nightingale | S_VLDL_CE | NA | G | A | 0.02462 | 0.004077 | 5.10E-09 |
| rs630014 | miR-10b-5p | trans | 2.46E-16 | Nightingale | M_VLDL_P | NA | G | A | 0.023825 | 0.003966 | 4.30E-09 |
| rs630014 | let-7b-5p | trans | 4.92E-16 | Nightingale | XL_VLDL_T | NA | G | A | -0.02212 | 0.004004 | 4.20E-08 |

| | | | | | | | | | | | |
|---|---|---|---|---|---|---|---|---|---|---|---|
| rs630014 | let-7b-5p | trans | 4.92E-16 | Nightingale | S_VLDL_CE | NA | G | A | 0.023822 | 0.004127 | 7.60E-09 |
| rs630014 | let-7b-5p | trans | 4.92E-16 | Nightingale | S_VLDL_FC | NA | G | A | 0.023102 | 0.004133 | 4.30E-08 |
| rs630014 | let-7b-5p | trans | 4.92E-16 | Nightingale | L_VLDL_CE | NA | G | A | 0.024915 | 0.003954 | 2.20E-10 |
| rs630014 | let-7b-5p | trans | 4.92E-16 | Nightingale | XL_VLDL_P | NA | G | A | 0.023649 | 0.004138 | 1.20E-08 |
| rs630014 | let-7b-5p | trans | 4.92E-16 | Nightingale | M_VLDL_F | NA | G | A | 0.022041 | 0.003932 | 3.60E-08 |
| rs630014 | let-7b-5p | trans | 4.92E-16 | Nightingale | L_VLDL_FC | NA | G | A | 0.029769 | 0.004093 | 1.90E-13 |
| rs630014 | let-7b-5p | trans | 4.92E-16 | Nightingale | S_VLDL_C | NA | G | A | 0.023768 | 0.004132 | 1.10E-08 |
| rs630014 | let-7b-5p | trans | 4.92E-16 | Nightingale | S_LDL_C_p | NA | G | A | 0.023635 | 0.004148 | 4.70E-09 |
| rs630014 | let-7b-5p | trans | 4.92E-16 | Nightingale | L_VLDL_C_ | NA | G | A | 0.028245 | 0.004036 | 1.50E-12 |
| rs630014 | let-7b-5p | trans | 4.92E-16 | Nightingale | L_VLDL_TG | NA | G | A | -0.03063 | 0.004113 | 4.80E-14 |
| rs630014 | let-7b-5p | trans | 4.92E-16 | Nightingale | S_VLDL_CE | NA | G | A | 0.02462 | 0.004077 | 5.10E-09 |
| rs630014 | let-7b-5p | trans | 4.92E-16 | Nightingale | M_VLDL_P | NA | G | A | 0.023825 | 0.003966 | 4.30E-09 |
| rs630510 | miR-126-3p | trans | 3.11E-19 | Nightingale | M_VLDL_P | NA | G | A | 0.023826 | 0.003966 | 4.30E-09 |
| rs630510 | miR-126-3p | trans | 3.11E-19 | Nightingale | S_VLDL_FC | NA | G | A | 0.023102 | 0.004133 | 4.30E-08 |
| rs630510 | miR-126-3p | trans | 3.11E-19 | Nightingale | S_VLDL_C | NA | G | A | 0.023768 | 0.004132 | 1.10E-08 |
| rs630510 | miR-126-3p | trans | 3.11E-19 | Nightingale | S_VLDL_CE | NA | G | A | 0.02462 | 0.004077 | 5.00E-09 |
| rs630510 | miR-126-3p | trans | 3.11E-19 | Nightingale | L_VLDL_TG | NA | G | A | -0.03063 | 0.004113 | 4.80E-14 |
| rs630510 | miR-126-3p | trans | 3.11E-19 | Nightingale | XL_VLDL_P | NA | G | A | 0.023649 | 0.004138 | 1.20E-08 |
| rs630510 | miR-126-3p | trans | 3.11E-19 | Nightingale | XL_VLDL_T | NA | G | A | -0.02212 | 0.004004 | 4.20E-08 |
| rs630510 | miR-126-3p | trans | 3.11E-19 | Nightingale | L_VLDL_FC | NA | G | A | 0.02977 | 0.004093 | 1.90E-13 |
| rs630510 | miR-126-3p | trans | 3.11E-19 | Nightingale | S_VLDL_CE | NA | G | A | 0.023822 | 0.004127 | 7.60E-09 |
| rs630510 | miR-126-3p | trans | 3.11E-19 | Nightingale | L_VLDL_C_ | NA | G | A | 0.028245 | 0.004036 | 1.50E-12 |
| rs630510 | miR-126-3p | trans | 3.11E-19 | Nightingale | M_VLDL_F | NA | G | A | 0.022041 | 0.003932 | 3.60E-08 |
| rs630510 | miR-126-3p | trans | 3.11E-19 | Nightingale | L_VLDL_CE | NA | G | A | 0.024915 | 0.003954 | 2.20E-10 |
| rs630510 | miR-126-3p | trans | 3.11E-19 | Nightingale | S_LDL_C_p | NA | G | A | 0.023635 | 0.004148 | 4.70E-09 |
| rs630510 | miR-10b-5p | trans | 2.40E-16 | Nightingale | M_VLDL_P | NA | G | A | 0.023826 | 0.003966 | 4.30E-09 |
| rs630510 | miR-10b-5p | trans | 2.40E-16 | Nightingale | S_VLDL_FC | NA | G | A | 0.023102 | 0.004133 | 4.30E-08 |
| rs630510 | miR-10b-5p | trans | 2.40E-16 | Nightingale | S_VLDL_C | NA | G | A | 0.023768 | 0.004132 | 1.10E-08 |
| rs630510 | miR-10b-5p | trans | 2.40E-16 | Nightingale | S_VLDL_CE | NA | G | A | 0.02462 | 0.004077 | 5.00E-09 |
| rs630510 | miR-10b-5p | trans | 2.40E-16 | Nightingale | L_VLDL_TG | NA | G | A | -0.03063 | 0.004113 | 4.80E-14 |
| rs630510 | miR-10b-5p | trans | 2.40E-16 | Nightingale | XL_VLDL_P | NA | G | A | 0.023649 | 0.004138 | 1.20E-08 |
| rs630510 | miR-10b-5p | trans | 2.40E-16 | Nightingale | XL_VLDL_T | NA | G | A | -0.02212 | 0.004004 | 4.20E-08 |
| rs630510 | miR-10b-5p | trans | 2.40E-16 | Nightingale | L_VLDL_FC | NA | G | A | 0.02977 | 0.004093 | 1.90E-13 |
| rs630510 | miR-10b-5p | trans | 2.40E-16 | Nightingale | S_VLDL_CE | NA | G | A | 0.023822 | 0.004127 | 7.60E-09 |
| rs630510 | miR-10b-5p | trans | 2.40E-16 | Nightingale | L_VLDL_C_ | NA | G | A | 0.028245 | 0.004036 | 1.50E-12 |
| rs630510 | miR-10b-5p | trans | 2.40E-16 | Nightingale | M_VLDL_F | NA | G | A | 0.022041 | 0.003932 | 3.60E-08 |
| rs630510 | miR-10b-5p | trans | 2.40E-16 | Nightingale | L_VLDL_CE | NA | G | A | 0.024915 | 0.003954 | 2.20E-10 |
| rs630510 | miR-10b-5p | trans | 2.40E-16 | Nightingale | S_LDL_C_p | NA | G | A | 0.023635 | 0.004148 | 4.70E-09 |
| rs630510 | let-7b-5p | trans | 4.85E-16 | Nightingale | M_VLDL_P | NA | G | A | 0.023826 | 0.003966 | 4.30E-09 |
| rs630510 | let-7b-5p | trans | 4.85E-16 | Nightingale | S_VLDL_FC | NA | G | A | 0.023102 | 0.004133 | 4.30E-08 |
| rs630510 | let-7b-5p | trans | 4.85E-16 | Nightingale | S_VLDL_C | NA | G | A | 0.023768 | 0.004132 | 1.10E-08 |
| rs630510 | let-7b-5p | trans | 4.85E-16 | Nightingale | S_VLDL_CE | NA | G | A | 0.02462 | 0.004077 | 5.00E-09 |
| rs630510 | let-7b-5p | trans | 4.85E-16 | Nightingale | L_VLDL_TG | NA | G | A | -0.03063 | 0.004113 | 4.80E-14 |
| rs630510 | let-7b-5p | trans | 4.85E-16 | Nightingale | XL_VLDL_P | NA | G | A | 0.023649 | 0.004138 | 1.20E-08 |
| rs630510 | let-7b-5p | trans | 4.85E-16 | Nightingale | XL_VLDL_T | NA | G | A | -0.02212 | 0.004004 | 4.20E-08 |
| rs630510 | let-7b-5p | trans | 4.85E-16 | Nightingale | L_VLDL_FC | NA | G | A | 0.02977 | 0.004093 | 1.90E-13 |
| rs630510 | let-7b-5p | trans | 4.85E-16 | Nightingale | S_VLDL_CE | NA | G | A | 0.023822 | 0.004127 | 7.60E-09 |
| rs630510 | let-7b-5p | trans | 4.85E-16 | Nightingale | L_VLDL_C_ | NA | G | A | 0.028245 | 0.004036 | 1.50E-12 |
| rs630510 | let-7b-5p | trans | 4.85E-16 | Nightingale | M_VLDL_F | NA | G | A | 0.022041 | 0.003932 | 3.60E-08 |
| rs630510 | let-7b-5p | trans | 4.85E-16 | Nightingale | L_VLDL_CE | NA | G | A | 0.024915 | 0.003954 | 2.20E-10 |
| rs630510 | let-7b-5p | trans | 4.85E-16 | Nightingale | S_LDL_C_p | NA | G | A | 0.023635 | 0.004148 | 4.70E-09 |
| rs643434 | miR-126-3p | trans | 1.18E-37 | Nightingale | XXL_VLDL_ | NA | A | G | -0.03445 | 0.004384 | 6.10E-16 |
| rs643434 | miR-126-3p | trans | 1.18E-37 | Nightingale | Sphingomy | NA | A | G | 0.025961 | 0.004064 | 3.00E-10 |
| rs643434 | miR-126-3p | trans | 1.18E-37 | Nightingale | Omega_6 | NA | A | G | 0.024274 | 0.004245 | 2.60E-09 |
| rs643434 | miR-126-3p | trans | 1.18E-37 | Nightingale | M_VLDL_C | NA | A | G | 0.031313 | 0.004084 | 3.50E-14 |
| rs643434 | miR-126-3p | trans | 1.18E-37 | Nightingale | M_VLDL_C | NA | A | G | 0.037416 | 0.004335 | 4.70E-18 |
| rs643434 | miR-126-3p | trans | 1.18E-37 | Nightingale | GlycA | NA | A | G | 0.025332 | 0.004328 | 5.00E-09 |
| rs643434 | miR-126-3p | trans | 1.18E-37 | Nightingale | L_VLDL_CE | NA | A | G | 0.044018 | 0.00416 | 9.50E-27 |
| rs643434 | miR-126-3p | trans | 1.18E-37 | Nightingale | IDL_PL | NA | A | G | 0.033641 | 0.004195 | 1.80E-15 |
| rs643434 | miR-126-3p | trans | 1.18E-37 | Nightingale | M_VLDL_F | NA | A | G | 0.032595 | 0.004346 | 3.50E-14 |
| rs643434 | miR-126-3p | trans | 1.18E-37 | Nightingale | non_HDL_( | NA | A | G | 0.034208 | 0.004323 | 2.20E-15 |
| rs643434 | miR-126-3p | trans | 1.18E-37 | Nightingale | XS_VLDL_L | NA | A | G | 0.033078 | 0.004273 | 1.20E-15 |
| rs643434 | miR-126-3p | trans | 1.18E-37 | Nightingale | LDL_L | NA | A | G | 0.03152 | 0.004332 | 5.40E-13 |

| | | | | | | | | | | | |
|---|---|---|---|---|---|---|---|---|---|---|---|
| rs643434 | miR-126-3p | trans | 1.18E-37 | Nightingale | S_LDL_PL_ | NA | A | G | -0.0281 | 0.004339 | 4.70E-12 |
| rs643434 | miR-126-3p | trans | 1.18E-37 | Nightingale | LDL_FC | NA | A | G | 0.031592 | 0.004301 | 6.20E-13 |
| rs643434 | miR-126-3p | trans | 1.18E-37 | Nightingale | ApoB | NA | A | G | 0.032994 | 0.004341 | 3.10E-14 |
| rs643434 | miR-126-3p | trans | 1.18E-37 | Nightingale | L_VLDL_CE | NA | A | G | 0.022834 | 0.004296 | 2.70E-08 |
| rs643434 | miR-126-3p | trans | 1.18E-37 | Nightingale | S_VLDL_CE | NA | A | G | 0.037357 | 0.004341 | 1.60E-18 |
| rs643434 | miR-126-3p | trans | 1.18E-37 | Nightingale | S_HDL_P | NA | A | G | 0.027834 | 0.004348 | 2.60E-10 |
| rs643434 | miR-126-3p | trans | 1.18E-37 | Nightingale | L_LDL_L | NA | A | G | 0.031581 | 0.004305 | 3.00E-13 |
| rs643434 | miR-126-3p | trans | 1.18E-37 | Nightingale | IDL_CE | NA | A | G | 0.032158 | 0.004197 | 3.80E-14 |
| rs643434 | miR-126-3p | trans | 1.18E-37 | Nightingale | L_LDL_CE | NA | A | G | 0.029584 | 0.004308 | 9.10E-12 |
| rs643434 | miR-126-3p | trans | 1.18E-37 | Nightingale | XL_VLDL_C | NA | A | G | 0.034537 | 0.00417 | 1.70E-16 |
| rs643434 | miR-126-3p | trans | 1.18E-37 | Nightingale | XS_VLDL_P | NA | A | G | 0.035851 | 0.004289 | 9.10E-18 |
| rs643434 | miR-126-3p | trans | 1.18E-37 | Nightingale | LDL_P | NA | A | G | 0.030723 | 0.004344 | 2.00E-12 |
| rs643434 | miR-126-3p | trans | 1.18E-37 | Nightingale | L_VLDL_C_ | NA | A | G | 0.049148 | 0.004246 | 1.20E-31 |
| rs643434 | miR-126-3p | trans | 1.18E-37 | Nightingale | M_VLDL_C | NA | A | G | 0.039267 | 0.004308 | 1.20E-19 |
| rs643434 | miR-126-3p | trans | 1.18E-37 | Nightingale | M_LDL_P | NA | A | G | 0.034038 | 0.004359 | 7.10E-15 |
| rs643434 | miR-126-3p | trans | 1.18E-37 | Nightingale | M_LDL_CE | NA | A | G | 0.031258 | 0.004353 | 8.10E-13 |
| rs643434 | miR-126-3p | trans | 1.18E-37 | Nightingale | XXL_VLDL_ | NA | A | G | 0.031806 | 0.00438 | 2.10E-13 |
| rs643434 | miR-126-3p | trans | 1.18E-37 | Nightingale | XXL_VLDL_ | NA | A | G | 0.033426 | 0.004356 | 9.00E-15 |
| rs643434 | miR-126-3p | trans | 1.18E-37 | Nightingale | XL_HDL_PL | NA | A | G | -0.02176 | 0.003994 | 4.90E-08 |
| rs643434 | miR-126-3p | trans | 1.18E-37 | Nightingale | S_LDL_C | NA | A | G | 0.029751 | 0.004351 | 1.50E-11 |
| rs643434 | miR-126-3p | trans | 1.18E-37 | Nightingale | S_VLDL_P | NA | A | G | 0.023452 | 0.004293 | 9.10E-09 |
| rs643434 | miR-126-3p | trans | 1.18E-37 | Nightingale | XS_VLDL_C | NA | A | G | 0.036711 | 0.00422 | 2.10E-18 |
| rs643434 | miR-126-3p | trans | 1.18E-37 | Nightingale | S_HDL_C | NA | A | G | 0.025404 | 0.00434 | 9.10E-09 |
| rs643434 | miR-126-3p | trans | 1.18E-37 | Nightingale | VLDL_FC | NA | A | G | 0.024523 | 0.004315 | 3.30E-09 |
| rs643434 | miR-126-3p | trans | 1.18E-37 | Nightingale | S_VLDL_FC | NA | A | G | 0.027147 | 0.004154 | 1.20E-10 |
| rs643434 | miR-126-3p | trans | 1.18E-37 | Nightingale | S_VLDL_TG | NA | A | G | -0.03596 | 0.004208 | 2.00E-17 |
| rs643434 | miR-126-3p | trans | 1.18E-37 | Nightingale | M_LDL_PL | NA | A | G | 0.031218 | 0.004353 | 9.40E-13 |
| rs643434 | miR-126-3p | trans | 1.18E-37 | Nightingale | L_LDL_P | NA | A | G | 0.028395 | 0.004332 | 7.00E-11 |
| rs643434 | miR-126-3p | trans | 1.18E-37 | Nightingale | ApoB_by_A | NA | A | G | 0.025706 | 0.004281 | 2.00E-09 |
| rs643434 | miR-126-3p | trans | 1.18E-37 | Nightingale | Remnant_( | NA | A | G | 0.035917 | 0.004319 | 4.70E-17 |
| rs643434 | miR-126-3p | trans | 1.18E-37 | Nightingale | XXL_VLDL_ | NA | A | G | 0.029234 | 0.004328 | 5.40E-12 |
| rs643434 | miR-126-3p | trans | 1.18E-37 | Nightingale | LA | NA | A | G | 0.023396 | 0.004263 | 1.30E-08 |
| rs643434 | miR-126-3p | trans | 1.18E-37 | Nightingale | M_VLDL_F | NA | A | G | 0.03934 | 0.004136 | 2.40E-21 |
| rs643434 | miR-126-3p | trans | 1.18E-37 | Nightingale | M_VLDL_P | NA | A | G | 0.028838 | 0.00434 | 1.20E-11 |
| rs643434 | miR-126-3p | trans | 1.18E-37 | Nightingale | LDL_PL | NA | A | G | 0.033367 | 0.004337 | 2.80E-14 |
| rs643434 | miR-126-3p | trans | 1.18E-37 | Nightingale | IDL_L | NA | A | G | 0.033725 | 0.004203 | 2.10E-15 |
| rs643434 | miR-126-3p | trans | 1.18E-37 | Nightingale | M_VLDL_P | NA | A | G | 0.042071 | 0.004172 | 6.90E-24 |
| rs643434 | miR-126-3p | trans | 1.18E-37 | Nightingale | IDL_C | NA | A | G | 0.033366 | 0.004198 | 4.40E-15 |
| rs643434 | miR-126-3p | trans | 1.18E-37 | Nightingale | Total_L | NA | A | G | 0.025099 | 0.004276 | 1.30E-09 |
| rs643434 | miR-126-3p | trans | 1.18E-37 | Nightingale | M_LDL_FC | NA | A | G | 0.032319 | 0.004342 | 2.20E-13 |
| rs643434 | miR-126-3p | trans | 1.18E-37 | Nightingale | S_VLDL_PL | NA | A | G | 0.032104 | 0.004339 | 4.40E-14 |
| rs643434 | miR-126-3p | trans | 1.18E-37 | Nightingale | XL_VLDL_P | NA | A | G | 0.039162 | 0.004353 | 6.90E-20 |
| rs643434 | miR-126-3p | trans | 1.18E-37 | Nightingale | M_LDL_C_ | NA | A | G | 0.026601 | 0.00435 | 1.60E-09 |
| rs643434 | miR-126-3p | trans | 1.18E-37 | Nightingale | XS_VLDL_F | NA | A | G | 0.035949 | 0.004272 | 6.80E-18 |
| rs643434 | miR-126-3p | trans | 1.18E-37 | Nightingale | S_VLDL_C_ | NA | A | G | 0.039702 | 0.004231 | 9.10E-21 |
| rs643434 | miR-126-3p | trans | 1.18E-37 | Nightingale | L_LDL_C | NA | A | G | 0.030283 | 0.004298 | 2.80E-12 |
| rs643434 | miR-126-3p | trans | 1.18E-37 | Nightingale | XXL_VLDL_ | NA | A | G | 0.023497 | 0.004375 | 5.50E-09 |
| rs643434 | miR-126-3p | trans | 1.18E-37 | Nightingale | S_LDL_P | NA | A | G | 0.029007 | 0.004351 | 2.80E-11 |
| rs643434 | miR-126-3p | trans | 1.18E-37 | Nightingale | S_LDL_L | NA | A | G | 0.026945 | 0.004346 | 1.10E-09 |
| rs643434 | miR-126-3p | trans | 1.18E-37 | Nightingale | M_LDL_C | NA | A | G | 0.032137 | 0.004355 | 2.20E-13 |
| rs643434 | miR-126-3p | trans | 1.18E-37 | Nightingale | LDL_CE | NA | A | G | 0.030677 | 0.004336 | 1.90E-12 |
| rs643434 | miR-126-3p | trans | 1.18E-37 | Nightingale | M_LDL_TG | NA | A | G | -0.02514 | 0.004293 | 8.20E-09 |
| rs643434 | miR-126-3p | trans | 1.18E-37 | Nightingale | XS_VLDL_P | NA | A | G | 0.031727 | 0.004286 | 1.80E-14 |
| rs643434 | miR-126-3p | trans | 1.18E-37 | Nightingale | L_LDL_FC | NA | A | G | 0.031521 | 0.004269 | 3.80E-13 |
| rs643434 | miR-126-3p | trans | 1.18E-37 | Nightingale | VLDL_C | NA | A | G | 0.031139 | 0.004341 | 1.90E-13 |
| rs643434 | miR-126-3p | trans | 1.18E-37 | Nightingale | XS_VLDL_C | NA | A | G | 0.036463 | 0.004195 | 3.70E-18 |
| rs643434 | miR-126-3p | trans | 1.18E-37 | Nightingale | S_LDL_FC | NA | A | G | 0.027234 | 0.00434 | 1.30E-09 |
| rs643434 | miR-126-3p | trans | 1.18E-37 | Nightingale | S_LDL_C_p | NA | A | G | 0.041888 | 0.004364 | 5.30E-22 |
| rs643434 | miR-126-3p | trans | 1.18E-37 | Nightingale | IDL_P | NA | A | G | 0.039091 | 0.004301 | 1.70E-19 |
| rs643434 | miR-126-3p | trans | 1.18E-37 | Nightingale | Total_CE | NA | A | G | 0.029456 | 0.004193 | 1.60E-12 |
| rs643434 | miR-126-3p | trans | 1.18E-37 | Nightingale | XS_VLDL_F | NA | A | G | 0.029136 | 0.004316 | 3.10E-11 |
| rs643434 | miR-126-3p | trans | 1.18E-37 | Nightingale | S_VLDL_L | NA | A | G | 0.024562 | 0.004305 | 2.30E-09 |
| rs643434 | miR-126-3p | trans | 1.18E-37 | Nightingale | S_LDL_CE_ | NA | A | G | 0.0289 | 0.00434 | 1.00E-12 |

| | | | | | | | | | | | |
|---|---|---|---|---|---|---|---|---|---|---|---|
| rs643434 | miR-126-3p | trans | 1.18E-37 | Nightingale | XL_VLDL_C | NA | A | G | 0.035551 | 0.004168 | 1.90E-17 |
| rs643434 | miR-126-3p | trans | 1.18E-37 | Nightingale | S_LDL_CE | NA | A | G | 0.0295 | 0.004349 | 1.40E-11 |
| rs643434 | miR-126-3p | trans | 1.18E-37 | Nightingale | Total_PL | NA | A | G | 0.023087 | 0.004139 | 6.80E-09 |
| rs643434 | miR-126-3p | trans | 1.18E-37 | Nightingale | Citrate | NA | A | G | 0.024643 | 0.004319 | 6.40E-09 |
| rs643434 | miR-126-3p | trans | 1.18E-37 | Nightingale | VLDL_P | NA | A | G | 0.026629 | 0.004324 | 1.80E-10 |
| rs643434 | miR-126-3p | trans | 1.18E-37 | Nightingale | S_HDL_L | NA | A | G | 0.024009 | 0.004332 | 2.60E-08 |
| rs643434 | miR-126-3p | trans | 1.18E-37 | Nightingale | IDL_FC | NA | A | G | 0.036015 | 0.004208 | 3.00E-17 |
| rs643434 | miR-126-3p | trans | 1.18E-37 | Nightingale | M_VLDL_T | NA | A | G | -0.0362 | 0.004108 | 1.70E-18 |
| rs643434 | miR-126-3p | trans | 1.18E-37 | Nightingale | M_LDL_L | NA | A | G | 0.031433 | 0.004353 | 6.60E-13 |
| rs643434 | miR-126-3p | trans | 1.18E-37 | Nightingale | Total_C | NA | A | G | 0.030215 | 0.004208 | 4.40E-13 |
| rs643434 | miR-126-3p | trans | 1.18E-37 | Nightingale | S_VLDL_FC | NA | A | G | 0.036708 | 0.004348 | 1.60E-17 |
| rs643434 | miR-126-3p | trans | 1.18E-37 | Nightingale | L_VLDL_FC | NA | A | G | 0.046831 | 0.004305 | 5.60E-28 |
| rs643434 | miR-126-3p | trans | 1.18E-37 | Nightingale | L_VLDL_PL | NA | A | G | 0.021642 | 0.004222 | 4.60E-08 |
| rs643434 | miR-126-3p | trans | 1.18E-37 | Nightingale | Clinical_LD | NA | A | G | 0.03461 | 0.004322 | 2.50E-15 |
| rs643434 | miR-126-3p | trans | 1.18E-37 | Nightingale | M_VLDL_P | NA | A | G | 0.028009 | 0.004336 | 4.20E-11 |
| rs643434 | miR-126-3p | trans | 1.18E-37 | Nightingale | Total_FC | NA | A | G | 0.031463 | 0.004247 | 4.00E-14 |
| rs643434 | miR-126-3p | trans | 1.18E-37 | Nightingale | S_VLDL_CE | NA | A | G | 0.045098 | 0.004289 | 1.70E-25 |
| rs643434 | miR-126-3p | trans | 1.18E-37 | Nightingale | LDL_C | NA | A | G | 0.031242 | 0.004328 | 8.80E-13 |
| rs643434 | miR-126-3p | trans | 1.18E-37 | Nightingale | L_LDL_PL | NA | A | G | 0.03597 | 0.004314 | 1.20E-16 |
| rs643434 | miR-126-3p | trans | 1.18E-37 | Nightingale | S_VLDL_C | NA | A | G | 0.037408 | 0.004347 | 2.50E-18 |
| rs643434 | miR-126-3p | trans | 1.18E-37 | Nightingale | S_VLDL_PL | NA | A | G | 0.025727 | 0.004162 | 1.50E-09 |
| rs643434 | miR-126-3p | trans | 1.18E-37 | Nightingale | XL_VLDL_T | NA | A | G | -0.0415 | 0.004213 | 5.60E-23 |
| rs643434 | miR-126-3p | trans | 1.18E-37 | Nightingale | L_VLDL_TG | NA | A | G | -0.04918 | 0.004326 | 7.90E-31 |
| rs643434 | miR-126-3p | trans | 1.18E-37 | Nightingale | VLDL_CE | NA | A | G | 0.03522 | 0.004348 | 1.90E-16 |
| rs643434 | miR-126-3p | trans | 1.18E-37 | Nightingale | M_VLDL_C | NA | A | G | 0.03381 | 0.004095 | 2.90E-16 |
| rs643434 | miR-126-3p | trans | 1.18E-37 | Nightingale | XL_VLDL_F | NA | A | G | 0.035154 | 0.004214 | 2.00E-17 |
| rs643434 | miR-126-3p | trans | 1.18E-37 | Nightingale | S_HDL_CE | NA | A | G | 0.025707 | 0.004354 | 6.70E-09 |
| rs643434 | miR-10b-5p | trans | 1.64E-34 | Nightingale | XXL_VLDL_ | NA | A | G | -0.03445 | 0.004384 | 6.10E-16 |
| rs643434 | miR-10b-5p | trans | 1.64E-34 | Nightingale | Sphingomy | NA | A | G | 0.025961 | 0.004064 | 3.00E-10 |
| rs643434 | miR-10b-5p | trans | 1.64E-34 | Nightingale | Omega_6 | NA | A | G | 0.024274 | 0.004245 | 2.60E-09 |
| rs643434 | miR-10b-5p | trans | 1.64E-34 | Nightingale | M_VLDL_C | NA | A | G | 0.031313 | 0.004084 | 3.50E-14 |
| rs643434 | miR-10b-5p | trans | 1.64E-34 | Nightingale | M_VLDL_C | NA | A | G | 0.037416 | 0.004335 | 4.70E-18 |
| rs643434 | miR-10b-5p | trans | 1.64E-34 | Nightingale | GlycA | NA | A | G | 0.025332 | 0.004328 | 5.00E-09 |
| rs643434 | miR-10b-5p | trans | 1.64E-34 | Nightingale | L_VLDL_CE | NA | A | G | 0.044018 | 0.00416 | 9.50E-27 |
| rs643434 | miR-10b-5p | trans | 1.64E-34 | Nightingale | IDL_PL | NA | A | G | 0.033641 | 0.004195 | 1.80E-15 |
| rs643434 | miR-10b-5p | trans | 1.64E-34 | Nightingale | M_VLDL_F | NA | A | G | 0.032595 | 0.004346 | 3.50E-14 |
| rs643434 | miR-10b-5p | trans | 1.64E-34 | Nightingale | non_HDL_( | NA | A | G | 0.034208 | 0.004323 | 2.20E-15 |
| rs643434 | miR-10b-5p | trans | 1.64E-34 | Nightingale | XS_VLDL_L | NA | A | G | 0.033078 | 0.004273 | 1.20E-15 |
| rs643434 | miR-10b-5p | trans | 1.64E-34 | Nightingale | LDL_L | NA | A | G | 0.03152 | 0.004332 | 5.40E-13 |
| rs643434 | miR-10b-5p | trans | 1.64E-34 | Nightingale | S_LDL_PL_ | NA | A | G | -0.0281 | 0.004339 | 4.70E-12 |
| rs643434 | miR-10b-5p | trans | 1.64E-34 | Nightingale | LDL_FC | NA | A | G | 0.031592 | 0.004301 | 6.20E-13 |
| rs643434 | miR-10b-5p | trans | 1.64E-34 | Nightingale | ApoB | NA | A | G | 0.032994 | 0.004341 | 3.10E-14 |
| rs643434 | miR-10b-5p | trans | 1.64E-34 | Nightingale | L_VLDL_CE | NA | A | G | 0.022834 | 0.004296 | 2.70E-08 |
| rs643434 | miR-10b-5p | trans | 1.64E-34 | Nightingale | S_VLDL_CE | NA | A | G | 0.037357 | 0.004341 | 1.60E-18 |
| rs643434 | miR-10b-5p | trans | 1.64E-34 | Nightingale | S_HDL_P | NA | A | G | 0.027834 | 0.004348 | 2.60E-10 |
| rs643434 | miR-10b-5p | trans | 1.64E-34 | Nightingale | L_LDL_L | NA | A | G | 0.031581 | 0.004305 | 3.00E-13 |
| rs643434 | miR-10b-5p | trans | 1.64E-34 | Nightingale | IDL_CE | NA | A | G | 0.032158 | 0.004197 | 3.80E-14 |
| rs643434 | miR-10b-5p | trans | 1.64E-34 | Nightingale | L_LDL_CE | NA | A | G | 0.029584 | 0.004308 | 9.10E-12 |
| rs643434 | miR-10b-5p | trans | 1.64E-34 | Nightingale | XL_VLDL_C | NA | A | G | 0.034537 | 0.00417 | 1.70E-16 |
| rs643434 | miR-10b-5p | trans | 1.64E-34 | Nightingale | XS_VLDL_P | NA | A | G | 0.035851 | 0.004289 | 9.10E-18 |
| rs643434 | miR-10b-5p | trans | 1.64E-34 | Nightingale | LDL_P | NA | A | G | 0.030723 | 0.004344 | 2.00E-12 |
| rs643434 | miR-10b-5p | trans | 1.64E-34 | Nightingale | L_VLDL_C_ | NA | A | G | 0.049148 | 0.004246 | 1.20E-31 |
| rs643434 | miR-10b-5p | trans | 1.64E-34 | Nightingale | M_VLDL_C | NA | A | G | 0.039267 | 0.004308 | 1.20E-19 |
| rs643434 | miR-10b-5p | trans | 1.64E-34 | Nightingale | M_LDL_P | NA | A | G | 0.034038 | 0.004359 | 7.10E-15 |
| rs643434 | miR-10b-5p | trans | 1.64E-34 | Nightingale | M_LDL_CE | NA | A | G | 0.031258 | 0.004353 | 8.10E-13 |
| rs643434 | miR-10b-5p | trans | 1.64E-34 | Nightingale | XXL_VLDL_ | NA | A | G | 0.031806 | 0.00438 | 2.10E-13 |
| rs643434 | miR-10b-5p | trans | 1.64E-34 | Nightingale | XXL_VLDL_ | NA | A | G | 0.033426 | 0.004356 | 9.00E-15 |
| rs643434 | miR-10b-5p | trans | 1.64E-34 | Nightingale | XL_HDL_PL | NA | A | G | -0.02176 | 0.003994 | 4.90E-08 |
| rs643434 | miR-10b-5p | trans | 1.64E-34 | Nightingale | S_LDL_C | NA | A | G | 0.029751 | 0.004351 | 1.50E-11 |
| rs643434 | miR-10b-5p | trans | 1.64E-34 | Nightingale | S_VLDL_P | NA | A | G | 0.023452 | 0.004293 | 9.10E-09 |
| rs643434 | miR-10b-5p | trans | 1.64E-34 | Nightingale | XS_VLDL_C | NA | A | G | 0.036711 | 0.00422 | 2.10E-18 |
| rs643434 | miR-10b-5p | trans | 1.64E-34 | Nightingale | S_HDL_C | NA | A | G | 0.025404 | 0.00434 | 9.10E-09 |
| rs643434 | miR-10b-5p | trans | 1.64E-34 | Nightingale | VLDL_FC | NA | A | G | 0.024523 | 0.004315 | 3.30E-09 |

| | | | | | | | | | | | |
|---|---|---|---|---|---|---|---|---|---|---|---|
| rs643434 | miR-10b-5p | trans | 1.64E-34 | Nightingale | S_VLDL_FC | NA | A | G | 0.027147 | 0.004154 | 1.20E-10 |
| rs643434 | miR-10b-5p | trans | 1.64E-34 | Nightingale | S_VLDL_TG | NA | A | G | -0.03596 | 0.004208 | 2.00E-17 |
| rs643434 | miR-10b-5p | trans | 1.64E-34 | Nightingale | M_LDL_PL | NA | A | G | 0.031218 | 0.004353 | 9.40E-13 |
| rs643434 | miR-10b-5p | trans | 1.64E-34 | Nightingale | L_LDL_P | NA | A | G | 0.028395 | 0.004332 | 7.00E-11 |
| rs643434 | miR-10b-5p | trans | 1.64E-34 | Nightingale | ApoB_by_A | NA | A | G | 0.025706 | 0.004281 | 2.00E-09 |
| rs643434 | miR-10b-5p | trans | 1.64E-34 | Nightingale | Remnant_C | NA | A | G | 0.035917 | 0.004319 | 4.70E-17 |
| rs643434 | miR-10b-5p | trans | 1.64E-34 | Nightingale | XXL_VLDL_ | NA | A | G | 0.029234 | 0.004328 | 5.40E-12 |
| rs643434 | miR-10b-5p | trans | 1.64E-34 | Nightingale | LA | NA | A | G | 0.023396 | 0.004263 | 1.30E-08 |
| rs643434 | miR-10b-5p | trans | 1.64E-34 | Nightingale | M_VLDL_F | NA | A | G | 0.03934 | 0.004136 | 2.40E-21 |
| rs643434 | miR-10b-5p | trans | 1.64E-34 | Nightingale | M_VLDL_P | NA | A | G | 0.028838 | 0.00434 | 1.20E-11 |
| rs643434 | miR-10b-5p | trans | 1.64E-34 | Nightingale | LDL_PL | NA | A | G | 0.033367 | 0.004337 | 2.80E-14 |
| rs643434 | miR-10b-5p | trans | 1.64E-34 | Nightingale | IDL_L | NA | A | G | 0.033725 | 0.004203 | 2.10E-15 |
| rs643434 | miR-10b-5p | trans | 1.64E-34 | Nightingale | M_VLDL_P | NA | A | G | 0.042071 | 0.004172 | 6.90E-24 |
| rs643434 | miR-10b-5p | trans | 1.64E-34 | Nightingale | IDL_C | NA | A | G | 0.033366 | 0.004198 | 4.40E-15 |
| rs643434 | miR-10b-5p | trans | 1.64E-34 | Nightingale | Total_L | NA | A | G | 0.025099 | 0.004276 | 1.30E-09 |
| rs643434 | miR-10b-5p | trans | 1.64E-34 | Nightingale | M_LDL_FC | NA | A | G | 0.032319 | 0.004342 | 2.20E-13 |
| rs643434 | miR-10b-5p | trans | 1.64E-34 | Nightingale | S_VLDL_PL | NA | A | G | 0.032104 | 0.004339 | 4.40E-14 |
| rs643434 | miR-10b-5p | trans | 1.64E-34 | Nightingale | XL_VLDL_P | NA | A | G | 0.039162 | 0.004353 | 6.90E-20 |
| rs643434 | miR-10b-5p | trans | 1.64E-34 | Nightingale | M_LDL_C_ | NA | A | G | 0.026601 | 0.00435 | 1.60E-09 |
| rs643434 | miR-10b-5p | trans | 1.64E-34 | Nightingale | XS_VLDL_F | NA | A | G | 0.035949 | 0.004272 | 6.80E-18 |
| rs643434 | miR-10b-5p | trans | 1.64E-34 | Nightingale | S_VLDL_C_ | NA | A | G | 0.039702 | 0.004231 | 9.10E-21 |
| rs643434 | miR-10b-5p | trans | 1.64E-34 | Nightingale | L_LDL_C | NA | A | G | 0.030283 | 0.004298 | 2.80E-12 |
| rs643434 | miR-10b-5p | trans | 1.64E-34 | Nightingale | XXL_VLDL_ | NA | A | G | 0.023497 | 0.004375 | 5.50E-09 |
| rs643434 | miR-10b-5p | trans | 1.64E-34 | Nightingale | S_LDL_P | NA | A | G | 0.029007 | 0.004351 | 2.80E-11 |
| rs643434 | miR-10b-5p | trans | 1.64E-34 | Nightingale | S_LDL_L | NA | A | G | 0.026945 | 0.004346 | 1.10E-09 |
| rs643434 | miR-10b-5p | trans | 1.64E-34 | Nightingale | M_LDL_C | NA | A | G | 0.032137 | 0.004355 | 2.20E-13 |
| rs643434 | miR-10b-5p | trans | 1.64E-34 | Nightingale | LDL_CE | NA | A | G | 0.030677 | 0.004336 | 1.90E-12 |
| rs643434 | miR-10b-5p | trans | 1.64E-34 | Nightingale | M_LDL_TG | NA | A | G | -0.02514 | 0.004293 | 8.20E-09 |
| rs643434 | miR-10b-5p | trans | 1.64E-34 | Nightingale | XS_VLDL_P | NA | A | G | 0.031727 | 0.004286 | 1.80E-14 |
| rs643434 | miR-10b-5p | trans | 1.64E-34 | Nightingale | L_LDL_FC | NA | A | G | 0.031521 | 0.004269 | 3.80E-13 |
| rs643434 | miR-10b-5p | trans | 1.64E-34 | Nightingale | VLDL_C | NA | A | G | 0.031139 | 0.004341 | 1.90E-13 |
| rs643434 | miR-10b-5p | trans | 1.64E-34 | Nightingale | XS_VLDL_C | NA | A | G | 0.036463 | 0.004195 | 3.70E-18 |
| rs643434 | miR-10b-5p | trans | 1.64E-34 | Nightingale | S_LDL_FC | NA | A | G | 0.027234 | 0.00434 | 1.30E-09 |
| rs643434 | miR-10b-5p | trans | 1.64E-34 | Nightingale | S_LDL_C_p | NA | A | G | 0.041888 | 0.004364 | 5.30E-22 |
| rs643434 | miR-10b-5p | trans | 1.64E-34 | Nightingale | IDL_P | NA | A | G | 0.039091 | 0.004301 | 1.70E-19 |
| rs643434 | miR-10b-5p | trans | 1.64E-34 | Nightingale | Total_CE | NA | A | G | 0.029456 | 0.004193 | 1.60E-12 |
| rs643434 | miR-10b-5p | trans | 1.64E-34 | Nightingale | XS_VLDL_F | NA | A | G | 0.029136 | 0.004316 | 3.10E-11 |
| rs643434 | miR-10b-5p | trans | 1.64E-34 | Nightingale | S_VLDL_L | NA | A | G | 0.024562 | 0.004305 | 2.30E-09 |
| rs643434 | miR-10b-5p | trans | 1.64E-34 | Nightingale | S_LDL_CE_ | NA | A | G | 0.0289 | 0.00434 | 1.00E-12 |
| rs643434 | miR-10b-5p | trans | 1.64E-34 | Nightingale | XL_VLDL_C | NA | A | G | 0.035551 | 0.004168 | 1.90E-17 |
| rs643434 | miR-10b-5p | trans | 1.64E-34 | Nightingale | S_LDL_CE | NA | A | G | 0.0295 | 0.004349 | 1.40E-11 |
| rs643434 | miR-10b-5p | trans | 1.64E-34 | Nightingale | Total_PL | NA | A | G | 0.023087 | 0.004139 | 6.80E-09 |
| rs643434 | miR-10b-5p | trans | 1.64E-34 | Nightingale | Citrate | NA | A | G | 0.024643 | 0.004319 | 6.40E-09 |
| rs643434 | miR-10b-5p | trans | 1.64E-34 | Nightingale | VLDL_P | NA | A | G | 0.026629 | 0.004324 | 1.80E-10 |
| rs643434 | miR-10b-5p | trans | 1.64E-34 | Nightingale | S_HDL_L | NA | A | G | 0.024009 | 0.004332 | 2.60E-08 |
| rs643434 | miR-10b-5p | trans | 1.64E-34 | Nightingale | IDL_FC | NA | A | G | 0.036015 | 0.004208 | 3.00E-17 |
| rs643434 | miR-10b-5p | trans | 1.64E-34 | Nightingale | M_VLDL_T | NA | A | G | -0.0362 | 0.004108 | 1.70E-18 |
| rs643434 | miR-10b-5p | trans | 1.64E-34 | Nightingale | M_LDL_L | NA | A | G | 0.031433 | 0.004353 | 6.60E-13 |
| rs643434 | miR-10b-5p | trans | 1.64E-34 | Nightingale | Total_C | NA | A | G | 0.030215 | 0.004208 | 4.40E-13 |
| rs643434 | miR-10b-5p | trans | 1.64E-34 | Nightingale | S_VLDL_FC | NA | A | G | 0.036708 | 0.004348 | 1.60E-17 |
| rs643434 | miR-10b-5p | trans | 1.64E-34 | Nightingale | L_VLDL_FC | NA | A | G | 0.046831 | 0.004305 | 5.60E-28 |
| rs643434 | miR-10b-5p | trans | 1.64E-34 | Nightingale | L_VLDL_PL | NA | A | G | 0.021642 | 0.004222 | 4.60E-08 |
| rs643434 | miR-10b-5p | trans | 1.64E-34 | Nightingale | Clinical_LD | NA | A | G | 0.03461 | 0.004322 | 2.50E-15 |
| rs643434 | miR-10b-5p | trans | 1.64E-34 | Nightingale | M_VLDL_P | NA | A | G | 0.028009 | 0.004336 | 4.20E-11 |
| rs643434 | miR-10b-5p | trans | 1.64E-34 | Nightingale | Total_FC | NA | A | G | 0.031463 | 0.004247 | 4.00E-14 |
| rs643434 | miR-10b-5p | trans | 1.64E-34 | Nightingale | S_VLDL_CE | NA | A | G | 0.045098 | 0.004289 | 1.70E-25 |
| rs643434 | miR-10b-5p | trans | 1.64E-34 | Nightingale | LDL_C | NA | A | G | 0.031242 | 0.004328 | 8.80E-13 |
| rs643434 | miR-10b-5p | trans | 1.64E-34 | Nightingale | L_LDL_PL | NA | A | G | 0.03597 | 0.004314 | 1.20E-16 |
| rs643434 | miR-10b-5p | trans | 1.64E-34 | Nightingale | S_VLDL_C | NA | A | G | 0.037408 | 0.004347 | 2.50E-18 |
| rs643434 | miR-10b-5p | trans | 1.64E-34 | Nightingale | S_VLDL_PL | NA | A | G | 0.025727 | 0.004162 | 1.50E-09 |
| rs643434 | miR-10b-5p | trans | 1.64E-34 | Nightingale | XL_VLDL_T | NA | A | G | -0.0415 | 0.004213 | 5.60E-23 |
| rs643434 | miR-10b-5p | trans | 1.64E-34 | Nightingale | L_VLDL_TG | NA | A | G | -0.04918 | 0.004326 | 7.90E-31 |
| rs643434 | miR-10b-5p | trans | 1.64E-34 | Nightingale | VLDL_CE | NA | A | G | 0.03522 | 0.004348 | 1.90E-16 |

| | | | | | | | | | | | |
|---|---|---|---|---|---|---|---|---|---|---|---|
| rs643434 | miR-10b-5p | trans | 1.64E-34 | Nightingale | M_VLDL_C | NA | A | G | 0.03381 | 0.004095 | 2.90E-16 |
| rs643434 | miR-10b-5p | trans | 1.64E-34 | Nightingale | XL_VLDL_F | NA | A | G | 0.035154 | 0.004214 | 2.00E-17 |
| rs643434 | miR-10b-5p | trans | 1.64E-34 | Nightingale | S_HDL_CE | NA | A | G | 0.025707 | 0.004354 | 6.70E-09 |
| rs643434 | let-7b-5p | trans | 1.70E-26 | Nightingale | XXL_VLDL_ | NA | A | G | -0.03445 | 0.004384 | 6.10E-16 |
| rs643434 | let-7b-5p | trans | 1.70E-26 | Nightingale | Sphingomy | NA | A | G | 0.025961 | 0.004064 | 3.00E-10 |
| rs643434 | let-7b-5p | trans | 1.70E-26 | Nightingale | Omega_6 | NA | A | G | 0.024274 | 0.004245 | 2.60E-09 |
| rs643434 | let-7b-5p | trans | 1.70E-26 | Nightingale | M_VLDL_C | NA | A | G | 0.031313 | 0.004084 | 3.50E-14 |
| rs643434 | let-7b-5p | trans | 1.70E-26 | Nightingale | M_VLDL_C | NA | A | G | 0.037416 | 0.004335 | 4.70E-18 |
| rs643434 | let-7b-5p | trans | 1.70E-26 | Nightingale | GlycA | NA | A | G | 0.025332 | 0.004328 | 5.00E-09 |
| rs643434 | let-7b-5p | trans | 1.70E-26 | Nightingale | L_VLDL_CE | NA | A | G | 0.044018 | 0.00416 | 9.50E-27 |
| rs643434 | let-7b-5p | trans | 1.70E-26 | Nightingale | IDL_PL | NA | A | G | 0.033641 | 0.004195 | 1.80E-15 |
| rs643434 | let-7b-5p | trans | 1.70E-26 | Nightingale | M_VLDL_F | NA | A | G | 0.032595 | 0.004346 | 3.50E-14 |
| rs643434 | let-7b-5p | trans | 1.70E-26 | Nightingale | non_HDL_( | NA | A | G | 0.034208 | 0.004323 | 2.20E-15 |
| rs643434 | let-7b-5p | trans | 1.70E-26 | Nightingale | XS_VLDL_L | NA | A | G | 0.033078 | 0.004273 | 1.20E-15 |
| rs643434 | let-7b-5p | trans | 1.70E-26 | Nightingale | LDL_L | NA | A | G | 0.03152 | 0.004332 | 5.40E-13 |
| rs643434 | let-7b-5p | trans | 1.70E-26 | Nightingale | S_LDL_PL_ | NA | A | G | -0.0281 | 0.004339 | 4.70E-12 |
| rs643434 | let-7b-5p | trans | 1.70E-26 | Nightingale | LDL_FC | NA | A | G | 0.031592 | 0.004301 | 6.20E-13 |
| rs643434 | let-7b-5p | trans | 1.70E-26 | Nightingale | ApoB | NA | A | G | 0.032994 | 0.004341 | 3.10E-14 |
| rs643434 | let-7b-5p | trans | 1.70E-26 | Nightingale | L_VLDL_CE | NA | A | G | 0.022834 | 0.004296 | 2.70E-08 |
| rs643434 | let-7b-5p | trans | 1.70E-26 | Nightingale | S_VLDL_CE | NA | A | G | 0.037357 | 0.004341 | 1.60E-18 |
| rs643434 | let-7b-5p | trans | 1.70E-26 | Nightingale | S_HDL_P | NA | A | G | 0.027834 | 0.004348 | 2.60E-10 |
| rs643434 | let-7b-5p | trans | 1.70E-26 | Nightingale | L_LDL_L | NA | A | G | 0.031581 | 0.004305 | 3.00E-13 |
| rs643434 | let-7b-5p | trans | 1.70E-26 | Nightingale | IDL_CE | NA | A | G | 0.032158 | 0.004197 | 3.80E-14 |
| rs643434 | let-7b-5p | trans | 1.70E-26 | Nightingale | L_LDL_CE | NA | A | G | 0.029584 | 0.004308 | 9.10E-12 |
| rs643434 | let-7b-5p | trans | 1.70E-26 | Nightingale | XL_VLDL_C | NA | A | G | 0.034537 | 0.00417 | 1.70E-16 |
| rs643434 | let-7b-5p | trans | 1.70E-26 | Nightingale | XS_VLDL_P | NA | A | G | 0.035851 | 0.004289 | 9.10E-18 |
| rs643434 | let-7b-5p | trans | 1.70E-26 | Nightingale | LDL_P | NA | A | G | 0.030723 | 0.004344 | 2.00E-12 |
| rs643434 | let-7b-5p | trans | 1.70E-26 | Nightingale | L_VLDL_C_ | NA | A | G | 0.049148 | 0.004246 | 1.20E-31 |
| rs643434 | let-7b-5p | trans | 1.70E-26 | Nightingale | M_VLDL_C | NA | A | G | 0.039267 | 0.004308 | 1.20E-19 |
| rs643434 | let-7b-5p | trans | 1.70E-26 | Nightingale | M_LDL_P | NA | A | G | 0.034038 | 0.004359 | 7.10E-15 |
| rs643434 | let-7b-5p | trans | 1.70E-26 | Nightingale | M_LDL_CE | NA | A | G | 0.031258 | 0.004353 | 8.10E-13 |
| rs643434 | let-7b-5p | trans | 1.70E-26 | Nightingale | XXL_VLDL_ | NA | A | G | 0.031806 | 0.00438 | 2.10E-13 |
| rs643434 | let-7b-5p | trans | 1.70E-26 | Nightingale | XXL_VLDL_ | NA | A | G | 0.033426 | 0.004356 | 9.00E-15 |
| rs643434 | let-7b-5p | trans | 1.70E-26 | Nightingale | XL_HDL_PL | NA | A | G | -0.02176 | 0.003994 | 4.90E-08 |
| rs643434 | let-7b-5p | trans | 1.70E-26 | Nightingale | S_LDL_C | NA | A | G | 0.029751 | 0.004351 | 1.50E-11 |
| rs643434 | let-7b-5p | trans | 1.70E-26 | Nightingale | S_VLDL_P | NA | A | G | 0.023452 | 0.004293 | 9.10E-09 |
| rs643434 | let-7b-5p | trans | 1.70E-26 | Nightingale | XS_VLDL_C | NA | A | G | 0.036711 | 0.00422 | 2.10E-18 |
| rs643434 | let-7b-5p | trans | 1.70E-26 | Nightingale | S_HDL_C | NA | A | G | 0.025404 | 0.00434 | 9.10E-09 |
| rs643434 | let-7b-5p | trans | 1.70E-26 | Nightingale | VLDL_FC | NA | A | G | 0.024523 | 0.004315 | 3.30E-09 |
| rs643434 | let-7b-5p | trans | 1.70E-26 | Nightingale | S_VLDL_FC | NA | A | G | 0.027147 | 0.004154 | 1.20E-10 |
| rs643434 | let-7b-5p | trans | 1.70E-26 | Nightingale | S_VLDL_TG | NA | A | G | -0.03596 | 0.004208 | 2.00E-17 |
| rs643434 | let-7b-5p | trans | 1.70E-26 | Nightingale | M_LDL_PL | NA | A | G | 0.031218 | 0.004353 | 9.40E-13 |
| rs643434 | let-7b-5p | trans | 1.70E-26 | Nightingale | L_LDL_P | NA | A | G | 0.028395 | 0.004332 | 7.00E-11 |
| rs643434 | let-7b-5p | trans | 1.70E-26 | Nightingale | ApoB_by_A | NA | A | G | 0.025706 | 0.004281 | 2.00E-09 |
| rs643434 | let-7b-5p | trans | 1.70E-26 | Nightingale | Remnant_( | NA | A | G | 0.035917 | 0.004319 | 4.70E-17 |
| rs643434 | let-7b-5p | trans | 1.70E-26 | Nightingale | XXL_VLDL_ | NA | A | G | 0.029234 | 0.004328 | 5.40E-12 |
| rs643434 | let-7b-5p | trans | 1.70E-26 | Nightingale | LA | NA | A | G | 0.023396 | 0.004263 | 1.30E-08 |
| rs643434 | let-7b-5p | trans | 1.70E-26 | Nightingale | M_VLDL_F | NA | A | G | 0.03934 | 0.004136 | 2.40E-21 |
| rs643434 | let-7b-5p | trans | 1.70E-26 | Nightingale | M_VLDL_P | NA | A | G | 0.028838 | 0.00434 | 1.20E-11 |
| rs643434 | let-7b-5p | trans | 1.70E-26 | Nightingale | LDL_PL | NA | A | G | 0.033367 | 0.004337 | 2.80E-14 |
| rs643434 | let-7b-5p | trans | 1.70E-26 | Nightingale | IDL_L | NA | A | G | 0.033725 | 0.004203 | 2.10E-15 |
| rs643434 | let-7b-5p | trans | 1.70E-26 | Nightingale | M_VLDL_P | NA | A | G | 0.042071 | 0.004172 | 6.90E-24 |
| rs643434 | let-7b-5p | trans | 1.70E-26 | Nightingale | IDL_C | NA | A | G | 0.033366 | 0.004198 | 4.40E-15 |
| rs643434 | let-7b-5p | trans | 1.70E-26 | Nightingale | Total_L | NA | A | G | 0.025099 | 0.004276 | 1.30E-09 |
| rs643434 | let-7b-5p | trans | 1.70E-26 | Nightingale | M_LDL_FC | NA | A | G | 0.032319 | 0.004342 | 2.20E-13 |
| rs643434 | let-7b-5p | trans | 1.70E-26 | Nightingale | S_VLDL_PL | NA | A | G | 0.032104 | 0.004339 | 4.40E-14 |
| rs643434 | let-7b-5p | trans | 1.70E-26 | Nightingale | XL_VLDL_P | NA | A | G | 0.039162 | 0.004353 | 6.90E-20 |
| rs643434 | let-7b-5p | trans | 1.70E-26 | Nightingale | M_LDL_C_ | NA | A | G | 0.026601 | 0.00435 | 1.60E-09 |
| rs643434 | let-7b-5p | trans | 1.70E-26 | Nightingale | XS_VLDL_F | NA | A | G | 0.035949 | 0.004272 | 6.80E-18 |
| rs643434 | let-7b-5p | trans | 1.70E-26 | Nightingale | S_VLDL_C_ | NA | A | G | 0.039702 | 0.004231 | 9.10E-21 |
| rs643434 | let-7b-5p | trans | 1.70E-26 | Nightingale | L_LDL_C | NA | A | G | 0.030283 | 0.004298 | 2.80E-12 |
| rs643434 | let-7b-5p | trans | 1.70E-26 | Nightingale | XXL_VLDL_ | NA | A | G | 0.023497 | 0.004375 | 5.50E-09 |
| rs643434 | let-7b-5p | trans | 1.70E-26 | Nightingale | S_LDL_P | NA | A | G | 0.029007 | 0.004351 | 2.80E-11 |

| | | | | | | | | | | | |
|---|---|---|---|---|---|---|---|---|---|---|---|
| rs643434 | let-7b-5p | trans | 1.70E-26 | Nightingale | S_LDL_L | NA | A | G | 0.026945 | 0.004346 | 1.10E-09 |
| rs643434 | let-7b-5p | trans | 1.70E-26 | Nightingale | M_LDL_C | NA | A | G | 0.032137 | 0.004355 | 2.20E-13 |
| rs643434 | let-7b-5p | trans | 1.70E-26 | Nightingale | LDL_CE | NA | A | G | 0.030677 | 0.004336 | 1.90E-12 |
| rs643434 | let-7b-5p | trans | 1.70E-26 | Nightingale | M_LDL_TG | NA | A | G | -0.02514 | 0.004293 | 8.20E-09 |
| rs643434 | let-7b-5p | trans | 1.70E-26 | Nightingale | XS_VLDL_P | NA | A | G | 0.031727 | 0.004286 | 1.80E-14 |
| rs643434 | let-7b-5p | trans | 1.70E-26 | Nightingale | L_LDL_FC | NA | A | G | 0.031521 | 0.004269 | 3.80E-13 |
| rs643434 | let-7b-5p | trans | 1.70E-26 | Nightingale | VLDL_C | NA | A | G | 0.031139 | 0.004341 | 1.90E-13 |
| rs643434 | let-7b-5p | trans | 1.70E-26 | Nightingale | XS_VLDL_C | NA | A | G | 0.036463 | 0.004195 | 3.70E-18 |
| rs643434 | let-7b-5p | trans | 1.70E-26 | Nightingale | S_LDL_FC | NA | A | G | 0.027234 | 0.00434 | 1.30E-09 |
| rs643434 | let-7b-5p | trans | 1.70E-26 | Nightingale | S_LDL_C_p | NA | A | G | 0.041888 | 0.004364 | 5.30E-22 |
| rs643434 | let-7b-5p | trans | 1.70E-26 | Nightingale | IDL_P | NA | A | G | 0.039091 | 0.004301 | 1.70E-19 |
| rs643434 | let-7b-5p | trans | 1.70E-26 | Nightingale | Total_CE | NA | A | G | 0.029456 | 0.004193 | 1.60E-12 |
| rs643434 | let-7b-5p | trans | 1.70E-26 | Nightingale | XS_VLDL_F | NA | A | G | 0.029136 | 0.004316 | 3.10E-11 |
| rs643434 | let-7b-5p | trans | 1.70E-26 | Nightingale | S_VLDL_L | NA | A | G | 0.024562 | 0.004305 | 2.30E-09 |
| rs643434 | let-7b-5p | trans | 1.70E-26 | Nightingale | S_LDL_CE_ | NA | A | G | 0.0289 | 0.00434 | 1.00E-12 |
| rs643434 | let-7b-5p | trans | 1.70E-26 | Nightingale | XL_VLDL_C | NA | A | G | 0.035551 | 0.004168 | 1.90E-17 |
| rs643434 | let-7b-5p | trans | 1.70E-26 | Nightingale | S_LDL_CE | NA | A | G | 0.0295 | 0.004349 | 1.40E-11 |
| rs643434 | let-7b-5p | trans | 1.70E-26 | Nightingale | Total_PL | NA | A | G | 0.023087 | 0.004139 | 6.80E-09 |
| rs643434 | let-7b-5p | trans | 1.70E-26 | Nightingale | Citrate | NA | A | G | 0.024643 | 0.004319 | 6.40E-09 |
| rs643434 | let-7b-5p | trans | 1.70E-26 | Nightingale | VLDL_P | NA | A | G | 0.026629 | 0.004324 | 1.80E-10 |
| rs643434 | let-7b-5p | trans | 1.70E-26 | Nightingale | S_HDL_L | NA | A | G | 0.024009 | 0.004332 | 2.60E-08 |
| rs643434 | let-7b-5p | trans | 1.70E-26 | Nightingale | IDL_FC | NA | A | G | 0.036015 | 0.004208 | 3.00E-17 |
| rs643434 | let-7b-5p | trans | 1.70E-26 | Nightingale | M_VLDL_T | NA | A | G | -0.0362 | 0.004108 | 1.70E-18 |
| rs643434 | let-7b-5p | trans | 1.70E-26 | Nightingale | M_LDL_L | NA | A | G | 0.031433 | 0.004353 | 6.60E-13 |
| rs643434 | let-7b-5p | trans | 1.70E-26 | Nightingale | Total_C | NA | A | G | 0.030215 | 0.004208 | 4.40E-13 |
| rs643434 | let-7b-5p | trans | 1.70E-26 | Nightingale | S_VLDL_FC | NA | A | G | 0.036708 | 0.004348 | 1.60E-17 |
| rs643434 | let-7b-5p | trans | 1.70E-26 | Nightingale | L_VLDL_FC | NA | A | G | 0.046831 | 0.004305 | 5.60E-28 |
| rs643434 | let-7b-5p | trans | 1.70E-26 | Nightingale | L_VLDL_PL | NA | A | G | 0.021642 | 0.004222 | 4.60E-08 |
| rs643434 | let-7b-5p | trans | 1.70E-26 | Nightingale | Clinical_LD | NA | A | G | 0.03461 | 0.004322 | 2.50E-15 |
| rs643434 | let-7b-5p | trans | 1.70E-26 | Nightingale | M_VLDL_P | NA | A | G | 0.028009 | 0.004336 | 4.20E-11 |
| rs643434 | let-7b-5p | trans | 1.70E-26 | Nightingale | Total_FC | NA | A | G | 0.031463 | 0.004247 | 4.00E-14 |
| rs643434 | let-7b-5p | trans | 1.70E-26 | Nightingale | S_VLDL_CE | NA | A | G | 0.045098 | 0.004289 | 1.70E-25 |
| rs643434 | let-7b-5p | trans | 1.70E-26 | Nightingale | LDL_C | NA | A | G | 0.031242 | 0.004328 | 8.80E-13 |
| rs643434 | let-7b-5p | trans | 1.70E-26 | Nightingale | L_LDL_PL | NA | A | G | 0.03597 | 0.004314 | 1.20E-16 |
| rs643434 | let-7b-5p | trans | 1.70E-26 | Nightingale | S_VLDL_C | NA | A | G | 0.037408 | 0.004347 | 2.50E-18 |
| rs643434 | let-7b-5p | trans | 1.70E-26 | Nightingale | S_VLDL_PL | NA | A | G | 0.025727 | 0.004162 | 1.50E-09 |
| rs643434 | let-7b-5p | trans | 1.70E-26 | Nightingale | XL_VLDL_T | NA | A | G | -0.0415 | 0.004213 | 5.60E-23 |
| rs643434 | let-7b-5p | trans | 1.70E-26 | Nightingale | L_VLDL_TG | NA | A | G | -0.04918 | 0.004326 | 7.90E-31 |
| rs643434 | let-7b-5p | trans | 1.70E-26 | Nightingale | VLDL_CE | NA | A | G | 0.03522 | 0.004348 | 1.90E-16 |
| rs643434 | let-7b-5p | trans | 1.70E-26 | Nightingale | M_VLDL_C | NA | A | G | 0.03381 | 0.004095 | 2.90E-16 |
| rs643434 | let-7b-5p | trans | 1.70E-26 | Nightingale | XL_VLDL_F | NA | A | G | 0.035154 | 0.004214 | 2.00E-17 |
| rs643434 | let-7b-5p | trans | 1.70E-26 | Nightingale | S_HDL_CE | NA | A | G | 0.025707 | 0.004354 | 6.70E-09 |
| rs643434 | miR-139-5p | trans | 1.31E-40 | Nightingale | XXL_VLDL_ | NA | A | G | -0.03445 | 0.004384 | 6.10E-16 |
| rs643434 | miR-139-5p | trans | 1.31E-40 | Nightingale | Sphingomy | NA | A | G | 0.025961 | 0.004064 | 3.00E-10 |
| rs643434 | miR-139-5p | trans | 1.31E-40 | Nightingale | Omega_6 | NA | A | G | 0.024274 | 0.004245 | 2.60E-09 |
| rs643434 | miR-139-5p | trans | 1.31E-40 | Nightingale | M_VLDL_C | NA | A | G | 0.031313 | 0.004084 | 3.50E-14 |
| rs643434 | miR-139-5p | trans | 1.31E-40 | Nightingale | M_VLDL_C | NA | A | G | 0.037416 | 0.004335 | 4.70E-18 |
| rs643434 | miR-139-5p | trans | 1.31E-40 | Nightingale | GlycA | NA | A | G | 0.025332 | 0.004328 | 5.00E-09 |
| rs643434 | miR-139-5p | trans | 1.31E-40 | Nightingale | L_VLDL_CE | NA | A | G | 0.044018 | 0.00416 | 9.50E-27 |
| rs643434 | miR-139-5p | trans | 1.31E-40 | Nightingale | IDL_PL | NA | A | G | 0.033641 | 0.004195 | 1.80E-15 |
| rs643434 | miR-139-5p | trans | 1.31E-40 | Nightingale | M_VLDL_F | NA | A | G | 0.032595 | 0.004346 | 3.50E-14 |
| rs643434 | miR-139-5p | trans | 1.31E-40 | Nightingale | non_HDL_( | NA | A | G | 0.034208 | 0.004323 | 2.20E-15 |
| rs643434 | miR-139-5p | trans | 1.31E-40 | Nightingale | XS_VLDL_L | NA | A | G | 0.033078 | 0.004273 | 1.20E-15 |
| rs643434 | miR-139-5p | trans | 1.31E-40 | Nightingale | LDL_L | NA | A | G | 0.03152 | 0.004332 | 5.40E-13 |
| rs643434 | miR-139-5p | trans | 1.31E-40 | Nightingale | S_LDL_PL_ | NA | A | G | -0.0281 | 0.004339 | 4.70E-12 |
| rs643434 | miR-139-5p | trans | 1.31E-40 | Nightingale | LDL_FC | NA | A | G | 0.031592 | 0.004301 | 6.20E-13 |
| rs643434 | miR-139-5p | trans | 1.31E-40 | Nightingale | ApoB | NA | A | G | 0.032994 | 0.004341 | 3.10E-14 |
| rs643434 | miR-139-5p | trans | 1.31E-40 | Nightingale | L_VLDL_CE | NA | A | G | 0.022834 | 0.004296 | 2.70E-08 |
| rs643434 | miR-139-5p | trans | 1.31E-40 | Nightingale | S_VLDL_CE | NA | A | G | 0.037357 | 0.004341 | 1.60E-18 |
| rs643434 | miR-139-5p | trans | 1.31E-40 | Nightingale | S_HDL_P | NA | A | G | 0.027834 | 0.004348 | 2.60E-10 |
| rs643434 | miR-139-5p | trans | 1.31E-40 | Nightingale | L_LDL_L | NA | A | G | 0.031581 | 0.004305 | 3.00E-13 |
| rs643434 | miR-139-5p | trans | 1.31E-40 | Nightingale | IDL_CE | NA | A | G | 0.032158 | 0.004197 | 3.80E-14 |
| rs643434 | miR-139-5p | trans | 1.31E-40 | Nightingale | L_LDL_CE | NA | A | G | 0.029584 | 0.004308 | 9.10E-12 |

| | | | | | | | | | | | |
|---|---|---|---|---|---|---|---|---|---|---|---|
| rs643434 | miR-139-5p | trans | 1.31E-40 | Nightingale | XL_VLDL_C | NA | A | G | 0.034537 | 0.00417 | 1.70E-16 |
| rs643434 | miR-139-5p | trans | 1.31E-40 | Nightingale | XS_VLDL_P | NA | A | G | 0.035851 | 0.004289 | 9.10E-18 |
| rs643434 | miR-139-5p | trans | 1.31E-40 | Nightingale | LDL_P | NA | A | G | 0.030723 | 0.004344 | 2.00E-12 |
| rs643434 | miR-139-5p | trans | 1.31E-40 | Nightingale | L_VLDL_C_ | NA | A | G | 0.049148 | 0.004246 | 1.20E-31 |
| rs643434 | miR-139-5p | trans | 1.31E-40 | Nightingale | M_VLDL_C | NA | A | G | 0.039267 | 0.004308 | 1.20E-19 |
| rs643434 | miR-139-5p | trans | 1.31E-40 | Nightingale | M_LDL_P | NA | A | G | 0.034038 | 0.004359 | 7.10E-15 |
| rs643434 | miR-139-5p | trans | 1.31E-40 | Nightingale | M_LDL_CE | NA | A | G | 0.031258 | 0.004353 | 8.10E-13 |
| rs643434 | miR-139-5p | trans | 1.31E-40 | Nightingale | XXL_VLDL_ | NA | A | G | 0.031806 | 0.00438 | 2.10E-13 |
| rs643434 | miR-139-5p | trans | 1.31E-40 | Nightingale | XXL_VLDL_ | NA | A | G | 0.033426 | 0.004356 | 9.00E-15 |
| rs643434 | miR-139-5p | trans | 1.31E-40 | Nightingale | XL_HDL_PL | NA | A | G | -0.02176 | 0.003994 | 4.90E-08 |
| rs643434 | miR-139-5p | trans | 1.31E-40 | Nightingale | S_LDL_C | NA | A | G | 0.029751 | 0.004351 | 1.50E-11 |
| rs643434 | miR-139-5p | trans | 1.31E-40 | Nightingale | S_VLDL_P | NA | A | G | 0.023452 | 0.004293 | 9.10E-09 |
| rs643434 | miR-139-5p | trans | 1.31E-40 | Nightingale | XS_VLDL_C | NA | A | G | 0.036711 | 0.00422 | 2.10E-18 |
| rs643434 | miR-139-5p | trans | 1.31E-40 | Nightingale | S_HDL_C | NA | A | G | 0.025404 | 0.00434 | 9.10E-09 |
| rs643434 | miR-139-5p | trans | 1.31E-40 | Nightingale | VLDL_FC | NA | A | G | 0.024523 | 0.004315 | 3.30E-09 |
| rs643434 | miR-139-5p | trans | 1.31E-40 | Nightingale | S_VLDL_FC | NA | A | G | 0.027147 | 0.004154 | 1.20E-10 |
| rs643434 | miR-139-5p | trans | 1.31E-40 | Nightingale | S_VLDL_TG | NA | A | G | -0.03596 | 0.004208 | 2.00E-17 |
| rs643434 | miR-139-5p | trans | 1.31E-40 | Nightingale | M_LDL_PL | NA | A | G | 0.031218 | 0.004353 | 9.40E-13 |
| rs643434 | miR-139-5p | trans | 1.31E-40 | Nightingale | L_LDL_P | NA | A | G | 0.028395 | 0.004332 | 7.00E-11 |
| rs643434 | miR-139-5p | trans | 1.31E-40 | Nightingale | ApoB_by_A | NA | A | G | 0.025706 | 0.004281 | 2.00E-09 |
| rs643434 | miR-139-5p | trans | 1.31E-40 | Nightingale | Remnant_C | NA | A | G | 0.035917 | 0.004319 | 4.70E-17 |
| rs643434 | miR-139-5p | trans | 1.31E-40 | Nightingale | XXL_VLDL_ | NA | A | G | 0.029234 | 0.004328 | 5.40E-12 |
| rs643434 | miR-139-5p | trans | 1.31E-40 | Nightingale | LA | NA | A | G | 0.023396 | 0.004263 | 1.30E-08 |
| rs643434 | miR-139-5p | trans | 1.31E-40 | Nightingale | M_VLDL_FC | NA | A | G | 0.03934 | 0.004136 | 2.40E-21 |
| rs643434 | miR-139-5p | trans | 1.31E-40 | Nightingale | M_VLDL_P | NA | A | G | 0.028838 | 0.00434 | 1.20E-11 |
| rs643434 | miR-139-5p | trans | 1.31E-40 | Nightingale | LDL_PL | NA | A | G | 0.033367 | 0.004337 | 2.80E-14 |
| rs643434 | miR-139-5p | trans | 1.31E-40 | Nightingale | IDL_L | NA | A | G | 0.033725 | 0.004203 | 2.10E-15 |
| rs643434 | miR-139-5p | trans | 1.31E-40 | Nightingale | M_VLDL_P | NA | A | G | 0.042071 | 0.004172 | 6.90E-24 |
| rs643434 | miR-139-5p | trans | 1.31E-40 | Nightingale | IDL_C | NA | A | G | 0.033366 | 0.004198 | 4.40E-15 |
| rs643434 | miR-139-5p | trans | 1.31E-40 | Nightingale | Total_L | NA | A | G | 0.025099 | 0.004276 | 1.30E-09 |
| rs643434 | miR-139-5p | trans | 1.31E-40 | Nightingale | M_LDL_FC | NA | A | G | 0.032319 | 0.004342 | 2.20E-13 |
| rs643434 | miR-139-5p | trans | 1.31E-40 | Nightingale | S_VLDL_PL | NA | A | G | 0.032104 | 0.004339 | 4.40E-14 |
| rs643434 | miR-139-5p | trans | 1.31E-40 | Nightingale | XL_VLDL_P | NA | A | G | 0.039162 | 0.004353 | 6.90E-20 |
| rs643434 | miR-139-5p | trans | 1.31E-40 | Nightingale | M_LDL_C_p | NA | A | G | 0.026601 | 0.00435 | 1.60E-09 |
| rs643434 | miR-139-5p | trans | 1.31E-40 | Nightingale | XS_VLDL_F | NA | A | G | 0.035949 | 0.004272 | 6.80E-18 |
| rs643434 | miR-139-5p | trans | 1.31E-40 | Nightingale | S_VLDL_C_ | NA | A | G | 0.039702 | 0.004231 | 9.10E-21 |
| rs643434 | miR-139-5p | trans | 1.31E-40 | Nightingale | L_LDL_C | NA | A | G | 0.030283 | 0.004298 | 2.80E-12 |
| rs643434 | miR-139-5p | trans | 1.31E-40 | Nightingale | XXL_VLDL_ | NA | A | G | 0.023497 | 0.004375 | 5.50E-09 |
| rs643434 | miR-139-5p | trans | 1.31E-40 | Nightingale | S_LDL_P | NA | A | G | 0.029007 | 0.004351 | 2.80E-11 |
| rs643434 | miR-139-5p | trans | 1.31E-40 | Nightingale | S_LDL_L | NA | A | G | 0.026945 | 0.004346 | 1.10E-09 |
| rs643434 | miR-139-5p | trans | 1.31E-40 | Nightingale | M_LDL_C | NA | A | G | 0.032137 | 0.004355 | 2.20E-13 |
| rs643434 | miR-139-5p | trans | 1.31E-40 | Nightingale | LDL_CE | NA | A | G | 0.030677 | 0.004336 | 1.90E-12 |
| rs643434 | miR-139-5p | trans | 1.31E-40 | Nightingale | M_LDL_TG | NA | A | G | -0.02514 | 0.004293 | 8.20E-09 |
| rs643434 | miR-139-5p | trans | 1.31E-40 | Nightingale | XS_VLDL_P | NA | A | G | 0.031727 | 0.004286 | 1.80E-14 |
| rs643434 | miR-139-5p | trans | 1.31E-40 | Nightingale | L_LDL_FC | NA | A | G | 0.031521 | 0.004269 | 3.80E-13 |
| rs643434 | miR-139-5p | trans | 1.31E-40 | Nightingale | VLDL_C | NA | A | G | 0.031139 | 0.004341 | 1.90E-13 |
| rs643434 | miR-139-5p | trans | 1.31E-40 | Nightingale | XS_VLDL_C | NA | A | G | 0.036463 | 0.004195 | 3.70E-18 |
| rs643434 | miR-139-5p | trans | 1.31E-40 | Nightingale | S_LDL_FC | NA | A | G | 0.027234 | 0.00434 | 1.30E-09 |
| rs643434 | miR-139-5p | trans | 1.31E-40 | Nightingale | S_LDL_C_p | NA | A | G | 0.041888 | 0.004364 | 5.30E-22 |
| rs643434 | miR-139-5p | trans | 1.31E-40 | Nightingale | IDL_P | NA | A | G | 0.039091 | 0.004301 | 1.70E-19 |
| rs643434 | miR-139-5p | trans | 1.31E-40 | Nightingale | Total_CE | NA | A | G | 0.029456 | 0.004193 | 1.60E-12 |
| rs643434 | miR-139-5p | trans | 1.31E-40 | Nightingale | XS_VLDL_F | NA | A | G | 0.029136 | 0.004316 | 3.10E-11 |
| rs643434 | miR-139-5p | trans | 1.31E-40 | Nightingale | S_VLDL_L | NA | A | G | 0.024562 | 0.004305 | 2.30E-09 |
| rs643434 | miR-139-5p | trans | 1.31E-40 | Nightingale | S_LDL_CE_ | NA | A | G | 0.0289 | 0.00434 | 1.00E-12 |
| rs643434 | miR-139-5p | trans | 1.31E-40 | Nightingale | XL_VLDL_C | NA | A | G | 0.035551 | 0.004168 | 1.90E-17 |
| rs643434 | miR-139-5p | trans | 1.31E-40 | Nightingale | S_LDL_CE | NA | A | G | 0.0295 | 0.004349 | 1.40E-11 |
| rs643434 | miR-139-5p | trans | 1.31E-40 | Nightingale | Total_PL | NA | A | G | 0.023087 | 0.004139 | 6.80E-09 |
| rs643434 | miR-139-5p | trans | 1.31E-40 | Nightingale | Citrate | NA | A | G | 0.024643 | 0.004319 | 6.40E-09 |
| rs643434 | miR-139-5p | trans | 1.31E-40 | Nightingale | VLDL_P | NA | A | G | 0.026629 | 0.004324 | 1.80E-10 |
| rs643434 | miR-139-5p | trans | 1.31E-40 | Nightingale | S_HDL_L | NA | A | G | 0.024009 | 0.004332 | 2.60E-08 |
| rs643434 | miR-139-5p | trans | 1.31E-40 | Nightingale | IDL_FC | NA | A | G | 0.036015 | 0.004208 | 3.00E-17 |
| rs643434 | miR-139-5p | trans | 1.31E-40 | Nightingale | M_VLDL_T | NA | A | G | -0.0362 | 0.004108 | 1.70E-18 |
| rs643434 | miR-139-5p | trans | 1.31E-40 | Nightingale | M_LDL_L | NA | A | G | 0.031433 | 0.004353 | 6.60E-13 |

| | | | | | | | | | | | |
|---|---|---|---|---|---|---|---|---|---|---|---|
| rs643434 | miR-139-5p | trans | 1.31E-40 | Nightingale | Total_C | NA | A | G | 0.030215 | 0.004208 | 4.40E-13 |
| rs643434 | miR-139-5p | trans | 1.31E-40 | Nightingale | S_VLDL_FC | NA | A | G | 0.036708 | 0.004348 | 1.60E-17 |
| rs643434 | miR-139-5p | trans | 1.31E-40 | Nightingale | L_VLDL_FC | NA | A | G | 0.046831 | 0.004305 | 5.60E-28 |
| rs643434 | miR-139-5p | trans | 1.31E-40 | Nightingale | L_VLDL_PL | NA | A | G | 0.021642 | 0.004222 | 4.60E-08 |
| rs643434 | miR-139-5p | trans | 1.31E-40 | Nightingale | Clinical_LD | NA | A | G | 0.03461 | 0.004322 | 2.50E-15 |
| rs643434 | miR-139-5p | trans | 1.31E-40 | Nightingale | M_VLDL_P | NA | A | G | 0.028009 | 0.004336 | 4.20E-11 |
| rs643434 | miR-139-5p | trans | 1.31E-40 | Nightingale | Total_FC | NA | A | G | 0.031463 | 0.004247 | 4.00E-14 |
| rs643434 | miR-139-5p | trans | 1.31E-40 | Nightingale | S_VLDL_CE | NA | A | G | 0.045098 | 0.004289 | 1.70E-25 |
| rs643434 | miR-139-5p | trans | 1.31E-40 | Nightingale | LDL_C | NA | A | G | 0.031242 | 0.004328 | 8.80E-13 |
| rs643434 | miR-139-5p | trans | 1.31E-40 | Nightingale | L_LDL_PL | NA | A | G | 0.03597 | 0.004314 | 1.20E-16 |
| rs643434 | miR-139-5p | trans | 1.31E-40 | Nightingale | S_VLDL_C | NA | A | G | 0.037408 | 0.004347 | 2.50E-18 |
| rs643434 | miR-139-5p | trans | 1.31E-40 | Nightingale | S_VLDL_PL | NA | A | G | 0.025727 | 0.004162 | 1.50E-09 |
| rs643434 | miR-139-5p | trans | 1.31E-40 | Nightingale | XL_VLDL_T | NA | A | G | -0.0415 | 0.004213 | 5.60E-23 |
| rs643434 | miR-139-5p | trans | 1.31E-40 | Nightingale | L_VLDL_TG | NA | A | G | -0.04918 | 0.004326 | 7.90E-31 |
| rs643434 | miR-139-5p | trans | 1.31E-40 | Nightingale | VLDL_CE | NA | A | G | 0.03522 | 0.004348 | 1.90E-16 |
| rs643434 | miR-139-5p | trans | 1.31E-40 | Nightingale | M_VLDL_C | NA | A | G | 0.03381 | 0.004095 | 2.90E-16 |
| rs643434 | miR-139-5p | trans | 1.31E-40 | Nightingale | XL_VLDL_F | NA | A | G | 0.035154 | 0.004214 | 2.00E-17 |
| rs643434 | miR-139-5p | trans | 1.31E-40 | Nightingale | S_HDL_CE | NA | A | G | 0.025707 | 0.004354 | 6.70E-09 |
| rs643434 | let-7a-5p | trans | 9.18E-13 | Nightingale | XXL_VLDL_ | NA | A | G | -0.03445 | 0.004384 | 6.10E-16 |
| rs643434 | let-7a-5p | trans | 9.18E-13 | Nightingale | Sphingomy | NA | A | G | 0.025961 | 0.004064 | 3.00E-10 |
| rs643434 | let-7a-5p | trans | 9.18E-13 | Nightingale | Omega_6 | NA | A | G | 0.024274 | 0.004245 | 2.60E-09 |
| rs643434 | let-7a-5p | trans | 9.18E-13 | Nightingale | M_VLDL_C | NA | A | G | 0.031313 | 0.004084 | 3.50E-14 |
| rs643434 | let-7a-5p | trans | 9.18E-13 | Nightingale | M_VLDL_C | NA | A | G | 0.037416 | 0.004335 | 4.70E-18 |
| rs643434 | let-7a-5p | trans | 9.18E-13 | Nightingale | GlycA | NA | A | G | 0.025332 | 0.004328 | 5.00E-09 |
| rs643434 | let-7a-5p | trans | 9.18E-13 | Nightingale | L_VLDL_CE | NA | A | G | 0.044018 | 0.00416 | 9.50E-27 |
| rs643434 | let-7a-5p | trans | 9.18E-13 | Nightingale | IDL_PL | NA | A | G | 0.033641 | 0.004195 | 1.80E-15 |
| rs643434 | let-7a-5p | trans | 9.18E-13 | Nightingale | M_VLDL_F | NA | A | G | 0.032595 | 0.004346 | 3.50E-14 |
| rs643434 | let-7a-5p | trans | 9.18E-13 | Nightingale | non_HDL_( | NA | A | G | 0.034208 | 0.004323 | 2.20E-15 |
| rs643434 | let-7a-5p | trans | 9.18E-13 | Nightingale | XS_VLDL_L | NA | A | G | 0.033078 | 0.004273 | 1.20E-15 |
| rs643434 | let-7a-5p | trans | 9.18E-13 | Nightingale | LDL_L | NA | A | G | 0.03152 | 0.004332 | 5.40E-13 |
| rs643434 | let-7a-5p | trans | 9.18E-13 | Nightingale | S_LDL_PL_ | NA | A | G | -0.0281 | 0.004339 | 4.70E-12 |
| rs643434 | let-7a-5p | trans | 9.18E-13 | Nightingale | LDL_FC | NA | A | G | 0.031592 | 0.004301 | 6.20E-13 |
| rs643434 | let-7a-5p | trans | 9.18E-13 | Nightingale | ApoB | NA | A | G | 0.032994 | 0.004341 | 3.10E-14 |
| rs643434 | let-7a-5p | trans | 9.18E-13 | Nightingale | L_VLDL_CE | NA | A | G | 0.022834 | 0.004296 | 2.70E-08 |
| rs643434 | let-7a-5p | trans | 9.18E-13 | Nightingale | S_VLDL_CE | NA | A | G | 0.037357 | 0.004341 | 1.60E-18 |
| rs643434 | let-7a-5p | trans | 9.18E-13 | Nightingale | S_HDL_P | NA | A | G | 0.027834 | 0.004348 | 2.60E-10 |
| rs643434 | let-7a-5p | trans | 9.18E-13 | Nightingale | L_LDL_L | NA | A | G | 0.031581 | 0.004305 | 3.00E-13 |
| rs643434 | let-7a-5p | trans | 9.18E-13 | Nightingale | IDL_CE | NA | A | G | 0.032158 | 0.004197 | 3.80E-14 |
| rs643434 | let-7a-5p | trans | 9.18E-13 | Nightingale | L_LDL_CE | NA | A | G | 0.029584 | 0.004308 | 9.10E-12 |
| rs643434 | let-7a-5p | trans | 9.18E-13 | Nightingale | XL_VLDL_C | NA | A | G | 0.034537 | 0.00417 | 1.70E-16 |
| rs643434 | let-7a-5p | trans | 9.18E-13 | Nightingale | XS_VLDL_P | NA | A | G | 0.035851 | 0.004289 | 9.10E-18 |
| rs643434 | let-7a-5p | trans | 9.18E-13 | Nightingale | LDL_P | NA | A | G | 0.030723 | 0.004344 | 2.00E-12 |
| rs643434 | let-7a-5p | trans | 9.18E-13 | Nightingale | L_VLDL_C_ | NA | A | G | 0.049148 | 0.004246 | 1.20E-31 |
| rs643434 | let-7a-5p | trans | 9.18E-13 | Nightingale | M_VLDL_C | NA | A | G | 0.039267 | 0.004308 | 1.20E-19 |
| rs643434 | let-7a-5p | trans | 9.18E-13 | Nightingale | M_LDL_P | NA | A | G | 0.034038 | 0.004359 | 7.10E-15 |
| rs643434 | let-7a-5p | trans | 9.18E-13 | Nightingale | M_LDL_CE | NA | A | G | 0.031258 | 0.004353 | 8.10E-13 |
| rs643434 | let-7a-5p | trans | 9.18E-13 | Nightingale | XXL_VLDL_ | NA | A | G | 0.031806 | 0.00438 | 2.10E-13 |
| rs643434 | let-7a-5p | trans | 9.18E-13 | Nightingale | XXL_VLDL_ | NA | A | G | 0.033426 | 0.004356 | 9.00E-15 |
| rs643434 | let-7a-5p | trans | 9.18E-13 | Nightingale | XL_HDL_PL | NA | A | G | -0.02176 | 0.003994 | 4.90E-08 |
| rs643434 | let-7a-5p | trans | 9.18E-13 | Nightingale | S_LDL_C | NA | A | G | 0.029751 | 0.004351 | 1.50E-11 |
| rs643434 | let-7a-5p | trans | 9.18E-13 | Nightingale | S_VLDL_P | NA | A | G | 0.023452 | 0.004293 | 9.10E-09 |
| rs643434 | let-7a-5p | trans | 9.18E-13 | Nightingale | XS_VLDL_C | NA | A | G | 0.036711 | 0.00422 | 2.10E-18 |
| rs643434 | let-7a-5p | trans | 9.18E-13 | Nightingale | S_HDL_C | NA | A | G | 0.025404 | 0.00434 | 9.10E-09 |
| rs643434 | let-7a-5p | trans | 9.18E-13 | Nightingale | VLDL_FC | NA | A | G | 0.024523 | 0.004315 | 3.30E-09 |
| rs643434 | let-7a-5p | trans | 9.18E-13 | Nightingale | S_VLDL_FC | NA | A | G | 0.027147 | 0.004154 | 1.20E-10 |
| rs643434 | let-7a-5p | trans | 9.18E-13 | Nightingale | S_VLDL_TG | NA | A | G | -0.03596 | 0.004208 | 2.00E-17 |
| rs643434 | let-7a-5p | trans | 9.18E-13 | Nightingale | M_LDL_PL | NA | A | G | 0.031218 | 0.004353 | 9.40E-13 |
| rs643434 | let-7a-5p | trans | 9.18E-13 | Nightingale | L_LDL_P | NA | A | G | 0.028395 | 0.004332 | 7.00E-11 |
| rs643434 | let-7a-5p | trans | 9.18E-13 | Nightingale | ApoB_by_A | NA | A | G | 0.025706 | 0.004281 | 2.00E-09 |
| rs643434 | let-7a-5p | trans | 9.18E-13 | Nightingale | Remnant_( | NA | A | G | 0.035917 | 0.004319 | 4.70E-17 |
| rs643434 | let-7a-5p | trans | 9.18E-13 | Nightingale | XXL_VLDL_ | NA | A | G | 0.029234 | 0.004328 | 5.40E-12 |
| rs643434 | let-7a-5p | trans | 9.18E-13 | Nightingale | LA | NA | A | G | 0.023396 | 0.004263 | 1.30E-08 |
| rs643434 | let-7a-5p | trans | 9.18E-13 | Nightingale | M_VLDL_F | NA | A | G | 0.03934 | 0.004136 | 2.40E-21 |

| | | | | | | | | | | | |
|---|---|---|---|---|---|---|---|---|---|---|---|
| rs643434 | let-7a-5p | trans | 9.18E-13 | Nightingale | M_VLDL_P | NA | A | G | 0.028838 | 0.00434 | 1.20E-11 |
| rs643434 | let-7a-5p | trans | 9.18E-13 | Nightingale | LDL_PL | NA | A | G | 0.033367 | 0.004337 | 2.80E-14 |
| rs643434 | let-7a-5p | trans | 9.18E-13 | Nightingale | IDL_L | NA | A | G | 0.033725 | 0.004203 | 2.10E-15 |
| rs643434 | let-7a-5p | trans | 9.18E-13 | Nightingale | M_VLDL_P | NA | A | G | 0.042071 | 0.004172 | 6.90E-24 |
| rs643434 | let-7a-5p | trans | 9.18E-13 | Nightingale | IDL_C | NA | A | G | 0.033366 | 0.004198 | 4.40E-15 |
| rs643434 | let-7a-5p | trans | 9.18E-13 | Nightingale | Total_L | NA | A | G | 0.025099 | 0.004276 | 1.30E-09 |
| rs643434 | let-7a-5p | trans | 9.18E-13 | Nightingale | M_LDL_FC | NA | A | G | 0.032319 | 0.004342 | 2.20E-13 |
| rs643434 | let-7a-5p | trans | 9.18E-13 | Nightingale | S_VLDL_PL | NA | A | G | 0.032104 | 0.004339 | 4.40E-14 |
| rs643434 | let-7a-5p | trans | 9.18E-13 | Nightingale | XL_VLDL_P | NA | A | G | 0.039162 | 0.004353 | 6.90E-20 |
| rs643434 | let-7a-5p | trans | 9.18E-13 | Nightingale | M_LDL_C_ | NA | A | G | 0.026601 | 0.00435 | 1.60E-09 |
| rs643434 | let-7a-5p | trans | 9.18E-13 | Nightingale | XS_VLDL_F | NA | A | G | 0.035949 | 0.004272 | 6.80E-18 |
| rs643434 | let-7a-5p | trans | 9.18E-13 | Nightingale | S_VLDL_C_ | NA | A | G | 0.039702 | 0.004231 | 9.10E-21 |
| rs643434 | let-7a-5p | trans | 9.18E-13 | Nightingale | L_LDL_C | NA | A | G | 0.030283 | 0.004298 | 2.80E-12 |
| rs643434 | let-7a-5p | trans | 9.18E-13 | Nightingale | XXL_VLDL_ | NA | A | G | 0.023497 | 0.004375 | 5.50E-09 |
| rs643434 | let-7a-5p | trans | 9.18E-13 | Nightingale | S_LDL_P | NA | A | G | 0.029007 | 0.004351 | 2.80E-11 |
| rs643434 | let-7a-5p | trans | 9.18E-13 | Nightingale | S_LDL_L | NA | A | G | 0.026945 | 0.004346 | 1.10E-09 |
| rs643434 | let-7a-5p | trans | 9.18E-13 | Nightingale | M_LDL_C | NA | A | G | 0.032137 | 0.004355 | 2.20E-13 |
| rs643434 | let-7a-5p | trans | 9.18E-13 | Nightingale | LDL_CE | NA | A | G | 0.030677 | 0.004336 | 1.90E-12 |
| rs643434 | let-7a-5p | trans | 9.18E-13 | Nightingale | M_LDL_TG | NA | A | G | -0.02514 | 0.004293 | 8.20E-09 |
| rs643434 | let-7a-5p | trans | 9.18E-13 | Nightingale | XS_VLDL_P | NA | A | G | 0.031727 | 0.004286 | 1.80E-14 |
| rs643434 | let-7a-5p | trans | 9.18E-13 | Nightingale | L_LDL_FC | NA | A | G | 0.031521 | 0.004269 | 3.80E-13 |
| rs643434 | let-7a-5p | trans | 9.18E-13 | Nightingale | VLDL_C | NA | A | G | 0.031139 | 0.004341 | 1.90E-13 |
| rs643434 | let-7a-5p | trans | 9.18E-13 | Nightingale | XS_VLDL_C | NA | A | G | 0.036463 | 0.004195 | 3.70E-18 |
| rs643434 | let-7a-5p | trans | 9.18E-13 | Nightingale | S_LDL_FC | NA | A | G | 0.027234 | 0.00434 | 1.30E-09 |
| rs643434 | let-7a-5p | trans | 9.18E-13 | Nightingale | S_LDL_C_p | NA | A | G | 0.041888 | 0.004364 | 5.30E-22 |
| rs643434 | let-7a-5p | trans | 9.18E-13 | Nightingale | IDL_P | NA | A | G | 0.039091 | 0.004301 | 1.70E-19 |
| rs643434 | let-7a-5p | trans | 9.18E-13 | Nightingale | Total_CE | NA | A | G | 0.029456 | 0.004193 | 1.60E-12 |
| rs643434 | let-7a-5p | trans | 9.18E-13 | Nightingale | XS_VLDL_F | NA | A | G | 0.029136 | 0.004316 | 3.10E-11 |
| rs643434 | let-7a-5p | trans | 9.18E-13 | Nightingale | S_VLDL_L | NA | A | G | 0.024562 | 0.004305 | 2.30E-09 |
| rs643434 | let-7a-5p | trans | 9.18E-13 | Nightingale | S_LDL_CE_ | NA | A | G | 0.0289 | 0.00434 | 1.00E-12 |
| rs643434 | let-7a-5p | trans | 9.18E-13 | Nightingale | XL_VLDL_C | NA | A | G | 0.035551 | 0.004168 | 1.90E-17 |
| rs643434 | let-7a-5p | trans | 9.18E-13 | Nightingale | S_LDL_CE | NA | A | G | 0.0295 | 0.004349 | 1.40E-11 |
| rs643434 | let-7a-5p | trans | 9.18E-13 | Nightingale | Total_PL | NA | A | G | 0.023087 | 0.004139 | 6.80E-09 |
| rs643434 | let-7a-5p | trans | 9.18E-13 | Nightingale | Citrate | NA | A | G | 0.024643 | 0.004319 | 6.40E-09 |
| rs643434 | let-7a-5p | trans | 9.18E-13 | Nightingale | VLDL_P | NA | A | G | 0.026629 | 0.004324 | 1.80E-10 |
| rs643434 | let-7a-5p | trans | 9.18E-13 | Nightingale | S_HDL_L | NA | A | G | 0.024009 | 0.004332 | 2.60E-08 |
| rs643434 | let-7a-5p | trans | 9.18E-13 | Nightingale | IDL_FC | NA | A | G | 0.036015 | 0.004208 | 3.00E-17 |
| rs643434 | let-7a-5p | trans | 9.18E-13 | Nightingale | M_VLDL_T | NA | A | G | -0.0362 | 0.004108 | 1.70E-18 |
| rs643434 | let-7a-5p | trans | 9.18E-13 | Nightingale | M_LDL_L | NA | A | G | 0.031433 | 0.004353 | 6.60E-13 |
| rs643434 | let-7a-5p | trans | 9.18E-13 | Nightingale | Total_C | NA | A | G | 0.030215 | 0.004208 | 4.40E-13 |
| rs643434 | let-7a-5p | trans | 9.18E-13 | Nightingale | S_VLDL_FC | NA | A | G | 0.036708 | 0.004348 | 1.60E-17 |
| rs643434 | let-7a-5p | trans | 9.18E-13 | Nightingale | L_VLDL_FC | NA | A | G | 0.046831 | 0.004305 | 5.60E-28 |
| rs643434 | let-7a-5p | trans | 9.18E-13 | Nightingale | L_VLDL_PL | NA | A | G | 0.021642 | 0.004222 | 4.60E-08 |
| rs643434 | let-7a-5p | trans | 9.18E-13 | Nightingale | Clinical_LD | NA | A | G | 0.03461 | 0.004322 | 2.50E-15 |
| rs643434 | let-7a-5p | trans | 9.18E-13 | Nightingale | M_VLDL_P | NA | A | G | 0.028009 | 0.004336 | 4.20E-11 |
| rs643434 | let-7a-5p | trans | 9.18E-13 | Nightingale | Total_FC | NA | A | G | 0.031463 | 0.004247 | 4.00E-14 |
| rs643434 | let-7a-5p | trans | 9.18E-13 | Nightingale | S_VLDL_CE | NA | A | G | 0.045098 | 0.004289 | 1.70E-25 |
| rs643434 | let-7a-5p | trans | 9.18E-13 | Nightingale | LDL_C | NA | A | G | 0.031242 | 0.004328 | 8.80E-13 |
| rs643434 | let-7a-5p | trans | 9.18E-13 | Nightingale | L_LDL_PL | NA | A | G | 0.03597 | 0.004314 | 1.20E-16 |
| rs643434 | let-7a-5p | trans | 9.18E-13 | Nightingale | S_VLDL_C | NA | A | G | 0.037408 | 0.004347 | 2.50E-18 |
| rs643434 | let-7a-5p | trans | 9.18E-13 | Nightingale | S_VLDL_PL | NA | A | G | 0.025727 | 0.004162 | 1.50E-09 |
| rs643434 | let-7a-5p | trans | 9.18E-13 | Nightingale | XL_VLDL_T | NA | A | G | -0.0415 | 0.004213 | 5.60E-23 |
| rs643434 | let-7a-5p | trans | 9.18E-13 | Nightingale | L_VLDL_TG | NA | A | G | -0.04918 | 0.004326 | 7.90E-31 |
| rs643434 | let-7a-5p | trans | 9.18E-13 | Nightingale | VLDL_CE | NA | A | G | 0.03522 | 0.004348 | 1.90E-16 |
| rs643434 | let-7a-5p | trans | 9.18E-13 | Nightingale | M_VLDL_C | NA | A | G | 0.03381 | 0.004095 | 2.90E-16 |
| rs643434 | let-7a-5p | trans | 9.18E-13 | Nightingale | XL_VLDL_F | NA | A | G | 0.035154 | 0.004214 | 2.00E-17 |
| rs643434 | let-7a-5p | trans | 9.18E-13 | Nightingale | S_HDL_CE | NA | A | G | 0.025707 | 0.004354 | 6.70E-09 |
| rs643434 | let-7c-5p | trans | 3.20E-23 | Nightingale | XXL_VLDL_ | NA | A | G | -0.03445 | 0.004384 | 6.10E-16 |
| rs643434 | let-7c-5p | trans | 3.20E-23 | Nightingale | Sphingomy | NA | A | G | 0.025961 | 0.004064 | 3.00E-10 |
| rs643434 | let-7c-5p | trans | 3.20E-23 | Nightingale | Omega_6 | NA | A | G | 0.024274 | 0.004245 | 2.60E-09 |
| rs643434 | let-7c-5p | trans | 3.20E-23 | Nightingale | M_VLDL_C | NA | A | G | 0.031313 | 0.004084 | 3.50E-14 |
| rs643434 | let-7c-5p | trans | 3.20E-23 | Nightingale | M_VLDL_C | NA | A | G | 0.037416 | 0.004335 | 4.70E-18 |
| rs643434 | let-7c-5p | trans | 3.20E-23 | Nightingale | GlycA | NA | A | G | 0.025332 | 0.004328 | 5.00E-09 |

| | | | | | | | | | | | |
|---|---|---|---|---|---|---|---|---|---|---|---|
| rs643434 | let-7c-5p | trans | 3.20E-23 | Nightingale | L_VLDL_CE | NA | A | G | 0.044018 | 0.00416 | 9.50E-27 |
| rs643434 | let-7c-5p | trans | 3.20E-23 | Nightingale | IDL_PL | NA | A | G | 0.033641 | 0.004195 | 1.80E-15 |
| rs643434 | let-7c-5p | trans | 3.20E-23 | Nightingale | M_VLDL_F | NA | A | G | 0.032595 | 0.004346 | 3.50E-14 |
| rs643434 | let-7c-5p | trans | 3.20E-23 | Nightingale | non_HDL_( | NA | A | G | 0.034208 | 0.004323 | 2.20E-15 |
| rs643434 | let-7c-5p | trans | 3.20E-23 | Nightingale | XS_VLDL_L | NA | A | G | 0.033078 | 0.004273 | 1.20E-15 |
| rs643434 | let-7c-5p | trans | 3.20E-23 | Nightingale | LDL_L | NA | A | G | 0.03152 | 0.004332 | 5.40E-13 |
| rs643434 | let-7c-5p | trans | 3.20E-23 | Nightingale | S_LDL_PL_ | NA | A | G | -0.0281 | 0.004339 | 4.70E-12 |
| rs643434 | let-7c-5p | trans | 3.20E-23 | Nightingale | LDL_FC | NA | A | G | 0.031592 | 0.004301 | 6.20E-13 |
| rs643434 | let-7c-5p | trans | 3.20E-23 | Nightingale | ApoB | NA | A | G | 0.032994 | 0.004341 | 3.10E-14 |
| rs643434 | let-7c-5p | trans | 3.20E-23 | Nightingale | L_VLDL_CE | NA | A | G | 0.022834 | 0.004296 | 2.70E-08 |
| rs643434 | let-7c-5p | trans | 3.20E-23 | Nightingale | S_VLDL_CE | NA | A | G | 0.037357 | 0.004341 | 1.60E-18 |
| rs643434 | let-7c-5p | trans | 3.20E-23 | Nightingale | S_HDL_P | NA | A | G | 0.027834 | 0.004348 | 2.60E-10 |
| rs643434 | let-7c-5p | trans | 3.20E-23 | Nightingale | L_LDL_L | NA | A | G | 0.031581 | 0.004305 | 3.00E-13 |
| rs643434 | let-7c-5p | trans | 3.20E-23 | Nightingale | IDL_CE | NA | A | G | 0.032158 | 0.004197 | 3.80E-14 |
| rs643434 | let-7c-5p | trans | 3.20E-23 | Nightingale | L_LDL_CE | NA | A | G | 0.029584 | 0.004308 | 9.10E-12 |
| rs643434 | let-7c-5p | trans | 3.20E-23 | Nightingale | XL_VLDL_C | NA | A | G | 0.034537 | 0.00417 | 1.70E-16 |
| rs643434 | let-7c-5p | trans | 3.20E-23 | Nightingale | XS_VLDL_P | NA | A | G | 0.035851 | 0.004289 | 9.10E-18 |
| rs643434 | let-7c-5p | trans | 3.20E-23 | Nightingale | LDL_P | NA | A | G | 0.030723 | 0.004344 | 2.00E-12 |
| rs643434 | let-7c-5p | trans | 3.20E-23 | Nightingale | L_VLDL_C_ | NA | A | G | 0.049148 | 0.004246 | 1.20E-31 |
| rs643434 | let-7c-5p | trans | 3.20E-23 | Nightingale | M_VLDL_C | NA | A | G | 0.039267 | 0.004308 | 1.20E-19 |
| rs643434 | let-7c-5p | trans | 3.20E-23 | Nightingale | M_LDL_P | NA | A | G | 0.034038 | 0.004359 | 7.10E-15 |
| rs643434 | let-7c-5p | trans | 3.20E-23 | Nightingale | M_LDL_CE | NA | A | G | 0.031258 | 0.004353 | 8.10E-13 |
| rs643434 | let-7c-5p | trans | 3.20E-23 | Nightingale | XXL_VLDL_ | NA | A | G | 0.031806 | 0.00438 | 2.10E-13 |
| rs643434 | let-7c-5p | trans | 3.20E-23 | Nightingale | XXL_VLDL_ | NA | A | G | 0.033426 | 0.004356 | 9.00E-15 |
| rs643434 | let-7c-5p | trans | 3.20E-23 | Nightingale | XL_HDL_PL | NA | A | G | -0.02176 | 0.003994 | 4.90E-08 |
| rs643434 | let-7c-5p | trans | 3.20E-23 | Nightingale | S_LDL_C | NA | A | G | 0.029751 | 0.004351 | 1.50E-11 |
| rs643434 | let-7c-5p | trans | 3.20E-23 | Nightingale | S_VLDL_P | NA | A | G | 0.023452 | 0.004293 | 9.10E-09 |
| rs643434 | let-7c-5p | trans | 3.20E-23 | Nightingale | XS_VLDL_C | NA | A | G | 0.036711 | 0.00422 | 2.10E-18 |
| rs643434 | let-7c-5p | trans | 3.20E-23 | Nightingale | S_HDL_C | NA | A | G | 0.025404 | 0.00434 | 9.10E-09 |
| rs643434 | let-7c-5p | trans | 3.20E-23 | Nightingale | VLDL_FC | NA | A | G | 0.024523 | 0.004315 | 3.30E-09 |
| rs643434 | let-7c-5p | trans | 3.20E-23 | Nightingale | S_VLDL_FC | NA | A | G | 0.027147 | 0.004154 | 1.20E-10 |
| rs643434 | let-7c-5p | trans | 3.20E-23 | Nightingale | S_VLDL_TG | NA | A | G | -0.03596 | 0.004208 | 2.00E-17 |
| rs643434 | let-7c-5p | trans | 3.20E-23 | Nightingale | M_LDL_PL | NA | A | G | 0.031218 | 0.004353 | 9.40E-13 |
| rs643434 | let-7c-5p | trans | 3.20E-23 | Nightingale | L_LDL_P | NA | A | G | 0.028395 | 0.004332 | 7.00E-11 |
| rs643434 | let-7c-5p | trans | 3.20E-23 | Nightingale | ApoB_by_A | NA | A | G | 0.025706 | 0.004281 | 2.00E-09 |
| rs643434 | let-7c-5p | trans | 3.20E-23 | Nightingale | Remnant_( | NA | A | G | 0.035917 | 0.004319 | 4.70E-17 |
| rs643434 | let-7c-5p | trans | 3.20E-23 | Nightingale | XXL_VLDL_ | NA | A | G | 0.029234 | 0.004328 | 5.40E-12 |
| rs643434 | let-7c-5p | trans | 3.20E-23 | Nightingale | LA | NA | A | G | 0.023396 | 0.004263 | 1.30E-08 |
| rs643434 | let-7c-5p | trans | 3.20E-23 | Nightingale | M_VLDL_F | NA | A | G | 0.03934 | 0.004136 | 2.40E-21 |
| rs643434 | let-7c-5p | trans | 3.20E-23 | Nightingale | M_VLDL_P | NA | A | G | 0.028838 | 0.00434 | 1.20E-11 |
| rs643434 | let-7c-5p | trans | 3.20E-23 | Nightingale | LDL_PL | NA | A | G | 0.033367 | 0.004337 | 2.80E-14 |
| rs643434 | let-7c-5p | trans | 3.20E-23 | Nightingale | IDL_L | NA | A | G | 0.033725 | 0.004203 | 2.10E-15 |
| rs643434 | let-7c-5p | trans | 3.20E-23 | Nightingale | M_VLDL_P | NA | A | G | 0.042071 | 0.004172 | 6.90E-24 |
| rs643434 | let-7c-5p | trans | 3.20E-23 | Nightingale | IDL_C | NA | A | G | 0.033366 | 0.004198 | 4.40E-15 |
| rs643434 | let-7c-5p | trans | 3.20E-23 | Nightingale | Total_L | NA | A | G | 0.025099 | 0.004276 | 1.30E-09 |
| rs643434 | let-7c-5p | trans | 3.20E-23 | Nightingale | M_LDL_FC | NA | A | G | 0.032319 | 0.004342 | 2.20E-13 |
| rs643434 | let-7c-5p | trans | 3.20E-23 | Nightingale | S_VLDL_PL | NA | A | G | 0.032104 | 0.004339 | 4.40E-14 |
| rs643434 | let-7c-5p | trans | 3.20E-23 | Nightingale | XL_VLDL_P | NA | A | G | 0.039162 | 0.004353 | 6.90E-20 |
| rs643434 | let-7c-5p | trans | 3.20E-23 | Nightingale | M_LDL_C_ | NA | A | G | 0.026601 | 0.00435 | 1.60E-09 |
| rs643434 | let-7c-5p | trans | 3.20E-23 | Nightingale | XS_VLDL_F | NA | A | G | 0.035949 | 0.004272 | 6.80E-18 |
| rs643434 | let-7c-5p | trans | 3.20E-23 | Nightingale | S_VLDL_C_ | NA | A | G | 0.039702 | 0.004231 | 9.10E-21 |
| rs643434 | let-7c-5p | trans | 3.20E-23 | Nightingale | L_LDL_C | NA | A | G | 0.030283 | 0.004298 | 2.80E-12 |
| rs643434 | let-7c-5p | trans | 3.20E-23 | Nightingale | XXL_VLDL_ | NA | A | G | 0.023497 | 0.004375 | 5.50E-09 |
| rs643434 | let-7c-5p | trans | 3.20E-23 | Nightingale | S_LDL_P | NA | A | G | 0.029007 | 0.004351 | 2.80E-11 |
| rs643434 | let-7c-5p | trans | 3.20E-23 | Nightingale | S_LDL_L | NA | A | G | 0.026945 | 0.004346 | 1.10E-09 |
| rs643434 | let-7c-5p | trans | 3.20E-23 | Nightingale | M_LDL_C | NA | A | G | 0.032137 | 0.004355 | 2.20E-13 |
| rs643434 | let-7c-5p | trans | 3.20E-23 | Nightingale | LDL_CE | NA | A | G | 0.030677 | 0.004336 | 1.90E-12 |
| rs643434 | let-7c-5p | trans | 3.20E-23 | Nightingale | M_LDL_TG | NA | A | G | -0.02514 | 0.004293 | 8.20E-09 |
| rs643434 | let-7c-5p | trans | 3.20E-23 | Nightingale | XS_VLDL_P | NA | A | G | 0.031727 | 0.004286 | 1.80E-14 |
| rs643434 | let-7c-5p | trans | 3.20E-23 | Nightingale | L_LDL_FC | NA | A | G | 0.031521 | 0.004269 | 3.80E-13 |
| rs643434 | let-7c-5p | trans | 3.20E-23 | Nightingale | VLDL_C | NA | A | G | 0.031139 | 0.004341 | 1.90E-13 |
| rs643434 | let-7c-5p | trans | 3.20E-23 | Nightingale | XS_VLDL_C | NA | A | G | 0.036463 | 0.004195 | 3.70E-18 |
| rs643434 | let-7c-5p | trans | 3.20E-23 | Nightingale | S_LDL_FC | NA | A | G | 0.027234 | 0.00434 | 1.30E-09 |

| | | | | | | | | | | | |
|---|---|---|---|---|---|---|---|---|---|---|---|
| rs643434 | let-7c-5p | trans | 3.20E-23 | Nightingale | S_LDL_C_p | NA | A | G | 0.041888 | 0.004364 | 5.30E-22 |
| rs643434 | let-7c-5p | trans | 3.20E-23 | Nightingale | IDL_P | NA | A | G | 0.039091 | 0.004301 | 1.70E-19 |
| rs643434 | let-7c-5p | trans | 3.20E-23 | Nightingale | Total_CE | NA | A | G | 0.029456 | 0.004193 | 1.60E-12 |
| rs643434 | let-7c-5p | trans | 3.20E-23 | Nightingale | XS_VLDL_F | NA | A | G | 0.029136 | 0.004316 | 3.10E-11 |
| rs643434 | let-7c-5p | trans | 3.20E-23 | Nightingale | S_VLDL_L | NA | A | G | 0.024562 | 0.004305 | 2.30E-09 |
| rs643434 | let-7c-5p | trans | 3.20E-23 | Nightingale | S_LDL_CE_ | NA | A | G | 0.0289 | 0.00434 | 1.00E-12 |
| rs643434 | let-7c-5p | trans | 3.20E-23 | Nightingale | XL_VLDL_C | NA | A | G | 0.035551 | 0.004168 | 1.90E-17 |
| rs643434 | let-7c-5p | trans | 3.20E-23 | Nightingale | S_LDL_CE | NA | A | G | 0.0295 | 0.004349 | 1.40E-11 |
| rs643434 | let-7c-5p | trans | 3.20E-23 | Nightingale | Total_PL | NA | A | G | 0.023087 | 0.004139 | 6.80E-09 |
| rs643434 | let-7c-5p | trans | 3.20E-23 | Nightingale | Citrate | NA | A | G | 0.024643 | 0.004319 | 6.40E-09 |
| rs643434 | let-7c-5p | trans | 3.20E-23 | Nightingale | VLDL_P | NA | A | G | 0.026629 | 0.004324 | 1.80E-10 |
| rs643434 | let-7c-5p | trans | 3.20E-23 | Nightingale | S_HDL_L | NA | A | G | 0.024009 | 0.004332 | 2.60E-08 |
| rs643434 | let-7c-5p | trans | 3.20E-23 | Nightingale | IDL_FC | NA | A | G | 0.036015 | 0.004208 | 3.00E-17 |
| rs643434 | let-7c-5p | trans | 3.20E-23 | Nightingale | M_VLDL_T | NA | A | G | -0.0362 | 0.004108 | 1.70E-18 |
| rs643434 | let-7c-5p | trans | 3.20E-23 | Nightingale | M_LDL_L | NA | A | G | 0.031433 | 0.004353 | 6.60E-13 |
| rs643434 | let-7c-5p | trans | 3.20E-23 | Nightingale | Total_C | NA | A | G | 0.030215 | 0.004208 | 4.40E-13 |
| rs643434 | let-7c-5p | trans | 3.20E-23 | Nightingale | S_VLDL_FC | NA | A | G | 0.036708 | 0.004348 | 1.60E-17 |
| rs643434 | let-7c-5p | trans | 3.20E-23 | Nightingale | L_VLDL_FC | NA | A | G | 0.046831 | 0.004305 | 5.60E-28 |
| rs643434 | let-7c-5p | trans | 3.20E-23 | Nightingale | L_VLDL_PL | NA | A | G | 0.021642 | 0.004222 | 4.60E-08 |
| rs643434 | let-7c-5p | trans | 3.20E-23 | Nightingale | Clinical_LD | NA | A | G | 0.03461 | 0.004322 | 2.50E-15 |
| rs643434 | let-7c-5p | trans | 3.20E-23 | Nightingale | M_VLDL_P | NA | A | G | 0.028009 | 0.004336 | 4.20E-11 |
| rs643434 | let-7c-5p | trans | 3.20E-23 | Nightingale | Total_FC | NA | A | G | 0.031463 | 0.004247 | 4.00E-14 |
| rs643434 | let-7c-5p | trans | 3.20E-23 | Nightingale | S_VLDL_CE | NA | A | G | 0.045098 | 0.004289 | 1.70E-25 |
| rs643434 | let-7c-5p | trans | 3.20E-23 | Nightingale | LDL_C | NA | A | G | 0.031242 | 0.004328 | 8.80E-13 |
| rs643434 | let-7c-5p | trans | 3.20E-23 | Nightingale | L_LDL_PL | NA | A | G | 0.03597 | 0.004314 | 1.20E-16 |
| rs643434 | let-7c-5p | trans | 3.20E-23 | Nightingale | S_VLDL_C | NA | A | G | 0.037408 | 0.004347 | 2.50E-18 |
| rs643434 | let-7c-5p | trans | 3.20E-23 | Nightingale | S_VLDL_PL | NA | A | G | 0.025727 | 0.004162 | 1.50E-09 |
| rs643434 | let-7c-5p | trans | 3.20E-23 | Nightingale | XL_VLDL_T | NA | A | G | -0.0415 | 0.004213 | 5.60E-23 |
| rs643434 | let-7c-5p | trans | 3.20E-23 | Nightingale | L_VLDL_TG | NA | A | G | -0.04918 | 0.004326 | 7.90E-31 |
| rs643434 | let-7c-5p | trans | 3.20E-23 | Nightingale | VLDL_CE | NA | A | G | 0.03522 | 0.004348 | 1.90E-16 |
| rs643434 | let-7c-5p | trans | 3.20E-23 | Nightingale | M_VLDL_C | NA | A | G | 0.03381 | 0.004095 | 2.90E-16 |
| rs643434 | let-7c-5p | trans | 3.20E-23 | Nightingale | XL_VLDL_F | NA | A | G | 0.035154 | 0.004214 | 2.00E-17 |
| rs643434 | let-7c-5p | trans | 3.20E-23 | Nightingale | S_HDL_CE | NA | A | G | 0.025707 | 0.004354 | 6.70E-09 |
| rs644234 | miR-10b-5p | trans | 1.46E-34 | Nightingale | S_LDL_L | NA | G | T | 0.026888 | 0.004346 | 1.20E-09 |
| rs644234 | miR-10b-5p | trans | 1.46E-34 | Nightingale | LDL_P | NA | G | T | 0.030662 | 0.004344 | 2.10E-12 |
| rs644234 | miR-10b-5p | trans | 1.46E-34 | Nightingale | M_VLDL_C | NA | G | T | 0.039209 | 0.004307 | 1.30E-19 |
| rs644234 | miR-10b-5p | trans | 1.46E-34 | Nightingale | non_HDL_( | NA | G | T | 0.03414 | 0.004323 | 2.40E-15 |
| rs644234 | miR-10b-5p | trans | 1.46E-34 | Nightingale | XXL_VLDL_ | NA | G | T | -0.03443 | 0.004384 | 6.20E-16 |
| rs644234 | miR-10b-5p | trans | 1.46E-34 | Nightingale | S_VLDL_C_ | NA | G | T | 0.039745 | 0.00423 | 7.90E-21 |
| rs644234 | miR-10b-5p | trans | 1.46E-34 | Nightingale | M_VLDL_C | NA | G | T | 0.031362 | 0.004084 | 3.10E-14 |
| rs644234 | miR-10b-5p | trans | 1.46E-34 | Nightingale | S_VLDL_L | NA | G | T | 0.024494 | 0.004305 | 2.50E-09 |
| rs644234 | miR-10b-5p | trans | 1.46E-34 | Nightingale | L_LDL_FC | NA | G | T | 0.031476 | 0.004268 | 4.00E-13 |
| rs644234 | miR-10b-5p | trans | 1.46E-34 | Nightingale | L_LDL_L | NA | G | T | 0.031526 | 0.004304 | 3.20E-13 |
| rs644234 | miR-10b-5p | trans | 1.46E-34 | Nightingale | L_VLDL_C_ | NA | G | T | 0.049243 | 0.004245 | 8.50E-32 |
| rs644234 | miR-10b-5p | trans | 1.46E-34 | Nightingale | M_LDL_C | NA | G | T | 0.032088 | 0.004355 | 2.30E-13 |
| rs644234 | miR-10b-5p | trans | 1.46E-34 | Nightingale | S_VLDL_PL | NA | G | T | 0.025736 | 0.004162 | 1.40E-09 |
| rs644234 | miR-10b-5p | trans | 1.46E-34 | Nightingale | XS_VLDL_F | NA | G | T | 0.029149 | 0.004316 | 3.00E-11 |
| rs644234 | miR-10b-5p | trans | 1.46E-34 | Nightingale | Clinical_LD | NA | G | T | 0.034557 | 0.004322 | 2.70E-15 |
| rs644234 | miR-10b-5p | trans | 1.46E-34 | Nightingale | S_LDL_PL_ | NA | G | T | -0.02807 | 0.004339 | 5.00E-12 |
| rs644234 | miR-10b-5p | trans | 1.46E-34 | Nightingale | S_HDL_C | NA | G | T | 0.025378 | 0.004339 | 9.40E-09 |
| rs644234 | miR-10b-5p | trans | 1.46E-34 | Nightingale | XL_VLDL_F | NA | G | T | 0.03523 | 0.004214 | 1.70E-17 |
| rs644234 | miR-10b-5p | trans | 1.46E-34 | Nightingale | L_LDL_C | NA | G | T | 0.030226 | 0.004298 | 3.00E-12 |
| rs644234 | miR-10b-5p | trans | 1.46E-34 | Nightingale | IDL_C | NA | G | T | 0.033306 | 0.004198 | 4.80E-15 |
| rs644234 | miR-10b-5p | trans | 1.46E-34 | Nightingale | S_LDL_CE_ | NA | G | T | 0.02885 | 0.004339 | 1.10E-12 |
| rs644234 | miR-10b-5p | trans | 1.46E-34 | Nightingale | L_VLDL_CE | NA | G | T | 0.022755 | 0.004296 | 2.90E-08 |
| rs644234 | miR-10b-5p | trans | 1.46E-34 | Nightingale | S_VLDL_C | NA | G | T | 0.037356 | 0.004346 | 2.60E-18 |
| rs644234 | miR-10b-5p | trans | 1.46E-34 | Nightingale | S_VLDL_PL | NA | G | T | 0.032035 | 0.004339 | 4.90E-14 |
| rs644234 | miR-10b-5p | trans | 1.46E-34 | Nightingale | Total_C | NA | G | T | 0.030145 | 0.004208 | 4.90E-13 |
| rs644234 | miR-10b-5p | trans | 1.46E-34 | Nightingale | M_VLDL_F | NA | G | T | 0.032516 | 0.004346 | 3.90E-14 |
| rs644234 | miR-10b-5p | trans | 1.46E-34 | Nightingale | IDL_PL | NA | G | T | 0.033582 | 0.004195 | 1.90E-15 |
| rs644234 | miR-10b-5p | trans | 1.46E-34 | Nightingale | S_LDL_CE | NA | G | T | 0.029439 | 0.004349 | 1.50E-11 |
| rs644234 | miR-10b-5p | trans | 1.46E-34 | Nightingale | M_LDL_CE | NA | G | T | 0.031208 | 0.004353 | 8.60E-13 |
| rs644234 | miR-10b-5p | trans | 1.46E-34 | Nightingale | S_LDL_C | NA | G | T | 0.029694 | 0.004351 | 1.60E-11 |

| | | | | | | | | | | | |
|---|---|---|---|---|---|---|---|---|---|---|---|
| rs644234 | miR-10b-5p | trans | 1.46E-34 | Nightingale | XXL_VLDL_ | NA | G | T | 0.031794 | 0.00438 | 2.10E-13 |
| rs644234 | miR-10b-5p | trans | 1.46E-34 | Nightingale | LDL_L | NA | G | T | 0.031466 | 0.004332 | 5.80E-13 |
| rs644234 | miR-10b-5p | trans | 1.46E-34 | Nightingale | XL_VLDL_P | NA | G | T | 0.039123 | 0.004353 | 7.10E-20 |
| rs644234 | miR-10b-5p | trans | 1.46E-34 | Nightingale | S_VLDL_FC | NA | G | T | 0.027162 | 0.004154 | 1.20E-10 |
| rs644234 | miR-10b-5p | trans | 1.46E-34 | Nightingale | L_VLDL_FC | NA | G | T | 0.046949 | 0.004305 | 3.90E-28 |
| rs644234 | miR-10b-5p | trans | 1.46E-34 | Nightingale | IDL_L | NA | G | T | 0.033664 | 0.004203 | 2.30E-15 |
| rs644234 | miR-10b-5p | trans | 1.46E-34 | Nightingale | XL_VLDL_C | NA | G | T | 0.03459 | 0.00417 | 1.50E-16 |
| rs644234 | miR-10b-5p | trans | 1.46E-34 | Nightingale | M_VLDL_C | NA | G | T | 0.033859 | 0.004094 | 2.60E-16 |
| rs644234 | miR-10b-5p | trans | 1.46E-34 | Nightingale | S_HDL_CE | NA | G | T | 0.025685 | 0.004354 | 7.00E-09 |
| rs644234 | miR-10b-5p | trans | 1.46E-34 | Nightingale | M_VLDL_P | NA | G | T | 0.027925 | 0.004336 | 4.70E-11 |
| rs644234 | miR-10b-5p | trans | 1.46E-34 | Nightingale | S_VLDL_CE | NA | G | T | 0.045161 | 0.004288 | 1.40E-25 |
| rs644234 | miR-10b-5p | trans | 1.46E-34 | Nightingale | L_VLDL_CE | NA | G | T | 0.044093 | 0.00416 | 7.40E-27 |
| rs644234 | miR-10b-5p | trans | 1.46E-34 | Nightingale | VLDL_C | NA | G | T | 0.031063 | 0.004341 | 2.10E-13 |
| rs644234 | miR-10b-5p | trans | 1.46E-34 | Nightingale | S_LDL_P | NA | G | T | 0.028954 | 0.004351 | 3.00E-11 |
| rs644234 | miR-10b-5p | trans | 1.46E-34 | Nightingale | LDL_CE | NA | G | T | 0.030617 | 0.004335 | 2.10E-12 |
| rs644234 | miR-10b-5p | trans | 1.46E-34 | Nightingale | L_LDL_PL | NA | G | T | 0.03592 | 0.004314 | 1.30E-16 |
| rs644234 | miR-10b-5p | trans | 1.46E-34 | Nightingale | Omega_6 | NA | G | T | 0.024169 | 0.004245 | 3.10E-09 |
| rs644234 | miR-10b-5p | trans | 1.46E-34 | Nightingale | XXL_VLDL_ | NA | G | T | 0.029285 | 0.004328 | 5.00E-12 |
| rs644234 | miR-10b-5p | trans | 1.46E-34 | Nightingale | XS_VLDL_P | NA | G | T | 0.035793 | 0.004289 | 9.90E-18 |
| rs644234 | miR-10b-5p | trans | 1.46E-34 | Nightingale | LDL_FC | NA | G | T | 0.031549 | 0.0043 | 6.50E-13 |
| rs644234 | miR-10b-5p | trans | 1.46E-34 | Nightingale | IDL_P | NA | G | T | 0.039024 | 0.004301 | 1.90E-19 |
| rs644234 | miR-10b-5p | trans | 1.46E-34 | Nightingale | M_LDL_PL | NA | G | T | 0.031167 | 0.004353 | 1.00E-12 |
| rs644234 | miR-10b-5p | trans | 1.46E-34 | Nightingale | ApoB_by_A | NA | G | T | 0.025671 | 0.004281 | 2.10E-09 |
| rs644234 | miR-10b-5p | trans | 1.46E-34 | Nightingale | Sphingomy | NA | G | T | 0.025922 | 0.004063 | 3.20E-10 |
| rs644234 | miR-10b-5p | trans | 1.46E-34 | Nightingale | XL_VLDL_C | NA | G | T | 0.035608 | 0.004168 | 1.70E-17 |
| rs644234 | miR-10b-5p | trans | 1.46E-34 | Nightingale | M_LDL_FC | NA | G | T | 0.032279 | 0.004341 | 2.40E-13 |
| rs644234 | miR-10b-5p | trans | 1.46E-34 | Nightingale | VLDL_P | NA | G | T | 0.026555 | 0.004324 | 2.00E-10 |
| rs644234 | miR-10b-5p | trans | 1.46E-34 | Nightingale | Total_PL | NA | G | T | 0.023006 | 0.004139 | 7.70E-09 |
| rs644234 | miR-10b-5p | trans | 1.46E-34 | Nightingale | S_VLDL_CE | NA | G | T | 0.037313 | 0.004341 | 1.70E-18 |
| rs644234 | miR-10b-5p | trans | 1.46E-34 | Nightingale | M_VLDL_C | NA | G | T | 0.037346 | 0.004335 | 5.30E-18 |
| rs644234 | miR-10b-5p | trans | 1.46E-34 | Nightingale | LDL_C | NA | G | T | 0.031186 | 0.004328 | 9.50E-13 |
| rs644234 | miR-10b-5p | trans | 1.46E-34 | Nightingale | XS_VLDL_L | NA | G | T | 0.033016 | 0.004273 | 1.30E-15 |
| rs644234 | miR-10b-5p | trans | 1.46E-34 | Nightingale | XS_VLDL_P | NA | G | T | 0.031667 | 0.004286 | 1.90E-14 |
| rs644234 | miR-10b-5p | trans | 1.46E-34 | Nightingale | S_LDL_FC | NA | G | T | 0.027196 | 0.00434 | 1.30E-09 |
| rs644234 | miR-10b-5p | trans | 1.46E-34 | Nightingale | GlycA | NA | G | T | 0.025325 | 0.004328 | 4.90E-09 |
| rs644234 | miR-10b-5p | trans | 1.46E-34 | Nightingale | M_VLDL_F | NA | G | T | 0.039386 | 0.004135 | 2.10E-21 |
| rs644234 | miR-10b-5p | trans | 1.46E-34 | Nightingale | Total_L | NA | G | T | 0.025011 | 0.004276 | 1.50E-09 |
| rs644234 | miR-10b-5p | trans | 1.46E-34 | Nightingale | XS_VLDL_C | NA | G | T | 0.036401 | 0.004195 | 4.00E-18 |
| rs644234 | miR-10b-5p | trans | 1.46E-34 | Nightingale | LA | NA | G | T | 0.023278 | 0.004263 | 1.60E-08 |
| rs644234 | miR-10b-5p | trans | 1.46E-34 | Nightingale | Total_FC | NA | G | T | 0.031385 | 0.004246 | 4.60E-14 |
| rs644234 | miR-10b-5p | trans | 1.46E-34 | Nightingale | XXL_VLDL_ | NA | G | T | 0.023513 | 0.004375 | 5.40E-09 |
| rs644234 | miR-10b-5p | trans | 1.46E-34 | Nightingale | VLDL_FC | NA | G | T | 0.024444 | 0.004315 | 3.60E-09 |
| rs644234 | miR-10b-5p | trans | 1.46E-34 | Nightingale | L_VLDL_TG | NA | G | T | -0.04925 | 0.004326 | 6.10E-31 |
| rs644234 | miR-10b-5p | trans | 1.46E-34 | Nightingale | S_VLDL_P | NA | G | T | 0.023388 | 0.004292 | 9.80E-09 |
| rs644234 | miR-10b-5p | trans | 1.46E-34 | Nightingale | Total_CE | NA | G | T | 0.029391 | 0.004193 | 1.80E-12 |
| rs644234 | miR-10b-5p | trans | 1.46E-34 | Nightingale | Citrate | NA | G | T | 0.024692 | 0.004318 | 6.00E-09 |
| rs644234 | miR-10b-5p | trans | 1.46E-34 | Nightingale | IDL_FC | NA | G | T | 0.035982 | 0.004208 | 3.10E-17 |
| rs644234 | miR-10b-5p | trans | 1.46E-34 | Nightingale | IDL_CE | NA | G | T | 0.032089 | 0.004197 | 4.30E-14 |
| rs644234 | miR-10b-5p | trans | 1.46E-34 | Nightingale | M_LDL_L | NA | G | T | 0.031384 | 0.004353 | 7.10E-13 |
| rs644234 | miR-10b-5p | trans | 1.46E-34 | Nightingale | L_LDL_P | NA | G | T | 0.028325 | 0.004332 | 7.70E-11 |
| rs644234 | miR-10b-5p | trans | 1.46E-34 | Nightingale | ApoB | NA | G | T | 0.032929 | 0.004341 | 3.40E-14 |
| rs644234 | miR-10b-5p | trans | 1.46E-34 | Nightingale | L_LDL_CE | NA | G | T | 0.029522 | 0.004308 | 9.80E-12 |
| rs644234 | miR-10b-5p | trans | 1.46E-34 | Nightingale | XL_VLDL_T | NA | G | T | -0.04156 | 0.004213 | 4.70E-23 |
| rs644234 | miR-10b-5p | trans | 1.46E-34 | Nightingale | M_VLDL_T | NA | G | T | -0.03625 | 0.004108 | 1.50E-18 |
| rs644234 | miR-10b-5p | trans | 1.46E-34 | Nightingale | S_HDL_P | NA | G | T | 0.027802 | 0.004347 | 2.70E-10 |
| rs644234 | miR-10b-5p | trans | 1.46E-34 | Nightingale | M_LDL_TG | NA | G | T | -0.02511 | 0.004293 | 8.50E-09 |
| rs644234 | miR-10b-5p | trans | 1.46E-34 | Nightingale | M_LDL_C_ | NA | G | T | 0.026584 | 0.00435 | 1.60E-09 |
| rs644234 | miR-10b-5p | trans | 1.46E-34 | Nightingale | M_VLDL_P | NA | G | T | 0.042108 | 0.004172 | 6.00E-24 |
| rs644234 | miR-10b-5p | trans | 1.46E-34 | Nightingale | M_VLDL_P | NA | G | T | 0.028752 | 0.00434 | 1.40E-11 |
| rs644234 | miR-10b-5p | trans | 1.46E-34 | Nightingale | S_HDL_L | NA | G | T | 0.023966 | 0.004332 | 2.70E-08 |
| rs644234 | miR-10b-5p | trans | 1.46E-34 | Nightingale | S_VLDL_FC | NA | G | T | 0.036644 | 0.004348 | 1.70E-17 |
| rs644234 | miR-10b-5p | trans | 1.46E-34 | Nightingale | XL_HDL_PL | NA | G | T | -0.02179 | 0.003994 | 4.70E-08 |
| rs644234 | miR-10b-5p | trans | 1.46E-34 | Nightingale | L_VLDL_PL | NA | G | T | 0.021654 | 0.004222 | 4.40E-08 |

| | | | | | | | | | | | |
|---|---|---|---|---|---|---|---|---|---|---|---|
| rs644234 | miR-10b-5p | trans | 1.46E-34 | Nightingale | M_LDL_P | NA | G | T | 0.033998 | 0.004359 | 7.50E-15 |
| rs644234 | miR-10b-5p | trans | 1.46E-34 | Nightingale | S_VLDL_TG | NA | G | T | -0.03599 | 0.004208 | 1.80E-17 |
| rs644234 | miR-10b-5p | trans | 1.46E-34 | Nightingale | XS_VLDL_F | NA | G | T | 0.035893 | 0.004271 | 7.40E-18 |
| rs644234 | miR-10b-5p | trans | 1.46E-34 | Nightingale | LDL_PL | NA | G | T | 0.033316 | 0.004337 | 3.00E-14 |
| rs644234 | miR-10b-5p | trans | 1.46E-34 | Nightingale | XXL_VLDL_ | NA | G | T | 0.033445 | 0.004356 | 8.70E-15 |
| rs644234 | miR-10b-5p | trans | 1.46E-34 | Nightingale | XS_VLDL_C | NA | G | T | 0.03665 | 0.004219 | 2.30E-18 |
| rs644234 | miR-10b-5p | trans | 1.46E-34 | Nightingale | Remnant_( | NA | G | T | 0.03584 | 0.004319 | 5.30E-17 |
| rs644234 | miR-10b-5p | trans | 1.46E-34 | Nightingale | S_LDL_C_p | NA | G | T | 0.041866 | 0.004364 | 5.50E-22 |
| rs644234 | miR-10b-5p | trans | 1.46E-34 | Nightingale | VLDL_CE | NA | G | T | 0.035147 | 0.004348 | 2.10E-16 |
| rs644234 | miR-126-3p | trans | 1.03E-37 | Nightingale | S_LDL_L | NA | G | T | 0.026888 | 0.004346 | 1.20E-09 |
| rs644234 | miR-126-3p | trans | 1.03E-37 | Nightingale | LDL_P | NA | G | T | 0.030662 | 0.004344 | 2.10E-12 |
| rs644234 | miR-126-3p | trans | 1.03E-37 | Nightingale | M_VLDL_C | NA | G | T | 0.039209 | 0.004307 | 1.30E-19 |
| rs644234 | miR-126-3p | trans | 1.03E-37 | Nightingale | non_HDL_( | NA | G | T | 0.03414 | 0.004323 | 2.40E-15 |
| rs644234 | miR-126-3p | trans | 1.03E-37 | Nightingale | XXL_VLDL_ | NA | G | T | -0.03443 | 0.004384 | 6.20E-16 |
| rs644234 | miR-126-3p | trans | 1.03E-37 | Nightingale | S_VLDL_C_ | NA | G | T | 0.039745 | 0.00423 | 7.90E-21 |
| rs644234 | miR-126-3p | trans | 1.03E-37 | Nightingale | M_VLDL_C | NA | G | T | 0.031362 | 0.004084 | 3.10E-14 |
| rs644234 | miR-126-3p | trans | 1.03E-37 | Nightingale | S_VLDL_L | NA | G | T | 0.024494 | 0.004305 | 2.50E-09 |
| rs644234 | miR-126-3p | trans | 1.03E-37 | Nightingale | L_LDL_FC | NA | G | T | 0.031476 | 0.004268 | 4.00E-13 |
| rs644234 | miR-126-3p | trans | 1.03E-37 | Nightingale | L_LDL_L | NA | G | T | 0.031526 | 0.004304 | 3.20E-13 |
| rs644234 | miR-126-3p | trans | 1.03E-37 | Nightingale | L_VLDL_C_ | NA | G | T | 0.049243 | 0.004245 | 8.50E-32 |
| rs644234 | miR-126-3p | trans | 1.03E-37 | Nightingale | M_LDL_C | NA | G | T | 0.032088 | 0.004355 | 2.30E-13 |
| rs644234 | miR-126-3p | trans | 1.03E-37 | Nightingale | S_VLDL_PL | NA | G | T | 0.025736 | 0.004162 | 1.40E-09 |
| rs644234 | miR-126-3p | trans | 1.03E-37 | Nightingale | XS_VLDL_F | NA | G | T | 0.029149 | 0.004316 | 3.00E-11 |
| rs644234 | miR-126-3p | trans | 1.03E-37 | Nightingale | Clinical_LD | NA | G | T | 0.034557 | 0.004322 | 2.70E-15 |
| rs644234 | miR-126-3p | trans | 1.03E-37 | Nightingale | S_LDL_PL_ | NA | G | T | -0.02807 | 0.004339 | 5.00E-12 |
| rs644234 | miR-126-3p | trans | 1.03E-37 | Nightingale | S_HDL_C | NA | G | T | 0.025378 | 0.004339 | 9.40E-09 |
| rs644234 | miR-126-3p | trans | 1.03E-37 | Nightingale | XL_VLDL_F | NA | G | T | 0.03523 | 0.004214 | 1.70E-17 |
| rs644234 | miR-126-3p | trans | 1.03E-37 | Nightingale | L_LDL_C | NA | G | T | 0.030226 | 0.004298 | 3.00E-12 |
| rs644234 | miR-126-3p | trans | 1.03E-37 | Nightingale | IDL_C | NA | G | T | 0.033306 | 0.004198 | 4.80E-15 |
| rs644234 | miR-126-3p | trans | 1.03E-37 | Nightingale | S_LDL_CE_ | NA | G | T | 0.02885 | 0.004339 | 1.10E-12 |
| rs644234 | miR-126-3p | trans | 1.03E-37 | Nightingale | L_VLDL_CE | NA | G | T | 0.022755 | 0.004296 | 2.90E-08 |
| rs644234 | miR-126-3p | trans | 1.03E-37 | Nightingale | S_VLDL_C | NA | G | T | 0.037356 | 0.004346 | 2.60E-18 |
| rs644234 | miR-126-3p | trans | 1.03E-37 | Nightingale | S_VLDL_PL | NA | G | T | 0.032035 | 0.004339 | 4.90E-14 |
| rs644234 | miR-126-3p | trans | 1.03E-37 | Nightingale | Total_C | NA | G | T | 0.030145 | 0.004208 | 4.90E-13 |
| rs644234 | miR-126-3p | trans | 1.03E-37 | Nightingale | M_VLDL_F | NA | G | T | 0.032516 | 0.004346 | 3.90E-14 |
| rs644234 | miR-126-3p | trans | 1.03E-37 | Nightingale | IDL_PL | NA | G | T | 0.033582 | 0.004195 | 1.90E-15 |
| rs644234 | miR-126-3p | trans | 1.03E-37 | Nightingale | S_LDL_CE | NA | G | T | 0.029439 | 0.004349 | 1.50E-11 |
| rs644234 | miR-126-3p | trans | 1.03E-37 | Nightingale | M_LDL_CE | NA | G | T | 0.031208 | 0.004353 | 8.60E-13 |
| rs644234 | miR-126-3p | trans | 1.03E-37 | Nightingale | S_LDL_C | NA | G | T | 0.029694 | 0.004351 | 1.60E-11 |
| rs644234 | miR-126-3p | trans | 1.03E-37 | Nightingale | XXL_VLDL_ | NA | G | T | 0.031794 | 0.00438 | 2.10E-13 |
| rs644234 | miR-126-3p | trans | 1.03E-37 | Nightingale | LDL_L | NA | G | T | 0.031466 | 0.004332 | 5.80E-13 |
| rs644234 | miR-126-3p | trans | 1.03E-37 | Nightingale | XL_VLDL_P | NA | G | T | 0.039123 | 0.004353 | 7.10E-20 |
| rs644234 | miR-126-3p | trans | 1.03E-37 | Nightingale | S_VLDL_FC | NA | G | T | 0.027162 | 0.004154 | 1.20E-10 |
| rs644234 | miR-126-3p | trans | 1.03E-37 | Nightingale | L_VLDL_FC | NA | G | T | 0.046949 | 0.004305 | 3.90E-28 |
| rs644234 | miR-126-3p | trans | 1.03E-37 | Nightingale | IDL_L | NA | G | T | 0.033664 | 0.004203 | 2.30E-15 |
| rs644234 | miR-126-3p | trans | 1.03E-37 | Nightingale | XL_VLDL_C | NA | G | T | 0.03459 | 0.00417 | 1.50E-16 |
| rs644234 | miR-126-3p | trans | 1.03E-37 | Nightingale | M_VLDL_C | NA | G | T | 0.033859 | 0.004094 | 2.60E-16 |
| rs644234 | miR-126-3p | trans | 1.03E-37 | Nightingale | S_HDL_CE | NA | G | T | 0.025685 | 0.004354 | 7.00E-09 |
| rs644234 | miR-126-3p | trans | 1.03E-37 | Nightingale | M_VLDL_P | NA | G | T | 0.027925 | 0.004336 | 4.70E-11 |
| rs644234 | miR-126-3p | trans | 1.03E-37 | Nightingale | S_VLDL_CE | NA | G | T | 0.045161 | 0.004288 | 1.40E-25 |
| rs644234 | miR-126-3p | trans | 1.03E-37 | Nightingale | L_VLDL_CE | NA | G | T | 0.044093 | 0.00416 | 7.40E-27 |
| rs644234 | miR-126-3p | trans | 1.03E-37 | Nightingale | VLDL_C | NA | G | T | 0.031063 | 0.004341 | 2.10E-13 |
| rs644234 | miR-126-3p | trans | 1.03E-37 | Nightingale | S_LDL_P | NA | G | T | 0.028954 | 0.004351 | 3.00E-11 |
| rs644234 | miR-126-3p | trans | 1.03E-37 | Nightingale | LDL_CE | NA | G | T | 0.030617 | 0.004335 | 2.10E-12 |
| rs644234 | miR-126-3p | trans | 1.03E-37 | Nightingale | L_LDL_PL | NA | G | T | 0.03592 | 0.004314 | 1.30E-16 |
| rs644234 | miR-126-3p | trans | 1.03E-37 | Nightingale | Omega_6 | NA | G | T | 0.024169 | 0.004245 | 3.10E-09 |
| rs644234 | miR-126-3p | trans | 1.03E-37 | Nightingale | XXL_VLDL_ | NA | G | T | 0.029285 | 0.004328 | 5.00E-12 |
| rs644234 | miR-126-3p | trans | 1.03E-37 | Nightingale | XS_VLDL_P | NA | G | T | 0.035793 | 0.004289 | 9.90E-18 |
| rs644234 | miR-126-3p | trans | 1.03E-37 | Nightingale | LDL_FC | NA | G | T | 0.031549 | 0.0043 | 6.50E-13 |
| rs644234 | miR-126-3p | trans | 1.03E-37 | Nightingale | IDL_P | NA | G | T | 0.039024 | 0.004301 | 1.90E-19 |
| rs644234 | miR-126-3p | trans | 1.03E-37 | Nightingale | M_LDL_PL | NA | G | T | 0.031167 | 0.004353 | 1.00E-12 |
| rs644234 | miR-126-3p | trans | 1.03E-37 | Nightingale | ApoB_by_A | NA | G | T | 0.025671 | 0.004281 | 2.10E-09 |
| rs644234 | miR-126-3p | trans | 1.03E-37 | Nightingale | Sphingomy | NA | G | T | 0.025922 | 0.004063 | 3.20E-10 |

| | | | | | | | | | | |
|---|---|---|---|---|---|---|---|---|---|---|
| rs644234 | miR-126-3p | trans | 1.03E-37 | Nightingale | XL_VLDL_C | NA | G | T | 0.035608 | 0.004168 | 1.70E-17 |
| rs644234 | miR-126-3p | trans | 1.03E-37 | Nightingale | M_LDL_FC | NA | G | T | 0.032279 | 0.004341 | 2.40E-13 |
| rs644234 | miR-126-3p | trans | 1.03E-37 | Nightingale | VLDL_P | NA | G | T | 0.026555 | 0.004324 | 2.00E-10 |
| rs644234 | miR-126-3p | trans | 1.03E-37 | Nightingale | Total_PL | NA | G | T | 0.023006 | 0.004139 | 7.70E-09 |
| rs644234 | miR-126-3p | trans | 1.03E-37 | Nightingale | S_VLDL_CE | NA | G | T | 0.037313 | 0.004341 | 1.70E-18 |
| rs644234 | miR-126-3p | trans | 1.03E-37 | Nightingale | M_VLDL_C | NA | G | T | 0.037346 | 0.004335 | 5.30E-18 |
| rs644234 | miR-126-3p | trans | 1.03E-37 | Nightingale | LDL_C | NA | G | T | 0.031186 | 0.004328 | 9.50E-13 |
| rs644234 | miR-126-3p | trans | 1.03E-37 | Nightingale | XS_VLDL_L | NA | G | T | 0.033016 | 0.004273 | 1.30E-15 |
| rs644234 | miR-126-3p | trans | 1.03E-37 | Nightingale | XS_VLDL_P | NA | G | T | 0.031667 | 0.004286 | 1.90E-14 |
| rs644234 | miR-126-3p | trans | 1.03E-37 | Nightingale | S_LDL_FC | NA | G | T | 0.027196 | 0.00434 | 1.30E-09 |
| rs644234 | miR-126-3p | trans | 1.03E-37 | Nightingale | GlycA | NA | G | T | 0.025325 | 0.004328 | 4.90E-09 |
| rs644234 | miR-126-3p | trans | 1.03E-37 | Nightingale | M_VLDL_F | NA | G | T | 0.039386 | 0.004135 | 2.10E-21 |
| rs644234 | miR-126-3p | trans | 1.03E-37 | Nightingale | Total_L | NA | G | T | 0.025011 | 0.004276 | 1.50E-09 |
| rs644234 | miR-126-3p | trans | 1.03E-37 | Nightingale | XS_VLDL_C | NA | G | T | 0.036401 | 0.004195 | 4.00E-18 |
| rs644234 | miR-126-3p | trans | 1.03E-37 | Nightingale | LA | NA | G | T | 0.023278 | 0.004263 | 1.60E-08 |
| rs644234 | miR-126-3p | trans | 1.03E-37 | Nightingale | Total_FC | NA | G | T | 0.031385 | 0.004246 | 4.60E-14 |
| rs644234 | miR-126-3p | trans | 1.03E-37 | Nightingale | XXL_VLDL_ | NA | G | T | 0.023513 | 0.004375 | 5.40E-09 |
| rs644234 | miR-126-3p | trans | 1.03E-37 | Nightingale | VLDL_FC | NA | G | T | 0.024444 | 0.004315 | 3.60E-09 |
| rs644234 | miR-126-3p | trans | 1.03E-37 | Nightingale | L_VLDL_TG | NA | G | T | -0.04925 | 0.004326 | 6.10E-31 |
| rs644234 | miR-126-3p | trans | 1.03E-37 | Nightingale | S_VLDL_P | NA | G | T | 0.023388 | 0.004292 | 9.80E-09 |
| rs644234 | miR-126-3p | trans | 1.03E-37 | Nightingale | Total_CE | NA | G | T | 0.029391 | 0.004193 | 1.80E-12 |
| rs644234 | miR-126-3p | trans | 1.03E-37 | Nightingale | Citrate | NA | G | T | 0.024692 | 0.004318 | 6.00E-09 |
| rs644234 | miR-126-3p | trans | 1.03E-37 | Nightingale | IDL_FC | NA | G | T | 0.035982 | 0.004208 | 3.10E-17 |
| rs644234 | miR-126-3p | trans | 1.03E-37 | Nightingale | IDL_CE | NA | G | T | 0.032089 | 0.004197 | 4.30E-14 |
| rs644234 | miR-126-3p | trans | 1.03E-37 | Nightingale | M_LDL_L | NA | G | T | 0.031384 | 0.004353 | 7.10E-13 |
| rs644234 | miR-126-3p | trans | 1.03E-37 | Nightingale | L_LDL_P | NA | G | T | 0.028325 | 0.004332 | 7.70E-11 |
| rs644234 | miR-126-3p | trans | 1.03E-37 | Nightingale | ApoB | NA | G | T | 0.032929 | 0.004341 | 3.40E-14 |
| rs644234 | miR-126-3p | trans | 1.03E-37 | Nightingale | L_LDL_CE | NA | G | T | 0.029522 | 0.004308 | 9.80E-12 |
| rs644234 | miR-126-3p | trans | 1.03E-37 | Nightingale | XL_VLDL_T | NA | G | T | -0.04156 | 0.004213 | 4.70E-23 |
| rs644234 | miR-126-3p | trans | 1.03E-37 | Nightingale | M_VLDL_T | NA | G | T | -0.03625 | 0.004108 | 1.50E-18 |
| rs644234 | miR-126-3p | trans | 1.03E-37 | Nightingale | S_HDL_P | NA | G | T | 0.027802 | 0.004347 | 2.70E-10 |
| rs644234 | miR-126-3p | trans | 1.03E-37 | Nightingale | M_LDL_TG | NA | G | T | -0.02511 | 0.004293 | 8.50E-09 |
| rs644234 | miR-126-3p | trans | 1.03E-37 | Nightingale | M_LDL_C_ | NA | G | T | 0.026584 | 0.00435 | 1.60E-09 |
| rs644234 | miR-126-3p | trans | 1.03E-37 | Nightingale | M_VLDL_P | NA | G | T | 0.042108 | 0.004172 | 6.00E-24 |
| rs644234 | miR-126-3p | trans | 1.03E-37 | Nightingale | M_VLDL_P | NA | G | T | 0.028752 | 0.00434 | 1.40E-11 |
| rs644234 | miR-126-3p | trans | 1.03E-37 | Nightingale | S_HDL_L | NA | G | T | 0.023966 | 0.004332 | 2.70E-08 |
| rs644234 | miR-126-3p | trans | 1.03E-37 | Nightingale | S_VLDL_FC | NA | G | T | 0.036644 | 0.004348 | 1.70E-17 |
| rs644234 | miR-126-3p | trans | 1.03E-37 | Nightingale | XL_HDL_PL | NA | G | T | -0.02179 | 0.003994 | 4.70E-08 |
| rs644234 | miR-126-3p | trans | 1.03E-37 | Nightingale | L_VLDL_PL | NA | G | T | 0.021654 | 0.004222 | 4.40E-08 |
| rs644234 | miR-126-3p | trans | 1.03E-37 | Nightingale | M_LDL_P | NA | G | T | 0.033998 | 0.004359 | 7.50E-15 |
| rs644234 | miR-126-3p | trans | 1.03E-37 | Nightingale | S_VLDL_TG | NA | G | T | -0.03599 | 0.004208 | 1.80E-17 |
| rs644234 | miR-126-3p | trans | 1.03E-37 | Nightingale | XS_VLDL_F | NA | G | T | 0.035893 | 0.004271 | 7.40E-18 |
| rs644234 | miR-126-3p | trans | 1.03E-37 | Nightingale | LDL_PL | NA | G | T | 0.033316 | 0.004337 | 3.00E-14 |
| rs644234 | miR-126-3p | trans | 1.03E-37 | Nightingale | XXL_VLDL_ | NA | G | T | 0.033445 | 0.004356 | 8.70E-15 |
| rs644234 | miR-126-3p | trans | 1.03E-37 | Nightingale | XS_VLDL_C | NA | G | T | 0.03665 | 0.004219 | 2.30E-18 |
| rs644234 | miR-126-3p | trans | 1.03E-37 | Nightingale | Remnant_( | NA | G | T | 0.03584 | 0.004319 | 5.30E-17 |
| rs644234 | miR-126-3p | trans | 1.03E-37 | Nightingale | S_LDL_C_p | NA | G | T | 0.041866 | 0.004364 | 5.50E-22 |
| rs644234 | miR-126-3p | trans | 1.03E-37 | Nightingale | VLDL_CE | NA | G | T | 0.035147 | 0.004348 | 2.10E-16 |
| rs644234 | let-7a-5p | trans | 8.38E-13 | Nightingale | S_LDL_L | NA | G | T | 0.026888 | 0.004346 | 1.20E-09 |
| rs644234 | let-7a-5p | trans | 8.38E-13 | Nightingale | LDL_P | NA | G | T | 0.030662 | 0.004344 | 2.10E-12 |
| rs644234 | let-7a-5p | trans | 8.38E-13 | Nightingale | M_VLDL_C | NA | G | T | 0.039209 | 0.004307 | 1.30E-19 |
| rs644234 | let-7a-5p | trans | 8.38E-13 | Nightingale | non_HDL_( | NA | G | T | 0.03414 | 0.004323 | 2.40E-15 |
| rs644234 | let-7a-5p | trans | 8.38E-13 | Nightingale | XXL_VLDL_ | NA | G | T | -0.03443 | 0.004384 | 6.20E-16 |
| rs644234 | let-7a-5p | trans | 8.38E-13 | Nightingale | S_VLDL_C_ | NA | G | T | 0.039745 | 0.00423 | 7.90E-21 |
| rs644234 | let-7a-5p | trans | 8.38E-13 | Nightingale | M_VLDL_C | NA | G | T | 0.031362 | 0.004084 | 3.10E-14 |
| rs644234 | let-7a-5p | trans | 8.38E-13 | Nightingale | S_VLDL_L | NA | G | T | 0.024494 | 0.004305 | 2.50E-09 |
| rs644234 | let-7a-5p | trans | 8.38E-13 | Nightingale | L_LDL_FC | NA | G | T | 0.031476 | 0.004268 | 4.00E-13 |
| rs644234 | let-7a-5p | trans | 8.38E-13 | Nightingale | L_LDL_L | NA | G | T | 0.031526 | 0.004304 | 3.20E-13 |
| rs644234 | let-7a-5p | trans | 8.38E-13 | Nightingale | L_VLDL_C_ | NA | G | T | 0.049243 | 0.004245 | 8.50E-32 |
| rs644234 | let-7a-5p | trans | 8.38E-13 | Nightingale | M_LDL_C | NA | G | T | 0.032088 | 0.004355 | 2.30E-13 |
| rs644234 | let-7a-5p | trans | 8.38E-13 | Nightingale | S_VLDL_PL | NA | G | T | 0.025736 | 0.004162 | 1.40E-09 |
| rs644234 | let-7a-5p | trans | 8.38E-13 | Nightingale | XS_VLDL_F | NA | G | T | 0.029149 | 0.004316 | 3.00E-11 |
| rs644234 | let-7a-5p | trans | 8.38E-13 | Nightingale | Clinical_LD | NA | G | T | 0.034557 | 0.004322 | 2.70E-15 |

| | | | | | | | | | | | |
|---|---|---|---|---|---|---|---|---|---|---|---|
| rs644234 | let-7a-5p | trans | 8.38E-13 | Nightingale | S_LDL_PL_ | NA | G | T | -0.02807 | 0.004339 | 5.00E-12 |
| rs644234 | let-7a-5p | trans | 8.38E-13 | Nightingale | S_HDL_C | NA | G | T | 0.025378 | 0.004339 | 9.40E-09 |
| rs644234 | let-7a-5p | trans | 8.38E-13 | Nightingale | XL_VLDL_F | NA | G | T | 0.03523 | 0.004214 | 1.70E-17 |
| rs644234 | let-7a-5p | trans | 8.38E-13 | Nightingale | L_LDL_C | NA | G | T | 0.030226 | 0.004298 | 3.00E-12 |
| rs644234 | let-7a-5p | trans | 8.38E-13 | Nightingale | IDL_C | NA | G | T | 0.033306 | 0.004198 | 4.80E-15 |
| rs644234 | let-7a-5p | trans | 8.38E-13 | Nightingale | S_LDL_CE_ | NA | G | T | 0.02885 | 0.004339 | 1.10E-12 |
| rs644234 | let-7a-5p | trans | 8.38E-13 | Nightingale | L_VLDL_CE | NA | G | T | 0.022755 | 0.004296 | 2.90E-08 |
| rs644234 | let-7a-5p | trans | 8.38E-13 | Nightingale | S_VLDL_C | NA | G | T | 0.037356 | 0.004346 | 2.60E-18 |
| rs644234 | let-7a-5p | trans | 8.38E-13 | Nightingale | S_VLDL_PL | NA | G | T | 0.032035 | 0.004339 | 4.90E-14 |
| rs644234 | let-7a-5p | trans | 8.38E-13 | Nightingale | Total_C | NA | G | T | 0.030145 | 0.004208 | 4.90E-13 |
| rs644234 | let-7a-5p | trans | 8.38E-13 | Nightingale | M_VLDL_F | NA | G | T | 0.032516 | 0.004346 | 3.90E-14 |
| rs644234 | let-7a-5p | trans | 8.38E-13 | Nightingale | IDL_PL | NA | G | T | 0.033582 | 0.004195 | 1.90E-15 |
| rs644234 | let-7a-5p | trans | 8.38E-13 | Nightingale | S_LDL_CE | NA | G | T | 0.029439 | 0.004349 | 1.50E-11 |
| rs644234 | let-7a-5p | trans | 8.38E-13 | Nightingale | M_LDL_CE | NA | G | T | 0.031208 | 0.004353 | 8.60E-13 |
| rs644234 | let-7a-5p | trans | 8.38E-13 | Nightingale | S_LDL_C | NA | G | T | 0.029694 | 0.004351 | 1.60E-11 |
| rs644234 | let-7a-5p | trans | 8.38E-13 | Nightingale | XXL_VLDL_ | NA | G | T | 0.031794 | 0.00438 | 2.10E-13 |
| rs644234 | let-7a-5p | trans | 8.38E-13 | Nightingale | LDL_L | NA | G | T | 0.031466 | 0.004332 | 5.80E-13 |
| rs644234 | let-7a-5p | trans | 8.38E-13 | Nightingale | XL_VLDL_P | NA | G | T | 0.039123 | 0.004353 | 7.10E-20 |
| rs644234 | let-7a-5p | trans | 8.38E-13 | Nightingale | S_VLDL_FC | NA | G | T | 0.027162 | 0.004154 | 1.20E-10 |
| rs644234 | let-7a-5p | trans | 8.38E-13 | Nightingale | L_VLDL_FC | NA | G | T | 0.046949 | 0.004305 | 3.90E-28 |
| rs644234 | let-7a-5p | trans | 8.38E-13 | Nightingale | IDL_L | NA | G | T | 0.033664 | 0.004203 | 2.30E-15 |
| rs644234 | let-7a-5p | trans | 8.38E-13 | Nightingale | XL_VLDL_C | NA | G | T | 0.03459 | 0.00417 | 1.50E-16 |
| rs644234 | let-7a-5p | trans | 8.38E-13 | Nightingale | M_VLDL_C | NA | G | T | 0.033859 | 0.004094 | 2.60E-16 |
| rs644234 | let-7a-5p | trans | 8.38E-13 | Nightingale | S_HDL_CE | NA | G | T | 0.025685 | 0.004354 | 7.00E-09 |
| rs644234 | let-7a-5p | trans | 8.38E-13 | Nightingale | M_VLDL_P | NA | G | T | 0.027925 | 0.004336 | 4.70E-11 |
| rs644234 | let-7a-5p | trans | 8.38E-13 | Nightingale | S_VLDL_CE | NA | G | T | 0.045161 | 0.004288 | 1.40E-25 |
| rs644234 | let-7a-5p | trans | 8.38E-13 | Nightingale | L_VLDL_CE | NA | G | T | 0.044093 | 0.00416 | 7.40E-27 |
| rs644234 | let-7a-5p | trans | 8.38E-13 | Nightingale | VLDL_C | NA | G | T | 0.031063 | 0.004341 | 2.10E-13 |
| rs644234 | let-7a-5p | trans | 8.38E-13 | Nightingale | S_LDL_P | NA | G | T | 0.028954 | 0.004351 | 3.00E-11 |
| rs644234 | let-7a-5p | trans | 8.38E-13 | Nightingale | LDL_CE | NA | G | T | 0.030617 | 0.004335 | 2.10E-12 |
| rs644234 | let-7a-5p | trans | 8.38E-13 | Nightingale | L_LDL_PL | NA | G | T | 0.03592 | 0.004314 | 1.30E-16 |
| rs644234 | let-7a-5p | trans | 8.38E-13 | Nightingale | Omega_6 | NA | G | T | 0.024169 | 0.004245 | 3.10E-09 |
| rs644234 | let-7a-5p | trans | 8.38E-13 | Nightingale | XXL_VLDL_ | NA | G | T | 0.029285 | 0.004328 | 5.00E-12 |
| rs644234 | let-7a-5p | trans | 8.38E-13 | Nightingale | XS_VLDL_P | NA | G | T | 0.035793 | 0.004289 | 9.90E-18 |
| rs644234 | let-7a-5p | trans | 8.38E-13 | Nightingale | LDL_FC | NA | G | T | 0.031549 | 0.0043 | 6.50E-13 |
| rs644234 | let-7a-5p | trans | 8.38E-13 | Nightingale | IDL_P | NA | G | T | 0.039024 | 0.004301 | 1.90E-19 |
| rs644234 | let-7a-5p | trans | 8.38E-13 | Nightingale | M_LDL_PL | NA | G | T | 0.031167 | 0.004353 | 1.00E-12 |
| rs644234 | let-7a-5p | trans | 8.38E-13 | Nightingale | ApoB_by_ | NA | G | T | 0.025671 | 0.004281 | 2.10E-09 |
| rs644234 | let-7a-5p | trans | 8.38E-13 | Nightingale | Sphingomy | NA | G | T | 0.025922 | 0.004063 | 3.20E-10 |
| rs644234 | let-7a-5p | trans | 8.38E-13 | Nightingale | XL_VLDL_C | NA | G | T | 0.035608 | 0.004168 | 1.70E-17 |
| rs644234 | let-7a-5p | trans | 8.38E-13 | Nightingale | M_LDL_FC | NA | G | T | 0.032279 | 0.004341 | 2.40E-13 |
| rs644234 | let-7a-5p | trans | 8.38E-13 | Nightingale | VLDL_P | NA | G | T | 0.026555 | 0.004324 | 2.00E-10 |
| rs644234 | let-7a-5p | trans | 8.38E-13 | Nightingale | Total_PL | NA | G | T | 0.023006 | 0.004139 | 7.70E-09 |
| rs644234 | let-7a-5p | trans | 8.38E-13 | Nightingale | S_VLDL_CE | NA | G | T | 0.037313 | 0.004341 | 1.70E-18 |
| rs644234 | let-7a-5p | trans | 8.38E-13 | Nightingale | M_VLDL_C | NA | G | T | 0.037346 | 0.004335 | 5.30E-18 |
| rs644234 | let-7a-5p | trans | 8.38E-13 | Nightingale | LDL_C | NA | G | T | 0.031186 | 0.004328 | 9.50E-13 |
| rs644234 | let-7a-5p | trans | 8.38E-13 | Nightingale | XS_VLDL_L | NA | G | T | 0.033016 | 0.004273 | 1.30E-15 |
| rs644234 | let-7a-5p | trans | 8.38E-13 | Nightingale | XS_VLDL_P | NA | G | T | 0.031667 | 0.004286 | 1.90E-14 |
| rs644234 | let-7a-5p | trans | 8.38E-13 | Nightingale | S_LDL_FC | NA | G | T | 0.027196 | 0.00434 | 1.30E-09 |
| rs644234 | let-7a-5p | trans | 8.38E-13 | Nightingale | GlycA | NA | G | T | 0.025325 | 0.004328 | 4.90E-09 |
| rs644234 | let-7a-5p | trans | 8.38E-13 | Nightingale | M_VLDL_F | NA | G | T | 0.039386 | 0.004135 | 2.10E-21 |
| rs644234 | let-7a-5p | trans | 8.38E-13 | Nightingale | Total_L | NA | G | T | 0.025011 | 0.004276 | 1.50E-09 |
| rs644234 | let-7a-5p | trans | 8.38E-13 | Nightingale | XS_VLDL_C | NA | G | T | 0.036401 | 0.004195 | 4.00E-18 |
| rs644234 | let-7a-5p | trans | 8.38E-13 | Nightingale | LA | NA | G | T | 0.023278 | 0.004263 | 1.60E-08 |
| rs644234 | let-7a-5p | trans | 8.38E-13 | Nightingale | Total_FC | NA | G | T | 0.031385 | 0.004246 | 4.60E-14 |
| rs644234 | let-7a-5p | trans | 8.38E-13 | Nightingale | XXL_VLDL_ | NA | G | T | 0.023513 | 0.004375 | 5.40E-09 |
| rs644234 | let-7a-5p | trans | 8.38E-13 | Nightingale | VLDL_FC | NA | G | T | 0.024444 | 0.004315 | 3.60E-09 |
| rs644234 | let-7a-5p | trans | 8.38E-13 | Nightingale | L_VLDL_TG | NA | G | T | -0.04925 | 0.004326 | 6.10E-31 |
| rs644234 | let-7a-5p | trans | 8.38E-13 | Nightingale | S_VLDL_P | NA | G | T | 0.023388 | 0.004292 | 9.80E-09 |
| rs644234 | let-7a-5p | trans | 8.38E-13 | Nightingale | Total_CE | NA | G | T | 0.029391 | 0.004193 | 1.80E-12 |
| rs644234 | let-7a-5p | trans | 8.38E-13 | Nightingale | Citrate | NA | G | T | 0.024692 | 0.004318 | 6.00E-09 |
| rs644234 | let-7a-5p | trans | 8.38E-13 | Nightingale | IDL_FC | NA | G | T | 0.035982 | 0.004208 | 3.10E-17 |
| rs644234 | let-7a-5p | trans | 8.38E-13 | Nightingale | IDL_CE | NA | G | T | 0.032089 | 0.004197 | 4.30E-14 |

| | | | | | | | | | | | |
|---|---|---|---|---|---|---|---|---|---|---|---|
| rs644234 | let-7a-5p | trans | 8.38E-13 | Nightingale | M_LDL_L | NA | G | T | 0.031384 | 0.004353 | 7.10E-13 |
| rs644234 | let-7a-5p | trans | 8.38E-13 | Nightingale | L_LDL_P | NA | G | T | 0.028325 | 0.004332 | 7.70E-11 |
| rs644234 | let-7a-5p | trans | 8.38E-13 | Nightingale | ApoB | NA | G | T | 0.032929 | 0.004341 | 3.40E-14 |
| rs644234 | let-7a-5p | trans | 8.38E-13 | Nightingale | L_LDL_CE | NA | G | T | 0.029522 | 0.004308 | 9.80E-12 |
| rs644234 | let-7a-5p | trans | 8.38E-13 | Nightingale | XL_VLDL_T | NA | G | T | -0.04156 | 0.004213 | 4.70E-23 |
| rs644234 | let-7a-5p | trans | 8.38E-13 | Nightingale | M_VLDL_T | NA | G | T | -0.03625 | 0.004108 | 1.50E-18 |
| rs644234 | let-7a-5p | trans | 8.38E-13 | Nightingale | S_HDL_P | NA | G | T | 0.027802 | 0.004347 | 2.70E-10 |
| rs644234 | let-7a-5p | trans | 8.38E-13 | Nightingale | M_LDL_TG | NA | G | T | -0.02511 | 0.004293 | 8.50E-09 |
| rs644234 | let-7a-5p | trans | 8.38E-13 | Nightingale | M_LDL_C_ | NA | G | T | 0.026584 | 0.00435 | 1.60E-09 |
| rs644234 | let-7a-5p | trans | 8.38E-13 | Nightingale | M_VLDL_P | NA | G | T | 0.042108 | 0.004172 | 6.00E-24 |
| rs644234 | let-7a-5p | trans | 8.38E-13 | Nightingale | M_VLDL_P | NA | G | T | 0.028752 | 0.00434 | 1.40E-11 |
| rs644234 | let-7a-5p | trans | 8.38E-13 | Nightingale | S_HDL_L | NA | G | T | 0.023966 | 0.004332 | 2.70E-08 |
| rs644234 | let-7a-5p | trans | 8.38E-13 | Nightingale | S_VLDL_FC | NA | G | T | 0.036644 | 0.004348 | 1.70E-17 |
| rs644234 | let-7a-5p | trans | 8.38E-13 | Nightingale | XL_HDL_PL | NA | G | T | -0.02179 | 0.003994 | 4.70E-08 |
| rs644234 | let-7a-5p | trans | 8.38E-13 | Nightingale | L_VLDL_PL | NA | G | T | 0.021654 | 0.004222 | 4.40E-08 |
| rs644234 | let-7a-5p | trans | 8.38E-13 | Nightingale | M_LDL_P | NA | G | T | 0.033998 | 0.004359 | 7.50E-15 |
| rs644234 | let-7a-5p | trans | 8.38E-13 | Nightingale | S_VLDL_TG | NA | G | T | -0.03599 | 0.004208 | 1.80E-17 |
| rs644234 | let-7a-5p | trans | 8.38E-13 | Nightingale | XS_VLDL_F | NA | G | T | 0.035893 | 0.004271 | 7.40E-18 |
| rs644234 | let-7a-5p | trans | 8.38E-13 | Nightingale | LDL_PL | NA | G | T | 0.033316 | 0.004337 | 3.00E-14 |
| rs644234 | let-7a-5p | trans | 8.38E-13 | Nightingale | XXL_VLDL_ | NA | G | T | 0.033445 | 0.004356 | 8.70E-15 |
| rs644234 | let-7a-5p | trans | 8.38E-13 | Nightingale | XS_VLDL_C | NA | G | T | 0.03665 | 0.004219 | 2.30E-18 |
| rs644234 | let-7a-5p | trans | 8.38E-13 | Nightingale | Remnant_( | NA | G | T | 0.03584 | 0.004319 | 5.30E-17 |
| rs644234 | let-7a-5p | trans | 8.38E-13 | Nightingale | S_LDL_C_p | NA | G | T | 0.041866 | 0.004364 | 5.50E-22 |
| rs644234 | let-7a-5p | trans | 8.38E-13 | Nightingale | VLDL_CE | NA | G | T | 0.035147 | 0.004348 | 2.10E-16 |
| rs644234 | let-7b-5p | trans | 1.49E-26 | Nightingale | S_LDL_L | NA | G | T | 0.026888 | 0.004346 | 1.20E-09 |
| rs644234 | let-7b-5p | trans | 1.49E-26 | Nightingale | LDL_P | NA | G | T | 0.030662 | 0.004344 | 2.10E-12 |
| rs644234 | let-7b-5p | trans | 1.49E-26 | Nightingale | M_VLDL_C | NA | G | T | 0.039209 | 0.004307 | 1.30E-19 |
| rs644234 | let-7b-5p | trans | 1.49E-26 | Nightingale | non_HDL_( | NA | G | T | 0.03414 | 0.004323 | 2.40E-15 |
| rs644234 | let-7b-5p | trans | 1.49E-26 | Nightingale | XXL_VLDL_ | NA | G | T | -0.03443 | 0.004384 | 6.20E-16 |
| rs644234 | let-7b-5p | trans | 1.49E-26 | Nightingale | S_VLDL_C_ | NA | G | T | 0.039745 | 0.00423 | 7.90E-21 |
| rs644234 | let-7b-5p | trans | 1.49E-26 | Nightingale | M_VLDL_C | NA | G | T | 0.031362 | 0.004084 | 3.10E-14 |
| rs644234 | let-7b-5p | trans | 1.49E-26 | Nightingale | S_VLDL_L | NA | G | T | 0.024494 | 0.004305 | 2.50E-09 |
| rs644234 | let-7b-5p | trans | 1.49E-26 | Nightingale | L_LDL_FC | NA | G | T | 0.031476 | 0.004268 | 4.00E-13 |
| rs644234 | let-7b-5p | trans | 1.49E-26 | Nightingale | L_LDL_L | NA | G | T | 0.031526 | 0.004304 | 3.20E-13 |
| rs644234 | let-7b-5p | trans | 1.49E-26 | Nightingale | L_VLDL_C_ | NA | G | T | 0.049243 | 0.004245 | 8.50E-32 |
| rs644234 | let-7b-5p | trans | 1.49E-26 | Nightingale | M_LDL_C | NA | G | T | 0.032088 | 0.004355 | 2.30E-13 |
| rs644234 | let-7b-5p | trans | 1.49E-26 | Nightingale | S_VLDL_PL | NA | G | T | 0.025736 | 0.004162 | 1.40E-09 |
| rs644234 | let-7b-5p | trans | 1.49E-26 | Nightingale | XS_VLDL_F | NA | G | T | 0.029149 | 0.004316 | 3.00E-11 |
| rs644234 | let-7b-5p | trans | 1.49E-26 | Nightingale | Clinical_LD | NA | G | T | 0.034557 | 0.004322 | 2.70E-15 |
| rs644234 | let-7b-5p | trans | 1.49E-26 | Nightingale | S_LDL_PL_ | NA | G | T | -0.02807 | 0.004339 | 5.00E-12 |
| rs644234 | let-7b-5p | trans | 1.49E-26 | Nightingale | S_HDL_C | NA | G | T | 0.025378 | 0.004339 | 9.40E-09 |
| rs644234 | let-7b-5p | trans | 1.49E-26 | Nightingale | XL_VLDL_F | NA | G | T | 0.03523 | 0.004214 | 1.70E-17 |
| rs644234 | let-7b-5p | trans | 1.49E-26 | Nightingale | L_LDL_C | NA | G | T | 0.030226 | 0.004298 | 3.00E-12 |
| rs644234 | let-7b-5p | trans | 1.49E-26 | Nightingale | IDL_C | NA | G | T | 0.033306 | 0.004198 | 4.80E-15 |
| rs644234 | let-7b-5p | trans | 1.49E-26 | Nightingale | S_LDL_CE_ | NA | G | T | 0.02885 | 0.004339 | 1.10E-12 |
| rs644234 | let-7b-5p | trans | 1.49E-26 | Nightingale | L_VLDL_CE | NA | G | T | 0.022755 | 0.004296 | 2.90E-08 |
| rs644234 | let-7b-5p | trans | 1.49E-26 | Nightingale | S_VLDL_C | NA | G | T | 0.037356 | 0.004346 | 2.60E-18 |
| rs644234 | let-7b-5p | trans | 1.49E-26 | Nightingale | S_VLDL_PL | NA | G | T | 0.032035 | 0.004339 | 4.90E-14 |
| rs644234 | let-7b-5p | trans | 1.49E-26 | Nightingale | Total_C | NA | G | T | 0.030145 | 0.004208 | 4.90E-13 |
| rs644234 | let-7b-5p | trans | 1.49E-26 | Nightingale | M_VLDL_F | NA | G | T | 0.032516 | 0.004346 | 3.90E-14 |
| rs644234 | let-7b-5p | trans | 1.49E-26 | Nightingale | IDL_PL | NA | G | T | 0.033582 | 0.004195 | 1.90E-15 |
| rs644234 | let-7b-5p | trans | 1.49E-26 | Nightingale | S_LDL_CE | NA | G | T | 0.029439 | 0.004349 | 1.50E-11 |
| rs644234 | let-7b-5p | trans | 1.49E-26 | Nightingale | M_LDL_CE | NA | G | T | 0.031208 | 0.004353 | 8.60E-13 |
| rs644234 | let-7b-5p | trans | 1.49E-26 | Nightingale | S_LDL_C | NA | G | T | 0.029694 | 0.004351 | 1.60E-11 |
| rs644234 | let-7b-5p | trans | 1.49E-26 | Nightingale | XXL_VLDL_ | NA | G | T | 0.031794 | 0.00438 | 2.10E-13 |
| rs644234 | let-7b-5p | trans | 1.49E-26 | Nightingale | LDL_L | NA | G | T | 0.031466 | 0.004332 | 5.80E-13 |
| rs644234 | let-7b-5p | trans | 1.49E-26 | Nightingale | XL_VLDL_P | NA | G | T | 0.039123 | 0.004353 | 7.10E-20 |
| rs644234 | let-7b-5p | trans | 1.49E-26 | Nightingale | S_VLDL_FC | NA | G | T | 0.027162 | 0.004154 | 1.20E-10 |
| rs644234 | let-7b-5p | trans | 1.49E-26 | Nightingale | L_VLDL_FC | NA | G | T | 0.046949 | 0.004305 | 3.90E-28 |
| rs644234 | let-7b-5p | trans | 1.49E-26 | Nightingale | IDL_L | NA | G | T | 0.033664 | 0.004203 | 2.30E-15 |
| rs644234 | let-7b-5p | trans | 1.49E-26 | Nightingale | XL_VLDL_C | NA | G | T | 0.03459 | 0.00417 | 1.50E-16 |
| rs644234 | let-7b-5p | trans | 1.49E-26 | Nightingale | M_VLDL_C | NA | G | T | 0.033859 | 0.004094 | 2.60E-16 |
| rs644234 | let-7b-5p | trans | 1.49E-26 | Nightingale | S_HDL_CE | NA | G | T | 0.025685 | 0.004354 | 7.00E-09 |

| | | | | | | | | | | | |
|---|---|---|---|---|---|---|---|---|---|---|---|
| rs644234 | let-7b-5p | trans | 1.49E-26 | Nightingale | M_VLDL_P | NA | G | T | 0.027925 | 0.004336 | 4.70E-11 |
| rs644234 | let-7b-5p | trans | 1.49E-26 | Nightingale | S_VLDL_CE | NA | G | T | 0.045161 | 0.004288 | 1.40E-25 |
| rs644234 | let-7b-5p | trans | 1.49E-26 | Nightingale | L_VLDL_CE | NA | G | T | 0.044093 | 0.00416 | 7.40E-27 |
| rs644234 | let-7b-5p | trans | 1.49E-26 | Nightingale | VLDL_C | NA | G | T | 0.031063 | 0.004341 | 2.10E-13 |
| rs644234 | let-7b-5p | trans | 1.49E-26 | Nightingale | S_LDL_P | NA | G | T | 0.028954 | 0.004351 | 3.00E-11 |
| rs644234 | let-7b-5p | trans | 1.49E-26 | Nightingale | LDL_CE | NA | G | T | 0.030617 | 0.004335 | 2.10E-12 |
| rs644234 | let-7b-5p | trans | 1.49E-26 | Nightingale | L_LDL_PL | NA | G | T | 0.03592 | 0.004314 | 1.30E-16 |
| rs644234 | let-7b-5p | trans | 1.49E-26 | Nightingale | Omega_6 | NA | G | T | 0.024169 | 0.004245 | 3.10E-09 |
| rs644234 | let-7b-5p | trans | 1.49E-26 | Nightingale | XXL_VLDL_ | NA | G | T | 0.029285 | 0.004328 | 5.00E-12 |
| rs644234 | let-7b-5p | trans | 1.49E-26 | Nightingale | XS_VLDL_P | NA | G | T | 0.035793 | 0.004289 | 9.90E-18 |
| rs644234 | let-7b-5p | trans | 1.49E-26 | Nightingale | LDL_FC | NA | G | T | 0.031549 | 0.0043 | 6.50E-13 |
| rs644234 | let-7b-5p | trans | 1.49E-26 | Nightingale | IDL_P | NA | G | T | 0.039024 | 0.004301 | 1.90E-19 |
| rs644234 | let-7b-5p | trans | 1.49E-26 | Nightingale | M_LDL_PL | NA | G | T | 0.031167 | 0.004353 | 1.00E-12 |
| rs644234 | let-7b-5p | trans | 1.49E-26 | Nightingale | ApoB_by_A | NA | G | T | 0.025671 | 0.004281 | 2.10E-09 |
| rs644234 | let-7b-5p | trans | 1.49E-26 | Nightingale | Sphingomy | NA | G | T | 0.025922 | 0.004063 | 3.20E-10 |
| rs644234 | let-7b-5p | trans | 1.49E-26 | Nightingale | XL_VLDL_C | NA | G | T | 0.035608 | 0.004168 | 1.70E-17 |
| rs644234 | let-7b-5p | trans | 1.49E-26 | Nightingale | M_LDL_FC | NA | G | T | 0.032279 | 0.004341 | 2.40E-13 |
| rs644234 | let-7b-5p | trans | 1.49E-26 | Nightingale | VLDL_P | NA | G | T | 0.026555 | 0.004324 | 2.00E-10 |
| rs644234 | let-7b-5p | trans | 1.49E-26 | Nightingale | Total_PL | NA | G | T | 0.023006 | 0.004139 | 7.70E-09 |
| rs644234 | let-7b-5p | trans | 1.49E-26 | Nightingale | S_VLDL_CE | NA | G | T | 0.037313 | 0.004341 | 1.70E-18 |
| rs644234 | let-7b-5p | trans | 1.49E-26 | Nightingale | M_VLDL_C | NA | G | T | 0.037346 | 0.004335 | 5.30E-18 |
| rs644234 | let-7b-5p | trans | 1.49E-26 | Nightingale | LDL_C | NA | G | T | 0.031186 | 0.004328 | 9.50E-13 |
| rs644234 | let-7b-5p | trans | 1.49E-26 | Nightingale | XS_VLDL_L | NA | G | T | 0.033016 | 0.004273 | 1.30E-15 |
| rs644234 | let-7b-5p | trans | 1.49E-26 | Nightingale | XS_VLDL_P | NA | G | T | 0.031667 | 0.004286 | 1.90E-14 |
| rs644234 | let-7b-5p | trans | 1.49E-26 | Nightingale | S_LDL_FC | NA | G | T | 0.027196 | 0.00434 | 1.30E-09 |
| rs644234 | let-7b-5p | trans | 1.49E-26 | Nightingale | GlycA | NA | G | T | 0.025325 | 0.004328 | 4.90E-09 |
| rs644234 | let-7b-5p | trans | 1.49E-26 | Nightingale | M_VLDL_F | NA | G | T | 0.039386 | 0.004135 | 2.10E-21 |
| rs644234 | let-7b-5p | trans | 1.49E-26 | Nightingale | Total_L | NA | G | T | 0.025011 | 0.004276 | 1.50E-09 |
| rs644234 | let-7b-5p | trans | 1.49E-26 | Nightingale | XS_VLDL_C | NA | G | T | 0.036401 | 0.004195 | 4.00E-18 |
| rs644234 | let-7b-5p | trans | 1.49E-26 | Nightingale | LA | NA | G | T | 0.023278 | 0.004263 | 1.60E-08 |
| rs644234 | let-7b-5p | trans | 1.49E-26 | Nightingale | Total_FC | NA | G | T | 0.031385 | 0.004246 | 4.60E-14 |
| rs644234 | let-7b-5p | trans | 1.49E-26 | Nightingale | XXL_VLDL_ | NA | G | T | 0.023513 | 0.004375 | 5.40E-09 |
| rs644234 | let-7b-5p | trans | 1.49E-26 | Nightingale | VLDL_FC | NA | G | T | 0.024444 | 0.004315 | 3.60E-09 |
| rs644234 | let-7b-5p | trans | 1.49E-26 | Nightingale | L_VLDL_TG | NA | G | T | -0.04925 | 0.004326 | 6.10E-31 |
| rs644234 | let-7b-5p | trans | 1.49E-26 | Nightingale | S_VLDL_P | NA | G | T | 0.023388 | 0.004292 | 9.80E-09 |
| rs644234 | let-7b-5p | trans | 1.49E-26 | Nightingale | Total_CE | NA | G | T | 0.029391 | 0.004193 | 1.80E-12 |
| rs644234 | let-7b-5p | trans | 1.49E-26 | Nightingale | Citrate | NA | G | T | 0.024692 | 0.004318 | 6.00E-09 |
| rs644234 | let-7b-5p | trans | 1.49E-26 | Nightingale | IDL_FC | NA | G | T | 0.035982 | 0.004208 | 3.10E-17 |
| rs644234 | let-7b-5p | trans | 1.49E-26 | Nightingale | IDL_CE | NA | G | T | 0.032089 | 0.004197 | 4.30E-14 |
| rs644234 | let-7b-5p | trans | 1.49E-26 | Nightingale | M_LDL_L | NA | G | T | 0.031384 | 0.004353 | 7.10E-13 |
| rs644234 | let-7b-5p | trans | 1.49E-26 | Nightingale | L_LDL_P | NA | G | T | 0.028325 | 0.004332 | 7.70E-11 |
| rs644234 | let-7b-5p | trans | 1.49E-26 | Nightingale | ApoB | NA | G | T | 0.032929 | 0.004341 | 3.40E-14 |
| rs644234 | let-7b-5p | trans | 1.49E-26 | Nightingale | L_LDL_CE | NA | G | T | 0.029522 | 0.004308 | 9.80E-12 |
| rs644234 | let-7b-5p | trans | 1.49E-26 | Nightingale | XL_VLDL_T | NA | G | T | -0.04156 | 0.004213 | 4.70E-23 |
| rs644234 | let-7b-5p | trans | 1.49E-26 | Nightingale | M_VLDL_T | NA | G | T | -0.03625 | 0.004108 | 1.50E-18 |
| rs644234 | let-7b-5p | trans | 1.49E-26 | Nightingale | S_HDL_P | NA | G | T | 0.027802 | 0.004347 | 2.70E-10 |
| rs644234 | let-7b-5p | trans | 1.49E-26 | Nightingale | M_LDL_TG | NA | G | T | -0.02511 | 0.004293 | 8.50E-09 |
| rs644234 | let-7b-5p | trans | 1.49E-26 | Nightingale | M_LDL_C_ | NA | G | T | 0.026584 | 0.00435 | 1.60E-09 |
| rs644234 | let-7b-5p | trans | 1.49E-26 | Nightingale | M_VLDL_P | NA | G | T | 0.042108 | 0.004172 | 6.00E-24 |
| rs644234 | let-7b-5p | trans | 1.49E-26 | Nightingale | M_VLDL_P | NA | G | T | 0.028752 | 0.00434 | 1.40E-11 |
| rs644234 | let-7b-5p | trans | 1.49E-26 | Nightingale | S_HDL_L | NA | G | T | 0.023966 | 0.004332 | 2.70E-08 |
| rs644234 | let-7b-5p | trans | 1.49E-26 | Nightingale | S_VLDL_FC | NA | G | T | 0.036644 | 0.004348 | 1.70E-17 |
| rs644234 | let-7b-5p | trans | 1.49E-26 | Nightingale | XL_HDL_PL | NA | G | T | -0.02179 | 0.003994 | 4.70E-08 |
| rs644234 | let-7b-5p | trans | 1.49E-26 | Nightingale | L_VLDL_PL | NA | G | T | 0.021654 | 0.004222 | 4.40E-08 |
| rs644234 | let-7b-5p | trans | 1.49E-26 | Nightingale | M_LDL_P | NA | G | T | 0.033998 | 0.004359 | 7.50E-15 |
| rs644234 | let-7b-5p | trans | 1.49E-26 | Nightingale | S_VLDL_TG | NA | G | T | -0.03599 | 0.004208 | 1.80E-17 |
| rs644234 | let-7b-5p | trans | 1.49E-26 | Nightingale | XS_VLDL_F | NA | G | T | 0.035893 | 0.004271 | 7.40E-18 |
| rs644234 | let-7b-5p | trans | 1.49E-26 | Nightingale | LDL_PL | NA | G | T | 0.033316 | 0.004337 | 3.00E-14 |
| rs644234 | let-7b-5p | trans | 1.49E-26 | Nightingale | XXL_VLDL_ | NA | G | T | 0.033445 | 0.004356 | 8.70E-15 |
| rs644234 | let-7b-5p | trans | 1.49E-26 | Nightingale | XS_VLDL_C | NA | G | T | 0.03665 | 0.004219 | 2.30E-18 |
| rs644234 | let-7b-5p | trans | 1.49E-26 | Nightingale | Remnant_( | NA | G | T | 0.03584 | 0.004319 | 5.30E-17 |
| rs644234 | let-7b-5p | trans | 1.49E-26 | Nightingale | S_LDL_C_p | NA | G | T | 0.041866 | 0.004364 | 5.50E-22 |
| rs644234 | let-7b-5p | trans | 1.49E-26 | Nightingale | VLDL_CE | NA | G | T | 0.035147 | 0.004348 | 2.10E-16 |

| | | | | | | | | | | | |
|---|---|---|---|---|---|---|---|---|---|---|---|
| rs644234 | let-7c-5p | trans | 2.80E-23 | Nightingale | S_LDL_L | NA | G | T | 0.026888 | 0.004346 | 1.20E-09 |
| rs644234 | let-7c-5p | trans | 2.80E-23 | Nightingale | LDL_P | NA | G | T | 0.030662 | 0.004344 | 2.10E-12 |
| rs644234 | let-7c-5p | trans | 2.80E-23 | Nightingale | M_VLDL_C | NA | G | T | 0.039209 | 0.004307 | 1.30E-19 |
| rs644234 | let-7c-5p | trans | 2.80E-23 | Nightingale | non_HDL_( | NA | G | T | 0.03414 | 0.004323 | 2.40E-15 |
| rs644234 | let-7c-5p | trans | 2.80E-23 | Nightingale | XXL_VLDL_ | NA | G | T | -0.03443 | 0.004384 | 6.20E-16 |
| rs644234 | let-7c-5p | trans | 2.80E-23 | Nightingale | S_VLDL_C_ | NA | G | T | 0.039745 | 0.00423 | 7.90E-21 |
| rs644234 | let-7c-5p | trans | 2.80E-23 | Nightingale | M_VLDL_C | NA | G | T | 0.031362 | 0.004084 | 3.10E-14 |
| rs644234 | let-7c-5p | trans | 2.80E-23 | Nightingale | S_VLDL_L | NA | G | T | 0.024494 | 0.004305 | 2.50E-09 |
| rs644234 | let-7c-5p | trans | 2.80E-23 | Nightingale | L_LDL_FC | NA | G | T | 0.031476 | 0.004268 | 4.00E-13 |
| rs644234 | let-7c-5p | trans | 2.80E-23 | Nightingale | L_LDL_L | NA | G | T | 0.031526 | 0.004304 | 3.20E-13 |
| rs644234 | let-7c-5p | trans | 2.80E-23 | Nightingale | L_VLDL_C_ | NA | G | T | 0.049243 | 0.004245 | 8.50E-32 |
| rs644234 | let-7c-5p | trans | 2.80E-23 | Nightingale | M_LDL_C | NA | G | T | 0.032088 | 0.004355 | 2.30E-13 |
| rs644234 | let-7c-5p | trans | 2.80E-23 | Nightingale | S_VLDL_PL | NA | G | T | 0.025736 | 0.004162 | 1.40E-09 |
| rs644234 | let-7c-5p | trans | 2.80E-23 | Nightingale | XS_VLDL_F | NA | G | T | 0.029149 | 0.004316 | 3.00E-11 |
| rs644234 | let-7c-5p | trans | 2.80E-23 | Nightingale | Clinical_LD | NA | G | T | 0.034557 | 0.004322 | 2.70E-15 |
| rs644234 | let-7c-5p | trans | 2.80E-23 | Nightingale | S_LDL_PL_ | NA | G | T | -0.02807 | 0.004339 | 5.00E-12 |
| rs644234 | let-7c-5p | trans | 2.80E-23 | Nightingale | S_HDL_C | NA | G | T | 0.025378 | 0.004339 | 9.40E-09 |
| rs644234 | let-7c-5p | trans | 2.80E-23 | Nightingale | XL_VLDL_F | NA | G | T | 0.03523 | 0.004214 | 1.70E-17 |
| rs644234 | let-7c-5p | trans | 2.80E-23 | Nightingale | L_LDL_C | NA | G | T | 0.030226 | 0.004298 | 3.00E-12 |
| rs644234 | let-7c-5p | trans | 2.80E-23 | Nightingale | IDL_C | NA | G | T | 0.033306 | 0.004198 | 4.80E-15 |
| rs644234 | let-7c-5p | trans | 2.80E-23 | Nightingale | S_LDL_CE_ | NA | G | T | 0.02885 | 0.004339 | 1.10E-12 |
| rs644234 | let-7c-5p | trans | 2.80E-23 | Nightingale | L_VLDL_CE | NA | G | T | 0.022755 | 0.004296 | 2.90E-08 |
| rs644234 | let-7c-5p | trans | 2.80E-23 | Nightingale | S_VLDL_C | NA | G | T | 0.037356 | 0.004346 | 2.60E-18 |
| rs644234 | let-7c-5p | trans | 2.80E-23 | Nightingale | S_VLDL_PL | NA | G | T | 0.032035 | 0.004339 | 4.90E-14 |
| rs644234 | let-7c-5p | trans | 2.80E-23 | Nightingale | Total_C | NA | G | T | 0.030145 | 0.004208 | 4.90E-13 |
| rs644234 | let-7c-5p | trans | 2.80E-23 | Nightingale | M_VLDL_F | NA | G | T | 0.032516 | 0.004346 | 3.90E-14 |
| rs644234 | let-7c-5p | trans | 2.80E-23 | Nightingale | IDL_PL | NA | G | T | 0.033582 | 0.004195 | 1.90E-15 |
| rs644234 | let-7c-5p | trans | 2.80E-23 | Nightingale | S_LDL_CE | NA | G | T | 0.029439 | 0.004349 | 1.50E-11 |
| rs644234 | let-7c-5p | trans | 2.80E-23 | Nightingale | M_LDL_CE | NA | G | T | 0.031208 | 0.004353 | 8.60E-13 |
| rs644234 | let-7c-5p | trans | 2.80E-23 | Nightingale | S_LDL_C | NA | G | T | 0.029694 | 0.004351 | 1.60E-11 |
| rs644234 | let-7c-5p | trans | 2.80E-23 | Nightingale | XXL_VLDL_ | NA | G | T | 0.031794 | 0.00438 | 2.10E-13 |
| rs644234 | let-7c-5p | trans | 2.80E-23 | Nightingale | LDL_L | NA | G | T | 0.031466 | 0.004332 | 5.80E-13 |
| rs644234 | let-7c-5p | trans | 2.80E-23 | Nightingale | XL_VLDL_P | NA | G | T | 0.039123 | 0.004353 | 7.10E-20 |
| rs644234 | let-7c-5p | trans | 2.80E-23 | Nightingale | S_VLDL_FC | NA | G | T | 0.027162 | 0.004154 | 1.20E-10 |
| rs644234 | let-7c-5p | trans | 2.80E-23 | Nightingale | L_VLDL_FC | NA | G | T | 0.046949 | 0.004305 | 3.90E-28 |
| rs644234 | let-7c-5p | trans | 2.80E-23 | Nightingale | IDL_L | NA | G | T | 0.033664 | 0.004203 | 2.30E-15 |
| rs644234 | let-7c-5p | trans | 2.80E-23 | Nightingale | XL_VLDL_C | NA | G | T | 0.03459 | 0.00417 | 1.50E-16 |
| rs644234 | let-7c-5p | trans | 2.80E-23 | Nightingale | M_VLDL_C | NA | G | T | 0.033859 | 0.004094 | 2.60E-16 |
| rs644234 | let-7c-5p | trans | 2.80E-23 | Nightingale | S_HDL_CE | NA | G | T | 0.025685 | 0.004354 | 7.00E-09 |
| rs644234 | let-7c-5p | trans | 2.80E-23 | Nightingale | M_VLDL_P | NA | G | T | 0.027925 | 0.004336 | 4.70E-11 |
| rs644234 | let-7c-5p | trans | 2.80E-23 | Nightingale | S_VLDL_CE | NA | G | T | 0.045161 | 0.004288 | 1.40E-25 |
| rs644234 | let-7c-5p | trans | 2.80E-23 | Nightingale | L_VLDL_CE | NA | G | T | 0.044093 | 0.00416 | 7.40E-27 |
| rs644234 | let-7c-5p | trans | 2.80E-23 | Nightingale | VLDL_C | NA | G | T | 0.031063 | 0.004341 | 2.10E-13 |
| rs644234 | let-7c-5p | trans | 2.80E-23 | Nightingale | S_LDL_P | NA | G | T | 0.028954 | 0.004351 | 3.00E-11 |
| rs644234 | let-7c-5p | trans | 2.80E-23 | Nightingale | LDL_CE | NA | G | T | 0.030617 | 0.004335 | 2.10E-12 |
| rs644234 | let-7c-5p | trans | 2.80E-23 | Nightingale | L_LDL_PL | NA | G | T | 0.03592 | 0.004314 | 1.30E-16 |
| rs644234 | let-7c-5p | trans | 2.80E-23 | Nightingale | Omega_6 | NA | G | T | 0.024169 | 0.004245 | 3.10E-09 |
| rs644234 | let-7c-5p | trans | 2.80E-23 | Nightingale | XXL_VLDL_ | NA | G | T | 0.029285 | 0.004328 | 5.00E-12 |
| rs644234 | let-7c-5p | trans | 2.80E-23 | Nightingale | XS_VLDL_P | NA | G | T | 0.035793 | 0.004289 | 9.90E-18 |
| rs644234 | let-7c-5p | trans | 2.80E-23 | Nightingale | LDL_FC | NA | G | T | 0.031549 | 0.0043 | 6.50E-13 |
| rs644234 | let-7c-5p | trans | 2.80E-23 | Nightingale | IDL_P | NA | G | T | 0.039024 | 0.004301 | 1.90E-19 |
| rs644234 | let-7c-5p | trans | 2.80E-23 | Nightingale | M_LDL_PL | NA | G | T | 0.031167 | 0.004353 | 1.00E-12 |
| rs644234 | let-7c-5p | trans | 2.80E-23 | Nightingale | ApoB_by_/ | NA | G | T | 0.025671 | 0.004281 | 2.10E-09 |
| rs644234 | let-7c-5p | trans | 2.80E-23 | Nightingale | Sphingomy | NA | G | T | 0.025922 | 0.004063 | 3.20E-10 |
| rs644234 | let-7c-5p | trans | 2.80E-23 | Nightingale | XL_VLDL_C | NA | G | T | 0.035608 | 0.004168 | 1.70E-17 |
| rs644234 | let-7c-5p | trans | 2.80E-23 | Nightingale | M_LDL_FC | NA | G | T | 0.032279 | 0.004341 | 2.40E-13 |
| rs644234 | let-7c-5p | trans | 2.80E-23 | Nightingale | VLDL_P | NA | G | T | 0.026555 | 0.004324 | 2.00E-10 |
| rs644234 | let-7c-5p | trans | 2.80E-23 | Nightingale | Total_PL | NA | G | T | 0.023006 | 0.004139 | 7.70E-09 |
| rs644234 | let-7c-5p | trans | 2.80E-23 | Nightingale | S_VLDL_CE | NA | G | T | 0.037313 | 0.004341 | 1.70E-18 |
| rs644234 | let-7c-5p | trans | 2.80E-23 | Nightingale | M_VLDL_C | NA | G | T | 0.037346 | 0.004335 | 5.30E-18 |
| rs644234 | let-7c-5p | trans | 2.80E-23 | Nightingale | LDL_C | NA | G | T | 0.031186 | 0.004328 | 9.50E-13 |
| rs644234 | let-7c-5p | trans | 2.80E-23 | Nightingale | XS_VLDL_L | NA | G | T | 0.033016 | 0.004273 | 1.30E-15 |
| rs644234 | let-7c-5p | trans | 2.80E-23 | Nightingale | XS_VLDL_P | NA | G | T | 0.031667 | 0.004286 | 1.90E-14 |

| | | | | | | | | | | | |
|---|---|---|---|---|---|---|---|---|---|---|---|
| rs644234 | let-7c-5p | trans | 2.80E-23 | Nightingale | S_LDL_FC | NA | G | T | 0.027196 | 0.00434 | 1.30E-09 |
| rs644234 | let-7c-5p | trans | 2.80E-23 | Nightingale | GlycA | NA | G | T | 0.025325 | 0.004328 | 4.90E-09 |
| rs644234 | let-7c-5p | trans | 2.80E-23 | Nightingale | M_VLDL_F | NA | G | T | 0.039386 | 0.004135 | 2.10E-21 |
| rs644234 | let-7c-5p | trans | 2.80E-23 | Nightingale | Total_L | NA | G | T | 0.025011 | 0.004276 | 1.50E-09 |
| rs644234 | let-7c-5p | trans | 2.80E-23 | Nightingale | XS_VLDL_C | NA | G | T | 0.036401 | 0.004195 | 4.00E-18 |
| rs644234 | let-7c-5p | trans | 2.80E-23 | Nightingale | LA | NA | G | T | 0.023278 | 0.004263 | 1.60E-08 |
| rs644234 | let-7c-5p | trans | 2.80E-23 | Nightingale | Total_FC | NA | G | T | 0.031385 | 0.004246 | 4.60E-14 |
| rs644234 | let-7c-5p | trans | 2.80E-23 | Nightingale | XXL_VLDL_ | NA | G | T | 0.023513 | 0.004375 | 5.40E-09 |
| rs644234 | let-7c-5p | trans | 2.80E-23 | Nightingale | VLDL_FC | NA | G | T | 0.024444 | 0.004315 | 3.60E-09 |
| rs644234 | let-7c-5p | trans | 2.80E-23 | Nightingale | L_VLDL_TG | NA | G | T | -0.04925 | 0.004326 | 6.10E-31 |
| rs644234 | let-7c-5p | trans | 2.80E-23 | Nightingale | S_VLDL_P | NA | G | T | 0.023388 | 0.004292 | 9.80E-09 |
| rs644234 | let-7c-5p | trans | 2.80E-23 | Nightingale | Total_CE | NA | G | T | 0.029391 | 0.004193 | 1.80E-12 |
| rs644234 | let-7c-5p | trans | 2.80E-23 | Nightingale | Citrate | NA | G | T | 0.024692 | 0.004318 | 6.00E-09 |
| rs644234 | let-7c-5p | trans | 2.80E-23 | Nightingale | IDL_FC | NA | G | T | 0.035982 | 0.004208 | 3.10E-17 |
| rs644234 | let-7c-5p | trans | 2.80E-23 | Nightingale | IDL_CE | NA | G | T | 0.032089 | 0.004197 | 4.30E-14 |
| rs644234 | let-7c-5p | trans | 2.80E-23 | Nightingale | M_LDL_L | NA | G | T | 0.031384 | 0.004353 | 7.10E-13 |
| rs644234 | let-7c-5p | trans | 2.80E-23 | Nightingale | L_LDL_P | NA | G | T | 0.028325 | 0.004332 | 7.70E-11 |
| rs644234 | let-7c-5p | trans | 2.80E-23 | Nightingale | ApoB | NA | G | T | 0.032929 | 0.004341 | 3.40E-14 |
| rs644234 | let-7c-5p | trans | 2.80E-23 | Nightingale | L_LDL_CE | NA | G | T | 0.029522 | 0.004308 | 9.80E-12 |
| rs644234 | let-7c-5p | trans | 2.80E-23 | Nightingale | XL_VLDL_T | NA | G | T | -0.04156 | 0.004213 | 4.70E-23 |
| rs644234 | let-7c-5p | trans | 2.80E-23 | Nightingale | M_VLDL_T | NA | G | T | -0.03625 | 0.004108 | 1.50E-18 |
| rs644234 | let-7c-5p | trans | 2.80E-23 | Nightingale | S_HDL_P | NA | G | T | 0.027802 | 0.004347 | 2.70E-10 |
| rs644234 | let-7c-5p | trans | 2.80E-23 | Nightingale | M_LDL_TG | NA | G | T | -0.02511 | 0.004293 | 8.50E-09 |
| rs644234 | let-7c-5p | trans | 2.80E-23 | Nightingale | M_LDL_C_ | NA | G | T | 0.026584 | 0.00435 | 1.60E-09 |
| rs644234 | let-7c-5p | trans | 2.80E-23 | Nightingale | M_VLDL_P | NA | G | T | 0.042108 | 0.004172 | 6.00E-24 |
| rs644234 | let-7c-5p | trans | 2.80E-23 | Nightingale | M_VLDL_P | NA | G | T | 0.028752 | 0.00434 | 1.40E-11 |
| rs644234 | let-7c-5p | trans | 2.80E-23 | Nightingale | S_HDL_L | NA | G | T | 0.023966 | 0.004332 | 2.70E-08 |
| rs644234 | let-7c-5p | trans | 2.80E-23 | Nightingale | S_VLDL_FC | NA | G | T | 0.036644 | 0.004348 | 1.70E-17 |
| rs644234 | let-7c-5p | trans | 2.80E-23 | Nightingale | XL_HDL_PL | NA | G | T | -0.02179 | 0.003994 | 4.70E-08 |
| rs644234 | let-7c-5p | trans | 2.80E-23 | Nightingale | L_VLDL_PL | NA | G | T | 0.021654 | 0.004222 | 4.40E-08 |
| rs644234 | let-7c-5p | trans | 2.80E-23 | Nightingale | M_LDL_P | NA | G | T | 0.033998 | 0.004359 | 7.50E-15 |
| rs644234 | let-7c-5p | trans | 2.80E-23 | Nightingale | S_VLDL_TG | NA | G | T | -0.03599 | 0.004208 | 1.80E-17 |
| rs644234 | let-7c-5p | trans | 2.80E-23 | Nightingale | XS_VLDL_F | NA | G | T | 0.035893 | 0.004271 | 7.40E-18 |
| rs644234 | let-7c-5p | trans | 2.80E-23 | Nightingale | LDL_PL | NA | G | T | 0.033316 | 0.004337 | 3.00E-14 |
| rs644234 | let-7c-5p | trans | 2.80E-23 | Nightingale | XXL_VLDL_ | NA | G | T | 0.033445 | 0.004356 | 8.70E-15 |
| rs644234 | let-7c-5p | trans | 2.80E-23 | Nightingale | XS_VLDL_C | NA | G | T | 0.03665 | 0.004219 | 2.30E-18 |
| rs644234 | let-7c-5p | trans | 2.80E-23 | Nightingale | Remnant_( | NA | G | T | 0.03584 | 0.004319 | 5.30E-17 |
| rs644234 | let-7c-5p | trans | 2.80E-23 | Nightingale | S_LDL_C_p | NA | G | T | 0.041866 | 0.004364 | 5.50E-22 |
| rs644234 | let-7c-5p | trans | 2.80E-23 | Nightingale | VLDL_CE | NA | G | T | 0.035147 | 0.004348 | 2.10E-16 |
| rs644234 | miR-139-5p | trans | 1.17E-40 | Nightingale | S_LDL_L | NA | G | T | 0.026888 | 0.004346 | 1.20E-09 |
| rs644234 | miR-139-5p | trans | 1.17E-40 | Nightingale | LDL_P | NA | G | T | 0.030662 | 0.004344 | 2.10E-12 |
| rs644234 | miR-139-5p | trans | 1.17E-40 | Nightingale | M_VLDL_C | NA | G | T | 0.039209 | 0.004307 | 1.30E-19 |
| rs644234 | miR-139-5p | trans | 1.17E-40 | Nightingale | non_HDL_( | NA | G | T | 0.03414 | 0.004323 | 2.40E-15 |
| rs644234 | miR-139-5p | trans | 1.17E-40 | Nightingale | XXL_VLDL_ | NA | G | T | -0.03443 | 0.004384 | 6.20E-16 |
| rs644234 | miR-139-5p | trans | 1.17E-40 | Nightingale | S_VLDL_C_ | NA | G | T | 0.039745 | 0.00423 | 7.90E-21 |
| rs644234 | miR-139-5p | trans | 1.17E-40 | Nightingale | M_VLDL_C | NA | G | T | 0.031362 | 0.004084 | 3.10E-14 |
| rs644234 | miR-139-5p | trans | 1.17E-40 | Nightingale | S_VLDL_L | NA | G | T | 0.024494 | 0.004305 | 2.50E-09 |
| rs644234 | miR-139-5p | trans | 1.17E-40 | Nightingale | L_LDL_FC | NA | G | T | 0.031476 | 0.004268 | 4.00E-13 |
| rs644234 | miR-139-5p | trans | 1.17E-40 | Nightingale | L_LDL_L | NA | G | T | 0.031526 | 0.004304 | 3.20E-13 |
| rs644234 | miR-139-5p | trans | 1.17E-40 | Nightingale | L_VLDL_C_ | NA | G | T | 0.049243 | 0.004245 | 8.50E-32 |
| rs644234 | miR-139-5p | trans | 1.17E-40 | Nightingale | M_LDL_C | NA | G | T | 0.032088 | 0.004355 | 2.30E-13 |
| rs644234 | miR-139-5p | trans | 1.17E-40 | Nightingale | S_VLDL_PL | NA | G | T | 0.025736 | 0.004162 | 1.40E-09 |
| rs644234 | miR-139-5p | trans | 1.17E-40 | Nightingale | XS_VLDL_F | NA | G | T | 0.029149 | 0.004316 | 3.00E-11 |
| rs644234 | miR-139-5p | trans | 1.17E-40 | Nightingale | Clinical_LD | NA | G | T | 0.034557 | 0.004322 | 2.70E-15 |
| rs644234 | miR-139-5p | trans | 1.17E-40 | Nightingale | S_LDL_PL_ | NA | G | T | -0.02807 | 0.004339 | 5.00E-12 |
| rs644234 | miR-139-5p | trans | 1.17E-40 | Nightingale | S_HDL_C | NA | G | T | 0.025378 | 0.004339 | 9.40E-09 |
| rs644234 | miR-139-5p | trans | 1.17E-40 | Nightingale | XL_VLDL_F | NA | G | T | 0.03523 | 0.004214 | 1.70E-17 |
| rs644234 | miR-139-5p | trans | 1.17E-40 | Nightingale | L_LDL_C | NA | G | T | 0.030226 | 0.004298 | 3.00E-12 |
| rs644234 | miR-139-5p | trans | 1.17E-40 | Nightingale | IDL_C | NA | G | T | 0.033306 | 0.004198 | 4.80E-15 |
| rs644234 | miR-139-5p | trans | 1.17E-40 | Nightingale | S_LDL_CE_ | NA | G | T | 0.02885 | 0.004339 | 1.10E-12 |
| rs644234 | miR-139-5p | trans | 1.17E-40 | Nightingale | L_VLDL_CE | NA | G | T | 0.022755 | 0.004296 | 2.90E-08 |
| rs644234 | miR-139-5p | trans | 1.17E-40 | Nightingale | S_VLDL_C | NA | G | T | 0.037356 | 0.004346 | 2.60E-18 |
| rs644234 | miR-139-5p | trans | 1.17E-40 | Nightingale | S_VLDL_PL | NA | G | T | 0.032035 | 0.004339 | 4.90E-14 |

| | | | | | | | | | | | |
|---|---|---|---|---|---|---|---|---|---|---|---|
| rs644234 | miR-139-5p | trans | 1.17E-40 | Nightingale | Total_C | NA | G | T | 0.030145 | 0.004208 | 4.90E-13 |
| rs644234 | miR-139-5p | trans | 1.17E-40 | Nightingale | M_VLDL_F | NA | G | T | 0.032516 | 0.004346 | 3.90E-14 |
| rs644234 | miR-139-5p | trans | 1.17E-40 | Nightingale | IDL_PL | NA | G | T | 0.033582 | 0.004195 | 1.90E-15 |
| rs644234 | miR-139-5p | trans | 1.17E-40 | Nightingale | S_LDL_CE | NA | G | T | 0.029439 | 0.004349 | 1.50E-11 |
| rs644234 | miR-139-5p | trans | 1.17E-40 | Nightingale | M_LDL_CE | NA | G | T | 0.031208 | 0.004353 | 8.60E-13 |
| rs644234 | miR-139-5p | trans | 1.17E-40 | Nightingale | S_LDL_C | NA | G | T | 0.029694 | 0.004351 | 1.60E-11 |
| rs644234 | miR-139-5p | trans | 1.17E-40 | Nightingale | XXL_VLDL_ | NA | G | T | 0.031794 | 0.00438 | 2.10E-13 |
| rs644234 | miR-139-5p | trans | 1.17E-40 | Nightingale | LDL_L | NA | G | T | 0.031466 | 0.004332 | 5.80E-13 |
| rs644234 | miR-139-5p | trans | 1.17E-40 | Nightingale | XL_VLDL_P | NA | G | T | 0.039123 | 0.004353 | 7.10E-20 |
| rs644234 | miR-139-5p | trans | 1.17E-40 | Nightingale | S_VLDL_FC | NA | G | T | 0.027162 | 0.004154 | 1.20E-10 |
| rs644234 | miR-139-5p | trans | 1.17E-40 | Nightingale | L_VLDL_FC | NA | G | T | 0.046949 | 0.004305 | 3.90E-28 |
| rs644234 | miR-139-5p | trans | 1.17E-40 | Nightingale | IDL_L | NA | G | T | 0.033664 | 0.004203 | 2.30E-15 |
| rs644234 | miR-139-5p | trans | 1.17E-40 | Nightingale | XL_VLDL_C | NA | G | T | 0.03459 | 0.00417 | 1.50E-16 |
| rs644234 | miR-139-5p | trans | 1.17E-40 | Nightingale | M_VLDL_C | NA | G | T | 0.033859 | 0.004094 | 2.60E-16 |
| rs644234 | miR-139-5p | trans | 1.17E-40 | Nightingale | S_HDL_CE | NA | G | T | 0.025685 | 0.004354 | 7.00E-09 |
| rs644234 | miR-139-5p | trans | 1.17E-40 | Nightingale | M_VLDL_P | NA | G | T | 0.027925 | 0.004336 | 4.70E-11 |
| rs644234 | miR-139-5p | trans | 1.17E-40 | Nightingale | S_VLDL_CE | NA | G | T | 0.045161 | 0.004288 | 1.40E-25 |
| rs644234 | miR-139-5p | trans | 1.17E-40 | Nightingale | L_VLDL_CE | NA | G | T | 0.044093 | 0.00416 | 7.40E-27 |
| rs644234 | miR-139-5p | trans | 1.17E-40 | Nightingale | VLDL_C | NA | G | T | 0.031063 | 0.004341 | 2.10E-13 |
| rs644234 | miR-139-5p | trans | 1.17E-40 | Nightingale | S_LDL_P | NA | G | T | 0.028954 | 0.004351 | 3.00E-11 |
| rs644234 | miR-139-5p | trans | 1.17E-40 | Nightingale | LDL_CE | NA | G | T | 0.030617 | 0.004335 | 2.10E-12 |
| rs644234 | miR-139-5p | trans | 1.17E-40 | Nightingale | L_LDL_PL | NA | G | T | 0.03592 | 0.004314 | 1.30E-16 |
| rs644234 | miR-139-5p | trans | 1.17E-40 | Nightingale | Omega_6 | NA | G | T | 0.024169 | 0.004245 | 3.10E-09 |
| rs644234 | miR-139-5p | trans | 1.17E-40 | Nightingale | XXL_VLDL_ | NA | G | T | 0.029285 | 0.004328 | 5.00E-12 |
| rs644234 | miR-139-5p | trans | 1.17E-40 | Nightingale | XS_VLDL_P | NA | G | T | 0.035793 | 0.004289 | 9.90E-18 |
| rs644234 | miR-139-5p | trans | 1.17E-40 | Nightingale | LDL_FC | NA | G | T | 0.031549 | 0.0043 | 6.50E-13 |
| rs644234 | miR-139-5p | trans | 1.17E-40 | Nightingale | IDL_P | NA | G | T | 0.039024 | 0.004301 | 1.90E-19 |
| rs644234 | miR-139-5p | trans | 1.17E-40 | Nightingale | M_LDL_PL | NA | G | T | 0.031167 | 0.004353 | 1.00E-12 |
| rs644234 | miR-139-5p | trans | 1.17E-40 | Nightingale | ApoB_by_/ | NA | G | T | 0.025671 | 0.004281 | 2.10E-09 |
| rs644234 | miR-139-5p | trans | 1.17E-40 | Nightingale | Sphingomy | NA | G | T | 0.025922 | 0.004063 | 3.20E-10 |
| rs644234 | miR-139-5p | trans | 1.17E-40 | Nightingale | XL_VLDL_C | NA | G | T | 0.035608 | 0.004168 | 1.70E-17 |
| rs644234 | miR-139-5p | trans | 1.17E-40 | Nightingale | M_LDL_FC | NA | G | T | 0.032279 | 0.004341 | 2.40E-13 |
| rs644234 | miR-139-5p | trans | 1.17E-40 | Nightingale | VLDL_P | NA | G | T | 0.026555 | 0.004324 | 2.00E-10 |
| rs644234 | miR-139-5p | trans | 1.17E-40 | Nightingale | Total_PL | NA | G | T | 0.023006 | 0.004139 | 7.70E-09 |
| rs644234 | miR-139-5p | trans | 1.17E-40 | Nightingale | S_VLDL_CE | NA | G | T | 0.037313 | 0.004341 | 1.70E-18 |
| rs644234 | miR-139-5p | trans | 1.17E-40 | Nightingale | M_VLDL_C | NA | G | T | 0.037346 | 0.004335 | 5.30E-18 |
| rs644234 | miR-139-5p | trans | 1.17E-40 | Nightingale | LDL_C | NA | G | T | 0.031186 | 0.004328 | 9.50E-13 |
| rs644234 | miR-139-5p | trans | 1.17E-40 | Nightingale | XS_VLDL_L | NA | G | T | 0.033016 | 0.004273 | 1.30E-15 |
| rs644234 | miR-139-5p | trans | 1.17E-40 | Nightingale | XS_VLDL_P | NA | G | T | 0.031667 | 0.004286 | 1.90E-14 |
| rs644234 | miR-139-5p | trans | 1.17E-40 | Nightingale | S_LDL_FC | NA | G | T | 0.027196 | 0.00434 | 1.30E-09 |
| rs644234 | miR-139-5p | trans | 1.17E-40 | Nightingale | GlycA | NA | G | T | 0.025325 | 0.004328 | 4.90E-09 |
| rs644234 | miR-139-5p | trans | 1.17E-40 | Nightingale | M_VLDL_F | NA | G | T | 0.039386 | 0.004135 | 2.10E-21 |
| rs644234 | miR-139-5p | trans | 1.17E-40 | Nightingale | Total_L | NA | G | T | 0.025011 | 0.004276 | 1.50E-09 |
| rs644234 | miR-139-5p | trans | 1.17E-40 | Nightingale | XS_VLDL_C | NA | G | T | 0.036401 | 0.004195 | 4.00E-18 |
| rs644234 | miR-139-5p | trans | 1.17E-40 | Nightingale | LA | NA | G | T | 0.023278 | 0.004263 | 1.60E-08 |
| rs644234 | miR-139-5p | trans | 1.17E-40 | Nightingale | Total_FC | NA | G | T | 0.031385 | 0.004246 | 4.60E-14 |
| rs644234 | miR-139-5p | trans | 1.17E-40 | Nightingale | XXL_VLDL_ | NA | G | T | 0.023513 | 0.004375 | 5.40E-09 |
| rs644234 | miR-139-5p | trans | 1.17E-40 | Nightingale | VLDL_FC | NA | G | T | 0.024444 | 0.004315 | 3.60E-09 |
| rs644234 | miR-139-5p | trans | 1.17E-40 | Nightingale | L_VLDL_TG | NA | G | T | -0.04925 | 0.004326 | 6.10E-31 |
| rs644234 | miR-139-5p | trans | 1.17E-40 | Nightingale | S_VLDL_P | NA | G | T | 0.023388 | 0.004292 | 9.80E-09 |
| rs644234 | miR-139-5p | trans | 1.17E-40 | Nightingale | Total_CE | NA | G | T | 0.029391 | 0.004193 | 1.80E-12 |
| rs644234 | miR-139-5p | trans | 1.17E-40 | Nightingale | Citrate | NA | G | T | 0.024692 | 0.004318 | 6.00E-09 |
| rs644234 | miR-139-5p | trans | 1.17E-40 | Nightingale | IDL_FC | NA | G | T | 0.035982 | 0.004208 | 3.10E-17 |
| rs644234 | miR-139-5p | trans | 1.17E-40 | Nightingale | IDL_CE | NA | G | T | 0.032089 | 0.004197 | 4.30E-14 |
| rs644234 | miR-139-5p | trans | 1.17E-40 | Nightingale | M_LDL_L | NA | G | T | 0.031384 | 0.004353 | 7.10E-13 |
| rs644234 | miR-139-5p | trans | 1.17E-40 | Nightingale | L_LDL_P | NA | G | T | 0.028325 | 0.004332 | 7.70E-11 |
| rs644234 | miR-139-5p | trans | 1.17E-40 | Nightingale | ApoB | NA | G | T | 0.032929 | 0.004341 | 3.40E-14 |
| rs644234 | miR-139-5p | trans | 1.17E-40 | Nightingale | L_LDL_CE | NA | G | T | 0.029522 | 0.004308 | 9.80E-12 |
| rs644234 | miR-139-5p | trans | 1.17E-40 | Nightingale | XL_VLDL_T | NA | G | T | -0.04156 | 0.004213 | 4.70E-23 |
| rs644234 | miR-139-5p | trans | 1.17E-40 | Nightingale | M_VLDL_T | NA | G | T | -0.03625 | 0.004108 | 1.50E-18 |
| rs644234 | miR-139-5p | trans | 1.17E-40 | Nightingale | S_HDL_P | NA | G | T | 0.027802 | 0.004347 | 2.70E-10 |
| rs644234 | miR-139-5p | trans | 1.17E-40 | Nightingale | M_LDL_TG | NA | G | T | -0.02511 | 0.004293 | 8.50E-09 |
| rs644234 | miR-139-5p | trans | 1.17E-40 | Nightingale | M_LDL_C_ | NA | G | T | 0.026584 | 0.00435 | 1.60E-09 |

| | | | | | | | | | | | |
|---|---|---|---|---|---|---|---|---|---|---|---|
| rs644234 | miR-139-5p | trans | 1.17E-40 | Nightingale | M_VLDL_P | NA | G | T | 0.042108 | 0.004172 | 6.00E-24 |
| rs644234 | miR-139-5p | trans | 1.17E-40 | Nightingale | M_VLDL_P | NA | G | T | 0.028752 | 0.00434 | 1.40E-11 |
| rs644234 | miR-139-5p | trans | 1.17E-40 | Nightingale | S_HDL_L | NA | G | T | 0.023966 | 0.004332 | 2.70E-08 |
| rs644234 | miR-139-5p | trans | 1.17E-40 | Nightingale | S_VLDL_FC | NA | G | T | 0.036644 | 0.004348 | 1.70E-17 |
| rs644234 | miR-139-5p | trans | 1.17E-40 | Nightingale | XL_HDL_PL | NA | G | T | -0.02179 | 0.003994 | 4.70E-08 |
| rs644234 | miR-139-5p | trans | 1.17E-40 | Nightingale | L_VLDL_PL | NA | G | T | 0.021654 | 0.004222 | 4.40E-08 |
| rs644234 | miR-139-5p | trans | 1.17E-40 | Nightingale | M_LDL_P | NA | G | T | 0.033998 | 0.004359 | 7.50E-15 |
| rs644234 | miR-139-5p | trans | 1.17E-40 | Nightingale | S_VLDL_TG | NA | G | T | -0.03599 | 0.004208 | 1.80E-17 |
| rs644234 | miR-139-5p | trans | 1.17E-40 | Nightingale | XS_VLDL_F | NA | G | T | 0.035893 | 0.004271 | 7.40E-18 |
| rs644234 | miR-139-5p | trans | 1.17E-40 | Nightingale | LDL_PL | NA | G | T | 0.033316 | 0.004337 | 3.00E-14 |
| rs644234 | miR-139-5p | trans | 1.17E-40 | Nightingale | XXL_VLDL_ | NA | G | T | 0.033445 | 0.004356 | 8.70E-15 |
| rs644234 | miR-139-5p | trans | 1.17E-40 | Nightingale | XS_VLDL_C | NA | G | T | 0.03665 | 0.004219 | 2.30E-18 |
| rs644234 | miR-139-5p | trans | 1.17E-40 | Nightingale | Remnant_( | NA | G | T | 0.03584 | 0.004319 | 5.30E-17 |
| rs644234 | miR-139-5p | trans | 1.17E-40 | Nightingale | S_LDL_C_p | NA | G | T | 0.041866 | 0.004364 | 5.50E-22 |
| rs644234 | miR-139-5p | trans | 1.17E-40 | Nightingale | VLDL_CE | NA | G | T | 0.035147 | 0.004348 | 2.10E-16 |
| rs645982 | miR-10b-5p | trans | 1.29E-19 | Nightingale | M_VLDL_T | NA | G | A | -0.02249 | 0.00392 | 2.80E-09 |
| rs645982 | miR-10b-5p | trans | 1.29E-19 | Nightingale | L_VLDL_CE | NA | G | A | 0.026433 | 0.00397 | 1.10E-11 |
| rs645982 | miR-10b-5p | trans | 1.29E-19 | Nightingale | S_VLDL_C_ | NA | G | A | 0.023398 | 0.004037 | 3.10E-09 |
| rs645982 | miR-10b-5p | trans | 1.29E-19 | Nightingale | S_VLDL_TG | NA | G | A | -0.02158 | 0.004016 | 3.50E-08 |
| rs645982 | miR-10b-5p | trans | 1.29E-19 | Nightingale | M_VLDL_C | NA | G | A | 0.02149 | 0.003908 | 1.00E-08 |
| rs645982 | miR-10b-5p | trans | 1.29E-19 | Nightingale | L_VLDL_FC | NA | G | A | 0.023839 | 0.004109 | 5.60E-09 |
| rs645982 | miR-10b-5p | trans | 1.29E-19 | Nightingale | XL_VLDL_F | NA | G | A | 0.021903 | 0.004021 | 1.50E-08 |
| rs645982 | miR-10b-5p | trans | 1.29E-19 | Nightingale | M_VLDL_F | NA | G | A | 0.024804 | 0.003947 | 1.30E-10 |
| rs645982 | miR-10b-5p | trans | 1.29E-19 | Nightingale | L_VLDL_C_ | NA | G | A | 0.028271 | 0.004052 | 1.50E-12 |
| rs645982 | miR-10b-5p | trans | 1.29E-19 | Nightingale | XL_VLDL_C | NA | G | A | 0.022584 | 0.003977 | 4.60E-09 |
| rs645982 | miR-10b-5p | trans | 1.29E-19 | Nightingale | XL_VLDL_C | NA | G | A | 0.022127 | 0.003979 | 8.60E-09 |
| rs645982 | miR-10b-5p | trans | 1.29E-19 | Nightingale | S_VLDL_CE | NA | G | A | 0.025444 | 0.004093 | 3.10E-10 |
| rs645982 | miR-10b-5p | trans | 1.29E-19 | Nightingale | IDL_P | NA | G | A | 0.023241 | 0.004105 | 1.70E-08 |
| rs645982 | miR-10b-5p | trans | 1.29E-19 | Nightingale | XL_VLDL_T | NA | G | A | -0.02541 | 0.00402 | 8.10E-11 |
| rs645982 | miR-10b-5p | trans | 1.29E-19 | Nightingale | L_VLDL_TG | NA | G | A | -0.0255 | 0.004129 | 5.60E-10 |
| rs645982 | miR-10b-5p | trans | 1.29E-19 | Nightingale | S_LDL_C_p | NA | G | A | 0.027055 | 0.004165 | 8.40E-11 |
| rs645982 | miR-10b-5p | trans | 1.29E-19 | Nightingale | M_VLDL_P | NA | G | A | 0.024568 | 0.003981 | 3.80E-10 |
| rs645982 | miR-10b-5p | trans | 1.29E-19 | Nightingale | IDL_FC | NA | G | A | 0.022239 | 0.004016 | 3.40E-08 |
| rs647800 | miR-10b-5p | trans | 5.81E-20 | Nightingale | S_VLDL_CE | NA | G | A | 0.025327 | 0.004106 | 4.00E-10 |
| rs647800 | miR-10b-5p | trans | 5.81E-20 | Nightingale | XL_VLDL_C | NA | G | A | 0.02151 | 0.00399 | 1.80E-08 |
| rs647800 | miR-10b-5p | trans | 5.81E-20 | Nightingale | L_VLDL_CE | NA | G | A | 0.025076 | 0.003983 | 8.30E-11 |
| rs647800 | miR-10b-5p | trans | 5.81E-20 | Nightingale | IDL_P | NA | G | A | 0.023764 | 0.004119 | 8.30E-09 |
| rs647800 | miR-10b-5p | trans | 5.81E-20 | Nightingale | XL_VLDL_C | NA | G | A | 0.020884 | 0.003992 | 4.30E-08 |
| rs647800 | miR-10b-5p | trans | 5.81E-20 | Nightingale | M_VLDL_P | NA | G | A | 0.024312 | 0.003995 | 5.00E-10 |
| rs647800 | miR-10b-5p | trans | 5.81E-20 | Nightingale | L_VLDL_FC | NA | G | A | 0.023999 | 0.004122 | 5.20E-09 |
| rs647800 | miR-10b-5p | trans | 5.81E-20 | Nightingale | L_VLDL_TG | NA | G | A | -0.02549 | 0.004142 | 5.60E-10 |
| rs647800 | miR-10b-5p | trans | 5.81E-20 | Nightingale | M_VLDL_F | NA | G | A | 0.02381 | 0.00396 | 5.80E-10 |
| rs647800 | miR-10b-5p | trans | 5.81E-20 | Nightingale | S_LDL_C_p | NA | G | A | 0.025365 | 0.004178 | 1.90E-09 |
| rs647800 | miR-10b-5p | trans | 5.81E-20 | Nightingale | S_VLDL_C_ | NA | G | A | 0.022859 | 0.004051 | 6.20E-09 |
| rs647800 | miR-10b-5p | trans | 5.81E-20 | Nightingale | M_VLDL_C | NA | G | A | 0.020588 | 0.003921 | 3.30E-08 |
| rs647800 | miR-10b-5p | trans | 5.81E-20 | Nightingale | M_VLDL_T | NA | G | A | -0.0217 | 0.003933 | 7.80E-09 |
| rs647800 | miR-10b-5p | trans | 5.81E-20 | Nightingale | XS_VLDL_C | NA | G | A | 0.021894 | 0.00404 | 4.70E-08 |
| rs647800 | miR-10b-5p | trans | 5.81E-20 | Nightingale | XL_VLDL_F | NA | G | A | 0.021414 | 0.004034 | 2.30E-08 |
| rs647800 | miR-10b-5p | trans | 5.81E-20 | Nightingale | XL_VLDL_T | NA | G | A | -0.02478 | 0.004033 | 2.00E-10 |
| rs647800 | miR-10b-5p | trans | 5.81E-20 | Nightingale | L_VLDL_C_ | NA | G | A | 0.027343 | 0.004065 | 7.20E-12 |
| rs649129 | let-7a-5p | trans | 1.48E-12 | Nightingale | M_LDL_L | NA | T | C | 0.051759 | 0.005111 | 6.60E-25 |
| rs649129 | let-7a-5p | trans | 1.48E-12 | Nightingale | S_HDL_FC | NA | T | C | 0.035785 | 0.00501 | 2.10E-13 |
| rs649129 | let-7a-5p | trans | 1.48E-12 | Nightingale | M_VLDL_C | NA | T | C | 0.056026 | 0.00509 | 9.10E-30 |
| rs649129 | let-7a-5p | trans | 1.48E-12 | Nightingale | M_LDL_C_ | NA | T | C | 0.037633 | 0.005107 | 1.40E-13 |
| rs649129 | let-7a-5p | trans | 1.48E-12 | Nightingale | S_HDL_CE | NA | T | C | 0.045905 | 0.005112 | 3.70E-19 |
| rs649129 | let-7a-5p | trans | 1.48E-12 | Nightingale | Remnant_( | NA | T | C | 0.053675 | 0.005071 | 4.00E-28 |
| rs649129 | let-7a-5p | trans | 1.48E-12 | Nightingale | HDL_size | NA | T | C | -0.03089 | 0.004525 | 3.00E-12 |
| rs649129 | let-7a-5p | trans | 1.48E-12 | Nightingale | Pyruvate | NA | T | C | 0.030064 | 0.005112 | 4.10E-09 |
| rs649129 | let-7a-5p | trans | 1.48E-12 | Nightingale | M_VLDL_P | NA | T | C | 0.041826 | 0.005096 | 7.80E-18 |
| rs649129 | let-7a-5p | trans | 1.48E-12 | Nightingale | L_VLDL_C_ | NA | T | C | 0.072931 | 0.004985 | 1.90E-49 |
| rs649129 | let-7a-5p | trans | 1.48E-12 | Nightingale | ApoB_by_/ | NA | T | C | 0.038436 | 0.005027 | 5.80E-15 |
| rs649129 | let-7a-5p | trans | 1.48E-12 | Nightingale | S_VLDL_L | NA | T | C | 0.03374 | 0.005054 | 7.60E-13 |
| rs649129 | let-7a-5p | trans | 1.48E-12 | Nightingale | XXL_VLDL_ | NA | T | C | 0.051604 | 0.005117 | 3.20E-24 |

| | | | | | | | | | | | |
|---|---|---|---|---|---|---|---|---|---|---|---|
| rs649129 | let-7a-5p | trans | 1.48E-12 | Nightingale | S_LDL_PL | NA | T | C | 0.038147 | 0.005092 | 3.30E-14 |
| rs649129 | let-7a-5p | trans | 1.48E-12 | Nightingale | S_VLDL_CE | NA | T | C | 0.053637 | 0.005097 | 6.20E-28 |
| rs649129 | let-7a-5p | trans | 1.48E-12 | Nightingale | IDL_CE | NA | T | C | 0.052641 | 0.004928 | 3.40E-27 |
| rs649129 | let-7a-5p | trans | 1.48E-12 | Nightingale | VLDL_CE | NA | T | C | 0.049788 | 0.005105 | 3.30E-24 |
| rs649129 | let-7a-5p | trans | 1.48E-12 | Nightingale | M_LDL_FC | NA | T | C | 0.056788 | 0.005097 | 3.80E-29 |
| rs649129 | let-7a-5p | trans | 1.48E-12 | Nightingale | M_VLDL_P | NA | T | C | 0.040449 | 0.005091 | 8.30E-17 |
| rs649129 | let-7a-5p | trans | 1.48E-12 | Nightingale | L_VLDL_FC | NA | T | C | 0.06918 | 0.005055 | 3.20E-43 |
| rs649129 | let-7a-5p | trans | 1.48E-12 | Nightingale | Total_P | NA | T | C | 0.030298 | 0.004804 | 6.30E-11 |
| rs649129 | let-7a-5p | trans | 1.48E-12 | Nightingale | M_VLDL_C | NA | T | C | 0.059663 | 0.005057 | 3.90E-33 |
| rs649129 | let-7a-5p | trans | 1.48E-12 | Nightingale | LDL_L | NA | T | C | 0.053444 | 0.005087 | 1.40E-26 |
| rs649129 | let-7a-5p | trans | 1.48E-12 | Nightingale | VLDL_PL | NA | T | C | 0.025811 | 0.005043 | 2.40E-08 |
| rs649129 | let-7a-5p | trans | 1.48E-12 | Nightingale | S_LDL_C | NA | T | C | 0.048975 | 0.005109 | 2.00E-22 |
| rs649129 | let-7a-5p | trans | 1.48E-12 | Nightingale | L_VLDL_CE | NA | T | C | 0.065497 | 0.004884 | 6.20E-42 |
| rs649129 | let-7a-5p | trans | 1.48E-12 | Nightingale | IDL_L | NA | T | C | 0.053788 | 0.004935 | 1.40E-28 |
| rs649129 | let-7a-5p | trans | 1.48E-12 | Nightingale | Clinical_LD | NA | T | C | 0.057836 | 0.005074 | 1.10E-30 |
| rs649129 | let-7a-5p | trans | 1.48E-12 | Nightingale | LDL_C | NA | T | C | 0.053645 | 0.005082 | 9.90E-27 |
| rs649129 | let-7a-5p | trans | 1.48E-12 | Nightingale | IDL_C | NA | T | C | 0.054101 | 0.004929 | 1.20E-28 |
| rs649129 | let-7a-5p | trans | 1.48E-12 | Nightingale | L_HDL_TG_ | NA | T | C | 0.024039 | 0.004822 | 1.90E-08 |
| rs649129 | let-7a-5p | trans | 1.48E-12 | Nightingale | LA | NA | T | C | 0.0377 | 0.005005 | 2.10E-15 |
| rs649129 | let-7a-5p | trans | 1.48E-12 | Nightingale | S_LDL_P | NA | T | C | 0.044892 | 0.005109 | 1.70E-19 |
| rs649129 | let-7a-5p | trans | 1.48E-12 | Nightingale | XXL_VLDL_ | NA | T | C | 0.033222 | 0.005139 | 3.50E-12 |
| rs649129 | let-7a-5p | trans | 1.48E-12 | Nightingale | M_VLDL_T | NA | T | C | -0.05565 | 0.004823 | 1.80E-30 |
| rs649129 | let-7a-5p | trans | 1.48E-12 | Nightingale | M_LDL_CE | NA | T | C | 0.04991 | 0.005111 | 1.90E-23 |
| rs649129 | let-7a-5p | trans | 1.48E-12 | Nightingale | L_LDL_L | NA | T | C | 0.054447 | 0.005054 | 6.20E-28 |
| rs649129 | let-7a-5p | trans | 1.48E-12 | Nightingale | S_VLDL_FC | NA | T | C | 0.054336 | 0.005105 | 5.10E-28 |
| rs649129 | let-7a-5p | trans | 1.48E-12 | Nightingale | IDL_C_pct | NA | T | C | 0.03258 | 0.004986 | 3.60E-10 |
| rs649129 | let-7a-5p | trans | 1.48E-12 | Nightingale | S_LDL_PL_ | NA | T | C | -0.04003 | 0.005094 | 5.10E-17 |
| rs649129 | let-7a-5p | trans | 1.48E-12 | Nightingale | S_HDL_C | NA | T | C | 0.045127 | 0.005095 | 8.00E-19 |
| rs649129 | let-7a-5p | trans | 1.48E-12 | Nightingale | S_VLDL_P | NA | T | C | 0.031863 | 0.00504 | 8.10E-12 |
| rs649129 | let-7a-5p | trans | 1.48E-12 | Nightingale | Phosphogly | NA | T | C | 0.027915 | 0.004834 | 9.60E-10 |
| rs649129 | let-7a-5p | trans | 1.48E-12 | Nightingale | ApoB | NA | T | C | 0.051073 | 0.005097 | 8.20E-25 |
| rs649129 | let-7a-5p | trans | 1.48E-12 | Nightingale | S_VLDL_FC | NA | T | C | 0.045731 | 0.004877 | 1.40E-20 |
| rs649129 | let-7a-5p | trans | 1.48E-12 | Nightingale | XS_VLDL_P | NA | T | C | 0.041856 | 0.005033 | 4.60E-19 |
| rs649129 | let-7a-5p | trans | 1.48E-12 | Nightingale | Total_PL | NA | T | C | 0.034937 | 0.00486 | 3.10E-14 |
| rs649129 | let-7a-5p | trans | 1.48E-12 | Nightingale | L_LDL_TG_ | NA | T | C | -0.03492 | 0.005006 | 9.60E-12 |
| rs649129 | let-7a-5p | trans | 1.48E-12 | Nightingale | S_HDL_PL | NA | T | C | 0.03281 | 0.00506 | 2.30E-11 |
| rs649129 | let-7a-5p | trans | 1.48E-12 | Nightingale | XL_VLDL_P | NA | T | C | 0.052761 | 0.005112 | 2.60E-26 |
| rs649129 | let-7a-5p | trans | 1.48E-12 | Nightingale | XS_VLDL_C | NA | T | C | 0.034575 | 0.004816 | 3.00E-12 |
| rs649129 | let-7a-5p | trans | 1.48E-12 | Nightingale | M_VLDL_C | NA | T | C | 0.049074 | 0.004795 | 4.20E-24 |
| rs649129 | let-7a-5p | trans | 1.48E-12 | Nightingale | L_LDL_P | NA | T | C | 0.046154 | 0.005087 | 1.60E-20 |
| rs649129 | let-7a-5p | trans | 1.48E-12 | Nightingale | XL_HDL_L | NA | T | C | -0.02498 | 0.00459 | 1.30E-08 |
| rs649129 | let-7a-5p | trans | 1.48E-12 | Nightingale | L_VLDL_TG | NA | T | C | -0.0723 | 0.00508 | 1.20E-47 |
| rs649129 | let-7a-5p | trans | 1.48E-12 | Nightingale | XL_HDL_C_ | NA | T | C | 0.033715 | 0.004874 | 2.30E-11 |
| rs649129 | let-7a-5p | trans | 1.48E-12 | Nightingale | LDL_P | NA | T | C | 0.048505 | 0.0051 | 2.00E-22 |
| rs649129 | let-7a-5p | trans | 1.48E-12 | Nightingale | XS_VLDL_P | NA | T | C | 0.049752 | 0.005036 | 1.20E-25 |
| rs649129 | let-7a-5p | trans | 1.48E-12 | Nightingale | XL_VLDL_F | NA | T | C | 0.050654 | 0.004948 | 3.70E-25 |
| rs649129 | let-7a-5p | trans | 1.48E-12 | Nightingale | S_VLDL_C | NA | T | C | 0.054311 | 0.005103 | 2.80E-28 |
| rs649129 | let-7a-5p | trans | 1.48E-12 | Nightingale | XL_VLDL_C | NA | T | C | 0.054551 | 0.004896 | 2.20E-28 |
| rs649129 | let-7a-5p | trans | 1.48E-12 | Nightingale | XS_VLDL_T | NA | T | C | -0.0371 | 0.004853 | 6.70E-14 |
| rs649129 | let-7a-5p | trans | 1.48E-12 | Nightingale | S_LDL_CE_ | NA | T | C | 0.039688 | 0.005095 | 5.70E-17 |
| rs649129 | let-7a-5p | trans | 1.48E-12 | Nightingale | Cholines | NA | T | C | 0.030997 | 0.0048 | 1.00E-11 |
| rs649129 | let-7a-5p | trans | 1.48E-12 | Nightingale | M_LDL_TG | NA | T | C | -0.04553 | 0.005041 | 3.70E-19 |
| rs649129 | let-7a-5p | trans | 1.48E-12 | Nightingale | S_VLDL_CE | NA | T | C | 0.06893 | 0.005035 | 2.20E-42 |
| rs649129 | let-7a-5p | trans | 1.48E-12 | Nightingale | M_LDL_C | NA | T | C | 0.052736 | 0.005113 | 1.10E-25 |
| rs649129 | let-7a-5p | trans | 1.48E-12 | Nightingale | S_VLDL_PL | NA | T | C | 0.046421 | 0.005094 | 1.80E-21 |
| rs649129 | let-7a-5p | trans | 1.48E-12 | Nightingale | M_VLDL_F | NA | T | C | 0.059402 | 0.004856 | 2.80E-34 |
| rs649129 | let-7a-5p | trans | 1.48E-12 | Nightingale | XS_VLDL_F | NA | T | C | 0.044222 | 0.005067 | 1.20E-17 |
| rs649129 | let-7a-5p | trans | 1.48E-12 | Nightingale | S_LDL_TG_ | NA | T | C | -0.03709 | 0.004949 | 3.70E-13 |
| rs649129 | let-7a-5p | trans | 1.48E-12 | Nightingale | XS_VLDL_C | NA | T | C | 0.037225 | 0.004841 | 5.30E-14 |
| rs649129 | let-7a-5p | trans | 1.48E-12 | Nightingale | M_VLDL_C_ | NA | T | C | 0.052426 | 0.004807 | 3.20E-27 |
| rs649129 | let-7a-5p | trans | 1.48E-12 | Nightingale | Total_C | NA | T | C | 0.04944 | 0.004941 | 7.20E-25 |
| rs649129 | let-7a-5p | trans | 1.48E-12 | Nightingale | M_LDL_PL | NA | T | C | 0.052914 | 0.005111 | 8.10E-26 |
| rs649129 | let-7a-5p | trans | 1.48E-12 | Nightingale | XL_HDL_PL | NA | T | C | -0.03729 | 0.004689 | 1.80E-15 |

| | | | | | | | | | | | |
|---|---|---|---|---|---|---|---|---|---|---|---|
| rs649129 | let-7a-5p | trans | 1.48E-12 | Nightingale | VLDL_C | NA | T | C | 0.043038 | 0.005097 | 6.90E-19 |
| rs649129 | let-7a-5p | trans | 1.48E-12 | Nightingale | S_LDL_C_p | NA | T | C | 0.066081 | 0.005124 | 1.30E-38 |
| rs649129 | let-7a-5p | trans | 1.48E-12 | Nightingale | S_HDL_PL_ | NA | T | C | -0.0287 | 0.004934 | 1.60E-08 |
| rs649129 | let-7a-5p | trans | 1.48E-12 | Nightingale | PUFA | NA | T | C | 0.032829 | 0.004978 | 4.10E-12 |
| rs649129 | let-7a-5p | trans | 1.48E-12 | Nightingale | LDL_CE | NA | T | C | 0.052086 | 0.00509 | 2.20E-25 |
| rs649129 | let-7a-5p | trans | 1.48E-12 | Nightingale | L_VLDL_PL | NA | T | C | 0.027864 | 0.004958 | 6.90E-10 |
| rs649129 | let-7a-5p | trans | 1.48E-12 | Nightingale | XXL_VLDL_ | NA | T | C | 0.041451 | 0.005085 | 1.40E-16 |
| rs649129 | let-7a-5p | trans | 1.48E-12 | Nightingale | non_HDL_( | NA | T | C | 0.05472 | 0.005076 | 2.20E-28 |
| rs649129 | let-7a-5p | trans | 1.48E-12 | Nightingale | S_VLDL_TG | NA | T | C | -0.05782 | 0.00494 | 1.20E-31 |
| rs649129 | let-7a-5p | trans | 1.48E-12 | Nightingale | IDL_PL | NA | T | C | 0.052831 | 0.004925 | 4.10E-28 |
| rs649129 | let-7a-5p | trans | 1.48E-12 | Nightingale | VLDL_P | NA | T | C | 0.036367 | 0.005077 | 2.60E-14 |
| rs649129 | let-7a-5p | trans | 1.48E-12 | Nightingale | XL_HDL_PL | NA | T | C | -0.02773 | 0.004591 | 5.90E-10 |
| rs649129 | let-7a-5p | trans | 1.48E-12 | Nightingale | M_VLDL_P | NA | T | C | 0.063527 | 0.004898 | 1.10E-38 |
| rs649129 | let-7a-5p | trans | 1.48E-12 | Nightingale | LDL_FC | NA | T | C | 0.055859 | 0.005049 | 1.30E-28 |
| rs649129 | let-7a-5p | trans | 1.48E-12 | Nightingale | IDL_FC_pct | NA | T | C | 0.033568 | 0.005064 | 7.50E-11 |
| rs649129 | let-7a-5p | trans | 1.48E-12 | Nightingale | L_LDL_CE | NA | T | C | 0.052051 | 0.005058 | 1.30E-25 |
| rs649129 | let-7a-5p | trans | 1.48E-12 | Nightingale | XS_VLDL_C | NA | T | C | 0.052439 | 0.004954 | 3.00E-28 |
| rs649129 | let-7a-5p | trans | 1.48E-12 | Nightingale | S_VLDL_PL | NA | T | C | 0.043676 | 0.004887 | 8.10E-19 |
| rs649129 | let-7a-5p | trans | 1.48E-12 | Nightingale | VLDL_FC | NA | T | C | 0.03243 | 0.005066 | 7.10E-12 |
| rs649129 | let-7a-5p | trans | 1.48E-12 | Nightingale | XS_VLDL_F | NA | T | C | 0.04923 | 0.005015 | 2.60E-25 |
| rs649129 | let-7a-5p | trans | 1.48E-12 | Nightingale | Total_CE | NA | T | C | 0.049106 | 0.004923 | 1.40E-24 |
| rs649129 | let-7a-5p | trans | 1.48E-12 | Nightingale | S_VLDL_C_ | NA | T | C | 0.062755 | 0.004967 | 1.10E-36 |
| rs649129 | let-7a-5p | trans | 1.48E-12 | Nightingale | IDL_P | NA | T | C | 0.059758 | 0.00505 | 4.30E-33 |
| rs649129 | let-7a-5p | trans | 1.48E-12 | Nightingale | Total_FC | NA | T | C | 0.049272 | 0.004986 | 5.60E-25 |
| rs649129 | let-7a-5p | trans | 1.48E-12 | Nightingale | LDL_PL | NA | T | C | 0.055857 | 0.005092 | 1.20E-28 |
| rs649129 | let-7a-5p | trans | 1.48E-12 | Nightingale | XL_VLDL_C | NA | T | C | 0.055011 | 0.004894 | 5.60E-29 |
| rs649129 | let-7a-5p | trans | 1.48E-12 | Nightingale | S_LDL_CE | NA | T | C | 0.047148 | 0.005106 | 3.20E-21 |
| rs649129 | let-7a-5p | trans | 1.48E-12 | Nightingale | Total_L | NA | T | C | 0.038384 | 0.005021 | 1.20E-15 |
| rs649129 | let-7a-5p | trans | 1.48E-12 | Nightingale | Sphingomy | NA | T | C | 0.040605 | 0.004771 | 1.30E-17 |
| rs649129 | let-7a-5p | trans | 1.48E-12 | Nightingale | L_VLDL_CE | NA | T | C | 0.030309 | 0.005044 | 1.00E-10 |
| rs649129 | let-7a-5p | trans | 1.48E-12 | Nightingale | XXL_VLDL_ | NA | T | C | 0.050813 | 0.005145 | 1.80E-23 |
| rs649129 | let-7a-5p | trans | 1.48E-12 | Nightingale | S_LDL_L | NA | T | C | 0.044422 | 0.005103 | 8.20E-19 |
| rs649129 | let-7a-5p | trans | 1.48E-12 | Nightingale | M_VLDL_F | NA | T | C | 0.047627 | 0.005103 | 3.80E-22 |
| rs649129 | let-7a-5p | trans | 1.48E-12 | Nightingale | L_LDL_FC | NA | T | C | 0.055728 | 0.005012 | 4.60E-29 |
| rs649129 | let-7a-5p | trans | 1.48E-12 | Nightingale | XXL_VLDL_ | NA | T | C | -0.05327 | 0.00515 | 8.30E-26 |
| rs649129 | let-7a-5p | trans | 1.48E-12 | Nightingale | IDL_TG_pc | NA | T | C | -0.03301 | 0.004934 | 7.00E-11 |
| rs649129 | let-7a-5p | trans | 1.48E-12 | Nightingale | XS_VLDL_L | NA | T | C | 0.045001 | 0.005017 | 9.20E-22 |
| rs649129 | let-7a-5p | trans | 1.48E-12 | Nightingale | L_LDL_PL | NA | T | C | 0.059608 | 0.005065 | 8.20E-33 |
| rs649129 | let-7a-5p | trans | 1.48E-12 | Nightingale | S_HDL_L | NA | T | C | 0.038495 | 0.005086 | 1.30E-14 |
| rs649129 | let-7a-5p | trans | 1.48E-12 | Nightingale | Phosphatic | NA | T | C | 0.02716 | 0.004782 | 1.80E-09 |
| rs649129 | let-7a-5p | trans | 1.48E-12 | Nightingale | L_LDL_C | NA | T | C | 0.053389 | 0.005046 | 7.00E-27 |
| rs649129 | let-7a-5p | trans | 1.48E-12 | Nightingale | XL_VLDL_T | NA | T | C | -0.06153 | 0.004947 | 1.10E-35 |
| rs649129 | let-7a-5p | trans | 1.48E-12 | Nightingale | S_HDL_P | NA | T | C | 0.048101 | 0.005104 | 4.00E-21 |
| rs649129 | let-7a-5p | trans | 1.48E-12 | Nightingale | Citrate | NA | T | C | 0.027695 | 0.005071 | 1.70E-08 |
| rs649129 | let-7a-5p | trans | 1.48E-12 | Nightingale | XL_VLDL_C | NA | T | C | 0.029049 | 0.005025 | 4.30E-10 |
| rs649129 | let-7a-5p | trans | 1.48E-12 | Nightingale | Omega_6 | NA | T | C | 0.03716 | 0.004984 | 3.70E-15 |
| rs649129 | let-7a-5p | trans | 1.48E-12 | Nightingale | M_LDL_P | NA | T | C | 0.051271 | 0.005118 | 1.50E-24 |
| rs649129 | let-7a-5p | trans | 1.48E-12 | Nightingale | S_LDL_FC | NA | T | C | 0.048996 | 0.005096 | 7.80E-22 |
| rs649129 | let-7a-5p | trans | 1.48E-12 | Nightingale | M_VLDL_L | NA | T | C | 0.030569 | 0.005069 | 1.20E-10 |
| rs649129 | let-7a-5p | trans | 1.48E-12 | Nightingale | XS_VLDL_C | NA | T | C | 0.052973 | 0.004925 | 6.90E-29 |
| rs649129 | let-7a-5p | trans | 1.48E-12 | Nightingale | IDL_FC | NA | T | C | 0.056993 | 0.004941 | 1.40E-31 |
| rs649129 | miR-10b-5 | trans | 2.54E-34 | Nightingale | M_LDL_L | NA | T | C | 0.051759 | 0.005111 | 6.60E-25 |
| rs649129 | miR-10b-5 | trans | 2.54E-34 | Nightingale | S_HDL_FC | NA | T | C | 0.035785 | 0.00501 | 2.10E-13 |
| rs649129 | miR-10b-5 | trans | 2.54E-34 | Nightingale | M_VLDL_C | NA | T | C | 0.056026 | 0.00509 | 9.10E-30 |
| rs649129 | miR-10b-5 | trans | 2.54E-34 | Nightingale | M_LDL_C_ | NA | T | C | 0.037633 | 0.005107 | 1.40E-13 |
| rs649129 | miR-10b-5 | trans | 2.54E-34 | Nightingale | S_HDL_CE | NA | T | C | 0.045905 | 0.005112 | 3.70E-19 |
| rs649129 | miR-10b-5 | trans | 2.54E-34 | Nightingale | Remnant_( | NA | T | C | 0.053675 | 0.005071 | 4.00E-28 |
| rs649129 | miR-10b-5 | trans | 2.54E-34 | Nightingale | HDL_size | NA | T | C | -0.03089 | 0.004525 | 3.00E-12 |
| rs649129 | miR-10b-5 | trans | 2.54E-34 | Nightingale | Pyruvate | NA | T | C | 0.030064 | 0.005112 | 4.10E-09 |
| rs649129 | miR-10b-5 | trans | 2.54E-34 | Nightingale | M_VLDL_P | NA | T | C | 0.041826 | 0.005096 | 7.80E-18 |
| rs649129 | miR-10b-5 | trans | 2.54E-34 | Nightingale | L_VLDL_C_ | NA | T | C | 0.072931 | 0.004985 | 1.90E-49 |
| rs649129 | miR-10b-5 | trans | 2.54E-34 | Nightingale | ApoB_by_/ | NA | T | C | 0.038436 | 0.005027 | 5.80E-15 |
| rs649129 | miR-10b-5 | trans | 2.54E-34 | Nightingale | S_VLDL_L | NA | T | C | 0.03374 | 0.005054 | 7.60E-13 |

| | | | | | | | | | | | |
|---|---|---|---|---|---|---|---|---|---|---|---|
| rs649129 | miR-10b-5 | trans | 2.54E-34 | Nightingale | XXL_VLDL_ | NA | T | C | 0.051604 | 0.005117 | 3.20E-24 |
| rs649129 | miR-10b-5 | trans | 2.54E-34 | Nightingale | S_LDL_PL | NA | T | C | 0.038147 | 0.005092 | 3.30E-14 |
| rs649129 | miR-10b-5 | trans | 2.54E-34 | Nightingale | S_VLDL_CE | NA | T | C | 0.053637 | 0.005097 | 6.20E-28 |
| rs649129 | miR-10b-5 | trans | 2.54E-34 | Nightingale | IDL_CE | NA | T | C | 0.052641 | 0.004928 | 3.40E-27 |
| rs649129 | miR-10b-5 | trans | 2.54E-34 | Nightingale | VLDL_CE | NA | T | C | 0.049788 | 0.005105 | 3.30E-24 |
| rs649129 | miR-10b-5 | trans | 2.54E-34 | Nightingale | M_LDL_FC | NA | T | C | 0.056788 | 0.005097 | 3.80E-29 |
| rs649129 | miR-10b-5 | trans | 2.54E-34 | Nightingale | M_VLDL_P | NA | T | C | 0.040449 | 0.005091 | 8.30E-17 |
| rs649129 | miR-10b-5 | trans | 2.54E-34 | Nightingale | L_VLDL_FC | NA | T | C | 0.06918 | 0.005055 | 3.20E-43 |
| rs649129 | miR-10b-5 | trans | 2.54E-34 | Nightingale | Total_P | NA | T | C | 0.030298 | 0.004804 | 6.30E-11 |
| rs649129 | miR-10b-5 | trans | 2.54E-34 | Nightingale | M_VLDL_C | NA | T | C | 0.059663 | 0.005057 | 3.90E-33 |
| rs649129 | miR-10b-5 | trans | 2.54E-34 | Nightingale | LDL_L | NA | T | C | 0.053444 | 0.005087 | 1.40E-26 |
| rs649129 | miR-10b-5 | trans | 2.54E-34 | Nightingale | VLDL_PL | NA | T | C | 0.025811 | 0.005043 | 2.40E-08 |
| rs649129 | miR-10b-5 | trans | 2.54E-34 | Nightingale | S_LDL_C | NA | T | C | 0.048975 | 0.005109 | 2.00E-22 |
| rs649129 | miR-10b-5 | trans | 2.54E-34 | Nightingale | L_VLDL_CE | NA | T | C | 0.065497 | 0.004884 | 6.20E-42 |
| rs649129 | miR-10b-5 | trans | 2.54E-34 | Nightingale | IDL_L | NA | T | C | 0.053788 | 0.004935 | 1.40E-28 |
| rs649129 | miR-10b-5 | trans | 2.54E-34 | Nightingale | Clinical_LD | NA | T | C | 0.057836 | 0.005074 | 1.10E-30 |
| rs649129 | miR-10b-5 | trans | 2.54E-34 | Nightingale | LDL_C | NA | T | C | 0.053645 | 0.005082 | 9.90E-27 |
| rs649129 | miR-10b-5 | trans | 2.54E-34 | Nightingale | IDL_C | NA | T | C | 0.054101 | 0.004929 | 1.20E-28 |
| rs649129 | miR-10b-5 | trans | 2.54E-34 | Nightingale | L_HDL_TG_ | NA | T | C | 0.024039 | 0.004822 | 1.90E-08 |
| rs649129 | miR-10b-5 | trans | 2.54E-34 | Nightingale | LA | NA | T | C | 0.0377 | 0.005005 | 2.10E-15 |
| rs649129 | miR-10b-5 | trans | 2.54E-34 | Nightingale | S_LDL_P | NA | T | C | 0.044892 | 0.005109 | 1.70E-19 |
| rs649129 | miR-10b-5 | trans | 2.54E-34 | Nightingale | XXL_VLDL_ | NA | T | C | 0.033222 | 0.005139 | 3.50E-12 |
| rs649129 | miR-10b-5 | trans | 2.54E-34 | Nightingale | M_VLDL_T | NA | T | C | -0.05565 | 0.004823 | 1.80E-30 |
| rs649129 | miR-10b-5 | trans | 2.54E-34 | Nightingale | M_LDL_CE | NA | T | C | 0.04991 | 0.005111 | 1.90E-23 |
| rs649129 | miR-10b-5 | trans | 2.54E-34 | Nightingale | L_LDL_L | NA | T | C | 0.054447 | 0.005054 | 6.20E-28 |
| rs649129 | miR-10b-5 | trans | 2.54E-34 | Nightingale | S_VLDL_FC | NA | T | C | 0.054336 | 0.005105 | 5.10E-28 |
| rs649129 | miR-10b-5 | trans | 2.54E-34 | Nightingale | IDL_C_pct | NA | T | C | 0.03258 | 0.004986 | 3.60E-10 |
| rs649129 | miR-10b-5 | trans | 2.54E-34 | Nightingale | S_LDL_PL_ | NA | T | C | -0.04003 | 0.005094 | 5.10E-17 |
| rs649129 | miR-10b-5 | trans | 2.54E-34 | Nightingale | S_HDL_C | NA | T | C | 0.045127 | 0.005095 | 8.00E-19 |
| rs649129 | miR-10b-5 | trans | 2.54E-34 | Nightingale | S_VLDL_P | NA | T | C | 0.031863 | 0.00504 | 8.10E-12 |
| rs649129 | miR-10b-5 | trans | 2.54E-34 | Nightingale | Phosphogl | NA | T | C | 0.027915 | 0.004834 | 9.60E-10 |
| rs649129 | miR-10b-5 | trans | 2.54E-34 | Nightingale | ApoB | NA | T | C | 0.051073 | 0.005097 | 8.20E-25 |
| rs649129 | miR-10b-5 | trans | 2.54E-34 | Nightingale | S_VLDL_FC | NA | T | C | 0.045731 | 0.004877 | 1.40E-20 |
| rs649129 | miR-10b-5 | trans | 2.54E-34 | Nightingale | XS_VLDL_P | NA | T | C | 0.041856 | 0.005033 | 4.60E-19 |
| rs649129 | miR-10b-5 | trans | 2.54E-34 | Nightingale | Total_PL | NA | T | C | 0.034937 | 0.00486 | 3.10E-14 |
| rs649129 | miR-10b-5 | trans | 2.54E-34 | Nightingale | L_LDL_TG_ | NA | T | C | -0.03492 | 0.005006 | 9.60E-12 |
| rs649129 | miR-10b-5 | trans | 2.54E-34 | Nightingale | S_HDL_PL | NA | T | C | 0.03281 | 0.00506 | 2.30E-11 |
| rs649129 | miR-10b-5 | trans | 2.54E-34 | Nightingale | XL_VLDL_P | NA | T | C | 0.052761 | 0.005112 | 2.60E-26 |
| rs649129 | miR-10b-5 | trans | 2.54E-34 | Nightingale | XS_VLDL_C | NA | T | C | 0.034575 | 0.004816 | 3.00E-12 |
| rs649129 | miR-10b-5 | trans | 2.54E-34 | Nightingale | M_VLDL_C | NA | T | C | 0.049074 | 0.004795 | 4.20E-24 |
| rs649129 | miR-10b-5 | trans | 2.54E-34 | Nightingale | L_LDL_P | NA | T | C | 0.046154 | 0.005087 | 1.60E-20 |
| rs649129 | miR-10b-5 | trans | 2.54E-34 | Nightingale | XL_HDL_L | NA | T | C | -0.02498 | 0.00459 | 1.30E-08 |
| rs649129 | miR-10b-5 | trans | 2.54E-34 | Nightingale | L_VLDL_TG | NA | T | C | -0.0723 | 0.00508 | 1.20E-47 |
| rs649129 | miR-10b-5 | trans | 2.54E-34 | Nightingale | XL_HDL_C_ | NA | T | C | 0.033715 | 0.004874 | 2.30E-11 |
| rs649129 | miR-10b-5 | trans | 2.54E-34 | Nightingale | LDL_P | NA | T | C | 0.048505 | 0.0051 | 2.00E-22 |
| rs649129 | miR-10b-5 | trans | 2.54E-34 | Nightingale | XS_VLDL_P | NA | T | C | 0.049752 | 0.005036 | 1.20E-25 |
| rs649129 | miR-10b-5 | trans | 2.54E-34 | Nightingale | XL_VLDL_F | NA | T | C | 0.050654 | 0.004948 | 3.70E-25 |
| rs649129 | miR-10b-5 | trans | 2.54E-34 | Nightingale | S_VLDL_C | NA | T | C | 0.054311 | 0.005103 | 2.80E-28 |
| rs649129 | miR-10b-5 | trans | 2.54E-34 | Nightingale | XL_VLDL_C | NA | T | C | 0.054551 | 0.004896 | 2.20E-28 |
| rs649129 | miR-10b-5 | trans | 2.54E-34 | Nightingale | XS_VLDL_T | NA | T | C | -0.0371 | 0.004853 | 6.70E-14 |
| rs649129 | miR-10b-5 | trans | 2.54E-34 | Nightingale | S_LDL_CE_ | NA | T | C | 0.039688 | 0.005095 | 5.70E-17 |
| rs649129 | miR-10b-5 | trans | 2.54E-34 | Nightingale | Cholines | NA | T | C | 0.030997 | 0.0048 | 1.00E-11 |
| rs649129 | miR-10b-5 | trans | 2.54E-34 | Nightingale | M_LDL_TG | NA | T | C | -0.04553 | 0.005041 | 3.70E-19 |
| rs649129 | miR-10b-5 | trans | 2.54E-34 | Nightingale | S_VLDL_CE | NA | T | C | 0.06893 | 0.005035 | 2.20E-42 |
| rs649129 | miR-10b-5 | trans | 2.54E-34 | Nightingale | M_LDL_C | NA | T | C | 0.052736 | 0.005113 | 1.10E-25 |
| rs649129 | miR-10b-5 | trans | 2.54E-34 | Nightingale | S_VLDL_PL | NA | T | C | 0.046421 | 0.005094 | 1.80E-21 |
| rs649129 | miR-10b-5 | trans | 2.54E-34 | Nightingale | M_VLDL_F | NA | T | C | 0.059402 | 0.004856 | 2.80E-34 |
| rs649129 | miR-10b-5 | trans | 2.54E-34 | Nightingale | XS_VLDL_F | NA | T | C | 0.044222 | 0.005067 | 1.20E-17 |
| rs649129 | miR-10b-5 | trans | 2.54E-34 | Nightingale | S_LDL_TG_ | NA | T | C | -0.03709 | 0.004949 | 3.70E-13 |
| rs649129 | miR-10b-5 | trans | 2.54E-34 | Nightingale | XS_VLDL_C | NA | T | C | 0.037225 | 0.004841 | 5.30E-14 |
| rs649129 | miR-10b-5 | trans | 2.54E-34 | Nightingale | M_VLDL_C | NA | T | C | 0.052426 | 0.004807 | 3.20E-27 |
| rs649129 | miR-10b-5 | trans | 2.54E-34 | Nightingale | Total_C | NA | T | C | 0.04944 | 0.004941 | 7.20E-25 |
| rs649129 | miR-10b-5 | trans | 2.54E-34 | Nightingale | M_LDL_PL | NA | T | C | 0.052914 | 0.005111 | 8.10E-26 |

| | | | | | | | | | | | |
|---|---|---|---|---|---|---|---|---|---|---|---|
| rs649129 | miR-10b-5| trans | 2.54E-34 | Nightingale | XL_HDL_PL | NA | T | C | -0.03729 | 0.004689 | 1.80E-15 |
| rs649129 | miR-10b-5| trans | 2.54E-34 | Nightingale | VLDL_C | NA | T | C | 0.043038 | 0.005097 | 6.90E-19 |
| rs649129 | miR-10b-5| trans | 2.54E-34 | Nightingale | S_LDL_C_p | NA | T | C | 0.066081 | 0.005124 | 1.30E-38 |
| rs649129 | miR-10b-5| trans | 2.54E-34 | Nightingale | S_HDL_PL_ | NA | T | C | -0.0287 | 0.004934 | 1.60E-08 |
| rs649129 | miR-10b-5| trans | 2.54E-34 | Nightingale | PUFA | NA | T | C | 0.032829 | 0.004978 | 4.10E-12 |
| rs649129 | miR-10b-5| trans | 2.54E-34 | Nightingale | LDL_CE | NA | T | C | 0.052086 | 0.00509 | 2.20E-25 |
| rs649129 | miR-10b-5| trans | 2.54E-34 | Nightingale | L_VLDL_PL | NA | T | C | 0.027864 | 0.004958 | 6.90E-10 |
| rs649129 | miR-10b-5| trans | 2.54E-34 | Nightingale | XXL_VLDL_ | NA | T | C | 0.041451 | 0.005085 | 1.40E-16 |
| rs649129 | miR-10b-5| trans | 2.54E-34 | Nightingale | non_HDL_( | NA | T | C | 0.05472 | 0.005076 | 2.20E-28 |
| rs649129 | miR-10b-5| trans | 2.54E-34 | Nightingale | S_VLDL_TG | NA | T | C | -0.05782 | 0.00494 | 1.20E-31 |
| rs649129 | miR-10b-5| trans | 2.54E-34 | Nightingale | IDL_PL | NA | T | C | 0.052831 | 0.004925 | 4.10E-28 |
| rs649129 | miR-10b-5| trans | 2.54E-34 | Nightingale | VLDL_P | NA | T | C | 0.036367 | 0.005077 | 2.60E-14 |
| rs649129 | miR-10b-5| trans | 2.54E-34 | Nightingale | XL_HDL_PL | NA | T | C | -0.02773 | 0.004591 | 5.90E-10 |
| rs649129 | miR-10b-5| trans | 2.54E-34 | Nightingale | M_VLDL_P | NA | T | C | 0.063527 | 0.004898 | 1.10E-38 |
| rs649129 | miR-10b-5| trans | 2.54E-34 | Nightingale | LDL_FC | NA | T | C | 0.055859 | 0.005049 | 1.30E-28 |
| rs649129 | miR-10b-5| trans | 2.54E-34 | Nightingale | IDL_FC_pct | NA | T | C | 0.033568 | 0.005064 | 7.50E-11 |
| rs649129 | miR-10b-5| trans | 2.54E-34 | Nightingale | L_LDL_CE | NA | T | C | 0.052051 | 0.005058 | 1.30E-25 |
| rs649129 | miR-10b-5| trans | 2.54E-34 | Nightingale | XS_VLDL_C | NA | T | C | 0.052439 | 0.004954 | 3.00E-28 |
| rs649129 | miR-10b-5| trans | 2.54E-34 | Nightingale | S_VLDL_PL | NA | T | C | 0.043676 | 0.004887 | 8.10E-19 |
| rs649129 | miR-10b-5| trans | 2.54E-34 | Nightingale | VLDL_FC | NA | T | C | 0.03243 | 0.005066 | 7.10E-12 |
| rs649129 | miR-10b-5| trans | 2.54E-34 | Nightingale | XS_VLDL_F | NA | T | C | 0.04923 | 0.005015 | 2.60E-25 |
| rs649129 | miR-10b-5| trans | 2.54E-34 | Nightingale | Total_CE | NA | T | C | 0.049106 | 0.004923 | 1.40E-24 |
| rs649129 | miR-10b-5| trans | 2.54E-34 | Nightingale | S_VLDL_C_ | NA | T | C | 0.062755 | 0.004967 | 1.10E-36 |
| rs649129 | miR-10b-5| trans | 2.54E-34 | Nightingale | IDL_P | NA | T | C | 0.059758 | 0.00505 | 4.30E-33 |
| rs649129 | miR-10b-5| trans | 2.54E-34 | Nightingale | Total_FC | NA | T | C | 0.049272 | 0.004986 | 5.60E-25 |
| rs649129 | miR-10b-5| trans | 2.54E-34 | Nightingale | LDL_PL | NA | T | C | 0.055857 | 0.005092 | 1.20E-28 |
| rs649129 | miR-10b-5| trans | 2.54E-34 | Nightingale | XL_VLDL_C | NA | T | C | 0.055011 | 0.004894 | 5.60E-29 |
| rs649129 | miR-10b-5| trans | 2.54E-34 | Nightingale | S_LDL_CE | NA | T | C | 0.047148 | 0.005106 | 3.20E-21 |
| rs649129 | miR-10b-5| trans | 2.54E-34 | Nightingale | Total_L | NA | T | C | 0.038384 | 0.005021 | 1.20E-15 |
| rs649129 | miR-10b-5| trans | 2.54E-34 | Nightingale | Sphingomy | NA | T | C | 0.040605 | 0.004771 | 1.30E-17 |
| rs649129 | miR-10b-5| trans | 2.54E-34 | Nightingale | L_VLDL_CE | NA | T | C | 0.030309 | 0.005044 | 1.00E-10 |
| rs649129 | miR-10b-5| trans | 2.54E-34 | Nightingale | XXL_VLDL_ | NA | T | C | 0.050813 | 0.005145 | 1.80E-23 |
| rs649129 | miR-10b-5| trans | 2.54E-34 | Nightingale | S_LDL_L | NA | T | C | 0.044422 | 0.005103 | 8.20E-19 |
| rs649129 | miR-10b-5| trans | 2.54E-34 | Nightingale | M_VLDL_F | NA | T | C | 0.047627 | 0.005103 | 3.80E-22 |
| rs649129 | miR-10b-5| trans | 2.54E-34 | Nightingale | L_LDL_FC | NA | T | C | 0.055728 | 0.005012 | 4.60E-29 |
| rs649129 | miR-10b-5| trans | 2.54E-34 | Nightingale | XXL_VLDL_ | NA | T | C | -0.05327 | 0.00515 | 8.30E-26 |
| rs649129 | miR-10b-5| trans | 2.54E-34 | Nightingale | IDL_TG_pc | NA | T | C | -0.03301 | 0.004934 | 7.00E-11 |
| rs649129 | miR-10b-5| trans | 2.54E-34 | Nightingale | XS_VLDL_L | NA | T | C | 0.045001 | 0.005017 | 9.20E-22 |
| rs649129 | miR-10b-5| trans | 2.54E-34 | Nightingale | L_LDL_PL | NA | T | C | 0.059608 | 0.005065 | 8.20E-33 |
| rs649129 | miR-10b-5| trans | 2.54E-34 | Nightingale | S_HDL_L | NA | T | C | 0.038495 | 0.005086 | 1.30E-14 |
| rs649129 | miR-10b-5| trans | 2.54E-34 | Nightingale | Phosphatic | NA | T | C | 0.02716 | 0.004782 | 1.80E-09 |
| rs649129 | miR-10b-5| trans | 2.54E-34 | Nightingale | L_LDL_C | NA | T | C | 0.053389 | 0.005046 | 7.00E-27 |
| rs649129 | miR-10b-5| trans | 2.54E-34 | Nightingale | XL_VLDL_T | NA | T | C | -0.06153 | 0.004947 | 1.10E-35 |
| rs649129 | miR-10b-5| trans | 2.54E-34 | Nightingale | S_HDL_P | NA | T | C | 0.048101 | 0.005104 | 4.00E-21 |
| rs649129 | miR-10b-5| trans | 2.54E-34 | Nightingale | Citrate | NA | T | C | 0.027695 | 0.005071 | 1.70E-08 |
| rs649129 | miR-10b-5| trans | 2.54E-34 | Nightingale | XL_VLDL_C | NA | T | C | 0.029049 | 0.005025 | 4.30E-10 |
| rs649129 | miR-10b-5| trans | 2.54E-34 | Nightingale | Omega_6 | NA | T | C | 0.03716 | 0.004984 | 3.70E-15 |
| rs649129 | miR-10b-5| trans | 2.54E-34 | Nightingale | M_LDL_P | NA | T | C | 0.051271 | 0.005118 | 1.50E-24 |
| rs649129 | miR-10b-5| trans | 2.54E-34 | Nightingale | S_LDL_FC | NA | T | C | 0.048996 | 0.005096 | 7.80E-22 |
| rs649129 | miR-10b-5| trans | 2.54E-34 | Nightingale | M_VLDL_L | NA | T | C | 0.030569 | 0.005069 | 1.20E-10 |
| rs649129 | miR-10b-5| trans | 2.54E-34 | Nightingale | XS_VLDL_C | NA | T | C | 0.052973 | 0.004925 | 6.90E-29 |
| rs649129 | miR-10b-5| trans | 2.54E-34 | Nightingale | IDL_FC | NA | T | C | 0.056993 | 0.004941 | 1.40E-31 |
| rs649129 | miR-126-3| trans | 1.95E-37 | Nightingale | M_LDL_L | NA | T | C | 0.051759 | 0.005111 | 6.60E-25 |
| rs649129 | miR-126-3| trans | 1.95E-37 | Nightingale | S_HDL_FC | NA | T | C | 0.035785 | 0.00501 | 2.10E-13 |
| rs649129 | miR-126-3| trans | 1.95E-37 | Nightingale | M_VLDL_C | NA | T | C | 0.056026 | 0.00509 | 9.10E-30 |
| rs649129 | miR-126-3| trans | 1.95E-37 | Nightingale | M_LDL_C_ | NA | T | C | 0.037633 | 0.005107 | 1.40E-13 |
| rs649129 | miR-126-3| trans | 1.95E-37 | Nightingale | S_HDL_CE | NA | T | C | 0.045905 | 0.005112 | 3.70E-19 |
| rs649129 | miR-126-3| trans | 1.95E-37 | Nightingale | Remnant_( | NA | T | C | 0.053675 | 0.005071 | 4.00E-28 |
| rs649129 | miR-126-3| trans | 1.95E-37 | Nightingale | HDL_size | NA | T | C | -0.03089 | 0.004525 | 3.00E-12 |
| rs649129 | miR-126-3| trans | 1.95E-37 | Nightingale | Pyruvate | NA | T | C | 0.030064 | 0.005112 | 4.10E-09 |
| rs649129 | miR-126-3| trans | 1.95E-37 | Nightingale | M_VLDL_P | NA | T | C | 0.041826 | 0.005096 | 7.80E-18 |
| rs649129 | miR-126-3| trans | 1.95E-37 | Nightingale | L_VLDL_C_ | NA | T | C | 0.072931 | 0.004985 | 1.90E-49 |
| rs649129 | miR-126-3| trans | 1.95E-37 | Nightingale | ApoB_by_/ | NA | T | C | 0.038436 | 0.005027 | 5.80E-15 |

| | | | | | | | | | | | |
|---|---|---|---|---|---|---|---|---|---|---|---|
| rs649129 | miR-126-3p | trans | 1.95E-37 | Nightingale | S_VLDL_L | NA | T | C | 0.03374 | 0.005054 | 7.60E-13 |
| rs649129 | miR-126-3p | trans | 1.95E-37 | Nightingale | XXL_VLDL_ | NA | T | C | 0.051604 | 0.005117 | 3.20E-24 |
| rs649129 | miR-126-3p | trans | 1.95E-37 | Nightingale | S_LDL_PL | NA | T | C | 0.038147 | 0.005092 | 3.30E-14 |
| rs649129 | miR-126-3p | trans | 1.95E-37 | Nightingale | S_VLDL_CE | NA | T | C | 0.053637 | 0.005097 | 6.20E-28 |
| rs649129 | miR-126-3p | trans | 1.95E-37 | Nightingale | IDL_CE | NA | T | C | 0.052641 | 0.004928 | 3.40E-27 |
| rs649129 | miR-126-3p | trans | 1.95E-37 | Nightingale | VLDL_CE | NA | T | C | 0.049788 | 0.005105 | 3.30E-24 |
| rs649129 | miR-126-3p | trans | 1.95E-37 | Nightingale | M_LDL_FC | NA | T | C | 0.056788 | 0.005097 | 3.80E-29 |
| rs649129 | miR-126-3p | trans | 1.95E-37 | Nightingale | M_VLDL_P | NA | T | C | 0.040449 | 0.005091 | 8.30E-17 |
| rs649129 | miR-126-3p | trans | 1.95E-37 | Nightingale | L_VLDL_FC | NA | T | C | 0.06918 | 0.005055 | 3.20E-43 |
| rs649129 | miR-126-3p | trans | 1.95E-37 | Nightingale | Total_P | NA | T | C | 0.030298 | 0.004804 | 6.30E-11 |
| rs649129 | miR-126-3p | trans | 1.95E-37 | Nightingale | M_VLDL_C | NA | T | C | 0.059663 | 0.005057 | 3.90E-33 |
| rs649129 | miR-126-3p | trans | 1.95E-37 | Nightingale | LDL_L | NA | T | C | 0.053444 | 0.005087 | 1.40E-26 |
| rs649129 | miR-126-3p | trans | 1.95E-37 | Nightingale | VLDL_PL | NA | T | C | 0.025811 | 0.005043 | 2.40E-08 |
| rs649129 | miR-126-3p | trans | 1.95E-37 | Nightingale | S_LDL_C | NA | T | C | 0.048975 | 0.005109 | 2.00E-22 |
| rs649129 | miR-126-3p | trans | 1.95E-37 | Nightingale | L_VLDL_CE | NA | T | C | 0.065497 | 0.004884 | 6.20E-42 |
| rs649129 | miR-126-3p | trans | 1.95E-37 | Nightingale | IDL_L | NA | T | C | 0.053788 | 0.004935 | 1.40E-28 |
| rs649129 | miR-126-3p | trans | 1.95E-37 | Nightingale | Clinical_LD | NA | T | C | 0.057836 | 0.005074 | 1.10E-30 |
| rs649129 | miR-126-3p | trans | 1.95E-37 | Nightingale | LDL_C | NA | T | C | 0.053645 | 0.005082 | 9.90E-27 |
| rs649129 | miR-126-3p | trans | 1.95E-37 | Nightingale | IDL_C | NA | T | C | 0.054101 | 0.004929 | 1.20E-28 |
| rs649129 | miR-126-3p | trans | 1.95E-37 | Nightingale | L_HDL_TG_ | NA | T | C | 0.024039 | 0.004822 | 1.90E-08 |
| rs649129 | miR-126-3p | trans | 1.95E-37 | Nightingale | LA | NA | T | C | 0.0377 | 0.005005 | 2.10E-15 |
| rs649129 | miR-126-3p | trans | 1.95E-37 | Nightingale | S_LDL_P | NA | T | C | 0.044892 | 0.005109 | 1.70E-19 |
| rs649129 | miR-126-3p | trans | 1.95E-37 | Nightingale | XXL_VLDL_ | NA | T | C | 0.033222 | 0.005139 | 3.50E-12 |
| rs649129 | miR-126-3p | trans | 1.95E-37 | Nightingale | M_VLDL_T | NA | T | C | -0.05565 | 0.004823 | 1.80E-30 |
| rs649129 | miR-126-3p | trans | 1.95E-37 | Nightingale | M_LDL_CE | NA | T | C | 0.04991 | 0.005111 | 1.90E-23 |
| rs649129 | miR-126-3p | trans | 1.95E-37 | Nightingale | L_LDL_L | NA | T | C | 0.054447 | 0.005054 | 6.20E-28 |
| rs649129 | miR-126-3p | trans | 1.95E-37 | Nightingale | S_VLDL_FC | NA | T | C | 0.054336 | 0.005105 | 5.10E-28 |
| rs649129 | miR-126-3p | trans | 1.95E-37 | Nightingale | IDL_C_pct | NA | T | C | 0.03258 | 0.004986 | 3.60E-10 |
| rs649129 | miR-126-3p | trans | 1.95E-37 | Nightingale | S_LDL_PL_ | NA | T | C | -0.04003 | 0.005094 | 5.10E-17 |
| rs649129 | miR-126-3p | trans | 1.95E-37 | Nightingale | S_HDL_C | NA | T | C | 0.045127 | 0.005095 | 8.00E-19 |
| rs649129 | miR-126-3p | trans | 1.95E-37 | Nightingale | S_VLDL_P | NA | T | C | 0.031863 | 0.00504 | 8.10E-12 |
| rs649129 | miR-126-3p | trans | 1.95E-37 | Nightingale | Phosphogly | NA | T | C | 0.027915 | 0.004834 | 9.60E-10 |
| rs649129 | miR-126-3p | trans | 1.95E-37 | Nightingale | ApoB | NA | T | C | 0.051073 | 0.005097 | 8.20E-25 |
| rs649129 | miR-126-3p | trans | 1.95E-37 | Nightingale | S_VLDL_FC | NA | T | C | 0.045731 | 0.004877 | 1.40E-20 |
| rs649129 | miR-126-3p | trans | 1.95E-37 | Nightingale | XS_VLDL_P | NA | T | C | 0.041856 | 0.005033 | 4.60E-19 |
| rs649129 | miR-126-3p | trans | 1.95E-37 | Nightingale | Total_PL | NA | T | C | 0.034937 | 0.00486 | 3.10E-14 |
| rs649129 | miR-126-3p | trans | 1.95E-37 | Nightingale | L_LDL_TG_ | NA | T | C | -0.03492 | 0.005006 | 9.60E-12 |
| rs649129 | miR-126-3p | trans | 1.95E-37 | Nightingale | S_HDL_PL | NA | T | C | 0.03281 | 0.00506 | 2.30E-11 |
| rs649129 | miR-126-3p | trans | 1.95E-37 | Nightingale | XL_VLDL_P | NA | T | C | 0.052761 | 0.005112 | 2.60E-26 |
| rs649129 | miR-126-3p | trans | 1.95E-37 | Nightingale | XS_VLDL_C | NA | T | C | 0.034575 | 0.004816 | 3.00E-12 |
| rs649129 | miR-126-3p | trans | 1.95E-37 | Nightingale | M_VLDL_C | NA | T | C | 0.049074 | 0.004795 | 4.20E-24 |
| rs649129 | miR-126-3p | trans | 1.95E-37 | Nightingale | L_LDL_P | NA | T | C | 0.046154 | 0.005087 | 1.60E-20 |
| rs649129 | miR-126-3p | trans | 1.95E-37 | Nightingale | XL_HDL_L | NA | T | C | -0.02498 | 0.00459 | 1.30E-08 |
| rs649129 | miR-126-3p | trans | 1.95E-37 | Nightingale | L_VLDL_TG | NA | T | C | -0.0723 | 0.00508 | 1.20E-47 |
| rs649129 | miR-126-3p | trans | 1.95E-37 | Nightingale | XL_HDL_C_ | NA | T | C | 0.033715 | 0.004874 | 2.30E-11 |
| rs649129 | miR-126-3p | trans | 1.95E-37 | Nightingale | LDL_P | NA | T | C | 0.048505 | 0.0051 | 2.00E-22 |
| rs649129 | miR-126-3p | trans | 1.95E-37 | Nightingale | XS_VLDL_P | NA | T | C | 0.049752 | 0.005036 | 1.20E-25 |
| rs649129 | miR-126-3p | trans | 1.95E-37 | Nightingale | XL_VLDL_F | NA | T | C | 0.050654 | 0.004948 | 3.70E-25 |
| rs649129 | miR-126-3p | trans | 1.95E-37 | Nightingale | S_VLDL_C | NA | T | C | 0.054311 | 0.005103 | 2.80E-28 |
| rs649129 | miR-126-3p | trans | 1.95E-37 | Nightingale | XL_VLDL_C | NA | T | C | 0.054551 | 0.004896 | 2.20E-28 |
| rs649129 | miR-126-3p | trans | 1.95E-37 | Nightingale | XS_VLDL_T | NA | T | C | -0.0371 | 0.004853 | 6.70E-14 |
| rs649129 | miR-126-3p | trans | 1.95E-37 | Nightingale | S_LDL_CE_ | NA | T | C | 0.039688 | 0.005095 | 5.70E-17 |
| rs649129 | miR-126-3p | trans | 1.95E-37 | Nightingale | Cholines | NA | T | C | 0.030997 | 0.0048 | 1.00E-11 |
| rs649129 | miR-126-3p | trans | 1.95E-37 | Nightingale | M_LDL_TG | NA | T | C | -0.04553 | 0.005041 | 3.70E-19 |
| rs649129 | miR-126-3p | trans | 1.95E-37 | Nightingale | S_VLDL_CE | NA | T | C | 0.06893 | 0.005035 | 2.20E-42 |
| rs649129 | miR-126-3p | trans | 1.95E-37 | Nightingale | M_LDL_C | NA | T | C | 0.052736 | 0.005113 | 1.10E-25 |
| rs649129 | miR-126-3p | trans | 1.95E-37 | Nightingale | S_VLDL_PL | NA | T | C | 0.046421 | 0.005094 | 1.80E-21 |
| rs649129 | miR-126-3p | trans | 1.95E-37 | Nightingale | M_VLDL_F | NA | T | C | 0.059402 | 0.004856 | 2.80E-34 |
| rs649129 | miR-126-3p | trans | 1.95E-37 | Nightingale | XS_VLDL_F | NA | T | C | 0.044222 | 0.005067 | 1.20E-17 |
| rs649129 | miR-126-3p | trans | 1.95E-37 | Nightingale | S_LDL_TG_ | NA | T | C | -0.03709 | 0.004949 | 3.70E-13 |
| rs649129 | miR-126-3p | trans | 1.95E-37 | Nightingale | XS_VLDL_C | NA | T | C | 0.037225 | 0.004841 | 5.30E-14 |
| rs649129 | miR-126-3p | trans | 1.95E-37 | Nightingale | M_VLDL_C | NA | T | C | 0.052426 | 0.004807 | 3.20E-27 |
| rs649129 | miR-126-3p | trans | 1.95E-37 | Nightingale | Total_C | NA | T | C | 0.04944 | 0.004941 | 7.20E-25 |

| | | | | | | | | | | | |
|---|---|---|---|---|---|---|---|---|---|---|---|
| rs649129 | miR-126-3p | trans | 1.95E-37 | Nightingale | M_LDL_PL | NA | T | C | 0.052914 | 0.005111 | 8.10E-26 |
| rs649129 | miR-126-3p | trans | 1.95E-37 | Nightingale | XL_HDL_PL | NA | T | C | -0.03729 | 0.004689 | 1.80E-15 |
| rs649129 | miR-126-3p | trans | 1.95E-37 | Nightingale | VLDL_C | NA | T | C | 0.043038 | 0.005097 | 6.90E-19 |
| rs649129 | miR-126-3p | trans | 1.95E-37 | Nightingale | S_LDL_C_p | NA | T | C | 0.066081 | 0.005124 | 1.30E-38 |
| rs649129 | miR-126-3p | trans | 1.95E-37 | Nightingale | S_HDL_PL_ | NA | T | C | -0.0287 | 0.004934 | 1.60E-08 |
| rs649129 | miR-126-3p | trans | 1.95E-37 | Nightingale | PUFA | NA | T | C | 0.032829 | 0.004978 | 4.10E-12 |
| rs649129 | miR-126-3p | trans | 1.95E-37 | Nightingale | LDL_CE | NA | T | C | 0.052086 | 0.00509 | 2.20E-25 |
| rs649129 | miR-126-3p | trans | 1.95E-37 | Nightingale | L_VLDL_PL | NA | T | C | 0.027864 | 0.004958 | 6.90E-10 |
| rs649129 | miR-126-3p | trans | 1.95E-37 | Nightingale | XXL_VLDL_ | NA | T | C | 0.041451 | 0.005085 | 1.40E-16 |
| rs649129 | miR-126-3p | trans | 1.95E-37 | Nightingale | non_HDL_( | NA | T | C | 0.05472 | 0.005076 | 2.20E-28 |
| rs649129 | miR-126-3p | trans | 1.95E-37 | Nightingale | S_VLDL_TG | NA | T | C | -0.05782 | 0.00494 | 1.20E-31 |
| rs649129 | miR-126-3p | trans | 1.95E-37 | Nightingale | IDL_PL | NA | T | C | 0.052831 | 0.004925 | 4.10E-28 |
| rs649129 | miR-126-3p | trans | 1.95E-37 | Nightingale | VLDL_P | NA | T | C | 0.036367 | 0.005077 | 2.60E-14 |
| rs649129 | miR-126-3p | trans | 1.95E-37 | Nightingale | XL_HDL_PL | NA | T | C | -0.02773 | 0.004591 | 5.90E-10 |
| rs649129 | miR-126-3p | trans | 1.95E-37 | Nightingale | M_VLDL_P | NA | T | C | 0.063527 | 0.004898 | 1.10E-38 |
| rs649129 | miR-126-3p | trans | 1.95E-37 | Nightingale | LDL_FC | NA | T | C | 0.055859 | 0.005049 | 1.30E-28 |
| rs649129 | miR-126-3p | trans | 1.95E-37 | Nightingale | IDL_FC_pct | NA | T | C | 0.033568 | 0.005064 | 7.50E-11 |
| rs649129 | miR-126-3p | trans | 1.95E-37 | Nightingale | L_LDL_CE | NA | T | C | 0.052051 | 0.005058 | 1.30E-25 |
| rs649129 | miR-126-3p | trans | 1.95E-37 | Nightingale | XS_VLDL_C | NA | T | C | 0.052439 | 0.004954 | 3.00E-28 |
| rs649129 | miR-126-3p | trans | 1.95E-37 | Nightingale | S_VLDL_PL | NA | T | C | 0.043676 | 0.004887 | 8.10E-19 |
| rs649129 | miR-126-3p | trans | 1.95E-37 | Nightingale | VLDL_FC | NA | T | C | 0.03243 | 0.005066 | 7.10E-12 |
| rs649129 | miR-126-3p | trans | 1.95E-37 | Nightingale | XS_VLDL_F | NA | T | C | 0.04923 | 0.005015 | 2.60E-25 |
| rs649129 | miR-126-3p | trans | 1.95E-37 | Nightingale | Total_CE | NA | T | C | 0.049106 | 0.004923 | 1.40E-24 |
| rs649129 | miR-126-3p | trans | 1.95E-37 | Nightingale | S_VLDL_C_ | NA | T | C | 0.062755 | 0.004967 | 1.10E-36 |
| rs649129 | miR-126-3p | trans | 1.95E-37 | Nightingale | IDL_P | NA | T | C | 0.059758 | 0.00505 | 4.30E-33 |
| rs649129 | miR-126-3p | trans | 1.95E-37 | Nightingale | Total_FC | NA | T | C | 0.049272 | 0.004986 | 5.60E-25 |
| rs649129 | miR-126-3p | trans | 1.95E-37 | Nightingale | LDL_PL | NA | T | C | 0.055857 | 0.005092 | 1.20E-28 |
| rs649129 | miR-126-3p | trans | 1.95E-37 | Nightingale | XL_VLDL_C | NA | T | C | 0.055011 | 0.004894 | 5.60E-29 |
| rs649129 | miR-126-3p | trans | 1.95E-37 | Nightingale | S_LDL_CE | NA | T | C | 0.047148 | 0.005106 | 3.20E-21 |
| rs649129 | miR-126-3p | trans | 1.95E-37 | Nightingale | Total_L | NA | T | C | 0.038384 | 0.005021 | 1.20E-15 |
| rs649129 | miR-126-3p | trans | 1.95E-37 | Nightingale | Sphingomy | NA | T | C | 0.040605 | 0.004771 | 1.30E-17 |
| rs649129 | miR-126-3p | trans | 1.95E-37 | Nightingale | L_VLDL_CE | NA | T | C | 0.030309 | 0.005044 | 1.00E-10 |
| rs649129 | miR-126-3p | trans | 1.95E-37 | Nightingale | XXL_VLDL_ | NA | T | C | 0.050813 | 0.005145 | 1.80E-23 |
| rs649129 | miR-126-3p | trans | 1.95E-37 | Nightingale | S_LDL_L | NA | T | C | 0.044422 | 0.005103 | 8.20E-19 |
| rs649129 | miR-126-3p | trans | 1.95E-37 | Nightingale | M_VLDL_F | NA | T | C | 0.047627 | 0.005103 | 3.80E-22 |
| rs649129 | miR-126-3p | trans | 1.95E-37 | Nightingale | L_LDL_FC | NA | T | C | 0.055728 | 0.005012 | 4.60E-29 |
| rs649129 | miR-126-3p | trans | 1.95E-37 | Nightingale | XXL_VLDL_ | NA | T | C | -0.05327 | 0.00515 | 8.30E-26 |
| rs649129 | miR-126-3p | trans | 1.95E-37 | Nightingale | IDL_TG_pct | NA | T | C | -0.03301 | 0.004934 | 7.00E-11 |
| rs649129 | miR-126-3p | trans | 1.95E-37 | Nightingale | XS_VLDL_L | NA | T | C | 0.045001 | 0.005017 | 9.20E-22 |
| rs649129 | miR-126-3p | trans | 1.95E-37 | Nightingale | L_LDL_PL | NA | T | C | 0.059608 | 0.005065 | 8.20E-33 |
| rs649129 | miR-126-3p | trans | 1.95E-37 | Nightingale | S_HDL_L | NA | T | C | 0.038495 | 0.005086 | 1.30E-14 |
| rs649129 | miR-126-3p | trans | 1.95E-37 | Nightingale | Phosphatic | NA | T | C | 0.02716 | 0.004782 | 1.80E-09 |
| rs649129 | miR-126-3p | trans | 1.95E-37 | Nightingale | L_LDL_C | NA | T | C | 0.053389 | 0.005046 | 7.00E-27 |
| rs649129 | miR-126-3p | trans | 1.95E-37 | Nightingale | XL_VLDL_T | NA | T | C | -0.06153 | 0.004947 | 1.10E-35 |
| rs649129 | miR-126-3p | trans | 1.95E-37 | Nightingale | S_HDL_P | NA | T | C | 0.048101 | 0.005104 | 4.00E-21 |
| rs649129 | miR-126-3p | trans | 1.95E-37 | Nightingale | Citrate | NA | T | C | 0.027695 | 0.005071 | 1.70E-08 |
| rs649129 | miR-126-3p | trans | 1.95E-37 | Nightingale | XL_VLDL_C | NA | T | C | 0.029049 | 0.005025 | 4.30E-10 |
| rs649129 | miR-126-3p | trans | 1.95E-37 | Nightingale | Omega_6 | NA | T | C | 0.03716 | 0.004984 | 3.70E-15 |
| rs649129 | miR-126-3p | trans | 1.95E-37 | Nightingale | M_LDL_P | NA | T | C | 0.051271 | 0.005118 | 1.50E-24 |
| rs649129 | miR-126-3p | trans | 1.95E-37 | Nightingale | S_LDL_FC | NA | T | C | 0.048996 | 0.005096 | 7.80E-22 |
| rs649129 | miR-126-3p | trans | 1.95E-37 | Nightingale | M_VLDL_L | NA | T | C | 0.030569 | 0.005069 | 1.20E-10 |
| rs649129 | miR-126-3p | trans | 1.95E-37 | Nightingale | XS_VLDL_C | NA | T | C | 0.052973 | 0.004925 | 6.90E-29 |
| rs649129 | miR-126-3p | trans | 1.95E-37 | Nightingale | IDL_FC | NA | T | C | 0.056993 | 0.004941 | 1.40E-31 |
| rs649129 | miR-139-5p | trans | 1.34E-34 | Nightingale | M_LDL_L | NA | T | C | 0.051759 | 0.005111 | 6.60E-25 |
| rs649129 | miR-139-5p | trans | 1.34E-34 | Nightingale | S_HDL_FC | NA | T | C | 0.035785 | 0.00501 | 2.10E-13 |
| rs649129 | miR-139-5p | trans | 1.34E-34 | Nightingale | M_VLDL_C | NA | T | C | 0.056026 | 0.00509 | 9.10E-30 |
| rs649129 | miR-139-5p | trans | 1.34E-34 | Nightingale | M_LDL_C_ | NA | T | C | 0.037633 | 0.005107 | 1.40E-13 |
| rs649129 | miR-139-5p | trans | 1.34E-34 | Nightingale | S_HDL_CE | NA | T | C | 0.045905 | 0.005112 | 3.70E-19 |
| rs649129 | miR-139-5p | trans | 1.34E-34 | Nightingale | Remnant_( | NA | T | C | 0.053675 | 0.005071 | 4.00E-28 |
| rs649129 | miR-139-5p | trans | 1.34E-34 | Nightingale | HDL_size | NA | T | C | -0.03089 | 0.004525 | 3.00E-12 |
| rs649129 | miR-139-5p | trans | 1.34E-34 | Nightingale | Pyruvate | NA | T | C | 0.030064 | 0.005112 | 4.10E-09 |
| rs649129 | miR-139-5p | trans | 1.34E-34 | Nightingale | M_VLDL_P | NA | T | C | 0.041826 | 0.005096 | 7.80E-18 |
| rs649129 | miR-139-5p | trans | 1.34E-34 | Nightingale | L_VLDL_C_ | NA | T | C | 0.072931 | 0.004985 | 1.90E-49 |

| | | | | | | | | | | | |
|---|---|---|---|---|---|---|---|---|---|---|---|
| rs649129 | miR-139-5p | trans | 1.34E-34 | Nightingale | ApoB_by_A | NA | T | C | 0.038436 | 0.005027 | 5.80E-15 |
| rs649129 | miR-139-5p | trans | 1.34E-34 | Nightingale | S_VLDL_L | NA | T | C | 0.03374 | 0.005054 | 7.60E-13 |
| rs649129 | miR-139-5p | trans | 1.34E-34 | Nightingale | XXL_VLDL_ | NA | T | C | 0.051604 | 0.005117 | 3.20E-24 |
| rs649129 | miR-139-5p | trans | 1.34E-34 | Nightingale | S_LDL_PL | NA | T | C | 0.038147 | 0.005092 | 3.30E-14 |
| rs649129 | miR-139-5p | trans | 1.34E-34 | Nightingale | S_VLDL_CE | NA | T | C | 0.053637 | 0.005097 | 6.20E-28 |
| rs649129 | miR-139-5p | trans | 1.34E-34 | Nightingale | IDL_CE | NA | T | C | 0.052641 | 0.004928 | 3.40E-27 |
| rs649129 | miR-139-5p | trans | 1.34E-34 | Nightingale | VLDL_CE | NA | T | C | 0.049788 | 0.005105 | 3.30E-24 |
| rs649129 | miR-139-5p | trans | 1.34E-34 | Nightingale | M_LDL_FC | NA | T | C | 0.056788 | 0.005097 | 3.80E-29 |
| rs649129 | miR-139-5p | trans | 1.34E-34 | Nightingale | M_VLDL_P | NA | T | C | 0.040449 | 0.005091 | 8.30E-17 |
| rs649129 | miR-139-5p | trans | 1.34E-34 | Nightingale | L_VLDL_FC | NA | T | C | 0.06918 | 0.005055 | 3.20E-43 |
| rs649129 | miR-139-5p | trans | 1.34E-34 | Nightingale | Total_P | NA | T | C | 0.030298 | 0.004804 | 6.30E-11 |
| rs649129 | miR-139-5p | trans | 1.34E-34 | Nightingale | M_VLDL_C | NA | T | C | 0.059663 | 0.005057 | 3.90E-33 |
| rs649129 | miR-139-5p | trans | 1.34E-34 | Nightingale | LDL_L | NA | T | C | 0.053444 | 0.005087 | 1.40E-26 |
| rs649129 | miR-139-5p | trans | 1.34E-34 | Nightingale | VLDL_PL | NA | T | C | 0.025811 | 0.005043 | 2.40E-08 |
| rs649129 | miR-139-5p | trans | 1.34E-34 | Nightingale | S_LDL_C | NA | T | C | 0.048975 | 0.005109 | 2.00E-22 |
| rs649129 | miR-139-5p | trans | 1.34E-34 | Nightingale | L_VLDL_CE | NA | T | C | 0.065497 | 0.004884 | 6.20E-42 |
| rs649129 | miR-139-5p | trans | 1.34E-34 | Nightingale | IDL_L | NA | T | C | 0.053788 | 0.004935 | 1.40E-28 |
| rs649129 | miR-139-5p | trans | 1.34E-34 | Nightingale | Clinical_LD | NA | T | C | 0.057836 | 0.005074 | 1.10E-30 |
| rs649129 | miR-139-5p | trans | 1.34E-34 | Nightingale | LDL_C | NA | T | C | 0.053645 | 0.005082 | 9.90E-27 |
| rs649129 | miR-139-5p | trans | 1.34E-34 | Nightingale | IDL_C | NA | T | C | 0.054101 | 0.004929 | 1.20E-28 |
| rs649129 | miR-139-5p | trans | 1.34E-34 | Nightingale | L_HDL_TG_ | NA | T | C | 0.024039 | 0.004822 | 1.90E-08 |
| rs649129 | miR-139-5p | trans | 1.34E-34 | Nightingale | LA | NA | T | C | 0.0377 | 0.005005 | 2.10E-15 |
| rs649129 | miR-139-5p | trans | 1.34E-34 | Nightingale | S_LDL_P | NA | T | C | 0.044892 | 0.005109 | 1.70E-19 |
| rs649129 | miR-139-5p | trans | 1.34E-34 | Nightingale | XXL_VLDL_ | NA | T | C | 0.033222 | 0.005139 | 3.50E-12 |
| rs649129 | miR-139-5p | trans | 1.34E-34 | Nightingale | M_VLDL_T | NA | T | C | -0.05565 | 0.004823 | 1.80E-30 |
| rs649129 | miR-139-5p | trans | 1.34E-34 | Nightingale | M_LDL_CE | NA | T | C | 0.04991 | 0.005111 | 1.90E-23 |
| rs649129 | miR-139-5p | trans | 1.34E-34 | Nightingale | L_LDL_L | NA | T | C | 0.054447 | 0.005054 | 6.20E-28 |
| rs649129 | miR-139-5p | trans | 1.34E-34 | Nightingale | S_VLDL_FC | NA | T | C | 0.054336 | 0.005105 | 5.10E-28 |
| rs649129 | miR-139-5p | trans | 1.34E-34 | Nightingale | IDL_C_pct | NA | T | C | 0.03258 | 0.004986 | 3.60E-10 |
| rs649129 | miR-139-5p | trans | 1.34E-34 | Nightingale | S_LDL_PL_ | NA | T | C | -0.04003 | 0.005094 | 5.10E-17 |
| rs649129 | miR-139-5p | trans | 1.34E-34 | Nightingale | S_HDL_C | NA | T | C | 0.045127 | 0.005095 | 8.00E-19 |
| rs649129 | miR-139-5p | trans | 1.34E-34 | Nightingale | S_VLDL_P | NA | T | C | 0.031863 | 0.00504 | 8.10E-12 |
| rs649129 | miR-139-5p | trans | 1.34E-34 | Nightingale | Phosphogly | NA | T | C | 0.027915 | 0.004834 | 9.60E-10 |
| rs649129 | miR-139-5p | trans | 1.34E-34 | Nightingale | ApoB | NA | T | C | 0.051073 | 0.005097 | 8.20E-25 |
| rs649129 | miR-139-5p | trans | 1.34E-34 | Nightingale | S_VLDL_FC | NA | T | C | 0.045731 | 0.004877 | 1.40E-20 |
| rs649129 | miR-139-5p | trans | 1.34E-34 | Nightingale | XS_VLDL_P | NA | T | C | 0.041856 | 0.005033 | 4.60E-19 |
| rs649129 | miR-139-5p | trans | 1.34E-34 | Nightingale | Total_PL | NA | T | C | 0.034937 | 0.00486 | 3.10E-14 |
| rs649129 | miR-139-5p | trans | 1.34E-34 | Nightingale | L_LDL_TG_ | NA | T | C | -0.03492 | 0.005006 | 9.60E-12 |
| rs649129 | miR-139-5p | trans | 1.34E-34 | Nightingale | S_HDL_PL | NA | T | C | 0.03281 | 0.00506 | 2.30E-11 |
| rs649129 | miR-139-5p | trans | 1.34E-34 | Nightingale | XL_VLDL_P | NA | T | C | 0.052761 | 0.005112 | 2.60E-26 |
| rs649129 | miR-139-5p | trans | 1.34E-34 | Nightingale | XS_VLDL_C | NA | T | C | 0.034575 | 0.004816 | 3.00E-12 |
| rs649129 | miR-139-5p | trans | 1.34E-34 | Nightingale | M_VLDL_C | NA | T | C | 0.049074 | 0.004795 | 4.20E-24 |
| rs649129 | miR-139-5p | trans | 1.34E-34 | Nightingale | L_LDL_P | NA | T | C | 0.046154 | 0.005087 | 1.60E-20 |
| rs649129 | miR-139-5p | trans | 1.34E-34 | Nightingale | XL_HDL_L | NA | T | C | -0.02498 | 0.00459 | 1.30E-08 |
| rs649129 | miR-139-5p | trans | 1.34E-34 | Nightingale | L_VLDL_TG | NA | T | C | -0.0723 | 0.00508 | 1.20E-47 |
| rs649129 | miR-139-5p | trans | 1.34E-34 | Nightingale | XL_HDL_C_ | NA | T | C | 0.033715 | 0.004874 | 2.30E-11 |
| rs649129 | miR-139-5p | trans | 1.34E-34 | Nightingale | LDL_P | NA | T | C | 0.048505 | 0.0051 | 2.00E-22 |
| rs649129 | miR-139-5p | trans | 1.34E-34 | Nightingale | XS_VLDL_P | NA | T | C | 0.049752 | 0.005036 | 1.20E-25 |
| rs649129 | miR-139-5p | trans | 1.34E-34 | Nightingale | XL_VLDL_F | NA | T | C | 0.050654 | 0.004948 | 3.70E-25 |
| rs649129 | miR-139-5p | trans | 1.34E-34 | Nightingale | S_VLDL_C | NA | T | C | 0.054311 | 0.005103 | 2.80E-28 |
| rs649129 | miR-139-5p | trans | 1.34E-34 | Nightingale | XL_VLDL_C | NA | T | C | 0.054551 | 0.004896 | 2.20E-28 |
| rs649129 | miR-139-5p | trans | 1.34E-34 | Nightingale | XS_VLDL_T | NA | T | C | -0.0371 | 0.004853 | 6.70E-14 |
| rs649129 | miR-139-5p | trans | 1.34E-34 | Nightingale | S_LDL_CE_ | NA | T | C | 0.039688 | 0.005095 | 5.70E-17 |
| rs649129 | miR-139-5p | trans | 1.34E-34 | Nightingale | Cholines | NA | T | C | 0.030997 | 0.0048 | 1.00E-11 |
| rs649129 | miR-139-5p | trans | 1.34E-34 | Nightingale | M_LDL_TG | NA | T | C | -0.04553 | 0.005041 | 3.70E-19 |
| rs649129 | miR-139-5p | trans | 1.34E-34 | Nightingale | S_VLDL_CE | NA | T | C | 0.06893 | 0.005035 | 2.20E-42 |
| rs649129 | miR-139-5p | trans | 1.34E-34 | Nightingale | M_LDL_C | NA | T | C | 0.052736 | 0.005113 | 1.10E-25 |
| rs649129 | miR-139-5p | trans | 1.34E-34 | Nightingale | S_VLDL_PL | NA | T | C | 0.046421 | 0.005094 | 1.80E-21 |
| rs649129 | miR-139-5p | trans | 1.34E-34 | Nightingale | M_VLDL_F | NA | T | C | 0.059402 | 0.004856 | 2.80E-34 |
| rs649129 | miR-139-5p | trans | 1.34E-34 | Nightingale | XS_VLDL_F | NA | T | C | 0.044222 | 0.005067 | 1.20E-17 |
| rs649129 | miR-139-5p | trans | 1.34E-34 | Nightingale | S_LDL_TG_ | NA | T | C | -0.03709 | 0.004949 | 3.70E-13 |
| rs649129 | miR-139-5p | trans | 1.34E-34 | Nightingale | XS_VLDL_C | NA | T | C | 0.037225 | 0.004841 | 5.30E-14 |
| rs649129 | miR-139-5p | trans | 1.34E-34 | Nightingale | M_VLDL_C | NA | T | C | 0.052426 | 0.004807 | 3.20E-27 |

| | | | | | | | | | | | |
|---|---|---|---|---|---|---|---|---|---|---|---|
| rs649129 | miR-139-5p | trans | 1.34E-34 | Nightingale | Total_C | NA | T | C | 0.04944 | 0.004941 | 7.20E-25 |
| rs649129 | miR-139-5p | trans | 1.34E-34 | Nightingale | M_LDL_PL | NA | T | C | 0.052914 | 0.005111 | 8.10E-26 |
| rs649129 | miR-139-5p | trans | 1.34E-34 | Nightingale | XL_HDL_PL | NA | T | C | -0.03729 | 0.004689 | 1.80E-15 |
| rs649129 | miR-139-5p | trans | 1.34E-34 | Nightingale | VLDL_C | NA | T | C | 0.043038 | 0.005097 | 6.90E-19 |
| rs649129 | miR-139-5p | trans | 1.34E-34 | Nightingale | S_LDL_C_p | NA | T | C | 0.066081 | 0.005124 | 1.30E-38 |
| rs649129 | miR-139-5p | trans | 1.34E-34 | Nightingale | S_HDL_PL_ | NA | T | C | -0.0287 | 0.004934 | 1.60E-08 |
| rs649129 | miR-139-5p | trans | 1.34E-34 | Nightingale | PUFA | NA | T | C | 0.032829 | 0.004978 | 4.10E-12 |
| rs649129 | miR-139-5p | trans | 1.34E-34 | Nightingale | LDL_CE | NA | T | C | 0.052086 | 0.00509 | 2.20E-25 |
| rs649129 | miR-139-5p | trans | 1.34E-34 | Nightingale | L_VLDL_PL | NA | T | C | 0.027864 | 0.004958 | 6.90E-10 |
| rs649129 | miR-139-5p | trans | 1.34E-34 | Nightingale | XXL_VLDL_ | NA | T | C | 0.041451 | 0.005085 | 1.40E-16 |
| rs649129 | miR-139-5p | trans | 1.34E-34 | Nightingale | non_HDL_( | NA | T | C | 0.05472 | 0.005076 | 2.20E-28 |
| rs649129 | miR-139-5p | trans | 1.34E-34 | Nightingale | S_VLDL_TG | NA | T | C | -0.05782 | 0.00494 | 1.20E-31 |
| rs649129 | miR-139-5p | trans | 1.34E-34 | Nightingale | IDL_PL | NA | T | C | 0.052831 | 0.004925 | 4.10E-28 |
| rs649129 | miR-139-5p | trans | 1.34E-34 | Nightingale | VLDL_P | NA | T | C | 0.036367 | 0.005077 | 2.60E-14 |
| rs649129 | miR-139-5p | trans | 1.34E-34 | Nightingale | XL_HDL_PL | NA | T | C | -0.02773 | 0.004591 | 5.90E-10 |
| rs649129 | miR-139-5p | trans | 1.34E-34 | Nightingale | M_VLDL_P | NA | T | C | 0.063527 | 0.004898 | 1.10E-38 |
| rs649129 | miR-139-5p | trans | 1.34E-34 | Nightingale | LDL_FC | NA | T | C | 0.055859 | 0.005049 | 1.30E-28 |
| rs649129 | miR-139-5p | trans | 1.34E-34 | Nightingale | IDL_FC_pct | NA | T | C | 0.033568 | 0.005064 | 7.50E-11 |
| rs649129 | miR-139-5p | trans | 1.34E-34 | Nightingale | L_LDL_CE | NA | T | C | 0.052051 | 0.005058 | 1.30E-25 |
| rs649129 | miR-139-5p | trans | 1.34E-34 | Nightingale | XS_VLDL_C | NA | T | C | 0.052439 | 0.004954 | 3.00E-28 |
| rs649129 | miR-139-5p | trans | 1.34E-34 | Nightingale | S_VLDL_PL | NA | T | C | 0.043676 | 0.004887 | 8.10E-19 |
| rs649129 | miR-139-5p | trans | 1.34E-34 | Nightingale | VLDL_FC | NA | T | C | 0.03243 | 0.005066 | 7.10E-12 |
| rs649129 | miR-139-5p | trans | 1.34E-34 | Nightingale | XS_VLDL_F | NA | T | C | 0.04923 | 0.005015 | 2.60E-25 |
| rs649129 | miR-139-5p | trans | 1.34E-34 | Nightingale | Total_CE | NA | T | C | 0.049106 | 0.004923 | 1.40E-24 |
| rs649129 | miR-139-5p | trans | 1.34E-34 | Nightingale | S_VLDL_C_ | NA | T | C | 0.062755 | 0.004967 | 1.10E-36 |
| rs649129 | miR-139-5p | trans | 1.34E-34 | Nightingale | IDL_P | NA | T | C | 0.059758 | 0.00505 | 4.30E-33 |
| rs649129 | miR-139-5p | trans | 1.34E-34 | Nightingale | Total_FC | NA | T | C | 0.049272 | 0.004986 | 5.60E-25 |
| rs649129 | miR-139-5p | trans | 1.34E-34 | Nightingale | LDL_PL | NA | T | C | 0.055857 | 0.005092 | 1.20E-28 |
| rs649129 | miR-139-5p | trans | 1.34E-34 | Nightingale | XL_VLDL_C | NA | T | C | 0.055011 | 0.004894 | 5.60E-29 |
| rs649129 | miR-139-5p | trans | 1.34E-34 | Nightingale | S_LDL_CE | NA | T | C | 0.047148 | 0.005106 | 3.20E-21 |
| rs649129 | miR-139-5p | trans | 1.34E-34 | Nightingale | Total_L | NA | T | C | 0.038384 | 0.005021 | 1.20E-15 |
| rs649129 | miR-139-5p | trans | 1.34E-34 | Nightingale | Sphingomy | NA | T | C | 0.040605 | 0.004771 | 1.30E-17 |
| rs649129 | miR-139-5p | trans | 1.34E-34 | Nightingale | L_VLDL_CE | NA | T | C | 0.030309 | 0.005044 | 1.00E-10 |
| rs649129 | miR-139-5p | trans | 1.34E-34 | Nightingale | XXL_VLDL_ | NA | T | C | 0.050813 | 0.005145 | 1.80E-23 |
| rs649129 | miR-139-5p | trans | 1.34E-34 | Nightingale | S_LDL_L | NA | T | C | 0.044422 | 0.005103 | 8.20E-19 |
| rs649129 | miR-139-5p | trans | 1.34E-34 | Nightingale | M_VLDL_F | NA | T | C | 0.047627 | 0.005103 | 3.80E-22 |
| rs649129 | miR-139-5p | trans | 1.34E-34 | Nightingale | L_LDL_FC | NA | T | C | 0.055728 | 0.005012 | 4.60E-29 |
| rs649129 | miR-139-5p | trans | 1.34E-34 | Nightingale | XXL_VLDL_ | NA | T | C | -0.05327 | 0.00515 | 8.30E-26 |
| rs649129 | miR-139-5p | trans | 1.34E-34 | Nightingale | IDL_TG_pc | NA | T | C | -0.03301 | 0.004934 | 7.00E-11 |
| rs649129 | miR-139-5p | trans | 1.34E-34 | Nightingale | XS_VLDL_L | NA | T | C | 0.045001 | 0.005017 | 9.20E-22 |
| rs649129 | miR-139-5p | trans | 1.34E-34 | Nightingale | L_LDL_PL | NA | T | C | 0.059608 | 0.005065 | 8.20E-33 |
| rs649129 | miR-139-5p | trans | 1.34E-34 | Nightingale | S_HDL_L | NA | T | C | 0.038495 | 0.005086 | 1.30E-14 |
| rs649129 | miR-139-5p | trans | 1.34E-34 | Nightingale | Phosphatic | NA | T | C | 0.02716 | 0.004782 | 1.80E-09 |
| rs649129 | miR-139-5p | trans | 1.34E-34 | Nightingale | L_LDL_C | NA | T | C | 0.053389 | 0.005046 | 7.00E-27 |
| rs649129 | miR-139-5p | trans | 1.34E-34 | Nightingale | XL_VLDL_T | NA | T | C | -0.06153 | 0.004947 | 1.10E-35 |
| rs649129 | miR-139-5p | trans | 1.34E-34 | Nightingale | S_HDL_P | NA | T | C | 0.048101 | 0.005104 | 4.00E-21 |
| rs649129 | miR-139-5p | trans | 1.34E-34 | Nightingale | Citrate | NA | T | C | 0.027695 | 0.005071 | 1.70E-08 |
| rs649129 | miR-139-5p | trans | 1.34E-34 | Nightingale | XL_VLDL_C | NA | T | C | 0.029049 | 0.005025 | 4.30E-10 |
| rs649129 | miR-139-5p | trans | 1.34E-34 | Nightingale | Omega_6 | NA | T | C | 0.03716 | 0.004984 | 3.70E-15 |
| rs649129 | miR-139-5p | trans | 1.34E-34 | Nightingale | M_LDL_P | NA | T | C | 0.051271 | 0.005118 | 1.50E-24 |
| rs649129 | miR-139-5p | trans | 1.34E-34 | Nightingale | S_LDL_FC | NA | T | C | 0.048996 | 0.005096 | 7.80E-22 |
| rs649129 | miR-139-5p | trans | 1.34E-34 | Nightingale | M_VLDL_L | NA | T | C | 0.030569 | 0.005069 | 1.20E-10 |
| rs649129 | miR-139-5p | trans | 1.34E-34 | Nightingale | XS_VLDL_C | NA | T | C | 0.052973 | 0.004925 | 6.90E-29 |
| rs649129 | miR-139-5p | trans | 1.34E-34 | Nightingale | IDL_FC | NA | T | C | 0.056993 | 0.004941 | 1.40E-31 |
| rs659104 | miR-10b-5p | trans | 1.28E-19 | Nightingale | L_VLDL_CE | NA | G | T | 0.026375 | 0.003969 | 1.20E-11 |
| rs659104 | miR-10b-5p | trans | 1.28E-19 | Nightingale | M_VLDL_C | NA | G | T | 0.021453 | 0.003907 | 1.10E-08 |
| rs659104 | miR-10b-5p | trans | 1.28E-19 | Nightingale | XL_VLDL_F | NA | G | T | 0.021895 | 0.00402 | 1.50E-08 |
| rs659104 | miR-10b-5p | trans | 1.28E-19 | Nightingale | L_VLDL_C_ | NA | G | T | 0.0282 | 0.004051 | 1.70E-12 |
| rs659104 | miR-10b-5p | trans | 1.28E-19 | Nightingale | L_VLDL_TG | NA | G | T | -0.02542 | 0.004128 | 6.30E-10 |
| rs659104 | miR-10b-5p | trans | 1.28E-19 | Nightingale | M_VLDL_T | NA | G | T | -0.02245 | 0.00392 | 3.00E-09 |
| rs659104 | miR-10b-5p | trans | 1.28E-19 | Nightingale | S_VLDL_C_ | NA | G | T | 0.023334 | 0.004037 | 3.50E-09 |
| rs659104 | miR-10b-5p | trans | 1.28E-19 | Nightingale | XL_VLDL_C | NA | G | T | 0.022082 | 0.003979 | 9.10E-09 |
| rs659104 | miR-10b-5p | trans | 1.28E-19 | Nightingale | L_VLDL_FC | NA | G | T | 0.023784 | 0.004108 | 6.10E-09 |

| | | | | | | | | | | | |
|---|---|---|---|---|---|---|---|---|---|---|---|
| rs659104 | miR-10b-5p | trans | 1.28E-19 | Nightingale | IDL_P | NA | G | T | 0.023178 | 0.004104 | 1.90E-08 |
| rs659104 | miR-10b-5p | trans | 1.28E-19 | Nightingale | XL_VLDL_C | NA | G | T | 0.022545 | 0.003976 | 4.90E-09 |
| rs659104 | miR-10b-5p | trans | 1.28E-19 | Nightingale | XL_VLDL_T | NA | G | T | -0.02536 | 0.00402 | 8.90E-11 |
| rs659104 | miR-10b-5p | trans | 1.28E-19 | Nightingale | IDL_FC | NA | G | T | 0.022179 | 0.004016 | 3.70E-08 |
| rs659104 | miR-10b-5p | trans | 1.28E-19 | Nightingale | S_VLDL_CE | NA | G | T | 0.025373 | 0.004092 | 3.50E-10 |
| rs659104 | miR-10b-5p | trans | 1.28E-19 | Nightingale | M_VLDL_F | NA | G | T | 0.024761 | 0.003946 | 1.50E-10 |
| rs659104 | miR-10b-5p | trans | 1.28E-19 | Nightingale | M_VLDL_P | NA | G | T | 0.02453 | 0.003981 | 4.10E-10 |
| rs659104 | miR-10b-5p | trans | 1.28E-19 | Nightingale | S_LDL_C_p | NA | G | T | 0.026925 | 0.004164 | 1.00E-10 |
| rs659104 | miR-10b-5p | trans | 1.28E-19 | Nightingale | S_VLDL_TG | NA | G | T | -0.02152 | 0.004015 | 3.90E-08 |
| rs660340 | miR-10b-5p | trans | 1.23E-19 | Nightingale | L_VLDL_C_ | NA | G | A | 0.028216 | 0.004051 | 1.60E-12 |
| rs660340 | miR-10b-5p | trans | 1.23E-19 | Nightingale | XL_VLDL_C | NA | G | A | 0.02257 | 0.003977 | 4.70E-09 |
| rs660340 | miR-10b-5p | trans | 1.23E-19 | Nightingale | S_VLDL_CE | NA | G | A | 0.025432 | 0.004092 | 3.10E-10 |
| rs660340 | miR-10b-5p | trans | 1.23E-19 | Nightingale | S_LDL_C_p | NA | G | A | 0.0269 | 0.004164 | 1.10E-10 |
| rs660340 | miR-10b-5p | trans | 1.23E-19 | Nightingale | S_VLDL_C_ | NA | G | A | 0.023361 | 0.004037 | 3.30E-09 |
| rs660340 | miR-10b-5p | trans | 1.23E-19 | Nightingale | M_VLDL_P | NA | G | A | 0.024507 | 0.003981 | 4.20E-10 |
| rs660340 | miR-10b-5p | trans | 1.23E-19 | Nightingale | IDL_FC | NA | G | A | 0.022146 | 0.004016 | 3.90E-08 |
| rs660340 | miR-10b-5p | trans | 1.23E-19 | Nightingale | XL_VLDL_T | NA | G | A | -0.02538 | 0.00402 | 8.70E-11 |
| rs660340 | miR-10b-5p | trans | 1.23E-19 | Nightingale | L_VLDL_FC | NA | G | A | 0.023814 | 0.004108 | 5.90E-09 |
| rs660340 | miR-10b-5p | trans | 1.23E-19 | Nightingale | XL_VLDL_F | NA | G | A | 0.021915 | 0.00402 | 1.50E-08 |
| rs660340 | miR-10b-5p | trans | 1.23E-19 | Nightingale | L_VLDL_TG | NA | G | A | -0.02545 | 0.004128 | 6.00E-10 |
| rs660340 | miR-10b-5p | trans | 1.23E-19 | Nightingale | S_VLDL_TG | NA | G | A | -0.02153 | 0.004015 | 3.70E-08 |
| rs660340 | miR-10b-5p | trans | 1.23E-19 | Nightingale | M_VLDL_T | NA | G | A | -0.02245 | 0.00392 | 3.00E-09 |
| rs660340 | miR-10b-5p | trans | 1.23E-19 | Nightingale | M_VLDL_C | NA | G | A | 0.021456 | 0.003907 | 1.10E-08 |
| rs660340 | miR-10b-5p | trans | 1.23E-19 | Nightingale | IDL_P | NA | G | A | 0.023172 | 0.004105 | 1.90E-08 |
| rs660340 | miR-10b-5p | trans | 1.23E-19 | Nightingale | L_VLDL_CE | NA | G | A | 0.026392 | 0.003969 | 1.10E-11 |
| rs660340 | miR-10b-5p | trans | 1.23E-19 | Nightingale | M_VLDL_F | NA | G | A | 0.024758 | 0.003946 | 1.40E-10 |
| rs660340 | miR-10b-5p | trans | 1.23E-19 | Nightingale | XL_VLDL_C | NA | G | A | 0.022109 | 0.003979 | 8.80E-09 |
| rs674302 | let-7c-5p | trans | 2.28E-23 | Nightingale | Remnant_( | NA | A | T | 0.036854 | 0.004396 | 3.70E-17 |
| rs674302 | let-7c-5p | trans | 2.28E-23 | Nightingale | L_LDL_PL | NA | A | T | 0.037956 | 0.004391 | 9.50E-18 |
| rs674302 | let-7c-5p | trans | 2.28E-23 | Nightingale | S_VLDL_CE | NA | A | T | 0.038162 | 0.004419 | 1.60E-18 |
| rs674302 | let-7c-5p | trans | 2.28E-23 | Nightingale | LDL_L | NA | A | T | 0.033263 | 0.00441 | 7.70E-14 |
| rs674302 | let-7c-5p | trans | 2.28E-23 | Nightingale | S_LDL_C | NA | A | T | 0.0309 | 0.004429 | 5.70E-12 |
| rs674302 | let-7c-5p | trans | 2.28E-23 | Nightingale | M_VLDL_F | NA | A | T | 0.041986 | 0.00421 | 4.20E-23 |
| rs674302 | let-7c-5p | trans | 2.28E-23 | Nightingale | IDL_L | NA | A | T | 0.03527 | 0.004278 | 4.90E-16 |
| rs674302 | let-7c-5p | trans | 2.28E-23 | Nightingale | S_LDL_FC | NA | A | T | 0.028528 | 0.004418 | 4.40E-10 |
| rs674302 | let-7c-5p | trans | 2.28E-23 | Nightingale | M_LDL_CE | NA | A | T | 0.032929 | 0.004431 | 1.00E-13 |
| rs674302 | let-7c-5p | trans | 2.28E-23 | Nightingale | S_LDL_CE | NA | A | T | 0.030504 | 0.004427 | 6.10E-12 |
| rs674302 | let-7c-5p | trans | 2.28E-23 | Nightingale | XXL_VLDL_ | NA | A | T | 0.030728 | 0.004406 | 1.30E-12 |
| rs674302 | let-7c-5p | trans | 2.28E-23 | Nightingale | M_LDL_C_ | NA | A | T | 0.031102 | 0.004428 | 3.20E-12 |
| rs674302 | let-7c-5p | trans | 2.28E-23 | Nightingale | Total_PL | NA | A | T | 0.024938 | 0.004213 | 1.10E-09 |
| rs674302 | let-7c-5p | trans | 2.28E-23 | Nightingale | Omega_6 | NA | A | T | 0.025874 | 0.004321 | 5.90E-10 |
| rs674302 | let-7c-5p | trans | 2.28E-23 | Nightingale | IDL_C | NA | A | T | 0.034959 | 0.004273 | 9.50E-16 |
| rs674302 | let-7c-5p | trans | 2.28E-23 | Nightingale | M_LDL_TG | NA | A | T | -0.02754 | 0.00437 | 3.60E-10 |
| rs674302 | let-7c-5p | trans | 2.28E-23 | Nightingale | IDL_P | NA | A | T | 0.04001 | 0.004378 | 1.30E-19 |
| rs674302 | let-7c-5p | trans | 2.28E-23 | Nightingale | L_LDL_CE | NA | A | T | 0.031522 | 0.004385 | 1.00E-12 |
| rs674302 | let-7c-5p | trans | 2.28E-23 | Nightingale | S_HDL_CE | NA | A | T | 0.028597 | 0.004432 | 1.40E-10 |
| rs674302 | let-7c-5p | trans | 2.28E-23 | Nightingale | IDL_CE | NA | A | T | 0.033517 | 0.004272 | 1.30E-14 |
| rs674302 | let-7c-5p | trans | 2.28E-23 | Nightingale | M_LDL_FC | NA | A | T | 0.034304 | 0.004419 | 2.00E-14 |
| rs674302 | let-7c-5p | trans | 2.28E-23 | Nightingale | M_VLDL_P | NA | A | T | 0.04397 | 0.004246 | 6.90E-25 |
| rs674302 | let-7c-5p | trans | 2.28E-23 | Nightingale | L_VLDL_TG | NA | A | T | -0.05103 | 0.004404 | 7.30E-32 |
| rs674302 | let-7c-5p | trans | 2.28E-23 | Nightingale | M_VLDL_P | NA | A | T | 0.028266 | 0.004414 | 6.40E-11 |
| rs674302 | let-7c-5p | trans | 2.28E-23 | Nightingale | LDL_P | NA | A | T | 0.031748 | 0.004421 | 1.10E-12 |
| rs674302 | let-7c-5p | trans | 2.28E-23 | Nightingale | M_VLDL_T | NA | A | T | -0.03848 | 0.004181 | 8.00E-20 |
| rs674302 | let-7c-5p | trans | 2.28E-23 | Nightingale | LDL_PL | NA | A | T | 0.034951 | 0.004414 | 4.90E-15 |
| rs674302 | let-7c-5p | trans | 2.28E-23 | Nightingale | S_HDL_L | NA | A | T | 0.02629 | 0.004409 | 1.60E-09 |
| rs674302 | let-7c-5p | trans | 2.28E-23 | Nightingale | S_HDL_C | NA | A | T | 0.028386 | 0.004417 | 1.80E-10 |
| rs674302 | let-7c-5p | trans | 2.28E-23 | Nightingale | S_HDL_FC | NA | A | T | 0.023467 | 0.004344 | 4.60E-08 |
| rs674302 | let-7c-5p | trans | 2.28E-23 | Nightingale | S_VLDL_C | NA | A | T | 0.038348 | 0.004424 | 1.80E-18 |
| rs674302 | let-7c-5p | trans | 2.28E-23 | Nightingale | LDL_C | NA | A | T | 0.033151 | 0.004405 | 9.30E-14 |
| rs674302 | let-7c-5p | trans | 2.28E-23 | Nightingale | M_VLDL_C | NA | A | T | 0.040808 | 0.004384 | 2.40E-20 |
| rs674302 | let-7c-5p | trans | 2.28E-23 | Nightingale | S_LDL_C_p | NA | A | T | 0.045309 | 0.004442 | 6.20E-25 |
| rs674302 | let-7c-5p | trans | 2.28E-23 | Nightingale | XXL_VLDL_ | NA | A | T | 0.033058 | 0.004458 | 9.70E-14 |
| rs674302 | let-7c-5p | trans | 2.28E-23 | Nightingale | IDL_PL | NA | A | T | 0.035323 | 0.00427 | 3.50E-16 |

| | | | | | | | | | | | |
|---|---|---|---|---|---|---|---|---|---|---|---|
| rs674302 | let-7c-5p | trans | 2.28E-23 | Nightingale | L_LDL_C | NA | A | T | 0.032278 | 0.004375 | 2.70E-13 |
| rs674302 | let-7c-5p | trans | 2.28E-23 | Nightingale | ApoB_by_A | NA | A | T | 0.024989 | 0.004358 | 1.40E-08 |
| rs674302 | let-7c-5p | trans | 2.28E-23 | Nightingale | IDL_FC_pct | NA | A | T | 0.025694 | 0.00439 | 1.20E-08 |
| rs674302 | let-7c-5p | trans | 2.28E-23 | Nightingale | Total_FC | NA | A | T | 0.033246 | 0.004322 | 6.90E-15 |
| rs674302 | let-7c-5p | trans | 2.28E-23 | Nightingale | S_LDL_CE_ | NA | A | T | 0.030904 | 0.004417 | 5.30E-14 |
| rs674302 | let-7c-5p | trans | 2.28E-23 | Nightingale | S_LDL_P | NA | A | T | 0.029552 | 0.004429 | 3.30E-11 |
| rs674302 | let-7c-5p | trans | 2.28E-23 | Nightingale | PUFA | NA | A | T | 0.023697 | 0.004316 | 1.50E-08 |
| rs674302 | let-7c-5p | trans | 2.28E-23 | Nightingale | ApoB | NA | A | T | 0.033904 | 0.004419 | 2.20E-14 |
| rs674302 | let-7c-5p | trans | 2.28E-23 | Nightingale | S_VLDL_C_ | NA | A | T | 0.04182 | 0.004306 | 6.00E-22 |
| rs674302 | let-7c-5p | trans | 2.28E-23 | Nightingale | M_VLDL_C | NA | A | T | 0.038673 | 0.004412 | 2.00E-18 |
| rs674302 | let-7c-5p | trans | 2.28E-23 | Nightingale | L_LDL_FC | NA | A | T | 0.033591 | 0.004345 | 3.00E-14 |
| rs674302 | let-7c-5p | trans | 2.28E-23 | Nightingale | S_VLDL_L | NA | A | T | 0.024659 | 0.004382 | 3.80E-09 |
| rs674302 | let-7c-5p | trans | 2.28E-23 | Nightingale | XL_VLDL_C | NA | A | T | 0.038131 | 0.004243 | 5.50E-19 |
| rs674302 | let-7c-5p | trans | 2.28E-23 | Nightingale | S_LDL_PL_ | NA | A | T | -0.03005 | 0.004416 | 2.80E-13 |
| rs674302 | let-7c-5p | trans | 2.28E-23 | Nightingale | S_HDL_PL | NA | A | T | 0.023787 | 0.004386 | 3.30E-08 |
| rs674302 | let-7c-5p | trans | 2.28E-23 | Nightingale | XXL_VLDL_ | NA | A | T | -0.03564 | 0.004463 | 2.50E-16 |
| rs674302 | let-7c-5p | trans | 2.28E-23 | Nightingale | XS_VLDL_C | NA | A | T | 0.038001 | 0.004295 | 1.30E-18 |
| rs674302 | let-7c-5p | trans | 2.28E-23 | Nightingale | LDL_FC | NA | A | T | 0.033595 | 0.004377 | 5.50E-14 |
| rs674302 | let-7c-5p | trans | 2.28E-23 | Nightingale | XS_VLDL_L | NA | A | T | 0.033833 | 0.004349 | 2.00E-15 |
| rs674302 | let-7c-5p | trans | 2.28E-23 | Nightingale | S_VLDL_TG | NA | A | T | -0.03806 | 0.004283 | 1.60E-18 |
| rs674302 | let-7c-5p | trans | 2.28E-23 | Nightingale | Total_CE | NA | A | T | 0.031729 | 0.004268 | 1.00E-13 |
| rs674302 | let-7c-5p | trans | 2.28E-23 | Nightingale | GlycA | NA | A | T | 0.027865 | 0.004406 | 1.80E-10 |
| rs674302 | let-7c-5p | trans | 2.28E-23 | Nightingale | S_VLDL_PL | NA | A | T | 0.027671 | 0.004237 | 1.90E-10 |
| rs674302 | let-7c-5p | trans | 2.28E-23 | Nightingale | LDL_CE | NA | A | T | 0.032508 | 0.004413 | 2.30E-13 |
| rs674302 | let-7c-5p | trans | 2.28E-23 | Nightingale | M_LDL_L | NA | A | T | 0.032989 | 0.004431 | 1.10E-13 |
| rs674302 | let-7c-5p | trans | 2.28E-23 | Nightingale | VLDL_FC | NA | A | T | 0.02414 | 0.004392 | 1.10E-08 |
| rs674302 | let-7c-5p | trans | 2.28E-23 | Nightingale | Cholines | NA | A | T | 0.022301 | 0.004161 | 4.90E-08 |
| rs674302 | let-7c-5p | trans | 2.28E-23 | Nightingale | L_LDL_L | NA | A | T | 0.033512 | 0.004381 | 3.20E-14 |
| rs674302 | let-7c-5p | trans | 2.28E-23 | Nightingale | XL_VLDL_F | NA | A | T | 0.037122 | 0.004289 | 2.30E-18 |
| rs674302 | let-7c-5p | trans | 2.28E-23 | Nightingale | XS_VLDL_P | NA | A | T | 0.032126 | 0.004363 | 5.20E-14 |
| rs674302 | let-7c-5p | trans | 2.28E-23 | Nightingale | VLDL_P | NA | A | T | 0.026627 | 0.004401 | 4.40E-10 |
| rs674302 | let-7c-5p | trans | 2.28E-23 | Nightingale | S_VLDL_FC | NA | A | T | 0.029296 | 0.004228 | 1.10E-11 |
| rs674302 | let-7c-5p | trans | 2.28E-23 | Nightingale | L_VLDL_FC | NA | A | T | 0.048437 | 0.004382 | 6.60E-29 |
| rs674302 | let-7c-5p | trans | 2.28E-23 | Nightingale | Total_L | NA | A | T | 0.026349 | 0.004353 | 4.60E-10 |
| rs674302 | let-7c-5p | trans | 2.28E-23 | Nightingale | XL_HDL_PL | NA | A | T | -0.02269 | 0.004065 | 1.20E-08 |
| rs674302 | let-7c-5p | trans | 2.28E-23 | Nightingale | XL_VLDL_C | NA | A | T | 0.037253 | 0.004245 | 3.80E-18 |
| rs674302 | let-7c-5p | trans | 2.28E-23 | Nightingale | S_VLDL_PL | NA | A | T | 0.032753 | 0.004416 | 4.20E-14 |
| rs674302 | let-7c-5p | trans | 2.28E-23 | Nightingale | Total_C | NA | A | T | 0.032359 | 0.004283 | 3.40E-14 |
| rs674302 | let-7c-5p | trans | 2.28E-23 | Nightingale | M_LDL_PL | NA | A | T | 0.032425 | 0.004431 | 2.70E-13 |
| rs674302 | let-7c-5p | trans | 2.28E-23 | Nightingale | L_VLDL_C_ | NA | A | T | 0.052079 | 0.004322 | 5.60E-34 |
| rs674302 | let-7c-5p | trans | 2.28E-23 | Nightingale | XXL_VLDL_ | NA | A | T | 0.034734 | 0.004434 | 3.40E-15 |
| rs674302 | let-7c-5p | trans | 2.28E-23 | Nightingale | M_VLDL_C | NA | A | T | 0.036135 | 0.004168 | 1.40E-17 |
| rs674302 | let-7c-5p | trans | 2.28E-23 | Nightingale | M_VLDL_P | NA | A | T | 0.029105 | 0.004418 | 2.10E-11 |
| rs674302 | let-7c-5p | trans | 2.28E-23 | Nightingale | Sphingomy | NA | A | T | 0.028962 | 0.004136 | 7.60E-12 |
| rs674302 | let-7c-5p | trans | 2.28E-23 | Nightingale | M_LDL_C | NA | A | T | 0.033944 | 0.004433 | 2.30E-14 |
| rs674302 | let-7c-5p | trans | 2.28E-23 | Nightingale | M_VLDL_F | NA | A | T | 0.033342 | 0.004424 | 3.20E-14 |
| rs674302 | let-7c-5p | trans | 2.28E-23 | Nightingale | S_VLDL_CE | NA | A | T | 0.046964 | 0.004365 | 1.40E-26 |
| rs674302 | let-7c-5p | trans | 2.28E-23 | Nightingale | XS_VLDL_P | NA | A | T | 0.036716 | 0.004366 | 1.30E-17 |
| rs674302 | let-7c-5p | trans | 2.28E-23 | Nightingale | S_LDL_L | NA | A | T | 0.027702 | 0.004424 | 8.50E-10 |
| rs674302 | let-7c-5p | trans | 2.28E-23 | Nightingale | L_LDL_P | NA | A | T | 0.029119 | 0.00441 | 6.60E-11 |
| rs674302 | let-7c-5p | trans | 2.28E-23 | Nightingale | S_HDL_P | NA | A | T | 0.03058 | 0.004425 | 5.40E-12 |
| rs674302 | let-7c-5p | trans | 2.28E-23 | Nightingale | XL_VLDL_T | NA | A | T | -0.04377 | 0.004289 | 2.50E-24 |
| rs674302 | let-7c-5p | trans | 2.28E-23 | Nightingale | XL_VLDL_P | NA | A | T | 0.038723 | 0.004431 | 6.70E-19 |
| rs674302 | let-7c-5p | trans | 2.28E-23 | Nightingale | non_HDL_( | NA | A | T | 0.035666 | 0.004401 | 5.30E-16 |
| rs674302 | let-7c-5p | trans | 2.28E-23 | Nightingale | XS_VLDL_F | NA | A | T | 0.03137 | 0.004393 | 2.60E-12 |
| rs674302 | let-7c-5p | trans | 2.28E-23 | Nightingale | S_VLDL_P | NA | A | T | 0.023462 | 0.004369 | 1.60E-08 |
| rs674302 | let-7c-5p | trans | 2.28E-23 | Nightingale | XS_VLDL_F | NA | A | T | 0.037073 | 0.004348 | 6.40E-18 |
| rs674302 | let-7c-5p | trans | 2.28E-23 | Nightingale | Clinical_LD | NA | A | T | 0.036436 | 0.004399 | 2.90E-16 |
| rs674302 | let-7c-5p | trans | 2.28E-23 | Nightingale | IDL_FC | NA | A | T | 0.03824 | 0.004284 | 1.50E-18 |
| rs674302 | let-7c-5p | trans | 2.28E-23 | Nightingale | XS_VLDL_C | NA | A | T | 0.037807 | 0.00427 | 2.10E-18 |
| rs674302 | let-7c-5p | trans | 2.28E-23 | Nightingale | LA | NA | A | T | 0.02494 | 0.004339 | 3.50E-09 |
| rs674302 | let-7c-5p | trans | 2.28E-23 | Nightingale | M_VLDL_C | NA | A | T | 0.033562 | 0.004157 | 2.10E-15 |
| rs674302 | let-7c-5p | trans | 2.28E-23 | Nightingale | VLDL_CE | NA | A | T | 0.035764 | 0.004426 | 3.10E-16 |

| | | | | | | | | | | | |
|---|---|---|---|---|---|---|---|---|---|---|---|
| rs674302 | let-7c-5p | trans | 2.28E-23 | Nightingale | S_VLDL_FC | NA | A | T | 0.037872 | 0.004425 | 7.50E-18 |
| rs674302 | let-7c-5p | trans | 2.28E-23 | Nightingale | L_VLDL_CE | NA | A | T | 0.047276 | 0.004234 | 2.90E-29 |
| rs674302 | let-7c-5p | trans | 2.28E-23 | Nightingale | XXL_VLDL_ | NA | A | T | 0.025014 | 0.004453 | 1.50E-09 |
| rs674302 | let-7c-5p | trans | 2.28E-23 | Nightingale | VLDL_C | NA | A | T | 0.031281 | 0.004419 | 5.00E-13 |
| rs674302 | let-7c-5p | trans | 2.28E-23 | Nightingale | Citrate | NA | A | T | 0.026836 | 0.004396 | 7.10E-10 |
| rs674302 | let-7c-5p | trans | 2.28E-23 | Nightingale | M_LDL_P | NA | A | T | 0.035851 | 0.004437 | 7.20E-16 |
| rs674302 | miR-126-3p | trans | 2.03E-39 | Nightingale | Remnant_( | NA | A | T | 0.036854 | 0.004396 | 3.70E-17 |
| rs674302 | miR-126-3p | trans | 2.03E-39 | Nightingale | L_LDL_PL | NA | A | T | 0.037956 | 0.004391 | 9.50E-18 |
| rs674302 | miR-126-3p | trans | 2.03E-39 | Nightingale | S_VLDL_CE | NA | A | T | 0.038162 | 0.004419 | 1.60E-18 |
| rs674302 | miR-126-3p | trans | 2.03E-39 | Nightingale | LDL_L | NA | A | T | 0.033263 | 0.00441 | 7.70E-14 |
| rs674302 | miR-126-3p | trans | 2.03E-39 | Nightingale | S_LDL_C | NA | A | T | 0.0309 | 0.004429 | 5.70E-12 |
| rs674302 | miR-126-3p | trans | 2.03E-39 | Nightingale | M_VLDL_F | NA | A | T | 0.041986 | 0.00421 | 4.20E-23 |
| rs674302 | miR-126-3p | trans | 2.03E-39 | Nightingale | IDL_L | NA | A | T | 0.03527 | 0.004278 | 4.90E-16 |
| rs674302 | miR-126-3p | trans | 2.03E-39 | Nightingale | S_LDL_FC | NA | A | T | 0.028528 | 0.004418 | 4.40E-10 |
| rs674302 | miR-126-3p | trans | 2.03E-39 | Nightingale | M_LDL_CE | NA | A | T | 0.032929 | 0.004431 | 1.00E-13 |
| rs674302 | miR-126-3p | trans | 2.03E-39 | Nightingale | S_LDL_CE | NA | A | T | 0.030504 | 0.004427 | 6.10E-12 |
| rs674302 | miR-126-3p | trans | 2.03E-39 | Nightingale | XXL_VLDL_ | NA | A | T | 0.030728 | 0.004406 | 1.30E-12 |
| rs674302 | miR-126-3p | trans | 2.03E-39 | Nightingale | M_LDL_C_ | NA | A | T | 0.031102 | 0.004428 | 3.20E-12 |
| rs674302 | miR-126-3p | trans | 2.03E-39 | Nightingale | Total_PL | NA | A | T | 0.024938 | 0.004213 | 1.10E-09 |
| rs674302 | miR-126-3p | trans | 2.03E-39 | Nightingale | Omega_6 | NA | A | T | 0.025874 | 0.004321 | 5.90E-10 |
| rs674302 | miR-126-3p | trans | 2.03E-39 | Nightingale | IDL_C | NA | A | T | 0.034959 | 0.004273 | 9.50E-16 |
| rs674302 | miR-126-3p | trans | 2.03E-39 | Nightingale | M_LDL_TG | NA | A | T | -0.02754 | 0.00437 | 3.60E-10 |
| rs674302 | miR-126-3p | trans | 2.03E-39 | Nightingale | IDL_P | NA | A | T | 0.04001 | 0.004378 | 1.30E-19 |
| rs674302 | miR-126-3p | trans | 2.03E-39 | Nightingale | L_LDL_CE | NA | A | T | 0.031522 | 0.004385 | 1.00E-12 |
| rs674302 | miR-126-3p | trans | 2.03E-39 | Nightingale | S_HDL_CE | NA | A | T | 0.028597 | 0.004432 | 1.40E-10 |
| rs674302 | miR-126-3p | trans | 2.03E-39 | Nightingale | IDL_CE | NA | A | T | 0.033517 | 0.004272 | 1.30E-14 |
| rs674302 | miR-126-3p | trans | 2.03E-39 | Nightingale | M_LDL_FC | NA | A | T | 0.034304 | 0.004419 | 2.00E-14 |
| rs674302 | miR-126-3p | trans | 2.03E-39 | Nightingale | M_VLDL_P | NA | A | T | 0.04397 | 0.004246 | 6.90E-25 |
| rs674302 | miR-126-3p | trans | 2.03E-39 | Nightingale | L_VLDL_TG | NA | A | T | -0.05103 | 0.004404 | 7.30E-32 |
| rs674302 | miR-126-3p | trans | 2.03E-39 | Nightingale | M_VLDL_P | NA | A | T | 0.028266 | 0.004414 | 6.40E-11 |
| rs674302 | miR-126-3p | trans | 2.03E-39 | Nightingale | LDL_P | NA | A | T | 0.031748 | 0.004421 | 1.10E-12 |
| rs674302 | miR-126-3p | trans | 2.03E-39 | Nightingale | M_VLDL_T | NA | A | T | -0.03848 | 0.004181 | 8.00E-20 |
| rs674302 | miR-126-3p | trans | 2.03E-39 | Nightingale | LDL_PL | NA | A | T | 0.034951 | 0.004414 | 4.90E-15 |
| rs674302 | miR-126-3p | trans | 2.03E-39 | Nightingale | S_HDL_L | NA | A | T | 0.02629 | 0.004409 | 1.60E-09 |
| rs674302 | miR-126-3p | trans | 2.03E-39 | Nightingale | S_HDL_C | NA | A | T | 0.028386 | 0.004417 | 1.80E-10 |
| rs674302 | miR-126-3p | trans | 2.03E-39 | Nightingale | S_HDL_FC | NA | A | T | 0.023467 | 0.004344 | 4.60E-08 |
| rs674302 | miR-126-3p | trans | 2.03E-39 | Nightingale | S_VLDL_C | NA | A | T | 0.038348 | 0.004424 | 1.80E-18 |
| rs674302 | miR-126-3p | trans | 2.03E-39 | Nightingale | LDL_C | NA | A | T | 0.033151 | 0.004405 | 9.30E-14 |
| rs674302 | miR-126-3p | trans | 2.03E-39 | Nightingale | M_VLDL_C | NA | A | T | 0.040808 | 0.004384 | 2.40E-20 |
| rs674302 | miR-126-3p | trans | 2.03E-39 | Nightingale | S_LDL_C_p | NA | A | T | 0.045309 | 0.004442 | 6.20E-25 |
| rs674302 | miR-126-3p | trans | 2.03E-39 | Nightingale | XXL_VLDL_ | NA | A | T | 0.033058 | 0.004458 | 9.70E-14 |
| rs674302 | miR-126-3p | trans | 2.03E-39 | Nightingale | IDL_PL | NA | A | T | 0.035323 | 0.00427 | 3.50E-16 |
| rs674302 | miR-126-3p | trans | 2.03E-39 | Nightingale | L_LDL_C | NA | A | T | 0.032278 | 0.004375 | 2.70E-13 |
| rs674302 | miR-126-3p | trans | 2.03E-39 | Nightingale | ApoB_by_A | NA | A | T | 0.024989 | 0.004358 | 1.40E-08 |
| rs674302 | miR-126-3p | trans | 2.03E-39 | Nightingale | IDL_FC_pct | NA | A | T | 0.025694 | 0.00439 | 1.20E-08 |
| rs674302 | miR-126-3p | trans | 2.03E-39 | Nightingale | Total_FC | NA | A | T | 0.033246 | 0.004322 | 6.90E-15 |
| rs674302 | miR-126-3p | trans | 2.03E-39 | Nightingale | S_LDL_CE_ | NA | A | T | 0.030904 | 0.004417 | 5.30E-14 |
| rs674302 | miR-126-3p | trans | 2.03E-39 | Nightingale | S_LDL_P | NA | A | T | 0.029552 | 0.004429 | 3.30E-11 |
| rs674302 | miR-126-3p | trans | 2.03E-39 | Nightingale | PUFA | NA | A | T | 0.023697 | 0.004316 | 1.50E-08 |
| rs674302 | miR-126-3p | trans | 2.03E-39 | Nightingale | ApoB | NA | A | T | 0.033904 | 0.004419 | 2.20E-14 |
| rs674302 | miR-126-3p | trans | 2.03E-39 | Nightingale | S_VLDL_C_ | NA | A | T | 0.04182 | 0.004306 | 6.00E-22 |
| rs674302 | miR-126-3p | trans | 2.03E-39 | Nightingale | M_VLDL_C | NA | A | T | 0.038673 | 0.004412 | 2.00E-18 |
| rs674302 | miR-126-3p | trans | 2.03E-39 | Nightingale | L_LDL_FC | NA | A | T | 0.033591 | 0.004345 | 3.00E-14 |
| rs674302 | miR-126-3p | trans | 2.03E-39 | Nightingale | S_VLDL_L | NA | A | T | 0.024659 | 0.004382 | 3.80E-09 |
| rs674302 | miR-126-3p | trans | 2.03E-39 | Nightingale | XL_VLDL_C | NA | A | T | 0.038131 | 0.004243 | 5.50E-19 |
| rs674302 | miR-126-3p | trans | 2.03E-39 | Nightingale | S_LDL_PL_ | NA | A | T | -0.03005 | 0.004416 | 2.80E-13 |
| rs674302 | miR-126-3p | trans | 2.03E-39 | Nightingale | S_HDL_PL | NA | A | T | 0.023787 | 0.004386 | 3.30E-08 |
| rs674302 | miR-126-3p | trans | 2.03E-39 | Nightingale | XXL_VLDL_ | NA | A | T | -0.03564 | 0.004463 | 2.50E-16 |
| rs674302 | miR-126-3p | trans | 2.03E-39 | Nightingale | XS_VLDL_C | NA | A | T | 0.038001 | 0.004295 | 1.30E-18 |
| rs674302 | miR-126-3p | trans | 2.03E-39 | Nightingale | LDL_FC | NA | A | T | 0.033595 | 0.004377 | 5.50E-14 |
| rs674302 | miR-126-3p | trans | 2.03E-39 | Nightingale | XS_VLDL_L | NA | A | T | 0.033833 | 0.004349 | 2.00E-15 |
| rs674302 | miR-126-3p | trans | 2.03E-39 | Nightingale | S_VLDL_TG | NA | A | T | -0.03806 | 0.004283 | 1.60E-18 |
| rs674302 | miR-126-3p | trans | 2.03E-39 | Nightingale | Total_CE | NA | A | T | 0.031729 | 0.004268 | 1.00E-13 |

| | | | | | | | | | | | |
|---|---|---|---|---|---|---|---|---|---|---|---|
| rs674302 | miR-126-3p | trans | 2.03E-39 | Nightingale | GlycA | NA | A | T | 0.027865 | 0.004406 | 1.80E-10 |
| rs674302 | miR-126-3p | trans | 2.03E-39 | Nightingale | S_VLDL_PL | NA | A | T | 0.027671 | 0.004237 | 1.90E-10 |
| rs674302 | miR-126-3p | trans | 2.03E-39 | Nightingale | LDL_CE | NA | A | T | 0.032508 | 0.004413 | 2.30E-13 |
| rs674302 | miR-126-3p | trans | 2.03E-39 | Nightingale | M_LDL_L | NA | A | T | 0.032989 | 0.004431 | 1.10E-13 |
| rs674302 | miR-126-3p | trans | 2.03E-39 | Nightingale | VLDL_FC | NA | A | T | 0.02414 | 0.004392 | 1.10E-08 |
| rs674302 | miR-126-3p | trans | 2.03E-39 | Nightingale | Cholines | NA | A | T | 0.022301 | 0.004161 | 4.90E-08 |
| rs674302 | miR-126-3p | trans | 2.03E-39 | Nightingale | L_LDL_L | NA | A | T | 0.033512 | 0.004381 | 3.20E-14 |
| rs674302 | miR-126-3p | trans | 2.03E-39 | Nightingale | XL_VLDL_F | NA | A | T | 0.037122 | 0.004289 | 2.30E-18 |
| rs674302 | miR-126-3p | trans | 2.03E-39 | Nightingale | XS_VLDL_P | NA | A | T | 0.032126 | 0.004363 | 5.20E-14 |
| rs674302 | miR-126-3p | trans | 2.03E-39 | Nightingale | VLDL_P | NA | A | T | 0.026627 | 0.004401 | 4.40E-10 |
| rs674302 | miR-126-3p | trans | 2.03E-39 | Nightingale | S_VLDL_FC | NA | A | T | 0.029296 | 0.004228 | 1.10E-11 |
| rs674302 | miR-126-3p | trans | 2.03E-39 | Nightingale | L_VLDL_FC | NA | A | T | 0.048437 | 0.004382 | 6.60E-29 |
| rs674302 | miR-126-3p | trans | 2.03E-39 | Nightingale | Total_L | NA | A | T | 0.026349 | 0.004353 | 4.60E-10 |
| rs674302 | miR-126-3p | trans | 2.03E-39 | Nightingale | XL_HDL_PL | NA | A | T | -0.02269 | 0.004065 | 1.20E-08 |
| rs674302 | miR-126-3p | trans | 2.03E-39 | Nightingale | XL_VLDL_C | NA | A | T | 0.037253 | 0.004245 | 3.80E-18 |
| rs674302 | miR-126-3p | trans | 2.03E-39 | Nightingale | S_VLDL_PL | NA | A | T | 0.032753 | 0.004416 | 4.20E-14 |
| rs674302 | miR-126-3p | trans | 2.03E-39 | Nightingale | Total_C | NA | A | T | 0.032359 | 0.004283 | 3.40E-14 |
| rs674302 | miR-126-3p | trans | 2.03E-39 | Nightingale | M_LDL_PL | NA | A | T | 0.032425 | 0.004431 | 2.70E-13 |
| rs674302 | miR-126-3p | trans | 2.03E-39 | Nightingale | L_VLDL_C_ | NA | A | T | 0.052079 | 0.004322 | 5.60E-34 |
| rs674302 | miR-126-3p | trans | 2.03E-39 | Nightingale | XXL_VLDL_ | NA | A | T | 0.034734 | 0.004434 | 3.40E-15 |
| rs674302 | miR-126-3p | trans | 2.03E-39 | Nightingale | M_VLDL_C | NA | A | T | 0.036135 | 0.004168 | 1.40E-17 |
| rs674302 | miR-126-3p | trans | 2.03E-39 | Nightingale | M_VLDL_P | NA | A | T | 0.029105 | 0.004418 | 2.10E-11 |
| rs674302 | miR-126-3p | trans | 2.03E-39 | Nightingale | Sphingomy | NA | A | T | 0.028962 | 0.004136 | 7.60E-12 |
| rs674302 | miR-126-3p | trans | 2.03E-39 | Nightingale | M_LDL_C | NA | A | T | 0.033944 | 0.004433 | 2.30E-14 |
| rs674302 | miR-126-3p | trans | 2.03E-39 | Nightingale | M_VLDL_F | NA | A | T | 0.033342 | 0.004424 | 3.20E-14 |
| rs674302 | miR-126-3p | trans | 2.03E-39 | Nightingale | S_VLDL_CE | NA | A | T | 0.046964 | 0.004365 | 1.40E-26 |
| rs674302 | miR-126-3p | trans | 2.03E-39 | Nightingale | XS_VLDL_P | NA | A | T | 0.036716 | 0.004366 | 1.30E-17 |
| rs674302 | miR-126-3p | trans | 2.03E-39 | Nightingale | S_LDL_L | NA | A | T | 0.027702 | 0.004424 | 8.50E-10 |
| rs674302 | miR-126-3p | trans | 2.03E-39 | Nightingale | L_LDL_P | NA | A | T | 0.029119 | 0.00441 | 6.60E-11 |
| rs674302 | miR-126-3p | trans | 2.03E-39 | Nightingale | S_HDL_P | NA | A | T | 0.03058 | 0.004425 | 5.40E-12 |
| rs674302 | miR-126-3p | trans | 2.03E-39 | Nightingale | XL_VLDL_T | NA | A | T | -0.04377 | 0.004289 | 2.50E-24 |
| rs674302 | miR-126-3p | trans | 2.03E-39 | Nightingale | XL_VLDL_P | NA | A | T | 0.038723 | 0.004431 | 6.70E-19 |
| rs674302 | miR-126-3p | trans | 2.03E-39 | Nightingale | non_HDL_( | NA | A | T | 0.035666 | 0.004401 | 5.30E-16 |
| rs674302 | miR-126-3p | trans | 2.03E-39 | Nightingale | XS_VLDL_F | NA | A | T | 0.03137 | 0.004393 | 2.60E-12 |
| rs674302 | miR-126-3p | trans | 2.03E-39 | Nightingale | S_VLDL_P | NA | A | T | 0.023462 | 0.004369 | 1.60E-08 |
| rs674302 | miR-126-3p | trans | 2.03E-39 | Nightingale | XS_VLDL_F | NA | A | T | 0.037073 | 0.004348 | 6.40E-18 |
| rs674302 | miR-126-3p | trans | 2.03E-39 | Nightingale | Clinical_LD | NA | A | T | 0.036436 | 0.004399 | 2.90E-16 |
| rs674302 | miR-126-3p | trans | 2.03E-39 | Nightingale | IDL_FC | NA | A | T | 0.03824 | 0.004284 | 1.50E-18 |
| rs674302 | miR-126-3p | trans | 2.03E-39 | Nightingale | XS_VLDL_C | NA | A | T | 0.037807 | 0.00427 | 2.10E-18 |
| rs674302 | miR-126-3p | trans | 2.03E-39 | Nightingale | LA | NA | A | T | 0.02494 | 0.004339 | 3.50E-09 |
| rs674302 | miR-126-3p | trans | 2.03E-39 | Nightingale | M_VLDL_C | NA | A | T | 0.033562 | 0.004157 | 2.10E-15 |
| rs674302 | miR-126-3p | trans | 2.03E-39 | Nightingale | VLDL_CE | NA | A | T | 0.035764 | 0.004426 | 3.10E-16 |
| rs674302 | miR-126-3p | trans | 2.03E-39 | Nightingale | S_VLDL_FC | NA | A | T | 0.037872 | 0.004425 | 7.50E-18 |
| rs674302 | miR-126-3p | trans | 2.03E-39 | Nightingale | L_VLDL_CE | NA | A | T | 0.047276 | 0.004234 | 2.90E-29 |
| rs674302 | miR-126-3p | trans | 2.03E-39 | Nightingale | XXL_VLDL_ | NA | A | T | 0.025014 | 0.004453 | 1.50E-09 |
| rs674302 | miR-126-3p | trans | 2.03E-39 | Nightingale | VLDL_C | NA | A | T | 0.031281 | 0.004419 | 5.00E-13 |
| rs674302 | miR-126-3p | trans | 2.03E-39 | Nightingale | Citrate | NA | A | T | 0.026836 | 0.004396 | 7.10E-10 |
| rs674302 | miR-126-3p | trans | 2.03E-39 | Nightingale | M_LDL_P | NA | A | T | 0.035851 | 0.004437 | 7.20E-16 |
| rs674302 | miR-139-5p | trans | 6.05E-42 | Nightingale | Remnant_( | NA | A | T | 0.036854 | 0.004396 | 3.70E-17 |
| rs674302 | miR-139-5p | trans | 6.05E-42 | Nightingale | L_LDL_PL | NA | A | T | 0.037956 | 0.004391 | 9.50E-18 |
| rs674302 | miR-139-5p | trans | 6.05E-42 | Nightingale | S_VLDL_CE | NA | A | T | 0.038162 | 0.004419 | 1.60E-18 |
| rs674302 | miR-139-5p | trans | 6.05E-42 | Nightingale | LDL_L | NA | A | T | 0.033263 | 0.00441 | 7.70E-14 |
| rs674302 | miR-139-5p | trans | 6.05E-42 | Nightingale | S_LDL_C | NA | A | T | 0.0309 | 0.004429 | 5.70E-12 |
| rs674302 | miR-139-5p | trans | 6.05E-42 | Nightingale | M_VLDL_F | NA | A | T | 0.041986 | 0.00421 | 4.20E-23 |
| rs674302 | miR-139-5p | trans | 6.05E-42 | Nightingale | IDL_L | NA | A | T | 0.03527 | 0.004278 | 4.90E-16 |
| rs674302 | miR-139-5p | trans | 6.05E-42 | Nightingale | S_LDL_FC | NA | A | T | 0.028528 | 0.004418 | 4.40E-10 |
| rs674302 | miR-139-5p | trans | 6.05E-42 | Nightingale | M_LDL_CE | NA | A | T | 0.032929 | 0.004431 | 1.00E-13 |
| rs674302 | miR-139-5p | trans | 6.05E-42 | Nightingale | S_LDL_CE | NA | A | T | 0.030504 | 0.004427 | 6.10E-12 |
| rs674302 | miR-139-5p | trans | 6.05E-42 | Nightingale | XXL_VLDL_ | NA | A | T | 0.030728 | 0.004406 | 1.30E-12 |
| rs674302 | miR-139-5p | trans | 6.05E-42 | Nightingale | M_LDL_C_ | NA | A | T | 0.031102 | 0.004428 | 3.20E-12 |
| rs674302 | miR-139-5p | trans | 6.05E-42 | Nightingale | Total_PL | NA | A | T | 0.024938 | 0.004213 | 1.10E-09 |
| rs674302 | miR-139-5p | trans | 6.05E-42 | Nightingale | Omega_6 | NA | A | T | 0.025874 | 0.004321 | 5.90E-10 |
| rs674302 | miR-139-5p | trans | 6.05E-42 | Nightingale | IDL_C | NA | A | T | 0.034959 | 0.004273 | 9.50E-16 |

| | | | | | | | | | | | |
|---|---|---|---|---|---|---|---|---|---|---|---|
| rs674302 | miR-139-5p | trans | 6.05E-42 | Nightingale | M_LDL_TG | NA | A | T | -0.02754 | 0.00437 | 3.60E-10 |
| rs674302 | miR-139-5p | trans | 6.05E-42 | Nightingale | IDL_P | NA | A | T | 0.04001 | 0.004378 | 1.30E-19 |
| rs674302 | miR-139-5p | trans | 6.05E-42 | Nightingale | L_LDL_CE | NA | A | T | 0.031522 | 0.004385 | 1.00E-12 |
| rs674302 | miR-139-5p | trans | 6.05E-42 | Nightingale | S_HDL_CE | NA | A | T | 0.028597 | 0.004432 | 1.40E-10 |
| rs674302 | miR-139-5p | trans | 6.05E-42 | Nightingale | IDL_CE | NA | A | T | 0.033517 | 0.004272 | 1.30E-14 |
| rs674302 | miR-139-5p | trans | 6.05E-42 | Nightingale | M_LDL_FC | NA | A | T | 0.034304 | 0.004419 | 2.00E-14 |
| rs674302 | miR-139-5p | trans | 6.05E-42 | Nightingale | M_VLDL_P | NA | A | T | 0.04397 | 0.004246 | 6.90E-25 |
| rs674302 | miR-139-5p | trans | 6.05E-42 | Nightingale | L_VLDL_TG | NA | A | T | -0.05103 | 0.004404 | 7.30E-32 |
| rs674302 | miR-139-5p | trans | 6.05E-42 | Nightingale | M_VLDL_P | NA | A | T | 0.028266 | 0.004414 | 6.40E-11 |
| rs674302 | miR-139-5p | trans | 6.05E-42 | Nightingale | LDL_P | NA | A | T | 0.031748 | 0.004421 | 1.10E-12 |
| rs674302 | miR-139-5p | trans | 6.05E-42 | Nightingale | M_VLDL_T | NA | A | T | -0.03848 | 0.004181 | 8.00E-20 |
| rs674302 | miR-139-5p | trans | 6.05E-42 | Nightingale | LDL_PL | NA | A | T | 0.034951 | 0.004414 | 4.90E-15 |
| rs674302 | miR-139-5p | trans | 6.05E-42 | Nightingale | S_HDL_L | NA | A | T | 0.02629 | 0.004409 | 1.60E-09 |
| rs674302 | miR-139-5p | trans | 6.05E-42 | Nightingale | S_HDL_C | NA | A | T | 0.028386 | 0.004417 | 1.80E-10 |
| rs674302 | miR-139-5p | trans | 6.05E-42 | Nightingale | S_HDL_FC | NA | A | T | 0.023467 | 0.004344 | 4.60E-08 |
| rs674302 | miR-139-5p | trans | 6.05E-42 | Nightingale | S_VLDL_C | NA | A | T | 0.038348 | 0.004424 | 1.80E-18 |
| rs674302 | miR-139-5p | trans | 6.05E-42 | Nightingale | LDL_C | NA | A | T | 0.033151 | 0.004405 | 9.30E-14 |
| rs674302 | miR-139-5p | trans | 6.05E-42 | Nightingale | M_VLDL_C | NA | A | T | 0.040808 | 0.004384 | 2.40E-20 |
| rs674302 | miR-139-5p | trans | 6.05E-42 | Nightingale | S_LDL_C_p | NA | A | T | 0.045309 | 0.004442 | 6.20E-25 |
| rs674302 | miR-139-5p | trans | 6.05E-42 | Nightingale | XXL_VLDL_ | NA | A | T | 0.033058 | 0.004458 | 9.70E-14 |
| rs674302 | miR-139-5p | trans | 6.05E-42 | Nightingale | IDL_PL | NA | A | T | 0.035323 | 0.00427 | 3.50E-16 |
| rs674302 | miR-139-5p | trans | 6.05E-42 | Nightingale | L_LDL_C | NA | A | T | 0.032278 | 0.004375 | 2.70E-13 |
| rs674302 | miR-139-5p | trans | 6.05E-42 | Nightingale | ApoB_by_A | NA | A | T | 0.024989 | 0.004358 | 1.40E-08 |
| rs674302 | miR-139-5p | trans | 6.05E-42 | Nightingale | IDL_FC_pct | NA | A | T | 0.025694 | 0.00439 | 1.20E-08 |
| rs674302 | miR-139-5p | trans | 6.05E-42 | Nightingale | Total_FC | NA | A | T | 0.033246 | 0.004322 | 6.90E-15 |
| rs674302 | miR-139-5p | trans | 6.05E-42 | Nightingale | S_LDL_CE_ | NA | A | T | 0.030904 | 0.004417 | 5.30E-14 |
| rs674302 | miR-139-5p | trans | 6.05E-42 | Nightingale | S_LDL_P | NA | A | T | 0.029552 | 0.004429 | 3.30E-11 |
| rs674302 | miR-139-5p | trans | 6.05E-42 | Nightingale | PUFA | NA | A | T | 0.023697 | 0.004316 | 1.50E-08 |
| rs674302 | miR-139-5p | trans | 6.05E-42 | Nightingale | ApoB | NA | A | T | 0.033904 | 0.004419 | 2.20E-14 |
| rs674302 | miR-139-5p | trans | 6.05E-42 | Nightingale | S_VLDL_C_ | NA | A | T | 0.04182 | 0.004306 | 6.00E-22 |
| rs674302 | miR-139-5p | trans | 6.05E-42 | Nightingale | M_VLDL_C | NA | A | T | 0.038673 | 0.004412 | 2.00E-18 |
| rs674302 | miR-139-5p | trans | 6.05E-42 | Nightingale | L_LDL_FC | NA | A | T | 0.033591 | 0.004345 | 3.00E-14 |
| rs674302 | miR-139-5p | trans | 6.05E-42 | Nightingale | S_VLDL_L | NA | A | T | 0.024659 | 0.004382 | 3.80E-09 |
| rs674302 | miR-139-5p | trans | 6.05E-42 | Nightingale | XL_VLDL_C | NA | A | T | 0.038131 | 0.004243 | 5.50E-19 |
| rs674302 | miR-139-5p | trans | 6.05E-42 | Nightingale | S_LDL_PL_ | NA | A | T | -0.03005 | 0.004416 | 2.80E-13 |
| rs674302 | miR-139-5p | trans | 6.05E-42 | Nightingale | S_HDL_PL | NA | A | T | 0.023787 | 0.004386 | 3.30E-08 |
| rs674302 | miR-139-5p | trans | 6.05E-42 | Nightingale | XXL_VLDL_ | NA | A | T | -0.03564 | 0.004463 | 2.50E-16 |
| rs674302 | miR-139-5p | trans | 6.05E-42 | Nightingale | XS_VLDL_C | NA | A | T | 0.038001 | 0.004295 | 1.30E-18 |
| rs674302 | miR-139-5p | trans | 6.05E-42 | Nightingale | LDL_FC | NA | A | T | 0.033595 | 0.004377 | 5.50E-14 |
| rs674302 | miR-139-5p | trans | 6.05E-42 | Nightingale | XS_VLDL_L | NA | A | T | 0.033833 | 0.004349 | 2.00E-15 |
| rs674302 | miR-139-5p | trans | 6.05E-42 | Nightingale | S_VLDL_TG | NA | A | T | -0.03806 | 0.004283 | 1.60E-18 |
| rs674302 | miR-139-5p | trans | 6.05E-42 | Nightingale | Total_CE | NA | A | T | 0.031729 | 0.004268 | 1.00E-13 |
| rs674302 | miR-139-5p | trans | 6.05E-42 | Nightingale | GlycA | NA | A | T | 0.027865 | 0.004406 | 1.80E-10 |
| rs674302 | miR-139-5p | trans | 6.05E-42 | Nightingale | S_VLDL_PL | NA | A | T | 0.027671 | 0.004237 | 1.90E-10 |
| rs674302 | miR-139-5p | trans | 6.05E-42 | Nightingale | LDL_CE | NA | A | T | 0.032508 | 0.004413 | 2.30E-13 |
| rs674302 | miR-139-5p | trans | 6.05E-42 | Nightingale | M_LDL_L | NA | A | T | 0.032989 | 0.004431 | 1.10E-13 |
| rs674302 | miR-139-5p | trans | 6.05E-42 | Nightingale | VLDL_FC | NA | A | T | 0.02414 | 0.004392 | 1.10E-08 |
| rs674302 | miR-139-5p | trans | 6.05E-42 | Nightingale | Cholines | NA | A | T | 0.022301 | 0.004161 | 4.90E-08 |
| rs674302 | miR-139-5p | trans | 6.05E-42 | Nightingale | L_LDL_L | NA | A | T | 0.033512 | 0.004381 | 3.20E-14 |
| rs674302 | miR-139-5p | trans | 6.05E-42 | Nightingale | XL_VLDL_F | NA | A | T | 0.037122 | 0.004289 | 2.30E-18 |
| rs674302 | miR-139-5p | trans | 6.05E-42 | Nightingale | XS_VLDL_P | NA | A | T | 0.032126 | 0.004363 | 5.20E-14 |
| rs674302 | miR-139-5p | trans | 6.05E-42 | Nightingale | VLDL_P | NA | A | T | 0.026627 | 0.004401 | 4.40E-10 |
| rs674302 | miR-139-5p | trans | 6.05E-42 | Nightingale | S_VLDL_FC | NA | A | T | 0.029296 | 0.004228 | 1.10E-11 |
| rs674302 | miR-139-5p | trans | 6.05E-42 | Nightingale | L_VLDL_FC | NA | A | T | 0.048437 | 0.004382 | 6.60E-29 |
| rs674302 | miR-139-5p | trans | 6.05E-42 | Nightingale | Total_L | NA | A | T | 0.026349 | 0.004353 | 4.60E-10 |
| rs674302 | miR-139-5p | trans | 6.05E-42 | Nightingale | XL_HDL_PL | NA | A | T | -0.02269 | 0.004065 | 1.20E-08 |
| rs674302 | miR-139-5p | trans | 6.05E-42 | Nightingale | XL_VLDL_C | NA | A | T | 0.037253 | 0.004245 | 3.80E-18 |
| rs674302 | miR-139-5p | trans | 6.05E-42 | Nightingale | S_VLDL_PL | NA | A | T | 0.032753 | 0.004416 | 4.20E-14 |
| rs674302 | miR-139-5p | trans | 6.05E-42 | Nightingale | Total_C | NA | A | T | 0.032359 | 0.004283 | 3.40E-14 |
| rs674302 | miR-139-5p | trans | 6.05E-42 | Nightingale | M_LDL_PL | NA | A | T | 0.032425 | 0.004431 | 2.70E-13 |
| rs674302 | miR-139-5p | trans | 6.05E-42 | Nightingale | L_VLDL_C_ | NA | A | T | 0.052079 | 0.004322 | 5.60E-34 |
| rs674302 | miR-139-5p | trans | 6.05E-42 | Nightingale | XXL_VLDL_ | NA | A | T | 0.034734 | 0.004434 | 3.40E-15 |
| rs674302 | miR-139-5p | trans | 6.05E-42 | Nightingale | M_VLDL_C | NA | A | T | 0.036135 | 0.004168 | 1.40E-17 |

| | | | | | | | | | | | |
|---|---|---|---|---|---|---|---|---|---|---|---|
| rs674302 | miR-139-5p | trans | 6.05E-42 | Nightingale | M_VLDL_P | NA | A | T | 0.029105 | 0.004418 | 2.10E-11 |
| rs674302 | miR-139-5p | trans | 6.05E-42 | Nightingale | Sphingomy | NA | A | T | 0.028962 | 0.004136 | 7.60E-12 |
| rs674302 | miR-139-5p | trans | 6.05E-42 | Nightingale | M_LDL_C | NA | A | T | 0.033944 | 0.004433 | 2.30E-14 |
| rs674302 | miR-139-5p | trans | 6.05E-42 | Nightingale | M_VLDL_F | NA | A | T | 0.033342 | 0.004424 | 3.20E-14 |
| rs674302 | miR-139-5p | trans | 6.05E-42 | Nightingale | S_VLDL_CE | NA | A | T | 0.046964 | 0.004365 | 1.40E-26 |
| rs674302 | miR-139-5p | trans | 6.05E-42 | Nightingale | XS_VLDL_P | NA | A | T | 0.036716 | 0.004366 | 1.30E-17 |
| rs674302 | miR-139-5p | trans | 6.05E-42 | Nightingale | S_LDL_L | NA | A | T | 0.027702 | 0.004424 | 8.50E-10 |
| rs674302 | miR-139-5p | trans | 6.05E-42 | Nightingale | L_LDL_P | NA | A | T | 0.029119 | 0.00441 | 6.60E-11 |
| rs674302 | miR-139-5p | trans | 6.05E-42 | Nightingale | S_HDL_P | NA | A | T | 0.03058 | 0.004425 | 5.40E-12 |
| rs674302 | miR-139-5p | trans | 6.05E-42 | Nightingale | XL_VLDL_T | NA | A | T | -0.04377 | 0.004289 | 2.50E-24 |
| rs674302 | miR-139-5p | trans | 6.05E-42 | Nightingale | XL_VLDL_P | NA | A | T | 0.038723 | 0.004431 | 6.70E-19 |
| rs674302 | miR-139-5p | trans | 6.05E-42 | Nightingale | non_HDL_( | NA | A | T | 0.035666 | 0.004401 | 5.30E-16 |
| rs674302 | miR-139-5p | trans | 6.05E-42 | Nightingale | XS_VLDL_F | NA | A | T | 0.03137 | 0.004393 | 2.60E-12 |
| rs674302 | miR-139-5p | trans | 6.05E-42 | Nightingale | S_VLDL_P | NA | A | T | 0.023462 | 0.004369 | 1.60E-08 |
| rs674302 | miR-139-5p | trans | 6.05E-42 | Nightingale | XS_VLDL_F | NA | A | T | 0.037073 | 0.004348 | 6.40E-18 |
| rs674302 | miR-139-5p | trans | 6.05E-42 | Nightingale | Clinical_LD | NA | A | T | 0.036436 | 0.004399 | 2.90E-16 |
| rs674302 | miR-139-5p | trans | 6.05E-42 | Nightingale | IDL_FC | NA | A | T | 0.03824 | 0.004284 | 1.50E-18 |
| rs674302 | miR-139-5p | trans | 6.05E-42 | Nightingale | XS_VLDL_C | NA | A | T | 0.037807 | 0.00427 | 2.10E-18 |
| rs674302 | miR-139-5p | trans | 6.05E-42 | Nightingale | LA | NA | A | T | 0.02494 | 0.004339 | 3.50E-09 |
| rs674302 | miR-139-5p | trans | 6.05E-42 | Nightingale | M_VLDL_C | NA | A | T | 0.033562 | 0.004157 | 2.10E-15 |
| rs674302 | miR-139-5p | trans | 6.05E-42 | Nightingale | VLDL_CE | NA | A | T | 0.035764 | 0.004426 | 3.10E-16 |
| rs674302 | miR-139-5p | trans | 6.05E-42 | Nightingale | S_VLDL_FC | NA | A | T | 0.037872 | 0.004425 | 7.50E-18 |
| rs674302 | miR-139-5p | trans | 6.05E-42 | Nightingale | L_VLDL_CE | NA | A | T | 0.047276 | 0.004234 | 2.90E-29 |
| rs674302 | miR-139-5p | trans | 6.05E-42 | Nightingale | XXL_VLDL_ | NA | A | T | 0.025014 | 0.004453 | 1.50E-09 |
| rs674302 | miR-139-5p | trans | 6.05E-42 | Nightingale | VLDL_C | NA | A | T | 0.031281 | 0.004419 | 5.00E-13 |
| rs674302 | miR-139-5p | trans | 6.05E-42 | Nightingale | Citrate | NA | A | T | 0.026836 | 0.004396 | 7.10E-10 |
| rs674302 | miR-139-5p | trans | 6.05E-42 | Nightingale | M_LDL_P | NA | A | T | 0.035851 | 0.004437 | 7.20E-16 |
| rs674302 | miR-10b-5p | trans | 2.75E-35 | Nightingale | Remnant_( | NA | A | T | 0.036854 | 0.004396 | 3.70E-17 |
| rs674302 | miR-10b-5p | trans | 2.75E-35 | Nightingale | L_LDL_PL | NA | A | T | 0.037956 | 0.004391 | 9.50E-18 |
| rs674302 | miR-10b-5p | trans | 2.75E-35 | Nightingale | S_VLDL_CE | NA | A | T | 0.038162 | 0.004419 | 1.60E-18 |
| rs674302 | miR-10b-5p | trans | 2.75E-35 | Nightingale | LDL_L | NA | A | T | 0.033263 | 0.00441 | 7.70E-14 |
| rs674302 | miR-10b-5p | trans | 2.75E-35 | Nightingale | S_LDL_C | NA | A | T | 0.0309 | 0.004429 | 5.70E-12 |
| rs674302 | miR-10b-5p | trans | 2.75E-35 | Nightingale | M_VLDL_F | NA | A | T | 0.041986 | 0.00421 | 4.20E-23 |
| rs674302 | miR-10b-5p | trans | 2.75E-35 | Nightingale | IDL_L | NA | A | T | 0.03527 | 0.004278 | 4.90E-16 |
| rs674302 | miR-10b-5p | trans | 2.75E-35 | Nightingale | S_LDL_FC | NA | A | T | 0.028528 | 0.004418 | 4.40E-10 |
| rs674302 | miR-10b-5p | trans | 2.75E-35 | Nightingale | M_LDL_CE | NA | A | T | 0.032929 | 0.004431 | 1.00E-13 |
| rs674302 | miR-10b-5p | trans | 2.75E-35 | Nightingale | S_LDL_CE | NA | A | T | 0.030504 | 0.004427 | 6.10E-12 |
| rs674302 | miR-10b-5p | trans | 2.75E-35 | Nightingale | XXL_VLDL_ | NA | A | T | 0.030728 | 0.004406 | 1.30E-12 |
| rs674302 | miR-10b-5p | trans | 2.75E-35 | Nightingale | M_LDL_C_ | NA | A | T | 0.031102 | 0.004428 | 3.20E-12 |
| rs674302 | miR-10b-5p | trans | 2.75E-35 | Nightingale | Total_PL | NA | A | T | 0.024938 | 0.004213 | 1.10E-09 |
| rs674302 | miR-10b-5p | trans | 2.75E-35 | Nightingale | Omega_6 | NA | A | T | 0.025874 | 0.004321 | 5.90E-10 |
| rs674302 | miR-10b-5p | trans | 2.75E-35 | Nightingale | IDL_C | NA | A | T | 0.034959 | 0.004273 | 9.50E-16 |
| rs674302 | miR-10b-5p | trans | 2.75E-35 | Nightingale | M_LDL_TG | NA | A | T | -0.02754 | 0.00437 | 3.60E-10 |
| rs674302 | miR-10b-5p | trans | 2.75E-35 | Nightingale | IDL_P | NA | A | T | 0.04001 | 0.004378 | 1.30E-19 |
| rs674302 | miR-10b-5p | trans | 2.75E-35 | Nightingale | L_LDL_CE | NA | A | T | 0.031522 | 0.004385 | 1.00E-12 |
| rs674302 | miR-10b-5p | trans | 2.75E-35 | Nightingale | S_HDL_CE | NA | A | T | 0.028597 | 0.004432 | 1.40E-10 |
| rs674302 | miR-10b-5p | trans | 2.75E-35 | Nightingale | IDL_CE | NA | A | T | 0.033517 | 0.004272 | 1.30E-14 |
| rs674302 | miR-10b-5p | trans | 2.75E-35 | Nightingale | M_LDL_FC | NA | A | T | 0.034304 | 0.004419 | 2.00E-14 |
| rs674302 | miR-10b-5p | trans | 2.75E-35 | Nightingale | M_VLDL_P | NA | A | T | 0.04397 | 0.004246 | 6.90E-25 |
| rs674302 | miR-10b-5p | trans | 2.75E-35 | Nightingale | L_VLDL_TG | NA | A | T | -0.05103 | 0.004404 | 7.30E-32 |
| rs674302 | miR-10b-5p | trans | 2.75E-35 | Nightingale | M_VLDL_P | NA | A | T | 0.028266 | 0.004414 | 6.40E-11 |
| rs674302 | miR-10b-5p | trans | 2.75E-35 | Nightingale | LDL_P | NA | A | T | 0.031748 | 0.004421 | 1.10E-12 |
| rs674302 | miR-10b-5p | trans | 2.75E-35 | Nightingale | M_VLDL_T | NA | A | T | -0.03848 | 0.004181 | 8.00E-20 |
| rs674302 | miR-10b-5p | trans | 2.75E-35 | Nightingale | LDL_PL | NA | A | T | 0.034951 | 0.004414 | 4.90E-15 |
| rs674302 | miR-10b-5p | trans | 2.75E-35 | Nightingale | S_HDL_L | NA | A | T | 0.02629 | 0.004409 | 1.60E-09 |
| rs674302 | miR-10b-5p | trans | 2.75E-35 | Nightingale | S_HDL_C | NA | A | T | 0.028386 | 0.004417 | 1.80E-10 |
| rs674302 | miR-10b-5p | trans | 2.75E-35 | Nightingale | S_HDL_FC | NA | A | T | 0.023467 | 0.004344 | 4.60E-08 |
| rs674302 | miR-10b-5p | trans | 2.75E-35 | Nightingale | S_VLDL_C | NA | A | T | 0.038348 | 0.004424 | 1.80E-18 |
| rs674302 | miR-10b-5p | trans | 2.75E-35 | Nightingale | LDL_C | NA | A | T | 0.033151 | 0.004405 | 9.30E-14 |
| rs674302 | miR-10b-5p | trans | 2.75E-35 | Nightingale | M_VLDL_C | NA | A | T | 0.040808 | 0.004384 | 2.40E-20 |
| rs674302 | miR-10b-5p | trans | 2.75E-35 | Nightingale | S_LDL_C_p | NA | A | T | 0.045309 | 0.004442 | 6.20E-25 |
| rs674302 | miR-10b-5p | trans | 2.75E-35 | Nightingale | XXL_VLDL_ | NA | A | T | 0.033058 | 0.004458 | 9.70E-14 |
| rs674302 | miR-10b-5p | trans | 2.75E-35 | Nightingale | IDL_PL | NA | A | T | 0.035323 | 0.00427 | 3.50E-16 |

| | | | | | | | | | | | |
|---|---|---|---|---|---|---|---|---|---|---|---|
| rs674302 | miR-10b-5p | trans | 2.75E-35 | Nightingale | L_LDL_C | NA | A | T | 0.032278 | 0.004375 | 2.70E-13 |
| rs674302 | miR-10b-5p | trans | 2.75E-35 | Nightingale | ApoB_by_A | NA | A | T | 0.024989 | 0.004358 | 1.40E-08 |
| rs674302 | miR-10b-5p | trans | 2.75E-35 | Nightingale | IDL_FC_pct | NA | A | T | 0.025694 | 0.00439 | 1.20E-08 |
| rs674302 | miR-10b-5p | trans | 2.75E-35 | Nightingale | Total_FC | NA | A | T | 0.033246 | 0.004322 | 6.90E-15 |
| rs674302 | miR-10b-5p | trans | 2.75E-35 | Nightingale | S_LDL_CE_ | NA | A | T | 0.030904 | 0.004417 | 5.30E-14 |
| rs674302 | miR-10b-5p | trans | 2.75E-35 | Nightingale | S_LDL_P | NA | A | T | 0.029552 | 0.004429 | 3.30E-11 |
| rs674302 | miR-10b-5p | trans | 2.75E-35 | Nightingale | PUFA | NA | A | T | 0.023697 | 0.004316 | 1.50E-08 |
| rs674302 | miR-10b-5p | trans | 2.75E-35 | Nightingale | ApoB | NA | A | T | 0.033904 | 0.004419 | 2.20E-14 |
| rs674302 | miR-10b-5p | trans | 2.75E-35 | Nightingale | S_VLDL_C_ | NA | A | T | 0.04182 | 0.004306 | 6.00E-22 |
| rs674302 | miR-10b-5p | trans | 2.75E-35 | Nightingale | M_VLDL_C | NA | A | T | 0.038673 | 0.004412 | 2.00E-18 |
| rs674302 | miR-10b-5p | trans | 2.75E-35 | Nightingale | L_LDL_FC | NA | A | T | 0.033591 | 0.004345 | 3.00E-14 |
| rs674302 | miR-10b-5p | trans | 2.75E-35 | Nightingale | S_VLDL_L | NA | A | T | 0.024659 | 0.004382 | 3.80E-09 |
| rs674302 | miR-10b-5p | trans | 2.75E-35 | Nightingale | XL_VLDL_C | NA | A | T | 0.038131 | 0.004243 | 5.50E-19 |
| rs674302 | miR-10b-5p | trans | 2.75E-35 | Nightingale | S_LDL_PL_ | NA | A | T | -0.03005 | 0.004416 | 2.80E-13 |
| rs674302 | miR-10b-5p | trans | 2.75E-35 | Nightingale | S_HDL_PL | NA | A | T | 0.023787 | 0.004386 | 3.30E-08 |
| rs674302 | miR-10b-5p | trans | 2.75E-35 | Nightingale | XXL_VLDL_ | NA | A | T | -0.03564 | 0.004463 | 2.50E-16 |
| rs674302 | miR-10b-5p | trans | 2.75E-35 | Nightingale | XS_VLDL_C | NA | A | T | 0.038001 | 0.004295 | 1.30E-18 |
| rs674302 | miR-10b-5p | trans | 2.75E-35 | Nightingale | LDL_FC | NA | A | T | 0.033595 | 0.004377 | 5.50E-14 |
| rs674302 | miR-10b-5p | trans | 2.75E-35 | Nightingale | XS_VLDL_L | NA | A | T | 0.033833 | 0.004349 | 2.00E-15 |
| rs674302 | miR-10b-5p | trans | 2.75E-35 | Nightingale | S_VLDL_TG | NA | A | T | -0.03806 | 0.004283 | 1.60E-18 |
| rs674302 | miR-10b-5p | trans | 2.75E-35 | Nightingale | Total_CE | NA | A | T | 0.031729 | 0.004268 | 1.00E-13 |
| rs674302 | miR-10b-5p | trans | 2.75E-35 | Nightingale | GlycA | NA | A | T | 0.027865 | 0.004406 | 1.80E-10 |
| rs674302 | miR-10b-5p | trans | 2.75E-35 | Nightingale | S_VLDL_PL | NA | A | T | 0.027671 | 0.004237 | 1.90E-10 |
| rs674302 | miR-10b-5p | trans | 2.75E-35 | Nightingale | LDL_CE | NA | A | T | 0.032508 | 0.004413 | 2.30E-13 |
| rs674302 | miR-10b-5p | trans | 2.75E-35 | Nightingale | M_LDL_L | NA | A | T | 0.032989 | 0.004431 | 1.10E-13 |
| rs674302 | miR-10b-5p | trans | 2.75E-35 | Nightingale | VLDL_FC | NA | A | T | 0.02414 | 0.004392 | 1.10E-08 |
| rs674302 | miR-10b-5p | trans | 2.75E-35 | Nightingale | Cholines | NA | A | T | 0.022301 | 0.004161 | 4.90E-08 |
| rs674302 | miR-10b-5p | trans | 2.75E-35 | Nightingale | L_LDL_L | NA | A | T | 0.033512 | 0.004381 | 3.20E-14 |
| rs674302 | miR-10b-5p | trans | 2.75E-35 | Nightingale | XL_VLDL_F | NA | A | T | 0.037122 | 0.004289 | 2.30E-18 |
| rs674302 | miR-10b-5p | trans | 2.75E-35 | Nightingale | XS_VLDL_P | NA | A | T | 0.032126 | 0.004363 | 5.20E-14 |
| rs674302 | miR-10b-5p | trans | 2.75E-35 | Nightingale | VLDL_P | NA | A | T | 0.026627 | 0.004401 | 4.40E-10 |
| rs674302 | miR-10b-5p | trans | 2.75E-35 | Nightingale | S_VLDL_FC | NA | A | T | 0.029296 | 0.004228 | 1.10E-11 |
| rs674302 | miR-10b-5p | trans | 2.75E-35 | Nightingale | L_VLDL_FC | NA | A | T | 0.048437 | 0.004382 | 6.60E-29 |
| rs674302 | miR-10b-5p | trans | 2.75E-35 | Nightingale | Total_L | NA | A | T | 0.026349 | 0.004353 | 4.60E-10 |
| rs674302 | miR-10b-5p | trans | 2.75E-35 | Nightingale | XL_HDL_PL | NA | A | T | -0.02269 | 0.004065 | 1.20E-08 |
| rs674302 | miR-10b-5p | trans | 2.75E-35 | Nightingale | XL_VLDL_C | NA | A | T | 0.037253 | 0.004245 | 3.80E-18 |
| rs674302 | miR-10b-5p | trans | 2.75E-35 | Nightingale | S_VLDL_PL | NA | A | T | 0.032753 | 0.004416 | 4.20E-14 |
| rs674302 | miR-10b-5p | trans | 2.75E-35 | Nightingale | Total_C | NA | A | T | 0.032359 | 0.004283 | 3.40E-14 |
| rs674302 | miR-10b-5p | trans | 2.75E-35 | Nightingale | M_LDL_PL | NA | A | T | 0.032425 | 0.004431 | 2.70E-13 |
| rs674302 | miR-10b-5p | trans | 2.75E-35 | Nightingale | L_VLDL_C_ | NA | A | T | 0.052079 | 0.004322 | 5.60E-34 |
| rs674302 | miR-10b-5p | trans | 2.75E-35 | Nightingale | XXL_VLDL_ | NA | A | T | 0.034734 | 0.004434 | 3.40E-15 |
| rs674302 | miR-10b-5p | trans | 2.75E-35 | Nightingale | M_VLDL_C | NA | A | T | 0.036135 | 0.004168 | 1.40E-17 |
| rs674302 | miR-10b-5p | trans | 2.75E-35 | Nightingale | M_VLDL_P | NA | A | T | 0.029105 | 0.004418 | 2.10E-11 |
| rs674302 | miR-10b-5p | trans | 2.75E-35 | Nightingale | Sphingomy | NA | A | T | 0.028962 | 0.004136 | 7.60E-12 |
| rs674302 | miR-10b-5p | trans | 2.75E-35 | Nightingale | M_LDL_C | NA | A | T | 0.033944 | 0.004433 | 2.30E-14 |
| rs674302 | miR-10b-5p | trans | 2.75E-35 | Nightingale | M_VLDL_F | NA | A | T | 0.033342 | 0.004424 | 3.20E-14 |
| rs674302 | miR-10b-5p | trans | 2.75E-35 | Nightingale | S_VLDL_CE | NA | A | T | 0.046964 | 0.004365 | 1.40E-26 |
| rs674302 | miR-10b-5p | trans | 2.75E-35 | Nightingale | XS_VLDL_P | NA | A | T | 0.036716 | 0.004366 | 1.30E-17 |
| rs674302 | miR-10b-5p | trans | 2.75E-35 | Nightingale | S_LDL_L | NA | A | T | 0.027702 | 0.004424 | 8.50E-10 |
| rs674302 | miR-10b-5p | trans | 2.75E-35 | Nightingale | L_LDL_P | NA | A | T | 0.029119 | 0.00441 | 6.60E-11 |
| rs674302 | miR-10b-5p | trans | 2.75E-35 | Nightingale | S_HDL_P | NA | A | T | 0.03058 | 0.004425 | 5.40E-12 |
| rs674302 | miR-10b-5p | trans | 2.75E-35 | Nightingale | XL_VLDL_T | NA | A | T | -0.04377 | 0.004289 | 2.50E-24 |
| rs674302 | miR-10b-5p | trans | 2.75E-35 | Nightingale | XL_VLDL_P | NA | A | T | 0.038723 | 0.004431 | 6.70E-19 |
| rs674302 | miR-10b-5p | trans | 2.75E-35 | Nightingale | non_HDL_( | NA | A | T | 0.035666 | 0.004401 | 5.30E-16 |
| rs674302 | miR-10b-5p | trans | 2.75E-35 | Nightingale | XS_VLDL_F | NA | A | T | 0.03137 | 0.004393 | 2.60E-12 |
| rs674302 | miR-10b-5p | trans | 2.75E-35 | Nightingale | S_VLDL_P | NA | A | T | 0.023462 | 0.004369 | 1.60E-08 |
| rs674302 | miR-10b-5p | trans | 2.75E-35 | Nightingale | XS_VLDL_F | NA | A | T | 0.037073 | 0.004348 | 6.40E-18 |
| rs674302 | miR-10b-5p | trans | 2.75E-35 | Nightingale | Clinical_LD | NA | A | T | 0.036436 | 0.004399 | 2.90E-16 |
| rs674302 | miR-10b-5p | trans | 2.75E-35 | Nightingale | IDL_FC | NA | A | T | 0.03824 | 0.004284 | 1.50E-18 |
| rs674302 | miR-10b-5p | trans | 2.75E-35 | Nightingale | XS_VLDL_C | NA | A | T | 0.037807 | 0.00427 | 2.10E-18 |
| rs674302 | miR-10b-5p | trans | 2.75E-35 | Nightingale | LA | NA | A | T | 0.02494 | 0.004339 | 3.50E-09 |
| rs674302 | miR-10b-5p | trans | 2.75E-35 | Nightingale | M_VLDL_C | NA | A | T | 0.033562 | 0.004157 | 2.10E-15 |
| rs674302 | miR-10b-5p | trans | 2.75E-35 | Nightingale | VLDL_CE | NA | A | T | 0.035764 | 0.004426 | 3.10E-16 |

| | | | | | | | | | | | |
|---|---|---|---|---|---|---|---|---|---|---|---|
| rs674302 | miR-10b-5p | trans | 2.75E-35 | Nightingale | S_VLDL_FC | NA | A | T | 0.037872 | 0.004425 | 7.50E-18 |
| rs674302 | miR-10b-5p | trans | 2.75E-35 | Nightingale | L_VLDL_CE | NA | A | T | 0.047276 | 0.004234 | 2.90E-29 |
| rs674302 | miR-10b-5p | trans | 2.75E-35 | Nightingale | XXL_VLDL_ | NA | A | T | 0.025014 | 0.004453 | 1.50E-09 |
| rs674302 | miR-10b-5p | trans | 2.75E-35 | Nightingale | VLDL_C | NA | A | T | 0.031281 | 0.004419 | 5.00E-13 |
| rs674302 | miR-10b-5p | trans | 2.75E-35 | Nightingale | Citrate | NA | A | T | 0.026836 | 0.004396 | 7.10E-10 |
| rs674302 | miR-10b-5p | trans | 2.75E-35 | Nightingale | M_LDL_P | NA | A | T | 0.035851 | 0.004437 | 7.20E-16 |
| rs674302 | let-7b-5p | trans | 7.63E-27 | Nightingale | Remnant_( | NA | A | T | 0.036854 | 0.004396 | 3.70E-17 |
| rs674302 | let-7b-5p | trans | 7.63E-27 | Nightingale | L_LDL_PL | NA | A | T | 0.037956 | 0.004391 | 9.50E-18 |
| rs674302 | let-7b-5p | trans | 7.63E-27 | Nightingale | S_VLDL_CE | NA | A | T | 0.038162 | 0.004419 | 1.60E-18 |
| rs674302 | let-7b-5p | trans | 7.63E-27 | Nightingale | LDL_L | NA | A | T | 0.033263 | 0.00441 | 7.70E-14 |
| rs674302 | let-7b-5p | trans | 7.63E-27 | Nightingale | S_LDL_C | NA | A | T | 0.0309 | 0.004429 | 5.70E-12 |
| rs674302 | let-7b-5p | trans | 7.63E-27 | Nightingale | M_VLDL_F | NA | A | T | 0.041986 | 0.00421 | 4.20E-23 |
| rs674302 | let-7b-5p | trans | 7.63E-27 | Nightingale | IDL_L | NA | A | T | 0.03527 | 0.004278 | 4.90E-16 |
| rs674302 | let-7b-5p | trans | 7.63E-27 | Nightingale | S_LDL_FC | NA | A | T | 0.028528 | 0.004418 | 4.40E-10 |
| rs674302 | let-7b-5p | trans | 7.63E-27 | Nightingale | M_LDL_CE | NA | A | T | 0.032929 | 0.004431 | 1.00E-13 |
| rs674302 | let-7b-5p | trans | 7.63E-27 | Nightingale | S_LDL_CE | NA | A | T | 0.030504 | 0.004427 | 6.10E-12 |
| rs674302 | let-7b-5p | trans | 7.63E-27 | Nightingale | XXL_VLDL_ | NA | A | T | 0.030728 | 0.004406 | 1.30E-12 |
| rs674302 | let-7b-5p | trans | 7.63E-27 | Nightingale | M_LDL_C_ | NA | A | T | 0.031102 | 0.004428 | 3.20E-12 |
| rs674302 | let-7b-5p | trans | 7.63E-27 | Nightingale | Total_PL | NA | A | T | 0.024938 | 0.004213 | 1.10E-09 |
| rs674302 | let-7b-5p | trans | 7.63E-27 | Nightingale | Omega_6 | NA | A | T | 0.025874 | 0.004321 | 5.90E-10 |
| rs674302 | let-7b-5p | trans | 7.63E-27 | Nightingale | IDL_C | NA | A | T | 0.034959 | 0.004273 | 9.50E-16 |
| rs674302 | let-7b-5p | trans | 7.63E-27 | Nightingale | M_LDL_TG | NA | A | T | -0.02754 | 0.00437 | 3.60E-10 |
| rs674302 | let-7b-5p | trans | 7.63E-27 | Nightingale | IDL_P | NA | A | T | 0.04001 | 0.004378 | 1.30E-19 |
| rs674302 | let-7b-5p | trans | 7.63E-27 | Nightingale | L_LDL_CE | NA | A | T | 0.031522 | 0.004385 | 1.00E-12 |
| rs674302 | let-7b-5p | trans | 7.63E-27 | Nightingale | S_HDL_CE | NA | A | T | 0.028597 | 0.004432 | 1.40E-10 |
| rs674302 | let-7b-5p | trans | 7.63E-27 | Nightingale | IDL_CE | NA | A | T | 0.033517 | 0.004272 | 1.30E-14 |
| rs674302 | let-7b-5p | trans | 7.63E-27 | Nightingale | M_LDL_FC | NA | A | T | 0.034304 | 0.004419 | 2.00E-14 |
| rs674302 | let-7b-5p | trans | 7.63E-27 | Nightingale | M_VLDL_P | NA | A | T | 0.04397 | 0.004246 | 6.90E-25 |
| rs674302 | let-7b-5p | trans | 7.63E-27 | Nightingale | L_VLDL_TG | NA | A | T | -0.05103 | 0.004404 | 7.30E-32 |
| rs674302 | let-7b-5p | trans | 7.63E-27 | Nightingale | M_VLDL_P | NA | A | T | 0.028266 | 0.004414 | 6.40E-11 |
| rs674302 | let-7b-5p | trans | 7.63E-27 | Nightingale | LDL_P | NA | A | T | 0.031748 | 0.004421 | 1.10E-12 |
| rs674302 | let-7b-5p | trans | 7.63E-27 | Nightingale | M_VLDL_T | NA | A | T | -0.03848 | 0.004181 | 8.00E-20 |
| rs674302 | let-7b-5p | trans | 7.63E-27 | Nightingale | LDL_PL | NA | A | T | 0.034951 | 0.004414 | 4.90E-15 |
| rs674302 | let-7b-5p | trans | 7.63E-27 | Nightingale | S_HDL_L | NA | A | T | 0.02629 | 0.004409 | 1.60E-09 |
| rs674302 | let-7b-5p | trans | 7.63E-27 | Nightingale | S_HDL_C | NA | A | T | 0.028386 | 0.004417 | 1.80E-10 |
| rs674302 | let-7b-5p | trans | 7.63E-27 | Nightingale | S_HDL_FC | NA | A | T | 0.023467 | 0.004344 | 4.60E-08 |
| rs674302 | let-7b-5p | trans | 7.63E-27 | Nightingale | S_VLDL_C | NA | A | T | 0.038348 | 0.004424 | 1.80E-18 |
| rs674302 | let-7b-5p | trans | 7.63E-27 | Nightingale | LDL_C | NA | A | T | 0.033151 | 0.004405 | 9.30E-14 |
| rs674302 | let-7b-5p | trans | 7.63E-27 | Nightingale | M_VLDL_C | NA | A | T | 0.040808 | 0.004384 | 2.40E-20 |
| rs674302 | let-7b-5p | trans | 7.63E-27 | Nightingale | S_LDL_C_p | NA | A | T | 0.045309 | 0.004442 | 6.20E-25 |
| rs674302 | let-7b-5p | trans | 7.63E-27 | Nightingale | XXL_VLDL_ | NA | A | T | 0.033058 | 0.004458 | 9.70E-14 |
| rs674302 | let-7b-5p | trans | 7.63E-27 | Nightingale | IDL_PL | NA | A | T | 0.035323 | 0.00427 | 3.50E-16 |
| rs674302 | let-7b-5p | trans | 7.63E-27 | Nightingale | L_LDL_C | NA | A | T | 0.032278 | 0.004375 | 2.70E-13 |
| rs674302 | let-7b-5p | trans | 7.63E-27 | Nightingale | ApoB_by_/ | NA | A | T | 0.024989 | 0.004358 | 1.40E-08 |
| rs674302 | let-7b-5p | trans | 7.63E-27 | Nightingale | IDL_FC_pct | NA | A | T | 0.025694 | 0.00439 | 1.20E-08 |
| rs674302 | let-7b-5p | trans | 7.63E-27 | Nightingale | Total_FC | NA | A | T | 0.033246 | 0.004322 | 6.90E-15 |
| rs674302 | let-7b-5p | trans | 7.63E-27 | Nightingale | S_LDL_CE_ | NA | A | T | 0.030904 | 0.004417 | 5.30E-14 |
| rs674302 | let-7b-5p | trans | 7.63E-27 | Nightingale | S_LDL_P | NA | A | T | 0.029552 | 0.004429 | 3.30E-11 |
| rs674302 | let-7b-5p | trans | 7.63E-27 | Nightingale | PUFA | NA | A | T | 0.023697 | 0.004316 | 1.50E-08 |
| rs674302 | let-7b-5p | trans | 7.63E-27 | Nightingale | ApoB | NA | A | T | 0.033904 | 0.004419 | 2.20E-14 |
| rs674302 | let-7b-5p | trans | 7.63E-27 | Nightingale | S_VLDL_C_ | NA | A | T | 0.04182 | 0.004306 | 6.00E-22 |
| rs674302 | let-7b-5p | trans | 7.63E-27 | Nightingale | M_VLDL_C | NA | A | T | 0.038673 | 0.004412 | 2.00E-18 |
| rs674302 | let-7b-5p | trans | 7.63E-27 | Nightingale | L_LDL_FC | NA | A | T | 0.033591 | 0.004345 | 3.00E-14 |
| rs674302 | let-7b-5p | trans | 7.63E-27 | Nightingale | S_VLDL_L | NA | A | T | 0.024659 | 0.004382 | 3.80E-09 |
| rs674302 | let-7b-5p | trans | 7.63E-27 | Nightingale | XL_VLDL_C | NA | A | T | 0.038131 | 0.004243 | 5.50E-19 |
| rs674302 | let-7b-5p | trans | 7.63E-27 | Nightingale | S_LDL_PL_ | NA | A | T | -0.03005 | 0.004416 | 2.80E-13 |
| rs674302 | let-7b-5p | trans | 7.63E-27 | Nightingale | S_HDL_PL | NA | A | T | 0.023787 | 0.004386 | 3.30E-08 |
| rs674302 | let-7b-5p | trans | 7.63E-27 | Nightingale | XXL_VLDL_ | NA | A | T | -0.03564 | 0.004463 | 2.50E-16 |
| rs674302 | let-7b-5p | trans | 7.63E-27 | Nightingale | XS_VLDL_C | NA | A | T | 0.038001 | 0.004295 | 1.30E-18 |
| rs674302 | let-7b-5p | trans | 7.63E-27 | Nightingale | LDL_FC | NA | A | T | 0.033595 | 0.004377 | 5.50E-14 |
| rs674302 | let-7b-5p | trans | 7.63E-27 | Nightingale | XS_VLDL_L | NA | A | T | 0.033833 | 0.004349 | 2.00E-15 |
| rs674302 | let-7b-5p | trans | 7.63E-27 | Nightingale | S_VLDL_TG | NA | A | T | -0.03806 | 0.004283 | 1.60E-18 |
| rs674302 | let-7b-5p | trans | 7.63E-27 | Nightingale | Total_CE | NA | A | T | 0.031729 | 0.004268 | 1.00E-13 |

| | | | | | | | | | | | |
|---|---|---|---|---|---|---|---|---|---|---|---|
| rs674302 | let-7b-5p | trans | 7.63E-27 | Nightingale | GlycA | NA | A | T | 0.027865 | 0.004406 | 1.80E-10 |
| rs674302 | let-7b-5p | trans | 7.63E-27 | Nightingale | S_VLDL_PL | NA | A | T | 0.027671 | 0.004237 | 1.90E-10 |
| rs674302 | let-7b-5p | trans | 7.63E-27 | Nightingale | LDL_CE | NA | A | T | 0.032508 | 0.004413 | 2.30E-13 |
| rs674302 | let-7b-5p | trans | 7.63E-27 | Nightingale | M_LDL_L | NA | A | T | 0.032989 | 0.004431 | 1.10E-13 |
| rs674302 | let-7b-5p | trans | 7.63E-27 | Nightingale | VLDL_FC | NA | A | T | 0.02414 | 0.004392 | 1.10E-08 |
| rs674302 | let-7b-5p | trans | 7.63E-27 | Nightingale | Cholines | NA | A | T | 0.022301 | 0.004161 | 4.90E-08 |
| rs674302 | let-7b-5p | trans | 7.63E-27 | Nightingale | L_LDL_L | NA | A | T | 0.033512 | 0.004381 | 3.20E-14 |
| rs674302 | let-7b-5p | trans | 7.63E-27 | Nightingale | XL_VLDL_F | NA | A | T | 0.037122 | 0.004289 | 2.30E-18 |
| rs674302 | let-7b-5p | trans | 7.63E-27 | Nightingale | XS_VLDL_P | NA | A | T | 0.032126 | 0.004363 | 5.20E-14 |
| rs674302 | let-7b-5p | trans | 7.63E-27 | Nightingale | VLDL_P | NA | A | T | 0.026627 | 0.004401 | 4.40E-10 |
| rs674302 | let-7b-5p | trans | 7.63E-27 | Nightingale | S_VLDL_FC | NA | A | T | 0.029296 | 0.004228 | 1.10E-11 |
| rs674302 | let-7b-5p | trans | 7.63E-27 | Nightingale | L_VLDL_FC | NA | A | T | 0.048437 | 0.004382 | 6.60E-29 |
| rs674302 | let-7b-5p | trans | 7.63E-27 | Nightingale | Total_L | NA | A | T | 0.026349 | 0.004353 | 4.60E-10 |
| rs674302 | let-7b-5p | trans | 7.63E-27 | Nightingale | XL_HDL_PL | NA | A | T | -0.02269 | 0.004065 | 1.20E-08 |
| rs674302 | let-7b-5p | trans | 7.63E-27 | Nightingale | XL_VLDL_C | NA | A | T | 0.037253 | 0.004245 | 3.80E-18 |
| rs674302 | let-7b-5p | trans | 7.63E-27 | Nightingale | S_VLDL_PL | NA | A | T | 0.032753 | 0.004416 | 4.20E-14 |
| rs674302 | let-7b-5p | trans | 7.63E-27 | Nightingale | Total_C | NA | A | T | 0.032359 | 0.004283 | 3.40E-14 |
| rs674302 | let-7b-5p | trans | 7.63E-27 | Nightingale | M_LDL_PL | NA | A | T | 0.032425 | 0.004431 | 2.70E-13 |
| rs674302 | let-7b-5p | trans | 7.63E-27 | Nightingale | L_VLDL_C_ | NA | A | T | 0.052079 | 0.004322 | 5.60E-34 |
| rs674302 | let-7b-5p | trans | 7.63E-27 | Nightingale | XXL_VLDL_ | NA | A | T | 0.034734 | 0.004434 | 3.40E-15 |
| rs674302 | let-7b-5p | trans | 7.63E-27 | Nightingale | M_VLDL_C | NA | A | T | 0.036135 | 0.004168 | 1.40E-17 |
| rs674302 | let-7b-5p | trans | 7.63E-27 | Nightingale | M_VLDL_P | NA | A | T | 0.029105 | 0.004418 | 2.10E-11 |
| rs674302 | let-7b-5p | trans | 7.63E-27 | Nightingale | Sphingomy | NA | A | T | 0.028962 | 0.004136 | 7.60E-12 |
| rs674302 | let-7b-5p | trans | 7.63E-27 | Nightingale | M_LDL_C | NA | A | T | 0.033944 | 0.004433 | 2.30E-14 |
| rs674302 | let-7b-5p | trans | 7.63E-27 | Nightingale | M_VLDL_F | NA | A | T | 0.033342 | 0.004424 | 3.20E-14 |
| rs674302 | let-7b-5p | trans | 7.63E-27 | Nightingale | S_VLDL_CE | NA | A | T | 0.046964 | 0.004365 | 1.40E-26 |
| rs674302 | let-7b-5p | trans | 7.63E-27 | Nightingale | XS_VLDL_P | NA | A | T | 0.036716 | 0.004366 | 1.30E-17 |
| rs674302 | let-7b-5p | trans | 7.63E-27 | Nightingale | S_LDL_L | NA | A | T | 0.027702 | 0.004424 | 8.50E-10 |
| rs674302 | let-7b-5p | trans | 7.63E-27 | Nightingale | L_LDL_P | NA | A | T | 0.029119 | 0.00441 | 6.60E-11 |
| rs674302 | let-7b-5p | trans | 7.63E-27 | Nightingale | S_HDL_P | NA | A | T | 0.03058 | 0.004425 | 5.40E-12 |
| rs674302 | let-7b-5p | trans | 7.63E-27 | Nightingale | XL_VLDL_T | NA | A | T | -0.04377 | 0.004289 | 2.50E-24 |
| rs674302 | let-7b-5p | trans | 7.63E-27 | Nightingale | XL_VLDL_P | NA | A | T | 0.038723 | 0.004431 | 6.70E-19 |
| rs674302 | let-7b-5p | trans | 7.63E-27 | Nightingale | non_HDL_( | NA | A | T | 0.035666 | 0.004401 | 5.30E-16 |
| rs674302 | let-7b-5p | trans | 7.63E-27 | Nightingale | XS_VLDL_F | NA | A | T | 0.03137 | 0.004393 | 2.60E-12 |
| rs674302 | let-7b-5p | trans | 7.63E-27 | Nightingale | S_VLDL_P | NA | A | T | 0.023462 | 0.004369 | 1.60E-08 |
| rs674302 | let-7b-5p | trans | 7.63E-27 | Nightingale | XS_VLDL_F | NA | A | T | 0.037073 | 0.004348 | 6.40E-18 |
| rs674302 | let-7b-5p | trans | 7.63E-27 | Nightingale | Clinical_LD | NA | A | T | 0.036436 | 0.004399 | 2.90E-16 |
| rs674302 | let-7b-5p | trans | 7.63E-27 | Nightingale | IDL_FC | NA | A | T | 0.03824 | 0.004284 | 1.50E-18 |
| rs674302 | let-7b-5p | trans | 7.63E-27 | Nightingale | XS_VLDL_C | NA | A | T | 0.037807 | 0.00427 | 2.10E-18 |
| rs674302 | let-7b-5p | trans | 7.63E-27 | Nightingale | LA | NA | A | T | 0.02494 | 0.004339 | 3.50E-09 |
| rs674302 | let-7b-5p | trans | 7.63E-27 | Nightingale | M_VLDL_C | NA | A | T | 0.033562 | 0.004157 | 2.10E-15 |
| rs674302 | let-7b-5p | trans | 7.63E-27 | Nightingale | VLDL_CE | NA | A | T | 0.035764 | 0.004426 | 3.10E-16 |
| rs674302 | let-7b-5p | trans | 7.63E-27 | Nightingale | S_VLDL_FC | NA | A | T | 0.037872 | 0.004425 | 7.50E-18 |
| rs674302 | let-7b-5p | trans | 7.63E-27 | Nightingale | L_VLDL_CE | NA | A | T | 0.047276 | 0.004234 | 2.90E-29 |
| rs674302 | let-7b-5p | trans | 7.63E-27 | Nightingale | XXL_VLDL_ | NA | A | T | 0.025014 | 0.004453 | 1.50E-09 |
| rs674302 | let-7b-5p | trans | 7.63E-27 | Nightingale | VLDL_C | NA | A | T | 0.031281 | 0.004419 | 5.00E-13 |
| rs674302 | let-7b-5p | trans | 7.63E-27 | Nightingale | Citrate | NA | A | T | 0.026836 | 0.004396 | 7.10E-10 |
| rs674302 | let-7b-5p | trans | 7.63E-27 | Nightingale | M_LDL_P | NA | A | T | 0.035851 | 0.004437 | 7.20E-16 |
| rs674302 | miR-195-5p | trans | 1.85E-17 | Nightingale | Remnant_( | NA | A | T | 0.036854 | 0.004396 | 3.70E-17 |
| rs674302 | miR-195-5p | trans | 1.85E-17 | Nightingale | L_LDL_PL | NA | A | T | 0.037956 | 0.004391 | 9.50E-18 |
| rs674302 | miR-195-5p | trans | 1.85E-17 | Nightingale | S_VLDL_CE | NA | A | T | 0.038162 | 0.004419 | 1.60E-18 |
| rs674302 | miR-195-5p | trans | 1.85E-17 | Nightingale | LDL_L | NA | A | T | 0.033263 | 0.00441 | 7.70E-14 |
| rs674302 | miR-195-5p | trans | 1.85E-17 | Nightingale | S_LDL_C | NA | A | T | 0.0309 | 0.004429 | 5.70E-12 |
| rs674302 | miR-195-5p | trans | 1.85E-17 | Nightingale | M_VLDL_F | NA | A | T | 0.041986 | 0.00421 | 4.20E-23 |
| rs674302 | miR-195-5p | trans | 1.85E-17 | Nightingale | IDL_L | NA | A | T | 0.03527 | 0.004278 | 4.90E-16 |
| rs674302 | miR-195-5p | trans | 1.85E-17 | Nightingale | S_LDL_FC | NA | A | T | 0.028528 | 0.004418 | 4.40E-10 |
| rs674302 | miR-195-5p | trans | 1.85E-17 | Nightingale | M_LDL_CE | NA | A | T | 0.032929 | 0.004431 | 1.00E-13 |
| rs674302 | miR-195-5p | trans | 1.85E-17 | Nightingale | S_LDL_CE | NA | A | T | 0.030504 | 0.004427 | 6.10E-12 |
| rs674302 | miR-195-5p | trans | 1.85E-17 | Nightingale | XXL_VLDL_ | NA | A | T | 0.030728 | 0.004406 | 1.30E-12 |
| rs674302 | miR-195-5p | trans | 1.85E-17 | Nightingale | M_LDL_C_ | NA | A | T | 0.031102 | 0.004428 | 3.20E-12 |
| rs674302 | miR-195-5p | trans | 1.85E-17 | Nightingale | Total_PL | NA | A | T | 0.024938 | 0.004213 | 1.10E-09 |
| rs674302 | miR-195-5p | trans | 1.85E-17 | Nightingale | Omega_6 | NA | A | T | 0.025874 | 0.004321 | 5.90E-10 |
| rs674302 | miR-195-5p | trans | 1.85E-17 | Nightingale | IDL_C | NA | A | T | 0.034959 | 0.004273 | 9.50E-16 |

| | | | | | | | | | | | |
|---|---|---|---|---|---|---|---|---|---|---|---|
| rs674302 | miR-195-5¦ | trans | 1.85E-17 | Nightingale | M_LDL_TG | NA | A | T | -0.02754 | 0.00437 | 3.60E-10 |
| rs674302 | miR-195-5¦ | trans | 1.85E-17 | Nightingale | IDL_P | NA | A | T | 0.04001 | 0.004378 | 1.30E-19 |
| rs674302 | miR-195-5¦ | trans | 1.85E-17 | Nightingale | L_LDL_CE | NA | A | T | 0.031522 | 0.004385 | 1.00E-12 |
| rs674302 | miR-195-5¦ | trans | 1.85E-17 | Nightingale | S_HDL_CE | NA | A | T | 0.028597 | 0.004432 | 1.40E-10 |
| rs674302 | miR-195-5¦ | trans | 1.85E-17 | Nightingale | IDL_CE | NA | A | T | 0.033517 | 0.004272 | 1.30E-14 |
| rs674302 | miR-195-5¦ | trans | 1.85E-17 | Nightingale | M_LDL_FC | NA | A | T | 0.034304 | 0.004419 | 2.00E-14 |
| rs674302 | miR-195-5¦ | trans | 1.85E-17 | Nightingale | M_VLDL_P | NA | A | T | 0.04397 | 0.004246 | 6.90E-25 |
| rs674302 | miR-195-5¦ | trans | 1.85E-17 | Nightingale | L_VLDL_TG | NA | A | T | -0.05103 | 0.004404 | 7.30E-32 |
| rs674302 | miR-195-5¦ | trans | 1.85E-17 | Nightingale | M_VLDL_P | NA | A | T | 0.028266 | 0.004414 | 6.40E-11 |
| rs674302 | miR-195-5¦ | trans | 1.85E-17 | Nightingale | LDL_P | NA | A | T | 0.031748 | 0.004421 | 1.10E-12 |
| rs674302 | miR-195-5¦ | trans | 1.85E-17 | Nightingale | M_VLDL_T | NA | A | T | -0.03848 | 0.004181 | 8.00E-20 |
| rs674302 | miR-195-5¦ | trans | 1.85E-17 | Nightingale | LDL_PL | NA | A | T | 0.034951 | 0.004414 | 4.90E-15 |
| rs674302 | miR-195-5¦ | trans | 1.85E-17 | Nightingale | S_HDL_L | NA | A | T | 0.02629 | 0.004409 | 1.60E-09 |
| rs674302 | miR-195-5¦ | trans | 1.85E-17 | Nightingale | S_HDL_C | NA | A | T | 0.028386 | 0.004417 | 1.80E-10 |
| rs674302 | miR-195-5¦ | trans | 1.85E-17 | Nightingale | S_HDL_FC | NA | A | T | 0.023467 | 0.004344 | 4.60E-08 |
| rs674302 | miR-195-5¦ | trans | 1.85E-17 | Nightingale | S_VLDL_C | NA | A | T | 0.038348 | 0.004424 | 1.80E-18 |
| rs674302 | miR-195-5¦ | trans | 1.85E-17 | Nightingale | LDL_C | NA | A | T | 0.033151 | 0.004405 | 9.30E-14 |
| rs674302 | miR-195-5¦ | trans | 1.85E-17 | Nightingale | M_VLDL_C | NA | A | T | 0.040808 | 0.004384 | 2.40E-20 |
| rs674302 | miR-195-5¦ | trans | 1.85E-17 | Nightingale | S_LDL_C_p | NA | A | T | 0.045309 | 0.004442 | 6.20E-25 |
| rs674302 | miR-195-5¦ | trans | 1.85E-17 | Nightingale | XXL_VLDL_ | NA | A | T | 0.033058 | 0.004458 | 9.70E-14 |
| rs674302 | miR-195-5¦ | trans | 1.85E-17 | Nightingale | IDL_PL | NA | A | T | 0.035323 | 0.00427 | 3.50E-16 |
| rs674302 | miR-195-5¦ | trans | 1.85E-17 | Nightingale | L_LDL_C | NA | A | T | 0.032278 | 0.004375 | 2.70E-13 |
| rs674302 | miR-195-5¦ | trans | 1.85E-17 | Nightingale | ApoB_by_A | NA | A | T | 0.024989 | 0.004358 | 1.40E-08 |
| rs674302 | miR-195-5¦ | trans | 1.85E-17 | Nightingale | IDL_FC_pct | NA | A | T | 0.025694 | 0.00439 | 1.20E-08 |
| rs674302 | miR-195-5¦ | trans | 1.85E-17 | Nightingale | Total_FC | NA | A | T | 0.033246 | 0.004322 | 6.90E-15 |
| rs674302 | miR-195-5¦ | trans | 1.85E-17 | Nightingale | S_LDL_CE_ | NA | A | T | 0.030904 | 0.004417 | 5.30E-14 |
| rs674302 | miR-195-5¦ | trans | 1.85E-17 | Nightingale | S_LDL_P | NA | A | T | 0.029552 | 0.004429 | 3.30E-11 |
| rs674302 | miR-195-5¦ | trans | 1.85E-17 | Nightingale | PUFA | NA | A | T | 0.023697 | 0.004316 | 1.50E-08 |
| rs674302 | miR-195-5¦ | trans | 1.85E-17 | Nightingale | ApoB | NA | A | T | 0.033904 | 0.004419 | 2.20E-14 |
| rs674302 | miR-195-5¦ | trans | 1.85E-17 | Nightingale | S_VLDL_C_ | NA | A | T | 0.04182 | 0.004306 | 6.00E-22 |
| rs674302 | miR-195-5¦ | trans | 1.85E-17 | Nightingale | M_VLDL_C | NA | A | T | 0.038673 | 0.004412 | 2.00E-18 |
| rs674302 | miR-195-5¦ | trans | 1.85E-17 | Nightingale | L_LDL_FC | NA | A | T | 0.033591 | 0.004345 | 3.00E-14 |
| rs674302 | miR-195-5¦ | trans | 1.85E-17 | Nightingale | S_VLDL_L | NA | A | T | 0.024659 | 0.004382 | 3.80E-09 |
| rs674302 | miR-195-5¦ | trans | 1.85E-17 | Nightingale | XL_VLDL_C | NA | A | T | 0.038131 | 0.004243 | 5.50E-19 |
| rs674302 | miR-195-5¦ | trans | 1.85E-17 | Nightingale | S_LDL_PL_ | NA | A | T | -0.03005 | 0.004416 | 2.80E-13 |
| rs674302 | miR-195-5¦ | trans | 1.85E-17 | Nightingale | S_HDL_PL | NA | A | T | 0.023787 | 0.004386 | 3.30E-08 |
| rs674302 | miR-195-5¦ | trans | 1.85E-17 | Nightingale | XXL_VLDL_ | NA | A | T | -0.03564 | 0.004463 | 2.50E-16 |
| rs674302 | miR-195-5¦ | trans | 1.85E-17 | Nightingale | XS_VLDL_C | NA | A | T | 0.038001 | 0.004295 | 1.30E-18 |
| rs674302 | miR-195-5¦ | trans | 1.85E-17 | Nightingale | LDL_FC | NA | A | T | 0.033595 | 0.004377 | 5.50E-14 |
| rs674302 | miR-195-5¦ | trans | 1.85E-17 | Nightingale | XS_VLDL_L | NA | A | T | 0.033833 | 0.004349 | 2.00E-15 |
| rs674302 | miR-195-5¦ | trans | 1.85E-17 | Nightingale | S_VLDL_TG | NA | A | T | -0.03806 | 0.004283 | 1.60E-18 |
| rs674302 | miR-195-5¦ | trans | 1.85E-17 | Nightingale | Total_CE | NA | A | T | 0.031729 | 0.004268 | 1.00E-13 |
| rs674302 | miR-195-5¦ | trans | 1.85E-17 | Nightingale | GlycA | NA | A | T | 0.027865 | 0.004406 | 1.80E-10 |
| rs674302 | miR-195-5¦ | trans | 1.85E-17 | Nightingale | S_VLDL_PL | NA | A | T | 0.027671 | 0.004237 | 1.90E-10 |
| rs674302 | miR-195-5¦ | trans | 1.85E-17 | Nightingale | LDL_CE | NA | A | T | 0.032508 | 0.004413 | 2.30E-13 |
| rs674302 | miR-195-5¦ | trans | 1.85E-17 | Nightingale | M_LDL_L | NA | A | T | 0.032989 | 0.004431 | 1.10E-13 |
| rs674302 | miR-195-5¦ | trans | 1.85E-17 | Nightingale | VLDL_FC | NA | A | T | 0.02414 | 0.004392 | 1.10E-08 |
| rs674302 | miR-195-5¦ | trans | 1.85E-17 | Nightingale | Cholines | NA | A | T | 0.022301 | 0.004161 | 4.90E-08 |
| rs674302 | miR-195-5¦ | trans | 1.85E-17 | Nightingale | L_LDL_L | NA | A | T | 0.033512 | 0.004381 | 3.20E-14 |
| rs674302 | miR-195-5¦ | trans | 1.85E-17 | Nightingale | XL_VLDL_F | NA | A | T | 0.037122 | 0.004289 | 2.30E-18 |
| rs674302 | miR-195-5¦ | trans | 1.85E-17 | Nightingale | XS_VLDL_P | NA | A | T | 0.032126 | 0.004363 | 5.20E-14 |
| rs674302 | miR-195-5¦ | trans | 1.85E-17 | Nightingale | VLDL_P | NA | A | T | 0.026627 | 0.004401 | 4.40E-10 |
| rs674302 | miR-195-5¦ | trans | 1.85E-17 | Nightingale | S_VLDL_FC | NA | A | T | 0.029296 | 0.004228 | 1.10E-11 |
| rs674302 | miR-195-5¦ | trans | 1.85E-17 | Nightingale | L_VLDL_FC | NA | A | T | 0.048437 | 0.004382 | 6.60E-29 |
| rs674302 | miR-195-5¦ | trans | 1.85E-17 | Nightingale | Total_L | NA | A | T | 0.026349 | 0.004353 | 4.60E-10 |
| rs674302 | miR-195-5¦ | trans | 1.85E-17 | Nightingale | XL_HDL_PL | NA | A | T | -0.02269 | 0.004065 | 1.20E-08 |
| rs674302 | miR-195-5¦ | trans | 1.85E-17 | Nightingale | XL_VLDL_C | NA | A | T | 0.037253 | 0.004245 | 3.80E-18 |
| rs674302 | miR-195-5¦ | trans | 1.85E-17 | Nightingale | S_VLDL_PL | NA | A | T | 0.032753 | 0.004416 | 4.20E-14 |
| rs674302 | miR-195-5¦ | trans | 1.85E-17 | Nightingale | Total_C | NA | A | T | 0.032359 | 0.004283 | 3.40E-14 |
| rs674302 | miR-195-5¦ | trans | 1.85E-17 | Nightingale | M_LDL_PL | NA | A | T | 0.032425 | 0.004431 | 2.70E-13 |
| rs674302 | miR-195-5¦ | trans | 1.85E-17 | Nightingale | L_VLDL_C_ | NA | A | T | 0.052079 | 0.004322 | 5.60E-34 |
| rs674302 | miR-195-5¦ | trans | 1.85E-17 | Nightingale | XXL_VLDL_ | NA | A | T | 0.034734 | 0.004434 | 3.40E-15 |
| rs674302 | miR-195-5¦ | trans | 1.85E-17 | Nightingale | M_VLDL_C | NA | A | T | 0.036135 | 0.004168 | 1.40E-17 |

| | | | | | | | | | | | |
|---|---|---|---|---|---|---|---|---|---|---|---|
| rs674302 | miR-195-5p | trans | 1.85E-17 | Nightingale | M_VLDL_P | NA | A | T | 0.029105 | 0.004418 | 2.10E-11 |
| rs674302 | miR-195-5p | trans | 1.85E-17 | Nightingale | Sphingomy | NA | A | T | 0.028962 | 0.004136 | 7.60E-12 |
| rs674302 | miR-195-5p | trans | 1.85E-17 | Nightingale | M_LDL_C | NA | A | T | 0.033944 | 0.004433 | 2.30E-14 |
| rs674302 | miR-195-5p | trans | 1.85E-17 | Nightingale | M_VLDL_F | NA | A | T | 0.033342 | 0.004424 | 3.20E-14 |
| rs674302 | miR-195-5p | trans | 1.85E-17 | Nightingale | S_VLDL_CE | NA | A | T | 0.046964 | 0.004365 | 1.40E-26 |
| rs674302 | miR-195-5p | trans | 1.85E-17 | Nightingale | XS_VLDL_P | NA | A | T | 0.036716 | 0.004366 | 1.30E-17 |
| rs674302 | miR-195-5p | trans | 1.85E-17 | Nightingale | S_LDL_L | NA | A | T | 0.027702 | 0.004424 | 8.50E-10 |
| rs674302 | miR-195-5p | trans | 1.85E-17 | Nightingale | L_LDL_P | NA | A | T | 0.029119 | 0.00441 | 6.60E-11 |
| rs674302 | miR-195-5p | trans | 1.85E-17 | Nightingale | S_HDL_P | NA | A | T | 0.03058 | 0.004425 | 5.40E-12 |
| rs674302 | miR-195-5p | trans | 1.85E-17 | Nightingale | XL_VLDL_T | NA | A | T | -0.04377 | 0.004289 | 2.50E-24 |
| rs674302 | miR-195-5p | trans | 1.85E-17 | Nightingale | XL_VLDL_P | NA | A | T | 0.038723 | 0.004431 | 6.70E-19 |
| rs674302 | miR-195-5p | trans | 1.85E-17 | Nightingale | non_HDL_( | NA | A | T | 0.035666 | 0.004401 | 5.30E-16 |
| rs674302 | miR-195-5p | trans | 1.85E-17 | Nightingale | XS_VLDL_F | NA | A | T | 0.03137 | 0.004393 | 2.60E-12 |
| rs674302 | miR-195-5p | trans | 1.85E-17 | Nightingale | S_VLDL_P | NA | A | T | 0.023462 | 0.004369 | 1.60E-08 |
| rs674302 | miR-195-5p | trans | 1.85E-17 | Nightingale | XS_VLDL_F | NA | A | T | 0.037073 | 0.004348 | 6.40E-18 |
| rs674302 | miR-195-5p | trans | 1.85E-17 | Nightingale | Clinical_LD | NA | A | T | 0.036436 | 0.004399 | 2.90E-16 |
| rs674302 | miR-195-5p | trans | 1.85E-17 | Nightingale | IDL_FC | NA | A | T | 0.03824 | 0.004284 | 1.50E-18 |
| rs674302 | miR-195-5p | trans | 1.85E-17 | Nightingale | XS_VLDL_C | NA | A | T | 0.037807 | 0.00427 | 2.10E-18 |
| rs674302 | miR-195-5p | trans | 1.85E-17 | Nightingale | LA | NA | A | T | 0.02494 | 0.004339 | 3.50E-09 |
| rs674302 | miR-195-5p | trans | 1.85E-17 | Nightingale | M_VLDL_C | NA | A | T | 0.033562 | 0.004157 | 2.10E-15 |
| rs674302 | miR-195-5p | trans | 1.85E-17 | Nightingale | VLDL_CE | NA | A | T | 0.035764 | 0.004426 | 3.10E-16 |
| rs674302 | miR-195-5p | trans | 1.85E-17 | Nightingale | S_VLDL_FC | NA | A | T | 0.037872 | 0.004425 | 7.50E-18 |
| rs674302 | miR-195-5p | trans | 1.85E-17 | Nightingale | L_VLDL_CE | NA | A | T | 0.047276 | 0.004234 | 2.90E-29 |
| rs674302 | miR-195-5p | trans | 1.85E-17 | Nightingale | XXL_VLDL_ | NA | A | T | 0.025014 | 0.004453 | 1.50E-09 |
| rs674302 | miR-195-5p | trans | 1.85E-17 | Nightingale | VLDL_C | NA | A | T | 0.031281 | 0.004419 | 5.00E-13 |
| rs674302 | miR-195-5p | trans | 1.85E-17 | Nightingale | Citrate | NA | A | T | 0.026836 | 0.004396 | 7.10E-10 |
| rs674302 | miR-195-5p | trans | 1.85E-17 | Nightingale | M_LDL_P | NA | A | T | 0.035851 | 0.004437 | 7.20E-16 |
| rs676457 | let-7b-5p | trans | 9.10E-27 | Nightingale | Remnant_( | NA | T | A | 0.03685 | 0.004396 | 3.70E-17 |
| rs676457 | let-7b-5p | trans | 9.10E-27 | Nightingale | S_HDL_PL | NA | T | A | 0.023785 | 0.004387 | 3.30E-08 |
| rs676457 | let-7b-5p | trans | 9.10E-27 | Nightingale | XL_HDL_PL | NA | T | A | -0.02267 | 0.004065 | 1.30E-08 |
| rs676457 | let-7b-5p | trans | 9.10E-27 | Nightingale | Total_C | NA | T | A | 0.032362 | 0.004284 | 3.40E-14 |
| rs676457 | let-7b-5p | trans | 9.10E-27 | Nightingale | LDL_P | NA | T | A | 0.031743 | 0.004422 | 1.10E-12 |
| rs676457 | let-7b-5p | trans | 9.10E-27 | Nightingale | M_LDL_P | NA | T | A | 0.035846 | 0.004437 | 7.30E-16 |
| rs676457 | let-7b-5p | trans | 9.10E-27 | Nightingale | S_HDL_C | NA | T | A | 0.028388 | 0.004417 | 1.80E-10 |
| rs676457 | let-7b-5p | trans | 9.10E-27 | Nightingale | M_LDL_FC | NA | T | A | 0.034303 | 0.004419 | 2.00E-14 |
| rs676457 | let-7b-5p | trans | 9.10E-27 | Nightingale | M_LDL_C_ | NA | T | A | 0.031099 | 0.004428 | 3.30E-12 |
| rs676457 | let-7b-5p | trans | 9.10E-27 | Nightingale | IDL_FC_pct | NA | T | A | 0.025704 | 0.004391 | 1.20E-08 |
| rs676457 | let-7b-5p | trans | 9.10E-27 | Nightingale | XL_VLDL_T | NA | T | A | -0.04378 | 0.004289 | 2.50E-24 |
| rs676457 | let-7b-5p | trans | 9.10E-27 | Nightingale | XS_VLDL_P | NA | T | A | 0.036716 | 0.004366 | 1.30E-17 |
| rs676457 | let-7b-5p | trans | 9.10E-27 | Nightingale | S_LDL_L | NA | T | A | 0.027696 | 0.004424 | 8.50E-10 |
| rs676457 | let-7b-5p | trans | 9.10E-27 | Nightingale | M_VLDL_F | NA | T | A | 0.041996 | 0.00421 | 4.10E-23 |
| rs676457 | let-7b-5p | trans | 9.10E-27 | Nightingale | LDL_C | NA | T | A | 0.033149 | 0.004406 | 9.40E-14 |
| rs676457 | let-7b-5p | trans | 9.10E-27 | Nightingale | XXL_VLDL_ | NA | T | A | 0.025022 | 0.004454 | 1.50E-09 |
| rs676457 | let-7b-5p | trans | 9.10E-27 | Nightingale | XXL_VLDL_ | NA | T | A | -0.03565 | 0.004463 | 2.50E-16 |
| rs676457 | let-7b-5p | trans | 9.10E-27 | Nightingale | L_LDL_L | NA | T | A | 0.033511 | 0.004382 | 3.20E-14 |
| rs676457 | let-7b-5p | trans | 9.10E-27 | Nightingale | IDL_L | NA | T | A | 0.035272 | 0.004278 | 4.90E-16 |
| rs676457 | let-7b-5p | trans | 9.10E-27 | Nightingale | M_VLDL_C | NA | T | A | 0.036148 | 0.004168 | 1.30E-17 |
| rs676457 | let-7b-5p | trans | 9.10E-27 | Nightingale | L_LDL_P | NA | T | A | 0.029115 | 0.00441 | 6.60E-11 |
| rs676457 | let-7b-5p | trans | 9.10E-27 | Nightingale | IDL_C | NA | T | A | 0.034961 | 0.004274 | 9.50E-16 |
| rs676457 | let-7b-5p | trans | 9.10E-27 | Nightingale | M_VLDL_P | NA | T | A | 0.043977 | 0.004247 | 6.90E-25 |
| rs676457 | let-7b-5p | trans | 9.10E-27 | Nightingale | ApoB_by_/ | NA | T | A | 0.024978 | 0.004358 | 1.40E-08 |
| rs676457 | let-7b-5p | trans | 9.10E-27 | Nightingale | Total_CE | NA | T | A | 0.031734 | 0.004268 | 9.90E-14 |
| rs676457 | let-7b-5p | trans | 9.10E-27 | Nightingale | LDL_L | NA | T | A | 0.033261 | 0.00441 | 7.80E-14 |
| rs676457 | let-7b-5p | trans | 9.10E-27 | Nightingale | S_HDL_L | NA | T | A | 0.026288 | 0.00441 | 1.60E-09 |
| rs676457 | let-7b-5p | trans | 9.10E-27 | Nightingale | IDL_FC | NA | T | A | 0.038245 | 0.004284 | 1.50E-18 |
| rs676457 | let-7b-5p | trans | 9.10E-27 | Nightingale | L_LDL_FC | NA | T | A | 0.033594 | 0.004345 | 3.00E-14 |
| rs676457 | let-7b-5p | trans | 9.10E-27 | Nightingale | S_HDL_P | NA | T | A | 0.03058 | 0.004425 | 5.40E-12 |
| rs676457 | let-7b-5p | trans | 9.10E-27 | Nightingale | Sphingomy | NA | T | A | 0.028973 | 0.004137 | 7.50E-12 |
| rs676457 | let-7b-5p | trans | 9.10E-27 | Nightingale | M_LDL_TG | NA | T | A | -0.02753 | 0.00437 | 3.60E-10 |
| rs676457 | let-7b-5p | trans | 9.10E-27 | Nightingale | XS_VLDL_C | NA | T | A | 0.03781 | 0.00427 | 2.10E-18 |
| rs676457 | let-7b-5p | trans | 9.10E-27 | Nightingale | M_VLDL_C | NA | T | A | 0.033576 | 0.004157 | 2.10E-15 |
| rs676457 | let-7b-5p | trans | 9.10E-27 | Nightingale | S_LDL_P | NA | T | A | 0.029546 | 0.004429 | 3.30E-11 |
| rs676457 | let-7b-5p | trans | 9.10E-27 | Nightingale | M_VLDL_P | NA | T | A | 0.028256 | 0.004414 | 6.50E-11 |

| | | | | | | | | | | | |
|---|---|---|---|---|---|---|---|---|---|---|---|
| rs676457 | let-7b-5p | trans | 9.10E-27 | Nightingale | S_LDL_C_p | NA | T | A | 0.045305 | 0.004442 | 6.40E-25 |
| rs676457 | let-7b-5p | trans | 9.10E-27 | Nightingale | S_VLDL_TG | NA | T | A | -0.03807 | 0.004283 | 1.60E-18 |
| rs676457 | let-7b-5p | trans | 9.10E-27 | Nightingale | M_LDL_CE | NA | T | A | 0.032923 | 0.004431 | 1.00E-13 |
| rs676457 | let-7b-5p | trans | 9.10E-27 | Nightingale | LDL_CE | NA | T | A | 0.032505 | 0.004413 | 2.30E-13 |
| rs676457 | let-7b-5p | trans | 9.10E-27 | Nightingale | GlycA | NA | T | A | 0.027864 | 0.004406 | 1.80E-10 |
| rs676457 | let-7b-5p | trans | 9.10E-27 | Nightingale | LDL_PL | NA | T | A | 0.034949 | 0.004415 | 4.90E-15 |
| rs676457 | let-7b-5p | trans | 9.10E-27 | Nightingale | L_VLDL_CE | NA | T | A | 0.04729 | 0.004234 | 2.80E-29 |
| rs676457 | let-7b-5p | trans | 9.10E-27 | Nightingale | L_VLDL_TG | NA | T | A | -0.05103 | 0.004404 | 7.20E-32 |
| rs676457 | let-7b-5p | trans | 9.10E-27 | Nightingale | Total_L | NA | T | A | 0.026347 | 0.004353 | 4.60E-10 |
| rs676457 | let-7b-5p | trans | 9.10E-27 | Nightingale | S_HDL_FC | NA | T | A | 0.023471 | 0.004344 | 4.60E-08 |
| rs676457 | let-7b-5p | trans | 9.10E-27 | Nightingale | S_VLDL_C_ | NA | T | A | 0.041827 | 0.004306 | 5.90E-22 |
| rs676457 | let-7b-5p | trans | 9.10E-27 | Nightingale | ApoB | NA | T | A | 0.033899 | 0.004419 | 2.20E-14 |
| rs676457 | let-7b-5p | trans | 9.10E-27 | Nightingale | Cholines | NA | T | A | 0.022307 | 0.004161 | 4.90E-08 |
| rs676457 | let-7b-5p | trans | 9.10E-27 | Nightingale | M_VLDL_C | NA | T | A | 0.03867 | 0.004413 | 2.00E-18 |
| rs676457 | let-7b-5p | trans | 9.10E-27 | Nightingale | Total_FC | NA | T | A | 0.033247 | 0.004323 | 6.90E-15 |
| rs676457 | let-7b-5p | trans | 9.10E-27 | Nightingale | S_LDL_CE | NA | T | A | 0.030497 | 0.004427 | 6.10E-12 |
| rs676457 | let-7b-5p | trans | 9.10E-27 | Nightingale | S_VLDL_CE | NA | T | A | 0.038156 | 0.004419 | 1.60E-18 |
| rs676457 | let-7b-5p | trans | 9.10E-27 | Nightingale | XS_VLDL_F | NA | T | A | 0.037075 | 0.004348 | 6.40E-18 |
| rs676457 | let-7b-5p | trans | 9.10E-27 | Nightingale | S_VLDL_C | NA | T | A | 0.038342 | 0.004424 | 1.80E-18 |
| rs676457 | let-7b-5p | trans | 9.10E-27 | Nightingale | XXL_VLDL_ | NA | T | A | 0.030738 | 0.004406 | 1.30E-12 |
| rs676457 | let-7b-5p | trans | 9.10E-27 | Nightingale | L_VLDL_C_ | NA | T | A | 0.05209 | 0.004322 | 5.50E-34 |
| rs676457 | let-7b-5p | trans | 9.10E-27 | Nightingale | XXL_VLDL_ | NA | T | A | 0.034746 | 0.004434 | 3.30E-15 |
| rs676457 | let-7b-5p | trans | 9.10E-27 | Nightingale | Omega_6 | NA | T | A | 0.025869 | 0.004321 | 6.00E-10 |
| rs676457 | let-7b-5p | trans | 9.10E-27 | Nightingale | S_VLDL_L | NA | T | A | 0.02465 | 0.004382 | 3.80E-09 |
| rs676457 | let-7b-5p | trans | 9.10E-27 | Nightingale | XL_VLDL_C | NA | T | A | 0.037267 | 0.004245 | 3.70E-18 |
| rs676457 | let-7b-5p | trans | 9.10E-27 | Nightingale | M_LDL_L | NA | T | A | 0.032983 | 0.004431 | 1.10E-13 |
| rs676457 | let-7b-5p | trans | 9.10E-27 | Nightingale | XL_VLDL_C | NA | T | A | 0.038144 | 0.004243 | 5.40E-19 |
| rs676457 | let-7b-5p | trans | 9.10E-27 | Nightingale | PUFA | NA | T | A | 0.023692 | 0.004316 | 1.50E-08 |
| rs676457 | let-7b-5p | trans | 9.10E-27 | Nightingale | Clinical_LD | NA | T | A | 0.036434 | 0.004399 | 2.90E-16 |
| rs676457 | let-7b-5p | trans | 9.10E-27 | Nightingale | non_HDL_( | NA | T | A | 0.035663 | 0.004401 | 5.30E-16 |
| rs676457 | let-7b-5p | trans | 9.10E-27 | Nightingale | VLDL_P | NA | T | A | 0.026619 | 0.004401 | 4.40E-10 |
| rs676457 | let-7b-5p | trans | 9.10E-27 | Nightingale | S_LDL_C | NA | T | A | 0.030895 | 0.004429 | 5.80E-12 |
| rs676457 | let-7b-5p | trans | 9.10E-27 | Nightingale | S_HDL_CE | NA | T | A | 0.028598 | 0.004432 | 1.40E-10 |
| rs676457 | let-7b-5p | trans | 9.10E-27 | Nightingale | S_LDL_PL_ | NA | T | A | -0.03004 | 0.004417 | 2.80E-13 |
| rs676457 | let-7b-5p | trans | 9.10E-27 | Nightingale | Total_PL | NA | T | A | 0.024941 | 0.004214 | 1.10E-09 |
| rs676457 | let-7b-5p | trans | 9.10E-27 | Nightingale | L_LDL_CE | NA | T | A | 0.031521 | 0.004385 | 1.00E-12 |
| rs676457 | let-7b-5p | trans | 9.10E-27 | Nightingale | M_VLDL_T | NA | T | A | -0.03849 | 0.004181 | 7.90E-20 |
| rs676457 | let-7b-5p | trans | 9.10E-27 | Nightingale | IDL_PL | NA | T | A | 0.035325 | 0.00427 | 3.50E-16 |
| rs676457 | let-7b-5p | trans | 9.10E-27 | Nightingale | XL_VLDL_P | NA | T | A | 0.038714 | 0.004432 | 6.80E-19 |
| rs676457 | let-7b-5p | trans | 9.10E-27 | Nightingale | XS_VLDL_P | NA | T | A | 0.032125 | 0.004363 | 5.20E-14 |
| rs676457 | let-7b-5p | trans | 9.10E-27 | Nightingale | IDL_CE | NA | T | A | 0.033519 | 0.004272 | 1.30E-14 |
| rs676457 | let-7b-5p | trans | 9.10E-27 | Nightingale | L_LDL_PL | NA | T | A | 0.037956 | 0.004391 | 9.50E-18 |
| rs676457 | let-7b-5p | trans | 9.10E-27 | Nightingale | S_VLDL_PL | NA | T | A | 0.027677 | 0.004237 | 1.90E-10 |
| rs676457 | let-7b-5p | trans | 9.10E-27 | Nightingale | XS_VLDL_C | NA | T | A | 0.038003 | 0.004295 | 1.30E-18 |
| rs676457 | let-7b-5p | trans | 9.10E-27 | Nightingale | M_VLDL_P | NA | T | A | 0.029096 | 0.004418 | 2.10E-11 |
| rs676457 | let-7b-5p | trans | 9.10E-27 | Nightingale | S_VLDL_PL | NA | T | A | 0.032746 | 0.004417 | 4.30E-14 |
| rs676457 | let-7b-5p | trans | 9.10E-27 | Nightingale | LDL_FC | NA | T | A | 0.033596 | 0.004378 | 5.50E-14 |
| rs676457 | let-7b-5p | trans | 9.10E-27 | Nightingale | S_VLDL_FC | NA | T | A | 0.029304 | 0.004228 | 1.10E-11 |
| rs676457 | let-7b-5p | trans | 9.10E-27 | Nightingale | L_VLDL_FC | NA | T | A | 0.048437 | 0.004382 | 6.60E-29 |
| rs676457 | let-7b-5p | trans | 9.10E-27 | Nightingale | Citrate | NA | T | A | 0.026851 | 0.004396 | 6.90E-10 |
| rs676457 | let-7b-5p | trans | 9.10E-27 | Nightingale | M_LDL_C | NA | T | A | 0.033939 | 0.004433 | 2.30E-14 |
| rs676457 | let-7b-5p | trans | 9.10E-27 | Nightingale | XS_VLDL_L | NA | T | A | 0.033833 | 0.00435 | 2.00E-15 |
| rs676457 | let-7b-5p | trans | 9.10E-27 | Nightingale | S_VLDL_FC | NA | T | A | 0.037867 | 0.004426 | 7.60E-18 |
| rs676457 | let-7b-5p | trans | 9.10E-27 | Nightingale | M_LDL_PL | NA | T | A | 0.032419 | 0.004431 | 2.80E-13 |
| rs676457 | let-7b-5p | trans | 9.10E-27 | Nightingale | VLDL_FC | NA | T | A | 0.024128 | 0.004392 | 1.10E-08 |
| rs676457 | let-7b-5p | trans | 9.10E-27 | Nightingale | M_VLDL_F | NA | T | A | 0.033335 | 0.004424 | 3.20E-14 |
| rs676457 | let-7b-5p | trans | 9.10E-27 | Nightingale | VLDL_C | NA | T | A | 0.031272 | 0.004419 | 5.00E-13 |
| rs676457 | let-7b-5p | trans | 9.10E-27 | Nightingale | M_VLDL_C | NA | T | A | 0.040809 | 0.004385 | 2.40E-20 |
| rs676457 | let-7b-5p | trans | 9.10E-27 | Nightingale | IDL_P | NA | T | A | 0.040008 | 0.004378 | 1.30E-19 |
| rs676457 | let-7b-5p | trans | 9.10E-27 | Nightingale | LA | NA | T | A | 0.024934 | 0.004339 | 3.60E-09 |
| rs676457 | let-7b-5p | trans | 9.10E-27 | Nightingale | VLDL_CE | NA | T | A | 0.035758 | 0.004426 | 3.10E-16 |
| rs676457 | let-7b-5p | trans | 9.10E-27 | Nightingale | S_LDL_FC | NA | T | A | 0.028528 | 0.004418 | 4.40E-10 |
| rs676457 | let-7b-5p | trans | 9.10E-27 | Nightingale | S_VLDL_CE | NA | T | A | 0.04697 | 0.004365 | 1.40E-26 |

| | | | | | | | | | | | |
|---|---|---|---|---|---|---|---|---|---|---|---|
| rs676457 | let-7b-5p | trans | 9.10E-27 | Nightingale | S_VLDL_P | NA | T | A | 0.023453 | 0.004369 | 1.70E-08 |
| rs676457 | let-7b-5p | trans | 9.10E-27 | Nightingale | XXL_VLDL_ | NA | T | A | 0.033069 | 0.004459 | 9.60E-14 |
| rs676457 | let-7b-5p | trans | 9.10E-27 | Nightingale | L_LDL_C | NA | T | A | 0.032277 | 0.004375 | 2.70E-13 |
| rs676457 | let-7b-5p | trans | 9.10E-27 | Nightingale | XL_VLDL_F | NA | T | A | 0.037129 | 0.00429 | 2.30E-18 |
| rs676457 | let-7b-5p | trans | 9.10E-27 | Nightingale | S_LDL_CE_ | NA | T | A | 0.030893 | 0.004417 | 5.50E-14 |
| rs676457 | let-7b-5p | trans | 9.10E-27 | Nightingale | XS_VLDL_F | NA | T | A | 0.031376 | 0.004393 | 2.60E-12 |
| rs676457 | let-7c-5p | trans | 2.46E-23 | Nightingale | Remnant_( | NA | T | A | 0.03685 | 0.004396 | 3.70E-17 |
| rs676457 | let-7c-5p | trans | 2.46E-23 | Nightingale | S_HDL_PL | NA | T | A | 0.023785 | 0.004387 | 3.30E-08 |
| rs676457 | let-7c-5p | trans | 2.46E-23 | Nightingale | XL_HDL_PL | NA | T | A | -0.02267 | 0.004065 | 1.30E-08 |
| rs676457 | let-7c-5p | trans | 2.46E-23 | Nightingale | Total_C | NA | T | A | 0.032362 | 0.004284 | 3.40E-14 |
| rs676457 | let-7c-5p | trans | 2.46E-23 | Nightingale | LDL_P | NA | T | A | 0.031743 | 0.004422 | 1.10E-12 |
| rs676457 | let-7c-5p | trans | 2.46E-23 | Nightingale | M_LDL_P | NA | T | A | 0.035846 | 0.004437 | 7.30E-16 |
| rs676457 | let-7c-5p | trans | 2.46E-23 | Nightingale | S_HDL_C | NA | T | A | 0.028388 | 0.004417 | 1.80E-10 |
| rs676457 | let-7c-5p | trans | 2.46E-23 | Nightingale | M_LDL_FC | NA | T | A | 0.034303 | 0.004419 | 2.00E-14 |
| rs676457 | let-7c-5p | trans | 2.46E-23 | Nightingale | M_LDL_C_ | NA | T | A | 0.031099 | 0.004428 | 3.30E-12 |
| rs676457 | let-7c-5p | trans | 2.46E-23 | Nightingale | IDL_FC_pct | NA | T | A | 0.025704 | 0.004391 | 1.20E-08 |
| rs676457 | let-7c-5p | trans | 2.46E-23 | Nightingale | XL_VLDL_T | NA | T | A | -0.04378 | 0.004289 | 2.50E-24 |
| rs676457 | let-7c-5p | trans | 2.46E-23 | Nightingale | XS_VLDL_P | NA | T | A | 0.036716 | 0.004366 | 1.30E-17 |
| rs676457 | let-7c-5p | trans | 2.46E-23 | Nightingale | S_LDL_L | NA | T | A | 0.027696 | 0.004424 | 8.50E-10 |
| rs676457 | let-7c-5p | trans | 2.46E-23 | Nightingale | M_VLDL_F | NA | T | A | 0.041996 | 0.00421 | 4.10E-23 |
| rs676457 | let-7c-5p | trans | 2.46E-23 | Nightingale | LDL_C | NA | T | A | 0.033149 | 0.004406 | 9.40E-14 |
| rs676457 | let-7c-5p | trans | 2.46E-23 | Nightingale | XXL_VLDL_ | NA | T | A | 0.025022 | 0.004454 | 1.50E-09 |
| rs676457 | let-7c-5p | trans | 2.46E-23 | Nightingale | XXL_VLDL_ | NA | T | A | -0.03565 | 0.004463 | 2.50E-16 |
| rs676457 | let-7c-5p | trans | 2.46E-23 | Nightingale | L_LDL_L | NA | T | A | 0.033511 | 0.004382 | 3.20E-14 |
| rs676457 | let-7c-5p | trans | 2.46E-23 | Nightingale | IDL_L | NA | T | A | 0.035272 | 0.004278 | 4.90E-16 |
| rs676457 | let-7c-5p | trans | 2.46E-23 | Nightingale | M_VLDL_C | NA | T | A | 0.036148 | 0.004168 | 1.30E-17 |
| rs676457 | let-7c-5p | trans | 2.46E-23 | Nightingale | L_LDL_P | NA | T | A | 0.029115 | 0.00441 | 6.60E-11 |
| rs676457 | let-7c-5p | trans | 2.46E-23 | Nightingale | IDL_C | NA | T | A | 0.034961 | 0.004274 | 9.50E-16 |
| rs676457 | let-7c-5p | trans | 2.46E-23 | Nightingale | M_VLDL_P | NA | T | A | 0.043977 | 0.004247 | 6.90E-25 |
| rs676457 | let-7c-5p | trans | 2.46E-23 | Nightingale | ApoB_by_/ | NA | T | A | 0.024978 | 0.004358 | 1.40E-08 |
| rs676457 | let-7c-5p | trans | 2.46E-23 | Nightingale | Total_CE | NA | T | A | 0.031734 | 0.004268 | 9.90E-14 |
| rs676457 | let-7c-5p | trans | 2.46E-23 | Nightingale | LDL_L | NA | T | A | 0.033261 | 0.00441 | 7.80E-14 |
| rs676457 | let-7c-5p | trans | 2.46E-23 | Nightingale | S_HDL_L | NA | T | A | 0.026288 | 0.00441 | 1.60E-09 |
| rs676457 | let-7c-5p | trans | 2.46E-23 | Nightingale | IDL_FC | NA | T | A | 0.038245 | 0.004284 | 1.50E-18 |
| rs676457 | let-7c-5p | trans | 2.46E-23 | Nightingale | L_LDL_FC | NA | T | A | 0.033594 | 0.004345 | 3.00E-14 |
| rs676457 | let-7c-5p | trans | 2.46E-23 | Nightingale | S_HDL_P | NA | T | A | 0.03058 | 0.004425 | 5.40E-12 |
| rs676457 | let-7c-5p | trans | 2.46E-23 | Nightingale | Sphingomy | NA | T | A | 0.028973 | 0.004137 | 7.50E-12 |
| rs676457 | let-7c-5p | trans | 2.46E-23 | Nightingale | M_LDL_TG | NA | T | A | -0.02753 | 0.00437 | 3.60E-10 |
| rs676457 | let-7c-5p | trans | 2.46E-23 | Nightingale | XS_VLDL_C | NA | T | A | 0.03781 | 0.00427 | 2.10E-18 |
| rs676457 | let-7c-5p | trans | 2.46E-23 | Nightingale | M_VLDL_C | NA | T | A | 0.033576 | 0.004157 | 2.10E-15 |
| rs676457 | let-7c-5p | trans | 2.46E-23 | Nightingale | S_LDL_P | NA | T | A | 0.029546 | 0.004429 | 3.30E-11 |
| rs676457 | let-7c-5p | trans | 2.46E-23 | Nightingale | M_VLDL_P | NA | T | A | 0.028256 | 0.004414 | 6.50E-11 |
| rs676457 | let-7c-5p | trans | 2.46E-23 | Nightingale | S_LDL_C_p | NA | T | A | 0.045305 | 0.004442 | 6.40E-25 |
| rs676457 | let-7c-5p | trans | 2.46E-23 | Nightingale | S_VLDL_TG | NA | T | A | -0.03807 | 0.004283 | 1.60E-18 |
| rs676457 | let-7c-5p | trans | 2.46E-23 | Nightingale | M_LDL_CE | NA | T | A | 0.032923 | 0.004431 | 1.00E-13 |
| rs676457 | let-7c-5p | trans | 2.46E-23 | Nightingale | LDL_CE | NA | T | A | 0.032505 | 0.004413 | 2.30E-13 |
| rs676457 | let-7c-5p | trans | 2.46E-23 | Nightingale | GlycA | NA | T | A | 0.027864 | 0.004406 | 1.80E-10 |
| rs676457 | let-7c-5p | trans | 2.46E-23 | Nightingale | LDL_PL | NA | T | A | 0.034949 | 0.004415 | 4.90E-15 |
| rs676457 | let-7c-5p | trans | 2.46E-23 | Nightingale | L_VLDL_CE | NA | T | A | 0.04729 | 0.004234 | 2.80E-29 |
| rs676457 | let-7c-5p | trans | 2.46E-23 | Nightingale | L_VLDL_TG | NA | T | A | -0.05103 | 0.004404 | 7.20E-32 |
| rs676457 | let-7c-5p | trans | 2.46E-23 | Nightingale | Total_L | NA | T | A | 0.026347 | 0.004353 | 4.60E-10 |
| rs676457 | let-7c-5p | trans | 2.46E-23 | Nightingale | S_HDL_FC | NA | T | A | 0.023471 | 0.004344 | 4.60E-08 |
| rs676457 | let-7c-5p | trans | 2.46E-23 | Nightingale | S_VLDL_C_ | NA | T | A | 0.041827 | 0.004306 | 5.90E-22 |
| rs676457 | let-7c-5p | trans | 2.46E-23 | Nightingale | ApoB | NA | T | A | 0.033899 | 0.004419 | 2.20E-14 |
| rs676457 | let-7c-5p | trans | 2.46E-23 | Nightingale | Cholines | NA | T | A | 0.022307 | 0.004161 | 4.90E-08 |
| rs676457 | let-7c-5p | trans | 2.46E-23 | Nightingale | M_VLDL_C | NA | T | A | 0.03867 | 0.004413 | 2.00E-18 |
| rs676457 | let-7c-5p | trans | 2.46E-23 | Nightingale | Total_FC | NA | T | A | 0.033247 | 0.004323 | 6.90E-15 |
| rs676457 | let-7c-5p | trans | 2.46E-23 | Nightingale | S_LDL_CE | NA | T | A | 0.030497 | 0.004427 | 6.10E-12 |
| rs676457 | let-7c-5p | trans | 2.46E-23 | Nightingale | S_VLDL_CE | NA | T | A | 0.038156 | 0.004419 | 1.60E-18 |
| rs676457 | let-7c-5p | trans | 2.46E-23 | Nightingale | XS_VLDL_F | NA | T | A | 0.037075 | 0.004348 | 6.40E-18 |
| rs676457 | let-7c-5p | trans | 2.46E-23 | Nightingale | S_VLDL_C | NA | T | A | 0.038342 | 0.004424 | 1.80E-18 |
| rs676457 | let-7c-5p | trans | 2.46E-23 | Nightingale | XXL_VLDL_ | NA | T | A | 0.030738 | 0.004406 | 1.30E-12 |
| rs676457 | let-7c-5p | trans | 2.46E-23 | Nightingale | L_VLDL_C_ | NA | T | A | 0.05209 | 0.004322 | 5.50E-34 |

| | | | | | | | | | | | |
|---|---|---|---|---|---|---|---|---|---|---|---|
| rs676457 | let-7c-5p | trans | 2.46E-23 | Nightingale | XXL_VLDL_ | NA | T | A | 0.034746 | 0.004434 | 3.30E-15 |
| rs676457 | let-7c-5p | trans | 2.46E-23 | Nightingale | Omega_6 | NA | T | A | 0.025869 | 0.004321 | 6.00E-10 |
| rs676457 | let-7c-5p | trans | 2.46E-23 | Nightingale | S_VLDL_L | NA | T | A | 0.02465 | 0.004382 | 3.80E-09 |
| rs676457 | let-7c-5p | trans | 2.46E-23 | Nightingale | XL_VLDL_C | NA | T | A | 0.037267 | 0.004245 | 3.70E-18 |
| rs676457 | let-7c-5p | trans | 2.46E-23 | Nightingale | M_LDL_L | NA | T | A | 0.032983 | 0.004431 | 1.10E-13 |
| rs676457 | let-7c-5p | trans | 2.46E-23 | Nightingale | XL_VLDL_C | NA | T | A | 0.038144 | 0.004243 | 5.40E-19 |
| rs676457 | let-7c-5p | trans | 2.46E-23 | Nightingale | PUFA | NA | T | A | 0.023692 | 0.004316 | 1.50E-08 |
| rs676457 | let-7c-5p | trans | 2.46E-23 | Nightingale | Clinical_LD | NA | T | A | 0.036434 | 0.004399 | 2.90E-16 |
| rs676457 | let-7c-5p | trans | 2.46E-23 | Nightingale | non_HDL_( | NA | T | A | 0.035663 | 0.004401 | 5.30E-16 |
| rs676457 | let-7c-5p | trans | 2.46E-23 | Nightingale | VLDL_P | NA | T | A | 0.026619 | 0.004401 | 4.40E-10 |
| rs676457 | let-7c-5p | trans | 2.46E-23 | Nightingale | S_LDL_C | NA | T | A | 0.030895 | 0.004429 | 5.80E-12 |
| rs676457 | let-7c-5p | trans | 2.46E-23 | Nightingale | S_HDL_CE | NA | T | A | 0.028598 | 0.004432 | 1.40E-10 |
| rs676457 | let-7c-5p | trans | 2.46E-23 | Nightingale | S_LDL_PL_ | NA | T | A | -0.03004 | 0.004417 | 2.80E-13 |
| rs676457 | let-7c-5p | trans | 2.46E-23 | Nightingale | Total_PL | NA | T | A | 0.024941 | 0.004214 | 1.10E-09 |
| rs676457 | let-7c-5p | trans | 2.46E-23 | Nightingale | L_LDL_CE | NA | T | A | 0.031521 | 0.004385 | 1.00E-12 |
| rs676457 | let-7c-5p | trans | 2.46E-23 | Nightingale | M_VLDL_T | NA | T | A | -0.03849 | 0.004181 | 7.90E-20 |
| rs676457 | let-7c-5p | trans | 2.46E-23 | Nightingale | IDL_PL | NA | T | A | 0.035325 | 0.00427 | 3.50E-16 |
| rs676457 | let-7c-5p | trans | 2.46E-23 | Nightingale | XL_VLDL_P | NA | T | A | 0.038714 | 0.004432 | 6.80E-19 |
| rs676457 | let-7c-5p | trans | 2.46E-23 | Nightingale | XS_VLDL_P | NA | T | A | 0.032125 | 0.004363 | 5.20E-14 |
| rs676457 | let-7c-5p | trans | 2.46E-23 | Nightingale | IDL_CE | NA | T | A | 0.033519 | 0.004272 | 1.30E-14 |
| rs676457 | let-7c-5p | trans | 2.46E-23 | Nightingale | L_LDL_PL | NA | T | A | 0.037956 | 0.004391 | 9.50E-18 |
| rs676457 | let-7c-5p | trans | 2.46E-23 | Nightingale | S_VLDL_PL | NA | T | A | 0.027677 | 0.004237 | 1.90E-10 |
| rs676457 | let-7c-5p | trans | 2.46E-23 | Nightingale | XS_VLDL_C | NA | T | A | 0.038003 | 0.004295 | 1.30E-18 |
| rs676457 | let-7c-5p | trans | 2.46E-23 | Nightingale | M_VLDL_P | NA | T | A | 0.029096 | 0.004418 | 2.10E-11 |
| rs676457 | let-7c-5p | trans | 2.46E-23 | Nightingale | S_VLDL_PL | NA | T | A | 0.032746 | 0.004417 | 4.30E-14 |
| rs676457 | let-7c-5p | trans | 2.46E-23 | Nightingale | LDL_FC | NA | T | A | 0.033596 | 0.004378 | 5.50E-14 |
| rs676457 | let-7c-5p | trans | 2.46E-23 | Nightingale | S_VLDL_FC | NA | T | A | 0.029304 | 0.004228 | 1.10E-11 |
| rs676457 | let-7c-5p | trans | 2.46E-23 | Nightingale | L_VLDL_FC | NA | T | A | 0.048437 | 0.004382 | 6.60E-29 |
| rs676457 | let-7c-5p | trans | 2.46E-23 | Nightingale | Citrate | NA | T | A | 0.026851 | 0.004396 | 6.90E-10 |
| rs676457 | let-7c-5p | trans | 2.46E-23 | Nightingale | M_LDL_C | NA | T | A | 0.033939 | 0.004433 | 2.30E-14 |
| rs676457 | let-7c-5p | trans | 2.46E-23 | Nightingale | XS_VLDL_L | NA | T | A | 0.033833 | 0.00435 | 2.00E-15 |
| rs676457 | let-7c-5p | trans | 2.46E-23 | Nightingale | S_VLDL_FC | NA | T | A | 0.037867 | 0.004426 | 7.60E-18 |
| rs676457 | let-7c-5p | trans | 2.46E-23 | Nightingale | M_LDL_PL | NA | T | A | 0.032419 | 0.004431 | 2.80E-13 |
| rs676457 | let-7c-5p | trans | 2.46E-23 | Nightingale | VLDL_FC | NA | T | A | 0.024128 | 0.004392 | 1.10E-08 |
| rs676457 | let-7c-5p | trans | 2.46E-23 | Nightingale | M_VLDL_F | NA | T | A | 0.033335 | 0.004424 | 3.20E-14 |
| rs676457 | let-7c-5p | trans | 2.46E-23 | Nightingale | VLDL_C | NA | T | A | 0.031272 | 0.004419 | 5.00E-13 |
| rs676457 | let-7c-5p | trans | 2.46E-23 | Nightingale | M_VLDL_C | NA | T | A | 0.040809 | 0.004385 | 2.40E-20 |
| rs676457 | let-7c-5p | trans | 2.46E-23 | Nightingale | IDL_P | NA | T | A | 0.040008 | 0.004378 | 1.30E-19 |
| rs676457 | let-7c-5p | trans | 2.46E-23 | Nightingale | LA | NA | T | A | 0.024934 | 0.004339 | 3.60E-09 |
| rs676457 | let-7c-5p | trans | 2.46E-23 | Nightingale | VLDL_CE | NA | T | A | 0.035758 | 0.004426 | 3.10E-16 |
| rs676457 | let-7c-5p | trans | 2.46E-23 | Nightingale | S_LDL_FC | NA | T | A | 0.028528 | 0.004418 | 4.40E-10 |
| rs676457 | let-7c-5p | trans | 2.46E-23 | Nightingale | S_VLDL_CE | NA | T | A | 0.04697 | 0.004365 | 1.40E-26 |
| rs676457 | let-7c-5p | trans | 2.46E-23 | Nightingale | S_VLDL_P | NA | T | A | 0.023453 | 0.004369 | 1.70E-08 |
| rs676457 | let-7c-5p | trans | 2.46E-23 | Nightingale | XXL_VLDL_ | NA | T | A | 0.033069 | 0.004459 | 9.60E-14 |
| rs676457 | let-7c-5p | trans | 2.46E-23 | Nightingale | L_LDL_C | NA | T | A | 0.032277 | 0.004375 | 2.70E-13 |
| rs676457 | let-7c-5p | trans | 2.46E-23 | Nightingale | XL_VLDL_F | NA | T | A | 0.037129 | 0.00429 | 2.30E-18 |
| rs676457 | let-7c-5p | trans | 2.46E-23 | Nightingale | S_LDL_CE_ | NA | T | A | 0.030893 | 0.004417 | 5.50E-14 |
| rs676457 | let-7c-5p | trans | 2.46E-23 | Nightingale | XS_VLDL_F | NA | T | A | 0.031376 | 0.004393 | 2.60E-12 |
| rs676457 | miR-139-5p | trans | 6.87E-42 | Nightingale | Remnant_( | NA | T | A | 0.03685 | 0.004396 | 3.70E-17 |
| rs676457 | miR-139-5p | trans | 6.87E-42 | Nightingale | S_HDL_PL | NA | T | A | 0.023785 | 0.004387 | 3.30E-08 |
| rs676457 | miR-139-5p | trans | 6.87E-42 | Nightingale | XL_HDL_PL | NA | T | A | -0.02267 | 0.004065 | 1.30E-08 |
| rs676457 | miR-139-5p | trans | 6.87E-42 | Nightingale | Total_C | NA | T | A | 0.032362 | 0.004284 | 3.40E-14 |
| rs676457 | miR-139-5p | trans | 6.87E-42 | Nightingale | LDL_P | NA | T | A | 0.031743 | 0.004422 | 1.10E-12 |
| rs676457 | miR-139-5p | trans | 6.87E-42 | Nightingale | M_LDL_P | NA | T | A | 0.035846 | 0.004437 | 7.30E-16 |
| rs676457 | miR-139-5p | trans | 6.87E-42 | Nightingale | S_HDL_C | NA | T | A | 0.028388 | 0.004417 | 1.80E-10 |
| rs676457 | miR-139-5p | trans | 6.87E-42 | Nightingale | M_LDL_FC | NA | T | A | 0.034303 | 0.004419 | 2.00E-14 |
| rs676457 | miR-139-5p | trans | 6.87E-42 | Nightingale | M_LDL_C_ | NA | T | A | 0.031099 | 0.004428 | 3.30E-12 |
| rs676457 | miR-139-5p | trans | 6.87E-42 | Nightingale | IDL_FC_pct | NA | T | A | 0.025704 | 0.004391 | 1.20E-08 |
| rs676457 | miR-139-5p | trans | 6.87E-42 | Nightingale | XL_VLDL_T | NA | T | A | -0.04378 | 0.004289 | 2.50E-24 |
| rs676457 | miR-139-5p | trans | 6.87E-42 | Nightingale | XS_VLDL_P | NA | T | A | 0.036716 | 0.004366 | 1.30E-17 |
| rs676457 | miR-139-5p | trans | 6.87E-42 | Nightingale | S_LDL_L | NA | T | A | 0.027696 | 0.004424 | 8.50E-10 |
| rs676457 | miR-139-5p | trans | 6.87E-42 | Nightingale | M_VLDL_F | NA | T | A | 0.041996 | 0.00421 | 4.10E-23 |
| rs676457 | miR-139-5p | trans | 6.87E-42 | Nightingale | LDL_C | NA | T | A | 0.033149 | 0.004406 | 9.40E-14 |

| | | | | | | | | | | | |
|---|---|---|---|---|---|---|---|---|---|---|---|
| rs676457 | miR-139-5p | trans | 6.87E-42 | Nightingale | XXL_VLDL_ | NA | T | A | 0.025022 | 0.004454 | 1.50E-09 |
| rs676457 | miR-139-5p | trans | 6.87E-42 | Nightingale | XXL_VLDL_ | NA | T | A | -0.03565 | 0.004463 | 2.50E-16 |
| rs676457 | miR-139-5p | trans | 6.87E-42 | Nightingale | L_LDL_L | NA | T | A | 0.033511 | 0.004382 | 3.20E-14 |
| rs676457 | miR-139-5p | trans | 6.87E-42 | Nightingale | IDL_L | NA | T | A | 0.035272 | 0.004278 | 4.90E-16 |
| rs676457 | miR-139-5p | trans | 6.87E-42 | Nightingale | M_VLDL_C | NA | T | A | 0.036148 | 0.004168 | 1.30E-17 |
| rs676457 | miR-139-5p | trans | 6.87E-42 | Nightingale | L_LDL_P | NA | T | A | 0.029115 | 0.00441 | 6.60E-11 |
| rs676457 | miR-139-5p | trans | 6.87E-42 | Nightingale | IDL_C | NA | T | A | 0.034961 | 0.004274 | 9.50E-16 |
| rs676457 | miR-139-5p | trans | 6.87E-42 | Nightingale | M_VLDL_P | NA | T | A | 0.043977 | 0.004247 | 6.90E-25 |
| rs676457 | miR-139-5p | trans | 6.87E-42 | Nightingale | ApoB_by_A | NA | T | A | 0.024978 | 0.004358 | 1.40E-08 |
| rs676457 | miR-139-5p | trans | 6.87E-42 | Nightingale | Total_CE | NA | T | A | 0.031734 | 0.004268 | 9.90E-14 |
| rs676457 | miR-139-5p | trans | 6.87E-42 | Nightingale | LDL_L | NA | T | A | 0.033261 | 0.00441 | 7.80E-14 |
| rs676457 | miR-139-5p | trans | 6.87E-42 | Nightingale | S_HDL_L | NA | T | A | 0.026288 | 0.00441 | 1.60E-09 |
| rs676457 | miR-139-5p | trans | 6.87E-42 | Nightingale | IDL_FC | NA | T | A | 0.038245 | 0.004284 | 1.50E-18 |
| rs676457 | miR-139-5p | trans | 6.87E-42 | Nightingale | L_LDL_FC | NA | T | A | 0.033594 | 0.004345 | 3.00E-14 |
| rs676457 | miR-139-5p | trans | 6.87E-42 | Nightingale | S_HDL_P | NA | T | A | 0.03058 | 0.004425 | 5.40E-12 |
| rs676457 | miR-139-5p | trans | 6.87E-42 | Nightingale | Sphingomy | NA | T | A | 0.028973 | 0.004137 | 7.50E-12 |
| rs676457 | miR-139-5p | trans | 6.87E-42 | Nightingale | M_LDL_TG | NA | T | A | -0.02753 | 0.00437 | 3.60E-10 |
| rs676457 | miR-139-5p | trans | 6.87E-42 | Nightingale | XS_VLDL_C | NA | T | A | 0.03781 | 0.00427 | 2.10E-18 |
| rs676457 | miR-139-5p | trans | 6.87E-42 | Nightingale | M_VLDL_C | NA | T | A | 0.033576 | 0.004157 | 2.10E-15 |
| rs676457 | miR-139-5p | trans | 6.87E-42 | Nightingale | S_LDL_P | NA | T | A | 0.029546 | 0.004429 | 3.30E-11 |
| rs676457 | miR-139-5p | trans | 6.87E-42 | Nightingale | M_VLDL_P | NA | T | A | 0.028256 | 0.004414 | 6.50E-11 |
| rs676457 | miR-139-5p | trans | 6.87E-42 | Nightingale | S_LDL_C_p | NA | T | A | 0.045305 | 0.004442 | 6.40E-25 |
| rs676457 | miR-139-5p | trans | 6.87E-42 | Nightingale | S_VLDL_TG | NA | T | A | -0.03807 | 0.004283 | 1.60E-18 |
| rs676457 | miR-139-5p | trans | 6.87E-42 | Nightingale | M_LDL_CE | NA | T | A | 0.032923 | 0.004431 | 1.00E-13 |
| rs676457 | miR-139-5p | trans | 6.87E-42 | Nightingale | LDL_CE | NA | T | A | 0.032505 | 0.004413 | 2.30E-13 |
| rs676457 | miR-139-5p | trans | 6.87E-42 | Nightingale | GlycA | NA | T | A | 0.027864 | 0.004406 | 1.80E-10 |
| rs676457 | miR-139-5p | trans | 6.87E-42 | Nightingale | LDL_PL | NA | T | A | 0.034949 | 0.004415 | 4.90E-15 |
| rs676457 | miR-139-5p | trans | 6.87E-42 | Nightingale | L_VLDL_CE | NA | T | A | 0.04729 | 0.004234 | 2.80E-29 |
| rs676457 | miR-139-5p | trans | 6.87E-42 | Nightingale | L_VLDL_TG | NA | T | A | -0.05103 | 0.004404 | 7.20E-32 |
| rs676457 | miR-139-5p | trans | 6.87E-42 | Nightingale | Total_L | NA | T | A | 0.026347 | 0.004353 | 4.60E-10 |
| rs676457 | miR-139-5p | trans | 6.87E-42 | Nightingale | S_HDL_FC | NA | T | A | 0.023471 | 0.004344 | 4.60E-08 |
| rs676457 | miR-139-5p | trans | 6.87E-42 | Nightingale | S_VLDL_C_ | NA | T | A | 0.041827 | 0.004306 | 5.90E-22 |
| rs676457 | miR-139-5p | trans | 6.87E-42 | Nightingale | ApoB | NA | T | A | 0.033899 | 0.004419 | 2.20E-14 |
| rs676457 | miR-139-5p | trans | 6.87E-42 | Nightingale | Cholines | NA | T | A | 0.022307 | 0.004161 | 4.90E-08 |
| rs676457 | miR-139-5p | trans | 6.87E-42 | Nightingale | M_VLDL_C | NA | T | A | 0.03867 | 0.004413 | 2.00E-18 |
| rs676457 | miR-139-5p | trans | 6.87E-42 | Nightingale | Total_FC | NA | T | A | 0.033247 | 0.004323 | 6.90E-15 |
| rs676457 | miR-139-5p | trans | 6.87E-42 | Nightingale | S_LDL_CE | NA | T | A | 0.030497 | 0.004427 | 6.10E-12 |
| rs676457 | miR-139-5p | trans | 6.87E-42 | Nightingale | S_VLDL_CE | NA | T | A | 0.038156 | 0.004419 | 1.60E-18 |
| rs676457 | miR-139-5p | trans | 6.87E-42 | Nightingale | XS_VLDL_F | NA | T | A | 0.037075 | 0.004348 | 6.40E-18 |
| rs676457 | miR-139-5p | trans | 6.87E-42 | Nightingale | S_VLDL_C | NA | T | A | 0.038342 | 0.004424 | 1.80E-18 |
| rs676457 | miR-139-5p | trans | 6.87E-42 | Nightingale | XXL_VLDL_ | NA | T | A | 0.030738 | 0.004406 | 1.30E-12 |
| rs676457 | miR-139-5p | trans | 6.87E-42 | Nightingale | L_VLDL_C_ | NA | T | A | 0.05209 | 0.004322 | 5.50E-34 |
| rs676457 | miR-139-5p | trans | 6.87E-42 | Nightingale | XXL_VLDL_ | NA | T | A | 0.034746 | 0.004434 | 3.30E-15 |
| rs676457 | miR-139-5p | trans | 6.87E-42 | Nightingale | Omega_6 | NA | T | A | 0.025869 | 0.004321 | 6.00E-10 |
| rs676457 | miR-139-5p | trans | 6.87E-42 | Nightingale | S_VLDL_L | NA | T | A | 0.02465 | 0.004382 | 3.80E-09 |
| rs676457 | miR-139-5p | trans | 6.87E-42 | Nightingale | XL_VLDL_C | NA | T | A | 0.037267 | 0.004245 | 3.70E-18 |
| rs676457 | miR-139-5p | trans | 6.87E-42 | Nightingale | M_LDL_L | NA | T | A | 0.032983 | 0.004431 | 1.10E-13 |
| rs676457 | miR-139-5p | trans | 6.87E-42 | Nightingale | XL_VLDL_C | NA | T | A | 0.038144 | 0.004243 | 5.40E-19 |
| rs676457 | miR-139-5p | trans | 6.87E-42 | Nightingale | PUFA | NA | T | A | 0.023692 | 0.004316 | 1.50E-08 |
| rs676457 | miR-139-5p | trans | 6.87E-42 | Nightingale | Clinical_LD | NA | T | A | 0.036434 | 0.004399 | 2.90E-16 |
| rs676457 | miR-139-5p | trans | 6.87E-42 | Nightingale | non_HDL_( | NA | T | A | 0.035663 | 0.004401 | 5.30E-16 |
| rs676457 | miR-139-5p | trans | 6.87E-42 | Nightingale | VLDL_P | NA | T | A | 0.026619 | 0.004401 | 4.40E-10 |
| rs676457 | miR-139-5p | trans | 6.87E-42 | Nightingale | S_LDL_C | NA | T | A | 0.030895 | 0.004429 | 5.80E-12 |
| rs676457 | miR-139-5p | trans | 6.87E-42 | Nightingale | S_HDL_CE | NA | T | A | 0.028598 | 0.004432 | 1.40E-10 |
| rs676457 | miR-139-5p | trans | 6.87E-42 | Nightingale | S_LDL_PL_ | NA | T | A | -0.03004 | 0.004417 | 2.80E-13 |
| rs676457 | miR-139-5p | trans | 6.87E-42 | Nightingale | Total_PL | NA | T | A | 0.024941 | 0.004214 | 1.10E-09 |
| rs676457 | miR-139-5p | trans | 6.87E-42 | Nightingale | L_LDL_CE | NA | T | A | 0.031521 | 0.004385 | 1.00E-12 |
| rs676457 | miR-139-5p | trans | 6.87E-42 | Nightingale | M_VLDL_T | NA | T | A | -0.03849 | 0.004181 | 7.90E-20 |
| rs676457 | miR-139-5p | trans | 6.87E-42 | Nightingale | IDL_PL | NA | T | A | 0.035325 | 0.00427 | 3.50E-16 |
| rs676457 | miR-139-5p | trans | 6.87E-42 | Nightingale | XL_VLDL_P | NA | T | A | 0.038714 | 0.004432 | 6.80E-19 |
| rs676457 | miR-139-5p | trans | 6.87E-42 | Nightingale | XS_VLDL_P | NA | T | A | 0.032125 | 0.004363 | 5.20E-14 |
| rs676457 | miR-139-5p | trans | 6.87E-42 | Nightingale | IDL_CE | NA | T | A | 0.033519 | 0.004272 | 1.30E-14 |
| rs676457 | miR-139-5p | trans | 6.87E-42 | Nightingale | L_LDL_PL | NA | T | A | 0.037956 | 0.004391 | 9.50E-18 |

| | | | | | | | | | | | |
|---|---|---|---|---|---|---|---|---|---|---|---|
| rs676457 | miR-139-5p | trans | 6.87E-42 | Nightingale | S_VLDL_PL | NA | T | A | 0.027677 | 0.004237 | 1.90E-10 |
| rs676457 | miR-139-5p | trans | 6.87E-42 | Nightingale | XS_VLDL_C | NA | T | A | 0.038003 | 0.004295 | 1.30E-18 |
| rs676457 | miR-139-5p | trans | 6.87E-42 | Nightingale | M_VLDL_P | NA | T | A | 0.029096 | 0.004418 | 2.10E-11 |
| rs676457 | miR-139-5p | trans | 6.87E-42 | Nightingale | S_VLDL_PL | NA | T | A | 0.032746 | 0.004417 | 4.30E-14 |
| rs676457 | miR-139-5p | trans | 6.87E-42 | Nightingale | LDL_FC | NA | T | A | 0.033596 | 0.004378 | 5.50E-14 |
| rs676457 | miR-139-5p | trans | 6.87E-42 | Nightingale | S_VLDL_FC | NA | T | A | 0.029304 | 0.004228 | 1.10E-11 |
| rs676457 | miR-139-5p | trans | 6.87E-42 | Nightingale | L_VLDL_FC | NA | T | A | 0.048437 | 0.004382 | 6.60E-29 |
| rs676457 | miR-139-5p | trans | 6.87E-42 | Nightingale | Citrate | NA | T | A | 0.026851 | 0.004396 | 6.90E-10 |
| rs676457 | miR-139-5p | trans | 6.87E-42 | Nightingale | M_LDL_C | NA | T | A | 0.033939 | 0.004433 | 2.30E-14 |
| rs676457 | miR-139-5p | trans | 6.87E-42 | Nightingale | XS_VLDL_L | NA | T | A | 0.033833 | 0.00435 | 2.00E-15 |
| rs676457 | miR-139-5p | trans | 6.87E-42 | Nightingale | S_VLDL_FC | NA | T | A | 0.037867 | 0.004426 | 7.60E-18 |
| rs676457 | miR-139-5p | trans | 6.87E-42 | Nightingale | M_LDL_PL | NA | T | A | 0.032419 | 0.004431 | 2.80E-13 |
| rs676457 | miR-139-5p | trans | 6.87E-42 | Nightingale | VLDL_FC | NA | T | A | 0.024128 | 0.004392 | 1.10E-08 |
| rs676457 | miR-139-5p | trans | 6.87E-42 | Nightingale | M_VLDL_F | NA | T | A | 0.033335 | 0.004424 | 3.20E-14 |
| rs676457 | miR-139-5p | trans | 6.87E-42 | Nightingale | VLDL_C | NA | T | A | 0.031272 | 0.004419 | 5.00E-13 |
| rs676457 | miR-139-5p | trans | 6.87E-42 | Nightingale | M_VLDL_C | NA | T | A | 0.040809 | 0.004385 | 2.40E-20 |
| rs676457 | miR-139-5p | trans | 6.87E-42 | Nightingale | IDL_P | NA | T | A | 0.040008 | 0.004378 | 1.30E-19 |
| rs676457 | miR-139-5p | trans | 6.87E-42 | Nightingale | LA | NA | T | A | 0.024934 | 0.004339 | 3.60E-09 |
| rs676457 | miR-139-5p | trans | 6.87E-42 | Nightingale | VLDL_CE | NA | T | A | 0.035758 | 0.004426 | 3.10E-16 |
| rs676457 | miR-139-5p | trans | 6.87E-42 | Nightingale | S_LDL_FC | NA | T | A | 0.028528 | 0.004418 | 4.40E-10 |
| rs676457 | miR-139-5p | trans | 6.87E-42 | Nightingale | S_VLDL_CE | NA | T | A | 0.04697 | 0.004365 | 1.40E-26 |
| rs676457 | miR-139-5p | trans | 6.87E-42 | Nightingale | S_VLDL_P | NA | T | A | 0.023453 | 0.004369 | 1.70E-08 |
| rs676457 | miR-139-5p | trans | 6.87E-42 | Nightingale | XXL_VLDL_ | NA | T | A | 0.033069 | 0.004459 | 9.60E-14 |
| rs676457 | miR-139-5p | trans | 6.87E-42 | Nightingale | L_LDL_C | NA | T | A | 0.032277 | 0.004375 | 2.70E-13 |
| rs676457 | miR-139-5p | trans | 6.87E-42 | Nightingale | XL_VLDL_F | NA | T | A | 0.037129 | 0.00429 | 2.30E-18 |
| rs676457 | miR-139-5p | trans | 6.87E-42 | Nightingale | S_LDL_CE_ | NA | T | A | 0.030893 | 0.004417 | 5.50E-14 |
| rs676457 | miR-139-5p | trans | 6.87E-42 | Nightingale | XS_VLDL_F | NA | T | A | 0.031376 | 0.004393 | 2.60E-12 |
| rs676457 | miR-10b-5p | trans | 3.04E-35 | Nightingale | Remnant_( | NA | T | A | 0.03685 | 0.004396 | 3.70E-17 |
| rs676457 | miR-10b-5p | trans | 3.04E-35 | Nightingale | S_HDL_PL | NA | T | A | 0.023785 | 0.004387 | 3.30E-08 |
| rs676457 | miR-10b-5p | trans | 3.04E-35 | Nightingale | XL_HDL_PL | NA | T | A | -0.02267 | 0.004065 | 1.30E-08 |
| rs676457 | miR-10b-5p | trans | 3.04E-35 | Nightingale | Total_C | NA | T | A | 0.032362 | 0.004284 | 3.40E-14 |
| rs676457 | miR-10b-5p | trans | 3.04E-35 | Nightingale | LDL_P | NA | T | A | 0.031743 | 0.004422 | 1.10E-12 |
| rs676457 | miR-10b-5p | trans | 3.04E-35 | Nightingale | M_LDL_P | NA | T | A | 0.035846 | 0.004437 | 7.30E-16 |
| rs676457 | miR-10b-5p | trans | 3.04E-35 | Nightingale | S_HDL_C | NA | T | A | 0.028388 | 0.004417 | 1.80E-10 |
| rs676457 | miR-10b-5p | trans | 3.04E-35 | Nightingale | M_LDL_FC | NA | T | A | 0.034303 | 0.004419 | 2.00E-14 |
| rs676457 | miR-10b-5p | trans | 3.04E-35 | Nightingale | M_LDL_C_ | NA | T | A | 0.031099 | 0.004428 | 3.30E-12 |
| rs676457 | miR-10b-5p | trans | 3.04E-35 | Nightingale | IDL_FC_pct | NA | T | A | 0.025704 | 0.004391 | 1.20E-08 |
| rs676457 | miR-10b-5p | trans | 3.04E-35 | Nightingale | XL_VLDL_T | NA | T | A | -0.04378 | 0.004289 | 2.50E-24 |
| rs676457 | miR-10b-5p | trans | 3.04E-35 | Nightingale | XS_VLDL_P | NA | T | A | 0.036716 | 0.004366 | 1.30E-17 |
| rs676457 | miR-10b-5p | trans | 3.04E-35 | Nightingale | S_LDL_L | NA | T | A | 0.027696 | 0.004424 | 8.50E-10 |
| rs676457 | miR-10b-5p | trans | 3.04E-35 | Nightingale | M_VLDL_F | NA | T | A | 0.041996 | 0.00421 | 4.10E-23 |
| rs676457 | miR-10b-5p | trans | 3.04E-35 | Nightingale | LDL_C | NA | T | A | 0.033149 | 0.004406 | 9.40E-14 |
| rs676457 | miR-10b-5p | trans | 3.04E-35 | Nightingale | XXL_VLDL_ | NA | T | A | 0.025022 | 0.004454 | 1.50E-09 |
| rs676457 | miR-10b-5p | trans | 3.04E-35 | Nightingale | XXL_VLDL_ | NA | T | A | -0.03565 | 0.004463 | 2.50E-16 |
| rs676457 | miR-10b-5p | trans | 3.04E-35 | Nightingale | L_LDL_L | NA | T | A | 0.033511 | 0.004382 | 3.20E-14 |
| rs676457 | miR-10b-5p | trans | 3.04E-35 | Nightingale | IDL_L | NA | T | A | 0.035272 | 0.004278 | 4.90E-16 |
| rs676457 | miR-10b-5p | trans | 3.04E-35 | Nightingale | M_VLDL_C | NA | T | A | 0.036148 | 0.004168 | 1.30E-17 |
| rs676457 | miR-10b-5p | trans | 3.04E-35 | Nightingale | L_LDL_P | NA | T | A | 0.029115 | 0.00441 | 6.60E-11 |
| rs676457 | miR-10b-5p | trans | 3.04E-35 | Nightingale | IDL_C | NA | T | A | 0.034961 | 0.004274 | 9.50E-16 |
| rs676457 | miR-10b-5p | trans | 3.04E-35 | Nightingale | M_VLDL_P | NA | T | A | 0.043977 | 0.004247 | 6.90E-25 |
| rs676457 | miR-10b-5p | trans | 3.04E-35 | Nightingale | ApoB_by_A | NA | T | A | 0.024978 | 0.004358 | 1.40E-08 |
| rs676457 | miR-10b-5p | trans | 3.04E-35 | Nightingale | Total_CE | NA | T | A | 0.031734 | 0.004268 | 9.90E-14 |
| rs676457 | miR-10b-5p | trans | 3.04E-35 | Nightingale | LDL_L | NA | T | A | 0.033261 | 0.00441 | 7.80E-14 |
| rs676457 | miR-10b-5p | trans | 3.04E-35 | Nightingale | S_HDL_L | NA | T | A | 0.026288 | 0.00441 | 1.60E-09 |
| rs676457 | miR-10b-5p | trans | 3.04E-35 | Nightingale | IDL_FC | NA | T | A | 0.038245 | 0.004284 | 1.50E-18 |
| rs676457 | miR-10b-5p | trans | 3.04E-35 | Nightingale | L_LDL_FC | NA | T | A | 0.033594 | 0.004345 | 3.00E-14 |
| rs676457 | miR-10b-5p | trans | 3.04E-35 | Nightingale | S_HDL_P | NA | T | A | 0.03058 | 0.004425 | 5.40E-12 |
| rs676457 | miR-10b-5p | trans | 3.04E-35 | Nightingale | Sphingomy | NA | T | A | 0.028973 | 0.004137 | 7.50E-12 |
| rs676457 | miR-10b-5p | trans | 3.04E-35 | Nightingale | M_LDL_TG | NA | T | A | -0.02753 | 0.00437 | 3.60E-10 |
| rs676457 | miR-10b-5p | trans | 3.04E-35 | Nightingale | XS_VLDL_C | NA | T | A | 0.03781 | 0.00427 | 2.10E-18 |
| rs676457 | miR-10b-5p | trans | 3.04E-35 | Nightingale | M_VLDL_C | NA | T | A | 0.033576 | 0.004157 | 2.10E-15 |
| rs676457 | miR-10b-5p | trans | 3.04E-35 | Nightingale | S_LDL_P | NA | T | A | 0.029546 | 0.004429 | 3.30E-11 |
| rs676457 | miR-10b-5p | trans | 3.04E-35 | Nightingale | M_VLDL_P | NA | T | A | 0.028256 | 0.004414 | 6.50E-11 |

| | | | | | | | | | | | |
|---|---|---|---|---|---|---|---|---|---|---|---|
| rs676457 | miR-10b-5p | trans | 3.04E-35 | Nightingale | S_LDL_C_p | NA | T | A | 0.045305 | 0.004442 | 6.40E-25 |
| rs676457 | miR-10b-5p | trans | 3.04E-35 | Nightingale | S_VLDL_TG | NA | T | A | -0.03807 | 0.004283 | 1.60E-18 |
| rs676457 | miR-10b-5p | trans | 3.04E-35 | Nightingale | M_LDL_CE | NA | T | A | 0.032923 | 0.004431 | 1.00E-13 |
| rs676457 | miR-10b-5p | trans | 3.04E-35 | Nightingale | LDL_CE | NA | T | A | 0.032505 | 0.004413 | 2.30E-13 |
| rs676457 | miR-10b-5p | trans | 3.04E-35 | Nightingale | GlycA | NA | T | A | 0.027864 | 0.004406 | 1.80E-10 |
| rs676457 | miR-10b-5p | trans | 3.04E-35 | Nightingale | LDL_PL | NA | T | A | 0.034949 | 0.004415 | 4.90E-15 |
| rs676457 | miR-10b-5p | trans | 3.04E-35 | Nightingale | L_VLDL_CE | NA | T | A | 0.04729 | 0.004234 | 2.80E-29 |
| rs676457 | miR-10b-5p | trans | 3.04E-35 | Nightingale | L_VLDL_TG | NA | T | A | -0.05103 | 0.004404 | 7.20E-32 |
| rs676457 | miR-10b-5p | trans | 3.04E-35 | Nightingale | Total_L | NA | T | A | 0.026347 | 0.004353 | 4.60E-10 |
| rs676457 | miR-10b-5p | trans | 3.04E-35 | Nightingale | S_HDL_FC | NA | T | A | 0.023471 | 0.004344 | 4.60E-08 |
| rs676457 | miR-10b-5p | trans | 3.04E-35 | Nightingale | S_VLDL_C_ | NA | T | A | 0.041827 | 0.004306 | 5.90E-22 |
| rs676457 | miR-10b-5p | trans | 3.04E-35 | Nightingale | ApoB | NA | T | A | 0.033899 | 0.004419 | 2.20E-14 |
| rs676457 | miR-10b-5p | trans | 3.04E-35 | Nightingale | Cholines | NA | T | A | 0.022307 | 0.004161 | 4.90E-08 |
| rs676457 | miR-10b-5p | trans | 3.04E-35 | Nightingale | M_VLDL_C | NA | T | A | 0.03867 | 0.004413 | 2.00E-18 |
| rs676457 | miR-10b-5p | trans | 3.04E-35 | Nightingale | Total_FC | NA | T | A | 0.033247 | 0.004323 | 6.90E-15 |
| rs676457 | miR-10b-5p | trans | 3.04E-35 | Nightingale | S_LDL_CE | NA | T | A | 0.030497 | 0.004427 | 6.10E-12 |
| rs676457 | miR-10b-5p | trans | 3.04E-35 | Nightingale | S_VLDL_CE | NA | T | A | 0.038156 | 0.004419 | 1.60E-18 |
| rs676457 | miR-10b-5p | trans | 3.04E-35 | Nightingale | XS_VLDL_F | NA | T | A | 0.037075 | 0.004348 | 6.40E-18 |
| rs676457 | miR-10b-5p | trans | 3.04E-35 | Nightingale | S_VLDL_C | NA | T | A | 0.038342 | 0.004424 | 1.80E-18 |
| rs676457 | miR-10b-5p | trans | 3.04E-35 | Nightingale | XXL_VLDL_ | NA | T | A | 0.030738 | 0.004406 | 1.30E-12 |
| rs676457 | miR-10b-5p | trans | 3.04E-35 | Nightingale | L_VLDL_C_ | NA | T | A | 0.05209 | 0.004322 | 5.50E-34 |
| rs676457 | miR-10b-5p | trans | 3.04E-35 | Nightingale | XXL_VLDL_ | NA | T | A | 0.034746 | 0.004434 | 3.30E-15 |
| rs676457 | miR-10b-5p | trans | 3.04E-35 | Nightingale | Omega_6 | NA | T | A | 0.025869 | 0.004321 | 6.00E-10 |
| rs676457 | miR-10b-5p | trans | 3.04E-35 | Nightingale | S_VLDL_L | NA | T | A | 0.02465 | 0.004382 | 3.80E-09 |
| rs676457 | miR-10b-5p | trans | 3.04E-35 | Nightingale | XL_VLDL_C | NA | T | A | 0.037267 | 0.004245 | 3.70E-18 |
| rs676457 | miR-10b-5p | trans | 3.04E-35 | Nightingale | M_LDL_L | NA | T | A | 0.032983 | 0.004431 | 1.10E-13 |
| rs676457 | miR-10b-5p | trans | 3.04E-35 | Nightingale | XL_VLDL_C | NA | T | A | 0.038144 | 0.004243 | 5.40E-19 |
| rs676457 | miR-10b-5p | trans | 3.04E-35 | Nightingale | PUFA | NA | T | A | 0.023692 | 0.004316 | 1.50E-08 |
| rs676457 | miR-10b-5p | trans | 3.04E-35 | Nightingale | Clinical_LD | NA | T | A | 0.036434 | 0.004399 | 2.90E-16 |
| rs676457 | miR-10b-5p | trans | 3.04E-35 | Nightingale | non_HDL_( | NA | T | A | 0.035663 | 0.004401 | 5.30E-16 |
| rs676457 | miR-10b-5p | trans | 3.04E-35 | Nightingale | VLDL_P | NA | T | A | 0.026619 | 0.004401 | 4.40E-10 |
| rs676457 | miR-10b-5p | trans | 3.04E-35 | Nightingale | S_LDL_C | NA | T | A | 0.030895 | 0.004429 | 5.80E-12 |
| rs676457 | miR-10b-5p | trans | 3.04E-35 | Nightingale | S_HDL_CE | NA | T | A | 0.028598 | 0.004432 | 1.40E-10 |
| rs676457 | miR-10b-5p | trans | 3.04E-35 | Nightingale | S_LDL_PL_ | NA | T | A | -0.03004 | 0.004417 | 2.80E-13 |
| rs676457 | miR-10b-5p | trans | 3.04E-35 | Nightingale | Total_PL | NA | T | A | 0.024941 | 0.004214 | 1.10E-09 |
| rs676457 | miR-10b-5p | trans | 3.04E-35 | Nightingale | L_LDL_CE | NA | T | A | 0.031521 | 0.004385 | 1.00E-12 |
| rs676457 | miR-10b-5p | trans | 3.04E-35 | Nightingale | M_VLDL_T | NA | T | A | -0.03849 | 0.004181 | 7.90E-20 |
| rs676457 | miR-10b-5p | trans | 3.04E-35 | Nightingale | IDL_PL | NA | T | A | 0.035325 | 0.00427 | 3.50E-16 |
| rs676457 | miR-10b-5p | trans | 3.04E-35 | Nightingale | XL_VLDL_P | NA | T | A | 0.038714 | 0.004432 | 6.80E-19 |
| rs676457 | miR-10b-5p | trans | 3.04E-35 | Nightingale | XS_VLDL_P | NA | T | A | 0.032125 | 0.004363 | 5.20E-14 |
| rs676457 | miR-10b-5p | trans | 3.04E-35 | Nightingale | IDL_CE | NA | T | A | 0.033519 | 0.004272 | 1.30E-14 |
| rs676457 | miR-10b-5p | trans | 3.04E-35 | Nightingale | L_LDL_PL | NA | T | A | 0.037956 | 0.004391 | 9.50E-18 |
| rs676457 | miR-10b-5p | trans | 3.04E-35 | Nightingale | S_VLDL_PL | NA | T | A | 0.027677 | 0.004237 | 1.90E-10 |
| rs676457 | miR-10b-5p | trans | 3.04E-35 | Nightingale | XS_VLDL_C | NA | T | A | 0.038003 | 0.004295 | 1.30E-18 |
| rs676457 | miR-10b-5p | trans | 3.04E-35 | Nightingale | M_VLDL_P | NA | T | A | 0.029096 | 0.004418 | 2.10E-11 |
| rs676457 | miR-10b-5p | trans | 3.04E-35 | Nightingale | S_VLDL_PL | NA | T | A | 0.032746 | 0.004417 | 4.30E-14 |
| rs676457 | miR-10b-5p | trans | 3.04E-35 | Nightingale | LDL_FC | NA | T | A | 0.033596 | 0.004378 | 5.50E-14 |
| rs676457 | miR-10b-5p | trans | 3.04E-35 | Nightingale | S_VLDL_FC | NA | T | A | 0.029304 | 0.004228 | 1.10E-11 |
| rs676457 | miR-10b-5p | trans | 3.04E-35 | Nightingale | L_VLDL_FC | NA | T | A | 0.048437 | 0.004382 | 6.60E-29 |
| rs676457 | miR-10b-5p | trans | 3.04E-35 | Nightingale | Citrate | NA | T | A | 0.026851 | 0.004396 | 6.90E-10 |
| rs676457 | miR-10b-5p | trans | 3.04E-35 | Nightingale | M_LDL_C | NA | T | A | 0.033939 | 0.004433 | 2.30E-14 |
| rs676457 | miR-10b-5p | trans | 3.04E-35 | Nightingale | XS_VLDL_L | NA | T | A | 0.033833 | 0.00435 | 2.00E-15 |
| rs676457 | miR-10b-5p | trans | 3.04E-35 | Nightingale | S_VLDL_FC | NA | T | A | 0.037867 | 0.004426 | 7.60E-18 |
| rs676457 | miR-10b-5p | trans | 3.04E-35 | Nightingale | M_LDL_PL | NA | T | A | 0.032419 | 0.004431 | 2.80E-13 |
| rs676457 | miR-10b-5p | trans | 3.04E-35 | Nightingale | VLDL_FC | NA | T | A | 0.024128 | 0.004392 | 1.10E-08 |
| rs676457 | miR-10b-5p | trans | 3.04E-35 | Nightingale | M_VLDL_F | NA | T | A | 0.033335 | 0.004424 | 3.20E-14 |
| rs676457 | miR-10b-5p | trans | 3.04E-35 | Nightingale | VLDL_C | NA | T | A | 0.031272 | 0.004419 | 5.00E-13 |
| rs676457 | miR-10b-5p | trans | 3.04E-35 | Nightingale | M_VLDL_C | NA | T | A | 0.040809 | 0.004385 | 2.40E-20 |
| rs676457 | miR-10b-5p | trans | 3.04E-35 | Nightingale | IDL_P | NA | T | A | 0.040008 | 0.004378 | 1.30E-19 |
| rs676457 | miR-10b-5p | trans | 3.04E-35 | Nightingale | LA | NA | T | A | 0.024934 | 0.004339 | 3.60E-09 |
| rs676457 | miR-10b-5p | trans | 3.04E-35 | Nightingale | VLDL_CE | NA | T | A | 0.035758 | 0.004426 | 3.10E-16 |
| rs676457 | miR-10b-5p | trans | 3.04E-35 | Nightingale | S_LDL_FC | NA | T | A | 0.028528 | 0.004418 | 4.40E-10 |
| rs676457 | miR-10b-5p | trans | 3.04E-35 | Nightingale | S_VLDL_CE | NA | T | A | 0.04697 | 0.004365 | 1.40E-26 |

| | | | | | | | | | | | |
|---|---|---|---|---|---|---|---|---|---|---|---|
| rs676457 | miR-10b-5p | trans | 3.04E-35 | Nightingale | S_VLDL_P | NA | T | A | 0.023453 | 0.004369 | 1.70E-08 |
| rs676457 | miR-10b-5p | trans | 3.04E-35 | Nightingale | XXL_VLDL_ | NA | T | A | 0.033069 | 0.004459 | 9.60E-14 |
| rs676457 | miR-10b-5p | trans | 3.04E-35 | Nightingale | L_LDL_C | NA | T | A | 0.032277 | 0.004375 | 2.70E-13 |
| rs676457 | miR-10b-5p | trans | 3.04E-35 | Nightingale | XL_VLDL_F | NA | T | A | 0.037129 | 0.00429 | 2.30E-18 |
| rs676457 | miR-10b-5p | trans | 3.04E-35 | Nightingale | S_LDL_CE_ | NA | T | A | 0.030893 | 0.004417 | 5.50E-14 |
| rs676457 | miR-10b-5p | trans | 3.04E-35 | Nightingale | XS_VLDL_F | NA | T | A | 0.031376 | 0.004393 | 2.60E-12 |
| rs676457 | miR-126-3p | trans | 2.46E-39 | Nightingale | Remnant_( | NA | T | A | 0.03685 | 0.004396 | 3.70E-17 |
| rs676457 | miR-126-3p | trans | 2.46E-39 | Nightingale | S_HDL_PL | NA | T | A | 0.023785 | 0.004387 | 3.30E-08 |
| rs676457 | miR-126-3p | trans | 2.46E-39 | Nightingale | XL_HDL_PL | NA | T | A | -0.02267 | 0.004065 | 1.30E-08 |
| rs676457 | miR-126-3p | trans | 2.46E-39 | Nightingale | Total_C | NA | T | A | 0.032362 | 0.004284 | 3.40E-14 |
| rs676457 | miR-126-3p | trans | 2.46E-39 | Nightingale | LDL_P | NA | T | A | 0.031743 | 0.004422 | 1.10E-12 |
| rs676457 | miR-126-3p | trans | 2.46E-39 | Nightingale | M_LDL_P | NA | T | A | 0.035846 | 0.004437 | 7.30E-16 |
| rs676457 | miR-126-3p | trans | 2.46E-39 | Nightingale | S_HDL_C | NA | T | A | 0.028388 | 0.004417 | 1.80E-10 |
| rs676457 | miR-126-3p | trans | 2.46E-39 | Nightingale | M_LDL_FC | NA | T | A | 0.034303 | 0.004419 | 2.00E-14 |
| rs676457 | miR-126-3p | trans | 2.46E-39 | Nightingale | M_LDL_C_ | NA | T | A | 0.031099 | 0.004428 | 3.30E-12 |
| rs676457 | miR-126-3p | trans | 2.46E-39 | Nightingale | IDL_FC_pct | NA | T | A | 0.025704 | 0.004391 | 1.20E-08 |
| rs676457 | miR-126-3p | trans | 2.46E-39 | Nightingale | XL_VLDL_T | NA | T | A | -0.04378 | 0.004289 | 2.50E-24 |
| rs676457 | miR-126-3p | trans | 2.46E-39 | Nightingale | XS_VLDL_P | NA | T | A | 0.036716 | 0.004366 | 1.30E-17 |
| rs676457 | miR-126-3p | trans | 2.46E-39 | Nightingale | S_LDL_L | NA | T | A | 0.027696 | 0.004424 | 8.50E-10 |
| rs676457 | miR-126-3p | trans | 2.46E-39 | Nightingale | M_VLDL_F | NA | T | A | 0.041996 | 0.00421 | 4.10E-23 |
| rs676457 | miR-126-3p | trans | 2.46E-39 | Nightingale | LDL_C | NA | T | A | 0.033149 | 0.004406 | 9.40E-14 |
| rs676457 | miR-126-3p | trans | 2.46E-39 | Nightingale | XXL_VLDL_ | NA | T | A | 0.025022 | 0.004454 | 1.50E-09 |
| rs676457 | miR-126-3p | trans | 2.46E-39 | Nightingale | XXL_VLDL_ | NA | T | A | -0.03565 | 0.004463 | 2.50E-16 |
| rs676457 | miR-126-3p | trans | 2.46E-39 | Nightingale | L_LDL_L | NA | T | A | 0.033511 | 0.004382 | 3.20E-14 |
| rs676457 | miR-126-3p | trans | 2.46E-39 | Nightingale | IDL_L | NA | T | A | 0.035272 | 0.004278 | 4.90E-16 |
| rs676457 | miR-126-3p | trans | 2.46E-39 | Nightingale | M_VLDL_C | NA | T | A | 0.036148 | 0.004168 | 1.30E-17 |
| rs676457 | miR-126-3p | trans | 2.46E-39 | Nightingale | L_LDL_P | NA | T | A | 0.029115 | 0.00441 | 6.60E-11 |
| rs676457 | miR-126-3p | trans | 2.46E-39 | Nightingale | IDL_C | NA | T | A | 0.034961 | 0.004274 | 9.50E-16 |
| rs676457 | miR-126-3p | trans | 2.46E-39 | Nightingale | M_VLDL_P | NA | T | A | 0.043977 | 0.004247 | 6.90E-25 |
| rs676457 | miR-126-3p | trans | 2.46E-39 | Nightingale | ApoB_by_/ | NA | T | A | 0.024978 | 0.004358 | 1.40E-08 |
| rs676457 | miR-126-3p | trans | 2.46E-39 | Nightingale | Total_CE | NA | T | A | 0.031734 | 0.004268 | 9.90E-14 |
| rs676457 | miR-126-3p | trans | 2.46E-39 | Nightingale | LDL_L | NA | T | A | 0.033261 | 0.00441 | 7.80E-14 |
| rs676457 | miR-126-3p | trans | 2.46E-39 | Nightingale | S_HDL_L | NA | T | A | 0.026288 | 0.00441 | 1.60E-09 |
| rs676457 | miR-126-3p | trans | 2.46E-39 | Nightingale | IDL_FC | NA | T | A | 0.038245 | 0.004284 | 1.50E-18 |
| rs676457 | miR-126-3p | trans | 2.46E-39 | Nightingale | L_LDL_FC | NA | T | A | 0.033594 | 0.004345 | 3.00E-14 |
| rs676457 | miR-126-3p | trans | 2.46E-39 | Nightingale | S_HDL_P | NA | T | A | 0.03058 | 0.004425 | 5.40E-12 |
| rs676457 | miR-126-3p | trans | 2.46E-39 | Nightingale | Sphingomy | NA | T | A | 0.028973 | 0.004137 | 7.50E-12 |
| rs676457 | miR-126-3p | trans | 2.46E-39 | Nightingale | M_LDL_TG | NA | T | A | -0.02753 | 0.00437 | 3.60E-10 |
| rs676457 | miR-126-3p | trans | 2.46E-39 | Nightingale | XS_VLDL_C | NA | T | A | 0.03781 | 0.00427 | 2.10E-18 |
| rs676457 | miR-126-3p | trans | 2.46E-39 | Nightingale | M_VLDL_C | NA | T | A | 0.033576 | 0.004157 | 2.10E-15 |
| rs676457 | miR-126-3p | trans | 2.46E-39 | Nightingale | S_LDL_P | NA | T | A | 0.029546 | 0.004429 | 3.30E-11 |
| rs676457 | miR-126-3p | trans | 2.46E-39 | Nightingale | M_VLDL_P | NA | T | A | 0.028256 | 0.004414 | 6.50E-11 |
| rs676457 | miR-126-3p | trans | 2.46E-39 | Nightingale | S_LDL_C_p | NA | T | A | 0.045305 | 0.004442 | 6.40E-25 |
| rs676457 | miR-126-3p | trans | 2.46E-39 | Nightingale | S_VLDL_TG | NA | T | A | -0.03807 | 0.004283 | 1.60E-18 |
| rs676457 | miR-126-3p | trans | 2.46E-39 | Nightingale | M_LDL_CE | NA | T | A | 0.032923 | 0.004431 | 1.00E-13 |
| rs676457 | miR-126-3p | trans | 2.46E-39 | Nightingale | LDL_CE | NA | T | A | 0.032505 | 0.004413 | 2.30E-13 |
| rs676457 | miR-126-3p | trans | 2.46E-39 | Nightingale | GlycA | NA | T | A | 0.027864 | 0.004406 | 1.80E-10 |
| rs676457 | miR-126-3p | trans | 2.46E-39 | Nightingale | LDL_PL | NA | T | A | 0.034949 | 0.004415 | 4.90E-15 |
| rs676457 | miR-126-3p | trans | 2.46E-39 | Nightingale | L_VLDL_CE | NA | T | A | 0.04729 | 0.004234 | 2.80E-29 |
| rs676457 | miR-126-3p | trans | 2.46E-39 | Nightingale | L_VLDL_TG | NA | T | A | -0.05103 | 0.004404 | 7.20E-32 |
| rs676457 | miR-126-3p | trans | 2.46E-39 | Nightingale | Total_L | NA | T | A | 0.026347 | 0.004353 | 4.60E-10 |
| rs676457 | miR-126-3p | trans | 2.46E-39 | Nightingale | S_HDL_FC | NA | T | A | 0.023471 | 0.004344 | 4.60E-08 |
| rs676457 | miR-126-3p | trans | 2.46E-39 | Nightingale | S_VLDL_C_ | NA | T | A | 0.041827 | 0.004306 | 5.90E-22 |
| rs676457 | miR-126-3p | trans | 2.46E-39 | Nightingale | ApoB | NA | T | A | 0.033899 | 0.004419 | 2.20E-14 |
| rs676457 | miR-126-3p | trans | 2.46E-39 | Nightingale | Cholines | NA | T | A | 0.022307 | 0.004161 | 4.90E-08 |
| rs676457 | miR-126-3p | trans | 2.46E-39 | Nightingale | M_VLDL_C | NA | T | A | 0.03867 | 0.004413 | 2.00E-18 |
| rs676457 | miR-126-3p | trans | 2.46E-39 | Nightingale | Total_FC | NA | T | A | 0.033247 | 0.004323 | 6.90E-15 |
| rs676457 | miR-126-3p | trans | 2.46E-39 | Nightingale | S_LDL_CE | NA | T | A | 0.030497 | 0.004427 | 6.10E-12 |
| rs676457 | miR-126-3p | trans | 2.46E-39 | Nightingale | S_VLDL_CE | NA | T | A | 0.038156 | 0.004419 | 1.60E-18 |
| rs676457 | miR-126-3p | trans | 2.46E-39 | Nightingale | XS_VLDL_F | NA | T | A | 0.037075 | 0.004348 | 6.40E-18 |
| rs676457 | miR-126-3p | trans | 2.46E-39 | Nightingale | S_VLDL_C | NA | T | A | 0.038342 | 0.004424 | 1.80E-18 |
| rs676457 | miR-126-3p | trans | 2.46E-39 | Nightingale | XXL_VLDL_ | NA | T | A | 0.030738 | 0.004406 | 1.30E-12 |
| rs676457 | miR-126-3p | trans | 2.46E-39 | Nightingale | L_VLDL_C_ | NA | T | A | 0.05209 | 0.004322 | 5.50E-34 |

| | | | | | | | | | | | |
|---|---|---|---|---|---|---|---|---|---|---|---|
| rs676457 | miR-126-3p | trans | 2.46E-39 | Nightingale | XXL_VLDL_ | NA | T | A | 0.034746 | 0.004434 | 3.30E-15 |
| rs676457 | miR-126-3p | trans | 2.46E-39 | Nightingale | Omega_6 | NA | T | A | 0.025869 | 0.004321 | 6.00E-10 |
| rs676457 | miR-126-3p | trans | 2.46E-39 | Nightingale | S_VLDL_L | NA | T | A | 0.02465 | 0.004382 | 3.80E-09 |
| rs676457 | miR-126-3p | trans | 2.46E-39 | Nightingale | XL_VLDL_C | NA | T | A | 0.037267 | 0.004245 | 3.70E-18 |
| rs676457 | miR-126-3p | trans | 2.46E-39 | Nightingale | M_LDL_L | NA | T | A | 0.032983 | 0.004431 | 1.10E-13 |
| rs676457 | miR-126-3p | trans | 2.46E-39 | Nightingale | XL_VLDL_C | NA | T | A | 0.038144 | 0.004243 | 5.40E-19 |
| rs676457 | miR-126-3p | trans | 2.46E-39 | Nightingale | PUFA | NA | T | A | 0.023692 | 0.004316 | 1.50E-08 |
| rs676457 | miR-126-3p | trans | 2.46E-39 | Nightingale | Clinical_LD | NA | T | A | 0.036434 | 0.004399 | 2.90E-16 |
| rs676457 | miR-126-3p | trans | 2.46E-39 | Nightingale | non_HDL_( | NA | T | A | 0.035663 | 0.004401 | 5.30E-16 |
| rs676457 | miR-126-3p | trans | 2.46E-39 | Nightingale | VLDL_P | NA | T | A | 0.026619 | 0.004401 | 4.40E-10 |
| rs676457 | miR-126-3p | trans | 2.46E-39 | Nightingale | S_LDL_C | NA | T | A | 0.030895 | 0.004429 | 5.80E-12 |
| rs676457 | miR-126-3p | trans | 2.46E-39 | Nightingale | S_HDL_CE | NA | T | A | 0.028598 | 0.004432 | 1.40E-10 |
| rs676457 | miR-126-3p | trans | 2.46E-39 | Nightingale | S_LDL_PL_ | NA | T | A | -0.03004 | 0.004417 | 2.80E-13 |
| rs676457 | miR-126-3p | trans | 2.46E-39 | Nightingale | Total_PL | NA | T | A | 0.024941 | 0.004214 | 1.10E-09 |
| rs676457 | miR-126-3p | trans | 2.46E-39 | Nightingale | L_LDL_CE | NA | T | A | 0.031521 | 0.004385 | 1.00E-12 |
| rs676457 | miR-126-3p | trans | 2.46E-39 | Nightingale | M_VLDL_T | NA | T | A | -0.03849 | 0.004181 | 7.90E-20 |
| rs676457 | miR-126-3p | trans | 2.46E-39 | Nightingale | IDL_PL | NA | T | A | 0.035325 | 0.00427 | 3.50E-16 |
| rs676457 | miR-126-3p | trans | 2.46E-39 | Nightingale | XL_VLDL_P | NA | T | A | 0.038714 | 0.004432 | 6.80E-19 |
| rs676457 | miR-126-3p | trans | 2.46E-39 | Nightingale | XS_VLDL_P | NA | T | A | 0.032125 | 0.004363 | 5.20E-14 |
| rs676457 | miR-126-3p | trans | 2.46E-39 | Nightingale | IDL_CE | NA | T | A | 0.033519 | 0.004272 | 1.30E-14 |
| rs676457 | miR-126-3p | trans | 2.46E-39 | Nightingale | L_LDL_PL | NA | T | A | 0.037956 | 0.004391 | 9.50E-18 |
| rs676457 | miR-126-3p | trans | 2.46E-39 | Nightingale | S_VLDL_PL | NA | T | A | 0.027677 | 0.004237 | 1.90E-10 |
| rs676457 | miR-126-3p | trans | 2.46E-39 | Nightingale | XS_VLDL_C | NA | T | A | 0.038003 | 0.004295 | 1.30E-18 |
| rs676457 | miR-126-3p | trans | 2.46E-39 | Nightingale | M_VLDL_P | NA | T | A | 0.029096 | 0.004418 | 2.10E-11 |
| rs676457 | miR-126-3p | trans | 2.46E-39 | Nightingale | S_VLDL_PL | NA | T | A | 0.032746 | 0.004417 | 4.30E-14 |
| rs676457 | miR-126-3p | trans | 2.46E-39 | Nightingale | LDL_FC | NA | T | A | 0.033596 | 0.004378 | 5.50E-14 |
| rs676457 | miR-126-3p | trans | 2.46E-39 | Nightingale | S_VLDL_FC | NA | T | A | 0.029304 | 0.004228 | 1.10E-11 |
| rs676457 | miR-126-3p | trans | 2.46E-39 | Nightingale | L_VLDL_FC | NA | T | A | 0.048437 | 0.004382 | 6.60E-29 |
| rs676457 | miR-126-3p | trans | 2.46E-39 | Nightingale | Citrate | NA | T | A | 0.026851 | 0.004396 | 6.90E-10 |
| rs676457 | miR-126-3p | trans | 2.46E-39 | Nightingale | M_LDL_C | NA | T | A | 0.033939 | 0.004433 | 2.30E-14 |
| rs676457 | miR-126-3p | trans | 2.46E-39 | Nightingale | XS_VLDL_L | NA | T | A | 0.033833 | 0.00435 | 2.00E-15 |
| rs676457 | miR-126-3p | trans | 2.46E-39 | Nightingale | S_VLDL_FC | NA | T | A | 0.037867 | 0.004426 | 7.60E-18 |
| rs676457 | miR-126-3p | trans | 2.46E-39 | Nightingale | M_LDL_PL | NA | T | A | 0.032419 | 0.004431 | 2.80E-13 |
| rs676457 | miR-126-3p | trans | 2.46E-39 | Nightingale | VLDL_FC | NA | T | A | 0.024128 | 0.004392 | 1.10E-08 |
| rs676457 | miR-126-3p | trans | 2.46E-39 | Nightingale | M_VLDL_F | NA | T | A | 0.033335 | 0.004424 | 3.20E-14 |
| rs676457 | miR-126-3p | trans | 2.46E-39 | Nightingale | VLDL_C | NA | T | A | 0.031272 | 0.004419 | 5.00E-13 |
| rs676457 | miR-126-3p | trans | 2.46E-39 | Nightingale | M_VLDL_C | NA | T | A | 0.040809 | 0.004385 | 2.40E-20 |
| rs676457 | miR-126-3p | trans | 2.46E-39 | Nightingale | IDL_P | NA | T | A | 0.040008 | 0.004378 | 1.30E-19 |
| rs676457 | miR-126-3p | trans | 2.46E-39 | Nightingale | LA | NA | T | A | 0.024934 | 0.004339 | 3.60E-09 |
| rs676457 | miR-126-3p | trans | 2.46E-39 | Nightingale | VLDL_CE | NA | T | A | 0.035758 | 0.004426 | 3.10E-16 |
| rs676457 | miR-126-3p | trans | 2.46E-39 | Nightingale | S_LDL_FC | NA | T | A | 0.028528 | 0.004418 | 4.40E-10 |
| rs676457 | miR-126-3p | trans | 2.46E-39 | Nightingale | S_VLDL_CE | NA | T | A | 0.04697 | 0.004365 | 1.40E-26 |
| rs676457 | miR-126-3p | trans | 2.46E-39 | Nightingale | S_VLDL_P | NA | T | A | 0.023453 | 0.004369 | 1.70E-08 |
| rs676457 | miR-126-3p | trans | 2.46E-39 | Nightingale | XXL_VLDL_ | NA | T | A | 0.033069 | 0.004459 | 9.60E-14 |
| rs676457 | miR-126-3p | trans | 2.46E-39 | Nightingale | L_LDL_C | NA | T | A | 0.032277 | 0.004375 | 2.70E-13 |
| rs676457 | miR-126-3p | trans | 2.46E-39 | Nightingale | XL_VLDL_F | NA | T | A | 0.037129 | 0.00429 | 2.30E-18 |
| rs676457 | miR-126-3p | trans | 2.46E-39 | Nightingale | S_LDL_CE_ | NA | T | A | 0.030893 | 0.004417 | 5.50E-14 |
| rs676457 | miR-126-3p | trans | 2.46E-39 | Nightingale | XS_VLDL_F | NA | T | A | 0.031376 | 0.004393 | 2.60E-12 |
| rs676457 | miR-195-5p | trans | 2.12E-17 | Nightingale | Remnant_( | NA | T | A | 0.03685 | 0.004396 | 3.70E-17 |
| rs676457 | miR-195-5p | trans | 2.12E-17 | Nightingale | S_HDL_PL | NA | T | A | 0.023785 | 0.004387 | 3.30E-08 |
| rs676457 | miR-195-5p | trans | 2.12E-17 | Nightingale | XL_HDL_PL | NA | T | A | -0.02267 | 0.004065 | 1.30E-08 |
| rs676457 | miR-195-5p | trans | 2.12E-17 | Nightingale | Total_C | NA | T | A | 0.032362 | 0.004284 | 3.40E-14 |
| rs676457 | miR-195-5p | trans | 2.12E-17 | Nightingale | LDL_P | NA | T | A | 0.031743 | 0.004422 | 1.10E-12 |
| rs676457 | miR-195-5p | trans | 2.12E-17 | Nightingale | M_LDL_P | NA | T | A | 0.035846 | 0.004437 | 7.30E-16 |
| rs676457 | miR-195-5p | trans | 2.12E-17 | Nightingale | S_HDL_C | NA | T | A | 0.028388 | 0.004417 | 1.80E-10 |
| rs676457 | miR-195-5p | trans | 2.12E-17 | Nightingale | M_LDL_FC | NA | T | A | 0.034303 | 0.004419 | 2.00E-14 |
| rs676457 | miR-195-5p | trans | 2.12E-17 | Nightingale | M_LDL_C_ | NA | T | A | 0.031099 | 0.004428 | 3.30E-12 |
| rs676457 | miR-195-5p | trans | 2.12E-17 | Nightingale | IDL_FC_pct | NA | T | A | 0.025704 | 0.004391 | 1.20E-08 |
| rs676457 | miR-195-5p | trans | 2.12E-17 | Nightingale | XL_VLDL_T | NA | T | A | -0.04378 | 0.004289 | 2.50E-24 |
| rs676457 | miR-195-5p | trans | 2.12E-17 | Nightingale | XS_VLDL_P | NA | T | A | 0.036716 | 0.004366 | 1.30E-17 |
| rs676457 | miR-195-5p | trans | 2.12E-17 | Nightingale | S_LDL_L | NA | T | A | 0.027696 | 0.004424 | 8.50E-10 |
| rs676457 | miR-195-5p | trans | 2.12E-17 | Nightingale | M_VLDL_F | NA | T | A | 0.041996 | 0.00421 | 4.10E-23 |
| rs676457 | miR-195-5p | trans | 2.12E-17 | Nightingale | LDL_C | NA | T | A | 0.033149 | 0.004406 | 9.40E-14 |

| | | | | | | | | | | | |
|---|---|---|---|---|---|---|---|---|---|---|---|
| rs676457 | miR-195-5p | trans | 2.12E-17 | Nightingale | XXL_VLDL_ | NA | T | A | 0.025022 | 0.004454 | 1.50E-09 |
| rs676457 | miR-195-5p | trans | 2.12E-17 | Nightingale | XXL_VLDL_ | NA | T | A | -0.03565 | 0.004463 | 2.50E-16 |
| rs676457 | miR-195-5p | trans | 2.12E-17 | Nightingale | L_LDL_L | NA | T | A | 0.033511 | 0.004382 | 3.20E-14 |
| rs676457 | miR-195-5p | trans | 2.12E-17 | Nightingale | IDL_L | NA | T | A | 0.035272 | 0.004278 | 4.90E-16 |
| rs676457 | miR-195-5p | trans | 2.12E-17 | Nightingale | M_VLDL_C | NA | T | A | 0.036148 | 0.004168 | 1.30E-17 |
| rs676457 | miR-195-5p | trans | 2.12E-17 | Nightingale | L_LDL_P | NA | T | A | 0.029115 | 0.00441 | 6.60E-11 |
| rs676457 | miR-195-5p | trans | 2.12E-17 | Nightingale | IDL_C | NA | T | A | 0.034961 | 0.004274 | 9.50E-16 |
| rs676457 | miR-195-5p | trans | 2.12E-17 | Nightingale | M_VLDL_P | NA | T | A | 0.043977 | 0.004247 | 6.90E-25 |
| rs676457 | miR-195-5p | trans | 2.12E-17 | Nightingale | ApoB_by_A | NA | T | A | 0.024978 | 0.004358 | 1.40E-08 |
| rs676457 | miR-195-5p | trans | 2.12E-17 | Nightingale | Total_CE | NA | T | A | 0.031734 | 0.004268 | 9.90E-14 |
| rs676457 | miR-195-5p | trans | 2.12E-17 | Nightingale | LDL_L | NA | T | A | 0.033261 | 0.00441 | 7.80E-14 |
| rs676457 | miR-195-5p | trans | 2.12E-17 | Nightingale | S_HDL_L | NA | T | A | 0.026288 | 0.00441 | 1.60E-09 |
| rs676457 | miR-195-5p | trans | 2.12E-17 | Nightingale | IDL_FC | NA | T | A | 0.038245 | 0.004284 | 1.50E-18 |
| rs676457 | miR-195-5p | trans | 2.12E-17 | Nightingale | L_LDL_FC | NA | T | A | 0.033594 | 0.004345 | 3.00E-14 |
| rs676457 | miR-195-5p | trans | 2.12E-17 | Nightingale | S_HDL_P | NA | T | A | 0.03058 | 0.004425 | 5.40E-12 |
| rs676457 | miR-195-5p | trans | 2.12E-17 | Nightingale | Sphingomy | NA | T | A | 0.028973 | 0.004137 | 7.50E-12 |
| rs676457 | miR-195-5p | trans | 2.12E-17 | Nightingale | M_LDL_TG | NA | T | A | -0.02753 | 0.00437 | 3.60E-10 |
| rs676457 | miR-195-5p | trans | 2.12E-17 | Nightingale | XS_VLDL_C | NA | T | A | 0.03781 | 0.00427 | 2.10E-18 |
| rs676457 | miR-195-5p | trans | 2.12E-17 | Nightingale | M_VLDL_C | NA | T | A | 0.033576 | 0.004157 | 2.10E-15 |
| rs676457 | miR-195-5p | trans | 2.12E-17 | Nightingale | S_LDL_P | NA | T | A | 0.029546 | 0.004429 | 3.30E-11 |
| rs676457 | miR-195-5p | trans | 2.12E-17 | Nightingale | M_VLDL_P | NA | T | A | 0.028256 | 0.004414 | 6.50E-11 |
| rs676457 | miR-195-5p | trans | 2.12E-17 | Nightingale | S_LDL_C_p | NA | T | A | 0.045305 | 0.004442 | 6.40E-25 |
| rs676457 | miR-195-5p | trans | 2.12E-17 | Nightingale | S_VLDL_TG | NA | T | A | -0.03807 | 0.004283 | 1.60E-18 |
| rs676457 | miR-195-5p | trans | 2.12E-17 | Nightingale | M_LDL_CE | NA | T | A | 0.032923 | 0.004431 | 1.00E-13 |
| rs676457 | miR-195-5p | trans | 2.12E-17 | Nightingale | LDL_CE | NA | T | A | 0.032505 | 0.004413 | 2.30E-13 |
| rs676457 | miR-195-5p | trans | 2.12E-17 | Nightingale | GlycA | NA | T | A | 0.027864 | 0.004406 | 1.80E-10 |
| rs676457 | miR-195-5p | trans | 2.12E-17 | Nightingale | LDL_PL | NA | T | A | 0.034949 | 0.004415 | 4.90E-15 |
| rs676457 | miR-195-5p | trans | 2.12E-17 | Nightingale | L_VLDL_CE | NA | T | A | 0.04729 | 0.004234 | 2.80E-29 |
| rs676457 | miR-195-5p | trans | 2.12E-17 | Nightingale | L_VLDL_TG | NA | T | A | -0.05103 | 0.004404 | 7.20E-32 |
| rs676457 | miR-195-5p | trans | 2.12E-17 | Nightingale | Total_L | NA | T | A | 0.026347 | 0.004353 | 4.60E-10 |
| rs676457 | miR-195-5p | trans | 2.12E-17 | Nightingale | S_HDL_FC | NA | T | A | 0.023471 | 0.004344 | 4.60E-08 |
| rs676457 | miR-195-5p | trans | 2.12E-17 | Nightingale | S_VLDL_C_ | NA | T | A | 0.041827 | 0.004306 | 5.90E-22 |
| rs676457 | miR-195-5p | trans | 2.12E-17 | Nightingale | ApoB | NA | T | A | 0.033899 | 0.004419 | 2.20E-14 |
| rs676457 | miR-195-5p | trans | 2.12E-17 | Nightingale | Cholines | NA | T | A | 0.022307 | 0.004161 | 4.90E-08 |
| rs676457 | miR-195-5p | trans | 2.12E-17 | Nightingale | M_VLDL_C | NA | T | A | 0.03867 | 0.004413 | 2.00E-18 |
| rs676457 | miR-195-5p | trans | 2.12E-17 | Nightingale | Total_FC | NA | T | A | 0.033247 | 0.004323 | 6.90E-15 |
| rs676457 | miR-195-5p | trans | 2.12E-17 | Nightingale | S_LDL_CE | NA | T | A | 0.030497 | 0.004427 | 6.10E-12 |
| rs676457 | miR-195-5p | trans | 2.12E-17 | Nightingale | S_VLDL_CE | NA | T | A | 0.038156 | 0.004419 | 1.60E-18 |
| rs676457 | miR-195-5p | trans | 2.12E-17 | Nightingale | XS_VLDL_F | NA | T | A | 0.037075 | 0.004348 | 6.40E-18 |
| rs676457 | miR-195-5p | trans | 2.12E-17 | Nightingale | S_VLDL_C | NA | T | A | 0.038342 | 0.004424 | 1.80E-18 |
| rs676457 | miR-195-5p | trans | 2.12E-17 | Nightingale | XXL_VLDL_ | NA | T | A | 0.030738 | 0.004406 | 1.30E-12 |
| rs676457 | miR-195-5p | trans | 2.12E-17 | Nightingale | L_VLDL_C_ | NA | T | A | 0.05209 | 0.004322 | 5.50E-34 |
| rs676457 | miR-195-5p | trans | 2.12E-17 | Nightingale | XXL_VLDL_ | NA | T | A | 0.034746 | 0.004434 | 3.30E-15 |
| rs676457 | miR-195-5p | trans | 2.12E-17 | Nightingale | Omega_6 | NA | T | A | 0.025869 | 0.004321 | 6.00E-10 |
| rs676457 | miR-195-5p | trans | 2.12E-17 | Nightingale | S_VLDL_L | NA | T | A | 0.02465 | 0.004382 | 3.80E-09 |
| rs676457 | miR-195-5p | trans | 2.12E-17 | Nightingale | XL_VLDL_C | NA | T | A | 0.037267 | 0.004245 | 3.70E-18 |
| rs676457 | miR-195-5p | trans | 2.12E-17 | Nightingale | M_LDL_L | NA | T | A | 0.032983 | 0.004431 | 1.10E-13 |
| rs676457 | miR-195-5p | trans | 2.12E-17 | Nightingale | XL_VLDL_C | NA | T | A | 0.038144 | 0.004243 | 5.40E-19 |
| rs676457 | miR-195-5p | trans | 2.12E-17 | Nightingale | PUFA | NA | T | A | 0.023692 | 0.004316 | 1.50E-08 |
| rs676457 | miR-195-5p | trans | 2.12E-17 | Nightingale | Clinical_LD | NA | T | A | 0.036434 | 0.004399 | 2.90E-16 |
| rs676457 | miR-195-5p | trans | 2.12E-17 | Nightingale | non_HDL_( | NA | T | A | 0.035663 | 0.004401 | 5.30E-16 |
| rs676457 | miR-195-5p | trans | 2.12E-17 | Nightingale | VLDL_P | NA | T | A | 0.026619 | 0.004401 | 4.40E-10 |
| rs676457 | miR-195-5p | trans | 2.12E-17 | Nightingale | S_LDL_C | NA | T | A | 0.030895 | 0.004429 | 5.80E-12 |
| rs676457 | miR-195-5p | trans | 2.12E-17 | Nightingale | S_HDL_CE | NA | T | A | 0.028598 | 0.004432 | 1.40E-10 |
| rs676457 | miR-195-5p | trans | 2.12E-17 | Nightingale | S_LDL_PL_ | NA | T | A | -0.03004 | 0.004417 | 2.80E-13 |
| rs676457 | miR-195-5p | trans | 2.12E-17 | Nightingale | Total_PL | NA | T | A | 0.024941 | 0.004214 | 1.10E-09 |
| rs676457 | miR-195-5p | trans | 2.12E-17 | Nightingale | L_LDL_CE | NA | T | A | 0.031521 | 0.004385 | 1.00E-12 |
| rs676457 | miR-195-5p | trans | 2.12E-17 | Nightingale | M_VLDL_T | NA | T | A | -0.03849 | 0.004181 | 7.90E-20 |
| rs676457 | miR-195-5p | trans | 2.12E-17 | Nightingale | IDL_PL | NA | T | A | 0.035325 | 0.00427 | 3.50E-16 |
| rs676457 | miR-195-5p | trans | 2.12E-17 | Nightingale | XL_VLDL_P | NA | T | A | 0.038714 | 0.004432 | 6.80E-19 |
| rs676457 | miR-195-5p | trans | 2.12E-17 | Nightingale | XS_VLDL_P | NA | T | A | 0.032125 | 0.004363 | 5.20E-14 |
| rs676457 | miR-195-5p | trans | 2.12E-17 | Nightingale | IDL_CE | NA | T | A | 0.033519 | 0.004272 | 1.30E-14 |
| rs676457 | miR-195-5p | trans | 2.12E-17 | Nightingale | L_LDL_PL | NA | T | A | 0.037956 | 0.004391 | 9.50E-18 |

| | | | | | | | | | | | |
|---|---|---|---|---|---|---|---|---|---|---|---|
| rs676457 | miR-195-5p | trans | 2.12E-17 | Nightingale | S_VLDL_PL | NA | T | A | 0.027677 | 0.004237 | 1.90E-10 |
| rs676457 | miR-195-5p | trans | 2.12E-17 | Nightingale | XS_VLDL_C | NA | T | A | 0.038003 | 0.004295 | 1.30E-18 |
| rs676457 | miR-195-5p | trans | 2.12E-17 | Nightingale | M_VLDL_P | NA | T | A | 0.029096 | 0.004418 | 2.10E-11 |
| rs676457 | miR-195-5p | trans | 2.12E-17 | Nightingale | S_VLDL_PL | NA | T | A | 0.032746 | 0.004417 | 4.30E-14 |
| rs676457 | miR-195-5p | trans | 2.12E-17 | Nightingale | LDL_FC | NA | T | A | 0.033596 | 0.004378 | 5.50E-14 |
| rs676457 | miR-195-5p | trans | 2.12E-17 | Nightingale | S_VLDL_FC | NA | T | A | 0.029304 | 0.004228 | 1.10E-11 |
| rs676457 | miR-195-5p | trans | 2.12E-17 | Nightingale | L_VLDL_FC | NA | T | A | 0.048437 | 0.004382 | 6.60E-29 |
| rs676457 | miR-195-5p | trans | 2.12E-17 | Nightingale | Citrate | NA | T | A | 0.026851 | 0.004396 | 6.90E-10 |
| rs676457 | miR-195-5p | trans | 2.12E-17 | Nightingale | M_LDL_C | NA | T | A | 0.033939 | 0.004433 | 2.30E-14 |
| rs676457 | miR-195-5p | trans | 2.12E-17 | Nightingale | XS_VLDL_L | NA | T | A | 0.033833 | 0.00435 | 2.00E-15 |
| rs676457 | miR-195-5p | trans | 2.12E-17 | Nightingale | S_VLDL_FC | NA | T | A | 0.037867 | 0.004426 | 7.60E-18 |
| rs676457 | miR-195-5p | trans | 2.12E-17 | Nightingale | M_LDL_PL | NA | T | A | 0.032419 | 0.004431 | 2.80E-13 |
| rs676457 | miR-195-5p | trans | 2.12E-17 | Nightingale | VLDL_FC | NA | T | A | 0.024128 | 0.004392 | 1.10E-08 |
| rs676457 | miR-195-5p | trans | 2.12E-17 | Nightingale | M_VLDL_F | NA | T | A | 0.033335 | 0.004424 | 3.20E-14 |
| rs676457 | miR-195-5p | trans | 2.12E-17 | Nightingale | VLDL_C | NA | T | A | 0.031272 | 0.004419 | 5.00E-13 |
| rs676457 | miR-195-5p | trans | 2.12E-17 | Nightingale | M_VLDL_C | NA | T | A | 0.040809 | 0.004385 | 2.40E-20 |
| rs676457 | miR-195-5p | trans | 2.12E-17 | Nightingale | IDL_P | NA | T | A | 0.040008 | 0.004378 | 1.30E-19 |
| rs676457 | miR-195-5p | trans | 2.12E-17 | Nightingale | LA | NA | T | A | 0.024934 | 0.004339 | 3.60E-09 |
| rs676457 | miR-195-5p | trans | 2.12E-17 | Nightingale | VLDL_CE | NA | T | A | 0.035758 | 0.004426 | 3.10E-16 |
| rs676457 | miR-195-5p | trans | 2.12E-17 | Nightingale | S_LDL_FC | NA | T | A | 0.028528 | 0.004418 | 4.40E-10 |
| rs676457 | miR-195-5p | trans | 2.12E-17 | Nightingale | S_VLDL_CE | NA | T | A | 0.04697 | 0.004365 | 1.40E-26 |
| rs676457 | miR-195-5p | trans | 2.12E-17 | Nightingale | S_VLDL_P | NA | T | A | 0.023453 | 0.004369 | 1.70E-08 |
| rs676457 | miR-195-5p | trans | 2.12E-17 | Nightingale | XXL_VLDL_ | NA | T | A | 0.033069 | 0.004459 | 9.60E-14 |
| rs676457 | miR-195-5p | trans | 2.12E-17 | Nightingale | L_LDL_C | NA | T | A | 0.032277 | 0.004375 | 2.70E-13 |
| rs676457 | miR-195-5p | trans | 2.12E-17 | Nightingale | XL_VLDL_F | NA | T | A | 0.037129 | 0.00429 | 2.30E-18 |
| rs676457 | miR-195-5p | trans | 2.12E-17 | Nightingale | S_LDL_CE_ | NA | T | A | 0.030893 | 0.004417 | 5.50E-14 |
| rs676457 | miR-195-5p | trans | 2.12E-17 | Nightingale | XS_VLDL_F | NA | T | A | 0.031376 | 0.004393 | 2.60E-12 |
| rs676996 | let-7c-5p | trans | 4.76E-23 | Nightingale | Remnant_( | NA | G | T | 0.036852 | 0.004396 | 3.70E-17 |
| rs676996 | let-7c-5p | trans | 4.76E-23 | Nightingale | ApoB | NA | G | T | 0.033901 | 0.004419 | 2.20E-14 |
| rs676996 | let-7c-5p | trans | 4.76E-23 | Nightingale | S_LDL_P | NA | G | T | 0.029548 | 0.004429 | 3.30E-11 |
| rs676996 | let-7c-5p | trans | 4.76E-23 | Nightingale | S_HDL_PL | NA | G | T | 0.023785 | 0.004387 | 3.30E-08 |
| rs676996 | let-7c-5p | trans | 4.76E-23 | Nightingale | XL_VLDL_P | NA | G | T | 0.038714 | 0.004432 | 6.80E-19 |
| rs676996 | let-7c-5p | trans | 4.76E-23 | Nightingale | Total_CE | NA | G | T | 0.031735 | 0.004268 | 9.90E-14 |
| rs676996 | let-7c-5p | trans | 4.76E-23 | Nightingale | ApoB_by_A | NA | G | T | 0.02498 | 0.004358 | 1.40E-08 |
| rs676996 | let-7c-5p | trans | 4.76E-23 | Nightingale | M_LDL_PL | NA | G | T | 0.032421 | 0.004431 | 2.80E-13 |
| rs676996 | let-7c-5p | trans | 4.76E-23 | Nightingale | S_LDL_PL_ | NA | G | T | -0.03004 | 0.004417 | 2.80E-13 |
| rs676996 | let-7c-5p | trans | 4.76E-23 | Nightingale | M_VLDL_P | NA | G | T | 0.028258 | 0.004414 | 6.50E-11 |
| rs676996 | let-7c-5p | trans | 4.76E-23 | Nightingale | Citrate | NA | G | T | 0.02685 | 0.004396 | 6.90E-10 |
| rs676996 | let-7c-5p | trans | 4.76E-23 | Nightingale | S_LDL_C_p | NA | G | T | 0.045304 | 0.004442 | 6.40E-25 |
| rs676996 | let-7c-5p | trans | 4.76E-23 | Nightingale | L_LDL_FC | NA | G | T | 0.033595 | 0.004345 | 3.00E-14 |
| rs676996 | let-7c-5p | trans | 4.76E-23 | Nightingale | IDL_CE | NA | G | T | 0.03352 | 0.004272 | 1.30E-14 |
| rs676996 | let-7c-5p | trans | 4.76E-23 | Nightingale | M_VLDL_C | NA | G | T | 0.033575 | 0.004157 | 2.10E-15 |
| rs676996 | let-7c-5p | trans | 4.76E-23 | Nightingale | S_VLDL_TG | NA | G | T | -0.03807 | 0.004283 | 1.60E-18 |
| rs676996 | let-7c-5p | trans | 4.76E-23 | Nightingale | Sphingomy | NA | G | T | 0.028974 | 0.004137 | 7.40E-12 |
| rs676996 | let-7c-5p | trans | 4.76E-23 | Nightingale | IDL_FC_pct | NA | G | T | 0.025703 | 0.004391 | 1.20E-08 |
| rs676996 | let-7c-5p | trans | 4.76E-23 | Nightingale | S_VLDL_L | NA | G | T | 0.024651 | 0.004382 | 3.80E-09 |
| rs676996 | let-7c-5p | trans | 4.76E-23 | Nightingale | XXL_VLDL_ | NA | G | T | -0.03564 | 0.004463 | 2.50E-16 |
| rs676996 | let-7c-5p | trans | 4.76E-23 | Nightingale | M_VLDL_F | NA | G | T | 0.041995 | 0.00421 | 4.10E-23 |
| rs676996 | let-7c-5p | trans | 4.76E-23 | Nightingale | Omega_6 | NA | G | T | 0.025871 | 0.004321 | 6.00E-10 |
| rs676996 | let-7c-5p | trans | 4.76E-23 | Nightingale | XL_VLDL_T | NA | G | T | -0.04377 | 0.004289 | 2.50E-24 |
| rs676996 | let-7c-5p | trans | 4.76E-23 | Nightingale | M_VLDL_F | NA | G | T | 0.033337 | 0.004424 | 3.20E-14 |
| rs676996 | let-7c-5p | trans | 4.76E-23 | Nightingale | XXL_VLDL_ | NA | G | T | 0.030734 | 0.004406 | 1.30E-12 |
| rs676996 | let-7c-5p | trans | 4.76E-23 | Nightingale | M_VLDL_P | NA | G | T | 0.029098 | 0.004418 | 2.10E-11 |
| rs676996 | let-7c-5p | trans | 4.76E-23 | Nightingale | M_LDL_TG | NA | G | T | -0.02753 | 0.00437 | 3.60E-10 |
| rs676996 | let-7c-5p | trans | 4.76E-23 | Nightingale | S_LDL_FC | NA | G | T | 0.02853 | 0.004418 | 4.40E-10 |
| rs676996 | let-7c-5p | trans | 4.76E-23 | Nightingale | XS_VLDL_L | NA | G | T | 0.033835 | 0.00435 | 2.00E-15 |
| rs676996 | let-7c-5p | trans | 4.76E-23 | Nightingale | M_LDL_L | NA | G | T | 0.032985 | 0.004431 | 1.10E-13 |
| rs676996 | let-7c-5p | trans | 4.76E-23 | Nightingale | LDL_PL | NA | G | T | 0.03495 | 0.004415 | 4.90E-15 |
| rs676996 | let-7c-5p | trans | 4.76E-23 | Nightingale | XS_VLDL_P | NA | G | T | 0.032127 | 0.004363 | 5.20E-14 |
| rs676996 | let-7c-5p | trans | 4.76E-23 | Nightingale | XS_VLDL_P | NA | G | T | 0.036717 | 0.004366 | 1.30E-17 |
| rs676996 | let-7c-5p | trans | 4.76E-23 | Nightingale | M_VLDL_C | NA | G | T | 0.04081 | 0.004385 | 2.40E-20 |
| rs676996 | let-7c-5p | trans | 4.76E-23 | Nightingale | XS_VLDL_C | NA | G | T | 0.038004 | 0.004295 | 1.30E-18 |
| rs676996 | let-7c-5p | trans | 4.76E-23 | Nightingale | S_VLDL_CE | NA | G | T | 0.038157 | 0.004419 | 1.60E-18 |

| | | | | | | | | | | | |
|---|---|---|---|---|---|---|---|---|---|---|---|
| rs676996 | let-7c-5p | trans | 4.76E-23 | Nightingale | Total_FC | NA | G | T | 0.033248 | 0.004323 | 6.90E-15 |
| rs676996 | let-7c-5p | trans | 4.76E-23 | Nightingale | LDL_C | NA | G | T | 0.03315 | 0.004406 | 9.30E-14 |
| rs676996 | let-7c-5p | trans | 4.76E-23 | Nightingale | S_LDL_C | NA | G | T | 0.030897 | 0.004429 | 5.80E-12 |
| rs676996 | let-7c-5p | trans | 4.76E-23 | Nightingale | L_LDL_PL | NA | G | T | 0.037957 | 0.004391 | 9.50E-18 |
| rs676996 | let-7c-5p | trans | 4.76E-23 | Nightingale | XXL_VLDL_ | NA | G | T | 0.034743 | 0.004434 | 3.30E-15 |
| rs676996 | let-7c-5p | trans | 4.76E-23 | Nightingale | non_HDL_( | NA | G | T | 0.035665 | 0.004401 | 5.30E-16 |
| rs676996 | let-7c-5p | trans | 4.76E-23 | Nightingale | S_LDL_CE_ | NA | G | T | 0.030893 | 0.004417 | 5.50E-14 |
| rs676996 | let-7c-5p | trans | 4.76E-23 | Nightingale | XXL_VLDL_ | NA | G | T | 0.025017 | 0.004454 | 1.50E-09 |
| rs676996 | let-7c-5p | trans | 4.76E-23 | Nightingale | S_LDL_L | NA | G | T | 0.027698 | 0.004424 | 8.50E-10 |
| rs676996 | let-7c-5p | trans | 4.76E-23 | Nightingale | S_VLDL_C_ | NA | G | T | 0.041827 | 0.004306 | 5.90E-22 |
| rs676996 | let-7c-5p | trans | 4.76E-23 | Nightingale | XS_VLDL_C | NA | G | T | 0.037811 | 0.00427 | 2.10E-18 |
| rs676996 | let-7c-5p | trans | 4.76E-23 | Nightingale | M_LDL_CE | NA | G | T | 0.032924 | 0.004431 | 1.00E-13 |
| rs676996 | let-7c-5p | trans | 4.76E-23 | Nightingale | XL_VLDL_C | NA | G | T | 0.037265 | 0.004245 | 3.70E-18 |
| rs676996 | let-7c-5p | trans | 4.76E-23 | Nightingale | XL_VLDL_C | NA | G | T | 0.038142 | 0.004243 | 5.40E-19 |
| rs676996 | let-7c-5p | trans | 4.76E-23 | Nightingale | S_HDL_P | NA | G | T | 0.030579 | 0.004425 | 5.40E-12 |
| rs676996 | let-7c-5p | trans | 4.76E-23 | Nightingale | XL_VLDL_F | NA | G | T | 0.037128 | 0.00429 | 2.30E-18 |
| rs676996 | let-7c-5p | trans | 4.76E-23 | Nightingale | S_HDL_C | NA | G | T | 0.028387 | 0.004417 | 1.80E-10 |
| rs676996 | let-7c-5p | trans | 4.76E-23 | Nightingale | M_VLDL_T | NA | G | T | -0.03849 | 0.004181 | 7.90E-20 |
| rs676996 | let-7c-5p | trans | 4.76E-23 | Nightingale | Total_C | NA | G | T | 0.032363 | 0.004284 | 3.40E-14 |
| rs676996 | let-7c-5p | trans | 4.76E-23 | Nightingale | GlycA | NA | G | T | 0.027865 | 0.004406 | 1.80E-10 |
| rs676996 | let-7c-5p | trans | 4.76E-23 | Nightingale | S_VLDL_FC | NA | G | T | 0.037868 | 0.004426 | 7.60E-18 |
| rs676996 | let-7c-5p | trans | 4.76E-23 | Nightingale | VLDL_FC | NA | G | T | 0.02413 | 0.004392 | 1.10E-08 |
| rs676996 | let-7c-5p | trans | 4.76E-23 | Nightingale | S_HDL_L | NA | G | T | 0.026288 | 0.00441 | 1.60E-09 |
| rs676996 | let-7c-5p | trans | 4.76E-23 | Nightingale | XL_HDL_PL | NA | G | T | -0.02267 | 0.004065 | 1.30E-08 |
| rs676996 | let-7c-5p | trans | 4.76E-23 | Nightingale | S_LDL_CE | NA | G | T | 0.030499 | 0.004427 | 6.10E-12 |
| rs676996 | let-7c-5p | trans | 4.76E-23 | Nightingale | XS_VLDL_F | NA | G | T | 0.037076 | 0.004348 | 6.40E-18 |
| rs676996 | let-7c-5p | trans | 4.76E-23 | Nightingale | L_LDL_CE | NA | G | T | 0.031522 | 0.004385 | 1.00E-12 |
| rs676996 | let-7c-5p | trans | 4.76E-23 | Nightingale | VLDL_CE | NA | G | T | 0.03576 | 0.004426 | 3.10E-16 |
| rs676996 | let-7c-5p | trans | 4.76E-23 | Nightingale | LDL_P | NA | G | T | 0.031745 | 0.004422 | 1.10E-12 |
| rs676996 | let-7c-5p | trans | 4.76E-23 | Nightingale | Total_L | NA | G | T | 0.026348 | 0.004353 | 4.60E-10 |
| rs676996 | let-7c-5p | trans | 4.76E-23 | Nightingale | LDL_L | NA | G | T | 0.033262 | 0.00441 | 7.80E-14 |
| rs676996 | let-7c-5p | trans | 4.76E-23 | Nightingale | M_LDL_P | NA | G | T | 0.035848 | 0.004437 | 7.30E-16 |
| rs676996 | let-7c-5p | trans | 4.76E-23 | Nightingale | XS_VLDL_F | NA | G | T | 0.031377 | 0.004393 | 2.60E-12 |
| rs676996 | let-7c-5p | trans | 4.76E-23 | Nightingale | IDL_PL | NA | G | T | 0.035327 | 0.00427 | 3.50E-16 |
| rs676996 | let-7c-5p | trans | 4.76E-23 | Nightingale | L_LDL_P | NA | G | T | 0.029116 | 0.00441 | 6.60E-11 |
| rs676996 | let-7c-5p | trans | 4.76E-23 | Nightingale | S_HDL_CE | NA | G | T | 0.028597 | 0.004432 | 1.40E-10 |
| rs676996 | let-7c-5p | trans | 4.76E-23 | Nightingale | IDL_P | NA | G | T | 0.040009 | 0.004378 | 1.30E-19 |
| rs676996 | let-7c-5p | trans | 4.76E-23 | Nightingale | IDL_C | NA | G | T | 0.034962 | 0.004274 | 9.40E-16 |
| rs676996 | let-7c-5p | trans | 4.76E-23 | Nightingale | LDL_FC | NA | G | T | 0.033597 | 0.004378 | 5.50E-14 |
| rs676996 | let-7c-5p | trans | 4.76E-23 | Nightingale | VLDL_C | NA | G | T | 0.031274 | 0.004419 | 5.00E-13 |
| rs676996 | let-7c-5p | trans | 4.76E-23 | Nightingale | L_LDL_C | NA | G | T | 0.032279 | 0.004375 | 2.70E-13 |
| rs676996 | let-7c-5p | trans | 4.76E-23 | Nightingale | S_VLDL_PL | NA | G | T | 0.032747 | 0.004417 | 4.30E-14 |
| rs676996 | let-7c-5p | trans | 4.76E-23 | Nightingale | M_LDL_C | NA | G | T | 0.03394 | 0.004433 | 2.30E-14 |
| rs676996 | let-7c-5p | trans | 4.76E-23 | Nightingale | Total_PL | NA | G | T | 0.024943 | 0.004214 | 1.10E-09 |
| rs676996 | let-7c-5p | trans | 4.76E-23 | Nightingale | S_VLDL_CE | NA | G | T | 0.046969 | 0.004365 | 1.40E-26 |
| rs676996 | let-7c-5p | trans | 4.76E-23 | Nightingale | S_VLDL_FC | NA | G | T | 0.029304 | 0.004228 | 1.10E-11 |
| rs676996 | let-7c-5p | trans | 4.76E-23 | Nightingale | M_LDL_FC | NA | G | T | 0.034304 | 0.004419 | 2.00E-14 |
| rs676996 | let-7c-5p | trans | 4.76E-23 | Nightingale | M_LDL_C_ | NA | G | T | 0.031099 | 0.004428 | 3.30E-12 |
| rs676996 | let-7c-5p | trans | 4.76E-23 | Nightingale | M_VLDL_P | NA | G | T | 0.043976 | 0.004247 | 6.90E-25 |
| rs676996 | let-7c-5p | trans | 4.76E-23 | Nightingale | Clinical_LD | NA | G | T | 0.036436 | 0.004399 | 2.90E-16 |
| rs676996 | let-7c-5p | trans | 4.76E-23 | Nightingale | M_VLDL_C | NA | G | T | 0.038672 | 0.004413 | 2.00E-18 |
| rs676996 | let-7c-5p | trans | 4.76E-23 | Nightingale | IDL_L | NA | G | T | 0.035274 | 0.004278 | 4.90E-16 |
| rs676996 | let-7c-5p | trans | 4.76E-23 | Nightingale | IDL_FC | NA | G | T | 0.038246 | 0.004284 | 1.50E-18 |
| rs676996 | let-7c-5p | trans | 4.76E-23 | Nightingale | S_VLDL_C | NA | G | T | 0.038343 | 0.004424 | 1.80E-18 |
| rs676996 | let-7c-5p | trans | 4.76E-23 | Nightingale | XXL_VLDL_ | NA | G | T | 0.033067 | 0.004459 | 9.60E-14 |
| rs676996 | let-7c-5p | trans | 4.76E-23 | Nightingale | Cholines | NA | G | T | 0.022308 | 0.004161 | 4.90E-08 |
| rs676996 | let-7c-5p | trans | 4.76E-23 | Nightingale | S_VLDL_PL | NA | G | T | 0.027677 | 0.004237 | 1.90E-10 |
| rs676996 | let-7c-5p | trans | 4.76E-23 | Nightingale | LDL_CE | NA | G | T | 0.032506 | 0.004413 | 2.30E-13 |
| rs676996 | let-7c-5p | trans | 4.76E-23 | Nightingale | L_LDL_L | NA | G | T | 0.033512 | 0.004382 | 3.20E-14 |
| rs676996 | let-7c-5p | trans | 4.76E-23 | Nightingale | L_VLDL_CE | NA | G | T | 0.047288 | 0.004234 | 2.80E-29 |
| rs676996 | let-7c-5p | trans | 4.76E-23 | Nightingale | L_VLDL_C_ | NA | G | T | 0.052087 | 0.004322 | 5.50E-34 |
| rs676996 | let-7c-5p | trans | 4.76E-23 | Nightingale | LA | NA | G | T | 0.024935 | 0.004339 | 3.50E-09 |
| rs676996 | let-7c-5p | trans | 4.76E-23 | Nightingale | S_VLDL_P | NA | G | T | 0.023454 | 0.004369 | 1.70E-08 |

| | | | | | | | | | | | |
|---|---|---|---|---|---|---|---|---|---|---|---|
| rs676996 | let-7c-5p | trans | 4.76E-23 | Nightingale | VLDL_P | NA | G | T | 0.02662 | 0.004401 | 4.40E-10 |
| rs676996 | let-7c-5p | trans | 4.76E-23 | Nightingale | PUFA | NA | G | T | 0.023693 | 0.004316 | 1.50E-08 |
| rs676996 | let-7c-5p | trans | 4.76E-23 | Nightingale | L_VLDL_FC | NA | G | T | 0.048434 | 0.004382 | 6.70E-29 |
| rs676996 | let-7c-5p | trans | 4.76E-23 | Nightingale | S_HDL_FC | NA | G | T | 0.023471 | 0.004344 | 4.60E-08 |
| rs676996 | let-7c-5p | trans | 4.76E-23 | Nightingale | M_VLDL_C | NA | G | T | 0.036147 | 0.004168 | 1.30E-17 |
| rs676996 | let-7c-5p | trans | 4.76E-23 | Nightingale | L_VLDL_TG | NA | G | T | -0.05103 | 0.004404 | 7.30E-32 |
| rs676996 | let-7b-5p | trans | 1.96E-26 | Nightingale | Remnant_( | NA | G | T | 0.036852 | 0.004396 | 3.70E-17 |
| rs676996 | let-7b-5p | trans | 1.96E-26 | Nightingale | ApoB | NA | G | T | 0.033901 | 0.004419 | 2.20E-14 |
| rs676996 | let-7b-5p | trans | 1.96E-26 | Nightingale | S_LDL_P | NA | G | T | 0.029548 | 0.004429 | 3.30E-11 |
| rs676996 | let-7b-5p | trans | 1.96E-26 | Nightingale | S_HDL_PL | NA | G | T | 0.023785 | 0.004387 | 3.30E-08 |
| rs676996 | let-7b-5p | trans | 1.96E-26 | Nightingale | XL_VLDL_P | NA | G | T | 0.038714 | 0.004432 | 6.80E-19 |
| rs676996 | let-7b-5p | trans | 1.96E-26 | Nightingale | Total_CE | NA | G | T | 0.031735 | 0.004268 | 9.90E-14 |
| rs676996 | let-7b-5p | trans | 1.96E-26 | Nightingale | ApoB_by_A | NA | G | T | 0.02498 | 0.004358 | 1.40E-08 |
| rs676996 | let-7b-5p | trans | 1.96E-26 | Nightingale | M_LDL_PL | NA | G | T | 0.032421 | 0.004431 | 2.80E-13 |
| rs676996 | let-7b-5p | trans | 1.96E-26 | Nightingale | S_LDL_PL_ | NA | G | T | -0.03004 | 0.004417 | 2.80E-13 |
| rs676996 | let-7b-5p | trans | 1.96E-26 | Nightingale | M_VLDL_P | NA | G | T | 0.028258 | 0.004414 | 6.50E-11 |
| rs676996 | let-7b-5p | trans | 1.96E-26 | Nightingale | Citrate | NA | G | T | 0.02685 | 0.004396 | 6.90E-10 |
| rs676996 | let-7b-5p | trans | 1.96E-26 | Nightingale | S_LDL_C_p | NA | G | T | 0.045304 | 0.004442 | 6.40E-25 |
| rs676996 | let-7b-5p | trans | 1.96E-26 | Nightingale | L_LDL_FC | NA | G | T | 0.033595 | 0.004345 | 3.00E-14 |
| rs676996 | let-7b-5p | trans | 1.96E-26 | Nightingale | IDL_CE | NA | G | T | 0.03352 | 0.004272 | 1.30E-14 |
| rs676996 | let-7b-5p | trans | 1.96E-26 | Nightingale | M_VLDL_C | NA | G | T | 0.033575 | 0.004157 | 2.10E-15 |
| rs676996 | let-7b-5p | trans | 1.96E-26 | Nightingale | S_VLDL_TG | NA | G | T | -0.03807 | 0.004283 | 1.60E-18 |
| rs676996 | let-7b-5p | trans | 1.96E-26 | Nightingale | Sphingomy | NA | G | T | 0.028974 | 0.004137 | 7.40E-12 |
| rs676996 | let-7b-5p | trans | 1.96E-26 | Nightingale | IDL_FC_pct | NA | G | T | 0.025703 | 0.004391 | 1.20E-08 |
| rs676996 | let-7b-5p | trans | 1.96E-26 | Nightingale | S_VLDL_L | NA | G | T | 0.024651 | 0.004382 | 3.80E-09 |
| rs676996 | let-7b-5p | trans | 1.96E-26 | Nightingale | XXL_VLDL_ | NA | G | T | -0.03564 | 0.004463 | 2.50E-16 |
| rs676996 | let-7b-5p | trans | 1.96E-26 | Nightingale | M_VLDL_F( | NA | G | T | 0.041995 | 0.00421 | 4.10E-23 |
| rs676996 | let-7b-5p | trans | 1.96E-26 | Nightingale | Omega_6 | NA | G | T | 0.025871 | 0.004321 | 6.00E-10 |
| rs676996 | let-7b-5p | trans | 1.96E-26 | Nightingale | XL_VLDL_T | NA | G | T | -0.04377 | 0.004289 | 2.50E-24 |
| rs676996 | let-7b-5p | trans | 1.96E-26 | Nightingale | M_VLDL_F( | NA | G | T | 0.033337 | 0.004424 | 3.20E-14 |
| rs676996 | let-7b-5p | trans | 1.96E-26 | Nightingale | XXL_VLDL_ | NA | G | T | 0.030734 | 0.004406 | 1.30E-12 |
| rs676996 | let-7b-5p | trans | 1.96E-26 | Nightingale | M_VLDL_P | NA | G | T | 0.029098 | 0.004418 | 2.10E-11 |
| rs676996 | let-7b-5p | trans | 1.96E-26 | Nightingale | M_LDL_TG | NA | G | T | -0.02753 | 0.00437 | 3.60E-10 |
| rs676996 | let-7b-5p | trans | 1.96E-26 | Nightingale | S_LDL_FC | NA | G | T | 0.02853 | 0.004418 | 4.40E-10 |
| rs676996 | let-7b-5p | trans | 1.96E-26 | Nightingale | XS_VLDL_L | NA | G | T | 0.033835 | 0.00435 | 2.00E-15 |
| rs676996 | let-7b-5p | trans | 1.96E-26 | Nightingale | M_LDL_L | NA | G | T | 0.032985 | 0.004431 | 1.10E-13 |
| rs676996 | let-7b-5p | trans | 1.96E-26 | Nightingale | LDL_PL | NA | G | T | 0.03495 | 0.004415 | 4.90E-15 |
| rs676996 | let-7b-5p | trans | 1.96E-26 | Nightingale | XS_VLDL_P | NA | G | T | 0.032127 | 0.004363 | 5.20E-14 |
| rs676996 | let-7b-5p | trans | 1.96E-26 | Nightingale | XS_VLDL_P | NA | G | T | 0.036717 | 0.004366 | 1.30E-17 |
| rs676996 | let-7b-5p | trans | 1.96E-26 | Nightingale | M_VLDL_C | NA | G | T | 0.04081 | 0.004385 | 2.40E-20 |
| rs676996 | let-7b-5p | trans | 1.96E-26 | Nightingale | XS_VLDL_C | NA | G | T | 0.038004 | 0.004295 | 1.30E-18 |
| rs676996 | let-7b-5p | trans | 1.96E-26 | Nightingale | S_VLDL_CE | NA | G | T | 0.038157 | 0.004419 | 1.60E-18 |
| rs676996 | let-7b-5p | trans | 1.96E-26 | Nightingale | Total_FC | NA | G | T | 0.033248 | 0.004323 | 6.90E-15 |
| rs676996 | let-7b-5p | trans | 1.96E-26 | Nightingale | LDL_C | NA | G | T | 0.03315 | 0.004406 | 9.30E-14 |
| rs676996 | let-7b-5p | trans | 1.96E-26 | Nightingale | S_LDL_C | NA | G | T | 0.030897 | 0.004429 | 5.80E-12 |
| rs676996 | let-7b-5p | trans | 1.96E-26 | Nightingale | L_LDL_PL | NA | G | T | 0.037957 | 0.004391 | 9.50E-18 |
| rs676996 | let-7b-5p | trans | 1.96E-26 | Nightingale | XXL_VLDL_ | NA | G | T | 0.034743 | 0.004434 | 3.30E-15 |
| rs676996 | let-7b-5p | trans | 1.96E-26 | Nightingale | non_HDL_( | NA | G | T | 0.035665 | 0.004401 | 5.30E-16 |
| rs676996 | let-7b-5p | trans | 1.96E-26 | Nightingale | S_LDL_CE_ | NA | G | T | 0.030893 | 0.004417 | 5.50E-14 |
| rs676996 | let-7b-5p | trans | 1.96E-26 | Nightingale | XXL_VLDL_ | NA | G | T | 0.025017 | 0.004454 | 1.50E-09 |
| rs676996 | let-7b-5p | trans | 1.96E-26 | Nightingale | S_LDL_L | NA | G | T | 0.027698 | 0.004424 | 8.50E-10 |
| rs676996 | let-7b-5p | trans | 1.96E-26 | Nightingale | S_VLDL_C_ | NA | G | T | 0.041827 | 0.004306 | 5.90E-22 |
| rs676996 | let-7b-5p | trans | 1.96E-26 | Nightingale | XS_VLDL_C | NA | G | T | 0.037811 | 0.00427 | 2.10E-18 |
| rs676996 | let-7b-5p | trans | 1.96E-26 | Nightingale | M_LDL_CE | NA | G | T | 0.032924 | 0.004431 | 1.00E-13 |
| rs676996 | let-7b-5p | trans | 1.96E-26 | Nightingale | XL_VLDL_C | NA | G | T | 0.037265 | 0.004245 | 3.70E-18 |
| rs676996 | let-7b-5p | trans | 1.96E-26 | Nightingale | XL_VLDL_C | NA | G | T | 0.038142 | 0.004243 | 5.40E-19 |
| rs676996 | let-7b-5p | trans | 1.96E-26 | Nightingale | S_HDL_P | NA | G | T | 0.030579 | 0.004425 | 5.40E-12 |
| rs676996 | let-7b-5p | trans | 1.96E-26 | Nightingale | XL_VLDL_F | NA | G | T | 0.037128 | 0.00429 | 2.30E-18 |
| rs676996 | let-7b-5p | trans | 1.96E-26 | Nightingale | S_HDL_C | NA | G | T | 0.028387 | 0.004417 | 1.80E-10 |
| rs676996 | let-7b-5p | trans | 1.96E-26 | Nightingale | M_VLDL_T( | NA | G | T | -0.03849 | 0.004181 | 7.90E-20 |
| rs676996 | let-7b-5p | trans | 1.96E-26 | Nightingale | Total_C | NA | G | T | 0.032363 | 0.004284 | 3.40E-14 |
| rs676996 | let-7b-5p | trans | 1.96E-26 | Nightingale | GlycA | NA | G | T | 0.027865 | 0.004406 | 1.80E-10 |
| rs676996 | let-7b-5p | trans | 1.96E-26 | Nightingale | S_VLDL_FC | NA | G | T | 0.037868 | 0.004426 | 7.60E-18 |

| | | | | | | | | | | | |
|---|---|---|---|---|---|---|---|---|---|---|---|
| rs676996 | let-7b-5p | trans | 1.96E-26 | Nightingale | VLDL_FC | NA | G | T | 0.02413 | 0.004392 | 1.10E-08 |
| rs676996 | let-7b-5p | trans | 1.96E-26 | Nightingale | S_HDL_L | NA | G | T | 0.026288 | 0.00441 | 1.60E-09 |
| rs676996 | let-7b-5p | trans | 1.96E-26 | Nightingale | XL_HDL_PL | NA | G | T | -0.02267 | 0.004065 | 1.30E-08 |
| rs676996 | let-7b-5p | trans | 1.96E-26 | Nightingale | S_LDL_CE | NA | G | T | 0.030499 | 0.004427 | 6.10E-12 |
| rs676996 | let-7b-5p | trans | 1.96E-26 | Nightingale | XS_VLDL_F | NA | G | T | 0.037076 | 0.004348 | 6.40E-18 |
| rs676996 | let-7b-5p | trans | 1.96E-26 | Nightingale | L_LDL_CE | NA | G | T | 0.031522 | 0.004385 | 1.00E-12 |
| rs676996 | let-7b-5p | trans | 1.96E-26 | Nightingale | VLDL_CE | NA | G | T | 0.03576 | 0.004426 | 3.10E-16 |
| rs676996 | let-7b-5p | trans | 1.96E-26 | Nightingale | LDL_P | NA | G | T | 0.031745 | 0.004422 | 1.10E-12 |
| rs676996 | let-7b-5p | trans | 1.96E-26 | Nightingale | Total_L | NA | G | T | 0.026348 | 0.004353 | 4.60E-10 |
| rs676996 | let-7b-5p | trans | 1.96E-26 | Nightingale | LDL_L | NA | G | T | 0.033262 | 0.00441 | 7.80E-14 |
| rs676996 | let-7b-5p | trans | 1.96E-26 | Nightingale | M_LDL_P | NA | G | T | 0.035848 | 0.004437 | 7.30E-16 |
| rs676996 | let-7b-5p | trans | 1.96E-26 | Nightingale | XS_VLDL_F | NA | G | T | 0.031377 | 0.004393 | 2.60E-12 |
| rs676996 | let-7b-5p | trans | 1.96E-26 | Nightingale | IDL_PL | NA | G | T | 0.035327 | 0.00427 | 3.50E-16 |
| rs676996 | let-7b-5p | trans | 1.96E-26 | Nightingale | L_LDL_P | NA | G | T | 0.029116 | 0.00441 | 6.60E-11 |
| rs676996 | let-7b-5p | trans | 1.96E-26 | Nightingale | S_HDL_CE | NA | G | T | 0.028597 | 0.004432 | 1.40E-10 |
| rs676996 | let-7b-5p | trans | 1.96E-26 | Nightingale | IDL_P | NA | G | T | 0.040009 | 0.004378 | 1.30E-19 |
| rs676996 | let-7b-5p | trans | 1.96E-26 | Nightingale | IDL_C | NA | G | T | 0.034962 | 0.004274 | 9.40E-16 |
| rs676996 | let-7b-5p | trans | 1.96E-26 | Nightingale | LDL_FC | NA | G | T | 0.033597 | 0.004378 | 5.50E-14 |
| rs676996 | let-7b-5p | trans | 1.96E-26 | Nightingale | VLDL_C | NA | G | T | 0.031274 | 0.004419 | 5.00E-13 |
| rs676996 | let-7b-5p | trans | 1.96E-26 | Nightingale | L_LDL_C | NA | G | T | 0.032279 | 0.004375 | 2.70E-13 |
| rs676996 | let-7b-5p | trans | 1.96E-26 | Nightingale | S_VLDL_PL | NA | G | T | 0.032747 | 0.004417 | 4.30E-14 |
| rs676996 | let-7b-5p | trans | 1.96E-26 | Nightingale | M_LDL_C | NA | G | T | 0.03394 | 0.004433 | 2.30E-14 |
| rs676996 | let-7b-5p | trans | 1.96E-26 | Nightingale | Total_PL | NA | G | T | 0.024943 | 0.004214 | 1.10E-09 |
| rs676996 | let-7b-5p | trans | 1.96E-26 | Nightingale | S_VLDL_CE | NA | G | T | 0.046969 | 0.004365 | 1.40E-26 |
| rs676996 | let-7b-5p | trans | 1.96E-26 | Nightingale | S_VLDL_FC | NA | G | T | 0.029304 | 0.004228 | 1.10E-11 |
| rs676996 | let-7b-5p | trans | 1.96E-26 | Nightingale | M_LDL_FC | NA | G | T | 0.034304 | 0.004419 | 2.00E-14 |
| rs676996 | let-7b-5p | trans | 1.96E-26 | Nightingale | M_LDL_C_ | NA | G | T | 0.031099 | 0.004428 | 3.30E-12 |
| rs676996 | let-7b-5p | trans | 1.96E-26 | Nightingale | M_VLDL_P | NA | G | T | 0.043976 | 0.004247 | 6.90E-25 |
| rs676996 | let-7b-5p | trans | 1.96E-26 | Nightingale | Clinical_LD | NA | G | T | 0.036436 | 0.004399 | 2.90E-16 |
| rs676996 | let-7b-5p | trans | 1.96E-26 | Nightingale | M_VLDL_C | NA | G | T | 0.038672 | 0.004413 | 2.00E-18 |
| rs676996 | let-7b-5p | trans | 1.96E-26 | Nightingale | IDL_L | NA | G | T | 0.035274 | 0.004278 | 4.90E-16 |
| rs676996 | let-7b-5p | trans | 1.96E-26 | Nightingale | IDL_FC | NA | G | T | 0.038246 | 0.004284 | 1.50E-18 |
| rs676996 | let-7b-5p | trans | 1.96E-26 | Nightingale | S_VLDL_C | NA | G | T | 0.038343 | 0.004424 | 1.80E-18 |
| rs676996 | let-7b-5p | trans | 1.96E-26 | Nightingale | XXL_VLDL_ | NA | G | T | 0.033067 | 0.004459 | 9.60E-14 |
| rs676996 | let-7b-5p | trans | 1.96E-26 | Nightingale | Cholines | NA | G | T | 0.022308 | 0.004161 | 4.90E-08 |
| rs676996 | let-7b-5p | trans | 1.96E-26 | Nightingale | S_VLDL_PL | NA | G | T | 0.027677 | 0.004237 | 1.90E-10 |
| rs676996 | let-7b-5p | trans | 1.96E-26 | Nightingale | LDL_CE | NA | G | T | 0.032506 | 0.004413 | 2.30E-13 |
| rs676996 | let-7b-5p | trans | 1.96E-26 | Nightingale | L_LDL_L | NA | G | T | 0.033512 | 0.004382 | 3.20E-14 |
| rs676996 | let-7b-5p | trans | 1.96E-26 | Nightingale | L_VLDL_CE | NA | G | T | 0.047288 | 0.004234 | 2.80E-29 |
| rs676996 | let-7b-5p | trans | 1.96E-26 | Nightingale | L_VLDL_C_ | NA | G | T | 0.052087 | 0.004322 | 5.50E-34 |
| rs676996 | let-7b-5p | trans | 1.96E-26 | Nightingale | LA | NA | G | T | 0.024935 | 0.004339 | 3.50E-09 |
| rs676996 | let-7b-5p | trans | 1.96E-26 | Nightingale | S_VLDL_P | NA | G | T | 0.023454 | 0.004369 | 1.70E-08 |
| rs676996 | let-7b-5p | trans | 1.96E-26 | Nightingale | VLDL_P | NA | G | T | 0.02662 | 0.004401 | 4.40E-10 |
| rs676996 | let-7b-5p | trans | 1.96E-26 | Nightingale | PUFA | NA | G | T | 0.023693 | 0.004316 | 1.50E-08 |
| rs676996 | let-7b-5p | trans | 1.96E-26 | Nightingale | L_VLDL_FC | NA | G | T | 0.048434 | 0.004382 | 6.70E-29 |
| rs676996 | let-7b-5p | trans | 1.96E-26 | Nightingale | S_HDL_FC | NA | G | T | 0.023471 | 0.004344 | 4.60E-08 |
| rs676996 | let-7b-5p | trans | 1.96E-26 | Nightingale | M_VLDL_C | NA | G | T | 0.036147 | 0.004168 | 1.30E-17 |
| rs676996 | let-7b-5p | trans | 1.96E-26 | Nightingale | L_VLDL_TG | NA | G | T | -0.05103 | 0.004404 | 7.30E-32 |
| rs676996 | miR-10b-5p | trans | 8.97E-35 | Nightingale | Remnant_( | NA | G | T | 0.036852 | 0.004396 | 3.70E-17 |
| rs676996 | miR-10b-5p | trans | 8.97E-35 | Nightingale | ApoB | NA | G | T | 0.033901 | 0.004419 | 2.20E-14 |
| rs676996 | miR-10b-5p | trans | 8.97E-35 | Nightingale | S_LDL_P | NA | G | T | 0.029548 | 0.004429 | 3.30E-11 |
| rs676996 | miR-10b-5p | trans | 8.97E-35 | Nightingale | S_HDL_PL | NA | G | T | 0.023785 | 0.004387 | 3.30E-08 |
| rs676996 | miR-10b-5p | trans | 8.97E-35 | Nightingale | XL_VLDL_P | NA | G | T | 0.038714 | 0.004432 | 6.80E-19 |
| rs676996 | miR-10b-5p | trans | 8.97E-35 | Nightingale | Total_CE | NA | G | T | 0.031735 | 0.004268 | 9.90E-14 |
| rs676996 | miR-10b-5p | trans | 8.97E-35 | Nightingale | ApoB_by_/ | NA | G | T | 0.02498 | 0.004358 | 1.40E-08 |
| rs676996 | miR-10b-5p | trans | 8.97E-35 | Nightingale | M_LDL_PL | NA | G | T | 0.032421 | 0.004431 | 2.80E-13 |
| rs676996 | miR-10b-5p | trans | 8.97E-35 | Nightingale | S_LDL_PL_ | NA | G | T | -0.03004 | 0.004417 | 2.80E-13 |
| rs676996 | miR-10b-5p | trans | 8.97E-35 | Nightingale | M_VLDL_P | NA | G | T | 0.028258 | 0.004414 | 6.50E-11 |
| rs676996 | miR-10b-5p | trans | 8.97E-35 | Nightingale | Citrate | NA | G | T | 0.02685 | 0.004396 | 6.90E-10 |
| rs676996 | miR-10b-5p | trans | 8.97E-35 | Nightingale | S_LDL_C_p | NA | G | T | 0.045304 | 0.004442 | 6.40E-25 |
| rs676996 | miR-10b-5p | trans | 8.97E-35 | Nightingale | L_LDL_FC | NA | G | T | 0.033595 | 0.004345 | 3.00E-14 |
| rs676996 | miR-10b-5p | trans | 8.97E-35 | Nightingale | IDL_CE | NA | G | T | 0.03352 | 0.004272 | 1.30E-14 |
| rs676996 | miR-10b-5p | trans | 8.97E-35 | Nightingale | M_VLDL_C | NA | G | T | 0.033575 | 0.004157 | 2.10E-15 |

| | | | | | | | | | | | |
|---|---|---|---|---|---|---|---|---|---|---|---|
| rs676996 | miR-10b-5p | trans | 8.97E-35 | Nightingale | S_VLDL_TG | NA | G | T | -0.03807 | 0.004283 | 1.60E-18 |
| rs676996 | miR-10b-5p | trans | 8.97E-35 | Nightingale | Sphingomy | NA | G | T | 0.028974 | 0.004137 | 7.40E-12 |
| rs676996 | miR-10b-5p | trans | 8.97E-35 | Nightingale | IDL_FC_pct | NA | G | T | 0.025703 | 0.004391 | 1.20E-08 |
| rs676996 | miR-10b-5p | trans | 8.97E-35 | Nightingale | S_VLDL_L | NA | G | T | 0.024651 | 0.004382 | 3.80E-09 |
| rs676996 | miR-10b-5p | trans | 8.97E-35 | Nightingale | XXL_VLDL_ | NA | G | T | -0.03564 | 0.004463 | 2.50E-16 |
| rs676996 | miR-10b-5p | trans | 8.97E-35 | Nightingale | M_VLDL_F | NA | G | T | 0.041995 | 0.00421 | 4.10E-23 |
| rs676996 | miR-10b-5p | trans | 8.97E-35 | Nightingale | Omega_6 | NA | G | T | 0.025871 | 0.004321 | 6.00E-10 |
| rs676996 | miR-10b-5p | trans | 8.97E-35 | Nightingale | XL_VLDL_T | NA | G | T | -0.04377 | 0.004289 | 2.50E-24 |
| rs676996 | miR-10b-5p | trans | 8.97E-35 | Nightingale | M_VLDL_F | NA | G | T | 0.033337 | 0.004424 | 3.20E-14 |
| rs676996 | miR-10b-5p | trans | 8.97E-35 | Nightingale | XXL_VLDL_ | NA | G | T | 0.030734 | 0.004406 | 1.30E-12 |
| rs676996 | miR-10b-5p | trans | 8.97E-35 | Nightingale | M_VLDL_P | NA | G | T | 0.029098 | 0.004418 | 2.10E-11 |
| rs676996 | miR-10b-5p | trans | 8.97E-35 | Nightingale | M_LDL_TG | NA | G | T | -0.02753 | 0.00437 | 3.60E-10 |
| rs676996 | miR-10b-5p | trans | 8.97E-35 | Nightingale | S_LDL_FC | NA | G | T | 0.02853 | 0.004418 | 4.40E-10 |
| rs676996 | miR-10b-5p | trans | 8.97E-35 | Nightingale | XS_VLDL_L | NA | G | T | 0.033835 | 0.00435 | 2.00E-15 |
| rs676996 | miR-10b-5p | trans | 8.97E-35 | Nightingale | M_LDL_L | NA | G | T | 0.032985 | 0.004431 | 1.10E-13 |
| rs676996 | miR-10b-5p | trans | 8.97E-35 | Nightingale | LDL_PL | NA | G | T | 0.03495 | 0.004415 | 4.90E-15 |
| rs676996 | miR-10b-5p | trans | 8.97E-35 | Nightingale | XS_VLDL_P | NA | G | T | 0.032127 | 0.004363 | 5.20E-14 |
| rs676996 | miR-10b-5p | trans | 8.97E-35 | Nightingale | XS_VLDL_P | NA | G | T | 0.036717 | 0.004366 | 1.30E-17 |
| rs676996 | miR-10b-5p | trans | 8.97E-35 | Nightingale | M_VLDL_C | NA | G | T | 0.04081 | 0.004385 | 2.40E-20 |
| rs676996 | miR-10b-5p | trans | 8.97E-35 | Nightingale | XS_VLDL_C | NA | G | T | 0.038004 | 0.004295 | 1.30E-18 |
| rs676996 | miR-10b-5p | trans | 8.97E-35 | Nightingale | S_VLDL_CE | NA | G | T | 0.038157 | 0.004419 | 1.60E-18 |
| rs676996 | miR-10b-5p | trans | 8.97E-35 | Nightingale | Total_FC | NA | G | T | 0.033248 | 0.004323 | 6.90E-15 |
| rs676996 | miR-10b-5p | trans | 8.97E-35 | Nightingale | LDL_C | NA | G | T | 0.03315 | 0.004406 | 9.30E-14 |
| rs676996 | miR-10b-5p | trans | 8.97E-35 | Nightingale | S_LDL_C | NA | G | T | 0.030897 | 0.004429 | 5.80E-12 |
| rs676996 | miR-10b-5p | trans | 8.97E-35 | Nightingale | L_LDL_PL | NA | G | T | 0.037957 | 0.004391 | 9.50E-18 |
| rs676996 | miR-10b-5p | trans | 8.97E-35 | Nightingale | XXL_VLDL_ | NA | G | T | 0.034743 | 0.004434 | 3.30E-15 |
| rs676996 | miR-10b-5p | trans | 8.97E-35 | Nightingale | non_HDL_( | NA | G | T | 0.035665 | 0.004401 | 5.30E-16 |
| rs676996 | miR-10b-5p | trans | 8.97E-35 | Nightingale | S_LDL_CE_ | NA | G | T | 0.030893 | 0.004417 | 5.50E-14 |
| rs676996 | miR-10b-5p | trans | 8.97E-35 | Nightingale | XXL_VLDL_ | NA | G | T | 0.025017 | 0.004454 | 1.50E-09 |
| rs676996 | miR-10b-5p | trans | 8.97E-35 | Nightingale | S_LDL_L | NA | G | T | 0.027698 | 0.004424 | 8.50E-10 |
| rs676996 | miR-10b-5p | trans | 8.97E-35 | Nightingale | S_VLDL_C_ | NA | G | T | 0.041827 | 0.004306 | 5.90E-22 |
| rs676996 | miR-10b-5p | trans | 8.97E-35 | Nightingale | XS_VLDL_C | NA | G | T | 0.037811 | 0.00427 | 2.10E-18 |
| rs676996 | miR-10b-5p | trans | 8.97E-35 | Nightingale | M_LDL_CE | NA | G | T | 0.032924 | 0.004431 | 1.00E-13 |
| rs676996 | miR-10b-5p | trans | 8.97E-35 | Nightingale | XL_VLDL_C | NA | G | T | 0.037265 | 0.004245 | 3.70E-18 |
| rs676996 | miR-10b-5p | trans | 8.97E-35 | Nightingale | XL_VLDL_C | NA | G | T | 0.038142 | 0.004243 | 5.40E-19 |
| rs676996 | miR-10b-5p | trans | 8.97E-35 | Nightingale | S_HDL_P | NA | G | T | 0.030579 | 0.004425 | 5.40E-12 |
| rs676996 | miR-10b-5p | trans | 8.97E-35 | Nightingale | XL_VLDL_F | NA | G | T | 0.037128 | 0.00429 | 2.30E-18 |
| rs676996 | miR-10b-5p | trans | 8.97E-35 | Nightingale | S_HDL_C | NA | G | T | 0.028387 | 0.004417 | 1.80E-10 |
| rs676996 | miR-10b-5p | trans | 8.97E-35 | Nightingale | M_VLDL_T | NA | G | T | -0.03849 | 0.004181 | 7.90E-20 |
| rs676996 | miR-10b-5p | trans | 8.97E-35 | Nightingale | Total_C | NA | G | T | 0.032363 | 0.004284 | 3.40E-14 |
| rs676996 | miR-10b-5p | trans | 8.97E-35 | Nightingale | GlycA | NA | G | T | 0.027865 | 0.004406 | 1.80E-10 |
| rs676996 | miR-10b-5p | trans | 8.97E-35 | Nightingale | S_VLDL_FC | NA | G | T | 0.037868 | 0.004426 | 7.60E-18 |
| rs676996 | miR-10b-5p | trans | 8.97E-35 | Nightingale | VLDL_FC | NA | G | T | 0.02413 | 0.004392 | 1.10E-08 |
| rs676996 | miR-10b-5p | trans | 8.97E-35 | Nightingale | S_HDL_L | NA | G | T | 0.026288 | 0.00441 | 1.60E-09 |
| rs676996 | miR-10b-5p | trans | 8.97E-35 | Nightingale | XL_HDL_PL | NA | G | T | -0.02267 | 0.004065 | 1.30E-08 |
| rs676996 | miR-10b-5p | trans | 8.97E-35 | Nightingale | S_LDL_CE | NA | G | T | 0.030499 | 0.004427 | 6.10E-12 |
| rs676996 | miR-10b-5p | trans | 8.97E-35 | Nightingale | XS_VLDL_F | NA | G | T | 0.037076 | 0.004348 | 6.40E-18 |
| rs676996 | miR-10b-5p | trans | 8.97E-35 | Nightingale | L_LDL_CE | NA | G | T | 0.031522 | 0.004385 | 1.00E-12 |
| rs676996 | miR-10b-5p | trans | 8.97E-35 | Nightingale | VLDL_CE | NA | G | T | 0.03576 | 0.004426 | 3.10E-16 |
| rs676996 | miR-10b-5p | trans | 8.97E-35 | Nightingale | LDL_P | NA | G | T | 0.031745 | 0.004422 | 1.10E-12 |
| rs676996 | miR-10b-5p | trans | 8.97E-35 | Nightingale | Total_L | NA | G | T | 0.026348 | 0.004353 | 4.60E-10 |
| rs676996 | miR-10b-5p | trans | 8.97E-35 | Nightingale | LDL_L | NA | G | T | 0.033262 | 0.00441 | 7.80E-14 |
| rs676996 | miR-10b-5p | trans | 8.97E-35 | Nightingale | M_LDL_P | NA | G | T | 0.035848 | 0.004437 | 7.30E-16 |
| rs676996 | miR-10b-5p | trans | 8.97E-35 | Nightingale | XS_VLDL_F | NA | G | T | 0.031377 | 0.004393 | 2.60E-12 |
| rs676996 | miR-10b-5p | trans | 8.97E-35 | Nightingale | IDL_PL | NA | G | T | 0.035327 | 0.00427 | 3.50E-16 |
| rs676996 | miR-10b-5p | trans | 8.97E-35 | Nightingale | L_LDL_P | NA | G | T | 0.029116 | 0.00441 | 6.60E-11 |
| rs676996 | miR-10b-5p | trans | 8.97E-35 | Nightingale | S_HDL_CE | NA | G | T | 0.028597 | 0.004432 | 1.40E-10 |
| rs676996 | miR-10b-5p | trans | 8.97E-35 | Nightingale | IDL_P | NA | G | T | 0.040009 | 0.004378 | 1.30E-19 |
| rs676996 | miR-10b-5p | trans | 8.97E-35 | Nightingale | IDL_C | NA | G | T | 0.034962 | 0.004274 | 9.40E-16 |
| rs676996 | miR-10b-5p | trans | 8.97E-35 | Nightingale | LDL_FC | NA | G | T | 0.033597 | 0.004378 | 5.50E-14 |
| rs676996 | miR-10b-5p | trans | 8.97E-35 | Nightingale | VLDL_C | NA | G | T | 0.031274 | 0.004419 | 5.00E-13 |
| rs676996 | miR-10b-5p | trans | 8.97E-35 | Nightingale | L_LDL_C | NA | G | T | 0.032279 | 0.004375 | 2.70E-13 |
| rs676996 | miR-10b-5p | trans | 8.97E-35 | Nightingale | S_VLDL_PL | NA | G | T | 0.032747 | 0.004417 | 4.30E-14 |

| | | | | | | | | | | | |
|---|---|---|---|---|---|---|---|---|---|---|---|
| rs676996 | miR-10b-5p | trans | 8.97E-35 | Nightingale | M_LDL_C | NA | G | T | 0.03394 | 0.004433 | 2.30E-14 |
| rs676996 | miR-10b-5p | trans | 8.97E-35 | Nightingale | Total_PL | NA | G | T | 0.024943 | 0.004214 | 1.10E-09 |
| rs676996 | miR-10b-5p | trans | 8.97E-35 | Nightingale | S_VLDL_CE | NA | G | T | 0.046969 | 0.004365 | 1.40E-26 |
| rs676996 | miR-10b-5p | trans | 8.97E-35 | Nightingale | S_VLDL_FC | NA | G | T | 0.029304 | 0.004228 | 1.10E-11 |
| rs676996 | miR-10b-5p | trans | 8.97E-35 | Nightingale | M_LDL_FC | NA | G | T | 0.034304 | 0.004419 | 2.00E-14 |
| rs676996 | miR-10b-5p | trans | 8.97E-35 | Nightingale | M_LDL_C_ | NA | G | T | 0.031099 | 0.004428 | 3.30E-12 |
| rs676996 | miR-10b-5p | trans | 8.97E-35 | Nightingale | M_VLDL_P | NA | G | T | 0.043976 | 0.004247 | 6.90E-25 |
| rs676996 | miR-10b-5p | trans | 8.97E-35 | Nightingale | Clinical_LD | NA | G | T | 0.036436 | 0.004399 | 2.90E-16 |
| rs676996 | miR-10b-5p | trans | 8.97E-35 | Nightingale | M_VLDL_C | NA | G | T | 0.038672 | 0.004413 | 2.00E-18 |
| rs676996 | miR-10b-5p | trans | 8.97E-35 | Nightingale | IDL_L | NA | G | T | 0.035274 | 0.004278 | 4.90E-16 |
| rs676996 | miR-10b-5p | trans | 8.97E-35 | Nightingale | IDL_FC | NA | G | T | 0.038246 | 0.004284 | 1.50E-18 |
| rs676996 | miR-10b-5p | trans | 8.97E-35 | Nightingale | S_VLDL_C | NA | G | T | 0.038343 | 0.004424 | 1.80E-18 |
| rs676996 | miR-10b-5p | trans | 8.97E-35 | Nightingale | XXL_VLDL_ | NA | G | T | 0.033067 | 0.004459 | 9.60E-14 |
| rs676996 | miR-10b-5p | trans | 8.97E-35 | Nightingale | Cholines | NA | G | T | 0.022308 | 0.004161 | 4.90E-08 |
| rs676996 | miR-10b-5p | trans | 8.97E-35 | Nightingale | S_VLDL_PL | NA | G | T | 0.027677 | 0.004237 | 1.90E-10 |
| rs676996 | miR-10b-5p | trans | 8.97E-35 | Nightingale | LDL_CE | NA | G | T | 0.032506 | 0.004413 | 2.30E-13 |
| rs676996 | miR-10b-5p | trans | 8.97E-35 | Nightingale | L_LDL_L | NA | G | T | 0.033512 | 0.004382 | 3.20E-14 |
| rs676996 | miR-10b-5p | trans | 8.97E-35 | Nightingale | L_VLDL_CE | NA | G | T | 0.047288 | 0.004234 | 2.80E-29 |
| rs676996 | miR-10b-5p | trans | 8.97E-35 | Nightingale | L_VLDL_C_ | NA | G | T | 0.052087 | 0.004322 | 5.50E-34 |
| rs676996 | miR-10b-5p | trans | 8.97E-35 | Nightingale | LA | NA | G | T | 0.024935 | 0.004339 | 3.50E-09 |
| rs676996 | miR-10b-5p | trans | 8.97E-35 | Nightingale | S_VLDL_P | NA | G | T | 0.023454 | 0.004369 | 1.70E-08 |
| rs676996 | miR-10b-5p | trans | 8.97E-35 | Nightingale | VLDL_P | NA | G | T | 0.02662 | 0.004401 | 4.40E-10 |
| rs676996 | miR-10b-5p | trans | 8.97E-35 | Nightingale | PUFA | NA | G | T | 0.023693 | 0.004316 | 1.50E-08 |
| rs676996 | miR-10b-5p | trans | 8.97E-35 | Nightingale | L_VLDL_FC | NA | G | T | 0.048434 | 0.004382 | 6.70E-29 |
| rs676996 | miR-10b-5p | trans | 8.97E-35 | Nightingale | S_HDL_FC | NA | G | T | 0.023471 | 0.004344 | 4.60E-08 |
| rs676996 | miR-10b-5p | trans | 8.97E-35 | Nightingale | M_VLDL_C | NA | G | T | 0.036147 | 0.004168 | 1.30E-17 |
| rs676996 | miR-10b-5p | trans | 8.97E-35 | Nightingale | L_VLDL_TG | NA | G | T | -0.05103 | 0.004404 | 7.30E-32 |
| rs676996 | miR-195-5p | trans | 3.08E-17 | Nightingale | Remnant_( | NA | G | T | 0.036852 | 0.004396 | 3.70E-17 |
| rs676996 | miR-195-5p | trans | 3.08E-17 | Nightingale | ApoB | NA | G | T | 0.033901 | 0.004419 | 2.20E-14 |
| rs676996 | miR-195-5p | trans | 3.08E-17 | Nightingale | S_LDL_P | NA | G | T | 0.029548 | 0.004429 | 3.30E-11 |
| rs676996 | miR-195-5p | trans | 3.08E-17 | Nightingale | S_HDL_PL | NA | G | T | 0.023785 | 0.004387 | 3.30E-08 |
| rs676996 | miR-195-5p | trans | 3.08E-17 | Nightingale | XL_VLDL_P | NA | G | T | 0.038714 | 0.004432 | 6.80E-19 |
| rs676996 | miR-195-5p | trans | 3.08E-17 | Nightingale | Total_CE | NA | G | T | 0.031735 | 0.004268 | 9.90E-14 |
| rs676996 | miR-195-5p | trans | 3.08E-17 | Nightingale | ApoB_by_A | NA | G | T | 0.02498 | 0.004358 | 1.40E-08 |
| rs676996 | miR-195-5p | trans | 3.08E-17 | Nightingale | M_LDL_PL | NA | G | T | 0.032421 | 0.004431 | 2.80E-13 |
| rs676996 | miR-195-5p | trans | 3.08E-17 | Nightingale | S_LDL_PL_ | NA | G | T | -0.03004 | 0.004417 | 2.80E-13 |
| rs676996 | miR-195-5p | trans | 3.08E-17 | Nightingale | M_VLDL_P | NA | G | T | 0.028258 | 0.004414 | 6.50E-11 |
| rs676996 | miR-195-5p | trans | 3.08E-17 | Nightingale | Citrate | NA | G | T | 0.02685 | 0.004396 | 6.90E-10 |
| rs676996 | miR-195-5p | trans | 3.08E-17 | Nightingale | S_LDL_C_p | NA | G | T | 0.045304 | 0.004442 | 6.40E-25 |
| rs676996 | miR-195-5p | trans | 3.08E-17 | Nightingale | L_LDL_FC | NA | G | T | 0.033595 | 0.004345 | 3.00E-14 |
| rs676996 | miR-195-5p | trans | 3.08E-17 | Nightingale | IDL_CE | NA | G | T | 0.03352 | 0.004272 | 1.30E-14 |
| rs676996 | miR-195-5p | trans | 3.08E-17 | Nightingale | M_VLDL_C | NA | G | T | 0.033575 | 0.004157 | 2.10E-15 |
| rs676996 | miR-195-5p | trans | 3.08E-17 | Nightingale | S_VLDL_TG | NA | G | T | -0.03807 | 0.004283 | 1.60E-18 |
| rs676996 | miR-195-5p | trans | 3.08E-17 | Nightingale | Sphingomy | NA | G | T | 0.028974 | 0.004137 | 7.40E-12 |
| rs676996 | miR-195-5p | trans | 3.08E-17 | Nightingale | IDL_FC_pct | NA | G | T | 0.025703 | 0.004391 | 1.20E-08 |
| rs676996 | miR-195-5p | trans | 3.08E-17 | Nightingale | S_VLDL_L | NA | G | T | 0.024651 | 0.004382 | 3.80E-09 |
| rs676996 | miR-195-5p | trans | 3.08E-17 | Nightingale | XXL_VLDL_ | NA | G | T | -0.03564 | 0.004463 | 2.50E-16 |
| rs676996 | miR-195-5p | trans | 3.08E-17 | Nightingale | M_VLDL_F | NA | G | T | 0.041995 | 0.00421 | 4.10E-23 |
| rs676996 | miR-195-5p | trans | 3.08E-17 | Nightingale | Omega_6 | NA | G | T | 0.025871 | 0.004321 | 6.00E-10 |
| rs676996 | miR-195-5p | trans | 3.08E-17 | Nightingale | XL_VLDL_T | NA | G | T | -0.04377 | 0.004289 | 2.50E-24 |
| rs676996 | miR-195-5p | trans | 3.08E-17 | Nightingale | M_VLDL_F | NA | G | T | 0.033337 | 0.004424 | 3.20E-14 |
| rs676996 | miR-195-5p | trans | 3.08E-17 | Nightingale | XXL_VLDL_ | NA | G | T | 0.030734 | 0.004406 | 1.30E-12 |
| rs676996 | miR-195-5p | trans | 3.08E-17 | Nightingale | M_VLDL_P | NA | G | T | 0.029098 | 0.004418 | 2.10E-11 |
| rs676996 | miR-195-5p | trans | 3.08E-17 | Nightingale | M_LDL_TG | NA | G | T | -0.02753 | 0.00437 | 3.60E-10 |
| rs676996 | miR-195-5p | trans | 3.08E-17 | Nightingale | S_LDL_FC | NA | G | T | 0.02853 | 0.004418 | 4.40E-10 |
| rs676996 | miR-195-5p | trans | 3.08E-17 | Nightingale | XS_VLDL_L | NA | G | T | 0.033835 | 0.00435 | 2.00E-15 |
| rs676996 | miR-195-5p | trans | 3.08E-17 | Nightingale | M_LDL_L | NA | G | T | 0.032985 | 0.004431 | 1.10E-13 |
| rs676996 | miR-195-5p | trans | 3.08E-17 | Nightingale | LDL_PL | NA | G | T | 0.03495 | 0.004415 | 4.90E-15 |
| rs676996 | miR-195-5p | trans | 3.08E-17 | Nightingale | XS_VLDL_P | NA | G | T | 0.032127 | 0.004363 | 5.20E-14 |
| rs676996 | miR-195-5p | trans | 3.08E-17 | Nightingale | XS_VLDL_P | NA | G | T | 0.036717 | 0.004366 | 1.30E-17 |
| rs676996 | miR-195-5p | trans | 3.08E-17 | Nightingale | M_VLDL_C | NA | G | T | 0.04081 | 0.004385 | 2.40E-20 |
| rs676996 | miR-195-5p | trans | 3.08E-17 | Nightingale | XS_VLDL_C | NA | G | T | 0.038004 | 0.004295 | 1.30E-18 |
| rs676996 | miR-195-5p | trans | 3.08E-17 | Nightingale | S_VLDL_CE | NA | G | T | 0.038157 | 0.004419 | 1.60E-18 |

| | | | | | | | | | | | |
|---|---|---|---|---|---|---|---|---|---|---|---|
| rs676996 | miR-195-5p | trans | 3.08E-17 | Nightingale | Total_FC | NA | G | T | 0.033248 | 0.004323 | 6.90E-15 |
| rs676996 | miR-195-5p | trans | 3.08E-17 | Nightingale | LDL_C | NA | G | T | 0.03315 | 0.004406 | 9.30E-14 |
| rs676996 | miR-195-5p | trans | 3.08E-17 | Nightingale | S_LDL_C | NA | G | T | 0.030897 | 0.004429 | 5.80E-12 |
| rs676996 | miR-195-5p | trans | 3.08E-17 | Nightingale | L_LDL_PL | NA | G | T | 0.037957 | 0.004391 | 9.50E-18 |
| rs676996 | miR-195-5p | trans | 3.08E-17 | Nightingale | XXL_VLDL_ | NA | G | T | 0.034743 | 0.004434 | 3.30E-15 |
| rs676996 | miR-195-5p | trans | 3.08E-17 | Nightingale | non_HDL_( | NA | G | T | 0.035665 | 0.004401 | 5.30E-16 |
| rs676996 | miR-195-5p | trans | 3.08E-17 | Nightingale | S_LDL_CE_ | NA | G | T | 0.030893 | 0.004417 | 5.50E-14 |
| rs676996 | miR-195-5p | trans | 3.08E-17 | Nightingale | XXL_VLDL_ | NA | G | T | 0.025017 | 0.004454 | 1.50E-09 |
| rs676996 | miR-195-5p | trans | 3.08E-17 | Nightingale | S_LDL_L | NA | G | T | 0.027698 | 0.004424 | 8.50E-10 |
| rs676996 | miR-195-5p | trans | 3.08E-17 | Nightingale | S_VLDL_C_ | NA | G | T | 0.041827 | 0.004306 | 5.90E-22 |
| rs676996 | miR-195-5p | trans | 3.08E-17 | Nightingale | XS_VLDL_C | NA | G | T | 0.037811 | 0.00427 | 2.10E-18 |
| rs676996 | miR-195-5p | trans | 3.08E-17 | Nightingale | M_LDL_CE | NA | G | T | 0.032924 | 0.004431 | 1.00E-13 |
| rs676996 | miR-195-5p | trans | 3.08E-17 | Nightingale | XL_VLDL_C | NA | G | T | 0.037265 | 0.004245 | 3.70E-18 |
| rs676996 | miR-195-5p | trans | 3.08E-17 | Nightingale | XL_VLDL_C | NA | G | T | 0.038142 | 0.004243 | 5.40E-19 |
| rs676996 | miR-195-5p | trans | 3.08E-17 | Nightingale | S_HDL_P | NA | G | T | 0.030579 | 0.004425 | 5.40E-12 |
| rs676996 | miR-195-5p | trans | 3.08E-17 | Nightingale | XL_VLDL_F | NA | G | T | 0.037128 | 0.00429 | 2.30E-18 |
| rs676996 | miR-195-5p | trans | 3.08E-17 | Nightingale | S_HDL_C | NA | G | T | 0.028387 | 0.004417 | 1.80E-10 |
| rs676996 | miR-195-5p | trans | 3.08E-17 | Nightingale | M_VLDL_T | NA | G | T | -0.03849 | 0.004181 | 7.90E-20 |
| rs676996 | miR-195-5p | trans | 3.08E-17 | Nightingale | Total_C | NA | G | T | 0.032363 | 0.004284 | 3.40E-14 |
| rs676996 | miR-195-5p | trans | 3.08E-17 | Nightingale | GlycA | NA | G | T | 0.027865 | 0.004406 | 1.80E-10 |
| rs676996 | miR-195-5p | trans | 3.08E-17 | Nightingale | S_VLDL_FC | NA | G | T | 0.037868 | 0.004426 | 7.60E-18 |
| rs676996 | miR-195-5p | trans | 3.08E-17 | Nightingale | VLDL_FC | NA | G | T | 0.02413 | 0.004392 | 1.10E-08 |
| rs676996 | miR-195-5p | trans | 3.08E-17 | Nightingale | S_HDL_L | NA | G | T | 0.026288 | 0.00441 | 1.60E-09 |
| rs676996 | miR-195-5p | trans | 3.08E-17 | Nightingale | XL_HDL_PL | NA | G | T | -0.02267 | 0.004065 | 1.30E-08 |
| rs676996 | miR-195-5p | trans | 3.08E-17 | Nightingale | S_LDL_CE | NA | G | T | 0.030499 | 0.004427 | 6.10E-12 |
| rs676996 | miR-195-5p | trans | 3.08E-17 | Nightingale | XS_VLDL_F | NA | G | T | 0.037076 | 0.004348 | 6.40E-18 |
| rs676996 | miR-195-5p | trans | 3.08E-17 | Nightingale | L_LDL_CE | NA | G | T | 0.031522 | 0.004385 | 1.00E-12 |
| rs676996 | miR-195-5p | trans | 3.08E-17 | Nightingale | VLDL_CE | NA | G | T | 0.03576 | 0.004426 | 3.10E-16 |
| rs676996 | miR-195-5p | trans | 3.08E-17 | Nightingale | LDL_P | NA | G | T | 0.031745 | 0.004422 | 1.10E-12 |
| rs676996 | miR-195-5p | trans | 3.08E-17 | Nightingale | Total_L | NA | G | T | 0.026348 | 0.004353 | 4.60E-10 |
| rs676996 | miR-195-5p | trans | 3.08E-17 | Nightingale | LDL_L | NA | G | T | 0.033262 | 0.00441 | 7.80E-14 |
| rs676996 | miR-195-5p | trans | 3.08E-17 | Nightingale | M_LDL_P | NA | G | T | 0.035848 | 0.004437 | 7.30E-16 |
| rs676996 | miR-195-5p | trans | 3.08E-17 | Nightingale | XS_VLDL_F | NA | G | T | 0.031377 | 0.004393 | 2.60E-12 |
| rs676996 | miR-195-5p | trans | 3.08E-17 | Nightingale | IDL_PL | NA | G | T | 0.035327 | 0.00427 | 3.50E-16 |
| rs676996 | miR-195-5p | trans | 3.08E-17 | Nightingale | L_LDL_P | NA | G | T | 0.029116 | 0.00441 | 6.60E-11 |
| rs676996 | miR-195-5p | trans | 3.08E-17 | Nightingale | S_HDL_CE | NA | G | T | 0.028597 | 0.004432 | 1.40E-10 |
| rs676996 | miR-195-5p | trans | 3.08E-17 | Nightingale | IDL_P | NA | G | T | 0.040009 | 0.004378 | 1.30E-19 |
| rs676996 | miR-195-5p | trans | 3.08E-17 | Nightingale | IDL_C | NA | G | T | 0.034962 | 0.004274 | 9.40E-16 |
| rs676996 | miR-195-5p | trans | 3.08E-17 | Nightingale | LDL_FC | NA | G | T | 0.033597 | 0.004378 | 5.50E-14 |
| rs676996 | miR-195-5p | trans | 3.08E-17 | Nightingale | VLDL_C | NA | G | T | 0.031274 | 0.004419 | 5.00E-13 |
| rs676996 | miR-195-5p | trans | 3.08E-17 | Nightingale | L_LDL_C | NA | G | T | 0.032279 | 0.004375 | 2.70E-13 |
| rs676996 | miR-195-5p | trans | 3.08E-17 | Nightingale | S_VLDL_PL | NA | G | T | 0.032747 | 0.004417 | 4.30E-14 |
| rs676996 | miR-195-5p | trans | 3.08E-17 | Nightingale | M_LDL_C | NA | G | T | 0.03394 | 0.004433 | 2.30E-14 |
| rs676996 | miR-195-5p | trans | 3.08E-17 | Nightingale | Total_PL | NA | G | T | 0.024943 | 0.004214 | 1.10E-09 |
| rs676996 | miR-195-5p | trans | 3.08E-17 | Nightingale | S_VLDL_CE | NA | G | T | 0.046969 | 0.004365 | 1.40E-26 |
| rs676996 | miR-195-5p | trans | 3.08E-17 | Nightingale | S_VLDL_FC | NA | G | T | 0.029304 | 0.004228 | 1.10E-11 |
| rs676996 | miR-195-5p | trans | 3.08E-17 | Nightingale | M_LDL_FC | NA | G | T | 0.034304 | 0.004419 | 2.00E-14 |
| rs676996 | miR-195-5p | trans | 3.08E-17 | Nightingale | M_LDL_C_ | NA | G | T | 0.031099 | 0.004428 | 3.30E-12 |
| rs676996 | miR-195-5p | trans | 3.08E-17 | Nightingale | M_VLDL_P | NA | G | T | 0.043976 | 0.004247 | 6.90E-25 |
| rs676996 | miR-195-5p | trans | 3.08E-17 | Nightingale | Clinical_LD | NA | G | T | 0.036436 | 0.004399 | 2.90E-16 |
| rs676996 | miR-195-5p | trans | 3.08E-17 | Nightingale | M_VLDL_C | NA | G | T | 0.038672 | 0.004413 | 2.00E-18 |
| rs676996 | miR-195-5p | trans | 3.08E-17 | Nightingale | IDL_L | NA | G | T | 0.035274 | 0.004278 | 4.90E-16 |
| rs676996 | miR-195-5p | trans | 3.08E-17 | Nightingale | IDL_FC | NA | G | T | 0.038246 | 0.004284 | 1.50E-18 |
| rs676996 | miR-195-5p | trans | 3.08E-17 | Nightingale | S_VLDL_C | NA | G | T | 0.038343 | 0.004424 | 1.80E-18 |
| rs676996 | miR-195-5p | trans | 3.08E-17 | Nightingale | XXL_VLDL_ | NA | G | T | 0.033067 | 0.004459 | 9.60E-14 |
| rs676996 | miR-195-5p | trans | 3.08E-17 | Nightingale | Cholines | NA | G | T | 0.022308 | 0.004161 | 4.90E-08 |
| rs676996 | miR-195-5p | trans | 3.08E-17 | Nightingale | S_VLDL_PL | NA | G | T | 0.027677 | 0.004237 | 1.90E-10 |
| rs676996 | miR-195-5p | trans | 3.08E-17 | Nightingale | LDL_CE | NA | G | T | 0.032506 | 0.004413 | 2.30E-13 |
| rs676996 | miR-195-5p | trans | 3.08E-17 | Nightingale | L_LDL_L | NA | G | T | 0.033512 | 0.004382 | 3.20E-14 |
| rs676996 | miR-195-5p | trans | 3.08E-17 | Nightingale | L_VLDL_CE | NA | G | T | 0.047288 | 0.004234 | 2.80E-29 |
| rs676996 | miR-195-5p | trans | 3.08E-17 | Nightingale | L_VLDL_C_ | NA | G | T | 0.052087 | 0.004322 | 5.50E-34 |
| rs676996 | miR-195-5p | trans | 3.08E-17 | Nightingale | LA | NA | G | T | 0.024935 | 0.004339 | 3.50E-09 |
| rs676996 | miR-195-5p | trans | 3.08E-17 | Nightingale | S_VLDL_P | NA | G | T | 0.023454 | 0.004369 | 1.70E-08 |

| | | | | | | | | | | | |
|---|---|---|---|---|---|---|---|---|---|---|---|
| rs676996 | miR-195-5p | trans | 3.08E-17 | Nightingale | VLDL_P | NA | G | T | 0.02662 | 0.004401 | 4.40E-10 |
| rs676996 | miR-195-5p | trans | 3.08E-17 | Nightingale | PUFA | NA | G | T | 0.023693 | 0.004316 | 1.50E-08 |
| rs676996 | miR-195-5p | trans | 3.08E-17 | Nightingale | L_VLDL_FC | NA | G | T | 0.048434 | 0.004382 | 6.70E-29 |
| rs676996 | miR-195-5p | trans | 3.08E-17 | Nightingale | S_HDL_FC | NA | G | T | 0.023471 | 0.004344 | 4.60E-08 |
| rs676996 | miR-195-5p | trans | 3.08E-17 | Nightingale | M_VLDL_C | NA | G | T | 0.036147 | 0.004168 | 1.30E-17 |
| rs676996 | miR-195-5p | trans | 3.08E-17 | Nightingale | L_VLDL_TG | NA | G | T | -0.05103 | 0.004404 | 7.30E-32 |
| rs676996 | miR-126-3p | trans | 4.67E-39 | Nightingale | Remnant_( | NA | G | T | 0.036852 | 0.004396 | 3.70E-17 |
| rs676996 | miR-126-3p | trans | 4.67E-39 | Nightingale | ApoB | NA | G | T | 0.033901 | 0.004419 | 2.20E-14 |
| rs676996 | miR-126-3p | trans | 4.67E-39 | Nightingale | S_LDL_P | NA | G | T | 0.029548 | 0.004429 | 3.30E-11 |
| rs676996 | miR-126-3p | trans | 4.67E-39 | Nightingale | S_HDL_PL | NA | G | T | 0.023785 | 0.004387 | 3.30E-08 |
| rs676996 | miR-126-3p | trans | 4.67E-39 | Nightingale | XL_VLDL_P | NA | G | T | 0.038714 | 0.004432 | 6.80E-19 |
| rs676996 | miR-126-3p | trans | 4.67E-39 | Nightingale | Total_CE | NA | G | T | 0.031735 | 0.004268 | 9.90E-14 |
| rs676996 | miR-126-3p | trans | 4.67E-39 | Nightingale | ApoB_by_A | NA | G | T | 0.02498 | 0.004358 | 1.40E-08 |
| rs676996 | miR-126-3p | trans | 4.67E-39 | Nightingale | M_LDL_PL | NA | G | T | 0.032421 | 0.004431 | 2.80E-13 |
| rs676996 | miR-126-3p | trans | 4.67E-39 | Nightingale | S_LDL_PL_ | NA | G | T | -0.03004 | 0.004417 | 2.80E-13 |
| rs676996 | miR-126-3p | trans | 4.67E-39 | Nightingale | M_VLDL_P | NA | G | T | 0.028258 | 0.004414 | 6.50E-11 |
| rs676996 | miR-126-3p | trans | 4.67E-39 | Nightingale | Citrate | NA | G | T | 0.02685 | 0.004396 | 6.90E-10 |
| rs676996 | miR-126-3p | trans | 4.67E-39 | Nightingale | S_LDL_C_p | NA | G | T | 0.045304 | 0.004442 | 6.40E-25 |
| rs676996 | miR-126-3p | trans | 4.67E-39 | Nightingale | L_LDL_FC | NA | G | T | 0.033595 | 0.004345 | 3.00E-14 |
| rs676996 | miR-126-3p | trans | 4.67E-39 | Nightingale | IDL_CE | NA | G | T | 0.03352 | 0.004272 | 1.30E-14 |
| rs676996 | miR-126-3p | trans | 4.67E-39 | Nightingale | M_VLDL_C | NA | G | T | 0.033575 | 0.004157 | 2.10E-15 |
| rs676996 | miR-126-3p | trans | 4.67E-39 | Nightingale | S_VLDL_TG | NA | G | T | -0.03807 | 0.004283 | 1.60E-18 |
| rs676996 | miR-126-3p | trans | 4.67E-39 | Nightingale | Sphingomy | NA | G | T | 0.028974 | 0.004137 | 7.40E-12 |
| rs676996 | miR-126-3p | trans | 4.67E-39 | Nightingale | IDL_FC_pct | NA | G | T | 0.025703 | 0.004391 | 1.20E-08 |
| rs676996 | miR-126-3p | trans | 4.67E-39 | Nightingale | S_VLDL_L | NA | G | T | 0.024651 | 0.004382 | 3.80E-09 |
| rs676996 | miR-126-3p | trans | 4.67E-39 | Nightingale | XXL_VLDL_ | NA | G | T | -0.03564 | 0.004463 | 2.50E-16 |
| rs676996 | miR-126-3p | trans | 4.67E-39 | Nightingale | M_VLDL_F( | NA | G | T | 0.041995 | 0.00421 | 4.10E-23 |
| rs676996 | miR-126-3p | trans | 4.67E-39 | Nightingale | Omega_6 | NA | G | T | 0.025871 | 0.004321 | 6.00E-10 |
| rs676996 | miR-126-3p | trans | 4.67E-39 | Nightingale | XL_VLDL_T | NA | G | T | -0.04377 | 0.004289 | 2.50E-24 |
| rs676996 | miR-126-3p | trans | 4.67E-39 | Nightingale | M_VLDL_F( | NA | G | T | 0.033337 | 0.004424 | 3.20E-14 |
| rs676996 | miR-126-3p | trans | 4.67E-39 | Nightingale | XXL_VLDL_ | NA | G | T | 0.030734 | 0.004406 | 1.30E-12 |
| rs676996 | miR-126-3p | trans | 4.67E-39 | Nightingale | M_VLDL_P | NA | G | T | 0.029098 | 0.004418 | 2.10E-11 |
| rs676996 | miR-126-3p | trans | 4.67E-39 | Nightingale | M_LDL_TG | NA | G | T | -0.02753 | 0.00437 | 3.60E-10 |
| rs676996 | miR-126-3p | trans | 4.67E-39 | Nightingale | S_LDL_FC | NA | G | T | 0.02853 | 0.004418 | 4.40E-10 |
| rs676996 | miR-126-3p | trans | 4.67E-39 | Nightingale | XS_VLDL_L | NA | G | T | 0.033835 | 0.00435 | 2.00E-15 |
| rs676996 | miR-126-3p | trans | 4.67E-39 | Nightingale | M_LDL_L | NA | G | T | 0.032985 | 0.004431 | 1.10E-13 |
| rs676996 | miR-126-3p | trans | 4.67E-39 | Nightingale | LDL_PL | NA | G | T | 0.03495 | 0.004415 | 4.90E-15 |
| rs676996 | miR-126-3p | trans | 4.67E-39 | Nightingale | XS_VLDL_P | NA | G | T | 0.032127 | 0.004363 | 5.20E-14 |
| rs676996 | miR-126-3p | trans | 4.67E-39 | Nightingale | XS_VLDL_P | NA | G | T | 0.036717 | 0.004366 | 1.30E-17 |
| rs676996 | miR-126-3p | trans | 4.67E-39 | Nightingale | M_VLDL_C | NA | G | T | 0.04081 | 0.004385 | 2.40E-20 |
| rs676996 | miR-126-3p | trans | 4.67E-39 | Nightingale | XS_VLDL_C | NA | G | T | 0.038004 | 0.004295 | 1.30E-18 |
| rs676996 | miR-126-3p | trans | 4.67E-39 | Nightingale | S_VLDL_CE | NA | G | T | 0.038157 | 0.004419 | 1.60E-18 |
| rs676996 | miR-126-3p | trans | 4.67E-39 | Nightingale | Total_FC | NA | G | T | 0.033248 | 0.004323 | 6.90E-15 |
| rs676996 | miR-126-3p | trans | 4.67E-39 | Nightingale | LDL_C | NA | G | T | 0.03315 | 0.004406 | 9.30E-14 |
| rs676996 | miR-126-3p | trans | 4.67E-39 | Nightingale | S_LDL_C | NA | G | T | 0.030897 | 0.004429 | 5.80E-12 |
| rs676996 | miR-126-3p | trans | 4.67E-39 | Nightingale | L_LDL_PL | NA | G | T | 0.037957 | 0.004391 | 9.50E-18 |
| rs676996 | miR-126-3p | trans | 4.67E-39 | Nightingale | XXL_VLDL_ | NA | G | T | 0.034743 | 0.004434 | 3.30E-15 |
| rs676996 | miR-126-3p | trans | 4.67E-39 | Nightingale | non_HDL_( | NA | G | T | 0.035665 | 0.004401 | 5.30E-16 |
| rs676996 | miR-126-3p | trans | 4.67E-39 | Nightingale | S_LDL_CE_ | NA | G | T | 0.030893 | 0.004417 | 5.50E-14 |
| rs676996 | miR-126-3p | trans | 4.67E-39 | Nightingale | XXL_VLDL_ | NA | G | T | 0.025017 | 0.004454 | 1.50E-09 |
| rs676996 | miR-126-3p | trans | 4.67E-39 | Nightingale | S_LDL_L | NA | G | T | 0.027698 | 0.004424 | 8.50E-10 |
| rs676996 | miR-126-3p | trans | 4.67E-39 | Nightingale | S_VLDL_C_ | NA | G | T | 0.041827 | 0.004306 | 5.90E-22 |
| rs676996 | miR-126-3p | trans | 4.67E-39 | Nightingale | XS_VLDL_C | NA | G | T | 0.037811 | 0.00427 | 2.10E-18 |
| rs676996 | miR-126-3p | trans | 4.67E-39 | Nightingale | M_LDL_CE | NA | G | T | 0.032924 | 0.004431 | 1.00E-13 |
| rs676996 | miR-126-3p | trans | 4.67E-39 | Nightingale | XL_VLDL_C | NA | G | T | 0.037265 | 0.004245 | 3.70E-18 |
| rs676996 | miR-126-3p | trans | 4.67E-39 | Nightingale | XL_VLDL_C | NA | G | T | 0.038142 | 0.004243 | 5.40E-19 |
| rs676996 | miR-126-3p | trans | 4.67E-39 | Nightingale | S_HDL_P | NA | G | T | 0.030579 | 0.004425 | 5.40E-12 |
| rs676996 | miR-126-3p | trans | 4.67E-39 | Nightingale | XL_VLDL_F | NA | G | T | 0.037128 | 0.00429 | 2.30E-18 |
| rs676996 | miR-126-3p | trans | 4.67E-39 | Nightingale | S_HDL_C | NA | G | T | 0.028387 | 0.004417 | 1.80E-10 |
| rs676996 | miR-126-3p | trans | 4.67E-39 | Nightingale | M_VLDL_T( | NA | G | T | -0.03849 | 0.004181 | 7.90E-20 |
| rs676996 | miR-126-3p | trans | 4.67E-39 | Nightingale | Total_C | NA | G | T | 0.032363 | 0.004284 | 3.40E-14 |
| rs676996 | miR-126-3p | trans | 4.67E-39 | Nightingale | GlycA | NA | G | T | 0.027865 | 0.004406 | 1.80E-10 |
| rs676996 | miR-126-3p | trans | 4.67E-39 | Nightingale | S_VLDL_FC | NA | G | T | 0.037868 | 0.004426 | 7.60E-18 |

| | | | | | | | | | | | |
|---|---|---|---|---|---|---|---|---|---|---|---|
| rs676996 | miR-126-3p | trans | 4.67E-39 | Nightingale | VLDL_FC | NA | G | T | 0.02413 | 0.004392 | 1.10E-08 |
| rs676996 | miR-126-3p | trans | 4.67E-39 | Nightingale | S_HDL_L | NA | G | T | 0.026288 | 0.00441 | 1.60E-09 |
| rs676996 | miR-126-3p | trans | 4.67E-39 | Nightingale | XL_HDL_PL | NA | G | T | -0.02267 | 0.004065 | 1.30E-08 |
| rs676996 | miR-126-3p | trans | 4.67E-39 | Nightingale | S_LDL_CE | NA | G | T | 0.030499 | 0.004427 | 6.10E-12 |
| rs676996 | miR-126-3p | trans | 4.67E-39 | Nightingale | XS_VLDL_F | NA | G | T | 0.037076 | 0.004348 | 6.40E-18 |
| rs676996 | miR-126-3p | trans | 4.67E-39 | Nightingale | L_LDL_CE | NA | G | T | 0.031522 | 0.004385 | 1.00E-12 |
| rs676996 | miR-126-3p | trans | 4.67E-39 | Nightingale | VLDL_CE | NA | G | T | 0.03576 | 0.004426 | 3.10E-16 |
| rs676996 | miR-126-3p | trans | 4.67E-39 | Nightingale | LDL_P | NA | G | T | 0.031745 | 0.004422 | 1.10E-12 |
| rs676996 | miR-126-3p | trans | 4.67E-39 | Nightingale | Total_L | NA | G | T | 0.026348 | 0.004353 | 4.60E-10 |
| rs676996 | miR-126-3p | trans | 4.67E-39 | Nightingale | LDL_L | NA | G | T | 0.033262 | 0.00441 | 7.80E-14 |
| rs676996 | miR-126-3p | trans | 4.67E-39 | Nightingale | M_LDL_P | NA | G | T | 0.035848 | 0.004437 | 7.30E-16 |
| rs676996 | miR-126-3p | trans | 4.67E-39 | Nightingale | XS_VLDL_F | NA | G | T | 0.031377 | 0.004393 | 2.60E-12 |
| rs676996 | miR-126-3p | trans | 4.67E-39 | Nightingale | IDL_PL | NA | G | T | 0.035327 | 0.00427 | 3.50E-16 |
| rs676996 | miR-126-3p | trans | 4.67E-39 | Nightingale | L_LDL_P | NA | G | T | 0.029116 | 0.00441 | 6.60E-11 |
| rs676996 | miR-126-3p | trans | 4.67E-39 | Nightingale | S_HDL_CE | NA | G | T | 0.028597 | 0.004432 | 1.40E-10 |
| rs676996 | miR-126-3p | trans | 4.67E-39 | Nightingale | IDL_P | NA | G | T | 0.040009 | 0.004378 | 1.30E-19 |
| rs676996 | miR-126-3p | trans | 4.67E-39 | Nightingale | IDL_C | NA | G | T | 0.034962 | 0.004274 | 9.40E-16 |
| rs676996 | miR-126-3p | trans | 4.67E-39 | Nightingale | LDL_FC | NA | G | T | 0.033597 | 0.004378 | 5.50E-14 |
| rs676996 | miR-126-3p | trans | 4.67E-39 | Nightingale | VLDL_C | NA | G | T | 0.031274 | 0.004419 | 5.00E-13 |
| rs676996 | miR-126-3p | trans | 4.67E-39 | Nightingale | L_LDL_C | NA | G | T | 0.032279 | 0.004375 | 2.70E-13 |
| rs676996 | miR-126-3p | trans | 4.67E-39 | Nightingale | S_VLDL_PL | NA | G | T | 0.032747 | 0.004417 | 4.30E-14 |
| rs676996 | miR-126-3p | trans | 4.67E-39 | Nightingale | M_LDL_C | NA | G | T | 0.03394 | 0.004433 | 2.30E-14 |
| rs676996 | miR-126-3p | trans | 4.67E-39 | Nightingale | Total_PL | NA | G | T | 0.024943 | 0.004214 | 1.10E-09 |
| rs676996 | miR-126-3p | trans | 4.67E-39 | Nightingale | S_VLDL_CE | NA | G | T | 0.046969 | 0.004365 | 1.40E-26 |
| rs676996 | miR-126-3p | trans | 4.67E-39 | Nightingale | S_VLDL_FC | NA | G | T | 0.029304 | 0.004228 | 1.10E-11 |
| rs676996 | miR-126-3p | trans | 4.67E-39 | Nightingale | M_LDL_FC | NA | G | T | 0.034304 | 0.004419 | 2.00E-14 |
| rs676996 | miR-126-3p | trans | 4.67E-39 | Nightingale | M_LDL_C_ | NA | G | T | 0.031099 | 0.004428 | 3.30E-12 |
| rs676996 | miR-126-3p | trans | 4.67E-39 | Nightingale | M_VLDL_P | NA | G | T | 0.043976 | 0.004247 | 6.90E-25 |
| rs676996 | miR-126-3p | trans | 4.67E-39 | Nightingale | Clinical_LD | NA | G | T | 0.036436 | 0.004399 | 2.90E-16 |
| rs676996 | miR-126-3p | trans | 4.67E-39 | Nightingale | M_VLDL_C | NA | G | T | 0.038672 | 0.004413 | 2.00E-18 |
| rs676996 | miR-126-3p | trans | 4.67E-39 | Nightingale | IDL_L | NA | G | T | 0.035274 | 0.004278 | 4.90E-16 |
| rs676996 | miR-126-3p | trans | 4.67E-39 | Nightingale | IDL_FC | NA | G | T | 0.038246 | 0.004284 | 1.50E-18 |
| rs676996 | miR-126-3p | trans | 4.67E-39 | Nightingale | S_VLDL_C | NA | G | T | 0.038343 | 0.004424 | 1.80E-18 |
| rs676996 | miR-126-3p | trans | 4.67E-39 | Nightingale | XXL_VLDL_ | NA | G | T | 0.033067 | 0.004459 | 9.60E-14 |
| rs676996 | miR-126-3p | trans | 4.67E-39 | Nightingale | Cholines | NA | G | T | 0.022308 | 0.004161 | 4.90E-08 |
| rs676996 | miR-126-3p | trans | 4.67E-39 | Nightingale | S_VLDL_PL | NA | G | T | 0.027677 | 0.004237 | 1.90E-10 |
| rs676996 | miR-126-3p | trans | 4.67E-39 | Nightingale | LDL_CE | NA | G | T | 0.032506 | 0.004413 | 2.30E-13 |
| rs676996 | miR-126-3p | trans | 4.67E-39 | Nightingale | L_LDL_L | NA | G | T | 0.033512 | 0.004382 | 3.20E-14 |
| rs676996 | miR-126-3p | trans | 4.67E-39 | Nightingale | L_VLDL_CE | NA | G | T | 0.047288 | 0.004234 | 2.80E-29 |
| rs676996 | miR-126-3p | trans | 4.67E-39 | Nightingale | L_VLDL_C_ | NA | G | T | 0.052087 | 0.004322 | 5.50E-34 |
| rs676996 | miR-126-3p | trans | 4.67E-39 | Nightingale | LA | NA | G | T | 0.024935 | 0.004339 | 3.50E-09 |
| rs676996 | miR-126-3p | trans | 4.67E-39 | Nightingale | S_VLDL_P | NA | G | T | 0.023454 | 0.004369 | 1.70E-08 |
| rs676996 | miR-126-3p | trans | 4.67E-39 | Nightingale | VLDL_P | NA | G | T | 0.02662 | 0.004401 | 4.40E-10 |
| rs676996 | miR-126-3p | trans | 4.67E-39 | Nightingale | PUFA | NA | G | T | 0.023693 | 0.004316 | 1.50E-08 |
| rs676996 | miR-126-3p | trans | 4.67E-39 | Nightingale | L_VLDL_FC | NA | G | T | 0.048434 | 0.004382 | 6.70E-29 |
| rs676996 | miR-126-3p | trans | 4.67E-39 | Nightingale | S_HDL_FC | NA | G | T | 0.023471 | 0.004344 | 4.60E-08 |
| rs676996 | miR-126-3p | trans | 4.67E-39 | Nightingale | M_VLDL_C | NA | G | T | 0.036147 | 0.004168 | 1.30E-17 |
| rs676996 | miR-126-3p | trans | 4.67E-39 | Nightingale | L_VLDL_TG | NA | G | T | -0.05103 | 0.004404 | 7.30E-32 |
| rs676996 | miR-139-5p | trans | 3.57E-41 | Nightingale | Remnant_( | NA | G | T | 0.036852 | 0.004396 | 3.70E-17 |
| rs676996 | miR-139-5p | trans | 3.57E-41 | Nightingale | ApoB | NA | G | T | 0.033901 | 0.004419 | 2.20E-14 |
| rs676996 | miR-139-5p | trans | 3.57E-41 | Nightingale | S_LDL_P | NA | G | T | 0.029548 | 0.004429 | 3.30E-11 |
| rs676996 | miR-139-5p | trans | 3.57E-41 | Nightingale | S_HDL_PL | NA | G | T | 0.023785 | 0.004387 | 3.30E-08 |
| rs676996 | miR-139-5p | trans | 3.57E-41 | Nightingale | XL_VLDL_P | NA | G | T | 0.038714 | 0.004432 | 6.80E-19 |
| rs676996 | miR-139-5p | trans | 3.57E-41 | Nightingale | Total_CE | NA | G | T | 0.031735 | 0.004268 | 9.90E-14 |
| rs676996 | miR-139-5p | trans | 3.57E-41 | Nightingale | ApoB_by_A | NA | G | T | 0.02498 | 0.004358 | 1.40E-08 |
| rs676996 | miR-139-5p | trans | 3.57E-41 | Nightingale | M_LDL_PL | NA | G | T | 0.032421 | 0.004431 | 2.80E-13 |
| rs676996 | miR-139-5p | trans | 3.57E-41 | Nightingale | S_LDL_PL_ | NA | G | T | -0.03004 | 0.004417 | 2.80E-13 |
| rs676996 | miR-139-5p | trans | 3.57E-41 | Nightingale | M_VLDL_P | NA | G | T | 0.028258 | 0.004414 | 6.50E-11 |
| rs676996 | miR-139-5p | trans | 3.57E-41 | Nightingale | Citrate | NA | G | T | 0.02685 | 0.004396 | 6.90E-10 |
| rs676996 | miR-139-5p | trans | 3.57E-41 | Nightingale | S_LDL_C_p | NA | G | T | 0.045304 | 0.004442 | 6.40E-25 |
| rs676996 | miR-139-5p | trans | 3.57E-41 | Nightingale | L_LDL_FC | NA | G | T | 0.033595 | 0.004345 | 3.00E-14 |
| rs676996 | miR-139-5p | trans | 3.57E-41 | Nightingale | IDL_CE | NA | G | T | 0.03352 | 0.004272 | 1.30E-14 |
| rs676996 | miR-139-5p | trans | 3.57E-41 | Nightingale | M_VLDL_C | NA | G | T | 0.033575 | 0.004157 | 2.10E-15 |

| | | | | | | | | | | | |
|---|---|---|---|---|---|---|---|---|---|---|---|
| rs676996 | miR-139-5p | trans | 3.57E-41 | Nightingale | S_VLDL_TG | NA | G | T | -0.03807 | 0.004283 | 1.60E-18 |
| rs676996 | miR-139-5p | trans | 3.57E-41 | Nightingale | Sphingomy | NA | G | T | 0.028974 | 0.004137 | 7.40E-12 |
| rs676996 | miR-139-5p | trans | 3.57E-41 | Nightingale | IDL_FC_pct | NA | G | T | 0.025703 | 0.004391 | 1.20E-08 |
| rs676996 | miR-139-5p | trans | 3.57E-41 | Nightingale | S_VLDL_L | NA | G | T | 0.024651 | 0.004382 | 3.80E-09 |
| rs676996 | miR-139-5p | trans | 3.57E-41 | Nightingale | XXL_VLDL_ | NA | G | T | -0.03564 | 0.004463 | 2.50E-16 |
| rs676996 | miR-139-5p | trans | 3.57E-41 | Nightingale | M_VLDL_F | NA | G | T | 0.041995 | 0.00421 | 4.10E-23 |
| rs676996 | miR-139-5p | trans | 3.57E-41 | Nightingale | Omega_6 | NA | G | T | 0.025871 | 0.004321 | 6.00E-10 |
| rs676996 | miR-139-5p | trans | 3.57E-41 | Nightingale | XL_VLDL_T | NA | G | T | -0.04377 | 0.004289 | 2.50E-24 |
| rs676996 | miR-139-5p | trans | 3.57E-41 | Nightingale | M_VLDL_F | NA | G | T | 0.033337 | 0.004424 | 3.20E-14 |
| rs676996 | miR-139-5p | trans | 3.57E-41 | Nightingale | XXL_VLDL_ | NA | G | T | 0.030734 | 0.004406 | 1.30E-12 |
| rs676996 | miR-139-5p | trans | 3.57E-41 | Nightingale | M_VLDL_P | NA | G | T | 0.029098 | 0.004418 | 2.10E-11 |
| rs676996 | miR-139-5p | trans | 3.57E-41 | Nightingale | M_LDL_TG | NA | G | T | -0.02753 | 0.00437 | 3.60E-10 |
| rs676996 | miR-139-5p | trans | 3.57E-41 | Nightingale | S_LDL_FC | NA | G | T | 0.02853 | 0.004418 | 4.40E-10 |
| rs676996 | miR-139-5p | trans | 3.57E-41 | Nightingale | XS_VLDL_L | NA | G | T | 0.033835 | 0.00435 | 2.00E-15 |
| rs676996 | miR-139-5p | trans | 3.57E-41 | Nightingale | M_LDL_L | NA | G | T | 0.032985 | 0.004431 | 1.10E-13 |
| rs676996 | miR-139-5p | trans | 3.57E-41 | Nightingale | LDL_PL | NA | G | T | 0.03495 | 0.004415 | 4.90E-15 |
| rs676996 | miR-139-5p | trans | 3.57E-41 | Nightingale | XS_VLDL_P | NA | G | T | 0.032127 | 0.004363 | 5.20E-14 |
| rs676996 | miR-139-5p | trans | 3.57E-41 | Nightingale | XS_VLDL_P | NA | G | T | 0.036717 | 0.004366 | 1.30E-17 |
| rs676996 | miR-139-5p | trans | 3.57E-41 | Nightingale | M_VLDL_C | NA | G | T | 0.04081 | 0.004385 | 2.40E-20 |
| rs676996 | miR-139-5p | trans | 3.57E-41 | Nightingale | XS_VLDL_C | NA | G | T | 0.038004 | 0.004295 | 1.30E-18 |
| rs676996 | miR-139-5p | trans | 3.57E-41 | Nightingale | S_VLDL_CE | NA | G | T | 0.038157 | 0.004419 | 1.60E-18 |
| rs676996 | miR-139-5p | trans | 3.57E-41 | Nightingale | Total_FC | NA | G | T | 0.033248 | 0.004323 | 6.90E-15 |
| rs676996 | miR-139-5p | trans | 3.57E-41 | Nightingale | LDL_C | NA | G | T | 0.03315 | 0.004406 | 9.30E-14 |
| rs676996 | miR-139-5p | trans | 3.57E-41 | Nightingale | S_LDL_C | NA | G | T | 0.030897 | 0.004429 | 5.80E-12 |
| rs676996 | miR-139-5p | trans | 3.57E-41 | Nightingale | L_LDL_PL | NA | G | T | 0.037957 | 0.004391 | 9.50E-18 |
| rs676996 | miR-139-5p | trans | 3.57E-41 | Nightingale | XXL_VLDL_ | NA | G | T | 0.034743 | 0.004434 | 3.30E-15 |
| rs676996 | miR-139-5p | trans | 3.57E-41 | Nightingale | non_HDL_( | NA | G | T | 0.035665 | 0.004401 | 5.30E-16 |
| rs676996 | miR-139-5p | trans | 3.57E-41 | Nightingale | S_LDL_CE_ | NA | G | T | 0.030893 | 0.004417 | 5.50E-14 |
| rs676996 | miR-139-5p | trans | 3.57E-41 | Nightingale | XXL_VLDL_ | NA | G | T | 0.025017 | 0.004454 | 1.50E-09 |
| rs676996 | miR-139-5p | trans | 3.57E-41 | Nightingale | S_LDL_L | NA | G | T | 0.027698 | 0.004424 | 8.50E-10 |
| rs676996 | miR-139-5p | trans | 3.57E-41 | Nightingale | S_VLDL_C_ | NA | G | T | 0.041827 | 0.004306 | 5.90E-22 |
| rs676996 | miR-139-5p | trans | 3.57E-41 | Nightingale | XS_VLDL_C | NA | G | T | 0.037811 | 0.00427 | 2.10E-18 |
| rs676996 | miR-139-5p | trans | 3.57E-41 | Nightingale | M_LDL_CE | NA | G | T | 0.032924 | 0.004431 | 1.00E-13 |
| rs676996 | miR-139-5p | trans | 3.57E-41 | Nightingale | XL_VLDL_C | NA | G | T | 0.037265 | 0.004245 | 3.70E-18 |
| rs676996 | miR-139-5p | trans | 3.57E-41 | Nightingale | XL_VLDL_C | NA | G | T | 0.038142 | 0.004243 | 5.40E-19 |
| rs676996 | miR-139-5p | trans | 3.57E-41 | Nightingale | S_HDL_P | NA | G | T | 0.030579 | 0.004425 | 5.40E-12 |
| rs676996 | miR-139-5p | trans | 3.57E-41 | Nightingale | XL_VLDL_F | NA | G | T | 0.037128 | 0.00429 | 2.30E-18 |
| rs676996 | miR-139-5p | trans | 3.57E-41 | Nightingale | S_HDL_C | NA | G | T | 0.028387 | 0.004417 | 1.80E-10 |
| rs676996 | miR-139-5p | trans | 3.57E-41 | Nightingale | M_VLDL_T | NA | G | T | -0.03849 | 0.004181 | 7.90E-20 |
| rs676996 | miR-139-5p | trans | 3.57E-41 | Nightingale | Total_C | NA | G | T | 0.032363 | 0.004284 | 3.40E-14 |
| rs676996 | miR-139-5p | trans | 3.57E-41 | Nightingale | GlycA | NA | G | T | 0.027865 | 0.004406 | 1.80E-10 |
| rs676996 | miR-139-5p | trans | 3.57E-41 | Nightingale | S_VLDL_FC | NA | G | T | 0.037868 | 0.004426 | 7.60E-18 |
| rs676996 | miR-139-5p | trans | 3.57E-41 | Nightingale | VLDL_FC | NA | G | T | 0.02413 | 0.004392 | 1.10E-08 |
| rs676996 | miR-139-5p | trans | 3.57E-41 | Nightingale | S_HDL_L | NA | G | T | 0.026288 | 0.00441 | 1.60E-09 |
| rs676996 | miR-139-5p | trans | 3.57E-41 | Nightingale | XL_HDL_PL | NA | G | T | -0.02267 | 0.004065 | 1.30E-08 |
| rs676996 | miR-139-5p | trans | 3.57E-41 | Nightingale | S_LDL_CE | NA | G | T | 0.030499 | 0.004427 | 6.10E-12 |
| rs676996 | miR-139-5p | trans | 3.57E-41 | Nightingale | XS_VLDL_F | NA | G | T | 0.037076 | 0.004348 | 6.40E-18 |
| rs676996 | miR-139-5p | trans | 3.57E-41 | Nightingale | L_LDL_CE | NA | G | T | 0.031522 | 0.004385 | 1.00E-12 |
| rs676996 | miR-139-5p | trans | 3.57E-41 | Nightingale | VLDL_CE | NA | G | T | 0.03576 | 0.004426 | 3.10E-16 |
| rs676996 | miR-139-5p | trans | 3.57E-41 | Nightingale | LDL_P | NA | G | T | 0.031745 | 0.004422 | 1.10E-12 |
| rs676996 | miR-139-5p | trans | 3.57E-41 | Nightingale | Total_L | NA | G | T | 0.026348 | 0.004353 | 4.60E-10 |
| rs676996 | miR-139-5p | trans | 3.57E-41 | Nightingale | LDL_L | NA | G | T | 0.033262 | 0.00441 | 7.80E-14 |
| rs676996 | miR-139-5p | trans | 3.57E-41 | Nightingale | M_LDL_P | NA | G | T | 0.035848 | 0.004437 | 7.30E-16 |
| rs676996 | miR-139-5p | trans | 3.57E-41 | Nightingale | XS_VLDL_F | NA | G | T | 0.031377 | 0.004393 | 2.60E-12 |
| rs676996 | miR-139-5p | trans | 3.57E-41 | Nightingale | IDL_PL | NA | G | T | 0.035327 | 0.00427 | 3.50E-16 |
| rs676996 | miR-139-5p | trans | 3.57E-41 | Nightingale | L_LDL_P | NA | G | T | 0.029116 | 0.00441 | 6.60E-11 |
| rs676996 | miR-139-5p | trans | 3.57E-41 | Nightingale | S_HDL_CE | NA | G | T | 0.028597 | 0.004432 | 1.40E-10 |
| rs676996 | miR-139-5p | trans | 3.57E-41 | Nightingale | IDL_P | NA | G | T | 0.040009 | 0.004378 | 1.30E-19 |
| rs676996 | miR-139-5p | trans | 3.57E-41 | Nightingale | IDL_C | NA | G | T | 0.034962 | 0.004274 | 9.40E-16 |
| rs676996 | miR-139-5p | trans | 3.57E-41 | Nightingale | LDL_FC | NA | G | T | 0.033597 | 0.004378 | 5.50E-14 |
| rs676996 | miR-139-5p | trans | 3.57E-41 | Nightingale | VLDL_C | NA | G | T | 0.031274 | 0.004419 | 5.00E-13 |
| rs676996 | miR-139-5p | trans | 3.57E-41 | Nightingale | L_LDL_C | NA | G | T | 0.032279 | 0.004375 | 2.70E-13 |
| rs676996 | miR-139-5p | trans | 3.57E-41 | Nightingale | S_VLDL_PL | NA | G | T | 0.032747 | 0.004417 | 4.30E-14 |

| | | | | | | | | | | | |
|---|---|---|---|---|---|---|---|---|---|---|---|
| rs676996 | miR-139-5p | trans | 3.57E-41 | Nightingale | M_LDL_C | NA | G | T | 0.03394 | 0.004433 | 2.30E-14 |
| rs676996 | miR-139-5p | trans | 3.57E-41 | Nightingale | Total_PL | NA | G | T | 0.024943 | 0.004214 | 1.10E-09 |
| rs676996 | miR-139-5p | trans | 3.57E-41 | Nightingale | S_VLDL_CE | NA | G | T | 0.046969 | 0.004365 | 1.40E-26 |
| rs676996 | miR-139-5p | trans | 3.57E-41 | Nightingale | S_VLDL_FC | NA | G | T | 0.029304 | 0.004228 | 1.10E-11 |
| rs676996 | miR-139-5p | trans | 3.57E-41 | Nightingale | M_LDL_FC | NA | G | T | 0.034304 | 0.004419 | 2.00E-14 |
| rs676996 | miR-139-5p | trans | 3.57E-41 | Nightingale | M_LDL_C_ | NA | G | T | 0.031099 | 0.004428 | 3.30E-12 |
| rs676996 | miR-139-5p | trans | 3.57E-41 | Nightingale | M_VLDL_P | NA | G | T | 0.043976 | 0.004247 | 6.90E-25 |
| rs676996 | miR-139-5p | trans | 3.57E-41 | Nightingale | Clinical_LD | NA | G | T | 0.036436 | 0.004399 | 2.90E-16 |
| rs676996 | miR-139-5p | trans | 3.57E-41 | Nightingale | M_VLDL_C | NA | G | T | 0.038672 | 0.004413 | 2.00E-18 |
| rs676996 | miR-139-5p | trans | 3.57E-41 | Nightingale | IDL_L | NA | G | T | 0.035274 | 0.004278 | 4.90E-16 |
| rs676996 | miR-139-5p | trans | 3.57E-41 | Nightingale | IDL_FC | NA | G | T | 0.038246 | 0.004284 | 1.50E-18 |
| rs676996 | miR-139-5p | trans | 3.57E-41 | Nightingale | S_VLDL_C | NA | G | T | 0.038343 | 0.004424 | 1.80E-18 |
| rs676996 | miR-139-5p | trans | 3.57E-41 | Nightingale | XXL_VLDL_ | NA | G | T | 0.033067 | 0.004459 | 9.60E-14 |
| rs676996 | miR-139-5p | trans | 3.57E-41 | Nightingale | Cholines | NA | G | T | 0.022308 | 0.004161 | 4.90E-08 |
| rs676996 | miR-139-5p | trans | 3.57E-41 | Nightingale | S_VLDL_PL | NA | G | T | 0.027677 | 0.004237 | 1.90E-10 |
| rs676996 | miR-139-5p | trans | 3.57E-41 | Nightingale | LDL_CE | NA | G | T | 0.032506 | 0.004413 | 2.30E-13 |
| rs676996 | miR-139-5p | trans | 3.57E-41 | Nightingale | L_LDL_L | NA | G | T | 0.033512 | 0.004382 | 3.20E-14 |
| rs676996 | miR-139-5p | trans | 3.57E-41 | Nightingale | L_VLDL_CE | NA | G | T | 0.047288 | 0.004234 | 2.80E-29 |
| rs676996 | miR-139-5p | trans | 3.57E-41 | Nightingale | L_VLDL_C_ | NA | G | T | 0.052087 | 0.004322 | 5.50E-34 |
| rs676996 | miR-139-5p | trans | 3.57E-41 | Nightingale | LA | NA | G | T | 0.024935 | 0.004339 | 3.50E-09 |
| rs676996 | miR-139-5p | trans | 3.57E-41 | Nightingale | S_VLDL_P | NA | G | T | 0.023454 | 0.004369 | 1.70E-08 |
| rs676996 | miR-139-5p | trans | 3.57E-41 | Nightingale | VLDL_P | NA | G | T | 0.02662 | 0.004401 | 4.40E-10 |
| rs676996 | miR-139-5p | trans | 3.57E-41 | Nightingale | PUFA | NA | G | T | 0.023693 | 0.004316 | 1.50E-08 |
| rs676996 | miR-139-5p | trans | 3.57E-41 | Nightingale | L_VLDL_FC | NA | G | T | 0.048434 | 0.004382 | 6.70E-29 |
| rs676996 | miR-139-5p | trans | 3.57E-41 | Nightingale | S_HDL_FC | NA | G | T | 0.023471 | 0.004344 | 4.60E-08 |
| rs676996 | miR-139-5p | trans | 3.57E-41 | Nightingale | M_VLDL_C | NA | G | T | 0.036147 | 0.004168 | 1.30E-17 |
| rs676996 | miR-139-5p | trans | 3.57E-41 | Nightingale | L_VLDL_TG | NA | G | T | -0.05103 | 0.004404 | 7.30E-32 |
| rs677355 | miR-126-3p | trans | 1.41E-38 | Nightingale | Citrate | NA | A | G | 0.026829 | 0.004397 | 7.20E-10 |
| rs677355 | miR-126-3p | trans | 1.41E-38 | Nightingale | ApoB_by_A | NA | A | G | 0.024942 | 0.004359 | 1.50E-08 |
| rs677355 | miR-126-3p | trans | 1.41E-38 | Nightingale | S_LDL_P | NA | A | G | 0.029497 | 0.00443 | 3.70E-11 |
| rs677355 | miR-126-3p | trans | 1.41E-38 | Nightingale | IDL_CE | NA | A | G | 0.033479 | 0.004273 | 1.50E-14 |
| rs677355 | miR-126-3p | trans | 1.41E-38 | Nightingale | M_LDL_FC | NA | A | G | 0.034256 | 0.00442 | 2.30E-14 |
| rs677355 | miR-126-3p | trans | 1.41E-38 | Nightingale | S_LDL_C_p | NA | A | G | 0.045307 | 0.004443 | 6.50E-25 |
| rs677355 | miR-126-3p | trans | 1.41E-38 | Nightingale | S_LDL_C | NA | A | G | 0.030835 | 0.00443 | 6.50E-12 |
| rs677355 | miR-126-3p | trans | 1.41E-38 | Nightingale | Total_PL | NA | A | G | 0.024874 | 0.004214 | 1.20E-09 |
| rs677355 | miR-126-3p | trans | 1.41E-38 | Nightingale | XS_VLDL_P | NA | A | G | 0.036669 | 0.004367 | 1.50E-17 |
| rs677355 | miR-126-3p | trans | 1.41E-38 | Nightingale | XS_VLDL_P | NA | A | G | 0.032072 | 0.004364 | 5.90E-14 |
| rs677355 | miR-126-3p | trans | 1.41E-38 | Nightingale | S_VLDL_FC | NA | A | G | 0.029355 | 0.004229 | 1.10E-11 |
| rs677355 | miR-126-3p | trans | 1.41E-38 | Nightingale | Remnant_( | NA | A | G | 0.036793 | 0.004397 | 4.30E-17 |
| rs677355 | miR-126-3p | trans | 1.41E-38 | Nightingale | Total_FC | NA | A | G | 0.033188 | 0.004324 | 8.00E-15 |
| rs677355 | miR-126-3p | trans | 1.41E-38 | Nightingale | IDL_PL | NA | A | G | 0.035285 | 0.004271 | 3.90E-16 |
| rs677355 | miR-126-3p | trans | 1.41E-38 | Nightingale | M_LDL_CE | NA | A | G | 0.032853 | 0.004432 | 1.20E-13 |
| rs677355 | miR-126-3p | trans | 1.41E-38 | Nightingale | S_HDL_PL | NA | A | G | 0.023722 | 0.004387 | 3.60E-08 |
| rs677355 | miR-126-3p | trans | 1.41E-38 | Nightingale | S_LDL_CE_ | NA | A | G | 0.030847 | 0.004418 | 6.00E-14 |
| rs677355 | miR-126-3p | trans | 1.41E-38 | Nightingale | ApoB | NA | A | G | 0.033845 | 0.00442 | 2.50E-14 |
| rs677355 | miR-126-3p | trans | 1.41E-38 | Nightingale | Total_C | NA | A | G | 0.032308 | 0.004285 | 3.90E-14 |
| rs677355 | miR-126-3p | trans | 1.41E-38 | Nightingale | XL_HDL_PL | NA | A | G | -0.02264 | 0.004066 | 1.30E-08 |
| rs677355 | miR-126-3p | trans | 1.41E-38 | Nightingale | L_LDL_FC | NA | A | G | 0.033559 | 0.004346 | 3.30E-14 |
| rs677355 | miR-126-3p | trans | 1.41E-38 | Nightingale | S_HDL_CE | NA | A | G | 0.028557 | 0.004433 | 1.50E-10 |
| rs677355 | miR-126-3p | trans | 1.41E-38 | Nightingale | XL_VLDL_C | NA | A | G | 0.038234 | 0.004244 | 4.60E-19 |
| rs677355 | miR-126-3p | trans | 1.41E-38 | Nightingale | S_VLDL_CE | NA | A | G | 0.038101 | 0.00442 | 1.90E-18 |
| rs677355 | miR-126-3p | trans | 1.41E-38 | Nightingale | M_LDL_P | NA | A | G | 0.035797 | 0.004438 | 8.20E-16 |
| rs677355 | miR-126-3p | trans | 1.41E-38 | Nightingale | M_VLDL_C | NA | A | G | 0.040775 | 0.004386 | 2.70E-20 |
| rs677355 | miR-126-3p | trans | 1.41E-38 | Nightingale | L_LDL_C | NA | A | G | 0.032229 | 0.004376 | 3.10E-13 |
| rs677355 | miR-126-3p | trans | 1.41E-38 | Nightingale | M_VLDL_P | NA | A | G | 0.029028 | 0.004419 | 2.40E-11 |
| rs677355 | miR-126-3p | trans | 1.41E-38 | Nightingale | M_LDL_C_ | NA | A | G | 0.031084 | 0.004429 | 3.40E-12 |
| rs677355 | miR-126-3p | trans | 1.41E-38 | Nightingale | M_VLDL_F | NA | A | G | 0.033273 | 0.004425 | 3.70E-14 |
| rs677355 | miR-126-3p | trans | 1.41E-38 | Nightingale | M_VLDL_P | NA | A | G | 0.028187 | 0.004415 | 7.40E-11 |
| rs677355 | miR-126-3p | trans | 1.41E-38 | Nightingale | XXL_VLDL_ | NA | A | G | 0.025023 | 0.004455 | 1.50E-09 |
| rs677355 | miR-126-3p | trans | 1.41E-38 | Nightingale | M_LDL_TG | NA | A | G | -0.02757 | 0.004371 | 3.50E-10 |
| rs677355 | miR-126-3p | trans | 1.41E-38 | Nightingale | L_LDL_CE | NA | A | G | 0.031469 | 0.004386 | 1.20E-12 |
| rs677355 | miR-126-3p | trans | 1.41E-38 | Nightingale | M_VLDL_C | NA | A | G | 0.033643 | 0.004158 | 1.90E-15 |
| rs677355 | miR-126-3p | trans | 1.41E-38 | Nightingale | XS_VLDL_C | NA | A | G | 0.037777 | 0.004271 | 2.40E-18 |

| | | | | | | | | | | | |
|---|---|---|---|---|---|---|---|---|---|---|---|
| rs677355 | miR-126-3p | trans | 1.41E-38 | Nightingale | M_VLDL_P | NA | A | G | 0.044027 | 0.004248 | 6.50E-25 |
| rs677355 | miR-126-3p | trans | 1.41E-38 | Nightingale | XS_VLDL_F | NA | A | G | 0.037025 | 0.004349 | 7.40E-18 |
| rs677355 | miR-126-3p | trans | 1.41E-38 | Nightingale | XS_VLDL_F | NA | A | G | 0.031383 | 0.004394 | 2.60E-12 |
| rs677355 | miR-126-3p | trans | 1.41E-38 | Nightingale | XXL_VLDL_ | NA | A | G | -0.03569 | 0.004464 | 2.30E-16 |
| rs677355 | miR-126-3p | trans | 1.41E-38 | Nightingale | Total_CE | NA | A | G | 0.031682 | 0.004269 | 1.10E-13 |
| rs677355 | miR-126-3p | trans | 1.41E-38 | Nightingale | LDL_PL | NA | A | G | 0.034895 | 0.004416 | 5.60E-15 |
| rs677355 | miR-126-3p | trans | 1.41E-38 | Nightingale | S_VLDL_TG | NA | A | G | -0.03812 | 0.004284 | 1.50E-18 |
| rs677355 | miR-126-3p | trans | 1.41E-38 | Nightingale | XL_VLDL_C | NA | A | G | 0.037355 | 0.004246 | 3.10E-18 |
| rs677355 | miR-126-3p | trans | 1.41E-38 | Nightingale | M_VLDL_C | NA | A | G | 0.036213 | 0.004169 | 1.20E-17 |
| rs677355 | miR-126-3p | trans | 1.41E-38 | Nightingale | LDL_P | NA | A | G | 0.031689 | 0.004423 | 1.20E-12 |
| rs677355 | miR-126-3p | trans | 1.41E-38 | Nightingale | XL_VLDL_T | NA | A | G | -0.04386 | 0.00429 | 2.10E-24 |
| rs677355 | miR-126-3p | trans | 1.41E-38 | Nightingale | S_LDL_PL_ | NA | A | G | -0.03 | 0.004418 | 3.00E-13 |
| rs677355 | miR-126-3p | trans | 1.41E-38 | Nightingale | S_LDL_CE | NA | A | G | 0.030433 | 0.004428 | 7.00E-12 |
| rs677355 | miR-126-3p | trans | 1.41E-38 | Nightingale | XXL_VLDL_ | NA | A | G | 0.034787 | 0.004435 | 3.10E-15 |
| rs677355 | miR-126-3p | trans | 1.41E-38 | Nightingale | S_VLDL_C | NA | A | G | 0.038286 | 0.004425 | 2.10E-18 |
| rs677355 | miR-126-3p | trans | 1.41E-38 | Nightingale | LDL_CE | NA | A | G | 0.032446 | 0.004414 | 2.60E-13 |
| rs677355 | miR-126-3p | trans | 1.41E-38 | Nightingale | M_LDL_L | NA | A | G | 0.032919 | 0.004432 | 1.20E-13 |
| rs677355 | miR-126-3p | trans | 1.41E-38 | Nightingale | IDL_FC | NA | A | G | 0.038214 | 0.004285 | 1.70E-18 |
| rs677355 | miR-126-3p | trans | 1.41E-38 | Nightingale | S_LDL_L | NA | A | G | 0.027634 | 0.004425 | 9.50E-10 |
| rs677355 | miR-126-3p | trans | 1.41E-38 | Nightingale | S_VLDL_L | NA | A | G | 0.024574 | 0.004383 | 4.30E-09 |
| rs677355 | miR-126-3p | trans | 1.41E-38 | Nightingale | VLDL_FC | NA | A | G | 0.024053 | 0.004393 | 1.30E-08 |
| rs677355 | miR-126-3p | trans | 1.41E-38 | Nightingale | S_HDL_L | NA | A | G | 0.026224 | 0.004411 | 1.80E-09 |
| rs677355 | miR-126-3p | trans | 1.41E-38 | Nightingale | M_LDL_PL | NA | A | G | 0.03236 | 0.004432 | 3.20E-13 |
| rs677355 | miR-126-3p | trans | 1.41E-38 | Nightingale | XXL_VLDL_ | NA | A | G | 0.030793 | 0.004407 | 1.20E-12 |
| rs677355 | miR-126-3p | trans | 1.41E-38 | Nightingale | Total_L | NA | A | G | 0.026271 | 0.004354 | 5.30E-10 |
| rs677355 | miR-126-3p | trans | 1.41E-38 | Nightingale | M_VLDL_C | NA | A | G | 0.038623 | 0.004414 | 2.30E-18 |
| rs677355 | miR-126-3p | trans | 1.41E-38 | Nightingale | S_VLDL_C_ | NA | A | G | 0.041886 | 0.004307 | 5.40E-22 |
| rs677355 | miR-126-3p | trans | 1.41E-38 | Nightingale | IDL_C | NA | A | G | 0.034923 | 0.004274 | 1.10E-15 |
| rs677355 | miR-126-3p | trans | 1.41E-38 | Nightingale | XXL_VLDL_ | NA | A | G | 0.033101 | 0.00446 | 9.20E-14 |
| rs677355 | miR-126-3p | trans | 1.41E-38 | Nightingale | L_VLDL_C_ | NA | A | G | 0.052158 | 0.004323 | 4.80E-34 |
| rs677355 | miR-126-3p | trans | 1.41E-38 | Nightingale | VLDL_C | NA | A | G | 0.031206 | 0.00442 | 5.80E-13 |
| rs677355 | miR-126-3p | trans | 1.41E-38 | Nightingale | Omega_6 | NA | A | G | 0.025797 | 0.004322 | 6.80E-10 |
| rs677355 | miR-126-3p | trans | 1.41E-38 | Nightingale | XS_VLDL_L | NA | A | G | 0.033781 | 0.004351 | 2.30E-15 |
| rs677355 | miR-126-3p | trans | 1.41E-38 | Nightingale | Sphingomy | NA | A | G | 0.028914 | 0.004137 | 8.50E-12 |
| rs677355 | miR-126-3p | trans | 1.41E-38 | Nightingale | LDL_FC | NA | A | G | 0.033557 | 0.004379 | 6.00E-14 |
| rs677355 | miR-126-3p | trans | 1.41E-38 | Nightingale | LDL_C | NA | A | G | 0.033095 | 0.004407 | 1.10E-13 |
| rs677355 | miR-126-3p | trans | 1.41E-38 | Nightingale | S_VLDL_FC | NA | A | G | 0.03781 | 0.004427 | 8.80E-18 |
| rs677355 | miR-126-3p | trans | 1.41E-38 | Nightingale | S_HDL_C | NA | A | G | 0.028339 | 0.004418 | 1.90E-10 |
| rs677355 | miR-126-3p | trans | 1.41E-38 | Nightingale | L_LDL_L | NA | A | G | 0.03346 | 0.004383 | 3.60E-14 |
| rs677355 | miR-126-3p | trans | 1.41E-38 | Nightingale | S_VLDL_CE | NA | A | G | 0.047027 | 0.004366 | 1.30E-26 |
| rs677355 | miR-126-3p | trans | 1.41E-38 | Nightingale | PUFA | NA | A | G | 0.023614 | 0.004317 | 1.70E-08 |
| rs677355 | miR-126-3p | trans | 1.41E-38 | Nightingale | S_VLDL_P | NA | A | G | 0.023377 | 0.00437 | 1.90E-08 |
| rs677355 | miR-126-3p | trans | 1.41E-38 | Nightingale | M_VLDL_F | NA | A | G | 0.042055 | 0.004211 | 3.70E-23 |
| rs677355 | miR-126-3p | trans | 1.41E-38 | Nightingale | S_LDL_FC | NA | A | G | 0.028483 | 0.004419 | 4.80E-10 |
| rs677355 | miR-126-3p | trans | 1.41E-38 | Nightingale | IDL_P | NA | A | G | 0.039974 | 0.004379 | 1.40E-19 |
| rs677355 | miR-126-3p | trans | 1.41E-38 | Nightingale | IDL_FC_pct | NA | A | G | 0.025744 | 0.004392 | 1.10E-08 |
| rs677355 | miR-126-3p | trans | 1.41E-38 | Nightingale | M_VLDL_T | NA | A | G | -0.03856 | 0.004182 | 7.10E-20 |
| rs677355 | miR-126-3p | trans | 1.41E-38 | Nightingale | S_HDL_P | NA | A | G | 0.030523 | 0.004426 | 6.00E-12 |
| rs677355 | miR-126-3p | trans | 1.41E-38 | Nightingale | XS_VLDL_C | NA | A | G | 0.037965 | 0.004296 | 1.50E-18 |
| rs677355 | miR-126-3p | trans | 1.41E-38 | Nightingale | Clinical_LD | NA | A | G | 0.036386 | 0.0044 | 3.20E-16 |
| rs677355 | miR-126-3p | trans | 1.41E-38 | Nightingale | LA | NA | A | G | 0.024871 | 0.00434 | 4.00E-09 |
| rs677355 | miR-126-3p | trans | 1.41E-38 | Nightingale | M_LDL_C | NA | A | G | 0.033874 | 0.004434 | 2.60E-14 |
| rs677355 | miR-126-3p | trans | 1.41E-38 | Nightingale | LDL_L | NA | A | G | 0.033204 | 0.004411 | 8.80E-14 |
| rs677355 | miR-126-3p | trans | 1.41E-38 | Nightingale | S_VLDL_PL | NA | A | G | 0.03268 | 0.004418 | 5.00E-14 |
| rs677355 | miR-126-3p | trans | 1.41E-38 | Nightingale | non_HDL_( | NA | A | G | 0.035607 | 0.004402 | 6.10E-16 |
| rs677355 | miR-126-3p | trans | 1.41E-38 | Nightingale | L_VLDL_FC | NA | A | G | 0.048463 | 0.004383 | 6.40E-29 |
| rs677355 | miR-126-3p | trans | 1.41E-38 | Nightingale | L_VLDL_TG | NA | A | G | -0.05108 | 0.004405 | 6.90E-32 |
| rs677355 | miR-126-3p | trans | 1.41E-38 | Nightingale | XL_VLDL_P | NA | A | G | 0.038718 | 0.004433 | 7.00E-19 |
| rs677355 | miR-126-3p | trans | 1.41E-38 | Nightingale | GlycA | NA | A | G | 0.02771 | 0.004407 | 2.30E-10 |
| rs677355 | miR-126-3p | trans | 1.41E-38 | Nightingale | S_VLDL_PL | NA | A | G | 0.027729 | 0.004238 | 1.80E-10 |
| rs677355 | miR-126-3p | trans | 1.41E-38 | Nightingale | VLDL_CE | NA | A | G | 0.035699 | 0.004427 | 3.60E-16 |
| rs677355 | miR-126-3p | trans | 1.41E-38 | Nightingale | IDL_L | NA | A | G | 0.03523 | 0.004279 | 5.50E-16 |
| rs677355 | miR-126-3p | trans | 1.41E-38 | Nightingale | L_LDL_PL | NA | A | G | 0.037906 | 0.004392 | 1.10E-17 |

| | | | | | | | | | | | |
|---|---|---|---|---|---|---|---|---|---|---|---|
| rs677355 | miR-126-3 | trans | 1.41E-38 | Nightingale | L_LDL_P | NA | A | G | 0.029058 | 0.004411 | 7.40E-11 |
| rs677355 | miR-126-3 | trans | 1.41E-38 | Nightingale | VLDL_P | NA | A | G | 0.026547 | 0.004402 | 5.00E-10 |
| rs677355 | miR-126-3 | trans | 1.41E-38 | Nightingale | L_VLDL_CE | NA | A | G | 0.047363 | 0.004235 | 2.40E-29 |
| rs677355 | miR-126-3 | trans | 1.41E-38 | Nightingale | XL_VLDL_F | NA | A | G | 0.037212 | 0.00429 | 2.00E-18 |
| rs677355 | let-7b-5p | trans | 2.73E-26 | Nightingale | Citrate | NA | A | G | 0.026829 | 0.004397 | 7.20E-10 |
| rs677355 | let-7b-5p | trans | 2.73E-26 | Nightingale | ApoB_by_A | NA | A | G | 0.024942 | 0.004359 | 1.50E-08 |
| rs677355 | let-7b-5p | trans | 2.73E-26 | Nightingale | S_LDL_P | NA | A | G | 0.029497 | 0.00443 | 3.70E-11 |
| rs677355 | let-7b-5p | trans | 2.73E-26 | Nightingale | IDL_CE | NA | A | G | 0.033479 | 0.004273 | 1.50E-14 |
| rs677355 | let-7b-5p | trans | 2.73E-26 | Nightingale | M_LDL_FC | NA | A | G | 0.034256 | 0.00442 | 2.30E-14 |
| rs677355 | let-7b-5p | trans | 2.73E-26 | Nightingale | S_LDL_C_p | NA | A | G | 0.045307 | 0.004443 | 6.50E-25 |
| rs677355 | let-7b-5p | trans | 2.73E-26 | Nightingale | S_LDL_C | NA | A | G | 0.030835 | 0.00443 | 6.50E-12 |
| rs677355 | let-7b-5p | trans | 2.73E-26 | Nightingale | Total_PL | NA | A | G | 0.024874 | 0.004214 | 1.20E-09 |
| rs677355 | let-7b-5p | trans | 2.73E-26 | Nightingale | XS_VLDL_P | NA | A | G | 0.036669 | 0.004367 | 1.50E-17 |
| rs677355 | let-7b-5p | trans | 2.73E-26 | Nightingale | XS_VLDL_P | NA | A | G | 0.032072 | 0.004364 | 5.90E-14 |
| rs677355 | let-7b-5p | trans | 2.73E-26 | Nightingale | S_VLDL_FC | NA | A | G | 0.029355 | 0.004229 | 1.10E-11 |
| rs677355 | let-7b-5p | trans | 2.73E-26 | Nightingale | Remnant_( | NA | A | G | 0.036793 | 0.004397 | 4.30E-17 |
| rs677355 | let-7b-5p | trans | 2.73E-26 | Nightingale | Total_FC | NA | A | G | 0.033188 | 0.004324 | 8.00E-15 |
| rs677355 | let-7b-5p | trans | 2.73E-26 | Nightingale | IDL_PL | NA | A | G | 0.035285 | 0.004271 | 3.90E-16 |
| rs677355 | let-7b-5p | trans | 2.73E-26 | Nightingale | M_LDL_CE | NA | A | G | 0.032853 | 0.004432 | 1.20E-13 |
| rs677355 | let-7b-5p | trans | 2.73E-26 | Nightingale | S_HDL_PL | NA | A | G | 0.023722 | 0.004387 | 3.60E-08 |
| rs677355 | let-7b-5p | trans | 2.73E-26 | Nightingale | S_LDL_CE_ | NA | A | G | 0.030847 | 0.004418 | 6.00E-14 |
| rs677355 | let-7b-5p | trans | 2.73E-26 | Nightingale | ApoB | NA | A | G | 0.033845 | 0.00442 | 2.50E-14 |
| rs677355 | let-7b-5p | trans | 2.73E-26 | Nightingale | Total_C | NA | A | G | 0.032308 | 0.004285 | 3.90E-14 |
| rs677355 | let-7b-5p | trans | 2.73E-26 | Nightingale | XL_HDL_PL | NA | A | G | -0.02264 | 0.004066 | 1.30E-08 |
| rs677355 | let-7b-5p | trans | 2.73E-26 | Nightingale | L_LDL_FC | NA | A | G | 0.033559 | 0.004346 | 3.30E-14 |
| rs677355 | let-7b-5p | trans | 2.73E-26 | Nightingale | S_HDL_CE | NA | A | G | 0.028557 | 0.004433 | 1.50E-10 |
| rs677355 | let-7b-5p | trans | 2.73E-26 | Nightingale | XL_VLDL_C | NA | A | G | 0.038234 | 0.004244 | 4.60E-19 |
| rs677355 | let-7b-5p | trans | 2.73E-26 | Nightingale | S_VLDL_CE | NA | A | G | 0.038101 | 0.00442 | 1.90E-18 |
| rs677355 | let-7b-5p | trans | 2.73E-26 | Nightingale | M_LDL_P | NA | A | G | 0.035797 | 0.004438 | 8.20E-16 |
| rs677355 | let-7b-5p | trans | 2.73E-26 | Nightingale | M_VLDL_C | NA | A | G | 0.040775 | 0.004386 | 2.70E-20 |
| rs677355 | let-7b-5p | trans | 2.73E-26 | Nightingale | L_LDL_C | NA | A | G | 0.032229 | 0.004376 | 3.10E-13 |
| rs677355 | let-7b-5p | trans | 2.73E-26 | Nightingale | M_VLDL_P | NA | A | G | 0.029028 | 0.004419 | 2.40E-11 |
| rs677355 | let-7b-5p | trans | 2.73E-26 | Nightingale | M_LDL_C_ | NA | A | G | 0.031084 | 0.004429 | 3.40E-12 |
| rs677355 | let-7b-5p | trans | 2.73E-26 | Nightingale | M_VLDL_F | NA | A | G | 0.033273 | 0.004425 | 3.70E-14 |
| rs677355 | let-7b-5p | trans | 2.73E-26 | Nightingale | M_VLDL_P | NA | A | G | 0.028187 | 0.004415 | 7.40E-11 |
| rs677355 | let-7b-5p | trans | 2.73E-26 | Nightingale | XXL_VLDL_ | NA | A | G | 0.025023 | 0.004455 | 1.50E-09 |
| rs677355 | let-7b-5p | trans | 2.73E-26 | Nightingale | M_LDL_TG | NA | A | G | -0.02757 | 0.004371 | 3.50E-10 |
| rs677355 | let-7b-5p | trans | 2.73E-26 | Nightingale | L_LDL_CE | NA | A | G | 0.031469 | 0.004386 | 1.20E-12 |
| rs677355 | let-7b-5p | trans | 2.73E-26 | Nightingale | M_VLDL_C | NA | A | G | 0.033643 | 0.004158 | 1.90E-15 |
| rs677355 | let-7b-5p | trans | 2.73E-26 | Nightingale | XS_VLDL_C | NA | A | G | 0.037777 | 0.004271 | 2.40E-18 |
| rs677355 | let-7b-5p | trans | 2.73E-26 | Nightingale | M_VLDL_P | NA | A | G | 0.044027 | 0.004248 | 6.50E-25 |
| rs677355 | let-7b-5p | trans | 2.73E-26 | Nightingale | XS_VLDL_F | NA | A | G | 0.037025 | 0.004349 | 7.40E-18 |
| rs677355 | let-7b-5p | trans | 2.73E-26 | Nightingale | XS_VLDL_F | NA | A | G | 0.031383 | 0.004394 | 2.60E-12 |
| rs677355 | let-7b-5p | trans | 2.73E-26 | Nightingale | XXL_VLDL_ | NA | A | G | -0.03569 | 0.004464 | 2.30E-16 |
| rs677355 | let-7b-5p | trans | 2.73E-26 | Nightingale | Total_CE | NA | A | G | 0.031682 | 0.004269 | 1.10E-13 |
| rs677355 | let-7b-5p | trans | 2.73E-26 | Nightingale | LDL_PL | NA | A | G | 0.034895 | 0.004416 | 5.60E-15 |
| rs677355 | let-7b-5p | trans | 2.73E-26 | Nightingale | S_VLDL_TG | NA | A | G | -0.03812 | 0.004284 | 1.50E-18 |
| rs677355 | let-7b-5p | trans | 2.73E-26 | Nightingale | XL_VLDL_C | NA | A | G | 0.037355 | 0.004246 | 3.10E-18 |
| rs677355 | let-7b-5p | trans | 2.73E-26 | Nightingale | M_VLDL_C | NA | A | G | 0.036213 | 0.004169 | 1.20E-17 |
| rs677355 | let-7b-5p | trans | 2.73E-26 | Nightingale | LDL_P | NA | A | G | 0.031689 | 0.004423 | 1.20E-12 |
| rs677355 | let-7b-5p | trans | 2.73E-26 | Nightingale | XL_VLDL_T | NA | A | G | -0.04386 | 0.00429 | 2.10E-24 |
| rs677355 | let-7b-5p | trans | 2.73E-26 | Nightingale | S_LDL_PL_ | NA | A | G | -0.03 | 0.004418 | 3.00E-13 |
| rs677355 | let-7b-5p | trans | 2.73E-26 | Nightingale | S_LDL_CE | NA | A | G | 0.030433 | 0.004428 | 7.00E-12 |
| rs677355 | let-7b-5p | trans | 2.73E-26 | Nightingale | XXL_VLDL_ | NA | A | G | 0.034787 | 0.004435 | 3.10E-15 |
| rs677355 | let-7b-5p | trans | 2.73E-26 | Nightingale | S_VLDL_C | NA | A | G | 0.038286 | 0.004425 | 2.10E-18 |
| rs677355 | let-7b-5p | trans | 2.73E-26 | Nightingale | LDL_CE | NA | A | G | 0.032446 | 0.004414 | 2.60E-13 |
| rs677355 | let-7b-5p | trans | 2.73E-26 | Nightingale | M_LDL_L | NA | A | G | 0.032919 | 0.004432 | 1.20E-13 |
| rs677355 | let-7b-5p | trans | 2.73E-26 | Nightingale | IDL_FC | NA | A | G | 0.038214 | 0.004285 | 1.70E-18 |
| rs677355 | let-7b-5p | trans | 2.73E-26 | Nightingale | S_LDL_L | NA | A | G | 0.027634 | 0.004425 | 9.50E-10 |
| rs677355 | let-7b-5p | trans | 2.73E-26 | Nightingale | S_VLDL_L | NA | A | G | 0.024574 | 0.004383 | 4.30E-09 |
| rs677355 | let-7b-5p | trans | 2.73E-26 | Nightingale | VLDL_FC | NA | A | G | 0.024053 | 0.004393 | 1.30E-08 |
| rs677355 | let-7b-5p | trans | 2.73E-26 | Nightingale | S_HDL_L | NA | A | G | 0.026224 | 0.004411 | 1.80E-09 |
| rs677355 | let-7b-5p | trans | 2.73E-26 | Nightingale | M_LDL_PL | NA | A | G | 0.03236 | 0.004432 | 3.20E-13 |

| | | | | | | | | | | | |
|---|---|---|---|---|---|---|---|---|---|---|---|
| rs677355 | let-7b-5p | trans | 2.73E-26 | Nightingale | XXL_VLDL_ | NA | A | G | 0.030793 | 0.004407 | 1.20E-12 |
| rs677355 | let-7b-5p | trans | 2.73E-26 | Nightingale | Total_L | NA | A | G | 0.026271 | 0.004354 | 5.30E-10 |
| rs677355 | let-7b-5p | trans | 2.73E-26 | Nightingale | M_VLDL_C | NA | A | G | 0.038623 | 0.004414 | 2.30E-18 |
| rs677355 | let-7b-5p | trans | 2.73E-26 | Nightingale | S_VLDL_C_ | NA | A | G | 0.041886 | 0.004307 | 5.40E-22 |
| rs677355 | let-7b-5p | trans | 2.73E-26 | Nightingale | IDL_C | NA | A | G | 0.034923 | 0.004274 | 1.10E-15 |
| rs677355 | let-7b-5p | trans | 2.73E-26 | Nightingale | XXL_VLDL_ | NA | A | G | 0.033101 | 0.00446 | 9.20E-14 |
| rs677355 | let-7b-5p | trans | 2.73E-26 | Nightingale | L_VLDL_C_ | NA | A | G | 0.052158 | 0.004323 | 4.80E-34 |
| rs677355 | let-7b-5p | trans | 2.73E-26 | Nightingale | VLDL_C | NA | A | G | 0.031206 | 0.00442 | 5.80E-13 |
| rs677355 | let-7b-5p | trans | 2.73E-26 | Nightingale | Omega_6 | NA | A | G | 0.025797 | 0.004322 | 6.80E-10 |
| rs677355 | let-7b-5p | trans | 2.73E-26 | Nightingale | XS_VLDL_L | NA | A | G | 0.033781 | 0.004351 | 2.30E-15 |
| rs677355 | let-7b-5p | trans | 2.73E-26 | Nightingale | Sphingomy | NA | A | G | 0.028914 | 0.004137 | 8.50E-12 |
| rs677355 | let-7b-5p | trans | 2.73E-26 | Nightingale | LDL_FC | NA | A | G | 0.033557 | 0.004379 | 6.00E-14 |
| rs677355 | let-7b-5p | trans | 2.73E-26 | Nightingale | LDL_C | NA | A | G | 0.033095 | 0.004407 | 1.10E-13 |
| rs677355 | let-7b-5p | trans | 2.73E-26 | Nightingale | S_VLDL_FC | NA | A | G | 0.03781 | 0.004427 | 8.80E-18 |
| rs677355 | let-7b-5p | trans | 2.73E-26 | Nightingale | S_HDL_C | NA | A | G | 0.028339 | 0.004418 | 1.90E-10 |
| rs677355 | let-7b-5p | trans | 2.73E-26 | Nightingale | L_LDL_L | NA | A | G | 0.03346 | 0.004383 | 3.60E-14 |
| rs677355 | let-7b-5p | trans | 2.73E-26 | Nightingale | S_VLDL_CE | NA | A | G | 0.047027 | 0.004366 | 1.30E-26 |
| rs677355 | let-7b-5p | trans | 2.73E-26 | Nightingale | PUFA | NA | A | G | 0.023614 | 0.004317 | 1.70E-08 |
| rs677355 | let-7b-5p | trans | 2.73E-26 | Nightingale | S_VLDL_P | NA | A | G | 0.023377 | 0.00437 | 1.90E-08 |
| rs677355 | let-7b-5p | trans | 2.73E-26 | Nightingale | M_VLDL_F | NA | A | G | 0.042055 | 0.004211 | 3.70E-23 |
| rs677355 | let-7b-5p | trans | 2.73E-26 | Nightingale | S_LDL_FC | NA | A | G | 0.028483 | 0.004419 | 4.80E-10 |
| rs677355 | let-7b-5p | trans | 2.73E-26 | Nightingale | IDL_P | NA | A | G | 0.039974 | 0.004379 | 1.40E-19 |
| rs677355 | let-7b-5p | trans | 2.73E-26 | Nightingale | IDL_FC_pct | NA | A | G | 0.025744 | 0.004392 | 1.10E-08 |
| rs677355 | let-7b-5p | trans | 2.73E-26 | Nightingale | M_VLDL_T | NA | A | G | -0.03856 | 0.004182 | 7.10E-20 |
| rs677355 | let-7b-5p | trans | 2.73E-26 | Nightingale | S_HDL_P | NA | A | G | 0.030523 | 0.004426 | 6.00E-12 |
| rs677355 | let-7b-5p | trans | 2.73E-26 | Nightingale | XS_VLDL_C | NA | A | G | 0.037965 | 0.004296 | 1.50E-18 |
| rs677355 | let-7b-5p | trans | 2.73E-26 | Nightingale | Clinical_LD | NA | A | G | 0.036386 | 0.0044 | 3.20E-16 |
| rs677355 | let-7b-5p | trans | 2.73E-26 | Nightingale | LA | NA | A | G | 0.024871 | 0.00434 | 4.00E-09 |
| rs677355 | let-7b-5p | trans | 2.73E-26 | Nightingale | M_LDL_C | NA | A | G | 0.033874 | 0.004434 | 2.60E-14 |
| rs677355 | let-7b-5p | trans | 2.73E-26 | Nightingale | LDL_L | NA | A | G | 0.033204 | 0.004411 | 8.80E-14 |
| rs677355 | let-7b-5p | trans | 2.73E-26 | Nightingale | S_VLDL_PL | NA | A | G | 0.03268 | 0.004418 | 5.00E-14 |
| rs677355 | let-7b-5p | trans | 2.73E-26 | Nightingale | non_HDL_( | NA | A | G | 0.035607 | 0.004402 | 6.10E-16 |
| rs677355 | let-7b-5p | trans | 2.73E-26 | Nightingale | L_VLDL_FC | NA | A | G | 0.048463 | 0.004383 | 6.40E-29 |
| rs677355 | let-7b-5p | trans | 2.73E-26 | Nightingale | L_VLDL_TG | NA | A | G | -0.05108 | 0.004405 | 6.90E-32 |
| rs677355 | let-7b-5p | trans | 2.73E-26 | Nightingale | XL_VLDL_P | NA | A | G | 0.038718 | 0.004433 | 7.00E-19 |
| rs677355 | let-7b-5p | trans | 2.73E-26 | Nightingale | GlycA | NA | A | G | 0.02771 | 0.004407 | 2.30E-10 |
| rs677355 | let-7b-5p | trans | 2.73E-26 | Nightingale | S_VLDL_PL | NA | A | G | 0.027729 | 0.004238 | 1.80E-10 |
| rs677355 | let-7b-5p | trans | 2.73E-26 | Nightingale | VLDL_CE | NA | A | G | 0.035699 | 0.004427 | 3.60E-16 |
| rs677355 | let-7b-5p | trans | 2.73E-26 | Nightingale | IDL_L | NA | A | G | 0.03523 | 0.004279 | 5.50E-16 |
| rs677355 | let-7b-5p | trans | 2.73E-26 | Nightingale | L_LDL_PL | NA | A | G | 0.037906 | 0.004392 | 1.10E-17 |
| rs677355 | let-7b-5p | trans | 2.73E-26 | Nightingale | L_LDL_P | NA | A | G | 0.029058 | 0.004411 | 7.40E-11 |
| rs677355 | let-7b-5p | trans | 2.73E-26 | Nightingale | VLDL_P | NA | A | G | 0.026547 | 0.004402 | 5.00E-10 |
| rs677355 | let-7b-5p | trans | 2.73E-26 | Nightingale | L_VLDL_CE | NA | A | G | 0.047363 | 0.004235 | 2.40E-29 |
| rs677355 | let-7b-5p | trans | 2.73E-26 | Nightingale | XL_VLDL_F | NA | A | G | 0.037212 | 0.00429 | 2.00E-18 |
| rs677355 | miR-139-5p | trans | 1.01E-40 | Nightingale | Citrate | NA | A | G | 0.026829 | 0.004397 | 7.20E-10 |
| rs677355 | miR-139-5p | trans | 1.01E-40 | Nightingale | ApoB_by_A | NA | A | G | 0.024942 | 0.004359 | 1.50E-08 |
| rs677355 | miR-139-5p | trans | 1.01E-40 | Nightingale | S_LDL_P | NA | A | G | 0.029497 | 0.00443 | 3.70E-11 |
| rs677355 | miR-139-5p | trans | 1.01E-40 | Nightingale | IDL_CE | NA | A | G | 0.033479 | 0.004273 | 1.50E-14 |
| rs677355 | miR-139-5p | trans | 1.01E-40 | Nightingale | M_LDL_FC | NA | A | G | 0.034256 | 0.00442 | 2.30E-14 |
| rs677355 | miR-139-5p | trans | 1.01E-40 | Nightingale | S_LDL_C_p | NA | A | G | 0.045307 | 0.004443 | 6.50E-25 |
| rs677355 | miR-139-5p | trans | 1.01E-40 | Nightingale | S_LDL_C | NA | A | G | 0.030835 | 0.00443 | 6.50E-12 |
| rs677355 | miR-139-5p | trans | 1.01E-40 | Nightingale | Total_PL | NA | A | G | 0.024874 | 0.004214 | 1.20E-09 |
| rs677355 | miR-139-5p | trans | 1.01E-40 | Nightingale | XS_VLDL_P | NA | A | G | 0.036669 | 0.004367 | 1.50E-17 |
| rs677355 | miR-139-5p | trans | 1.01E-40 | Nightingale | XS_VLDL_P | NA | A | G | 0.032072 | 0.004364 | 5.90E-14 |
| rs677355 | miR-139-5p | trans | 1.01E-40 | Nightingale | S_VLDL_FC | NA | A | G | 0.029355 | 0.004229 | 1.10E-11 |
| rs677355 | miR-139-5p | trans | 1.01E-40 | Nightingale | Remnant_( | NA | A | G | 0.036793 | 0.004397 | 4.30E-17 |
| rs677355 | miR-139-5p | trans | 1.01E-40 | Nightingale | Total_FC | NA | A | G | 0.033188 | 0.004324 | 8.00E-15 |
| rs677355 | miR-139-5p | trans | 1.01E-40 | Nightingale | IDL_PL | NA | A | G | 0.035285 | 0.004271 | 3.90E-16 |
| rs677355 | miR-139-5p | trans | 1.01E-40 | Nightingale | M_LDL_CE | NA | A | G | 0.032853 | 0.004432 | 1.20E-13 |
| rs677355 | miR-139-5p | trans | 1.01E-40 | Nightingale | S_HDL_PL | NA | A | G | 0.023722 | 0.004387 | 3.60E-08 |
| rs677355 | miR-139-5p | trans | 1.01E-40 | Nightingale | S_LDL_CE_ | NA | A | G | 0.030847 | 0.004418 | 6.00E-14 |
| rs677355 | miR-139-5p | trans | 1.01E-40 | Nightingale | ApoB | NA | A | G | 0.033845 | 0.00442 | 2.50E-14 |
| rs677355 | miR-139-5p | trans | 1.01E-40 | Nightingale | Total_C | NA | A | G | 0.032308 | 0.004285 | 3.90E-14 |

| | | | | | | | | | | | |
|---|---|---|---|---|---|---|---|---|---|---|---|
| rs677355 | miR-139-5p | trans | 1.01E-40 | Nightingale | XL_HDL_PL | NA | A | G | -0.02264 | 0.004066 | 1.30E-08 |
| rs677355 | miR-139-5p | trans | 1.01E-40 | Nightingale | L_LDL_FC | NA | A | G | 0.033559 | 0.004346 | 3.30E-14 |
| rs677355 | miR-139-5p | trans | 1.01E-40 | Nightingale | S_HDL_CE | NA | A | G | 0.028557 | 0.004433 | 1.50E-10 |
| rs677355 | miR-139-5p | trans | 1.01E-40 | Nightingale | XL_VLDL_C | NA | A | G | 0.038234 | 0.004244 | 4.60E-19 |
| rs677355 | miR-139-5p | trans | 1.01E-40 | Nightingale | S_VLDL_CE | NA | A | G | 0.038101 | 0.00442 | 1.90E-18 |
| rs677355 | miR-139-5p | trans | 1.01E-40 | Nightingale | M_LDL_P | NA | A | G | 0.035797 | 0.004438 | 8.20E-16 |
| rs677355 | miR-139-5p | trans | 1.01E-40 | Nightingale | M_VLDL_C | NA | A | G | 0.040775 | 0.004386 | 2.70E-20 |
| rs677355 | miR-139-5p | trans | 1.01E-40 | Nightingale | L_LDL_C | NA | A | G | 0.032229 | 0.004376 | 3.10E-13 |
| rs677355 | miR-139-5p | trans | 1.01E-40 | Nightingale | M_VLDL_P | NA | A | G | 0.029028 | 0.004419 | 2.40E-11 |
| rs677355 | miR-139-5p | trans | 1.01E-40 | Nightingale | M_LDL_C_ | NA | A | G | 0.031084 | 0.004429 | 3.40E-12 |
| rs677355 | miR-139-5p | trans | 1.01E-40 | Nightingale | M_VLDL_F | NA | A | G | 0.033273 | 0.004425 | 3.70E-14 |
| rs677355 | miR-139-5p | trans | 1.01E-40 | Nightingale | M_VLDL_P | NA | A | G | 0.028187 | 0.004415 | 7.40E-11 |
| rs677355 | miR-139-5p | trans | 1.01E-40 | Nightingale | XXL_VLDL_ | NA | A | G | 0.025023 | 0.004455 | 1.50E-09 |
| rs677355 | miR-139-5p | trans | 1.01E-40 | Nightingale | M_LDL_TG | NA | A | G | -0.02757 | 0.004371 | 3.50E-10 |
| rs677355 | miR-139-5p | trans | 1.01E-40 | Nightingale | L_LDL_CE | NA | A | G | 0.031469 | 0.004386 | 1.20E-12 |
| rs677355 | miR-139-5p | trans | 1.01E-40 | Nightingale | M_VLDL_C | NA | A | G | 0.033643 | 0.004158 | 1.90E-15 |
| rs677355 | miR-139-5p | trans | 1.01E-40 | Nightingale | XS_VLDL_C | NA | A | G | 0.037777 | 0.004271 | 2.40E-18 |
| rs677355 | miR-139-5p | trans | 1.01E-40 | Nightingale | M_VLDL_P | NA | A | G | 0.044027 | 0.004248 | 6.50E-25 |
| rs677355 | miR-139-5p | trans | 1.01E-40 | Nightingale | XS_VLDL_F | NA | A | G | 0.037025 | 0.004349 | 7.40E-18 |
| rs677355 | miR-139-5p | trans | 1.01E-40 | Nightingale | XS_VLDL_F | NA | A | G | 0.031383 | 0.004394 | 2.60E-12 |
| rs677355 | miR-139-5p | trans | 1.01E-40 | Nightingale | XXL_VLDL_ | NA | A | G | -0.03569 | 0.004464 | 2.30E-16 |
| rs677355 | miR-139-5p | trans | 1.01E-40 | Nightingale | Total_CE | NA | A | G | 0.031682 | 0.004269 | 1.10E-13 |
| rs677355 | miR-139-5p | trans | 1.01E-40 | Nightingale | LDL_PL | NA | A | G | 0.034895 | 0.004416 | 5.60E-15 |
| rs677355 | miR-139-5p | trans | 1.01E-40 | Nightingale | S_VLDL_TG | NA | A | G | -0.03812 | 0.004284 | 1.50E-18 |
| rs677355 | miR-139-5p | trans | 1.01E-40 | Nightingale | XL_VLDL_C | NA | A | G | 0.037355 | 0.004246 | 3.10E-18 |
| rs677355 | miR-139-5p | trans | 1.01E-40 | Nightingale | M_VLDL_C | NA | A | G | 0.036213 | 0.004169 | 1.20E-17 |
| rs677355 | miR-139-5p | trans | 1.01E-40 | Nightingale | LDL_P | NA | A | G | 0.031689 | 0.004423 | 1.20E-12 |
| rs677355 | miR-139-5p | trans | 1.01E-40 | Nightingale | XL_VLDL_T | NA | A | G | -0.04386 | 0.00429 | 2.10E-24 |
| rs677355 | miR-139-5p | trans | 1.01E-40 | Nightingale | S_LDL_PL_ | NA | A | G | -0.03 | 0.004418 | 3.00E-13 |
| rs677355 | miR-139-5p | trans | 1.01E-40 | Nightingale | S_LDL_CE | NA | A | G | 0.030433 | 0.004428 | 7.00E-12 |
| rs677355 | miR-139-5p | trans | 1.01E-40 | Nightingale | XXL_VLDL_ | NA | A | G | 0.034787 | 0.004435 | 3.10E-15 |
| rs677355 | miR-139-5p | trans | 1.01E-40 | Nightingale | S_VLDL_C | NA | A | G | 0.038286 | 0.004425 | 2.10E-18 |
| rs677355 | miR-139-5p | trans | 1.01E-40 | Nightingale | LDL_CE | NA | A | G | 0.032446 | 0.004414 | 2.60E-13 |
| rs677355 | miR-139-5p | trans | 1.01E-40 | Nightingale | M_LDL_L | NA | A | G | 0.032919 | 0.004432 | 1.20E-13 |
| rs677355 | miR-139-5p | trans | 1.01E-40 | Nightingale | IDL_FC | NA | A | G | 0.038214 | 0.004285 | 1.70E-18 |
| rs677355 | miR-139-5p | trans | 1.01E-40 | Nightingale | S_LDL_L | NA | A | G | 0.027634 | 0.004425 | 9.50E-10 |
| rs677355 | miR-139-5p | trans | 1.01E-40 | Nightingale | S_VLDL_L | NA | A | G | 0.024574 | 0.004383 | 4.30E-09 |
| rs677355 | miR-139-5p | trans | 1.01E-40 | Nightingale | VLDL_FC | NA | A | G | 0.024053 | 0.004393 | 1.30E-08 |
| rs677355 | miR-139-5p | trans | 1.01E-40 | Nightingale | S_HDL_L | NA | A | G | 0.026224 | 0.004411 | 1.80E-09 |
| rs677355 | miR-139-5p | trans | 1.01E-40 | Nightingale | M_LDL_PL | NA | A | G | 0.03236 | 0.004432 | 3.20E-13 |
| rs677355 | miR-139-5p | trans | 1.01E-40 | Nightingale | XXL_VLDL_ | NA | A | G | 0.030793 | 0.004407 | 1.20E-12 |
| rs677355 | miR-139-5p | trans | 1.01E-40 | Nightingale | Total_L | NA | A | G | 0.026271 | 0.004354 | 5.30E-10 |
| rs677355 | miR-139-5p | trans | 1.01E-40 | Nightingale | M_VLDL_C | NA | A | G | 0.038623 | 0.004414 | 2.30E-18 |
| rs677355 | miR-139-5p | trans | 1.01E-40 | Nightingale | S_VLDL_C_ | NA | A | G | 0.041886 | 0.004307 | 5.40E-22 |
| rs677355 | miR-139-5p | trans | 1.01E-40 | Nightingale | IDL_C | NA | A | G | 0.034923 | 0.004274 | 1.10E-15 |
| rs677355 | miR-139-5p | trans | 1.01E-40 | Nightingale | XXL_VLDL_ | NA | A | G | 0.033101 | 0.00446 | 9.20E-14 |
| rs677355 | miR-139-5p | trans | 1.01E-40 | Nightingale | L_VLDL_C_ | NA | A | G | 0.052158 | 0.004323 | 4.80E-34 |
| rs677355 | miR-139-5p | trans | 1.01E-40 | Nightingale | VLDL_C | NA | A | G | 0.031206 | 0.00442 | 5.80E-13 |
| rs677355 | miR-139-5p | trans | 1.01E-40 | Nightingale | Omega_6 | NA | A | G | 0.025797 | 0.004322 | 6.80E-10 |
| rs677355 | miR-139-5p | trans | 1.01E-40 | Nightingale | XS_VLDL_L | NA | A | G | 0.033781 | 0.004351 | 2.30E-15 |
| rs677355 | miR-139-5p | trans | 1.01E-40 | Nightingale | Sphingomy | NA | A | G | 0.028914 | 0.004137 | 8.50E-12 |
| rs677355 | miR-139-5p | trans | 1.01E-40 | Nightingale | LDL_FC | NA | A | G | 0.033557 | 0.004379 | 6.00E-14 |
| rs677355 | miR-139-5p | trans | 1.01E-40 | Nightingale | LDL_C | NA | A | G | 0.033095 | 0.004407 | 1.10E-13 |
| rs677355 | miR-139-5p | trans | 1.01E-40 | Nightingale | S_VLDL_FC | NA | A | G | 0.03781 | 0.004427 | 8.80E-18 |
| rs677355 | miR-139-5p | trans | 1.01E-40 | Nightingale | S_HDL_C | NA | A | G | 0.028339 | 0.004418 | 1.90E-10 |
| rs677355 | miR-139-5p | trans | 1.01E-40 | Nightingale | L_LDL_L | NA | A | G | 0.03346 | 0.004383 | 3.60E-14 |
| rs677355 | miR-139-5p | trans | 1.01E-40 | Nightingale | S_VLDL_CE | NA | A | G | 0.047027 | 0.004366 | 1.30E-26 |
| rs677355 | miR-139-5p | trans | 1.01E-40 | Nightingale | PUFA | NA | A | G | 0.023614 | 0.004317 | 1.70E-08 |
| rs677355 | miR-139-5p | trans | 1.01E-40 | Nightingale | S_VLDL_P | NA | A | G | 0.023377 | 0.00437 | 1.90E-08 |
| rs677355 | miR-139-5p | trans | 1.01E-40 | Nightingale | M_VLDL_F | NA | A | G | 0.042055 | 0.004211 | 3.70E-23 |
| rs677355 | miR-139-5p | trans | 1.01E-40 | Nightingale | S_LDL_FC | NA | A | G | 0.028483 | 0.004419 | 4.80E-10 |
| rs677355 | miR-139-5p | trans | 1.01E-40 | Nightingale | IDL_P | NA | A | G | 0.039974 | 0.004379 | 1.40E-19 |
| rs677355 | miR-139-5p | trans | 1.01E-40 | Nightingale | IDL_FC_pct | NA | A | G | 0.025744 | 0.004392 | 1.10E-08 |

| | | | | | | | | | | | |
|---|---|---|---|---|---|---|---|---|---|---|---|
| rs677355 | miR-139-5ן | trans | 1.01E-40 | Nightingale | M_VLDL_T | NA | A | G | -0.03856 | 0.004182 | 7.10E-20 |
| rs677355 | miR-139-5ן | trans | 1.01E-40 | Nightingale | S_HDL_P | NA | A | G | 0.030523 | 0.004426 | 6.00E-12 |
| rs677355 | miR-139-5ן | trans | 1.01E-40 | Nightingale | XS_VLDL_C | NA | A | G | 0.037965 | 0.004296 | 1.50E-18 |
| rs677355 | miR-139-5ן | trans | 1.01E-40 | Nightingale | Clinical_LD | NA | A | G | 0.036386 | 0.0044 | 3.20E-16 |
| rs677355 | miR-139-5ן | trans | 1.01E-40 | Nightingale | LA | NA | A | G | 0.024871 | 0.00434 | 4.00E-09 |
| rs677355 | miR-139-5ן | trans | 1.01E-40 | Nightingale | M_LDL_C | NA | A | G | 0.033874 | 0.004434 | 2.60E-14 |
| rs677355 | miR-139-5ן | trans | 1.01E-40 | Nightingale | LDL_L | NA | A | G | 0.033204 | 0.004411 | 8.80E-14 |
| rs677355 | miR-139-5ן | trans | 1.01E-40 | Nightingale | S_VLDL_PL | NA | A | G | 0.03268 | 0.004418 | 5.00E-14 |
| rs677355 | miR-139-5ן | trans | 1.01E-40 | Nightingale | non_HDL_( | NA | A | G | 0.035607 | 0.004402 | 6.10E-16 |
| rs677355 | miR-139-5ן | trans | 1.01E-40 | Nightingale | L_VLDL_FC | NA | A | G | 0.048463 | 0.004383 | 6.40E-29 |
| rs677355 | miR-139-5ן | trans | 1.01E-40 | Nightingale | L_VLDL_TG | NA | A | G | -0.05108 | 0.004405 | 6.90E-32 |
| rs677355 | miR-139-5ן | trans | 1.01E-40 | Nightingale | XL_VLDL_P | NA | A | G | 0.038718 | 0.004433 | 7.00E-19 |
| rs677355 | miR-139-5ן | trans | 1.01E-40 | Nightingale | GlycA | NA | A | G | 0.02771 | 0.004407 | 2.30E-10 |
| rs677355 | miR-139-5ן | trans | 1.01E-40 | Nightingale | S_VLDL_PL | NA | A | G | 0.027729 | 0.004238 | 1.80E-10 |
| rs677355 | miR-139-5ן | trans | 1.01E-40 | Nightingale | VLDL_CE | NA | A | G | 0.035699 | 0.004427 | 3.60E-16 |
| rs677355 | miR-139-5ן | trans | 1.01E-40 | Nightingale | IDL_L | NA | A | G | 0.03523 | 0.004279 | 5.50E-16 |
| rs677355 | miR-139-5ן | trans | 1.01E-40 | Nightingale | L_LDL_PL | NA | A | G | 0.037906 | 0.004392 | 1.10E-17 |
| rs677355 | miR-139-5ן | trans | 1.01E-40 | Nightingale | L_LDL_P | NA | A | G | 0.029058 | 0.004411 | 7.40E-11 |
| rs677355 | miR-139-5ן | trans | 1.01E-40 | Nightingale | VLDL_P | NA | A | G | 0.026547 | 0.004402 | 5.00E-10 |
| rs677355 | miR-139-5ן | trans | 1.01E-40 | Nightingale | L_VLDL_CE | NA | A | G | 0.047363 | 0.004235 | 2.40E-29 |
| rs677355 | miR-139-5ן | trans | 1.01E-40 | Nightingale | XL_VLDL_F | NA | A | G | 0.037212 | 0.00429 | 2.00E-18 |
| rs677355 | miR-195-5ן | trans | 5.38E-17 | Nightingale | Citrate | NA | A | G | 0.026829 | 0.004397 | 7.20E-10 |
| rs677355 | miR-195-5ן | trans | 5.38E-17 | Nightingale | ApoB_by_/ | NA | A | G | 0.024942 | 0.004359 | 1.50E-08 |
| rs677355 | miR-195-5ן | trans | 5.38E-17 | Nightingale | S_LDL_P | NA | A | G | 0.029497 | 0.00443 | 3.70E-11 |
| rs677355 | miR-195-5ן | trans | 5.38E-17 | Nightingale | IDL_CE | NA | A | G | 0.033479 | 0.004273 | 1.50E-14 |
| rs677355 | miR-195-5ן | trans | 5.38E-17 | Nightingale | M_LDL_FC | NA | A | G | 0.034256 | 0.00442 | 2.30E-14 |
| rs677355 | miR-195-5ן | trans | 5.38E-17 | Nightingale | S_LDL_C_p | NA | A | G | 0.045307 | 0.004443 | 6.50E-25 |
| rs677355 | miR-195-5ן | trans | 5.38E-17 | Nightingale | S_LDL_C | NA | A | G | 0.030835 | 0.00443 | 6.50E-12 |
| rs677355 | miR-195-5ן | trans | 5.38E-17 | Nightingale | Total_PL | NA | A | G | 0.024874 | 0.004214 | 1.20E-09 |
| rs677355 | miR-195-5ן | trans | 5.38E-17 | Nightingale | XS_VLDL_P | NA | A | G | 0.036669 | 0.004367 | 1.50E-17 |
| rs677355 | miR-195-5ן | trans | 5.38E-17 | Nightingale | XS_VLDL_P | NA | A | G | 0.032072 | 0.004364 | 5.90E-14 |
| rs677355 | miR-195-5ן | trans | 5.38E-17 | Nightingale | S_VLDL_FC | NA | A | G | 0.029355 | 0.004229 | 1.10E-11 |
| rs677355 | miR-195-5ן | trans | 5.38E-17 | Nightingale | Remnant_( | NA | A | G | 0.036793 | 0.004397 | 4.30E-17 |
| rs677355 | miR-195-5ן | trans | 5.38E-17 | Nightingale | Total_FC | NA | A | G | 0.033188 | 0.004324 | 8.00E-15 |
| rs677355 | miR-195-5ן | trans | 5.38E-17 | Nightingale | IDL_PL | NA | A | G | 0.035285 | 0.004271 | 3.90E-16 |
| rs677355 | miR-195-5ן | trans | 5.38E-17 | Nightingale | M_LDL_CE | NA | A | G | 0.032853 | 0.004432 | 1.20E-13 |
| rs677355 | miR-195-5ן | trans | 5.38E-17 | Nightingale | S_HDL_PL | NA | A | G | 0.023722 | 0.004387 | 3.60E-08 |
| rs677355 | miR-195-5ן | trans | 5.38E-17 | Nightingale | S_LDL_CE_ | NA | A | G | 0.030847 | 0.004418 | 6.00E-14 |
| rs677355 | miR-195-5ן | trans | 5.38E-17 | Nightingale | ApoB | NA | A | G | 0.033845 | 0.00442 | 2.50E-14 |
| rs677355 | miR-195-5ן | trans | 5.38E-17 | Nightingale | Total_C | NA | A | G | 0.032308 | 0.004285 | 3.90E-14 |
| rs677355 | miR-195-5ן | trans | 5.38E-17 | Nightingale | XL_HDL_PL | NA | A | G | -0.02264 | 0.004066 | 1.30E-08 |
| rs677355 | miR-195-5ן | trans | 5.38E-17 | Nightingale | L_LDL_FC | NA | A | G | 0.033559 | 0.004346 | 3.30E-14 |
| rs677355 | miR-195-5ן | trans | 5.38E-17 | Nightingale | S_HDL_CE | NA | A | G | 0.028557 | 0.004433 | 1.50E-10 |
| rs677355 | miR-195-5ן | trans | 5.38E-17 | Nightingale | XL_VLDL_C | NA | A | G | 0.038234 | 0.004244 | 4.60E-19 |
| rs677355 | miR-195-5ן | trans | 5.38E-17 | Nightingale | S_VLDL_CE | NA | A | G | 0.038101 | 0.00442 | 1.90E-18 |
| rs677355 | miR-195-5ן | trans | 5.38E-17 | Nightingale | M_LDL_P | NA | A | G | 0.035797 | 0.004438 | 8.20E-16 |
| rs677355 | miR-195-5ן | trans | 5.38E-17 | Nightingale | M_VLDL_C | NA | A | G | 0.040775 | 0.004386 | 2.70E-20 |
| rs677355 | miR-195-5ן | trans | 5.38E-17 | Nightingale | L_LDL_C | NA | A | G | 0.032229 | 0.004376 | 3.10E-13 |
| rs677355 | miR-195-5ן | trans | 5.38E-17 | Nightingale | M_VLDL_P | NA | A | G | 0.029028 | 0.004419 | 2.40E-11 |
| rs677355 | miR-195-5ן | trans | 5.38E-17 | Nightingale | M_LDL_C_ן | NA | A | G | 0.031084 | 0.004429 | 3.40E-12 |
| rs677355 | miR-195-5ן | trans | 5.38E-17 | Nightingale | M_VLDL_F | NA | A | G | 0.033273 | 0.004425 | 3.70E-14 |
| rs677355 | miR-195-5ן | trans | 5.38E-17 | Nightingale | M_VLDL_P | NA | A | G | 0.028187 | 0.004415 | 7.40E-11 |
| rs677355 | miR-195-5ן | trans | 5.38E-17 | Nightingale | XXL_VLDL_ | NA | A | G | 0.025023 | 0.004455 | 1.50E-09 |
| rs677355 | miR-195-5ן | trans | 5.38E-17 | Nightingale | M_LDL_TG | NA | A | G | -0.02757 | 0.004371 | 3.50E-10 |
| rs677355 | miR-195-5ן | trans | 5.38E-17 | Nightingale | L_LDL_CE | NA | A | G | 0.031469 | 0.004386 | 1.20E-12 |
| rs677355 | miR-195-5ן | trans | 5.38E-17 | Nightingale | M_VLDL_C | NA | A | G | 0.033643 | 0.004158 | 1.90E-15 |
| rs677355 | miR-195-5ן | trans | 5.38E-17 | Nightingale | XS_VLDL_C | NA | A | G | 0.037777 | 0.004271 | 2.40E-18 |
| rs677355 | miR-195-5ן | trans | 5.38E-17 | Nightingale | M_VLDL_P | NA | A | G | 0.044027 | 0.004248 | 6.50E-25 |
| rs677355 | miR-195-5ן | trans | 5.38E-17 | Nightingale | XS_VLDL_F | NA | A | G | 0.037025 | 0.004349 | 7.40E-18 |
| rs677355 | miR-195-5ן | trans | 5.38E-17 | Nightingale | XS_VLDL_F | NA | A | G | 0.031383 | 0.004394 | 2.60E-12 |
| rs677355 | miR-195-5ן | trans | 5.38E-17 | Nightingale | XXL_VLDL_ | NA | A | G | -0.03569 | 0.004464 | 2.30E-16 |
| rs677355 | miR-195-5ן | trans | 5.38E-17 | Nightingale | Total_CE | NA | A | G | 0.031682 | 0.004269 | 1.10E-13 |
| rs677355 | miR-195-5ן | trans | 5.38E-17 | Nightingale | LDL_PL | NA | A | G | 0.034895 | 0.004416 | 5.60E-15 |

| | | | | | | | | | | | | |
|---|---|---|---|---|---|---|---|---|---|---|---|---|
| rs677355 | miR-195-5p | trans | | 5.38E-17 | Nightingale | S_VLDL_TG | NA | A | G | -0.03812 | 0.004284 | 1.50E-18 |
| rs677355 | miR-195-5p | trans | | 5.38E-17 | Nightingale | XL_VLDL_C | NA | A | G | 0.037355 | 0.004246 | 3.10E-18 |
| rs677355 | miR-195-5p | trans | | 5.38E-17 | Nightingale | M_VLDL_C | NA | A | G | 0.036213 | 0.004169 | 1.20E-17 |
| rs677355 | miR-195-5p | trans | | 5.38E-17 | Nightingale | LDL_P | NA | A | G | 0.031689 | 0.004423 | 1.20E-12 |
| rs677355 | miR-195-5p | trans | | 5.38E-17 | Nightingale | XL_VLDL_T | NA | A | G | -0.04386 | 0.00429 | 2.10E-24 |
| rs677355 | miR-195-5p | trans | | 5.38E-17 | Nightingale | S_LDL_PL_ | NA | A | G | -0.03 | 0.004418 | 3.00E-13 |
| rs677355 | miR-195-5p | trans | | 5.38E-17 | Nightingale | S_LDL_CE | NA | A | G | 0.030433 | 0.004428 | 7.00E-12 |
| rs677355 | miR-195-5p | trans | | 5.38E-17 | Nightingale | XXL_VLDL_ | NA | A | G | 0.034787 | 0.004435 | 3.10E-15 |
| rs677355 | miR-195-5p | trans | | 5.38E-17 | Nightingale | S_VLDL_C | NA | A | G | 0.038286 | 0.004425 | 2.10E-18 |
| rs677355 | miR-195-5p | trans | | 5.38E-17 | Nightingale | LDL_CE | NA | A | G | 0.032446 | 0.004414 | 2.60E-13 |
| rs677355 | miR-195-5p | trans | | 5.38E-17 | Nightingale | M_LDL_L | NA | A | G | 0.032919 | 0.004432 | 1.20E-13 |
| rs677355 | miR-195-5p | trans | | 5.38E-17 | Nightingale | IDL_FC | NA | A | G | 0.038214 | 0.004285 | 1.70E-18 |
| rs677355 | miR-195-5p | trans | | 5.38E-17 | Nightingale | S_LDL_L | NA | A | G | 0.027634 | 0.004425 | 9.50E-10 |
| rs677355 | miR-195-5p | trans | | 5.38E-17 | Nightingale | S_VLDL_L | NA | A | G | 0.024574 | 0.004383 | 4.30E-09 |
| rs677355 | miR-195-5p | trans | | 5.38E-17 | Nightingale | VLDL_FC | NA | A | G | 0.024053 | 0.004393 | 1.30E-08 |
| rs677355 | miR-195-5p | trans | | 5.38E-17 | Nightingale | S_HDL_L | NA | A | G | 0.026224 | 0.004411 | 1.80E-09 |
| rs677355 | miR-195-5p | trans | | 5.38E-17 | Nightingale | M_LDL_PL | NA | A | G | 0.03236 | 0.004432 | 3.20E-13 |
| rs677355 | miR-195-5p | trans | | 5.38E-17 | Nightingale | XXL_VLDL_ | NA | A | G | 0.030793 | 0.004407 | 1.20E-12 |
| rs677355 | miR-195-5p | trans | | 5.38E-17 | Nightingale | Total_L | NA | A | G | 0.026271 | 0.004354 | 5.30E-10 |
| rs677355 | miR-195-5p | trans | | 5.38E-17 | Nightingale | M_VLDL_C | NA | A | G | 0.038623 | 0.004414 | 2.30E-18 |
| rs677355 | miR-195-5p | trans | | 5.38E-17 | Nightingale | S_VLDL_C_ | NA | A | G | 0.041886 | 0.004307 | 5.40E-22 |
| rs677355 | miR-195-5p | trans | | 5.38E-17 | Nightingale | IDL_C | NA | A | G | 0.034923 | 0.004274 | 1.10E-15 |
| rs677355 | miR-195-5p | trans | | 5.38E-17 | Nightingale | XXL_VLDL_ | NA | A | G | 0.033101 | 0.00446 | 9.20E-14 |
| rs677355 | miR-195-5p | trans | | 5.38E-17 | Nightingale | L_VLDL_C_ | NA | A | G | 0.052158 | 0.004323 | 4.80E-34 |
| rs677355 | miR-195-5p | trans | | 5.38E-17 | Nightingale | VLDL_C | NA | A | G | 0.031206 | 0.00442 | 5.80E-13 |
| rs677355 | miR-195-5p | trans | | 5.38E-17 | Nightingale | Omega_6 | NA | A | G | 0.025797 | 0.004322 | 6.80E-10 |
| rs677355 | miR-195-5p | trans | | 5.38E-17 | Nightingale | XS_VLDL_L | NA | A | G | 0.033781 | 0.004351 | 2.30E-15 |
| rs677355 | miR-195-5p | trans | | 5.38E-17 | Nightingale | Sphingomy | NA | A | G | 0.028914 | 0.004137 | 8.50E-12 |
| rs677355 | miR-195-5p | trans | | 5.38E-17 | Nightingale | LDL_FC | NA | A | G | 0.033557 | 0.004379 | 6.00E-14 |
| rs677355 | miR-195-5p | trans | | 5.38E-17 | Nightingale | LDL_C | NA | A | G | 0.033095 | 0.004407 | 1.10E-13 |
| rs677355 | miR-195-5p | trans | | 5.38E-17 | Nightingale | S_VLDL_FC | NA | A | G | 0.03781 | 0.004427 | 8.80E-18 |
| rs677355 | miR-195-5p | trans | | 5.38E-17 | Nightingale | S_HDL_C | NA | A | G | 0.028339 | 0.004418 | 1.90E-10 |
| rs677355 | miR-195-5p | trans | | 5.38E-17 | Nightingale | L_LDL_L | NA | A | G | 0.03346 | 0.004383 | 3.60E-14 |
| rs677355 | miR-195-5p | trans | | 5.38E-17 | Nightingale | S_VLDL_CE | NA | A | G | 0.047027 | 0.004366 | 1.30E-26 |
| rs677355 | miR-195-5p | trans | | 5.38E-17 | Nightingale | PUFA | NA | A | G | 0.023614 | 0.004317 | 1.70E-08 |
| rs677355 | miR-195-5p | trans | | 5.38E-17 | Nightingale | S_VLDL_P | NA | A | G | 0.023377 | 0.00437 | 1.90E-08 |
| rs677355 | miR-195-5p | trans | | 5.38E-17 | Nightingale | M_VLDL_F | NA | A | G | 0.042055 | 0.004211 | 3.70E-23 |
| rs677355 | miR-195-5p | trans | | 5.38E-17 | Nightingale | S_LDL_FC | NA | A | G | 0.028483 | 0.004419 | 4.80E-10 |
| rs677355 | miR-195-5p | trans | | 5.38E-17 | Nightingale | IDL_P | NA | A | G | 0.039974 | 0.004379 | 1.40E-19 |
| rs677355 | miR-195-5p | trans | | 5.38E-17 | Nightingale | IDL_FC_pct | NA | A | G | 0.025744 | 0.004392 | 1.10E-08 |
| rs677355 | miR-195-5p | trans | | 5.38E-17 | Nightingale | M_VLDL_T | NA | A | G | -0.03856 | 0.004182 | 7.10E-20 |
| rs677355 | miR-195-5p | trans | | 5.38E-17 | Nightingale | S_HDL_P | NA | A | G | 0.030523 | 0.004426 | 6.00E-12 |
| rs677355 | miR-195-5p | trans | | 5.38E-17 | Nightingale | XS_VLDL_C | NA | A | G | 0.037965 | 0.004296 | 1.50E-18 |
| rs677355 | miR-195-5p | trans | | 5.38E-17 | Nightingale | Clinical_LD | NA | A | G | 0.036386 | 0.0044 | 3.20E-16 |
| rs677355 | miR-195-5p | trans | | 5.38E-17 | Nightingale | LA | NA | A | G | 0.024871 | 0.00434 | 4.00E-09 |
| rs677355 | miR-195-5p | trans | | 5.38E-17 | Nightingale | M_LDL_C | NA | A | G | 0.033874 | 0.004434 | 2.60E-14 |
| rs677355 | miR-195-5p | trans | | 5.38E-17 | Nightingale | LDL_L | NA | A | G | 0.033204 | 0.004411 | 8.80E-14 |
| rs677355 | miR-195-5p | trans | | 5.38E-17 | Nightingale | S_VLDL_PL | NA | A | G | 0.03268 | 0.004418 | 5.00E-14 |
| rs677355 | miR-195-5p | trans | | 5.38E-17 | Nightingale | non_HDL_( | NA | A | G | 0.035607 | 0.004402 | 6.10E-16 |
| rs677355 | miR-195-5p | trans | | 5.38E-17 | Nightingale | L_VLDL_FC | NA | A | G | 0.048463 | 0.004383 | 6.40E-29 |
| rs677355 | miR-195-5p | trans | | 5.38E-17 | Nightingale | L_VLDL_TG | NA | A | G | -0.05108 | 0.004405 | 6.90E-32 |
| rs677355 | miR-195-5p | trans | | 5.38E-17 | Nightingale | XL_VLDL_P | NA | A | G | 0.038718 | 0.004433 | 7.00E-19 |
| rs677355 | miR-195-5p | trans | | 5.38E-17 | Nightingale | GlycA | NA | A | G | 0.02771 | 0.004407 | 2.30E-10 |
| rs677355 | miR-195-5p | trans | | 5.38E-17 | Nightingale | S_VLDL_PL | NA | A | G | 0.027729 | 0.004238 | 1.80E-10 |
| rs677355 | miR-195-5p | trans | | 5.38E-17 | Nightingale | VLDL_CE | NA | A | G | 0.035699 | 0.004427 | 3.60E-16 |
| rs677355 | miR-195-5p | trans | | 5.38E-17 | Nightingale | IDL_L | NA | A | G | 0.03523 | 0.004279 | 5.50E-16 |
| rs677355 | miR-195-5p | trans | | 5.38E-17 | Nightingale | L_LDL_PL | NA | A | G | 0.037906 | 0.004392 | 1.10E-17 |
| rs677355 | miR-195-5p | trans | | 5.38E-17 | Nightingale | L_LDL_P | NA | A | G | 0.029058 | 0.004411 | 7.40E-11 |
| rs677355 | miR-195-5p | trans | | 5.38E-17 | Nightingale | VLDL_P | NA | A | G | 0.026547 | 0.004402 | 5.00E-10 |
| rs677355 | miR-195-5p | trans | | 5.38E-17 | Nightingale | L_VLDL_CE | NA | A | G | 0.047363 | 0.004235 | 2.40E-29 |
| rs677355 | miR-195-5p | trans | | 5.38E-17 | Nightingale | XL_VLDL_F | NA | A | G | 0.037212 | 0.00429 | 2.00E-18 |
| rs677355 | miR-10b-5p | trans | | 1.02E-34 | Nightingale | Citrate | NA | A | G | 0.026829 | 0.004397 | 7.20E-10 |
| rs677355 | miR-10b-5p | trans | | 1.02E-34 | Nightingale | ApoB_by_A | NA | A | G | 0.024942 | 0.004359 | 1.50E-08 |

| | | | | | | | | | | | |
|---|---|---|---|---|---|---|---|---|---|---|---|
| rs677355 | miR-10b-5p | trans | 1.02E-34 | Nightingale | S_LDL_P | NA | A | G | 0.029497 | 0.00443 | 3.70E-11 |
| rs677355 | miR-10b-5p | trans | 1.02E-34 | Nightingale | IDL_CE | NA | A | G | 0.033479 | 0.004273 | 1.50E-14 |
| rs677355 | miR-10b-5p | trans | 1.02E-34 | Nightingale | M_LDL_FC | NA | A | G | 0.034256 | 0.00442 | 2.30E-14 |
| rs677355 | miR-10b-5p | trans | 1.02E-34 | Nightingale | S_LDL_C_p | NA | A | G | 0.045307 | 0.004443 | 6.50E-25 |
| rs677355 | miR-10b-5p | trans | 1.02E-34 | Nightingale | S_LDL_C | NA | A | G | 0.030835 | 0.00443 | 6.50E-12 |
| rs677355 | miR-10b-5p | trans | 1.02E-34 | Nightingale | Total_PL | NA | A | G | 0.024874 | 0.004214 | 1.20E-09 |
| rs677355 | miR-10b-5p | trans | 1.02E-34 | Nightingale | XS_VLDL_P | NA | A | G | 0.036669 | 0.004367 | 1.50E-17 |
| rs677355 | miR-10b-5p | trans | 1.02E-34 | Nightingale | XS_VLDL_P | NA | A | G | 0.032072 | 0.004364 | 5.90E-14 |
| rs677355 | miR-10b-5p | trans | 1.02E-34 | Nightingale | S_VLDL_FC | NA | A | G | 0.029355 | 0.004229 | 1.10E-11 |
| rs677355 | miR-10b-5p | trans | 1.02E-34 | Nightingale | Remnant_( | NA | A | G | 0.036793 | 0.004397 | 4.30E-17 |
| rs677355 | miR-10b-5p | trans | 1.02E-34 | Nightingale | Total_FC | NA | A | G | 0.033188 | 0.004324 | 8.00E-15 |
| rs677355 | miR-10b-5p | trans | 1.02E-34 | Nightingale | IDL_PL | NA | A | G | 0.035285 | 0.004271 | 3.90E-16 |
| rs677355 | miR-10b-5p | trans | 1.02E-34 | Nightingale | M_LDL_CE | NA | A | G | 0.032853 | 0.004432 | 1.20E-13 |
| rs677355 | miR-10b-5p | trans | 1.02E-34 | Nightingale | S_HDL_PL | NA | A | G | 0.023722 | 0.004387 | 3.60E-08 |
| rs677355 | miR-10b-5p | trans | 1.02E-34 | Nightingale | S_LDL_CE_ | NA | A | G | 0.030847 | 0.004418 | 6.00E-14 |
| rs677355 | miR-10b-5p | trans | 1.02E-34 | Nightingale | ApoB | NA | A | G | 0.033845 | 0.00442 | 2.50E-14 |
| rs677355 | miR-10b-5p | trans | 1.02E-34 | Nightingale | Total_C | NA | A | G | 0.032308 | 0.004285 | 3.90E-14 |
| rs677355 | miR-10b-5p | trans | 1.02E-34 | Nightingale | XL_HDL_PL | NA | A | G | -0.02264 | 0.004066 | 1.30E-08 |
| rs677355 | miR-10b-5p | trans | 1.02E-34 | Nightingale | L_LDL_FC | NA | A | G | 0.033559 | 0.004346 | 3.30E-14 |
| rs677355 | miR-10b-5p | trans | 1.02E-34 | Nightingale | S_HDL_CE | NA | A | G | 0.028557 | 0.004433 | 1.50E-10 |
| rs677355 | miR-10b-5p | trans | 1.02E-34 | Nightingale | XL_VLDL_C | NA | A | G | 0.038234 | 0.004244 | 4.60E-19 |
| rs677355 | miR-10b-5p | trans | 1.02E-34 | Nightingale | S_VLDL_CE | NA | A | G | 0.038101 | 0.00442 | 1.90E-18 |
| rs677355 | miR-10b-5p | trans | 1.02E-34 | Nightingale | M_LDL_P | NA | A | G | 0.035797 | 0.004438 | 8.20E-16 |
| rs677355 | miR-10b-5p | trans | 1.02E-34 | Nightingale | M_VLDL_C | NA | A | G | 0.040775 | 0.004386 | 2.70E-20 |
| rs677355 | miR-10b-5p | trans | 1.02E-34 | Nightingale | L_LDL_C | NA | A | G | 0.032229 | 0.004376 | 3.10E-13 |
| rs677355 | miR-10b-5p | trans | 1.02E-34 | Nightingale | M_VLDL_P | NA | A | G | 0.029028 | 0.004419 | 2.40E-11 |
| rs677355 | miR-10b-5p | trans | 1.02E-34 | Nightingale | M_LDL_C_p | NA | A | G | 0.031084 | 0.004429 | 3.40E-12 |
| rs677355 | miR-10b-5p | trans | 1.02E-34 | Nightingale | M_VLDL_F | NA | A | G | 0.033273 | 0.004425 | 3.70E-14 |
| rs677355 | miR-10b-5p | trans | 1.02E-34 | Nightingale | M_VLDL_P | NA | A | G | 0.028187 | 0.004415 | 7.40E-11 |
| rs677355 | miR-10b-5p | trans | 1.02E-34 | Nightingale | XXL_VLDL_ | NA | A | G | 0.025023 | 0.004455 | 1.50E-09 |
| rs677355 | miR-10b-5p | trans | 1.02E-34 | Nightingale | M_LDL_TG | NA | A | G | -0.02757 | 0.004371 | 3.50E-10 |
| rs677355 | miR-10b-5p | trans | 1.02E-34 | Nightingale | L_LDL_CE | NA | A | G | 0.031469 | 0.004386 | 1.20E-12 |
| rs677355 | miR-10b-5p | trans | 1.02E-34 | Nightingale | M_VLDL_C | NA | A | G | 0.033643 | 0.004158 | 1.90E-15 |
| rs677355 | miR-10b-5p | trans | 1.02E-34 | Nightingale | XS_VLDL_C | NA | A | G | 0.037777 | 0.004271 | 2.40E-18 |
| rs677355 | miR-10b-5p | trans | 1.02E-34 | Nightingale | M_VLDL_P | NA | A | G | 0.044027 | 0.004248 | 6.50E-25 |
| rs677355 | miR-10b-5p | trans | 1.02E-34 | Nightingale | XS_VLDL_F | NA | A | G | 0.037025 | 0.004349 | 7.40E-18 |
| rs677355 | miR-10b-5p | trans | 1.02E-34 | Nightingale | XS_VLDL_F | NA | A | G | 0.031383 | 0.004394 | 2.60E-12 |
| rs677355 | miR-10b-5p | trans | 1.02E-34 | Nightingale | XXL_VLDL_ | NA | A | G | -0.03569 | 0.004464 | 2.30E-16 |
| rs677355 | miR-10b-5p | trans | 1.02E-34 | Nightingale | Total_CE | NA | A | G | 0.031682 | 0.004269 | 1.10E-13 |
| rs677355 | miR-10b-5p | trans | 1.02E-34 | Nightingale | LDL_PL | NA | A | G | 0.034895 | 0.004416 | 5.60E-15 |
| rs677355 | miR-10b-5p | trans | 1.02E-34 | Nightingale | S_VLDL_TG | NA | A | G | -0.03812 | 0.004284 | 1.50E-18 |
| rs677355 | miR-10b-5p | trans | 1.02E-34 | Nightingale | XL_VLDL_C | NA | A | G | 0.037355 | 0.004246 | 3.10E-18 |
| rs677355 | miR-10b-5p | trans | 1.02E-34 | Nightingale | M_VLDL_C | NA | A | G | 0.036213 | 0.004169 | 1.20E-17 |
| rs677355 | miR-10b-5p | trans | 1.02E-34 | Nightingale | LDL_P | NA | A | G | 0.031689 | 0.004423 | 1.20E-12 |
| rs677355 | miR-10b-5p | trans | 1.02E-34 | Nightingale | XL_VLDL_T | NA | A | G | -0.04386 | 0.00429 | 2.10E-24 |
| rs677355 | miR-10b-5p | trans | 1.02E-34 | Nightingale | S_LDL_PL_ | NA | A | G | -0.03 | 0.004418 | 3.00E-13 |
| rs677355 | miR-10b-5p | trans | 1.02E-34 | Nightingale | S_LDL_CE | NA | A | G | 0.030433 | 0.004428 | 7.00E-12 |
| rs677355 | miR-10b-5p | trans | 1.02E-34 | Nightingale | XXL_VLDL_ | NA | A | G | 0.034787 | 0.004435 | 3.10E-15 |
| rs677355 | miR-10b-5p | trans | 1.02E-34 | Nightingale | S_VLDL_C | NA | A | G | 0.038286 | 0.004425 | 2.10E-18 |
| rs677355 | miR-10b-5p | trans | 1.02E-34 | Nightingale | LDL_CE | NA | A | G | 0.032446 | 0.004414 | 2.60E-13 |
| rs677355 | miR-10b-5p | trans | 1.02E-34 | Nightingale | M_LDL_L | NA | A | G | 0.032919 | 0.004432 | 1.20E-13 |
| rs677355 | miR-10b-5p | trans | 1.02E-34 | Nightingale | IDL_FC | NA | A | G | 0.038214 | 0.004285 | 1.70E-18 |
| rs677355 | miR-10b-5p | trans | 1.02E-34 | Nightingale | S_LDL_L | NA | A | G | 0.027634 | 0.004425 | 9.50E-10 |
| rs677355 | miR-10b-5p | trans | 1.02E-34 | Nightingale | S_VLDL_L | NA | A | G | 0.024574 | 0.004383 | 4.30E-09 |
| rs677355 | miR-10b-5p | trans | 1.02E-34 | Nightingale | VLDL_FC | NA | A | G | 0.024053 | 0.004393 | 1.30E-08 |
| rs677355 | miR-10b-5p | trans | 1.02E-34 | Nightingale | S_HDL_L | NA | A | G | 0.026224 | 0.004411 | 1.80E-09 |
| rs677355 | miR-10b-5p | trans | 1.02E-34 | Nightingale | M_LDL_PL | NA | A | G | 0.03236 | 0.004432 | 3.20E-13 |
| rs677355 | miR-10b-5p | trans | 1.02E-34 | Nightingale | XXL_VLDL_ | NA | A | G | 0.030793 | 0.004407 | 1.20E-12 |
| rs677355 | miR-10b-5p | trans | 1.02E-34 | Nightingale | Total_L | NA | A | G | 0.026271 | 0.004354 | 5.30E-10 |
| rs677355 | miR-10b-5p | trans | 1.02E-34 | Nightingale | M_VLDL_C | NA | A | G | 0.038623 | 0.004414 | 2.30E-18 |
| rs677355 | miR-10b-5p | trans | 1.02E-34 | Nightingale | S_VLDL_C_ | NA | A | G | 0.041886 | 0.004307 | 5.40E-22 |
| rs677355 | miR-10b-5p | trans | 1.02E-34 | Nightingale | IDL_C | NA | A | G | 0.034923 | 0.004274 | 1.10E-15 |
| rs677355 | miR-10b-5p | trans | 1.02E-34 | Nightingale | XXL_VLDL_ | NA | A | G | 0.033101 | 0.00446 | 9.20E-14 |

| | | | | | | | | | | | |
|---|---|---|---|---|---|---|---|---|---|---|---|
| rs677355 | miR-10b-5| | trans | 1.02E-34 | Nightingale | L_VLDL_C_ | NA | A | G | 0.052158 | 0.004323 | 4.80E-34 |
| rs677355 | miR-10b-5| | trans | 1.02E-34 | Nightingale | VLDL_C | NA | A | G | 0.031206 | 0.00442 | 5.80E-13 |
| rs677355 | miR-10b-5| | trans | 1.02E-34 | Nightingale | Omega_6 | NA | A | G | 0.025797 | 0.004322 | 6.80E-10 |
| rs677355 | miR-10b-5| | trans | 1.02E-34 | Nightingale | XS_VLDL_L | NA | A | G | 0.033781 | 0.004351 | 2.30E-15 |
| rs677355 | miR-10b-5| | trans | 1.02E-34 | Nightingale | Sphingomy | NA | A | G | 0.028914 | 0.004137 | 8.50E-12 |
| rs677355 | miR-10b-5| | trans | 1.02E-34 | Nightingale | LDL_FC | NA | A | G | 0.033557 | 0.004379 | 6.00E-14 |
| rs677355 | miR-10b-5| | trans | 1.02E-34 | Nightingale | LDL_C | NA | A | G | 0.033095 | 0.004407 | 1.10E-13 |
| rs677355 | miR-10b-5| | trans | 1.02E-34 | Nightingale | S_VLDL_FC | NA | A | G | 0.03781 | 0.004427 | 8.80E-18 |
| rs677355 | miR-10b-5| | trans | 1.02E-34 | Nightingale | S_HDL_C | NA | A | G | 0.028339 | 0.004418 | 1.90E-10 |
| rs677355 | miR-10b-5| | trans | 1.02E-34 | Nightingale | L_LDL_L | NA | A | G | 0.03346 | 0.004383 | 3.60E-14 |
| rs677355 | miR-10b-5| | trans | 1.02E-34 | Nightingale | S_VLDL_CE | NA | A | G | 0.047027 | 0.004366 | 1.30E-26 |
| rs677355 | miR-10b-5| | trans | 1.02E-34 | Nightingale | PUFA | NA | A | G | 0.023614 | 0.004317 | 1.70E-08 |
| rs677355 | miR-10b-5| | trans | 1.02E-34 | Nightingale | S_VLDL_P | NA | A | G | 0.023377 | 0.00437 | 1.90E-08 |
| rs677355 | miR-10b-5| | trans | 1.02E-34 | Nightingale | M_VLDL_F | NA | A | G | 0.042055 | 0.004211 | 3.70E-23 |
| rs677355 | miR-10b-5| | trans | 1.02E-34 | Nightingale | S_LDL_FC | NA | A | G | 0.028483 | 0.004419 | 4.80E-10 |
| rs677355 | miR-10b-5| | trans | 1.02E-34 | Nightingale | IDL_P | NA | A | G | 0.039974 | 0.004379 | 1.40E-19 |
| rs677355 | miR-10b-5| | trans | 1.02E-34 | Nightingale | IDL_FC_pct | NA | A | G | 0.025744 | 0.004392 | 1.10E-08 |
| rs677355 | miR-10b-5| | trans | 1.02E-34 | Nightingale | M_VLDL_T | NA | A | G | -0.03856 | 0.004182 | 7.10E-20 |
| rs677355 | miR-10b-5| | trans | 1.02E-34 | Nightingale | S_HDL_P | NA | A | G | 0.030523 | 0.004426 | 6.00E-12 |
| rs677355 | miR-10b-5| | trans | 1.02E-34 | Nightingale | XS_VLDL_C | NA | A | G | 0.037965 | 0.004296 | 1.50E-18 |
| rs677355 | miR-10b-5| | trans | 1.02E-34 | Nightingale | Clinical_LD | NA | A | G | 0.036386 | 0.0044 | 3.20E-16 |
| rs677355 | miR-10b-5| | trans | 1.02E-34 | Nightingale | LA | NA | A | G | 0.024871 | 0.00434 | 4.00E-09 |
| rs677355 | miR-10b-5| | trans | 1.02E-34 | Nightingale | M_LDL_C | NA | A | G | 0.033874 | 0.004434 | 2.60E-14 |
| rs677355 | miR-10b-5| | trans | 1.02E-34 | Nightingale | LDL_L | NA | A | G | 0.033204 | 0.004411 | 8.80E-14 |
| rs677355 | miR-10b-5| | trans | 1.02E-34 | Nightingale | S_VLDL_PL | NA | A | G | 0.03268 | 0.004418 | 5.00E-14 |
| rs677355 | miR-10b-5| | trans | 1.02E-34 | Nightingale | non_HDL_( | NA | A | G | 0.035607 | 0.004402 | 6.10E-16 |
| rs677355 | miR-10b-5| | trans | 1.02E-34 | Nightingale | L_VLDL_FC | NA | A | G | 0.048463 | 0.004383 | 6.40E-29 |
| rs677355 | miR-10b-5| | trans | 1.02E-34 | Nightingale | L_VLDL_TG | NA | A | G | -0.05108 | 0.004405 | 6.90E-32 |
| rs677355 | miR-10b-5| | trans | 1.02E-34 | Nightingale | XL_VLDL_P | NA | A | G | 0.038718 | 0.004433 | 7.00E-19 |
| rs677355 | miR-10b-5| | trans | 1.02E-34 | Nightingale | GlycA | NA | A | G | 0.02771 | 0.004407 | 2.30E-10 |
| rs677355 | miR-10b-5| | trans | 1.02E-34 | Nightingale | S_VLDL_PL | NA | A | G | 0.027729 | 0.004238 | 1.80E-10 |
| rs677355 | miR-10b-5| | trans | 1.02E-34 | Nightingale | VLDL_CE | NA | A | G | 0.035699 | 0.004427 | 3.60E-16 |
| rs677355 | miR-10b-5| | trans | 1.02E-34 | Nightingale | IDL_L | NA | A | G | 0.03523 | 0.004279 | 5.50E-16 |
| rs677355 | miR-10b-5| | trans | 1.02E-34 | Nightingale | L_LDL_PL | NA | A | G | 0.037906 | 0.004392 | 1.10E-17 |
| rs677355 | miR-10b-5| | trans | 1.02E-34 | Nightingale | L_LDL_P | NA | A | G | 0.029058 | 0.004411 | 7.40E-11 |
| rs677355 | miR-10b-5| | trans | 1.02E-34 | Nightingale | VLDL_P | NA | A | G | 0.026547 | 0.004402 | 5.00E-10 |
| rs677355 | miR-10b-5| | trans | 1.02E-34 | Nightingale | L_VLDL_CE | NA | A | G | 0.047363 | 0.004235 | 2.40E-29 |
| rs677355 | miR-10b-5| | trans | 1.02E-34 | Nightingale | XL_VLDL_F | NA | A | G | 0.037212 | 0.00429 | 2.00E-18 |
| rs677355 | let-7c-5p | trans | 5.81E-23 | Nightingale | Citrate | NA | A | G | 0.026829 | 0.004397 | 7.20E-10 |
| rs677355 | let-7c-5p | trans | 5.81E-23 | Nightingale | ApoB_by_/ | NA | A | G | 0.024942 | 0.004359 | 1.50E-08 |
| rs677355 | let-7c-5p | trans | 5.81E-23 | Nightingale | S_LDL_P | NA | A | G | 0.029497 | 0.00443 | 3.70E-11 |
| rs677355 | let-7c-5p | trans | 5.81E-23 | Nightingale | IDL_CE | NA | A | G | 0.033479 | 0.004273 | 1.50E-14 |
| rs677355 | let-7c-5p | trans | 5.81E-23 | Nightingale | M_LDL_FC | NA | A | G | 0.034256 | 0.00442 | 2.30E-14 |
| rs677355 | let-7c-5p | trans | 5.81E-23 | Nightingale | S_LDL_C_p | NA | A | G | 0.045307 | 0.004443 | 6.50E-25 |
| rs677355 | let-7c-5p | trans | 5.81E-23 | Nightingale | S_LDL_C | NA | A | G | 0.030835 | 0.00443 | 6.50E-12 |
| rs677355 | let-7c-5p | trans | 5.81E-23 | Nightingale | Total_PL | NA | A | G | 0.024874 | 0.004214 | 1.20E-09 |
| rs677355 | let-7c-5p | trans | 5.81E-23 | Nightingale | XS_VLDL_P | NA | A | G | 0.036669 | 0.004367 | 1.50E-17 |
| rs677355 | let-7c-5p | trans | 5.81E-23 | Nightingale | XS_VLDL_P | NA | A | G | 0.032072 | 0.004364 | 5.90E-14 |
| rs677355 | let-7c-5p | trans | 5.81E-23 | Nightingale | S_VLDL_FC | NA | A | G | 0.029355 | 0.004229 | 1.10E-11 |
| rs677355 | let-7c-5p | trans | 5.81E-23 | Nightingale | Remnant_( | NA | A | G | 0.036793 | 0.004397 | 4.30E-17 |
| rs677355 | let-7c-5p | trans | 5.81E-23 | Nightingale | Total_FC | NA | A | G | 0.033188 | 0.004324 | 8.00E-15 |
| rs677355 | let-7c-5p | trans | 5.81E-23 | Nightingale | IDL_PL | NA | A | G | 0.035285 | 0.004271 | 3.90E-16 |
| rs677355 | let-7c-5p | trans | 5.81E-23 | Nightingale | M_LDL_CE | NA | A | G | 0.032853 | 0.004432 | 1.20E-13 |
| rs677355 | let-7c-5p | trans | 5.81E-23 | Nightingale | S_HDL_PL | NA | A | G | 0.023722 | 0.004387 | 3.60E-08 |
| rs677355 | let-7c-5p | trans | 5.81E-23 | Nightingale | S_LDL_CE_ | NA | A | G | 0.030847 | 0.004418 | 6.00E-14 |
| rs677355 | let-7c-5p | trans | 5.81E-23 | Nightingale | ApoB | NA | A | G | 0.033845 | 0.00442 | 2.50E-14 |
| rs677355 | let-7c-5p | trans | 5.81E-23 | Nightingale | Total_C | NA | A | G | 0.032308 | 0.004285 | 3.90E-14 |
| rs677355 | let-7c-5p | trans | 5.81E-23 | Nightingale | XL_HDL_PL | NA | A | G | -0.02264 | 0.004066 | 1.30E-08 |
| rs677355 | let-7c-5p | trans | 5.81E-23 | Nightingale | L_LDL_FC | NA | A | G | 0.033559 | 0.004346 | 3.30E-14 |
| rs677355 | let-7c-5p | trans | 5.81E-23 | Nightingale | S_HDL_CE | NA | A | G | 0.028557 | 0.004433 | 1.50E-10 |
| rs677355 | let-7c-5p | trans | 5.81E-23 | Nightingale | XL_VLDL_C | NA | A | G | 0.038234 | 0.004244 | 4.60E-19 |
| rs677355 | let-7c-5p | trans | 5.81E-23 | Nightingale | S_VLDL_CE | NA | A | G | 0.038101 | 0.00442 | 1.90E-18 |
| rs677355 | let-7c-5p | trans | 5.81E-23 | Nightingale | M_LDL_P | NA | A | G | 0.035797 | 0.004438 | 8.20E-16 |

| | | | | | | | | | | | |
|---|---|---|---|---|---|---|---|---|---|---|---|
| rs677355 | let-7c-5p | trans | 5.81E-23 | Nightingale | M_VLDL_C | NA | A | G | 0.040775 | 0.004386 | 2.70E-20 |
| rs677355 | let-7c-5p | trans | 5.81E-23 | Nightingale | L_LDL_C | NA | A | G | 0.032229 | 0.004376 | 3.10E-13 |
| rs677355 | let-7c-5p | trans | 5.81E-23 | Nightingale | M_VLDL_P | NA | A | G | 0.029028 | 0.004419 | 2.40E-11 |
| rs677355 | let-7c-5p | trans | 5.81E-23 | Nightingale | M_LDL_C_ | NA | A | G | 0.031084 | 0.004429 | 3.40E-12 |
| rs677355 | let-7c-5p | trans | 5.81E-23 | Nightingale | M_VLDL_F | NA | A | G | 0.033273 | 0.004425 | 3.70E-14 |
| rs677355 | let-7c-5p | trans | 5.81E-23 | Nightingale | M_VLDL_P | NA | A | G | 0.028187 | 0.004415 | 7.40E-11 |
| rs677355 | let-7c-5p | trans | 5.81E-23 | Nightingale | XXL_VLDL_ | NA | A | G | 0.025023 | 0.004455 | 1.50E-09 |
| rs677355 | let-7c-5p | trans | 5.81E-23 | Nightingale | M_LDL_TG | NA | A | G | -0.02757 | 0.004371 | 3.50E-10 |
| rs677355 | let-7c-5p | trans | 5.81E-23 | Nightingale | L_LDL_CE | NA | A | G | 0.031469 | 0.004386 | 1.20E-12 |
| rs677355 | let-7c-5p | trans | 5.81E-23 | Nightingale | M_VLDL_C | NA | A | G | 0.033643 | 0.004158 | 1.90E-15 |
| rs677355 | let-7c-5p | trans | 5.81E-23 | Nightingale | XS_VLDL_C | NA | A | G | 0.037777 | 0.004271 | 2.40E-18 |
| rs677355 | let-7c-5p | trans | 5.81E-23 | Nightingale | M_VLDL_P | NA | A | G | 0.044027 | 0.004248 | 6.50E-25 |
| rs677355 | let-7c-5p | trans | 5.81E-23 | Nightingale | XS_VLDL_F | NA | A | G | 0.037025 | 0.004349 | 7.40E-18 |
| rs677355 | let-7c-5p | trans | 5.81E-23 | Nightingale | XS_VLDL_F | NA | A | G | 0.031383 | 0.004394 | 2.60E-12 |
| rs677355 | let-7c-5p | trans | 5.81E-23 | Nightingale | XXL_VLDL_ | NA | A | G | -0.03569 | 0.004464 | 2.30E-16 |
| rs677355 | let-7c-5p | trans | 5.81E-23 | Nightingale | Total_CE | NA | A | G | 0.031682 | 0.004269 | 1.10E-13 |
| rs677355 | let-7c-5p | trans | 5.81E-23 | Nightingale | LDL_PL | NA | A | G | 0.034895 | 0.004416 | 5.60E-15 |
| rs677355 | let-7c-5p | trans | 5.81E-23 | Nightingale | S_VLDL_TG | NA | A | G | -0.03812 | 0.004284 | 1.50E-18 |
| rs677355 | let-7c-5p | trans | 5.81E-23 | Nightingale | XL_VLDL_C | NA | A | G | 0.037355 | 0.004246 | 3.10E-18 |
| rs677355 | let-7c-5p | trans | 5.81E-23 | Nightingale | M_VLDL_C | NA | A | G | 0.036213 | 0.004169 | 1.20E-17 |
| rs677355 | let-7c-5p | trans | 5.81E-23 | Nightingale | LDL_P | NA | A | G | 0.031689 | 0.004423 | 1.20E-12 |
| rs677355 | let-7c-5p | trans | 5.81E-23 | Nightingale | XL_VLDL_T | NA | A | G | -0.04386 | 0.00429 | 2.10E-24 |
| rs677355 | let-7c-5p | trans | 5.81E-23 | Nightingale | S_LDL_PL_ | NA | A | G | -0.03 | 0.004418 | 3.00E-13 |
| rs677355 | let-7c-5p | trans | 5.81E-23 | Nightingale | S_LDL_CE | NA | A | G | 0.030433 | 0.004428 | 7.00E-12 |
| rs677355 | let-7c-5p | trans | 5.81E-23 | Nightingale | XXL_VLDL_ | NA | A | G | 0.034787 | 0.004435 | 3.10E-15 |
| rs677355 | let-7c-5p | trans | 5.81E-23 | Nightingale | S_VLDL_C | NA | A | G | 0.038286 | 0.004425 | 2.10E-18 |
| rs677355 | let-7c-5p | trans | 5.81E-23 | Nightingale | LDL_CE | NA | A | G | 0.032446 | 0.004414 | 2.60E-13 |
| rs677355 | let-7c-5p | trans | 5.81E-23 | Nightingale | M_LDL_L | NA | A | G | 0.032919 | 0.004432 | 1.20E-13 |
| rs677355 | let-7c-5p | trans | 5.81E-23 | Nightingale | IDL_FC | NA | A | G | 0.038214 | 0.004285 | 1.70E-18 |
| rs677355 | let-7c-5p | trans | 5.81E-23 | Nightingale | S_LDL_L | NA | A | G | 0.027634 | 0.004425 | 9.50E-10 |
| rs677355 | let-7c-5p | trans | 5.81E-23 | Nightingale | S_VLDL_L | NA | A | G | 0.024574 | 0.004383 | 4.30E-09 |
| rs677355 | let-7c-5p | trans | 5.81E-23 | Nightingale | VLDL_FC | NA | A | G | 0.024053 | 0.004393 | 1.30E-08 |
| rs677355 | let-7c-5p | trans | 5.81E-23 | Nightingale | S_HDL_L | NA | A | G | 0.026224 | 0.004411 | 1.80E-09 |
| rs677355 | let-7c-5p | trans | 5.81E-23 | Nightingale | M_LDL_PL | NA | A | G | 0.03236 | 0.004432 | 3.20E-13 |
| rs677355 | let-7c-5p | trans | 5.81E-23 | Nightingale | XXL_VLDL_ | NA | A | G | 0.030793 | 0.004407 | 1.20E-12 |
| rs677355 | let-7c-5p | trans | 5.81E-23 | Nightingale | Total_L | NA | A | G | 0.026271 | 0.004354 | 5.30E-10 |
| rs677355 | let-7c-5p | trans | 5.81E-23 | Nightingale | M_VLDL_C | NA | A | G | 0.038623 | 0.004414 | 2.30E-18 |
| rs677355 | let-7c-5p | trans | 5.81E-23 | Nightingale | S_VLDL_C_ | NA | A | G | 0.041886 | 0.004307 | 5.40E-22 |
| rs677355 | let-7c-5p | trans | 5.81E-23 | Nightingale | IDL_C | NA | A | G | 0.034923 | 0.004274 | 1.10E-15 |
| rs677355 | let-7c-5p | trans | 5.81E-23 | Nightingale | XXL_VLDL_ | NA | A | G | 0.033101 | 0.00446 | 9.20E-14 |
| rs677355 | let-7c-5p | trans | 5.81E-23 | Nightingale | L_VLDL_C_ | NA | A | G | 0.052158 | 0.004323 | 4.80E-34 |
| rs677355 | let-7c-5p | trans | 5.81E-23 | Nightingale | VLDL_C | NA | A | G | 0.031206 | 0.00442 | 5.80E-13 |
| rs677355 | let-7c-5p | trans | 5.81E-23 | Nightingale | Omega_6 | NA | A | G | 0.025797 | 0.004322 | 6.80E-10 |
| rs677355 | let-7c-5p | trans | 5.81E-23 | Nightingale | XS_VLDL_L | NA | A | G | 0.033781 | 0.004351 | 2.30E-15 |
| rs677355 | let-7c-5p | trans | 5.81E-23 | Nightingale | Sphingomy | NA | A | G | 0.028914 | 0.004137 | 8.50E-12 |
| rs677355 | let-7c-5p | trans | 5.81E-23 | Nightingale | LDL_FC | NA | A | G | 0.033557 | 0.004379 | 6.00E-14 |
| rs677355 | let-7c-5p | trans | 5.81E-23 | Nightingale | LDL_C | NA | A | G | 0.033095 | 0.004407 | 1.10E-13 |
| rs677355 | let-7c-5p | trans | 5.81E-23 | Nightingale | S_VLDL_FC | NA | A | G | 0.03781 | 0.004427 | 8.80E-18 |
| rs677355 | let-7c-5p | trans | 5.81E-23 | Nightingale | S_HDL_C | NA | A | G | 0.028339 | 0.004418 | 1.90E-10 |
| rs677355 | let-7c-5p | trans | 5.81E-23 | Nightingale | L_LDL_L | NA | A | G | 0.03346 | 0.004383 | 3.60E-14 |
| rs677355 | let-7c-5p | trans | 5.81E-23 | Nightingale | S_VLDL_CE | NA | A | G | 0.047027 | 0.004366 | 1.30E-26 |
| rs677355 | let-7c-5p | trans | 5.81E-23 | Nightingale | PUFA | NA | A | G | 0.023614 | 0.004317 | 1.70E-08 |
| rs677355 | let-7c-5p | trans | 5.81E-23 | Nightingale | S_VLDL_P | NA | A | G | 0.023377 | 0.00437 | 1.90E-08 |
| rs677355 | let-7c-5p | trans | 5.81E-23 | Nightingale | M_VLDL_F | NA | A | G | 0.042055 | 0.004211 | 3.70E-23 |
| rs677355 | let-7c-5p | trans | 5.81E-23 | Nightingale | S_LDL_FC | NA | A | G | 0.028483 | 0.004419 | 4.80E-10 |
| rs677355 | let-7c-5p | trans | 5.81E-23 | Nightingale | IDL_P | NA | A | G | 0.039974 | 0.004379 | 1.40E-19 |
| rs677355 | let-7c-5p | trans | 5.81E-23 | Nightingale | IDL_FC_pct | NA | A | G | 0.025744 | 0.004392 | 1.10E-08 |
| rs677355 | let-7c-5p | trans | 5.81E-23 | Nightingale | M_VLDL_T | NA | A | G | -0.03856 | 0.004182 | 7.10E-20 |
| rs677355 | let-7c-5p | trans | 5.81E-23 | Nightingale | S_HDL_P | NA | A | G | 0.030523 | 0.004426 | 6.00E-12 |
| rs677355 | let-7c-5p | trans | 5.81E-23 | Nightingale | XS_VLDL_C | NA | A | G | 0.037965 | 0.004296 | 1.50E-18 |
| rs677355 | let-7c-5p | trans | 5.81E-23 | Nightingale | Clinical_LD | NA | A | G | 0.036386 | 0.0044 | 3.20E-16 |
| rs677355 | let-7c-5p | trans | 5.81E-23 | Nightingale | LA | NA | A | G | 0.024871 | 0.00434 | 4.00E-09 |
| rs677355 | let-7c-5p | trans | 5.81E-23 | Nightingale | M_LDL_C | NA | A | G | 0.033874 | 0.004434 | 2.60E-14 |

| | | | | | | | | | | | |
|---|---|---|---|---|---|---|---|---|---|---|---|
| rs677355 | let-7c-5p | trans | 5.81E-23 | Nightingale | LDL_L | NA | A | G | 0.033204 | 0.004411 | 8.80E-14 |
| rs677355 | let-7c-5p | trans | 5.81E-23 | Nightingale | S_VLDL_PL | NA | A | G | 0.03268 | 0.004418 | 5.00E-14 |
| rs677355 | let-7c-5p | trans | 5.81E-23 | Nightingale | non_HDL_( | NA | A | G | 0.035607 | 0.004402 | 6.10E-16 |
| rs677355 | let-7c-5p | trans | 5.81E-23 | Nightingale | L_VLDL_FC | NA | A | G | 0.048463 | 0.004383 | 6.40E-29 |
| rs677355 | let-7c-5p | trans | 5.81E-23 | Nightingale | L_VLDL_TG | NA | A | G | -0.05108 | 0.004405 | 6.90E-32 |
| rs677355 | let-7c-5p | trans | 5.81E-23 | Nightingale | XL_VLDL_P | NA | A | G | 0.038718 | 0.004433 | 7.00E-19 |
| rs677355 | let-7c-5p | trans | 5.81E-23 | Nightingale | GlycA | NA | A | G | 0.02771 | 0.004407 | 2.30E-10 |
| rs677355 | let-7c-5p | trans | 5.81E-23 | Nightingale | S_VLDL_PL | NA | A | G | 0.027729 | 0.004238 | 1.80E-10 |
| rs677355 | let-7c-5p | trans | 5.81E-23 | Nightingale | VLDL_CE | NA | A | G | 0.035699 | 0.004427 | 3.60E-16 |
| rs677355 | let-7c-5p | trans | 5.81E-23 | Nightingale | IDL_L | NA | A | G | 0.03523 | 0.004279 | 5.50E-16 |
| rs677355 | let-7c-5p | trans | 5.81E-23 | Nightingale | L_LDL_PL | NA | A | G | 0.037906 | 0.004392 | 1.10E-17 |
| rs677355 | let-7c-5p | trans | 5.81E-23 | Nightingale | L_LDL_P | NA | A | G | 0.029058 | 0.004411 | 7.40E-11 |
| rs677355 | let-7c-5p | trans | 5.81E-23 | Nightingale | VLDL_P | NA | A | G | 0.026547 | 0.004402 | 5.00E-10 |
| rs677355 | let-7c-5p | trans | 5.81E-23 | Nightingale | L_VLDL_CE | NA | A | G | 0.047363 | 0.004235 | 2.40E-29 |
| rs677355 | let-7c-5p | trans | 5.81E-23 | Nightingale | XL_VLDL_F | NA | A | G | 0.037212 | 0.00429 | 2.00E-18 |
| rs687289 | let-7c-5p | trans | 5.91E-24 | Nightingale | XS_VLDL_C | NA | A | G | 0.03768 | 0.004274 | 2.70E-18 |
| rs687289 | let-7c-5p | trans | 5.91E-24 | Nightingale | ApoB | NA | A | G | 0.03375 | 0.004423 | 2.90E-14 |
| rs687289 | let-7c-5p | trans | 5.91E-24 | Nightingale | LDL_C | NA | A | G | 0.032964 | 0.004409 | 1.20E-13 |
| rs687289 | let-7c-5p | trans | 5.91E-24 | Nightingale | S_VLDL_PL | NA | A | G | 0.032601 | 0.00442 | 5.80E-14 |
| rs687289 | let-7c-5p | trans | 5.91E-24 | Nightingale | M_LDL_C_ | NA | A | G | 0.031111 | 0.004432 | 3.10E-12 |
| rs687289 | let-7c-5p | trans | 5.91E-24 | Nightingale | L_VLDL_TG | NA | A | G | -0.05139 | 0.004408 | 3.50E-32 |
| rs687289 | let-7c-5p | trans | 5.91E-24 | Nightingale | XL_VLDL_C | NA | A | G | 0.037793 | 0.004249 | 1.10E-18 |
| rs687289 | let-7c-5p | trans | 5.91E-24 | Nightingale | S_VLDL_L | NA | A | G | 0.024476 | 0.004386 | 5.10E-09 |
| rs687289 | let-7c-5p | trans | 5.91E-24 | Nightingale | S_VLDL_P | NA | A | G | 0.023298 | 0.004373 | 2.10E-08 |
| rs687289 | let-7c-5p | trans | 5.91E-24 | Nightingale | M_VLDL_C | NA | A | G | 0.036468 | 0.004171 | 5.90E-18 |
| rs687289 | let-7c-5p | trans | 5.91E-24 | Nightingale | S_LDL_C | NA | A | G | 0.030716 | 0.004433 | 7.50E-12 |
| rs687289 | let-7c-5p | trans | 5.91E-24 | Nightingale | LDL_L | NA | A | G | 0.033082 | 0.004414 | 1.00E-13 |
| rs687289 | let-7c-5p | trans | 5.91E-24 | Nightingale | M_VLDL_C | NA | A | G | 0.033905 | 0.004161 | 9.50E-16 |
| rs687289 | let-7c-5p | trans | 5.91E-24 | Nightingale | S_HDL_P | NA | A | G | 0.030278 | 0.004429 | 9.00E-12 |
| rs687289 | let-7c-5p | trans | 5.91E-24 | Nightingale | M_LDL_L | NA | A | G | 0.032826 | 0.004435 | 1.40E-13 |
| rs687289 | let-7c-5p | trans | 5.91E-24 | Nightingale | M_VLDL_F | NA | A | G | 0.042299 | 0.004213 | 1.80E-23 |
| rs687289 | let-7c-5p | trans | 5.91E-24 | Nightingale | GlycA | NA | A | G | 0.028099 | 0.00441 | 1.30E-10 |
| rs687289 | let-7c-5p | trans | 5.91E-24 | Nightingale | S_LDL_CE | NA | A | G | 0.030317 | 0.004431 | 8.30E-12 |
| rs687289 | let-7c-5p | trans | 5.91E-24 | Nightingale | M_VLDL_T | NA | A | G | -0.03881 | 0.004185 | 3.40E-20 |
| rs687289 | let-7c-5p | trans | 5.91E-24 | Nightingale | S_VLDL_C_ | NA | A | G | 0.042052 | 0.00431 | 3.20E-22 |
| rs687289 | let-7c-5p | trans | 5.91E-24 | Nightingale | M_LDL_C | NA | A | G | 0.033786 | 0.004437 | 3.00E-14 |
| rs687289 | let-7c-5p | trans | 5.91E-24 | Nightingale | M_VLDL_C | NA | A | G | 0.038509 | 0.004416 | 2.60E-18 |
| rs687289 | let-7c-5p | trans | 5.91E-24 | Nightingale | XL_VLDL_F | NA | A | G | 0.037531 | 0.004293 | 1.00E-18 |
| rs687289 | let-7c-5p | trans | 5.91E-24 | Nightingale | M_VLDL_C | NA | A | G | 0.040645 | 0.004388 | 3.20E-20 |
| rs687289 | let-7c-5p | trans | 5.91E-24 | Nightingale | S_LDL_PL_ | NA | A | G | -0.02989 | 0.00442 | 4.10E-13 |
| rs687289 | let-7c-5p | trans | 5.91E-24 | Nightingale | IDL_CE | NA | A | G | 0.033304 | 0.004276 | 1.80E-14 |
| rs687289 | let-7c-5p | trans | 5.91E-24 | Nightingale | Omega_6 | NA | A | G | 0.025429 | 0.004325 | 1.10E-09 |
| rs687289 | let-7c-5p | trans | 5.91E-24 | Nightingale | XL_VLDL_T | NA | A | G | -0.04438 | 0.004292 | 5.40E-25 |
| rs687289 | let-7c-5p | trans | 5.91E-24 | Nightingale | LDL_CE | NA | A | G | 0.032325 | 0.004417 | 3.10E-13 |
| rs687289 | let-7c-5p | trans | 5.91E-24 | Nightingale | XXL_VLDL_ | NA | A | G | 0.035053 | 0.004438 | 2.10E-15 |
| rs687289 | let-7c-5p | trans | 5.91E-24 | Nightingale | XS_VLDL_P | NA | A | G | 0.036638 | 0.00437 | 1.60E-17 |
| rs687289 | let-7c-5p | trans | 5.91E-24 | Nightingale | S_LDL_L | NA | A | G | 0.027511 | 0.004428 | 1.10E-09 |
| rs687289 | let-7c-5p | trans | 5.91E-24 | Nightingale | L_LDL_C | NA | A | G | 0.032081 | 0.004379 | 3.60E-13 |
| rs687289 | let-7c-5p | trans | 5.91E-24 | Nightingale | M_LDL_TG | NA | A | G | -0.02742 | 0.004374 | 4.00E-10 |
| rs687289 | let-7c-5p | trans | 5.91E-24 | Nightingale | S_VLDL_C | NA | A | G | 0.038239 | 0.004428 | 2.40E-18 |
| rs687289 | let-7c-5p | trans | 5.91E-24 | Nightingale | M_VLDL_P | NA | A | G | 0.028859 | 0.004422 | 3.10E-11 |
| rs687289 | let-7c-5p | trans | 5.91E-24 | Nightingale | L_LDL_P | NA | A | G | 0.028943 | 0.004414 | 8.30E-11 |
| rs687289 | let-7c-5p | trans | 5.91E-24 | Nightingale | M_LDL_P | NA | A | G | 0.035821 | 0.004441 | 7.50E-16 |
| rs687289 | let-7c-5p | trans | 5.91E-24 | Nightingale | VLDL_CE | NA | A | G | 0.03558 | 0.00443 | 4.60E-16 |
| rs687289 | let-7c-5p | trans | 5.91E-24 | Nightingale | XL_VLDL_P | NA | A | G | 0.038789 | 0.004435 | 6.40E-19 |
| rs687289 | let-7c-5p | trans | 5.91E-24 | Nightingale | S_LDL_P | NA | A | G | 0.029429 | 0.004433 | 4.00E-11 |
| rs687289 | let-7c-5p | trans | 5.91E-24 | Nightingale | XS_VLDL_F | NA | A | G | 0.037022 | 0.004352 | 7.50E-18 |
| rs687289 | let-7c-5p | trans | 5.91E-24 | Nightingale | M_VLDL_P | NA | A | G | 0.028023 | 0.004418 | 9.80E-11 |
| rs687289 | let-7c-5p | trans | 5.91E-24 | Nightingale | LDL_P | NA | A | G | 0.031612 | 0.004425 | 1.30E-12 |
| rs687289 | let-7c-5p | trans | 5.91E-24 | Nightingale | L_VLDL_C_ | NA | A | G | 0.052543 | 0.004325 | 1.80E-34 |
| rs687289 | let-7c-5p | trans | 5.91E-24 | Nightingale | L_LDL_L | NA | A | G | 0.033323 | 0.004385 | 4.20E-14 |
| rs687289 | let-7c-5p | trans | 5.91E-24 | Nightingale | LDL_PL | NA | A | G | 0.034755 | 0.004418 | 6.70E-15 |
| rs687289 | let-7c-5p | trans | 5.91E-24 | Nightingale | XS_VLDL_C | NA | A | G | 0.037903 | 0.004299 | 1.70E-18 |

| | | | | | | | | | | |
|---|---|---|---|---|---|---|---|---|---|---|
| rs687289 | let-7c-5p | trans | 5.91E-24 | Nightingale | M_LDL_CE | NA | A | G | 0.032783 | 0.004435 | 1.30E-13 |
| rs687289 | let-7c-5p | trans | 5.91E-24 | Nightingale | LDL_FC | NA | A | G | 0.033395 | 0.004381 | 7.50E-14 |
| rs687289 | let-7c-5p | trans | 5.91E-24 | Nightingale | S_HDL_C | NA | A | G | 0.028109 | 0.004421 | 2.80E-10 |
| rs687289 | let-7c-5p | trans | 5.91E-24 | Nightingale | IDL_P | NA | A | G | 0.03981 | 0.004382 | 1.80E-19 |
| rs687289 | let-7c-5p | trans | 5.91E-24 | Nightingale | XS_VLDL_P | NA | A | G | 0.032077 | 0.004367 | 5.90E-14 |
| rs687289 | let-7c-5p | trans | 5.91E-24 | Nightingale | XL_VLDL_C | NA | A | G | 0.038665 | 0.004247 | 1.60E-19 |
| rs687289 | let-7c-5p | trans | 5.91E-24 | Nightingale | Remnant_( | NA | A | G | 0.036652 | 0.0044 | 5.60E-17 |
| rs687289 | let-7c-5p | trans | 5.91E-24 | Nightingale | L_LDL_CE | NA | A | G | 0.031327 | 0.004389 | 1.40E-12 |
| rs687289 | let-7c-5p | trans | 5.91E-24 | Nightingale | S_HDL_L | NA | A | G | 0.026 | 0.004413 | 2.50E-09 |
| rs687289 | let-7c-5p | trans | 5.91E-24 | Nightingale | Total_PL | NA | A | G | 0.02466 | 0.004217 | 1.60E-09 |
| rs687289 | let-7c-5p | trans | 5.91E-24 | Nightingale | IDL_PL | NA | A | G | 0.035187 | 0.004274 | 4.50E-16 |
| rs687289 | let-7c-5p | trans | 5.91E-24 | Nightingale | XXL_VLDL_ | NA | A | G | 0.02523 | 0.004458 | 1.10E-09 |
| rs687289 | let-7c-5p | trans | 5.91E-24 | Nightingale | ApoB_by_/ | NA | A | G | 0.024977 | 0.004362 | 1.30E-08 |
| rs687289 | let-7c-5p | trans | 5.91E-24 | Nightingale | S_HDL_CE | NA | A | G | 0.028306 | 0.004436 | 2.20E-10 |
| rs687289 | let-7c-5p | trans | 5.91E-24 | Nightingale | Total_CE | NA | A | G | 0.031498 | 0.004272 | 1.40E-13 |
| rs687289 | let-7c-5p | trans | 5.91E-24 | Nightingale | Total_FC | NA | A | G | 0.033014 | 0.004326 | 1.00E-14 |
| rs687289 | let-7c-5p | trans | 5.91E-24 | Nightingale | S_VLDL_CE | NA | A | G | 0.047288 | 0.004369 | 6.20E-27 |
| rs687289 | let-7c-5p | trans | 5.91E-24 | Nightingale | M_VLDL_F | NA | A | G | 0.033132 | 0.004428 | 4.80E-14 |
| rs687289 | let-7c-5p | trans | 5.91E-24 | Nightingale | IDL_C | NA | A | G | 0.034761 | 0.004277 | 1.30E-15 |
| rs687289 | let-7c-5p | trans | 5.91E-24 | Nightingale | non_HDL_( | NA | A | G | 0.03547 | 0.004405 | 7.40E-16 |
| rs687289 | let-7c-5p | trans | 5.91E-24 | Nightingale | S_LDL_C_p | NA | A | G | 0.045227 | 0.004446 | 8.10E-25 |
| rs687289 | let-7c-5p | trans | 5.91E-24 | Nightingale | XS_VLDL_F | NA | A | G | 0.031381 | 0.004397 | 2.60E-12 |
| rs687289 | let-7c-5p | trans | 5.91E-24 | Nightingale | VLDL_P | NA | A | G | 0.026418 | 0.004405 | 6.20E-10 |
| rs687289 | let-7c-5p | trans | 5.91E-24 | Nightingale | M_VLDL_P | NA | A | G | 0.044251 | 0.00425 | 3.40E-25 |
| rs687289 | let-7c-5p | trans | 5.91E-24 | Nightingale | L_VLDL_FC | NA | A | G | 0.048909 | 0.004386 | 2.40E-29 |
| rs687289 | let-7c-5p | trans | 5.91E-24 | Nightingale | Total_C | NA | A | G | 0.032124 | 0.004287 | 4.80E-14 |
| rs687289 | let-7c-5p | trans | 5.91E-24 | Nightingale | L_LDL_FC | NA | A | G | 0.033386 | 0.004349 | 4.10E-14 |
| rs687289 | let-7c-5p | trans | 5.91E-24 | Nightingale | S_HDL_PL | NA | A | G | 0.023499 | 0.00439 | 5.00E-08 |
| rs687289 | let-7c-5p | trans | 5.91E-24 | Nightingale | M_LDL_FC | NA | A | G | 0.034117 | 0.004423 | 2.80E-14 |
| rs687289 | let-7c-5p | trans | 5.91E-24 | Nightingale | S_VLDL_TG | NA | A | G | -0.03827 | 0.004287 | 9.50E-19 |
| rs687289 | let-7c-5p | trans | 5.91E-24 | Nightingale | XXL_VLDL_ | NA | A | G | 0.033249 | 0.004463 | 7.70E-14 |
| rs687289 | let-7c-5p | trans | 5.91E-24 | Nightingale | S_LDL_FC | NA | A | G | 0.028345 | 0.004422 | 5.60E-10 |
| rs687289 | let-7c-5p | trans | 5.91E-24 | Nightingale | L_VLDL_CE | NA | A | G | 0.047672 | 0.004238 | 1.00E-29 |
| rs687289 | let-7c-5p | trans | 5.91E-24 | Nightingale | Citrate | NA | A | G | 0.026536 | 0.0044 | 1.10E-09 |
| rs687289 | let-7c-5p | trans | 5.91E-24 | Nightingale | M_LDL_PL | NA | A | G | 0.032235 | 0.004435 | 3.90E-13 |
| rs687289 | let-7c-5p | trans | 5.91E-24 | Nightingale | Total_L | NA | A | G | 0.026033 | 0.004357 | 7.20E-10 |
| rs687289 | let-7c-5p | trans | 5.91E-24 | Nightingale | VLDL_C | NA | A | G | 0.031062 | 0.004422 | 7.60E-13 |
| rs687289 | let-7c-5p | trans | 5.91E-24 | Nightingale | XS_VLDL_L | NA | A | G | 0.033757 | 0.004353 | 2.40E-15 |
| rs687289 | let-7c-5p | trans | 5.91E-24 | Nightingale | IDL_FC | NA | A | G | 0.038096 | 0.004287 | 2.00E-18 |
| rs687289 | let-7c-5p | trans | 5.91E-24 | Nightingale | S_VLDL_CE | NA | A | G | 0.038069 | 0.004423 | 2.20E-18 |
| rs687289 | let-7c-5p | trans | 5.91E-24 | Nightingale | PUFA | NA | A | G | 0.023493 | 0.00432 | 2.00E-08 |
| rs687289 | let-7c-5p | trans | 5.91E-24 | Nightingale | XXL_VLDL_ | NA | A | G | -0.03594 | 0.004467 | 1.60E-16 |
| rs687289 | let-7c-5p | trans | 5.91E-24 | Nightingale | S_VLDL_PL | NA | A | G | 0.027844 | 0.004241 | 1.40E-10 |
| rs687289 | let-7c-5p | trans | 5.91E-24 | Nightingale | Clinical_LD | NA | A | G | 0.036238 | 0.004403 | 4.00E-16 |
| rs687289 | let-7c-5p | trans | 5.91E-24 | Nightingale | S_LDL_CE_ | NA | A | G | 0.03078 | 0.004421 | 7.20E-14 |
| rs687289 | let-7c-5p | trans | 5.91E-24 | Nightingale | Sphingomy | NA | A | G | 0.028834 | 0.00414 | 9.00E-12 |
| rs687289 | let-7c-5p | trans | 5.91E-24 | Nightingale | S_VLDL_FC | NA | A | G | 0.029424 | 0.004232 | 8.60E-12 |
| rs687289 | let-7c-5p | trans | 5.91E-24 | Nightingale | S_VLDL_FC | NA | A | G | 0.037731 | 0.004429 | 1.00E-17 |
| rs687289 | let-7c-5p | trans | 5.91E-24 | Nightingale | XL_HDL_PL | NA | A | G | -0.02251 | 0.004069 | 1.90E-08 |
| rs687289 | let-7c-5p | trans | 5.91E-24 | Nightingale | IDL_L | NA | A | G | 0.035103 | 0.004282 | 6.50E-16 |
| rs687289 | let-7c-5p | trans | 5.91E-24 | Nightingale | L_LDL_PL | NA | A | G | 0.037753 | 0.004395 | 1.30E-17 |
| rs687289 | let-7c-5p | trans | 5.91E-24 | Nightingale | XXL_VLDL_ | NA | A | G | 0.031237 | 0.00441 | 6.10E-13 |
| rs687289 | let-7c-5p | trans | 5.91E-24 | Nightingale | LA | NA | A | G | 0.024416 | 0.004343 | 7.10E-09 |
| rs687289 | let-7c-5p | trans | 5.91E-24 | Nightingale | IDL_FC_pct | NA | A | G | 0.025842 | 0.004394 | 9.80E-09 |
| rs687289 | let-7c-5p | trans | 5.91E-24 | Nightingale | VLDL_FC | NA | A | G | 0.02388 | 0.004396 | 1.60E-08 |
| rs687289 | let-7b-5p | trans | 8.61E-28 | Nightingale | XS_VLDL_C | NA | A | G | 0.03768 | 0.004274 | 2.70E-18 |
| rs687289 | let-7b-5p | trans | 8.61E-28 | Nightingale | ApoB | NA | A | G | 0.03375 | 0.004423 | 2.90E-14 |
| rs687289 | let-7b-5p | trans | 8.61E-28 | Nightingale | LDL_C | NA | A | G | 0.032964 | 0.004409 | 1.20E-13 |
| rs687289 | let-7b-5p | trans | 8.61E-28 | Nightingale | S_VLDL_PL | NA | A | G | 0.032601 | 0.00442 | 5.80E-14 |
| rs687289 | let-7b-5p | trans | 8.61E-28 | Nightingale | M_LDL_C_ | NA | A | G | 0.031111 | 0.004432 | 3.10E-12 |
| rs687289 | let-7b-5p | trans | 8.61E-28 | Nightingale | L_VLDL_TG | NA | A | G | -0.05139 | 0.004408 | 3.50E-32 |
| rs687289 | let-7b-5p | trans | 8.61E-28 | Nightingale | XL_VLDL_C | NA | A | G | 0.037793 | 0.004249 | 1.10E-18 |
| rs687289 | let-7b-5p | trans | 8.61E-28 | Nightingale | S_VLDL_L | NA | A | G | 0.024476 | 0.004386 | 5.10E-09 |

| | | | | | | | | | | | |
|---|---|---|---|---|---|---|---|---|---|---|---|
| rs687289 | let-7b-5p | trans | 8.61E-28 | Nightingale | S_VLDL_P | NA | A | G | 0.023298 | 0.004373 | 2.10E-08 |
| rs687289 | let-7b-5p | trans | 8.61E-28 | Nightingale | M_VLDL_C | NA | A | G | 0.036468 | 0.004171 | 5.90E-18 |
| rs687289 | let-7b-5p | trans | 8.61E-28 | Nightingale | S_LDL_C | NA | A | G | 0.030716 | 0.004433 | 7.50E-12 |
| rs687289 | let-7b-5p | trans | 8.61E-28 | Nightingale | LDL_L | NA | A | G | 0.033082 | 0.004414 | 1.00E-13 |
| rs687289 | let-7b-5p | trans | 8.61E-28 | Nightingale | M_VLDL_C | NA | A | G | 0.033905 | 0.004161 | 9.50E-16 |
| rs687289 | let-7b-5p | trans | 8.61E-28 | Nightingale | S_HDL_P | NA | A | G | 0.030278 | 0.004429 | 9.00E-12 |
| rs687289 | let-7b-5p | trans | 8.61E-28 | Nightingale | M_LDL_L | NA | A | G | 0.032826 | 0.004435 | 1.40E-13 |
| rs687289 | let-7b-5p | trans | 8.61E-28 | Nightingale | M_VLDL_F | NA | A | G | 0.042299 | 0.004213 | 1.80E-23 |
| rs687289 | let-7b-5p | trans | 8.61E-28 | Nightingale | GlycA | NA | A | G | 0.028099 | 0.00441 | 1.30E-10 |
| rs687289 | let-7b-5p | trans | 8.61E-28 | Nightingale | S_LDL_CE | NA | A | G | 0.030317 | 0.004431 | 8.30E-12 |
| rs687289 | let-7b-5p | trans | 8.61E-28 | Nightingale | M_VLDL_T | NA | A | G | -0.03881 | 0.004185 | 3.40E-20 |
| rs687289 | let-7b-5p | trans | 8.61E-28 | Nightingale | S_VLDL_C_ | NA | A | G | 0.042052 | 0.00431 | 3.20E-22 |
| rs687289 | let-7b-5p | trans | 8.61E-28 | Nightingale | M_LDL_C | NA | A | G | 0.033786 | 0.004437 | 3.00E-14 |
| rs687289 | let-7b-5p | trans | 8.61E-28 | Nightingale | M_VLDL_C | NA | A | G | 0.038509 | 0.004416 | 2.60E-18 |
| rs687289 | let-7b-5p | trans | 8.61E-28 | Nightingale | XL_VLDL_F | NA | A | G | 0.037531 | 0.004293 | 1.00E-18 |
| rs687289 | let-7b-5p | trans | 8.61E-28 | Nightingale | M_VLDL_C | NA | A | G | 0.040645 | 0.004388 | 3.20E-20 |
| rs687289 | let-7b-5p | trans | 8.61E-28 | Nightingale | S_LDL_PL_ | NA | A | G | -0.02989 | 0.00442 | 4.10E-13 |
| rs687289 | let-7b-5p | trans | 8.61E-28 | Nightingale | IDL_CE | NA | A | G | 0.033304 | 0.004276 | 1.80E-14 |
| rs687289 | let-7b-5p | trans | 8.61E-28 | Nightingale | Omega_6 | NA | A | G | 0.025429 | 0.004325 | 1.10E-09 |
| rs687289 | let-7b-5p | trans | 8.61E-28 | Nightingale | XL_VLDL_T | NA | A | G | -0.04438 | 0.004292 | 5.40E-25 |
| rs687289 | let-7b-5p | trans | 8.61E-28 | Nightingale | LDL_CE | NA | A | G | 0.032325 | 0.004417 | 3.10E-13 |
| rs687289 | let-7b-5p | trans | 8.61E-28 | Nightingale | XXL_VLDL_ | NA | A | G | 0.035053 | 0.004438 | 2.10E-15 |
| rs687289 | let-7b-5p | trans | 8.61E-28 | Nightingale | XS_VLDL_P | NA | A | G | 0.036638 | 0.00437 | 1.60E-17 |
| rs687289 | let-7b-5p | trans | 8.61E-28 | Nightingale | S_LDL_L | NA | A | G | 0.027511 | 0.004428 | 1.10E-09 |
| rs687289 | let-7b-5p | trans | 8.61E-28 | Nightingale | L_LDL_C | NA | A | G | 0.032081 | 0.004379 | 3.60E-13 |
| rs687289 | let-7b-5p | trans | 8.61E-28 | Nightingale | M_LDL_TG | NA | A | G | -0.02742 | 0.004374 | 4.00E-10 |
| rs687289 | let-7b-5p | trans | 8.61E-28 | Nightingale | S_VLDL_C | NA | A | G | 0.038239 | 0.004428 | 2.40E-18 |
| rs687289 | let-7b-5p | trans | 8.61E-28 | Nightingale | M_VLDL_P | NA | A | G | 0.028859 | 0.004422 | 3.10E-11 |
| rs687289 | let-7b-5p | trans | 8.61E-28 | Nightingale | L_LDL_P | NA | A | G | 0.028943 | 0.004414 | 8.30E-11 |
| rs687289 | let-7b-5p | trans | 8.61E-28 | Nightingale | M_LDL_P | NA | A | G | 0.035821 | 0.004441 | 7.50E-16 |
| rs687289 | let-7b-5p | trans | 8.61E-28 | Nightingale | VLDL_CE | NA | A | G | 0.03558 | 0.00443 | 4.60E-16 |
| rs687289 | let-7b-5p | trans | 8.61E-28 | Nightingale | XL_VLDL_P | NA | A | G | 0.038789 | 0.004435 | 6.40E-19 |
| rs687289 | let-7b-5p | trans | 8.61E-28 | Nightingale | S_LDL_P | NA | A | G | 0.029429 | 0.004433 | 4.00E-11 |
| rs687289 | let-7b-5p | trans | 8.61E-28 | Nightingale | XS_VLDL_F | NA | A | G | 0.037022 | 0.004352 | 7.50E-18 |
| rs687289 | let-7b-5p | trans | 8.61E-28 | Nightingale | M_VLDL_P | NA | A | G | 0.028023 | 0.004418 | 9.80E-11 |
| rs687289 | let-7b-5p | trans | 8.61E-28 | Nightingale | LDL_P | NA | A | G | 0.031612 | 0.004425 | 1.30E-12 |
| rs687289 | let-7b-5p | trans | 8.61E-28 | Nightingale | L_VLDL_C_ | NA | A | G | 0.052543 | 0.004325 | 1.80E-34 |
| rs687289 | let-7b-5p | trans | 8.61E-28 | Nightingale | L_LDL_L | NA | A | G | 0.033323 | 0.004385 | 4.20E-14 |
| rs687289 | let-7b-5p | trans | 8.61E-28 | Nightingale | LDL_PL | NA | A | G | 0.034755 | 0.004418 | 6.70E-15 |
| rs687289 | let-7b-5p | trans | 8.61E-28 | Nightingale | XS_VLDL_C | NA | A | G | 0.037903 | 0.004299 | 1.70E-18 |
| rs687289 | let-7b-5p | trans | 8.61E-28 | Nightingale | M_LDL_CE | NA | A | G | 0.032783 | 0.004435 | 1.30E-13 |
| rs687289 | let-7b-5p | trans | 8.61E-28 | Nightingale | LDL_FC | NA | A | G | 0.033395 | 0.004381 | 7.50E-14 |
| rs687289 | let-7b-5p | trans | 8.61E-28 | Nightingale | S_HDL_C | NA | A | G | 0.028109 | 0.004421 | 2.80E-10 |
| rs687289 | let-7b-5p | trans | 8.61E-28 | Nightingale | IDL_P | NA | A | G | 0.03981 | 0.004382 | 1.80E-19 |
| rs687289 | let-7b-5p | trans | 8.61E-28 | Nightingale | XS_VLDL_P | NA | A | G | 0.032077 | 0.004367 | 5.90E-14 |
| rs687289 | let-7b-5p | trans | 8.61E-28 | Nightingale | XL_VLDL_C | NA | A | G | 0.038665 | 0.004247 | 1.60E-19 |
| rs687289 | let-7b-5p | trans | 8.61E-28 | Nightingale | Remnant_( | NA | A | G | 0.036652 | 0.0044 | 5.60E-17 |
| rs687289 | let-7b-5p | trans | 8.61E-28 | Nightingale | L_LDL_CE | NA | A | G | 0.031327 | 0.004389 | 1.40E-12 |
| rs687289 | let-7b-5p | trans | 8.61E-28 | Nightingale | S_HDL_L | NA | A | G | 0.026 | 0.004413 | 2.50E-09 |
| rs687289 | let-7b-5p | trans | 8.61E-28 | Nightingale | Total_PL | NA | A | G | 0.02466 | 0.004217 | 1.60E-09 |
| rs687289 | let-7b-5p | trans | 8.61E-28 | Nightingale | IDL_PL | NA | A | G | 0.035187 | 0.004274 | 4.50E-16 |
| rs687289 | let-7b-5p | trans | 8.61E-28 | Nightingale | XXL_VLDL_ | NA | A | G | 0.02523 | 0.004458 | 1.10E-09 |
| rs687289 | let-7b-5p | trans | 8.61E-28 | Nightingale | ApoB_by_/ | NA | A | G | 0.024977 | 0.004362 | 1.30E-08 |
| rs687289 | let-7b-5p | trans | 8.61E-28 | Nightingale | S_HDL_CE | NA | A | G | 0.028306 | 0.004436 | 2.20E-10 |
| rs687289 | let-7b-5p | trans | 8.61E-28 | Nightingale | Total_CE | NA | A | G | 0.031498 | 0.004272 | 1.40E-13 |
| rs687289 | let-7b-5p | trans | 8.61E-28 | Nightingale | Total_FC | NA | A | G | 0.033014 | 0.004326 | 1.00E-14 |
| rs687289 | let-7b-5p | trans | 8.61E-28 | Nightingale | S_VLDL_CE | NA | A | G | 0.047288 | 0.004369 | 6.20E-27 |
| rs687289 | let-7b-5p | trans | 8.61E-28 | Nightingale | M_VLDL_F | NA | A | G | 0.033132 | 0.004428 | 4.80E-14 |
| rs687289 | let-7b-5p | trans | 8.61E-28 | Nightingale | IDL_C | NA | A | G | 0.034761 | 0.004277 | 1.30E-15 |
| rs687289 | let-7b-5p | trans | 8.61E-28 | Nightingale | non_HDL_( | NA | A | G | 0.03547 | 0.004405 | 7.40E-16 |
| rs687289 | let-7b-5p | trans | 8.61E-28 | Nightingale | S_LDL_C_p | NA | A | G | 0.045227 | 0.004446 | 8.10E-25 |
| rs687289 | let-7b-5p | trans | 8.61E-28 | Nightingale | XS_VLDL_F | NA | A | G | 0.031381 | 0.004397 | 2.60E-12 |
| rs687289 | let-7b-5p | trans | 8.61E-28 | Nightingale | VLDL_P | NA | A | G | 0.026418 | 0.004405 | 6.20E-10 |

| | | | | | | | | | | | |
|---|---|---|---|---|---|---|---|---|---|---|---|
| rs687289 | let-7b-5p | trans | 8.61E-28 | Nightingale | M_VLDL_P | NA | A | G | 0.044251 | 0.00425 | 3.40E-25 |
| rs687289 | let-7b-5p | trans | 8.61E-28 | Nightingale | L_VLDL_FC | NA | A | G | 0.048909 | 0.004386 | 2.40E-29 |
| rs687289 | let-7b-5p | trans | 8.61E-28 | Nightingale | Total_C | NA | A | G | 0.032124 | 0.004287 | 4.80E-14 |
| rs687289 | let-7b-5p | trans | 8.61E-28 | Nightingale | L_LDL_FC | NA | A | G | 0.033386 | 0.004349 | 4.10E-14 |
| rs687289 | let-7b-5p | trans | 8.61E-28 | Nightingale | S_HDL_PL | NA | A | G | 0.023499 | 0.00439 | 5.00E-08 |
| rs687289 | let-7b-5p | trans | 8.61E-28 | Nightingale | M_LDL_FC | NA | A | G | 0.034117 | 0.004423 | 2.80E-14 |
| rs687289 | let-7b-5p | trans | 8.61E-28 | Nightingale | S_VLDL_TG | NA | A | G | -0.03827 | 0.004287 | 9.50E-19 |
| rs687289 | let-7b-5p | trans | 8.61E-28 | Nightingale | XXL_VLDL_ | NA | A | G | 0.033249 | 0.004463 | 7.70E-14 |
| rs687289 | let-7b-5p | trans | 8.61E-28 | Nightingale | S_LDL_FC | NA | A | G | 0.028345 | 0.004422 | 5.60E-10 |
| rs687289 | let-7b-5p | trans | 8.61E-28 | Nightingale | L_VLDL_CE | NA | A | G | 0.047672 | 0.004238 | 1.00E-29 |
| rs687289 | let-7b-5p | trans | 8.61E-28 | Nightingale | Citrate | NA | A | G | 0.026536 | 0.0044 | 1.10E-09 |
| rs687289 | let-7b-5p | trans | 8.61E-28 | Nightingale | M_LDL_PL | NA | A | G | 0.032235 | 0.004435 | 3.90E-13 |
| rs687289 | let-7b-5p | trans | 8.61E-28 | Nightingale | Total_L | NA | A | G | 0.026033 | 0.004357 | 7.20E-10 |
| rs687289 | let-7b-5p | trans | 8.61E-28 | Nightingale | VLDL_C | NA | A | G | 0.031062 | 0.004422 | 7.60E-13 |
| rs687289 | let-7b-5p | trans | 8.61E-28 | Nightingale | XS_VLDL_L | NA | A | G | 0.033757 | 0.004353 | 2.40E-15 |
| rs687289 | let-7b-5p | trans | 8.61E-28 | Nightingale | IDL_FC | NA | A | G | 0.038096 | 0.004287 | 2.00E-18 |
| rs687289 | let-7b-5p | trans | 8.61E-28 | Nightingale | S_VLDL_CE | NA | A | G | 0.038069 | 0.004423 | 2.20E-18 |
| rs687289 | let-7b-5p | trans | 8.61E-28 | Nightingale | PUFA | NA | A | G | 0.023493 | 0.00432 | 2.00E-08 |
| rs687289 | let-7b-5p | trans | 8.61E-28 | Nightingale | XXL_VLDL_ | NA | A | G | -0.03594 | 0.004467 | 1.60E-16 |
| rs687289 | let-7b-5p | trans | 8.61E-28 | Nightingale | S_VLDL_PL | NA | A | G | 0.027844 | 0.004241 | 1.40E-10 |
| rs687289 | let-7b-5p | trans | 8.61E-28 | Nightingale | Clinical_LD | NA | A | G | 0.036238 | 0.004403 | 4.00E-16 |
| rs687289 | let-7b-5p | trans | 8.61E-28 | Nightingale | S_LDL_CE_ | NA | A | G | 0.03078 | 0.004421 | 7.20E-14 |
| rs687289 | let-7b-5p | trans | 8.61E-28 | Nightingale | Sphingomy | NA | A | G | 0.028834 | 0.00414 | 9.00E-12 |
| rs687289 | let-7b-5p | trans | 8.61E-28 | Nightingale | S_VLDL_FC | NA | A | G | 0.029424 | 0.004232 | 8.60E-12 |
| rs687289 | let-7b-5p | trans | 8.61E-28 | Nightingale | S_VLDL_FC | NA | A | G | 0.037731 | 0.004429 | 1.00E-17 |
| rs687289 | let-7b-5p | trans | 8.61E-28 | Nightingale | XL_HDL_PL | NA | A | G | -0.02251 | 0.004069 | 1.90E-08 |
| rs687289 | let-7b-5p | trans | 8.61E-28 | Nightingale | IDL_L | NA | A | G | 0.035103 | 0.004282 | 6.50E-16 |
| rs687289 | let-7b-5p | trans | 8.61E-28 | Nightingale | L_LDL_PL | NA | A | G | 0.037753 | 0.004395 | 1.30E-17 |
| rs687289 | let-7b-5p | trans | 8.61E-28 | Nightingale | XXL_VLDL_ | NA | A | G | 0.031237 | 0.00441 | 6.10E-13 |
| rs687289 | let-7b-5p | trans | 8.61E-28 | Nightingale | LA | NA | A | G | 0.024416 | 0.004343 | 7.10E-09 |
| rs687289 | let-7b-5p | trans | 8.61E-28 | Nightingale | IDL_FC_pct | NA | A | G | 0.025842 | 0.004394 | 9.80E-09 |
| rs687289 | let-7b-5p | trans | 8.61E-28 | Nightingale | VLDL_FC | NA | A | G | 0.02388 | 0.004396 | 1.60E-08 |
| rs687289 | miR-139-5p | trans | 2.87E-43 | Nightingale | XS_VLDL_C | NA | A | G | 0.03768 | 0.004274 | 2.70E-18 |
| rs687289 | miR-139-5p | trans | 2.87E-43 | Nightingale | ApoB | NA | A | G | 0.03375 | 0.004423 | 2.90E-14 |
| rs687289 | miR-139-5p | trans | 2.87E-43 | Nightingale | LDL_C | NA | A | G | 0.032964 | 0.004409 | 1.20E-13 |
| rs687289 | miR-139-5p | trans | 2.87E-43 | Nightingale | S_VLDL_PL | NA | A | G | 0.032601 | 0.00442 | 5.80E-14 |
| rs687289 | miR-139-5p | trans | 2.87E-43 | Nightingale | M_LDL_C_ | NA | A | G | 0.031111 | 0.004432 | 3.10E-12 |
| rs687289 | miR-139-5p | trans | 2.87E-43 | Nightingale | L_VLDL_TG | NA | A | G | -0.05139 | 0.004408 | 3.50E-32 |
| rs687289 | miR-139-5p | trans | 2.87E-43 | Nightingale | XL_VLDL_C | NA | A | G | 0.037793 | 0.004249 | 1.10E-18 |
| rs687289 | miR-139-5p | trans | 2.87E-43 | Nightingale | S_VLDL_L | NA | A | G | 0.024476 | 0.004386 | 5.10E-09 |
| rs687289 | miR-139-5p | trans | 2.87E-43 | Nightingale | S_VLDL_P | NA | A | G | 0.023298 | 0.004373 | 2.10E-08 |
| rs687289 | miR-139-5p | trans | 2.87E-43 | Nightingale | M_VLDL_C | NA | A | G | 0.036468 | 0.004171 | 5.90E-18 |
| rs687289 | miR-139-5p | trans | 2.87E-43 | Nightingale | S_LDL_C | NA | A | G | 0.030716 | 0.004433 | 7.50E-12 |
| rs687289 | miR-139-5p | trans | 2.87E-43 | Nightingale | LDL_L | NA | A | G | 0.033082 | 0.004414 | 1.00E-13 |
| rs687289 | miR-139-5p | trans | 2.87E-43 | Nightingale | M_VLDL_C | NA | A | G | 0.033905 | 0.004161 | 9.50E-16 |
| rs687289 | miR-139-5p | trans | 2.87E-43 | Nightingale | S_HDL_P | NA | A | G | 0.030278 | 0.004429 | 9.00E-12 |
| rs687289 | miR-139-5p | trans | 2.87E-43 | Nightingale | M_LDL_L | NA | A | G | 0.032826 | 0.004435 | 1.40E-13 |
| rs687289 | miR-139-5p | trans | 2.87E-43 | Nightingale | M_VLDL_F | NA | A | G | 0.042299 | 0.004213 | 1.80E-23 |
| rs687289 | miR-139-5p | trans | 2.87E-43 | Nightingale | GlycA | NA | A | G | 0.028099 | 0.00441 | 1.30E-10 |
| rs687289 | miR-139-5p | trans | 2.87E-43 | Nightingale | S_LDL_CE | NA | A | G | 0.030317 | 0.004431 | 8.30E-12 |
| rs687289 | miR-139-5p | trans | 2.87E-43 | Nightingale | M_VLDL_T | NA | A | G | -0.03881 | 0.004185 | 3.40E-20 |
| rs687289 | miR-139-5p | trans | 2.87E-43 | Nightingale | S_VLDL_C_ | NA | A | G | 0.042052 | 0.00431 | 3.20E-22 |
| rs687289 | miR-139-5p | trans | 2.87E-43 | Nightingale | M_LDL_C | NA | A | G | 0.033786 | 0.004437 | 3.00E-14 |
| rs687289 | miR-139-5p | trans | 2.87E-43 | Nightingale | M_VLDL_C | NA | A | G | 0.038509 | 0.004416 | 2.60E-18 |
| rs687289 | miR-139-5p | trans | 2.87E-43 | Nightingale | XL_VLDL_F | NA | A | G | 0.037531 | 0.004293 | 1.00E-18 |
| rs687289 | miR-139-5p | trans | 2.87E-43 | Nightingale | M_VLDL_C | NA | A | G | 0.040645 | 0.004388 | 3.20E-20 |
| rs687289 | miR-139-5p | trans | 2.87E-43 | Nightingale | S_LDL_PL_ | NA | A | G | -0.02989 | 0.00442 | 4.10E-13 |
| rs687289 | miR-139-5p | trans | 2.87E-43 | Nightingale | IDL_CE | NA | A | G | 0.033304 | 0.004276 | 1.80E-14 |
| rs687289 | miR-139-5p | trans | 2.87E-43 | Nightingale | Omega_6 | NA | A | G | 0.025429 | 0.004325 | 1.10E-09 |
| rs687289 | miR-139-5p | trans | 2.87E-43 | Nightingale | XL_VLDL_T | NA | A | G | -0.04438 | 0.004292 | 5.40E-25 |
| rs687289 | miR-139-5p | trans | 2.87E-43 | Nightingale | LDL_CE | NA | A | G | 0.032325 | 0.004417 | 3.10E-13 |
| rs687289 | miR-139-5p | trans | 2.87E-43 | Nightingale | XXL_VLDL_ | NA | A | G | 0.035053 | 0.004438 | 2.10E-15 |
| rs687289 | miR-139-5p | trans | 2.87E-43 | Nightingale | XS_VLDL_P | NA | A | G | 0.036638 | 0.00437 | 1.60E-17 |

| | | | | | | | | | | | |
|---|---|---|---|---|---|---|---|---|---|---|---|
| rs687289 | miR-139-5p | trans | 2.87E-43 | Nightingale | S_LDL_L | NA | A | G | 0.027511 | 0.004428 | 1.10E-09 |
| rs687289 | miR-139-5p | trans | 2.87E-43 | Nightingale | L_LDL_C | NA | A | G | 0.032081 | 0.004379 | 3.60E-13 |
| rs687289 | miR-139-5p | trans | 2.87E-43 | Nightingale | M_LDL_TG | NA | A | G | -0.02742 | 0.004374 | 4.00E-10 |
| rs687289 | miR-139-5p | trans | 2.87E-43 | Nightingale | S_VLDL_C | NA | A | G | 0.038239 | 0.004428 | 2.40E-18 |
| rs687289 | miR-139-5p | trans | 2.87E-43 | Nightingale | M_VLDL_P | NA | A | G | 0.028859 | 0.004422 | 3.10E-11 |
| rs687289 | miR-139-5p | trans | 2.87E-43 | Nightingale | L_LDL_P | NA | A | G | 0.028943 | 0.004414 | 8.30E-11 |
| rs687289 | miR-139-5p | trans | 2.87E-43 | Nightingale | M_LDL_P | NA | A | G | 0.035821 | 0.004441 | 7.50E-16 |
| rs687289 | miR-139-5p | trans | 2.87E-43 | Nightingale | VLDL_CE | NA | A | G | 0.03558 | 0.00443 | 4.60E-16 |
| rs687289 | miR-139-5p | trans | 2.87E-43 | Nightingale | XL_VLDL_P | NA | A | G | 0.038789 | 0.004435 | 6.40E-19 |
| rs687289 | miR-139-5p | trans | 2.87E-43 | Nightingale | S_LDL_P | NA | A | G | 0.029429 | 0.004433 | 4.00E-11 |
| rs687289 | miR-139-5p | trans | 2.87E-43 | Nightingale | XS_VLDL_F | NA | A | G | 0.037022 | 0.004352 | 7.50E-18 |
| rs687289 | miR-139-5p | trans | 2.87E-43 | Nightingale | M_VLDL_P | NA | A | G | 0.028023 | 0.004418 | 9.80E-11 |
| rs687289 | miR-139-5p | trans | 2.87E-43 | Nightingale | LDL_P | NA | A | G | 0.031612 | 0.004425 | 1.30E-12 |
| rs687289 | miR-139-5p | trans | 2.87E-43 | Nightingale | L_VLDL_C_ | NA | A | G | 0.052543 | 0.004325 | 1.80E-34 |
| rs687289 | miR-139-5p | trans | 2.87E-43 | Nightingale | L_LDL_L | NA | A | G | 0.033323 | 0.004385 | 4.20E-14 |
| rs687289 | miR-139-5p | trans | 2.87E-43 | Nightingale | LDL_PL | NA | A | G | 0.034755 | 0.004418 | 6.70E-15 |
| rs687289 | miR-139-5p | trans | 2.87E-43 | Nightingale | XS_VLDL_C | NA | A | G | 0.037903 | 0.004299 | 1.70E-18 |
| rs687289 | miR-139-5p | trans | 2.87E-43 | Nightingale | M_LDL_CE | NA | A | G | 0.032783 | 0.004435 | 1.30E-13 |
| rs687289 | miR-139-5p | trans | 2.87E-43 | Nightingale | LDL_FC | NA | A | G | 0.033395 | 0.004381 | 7.50E-14 |
| rs687289 | miR-139-5p | trans | 2.87E-43 | Nightingale | S_HDL_C | NA | A | G | 0.028109 | 0.004421 | 2.80E-10 |
| rs687289 | miR-139-5p | trans | 2.87E-43 | Nightingale | IDL_P | NA | A | G | 0.03981 | 0.004382 | 1.80E-19 |
| rs687289 | miR-139-5p | trans | 2.87E-43 | Nightingale | XS_VLDL_P | NA | A | G | 0.032077 | 0.004367 | 5.90E-14 |
| rs687289 | miR-139-5p | trans | 2.87E-43 | Nightingale | XL_VLDL_C | NA | A | G | 0.038665 | 0.004247 | 1.60E-19 |
| rs687289 | miR-139-5p | trans | 2.87E-43 | Nightingale | Remnant_( | NA | A | G | 0.036652 | 0.0044 | 5.60E-17 |
| rs687289 | miR-139-5p | trans | 2.87E-43 | Nightingale | L_LDL_CE | NA | A | G | 0.031327 | 0.004389 | 1.40E-12 |
| rs687289 | miR-139-5p | trans | 2.87E-43 | Nightingale | S_HDL_L | NA | A | G | 0.026 | 0.004413 | 2.50E-09 |
| rs687289 | miR-139-5p | trans | 2.87E-43 | Nightingale | Total_PL | NA | A | G | 0.02466 | 0.004217 | 1.60E-09 |
| rs687289 | miR-139-5p | trans | 2.87E-43 | Nightingale | IDL_PL | NA | A | G | 0.035187 | 0.004274 | 4.50E-16 |
| rs687289 | miR-139-5p | trans | 2.87E-43 | Nightingale | XXL_VLDL_ | NA | A | G | 0.02523 | 0.004458 | 1.10E-09 |
| rs687289 | miR-139-5p | trans | 2.87E-43 | Nightingale | ApoB_by_A | NA | A | G | 0.024977 | 0.004362 | 1.30E-08 |
| rs687289 | miR-139-5p | trans | 2.87E-43 | Nightingale | S_HDL_CE | NA | A | G | 0.028306 | 0.004436 | 2.20E-10 |
| rs687289 | miR-139-5p | trans | 2.87E-43 | Nightingale | Total_CE | NA | A | G | 0.031498 | 0.004272 | 1.40E-13 |
| rs687289 | miR-139-5p | trans | 2.87E-43 | Nightingale | Total_FC | NA | A | G | 0.033014 | 0.004326 | 1.00E-14 |
| rs687289 | miR-139-5p | trans | 2.87E-43 | Nightingale | S_VLDL_CE | NA | A | G | 0.047288 | 0.004369 | 6.20E-27 |
| rs687289 | miR-139-5p | trans | 2.87E-43 | Nightingale | M_VLDL_F | NA | A | G | 0.033132 | 0.004428 | 4.80E-14 |
| rs687289 | miR-139-5p | trans | 2.87E-43 | Nightingale | IDL_C | NA | A | G | 0.034761 | 0.004277 | 1.30E-15 |
| rs687289 | miR-139-5p | trans | 2.87E-43 | Nightingale | non_HDL_( | NA | A | G | 0.03547 | 0.004405 | 7.40E-16 |
| rs687289 | miR-139-5p | trans | 2.87E-43 | Nightingale | S_LDL_C_p | NA | A | G | 0.045227 | 0.004446 | 8.10E-25 |
| rs687289 | miR-139-5p | trans | 2.87E-43 | Nightingale | XS_VLDL_F | NA | A | G | 0.031381 | 0.004397 | 2.60E-12 |
| rs687289 | miR-139-5p | trans | 2.87E-43 | Nightingale | VLDL_P | NA | A | G | 0.026418 | 0.004405 | 6.20E-10 |
| rs687289 | miR-139-5p | trans | 2.87E-43 | Nightingale | M_VLDL_P | NA | A | G | 0.044251 | 0.00425 | 3.40E-25 |
| rs687289 | miR-139-5p | trans | 2.87E-43 | Nightingale | L_VLDL_FC | NA | A | G | 0.048909 | 0.004386 | 2.40E-29 |
| rs687289 | miR-139-5p | trans | 2.87E-43 | Nightingale | Total_C | NA | A | G | 0.032124 | 0.004287 | 4.80E-14 |
| rs687289 | miR-139-5p | trans | 2.87E-43 | Nightingale | L_LDL_FC | NA | A | G | 0.033386 | 0.004349 | 4.10E-14 |
| rs687289 | miR-139-5p | trans | 2.87E-43 | Nightingale | S_HDL_PL | NA | A | G | 0.023499 | 0.00439 | 5.00E-08 |
| rs687289 | miR-139-5p | trans | 2.87E-43 | Nightingale | M_LDL_FC | NA | A | G | 0.034117 | 0.004423 | 2.80E-14 |
| rs687289 | miR-139-5p | trans | 2.87E-43 | Nightingale | S_VLDL_TG | NA | A | G | -0.03827 | 0.004287 | 9.50E-19 |
| rs687289 | miR-139-5p | trans | 2.87E-43 | Nightingale | XXL_VLDL_ | NA | A | G | 0.033249 | 0.004463 | 7.70E-14 |
| rs687289 | miR-139-5p | trans | 2.87E-43 | Nightingale | S_LDL_FC | NA | A | G | 0.028345 | 0.004422 | 5.60E-10 |
| rs687289 | miR-139-5p | trans | 2.87E-43 | Nightingale | L_VLDL_CE | NA | A | G | 0.047672 | 0.004238 | 1.00E-29 |
| rs687289 | miR-139-5p | trans | 2.87E-43 | Nightingale | Citrate | NA | A | G | 0.026536 | 0.0044 | 1.10E-09 |
| rs687289 | miR-139-5p | trans | 2.87E-43 | Nightingale | M_LDL_PL | NA | A | G | 0.032235 | 0.004435 | 3.90E-13 |
| rs687289 | miR-139-5p | trans | 2.87E-43 | Nightingale | Total_L | NA | A | G | 0.026033 | 0.004357 | 7.20E-10 |
| rs687289 | miR-139-5p | trans | 2.87E-43 | Nightingale | VLDL_C | NA | A | G | 0.031062 | 0.004422 | 7.60E-13 |
| rs687289 | miR-139-5p | trans | 2.87E-43 | Nightingale | XS_VLDL_L | NA | A | G | 0.033757 | 0.004353 | 2.40E-15 |
| rs687289 | miR-139-5p | trans | 2.87E-43 | Nightingale | IDL_FC | NA | A | G | 0.038096 | 0.004287 | 2.00E-18 |
| rs687289 | miR-139-5p | trans | 2.87E-43 | Nightingale | S_VLDL_CE | NA | A | G | 0.038069 | 0.004423 | 2.20E-18 |
| rs687289 | miR-139-5p | trans | 2.87E-43 | Nightingale | PUFA | NA | A | G | 0.023493 | 0.00432 | 2.00E-08 |
| rs687289 | miR-139-5p | trans | 2.87E-43 | Nightingale | XXL_VLDL_ | NA | A | G | -0.03594 | 0.004467 | 1.60E-16 |
| rs687289 | miR-139-5p | trans | 2.87E-43 | Nightingale | S_VLDL_PL | NA | A | G | 0.027844 | 0.004241 | 1.40E-10 |
| rs687289 | miR-139-5p | trans | 2.87E-43 | Nightingale | Clinical_LD | NA | A | G | 0.036238 | 0.004403 | 4.00E-16 |
| rs687289 | miR-139-5p | trans | 2.87E-43 | Nightingale | S_LDL_CE_ | NA | A | G | 0.03078 | 0.004421 | 7.20E-14 |
| rs687289 | miR-139-5p | trans | 2.87E-43 | Nightingale | Sphingomy | NA | A | G | 0.028834 | 0.00414 | 9.00E-12 |

| | | | | | | | | | | | |
|---|---|---|---|---|---|---|---|---|---|---|---|
| rs687289 | miR-139-5p | trans | 2.87E-43 | Nightingale | S_VLDL_FC | NA | A | G | 0.029424 | 0.004232 | 8.60E-12 |
| rs687289 | miR-139-5p | trans | 2.87E-43 | Nightingale | S_VLDL_FC | NA | A | G | 0.037731 | 0.004429 | 1.00E-17 |
| rs687289 | miR-139-5p | trans | 2.87E-43 | Nightingale | XL_HDL_PL | NA | A | G | -0.02251 | 0.004069 | 1.90E-08 |
| rs687289 | miR-139-5p | trans | 2.87E-43 | Nightingale | IDL_L | NA | A | G | 0.035103 | 0.004282 | 6.50E-16 |
| rs687289 | miR-139-5p | trans | 2.87E-43 | Nightingale | L_LDL_PL | NA | A | G | 0.037753 | 0.004395 | 1.30E-17 |
| rs687289 | miR-139-5p | trans | 2.87E-43 | Nightingale | XXL_VLDL_ | NA | A | G | 0.031237 | 0.00441 | 6.10E-13 |
| rs687289 | miR-139-5p | trans | 2.87E-43 | Nightingale | LA | NA | A | G | 0.024416 | 0.004343 | 7.10E-09 |
| rs687289 | miR-139-5p | trans | 2.87E-43 | Nightingale | IDL_FC_pct | NA | A | G | 0.025842 | 0.004394 | 9.80E-09 |
| rs687289 | miR-139-5p | trans | 2.87E-43 | Nightingale | VLDL_FC | NA | A | G | 0.02388 | 0.004396 | 1.60E-08 |
| rs687289 | miR-195-5p | trans | 1.08E-17 | Nightingale | XS_VLDL_C | NA | A | G | 0.03768 | 0.004274 | 2.70E-18 |
| rs687289 | miR-195-5p | trans | 1.08E-17 | Nightingale | ApoB | NA | A | G | 0.03375 | 0.004423 | 2.90E-14 |
| rs687289 | miR-195-5p | trans | 1.08E-17 | Nightingale | LDL_C | NA | A | G | 0.032964 | 0.004409 | 1.20E-13 |
| rs687289 | miR-195-5p | trans | 1.08E-17 | Nightingale | S_VLDL_PL | NA | A | G | 0.032601 | 0.00442 | 5.80E-14 |
| rs687289 | miR-195-5p | trans | 1.08E-17 | Nightingale | M_LDL_C_p | NA | A | G | 0.031111 | 0.004432 | 3.10E-12 |
| rs687289 | miR-195-5p | trans | 1.08E-17 | Nightingale | L_VLDL_TG | NA | A | G | -0.05139 | 0.004408 | 3.50E-32 |
| rs687289 | miR-195-5p | trans | 1.08E-17 | Nightingale | XL_VLDL_C | NA | A | G | 0.037793 | 0.004249 | 1.10E-18 |
| rs687289 | miR-195-5p | trans | 1.08E-17 | Nightingale | S_VLDL_L | NA | A | G | 0.024476 | 0.004386 | 5.10E-09 |
| rs687289 | miR-195-5p | trans | 1.08E-17 | Nightingale | S_VLDL_P | NA | A | G | 0.023298 | 0.004373 | 2.10E-08 |
| rs687289 | miR-195-5p | trans | 1.08E-17 | Nightingale | M_VLDL_C | NA | A | G | 0.036468 | 0.004171 | 5.90E-18 |
| rs687289 | miR-195-5p | trans | 1.08E-17 | Nightingale | S_LDL_C | NA | A | G | 0.030716 | 0.004433 | 7.50E-12 |
| rs687289 | miR-195-5p | trans | 1.08E-17 | Nightingale | LDL_L | NA | A | G | 0.033082 | 0.004414 | 1.00E-13 |
| rs687289 | miR-195-5p | trans | 1.08E-17 | Nightingale | M_VLDL_C | NA | A | G | 0.033905 | 0.004161 | 9.50E-16 |
| rs687289 | miR-195-5p | trans | 1.08E-17 | Nightingale | S_HDL_P | NA | A | G | 0.030278 | 0.004429 | 9.00E-12 |
| rs687289 | miR-195-5p | trans | 1.08E-17 | Nightingale | M_LDL_L | NA | A | G | 0.032826 | 0.004435 | 1.40E-13 |
| rs687289 | miR-195-5p | trans | 1.08E-17 | Nightingale | M_VLDL_F | NA | A | G | 0.042299 | 0.004213 | 1.80E-23 |
| rs687289 | miR-195-5p | trans | 1.08E-17 | Nightingale | GlycA | NA | A | G | 0.028099 | 0.00441 | 1.30E-10 |
| rs687289 | miR-195-5p | trans | 1.08E-17 | Nightingale | S_LDL_CE | NA | A | G | 0.030317 | 0.004431 | 8.30E-12 |
| rs687289 | miR-195-5p | trans | 1.08E-17 | Nightingale | M_VLDL_T | NA | A | G | -0.03881 | 0.004185 | 3.40E-20 |
| rs687289 | miR-195-5p | trans | 1.08E-17 | Nightingale | S_VLDL_C_ | NA | A | G | 0.042052 | 0.00431 | 3.20E-22 |
| rs687289 | miR-195-5p | trans | 1.08E-17 | Nightingale | M_LDL_C | NA | A | G | 0.033786 | 0.004437 | 3.00E-14 |
| rs687289 | miR-195-5p | trans | 1.08E-17 | Nightingale | M_VLDL_C | NA | A | G | 0.038509 | 0.004416 | 2.60E-18 |
| rs687289 | miR-195-5p | trans | 1.08E-17 | Nightingale | XL_VLDL_F | NA | A | G | 0.037531 | 0.004293 | 1.00E-18 |
| rs687289 | miR-195-5p | trans | 1.08E-17 | Nightingale | M_VLDL_C | NA | A | G | 0.040645 | 0.004388 | 3.20E-20 |
| rs687289 | miR-195-5p | trans | 1.08E-17 | Nightingale | S_LDL_PL_ | NA | A | G | -0.02989 | 0.00442 | 4.10E-13 |
| rs687289 | miR-195-5p | trans | 1.08E-17 | Nightingale | IDL_CE | NA | A | G | 0.033304 | 0.004276 | 1.80E-14 |
| rs687289 | miR-195-5p | trans | 1.08E-17 | Nightingale | Omega_6 | NA | A | G | 0.025429 | 0.004325 | 1.10E-09 |
| rs687289 | miR-195-5p | trans | 1.08E-17 | Nightingale | XL_VLDL_T | NA | A | G | -0.04438 | 0.004292 | 5.40E-25 |
| rs687289 | miR-195-5p | trans | 1.08E-17 | Nightingale | LDL_CE | NA | A | G | 0.032325 | 0.004417 | 3.10E-13 |
| rs687289 | miR-195-5p | trans | 1.08E-17 | Nightingale | XXL_VLDL_ | NA | A | G | 0.035053 | 0.004438 | 2.10E-15 |
| rs687289 | miR-195-5p | trans | 1.08E-17 | Nightingale | XS_VLDL_P | NA | A | G | 0.036638 | 0.00437 | 1.60E-17 |
| rs687289 | miR-195-5p | trans | 1.08E-17 | Nightingale | S_LDL_L | NA | A | G | 0.027511 | 0.004428 | 1.10E-09 |
| rs687289 | miR-195-5p | trans | 1.08E-17 | Nightingale | L_LDL_C | NA | A | G | 0.032081 | 0.004379 | 3.60E-13 |
| rs687289 | miR-195-5p | trans | 1.08E-17 | Nightingale | M_LDL_TG | NA | A | G | -0.02742 | 0.004374 | 4.00E-10 |
| rs687289 | miR-195-5p | trans | 1.08E-17 | Nightingale | S_VLDL_C | NA | A | G | 0.038239 | 0.004428 | 2.40E-18 |
| rs687289 | miR-195-5p | trans | 1.08E-17 | Nightingale | M_VLDL_P | NA | A | G | 0.028859 | 0.004422 | 3.10E-11 |
| rs687289 | miR-195-5p | trans | 1.08E-17 | Nightingale | L_LDL_P | NA | A | G | 0.028943 | 0.004414 | 8.30E-11 |
| rs687289 | miR-195-5p | trans | 1.08E-17 | Nightingale | M_LDL_P | NA | A | G | 0.035821 | 0.004441 | 7.50E-16 |
| rs687289 | miR-195-5p | trans | 1.08E-17 | Nightingale | VLDL_CE | NA | A | G | 0.03558 | 0.00443 | 4.60E-16 |
| rs687289 | miR-195-5p | trans | 1.08E-17 | Nightingale | XL_VLDL_P | NA | A | G | 0.038789 | 0.004435 | 6.40E-19 |
| rs687289 | miR-195-5p | trans | 1.08E-17 | Nightingale | S_LDL_P | NA | A | G | 0.029429 | 0.004433 | 4.00E-11 |
| rs687289 | miR-195-5p | trans | 1.08E-17 | Nightingale | XS_VLDL_F | NA | A | G | 0.037022 | 0.004352 | 7.50E-18 |
| rs687289 | miR-195-5p | trans | 1.08E-17 | Nightingale | M_VLDL_P | NA | A | G | 0.028023 | 0.004418 | 9.80E-11 |
| rs687289 | miR-195-5p | trans | 1.08E-17 | Nightingale | LDL_P | NA | A | G | 0.031612 | 0.004425 | 1.30E-12 |
| rs687289 | miR-195-5p | trans | 1.08E-17 | Nightingale | L_VLDL_C_ | NA | A | G | 0.052543 | 0.004325 | 1.80E-34 |
| rs687289 | miR-195-5p | trans | 1.08E-17 | Nightingale | L_LDL_L | NA | A | G | 0.033323 | 0.004385 | 4.20E-14 |
| rs687289 | miR-195-5p | trans | 1.08E-17 | Nightingale | LDL_PL | NA | A | G | 0.034755 | 0.004418 | 6.70E-15 |
| rs687289 | miR-195-5p | trans | 1.08E-17 | Nightingale | XS_VLDL_C | NA | A | G | 0.037903 | 0.004299 | 1.70E-18 |
| rs687289 | miR-195-5p | trans | 1.08E-17 | Nightingale | M_LDL_CE | NA | A | G | 0.032783 | 0.004435 | 1.30E-13 |
| rs687289 | miR-195-5p | trans | 1.08E-17 | Nightingale | LDL_FC | NA | A | G | 0.033395 | 0.004381 | 7.50E-14 |
| rs687289 | miR-195-5p | trans | 1.08E-17 | Nightingale | S_HDL_C | NA | A | G | 0.028109 | 0.004421 | 2.80E-10 |
| rs687289 | miR-195-5p | trans | 1.08E-17 | Nightingale | IDL_P | NA | A | G | 0.03981 | 0.004382 | 1.80E-19 |
| rs687289 | miR-195-5p | trans | 1.08E-17 | Nightingale | XS_VLDL_P | NA | A | G | 0.032077 | 0.004367 | 5.90E-14 |
| rs687289 | miR-195-5p | trans | 1.08E-17 | Nightingale | XL_VLDL_C | NA | A | G | 0.038665 | 0.004247 | 1.60E-19 |

| | | | | | | | | | | | |
|---|---|---|---|---|---|---|---|---|---|---|---|
| rs687289 | miR-195-5p | trans | 1.08E-17 | Nightingale | Remnant_( | NA | A | G | 0.036652 | 0.0044 | 5.60E-17 |
| rs687289 | miR-195-5p | trans | 1.08E-17 | Nightingale | L_LDL_CE | NA | A | G | 0.031327 | 0.004389 | 1.40E-12 |
| rs687289 | miR-195-5p | trans | 1.08E-17 | Nightingale | S_HDL_L | NA | A | G | 0.026 | 0.004413 | 2.50E-09 |
| rs687289 | miR-195-5p | trans | 1.08E-17 | Nightingale | Total_PL | NA | A | G | 0.02466 | 0.004217 | 1.60E-09 |
| rs687289 | miR-195-5p | trans | 1.08E-17 | Nightingale | IDL_PL | NA | A | G | 0.035187 | 0.004274 | 4.50E-16 |
| rs687289 | miR-195-5p | trans | 1.08E-17 | Nightingale | XXL_VLDL_ | NA | A | G | 0.02523 | 0.004458 | 1.10E-09 |
| rs687289 | miR-195-5p | trans | 1.08E-17 | Nightingale | ApoB_by_A | NA | A | G | 0.024977 | 0.004362 | 1.30E-08 |
| rs687289 | miR-195-5p | trans | 1.08E-17 | Nightingale | S_HDL_CE | NA | A | G | 0.028306 | 0.004436 | 2.20E-10 |
| rs687289 | miR-195-5p | trans | 1.08E-17 | Nightingale | Total_CE | NA | A | G | 0.031498 | 0.004272 | 1.40E-13 |
| rs687289 | miR-195-5p | trans | 1.08E-17 | Nightingale | Total_FC | NA | A | G | 0.033014 | 0.004326 | 1.00E-14 |
| rs687289 | miR-195-5p | trans | 1.08E-17 | Nightingale | S_VLDL_CE | NA | A | G | 0.047288 | 0.004369 | 6.20E-27 |
| rs687289 | miR-195-5p | trans | 1.08E-17 | Nightingale | M_VLDL_F( | NA | A | G | 0.033132 | 0.004428 | 4.80E-14 |
| rs687289 | miR-195-5p | trans | 1.08E-17 | Nightingale | IDL_C | NA | A | G | 0.034761 | 0.004277 | 1.30E-15 |
| rs687289 | miR-195-5p | trans | 1.08E-17 | Nightingale | non_HDL_( | NA | A | G | 0.03547 | 0.004405 | 7.40E-16 |
| rs687289 | miR-195-5p | trans | 1.08E-17 | Nightingale | S_LDL_C_p | NA | A | G | 0.045227 | 0.004446 | 8.10E-25 |
| rs687289 | miR-195-5p | trans | 1.08E-17 | Nightingale | XS_VLDL_F | NA | A | G | 0.031381 | 0.004397 | 2.60E-12 |
| rs687289 | miR-195-5p | trans | 1.08E-17 | Nightingale | VLDL_P | NA | A | G | 0.026418 | 0.004405 | 6.20E-10 |
| rs687289 | miR-195-5p | trans | 1.08E-17 | Nightingale | M_VLDL_P | NA | A | G | 0.044251 | 0.00425 | 3.40E-25 |
| rs687289 | miR-195-5p | trans | 1.08E-17 | Nightingale | L_VLDL_FC | NA | A | G | 0.048909 | 0.004386 | 2.40E-29 |
| rs687289 | miR-195-5p | trans | 1.08E-17 | Nightingale | Total_C | NA | A | G | 0.032124 | 0.004287 | 4.80E-14 |
| rs687289 | miR-195-5p | trans | 1.08E-17 | Nightingale | L_LDL_FC | NA | A | G | 0.033386 | 0.004349 | 4.10E-14 |
| rs687289 | miR-195-5p | trans | 1.08E-17 | Nightingale | S_HDL_PL | NA | A | G | 0.023499 | 0.00439 | 5.00E-08 |
| rs687289 | miR-195-5p | trans | 1.08E-17 | Nightingale | M_LDL_FC | NA | A | G | 0.034117 | 0.004423 | 2.80E-14 |
| rs687289 | miR-195-5p | trans | 1.08E-17 | Nightingale | S_VLDL_TG | NA | A | G | -0.03827 | 0.004287 | 9.50E-19 |
| rs687289 | miR-195-5p | trans | 1.08E-17 | Nightingale | XXL_VLDL_ | NA | A | G | 0.033249 | 0.004463 | 7.70E-14 |
| rs687289 | miR-195-5p | trans | 1.08E-17 | Nightingale | S_LDL_FC | NA | A | G | 0.028345 | 0.004422 | 5.60E-10 |
| rs687289 | miR-195-5p | trans | 1.08E-17 | Nightingale | L_VLDL_CE | NA | A | G | 0.047672 | 0.004238 | 1.00E-29 |
| rs687289 | miR-195-5p | trans | 1.08E-17 | Nightingale | Citrate | NA | A | G | 0.026536 | 0.0044 | 1.10E-09 |
| rs687289 | miR-195-5p | trans | 1.08E-17 | Nightingale | M_LDL_PL | NA | A | G | 0.032235 | 0.004435 | 3.90E-13 |
| rs687289 | miR-195-5p | trans | 1.08E-17 | Nightingale | Total_L | NA | A | G | 0.026033 | 0.004357 | 7.20E-10 |
| rs687289 | miR-195-5p | trans | 1.08E-17 | Nightingale | VLDL_C | NA | A | G | 0.031062 | 0.004422 | 7.60E-13 |
| rs687289 | miR-195-5p | trans | 1.08E-17 | Nightingale | XS_VLDL_L | NA | A | G | 0.033757 | 0.004353 | 2.40E-15 |
| rs687289 | miR-195-5p | trans | 1.08E-17 | Nightingale | IDL_FC | NA | A | G | 0.038096 | 0.004287 | 2.00E-18 |
| rs687289 | miR-195-5p | trans | 1.08E-17 | Nightingale | S_VLDL_CE | NA | A | G | 0.038069 | 0.004423 | 2.20E-18 |
| rs687289 | miR-195-5p | trans | 1.08E-17 | Nightingale | PUFA | NA | A | G | 0.023493 | 0.00432 | 2.00E-08 |
| rs687289 | miR-195-5p | trans | 1.08E-17 | Nightingale | XXL_VLDL_ | NA | A | G | -0.03594 | 0.004467 | 1.60E-16 |
| rs687289 | miR-195-5p | trans | 1.08E-17 | Nightingale | S_VLDL_PL | NA | A | G | 0.027844 | 0.004241 | 1.40E-10 |
| rs687289 | miR-195-5p | trans | 1.08E-17 | Nightingale | Clinical_LD | NA | A | G | 0.036238 | 0.004403 | 4.00E-16 |
| rs687289 | miR-195-5p | trans | 1.08E-17 | Nightingale | S_LDL_CE_ | NA | A | G | 0.03078 | 0.004421 | 7.20E-14 |
| rs687289 | miR-195-5p | trans | 1.08E-17 | Nightingale | Sphingomy | NA | A | G | 0.028834 | 0.00414 | 9.00E-12 |
| rs687289 | miR-195-5p | trans | 1.08E-17 | Nightingale | S_VLDL_FC | NA | A | G | 0.029424 | 0.004232 | 8.60E-12 |
| rs687289 | miR-195-5p | trans | 1.08E-17 | Nightingale | S_VLDL_FC | NA | A | G | 0.037731 | 0.004429 | 1.00E-17 |
| rs687289 | miR-195-5p | trans | 1.08E-17 | Nightingale | XL_HDL_PL | NA | A | G | -0.02251 | 0.004069 | 1.90E-08 |
| rs687289 | miR-195-5p | trans | 1.08E-17 | Nightingale | IDL_L | NA | A | G | 0.035103 | 0.004282 | 6.50E-16 |
| rs687289 | miR-195-5p | trans | 1.08E-17 | Nightingale | L_LDL_PL | NA | A | G | 0.037753 | 0.004395 | 1.30E-17 |
| rs687289 | miR-195-5p | trans | 1.08E-17 | Nightingale | XXL_VLDL_ | NA | A | G | 0.031237 | 0.00441 | 6.10E-13 |
| rs687289 | miR-195-5p | trans | 1.08E-17 | Nightingale | LA | NA | A | G | 0.024416 | 0.004343 | 7.10E-09 |
| rs687289 | miR-195-5p | trans | 1.08E-17 | Nightingale | IDL_FC_pct | NA | A | G | 0.025842 | 0.004394 | 9.80E-09 |
| rs687289 | miR-195-5p | trans | 1.08E-17 | Nightingale | VLDL_FC | NA | A | G | 0.02388 | 0.004396 | 1.60E-08 |
| rs687289 | miR-10b-5p | trans | 5.80E-37 | Nightingale | XS_VLDL_C | NA | A | G | 0.03768 | 0.004274 | 2.70E-18 |
| rs687289 | miR-10b-5p | trans | 5.80E-37 | Nightingale | ApoB | NA | A | G | 0.03375 | 0.004423 | 2.90E-14 |
| rs687289 | miR-10b-5p | trans | 5.80E-37 | Nightingale | LDL_C | NA | A | G | 0.032964 | 0.004409 | 1.20E-13 |
| rs687289 | miR-10b-5p | trans | 5.80E-37 | Nightingale | S_VLDL_PL | NA | A | G | 0.032601 | 0.00442 | 5.80E-14 |
| rs687289 | miR-10b-5p | trans | 5.80E-37 | Nightingale | M_LDL_C_p | NA | A | G | 0.031111 | 0.004432 | 3.10E-12 |
| rs687289 | miR-10b-5p | trans | 5.80E-37 | Nightingale | L_VLDL_TG | NA | A | G | -0.05139 | 0.004408 | 3.50E-32 |
| rs687289 | miR-10b-5p | trans | 5.80E-37 | Nightingale | XL_VLDL_C | NA | A | G | 0.037793 | 0.004249 | 1.10E-18 |
| rs687289 | miR-10b-5p | trans | 5.80E-37 | Nightingale | S_VLDL_L | NA | A | G | 0.024476 | 0.004386 | 5.10E-09 |
| rs687289 | miR-10b-5p | trans | 5.80E-37 | Nightingale | S_VLDL_P | NA | A | G | 0.023298 | 0.004373 | 2.10E-08 |
| rs687289 | miR-10b-5p | trans | 5.80E-37 | Nightingale | M_VLDL_C | NA | A | G | 0.036468 | 0.004171 | 5.90E-18 |
| rs687289 | miR-10b-5p | trans | 5.80E-37 | Nightingale | S_LDL_C | NA | A | G | 0.030716 | 0.004433 | 7.50E-12 |
| rs687289 | miR-10b-5p | trans | 5.80E-37 | Nightingale | LDL_L | NA | A | G | 0.033082 | 0.004414 | 1.00E-13 |
| rs687289 | miR-10b-5p | trans | 5.80E-37 | Nightingale | M_VLDL_C | NA | A | G | 0.033905 | 0.004161 | 9.50E-16 |
| rs687289 | miR-10b-5p | trans | 5.80E-37 | Nightingale | S_HDL_P | NA | A | G | 0.030278 | 0.004429 | 9.00E-12 |

| | | | | | | | | | | | |
|---|---|---|---|---|---|---|---|---|---|---|---|
| rs687289 | miR-10b-5p | trans | 5.80E-37 | Nightingale | M_LDL_L | NA | A | G | 0.032826 | 0.004435 | 1.40E-13 |
| rs687289 | miR-10b-5p | trans | 5.80E-37 | Nightingale | M_VLDL_F | NA | A | G | 0.042299 | 0.004213 | 1.80E-23 |
| rs687289 | miR-10b-5p | trans | 5.80E-37 | Nightingale | GlycA | NA | A | G | 0.028099 | 0.00441 | 1.30E-10 |
| rs687289 | miR-10b-5p | trans | 5.80E-37 | Nightingale | S_LDL_CE | NA | A | G | 0.030317 | 0.004431 | 8.30E-12 |
| rs687289 | miR-10b-5p | trans | 5.80E-37 | Nightingale | M_VLDL_T | NA | A | G | -0.03881 | 0.004185 | 3.40E-20 |
| rs687289 | miR-10b-5p | trans | 5.80E-37 | Nightingale | S_VLDL_C_ | NA | A | G | 0.042052 | 0.00431 | 3.20E-22 |
| rs687289 | miR-10b-5p | trans | 5.80E-37 | Nightingale | M_LDL_C | NA | A | G | 0.033786 | 0.004437 | 3.00E-14 |
| rs687289 | miR-10b-5p | trans | 5.80E-37 | Nightingale | M_VLDL_C | NA | A | G | 0.038509 | 0.004416 | 2.60E-18 |
| rs687289 | miR-10b-5p | trans | 5.80E-37 | Nightingale | XL_VLDL_F | NA | A | G | 0.037531 | 0.004293 | 1.00E-18 |
| rs687289 | miR-10b-5p | trans | 5.80E-37 | Nightingale | M_VLDL_C | NA | A | G | 0.040645 | 0.004388 | 3.20E-20 |
| rs687289 | miR-10b-5p | trans | 5.80E-37 | Nightingale | S_LDL_PL_ | NA | A | G | -0.02989 | 0.00442 | 4.10E-13 |
| rs687289 | miR-10b-5p | trans | 5.80E-37 | Nightingale | IDL_CE | NA | A | G | 0.033304 | 0.004276 | 1.80E-14 |
| rs687289 | miR-10b-5p | trans | 5.80E-37 | Nightingale | Omega_6 | NA | A | G | 0.025429 | 0.004325 | 1.10E-09 |
| rs687289 | miR-10b-5p | trans | 5.80E-37 | Nightingale | XL_VLDL_T | NA | A | G | -0.04438 | 0.004292 | 5.40E-25 |
| rs687289 | miR-10b-5p | trans | 5.80E-37 | Nightingale | LDL_CE | NA | A | G | 0.032325 | 0.004417 | 3.10E-13 |
| rs687289 | miR-10b-5p | trans | 5.80E-37 | Nightingale | XXL_VLDL_ | NA | A | G | 0.035053 | 0.004438 | 2.10E-15 |
| rs687289 | miR-10b-5p | trans | 5.80E-37 | Nightingale | XS_VLDL_P | NA | A | G | 0.036638 | 0.00437 | 1.60E-17 |
| rs687289 | miR-10b-5p | trans | 5.80E-37 | Nightingale | S_LDL_L | NA | A | G | 0.027511 | 0.004428 | 1.10E-09 |
| rs687289 | miR-10b-5p | trans | 5.80E-37 | Nightingale | L_LDL_C | NA | A | G | 0.032081 | 0.004379 | 3.60E-13 |
| rs687289 | miR-10b-5p | trans | 5.80E-37 | Nightingale | M_LDL_TG | NA | A | G | -0.02742 | 0.004374 | 4.00E-10 |
| rs687289 | miR-10b-5p | trans | 5.80E-37 | Nightingale | S_VLDL_C | NA | A | G | 0.038239 | 0.004428 | 2.40E-18 |
| rs687289 | miR-10b-5p | trans | 5.80E-37 | Nightingale | M_VLDL_P | NA | A | G | 0.028859 | 0.004422 | 3.10E-11 |
| rs687289 | miR-10b-5p | trans | 5.80E-37 | Nightingale | L_LDL_P | NA | A | G | 0.028943 | 0.004414 | 8.30E-11 |
| rs687289 | miR-10b-5p | trans | 5.80E-37 | Nightingale | M_LDL_P | NA | A | G | 0.035821 | 0.004441 | 7.50E-16 |
| rs687289 | miR-10b-5p | trans | 5.80E-37 | Nightingale | VLDL_CE | NA | A | G | 0.03558 | 0.00443 | 4.60E-16 |
| rs687289 | miR-10b-5p | trans | 5.80E-37 | Nightingale | XL_VLDL_P | NA | A | G | 0.038789 | 0.004435 | 6.40E-19 |
| rs687289 | miR-10b-5p | trans | 5.80E-37 | Nightingale | S_LDL_P | NA | A | G | 0.029429 | 0.004433 | 4.00E-11 |
| rs687289 | miR-10b-5p | trans | 5.80E-37 | Nightingale | XS_VLDL_F | NA | A | G | 0.037022 | 0.004352 | 7.50E-18 |
| rs687289 | miR-10b-5p | trans | 5.80E-37 | Nightingale | M_VLDL_P | NA | A | G | 0.028023 | 0.004418 | 9.80E-11 |
| rs687289 | miR-10b-5p | trans | 5.80E-37 | Nightingale | LDL_P | NA | A | G | 0.031612 | 0.004425 | 1.30E-12 |
| rs687289 | miR-10b-5p | trans | 5.80E-37 | Nightingale | L_VLDL_C_ | NA | A | G | 0.052543 | 0.004325 | 1.80E-34 |
| rs687289 | miR-10b-5p | trans | 5.80E-37 | Nightingale | L_LDL_L | NA | A | G | 0.033323 | 0.004385 | 4.20E-14 |
| rs687289 | miR-10b-5p | trans | 5.80E-37 | Nightingale | LDL_PL | NA | A | G | 0.034755 | 0.004418 | 6.70E-15 |
| rs687289 | miR-10b-5p | trans | 5.80E-37 | Nightingale | XS_VLDL_C | NA | A | G | 0.037903 | 0.004299 | 1.70E-18 |
| rs687289 | miR-10b-5p | trans | 5.80E-37 | Nightingale | M_LDL_CE | NA | A | G | 0.032783 | 0.004435 | 1.30E-13 |
| rs687289 | miR-10b-5p | trans | 5.80E-37 | Nightingale | LDL_FC | NA | A | G | 0.033395 | 0.004381 | 7.50E-14 |
| rs687289 | miR-10b-5p | trans | 5.80E-37 | Nightingale | S_HDL_C | NA | A | G | 0.028109 | 0.004421 | 2.80E-10 |
| rs687289 | miR-10b-5p | trans | 5.80E-37 | Nightingale | IDL_P | NA | A | G | 0.03981 | 0.004382 | 1.80E-19 |
| rs687289 | miR-10b-5p | trans | 5.80E-37 | Nightingale | XS_VLDL_P | NA | A | G | 0.032077 | 0.004367 | 5.90E-14 |
| rs687289 | miR-10b-5p | trans | 5.80E-37 | Nightingale | XL_VLDL_C | NA | A | G | 0.038665 | 0.004247 | 1.60E-19 |
| rs687289 | miR-10b-5p | trans | 5.80E-37 | Nightingale | Remnant_( | NA | A | G | 0.036652 | 0.0044 | 5.60E-17 |
| rs687289 | miR-10b-5p | trans | 5.80E-37 | Nightingale | L_LDL_CE | NA | A | G | 0.031327 | 0.004389 | 1.40E-12 |
| rs687289 | miR-10b-5p | trans | 5.80E-37 | Nightingale | S_HDL_L | NA | A | G | 0.026 | 0.004413 | 2.50E-09 |
| rs687289 | miR-10b-5p | trans | 5.80E-37 | Nightingale | Total_PL | NA | A | G | 0.02466 | 0.004217 | 1.60E-09 |
| rs687289 | miR-10b-5p | trans | 5.80E-37 | Nightingale | IDL_PL | NA | A | G | 0.035187 | 0.004274 | 4.50E-16 |
| rs687289 | miR-10b-5p | trans | 5.80E-37 | Nightingale | XXL_VLDL_ | NA | A | G | 0.02523 | 0.004458 | 1.10E-09 |
| rs687289 | miR-10b-5p | trans | 5.80E-37 | Nightingale | ApoB_by_A | NA | A | G | 0.024977 | 0.004362 | 1.30E-08 |
| rs687289 | miR-10b-5p | trans | 5.80E-37 | Nightingale | S_HDL_CE | NA | A | G | 0.028306 | 0.004436 | 2.20E-10 |
| rs687289 | miR-10b-5p | trans | 5.80E-37 | Nightingale | Total_CE | NA | A | G | 0.031498 | 0.004272 | 1.40E-13 |
| rs687289 | miR-10b-5p | trans | 5.80E-37 | Nightingale | Total_FC | NA | A | G | 0.033014 | 0.004326 | 1.00E-14 |
| rs687289 | miR-10b-5p | trans | 5.80E-37 | Nightingale | S_VLDL_CE | NA | A | G | 0.047288 | 0.004369 | 6.20E-27 |
| rs687289 | miR-10b-5p | trans | 5.80E-37 | Nightingale | M_VLDL_F | NA | A | G | 0.033132 | 0.004428 | 4.80E-14 |
| rs687289 | miR-10b-5p | trans | 5.80E-37 | Nightingale | IDL_C | NA | A | G | 0.034761 | 0.004277 | 1.30E-15 |
| rs687289 | miR-10b-5p | trans | 5.80E-37 | Nightingale | non_HDL_( | NA | A | G | 0.03547 | 0.004405 | 7.40E-16 |
| rs687289 | miR-10b-5p | trans | 5.80E-37 | Nightingale | S_LDL_C_p | NA | A | G | 0.045227 | 0.004446 | 8.10E-25 |
| rs687289 | miR-10b-5p | trans | 5.80E-37 | Nightingale | XS_VLDL_F | NA | A | G | 0.031381 | 0.004397 | 2.60E-12 |
| rs687289 | miR-10b-5p | trans | 5.80E-37 | Nightingale | VLDL_P | NA | A | G | 0.026418 | 0.004405 | 6.20E-10 |
| rs687289 | miR-10b-5p | trans | 5.80E-37 | Nightingale | M_VLDL_P | NA | A | G | 0.044251 | 0.00425 | 3.40E-25 |
| rs687289 | miR-10b-5p | trans | 5.80E-37 | Nightingale | L_VLDL_FC | NA | A | G | 0.048909 | 0.004386 | 2.40E-29 |
| rs687289 | miR-10b-5p | trans | 5.80E-37 | Nightingale | Total_C | NA | A | G | 0.032124 | 0.004287 | 4.80E-14 |
| rs687289 | miR-10b-5p | trans | 5.80E-37 | Nightingale | L_LDL_FC | NA | A | G | 0.033386 | 0.004349 | 4.10E-14 |
| rs687289 | miR-10b-5p | trans | 5.80E-37 | Nightingale | S_HDL_PL | NA | A | G | 0.023499 | 0.00439 | 5.00E-08 |
| rs687289 | miR-10b-5p | trans | 5.80E-37 | Nightingale | M_LDL_FC | NA | A | G | 0.034117 | 0.004423 | 2.80E-14 |

| | | | | | | | | | | | |
|---|---|---|---|---|---|---|---|---|---|---|---|
| rs687289 | miR-10b-5p | trans | 5.80E-37 | Nightingale | S_VLDL_TG | NA | A | G | -0.03827 | 0.004287 | 9.50E-19 |
| rs687289 | miR-10b-5p | trans | 5.80E-37 | Nightingale | XXL_VLDL_ | NA | A | G | 0.033249 | 0.004463 | 7.70E-14 |
| rs687289 | miR-10b-5p | trans | 5.80E-37 | Nightingale | S_LDL_FC | NA | A | G | 0.028345 | 0.004422 | 5.60E-10 |
| rs687289 | miR-10b-5p | trans | 5.80E-37 | Nightingale | L_VLDL_CE | NA | A | G | 0.047672 | 0.004238 | 1.00E-29 |
| rs687289 | miR-10b-5p | trans | 5.80E-37 | Nightingale | Citrate | NA | A | G | 0.026536 | 0.0044 | 1.10E-09 |
| rs687289 | miR-10b-5p | trans | 5.80E-37 | Nightingale | M_LDL_PL | NA | A | G | 0.032235 | 0.004435 | 3.90E-13 |
| rs687289 | miR-10b-5p | trans | 5.80E-37 | Nightingale | Total_L | NA | A | G | 0.026033 | 0.004357 | 7.20E-10 |
| rs687289 | miR-10b-5p | trans | 5.80E-37 | Nightingale | VLDL_C | NA | A | G | 0.031062 | 0.004422 | 7.60E-13 |
| rs687289 | miR-10b-5p | trans | 5.80E-37 | Nightingale | XS_VLDL_L | NA | A | G | 0.033757 | 0.004353 | 2.40E-15 |
| rs687289 | miR-10b-5p | trans | 5.80E-37 | Nightingale | IDL_FC | NA | A | G | 0.038096 | 0.004287 | 2.00E-18 |
| rs687289 | miR-10b-5p | trans | 5.80E-37 | Nightingale | S_VLDL_CE | NA | A | G | 0.038069 | 0.004423 | 2.20E-18 |
| rs687289 | miR-10b-5p | trans | 5.80E-37 | Nightingale | PUFA | NA | A | G | 0.023493 | 0.00432 | 2.00E-08 |
| rs687289 | miR-10b-5p | trans | 5.80E-37 | Nightingale | XXL_VLDL_ | NA | A | G | -0.03594 | 0.004467 | 1.60E-16 |
| rs687289 | miR-10b-5p | trans | 5.80E-37 | Nightingale | S_VLDL_PL | NA | A | G | 0.027844 | 0.004241 | 1.40E-10 |
| rs687289 | miR-10b-5p | trans | 5.80E-37 | Nightingale | Clinical_LD | NA | A | G | 0.036238 | 0.004403 | 4.00E-16 |
| rs687289 | miR-10b-5p | trans | 5.80E-37 | Nightingale | S_LDL_CE_ | NA | A | G | 0.03078 | 0.004421 | 7.20E-14 |
| rs687289 | miR-10b-5p | trans | 5.80E-37 | Nightingale | Sphingomy | NA | A | G | 0.028834 | 0.00414 | 9.00E-12 |
| rs687289 | miR-10b-5p | trans | 5.80E-37 | Nightingale | S_VLDL_FC | NA | A | G | 0.029424 | 0.004232 | 8.60E-12 |
| rs687289 | miR-10b-5p | trans | 5.80E-37 | Nightingale | S_VLDL_FC | NA | A | G | 0.037731 | 0.004429 | 1.00E-17 |
| rs687289 | miR-10b-5p | trans | 5.80E-37 | Nightingale | XL_HDL_PL | NA | A | G | -0.02251 | 0.004069 | 1.90E-08 |
| rs687289 | miR-10b-5p | trans | 5.80E-37 | Nightingale | IDL_L | NA | A | G | 0.035103 | 0.004282 | 6.50E-16 |
| rs687289 | miR-10b-5p | trans | 5.80E-37 | Nightingale | L_LDL_PL | NA | A | G | 0.037753 | 0.004395 | 1.30E-17 |
| rs687289 | miR-10b-5p | trans | 5.80E-37 | Nightingale | XXL_VLDL_ | NA | A | G | 0.031237 | 0.00441 | 6.10E-13 |
| rs687289 | miR-10b-5p | trans | 5.80E-37 | Nightingale | LA | NA | A | G | 0.024416 | 0.004343 | 7.10E-09 |
| rs687289 | miR-10b-5p | trans | 5.80E-37 | Nightingale | IDL_FC_pct | NA | A | G | 0.025842 | 0.004394 | 9.80E-09 |
| rs687289 | miR-10b-5p | trans | 5.80E-37 | Nightingale | VLDL_FC | NA | A | G | 0.02388 | 0.004396 | 1.60E-08 |
| rs687289 | miR-126-3p | trans | 3.06E-41 | Nightingale | XS_VLDL_C | NA | A | G | 0.03768 | 0.004274 | 2.70E-18 |
| rs687289 | miR-126-3p | trans | 3.06E-41 | Nightingale | ApoB | NA | A | G | 0.03375 | 0.004423 | 2.90E-14 |
| rs687289 | miR-126-3p | trans | 3.06E-41 | Nightingale | LDL_C | NA | A | G | 0.032964 | 0.004409 | 1.20E-13 |
| rs687289 | miR-126-3p | trans | 3.06E-41 | Nightingale | S_VLDL_PL | NA | A | G | 0.032601 | 0.00442 | 5.80E-14 |
| rs687289 | miR-126-3p | trans | 3.06E-41 | Nightingale | M_LDL_C_ | NA | A | G | 0.031111 | 0.004432 | 3.10E-12 |
| rs687289 | miR-126-3p | trans | 3.06E-41 | Nightingale | L_VLDL_TG | NA | A | G | -0.05139 | 0.004408 | 3.50E-32 |
| rs687289 | miR-126-3p | trans | 3.06E-41 | Nightingale | XL_VLDL_C | NA | A | G | 0.037793 | 0.004249 | 1.10E-18 |
| rs687289 | miR-126-3p | trans | 3.06E-41 | Nightingale | S_VLDL_L | NA | A | G | 0.024476 | 0.004386 | 5.10E-09 |
| rs687289 | miR-126-3p | trans | 3.06E-41 | Nightingale | S_VLDL_P | NA | A | G | 0.023298 | 0.004373 | 2.10E-08 |
| rs687289 | miR-126-3p | trans | 3.06E-41 | Nightingale | M_VLDL_C | NA | A | G | 0.036468 | 0.004171 | 5.90E-18 |
| rs687289 | miR-126-3p | trans | 3.06E-41 | Nightingale | S_LDL_C | NA | A | G | 0.030716 | 0.004433 | 7.50E-12 |
| rs687289 | miR-126-3p | trans | 3.06E-41 | Nightingale | LDL_L | NA | A | G | 0.033082 | 0.004414 | 1.00E-13 |
| rs687289 | miR-126-3p | trans | 3.06E-41 | Nightingale | M_VLDL_C | NA | A | G | 0.033905 | 0.004161 | 9.50E-16 |
| rs687289 | miR-126-3p | trans | 3.06E-41 | Nightingale | S_HDL_P | NA | A | G | 0.030278 | 0.004429 | 9.00E-12 |
| rs687289 | miR-126-3p | trans | 3.06E-41 | Nightingale | M_LDL_L | NA | A | G | 0.032826 | 0.004435 | 1.40E-13 |
| rs687289 | miR-126-3p | trans | 3.06E-41 | Nightingale | M_VLDL_F | NA | A | G | 0.042299 | 0.004213 | 1.80E-23 |
| rs687289 | miR-126-3p | trans | 3.06E-41 | Nightingale | GlycA | NA | A | G | 0.028099 | 0.00441 | 1.30E-10 |
| rs687289 | miR-126-3p | trans | 3.06E-41 | Nightingale | S_LDL_CE | NA | A | G | 0.030317 | 0.004431 | 8.30E-12 |
| rs687289 | miR-126-3p | trans | 3.06E-41 | Nightingale | M_VLDL_T | NA | A | G | -0.03881 | 0.004185 | 3.40E-20 |
| rs687289 | miR-126-3p | trans | 3.06E-41 | Nightingale | S_VLDL_C_ | NA | A | G | 0.042052 | 0.00431 | 3.20E-22 |
| rs687289 | miR-126-3p | trans | 3.06E-41 | Nightingale | M_LDL_C | NA | A | G | 0.033786 | 0.004437 | 3.00E-14 |
| rs687289 | miR-126-3p | trans | 3.06E-41 | Nightingale | M_VLDL_C | NA | A | G | 0.038509 | 0.004416 | 2.60E-18 |
| rs687289 | miR-126-3p | trans | 3.06E-41 | Nightingale | XL_VLDL_F | NA | A | G | 0.037531 | 0.004293 | 1.00E-18 |
| rs687289 | miR-126-3p | trans | 3.06E-41 | Nightingale | M_VLDL_C | NA | A | G | 0.040645 | 0.004388 | 3.20E-20 |
| rs687289 | miR-126-3p | trans | 3.06E-41 | Nightingale | S_LDL_PL_ | NA | A | G | -0.02989 | 0.00442 | 4.10E-13 |
| rs687289 | miR-126-3p | trans | 3.06E-41 | Nightingale | IDL_CE | NA | A | G | 0.033304 | 0.004276 | 1.80E-14 |
| rs687289 | miR-126-3p | trans | 3.06E-41 | Nightingale | Omega_6 | NA | A | G | 0.025429 | 0.004325 | 1.10E-09 |
| rs687289 | miR-126-3p | trans | 3.06E-41 | Nightingale | XL_VLDL_T | NA | A | G | -0.04438 | 0.004292 | 5.40E-25 |
| rs687289 | miR-126-3p | trans | 3.06E-41 | Nightingale | LDL_CE | NA | A | G | 0.032325 | 0.004417 | 3.10E-13 |
| rs687289 | miR-126-3p | trans | 3.06E-41 | Nightingale | XXL_VLDL_ | NA | A | G | 0.035053 | 0.004438 | 2.10E-15 |
| rs687289 | miR-126-3p | trans | 3.06E-41 | Nightingale | XS_VLDL_P | NA | A | G | 0.036638 | 0.00437 | 1.60E-17 |
| rs687289 | miR-126-3p | trans | 3.06E-41 | Nightingale | S_LDL_L | NA | A | G | 0.027511 | 0.004428 | 1.10E-09 |
| rs687289 | miR-126-3p | trans | 3.06E-41 | Nightingale | L_LDL_C | NA | A | G | 0.032081 | 0.004379 | 3.60E-13 |
| rs687289 | miR-126-3p | trans | 3.06E-41 | Nightingale | M_LDL_TG | NA | A | G | -0.02742 | 0.004374 | 4.00E-10 |
| rs687289 | miR-126-3p | trans | 3.06E-41 | Nightingale | S_VLDL_C | NA | A | G | 0.038239 | 0.004428 | 2.40E-18 |
| rs687289 | miR-126-3p | trans | 3.06E-41 | Nightingale | M_VLDL_P | NA | A | G | 0.028859 | 0.004422 | 3.10E-11 |
| rs687289 | miR-126-3p | trans | 3.06E-41 | Nightingale | L_LDL_P | NA | A | G | 0.028943 | 0.004414 | 8.30E-11 |

| | | | | | | | | | | | |
|---|---|---|---|---|---|---|---|---|---|---|---|
| rs687289 | miR-126-3p | trans | 3.06E-41 | Nightingale | M_LDL_P | NA | A | G | 0.035821 | 0.004441 | 7.50E-16 |
| rs687289 | miR-126-3p | trans | 3.06E-41 | Nightingale | VLDL_CE | NA | A | G | 0.03558 | 0.00443 | 4.60E-16 |
| rs687289 | miR-126-3p | trans | 3.06E-41 | Nightingale | XL_VLDL_P | NA | A | G | 0.038789 | 0.004435 | 6.40E-19 |
| rs687289 | miR-126-3p | trans | 3.06E-41 | Nightingale | S_LDL_P | NA | A | G | 0.029429 | 0.004433 | 4.00E-11 |
| rs687289 | miR-126-3p | trans | 3.06E-41 | Nightingale | XS_VLDL_F | NA | A | G | 0.037022 | 0.004352 | 7.50E-18 |
| rs687289 | miR-126-3p | trans | 3.06E-41 | Nightingale | M_VLDL_P | NA | A | G | 0.028023 | 0.004418 | 9.80E-11 |
| rs687289 | miR-126-3p | trans | 3.06E-41 | Nightingale | LDL_P | NA | A | G | 0.031612 | 0.004425 | 1.30E-12 |
| rs687289 | miR-126-3p | trans | 3.06E-41 | Nightingale | L_VLDL_C_ | NA | A | G | 0.052543 | 0.004325 | 1.80E-34 |
| rs687289 | miR-126-3p | trans | 3.06E-41 | Nightingale | L_LDL_L | NA | A | G | 0.033323 | 0.004385 | 4.20E-14 |
| rs687289 | miR-126-3p | trans | 3.06E-41 | Nightingale | LDL_PL | NA | A | G | 0.034755 | 0.004418 | 6.70E-15 |
| rs687289 | miR-126-3p | trans | 3.06E-41 | Nightingale | XS_VLDL_C | NA | A | G | 0.037903 | 0.004299 | 1.70E-18 |
| rs687289 | miR-126-3p | trans | 3.06E-41 | Nightingale | M_LDL_CE | NA | A | G | 0.032783 | 0.004435 | 1.30E-13 |
| rs687289 | miR-126-3p | trans | 3.06E-41 | Nightingale | LDL_FC | NA | A | G | 0.033395 | 0.004381 | 7.50E-14 |
| rs687289 | miR-126-3p | trans | 3.06E-41 | Nightingale | S_HDL_C | NA | A | G | 0.028109 | 0.004421 | 2.80E-10 |
| rs687289 | miR-126-3p | trans | 3.06E-41 | Nightingale | IDL_P | NA | A | G | 0.03981 | 0.004382 | 1.80E-19 |
| rs687289 | miR-126-3p | trans | 3.06E-41 | Nightingale | XS_VLDL_P | NA | A | G | 0.032077 | 0.004367 | 5.90E-14 |
| rs687289 | miR-126-3p | trans | 3.06E-41 | Nightingale | XL_VLDL_C | NA | A | G | 0.038665 | 0.004247 | 1.60E-19 |
| rs687289 | miR-126-3p | trans | 3.06E-41 | Nightingale | Remnant_( | NA | A | G | 0.036652 | 0.0044 | 5.60E-17 |
| rs687289 | miR-126-3p | trans | 3.06E-41 | Nightingale | L_LDL_CE | NA | A | G | 0.031327 | 0.004389 | 1.40E-12 |
| rs687289 | miR-126-3p | trans | 3.06E-41 | Nightingale | S_HDL_L | NA | A | G | 0.026 | 0.004413 | 2.50E-09 |
| rs687289 | miR-126-3p | trans | 3.06E-41 | Nightingale | Total_PL | NA | A | G | 0.02466 | 0.004217 | 1.60E-09 |
| rs687289 | miR-126-3p | trans | 3.06E-41 | Nightingale | IDL_PL | NA | A | G | 0.035187 | 0.004274 | 4.50E-16 |
| rs687289 | miR-126-3p | trans | 3.06E-41 | Nightingale | XXL_VLDL_ | NA | A | G | 0.02523 | 0.004458 | 1.10E-09 |
| rs687289 | miR-126-3p | trans | 3.06E-41 | Nightingale | ApoB_by_A | NA | A | G | 0.024977 | 0.004362 | 1.30E-08 |
| rs687289 | miR-126-3p | trans | 3.06E-41 | Nightingale | S_HDL_CE | NA | A | G | 0.028306 | 0.004436 | 2.20E-10 |
| rs687289 | miR-126-3p | trans | 3.06E-41 | Nightingale | Total_CE | NA | A | G | 0.031498 | 0.004272 | 1.40E-13 |
| rs687289 | miR-126-3p | trans | 3.06E-41 | Nightingale | Total_FC | NA | A | G | 0.033014 | 0.004326 | 1.00E-14 |
| rs687289 | miR-126-3p | trans | 3.06E-41 | Nightingale | S_VLDL_CE | NA | A | G | 0.047288 | 0.004369 | 6.20E-27 |
| rs687289 | miR-126-3p | trans | 3.06E-41 | Nightingale | M_VLDL_F | NA | A | G | 0.033132 | 0.004428 | 4.80E-14 |
| rs687289 | miR-126-3p | trans | 3.06E-41 | Nightingale | IDL_C | NA | A | G | 0.034761 | 0.004277 | 1.30E-15 |
| rs687289 | miR-126-3p | trans | 3.06E-41 | Nightingale | non_HDL_( | NA | A | G | 0.03547 | 0.004405 | 7.40E-16 |
| rs687289 | miR-126-3p | trans | 3.06E-41 | Nightingale | S_LDL_C_p | NA | A | G | 0.045227 | 0.004446 | 8.10E-25 |
| rs687289 | miR-126-3p | trans | 3.06E-41 | Nightingale | XS_VLDL_F | NA | A | G | 0.031381 | 0.004397 | 2.60E-12 |
| rs687289 | miR-126-3p | trans | 3.06E-41 | Nightingale | VLDL_P | NA | A | G | 0.026418 | 0.004405 | 6.20E-10 |
| rs687289 | miR-126-3p | trans | 3.06E-41 | Nightingale | M_VLDL_P | NA | A | G | 0.044251 | 0.00425 | 3.40E-25 |
| rs687289 | miR-126-3p | trans | 3.06E-41 | Nightingale | L_VLDL_FC | NA | A | G | 0.048909 | 0.004386 | 2.40E-29 |
| rs687289 | miR-126-3p | trans | 3.06E-41 | Nightingale | Total_C | NA | A | G | 0.032124 | 0.004287 | 4.80E-14 |
| rs687289 | miR-126-3p | trans | 3.06E-41 | Nightingale | L_LDL_FC | NA | A | G | 0.033386 | 0.004349 | 4.10E-14 |
| rs687289 | miR-126-3p | trans | 3.06E-41 | Nightingale | S_HDL_PL | NA | A | G | 0.023499 | 0.00439 | 5.00E-08 |
| rs687289 | miR-126-3p | trans | 3.06E-41 | Nightingale | M_LDL_FC | NA | A | G | 0.034117 | 0.004423 | 2.80E-14 |
| rs687289 | miR-126-3p | trans | 3.06E-41 | Nightingale | S_VLDL_TG | NA | A | G | -0.03827 | 0.004287 | 9.50E-19 |
| rs687289 | miR-126-3p | trans | 3.06E-41 | Nightingale | XXL_VLDL_ | NA | A | G | 0.033249 | 0.004463 | 7.70E-14 |
| rs687289 | miR-126-3p | trans | 3.06E-41 | Nightingale | S_LDL_FC | NA | A | G | 0.028345 | 0.004422 | 5.60E-10 |
| rs687289 | miR-126-3p | trans | 3.06E-41 | Nightingale | L_VLDL_CE | NA | A | G | 0.047672 | 0.004238 | 1.00E-29 |
| rs687289 | miR-126-3p | trans | 3.06E-41 | Nightingale | Citrate | NA | A | G | 0.026536 | 0.0044 | 1.10E-09 |
| rs687289 | miR-126-3p | trans | 3.06E-41 | Nightingale | M_LDL_PL | NA | A | G | 0.032235 | 0.004435 | 3.90E-13 |
| rs687289 | miR-126-3p | trans | 3.06E-41 | Nightingale | Total_L | NA | A | G | 0.026033 | 0.004357 | 7.20E-10 |
| rs687289 | miR-126-3p | trans | 3.06E-41 | Nightingale | VLDL_C | NA | A | G | 0.031062 | 0.004422 | 7.60E-13 |
| rs687289 | miR-126-3p | trans | 3.06E-41 | Nightingale | XS_VLDL_L | NA | A | G | 0.033757 | 0.004353 | 2.40E-15 |
| rs687289 | miR-126-3p | trans | 3.06E-41 | Nightingale | IDL_FC | NA | A | G | 0.038096 | 0.004287 | 2.00E-18 |
| rs687289 | miR-126-3p | trans | 3.06E-41 | Nightingale | S_VLDL_CE | NA | A | G | 0.038069 | 0.004423 | 2.20E-18 |
| rs687289 | miR-126-3p | trans | 3.06E-41 | Nightingale | PUFA | NA | A | G | 0.023493 | 0.00432 | 2.00E-08 |
| rs687289 | miR-126-3p | trans | 3.06E-41 | Nightingale | XXL_VLDL_ | NA | A | G | -0.03594 | 0.004467 | 1.60E-16 |
| rs687289 | miR-126-3p | trans | 3.06E-41 | Nightingale | S_VLDL_PL | NA | A | G | 0.027844 | 0.004241 | 1.40E-10 |
| rs687289 | miR-126-3p | trans | 3.06E-41 | Nightingale | Clinical_LD | NA | A | G | 0.036238 | 0.004403 | 4.00E-16 |
| rs687289 | miR-126-3p | trans | 3.06E-41 | Nightingale | S_LDL_CE_ | NA | A | G | 0.03078 | 0.004421 | 7.20E-14 |
| rs687289 | miR-126-3p | trans | 3.06E-41 | Nightingale | Sphingomy | NA | A | G | 0.028834 | 0.00414 | 9.00E-12 |
| rs687289 | miR-126-3p | trans | 3.06E-41 | Nightingale | S_VLDL_FC | NA | A | G | 0.029424 | 0.004232 | 8.60E-12 |
| rs687289 | miR-126-3p | trans | 3.06E-41 | Nightingale | S_VLDL_FC | NA | A | G | 0.037731 | 0.004429 | 1.00E-17 |
| rs687289 | miR-126-3p | trans | 3.06E-41 | Nightingale | XL_HDL_PL | NA | A | G | -0.02251 | 0.004069 | 1.90E-08 |
| rs687289 | miR-126-3p | trans | 3.06E-41 | Nightingale | IDL_L | NA | A | G | 0.035103 | 0.004282 | 6.50E-16 |
| rs687289 | miR-126-3p | trans | 3.06E-41 | Nightingale | L_LDL_PL | NA | A | G | 0.037753 | 0.004395 | 1.30E-17 |
| rs687289 | miR-126-3p | trans | 3.06E-41 | Nightingale | XXL_VLDL_ | NA | A | G | 0.031237 | 0.00441 | 6.10E-13 |

| | | | | | | | | | | | |
|---|---|---|---|---|---|---|---|---|---|---|---|
| rs687289 | miR-126-3p | trans | 3.06E-41 | Nightingale | LA | NA | A | G | 0.024416 | 0.004343 | 7.10E-09 |
| rs687289 | miR-126-3p | trans | 3.06E-41 | Nightingale | IDL_FC_pct | NA | A | G | 0.025842 | 0.004394 | 9.80E-09 |
| rs687289 | miR-126-3p | trans | 3.06E-41 | Nightingale | VLDL_FC | NA | A | G | 0.02388 | 0.004396 | 1.60E-08 |
| rs7036642 | miR-10a-5p | trans | 1.52E-13 | Nightingale | Clinical_LD | NA | A | G | -0.02345 | 0.004163 | 1.60E-08 |
| rs7036642 | miR-10a-5p | trans | 1.52E-13 | Nightingale | IDL_PL | NA | A | G | -0.02247 | 0.00404 | 2.00E-08 |
| rs7036642 | miR-10a-5p | trans | 1.52E-13 | Nightingale | M_VLDL_C | NA | A | G | -0.02365 | 0.003944 | 1.30E-08 |
| rs7036642 | miR-10a-5p | trans | 1.52E-13 | Nightingale | S_VLDL_CE | NA | A | G | -0.02316 | 0.004181 | 6.50E-09 |
| rs7036642 | miR-10a-5p | trans | 1.52E-13 | Nightingale | XL_VLDL_C | NA | A | G | -0.02594 | 0.004014 | 7.80E-10 |
| rs7036642 | miR-10a-5p | trans | 1.52E-13 | Nightingale | M_LDL_P | NA | A | G | -0.02333 | 0.004199 | 1.40E-08 |
| rs7036642 | miR-10a-5p | trans | 1.52E-13 | Nightingale | XL_VLDL_C | NA | A | G | -0.02532 | 0.004017 | 2.60E-09 |
| rs7036642 | miR-10a-5p | trans | 1.52E-13 | Nightingale | XXL_VLDL_ | NA | A | G | -0.02525 | 0.004194 | 1.80E-09 |
| rs7036642 | miR-10a-5p | trans | 1.52E-13 | Nightingale | XS_VLDL_C | NA | A | G | -0.02471 | 0.00404 | 3.50E-10 |
| rs7036642 | miR-10a-5p | trans | 1.52E-13 | Nightingale | XXL_VLDL_ | NA | A | G | -0.02435 | 0.004217 | 7.80E-09 |
| rs7036642 | miR-10a-5p | trans | 1.52E-13 | Nightingale | S_VLDL_CE | NA | A | G | -0.03238 | 0.004131 | 8.20E-15 |
| rs7036642 | miR-10a-5p | trans | 1.52E-13 | Nightingale | L_LDL_PL | NA | A | G | -0.02441 | 0.004155 | 3.10E-09 |
| rs7036642 | miR-10a-5p | trans | 1.52E-13 | Nightingale | M_VLDL_P | NA | A | G | -0.0288 | 0.004018 | 1.30E-12 |
| rs7036642 | miR-10a-5p | trans | 1.52E-13 | Nightingale | IDL_P | NA | A | G | -0.02514 | 0.004143 | 7.80E-10 |
| rs7036642 | miR-10a-5p | trans | 1.52E-13 | Nightingale | XS_VLDL_C | NA | A | G | -0.02428 | 0.004064 | 7.60E-10 |
| rs7036642 | miR-10a-5p | trans | 1.52E-13 | Nightingale | S_VLDL_C_ | NA | A | G | -0.0285 | 0.004075 | 1.00E-11 |
| rs7036642 | miR-10a-5p | trans | 1.52E-13 | Nightingale | Remnant_( | NA | A | G | -0.02289 | 0.00416 | 9.30E-09 |
| rs7036642 | miR-10a-5p | trans | 1.52E-13 | Nightingale | XS_VLDL_P | NA | A | G | -0.02209 | 0.004131 | 1.30E-08 |
| rs7036642 | miR-10a-5p | trans | 1.52E-13 | Nightingale | M_VLDL_C | NA | A | G | -0.02433 | 0.004175 | 1.70E-09 |
| rs7036642 | miR-10a-5p | trans | 1.52E-13 | Nightingale | L_VLDL_C_ | NA | A | G | -0.03516 | 0.004089 | 8.10E-18 |
| rs7036642 | miR-10a-5p | trans | 1.52E-13 | Nightingale | S_VLDL_FC | NA | A | G | -0.02292 | 0.004188 | 1.50E-08 |
| rs7036642 | miR-10a-5p | trans | 1.52E-13 | Nightingale | L_VLDL_TG | NA | A | G | 0.033443 | 0.004167 | 2.40E-16 |
| rs7036642 | miR-10a-5p | trans | 1.52E-13 | Nightingale | XS_VLDL_F | NA | A | G | -0.02238 | 0.004114 | 9.70E-09 |
| rs7036642 | miR-10a-5p | trans | 1.52E-13 | Nightingale | M_VLDL_C | NA | A | G | -0.02653 | 0.004149 | 8.70E-11 |
| rs7036642 | miR-10a-5p | trans | 1.52E-13 | Nightingale | L_VLDL_CE | NA | A | G | -0.03188 | 0.004006 | 3.50E-15 |
| rs7036642 | miR-10a-5p | trans | 1.52E-13 | Nightingale | IDL_FC | NA | A | G | -0.02519 | 0.004053 | 4.90E-10 |
| rs7036642 | miR-10a-5p | trans | 1.52E-13 | Nightingale | M_VLDL_F | NA | A | G | -0.02683 | 0.003983 | 6.40E-11 |
| rs7036642 | miR-10a-5p | trans | 1.52E-13 | Nightingale | IDL_CE | NA | A | G | -0.02213 | 0.004042 | 4.00E-08 |
| rs7036642 | miR-10a-5p | trans | 1.52E-13 | Nightingale | IDL_C | NA | A | G | -0.02309 | 0.004044 | 1.10E-08 |
| rs7036642 | miR-10a-5p | trans | 1.52E-13 | Nightingale | L_VLDL_FC | NA | A | G | -0.03157 | 0.004147 | 1.40E-14 |
| rs7036642 | miR-10a-5p | trans | 1.52E-13 | Nightingale | S_LDL_C_p | NA | A | G | -0.0296 | 0.004203 | 1.40E-12 |
| rs7036642 | miR-10a-5p | trans | 1.52E-13 | Nightingale | XXL_VLDL_ | NA | A | G | 0.025977 | 0.004221 | 3.70E-10 |
| rs7036642 | miR-10a-5p | trans | 1.52E-13 | Nightingale | S_VLDL_C | NA | A | G | -0.02326 | 0.004186 | 6.90E-09 |
| rs7036642 | miR-10a-5p | trans | 1.52E-13 | Nightingale | XL_VLDL_P | NA | A | G | -0.02451 | 0.004193 | 1.40E-09 |
| rs7036642 | miR-10a-5p | trans | 1.52E-13 | Nightingale | VLDL_CE | NA | A | G | -0.02175 | 0.004188 | 4.80E-08 |
| rs7036642 | miR-10a-5p | trans | 1.52E-13 | Nightingale | IDL_L | NA | A | G | -0.02279 | 0.004048 | 1.60E-08 |
| rs7036642 | miR-10a-5p | trans | 1.52E-13 | Nightingale | S_VLDL_TG | NA | A | G | 0.026003 | 0.004053 | 5.70E-10 |
| rs7036642 | miR-10a-5p | trans | 1.52E-13 | Nightingale | M_VLDL_T | NA | A | G | 0.025138 | 0.003957 | 1.10E-09 |
| rs7036642 | miR-10a-5p | trans | 1.52E-13 | Nightingale | XL_VLDL_T | NA | A | G | 0.029657 | 0.004058 | 1.20E-12 |
| rs7036642 | miR-10a-5p | trans | 1.52E-13 | Nightingale | XL_VLDL_F | NA | A | G | -0.02503 | 0.004059 | 1.10E-09 |
| rs7036642 | miR-126-3p | trans | 9.29E-19 | Nightingale | Clinical_LD | NA | A | G | -0.02345 | 0.004163 | 1.60E-08 |
| rs7036642 | miR-126-3p | trans | 9.29E-19 | Nightingale | IDL_PL | NA | A | G | -0.02247 | 0.00404 | 2.00E-08 |
| rs7036642 | miR-126-3p | trans | 9.29E-19 | Nightingale | M_VLDL_C | NA | A | G | -0.02365 | 0.003944 | 1.30E-08 |
| rs7036642 | miR-126-3p | trans | 9.29E-19 | Nightingale | S_VLDL_CE | NA | A | G | -0.02316 | 0.004181 | 6.50E-09 |
| rs7036642 | miR-126-3p | trans | 9.29E-19 | Nightingale | XL_VLDL_C | NA | A | G | -0.02594 | 0.004014 | 7.80E-10 |
| rs7036642 | miR-126-3p | trans | 9.29E-19 | Nightingale | M_LDL_P | NA | A | G | -0.02333 | 0.004199 | 1.40E-08 |
| rs7036642 | miR-126-3p | trans | 9.29E-19 | Nightingale | XL_VLDL_C | NA | A | G | -0.02532 | 0.004017 | 2.60E-09 |
| rs7036642 | miR-126-3p | trans | 9.29E-19 | Nightingale | XXL_VLDL_ | NA | A | G | -0.02525 | 0.004194 | 1.80E-09 |
| rs7036642 | miR-126-3p | trans | 9.29E-19 | Nightingale | XS_VLDL_C | NA | A | G | -0.02471 | 0.00404 | 3.50E-10 |
| rs7036642 | miR-126-3p | trans | 9.29E-19 | Nightingale | XXL_VLDL_ | NA | A | G | -0.02435 | 0.004217 | 7.80E-09 |
| rs7036642 | miR-126-3p | trans | 9.29E-19 | Nightingale | S_VLDL_CE | NA | A | G | -0.03238 | 0.004131 | 8.20E-15 |
| rs7036642 | miR-126-3p | trans | 9.29E-19 | Nightingale | L_LDL_PL | NA | A | G | -0.02441 | 0.004155 | 3.10E-09 |
| rs7036642 | miR-126-3p | trans | 9.29E-19 | Nightingale | M_VLDL_P | NA | A | G | -0.0288 | 0.004018 | 1.30E-12 |
| rs7036642 | miR-126-3p | trans | 9.29E-19 | Nightingale | IDL_P | NA | A | G | -0.02514 | 0.004143 | 7.80E-10 |
| rs7036642 | miR-126-3p | trans | 9.29E-19 | Nightingale | XS_VLDL_C | NA | A | G | -0.02428 | 0.004064 | 7.60E-10 |
| rs7036642 | miR-126-3p | trans | 9.29E-19 | Nightingale | S_VLDL_C_ | NA | A | G | -0.0285 | 0.004075 | 1.00E-11 |
| rs7036642 | miR-126-3p | trans | 9.29E-19 | Nightingale | Remnant_( | NA | A | G | -0.02289 | 0.00416 | 9.30E-09 |
| rs7036642 | miR-126-3p | trans | 9.29E-19 | Nightingale | XS_VLDL_P | NA | A | G | -0.02209 | 0.004131 | 1.30E-08 |
| rs7036642 | miR-126-3p | trans | 9.29E-19 | Nightingale | M_VLDL_C | NA | A | G | -0.02433 | 0.004175 | 1.70E-09 |
| rs7036642 | miR-126-3p | trans | 9.29E-19 | Nightingale | L_VLDL_C_ | NA | A | G | -0.03516 | 0.004089 | 8.10E-18 |

| | | | | | | | | | | | |
|---|---|---|---|---|---|---|---|---|---|---|---|
| rs7036642 | miR-126-3p | trans | 9.29E-19 | Nightingale | S_VLDL_FC | NA | A | G | -0.02292 | 0.004188 | 1.50E-08 |
| rs7036642 | miR-126-3p | trans | 9.29E-19 | Nightingale | L_VLDL_TG | NA | A | G | 0.033443 | 0.004167 | 2.40E-16 |
| rs7036642 | miR-126-3p | trans | 9.29E-19 | Nightingale | XS_VLDL_F | NA | A | G | -0.02238 | 0.004114 | 9.70E-09 |
| rs7036642 | miR-126-3p | trans | 9.29E-19 | Nightingale | M_VLDL_C | NA | A | G | -0.02653 | 0.004149 | 8.70E-11 |
| rs7036642 | miR-126-3p | trans | 9.29E-19 | Nightingale | L_VLDL_CE | NA | A | G | -0.03188 | 0.004006 | 3.50E-15 |
| rs7036642 | miR-126-3p | trans | 9.29E-19 | Nightingale | IDL_FC | NA | A | G | -0.02519 | 0.004053 | 4.90E-10 |
| rs7036642 | miR-126-3p | trans | 9.29E-19 | Nightingale | M_VLDL_F | NA | A | G | -0.02683 | 0.003983 | 6.40E-11 |
| rs7036642 | miR-126-3p | trans | 9.29E-19 | Nightingale | IDL_CE | NA | A | G | -0.02213 | 0.004042 | 4.00E-08 |
| rs7036642 | miR-126-3p | trans | 9.29E-19 | Nightingale | IDL_C | NA | A | G | -0.02309 | 0.004044 | 1.10E-08 |
| rs7036642 | miR-126-3p | trans | 9.29E-19 | Nightingale | L_VLDL_FC | NA | A | G | -0.03157 | 0.004147 | 1.40E-14 |
| rs7036642 | miR-126-3p | trans | 9.29E-19 | Nightingale | S_LDL_C_p | NA | A | G | -0.0296 | 0.004203 | 1.40E-12 |
| rs7036642 | miR-126-3p | trans | 9.29E-19 | Nightingale | XXL_VLDL_ | NA | A | G | 0.025977 | 0.004221 | 3.70E-10 |
| rs7036642 | miR-126-3p | trans | 9.29E-19 | Nightingale | S_VLDL_C | NA | A | G | -0.02326 | 0.004186 | 6.90E-09 |
| rs7036642 | miR-126-3p | trans | 9.29E-19 | Nightingale | XL_VLDL_P | NA | A | G | -0.02451 | 0.004193 | 1.40E-09 |
| rs7036642 | miR-126-3p | trans | 9.29E-19 | Nightingale | VLDL_CE | NA | A | G | -0.02175 | 0.004188 | 4.80E-08 |
| rs7036642 | miR-126-3p | trans | 9.29E-19 | Nightingale | IDL_L | NA | A | G | -0.02279 | 0.004048 | 1.60E-08 |
| rs7036642 | miR-126-3p | trans | 9.29E-19 | Nightingale | S_VLDL_TG | NA | A | G | 0.026003 | 0.004053 | 5.70E-10 |
| rs7036642 | miR-126-3p | trans | 9.29E-19 | Nightingale | M_VLDL_T | NA | A | G | 0.025138 | 0.003957 | 1.10E-09 |
| rs7036642 | miR-126-3p | trans | 9.29E-19 | Nightingale | XL_VLDL_T | NA | A | G | 0.029657 | 0.004058 | 1.20E-12 |
| rs7036642 | miR-126-3p | trans | 9.29E-19 | Nightingale | XL_VLDL_F | NA | A | G | -0.02503 | 0.004059 | 1.10E-09 |
| rs7046674 | miR-10a-5p | trans | 9.50E-14 | Nightingale | Clinical_LD | NA | T | C | -0.02355 | 0.004163 | 1.30E-08 |
| rs7046674 | miR-10a-5p | trans | 9.50E-14 | Nightingale | VLDL_CE | NA | T | C | -0.02189 | 0.004188 | 3.80E-08 |
| rs7046674 | miR-10a-5p | trans | 9.50E-14 | Nightingale | L_LDL_PL | NA | T | C | -0.02451 | 0.004155 | 2.50E-09 |
| rs7046674 | miR-10a-5p | trans | 9.50E-14 | Nightingale | XL_VLDL_C | NA | T | C | -0.02581 | 0.004015 | 9.30E-10 |
| rs7046674 | miR-10a-5p | trans | 9.50E-14 | Nightingale | M_VLDL_C | NA | T | C | -0.02665 | 0.004149 | 6.70E-11 |
| rs7046674 | miR-10a-5p | trans | 9.50E-14 | Nightingale | XL_VLDL_F | NA | T | C | -0.02491 | 0.004059 | 1.20E-09 |
| rs7046674 | miR-10a-5p | trans | 9.50E-14 | Nightingale | XL_VLDL_T | NA | T | C | 0.029538 | 0.004058 | 1.50E-12 |
| rs7046674 | miR-10a-5p | trans | 9.50E-14 | Nightingale | IDL_P | NA | T | C | -0.02532 | 0.004143 | 5.70E-10 |
| rs7046674 | miR-10a-5p | trans | 9.50E-14 | Nightingale | XS_VLDL_C | NA | T | C | -0.02442 | 0.004064 | 6.00E-10 |
| rs7046674 | miR-10a-5p | trans | 9.50E-14 | Nightingale | IDL_L | NA | T | C | -0.0229 | 0.004048 | 1.40E-08 |
| rs7046674 | miR-10a-5p | trans | 9.50E-14 | Nightingale | Remnant_( | NA | T | C | -0.02302 | 0.00416 | 7.30E-09 |
| rs7046674 | miR-10a-5p | trans | 9.50E-14 | Nightingale | XL_VLDL_C | NA | T | C | -0.02519 | 0.004017 | 3.00E-09 |
| rs7046674 | miR-10a-5p | trans | 9.50E-14 | Nightingale | S_VLDL_C | NA | T | C | -0.0234 | 0.004187 | 5.40E-09 |
| rs7046674 | miR-10a-5p | trans | 9.50E-14 | Nightingale | S_VLDL_C_ | NA | T | C | -0.02853 | 0.004075 | 8.90E-12 |
| rs7046674 | miR-10a-5p | trans | 9.50E-14 | Nightingale | XXL_VLDL_ | NA | T | C | -0.02444 | 0.004217 | 6.70E-09 |
| rs7046674 | miR-10a-5p | trans | 9.50E-14 | Nightingale | IDL_PL | NA | T | C | -0.02256 | 0.004041 | 1.70E-08 |
| rs7046674 | miR-10a-5p | trans | 9.50E-14 | Nightingale | XXL_VLDL_ | NA | T | C | -0.02528 | 0.004194 | 1.80E-09 |
| rs7046674 | miR-10a-5p | trans | 9.50E-14 | Nightingale | M_VLDL_T | NA | T | C | 0.025093 | 0.003957 | 1.20E-09 |
| rs7046674 | miR-10a-5p | trans | 9.50E-14 | Nightingale | S_LDL_C_p | NA | T | C | -0.0297 | 0.004203 | 1.20E-12 |
| rs7046674 | miR-10a-5p | trans | 9.50E-14 | Nightingale | S_VLDL_CE | NA | T | C | -0.03245 | 0.004131 | 6.70E-15 |
| rs7046674 | miR-10a-5p | trans | 9.50E-14 | Nightingale | S_VLDL_CE | NA | T | C | -0.02331 | 0.004182 | 5.00E-09 |
| rs7046674 | miR-10a-5p | trans | 9.50E-14 | Nightingale | L_VLDL_FC | NA | T | C | -0.03163 | 0.004147 | 1.30E-14 |
| rs7046674 | miR-10a-5p | trans | 9.50E-14 | Nightingale | L_VLDL_CE | NA | T | C | -0.03189 | 0.004007 | 3.50E-15 |
| rs7046674 | miR-10a-5p | trans | 9.50E-14 | Nightingale | XXL_VLDL_ | NA | T | C | 0.026027 | 0.004222 | 3.40E-10 |
| rs7046674 | miR-10a-5p | trans | 9.50E-14 | Nightingale | non_HDL_( | NA | T | C | -0.02236 | 0.004164 | 4.20E-08 |
| rs7046674 | miR-10a-5p | trans | 9.50E-14 | Nightingale | XL_VLDL_P | NA | T | C | -0.02456 | 0.004193 | 1.30E-09 |
| rs7046674 | miR-10a-5p | trans | 9.50E-14 | Nightingale | S_VLDL_FC | NA | T | C | -0.02306 | 0.004188 | 1.20E-08 |
| rs7046674 | miR-10a-5p | trans | 9.50E-14 | Nightingale | IDL_C | NA | T | C | -0.0232 | 0.004044 | 8.90E-09 |
| rs7046674 | miR-10a-5p | trans | 9.50E-14 | Nightingale | XS_VLDL_P | NA | T | C | -0.02224 | 0.004132 | 1.00E-08 |
| rs7046674 | miR-10a-5p | trans | 9.50E-14 | Nightingale | M_VLDL_C | NA | T | C | -0.02445 | 0.004175 | 1.30E-09 |
| rs7046674 | miR-10a-5p | trans | 9.50E-14 | Nightingale | M_VLDL_C | NA | T | C | -0.02359 | 0.003944 | 1.40E-08 |
| rs7046674 | miR-10a-5p | trans | 9.50E-14 | Nightingale | IDL_CE | NA | T | C | -0.02226 | 0.004043 | 3.20E-08 |
| rs7046674 | miR-10a-5p | trans | 9.50E-14 | Nightingale | M_VLDL_F | NA | T | C | -0.0268 | 0.003984 | 6.60E-11 |
| rs7046674 | miR-10a-5p | trans | 9.50E-14 | Nightingale | XS_VLDL_C | NA | T | C | -0.02485 | 0.004041 | 2.80E-10 |
| rs7046674 | miR-10a-5p | trans | 9.50E-14 | Nightingale | L_VLDL_TG | NA | T | C | 0.03358 | 0.004167 | 1.80E-16 |
| rs7046674 | miR-10a-5p | trans | 9.50E-14 | Nightingale | L_VLDL_C_ | NA | T | C | -0.03517 | 0.004089 | 8.00E-18 |
| rs7046674 | miR-10a-5p | trans | 9.50E-14 | Nightingale | XS_VLDL_F | NA | T | C | -0.02251 | 0.004114 | 7.80E-09 |
| rs7046674 | miR-10a-5p | trans | 9.50E-14 | Nightingale | S_VLDL_TG | NA | T | C | 0.026025 | 0.004053 | 5.20E-10 |
| rs7046674 | miR-10a-5p | trans | 9.50E-14 | Nightingale | M_VLDL_P | NA | T | C | -0.02881 | 0.004018 | 1.30E-12 |
| rs7046674 | miR-10a-5p | trans | 9.50E-14 | Nightingale | IDL_FC | NA | T | C | -0.02526 | 0.004054 | 4.20E-10 |
| rs7046674 | miR-10a-5p | trans | 9.50E-14 | Nightingale | M_LDL_P | NA | T | C | -0.02341 | 0.004199 | 1.20E-08 |
| rs7046674 | miR-126-3p | trans | 6.62E-19 | Nightingale | Clinical_LD | NA | T | C | -0.02355 | 0.004163 | 1.30E-08 |
| rs7046674 | miR-126-3p | trans | 6.62E-19 | Nightingale | VLDL_CE | NA | T | C | -0.02189 | 0.004188 | 3.80E-08 |

| | | | | | | | | | | | |
|---|---|---|---|---|---|---|---|---|---|---|---|
| rs7046674 | miR-126-3p | trans | 6.62E-19 | Nightingale | L_LDL_PL | NA | T | C | -0.02451 | 0.004155 | 2.50E-09 |
| rs7046674 | miR-126-3p | trans | 6.62E-19 | Nightingale | XL_VLDL_C | NA | T | C | -0.02581 | 0.004015 | 9.30E-10 |
| rs7046674 | miR-126-3p | trans | 6.62E-19 | Nightingale | M_VLDL_C | NA | T | C | -0.02665 | 0.004149 | 6.70E-11 |
| rs7046674 | miR-126-3p | trans | 6.62E-19 | Nightingale | XL_VLDL_F | NA | T | C | -0.02491 | 0.004059 | 1.20E-09 |
| rs7046674 | miR-126-3p | trans | 6.62E-19 | Nightingale | XL_VLDL_T | NA | T | C | 0.029538 | 0.004058 | 1.50E-12 |
| rs7046674 | miR-126-3p | trans | 6.62E-19 | Nightingale | IDL_P | NA | T | C | -0.02532 | 0.004143 | 5.70E-10 |
| rs7046674 | miR-126-3p | trans | 6.62E-19 | Nightingale | XS_VLDL_C | NA | T | C | -0.02442 | 0.004064 | 6.00E-10 |
| rs7046674 | miR-126-3p | trans | 6.62E-19 | Nightingale | IDL_L | NA | T | C | -0.0229 | 0.004048 | 1.40E-08 |
| rs7046674 | miR-126-3p | trans | 6.62E-19 | Nightingale | Remnant_( | NA | T | C | -0.02302 | 0.00416 | 7.30E-09 |
| rs7046674 | miR-126-3p | trans | 6.62E-19 | Nightingale | XL_VLDL_C | NA | T | C | -0.02519 | 0.004017 | 3.00E-09 |
| rs7046674 | miR-126-3p | trans | 6.62E-19 | Nightingale | S_VLDL_C | NA | T | C | -0.0234 | 0.004187 | 5.40E-09 |
| rs7046674 | miR-126-3p | trans | 6.62E-19 | Nightingale | S_VLDL_C_ | NA | T | C | -0.02853 | 0.004075 | 8.90E-12 |
| rs7046674 | miR-126-3p | trans | 6.62E-19 | Nightingale | XXL_VLDL_ | NA | T | C | -0.02444 | 0.004217 | 6.70E-09 |
| rs7046674 | miR-126-3p | trans | 6.62E-19 | Nightingale | IDL_PL | NA | T | C | -0.02256 | 0.004041 | 1.70E-08 |
| rs7046674 | miR-126-3p | trans | 6.62E-19 | Nightingale | XXL_VLDL_ | NA | T | C | -0.02528 | 0.004194 | 1.80E-09 |
| rs7046674 | miR-126-3p | trans | 6.62E-19 | Nightingale | M_VLDL_T | NA | T | C | 0.025093 | 0.003957 | 1.20E-09 |
| rs7046674 | miR-126-3p | trans | 6.62E-19 | Nightingale | S_LDL_C_p | NA | T | C | -0.0297 | 0.004203 | 1.20E-12 |
| rs7046674 | miR-126-3p | trans | 6.62E-19 | Nightingale | S_VLDL_CE | NA | T | C | -0.03245 | 0.004131 | 6.70E-15 |
| rs7046674 | miR-126-3p | trans | 6.62E-19 | Nightingale | S_VLDL_CE | NA | T | C | -0.02331 | 0.004182 | 5.00E-09 |
| rs7046674 | miR-126-3p | trans | 6.62E-19 | Nightingale | L_VLDL_FC | NA | T | C | -0.03163 | 0.004147 | 1.30E-14 |
| rs7046674 | miR-126-3p | trans | 6.62E-19 | Nightingale | L_VLDL_CE | NA | T | C | -0.03189 | 0.004007 | 3.50E-15 |
| rs7046674 | miR-126-3p | trans | 6.62E-19 | Nightingale | XXL_VLDL_ | NA | T | C | 0.026027 | 0.004222 | 3.40E-10 |
| rs7046674 | miR-126-3p | trans | 6.62E-19 | Nightingale | non_HDL_( | NA | T | C | -0.02236 | 0.004164 | 4.20E-08 |
| rs7046674 | miR-126-3p | trans | 6.62E-19 | Nightingale | XL_VLDL_P | NA | T | C | -0.02456 | 0.004193 | 1.30E-09 |
| rs7046674 | miR-126-3p | trans | 6.62E-19 | Nightingale | S_VLDL_FC | NA | T | C | -0.02306 | 0.004188 | 1.20E-08 |
| rs7046674 | miR-126-3p | trans | 6.62E-19 | Nightingale | IDL_C | NA | T | C | -0.0232 | 0.004044 | 8.90E-09 |
| rs7046674 | miR-126-3p | trans | 6.62E-19 | Nightingale | XS_VLDL_P | NA | T | C | -0.02224 | 0.004132 | 1.00E-08 |
| rs7046674 | miR-126-3p | trans | 6.62E-19 | Nightingale | M_VLDL_C | NA | T | C | -0.02445 | 0.004175 | 1.30E-09 |
| rs7046674 | miR-126-3p | trans | 6.62E-19 | Nightingale | M_VLDL_C | NA | T | C | -0.02359 | 0.003944 | 1.40E-08 |
| rs7046674 | miR-126-3p | trans | 6.62E-19 | Nightingale | IDL_CE | NA | T | C | -0.02226 | 0.004043 | 3.20E-08 |
| rs7046674 | miR-126-3p | trans | 6.62E-19 | Nightingale | M_VLDL_F | NA | T | C | -0.0268 | 0.003984 | 6.60E-11 |
| rs7046674 | miR-126-3p | trans | 6.62E-19 | Nightingale | XS_VLDL_C | NA | T | C | -0.02485 | 0.004041 | 2.80E-10 |
| rs7046674 | miR-126-3p | trans | 6.62E-19 | Nightingale | L_VLDL_TG | NA | T | C | 0.03358 | 0.004167 | 1.80E-16 |
| rs7046674 | miR-126-3p | trans | 6.62E-19 | Nightingale | L_VLDL_C_ | NA | T | C | -0.03517 | 0.004089 | 8.00E-18 |
| rs7046674 | miR-126-3p | trans | 6.62E-19 | Nightingale | XS_VLDL_F | NA | T | C | -0.02251 | 0.004114 | 7.80E-09 |
| rs7046674 | miR-126-3p | trans | 6.62E-19 | Nightingale | S_VLDL_TG | NA | T | C | 0.026025 | 0.004053 | 5.20E-10 |
| rs7046674 | miR-126-3p | trans | 6.62E-19 | Nightingale | M_VLDL_P | NA | T | C | -0.02881 | 0.004018 | 1.30E-12 |
| rs7046674 | miR-126-3p | trans | 6.62E-19 | Nightingale | IDL_FC | NA | T | C | -0.02526 | 0.004054 | 4.20E-10 |
| rs7046674 | miR-126-3p | trans | 6.62E-19 | Nightingale | M_LDL_P | NA | T | C | -0.02341 | 0.004199 | 1.20E-08 |
| rs7832219 | miR-143-3p | trans | 4.55E-10 | Nightingale | Lactate | NA | C | T | -0.02899 | 0.004615 | 3.30E-10 |
| rs7832219 | miR-143-3p | trans | 4.55E-10 | Nightingale | Pyruvate | NA | C | T | -0.03346 | 0.004616 | 4.20E-13 |
| rs7832219 | miR-145-5p | trans | 7.16E-08 | Nightingale | Lactate | NA | C | T | -0.02899 | 0.004615 | 3.30E-10 |
| rs7832219 | miR-145-5p | trans | 7.16E-08 | Nightingale | Pyruvate | NA | C | T | -0.03346 | 0.004616 | 4.20E-13 |
| rs7873635 | miR-10a-5p | trans | 2.32E-12 | Nightingale | L_VLDL_C_ | NA | C | T | 0.035379 | 0.004093 | 4.10E-18 |
| rs7873635 | miR-10a-5p | trans | 2.32E-12 | Nightingale | L_VLDL_CE | NA | C | T | 0.032184 | 0.004011 | 1.50E-15 |
| rs7873635 | miR-10a-5p | trans | 2.32E-12 | Nightingale | XXL_VLDL_ | NA | C | T | 0.024544 | 0.004221 | 5.30E-09 |
| rs7873635 | miR-10a-5p | trans | 2.32E-12 | Nightingale | S_VLDL_TG | NA | C | T | -0.02591 | 0.004057 | 5.90E-10 |
| rs7873635 | miR-10a-5p | trans | 2.32E-12 | Nightingale | S_VLDL_CE | NA | C | T | 0.032025 | 0.004135 | 1.20E-14 |
| rs7873635 | miR-10a-5p | trans | 2.32E-12 | Nightingale | M_VLDL_C | NA | C | T | 0.025351 | 0.004153 | 5.30E-10 |
| rs7873635 | miR-10a-5p | trans | 2.32E-12 | Nightingale | XXL_VLDL_ | NA | C | T | -0.02625 | 0.004225 | 2.00E-10 |
| rs7873635 | miR-10a-5p | trans | 2.32E-12 | Nightingale | S_VLDL_CE | NA | C | T | 0.021726 | 0.004186 | 4.30E-08 |
| rs7873635 | miR-10a-5p | trans | 2.32E-12 | Nightingale | S_VLDL_C_ | NA | C | T | 0.028317 | 0.004079 | 1.10E-11 |
| rs7873635 | miR-10a-5p | trans | 2.32E-12 | Nightingale | L_VLDL_FC | NA | C | T | 0.031291 | 0.004151 | 1.80E-14 |
| rs7873635 | miR-10a-5p | trans | 2.32E-12 | Nightingale | XXL_VLDL_ | NA | C | T | 0.025558 | 0.004198 | 1.00E-09 |
| rs7873635 | miR-10a-5p | trans | 2.32E-12 | Nightingale | M_VLDL_P | NA | C | T | 0.028802 | 0.004022 | 1.00E-12 |
| rs7873635 | miR-10a-5p | trans | 2.32E-12 | Nightingale | IDL_FC | NA | C | T | 0.02448 | 0.004058 | 1.50E-09 |
| rs7873635 | miR-10a-5p | trans | 2.32E-12 | Nightingale | Remnant_( | NA | C | T | 0.021758 | 0.004164 | 4.70E-08 |
| rs7873635 | miR-10a-5p | trans | 2.32E-12 | Nightingale | XL_VLDL_C | NA | C | T | 0.026244 | 0.004019 | 4.30E-10 |
| rs7873635 | miR-10a-5p | trans | 2.32E-12 | Nightingale | XL_VLDL_F | NA | C | T | 0.025169 | 0.004063 | 7.80E-10 |
| rs7873635 | miR-10a-5p | trans | 2.32E-12 | Nightingale | M_VLDL_C | NA | C | T | 0.023103 | 0.004179 | 9.50E-09 |
| rs7873635 | miR-10a-5p | trans | 2.32E-12 | Nightingale | M_VLDL_T | NA | C | T | -0.02539 | 0.003961 | 6.60E-10 |
| rs7873635 | miR-10a-5p | trans | 2.32E-12 | Nightingale | M_VLDL_F | NA | C | T | 0.026997 | 0.003987 | 4.00E-11 |
| rs7873635 | miR-10a-5p | trans | 2.32E-12 | Nightingale | IDL_C | NA | C | T | 0.022431 | 0.004048 | 2.80E-08 |

| | | | | | | | | | | | |
|---|---|---|---|---|---|---|---|---|---|---|---|
| rs7873635 | miR-10a-5p | trans | 2.32E-12 | Nightingale | L_VLDL_TG | NA | C | T | -0.0331 | 0.004171 | 3.40E-16 |
| rs7873635 | miR-10a-5p | trans | 2.32E-12 | Nightingale | IDL_L | NA | C | T | 0.022016 | 0.004052 | 4.90E-08 |
| rs7873635 | miR-10a-5p | trans | 2.32E-12 | Nightingale | XS_VLDL_C | NA | C | T | 0.023605 | 0.004045 | 1.90E-09 |
| rs7873635 | miR-10a-5p | trans | 2.32E-12 | Nightingale | XL_VLDL_T | NA | C | T | -0.0299 | 0.004062 | 7.00E-13 |
| rs7873635 | miR-10a-5p | trans | 2.32E-12 | Nightingale | L_LDL_PL | NA | C | T | 0.023563 | 0.004159 | 1.10E-08 |
| rs7873635 | miR-10a-5p | trans | 2.32E-12 | Nightingale | XL_VLDL_P | NA | C | T | 0.023547 | 0.004197 | 5.00E-09 |
| rs7873635 | miR-10a-5p | trans | 2.32E-12 | Nightingale | M_LDL_P | NA | C | T | 0.022525 | 0.004203 | 4.40E-08 |
| rs7873635 | miR-10a-5p | trans | 2.32E-12 | Nightingale | IDL_P | NA | C | T | 0.024159 | 0.004147 | 3.20E-09 |
| rs7873635 | miR-10a-5p | trans | 2.32E-12 | Nightingale | S_VLDL_C | NA | C | T | 0.021871 | 0.004191 | 4.40E-08 |
| rs7873635 | miR-10a-5p | trans | 2.32E-12 | Nightingale | XXL_VLDL_ | NA | C | T | 0.023062 | 0.004171 | 3.20E-08 |
| rs7873635 | miR-10a-5p | trans | 2.32E-12 | Nightingale | M_VLDL_C | NA | C | T | 0.023938 | 0.003948 | 7.30E-09 |
| rs7873635 | miR-10a-5p | trans | 2.32E-12 | Nightingale | XS_VLDL_C | NA | C | T | 0.023128 | 0.004068 | 4.20E-09 |
| rs7873635 | miR-10a-5p | trans | 2.32E-12 | Nightingale | S_LDL_C_p | NA | C | T | 0.029512 | 0.004207 | 1.70E-12 |
| rs7873635 | miR-10a-5p | trans | 2.32E-12 | Nightingale | XL_VLDL_C | NA | C | T | 0.025622 | 0.004021 | 1.40E-09 |
| rs7873635 | miR-126-3p | trans | 1.04E-17 | Nightingale | L_VLDL_C_ | NA | C | T | 0.035379 | 0.004093 | 4.10E-18 |
| rs7873635 | miR-126-3p | trans | 1.04E-17 | Nightingale | L_VLDL_CE | NA | C | T | 0.032184 | 0.004011 | 1.50E-15 |
| rs7873635 | miR-126-3p | trans | 1.04E-17 | Nightingale | XXL_VLDL_ | NA | C | T | 0.024544 | 0.004221 | 5.30E-09 |
| rs7873635 | miR-126-3p | trans | 1.04E-17 | Nightingale | S_VLDL_TG | NA | C | T | -0.02591 | 0.004057 | 5.90E-10 |
| rs7873635 | miR-126-3p | trans | 1.04E-17 | Nightingale | S_VLDL_CE | NA | C | T | 0.032025 | 0.004135 | 1.20E-14 |
| rs7873635 | miR-126-3p | trans | 1.04E-17 | Nightingale | M_VLDL_C | NA | C | T | 0.025351 | 0.004153 | 5.30E-10 |
| rs7873635 | miR-126-3p | trans | 1.04E-17 | Nightingale | XXL_VLDL_ | NA | C | T | -0.02625 | 0.004225 | 2.00E-10 |
| rs7873635 | miR-126-3p | trans | 1.04E-17 | Nightingale | S_VLDL_CE | NA | C | T | 0.021726 | 0.004186 | 4.30E-08 |
| rs7873635 | miR-126-3p | trans | 1.04E-17 | Nightingale | S_VLDL_C_ | NA | C | T | 0.028317 | 0.004079 | 1.10E-11 |
| rs7873635 | miR-126-3p | trans | 1.04E-17 | Nightingale | L_VLDL_FC | NA | C | T | 0.031291 | 0.004151 | 1.80E-14 |
| rs7873635 | miR-126-3p | trans | 1.04E-17 | Nightingale | XXL_VLDL_ | NA | C | T | 0.025558 | 0.004198 | 1.00E-09 |
| rs7873635 | miR-126-3p | trans | 1.04E-17 | Nightingale | M_VLDL_P | NA | C | T | 0.028802 | 0.004022 | 1.00E-12 |
| rs7873635 | miR-126-3p | trans | 1.04E-17 | Nightingale | IDL_FC | NA | C | T | 0.02448 | 0.004058 | 1.50E-09 |
| rs7873635 | miR-126-3p | trans | 1.04E-17 | Nightingale | Remnant_( | NA | C | T | 0.021758 | 0.004164 | 4.70E-08 |
| rs7873635 | miR-126-3p | trans | 1.04E-17 | Nightingale | XL_VLDL_C | NA | C | T | 0.026244 | 0.004019 | 4.30E-10 |
| rs7873635 | miR-126-3p | trans | 1.04E-17 | Nightingale | XL_VLDL_F | NA | C | T | 0.025169 | 0.004063 | 7.80E-10 |
| rs7873635 | miR-126-3p | trans | 1.04E-17 | Nightingale | M_VLDL_C | NA | C | T | 0.023103 | 0.004179 | 9.50E-09 |
| rs7873635 | miR-126-3p | trans | 1.04E-17 | Nightingale | M_VLDL_T | NA | C | T | -0.02539 | 0.003961 | 6.60E-10 |
| rs7873635 | miR-126-3p | trans | 1.04E-17 | Nightingale | M_VLDL_F | NA | C | T | 0.026997 | 0.003987 | 4.00E-11 |
| rs7873635 | miR-126-3p | trans | 1.04E-17 | Nightingale | IDL_C | NA | C | T | 0.022431 | 0.004048 | 2.80E-08 |
| rs7873635 | miR-126-3p | trans | 1.04E-17 | Nightingale | L_VLDL_TG | NA | C | T | -0.0331 | 0.004171 | 3.40E-16 |
| rs7873635 | miR-126-3p | trans | 1.04E-17 | Nightingale | IDL_L | NA | C | T | 0.022016 | 0.004052 | 4.90E-08 |
| rs7873635 | miR-126-3p | trans | 1.04E-17 | Nightingale | XS_VLDL_C | NA | C | T | 0.023605 | 0.004045 | 1.90E-09 |
| rs7873635 | miR-126-3p | trans | 1.04E-17 | Nightingale | XL_VLDL_T | NA | C | T | -0.0299 | 0.004062 | 7.00E-13 |
| rs7873635 | miR-126-3p | trans | 1.04E-17 | Nightingale | L_LDL_PL | NA | C | T | 0.023563 | 0.004159 | 1.10E-08 |
| rs7873635 | miR-126-3p | trans | 1.04E-17 | Nightingale | XL_VLDL_P | NA | C | T | 0.023547 | 0.004197 | 5.00E-09 |
| rs7873635 | miR-126-3p | trans | 1.04E-17 | Nightingale | M_LDL_P | NA | C | T | 0.022525 | 0.004203 | 4.40E-08 |
| rs7873635 | miR-126-3p | trans | 1.04E-17 | Nightingale | IDL_P | NA | C | T | 0.024159 | 0.004147 | 3.20E-09 |
| rs7873635 | miR-126-3p | trans | 1.04E-17 | Nightingale | S_VLDL_C | NA | C | T | 0.021871 | 0.004191 | 4.40E-08 |
| rs7873635 | miR-126-3p | trans | 1.04E-17 | Nightingale | XXL_VLDL_ | NA | C | T | 0.023062 | 0.004171 | 3.20E-08 |
| rs7873635 | miR-126-3p | trans | 1.04E-17 | Nightingale | M_VLDL_C | NA | C | T | 0.023938 | 0.003948 | 7.30E-09 |
| rs7873635 | miR-126-3p | trans | 1.04E-17 | Nightingale | XS_VLDL_C | NA | C | T | 0.023128 | 0.004068 | 4.20E-09 |
| rs7873635 | miR-126-3p | trans | 1.04E-17 | Nightingale | S_LDL_C_p | NA | C | T | 0.029512 | 0.004207 | 1.70E-12 |
| rs7873635 | miR-126-3p | trans | 1.04E-17 | Nightingale | XL_VLDL_C | NA | C | T | 0.025622 | 0.004021 | 1.40E-09 |
| rs8176649 | miR-126-3p | trans | 7.78E-19 | Nightingale | XXL_VLDL_ | NA | A | G | -0.02534 | 0.004195 | 1.70E-09 |
| rs8176649 | miR-126-3p | trans | 7.78E-19 | Nightingale | VLDL_CE | NA | A | G | -0.02181 | 0.004189 | 4.30E-08 |
| rs8176649 | miR-126-3p | trans | 7.78E-19 | Nightingale | XS_VLDL_C | NA | A | G | -0.0244 | 0.004065 | 6.40E-10 |
| rs8176649 | miR-126-3p | trans | 7.78E-19 | Nightingale | XXL_VLDL_ | NA | A | G | 0.026063 | 0.004222 | 3.50E-10 |
| rs8176649 | miR-126-3p | trans | 7.78E-19 | Nightingale | XL_VLDL_T | NA | A | G | 0.029644 | 0.004059 | 1.30E-12 |
| rs8176649 | miR-126-3p | trans | 7.78E-19 | Nightingale | S_VLDL_TG | NA | A | G | 0.026074 | 0.004053 | 4.90E-10 |
| rs8176649 | miR-126-3p | trans | 7.78E-19 | Nightingale | IDL_L | NA | A | G | -0.02287 | 0.004049 | 1.40E-08 |
| rs8176649 | miR-126-3p | trans | 7.78E-19 | Nightingale | IDL_PL | NA | A | G | -0.02254 | 0.004041 | 1.70E-08 |
| rs8176649 | miR-126-3p | trans | 7.78E-19 | Nightingale | IDL_CE | NA | A | G | -0.02223 | 0.004043 | 3.40E-08 |
| rs8176649 | miR-126-3p | trans | 7.78E-19 | Nightingale | L_VLDL_TG | NA | A | G | 0.033649 | 0.004168 | 1.70E-16 |
| rs8176649 | miR-126-3p | trans | 7.78E-19 | Nightingale | XXL_VLDL_ | NA | A | G | -0.0245 | 0.004218 | 6.50E-09 |
| rs8176649 | miR-126-3p | trans | 7.78E-19 | Nightingale | M_LDL_P | NA | A | G | -0.02333 | 0.004199 | 1.40E-08 |
| rs8176649 | miR-126-3p | trans | 7.78E-19 | Nightingale | IDL_C | NA | A | G | -0.02316 | 0.004044 | 9.40E-09 |
| rs8176649 | miR-126-3p | trans | 7.78E-19 | Nightingale | XS_VLDL_C | NA | A | G | -0.02483 | 0.004041 | 3.00E-10 |
| rs8176649 | miR-126-3p | trans | 7.78E-19 | Nightingale | M_VLDL_C | NA | A | G | -0.02367 | 0.003944 | 1.20E-08 |

| | | | | | | | | | | | |
|---|---|---|---|---|---|---|---|---|---|---|---|
| rs8176649 | miR-126-3p | trans | 7.78E-19 | Nightingale | L_VLDL_C_ | NA | A | G | -0.03529 | 0.00409 | 6.50E-18 |
| rs8176649 | miR-126-3p | trans | 7.78E-19 | Nightingale | IDL_FC | NA | A | G | -0.02522 | 0.004054 | 4.50E-10 |
| rs8176649 | miR-126-3p | trans | 7.78E-19 | Nightingale | XS_VLDL_F | NA | A | G | -0.02249 | 0.004115 | 8.20E-09 |
| rs8176649 | miR-126-3p | trans | 7.78E-19 | Nightingale | S_VLDL_CE | NA | A | G | -0.03253 | 0.004131 | 6.00E-15 |
| rs8176649 | miR-126-3p | trans | 7.78E-19 | Nightingale | XL_VLDL_F | NA | A | G | -0.02504 | 0.004059 | 1.10E-09 |
| rs8176649 | miR-126-3p | trans | 7.78E-19 | Nightingale | L_VLDL_CE | NA | A | G | -0.03201 | 0.004007 | 2.90E-15 |
| rs8176649 | miR-126-3p | trans | 7.78E-19 | Nightingale | S_VLDL_CE | NA | A | G | -0.02324 | 0.004182 | 5.60E-09 |
| rs8176649 | miR-126-3p | trans | 7.78E-19 | Nightingale | M_VLDL_C | NA | A | G | -0.02656 | 0.004149 | 7.90E-11 |
| rs8176649 | miR-126-3p | trans | 7.78E-19 | Nightingale | M_VLDL_P | NA | A | G | -0.0289 | 0.004019 | 1.10E-12 |
| rs8176649 | miR-126-3p | trans | 7.78E-19 | Nightingale | Clinical_LD | NA | A | G | -0.02347 | 0.004163 | 1.50E-08 |
| rs8176649 | miR-126-3p | trans | 7.78E-19 | Nightingale | XS_VLDL_P | NA | A | G | -0.02223 | 0.004132 | 1.10E-08 |
| rs8176649 | miR-126-3p | trans | 7.78E-19 | Nightingale | S_VLDL_C_ | NA | A | G | -0.0286 | 0.004075 | 8.30E-12 |
| rs8176649 | miR-126-3p | trans | 7.78E-19 | Nightingale | S_VLDL_FC | NA | A | G | -0.02296 | 0.004188 | 1.30E-08 |
| rs8176649 | miR-126-3p | trans | 7.78E-19 | Nightingale | M_VLDL_F | NA | A | G | -0.02687 | 0.003984 | 5.90E-11 |
| rs8176649 | miR-126-3p | trans | 7.78E-19 | Nightingale | L_VLDL_FC | NA | A | G | -0.0317 | 0.004147 | 1.20E-14 |
| rs8176649 | miR-126-3p | trans | 7.78E-19 | Nightingale | XL_VLDL_C | NA | A | G | -0.02593 | 0.004015 | 7.90E-10 |
| rs8176649 | miR-126-3p | trans | 7.78E-19 | Nightingale | XL_VLDL_C | NA | A | G | -0.0253 | 0.004017 | 2.60E-09 |
| rs8176649 | miR-126-3p | trans | 7.78E-19 | Nightingale | M_VLDL_C | NA | A | G | -0.02435 | 0.004176 | 1.50E-09 |
| rs8176649 | miR-126-3p | trans | 7.78E-19 | Nightingale | non_HDL_( | NA | A | G | -0.02227 | 0.004165 | 4.60E-08 |
| rs8176649 | miR-126-3p | trans | 7.78E-19 | Nightingale | S_VLDL_C | NA | A | G | -0.02332 | 0.004187 | 6.10E-09 |
| rs8176649 | miR-126-3p | trans | 7.78E-19 | Nightingale | Remnant_( | NA | A | G | -0.02295 | 0.00416 | 8.10E-09 |
| rs8176649 | miR-126-3p | trans | 7.78E-19 | Nightingale | M_VLDL_T | NA | A | G | 0.025177 | 0.003957 | 1.10E-09 |
| rs8176649 | miR-126-3p | trans | 7.78E-19 | Nightingale | L_LDL_PL | NA | A | G | -0.02444 | 0.004156 | 2.80E-09 |
| rs8176649 | miR-126-3p | trans | 7.78E-19 | Nightingale | IDL_P | NA | A | G | -0.0253 | 0.004144 | 5.90E-10 |
| rs8176649 | miR-126-3p | trans | 7.78E-19 | Nightingale | XL_VLDL_P | NA | A | G | -0.02452 | 0.004194 | 1.40E-09 |
| rs8176649 | miR-126-3p | trans | 7.78E-19 | Nightingale | S_LDL_C_p | NA | A | G | -0.02953 | 0.004204 | 1.60E-12 |
| rs8176649 | miR-10a-5p | trans | 9.35E-14 | Nightingale | XXL_VLDL_ | NA | A | G | -0.02534 | 0.004195 | 1.70E-09 |
| rs8176649 | miR-10a-5p | trans | 9.35E-14 | Nightingale | VLDL_CE | NA | A | G | -0.02181 | 0.004189 | 4.30E-08 |
| rs8176649 | miR-10a-5p | trans | 9.35E-14 | Nightingale | XS_VLDL_C | NA | A | G | -0.0244 | 0.004065 | 6.40E-10 |
| rs8176649 | miR-10a-5p | trans | 9.35E-14 | Nightingale | XXL_VLDL_ | NA | A | G | 0.026063 | 0.004222 | 3.50E-10 |
| rs8176649 | miR-10a-5p | trans | 9.35E-14 | Nightingale | XL_VLDL_T | NA | A | G | 0.029644 | 0.004059 | 1.30E-12 |
| rs8176649 | miR-10a-5p | trans | 9.35E-14 | Nightingale | S_VLDL_TG | NA | A | G | 0.026074 | 0.004053 | 4.90E-10 |
| rs8176649 | miR-10a-5p | trans | 9.35E-14 | Nightingale | IDL_L | NA | A | G | -0.02287 | 0.004049 | 1.40E-08 |
| rs8176649 | miR-10a-5p | trans | 9.35E-14 | Nightingale | IDL_PL | NA | A | G | -0.02254 | 0.004041 | 1.70E-08 |
| rs8176649 | miR-10a-5p | trans | 9.35E-14 | Nightingale | IDL_CE | NA | A | G | -0.02223 | 0.004043 | 3.40E-08 |
| rs8176649 | miR-10a-5p | trans | 9.35E-14 | Nightingale | L_VLDL_TG | NA | A | G | 0.033649 | 0.004168 | 1.70E-16 |
| rs8176649 | miR-10a-5p | trans | 9.35E-14 | Nightingale | XXL_VLDL_ | NA | A | G | -0.0245 | 0.004218 | 6.50E-09 |
| rs8176649 | miR-10a-5p | trans | 9.35E-14 | Nightingale | M_LDL_P | NA | A | G | -0.02333 | 0.004199 | 1.40E-08 |
| rs8176649 | miR-10a-5p | trans | 9.35E-14 | Nightingale | IDL_C | NA | A | G | -0.02316 | 0.004044 | 9.40E-09 |
| rs8176649 | miR-10a-5p | trans | 9.35E-14 | Nightingale | XS_VLDL_C | NA | A | G | -0.02483 | 0.004041 | 3.00E-10 |
| rs8176649 | miR-10a-5p | trans | 9.35E-14 | Nightingale | M_VLDL_C | NA | A | G | -0.02367 | 0.003944 | 1.20E-08 |
| rs8176649 | miR-10a-5p | trans | 9.35E-14 | Nightingale | L_VLDL_C_ | NA | A | G | -0.03529 | 0.00409 | 6.50E-18 |
| rs8176649 | miR-10a-5p | trans | 9.35E-14 | Nightingale | IDL_FC | NA | A | G | -0.02522 | 0.004054 | 4.50E-10 |
| rs8176649 | miR-10a-5p | trans | 9.35E-14 | Nightingale | XS_VLDL_F | NA | A | G | -0.02249 | 0.004115 | 8.20E-09 |
| rs8176649 | miR-10a-5p | trans | 9.35E-14 | Nightingale | S_VLDL_CE | NA | A | G | -0.03253 | 0.004131 | 6.00E-15 |
| rs8176649 | miR-10a-5p | trans | 9.35E-14 | Nightingale | XL_VLDL_F | NA | A | G | -0.02504 | 0.004059 | 1.10E-09 |
| rs8176649 | miR-10a-5p | trans | 9.35E-14 | Nightingale | L_VLDL_CE | NA | A | G | -0.03201 | 0.004007 | 2.90E-15 |
| rs8176649 | miR-10a-5p | trans | 9.35E-14 | Nightingale | S_VLDL_CE | NA | A | G | -0.02324 | 0.004182 | 5.60E-09 |
| rs8176649 | miR-10a-5p | trans | 9.35E-14 | Nightingale | M_VLDL_C | NA | A | G | -0.02656 | 0.004149 | 7.90E-11 |
| rs8176649 | miR-10a-5p | trans | 9.35E-14 | Nightingale | M_VLDL_P | NA | A | G | -0.0289 | 0.004019 | 1.10E-12 |
| rs8176649 | miR-10a-5p | trans | 9.35E-14 | Nightingale | Clinical_LD | NA | A | G | -0.02347 | 0.004163 | 1.50E-08 |
| rs8176649 | miR-10a-5p | trans | 9.35E-14 | Nightingale | XS_VLDL_P | NA | A | G | -0.02223 | 0.004132 | 1.10E-08 |
| rs8176649 | miR-10a-5p | trans | 9.35E-14 | Nightingale | S_VLDL_C_ | NA | A | G | -0.0286 | 0.004075 | 8.30E-12 |
| rs8176649 | miR-10a-5p | trans | 9.35E-14 | Nightingale | S_VLDL_FC | NA | A | G | -0.02296 | 0.004188 | 1.30E-08 |
| rs8176649 | miR-10a-5p | trans | 9.35E-14 | Nightingale | M_VLDL_F | NA | A | G | -0.02687 | 0.003984 | 5.90E-11 |
| rs8176649 | miR-10a-5p | trans | 9.35E-14 | Nightingale | L_VLDL_FC | NA | A | G | -0.0317 | 0.004147 | 1.20E-14 |
| rs8176649 | miR-10a-5p | trans | 9.35E-14 | Nightingale | XL_VLDL_C | NA | A | G | -0.02593 | 0.004015 | 7.90E-10 |
| rs8176649 | miR-10a-5p | trans | 9.35E-14 | Nightingale | XL_VLDL_C | NA | A | G | -0.0253 | 0.004017 | 2.60E-09 |
| rs8176649 | miR-10a-5p | trans | 9.35E-14 | Nightingale | M_VLDL_C | NA | A | G | -0.02435 | 0.004176 | 1.50E-09 |
| rs8176649 | miR-10a-5p | trans | 9.35E-14 | Nightingale | non_HDL_( | NA | A | G | -0.02227 | 0.004165 | 4.60E-08 |
| rs8176649 | miR-10a-5p | trans | 9.35E-14 | Nightingale | S_VLDL_C | NA | A | G | -0.02332 | 0.004187 | 6.10E-09 |
| rs8176649 | miR-10a-5p | trans | 9.35E-14 | Nightingale | Remnant_( | NA | A | G | -0.02295 | 0.00416 | 8.10E-09 |
| rs8176649 | miR-10a-5p | trans | 9.35E-14 | Nightingale | M_VLDL_T | NA | A | G | 0.025177 | 0.003957 | 1.10E-09 |

| | | | | | | | | | | | |
|---|---|---|---|---|---|---|---|---|---|---|---|
| rs8176649 | miR-10a-5p | trans | 9.35E-14 | Nightingale | L_LDL_PL | NA | A | G | -0.02444 | 0.004156 | 2.80E-09 |
| rs8176649 | miR-10a-5p | trans | 9.35E-14 | Nightingale | IDL_P | NA | A | G | -0.0253 | 0.004144 | 5.90E-10 |
| rs8176649 | miR-10a-5p | trans | 9.35E-14 | Nightingale | XL_VLDL_P | NA | A | G | -0.02452 | 0.004194 | 1.40E-09 |
| rs8176649 | miR-10a-5p | trans | 9.35E-14 | Nightingale | S_LDL_C_p | NA | A | G | -0.02953 | 0.004204 | 1.60E-12 |
| rs8176663 | let-7b-5p | trans | 1.07E-26 | Nightingale | L_LDL_P | NA | C | T | 0.029177 | 0.00441 | 6.20E-11 |
| rs8176663 | let-7b-5p | trans | 1.07E-26 | Nightingale | S_VLDL_C | NA | C | T | 0.038373 | 0.004424 | 1.80E-18 |
| rs8176663 | let-7b-5p | trans | 1.07E-26 | Nightingale | L_LDL_CE | NA | C | T | 0.031569 | 0.004385 | 9.80E-13 |
| rs8176663 | let-7b-5p | trans | 1.07E-26 | Nightingale | LDL_CE | NA | C | T | 0.032556 | 0.004413 | 2.20E-13 |
| rs8176663 | let-7b-5p | trans | 1.07E-26 | Nightingale | XL_VLDL_F | NA | C | T | 0.037195 | 0.00429 | 2.00E-18 |
| rs8176663 | let-7b-5p | trans | 1.07E-26 | Nightingale | S_HDL_L | NA | C | T | 0.026297 | 0.00441 | 1.50E-09 |
| rs8176663 | let-7b-5p | trans | 1.07E-26 | Nightingale | S_LDL_C | NA | C | T | 0.030946 | 0.004429 | 5.50E-12 |
| rs8176663 | let-7b-5p | trans | 1.07E-26 | Nightingale | PUFA | NA | C | T | 0.023713 | 0.004316 | 1.40E-08 |
| rs8176663 | let-7b-5p | trans | 1.07E-26 | Nightingale | L_VLDL_TG | NA | C | T | -0.05114 | 0.004404 | 5.30E-32 |
| rs8176663 | let-7b-5p | trans | 1.07E-26 | Nightingale | IDL_FC | NA | C | T | 0.038254 | 0.004284 | 1.50E-18 |
| rs8176663 | let-7b-5p | trans | 1.07E-26 | Nightingale | L_LDL_PL | NA | C | T | 0.037992 | 0.004391 | 9.00E-18 |
| rs8176663 | let-7b-5p | trans | 1.07E-26 | Nightingale | S_VLDL_L | NA | C | T | 0.024668 | 0.004382 | 3.70E-09 |
| rs8176663 | let-7b-5p | trans | 1.07E-26 | Nightingale | M_VLDL_P | NA | C | T | 0.044056 | 0.004247 | 5.50E-25 |
| rs8176663 | let-7b-5p | trans | 1.07E-26 | Nightingale | S_LDL_L | NA | C | T | 0.027745 | 0.004424 | 8.20E-10 |
| rs8176663 | let-7b-5p | trans | 1.07E-26 | Nightingale | S_LDL_FC | NA | C | T | 0.028568 | 0.004418 | 4.20E-10 |
| rs8176663 | let-7b-5p | trans | 1.07E-26 | Nightingale | XL_VLDL_C | NA | C | T | 0.038222 | 0.004243 | 4.70E-19 |
| rs8176663 | let-7b-5p | trans | 1.07E-26 | Nightingale | Sphingomy | NA | C | T | 0.029016 | 0.004136 | 6.80E-12 |
| rs8176663 | let-7b-5p | trans | 1.07E-26 | Nightingale | IDL_C | NA | C | T | 0.034955 | 0.004273 | 9.40E-16 |
| rs8176663 | let-7b-5p | trans | 1.07E-26 | Nightingale | IDL_FC_pct | NA | C | T | 0.025773 | 0.004391 | 1.10E-08 |
| rs8176663 | let-7b-5p | trans | 1.07E-26 | Nightingale | GlycA | NA | C | T | 0.028013 | 0.004406 | 1.40E-10 |
| rs8176663 | let-7b-5p | trans | 1.07E-26 | Nightingale | Remnant_( | NA | C | T | 0.036867 | 0.004396 | 3.70E-17 |
| rs8176663 | let-7b-5p | trans | 1.07E-26 | Nightingale | S_VLDL_TG | NA | C | T | -0.03813 | 0.004283 | 1.40E-18 |
| rs8176663 | let-7b-5p | trans | 1.07E-26 | Nightingale | XS_VLDL_C | NA | C | T | 0.037794 | 0.00427 | 2.10E-18 |
| rs8176663 | let-7b-5p | trans | 1.07E-26 | Nightingale | S_VLDL_PL | NA | C | T | 0.032777 | 0.004417 | 4.20E-14 |
| rs8176663 | let-7b-5p | trans | 1.07E-26 | Nightingale | IDL_L | NA | C | T | 0.035268 | 0.004278 | 4.80E-16 |
| rs8176663 | let-7b-5p | trans | 1.07E-26 | Nightingale | Cholines | NA | C | T | 0.022288 | 0.004161 | 4.90E-08 |
| rs8176663 | let-7b-5p | trans | 1.07E-26 | Nightingale | M_VLDL_C | NA | C | T | 0.036205 | 0.004168 | 1.20E-17 |
| rs8176663 | let-7b-5p | trans | 1.07E-26 | Nightingale | LDL_PL | NA | C | T | 0.034991 | 0.004415 | 4.70E-15 |
| rs8176663 | let-7b-5p | trans | 1.07E-26 | Nightingale | XXL_VLDL_ | NA | C | T | 0.033119 | 0.004459 | 9.00E-14 |
| rs8176663 | let-7b-5p | trans | 1.07E-26 | Nightingale | LDL_L | NA | C | T | 0.033306 | 0.00441 | 7.40E-14 |
| rs8176663 | let-7b-5p | trans | 1.07E-26 | Nightingale | L_VLDL_C_ | NA | C | T | 0.052192 | 0.004322 | 4.00E-34 |
| rs8176663 | let-7b-5p | trans | 1.07E-26 | Nightingale | S_VLDL_PL | NA | C | T | 0.02774 | 0.004237 | 1.80E-10 |
| rs8176663 | let-7b-5p | trans | 1.07E-26 | Nightingale | M_LDL_L | NA | C | T | 0.033035 | 0.004431 | 1.00E-13 |
| rs8176663 | let-7b-5p | trans | 1.07E-26 | Nightingale | XS_VLDL_C | NA | C | T | 0.037989 | 0.004295 | 1.30E-18 |
| rs8176663 | let-7b-5p | trans | 1.07E-26 | Nightingale | LA | NA | C | T | 0.024903 | 0.004339 | 3.70E-09 |
| rs8176663 | let-7b-5p | trans | 1.07E-26 | Nightingale | LDL_C | NA | C | T | 0.033196 | 0.004406 | 8.80E-14 |
| rs8176663 | let-7b-5p | trans | 1.07E-26 | Nightingale | S_VLDL_CE | NA | C | T | 0.047034 | 0.004365 | 1.20E-26 |
| rs8176663 | let-7b-5p | trans | 1.07E-26 | Nightingale | S_HDL_P | NA | C | T | 0.030615 | 0.004425 | 5.10E-12 |
| rs8176663 | let-7b-5p | trans | 1.07E-26 | Nightingale | S_HDL_FC | NA | C | T | 0.02345 | 0.004344 | 4.60E-08 |
| rs8176663 | let-7b-5p | trans | 1.07E-26 | Nightingale | Total_FC | NA | C | T | 0.033253 | 0.004323 | 6.80E-15 |
| rs8176663 | let-7b-5p | trans | 1.07E-26 | Nightingale | Clinical_LD | NA | C | T | 0.036477 | 0.004399 | 2.70E-16 |
| rs8176663 | let-7b-5p | trans | 1.07E-26 | Nightingale | XXL_VLDL_ | NA | C | T | 0.025004 | 0.004454 | 1.50E-09 |
| rs8176663 | let-7b-5p | trans | 1.07E-26 | Nightingale | XS_VLDL_F | NA | C | T | 0.031441 | 0.004393 | 2.40E-12 |
| rs8176663 | let-7b-5p | trans | 1.07E-26 | Nightingale | XL_VLDL_C | NA | C | T | 0.037351 | 0.004245 | 3.10E-18 |
| rs8176663 | let-7b-5p | trans | 1.07E-26 | Nightingale | ApoB_by_/ | NA | C | T | 0.025055 | 0.004358 | 1.30E-08 |
| rs8176663 | let-7b-5p | trans | 1.07E-26 | Nightingale | VLDL_CE | NA | C | T | 0.035786 | 0.004426 | 3.00E-16 |
| rs8176663 | let-7b-5p | trans | 1.07E-26 | Nightingale | Total_C | NA | C | T | 0.032372 | 0.004284 | 3.30E-14 |
| rs8176663 | let-7b-5p | trans | 1.07E-26 | Nightingale | L_LDL_FC | NA | C | T | 0.033627 | 0.004345 | 2.90E-14 |
| rs8176663 | let-7b-5p | trans | 1.07E-26 | Nightingale | Total_L | NA | C | T | 0.026346 | 0.004353 | 4.60E-10 |
| rs8176663 | let-7b-5p | trans | 1.07E-26 | Nightingale | S_VLDL_FC | NA | C | T | 0.037905 | 0.004426 | 7.30E-18 |
| rs8176663 | let-7b-5p | trans | 1.07E-26 | Nightingale | IDL_CE | NA | C | T | 0.033507 | 0.004272 | 1.30E-14 |
| rs8176663 | let-7b-5p | trans | 1.07E-26 | Nightingale | S_VLDL_FC | NA | C | T | 0.029358 | 0.004228 | 1.10E-11 |
| rs8176663 | let-7b-5p | trans | 1.07E-26 | Nightingale | VLDL_FC | NA | C | T | 0.024159 | 0.004392 | 1.10E-08 |
| rs8176663 | let-7b-5p | trans | 1.07E-26 | Nightingale | Omega_6 | NA | C | T | 0.025889 | 0.004321 | 5.80E-10 |
| rs8176663 | let-7b-5p | trans | 1.07E-26 | Nightingale | L_LDL_L | NA | C | T | 0.033551 | 0.004382 | 3.00E-14 |
| rs8176663 | let-7b-5p | trans | 1.07E-26 | Nightingale | Total_CE | NA | C | T | 0.031745 | 0.004268 | 9.70E-14 |
| rs8176663 | let-7b-5p | trans | 1.07E-26 | Nightingale | S_VLDL_P | NA | C | T | 0.023465 | 0.004369 | 1.70E-08 |
| rs8176663 | let-7b-5p | trans | 1.07E-26 | Nightingale | S_LDL_P | NA | C | T | 0.029589 | 0.004429 | 3.20E-11 |
| rs8176663 | let-7b-5p | trans | 1.07E-26 | Nightingale | M_LDL_PL | NA | C | T | 0.032471 | 0.004431 | 2.60E-13 |

| | | | | | | | | | | | |
|---|---|---|---|---|---|---|---|---|---|---|---|
| rs8176663 | let-7b-5p | trans | 1.07E-26 | Nightingale | M_LDL_C_ | NA | C | T | 0.031165 | 0.004428 | 2.90E-12 |
| rs8176663 | let-7b-5p | trans | 1.07E-26 | Nightingale | L_LDL_C | NA | C | T | 0.032322 | 0.004375 | 2.60E-13 |
| rs8176663 | let-7b-5p | trans | 1.07E-26 | Nightingale | M_VLDL_F | NA | C | T | 0.033372 | 0.004424 | 3.10E-14 |
| rs8176663 | let-7b-5p | trans | 1.07E-26 | Nightingale | IDL_P | NA | C | T | 0.039988 | 0.004378 | 1.30E-19 |
| rs8176663 | let-7b-5p | trans | 1.07E-26 | Nightingale | XXL_VLDL_ | NA | C | T | 0.034827 | 0.004434 | 2.90E-15 |
| rs8176663 | let-7b-5p | trans | 1.07E-26 | Nightingale | XS_VLDL_F | NA | C | T | 0.037065 | 0.004348 | 6.50E-18 |
| rs8176663 | let-7b-5p | trans | 1.07E-26 | Nightingale | M_VLDL_C | NA | C | T | 0.033628 | 0.004157 | 1.90E-15 |
| rs8176663 | let-7b-5p | trans | 1.07E-26 | Nightingale | M_VLDL_T | NA | C | T | -0.03856 | 0.004181 | 6.80E-20 |
| rs8176663 | let-7b-5p | trans | 1.07E-26 | Nightingale | S_VLDL_CE | NA | C | T | 0.038182 | 0.004419 | 1.60E-18 |
| rs8176663 | let-7b-5p | trans | 1.07E-26 | Nightingale | IDL_PL | NA | C | T | 0.035341 | 0.00427 | 3.30E-16 |
| rs8176663 | let-7b-5p | trans | 1.07E-26 | Nightingale | XXL_VLDL_ | NA | C | T | 0.030847 | 0.004406 | 1.10E-12 |
| rs8176663 | let-7b-5p | trans | 1.07E-26 | Nightingale | S_HDL_CE | NA | C | T | 0.02864 | 0.004432 | 1.30E-10 |
| rs8176663 | let-7b-5p | trans | 1.07E-26 | Nightingale | M_VLDL_C | NA | C | T | 0.038711 | 0.004413 | 1.90E-18 |
| rs8176663 | let-7b-5p | trans | 1.07E-26 | Nightingale | S_VLDL_C_ | NA | C | T | 0.041889 | 0.004306 | 5.20E-22 |
| rs8176663 | let-7b-5p | trans | 1.07E-26 | Nightingale | VLDL_C | NA | C | T | 0.031302 | 0.004419 | 4.90E-13 |
| rs8176663 | let-7b-5p | trans | 1.07E-26 | Nightingale | L_VLDL_CE | NA | C | T | 0.047336 | 0.004234 | 2.40E-29 |
| rs8176663 | let-7b-5p | trans | 1.07E-26 | Nightingale | Total_PL | NA | C | T | 0.024922 | 0.004213 | 1.10E-09 |
| rs8176663 | let-7b-5p | trans | 1.07E-26 | Nightingale | S_LDL_C_p | NA | C | T | 0.045366 | 0.004442 | 5.40E-25 |
| rs8176663 | let-7b-5p | trans | 1.07E-26 | Nightingale | XS_VLDL_P | NA | C | T | 0.036698 | 0.004366 | 1.40E-17 |
| rs8176663 | let-7b-5p | trans | 1.07E-26 | Nightingale | VLDL_P | NA | C | T | 0.026633 | 0.004401 | 4.40E-10 |
| rs8176663 | let-7b-5p | trans | 1.07E-26 | Nightingale | M_LDL_TG | NA | C | T | -0.02763 | 0.00437 | 3.20E-10 |
| rs8176663 | let-7b-5p | trans | 1.07E-26 | Nightingale | non_HDL_( | NA | C | T | 0.035698 | 0.004401 | 5.10E-16 |
| rs8176663 | let-7b-5p | trans | 1.07E-26 | Nightingale | XS_VLDL_L | NA | C | T | 0.033813 | 0.00435 | 2.10E-15 |
| rs8176663 | let-7b-5p | trans | 1.07E-26 | Nightingale | S_LDL_CE_ | NA | C | T | 0.030946 | 0.004417 | 4.90E-14 |
| rs8176663 | let-7b-5p | trans | 1.07E-26 | Nightingale | M_VLDL_F | NA | C | T | 0.042064 | 0.00421 | 3.50E-23 |
| rs8176663 | let-7b-5p | trans | 1.07E-26 | Nightingale | M_LDL_FC | NA | C | T | 0.034352 | 0.004419 | 1.90E-14 |
| rs8176663 | let-7b-5p | trans | 1.07E-26 | Nightingale | M_LDL_C | NA | C | T | 0.033991 | 0.004433 | 2.10E-14 |
| rs8176663 | let-7b-5p | trans | 1.07E-26 | Nightingale | XS_VLDL_P | NA | C | T | 0.032112 | 0.004363 | 5.40E-14 |
| rs8176663 | let-7b-5p | trans | 1.07E-26 | Nightingale | ApoB | NA | C | T | 0.033939 | 0.004419 | 2.10E-14 |
| rs8176663 | let-7b-5p | trans | 1.07E-26 | Nightingale | L_VLDL_FC | NA | C | T | 0.048588 | 0.004382 | 4.50E-29 |
| rs8176663 | let-7b-5p | trans | 1.07E-26 | Nightingale | S_HDL_C | NA | C | T | 0.028415 | 0.004417 | 1.70E-10 |
| rs8176663 | let-7b-5p | trans | 1.07E-26 | Nightingale | XL_VLDL_T | NA | C | T | -0.04392 | 0.004289 | 1.70E-24 |
| rs8176663 | let-7b-5p | trans | 1.07E-26 | Nightingale | S_LDL_CE | NA | C | T | 0.030551 | 0.004427 | 5.80E-12 |
| rs8176663 | let-7b-5p | trans | 1.07E-26 | Nightingale | LDL_P | NA | C | T | 0.031801 | 0.004422 | 1.00E-12 |
| rs8176663 | let-7b-5p | trans | 1.07E-26 | Nightingale | XXL_VLDL_ | NA | C | T | -0.03573 | 0.004463 | 2.20E-16 |
| rs8176663 | let-7b-5p | trans | 1.07E-26 | Nightingale | XL_HDL_PL | NA | C | T | -0.02277 | 0.004065 | 1.10E-08 |
| rs8176663 | let-7b-5p | trans | 1.07E-26 | Nightingale | XL_VLDL_P | NA | C | T | 0.038847 | 0.004432 | 5.10E-19 |
| rs8176663 | let-7b-5p | trans | 1.07E-26 | Nightingale | M_VLDL_C | NA | C | T | 0.040844 | 0.004385 | 2.30E-20 |
| rs8176663 | let-7b-5p | trans | 1.07E-26 | Nightingale | M_LDL_P | NA | C | T | 0.035895 | 0.004437 | 6.80E-16 |
| rs8176663 | let-7b-5p | trans | 1.07E-26 | Nightingale | M_VLDL_P | NA | C | T | 0.029128 | 0.004418 | 2.00E-11 |
| rs8176663 | let-7b-5p | trans | 1.07E-26 | Nightingale | M_VLDL_P | NA | C | T | 0.028296 | 0.004414 | 6.30E-11 |
| rs8176663 | let-7b-5p | trans | 1.07E-26 | Nightingale | LDL_FC | NA | C | T | 0.033635 | 0.004378 | 5.20E-14 |
| rs8176663 | let-7b-5p | trans | 1.07E-26 | Nightingale | S_HDL_PL | NA | C | T | 0.023784 | 0.004386 | 3.20E-08 |
| rs8176663 | let-7b-5p | trans | 1.07E-26 | Nightingale | S_LDL_PL_ | NA | C | T | -0.03006 | 0.004417 | 2.60E-13 |
| rs8176663 | let-7b-5p | trans | 1.07E-26 | Nightingale | Citrate | NA | C | T | 0.026829 | 0.004396 | 7.30E-10 |
| rs8176663 | let-7b-5p | trans | 1.07E-26 | Nightingale | M_LDL_CE | NA | C | T | 0.032977 | 0.004431 | 9.60E-14 |
| rs8176663 | miR-10b-5p | trans | 3.40E-35 | Nightingale | L_LDL_P | NA | C | T | 0.029177 | 0.00441 | 6.20E-11 |
| rs8176663 | miR-10b-5p | trans | 3.40E-35 | Nightingale | S_VLDL_C | NA | C | T | 0.038373 | 0.004424 | 1.80E-18 |
| rs8176663 | miR-10b-5p | trans | 3.40E-35 | Nightingale | L_LDL_CE | NA | C | T | 0.031569 | 0.004385 | 9.80E-13 |
| rs8176663 | miR-10b-5p | trans | 3.40E-35 | Nightingale | LDL_CE | NA | C | T | 0.032556 | 0.004413 | 2.20E-13 |
| rs8176663 | miR-10b-5p | trans | 3.40E-35 | Nightingale | XL_VLDL_F | NA | C | T | 0.037195 | 0.00429 | 2.00E-18 |
| rs8176663 | miR-10b-5p | trans | 3.40E-35 | Nightingale | S_HDL_L | NA | C | T | 0.026297 | 0.00441 | 1.50E-09 |
| rs8176663 | miR-10b-5p | trans | 3.40E-35 | Nightingale | S_LDL_C | NA | C | T | 0.030946 | 0.004429 | 5.50E-12 |
| rs8176663 | miR-10b-5p | trans | 3.40E-35 | Nightingale | PUFA | NA | C | T | 0.023713 | 0.004316 | 1.40E-08 |
| rs8176663 | miR-10b-5p | trans | 3.40E-35 | Nightingale | L_VLDL_TG | NA | C | T | -0.05114 | 0.004404 | 5.30E-32 |
| rs8176663 | miR-10b-5p | trans | 3.40E-35 | Nightingale | IDL_FC | NA | C | T | 0.038254 | 0.004284 | 1.50E-18 |
| rs8176663 | miR-10b-5p | trans | 3.40E-35 | Nightingale | L_LDL_PL | NA | C | T | 0.037992 | 0.004391 | 9.00E-18 |
| rs8176663 | miR-10b-5p | trans | 3.40E-35 | Nightingale | S_VLDL_L | NA | C | T | 0.024668 | 0.004382 | 3.70E-09 |
| rs8176663 | miR-10b-5p | trans | 3.40E-35 | Nightingale | M_VLDL_P | NA | C | T | 0.044056 | 0.004247 | 5.50E-25 |
| rs8176663 | miR-10b-5p | trans | 3.40E-35 | Nightingale | S_LDL_L | NA | C | T | 0.027745 | 0.004424 | 8.20E-10 |
| rs8176663 | miR-10b-5p | trans | 3.40E-35 | Nightingale | S_LDL_FC | NA | C | T | 0.028568 | 0.004418 | 4.20E-10 |
| rs8176663 | miR-10b-5p | trans | 3.40E-35 | Nightingale | XL_VLDL_C | NA | C | T | 0.038222 | 0.004243 | 4.70E-19 |
| rs8176663 | miR-10b-5p | trans | 3.40E-35 | Nightingale | Sphingomy | NA | C | T | 0.029016 | 0.004136 | 6.80E-12 |

| | | | | | | | | | | | |
|---|---|---|---|---|---|---|---|---|---|---|---|
| rs8176663 | miR-10b-5p | trans | 3.40E-35 | Nightingale | IDL_C | NA | C | T | 0.034955 | 0.004273 | 9.40E-16 |
| rs8176663 | miR-10b-5p | trans | 3.40E-35 | Nightingale | IDL_FC_pct | NA | C | T | 0.025773 | 0.004391 | 1.10E-08 |
| rs8176663 | miR-10b-5p | trans | 3.40E-35 | Nightingale | GlycA | NA | C | T | 0.028013 | 0.004406 | 1.40E-10 |
| rs8176663 | miR-10b-5p | trans | 3.40E-35 | Nightingale | Remnant_( | NA | C | T | 0.036867 | 0.004396 | 3.70E-17 |
| rs8176663 | miR-10b-5p | trans | 3.40E-35 | Nightingale | S_VLDL_TG | NA | C | T | -0.03813 | 0.004283 | 1.40E-18 |
| rs8176663 | miR-10b-5p | trans | 3.40E-35 | Nightingale | XS_VLDL_C | NA | C | T | 0.037794 | 0.00427 | 2.10E-18 |
| rs8176663 | miR-10b-5p | trans | 3.40E-35 | Nightingale | S_VLDL_PL | NA | C | T | 0.032777 | 0.004417 | 4.20E-14 |
| rs8176663 | miR-10b-5p | trans | 3.40E-35 | Nightingale | IDL_L | NA | C | T | 0.035268 | 0.004278 | 4.80E-16 |
| rs8176663 | miR-10b-5p | trans | 3.40E-35 | Nightingale | Cholines | NA | C | T | 0.022288 | 0.004161 | 4.90E-08 |
| rs8176663 | miR-10b-5p | trans | 3.40E-35 | Nightingale | M_VLDL_C | NA | C | T | 0.036205 | 0.004168 | 1.20E-17 |
| rs8176663 | miR-10b-5p | trans | 3.40E-35 | Nightingale | LDL_PL | NA | C | T | 0.034991 | 0.004415 | 4.70E-15 |
| rs8176663 | miR-10b-5p | trans | 3.40E-35 | Nightingale | XXL_VLDL_ | NA | C | T | 0.033119 | 0.004459 | 9.00E-14 |
| rs8176663 | miR-10b-5p | trans | 3.40E-35 | Nightingale | LDL_L | NA | C | T | 0.033306 | 0.00441 | 7.40E-14 |
| rs8176663 | miR-10b-5p | trans | 3.40E-35 | Nightingale | L_VLDL_C_ | NA | C | T | 0.052192 | 0.004322 | 4.00E-34 |
| rs8176663 | miR-10b-5p | trans | 3.40E-35 | Nightingale | S_VLDL_PL | NA | C | T | 0.02774 | 0.004237 | 1.80E-10 |
| rs8176663 | miR-10b-5p | trans | 3.40E-35 | Nightingale | M_LDL_L | NA | C | T | 0.033035 | 0.004431 | 1.00E-13 |
| rs8176663 | miR-10b-5p | trans | 3.40E-35 | Nightingale | XS_VLDL_C | NA | C | T | 0.037989 | 0.004295 | 1.30E-18 |
| rs8176663 | miR-10b-5p | trans | 3.40E-35 | Nightingale | LA | NA | C | T | 0.024903 | 0.004339 | 3.70E-09 |
| rs8176663 | miR-10b-5p | trans | 3.40E-35 | Nightingale | LDL_C | NA | C | T | 0.033196 | 0.004406 | 8.80E-14 |
| rs8176663 | miR-10b-5p | trans | 3.40E-35 | Nightingale | S_VLDL_CE | NA | C | T | 0.047034 | 0.004365 | 1.20E-26 |
| rs8176663 | miR-10b-5p | trans | 3.40E-35 | Nightingale | S_HDL_P | NA | C | T | 0.030615 | 0.004425 | 5.10E-12 |
| rs8176663 | miR-10b-5p | trans | 3.40E-35 | Nightingale | S_HDL_FC | NA | C | T | 0.02345 | 0.004344 | 4.60E-08 |
| rs8176663 | miR-10b-5p | trans | 3.40E-35 | Nightingale | Total_FC | NA | C | T | 0.033253 | 0.004323 | 6.80E-15 |
| rs8176663 | miR-10b-5p | trans | 3.40E-35 | Nightingale | Clinical_LD | NA | C | T | 0.036477 | 0.004399 | 2.70E-16 |
| rs8176663 | miR-10b-5p | trans | 3.40E-35 | Nightingale | XXL_VLDL_ | NA | C | T | 0.025004 | 0.004454 | 1.50E-09 |
| rs8176663 | miR-10b-5p | trans | 3.40E-35 | Nightingale | XS_VLDL_F | NA | C | T | 0.031441 | 0.004393 | 2.40E-12 |
| rs8176663 | miR-10b-5p | trans | 3.40E-35 | Nightingale | XL_VLDL_C | NA | C | T | 0.037351 | 0.004245 | 3.10E-18 |
| rs8176663 | miR-10b-5p | trans | 3.40E-35 | Nightingale | ApoB_by_A | NA | C | T | 0.025055 | 0.004358 | 1.30E-08 |
| rs8176663 | miR-10b-5p | trans | 3.40E-35 | Nightingale | VLDL_CE | NA | C | T | 0.035786 | 0.004426 | 3.00E-16 |
| rs8176663 | miR-10b-5p | trans | 3.40E-35 | Nightingale | Total_C | NA | C | T | 0.032372 | 0.004284 | 3.30E-14 |
| rs8176663 | miR-10b-5p | trans | 3.40E-35 | Nightingale | L_LDL_FC | NA | C | T | 0.033627 | 0.004345 | 2.90E-14 |
| rs8176663 | miR-10b-5p | trans | 3.40E-35 | Nightingale | Total_L | NA | C | T | 0.026346 | 0.004353 | 4.60E-10 |
| rs8176663 | miR-10b-5p | trans | 3.40E-35 | Nightingale | S_VLDL_FC | NA | C | T | 0.037905 | 0.004426 | 7.30E-18 |
| rs8176663 | miR-10b-5p | trans | 3.40E-35 | Nightingale | IDL_CE | NA | C | T | 0.033507 | 0.004272 | 1.30E-14 |
| rs8176663 | miR-10b-5p | trans | 3.40E-35 | Nightingale | S_VLDL_FC | NA | C | T | 0.029358 | 0.004228 | 1.10E-11 |
| rs8176663 | miR-10b-5p | trans | 3.40E-35 | Nightingale | VLDL_FC | NA | C | T | 0.024159 | 0.004392 | 1.10E-08 |
| rs8176663 | miR-10b-5p | trans | 3.40E-35 | Nightingale | Omega_6 | NA | C | T | 0.025889 | 0.004321 | 5.80E-10 |
| rs8176663 | miR-10b-5p | trans | 3.40E-35 | Nightingale | L_LDL_L | NA | C | T | 0.033551 | 0.004382 | 3.00E-14 |
| rs8176663 | miR-10b-5p | trans | 3.40E-35 | Nightingale | Total_CE | NA | C | T | 0.031745 | 0.004268 | 9.70E-14 |
| rs8176663 | miR-10b-5p | trans | 3.40E-35 | Nightingale | S_VLDL_P | NA | C | T | 0.023465 | 0.004369 | 1.70E-08 |
| rs8176663 | miR-10b-5p | trans | 3.40E-35 | Nightingale | S_LDL_P | NA | C | T | 0.029589 | 0.004429 | 3.20E-11 |
| rs8176663 | miR-10b-5p | trans | 3.40E-35 | Nightingale | M_LDL_PL | NA | C | T | 0.032471 | 0.004431 | 2.60E-13 |
| rs8176663 | miR-10b-5p | trans | 3.40E-35 | Nightingale | M_LDL_C_p | NA | C | T | 0.031165 | 0.004428 | 2.90E-12 |
| rs8176663 | miR-10b-5p | trans | 3.40E-35 | Nightingale | L_LDL_C | NA | C | T | 0.032322 | 0.004375 | 2.60E-13 |
| rs8176663 | miR-10b-5p | trans | 3.40E-35 | Nightingale | M_VLDL_F | NA | C | T | 0.033372 | 0.004424 | 3.10E-14 |
| rs8176663 | miR-10b-5p | trans | 3.40E-35 | Nightingale | IDL_P | NA | C | T | 0.039988 | 0.004378 | 1.30E-19 |
| rs8176663 | miR-10b-5p | trans | 3.40E-35 | Nightingale | XXL_VLDL_ | NA | C | T | 0.034827 | 0.004434 | 2.90E-15 |
| rs8176663 | miR-10b-5p | trans | 3.40E-35 | Nightingale | XS_VLDL_F | NA | C | T | 0.037065 | 0.004348 | 6.50E-18 |
| rs8176663 | miR-10b-5p | trans | 3.40E-35 | Nightingale | M_VLDL_C | NA | C | T | 0.033628 | 0.004157 | 1.90E-15 |
| rs8176663 | miR-10b-5p | trans | 3.40E-35 | Nightingale | M_VLDL_T | NA | C | T | -0.03856 | 0.004181 | 6.80E-20 |
| rs8176663 | miR-10b-5p | trans | 3.40E-35 | Nightingale | S_VLDL_CE | NA | C | T | 0.038182 | 0.004419 | 1.60E-18 |
| rs8176663 | miR-10b-5p | trans | 3.40E-35 | Nightingale | IDL_PL | NA | C | T | 0.035341 | 0.00427 | 3.30E-16 |
| rs8176663 | miR-10b-5p | trans | 3.40E-35 | Nightingale | XXL_VLDL_ | NA | C | T | 0.030847 | 0.004406 | 1.10E-12 |
| rs8176663 | miR-10b-5p | trans | 3.40E-35 | Nightingale | S_HDL_CE | NA | C | T | 0.02864 | 0.004432 | 1.30E-10 |
| rs8176663 | miR-10b-5p | trans | 3.40E-35 | Nightingale | M_VLDL_C | NA | C | T | 0.038711 | 0.004413 | 1.90E-18 |
| rs8176663 | miR-10b-5p | trans | 3.40E-35 | Nightingale | S_VLDL_C_ | NA | C | T | 0.041889 | 0.004306 | 5.20E-22 |
| rs8176663 | miR-10b-5p | trans | 3.40E-35 | Nightingale | VLDL_C | NA | C | T | 0.031302 | 0.004419 | 4.90E-13 |
| rs8176663 | miR-10b-5p | trans | 3.40E-35 | Nightingale | L_VLDL_CE | NA | C | T | 0.047336 | 0.004234 | 2.40E-29 |
| rs8176663 | miR-10b-5p | trans | 3.40E-35 | Nightingale | Total_PL | NA | C | T | 0.024922 | 0.004213 | 1.10E-09 |
| rs8176663 | miR-10b-5p | trans | 3.40E-35 | Nightingale | S_LDL_C_p | NA | C | T | 0.045366 | 0.004442 | 5.40E-25 |
| rs8176663 | miR-10b-5p | trans | 3.40E-35 | Nightingale | XS_VLDL_P | NA | C | T | 0.036698 | 0.004366 | 1.40E-17 |
| rs8176663 | miR-10b-5p | trans | 3.40E-35 | Nightingale | VLDL_P | NA | C | T | 0.026633 | 0.004401 | 4.40E-10 |
| rs8176663 | miR-10b-5p | trans | 3.40E-35 | Nightingale | M_LDL_TG | NA | C | T | -0.02763 | 0.00437 | 3.20E-10 |

| | | | | | | | | | | | |
|---|---|---|---|---|---|---|---|---|---|---|---|
| rs8176663 | miR-10b-5p | trans | 3.40E-35 | Nightingale | non_HDL_( | NA | C | T | 0.035698 | 0.004401 | 5.10E-16 |
| rs8176663 | miR-10b-5p | trans | 3.40E-35 | Nightingale | XS_VLDL_L | NA | C | T | 0.033813 | 0.00435 | 2.10E-15 |
| rs8176663 | miR-10b-5p | trans | 3.40E-35 | Nightingale | S_LDL_CE_ | NA | C | T | 0.030946 | 0.004417 | 4.90E-14 |
| rs8176663 | miR-10b-5p | trans | 3.40E-35 | Nightingale | M_VLDL_F | NA | C | T | 0.042064 | 0.00421 | 3.50E-23 |
| rs8176663 | miR-10b-5p | trans | 3.40E-35 | Nightingale | M_LDL_FC | NA | C | T | 0.034352 | 0.004419 | 1.90E-14 |
| rs8176663 | miR-10b-5p | trans | 3.40E-35 | Nightingale | M_LDL_C | NA | C | T | 0.033991 | 0.004433 | 2.10E-14 |
| rs8176663 | miR-10b-5p | trans | 3.40E-35 | Nightingale | XS_VLDL_P | NA | C | T | 0.032112 | 0.004363 | 5.40E-14 |
| rs8176663 | miR-10b-5p | trans | 3.40E-35 | Nightingale | ApoB | NA | C | T | 0.033939 | 0.004419 | 2.10E-14 |
| rs8176663 | miR-10b-5p | trans | 3.40E-35 | Nightingale | L_VLDL_FC | NA | C | T | 0.048588 | 0.004382 | 4.50E-29 |
| rs8176663 | miR-10b-5p | trans | 3.40E-35 | Nightingale | S_HDL_C | NA | C | T | 0.028415 | 0.004417 | 1.70E-10 |
| rs8176663 | miR-10b-5p | trans | 3.40E-35 | Nightingale | XL_VLDL_T | NA | C | T | -0.04392 | 0.004289 | 1.70E-24 |
| rs8176663 | miR-10b-5p | trans | 3.40E-35 | Nightingale | S_LDL_CE | NA | C | T | 0.030551 | 0.004427 | 5.80E-12 |
| rs8176663 | miR-10b-5p | trans | 3.40E-35 | Nightingale | LDL_P | NA | C | T | 0.031801 | 0.004422 | 1.00E-12 |
| rs8176663 | miR-10b-5p | trans | 3.40E-35 | Nightingale | XXL_VLDL_ | NA | C | T | -0.03573 | 0.004463 | 2.20E-16 |
| rs8176663 | miR-10b-5p | trans | 3.40E-35 | Nightingale | XL_HDL_PL | NA | C | T | -0.02277 | 0.004065 | 1.10E-08 |
| rs8176663 | miR-10b-5p | trans | 3.40E-35 | Nightingale | XL_VLDL_P | NA | C | T | 0.038847 | 0.004432 | 5.10E-19 |
| rs8176663 | miR-10b-5p | trans | 3.40E-35 | Nightingale | M_VLDL_C | NA | C | T | 0.040844 | 0.004385 | 2.30E-20 |
| rs8176663 | miR-10b-5p | trans | 3.40E-35 | Nightingale | M_LDL_P | NA | C | T | 0.035895 | 0.004437 | 6.80E-16 |
| rs8176663 | miR-10b-5p | trans | 3.40E-35 | Nightingale | M_VLDL_P | NA | C | T | 0.029128 | 0.004418 | 2.00E-11 |
| rs8176663 | miR-10b-5p | trans | 3.40E-35 | Nightingale | M_VLDL_P | NA | C | T | 0.028296 | 0.004414 | 6.30E-11 |
| rs8176663 | miR-10b-5p | trans | 3.40E-35 | Nightingale | LDL_FC | NA | C | T | 0.033635 | 0.004378 | 5.20E-14 |
| rs8176663 | miR-10b-5p | trans | 3.40E-35 | Nightingale | S_HDL_PL | NA | C | T | 0.023784 | 0.004386 | 3.20E-08 |
| rs8176663 | miR-10b-5p | trans | 3.40E-35 | Nightingale | S_LDL_PL_ | NA | C | T | -0.03006 | 0.004417 | 2.60E-13 |
| rs8176663 | miR-10b-5p | trans | 3.40E-35 | Nightingale | Citrate | NA | C | T | 0.026829 | 0.004396 | 7.30E-10 |
| rs8176663 | miR-10b-5p | trans | 3.40E-35 | Nightingale | M_LDL_CE | NA | C | T | 0.032977 | 0.004431 | 9.60E-14 |
| rs8176663 | miR-195-5p | trans | 2.48E-17 | Nightingale | L_LDL_P | NA | C | T | 0.029177 | 0.00441 | 6.20E-11 |
| rs8176663 | miR-195-5p | trans | 2.48E-17 | Nightingale | S_VLDL_C | NA | C | T | 0.038373 | 0.004424 | 1.80E-18 |
| rs8176663 | miR-195-5p | trans | 2.48E-17 | Nightingale | L_LDL_CE | NA | C | T | 0.031569 | 0.004385 | 9.80E-13 |
| rs8176663 | miR-195-5p | trans | 2.48E-17 | Nightingale | LDL_CE | NA | C | T | 0.032556 | 0.004413 | 2.20E-13 |
| rs8176663 | miR-195-5p | trans | 2.48E-17 | Nightingale | XL_VLDL_F | NA | C | T | 0.037195 | 0.00429 | 2.00E-18 |
| rs8176663 | miR-195-5p | trans | 2.48E-17 | Nightingale | S_HDL_L | NA | C | T | 0.026297 | 0.00441 | 1.50E-09 |
| rs8176663 | miR-195-5p | trans | 2.48E-17 | Nightingale | S_LDL_C | NA | C | T | 0.030946 | 0.004429 | 5.50E-12 |
| rs8176663 | miR-195-5p | trans | 2.48E-17 | Nightingale | PUFA | NA | C | T | 0.023713 | 0.004316 | 1.40E-08 |
| rs8176663 | miR-195-5p | trans | 2.48E-17 | Nightingale | L_VLDL_TG | NA | C | T | -0.05114 | 0.004404 | 5.30E-32 |
| rs8176663 | miR-195-5p | trans | 2.48E-17 | Nightingale | IDL_FC | NA | C | T | 0.038254 | 0.004284 | 1.50E-18 |
| rs8176663 | miR-195-5p | trans | 2.48E-17 | Nightingale | L_LDL_PL | NA | C | T | 0.037992 | 0.004391 | 9.00E-18 |
| rs8176663 | miR-195-5p | trans | 2.48E-17 | Nightingale | S_VLDL_L | NA | C | T | 0.024668 | 0.004382 | 3.70E-09 |
| rs8176663 | miR-195-5p | trans | 2.48E-17 | Nightingale | M_VLDL_P | NA | C | T | 0.044056 | 0.004247 | 5.50E-25 |
| rs8176663 | miR-195-5p | trans | 2.48E-17 | Nightingale | S_LDL_L | NA | C | T | 0.027745 | 0.004424 | 8.20E-10 |
| rs8176663 | miR-195-5p | trans | 2.48E-17 | Nightingale | S_LDL_FC | NA | C | T | 0.028568 | 0.004418 | 4.20E-10 |
| rs8176663 | miR-195-5p | trans | 2.48E-17 | Nightingale | XL_VLDL_C | NA | C | T | 0.038222 | 0.004243 | 4.70E-19 |
| rs8176663 | miR-195-5p | trans | 2.48E-17 | Nightingale | Sphingomy | NA | C | T | 0.029016 | 0.004136 | 6.80E-12 |
| rs8176663 | miR-195-5p | trans | 2.48E-17 | Nightingale | IDL_C | NA | C | T | 0.034955 | 0.004273 | 9.40E-16 |
| rs8176663 | miR-195-5p | trans | 2.48E-17 | Nightingale | IDL_FC_pct | NA | C | T | 0.025773 | 0.004391 | 1.10E-08 |
| rs8176663 | miR-195-5p | trans | 2.48E-17 | Nightingale | GlycA | NA | C | T | 0.028013 | 0.004406 | 1.40E-10 |
| rs8176663 | miR-195-5p | trans | 2.48E-17 | Nightingale | Remnant_( | NA | C | T | 0.036867 | 0.004396 | 3.70E-17 |
| rs8176663 | miR-195-5p | trans | 2.48E-17 | Nightingale | S_VLDL_TG | NA | C | T | -0.03813 | 0.004283 | 1.40E-18 |
| rs8176663 | miR-195-5p | trans | 2.48E-17 | Nightingale | XS_VLDL_C | NA | C | T | 0.037794 | 0.00427 | 2.10E-18 |
| rs8176663 | miR-195-5p | trans | 2.48E-17 | Nightingale | S_VLDL_PL | NA | C | T | 0.032777 | 0.004417 | 4.20E-14 |
| rs8176663 | miR-195-5p | trans | 2.48E-17 | Nightingale | IDL_L | NA | C | T | 0.035268 | 0.004278 | 4.80E-16 |
| rs8176663 | miR-195-5p | trans | 2.48E-17 | Nightingale | Cholines | NA | C | T | 0.022288 | 0.004161 | 4.90E-08 |
| rs8176663 | miR-195-5p | trans | 2.48E-17 | Nightingale | M_VLDL_C | NA | C | T | 0.036205 | 0.004168 | 1.20E-17 |
| rs8176663 | miR-195-5p | trans | 2.48E-17 | Nightingale | LDL_PL | NA | C | T | 0.034991 | 0.004415 | 4.70E-15 |
| rs8176663 | miR-195-5p | trans | 2.48E-17 | Nightingale | XXL_VLDL_ | NA | C | T | 0.033119 | 0.004459 | 9.00E-14 |
| rs8176663 | miR-195-5p | trans | 2.48E-17 | Nightingale | LDL_L | NA | C | T | 0.033306 | 0.00441 | 7.40E-14 |
| rs8176663 | miR-195-5p | trans | 2.48E-17 | Nightingale | L_VLDL_C_ | NA | C | T | 0.052192 | 0.004322 | 4.00E-34 |
| rs8176663 | miR-195-5p | trans | 2.48E-17 | Nightingale | S_VLDL_PL | NA | C | T | 0.02774 | 0.004237 | 1.80E-10 |
| rs8176663 | miR-195-5p | trans | 2.48E-17 | Nightingale | M_LDL_L | NA | C | T | 0.033035 | 0.004431 | 1.00E-13 |
| rs8176663 | miR-195-5p | trans | 2.48E-17 | Nightingale | XS_VLDL_C | NA | C | T | 0.037989 | 0.004295 | 1.30E-18 |
| rs8176663 | miR-195-5p | trans | 2.48E-17 | Nightingale | LA | NA | C | T | 0.024903 | 0.004339 | 3.70E-09 |
| rs8176663 | miR-195-5p | trans | 2.48E-17 | Nightingale | LDL_C | NA | C | T | 0.033196 | 0.004406 | 8.80E-14 |
| rs8176663 | miR-195-5p | trans | 2.48E-17 | Nightingale | S_VLDL_CE | NA | C | T | 0.047034 | 0.004365 | 1.20E-26 |
| rs8176663 | miR-195-5p | trans | 2.48E-17 | Nightingale | S_HDL_P | NA | C | T | 0.030615 | 0.004425 | 5.10E-12 |

| | | | | | | | | | | | |
|---|---|---|---|---|---|---|---|---|---|---|---|
| rs8176663 | miR-195-5p | trans | 2.48E-17 | Nightingale | S_HDL_FC | NA | C | T | 0.02345 | 0.004344 | 4.60E-08 |
| rs8176663 | miR-195-5p | trans | 2.48E-17 | Nightingale | Total_FC | NA | C | T | 0.033253 | 0.004323 | 6.80E-15 |
| rs8176663 | miR-195-5p | trans | 2.48E-17 | Nightingale | Clinical_LD | NA | C | T | 0.036477 | 0.004399 | 2.70E-16 |
| rs8176663 | miR-195-5p | trans | 2.48E-17 | Nightingale | XXL_VLDL_ | NA | C | T | 0.025004 | 0.004454 | 1.50E-09 |
| rs8176663 | miR-195-5p | trans | 2.48E-17 | Nightingale | XS_VLDL_F | NA | C | T | 0.031441 | 0.004393 | 2.40E-12 |
| rs8176663 | miR-195-5p | trans | 2.48E-17 | Nightingale | XL_VLDL_C | NA | C | T | 0.037351 | 0.004245 | 3.10E-18 |
| rs8176663 | miR-195-5p | trans | 2.48E-17 | Nightingale | ApoB_by_A | NA | C | T | 0.025055 | 0.004358 | 1.30E-08 |
| rs8176663 | miR-195-5p | trans | 2.48E-17 | Nightingale | VLDL_CE | NA | C | T | 0.035786 | 0.004426 | 3.00E-16 |
| rs8176663 | miR-195-5p | trans | 2.48E-17 | Nightingale | Total_C | NA | C | T | 0.032372 | 0.004284 | 3.30E-14 |
| rs8176663 | miR-195-5p | trans | 2.48E-17 | Nightingale | L_LDL_FC | NA | C | T | 0.033627 | 0.004345 | 2.90E-14 |
| rs8176663 | miR-195-5p | trans | 2.48E-17 | Nightingale | Total_L | NA | C | T | 0.026346 | 0.004353 | 4.60E-10 |
| rs8176663 | miR-195-5p | trans | 2.48E-17 | Nightingale | S_VLDL_FC | NA | C | T | 0.037905 | 0.004426 | 7.30E-18 |
| rs8176663 | miR-195-5p | trans | 2.48E-17 | Nightingale | IDL_CE | NA | C | T | 0.033507 | 0.004272 | 1.30E-14 |
| rs8176663 | miR-195-5p | trans | 2.48E-17 | Nightingale | S_VLDL_FC | NA | C | T | 0.029358 | 0.004228 | 1.10E-11 |
| rs8176663 | miR-195-5p | trans | 2.48E-17 | Nightingale | VLDL_FC | NA | C | T | 0.024159 | 0.004392 | 1.10E-08 |
| rs8176663 | miR-195-5p | trans | 2.48E-17 | Nightingale | Omega_6 | NA | C | T | 0.025889 | 0.004321 | 5.80E-10 |
| rs8176663 | miR-195-5p | trans | 2.48E-17 | Nightingale | L_LDL_L | NA | C | T | 0.033551 | 0.004382 | 3.00E-14 |
| rs8176663 | miR-195-5p | trans | 2.48E-17 | Nightingale | Total_CE | NA | C | T | 0.031745 | 0.004268 | 9.70E-14 |
| rs8176663 | miR-195-5p | trans | 2.48E-17 | Nightingale | S_VLDL_P | NA | C | T | 0.023465 | 0.004369 | 1.70E-08 |
| rs8176663 | miR-195-5p | trans | 2.48E-17 | Nightingale | S_LDL_P | NA | C | T | 0.029589 | 0.004429 | 3.20E-11 |
| rs8176663 | miR-195-5p | trans | 2.48E-17 | Nightingale | M_LDL_PL | NA | C | T | 0.032471 | 0.004431 | 2.60E-13 |
| rs8176663 | miR-195-5p | trans | 2.48E-17 | Nightingale | M_LDL_C_p | NA | C | T | 0.031165 | 0.004428 | 2.90E-12 |
| rs8176663 | miR-195-5p | trans | 2.48E-17 | Nightingale | L_LDL_C | NA | C | T | 0.032322 | 0.004375 | 2.60E-13 |
| rs8176663 | miR-195-5p | trans | 2.48E-17 | Nightingale | M_VLDL_F | NA | C | T | 0.033372 | 0.004424 | 3.10E-14 |
| rs8176663 | miR-195-5p | trans | 2.48E-17 | Nightingale | IDL_P | NA | C | T | 0.039988 | 0.004378 | 1.30E-19 |
| rs8176663 | miR-195-5p | trans | 2.48E-17 | Nightingale | XXL_VLDL_ | NA | C | T | 0.034827 | 0.004434 | 2.90E-15 |
| rs8176663 | miR-195-5p | trans | 2.48E-17 | Nightingale | XS_VLDL_F | NA | C | T | 0.037065 | 0.004348 | 6.50E-18 |
| rs8176663 | miR-195-5p | trans | 2.48E-17 | Nightingale | M_VLDL_C | NA | C | T | 0.033628 | 0.004157 | 1.90E-15 |
| rs8176663 | miR-195-5p | trans | 2.48E-17 | Nightingale | M_VLDL_T | NA | C | T | -0.03856 | 0.004181 | 6.80E-20 |
| rs8176663 | miR-195-5p | trans | 2.48E-17 | Nightingale | S_VLDL_CE | NA | C | T | 0.038182 | 0.004419 | 1.60E-18 |
| rs8176663 | miR-195-5p | trans | 2.48E-17 | Nightingale | IDL_PL | NA | C | T | 0.035341 | 0.00427 | 3.30E-16 |
| rs8176663 | miR-195-5p | trans | 2.48E-17 | Nightingale | XXL_VLDL_ | NA | C | T | 0.030847 | 0.004406 | 1.10E-12 |
| rs8176663 | miR-195-5p | trans | 2.48E-17 | Nightingale | S_HDL_CE | NA | C | T | 0.02864 | 0.004432 | 1.30E-10 |
| rs8176663 | miR-195-5p | trans | 2.48E-17 | Nightingale | M_VLDL_C | NA | C | T | 0.038711 | 0.004413 | 1.90E-18 |
| rs8176663 | miR-195-5p | trans | 2.48E-17 | Nightingale | S_VLDL_C_ | NA | C | T | 0.041889 | 0.004306 | 5.20E-22 |
| rs8176663 | miR-195-5p | trans | 2.48E-17 | Nightingale | VLDL_C | NA | C | T | 0.031302 | 0.004419 | 4.90E-13 |
| rs8176663 | miR-195-5p | trans | 2.48E-17 | Nightingale | L_VLDL_CE | NA | C | T | 0.047336 | 0.004234 | 2.40E-29 |
| rs8176663 | miR-195-5p | trans | 2.48E-17 | Nightingale | Total_PL | NA | C | T | 0.024922 | 0.004213 | 1.10E-09 |
| rs8176663 | miR-195-5p | trans | 2.48E-17 | Nightingale | S_LDL_C_p | NA | C | T | 0.045366 | 0.004442 | 5.40E-25 |
| rs8176663 | miR-195-5p | trans | 2.48E-17 | Nightingale | XS_VLDL_P | NA | C | T | 0.036698 | 0.004366 | 1.40E-17 |
| rs8176663 | miR-195-5p | trans | 2.48E-17 | Nightingale | VLDL_P | NA | C | T | 0.026633 | 0.004401 | 4.40E-10 |
| rs8176663 | miR-195-5p | trans | 2.48E-17 | Nightingale | M_LDL_TG | NA | C | T | -0.02763 | 0.00437 | 3.20E-10 |
| rs8176663 | miR-195-5p | trans | 2.48E-17 | Nightingale | non_HDL_( | NA | C | T | 0.035698 | 0.004401 | 5.10E-16 |
| rs8176663 | miR-195-5p | trans | 2.48E-17 | Nightingale | XS_VLDL_L | NA | C | T | 0.033813 | 0.00435 | 2.10E-15 |
| rs8176663 | miR-195-5p | trans | 2.48E-17 | Nightingale | S_LDL_CE_ | NA | C | T | 0.030946 | 0.004417 | 4.90E-14 |
| rs8176663 | miR-195-5p | trans | 2.48E-17 | Nightingale | M_VLDL_F | NA | C | T | 0.042064 | 0.00421 | 3.50E-23 |
| rs8176663 | miR-195-5p | trans | 2.48E-17 | Nightingale | M_LDL_FC | NA | C | T | 0.034352 | 0.004419 | 1.90E-14 |
| rs8176663 | miR-195-5p | trans | 2.48E-17 | Nightingale | M_LDL_C | NA | C | T | 0.033991 | 0.004433 | 2.10E-14 |
| rs8176663 | miR-195-5p | trans | 2.48E-17 | Nightingale | XS_VLDL_P | NA | C | T | 0.032112 | 0.004363 | 5.40E-14 |
| rs8176663 | miR-195-5p | trans | 2.48E-17 | Nightingale | ApoB | NA | C | T | 0.033939 | 0.004419 | 2.10E-14 |
| rs8176663 | miR-195-5p | trans | 2.48E-17 | Nightingale | L_VLDL_FC | NA | C | T | 0.048588 | 0.004382 | 4.50E-29 |
| rs8176663 | miR-195-5p | trans | 2.48E-17 | Nightingale | S_HDL_C | NA | C | T | 0.028415 | 0.004417 | 1.70E-10 |
| rs8176663 | miR-195-5p | trans | 2.48E-17 | Nightingale | XL_VLDL_T | NA | C | T | -0.04392 | 0.004289 | 1.70E-24 |
| rs8176663 | miR-195-5p | trans | 2.48E-17 | Nightingale | S_LDL_CE | NA | C | T | 0.030551 | 0.004427 | 5.80E-12 |
| rs8176663 | miR-195-5p | trans | 2.48E-17 | Nightingale | LDL_P | NA | C | T | 0.031801 | 0.004422 | 1.00E-12 |
| rs8176663 | miR-195-5p | trans | 2.48E-17 | Nightingale | XXL_VLDL_ | NA | C | T | -0.03573 | 0.004463 | 2.20E-16 |
| rs8176663 | miR-195-5p | trans | 2.48E-17 | Nightingale | XL_HDL_PL | NA | C | T | -0.02277 | 0.004065 | 1.10E-08 |
| rs8176663 | miR-195-5p | trans | 2.48E-17 | Nightingale | XL_VLDL_P | NA | C | T | 0.038847 | 0.004432 | 5.10E-19 |
| rs8176663 | miR-195-5p | trans | 2.48E-17 | Nightingale | M_VLDL_C | NA | C | T | 0.040844 | 0.004385 | 2.30E-20 |
| rs8176663 | miR-195-5p | trans | 2.48E-17 | Nightingale | M_LDL_P | NA | C | T | 0.035895 | 0.004437 | 6.80E-16 |
| rs8176663 | miR-195-5p | trans | 2.48E-17 | Nightingale | M_VLDL_P | NA | C | T | 0.029128 | 0.004418 | 2.00E-11 |
| rs8176663 | miR-195-5p | trans | 2.48E-17 | Nightingale | M_VLDL_P | NA | C | T | 0.028296 | 0.004414 | 6.30E-11 |
| rs8176663 | miR-195-5p | trans | 2.48E-17 | Nightingale | LDL_FC | NA | C | T | 0.033635 | 0.004378 | 5.20E-14 |

| | | | | | | | | | | | |
|---|---|---|---|---|---|---|---|---|---|---|---|
| rs8176663 | miR-195-5p | trans | 2.48E-17 | Nightingale | S_HDL_PL | NA | C | T | 0.023784 | 0.004386 | 3.20E-08 |
| rs8176663 | miR-195-5p | trans | 2.48E-17 | Nightingale | S_LDL_PL_ | NA | C | T | -0.03006 | 0.004417 | 2.60E-13 |
| rs8176663 | miR-195-5p | trans | 2.48E-17 | Nightingale | Citrate | NA | C | T | 0.026829 | 0.004396 | 7.30E-10 |
| rs8176663 | miR-195-5p | trans | 2.48E-17 | Nightingale | M_LDL_CE | NA | C | T | 0.032977 | 0.004431 | 9.60E-14 |
| rs8176663 | miR-126-3p | trans | 2.73E-39 | Nightingale | L_LDL_P | NA | C | T | 0.029177 | 0.00441 | 6.20E-11 |
| rs8176663 | miR-126-3p | trans | 2.73E-39 | Nightingale | S_VLDL_C | NA | C | T | 0.038373 | 0.004424 | 1.80E-18 |
| rs8176663 | miR-126-3p | trans | 2.73E-39 | Nightingale | L_LDL_CE | NA | C | T | 0.031569 | 0.004385 | 9.80E-13 |
| rs8176663 | miR-126-3p | trans | 2.73E-39 | Nightingale | LDL_CE | NA | C | T | 0.032556 | 0.004413 | 2.20E-13 |
| rs8176663 | miR-126-3p | trans | 2.73E-39 | Nightingale | XL_VLDL_F | NA | C | T | 0.037195 | 0.00429 | 2.00E-18 |
| rs8176663 | miR-126-3p | trans | 2.73E-39 | Nightingale | S_HDL_L | NA | C | T | 0.026297 | 0.00441 | 1.50E-09 |
| rs8176663 | miR-126-3p | trans | 2.73E-39 | Nightingale | S_LDL_C | NA | C | T | 0.030946 | 0.004429 | 5.50E-12 |
| rs8176663 | miR-126-3p | trans | 2.73E-39 | Nightingale | PUFA | NA | C | T | 0.023713 | 0.004316 | 1.40E-08 |
| rs8176663 | miR-126-3p | trans | 2.73E-39 | Nightingale | L_VLDL_TG | NA | C | T | -0.05114 | 0.004404 | 5.30E-32 |
| rs8176663 | miR-126-3p | trans | 2.73E-39 | Nightingale | IDL_FC | NA | C | T | 0.038254 | 0.004284 | 1.50E-18 |
| rs8176663 | miR-126-3p | trans | 2.73E-39 | Nightingale | L_LDL_PL | NA | C | T | 0.037992 | 0.004391 | 9.00E-18 |
| rs8176663 | miR-126-3p | trans | 2.73E-39 | Nightingale | S_VLDL_L | NA | C | T | 0.024668 | 0.004382 | 3.70E-09 |
| rs8176663 | miR-126-3p | trans | 2.73E-39 | Nightingale | M_VLDL_P | NA | C | T | 0.044056 | 0.004247 | 5.50E-25 |
| rs8176663 | miR-126-3p | trans | 2.73E-39 | Nightingale | S_LDL_L | NA | C | T | 0.027745 | 0.004424 | 8.20E-10 |
| rs8176663 | miR-126-3p | trans | 2.73E-39 | Nightingale | S_LDL_FC | NA | C | T | 0.028568 | 0.004418 | 4.20E-10 |
| rs8176663 | miR-126-3p | trans | 2.73E-39 | Nightingale | XL_VLDL_C | NA | C | T | 0.038222 | 0.004243 | 4.70E-19 |
| rs8176663 | miR-126-3p | trans | 2.73E-39 | Nightingale | Sphingomy | NA | C | T | 0.029016 | 0.004136 | 6.80E-12 |
| rs8176663 | miR-126-3p | trans | 2.73E-39 | Nightingale | IDL_C | NA | C | T | 0.034955 | 0.004273 | 9.40E-16 |
| rs8176663 | miR-126-3p | trans | 2.73E-39 | Nightingale | IDL_FC_pct | NA | C | T | 0.025773 | 0.004391 | 1.10E-08 |
| rs8176663 | miR-126-3p | trans | 2.73E-39 | Nightingale | GlycA | NA | C | T | 0.028013 | 0.004406 | 1.40E-10 |
| rs8176663 | miR-126-3p | trans | 2.73E-39 | Nightingale | Remnant_( | NA | C | T | 0.036867 | 0.004396 | 3.70E-17 |
| rs8176663 | miR-126-3p | trans | 2.73E-39 | Nightingale | S_VLDL_TG | NA | C | T | -0.03813 | 0.004283 | 1.40E-18 |
| rs8176663 | miR-126-3p | trans | 2.73E-39 | Nightingale | XS_VLDL_C | NA | C | T | 0.037794 | 0.00427 | 2.10E-18 |
| rs8176663 | miR-126-3p | trans | 2.73E-39 | Nightingale | S_VLDL_PL | NA | C | T | 0.032777 | 0.004417 | 4.20E-14 |
| rs8176663 | miR-126-3p | trans | 2.73E-39 | Nightingale | IDL_L | NA | C | T | 0.035268 | 0.004278 | 4.80E-16 |
| rs8176663 | miR-126-3p | trans | 2.73E-39 | Nightingale | Cholines | NA | C | T | 0.022288 | 0.004161 | 4.90E-08 |
| rs8176663 | miR-126-3p | trans | 2.73E-39 | Nightingale | M_VLDL_C | NA | C | T | 0.036205 | 0.004168 | 1.20E-17 |
| rs8176663 | miR-126-3p | trans | 2.73E-39 | Nightingale | LDL_PL | NA | C | T | 0.034991 | 0.004415 | 4.70E-15 |
| rs8176663 | miR-126-3p | trans | 2.73E-39 | Nightingale | XXL_VLDL_ | NA | C | T | 0.033119 | 0.004459 | 9.00E-14 |
| rs8176663 | miR-126-3p | trans | 2.73E-39 | Nightingale | LDL_L | NA | C | T | 0.033306 | 0.00441 | 7.40E-14 |
| rs8176663 | miR-126-3p | trans | 2.73E-39 | Nightingale | L_VLDL_C_ | NA | C | T | 0.052192 | 0.004322 | 4.00E-34 |
| rs8176663 | miR-126-3p | trans | 2.73E-39 | Nightingale | S_VLDL_PL | NA | C | T | 0.02774 | 0.004237 | 1.80E-10 |
| rs8176663 | miR-126-3p | trans | 2.73E-39 | Nightingale | M_LDL_L | NA | C | T | 0.033035 | 0.004431 | 1.00E-13 |
| rs8176663 | miR-126-3p | trans | 2.73E-39 | Nightingale | XS_VLDL_C | NA | C | T | 0.037989 | 0.004295 | 1.30E-18 |
| rs8176663 | miR-126-3p | trans | 2.73E-39 | Nightingale | LA | NA | C | T | 0.024903 | 0.004339 | 3.70E-09 |
| rs8176663 | miR-126-3p | trans | 2.73E-39 | Nightingale | LDL_C | NA | C | T | 0.033196 | 0.004406 | 8.80E-14 |
| rs8176663 | miR-126-3p | trans | 2.73E-39 | Nightingale | S_VLDL_CE | NA | C | T | 0.047034 | 0.004365 | 1.20E-26 |
| rs8176663 | miR-126-3p | trans | 2.73E-39 | Nightingale | S_HDL_P | NA | C | T | 0.030615 | 0.004425 | 5.10E-12 |
| rs8176663 | miR-126-3p | trans | 2.73E-39 | Nightingale | S_HDL_FC | NA | C | T | 0.02345 | 0.004344 | 4.60E-08 |
| rs8176663 | miR-126-3p | trans | 2.73E-39 | Nightingale | Total_FC | NA | C | T | 0.033253 | 0.004323 | 6.80E-15 |
| rs8176663 | miR-126-3p | trans | 2.73E-39 | Nightingale | Clinical_LD | NA | C | T | 0.036477 | 0.004399 | 2.70E-16 |
| rs8176663 | miR-126-3p | trans | 2.73E-39 | Nightingale | XXL_VLDL_ | NA | C | T | 0.025004 | 0.004454 | 1.50E-09 |
| rs8176663 | miR-126-3p | trans | 2.73E-39 | Nightingale | XS_VLDL_F | NA | C | T | 0.031441 | 0.004393 | 2.40E-12 |
| rs8176663 | miR-126-3p | trans | 2.73E-39 | Nightingale | XL_VLDL_C | NA | C | T | 0.037351 | 0.004245 | 3.10E-18 |
| rs8176663 | miR-126-3p | trans | 2.73E-39 | Nightingale | ApoB_by_A | NA | C | T | 0.025055 | 0.004358 | 1.30E-08 |
| rs8176663 | miR-126-3p | trans | 2.73E-39 | Nightingale | VLDL_CE | NA | C | T | 0.035786 | 0.004426 | 3.00E-16 |
| rs8176663 | miR-126-3p | trans | 2.73E-39 | Nightingale | Total_C | NA | C | T | 0.032372 | 0.004284 | 3.30E-14 |
| rs8176663 | miR-126-3p | trans | 2.73E-39 | Nightingale | L_LDL_FC | NA | C | T | 0.033627 | 0.004345 | 2.90E-14 |
| rs8176663 | miR-126-3p | trans | 2.73E-39 | Nightingale | Total_L | NA | C | T | 0.026346 | 0.004353 | 4.60E-10 |
| rs8176663 | miR-126-3p | trans | 2.73E-39 | Nightingale | S_VLDL_FC | NA | C | T | 0.037905 | 0.004426 | 7.30E-18 |
| rs8176663 | miR-126-3p | trans | 2.73E-39 | Nightingale | IDL_CE | NA | C | T | 0.033507 | 0.004272 | 1.30E-14 |
| rs8176663 | miR-126-3p | trans | 2.73E-39 | Nightingale | S_VLDL_FC | NA | C | T | 0.029358 | 0.004228 | 1.10E-11 |
| rs8176663 | miR-126-3p | trans | 2.73E-39 | Nightingale | VLDL_FC | NA | C | T | 0.024159 | 0.004392 | 1.10E-08 |
| rs8176663 | miR-126-3p | trans | 2.73E-39 | Nightingale | Omega_6 | NA | C | T | 0.025889 | 0.004321 | 5.80E-10 |
| rs8176663 | miR-126-3p | trans | 2.73E-39 | Nightingale | L_LDL_L | NA | C | T | 0.033551 | 0.004382 | 3.00E-14 |
| rs8176663 | miR-126-3p | trans | 2.73E-39 | Nightingale | Total_CE | NA | C | T | 0.031745 | 0.004268 | 9.70E-14 |
| rs8176663 | miR-126-3p | trans | 2.73E-39 | Nightingale | S_VLDL_P | NA | C | T | 0.023465 | 0.004369 | 1.70E-08 |
| rs8176663 | miR-126-3p | trans | 2.73E-39 | Nightingale | S_LDL_P | NA | C | T | 0.029589 | 0.004429 | 3.20E-11 |
| rs8176663 | miR-126-3p | trans | 2.73E-39 | Nightingale | M_LDL_PL | NA | C | T | 0.032471 | 0.004431 | 2.60E-13 |

| | | | | | | | | | | | |
|---|---|---|---|---|---|---|---|---|---|---|---|
| rs8176663 | miR-126-3p | trans | 2.73E-39 | Nightingale | M_LDL_C_p | NA | C | T | 0.031165 | 0.004428 | 2.90E-12 |
| rs8176663 | miR-126-3p | trans | 2.73E-39 | Nightingale | L_LDL_C | NA | C | T | 0.032322 | 0.004375 | 2.60E-13 |
| rs8176663 | miR-126-3p | trans | 2.73E-39 | Nightingale | M_VLDL_F | NA | C | T | 0.033372 | 0.004424 | 3.10E-14 |
| rs8176663 | miR-126-3p | trans | 2.73E-39 | Nightingale | IDL_P | NA | C | T | 0.039988 | 0.004378 | 1.30E-19 |
| rs8176663 | miR-126-3p | trans | 2.73E-39 | Nightingale | XXL_VLDL_ | NA | C | T | 0.034827 | 0.004434 | 2.90E-15 |
| rs8176663 | miR-126-3p | trans | 2.73E-39 | Nightingale | XS_VLDL_F | NA | C | T | 0.037065 | 0.004348 | 6.50E-18 |
| rs8176663 | miR-126-3p | trans | 2.73E-39 | Nightingale | M_VLDL_C | NA | C | T | 0.033628 | 0.004157 | 1.90E-15 |
| rs8176663 | miR-126-3p | trans | 2.73E-39 | Nightingale | M_VLDL_T | NA | C | T | -0.03856 | 0.004181 | 6.80E-20 |
| rs8176663 | miR-126-3p | trans | 2.73E-39 | Nightingale | S_VLDL_CE | NA | C | T | 0.038182 | 0.004419 | 1.60E-18 |
| rs8176663 | miR-126-3p | trans | 2.73E-39 | Nightingale | IDL_PL | NA | C | T | 0.035341 | 0.00427 | 3.30E-16 |
| rs8176663 | miR-126-3p | trans | 2.73E-39 | Nightingale | XXL_VLDL_ | NA | C | T | 0.030847 | 0.004406 | 1.10E-12 |
| rs8176663 | miR-126-3p | trans | 2.73E-39 | Nightingale | S_HDL_CE | NA | C | T | 0.02864 | 0.004432 | 1.30E-10 |
| rs8176663 | miR-126-3p | trans | 2.73E-39 | Nightingale | M_VLDL_C | NA | C | T | 0.038711 | 0.004413 | 1.90E-18 |
| rs8176663 | miR-126-3p | trans | 2.73E-39 | Nightingale | S_VLDL_C_ | NA | C | T | 0.041889 | 0.004306 | 5.20E-22 |
| rs8176663 | miR-126-3p | trans | 2.73E-39 | Nightingale | VLDL_C | NA | C | T | 0.031302 | 0.004419 | 4.90E-13 |
| rs8176663 | miR-126-3p | trans | 2.73E-39 | Nightingale | L_VLDL_CE | NA | C | T | 0.047336 | 0.004234 | 2.40E-29 |
| rs8176663 | miR-126-3p | trans | 2.73E-39 | Nightingale | Total_PL | NA | C | T | 0.024922 | 0.004213 | 1.10E-09 |
| rs8176663 | miR-126-3p | trans | 2.73E-39 | Nightingale | S_LDL_C_p | NA | C | T | 0.045366 | 0.004442 | 5.40E-25 |
| rs8176663 | miR-126-3p | trans | 2.73E-39 | Nightingale | XS_VLDL_P | NA | C | T | 0.036698 | 0.004366 | 1.40E-17 |
| rs8176663 | miR-126-3p | trans | 2.73E-39 | Nightingale | VLDL_P | NA | C | T | 0.026633 | 0.004401 | 4.40E-10 |
| rs8176663 | miR-126-3p | trans | 2.73E-39 | Nightingale | M_LDL_TG | NA | C | T | -0.02763 | 0.00437 | 3.20E-10 |
| rs8176663 | miR-126-3p | trans | 2.73E-39 | Nightingale | non_HDL_( | NA | C | T | 0.035698 | 0.004401 | 5.10E-16 |
| rs8176663 | miR-126-3p | trans | 2.73E-39 | Nightingale | XS_VLDL_L | NA | C | T | 0.033813 | 0.00435 | 2.10E-15 |
| rs8176663 | miR-126-3p | trans | 2.73E-39 | Nightingale | S_LDL_CE_ | NA | C | T | 0.030946 | 0.004417 | 4.90E-14 |
| rs8176663 | miR-126-3p | trans | 2.73E-39 | Nightingale | M_VLDL_F | NA | C | T | 0.042064 | 0.00421 | 3.50E-23 |
| rs8176663 | miR-126-3p | trans | 2.73E-39 | Nightingale | M_LDL_FC | NA | C | T | 0.034352 | 0.004419 | 1.90E-14 |
| rs8176663 | miR-126-3p | trans | 2.73E-39 | Nightingale | M_LDL_C | NA | C | T | 0.033991 | 0.004433 | 2.10E-14 |
| rs8176663 | miR-126-3p | trans | 2.73E-39 | Nightingale | XS_VLDL_P | NA | C | T | 0.032112 | 0.004363 | 5.40E-14 |
| rs8176663 | miR-126-3p | trans | 2.73E-39 | Nightingale | ApoB | NA | C | T | 0.033939 | 0.004419 | 2.10E-14 |
| rs8176663 | miR-126-3p | trans | 2.73E-39 | Nightingale | L_VLDL_FC | NA | C | T | 0.048588 | 0.004382 | 4.50E-29 |
| rs8176663 | miR-126-3p | trans | 2.73E-39 | Nightingale | S_HDL_C | NA | C | T | 0.028415 | 0.004417 | 1.70E-10 |
| rs8176663 | miR-126-3p | trans | 2.73E-39 | Nightingale | XL_VLDL_T | NA | C | T | -0.04392 | 0.004289 | 1.70E-24 |
| rs8176663 | miR-126-3p | trans | 2.73E-39 | Nightingale | S_LDL_CE | NA | C | T | 0.030551 | 0.004427 | 5.80E-12 |
| rs8176663 | miR-126-3p | trans | 2.73E-39 | Nightingale | LDL_P | NA | C | T | 0.031801 | 0.004422 | 1.00E-12 |
| rs8176663 | miR-126-3p | trans | 2.73E-39 | Nightingale | XXL_VLDL_ | NA | C | T | -0.03573 | 0.004463 | 2.20E-16 |
| rs8176663 | miR-126-3p | trans | 2.73E-39 | Nightingale | XL_HDL_PL | NA | C | T | -0.02277 | 0.004065 | 1.10E-08 |
| rs8176663 | miR-126-3p | trans | 2.73E-39 | Nightingale | XL_VLDL_P | NA | C | T | 0.038847 | 0.004432 | 5.10E-19 |
| rs8176663 | miR-126-3p | trans | 2.73E-39 | Nightingale | M_VLDL_C | NA | C | T | 0.040844 | 0.004385 | 2.30E-20 |
| rs8176663 | miR-126-3p | trans | 2.73E-39 | Nightingale | M_LDL_P | NA | C | T | 0.035895 | 0.004437 | 6.80E-16 |
| rs8176663 | miR-126-3p | trans | 2.73E-39 | Nightingale | M_VLDL_P | NA | C | T | 0.029128 | 0.004418 | 2.00E-11 |
| rs8176663 | miR-126-3p | trans | 2.73E-39 | Nightingale | M_VLDL_P | NA | C | T | 0.028296 | 0.004414 | 6.30E-11 |
| rs8176663 | miR-126-3p | trans | 2.73E-39 | Nightingale | LDL_FC | NA | C | T | 0.033635 | 0.004378 | 5.20E-14 |
| rs8176663 | miR-126-3p | trans | 2.73E-39 | Nightingale | S_HDL_PL | NA | C | T | 0.023784 | 0.004386 | 3.20E-08 |
| rs8176663 | miR-126-3p | trans | 2.73E-39 | Nightingale | S_LDL_PL_ | NA | C | T | -0.03006 | 0.004417 | 2.60E-13 |
| rs8176663 | miR-126-3p | trans | 2.73E-39 | Nightingale | Citrate | NA | C | T | 0.026829 | 0.004396 | 7.30E-10 |
| rs8176663 | miR-126-3p | trans | 2.73E-39 | Nightingale | M_LDL_CE | NA | C | T | 0.032977 | 0.004431 | 9.60E-14 |
| rs8176663 | miR-139-5p | trans | 7.92E-42 | Nightingale | L_LDL_P | NA | C | T | 0.029177 | 0.00441 | 6.20E-11 |
| rs8176663 | miR-139-5p | trans | 7.92E-42 | Nightingale | S_VLDL_C | NA | C | T | 0.038373 | 0.004424 | 1.80E-18 |
| rs8176663 | miR-139-5p | trans | 7.92E-42 | Nightingale | L_LDL_CE | NA | C | T | 0.031569 | 0.004385 | 9.80E-13 |
| rs8176663 | miR-139-5p | trans | 7.92E-42 | Nightingale | LDL_CE | NA | C | T | 0.032556 | 0.004413 | 2.20E-13 |
| rs8176663 | miR-139-5p | trans | 7.92E-42 | Nightingale | XL_VLDL_F | NA | C | T | 0.037195 | 0.00429 | 2.00E-18 |
| rs8176663 | miR-139-5p | trans | 7.92E-42 | Nightingale | S_HDL_L | NA | C | T | 0.026297 | 0.00441 | 1.50E-09 |
| rs8176663 | miR-139-5p | trans | 7.92E-42 | Nightingale | S_LDL_C | NA | C | T | 0.030946 | 0.004429 | 5.50E-12 |
| rs8176663 | miR-139-5p | trans | 7.92E-42 | Nightingale | PUFA | NA | C | T | 0.023713 | 0.004316 | 1.40E-08 |
| rs8176663 | miR-139-5p | trans | 7.92E-42 | Nightingale | L_VLDL_TG | NA | C | T | -0.05114 | 0.004404 | 5.30E-32 |
| rs8176663 | miR-139-5p | trans | 7.92E-42 | Nightingale | IDL_FC | NA | C | T | 0.038254 | 0.004284 | 1.50E-18 |
| rs8176663 | miR-139-5p | trans | 7.92E-42 | Nightingale | L_LDL_PL | NA | C | T | 0.037992 | 0.004391 | 9.00E-18 |
| rs8176663 | miR-139-5p | trans | 7.92E-42 | Nightingale | S_VLDL_L | NA | C | T | 0.024668 | 0.004382 | 3.70E-09 |
| rs8176663 | miR-139-5p | trans | 7.92E-42 | Nightingale | M_VLDL_P | NA | C | T | 0.044056 | 0.004247 | 5.50E-25 |
| rs8176663 | miR-139-5p | trans | 7.92E-42 | Nightingale | S_LDL_L | NA | C | T | 0.027745 | 0.004424 | 8.20E-10 |
| rs8176663 | miR-139-5p | trans | 7.92E-42 | Nightingale | S_LDL_FC | NA | C | T | 0.028568 | 0.004418 | 4.20E-10 |
| rs8176663 | miR-139-5p | trans | 7.92E-42 | Nightingale | XL_VLDL_C | NA | C | T | 0.038222 | 0.004243 | 4.70E-19 |
| rs8176663 | miR-139-5p | trans | 7.92E-42 | Nightingale | Sphingomy | NA | C | T | 0.029016 | 0.004136 | 6.80E-12 |

| | | | | | | | | | | | |
|---|---|---|---|---|---|---|---|---|---|---|---|
| rs8176663 | miR-139-5p | trans | 7.92E-42 | Nightingale | IDL_C | NA | C | T | 0.034955 | 0.004273 | 9.40E-16 |
| rs8176663 | miR-139-5p | trans | 7.92E-42 | Nightingale | IDL_FC_pct | NA | C | T | 0.025773 | 0.004391 | 1.10E-08 |
| rs8176663 | miR-139-5p | trans | 7.92E-42 | Nightingale | GlycA | NA | C | T | 0.028013 | 0.004406 | 1.40E-10 |
| rs8176663 | miR-139-5p | trans | 7.92E-42 | Nightingale | Remnant_( | NA | C | T | 0.036867 | 0.004396 | 3.70E-17 |
| rs8176663 | miR-139-5p | trans | 7.92E-42 | Nightingale | S_VLDL_TG | NA | C | T | -0.03813 | 0.004283 | 1.40E-18 |
| rs8176663 | miR-139-5p | trans | 7.92E-42 | Nightingale | XS_VLDL_C | NA | C | T | 0.037794 | 0.00427 | 2.10E-18 |
| rs8176663 | miR-139-5p | trans | 7.92E-42 | Nightingale | S_VLDL_PL | NA | C | T | 0.032777 | 0.004417 | 4.20E-14 |
| rs8176663 | miR-139-5p | trans | 7.92E-42 | Nightingale | IDL_L | NA | C | T | 0.035268 | 0.004278 | 4.80E-16 |
| rs8176663 | miR-139-5p | trans | 7.92E-42 | Nightingale | Cholines | NA | C | T | 0.022288 | 0.004161 | 4.90E-08 |
| rs8176663 | miR-139-5p | trans | 7.92E-42 | Nightingale | M_VLDL_C | NA | C | T | 0.036205 | 0.004168 | 1.20E-17 |
| rs8176663 | miR-139-5p | trans | 7.92E-42 | Nightingale | LDL_PL | NA | C | T | 0.034991 | 0.004415 | 4.70E-15 |
| rs8176663 | miR-139-5p | trans | 7.92E-42 | Nightingale | XXL_VLDL_ | NA | C | T | 0.033119 | 0.004459 | 9.00E-14 |
| rs8176663 | miR-139-5p | trans | 7.92E-42 | Nightingale | LDL_L | NA | C | T | 0.033306 | 0.00441 | 7.40E-14 |
| rs8176663 | miR-139-5p | trans | 7.92E-42 | Nightingale | L_VLDL_C_ | NA | C | T | 0.052192 | 0.004322 | 4.00E-34 |
| rs8176663 | miR-139-5p | trans | 7.92E-42 | Nightingale | S_VLDL_PL | NA | C | T | 0.02774 | 0.004237 | 1.80E-10 |
| rs8176663 | miR-139-5p | trans | 7.92E-42 | Nightingale | M_LDL_L | NA | C | T | 0.033035 | 0.004431 | 1.00E-13 |
| rs8176663 | miR-139-5p | trans | 7.92E-42 | Nightingale | XS_VLDL_C | NA | C | T | 0.037989 | 0.004295 | 1.30E-18 |
| rs8176663 | miR-139-5p | trans | 7.92E-42 | Nightingale | LA | NA | C | T | 0.024903 | 0.004339 | 3.70E-09 |
| rs8176663 | miR-139-5p | trans | 7.92E-42 | Nightingale | LDL_C | NA | C | T | 0.033196 | 0.004406 | 8.80E-14 |
| rs8176663 | miR-139-5p | trans | 7.92E-42 | Nightingale | S_VLDL_CE | NA | C | T | 0.047034 | 0.004365 | 1.20E-26 |
| rs8176663 | miR-139-5p | trans | 7.92E-42 | Nightingale | S_HDL_P | NA | C | T | 0.030615 | 0.004425 | 5.10E-12 |
| rs8176663 | miR-139-5p | trans | 7.92E-42 | Nightingale | S_HDL_FC | NA | C | T | 0.02345 | 0.004344 | 4.60E-08 |
| rs8176663 | miR-139-5p | trans | 7.92E-42 | Nightingale | Total_FC | NA | C | T | 0.033253 | 0.004323 | 6.80E-15 |
| rs8176663 | miR-139-5p | trans | 7.92E-42 | Nightingale | Clinical_LD | NA | C | T | 0.036477 | 0.004399 | 2.70E-16 |
| rs8176663 | miR-139-5p | trans | 7.92E-42 | Nightingale | XXL_VLDL_ | NA | C | T | 0.025004 | 0.004454 | 1.50E-09 |
| rs8176663 | miR-139-5p | trans | 7.92E-42 | Nightingale | XS_VLDL_F | NA | C | T | 0.031441 | 0.004393 | 2.40E-12 |
| rs8176663 | miR-139-5p | trans | 7.92E-42 | Nightingale | XL_VLDL_C | NA | C | T | 0.037351 | 0.004245 | 3.10E-18 |
| rs8176663 | miR-139-5p | trans | 7.92E-42 | Nightingale | ApoB_by_A | NA | C | T | 0.025055 | 0.004358 | 1.30E-08 |
| rs8176663 | miR-139-5p | trans | 7.92E-42 | Nightingale | VLDL_CE | NA | C | T | 0.035786 | 0.004426 | 3.00E-16 |
| rs8176663 | miR-139-5p | trans | 7.92E-42 | Nightingale | Total_C | NA | C | T | 0.032372 | 0.004284 | 3.30E-14 |
| rs8176663 | miR-139-5p | trans | 7.92E-42 | Nightingale | L_LDL_FC | NA | C | T | 0.033627 | 0.004345 | 2.90E-14 |
| rs8176663 | miR-139-5p | trans | 7.92E-42 | Nightingale | Total_L | NA | C | T | 0.026346 | 0.004353 | 4.60E-10 |
| rs8176663 | miR-139-5p | trans | 7.92E-42 | Nightingale | S_VLDL_FC | NA | C | T | 0.037905 | 0.004426 | 7.30E-18 |
| rs8176663 | miR-139-5p | trans | 7.92E-42 | Nightingale | IDL_CE | NA | C | T | 0.033507 | 0.004272 | 1.30E-14 |
| rs8176663 | miR-139-5p | trans | 7.92E-42 | Nightingale | S_VLDL_FC | NA | C | T | 0.029358 | 0.004228 | 1.10E-11 |
| rs8176663 | miR-139-5p | trans | 7.92E-42 | Nightingale | VLDL_FC | NA | C | T | 0.024159 | 0.004392 | 1.10E-08 |
| rs8176663 | miR-139-5p | trans | 7.92E-42 | Nightingale | Omega_6 | NA | C | T | 0.025889 | 0.004321 | 5.80E-10 |
| rs8176663 | miR-139-5p | trans | 7.92E-42 | Nightingale | L_LDL_L | NA | C | T | 0.033551 | 0.004382 | 3.00E-14 |
| rs8176663 | miR-139-5p | trans | 7.92E-42 | Nightingale | Total_CE | NA | C | T | 0.031745 | 0.004268 | 9.70E-14 |
| rs8176663 | miR-139-5p | trans | 7.92E-42 | Nightingale | S_VLDL_P | NA | C | T | 0.023465 | 0.004369 | 1.70E-08 |
| rs8176663 | miR-139-5p | trans | 7.92E-42 | Nightingale | S_LDL_P | NA | C | T | 0.029589 | 0.004429 | 3.20E-11 |
| rs8176663 | miR-139-5p | trans | 7.92E-42 | Nightingale | M_LDL_PL | NA | C | T | 0.032471 | 0.004431 | 2.60E-13 |
| rs8176663 | miR-139-5p | trans | 7.92E-42 | Nightingale | M_LDL_C_p | NA | C | T | 0.031165 | 0.004428 | 2.90E-12 |
| rs8176663 | miR-139-5p | trans | 7.92E-42 | Nightingale | L_LDL_C | NA | C | T | 0.032322 | 0.004375 | 2.60E-13 |
| rs8176663 | miR-139-5p | trans | 7.92E-42 | Nightingale | M_VLDL_F( | NA | C | T | 0.033372 | 0.004424 | 3.10E-14 |
| rs8176663 | miR-139-5p | trans | 7.92E-42 | Nightingale | IDL_P | NA | C | T | 0.039988 | 0.004378 | 1.30E-19 |
| rs8176663 | miR-139-5p | trans | 7.92E-42 | Nightingale | XXL_VLDL_ | NA | C | T | 0.034827 | 0.004434 | 2.90E-15 |
| rs8176663 | miR-139-5p | trans | 7.92E-42 | Nightingale | XS_VLDL_F | NA | C | T | 0.037065 | 0.004348 | 6.50E-18 |
| rs8176663 | miR-139-5p | trans | 7.92E-42 | Nightingale | M_VLDL_C | NA | C | T | 0.033628 | 0.004157 | 1.90E-15 |
| rs8176663 | miR-139-5p | trans | 7.92E-42 | Nightingale | M_VLDL_T( | NA | C | T | -0.03856 | 0.004181 | 6.80E-20 |
| rs8176663 | miR-139-5p | trans | 7.92E-42 | Nightingale | S_VLDL_CE | NA | C | T | 0.038182 | 0.004419 | 1.60E-18 |
| rs8176663 | miR-139-5p | trans | 7.92E-42 | Nightingale | IDL_PL | NA | C | T | 0.035341 | 0.00427 | 3.30E-16 |
| rs8176663 | miR-139-5p | trans | 7.92E-42 | Nightingale | XXL_VLDL_ | NA | C | T | 0.030847 | 0.004406 | 1.10E-12 |
| rs8176663 | miR-139-5p | trans | 7.92E-42 | Nightingale | S_HDL_CE | NA | C | T | 0.02864 | 0.004432 | 1.30E-10 |
| rs8176663 | miR-139-5p | trans | 7.92E-42 | Nightingale | M_VLDL_C | NA | C | T | 0.038711 | 0.004413 | 1.90E-18 |
| rs8176663 | miR-139-5p | trans | 7.92E-42 | Nightingale | S_VLDL_C_ | NA | C | T | 0.041889 | 0.004306 | 5.20E-22 |
| rs8176663 | miR-139-5p | trans | 7.92E-42 | Nightingale | VLDL_C | NA | C | T | 0.031302 | 0.004419 | 4.90E-13 |
| rs8176663 | miR-139-5p | trans | 7.92E-42 | Nightingale | L_VLDL_CE | NA | C | T | 0.047336 | 0.004234 | 2.40E-29 |
| rs8176663 | miR-139-5p | trans | 7.92E-42 | Nightingale | Total_PL | NA | C | T | 0.024922 | 0.004213 | 1.10E-09 |
| rs8176663 | miR-139-5p | trans | 7.92E-42 | Nightingale | S_LDL_C_p | NA | C | T | 0.045366 | 0.004442 | 5.40E-25 |
| rs8176663 | miR-139-5p | trans | 7.92E-42 | Nightingale | XS_VLDL_P | NA | C | T | 0.036698 | 0.004366 | 1.40E-17 |
| rs8176663 | miR-139-5p | trans | 7.92E-42 | Nightingale | VLDL_P | NA | C | T | 0.026633 | 0.004401 | 4.40E-10 |
| rs8176663 | miR-139-5p | trans | 7.92E-42 | Nightingale | M_LDL_TG | NA | C | T | -0.02763 | 0.00437 | 3.20E-10 |

| | | | | | | | | | | | | |
|---|---|---|---|---|---|---|---|---|---|---|---|---|
| rs8176663 | miR-139-5| | trans | 7.92E-42 | Nightingale | non_HDL_( | NA | C | T | 0.035698 | 0.004401 | 5.10E-16 |
| rs8176663 | miR-139-5| | trans | 7.92E-42 | Nightingale | XS_VLDL_L | NA | C | T | 0.033813 | 0.00435 | 2.10E-15 |
| rs8176663 | miR-139-5| | trans | 7.92E-42 | Nightingale | S_LDL_CE_ | NA | C | T | 0.030946 | 0.004417 | 4.90E-14 |
| rs8176663 | miR-139-5| | trans | 7.92E-42 | Nightingale | M_VLDL_F | NA | C | T | 0.042064 | 0.00421 | 3.50E-23 |
| rs8176663 | miR-139-5| | trans | 7.92E-42 | Nightingale | M_LDL_FC | NA | C | T | 0.034352 | 0.004419 | 1.90E-14 |
| rs8176663 | miR-139-5| | trans | 7.92E-42 | Nightingale | M_LDL_C | NA | C | T | 0.033991 | 0.004433 | 2.10E-14 |
| rs8176663 | miR-139-5| | trans | 7.92E-42 | Nightingale | XS_VLDL_P | NA | C | T | 0.032112 | 0.004363 | 5.40E-14 |
| rs8176663 | miR-139-5| | trans | 7.92E-42 | Nightingale | ApoB | NA | C | T | 0.033939 | 0.004419 | 2.10E-14 |
| rs8176663 | miR-139-5| | trans | 7.92E-42 | Nightingale | L_VLDL_FC | NA | C | T | 0.048588 | 0.004382 | 4.50E-29 |
| rs8176663 | miR-139-5| | trans | 7.92E-42 | Nightingale | S_HDL_C | NA | C | T | 0.028415 | 0.004417 | 1.70E-10 |
| rs8176663 | miR-139-5| | trans | 7.92E-42 | Nightingale | XL_VLDL_T | NA | C | T | -0.04392 | 0.004289 | 1.70E-24 |
| rs8176663 | miR-139-5| | trans | 7.92E-42 | Nightingale | S_LDL_CE | NA | C | T | 0.030551 | 0.004427 | 5.80E-12 |
| rs8176663 | miR-139-5| | trans | 7.92E-42 | Nightingale | LDL_P | NA | C | T | 0.031801 | 0.004422 | 1.00E-12 |
| rs8176663 | miR-139-5| | trans | 7.92E-42 | Nightingale | XXL_VLDL_ | NA | C | T | -0.03573 | 0.004463 | 2.20E-16 |
| rs8176663 | miR-139-5| | trans | 7.92E-42 | Nightingale | XL_HDL_PL | NA | C | T | -0.02277 | 0.004065 | 1.10E-08 |
| rs8176663 | miR-139-5| | trans | 7.92E-42 | Nightingale | XL_VLDL_P | NA | C | T | 0.038847 | 0.004432 | 5.10E-19 |
| rs8176663 | miR-139-5| | trans | 7.92E-42 | Nightingale | M_VLDL_C | NA | C | T | 0.040844 | 0.004385 | 2.30E-20 |
| rs8176663 | miR-139-5| | trans | 7.92E-42 | Nightingale | M_LDL_P | NA | C | T | 0.035895 | 0.004437 | 6.80E-16 |
| rs8176663 | miR-139-5| | trans | 7.92E-42 | Nightingale | M_VLDL_P | NA | C | T | 0.029128 | 0.004418 | 2.00E-11 |
| rs8176663 | miR-139-5| | trans | 7.92E-42 | Nightingale | M_VLDL_P | NA | C | T | 0.028296 | 0.004414 | 6.30E-11 |
| rs8176663 | miR-139-5| | trans | 7.92E-42 | Nightingale | LDL_FC | NA | C | T | 0.033635 | 0.004378 | 5.20E-14 |
| rs8176663 | miR-139-5| | trans | 7.92E-42 | Nightingale | S_HDL_PL | NA | C | T | 0.023784 | 0.004386 | 3.20E-08 |
| rs8176663 | miR-139-5| | trans | 7.92E-42 | Nightingale | S_LDL_PL_ | NA | C | T | -0.03006 | 0.004417 | 2.60E-13 |
| rs8176663 | miR-139-5| | trans | 7.92E-42 | Nightingale | Citrate | NA | C | T | 0.026829 | 0.004396 | 7.30E-10 |
| rs8176663 | miR-139-5| | trans | 7.92E-42 | Nightingale | M_LDL_CE | NA | C | T | 0.032977 | 0.004431 | 9.60E-14 |
| rs8176663 | let-7c-5p | trans | 2.74E-23 | Nightingale | L_LDL_P | NA | C | T | 0.029177 | 0.00441 | 6.20E-11 |
| rs8176663 | let-7c-5p | trans | 2.74E-23 | Nightingale | S_VLDL_C | NA | C | T | 0.038373 | 0.004424 | 1.80E-18 |
| rs8176663 | let-7c-5p | trans | 2.74E-23 | Nightingale | L_LDL_CE | NA | C | T | 0.031569 | 0.004385 | 9.80E-13 |
| rs8176663 | let-7c-5p | trans | 2.74E-23 | Nightingale | LDL_CE | NA | C | T | 0.032556 | 0.004413 | 2.20E-13 |
| rs8176663 | let-7c-5p | trans | 2.74E-23 | Nightingale | XL_VLDL_F | NA | C | T | 0.037195 | 0.00429 | 2.00E-18 |
| rs8176663 | let-7c-5p | trans | 2.74E-23 | Nightingale | S_HDL_L | NA | C | T | 0.026297 | 0.00441 | 1.50E-09 |
| rs8176663 | let-7c-5p | trans | 2.74E-23 | Nightingale | S_LDL_C | NA | C | T | 0.030946 | 0.004429 | 5.50E-12 |
| rs8176663 | let-7c-5p | trans | 2.74E-23 | Nightingale | PUFA | NA | C | T | 0.023713 | 0.004316 | 1.40E-08 |
| rs8176663 | let-7c-5p | trans | 2.74E-23 | Nightingale | L_VLDL_TG | NA | C | T | -0.05114 | 0.004404 | 5.30E-32 |
| rs8176663 | let-7c-5p | trans | 2.74E-23 | Nightingale | IDL_FC | NA | C | T | 0.038254 | 0.004284 | 1.50E-18 |
| rs8176663 | let-7c-5p | trans | 2.74E-23 | Nightingale | L_LDL_PL | NA | C | T | 0.037992 | 0.004391 | 9.00E-18 |
| rs8176663 | let-7c-5p | trans | 2.74E-23 | Nightingale | S_VLDL_L | NA | C | T | 0.024668 | 0.004382 | 3.70E-09 |
| rs8176663 | let-7c-5p | trans | 2.74E-23 | Nightingale | M_VLDL_P | NA | C | T | 0.044056 | 0.004247 | 5.50E-25 |
| rs8176663 | let-7c-5p | trans | 2.74E-23 | Nightingale | S_LDL_L | NA | C | T | 0.027745 | 0.004424 | 8.20E-10 |
| rs8176663 | let-7c-5p | trans | 2.74E-23 | Nightingale | S_LDL_FC | NA | C | T | 0.028568 | 0.004418 | 4.20E-10 |
| rs8176663 | let-7c-5p | trans | 2.74E-23 | Nightingale | XL_VLDL_C | NA | C | T | 0.038222 | 0.004243 | 4.70E-19 |
| rs8176663 | let-7c-5p | trans | 2.74E-23 | Nightingale | Sphingomy | NA | C | T | 0.029016 | 0.004136 | 6.80E-12 |
| rs8176663 | let-7c-5p | trans | 2.74E-23 | Nightingale | IDL_C | NA | C | T | 0.034955 | 0.004273 | 9.40E-16 |
| rs8176663 | let-7c-5p | trans | 2.74E-23 | Nightingale | IDL_FC_pct | NA | C | T | 0.025773 | 0.004391 | 1.10E-08 |
| rs8176663 | let-7c-5p | trans | 2.74E-23 | Nightingale | GlycA | NA | C | T | 0.028013 | 0.004406 | 1.40E-10 |
| rs8176663 | let-7c-5p | trans | 2.74E-23 | Nightingale | Remnant_( | NA | C | T | 0.036867 | 0.004396 | 3.70E-17 |
| rs8176663 | let-7c-5p | trans | 2.74E-23 | Nightingale | S_VLDL_TG | NA | C | T | -0.03813 | 0.004283 | 1.40E-18 |
| rs8176663 | let-7c-5p | trans | 2.74E-23 | Nightingale | XS_VLDL_C | NA | C | T | 0.037794 | 0.00427 | 2.10E-18 |
| rs8176663 | let-7c-5p | trans | 2.74E-23 | Nightingale | S_VLDL_PL | NA | C | T | 0.032777 | 0.004417 | 4.20E-14 |
| rs8176663 | let-7c-5p | trans | 2.74E-23 | Nightingale | IDL_L | NA | C | T | 0.035268 | 0.004278 | 4.80E-16 |
| rs8176663 | let-7c-5p | trans | 2.74E-23 | Nightingale | Cholines | NA | C | T | 0.022288 | 0.004161 | 4.90E-08 |
| rs8176663 | let-7c-5p | trans | 2.74E-23 | Nightingale | M_VLDL_C | NA | C | T | 0.036205 | 0.004168 | 1.20E-17 |
| rs8176663 | let-7c-5p | trans | 2.74E-23 | Nightingale | LDL_PL | NA | C | T | 0.034991 | 0.004415 | 4.70E-15 |
| rs8176663 | let-7c-5p | trans | 2.74E-23 | Nightingale | XXL_VLDL_ | NA | C | T | 0.033119 | 0.004459 | 9.00E-14 |
| rs8176663 | let-7c-5p | trans | 2.74E-23 | Nightingale | LDL_L | NA | C | T | 0.033306 | 0.00441 | 7.40E-14 |
| rs8176663 | let-7c-5p | trans | 2.74E-23 | Nightingale | L_VLDL_C_ | NA | C | T | 0.052192 | 0.004322 | 4.00E-34 |
| rs8176663 | let-7c-5p | trans | 2.74E-23 | Nightingale | S_VLDL_PL | NA | C | T | 0.02774 | 0.004237 | 1.80E-10 |
| rs8176663 | let-7c-5p | trans | 2.74E-23 | Nightingale | M_LDL_L | NA | C | T | 0.033035 | 0.004431 | 1.00E-13 |
| rs8176663 | let-7c-5p | trans | 2.74E-23 | Nightingale | XS_VLDL_C | NA | C | T | 0.037989 | 0.004295 | 1.30E-18 |
| rs8176663 | let-7c-5p | trans | 2.74E-23 | Nightingale | LA | NA | C | T | 0.024903 | 0.004339 | 3.70E-09 |
| rs8176663 | let-7c-5p | trans | 2.74E-23 | Nightingale | LDL_C | NA | C | T | 0.033196 | 0.004406 | 8.80E-14 |
| rs8176663 | let-7c-5p | trans | 2.74E-23 | Nightingale | S_VLDL_CE | NA | C | T | 0.047034 | 0.004365 | 1.20E-26 |
| rs8176663 | let-7c-5p | trans | 2.74E-23 | Nightingale | S_HDL_P | NA | C | T | 0.030615 | 0.004425 | 5.10E-12 |

| | | | | | | | | | | | |
|---|---|---|---|---|---|---|---|---|---|---|---|
| rs8176663 | let-7c-5p | trans | 2.74E-23 | Nightingale | S_HDL_FC | NA | C | T | 0.02345 | 0.004344 | 4.60E-08 |
| rs8176663 | let-7c-5p | trans | 2.74E-23 | Nightingale | Total_FC | NA | C | T | 0.033253 | 0.004323 | 6.80E-15 |
| rs8176663 | let-7c-5p | trans | 2.74E-23 | Nightingale | Clinical_LD | NA | C | T | 0.036477 | 0.004399 | 2.70E-16 |
| rs8176663 | let-7c-5p | trans | 2.74E-23 | Nightingale | XXL_VLDL_ | NA | C | T | 0.025004 | 0.004454 | 1.50E-09 |
| rs8176663 | let-7c-5p | trans | 2.74E-23 | Nightingale | XS_VLDL_F | NA | C | T | 0.031441 | 0.004393 | 2.40E-12 |
| rs8176663 | let-7c-5p | trans | 2.74E-23 | Nightingale | XL_VLDL_C | NA | C | T | 0.037351 | 0.004245 | 3.10E-18 |
| rs8176663 | let-7c-5p | trans | 2.74E-23 | Nightingale | ApoB_by_A | NA | C | T | 0.025055 | 0.004358 | 1.30E-08 |
| rs8176663 | let-7c-5p | trans | 2.74E-23 | Nightingale | VLDL_CE | NA | C | T | 0.035786 | 0.004426 | 3.00E-16 |
| rs8176663 | let-7c-5p | trans | 2.74E-23 | Nightingale | Total_C | NA | C | T | 0.032372 | 0.004284 | 3.30E-14 |
| rs8176663 | let-7c-5p | trans | 2.74E-23 | Nightingale | L_LDL_FC | NA | C | T | 0.033627 | 0.004345 | 2.90E-14 |
| rs8176663 | let-7c-5p | trans | 2.74E-23 | Nightingale | Total_L | NA | C | T | 0.026346 | 0.004353 | 4.60E-10 |
| rs8176663 | let-7c-5p | trans | 2.74E-23 | Nightingale | S_VLDL_FC | NA | C | T | 0.037905 | 0.004426 | 7.30E-18 |
| rs8176663 | let-7c-5p | trans | 2.74E-23 | Nightingale | IDL_CE | NA | C | T | 0.033507 | 0.004272 | 1.30E-14 |
| rs8176663 | let-7c-5p | trans | 2.74E-23 | Nightingale | S_VLDL_FC | NA | C | T | 0.029358 | 0.004228 | 1.10E-11 |
| rs8176663 | let-7c-5p | trans | 2.74E-23 | Nightingale | VLDL_FC | NA | C | T | 0.024159 | 0.004392 | 1.10E-08 |
| rs8176663 | let-7c-5p | trans | 2.74E-23 | Nightingale | Omega_6 | NA | C | T | 0.025889 | 0.004321 | 5.80E-10 |
| rs8176663 | let-7c-5p | trans | 2.74E-23 | Nightingale | L_LDL_L | NA | C | T | 0.033551 | 0.004382 | 3.00E-14 |
| rs8176663 | let-7c-5p | trans | 2.74E-23 | Nightingale | Total_CE | NA | C | T | 0.031745 | 0.004268 | 9.70E-14 |
| rs8176663 | let-7c-5p | trans | 2.74E-23 | Nightingale | S_VLDL_P | NA | C | T | 0.023465 | 0.004369 | 1.70E-08 |
| rs8176663 | let-7c-5p | trans | 2.74E-23 | Nightingale | S_LDL_P | NA | C | T | 0.029589 | 0.004429 | 3.20E-11 |
| rs8176663 | let-7c-5p | trans | 2.74E-23 | Nightingale | M_LDL_PL | NA | C | T | 0.032471 | 0.004431 | 2.60E-13 |
| rs8176663 | let-7c-5p | trans | 2.74E-23 | Nightingale | M_LDL_C_ | NA | C | T | 0.031165 | 0.004428 | 2.90E-12 |
| rs8176663 | let-7c-5p | trans | 2.74E-23 | Nightingale | L_LDL_C | NA | C | T | 0.032322 | 0.004375 | 2.60E-13 |
| rs8176663 | let-7c-5p | trans | 2.74E-23 | Nightingale | M_VLDL_F | NA | C | T | 0.033372 | 0.004424 | 3.10E-14 |
| rs8176663 | let-7c-5p | trans | 2.74E-23 | Nightingale | IDL_P | NA | C | T | 0.039988 | 0.004378 | 1.30E-19 |
| rs8176663 | let-7c-5p | trans | 2.74E-23 | Nightingale | XXL_VLDL_ | NA | C | T | 0.034827 | 0.004434 | 2.90E-15 |
| rs8176663 | let-7c-5p | trans | 2.74E-23 | Nightingale | XS_VLDL_F | NA | C | T | 0.037065 | 0.004348 | 6.50E-18 |
| rs8176663 | let-7c-5p | trans | 2.74E-23 | Nightingale | M_VLDL_C | NA | C | T | 0.033628 | 0.004157 | 1.90E-15 |
| rs8176663 | let-7c-5p | trans | 2.74E-23 | Nightingale | M_VLDL_T | NA | C | T | -0.03856 | 0.004181 | 6.80E-20 |
| rs8176663 | let-7c-5p | trans | 2.74E-23 | Nightingale | S_VLDL_CE | NA | C | T | 0.038182 | 0.004419 | 1.60E-18 |
| rs8176663 | let-7c-5p | trans | 2.74E-23 | Nightingale | IDL_PL | NA | C | T | 0.035341 | 0.00427 | 3.30E-16 |
| rs8176663 | let-7c-5p | trans | 2.74E-23 | Nightingale | XXL_VLDL_ | NA | C | T | 0.030847 | 0.004406 | 1.10E-12 |
| rs8176663 | let-7c-5p | trans | 2.74E-23 | Nightingale | S_HDL_CE | NA | C | T | 0.02864 | 0.004432 | 1.30E-10 |
| rs8176663 | let-7c-5p | trans | 2.74E-23 | Nightingale | M_VLDL_C | NA | C | T | 0.038711 | 0.004413 | 1.90E-18 |
| rs8176663 | let-7c-5p | trans | 2.74E-23 | Nightingale | S_VLDL_C_ | NA | C | T | 0.041889 | 0.004306 | 5.20E-22 |
| rs8176663 | let-7c-5p | trans | 2.74E-23 | Nightingale | VLDL_C | NA | C | T | 0.031302 | 0.004419 | 4.90E-13 |
| rs8176663 | let-7c-5p | trans | 2.74E-23 | Nightingale | L_VLDL_CE | NA | C | T | 0.047336 | 0.004234 | 2.40E-29 |
| rs8176663 | let-7c-5p | trans | 2.74E-23 | Nightingale | Total_PL | NA | C | T | 0.024922 | 0.004213 | 1.10E-09 |
| rs8176663 | let-7c-5p | trans | 2.74E-23 | Nightingale | S_LDL_C_p | NA | C | T | 0.045366 | 0.004442 | 5.40E-25 |
| rs8176663 | let-7c-5p | trans | 2.74E-23 | Nightingale | XS_VLDL_P | NA | C | T | 0.036698 | 0.004366 | 1.40E-17 |
| rs8176663 | let-7c-5p | trans | 2.74E-23 | Nightingale | VLDL_P | NA | C | T | 0.026633 | 0.004401 | 4.40E-10 |
| rs8176663 | let-7c-5p | trans | 2.74E-23 | Nightingale | M_LDL_TG | NA | C | T | -0.02763 | 0.00437 | 3.20E-10 |
| rs8176663 | let-7c-5p | trans | 2.74E-23 | Nightingale | non_HDL_( | NA | C | T | 0.035698 | 0.004401 | 5.10E-16 |
| rs8176663 | let-7c-5p | trans | 2.74E-23 | Nightingale | XS_VLDL_L | NA | C | T | 0.033813 | 0.00435 | 2.10E-15 |
| rs8176663 | let-7c-5p | trans | 2.74E-23 | Nightingale | S_LDL_CE_ | NA | C | T | 0.030946 | 0.004417 | 4.90E-14 |
| rs8176663 | let-7c-5p | trans | 2.74E-23 | Nightingale | M_VLDL_F | NA | C | T | 0.042064 | 0.00421 | 3.50E-23 |
| rs8176663 | let-7c-5p | trans | 2.74E-23 | Nightingale | M_LDL_FC | NA | C | T | 0.034352 | 0.004419 | 1.90E-14 |
| rs8176663 | let-7c-5p | trans | 2.74E-23 | Nightingale | M_LDL_C | NA | C | T | 0.033991 | 0.004433 | 2.10E-14 |
| rs8176663 | let-7c-5p | trans | 2.74E-23 | Nightingale | XS_VLDL_P | NA | C | T | 0.032112 | 0.004363 | 5.40E-14 |
| rs8176663 | let-7c-5p | trans | 2.74E-23 | Nightingale | ApoB | NA | C | T | 0.033939 | 0.004419 | 2.10E-14 |
| rs8176663 | let-7c-5p | trans | 2.74E-23 | Nightingale | L_VLDL_FC | NA | C | T | 0.048588 | 0.004382 | 4.50E-29 |
| rs8176663 | let-7c-5p | trans | 2.74E-23 | Nightingale | S_HDL_C | NA | C | T | 0.028415 | 0.004417 | 1.70E-10 |
| rs8176663 | let-7c-5p | trans | 2.74E-23 | Nightingale | XL_VLDL_T | NA | C | T | -0.04392 | 0.004289 | 1.70E-24 |
| rs8176663 | let-7c-5p | trans | 2.74E-23 | Nightingale | S_LDL_CE | NA | C | T | 0.030551 | 0.004427 | 5.80E-12 |
| rs8176663 | let-7c-5p | trans | 2.74E-23 | Nightingale | LDL_P | NA | C | T | 0.031801 | 0.004422 | 1.00E-12 |
| rs8176663 | let-7c-5p | trans | 2.74E-23 | Nightingale | XXL_VLDL_ | NA | C | T | -0.03573 | 0.004463 | 2.20E-16 |
| rs8176663 | let-7c-5p | trans | 2.74E-23 | Nightingale | XL_HDL_PL | NA | C | T | -0.02277 | 0.004065 | 1.10E-08 |
| rs8176663 | let-7c-5p | trans | 2.74E-23 | Nightingale | XL_VLDL_P | NA | C | T | 0.038847 | 0.004432 | 5.10E-19 |
| rs8176663 | let-7c-5p | trans | 2.74E-23 | Nightingale | M_VLDL_C | NA | C | T | 0.040844 | 0.004385 | 2.30E-20 |
| rs8176663 | let-7c-5p | trans | 2.74E-23 | Nightingale | M_LDL_P | NA | C | T | 0.035895 | 0.004437 | 6.80E-16 |
| rs8176663 | let-7c-5p | trans | 2.74E-23 | Nightingale | M_VLDL_P | NA | C | T | 0.029128 | 0.004418 | 2.00E-11 |
| rs8176663 | let-7c-5p | trans | 2.74E-23 | Nightingale | M_VLDL_P | NA | C | T | 0.028296 | 0.004414 | 6.30E-11 |
| rs8176663 | let-7c-5p | trans | 2.74E-23 | Nightingale | LDL_FC | NA | C | T | 0.033635 | 0.004378 | 5.20E-14 |

| | | | | | | | | | | | |
|---|---|---|---|---|---|---|---|---|---|---|---|
| rs8176663 | let-7c-5p | trans | 2.74E-23 | Nightingale | S_HDL_PL | NA | C | T | 0.023784 | 0.004386 | 3.20E-08 |
| rs8176663 | let-7c-5p | trans | 2.74E-23 | Nightingale | S_LDL_PL_ | NA | C | T | -0.03006 | 0.004417 | 2.60E-13 |
| rs8176663 | let-7c-5p | trans | 2.74E-23 | Nightingale | Citrate | NA | C | T | 0.026829 | 0.004396 | 7.30E-10 |
| rs8176663 | let-7c-5p | trans | 2.74E-23 | Nightingale | M_LDL_CE | NA | C | T | 0.032977 | 0.004431 | 9.60E-14 |
| rs8176668 | miR-126-3p | trans | 8.74E-19 | Nightingale | L_VLDL_FC | NA | T | A | -0.03165 | 0.004146 | 1.20E-14 |
| rs8176668 | miR-126-3p | trans | 8.74E-19 | Nightingale | XL_VLDL_F | NA | T | A | -0.02519 | 0.004058 | 8.00E-10 |
| rs8176668 | miR-126-3p | trans | 8.74E-19 | Nightingale | XS_VLDL_F | NA | T | A | -0.02234 | 0.004114 | 1.00E-08 |
| rs8176668 | miR-126-3p | trans | 8.74E-19 | Nightingale | S_VLDL_C_ | NA | T | A | -0.02856 | 0.004074 | 8.70E-12 |
| rs8176668 | miR-126-3p | trans | 8.74E-19 | Nightingale | XS_VLDL_C | NA | T | A | -0.02424 | 0.004064 | 8.20E-10 |
| rs8176668 | miR-126-3p | trans | 8.74E-19 | Nightingale | IDL_PL | NA | T | A | -0.02241 | 0.00404 | 2.20E-08 |
| rs8176668 | miR-126-3p | trans | 8.74E-19 | Nightingale | S_VLDL_TG | NA | T | A | 0.026056 | 0.004052 | 5.10E-10 |
| rs8176668 | miR-126-3p | trans | 8.74E-19 | Nightingale | XS_VLDL_P | NA | T | A | -0.02205 | 0.004131 | 1.40E-08 |
| rs8176668 | miR-126-3p | trans | 8.74E-19 | Nightingale | XL_VLDL_C | NA | T | A | -0.02539 | 0.004016 | 2.30E-09 |
| rs8176668 | miR-126-3p | trans | 8.74E-19 | Nightingale | IDL_P | NA | T | A | -0.0251 | 0.004143 | 8.40E-10 |
| rs8176668 | miR-126-3p | trans | 8.74E-19 | Nightingale | XL_VLDL_T | NA | T | A | 0.029823 | 0.004058 | 8.90E-13 |
| rs8176668 | miR-126-3p | trans | 8.74E-19 | Nightingale | XL_VLDL_P | NA | T | A | -0.02461 | 0.004193 | 1.20E-09 |
| rs8176668 | miR-126-3p | trans | 8.74E-19 | Nightingale | XXL_VLDL_ | NA | T | A | -0.02535 | 0.004194 | 1.60E-09 |
| rs8176668 | miR-126-3p | trans | 8.74E-19 | Nightingale | IDL_C | NA | T | A | -0.02302 | 0.004043 | 1.20E-08 |
| rs8176668 | miR-126-3p | trans | 8.74E-19 | Nightingale | Remnant_( | NA | T | A | -0.02282 | 0.004159 | 1.10E-08 |
| rs8176668 | miR-126-3p | trans | 8.74E-19 | Nightingale | M_LDL_P | NA | T | A | -0.02329 | 0.004198 | 1.50E-08 |
| rs8176668 | miR-126-3p | trans | 8.74E-19 | Nightingale | S_LDL_C_p | NA | T | A | -0.02958 | 0.004203 | 1.40E-12 |
| rs8176668 | miR-126-3p | trans | 8.74E-19 | Nightingale | IDL_CE | NA | T | A | -0.02206 | 0.004042 | 4.50E-08 |
| rs8176668 | miR-126-3p | trans | 8.74E-19 | Nightingale | L_VLDL_CE | NA | T | A | -0.03202 | 0.004006 | 2.60E-15 |
| rs8176668 | miR-126-3p | trans | 8.74E-19 | Nightingale | M_VLDL_C | NA | T | A | -0.02429 | 0.004175 | 1.80E-09 |
| rs8176668 | miR-126-3p | trans | 8.74E-19 | Nightingale | M_VLDL_F | NA | T | A | -0.02691 | 0.003983 | 5.50E-11 |
| rs8176668 | miR-126-3p | trans | 8.74E-19 | Nightingale | IDL_FC | NA | T | A | -0.02513 | 0.004053 | 5.50E-10 |
| rs8176668 | miR-126-3p | trans | 8.74E-19 | Nightingale | XL_VLDL_C | NA | T | A | -0.02604 | 0.004014 | 6.70E-10 |
| rs8176668 | miR-126-3p | trans | 8.74E-19 | Nightingale | S_VLDL_FC | NA | T | A | -0.02288 | 0.004187 | 1.60E-08 |
| rs8176668 | miR-126-3p | trans | 8.74E-19 | Nightingale | L_VLDL_C_ | NA | T | A | -0.03531 | 0.004089 | 5.70E-18 |
| rs8176668 | miR-126-3p | trans | 8.74E-19 | Nightingale | IDL_L | NA | T | A | -0.02272 | 0.004048 | 1.90E-08 |
| rs8176668 | miR-126-3p | trans | 8.74E-19 | Nightingale | L_VLDL_TG | NA | T | A | 0.03357 | 0.004167 | 1.80E-16 |
| rs8176668 | miR-126-3p | trans | 8.74E-19 | Nightingale | L_LDL_PL | NA | T | A | -0.02433 | 0.004155 | 3.60E-09 |
| rs8176668 | miR-126-3p | trans | 8.74E-19 | Nightingale | S_VLDL_CE | NA | T | A | -0.03248 | 0.00413 | 6.60E-15 |
| rs8176668 | miR-126-3p | trans | 8.74E-19 | Nightingale | M_VLDL_T | NA | T | A | 0.025206 | 0.003956 | 1.00E-09 |
| rs8176668 | miR-126-3p | trans | 8.74E-19 | Nightingale | XS_VLDL_C | NA | T | A | -0.02466 | 0.00404 | 3.80E-10 |
| rs8176668 | miR-126-3p | trans | 8.74E-19 | Nightingale | M_VLDL_C | NA | T | A | -0.02651 | 0.004149 | 9.10E-11 |
| rs8176668 | miR-126-3p | trans | 8.74E-19 | Nightingale | Clinical_LD | NA | T | A | -0.02337 | 0.004162 | 1.80E-08 |
| rs8176668 | miR-126-3p | trans | 8.74E-19 | Nightingale | M_VLDL_C | NA | T | A | -0.02371 | 0.003943 | 1.20E-08 |
| rs8176668 | miR-126-3p | trans | 8.74E-19 | Nightingale | S_VLDL_CE | NA | T | A | -0.02313 | 0.004181 | 6.70E-09 |
| rs8176668 | miR-126-3p | trans | 8.74E-19 | Nightingale | XXL_VLDL_ | NA | T | A | -0.02444 | 0.004217 | 6.80E-09 |
| rs8176668 | miR-126-3p | trans | 8.74E-19 | Nightingale | XXL_VLDL_ | NA | T | A | 0.02607 | 0.004221 | 3.10E-10 |
| rs8176668 | miR-126-3p | trans | 8.74E-19 | Nightingale | S_VLDL_C | NA | T | A | -0.02322 | 0.004186 | 7.30E-09 |
| rs8176668 | miR-126-3p | trans | 8.74E-19 | Nightingale | M_VLDL_P | NA | T | A | -0.02888 | 0.004018 | 1.10E-12 |
| rs8176668 | miR-10a-5p | trans | 1.84E-13 | Nightingale | L_VLDL_FC | NA | T | A | -0.03165 | 0.004146 | 1.20E-14 |
| rs8176668 | miR-10a-5p | trans | 1.84E-13 | Nightingale | XL_VLDL_F | NA | T | A | -0.02519 | 0.004058 | 8.00E-10 |
| rs8176668 | miR-10a-5p | trans | 1.84E-13 | Nightingale | XS_VLDL_F | NA | T | A | -0.02234 | 0.004114 | 1.00E-08 |
| rs8176668 | miR-10a-5p | trans | 1.84E-13 | Nightingale | S_VLDL_C_ | NA | T | A | -0.02856 | 0.004074 | 8.70E-12 |
| rs8176668 | miR-10a-5p | trans | 1.84E-13 | Nightingale | XS_VLDL_C | NA | T | A | -0.02424 | 0.004064 | 8.20E-10 |
| rs8176668 | miR-10a-5p | trans | 1.84E-13 | Nightingale | IDL_PL | NA | T | A | -0.02241 | 0.00404 | 2.20E-08 |
| rs8176668 | miR-10a-5p | trans | 1.84E-13 | Nightingale | S_VLDL_TG | NA | T | A | 0.026056 | 0.004052 | 5.10E-10 |
| rs8176668 | miR-10a-5p | trans | 1.84E-13 | Nightingale | XS_VLDL_P | NA | T | A | -0.02205 | 0.004131 | 1.40E-08 |
| rs8176668 | miR-10a-5p | trans | 1.84E-13 | Nightingale | XL_VLDL_C | NA | T | A | -0.02539 | 0.004016 | 2.30E-09 |
| rs8176668 | miR-10a-5p | trans | 1.84E-13 | Nightingale | IDL_P | NA | T | A | -0.0251 | 0.004143 | 8.40E-10 |
| rs8176668 | miR-10a-5p | trans | 1.84E-13 | Nightingale | XL_VLDL_T | NA | T | A | 0.029823 | 0.004058 | 8.90E-13 |
| rs8176668 | miR-10a-5p | trans | 1.84E-13 | Nightingale | XL_VLDL_P | NA | T | A | -0.02461 | 0.004193 | 1.20E-09 |
| rs8176668 | miR-10a-5p | trans | 1.84E-13 | Nightingale | XXL_VLDL_ | NA | T | A | -0.02535 | 0.004194 | 1.60E-09 |
| rs8176668 | miR-10a-5p | trans | 1.84E-13 | Nightingale | IDL_C | NA | T | A | -0.02302 | 0.004043 | 1.20E-08 |
| rs8176668 | miR-10a-5p | trans | 1.84E-13 | Nightingale | Remnant_( | NA | T | A | -0.02282 | 0.004159 | 1.10E-08 |
| rs8176668 | miR-10a-5p | trans | 1.84E-13 | Nightingale | M_LDL_P | NA | T | A | -0.02329 | 0.004198 | 1.50E-08 |
| rs8176668 | miR-10a-5p | trans | 1.84E-13 | Nightingale | S_LDL_C_p | NA | T | A | -0.02958 | 0.004203 | 1.40E-12 |
| rs8176668 | miR-10a-5p | trans | 1.84E-13 | Nightingale | IDL_CE | NA | T | A | -0.02206 | 0.004042 | 4.50E-08 |
| rs8176668 | miR-10a-5p | trans | 1.84E-13 | Nightingale | L_VLDL_CE | NA | T | A | -0.03202 | 0.004006 | 2.60E-15 |
| rs8176668 | miR-10a-5p | trans | 1.84E-13 | Nightingale | M_VLDL_C | NA | T | A | -0.02429 | 0.004175 | 1.80E-09 |

| | | | | | | | | | | | |
|---|---|---|---|---|---|---|---|---|---|---|---|
| rs8176668 | miR-10a-5p | trans | 1.84E-13 | Nightingale | M_VLDL_F | NA | T | A | -0.02691 | 0.003983 | 5.50E-11 |
| rs8176668 | miR-10a-5p | trans | 1.84E-13 | Nightingale | IDL_FC | NA | T | A | -0.02513 | 0.004053 | 5.50E-10 |
| rs8176668 | miR-10a-5p | trans | 1.84E-13 | Nightingale | XL_VLDL_C | NA | T | A | -0.02604 | 0.004014 | 6.70E-10 |
| rs8176668 | miR-10a-5p | trans | 1.84E-13 | Nightingale | S_VLDL_FC | NA | T | A | -0.02288 | 0.004187 | 1.60E-08 |
| rs8176668 | miR-10a-5p | trans | 1.84E-13 | Nightingale | L_VLDL_C_ | NA | T | A | -0.03531 | 0.004089 | 5.70E-18 |
| rs8176668 | miR-10a-5p | trans | 1.84E-13 | Nightingale | IDL_L | NA | T | A | -0.02272 | 0.004048 | 1.90E-08 |
| rs8176668 | miR-10a-5p | trans | 1.84E-13 | Nightingale | L_VLDL_TG | NA | T | A | 0.03357 | 0.004167 | 1.80E-16 |
| rs8176668 | miR-10a-5p | trans | 1.84E-13 | Nightingale | L_LDL_PL | NA | T | A | -0.02433 | 0.004155 | 3.60E-09 |
| rs8176668 | miR-10a-5p | trans | 1.84E-13 | Nightingale | S_VLDL_CE | NA | T | A | -0.03248 | 0.00413 | 6.60E-15 |
| rs8176668 | miR-10a-5p | trans | 1.84E-13 | Nightingale | M_VLDL_T | NA | T | A | 0.025206 | 0.003956 | 1.00E-09 |
| rs8176668 | miR-10a-5p | trans | 1.84E-13 | Nightingale | XS_VLDL_C | NA | T | A | -0.02466 | 0.00404 | 3.80E-10 |
| rs8176668 | miR-10a-5p | trans | 1.84E-13 | Nightingale | M_VLDL_C | NA | T | A | -0.02651 | 0.004149 | 9.10E-11 |
| rs8176668 | miR-10a-5p | trans | 1.84E-13 | Nightingale | Clinical_LD | NA | T | A | -0.02337 | 0.004162 | 1.80E-08 |
| rs8176668 | miR-10a-5p | trans | 1.84E-13 | Nightingale | M_VLDL_C | NA | T | A | -0.02371 | 0.003943 | 1.20E-08 |
| rs8176668 | miR-10a-5p | trans | 1.84E-13 | Nightingale | S_VLDL_CE | NA | T | A | -0.02313 | 0.004181 | 6.70E-09 |
| rs8176668 | miR-10a-5p | trans | 1.84E-13 | Nightingale | XXL_VLDL_ | NA | T | A | -0.02444 | 0.004217 | 6.80E-09 |
| rs8176668 | miR-10a-5p | trans | 1.84E-13 | Nightingale | XXL_VLDL_ | NA | T | A | 0.02607 | 0.004221 | 3.10E-10 |
| rs8176668 | miR-10a-5p | trans | 1.84E-13 | Nightingale | S_VLDL_C | NA | T | A | -0.02322 | 0.004186 | 7.30E-09 |
| rs8176668 | miR-10a-5p | trans | 1.84E-13 | Nightingale | M_VLDL_P | NA | T | A | -0.02888 | 0.004018 | 1.10E-12 |
| rs8176681 | miR-126-3p | trans | 2.18E-18 | Nightingale | S_VLDL_C | NA | C | T | -0.02257 | 0.004162 | 2.00E-08 |
| rs8176681 | miR-126-3p | trans | 2.18E-18 | Nightingale | XXL_VLDL_ | NA | C | T | -0.02484 | 0.004192 | 4.30E-09 |
| rs8176681 | miR-126-3p | trans | 2.18E-18 | Nightingale | M_LDL_C_ | NA | C | T | -0.02516 | 0.004165 | 1.60E-09 |
| rs8176681 | miR-126-3p | trans | 2.18E-18 | Nightingale | M_VLDL_P | NA | C | T | -0.02923 | 0.003995 | 5.90E-13 |
| rs8176681 | miR-126-3p | trans | 2.18E-18 | Nightingale | IDL_FC | NA | C | T | -0.02577 | 0.00403 | 1.70E-10 |
| rs8176681 | miR-126-3p | trans | 2.18E-18 | Nightingale | S_VLDL_FC | NA | C | T | -0.02243 | 0.004163 | 3.10E-08 |
| rs8176681 | miR-126-3p | trans | 2.18E-18 | Nightingale | IDL_P | NA | C | T | -0.0244 | 0.004119 | 2.20E-09 |
| rs8176681 | miR-126-3p | trans | 2.18E-18 | Nightingale | XL_VLDL_C | NA | C | T | -0.02692 | 0.003993 | 1.90E-10 |
| rs8176681 | miR-126-3p | trans | 2.18E-18 | Nightingale | M_LDL_P | NA | C | T | -0.02364 | 0.004174 | 7.20E-09 |
| rs8176681 | miR-126-3p | trans | 2.18E-18 | Nightingale | M_VLDL_C | NA | C | T | -0.0263 | 0.004124 | 1.10E-10 |
| rs8176681 | miR-126-3p | trans | 2.18E-18 | Nightingale | L_VLDL_FC | NA | C | T | -0.03174 | 0.004122 | 7.80E-15 |
| rs8176681 | miR-126-3p | trans | 2.18E-18 | Nightingale | XS_VLDL_C | NA | C | T | -0.02448 | 0.004017 | 7.60E-10 |
| rs8176681 | miR-126-3p | trans | 2.18E-18 | Nightingale | Clinical_LD | NA | C | T | -0.0237 | 0.004138 | 9.20E-09 |
| rs8176681 | miR-126-3p | trans | 2.18E-18 | Nightingale | XS_VLDL_P | NA | C | T | -0.02149 | 0.004107 | 5.00E-08 |
| rs8176681 | miR-126-3p | trans | 2.18E-18 | Nightingale | XXL_VLDL_ | NA | C | T | 0.026529 | 0.004196 | 1.50E-10 |
| rs8176681 | miR-126-3p | trans | 2.18E-18 | Nightingale | L_LDL_FC | NA | C | T | -0.02249 | 0.004087 | 4.40E-08 |
| rs8176681 | miR-126-3p | trans | 2.18E-18 | Nightingale | M_VLDL_T | NA | C | T | 0.026094 | 0.003933 | 2.40E-10 |
| rs8176681 | miR-126-3p | trans | 2.18E-18 | Nightingale | M_VLDL_F | NA | C | T | -0.02797 | 0.00396 | 8.50E-12 |
| rs8176681 | miR-126-3p | trans | 2.18E-18 | Nightingale | XXL_VLDL_ | NA | C | T | -0.02343 | 0.004143 | 2.00E-08 |
| rs8176681 | miR-126-3p | trans | 2.18E-18 | Nightingale | IDL_C | NA | C | T | -0.02322 | 0.00402 | 9.10E-09 |
| rs8176681 | miR-126-3p | trans | 2.18E-18 | Nightingale | XL_VLDL_C | NA | C | T | -0.02738 | 0.003991 | 7.40E-11 |
| rs8176681 | miR-126-3p | trans | 2.18E-18 | Nightingale | XL_VLDL_F | NA | C | T | -0.02581 | 0.004035 | 3.70E-10 |
| rs8176681 | miR-126-3p | trans | 2.18E-18 | Nightingale | S_VLDL_CE | NA | C | T | -0.02237 | 0.004157 | 2.30E-08 |
| rs8176681 | miR-126-3p | trans | 2.18E-18 | Nightingale | L_VLDL_CE | NA | C | T | -0.03355 | 0.003983 | 1.20E-16 |
| rs8176681 | miR-126-3p | trans | 2.18E-18 | Nightingale | M_VLDL_C | NA | C | T | -0.02328 | 0.00391 | 2.20E-08 |
| rs8176681 | miR-126-3p | trans | 2.18E-18 | Nightingale | S_VLDL_C_ | NA | C | T | -0.02914 | 0.004051 | 2.90E-12 |
| rs8176681 | miR-126-3p | trans | 2.18E-18 | Nightingale | IDL_L | NA | C | T | -0.02289 | 0.004024 | 1.50E-08 |
| rs8176681 | miR-126-3p | trans | 2.18E-18 | Nightingale | L_VLDL_C_ | NA | C | T | -0.0365 | 0.004065 | 3.90E-19 |
| rs8176681 | miR-126-3p | trans | 2.18E-18 | Nightingale | IDL_PL | NA | C | T | -0.02269 | 0.004017 | 1.60E-08 |
| rs8176681 | miR-126-3p | trans | 2.18E-18 | Nightingale | XL_VLDL_P | NA | C | T | -0.02288 | 0.004169 | 1.30E-08 |
| rs8176681 | miR-126-3p | trans | 2.18E-18 | Nightingale | S_VLDL_TG | NA | C | T | 0.026753 | 0.004029 | 1.60E-10 |
| rs8176681 | miR-126-3p | trans | 2.18E-18 | Nightingale | M_VLDL_C | NA | C | T | -0.02388 | 0.004151 | 3.30E-09 |
| rs8176681 | miR-126-3p | trans | 2.18E-18 | Nightingale | S_VLDL_CE | NA | C | T | -0.03268 | 0.004106 | 3.20E-15 |
| rs8176681 | miR-126-3p | trans | 2.18E-18 | Nightingale | M_VLDL_C | NA | C | T | -0.02471 | 0.003921 | 2.50E-09 |
| rs8176681 | miR-126-3p | trans | 2.18E-18 | Nightingale | XS_VLDL_F | NA | C | T | -0.02199 | 0.00409 | 2.80E-08 |
| rs8176681 | miR-126-3p | trans | 2.18E-18 | Nightingale | XS_VLDL_C | NA | C | T | -0.024 | 0.00404 | 1.80E-09 |
| rs8176681 | miR-126-3p | trans | 2.18E-18 | Nightingale | L_LDL_PL | NA | C | T | -0.02478 | 0.004131 | 1.50E-09 |
| rs8176681 | miR-126-3p | trans | 2.18E-18 | Nightingale | S_LDL_C_p | NA | C | T | -0.03119 | 0.004178 | 3.70E-14 |
| rs8176681 | miR-126-3p | trans | 2.18E-18 | Nightingale | L_VLDL_TG | NA | C | T | 0.033794 | 0.004142 | 1.10E-16 |
| rs8176681 | miR-126-3p | trans | 2.18E-18 | Nightingale | Remnant_( | NA | C | T | -0.02226 | 0.004135 | 2.50E-08 |
| rs8176681 | miR-126-3p | trans | 2.18E-18 | Nightingale | XL_VLDL_T | NA | C | T | 0.030651 | 0.004034 | 1.80E-13 |
| rs8176681 | miR-126-3p | trans | 2.18E-18 | Nightingale | XXL_VLDL_ | NA | C | T | -0.02579 | 0.004169 | 8.40E-10 |
| rs8176681 | miR-126-3p | trans | 2.18E-18 | Nightingale | IDL_CE | NA | C | T | -0.02212 | 0.004019 | 4.30E-08 |
| rs8176681 | miR-10a-5p | trans | 9.94E-13 | Nightingale | S_VLDL_C | NA | C | T | -0.02257 | 0.004162 | 2.00E-08 |

| | | | | | | | | | | | |
|---|---|---|---|---|---|---|---|---|---|---|---|
| rs8176681 | miR-10a-5p | trans | 9.94E-13 | Nightingale | XXL_VLDL_ | NA | C | T | -0.02484 | 0.004192 | 4.30E-09 |
| rs8176681 | miR-10a-5p | trans | 9.94E-13 | Nightingale | M_LDL_C_ | NA | C | T | -0.02516 | 0.004165 | 1.60E-09 |
| rs8176681 | miR-10a-5p | trans | 9.94E-13 | Nightingale | M_VLDL_P | NA | C | T | -0.02923 | 0.003995 | 5.90E-13 |
| rs8176681 | miR-10a-5p | trans | 9.94E-13 | Nightingale | IDL_FC | NA | C | T | -0.02577 | 0.00403 | 1.70E-10 |
| rs8176681 | miR-10a-5p | trans | 9.94E-13 | Nightingale | S_VLDL_FC | NA | C | T | -0.02243 | 0.004163 | 3.10E-08 |
| rs8176681 | miR-10a-5p | trans | 9.94E-13 | Nightingale | IDL_P | NA | C | T | -0.0244 | 0.004119 | 2.20E-09 |
| rs8176681 | miR-10a-5p | trans | 9.94E-13 | Nightingale | XL_VLDL_C | NA | C | T | -0.02692 | 0.003993 | 1.90E-10 |
| rs8176681 | miR-10a-5p | trans | 9.94E-13 | Nightingale | M_LDL_P | NA | C | T | -0.02364 | 0.004174 | 7.20E-09 |
| rs8176681 | miR-10a-5p | trans | 9.94E-13 | Nightingale | M_VLDL_C | NA | C | T | -0.0263 | 0.004124 | 1.10E-10 |
| rs8176681 | miR-10a-5p | trans | 9.94E-13 | Nightingale | L_VLDL_FC | NA | C | T | -0.03174 | 0.004122 | 7.80E-15 |
| rs8176681 | miR-10a-5p | trans | 9.94E-13 | Nightingale | XS_VLDL_C | NA | C | T | -0.02448 | 0.004017 | 7.60E-10 |
| rs8176681 | miR-10a-5p | trans | 9.94E-13 | Nightingale | Clinical_LD | NA | C | T | -0.0237 | 0.004138 | 9.20E-09 |
| rs8176681 | miR-10a-5p | trans | 9.94E-13 | Nightingale | XS_VLDL_P | NA | C | T | -0.02149 | 0.004107 | 5.00E-08 |
| rs8176681 | miR-10a-5p | trans | 9.94E-13 | Nightingale | XXL_VLDL_ | NA | C | T | 0.026529 | 0.004196 | 1.50E-10 |
| rs8176681 | miR-10a-5p | trans | 9.94E-13 | Nightingale | L_LDL_FC | NA | C | T | -0.02249 | 0.004087 | 4.40E-08 |
| rs8176681 | miR-10a-5p | trans | 9.94E-13 | Nightingale | M_VLDL_T | NA | C | T | 0.026094 | 0.003933 | 2.40E-10 |
| rs8176681 | miR-10a-5p | trans | 9.94E-13 | Nightingale | M_VLDL_F | NA | C | T | -0.02797 | 0.00396 | 8.50E-12 |
| rs8176681 | miR-10a-5p | trans | 9.94E-13 | Nightingale | XXL_VLDL_ | NA | C | T | -0.02343 | 0.004143 | 2.00E-08 |
| rs8176681 | miR-10a-5p | trans | 9.94E-13 | Nightingale | IDL_C | NA | C | T | -0.02322 | 0.00402 | 9.10E-09 |
| rs8176681 | miR-10a-5p | trans | 9.94E-13 | Nightingale | XL_VLDL_C | NA | C | T | -0.02738 | 0.003991 | 7.40E-11 |
| rs8176681 | miR-10a-5p | trans | 9.94E-13 | Nightingale | XL_VLDL_F | NA | C | T | -0.02581 | 0.004035 | 3.70E-10 |
| rs8176681 | miR-10a-5p | trans | 9.94E-13 | Nightingale | S_VLDL_CE | NA | C | T | -0.02237 | 0.004157 | 2.30E-08 |
| rs8176681 | miR-10a-5p | trans | 9.94E-13 | Nightingale | L_VLDL_CE | NA | C | T | -0.03355 | 0.003983 | 1.20E-16 |
| rs8176681 | miR-10a-5p | trans | 9.94E-13 | Nightingale | M_VLDL_C | NA | C | T | -0.02328 | 0.00391 | 2.20E-08 |
| rs8176681 | miR-10a-5p | trans | 9.94E-13 | Nightingale | S_VLDL_C_ | NA | C | T | -0.02914 | 0.004051 | 2.90E-12 |
| rs8176681 | miR-10a-5p | trans | 9.94E-13 | Nightingale | IDL_L | NA | C | T | -0.02289 | 0.004024 | 1.50E-08 |
| rs8176681 | miR-10a-5p | trans | 9.94E-13 | Nightingale | L_VLDL_C_ | NA | C | T | -0.0365 | 0.004065 | 3.90E-19 |
| rs8176681 | miR-10a-5p | trans | 9.94E-13 | Nightingale | IDL_PL | NA | C | T | -0.02269 | 0.004017 | 1.60E-08 |
| rs8176681 | miR-10a-5p | trans | 9.94E-13 | Nightingale | XL_VLDL_P | NA | C | T | -0.02288 | 0.004169 | 1.30E-08 |
| rs8176681 | miR-10a-5p | trans | 9.94E-13 | Nightingale | S_VLDL_TG | NA | C | T | 0.026753 | 0.004029 | 1.60E-10 |
| rs8176681 | miR-10a-5p | trans | 9.94E-13 | Nightingale | M_VLDL_C | NA | C | T | -0.02388 | 0.004151 | 3.30E-09 |
| rs8176681 | miR-10a-5p | trans | 9.94E-13 | Nightingale | S_VLDL_CE | NA | C | T | -0.03268 | 0.004106 | 3.20E-15 |
| rs8176681 | miR-10a-5p | trans | 9.94E-13 | Nightingale | M_VLDL_C | NA | C | T | -0.02471 | 0.003921 | 2.50E-09 |
| rs8176681 | miR-10a-5p | trans | 9.94E-13 | Nightingale | XS_VLDL_F | NA | C | T | -0.02199 | 0.00409 | 2.80E-08 |
| rs8176681 | miR-10a-5p | trans | 9.94E-13 | Nightingale | XS_VLDL_C | NA | C | T | -0.024 | 0.00404 | 1.80E-09 |
| rs8176681 | miR-10a-5p | trans | 9.94E-13 | Nightingale | L_LDL_PL | NA | C | T | -0.02478 | 0.004131 | 1.50E-09 |
| rs8176681 | miR-10a-5p | trans | 9.94E-13 | Nightingale | S_LDL_C_p | NA | C | T | -0.03119 | 0.004178 | 3.70E-14 |
| rs8176681 | miR-10a-5p | trans | 9.94E-13 | Nightingale | L_VLDL_TG | NA | C | T | 0.033794 | 0.004142 | 1.10E-16 |
| rs8176681 | miR-10a-5p | trans | 9.94E-13 | Nightingale | Remnant_( | NA | C | T | -0.02226 | 0.004135 | 2.50E-08 |
| rs8176681 | miR-10a-5p | trans | 9.94E-13 | Nightingale | XL_VLDL_T | NA | C | T | 0.030651 | 0.004034 | 1.80E-13 |
| rs8176681 | miR-10a-5p | trans | 9.94E-13 | Nightingale | XXL_VLDL_ | NA | C | T | -0.02579 | 0.004169 | 8.40E-10 |
| rs8176681 | miR-10a-5p | trans | 9.94E-13 | Nightingale | IDL_CE | NA | C | T | -0.02212 | 0.004019 | 4.30E-08 |
| rs8176682 | miR-10a-5p | trans | 2.24E-13 | Nightingale | XL_VLDL_C | NA | T | C | -0.02622 | 0.004016 | 5.90E-10 |
| rs8176682 | miR-10a-5p | trans | 2.24E-13 | Nightingale | IDL_FC | NA | T | C | -0.02532 | 0.004053 | 3.80E-10 |
| rs8176682 | miR-10a-5p | trans | 2.24E-13 | Nightingale | M_VLDL_C | NA | T | C | -0.02292 | 0.003933 | 3.50E-08 |
| rs8176682 | miR-10a-5p | trans | 2.24E-13 | Nightingale | S_VLDL_CE | NA | T | C | -0.03305 | 0.00413 | 2.10E-15 |
| rs8176682 | miR-10a-5p | trans | 2.24E-13 | Nightingale | IDL_L | NA | T | C | -0.02283 | 0.004047 | 1.50E-08 |
| rs8176682 | miR-10a-5p | trans | 2.24E-13 | Nightingale | XL_VLDL_P | NA | T | C | -0.02472 | 0.004192 | 1.10E-09 |
| rs8176682 | miR-10a-5p | trans | 2.24E-13 | Nightingale | XL_VLDL_F | NA | T | C | -0.02592 | 0.004058 | 2.60E-10 |
| rs8176682 | miR-10a-5p | trans | 2.24E-13 | Nightingale | XXL_VLDL_ | NA | T | C | 0.026931 | 0.00422 | 8.70E-11 |
| rs8176682 | miR-10a-5p | trans | 2.24E-13 | Nightingale | XS_VLDL_P | NA | T | C | -0.02185 | 0.00413 | 1.90E-08 |
| rs8176682 | miR-10a-5p | trans | 2.24E-13 | Nightingale | S_VLDL_C_ | NA | T | C | -0.02915 | 0.004074 | 2.90E-12 |
| rs8176682 | miR-10a-5p | trans | 2.24E-13 | Nightingale | S_VLDL_C | NA | T | C | -0.02288 | 0.004185 | 1.20E-08 |
| rs8176682 | miR-10a-5p | trans | 2.24E-13 | Nightingale | XXL_VLDL_ | NA | T | C | -0.02517 | 0.004216 | 2.40E-09 |
| rs8176682 | miR-10a-5p | trans | 2.24E-13 | Nightingale | L_VLDL_TG | NA | T | C | 0.034082 | 0.004166 | 7.00E-17 |
| rs8176682 | miR-10a-5p | trans | 2.24E-13 | Nightingale | M_VLDL_P | NA | T | C | -0.02953 | 0.004017 | 3.20E-13 |
| rs8176682 | miR-10a-5p | trans | 2.24E-13 | Nightingale | M_VLDL_T | NA | T | C | 0.025898 | 0.003956 | 3.00E-10 |
| rs8176682 | miR-10a-5p | trans | 2.24E-13 | Nightingale | Clinical_LD | NA | T | C | -0.0234 | 0.004162 | 1.60E-08 |
| rs8176682 | miR-10a-5p | trans | 2.24E-13 | Nightingale | XS_VLDL_C | NA | T | C | -0.02416 | 0.004063 | 8.90E-10 |
| rs8176682 | miR-10a-5p | trans | 2.24E-13 | Nightingale | IDL_P | NA | T | C | -0.02509 | 0.004142 | 7.90E-10 |
| rs8176682 | miR-10a-5p | trans | 2.24E-13 | Nightingale | L_VLDL_C_ | NA | T | C | -0.03612 | 0.004088 | 1.00E-18 |
| rs8176682 | miR-10a-5p | trans | 2.24E-13 | Nightingale | M_VLDL_C | NA | T | C | -0.02641 | 0.004148 | 9.70E-11 |
| rs8176682 | miR-10a-5p | trans | 2.24E-13 | Nightingale | M_VLDL_C | NA | T | C | -0.02407 | 0.004174 | 2.30E-09 |

| | | | | | | | | | | | |
|---|---|---|---|---|---|---|---|---|---|---|---|
| rs8176682 | miR-10a-5p | trans | 2.24E-13 | Nightingale | M_LDL_P | NA | T | C | -0.02326 | 0.004198 | 1.50E-08 |
| rs8176682 | miR-10a-5p | trans | 2.24E-13 | Nightingale | M_VLDL_F | NA | T | C | -0.02759 | 0.003982 | 1.60E-11 |
| rs8176682 | miR-10a-5p | trans | 2.24E-13 | Nightingale | XS_VLDL_F | NA | T | C | -0.02215 | 0.004113 | 1.30E-08 |
| rs8176682 | miR-10a-5p | trans | 2.24E-13 | Nightingale | XL_VLDL_C | NA | T | C | -0.02687 | 0.004014 | 1.60E-10 |
| rs8176682 | miR-10a-5p | trans | 2.24E-13 | Nightingale | XXL_VLDL_ | NA | T | C | -0.02345 | 0.004167 | 2.10E-08 |
| rs8176682 | miR-10a-5p | trans | 2.24E-13 | Nightingale | S_VLDL_TG | NA | T | C | 0.026645 | 0.004052 | 1.80E-10 |
| rs8176682 | miR-10a-5p | trans | 2.24E-13 | Nightingale | L_VLDL_CE | NA | T | C | -0.03277 | 0.004006 | 5.60E-16 |
| rs8176682 | miR-10a-5p | trans | 2.24E-13 | Nightingale | IDL_C | NA | T | C | -0.02318 | 0.004043 | 8.90E-09 |
| rs8176682 | miR-10a-5p | trans | 2.24E-13 | Nightingale | IDL_PL | NA | T | C | -0.02245 | 0.004039 | 1.90E-08 |
| rs8176682 | miR-10a-5p | trans | 2.24E-13 | Nightingale | XS_VLDL_C | NA | T | C | -0.02464 | 0.00404 | 3.80E-10 |
| rs8176682 | miR-10a-5p | trans | 2.24E-13 | Nightingale | L_LDL_PL | NA | T | C | -0.02438 | 0.004154 | 3.00E-09 |
| rs8176682 | miR-10a-5p | trans | 2.24E-13 | Nightingale | Remnant_( | NA | T | C | -0.02265 | 0.004159 | 1.30E-08 |
| rs8176682 | miR-10a-5p | trans | 2.24E-13 | Nightingale | XXL_VLDL_ | NA | T | C | -0.02619 | 0.004193 | 4.70E-10 |
| rs8176682 | miR-10a-5p | trans | 2.24E-13 | Nightingale | L_VLDL_FC | NA | T | C | -0.03213 | 0.004146 | 5.50E-15 |
| rs8176682 | miR-10a-5p | trans | 2.24E-13 | Nightingale | M_VLDL_C | NA | T | C | -0.02439 | 0.003943 | 3.80E-09 |
| rs8176682 | miR-10a-5p | trans | 2.24E-13 | Nightingale | IDL_CE | NA | T | C | -0.02222 | 0.004041 | 3.40E-08 |
| rs8176682 | miR-10a-5p | trans | 2.24E-13 | Nightingale | XL_VLDL_T | NA | T | C | 0.030619 | 0.004057 | 2.00E-13 |
| rs8176682 | miR-10a-5p | trans | 2.24E-13 | Nightingale | S_VLDL_CE | NA | T | C | -0.02277 | 0.004181 | 1.20E-08 |
| rs8176682 | miR-10a-5p | trans | 2.24E-13 | Nightingale | S_VLDL_FC | NA | T | C | -0.02256 | 0.004187 | 2.50E-08 |
| rs8176682 | miR-10a-5p | trans | 2.24E-13 | Nightingale | S_LDL_C_p | NA | T | C | -0.02983 | 0.004202 | 9.00E-13 |
| rs8176682 | miR-126-3p | trans | 1.54E-18 | Nightingale | XL_VLDL_C | NA | T | C | -0.02622 | 0.004016 | 5.90E-10 |
| rs8176682 | miR-126-3p | trans | 1.54E-18 | Nightingale | IDL_FC | NA | T | C | -0.02532 | 0.004053 | 3.80E-10 |
| rs8176682 | miR-126-3p | trans | 1.54E-18 | Nightingale | M_VLDL_C | NA | T | C | -0.02292 | 0.003933 | 3.50E-08 |
| rs8176682 | miR-126-3p | trans | 1.54E-18 | Nightingale | S_VLDL_CE | NA | T | C | -0.03305 | 0.00413 | 2.10E-15 |
| rs8176682 | miR-126-3p | trans | 1.54E-18 | Nightingale | IDL_L | NA | T | C | -0.02283 | 0.004047 | 1.50E-08 |
| rs8176682 | miR-126-3p | trans | 1.54E-18 | Nightingale | XL_VLDL_P | NA | T | C | -0.02472 | 0.004192 | 1.10E-09 |
| rs8176682 | miR-126-3p | trans | 1.54E-18 | Nightingale | XL_VLDL_F | NA | T | C | -0.02592 | 0.004058 | 2.60E-10 |
| rs8176682 | miR-126-3p | trans | 1.54E-18 | Nightingale | XXL_VLDL_ | NA | T | C | 0.026931 | 0.00422 | 8.70E-11 |
| rs8176682 | miR-126-3p | trans | 1.54E-18 | Nightingale | XS_VLDL_P | NA | T | C | -0.02185 | 0.00413 | 1.90E-08 |
| rs8176682 | miR-126-3p | trans | 1.54E-18 | Nightingale | S_VLDL_C_ | NA | T | C | -0.02915 | 0.004074 | 2.90E-12 |
| rs8176682 | miR-126-3p | trans | 1.54E-18 | Nightingale | S_VLDL_C | NA | T | C | -0.02288 | 0.004185 | 1.20E-08 |
| rs8176682 | miR-126-3p | trans | 1.54E-18 | Nightingale | XXL_VLDL_ | NA | T | C | -0.02517 | 0.004216 | 2.40E-09 |
| rs8176682 | miR-126-3p | trans | 1.54E-18 | Nightingale | L_VLDL_TG | NA | T | C | 0.034082 | 0.004166 | 7.00E-17 |
| rs8176682 | miR-126-3p | trans | 1.54E-18 | Nightingale | M_VLDL_P | NA | T | C | -0.02953 | 0.004017 | 3.20E-13 |
| rs8176682 | miR-126-3p | trans | 1.54E-18 | Nightingale | M_VLDL_T | NA | T | C | 0.025898 | 0.003956 | 3.00E-10 |
| rs8176682 | miR-126-3p | trans | 1.54E-18 | Nightingale | Clinical_LD | NA | T | C | -0.0234 | 0.004162 | 1.60E-08 |
| rs8176682 | miR-126-3p | trans | 1.54E-18 | Nightingale | XS_VLDL_C | NA | T | C | -0.02416 | 0.004063 | 8.90E-10 |
| rs8176682 | miR-126-3p | trans | 1.54E-18 | Nightingale | IDL_P | NA | T | C | -0.02509 | 0.004142 | 7.90E-10 |
| rs8176682 | miR-126-3p | trans | 1.54E-18 | Nightingale | L_VLDL_C_ | NA | T | C | -0.03612 | 0.004088 | 1.00E-18 |
| rs8176682 | miR-126-3p | trans | 1.54E-18 | Nightingale | M_VLDL_C | NA | T | C | -0.02641 | 0.004148 | 9.70E-11 |
| rs8176682 | miR-126-3p | trans | 1.54E-18 | Nightingale | M_VLDL_C | NA | T | C | -0.02407 | 0.004174 | 2.30E-09 |
| rs8176682 | miR-126-3p | trans | 1.54E-18 | Nightingale | M_LDL_P | NA | T | C | -0.02326 | 0.004198 | 1.50E-08 |
| rs8176682 | miR-126-3p | trans | 1.54E-18 | Nightingale | M_VLDL_F | NA | T | C | -0.02759 | 0.003982 | 1.60E-11 |
| rs8176682 | miR-126-3p | trans | 1.54E-18 | Nightingale | XS_VLDL_F | NA | T | C | -0.02215 | 0.004113 | 1.30E-08 |
| rs8176682 | miR-126-3p | trans | 1.54E-18 | Nightingale | XL_VLDL_C | NA | T | C | -0.02687 | 0.004014 | 1.60E-10 |
| rs8176682 | miR-126-3p | trans | 1.54E-18 | Nightingale | XXL_VLDL_ | NA | T | C | -0.02345 | 0.004167 | 2.10E-08 |
| rs8176682 | miR-126-3p | trans | 1.54E-18 | Nightingale | S_VLDL_TG | NA | T | C | 0.026645 | 0.004052 | 1.80E-10 |
| rs8176682 | miR-126-3p | trans | 1.54E-18 | Nightingale | L_VLDL_CE | NA | T | C | -0.03277 | 0.004006 | 5.60E-16 |
| rs8176682 | miR-126-3p | trans | 1.54E-18 | Nightingale | IDL_C | NA | T | C | -0.02318 | 0.004043 | 8.90E-09 |
| rs8176682 | miR-126-3p | trans | 1.54E-18 | Nightingale | IDL_PL | NA | T | C | -0.02245 | 0.004039 | 1.90E-08 |
| rs8176682 | miR-126-3p | trans | 1.54E-18 | Nightingale | XS_VLDL_C | NA | T | C | -0.02464 | 0.00404 | 3.80E-10 |
| rs8176682 | miR-126-3p | trans | 1.54E-18 | Nightingale | L_LDL_PL | NA | T | C | -0.02438 | 0.004154 | 3.00E-09 |
| rs8176682 | miR-126-3p | trans | 1.54E-18 | Nightingale | Remnant_( | NA | T | C | -0.02265 | 0.004159 | 1.30E-08 |
| rs8176682 | miR-126-3p | trans | 1.54E-18 | Nightingale | XXL_VLDL_ | NA | T | C | -0.02619 | 0.004193 | 4.70E-10 |
| rs8176682 | miR-126-3p | trans | 1.54E-18 | Nightingale | L_VLDL_FC | NA | T | C | -0.03213 | 0.004146 | 5.50E-15 |
| rs8176682 | miR-126-3p | trans | 1.54E-18 | Nightingale | M_VLDL_C | NA | T | C | -0.02439 | 0.003943 | 3.80E-09 |
| rs8176682 | miR-126-3p | trans | 1.54E-18 | Nightingale | IDL_CE | NA | T | C | -0.02222 | 0.004041 | 3.40E-08 |
| rs8176682 | miR-126-3p | trans | 1.54E-18 | Nightingale | XL_VLDL_T | NA | T | C | 0.030619 | 0.004057 | 2.00E-13 |
| rs8176682 | miR-126-3p | trans | 1.54E-18 | Nightingale | S_VLDL_CE | NA | T | C | -0.02277 | 0.004181 | 1.20E-08 |
| rs8176682 | miR-126-3p | trans | 1.54E-18 | Nightingale | S_VLDL_FC | NA | T | C | -0.02256 | 0.004187 | 2.50E-08 |
| rs8176682 | miR-126-3p | trans | 1.54E-18 | Nightingale | S_LDL_C_p | NA | T | C | -0.02983 | 0.004202 | 9.00E-13 |
| rs8176690 | miR-10a-5p | trans | 1.35E-12 | Nightingale | Clinical_LD | NA | G | A | -0.0237 | 0.004137 | 8.80E-09 |
| rs8176690 | miR-10a-5p | trans | 1.35E-12 | Nightingale | IDL_PL | NA | G | A | -0.02263 | 0.004016 | 1.70E-08 |

| | | | | | | | | | | | |
|---|---|---|---|---|---|---|---|---|---|---|---|
| rs8176690 | miR-10a-5p | trans | 1.35E-12 | Nightingale | L_LDL_FC | NA | G | A | -0.02255 | 0.004086 | 3.90E-08 |
| rs8176690 | miR-10a-5p | trans | 1.35E-12 | Nightingale | S_VLDL_C_ | NA | G | A | -0.0293 | 0.00405 | 2.20E-12 |
| rs8176690 | miR-10a-5p | trans | 1.35E-12 | Nightingale | XS_VLDL_C | NA | G | A | -0.02388 | 0.004039 | 2.00E-09 |
| rs8176690 | miR-10a-5p | trans | 1.35E-12 | Nightingale | L_VLDL_CE | NA | G | A | -0.03364 | 0.003982 | 9.70E-17 |
| rs8176690 | miR-10a-5p | trans | 1.35E-12 | Nightingale | M_VLDL_F | NA | G | A | -0.02819 | 0.003959 | 5.60E-12 |
| rs8176690 | miR-10a-5p | trans | 1.35E-12 | Nightingale | IDL_C | NA | G | A | -0.02325 | 0.004019 | 8.20E-09 |
| rs8176690 | miR-10a-5p | trans | 1.35E-12 | Nightingale | S_LDL_C_p | NA | G | A | -0.03133 | 0.004178 | 2.70E-14 |
| rs8176690 | miR-10a-5p | trans | 1.35E-12 | Nightingale | XXL_VLDL_ | NA | G | A | 0.026524 | 0.004195 | 1.40E-10 |
| rs8176690 | miR-10a-5p | trans | 1.35E-12 | Nightingale | IDL_L | NA | G | A | -0.02286 | 0.004024 | 1.50E-08 |
| rs8176690 | miR-10a-5p | trans | 1.35E-12 | Nightingale | Remnant_( | NA | G | A | -0.02216 | 0.004134 | 2.80E-08 |
| rs8176690 | miR-10a-5p | trans | 1.35E-12 | Nightingale | XXL_VLDL_ | NA | G | A | -0.02341 | 0.004142 | 2.00E-08 |
| rs8176690 | miR-10a-5p | trans | 1.35E-12 | Nightingale | L_LDL_PL | NA | G | A | -0.02476 | 0.00413 | 1.40E-09 |
| rs8176690 | miR-10a-5p | trans | 1.35E-12 | Nightingale | M_VLDL_T | NA | G | A | 0.026294 | 0.003933 | 1.70E-10 |
| rs8176690 | miR-10a-5p | trans | 1.35E-12 | Nightingale | XXL_VLDL_ | NA | G | A | -0.02483 | 0.004191 | 4.30E-09 |
| rs8176690 | miR-10a-5p | trans | 1.35E-12 | Nightingale | XL_VLDL_P | NA | G | A | -0.0229 | 0.004168 | 1.20E-08 |
| rs8176690 | miR-10a-5p | trans | 1.35E-12 | Nightingale | IDL_CE | NA | G | A | -0.02213 | 0.004018 | 4.00E-08 |
| rs8176690 | miR-10a-5p | trans | 1.35E-12 | Nightingale | XL_VLDL_T | NA | G | A | 0.0308 | 0.004033 | 1.40E-13 |
| rs8176690 | miR-10a-5p | trans | 1.35E-12 | Nightingale | S_VLDL_CE | NA | G | A | -0.02217 | 0.004156 | 2.90E-08 |
| rs8176690 | miR-10a-5p | trans | 1.35E-12 | Nightingale | S_VLDL_C | NA | G | A | -0.02239 | 0.004161 | 2.50E-08 |
| rs8176690 | miR-10a-5p | trans | 1.35E-12 | Nightingale | M_LDL_C_ | NA | G | A | -0.02545 | 0.004164 | 1.10E-09 |
| rs8176690 | miR-10a-5p | trans | 1.35E-12 | Nightingale | XXL_VLDL_ | NA | G | A | -0.02579 | 0.004168 | 8.30E-10 |
| rs8176690 | miR-10a-5p | trans | 1.35E-12 | Nightingale | M_VLDL_C | NA | G | A | -0.02382 | 0.00415 | 3.50E-09 |
| rs8176690 | miR-10a-5p | trans | 1.35E-12 | Nightingale | L_VLDL_FC | NA | G | A | -0.03173 | 0.004121 | 7.50E-15 |
| rs8176690 | miR-10a-5p | trans | 1.35E-12 | Nightingale | S_VLDL_TG | NA | G | A | 0.026931 | 0.004028 | 1.20E-10 |
| rs8176690 | miR-10a-5p | trans | 1.35E-12 | Nightingale | XL_VLDL_F | NA | G | A | -0.02593 | 0.004034 | 3.10E-10 |
| rs8176690 | miR-10a-5p | trans | 1.35E-12 | Nightingale | XS_VLDL_C | NA | G | A | -0.02439 | 0.004016 | 8.10E-10 |
| rs8176690 | miR-10a-5p | trans | 1.35E-12 | Nightingale | M_VLDL_C | NA | G | A | -0.02627 | 0.004124 | 1.10E-10 |
| rs8176690 | miR-10a-5p | trans | 1.35E-12 | Nightingale | M_VLDL_C | NA | G | A | -0.02492 | 0.00392 | 1.80E-09 |
| rs8176690 | miR-10a-5p | trans | 1.35E-12 | Nightingale | XS_VLDL_F | NA | G | A | -0.0218 | 0.004089 | 3.40E-08 |
| rs8176690 | miR-10a-5p | trans | 1.35E-12 | Nightingale | IDL_P | NA | G | A | -0.02431 | 0.004118 | 2.30E-09 |
| rs8176690 | miR-10a-5p | trans | 1.35E-12 | Nightingale | S_VLDL_CE | NA | G | A | -0.03277 | 0.004106 | 2.60E-15 |
| rs8176690 | miR-10a-5p | trans | 1.35E-12 | Nightingale | M_LDL_P | NA | G | A | -0.02363 | 0.004173 | 7.10E-09 |
| rs8176690 | miR-10a-5p | trans | 1.35E-12 | Nightingale | IDL_FC | NA | G | A | -0.02583 | 0.004029 | 1.50E-10 |
| rs8176690 | miR-10a-5p | trans | 1.35E-12 | Nightingale | S_VLDL_FC | NA | G | A | -0.02229 | 0.004162 | 3.70E-08 |
| rs8176690 | miR-10a-5p | trans | 1.35E-12 | Nightingale | XL_VLDL_C | NA | G | A | -0.02752 | 0.00399 | 5.90E-11 |
| rs8176690 | miR-10a-5p | trans | 1.35E-12 | Nightingale | L_VLDL_C_ | NA | G | A | -0.03655 | 0.004064 | 3.30E-19 |
| rs8176690 | miR-10a-5p | trans | 1.35E-12 | Nightingale | L_VLDL_TG | NA | G | A | 0.033748 | 0.004142 | 1.10E-16 |
| rs8176690 | miR-10a-5p | trans | 1.35E-12 | Nightingale | XL_VLDL_C | NA | G | A | -0.02706 | 0.003992 | 1.60E-10 |
| rs8176690 | miR-10a-5p | trans | 1.35E-12 | Nightingale | M_VLDL_P | NA | G | A | -0.02937 | 0.003994 | 4.30E-13 |
| rs8176690 | miR-10a-5p | trans | 1.35E-12 | Nightingale | M_VLDL_C | NA | G | A | -0.02348 | 0.00391 | 1.70E-08 |
| rs8176690 | miR-126-3p | trans | 2.30E-18 | Nightingale | Clinical_LD | NA | G | A | -0.0237 | 0.004137 | 8.80E-09 |
| rs8176690 | miR-126-3p | trans | 2.30E-18 | Nightingale | IDL_PL | NA | G | A | -0.02263 | 0.004016 | 1.70E-08 |
| rs8176690 | miR-126-3p | trans | 2.30E-18 | Nightingale | L_LDL_FC | NA | G | A | -0.02255 | 0.004086 | 3.90E-08 |
| rs8176690 | miR-126-3p | trans | 2.30E-18 | Nightingale | S_VLDL_C_ | NA | G | A | -0.0293 | 0.00405 | 2.20E-12 |
| rs8176690 | miR-126-3p | trans | 2.30E-18 | Nightingale | XS_VLDL_C | NA | G | A | -0.02388 | 0.004039 | 2.00E-09 |
| rs8176690 | miR-126-3p | trans | 2.30E-18 | Nightingale | L_VLDL_CE | NA | G | A | -0.03364 | 0.003982 | 9.70E-17 |
| rs8176690 | miR-126-3p | trans | 2.30E-18 | Nightingale | M_VLDL_F | NA | G | A | -0.02819 | 0.003959 | 5.60E-12 |
| rs8176690 | miR-126-3p | trans | 2.30E-18 | Nightingale | IDL_C | NA | G | A | -0.02325 | 0.004019 | 8.20E-09 |
| rs8176690 | miR-126-3p | trans | 2.30E-18 | Nightingale | S_LDL_C_p | NA | G | A | -0.03133 | 0.004178 | 2.70E-14 |
| rs8176690 | miR-126-3p | trans | 2.30E-18 | Nightingale | XXL_VLDL_ | NA | G | A | 0.026524 | 0.004195 | 1.40E-10 |
| rs8176690 | miR-126-3p | trans | 2.30E-18 | Nightingale | IDL_L | NA | G | A | -0.02286 | 0.004024 | 1.50E-08 |
| rs8176690 | miR-126-3p | trans | 2.30E-18 | Nightingale | Remnant_( | NA | G | A | -0.02216 | 0.004134 | 2.80E-08 |
| rs8176690 | miR-126-3p | trans | 2.30E-18 | Nightingale | XXL_VLDL_ | NA | G | A | -0.02341 | 0.004142 | 2.00E-08 |
| rs8176690 | miR-126-3p | trans | 2.30E-18 | Nightingale | L_LDL_PL | NA | G | A | -0.02476 | 0.00413 | 1.40E-09 |
| rs8176690 | miR-126-3p | trans | 2.30E-18 | Nightingale | M_VLDL_T | NA | G | A | 0.026294 | 0.003933 | 1.70E-10 |
| rs8176690 | miR-126-3p | trans | 2.30E-18 | Nightingale | XXL_VLDL_ | NA | G | A | -0.02483 | 0.004191 | 4.30E-09 |
| rs8176690 | miR-126-3p | trans | 2.30E-18 | Nightingale | XL_VLDL_P | NA | G | A | -0.0229 | 0.004168 | 1.20E-08 |
| rs8176690 | miR-126-3p | trans | 2.30E-18 | Nightingale | IDL_CE | NA | G | A | -0.02213 | 0.004018 | 4.00E-08 |
| rs8176690 | miR-126-3p | trans | 2.30E-18 | Nightingale | XL_VLDL_T | NA | G | A | 0.0308 | 0.004033 | 1.40E-13 |
| rs8176690 | miR-126-3p | trans | 2.30E-18 | Nightingale | S_VLDL_CE | NA | G | A | -0.02217 | 0.004156 | 2.90E-08 |
| rs8176690 | miR-126-3p | trans | 2.30E-18 | Nightingale | S_VLDL_C | NA | G | A | -0.02239 | 0.004161 | 2.50E-08 |
| rs8176690 | miR-126-3p | trans | 2.30E-18 | Nightingale | M_LDL_C_ | NA | G | A | -0.02545 | 0.004164 | 1.10E-09 |
| rs8176690 | miR-126-3p | trans | 2.30E-18 | Nightingale | XXL_VLDL_ | NA | G | A | -0.02579 | 0.004168 | 8.30E-10 |

| | | | | | | | | | | | |
|---|---|---|---|---|---|---|---|---|---|---|---|
| rs8176690 | miR-126-3ｐ | trans | 2.30E-18 | Nightingale | M_VLDL_C | NA | G | A | -0.02382 | 0.00415 | 3.50E-09 |
| rs8176690 | miR-126-3ｐ | trans | 2.30E-18 | Nightingale | L_VLDL_FC | NA | G | A | -0.03173 | 0.004121 | 7.50E-15 |
| rs8176690 | miR-126-3ｐ | trans | 2.30E-18 | Nightingale | S_VLDL_TG | NA | G | A | 0.026931 | 0.004028 | 1.20E-10 |
| rs8176690 | miR-126-3ｐ | trans | 2.30E-18 | Nightingale | XL_VLDL_F | NA | G | A | -0.02593 | 0.004034 | 3.10E-10 |
| rs8176690 | miR-126-3ｐ | trans | 2.30E-18 | Nightingale | XS_VLDL_C | NA | G | A | -0.02439 | 0.004016 | 8.10E-10 |
| rs8176690 | miR-126-3ｐ | trans | 2.30E-18 | Nightingale | M_VLDL_C | NA | G | A | -0.02627 | 0.004124 | 1.10E-10 |
| rs8176690 | miR-126-3ｐ | trans | 2.30E-18 | Nightingale | M_VLDL_C | NA | G | A | -0.02492 | 0.00392 | 1.80E-09 |
| rs8176690 | miR-126-3ｐ | trans | 2.30E-18 | Nightingale | XS_VLDL_F | NA | G | A | -0.0218 | 0.004089 | 3.40E-08 |
| rs8176690 | miR-126-3ｐ | trans | 2.30E-18 | Nightingale | IDL_P | NA | G | A | -0.02431 | 0.004118 | 2.30E-09 |
| rs8176690 | miR-126-3ｐ | trans | 2.30E-18 | Nightingale | S_VLDL_CE | NA | G | A | -0.03277 | 0.004106 | 2.60E-15 |
| rs8176690 | miR-126-3ｐ | trans | 2.30E-18 | Nightingale | M_LDL_P | NA | G | A | -0.02363 | 0.004173 | 7.10E-09 |
| rs8176690 | miR-126-3ｐ | trans | 2.30E-18 | Nightingale | IDL_FC | NA | G | A | -0.02583 | 0.004029 | 1.50E-10 |
| rs8176690 | miR-126-3ｐ | trans | 2.30E-18 | Nightingale | S_VLDL_FC | NA | G | A | -0.02229 | 0.004162 | 3.70E-08 |
| rs8176690 | miR-126-3ｐ | trans | 2.30E-18 | Nightingale | XL_VLDL_C | NA | G | A | -0.02752 | 0.00399 | 5.90E-11 |
| rs8176690 | miR-126-3ｐ | trans | 2.30E-18 | Nightingale | L_VLDL_C_ | NA | G | A | -0.03655 | 0.004064 | 3.30E-19 |
| rs8176690 | miR-126-3ｐ | trans | 2.30E-18 | Nightingale | L_VLDL_TG | NA | G | A | 0.033748 | 0.004142 | 1.10E-16 |
| rs8176690 | miR-126-3ｐ | trans | 2.30E-18 | Nightingale | XL_VLDL_C | NA | G | A | -0.02706 | 0.003992 | 1.60E-10 |
| rs8176690 | miR-126-3ｐ | trans | 2.30E-18 | Nightingale | M_VLDL_P | NA | G | A | -0.02937 | 0.003994 | 4.30E-13 |
| rs8176690 | miR-126-3ｐ | trans | 2.30E-18 | Nightingale | M_VLDL_C | NA | G | A | -0.02348 | 0.00391 | 1.70E-08 |
| rs8176691 | miR-126-3ｐ | trans | 1.22E-18 | Nightingale | M_VLDL_T | NA | T | C | 0.025451 | 0.003957 | 6.30E-10 |
| rs8176691 | miR-126-3ｐ | trans | 1.22E-18 | Nightingale | S_VLDL_C_ | NA | T | C | -0.02862 | 0.004075 | 7.40E-12 |
| rs8176691 | miR-126-3ｐ | trans | 1.22E-18 | Nightingale | M_VLDL_P | NA | T | C | -0.02895 | 0.004018 | 9.00E-13 |
| rs8176691 | miR-126-3ｐ | trans | 1.22E-18 | Nightingale | XL_VLDL_C | NA | T | C | -0.02583 | 0.004017 | 1.10E-09 |
| rs8176691 | miR-126-3ｐ | trans | 1.22E-18 | Nightingale | S_VLDL_FC | NA | T | C | -0.02216 | 0.004188 | 4.20E-08 |
| rs8176691 | miR-126-3ｐ | trans | 1.22E-18 | Nightingale | S_VLDL_C | NA | T | C | -0.02244 | 0.004187 | 2.20E-08 |
| rs8176691 | miR-126-3ｐ | trans | 1.22E-18 | Nightingale | IDL_L | NA | T | C | -0.02235 | 0.004048 | 3.10E-08 |
| rs8176691 | miR-126-3ｐ | trans | 1.22E-18 | Nightingale | S_VLDL_TG | NA | T | C | 0.02616 | 0.004053 | 4.10E-10 |
| rs8176691 | miR-126-3ｐ | trans | 1.22E-18 | Nightingale | Clinical_LD | NA | T | C | -0.02298 | 0.004163 | 2.90E-08 |
| rs8176691 | miR-126-3ｐ | trans | 1.22E-18 | Nightingale | L_VLDL_CE | NA | T | C | -0.03232 | 0.004007 | 1.30E-15 |
| rs8176691 | miR-126-3ｐ | trans | 1.22E-18 | Nightingale | S_LDL_C_p | NA | T | C | -0.02968 | 0.004203 | 1.20E-12 |
| rs8176691 | miR-126-3ｐ | trans | 1.22E-18 | Nightingale | IDL_P | NA | T | C | -0.02456 | 0.004143 | 1.80E-09 |
| rs8176691 | miR-126-3ｐ | trans | 1.22E-18 | Nightingale | M_VLDL_C | NA | T | C | -0.02363 | 0.004175 | 4.50E-09 |
| rs8176691 | miR-126-3ｐ | trans | 1.22E-18 | Nightingale | Remnant_( | NA | T | C | -0.02223 | 0.00416 | 2.50E-08 |
| rs8176691 | miR-126-3ｐ | trans | 1.22E-18 | Nightingale | S_VLDL_CE | NA | T | C | -0.0324 | 0.004131 | 6.60E-15 |
| rs8176691 | miR-126-3ｐ | trans | 1.22E-18 | Nightingale | S_VLDL_CE | NA | T | C | -0.02232 | 0.004182 | 2.10E-08 |
| rs8176691 | miR-126-3ｐ | trans | 1.22E-18 | Nightingale | XL_VLDL_T | NA | T | C | 0.030151 | 0.004058 | 4.60E-13 |
| rs8176691 | miR-126-3ｐ | trans | 1.22E-18 | Nightingale | M_LDL_P | NA | T | C | -0.02284 | 0.004199 | 2.80E-08 |
| rs8176691 | miR-126-3ｐ | trans | 1.22E-18 | Nightingale | IDL_C | NA | T | C | -0.02274 | 0.004044 | 1.80E-08 |
| rs8176691 | miR-126-3ｐ | trans | 1.22E-18 | Nightingale | XXL_VLDL_ | NA | T | C | -0.02576 | 0.004194 | 8.30E-10 |
| rs8176691 | miR-126-3ｐ | trans | 1.22E-18 | Nightingale | XXL_VLDL_ | NA | T | C | -0.02482 | 0.004217 | 3.90E-09 |
| rs8176691 | miR-126-3ｐ | trans | 1.22E-18 | Nightingale | L_VLDL_FC | NA | T | C | -0.03146 | 0.004147 | 1.60E-14 |
| rs8176691 | miR-126-3ｐ | trans | 1.22E-18 | Nightingale | IDL_FC | NA | T | C | -0.0248 | 0.004054 | 9.30E-10 |
| rs8176691 | miR-126-3ｐ | trans | 1.22E-18 | Nightingale | L_VLDL_TG | NA | T | C | 0.033346 | 0.004167 | 2.60E-16 |
| rs8176691 | miR-126-3ｐ | trans | 1.22E-18 | Nightingale | IDL_PL | NA | T | C | -0.02192 | 0.004041 | 4.30E-08 |
| rs8176691 | miR-126-3ｐ | trans | 1.22E-18 | Nightingale | L_LDL_PL | NA | T | C | -0.02396 | 0.004155 | 5.80E-09 |
| rs8176691 | miR-126-3ｐ | trans | 1.22E-18 | Nightingale | XXL_VLDL_ | NA | T | C | -0.02308 | 0.004168 | 3.30E-08 |
| rs8176691 | miR-126-3ｐ | trans | 1.22E-18 | Nightingale | M_VLDL_C | NA | T | C | -0.02399 | 0.003944 | 7.10E-09 |
| rs8176691 | miR-126-3ｐ | trans | 1.22E-18 | Nightingale | XS_VLDL_F | NA | T | C | -0.02161 | 0.004114 | 2.80E-08 |
| rs8176691 | miR-126-3ｐ | trans | 1.22E-18 | Nightingale | XS_VLDL_C | NA | T | C | -0.02406 | 0.004041 | 9.60E-10 |
| rs8176691 | miR-126-3ｐ | trans | 1.22E-18 | Nightingale | M_VLDL_C | NA | T | C | -0.02588 | 0.004149 | 2.30E-10 |
| rs8176691 | miR-126-3ｐ | trans | 1.22E-18 | Nightingale | XXL_VLDL_ | NA | T | C | 0.026515 | 0.004221 | 1.60E-10 |
| rs8176691 | miR-126-3ｐ | trans | 1.22E-18 | Nightingale | XL_VLDL_F | NA | T | C | -0.02537 | 0.004059 | 6.20E-10 |
| rs8176691 | miR-126-3ｐ | trans | 1.22E-18 | Nightingale | XL_VLDL_P | NA | T | C | -0.02399 | 0.004193 | 3.00E-09 |
| rs8176691 | miR-126-3ｐ | trans | 1.22E-18 | Nightingale | XL_VLDL_C | NA | T | C | -0.02645 | 0.004015 | 3.20E-10 |
| rs8176691 | miR-126-3ｐ | trans | 1.22E-18 | Nightingale | L_VLDL_C_ | NA | T | C | -0.03556 | 0.004089 | 3.20E-18 |
| rs8176691 | miR-126-3ｐ | trans | 1.22E-18 | Nightingale | XS_VLDL_C | NA | T | C | -0.0236 | 0.004064 | 2.10E-09 |
| rs8176691 | miR-126-3ｐ | trans | 1.22E-18 | Nightingale | M_VLDL_F | NA | T | C | -0.02709 | 0.003984 | 3.70E-11 |
| rs8176691 | miR-126-3ｐ | trans | 1.22E-18 | Nightingale | XS_VLDL_P | NA | T | C | -0.02135 | 0.004132 | 3.70E-08 |
| rs8176691 | miR-10a-5ｐ | trans | 2.21E-13 | Nightingale | M_VLDL_T | NA | T | C | 0.025451 | 0.003957 | 6.30E-10 |
| rs8176691 | miR-10a-5ｐ | trans | 2.21E-13 | Nightingale | S_VLDL_C_ | NA | T | C | -0.02862 | 0.004075 | 7.40E-12 |
| rs8176691 | miR-10a-5ｐ | trans | 2.21E-13 | Nightingale | M_VLDL_P | NA | T | C | -0.02895 | 0.004018 | 9.00E-13 |
| rs8176691 | miR-10a-5ｐ | trans | 2.21E-13 | Nightingale | XL_VLDL_C | NA | T | C | -0.02583 | 0.004017 | 1.10E-09 |
| rs8176691 | miR-10a-5ｐ | trans | 2.21E-13 | Nightingale | S_VLDL_FC | NA | T | C | -0.02216 | 0.004188 | 4.20E-08 |

| | | | | | | | | | | | |
|---|---|---|---|---|---|---|---|---|---|---|---|
| rs8176691 | miR-10a-5p | trans | 2.21E-13 | Nightingale | S_VLDL_C | NA | T | C | -0.02244 | 0.004187 | 2.20E-08 |
| rs8176691 | miR-10a-5p | trans | 2.21E-13 | Nightingale | IDL_L | NA | T | C | -0.02235 | 0.004048 | 3.10E-08 |
| rs8176691 | miR-10a-5p | trans | 2.21E-13 | Nightingale | S_VLDL_TG | NA | T | C | 0.02616 | 0.004053 | 4.10E-10 |
| rs8176691 | miR-10a-5p | trans | 2.21E-13 | Nightingale | Clinical_LD | NA | T | C | -0.02298 | 0.004163 | 2.90E-08 |
| rs8176691 | miR-10a-5p | trans | 2.21E-13 | Nightingale | L_VLDL_CE | NA | T | C | -0.03232 | 0.004007 | 1.30E-15 |
| rs8176691 | miR-10a-5p | trans | 2.21E-13 | Nightingale | S_LDL_C_p | NA | T | C | -0.02968 | 0.004203 | 1.20E-12 |
| rs8176691 | miR-10a-5p | trans | 2.21E-13 | Nightingale | IDL_P | NA | T | C | -0.02456 | 0.004143 | 1.80E-09 |
| rs8176691 | miR-10a-5p | trans | 2.21E-13 | Nightingale | M_VLDL_C | NA | T | C | -0.02363 | 0.004175 | 4.50E-09 |
| rs8176691 | miR-10a-5p | trans | 2.21E-13 | Nightingale | Remnant_( | NA | T | C | -0.02223 | 0.00416 | 2.50E-08 |
| rs8176691 | miR-10a-5p | trans | 2.21E-13 | Nightingale | S_VLDL_CE | NA | T | C | -0.0324 | 0.004131 | 6.60E-15 |
| rs8176691 | miR-10a-5p | trans | 2.21E-13 | Nightingale | S_VLDL_CE | NA | T | C | -0.02232 | 0.004182 | 2.10E-08 |
| rs8176691 | miR-10a-5p | trans | 2.21E-13 | Nightingale | XL_VLDL_T | NA | T | C | 0.030151 | 0.004058 | 4.60E-13 |
| rs8176691 | miR-10a-5p | trans | 2.21E-13 | Nightingale | M_LDL_P | NA | T | C | -0.02284 | 0.004199 | 2.80E-08 |
| rs8176691 | miR-10a-5p | trans | 2.21E-13 | Nightingale | IDL_C | NA | T | C | -0.02274 | 0.004044 | 1.80E-08 |
| rs8176691 | miR-10a-5p | trans | 2.21E-13 | Nightingale | XXL_VLDL_ | NA | T | C | -0.02576 | 0.004194 | 8.30E-10 |
| rs8176691 | miR-10a-5p | trans | 2.21E-13 | Nightingale | XXL_VLDL_ | NA | T | C | -0.02482 | 0.004217 | 3.90E-09 |
| rs8176691 | miR-10a-5p | trans | 2.21E-13 | Nightingale | L_VLDL_FC | NA | T | C | -0.03146 | 0.004147 | 1.60E-14 |
| rs8176691 | miR-10a-5p | trans | 2.21E-13 | Nightingale | IDL_FC | NA | T | C | -0.0248 | 0.004054 | 9.30E-10 |
| rs8176691 | miR-10a-5p | trans | 2.21E-13 | Nightingale | L_VLDL_TG | NA | T | C | 0.033346 | 0.004167 | 2.60E-16 |
| rs8176691 | miR-10a-5p | trans | 2.21E-13 | Nightingale | IDL_PL | NA | T | C | -0.02192 | 0.004041 | 4.30E-08 |
| rs8176691 | miR-10a-5p | trans | 2.21E-13 | Nightingale | L_LDL_PL | NA | T | C | -0.02396 | 0.004155 | 5.80E-09 |
| rs8176691 | miR-10a-5p | trans | 2.21E-13 | Nightingale | XXL_VLDL_ | NA | T | C | -0.02308 | 0.004168 | 3.30E-08 |
| rs8176691 | miR-10a-5p | trans | 2.21E-13 | Nightingale | M_VLDL_C | NA | T | C | -0.02399 | 0.003944 | 7.10E-09 |
| rs8176691 | miR-10a-5p | trans | 2.21E-13 | Nightingale | XS_VLDL_F | NA | T | C | -0.02161 | 0.004114 | 2.80E-08 |
| rs8176691 | miR-10a-5p | trans | 2.21E-13 | Nightingale | XS_VLDL_C | NA | T | C | -0.02406 | 0.004041 | 9.60E-10 |
| rs8176691 | miR-10a-5p | trans | 2.21E-13 | Nightingale | M_VLDL_C | NA | T | C | -0.02588 | 0.004149 | 2.30E-10 |
| rs8176691 | miR-10a-5p | trans | 2.21E-13 | Nightingale | XXL_VLDL_ | NA | T | C | 0.026515 | 0.004221 | 1.60E-10 |
| rs8176691 | miR-10a-5p | trans | 2.21E-13 | Nightingale | XL_VLDL_F | NA | T | C | -0.02537 | 0.004059 | 6.20E-10 |
| rs8176691 | miR-10a-5p | trans | 2.21E-13 | Nightingale | XL_VLDL_P | NA | T | C | -0.02399 | 0.004193 | 3.00E-09 |
| rs8176691 | miR-10a-5p | trans | 2.21E-13 | Nightingale | XL_VLDL_C | NA | T | C | -0.02645 | 0.004015 | 3.20E-10 |
| rs8176691 | miR-10a-5p | trans | 2.21E-13 | Nightingale | L_VLDL_C_ | NA | T | C | -0.03556 | 0.004089 | 3.20E-18 |
| rs8176691 | miR-10a-5p | trans | 2.21E-13 | Nightingale | XS_VLDL_C | NA | T | C | -0.0236 | 0.004064 | 2.10E-09 |
| rs8176691 | miR-10a-5p | trans | 2.21E-13 | Nightingale | M_VLDL_F | NA | T | C | -0.02709 | 0.003984 | 3.70E-11 |
| rs8176691 | miR-10a-5p | trans | 2.21E-13 | Nightingale | XS_VLDL_P | NA | T | C | -0.02135 | 0.004132 | 3.70E-08 |
| rs8176702 | miR-10a-5p | trans | 2.18E-13 | Nightingale | M_VLDL_C | NA | A | G | -0.02338 | 0.004176 | 5.90E-09 |
| rs8176702 | miR-10a-5p | trans | 2.18E-13 | Nightingale | L_VLDL_CE | NA | A | G | -0.03194 | 0.004007 | 2.60E-15 |
| rs8176702 | miR-10a-5p | trans | 2.18E-13 | Nightingale | L_VLDL_C_ | NA | A | G | -0.03521 | 0.00409 | 6.30E-18 |
| rs8176702 | miR-10a-5p | trans | 2.18E-13 | Nightingale | S_VLDL_TG | NA | A | G | 0.025803 | 0.004054 | 6.90E-10 |
| rs8176702 | miR-10a-5p | trans | 2.18E-13 | Nightingale | L_VLDL_TG | NA | A | G | 0.033013 | 0.004168 | 4.70E-16 |
| rs8176702 | miR-10a-5p | trans | 2.18E-13 | Nightingale | XL_VLDL_T | NA | A | G | 0.029734 | 0.004059 | 9.30E-13 |
| rs8176702 | miR-10a-5p | trans | 2.18E-13 | Nightingale | M_VLDL_C | NA | A | G | -0.02556 | 0.00415 | 3.60E-10 |
| rs8176702 | miR-10a-5p | trans | 2.18E-13 | Nightingale | S_VLDL_CE | NA | A | G | -0.03205 | 0.004131 | 1.20E-14 |
| rs8176702 | miR-10a-5p | trans | 2.18E-13 | Nightingale | S_VLDL_C_ | NA | A | G | -0.02826 | 0.004075 | 1.30E-11 |
| rs8176702 | miR-10a-5p | trans | 2.18E-13 | Nightingale | S_LDL_C_p | NA | A | G | -0.02943 | 0.004204 | 2.00E-12 |
| rs8176702 | miR-10a-5p | trans | 2.18E-13 | Nightingale | XS_VLDL_C | NA | A | G | -0.02342 | 0.004065 | 2.60E-09 |
| rs8176702 | miR-10a-5p | trans | 2.18E-13 | Nightingale | S_VLDL_C | NA | A | G | -0.02231 | 0.004187 | 2.50E-08 |
| rs8176702 | miR-10a-5p | trans | 2.18E-13 | Nightingale | IDL_FC | NA | A | G | -0.02457 | 0.004054 | 1.30E-09 |
| rs8176702 | miR-10a-5p | trans | 2.18E-13 | Nightingale | M_VLDL_P | NA | A | G | -0.02863 | 0.004019 | 1.50E-12 |
| rs8176702 | miR-10a-5p | trans | 2.18E-13 | Nightingale | XXL_VLDL_ | NA | A | G | -0.02284 | 0.004168 | 4.50E-08 |
| rs8176702 | miR-10a-5p | trans | 2.18E-13 | Nightingale | Remnant_( | NA | A | G | -0.02207 | 0.00416 | 3.00E-08 |
| rs8176702 | miR-10a-5p | trans | 2.18E-13 | Nightingale | XXL_VLDL_ | NA | A | G | -0.02449 | 0.004218 | 6.10E-09 |
| rs8176702 | miR-10a-5p | trans | 2.18E-13 | Nightingale | IDL_P | NA | A | G | -0.02432 | 0.004144 | 2.50E-09 |
| rs8176702 | miR-10a-5p | trans | 2.18E-13 | Nightingale | XL_VLDL_C | NA | A | G | -0.02606 | 0.004015 | 5.60E-10 |
| rs8176702 | miR-10a-5p | trans | 2.18E-13 | Nightingale | M_VLDL_T | NA | A | G | 0.025136 | 0.003957 | 9.60E-10 |
| rs8176702 | miR-10a-5p | trans | 2.18E-13 | Nightingale | XL_VLDL_P | NA | A | G | -0.02377 | 0.004194 | 3.90E-09 |
| rs8176702 | miR-10a-5p | trans | 2.18E-13 | Nightingale | S_VLDL_CE | NA | A | G | -0.0222 | 0.004182 | 2.40E-08 |
| rs8176702 | miR-10a-5p | trans | 2.18E-13 | Nightingale | M_LDL_P | NA | A | G | -0.02275 | 0.004199 | 3.10E-08 |
| rs8176702 | miR-10a-5p | trans | 2.18E-13 | Nightingale | XS_VLDL_P | NA | A | G | -0.02127 | 0.004132 | 3.90E-08 |
| rs8176702 | miR-10a-5p | trans | 2.18E-13 | Nightingale | IDL_L | NA | A | G | -0.02215 | 0.004049 | 4.00E-08 |
| rs8176702 | miR-10a-5p | trans | 2.18E-13 | Nightingale | M_VLDL_F | NA | A | G | -0.02681 | 0.003984 | 5.50E-11 |
| rs8176702 | miR-10a-5p | trans | 2.18E-13 | Nightingale | XS_VLDL_C | NA | A | G | -0.02386 | 0.004041 | 1.20E-09 |
| rs8176702 | miR-10a-5p | trans | 2.18E-13 | Nightingale | IDL_C | NA | A | G | -0.02251 | 0.004044 | 2.50E-08 |
| rs8176702 | miR-10a-5p | trans | 2.18E-13 | Nightingale | L_VLDL_FC | NA | A | G | -0.0313 | 0.004147 | 2.00E-14 |

| | | | | | | | | | | | |
|---|---|---|---|---|---|---|---|---|---|---|---|
| rs8176702 | miR-10a-5p | trans | 2.18E-13 | Nightingale | XXL_VLDL_ | NA | A | G | -0.02544 | 0.004195 | 1.30E-09 |
| rs8176702 | miR-10a-5p | trans | 2.18E-13 | Nightingale | S_VLDL_FC | NA | A | G | -0.02201 | 0.004188 | 4.90E-08 |
| rs8176702 | miR-10a-5p | trans | 2.18E-13 | Nightingale | XXL_VLDL_ | NA | A | G | 0.026148 | 0.004222 | 2.60E-10 |
| rs8176702 | miR-10a-5p | trans | 2.18E-13 | Nightingale | XS_VLDL_F | NA | A | G | -0.02151 | 0.004115 | 3.10E-08 |
| rs8176702 | miR-10a-5p | trans | 2.18E-13 | Nightingale | XL_VLDL_C | NA | A | G | -0.02545 | 0.004017 | 1.80E-09 |
| rs8176702 | miR-10a-5p | trans | 2.18E-13 | Nightingale | L_LDL_PL | NA | A | G | -0.02376 | 0.004156 | 7.50E-09 |
| rs8176702 | miR-10a-5p | trans | 2.18E-13 | Nightingale | Clinical_LD | NA | A | G | -0.02277 | 0.004163 | 3.80E-08 |
| rs8176702 | miR-10a-5p | trans | 2.18E-13 | Nightingale | M_VLDL_C | NA | A | G | -0.02368 | 0.003944 | 1.00E-08 |
| rs8176702 | miR-10a-5p | trans | 2.18E-13 | Nightingale | XL_VLDL_F | NA | A | G | -0.025 | 0.004059 | 1.10E-09 |
| rs8176702 | miR-126-3p | trans | 1.25E-18 | Nightingale | M_VLDL_C | NA | A | G | -0.02338 | 0.004176 | 5.90E-09 |
| rs8176702 | miR-126-3p | trans | 1.25E-18 | Nightingale | L_VLDL_CE | NA | A | G | -0.03194 | 0.004007 | 2.60E-15 |
| rs8176702 | miR-126-3p | trans | 1.25E-18 | Nightingale | L_VLDL_C_ | NA | A | G | -0.03521 | 0.00409 | 6.30E-18 |
| rs8176702 | miR-126-3p | trans | 1.25E-18 | Nightingale | S_VLDL_TG | NA | A | G | 0.025803 | 0.004054 | 6.90E-10 |
| rs8176702 | miR-126-3p | trans | 1.25E-18 | Nightingale | L_VLDL_TG | NA | A | G | 0.033013 | 0.004168 | 4.70E-16 |
| rs8176702 | miR-126-3p | trans | 1.25E-18 | Nightingale | XL_VLDL_T | NA | A | G | 0.029734 | 0.004059 | 9.30E-13 |
| rs8176702 | miR-126-3p | trans | 1.25E-18 | Nightingale | M_VLDL_C | NA | A | G | -0.02556 | 0.00415 | 3.60E-10 |
| rs8176702 | miR-126-3p | trans | 1.25E-18 | Nightingale | S_VLDL_CE | NA | A | G | -0.03205 | 0.004131 | 1.20E-14 |
| rs8176702 | miR-126-3p | trans | 1.25E-18 | Nightingale | S_VLDL_C_ | NA | A | G | -0.02826 | 0.004075 | 1.30E-11 |
| rs8176702 | miR-126-3p | trans | 1.25E-18 | Nightingale | S_LDL_C_p | NA | A | G | -0.02943 | 0.004204 | 2.00E-12 |
| rs8176702 | miR-126-3p | trans | 1.25E-18 | Nightingale | XS_VLDL_C | NA | A | G | -0.02342 | 0.004065 | 2.60E-09 |
| rs8176702 | miR-126-3p | trans | 1.25E-18 | Nightingale | S_VLDL_C | NA | A | G | -0.02231 | 0.004187 | 2.50E-08 |
| rs8176702 | miR-126-3p | trans | 1.25E-18 | Nightingale | IDL_FC | NA | A | G | -0.02457 | 0.004054 | 1.30E-09 |
| rs8176702 | miR-126-3p | trans | 1.25E-18 | Nightingale | M_VLDL_P | NA | A | G | -0.02863 | 0.004019 | 1.50E-12 |
| rs8176702 | miR-126-3p | trans | 1.25E-18 | Nightingale | XXL_VLDL_ | NA | A | G | -0.02284 | 0.004168 | 4.50E-08 |
| rs8176702 | miR-126-3p | trans | 1.25E-18 | Nightingale | Remnant_C | NA | A | G | -0.02207 | 0.00416 | 3.00E-08 |
| rs8176702 | miR-126-3p | trans | 1.25E-18 | Nightingale | XXL_VLDL_ | NA | A | G | -0.02449 | 0.004218 | 6.10E-09 |
| rs8176702 | miR-126-3p | trans | 1.25E-18 | Nightingale | IDL_P | NA | A | G | -0.02432 | 0.004144 | 2.50E-09 |
| rs8176702 | miR-126-3p | trans | 1.25E-18 | Nightingale | XL_VLDL_C | NA | A | G | -0.02606 | 0.004015 | 5.60E-10 |
| rs8176702 | miR-126-3p | trans | 1.25E-18 | Nightingale | M_VLDL_T | NA | A | G | 0.025136 | 0.003957 | 9.60E-10 |
| rs8176702 | miR-126-3p | trans | 1.25E-18 | Nightingale | XL_VLDL_P | NA | A | G | -0.02377 | 0.004194 | 3.90E-09 |
| rs8176702 | miR-126-3p | trans | 1.25E-18 | Nightingale | S_VLDL_CE | NA | A | G | -0.0222 | 0.004182 | 2.40E-08 |
| rs8176702 | miR-126-3p | trans | 1.25E-18 | Nightingale | M_LDL_P | NA | A | G | -0.02275 | 0.004199 | 3.10E-08 |
| rs8176702 | miR-126-3p | trans | 1.25E-18 | Nightingale | XS_VLDL_P | NA | A | G | -0.02127 | 0.004132 | 3.90E-08 |
| rs8176702 | miR-126-3p | trans | 1.25E-18 | Nightingale | IDL_L | NA | A | G | -0.02215 | 0.004049 | 4.00E-08 |
| rs8176702 | miR-126-3p | trans | 1.25E-18 | Nightingale | M_VLDL_F | NA | A | G | -0.02681 | 0.003984 | 5.50E-11 |
| rs8176702 | miR-126-3p | trans | 1.25E-18 | Nightingale | XS_VLDL_C | NA | A | G | -0.02386 | 0.004041 | 1.20E-09 |
| rs8176702 | miR-126-3p | trans | 1.25E-18 | Nightingale | IDL_C | NA | A | G | -0.02251 | 0.004044 | 2.50E-08 |
| rs8176702 | miR-126-3p | trans | 1.25E-18 | Nightingale | L_VLDL_FC | NA | A | G | -0.0313 | 0.004147 | 2.00E-14 |
| rs8176702 | miR-126-3p | trans | 1.25E-18 | Nightingale | XXL_VLDL_ | NA | A | G | -0.02544 | 0.004195 | 1.30E-09 |
| rs8176702 | miR-126-3p | trans | 1.25E-18 | Nightingale | S_VLDL_FC | NA | A | G | -0.02201 | 0.004188 | 4.90E-08 |
| rs8176702 | miR-126-3p | trans | 1.25E-18 | Nightingale | XXL_VLDL_ | NA | A | G | 0.026148 | 0.004222 | 2.60E-10 |
| rs8176702 | miR-126-3p | trans | 1.25E-18 | Nightingale | XS_VLDL_F | NA | A | G | -0.02151 | 0.004115 | 3.10E-08 |
| rs8176702 | miR-126-3p | trans | 1.25E-18 | Nightingale | XL_VLDL_C | NA | A | G | -0.02545 | 0.004017 | 1.80E-09 |
| rs8176702 | miR-126-3p | trans | 1.25E-18 | Nightingale | L_LDL_PL | NA | A | G | -0.02376 | 0.004156 | 7.50E-09 |
| rs8176702 | miR-126-3p | trans | 1.25E-18 | Nightingale | Clinical_LD | NA | A | G | -0.02277 | 0.004163 | 3.80E-08 |
| rs8176702 | miR-126-3p | trans | 1.25E-18 | Nightingale | M_VLDL_C | NA | A | G | -0.02368 | 0.003944 | 1.00E-08 |
| rs8176702 | miR-126-3p | trans | 1.25E-18 | Nightingale | XL_VLDL_F | NA | A | G | -0.025 | 0.004059 | 1.10E-09 |

Table S19. GWAS traits associated with miR-eQTLs

| SNPS | CONTEXT | DISEASE.TR | PUBMEDID | P.VALUE | MAPPED_G | miRNA | miRNA_context | cismark | P_miR-eQTL |
|---|---|---|---|---|---|---|---|---|---|
| rs630510 | intron_variant | Blood prot | 28240269 | 2.00E-15 | ABO | let-7b-5p | intergenic | trans | 4.85E-16 |
| rs2519093 | intron_variant | Total chole | 29507422 | 3.00E-41 | ABO | let-7c-5p | intergenic | trans | 4.94E-22 |
| rs2519093 | intron_variant | Venous thr | 28373160 | 6.00E-26 | ABO | let-7c-5p | intergenic | trans | 4.94E-22 |
| rs2519093 | intron_variant | Venous thr | 22672568 | 8.00E-16 | ABO | let-7c-5p | intergenic | trans | 4.94E-22 |
| rs2519093 | intron_variant | Coronary a | 29212778 | 2.00E-14 | ABO | let-7c-5p | intergenic | trans | 4.94E-22 |
| rs2519093 | intron_variant | Red blood | 29403010 | 7.00E-45 | ABO | let-7c-5p | intergenic | trans | 4.94E-22 |
| rs2519093 | intron_variant | Allergy | 27182965 | 4.00E-08 | ABO | let-7c-5p | intergenic | trans | 4.94E-22 |
| rs2519093 | intron_variant | Coronary a | 26343387 | 1.00E-11 | ABO | let-7c-5p | intergenic | trans | 4.94E-22 |
| rs643434 | intron_variant | Inflammato | 22291609 | 9.00E-25 | ABO | let-7c-5p | intergenic | trans | 3.20E-23 |
| rs2519093 | intron_variant | Triglyceride | 32203549 | 1.00E-16 | ABO | let-7c-5p | intergenic | trans | 4.94E-22 |
| rs643434 | intron_variant | End-stage | 23381943 | 1.00E-49 | ABO | let-7c-5p | intergenic | trans | 3.20E-23 |
| rs1165522 | intron_variant | Thyroid stir | 25743335 | 2.00E-14 | ABO | miR-10b-5 | intragenic (HOXD4) | trans | 2.13E-34 |
| rs649129 | intergenic_variant | Soluble lev | 20167578 | 1.00E-15 | ABO - Y_RN | miR-10b-5 | intragenic (HOXD4) | trans | 2.54E-34 |
| rs532436 | intron_variant | Myocardial | 26343387 | 2.00E-17 | ABO | miR-10b-5 | intragenic (HOXD4) | trans | 5.20E-36 |
| rs1165522 | intron_variant | Non-album | 29403010 | 2.00E-08 | ABO | miR-10b-5 | intragenic (HOXD4) | trans | 2.13E-34 |
| rs649129 | intergenic_variant | Total chole | 29507422 | 1.00E-48 | ABO - Y_RN | miR-10b-5 | intragenic (HOXD4) | trans | 2.54E-34 |
| rs660340 | intron_variant | vWF levels | 30586737 | 9.9998886 | ABO | miR-10b-5 | intragenic (HOXD4) | trans | 1.23E-19 |
| rs600038 | regulatory_region_variant | Heart failu | 31919418 | 4.00E-09 | ABO - Y_RN | miR-10b-5 | intragenic (HOXD4) | trans | 2.32E-34 |
| rs529565 | intron_variant | Venous thr | 25772935 | 4.00E-75 | ABO | miR-10b-5 | intragenic (HOXD4) | trans | 4.06E-36 |
| rs649129 | intergenic_variant | Blood meta | 24816252 | 9.00E-37 | ABO - Y_RN | miR-10b-5 | intragenic (HOXD4) | trans | 2.54E-34 |
| rs529565 | intron_variant | Thrombosi | 26908601 | 7.00E-63 | ABO | miR-10b-5 | intragenic (HOXD4) | trans | 4.06E-36 |
| rs492488 | intron_variant | Blood prot | 30072576 | 7.00E-23 | ABO | miR-126-3 | intragenic (EGFL7) | trans | 2.58E-39 |
| rs687289 | intron_variant | Coagulatio | 23267103 | 1.00E-128 | ABO | miR-126-3 | intragenic (EGFL7) | trans | 3.06E-41 |
| rs687289 | intron_variant | Pancreatic | 25086665 | 2.00E-16 | ABO | miR-126-3 | intragenic (EGFL7) | trans | 3.06E-41 |
| rs687289 | intron_variant | Factor VIII | 30586737 | 0 | ABO | miR-126-3 | intragenic (EGFL7) | trans | 3.06E-41 |
| rs687289 | intron_variant | Urinary me | 24586186 | 2.00E-12 | ABO | miR-126-3 | intragenic (EGFL7) | trans | 3.06E-41 |
| rs687289 | intron_variant | Monocyte | 27863252 | 8.00E-26 | ABO | miR-126-3 | intragenic (EGFL7) | trans | 3.06E-41 |
| rs687289 | intron_variant | Activated p | 29403010 | 9.00E-68 | ABO | miR-126-3 | intragenic (EGFL7) | trans | 3.06E-41 |
| rs687289 | intron_variant | Blood prot | 28240269 | 8.00E-10 | ABO | miR-126-3 | intragenic (EGFL7) | trans | 3.06E-41 |
| rs644234 | intron_variant | Protein bio | 23056639 | 4.00E-47 | ABO | miR-126-3 | intragenic (EGFL7) | trans | 1.03E-37 |
| rs687289 | intron_variant | Blood prot | 27532455 | 1.00E-40 | ABO | miR-126-3 | intragenic (EGFL7) | trans | 3.06E-41 |
| rs687289 | intron_variant | Factor VIII | 20231535 | 4.9406564 | ABO | miR-126-3 | intragenic (EGFL7) | trans | 3.06E-41 |
| rs1089663 | intron_variant | Neuroticisr | 29255261 | 7.00E-11 | ZDHHC5 | miR-130a-3 | intergenic | cis | 2.02E-27 |
| rs1089663 | intron_variant | Neurocitici | 29500382 | 9.00E-11 | ZDHHC5 | miR-130a-3 | intergenic | cis | 2.02E-27 |
| rs687432 | intergenic_variant | Parkinson's | 27182965 | 2.00E-09 | OR5AZ1P - | miR-130a-3 | intergenic | cis | 0.00033 |
| rs7102271 | intergenic_variant | Blood prot | 30072576 | 3.00E-19 | SERPING1 - | miR-130a-3 | intergenic | cis | 2.50E-38 |
| rs1122908 | intergenic_variant | Blood prot | 30072576 | 0 | SERPING1 - | miR-130a-3 | intergenic | cis | 3.01E-38 |
| rs1160302 | intron_variant | Blood prot | 28240269 | 5.00E-96 | SERPING1 | miR-130a-3 | intergenic | cis | 6.00E-38 |
| rs9420 | non_coding_transcript_exo | Schizophre | 28991256 | 9.00E-09 | SELENOH | miR-130a-3 | intergenic | cis | 3.59E-27 |
| rs641325 | intergenic_variant | Worry too | 29500382 | 7.00E-10 | OR5BA1P - | miR-130a-3 | intergenic | cis | 2.30E-18 |
| rs2729354 | TF_binding_site_variant | Blood prot | 28240269 | 8.00E-14 | RPS4XP13 - | miR-130a-3 | intergenic | cis | 0.000809 |
| rs9420 | non_coding_transcript_exo | Schizophre | 26198764 | 2.00E-08 | SELENOH | miR-130a-3 | intergenic | cis | 3.59E-27 |
| rs3781885 | intron_variant | Worry too | 29500382 | 1.00E-11 | CTNND1 | miR-130a-3 | intergenic | cis | 2.31E-28 |
| rs674094 | intergenic_variant | Feeling hur | 29500382 | 3.00E-08 | OR5BA1P - | miR-130a-3 | intergenic | cis | 9.64E-18 |
| rs7117205 | intergenic_variant | Neurocitici | 29500382 | 6.00E-10 | OR5BA1P - | miR-130a-3 | intergenic | cis | 6.23E-24 |
| rs9420 | non_coding_transcript_exo | Schizophre | 25056061 | 2.00E-09 | SELENOH | miR-130a-3 | intergenic | cis | 3.59E-27 |
| rs673344 | intergenic_variant | Neuroticisr | 29255261 | 1.00E-10 | OR5BA1P - | miR-130a-3 | intergenic | cis | 1.53E-23 |
| rs6062278 | intron_variant | Brain regio | 31676860 | 1.00E-09 | MIR1-1HG- | miR-133a-3 | intragenic (MIB1) | cis | 5.28E-08 |
| rs6062278 | intron_variant | Brain regio | 31676860 | 1.00E-09 | MIR1-1HG- | miR-133a-3 | intragenic (MIR1-1HG) | cis | 5.28E-08 |
| rs7269123 | intron_variant | Atrial fibrill | 29892015 | 6.00E-09 | MIR1-1HG | miR-133a-3 | intragenic (MIB1) | cis | 9.75E-07 |
| rs7269123 | intron_variant | Atrial fibrill | 29892015 | 6.00E-09 | MIR1-1HG | miR-133a-3 | intragenic (MIR1-1HG) | cis | 9.75E-07 |
| rs495828 | intergenic_variant | Coronary a | 29212778 | 9.00E-12 | ABO - Y_RN | miR-139-5 | intragenic (PDE2A) | trans | 1.46E-34 |
| rs495828 | intergenic_variant | LDL cholest | 30698716 | 4.00E-86 | ABO - Y_RN | miR-139-5 | intragenic (PDE2A) | trans | 1.46E-34 |
| rs495828 | intergenic_variant | Hematocrit | 28017375 | 3.00E-09 | ABO - Y_RN | miR-139-5 | intragenic (PDE2A) | trans | 1.46E-34 |
| rs495828 | intergenic_variant | Hematolog | 20139978 | 6.00E-10 | ABO - Y_RN | miR-139-5 | intragenic (PDE2A) | trans | 1.46E-34 |
| rs550057 | intron_variant | Total chole | 29507422 | 6.00E-11 | ABO | miR-139-5 | intragenic (PDE2A) | trans | 5.52E-32 |
| rs491626 | intron_variant | Creatine ki | 29403010 | 1.00E-09 | ABO | miR-139-5 | intragenic (PDE2A) | trans | 8.32E-42 |
| rs550057 | intron_variant | LDL cholest | 25961943 | 6.00E-20 | ABO | miR-139-5 | intragenic (PDE2A) | trans | 5.52E-32 |
| rs550057 | intron_variant | Cytokine n | 31679650 | 3.00E-08 | ABO | miR-139-5 | intragenic (PDE2A) | trans | 5.52E-32 |
| rs495828 | intergenic_variant | Angiotensi | 20066004 | 3.00E-08 | ABO - Y_RN | miR-139-5 | intragenic (PDE2A) | trans | 1.46E-34 |
| rs495828 | intergenic_variant | Blood meta | 24816252 | 6.00E-34 | ABO - Y_RN | miR-139-5 | intragenic (PDE2A) | trans | 1.46E-34 |
| rs495828 | intergenic_variant | Venous thr | 22672568 | 3.00E-16 | ABO - Y_RN | miR-139-5 | intragenic (PDE2A) | trans | 1.46E-34 |
| rs2343592 | intron_variant | Platelet co | 29403010 | 2.00E-11 | ZFPM2-AS1 | miR-145-5 | intergenic | trans | 1.93E-08 |
| rs4719841 | intergenic_variant | Triglyceride | 28334899 | 9.00E-13 | MIR148A - | miR-148a-3 | intergenic | cis | 0.000239 |
| rs1036332 | intron_variant | Eosinophil | 27863252 | 1.00E-20 | LINC01221 | miR-181a-3 | intergenic | cis | 2.64E-07 |
| rs1434282 | non_coding_transcript_exo | Mean corp | 27863252 | 1.00E-39 | LINC01221 | miR-181a-3 | intergenic | cis | 2.00E-07 |
| rs1434282 | non_coding_transcript_exo | Mean corp | 29403010 | 1.00E-17 | LINC01221 | miR-181a-3 | intergenic | cis | 2.00E-07 |
| rs7529925 | intron_variant | Red blood | 23222517 | 8.00E-09 | LINC01221 | miR-181a-3 | intergenic | cis | 2.54E-07 |
| rs7529925 | intron_variant | Platelet co | 29403010 | 1.00E-13 | LINC01221 | miR-181a-3 | intergenic | cis | 2.54E-07 |

| | | | | | | | | | |
|---|---|---|---|---|---|---|---|---|---|
| rs6427756 | intron_variant | White bloo | 29403010 | 2.00E-08 | LINC01221 | miR-181a-5 | intragenic (NR6A1) | cis | 1.69E-37 |
| rs1223917 | non_coding_transcript_exo | Reticulocyt | 27863252 | 5.00E-13 | LINC01222 | miR-181a-5 | intragenic (NR6A1) | cis | 5.38E-30 |
| rs174535 | missense_variant | Red blood | 25500335 | 2.00E-12 | MYRF, TME | miR-1908-5 | intragenic (FADS1) | cis | 6.98E-09 |
| rs174530 | non_coding_transcript_exo | Serum tota | 29403010 | 1.00E-12 | TMEM258, | miR-1908-5 | intragenic (FADS1) | cis | 1.12E-08 |
| rs174549 | 5_prime_UTR_variant | Red blood | 25500335 | 4.00E-10 | FADS1, FAD | miR-1908-5 | intragenic (FADS1) | cis | 4.04E-08 |
| rs174533 | intron_variant | Total chole | 28334899 | 2.00E-09 | MYRF, TME | miR-1908-5 | intragenic (FADS1) | cis | 1.26E-08 |
| rs102274 | non_coding_transcript_exo | Cholesteryl | 31551469 | 2.00E-32 | TMEM258 | miR-1908-5 | intragenic (FADS1) | cis | 9.41E-09 |
| rs102274 | non_coding_transcript_exo | Serum met | 29545352 | 8.00E-21 | TMEM258 | miR-1908-5 | intragenic (FADS1) | cis | 9.41E-09 |
| rs174535 | missense_variant | Blood meta | 24816252 | 2.00E-94 | MYRF, TME | miR-1908-5 | intragenic (FADS1) | cis | 6.98E-09 |
| rs174537 | non_coding_transcript_exo | Crohn's dis | 26192919 | 2.00E-12 | MYRF, TME | miR-1908-5 | intragenic (FADS1) | cis | 9.41E-09 |
| rs174538 | 5_prime_UTR_variant | Plasma om | 21829377 | 5.00E-58 | TMEM258 | miR-1908-5 | intragenic (FADS1) | cis | 1.35E-07 |
| rs174535 | missense_variant | Plasma om | 21829377 | 1.00E-151 | MYRF, TME | miR-1908-5 | intragenic (FADS1) | cis | 6.98E-09 |
| rs174536 | non_coding_transcript_exo | Glycerophc | 26068415 | 5.00E-59 | MYRF, TME | miR-1908-5 | intragenic (FADS1) | cis | 1.31E-08 |
| rs174533 | intron_variant | Serum met | 29545352 | 5.00E-17 | MYRF, TME | miR-1908-5 | intragenic (FADS1) | cis | 1.26E-08 |
| rs174538 | 5_prime_UTR_variant | Blood meta | 24816252 | 9.00E-14 | TMEM258 | miR-1908-5 | intragenic (FADS1) | cis | 1.35E-07 |
| rs174556 | intron_variant | Blood meta | 24816252 | 2.00E-22 | FADS2, FAD | miR-1908-5 | intragenic (FADS1) | cis | 7.67E-08 |
| rs174533 | intron_variant | Colorectal | 30510241 | 1.00E-11 | MYRF, TME | miR-1908-5 | intragenic (FADS1) | cis | 1.26E-08 |
| rs174537 | non_coding_transcript_exo | Serum met | 29545352 | 2.00E-14 | MYRF, TME | miR-1908-5 | intragenic (FADS1) | cis | 9.41E-09 |
| rs174537 | non_coding_transcript_exo | Colorectal | 24836286 | 9.00E-21 | MYRF, TME | miR-1908-5 | intragenic (FADS1) | cis | 9.41E-09 |
| rs174545 | 3_prime_UTR_variant | Red blood | 25500335 | 8.00E-90 | FADS2, FAD | miR-1908-5 | intragenic (FADS1) | cis | 2.29E-08 |
| rs174535 | missense_variant | Chronic inf | 26974007 | 2.00E-11 | MYRF, TME | miR-1908-5 | intragenic (FADS1) | cis | 6.98E-09 |
| rs174535 | missense_variant | Trans fatty | 25646338 | 1.00E-13 | MYRF, TME | miR-1908-5 | intragenic (FADS1) | cis | 6.98E-09 |
| rs174548 | 5_prime_UTR_variant | Serum met | 29545352 | 1.00E-15 | FADS2, FAD | miR-1908-5 | intragenic (FADS1) | cis | 3.01E-08 |
| rs174548 | 5_prime_UTR_variant | Metabolite | 23823483 | 7.00E-16 | FADS2, FAD | miR-1908-5 | intragenic (FADS1) | cis | 3.01E-08 |
| rs174537 | non_coding_transcript_exo | Plasma om | 26584805 | 4.00E-164 | MYRF, TME | miR-1908-5 | intragenic (FADS1) | cis | 9.41E-09 |
| rs174530 | non_coding_transcript_exo | Metabolite | 31628463 | 1.00E-09 | TMEM258, | miR-1908-5 | intragenic (FADS1) | cis | 1.12E-08 |
| rs174538 | 5_prime_UTR_variant | Trans fatty | 25646338 | 1.00E-12 | TMEM258 | miR-1908-5 | intragenic (FADS1) | cis | 1.35E-07 |
| rs174548 | 5_prime_UTR_variant | Triglyceride | 20864672 | 5.00E-14 | FADS2, FAD | miR-1908-5 | intragenic (FADS1) | cis | 3.01E-08 |
| rs174548 | 5_prime_UTR_variant | Trans fatty | 25646338 | 5.00E-15 | FADS2, FAD | miR-1908-5 | intragenic (FADS1) | cis | 3.01E-08 |
| rs174545 | 3_prime_UTR_variant | Trans fatty | 25646338 | 7.00E-14 | FADS2, FAD | miR-1908-5 | intragenic (FADS1) | cis | 2.29E-08 |
| rs174545 | 3_prime_UTR_variant | Delta-6 des | 26584805 | 5.00E-37 | FADS2, FAD | miR-1908-5 | intragenic (FADS1) | cis | 2.29E-08 |
| rs174548 | 5_prime_UTR_variant | Hematolog | 23303382 | 4.00E-08 | FADS2, FAD | miR-1908-5 | intragenic (FADS1) | cis | 3.01E-08 |
| rs174448 | regulatory_region_variant | Plasma om | 21829377 | 7.00E-28 | FADS2 - FA | miR-1908-5 | intragenic (FADS1) | cis | 1.95E-05 |
| rs174549 | 5_prime_UTR_variant | Serum met | 29545352 | 1.00E-20 | FADS1, FAD | miR-1908-5 | intragenic (FADS1) | cis | 4.04E-08 |
| rs174549 | 5_prime_UTR_variant | Laryngeal s | 25194280 | 1.00E-20 | FADS1, FAD | miR-1908-5 | intragenic (FADS1) | cis | 4.04E-08 |
| rs174536 | non_coding_transcript_exo | Plasma om | 21829377 | 1.00E-63 | MYRF, TME | miR-1908-5 | intragenic (FADS1) | cis | 1.31E-08 |
| rs174536 | non_coding_transcript_exo | Resting hea | 27798624 | 2.00E-30 | MYRF, TME | miR-1908-5 | intragenic (FADS1) | cis | 1.31E-08 |
| rs174549 | 5_prime_UTR_variant | Phosphatid | 31551469 | 6.00E-09 | FADS1, FAD | miR-1908-5 | intragenic (FADS1) | cis | 4.04E-08 |
| rs174536 | non_coding_transcript_exo | Phosphatid | 31551469 | 7.00E-09 | MYRF, TME | miR-1908-5 | intragenic (FADS1) | cis | 1.31E-08 |
| rs174536 | non_coding_transcript_exo | Trans fatty | 25646338 | 1.00E-13 | MYRF, TME | miR-1908-5 | intragenic (FADS1) | cis | 1.31E-08 |
| rs174548 | 5_prime_UTR_variant | Plasma om | 26584805 | 7.00E-31 | FADS2, FAD | miR-1908-5 | intragenic (FADS1) | cis | 3.01E-08 |
| rs174567 | intron_variant | Serum met | 29545352 | 2.00E-13 | FADS1, FAD | miR-1908-5 | intragenic (FADS1) | cis | 1.32E-08 |
| rs174554 | intron_variant | Serum met | 29545352 | 3.00E-17 | FADS2, FAD | miR-1908-5 | intragenic (FADS1) | cis | 3.17E-08 |
| rs174555 | intron_variant | Trans fatty | 25646338 | 6.00E-15 | FADS1, FAD | miR-1908-5 | intragenic (FADS1) | cis | 7.43E-08 |
| rs174549 | 5_prime_UTR_variant | White bloo | 27863252 | 6.00E-11 | FADS1, FAD | miR-1908-5 | intragenic (FADS1) | cis | 4.04E-08 |
| rs174537 | non_coding_transcript_exo | Trans fatty | 25646338 | 8.00E-14 | MYRF, TME | miR-1908-5 | intragenic (FADS1) | cis | 9.41E-09 |
| rs174537 | non_coding_transcript_exo | Colorectal | 26965516 | 9.00E-21 | MYRF, TME | miR-1908-5 | intragenic (FADS1) | cis | 9.41E-09 |
| rs174556 | intron_variant | Trans fatty | 25646338 | 3.00E-14 | FADS2, FAD | miR-1908-5 | intragenic (FADS1) | cis | 7.67E-08 |
| rs174556 | intron_variant | Metabolite | 23823483 | 3.00E-16 | FADS2, FAD | miR-1908-5 | intragenic (FADS1) | cis | 7.67E-08 |
| rs174556 | intron_variant | Granulocyt | 27863252 | 2.00E-17 | FADS2, FAD | miR-1908-5 | intragenic (FADS1) | cis | 7.67E-08 |
| rs174578 | intron_variant | Blood meta | 24816252 | 6.00E-42 | FADS2 | miR-1908-5 | intragenic (FADS1) | cis | 2.55E-08 |
| rs174544 | 3_prime_UTR_variant | Phosphatid | 31551469 | 2.00E-16 | FADS1, FAD | miR-1908-5 | intragenic (FADS1) | cis | 2.30E-08 |
| rs174554 | intron_variant | Triglyceride | 29507422 | 4.00E-41 | FADS2, FAD | miR-1908-5 | intragenic (FADS1) | cis | 3.17E-08 |
| rs174554 | intron_variant | Cholestero | 25961943 | 5.00E-24 | FADS2, FAD | miR-1908-5 | intragenic (FADS1) | cis | 3.17E-08 |
| rs174561 | 5_prime_UTR_variant | Serum met | 29545352 | 3.00E-16 | FADS1, MIF | miR-1908-5 | intragenic (FADS1) | cis | 7.66E-08 |
| rs174562 | intron_variant | Serum met | 29545352 | 3.00E-21 | FADS2, FAD | miR-1908-5 | intragenic (FADS1) | cis | 4.92E-08 |
| rs174545 | 3_prime_UTR_variant | Serum met | 29545352 | 7.00E-21 | FADS2, FAD | miR-1908-5 | intragenic (FADS1) | cis | 2.29E-08 |
| rs174564 | intron_variant | Low densit | 32154731 | 6.00E-63 | FADS2, FAD | miR-1908-5 | intragenic (FADS1) | cis | 1.61E-08 |
| rs174564 | intron_variant | LDL cholest | 32203549 | 3.00E-48 | FADS2, FAD | miR-1908-5 | intragenic (FADS1) | cis | 1.61E-08 |
| rs174530 | non_coding_transcript_exo | Serum met | 29545352 | 2.00E-12 | TMEM258, | miR-1908-5 | intragenic (FADS1) | cis | 1.12E-08 |
| rs174548 | 5_prime_UTR_variant | Cholesteryl | 31551469 | 1.00E-08 | FADS2, FAD | miR-1908-5 | intragenic (FADS1) | cis | 3.01E-08 |
| rs174533 | intron_variant | Hemoglobi | 27863252 | 3.00E-14 | MYRF, TME | miR-1908-5 | intragenic (FADS1) | cis | 1.26E-08 |
| rs174566 | intron_variant | Asthma | 31619474 | 5.00E-09 | FADS1, FAD | miR-1908-5 | intragenic (FADS1) | cis | 1.58E-08 |
| rs174548 | 5_prime_UTR_variant | Sum eosinc | 27863252 | 4.00E-11 | FADS2, FAD | miR-1908-5 | intragenic (FADS1) | cis | 3.01E-08 |
| rs174560 | non_coding_transcript_exo | Serum met | 29545352 | 1.00E-17 | FADS2, FAD | miR-1908-5 | intragenic (FADS1) | cis | 4.91E-08 |
| rs174601 | non_coding_transcript_exo | Blood meta | 24816252 | 8.00E-16 | FADS2 | miR-1908-5 | intragenic (FADS1) | cis | 5.98E-09 |
| rs174568 | 3_prime_UTR_variant | Serum met | 29545352 | 4.00E-24 | FADS2, FAD | miR-1908-5 | intragenic (FADS1) | cis | 3.76E-08 |
| rs174574 | intron_variant | Trans fatty | 25646338 | 3.00E-14 | FADS2 | miR-1908-5 | intragenic (FADS1) | cis | 8.50E-09 |
| rs174574 | intron_variant | Phosphatid | 31551469 | 4.00E-12 | FADS2 | miR-1908-5 | intragenic (FADS1) | cis | 8.50E-09 |
| rs174574 | intron_variant | C-reactive | 27286809 | 8.00E-10 | FADS2 | miR-1908-5 | intragenic (FADS1) | cis | 8.50E-09 |
| rs174574 | intron_variant | Serum met | 29545352 | 2.00E-17 | FADS2 | miR-1908-5 | intragenic (FADS1) | cis | 8.50E-09 |

| | | | | | | | | | |
|---|---|---|---|---|---|---|---|---|---|
| rs174549 | 5_prime_UTR_variant | Fatty acid c | 30453627 | 1.00E-27 | FADS1, FAD | miR-1908-5 | intragenic (FADS1) | cis | 4.04E-08 |
| rs174549 | 5_prime_UTR_variant | Metabolite | 23281178 | 2.00E-30 | FADS1, FAD | miR-1908-5 | intragenic (FADS1) | cis | 4.04E-08 |
| rs174549 | 5_prime_UTR_variant | Trans fatty | 25646338 | 5.00E-15 | FADS1, FAD | miR-1908-5 | intragenic (FADS1) | cis | 4.04E-08 |
| rs174549 | 5_prime_UTR_variant | Heart rate | 23583979 | 1.00E-22 | FADS1, FAD | miR-1908-5 | intragenic (FADS1) | cis | 4.04E-08 |
| rs174567 | intron_variant | Phosphatid | 31551469 | 4.00E-30 | FADS1, FAD | miR-1908-5 | intragenic (FADS1) | cis | 1.32E-08 |
| rs174576 | intron_variant | Trans fatty | 25646338 | 3.00E-14 | FADS2 | miR-1908-5 | intragenic (FADS1) | cis | 3.13E-08 |
| rs174576 | intron_variant | Bipolar disc | 28115744 | 1.00E-10 | FADS2 | miR-1908-5 | intragenic (FADS1) | cis | 3.13E-08 |
| rs174577 | intron_variant | Triglyceride | 29507422 | 8.00E-39 | FADS2 | miR-1908-5 | intragenic (FADS1) | cis | 1.89E-08 |
| rs174537 | non_coding_transcript_exo | Glycerophc | 26068415 | 3.00E-19 | MYRF, TME | miR-1908-5 | intragenic (FADS1) | cis | 9.41E-09 |
| rs174555 | intron_variant | Plasma om | 24823311 | 5.00E-168 | FADS1, FAD | miR-1908-5 | intragenic (FADS1) | cis | 7.43E-08 |
| rs174555 | intron_variant | Sum neutrc | 27863252 | 7.00E-14 | FADS1, FAD | miR-1908-5 | intragenic (FADS1) | cis | 7.43E-08 |
| rs174556 | intron_variant | Breast milk | 29514873 | 5.00E-10 | FADS2, FAD | miR-1908-5 | intragenic (FADS1) | cis | 7.67E-08 |
| rs174577 | intron_variant | Plasma om | 26584805 | 4.00E-68 | FADS2 | miR-1908-5 | intragenic (FADS1) | cis | 1.89E-08 |
| rs174577 | intron_variant | Iron status | 25352340 | 2.00E-17 | FADS2 | miR-1908-5 | intragenic (FADS1) | cis | 1.89E-08 |
| rs174578 | intron_variant | Serum met | 29545352 | 9.00E-18 | FADS2 | miR-1908-5 | intragenic (FADS1) | cis | 2.55E-08 |
| rs2727271 | intron_variant | Trans fatty | 25646338 | 7.00E-09 | FADS2 | miR-1908-5 | intragenic (FADS1) | cis | 4.23E-05 |
| rs174559 | non_coding_transcript_exo | Red cell dis | 28957414 | 5.00E-44 | FADS1, FAD | miR-1908-5 | intragenic (FADS1) | cis | 5.55E-07 |
| rs174576 | intron_variant | Metabolite | 23823483 | 2.00E-10 | FADS2 | miR-1908-5 | intragenic (FADS1) | cis | 3.13E-08 |
| rs174578 | intron_variant | Trans fatty | 25646338 | 5.00E-14 | FADS2 | miR-1908-5 | intragenic (FADS1) | cis | 2.55E-08 |
| rs174580 | intron_variant | Phosphatid | 31551469 | 5.00E-22 | FADS2 | miR-1908-5 | intragenic (FADS1) | cis | 1.81E-08 |
| rs174580 | intron_variant | Serum met | 29545352 | 6.00E-21 | FADS2 | miR-1908-5 | intragenic (FADS1) | cis | 1.81E-08 |
| rs174545 | 3_prime_UTR_variant | Metabolite | 23823483 | 3.00E-61 | FADS2, FAD | miR-1908-5 | intragenic (FADS1) | cis | 2.29E-08 |
| rs174564 | intron_variant | Phosphatid | 31551469 | 4.00E-33 | FADS2, FAD | miR-1908-5 | intragenic (FADS1) | cis | 1.61E-08 |
| rs174592 | intron_variant | Serum met | 29545352 | 3.00E-13 | FADS2 | miR-1908-5 | intragenic (FADS1) | cis | 3.81E-09 |
| rs174592 | intron_variant | Triacylglyce | 31551469 | 2.00E-08 | FADS2 | miR-1908-5 | intragenic (FADS1) | cis | 3.81E-09 |
| rs174548 | 5_prime_UTR_variant | Blood meta | 24816252 | 1.00E-84 | FADS2, FAD | miR-1908-5 | intragenic (FADS1) | cis | 3.01E-08 |
| rs174566 | intron_variant | LDL cholest | 30698716 | 8.00E-49 | FADS1, FAD | miR-1908-5 | intragenic (FADS1) | cis | 1.58E-08 |
| rs174548 | 5_prime_UTR_variant | Serum met | 23093944 | 1.00E-21 | FADS2, FAD | miR-1908-5 | intragenic (FADS1) | cis | 3.01E-08 |
| rs174601 | non_coding_transcript_exo | Total chole | 29507422 | 1.00E-23 | FADS2 | miR-1908-5 | intragenic (FADS1) | cis | 5.98E-09 |
| rs174566 | intron_variant | Triglyceride | 32203549 | 2.00E-120 | FADS1, FAD | miR-1908-5 | intragenic (FADS1) | cis | 1.58E-08 |
| rs174601 | non_coding_transcript_exo | Phosphatid | 31551469 | 1.00E-15 | FADS2 | miR-1908-5 | intragenic (FADS1) | cis | 5.98E-09 |
| rs174601 | non_coding_transcript_exo | Liver enzyn | 22001757 | 3.00E-09 | FADS2 | miR-1908-5 | intragenic (FADS1) | cis | 5.98E-09 |
| rs28456 | intron_variant | Bipolar disc | 28115744 | 6.00E-09 | FADS1, FAD | miR-1908-5 | intragenic (FADS1) | cis | 6.85E-08 |
| rs28456 | intron_variant | Phosphatid | 31551469 | 1.00E-19 | FADS1, FAD | miR-1908-5 | intragenic (FADS1) | cis | 6.85E-08 |
| rs28456 | intron_variant | Serum met | 29545352 | 1.00E-18 | FADS1, FAD | miR-1908-5 | intragenic (FADS1) | cis | 6.85E-08 |
| rs2072113 | intron_variant | Fatty acid c | 30453627 | 4.00E-55 | FADS2 | miR-1908-5 | intragenic (FADS1) | cis | 3.35E-05 |
| rs2072114 | intron_variant | Phosphatid | 31551469 | 1.00E-15 | FADS2 | miR-1908-5 | intragenic (FADS1) | cis | 1.34E-05 |
| rs2072114 | intron_variant | Trans fatty | 25646338 | 3.00E-09 | FADS2 | miR-1908-5 | intragenic (FADS1) | cis | 1.34E-05 |
| rs174577 | intron_variant | QRS durati | 27659466 | 4.00E-11 | FADS2 | miR-1908-5 | intragenic (FADS1) | cis | 1.89E-08 |
| rs174574 | intron_variant | Plasma om | 21829377 | 4.00E-55 | FADS2 | miR-1908-5 | intragenic (FADS1) | cis | 8.50E-09 |
| rs174574 | intron_variant | Heel bone | 28869591 | 5.00E-09 | FADS2 | miR-1908-5 | intragenic (FADS1) | cis | 8.50E-09 |
| rs2727271 | intron_variant | Non-album | 29403010 | 2.00E-13 | FADS2 | miR-1908-5 | intragenic (FADS1) | cis | 4.23E-05 |
| rs174576 | intron_variant | Serum met | 29545352 | 1.00E-30 | FADS2 | miR-1908-5 | intragenic (FADS1) | cis | 3.13E-08 |
| rs174576 | intron_variant | Glycerophc | 26068415 | 1.00E-17 | FADS2 | miR-1908-5 | intragenic (FADS1) | cis | 3.13E-08 |
| rs174544 | 3_prime_UTR_variant | Monocyte | 27863252 | 3.00E-15 | FADS1, FAD | miR-1908-5 | intragenic (FADS1) | cis | 2.30E-08 |
| rs174560 | non_coding_transcript_exo | Phosphatid | 31551469 | 2.00E-08 | FADS2, FAD | miR-1908-5 | intragenic (FADS1) | cis | 4.91E-08 |
| rs174578 | intron_variant | Glycerophc | 26068415 | 2.00E-43 | FADS2 | miR-1908-5 | intragenic (FADS1) | cis | 2.55E-08 |
| rs2524299 | intron_variant | Trans fatty | 25646338 | 7.00E-09 | FADS2 | miR-1908-5 | intragenic (FADS1) | cis | 4.82E-05 |
| rs174555 | intron_variant | Glycerophc | 26068415 | 1.00E-35 | FADS1, FAD | miR-1908-5 | intragenic (FADS1) | cis | 7.43E-08 |
| rs174577 | intron_variant | Trans fatty | 25646338 | 4.00E-14 | FADS2 | miR-1908-5 | intragenic (FADS1) | cis | 1.89E-08 |
| rs174577 | intron_variant | P wave dur | 24850809 | 3.00E-08 | FADS2 | miR-1908-5 | intragenic (FADS1) | cis | 1.89E-08 |
| rs174601 | non_coding_transcript_exo | Gondoic ac | 28298293 | 1.00E-45 | FADS2 | miR-1908-5 | intragenic (FADS1) | cis | 5.98E-09 |
| rs2727270 | intron_variant | Trans fatty | 25646338 | 7.00E-09 | FADS2 | miR-1908-5 | intragenic (FADS1) | cis | 4.45E-05 |
| rs2727271 | intron_variant | Blood meta | 24816252 | 3.00E-11 | FADS2 | miR-1908-5 | intragenic (FADS1) | cis | 4.23E-05 |
| rs174566 | intron_variant | Serum met | 29545352 | 1.00E-22 | FADS1, FAD | miR-1908-5 | intragenic (FADS1) | cis | 1.58E-08 |
| rs99780 | intron_variant | Serum met | 29545352 | 2.00E-16 | FADS2, FAD | miR-1908-5 | intragenic (FADS1) | cis | 1.74E-08 |
| rs174578 | intron_variant | Plasma om | 26584805 | 2.00E-16 | FADS2 | miR-1908-5 | intragenic (FADS1) | cis | 2.55E-08 |
| rs174584 | intron_variant | Lysophospl | 31551469 | 3.00E-15 | FADS2 | miR-1908-5 | intragenic (FADS1) | cis | 1.39E-08 |
| rs174581 | intron_variant | Serum met | 29545352 | 3.00E-18 | FADS2 | miR-1908-5 | intragenic (FADS1) | cis | 1.68E-08 |
| rs174564 | intron_variant | Serum met | 29545352 | 1.00E-35 | FADS2, FAD | miR-1908-5 | intragenic (FADS1) | cis | 1.61E-08 |
| rs174601 | non_coding_transcript_exo | Trans fatty | 25646338 | 3.00E-13 | FADS2 | miR-1908-5 | intragenic (FADS1) | cis | 5.98E-09 |
| rs174592 | intron_variant | Male-patte | 30573740 | 7.00E-22 | FADS2 | miR-1908-5 | intragenic (FADS1) | cis | 3.81E-09 |
| rs174564 | intron_variant | Pulse press | 30578418 | 6.00E-10 | FADS2, FAD | miR-1908-5 | intragenic (FADS1) | cis | 1.61E-08 |
| rs2727270 | intron_variant | Plasma om | 24823311 | 3.00E-21 | FADS2 | miR-1908-5 | intragenic (FADS1) | cis | 4.45E-05 |
| rs174601 | non_coding_transcript_exo | Red blood | 25500335 | 3.00E-305 | FADS2 | miR-1908-5 | intragenic (FADS1) | cis | 5.98E-09 |
| rs612169 | intron_variant | Blood prote | 28240269 | 1.00E-20 | ABO | miR-195-5p | intergenic | trans | 2.56E-17 |
| rs612169 | intron_variant | Metabolic t | 21886157 | 9.00E-40 | ABO | miR-195-5p | intergenic | trans | 2.56E-17 |
| rs1541777 | intron_variant | Height | 25429064 | 2.00E-08 | TTLL4 | miR-26b-5p | intragenic (CTDSP1) | cis | 0.000663 |
| rs921968 | TF_binding_site_variant | Mean corp | 27863252 | 4.00E-12 | CTDSP1 - V | miR-26b-5p | intragenic (CTDSP1) | cis | 5.63E-05 |
| rs1541777 | intron_variant | Body mass | 30239722 | 2.00E-13 | TTLL4 | miR-26b-5p | intragenic (CTDSP1) | cis | 0.000663 |
| rs7595717 | regulatory_region_variant | Multiple sc | 24076602 | 1.00E-13 | CNRIP1 - PI | miR-320b | intragenic (NVL) | trans | 7.20E-05 |

| | | | | | | | | | |
|---|---|---|---|---|---|---|---|---|---|
| rs2847302: | regulatory_region_variant | Predicted v | 31501611 | 1.00E-09 | LINC00523 | miR-370-3p | intergenic | cis | 7.84E-06 |
| rs1555405 | regulatory_region_variant | Platelet cou | 27863252 | 9.00E-33 | LINC00523 | miR-433-3p | intragenic (RTL1) | cis | 4.59E-07 |
| rs1301107! | intergenic_variant | Mean corp | 27863252 | 2.00E-09 | PLEK - WDF | miR-4429 | intragenic (GREB1) | trans | 6.66E-14 |
| rs3434488 | intron_variant | Educationa | 27225129 | 9.00E-13 | RBM23 | miR-4707-3 | intragenic (HAUS4) | cis | 1.99E-10 |
| rs8005677 | intron_variant | Cognitive a | 29186694 | 9.00E-09 | RBM23 | miR-4707-3 | intragenic (HAUS4) | cis | 1.78E-10 |
| rs5589413: | intron_variant | Intelligence | 29326435 | 5.00E-10 | PRMT5 - H/ | miR-4707-3 | intragenic (HAUS4) | cis | 5.95E-12 |
| rs1243715: | non_coding_transcript_exo | Vertical cup | 31959993 | 2.00E-16 | RBM23 | miR-4707-3 | intragenic (HAUS4) | cis | 2.35E-11 |
| rs1214784! | intron_variant | Body mass | 30239722 | 5.00E-12 | LINC00523 | miR-494-3p | intergenic | cis | 1.61E-10 |
| rs1214784! | intron_variant | Body mass | 30595370 | 1.00E-11 | LINC00523 | miR-494-3p | intergenic | cis | 1.61E-10 |
| rs7149242 | intergenic_variant | Platelet cou | 22139419 | 3.00E-08 | LINC00523 | miR-494-3p | intergenic | cis | 3.33E-10 |
| rs2400940 | intergenic_variant | Diastolic bl | 31754133 | 4.00E-08 | DLK1 - MEC | miR-543 | intergenic | cis | 2.33E-06 |
| rs9324022 | intergenic_variant | Plateletcrit | 27863252 | 3.00E-38 | LINC00523 | miR-543 | intergenic | cis | 4.37E-10 |
| rs9635298 | regulatory_region_variant | Platelet cou | 29403010 | 2.00E-10 | LINC00523 | miR-543 | intergenic | cis | 4.07E-05 |
| rs1036190 | 3_prime_UTR_variant | Systolic blo | 30578418 | 2.00E-10 | SH3TC2 | miR-584-5p | intragenic (SH3TC2) | cis | 1.16E-05 |
| rs1115859: | intergenic_variant | IgG glycosy | 29535710 | 1.00E-17 | MIR4708 - | miR-625-5p | intragenic (FUT8) | cis | 2.48E-11 |
| rs1048378! | intergenic_variant | IgG glycosy | 29535710 | 1.00E-08 | FUT8 - NA | miR-625-5p | intragenic (FUT8) | cis | 2.84E-11 |
| rs867972 | intron_variant | IgG glycosy | 29535710 | 2.00E-17 | FUT8 | miR-625-5p | intragenic (FUT8) | cis | 1.90E-13 |
| rs1958560 | intron_variant | Menarche ( | 25231870 | 4.00E-08 | FUT8 | miR-625-5p | intragenic (FUT8) | cis | 5.40E-17 |
| rs7157006 | intergenic_variant | IgG glycosy | 29535710 | 3.00E-16 | MIR4708 - | miR-625-5p | intragenic (FUT8) | cis | 9.71E-25 |
| rs3742597 | intron_variant | N-glycan le | 21908519 | 1.00E-14 | FUT8 | miR-625-5p | intragenic (FUT8) | cis | 9.19E-09 |
| rs1162168( | intron_variant | IgG glycosy | 29535710 | 6.00E-11 | FUT8 | miR-625-5p | intragenic (FUT8) | cis | 2.44E-11 |
| rs2411822 | 5_prime_UTR_variant | IgG glycosy | 29535710 | 2.00E-13 | FUT8, FUT8 | miR-625-5p | intragenic (FUT8) | cis | 1.16E-13 |
| rs1184726: | intron_variant | IgG glycosy | 29535710 | 2.00E-19 | PTBP1P - M | miR-625-5p | intragenic (FUT8) | cis | 1.15E-13 |
| rs4902389 | intergenic_variant | Blood prote | 30072576 | 0 | PTBP1P - M | miR-625-5p | intragenic (FUT8) | cis | 5.70E-58 |
| rs7147636 | intron_variant | N-glycan le | 31163085 | 5.00E-37 | FUT8 | miR-625-5p | intragenic (FUT8) | cis | 6.53E-17 |
| rs1162282! | intron_variant | IgG glycosy | 29535710 | 3.00E-12 | FUT8 | miR-625-5p | intragenic (FUT8) | cis | 2.93E-13 |
| rs1115860: | intron_variant | IgG glycosy | 29535710 | 1.00E-10 | FUT8 | miR-625-5p | intragenic (FUT8) | cis | 7.53E-13 |
| rs7153297 | intron_variant | IgG glycosy | 29535710 | 5.00E-15 | FUT8 | miR-625-5p | intragenic (FUT8) | cis | 1.30E-13 |
| rs1115858! | intergenic_variant | Plateletcrit | 27863252 | 5.00E-12 | PTBP1P - M | miR-625-5p | intragenic (FUT8) | cis | 5.00E-59 |
| rs1162112: | regulatory_region_variant | N-glycan le | 21908519 | 2.00E-23 | MIR4708 - | miR-625-5p | intragenic (FUT8) | cis | 1.79E-24 |
| rs2149841 | intron_variant | IgG glycosy | 29535710 | 2.00E-12 | FUT8 | miR-625-5p | intragenic (FUT8) | cis | 9.11E-14 |
| rs4073416 | regulatory_region_variant | N-glycan le | 21908519 | 7.00E-15 | FUT8 - NCC | miR-625-5p | intragenic (FUT8) | cis | 2.29E-12 |
| rs1184726: | intron_variant | IgG glycosy | 23382691 | 7.00E-14 | PTBP1P - M | miR-625-5p | intragenic (FUT8) | cis | 1.15E-13 |
| rs8018278 | intron_variant | IgG glycosy | 29535710 | 1.00E-12 | FUT8 | miR-625-5p | intragenic (FUT8) | cis | 2.26E-13 |
| rs6573626 | intergenic_variant | IgG glycosy | 29535710 | 5.00E-10 | FUT8 - NA | miR-625-5p | intragenic (FUT8) | cis | 7.72E-14 |
| rs1289490: | intron_variant | IgG glycosy | 29535710 | 2.00E-13 | FUT8 | miR-625-5p | intragenic (FUT8) | cis | 6.95E-14 |

**Table S20. miR-eQTLs in chr9:136128546-136296530 and other omics**

| SNP | type | effect_a | P | ID | name |
|---|---|---|---|---|---|
| rs7873635 | pQTLs | T | 4.68E-233 | P16442 | Histo-blood group ABO system transferase |
| rs4962040 | pQTLs | A | 3.02E-231 | P16442 | Histo-blood group ABO system transferase |
| rs8176702 | pQTLs | A | 1.58E-233 | P16442 | Histo-blood group ABO system transferase |
| rs2073826 | pQTLs | T | 2.29E-270 | P16442 | Histo-blood group ABO system transferase |
| rs687289 | pQTLs | A | 0 | P16442 | Histo-blood group ABO system transferase |
| rs2073827 | pQTLs | C | 1.05E-269 | P16442 | Histo-blood group ABO system transferase |
| rs2073828 | pQTLs | A | 7.94E-269 | P16442 | Histo-blood group ABO system transferase |
| rs8176691 | pQTLs | T | 3.89E-233 | P16442 | Histo-blood group ABO system transferase |
| rs8176690 | pQTLs | A | 3.55E-271 | P16442 | Histo-blood group ABO system transferase |
| rs8176682 | pQTLs | T | 1.91E-233 | P16442 | Histo-blood group ABO system transferase |
| rs8176681 | pQTLs | T | 2.45E-270 | P16442 | Histo-blood group ABO system transferase |
| rs2519093 | pQTLs | T | 0 | P16442 | Histo-blood group ABO system transferase |
| rs644234 | pQTLs | T | 0 | P16442 | Histo-blood group ABO system transferase |
| rs643434 | pQTLs | A | 0 | P16442 | Histo-blood group ABO system transferase |
| rs543040 | pQTLs | A | 0 | P16442 | Histo-blood group ABO system transferase |
| rs613534 | pQTLs | A | 0 | P16442 | Histo-blood group ABO system transferase |
| rs543968 | pQTLs | T | 0 | P16442 | Histo-blood group ABO system transferase |
| rs544873 | pQTLs | A | 0 | P16442 | Histo-blood group ABO system transferase |
| rs545971 | pQTLs | T | 0 | P16442 | Histo-blood group ABO system transferase |
| rs612169 | pQTLs | A | 0 | P16442 | Histo-blood group ABO system transferase |
| rs8176668 | pQTLs | A | 1.74E-231 | P16442 | Histo-blood group ABO system transferase |
| rs8176663 | pQTLs | T | 0 | P16442 | Histo-blood group ABO system transferase |
| rs7036642 | pQTLs | A | 1.05E-231 | P16442 | Histo-blood group ABO system transferase |
| rs491626 | pQTLs | T | 0 | P16442 | Histo-blood group ABO system transferase |
| rs492488 | pQTLs | A | 0 | P16442 | Histo-blood group ABO system transferase |
| rs493246 | pQTLs | A | 0 | P16442 | Histo-blood group ABO system transferase |
| rs494242 | pQTLs | T | 0 | P16442 | Histo-blood group ABO system transferase |
| rs495203 | pQTLs | T | 0 | P16442 | Histo-blood group ABO system transferase |
| rs582118 | pQTLs | A | 0 | P16442 | Histo-blood group ABO system transferase |
| rs582094 | pQTLs | A | 0 | P16442 | Histo-blood group ABO system transferase |
| rs2769071 | pQTLs | A | 0 | P16442 | Histo-blood group ABO system transferase |
| rs677355 | pQTLs | A | 0 | P16442 | Histo-blood group ABO system transferase |
| rs676996 | pQTLs | T | 0 | P16442 | Histo-blood group ABO system transferase |
| rs676457 | pQTLs | A | 0 | P16442 | Histo-blood group ABO system transferase |
| rs550057 | pQTLs | T | 0 | P16442 | Histo-blood group ABO system transferase |
| rs674302 | pQTLs | A | 0 | P16442 | Histo-blood group ABO system transferase |
| rs7046674 | pQTLs | T | 2.57E-230 | P16442 | Histo-blood group ABO system transferase |
| rs554833 | pQTLs | T | 0 | P16442 | Histo-blood group ABO system transferase |
| rs8176649 | pQTLs | A | 3.80E-232 | P16442 | Histo-blood group ABO system transferase |
| rs660340 | pQTLs | A | 2.24E-291 | P16442 | Histo-blood group ABO system transferase |
| rs581107 | pQTLs | T | 4.27E-294 | P16442 | Histo-blood group ABO system transferase |
| rs659104 | pQTLs | T | 8.13E-294 | P16442 | Histo-blood group ABO system transferase |
| rs647800 | pQTLs | A | 1.00E-252 | P16442 | Histo-blood group ABO system transferase |
| rs473533 | pQTLs | T | 9.33E-294 | P16442 | Histo-blood group ABO system transferase |
| rs476410 | pQTLs | C | 3.80E-293 | P16442 | Histo-blood group ABO system transferase |
| rs645982 | pQTLs | A | 2.40E-292 | P16442 | Histo-blood group ABO system transferase |
| rs529565 | pQTLs | T | 0 | P16442 | Histo-blood group ABO system transferase |
| rs630510 | pQTLs | A | 5.6234156 | P16442 | Histo-blood group ABO system transferase |

| | | | | | |
|---|---|---|---|---|---|
| rs630014 | pQTLs | A | 4.1686938! | P16442 | Histo-blood group ABO system transferase |
| rs532436 | pQTLs | A | 0 | P16442 | Histo-blood group ABO system transferase |
| rs600038 | pQTLs | T | 0 | P16442 | Histo-blood group ABO system transferase |
| rs1124406: | pQTLs | T | 1.66E-238 | P16442 | Histo-blood group ABO system transferase |
| rs649129 | pQTLs | T | 2.2908676! | P16442 | Histo-blood group ABO system transferase |
| rs495828 | pQTLs | T | 0 | P16442 | Histo-blood group ABO system transferase |
| rs660340 | pQTLs | A | 9.33E-09 | Q76LX8 | A disintegrin and metalloproteinase with thrombospon |
| rs581107 | pQTLs | T | 7.76E-09 | Q76LX8 | A disintegrin and metalloproteinase with thrombospon |
| rs659104 | pQTLs | T | 7.76E-09 | Q76LX8 | A disintegrin and metalloproteinase with thrombospon |
| rs473533 | pQTLs | T | 9.12E-09 | Q76LX8 | A disintegrin and metalloproteinase with thrombospon |
| rs476410 | pQTLs | C | 7.24E-09 | Q76LX8 | A disintegrin and metalloproteinase with thrombospon |
| rs645982 | pQTLs | A | 8.13E-09 | Q76LX8 | A disintegrin and metalloproteinase with thrombospon |
| rs7873635 | pQTLs | T | 2.75E-13 | Q8IZF2 | Adhesion G protein-coupled receptor F5 |
| rs4962040 | pQTLs | A | 8.71E-14 | Q8IZF2 | Adhesion G protein-coupled receptor F5 |
| rs8176702 | pQTLs | A | 8.51E-14 | Q8IZF2 | Adhesion G protein-coupled receptor F5 |
| rs2073826 | pQTLs | T | 5.89E-15 | Q8IZF2 | Adhesion G protein-coupled receptor F5 |
| rs687289 | pQTLs | A | 1.70E-36 | Q8IZF2 | Adhesion G protein-coupled receptor F5 |
| rs2073827 | pQTLs | C | 5.75E-15 | Q8IZF2 | Adhesion G protein-coupled receptor F5 |
| rs2073828 | pQTLs | A | 6.76E-15 | Q8IZF2 | Adhesion G protein-coupled receptor F5 |
| rs8176691 | pQTLs | T | 8.32E-14 | Q8IZF2 | Adhesion G protein-coupled receptor F5 |
| rs8176690 | pQTLs | A | 3.63E-15 | Q8IZF2 | Adhesion G protein-coupled receptor F5 |
| rs8176682 | pQTLs | T | 1.29E-13 | Q8IZF2 | Adhesion G protein-coupled receptor F5 |
| rs8176681 | pQTLs | T | 2.24E-14 | Q8IZF2 | Adhesion G protein-coupled receptor F5 |
| rs2519093 | pQTLs | T | 1.38E-155 | Q8IZF2 | Adhesion G protein-coupled receptor F5 |
| rs644234 | pQTLs | T | 1.58E-32 | Q8IZF2 | Adhesion G protein-coupled receptor F5 |
| rs643434 | pQTLs | A | 2.14E-32 | Q8IZF2 | Adhesion G protein-coupled receptor F5 |
| rs543040 | pQTLs | A | 3.47E-36 | Q8IZF2 | Adhesion G protein-coupled receptor F5 |
| rs613534 | pQTLs | A | 1.91E-29 | Q8IZF2 | Adhesion G protein-coupled receptor F5 |
| rs543968 | pQTLs | T | 1.62E-29 | Q8IZF2 | Adhesion G protein-coupled receptor F5 |
| rs544873 | pQTLs | A | 1.78E-32 | Q8IZF2 | Adhesion G protein-coupled receptor F5 |
| rs545971 | pQTLs | T | 6.61E-36 | Q8IZF2 | Adhesion G protein-coupled receptor F5 |
| rs612169 | pQTLs | A | 6.46E-36 | Q8IZF2 | Adhesion G protein-coupled receptor F5 |
| rs8176668 | pQTLs | A | 2.45E-13 | Q8IZF2 | Adhesion G protein-coupled receptor F5 |
| rs8176663 | pQTLs | T | 4.79E-36 | Q8IZF2 | Adhesion G protein-coupled receptor F5 |
| rs7036642 | pQTLs | A | 4.79E-13 | Q8IZF2 | Adhesion G protein-coupled receptor F5 |
| rs491626 | pQTLs | T | 4.47E-36 | Q8IZF2 | Adhesion G protein-coupled receptor F5 |
| rs492488 | pQTLs | A | 6.17E-36 | Q8IZF2 | Adhesion G protein-coupled receptor F5 |
| rs493246 | pQTLs | A | 7.24E-36 | Q8IZF2 | Adhesion G protein-coupled receptor F5 |
| rs494242 | pQTLs | T | 2.00E-32 | Q8IZF2 | Adhesion G protein-coupled receptor F5 |
| rs495203 | pQTLs | T | 2.95E-36 | Q8IZF2 | Adhesion G protein-coupled receptor F5 |
| rs582118 | pQTLs | A | 1.15E-33 | Q8IZF2 | Adhesion G protein-coupled receptor F5 |
| rs582094 | pQTLs | A | 2.04E-34 | Q8IZF2 | Adhesion G protein-coupled receptor F5 |
| rs2769071 | pQTLs | A | 5.37E-36 | Q8IZF2 | Adhesion G protein-coupled receptor F5 |
| rs677355 | pQTLs | A | 3.39E-36 | Q8IZF2 | Adhesion G protein-coupled receptor F5 |
| rs676996 | pQTLs | T | 2.14E-34 | Q8IZF2 | Adhesion G protein-coupled receptor F5 |
| rs676457 | pQTLs | A | 1.45E-35 | Q8IZF2 | Adhesion G protein-coupled receptor F5 |
| rs550057 | pQTLs | T | 3.09E-122 | Q8IZF2 | Adhesion G protein-coupled receptor F5 |
| rs674302 | pQTLs | A | 3.09E-36 | Q8IZF2 | Adhesion G protein-coupled receptor F5 |
| rs7046674 | pQTLs | T | 2.88E-13 | Q8IZF2 | Adhesion G protein-coupled receptor F5 |
| rs554833 | pQTLs | T | 2.88E-36 | Q8IZF2 | Adhesion G protein-coupled receptor F5 |

| | | | | | |
|---|---|---|---|---|---|
| rs8176649 | pQTLs | A | 3.47E-13 | Q8IZF2 | Adhesion G protein-coupled receptor F5 |
| rs660340 | pQTLs | A | 7.41E-12 | Q8IZF2 | Adhesion G protein-coupled receptor F5 |
| rs581107 | pQTLs | T | 4.57E-12 | Q8IZF2 | Adhesion G protein-coupled receptor F5 |
| rs659104 | pQTLs | T | 3.98E-12 | Q8IZF2 | Adhesion G protein-coupled receptor F5 |
| rs647800 | pQTLs | A | 4.07E-11 | Q8IZF2 | Adhesion G protein-coupled receptor F5 |
| rs473533 | pQTLs | T | 3.89E-12 | Q8IZF2 | Adhesion G protein-coupled receptor F5 |
| rs476410 | pQTLs | C | 2.82E-12 | Q8IZF2 | Adhesion G protein-coupled receptor F5 |
| rs645982 | pQTLs | A | 4.57E-12 | Q8IZF2 | Adhesion G protein-coupled receptor F5 |
| rs529565 | pQTLs | T | 1.58E-38 | Q8IZF2 | Adhesion G protein-coupled receptor F5 |
| rs630510 | pQTLs | A | 1.86E-19 | Q8IZF2 | Adhesion G protein-coupled receptor F5 |
| rs630014 | pQTLs | A | 3.89E-19 | Q8IZF2 | Adhesion G protein-coupled receptor F5 |
| rs532436 | pQTLs | A | 1.23E-152 | Q8IZF2 | Adhesion G protein-coupled receptor F5 |
| rs600038 | pQTLs | T | 6.76E-128 | Q8IZF2 | Adhesion G protein-coupled receptor F5 |
| rs1124406: | pQTLs | T | 1.58E-77 | Q8IZF2 | Adhesion G protein-coupled receptor F5 |
| rs649129 | pQTLs | T | 1.74E-129 | Q8IZF2 | Adhesion G protein-coupled receptor F5 |
| rs495828 | pQTLs | T | 4.17E-127 | Q8IZF2 | Adhesion G protein-coupled receptor F5 |
| rs687289 | pQTLs | A | 1.10E-12 | P09923 | Intestinal-type alkaline phosphatase |
| rs2519093 | pQTLs | T | 1.23E-15 | P09923 | Intestinal-type alkaline phosphatase |
| rs644234 | pQTLs | T | 9.33E-11 | P09923 | Intestinal-type alkaline phosphatase |
| rs643434 | pQTLs | A | 1.58E-10 | P09923 | Intestinal-type alkaline phosphatase |
| rs543040 | pQTLs | A | 2.40E-12 | P09923 | Intestinal-type alkaline phosphatase |
| rs613534 | pQTLs | A | 2.82E-10 | P09923 | Intestinal-type alkaline phosphatase |
| rs543968 | pQTLs | T | 5.89E-10 | P09923 | Intestinal-type alkaline phosphatase |
| rs544873 | pQTLs | A | 1.70E-10 | P09923 | Intestinal-type alkaline phosphatase |
| rs545971 | pQTLs | T | 1.70E-12 | P09923 | Intestinal-type alkaline phosphatase |
| rs612169 | pQTLs | A | 1.38E-12 | P09923 | Intestinal-type alkaline phosphatase |
| rs8176663 | pQTLs | T | 1.05E-12 | P09923 | Intestinal-type alkaline phosphatase |
| rs491626 | pQTLs | T | 7.41E-13 | P09923 | Intestinal-type alkaline phosphatase |
| rs492488 | pQTLs | A | 9.33E-13 | P09923 | Intestinal-type alkaline phosphatase |
| rs493246 | pQTLs | A | 1.78E-12 | P09923 | Intestinal-type alkaline phosphatase |
| rs494242 | pQTLs | T | 1.95E-10 | P09923 | Intestinal-type alkaline phosphatase |
| rs495203 | pQTLs | T | 1.17E-12 | P09923 | Intestinal-type alkaline phosphatase |
| rs582118 | pQTLs | A | 4.07E-12 | P09923 | Intestinal-type alkaline phosphatase |
| rs582094 | pQTLs | A | 2.29E-12 | P09923 | Intestinal-type alkaline phosphatase |
| rs2769071 | pQTLs | A | 1.00E-12 | P09923 | Intestinal-type alkaline phosphatase |
| rs677355 | pQTLs | A | 1.17E-12 | P09923 | Intestinal-type alkaline phosphatase |
| rs676996 | pQTLs | T | 4.57E-13 | P09923 | Intestinal-type alkaline phosphatase |
| rs676457 | pQTLs | A | 1.00E-12 | P09923 | Intestinal-type alkaline phosphatase |
| rs550057 | pQTLs | T | 2.75E-19 | P09923 | Intestinal-type alkaline phosphatase |
| rs674302 | pQTLs | A | 1.23E-12 | P09923 | Intestinal-type alkaline phosphatase |
| rs554833 | pQTLs | T | 1.10E-12 | P09923 | Intestinal-type alkaline phosphatase |
| rs660340 | pQTLs | A | 3.31E-08 | P09923 | Intestinal-type alkaline phosphatase |
| rs581107 | pQTLs | T | 4.90E-08 | P09923 | Intestinal-type alkaline phosphatase |
| rs659104 | pQTLs | T | 4.27E-08 | P09923 | Intestinal-type alkaline phosphatase |
| rs473533 | pQTLs | T | 4.68E-08 | P09923 | Intestinal-type alkaline phosphatase |
| rs529565 | pQTLs | T | 7.08E-13 | P09923 | Intestinal-type alkaline phosphatase |
| rs532436 | pQTLs | A | 6.17E-16 | P09923 | Intestinal-type alkaline phosphatase |
| rs600038 | pQTLs | T | 6.31E-12 | P09923 | Intestinal-type alkaline phosphatase |
| rs1124406: | pQTLs | T | 9.12E-10 | P09923 | Intestinal-type alkaline phosphatase |
| rs649129 | pQTLs | T | 2.69E-12 | P09923 | Intestinal-type alkaline phosphatase |

| | | | | | |
|---|---|---|---|---|---|
| rs495828 | pQTLs | T | 4.68E-12 | P09923 | Intestinal-type alkaline phosphatase |
| rs7873635 | pQTLs | T | 2.63E-12 | Q9NY97 | N-acetyllactosaminide beta-1,3-N-acetylglucosaminyltr |
| rs4962040 | pQTLs | A | 5.25E-12 | Q9NY97 | N-acetyllactosaminide beta-1,3-N-acetylglucosaminyltr |
| rs8176702 | pQTLs | A | 4.47E-12 | Q9NY97 | N-acetyllactosaminide beta-1,3-N-acetylglucosaminyltr |
| rs2073826 | pQTLs | T | 2.04E-11 | Q9NY97 | N-acetyllactosaminide beta-1,3-N-acetylglucosaminyltr |
| rs687289 | pQTLs | A | 1.07E-20 | Q9NY97 | N-acetyllactosaminide beta-1,3-N-acetylglucosaminyltr |
| rs2073827 | pQTLs | C | 2.69E-11 | Q9NY97 | N-acetyllactosaminide beta-1,3-N-acetylglucosaminyltr |
| rs2073828 | pQTLs | A | 3.80E-11 | Q9NY97 | N-acetyllactosaminide beta-1,3-N-acetylglucosaminyltr |
| rs8176691 | pQTLs | T | 5.37E-12 | Q9NY97 | N-acetyllactosaminide beta-1,3-N-acetylglucosaminyltr |
| rs8176690 | pQTLs | A | 3.31E-11 | Q9NY97 | N-acetyllactosaminide beta-1,3-N-acetylglucosaminyltr |
| rs8176682 | pQTLs | T | 5.89E-12 | Q9NY97 | N-acetyllactosaminide beta-1,3-N-acetylglucosaminyltr |
| rs8176681 | pQTLs | T | 3.39E-11 | Q9NY97 | N-acetyllactosaminide beta-1,3-N-acetylglucosaminyltr |
| rs2519093 | pQTLs | T | 4.17E-23 | Q9NY97 | N-acetyllactosaminide beta-1,3-N-acetylglucosaminyltr |
| rs644234 | pQTLs | T | 5.25E-20 | Q9NY97 | N-acetyllactosaminide beta-1,3-N-acetylglucosaminyltr |
| rs643434 | pQTLs | A | 1.66E-20 | Q9NY97 | N-acetyllactosaminide beta-1,3-N-acetylglucosaminyltr |
| rs543040 | pQTLs | A | 8.32E-21 | Q9NY97 | N-acetyllactosaminide beta-1,3-N-acetylglucosaminyltr |
| rs613534 | pQTLs | A | 1.23E-18 | Q9NY97 | N-acetyllactosaminide beta-1,3-N-acetylglucosaminyltr |
| rs543968 | pQTLs | T | 1.12E-19 | Q9NY97 | N-acetyllactosaminide beta-1,3-N-acetylglucosaminyltr |
| rs544873 | pQTLs | A | 2.00E-20 | Q9NY97 | N-acetyllactosaminide beta-1,3-N-acetylglucosaminyltr |
| rs545971 | pQTLs | T | 4.07E-21 | Q9NY97 | N-acetyllactosaminide beta-1,3-N-acetylglucosaminyltr |
| rs612169 | pQTLs | A | 1.00E-20 | Q9NY97 | N-acetyllactosaminide beta-1,3-N-acetylglucosaminyltr |
| rs8176668 | pQTLs | A | 1.48E-11 | Q9NY97 | N-acetyllactosaminide beta-1,3-N-acetylglucosaminyltr |
| rs8176663 | pQTLs | T | 2.00E-20 | Q9NY97 | N-acetyllactosaminide beta-1,3-N-acetylglucosaminyltr |
| rs7036642 | pQTLs | A | 1.58E-11 | Q9NY97 | N-acetyllactosaminide beta-1,3-N-acetylglucosaminyltr |
| rs491626 | pQTLs | T | 1.70E-20 | Q9NY97 | N-acetyllactosaminide beta-1,3-N-acetylglucosaminyltr |
| rs492488 | pQTLs | A | 2.69E-20 | Q9NY97 | N-acetyllactosaminide beta-1,3-N-acetylglucosaminyltr |
| rs493246 | pQTLs | A | 1.58E-20 | Q9NY97 | N-acetyllactosaminide beta-1,3-N-acetylglucosaminyltr |
| rs494242 | pQTLs | T | 2.00E-20 | Q9NY97 | N-acetyllactosaminide beta-1,3-N-acetylglucosaminyltr |
| rs495203 | pQTLs | T | 1.91E-20 | Q9NY97 | N-acetyllactosaminide beta-1,3-N-acetylglucosaminyltr |
| rs582118 | pQTLs | A | 1.05E-19 | Q9NY97 | N-acetyllactosaminide beta-1,3-N-acetylglucosaminyltr |
| rs582094 | pQTLs | A | 2.95E-20 | Q9NY97 | N-acetyllactosaminide beta-1,3-N-acetylglucosaminyltr |
| rs2769071 | pQTLs | A | 4.37E-20 | Q9NY97 | N-acetyllactosaminide beta-1,3-N-acetylglucosaminyltr |
| rs677355 | pQTLs | A | 3.16E-20 | Q9NY97 | N-acetyllactosaminide beta-1,3-N-acetylglucosaminyltr |
| rs676996 | pQTLs | T | 8.51E-19 | Q9NY97 | N-acetyllactosaminide beta-1,3-N-acetylglucosaminyltr |
| rs676457 | pQTLs | A | 1.15E-19 | Q9NY97 | N-acetyllactosaminide beta-1,3-N-acetylglucosaminyltr |
| rs550057 | pQTLs | T | 6.46E-17 | Q9NY97 | N-acetyllactosaminide beta-1,3-N-acetylglucosaminyltr |
| rs674302 | pQTLs | A | 4.17E-20 | Q9NY97 | N-acetyllactosaminide beta-1,3-N-acetylglucosaminyltr |
| rs7046674 | pQTLs | T | 5.37E-11 | Q9NY97 | N-acetyllactosaminide beta-1,3-N-acetylglucosaminyltr |
| rs554833 | pQTLs | T | 5.62E-20 | Q9NY97 | N-acetyllactosaminide beta-1,3-N-acetylglucosaminyltr |
| rs8176649 | pQTLs | A | 3.16E-11 | Q9NY97 | N-acetyllactosaminide beta-1,3-N-acetylglucosaminyltr |
| rs529565 | pQTLs | T | 4.37E-20 | Q9NY97 | N-acetyllactosaminide beta-1,3-N-acetylglucosaminyltr |
| rs532436 | pQTLs | A | 1.70E-22 | Q9NY97 | N-acetyllactosaminide beta-1,3-N-acetylglucosaminyltr |
| rs600038 | pQTLs | T | 9.12E-21 | Q9NY97 | N-acetyllactosaminide beta-1,3-N-acetylglucosaminyltr |
| rs1124406: | pQTLs | T | 1.78E-15 | Q9NY97 | N-acetyllactosaminide beta-1,3-N-acetylglucosaminyltr |
| rs649129 | pQTLs | T | 4.37E-19 | Q9NY97 | N-acetyllactosaminide beta-1,3-N-acetylglucosaminyltr |
| rs495828 | pQTLs | T | 1.15E-20 | Q9NY97 | N-acetyllactosaminide beta-1,3-N-acetylglucosaminyltr |
| rs2519093 | pQTLs | T | 4.90E-14 | P15291 | Beta-1,4-galactosyltransferase 1 |
| rs644234 | pQTLs | T | 1.70E-08 | P15291 | Beta-1,4-galactosyltransferase 1 |
| rs643434 | pQTLs | A | 1.74E-08 | P15291 | Beta-1,4-galactosyltransferase 1 |
| rs543040 | pQTLs | A | 4.79E-08 | P15291 | Beta-1,4-galactosyltransferase 1 |

| | | | | | |
|---|---|---|---|---|---|
| rs543968 | pQTLs | T | 4.90E-08 | P15291 | Beta-1,4-galactosyltransferase 1 |
| rs544873 | pQTLs | A | 1.51E-08 | P15291 | Beta-1,4-galactosyltransferase 1 |
| rs545971 | pQTLs | T | 4.47E-08 | P15291 | Beta-1,4-galactosyltransferase 1 |
| rs494242 | pQTLs | T | 1.86E-08 | P15291 | Beta-1,4-galactosyltransferase 1 |
| rs550057 | pQTLs | T | 3.98E-10 | P15291 | Beta-1,4-galactosyltransferase 1 |
| rs532436 | pQTLs | A | 1.55E-13 | P15291 | Beta-1,4-galactosyltransferase 1 |
| rs600038 | pQTLs | T | 3.80E-14 | P15291 | Beta-1,4-galactosyltransferase 1 |
| rs1124406: | pQTLs | T | 4.79E-09 | P15291 | Beta-1,4-galactosyltransferase 1 |
| rs649129 | pQTLs | T | 1.07E-13 | P15291 | Beta-1,4-galactosyltransferase 1 |
| rs495828 | pQTLs | T | 5.75E-14 | P15291 | Beta-1,4-galactosyltransferase 1 |
| rs2519093 | pQTLs | T | 2.95E-08 | O60909 | Beta-1,4-galactosyltransferase 2 |
| rs550057 | pQTLs | T | 1.35E-08 | O60909 | Beta-1,4-galactosyltransferase 2 |
| rs532436 | pQTLs | A | 3.39E-08 | O60909 | Beta-1,4-galactosyltransferase 2 |
| rs600038 | pQTLs | T | 4.07E-08 | O60909 | Beta-1,4-galactosyltransferase 2 |
| rs1124406: | pQTLs | T | 2.88E-08 | O60909 | Beta-1,4-galactosyltransferase 2 |
| rs7873635 | pQTLs | T | 1.20E-21 | Q96EU7 | C1GALT1-specific chaperone 1 |
| rs4962040 | pQTLs | A | 1.02E-21 | Q96EU7 | C1GALT1-specific chaperone 1 |
| rs8176702 | pQTLs | A | 6.17E-22 | Q96EU7 | C1GALT1-specific chaperone 1 |
| rs2073826 | pQTLs | T | 2.14E-20 | Q96EU7 | C1GALT1-specific chaperone 1 |
| rs687289 | pQTLs | A | 8.71E-50 | Q96EU7 | C1GALT1-specific chaperone 1 |
| rs2073827 | pQTLs | C | 2.34E-20 | Q96EU7 | C1GALT1-specific chaperone 1 |
| rs2073828 | pQTLs | A | 2.88E-20 | Q96EU7 | C1GALT1-specific chaperone 1 |
| rs8176691 | pQTLs | T | 6.76E-22 | Q96EU7 | C1GALT1-specific chaperone 1 |
| rs8176690 | pQTLs | A | 4.27E-20 | Q96EU7 | C1GALT1-specific chaperone 1 |
| rs8176682 | pQTLs | T | 8.13E-22 | Q96EU7 | C1GALT1-specific chaperone 1 |
| rs8176681 | pQTLs | T | 4.27E-20 | Q96EU7 | C1GALT1-specific chaperone 1 |
| rs2519093 | pQTLs | T | 1.91E-89 | Q96EU7 | C1GALT1-specific chaperone 1 |
| rs644234 | pQTLs | T | 2.40E-50 | Q96EU7 | C1GALT1-specific chaperone 1 |
| rs643434 | pQTLs | A | 1.70E-49 | Q96EU7 | C1GALT1-specific chaperone 1 |
| rs543040 | pQTLs | A | 1.32E-49 | Q96EU7 | C1GALT1-specific chaperone 1 |
| rs613534 | pQTLs | A | 3.16E-47 | Q96EU7 | C1GALT1-specific chaperone 1 |
| rs543968 | pQTLs | T | 6.31E-47 | Q96EU7 | C1GALT1-specific chaperone 1 |
| rs544873 | pQTLs | A | 1.74E-49 | Q96EU7 | C1GALT1-specific chaperone 1 |
| rs545971 | pQTLs | T | 3.63E-49 | Q96EU7 | C1GALT1-specific chaperone 1 |
| rs612169 | pQTLs | A | 1.74E-49 | Q96EU7 | C1GALT1-specific chaperone 1 |
| rs8176668 | pQTLs | A | 1.51E-21 | Q96EU7 | C1GALT1-specific chaperone 1 |
| rs8176663 | pQTLs | T | 5.13E-49 | Q96EU7 | C1GALT1-specific chaperone 1 |
| rs7036642 | pQTLs | A | 7.08E-21 | Q96EU7 | C1GALT1-specific chaperone 1 |
| rs491626 | pQTLs | T | 1.66E-48 | Q96EU7 | C1GALT1-specific chaperone 1 |
| rs492488 | pQTLs | A | 1.66E-48 | Q96EU7 | C1GALT1-specific chaperone 1 |
| rs493246 | pQTLs | A | 1.02E-48 | Q96EU7 | C1GALT1-specific chaperone 1 |
| rs494242 | pQTLs | T | 7.08E-49 | Q96EU7 | C1GALT1-specific chaperone 1 |
| rs495203 | pQTLs | T | 3.09E-49 | Q96EU7 | C1GALT1-specific chaperone 1 |
| rs582118 | pQTLs | A | 2.40E-47 | Q96EU7 | C1GALT1-specific chaperone 1 |
| rs582094 | pQTLs | A | 4.27E-48 | Q96EU7 | C1GALT1-specific chaperone 1 |
| rs2769071 | pQTLs | A | 9.12E-49 | Q96EU7 | C1GALT1-specific chaperone 1 |
| rs677355 | pQTLs | A | 6.61E-49 | Q96EU7 | C1GALT1-specific chaperone 1 |
| rs676996 | pQTLs | T | 7.59E-45 | Q96EU7 | C1GALT1-specific chaperone 1 |
| rs676457 | pQTLs | A | 4.07E-49 | Q96EU7 | C1GALT1-specific chaperone 1 |
| rs550057 | pQTLs | T | 9.77E-77 | Q96EU7 | C1GALT1-specific chaperone 1 |

| | | | | | |
|---|---|---|---|---|---|
| rs674302 | pQTLs | A | 6.03E-49 | Q96EU7 | C1GALT1-specific chaperone 1 |
| rs7046674 | pQTLs | T | 8.71E-21 | Q96EU7 | C1GALT1-specific chaperone 1 |
| rs554833 | pQTLs | T | 3.39E-49 | Q96EU7 | C1GALT1-specific chaperone 1 |
| rs8176649 | pQTLs | A | 5.89E-21 | Q96EU7 | C1GALT1-specific chaperone 1 |
| rs660340 | pQTLs | A | 1.00E-17 | Q96EU7 | C1GALT1-specific chaperone 1 |
| rs581107 | pQTLs | T | 7.59E-18 | Q96EU7 | C1GALT1-specific chaperone 1 |
| rs659104 | pQTLs | T | 8.51E-18 | Q96EU7 | C1GALT1-specific chaperone 1 |
| rs647800 | pQTLs | A | 3.98E-19 | Q96EU7 | C1GALT1-specific chaperone 1 |
| rs473533 | pQTLs | T | 1.05E-17 | Q96EU7 | C1GALT1-specific chaperone 1 |
| rs476410 | pQTLs | C | 3.89E-18 | Q96EU7 | C1GALT1-specific chaperone 1 |
| rs645982 | pQTLs | A | 8.13E-18 | Q96EU7 | C1GALT1-specific chaperone 1 |
| rs529565 | pQTLs | T | 3.89E-48 | Q96EU7 | C1GALT1-specific chaperone 1 |
| rs630510 | pQTLs | A | 3.31E-19 | Q96EU7 | C1GALT1-specific chaperone 1 |
| rs630014 | pQTLs | A | 4.57E-19 | Q96EU7 | C1GALT1-specific chaperone 1 |
| rs532436 | pQTLs | A | 2.45E-87 | Q96EU7 | C1GALT1-specific chaperone 1 |
| rs600038 | pQTLs | T | 3.09E-80 | Q96EU7 | C1GALT1-specific chaperone 1 |
| rs1124406: | pQTLs | T | 2.40E-57 | Q96EU7 | C1GALT1-specific chaperone 1 |
| rs649129 | pQTLs | T | 2.45E-78 | Q96EU7 | C1GALT1-specific chaperone 1 |
| rs495828 | pQTLs | T | 5.75E-80 | Q96EU7 | C1GALT1-specific chaperone 1 |
| rs687289 | pQTLs | A | 1.48E-09 | O15444 | C-C motif chemokine 25 |
| rs644234 | pQTLs | T | 3.31E-09 | O15444 | C-C motif chemokine 25 |
| rs643434 | pQTLs | A | 3.80E-09 | O15444 | C-C motif chemokine 25 |
| rs543040 | pQTLs | A | 2.51E-09 | O15444 | C-C motif chemokine 25 |
| rs613534 | pQTLs | A | 1.51E-08 | O15444 | C-C motif chemokine 25 |
| rs543968 | pQTLs | T | 2.24E-08 | O15444 | C-C motif chemokine 25 |
| rs544873 | pQTLs | A | 3.98E-09 | O15444 | C-C motif chemokine 25 |
| rs545971 | pQTLs | T | 3.24E-09 | O15444 | C-C motif chemokine 25 |
| rs612169 | pQTLs | A | 1.74E-09 | O15444 | C-C motif chemokine 25 |
| rs8176663 | pQTLs | T | 1.35E-09 | O15444 | C-C motif chemokine 25 |
| rs491626 | pQTLs | T | 1.23E-09 | O15444 | C-C motif chemokine 25 |
| rs492488 | pQTLs | A | 1.12E-09 | O15444 | C-C motif chemokine 25 |
| rs493246 | pQTLs | A | 1.23E-09 | O15444 | C-C motif chemokine 25 |
| rs494242 | pQTLs | T | 2.75E-09 | O15444 | C-C motif chemokine 25 |
| rs495203 | pQTLs | T | 1.32E-09 | O15444 | C-C motif chemokine 25 |
| rs582118 | pQTLs | A | 7.94E-09 | O15444 | C-C motif chemokine 25 |
| rs582094 | pQTLs | A | 4.07E-09 | O15444 | C-C motif chemokine 25 |
| rs2769071 | pQTLs | A | 1.07E-09 | O15444 | C-C motif chemokine 25 |
| rs677355 | pQTLs | A | 1.55E-09 | O15444 | C-C motif chemokine 25 |
| rs676996 | pQTLs | T | 2.69E-09 | O15444 | C-C motif chemokine 25 |
| rs676457 | pQTLs | A | 1.12E-09 | O15444 | C-C motif chemokine 25 |
| rs550057 | pQTLs | T | 4.07E-12 | O15444 | C-C motif chemokine 25 |
| rs674302 | pQTLs | A | 1.45E-09 | O15444 | C-C motif chemokine 25 |
| rs554833 | pQTLs | T | 1.10E-09 | O15444 | C-C motif chemokine 25 |
| rs529565 | pQTLs | T | 2.95E-09 | O15444 | C-C motif chemokine 25 |
| rs687289 | pQTLs | A | 1.02E-09 | O15444 | C-C motif chemokine 25 |
| rs644234 | pQTLs | T | 1.78E-08 | O15444 | C-C motif chemokine 25 |
| rs643434 | pQTLs | A | 1.48E-08 | O15444 | C-C motif chemokine 25 |
| rs543040 | pQTLs | A | 3.55E-09 | O15444 | C-C motif chemokine 25 |
| rs613534 | pQTLs | A | 2.45E-08 | O15444 | C-C motif chemokine 25 |
| rs544873 | pQTLs | A | 1.45E-08 | O15444 | C-C motif chemokine 25 |

| | | | | | |
|---|---|---|---|---|---|
| rs545971 | pQTLs | T | 2.69E-09 | O15444 | C-C motif chemokine 25 |
| rs612169 | pQTLs | A | 1.66E-09 | O15444 | C-C motif chemokine 25 |
| rs8176663 | pQTLs | T | 1.20E-09 | O15444 | C-C motif chemokine 25 |
| rs491626 | pQTLs | T | 1.07E-09 | O15444 | C-C motif chemokine 25 |
| rs492488 | pQTLs | A | 1.05E-09 | O15444 | C-C motif chemokine 25 |
| rs493246 | pQTLs | A | 1.26E-09 | O15444 | C-C motif chemokine 25 |
| rs494242 | pQTLs | T | 1.23E-08 | O15444 | C-C motif chemokine 25 |
| rs495203 | pQTLs | T | 1.15E-09 | O15444 | C-C motif chemokine 25 |
| rs582118 | pQTLs | A | 8.71E-09 | O15444 | C-C motif chemokine 25 |
| rs582094 | pQTLs | A | 2.82E-09 | O15444 | C-C motif chemokine 25 |
| rs2769071 | pQTLs | A | 9.12E-10 | O15444 | C-C motif chemokine 25 |
| rs677355 | pQTLs | A | 9.55E-10 | O15444 | C-C motif chemokine 25 |
| rs676996 | pQTLs | T | 2.09E-09 | O15444 | C-C motif chemokine 25 |
| rs676457 | pQTLs | A | 7.41E-10 | O15444 | C-C motif chemokine 25 |
| rs550057 | pQTLs | T | 2.75E-12 | O15444 | C-C motif chemokine 25 |
| rs674302 | pQTLs | A | 1.17E-09 | O15444 | C-C motif chemokine 25 |
| rs554833 | pQTLs | T | 8.71E-10 | O15444 | C-C motif chemokine 25 |
| rs529565 | pQTLs | T | 2.09E-09 | O15444 | C-C motif chemokine 25 |
| rs2519093 | pQTLs | T | 1.48E-08 | P41217 | OX-2 membrane glycoprotein |
| rs532436 | pQTLs | A | 7.24E-09 | P41217 | OX-2 membrane glycoprotein |
| rs7873635 | pQTLs | T | 1.38E-78 | Q9NNX6 | CD209 antigen |
| rs4962040 | pQTLs | A | 7.76E-78 | Q9NNX6 | CD209 antigen |
| rs8176702 | pQTLs | A | 3.09E-78 | Q9NNX6 | CD209 antigen |
| rs2073826 | pQTLs | T | 6.76E-89 | Q9NNX6 | CD209 antigen |
| rs687289 | pQTLs | A | 1.74E-300 | Q9NNX6 | CD209 antigen |
| rs2073827 | pQTLs | C | 7.94E-89 | Q9NNX6 | CD209 antigen |
| rs2073828 | pQTLs | A | 8.71E-89 | Q9NNX6 | CD209 antigen |
| rs8176691 | pQTLs | T | 4.07E-78 | Q9NNX6 | CD209 antigen |
| rs8176690 | pQTLs | A | 4.37E-89 | Q9NNX6 | CD209 antigen |
| rs8176682 | pQTLs | T | 1.32E-78 | Q9NNX6 | CD209 antigen |
| rs8176681 | pQTLs | T | 4.47E-89 | Q9NNX6 | CD209 antigen |
| rs2519093 | pQTLs | T | 1.17E-171 | Q9NNX6 | CD209 antigen |
| rs644234 | pQTLs | T | 4.90E-251 | Q9NNX6 | CD209 antigen |
| rs643434 | pQTLs | A | 3.80E-253 | Q9NNX6 | CD209 antigen |
| rs543040 | pQTLs | A | 4.79E-300 | Q9NNX6 | CD209 antigen |
| rs613534 | pQTLs | A | 1.55E-242 | Q9NNX6 | CD209 antigen |
| rs543968 | pQTLs | T | 1.51E-241 | Q9NNX6 | CD209 antigen |
| rs544873 | pQTLs | A | 6.17E-253 | Q9NNX6 | CD209 antigen |
| rs545971 | pQTLs | T | 1.26E-300 | Q9NNX6 | CD209 antigen |
| rs612169 | pQTLs | A | 6.03E-300 | Q9NNX6 | CD209 antigen |
| rs8176668 | pQTLs | A | 5.13E-78 | Q9NNX6 | CD209 antigen |
| rs8176663 | pQTLs | T | 2.45E-299 | Q9NNX6 | CD209 antigen |
| rs7036642 | pQTLs | A | 4.37E-78 | Q9NNX6 | CD209 antigen |
| rs491626 | pQTLs | T | 1.86E-298 | Q9NNX6 | CD209 antigen |
| rs492488 | pQTLs | A | 6.31E-298 | Q9NNX6 | CD209 antigen |
| rs493246 | pQTLs | A | 2.34E-297 | Q9NNX6 | CD209 antigen |
| rs494242 | pQTLs | T | 7.41E-253 | Q9NNX6 | CD209 antigen |
| rs495203 | pQTLs | T | 2.24E-299 | Q9NNX6 | CD209 antigen |
| rs582118 | pQTLs | A | 4.27E-294 | Q9NNX6 | CD209 antigen |
| rs582094 | pQTLs | A | 9.12E-295 | Q9NNX6 | CD209 antigen |

| | | | | | |
|---|---|---|---|---|---|
| rs2769071 | pQTLs | A | 1.86E-297 | Q9NNX6 | CD209 antigen |
| rs677355 | pQTLs | A | 1.45E-297 | Q9NNX6 | CD209 antigen |
| rs676996 | pQTLs | T | 2.00E-286 | Q9NNX6 | CD209 antigen |
| rs676457 | pQTLs | A | 1.78E-295 | Q9NNX6 | CD209 antigen |
| rs550057 | pQTLs | T | 2.24E-159 | Q9NNX6 | CD209 antigen |
| rs674302 | pQTLs | A | 2.51E-298 | Q9NNX6 | CD209 antigen |
| rs7046674 | pQTLs | T | 3.24E-77 | Q9NNX6 | CD209 antigen |
| rs554833 | pQTLs | T | 4.47E-297 | Q9NNX6 | CD209 antigen |
| rs8176649 | pQTLs | A | 1.78E-76 | Q9NNX6 | CD209 antigen |
| rs660340 | pQTLs | A | 1.02E-98 | Q9NNX6 | CD209 antigen |
| rs581107 | pQTLs | T | 5.89E-99 | Q9NNX6 | CD209 antigen |
| rs659104 | pQTLs | T | 1.78E-98 | Q9NNX6 | CD209 antigen |
| rs647800 | pQTLs | A | 1.07E-86 | Q9NNX6 | CD209 antigen |
| rs473533 | pQTLs | T | 1.58E-98 | Q9NNX6 | CD209 antigen |
| rs476410 | pQTLs | C | 7.59E-98 | Q9NNX6 | CD209 antigen |
| rs645982 | pQTLs | A | 3.80E-98 | Q9NNX6 | CD209 antigen |
| rs529565 | pQTLs | T | 9.77E-301 | Q9NNX6 | CD209 antigen |
| rs630510 | pQTLs | A | 9.33E-109 | Q9NNX6 | CD209 antigen |
| rs630014 | pQTLs | A | 5.62E-109 | Q9NNX6 | CD209 antigen |
| rs532436 | pQTLs | A | 6.92E-171 | Q9NNX6 | CD209 antigen |
| rs600038 | pQTLs | T | 5.75E-129 | Q9NNX6 | CD209 antigen |
| rs1124406: | pQTLs | T | 2.45E-94 | Q9NNX6 | CD209 antigen |
| rs649129 | pQTLs | T | 2.57E-123 | Q9NNX6 | CD209 antigen |
| rs495828 | pQTLs | T | 1.58E-127 | Q9NNX6 | CD209 antigen |
| rs7873635 | pQTLs | T | 1.41E-08 | P33151 | Cadherin-5 |
| rs8176702 | pQTLs | A | 4.90E-08 | P33151 | Cadherin-5 |
| rs2073826 | pQTLs | T | 3.31E-09 | P33151 | Cadherin-5 |
| rs687289 | pQTLs | A | 2.00E-10 | P33151 | Cadherin-5 |
| rs2073827 | pQTLs | C | 5.01E-09 | P33151 | Cadherin-5 |
| rs2073828 | pQTLs | A | 5.62E-09 | P33151 | Cadherin-5 |
| rs8176690 | pQTLs | A | 2.34E-09 | P33151 | Cadherin-5 |
| rs8176682 | pQTLs | T | 2.51E-08 | P33151 | Cadherin-5 |
| rs8176681 | pQTLs | T | 1.86E-09 | P33151 | Cadherin-5 |
| rs2519093 | pQTLs | T | 3.39E-14 | P33151 | Cadherin-5 |
| rs644234 | pQTLs | T | 1.51E-08 | P33151 | Cadherin-5 |
| rs643434 | pQTLs | A | 1.32E-08 | P33151 | Cadherin-5 |
| rs543040 | pQTLs | A | 2.45E-10 | P33151 | Cadherin-5 |
| rs613534 | pQTLs | A | 1.17E-08 | P33151 | Cadherin-5 |
| rs543968 | pQTLs | T | 7.24E-09 | P33151 | Cadherin-5 |
| rs544873 | pQTLs | A | 1.45E-08 | P33151 | Cadherin-5 |
| rs545971 | pQTLs | T | 6.76E-11 | P33151 | Cadherin-5 |
| rs612169 | pQTLs | A | 1.41E-10 | P33151 | Cadherin-5 |
| rs8176663 | pQTLs | T | 2.29E-10 | P33151 | Cadherin-5 |
| rs7036642 | pQTLs | A | 4.79E-08 | P33151 | Cadherin-5 |
| rs491626 | pQTLs | T | 2.82E-10 | P33151 | Cadherin-5 |
| rs492488 | pQTLs | A | 2.51E-10 | P33151 | Cadherin-5 |
| rs493246 | pQTLs | A | 2.19E-10 | P33151 | Cadherin-5 |
| rs494242 | pQTLs | T | 1.48E-08 | P33151 | Cadherin-5 |
| rs495203 | pQTLs | T | 2.63E-10 | P33151 | Cadherin-5 |
| rs582118 | pQTLs | A | 4.90E-11 | P33151 | Cadherin-5 |

| | | | | | |
|---|---|---|---|---|---|
| rs582094 | pQTLs | A | 4.57E-11 | P33151 | Cadherin-5 |
| rs2769071 | pQTLs | A | 3.63E-10 | P33151 | Cadherin-5 |
| rs677355 | pQTLs | A | 2.63E-10 | P33151 | Cadherin-5 |
| rs676996 | pQTLs | T | 2.82E-10 | P33151 | Cadherin-5 |
| rs676457 | pQTLs | A | 5.13E-10 | P33151 | Cadherin-5 |
| rs550057 | pQTLs | T | 2.24E-12 | P33151 | Cadherin-5 |
| rs674302 | pQTLs | A | 2.95E-10 | P33151 | Cadherin-5 |
| rs554833 | pQTLs | T | 5.13E-10 | P33151 | Cadherin-5 |
| rs529565 | pQTLs | T | 1.38E-09 | P33151 | Cadherin-5 |
| rs532436 | pQTLs | A | 8.71E-14 | P33151 | Cadherin-5 |
| rs600038 | pQTLs | T | 1.26E-14 | P33151 | Cadherin-5 |
| rs649129 | pQTLs | T | 3.47E-15 | P33151 | Cadherin-5 |
| rs495828 | pQTLs | T | 5.75E-15 | P33151 | Cadherin-5 |
| rs687289 | pQTLs | A | 1.00E-11 | Q9NPF2 | Carbohydrate sulfotransferase 11 |
| rs2519093 | pQTLs | T | 2.24E-10 | Q9NPF2 | Carbohydrate sulfotransferase 11 |
| rs644234 | pQTLs | T | 2.88E-12 | Q9NPF2 | Carbohydrate sulfotransferase 11 |
| rs643434 | pQTLs | A | 4.27E-12 | Q9NPF2 | Carbohydrate sulfotransferase 11 |
| rs543040 | pQTLs | A | 1.51E-11 | Q9NPF2 | Carbohydrate sulfotransferase 11 |
| rs613534 | pQTLs | A | 3.24E-12 | Q9NPF2 | Carbohydrate sulfotransferase 11 |
| rs543968 | pQTLs | T | 5.13E-12 | Q9NPF2 | Carbohydrate sulfotransferase 11 |
| rs544873 | pQTLs | A | 4.17E-12 | Q9NPF2 | Carbohydrate sulfotransferase 11 |
| rs545971 | pQTLs | T | 1.78E-11 | Q9NPF2 | Carbohydrate sulfotransferase 11 |
| rs612169 | pQTLs | A | 1.55E-11 | Q9NPF2 | Carbohydrate sulfotransferase 11 |
| rs8176663 | pQTLs | T | 1.70E-11 | Q9NPF2 | Carbohydrate sulfotransferase 11 |
| rs491626 | pQTLs | T | 1.95E-11 | Q9NPF2 | Carbohydrate sulfotransferase 11 |
| rs492488 | pQTLs | A | 1.91E-11 | Q9NPF2 | Carbohydrate sulfotransferase 11 |
| rs493246 | pQTLs | A | 1.82E-11 | Q9NPF2 | Carbohydrate sulfotransferase 11 |
| rs494242 | pQTLs | T | 6.17E-12 | Q9NPF2 | Carbohydrate sulfotransferase 11 |
| rs495203 | pQTLs | T | 1.35E-11 | Q9NPF2 | Carbohydrate sulfotransferase 11 |
| rs582118 | pQTLs | A | 4.37E-11 | Q9NPF2 | Carbohydrate sulfotransferase 11 |
| rs582094 | pQTLs | A | 2.75E-11 | Q9NPF2 | Carbohydrate sulfotransferase 11 |
| rs2769071 | pQTLs | A | 2.14E-11 | Q9NPF2 | Carbohydrate sulfotransferase 11 |
| rs677355 | pQTLs | A | 1.62E-11 | Q9NPF2 | Carbohydrate sulfotransferase 11 |
| rs676996 | pQTLs | T | 1.51E-11 | Q9NPF2 | Carbohydrate sulfotransferase 11 |
| rs676457 | pQTLs | A | 1.07E-11 | Q9NPF2 | Carbohydrate sulfotransferase 11 |
| rs550057 | pQTLs | T | 1.10E-09 | Q9NPF2 | Carbohydrate sulfotransferase 11 |
| rs674302 | pQTLs | A | 1.70E-11 | Q9NPF2 | Carbohydrate sulfotransferase 11 |
| rs554833 | pQTLs | T | 1.70E-11 | Q9NPF2 | Carbohydrate sulfotransferase 11 |
| rs529565 | pQTLs | T | 1.35E-11 | Q9NPF2 | Carbohydrate sulfotransferase 11 |
| rs532436 | pQTLs | A | 4.90E-10 | Q9NPF2 | Carbohydrate sulfotransferase 11 |
| rs600038 | pQTLs | T | 5.89E-10 | Q9NPF2 | Carbohydrate sulfotransferase 11 |
| rs649129 | pQTLs | T | 1.20E-09 | Q9NPF2 | Carbohydrate sulfotransferase 11 |
| rs495828 | pQTLs | T | 2.57E-10 | Q9NPF2 | Carbohydrate sulfotransferase 11 |
| rs687289 | pQTLs | A | 3.24E-20 | Q7LFX5 | Carbohydrate sulfotransferase 15 |
| rs2519093 | pQTLs | T | 1.12E-18 | Q7LFX5 | Carbohydrate sulfotransferase 15 |
| rs644234 | pQTLs | T | 2.09E-17 | Q7LFX5 | Carbohydrate sulfotransferase 15 |
| rs643434 | pQTLs | A | 5.75E-18 | Q7LFX5 | Carbohydrate sulfotransferase 15 |
| rs543040 | pQTLs | A | 8.91E-20 | Q7LFX5 | Carbohydrate sulfotransferase 15 |
| rs613534 | pQTLs | A | 4.57E-17 | Q7LFX5 | Carbohydrate sulfotransferase 15 |
| rs543968 | pQTLs | T | 5.75E-17 | Q7LFX5 | Carbohydrate sulfotransferase 15 |

| | | | | | |
|---|---|---|---|---|---|
| rs544873 | pQTLs | A | 7.08E-18 | Q7LFX5 | Carbohydrate sulfotransferase 15 |
| rs545971 | pQTLs | T | 1.66E-19 | Q7LFX5 | Carbohydrate sulfotransferase 15 |
| rs612169 | pQTLs | A | 5.50E-20 | Q7LFX5 | Carbohydrate sulfotransferase 15 |
| rs8176663 | pQTLs | T | 2.88E-20 | Q7LFX5 | Carbohydrate sulfotransferase 15 |
| rs491626 | pQTLs | T | 2.40E-20 | Q7LFX5 | Carbohydrate sulfotransferase 15 |
| rs492488 | pQTLs | A | 2.40E-20 | Q7LFX5 | Carbohydrate sulfotransferase 15 |
| rs493246 | pQTLs | A | 2.14E-20 | Q7LFX5 | Carbohydrate sulfotransferase 15 |
| rs494242 | pQTLs | T | 3.80E-18 | Q7LFX5 | Carbohydrate sulfotransferase 15 |
| rs495203 | pQTLs | T | 2.14E-20 | Q7LFX5 | Carbohydrate sulfotransferase 15 |
| rs582118 | pQTLs | A | 2.00E-19 | Q7LFX5 | Carbohydrate sulfotransferase 15 |
| rs582094 | pQTLs | A | 2.00E-19 | Q7LFX5 | Carbohydrate sulfotransferase 15 |
| rs2769071 | pQTLs | A | 1.95E-20 | Q7LFX5 | Carbohydrate sulfotransferase 15 |
| rs677355 | pQTLs | A | 2.40E-20 | Q7LFX5 | Carbohydrate sulfotransferase 15 |
| rs676996 | pQTLs | T | 1.70E-19 | Q7LFX5 | Carbohydrate sulfotransferase 15 |
| rs676457 | pQTLs | A | 3.09E-20 | Q7LFX5 | Carbohydrate sulfotransferase 15 |
| rs550057 | pQTLs | T | 3.24E-25 | Q7LFX5 | Carbohydrate sulfotransferase 15 |
| rs674302 | pQTLs | A | 1.48E-20 | Q7LFX5 | Carbohydrate sulfotransferase 15 |
| rs554833 | pQTLs | T | 2.45E-20 | Q7LFX5 | Carbohydrate sulfotransferase 15 |
| rs660340 | pQTLs | A | 5.62E-10 | Q7LFX5 | Carbohydrate sulfotransferase 15 |
| rs581107 | pQTLs | T | 4.68E-10 | Q7LFX5 | Carbohydrate sulfotransferase 15 |
| rs659104 | pQTLs | T | 4.27E-10 | Q7LFX5 | Carbohydrate sulfotransferase 15 |
| rs647800 | pQTLs | A | 2.40E-09 | Q7LFX5 | Carbohydrate sulfotransferase 15 |
| rs473533 | pQTLs | T | 5.89E-10 | Q7LFX5 | Carbohydrate sulfotransferase 15 |
| rs476410 | pQTLs | C | 7.24E-10 | Q7LFX5 | Carbohydrate sulfotransferase 15 |
| rs645982 | pQTLs | A | 5.37E-10 | Q7LFX5 | Carbohydrate sulfotransferase 15 |
| rs529565 | pQTLs | T | 3.89E-20 | Q7LFX5 | Carbohydrate sulfotransferase 15 |
| rs630510 | pQTLs | A | 5.50E-09 | Q7LFX5 | Carbohydrate sulfotransferase 15 |
| rs630014 | pQTLs | A | 3.80E-09 | Q7LFX5 | Carbohydrate sulfotransferase 15 |
| rs532436 | pQTLs | A | 2.82E-19 | Q7LFX5 | Carbohydrate sulfotransferase 15 |
| rs600038 | pQTLs | T | 9.77E-16 | Q7LFX5 | Carbohydrate sulfotransferase 15 |
| rs1124406: | pQTLs | T | 4.17E-09 | Q7LFX5 | Carbohydrate sulfotransferase 15 |
| rs649129 | pQTLs | T | 6.03E-16 | Q7LFX5 | Carbohydrate sulfotransferase 15 |
| rs495828 | pQTLs | T | 4.27E-16 | Q7LFX5 | Carbohydrate sulfotransferase 15 |
| rs7873635 | pQTLs | T | 2.51E-08 | Q7LFX5 | Carbohydrate sulfotransferase 15 |
| rs2073826 | pQTLs | T | 4.17E-09 | Q7LFX5 | Carbohydrate sulfotransferase 15 |
| rs687289 | pQTLs | A | 1.58E-24 | Q7LFX5 | Carbohydrate sulfotransferase 15 |
| rs2073827 | pQTLs | C | 3.47E-09 | Q7LFX5 | Carbohydrate sulfotransferase 15 |
| rs2073828 | pQTLs | A | 3.63E-09 | Q7LFX5 | Carbohydrate sulfotransferase 15 |
| rs8176690 | pQTLs | A | 6.31E-09 | Q7LFX5 | Carbohydrate sulfotransferase 15 |
| rs8176681 | pQTLs | T | 4.90E-09 | Q7LFX5 | Carbohydrate sulfotransferase 15 |
| rs2519093 | pQTLs | T | 6.76E-23 | Q7LFX5 | Carbohydrate sulfotransferase 15 |
| rs644234 | pQTLs | T | 6.61E-21 | Q7LFX5 | Carbohydrate sulfotransferase 15 |
| rs643434 | pQTLs | A | 2.14E-21 | Q7LFX5 | Carbohydrate sulfotransferase 15 |
| rs543040 | pQTLs | A | 4.68E-24 | Q7LFX5 | Carbohydrate sulfotransferase 15 |
| rs613534 | pQTLs | A | 1.12E-20 | Q7LFX5 | Carbohydrate sulfotransferase 15 |
| rs543968 | pQTLs | T | 2.00E-20 | Q7LFX5 | Carbohydrate sulfotransferase 15 |
| rs544873 | pQTLs | A | 2.57E-21 | Q7LFX5 | Carbohydrate sulfotransferase 15 |
| rs545971 | pQTLs | T | 8.51E-24 | Q7LFX5 | Carbohydrate sulfotransferase 15 |
| rs612169 | pQTLs | A | 2.82E-24 | Q7LFX5 | Carbohydrate sulfotransferase 15 |
| rs8176668 | pQTLs | A | 3.89E-08 | Q7LFX5 | Carbohydrate sulfotransferase 15 |

| | | | | | |
|---|---|---|---|---|---|
| rs8176663 | pQTLs | T | 1.12E-24 | Q7LFX5 | Carbohydrate sulfotransferase 15 |
| rs7036642 | pQTLs | A | 3.02E-08 | Q7LFX5 | Carbohydrate sulfotransferase 15 |
| rs491626 | pQTLs | T | 1.12E-24 | Q7LFX5 | Carbohydrate sulfotransferase 15 |
| rs492488 | pQTLs | A | 1.05E-24 | Q7LFX5 | Carbohydrate sulfotransferase 15 |
| rs493246 | pQTLs | A | 1.00E-24 | Q7LFX5 | Carbohydrate sulfotransferase 15 |
| rs494242 | pQTLs | T | 1.32E-21 | Q7LFX5 | Carbohydrate sulfotransferase 15 |
| rs495203 | pQTLs | T | 8.91E-25 | Q7LFX5 | Carbohydrate sulfotransferase 15 |
| rs582118 | pQTLs | A | 1.15E-23 | Q7LFX5 | Carbohydrate sulfotransferase 15 |
| rs582094 | pQTLs | A | 9.77E-24 | Q7LFX5 | Carbohydrate sulfotransferase 15 |
| rs2769071 | pQTLs | A | 7.94E-25 | Q7LFX5 | Carbohydrate sulfotransferase 15 |
| rs677355 | pQTLs | A | 1.07E-24 | Q7LFX5 | Carbohydrate sulfotransferase 15 |
| rs676996 | pQTLs | T | 7.24E-24 | Q7LFX5 | Carbohydrate sulfotransferase 15 |
| rs676457 | pQTLs | A | 1.20E-24 | Q7LFX5 | Carbohydrate sulfotransferase 15 |
| rs550057 | pQTLs | T | 1.02E-30 | Q7LFX5 | Carbohydrate sulfotransferase 15 |
| rs674302 | pQTLs | A | 6.03E-25 | Q7LFX5 | Carbohydrate sulfotransferase 15 |
| rs7046674 | pQTLs | T | 1.58E-08 | Q7LFX5 | Carbohydrate sulfotransferase 15 |
| rs554833 | pQTLs | T | 1.12E-24 | Q7LFX5 | Carbohydrate sulfotransferase 15 |
| rs8176649 | pQTLs | A | 3.24E-08 | Q7LFX5 | Carbohydrate sulfotransferase 15 |
| rs660340 | pQTLs | A | 2.69E-12 | Q7LFX5 | Carbohydrate sulfotransferase 15 |
| rs581107 | pQTLs | T | 2.29E-12 | Q7LFX5 | Carbohydrate sulfotransferase 15 |
| rs659104 | pQTLs | T | 2.00E-12 | Q7LFX5 | Carbohydrate sulfotransferase 15 |
| rs647800 | pQTLs | A | 3.39E-11 | Q7LFX5 | Carbohydrate sulfotransferase 15 |
| rs473533 | pQTLs | T | 2.82E-12 | Q7LFX5 | Carbohydrate sulfotransferase 15 |
| rs476410 | pQTLs | C | 4.37E-12 | Q7LFX5 | Carbohydrate sulfotransferase 15 |
| rs645982 | pQTLs | A | 2.63E-12 | Q7LFX5 | Carbohydrate sulfotransferase 15 |
| rs529565 | pQTLs | T | 1.29E-24 | Q7LFX5 | Carbohydrate sulfotransferase 15 |
| rs630510 | pQTLs | A | 1.41E-10 | Q7LFX5 | Carbohydrate sulfotransferase 15 |
| rs630014 | pQTLs | A | 8.91E-11 | Q7LFX5 | Carbohydrate sulfotransferase 15 |
| rs532436 | pQTLs | A | 1.20E-23 | Q7LFX5 | Carbohydrate sulfotransferase 15 |
| rs600038 | pQTLs | T | 7.94E-19 | Q7LFX5 | Carbohydrate sulfotransferase 15 |
| rs1124406: | pQTLs | T | 1.58E-11 | Q7LFX5 | Carbohydrate sulfotransferase 15 |
| rs649129 | pQTLs | T | 5.01E-19 | Q7LFX5 | Carbohydrate sulfotransferase 15 |
| rs495828 | pQTLs | T | 2.40E-19 | Q7LFX5 | Carbohydrate sulfotransferase 15 |
| rs687289 | pQTLs | A | 2.04E-09 | Q9NZA1 | Chloride intracellular channel protein 5 |
| rs2519093 | pQTLs | T | 2.57E-09 | Q9NZA1 | Chloride intracellular channel protein 5 |
| rs643434 | pQTLs | A | 2.51E-08 | Q9NZA1 | Chloride intracellular channel protein 5 |
| rs543040 | pQTLs | A | 2.51E-09 | Q9NZA1 | Chloride intracellular channel protein 5 |
| rs613534 | pQTLs | A | 3.47E-08 | Q9NZA1 | Chloride intracellular channel protein 5 |
| rs543968 | pQTLs | T | 2.45E-08 | Q9NZA1 | Chloride intracellular channel protein 5 |
| rs544873 | pQTLs | A | 2.40E-08 | Q9NZA1 | Chloride intracellular channel protein 5 |
| rs545971 | pQTLs | T | 1.32E-09 | Q9NZA1 | Chloride intracellular channel protein 5 |
| rs612169 | pQTLs | A | 2.69E-09 | Q9NZA1 | Chloride intracellular channel protein 5 |
| rs8176663 | pQTLs | T | 4.57E-09 | Q9NZA1 | Chloride intracellular channel protein 5 |
| rs491626 | pQTLs | T | 3.72E-09 | Q9NZA1 | Chloride intracellular channel protein 5 |
| rs492488 | pQTLs | A | 5.50E-09 | Q9NZA1 | Chloride intracellular channel protein 5 |
| rs493246 | pQTLs | A | 5.62E-09 | Q9NZA1 | Chloride intracellular channel protein 5 |
| rs494242 | pQTLs | T | 2.69E-08 | Q9NZA1 | Chloride intracellular channel protein 5 |
| rs495203 | pQTLs | T | 3.98E-09 | Q9NZA1 | Chloride intracellular channel protein 5 |
| rs582118 | pQTLs | A | 1.17E-08 | Q9NZA1 | Chloride intracellular channel protein 5 |
| rs582094 | pQTLs | A | 5.89E-09 | Q9NZA1 | Chloride intracellular channel protein 5 |

| | | | | | |
|---|---|---|---|---|---|
| rs2769071 | pQTLs | A | 5.89E-09 | Q9NZA1 | Chloride intracellular channel protein 5 |
| rs677355 | pQTLs | A | 5.50E-09 | Q9NZA1 | Chloride intracellular channel protein 5 |
| rs676996 | pQTLs | T | 1.07E-08 | Q9NZA1 | Chloride intracellular channel protein 5 |
| rs676457 | pQTLs | A | 7.41E-09 | Q9NZA1 | Chloride intracellular channel protein 5 |
| rs674302 | pQTLs | A | 5.37E-09 | Q9NZA1 | Chloride intracellular channel protein 5 |
| rs554833 | pQTLs | T | 8.71E-09 | Q9NZA1 | Chloride intracellular channel protein 5 |
| rs529565 | pQTLs | T | 1.91E-09 | Q9NZA1 | Chloride intracellular channel protein 5 |
| rs532436 | pQTLs | A | 2.57E-09 | Q9NZA1 | Chloride intracellular channel protein 5 |
| rs687289 | pQTLs | A | 6.17E-12 | P00374 | Dihydrofolate reductase |
| rs644234 | pQTLs | T | 6.92E-11 | P00374 | Dihydrofolate reductase |
| rs643434 | pQTLs | A | 1.12E-10 | P00374 | Dihydrofolate reductase |
| rs543040 | pQTLs | A | 5.89E-12 | P00374 | Dihydrofolate reductase |
| rs613534 | pQTLs | A | 2.82E-10 | P00374 | Dihydrofolate reductase |
| rs543968 | pQTLs | T | 1.02E-09 | P00374 | Dihydrofolate reductase |
| rs544873 | pQTLs | A | 1.10E-10 | P00374 | Dihydrofolate reductase |
| rs545971 | pQTLs | T | 1.62E-11 | P00374 | Dihydrofolate reductase |
| rs612169 | pQTLs | A | 8.13E-12 | P00374 | Dihydrofolate reductase |
| rs8176663 | pQTLs | T | 4.07E-12 | P00374 | Dihydrofolate reductase |
| rs491626 | pQTLs | T | 6.31E-12 | P00374 | Dihydrofolate reductase |
| rs492488 | pQTLs | A | 4.68E-12 | P00374 | Dihydrofolate reductase |
| rs493246 | pQTLs | A | 4.47E-12 | P00374 | Dihydrofolate reductase |
| rs494242 | pQTLs | T | 9.77E-11 | P00374 | Dihydrofolate reductase |
| rs495203 | pQTLs | T | 4.68E-12 | P00374 | Dihydrofolate reductase |
| rs582118 | pQTLs | A | 1.66E-11 | P00374 | Dihydrofolate reductase |
| rs582094 | pQTLs | A | 1.86E-11 | P00374 | Dihydrofolate reductase |
| rs2769071 | pQTLs | A | 3.47E-12 | P00374 | Dihydrofolate reductase |
| rs677355 | pQTLs | A | 4.37E-12 | P00374 | Dihydrofolate reductase |
| rs676996 | pQTLs | T | 5.25E-12 | P00374 | Dihydrofolate reductase |
| rs676457 | pQTLs | A | 2.45E-12 | P00374 | Dihydrofolate reductase |
| rs674302 | pQTLs | A | 3.47E-12 | P00374 | Dihydrofolate reductase |
| rs554833 | pQTLs | T | 3.16E-12 | P00374 | Dihydrofolate reductase |
| rs529565 | pQTLs | T | 7.94E-12 | P00374 | Dihydrofolate reductase |
| rs2519093 | pQTLs | T | 1.82E-12 | Q9H4A9 | Dipeptidase 2 |
| rs550057 | pQTLs | T | 1.86E-09 | Q9H4A9 | Dipeptidase 2 |
| rs532436 | pQTLs | A | 2.57E-12 | Q9H4A9 | Dipeptidase 2 |
| rs600038 | pQTLs | T | 5.89E-10 | Q9H4A9 | Dipeptidase 2 |
| rs649129 | pQTLs | T | 1.23E-09 | Q9H4A9 | Dipeptidase 2 |
| rs495828 | pQTLs | T | 1.10E-09 | Q9H4A9 | Dipeptidase 2 |
| rs7873635 | pQTLs | T | 1.86E-14 | Q14126 | Desmoglein-2 |
| rs4962040 | pQTLs | A | 1.74E-14 | Q14126 | Desmoglein-2 |
| rs8176702 | pQTLs | A | 3.02E-14 | Q14126 | Desmoglein-2 |
| rs2073826 | pQTLs | T | 2.95E-15 | Q14126 | Desmoglein-2 |
| rs687289 | pQTLs | A | 2.00E-16 | Q14126 | Desmoglein-2 |
| rs2073827 | pQTLs | C | 8.71E-15 | Q14126 | Desmoglein-2 |
| rs2073828 | pQTLs | A | 8.51E-15 | Q14126 | Desmoglein-2 |
| rs8176691 | pQTLs | T | 3.16E-14 | Q14126 | Desmoglein-2 |
| rs8176690 | pQTLs | A | 4.27E-15 | Q14126 | Desmoglein-2 |
| rs8176682 | pQTLs | T | 1.91E-14 | Q14126 | Desmoglein-2 |
| rs8176681 | pQTLs | T | 6.76E-15 | Q14126 | Desmoglein-2 |
| rs2519093 | pQTLs | T | 4.68E-08 | Q14126 | Desmoglein-2 |

| | | | | | |
|---|---|---|---|---|---|
| rs644234 | pQTLs | T | 1.29E-14 | Q14126 | Desmoglein-2 |
| rs643434 | pQTLs | A | 2.63E-14 | Q14126 | Desmoglein-2 |
| rs543040 | pQTLs | A | 5.62E-16 | Q14126 | Desmoglein-2 |
| rs613534 | pQTLs | A | 5.75E-14 | Q14126 | Desmoglein-2 |
| rs543968 | pQTLs | T | 6.17E-14 | Q14126 | Desmoglein-2 |
| rs544873 | pQTLs | A | 3.24E-14 | Q14126 | Desmoglein-2 |
| rs545971 | pQTLs | T | 1.70E-16 | Q14126 | Desmoglein-2 |
| rs612169 | pQTLs | A | 4.68E-16 | Q14126 | Desmoglein-2 |
| rs8176668 | pQTLs | A | 1.29E-13 | Q14126 | Desmoglein-2 |
| rs8176663 | pQTLs | T | 8.91E-16 | Q14126 | Desmoglein-2 |
| rs7036642 | pQTLs | A | 1.20E-13 | Q14126 | Desmoglein-2 |
| rs491626 | pQTLs | T | 7.94E-16 | Q14126 | Desmoglein-2 |
| rs492488 | pQTLs | A | 9.77E-16 | Q14126 | Desmoglein-2 |
| rs493246 | pQTLs | A | 8.91E-16 | Q14126 | Desmoglein-2 |
| rs494242 | pQTLs | T | 2.40E-14 | Q14126 | Desmoglein-2 |
| rs495203 | pQTLs | T | 8.71E-16 | Q14126 | Desmoglein-2 |
| rs582118 | pQTLs | A | 4.07E-16 | Q14126 | Desmoglein-2 |
| rs582094 | pQTLs | A | 3.72E-16 | Q14126 | Desmoglein-2 |
| rs2769071 | pQTLs | A | 1.48E-15 | Q14126 | Desmoglein-2 |
| rs677355 | pQTLs | A | 7.94E-16 | Q14126 | Desmoglein-2 |
| rs676996 | pQTLs | T | 6.03E-16 | Q14126 | Desmoglein-2 |
| rs676457 | pQTLs | A | 1.48E-15 | Q14126 | Desmoglein-2 |
| rs674302 | pQTLs | A | 1.55E-15 | Q14126 | Desmoglein-2 |
| rs7046674 | pQTLs | T | 6.17E-13 | Q14126 | Desmoglein-2 |
| rs554833 | pQTLs | T | 1.91E-15 | Q14126 | Desmoglein-2 |
| rs8176649 | pQTLs | A | 1.17E-13 | Q14126 | Desmoglein-2 |
| rs529565 | pQTLs | T | 4.27E-16 | Q14126 | Desmoglein-2 |
| rs2519093 | pQTLs | T | 1.95E-13 | P17813 | Endoglin |
| rs550057 | pQTLs | T | 2.34E-12 | P17813 | Endoglin |
| rs532436 | pQTLs | A | 1.86E-13 | P17813 | Endoglin |
| rs600038 | pQTLs | T | 1.41E-10 | P17813 | Endoglin |
| rs1124406: | pQTLs | T | 8.51E-10 | P17813 | Endoglin |
| rs649129 | pQTLs | T | 9.77E-11 | P17813 | Endoglin |
| rs495828 | pQTLs | T | 8.91E-11 | P17813 | Endoglin |
| rs7873635 | pQTLs | T | 3.39E-20 | P00451 | Coagulation Factor VIII |
| rs4962040 | pQTLs | A | 1.38E-21 | P00451 | Coagulation Factor VIII |
| rs8176702 | pQTLs | A | 1.15E-21 | P00451 | Coagulation Factor VIII |
| rs2073826 | pQTLs | T | 5.62E-23 | P00451 | Coagulation Factor VIII |
| rs687289 | pQTLs | A | 1.20E-77 | P00451 | Coagulation Factor VIII |
| rs2073827 | pQTLs | C | 7.59E-23 | P00451 | Coagulation Factor VIII |
| rs2073828 | pQTLs | A | 9.33E-23 | P00451 | Coagulation Factor VIII |
| rs8176691 | pQTLs | T | 1.66E-21 | P00451 | Coagulation Factor VIII |
| rs8176690 | pQTLs | A | 2.00E-23 | P00451 | Coagulation Factor VIII |
| rs8176682 | pQTLs | T | 6.76E-22 | P00451 | Coagulation Factor VIII |
| rs8176681 | pQTLs | T | 1.29E-23 | P00451 | Coagulation Factor VIII |
| rs2519093 | pQTLs | T | 7.08E-65 | P00451 | Coagulation Factor VIII |
| rs644234 | pQTLs | T | 4.79E-71 | P00451 | Coagulation Factor VIII |
| rs643434 | pQTLs | A | 6.92E-71 | P00451 | Coagulation Factor VIII |
| rs543040 | pQTLs | A | 4.68E-78 | P00451 | Coagulation Factor VIII |
| rs613534 | pQTLs | A | 1.86E-69 | P00451 | Coagulation Factor VIII |

| | | | | | |
|---|---|---|---|---|---|
| rs543968 | pQTLs | T | 2.04E-69 | P00451 | Coagulation Factor VIII |
| rs544873 | pQTLs | A | 3.47E-71 | P00451 | Coagulation Factor VIII |
| rs545971 | pQTLs | T | 1.38E-77 | P00451 | Coagulation Factor VIII |
| rs612169 | pQTLs | A | 7.59E-78 | P00451 | Coagulation Factor VIII |
| rs8176668 | pQTLs | A | 1.15E-21 | P00451 | Coagulation Factor VIII |
| rs8176663 | pQTLs | T | 5.75E-78 | P00451 | Coagulation Factor VIII |
| rs7036642 | pQTLs | A | 3.24E-22 | P00451 | Coagulation Factor VIII |
| rs491626 | pQTLs | T | 3.02E-78 | P00451 | Coagulation Factor VIII |
| rs492488 | pQTLs | A | 5.37E-78 | P00451 | Coagulation Factor VIII |
| rs493246 | pQTLs | A | 3.98E-78 | P00451 | Coagulation Factor VIII |
| rs494242 | pQTLs | T | 2.63E-71 | P00451 | Coagulation Factor VIII |
| rs495203 | pQTLs | T | 5.25E-78 | P00451 | Coagulation Factor VIII |
| rs582118 | pQTLs | A | 8.91E-78 | P00451 | Coagulation Factor VIII |
| rs582094 | pQTLs | A | 3.47E-77 | P00451 | Coagulation Factor VIII |
| rs2769071 | pQTLs | A | 1.38E-77 | P00451 | Coagulation Factor VIII |
| rs677355 | pQTLs | A | 1.78E-77 | P00451 | Coagulation Factor VIII |
| rs676996 | pQTLs | T | 3.39E-77 | P00451 | Coagulation Factor VIII |
| rs676457 | pQTLs | A | 1.26E-78 | P00451 | Coagulation Factor VIII |
| rs550057 | pQTLs | T | 1.10E-45 | P00451 | Coagulation Factor VIII |
| rs674302 | pQTLs | A | 9.12E-78 | P00451 | Coagulation Factor VIII |
| rs7046674 | pQTLs | T | 9.33E-22 | P00451 | Coagulation Factor VIII |
| rs554833 | pQTLs | T | 8.13E-78 | P00451 | Coagulation Factor VIII |
| rs8176649 | pQTLs | A | 7.59E-23 | P00451 | Coagulation Factor VIII |
| rs660340 | pQTLs | A | 5.75E-31 | P00451 | Coagulation Factor VIII |
| rs581107 | pQTLs | T | 2.45E-31 | P00451 | Coagulation Factor VIII |
| rs659104 | pQTLs | T | 2.09E-31 | P00451 | Coagulation Factor VIII |
| rs647800 | pQTLs | A | 1.58E-29 | P00451 | Coagulation Factor VIII |
| rs473533 | pQTLs | T | 1.41E-31 | P00451 | Coagulation Factor VIII |
| rs476410 | pQTLs | C | 1.12E-31 | P00451 | Coagulation Factor VIII |
| rs645982 | pQTLs | A | 1.12E-31 | P00451 | Coagulation Factor VIII |
| rs529565 | pQTLs | T | 3.16E-78 | P00451 | Coagulation Factor VIII |
| rs630510 | pQTLs | A | 7.41E-36 | P00451 | Coagulation Factor VIII |
| rs630014 | pQTLs | A | 6.76E-36 | P00451 | Coagulation Factor VIII |
| rs532436 | pQTLs | A | 8.13E-66 | P00451 | Coagulation Factor VIII |
| rs600038 | pQTLs | T | 1.55E-54 | P00451 | Coagulation Factor VIII |
| rs1124406: | pQTLs | T | 9.12E-37 | P00451 | Coagulation Factor VIII |
| rs649129 | pQTLs | T | 3.02E-54 | P00451 | Coagulation Factor VIII |
| rs495828 | pQTLs | T | 1.86E-54 | P00451 | Coagulation Factor VIII |
| rs687289 | pQTLs | A | 2.82E-10 | Q8N128 | Protein FAM177A1 |
| rs2519093 | pQTLs | T | 6.46E-10 | Q8N128 | Protein FAM177A1 |
| rs644234 | pQTLs | T | 2.57E-09 | Q8N128 | Protein FAM177A1 |
| rs643434 | pQTLs | A | 2.34E-09 | Q8N128 | Protein FAM177A1 |
| rs543040 | pQTLs | A | 2.95E-10 | Q8N128 | Protein FAM177A1 |
| rs613534 | pQTLs | A | 1.41E-09 | Q8N128 | Protein FAM177A1 |
| rs543968 | pQTLs | T | 3.89E-09 | Q8N128 | Protein FAM177A1 |
| rs544873 | pQTLs | A | 2.34E-09 | Q8N128 | Protein FAM177A1 |
| rs545971 | pQTLs | T | 3.02E-10 | Q8N128 | Protein FAM177A1 |
| rs612169 | pQTLs | A | 2.75E-10 | Q8N128 | Protein FAM177A1 |
| rs8176663 | pQTLs | T | 1.66E-10 | Q8N128 | Protein FAM177A1 |
| rs491626 | pQTLs | T | 1.45E-10 | Q8N128 | Protein FAM177A1 |

| | | | | | |
|---|---|---|---|---|---|
| rs492488 | pQTLs | A | 1.32E-10 | Q8N128 | Protein FAM177A1 |
| rs493246 | pQTLs | A | 1.78E-10 | Q8N128 | Protein FAM177A1 |
| rs494242 | pQTLs | T | 2.09E-09 | Q8N128 | Protein FAM177A1 |
| rs495203 | pQTLs | T | 1.86E-10 | Q8N128 | Protein FAM177A1 |
| rs582118 | pQTLs | A | 2.88E-10 | Q8N128 | Protein FAM177A1 |
| rs582094 | pQTLs | A | 2.45E-10 | Q8N128 | Protein FAM177A1 |
| rs2769071 | pQTLs | A | 1.41E-10 | Q8N128 | Protein FAM177A1 |
| rs677355 | pQTLs | A | 1.95E-10 | Q8N128 | Protein FAM177A1 |
| rs676996 | pQTLs | T | 1.07E-09 | Q8N128 | Protein FAM177A1 |
| rs676457 | pQTLs | A | 8.71E-11 | Q8N128 | Protein FAM177A1 |
| rs550057 | pQTLs | T | 9.33E-19 | Q8N128 | Protein FAM177A1 |
| rs674302 | pQTLs | A | 1.91E-10 | Q8N128 | Protein FAM177A1 |
| rs554833 | pQTLs | T | 2.51E-10 | Q8N128 | Protein FAM177A1 |
| rs529565 | pQTLs | T | 1.02E-10 | Q8N128 | Protein FAM177A1 |
| rs532436 | pQTLs | A | 3.89E-10 | Q8N128 | Protein FAM177A1 |
| rs600038 | pQTLs | T | 1.02E-08 | Q8N128 | Protein FAM177A1 |
| rs649129 | pQTLs | T | 8.71E-09 | Q8N128 | Protein FAM177A1 |
| rs495828 | pQTLs | T | 1.35E-08 | Q8N128 | Protein FAM177A1 |
| rs7873635 | pQTLs | T | 4.37E-08 | O75063 | Glycosaminoglycan xylosylkinase |
| rs2519093 | pQTLs | T | 1.58E-16 | O75063 | Glycosaminoglycan xylosylkinase |
| rs550057 | pQTLs | T | 8.91E-12 | O75063 | Glycosaminoglycan xylosylkinase |
| rs532436 | pQTLs | A | 1.26E-16 | O75063 | Glycosaminoglycan xylosylkinase |
| rs600038 | pQTLs | T | 9.33E-16 | O75063 | Glycosaminoglycan xylosylkinase |
| rs1124406: | pQTLs | T | 6.17E-12 | O75063 | Glycosaminoglycan xylosylkinase |
| rs649129 | pQTLs | T | 7.08E-15 | O75063 | Glycosaminoglycan xylosylkinase |
| rs495828 | pQTLs | T | 1.51E-15 | O75063 | Glycosaminoglycan xylosylkinase |
| rs2519093 | pQTLs | T | 1.02E-10 | P58499 | Protein FAM3B |
| rs532436 | pQTLs | A | 1.70E-10 | P58499 | Protein FAM3B |
| rs600038 | pQTLs | T | 3.39E-09 | P58499 | Protein FAM3B |
| rs1124406: | pQTLs | T | 5.25E-09 | P58499 | Protein FAM3B |
| rs649129 | pQTLs | T | 1.29E-08 | P58499 | Protein FAM3B |
| rs495828 | pQTLs | T | 6.03E-09 | P58499 | Protein FAM3B |
| rs7873635 | pQTLs | T | 3.31E-10 | P58499 | Protein FAM3B |
| rs4962040 | pQTLs | A | 1.66E-10 | P58499 | Protein FAM3B |
| rs8176702 | pQTLs | A | 1.78E-10 | P58499 | Protein FAM3B |
| rs2073826 | pQTLs | T | 3.72E-10 | P58499 | Protein FAM3B |
| rs687289 | pQTLs | A | 2.40E-14 | P58499 | Protein FAM3B |
| rs2073827 | pQTLs | C | 3.39E-10 | P58499 | Protein FAM3B |
| rs2073828 | pQTLs | A | 3.31E-10 | P58499 | Protein FAM3B |
| rs8176691 | pQTLs | T | 1.95E-10 | P58499 | Protein FAM3B |
| rs8176690 | pQTLs | A | 3.47E-10 | P58499 | Protein FAM3B |
| rs8176682 | pQTLs | T | 8.51E-11 | P58499 | Protein FAM3B |
| rs8176681 | pQTLs | T | 4.79E-10 | P58499 | Protein FAM3B |
| rs2519093 | pQTLs | T | 3.09E-20 | P58499 | Protein FAM3B |
| rs644234 | pQTLs | T | 7.08E-14 | P58499 | Protein FAM3B |
| rs643434 | pQTLs | A | 1.15E-13 | P58499 | Protein FAM3B |
| rs543040 | pQTLs | A | 5.13E-14 | P58499 | Protein FAM3B |
| rs613534 | pQTLs | A | 6.17E-13 | P58499 | Protein FAM3B |
| rs543968 | pQTLs | T | 2.09E-13 | P58499 | Protein FAM3B |
| rs544873 | pQTLs | A | 1.74E-13 | P58499 | Protein FAM3B |

| | | | | | |
|---|---|---|---|---|---|
| rs545971 | pQTLs | T | 4.90E-14 | P58499 | Protein FAM3B |
| rs612169 | pQTLs | A | 1.15E-13 | P58499 | Protein FAM3B |
| rs8176668 | pQTLs | A | 1.74E-10 | P58499 | Protein FAM3B |
| rs8176663 | pQTLs | T | 1.74E-13 | P58499 | Protein FAM3B |
| rs7036642 | pQTLs | A | 2.69E-10 | P58499 | Protein FAM3B |
| rs491626 | pQTLs | T | 1.95E-13 | P58499 | Protein FAM3B |
| rs492488 | pQTLs | A | 2.75E-13 | P58499 | Protein FAM3B |
| rs493246 | pQTLs | A | 2.40E-13 | P58499 | Protein FAM3B |
| rs494242 | pQTLs | T | 1.41E-13 | P58499 | Protein FAM3B |
| rs495203 | pQTLs | T | 1.66E-13 | P58499 | Protein FAM3B |
| rs582118 | pQTLs | A | 2.75E-13 | P58499 | Protein FAM3B |
| rs582094 | pQTLs | A | 8.13E-14 | P58499 | Protein FAM3B |
| rs2769071 | pQTLs | A | 3.39E-13 | P58499 | Protein FAM3B |
| rs677355 | pQTLs | A | 3.72E-13 | P58499 | Protein FAM3B |
| rs676996 | pQTLs | T | 9.12E-13 | P58499 | Protein FAM3B |
| rs676457 | pQTLs | A | 6.17E-13 | P58499 | Protein FAM3B |
| rs550057 | pQTLs | T | 7.59E-16 | P58499 | Protein FAM3B |
| rs674302 | pQTLs | A | 2.69E-13 | P58499 | Protein FAM3B |
| rs7046674 | pQTLs | T | 6.31E-10 | P58499 | Protein FAM3B |
| rs554833 | pQTLs | T | 2.57E-13 | P58499 | Protein FAM3B |
| rs8176649 | pQTLs | A | 5.01E-10 | P58499 | Protein FAM3B |
| rs529565 | pQTLs | T | 2.63E-13 | P58499 | Protein FAM3B |
| rs532436 | pQTLs | A | 7.41E-20 | P58499 | Protein FAM3B |
| rs600038 | pQTLs | T | 3.89E-18 | P58499 | Protein FAM3B |
| rs1124406: | pQTLs | T | 8.32E-17 | P58499 | Protein FAM3B |
| rs649129 | pQTLs | T | 8.91E-17 | P58499 | Protein FAM3B |
| rs495828 | pQTLs | T | 4.68E-18 | P58499 | Protein FAM3B |
| rs7873635 | pQTLs | T | 1.55E-47 | Q96BQ1 | Protein FAM3D |
| rs4962040 | pQTLs | A | 1.32E-47 | Q96BQ1 | Protein FAM3D |
| rs8176702 | pQTLs | A | 1.70E-47 | Q96BQ1 | Protein FAM3D |
| rs2073826 | pQTLs | T | 1.07E-54 | Q96BQ1 | Protein FAM3D |
| rs687289 | pQTLs | A | 7.76E-140 | Q96BQ1 | Protein FAM3D |
| rs2073827 | pQTLs | C | 1.82E-54 | Q96BQ1 | Protein FAM3D |
| rs2073828 | pQTLs | A | 2.69E-54 | Q96BQ1 | Protein FAM3D |
| rs8176691 | pQTLs | T | 2.40E-47 | Q96BQ1 | Protein FAM3D |
| rs8176690 | pQTLs | A | 8.13E-55 | Q96BQ1 | Protein FAM3D |
| rs8176682 | pQTLs | T | 2.40E-47 | Q96BQ1 | Protein FAM3D |
| rs8176681 | pQTLs | T | 3.63E-54 | Q96BQ1 | Protein FAM3D |
| rs2519093 | pQTLs | T | 8.91E-123 | Q96BQ1 | Protein FAM3D |
| rs644234 | pQTLs | T | 1.95E-117 | Q96BQ1 | Protein FAM3D |
| rs643434 | pQTLs | A | 2.75E-118 | Q96BQ1 | Protein FAM3D |
| rs543040 | pQTLs | A | 1.91E-139 | Q96BQ1 | Protein FAM3D |
| rs613534 | pQTLs | A | 1.86E-110 | Q96BQ1 | Protein FAM3D |
| rs543968 | pQTLs | T | 2.04E-111 | Q96BQ1 | Protein FAM3D |
| rs544873 | pQTLs | A | 8.71E-118 | Q96BQ1 | Protein FAM3D |
| rs545971 | pQTLs | T | 1.58E-141 | Q96BQ1 | Protein FAM3D |
| rs612169 | pQTLs | A | 6.17E-139 | Q96BQ1 | Protein FAM3D |
| rs8176668 | pQTLs | A | 2.63E-47 | Q96BQ1 | Protein FAM3D |
| rs8176663 | pQTLs | T | 1.58E-139 | Q96BQ1 | Protein FAM3D |
| rs7036642 | pQTLs | A | 3.55E-47 | Q96BQ1 | Protein FAM3D |

| | | | | | |
|---|---|---|---|---|---|
| rs491626 | pQTLs | T | 8.91E-140 | Q96BQ1 | Protein FAM3D |
| rs492488 | pQTLs | A | 2.45E-139 | Q96BQ1 | Protein FAM3D |
| rs493246 | pQTLs | A | 4.68E-140 | Q96BQ1 | Protein FAM3D |
| rs494242 | pQTLs | T | 3.72E-118 | Q96BQ1 | Protein FAM3D |
| rs495203 | pQTLs | T | 6.03E-140 | Q96BQ1 | Protein FAM3D |
| rs582118 | pQTLs | A | 9.77E-140 | Q96BQ1 | Protein FAM3D |
| rs582094 | pQTLs | A | 2.34E-139 | Q96BQ1 | Protein FAM3D |
| rs2769071 | pQTLs | A | 3.24E-139 | Q96BQ1 | Protein FAM3D |
| rs677355 | pQTLs | A | 1.26E-139 | Q96BQ1 | Protein FAM3D |
| rs676996 | pQTLs | T | 4.79E-136 | Q96BQ1 | Protein FAM3D |
| rs676457 | pQTLs | A | 1.15E-139 | Q96BQ1 | Protein FAM3D |
| rs550057 | pQTLs | T | 9.55E-171 | Q96BQ1 | Protein FAM3D |
| rs674302 | pQTLs | A | 2.51E-139 | Q96BQ1 | Protein FAM3D |
| rs7046674 | pQTLs | T | 1.95E-46 | Q96BQ1 | Protein FAM3D |
| rs554833 | pQTLs | T | 3.89E-140 | Q96BQ1 | Protein FAM3D |
| rs8176649 | pQTLs | A | 4.17E-47 | Q96BQ1 | Protein FAM3D |
| rs660340 | pQTLs | A | 1.95E-56 | Q96BQ1 | Protein FAM3D |
| rs581107 | pQTLs | T | 3.89E-57 | Q96BQ1 | Protein FAM3D |
| rs659104 | pQTLs | T | 3.98E-57 | Q96BQ1 | Protein FAM3D |
| rs647800 | pQTLs | A | 2.04E-49 | Q96BQ1 | Protein FAM3D |
| rs473533 | pQTLs | T | 5.01E-57 | Q96BQ1 | Protein FAM3D |
| rs476410 | pQTLs | C | 2.45E-57 | Q96BQ1 | Protein FAM3D |
| rs645982 | pQTLs | A | 9.55E-57 | Q96BQ1 | Protein FAM3D |
| rs529565 | pQTLs | T | 2.95E-139 | Q96BQ1 | Protein FAM3D |
| rs630510 | pQTLs | A | 1.17E-46 | Q96BQ1 | Protein FAM3D |
| rs630014 | pQTLs | A | 1.23E-46 | Q96BQ1 | Protein FAM3D |
| rs532436 | pQTLs | A | 4.17E-122 | Q96BQ1 | Protein FAM3D |
| rs600038 | pQTLs | T | 2.95E-91 | Q96BQ1 | Protein FAM3D |
| rs1124406[illegible] | pQTLs | T | 4.79E-82 | Q96BQ1 | Protein FAM3D |
| rs649129 | pQTLs | T | 3.39E-88 | Q96BQ1 | Protein FAM3D |
| rs495828 | pQTLs | T | 4.79E-92 | Q96BQ1 | Protein FAM3D |
| rs7873635 | pQTLs | T | 5.75E-11 | P35916 | Vascular endothelial growth factor receptor 3 |
| rs4962040 | pQTLs | A | 6.76E-11 | P35916 | Vascular endothelial growth factor receptor 3 |
| rs8176702 | pQTLs | A | 3.02E-11 | P35916 | Vascular endothelial growth factor receptor 3 |
| rs2073826 | pQTLs | T | 2.88E-13 | P35916 | Vascular endothelial growth factor receptor 3 |
| rs687289 | pQTLs | A | 1.38E-33 | P35916 | Vascular endothelial growth factor receptor 3 |
| rs2073827 | pQTLs | C | 2.34E-13 | P35916 | Vascular endothelial growth factor receptor 3 |
| rs2073828 | pQTLs | A | 2.34E-13 | P35916 | Vascular endothelial growth factor receptor 3 |
| rs8176691 | pQTLs | T | 2.95E-11 | P35916 | Vascular endothelial growth factor receptor 3 |
| rs8176690 | pQTLs | A | 1.62E-12 | P35916 | Vascular endothelial growth factor receptor 3 |
| rs8176682 | pQTLs | T | 1.02E-10 | P35916 | Vascular endothelial growth factor receptor 3 |
| rs8176681 | pQTLs | T | 1.17E-12 | P35916 | Vascular endothelial growth factor receptor 3 |
| rs2519093 | pQTLs | T | 5.89E-80 | P35916 | Vascular endothelial growth factor receptor 3 |
| rs644234 | pQTLs | T | 1.41E-28 | P35916 | Vascular endothelial growth factor receptor 3 |
| rs643434 | pQTLs | A | 1.29E-28 | P35916 | Vascular endothelial growth factor receptor 3 |
| rs543040 | pQTLs | A | 1.95E-33 | P35916 | Vascular endothelial growth factor receptor 3 |
| rs613534 | pQTLs | A | 4.79E-27 | P35916 | Vascular endothelial growth factor receptor 3 |
| rs543968 | pQTLs | T | 6.76E-27 | P35916 | Vascular endothelial growth factor receptor 3 |
| rs544873 | pQTLs | A | 1.48E-28 | P35916 | Vascular endothelial growth factor receptor 3 |
| rs545971 | pQTLs | T | 8.71E-33 | P35916 | Vascular endothelial growth factor receptor 3 |

| | | | | | |
|---|---|---|---|---|---|
| rs612169 | pQTLs | A | 2.14E-33 | P35916 | Vascular endothelial growth factor receptor 3 |
| rs8176668 | pQTLs | A | 8.71E-11 | P35916 | Vascular endothelial growth factor receptor 3 |
| rs8176663 | pQTLs | T | 2.51E-33 | P35916 | Vascular endothelial growth factor receptor 3 |
| rs7036642 | pQTLs | A | 1.07E-10 | P35916 | Vascular endothelial growth factor receptor 3 |
| rs491626 | pQTLs | T | 4.47E-33 | P35916 | Vascular endothelial growth factor receptor 3 |
| rs492488 | pQTLs | A | 7.08E-33 | P35916 | Vascular endothelial growth factor receptor 3 |
| rs493246 | pQTLs | A | 7.59E-33 | P35916 | Vascular endothelial growth factor receptor 3 |
| rs494242 | pQTLs | T | 3.47E-28 | P35916 | Vascular endothelial growth factor receptor 3 |
| rs495203 | pQTLs | T | 2.82E-33 | P35916 | Vascular endothelial growth factor receptor 3 |
| rs582118 | pQTLs | A | 3.39E-32 | P35916 | Vascular endothelial growth factor receptor 3 |
| rs582094 | pQTLs | A | 5.50E-32 | P35916 | Vascular endothelial growth factor receptor 3 |
| rs2769071 | pQTLs | A | 6.61E-33 | P35916 | Vascular endothelial growth factor receptor 3 |
| rs677355 | pQTLs | A | 5.25E-33 | P35916 | Vascular endothelial growth factor receptor 3 |
| rs676996 | pQTLs | T | 2.88E-31 | P35916 | Vascular endothelial growth factor receptor 3 |
| rs676457 | pQTLs | A | 7.59E-33 | P35916 | Vascular endothelial growth factor receptor 3 |
| rs550057 | pQTLs | T | 3.09E-68 | P35916 | Vascular endothelial growth factor receptor 3 |
| rs674302 | pQTLs | A | 3.02E-33 | P35916 | Vascular endothelial growth factor receptor 3 |
| rs7046674 | pQTLs | T | 8.51E-11 | P35916 | Vascular endothelial growth factor receptor 3 |
| rs554833 | pQTLs | T | 2.63E-33 | P35916 | Vascular endothelial growth factor receptor 3 |
| rs8176649 | pQTLs | A | 8.32E-11 | P35916 | Vascular endothelial growth factor receptor 3 |
| rs660340 | pQTLs | A | 1.20E-14 | P35916 | Vascular endothelial growth factor receptor 3 |
| rs581107 | pQTLs | T | 1.38E-14 | P35916 | Vascular endothelial growth factor receptor 3 |
| rs659104 | pQTLs | T | 1.12E-14 | P35916 | Vascular endothelial growth factor receptor 3 |
| rs647800 | pQTLs | A | 1.35E-12 | P35916 | Vascular endothelial growth factor receptor 3 |
| rs473533 | pQTLs | T | 1.82E-14 | P35916 | Vascular endothelial growth factor receptor 3 |
| rs476410 | pQTLs | C | 2.00E-14 | P35916 | Vascular endothelial growth factor receptor 3 |
| rs645982 | pQTLs | A | 2.19E-14 | P35916 | Vascular endothelial growth factor receptor 3 |
| rs529565 | pQTLs | T | 3.98E-34 | P35916 | Vascular endothelial growth factor receptor 3 |
| rs630510 | pQTLs | A | 3.89E-16 | P35916 | Vascular endothelial growth factor receptor 3 |
| rs630014 | pQTLs | A | 3.89E-16 | P35916 | Vascular endothelial growth factor receptor 3 |
| rs532436 | pQTLs | A | 1.48E-77 | P35916 | Vascular endothelial growth factor receptor 3 |
| rs600038 | pQTLs | T | 4.57E-62 | P35916 | Vascular endothelial growth factor receptor 3 |
| rs1124406: | pQTLs | T | 5.75E-43 | P35916 | Vascular endothelial growth factor receptor 3 |
| rs649129 | pQTLs | T | 4.27E-62 | P35916 | Vascular endothelial growth factor receptor 3 |
| rs495828 | pQTLs | T | 3.16E-62 | P35916 | Vascular endothelial growth factor receptor 3 |
| rs7873635 | pQTLs | T | 1.20E-20 | O94923 | D-glucuronyl C5-epimerase |
| rs4962040 | pQTLs | A | 2.75E-20 | O94923 | D-glucuronyl C5-epimerase |
| rs8176702 | pQTLs | A | 1.45E-20 | O94923 | D-glucuronyl C5-epimerase |
| rs2073826 | pQTLs | T | 1.74E-21 | O94923 | D-glucuronyl C5-epimerase |
| rs687289 | pQTLs | A | 6.92E-47 | O94923 | D-glucuronyl C5-epimerase |
| rs2073827 | pQTLs | C | 1.70E-21 | O94923 | D-glucuronyl C5-epimerase |
| rs2073828 | pQTLs | A | 1.41E-21 | O94923 | D-glucuronyl C5-epimerase |
| rs8176691 | pQTLs | T | 1.51E-20 | O94923 | D-glucuronyl C5-epimerase |
| rs8176690 | pQTLs | A | 2.19E-21 | O94923 | D-glucuronyl C5-epimerase |
| rs8176682 | pQTLs | T | 1.48E-20 | O94923 | D-glucuronyl C5-epimerase |
| rs8176681 | pQTLs | T | 1.91E-21 | O94923 | D-glucuronyl C5-epimerase |
| rs2519093 | pQTLs | T | 1.07E-78 | O94923 | D-glucuronyl C5-epimerase |
| rs644234 | pQTLs | T | 4.68E-43 | O94923 | D-glucuronyl C5-epimerase |
| rs643434 | pQTLs | A | 8.71E-43 | O94923 | D-glucuronyl C5-epimerase |
| rs543040 | pQTLs | A | 1.95E-46 | O94923 | D-glucuronyl C5-epimerase |

| | | | | | |
|---|---|---|---|---|---|
| rs613534 | pQTLs | A | 5.01E-41 | O94923 | D-glucuronyl C5-epimerase |
| rs543968 | pQTLs | T | 7.94E-41 | O94923 | D-glucuronyl C5-epimerase |
| rs544873 | pQTLs | A | 1.35E-42 | O94923 | D-glucuronyl C5-epimerase |
| rs545971 | pQTLs | T | 3.72E-46 | O94923 | D-glucuronyl C5-epimerase |
| rs612169 | pQTLs | A | 1.55E-46 | O94923 | D-glucuronyl C5-epimerase |
| rs8176668 | pQTLs | A | 2.69E-20 | O94923 | D-glucuronyl C5-epimerase |
| rs8176663 | pQTLs | T | 2.04E-46 | O94923 | D-glucuronyl C5-epimerase |
| rs7036642 | pQTLs | A | 2.40E-20 | O94923 | D-glucuronyl C5-epimerase |
| rs491626 | pQTLs | T | 5.75E-46 | O94923 | D-glucuronyl C5-epimerase |
| rs492488 | pQTLs | A | 5.25E-46 | O94923 | D-glucuronyl C5-epimerase |
| rs493246 | pQTLs | A | 5.75E-46 | O94923 | D-glucuronyl C5-epimerase |
| rs494242 | pQTLs | T | 1.55E-42 | O94923 | D-glucuronyl C5-epimerase |
| rs495203 | pQTLs | T | 1.51E-46 | O94923 | D-glucuronyl C5-epimerase |
| rs582118 | pQTLs | A | 5.62E-44 | O94923 | D-glucuronyl C5-epimerase |
| rs582094 | pQTLs | A | 5.01E-45 | O94923 | D-glucuronyl C5-epimerase |
| rs2769071 | pQTLs | A | 3.63E-46 | O94923 | D-glucuronyl C5-epimerase |
| rs677355 | pQTLs | A | 3.63E-46 | O94923 | D-glucuronyl C5-epimerase |
| rs676996 | pQTLs | T | 2.14E-42 | O94923 | D-glucuronyl C5-epimerase |
| rs676457 | pQTLs | A | 2.29E-46 | O94923 | D-glucuronyl C5-epimerase |
| rs550057 | pQTLs | T | 1.23E-70 | O94923 | D-glucuronyl C5-epimerase |
| rs674302 | pQTLs | A | 2.19E-46 | O94923 | D-glucuronyl C5-epimerase |
| rs7046674 | pQTLs | T | 2.51E-20 | O94923 | D-glucuronyl C5-epimerase |
| rs554833 | pQTLs | T | 1.26E-46 | O94923 | D-glucuronyl C5-epimerase |
| rs8176649 | pQTLs | A | 9.55E-20 | O94923 | D-glucuronyl C5-epimerase |
| rs660340 | pQTLs | A | 1.95E-15 | O94923 | D-glucuronyl C5-epimerase |
| rs581107 | pQTLs | T | 1.10E-15 | O94923 | D-glucuronyl C5-epimerase |
| rs659104 | pQTLs | T | 1.26E-15 | O94923 | D-glucuronyl C5-epimerase |
| rs647800 | pQTLs | A | 6.92E-15 | O94923 | D-glucuronyl C5-epimerase |
| rs473533 | pQTLs | T | 1.86E-15 | O94923 | D-glucuronyl C5-epimerase |
| rs476410 | pQTLs | C | 1.12E-15 | O94923 | D-glucuronyl C5-epimerase |
| rs645982 | pQTLs | A | 1.66E-15 | O94923 | D-glucuronyl C5-epimerase |
| rs529565 | pQTLs | T | 6.61E-48 | O94923 | D-glucuronyl C5-epimerase |
| rs630510 | pQTLs | A | 2.09E-20 | O94923 | D-glucuronyl C5-epimerase |
| rs630014 | pQTLs | A | 2.09E-20 | O94923 | D-glucuronyl C5-epimerase |
| rs532436 | pQTLs | A | 3.09E-77 | O94923 | D-glucuronyl C5-epimerase |
| rs600038 | pQTLs | T | 2.00E-65 | O94923 | D-glucuronyl C5-epimerase |
| rs1124406: | pQTLs | T | 1.55E-42 | O94923 | D-glucuronyl C5-epimerase |
| rs649129 | pQTLs | T | 4.07E-64 | O94923 | D-glucuronyl C5-epimerase |
| rs495828 | pQTLs | T | 1.58E-66 | O94923 | D-glucuronyl C5-epimerase |
| rs687289 | pQTLs | A | 4.47E-09 | P15586 | N-acetylglucosamine-6-sulfatase |
| rs543040 | pQTLs | A | 9.55E-09 | P15586 | N-acetylglucosamine-6-sulfatase |
| rs543968 | pQTLs | T | 2.82E-08 | P15586 | N-acetylglucosamine-6-sulfatase |
| rs545971 | pQTLs | T | 4.57E-09 | P15586 | N-acetylglucosamine-6-sulfatase |
| rs612169 | pQTLs | A | 5.37E-09 | P15586 | N-acetylglucosamine-6-sulfatase |
| rs8176663 | pQTLs | T | 4.47E-09 | P15586 | N-acetylglucosamine-6-sulfatase |
| rs491626 | pQTLs | T | 2.95E-09 | P15586 | N-acetylglucosamine-6-sulfatase |
| rs492488 | pQTLs | A | 4.07E-09 | P15586 | N-acetylglucosamine-6-sulfatase |
| rs493246 | pQTLs | A | 4.79E-09 | P15586 | N-acetylglucosamine-6-sulfatase |
| rs495203 | pQTLs | T | 4.27E-09 | P15586 | N-acetylglucosamine-6-sulfatase |
| rs582118 | pQTLs | A | 4.57E-09 | P15586 | N-acetylglucosamine-6-sulfatase |

| | | | | | |
|---|---|---|---|---|---|
| rs582094 | pQTLs | A | 5.62E-09 | P15586 | N-acetylglucosamine-6-sulfatase |
| rs2769071 | pQTLs | A | 4.79E-09 | P15586 | N-acetylglucosamine-6-sulfatase |
| rs677355 | pQTLs | A | 5.75E-09 | P15586 | N-acetylglucosamine-6-sulfatase |
| rs676996 | pQTLs | T | 2.45E-09 | P15586 | N-acetylglucosamine-6-sulfatase |
| rs676457 | pQTLs | A | 4.37E-09 | P15586 | N-acetylglucosamine-6-sulfatase |
| rs550057 | pQTLs | T | 1.45E-09 | P15586 | N-acetylglucosamine-6-sulfatase |
| rs674302 | pQTLs | A | 4.90E-09 | P15586 | N-acetylglucosamine-6-sulfatase |
| rs554833 | pQTLs | T | 6.46E-09 | P15586 | N-acetylglucosamine-6-sulfatase |
| rs529565 | pQTLs | T | 7.24E-09 | P15586 | N-acetylglucosamine-6-sulfatase |
| rs7873635 | pQTLs | T | 3.09E-24 | Q8NBJ4 | Golgi membrane protein 1 |
| rs4962040 | pQTLs | A | 2.69E-24 | Q8NBJ4 | Golgi membrane protein 1 |
| rs8176702 | pQTLs | A | 3.31E-24 | Q8NBJ4 | Golgi membrane protein 1 |
| rs2073826 | pQTLs | T | 6.61E-30 | Q8NBJ4 | Golgi membrane protein 1 |
| rs687289 | pQTLs | A | 1.86E-88 | Q8NBJ4 | Golgi membrane protein 1 |
| rs2073827 | pQTLs | C | 5.37E-30 | Q8NBJ4 | Golgi membrane protein 1 |
| rs2073828 | pQTLs | A | 7.24E-30 | Q8NBJ4 | Golgi membrane protein 1 |
| rs8176691 | pQTLs | T | 3.39E-24 | Q8NBJ4 | Golgi membrane protein 1 |
| rs8176690 | pQTLs | A | 7.76E-30 | Q8NBJ4 | Golgi membrane protein 1 |
| rs8176682 | pQTLs | T | 3.24E-24 | Q8NBJ4 | Golgi membrane protein 1 |
| rs8176681 | pQTLs | T | 3.47E-30 | Q8NBJ4 | Golgi membrane protein 1 |
| rs2519093 | pQTLs | T | 5.01E-66 | Q8NBJ4 | Golgi membrane protein 1 |
| rs644234 | pQTLs | T | 5.01E-72 | Q8NBJ4 | Golgi membrane protein 1 |
| rs643434 | pQTLs | A | 1.86E-72 | Q8NBJ4 | Golgi membrane protein 1 |
| rs543040 | pQTLs | A | 3.98E-88 | Q8NBJ4 | Golgi membrane protein 1 |
| rs613534 | pQTLs | A | 1.26E-66 | Q8NBJ4 | Golgi membrane protein 1 |
| rs543968 | pQTLs | T | 1.02E-67 | Q8NBJ4 | Golgi membrane protein 1 |
| rs544873 | pQTLs | A | 3.63E-72 | Q8NBJ4 | Golgi membrane protein 1 |
| rs545971 | pQTLs | T | 8.91E-89 | Q8NBJ4 | Golgi membrane protein 1 |
| rs612169 | pQTLs | A | 2.95E-88 | Q8NBJ4 | Golgi membrane protein 1 |
| rs8176668 | pQTLs | A | 2.51E-24 | Q8NBJ4 | Golgi membrane protein 1 |
| rs8176663 | pQTLs | T | 7.24E-88 | Q8NBJ4 | Golgi membrane protein 1 |
| rs7036642 | pQTLs | A | 6.03E-24 | Q8NBJ4 | Golgi membrane protein 1 |
| rs491626 | pQTLs | T | 1.55E-87 | Q8NBJ4 | Golgi membrane protein 1 |
| rs492488 | pQTLs | A | 1.95E-87 | Q8NBJ4 | Golgi membrane protein 1 |
| rs493246 | pQTLs | A | 2.14E-87 | Q8NBJ4 | Golgi membrane protein 1 |
| rs494242 | pQTLs | T | 4.47E-72 | Q8NBJ4 | Golgi membrane protein 1 |
| rs495203 | pQTLs | T | 2.45E-88 | Q8NBJ4 | Golgi membrane protein 1 |
| rs582118 | pQTLs | A | 1.10E-86 | Q8NBJ4 | Golgi membrane protein 1 |
| rs582094 | pQTLs | A | 1.58E-86 | Q8NBJ4 | Golgi membrane protein 1 |
| rs2769071 | pQTLs | A | 1.02E-87 | Q8NBJ4 | Golgi membrane protein 1 |
| rs677355 | pQTLs | A | 1.82E-87 | Q8NBJ4 | Golgi membrane protein 1 |
| rs676996 | pQTLs | T | 5.50E-84 | Q8NBJ4 | Golgi membrane protein 1 |
| rs676457 | pQTLs | A | 1.91E-87 | Q8NBJ4 | Golgi membrane protein 1 |
| rs550057 | pQTLs | T | 3.55E-92 | Q8NBJ4 | Golgi membrane protein 1 |
| rs674302 | pQTLs | A | 6.76E-88 | Q8NBJ4 | Golgi membrane protein 1 |
| rs7046674 | pQTLs | T | 2.95E-24 | Q8NBJ4 | Golgi membrane protein 1 |
| rs554833 | pQTLs | T | 4.07E-88 | Q8NBJ4 | Golgi membrane protein 1 |
| rs8176649 | pQTLs | A | 2.09E-24 | Q8NBJ4 | Golgi membrane protein 1 |
| rs660340 | pQTLs | A | 6.17E-44 | Q8NBJ4 | Golgi membrane protein 1 |
| rs581107 | pQTLs | T | 3.80E-44 | Q8NBJ4 | Golgi membrane protein 1 |

| | | | | | |
|---|---|---|---|---|---|
| rs659104 | pQTLs | T | 4.90E-44 | Q8NBJ4 | Golgi membrane protein 1 |
| rs647800 | pQTLs | A | 4.90E-37 | Q8NBJ4 | Golgi membrane protein 1 |
| rs473533 | pQTLs | T | 6.17E-44 | Q8NBJ4 | Golgi membrane protein 1 |
| rs476410 | pQTLs | C | 3.47E-44 | Q8NBJ4 | Golgi membrane protein 1 |
| rs645982 | pQTLs | A | 4.68E-44 | Q8NBJ4 | Golgi membrane protein 1 |
| rs529565 | pQTLs | T | 1.12E-85 | Q8NBJ4 | Golgi membrane protein 1 |
| rs630510 | pQTLs | A | 3.80E-32 | Q8NBJ4 | Golgi membrane protein 1 |
| rs630014 | pQTLs | A | 4.79E-32 | Q8NBJ4 | Golgi membrane protein 1 |
| rs532436 | pQTLs | A | 8.91E-65 | Q8NBJ4 | Golgi membrane protein 1 |
| rs600038 | pQTLs | T | 4.27E-47 | Q8NBJ4 | Golgi membrane protein 1 |
| rs1124406: | pQTLs | T | 6.61E-47 | Q8NBJ4 | Golgi membrane protein 1 |
| rs649129 | pQTLs | T | 1.51E-45 | Q8NBJ4 | Golgi membrane protein 1 |
| rs495828 | pQTLs | T | 4.07E-48 | Q8NBJ4 | Golgi membrane protein 1 |
| rs687289 | pQTLs | A | 1.55E-11 | P05362 | Intercellular adhesion molecule 1 |
| rs2519093 | pQTLs | T | 3.55E-17 | P05362 | Intercellular adhesion molecule 1 |
| rs644234 | pQTLs | T | 3.63E-10 | P05362 | Intercellular adhesion molecule 1 |
| rs643434 | pQTLs | A | 7.08E-10 | P05362 | Intercellular adhesion molecule 1 |
| rs543040 | pQTLs | A | 9.33E-12 | P05362 | Intercellular adhesion molecule 1 |
| rs613534 | pQTLs | A | 3.39E-09 | P05362 | Intercellular adhesion molecule 1 |
| rs543968 | pQTLs | T | 3.89E-09 | P05362 | Intercellular adhesion molecule 1 |
| rs544873 | pQTLs | A | 6.31E-10 | P05362 | Intercellular adhesion molecule 1 |
| rs545971 | pQTLs | T | 1.70E-11 | P05362 | Intercellular adhesion molecule 1 |
| rs612169 | pQTLs | A | 2.09E-11 | P05362 | Intercellular adhesion molecule 1 |
| rs8176663 | pQTLs | T | 1.32E-11 | P05362 | Intercellular adhesion molecule 1 |
| rs491626 | pQTLs | T | 1.26E-11 | P05362 | Intercellular adhesion molecule 1 |
| rs492488 | pQTLs | A | 1.23E-11 | P05362 | Intercellular adhesion molecule 1 |
| rs493246 | pQTLs | A | 1.35E-11 | P05362 | Intercellular adhesion molecule 1 |
| rs494242 | pQTLs | T | 4.68E-10 | P05362 | Intercellular adhesion molecule 1 |
| rs495203 | pQTLs | T | 1.45E-11 | P05362 | Intercellular adhesion molecule 1 |
| rs582118 | pQTLs | A | 1.55E-11 | P05362 | Intercellular adhesion molecule 1 |
| rs582094 | pQTLs | A | 2.69E-11 | P05362 | Intercellular adhesion molecule 1 |
| rs2769071 | pQTLs | A | 1.26E-11 | P05362 | Intercellular adhesion molecule 1 |
| rs677355 | pQTLs | A | 1.48E-11 | P05362 | Intercellular adhesion molecule 1 |
| rs676996 | pQTLs | T | 1.02E-10 | P05362 | Intercellular adhesion molecule 1 |
| rs676457 | pQTLs | A | 2.45E-11 | P05362 | Intercellular adhesion molecule 1 |
| rs550057 | pQTLs | T | 9.33E-13 | P05362 | Intercellular adhesion molecule 1 |
| rs674302 | pQTLs | A | 1.07E-11 | P05362 | Intercellular adhesion molecule 1 |
| rs554833 | pQTLs | T | 7.59E-12 | P05362 | Intercellular adhesion molecule 1 |
| rs529565 | pQTLs | T | 1.29E-11 | P05362 | Intercellular adhesion molecule 1 |
| rs532436 | pQTLs | A | 3.31E-17 | P05362 | Intercellular adhesion molecule 1 |
| rs600038 | pQTLs | T | 8.71E-14 | P05362 | Intercellular adhesion molecule 1 |
| rs1124406: | pQTLs | T | 1.55E-08 | P05362 | Intercellular adhesion molecule 1 |
| rs649129 | pQTLs | T | 1.29E-13 | P05362 | Intercellular adhesion molecule 1 |
| rs495828 | pQTLs | T | 9.12E-14 | P05362 | Intercellular adhesion molecule 1 |
| rs687289 | pQTLs | A | 1.86E-09 | P13598 | Intercellular adhesion molecule 2 |
| rs2519093 | pQTLs | T | 8.32E-14 | P13598 | Intercellular adhesion molecule 2 |
| rs644234 | pQTLs | T | 4.17E-10 | P13598 | Intercellular adhesion molecule 2 |
| rs643434 | pQTLs | A | 3.31E-10 | P13598 | Intercellular adhesion molecule 2 |
| rs543040 | pQTLs | A | 7.24E-10 | P13598 | Intercellular adhesion molecule 2 |
| rs613534 | pQTLs | A | 2.04E-09 | P13598 | Intercellular adhesion molecule 2 |

| | | | | | |
|---|---|---|---|---|---|
| rs543968 | pQTLs | T | 3.72E-10 | P13598 | Intercellular adhesion molecule 2 |
| rs544873 | pQTLs | A | 3.16E-10 | P13598 | Intercellular adhesion molecule 2 |
| rs545971 | pQTLs | T | 1.58E-09 | P13598 | Intercellular adhesion molecule 2 |
| rs612169 | pQTLs | A | 9.55E-10 | P13598 | Intercellular adhesion molecule 2 |
| rs8176663 | pQTLs | T | 1.17E-09 | P13598 | Intercellular adhesion molecule 2 |
| rs491626 | pQTLs | T | 1.10E-09 | P13598 | Intercellular adhesion molecule 2 |
| rs492488 | pQTLs | A | 1.05E-09 | P13598 | Intercellular adhesion molecule 2 |
| rs493246 | pQTLs | A | 9.55E-10 | P13598 | Intercellular adhesion molecule 2 |
| rs494242 | pQTLs | T | 2.40E-10 | P13598 | Intercellular adhesion molecule 2 |
| rs495203 | pQTLs | T | 1.15E-09 | P13598 | Intercellular adhesion molecule 2 |
| rs582118 | pQTLs | A | 8.13E-10 | P13598 | Intercellular adhesion molecule 2 |
| rs582094 | pQTLs | A | 1.78E-09 | P13598 | Intercellular adhesion molecule 2 |
| rs2769071 | pQTLs | A | 1.35E-09 | P13598 | Intercellular adhesion molecule 2 |
| rs677355 | pQTLs | A | 1.35E-09 | P13598 | Intercellular adhesion molecule 2 |
| rs676996 | pQTLs | T | 1.07E-09 | P13598 | Intercellular adhesion molecule 2 |
| rs676457 | pQTLs | A | 1.41E-09 | P13598 | Intercellular adhesion molecule 2 |
| rs550057 | pQTLs | T | 1.02E-13 | P13598 | Intercellular adhesion molecule 2 |
| rs674302 | pQTLs | A | 9.12E-10 | P13598 | Intercellular adhesion molecule 2 |
| rs554833 | pQTLs | T | 1.41E-09 | P13598 | Intercellular adhesion molecule 2 |
| rs529565 | pQTLs | T | 1.86E-09 | P13598 | Intercellular adhesion molecule 2 |
| rs532436 | pQTLs | A | 1.38E-13 | P13598 | Intercellular adhesion molecule 2 |
| rs600038 | pQTLs | T | 1.02E-13 | P13598 | Intercellular adhesion molecule 2 |
| rs1124406: | pQTLs | T | 5.89E-13 | P13598 | Intercellular adhesion molecule 2 |
| rs649129 | pQTLs | T | 2.69E-13 | P13598 | Intercellular adhesion molecule 2 |
| rs495828 | pQTLs | T | 1.05E-13 | P13598 | Intercellular adhesion molecule 2 |
| rs2519093 | pQTLs | T | 2.29E-10 | Q14773 | Intercellular adhesion molecule 4 |
| rs532436 | pQTLs | A | 1.02E-10 | Q14773 | Intercellular adhesion molecule 4 |
| rs600038 | pQTLs | T | 9.55E-09 | Q14773 | Intercellular adhesion molecule 4 |
| rs649129 | pQTLs | T | 1.95E-08 | Q14773 | Intercellular adhesion molecule 4 |
| rs495828 | pQTLs | T | 1.91E-08 | Q14773 | Intercellular adhesion molecule 4 |
| rs687289 | pQTLs | A | 4.27E-10 | Q9UMF0 | Intercellular adhesion molecule 5 |
| rs2519093 | pQTLs | T | 1.58E-16 | Q9UMF0 | Intercellular adhesion molecule 5 |
| rs543040 | pQTLs | A | 6.31E-10 | Q9UMF0 | Intercellular adhesion molecule 5 |
| rs545971 | pQTLs | T | 8.32E-10 | Q9UMF0 | Intercellular adhesion molecule 5 |
| rs612169 | pQTLs | A | 4.27E-10 | Q9UMF0 | Intercellular adhesion molecule 5 |
| rs8176663 | pQTLs | T | 2.69E-10 | Q9UMF0 | Intercellular adhesion molecule 5 |
| rs491626 | pQTLs | T | 3.16E-10 | Q9UMF0 | Intercellular adhesion molecule 5 |
| rs492488 | pQTLs | A | 2.51E-10 | Q9UMF0 | Intercellular adhesion molecule 5 |
| rs493246 | pQTLs | A | 2.57E-10 | Q9UMF0 | Intercellular adhesion molecule 5 |
| rs495203 | pQTLs | T | 2.57E-10 | Q9UMF0 | Intercellular adhesion molecule 5 |
| rs582118 | pQTLs | A | 5.25E-10 | Q9UMF0 | Intercellular adhesion molecule 5 |
| rs582094 | pQTLs | A | 5.62E-10 | Q9UMF0 | Intercellular adhesion molecule 5 |
| rs2769071 | pQTLs | A | 2.19E-10 | Q9UMF0 | Intercellular adhesion molecule 5 |
| rs677355 | pQTLs | A | 2.51E-10 | Q9UMF0 | Intercellular adhesion molecule 5 |
| rs676996 | pQTLs | T | 5.75E-10 | Q9UMF0 | Intercellular adhesion molecule 5 |
| rs676457 | pQTLs | A | 1.26E-10 | Q9UMF0 | Intercellular adhesion molecule 5 |
| rs550057 | pQTLs | T | 2.51E-12 | Q9UMF0 | Intercellular adhesion molecule 5 |
| rs674302 | pQTLs | A | 2.19E-10 | Q9UMF0 | Intercellular adhesion molecule 5 |
| rs554833 | pQTLs | T | 2.29E-10 | Q9UMF0 | Intercellular adhesion molecule 5 |
| rs529565 | pQTLs | T | 4.27E-10 | Q9UMF0 | Intercellular adhesion molecule 5 |

| | | | | | |
|---|---|---|---|---|---|
| rs532436 | pQTLs | A | 8.51E-17 | Q9UMF0 | Intercellular adhesion molecule 5 |
| rs600038 | pQTLs | T | 1.48E-12 | Q9UMF0 | Intercellular adhesion molecule 5 |
| rs1124406: | pQTLs | T | 2.19E-11 | Q9UMF0 | Intercellular adhesion molecule 5 |
| rs649129 | pQTLs | T | 5.01E-13 | Q9UMF0 | Intercellular adhesion molecule 5 |
| rs495828 | pQTLs | T | 2.88E-13 | Q9UMF0 | Intercellular adhesion molecule 5 |
| rs687289 | pQTLs | A | 4.68E-22 | Q9UMF0 | Intercellular adhesion molecule 5 |
| rs2519093 | pQTLs | T | 8.32E-38 | Q9UMF0 | Intercellular adhesion molecule 5 |
| rs644234 | pQTLs | T | 2.09E-15 | Q9UMF0 | Intercellular adhesion molecule 5 |
| rs643434 | pQTLs | A | 1.29E-15 | Q9UMF0 | Intercellular adhesion molecule 5 |
| rs543040 | pQTLs | A | 1.05E-21 | Q9UMF0 | Intercellular adhesion molecule 5 |
| rs613534 | pQTLs | A | 2.63E-15 | Q9UMF0 | Intercellular adhesion molecule 5 |
| rs543968 | pQTLs | T | 3.02E-15 | Q9UMF0 | Intercellular adhesion molecule 5 |
| rs544873 | pQTLs | A | 1.23E-15 | Q9UMF0 | Intercellular adhesion molecule 5 |
| rs545971 | pQTLs | T | 1.58E-21 | Q9UMF0 | Intercellular adhesion molecule 5 |
| rs612169 | pQTLs | A | 9.55E-22 | Q9UMF0 | Intercellular adhesion molecule 5 |
| rs8176663 | pQTLs | T | 5.50E-22 | Q9UMF0 | Intercellular adhesion molecule 5 |
| rs491626 | pQTLs | T | 4.68E-22 | Q9UMF0 | Intercellular adhesion molecule 5 |
| rs492488 | pQTLs | A | 6.17E-22 | Q9UMF0 | Intercellular adhesion molecule 5 |
| rs493246 | pQTLs | A | 8.51E-22 | Q9UMF0 | Intercellular adhesion molecule 5 |
| rs494242 | pQTLs | T | 1.41E-15 | Q9UMF0 | Intercellular adhesion molecule 5 |
| rs495203 | pQTLs | T | 4.90E-22 | Q9UMF0 | Intercellular adhesion molecule 5 |
| rs582118 | pQTLs | A | 2.09E-21 | Q9UMF0 | Intercellular adhesion molecule 5 |
| rs582094 | pQTLs | A | 1.66E-21 | Q9UMF0 | Intercellular adhesion molecule 5 |
| rs2769071 | pQTLs | A | 5.37E-22 | Q9UMF0 | Intercellular adhesion molecule 5 |
| rs677355 | pQTLs | A | 6.46E-22 | Q9UMF0 | Intercellular adhesion molecule 5 |
| rs676996 | pQTLs | T | 2.51E-21 | Q9UMF0 | Intercellular adhesion molecule 5 |
| rs676457 | pQTLs | A | 1.26E-22 | Q9UMF0 | Intercellular adhesion molecule 5 |
| rs550057 | pQTLs | T | 1.51E-28 | Q9UMF0 | Intercellular adhesion molecule 5 |
| rs674302 | pQTLs | A | 5.50E-22 | Q9UMF0 | Intercellular adhesion molecule 5 |
| rs554833 | pQTLs | T | 7.41E-22 | Q9UMF0 | Intercellular adhesion molecule 5 |
| rs660340 | pQTLs | A | 2.45E-11 | Q9UMF0 | Intercellular adhesion molecule 5 |
| rs581107 | pQTLs | T | 2.24E-11 | Q9UMF0 | Intercellular adhesion molecule 5 |
| rs659104 | pQTLs | T | 1.91E-11 | Q9UMF0 | Intercellular adhesion molecule 5 |
| rs473533 | pQTLs | T | 3.16E-11 | Q9UMF0 | Intercellular adhesion molecule 5 |
| rs476410 | pQTLs | C | 2.57E-11 | Q9UMF0 | Intercellular adhesion molecule 5 |
| rs645982 | pQTLs | A | 1.82E-11 | Q9UMF0 | Intercellular adhesion molecule 5 |
| rs529565 | pQTLs | T | 4.07E-22 | Q9UMF0 | Intercellular adhesion molecule 5 |
| rs630510 | pQTLs | A | 6.76E-15 | Q9UMF0 | Intercellular adhesion molecule 5 |
| rs630014 | pQTLs | A | 1.12E-14 | Q9UMF0 | Intercellular adhesion molecule 5 |
| rs532436 | pQTLs | A | 1.51E-38 | Q9UMF0 | Intercellular adhesion molecule 5 |
| rs600038 | pQTLs | T | 3.80E-26 | Q9UMF0 | Intercellular adhesion molecule 5 |
| rs1124406: | pQTLs | T | 1.07E-25 | Q9UMF0 | Intercellular adhesion molecule 5 |
| rs649129 | pQTLs | T | 1.38E-26 | Q9UMF0 | Intercellular adhesion molecule 5 |
| rs495828 | pQTLs | T | 3.09E-27 | Q9UMF0 | Intercellular adhesion molecule 5 |
| rs2519093 | pQTLs | T | 7.94E-11 | P08069 | Insulin-like growth factor 1 receptor |
| rs550057 | pQTLs | T | 3.47E-09 | P08069 | Insulin-like growth factor 1 receptor |
| rs532436 | pQTLs | A | 6.17E-11 | P08069 | Insulin-like growth factor 1 receptor |
| rs600038 | pQTLs | T | 2.34E-09 | P08069 | Insulin-like growth factor 1 receptor |
| rs649129 | pQTLs | T | 8.13E-10 | P08069 | Insulin-like growth factor 1 receptor |
| rs495828 | pQTLs | T | 6.92E-10 | P08069 | Insulin-like growth factor 1 receptor |

| | | | | | |
|---|---|---|---|---|---|
| rs7873635 | pQTLs | T | 5.37E-15 | P26951 | Interleukin-3 receptor subunit alpha |
| rs4962040 | pQTLs | A | 1.41E-15 | P26951 | Interleukin-3 receptor subunit alpha |
| rs8176702 | pQTLs | A | 1.45E-15 | P26951 | Interleukin-3 receptor subunit alpha |
| rs2073826 | pQTLs | T | 1.29E-17 | P26951 | Interleukin-3 receptor subunit alpha |
| rs687289 | pQTLs | A | 3.31E-51 | P26951 | Interleukin-3 receptor subunit alpha |
| rs2073827 | pQTLs | C | 1.45E-17 | P26951 | Interleukin-3 receptor subunit alpha |
| rs2073828 | pQTLs | A | 1.38E-17 | P26951 | Interleukin-3 receptor subunit alpha |
| rs8176691 | pQTLs | T | 1.45E-15 | P26951 | Interleukin-3 receptor subunit alpha |
| rs8176690 | pQTLs | A | 3.72E-17 | P26951 | Interleukin-3 receptor subunit alpha |
| rs8176682 | pQTLs | T | 3.39E-15 | P26951 | Interleukin-3 receptor subunit alpha |
| rs8176681 | pQTLs | T | 3.98E-17 | P26951 | Interleukin-3 receptor subunit alpha |
| rs2519093 | pQTLs | T | 8.32E-198 | P26951 | Interleukin-3 receptor subunit alpha |
| rs644234 | pQTLs | T | 4.90E-44 | P26951 | Interleukin-3 receptor subunit alpha |
| rs643434 | pQTLs | A | 1.12E-44 | P26951 | Interleukin-3 receptor subunit alpha |
| rs543040 | pQTLs | A | 5.13E-51 | P26951 | Interleukin-3 receptor subunit alpha |
| rs613534 | pQTLs | A | 6.17E-41 | P26951 | Interleukin-3 receptor subunit alpha |
| rs543968 | pQTLs | T | 3.98E-40 | P26951 | Interleukin-3 receptor subunit alpha |
| rs544873 | pQTLs | A | 9.77E-45 | P26951 | Interleukin-3 receptor subunit alpha |
| rs545971 | pQTLs | T | 4.07E-50 | P26951 | Interleukin-3 receptor subunit alpha |
| rs612169 | pQTLs | A | 2.00E-50 | P26951 | Interleukin-3 receptor subunit alpha |
| rs8176668 | pQTLs | A | 4.27E-15 | P26951 | Interleukin-3 receptor subunit alpha |
| rs8176663 | pQTLs | T | 1.51E-50 | P26951 | Interleukin-3 receptor subunit alpha |
| rs7036642 | pQTLs | A | 5.25E-15 | P26951 | Interleukin-3 receptor subunit alpha |
| rs491626 | pQTLs | T | 1.00E-50 | P26951 | Interleukin-3 receptor subunit alpha |
| rs492488 | pQTLs | A | 2.40E-50 | P26951 | Interleukin-3 receptor subunit alpha |
| rs493246 | pQTLs | A | 2.57E-50 | P26951 | Interleukin-3 receptor subunit alpha |
| rs494242 | pQTLs | T | 1.91E-44 | P26951 | Interleukin-3 receptor subunit alpha |
| rs495203 | pQTLs | T | 1.35E-50 | P26951 | Interleukin-3 receptor subunit alpha |
| rs582118 | pQTLs | A | 2.40E-48 | P26951 | Interleukin-3 receptor subunit alpha |
| rs582094 | pQTLs | A | 4.47E-48 | P26951 | Interleukin-3 receptor subunit alpha |
| rs2769071 | pQTLs | A | 3.98E-50 | P26951 | Interleukin-3 receptor subunit alpha |
| rs677355 | pQTLs | A | 2.45E-50 | P26951 | Interleukin-3 receptor subunit alpha |
| rs676996 | pQTLs | T | 1.32E-46 | P26951 | Interleukin-3 receptor subunit alpha |
| rs676457 | pQTLs | A | 4.68E-50 | P26951 | Interleukin-3 receptor subunit alpha |
| rs550057 | pQTLs | T | 3.39E-169 | P26951 | Interleukin-3 receptor subunit alpha |
| rs674302 | pQTLs | A | 2.82E-50 | P26951 | Interleukin-3 receptor subunit alpha |
| rs7046674 | pQTLs | T | 8.32E-15 | P26951 | Interleukin-3 receptor subunit alpha |
| rs554833 | pQTLs | T | 3.02E-50 | P26951 | Interleukin-3 receptor subunit alpha |
| rs8176649 | pQTLs | A | 1.35E-14 | P26951 | Interleukin-3 receptor subunit alpha |
| rs660340 | pQTLs | A | 5.37E-26 | P26951 | Interleukin-3 receptor subunit alpha |
| rs581107 | pQTLs | T | 3.39E-26 | P26951 | Interleukin-3 receptor subunit alpha |
| rs659104 | pQTLs | T | 2.95E-26 | P26951 | Interleukin-3 receptor subunit alpha |
| rs647800 | pQTLs | A | 5.89E-24 | P26951 | Interleukin-3 receptor subunit alpha |
| rs473533 | pQTLs | T | 3.80E-26 | P26951 | Interleukin-3 receptor subunit alpha |
| rs476410 | pQTLs | C | 1.66E-26 | P26951 | Interleukin-3 receptor subunit alpha |
| rs645982 | pQTLs | A | 4.17E-26 | P26951 | Interleukin-3 receptor subunit alpha |
| rs529565 | pQTLs | T | 1.23E-52 | P26951 | Interleukin-3 receptor subunit alpha |
| rs630510 | pQTLs | A | 6.31E-20 | P26951 | Interleukin-3 receptor subunit alpha |
| rs630014 | pQTLs | A | 7.59E-20 | P26951 | Interleukin-3 receptor subunit alpha |
| rs532436 | pQTLs | A | 6.61E-195 | P26951 | Interleukin-3 receptor subunit alpha |

| | | | | | |
|---|---|---|---|---|---|
| rs600038 | pQTLs | T | 2.75E-159 | P26951 | Interleukin-3 receptor subunit alpha |
| rs1124406: | pQTLs | T | 4.27E-106 | P26951 | Interleukin-3 receptor subunit alpha |
| rs649129 | pQTLs | T | 2.45E-157 | P26951 | Interleukin-3 receptor subunit alpha |
| rs495828 | pQTLs | T | 1.51E-159 | P26951 | Interleukin-3 receptor subunit alpha |
| rs2519093 | pQTLs | T | 4.27E-12 | P40189 | Interleukin-6 receptor subunit beta |
| rs550057 | pQTLs | T | 9.33E-11 | P40189 | Interleukin-6 receptor subunit beta |
| rs532436 | pQTLs | A | 1.45E-12 | P40189 | Interleukin-6 receptor subunit beta |
| rs600038 | pQTLs | T | 1.32E-09 | P40189 | Interleukin-6 receptor subunit beta |
| rs649129 | pQTLs | T | 2.69E-10 | P40189 | Interleukin-6 receptor subunit beta |
| rs495828 | pQTLs | T | 5.89E-10 | P40189 | Interleukin-6 receptor subunit beta |
| rs7873635 | pQTLs | T | 7.41E-10 | P06213 | Insulin receptor |
| rs4962040 | pQTLs | A | 1.70E-09 | P06213 | Insulin receptor |
| rs8176702 | pQTLs | A | 7.76E-10 | P06213 | Insulin receptor |
| rs2073826 | pQTLs | T | 1.10E-12 | P06213 | Insulin receptor |
| rs687289 | pQTLs | A | 1.45E-35 | P06213 | Insulin receptor |
| rs2073827 | pQTLs | C | 1.07E-12 | P06213 | Insulin receptor |
| rs2073828 | pQTLs | A | 9.12E-13 | P06213 | Insulin receptor |
| rs8176691 | pQTLs | T | 7.41E-10 | P06213 | Insulin receptor |
| rs8176690 | pQTLs | A | 1.66E-12 | P06213 | Insulin receptor |
| rs8176682 | pQTLs | T | 1.17E-09 | P06213 | Insulin receptor |
| rs8176681 | pQTLs | T | 1.15E-12 | P06213 | Insulin receptor |
| rs2519093 | pQTLs | T | 6.92E-79 | P06213 | Insulin receptor |
| rs644234 | pQTLs | T | 5.89E-29 | P06213 | Insulin receptor |
| rs643434 | pQTLs | A | 7.59E-29 | P06213 | Insulin receptor |
| rs543040 | pQTLs | A | 4.68E-35 | P06213 | Insulin receptor |
| rs613534 | pQTLs | A | 1.48E-28 | P06213 | Insulin receptor |
| rs543968 | pQTLs | T | 1.82E-28 | P06213 | Insulin receptor |
| rs544873 | pQTLs | A | 5.37E-29 | P06213 | Insulin receptor |
| rs545971 | pQTLs | T | 2.04E-35 | P06213 | Insulin receptor |
| rs612169 | pQTLs | A | 1.26E-35 | P06213 | Insulin receptor |
| rs8176668 | pQTLs | A | 7.94E-10 | P06213 | Insulin receptor |
| rs8176663 | pQTLs | T | 3.47E-36 | P06213 | Insulin receptor |
| rs7036642 | pQTLs | A | 4.37E-10 | P06213 | Insulin receptor |
| rs491626 | pQTLs | T | 2.82E-36 | P06213 | Insulin receptor |
| rs492488 | pQTLs | A | 4.27E-36 | P06213 | Insulin receptor |
| rs493246 | pQTLs | A | 1.00E-35 | P06213 | Insulin receptor |
| rs494242 | pQTLs | T | 4.68E-29 | P06213 | Insulin receptor |
| rs495203 | pQTLs | T | 3.09E-36 | P06213 | Insulin receptor |
| rs582118 | pQTLs | A | 8.13E-34 | P06213 | Insulin receptor |
| rs582094 | pQTLs | A | 4.07E-34 | P06213 | Insulin receptor |
| rs2769071 | pQTLs | A | 2.82E-36 | P06213 | Insulin receptor |
| rs677355 | pQTLs | A | 4.27E-36 | P06213 | Insulin receptor |
| rs676996 | pQTLs | T | 3.55E-34 | P06213 | Insulin receptor |
| rs676457 | pQTLs | A | 1.32E-36 | P06213 | Insulin receptor |
| rs550057 | pQTLs | T | 1.12E-59 | P06213 | Insulin receptor |
| rs674302 | pQTLs | A | 2.14E-36 | P06213 | Insulin receptor |
| rs7046674 | pQTLs | T | 2.04E-10 | P06213 | Insulin receptor |
| rs554833 | pQTLs | T | 1.48E-36 | P06213 | Insulin receptor |
| rs8176649 | pQTLs | A | 2.82E-10 | P06213 | Insulin receptor |
| rs660340 | pQTLs | A | 1.26E-15 | P06213 | Insulin receptor |

| | | | | | |
|---|---|---|---|---|---|
| rs581107 | pQTLs | T | 1.26E-15 | P06213 | Insulin receptor |
| rs659104 | pQTLs | T | 1.20E-15 | P06213 | Insulin receptor |
| rs647800 | pQTLs | A | 1.55E-12 | P06213 | Insulin receptor |
| rs473533 | pQTLs | T | 1.12E-15 | P06213 | Insulin receptor |
| rs476410 | pQTLs | C | 2.00E-15 | P06213 | Insulin receptor |
| rs645982 | pQTLs | A | 1.74E-15 | P06213 | Insulin receptor |
| rs529565 | pQTLs | T | 1.15E-36 | P06213 | Insulin receptor |
| rs630510 | pQTLs | A | 6.31E-19 | P06213 | Insulin receptor |
| rs630014 | pQTLs | A | 8.51E-19 | P06213 | Insulin receptor |
| rs532436 | pQTLs | A | 6.61E-79 | P06213 | Insulin receptor |
| rs600038 | pQTLs | T | 1.10E-59 | P06213 | Insulin receptor |
| rs1124406: | pQTLs | T | 4.17E-42 | P06213 | Insulin receptor |
| rs649129 | pQTLs | T | 4.17E-60 | P06213 | Insulin receptor |
| rs495828 | pQTLs | T | 2.45E-60 | P06213 | Insulin receptor |
| rs687289 | pQTLs | A | 9.55E-09 | Q6UXK2 | Immunoglobulin superfamily containing leucine-rich re |
| rs2519093 | pQTLs | T | 9.12E-46 | Q6UXK2 | Immunoglobulin superfamily containing leucine-rich re |
| rs543040 | pQTLs | A | 9.55E-09 | Q6UXK2 | Immunoglobulin superfamily containing leucine-rich re |
| rs545971 | pQTLs | T | 1.86E-08 | Q6UXK2 | Immunoglobulin superfamily containing leucine-rich re |
| rs612169 | pQTLs | A | 9.12E-09 | Q6UXK2 | Immunoglobulin superfamily containing leucine-rich re |
| rs8176663 | pQTLs | T | 5.89E-09 | Q6UXK2 | Immunoglobulin superfamily containing leucine-rich re |
| rs491626 | pQTLs | T | 5.75E-09 | Q6UXK2 | Immunoglobulin superfamily containing leucine-rich re |
| rs492488 | pQTLs | A | 5.50E-09 | Q6UXK2 | Immunoglobulin superfamily containing leucine-rich re |
| rs493246 | pQTLs | A | 8.51E-09 | Q6UXK2 | Immunoglobulin superfamily containing leucine-rich re |
| rs495203 | pQTLs | T | 5.75E-09 | Q6UXK2 | Immunoglobulin superfamily containing leucine-rich re |
| rs582118 | pQTLs | A | 2.40E-08 | Q6UXK2 | Immunoglobulin superfamily containing leucine-rich re |
| rs582094 | pQTLs | A | 2.29E-08 | Q6UXK2 | Immunoglobulin superfamily containing leucine-rich re |
| rs2769071 | pQTLs | A | 5.13E-09 | Q6UXK2 | Immunoglobulin superfamily containing leucine-rich re |
| rs677355 | pQTLs | A | 7.41E-09 | Q6UXK2 | Immunoglobulin superfamily containing leucine-rich re |
| rs676996 | pQTLs | T | 1.41E-08 | Q6UXK2 | Immunoglobulin superfamily containing leucine-rich re |
| rs676457 | pQTLs | A | 4.79E-09 | Q6UXK2 | Immunoglobulin superfamily containing leucine-rich re |
| rs550057 | pQTLs | T | 2.45E-43 | Q6UXK2 | Immunoglobulin superfamily containing leucine-rich re |
| rs674302 | pQTLs | A | 5.62E-09 | Q6UXK2 | Immunoglobulin superfamily containing leucine-rich re |
| rs554833 | pQTLs | T | 3.47E-09 | Q6UXK2 | Immunoglobulin superfamily containing leucine-rich re |
| rs529565 | pQTLs | T | 7.94E-09 | Q6UXK2 | Immunoglobulin superfamily containing leucine-rich re |
| rs532436 | pQTLs | A | 1.86E-45 | Q6UXK2 | Immunoglobulin superfamily containing leucine-rich re |
| rs600038 | pQTLs | T | 3.31E-38 | Q6UXK2 | Immunoglobulin superfamily containing leucine-rich re |
| rs1124406: | pQTLs | T | 2.45E-25 | Q6UXK2 | Immunoglobulin superfamily containing leucine-rich re |
| rs649129 | pQTLs | T | 1.48E-39 | Q6UXK2 | Immunoglobulin superfamily containing leucine-rich re |
| rs495828 | pQTLs | T | 6.31E-39 | Q6UXK2 | Immunoglobulin superfamily containing leucine-rich re |
| rs2519093 | pQTLs | T | 3.55E-13 | P78504 | Protein jagged-1 |
| rs550057 | pQTLs | T | 8.32E-14 | P78504 | Protein jagged-1 |
| rs532436 | pQTLs | A | 3.89E-13 | P78504 | Protein jagged-1 |
| rs600038 | pQTLs | T | 2.51E-09 | P78504 | Protein jagged-1 |
| rs649129 | pQTLs | T | 1.02E-09 | P78504 | Protein jagged-1 |
| rs495828 | pQTLs | T | 1.10E-09 | P78504 | Protein jagged-1 |
| rs7873635 | pQTLs | T | 1.55E-16 | P35968 | Vascular endothelial growth factor receptor 2 |
| rs4962040 | pQTLs | A | 2.24E-16 | P35968 | Vascular endothelial growth factor receptor 2 |
| rs8176702 | pQTLs | A | 1.00E-16 | P35968 | Vascular endothelial growth factor receptor 2 |
| rs2073826 | pQTLs | T | 2.63E-19 | P35968 | Vascular endothelial growth factor receptor 2 |
| rs687289 | pQTLs | A | 3.47E-53 | P35968 | Vascular endothelial growth factor receptor 2 |

| | | | | | |
|---|---|---|---|---|---|
| rs2073827 | pQTLs | C | 1.48E-19 | P35968 | Vascular endothelial growth factor receptor 2 |
| rs2073828 | pQTLs | A | 1.00E-19 | P35968 | Vascular endothelial growth factor receptor 2 |
| rs8176691 | pQTLs | T | 1.32E-16 | P35968 | Vascular endothelial growth factor receptor 2 |
| rs8176690 | pQTLs | A | 1.45E-18 | P35968 | Vascular endothelial growth factor receptor 2 |
| rs8176682 | pQTLs | T | 6.03E-16 | P35968 | Vascular endothelial growth factor receptor 2 |
| rs8176681 | pQTLs | T | 2.82E-18 | P35968 | Vascular endothelial growth factor receptor 2 |
| rs2519093 | pQTLs | T | 4.90E-80 | P35968 | Vascular endothelial growth factor receptor 2 |
| rs644234 | pQTLs | T | 2.34E-45 | P35968 | Vascular endothelial growth factor receptor 2 |
| rs643434 | pQTLs | A | 2.24E-45 | P35968 | Vascular endothelial growth factor receptor 2 |
| rs543040 | pQTLs | A | 5.37E-52 | P35968 | Vascular endothelial growth factor receptor 2 |
| rs613534 | pQTLs | A | 5.62E-44 | P35968 | Vascular endothelial growth factor receptor 2 |
| rs543968 | pQTLs | T | 4.37E-42 | P35968 | Vascular endothelial growth factor receptor 2 |
| rs544873 | pQTLs | A | 2.75E-45 | P35968 | Vascular endothelial growth factor receptor 2 |
| rs545971 | pQTLs | T | 2.63E-52 | P35968 | Vascular endothelial growth factor receptor 2 |
| rs612169 | pQTLs | A | 3.24E-52 | P35968 | Vascular endothelial growth factor receptor 2 |
| rs8176668 | pQTLs | A | 2.69E-16 | P35968 | Vascular endothelial growth factor receptor 2 |
| rs8176663 | pQTLs | T | 1.35E-52 | P35968 | Vascular endothelial growth factor receptor 2 |
| rs7036642 | pQTLs | A | 3.72E-16 | P35968 | Vascular endothelial growth factor receptor 2 |
| rs491626 | pQTLs | T | 1.82E-52 | P35968 | Vascular endothelial growth factor receptor 2 |
| rs492488 | pQTLs | A | 2.51E-52 | P35968 | Vascular endothelial growth factor receptor 2 |
| rs493246 | pQTLs | A | 2.40E-52 | P35968 | Vascular endothelial growth factor receptor 2 |
| rs494242 | pQTLs | T | 1.78E-45 | P35968 | Vascular endothelial growth factor receptor 2 |
| rs495203 | pQTLs | T | 1.48E-52 | P35968 | Vascular endothelial growth factor receptor 2 |
| rs582118 | pQTLs | A | 2.51E-52 | P35968 | Vascular endothelial growth factor receptor 2 |
| rs582094 | pQTLs | A | 9.33E-52 | P35968 | Vascular endothelial growth factor receptor 2 |
| rs2769071 | pQTLs | A | 4.07E-52 | P35968 | Vascular endothelial growth factor receptor 2 |
| rs677355 | pQTLs | A | 1.26E-52 | P35968 | Vascular endothelial growth factor receptor 2 |
| rs676996 | pQTLs | T | 4.07E-51 | P35968 | Vascular endothelial growth factor receptor 2 |
| rs676457 | pQTLs | A | 6.61E-52 | P35968 | Vascular endothelial growth factor receptor 2 |
| rs550057 | pQTLs | T | 3.80E-67 | P35968 | Vascular endothelial growth factor receptor 2 |
| rs674302 | pQTLs | A | 9.77E-53 | P35968 | Vascular endothelial growth factor receptor 2 |
| rs7046674 | pQTLs | T | 3.80E-16 | P35968 | Vascular endothelial growth factor receptor 2 |
| rs554833 | pQTLs | T | 3.02E-52 | P35968 | Vascular endothelial growth factor receptor 2 |
| rs8176649 | pQTLs | A | 2.45E-16 | P35968 | Vascular endothelial growth factor receptor 2 |
| rs660340 | pQTLs | A | 7.94E-23 | P35968 | Vascular endothelial growth factor receptor 2 |
| rs581107 | pQTLs | T | 6.92E-23 | P35968 | Vascular endothelial growth factor receptor 2 |
| rs659104 | pQTLs | T | 5.75E-23 | P35968 | Vascular endothelial growth factor receptor 2 |
| rs647800 | pQTLs | A | 2.75E-20 | P35968 | Vascular endothelial growth factor receptor 2 |
| rs473533 | pQTLs | T | 5.75E-23 | P35968 | Vascular endothelial growth factor receptor 2 |
| rs476410 | pQTLs | C | 9.77E-23 | P35968 | Vascular endothelial growth factor receptor 2 |
| rs645982 | pQTLs | A | 9.33E-23 | P35968 | Vascular endothelial growth factor receptor 2 |
| rs529565 | pQTLs | T | 4.07E-53 | P35968 | Vascular endothelial growth factor receptor 2 |
| rs630510 | pQTLs | A | 1.23E-23 | P35968 | Vascular endothelial growth factor receptor 2 |
| rs630014 | pQTLs | A | 1.05E-23 | P35968 | Vascular endothelial growth factor receptor 2 |
| rs532436 | pQTLs | A | 1.82E-79 | P35968 | Vascular endothelial growth factor receptor 2 |
| rs600038 | pQTLs | T | 8.32E-63 | P35968 | Vascular endothelial growth factor receptor 2 |
| rs1124406[illegible] | pQTLs | T | 3.16E-46 | P35968 | Vascular endothelial growth factor receptor 2 |
| rs649129 | pQTLs | T | 4.17E-64 | P35968 | Vascular endothelial growth factor receptor 2 |
| rs495828 | pQTLs | T | 8.13E-64 | P35968 | Vascular endothelial growth factor receptor 2 |
| rs687289 | pQTLs | A | 2.45E-14 | O60870 | DNA/RNA-binding protein KIN17 |

| | | | | | |
|---|---|---|---|---|---|
| rs2519093 | pQTLs | T | 1.62E-23 | O60870 | DNA/RNA-binding protein KIN17 |
| rs644234 | pQTLs | T | 2.34E-11 | O60870 | DNA/RNA-binding protein KIN17 |
| rs643434 | pQTLs | A | 1.10E-11 | O60870 | DNA/RNA-binding protein KIN17 |
| rs543040 | pQTLs | A | 5.25E-14 | O60870 | DNA/RNA-binding protein KIN17 |
| rs613534 | pQTLs | A | 5.37E-11 | O60870 | DNA/RNA-binding protein KIN17 |
| rs543968 | pQTLs | T | 2.75E-12 | O60870 | DNA/RNA-binding protein KIN17 |
| rs544873 | pQTLs | A | 1.20E-11 | O60870 | DNA/RNA-binding protein KIN17 |
| rs545971 | pQTLs | T | 1.20E-14 | O60870 | DNA/RNA-binding protein KIN17 |
| rs612169 | pQTLs | A | 2.82E-14 | O60870 | DNA/RNA-binding protein KIN17 |
| rs8176663 | pQTLs | T | 5.50E-14 | O60870 | DNA/RNA-binding protein KIN17 |
| rs491626 | pQTLs | T | 2.95E-14 | O60870 | DNA/RNA-binding protein KIN17 |
| rs492488 | pQTLs | A | 4.27E-14 | O60870 | DNA/RNA-binding protein KIN17 |
| rs493246 | pQTLs | A | 5.75E-14 | O60870 | DNA/RNA-binding protein KIN17 |
| rs494242 | pQTLs | T | 1.05E-11 | O60870 | DNA/RNA-binding protein KIN17 |
| rs495203 | pQTLs | T | 4.07E-14 | O60870 | DNA/RNA-binding protein KIN17 |
| rs582118 | pQTLs | A | 1.86E-14 | O60870 | DNA/RNA-binding protein KIN17 |
| rs582094 | pQTLs | A | 3.24E-14 | O60870 | DNA/RNA-binding protein KIN17 |
| rs2769071 | pQTLs | A | 5.89E-14 | O60870 | DNA/RNA-binding protein KIN17 |
| rs677355 | pQTLs | A | 6.92E-14 | O60870 | DNA/RNA-binding protein KIN17 |
| rs676996 | pQTLs | T | 8.71E-14 | O60870 | DNA/RNA-binding protein KIN17 |
| rs676457 | pQTLs | A | 5.01E-14 | O60870 | DNA/RNA-binding protein KIN17 |
| rs550057 | pQTLs | T | 5.13E-23 | O60870 | DNA/RNA-binding protein KIN17 |
| rs674302 | pQTLs | A | 5.50E-14 | O60870 | DNA/RNA-binding protein KIN17 |
| rs554833 | pQTLs | T | 8.32E-14 | O60870 | DNA/RNA-binding protein KIN17 |
| rs660340 | pQTLs | A | 2.14E-09 | O60870 | DNA/RNA-binding protein KIN17 |
| rs581107 | pQTLs | T | 1.86E-09 | O60870 | DNA/RNA-binding protein KIN17 |
| rs659104 | pQTLs | T | 1.74E-09 | O60870 | DNA/RNA-binding protein KIN17 |
| rs473533 | pQTLs | T | 1.95E-09 | O60870 | DNA/RNA-binding protein KIN17 |
| rs476410 | pQTLs | C | 1.48E-09 | O60870 | DNA/RNA-binding protein KIN17 |
| rs645982 | pQTLs | A | 1.51E-09 | O60870 | DNA/RNA-binding protein KIN17 |
| rs529565 | pQTLs | T | 5.37E-14 | O60870 | DNA/RNA-binding protein KIN17 |
| rs532436 | pQTLs | A | 9.12E-23 | O60870 | DNA/RNA-binding protein KIN17 |
| rs600038 | pQTLs | T | 4.27E-17 | O60870 | DNA/RNA-binding protein KIN17 |
| rs1124406: | pQTLs | T | 1.20E-13 | O60870 | DNA/RNA-binding protein KIN17 |
| rs649129 | pQTLs | T | 4.47E-18 | O60870 | DNA/RNA-binding protein KIN17 |
| rs495828 | pQTLs | T | 6.76E-17 | O60870 | DNA/RNA-binding protein KIN17 |
| rs2519093 | pQTLs | T | 8.91E-11 | P32004 | Neural cell adhesion molecule L1 |
| rs550057 | pQTLs | T | 1.02E-08 | P32004 | Neural cell adhesion molecule L1 |
| rs532436 | pQTLs | A | 1.29E-10 | P32004 | Neural cell adhesion molecule L1 |
| rs600038 | pQTLs | T | 3.24E-10 | P32004 | Neural cell adhesion molecule L1 |
| rs649129 | pQTLs | T | 6.46E-11 | P32004 | Neural cell adhesion molecule L1 |
| rs495828 | pQTLs | T | 1.23E-10 | P32004 | Neural cell adhesion molecule L1 |
| rs2519093 | pQTLs | T | 2.88E-08 | P07942,P1: | Laminin |
| rs532436 | pQTLs | A | 2.63E-08 | P07942,P1: | Laminin |
| rs2519093 | pQTLs | T | 9.12E-09 | P09848 | Lactase-phlorizin hydrolase |
| rs550057 | pQTLs | T | 8.51E-09 | P09848 | Lactase-phlorizin hydrolase |
| rs532436 | pQTLs | A | 6.46E-09 | P09848 | Lactase-phlorizin hydrolase |
| rs2519093 | pQTLs | T | 7.59E-20 | P42702 | Leukemia inhibitory factor receptor |
| rs8176663 | pQTLs | T | 4.07E-08 | P42702 | Leukemia inhibitory factor receptor |
| rs492488 | pQTLs | A | 3.39E-08 | P42702 | Leukemia inhibitory factor receptor |

| | | | | | |
|---|---|---|---|---|---|
| rs493246 | pQTLs | A | 3.31E-08 | P42702 | Leukemia inhibitory factor receptor |
| rs495203 | pQTLs | T | 3.98E-08 | P42702 | Leukemia inhibitory factor receptor |
| rs2769071 | pQTLs | A | 3.09E-08 | P42702 | Leukemia inhibitory factor receptor |
| rs677355 | pQTLs | A | 3.55E-08 | P42702 | Leukemia inhibitory factor receptor |
| rs676457 | pQTLs | A | 2.57E-08 | P42702 | Leukemia inhibitory factor receptor |
| rs550057 | pQTLs | T | 2.34E-15 | P42702 | Leukemia inhibitory factor receptor |
| rs674302 | pQTLs | A | 2.95E-08 | P42702 | Leukemia inhibitory factor receptor |
| rs554833 | pQTLs | T | 2.82E-08 | P42702 | Leukemia inhibitory factor receptor |
| rs529565 | pQTLs | T | 4.07E-08 | P42702 | Leukemia inhibitory factor receptor |
| rs532436 | pQTLs | A | 3.47E-20 | P42702 | Leukemia inhibitory factor receptor |
| rs600038 | pQTLs | T | 1.78E-17 | P42702 | Leukemia inhibitory factor receptor |
| rs1124406: | pQTLs | T | 7.41E-11 | P42702 | Leukemia inhibitory factor receptor |
| rs649129 | pQTLs | T | 5.01E-18 | P42702 | Leukemia inhibitory factor receptor |
| rs495828 | pQTLs | T | 6.17E-18 | P42702 | Leukemia inhibitory factor receptor |
| rs2519093 | pQTLs | T | 7.41E-12 | O60476 | Mannosyl-oligosaccharide 1,2-alpha-mannosidase IB |
| rs532436 | pQTLs | A | 1.12E-11 | O60476 | Mannosyl-oligosaccharide 1,2-alpha-mannosidase IB |
| rs600038 | pQTLs | T | 1.23E-11 | O60476 | Mannosyl-oligosaccharide 1,2-alpha-mannosidase IB |
| rs649129 | pQTLs | T | 3.72E-11 | O60476 | Mannosyl-oligosaccharide 1,2-alpha-mannosidase IB |
| rs495828 | pQTLs | T | 8.32E-12 | O60476 | Mannosyl-oligosaccharide 1,2-alpha-mannosidase IB |
| rs2073826 | pQTLs | T | 4.90E-09 | P11226 | Mannose-binding protein C |
| rs687289 | pQTLs | A | 2.40E-18 | P11226 | Mannose-binding protein C |
| rs2073827 | pQTLs | C | 5.50E-09 | P11226 | Mannose-binding protein C |
| rs2073828 | pQTLs | A | 5.89E-09 | P11226 | Mannose-binding protein C |
| rs8176690 | pQTLs | A | 6.92E-09 | P11226 | Mannose-binding protein C |
| rs8176681 | pQTLs | T | 6.92E-09 | P11226 | Mannose-binding protein C |
| rs2519093 | pQTLs | T | 4.17E-14 | P11226 | Mannose-binding protein C |
| rs644234 | pQTLs | T | 1.20E-14 | P11226 | Mannose-binding protein C |
| rs643434 | pQTLs | A | 3.55E-15 | P11226 | Mannose-binding protein C |
| rs543040 | pQTLs | A | 1.23E-18 | P11226 | Mannose-binding protein C |
| rs613534 | pQTLs | A | 6.03E-15 | P11226 | Mannose-binding protein C |
| rs543968 | pQTLs | T | 1.95E-15 | P11226 | Mannose-binding protein C |
| rs544873 | pQTLs | A | 3.09E-15 | P11226 | Mannose-binding protein C |
| rs545971 | pQTLs | T | 5.37E-19 | P11226 | Mannose-binding protein C |
| rs612169 | pQTLs | A | 6.61E-19 | P11226 | Mannose-binding protein C |
| rs8176663 | pQTLs | T | 1.74E-18 | P11226 | Mannose-binding protein C |
| rs491626 | pQTLs | T | 1.70E-18 | P11226 | Mannose-binding protein C |
| rs492488 | pQTLs | A | 2.19E-18 | P11226 | Mannose-binding protein C |
| rs493246 | pQTLs | A | 2.00E-18 | P11226 | Mannose-binding protein C |
| rs494242 | pQTLs | T | 5.37E-15 | P11226 | Mannose-binding protein C |
| rs495203 | pQTLs | T | 1.55E-18 | P11226 | Mannose-binding protein C |
| rs582118 | pQTLs | A | 3.63E-18 | P11226 | Mannose-binding protein C |
| rs582094 | pQTLs | A | 9.55E-18 | P11226 | Mannose-binding protein C |
| rs2769071 | pQTLs | A | 3.02E-18 | P11226 | Mannose-binding protein C |
| rs677355 | pQTLs | A | 3.16E-18 | P11226 | Mannose-binding protein C |
| rs676996 | pQTLs | T | 5.89E-19 | P11226 | Mannose-binding protein C |
| rs676457 | pQTLs | A | 3.80E-18 | P11226 | Mannose-binding protein C |
| rs550057 | pQTLs | T | 2.95E-14 | P11226 | Mannose-binding protein C |
| rs674302 | pQTLs | A | 2.51E-18 | P11226 | Mannose-binding protein C |
| rs554833 | pQTLs | T | 4.47E-18 | P11226 | Mannose-binding protein C |
| rs529565 | pQTLs | T | 7.08E-18 | P11226 | Mannose-binding protein C |

| | | | | | |
|---|---|---|---|---|---|
| rs630510 | pQTLs | A | 2.57E-10 | P11226 | Mannose-binding protein C |
| rs630014 | pQTLs | A | 3.39E-10 | P11226 | Mannose-binding protein C |
| rs532436 | pQTLs | A | 9.55E-14 | P11226 | Mannose-binding protein C |
| rs600038 | pQTLs | T | 1.51E-09 | P11226 | Mannose-binding protein C |
| rs1124406: | pQTLs | T | 2.34E-11 | P11226 | Mannose-binding protein C |
| rs649129 | pQTLs | T | 2.19E-09 | P11226 | Mannose-binding protein C |
| rs495828 | pQTLs | T | 1.10E-09 | P11226 | Mannose-binding protein C |
| rs687289 | pQTLs | A | 4.79E-23 | P08581 | Hepatocyte growth factor receptor |
| rs2519093 | pQTLs | T | 4.57E-32 | P08581 | Hepatocyte growth factor receptor |
| rs644234 | pQTLs | T | 5.50E-18 | P08581 | Hepatocyte growth factor receptor |
| rs643434 | pQTLs | A | 3.02E-18 | P08581 | Hepatocyte growth factor receptor |
| rs543040 | pQTLs | A | 7.59E-23 | P08581 | Hepatocyte growth factor receptor |
| rs613534 | pQTLs | A | 6.61E-18 | P08581 | Hepatocyte growth factor receptor |
| rs543968 | pQTLs | T | 1.55E-17 | P08581 | Hepatocyte growth factor receptor |
| rs544873 | pQTLs | A | 3.16E-18 | P08581 | Hepatocyte growth factor receptor |
| rs545971 | pQTLs | T | 5.50E-23 | P08581 | Hepatocyte growth factor receptor |
| rs612169 | pQTLs | A | 5.75E-23 | P08581 | Hepatocyte growth factor receptor |
| rs8176663 | pQTLs | T | 1.74E-23 | P08581 | Hepatocyte growth factor receptor |
| rs491626 | pQTLs | T | 2.34E-23 | P08581 | Hepatocyte growth factor receptor |
| rs492488 | pQTLs | A | 1.78E-23 | P08581 | Hepatocyte growth factor receptor |
| rs493246 | pQTLs | A | 2.29E-23 | P08581 | Hepatocyte growth factor receptor |
| rs494242 | pQTLs | T | 2.04E-18 | P08581 | Hepatocyte growth factor receptor |
| rs495203 | pQTLs | T | 2.00E-23 | P08581 | Hepatocyte growth factor receptor |
| rs582118 | pQTLs | A | 1.86E-23 | P08581 | Hepatocyte growth factor receptor |
| rs582094 | pQTLs | A | 3.89E-23 | P08581 | Hepatocyte growth factor receptor |
| rs2769071 | pQTLs | A | 1.74E-23 | P08581 | Hepatocyte growth factor receptor |
| rs677355 | pQTLs | A | 2.29E-23 | P08581 | Hepatocyte growth factor receptor |
| rs676996 | pQTLs | T | 1.41E-23 | P08581 | Hepatocyte growth factor receptor |
| rs676457 | pQTLs | A | 1.29E-23 | P08581 | Hepatocyte growth factor receptor |
| rs550057 | pQTLs | T | 3.55E-33 | P08581 | Hepatocyte growth factor receptor |
| rs674302 | pQTLs | A | 1.38E-23 | P08581 | Hepatocyte growth factor receptor |
| rs554833 | pQTLs | T | 2.09E-23 | P08581 | Hepatocyte growth factor receptor |
| rs660340 | pQTLs | A | 5.37E-18 | P08581 | Hepatocyte growth factor receptor |
| rs581107 | pQTLs | T | 1.07E-17 | P08581 | Hepatocyte growth factor receptor |
| rs659104 | pQTLs | T | 1.05E-17 | P08581 | Hepatocyte growth factor receptor |
| rs647800 | pQTLs | A | 1.58E-14 | P08581 | Hepatocyte growth factor receptor |
| rs473533 | pQTLs | T | 1.15E-17 | P08581 | Hepatocyte growth factor receptor |
| rs476410 | pQTLs | C | 2.34E-17 | P08581 | Hepatocyte growth factor receptor |
| rs645982 | pQTLs | A | 1.62E-17 | P08581 | Hepatocyte growth factor receptor |
| rs529565 | pQTLs | T | 1.10E-24 | P08581 | Hepatocyte growth factor receptor |
| rs630510 | pQTLs | A | 5.25E-12 | P08581 | Hepatocyte growth factor receptor |
| rs630014 | pQTLs | A | 6.03E-12 | P08581 | Hepatocyte growth factor receptor |
| rs532436 | pQTLs | A | 2.24E-32 | P08581 | Hepatocyte growth factor receptor |
| rs600038 | pQTLs | T | 1.91E-23 | P08581 | Hepatocyte growth factor receptor |
| rs1124406: | pQTLs | T | 1.10E-15 | P08581 | Hepatocyte growth factor receptor |
| rs649129 | pQTLs | T | 2.57E-24 | P08581 | Hepatocyte growth factor receptor |
| rs495828 | pQTLs | T | 7.08E-24 | P08581 | Hepatocyte growth factor receptor |
| rs2519093 | pQTLs | T | 3.80E-11 | Q5VY43 | Platelet endothelial aggregation receptor 1 |
| rs550057 | pQTLs | T | 1.00E-11 | Q5VY43 | Platelet endothelial aggregation receptor 1 |
| rs532436 | pQTLs | A | 2.63E-11 | Q5VY43 | Platelet endothelial aggregation receptor 1 |

| | | | | | |
|---|---|---|---|---|---|
| rs600038 | pQTLs | T | 1.20E-08 | Q5VY43 | Platelet endothelial aggregation receptor 1 |
| rs649129 | pQTLs | T | 2.69E-09 | Q5VY43 | Platelet endothelial aggregation receptor 1 |
| rs495828 | pQTLs | T | 7.24E-09 | Q5VY43 | Platelet endothelial aggregation receptor 1 |
| rs630510 | pQTLs | A | 2.14E-08 | O15031 | Plexin-B2 |
| rs630014 | pQTLs | A | 2.88E-08 | O15031 | Plexin-B2 |
| rs7873635 | pQTLs | T | 8.91E-68 | Q6ZRP7 | Sulfhydryl oxidase 2 |
| rs4962040 | pQTLs | A | 1.58E-68 | Q6ZRP7 | Sulfhydryl oxidase 2 |
| rs8176702 | pQTLs | A | 1.17E-68 | Q6ZRP7 | Sulfhydryl oxidase 2 |
| rs2073826 | pQTLs | T | 3.31E-74 | Q6ZRP7 | Sulfhydryl oxidase 2 |
| rs687289 | pQTLs | A | 2.04E-241 | Q6ZRP7 | Sulfhydryl oxidase 2 |
| rs2073827 | pQTLs | C | 2.34E-74 | Q6ZRP7 | Sulfhydryl oxidase 2 |
| rs2073828 | pQTLs | A | 4.37E-74 | Q6ZRP7 | Sulfhydryl oxidase 2 |
| rs8176691 | pQTLs | T | 1.12E-68 | Q6ZRP7 | Sulfhydryl oxidase 2 |
| rs8176690 | pQTLs | A | 1.23E-74 | Q6ZRP7 | Sulfhydryl oxidase 2 |
| rs8176682 | pQTLs | T | 3.80E-68 | Q6ZRP7 | Sulfhydryl oxidase 2 |
| rs8176681 | pQTLs | T | 2.09E-73 | Q6ZRP7 | Sulfhydryl oxidase 2 |
| rs2519093 | pQTLs | T | 7.76E-257 | Q6ZRP7 | Sulfhydryl oxidase 2 |
| rs644234 | pQTLs | T | 6.17E-213 | Q6ZRP7 | Sulfhydryl oxidase 2 |
| rs643434 | pQTLs | A | 7.76E-211 | Q6ZRP7 | Sulfhydryl oxidase 2 |
| rs543040 | pQTLs | A | 2.63E-239 | Q6ZRP7 | Sulfhydryl oxidase 2 |
| rs613534 | pQTLs | A | 1.48E-198 | Q6ZRP7 | Sulfhydryl oxidase 2 |
| rs543968 | pQTLs | T | 1.20E-198 | Q6ZRP7 | Sulfhydryl oxidase 2 |
| rs544873 | pQTLs | A | 1.05E-210 | Q6ZRP7 | Sulfhydryl oxidase 2 |
| rs545971 | pQTLs | T | 8.51E-239 | Q6ZRP7 | Sulfhydryl oxidase 2 |
| rs612169 | pQTLs | A | 7.59E-239 | Q6ZRP7 | Sulfhydryl oxidase 2 |
| rs8176668 | pQTLs | A | 1.12E-67 | Q6ZRP7 | Sulfhydryl oxidase 2 |
| rs8176663 | pQTLs | T | 3.02E-238 | Q6ZRP7 | Sulfhydryl oxidase 2 |
| rs7036642 | pQTLs | A | 3.31E-67 | Q6ZRP7 | Sulfhydryl oxidase 2 |
| rs491626 | pQTLs | T | 6.03E-238 | Q6ZRP7 | Sulfhydryl oxidase 2 |
| rs492488 | pQTLs | A | 4.27E-237 | Q6ZRP7 | Sulfhydryl oxidase 2 |
| rs493246 | pQTLs | A | 4.17E-237 | Q6ZRP7 | Sulfhydryl oxidase 2 |
| rs494242 | pQTLs | T | 3.39E-210 | Q6ZRP7 | Sulfhydryl oxidase 2 |
| rs495203 | pQTLs | T | 4.68E-239 | Q6ZRP7 | Sulfhydryl oxidase 2 |
| rs582118 | pQTLs | A | 8.91E-233 | Q6ZRP7 | Sulfhydryl oxidase 2 |
| rs582094 | pQTLs | A | 6.17E-235 | Q6ZRP7 | Sulfhydryl oxidase 2 |
| rs2769071 | pQTLs | A | 7.41E-238 | Q6ZRP7 | Sulfhydryl oxidase 2 |
| rs677355 | pQTLs | A | 2.57E-238 | Q6ZRP7 | Sulfhydryl oxidase 2 |
| rs676996 | pQTLs | T | 3.55E-228 | Q6ZRP7 | Sulfhydryl oxidase 2 |
| rs676457 | pQTLs | A | 8.13E-237 | Q6ZRP7 | Sulfhydryl oxidase 2 |
| rs550057 | pQTLs | T | 3.8904514 | Q6ZRP7 | Sulfhydryl oxidase 2 |
| rs674302 | pQTLs | A | 6.61E-238 | Q6ZRP7 | Sulfhydryl oxidase 2 |
| rs7046674 | pQTLs | T | 1.58E-67 | Q6ZRP7 | Sulfhydryl oxidase 2 |
| rs554833 | pQTLs | T | 2.51E-238 | Q6ZRP7 | Sulfhydryl oxidase 2 |
| rs8176649 | pQTLs | A | 2.57E-67 | Q6ZRP7 | Sulfhydryl oxidase 2 |
| rs660340 | pQTLs | A | 2.63E-81 | Q6ZRP7 | Sulfhydryl oxidase 2 |
| rs581107 | pQTLs | T | 5.37E-82 | Q6ZRP7 | Sulfhydryl oxidase 2 |
| rs659104 | pQTLs | T | 2.95E-82 | Q6ZRP7 | Sulfhydryl oxidase 2 |
| rs647800 | pQTLs | A | 1.12E-75 | Q6ZRP7 | Sulfhydryl oxidase 2 |
| rs473533 | pQTLs | T | 8.71E-82 | Q6ZRP7 | Sulfhydryl oxidase 2 |
| rs476410 | pQTLs | C | 5.50E-83 | Q6ZRP7 | Sulfhydryl oxidase 2 |

| | | | | | |
|---|---|---|---|---|---|
| rs645982 | pQTLs | A | 4.47E-82 | Q6ZRP7 | Sulfhydryl oxidase 2 |
| rs529565 | pQTLs | T | 8.91E-245 | Q6ZRP7 | Sulfhydryl oxidase 2 |
| rs630510 | pQTLs | A | 4.47E-89 | Q6ZRP7 | Sulfhydryl oxidase 2 |
| rs630014 | pQTLs | A | 3.72E-88 | Q6ZRP7 | Sulfhydryl oxidase 2 |
| rs532436 | pQTLs | A | 3.63E-252 | Q6ZRP7 | Sulfhydryl oxidase 2 |
| rs600038 | pQTLs | T | 1.00E-198 | Q6ZRP7 | Sulfhydryl oxidase 2 |
| rs1124406: | pQTLs | T | 1.62E-124 | Q6ZRP7 | Sulfhydryl oxidase 2 |
| rs649129 | pQTLs | T | 1.86E-194 | Q6ZRP7 | Sulfhydryl oxidase 2 |
| rs495828 | pQTLs | T | 2.57E-200 | Q6ZRP7 | Sulfhydryl oxidase 2 |
| rs600038 | pQTLs | T | 2.75E-08 | Q8WVN6 | Secreted and transmembrane protein 1 |
| rs649129 | pQTLs | T | 2.14E-08 | Q8WVN6 | Secreted and transmembrane protein 1 |
| rs495828 | pQTLs | T | 2.45E-08 | Q8WVN6 | Secreted and transmembrane protein 1 |
| rs7873635 | pQTLs | T | 8.51E-57 | P16581 | E-selectin |
| rs4962040 | pQTLs | A | 9.77E-58 | P16581 | E-selectin |
| rs8176702 | pQTLs | A | 2.29E-58 | P16581 | E-selectin |
| rs2073826 | pQTLs | T | 1.45E-68 | P16581 | E-selectin |
| rs687289 | pQTLs | A | 2.24E-222 | P16581 | E-selectin |
| rs2073827 | pQTLs | C | 3.47E-68 | P16581 | E-selectin |
| rs2073828 | pQTLs | A | 4.57E-68 | P16581 | E-selectin |
| rs8176691 | pQTLs | T | 2.82E-58 | P16581 | E-selectin |
| rs8176690 | pQTLs | A | 5.13E-68 | P16581 | E-selectin |
| rs8176682 | pQTLs | T | 3.63E-58 | P16581 | E-selectin |
| rs8176681 | pQTLs | T | 1.70E-67 | P16581 | E-selectin |
| rs2519093 | pQTLs | T | 0 | P16581 | E-selectin |
| rs644234 | pQTLs | T | 3.24E-183 | P16581 | E-selectin |
| rs643434 | pQTLs | A | 6.46E-184 | P16581 | E-selectin |
| rs543040 | pQTLs | A | 2.34E-219 | P16581 | E-selectin |
| rs613534 | pQTLs | A | 9.77E-172 | P16581 | E-selectin |
| rs543968 | pQTLs | T | 9.12E-173 | P16581 | E-selectin |
| rs544873 | pQTLs | A | 9.33E-184 | P16581 | E-selectin |
| rs545971 | pQTLs | T | 8.32E-220 | P16581 | E-selectin |
| rs612169 | pQTLs | A | 1.10E-220 | P16581 | E-selectin |
| rs8176668 | pQTLs | A | 3.98E-57 | P16581 | E-selectin |
| rs8176663 | pQTLs | T | 6.76E-221 | P16581 | E-selectin |
| rs7036642 | pQTLs | A | 1.58E-57 | P16581 | E-selectin |
| rs491626 | pQTLs | T | 9.77E-221 | P16581 | E-selectin |
| rs492488 | pQTLs | A | 1.05E-220 | P16581 | E-selectin |
| rs493246 | pQTLs | A | 1.95E-220 | P16581 | E-selectin |
| rs494242 | pQTLs | T | 4.27E-183 | P16581 | E-selectin |
| rs495203 | pQTLs | T | 4.07E-221 | P16581 | E-selectin |
| rs582118 | pQTLs | A | 5.13E-213 | P16581 | E-selectin |
| rs582094 | pQTLs | A | 8.51E-213 | P16581 | E-selectin |
| rs2769071 | pQTLs | A | 5.75E-220 | P16581 | E-selectin |
| rs677355 | pQTLs | A | 6.61E-221 | P16581 | E-selectin |
| rs676996 | pQTLs | T | 2.14E-210 | P16581 | E-selectin |
| rs676457 | pQTLs | A | 2.34E-220 | P16581 | E-selectin |
| rs550057 | pQTLs | T | 7.41E-269 | P16581 | E-selectin |
| rs674302 | pQTLs | A | 6.03E-221 | P16581 | E-selectin |
| rs7046674 | pQTLs | T | 1.66E-57 | P16581 | E-selectin |
| rs554833 | pQTLs | T | 3.55E-220 | P16581 | E-selectin |

| | | | | | |
|---|---|---|---|---|---|
| rs8176649 | pQTLs | A | 7.94E-59 | P16581 | E-selectin |
| rs660340 | pQTLs | A | 1.35E-78 | P16581 | E-selectin |
| rs581107 | pQTLs | T | 3.63E-79 | P16581 | E-selectin |
| rs659104 | pQTLs | T | 2.57E-79 | P16581 | E-selectin |
| rs647800 | pQTLs | A | 3.16E-68 | P16581 | E-selectin |
| rs473533 | pQTLs | T | 3.89E-79 | P16581 | E-selectin |
| rs476410 | pQTLs | C | 9.77E-80 | P16581 | E-selectin |
| rs645982 | pQTLs | A | 1.66E-79 | P16581 | E-selectin |
| rs529565 | pQTLs | T | 6.03E-225 | P16581 | E-selectin |
| rs630510 | pQTLs | A | 1.51E-87 | P16581 | E-selectin |
| rs630014 | pQTLs | A | 1.07E-86 | P16581 | E-selectin |
| rs532436 | pQTLs | A | 0 | P16581 | E-selectin |
| rs600038 | pQTLs | T | 0 | P16581 | E-selectin |
| rs1124406: | pQTLs | T | 2.40E-216 | P16581 | E-selectin |
| rs649129 | pQTLs | T | 0 | P16581 | E-selectin |
| rs495828 | pQTLs | T | 0 | P16581 | E-selectin |
| rs2073826 | pQTLs | T | 7.59E-10 | P16109 | P-selectin |
| rs687289 | pQTLs | A | 2.88E-24 | P16109 | P-selectin |
| rs2073827 | pQTLs | C | 1.00E-09 | P16109 | P-selectin |
| rs2073828 | pQTLs | A | 1.35E-09 | P16109 | P-selectin |
| rs8176690 | pQTLs | A | 7.41E-10 | P16109 | P-selectin |
| rs8176682 | pQTLs | T | 2.63E-08 | P16109 | P-selectin |
| rs8176681 | pQTLs | T | 4.47E-10 | P16109 | P-selectin |
| rs2519093 | pQTLs | T | 4.27E-48 | P16109 | P-selectin |
| rs644234 | pQTLs | T | 3.02E-19 | P16109 | P-selectin |
| rs643434 | pQTLs | A | 8.32E-20 | P16109 | P-selectin |
| rs543040 | pQTLs | A | 1.02E-23 | P16109 | P-selectin |
| rs613534 | pQTLs | A | 4.07E-19 | P16109 | P-selectin |
| rs543968 | pQTLs | T | 8.71E-20 | P16109 | P-selectin |
| rs544873 | pQTLs | A | 6.76E-20 | P16109 | P-selectin |
| rs545971 | pQTLs | T | 1.48E-24 | P16109 | P-selectin |
| rs612169 | pQTLs | A | 3.98E-24 | P16109 | P-selectin |
| rs8176668 | pQTLs | A | 4.90E-08 | P16109 | P-selectin |
| rs8176663 | pQTLs | T | 6.31E-24 | P16109 | P-selectin |
| rs491626 | pQTLs | T | 2.09E-24 | P16109 | P-selectin |
| rs492488 | pQTLs | A | 4.27E-24 | P16109 | P-selectin |
| rs493246 | pQTLs | A | 4.57E-24 | P16109 | P-selectin |
| rs494242 | pQTLs | T | 5.50E-20 | P16109 | P-selectin |
| rs495203 | pQTLs | T | 4.37E-24 | P16109 | P-selectin |
| rs582118 | pQTLs | A | 5.75E-24 | P16109 | P-selectin |
| rs582094 | pQTLs | A | 2.24E-23 | P16109 | P-selectin |
| rs2769071 | pQTLs | A | 6.03E-24 | P16109 | P-selectin |
| rs677355 | pQTLs | A | 6.46E-24 | P16109 | P-selectin |
| rs676996 | pQTLs | T | 3.16E-23 | P16109 | P-selectin |
| rs676457 | pQTLs | A | 5.25E-24 | P16109 | P-selectin |
| rs550057 | pQTLs | T | 4.17E-36 | P16109 | P-selectin |
| rs674302 | pQTLs | A | 5.25E-24 | P16109 | P-selectin |
| rs554833 | pQTLs | T | 4.57E-24 | P16109 | P-selectin |
| rs8176649 | pQTLs | A | 3.80E-08 | P16109 | P-selectin |
| rs660340 | pQTLs | A | 1.62E-11 | P16109 | P-selectin |

| | | | | | |
|---|---|---|---|---|---|
| rs581107 | pQTLs | T | 1.32E-11 | P16109 | P-selectin |
| rs659104 | pQTLs | T | 1.20E-11 | P16109 | P-selectin |
| rs647800 | pQTLs | A | 1.38E-09 | P16109 | P-selectin |
| rs473533 | pQTLs | T | 1.58E-11 | P16109 | P-selectin |
| rs476410 | pQTLs | C | 7.08E-12 | P16109 | P-selectin |
| rs645982 | pQTLs | A | 8.51E-12 | P16109 | P-selectin |
| rs529565 | pQTLs | T | 3.89E-24 | P16109 | P-selectin |
| rs630510 | pQTLs | A | 2.51E-11 | P16109 | P-selectin |
| rs630014 | pQTLs | A | 6.03E-11 | P16109 | P-selectin |
| rs532436 | pQTLs | A | 2.51E-47 | P16109 | P-selectin |
| rs600038 | pQTLs | T | 2.45E-36 | P16109 | P-selectin |
| rs1124406: | pQTLs | T | 4.90E-27 | P16109 | P-selectin |
| rs649129 | pQTLs | T | 7.08E-37 | P16109 | P-selectin |
| rs495828 | pQTLs | T | 2.09E-35 | P16109 | P-selectin |
| rs2519093 | pQTLs | T | 1.38E-19 | Q9H2E6 | Semaphorin-6A |
| rs550057 | pQTLs | T | 6.92E-20 | Q9H2E6 | Semaphorin-6A |
| rs532436 | pQTLs | A | 6.17E-19 | Q9H2E6 | Semaphorin-6A |
| rs600038 | pQTLs | T | 3.63E-16 | Q9H2E6 | Semaphorin-6A |
| rs1124406: | pQTLs | T | 4.79E-10 | Q9H2E6 | Semaphorin-6A |
| rs649129 | pQTLs | T | 3.47E-17 | Q9H2E6 | Semaphorin-6A |
| rs495828 | pQTLs | T | 1.10E-16 | Q9H2E6 | Semaphorin-6A |
| rs687289 | pQTLs | A | 2.95E-09 | Q9NS62 | Thrombospondin type-1 domain-containing protein 1 |
| rs2519093 | pQTLs | T | 2.69E-19 | Q9NS62 | Thrombospondin type-1 domain-containing protein 1 |
| rs644234 | pQTLs | T | 1.78E-09 | Q9NS62 | Thrombospondin type-1 domain-containing protein 1 |
| rs643434 | pQTLs | A | 1.26E-09 | Q9NS62 | Thrombospondin type-1 domain-containing protein 1 |
| rs543040 | pQTLs | A | 9.77E-09 | Q9NS62 | Thrombospondin type-1 domain-containing protein 1 |
| rs613534 | pQTLs | A | 3.24E-09 | Q9NS62 | Thrombospondin type-1 domain-containing protein 1 |
| rs543968 | pQTLs | T | 6.31E-09 | Q9NS62 | Thrombospondin type-1 domain-containing protein 1 |
| rs544873 | pQTLs | A | 1.26E-09 | Q9NS62 | Thrombospondin type-1 domain-containing protein 1 |
| rs545971 | pQTLs | T | 5.75E-09 | Q9NS62 | Thrombospondin type-1 domain-containing protein 1 |
| rs612169 | pQTLs | A | 5.13E-09 | Q9NS62 | Thrombospondin type-1 domain-containing protein 1 |
| rs8176663 | pQTLs | T | 7.59E-09 | Q9NS62 | Thrombospondin type-1 domain-containing protein 1 |
| rs491626 | pQTLs | T | 6.31E-09 | Q9NS62 | Thrombospondin type-1 domain-containing protein 1 |
| rs492488 | pQTLs | A | 6.61E-09 | Q9NS62 | Thrombospondin type-1 domain-containing protein 1 |
| rs493246 | pQTLs | A | 5.25E-09 | Q9NS62 | Thrombospondin type-1 domain-containing protein 1 |
| rs494242 | pQTLs | T | 1.95E-09 | Q9NS62 | Thrombospondin type-1 domain-containing protein 1 |
| rs495203 | pQTLs | T | 6.61E-09 | Q9NS62 | Thrombospondin type-1 domain-containing protein 1 |
| rs582118 | pQTLs | A | 3.09E-09 | Q9NS62 | Thrombospondin type-1 domain-containing protein 1 |
| rs582094 | pQTLs | A | 7.24E-09 | Q9NS62 | Thrombospondin type-1 domain-containing protein 1 |
| rs2769071 | pQTLs | A | 7.41E-09 | Q9NS62 | Thrombospondin type-1 domain-containing protein 1 |
| rs677355 | pQTLs | A | 9.12E-09 | Q9NS62 | Thrombospondin type-1 domain-containing protein 1 |
| rs676996 | pQTLs | T | 1.91E-08 | Q9NS62 | Thrombospondin type-1 domain-containing protein 1 |
| rs676457 | pQTLs | A | 8.51E-09 | Q9NS62 | Thrombospondin type-1 domain-containing protein 1 |
| rs550057 | pQTLs | T | 2.14E-13 | Q9NS62 | Thrombospondin type-1 domain-containing protein 1 |
| rs674302 | pQTLs | A | 5.62E-09 | Q9NS62 | Thrombospondin type-1 domain-containing protein 1 |
| rs554833 | pQTLs | T | 7.76E-09 | Q9NS62 | Thrombospondin type-1 domain-containing protein 1 |
| rs529565 | pQTLs | T | 2.88E-08 | Q9NS62 | Thrombospondin type-1 domain-containing protein 1 |
| rs532436 | pQTLs | A | 4.57E-19 | Q9NS62 | Thrombospondin type-1 domain-containing protein 1 |
| rs600038 | pQTLs | T | 7.94E-19 | Q9NS62 | Thrombospondin type-1 domain-containing protein 1 |
| rs1124406: | pQTLs | T | 2.40E-11 | Q9NS62 | Thrombospondin type-1 domain-containing protein 1 |

| | | | | | |
|---|---|---|---|---|---|
| rs649129 | pQTLs | T | 3.63E-19 | Q9NS62 | Thrombospondin type-1 domain-containing protein 1 |
| rs495828 | pQTLs | T | 7.76E-19 | Q9NS62 | Thrombospondin type-1 domain-containing protein 1 |
| rs7873635 | pQTLs | T | 2.88E-27 | P35590 | Tyrosine-protein kinase receptor Tie-1, soluble |
| rs4962040 | pQTLs | A | 3.24E-27 | P35590 | Tyrosine-protein kinase receptor Tie-1, soluble |
| rs8176702 | pQTLs | A | 3.47E-27 | P35590 | Tyrosine-protein kinase receptor Tie-1, soluble |
| rs2073826 | pQTLs | T | 1.78E-29 | P35590 | Tyrosine-protein kinase receptor Tie-1, soluble |
| rs687289 | pQTLs | A | 3.02E-77 | P35590 | Tyrosine-protein kinase receptor Tie-1, soluble |
| rs2073827 | pQTLs | C | 3.72E-29 | P35590 | Tyrosine-protein kinase receptor Tie-1, soluble |
| rs2073828 | pQTLs | A | 4.79E-29 | P35590 | Tyrosine-protein kinase receptor Tie-1, soluble |
| rs8176691 | pQTLs | T | 3.98E-27 | P35590 | Tyrosine-protein kinase receptor Tie-1, soluble |
| rs8176690 | pQTLs | A | 4.47E-30 | P35590 | Tyrosine-protein kinase receptor Tie-1, soluble |
| rs8176682 | pQTLs | T | 4.79E-28 | P35590 | Tyrosine-protein kinase receptor Tie-1, soluble |
| rs8176681 | pQTLs | T | 3.55E-30 | P35590 | Tyrosine-protein kinase receptor Tie-1, soluble |
| rs2519093 | pQTLs | T | 4.27E-14 | P35590 | Tyrosine-protein kinase receptor Tie-1, soluble |
| rs644234 | pQTLs | T | 6.03E-69 | P35590 | Tyrosine-protein kinase receptor Tie-1, soluble |
| rs643434 | pQTLs | A | 2.57E-69 | P35590 | Tyrosine-protein kinase receptor Tie-1, soluble |
| rs543040 | pQTLs | A | 4.27E-77 | P35590 | Tyrosine-protein kinase receptor Tie-1, soluble |
| rs613534 | pQTLs | A | 2.82E-67 | P35590 | Tyrosine-protein kinase receptor Tie-1, soluble |
| rs543968 | pQTLs | T | 4.79E-68 | P35590 | Tyrosine-protein kinase receptor Tie-1, soluble |
| rs544873 | pQTLs | A | 2.29E-69 | P35590 | Tyrosine-protein kinase receptor Tie-1, soluble |
| rs545971 | pQTLs | T | 4.47E-78 | P35590 | Tyrosine-protein kinase receptor Tie-1, soluble |
| rs612169 | pQTLs | A | 8.32E-78 | P35590 | Tyrosine-protein kinase receptor Tie-1, soluble |
| rs8176668 | pQTLs | A | 2.51E-27 | P35590 | Tyrosine-protein kinase receptor Tie-1, soluble |
| rs8176663 | pQTLs | T | 2.95E-77 | P35590 | Tyrosine-protein kinase receptor Tie-1, soluble |
| rs7036642 | pQTLs | A | 1.48E-27 | P35590 | Tyrosine-protein kinase receptor Tie-1, soluble |
| rs491626 | pQTLs | T | 1.86E-77 | P35590 | Tyrosine-protein kinase receptor Tie-1, soluble |
| rs492488 | pQTLs | A | 2.19E-77 | P35590 | Tyrosine-protein kinase receptor Tie-1, soluble |
| rs493246 | pQTLs | A | 2.40E-77 | P35590 | Tyrosine-protein kinase receptor Tie-1, soluble |
| rs494242 | pQTLs | T | 2.63E-69 | P35590 | Tyrosine-protein kinase receptor Tie-1, soluble |
| rs495203 | pQTLs | T | 3.02E-77 | P35590 | Tyrosine-protein kinase receptor Tie-1, soluble |
| rs582118 | pQTLs | A | 3.89E-78 | P35590 | Tyrosine-protein kinase receptor Tie-1, soluble |
| rs582094 | pQTLs | A | 4.57E-78 | P35590 | Tyrosine-protein kinase receptor Tie-1, soluble |
| rs2769071 | pQTLs | A | 8.71E-77 | P35590 | Tyrosine-protein kinase receptor Tie-1, soluble |
| rs677355 | pQTLs | A | 7.59E-77 | P35590 | Tyrosine-protein kinase receptor Tie-1, soluble |
| rs676996 | pQTLs | T | 3.09E-74 | P35590 | Tyrosine-protein kinase receptor Tie-1, soluble |
| rs676457 | pQTLs | A | 3.39E-77 | P35590 | Tyrosine-protein kinase receptor Tie-1, soluble |
| rs550057 | pQTLs | T | 6.92E-10 | P35590 | Tyrosine-protein kinase receptor Tie-1, soluble |
| rs674302 | pQTLs | A | 5.13E-77 | P35590 | Tyrosine-protein kinase receptor Tie-1, soluble |
| rs7046674 | pQTLs | T | 7.24E-27 | P35590 | Tyrosine-protein kinase receptor Tie-1, soluble |
| rs554833 | pQTLs | T | 3.09E-76 | P35590 | Tyrosine-protein kinase receptor Tie-1, soluble |
| rs8176649 | pQTLs | A | 5.89E-28 | P35590 | Tyrosine-protein kinase receptor Tie-1, soluble |
| rs660340 | pQTLs | A | 8.13E-28 | P35590 | Tyrosine-protein kinase receptor Tie-1, soluble |
| rs581107 | pQTLs | T | 3.80E-28 | P35590 | Tyrosine-protein kinase receptor Tie-1, soluble |
| rs659104 | pQTLs | T | 4.57E-28 | P35590 | Tyrosine-protein kinase receptor Tie-1, soluble |
| rs647800 | pQTLs | A | 1.20E-25 | P35590 | Tyrosine-protein kinase receptor Tie-1, soluble |
| rs473533 | pQTLs | T | 2.69E-28 | P35590 | Tyrosine-protein kinase receptor Tie-1, soluble |
| rs476410 | pQTLs | C | 1.95E-28 | P35590 | Tyrosine-protein kinase receptor Tie-1, soluble |
| rs645982 | pQTLs | A | 1.32E-28 | P35590 | Tyrosine-protein kinase receptor Tie-1, soluble |
| rs529565 | pQTLs | T | 4.90E-76 | P35590 | Tyrosine-protein kinase receptor Tie-1, soluble |
| rs630510 | pQTLs | A | 6.76E-33 | P35590 | Tyrosine-protein kinase receptor Tie-1, soluble |

| | | | | | |
|---|---|---|---|---|---|
| rs630014 | pQTLs | A | 1.29E-32 | P35590 | Tyrosine-protein kinase receptor Tie-1, soluble |
| rs532436 | pQTLs | A | 2.19E-14 | P35590 | Tyrosine-protein kinase receptor Tie-1, soluble |
| rs600038 | pQTLs | T | 9.12E-11 | P35590 | Tyrosine-protein kinase receptor Tie-1, soluble |
| rs1124406: | pQTLs | T | 1.66E-08 | P35590 | Tyrosine-protein kinase receptor Tie-1, soluble |
| rs649129 | pQTLs | T | 7.59E-10 | P35590 | Tyrosine-protein kinase receptor Tie-1, soluble |
| rs495828 | pQTLs | T | 2.19E-10 | P35590 | Tyrosine-protein kinase receptor Tie-1, soluble |
| rs687289 | pQTLs | A | 2.00E-11 | O00206,Q9 | Toll-like receptor 4:Lymphocyte antigen 96 complex |
| rs2519093 | pQTLs | T | 6.46E-23 | O00206,Q9 | Toll-like receptor 4:Lymphocyte antigen 96 complex |
| rs644234 | pQTLs | T | 4.27E-09 | O00206,Q9 | Toll-like receptor 4:Lymphocyte antigen 96 complex |
| rs643434 | pQTLs | A | 2.69E-09 | O00206,Q9 | Toll-like receptor 4:Lymphocyte antigen 96 complex |
| rs543040 | pQTLs | A | 1.91E-11 | O00206,Q9 | Toll-like receptor 4:Lymphocyte antigen 96 complex |
| rs613534 | pQTLs | A | 1.15E-08 | O00206,Q9 | Toll-like receptor 4:Lymphocyte antigen 96 complex |
| rs543968 | pQTLs | T | 1.20E-08 | O00206,Q9 | Toll-like receptor 4:Lymphocyte antigen 96 complex |
| rs544873 | pQTLs | A | 2.24E-09 | O00206,Q9 | Toll-like receptor 4:Lymphocyte antigen 96 complex |
| rs545971 | pQTLs | T | 2.88E-11 | O00206,Q9 | Toll-like receptor 4:Lymphocyte antigen 96 complex |
| rs612169 | pQTLs | A | 1.51E-11 | O00206,Q9 | Toll-like receptor 4:Lymphocyte antigen 96 complex |
| rs8176663 | pQTLs | T | 7.59E-12 | O00206,Q9 | Toll-like receptor 4:Lymphocyte antigen 96 complex |
| rs491626 | pQTLs | T | 6.76E-12 | O00206,Q9 | Toll-like receptor 4:Lymphocyte antigen 96 complex |
| rs492488 | pQTLs | A | 5.75E-12 | O00206,Q9 | Toll-like receptor 4:Lymphocyte antigen 96 complex |
| rs493246 | pQTLs | A | 7.76E-12 | O00206,Q9 | Toll-like receptor 4:Lymphocyte antigen 96 complex |
| rs494242 | pQTLs | T | 1.51E-09 | O00206,Q9 | Toll-like receptor 4:Lymphocyte antigen 96 complex |
| rs495203 | pQTLs | T | 6.46E-12 | O00206,Q9 | Toll-like receptor 4:Lymphocyte antigen 96 complex |
| rs582118 | pQTLs | A | 1.82E-11 | O00206,Q9 | Toll-like receptor 4:Lymphocyte antigen 96 complex |
| rs582094 | pQTLs | A | 4.07E-11 | O00206,Q9 | Toll-like receptor 4:Lymphocyte antigen 96 complex |
| rs2769071 | pQTLs | A | 4.47E-12 | O00206,Q9 | Toll-like receptor 4:Lymphocyte antigen 96 complex |
| rs677355 | pQTLs | A | 6.03E-12 | O00206,Q9 | Toll-like receptor 4:Lymphocyte antigen 96 complex |
| rs676996 | pQTLs | T | 1.26E-11 | O00206,Q9 | Toll-like receptor 4:Lymphocyte antigen 96 complex |
| rs676457 | pQTLs | A | 3.89E-12 | O00206,Q9 | Toll-like receptor 4:Lymphocyte antigen 96 complex |
| rs550057 | pQTLs | T | 5.89E-18 | O00206,Q9 | Toll-like receptor 4:Lymphocyte antigen 96 complex |
| rs674302 | pQTLs | A | 4.57E-12 | O00206,Q9 | Toll-like receptor 4:Lymphocyte antigen 96 complex |
| rs554833 | pQTLs | T | 3.98E-12 | O00206,Q9 | Toll-like receptor 4:Lymphocyte antigen 96 complex |
| rs529565 | pQTLs | T | 5.62E-12 | O00206,Q9 | Toll-like receptor 4:Lymphocyte antigen 96 complex |
| rs532436 | pQTLs | A | 3.72E-23 | O00206,Q9 | Toll-like receptor 4:Lymphocyte antigen 96 complex |
| rs600038 | pQTLs | T | 5.89E-18 | O00206,Q9 | Toll-like receptor 4:Lymphocyte antigen 96 complex |
| rs1124406: | pQTLs | T | 1.48E-15 | O00206,Q9 | Toll-like receptor 4:Lymphocyte antigen 96 complex |
| rs649129 | pQTLs | T | 3.98E-18 | O00206,Q9 | Toll-like receptor 4:Lymphocyte antigen 96 complex |
| rs495828 | pQTLs | T | 3.72E-18 | O00206,Q9 | Toll-like receptor 4:Lymphocyte antigen 96 complex |
| rs687289 | pQTLs | A | 3.63E-14 | O60704 | Protein-tyrosine sulfotransferase 2 |
| rs2519093 | pQTLs | T | 8.13E-23 | O60704 | Protein-tyrosine sulfotransferase 2 |
| rs644234 | pQTLs | T | 1.55E-14 | O60704 | Protein-tyrosine sulfotransferase 2 |
| rs643434 | pQTLs | A | 1.23E-14 | O60704 | Protein-tyrosine sulfotransferase 2 |
| rs543040 | pQTLs | A | 1.23E-14 | O60704 | Protein-tyrosine sulfotransferase 2 |
| rs613534 | pQTLs | A | 1.48E-14 | O60704 | Protein-tyrosine sulfotransferase 2 |
| rs543968 | pQTLs | T | 2.24E-14 | O60704 | Protein-tyrosine sulfotransferase 2 |
| rs544873 | pQTLs | A | 9.33E-15 | O60704 | Protein-tyrosine sulfotransferase 2 |
| rs545971 | pQTLs | T | 3.98E-14 | O60704 | Protein-tyrosine sulfotransferase 2 |
| rs612169 | pQTLs | A | 2.24E-14 | O60704 | Protein-tyrosine sulfotransferase 2 |
| rs8176663 | pQTLs | T | 1.26E-14 | O60704 | Protein-tyrosine sulfotransferase 2 |
| rs491626 | pQTLs | T | 2.14E-14 | O60704 | Protein-tyrosine sulfotransferase 2 |
| rs492488 | pQTLs | A | 1.48E-14 | O60704 | Protein-tyrosine sulfotransferase 2 |

| | | | | | |
|---|---|---|---|---|---|
| rs493246 | pQTLs | A | 2.34E-14 | O60704 | Protein-tyrosine sulfotransferase 2 |
| rs494242 | pQTLs | T | 5.75E-15 | O60704 | Protein-tyrosine sulfotransferase 2 |
| rs495203 | pQTLs | T | 1.23E-14 | O60704 | Protein-tyrosine sulfotransferase 2 |
| rs582118 | pQTLs | A | 3.98E-14 | O60704 | Protein-tyrosine sulfotransferase 2 |
| rs582094 | pQTLs | A | 1.82E-14 | O60704 | Protein-tyrosine sulfotransferase 2 |
| rs2769071 | pQTLs | A | 9.55E-15 | O60704 | Protein-tyrosine sulfotransferase 2 |
| rs677355 | pQTLs | A | 1.23E-14 | O60704 | Protein-tyrosine sulfotransferase 2 |
| rs676996 | pQTLs | T | 3.02E-14 | O60704 | Protein-tyrosine sulfotransferase 2 |
| rs676457 | pQTLs | A | 3.98E-15 | O60704 | Protein-tyrosine sulfotransferase 2 |
| rs550057 | pQTLs | T | 2.14E-16 | O60704 | Protein-tyrosine sulfotransferase 2 |
| rs674302 | pQTLs | A | 1.45E-14 | O60704 | Protein-tyrosine sulfotransferase 2 |
| rs554833 | pQTLs | T | 4.90E-15 | O60704 | Protein-tyrosine sulfotransferase 2 |
| rs529565 | pQTLs | T | 1.48E-14 | O60704 | Protein-tyrosine sulfotransferase 2 |
| rs630510 | pQTLs | A | 6.17E-09 | O60704 | Protein-tyrosine sulfotransferase 2 |
| rs630014 | pQTLs | A | 4.57E-09 | O60704 | Protein-tyrosine sulfotransferase 2 |
| rs532436 | pQTLs | A | 2.45E-23 | O60704 | Protein-tyrosine sulfotransferase 2 |
| rs600038 | pQTLs | T | 3.16E-22 | O60704 | Protein-tyrosine sulfotransferase 2 |
| rs1124406 | pQTLs | T | 6.03E-13 | O60704 | Protein-tyrosine sulfotransferase 2 |
| rs649129 | pQTLs | T | 2.04E-21 | O60704 | Protein-tyrosine sulfotransferase 2 |
| rs495828 | pQTLs | T | 1.95E-22 | O60704 | Protein-tyrosine sulfotransferase 2 |
| rs649129 | pQTLs | T | 1.41E-08 | Q9NNX1 | Tuftelin |
| rs495828 | pQTLs | T | 4.37E-08 | Q9NNX1 | Tuftelin |
| rs7873635 | cardiovascu | t | 1.23E-187 | P16581 | E-selectin |
| rs7873635 | cardiovascu | t | 1.28E-08 | Q02763 | Angiopoietin-1 receptor |
| rs7873635 | cardiovascu | t | 3.45E-27 | P16284 | Platelet endothelial cell adhesion molecule |
| rs7873635 | cardiovascu | t | 1.31E-16 | P07204 | Thrombomodulin |
| rs4962040 | cardiovascu | a | 1.70E-08 | Q02763 | Angiopoietin-1 receptor |
| rs4962040 | cardiovascu | a | 3.87E-27 | P16284 | Platelet endothelial cell adhesion molecule |
| rs4962040 | cardiovascu | a | 1.15E-16 | P07204 | Thrombomodulin |
| rs4962040 | cardiovascu | a | 1.22E-188 | P16581 | E-selectin |
| rs8176702 | cardiovascu | a | 3.05E-189 | P16581 | E-selectin |
| rs8176702 | cardiovascu | a | 1.54E-08 | Q02763 | Angiopoietin-1 receptor |
| rs8176702 | cardiovascu | a | 1.53E-16 | P07204 | Thrombomodulin |
| rs8176702 | cardiovascu | a | 2.07E-27 | P16284 | Platelet endothelial cell adhesion molecule |
| rs2073826 | cardiovascu | t | 4.08E-17 | P07204 | Thrombomodulin |
| rs2073826 | cardiovascu | t | 2.53E-29 | P16284 | Platelet endothelial cell adhesion molecule |
| rs2073826 | cardiovascu | t | 1.62E-215 | P16581 | E-selectin |
| rs2073826 | cardiovascu | t | 3.45E-10 | Q02763 | Angiopoietin-1 receptor |
| rs687289 | cardiovascu | a | 0 | P16581 | E-selectin |
| rs687289 | cardiovascu | a | 2.85E-70 | P16284 | Platelet endothelial cell adhesion molecule |
| rs687289 | cardiovascu | a | 5.45E-11 | Q9GZV9 | Fibroblast growth factor 23 |
| rs687289 | cardiovascu | a | 2.04E-26 | Q02763 | Angiopoietin-1 receptor |
| rs687289 | cardiovascu | a | 2.66E-50 | P07204 | Thrombomodulin |
| rs2073827 | cardiovascu | c | 4.23E-10 | Q02763 | Angiopoietin-1 receptor |
| rs2073827 | cardiovascu | c | 1.34E-215 | P16581 | E-selectin |
| rs2073827 | cardiovascu | c | 4.78E-17 | P07204 | Thrombomodulin |
| rs2073827 | cardiovascu | c | 2.93E-29 | P16284 | Platelet endothelial cell adhesion molecule |
| rs2073828 | cardiovascu | a | 4.30E-10 | Q02763 | Angiopoietin-1 receptor |
| rs2073828 | cardiovascu | a | 2.63E-29 | P16284 | Platelet endothelial cell adhesion molecule |
| rs2073828 | cardiovascu | a | 4.85E-17 | P07204 | Thrombomodulin |

| | | | | | |
|---|---|---|---|---|---|
| rs2073828 | cardiovascı | a | 9.98E-216 | P16581 | E-selectin |
| rs8176691 | cardiovascı | t | 5.04E-190 | P16581 | E-selectin |
| rs8176691 | cardiovascı | t | 2.60E-27 | P16284 | Platelet endothelial cell adhesion molecule |
| rs8176691 | cardiovascı | t | 1.52E-08 | Q02763 | Angiopoietin-1 receptor |
| rs8176691 | cardiovascı | t | 6.28E-17 | P07204 | Thrombomodulin |
| rs8176690 | cardiovascı | a | 3.58E-10 | Q02763 | Angiopoietin-1 receptor |
| rs8176690 | cardiovascı | a | 4.96E-29 | P16284 | Platelet endothelial cell adhesion molecule |
| rs8176690 | cardiovascı | a | 5.60E-216 | P16581 | E-selectin |
| rs8176690 | cardiovascı | a | 1.17E-17 | P07204 | Thrombomodulin |
| rs8176682 | cardiovascı | t | 1.50E-26 | P16284 | Platelet endothelial cell adhesion molecule |
| rs8176682 | cardiovascı | t | 5.87E-17 | P07204 | Thrombomodulin |
| rs8176682 | cardiovascı | t | 2.03E-09 | Q02763 | Angiopoietin-1 receptor |
| rs8176682 | cardiovascı | t | 1.68E-196 | P16581 | E-selectin |
| rs8176681 | cardiovascı | t | 1.44E-17 | P07204 | Thrombomodulin |
| rs8176681 | cardiovascı | t | 2.93E-10 | Q02763 | Angiopoietin-1 receptor |
| rs8176681 | cardiovascı | t | 3.33E-216 | P16581 | E-selectin |
| rs8176681 | cardiovascı | t | 1.04E-28 | P16284 | Platelet endothelial cell adhesion molecule |
| rs2519093 | cardiovascı | t | 4.67E-09 | Q9GZV9 | Fibroblast growth factor 23 |
| rs2519093 | cardiovascı | t | 8.72E-09 | P78423 | Fractalkine |
| rs2519093 | cardiovascı | t | 1.01E-28 | Q02763 | Angiopoietin-1 receptor |
| rs2519093 | cardiovascı | t | 8.31E-57 | P13726 | Tissue factor |
| rs2519093 | cardiovascı | t | 2.39E-15 | P07204 | Thrombomodulin |
| rs2519093 | cardiovascı | t | 0 | P16284 | Platelet endothelial cell adhesion molecule |
| rs2519093 | cardiovascı | t | 0 | P16581 | E-selectin |
| rs2519093 | cardiovascı | t | 2.82E-10 | Q01638 | ST2 protein |
| rs644234 | cardiovascı | t | 4.44E-22 | Q02763 | Angiopoietin-1 receptor |
| rs644234 | cardiovascı | t | 0 | P16581 | E-selectin |
| rs644234 | cardiovascı | t | 4.80E-63 | P16284 | Platelet endothelial cell adhesion molecule |
| rs644234 | cardiovascı | t | 1.08E-48 | P07204 | Thrombomodulin |
| rs644234 | cardiovascı | t | 1.53E-10 | Q9GZV9 | Fibroblast growth factor 23 |
| rs643434 | cardiovascı | a | 3.36E-22 | Q02763 | Angiopoietin-1 receptor |
| rs643434 | cardiovascı | a | 2.13E-10 | Q9GZV9 | Fibroblast growth factor 23 |
| rs643434 | cardiovascı | a | 3.47E-63 | P16284 | Platelet endothelial cell adhesion molecule |
| rs643434 | cardiovascı | a | 1.48E-48 | P07204 | Thrombomodulin |
| rs643434 | cardiovascı | a | 0 | P16581 | E-selectin |
| rs543040 | cardiovascı | a | 0 | P16581 | E-selectin |
| rs543040 | cardiovascı | a | 1.55E-48 | P07204 | Thrombomodulin |
| rs543040 | cardiovascı | a | 9.90E-20 | Q02763 | Angiopoietin-1 receptor |
| rs543040 | cardiovascı | a | 2.30E-51 | P16284 | Platelet endothelial cell adhesion molecule |
| rs543040 | cardiovascı | a | 7.95E-09 | Q9GZV9 | Fibroblast growth factor 23 |
| rs613534 | cardiovascı | a | 6.09E-23 | Q02763 | Angiopoietin-1 receptor |
| rs613534 | cardiovascı | a | 0 | P16581 | E-selectin |
| rs613534 | cardiovascı | a | 3.03E-62 | P16284 | Platelet endothelial cell adhesion molecule |
| rs613534 | cardiovascı | a | 1.28E-48 | P07204 | Thrombomodulin |
| rs613534 | cardiovascı | a | 1.62E-10 | Q9GZV9 | Fibroblast growth factor 23 |
| rs543968 | cardiovascı | t | 1.28E-10 | Q9GZV9 | Fibroblast growth factor 23 |
| rs543968 | cardiovascı | t | 1.55E-48 | P07204 | Thrombomodulin |
| rs543968 | cardiovascı | t | 0 | P16581 | E-selectin |
| rs543968 | cardiovascı | t | 9.70E-23 | Q02763 | Angiopoietin-1 receptor |
| rs543968 | cardiovascı | t | 3.50E-62 | P16284 | Platelet endothelial cell adhesion molecule |

| | | | | | |
|---|---|---|---|---|---|
| rs544873 | cardiovascı | a | 2.23E-48 | P07204 | Thrombomodulin |
| rs544873 | cardiovascı | a | 1.81E-10 | Q9GZV9 | Fibroblast growth factor 23 |
| rs544873 | cardiovascı | a | 2.87E-63 | P16284 | Platelet endothelial cell adhesion molecule |
| rs544873 | cardiovascı | a | 0 | P16581 | E-selectin |
| rs544873 | cardiovascı | a | 2.63E-22 | Q02763 | Angiopoietin-1 receptor |
| rs545971 | cardiovascı | t | 8.13E-51 | P07204 | Thrombomodulin |
| rs545971 | cardiovascı | t | 1.07E-67 | P16284 | Platelet endothelial cell adhesion molecule |
| rs545971 | cardiovascı | t | 0 | P16581 | E-selectin |
| rs545971 | cardiovascı | t | 1.16E-25 | Q02763 | Angiopoietin-1 receptor |
| rs545971 | cardiovascı | t | 6.71E-11 | Q9GZV9 | Fibroblast growth factor 23 |
| rs612169 | cardiovascı | a | 0 | P16581 | E-selectin |
| rs612169 | cardiovascı | a | 9.43E-11 | Q9GZV9 | Fibroblast growth factor 23 |
| rs612169 | cardiovascı | a | 1.09E-67 | P16284 | Platelet endothelial cell adhesion molecule |
| rs612169 | cardiovascı | a | 4.57E-51 | P07204 | Thrombomodulin |
| rs612169 | cardiovascı | a | 8.79E-26 | Q02763 | Angiopoietin-1 receptor |
| rs8176668 | cardiovascı | a | 1.99E-190 | P16581 | E-selectin |
| rs8176668 | cardiovascı | a | 3.40E-17 | P07204 | Thrombomodulin |
| rs8176668 | cardiovascı | a | 1.67E-08 | Q02763 | Angiopoietin-1 receptor |
| rs8176668 | cardiovascı | a | 2.96E-27 | P16284 | Platelet endothelial cell adhesion molecule |
| rs8176663 | cardiovascı | t | 7.53E-51 | P07204 | Thrombomodulin |
| rs8176663 | cardiovascı | t | 7.37E-26 | Q02763 | Angiopoietin-1 receptor |
| rs8176663 | cardiovascı | t | 0 | P16581 | E-selectin |
| rs8176663 | cardiovascı | t | 8.93E-11 | Q9GZV9 | Fibroblast growth factor 23 |
| rs8176663 | cardiovascı | t | 3.05E-68 | P16284 | Platelet endothelial cell adhesion molecule |
| rs7036642 | cardiovascı | a | 4.10E-189 | P16581 | E-selectin |
| rs7036642 | cardiovascı | a | 1.45E-08 | Q02763 | Angiopoietin-1 receptor |
| rs7036642 | cardiovascı | a | 3.18E-27 | P16284 | Platelet endothelial cell adhesion molecule |
| rs7036642 | cardiovascı | a | 1.64E-17 | P07204 | Thrombomodulin |
| rs491626 | cardiovascı | t | 1.09E-25 | Q02763 | Angiopoietin-1 receptor |
| rs491626 | cardiovascı | t | 4.41E-68 | P16284 | Platelet endothelial cell adhesion molecule |
| rs491626 | cardiovascı | t | 0 | P16581 | E-selectin |
| rs491626 | cardiovascı | t | 8.45E-51 | P07204 | Thrombomodulin |
| rs491626 | cardiovascı | t | 9.42E-11 | Q9GZV9 | Fibroblast growth factor 23 |
| rs492488 | cardiovascı | a | 8.91E-51 | P07204 | Thrombomodulin |
| rs492488 | cardiovascı | a | 1.32E-25 | Q02763 | Angiopoietin-1 receptor |
| rs492488 | cardiovascı | a | 0 | P16581 | E-selectin |
| rs492488 | cardiovascı | a | 8.91E-11 | Q9GZV9 | Fibroblast growth factor 23 |
| rs492488 | cardiovascı | a | 3.08E-68 | P16284 | Platelet endothelial cell adhesion molecule |
| rs493246 | cardiovascı | a | 1.18E-25 | Q02763 | Angiopoietin-1 receptor |
| rs493246 | cardiovascı | a | 0 | P16581 | E-selectin |
| rs493246 | cardiovascı | a | 5.33E-68 | P16284 | Platelet endothelial cell adhesion molecule |
| rs493246 | cardiovascı | a | 1.09E-50 | P07204 | Thrombomodulin |
| rs493246 | cardiovascı | a | 9.57E-11 | Q9GZV9 | Fibroblast growth factor 23 |
| rs494242 | cardiovascı | t | 2.25E-63 | P16284 | Platelet endothelial cell adhesion molecule |
| rs494242 | cardiovascı | t | 2.43E-22 | Q02763 | Angiopoietin-1 receptor |
| rs494242 | cardiovascı | t | 2.04E-10 | Q9GZV9 | Fibroblast growth factor 23 |
| rs494242 | cardiovascı | t | 0 | P16581 | E-selectin |
| rs494242 | cardiovascı | t | 2.07E-48 | P07204 | Thrombomodulin |
| rs495203 | cardiovascı | t | 3.21E-68 | P16284 | Platelet endothelial cell adhesion molecule |
| rs495203 | cardiovascı | t | 0 | P16581 | E-selectin |

| | | | | | |
|---|---|---|---|---|---|
| rs495203 | cardiovascı | t | 9.09E-26 | Q02763 | Angiopoietin-1 receptor |
| rs495203 | cardiovascı | t | 8.70E-11 | Q9GZV9 | Fibroblast growth factor 23 |
| rs495203 | cardiovascı | t | 6.72E-51 | P07204 | Thrombomodulin |
| rs582118 | cardiovascı | a | 2.18E-68 | P16284 | Platelet endothelial cell adhesion molecule |
| rs582118 | cardiovascı | a | 1.39E-10 | Q9GZV9 | Fibroblast growth factor 23 |
| rs582118 | cardiovascı | a | 9.30E-26 | Q02763 | Angiopoietin-1 receptor |
| rs582118 | cardiovascı | a | 6.32E-51 | P07204 | Thrombomodulin |
| rs582118 | cardiovascı | a | 0 | P16581 | E-selectin |
| rs582094 | cardiovascı | a | 2.18E-68 | P16284 | Platelet endothelial cell adhesion molecule |
| rs582094 | cardiovascı | a | 2.09E-50 | P07204 | Thrombomodulin |
| rs582094 | cardiovascı | a | 7.58E-26 | Q02763 | Angiopoietin-1 receptor |
| rs582094 | cardiovascı | a | 0 | P16581 | E-selectin |
| rs582094 | cardiovascı | a | 8.86E-11 | Q9GZV9 | Fibroblast growth factor 23 |
| rs2769071 | cardiovascı | a | 4.00E-67 | P16284 | Platelet endothelial cell adhesion molecule |
| rs2769071 | cardiovascı | a | 6.41E-26 | Q02763 | Angiopoietin-1 receptor |
| rs2769071 | cardiovascı | a | 1.34E-10 | Q9GZV9 | Fibroblast growth factor 23 |
| rs2769071 | cardiovascı | a | 0 | P16581 | E-selectin |
| rs2769071 | cardiovascı | a | 4.69E-51 | P07204 | Thrombomodulin |
| rs677355 | cardiovascı | a | 1.71E-28 | Q02763 | Angiopoietin-1 receptor |
| rs677355 | cardiovascı | a | 2.35E-63 | P16284 | Platelet endothelial cell adhesion molecule |
| rs677355 | cardiovascı | a | 1.86E-10 | Q9GZV9 | Fibroblast growth factor 23 |
| rs677355 | cardiovascı | a | 3.06E-50 | P07204 | Thrombomodulin |
| rs677355 | cardiovascı | a | 0 | P16581 | E-selectin |
| rs676996 | cardiovascı | t | 3.20E-49 | P07204 | Thrombomodulin |
| rs676996 | cardiovascı | t | 0 | P16581 | E-selectin |
| rs676996 | cardiovascı | t | 2.04E-10 | Q9GZV9 | Fibroblast growth factor 23 |
| rs676996 | cardiovascı | t | 1.85E-28 | Q02763 | Angiopoietin-1 receptor |
| rs676996 | cardiovascı | t | 9.36E-63 | P16284 | Platelet endothelial cell adhesion molecule |
| rs676457 | cardiovascı | a | 2.20E-68 | P16284 | Platelet endothelial cell adhesion molecule |
| rs676457 | cardiovascı | a | 1.95E-10 | Q9GZV9 | Fibroblast growth factor 23 |
| rs676457 | cardiovascı | a | 2.16E-25 | Q02763 | Angiopoietin-1 receptor |
| rs676457 | cardiovascı | a | 1.92E-50 | P07204 | Thrombomodulin |
| rs676457 | cardiovascı | a | 0 | P16581 | E-selectin |
| rs550057 | cardiovascı | t | 1.16E-09 | Q9GZV9 | Fibroblast growth factor 23 |
| rs550057 | cardiovascı | t | 2.16E-252 | P16284 | Platelet endothelial cell adhesion molecule |
| rs550057 | cardiovascı | t | 2.49E-13 | P07204 | Thrombomodulin |
| rs550057 | cardiovascı | t | 0 | P16581 | E-selectin |
| rs550057 | cardiovascı | t | 8.56E-26 | Q02763 | Angiopoietin-1 receptor |
| rs550057 | cardiovascı | t | 2.75E-41 | P13726 | Tissue factor |
| rs674302 | cardiovascı | a | 0 | P16581 | E-selectin |
| rs674302 | cardiovascı | a | 8.79E-51 | P07204 | Thrombomodulin |
| rs674302 | cardiovascı | a | 7.81E-26 | Q02763 | Angiopoietin-1 receptor |
| rs674302 | cardiovascı | a | 1.01E-10 | Q9GZV9 | Fibroblast growth factor 23 |
| rs674302 | cardiovascı | a | 6.65E-68 | P16284 | Platelet endothelial cell adhesion molecule |
| rs7046674 | cardiovascı | t | 1.51E-27 | P16284 | Platelet endothelial cell adhesion molecule |
| rs7046674 | cardiovascı | t | 2.10E-189 | P16581 | E-selectin |
| rs7046674 | cardiovascı | t | 3.60E-17 | P07204 | Thrombomodulin |
| rs7046674 | cardiovascı | t | 1.67E-08 | Q02763 | Angiopoietin-1 receptor |
| rs554833 | cardiovascı | t | 1.16E-10 | Q9GZV9 | Fibroblast growth factor 23 |
| rs554833 | cardiovascı | t | 2.41E-68 | P16284 | Platelet endothelial cell adhesion molecule |

| | | | | | |
|---|---|---|---|---|---|
| rs554833 | cardiovascı | t | 1.09E-50 | P07204 | Thrombomodulin |
| rs554833 | cardiovascı | t | 1.03E-25 | Q02763 | Angiopoietin-1 receptor |
| rs554833 | cardiovascı | t | 0 | P16581 | E-selectin |
| rs8176649 | cardiovascı | a | 1.02E-27 | P16284 | Platelet endothelial cell adhesion molecule |
| rs8176649 | cardiovascı | a | 1.73E-08 | Q02763 | Angiopoietin-1 receptor |
| rs8176649 | cardiovascı | a | 7.83E-190 | P16581 | E-selectin |
| rs8176649 | cardiovascı | a | 3.46E-17 | P07204 | Thrombomodulin |
| rs660340 | cardiovascı | a | 1.29E-14 | Q02763 | Angiopoietin-1 receptor |
| rs660340 | cardiovascı | a | 3.43E-31 | P16284 | Platelet endothelial cell adhesion molecule |
| rs660340 | cardiovascı | a | 4.10E-23 | P07204 | Thrombomodulin |
| rs660340 | cardiovascı | a | 5.66E-271 | P16581 | E-selectin |
| rs581107 | cardiovascı | t | 3.90E-31 | P16284 | Platelet endothelial cell adhesion molecule |
| rs581107 | cardiovascı | t | 2.90E-23 | P07204 | Thrombomodulin |
| rs581107 | cardiovascı | t | 2.38E-271 | P16581 | E-selectin |
| rs581107 | cardiovascı | t | 6.99E-15 | Q02763 | Angiopoietin-1 receptor |
| rs659104 | cardiovascı | t | 3.11E-23 | P07204 | Thrombomodulin |
| rs659104 | cardiovascı | t | 8.27E-15 | Q02763 | Angiopoietin-1 receptor |
| rs659104 | cardiovascı | t | 1.13E-271 | P16581 | E-selectin |
| rs659104 | cardiovascı | t | 2.27E-31 | P16284 | Platelet endothelial cell adhesion molecule |
| rs647800 | cardiovascı | a | 2.51E-22 | P07204 | Thrombomodulin |
| rs647800 | cardiovascı | a | 1.11E-29 | P16284 | Platelet endothelial cell adhesion molecule |
| rs647800 | cardiovascı | a | 9.06E-13 | Q02763 | Angiopoietin-1 receptor |
| rs647800 | cardiovascı | a | 3.12E-242 | P16581 | E-selectin |
| rs473533 | cardiovascı | t | 5.34E-23 | P07204 | Thrombomodulin |
| rs473533 | cardiovascı | t | 2.16E-31 | P16284 | Platelet endothelial cell adhesion molecule |
| rs473533 | cardiovascı | t | 8.60E-15 | Q02763 | Angiopoietin-1 receptor |
| rs473533 | cardiovascı | t | 9.55E-272 | P16581 | E-selectin |
| rs476410 | cardiovascı | c | 3.21E-31 | P16284 | Platelet endothelial cell adhesion molecule |
| rs476410 | cardiovascı | c | 8.91E-15 | Q02763 | Angiopoietin-1 receptor |
| rs476410 | cardiovascı | c | 1.03E-271 | P16581 | E-selectin |
| rs476410 | cardiovascı | c | 4.82E-23 | P07204 | Thrombomodulin |
| rs645982 | cardiovascı | a | 3.85E-23 | P07204 | Thrombomodulin |
| rs645982 | cardiovascı | a | 3.97E-31 | P16284 | Platelet endothelial cell adhesion molecule |
| rs645982 | cardiovascı | a | 1.56E-271 | P16581 | E-selectin |
| rs645982 | cardiovascı | a | 8.93E-15 | Q02763 | Angiopoietin-1 receptor |
| rs1165522⁴ | cardiovascı | a | 2.10E-20 | Q02763 | Angiopoietin-1 receptor |
| rs1165522⁴ | cardiovascı | a | 0 | P16581 | E-selectin |
| rs1165522⁴ | cardiovascı | a | 3.98E-10 | Q9GZV9 | Fibroblast growth factor 23 |
| rs1165522⁴ | cardiovascı | a | 6.06E-64 | P16284 | Platelet endothelial cell adhesion molecule |
| rs1165522⁴ | cardiovascı | a | 7.66E-49 | P07204 | Thrombomodulin |
| rs529565 | cardiovascı | t | 1.19E-25 | Q02763 | Angiopoietin-1 receptor |
| rs529565 | cardiovascı | t | 7.52E-11 | Q9GZV9 | Fibroblast growth factor 23 |
| rs529565 | cardiovascı | t | 7.20E-51 | P07204 | Thrombomodulin |
| rs529565 | cardiovascı | t | 5.18E-69 | P16284 | Platelet endothelial cell adhesion molecule |
| rs529565 | cardiovascı | t | 0 | P16581 | E-selectin |
| rs630510 | cardiovascı | a | 4.38E-29 | P16284 | Platelet endothelial cell adhesion molecule |
| rs630510 | cardiovascı | a | 2.57E-28 | P07204 | Thrombomodulin |
| rs630510 | cardiovascı | a | 5.60E-13 | Q02763 | Angiopoietin-1 receptor |
| rs630510 | cardiovascı | a | 1.18E-301 | P16581 | E-selectin |
| rs630014 | cardiovascı | a | 1.52E-28 | P16284 | Platelet endothelial cell adhesion molecule |

| | | | | |
|---|---|---|---|---|
| rs630014 | cardiovascu a | 3.13E-13 | Q02763 | Angiopoietin-1 receptor |
| rs630014 | cardiovascu a | 5.31E-300 | P16581 | E-selectin |
| rs630014 | cardiovascu a | 8.01E-29 | P07204 | Thrombomodulin |
| rs532436 | cardiovascu a | 4.85E-28 | Q02763 | Angiopoietin-1 receptor |
| rs532436 | cardiovascu a | 2.77E-56 | P13726 | Tissue factor |
| rs532436 | cardiovascu a | 2.93E-09 | Q9GZV9 | Fibroblast growth factor 23 |
| rs532436 | cardiovascu a | 0 | P16284 | Platelet endothelial cell adhesion molecule |
| rs532436 | cardiovascu a | 3.63E-16 | P07204 | Thrombomodulin |
| rs532436 | cardiovascu a | 2.30E-10 | Q01638 | ST2 protein |
| rs532436 | cardiovascu a | 0 | P16581 | E-selectin |
| rs532436 | cardiovascu a | 1.57E-08 | P78423 | Fractalkine |
| rs600038 | cardiovascu t | 6.85E-09 | P21583 | Stem cell factor |
| rs600038 | cardiovascu t | 0 | P16581 | E-selectin |
| rs600038 | cardiovascu t | 2.91E-08 | P78423 | Fractalkine |
| rs600038 | cardiovascu t | 2.71E-08 | Q9GZV9 | Fibroblast growth factor 23 |
| rs600038 | cardiovascu t | 1.81E-30 | Q02763 | Angiopoietin-1 receptor |
| rs600038 | cardiovascu t | 9.28E-14 | P07204 | Thrombomodulin |
| rs600038 | cardiovascu t | 6.05E-53 | P13726 | Tissue factor |
| rs600038 | cardiovascu t | 0 | P16284 | Platelet endothelial cell adhesion molecule |
| rs600038 | cardiovascu t | 8.61E-10 | Q01638 | ST2 protein |
| rs1124406: | cardiovascu t | 0 | P16581 | E-selectin |
| rs1124406: | cardiovascu t | 3.94E-10 | P07204 | Thrombomodulin |
| rs1124406: | cardiovascu t | 7.62E-30 | P13726 | Tissue factor |
| rs1124406: | cardiovascu t | 3.24E-17 | Q02763 | Angiopoietin-1 receptor |
| rs1124406: | cardiovascu t | 4.32E-197 | P16284 | Platelet endothelial cell adhesion molecule |
| rs649129 | cardiovascu t | 1.31E-52 | P13726 | Tissue factor |
| rs649129 | cardiovascu t | 6.59E-31 | Q02763 | Angiopoietin-1 receptor |
| rs649129 | cardiovascu t | 0 | P16284 | Platelet endothelial cell adhesion molecule |
| rs649129 | cardiovascu t | 3.74E-13 | P07204 | Thrombomodulin |
| rs649129 | cardiovascu t | 4.02E-08 | P78423 | Fractalkine |
| rs649129 | cardiovascu t | 0 | P16581 | E-selectin |
| rs649129 | cardiovascu t | 1.17E-08 | P21583 | Stem cell factor |
| rs649129 | cardiovascu t | 3.76E-08 | Q9GZV9 | Fibroblast growth factor 23 |
| rs649129 | cardiovascu t | 7.34E-10 | Q01638 | ST2 protein |
| rs495828 | cardiovascu t | 0 | P16284 | Platelet endothelial cell adhesion molecule |
| rs495828 | cardiovascu t | 5.84E-09 | P21583 | Stem cell factor |
| rs495828 | cardiovascu t | 4.90E-52 | P13726 | Tissue factor |
| rs495828 | cardiovascu t | 0 | P16581 | E-selectin |
| rs495828 | cardiovascu t | 1.88E-09 | Q01638 | ST2 protein |
| rs495828 | cardiovascu t | 1.13E-13 | P07204 | Thrombomodulin |
| rs495828 | cardiovascu t | 3.31E-08 | P78423 | Fractalkine |
| rs495828 | cardiovascu t | 7.41E-31 | Q02763 | Angiopoietin-1 receptor |
| rs495828 | metabolite t | 1.46E-08 | M32740 | X-11423 |
| rs649129 | metabolite t | 3.02E-09 | M32740 | X-11423 |
| rs674302 | metabolite a | 3.56E-13 | M33801 | ADpSGEGDFXAEGGGVR* |
| rs643434 | metabolite a | 1.45E-13 | M33801 | ADpSGEGDFXAEGGGVR* |
| rs644234 | metabolite t | 1.43E-13 | M33801 | ADpSGEGDFXAEGGGVR* |
| rs529565 | metabolite t | 5.41E-13 | M33801 | ADpSGEGDFXAEGGGVR* |
| rs630014 | metabolite a | 3.53E-09 | M33801 | ADpSGEGDFXAEGGGVR* |
| rs495828 | metabolite t | 2.72E-18 | M33801 | ADpSGEGDFXAEGGGVR* |

| | | | | |
|---|---|---|---|---|
| rs612169 | metabolite a | 9.66E-14 | M33801 | ADpSGEGDFXAEGGGVR* |
| rs545971 | metabolite t | 3.33E-13 | M33801 | ADpSGEGDFXAEGGGVR* |
| rs687289 | metabolite a | 2.30E-13 | M33801 | ADpSGEGDFXAEGGGVR* |
| rs649129 | metabolite t | 2.09E-19 | M33801 | ADpSGEGDFXAEGGGVR* |
| rs495828 | metabolite t | 2.26E-09 | M36115 | X-14189 |
| rs649129 | metabolite t | 4.32E-09 | M36115 | X-14189 |
| rs495828 | metabolite t | 1.98E-08 | M36230 | X-14304 |
| rs649129 | metabolite t | 1.26E-08 | M36230 | X-14304 |
| rs2519093 | metabolite T | 9.10E-09 | ApoB | ApoB |
| rs550057 | metabolite T | 3.76E-08 | ApoB | ApoB |
| rs532436 | metabolite A | 1.22E-08 | ApoB | ApoB |
| rs600038 | metabolite C | 4.20E-08 | ApoB | ApoB |
| rs649129 | metabolite T | 3.23E-08 | ApoB | ApoB |
| rs495828 | metabolite T | 2.60E-08 | ApoB | ApoB |
| rs2519093 | metabolite T | 5.12E-10 | IDL.C | Total cholesterol in IDL |
| rs550057 | metabolite T | 9.72E-09 | IDL.C | Total cholesterol in IDL |
| rs532436 | metabolite A | 2.79E-10 | IDL.C | Total cholesterol in IDL |
| rs600038 | metabolite C | 6.84E-10 | IDL.C | Total cholesterol in IDL |
| rs649129 | metabolite T | 6.08E-10 | IDL.C | Total cholesterol in IDL |
| rs495828 | metabolite T | 4.46E-10 | IDL.C | Total cholesterol in IDL |
| rs2519093 | metabolite T | 2.47E-09 | IDL.FC | Free cholesterol in IDL |
| rs532436 | metabolite A | 1.01E-09 | IDL.FC | Free cholesterol in IDL |
| rs600038 | metabolite C | 5.42E-09 | IDL.FC | Free cholesterol in IDL |
| rs649129 | metabolite T | 4.36E-09 | IDL.FC | Free cholesterol in IDL |
| rs495828 | metabolite T | 2.84E-09 | IDL.FC | Free cholesterol in IDL |
| rs2519093 | metabolite T | 1.92E-10 | IDL.L | Total lipids in IDL |
| rs550057 | metabolite T | 2.46E-09 | IDL.L | Total lipids in IDL |
| rs532436 | metabolite A | 8.77E-11 | IDL.L | Total lipids in IDL |
| rs600038 | metabolite C | 3.93E-10 | IDL.L | Total lipids in IDL |
| rs649129 | metabolite T | 3.15E-10 | IDL.L | Total lipids in IDL |
| rs495828 | metabolite T | 2.51E-10 | IDL.L | Total lipids in IDL |
| rs2519093 | metabolite T | 2.87E-10 | IDL.P | Concentration of IDL particles |
| rs550057 | metabolite T | 2.74E-09 | IDL.P | Concentration of IDL particles |
| rs532436 | metabolite A | 1.21E-10 | IDL.P | Concentration of IDL particles |
| rs600038 | metabolite C | 7.10E-10 | IDL.P | Concentration of IDL particles |
| rs649129 | metabolite T | 5.54E-10 | IDL.P | Concentration of IDL particles |
| rs495828 | metabolite T | 4.50E-10 | IDL.P | Concentration of IDL particles |
| rs2519093 | metabolite T | 2.78E-09 | IDL.PL | Phospholipids in IDL |
| rs550057 | metabolite T | 2.51E-08 | IDL.PL | Phospholipids in IDL |
| rs532436 | metabolite A | 1.23E-09 | IDL.PL | Phospholipids in IDL |
| rs600038 | metabolite C | 1.08E-08 | IDL.PL | Phospholipids in IDL |
| rs649129 | metabolite T | 7.90E-09 | IDL.PL | Phospholipids in IDL |
| rs495828 | metabolite T | 5.14E-09 | IDL.PL | Phospholipids in IDL |
| rs2519093 | metabolite T | 2.70E-09 | L.LDL.C | Total cholesterol in large LDL |
| rs550057 | metabolite T | 4.00E-08 | L.LDL.C | Total cholesterol in large LDL |
| rs532436 | metabolite A | 2.27E-09 | L.LDL.C | Total cholesterol in large LDL |
| rs600038 | metabolite C | 3.44E-09 | L.LDL.C | Total cholesterol in large LDL |
| rs649129 | metabolite T | 3.02E-09 | L.LDL.C | Total cholesterol in large LDL |
| rs495828 | metabolite T | 1.61E-09 | L.LDL.C | Total cholesterol in large LDL |
| rs2519093 | metabolite T | 1.72E-08 | L.LDL.CE | Cholesterol esters in large LDL |

| | | | | |
|---|---|---|---|---|
| rs532436 | metabolite A | 1.48E-08 | L.LDL.CE | Cholesterol esters in large LDL |
| rs600038 | metabolite C | 3.57E-08 | L.LDL.CE | Cholesterol esters in large LDL |
| rs649129 | metabolite T | 3.31E-08 | L.LDL.CE | Cholesterol esters in large LDL |
| rs495828 | metabolite T | 1.80E-08 | L.LDL.CE | Cholesterol esters in large LDL |
| rs2519093 | metabolite T | 1.91E-09 | L.LDL.FC | Free cholesterol in large LDL |
| rs550057 | metabolite T | 1.82E-08 | L.LDL.FC | Free cholesterol in large LDL |
| rs532436 | metabolite A | 1.14E-09 | L.LDL.FC | Free cholesterol in large LDL |
| rs600038 | metabolite C | 1.12E-09 | L.LDL.FC | Free cholesterol in large LDL |
| rs649129 | metabolite T | 8.80E-10 | L.LDL.FC | Free cholesterol in large LDL |
| rs495828 | metabolite T | 4.67E-10 | L.LDL.FC | Free cholesterol in large LDL |
| rs2519093 | metabolite T | 7.67E-09 | L.LDL.L | Total lipids in large LDL |
| rs532436 | metabolite A | 5.39E-09 | L.LDL.L | Total lipids in large LDL |
| rs600038 | metabolite C | 1.52E-08 | L.LDL.L | Total lipids in large LDL |
| rs649129 | metabolite T | 1.28E-08 | L.LDL.L | Total lipids in large LDL |
| rs495828 | metabolite T | 7.09E-09 | L.LDL.L | Total lipids in large LDL |
| rs2519093 | metabolite T | 5.84E-09 | L.LDL.P | Concentration of large LDL particles |
| rs550057 | metabolite T | 4.88E-08 | L.LDL.P | Concentration of large LDL particles |
| rs532436 | metabolite A | 3.91E-09 | L.LDL.P | Concentration of large LDL particles |
| rs600038 | metabolite C | 1.22E-08 | L.LDL.P | Concentration of large LDL particles |
| rs649129 | metabolite T | 9.99E-09 | L.LDL.P | Concentration of large LDL particles |
| rs495828 | metabolite T | 5.88E-09 | L.LDL.P | Concentration of large LDL particles |
| rs2519093 | metabolite T | 9.21E-10 | L.LDL.PL | Phospholipids in large LDL |
| rs550057 | metabolite T | 6.16E-09 | L.LDL.PL | Phospholipids in large LDL |
| rs532436 | metabolite A | 6.03E-10 | L.LDL.PL | Phospholipids in large LDL |
| rs600038 | metabolite C | 1.30E-09 | L.LDL.PL | Phospholipids in large LDL |
| rs649129 | metabolite T | 9.88E-10 | L.LDL.PL | Phospholipids in large LDL |
| rs495828 | metabolite T | 5.63E-10 | L.LDL.PL | Phospholipids in large LDL |
| rs2519093 | metabolite T | 4.57E-08 | LDL.C | Total cholesterol in LDL |
| rs532436 | metabolite A | 3.79E-08 | LDL.C | Total cholesterol in LDL |
| rs495828 | metabolite T | 3.24E-08 | LDL.C | Total cholesterol in LDL |
| rs2519093 | metabolite T | 1.73E-09 | XS.VLDL.L | Total lipids in very small VLDL |
| rs550057 | metabolite T | 3.00E-09 | XS.VLDL.L | Total lipids in very small VLDL |
| rs532436 | metabolite A | 7.07E-10 | XS.VLDL.L | Total lipids in very small VLDL |
| rs600038 | metabolite C | 4.45E-09 | XS.VLDL.L | Total lipids in very small VLDL |
| rs649129 | metabolite T | 4.24E-09 | XS.VLDL.L | Total lipids in very small VLDL |
| rs495828 | metabolite T | 5.15E-09 | XS.VLDL.L | Total lipids in very small VLDL |
| rs2519093 | metabolite T | 1.16E-08 | XS.VLDL.P | Concentration of very small VLDL particles |
| rs550057 | metabolite T | 1.22E-08 | XS.VLDL.P | Concentration of very small VLDL particles |
| rs532436 | metabolite A | 4.92E-09 | XS.VLDL.P | Concentration of very small VLDL particles |
| rs600038 | metabolite C | 3.82E-08 | XS.VLDL.P | Concentration of very small VLDL particles |
| rs649129 | metabolite T | 3.59E-08 | XS.VLDL.P | Concentration of very small VLDL particles |
| rs495828 | metabolite T | 4.11E-08 | XS.VLDL.P | Concentration of very small VLDL particles |
| rs2519093 | metabolite T | 8.39E-10 | XS.VLDL.PL | Phospholipids in very small VLDL |
| rs550057 | metabolite T | 6.89E-09 | XS.VLDL.PL | Phospholipids in very small VLDL |
| rs532436 | metabolite A | 2.59E-10 | XS.VLDL.PL | Phospholipids in very small VLDL |
| rs600038 | metabolite C | 3.53E-09 | XS.VLDL.PL | Phospholipids in very small VLDL |
| rs649129 | metabolite T | 3.14E-09 | XS.VLDL.PL | Phospholipids in very small VLDL |
| rs495828 | metabolite T | 3.03E-09 | XS.VLDL.PL | Phospholipids in very small VLDL |
| rs687289 | metabolite A | 2.90E-14 | ApoB | ApoB |
| rs2519093 | metabolite T | 1.10E-26 | ApoB | ApoB |

| | | | | |
|---|---|---|---|---|
| rs644234 | metabolite G | 3.40E-14 | ApoB | ApoB |
| rs643434 | metabolite A | 3.10E-14 | ApoB | ApoB |
| rs543040 | metabolite T | 2.10E-14 | ApoB | ApoB |
| rs613534 | metabolite G | 3.40E-14 | ApoB | ApoB |
| rs543968 | metabolite C | 3.40E-14 | ApoB | ApoB |
| rs544873 | metabolite A | 3.40E-14 | ApoB | ApoB |
| rs545971 | metabolite T | 2.10E-14 | ApoB | ApoB |
| rs612169 | metabolite G | 2.70E-14 | ApoB | ApoB |
| rs8176663 | metabolite C | 2.10E-14 | ApoB | ApoB |
| rs491626 | metabolite T | 2.10E-14 | ApoB | ApoB |
| rs492488 | metabolite A | 2.10E-14 | ApoB | ApoB |
| rs493246 | metabolite A | 2.10E-14 | ApoB | ApoB |
| rs494242 | metabolite T | 3.30E-14 | ApoB | ApoB |
| rs495203 | metabolite T | 2.10E-14 | ApoB | ApoB |
| rs582118 | metabolite G | 2.20E-14 | ApoB | ApoB |
| rs582094 | metabolite T | 3.10E-14 | ApoB | ApoB |
| rs2769071 | metabolite G | 2.20E-14 | ApoB | ApoB |
| rs677355 | metabolite A | 2.50E-14 | ApoB | ApoB |
| rs676996 | metabolite G | 2.20E-14 | ApoB | ApoB |
| rs676457 | metabolite T | 2.20E-14 | ApoB | ApoB |
| rs550057 | metabolite T | 1.20E-22 | ApoB | ApoB |
| rs674302 | metabolite A | 2.20E-14 | ApoB | ApoB |
| rs554833 | metabolite T | 2.40E-14 | ApoB | ApoB |
| rs529565 | metabolite C | 3.90E-14 | ApoB | ApoB |
| rs532436 | metabolite A | 1.40E-26 | ApoB | ApoB |
| rs600038 | metabolite C | 1.40E-24 | ApoB | ApoB |
| rs1124406: | metabolite T | 2.80E-16 | ApoB | ApoB |
| rs649129 | metabolite T | 8.20E-25 | ApoB | ApoB |
| rs495828 | metabolite T | 3.80E-25 | ApoB | ApoB |
| rs2519093 | metabolite T | 9.30E-13 | HDL_size | HDL_size |
| rs550057 | metabolite T | 4.40E-10 | HDL_size | HDL_size |
| rs532436 | metabolite A | 1.70E-12 | HDL_size | HDL_size |
| rs600038 | metabolite C | 3.50E-12 | HDL_size | HDL_size |
| rs1124406: | metabolite T | 2.40E-10 | HDL_size | HDL_size |
| rs649129 | metabolite T | 3.00E-12 | HDL_size | HDL_size |
| rs495828 | metabolite T | 3.10E-12 | HDL_size | HDL_size |
| rs7873635 | metabolite C | 2.00E-10 | XXL_VLDL_ | XXL_VLDL_TG_pct |
| rs4962040 | metabolite G | 1.50E-10 | XXL_VLDL_ | XXL_VLDL_TG_pct |
| rs8176702 | metabolite A | 2.60E-10 | XXL_VLDL_ | XXL_VLDL_TG_pct |
| rs2073826 | metabolite T | 3.20E-10 | XXL_VLDL_ | XXL_VLDL_TG_pct |
| rs687289 | metabolite A | 1.60E-16 | XXL_VLDL_ | XXL_VLDL_TG_pct |
| rs2073827 | metabolite C | 2.40E-10 | XXL_VLDL_ | XXL_VLDL_TG_pct |
| rs2073828 | metabolite A | 2.50E-10 | XXL_VLDL_ | XXL_VLDL_TG_pct |
| rs8176691 | metabolite T | 1.60E-10 | XXL_VLDL_ | XXL_VLDL_TG_pct |
| rs8176690 | metabolite G | 1.40E-10 | XXL_VLDL_ | XXL_VLDL_TG_pct |
| rs8176682 | metabolite T | 8.70E-11 | XXL_VLDL_ | XXL_VLDL_TG_pct |
| rs8176681 | metabolite C | 1.50E-10 | XXL_VLDL_ | XXL_VLDL_TG_pct |
| rs2519093 | metabolite T | 5.10E-28 | XXL_VLDL_ | XXL_VLDL_TG_pct |
| rs644234 | metabolite G | 6.20E-16 | XXL_VLDL_ | XXL_VLDL_TG_pct |
| rs643434 | metabolite A | 6.10E-16 | XXL_VLDL_ | XXL_VLDL_TG_pct |

| | | | | |
|---|---|---|---|---|
| rs543040 | metabolite T | 3.10E-16 | XXL_VLDL_ | XXL_VLDL_TG_pct |
| rs613534 | metabolite G | 6.30E-16 | XXL_VLDL_ | XXL_VLDL_TG_pct |
| rs543968 | metabolite C | 6.30E-16 | XXL_VLDL_ | XXL_VLDL_TG_pct |
| rs544873 | metabolite A | 6.30E-16 | XXL_VLDL_ | XXL_VLDL_TG_pct |
| rs545971 | metabolite T | 2.20E-16 | XXL_VLDL_ | XXL_VLDL_TG_pct |
| rs612169 | metabolite G | 2.20E-16 | XXL_VLDL_ | XXL_VLDL_TG_pct |
| rs8176668 | metabolite T | 3.10E-10 | XXL_VLDL_ | XXL_VLDL_TG_pct |
| rs8176663 | metabolite C | 2.20E-16 | XXL_VLDL_ | XXL_VLDL_TG_pct |
| rs7036642 | metabolite A | 3.70E-10 | XXL_VLDL_ | XXL_VLDL_TG_pct |
| rs491626 | metabolite T | 2.10E-16 | XXL_VLDL_ | XXL_VLDL_TG_pct |
| rs492488 | metabolite A | 2.10E-16 | XXL_VLDL_ | XXL_VLDL_TG_pct |
| rs493246 | metabolite A | 2.10E-16 | XXL_VLDL_ | XXL_VLDL_TG_pct |
| rs494242 | metabolite T | 6.30E-16 | XXL_VLDL_ | XXL_VLDL_TG_pct |
| rs495203 | metabolite T | 2.10E-16 | XXL_VLDL_ | XXL_VLDL_TG_pct |
| rs582118 | metabolite G | 2.20E-16 | XXL_VLDL_ | XXL_VLDL_TG_pct |
| rs582094 | metabolite T | 2.70E-16 | XXL_VLDL_ | XXL_VLDL_TG_pct |
| rs2769071 | metabolite G | 2.70E-16 | XXL_VLDL_ | XXL_VLDL_TG_pct |
| rs677355 | metabolite A | 2.30E-16 | XXL_VLDL_ | XXL_VLDL_TG_pct |
| rs676996 | metabolite G | 2.50E-16 | XXL_VLDL_ | XXL_VLDL_TG_pct |
| rs676457 | metabolite T | 2.50E-16 | XXL_VLDL_ | XXL_VLDL_TG_pct |
| rs550057 | metabolite T | 2.80E-18 | XXL_VLDL_ | XXL_VLDL_TG_pct |
| rs674302 | metabolite A | 2.50E-16 | XXL_VLDL_ | XXL_VLDL_TG_pct |
| rs7046674 | metabolite T | 3.40E-10 | XXL_VLDL_ | XXL_VLDL_TG_pct |
| rs554833 | metabolite T | 2.80E-16 | XXL_VLDL_ | XXL_VLDL_TG_pct |
| rs8176649 | metabolite A | 3.50E-10 | XXL_VLDL_ | XXL_VLDL_TG_pct |
| rs529565 | metabolite C | 1.80E-16 | XXL_VLDL_ | XXL_VLDL_TG_pct |
| rs532436 | metabolite A | 5.80E-28 | XXL_VLDL_ | XXL_VLDL_TG_pct |
| rs600038 | metabolite C | 1.90E-25 | XXL_VLDL_ | XXL_VLDL_TG_pct |
| rs1124406: | metabolite T | 1.40E-20 | XXL_VLDL_ | XXL_VLDL_TG_pct |
| rs649129 | metabolite T | 8.30E-26 | XXL_VLDL_ | XXL_VLDL_TG_pct |
| rs495828 | metabolite T | 3.00E-25 | XXL_VLDL_ | XXL_VLDL_TG_pct |
| rs687289 | metabolite A | 1.10E-09 | XXL_VLDL_ | XXL_VLDL_PL_pct |
| rs2519093 | metabolite T | 2.00E-13 | XXL_VLDL_ | XXL_VLDL_PL_pct |
| rs644234 | metabolite G | 5.40E-09 | XXL_VLDL_ | XXL_VLDL_PL_pct |
| rs643434 | metabolite A | 5.50E-09 | XXL_VLDL_ | XXL_VLDL_PL_pct |
| rs543040 | metabolite T | 3.20E-09 | XXL_VLDL_ | XXL_VLDL_PL_pct |
| rs613534 | metabolite G | 5.40E-09 | XXL_VLDL_ | XXL_VLDL_PL_pct |
| rs543968 | metabolite C | 5.40E-09 | XXL_VLDL_ | XXL_VLDL_PL_pct |
| rs544873 | metabolite A | 5.40E-09 | XXL_VLDL_ | XXL_VLDL_PL_pct |
| rs545971 | metabolite T | 1.50E-09 | XXL_VLDL_ | XXL_VLDL_PL_pct |
| rs612169 | metabolite G | 1.90E-09 | XXL_VLDL_ | XXL_VLDL_PL_pct |
| rs8176663 | metabolite C | 1.50E-09 | XXL_VLDL_ | XXL_VLDL_PL_pct |
| rs491626 | metabolite T | 1.50E-09 | XXL_VLDL_ | XXL_VLDL_PL_pct |
| rs492488 | metabolite A | 1.50E-09 | XXL_VLDL_ | XXL_VLDL_PL_pct |
| rs493246 | metabolite A | 1.50E-09 | XXL_VLDL_ | XXL_VLDL_PL_pct |
| rs494242 | metabolite T | 5.40E-09 | XXL_VLDL_ | XXL_VLDL_PL_pct |
| rs495203 | metabolite T | 1.50E-09 | XXL_VLDL_ | XXL_VLDL_PL_pct |
| rs582118 | metabolite G | 1.50E-09 | XXL_VLDL_ | XXL_VLDL_PL_pct |
| rs582094 | metabolite T | 1.40E-09 | XXL_VLDL_ | XXL_VLDL_PL_pct |
| rs2769071 | metabolite G | 1.60E-09 | XXL_VLDL_ | XXL_VLDL_PL_pct |

| | | | | |
|---|---|---|---|---|
| rs677355 | metabolite A | 1.50E-09 | XXL_VLDL_ | XXL_VLDL_PL_pct |
| rs676996 | metabolite G | 1.50E-09 | XXL_VLDL_ | XXL_VLDL_PL_pct |
| rs676457 | metabolite T | 1.50E-09 | XXL_VLDL_ | XXL_VLDL_PL_pct |
| rs550057 | metabolite T | 7.10E-09 | XXL_VLDL_ | XXL_VLDL_PL_pct |
| rs674302 | metabolite A | 1.50E-09 | XXL_VLDL_ | XXL_VLDL_PL_pct |
| rs554833 | metabolite T | 1.70E-09 | XXL_VLDL_ | XXL_VLDL_PL_pct |
| rs529565 | metabolite C | 7.00E-10 | XXL_VLDL_ | XXL_VLDL_PL_pct |
| rs532436 | metabolite A | 1.70E-13 | XXL_VLDL_ | XXL_VLDL_PL_pct |
| rs600038 | metabolite C | 5.00E-12 | XXL_VLDL_ | XXL_VLDL_PL_pct |
| rs1124406: | metabolite T | 5.50E-12 | XXL_VLDL_ | XXL_VLDL_PL_pct |
| rs649129 | metabolite T | 3.50E-12 | XXL_VLDL_ | XXL_VLDL_PL_pct |
| rs495828 | metabolite T | 7.40E-12 | XXL_VLDL_ | XXL_VLDL_PL_pct |
| rs687289 | metabolite A | 1.00E-13 | LDL_L | LDL_L |
| rs2519093 | metabolite T | 6.80E-30 | LDL_L | LDL_L |
| rs644234 | metabolite G | 5.80E-13 | LDL_L | LDL_L |
| rs643434 | metabolite A | 5.40E-13 | LDL_L | LDL_L |
| rs543040 | metabolite T | 7.50E-14 | LDL_L | LDL_L |
| rs613534 | metabolite G | 5.80E-13 | LDL_L | LDL_L |
| rs543968 | metabolite C | 5.80E-13 | LDL_L | LDL_L |
| rs544873 | metabolite A | 5.80E-13 | LDL_L | LDL_L |
| rs545971 | metabolite T | 7.40E-14 | LDL_L | LDL_L |
| rs612169 | metabolite G | 9.30E-14 | LDL_L | LDL_L |
| rs8176663 | metabolite C | 7.40E-14 | LDL_L | LDL_L |
| rs491626 | metabolite T | 7.30E-14 | LDL_L | LDL_L |
| rs492488 | metabolite A | 7.30E-14 | LDL_L | LDL_L |
| rs493246 | metabolite A | 7.30E-14 | LDL_L | LDL_L |
| rs494242 | metabolite T | 5.60E-13 | LDL_L | LDL_L |
| rs495203 | metabolite T | 7.30E-14 | LDL_L | LDL_L |
| rs582118 | metabolite G | 7.60E-14 | LDL_L | LDL_L |
| rs582094 | metabolite T | 1.10E-13 | LDL_L | LDL_L |
| rs2769071 | metabolite G | 7.70E-14 | LDL_L | LDL_L |
| rs677355 | metabolite A | 8.80E-14 | LDL_L | LDL_L |
| rs676996 | metabolite G | 7.80E-14 | LDL_L | LDL_L |
| rs676457 | metabolite T | 7.80E-14 | LDL_L | LDL_L |
| rs550057 | metabolite T | 6.60E-24 | LDL_L | LDL_L |
| rs674302 | metabolite A | 7.70E-14 | LDL_L | LDL_L |
| rs554833 | metabolite T | 8.50E-14 | LDL_L | LDL_L |
| rs529565 | metabolite C | 1.30E-13 | LDL_L | LDL_L |
| rs532436 | metabolite A | 9.50E-30 | LDL_L | LDL_L |
| rs600038 | metabolite C | 2.30E-26 | LDL_L | LDL_L |
| rs1124406: | metabolite T | 2.10E-18 | LDL_L | LDL_L |
| rs649129 | metabolite T | 1.40E-26 | LDL_L | LDL_L |
| rs495828 | metabolite T | 9.10E-27 | LDL_L | LDL_L |
| rs2519093 | metabolite T | 4.80E-14 | Total_P | Total_P |
| rs550057 | metabolite T | 1.70E-09 | Total_P | Total_P |
| rs532436 | metabolite A | 5.50E-14 | Total_P | Total_P |
| rs600038 | metabolite C | 7.20E-11 | Total_P | Total_P |
| rs1124406: | metabolite T | 9.10E-10 | Total_P | Total_P |
| rs649129 | metabolite T | 6.30E-11 | Total_P | Total_P |
| rs495828 | metabolite T | 9.80E-11 | Total_P | Total_P |

| | | | | |
|---|---|---|---|---|
| rs687289 | metabolite A | 5.10E-09 | S_VLDL_L | S_VLDL_L |
| rs2519093 | metabolite T | 4.30E-13 | S_VLDL_L | S_VLDL_L |
| rs644234 | metabolite G | 2.50E-09 | S_VLDL_L | S_VLDL_L |
| rs643434 | metabolite A | 2.30E-09 | S_VLDL_L | S_VLDL_L |
| rs543040 | metabolite T | 3.10E-09 | S_VLDL_L | S_VLDL_L |
| rs613534 | metabolite G | 2.50E-09 | S_VLDL_L | S_VLDL_L |
| rs543968 | metabolite C | 2.50E-09 | S_VLDL_L | S_VLDL_L |
| rs544873 | metabolite A | 2.50E-09 | S_VLDL_L | S_VLDL_L |
| rs545971 | metabolite T | 3.70E-09 | S_VLDL_L | S_VLDL_L |
| rs612169 | metabolite G | 4.70E-09 | S_VLDL_L | S_VLDL_L |
| rs8176663 | metabolite C | 3.70E-09 | S_VLDL_L | S_VLDL_L |
| rs491626 | metabolite T | 3.70E-09 | S_VLDL_L | S_VLDL_L |
| rs492488 | metabolite A | 3.70E-09 | S_VLDL_L | S_VLDL_L |
| rs493246 | metabolite A | 3.70E-09 | S_VLDL_L | S_VLDL_L |
| rs494242 | metabolite T | 2.40E-09 | S_VLDL_L | S_VLDL_L |
| rs495203 | metabolite T | 3.70E-09 | S_VLDL_L | S_VLDL_L |
| rs582118 | metabolite G | 3.80E-09 | S_VLDL_L | S_VLDL_L |
| rs582094 | metabolite T | 4.70E-09 | S_VLDL_L | S_VLDL_L |
| rs2769071 | metabolite G | 3.70E-09 | S_VLDL_L | S_VLDL_L |
| rs677355 | metabolite A | 4.30E-09 | S_VLDL_L | S_VLDL_L |
| rs676996 | metabolite G | 3.80E-09 | S_VLDL_L | S_VLDL_L |
| rs676457 | metabolite T | 3.80E-09 | S_VLDL_L | S_VLDL_L |
| rs550057 | metabolite T | 3.50E-11 | S_VLDL_L | S_VLDL_L |
| rs674302 | metabolite A | 3.80E-09 | S_VLDL_L | S_VLDL_L |
| rs554833 | metabolite T | 4.00E-09 | S_VLDL_L | S_VLDL_L |
| rs529565 | metabolite C | 4.50E-09 | S_VLDL_L | S_VLDL_L |
| rs532436 | metabolite A | 4.90E-13 | S_VLDL_L | S_VLDL_L |
| rs600038 | metabolite C | 1.40E-12 | S_VLDL_L | S_VLDL_L |
| rs1124406: | metabolite T | 5.10E-09 | S_VLDL_L | S_VLDL_L |
| rs649129 | metabolite T | 7.60E-13 | S_VLDL_L | S_VLDL_L |
| rs495828 | metabolite T | 4.60E-13 | S_VLDL_L | S_VLDL_L |
| rs687289 | metabolite A | 6.20E-10 | VLDL_P | VLDL_P |
| rs2519093 | metabolite T | 2.10E-14 | VLDL_P | VLDL_P |
| rs644234 | metabolite G | 2.00E-10 | VLDL_P | VLDL_P |
| rs643434 | metabolite A | 1.80E-10 | VLDL_P | VLDL_P |
| rs543040 | metabolite T | 3.40E-10 | VLDL_P | VLDL_P |
| rs613534 | metabolite G | 2.00E-10 | VLDL_P | VLDL_P |
| rs543968 | metabolite C | 2.00E-10 | VLDL_P | VLDL_P |
| rs544873 | metabolite A | 2.00E-10 | VLDL_P | VLDL_P |
| rs545971 | metabolite T | 4.40E-10 | VLDL_P | VLDL_P |
| rs612169 | metabolite G | 5.60E-10 | VLDL_P | VLDL_P |
| rs8176663 | metabolite C | 4.40E-10 | VLDL_P | VLDL_P |
| rs491626 | metabolite T | 4.40E-10 | VLDL_P | VLDL_P |
| rs492488 | metabolite A | 4.40E-10 | VLDL_P | VLDL_P |
| rs493246 | metabolite A | 4.40E-10 | VLDL_P | VLDL_P |
| rs494242 | metabolite T | 1.90E-10 | VLDL_P | VLDL_P |
| rs495203 | metabolite T | 4.40E-10 | VLDL_P | VLDL_P |
| rs582118 | metabolite G | 4.50E-10 | VLDL_P | VLDL_P |
| rs582094 | metabolite T | 5.50E-10 | VLDL_P | VLDL_P |
| rs2769071 | metabolite G | 4.40E-10 | VLDL_P | VLDL_P |

| | | | | |
|---|---|---|---|---|
| rs677355 | metabolite A | 5.00E-10 | VLDL_P | VLDL_P |
| rs676996 | metabolite G | 4.40E-10 | VLDL_P | VLDL_P |
| rs676457 | metabolite T | 4.40E-10 | VLDL_P | VLDL_P |
| rs550057 | metabolite T | 1.00E-12 | VLDL_P | VLDL_P |
| rs674302 | metabolite A | 4.40E-10 | VLDL_P | VLDL_P |
| rs554833 | metabolite T | 4.70E-10 | VLDL_P | VLDL_P |
| rs529565 | metabolite C | 5.50E-10 | VLDL_P | VLDL_P |
| rs532436 | metabolite A | 2.40E-14 | VLDL_P | VLDL_P |
| rs600038 | metabolite C | 4.70E-14 | VLDL_P | VLDL_P |
| rs1124406: | metabolite T | 1.60E-09 | VLDL_P | VLDL_P |
| rs649129 | metabolite T | 2.60E-14 | VLDL_P | VLDL_P |
| rs495828 | metabolite T | 1.40E-14 | VLDL_P | VLDL_P |
| rs687289 | metabolite A | 7.60E-13 | VLDL_C | VLDL_C |
| rs2519093 | metabolite T | 2.40E-19 | VLDL_C | VLDL_C |
| rs644234 | metabolite G | 2.10E-13 | VLDL_C | VLDL_C |
| rs643434 | metabolite A | 1.90E-13 | VLDL_C | VLDL_C |
| rs543040 | metabolite T | 3.80E-13 | VLDL_C | VLDL_C |
| rs613534 | metabolite G | 2.10E-13 | VLDL_C | VLDL_C |
| rs543968 | metabolite C | 2.10E-13 | VLDL_C | VLDL_C |
| rs544873 | metabolite A | 2.10E-13 | VLDL_C | VLDL_C |
| rs545971 | metabolite T | 4.90E-13 | VLDL_C | VLDL_C |
| rs612169 | metabolite G | 6.50E-13 | VLDL_C | VLDL_C |
| rs8176663 | metabolite C | 4.90E-13 | VLDL_C | VLDL_C |
| rs491626 | metabolite T | 4.80E-13 | VLDL_C | VLDL_C |
| rs492488 | metabolite A | 4.80E-13 | VLDL_C | VLDL_C |
| rs493246 | metabolite A | 4.80E-13 | VLDL_C | VLDL_C |
| rs494242 | metabolite T | 2.10E-13 | VLDL_C | VLDL_C |
| rs495203 | metabolite T | 4.90E-13 | VLDL_C | VLDL_C |
| rs582118 | metabolite G | 5.00E-13 | VLDL_C | VLDL_C |
| rs582094 | metabolite T | 6.70E-13 | VLDL_C | VLDL_C |
| rs2769071 | metabolite G | 5.00E-13 | VLDL_C | VLDL_C |
| rs677355 | metabolite A | 5.80E-13 | VLDL_C | VLDL_C |
| rs676996 | metabolite G | 5.00E-13 | VLDL_C | VLDL_C |
| rs676457 | metabolite T | 5.00E-13 | VLDL_C | VLDL_C |
| rs550057 | metabolite T | 4.20E-17 | VLDL_C | VLDL_C |
| rs674302 | metabolite A | 5.00E-13 | VLDL_C | VLDL_C |
| rs554833 | metabolite T | 5.50E-13 | VLDL_C | VLDL_C |
| rs529565 | metabolite C | 6.40E-13 | VLDL_C | VLDL_C |
| rs532436 | metabolite A | 2.90E-19 | VLDL_C | VLDL_C |
| rs600038 | metabolite C | 1.20E-18 | VLDL_C | VLDL_C |
| rs1124406: | metabolite T | 1.50E-12 | VLDL_C | VLDL_C |
| rs649129 | metabolite T | 6.90E-19 | VLDL_C | VLDL_C |
| rs495828 | metabolite T | 3.50E-19 | VLDL_C | VLDL_C |
| rs2519093 | metabolite T | 2.50E-16 | XS_VLDL_T | XS_VLDL_TG_pct |
| rs550057 | metabolite T | 1.60E-14 | XS_VLDL_T | XS_VLDL_TG_pct |
| rs532436 | metabolite A | 3.40E-16 | XS_VLDL_T | XS_VLDL_TG_pct |
| rs600038 | metabolite C | 4.10E-14 | XS_VLDL_T | XS_VLDL_TG_pct |
| rs649129 | metabolite T | 6.70E-14 | XS_VLDL_T | XS_VLDL_TG_pct |
| rs495828 | metabolite T | 6.80E-14 | XS_VLDL_T | XS_VLDL_TG_pct |
| rs7873635 | metabolite C | 7.00E-13 | XL_VLDL_T | XL_VLDL_TG_pct |

| | | | | |
|---|---|---|---|---|
| rs4962040 | metabolite G | 6.90E-13 | XL_VLDL_T | XL_VLDL_TG_pct |
| rs8176702 | metabolite A | 9.30E-13 | XL_VLDL_T | XL_VLDL_TG_pct |
| rs2073826 | metabolite T | 5.00E-13 | XL_VLDL_T | XL_VLDL_TG_pct |
| rs687289 | metabolite A | 5.40E-25 | XL_VLDL_T | XL_VLDL_TG_pct |
| rs2073827 | metabolite C | 4.30E-13 | XL_VLDL_T | XL_VLDL_TG_pct |
| rs2073828 | metabolite A | 4.50E-13 | XL_VLDL_T | XL_VLDL_TG_pct |
| rs8176691 | metabolite T | 4.60E-13 | XL_VLDL_T | XL_VLDL_TG_pct |
| rs8176690 | metabolite G | 1.40E-13 | XL_VLDL_T | XL_VLDL_TG_pct |
| rs8176682 | metabolite T | 2.00E-13 | XL_VLDL_T | XL_VLDL_TG_pct |
| rs8176681 | metabolite C | 1.80E-13 | XL_VLDL_T | XL_VLDL_TG_pct |
| rs2519093 | metabolite T | 9.80E-41 | XL_VLDL_T | XL_VLDL_TG_pct |
| rs644234 | metabolite G | 4.70E-23 | XL_VLDL_T | XL_VLDL_TG_pct |
| rs643434 | metabolite A | 5.60E-23 | XL_VLDL_T | XL_VLDL_TG_pct |
| rs543040 | metabolite T | 3.50E-24 | XL_VLDL_T | XL_VLDL_TG_pct |
| rs613534 | metabolite G | 4.90E-23 | XL_VLDL_T | XL_VLDL_TG_pct |
| rs543968 | metabolite C | 4.90E-23 | XL_VLDL_T | XL_VLDL_TG_pct |
| rs544873 | metabolite A | 4.90E-23 | XL_VLDL_T | XL_VLDL_TG_pct |
| rs545971 | metabolite T | 1.70E-24 | XL_VLDL_T | XL_VLDL_TG_pct |
| rs612169 | metabolite G | 1.80E-24 | XL_VLDL_T | XL_VLDL_TG_pct |
| rs8176668 | metabolite T | 8.90E-13 | XL_VLDL_T | XL_VLDL_TG_pct |
| rs8176663 | metabolite C | 1.70E-24 | XL_VLDL_T | XL_VLDL_TG_pct |
| rs7036642 | metabolite A | 1.20E-12 | XL_VLDL_T | XL_VLDL_TG_pct |
| rs491626 | metabolite T | 1.70E-24 | XL_VLDL_T | XL_VLDL_TG_pct |
| rs492488 | metabolite A | 1.70E-24 | XL_VLDL_T | XL_VLDL_TG_pct |
| rs493246 | metabolite A | 1.70E-24 | XL_VLDL_T | XL_VLDL_TG_pct |
| rs494242 | metabolite T | 5.00E-23 | XL_VLDL_T | XL_VLDL_TG_pct |
| rs495203 | metabolite T | 1.70E-24 | XL_VLDL_T | XL_VLDL_TG_pct |
| rs582118 | metabolite G | 1.80E-24 | XL_VLDL_T | XL_VLDL_TG_pct |
| rs582094 | metabolite T | 2.20E-24 | XL_VLDL_T | XL_VLDL_TG_pct |
| rs2769071 | metabolite G | 3.40E-24 | XL_VLDL_T | XL_VLDL_TG_pct |
| rs677355 | metabolite A | 2.10E-24 | XL_VLDL_T | XL_VLDL_TG_pct |
| rs676996 | metabolite G | 2.50E-24 | XL_VLDL_T | XL_VLDL_TG_pct |
| rs676457 | metabolite T | 2.50E-24 | XL_VLDL_T | XL_VLDL_TG_pct |
| rs550057 | metabolite T | 3.30E-32 | XL_VLDL_T | XL_VLDL_TG_pct |
| rs674302 | metabolite A | 2.50E-24 | XL_VLDL_T | XL_VLDL_TG_pct |
| rs7046674 | metabolite T | 1.50E-12 | XL_VLDL_T | XL_VLDL_TG_pct |
| rs554833 | metabolite T | 2.60E-24 | XL_VLDL_T | XL_VLDL_TG_pct |
| rs8176649 | metabolite A | 1.30E-12 | XL_VLDL_T | XL_VLDL_TG_pct |
| rs660340 | metabolite G | 8.70E-11 | XL_VLDL_T | XL_VLDL_TG_pct |
| rs581107 | metabolite T | 8.80E-11 | XL_VLDL_T | XL_VLDL_TG_pct |
| rs659104 | metabolite G | 8.90E-11 | XL_VLDL_T | XL_VLDL_TG_pct |
| rs647800 | metabolite G | 2.00E-10 | XL_VLDL_T | XL_VLDL_TG_pct |
| rs473533 | metabolite C | 9.10E-11 | XL_VLDL_T | XL_VLDL_TG_pct |
| rs476410 | metabolite C | 1.00E-10 | XL_VLDL_T | XL_VLDL_TG_pct |
| rs645982 | metabolite G | 8.10E-11 | XL_VLDL_T | XL_VLDL_TG_pct |
| rs529565 | metabolite C | 3.70E-24 | XL_VLDL_T | XL_VLDL_TG_pct |
| rs630510 | metabolite G | 4.20E-08 | XL_VLDL_T | XL_VLDL_TG_pct |
| rs630014 | metabolite G | 4.20E-08 | XL_VLDL_T | XL_VLDL_TG_pct |
| rs532436 | metabolite A | 1.90E-40 | XL_VLDL_T | XL_VLDL_TG_pct |
| rs600038 | metabolite C | 1.30E-35 | XL_VLDL_T | XL_VLDL_TG_pct |

| | | | | |
|---|---|---|---|---|
| rs1124406: | metabolite T | 1.70E-24 | XL_VLDL_T | XL_VLDL_TG_pct |
| rs649129 | metabolite T | 1.10E-35 | XL_VLDL_T | XL_VLDL_TG_pct |
| rs495828 | metabolite T | 1.60E-35 | XL_VLDL_T | XL_VLDL_TG_pct |
| rs7873635 | metabolite C | 5.30E-09 | XXL_VLDL_ | XXL_VLDL_CE_pct |
| rs4962040 | metabolite G | 3.50E-09 | XXL_VLDL_ | XXL_VLDL_CE_pct |
| rs8176702 | metabolite A | 6.10E-09 | XXL_VLDL_ | XXL_VLDL_CE_pct |
| rs2073826 | metabolite T | 8.20E-09 | XXL_VLDL_ | XXL_VLDL_CE_pct |
| rs687289 | metabolite A | 7.70E-14 | XXL_VLDL_ | XXL_VLDL_CE_pct |
| rs2073827 | metabolite C | 6.80E-09 | XXL_VLDL_ | XXL_VLDL_CE_pct |
| rs2073828 | metabolite A | 7.20E-09 | XXL_VLDL_ | XXL_VLDL_CE_pct |
| rs8176691 | metabolite T | 3.90E-09 | XXL_VLDL_ | XXL_VLDL_CE_pct |
| rs8176690 | metabolite G | 4.30E-09 | XXL_VLDL_ | XXL_VLDL_CE_pct |
| rs8176682 | metabolite T | 2.40E-09 | XXL_VLDL_ | XXL_VLDL_CE_pct |
| rs8176681 | metabolite C | 4.30E-09 | XXL_VLDL_ | XXL_VLDL_CE_pct |
| rs2519093 | metabolite T | 1.00E-25 | XXL_VLDL_ | XXL_VLDL_CE_pct |
| rs644234 | metabolite G | 2.10E-13 | XXL_VLDL_ | XXL_VLDL_CE_pct |
| rs643434 | metabolite A | 2.10E-13 | XXL_VLDL_ | XXL_VLDL_CE_pct |
| rs543040 | metabolite T | 1.00E-13 | XXL_VLDL_ | XXL_VLDL_CE_pct |
| rs613534 | metabolite G | 2.10E-13 | XXL_VLDL_ | XXL_VLDL_CE_pct |
| rs543968 | metabolite C | 2.10E-13 | XXL_VLDL_ | XXL_VLDL_CE_pct |
| rs544873 | metabolite A | 2.10E-13 | XXL_VLDL_ | XXL_VLDL_CE_pct |
| rs545971 | metabolite T | 9.00E-14 | XXL_VLDL_ | XXL_VLDL_CE_pct |
| rs612169 | metabolite G | 9.30E-14 | XXL_VLDL_ | XXL_VLDL_CE_pct |
| rs8176668 | metabolite T | 6.80E-09 | XXL_VLDL_ | XXL_VLDL_CE_pct |
| rs8176663 | metabolite C | 9.00E-14 | XXL_VLDL_ | XXL_VLDL_CE_pct |
| rs7036642 | metabolite A | 7.80E-09 | XXL_VLDL_ | XXL_VLDL_CE_pct |
| rs491626 | metabolite T | 8.80E-14 | XXL_VLDL_ | XXL_VLDL_CE_pct |
| rs492488 | metabolite A | 8.80E-14 | XXL_VLDL_ | XXL_VLDL_CE_pct |
| rs493246 | metabolite A | 8.80E-14 | XXL_VLDL_ | XXL_VLDL_CE_pct |
| rs494242 | metabolite T | 2.10E-13 | XXL_VLDL_ | XXL_VLDL_CE_pct |
| rs495203 | metabolite T | 8.80E-14 | XXL_VLDL_ | XXL_VLDL_CE_pct |
| rs582118 | metabolite G | 9.10E-14 | XXL_VLDL_ | XXL_VLDL_CE_pct |
| rs582094 | metabolite T | 1.30E-13 | XXL_VLDL_ | XXL_VLDL_CE_pct |
| rs2769071 | metabolite G | 9.80E-14 | XXL_VLDL_ | XXL_VLDL_CE_pct |
| rs677355 | metabolite A | 9.20E-14 | XXL_VLDL_ | XXL_VLDL_CE_pct |
| rs676996 | metabolite G | 9.60E-14 | XXL_VLDL_ | XXL_VLDL_CE_pct |
| rs676457 | metabolite T | 9.60E-14 | XXL_VLDL_ | XXL_VLDL_CE_pct |
| rs550057 | metabolite T | 1.80E-17 | XXL_VLDL_ | XXL_VLDL_CE_pct |
| rs674302 | metabolite A | 9.70E-14 | XXL_VLDL_ | XXL_VLDL_CE_pct |
| rs7046674 | metabolite T | 6.70E-09 | XXL_VLDL_ | XXL_VLDL_CE_pct |
| rs554833 | metabolite T | 1.00E-13 | XXL_VLDL_ | XXL_VLDL_CE_pct |
| rs8176649 | metabolite A | 6.50E-09 | XXL_VLDL_ | XXL_VLDL_CE_pct |
| rs529565 | metabolite C | 1.10E-13 | XXL_VLDL_ | XXL_VLDL_CE_pct |
| rs532436 | metabolite A | 1.40E-25 | XXL_VLDL_ | XXL_VLDL_CE_pct |
| rs600038 | metabolite C | 4.30E-23 | XXL_VLDL_ | XXL_VLDL_CE_pct |
| rs1124406: | metabolite T | 4.30E-18 | XXL_VLDL_ | XXL_VLDL_CE_pct |
| rs649129 | metabolite T | 1.80E-23 | XXL_VLDL_ | XXL_VLDL_CE_pct |
| rs495828 | metabolite T | 4.00E-23 | XXL_VLDL_ | XXL_VLDL_CE_pct |
| rs7873635 | metabolite C | 3.20E-08 | XXL_VLDL_ | XXL_VLDL_FC_pct |
| rs4962040 | metabolite G | 2.80E-08 | XXL_VLDL_ | XXL_VLDL_FC_pct |

| | | | | |
|---|---|---|---|---|
| rs8176702 | metabolite A | 4.50E-08 | XXL_VLDL_ | XXL_VLDL_FC_pct |
| rs2073826 | metabolite T | 3.50E-08 | XXL_VLDL_ | XXL_VLDL_FC_pct |
| rs687289 | metabolite A | 6.10E-13 | XXL_VLDL_ | XXL_VLDL_FC_pct |
| rs2073827 | metabolite C | 2.70E-08 | XXL_VLDL_ | XXL_VLDL_FC_pct |
| rs2073828 | metabolite A | 2.90E-08 | XXL_VLDL_ | XXL_VLDL_FC_pct |
| rs8176691 | metabolite T | 3.30E-08 | XXL_VLDL_ | XXL_VLDL_FC_pct |
| rs8176690 | metabolite G | 2.00E-08 | XXL_VLDL_ | XXL_VLDL_FC_pct |
| rs8176682 | metabolite T | 2.10E-08 | XXL_VLDL_ | XXL_VLDL_FC_pct |
| rs8176681 | metabolite C | 2.00E-08 | XXL_VLDL_ | XXL_VLDL_FC_pct |
| rs2519093 | metabolite T | 1.50E-18 | XXL_VLDL_ | XXL_VLDL_FC_pct |
| rs644234 | metabolite G | 5.00E-12 | XXL_VLDL_ | XXL_VLDL_FC_pct |
| rs643434 | metabolite A | 5.40E-12 | XXL_VLDL_ | XXL_VLDL_FC_pct |
| rs543040 | metabolite T | 2.00E-12 | XXL_VLDL_ | XXL_VLDL_FC_pct |
| rs613534 | metabolite G | 5.10E-12 | XXL_VLDL_ | XXL_VLDL_FC_pct |
| rs543968 | metabolite C | 5.10E-12 | XXL_VLDL_ | XXL_VLDL_FC_pct |
| rs544873 | metabolite A | 5.10E-12 | XXL_VLDL_ | XXL_VLDL_FC_pct |
| rs545971 | metabolite T | 1.10E-12 | XXL_VLDL_ | XXL_VLDL_FC_pct |
| rs612169 | metabolite G | 9.30E-13 | XXL_VLDL_ | XXL_VLDL_FC_pct |
| rs8176663 | metabolite C | 1.10E-12 | XXL_VLDL_ | XXL_VLDL_FC_pct |
| rs491626 | metabolite T | 1.10E-12 | XXL_VLDL_ | XXL_VLDL_FC_pct |
| rs492488 | metabolite A | 1.10E-12 | XXL_VLDL_ | XXL_VLDL_FC_pct |
| rs493246 | metabolite A | 1.10E-12 | XXL_VLDL_ | XXL_VLDL_FC_pct |
| rs494242 | metabolite T | 5.20E-12 | XXL_VLDL_ | XXL_VLDL_FC_pct |
| rs495203 | metabolite T | 1.10E-12 | XXL_VLDL_ | XXL_VLDL_FC_pct |
| rs582118 | metabolite G | 1.10E-12 | XXL_VLDL_ | XXL_VLDL_FC_pct |
| rs582094 | metabolite T | 1.20E-12 | XXL_VLDL_ | XXL_VLDL_FC_pct |
| rs2769071 | metabolite G | 1.30E-12 | XXL_VLDL_ | XXL_VLDL_FC_pct |
| rs677355 | metabolite A | 1.20E-12 | XXL_VLDL_ | XXL_VLDL_FC_pct |
| rs676996 | metabolite G | 1.30E-12 | XXL_VLDL_ | XXL_VLDL_FC_pct |
| rs676457 | metabolite T | 1.30E-12 | XXL_VLDL_ | XXL_VLDL_FC_pct |
| rs550057 | metabolite T | 8.10E-13 | XXL_VLDL_ | XXL_VLDL_FC_pct |
| rs674302 | metabolite A | 1.30E-12 | XXL_VLDL_ | XXL_VLDL_FC_pct |
| rs554833 | metabolite T | 1.40E-12 | XXL_VLDL_ | XXL_VLDL_FC_pct |
| rs529565 | metabolite C | 9.30E-13 | XXL_VLDL_ | XXL_VLDL_FC_pct |
| rs532436 | metabolite A | 1.30E-18 | XXL_VLDL_ | XXL_VLDL_FC_pct |
| rs600038 | metabolite C | 1.00E-16 | XXL_VLDL_ | XXL_VLDL_FC_pct |
| rs1124406: | metabolite T | 6.10E-13 | XXL_VLDL_ | XXL_VLDL_FC_pct |
| rs649129 | metabolite T | 1.40E-16 | XXL_VLDL_ | XXL_VLDL_FC_pct |
| rs495828 | metabolite T | 3.40E-16 | XXL_VLDL_ | XXL_VLDL_FC_pct |
| rs7873635 | metabolite C | 1.00E-09 | XXL_VLDL_ | XXL_VLDL_C_pct |
| rs4962040 | metabolite G | 6.90E-10 | XXL_VLDL_ | XXL_VLDL_C_pct |
| rs8176702 | metabolite A | 1.30E-09 | XXL_VLDL_ | XXL_VLDL_C_pct |
| rs2073826 | metabolite T | 1.70E-09 | XXL_VLDL_ | XXL_VLDL_C_pct |
| rs687289 | metabolite A | 2.10E-15 | XXL_VLDL_ | XXL_VLDL_C_pct |
| rs2073827 | metabolite C | 1.30E-09 | XXL_VLDL_ | XXL_VLDL_C_pct |
| rs2073828 | metabolite A | 1.40E-09 | XXL_VLDL_ | XXL_VLDL_C_pct |
| rs8176691 | metabolite T | 8.30E-10 | XXL_VLDL_ | XXL_VLDL_C_pct |
| rs8176690 | metabolite G | 8.30E-10 | XXL_VLDL_ | XXL_VLDL_C_pct |
| rs8176682 | metabolite T | 4.70E-10 | XXL_VLDL_ | XXL_VLDL_C_pct |
| rs8176681 | metabolite C | 8.40E-10 | XXL_VLDL_ | XXL_VLDL_C_pct |

| | | | | | |
|---|---|---|---|---|---|
| rs2519093 | metabolite | T | 1.20E-26 | XXL_VLDL_ | XXL_VLDL_C_pct |
| rs644234 | metabolite | G | 8.70E-15 | XXL_VLDL_ | XXL_VLDL_C_pct |
| rs643434 | metabolite | A | 9.00E-15 | XXL_VLDL_ | XXL_VLDL_C_pct |
| rs543040 | metabolite | T | 4.10E-15 | XXL_VLDL_ | XXL_VLDL_C_pct |
| rs613534 | metabolite | G | 8.80E-15 | XXL_VLDL_ | XXL_VLDL_C_pct |
| rs543968 | metabolite | C | 8.80E-15 | XXL_VLDL_ | XXL_VLDL_C_pct |
| rs544873 | metabolite | A | 8.80E-15 | XXL_VLDL_ | XXL_VLDL_C_pct |
| rs545971 | metabolite | T | 2.90E-15 | XXL_VLDL_ | XXL_VLDL_C_pct |
| rs612169 | metabolite | G | 2.70E-15 | XXL_VLDL_ | XXL_VLDL_C_pct |
| rs8176668 | metabolite | T | 1.60E-09 | XXL_VLDL_ | XXL_VLDL_C_pct |
| rs8176663 | metabolite | C | 2.90E-15 | XXL_VLDL_ | XXL_VLDL_C_pct |
| rs7036642 | metabolite | A | 1.80E-09 | XXL_VLDL_ | XXL_VLDL_C_pct |
| rs491626 | metabolite | T | 2.90E-15 | XXL_VLDL_ | XXL_VLDL_C_pct |
| rs492488 | metabolite | A | 2.90E-15 | XXL_VLDL_ | XXL_VLDL_C_pct |
| rs493246 | metabolite | A | 2.90E-15 | XXL_VLDL_ | XXL_VLDL_C_pct |
| rs494242 | metabolite | T | 8.80E-15 | XXL_VLDL_ | XXL_VLDL_C_pct |
| rs495203 | metabolite | T | 2.90E-15 | XXL_VLDL_ | XXL_VLDL_C_pct |
| rs582118 | metabolite | G | 2.90E-15 | XXL_VLDL_ | XXL_VLDL_C_pct |
| rs582094 | metabolite | T | 4.10E-15 | XXL_VLDL_ | XXL_VLDL_C_pct |
| rs2769071 | metabolite | G | 3.40E-15 | XXL_VLDL_ | XXL_VLDL_C_pct |
| rs677355 | metabolite | A | 3.10E-15 | XXL_VLDL_ | XXL_VLDL_C_pct |
| rs676996 | metabolite | G | 3.30E-15 | XXL_VLDL_ | XXL_VLDL_C_pct |
| rs676457 | metabolite | T | 3.30E-15 | XXL_VLDL_ | XXL_VLDL_C_pct |
| rs550057 | metabolite | T | 1.60E-17 | XXL_VLDL_ | XXL_VLDL_C_pct |
| rs674302 | metabolite | A | 3.40E-15 | XXL_VLDL_ | XXL_VLDL_C_pct |
| rs7046674 | metabolite | T | 1.80E-09 | XXL_VLDL_ | XXL_VLDL_C_pct |
| rs554833 | metabolite | T | 3.60E-15 | XXL_VLDL_ | XXL_VLDL_C_pct |
| rs8176649 | metabolite | A | 1.70E-09 | XXL_VLDL_ | XXL_VLDL_C_pct |
| rs529565 | metabolite | C | 2.80E-15 | XXL_VLDL_ | XXL_VLDL_C_pct |
| rs532436 | metabolite | A | 1.50E-26 | XXL_VLDL_ | XXL_VLDL_C_pct |
| rs600038 | metabolite | C | 5.90E-24 | XXL_VLDL_ | XXL_VLDL_C_pct |
| rs1124406: | metabolite | T | 1.50E-18 | XXL_VLDL_ | XXL_VLDL_C_pct |
| rs649129 | metabolite | T | 3.20E-24 | XXL_VLDL_ | XXL_VLDL_C_pct |
| rs495828 | metabolite | T | 7.70E-24 | XXL_VLDL_ | XXL_VLDL_C_pct |
| rs7873635 | metabolite | C | 2.80E-08 | IDL_C | IDL_C |
| rs4962040 | metabolite | G | 2.10E-08 | IDL_C | IDL_C |
| rs8176702 | metabolite | A | 2.50E-08 | IDL_C | IDL_C |
| rs2073826 | metabolite | T | 1.90E-08 | IDL_C | IDL_C |
| rs687289 | metabolite | A | 1.30E-15 | IDL_C | IDL_C |
| rs2073827 | metabolite | C | 2.30E-08 | IDL_C | IDL_C |
| rs2073828 | metabolite | A | 2.30E-08 | IDL_C | IDL_C |
| rs8176691 | metabolite | T | 1.80E-08 | IDL_C | IDL_C |
| rs8176690 | metabolite | G | 8.20E-09 | IDL_C | IDL_C |
| rs8176682 | metabolite | T | 8.90E-09 | IDL_C | IDL_C |
| rs8176681 | metabolite | C | 9.10E-09 | IDL_C | IDL_C |
| rs2519093 | metabolite | T | 6.00E-32 | IDL_C | IDL_C |
| rs644234 | metabolite | G | 4.80E-15 | IDL_C | IDL_C |
| rs643434 | metabolite | A | 4.40E-15 | IDL_C | IDL_C |
| rs543040 | metabolite | T | 1.20E-15 | IDL_C | IDL_C |
| rs613534 | metabolite | G | 4.90E-15 | IDL_C | IDL_C |

| | | | | |
|---|---|---|---|---|
| rs543968 | metabolite C | 4.90E-15 | IDL_C | IDL_C |
| rs544873 | metabolite A | 4.90E-15 | IDL_C | IDL_C |
| rs545971 | metabolite T | 9.40E-16 | IDL_C | IDL_C |
| rs612169 | metabolite G | 1.20E-15 | IDL_C | IDL_C |
| rs8176668 | metabolite T | 1.20E-08 | IDL_C | IDL_C |
| rs8176663 | metabolite C | 9.40E-16 | IDL_C | IDL_C |
| rs7036642 | metabolite A | 1.10E-08 | IDL_C | IDL_C |
| rs491626 | metabolite T | 9.30E-16 | IDL_C | IDL_C |
| rs492488 | metabolite A | 9.20E-16 | IDL_C | IDL_C |
| rs493246 | metabolite A | 9.20E-16 | IDL_C | IDL_C |
| rs494242 | metabolite T | 4.70E-15 | IDL_C | IDL_C |
| rs495203 | metabolite T | 9.40E-16 | IDL_C | IDL_C |
| rs582118 | metabolite G | 9.70E-16 | IDL_C | IDL_C |
| rs582094 | metabolite T | 1.40E-15 | IDL_C | IDL_C |
| rs2769071 | metabolite G | 9.40E-16 | IDL_C | IDL_C |
| rs677355 | metabolite A | 1.10E-15 | IDL_C | IDL_C |
| rs676996 | metabolite G | 9.40E-16 | IDL_C | IDL_C |
| rs676457 | metabolite T | 9.50E-16 | IDL_C | IDL_C |
| rs550057 | metabolite T | 5.10E-26 | IDL_C | IDL_C |
| rs674302 | metabolite A | 9.50E-16 | IDL_C | IDL_C |
| rs7046674 | metabolite T | 8.90E-09 | IDL_C | IDL_C |
| rs554833 | metabolite T | 1.10E-15 | IDL_C | IDL_C |
| rs8176649 | metabolite A | 9.40E-09 | IDL_C | IDL_C |
| rs529565 | metabolite C | 3.00E-15 | IDL_C | IDL_C |
| rs532436 | metabolite A | 9.90E-32 | IDL_C | IDL_C |
| rs600038 | metabolite C | 1.40E-28 | IDL_C | IDL_C |
| rs1124406: | metabolite T | 3.60E-18 | IDL_C | IDL_C |
| rs649129 | metabolite T | 1.20E-28 | IDL_C | IDL_C |
| rs495828 | metabolite T | 6.00E-29 | IDL_C | IDL_C |
| rs687289 | metabolite A | 3.60E-13 | L_LDL_C | L_LDL_C |
| rs2519093 | metabolite T | 1.10E-30 | L_LDL_C | L_LDL_C |
| rs644234 | metabolite G | 3.00E-12 | L_LDL_C | L_LDL_C |
| rs643434 | metabolite A | 2.80E-12 | L_LDL_C | L_LDL_C |
| rs543040 | metabolite T | 2.70E-13 | L_LDL_C | L_LDL_C |
| rs613534 | metabolite G | 3.00E-12 | L_LDL_C | L_LDL_C |
| rs543968 | metabolite C | 3.00E-12 | L_LDL_C | L_LDL_C |
| rs544873 | metabolite A | 3.00E-12 | L_LDL_C | L_LDL_C |
| rs545971 | metabolite T | 2.60E-13 | L_LDL_C | L_LDL_C |
| rs612169 | metabolite G | 3.10E-13 | L_LDL_C | L_LDL_C |
| rs8176663 | metabolite C | 2.60E-13 | L_LDL_C | L_LDL_C |
| rs491626 | metabolite T | 2.60E-13 | L_LDL_C | L_LDL_C |
| rs492488 | metabolite A | 2.50E-13 | L_LDL_C | L_LDL_C |
| rs493246 | metabolite A | 2.50E-13 | L_LDL_C | L_LDL_C |
| rs494242 | metabolite T | 2.90E-12 | L_LDL_C | L_LDL_C |
| rs495203 | metabolite T | 2.60E-13 | L_LDL_C | L_LDL_C |
| rs582118 | metabolite G | 2.60E-13 | L_LDL_C | L_LDL_C |
| rs582094 | metabolite T | 4.00E-13 | L_LDL_C | L_LDL_C |
| rs2769071 | metabolite G | 2.70E-13 | L_LDL_C | L_LDL_C |
| rs677355 | metabolite A | 3.10E-13 | L_LDL_C | L_LDL_C |
| rs676996 | metabolite G | 2.70E-13 | L_LDL_C | L_LDL_C |

| | | | | |
|---|---|---|---|---|
| rs676457 | metabolite T | 2.70E-13 | L_LDL_C | L_LDL_C |
| rs550057 | metabolite T | 5.60E-24 | L_LDL_C | L_LDL_C |
| rs674302 | metabolite A | 2.70E-13 | L_LDL_C | L_LDL_C |
| rs554833 | metabolite T | 3.00E-13 | L_LDL_C | L_LDL_C |
| rs529565 | metabolite C | 4.80E-13 | L_LDL_C | L_LDL_C |
| rs532436 | metabolite A | 1.70E-30 | L_LDL_C | L_LDL_C |
| rs600038 | metabolite C | 1.10E-26 | L_LDL_C | L_LDL_C |
| rs1124406: | metabolite T | 2.70E-18 | L_LDL_C | L_LDL_C |
| rs649129 | metabolite T | 7.00E-27 | L_LDL_C | L_LDL_C |
| rs495828 | metabolite T | 5.20E-27 | L_LDL_C | L_LDL_C |
| rs687289 | metabolite A | 3.00E-14 | M_LDL_C | M_LDL_C |
| rs2519093 | metabolite T | 4.10E-29 | M_LDL_C | M_LDL_C |
| rs644234 | metabolite G | 2.30E-13 | M_LDL_C | M_LDL_C |
| rs643434 | metabolite A | 2.20E-13 | M_LDL_C | M_LDL_C |
| rs543040 | metabolite T | 2.10E-14 | M_LDL_C | M_LDL_C |
| rs613534 | metabolite G | 2.30E-13 | M_LDL_C | M_LDL_C |
| rs543968 | metabolite C | 2.30E-13 | M_LDL_C | M_LDL_C |
| rs544873 | metabolite A | 2.30E-13 | M_LDL_C | M_LDL_C |
| rs545971 | metabolite T | 2.10E-14 | M_LDL_C | M_LDL_C |
| rs612169 | metabolite G | 2.80E-14 | M_LDL_C | M_LDL_C |
| rs8176663 | metabolite C | 2.10E-14 | M_LDL_C | M_LDL_C |
| rs491626 | metabolite T | 2.10E-14 | M_LDL_C | M_LDL_C |
| rs492488 | metabolite A | 2.10E-14 | M_LDL_C | M_LDL_C |
| rs493246 | metabolite A | 2.10E-14 | M_LDL_C | M_LDL_C |
| rs494242 | metabolite T | 2.20E-13 | M_LDL_C | M_LDL_C |
| rs495203 | metabolite T | 2.10E-14 | M_LDL_C | M_LDL_C |
| rs582118 | metabolite G | 2.20E-14 | M_LDL_C | M_LDL_C |
| rs582094 | metabolite T | 3.20E-14 | M_LDL_C | M_LDL_C |
| rs2769071 | metabolite G | 2.30E-14 | M_LDL_C | M_LDL_C |
| rs677355 | metabolite A | 2.60E-14 | M_LDL_C | M_LDL_C |
| rs676996 | metabolite G | 2.30E-14 | M_LDL_C | M_LDL_C |
| rs676457 | metabolite T | 2.30E-14 | M_LDL_C | M_LDL_C |
| rs550057 | metabolite T | 1.00E-23 | M_LDL_C | M_LDL_C |
| rs674302 | metabolite A | 2.30E-14 | M_LDL_C | M_LDL_C |
| rs554833 | metabolite T | 2.40E-14 | M_LDL_C | M_LDL_C |
| rs529565 | metabolite C | 3.10E-14 | M_LDL_C | M_LDL_C |
| rs532436 | metabolite A | 5.20E-29 | M_LDL_C | M_LDL_C |
| rs600038 | metabolite C | 1.70E-25 | M_LDL_C | M_LDL_C |
| rs1124406: | metabolite T | 5.60E-18 | M_LDL_C | M_LDL_C |
| rs649129 | metabolite T | 1.10E-25 | M_LDL_C | M_LDL_C |
| rs495828 | metabolite T | 7.60E-26 | M_LDL_C | M_LDL_C |
| rs7873635 | metabolite C | 9.50E-09 | M_VLDL_C | M_VLDL_C |
| rs4962040 | metabolite G | 5.50E-09 | M_VLDL_C | M_VLDL_C |
| rs8176702 | metabolite A | 5.90E-09 | M_VLDL_C | M_VLDL_C |
| rs2073826 | metabolite T | 6.90E-09 | M_VLDL_C | M_VLDL_C |
| rs687289 | metabolite A | 2.60E-18 | M_VLDL_C | M_VLDL_C |
| rs2073827 | metabolite C | 7.80E-09 | M_VLDL_C | M_VLDL_C |
| rs2073828 | metabolite A | 8.00E-09 | M_VLDL_C | M_VLDL_C |
| rs8176691 | metabolite T | 4.50E-09 | M_VLDL_C | M_VLDL_C |
| rs8176690 | metabolite G | 3.50E-09 | M_VLDL_C | M_VLDL_C |

| | | | | |
|---|---|---|---|---|
| rs8176682 | metabolite T | 2.30E-09 | M_VLDL_C | M_VLDL_C |
| rs8176681 | metabolite C | 3.30E-09 | M_VLDL_C | M_VLDL_C |
| rs2519093 | metabolite T | 1.60E-32 | M_VLDL_C | M_VLDL_C |
| rs644234 | metabolite G | 5.30E-18 | M_VLDL_C | M_VLDL_C |
| rs643434 | metabolite A | 4.70E-18 | M_VLDL_C | M_VLDL_C |
| rs543040 | metabolite T | 1.90E-18 | M_VLDL_C | M_VLDL_C |
| rs613534 | metabolite G | 5.40E-18 | M_VLDL_C | M_VLDL_C |
| rs543968 | metabolite C | 5.40E-18 | M_VLDL_C | M_VLDL_C |
| rs544873 | metabolite A | 5.40E-18 | M_VLDL_C | M_VLDL_C |
| rs545971 | metabolite T | 1.90E-18 | M_VLDL_C | M_VLDL_C |
| rs612169 | metabolite G | 2.70E-18 | M_VLDL_C | M_VLDL_C |
| rs8176668 | metabolite T | 1.80E-09 | M_VLDL_C | M_VLDL_C |
| rs8176663 | metabolite C | 1.90E-18 | M_VLDL_C | M_VLDL_C |
| rs7036642 | metabolite A | 1.70E-09 | M_VLDL_C | M_VLDL_C |
| rs491626 | metabolite T | 1.90E-18 | M_VLDL_C | M_VLDL_C |
| rs492488 | metabolite A | 1.90E-18 | M_VLDL_C | M_VLDL_C |
| rs493246 | metabolite A | 1.90E-18 | M_VLDL_C | M_VLDL_C |
| rs494242 | metabolite T | 5.20E-18 | M_VLDL_C | M_VLDL_C |
| rs495203 | metabolite T | 1.90E-18 | M_VLDL_C | M_VLDL_C |
| rs582118 | metabolite G | 2.00E-18 | M_VLDL_C | M_VLDL_C |
| rs582094 | metabolite T | 3.20E-18 | M_VLDL_C | M_VLDL_C |
| rs2769071 | metabolite G | 2.00E-18 | M_VLDL_C | M_VLDL_C |
| rs677355 | metabolite A | 2.30E-18 | M_VLDL_C | M_VLDL_C |
| rs676996 | metabolite G | 2.00E-18 | M_VLDL_C | M_VLDL_C |
| rs676457 | metabolite T | 2.00E-18 | M_VLDL_C | M_VLDL_C |
| rs550057 | metabolite T | 1.60E-27 | M_VLDL_C | M_VLDL_C |
| rs674302 | metabolite A | 2.00E-18 | M_VLDL_C | M_VLDL_C |
| rs7046674 | metabolite T | 1.30E-09 | M_VLDL_C | M_VLDL_C |
| rs554833 | metabolite T | 2.20E-18 | M_VLDL_C | M_VLDL_C |
| rs8176649 | metabolite A | 1.50E-09 | M_VLDL_C | M_VLDL_C |
| rs529565 | metabolite C | 4.00E-18 | M_VLDL_C | M_VLDL_C |
| rs532436 | metabolite A | 2.10E-32 | M_VLDL_C | M_VLDL_C |
| rs600038 | metabolite C | 1.20E-29 | M_VLDL_C | M_VLDL_C |
| rs1124406: | metabolite T | 3.00E-19 | M_VLDL_C | M_VLDL_C |
| rs649129 | metabolite T | 9.10E-30 | M_VLDL_C | M_VLDL_C |
| rs495828 | metabolite T | 4.20E-30 | M_VLDL_C | M_VLDL_C |
| rs687289 | metabolite A | 2.80E-10 | S_HDL_C | S_HDL_C |
| rs2519093 | metabolite T | 2.20E-23 | S_HDL_C | S_HDL_C |
| rs644234 | metabolite G | 9.40E-09 | S_HDL_C | S_HDL_C |
| rs643434 | metabolite A | 9.10E-09 | S_HDL_C | S_HDL_C |
| rs543040 | metabolite T | 2.00E-10 | S_HDL_C | S_HDL_C |
| rs613534 | metabolite G | 9.40E-09 | S_HDL_C | S_HDL_C |
| rs543968 | metabolite C | 9.40E-09 | S_HDL_C | S_HDL_C |
| rs544873 | metabolite A | 9.40E-09 | S_HDL_C | S_HDL_C |
| rs545971 | metabolite T | 1.70E-10 | S_HDL_C | S_HDL_C |
| rs612169 | metabolite G | 1.80E-10 | S_HDL_C | S_HDL_C |
| rs8176663 | metabolite C | 1.70E-10 | S_HDL_C | S_HDL_C |
| rs491626 | metabolite T | 1.70E-10 | S_HDL_C | S_HDL_C |
| rs492488 | metabolite A | 1.70E-10 | S_HDL_C | S_HDL_C |
| rs493246 | metabolite A | 1.70E-10 | S_HDL_C | S_HDL_C |

| | | | | |
|---|---|---|---|---|
| rs494242 | metabolite T | 9.00E-09 | S_HDL_C | S_HDL_C |
| rs495203 | metabolite T | 1.70E-10 | S_HDL_C | S_HDL_C |
| rs582118 | metabolite G | 1.70E-10 | S_HDL_C | S_HDL_C |
| rs582094 | metabolite T | 2.50E-10 | S_HDL_C | S_HDL_C |
| rs2769071 | metabolite G | 1.70E-10 | S_HDL_C | S_HDL_C |
| rs677355 | metabolite A | 1.90E-10 | S_HDL_C | S_HDL_C |
| rs676996 | metabolite G | 1.80E-10 | S_HDL_C | S_HDL_C |
| rs676457 | metabolite T | 1.80E-10 | S_HDL_C | S_HDL_C |
| rs550057 | metabolite T | 1.40E-15 | S_HDL_C | S_HDL_C |
| rs674302 | metabolite A | 1.80E-10 | S_HDL_C | S_HDL_C |
| rs554833 | metabolite T | 1.80E-10 | S_HDL_C | S_HDL_C |
| rs529565 | metabolite C | 1.20E-10 | S_HDL_C | S_HDL_C |
| rs532436 | metabolite A | 3.50E-23 | S_HDL_C | S_HDL_C |
| rs600038 | metabolite C | 8.20E-19 | S_HDL_C | S_HDL_C |
| rs1124406: | metabolite T | 1.20E-17 | S_HDL_C | S_HDL_C |
| rs649129 | metabolite T | 8.00E-19 | S_HDL_C | S_HDL_C |
| rs495828 | metabolite T | 1.80E-18 | S_HDL_C | S_HDL_C |
| rs687289 | metabolite A | 7.50E-12 | S_LDL_C | S_LDL_C |
| rs2519093 | metabolite T | 1.20E-24 | S_LDL_C | S_LDL_C |
| rs644234 | metabolite G | 1.60E-11 | S_LDL_C | S_LDL_C |
| rs643434 | metabolite A | 1.50E-11 | S_LDL_C | S_LDL_C |
| rs543040 | metabolite T | 5.40E-12 | S_LDL_C | S_LDL_C |
| rs613534 | metabolite G | 1.60E-11 | S_LDL_C | S_LDL_C |
| rs543968 | metabolite C | 1.60E-11 | S_LDL_C | S_LDL_C |
| rs544873 | metabolite A | 1.60E-11 | S_LDL_C | S_LDL_C |
| rs545971 | metabolite T | 5.50E-12 | S_LDL_C | S_LDL_C |
| rs612169 | metabolite G | 6.90E-12 | S_LDL_C | S_LDL_C |
| rs8176663 | metabolite C | 5.50E-12 | S_LDL_C | S_LDL_C |
| rs491626 | metabolite T | 5.50E-12 | S_LDL_C | S_LDL_C |
| rs492488 | metabolite A | 5.50E-12 | S_LDL_C | S_LDL_C |
| rs493246 | metabolite A | 5.50E-12 | S_LDL_C | S_LDL_C |
| rs494242 | metabolite T | 1.50E-11 | S_LDL_C | S_LDL_C |
| rs495203 | metabolite T | 5.50E-12 | S_LDL_C | S_LDL_C |
| rs582118 | metabolite G | 5.60E-12 | S_LDL_C | S_LDL_C |
| rs582094 | metabolite T | 7.70E-12 | S_LDL_C | S_LDL_C |
| rs2769071 | metabolite G | 5.80E-12 | S_LDL_C | S_LDL_C |
| rs677355 | metabolite A | 6.50E-12 | S_LDL_C | S_LDL_C |
| rs676996 | metabolite G | 5.80E-12 | S_LDL_C | S_LDL_C |
| rs676457 | metabolite T | 5.80E-12 | S_LDL_C | S_LDL_C |
| rs550057 | metabolite T | 1.20E-20 | S_LDL_C | S_LDL_C |
| rs674302 | metabolite A | 5.70E-12 | S_LDL_C | S_LDL_C |
| rs554833 | metabolite T | 6.20E-12 | S_LDL_C | S_LDL_C |
| rs529565 | metabolite C | 8.70E-12 | S_LDL_C | S_LDL_C |
| rs532436 | metabolite A | 1.50E-24 | S_LDL_C | S_LDL_C |
| rs600038 | metabolite C | 3.30E-22 | S_LDL_C | S_LDL_C |
| rs1124406: | metabolite T | 5.10E-15 | S_LDL_C | S_LDL_C |
| rs649129 | metabolite T | 2.00E-22 | S_LDL_C | S_LDL_C |
| rs495828 | metabolite T | 1.10E-22 | S_LDL_C | S_LDL_C |
| rs7873635 | metabolite C | 4.40E-08 | S_VLDL_C | S_VLDL_C |
| rs4962040 | metabolite G | 2.50E-08 | S_VLDL_C | S_VLDL_C |

| | | | | |
|---|---|---|---|---|
| rs8176702 | metabolite A | 2.50E-08 | S_VLDL_C | S_VLDL_C |
| rs2073826 | metabolite T | 3.80E-08 | S_VLDL_C | S_VLDL_C |
| rs687289 | metabolite A | 2.40E-18 | S_VLDL_C | S_VLDL_C |
| rs2073827 | metabolite C | 4.50E-08 | S_VLDL_C | S_VLDL_C |
| rs2073828 | metabolite A | 4.60E-08 | S_VLDL_C | S_VLDL_C |
| rs8176691 | metabolite T | 2.20E-08 | S_VLDL_C | S_VLDL_C |
| rs8176690 | metabolite G | 2.50E-08 | S_VLDL_C | S_VLDL_C |
| rs8176682 | metabolite T | 1.20E-08 | S_VLDL_C | S_VLDL_C |
| rs8176681 | metabolite C | 2.00E-08 | S_VLDL_C | S_VLDL_C |
| rs2519093 | metabolite T | 2.80E-30 | S_VLDL_C | S_VLDL_C |
| rs644234 | metabolite G | 2.60E-18 | S_VLDL_C | S_VLDL_C |
| rs643434 | metabolite A | 2.50E-18 | S_VLDL_C | S_VLDL_C |
| rs543040 | metabolite T | 1.80E-18 | S_VLDL_C | S_VLDL_C |
| rs613534 | metabolite G | 2.60E-18 | S_VLDL_C | S_VLDL_C |
| rs543968 | metabolite C | 2.60E-18 | S_VLDL_C | S_VLDL_C |
| rs544873 | metabolite A | 2.60E-18 | S_VLDL_C | S_VLDL_C |
| rs545971 | metabolite T | 1.80E-18 | S_VLDL_C | S_VLDL_C |
| rs612169 | metabolite G | 2.70E-18 | S_VLDL_C | S_VLDL_C |
| rs8176668 | metabolite T | 7.30E-09 | S_VLDL_C | S_VLDL_C |
| rs8176663 | metabolite C | 1.80E-18 | S_VLDL_C | S_VLDL_C |
| rs7036642 | metabolite A | 6.90E-09 | S_VLDL_C | S_VLDL_C |
| rs491626 | metabolite T | 1.80E-18 | S_VLDL_C | S_VLDL_C |
| rs492488 | metabolite A | 1.80E-18 | S_VLDL_C | S_VLDL_C |
| rs493246 | metabolite A | 1.80E-18 | S_VLDL_C | S_VLDL_C |
| rs494242 | metabolite T | 2.50E-18 | S_VLDL_C | S_VLDL_C |
| rs495203 | metabolite T | 1.80E-18 | S_VLDL_C | S_VLDL_C |
| rs582118 | metabolite G | 1.80E-18 | S_VLDL_C | S_VLDL_C |
| rs582094 | metabolite T | 2.80E-18 | S_VLDL_C | S_VLDL_C |
| rs2769071 | metabolite G | 1.90E-18 | S_VLDL_C | S_VLDL_C |
| rs677355 | metabolite A | 2.10E-18 | S_VLDL_C | S_VLDL_C |
| rs676996 | metabolite G | 1.80E-18 | S_VLDL_C | S_VLDL_C |
| rs676457 | metabolite T | 1.80E-18 | S_VLDL_C | S_VLDL_C |
| rs550057 | metabolite T | 1.80E-25 | S_VLDL_C | S_VLDL_C |
| rs674302 | metabolite A | 1.80E-18 | S_VLDL_C | S_VLDL_C |
| rs7046674 | metabolite T | 5.40E-09 | S_VLDL_C | S_VLDL_C |
| rs554833 | metabolite T | 2.00E-18 | S_VLDL_C | S_VLDL_C |
| rs8176649 | metabolite A | 6.10E-09 | S_VLDL_C | S_VLDL_C |
| rs529565 | metabolite C | 2.60E-18 | S_VLDL_C | S_VLDL_C |
| rs630510 | metabolite G | 1.10E-08 | S_VLDL_C | S_VLDL_C |
| rs630014 | metabolite G | 1.10E-08 | S_VLDL_C | S_VLDL_C |
| rs532436 | metabolite A | 3.90E-30 | S_VLDL_C | S_VLDL_C |
| rs600038 | metabolite C | 5.10E-28 | S_VLDL_C | S_VLDL_C |
| rs1124406: | metabolite T | 1.50E-19 | S_VLDL_C | S_VLDL_C |
| rs649129 | metabolite T | 2.80E-28 | S_VLDL_C | S_VLDL_C |
| rs495828 | metabolite T | 1.40E-28 | S_VLDL_C | S_VLDL_C |
| rs7873635 | metabolite C | 4.20E-09 | XS_VLDL_C | XS_VLDL_C |
| rs4962040 | metabolite G | 2.60E-09 | XS_VLDL_C | XS_VLDL_C |
| rs8176702 | metabolite A | 2.60E-09 | XS_VLDL_C | XS_VLDL_C |
| rs2073826 | metabolite T | 4.30E-09 | XS_VLDL_C | XS_VLDL_C |
| rs687289 | metabolite A | 1.70E-18 | XS_VLDL_C | XS_VLDL_C |

| | | | | |
|---|---|---|---|---|
| rs2073827 | metabolite C | 5.60E-09 | XS_VLDL_C | XS_VLDL_C |
| rs2073828 | metabolite A | 5.60E-09 | XS_VLDL_C | XS_VLDL_C |
| rs8176691 | metabolite T | 2.10E-09 | XS_VLDL_C | XS_VLDL_C |
| rs8176690 | metabolite G | 2.00E-09 | XS_VLDL_C | XS_VLDL_C |
| rs8176682 | metabolite T | 8.90E-10 | XS_VLDL_C | XS_VLDL_C |
| rs8176681 | metabolite C | 1.80E-09 | XS_VLDL_C | XS_VLDL_C |
| rs2519093 | metabolite T | 1.80E-30 | XS_VLDL_C | XS_VLDL_C |
| rs644234 | metabolite G | 2.30E-18 | XS_VLDL_C | XS_VLDL_C |
| rs643434 | metabolite A | 2.10E-18 | XS_VLDL_C | XS_VLDL_C |
| rs543040 | metabolite T | 1.40E-18 | XS_VLDL_C | XS_VLDL_C |
| rs613534 | metabolite G | 2.40E-18 | XS_VLDL_C | XS_VLDL_C |
| rs543968 | metabolite C | 2.40E-18 | XS_VLDL_C | XS_VLDL_C |
| rs544873 | metabolite A | 2.40E-18 | XS_VLDL_C | XS_VLDL_C |
| rs545971 | metabolite T | 1.30E-18 | XS_VLDL_C | XS_VLDL_C |
| rs612169 | metabolite G | 1.80E-18 | XS_VLDL_C | XS_VLDL_C |
| rs8176668 | metabolite T | 8.20E-10 | XS_VLDL_C | XS_VLDL_C |
| rs8176663 | metabolite C | 1.30E-18 | XS_VLDL_C | XS_VLDL_C |
| rs7036642 | metabolite A | 7.60E-10 | XS_VLDL_C | XS_VLDL_C |
| rs491626 | metabolite T | 1.30E-18 | XS_VLDL_C | XS_VLDL_C |
| rs492488 | metabolite A | 1.30E-18 | XS_VLDL_C | XS_VLDL_C |
| rs493246 | metabolite A | 1.30E-18 | XS_VLDL_C | XS_VLDL_C |
| rs494242 | metabolite T | 2.30E-18 | XS_VLDL_C | XS_VLDL_C |
| rs495203 | metabolite T | 1.30E-18 | XS_VLDL_C | XS_VLDL_C |
| rs582118 | metabolite G | 1.40E-18 | XS_VLDL_C | XS_VLDL_C |
| rs582094 | metabolite T | 2.00E-18 | XS_VLDL_C | XS_VLDL_C |
| rs2769071 | metabolite G | 1.30E-18 | XS_VLDL_C | XS_VLDL_C |
| rs677355 | metabolite A | 1.50E-18 | XS_VLDL_C | XS_VLDL_C |
| rs676996 | metabolite G | 1.30E-18 | XS_VLDL_C | XS_VLDL_C |
| rs676457 | metabolite T | 1.30E-18 | XS_VLDL_C | XS_VLDL_C |
| rs550057 | metabolite T | 3.90E-25 | XS_VLDL_C | XS_VLDL_C |
| rs674302 | metabolite A | 1.30E-18 | XS_VLDL_C | XS_VLDL_C |
| rs7046674 | metabolite T | 6.00E-10 | XS_VLDL_C | XS_VLDL_C |
| rs554833 | metabolite T | 1.60E-18 | XS_VLDL_C | XS_VLDL_C |
| rs8176649 | metabolite A | 6.40E-10 | XS_VLDL_C | XS_VLDL_C |
| rs647800 | metabolite G | 4.70E-08 | XS_VLDL_C | XS_VLDL_C |
| rs529565 | metabolite C | 3.30E-18 | XS_VLDL_C | XS_VLDL_C |
| rs532436 | metabolite A | 2.50E-30 | XS_VLDL_C | XS_VLDL_C |
| rs600038 | metabolite C | 3.80E-28 | XS_VLDL_C | XS_VLDL_C |
| rs1124406: | metabolite T | 7.70E-18 | XS_VLDL_C | XS_VLDL_C |
| rs649129 | metabolite T | 3.00E-28 | XS_VLDL_C | XS_VLDL_C |
| rs495828 | metabolite T | 1.50E-28 | XS_VLDL_C | XS_VLDL_C |
| rs2519093 | metabolite T | 4.50E-12 | IDL_C_pct | IDL_C_pct |
| rs550057 | metabolite T | 3.40E-11 | IDL_C_pct | IDL_C_pct |
| rs532436 | metabolite A | 6.30E-12 | IDL_C_pct | IDL_C_pct |
| rs600038 | metabolite C | 2.60E-10 | IDL_C_pct | IDL_C_pct |
| rs649129 | metabolite T | 3.60E-10 | IDL_C_pct | IDL_C_pct |
| rs495828 | metabolite T | 3.20E-10 | IDL_C_pct | IDL_C_pct |
| rs2073826 | metabolite T | 2.00E-09 | M_LDL_C_ | M_LDL_C_pct |
| rs687289 | metabolite A | 3.10E-12 | M_LDL_C_ | M_LDL_C_pct |
| rs2073827 | metabolite C | 1.90E-09 | M_LDL_C_ | M_LDL_C_pct |

| | | | | |
|---|---|---|---|---|
| rs2073828 | metabolite A | 2.00E-09 | M_LDL_C_ | M_LDL_C_pct |
| rs8176690 | metabolite G | 1.10E-09 | M_LDL_C_ | M_LDL_C_pct |
| rs8176681 | metabolite C | 1.60E-09 | M_LDL_C_ | M_LDL_C_pct |
| rs2519093 | metabolite T | 3.80E-19 | M_LDL_C_ | M_LDL_C_pct |
| rs644234 | metabolite G | 1.60E-09 | M_LDL_C_ | M_LDL_C_pct |
| rs643434 | metabolite A | 1.60E-09 | M_LDL_C_ | M_LDL_C_pct |
| rs543040 | metabolite T | 2.60E-12 | M_LDL_C_ | M_LDL_C_pct |
| rs613534 | metabolite G | 1.60E-09 | M_LDL_C_ | M_LDL_C_pct |
| rs543968 | metabolite C | 1.60E-09 | M_LDL_C_ | M_LDL_C_pct |
| rs544873 | metabolite A | 1.60E-09 | M_LDL_C_ | M_LDL_C_pct |
| rs545971 | metabolite T | 2.90E-12 | M_LDL_C_ | M_LDL_C_pct |
| rs612169 | metabolite G | 3.10E-12 | M_LDL_C_ | M_LDL_C_pct |
| rs8176663 | metabolite C | 2.90E-12 | M_LDL_C_ | M_LDL_C_pct |
| rs491626 | metabolite T | 2.90E-12 | M_LDL_C_ | M_LDL_C_pct |
| rs492488 | metabolite A | 2.90E-12 | M_LDL_C_ | M_LDL_C_pct |
| rs493246 | metabolite A | 2.90E-12 | M_LDL_C_ | M_LDL_C_pct |
| rs494242 | metabolite T | 1.60E-09 | M_LDL_C_ | M_LDL_C_pct |
| rs495203 | metabolite T | 2.90E-12 | M_LDL_C_ | M_LDL_C_pct |
| rs582118 | metabolite G | 2.90E-12 | M_LDL_C_ | M_LDL_C_pct |
| rs582094 | metabolite T | 3.90E-12 | M_LDL_C_ | M_LDL_C_pct |
| rs2769071 | metabolite G | 3.20E-12 | M_LDL_C_ | M_LDL_C_pct |
| rs677355 | metabolite A | 3.40E-12 | M_LDL_C_ | M_LDL_C_pct |
| rs676996 | metabolite G | 3.30E-12 | M_LDL_C_ | M_LDL_C_pct |
| rs676457 | metabolite T | 3.30E-12 | M_LDL_C_ | M_LDL_C_pct |
| rs550057 | metabolite T | 4.30E-16 | M_LDL_C_ | M_LDL_C_pct |
| rs674302 | metabolite A | 3.20E-12 | M_LDL_C_ | M_LDL_C_pct |
| rs554833 | metabolite T | 3.10E-12 | M_LDL_C_ | M_LDL_C_pct |
| rs529565 | metabolite C | 4.00E-12 | M_LDL_C_ | M_LDL_C_pct |
| rs532436 | metabolite A | 4.30E-19 | M_LDL_C_ | M_LDL_C_pct |
| rs600038 | metabolite C | 7.20E-14 | M_LDL_C_ | M_LDL_C_pct |
| rs1124406 | metabolite T | 2.90E-09 | M_LDL_C_ | M_LDL_C_pct |
| rs649129 | metabolite T | 1.40E-13 | M_LDL_C_ | M_LDL_C_pct |
| rs495828 | metabolite T | 2.30E-13 | M_LDL_C_ | M_LDL_C_pct |
| rs7873635 | metabolite C | 7.30E-09 | M_VLDL_C | M_VLDL_C_pct |
| rs4962040 | metabolite G | 7.60E-09 | M_VLDL_C | M_VLDL_C_pct |
| rs8176702 | metabolite A | 1.00E-08 | M_VLDL_C | M_VLDL_C_pct |
| rs2073826 | metabolite T | 4.50E-09 | M_VLDL_C | M_VLDL_C_pct |
| rs687289 | metabolite A | 5.90E-18 | M_VLDL_C | M_VLDL_C_pct |
| rs2073827 | metabolite C | 4.60E-09 | M_VLDL_C | M_VLDL_C_pct |
| rs2073828 | metabolite A | 4.60E-09 | M_VLDL_C | M_VLDL_C_pct |
| rs8176691 | metabolite T | 7.10E-09 | M_VLDL_C | M_VLDL_C_pct |
| rs8176690 | metabolite G | 1.80E-09 | M_VLDL_C | M_VLDL_C_pct |
| rs8176682 | metabolite T | 3.80E-09 | M_VLDL_C | M_VLDL_C_pct |
| rs8176681 | metabolite C | 2.50E-09 | M_VLDL_C | M_VLDL_C_pct |
| rs2519093 | metabolite T | 1.20E-31 | M_VLDL_C | M_VLDL_C_pct |
| rs644234 | metabolite G | 2.60E-16 | M_VLDL_C | M_VLDL_C_pct |
| rs643434 | metabolite A | 2.90E-16 | M_VLDL_C | M_VLDL_C_pct |
| rs543040 | metabolite T | 2.10E-17 | M_VLDL_C | M_VLDL_C_pct |
| rs613534 | metabolite G | 2.70E-16 | M_VLDL_C | M_VLDL_C_pct |
| rs543968 | metabolite C | 2.70E-16 | M_VLDL_C | M_VLDL_C_pct |

| | | | | |
|---|---|---|---|---|
| rs544873 | metabolite A | 2.70E-16 | M_VLDL_C | M_VLDL_C_pct |
| rs545971 | metabolite T | 1.20E-17 | M_VLDL_C | M_VLDL_C_pct |
| rs612169 | metabolite G | 1.30E-17 | M_VLDL_C | M_VLDL_C_pct |
| rs8176668 | metabolite T | 1.20E-08 | M_VLDL_C | M_VLDL_C_pct |
| rs8176663 | metabolite C | 1.20E-17 | M_VLDL_C | M_VLDL_C_pct |
| rs7036642 | metabolite A | 1.30E-08 | M_VLDL_C | M_VLDL_C_pct |
| rs491626 | metabolite T | 1.20E-17 | M_VLDL_C | M_VLDL_C_pct |
| rs492488 | metabolite A | 1.20E-17 | M_VLDL_C | M_VLDL_C_pct |
| rs493246 | metabolite A | 1.20E-17 | M_VLDL_C | M_VLDL_C_pct |
| rs494242 | metabolite T | 2.70E-16 | M_VLDL_C | M_VLDL_C_pct |
| rs495203 | metabolite T | 1.20E-17 | M_VLDL_C | M_VLDL_C_pct |
| rs582118 | metabolite G | 1.20E-17 | M_VLDL_C | M_VLDL_C_pct |
| rs582094 | metabolite T | 1.50E-17 | M_VLDL_C | M_VLDL_C_pct |
| rs2769071 | metabolite G | 1.50E-17 | M_VLDL_C | M_VLDL_C_pct |
| rs677355 | metabolite A | 1.20E-17 | M_VLDL_C | M_VLDL_C_pct |
| rs676996 | metabolite G | 1.30E-17 | M_VLDL_C | M_VLDL_C_pct |
| rs676457 | metabolite T | 1.30E-17 | M_VLDL_C | M_VLDL_C_pct |
| rs550057 | metabolite T | 3.30E-26 | M_VLDL_C | M_VLDL_C_pct |
| rs674302 | metabolite A | 1.40E-17 | M_VLDL_C | M_VLDL_C_pct |
| rs7046674 | metabolite T | 1.40E-08 | M_VLDL_C | M_VLDL_C_pct |
| rs554833 | metabolite T | 1.40E-17 | M_VLDL_C | M_VLDL_C_pct |
| rs8176649 | metabolite A | 1.20E-08 | M_VLDL_C | M_VLDL_C_pct |
| rs660340 | metabolite G | 1.10E-08 | M_VLDL_C | M_VLDL_C_pct |
| rs581107 | metabolite T | 1.10E-08 | M_VLDL_C | M_VLDL_C_pct |
| rs659104 | metabolite G | 1.10E-08 | M_VLDL_C | M_VLDL_C_pct |
| rs647800 | metabolite G | 3.30E-08 | M_VLDL_C | M_VLDL_C_pct |
| rs473533 | metabolite C | 1.10E-08 | M_VLDL_C | M_VLDL_C_pct |
| rs476410 | metabolite C | 1.20E-08 | M_VLDL_C | M_VLDL_C_pct |
| rs645982 | metabolite G | 1.00E-08 | M_VLDL_C | M_VLDL_C_pct |
| rs529565 | metabolite C | 2.10E-17 | M_VLDL_C | M_VLDL_C_pct |
| rs532436 | metabolite A | 1.90E-31 | M_VLDL_C | M_VLDL_C_pct |
| rs600038 | metabolite C | 2.00E-27 | M_VLDL_C | M_VLDL_C_pct |
| rs1124406 | metabolite T | 1.20E-17 | M_VLDL_C | M_VLDL_C_pct |
| rs649129 | metabolite T | 3.20E-27 | M_VLDL_C | M_VLDL_C_pct |
| rs495828 | metabolite T | 2.80E-27 | M_VLDL_C | M_VLDL_C_pct |
| rs7873635 | metabolite C | 1.70E-12 | S_LDL_C_p | S_LDL_C_pct |
| rs4962040 | metabolite G | 1.30E-12 | S_LDL_C_p | S_LDL_C_pct |
| rs8176702 | metabolite A | 2.00E-12 | S_LDL_C_p | S_LDL_C_pct |
| rs2073826 | metabolite T | 5.80E-14 | S_LDL_C_p | S_LDL_C_pct |
| rs687289 | metabolite A | 8.10E-25 | S_LDL_C_p | S_LDL_C_pct |
| rs2073827 | metabolite C | 4.90E-14 | S_LDL_C_p | S_LDL_C_pct |
| rs2073828 | metabolite A | 5.10E-14 | S_LDL_C_p | S_LDL_C_pct |
| rs8176691 | metabolite T | 1.20E-12 | S_LDL_C_p | S_LDL_C_pct |
| rs8176690 | metabolite G | 2.70E-14 | S_LDL_C_p | S_LDL_C_pct |
| rs8176682 | metabolite T | 9.00E-13 | S_LDL_C_p | S_LDL_C_pct |
| rs8176681 | metabolite C | 3.70E-14 | S_LDL_C_p | S_LDL_C_pct |
| rs2519093 | metabolite T | 8.10E-47 | S_LDL_C_p | S_LDL_C_pct |
| rs644234 | metabolite G | 5.50E-22 | S_LDL_C_p | S_LDL_C_pct |
| rs643434 | metabolite A | 5.30E-22 | S_LDL_C_p | S_LDL_C_pct |
| rs543040 | metabolite T | 7.00E-25 | S_LDL_C_p | S_LDL_C_pct |

| | | | | |
|---|---|---|---|---|
| rs613534 | metabolite G | 5.60E-22 | S_LDL_C_p | S_LDL_C_pct |
| rs543968 | metabolite C | 5.60E-22 | S_LDL_C_p | S_LDL_C_pct |
| rs544873 | metabolite A | 5.60E-22 | S_LDL_C_p | S_LDL_C_pct |
| rs545971 | metabolite T | 5.40E-25 | S_LDL_C_p | S_LDL_C_pct |
| rs612169 | metabolite G | 8.20E-25 | S_LDL_C_p | S_LDL_C_pct |
| rs8176668 | metabolite T | 1.40E-12 | S_LDL_C_p | S_LDL_C_pct |
| rs8176663 | metabolite C | 5.40E-25 | S_LDL_C_p | S_LDL_C_pct |
| rs7036642 | metabolite A | 1.40E-12 | S_LDL_C_p | S_LDL_C_pct |
| rs491626 | metabolite T | 5.40E-25 | S_LDL_C_p | S_LDL_C_pct |
| rs492488 | metabolite A | 5.40E-25 | S_LDL_C_p | S_LDL_C_pct |
| rs493246 | metabolite A | 5.40E-25 | S_LDL_C_p | S_LDL_C_pct |
| rs494242 | metabolite T | 5.60E-22 | S_LDL_C_p | S_LDL_C_pct |
| rs495203 | metabolite T | 5.50E-25 | S_LDL_C_p | S_LDL_C_pct |
| rs582118 | metabolite G | 5.70E-25 | S_LDL_C_p | S_LDL_C_pct |
| rs582094 | metabolite T | 9.90E-25 | S_LDL_C_p | S_LDL_C_pct |
| rs2769071 | metabolite G | 7.30E-25 | S_LDL_C_p | S_LDL_C_pct |
| rs677355 | metabolite A | 6.50E-25 | S_LDL_C_p | S_LDL_C_pct |
| rs676996 | metabolite G | 6.40E-25 | S_LDL_C_p | S_LDL_C_pct |
| rs676457 | metabolite T | 6.40E-25 | S_LDL_C_p | S_LDL_C_pct |
| rs550057 | metabolite T | 3.20E-37 | S_LDL_C_p | S_LDL_C_pct |
| rs674302 | metabolite A | 6.20E-25 | S_LDL_C_p | S_LDL_C_pct |
| rs7046674 | metabolite T | 1.20E-12 | S_LDL_C_p | S_LDL_C_pct |
| rs554833 | metabolite T | 6.00E-25 | S_LDL_C_p | S_LDL_C_pct |
| rs8176649 | metabolite A | 1.60E-12 | S_LDL_C_p | S_LDL_C_pct |
| rs660340 | metabolite G | 1.10E-10 | S_LDL_C_p | S_LDL_C_pct |
| rs581107 | metabolite T | 8.80E-11 | S_LDL_C_p | S_LDL_C_pct |
| rs659104 | metabolite G | 1.00E-10 | S_LDL_C_p | S_LDL_C_pct |
| rs647800 | metabolite G | 1.90E-09 | S_LDL_C_p | S_LDL_C_pct |
| rs473533 | metabolite C | 1.10E-10 | S_LDL_C_p | S_LDL_C_pct |
| rs476410 | metabolite C | 1.10E-10 | S_LDL_C_p | S_LDL_C_pct |
| rs645982 | metabolite G | 8.40E-11 | S_LDL_C_p | S_LDL_C_pct |
| rs529565 | metabolite C | 4.00E-25 | S_LDL_C_p | S_LDL_C_pct |
| rs630510 | metabolite G | 4.70E-09 | S_LDL_C_p | S_LDL_C_pct |
| rs630014 | metabolite G | 4.70E-09 | S_LDL_C_p | S_LDL_C_pct |
| rs532436 | metabolite A | 2.10E-46 | S_LDL_C_p | S_LDL_C_pct |
| rs600038 | metabolite C | 1.80E-38 | S_LDL_C_p | S_LDL_C_pct |
| rs1124406: | metabolite T | 1.40E-28 | S_LDL_C_p | S_LDL_C_pct |
| rs649129 | metabolite T | 1.30E-38 | S_LDL_C_p | S_LDL_C_pct |
| rs495828 | metabolite T | 3.20E-38 | S_LDL_C_p | S_LDL_C_pct |
| rs7873635 | metabolite C | 1.10E-11 | S_VLDL_C_ | S_VLDL_C_pct |
| rs4962040 | metabolite G | 1.00E-11 | S_VLDL_C_ | S_VLDL_C_pct |
| rs8176702 | metabolite A | 1.30E-11 | S_VLDL_C_ | S_VLDL_C_pct |
| rs2073826 | metabolite T | 8.10E-12 | S_VLDL_C_ | S_VLDL_C_pct |
| rs687289 | metabolite A | 3.20E-22 | S_VLDL_C_ | S_VLDL_C_pct |
| rs2073827 | metabolite C | 7.80E-12 | S_VLDL_C_ | S_VLDL_C_pct |
| rs2073828 | metabolite A | 7.90E-12 | S_VLDL_C_ | S_VLDL_C_pct |
| rs8176691 | metabolite T | 7.40E-12 | S_VLDL_C_ | S_VLDL_C_pct |
| rs8176690 | metabolite G | 2.20E-12 | S_VLDL_C_ | S_VLDL_C_pct |
| rs8176682 | metabolite T | 2.90E-12 | S_VLDL_C_ | S_VLDL_C_pct |
| rs8176681 | metabolite C | 2.90E-12 | S_VLDL_C_ | S_VLDL_C_pct |

| | | | | | |
|---|---|---|---|---|---|
| rs2519093 | metabolite | T | 5.60E-42 | S_VLDL_C_ | S_VLDL_C_pct |
| rs644234 | metabolite | G | 7.90E-21 | S_VLDL_C_ | S_VLDL_C_pct |
| rs643434 | metabolite | A | 9.10E-21 | S_VLDL_C_ | S_VLDL_C_pct |
| rs543040 | metabolite | T | 1.10E-21 | S_VLDL_C_ | S_VLDL_C_pct |
| rs613534 | metabolite | G | 8.30E-21 | S_VLDL_C_ | S_VLDL_C_pct |
| rs543968 | metabolite | C | 8.30E-21 | S_VLDL_C_ | S_VLDL_C_pct |
| rs544873 | metabolite | A | 8.30E-21 | S_VLDL_C_ | S_VLDL_C_pct |
| rs545971 | metabolite | T | 5.20E-22 | S_VLDL_C_ | S_VLDL_C_pct |
| rs612169 | metabolite | G | 6.30E-22 | S_VLDL_C_ | S_VLDL_C_pct |
| rs8176668 | metabolite | T | 8.70E-12 | S_VLDL_C_ | S_VLDL_C_pct |
| rs8176663 | metabolite | C | 5.20E-22 | S_VLDL_C_ | S_VLDL_C_pct |
| rs7036642 | metabolite | A | 1.00E-11 | S_VLDL_C_ | S_VLDL_C_pct |
| rs491626 | metabolite | T | 5.20E-22 | S_VLDL_C_ | S_VLDL_C_pct |
| rs492488 | metabolite | A | 5.20E-22 | S_VLDL_C_ | S_VLDL_C_pct |
| rs493246 | metabolite | A | 5.20E-22 | S_VLDL_C_ | S_VLDL_C_pct |
| rs494242 | metabolite | T | 8.60E-21 | S_VLDL_C_ | S_VLDL_C_pct |
| rs495203 | metabolite | T | 5.20E-22 | S_VLDL_C_ | S_VLDL_C_pct |
| rs582118 | metabolite | G | 5.30E-22 | S_VLDL_C_ | S_VLDL_C_pct |
| rs582094 | metabolite | T | 8.50E-22 | S_VLDL_C_ | S_VLDL_C_pct |
| rs2769071 | metabolite | G | 6.50E-22 | S_VLDL_C_ | S_VLDL_C_pct |
| rs677355 | metabolite | A | 5.40E-22 | S_VLDL_C_ | S_VLDL_C_pct |
| rs676996 | metabolite | G | 5.90E-22 | S_VLDL_C_ | S_VLDL_C_pct |
| rs676457 | metabolite | T | 5.90E-22 | S_VLDL_C_ | S_VLDL_C_pct |
| rs550057 | metabolite | T | 6.60E-35 | S_VLDL_C_ | S_VLDL_C_pct |
| rs674302 | metabolite | A | 6.00E-22 | S_VLDL_C_ | S_VLDL_C_pct |
| rs7046674 | metabolite | T | 8.90E-12 | S_VLDL_C_ | S_VLDL_C_pct |
| rs554833 | metabolite | T | 6.50E-22 | S_VLDL_C_ | S_VLDL_C_pct |
| rs8176649 | metabolite | A | 8.30E-12 | S_VLDL_C_ | S_VLDL_C_pct |
| rs660340 | metabolite | G | 3.30E-09 | S_VLDL_C_ | S_VLDL_C_pct |
| rs581107 | metabolite | T | 3.20E-09 | S_VLDL_C_ | S_VLDL_C_pct |
| rs659104 | metabolite | G | 3.50E-09 | S_VLDL_C_ | S_VLDL_C_pct |
| rs647800 | metabolite | G | 6.20E-09 | S_VLDL_C_ | S_VLDL_C_pct |
| rs473533 | metabolite | C | 3.60E-09 | S_VLDL_C_ | S_VLDL_C_pct |
| rs476410 | metabolite | C | 3.60E-09 | S_VLDL_C_ | S_VLDL_C_pct |
| rs645982 | metabolite | G | 3.10E-09 | S_VLDL_C_ | S_VLDL_C_pct |
| rs529565 | metabolite | C | 1.10E-21 | S_VLDL_C_ | S_VLDL_C_pct |
| rs532436 | metabolite | A | 1.30E-41 | S_VLDL_C_ | S_VLDL_C_pct |
| rs600038 | metabolite | C | 6.40E-37 | S_VLDL_C_ | S_VLDL_C_pct |
| rs1124406: | metabolite | T | 1.20E-24 | S_VLDL_C_ | S_VLDL_C_pct |
| rs649129 | metabolite | T | 1.10E-36 | S_VLDL_C_ | S_VLDL_C_pct |
| rs495828 | metabolite | T | 1.10E-36 | S_VLDL_C_ | S_VLDL_C_pct |
| rs2519093 | metabolite | T | 7.70E-17 | XS_VLDL_C | XS_VLDL_C_pct |
| rs550057 | metabolite | T | 2.30E-14 | XS_VLDL_C | XS_VLDL_C_pct |
| rs532436 | metabolite | A | 1.10E-16 | XS_VLDL_C | XS_VLDL_C_pct |
| rs600038 | metabolite | C | 3.10E-14 | XS_VLDL_C | XS_VLDL_C_pct |
| rs1124406: | metabolite | T | 2.70E-08 | XS_VLDL_C | XS_VLDL_C_pct |
| rs649129 | metabolite | T | 5.30E-14 | XS_VLDL_C | XS_VLDL_C_pct |
| rs495828 | metabolite | T | 5.50E-14 | XS_VLDL_C | XS_VLDL_C_pct |
| rs687289 | metabolite | A | 1.80E-14 | IDL_CE | IDL_CE |
| rs8176690 | metabolite | G | 4.00E-08 | IDL_CE | IDL_CE |

| | | | | |
|---|---|---|---|---|
| rs8176682 | metabolite T | 3.40E-08 | IDL_CE | IDL_CE |
| rs8176681 | metabolite C | 4.30E-08 | IDL_CE | IDL_CE |
| rs2519093 | metabolite T | 4.70E-30 | IDL_CE | IDL_CE |
| rs644234 | metabolite G | 4.30E-14 | IDL_CE | IDL_CE |
| rs643434 | metabolite A | 3.80E-14 | IDL_CE | IDL_CE |
| rs543040 | metabolite T | 1.60E-14 | IDL_CE | IDL_CE |
| rs613534 | metabolite G | 4.30E-14 | IDL_CE | IDL_CE |
| rs543968 | metabolite C | 4.30E-14 | IDL_CE | IDL_CE |
| rs544873 | metabolite A | 4.30E-14 | IDL_CE | IDL_CE |
| rs545971 | metabolite T | 1.30E-14 | IDL_CE | IDL_CE |
| rs612169 | metabolite G | 1.60E-14 | IDL_CE | IDL_CE |
| rs8176668 | metabolite T | 4.50E-08 | IDL_CE | IDL_CE |
| rs8176663 | metabolite C | 1.30E-14 | IDL_CE | IDL_CE |
| rs7036642 | metabolite A | 4.00E-08 | IDL_CE | IDL_CE |
| rs491626 | metabolite T | 1.30E-14 | IDL_CE | IDL_CE |
| rs492488 | metabolite A | 1.30E-14 | IDL_CE | IDL_CE |
| rs493246 | metabolite A | 1.30E-14 | IDL_CE | IDL_CE |
| rs494242 | metabolite T | 4.20E-14 | IDL_CE | IDL_CE |
| rs495203 | metabolite T | 1.30E-14 | IDL_CE | IDL_CE |
| rs582118 | metabolite G | 1.40E-14 | IDL_CE | IDL_CE |
| rs582094 | metabolite T | 1.80E-14 | IDL_CE | IDL_CE |
| rs2769071 | metabolite G | 1.30E-14 | IDL_CE | IDL_CE |
| rs677355 | metabolite A | 1.50E-14 | IDL_CE | IDL_CE |
| rs676996 | metabolite G | 1.30E-14 | IDL_CE | IDL_CE |
| rs676457 | metabolite T | 1.30E-14 | IDL_CE | IDL_CE |
| rs550057 | metabolite T | 1.80E-24 | IDL_CE | IDL_CE |
| rs674302 | metabolite A | 1.30E-14 | IDL_CE | IDL_CE |
| rs7046674 | metabolite T | 3.20E-08 | IDL_CE | IDL_CE |
| rs554833 | metabolite T | 1.50E-14 | IDL_CE | IDL_CE |
| rs8176649 | metabolite A | 3.40E-08 | IDL_CE | IDL_CE |
| rs529565 | metabolite C | 4.10E-14 | IDL_CE | IDL_CE |
| rs532436 | metabolite A | 7.40E-30 | IDL_CE | IDL_CE |
| rs600038 | metabolite C | 4.00E-27 | IDL_CE | IDL_CE |
| rs1124406: | metabolite T | 4.10E-17 | IDL_CE | IDL_CE |
| rs649129 | metabolite T | 3.40E-27 | IDL_CE | IDL_CE |
| rs495828 | metabolite T | 1.60E-27 | IDL_CE | IDL_CE |
| rs687289 | metabolite A | 1.40E-12 | L_LDL_CE | L_LDL_CE |
| rs2519093 | metabolite T | 3.70E-29 | L_LDL_CE | L_LDL_CE |
| rs644234 | metabolite G | 9.80E-12 | L_LDL_CE | L_LDL_CE |
| rs643434 | metabolite A | 9.10E-12 | L_LDL_CE | L_LDL_CE |
| rs543040 | metabolite T | 1.00E-12 | L_LDL_CE | L_LDL_CE |
| rs613534 | metabolite G | 9.90E-12 | L_LDL_CE | L_LDL_CE |
| rs543968 | metabolite C | 9.90E-12 | L_LDL_CE | L_LDL_CE |
| rs544873 | metabolite A | 9.90E-12 | L_LDL_CE | L_LDL_CE |
| rs545971 | metabolite T | 9.80E-13 | L_LDL_CE | L_LDL_CE |
| rs612169 | metabolite G | 1.20E-12 | L_LDL_CE | L_LDL_CE |
| rs8176663 | metabolite C | 9.80E-13 | L_LDL_CE | L_LDL_CE |
| rs491626 | metabolite T | 9.70E-13 | L_LDL_CE | L_LDL_CE |
| rs492488 | metabolite A | 9.70E-13 | L_LDL_CE | L_LDL_CE |
| rs493246 | metabolite A | 9.70E-13 | L_LDL_CE | L_LDL_CE |

| | | | | |
|---|---|---|---|---|
| rs494242 | metabolite T | 9.50E-12 | L_LDL_CE | L_LDL_CE |
| rs495203 | metabolite T | 9.80E-13 | L_LDL_CE | L_LDL_CE |
| rs582118 | metabolite G | 1.00E-12 | L_LDL_CE | L_LDL_CE |
| rs582094 | metabolite T | 1.50E-12 | L_LDL_CE | L_LDL_CE |
| rs2769071 | metabolite G | 1.00E-12 | L_LDL_CE | L_LDL_CE |
| rs677355 | metabolite A | 1.20E-12 | L_LDL_CE | L_LDL_CE |
| rs676996 | metabolite G | 1.00E-12 | L_LDL_CE | L_LDL_CE |
| rs676457 | metabolite T | 1.00E-12 | L_LDL_CE | L_LDL_CE |
| rs550057 | metabolite T | 1.00E-22 | L_LDL_CE | L_LDL_CE |
| rs674302 | metabolite A | 1.00E-12 | L_LDL_CE | L_LDL_CE |
| rs554833 | metabolite T | 1.10E-12 | L_LDL_CE | L_LDL_CE |
| rs529565 | metabolite C | 1.70E-12 | L_LDL_CE | L_LDL_CE |
| rs532436 | metabolite A | 5.40E-29 | L_LDL_CE | L_LDL_CE |
| rs600038 | metabolite C | 2.00E-25 | L_LDL_CE | L_LDL_CE |
| rs1124406: | metabolite T | 1.20E-17 | L_LDL_CE | L_LDL_CE |
| rs649129 | metabolite T | 1.30E-25 | L_LDL_CE | L_LDL_CE |
| rs495828 | metabolite T | 9.70E-26 | L_LDL_CE | L_LDL_CE |
| rs2519093 | metabolite T | 1.30E-10 | L_VLDL_CE | L_VLDL_CE |
| rs644234 | metabolite G | 2.90E-08 | L_VLDL_CE | L_VLDL_CE |
| rs643434 | metabolite A | 2.70E-08 | L_VLDL_CE | L_VLDL_CE |
| rs613534 | metabolite G | 2.90E-08 | L_VLDL_CE | L_VLDL_CE |
| rs543968 | metabolite C | 2.90E-08 | L_VLDL_CE | L_VLDL_CE |
| rs544873 | metabolite A | 2.90E-08 | L_VLDL_CE | L_VLDL_CE |
| rs494242 | metabolite T | 2.80E-08 | L_VLDL_CE | L_VLDL_CE |
| rs550057 | metabolite T | 7.80E-10 | L_VLDL_CE | L_VLDL_CE |
| rs532436 | metabolite A | 1.40E-10 | L_VLDL_CE | L_VLDL_CE |
| rs600038 | metabolite C | 1.80E-10 | L_VLDL_CE | L_VLDL_CE |
| rs649129 | metabolite T | 1.00E-10 | L_VLDL_CE | L_VLDL_CE |
| rs495828 | metabolite T | 6.30E-11 | L_VLDL_CE | L_VLDL_CE |
| rs687289 | metabolite A | 3.10E-13 | LDL_CE | LDL_CE |
| rs2519093 | metabolite T | 9.20E-29 | LDL_CE | LDL_CE |
| rs644234 | metabolite G | 2.10E-12 | LDL_CE | LDL_CE |
| rs643434 | metabolite A | 1.90E-12 | LDL_CE | LDL_CE |
| rs543040 | metabolite T | 2.10E-13 | LDL_CE | LDL_CE |
| rs613534 | metabolite G | 2.10E-12 | LDL_CE | LDL_CE |
| rs543968 | metabolite C | 2.10E-12 | LDL_CE | LDL_CE |
| rs544873 | metabolite A | 2.10E-12 | LDL_CE | LDL_CE |
| rs545971 | metabolite T | 2.20E-13 | LDL_CE | LDL_CE |
| rs612169 | metabolite G | 2.70E-13 | LDL_CE | LDL_CE |
| rs8176663 | metabolite C | 2.20E-13 | LDL_CE | LDL_CE |
| rs491626 | metabolite T | 2.20E-13 | LDL_CE | LDL_CE |
| rs492488 | metabolite A | 2.20E-13 | LDL_CE | LDL_CE |
| rs493246 | metabolite A | 2.20E-13 | LDL_CE | LDL_CE |
| rs494242 | metabolite T | 2.00E-12 | LDL_CE | LDL_CE |
| rs495203 | metabolite T | 2.20E-13 | LDL_CE | LDL_CE |
| rs582118 | metabolite G | 2.20E-13 | LDL_CE | LDL_CE |
| rs582094 | metabolite T | 3.30E-13 | LDL_CE | LDL_CE |
| rs2769071 | metabolite G | 2.30E-13 | LDL_CE | LDL_CE |
| rs677355 | metabolite A | 2.60E-13 | LDL_CE | LDL_CE |
| rs676996 | metabolite G | 2.30E-13 | LDL_CE | LDL_CE |

| | | | | |
|---|---|---|---|---|
| rs676457 | metabolite T | 2.30E-13 | LDL_CE | LDL_CE |
| rs550057 | metabolite T | 6.60E-23 | LDL_CE | LDL_CE |
| rs674302 | metabolite A | 2.30E-13 | LDL_CE | LDL_CE |
| rs554833 | metabolite T | 2.50E-13 | LDL_CE | LDL_CE |
| rs529565 | metabolite C | 3.50E-13 | LDL_CE | LDL_CE |
| rs532436 | metabolite A | 1.20E-28 | LDL_CE | LDL_CE |
| rs600038 | metabolite C | 3.60E-25 | LDL_CE | LDL_CE |
| rs1124406: | metabolite T | 1.30E-17 | LDL_CE | LDL_CE |
| rs649129 | metabolite T | 2.20E-25 | LDL_CE | LDL_CE |
| rs495828 | metabolite T | 1.60E-25 | LDL_CE | LDL_CE |
| rs687289 | metabolite A | 1.30E-13 | M_LDL_CE | M_LDL_CE |
| rs2519093 | metabolite T | 1.90E-26 | M_LDL_CE | M_LDL_CE |
| rs644234 | metabolite G | 8.60E-13 | M_LDL_CE | M_LDL_CE |
| rs643434 | metabolite A | 8.10E-13 | M_LDL_CE | M_LDL_CE |
| rs543040 | metabolite T | 8.90E-14 | M_LDL_CE | M_LDL_CE |
| rs613534 | metabolite G | 8.50E-13 | M_LDL_CE | M_LDL_CE |
| rs543968 | metabolite C | 8.50E-13 | M_LDL_CE | M_LDL_CE |
| rs544873 | metabolite A | 8.50E-13 | M_LDL_CE | M_LDL_CE |
| rs545971 | metabolite T | 9.60E-14 | M_LDL_CE | M_LDL_CE |
| rs612169 | metabolite G | 1.20E-13 | M_LDL_CE | M_LDL_CE |
| rs8176663 | metabolite C | 9.60E-14 | M_LDL_CE | M_LDL_CE |
| rs491626 | metabolite T | 9.50E-14 | M_LDL_CE | M_LDL_CE |
| rs492488 | metabolite A | 9.50E-14 | M_LDL_CE | M_LDL_CE |
| rs493246 | metabolite A | 9.50E-14 | M_LDL_CE | M_LDL_CE |
| rs494242 | metabolite T | 8.20E-13 | M_LDL_CE | M_LDL_CE |
| rs495203 | metabolite T | 9.60E-14 | M_LDL_CE | M_LDL_CE |
| rs582118 | metabolite G | 9.90E-14 | M_LDL_CE | M_LDL_CE |
| rs582094 | metabolite T | 1.40E-13 | M_LDL_CE | M_LDL_CE |
| rs2769071 | metabolite G | 1.00E-13 | M_LDL_CE | M_LDL_CE |
| rs677355 | metabolite A | 1.20E-13 | M_LDL_CE | M_LDL_CE |
| rs676996 | metabolite G | 1.00E-13 | M_LDL_CE | M_LDL_CE |
| rs676457 | metabolite T | 1.00E-13 | M_LDL_CE | M_LDL_CE |
| rs550057 | metabolite T | 9.80E-22 | M_LDL_CE | M_LDL_CE |
| rs674302 | metabolite A | 1.00E-13 | M_LDL_CE | M_LDL_CE |
| rs554833 | metabolite T | 1.10E-13 | M_LDL_CE | M_LDL_CE |
| rs529565 | metabolite C | 1.20E-13 | M_LDL_CE | M_LDL_CE |
| rs532436 | metabolite A | 2.20E-26 | M_LDL_CE | M_LDL_CE |
| rs600038 | metabolite C | 3.20E-23 | M_LDL_CE | M_LDL_CE |
| rs1124406: | metabolite T | 1.50E-16 | M_LDL_CE | M_LDL_CE |
| rs649129 | metabolite T | 1.90E-23 | M_LDL_CE | M_LDL_CE |
| rs495828 | metabolite T | 1.40E-23 | M_LDL_CE | M_LDL_CE |
| rs7873635 | metabolite C | 5.30E-10 | M_VLDL_C | M_VLDL_CE |
| rs4962040 | metabolite G | 3.20E-10 | M_VLDL_C | M_VLDL_CE |
| rs8176702 | metabolite A | 3.60E-10 | M_VLDL_C | M_VLDL_CE |
| rs2073826 | metabolite T | 3.00E-10 | M_VLDL_C | M_VLDL_CE |
| rs687289 | metabolite A | 3.20E-20 | M_VLDL_C | M_VLDL_CE |
| rs2073827 | metabolite C | 3.20E-10 | M_VLDL_C | M_VLDL_CE |
| rs2073828 | metabolite A | 3.30E-10 | M_VLDL_C | M_VLDL_CE |
| rs8176691 | metabolite T | 2.30E-10 | M_VLDL_C | M_VLDL_CE |
| rs8176690 | metabolite G | 1.10E-10 | M_VLDL_C | M_VLDL_CE |

| | | | | |
|---|---|---|---|---|
| rs8176682 | metabolite T | 9.70E-11 | M_VLDL_C | M_VLDL_CE |
| rs8176681 | metabolite C | 1.10E-10 | M_VLDL_C | M_VLDL_CE |
| rs2519093 | metabolite T | 7.60E-37 | M_VLDL_C | M_VLDL_CE |
| rs644234 | metabolite G | 1.30E-19 | M_VLDL_C | M_VLDL_CE |
| rs643434 | metabolite A | 1.20E-19 | M_VLDL_C | M_VLDL_CE |
| rs543040 | metabolite T | 2.70E-20 | M_VLDL_C | M_VLDL_CE |
| rs613534 | metabolite G | 1.30E-19 | M_VLDL_C | M_VLDL_CE |
| rs543968 | metabolite C | 1.30E-19 | M_VLDL_C | M_VLDL_CE |
| rs544873 | metabolite A | 1.30E-19 | M_VLDL_C | M_VLDL_CE |
| rs545971 | metabolite T | 2.30E-20 | M_VLDL_C | M_VLDL_CE |
| rs612169 | metabolite G | 3.10E-20 | M_VLDL_C | M_VLDL_CE |
| rs8176668 | metabolite T | 9.10E-11 | M_VLDL_C | M_VLDL_CE |
| rs8176663 | metabolite C | 2.30E-20 | M_VLDL_C | M_VLDL_CE |
| rs7036642 | metabolite A | 8.70E-11 | M_VLDL_C | M_VLDL_CE |
| rs491626 | metabolite T | 2.30E-20 | M_VLDL_C | M_VLDL_CE |
| rs492488 | metabolite A | 2.30E-20 | M_VLDL_C | M_VLDL_CE |
| rs493246 | metabolite A | 2.30E-20 | M_VLDL_C | M_VLDL_CE |
| rs494242 | metabolite T | 1.30E-19 | M_VLDL_C | M_VLDL_CE |
| rs495203 | metabolite T | 2.30E-20 | M_VLDL_C | M_VLDL_CE |
| rs582118 | metabolite G | 2.40E-20 | M_VLDL_C | M_VLDL_CE |
| rs582094 | metabolite T | 3.90E-20 | M_VLDL_C | M_VLDL_CE |
| rs2769071 | metabolite G | 2.50E-20 | M_VLDL_C | M_VLDL_CE |
| rs677355 | metabolite A | 2.70E-20 | M_VLDL_C | M_VLDL_CE |
| rs676996 | metabolite G | 2.40E-20 | M_VLDL_C | M_VLDL_CE |
| rs676457 | metabolite T | 2.40E-20 | M_VLDL_C | M_VLDL_CE |
| rs550057 | metabolite T | 7.40E-31 | M_VLDL_C | M_VLDL_CE |
| rs674302 | metabolite A | 2.40E-20 | M_VLDL_C | M_VLDL_CE |
| rs7046674 | metabolite T | 6.70E-11 | M_VLDL_C | M_VLDL_CE |
| rs554833 | metabolite T | 2.60E-20 | M_VLDL_C | M_VLDL_CE |
| rs8176649 | metabolite A | 7.90E-11 | M_VLDL_C | M_VLDL_CE |
| rs529565 | metabolite C | 5.90E-20 | M_VLDL_C | M_VLDL_CE |
| rs532436 | metabolite A | 1.00E-36 | M_VLDL_C | M_VLDL_CE |
| rs600038 | metabolite C | 4.30E-33 | M_VLDL_C | M_VLDL_CE |
| rs1124406: | metabolite T | 2.00E-21 | M_VLDL_C | M_VLDL_CE |
| rs649129 | metabolite T | 3.90E-33 | M_VLDL_C | M_VLDL_CE |
| rs495828 | metabolite T | 2.00E-33 | M_VLDL_C | M_VLDL_CE |
| rs687289 | metabolite A | 2.20E-10 | S_HDL_CE | S_HDL_CE |
| rs2519093 | metabolite T | 7.30E-24 | S_HDL_CE | S_HDL_CE |
| rs644234 | metabolite G | 7.00E-09 | S_HDL_CE | S_HDL_CE |
| rs643434 | metabolite A | 6.70E-09 | S_HDL_CE | S_HDL_CE |
| rs543040 | metabolite T | 1.60E-10 | S_HDL_CE | S_HDL_CE |
| rs613534 | metabolite G | 7.00E-09 | S_HDL_CE | S_HDL_CE |
| rs543968 | metabolite C | 7.00E-09 | S_HDL_CE | S_HDL_CE |
| rs544873 | metabolite A | 7.00E-09 | S_HDL_CE | S_HDL_CE |
| rs545971 | metabolite T | 1.30E-10 | S_HDL_CE | S_HDL_CE |
| rs612169 | metabolite G | 1.40E-10 | S_HDL_CE | S_HDL_CE |
| rs8176663 | metabolite C | 1.30E-10 | S_HDL_CE | S_HDL_CE |
| rs491626 | metabolite T | 1.30E-10 | S_HDL_CE | S_HDL_CE |
| rs492488 | metabolite A | 1.30E-10 | S_HDL_CE | S_HDL_CE |
| rs493246 | metabolite A | 1.30E-10 | S_HDL_CE | S_HDL_CE |

| | | | | |
|---|---|---|---|---|
| rs494242 | metabolite T | 6.80E-09 | S_HDL_CE | S_HDL_CE |
| rs495203 | metabolite T | 1.30E-10 | S_HDL_CE | S_HDL_CE |
| rs582118 | metabolite G | 1.30E-10 | S_HDL_CE | S_HDL_CE |
| rs582094 | metabolite T | 1.90E-10 | S_HDL_CE | S_HDL_CE |
| rs2769071 | metabolite G | 1.40E-10 | S_HDL_CE | S_HDL_CE |
| rs677355 | metabolite A | 1.50E-10 | S_HDL_CE | S_HDL_CE |
| rs676996 | metabolite G | 1.40E-10 | S_HDL_CE | S_HDL_CE |
| rs676457 | metabolite T | 1.40E-10 | S_HDL_CE | S_HDL_CE |
| rs550057 | metabolite T | 6.00E-16 | S_HDL_CE | S_HDL_CE |
| rs674302 | metabolite A | 1.40E-10 | S_HDL_CE | S_HDL_CE |
| rs554833 | metabolite T | 1.40E-10 | S_HDL_CE | S_HDL_CE |
| rs529565 | metabolite C | 8.90E-11 | S_HDL_CE | S_HDL_CE |
| rs532436 | metabolite A | 1.20E-23 | S_HDL_CE | S_HDL_CE |
| rs600038 | metabolite C | 3.30E-19 | S_HDL_CE | S_HDL_CE |
| rs1124406: | metabolite T | 4.10E-18 | S_HDL_CE | S_HDL_CE |
| rs649129 | metabolite T | 3.70E-19 | S_HDL_CE | S_HDL_CE |
| rs495828 | metabolite T | 8.90E-19 | S_HDL_CE | S_HDL_CE |
| rs2519093 | metabolite T | 5.00E-08 | XS_VLDL_P | XS_VLDL_PL_pct |
| rs532436 | metabolite A | 5.00E-08 | XS_VLDL_P | XS_VLDL_PL_pct |
| rs687289 | metabolite A | 5.90E-14 | XS_VLDL_P | XS_VLDL_PL |
| rs2519093 | metabolite T | 2.50E-19 | XS_VLDL_P | XS_VLDL_PL |
| rs644234 | metabolite G | 1.90E-14 | XS_VLDL_P | XS_VLDL_PL |
| rs643434 | metabolite A | 1.80E-14 | XS_VLDL_P | XS_VLDL_PL |
| rs543040 | metabolite T | 4.80E-14 | XS_VLDL_P | XS_VLDL_PL |
| rs613534 | metabolite G | 2.00E-14 | XS_VLDL_P | XS_VLDL_PL |
| rs543968 | metabolite C | 2.00E-14 | XS_VLDL_P | XS_VLDL_PL |
| rs544873 | metabolite A | 2.00E-14 | XS_VLDL_P | XS_VLDL_PL |
| rs545971 | metabolite T | 5.40E-14 | XS_VLDL_P | XS_VLDL_PL |
| rs612169 | metabolite G | 6.80E-14 | XS_VLDL_P | XS_VLDL_PL |
| rs8176663 | metabolite C | 5.40E-14 | XS_VLDL_P | XS_VLDL_PL |
| rs491626 | metabolite T | 5.30E-14 | XS_VLDL_P | XS_VLDL_PL |
| rs492488 | metabolite A | 5.30E-14 | XS_VLDL_P | XS_VLDL_PL |
| rs493246 | metabolite A | 5.30E-14 | XS_VLDL_P | XS_VLDL_PL |
| rs494242 | metabolite T | 1.90E-14 | XS_VLDL_P | XS_VLDL_PL |
| rs495203 | metabolite T | 5.30E-14 | XS_VLDL_P | XS_VLDL_PL |
| rs582118 | metabolite G | 5.50E-14 | XS_VLDL_P | XS_VLDL_PL |
| rs582094 | metabolite T | 6.90E-14 | XS_VLDL_P | XS_VLDL_PL |
| rs2769071 | metabolite G | 4.90E-14 | XS_VLDL_P | XS_VLDL_PL |
| rs677355 | metabolite A | 5.90E-14 | XS_VLDL_P | XS_VLDL_PL |
| rs676996 | metabolite G | 5.20E-14 | XS_VLDL_P | XS_VLDL_PL |
| rs676457 | metabolite T | 5.20E-14 | XS_VLDL_P | XS_VLDL_PL |
| rs550057 | metabolite T | 2.30E-16 | XS_VLDL_P | XS_VLDL_PL |
| rs674302 | metabolite A | 5.20E-14 | XS_VLDL_P | XS_VLDL_PL |
| rs554833 | metabolite T | 6.00E-14 | XS_VLDL_P | XS_VLDL_PL |
| rs529565 | metabolite C | 8.00E-14 | XS_VLDL_P | XS_VLDL_PL |
| rs532436 | metabolite A | 3.00E-19 | XS_VLDL_P | XS_VLDL_PL |
| rs600038 | metabolite C | 8.10E-19 | XS_VLDL_P | XS_VLDL_PL |
| rs1124406: | metabolite T | 5.80E-12 | XS_VLDL_P | XS_VLDL_PL |
| rs649129 | metabolite T | 4.60E-19 | XS_VLDL_P | XS_VLDL_PL |
| rs495828 | metabolite T | 2.50E-19 | XS_VLDL_P | XS_VLDL_PL |

| | | | | |
|---|---|---|---|---|
| rs4962040 | metabolite G | 4.00E-08 | XS_VLDL_P | XS_VLDL_P |
| rs8176702 | metabolite A | 3.90E-08 | XS_VLDL_P | XS_VLDL_P |
| rs687289 | metabolite A | 1.60E-17 | XS_VLDL_P | XS_VLDL_P |
| rs8176691 | metabolite T | 3.70E-08 | XS_VLDL_P | XS_VLDL_P |
| rs8176682 | metabolite T | 1.90E-08 | XS_VLDL_P | XS_VLDL_P |
| rs8176681 | metabolite C | 5.00E-08 | XS_VLDL_P | XS_VLDL_P |
| rs2519093 | metabolite T | 8.60E-27 | XS_VLDL_P | XS_VLDL_P |
| rs644234 | metabolite G | 9.90E-18 | XS_VLDL_P | XS_VLDL_P |
| rs643434 | metabolite A | 9.10E-18 | XS_VLDL_P | XS_VLDL_P |
| rs543040 | metabolite T | 1.30E-17 | XS_VLDL_P | XS_VLDL_P |
| rs613534 | metabolite G | 1.00E-17 | XS_VLDL_P | XS_VLDL_P |
| rs543968 | metabolite C | 1.00E-17 | XS_VLDL_P | XS_VLDL_P |
| rs544873 | metabolite A | 1.00E-17 | XS_VLDL_P | XS_VLDL_P |
| rs545971 | metabolite T | 1.40E-17 | XS_VLDL_P | XS_VLDL_P |
| rs612169 | metabolite G | 2.00E-17 | XS_VLDL_P | XS_VLDL_P |
| rs8176668 | metabolite T | 1.40E-08 | XS_VLDL_P | XS_VLDL_P |
| rs8176663 | metabolite C | 1.40E-17 | XS_VLDL_P | XS_VLDL_P |
| rs7036642 | metabolite A | 1.30E-08 | XS_VLDL_P | XS_VLDL_P |
| rs491626 | metabolite T | 1.40E-17 | XS_VLDL_P | XS_VLDL_P |
| rs492488 | metabolite A | 1.40E-17 | XS_VLDL_P | XS_VLDL_P |
| rs493246 | metabolite A | 1.40E-17 | XS_VLDL_P | XS_VLDL_P |
| rs494242 | metabolite T | 9.60E-18 | XS_VLDL_P | XS_VLDL_P |
| rs495203 | metabolite T | 1.40E-17 | XS_VLDL_P | XS_VLDL_P |
| rs582118 | metabolite G | 1.40E-17 | XS_VLDL_P | XS_VLDL_P |
| rs582094 | metabolite T | 1.90E-17 | XS_VLDL_P | XS_VLDL_P |
| rs2769071 | metabolite G | 1.30E-17 | XS_VLDL_P | XS_VLDL_P |
| rs677355 | metabolite A | 1.50E-17 | XS_VLDL_P | XS_VLDL_P |
| rs676996 | metabolite G | 1.30E-17 | XS_VLDL_P | XS_VLDL_P |
| rs676457 | metabolite T | 1.30E-17 | XS_VLDL_P | XS_VLDL_P |
| rs550057 | metabolite T | 1.80E-22 | XS_VLDL_P | XS_VLDL_P |
| rs674302 | metabolite A | 1.30E-17 | XS_VLDL_P | XS_VLDL_P |
| rs7046674 | metabolite T | 1.00E-08 | XS_VLDL_P | XS_VLDL_P |
| rs554833 | metabolite T | 1.60E-17 | XS_VLDL_P | XS_VLDL_P |
| rs8176649 | metabolite A | 1.10E-08 | XS_VLDL_P | XS_VLDL_P |
| rs529565 | metabolite C | 2.20E-17 | XS_VLDL_P | XS_VLDL_P |
| rs532436 | metabolite A | 1.20E-26 | XS_VLDL_P | XS_VLDL_P |
| rs600038 | metabolite C | 2.10E-25 | XS_VLDL_P | XS_VLDL_P |
| rs1124406: | metabolite T | 8.80E-17 | XS_VLDL_P | XS_VLDL_P |
| rs649129 | metabolite T | 1.20E-25 | XS_VLDL_P | XS_VLDL_P |
| rs495828 | metabolite T | 5.90E-26 | XS_VLDL_P | XS_VLDL_P |
| rs687289 | metabolite A | 2.40E-15 | XS_VLDL_L | XS_VLDL_L |
| rs2519093 | metabolite T | 1.40E-22 | XS_VLDL_L | XS_VLDL_L |
| rs644234 | metabolite G | 1.30E-15 | XS_VLDL_L | XS_VLDL_L |
| rs643434 | metabolite A | 1.20E-15 | XS_VLDL_L | XS_VLDL_L |
| rs543040 | metabolite T | 1.80E-15 | XS_VLDL_L | XS_VLDL_L |
| rs613534 | metabolite G | 1.40E-15 | XS_VLDL_L | XS_VLDL_L |
| rs543968 | metabolite C | 1.40E-15 | XS_VLDL_L | XS_VLDL_L |
| rs544873 | metabolite A | 1.40E-15 | XS_VLDL_L | XS_VLDL_L |
| rs545971 | metabolite T | 2.10E-15 | XS_VLDL_L | XS_VLDL_L |
| rs612169 | metabolite G | 2.70E-15 | XS_VLDL_L | XS_VLDL_L |

| | | | | |
|---|---|---|---|---|
| rs8176663 | metabolite C | 2.10E-15 | XS_VLDL_L | XS_VLDL_L |
| rs491626 | metabolite T | 2.00E-15 | XS_VLDL_L | XS_VLDL_L |
| rs492488 | metabolite A | 2.00E-15 | XS_VLDL_L | XS_VLDL_L |
| rs493246 | metabolite A | 2.00E-15 | XS_VLDL_L | XS_VLDL_L |
| rs494242 | metabolite T | 1.30E-15 | XS_VLDL_L | XS_VLDL_L |
| rs495203 | metabolite T | 2.00E-15 | XS_VLDL_L | XS_VLDL_L |
| rs582118 | metabolite G | 2.10E-15 | XS_VLDL_L | XS_VLDL_L |
| rs582094 | metabolite T | 2.70E-15 | XS_VLDL_L | XS_VLDL_L |
| rs2769071 | metabolite G | 1.90E-15 | XS_VLDL_L | XS_VLDL_L |
| rs677355 | metabolite A | 2.30E-15 | XS_VLDL_L | XS_VLDL_L |
| rs676996 | metabolite G | 2.00E-15 | XS_VLDL_L | XS_VLDL_L |
| rs676457 | metabolite T | 2.00E-15 | XS_VLDL_L | XS_VLDL_L |
| rs550057 | metabolite T | 8.60E-19 | XS_VLDL_L | XS_VLDL_L |
| rs674302 | metabolite A | 2.00E-15 | XS_VLDL_L | XS_VLDL_L |
| rs554833 | metabolite T | 2.30E-15 | XS_VLDL_L | XS_VLDL_L |
| rs529565 | metabolite C | 3.30E-15 | XS_VLDL_L | XS_VLDL_L |
| rs532436 | metabolite A | 1.70E-22 | XS_VLDL_L | XS_VLDL_L |
| rs600038 | metabolite C | 1.60E-21 | XS_VLDL_L | XS_VLDL_L |
| rs1124406: | metabolite T | 5.20E-14 | XS_VLDL_L | XS_VLDL_L |
| rs649129 | metabolite T | 9.20E-22 | XS_VLDL_L | XS_VLDL_L |
| rs495828 | metabolite T | 4.90E-22 | XS_VLDL_L | XS_VLDL_L |
| rs687289 | metabolite A | 2.60E-12 | XS_VLDL_F | XS_VLDL_FC_pct |
| rs2519093 | metabolite T | 6.20E-21 | XS_VLDL_F | XS_VLDL_FC_pct |
| rs644234 | metabolite G | 3.00E-11 | XS_VLDL_F | XS_VLDL_FC_pct |
| rs643434 | metabolite A | 3.10E-11 | XS_VLDL_F | XS_VLDL_FC_pct |
| rs543040 | metabolite T | 3.50E-12 | XS_VLDL_F | XS_VLDL_FC_pct |
| rs613534 | metabolite G | 3.10E-11 | XS_VLDL_F | XS_VLDL_FC_pct |
| rs543968 | metabolite C | 3.10E-11 | XS_VLDL_F | XS_VLDL_FC_pct |
| rs544873 | metabolite A | 3.10E-11 | XS_VLDL_F | XS_VLDL_FC_pct |
| rs545971 | metabolite T | 2.40E-12 | XS_VLDL_F | XS_VLDL_FC_pct |
| rs612169 | metabolite G | 2.70E-12 | XS_VLDL_F | XS_VLDL_FC_pct |
| rs8176663 | metabolite C | 2.40E-12 | XS_VLDL_F | XS_VLDL_FC_pct |
| rs491626 | metabolite T | 2.40E-12 | XS_VLDL_F | XS_VLDL_FC_pct |
| rs492488 | metabolite A | 2.40E-12 | XS_VLDL_F | XS_VLDL_FC_pct |
| rs493246 | metabolite A | 2.40E-12 | XS_VLDL_F | XS_VLDL_FC_pct |
| rs494242 | metabolite T | 3.20E-11 | XS_VLDL_F | XS_VLDL_FC_pct |
| rs495203 | metabolite T | 2.50E-12 | XS_VLDL_F | XS_VLDL_FC_pct |
| rs582118 | metabolite G | 2.50E-12 | XS_VLDL_F | XS_VLDL_FC_pct |
| rs582094 | metabolite T | 3.70E-12 | XS_VLDL_F | XS_VLDL_FC_pct |
| rs2769071 | metabolite G | 2.50E-12 | XS_VLDL_F | XS_VLDL_FC_pct |
| rs677355 | metabolite A | 2.60E-12 | XS_VLDL_F | XS_VLDL_FC_pct |
| rs676996 | metabolite G | 2.60E-12 | XS_VLDL_F | XS_VLDL_FC_pct |
| rs676457 | metabolite T | 2.60E-12 | XS_VLDL_F | XS_VLDL_FC_pct |
| rs550057 | metabolite T | 1.40E-19 | XS_VLDL_F | XS_VLDL_FC_pct |
| rs674302 | metabolite A | 2.60E-12 | XS_VLDL_F | XS_VLDL_FC_pct |
| rs554833 | metabolite T | 2.50E-12 | XS_VLDL_F | XS_VLDL_FC_pct |
| rs529565 | metabolite C | 5.80E-12 | XS_VLDL_F | XS_VLDL_FC_pct |
| rs532436 | metabolite A | 9.70E-21 | XS_VLDL_F | XS_VLDL_FC_pct |
| rs600038 | metabolite C | 8.00E-18 | XS_VLDL_F | XS_VLDL_FC_pct |
| rs1124406: | metabolite T | 1.10E-10 | XS_VLDL_F | XS_VLDL_FC_pct |

| | | | | |
|---|---|---|---|---|
| rs649129 | metabolite T | 1.20E-17 | XS_VLDL_F | XS_VLDL_FC_pct |
| rs495828 | metabolite T | 1.70E-17 | XS_VLDL_F | XS_VLDL_FC_pct |
| rs4962040 | metabolite G | 3.10E-08 | XS_VLDL_F | XS_VLDL_FC |
| rs8176702 | metabolite A | 3.10E-08 | XS_VLDL_F | XS_VLDL_FC |
| rs687289 | metabolite A | 7.50E-18 | XS_VLDL_F | XS_VLDL_FC |
| rs8176691 | metabolite T | 2.80E-08 | XS_VLDL_F | XS_VLDL_FC |
| rs8176690 | metabolite G | 3.40E-08 | XS_VLDL_F | XS_VLDL_FC |
| rs8176682 | metabolite T | 1.30E-08 | XS_VLDL_F | XS_VLDL_FC |
| rs8176681 | metabolite C | 2.80E-08 | XS_VLDL_F | XS_VLDL_FC |
| rs2519093 | metabolite T | 8.50E-27 | XS_VLDL_F | XS_VLDL_FC |
| rs644234 | metabolite G | 7.40E-18 | XS_VLDL_F | XS_VLDL_FC |
| rs643434 | metabolite A | 6.80E-18 | XS_VLDL_F | XS_VLDL_FC |
| rs543040 | metabolite T | 6.40E-18 | XS_VLDL_F | XS_VLDL_FC |
| rs613534 | metabolite G | 7.50E-18 | XS_VLDL_F | XS_VLDL_FC |
| rs543968 | metabolite C | 7.50E-18 | XS_VLDL_F | XS_VLDL_FC |
| rs544873 | metabolite A | 7.50E-18 | XS_VLDL_F | XS_VLDL_FC |
| rs545971 | metabolite T | 6.50E-18 | XS_VLDL_F | XS_VLDL_FC |
| rs612169 | metabolite G | 8.90E-18 | XS_VLDL_F | XS_VLDL_FC |
| rs8176668 | metabolite T | 1.00E-08 | XS_VLDL_F | XS_VLDL_FC |
| rs8176663 | metabolite C | 6.50E-18 | XS_VLDL_F | XS_VLDL_FC |
| rs7036642 | metabolite A | 9.70E-09 | XS_VLDL_F | XS_VLDL_FC |
| rs491626 | metabolite T | 6.40E-18 | XS_VLDL_F | XS_VLDL_FC |
| rs492488 | metabolite A | 6.40E-18 | XS_VLDL_F | XS_VLDL_FC |
| rs493246 | metabolite A | 6.40E-18 | XS_VLDL_F | XS_VLDL_FC |
| rs494242 | metabolite T | 7.30E-18 | XS_VLDL_F | XS_VLDL_FC |
| rs495203 | metabolite T | 6.50E-18 | XS_VLDL_F | XS_VLDL_FC |
| rs582118 | metabolite G | 6.70E-18 | XS_VLDL_F | XS_VLDL_FC |
| rs582094 | metabolite T | 9.50E-18 | XS_VLDL_F | XS_VLDL_FC |
| rs2769071 | metabolite G | 6.00E-18 | XS_VLDL_F | XS_VLDL_FC |
| rs677355 | metabolite A | 7.40E-18 | XS_VLDL_F | XS_VLDL_FC |
| rs676996 | metabolite G | 6.40E-18 | XS_VLDL_F | XS_VLDL_FC |
| rs676457 | metabolite T | 6.40E-18 | XS_VLDL_F | XS_VLDL_FC |
| rs550057 | metabolite T | 1.60E-22 | XS_VLDL_F | XS_VLDL_FC |
| rs674302 | metabolite A | 6.40E-18 | XS_VLDL_F | XS_VLDL_FC |
| rs7046674 | metabolite T | 7.80E-09 | XS_VLDL_F | XS_VLDL_FC |
| rs554833 | metabolite T | 7.40E-18 | XS_VLDL_F | XS_VLDL_FC |
| rs8176649 | metabolite A | 8.20E-09 | XS_VLDL_F | XS_VLDL_FC |
| rs529565 | metabolite C | 1.20E-17 | XS_VLDL_F | XS_VLDL_FC |
| rs532436 | metabolite A | 1.10E-26 | XS_VLDL_F | XS_VLDL_FC |
| rs600038 | metabolite C | 4.20E-25 | XS_VLDL_F | XS_VLDL_FC |
| rs1124406: | metabolite T | 3.50E-16 | XS_VLDL_F | XS_VLDL_FC |
| rs649129 | metabolite T | 2.60E-25 | XS_VLDL_F | XS_VLDL_FC |
| rs495828 | metabolite T | 1.30E-25 | XS_VLDL_F | XS_VLDL_FC |
| rs2519093 | metabolite T | 1.00E-14 | XS_VLDL_C | XS_VLDL_CE_pct |
| rs550057 | metabolite T | 2.20E-12 | XS_VLDL_C | XS_VLDL_CE_pct |
| rs532436 | metabolite A | 1.40E-14 | XS_VLDL_C | XS_VLDL_CE_pct |
| rs600038 | metabolite C | 1.80E-12 | XS_VLDL_C | XS_VLDL_CE_pct |
| rs649129 | metabolite T | 3.00E-12 | XS_VLDL_C | XS_VLDL_CE_pct |
| rs495828 | metabolite T | 3.10E-12 | XS_VLDL_C | XS_VLDL_CE_pct |
| rs7873635 | metabolite C | 1.90E-09 | XS_VLDL_C | XS_VLDL_CE |

| | | | | |
|---|---|---|---|---|
| rs4962040 | metabolite G | 1.20E-09 | XS_VLDL_C | XS_VLDL_CE |
| rs8176702 | metabolite A | 1.20E-09 | XS_VLDL_C | XS_VLDL_CE |
| rs2073826 | metabolite T | 2.00E-09 | XS_VLDL_C | XS_VLDL_CE |
| rs687289 | metabolite A | 2.70E-18 | XS_VLDL_C | XS_VLDL_CE |
| rs2073827 | metabolite C | 2.50E-09 | XS_VLDL_C | XS_VLDL_CE |
| rs2073828 | metabolite A | 2.50E-09 | XS_VLDL_C | XS_VLDL_CE |
| rs8176691 | metabolite T | 9.60E-10 | XS_VLDL_C | XS_VLDL_CE |
| rs8176690 | metabolite G | 8.10E-10 | XS_VLDL_C | XS_VLDL_CE |
| rs8176682 | metabolite T | 3.80E-10 | XS_VLDL_C | XS_VLDL_CE |
| rs8176681 | metabolite C | 7.60E-10 | XS_VLDL_C | XS_VLDL_CE |
| rs2519093 | metabolite T | 2.40E-31 | XS_VLDL_C | XS_VLDL_CE |
| rs644234 | metabolite G | 4.00E-18 | XS_VLDL_C | XS_VLDL_CE |
| rs643434 | metabolite A | 3.70E-18 | XS_VLDL_C | XS_VLDL_CE |
| rs543040 | metabolite T | 2.30E-18 | XS_VLDL_C | XS_VLDL_CE |
| rs613534 | metabolite G | 4.10E-18 | XS_VLDL_C | XS_VLDL_CE |
| rs543968 | metabolite C | 4.10E-18 | XS_VLDL_C | XS_VLDL_CE |
| rs544873 | metabolite A | 4.10E-18 | XS_VLDL_C | XS_VLDL_CE |
| rs545971 | metabolite T | 2.10E-18 | XS_VLDL_C | XS_VLDL_CE |
| rs612169 | metabolite G | 2.80E-18 | XS_VLDL_C | XS_VLDL_CE |
| rs8176668 | metabolite T | 3.80E-10 | XS_VLDL_C | XS_VLDL_CE |
| rs8176663 | metabolite C | 2.10E-18 | XS_VLDL_C | XS_VLDL_CE |
| rs7036642 | metabolite A | 3.50E-10 | XS_VLDL_C | XS_VLDL_CE |
| rs491626 | metabolite T | 2.10E-18 | XS_VLDL_C | XS_VLDL_CE |
| rs492488 | metabolite A | 2.10E-18 | XS_VLDL_C | XS_VLDL_CE |
| rs493246 | metabolite A | 2.10E-18 | XS_VLDL_C | XS_VLDL_CE |
| rs494242 | metabolite T | 4.00E-18 | XS_VLDL_C | XS_VLDL_CE |
| rs495203 | metabolite T | 2.10E-18 | XS_VLDL_C | XS_VLDL_CE |
| rs582118 | metabolite G | 2.10E-18 | XS_VLDL_C | XS_VLDL_CE |
| rs582094 | metabolite T | 3.10E-18 | XS_VLDL_C | XS_VLDL_CE |
| rs2769071 | metabolite G | 2.00E-18 | XS_VLDL_C | XS_VLDL_CE |
| rs677355 | metabolite A | 2.40E-18 | XS_VLDL_C | XS_VLDL_CE |
| rs676996 | metabolite G | 2.10E-18 | XS_VLDL_C | XS_VLDL_CE |
| rs676457 | metabolite T | 2.10E-18 | XS_VLDL_C | XS_VLDL_CE |
| rs550057 | metabolite T | 1.30E-25 | XS_VLDL_C | XS_VLDL_CE |
| rs674302 | metabolite A | 2.10E-18 | XS_VLDL_C | XS_VLDL_CE |
| rs7046674 | metabolite T | 2.80E-10 | XS_VLDL_C | XS_VLDL_CE |
| rs554833 | metabolite T | 2.50E-18 | XS_VLDL_C | XS_VLDL_CE |
| rs8176649 | metabolite A | 3.00E-10 | XS_VLDL_C | XS_VLDL_CE |
| rs529565 | metabolite C | 5.70E-18 | XS_VLDL_C | XS_VLDL_CE |
| rs532436 | metabolite A | 3.40E-31 | XS_VLDL_C | XS_VLDL_CE |
| rs600038 | metabolite C | 7.70E-29 | XS_VLDL_C | XS_VLDL_CE |
| rs1124406: | metabolite T | 4.10E-18 | XS_VLDL_C | XS_VLDL_CE |
| rs649129 | metabolite T | 6.90E-29 | XS_VLDL_C | XS_VLDL_CE |
| rs495828 | metabolite T | 3.40E-29 | XS_VLDL_C | XS_VLDL_CE |
| rs7873635 | metabolite C | 5.00E-09 | XL_VLDL_P | XL_VLDL_PL_pct |
| rs4962040 | metabolite G | 4.90E-09 | XL_VLDL_P | XL_VLDL_PL_pct |
| rs8176702 | metabolite A | 3.90E-09 | XL_VLDL_P | XL_VLDL_PL_pct |
| rs2073826 | metabolite T | 3.20E-08 | XL_VLDL_P | XL_VLDL_PL_pct |
| rs687289 | metabolite A | 6.40E-19 | XL_VLDL_P | XL_VLDL_PL_pct |
| rs2073827 | metabolite C | 3.60E-08 | XL_VLDL_P | XL_VLDL_PL_pct |

| | | | | |
|---|---|---|---|---|
| rs2073828 | metabolite A | 3.70E-08 | XL_VLDL_P | XL_VLDL_PL_pct |
| rs8176691 | metabolite T | 3.00E-09 | XL_VLDL_P | XL_VLDL_PL_pct |
| rs8176690 | metabolite G | 1.20E-08 | XL_VLDL_P | XL_VLDL_PL_pct |
| rs8176682 | metabolite T | 1.10E-09 | XL_VLDL_P | XL_VLDL_PL_pct |
| rs8176681 | metabolite C | 1.30E-08 | XL_VLDL_P | XL_VLDL_PL_pct |
| rs2519093 | metabolite T | 2.80E-27 | XL_VLDL_P | XL_VLDL_PL_pct |
| rs644234 | metabolite G | 7.10E-20 | XL_VLDL_P | XL_VLDL_PL_pct |
| rs643434 | metabolite A | 6.90E-20 | XL_VLDL_P | XL_VLDL_PL_pct |
| rs543040 | metabolite T | 5.80E-19 | XL_VLDL_P | XL_VLDL_PL_pct |
| rs613534 | metabolite G | 7.40E-20 | XL_VLDL_P | XL_VLDL_PL_pct |
| rs543968 | metabolite C | 7.40E-20 | XL_VLDL_P | XL_VLDL_PL_pct |
| rs544873 | metabolite A | 7.40E-20 | XL_VLDL_P | XL_VLDL_PL_pct |
| rs545971 | metabolite T | 5.10E-19 | XL_VLDL_P | XL_VLDL_PL_pct |
| rs612169 | metabolite G | 7.90E-19 | XL_VLDL_P | XL_VLDL_PL_pct |
| rs8176668 | metabolite T | 1.20E-09 | XL_VLDL_P | XL_VLDL_PL_pct |
| rs8176663 | metabolite C | 5.10E-19 | XL_VLDL_P | XL_VLDL_PL_pct |
| rs7036642 | metabolite A | 1.40E-09 | XL_VLDL_P | XL_VLDL_PL_pct |
| rs491626 | metabolite T | 5.20E-19 | XL_VLDL_P | XL_VLDL_PL_pct |
| rs492488 | metabolite A | 5.20E-19 | XL_VLDL_P | XL_VLDL_PL_pct |
| rs493246 | metabolite A | 5.20E-19 | XL_VLDL_P | XL_VLDL_PL_pct |
| rs494242 | metabolite T | 7.70E-20 | XL_VLDL_P | XL_VLDL_PL_pct |
| rs495203 | metabolite T | 5.30E-19 | XL_VLDL_P | XL_VLDL_PL_pct |
| rs582118 | metabolite G | 5.30E-19 | XL_VLDL_P | XL_VLDL_PL_pct |
| rs582094 | metabolite T | 4.80E-19 | XL_VLDL_P | XL_VLDL_PL_pct |
| rs2769071 | metabolite G | 7.10E-19 | XL_VLDL_P | XL_VLDL_PL_pct |
| rs677355 | metabolite A | 7.00E-19 | XL_VLDL_P | XL_VLDL_PL_pct |
| rs676996 | metabolite G | 6.80E-19 | XL_VLDL_P | XL_VLDL_PL_pct |
| rs676457 | metabolite T | 6.80E-19 | XL_VLDL_P | XL_VLDL_PL_pct |
| rs550057 | metabolite T | 7.70E-24 | XL_VLDL_P | XL_VLDL_PL_pct |
| rs674302 | metabolite A | 6.70E-19 | XL_VLDL_P | XL_VLDL_PL_pct |
| rs7046674 | metabolite T | 1.30E-09 | XL_VLDL_P | XL_VLDL_PL_pct |
| rs554833 | metabolite T | 8.20E-19 | XL_VLDL_P | XL_VLDL_PL_pct |
| rs8176649 | metabolite A | 1.40E-09 | XL_VLDL_P | XL_VLDL_PL_pct |
| rs529565 | metabolite C | 1.50E-18 | XL_VLDL_P | XL_VLDL_PL_pct |
| rs630510 | metabolite G | 1.20E-08 | XL_VLDL_P | XL_VLDL_PL_pct |
| rs630014 | metabolite G | 1.20E-08 | XL_VLDL_P | XL_VLDL_PL_pct |
| rs532436 | metabolite A | 6.50E-27 | XL_VLDL_P | XL_VLDL_PL_pct |
| rs600038 | metabolite C | 2.40E-26 | XL_VLDL_P | XL_VLDL_PL_pct |
| rs1124406: | metabolite T | 1.30E-14 | XL_VLDL_P | XL_VLDL_PL_pct |
| rs649129 | metabolite T | 2.60E-26 | XL_VLDL_P | XL_VLDL_PL_pct |
| rs495828 | metabolite T | 1.40E-26 | XL_VLDL_P | XL_VLDL_PL_pct |
| rs7873635 | metabolite C | 7.80E-10 | XL_VLDL_F | XL_VLDL_FC_pct |
| rs4962040 | metabolite G | 8.50E-10 | XL_VLDL_F | XL_VLDL_FC_pct |
| rs8176702 | metabolite A | 1.10E-09 | XL_VLDL_F | XL_VLDL_FC_pct |
| rs2073826 | metabolite T | 9.30E-10 | XL_VLDL_F | XL_VLDL_FC_pct |
| rs687289 | metabolite A | 1.00E-18 | XL_VLDL_F | XL_VLDL_FC_pct |
| rs2073827 | metabolite C | 9.00E-10 | XL_VLDL_F | XL_VLDL_FC_pct |
| rs2073828 | metabolite A | 9.50E-10 | XL_VLDL_F | XL_VLDL_FC_pct |
| rs8176691 | metabolite T | 6.20E-10 | XL_VLDL_F | XL_VLDL_FC_pct |
| rs8176690 | metabolite G | 3.10E-10 | XL_VLDL_F | XL_VLDL_FC_pct |

| | | | | |
|---|---|---|---|---|
| rs8176682 | metabolite T | 2.60E-10 | XL_VLDL_F | XL_VLDL_FC_pct |
| rs8176681 | metabolite C | 3.70E-10 | XL_VLDL_F | XL_VLDL_FC_pct |
| rs2519093 | metabolite T | 2.50E-28 | XL_VLDL_F | XL_VLDL_FC_pct |
| rs644234 | metabolite G | 1.70E-17 | XL_VLDL_F | XL_VLDL_FC_pct |
| rs643434 | metabolite A | 2.00E-17 | XL_VLDL_F | XL_VLDL_FC_pct |
| rs543040 | metabolite T | 3.40E-18 | XL_VLDL_F | XL_VLDL_FC_pct |
| rs613534 | metabolite G | 1.80E-17 | XL_VLDL_F | XL_VLDL_FC_pct |
| rs543968 | metabolite C | 1.80E-17 | XL_VLDL_F | XL_VLDL_FC_pct |
| rs544873 | metabolite A | 1.80E-17 | XL_VLDL_F | XL_VLDL_FC_pct |
| rs545971 | metabolite T | 2.00E-18 | XL_VLDL_F | XL_VLDL_FC_pct |
| rs612169 | metabolite G | 2.00E-18 | XL_VLDL_F | XL_VLDL_FC_pct |
| rs8176668 | metabolite T | 8.00E-10 | XL_VLDL_F | XL_VLDL_FC_pct |
| rs8176663 | metabolite C | 2.00E-18 | XL_VLDL_F | XL_VLDL_FC_pct |
| rs7036642 | metabolite A | 1.10E-09 | XL_VLDL_F | XL_VLDL_FC_pct |
| rs491626 | metabolite T | 2.00E-18 | XL_VLDL_F | XL_VLDL_FC_pct |
| rs492488 | metabolite A | 2.00E-18 | XL_VLDL_F | XL_VLDL_FC_pct |
| rs493246 | metabolite A | 2.00E-18 | XL_VLDL_F | XL_VLDL_FC_pct |
| rs494242 | metabolite T | 1.80E-17 | XL_VLDL_F | XL_VLDL_FC_pct |
| rs495203 | metabolite T | 2.00E-18 | XL_VLDL_F | XL_VLDL_FC_pct |
| rs582118 | metabolite G | 2.00E-18 | XL_VLDL_F | XL_VLDL_FC_pct |
| rs582094 | metabolite T | 2.40E-18 | XL_VLDL_F | XL_VLDL_FC_pct |
| rs2769071 | metabolite G | 3.20E-18 | XL_VLDL_F | XL_VLDL_FC_pct |
| rs677355 | metabolite A | 2.00E-18 | XL_VLDL_F | XL_VLDL_FC_pct |
| rs676996 | metabolite G | 2.30E-18 | XL_VLDL_F | XL_VLDL_FC_pct |
| rs676457 | metabolite T | 2.30E-18 | XL_VLDL_F | XL_VLDL_FC_pct |
| rs550057 | metabolite T | 2.40E-23 | XL_VLDL_F | XL_VLDL_FC_pct |
| rs674302 | metabolite A | 2.30E-18 | XL_VLDL_F | XL_VLDL_FC_pct |
| rs7046674 | metabolite T | 1.20E-09 | XL_VLDL_F | XL_VLDL_FC_pct |
| rs554833 | metabolite T | 2.50E-18 | XL_VLDL_F | XL_VLDL_FC_pct |
| rs8176649 | metabolite A | 1.10E-09 | XL_VLDL_F | XL_VLDL_FC_pct |
| rs660340 | metabolite G | 1.50E-08 | XL_VLDL_F | XL_VLDL_FC_pct |
| rs581107 | metabolite T | 1.40E-08 | XL_VLDL_F | XL_VLDL_FC_pct |
| rs659104 | metabolite G | 1.50E-08 | XL_VLDL_F | XL_VLDL_FC_pct |
| rs647800 | metabolite G | 2.30E-08 | XL_VLDL_F | XL_VLDL_FC_pct |
| rs473533 | metabolite C | 1.50E-08 | XL_VLDL_F | XL_VLDL_FC_pct |
| rs476410 | metabolite C | 1.80E-08 | XL_VLDL_F | XL_VLDL_FC_pct |
| rs645982 | metabolite G | 1.50E-08 | XL_VLDL_F | XL_VLDL_FC_pct |
| rs529565 | metabolite C | 3.90E-18 | XL_VLDL_F | XL_VLDL_FC_pct |
| rs532436 | metabolite A | 2.60E-28 | XL_VLDL_F | XL_VLDL_FC_pct |
| rs600038 | metabolite C | 2.90E-25 | XL_VLDL_F | XL_VLDL_FC_pct |
| rs1124406: | metabolite T | 6.40E-16 | XL_VLDL_F | XL_VLDL_FC_pct |
| rs649129 | metabolite T | 3.70E-25 | XL_VLDL_F | XL_VLDL_FC_pct |
| rs495828 | metabolite T | 3.60E-25 | XL_VLDL_F | XL_VLDL_FC_pct |
| rs7873635 | metabolite C | 1.40E-09 | XL_VLDL_C | XL_VLDL_CE_pct |
| rs4962040 | metabolite G | 1.30E-09 | XL_VLDL_C | XL_VLDL_CE_pct |
| rs8176702 | metabolite A | 1.80E-09 | XL_VLDL_C | XL_VLDL_CE_pct |
| rs2073826 | metabolite T | 4.10E-10 | XL_VLDL_C | XL_VLDL_CE_pct |
| rs687289 | metabolite A | 1.10E-18 | XL_VLDL_C | XL_VLDL_CE_pct |
| rs2073827 | metabolite C | 3.40E-10 | XL_VLDL_C | XL_VLDL_CE_pct |
| rs2073828 | metabolite A | 3.50E-10 | XL_VLDL_C | XL_VLDL_CE_pct |

| | | | | | |
|---|---|---|---|---|---|
| rs8176691 | metabolite | T | 1.10E-09 | XL_VLDL_C | XL_VLDL_CE_pct |
| rs8176690 | metabolite | G | 1.60E-10 | XL_VLDL_C | XL_VLDL_CE_pct |
| rs8176682 | metabolite | T | 5.90E-10 | XL_VLDL_C | XL_VLDL_CE_pct |
| rs8176681 | metabolite | C | 1.90E-10 | XL_VLDL_C | XL_VLDL_CE_pct |
| rs2519093 | metabolite | T | 1.50E-33 | XL_VLDL_C | XL_VLDL_CE_pct |
| rs644234 | metabolite | G | 1.50E-16 | XL_VLDL_C | XL_VLDL_CE_pct |
| rs643434 | metabolite | A | 1.70E-16 | XL_VLDL_C | XL_VLDL_CE_pct |
| rs543040 | metabolite | T | 6.40E-18 | XL_VLDL_C | XL_VLDL_CE_pct |
| rs613534 | metabolite | G | 1.50E-16 | XL_VLDL_C | XL_VLDL_CE_pct |
| rs543968 | metabolite | C | 1.50E-16 | XL_VLDL_C | XL_VLDL_CE_pct |
| rs544873 | metabolite | A | 1.50E-16 | XL_VLDL_C | XL_VLDL_CE_pct |
| rs545971 | metabolite | T | 3.10E-18 | XL_VLDL_C | XL_VLDL_CE_pct |
| rs612169 | metabolite | G | 2.90E-18 | XL_VLDL_C | XL_VLDL_CE_pct |
| rs8176668 | metabolite | T | 2.30E-09 | XL_VLDL_C | XL_VLDL_CE_pct |
| rs8176663 | metabolite | C | 3.10E-18 | XL_VLDL_C | XL_VLDL_CE_pct |
| rs7036642 | metabolite | A | 2.60E-09 | XL_VLDL_C | XL_VLDL_CE_pct |
| rs491626 | metabolite | T | 3.10E-18 | XL_VLDL_C | XL_VLDL_CE_pct |
| rs492488 | metabolite | A | 3.10E-18 | XL_VLDL_C | XL_VLDL_CE_pct |
| rs493246 | metabolite | A | 3.10E-18 | XL_VLDL_C | XL_VLDL_CE_pct |
| rs494242 | metabolite | T | 1.60E-16 | XL_VLDL_C | XL_VLDL_CE_pct |
| rs495203 | metabolite | T | 3.10E-18 | XL_VLDL_C | XL_VLDL_CE_pct |
| rs582118 | metabolite | G | 3.10E-18 | XL_VLDL_C | XL_VLDL_CE_pct |
| rs582094 | metabolite | T | 4.70E-18 | XL_VLDL_C | XL_VLDL_CE_pct |
| rs2769071 | metabolite | G | 4.50E-18 | XL_VLDL_C | XL_VLDL_CE_pct |
| rs677355 | metabolite | A | 3.10E-18 | XL_VLDL_C | XL_VLDL_CE_pct |
| rs676996 | metabolite | G | 3.70E-18 | XL_VLDL_C | XL_VLDL_CE_pct |
| rs676457 | metabolite | T | 3.70E-18 | XL_VLDL_C | XL_VLDL_CE_pct |
| rs550057 | metabolite | T | 1.10E-25 | XL_VLDL_C | XL_VLDL_CE_pct |
| rs674302 | metabolite | A | 3.80E-18 | XL_VLDL_C | XL_VLDL_CE_pct |
| rs7046674 | metabolite | T | 3.00E-09 | XL_VLDL_C | XL_VLDL_CE_pct |
| rs554833 | metabolite | T | 3.80E-18 | XL_VLDL_C | XL_VLDL_CE_pct |
| rs8176649 | metabolite | A | 2.60E-09 | XL_VLDL_C | XL_VLDL_CE_pct |
| rs660340 | metabolite | G | 8.80E-09 | XL_VLDL_C | XL_VLDL_CE_pct |
| rs581107 | metabolite | T | 9.20E-09 | XL_VLDL_C | XL_VLDL_CE_pct |
| rs659104 | metabolite | G | 9.10E-09 | XL_VLDL_C | XL_VLDL_CE_pct |
| rs647800 | metabolite | G | 4.30E-08 | XL_VLDL_C | XL_VLDL_CE_pct |
| rs473533 | metabolite | C | 9.30E-09 | XL_VLDL_C | XL_VLDL_CE_pct |
| rs476410 | metabolite | C | 9.70E-09 | XL_VLDL_C | XL_VLDL_CE_pct |
| rs645982 | metabolite | G | 8.60E-09 | XL_VLDL_C | XL_VLDL_CE_pct |
| rs529565 | metabolite | C | 3.10E-18 | XL_VLDL_C | XL_VLDL_CE_pct |
| rs532436 | metabolite | A | 2.90E-33 | XL_VLDL_C | XL_VLDL_CE_pct |
| rs600038 | metabolite | C | 2.80E-28 | XL_VLDL_C | XL_VLDL_CE_pct |
| rs1124406: | metabolite | T | 1.20E-21 | XL_VLDL_C | XL_VLDL_CE_pct |
| rs649129 | metabolite | T | 2.20E-28 | XL_VLDL_C | XL_VLDL_CE_pct |
| rs495828 | metabolite | T | 4.50E-28 | XL_VLDL_C | XL_VLDL_CE_pct |
| rs2519093 | metabolite | T | 9.50E-10 | XL_VLDL_C | XL_VLDL_CE |
| rs550057 | metabolite | T | 2.80E-09 | XL_VLDL_C | XL_VLDL_CE |
| rs532436 | metabolite | A | 1.10E-09 | XL_VLDL_C | XL_VLDL_CE |
| rs600038 | metabolite | C | 8.00E-10 | XL_VLDL_C | XL_VLDL_CE |
| rs649129 | metabolite | T | 4.30E-10 | XL_VLDL_C | XL_VLDL_CE |

| | | | | |
|---|---|---|---|---|
| rs495828 | metabolite T | 2.80E-10 | XL_VLDL_C | XL_VLDL_CE |
| rs7873635 | metabolite C | 4.30E-10 | XL_VLDL_C | XL_VLDL_C_pct |
| rs4962040 | metabolite G | 4.00E-10 | XL_VLDL_C | XL_VLDL_C_pct |
| rs8176702 | metabolite A | 5.60E-10 | XL_VLDL_C | XL_VLDL_C_pct |
| rs2073826 | metabolite T | 1.70E-10 | XL_VLDL_C | XL_VLDL_C_pct |
| rs687289 | metabolite A | 1.60E-19 | XL_VLDL_C | XL_VLDL_C_pct |
| rs2073827 | metabolite C | 1.40E-10 | XL_VLDL_C | XL_VLDL_C_pct |
| rs2073828 | metabolite A | 1.50E-10 | XL_VLDL_C | XL_VLDL_C_pct |
| rs8176691 | metabolite T | 3.20E-10 | XL_VLDL_C | XL_VLDL_C_pct |
| rs8176690 | metabolite G | 5.90E-11 | XL_VLDL_C | XL_VLDL_C_pct |
| rs8176682 | metabolite T | 1.60E-10 | XL_VLDL_C | XL_VLDL_C_pct |
| rs8176681 | metabolite C | 7.40E-11 | XL_VLDL_C | XL_VLDL_C_pct |
| rs2519093 | metabolite T | 8.10E-34 | XL_VLDL_C | XL_VLDL_C_pct |
| rs644234 | metabolite G | 1.70E-17 | XL_VLDL_C | XL_VLDL_C_pct |
| rs643434 | metabolite A | 1.90E-17 | XL_VLDL_C | XL_VLDL_C_pct |
| rs543040 | metabolite T | 9.30E-19 | XL_VLDL_C | XL_VLDL_C_pct |
| rs613534 | metabolite G | 1.70E-17 | XL_VLDL_C | XL_VLDL_C_pct |
| rs543968 | metabolite C | 1.70E-17 | XL_VLDL_C | XL_VLDL_C_pct |
| rs544873 | metabolite A | 1.70E-17 | XL_VLDL_C | XL_VLDL_C_pct |
| rs545971 | metabolite T | 4.70E-19 | XL_VLDL_C | XL_VLDL_C_pct |
| rs612169 | metabolite G | 4.10E-19 | XL_VLDL_C | XL_VLDL_C_pct |
| rs8176668 | metabolite T | 6.70E-10 | XL_VLDL_C | XL_VLDL_C_pct |
| rs8176663 | metabolite C | 4.70E-19 | XL_VLDL_C | XL_VLDL_C_pct |
| rs7036642 | metabolite A | 7.80E-10 | XL_VLDL_C | XL_VLDL_C_pct |
| rs491626 | metabolite T | 4.60E-19 | XL_VLDL_C | XL_VLDL_C_pct |
| rs492488 | metabolite A | 4.60E-19 | XL_VLDL_C | XL_VLDL_C_pct |
| rs493246 | metabolite A | 4.60E-19 | XL_VLDL_C | XL_VLDL_C_pct |
| rs494242 | metabolite T | 1.80E-17 | XL_VLDL_C | XL_VLDL_C_pct |
| rs495203 | metabolite T | 4.60E-19 | XL_VLDL_C | XL_VLDL_C_pct |
| rs582118 | metabolite G | 4.60E-19 | XL_VLDL_C | XL_VLDL_C_pct |
| rs582094 | metabolite T | 6.90E-19 | XL_VLDL_C | XL_VLDL_C_pct |
| rs2769071 | metabolite G | 6.90E-19 | XL_VLDL_C | XL_VLDL_C_pct |
| rs677355 | metabolite A | 4.60E-19 | XL_VLDL_C | XL_VLDL_C_pct |
| rs676996 | metabolite G | 5.40E-19 | XL_VLDL_C | XL_VLDL_C_pct |
| rs676457 | metabolite T | 5.40E-19 | XL_VLDL_C | XL_VLDL_C_pct |
| rs550057 | metabolite T | 3.40E-26 | XL_VLDL_C | XL_VLDL_C_pct |
| rs674302 | metabolite A | 5.50E-19 | XL_VLDL_C | XL_VLDL_C_pct |
| rs7046674 | metabolite T | 9.30E-10 | XL_VLDL_C | XL_VLDL_C_pct |
| rs554833 | metabolite T | 5.70E-19 | XL_VLDL_C | XL_VLDL_C_pct |
| rs8176649 | metabolite A | 7.90E-10 | XL_VLDL_C | XL_VLDL_C_pct |
| rs660340 | metabolite G | 4.70E-09 | XL_VLDL_C | XL_VLDL_C_pct |
| rs581107 | metabolite T | 4.90E-09 | XL_VLDL_C | XL_VLDL_C_pct |
| rs659104 | metabolite G | 4.90E-09 | XL_VLDL_C | XL_VLDL_C_pct |
| rs647800 | metabolite G | 1.80E-08 | XL_VLDL_C | XL_VLDL_C_pct |
| rs473533 | metabolite C | 5.00E-09 | XL_VLDL_C | XL_VLDL_C_pct |
| rs476410 | metabolite C | 5.40E-09 | XL_VLDL_C | XL_VLDL_C_pct |
| rs645982 | metabolite G | 4.60E-09 | XL_VLDL_C | XL_VLDL_C_pct |
| rs529565 | metabolite C | 5.20E-19 | XL_VLDL_C | XL_VLDL_C_pct |
| rs532436 | metabolite A | 1.30E-33 | XL_VLDL_C | XL_VLDL_C_pct |
| rs600038 | metabolite C | 6.90E-29 | XL_VLDL_C | XL_VLDL_C_pct |

| | | | | |
|---|---|---|---|---|
| rs1124406: | metabolite T | 3.20E-21 | XL_VLDL_C | XL_VLDL_C_pct |
| rs649129 | metabolite T | 5.60E-29 | XL_VLDL_C | XL_VLDL_C_pct |
| rs495828 | metabolite T | 1.00E-28 | XL_VLDL_C | XL_VLDL_C_pct |
| rs687289 | metabolite A | 1.90E-08 | XL_HDL_PL | XL_HDL_PL_pct |
| rs2519093 | metabolite T | 9.60E-18 | XL_HDL_PL | XL_HDL_PL_pct |
| rs644234 | metabolite G | 4.70E-08 | XL_HDL_PL | XL_HDL_PL_pct |
| rs643434 | metabolite A | 4.90E-08 | XL_HDL_PL | XL_HDL_PL_pct |
| rs543040 | metabolite T | 1.20E-08 | XL_HDL_PL | XL_HDL_PL_pct |
| rs613534 | metabolite G | 4.60E-08 | XL_HDL_PL | XL_HDL_PL_pct |
| rs543968 | metabolite C | 4.60E-08 | XL_HDL_PL | XL_HDL_PL_pct |
| rs544873 | metabolite A | 4.60E-08 | XL_HDL_PL | XL_HDL_PL_pct |
| rs545971 | metabolite T | 1.10E-08 | XL_HDL_PL | XL_HDL_PL_pct |
| rs612169 | metabolite G | 1.40E-08 | XL_HDL_PL | XL_HDL_PL_pct |
| rs8176663 | metabolite C | 1.10E-08 | XL_HDL_PL | XL_HDL_PL_pct |
| rs491626 | metabolite T | 1.20E-08 | XL_HDL_PL | XL_HDL_PL_pct |
| rs492488 | metabolite A | 1.20E-08 | XL_HDL_PL | XL_HDL_PL_pct |
| rs493246 | metabolite A | 1.20E-08 | XL_HDL_PL | XL_HDL_PL_pct |
| rs494242 | metabolite T | 4.60E-08 | XL_HDL_PL | XL_HDL_PL_pct |
| rs495203 | metabolite T | 1.20E-08 | XL_HDL_PL | XL_HDL_PL_pct |
| rs582118 | metabolite G | 1.20E-08 | XL_HDL_PL | XL_HDL_PL_pct |
| rs582094 | metabolite T | 1.40E-08 | XL_HDL_PL | XL_HDL_PL_pct |
| rs2769071 | metabolite G | 1.40E-08 | XL_HDL_PL | XL_HDL_PL_pct |
| rs677355 | metabolite A | 1.30E-08 | XL_HDL_PL | XL_HDL_PL_pct |
| rs676996 | metabolite G | 1.30E-08 | XL_HDL_PL | XL_HDL_PL_pct |
| rs676457 | metabolite T | 1.30E-08 | XL_HDL_PL | XL_HDL_PL_pct |
| rs550057 | metabolite T | 1.10E-13 | XL_HDL_PL | XL_HDL_PL_pct |
| rs674302 | metabolite A | 1.20E-08 | XL_HDL_PL | XL_HDL_PL_pct |
| rs554833 | metabolite T | 1.20E-08 | XL_HDL_PL | XL_HDL_PL_pct |
| rs529565 | metabolite C | 1.10E-08 | XL_HDL_PL | XL_HDL_PL_pct |
| rs532436 | metabolite A | 1.70E-17 | XL_HDL_PL | XL_HDL_PL_pct |
| rs600038 | metabolite C | 2.50E-15 | XL_HDL_PL | XL_HDL_PL_pct |
| rs1124406: | metabolite T | 5.80E-14 | XL_HDL_PL | XL_HDL_PL_pct |
| rs649129 | metabolite T | 1.80E-15 | XL_HDL_PL | XL_HDL_PL_pct |
| rs495828 | metabolite T | 2.10E-15 | XL_HDL_PL | XL_HDL_PL_pct |
| rs2519093 | metabolite T | 4.90E-11 | XL_HDL_PL | XL_HDL_PL |
| rs550057 | metabolite T | 5.00E-08 | XL_HDL_PL | XL_HDL_PL |
| rs532436 | metabolite A | 6.90E-11 | XL_HDL_PL | XL_HDL_PL |
| rs600038 | metabolite C | 6.70E-10 | XL_HDL_PL | XL_HDL_PL |
| rs1124406: | metabolite T | 7.50E-10 | XL_HDL_PL | XL_HDL_PL |
| rs649129 | metabolite T | 5.90E-10 | XL_HDL_PL | XL_HDL_PL |
| rs495828 | metabolite T | 8.00E-10 | XL_HDL_PL | XL_HDL_PL |
| rs687289 | metabolite A | 1.00E-14 | Total_FC | Total_FC |
| rs2519093 | metabolite T | 7.90E-28 | Total_FC | Total_FC |
| rs644234 | metabolite G | 4.60E-14 | Total_FC | Total_FC |
| rs643434 | metabolite A | 4.00E-14 | Total_FC | Total_FC |
| rs543040 | metabolite T | 6.10E-15 | Total_FC | Total_FC |
| rs613534 | metabolite G | 4.70E-14 | Total_FC | Total_FC |
| rs543968 | metabolite C | 4.70E-14 | Total_FC | Total_FC |
| rs544873 | metabolite A | 4.70E-14 | Total_FC | Total_FC |
| rs545971 | metabolite T | 6.80E-15 | Total_FC | Total_FC |

| | | | | |
|---|---|---|---|---|
| rs612169 | metabolite G | 8.30E-15 | Total_FC | Total_FC |
| rs8176663 | metabolite C | 6.80E-15 | Total_FC | Total_FC |
| rs491626 | metabolite T | 6.70E-15 | Total_FC | Total_FC |
| rs492488 | metabolite A | 6.70E-15 | Total_FC | Total_FC |
| rs493246 | metabolite A | 6.70E-15 | Total_FC | Total_FC |
| rs494242 | metabolite T | 4.40E-14 | Total_FC | Total_FC |
| rs495203 | metabolite T | 6.70E-15 | Total_FC | Total_FC |
| rs582118 | metabolite G | 6.90E-15 | Total_FC | Total_FC |
| rs582094 | metabolite T | 1.00E-14 | Total_FC | Total_FC |
| rs2769071 | metabolite G | 6.50E-15 | Total_FC | Total_FC |
| rs677355 | metabolite A | 8.00E-15 | Total_FC | Total_FC |
| rs676996 | metabolite G | 6.90E-15 | Total_FC | Total_FC |
| rs676457 | metabolite T | 6.90E-15 | Total_FC | Total_FC |
| rs550057 | metabolite T | 8.00E-23 | Total_FC | Total_FC |
| rs674302 | metabolite A | 6.90E-15 | Total_FC | Total_FC |
| rs554833 | metabolite T | 7.70E-15 | Total_FC | Total_FC |
| rs529565 | metabolite C | 1.20E-14 | Total_FC | Total_FC |
| rs532436 | metabolite A | 1.00E-27 | Total_FC | Total_FC |
| rs600038 | metabolite C | 8.70E-25 | Total_FC | Total_FC |
| rs1124406: | metabolite T | 1.90E-16 | Total_FC | Total_FC |
| rs649129 | metabolite T | 5.60E-25 | Total_FC | Total_FC |
| rs495828 | metabolite T | 3.30E-25 | Total_FC | Total_FC |
| rs687289 | metabolite A | 5.00E-08 | S_HDL_PL | S_HDL_PL |
| rs2519093 | metabolite T | 1.40E-13 | S_HDL_PL | S_HDL_PL |
| rs543040 | metabolite T | 3.00E-08 | S_HDL_PL | S_HDL_PL |
| rs545971 | metabolite T | 3.20E-08 | S_HDL_PL | S_HDL_PL |
| rs612169 | metabolite G | 3.40E-08 | S_HDL_PL | S_HDL_PL |
| rs8176663 | metabolite C | 3.20E-08 | S_HDL_PL | S_HDL_PL |
| rs491626 | metabolite T | 3.20E-08 | S_HDL_PL | S_HDL_PL |
| rs492488 | metabolite A | 3.20E-08 | S_HDL_PL | S_HDL_PL |
| rs493246 | metabolite A | 3.20E-08 | S_HDL_PL | S_HDL_PL |
| rs495203 | metabolite T | 3.20E-08 | S_HDL_PL | S_HDL_PL |
| rs582118 | metabolite G | 3.30E-08 | S_HDL_PL | S_HDL_PL |
| rs582094 | metabolite T | 4.10E-08 | S_HDL_PL | S_HDL_PL |
| rs2769071 | metabolite G | 3.00E-08 | S_HDL_PL | S_HDL_PL |
| rs677355 | metabolite A | 3.60E-08 | S_HDL_PL | S_HDL_PL |
| rs676996 | metabolite G | 3.30E-08 | S_HDL_PL | S_HDL_PL |
| rs676457 | metabolite T | 3.30E-08 | S_HDL_PL | S_HDL_PL |
| rs550057 | metabolite T | 2.80E-09 | S_HDL_PL | S_HDL_PL |
| rs674302 | metabolite A | 3.30E-08 | S_HDL_PL | S_HDL_PL |
| rs554833 | metabolite T | 3.40E-08 | S_HDL_PL | S_HDL_PL |
| rs529565 | metabolite C | 1.90E-08 | S_HDL_PL | S_HDL_PL |
| rs532436 | metabolite A | 1.80E-13 | S_HDL_PL | S_HDL_PL |
| rs600038 | metabolite C | 2.80E-11 | S_HDL_PL | S_HDL_PL |
| rs1124406: | metabolite T | 1.70E-11 | S_HDL_PL | S_HDL_PL |
| rs649129 | metabolite T | 2.30E-11 | S_HDL_PL | S_HDL_PL |
| rs495828 | metabolite T | 3.90E-11 | S_HDL_PL | S_HDL_PL |
| rs687289 | metabolite A | 7.10E-09 | LA | LA |
| rs2519093 | metabolite T | 4.40E-17 | LA | LA |
| rs644234 | metabolite G | 1.60E-08 | LA | LA |

| | | | | |
|---|---|---|---|---|
| rs643434 | metabolite A | 1.30E-08 | LA | LA |
| rs543040 | metabolite T | 3.40E-09 | LA | LA |
| rs613534 | metabolite G | 1.60E-08 | LA | LA |
| rs543968 | metabolite C | 1.60E-08 | LA | LA |
| rs544873 | metabolite A | 1.60E-08 | LA | LA |
| rs545971 | metabolite T | 3.70E-09 | LA | LA |
| rs612169 | metabolite G | 4.00E-09 | LA | LA |
| rs8176663 | metabolite C | 3.70E-09 | LA | LA |
| rs491626 | metabolite T | 3.60E-09 | LA | LA |
| rs492488 | metabolite A | 3.60E-09 | LA | LA |
| rs493246 | metabolite A | 3.60E-09 | LA | LA |
| rs494242 | metabolite T | 1.50E-08 | LA | LA |
| rs495203 | metabolite T | 3.60E-09 | LA | LA |
| rs582118 | metabolite G | 3.70E-09 | LA | LA |
| rs582094 | metabolite T | 5.20E-09 | LA | LA |
| rs2769071 | metabolite G | 3.50E-09 | LA | LA |
| rs677355 | metabolite A | 4.00E-09 | LA | LA |
| rs676996 | metabolite G | 3.50E-09 | LA | LA |
| rs676457 | metabolite T | 3.60E-09 | LA | LA |
| rs550057 | metabolite T | 1.40E-13 | LA | LA |
| rs674302 | metabolite A | 3.50E-09 | LA | LA |
| rs554833 | metabolite T | 3.80E-09 | LA | LA |
| rs529565 | metabolite C | 3.90E-09 | LA | LA |
| rs532436 | metabolite A | 3.30E-17 | LA | LA |
| rs600038 | metabolite C | 3.30E-15 | LA | LA |
| rs1124406: | metabolite T | 3.90E-11 | LA | LA |
| rs649129 | metabolite T | 2.10E-15 | LA | LA |
| rs495828 | metabolite T | 1.40E-15 | LA | LA |
| rs687289 | metabolite A | 1.30E-08 | ApoB_by_A | ApoB_by_ApoA1 |
| rs2519093 | metabolite T | 9.90E-15 | ApoB_by_A | ApoB_by_ApoA1 |
| rs644234 | metabolite G | 2.10E-09 | ApoB_by_A | ApoB_by_ApoA1 |
| rs643434 | metabolite A | 2.00E-09 | ApoB_by_A | ApoB_by_ApoA1 |
| rs543040 | metabolite T | 1.30E-08 | ApoB_by_A | ApoB_by_ApoA1 |
| rs613534 | metabolite G | 2.00E-09 | ApoB_by_A | ApoB_by_ApoA1 |
| rs543968 | metabolite C | 2.00E-09 | ApoB_by_A | ApoB_by_ApoA1 |
| rs544873 | metabolite A | 2.00E-09 | ApoB_by_A | ApoB_by_ApoA1 |
| rs545971 | metabolite T | 1.30E-08 | ApoB_by_A | ApoB_by_ApoA1 |
| rs612169 | metabolite G | 1.60E-08 | ApoB_by_A | ApoB_by_ApoA1 |
| rs8176663 | metabolite C | 1.30E-08 | ApoB_by_A | ApoB_by_ApoA1 |
| rs491626 | metabolite T | 1.30E-08 | ApoB_by_A | ApoB_by_ApoA1 |
| rs492488 | metabolite A | 1.30E-08 | ApoB_by_A | ApoB_by_ApoA1 |
| rs493246 | metabolite A | 1.30E-08 | ApoB_by_A | ApoB_by_ApoA1 |
| rs494242 | metabolite T | 2.10E-09 | ApoB_by_A | ApoB_by_ApoA1 |
| rs495203 | metabolite T | 1.30E-08 | ApoB_by_A | ApoB_by_ApoA1 |
| rs582118 | metabolite G | 1.30E-08 | ApoB_by_A | ApoB_by_ApoA1 |
| rs582094 | metabolite T | 1.60E-08 | ApoB_by_A | ApoB_by_ApoA1 |
| rs2769071 | metabolite G | 1.40E-08 | ApoB_by_A | ApoB_by_ApoA1 |
| rs677355 | metabolite A | 1.50E-08 | ApoB_by_A | ApoB_by_ApoA1 |
| rs676996 | metabolite G | 1.40E-08 | ApoB_by_A | ApoB_by_ApoA1 |
| rs676457 | metabolite T | 1.40E-08 | ApoB_by_A | ApoB_by_ApoA1 |

| | | | | |
|---|---|---|---|---|
| rs550057 | metabolite T | 5.50E-14 | ApoB_by_/ | ApoB_by_ApoA1 |
| rs674302 | metabolite A | 1.40E-08 | ApoB_by_/ | ApoB_by_ApoA1 |
| rs554833 | metabolite T | 1.40E-08 | ApoB_by_/ | ApoB_by_ApoA1 |
| rs529565 | metabolite C | 2.40E-08 | ApoB_by_/ | ApoB_by_ApoA1 |
| rs532436 | metabolite A | 1.30E-14 | ApoB_by_/ | ApoB_by_ApoA1 |
| rs600038 | metabolite C | 8.20E-15 | ApoB_by_/ | ApoB_by_ApoA1 |
| rs1124406: | metabolite T | 4.90E-09 | ApoB_by_/ | ApoB_by_ApoA1 |
| rs649129 | metabolite T | 5.80E-15 | ApoB_by_/ | ApoB_by_ApoA1 |
| rs495828 | metabolite T | 2.60E-15 | ApoB_by_/ | ApoB_by_ApoA1 |
| rs687289 | metabolite A | 1.20E-13 | LDL_C | LDL_C |
| rs2519093 | metabolite T | 2.20E-30 | LDL_C | LDL_C |
| rs644234 | metabolite G | 9.50E-13 | LDL_C | LDL_C |
| rs643434 | metabolite A | 8.80E-13 | LDL_C | LDL_C |
| rs543040 | metabolite T | 9.10E-14 | LDL_C | LDL_C |
| rs613534 | metabolite G | 9.60E-13 | LDL_C | LDL_C |
| rs543968 | metabolite C | 9.60E-13 | LDL_C | LDL_C |
| rs544873 | metabolite A | 9.60E-13 | LDL_C | LDL_C |
| rs545971 | metabolite T | 8.80E-14 | LDL_C | LDL_C |
| rs612169 | metabolite G | 1.10E-13 | LDL_C | LDL_C |
| rs8176663 | metabolite C | 8.80E-14 | LDL_C | LDL_C |
| rs491626 | metabolite T | 8.70E-14 | LDL_C | LDL_C |
| rs492488 | metabolite A | 8.70E-14 | LDL_C | LDL_C |
| rs493246 | metabolite A | 8.70E-14 | LDL_C | LDL_C |
| rs494242 | metabolite T | 9.20E-13 | LDL_C | LDL_C |
| rs495203 | metabolite T | 8.80E-14 | LDL_C | LDL_C |
| rs582118 | metabolite G | 9.00E-14 | LDL_C | LDL_C |
| rs582094 | metabolite T | 1.40E-13 | LDL_C | LDL_C |
| rs2769071 | metabolite G | 9.30E-14 | LDL_C | LDL_C |
| rs677355 | metabolite A | 1.10E-13 | LDL_C | LDL_C |
| rs676996 | metabolite G | 9.30E-14 | LDL_C | LDL_C |
| rs676457 | metabolite T | 9.40E-14 | LDL_C | LDL_C |
| rs550057 | metabolite T | 3.90E-24 | LDL_C | LDL_C |
| rs674302 | metabolite A | 9.30E-14 | LDL_C | LDL_C |
| rs554833 | metabolite T | 1.00E-13 | LDL_C | LDL_C |
| rs529565 | metabolite C | 1.60E-13 | LDL_C | LDL_C |
| rs532436 | metabolite A | 3.10E-30 | LDL_C | LDL_C |
| rs600038 | metabolite C | 1.60E-26 | LDL_C | LDL_C |
| rs1124406: | metabolite T | 2.60E-18 | LDL_C | LDL_C |
| rs649129 | metabolite T | 9.90E-27 | LDL_C | LDL_C |
| rs495828 | metabolite T | 7.00E-27 | LDL_C | LDL_C |
| rs687289 | metabolite A | 2.50E-09 | S_HDL_L | S_HDL_L |
| rs2519093 | metabolite T | 1.60E-17 | S_HDL_L | S_HDL_L |
| rs644234 | metabolite G | 2.70E-08 | S_HDL_L | S_HDL_L |
| rs643434 | metabolite A | 2.60E-08 | S_HDL_L | S_HDL_L |
| rs543040 | metabolite T | 1.50E-09 | S_HDL_L | S_HDL_L |
| rs613534 | metabolite G | 2.70E-08 | S_HDL_L | S_HDL_L |
| rs543968 | metabolite C | 2.70E-08 | S_HDL_L | S_HDL_L |
| rs544873 | metabolite A | 2.70E-08 | S_HDL_L | S_HDL_L |
| rs545971 | metabolite T | 1.50E-09 | S_HDL_L | S_HDL_L |
| rs612169 | metabolite G | 1.60E-09 | S_HDL_L | S_HDL_L |

| | | | | |
|---|---|---|---|---|
| rs8176663 | metabolite C | 1.50E-09 | S_HDL_L | S_HDL_L |
| rs491626 | metabolite T | 1.50E-09 | S_HDL_L | S_HDL_L |
| rs492488 | metabolite A | 1.50E-09 | S_HDL_L | S_HDL_L |
| rs493246 | metabolite A | 1.50E-09 | S_HDL_L | S_HDL_L |
| rs494242 | metabolite T | 2.60E-08 | S_HDL_L | S_HDL_L |
| rs495203 | metabolite T | 1.50E-09 | S_HDL_L | S_HDL_L |
| rs582118 | metabolite G | 1.60E-09 | S_HDL_L | S_HDL_L |
| rs582094 | metabolite T | 2.10E-09 | S_HDL_L | S_HDL_L |
| rs2769071 | metabolite G | 1.50E-09 | S_HDL_L | S_HDL_L |
| rs677355 | metabolite A | 1.80E-09 | S_HDL_L | S_HDL_L |
| rs676996 | metabolite G | 1.60E-09 | S_HDL_L | S_HDL_L |
| rs676457 | metabolite T | 1.60E-09 | S_HDL_L | S_HDL_L |
| rs550057 | metabolite T | 5.20E-12 | S_HDL_L | S_HDL_L |
| rs674302 | metabolite A | 1.60E-09 | S_HDL_L | S_HDL_L |
| rs554833 | metabolite T | 1.60E-09 | S_HDL_L | S_HDL_L |
| rs529565 | metabolite C | 8.80E-10 | S_HDL_L | S_HDL_L |
| rs532436 | metabolite A | 2.20E-17 | S_HDL_L | S_HDL_L |
| rs600038 | metabolite C | 1.60E-14 | S_HDL_L | S_HDL_L |
| rs1124406: | metabolite T | 2.50E-14 | S_HDL_L | S_HDL_L |
| rs649129 | metabolite T | 1.30E-14 | S_HDL_L | S_HDL_L |
| rs495828 | metabolite T | 2.30E-14 | S_HDL_L | S_HDL_L |
| rs2519093 | metabolite T | 1.10E-10 | Pyruvate | Pyruvate |
| rs550057 | metabolite T | 6.40E-09 | Pyruvate | Pyruvate |
| rs532436 | metabolite A | 1.60E-10 | Pyruvate | Pyruvate |
| rs600038 | metabolite C | 3.60E-09 | Pyruvate | Pyruvate |
| rs1124406: | metabolite T | 5.00E-09 | Pyruvate | Pyruvate |
| rs649129 | metabolite T | 4.10E-09 | Pyruvate | Pyruvate |
| rs495828 | metabolite T | 5.60E-09 | Pyruvate | Pyruvate |
| rs2519093 | metabolite T | 1.90E-16 | S_HDL_FC | S_HDL_FC |
| rs543040 | metabolite T | 4.50E-08 | S_HDL_FC | S_HDL_FC |
| rs545971 | metabolite T | 4.60E-08 | S_HDL_FC | S_HDL_FC |
| rs612169 | metabolite G | 4.80E-08 | S_HDL_FC | S_HDL_FC |
| rs8176663 | metabolite C | 4.60E-08 | S_HDL_FC | S_HDL_FC |
| rs491626 | metabolite T | 4.50E-08 | S_HDL_FC | S_HDL_FC |
| rs492488 | metabolite A | 4.50E-08 | S_HDL_FC | S_HDL_FC |
| rs493246 | metabolite A | 4.50E-08 | S_HDL_FC | S_HDL_FC |
| rs495203 | metabolite T | 4.50E-08 | S_HDL_FC | S_HDL_FC |
| rs582118 | metabolite G | 4.60E-08 | S_HDL_FC | S_HDL_FC |
| rs2769071 | metabolite G | 4.30E-08 | S_HDL_FC | S_HDL_FC |
| rs676996 | metabolite G | 4.60E-08 | S_HDL_FC | S_HDL_FC |
| rs676457 | metabolite T | 4.60E-08 | S_HDL_FC | S_HDL_FC |
| rs550057 | metabolite T | 2.70E-11 | S_HDL_FC | S_HDL_FC |
| rs674302 | metabolite A | 4.60E-08 | S_HDL_FC | S_HDL_FC |
| rs554833 | metabolite T | 4.80E-08 | S_HDL_FC | S_HDL_FC |
| rs529565 | metabolite C | 3.60E-08 | S_HDL_FC | S_HDL_FC |
| rs532436 | metabolite A | 2.40E-16 | S_HDL_FC | S_HDL_FC |
| rs600038 | metabolite C | 3.10E-13 | S_HDL_FC | S_HDL_FC |
| rs1124406: | metabolite T | 2.30E-12 | S_HDL_FC | S_HDL_FC |
| rs649129 | metabolite T | 2.10E-13 | S_HDL_FC | S_HDL_FC |
| rs495828 | metabolite T | 3.00E-13 | S_HDL_FC | S_HDL_FC |

| | | | | |
|---|---|---|---|---|
| rs7873635 | metabolite C | 5.90E-10 | S_VLDL_TG | S_VLDL_TG_pct |
| rs4962040 | metabolite G | 5.20E-10 | S_VLDL_TG | S_VLDL_TG_pct |
| rs8176702 | metabolite A | 6.90E-10 | S_VLDL_TG | S_VLDL_TG_pct |
| rs2073826 | metabolite T | 4.00E-10 | S_VLDL_TG | S_VLDL_TG_pct |
| rs687289 | metabolite A | 9.50E-19 | S_VLDL_TG | S_VLDL_TG_pct |
| rs2073827 | metabolite C | 3.80E-10 | S_VLDL_TG | S_VLDL_TG_pct |
| rs2073828 | metabolite A | 3.90E-10 | S_VLDL_TG | S_VLDL_TG_pct |
| rs8176691 | metabolite T | 4.10E-10 | S_VLDL_TG | S_VLDL_TG_pct |
| rs8176690 | metabolite G | 1.20E-10 | S_VLDL_TG | S_VLDL_TG_pct |
| rs8176682 | metabolite T | 1.80E-10 | S_VLDL_TG | S_VLDL_TG_pct |
| rs8176681 | metabolite C | 1.60E-10 | S_VLDL_TG | S_VLDL_TG_pct |
| rs2519093 | metabolite T | 2.20E-36 | S_VLDL_TG | S_VLDL_TG_pct |
| rs644234 | metabolite G | 1.80E-17 | S_VLDL_TG | S_VLDL_TG_pct |
| rs643434 | metabolite A | 2.00E-17 | S_VLDL_TG | S_VLDL_TG_pct |
| rs543040 | metabolite T | 2.60E-18 | S_VLDL_TG | S_VLDL_TG_pct |
| rs613534 | metabolite G | 1.90E-17 | S_VLDL_TG | S_VLDL_TG_pct |
| rs543968 | metabolite C | 1.90E-17 | S_VLDL_TG | S_VLDL_TG_pct |
| rs544873 | metabolite A | 1.90E-17 | S_VLDL_TG | S_VLDL_TG_pct |
| rs545971 | metabolite T | 1.40E-18 | S_VLDL_TG | S_VLDL_TG_pct |
| rs612169 | metabolite G | 1.60E-18 | S_VLDL_TG | S_VLDL_TG_pct |
| rs8176668 | metabolite T | 5.10E-10 | S_VLDL_TG | S_VLDL_TG_pct |
| rs8176663 | metabolite C | 1.40E-18 | S_VLDL_TG | S_VLDL_TG_pct |
| rs7036642 | metabolite A | 5.70E-10 | S_VLDL_TG | S_VLDL_TG_pct |
| rs491626 | metabolite T | 1.40E-18 | S_VLDL_TG | S_VLDL_TG_pct |
| rs492488 | metabolite A | 1.40E-18 | S_VLDL_TG | S_VLDL_TG_pct |
| rs493246 | metabolite A | 1.40E-18 | S_VLDL_TG | S_VLDL_TG_pct |
| rs494242 | metabolite T | 1.90E-17 | S_VLDL_TG | S_VLDL_TG_pct |
| rs495203 | metabolite T | 1.40E-18 | S_VLDL_TG | S_VLDL_TG_pct |
| rs582118 | metabolite G | 1.50E-18 | S_VLDL_TG | S_VLDL_TG_pct |
| rs582094 | metabolite T | 2.20E-18 | S_VLDL_TG | S_VLDL_TG_pct |
| rs2769071 | metabolite G | 1.70E-18 | S_VLDL_TG | S_VLDL_TG_pct |
| rs677355 | metabolite A | 1.50E-18 | S_VLDL_TG | S_VLDL_TG_pct |
| rs676996 | metabolite G | 1.60E-18 | S_VLDL_TG | S_VLDL_TG_pct |
| rs676457 | metabolite T | 1.60E-18 | S_VLDL_TG | S_VLDL_TG_pct |
| rs550057 | metabolite T | 2.10E-30 | S_VLDL_TG | S_VLDL_TG_pct |
| rs674302 | metabolite A | 1.60E-18 | S_VLDL_TG | S_VLDL_TG_pct |
| rs7046674 | metabolite T | 5.20E-10 | S_VLDL_TG | S_VLDL_TG_pct |
| rs554833 | metabolite T | 1.70E-18 | S_VLDL_TG | S_VLDL_TG_pct |
| rs8176649 | metabolite A | 4.90E-10 | S_VLDL_TG | S_VLDL_TG_pct |
| rs660340 | metabolite G | 3.70E-08 | S_VLDL_TG | S_VLDL_TG_pct |
| rs581107 | metabolite T | 3.70E-08 | S_VLDL_TG | S_VLDL_TG_pct |
| rs659104 | metabolite G | 3.90E-08 | S_VLDL_TG | S_VLDL_TG_pct |
| rs473533 | metabolite C | 4.00E-08 | S_VLDL_TG | S_VLDL_TG_pct |
| rs476410 | metabolite C | 4.00E-08 | S_VLDL_TG | S_VLDL_TG_pct |
| rs645982 | metabolite G | 3.50E-08 | S_VLDL_TG | S_VLDL_TG_pct |
| rs529565 | metabolite C | 2.80E-18 | S_VLDL_TG | S_VLDL_TG_pct |
| rs532436 | metabolite A | 4.40E-36 | S_VLDL_TG | S_VLDL_TG_pct |
| rs600038 | metabolite C | 6.80E-32 | S_VLDL_TG | S_VLDL_TG_pct |
| rs1124406: | metabolite T | 7.20E-21 | S_VLDL_TG | S_VLDL_TG_pct |
| rs649129 | metabolite T | 1.20E-31 | S_VLDL_TG | S_VLDL_TG_pct |

| | | | | |
|---|---|---|---|---|
| rs495828 | metabolite T | 1.10E-31 | S_VLDL_TG | S_VLDL_TG_pct |
| rs2519093 | metabolite T | 2.30E-16 | S_LDL_TG_ | S_LDL_TG_pct |
| rs550057 | metabolite T | 2.30E-13 | S_LDL_TG_ | S_LDL_TG_pct |
| rs532436 | metabolite A | 3.00E-16 | S_LDL_TG_ | S_LDL_TG_pct |
| rs600038 | metabolite C | 2.20E-13 | S_LDL_TG_ | S_LDL_TG_pct |
| rs1124406 | metabolite T | 1.50E-08 | S_LDL_TG_ | S_LDL_TG_pct |
| rs649129 | metabolite T | 3.70E-13 | S_LDL_TG_ | S_LDL_TG_pct |
| rs495828 | metabolite T | 5.00E-13 | S_LDL_TG_ | S_LDL_TG_pct |
| rs7873635 | metabolite C | 6.60E-10 | M_VLDL_T | M_VLDL_TG_pct |
| rs4962040 | metabolite G | 6.90E-10 | M_VLDL_T | M_VLDL_TG_pct |
| rs8176702 | metabolite A | 9.60E-10 | M_VLDL_T | M_VLDL_TG_pct |
| rs2073826 | metabolite T | 4.80E-10 | M_VLDL_T | M_VLDL_TG_pct |
| rs687289 | metabolite A | 3.40E-20 | M_VLDL_T | M_VLDL_TG_pct |
| rs2073827 | metabolite C | 5.00E-10 | M_VLDL_T | M_VLDL_TG_pct |
| rs2073828 | metabolite A | 5.10E-10 | M_VLDL_T | M_VLDL_TG_pct |
| rs8176691 | metabolite T | 6.30E-10 | M_VLDL_T | M_VLDL_TG_pct |
| rs8176690 | metabolite G | 1.70E-10 | M_VLDL_T | M_VLDL_TG_pct |
| rs8176682 | metabolite T | 3.00E-10 | M_VLDL_T | M_VLDL_TG_pct |
| rs8176681 | metabolite C | 2.40E-10 | M_VLDL_T | M_VLDL_TG_pct |
| rs2519093 | metabolite T | 4.00E-35 | M_VLDL_T | M_VLDL_TG_pct |
| rs644234 | metabolite G | 1.50E-18 | M_VLDL_T | M_VLDL_TG_pct |
| rs643434 | metabolite A | 1.70E-18 | M_VLDL_T | M_VLDL_TG_pct |
| rs543040 | metabolite T | 1.20E-19 | M_VLDL_T | M_VLDL_TG_pct |
| rs613534 | metabolite G | 1.50E-18 | M_VLDL_T | M_VLDL_TG_pct |
| rs543968 | metabolite C | 1.50E-18 | M_VLDL_T | M_VLDL_TG_pct |
| rs544873 | metabolite A | 1.50E-18 | M_VLDL_T | M_VLDL_TG_pct |
| rs545971 | metabolite T | 6.80E-20 | M_VLDL_T | M_VLDL_TG_pct |
| rs612169 | metabolite G | 7.80E-20 | M_VLDL_T | M_VLDL_TG_pct |
| rs8176668 | metabolite T | 1.00E-09 | M_VLDL_T | M_VLDL_TG_pct |
| rs8176663 | metabolite C | 6.80E-20 | M_VLDL_T | M_VLDL_TG_pct |
| rs7036642 | metabolite A | 1.10E-09 | M_VLDL_T | M_VLDL_TG_pct |
| rs491626 | metabolite T | 6.70E-20 | M_VLDL_T | M_VLDL_TG_pct |
| rs492488 | metabolite A | 6.70E-20 | M_VLDL_T | M_VLDL_TG_pct |
| rs493246 | metabolite A | 6.70E-20 | M_VLDL_T | M_VLDL_TG_pct |
| rs494242 | metabolite T | 1.60E-18 | M_VLDL_T | M_VLDL_TG_pct |
| rs495203 | metabolite T | 6.80E-20 | M_VLDL_T | M_VLDL_TG_pct |
| rs582118 | metabolite G | 6.70E-20 | M_VLDL_T | M_VLDL_TG_pct |
| rs582094 | metabolite T | 8.80E-20 | M_VLDL_T | M_VLDL_TG_pct |
| rs2769071 | metabolite G | 9.20E-20 | M_VLDL_T | M_VLDL_TG_pct |
| rs677355 | metabolite A | 7.10E-20 | M_VLDL_T | M_VLDL_TG_pct |
| rs676996 | metabolite G | 7.90E-20 | M_VLDL_T | M_VLDL_TG_pct |
| rs676457 | metabolite T | 7.90E-20 | M_VLDL_T | M_VLDL_TG_pct |
| rs550057 | metabolite T | 4.10E-29 | M_VLDL_T | M_VLDL_TG_pct |
| rs674302 | metabolite A | 8.00E-20 | M_VLDL_T | M_VLDL_TG_pct |
| rs7046674 | metabolite T | 1.20E-09 | M_VLDL_T | M_VLDL_TG_pct |
| rs554833 | metabolite T | 8.80E-20 | M_VLDL_T | M_VLDL_TG_pct |
| rs8176649 | metabolite A | 1.10E-09 | M_VLDL_T | M_VLDL_TG_pct |
| rs660340 | metabolite G | 3.00E-09 | M_VLDL_T | M_VLDL_TG_pct |
| rs581107 | metabolite T | 3.10E-09 | M_VLDL_T | M_VLDL_TG_pct |
| rs659104 | metabolite G | 3.00E-09 | M_VLDL_T | M_VLDL_TG_pct |

| | | | | |
|---|---|---|---|---|
| rs647800 | metabolite G | 7.80E-09 | M_VLDL_T | M_VLDL_TG_pct |
| rs473533 | metabolite C | 3.10E-09 | M_VLDL_T | M_VLDL_TG_pct |
| rs476410 | metabolite C | 3.30E-09 | M_VLDL_T | M_VLDL_TG_pct |
| rs645982 | metabolite G | 2.80E-09 | M_VLDL_T | M_VLDL_TG_pct |
| rs529565 | metabolite C | 1.50E-19 | M_VLDL_T | M_VLDL_TG_pct |
| rs532436 | metabolite A | 7.40E-35 | M_VLDL_T | M_VLDL_TG_pct |
| rs600038 | metabolite C | 1.20E-30 | M_VLDL_T | M_VLDL_TG_pct |
| rs1124406 | metabolite T | 1.10E-19 | M_VLDL_T | M_VLDL_TG_pct |
| rs649129 | metabolite T | 1.80E-30 | M_VLDL_T | M_VLDL_TG_pct |
| rs495828 | metabolite T | 1.50E-30 | M_VLDL_T | M_VLDL_TG_pct |
| rs687289 | metabolite A | 4.00E-10 | M_LDL_TG | M_LDL_TG_pct |
| rs2519093 | metabolite T | 6.00E-24 | M_LDL_TG | M_LDL_TG_pct |
| rs644234 | metabolite G | 8.50E-09 | M_LDL_TG | M_LDL_TG_pct |
| rs643434 | metabolite A | 8.20E-09 | M_LDL_TG | M_LDL_TG_pct |
| rs543040 | metabolite T | 4.30E-10 | M_LDL_TG | M_LDL_TG_pct |
| rs613534 | metabolite G | 8.60E-09 | M_LDL_TG | M_LDL_TG_pct |
| rs543968 | metabolite C | 8.60E-09 | M_LDL_TG | M_LDL_TG_pct |
| rs544873 | metabolite A | 8.60E-09 | M_LDL_TG | M_LDL_TG_pct |
| rs545971 | metabolite T | 3.20E-10 | M_LDL_TG | M_LDL_TG_pct |
| rs612169 | metabolite G | 3.20E-10 | M_LDL_TG | M_LDL_TG_pct |
| rs8176663 | metabolite C | 3.20E-10 | M_LDL_TG | M_LDL_TG_pct |
| rs491626 | metabolite T | 3.20E-10 | M_LDL_TG | M_LDL_TG_pct |
| rs492488 | metabolite A | 3.20E-10 | M_LDL_TG | M_LDL_TG_pct |
| rs493246 | metabolite A | 3.20E-10 | M_LDL_TG | M_LDL_TG_pct |
| rs494242 | metabolite T | 8.80E-09 | M_LDL_TG | M_LDL_TG_pct |
| rs495203 | metabolite T | 3.20E-10 | M_LDL_TG | M_LDL_TG_pct |
| rs582118 | metabolite G | 3.20E-10 | M_LDL_TG | M_LDL_TG_pct |
| rs582094 | metabolite T | 4.70E-10 | M_LDL_TG | M_LDL_TG_pct |
| rs2769071 | metabolite G | 3.80E-10 | M_LDL_TG | M_LDL_TG_pct |
| rs677355 | metabolite A | 3.50E-10 | M_LDL_TG | M_LDL_TG_pct |
| rs676996 | metabolite G | 3.60E-10 | M_LDL_TG | M_LDL_TG_pct |
| rs676457 | metabolite T | 3.60E-10 | M_LDL_TG | M_LDL_TG_pct |
| rs550057 | metabolite T | 8.30E-20 | M_LDL_TG | M_LDL_TG_pct |
| rs674302 | metabolite A | 3.60E-10 | M_LDL_TG | M_LDL_TG_pct |
| rs554833 | metabolite T | 3.50E-10 | M_LDL_TG | M_LDL_TG_pct |
| rs529565 | metabolite C | 5.60E-10 | M_LDL_TG | M_LDL_TG_pct |
| rs532436 | metabolite A | 1.10E-23 | M_LDL_TG | M_LDL_TG_pct |
| rs600038 | metabolite C | 1.80E-19 | M_LDL_TG | M_LDL_TG_pct |
| rs1124406 | metabolite T | 2.40E-12 | M_LDL_TG | M_LDL_TG_pct |
| rs649129 | metabolite T | 3.70E-19 | M_LDL_TG | M_LDL_TG_pct |
| rs495828 | metabolite T | 5.60E-19 | M_LDL_TG | M_LDL_TG_pct |
| rs7873635 | metabolite C | 3.40E-16 | L_VLDL_TG | L_VLDL_TG_pct |
| rs4962040 | metabolite G | 4.10E-16 | L_VLDL_TG | L_VLDL_TG_pct |
| rs8176702 | metabolite A | 4.70E-16 | L_VLDL_TG | L_VLDL_TG_pct |
| rs2073826 | metabolite T | 4.10E-16 | L_VLDL_TG | L_VLDL_TG_pct |
| rs687289 | metabolite A | 3.50E-32 | L_VLDL_TG | L_VLDL_TG_pct |
| rs2073827 | metabolite C | 4.70E-16 | L_VLDL_TG | L_VLDL_TG_pct |
| rs2073828 | metabolite A | 5.00E-16 | L_VLDL_TG | L_VLDL_TG_pct |
| rs8176691 | metabolite T | 2.60E-16 | L_VLDL_TG | L_VLDL_TG_pct |
| rs8176690 | metabolite G | 1.10E-16 | L_VLDL_TG | L_VLDL_TG_pct |

| | | | | |
|---|---|---|---|---|
| rs8176682 | metabolite T | 7.00E-17 | L_VLDL_TG | L_VLDL_TG_pct |
| rs8176681 | metabolite C | 1.10E-16 | L_VLDL_TG | L_VLDL_TG_pct |
| rs2519093 | metabolite T | 4.90E-53 | L_VLDL_TG | L_VLDL_TG_pct |
| rs644234 | metabolite G | 6.10E-31 | L_VLDL_TG | L_VLDL_TG_pct |
| rs643434 | metabolite A | 7.90E-31 | L_VLDL_TG | L_VLDL_TG_pct |
| rs543040 | metabolite T | 1.20E-31 | L_VLDL_TG | L_VLDL_TG_pct |
| rs613534 | metabolite G | 6.40E-31 | L_VLDL_TG | L_VLDL_TG_pct |
| rs543968 | metabolite C | 6.40E-31 | L_VLDL_TG | L_VLDL_TG_pct |
| rs544873 | metabolite A | 6.40E-31 | L_VLDL_TG | L_VLDL_TG_pct |
| rs545971 | metabolite T | 5.30E-32 | L_VLDL_TG | L_VLDL_TG_pct |
| rs612169 | metabolite G | 9.20E-32 | L_VLDL_TG | L_VLDL_TG_pct |
| rs8176668 | metabolite T | 1.80E-16 | L_VLDL_TG | L_VLDL_TG_pct |
| rs8176663 | metabolite C | 5.30E-32 | L_VLDL_TG | L_VLDL_TG_pct |
| rs7036642 | metabolite A | 2.40E-16 | L_VLDL_TG | L_VLDL_TG_pct |
| rs491626 | metabolite T | 5.30E-32 | L_VLDL_TG | L_VLDL_TG_pct |
| rs492488 | metabolite A | 5.30E-32 | L_VLDL_TG | L_VLDL_TG_pct |
| rs493246 | metabolite A | 5.30E-32 | L_VLDL_TG | L_VLDL_TG_pct |
| rs494242 | metabolite T | 6.60E-31 | L_VLDL_TG | L_VLDL_TG_pct |
| rs495203 | metabolite T | 5.30E-32 | L_VLDL_TG | L_VLDL_TG_pct |
| rs582118 | metabolite G | 5.60E-32 | L_VLDL_TG | L_VLDL_TG_pct |
| rs582094 | metabolite T | 7.10E-32 | L_VLDL_TG | L_VLDL_TG_pct |
| rs2769071 | metabolite G | 9.70E-32 | L_VLDL_TG | L_VLDL_TG_pct |
| rs677355 | metabolite A | 6.90E-32 | L_VLDL_TG | L_VLDL_TG_pct |
| rs676996 | metabolite G | 7.30E-32 | L_VLDL_TG | L_VLDL_TG_pct |
| rs676457 | metabolite T | 7.20E-32 | L_VLDL_TG | L_VLDL_TG_pct |
| rs550057 | metabolite T | 4.90E-41 | L_VLDL_TG | L_VLDL_TG_pct |
| rs674302 | metabolite A | 7.30E-32 | L_VLDL_TG | L_VLDL_TG_pct |
| rs7046674 | metabolite T | 1.80E-16 | L_VLDL_TG | L_VLDL_TG_pct |
| rs554833 | metabolite T | 9.30E-32 | L_VLDL_TG | L_VLDL_TG_pct |
| rs8176649 | metabolite A | 1.70E-16 | L_VLDL_TG | L_VLDL_TG_pct |
| rs660340 | metabolite G | 6.00E-10 | L_VLDL_TG | L_VLDL_TG_pct |
| rs581107 | metabolite T | 5.80E-10 | L_VLDL_TG | L_VLDL_TG_pct |
| rs659104 | metabolite G | 6.30E-10 | L_VLDL_TG | L_VLDL_TG_pct |
| rs647800 | metabolite G | 5.60E-10 | L_VLDL_TG | L_VLDL_TG_pct |
| rs473533 | metabolite C | 6.60E-10 | L_VLDL_TG | L_VLDL_TG_pct |
| rs476410 | metabolite C | 5.90E-10 | L_VLDL_TG | L_VLDL_TG_pct |
| rs645982 | metabolite G | 5.60E-10 | L_VLDL_TG | L_VLDL_TG_pct |
| rs529565 | metabolite C | 1.60E-31 | L_VLDL_TG | L_VLDL_TG_pct |
| rs630510 | metabolite G | 4.80E-14 | L_VLDL_TG | L_VLDL_TG_pct |
| rs630014 | metabolite G | 4.80E-14 | L_VLDL_TG | L_VLDL_TG_pct |
| rs532436 | metabolite A | 2.00E-52 | L_VLDL_TG | L_VLDL_TG_pct |
| rs600038 | metabolite C | 1.80E-47 | L_VLDL_TG | L_VLDL_TG_pct |
| rs1124406: | metabolite T | 2.00E-35 | L_VLDL_TG | L_VLDL_TG_pct |
| rs649129 | metabolite T | 1.20E-47 | L_VLDL_TG | L_VLDL_TG_pct |
| rs495828 | metabolite T | 2.50E-47 | L_VLDL_TG | L_VLDL_TG_pct |
| rs2519093 | metabolite T | 5.50E-15 | L_LDL_TG_ | L_LDL_TG_pct |
| rs550057 | metabolite T | 1.40E-12 | L_LDL_TG_ | L_LDL_TG_pct |
| rs532436 | metabolite A | 8.10E-15 | L_LDL_TG_ | L_LDL_TG_pct |
| rs600038 | metabolite C | 5.10E-12 | L_LDL_TG_ | L_LDL_TG_pct |
| rs649129 | metabolite T | 9.60E-12 | L_LDL_TG_ | L_LDL_TG_pct |

| | | | | |
|---|---|---|---|---|
| rs495828 | metabolite T | 1.30E-11 | L_LDL_TG_ | L_LDL_TG_pct |
| rs600038 | metabolite C | 3.50E-08 | L_HDL_TG_ | L_HDL_TG_pct |
| rs649129 | metabolite T | 1.90E-08 | L_HDL_TG_ | L_HDL_TG_pct |
| rs495828 | metabolite T | 1.30E-08 | L_HDL_TG_ | L_HDL_TG_pct |
| rs2519093 | metabolite T | 2.10E-13 | IDL_TG_pct | IDL_TG_pct |
| rs550057 | metabolite T | 7.70E-12 | IDL_TG_pct | IDL_TG_pct |
| rs532436 | metabolite A | 3.10E-13 | IDL_TG_pct | IDL_TG_pct |
| rs600038 | metabolite C | 4.00E-11 | IDL_TG_pct | IDL_TG_pct |
| rs649129 | metabolite T | 7.00E-11 | IDL_TG_pct | IDL_TG_pct |
| rs495828 | metabolite T | 7.40E-11 | IDL_TG_pct | IDL_TG_pct |
| rs687289 | metabolite A | 1.60E-09 | Total_PL | Total_PL |
| rs2519093 | metabolite T | 4.00E-16 | Total_PL | Total_PL |
| rs644234 | metabolite G | 7.70E-09 | Total_PL | Total_PL |
| rs643434 | metabolite A | 6.80E-09 | Total_PL | Total_PL |
| rs543040 | metabolite T | 8.60E-10 | Total_PL | Total_PL |
| rs613534 | metabolite G | 7.80E-09 | Total_PL | Total_PL |
| rs543968 | metabolite C | 7.80E-09 | Total_PL | Total_PL |
| rs544873 | metabolite A | 7.80E-09 | Total_PL | Total_PL |
| rs545971 | metabolite T | 1.10E-09 | Total_PL | Total_PL |
| rs612169 | metabolite G | 1.10E-09 | Total_PL | Total_PL |
| rs8176663 | metabolite C | 1.10E-09 | Total_PL | Total_PL |
| rs491626 | metabolite T | 1.00E-09 | Total_PL | Total_PL |
| rs492488 | metabolite A | 1.00E-09 | Total_PL | Total_PL |
| rs493246 | metabolite A | 1.00E-09 | Total_PL | Total_PL |
| rs494242 | metabolite T | 7.30E-09 | Total_PL | Total_PL |
| rs495203 | metabolite T | 1.00E-09 | Total_PL | Total_PL |
| rs582118 | metabolite G | 1.10E-09 | Total_PL | Total_PL |
| rs582094 | metabolite T | 1.40E-09 | Total_PL | Total_PL |
| rs2769071 | metabolite G | 9.60E-10 | Total_PL | Total_PL |
| rs677355 | metabolite A | 1.20E-09 | Total_PL | Total_PL |
| rs676996 | metabolite G | 1.10E-09 | Total_PL | Total_PL |
| rs676457 | metabolite T | 1.10E-09 | Total_PL | Total_PL |
| rs550057 | metabolite T | 1.20E-12 | Total_PL | Total_PL |
| rs674302 | metabolite A | 1.10E-09 | Total_PL | Total_PL |
| rs554833 | metabolite T | 1.20E-09 | Total_PL | Total_PL |
| rs529565 | metabolite C | 1.20E-09 | Total_PL | Total_PL |
| rs532436 | metabolite A | 4.30E-16 | Total_PL | Total_PL |
| rs600038 | metabolite C | 4.60E-14 | Total_PL | Total_PL |
| rs1124406: | metabolite T | 1.90E-10 | Total_PL | Total_PL |
| rs649129 | metabolite T | 3.10E-14 | Total_PL | Total_PL |
| rs495828 | metabolite T | 2.70E-14 | Total_PL | Total_PL |
| rs2519093 | metabolite T | 1.80E-09 | XL_HDL_L | XL_HDL_L |
| rs532436 | metabolite A | 2.40E-09 | XL_HDL_L | XL_HDL_L |
| rs600038 | metabolite C | 1.40E-08 | XL_HDL_L | XL_HDL_L |
| rs1124406: | metabolite T | 1.30E-08 | XL_HDL_L | XL_HDL_L |
| rs649129 | metabolite T | 1.30E-08 | XL_HDL_L | XL_HDL_L |
| rs495828 | metabolite T | 1.70E-08 | XL_HDL_L | XL_HDL_L |
| rs550057 | metabolite T | 2.10E-08 | Glucose | Glucose |
| rs2519093 | metabolite T | 3.40E-11 | Phosphogl\ | Phosphoglyc |
| rs550057 | metabolite T | 3.00E-08 | Phosphogl\ | Phosphoglyc |

| | | | | |
|---|---|---|---|---|
| rs532436 | metabolite A | 3.60E-11 | Phosphogly | Phosphoglyc |
| rs600038 | metabolite C | 1.30E-09 | Phosphogly | Phosphoglyc |
| rs649129 | metabolite T | 9.60E-10 | Phosphogly | Phosphoglyc |
| rs495828 | metabolite T | 9.60E-10 | Phosphogly | Phosphoglyc |
| rs2519093 | metabolite T | 1.00E-13 | Cholines | Cholines |
| rs543040 | metabolite T | 4.00E-08 | Cholines | Cholines |
| rs545971 | metabolite T | 4.90E-08 | Cholines | Cholines |
| rs612169 | metabolite G | 4.90E-08 | Cholines | Cholines |
| rs8176663 | metabolite C | 4.90E-08 | Cholines | Cholines |
| rs491626 | metabolite T | 4.70E-08 | Cholines | Cholines |
| rs492488 | metabolite A | 4.70E-08 | Cholines | Cholines |
| rs493246 | metabolite A | 4.70E-08 | Cholines | Cholines |
| rs495203 | metabolite T | 4.70E-08 | Cholines | Cholines |
| rs582118 | metabolite G | 4.80E-08 | Cholines | Cholines |
| rs2769071 | metabolite G | 4.50E-08 | Cholines | Cholines |
| rs676996 | metabolite G | 4.90E-08 | Cholines | Cholines |
| rs676457 | metabolite T | 4.90E-08 | Cholines | Cholines |
| rs550057 | metabolite T | 3.90E-10 | Cholines | Cholines |
| rs674302 | metabolite A | 4.90E-08 | Cholines | Cholines |
| rs532436 | metabolite A | 1.10E-13 | Cholines | Cholines |
| rs600038 | metabolite C | 1.40E-11 | Cholines | Cholines |
| rs1124406: | metabolite T | 7.10E-09 | Cholines | Cholines |
| rs649129 | metabolite T | 1.00E-11 | Cholines | Cholines |
| rs495828 | metabolite T | 1.00E-11 | Cholines | Cholines |
| rs687289 | metabolite A | 9.00E-12 | Sphingomy | Sphingomyelins |
| rs2519093 | metabolite T | 2.20E-21 | Sphingomy | Sphingomyelins |
| rs644234 | metabolite G | 3.20E-10 | Sphingomy | Sphingomyelins |
| rs643434 | metabolite A | 3.00E-10 | Sphingomy | Sphingomyelins |
| rs543040 | metabolite T | 6.00E-12 | Sphingomy | Sphingomyelins |
| rs613534 | metabolite G | 3.30E-10 | Sphingomy | Sphingomyelins |
| rs543968 | metabolite C | 3.30E-10 | Sphingomy | Sphingomyelins |
| rs544873 | metabolite A | 3.30E-10 | Sphingomy | Sphingomyelins |
| rs545971 | metabolite T | 6.80E-12 | Sphingomy | Sphingomyelins |
| rs612169 | metabolite G | 7.10E-12 | Sphingomy | Sphingomyelins |
| rs8176663 | metabolite C | 6.80E-12 | Sphingomy | Sphingomyelins |
| rs491626 | metabolite T | 6.60E-12 | Sphingomy | Sphingomyelins |
| rs492488 | metabolite A | 6.60E-12 | Sphingomy | Sphingomyelins |
| rs493246 | metabolite A | 6.60E-12 | Sphingomy | Sphingomyelins |
| rs494242 | metabolite T | 3.10E-10 | Sphingomy | Sphingomyelins |
| rs495203 | metabolite T | 6.60E-12 | Sphingomy | Sphingomyelins |
| rs582118 | metabolite G | 6.70E-12 | Sphingomy | Sphingomyelins |
| rs582094 | metabolite T | 8.90E-12 | Sphingomy | Sphingomyelins |
| rs2769071 | metabolite G | 6.70E-12 | Sphingomy | Sphingomyelins |
| rs677355 | metabolite A | 8.50E-12 | Sphingomy | Sphingomyelins |
| rs676996 | metabolite G | 7.40E-12 | Sphingomy | Sphingomyelins |
| rs676457 | metabolite T | 7.50E-12 | Sphingomy | Sphingomyelins |
| rs550057 | metabolite T | 6.30E-17 | Sphingomy | Sphingomyelins |
| rs674302 | metabolite A | 7.60E-12 | Sphingomy | Sphingomyelins |
| rs554833 | metabolite T | 8.10E-12 | Sphingomy | Sphingomyelins |
| rs529565 | metabolite C | 1.30E-11 | Sphingomy | Sphingomyelins |

| | | | | |
|---|---|---|---|---|
| rs532436 | metabolite A | 3.70E-21 | Sphingomy | Sphingomyelins |
| rs600038 | metabolite C | 1.70E-17 | Sphingomy | Sphingomyelins |
| rs1124406: | metabolite T | 3.30E-11 | Sphingomy | Sphingomyelins |
| rs649129 | metabolite T | 1.30E-17 | Sphingomy | Sphingomyelins |
| rs495828 | metabolite T | 1.10E-17 | Sphingomy | Sphingomyelins |
| rs2519093 | metabolite T | 1.10E-09 | LA_pct | LA_pct |
| rs532436 | metabolite A | 6.90E-10 | LA_pct | LA_pct |
| rs2519093 | metabolite T | 6.70E-11 | Phosphatid | Phosphatidylc |
| rs532436 | metabolite A | 6.60E-11 | Phosphatid | Phosphatidylc |
| rs600038 | metabolite C | 2.30E-09 | Phosphatid | Phosphatidylc |
| rs649129 | metabolite T | 1.80E-09 | Phosphatid | Phosphatidylc |
| rs495828 | metabolite T | 1.70E-09 | Phosphatid | Phosphatidylc |
| rs687289 | metabolite A | 2.00E-08 | PUFA | PUFA |
| rs2519093 | metabolite T | 1.30E-13 | PUFA | PUFA |
| rs543040 | metabolite T | 1.30E-08 | PUFA | PUFA |
| rs545971 | metabolite T | 1.40E-08 | PUFA | PUFA |
| rs612169 | metabolite G | 1.60E-08 | PUFA | PUFA |
| rs8176663 | metabolite C | 1.40E-08 | PUFA | PUFA |
| rs491626 | metabolite T | 1.40E-08 | PUFA | PUFA |
| rs492488 | metabolite A | 1.40E-08 | PUFA | PUFA |
| rs493246 | metabolite A | 1.40E-08 | PUFA | PUFA |
| rs495203 | metabolite T | 1.40E-08 | PUFA | PUFA |
| rs582118 | metabolite G | 1.40E-08 | PUFA | PUFA |
| rs582094 | metabolite T | 1.80E-08 | PUFA | PUFA |
| rs2769071 | metabolite G | 1.40E-08 | PUFA | PUFA |
| rs677355 | metabolite A | 1.70E-08 | PUFA | PUFA |
| rs676996 | metabolite G | 1.50E-08 | PUFA | PUFA |
| rs676457 | metabolite T | 1.50E-08 | PUFA | PUFA |
| rs550057 | metabolite T | 2.60E-11 | PUFA | PUFA |
| rs674302 | metabolite A | 1.50E-08 | PUFA | PUFA |
| rs554833 | metabolite T | 1.60E-08 | PUFA | PUFA |
| rs529565 | metabolite C | 1.80E-08 | PUFA | PUFA |
| rs532436 | metabolite A | 1.50E-13 | PUFA | PUFA |
| rs600038 | metabolite C | 6.00E-12 | PUFA | PUFA |
| rs1124406: | metabolite T | 6.50E-09 | PUFA | PUFA |
| rs649129 | metabolite T | 4.10E-12 | PUFA | PUFA |
| rs495828 | metabolite T | 3.80E-12 | PUFA | PUFA |
| rs687289 | metabolite A | 1.10E-09 | Omega_6 | Omega_6 |
| rs2519093 | metabolite T | 5.40E-17 | Omega_6 | Omega_6 |
| rs644234 | metabolite G | 3.10E-09 | Omega_6 | Omega_6 |
| rs643434 | metabolite A | 2.60E-09 | Omega_6 | Omega_6 |
| rs543040 | metabolite T | 5.10E-10 | Omega_6 | Omega_6 |
| rs613534 | metabolite G | 3.10E-09 | Omega_6 | Omega_6 |
| rs543968 | metabolite C | 3.10E-09 | Omega_6 | Omega_6 |
| rs544873 | metabolite A | 3.10E-09 | Omega_6 | Omega_6 |
| rs545971 | metabolite T | 5.80E-10 | Omega_6 | Omega_6 |
| rs612169 | metabolite G | 6.30E-10 | Omega_6 | Omega_6 |
| rs8176663 | metabolite C | 5.80E-10 | Omega_6 | Omega_6 |
| rs491626 | metabolite T | 5.60E-10 | Omega_6 | Omega_6 |
| rs492488 | metabolite A | 5.60E-10 | Omega_6 | Omega_6 |

| | | | | |
|---|---|---|---|---|
| rs493246 | metabolite A | 5.60E-10 | Omega_6 | Omega_6 |
| rs494242 | metabolite T | 2.90E-09 | Omega_6 | Omega_6 |
| rs495203 | metabolite T | 5.60E-10 | Omega_6 | Omega_6 |
| rs582118 | metabolite G | 5.80E-10 | Omega_6 | Omega_6 |
| rs582094 | metabolite T | 8.00E-10 | Omega_6 | Omega_6 |
| rs2769071 | metabolite G | 5.60E-10 | Omega_6 | Omega_6 |
| rs677355 | metabolite A | 6.80E-10 | Omega_6 | Omega_6 |
| rs676996 | metabolite G | 6.00E-10 | Omega_6 | Omega_6 |
| rs676457 | metabolite T | 6.00E-10 | Omega_6 | Omega_6 |
| rs550057 | metabolite T | 1.30E-13 | Omega_6 | Omega_6 |
| rs674302 | metabolite A | 5.90E-10 | Omega_6 | Omega_6 |
| rs554833 | metabolite T | 6.50E-10 | Omega_6 | Omega_6 |
| rs529565 | metabolite C | 6.10E-10 | Omega_6 | Omega_6 |
| rs532436 | metabolite A | 5.30E-17 | Omega_6 | Omega_6 |
| rs600038 | metabolite C | 5.10E-15 | Omega_6 | Omega_6 |
| rs1124406: | metabolite T | 4.70E-11 | Omega_6 | Omega_6 |
| rs649129 | metabolite T | 3.70E-15 | Omega_6 | Omega_6 |
| rs495828 | metabolite T | 2.90E-15 | Omega_6 | Omega_6 |
| rs687289 | metabolite A | 8.30E-12 | S_LDL_CE | S_LDL_CE |
| rs2519093 | metabolite T | 4.00E-23 | S_LDL_CE | S_LDL_CE |
| rs644234 | metabolite G | 1.50E-11 | S_LDL_CE | S_LDL_CE |
| rs643434 | metabolite A | 1.40E-11 | S_LDL_CE | S_LDL_CE |
| rs543040 | metabolite T | 5.20E-12 | S_LDL_CE | S_LDL_CE |
| rs613534 | metabolite G | 1.50E-11 | S_LDL_CE | S_LDL_CE |
| rs543968 | metabolite C | 1.50E-11 | S_LDL_CE | S_LDL_CE |
| rs544873 | metabolite A | 1.50E-11 | S_LDL_CE | S_LDL_CE |
| rs545971 | metabolite T | 5.80E-12 | S_LDL_CE | S_LDL_CE |
| rs612169 | metabolite G | 7.20E-12 | S_LDL_CE | S_LDL_CE |
| rs8176663 | metabolite C | 5.80E-12 | S_LDL_CE | S_LDL_CE |
| rs491626 | metabolite T | 5.80E-12 | S_LDL_CE | S_LDL_CE |
| rs492488 | metabolite A | 5.80E-12 | S_LDL_CE | S_LDL_CE |
| rs493246 | metabolite A | 5.80E-12 | S_LDL_CE | S_LDL_CE |
| rs494242 | metabolite T | 1.40E-11 | S_LDL_CE | S_LDL_CE |
| rs495203 | metabolite T | 5.80E-12 | S_LDL_CE | S_LDL_CE |
| rs582118 | metabolite G | 6.00E-12 | S_LDL_CE | S_LDL_CE |
| rs582094 | metabolite T | 7.90E-12 | S_LDL_CE | S_LDL_CE |
| rs2769071 | metabolite G | 6.20E-12 | S_LDL_CE | S_LDL_CE |
| rs677355 | metabolite A | 7.00E-12 | S_LDL_CE | S_LDL_CE |
| rs676996 | metabolite G | 6.10E-12 | S_LDL_CE | S_LDL_CE |
| rs676457 | metabolite T | 6.10E-12 | S_LDL_CE | S_LDL_CE |
| rs550057 | metabolite T | 1.20E-19 | S_LDL_CE | S_LDL_CE |
| rs674302 | metabolite A | 6.10E-12 | S_LDL_CE | S_LDL_CE |
| rs554833 | metabolite T | 6.60E-12 | S_LDL_CE | S_LDL_CE |
| rs529565 | metabolite C | 7.50E-12 | S_LDL_CE | S_LDL_CE |
| rs532436 | metabolite A | 4.80E-23 | S_LDL_CE | S_LDL_CE |
| rs600038 | metabolite C | 5.90E-21 | S_LDL_CE | S_LDL_CE |
| rs1124406: | metabolite T | 1.40E-14 | S_LDL_CE | S_LDL_CE |
| rs649129 | metabolite T | 3.20E-21 | S_LDL_CE | S_LDL_CE |
| rs495828 | metabolite T | 1.80E-21 | S_LDL_CE | S_LDL_CE |
| rs7873635 | metabolite C | 4.30E-08 | S_VLDL_CE | S_VLDL_CE |

| | | | | |
|---|---|---|---|---|
| rs4962040 | metabolite G | 2.50E-08 | S_VLDL_CE | S_VLDL_CE |
| rs8176702 | metabolite A | 2.40E-08 | S_VLDL_CE | S_VLDL_CE |
| rs2073826 | metabolite T | 4.30E-08 | S_VLDL_CE | S_VLDL_CE |
| rs687289 | metabolite A | 2.20E-18 | S_VLDL_CE | S_VLDL_CE |
| rs8176691 | metabolite T | 2.10E-08 | S_VLDL_CE | S_VLDL_CE |
| rs8176690 | metabolite G | 2.90E-08 | S_VLDL_CE | S_VLDL_CE |
| rs8176682 | metabolite T | 1.20E-08 | S_VLDL_CE | S_VLDL_CE |
| rs8176681 | metabolite C | 2.30E-08 | S_VLDL_CE | S_VLDL_CE |
| rs2519093 | metabolite T | 1.10E-29 | S_VLDL_CE | S_VLDL_CE |
| rs644234 | metabolite G | 1.70E-18 | S_VLDL_CE | S_VLDL_CE |
| rs643434 | metabolite A | 1.60E-18 | S_VLDL_CE | S_VLDL_CE |
| rs543040 | metabolite T | 1.60E-18 | S_VLDL_CE | S_VLDL_CE |
| rs613534 | metabolite G | 1.70E-18 | S_VLDL_CE | S_VLDL_CE |
| rs543968 | metabolite C | 1.70E-18 | S_VLDL_CE | S_VLDL_CE |
| rs544873 | metabolite A | 1.70E-18 | S_VLDL_CE | S_VLDL_CE |
| rs545971 | metabolite T | 1.60E-18 | S_VLDL_CE | S_VLDL_CE |
| rs612169 | metabolite G | 2.50E-18 | S_VLDL_CE | S_VLDL_CE |
| rs8176668 | metabolite T | 6.70E-09 | S_VLDL_CE | S_VLDL_CE |
| rs8176663 | metabolite C | 1.60E-18 | S_VLDL_CE | S_VLDL_CE |
| rs7036642 | metabolite A | 6.50E-09 | S_VLDL_CE | S_VLDL_CE |
| rs491626 | metabolite T | 1.60E-18 | S_VLDL_CE | S_VLDL_CE |
| rs492488 | metabolite A | 1.60E-18 | S_VLDL_CE | S_VLDL_CE |
| rs493246 | metabolite A | 1.60E-18 | S_VLDL_CE | S_VLDL_CE |
| rs494242 | metabolite T | 1.70E-18 | S_VLDL_CE | S_VLDL_CE |
| rs495203 | metabolite T | 1.60E-18 | S_VLDL_CE | S_VLDL_CE |
| rs582118 | metabolite G | 1.70E-18 | S_VLDL_CE | S_VLDL_CE |
| rs582094 | metabolite T | 2.50E-18 | S_VLDL_CE | S_VLDL_CE |
| rs2769071 | metabolite G | 1.70E-18 | S_VLDL_CE | S_VLDL_CE |
| rs677355 | metabolite A | 1.90E-18 | S_VLDL_CE | S_VLDL_CE |
| rs676996 | metabolite G | 1.60E-18 | S_VLDL_CE | S_VLDL_CE |
| rs676457 | metabolite T | 1.60E-18 | S_VLDL_CE | S_VLDL_CE |
| rs550057 | metabolite T | 6.20E-25 | S_VLDL_CE | S_VLDL_CE |
| rs674302 | metabolite A | 1.60E-18 | S_VLDL_CE | S_VLDL_CE |
| rs7046674 | metabolite T | 5.00E-09 | S_VLDL_CE | S_VLDL_CE |
| rs554833 | metabolite T | 1.80E-18 | S_VLDL_CE | S_VLDL_CE |
| rs8176649 | metabolite A | 5.60E-09 | S_VLDL_CE | S_VLDL_CE |
| rs529565 | metabolite C | 2.20E-18 | S_VLDL_CE | S_VLDL_CE |
| rs630510 | metabolite G | 7.60E-09 | S_VLDL_CE | S_VLDL_CE |
| rs630014 | metabolite G | 7.60E-09 | S_VLDL_CE | S_VLDL_CE |
| rs532436 | metabolite A | 1.60E-29 | S_VLDL_CE | S_VLDL_CE |
| rs600038 | metabolite C | 1.20E-27 | S_VLDL_CE | S_VLDL_CE |
| rs1124406: | metabolite T | 1.40E-19 | S_VLDL_CE | S_VLDL_CE |
| rs649129 | metabolite T | 6.20E-28 | S_VLDL_CE | S_VLDL_CE |
| rs495828 | metabolite T | 3.30E-28 | S_VLDL_CE | S_VLDL_CE |
| rs687289 | metabolite A | 4.60E-16 | VLDL_CE | VLDL_CE |
| rs2519093 | metabolite T | 2.10E-25 | VLDL_CE | VLDL_CE |
| rs644234 | metabolite G | 2.10E-16 | VLDL_CE | VLDL_CE |
| rs643434 | metabolite A | 1.90E-16 | VLDL_CE | VLDL_CE |
| rs543040 | metabolite T | 2.50E-16 | VLDL_CE | VLDL_CE |
| rs613534 | metabolite G | 2.10E-16 | VLDL_CE | VLDL_CE |

| | | | | |
|---|---|---|---|---|
| rs543968 | metabolite C | 2.10E-16 | VLDL_CE | VLDL_CE |
| rs544873 | metabolite A | 2.10E-16 | VLDL_CE | VLDL_CE |
| rs545971 | metabolite T | 3.00E-16 | VLDL_CE | VLDL_CE |
| rs612169 | metabolite G | 4.20E-16 | VLDL_CE | VLDL_CE |
| rs8176663 | metabolite C | 3.00E-16 | VLDL_CE | VLDL_CE |
| rs7036642 | metabolite A | 4.80E-08 | VLDL_CE | VLDL_CE |
| rs491626 | metabolite T | 3.00E-16 | VLDL_CE | VLDL_CE |
| rs492488 | metabolite A | 3.00E-16 | VLDL_CE | VLDL_CE |
| rs493246 | metabolite A | 3.00E-16 | VLDL_CE | VLDL_CE |
| rs494242 | metabolite T | 2.00E-16 | VLDL_CE | VLDL_CE |
| rs495203 | metabolite T | 3.00E-16 | VLDL_CE | VLDL_CE |
| rs582118 | metabolite G | 3.10E-16 | VLDL_CE | VLDL_CE |
| rs582094 | metabolite T | 4.50E-16 | VLDL_CE | VLDL_CE |
| rs2769071 | metabolite G | 3.10E-16 | VLDL_CE | VLDL_CE |
| rs677355 | metabolite A | 3.60E-16 | VLDL_CE | VLDL_CE |
| rs676996 | metabolite G | 3.10E-16 | VLDL_CE | VLDL_CE |
| rs676457 | metabolite T | 3.10E-16 | VLDL_CE | VLDL_CE |
| rs550057 | metabolite T | 5.90E-22 | VLDL_CE | VLDL_CE |
| rs674302 | metabolite A | 3.10E-16 | VLDL_CE | VLDL_CE |
| rs7046674 | metabolite T | 3.80E-08 | VLDL_CE | VLDL_CE |
| rs554833 | metabolite T | 3.40E-16 | VLDL_CE | VLDL_CE |
| rs8176649 | metabolite A | 4.30E-08 | VLDL_CE | VLDL_CE |
| rs529565 | metabolite C | 4.50E-16 | VLDL_CE | VLDL_CE |
| rs532436 | metabolite A | 2.70E-25 | VLDL_CE | VLDL_CE |
| rs600038 | metabolite C | 5.70E-24 | VLDL_CE | VLDL_CE |
| rs1124406: | metabolite T | 3.80E-16 | VLDL_CE | VLDL_CE |
| rs649129 | metabolite T | 3.30E-24 | VLDL_CE | VLDL_CE |
| rs495828 | metabolite T | 1.60E-24 | VLDL_CE | VLDL_CE |
| rs687289 | metabolite A | 1.10E-09 | Citrate | Citrate |
| rs2519093 | metabolite T | 1.70E-09 | Citrate | Citrate |
| rs644234 | metabolite G | 6.00E-09 | Citrate | Citrate |
| rs643434 | metabolite A | 6.40E-09 | Citrate | Citrate |
| rs543040 | metabolite T | 5.00E-10 | Citrate | Citrate |
| rs613534 | metabolite G | 6.10E-09 | Citrate | Citrate |
| rs543968 | metabolite C | 6.10E-09 | Citrate | Citrate |
| rs544873 | metabolite A | 6.10E-09 | Citrate | Citrate |
| rs545971 | metabolite T | 7.30E-10 | Citrate | Citrate |
| rs612169 | metabolite G | 7.80E-10 | Citrate | Citrate |
| rs8176663 | metabolite C | 7.30E-10 | Citrate | Citrate |
| rs491626 | metabolite T | 7.40E-10 | Citrate | Citrate |
| rs492488 | metabolite A | 7.40E-10 | Citrate | Citrate |
| rs493246 | metabolite A | 7.40E-10 | Citrate | Citrate |
| rs494242 | metabolite T | 6.10E-09 | Citrate | Citrate |
| rs495203 | metabolite T | 7.30E-10 | Citrate | Citrate |
| rs582118 | metabolite G | 7.30E-10 | Citrate | Citrate |
| rs582094 | metabolite T | 7.90E-10 | Citrate | Citrate |
| rs2769071 | metabolite G | 7.90E-10 | Citrate | Citrate |
| rs677355 | metabolite A | 7.20E-10 | Citrate | Citrate |
| rs676996 | metabolite G | 6.90E-10 | Citrate | Citrate |
| rs676457 | metabolite T | 6.90E-10 | Citrate | Citrate |

| | | | | |
|---|---|---|---|---|
| rs550057 | metabolite T | 8.90E-09 | Citrate | Citrate |
| rs674302 | metabolite A | 7.10E-10 | Citrate | Citrate |
| rs554833 | metabolite T | 6.50E-10 | Citrate | Citrate |
| rs529565 | metabolite C | 2.10E-10 | Citrate | Citrate |
| rs532436 | metabolite A | 1.60E-09 | Citrate | Citrate |
| rs600038 | metabolite C | 2.60E-08 | Citrate | Citrate |
| rs649129 | metabolite T | 1.70E-08 | Citrate | Citrate |
| rs495828 | metabolite T | 2.00E-08 | Citrate | Citrate |
| rs4962040 | metabolite G | 3.40E-08 | Clinical_LD | Clinical_LDL_C |
| rs8176702 | metabolite A | 3.80E-08 | Clinical_LD | Clinical_LDL_C |
| rs2073826 | metabolite T | 1.80E-08 | Clinical_LD | Clinical_LDL_C |
| rs687289 | metabolite A | 4.00E-16 | Clinical_LD | Clinical_LDL_C |
| rs2073827 | metabolite C | 2.00E-08 | Clinical_LD | Clinical_LDL_C |
| rs2073828 | metabolite A | 2.00E-08 | Clinical_LD | Clinical_LDL_C |
| rs8176691 | metabolite T | 2.90E-08 | Clinical_LD | Clinical_LDL_C |
| rs8176690 | metabolite G | 8.80E-09 | Clinical_LD | Clinical_LDL_C |
| rs8176682 | metabolite T | 1.60E-08 | Clinical_LD | Clinical_LDL_C |
| rs8176681 | metabolite C | 9.20E-09 | Clinical_LD | Clinical_LDL_C |
| rs2519093 | metabolite T | 1.10E-34 | Clinical_LD | Clinical_LDL_C |
| rs644234 | metabolite G | 2.70E-15 | Clinical_LD | Clinical_LDL_C |
| rs643434 | metabolite A | 2.50E-15 | Clinical_LD | Clinical_LDL_C |
| rs543040 | metabolite T | 3.10E-16 | Clinical_LD | Clinical_LDL_C |
| rs613534 | metabolite G | 2.70E-15 | Clinical_LD | Clinical_LDL_C |
| rs543968 | metabolite C | 2.70E-15 | Clinical_LD | Clinical_LDL_C |
| rs544873 | metabolite A | 2.70E-15 | Clinical_LD | Clinical_LDL_C |
| rs545971 | metabolite T | 2.70E-16 | Clinical_LD | Clinical_LDL_C |
| rs612169 | metabolite G | 3.60E-16 | Clinical_LD | Clinical_LDL_C |
| rs8176668 | metabolite T | 1.80E-08 | Clinical_LD | Clinical_LDL_C |
| rs8176663 | metabolite C | 2.70E-16 | Clinical_LD | Clinical_LDL_C |
| rs7036642 | metabolite A | 1.60E-08 | Clinical_LD | Clinical_LDL_C |
| rs491626 | metabolite T | 2.70E-16 | Clinical_LD | Clinical_LDL_C |
| rs492488 | metabolite A | 2.70E-16 | Clinical_LD | Clinical_LDL_C |
| rs493246 | metabolite A | 2.70E-16 | Clinical_LD | Clinical_LDL_C |
| rs494242 | metabolite T | 2.70E-15 | Clinical_LD | Clinical_LDL_C |
| rs495203 | metabolite T | 2.70E-16 | Clinical_LD | Clinical_LDL_C |
| rs582118 | metabolite G | 2.80E-16 | Clinical_LD | Clinical_LDL_C |
| rs582094 | metabolite T | 4.30E-16 | Clinical_LD | Clinical_LDL_C |
| rs2769071 | metabolite G | 2.90E-16 | Clinical_LD | Clinical_LDL_C |
| rs677355 | metabolite A | 3.20E-16 | Clinical_LD | Clinical_LDL_C |
| rs676996 | metabolite G | 2.90E-16 | Clinical_LD | Clinical_LDL_C |
| rs676457 | metabolite T | 2.90E-16 | Clinical_LD | Clinical_LDL_C |
| rs550057 | metabolite T | 4.80E-28 | Clinical_LD | Clinical_LDL_C |
| rs674302 | metabolite A | 2.90E-16 | Clinical_LD | Clinical_LDL_C |
| rs7046674 | metabolite T | 1.30E-08 | Clinical_LD | Clinical_LDL_C |
| rs554833 | metabolite T | 3.20E-16 | Clinical_LD | Clinical_LDL_C |
| rs8176649 | metabolite A | 1.50E-08 | Clinical_LD | Clinical_LDL_C |
| rs529565 | metabolite C | 5.80E-16 | Clinical_LD | Clinical_LDL_C |
| rs532436 | metabolite A | 1.80E-34 | Clinical_LD | Clinical_LDL_C |
| rs600038 | metabolite C | 1.50E-30 | Clinical_LD | Clinical_LDL_C |
| rs1124406: | metabolite T | 1.60E-20 | Clinical_LD | Clinical_LDL_C |

| | | | | |
|---|---|---|---|---|
| rs649129 | metabolite T | 1.10E-30 | Clinical_LD | Clinical_LDL_C |
| rs495828 | metabolite T | 6.80E-31 | Clinical_LD | Clinical_LDL_C |
| rs687289 | metabolite A | 1.30E-10 | GlycA | GlycA |
| rs2519093 | metabolite T | 4.90E-09 | GlycA | GlycA |
| rs644234 | metabolite G | 4.90E-09 | GlycA | GlycA |
| rs643434 | metabolite A | 5.00E-09 | GlycA | GlycA |
| rs543040 | metabolite T | 8.90E-11 | GlycA | GlycA |
| rs613534 | metabolite G | 4.80E-09 | GlycA | GlycA |
| rs543968 | metabolite C | 4.80E-09 | GlycA | GlycA |
| rs544873 | metabolite A | 4.80E-09 | GlycA | GlycA |
| rs545971 | metabolite T | 1.40E-10 | GlycA | GlycA |
| rs612169 | metabolite G | 1.70E-10 | GlycA | GlycA |
| rs8176663 | metabolite C | 1.40E-10 | GlycA | GlycA |
| rs491626 | metabolite T | 1.40E-10 | GlycA | GlycA |
| rs492488 | metabolite A | 1.40E-10 | GlycA | GlycA |
| rs493246 | metabolite A | 1.40E-10 | GlycA | GlycA |
| rs494242 | metabolite T | 4.70E-09 | GlycA | GlycA |
| rs495203 | metabolite T | 1.40E-10 | GlycA | GlycA |
| rs582118 | metabolite G | 1.40E-10 | GlycA | GlycA |
| rs582094 | metabolite T | 1.60E-10 | GlycA | GlycA |
| rs2769071 | metabolite G | 1.50E-10 | GlycA | GlycA |
| rs677355 | metabolite A | 2.30E-10 | GlycA | GlycA |
| rs676996 | metabolite G | 1.80E-10 | GlycA | GlycA |
| rs676457 | metabolite T | 1.80E-10 | GlycA | GlycA |
| rs550057 | metabolite T | 1.00E-08 | GlycA | GlycA |
| rs674302 | metabolite A | 1.80E-10 | GlycA | GlycA |
| rs554833 | metabolite T | 1.80E-10 | GlycA | GlycA |
| rs529565 | metabolite C | 2.40E-10 | GlycA | GlycA |
| rs532436 | metabolite A | 6.60E-09 | GlycA | GlycA |
| rs2519093 | metabolite T | 3.00E-09 | HDL_P | HDL_P |
| rs532436 | metabolite A | 3.30E-09 | HDL_P | HDL_P |
| rs7873635 | metabolite C | 1.50E-09 | IDL_FC | IDL_FC |
| rs4962040 | metabolite G | 1.10E-09 | IDL_FC | IDL_FC |
| rs8176702 | metabolite A | 1.30E-09 | IDL_FC | IDL_FC |
| rs2073826 | metabolite T | 4.40E-10 | IDL_FC | IDL_FC |
| rs687289 | metabolite A | 2.00E-18 | IDL_FC | IDL_FC |
| rs2073827 | metabolite C | 5.20E-10 | IDL_FC | IDL_FC |
| rs2073828 | metabolite A | 5.30E-10 | IDL_FC | IDL_FC |
| rs8176691 | metabolite T | 9.30E-10 | IDL_FC | IDL_FC |
| rs8176690 | metabolite G | 1.50E-10 | IDL_FC | IDL_FC |
| rs8176682 | metabolite T | 3.80E-10 | IDL_FC | IDL_FC |
| rs8176681 | metabolite C | 1.70E-10 | IDL_FC | IDL_FC |
| rs2519093 | metabolite T | 4.20E-36 | IDL_FC | IDL_FC |
| rs644234 | metabolite G | 3.10E-17 | IDL_FC | IDL_FC |
| rs643434 | metabolite A | 3.00E-17 | IDL_FC | IDL_FC |
| rs543040 | metabolite T | 1.80E-18 | IDL_FC | IDL_FC |
| rs613534 | metabolite G | 3.20E-17 | IDL_FC | IDL_FC |
| rs543968 | metabolite C | 3.20E-17 | IDL_FC | IDL_FC |
| rs544873 | metabolite A | 3.20E-17 | IDL_FC | IDL_FC |
| rs545971 | metabolite T | 1.50E-18 | IDL_FC | IDL_FC |

| | | | | |
|---|---|---|---|---|
| rs612169 | metabolite G | 2.00E-18 | IDL_FC | IDL_FC |
| rs8176668 | metabolite T | 5.50E-10 | IDL_FC | IDL_FC |
| rs8176663 | metabolite C | 1.50E-18 | IDL_FC | IDL_FC |
| rs7036642 | metabolite A | 4.90E-10 | IDL_FC | IDL_FC |
| rs491626 | metabolite T | 1.40E-18 | IDL_FC | IDL_FC |
| rs492488 | metabolite A | 1.40E-18 | IDL_FC | IDL_FC |
| rs493246 | metabolite A | 1.40E-18 | IDL_FC | IDL_FC |
| rs494242 | metabolite T | 3.10E-17 | IDL_FC | IDL_FC |
| rs495203 | metabolite T | 1.40E-18 | IDL_FC | IDL_FC |
| rs582118 | metabolite G | 1.50E-18 | IDL_FC | IDL_FC |
| rs582094 | metabolite T | 2.20E-18 | IDL_FC | IDL_FC |
| rs2769071 | metabolite G | 1.50E-18 | IDL_FC | IDL_FC |
| rs677355 | metabolite A | 1.70E-18 | IDL_FC | IDL_FC |
| rs676996 | metabolite G | 1.50E-18 | IDL_FC | IDL_FC |
| rs676457 | metabolite T | 1.50E-18 | IDL_FC | IDL_FC |
| rs550057 | metabolite T | 1.70E-29 | IDL_FC | IDL_FC |
| rs674302 | metabolite A | 1.50E-18 | IDL_FC | IDL_FC |
| rs7046674 | metabolite T | 4.20E-10 | IDL_FC | IDL_FC |
| rs554833 | metabolite T | 1.80E-18 | IDL_FC | IDL_FC |
| rs8176649 | metabolite A | 4.50E-10 | IDL_FC | IDL_FC |
| rs660340 | metabolite G | 3.90E-08 | IDL_FC | IDL_FC |
| rs581107 | metabolite T | 3.40E-08 | IDL_FC | IDL_FC |
| rs659104 | metabolite G | 3.70E-08 | IDL_FC | IDL_FC |
| rs473533 | metabolite C | 3.80E-08 | IDL_FC | IDL_FC |
| rs476410 | metabolite C | 4.00E-08 | IDL_FC | IDL_FC |
| rs645982 | metabolite G | 3.40E-08 | IDL_FC | IDL_FC |
| rs529565 | metabolite C | 4.70E-18 | IDL_FC | IDL_FC |
| rs532436 | metabolite A | 7.70E-36 | IDL_FC | IDL_FC |
| rs600038 | metabolite C | 1.50E-31 | IDL_FC | IDL_FC |
| rs1124406 | metabolite T | 1.80E-20 | IDL_FC | IDL_FC |
| rs649129 | metabolite T | 1.40E-31 | IDL_FC | IDL_FC |
| rs495828 | metabolite T | 9.10E-32 | IDL_FC | IDL_FC |
| rs687289 | metabolite A | 9.80E-09 | IDL_FC_pct | IDL_FC_pct |
| rs2519093 | metabolite T | 1.40E-14 | IDL_FC_pct | IDL_FC_pct |
| rs543040 | metabolite T | 1.20E-08 | IDL_FC_pct | IDL_FC_pct |
| rs545971 | metabolite T | 1.10E-08 | IDL_FC_pct | IDL_FC_pct |
| rs612169 | metabolite G | 1.30E-08 | IDL_FC_pct | IDL_FC_pct |
| rs8176663 | metabolite C | 1.10E-08 | IDL_FC_pct | IDL_FC_pct |
| rs491626 | metabolite T | 1.10E-08 | IDL_FC_pct | IDL_FC_pct |
| rs492488 | metabolite A | 1.10E-08 | IDL_FC_pct | IDL_FC_pct |
| rs493246 | metabolite A | 1.10E-08 | IDL_FC_pct | IDL_FC_pct |
| rs495203 | metabolite T | 1.10E-08 | IDL_FC_pct | IDL_FC_pct |
| rs582118 | metabolite G | 1.10E-08 | IDL_FC_pct | IDL_FC_pct |
| rs582094 | metabolite T | 1.20E-08 | IDL_FC_pct | IDL_FC_pct |
| rs2769071 | metabolite G | 1.20E-08 | IDL_FC_pct | IDL_FC_pct |
| rs677355 | metabolite A | 1.10E-08 | IDL_FC_pct | IDL_FC_pct |
| rs676996 | metabolite G | 1.20E-08 | IDL_FC_pct | IDL_FC_pct |
| rs676457 | metabolite T | 1.20E-08 | IDL_FC_pct | IDL_FC_pct |
| rs550057 | metabolite T | 4.70E-13 | IDL_FC_pct | IDL_FC_pct |
| rs674302 | metabolite A | 1.20E-08 | IDL_FC_pct | IDL_FC_pct |

| | | | | |
|---|---|---|---|---|
| rs554833 | metabolite T | 1.10E-08 | IDL_FC_pct | IDL_FC_pct |
| rs529565 | metabolite C | 1.50E-08 | IDL_FC_pct | IDL_FC_pct |
| rs532436 | metabolite A | 2.10E-14 | IDL_FC_pct | IDL_FC_pct |
| rs600038 | metabolite C | 4.50E-11 | IDL_FC_pct | IDL_FC_pct |
| rs649129 | metabolite T | 7.50E-11 | IDL_FC_pct | IDL_FC_pct |
| rs495828 | metabolite T | 1.40E-10 | IDL_FC_pct | IDL_FC_pct |
| rs7873635 | metabolite C | 4.90E-08 | IDL_L | IDL_L |
| rs4962040 | metabolite G | 3.50E-08 | IDL_L | IDL_L |
| rs8176702 | metabolite A | 4.00E-08 | IDL_L | IDL_L |
| rs2073826 | metabolite T | 3.20E-08 | IDL_L | IDL_L |
| rs687289 | metabolite A | 6.50E-16 | IDL_L | IDL_L |
| rs2073827 | metabolite C | 3.90E-08 | IDL_L | IDL_L |
| rs2073828 | metabolite A | 3.90E-08 | IDL_L | IDL_L |
| rs8176691 | metabolite T | 3.10E-08 | IDL_L | IDL_L |
| rs8176690 | metabolite G | 1.50E-08 | IDL_L | IDL_L |
| rs8176682 | metabolite T | 1.50E-08 | IDL_L | IDL_L |
| rs8176681 | metabolite C | 1.50E-08 | IDL_L | IDL_L |
| rs2519093 | metabolite T | 1.10E-31 | IDL_L | IDL_L |
| rs644234 | metabolite G | 2.30E-15 | IDL_L | IDL_L |
| rs643434 | metabolite A | 2.10E-15 | IDL_L | IDL_L |
| rs543040 | metabolite T | 5.60E-16 | IDL_L | IDL_L |
| rs613534 | metabolite G | 2.30E-15 | IDL_L | IDL_L |
| rs543968 | metabolite C | 2.30E-15 | IDL_L | IDL_L |
| rs544873 | metabolite A | 2.30E-15 | IDL_L | IDL_L |
| rs545971 | metabolite T | 4.80E-16 | IDL_L | IDL_L |
| rs612169 | metabolite G | 6.10E-16 | IDL_L | IDL_L |
| rs8176668 | metabolite T | 1.90E-08 | IDL_L | IDL_L |
| rs8176663 | metabolite C | 4.80E-16 | IDL_L | IDL_L |
| rs7036642 | metabolite A | 1.60E-08 | IDL_L | IDL_L |
| rs491626 | metabolite T | 4.70E-16 | IDL_L | IDL_L |
| rs492488 | metabolite A | 4.70E-16 | IDL_L | IDL_L |
| rs493246 | metabolite A | 4.70E-16 | IDL_L | IDL_L |
| rs494242 | metabolite T | 2.20E-15 | IDL_L | IDL_L |
| rs495203 | metabolite T | 4.80E-16 | IDL_L | IDL_L |
| rs582118 | metabolite G | 5.00E-16 | IDL_L | IDL_L |
| rs582094 | metabolite T | 7.00E-16 | IDL_L | IDL_L |
| rs2769071 | metabolite G | 4.70E-16 | IDL_L | IDL_L |
| rs677355 | metabolite A | 5.50E-16 | IDL_L | IDL_L |
| rs676996 | metabolite G | 4.90E-16 | IDL_L | IDL_L |
| rs676457 | metabolite T | 4.90E-16 | IDL_L | IDL_L |
| rs550057 | metabolite T | 1.10E-25 | IDL_L | IDL_L |
| rs674302 | metabolite A | 4.90E-16 | IDL_L | IDL_L |
| rs7046674 | metabolite T | 1.40E-08 | IDL_L | IDL_L |
| rs554833 | metabolite T | 5.80E-16 | IDL_L | IDL_L |
| rs8176649 | metabolite A | 1.40E-08 | IDL_L | IDL_L |
| rs529565 | metabolite C | 1.30E-15 | IDL_L | IDL_L |
| rs532436 | metabolite A | 1.70E-31 | IDL_L | IDL_L |
| rs600038 | metabolite C | 1.90E-28 | IDL_L | IDL_L |
| rs1124406: | metabolite T | 1.80E-18 | IDL_L | IDL_L |
| rs649129 | metabolite T | 1.40E-28 | IDL_L | IDL_L |

| | | | | |
|---|---|---|---|---|
| rs495828 | metabolite T | 7.10E-29 | IDL_L | IDL_L |
| rs7873635 | metabolite C | 3.20E-09 | IDL_P | IDL_P |
| rs4962040 | metabolite G | 2.30E-09 | IDL_P | IDL_P |
| rs8176702 | metabolite A | 2.50E-09 | IDL_P | IDL_P |
| rs2073826 | metabolite T | 5.40E-09 | IDL_P | IDL_P |
| rs687289 | metabolite A | 1.80E-19 | IDL_P | IDL_P |
| rs2073827 | metabolite C | 6.60E-09 | IDL_P | IDL_P |
| rs2073828 | metabolite A | 6.50E-09 | IDL_P | IDL_P |
| rs8176691 | metabolite T | 1.80E-09 | IDL_P | IDL_P |
| rs8176690 | metabolite G | 2.30E-09 | IDL_P | IDL_P |
| rs8176682 | metabolite T | 7.90E-10 | IDL_P | IDL_P |
| rs8176681 | metabolite C | 2.20E-09 | IDL_P | IDL_P |
| rs2519093 | metabolite T | 1.10E-35 | IDL_P | IDL_P |
| rs644234 | metabolite G | 1.90E-19 | IDL_P | IDL_P |
| rs643434 | metabolite A | 1.70E-19 | IDL_P | IDL_P |
| rs543040 | metabolite T | 1.70E-19 | IDL_P | IDL_P |
| rs613534 | metabolite G | 1.80E-19 | IDL_P | IDL_P |
| rs543968 | metabolite C | 1.80E-19 | IDL_P | IDL_P |
| rs544873 | metabolite A | 1.80E-19 | IDL_P | IDL_P |
| rs545971 | metabolite T | 1.30E-19 | IDL_P | IDL_P |
| rs612169 | metabolite G | 1.90E-19 | IDL_P | IDL_P |
| rs8176668 | metabolite T | 8.40E-10 | IDL_P | IDL_P |
| rs8176663 | metabolite C | 1.30E-19 | IDL_P | IDL_P |
| rs7036642 | metabolite A | 7.80E-10 | IDL_P | IDL_P |
| rs491626 | metabolite T | 1.30E-19 | IDL_P | IDL_P |
| rs492488 | metabolite A | 1.30E-19 | IDL_P | IDL_P |
| rs493246 | metabolite A | 1.30E-19 | IDL_P | IDL_P |
| rs494242 | metabolite T | 1.80E-19 | IDL_P | IDL_P |
| rs495203 | metabolite T | 1.30E-19 | IDL_P | IDL_P |
| rs582118 | metabolite G | 1.40E-19 | IDL_P | IDL_P |
| rs582094 | metabolite T | 1.90E-19 | IDL_P | IDL_P |
| rs2769071 | metabolite G | 1.40E-19 | IDL_P | IDL_P |
| rs677355 | metabolite A | 1.40E-19 | IDL_P | IDL_P |
| rs676996 | metabolite G | 1.30E-19 | IDL_P | IDL_P |
| rs676457 | metabolite T | 1.30E-19 | IDL_P | IDL_P |
| rs550057 | metabolite T | 2.00E-30 | IDL_P | IDL_P |
| rs674302 | metabolite A | 1.30E-19 | IDL_P | IDL_P |
| rs7046674 | metabolite T | 5.70E-10 | IDL_P | IDL_P |
| rs554833 | metabolite T | 1.50E-19 | IDL_P | IDL_P |
| rs8176649 | metabolite A | 5.90E-10 | IDL_P | IDL_P |
| rs660340 | metabolite G | 1.90E-08 | IDL_P | IDL_P |
| rs581107 | metabolite T | 1.60E-08 | IDL_P | IDL_P |
| rs659104 | metabolite G | 1.90E-08 | IDL_P | IDL_P |
| rs647800 | metabolite G | 8.30E-09 | IDL_P | IDL_P |
| rs473533 | metabolite C | 2.00E-08 | IDL_P | IDL_P |
| rs476410 | metabolite C | 2.10E-08 | IDL_P | IDL_P |
| rs645982 | metabolite G | 1.70E-08 | IDL_P | IDL_P |
| rs529565 | metabolite C | 3.50E-19 | IDL_P | IDL_P |
| rs532436 | metabolite A | 1.90E-35 | IDL_P | IDL_P |
| rs600038 | metabolite C | 5.60E-33 | IDL_P | IDL_P |

| | | | | | |
|---|---|---|---|---|---|
| rs1124406: | metabolite | T | 3.40E-22 | IDL_P | IDL_P |
| rs649129 | metabolite | T | 4.30E-33 | IDL_P | IDL_P |
| rs495828 | metabolite | T | 1.60E-33 | IDL_P | IDL_P |
| rs4962040 | metabolite | G | 4.90E-08 | IDL_PL | IDL_PL |
| rs2073826 | metabolite | T | 3.70E-08 | IDL_PL | IDL_PL |
| rs687289 | metabolite | A | 4.50E-16 | IDL_PL | IDL_PL |
| rs2073827 | metabolite | C | 4.50E-08 | IDL_PL | IDL_PL |
| rs2073828 | metabolite | A | 4.50E-08 | IDL_PL | IDL_PL |
| rs8176691 | metabolite | T | 4.30E-08 | IDL_PL | IDL_PL |
| rs8176690 | metabolite | G | 1.70E-08 | IDL_PL | IDL_PL |
| rs8176682 | metabolite | T | 1.90E-08 | IDL_PL | IDL_PL |
| rs8176681 | metabolite | C | 1.60E-08 | IDL_PL | IDL_PL |
| rs2519093 | metabolite | T | 2.40E-31 | IDL_PL | IDL_PL |
| rs644234 | metabolite | G | 1.90E-15 | IDL_PL | IDL_PL |
| rs643434 | metabolite | A | 1.80E-15 | IDL_PL | IDL_PL |
| rs543040 | metabolite | T | 3.60E-16 | IDL_PL | IDL_PL |
| rs613534 | metabolite | G | 2.00E-15 | IDL_PL | IDL_PL |
| rs543968 | metabolite | C | 2.00E-15 | IDL_PL | IDL_PL |
| rs544873 | metabolite | A | 2.00E-15 | IDL_PL | IDL_PL |
| rs545971 | metabolite | T | 3.30E-16 | IDL_PL | IDL_PL |
| rs612169 | metabolite | G | 4.10E-16 | IDL_PL | IDL_PL |
| rs8176668 | metabolite | T | 2.20E-08 | IDL_PL | IDL_PL |
| rs8176663 | metabolite | C | 3.30E-16 | IDL_PL | IDL_PL |
| rs7036642 | metabolite | A | 2.00E-08 | IDL_PL | IDL_PL |
| rs491626 | metabolite | T | 3.30E-16 | IDL_PL | IDL_PL |
| rs492488 | metabolite | A | 3.30E-16 | IDL_PL | IDL_PL |
| rs493246 | metabolite | A | 3.30E-16 | IDL_PL | IDL_PL |
| rs494242 | metabolite | T | 1.90E-15 | IDL_PL | IDL_PL |
| rs495203 | metabolite | T | 3.30E-16 | IDL_PL | IDL_PL |
| rs582118 | metabolite | G | 3.40E-16 | IDL_PL | IDL_PL |
| rs582094 | metabolite | T | 4.90E-16 | IDL_PL | IDL_PL |
| rs2769071 | metabolite | G | 3.30E-16 | IDL_PL | IDL_PL |
| rs677355 | metabolite | A | 3.90E-16 | IDL_PL | IDL_PL |
| rs676996 | metabolite | G | 3.50E-16 | IDL_PL | IDL_PL |
| rs676457 | metabolite | T | 3.50E-16 | IDL_PL | IDL_PL |
| rs550057 | metabolite | T | 4.80E-25 | IDL_PL | IDL_PL |
| rs674302 | metabolite | A | 3.50E-16 | IDL_PL | IDL_PL |
| rs7046674 | metabolite | T | 1.70E-08 | IDL_PL | IDL_PL |
| rs554833 | metabolite | T | 4.10E-16 | IDL_PL | IDL_PL |
| rs8176649 | metabolite | A | 1.70E-08 | IDL_PL | IDL_PL |
| rs529565 | metabolite | C | 7.90E-16 | IDL_PL | IDL_PL |
| rs532436 | metabolite | A | 3.70E-31 | IDL_PL | IDL_PL |
| rs600038 | metabolite | C | 5.20E-28 | IDL_PL | IDL_PL |
| rs1124406: | metabolite | T | 5.00E-18 | IDL_PL | IDL_PL |
| rs649129 | metabolite | T | 4.10E-28 | IDL_PL | IDL_PL |
| rs495828 | metabolite | T | 2.50E-28 | IDL_PL | IDL_PL |
| rs7873635 | metabolite | C | 4.10E-18 | L_VLDL_C_ | L_VLDL_C_pct |
| rs4962040 | metabolite | G | 4.50E-18 | L_VLDL_C_ | L_VLDL_C_pct |
| rs8176702 | metabolite | A | 6.30E-18 | L_VLDL_C_ | L_VLDL_C_pct |
| rs2073826 | metabolite | T | 1.40E-18 | L_VLDL_C_ | L_VLDL_C_pct |

| | | | | |
|---|---|---|---|---|
| rs687289 | metabolite A | 1.80E-34 | L_VLDL_C_ | L_VLDL_C_pct |
| rs2073827 | metabolite C | 1.40E-18 | L_VLDL_C_ | L_VLDL_C_pct |
| rs2073828 | metabolite A | 1.50E-18 | L_VLDL_C_ | L_VLDL_C_pct |
| rs8176691 | metabolite T | 3.20E-18 | L_VLDL_C_ | L_VLDL_C_pct |
| rs8176690 | metabolite G | 3.30E-19 | L_VLDL_C_ | L_VLDL_C_pct |
| rs8176682 | metabolite T | 1.00E-18 | L_VLDL_C_ | L_VLDL_C_pct |
| rs8176681 | metabolite C | 3.90E-19 | L_VLDL_C_ | L_VLDL_C_pct |
| rs2519093 | metabolite T | 9.00E-57 | L_VLDL_C_ | L_VLDL_C_pct |
| rs644234 | metabolite G | 8.50E-32 | L_VLDL_C_ | L_VLDL_C_pct |
| rs643434 | metabolite A | 1.20E-31 | L_VLDL_C_ | L_VLDL_C_pct |
| rs543040 | metabolite T | 1.30E-33 | L_VLDL_C_ | L_VLDL_C_pct |
| rs613534 | metabolite G | 8.90E-32 | L_VLDL_C_ | L_VLDL_C_pct |
| rs543968 | metabolite C | 8.90E-32 | L_VLDL_C_ | L_VLDL_C_pct |
| rs544873 | metabolite A | 8.90E-32 | L_VLDL_C_ | L_VLDL_C_pct |
| rs545971 | metabolite T | 4.00E-34 | L_VLDL_C_ | L_VLDL_C_pct |
| rs612169 | metabolite G | 5.00E-34 | L_VLDL_C_ | L_VLDL_C_pct |
| rs8176668 | metabolite T | 5.70E-18 | L_VLDL_C_ | L_VLDL_C_pct |
| rs8176663 | metabolite C | 4.00E-34 | L_VLDL_C_ | L_VLDL_C_pct |
| rs7036642 | metabolite A | 8.10E-18 | L_VLDL_C_ | L_VLDL_C_pct |
| rs491626 | metabolite T | 3.90E-34 | L_VLDL_C_ | L_VLDL_C_pct |
| rs492488 | metabolite A | 3.90E-34 | L_VLDL_C_ | L_VLDL_C_pct |
| rs493246 | metabolite A | 3.90E-34 | L_VLDL_C_ | L_VLDL_C_pct |
| rs494242 | metabolite T | 9.30E-32 | L_VLDL_C_ | L_VLDL_C_pct |
| rs495203 | metabolite T | 4.00E-34 | L_VLDL_C_ | L_VLDL_C_pct |
| rs582118 | metabolite G | 4.00E-34 | L_VLDL_C_ | L_VLDL_C_pct |
| rs582094 | metabolite T | 4.90E-34 | L_VLDL_C_ | L_VLDL_C_pct |
| rs2769071 | metabolite G | 7.60E-34 | L_VLDL_C_ | L_VLDL_C_pct |
| rs677355 | metabolite A | 4.80E-34 | L_VLDL_C_ | L_VLDL_C_pct |
| rs676996 | metabolite G | 5.50E-34 | L_VLDL_C_ | L_VLDL_C_pct |
| rs676457 | metabolite T | 5.50E-34 | L_VLDL_C_ | L_VLDL_C_pct |
| rs550057 | metabolite T | 1.20E-45 | L_VLDL_C_ | L_VLDL_C_pct |
| rs674302 | metabolite A | 5.60E-34 | L_VLDL_C_ | L_VLDL_C_pct |
| rs7046674 | metabolite T | 8.00E-18 | L_VLDL_C_ | L_VLDL_C_pct |
| rs554833 | metabolite T | 6.60E-34 | L_VLDL_C_ | L_VLDL_C_pct |
| rs8176649 | metabolite A | 6.50E-18 | L_VLDL_C_ | L_VLDL_C_pct |
| rs660340 | metabolite G | 1.60E-12 | L_VLDL_C_ | L_VLDL_C_pct |
| rs581107 | metabolite T | 1.60E-12 | L_VLDL_C_ | L_VLDL_C_pct |
| rs659104 | metabolite G | 1.70E-12 | L_VLDL_C_ | L_VLDL_C_pct |
| rs647800 | metabolite G | 7.20E-12 | L_VLDL_C_ | L_VLDL_C_pct |
| rs473533 | metabolite C | 1.80E-12 | L_VLDL_C_ | L_VLDL_C_pct |
| rs476410 | metabolite C | 1.80E-12 | L_VLDL_C_ | L_VLDL_C_pct |
| rs645982 | metabolite G | 1.50E-12 | L_VLDL_C_ | L_VLDL_C_pct |
| rs529565 | metabolite C | 9.90E-34 | L_VLDL_C_ | L_VLDL_C_pct |
| rs630510 | metabolite G | 1.50E-12 | L_VLDL_C_ | L_VLDL_C_pct |
| rs630014 | metabolite G | 1.50E-12 | L_VLDL_C_ | L_VLDL_C_pct |
| rs532436 | metabolite A | 2.50E-56 | L_VLDL_C_ | L_VLDL_C_pct |
| rs600038 | metabolite C | 1.60E-49 | L_VLDL_C_ | L_VLDL_C_pct |
| rs1124406: | metabolite T | 3.40E-34 | L_VLDL_C_ | L_VLDL_C_pct |
| rs649129 | metabolite T | 1.90E-49 | L_VLDL_C_ | L_VLDL_C_pct |
| rs495828 | metabolite T | 3.00E-49 | L_VLDL_C_ | L_VLDL_C_pct |

| | | | | |
|---|---|---|---|---|
| rs2519093 | metabolite T | 1.00E-13 | XL_HDL_C_ | XL_HDL_C_pct |
| rs550057 | metabolite T | 1.90E-11 | XL_HDL_C_ | XL_HDL_C_pct |
| rs532436 | metabolite A | 1.70E-13 | XL_HDL_C_ | XL_HDL_C_pct |
| rs600038 | metabolite C | 2.10E-11 | XL_HDL_C_ | XL_HDL_C_pct |
| rs1124406: | metabolite T | 1.50E-10 | XL_HDL_C_ | XL_HDL_C_pct |
| rs649129 | metabolite T | 2.30E-11 | XL_HDL_C_ | XL_HDL_C_pct |
| rs495828 | metabolite T | 3.10E-11 | XL_HDL_C_ | XL_HDL_C_pct |
| rs2519093 | metabolite T | 3.60E-08 | XL_HDL_CE | XL_HDL_CE_pct |
| rs532436 | metabolite A | 4.20E-08 | XL_HDL_CE | XL_HDL_CE_pct |
| rs687289 | metabolite A | 6.70E-15 | LDL_PL | LDL_PL |
| rs2519093 | metabolite T | 5.60E-32 | LDL_PL | LDL_PL |
| rs644234 | metabolite G | 3.00E-14 | LDL_PL | LDL_PL |
| rs643434 | metabolite A | 2.80E-14 | LDL_PL | LDL_PL |
| rs543040 | metabolite T | 5.10E-15 | LDL_PL | LDL_PL |
| rs613534 | metabolite G | 3.00E-14 | LDL_PL | LDL_PL |
| rs543968 | metabolite C | 3.00E-14 | LDL_PL | LDL_PL |
| rs544873 | metabolite A | 3.00E-14 | LDL_PL | LDL_PL |
| rs545971 | metabolite T | 4.70E-15 | LDL_PL | LDL_PL |
| rs612169 | metabolite G | 6.10E-15 | LDL_PL | LDL_PL |
| rs8176663 | metabolite C | 4.70E-15 | LDL_PL | LDL_PL |
| rs491626 | metabolite T | 4.70E-15 | LDL_PL | LDL_PL |
| rs492488 | metabolite A | 4.70E-15 | LDL_PL | LDL_PL |
| rs493246 | metabolite A | 4.70E-15 | LDL_PL | LDL_PL |
| rs494242 | metabolite T | 2.90E-14 | LDL_PL | LDL_PL |
| rs495203 | metabolite T | 4.70E-15 | LDL_PL | LDL_PL |
| rs582118 | metabolite G | 4.90E-15 | LDL_PL | LDL_PL |
| rs582094 | metabolite T | 7.30E-15 | LDL_PL | LDL_PL |
| rs2769071 | metabolite G | 5.00E-15 | LDL_PL | LDL_PL |
| rs677355 | metabolite A | 5.60E-15 | LDL_PL | LDL_PL |
| rs676996 | metabolite G | 4.90E-15 | LDL_PL | LDL_PL |
| rs676457 | metabolite T | 4.90E-15 | LDL_PL | LDL_PL |
| rs550057 | metabolite T | 6.40E-26 | LDL_PL | LDL_PL |
| rs674302 | metabolite A | 4.90E-15 | LDL_PL | LDL_PL |
| rs554833 | metabolite T | 5.50E-15 | LDL_PL | LDL_PL |
| rs529565 | metabolite C | 9.20E-15 | LDL_PL | LDL_PL |
| rs532436 | metabolite A | 7.90E-32 | LDL_PL | LDL_PL |
| rs600038 | metabolite C | 1.90E-28 | LDL_PL | LDL_PL |
| rs1124406: | metabolite T | 1.80E-19 | LDL_PL | LDL_PL |
| rs649129 | metabolite T | 1.20E-28 | LDL_PL | LDL_PL |
| rs495828 | metabolite T | 7.10E-29 | LDL_PL | LDL_PL |
| rs7873635 | metabolite C | 4.00E-11 | M_VLDL_F( | M_VLDL_FC_pct |
| rs4962040 | metabolite G | 4.00E-11 | M_VLDL_F( | M_VLDL_FC_pct |
| rs8176702 | metabolite A | 5.50E-11 | M_VLDL_F( | M_VLDL_FC_pct |
| rs2073826 | metabolite T | 1.70E-11 | M_VLDL_F( | M_VLDL_FC_pct |
| rs687289 | metabolite A | 1.80E-23 | M_VLDL_F( | M_VLDL_FC_pct |
| rs2073827 | metabolite C | 2.00E-11 | M_VLDL_F( | M_VLDL_FC_pct |
| rs2073828 | metabolite A | 2.00E-11 | M_VLDL_F( | M_VLDL_FC_pct |
| rs8176691 | metabolite T | 3.70E-11 | M_VLDL_F( | M_VLDL_FC_pct |
| rs8176690 | metabolite G | 5.60E-12 | M_VLDL_F( | M_VLDL_FC_pct |
| rs8176682 | metabolite T | 1.60E-11 | M_VLDL_F( | M_VLDL_FC_pct |

| | | | | |
|---|---|---|---|---|
| rs8176681 | metabolite C | 8.50E-12 | M_VLDL_F( | M_VLDL_FC_pct |
| rs2519093 | metabolite T | 8.00E-40 | M_VLDL_F( | M_VLDL_FC_pct |
| rs644234 | metabolite G | 2.10E-21 | M_VLDL_F( | M_VLDL_FC_pct |
| rs643434 | metabolite A | 2.40E-21 | M_VLDL_F( | M_VLDL_FC_pct |
| rs543040 | metabolite T | 6.40E-23 | M_VLDL_F( | M_VLDL_FC_pct |
| rs613534 | metabolite G | 2.20E-21 | M_VLDL_F( | M_VLDL_FC_pct |
| rs543968 | metabolite C | 2.20E-21 | M_VLDL_F( | M_VLDL_FC_pct |
| rs544873 | metabolite A | 2.20E-21 | M_VLDL_F( | M_VLDL_FC_pct |
| rs545971 | metabolite T | 3.50E-23 | M_VLDL_F( | M_VLDL_FC_pct |
| rs612169 | metabolite G | 4.30E-23 | M_VLDL_F( | M_VLDL_FC_pct |
| rs8176668 | metabolite T | 5.50E-11 | M_VLDL_F( | M_VLDL_FC_pct |
| rs8176663 | metabolite C | 3.50E-23 | M_VLDL_F( | M_VLDL_FC_pct |
| rs7036642 | metabolite A | 6.40E-11 | M_VLDL_F( | M_VLDL_FC_pct |
| rs491626 | metabolite T | 3.40E-23 | M_VLDL_F( | M_VLDL_FC_pct |
| rs492488 | metabolite A | 3.40E-23 | M_VLDL_F( | M_VLDL_FC_pct |
| rs493246 | metabolite A | 3.40E-23 | M_VLDL_F( | M_VLDL_FC_pct |
| rs494242 | metabolite T | 2.20E-21 | M_VLDL_F( | M_VLDL_FC_pct |
| rs495203 | metabolite T | 3.50E-23 | M_VLDL_F( | M_VLDL_FC_pct |
| rs582118 | metabolite G | 3.40E-23 | M_VLDL_F( | M_VLDL_FC_pct |
| rs582094 | metabolite T | 4.50E-23 | M_VLDL_F( | M_VLDL_FC_pct |
| rs2769071 | metabolite G | 4.70E-23 | M_VLDL_F( | M_VLDL_FC_pct |
| rs677355 | metabolite A | 3.70E-23 | M_VLDL_F( | M_VLDL_FC_pct |
| rs676996 | metabolite G | 4.10E-23 | M_VLDL_F( | M_VLDL_FC_pct |
| rs676457 | metabolite T | 4.10E-23 | M_VLDL_F( | M_VLDL_FC_pct |
| rs550057 | metabolite T | 3.10E-33 | M_VLDL_F( | M_VLDL_FC_pct |
| rs674302 | metabolite A | 4.20E-23 | M_VLDL_F( | M_VLDL_FC_pct |
| rs7046674 | metabolite T | 6.60E-11 | M_VLDL_F( | M_VLDL_FC_pct |
| rs554833 | metabolite T | 4.60E-23 | M_VLDL_F( | M_VLDL_FC_pct |
| rs8176649 | metabolite A | 5.90E-11 | M_VLDL_F( | M_VLDL_FC_pct |
| rs660340 | metabolite G | 1.40E-10 | M_VLDL_F( | M_VLDL_FC_pct |
| rs581107 | metabolite T | 1.40E-10 | M_VLDL_F( | M_VLDL_FC_pct |
| rs659104 | metabolite G | 1.50E-10 | M_VLDL_F( | M_VLDL_FC_pct |
| rs647800 | metabolite G | 5.80E-10 | M_VLDL_F( | M_VLDL_FC_pct |
| rs473533 | metabolite C | 1.50E-10 | M_VLDL_F( | M_VLDL_FC_pct |
| rs476410 | metabolite C | 1.60E-10 | M_VLDL_F( | M_VLDL_FC_pct |
| rs645982 | metabolite G | 1.30E-10 | M_VLDL_F( | M_VLDL_FC_pct |
| rs529565 | metabolite C | 1.10E-22 | M_VLDL_F( | M_VLDL_FC_pct |
| rs630510 | metabolite G | 3.60E-08 | M_VLDL_F( | M_VLDL_FC_pct |
| rs630014 | metabolite G | 3.60E-08 | M_VLDL_F( | M_VLDL_FC_pct |
| rs532436 | metabolite A | 1.70E-39 | M_VLDL_F( | M_VLDL_FC_pct |
| rs600038 | metabolite C | 1.60E-34 | M_VLDL_F( | M_VLDL_FC_pct |
| rs1124406: | metabolite T | 1.90E-21 | M_VLDL_F( | M_VLDL_FC_pct |
| rs649129 | metabolite T | 2.80E-34 | M_VLDL_F( | M_VLDL_FC_pct |
| rs495828 | metabolite T | 2.10E-34 | M_VLDL_F( | M_VLDL_FC_pct |
| rs7873635 | metabolite C | 4.70E-08 | Remnant_( | Remnant_C |
| rs4962040 | metabolite G | 2.90E-08 | Remnant_( | Remnant_C |
| rs8176702 | metabolite A | 3.00E-08 | Remnant_( | Remnant_C |
| rs2073826 | metabolite T | 5.00E-08 | Remnant_( | Remnant_C |
| rs687289 | metabolite A | 5.60E-17 | Remnant_( | Remnant_C |
| rs8176691 | metabolite T | 2.50E-08 | Remnant_( | Remnant_C |

| | | | | |
|---|---|---|---|---|
| rs8176690 | metabolite G | 2.80E-08 | Remnant_( | Remnant_C |
| rs8176682 | metabolite T | 1.30E-08 | Remnant_( | Remnant_C |
| rs8176681 | metabolite C | 2.50E-08 | Remnant_( | Remnant_C |
| rs2519093 | metabolite T | 4.00E-30 | Remnant_( | Remnant_C |
| rs644234 | metabolite G | 5.30E-17 | Remnant_( | Remnant_C |
| rs643434 | metabolite A | 4.70E-17 | Remnant_( | Remnant_C |
| rs543040 | metabolite T | 3.40E-17 | Remnant_( | Remnant_C |
| rs613534 | metabolite G | 5.30E-17 | Remnant_( | Remnant_C |
| rs543968 | metabolite C | 5.30E-17 | Remnant_( | Remnant_C |
| rs544873 | metabolite A | 5.30E-17 | Remnant_( | Remnant_C |
| rs545971 | metabolite T | 3.70E-17 | Remnant_( | Remnant_C |
| rs612169 | metabolite G | 5.00E-17 | Remnant_( | Remnant_C |
| rs8176668 | metabolite T | 1.10E-08 | Remnant_( | Remnant_C |
| rs8176663 | metabolite C | 3.70E-17 | Remnant_( | Remnant_C |
| rs7036642 | metabolite A | 9.30E-09 | Remnant_( | Remnant_C |
| rs491626 | metabolite T | 3.60E-17 | Remnant_( | Remnant_C |
| rs492488 | metabolite A | 3.60E-17 | Remnant_( | Remnant_C |
| rs493246 | metabolite A | 3.60E-17 | Remnant_( | Remnant_C |
| rs494242 | metabolite T | 5.10E-17 | Remnant_( | Remnant_C |
| rs495203 | metabolite T | 3.70E-17 | Remnant_( | Remnant_C |
| rs582118 | metabolite G | 3.80E-17 | Remnant_( | Remnant_C |
| rs582094 | metabolite T | 5.60E-17 | Remnant_( | Remnant_C |
| rs2769071 | metabolite G | 3.70E-17 | Remnant_( | Remnant_C |
| rs677355 | metabolite A | 4.30E-17 | Remnant_( | Remnant_C |
| rs676996 | metabolite G | 3.70E-17 | Remnant_( | Remnant_C |
| rs676457 | metabolite T | 3.70E-17 | Remnant_( | Remnant_C |
| rs550057 | metabolite T | 1.40E-25 | Remnant_( | Remnant_C |
| rs674302 | metabolite A | 3.70E-17 | Remnant_( | Remnant_C |
| rs7046674 | metabolite T | 7.30E-09 | Remnant_( | Remnant_C |
| rs554833 | metabolite T | 4.30E-17 | Remnant_( | Remnant_C |
| rs8176649 | metabolite A | 8.10E-09 | Remnant_( | Remnant_C |
| rs529565 | metabolite C | 7.40E-17 | Remnant_( | Remnant_C |
| rs532436 | metabolite A | 5.50E-30 | Remnant_( | Remnant_C |
| rs600038 | metabolite C | 6.60E-28 | Remnant_( | Remnant_C |
| rs1124406: | metabolite T | 2.50E-18 | Remnant_( | Remnant_C |
| rs649129 | metabolite T | 4.00E-28 | Remnant_( | Remnant_C |
| rs495828 | metabolite T | 1.70E-28 | Remnant_( | Remnant_C |
| rs7873635 | metabolite C | 1.50E-15 | L_VLDL_CE | L_VLDL_CE_pct |
| rs4962040 | metabolite G | 1.70E-15 | L_VLDL_CE | L_VLDL_CE_pct |
| rs8176702 | metabolite A | 2.60E-15 | L_VLDL_CE | L_VLDL_CE_pct |
| rs2073826 | metabolite T | 3.30E-16 | L_VLDL_CE | L_VLDL_CE_pct |
| rs687289 | metabolite A | 1.00E-29 | L_VLDL_CE | L_VLDL_CE_pct |
| rs2073827 | metabolite C | 3.20E-16 | L_VLDL_CE | L_VLDL_CE_pct |
| rs2073828 | metabolite A | 3.40E-16 | L_VLDL_CE | L_VLDL_CE_pct |
| rs8176691 | metabolite T | 1.30E-15 | L_VLDL_CE | L_VLDL_CE_pct |
| rs8176690 | metabolite G | 9.70E-17 | L_VLDL_CE | L_VLDL_CE_pct |
| rs8176682 | metabolite T | 5.60E-16 | L_VLDL_CE | L_VLDL_CE_pct |
| rs8176681 | metabolite C | 1.20E-16 | L_VLDL_CE | L_VLDL_CE_pct |
| rs2519093 | metabolite T | 7.40E-49 | L_VLDL_CE | L_VLDL_CE_pct |
| rs644234 | metabolite G | 7.40E-27 | L_VLDL_CE | L_VLDL_CE_pct |

| | | | | |
|---|---|---|---|---|
| rs643434 | metabolite A | 9.50E-27 | L_VLDL_CE | L_VLDL_CE_pct |
| rs543040 | metabolite T | 7.50E-29 | L_VLDL_CE | L_VLDL_CE_pct |
| rs613534 | metabolite G | 7.90E-27 | L_VLDL_CE | L_VLDL_CE_pct |
| rs543968 | metabolite C | 7.90E-27 | L_VLDL_CE | L_VLDL_CE_pct |
| rs544873 | metabolite A | 7.90E-27 | L_VLDL_CE | L_VLDL_CE_pct |
| rs545971 | metabolite T | 2.40E-29 | L_VLDL_CE | L_VLDL_CE_pct |
| rs612169 | metabolite G | 2.50E-29 | L_VLDL_CE | L_VLDL_CE_pct |
| rs8176668 | metabolite T | 2.60E-15 | L_VLDL_CE | L_VLDL_CE_pct |
| rs8176663 | metabolite C | 2.40E-29 | L_VLDL_CE | L_VLDL_CE_pct |
| rs7036642 | metabolite A | 3.50E-15 | L_VLDL_CE | L_VLDL_CE_pct |
| rs491626 | metabolite T | 2.40E-29 | L_VLDL_CE | L_VLDL_CE_pct |
| rs492488 | metabolite A | 2.40E-29 | L_VLDL_CE | L_VLDL_CE_pct |
| rs493246 | metabolite A | 2.40E-29 | L_VLDL_CE | L_VLDL_CE_pct |
| rs494242 | metabolite T | 8.10E-27 | L_VLDL_CE | L_VLDL_CE_pct |
| rs495203 | metabolite T | 2.40E-29 | L_VLDL_CE | L_VLDL_CE_pct |
| rs582118 | metabolite G | 2.40E-29 | L_VLDL_CE | L_VLDL_CE_pct |
| rs582094 | metabolite T | 3.20E-29 | L_VLDL_CE | L_VLDL_CE_pct |
| rs2769071 | metabolite G | 3.80E-29 | L_VLDL_CE | L_VLDL_CE_pct |
| rs677355 | metabolite A | 2.40E-29 | L_VLDL_CE | L_VLDL_CE_pct |
| rs676996 | metabolite G | 2.80E-29 | L_VLDL_CE | L_VLDL_CE_pct |
| rs676457 | metabolite T | 2.80E-29 | L_VLDL_CE | L_VLDL_CE_pct |
| rs550057 | metabolite T | 2.60E-39 | L_VLDL_CE | L_VLDL_CE_pct |
| rs674302 | metabolite A | 2.90E-29 | L_VLDL_CE | L_VLDL_CE_pct |
| rs7046674 | metabolite T | 3.50E-15 | L_VLDL_CE | L_VLDL_CE_pct |
| rs554833 | metabolite T | 3.20E-29 | L_VLDL_CE | L_VLDL_CE_pct |
| rs8176649 | metabolite A | 2.90E-15 | L_VLDL_CE | L_VLDL_CE_pct |
| rs660340 | metabolite G | 1.10E-11 | L_VLDL_CE | L_VLDL_CE_pct |
| rs581107 | metabolite T | 1.20E-11 | L_VLDL_CE | L_VLDL_CE_pct |
| rs659104 | metabolite G | 1.20E-11 | L_VLDL_CE | L_VLDL_CE_pct |
| rs647800 | metabolite G | 8.30E-11 | L_VLDL_CE | L_VLDL_CE_pct |
| rs473533 | metabolite C | 1.20E-11 | L_VLDL_CE | L_VLDL_CE_pct |
| rs476410 | metabolite C | 1.30E-11 | L_VLDL_CE | L_VLDL_CE_pct |
| rs645982 | metabolite G | 1.10E-11 | L_VLDL_CE | L_VLDL_CE_pct |
| rs529565 | metabolite C | 5.10E-29 | L_VLDL_CE | L_VLDL_CE_pct |
| rs630510 | metabolite G | 2.20E-10 | L_VLDL_CE | L_VLDL_CE_pct |
| rs630014 | metabolite G | 2.20E-10 | L_VLDL_CE | L_VLDL_CE_pct |
| rs532436 | metabolite A | 1.30E-48 | L_VLDL_CE | L_VLDL_CE_pct |
| rs600038 | metabolite C | 4.90E-42 | L_VLDL_CE | L_VLDL_CE_pct |
| rs1124406: | metabolite T | 1.80E-27 | L_VLDL_CE | L_VLDL_CE_pct |
| rs649129 | metabolite T | 6.20E-42 | L_VLDL_CE | L_VLDL_CE_pct |
| rs495828 | metabolite T | 7.50E-42 | L_VLDL_CE | L_VLDL_CE_pct |
| rs687289 | metabolite A | 4.00E-11 | S_LDL_P | S_LDL_P |
| rs2519093 | metabolite T | 1.60E-20 | S_LDL_P | S_LDL_P |
| rs644234 | metabolite G | 3.00E-11 | S_LDL_P | S_LDL_P |
| rs643434 | metabolite A | 2.80E-11 | S_LDL_P | S_LDL_P |
| rs543040 | metabolite T | 2.90E-11 | S_LDL_P | S_LDL_P |
| rs613534 | metabolite G | 3.00E-11 | S_LDL_P | S_LDL_P |
| rs543968 | metabolite C | 3.00E-11 | S_LDL_P | S_LDL_P |
| rs544873 | metabolite A | 3.00E-11 | S_LDL_P | S_LDL_P |
| rs545971 | metabolite T | 3.20E-11 | S_LDL_P | S_LDL_P |

| | | | | |
|---|---|---|---|---|
| rs612169 | metabolite G | 4.20E-11 | S_LDL_P | S_LDL_P |
| rs8176663 | metabolite C | 3.20E-11 | S_LDL_P | S_LDL_P |
| rs491626 | metabolite T | 3.20E-11 | S_LDL_P | S_LDL_P |
| rs492488 | metabolite A | 3.20E-11 | S_LDL_P | S_LDL_P |
| rs493246 | metabolite A | 3.20E-11 | S_LDL_P | S_LDL_P |
| rs494242 | metabolite T | 2.90E-11 | S_LDL_P | S_LDL_P |
| rs495203 | metabolite T | 3.20E-11 | S_LDL_P | S_LDL_P |
| rs582118 | metabolite G | 3.30E-11 | S_LDL_P | S_LDL_P |
| rs582094 | metabolite T | 4.10E-11 | S_LDL_P | S_LDL_P |
| rs2769071 | metabolite G | 3.40E-11 | S_LDL_P | S_LDL_P |
| rs677355 | metabolite A | 3.70E-11 | S_LDL_P | S_LDL_P |
| rs676996 | metabolite G | 3.30E-11 | S_LDL_P | S_LDL_P |
| rs676457 | metabolite T | 3.30E-11 | S_LDL_P | S_LDL_P |
| rs550057 | metabolite T | 7.70E-18 | S_LDL_P | S_LDL_P |
| rs674302 | metabolite A | 3.30E-11 | S_LDL_P | S_LDL_P |
| rs554833 | metabolite T | 3.50E-11 | S_LDL_P | S_LDL_P |
| rs529565 | metabolite C | 4.60E-11 | S_LDL_P | S_LDL_P |
| rs532436 | metabolite A | 1.90E-20 | S_LDL_P | S_LDL_P |
| rs600038 | metabolite C | 3.00E-19 | S_LDL_P | S_LDL_P |
| rs1124406: | metabolite T | 4.30E-12 | S_LDL_P | S_LDL_P |
| rs649129 | metabolite T | 1.70E-19 | S_LDL_P | S_LDL_P |
| rs495828 | metabolite T | 7.80E-20 | S_LDL_P | S_LDL_P |
| rs687289 | metabolite A | 7.40E-16 | non_HDL_( | non_HDL_C |
| rs2519093 | metabolite T | 2.30E-31 | non_HDL_( | non_HDL_C |
| rs644234 | metabolite G | 2.40E-15 | non_HDL_( | non_HDL_C |
| rs643434 | metabolite A | 2.20E-15 | non_HDL_( | non_HDL_C |
| rs543040 | metabolite T | 5.00E-16 | non_HDL_( | non_HDL_C |
| rs613534 | metabolite G | 2.40E-15 | non_HDL_( | non_HDL_C |
| rs543968 | metabolite C | 2.40E-15 | non_HDL_( | non_HDL_C |
| rs544873 | metabolite A | 2.40E-15 | non_HDL_( | non_HDL_C |
| rs545971 | metabolite T | 5.10E-16 | non_HDL_( | non_HDL_C |
| rs612169 | metabolite G | 6.60E-16 | non_HDL_( | non_HDL_C |
| rs8176663 | metabolite C | 5.10E-16 | non_HDL_( | non_HDL_C |
| rs491626 | metabolite T | 5.00E-16 | non_HDL_( | non_HDL_C |
| rs492488 | metabolite A | 5.00E-16 | non_HDL_( | non_HDL_C |
| rs493246 | metabolite A | 5.00E-16 | non_HDL_( | non_HDL_C |
| rs494242 | metabolite T | 2.30E-15 | non_HDL_( | non_HDL_C |
| rs495203 | metabolite T | 5.00E-16 | non_HDL_( | non_HDL_C |
| rs582118 | metabolite G | 5.20E-16 | non_HDL_( | non_HDL_C |
| rs582094 | metabolite T | 7.90E-16 | non_HDL_( | non_HDL_C |
| rs2769071 | metabolite G | 5.30E-16 | non_HDL_( | non_HDL_C |
| rs677355 | metabolite A | 6.10E-16 | non_HDL_( | non_HDL_C |
| rs676996 | metabolite G | 5.30E-16 | non_HDL_( | non_HDL_C |
| rs676457 | metabolite T | 5.30E-16 | non_HDL_( | non_HDL_C |
| rs550057 | metabolite T | 9.80E-26 | non_HDL_( | non_HDL_C |
| rs674302 | metabolite A | 5.30E-16 | non_HDL_( | non_HDL_C |
| rs7046674 | metabolite T | 4.20E-08 | non_HDL_( | non_HDL_C |
| rs554833 | metabolite T | 6.00E-16 | non_HDL_( | non_HDL_C |
| rs8176649 | metabolite A | 4.60E-08 | non_HDL_( | non_HDL_C |
| rs529565 | metabolite C | 1.00E-15 | non_HDL_( | non_HDL_C |

| | | | | |
|---|---|---|---|---|
| rs532436 | metabolite A | 3.40E-31 | non_HDL_( | non_HDL_C |
| rs600038 | metabolite C | 3.60E-28 | non_HDL_( | non_HDL_C |
| rs1124406: | metabolite T | 4.50E-19 | non_HDL_( | non_HDL_C |
| rs649129 | metabolite T | 2.20E-28 | non_HDL_( | non_HDL_C |
| rs495828 | metabolite T | 1.20E-28 | non_HDL_( | non_HDL_C |
| rs4962040 | metabolite G | 4.70E-08 | S_VLDL_FC | S_VLDL_FC |
| rs8176702 | metabolite A | 4.90E-08 | S_VLDL_FC | S_VLDL_FC |
| rs687289 | metabolite A | 1.00E-17 | S_VLDL_FC | S_VLDL_FC |
| rs8176691 | metabolite T | 4.20E-08 | S_VLDL_FC | S_VLDL_FC |
| rs8176690 | metabolite G | 3.70E-08 | S_VLDL_FC | S_VLDL_FC |
| rs8176682 | metabolite T | 2.50E-08 | S_VLDL_FC | S_VLDL_FC |
| rs8176681 | metabolite C | 3.10E-08 | S_VLDL_FC | S_VLDL_FC |
| rs2519093 | metabolite T | 2.30E-30 | S_VLDL_FC | S_VLDL_FC |
| rs644234 | metabolite G | 1.70E-17 | S_VLDL_FC | S_VLDL_FC |
| rs643434 | metabolite A | 1.60E-17 | S_VLDL_FC | S_VLDL_FC |
| rs543040 | metabolite T | 7.30E-18 | S_VLDL_FC | S_VLDL_FC |
| rs613534 | metabolite G | 1.70E-17 | S_VLDL_FC | S_VLDL_FC |
| rs543968 | metabolite C | 1.70E-17 | S_VLDL_FC | S_VLDL_FC |
| rs544873 | metabolite A | 1.70E-17 | S_VLDL_FC | S_VLDL_FC |
| rs545971 | metabolite T | 7.30E-18 | S_VLDL_FC | S_VLDL_FC |
| rs612169 | metabolite G | 1.10E-17 | S_VLDL_FC | S_VLDL_FC |
| rs8176668 | metabolite T | 1.60E-08 | S_VLDL_FC | S_VLDL_FC |
| rs8176663 | metabolite C | 7.30E-18 | S_VLDL_FC | S_VLDL_FC |
| rs7036642 | metabolite A | 1.50E-08 | S_VLDL_FC | S_VLDL_FC |
| rs491626 | metabolite T | 7.30E-18 | S_VLDL_FC | S_VLDL_FC |
| rs492488 | metabolite A | 7.30E-18 | S_VLDL_FC | S_VLDL_FC |
| rs493246 | metabolite A | 7.30E-18 | S_VLDL_FC | S_VLDL_FC |
| rs494242 | metabolite T | 1.70E-17 | S_VLDL_FC | S_VLDL_FC |
| rs495203 | metabolite T | 7.30E-18 | S_VLDL_FC | S_VLDL_FC |
| rs582118 | metabolite G | 7.60E-18 | S_VLDL_FC | S_VLDL_FC |
| rs582094 | metabolite T | 1.20E-17 | S_VLDL_FC | S_VLDL_FC |
| rs2769071 | metabolite G | 7.60E-18 | S_VLDL_FC | S_VLDL_FC |
| rs677355 | metabolite A | 8.80E-18 | S_VLDL_FC | S_VLDL_FC |
| rs676996 | metabolite G | 7.60E-18 | S_VLDL_FC | S_VLDL_FC |
| rs676457 | metabolite T | 7.60E-18 | S_VLDL_FC | S_VLDL_FC |
| rs550057 | metabolite T | 1.40E-25 | S_VLDL_FC | S_VLDL_FC |
| rs674302 | metabolite A | 7.50E-18 | S_VLDL_FC | S_VLDL_FC |
| rs7046674 | metabolite T | 1.20E-08 | S_VLDL_FC | S_VLDL_FC |
| rs554833 | metabolite T | 8.10E-18 | S_VLDL_FC | S_VLDL_FC |
| rs8176649 | metabolite A | 1.30E-08 | S_VLDL_FC | S_VLDL_FC |
| rs529565 | metabolite C | 1.30E-17 | S_VLDL_FC | S_VLDL_FC |
| rs630510 | metabolite G | 4.30E-08 | S_VLDL_FC | S_VLDL_FC |
| rs630014 | metabolite G | 4.30E-08 | S_VLDL_FC | S_VLDL_FC |
| rs532436 | metabolite A | 3.00E-30 | S_VLDL_FC | S_VLDL_FC |
| rs600038 | metabolite C | 8.50E-28 | S_VLDL_FC | S_VLDL_FC |
| rs1124406: | metabolite T | 6.10E-19 | S_VLDL_FC | S_VLDL_FC |
| rs649129 | metabolite T | 5.10E-28 | S_VLDL_FC | S_VLDL_FC |
| rs495828 | metabolite T | 2.60E-28 | S_VLDL_FC | S_VLDL_FC |
| rs687289 | metabolite A | 5.60E-10 | S_LDL_FC | S_LDL_FC |
| rs2519093 | metabolite T | 1.90E-24 | S_LDL_FC | S_LDL_FC |

| | | | | |
|---|---|---|---|---|
| rs644234 | metabolite G | 1.30E-09 | S_LDL_FC | S_LDL_FC |
| rs643434 | metabolite A | 1.30E-09 | S_LDL_FC | S_LDL_FC |
| rs543040 | metabolite T | 5.20E-10 | S_LDL_FC | S_LDL_FC |
| rs613534 | metabolite G | 1.30E-09 | S_LDL_FC | S_LDL_FC |
| rs543968 | metabolite C | 1.30E-09 | S_LDL_FC | S_LDL_FC |
| rs544873 | metabolite A | 1.30E-09 | S_LDL_FC | S_LDL_FC |
| rs545971 | metabolite T | 4.20E-10 | S_LDL_FC | S_LDL_FC |
| rs612169 | metabolite G | 5.20E-10 | S_LDL_FC | S_LDL_FC |
| rs8176663 | metabolite C | 4.20E-10 | S_LDL_FC | S_LDL_FC |
| rs491626 | metabolite T | 4.20E-10 | S_LDL_FC | S_LDL_FC |
| rs492488 | metabolite A | 4.20E-10 | S_LDL_FC | S_LDL_FC |
| rs493246 | metabolite A | 4.20E-10 | S_LDL_FC | S_LDL_FC |
| rs494242 | metabolite T | 1.30E-09 | S_LDL_FC | S_LDL_FC |
| rs495203 | metabolite T | 4.30E-10 | S_LDL_FC | S_LDL_FC |
| rs582118 | metabolite G | 4.40E-10 | S_LDL_FC | S_LDL_FC |
| rs582094 | metabolite T | 6.00E-10 | S_LDL_FC | S_LDL_FC |
| rs2769071 | metabolite G | 4.40E-10 | S_LDL_FC | S_LDL_FC |
| rs677355 | metabolite A | 4.80E-10 | S_LDL_FC | S_LDL_FC |
| rs676996 | metabolite G | 4.40E-10 | S_LDL_FC | S_LDL_FC |
| rs676457 | metabolite T | 4.40E-10 | S_LDL_FC | S_LDL_FC |
| rs550057 | metabolite T | 8.00E-20 | S_LDL_FC | S_LDL_FC |
| rs674302 | metabolite A | 4.40E-10 | S_LDL_FC | S_LDL_FC |
| rs554833 | metabolite T | 4.50E-10 | S_LDL_FC | S_LDL_FC |
| rs529565 | metabolite C | 9.30E-10 | S_LDL_FC | S_LDL_FC |
| rs532436 | metabolite A | 2.40E-24 | S_LDL_FC | S_LDL_FC |
| rs600038 | metabolite C | 7.70E-22 | S_LDL_FC | S_LDL_FC |
| rs1124406: | metabolite T | 1.20E-13 | S_LDL_FC | S_LDL_FC |
| rs649129 | metabolite T | 7.80E-22 | S_LDL_FC | S_LDL_FC |
| rs495828 | metabolite T | 4.70E-22 | S_LDL_FC | S_LDL_FC |
| rs687289 | metabolite A | 9.00E-12 | S_HDL_P | S_HDL_P |
| rs2519093 | metabolite T | 1.10E-25 | S_HDL_P | S_HDL_P |
| rs644234 | metabolite G | 2.70E-10 | S_HDL_P | S_HDL_P |
| rs643434 | metabolite A | 2.60E-10 | S_HDL_P | S_HDL_P |
| rs543040 | metabolite T | 5.60E-12 | S_HDL_P | S_HDL_P |
| rs613534 | metabolite G | 2.70E-10 | S_HDL_P | S_HDL_P |
| rs543968 | metabolite C | 2.70E-10 | S_HDL_P | S_HDL_P |
| rs544873 | metabolite A | 2.70E-10 | S_HDL_P | S_HDL_P |
| rs545971 | metabolite T | 5.10E-12 | S_HDL_P | S_HDL_P |
| rs612169 | metabolite G | 5.70E-12 | S_HDL_P | S_HDL_P |
| rs8176663 | metabolite C | 5.10E-12 | S_HDL_P | S_HDL_P |
| rs491626 | metabolite T | 5.00E-12 | S_HDL_P | S_HDL_P |
| rs492488 | metabolite A | 5.00E-12 | S_HDL_P | S_HDL_P |
| rs493246 | metabolite A | 5.00E-12 | S_HDL_P | S_HDL_P |
| rs494242 | metabolite T | 2.60E-10 | S_HDL_P | S_HDL_P |
| rs495203 | metabolite T | 5.00E-12 | S_HDL_P | S_HDL_P |
| rs582118 | metabolite G | 5.20E-12 | S_HDL_P | S_HDL_P |
| rs582094 | metabolite T | 7.50E-12 | S_HDL_P | S_HDL_P |
| rs2769071 | metabolite G | 5.20E-12 | S_HDL_P | S_HDL_P |
| rs677355 | metabolite A | 6.00E-12 | S_HDL_P | S_HDL_P |
| rs676996 | metabolite G | 5.40E-12 | S_HDL_P | S_HDL_P |

| | | | | |
|---|---|---|---|---|
| rs676457 | metabolite T | 5.40E-12 | S_HDL_P | S_HDL_P |
| rs550057 | metabolite T | 1.30E-17 | S_HDL_P | S_HDL_P |
| rs674302 | metabolite A | 5.40E-12 | S_HDL_P | S_HDL_P |
| rs554833 | metabolite T | 5.40E-12 | S_HDL_P | S_HDL_P |
| rs529565 | metabolite C | 3.30E-12 | S_HDL_P | S_HDL_P |
| rs532436 | metabolite A | 1.70E-25 | S_HDL_P | S_HDL_P |
| rs600038 | metabolite C | 4.50E-21 | S_HDL_P | S_HDL_P |
| rs1124406: | metabolite T | 1.80E-19 | S_HDL_P | S_HDL_P |
| rs649129 | metabolite T | 4.00E-21 | S_HDL_P | S_HDL_P |
| rs495828 | metabolite T | 8.50E-21 | S_HDL_P | S_HDL_P |
| rs2073826 | metabolite T | 3.90E-08 | M_VLDL_C | M_VLDL_CE_pct |
| rs687289 | metabolite A | 9.50E-16 | M_VLDL_C | M_VLDL_CE_pct |
| rs2073827 | metabolite C | 3.80E-08 | M_VLDL_C | M_VLDL_CE_pct |
| rs2073828 | metabolite A | 3.80E-08 | M_VLDL_C | M_VLDL_CE_pct |
| rs8176690 | metabolite G | 1.70E-08 | M_VLDL_C | M_VLDL_CE_pct |
| rs8176682 | metabolite T | 3.50E-08 | M_VLDL_C | M_VLDL_CE_pct |
| rs8176681 | metabolite C | 2.20E-08 | M_VLDL_C | M_VLDL_CE_pct |
| rs2519093 | metabolite T | 3.90E-28 | M_VLDL_C | M_VLDL_CE_pct |
| rs644234 | metabolite G | 3.10E-14 | M_VLDL_C | M_VLDL_CE_pct |
| rs643434 | metabolite A | 3.50E-14 | M_VLDL_C | M_VLDL_CE_pct |
| rs543040 | metabolite T | 3.20E-15 | M_VLDL_C | M_VLDL_CE_pct |
| rs613534 | metabolite G | 3.30E-14 | M_VLDL_C | M_VLDL_CE_pct |
| rs543968 | metabolite C | 3.30E-14 | M_VLDL_C | M_VLDL_CE_pct |
| rs544873 | metabolite A | 3.30E-14 | M_VLDL_C | M_VLDL_CE_pct |
| rs545971 | metabolite T | 1.90E-15 | M_VLDL_C | M_VLDL_CE_pct |
| rs612169 | metabolite G | 2.00E-15 | M_VLDL_C | M_VLDL_CE_pct |
| rs8176663 | metabolite C | 1.90E-15 | M_VLDL_C | M_VLDL_CE_pct |
| rs491626 | metabolite T | 1.90E-15 | M_VLDL_C | M_VLDL_CE_pct |
| rs492488 | metabolite A | 1.90E-15 | M_VLDL_C | M_VLDL_CE_pct |
| rs493246 | metabolite A | 1.90E-15 | M_VLDL_C | M_VLDL_CE_pct |
| rs494242 | metabolite T | 3.30E-14 | M_VLDL_C | M_VLDL_CE_pct |
| rs495203 | metabolite T | 1.90E-15 | M_VLDL_C | M_VLDL_CE_pct |
| rs582118 | metabolite G | 1.90E-15 | M_VLDL_C | M_VLDL_CE_pct |
| rs582094 | metabolite T | 2.40E-15 | M_VLDL_C | M_VLDL_CE_pct |
| rs2769071 | metabolite G | 2.40E-15 | M_VLDL_C | M_VLDL_CE_pct |
| rs677355 | metabolite A | 1.90E-15 | M_VLDL_C | M_VLDL_CE_pct |
| rs676996 | metabolite G | 2.10E-15 | M_VLDL_C | M_VLDL_CE_pct |
| rs676457 | metabolite T | 2.10E-15 | M_VLDL_C | M_VLDL_CE_pct |
| rs550057 | metabolite T | 2.60E-23 | M_VLDL_C | M_VLDL_CE_pct |
| rs674302 | metabolite A | 2.10E-15 | M_VLDL_C | M_VLDL_CE_pct |
| rs554833 | metabolite T | 2.20E-15 | M_VLDL_C | M_VLDL_CE_pct |
| rs529565 | metabolite C | 2.80E-15 | M_VLDL_C | M_VLDL_CE_pct |
| rs532436 | metabolite A | 5.70E-28 | M_VLDL_C | M_VLDL_CE_pct |
| rs600038 | metabolite C | 2.70E-24 | M_VLDL_C | M_VLDL_CE_pct |
| rs1124406: | metabolite T | 5.10E-16 | M_VLDL_C | M_VLDL_CE_pct |
| rs649129 | metabolite T | 4.20E-24 | M_VLDL_C | M_VLDL_CE_pct |
| rs495828 | metabolite T | 4.00E-24 | M_VLDL_C | M_VLDL_CE_pct |
| rs687289 | metabolite A | 4.10E-13 | S_LDL_PL_ | S_LDL_PL_pct |
| rs2519093 | metabolite T | 8.80E-20 | S_LDL_PL_ | S_LDL_PL_pct |
| rs644234 | metabolite G | 5.00E-12 | S_LDL_PL_ | S_LDL_PL_pct |

| | | | | |
|---|---|---|---|---|
| rs643434 | metabolite A | 4.70E-12 | S_LDL_PL_ | S_LDL_PL_pct |
| rs543040 | metabolite T | 1.90E-13 | S_LDL_PL_ | S_LDL_PL_pct |
| rs613534 | metabolite G | 4.90E-12 | S_LDL_PL_ | S_LDL_PL_pct |
| rs543968 | metabolite C | 4.90E-12 | S_LDL_PL_ | S_LDL_PL_pct |
| rs544873 | metabolite A | 4.90E-12 | S_LDL_PL_ | S_LDL_PL_pct |
| rs545971 | metabolite T | 2.60E-13 | S_LDL_PL_ | S_LDL_PL_pct |
| rs612169 | metabolite G | 3.40E-13 | S_LDL_PL_ | S_LDL_PL_pct |
| rs8176663 | metabolite C | 2.60E-13 | S_LDL_PL_ | S_LDL_PL_pct |
| rs491626 | metabolite T | 2.60E-13 | S_LDL_PL_ | S_LDL_PL_pct |
| rs492488 | metabolite A | 2.60E-13 | S_LDL_PL_ | S_LDL_PL_pct |
| rs493246 | metabolite A | 2.60E-13 | S_LDL_PL_ | S_LDL_PL_pct |
| rs494242 | metabolite T | 4.70E-12 | S_LDL_PL_ | S_LDL_PL_pct |
| rs495203 | metabolite T | 2.60E-13 | S_LDL_PL_ | S_LDL_PL_pct |
| rs582118 | metabolite G | 2.70E-13 | S_LDL_PL_ | S_LDL_PL_pct |
| rs582094 | metabolite T | 3.30E-13 | S_LDL_PL_ | S_LDL_PL_pct |
| rs2769071 | metabolite G | 2.90E-13 | S_LDL_PL_ | S_LDL_PL_pct |
| rs677355 | metabolite A | 3.00E-13 | S_LDL_PL_ | S_LDL_PL_pct |
| rs676996 | metabolite G | 2.80E-13 | S_LDL_PL_ | S_LDL_PL_pct |
| rs676457 | metabolite T | 2.80E-13 | S_LDL_PL_ | S_LDL_PL_pct |
| rs550057 | metabolite T | 8.30E-16 | S_LDL_PL_ | S_LDL_PL_pct |
| rs674302 | metabolite A | 2.80E-13 | S_LDL_PL_ | S_LDL_PL_pct |
| rs554833 | metabolite T | 2.80E-13 | S_LDL_PL_ | S_LDL_PL_pct |
| rs529565 | metabolite C | 9.40E-14 | S_LDL_PL_ | S_LDL_PL_pct |
| rs532436 | metabolite A | 1.60E-19 | S_LDL_PL_ | S_LDL_PL_pct |
| rs600038 | metabolite C | 1.10E-16 | S_LDL_PL_ | S_LDL_PL_pct |
| rs1124406 | metabolite T | 1.90E-15 | S_LDL_PL_ | S_LDL_PL_pct |
| rs649129 | metabolite T | 5.10E-17 | S_LDL_PL_ | S_LDL_PL_pct |
| rs495828 | metabolite T | 7.80E-17 | S_LDL_PL_ | S_LDL_PL_pct |
| rs687289 | metabolite A | 1.40E-13 | Total_CE | Total_CE |
| rs2519093 | metabolite T | 2.40E-28 | Total_CE | Total_CE |
| rs644234 | metabolite G | 1.80E-12 | Total_CE | Total_CE |
| rs643434 | metabolite A | 1.60E-12 | Total_CE | Total_CE |
| rs543040 | metabolite T | 9.70E-14 | Total_CE | Total_CE |
| rs613534 | metabolite G | 1.80E-12 | Total_CE | Total_CE |
| rs543968 | metabolite C | 1.80E-12 | Total_CE | Total_CE |
| rs544873 | metabolite A | 1.80E-12 | Total_CE | Total_CE |
| rs545971 | metabolite T | 9.70E-14 | Total_CE | Total_CE |
| rs612169 | metabolite G | 1.10E-13 | Total_CE | Total_CE |
| rs8176663 | metabolite C | 9.70E-14 | Total_CE | Total_CE |
| rs491626 | metabolite T | 9.50E-14 | Total_CE | Total_CE |
| rs492488 | metabolite A | 9.50E-14 | Total_CE | Total_CE |
| rs493246 | metabolite A | 9.50E-14 | Total_CE | Total_CE |
| rs494242 | metabolite T | 1.80E-12 | Total_CE | Total_CE |
| rs495203 | metabolite T | 9.50E-14 | Total_CE | Total_CE |
| rs582118 | metabolite G | 9.80E-14 | Total_CE | Total_CE |
| rs582094 | metabolite T | 1.40E-13 | Total_CE | Total_CE |
| rs2769071 | metabolite G | 9.40E-14 | Total_CE | Total_CE |
| rs677355 | metabolite A | 1.10E-13 | Total_CE | Total_CE |
| rs676996 | metabolite G | 9.90E-14 | Total_CE | Total_CE |
| rs676457 | metabolite T | 9.90E-14 | Total_CE | Total_CE |

| | | | | |
|---|---|---|---|---|
| rs550057 | metabolite T | 3.60E-22 | Total_CE | Total_CE |
| rs674302 | metabolite A | 1.00E-13 | Total_CE | Total_CE |
| rs554833 | metabolite T | 1.10E-13 | Total_CE | Total_CE |
| rs529565 | metabolite C | 1.80E-13 | Total_CE | Total_CE |
| rs532436 | metabolite A | 3.40E-28 | Total_CE | Total_CE |
| rs600038 | metabolite C | 2.10E-24 | Total_CE | Total_CE |
| rs1124406: | metabolite T | 1.10E-16 | Total_CE | Total_CE |
| rs649129 | metabolite T | 1.40E-24 | Total_CE | Total_CE |
| rs495828 | metabolite T | 1.10E-24 | Total_CE | Total_CE |
| rs687289 | metabolite A | 4.10E-14 | L_LDL_FC | L_LDL_FC |
| rs8176690 | metabolite G | 3.90E-08 | L_LDL_FC | L_LDL_FC |
| rs8176681 | metabolite C | 4.40E-08 | L_LDL_FC | L_LDL_FC |
| rs2519093 | metabolite T | 2.40E-33 | L_LDL_FC | L_LDL_FC |
| rs644234 | metabolite G | 4.00E-13 | L_LDL_FC | L_LDL_FC |
| rs643434 | metabolite A | 3.80E-13 | L_LDL_FC | L_LDL_FC |
| rs543040 | metabolite T | 3.40E-14 | L_LDL_FC | L_LDL_FC |
| rs613534 | metabolite G | 4.10E-13 | L_LDL_FC | L_LDL_FC |
| rs543968 | metabolite C | 4.10E-13 | L_LDL_FC | L_LDL_FC |
| rs544873 | metabolite A | 4.10E-13 | L_LDL_FC | L_LDL_FC |
| rs545971 | metabolite T | 2.90E-14 | L_LDL_FC | L_LDL_FC |
| rs612169 | metabolite G | 3.50E-14 | L_LDL_FC | L_LDL_FC |
| rs8176663 | metabolite C | 2.90E-14 | L_LDL_FC | L_LDL_FC |
| rs491626 | metabolite T | 2.80E-14 | L_LDL_FC | L_LDL_FC |
| rs492488 | metabolite A | 2.80E-14 | L_LDL_FC | L_LDL_FC |
| rs493246 | metabolite A | 2.80E-14 | L_LDL_FC | L_LDL_FC |
| rs494242 | metabolite T | 4.00E-13 | L_LDL_FC | L_LDL_FC |
| rs495203 | metabolite T | 2.80E-14 | L_LDL_FC | L_LDL_FC |
| rs582118 | metabolite G | 2.90E-14 | L_LDL_FC | L_LDL_FC |
| rs582094 | metabolite T | 4.40E-14 | L_LDL_FC | L_LDL_FC |
| rs2769071 | metabolite G | 3.00E-14 | L_LDL_FC | L_LDL_FC |
| rs677355 | metabolite A | 3.30E-14 | L_LDL_FC | L_LDL_FC |
| rs676996 | metabolite G | 3.00E-14 | L_LDL_FC | L_LDL_FC |
| rs676457 | metabolite T | 3.00E-14 | L_LDL_FC | L_LDL_FC |
| rs550057 | metabolite T | 3.20E-26 | L_LDL_FC | L_LDL_FC |
| rs674302 | metabolite A | 3.00E-14 | L_LDL_FC | L_LDL_FC |
| rs554833 | metabolite T | 3.30E-14 | L_LDL_FC | L_LDL_FC |
| rs529565 | metabolite C | 6.50E-14 | L_LDL_FC | L_LDL_FC |
| rs532436 | metabolite A | 3.80E-33 | L_LDL_FC | L_LDL_FC |
| rs600038 | metabolite C | 5.90E-29 | L_LDL_FC | L_LDL_FC |
| rs1124406: | metabolite T | 2.60E-19 | L_LDL_FC | L_LDL_FC |
| rs649129 | metabolite T | 4.60E-29 | L_LDL_FC | L_LDL_FC |
| rs495828 | metabolite T | 3.50E-29 | L_LDL_FC | L_LDL_FC |
| rs687289 | metabolite A | 1.10E-09 | S_LDL_L | S_LDL_L |
| rs2519093 | metabolite T | 3.30E-20 | S_LDL_L | S_LDL_L |
| rs644234 | metabolite G | 1.20E-09 | S_LDL_L | S_LDL_L |
| rs643434 | metabolite A | 1.10E-09 | S_LDL_L | S_LDL_L |
| rs543040 | metabolite T | 7.80E-10 | S_LDL_L | S_LDL_L |
| rs613534 | metabolite G | 1.20E-09 | S_LDL_L | S_LDL_L |
| rs543968 | metabolite C | 1.20E-09 | S_LDL_L | S_LDL_L |
| rs544873 | metabolite A | 1.20E-09 | S_LDL_L | S_LDL_L |

| | | | | |
|---|---|---|---|---|
| rs545971 | metabolite T | 8.20E-10 | S_LDL_L | S_LDL_L |
| rs612169 | metabolite G | 9.90E-10 | S_LDL_L | S_LDL_L |
| rs8176663 | metabolite C | 8.20E-10 | S_LDL_L | S_LDL_L |
| rs491626 | metabolite T | 8.20E-10 | S_LDL_L | S_LDL_L |
| rs492488 | metabolite A | 8.20E-10 | S_LDL_L | S_LDL_L |
| rs493246 | metabolite A | 8.20E-10 | S_LDL_L | S_LDL_L |
| rs494242 | metabolite T | 1.20E-09 | S_LDL_L | S_LDL_L |
| rs495203 | metabolite T | 8.20E-10 | S_LDL_L | S_LDL_L |
| rs582118 | metabolite G | 8.40E-10 | S_LDL_L | S_LDL_L |
| rs582094 | metabolite T | 1.10E-09 | S_LDL_L | S_LDL_L |
| rs2769071 | metabolite G | 8.50E-10 | S_LDL_L | S_LDL_L |
| rs677355 | metabolite A | 9.50E-10 | S_LDL_L | S_LDL_L |
| rs676996 | metabolite G | 8.50E-10 | S_LDL_L | S_LDL_L |
| rs676457 | metabolite T | 8.50E-10 | S_LDL_L | S_LDL_L |
| rs550057 | metabolite T | 3.50E-17 | S_LDL_L | S_LDL_L |
| rs674302 | metabolite A | 8.50E-10 | S_LDL_L | S_LDL_L |
| rs554833 | metabolite T | 9.10E-10 | S_LDL_L | S_LDL_L |
| rs529565 | metabolite C | 1.30E-09 | S_LDL_L | S_LDL_L |
| rs532436 | metabolite A | 3.80E-20 | S_LDL_L | S_LDL_L |
| rs600038 | metabolite C | 1.30E-18 | S_LDL_L | S_LDL_L |
| rs1124406: | metabolite T | 2.30E-12 | S_LDL_L | S_LDL_L |
| rs649129 | metabolite T | 8.20E-19 | S_LDL_L | S_LDL_L |
| rs495828 | metabolite T | 4.30E-19 | S_LDL_L | S_LDL_L |
| rs2519093 | metabolite T | 1.00E-10 | M_VLDL_L | M_VLDL_L |
| rs550057 | metabolite T | 1.30E-09 | M_VLDL_L | M_VLDL_L |
| rs532436 | metabolite A | 1.10E-10 | M_VLDL_L | M_VLDL_L |
| rs600038 | metabolite C | 2.00E-10 | M_VLDL_L | M_VLDL_L |
| rs649129 | metabolite T | 1.20E-10 | M_VLDL_L | M_VLDL_L |
| rs495828 | metabolite T | 7.30E-11 | M_VLDL_L | M_VLDL_L |
| rs687289 | metabolite A | 1.40E-13 | M_LDL_L | M_LDL_L |
| rs2519093 | metabolite T | 6.90E-28 | M_LDL_L | M_LDL_L |
| rs644234 | metabolite G | 7.10E-13 | M_LDL_L | M_LDL_L |
| rs643434 | metabolite A | 6.60E-13 | M_LDL_L | M_LDL_L |
| rs543040 | metabolite T | 9.90E-14 | M_LDL_L | M_LDL_L |
| rs613534 | metabolite G | 7.00E-13 | M_LDL_L | M_LDL_L |
| rs543968 | metabolite C | 7.00E-13 | M_LDL_L | M_LDL_L |
| rs544873 | metabolite A | 7.00E-13 | M_LDL_L | M_LDL_L |
| rs545971 | metabolite T | 1.00E-13 | M_LDL_L | M_LDL_L |
| rs612169 | metabolite G | 1.30E-13 | M_LDL_L | M_LDL_L |
| rs8176663 | metabolite C | 1.00E-13 | M_LDL_L | M_LDL_L |
| rs491626 | metabolite T | 1.00E-13 | M_LDL_L | M_LDL_L |
| rs492488 | metabolite A | 1.00E-13 | M_LDL_L | M_LDL_L |
| rs493246 | metabolite A | 1.00E-13 | M_LDL_L | M_LDL_L |
| rs494242 | metabolite T | 6.70E-13 | M_LDL_L | M_LDL_L |
| rs495203 | metabolite T | 1.00E-13 | M_LDL_L | M_LDL_L |
| rs582118 | metabolite G | 1.00E-13 | M_LDL_L | M_LDL_L |
| rs582094 | metabolite T | 1.50E-13 | M_LDL_L | M_LDL_L |
| rs2769071 | metabolite G | 1.10E-13 | M_LDL_L | M_LDL_L |
| rs677355 | metabolite A | 1.20E-13 | M_LDL_L | M_LDL_L |
| rs676996 | metabolite G | 1.10E-13 | M_LDL_L | M_LDL_L |

| | | | | |
|---|---|---|---|---|
| rs676457 | metabolite T | 1.10E-13 | M_LDL_L | M_LDL_L |
| rs550057 | metabolite T | 8.50E-23 | M_LDL_L | M_LDL_L |
| rs674302 | metabolite A | 1.10E-13 | M_LDL_L | M_LDL_L |
| rs554833 | metabolite T | 1.10E-13 | M_LDL_L | M_LDL_L |
| rs529565 | metabolite C | 1.40E-13 | M_LDL_L | M_LDL_L |
| rs532436 | metabolite A | 8.90E-28 | M_LDL_L | M_LDL_L |
| rs600038 | metabolite C | 1.20E-24 | M_LDL_L | M_LDL_L |
| rs1124406: | metabolite T | 1.40E-17 | M_LDL_L | M_LDL_L |
| rs649129 | metabolite T | 6.60E-25 | M_LDL_L | M_LDL_L |
| rs495828 | metabolite T | 4.40E-25 | M_LDL_L | M_LDL_L |
| rs687289 | metabolite A | 7.20E-10 | Total_L | Total_L |
| rs2519093 | metabolite T | 4.30E-17 | Total_L | Total_L |
| rs644234 | metabolite G | 1.50E-09 | Total_L | Total_L |
| rs643434 | metabolite A | 1.30E-09 | Total_L | Total_L |
| rs543040 | metabolite T | 3.50E-10 | Total_L | Total_L |
| rs613534 | metabolite G | 1.50E-09 | Total_L | Total_L |
| rs543968 | metabolite C | 1.50E-09 | Total_L | Total_L |
| rs544873 | metabolite A | 1.50E-09 | Total_L | Total_L |
| rs545971 | metabolite T | 4.60E-10 | Total_L | Total_L |
| rs612169 | metabolite G | 5.10E-10 | Total_L | Total_L |
| rs8176663 | metabolite C | 4.60E-10 | Total_L | Total_L |
| rs491626 | metabolite T | 4.50E-10 | Total_L | Total_L |
| rs492488 | metabolite A | 4.50E-10 | Total_L | Total_L |
| rs493246 | metabolite A | 4.50E-10 | Total_L | Total_L |
| rs494242 | metabolite T | 1.40E-09 | Total_L | Total_L |
| rs495203 | metabolite T | 4.50E-10 | Total_L | Total_L |
| rs582118 | metabolite G | 4.60E-10 | Total_L | Total_L |
| rs582094 | metabolite T | 5.90E-10 | Total_L | Total_L |
| rs2769071 | metabolite G | 4.30E-10 | Total_L | Total_L |
| rs677355 | metabolite A | 5.30E-10 | Total_L | Total_L |
| rs676996 | metabolite G | 4.60E-10 | Total_L | Total_L |
| rs676457 | metabolite T | 4.60E-10 | Total_L | Total_L |
| rs550057 | metabolite T | 4.10E-14 | Total_L | Total_L |
| rs674302 | metabolite A | 4.60E-10 | Total_L | Total_L |
| rs554833 | metabolite T | 5.00E-10 | Total_L | Total_L |
| rs529565 | metabolite C | 5.60E-10 | Total_L | Total_L |
| rs532436 | metabolite A | 4.90E-17 | Total_L | Total_L |
| rs600038 | metabolite C | 2.10E-15 | Total_L | Total_L |
| rs1124406: | metabolite T | 4.00E-11 | Total_L | Total_L |
| rs649129 | metabolite T | 1.20E-15 | Total_L | Total_L |
| rs495828 | metabolite T | 7.90E-16 | Total_L | Total_L |
| rs687289 | metabolite A | 4.20E-14 | L_LDL_L | L_LDL_L |
| rs2519093 | metabolite T | 1.20E-31 | L_LDL_L | L_LDL_L |
| rs644234 | metabolite G | 3.20E-13 | L_LDL_L | L_LDL_L |
| rs643434 | metabolite A | 3.00E-13 | L_LDL_L | L_LDL_L |
| rs543040 | metabolite T | 3.20E-14 | L_LDL_L | L_LDL_L |
| rs613534 | metabolite G | 3.20E-13 | L_LDL_L | L_LDL_L |
| rs543968 | metabolite C | 3.20E-13 | L_LDL_L | L_LDL_L |
| rs544873 | metabolite A | 3.20E-13 | L_LDL_L | L_LDL_L |
| rs545971 | metabolite T | 3.00E-14 | L_LDL_L | L_LDL_L |

| | | | | |
|---|---|---|---|---|
| rs612169 | metabolite G | 3.70E-14 | L_LDL_L | L_LDL_L |
| rs8176663 | metabolite C | 3.00E-14 | L_LDL_L | L_LDL_L |
| rs491626 | metabolite T | 2.90E-14 | L_LDL_L | L_LDL_L |
| rs492488 | metabolite A | 2.90E-14 | L_LDL_L | L_LDL_L |
| rs493246 | metabolite A | 2.90E-14 | L_LDL_L | L_LDL_L |
| rs494242 | metabolite T | 3.10E-13 | L_LDL_L | L_LDL_L |
| rs495203 | metabolite T | 3.00E-14 | L_LDL_L | L_LDL_L |
| rs582118 | metabolite G | 3.10E-14 | L_LDL_L | L_LDL_L |
| rs582094 | metabolite T | 4.70E-14 | L_LDL_L | L_LDL_L |
| rs2769071 | metabolite G | 3.10E-14 | L_LDL_L | L_LDL_L |
| rs677355 | metabolite A | 3.60E-14 | L_LDL_L | L_LDL_L |
| rs676996 | metabolite G | 3.20E-14 | L_LDL_L | L_LDL_L |
| rs676457 | metabolite T | 3.20E-14 | L_LDL_L | L_LDL_L |
| rs550057 | metabolite T | 6.80E-25 | L_LDL_L | L_LDL_L |
| rs674302 | metabolite A | 3.20E-14 | L_LDL_L | L_LDL_L |
| rs554833 | metabolite T | 3.50E-14 | L_LDL_L | L_LDL_L |
| rs529565 | metabolite C | 5.60E-14 | L_LDL_L | L_LDL_L |
| rs532436 | metabolite A | 1.80E-31 | L_LDL_L | L_LDL_L |
| rs600038 | metabolite C | 1.00E-27 | L_LDL_L | L_LDL_L |
| rs1124406: | metabolite T | 2.60E-19 | L_LDL_L | L_LDL_L |
| rs649129 | metabolite T | 6.20E-28 | L_LDL_L | L_LDL_L |
| rs495828 | metabolite T | 4.40E-28 | L_LDL_L | L_LDL_L |
| rs687289 | metabolite A | 1.60E-08 | VLDL_FC | VLDL_FC |
| rs2519093 | metabolite T | 1.10E-11 | VLDL_FC | VLDL_FC |
| rs644234 | metabolite G | 3.60E-09 | VLDL_FC | VLDL_FC |
| rs643434 | metabolite A | 3.30E-09 | VLDL_FC | VLDL_FC |
| rs543040 | metabolite T | 8.20E-09 | VLDL_FC | VLDL_FC |
| rs613534 | metabolite G | 3.60E-09 | VLDL_FC | VLDL_FC |
| rs543968 | metabolite C | 3.60E-09 | VLDL_FC | VLDL_FC |
| rs544873 | metabolite A | 3.60E-09 | VLDL_FC | VLDL_FC |
| rs545971 | metabolite T | 1.10E-08 | VLDL_FC | VLDL_FC |
| rs612169 | metabolite G | 1.30E-08 | VLDL_FC | VLDL_FC |
| rs8176663 | metabolite C | 1.10E-08 | VLDL_FC | VLDL_FC |
| rs491626 | metabolite T | 1.10E-08 | VLDL_FC | VLDL_FC |
| rs492488 | metabolite A | 1.10E-08 | VLDL_FC | VLDL_FC |
| rs493246 | metabolite A | 1.10E-08 | VLDL_FC | VLDL_FC |
| rs494242 | metabolite T | 3.40E-09 | VLDL_FC | VLDL_FC |
| rs495203 | metabolite T | 1.10E-08 | VLDL_FC | VLDL_FC |
| rs582118 | metabolite G | 1.10E-08 | VLDL_FC | VLDL_FC |
| rs582094 | metabolite T | 1.30E-08 | VLDL_FC | VLDL_FC |
| rs2769071 | metabolite G | 1.10E-08 | VLDL_FC | VLDL_FC |
| rs677355 | metabolite A | 1.30E-08 | VLDL_FC | VLDL_FC |
| rs676996 | metabolite G | 1.10E-08 | VLDL_FC | VLDL_FC |
| rs676457 | metabolite T | 1.10E-08 | VLDL_FC | VLDL_FC |
| rs550057 | metabolite T | 9.50E-11 | VLDL_FC | VLDL_FC |
| rs674302 | metabolite A | 1.10E-08 | VLDL_FC | VLDL_FC |
| rs554833 | metabolite T | 1.20E-08 | VLDL_FC | VLDL_FC |
| rs529565 | metabolite C | 1.30E-08 | VLDL_FC | VLDL_FC |
| rs532436 | metabolite A | 1.30E-11 | VLDL_FC | VLDL_FC |
| rs600038 | metabolite C | 1.10E-11 | VLDL_FC | VLDL_FC |

| | | | | |
|---|---|---|---|---|
| rs649129 | metabolite T | 7.10E-12 | VLDL_FC | VLDL_FC |
| rs495828 | metabolite T | 3.90E-12 | VLDL_FC | VLDL_FC |
| rs687289 | metabolite A | 1.30E-12 | LDL_P | LDL_P |
| rs2519093 | metabolite T | 2.30E-24 | LDL_P | LDL_P |
| rs644234 | metabolite G | 2.10E-12 | LDL_P | LDL_P |
| rs643434 | metabolite A | 2.00E-12 | LDL_P | LDL_P |
| rs543040 | metabolite T | 9.70E-13 | LDL_P | LDL_P |
| rs613534 | metabolite G | 2.20E-12 | LDL_P | LDL_P |
| rs543968 | metabolite C | 2.20E-12 | LDL_P | LDL_P |
| rs544873 | metabolite A | 2.20E-12 | LDL_P | LDL_P |
| rs545971 | metabolite T | 1.00E-12 | LDL_P | LDL_P |
| rs612169 | metabolite G | 1.30E-12 | LDL_P | LDL_P |
| rs8176663 | metabolite C | 1.00E-12 | LDL_P | LDL_P |
| rs491626 | metabolite T | 1.00E-12 | LDL_P | LDL_P |
| rs492488 | metabolite A | 1.00E-12 | LDL_P | LDL_P |
| rs493246 | metabolite A | 1.00E-12 | LDL_P | LDL_P |
| rs494242 | metabolite T | 2.10E-12 | LDL_P | LDL_P |
| rs495203 | metabolite T | 1.00E-12 | LDL_P | LDL_P |
| rs582118 | metabolite G | 1.00E-12 | LDL_P | LDL_P |
| rs582094 | metabolite T | 1.50E-12 | LDL_P | LDL_P |
| rs2769071 | metabolite G | 1.10E-12 | LDL_P | LDL_P |
| rs677355 | metabolite A | 1.20E-12 | LDL_P | LDL_P |
| rs676996 | metabolite G | 1.10E-12 | LDL_P | LDL_P |
| rs676457 | metabolite T | 1.10E-12 | LDL_P | LDL_P |
| rs550057 | metabolite T | 1.40E-20 | LDL_P | LDL_P |
| rs674302 | metabolite A | 1.10E-12 | LDL_P | LDL_P |
| rs554833 | metabolite T | 1.20E-12 | LDL_P | LDL_P |
| rs529565 | metabolite C | 1.80E-12 | LDL_P | LDL_P |
| rs532436 | metabolite A | 2.90E-24 | LDL_P | LDL_P |
| rs600038 | metabolite C | 3.30E-22 | LDL_P | LDL_P |
| rs1124406: | metabolite T | 1.40E-14 | LDL_P | LDL_P |
| rs649129 | metabolite T | 2.00E-22 | LDL_P | LDL_P |
| rs495828 | metabolite T | 1.00E-22 | LDL_P | LDL_P |
| rs687289 | metabolite A | 7.20E-14 | S_LDL_CE_ | S_LDL_CE_pct |
| rs2519093 | metabolite T | 8.40E-20 | S_LDL_CE_ | S_LDL_CE_pct |
| rs644234 | metabolite G | 1.10E-12 | S_LDL_CE_ | S_LDL_CE_pct |
| rs643434 | metabolite A | 1.00E-12 | S_LDL_CE_ | S_LDL_CE_pct |
| rs543040 | metabolite T | 3.30E-14 | S_LDL_CE_ | S_LDL_CE_pct |
| rs613534 | metabolite G | 1.10E-12 | S_LDL_CE_ | S_LDL_CE_pct |
| rs543968 | metabolite C | 1.10E-12 | S_LDL_CE_ | S_LDL_CE_pct |
| rs544873 | metabolite A | 1.10E-12 | S_LDL_CE_ | S_LDL_CE_pct |
| rs545971 | metabolite T | 4.90E-14 | S_LDL_CE_ | S_LDL_CE_pct |
| rs612169 | metabolite G | 6.20E-14 | S_LDL_CE_ | S_LDL_CE_pct |
| rs8176663 | metabolite C | 4.90E-14 | S_LDL_CE_ | S_LDL_CE_pct |
| rs491626 | metabolite T | 4.80E-14 | S_LDL_CE_ | S_LDL_CE_pct |
| rs492488 | metabolite A | 4.80E-14 | S_LDL_CE_ | S_LDL_CE_pct |
| rs493246 | metabolite A | 4.80E-14 | S_LDL_CE_ | S_LDL_CE_pct |
| rs494242 | metabolite T | 1.00E-12 | S_LDL_CE_ | S_LDL_CE_pct |
| rs495203 | metabolite T | 4.90E-14 | S_LDL_CE_ | S_LDL_CE_pct |
| rs582118 | metabolite G | 5.00E-14 | S_LDL_CE_ | S_LDL_CE_pct |

| | | | | |
|---|---|---|---|---|
| rs582094 | metabolite T | 6.10E-14 | S_LDL_CE_ | S_LDL_CE_pct |
| rs2769071 | metabolite G | 5.50E-14 | S_LDL_CE_ | S_LDL_CE_pct |
| rs677355 | metabolite A | 6.00E-14 | S_LDL_CE_ | S_LDL_CE_pct |
| rs676996 | metabolite G | 5.50E-14 | S_LDL_CE_ | S_LDL_CE_pct |
| rs676457 | metabolite T | 5.50E-14 | S_LDL_CE_ | S_LDL_CE_pct |
| rs550057 | metabolite T | 2.00E-16 | S_LDL_CE_ | S_LDL_CE_pct |
| rs674302 | metabolite A | 5.30E-14 | S_LDL_CE_ | S_LDL_CE_pct |
| rs554833 | metabolite T | 5.70E-14 | S_LDL_CE_ | S_LDL_CE_pct |
| rs529565 | metabolite C | 1.80E-14 | S_LDL_CE_ | S_LDL_CE_pct |
| rs532436 | metabolite A | 1.40E-19 | S_LDL_CE_ | S_LDL_CE_pct |
| rs600038 | metabolite C | 1.30E-16 | S_LDL_CE_ | S_LDL_CE_pct |
| rs1124406: | metabolite T | 3.00E-15 | S_LDL_CE_ | S_LDL_CE_pct |
| rs649129 | metabolite T | 5.70E-17 | S_LDL_CE_ | S_LDL_CE_pct |
| rs495828 | metabolite T | 7.30E-17 | S_LDL_CE_ | S_LDL_CE_pct |
| rs7873635 | metabolite C | 1.20E-14 | S_VLDL_CE | S_VLDL_CE_pct |
| rs4962040 | metabolite G | 1.10E-14 | S_VLDL_CE | S_VLDL_CE_pct |
| rs8176702 | metabolite A | 1.20E-14 | S_VLDL_CE | S_VLDL_CE_pct |
| rs2073826 | metabolite T | 1.10E-14 | S_VLDL_CE | S_VLDL_CE_pct |
| rs687289 | metabolite A | 6.20E-27 | S_VLDL_CE | S_VLDL_CE_pct |
| rs2073827 | metabolite C | 1.10E-14 | S_VLDL_CE | S_VLDL_CE_pct |
| rs2073828 | metabolite A | 1.10E-14 | S_VLDL_CE | S_VLDL_CE_pct |
| rs8176691 | metabolite T | 6.60E-15 | S_VLDL_CE | S_VLDL_CE_pct |
| rs8176690 | metabolite G | 2.60E-15 | S_VLDL_CE | S_VLDL_CE_pct |
| rs8176682 | metabolite T | 2.10E-15 | S_VLDL_CE | S_VLDL_CE_pct |
| rs8176681 | metabolite C | 3.20E-15 | S_VLDL_CE | S_VLDL_CE_pct |
| rs2519093 | metabolite T | 7.60E-48 | S_VLDL_CE | S_VLDL_CE_pct |
| rs644234 | metabolite G | 1.40E-25 | S_VLDL_CE | S_VLDL_CE_pct |
| rs643434 | metabolite A | 1.70E-25 | S_VLDL_CE | S_VLDL_CE_pct |
| rs543040 | metabolite T | 2.90E-26 | S_VLDL_CE | S_VLDL_CE_pct |
| rs613534 | metabolite G | 1.40E-25 | S_VLDL_CE | S_VLDL_CE_pct |
| rs543968 | metabolite C | 1.40E-25 | S_VLDL_CE | S_VLDL_CE_pct |
| rs544873 | metabolite A | 1.40E-25 | S_VLDL_CE | S_VLDL_CE_pct |
| rs545971 | metabolite T | 1.20E-26 | S_VLDL_CE | S_VLDL_CE_pct |
| rs612169 | metabolite G | 1.70E-26 | S_VLDL_CE | S_VLDL_CE_pct |
| rs8176668 | metabolite T | 6.60E-15 | S_VLDL_CE | S_VLDL_CE_pct |
| rs8176663 | metabolite C | 1.20E-26 | S_VLDL_CE | S_VLDL_CE_pct |
| rs7036642 | metabolite A | 8.20E-15 | S_VLDL_CE | S_VLDL_CE_pct |
| rs491626 | metabolite T | 1.20E-26 | S_VLDL_CE | S_VLDL_CE_pct |
| rs492488 | metabolite A | 1.20E-26 | S_VLDL_CE | S_VLDL_CE_pct |
| rs493246 | metabolite A | 1.20E-26 | S_VLDL_CE | S_VLDL_CE_pct |
| rs494242 | metabolite T | 1.50E-25 | S_VLDL_CE | S_VLDL_CE_pct |
| rs495203 | metabolite T | 1.20E-26 | S_VLDL_CE | S_VLDL_CE_pct |
| rs582118 | metabolite G | 1.20E-26 | S_VLDL_CE | S_VLDL_CE_pct |
| rs582094 | metabolite T | 1.80E-26 | S_VLDL_CE | S_VLDL_CE_pct |
| rs2769071 | metabolite G | 1.60E-26 | S_VLDL_CE | S_VLDL_CE_pct |
| rs677355 | metabolite A | 1.30E-26 | S_VLDL_CE | S_VLDL_CE_pct |
| rs676996 | metabolite G | 1.40E-26 | S_VLDL_CE | S_VLDL_CE_pct |
| rs676457 | metabolite T | 1.40E-26 | S_VLDL_CE | S_VLDL_CE_pct |
| rs550057 | metabolite T | 2.00E-39 | S_VLDL_CE | S_VLDL_CE_pct |
| rs674302 | metabolite A | 1.40E-26 | S_VLDL_CE | S_VLDL_CE_pct |

| | | | | | |
|---|---|---|---|---|---|
| rs7046674 | metabolite | T | 6.70E-15 | S_VLDL_CE | S_VLDL_CE_pct |
| rs554833 | metabolite | T | 1.60E-26 | S_VLDL_CE | S_VLDL_CE_pct |
| rs8176649 | metabolite | A | 6.00E-15 | S_VLDL_CE | S_VLDL_CE_pct |
| rs660340 | metabolite | G | 3.10E-10 | S_VLDL_CE | S_VLDL_CE_pct |
| rs581107 | metabolite | T | 3.00E-10 | S_VLDL_CE | S_VLDL_CE_pct |
| rs659104 | metabolite | G | 3.50E-10 | S_VLDL_CE | S_VLDL_CE_pct |
| rs647800 | metabolite | G | 4.00E-10 | S_VLDL_CE | S_VLDL_CE_pct |
| rs473533 | metabolite | C | 3.70E-10 | S_VLDL_CE | S_VLDL_CE_pct |
| rs476410 | metabolite | C | 3.40E-10 | S_VLDL_CE | S_VLDL_CE_pct |
| rs645982 | metabolite | G | 3.10E-10 | S_VLDL_CE | S_VLDL_CE_pct |
| rs529565 | metabolite | C | 2.40E-26 | S_VLDL_CE | S_VLDL_CE_pct |
| rs630510 | metabolite | G | 5.00E-09 | S_VLDL_CE | S_VLDL_CE_pct |
| rs630014 | metabolite | G | 5.10E-09 | S_VLDL_CE | S_VLDL_CE_pct |
| rs532436 | metabolite | A | 2.10E-47 | S_VLDL_CE | S_VLDL_CE_pct |
| rs600038 | metabolite | C | 1.40E-42 | S_VLDL_CE | S_VLDL_CE_pct |
| rs1124406: | metabolite | T | 1.20E-29 | S_VLDL_CE | S_VLDL_CE_pct |
| rs649129 | metabolite | T | 2.20E-42 | S_VLDL_CE | S_VLDL_CE_pct |
| rs495828 | metabolite | T | 2.10E-42 | S_VLDL_CE | S_VLDL_CE_pct |
| rs600038 | metabolite | C | 3.80E-08 | VLDL_PL | VLDL_PL |
| rs649129 | metabolite | T | 2.40E-08 | VLDL_PL | VLDL_PL |
| rs495828 | metabolite | T | 1.40E-08 | VLDL_PL | VLDL_PL |
| rs687289 | metabolite | A | 2.80E-14 | M_LDL_FC | M_LDL_FC |
| rs2519093 | metabolite | T | 2.80E-33 | M_LDL_FC | M_LDL_FC |
| rs644234 | metabolite | G | 2.40E-13 | M_LDL_FC | M_LDL_FC |
| rs643434 | metabolite | A | 2.20E-13 | M_LDL_FC | M_LDL_FC |
| rs543040 | metabolite | T | 2.40E-14 | M_LDL_FC | M_LDL_FC |
| rs613534 | metabolite | G | 2.40E-13 | M_LDL_FC | M_LDL_FC |
| rs543968 | metabolite | C | 2.40E-13 | M_LDL_FC | M_LDL_FC |
| rs544873 | metabolite | A | 2.40E-13 | M_LDL_FC | M_LDL_FC |
| rs545971 | metabolite | T | 1.90E-14 | M_LDL_FC | M_LDL_FC |
| rs612169 | metabolite | G | 2.50E-14 | M_LDL_FC | M_LDL_FC |
| rs8176663 | metabolite | C | 1.90E-14 | M_LDL_FC | M_LDL_FC |
| rs491626 | metabolite | T | 1.90E-14 | M_LDL_FC | M_LDL_FC |
| rs492488 | metabolite | A | 1.90E-14 | M_LDL_FC | M_LDL_FC |
| rs493246 | metabolite | A | 1.90E-14 | M_LDL_FC | M_LDL_FC |
| rs494242 | metabolite | T | 2.30E-13 | M_LDL_FC | M_LDL_FC |
| rs495203 | metabolite | T | 1.90E-14 | M_LDL_FC | M_LDL_FC |
| rs582118 | metabolite | G | 2.00E-14 | M_LDL_FC | M_LDL_FC |
| rs582094 | metabolite | T | 3.00E-14 | M_LDL_FC | M_LDL_FC |
| rs2769071 | metabolite | G | 2.10E-14 | M_LDL_FC | M_LDL_FC |
| rs677355 | metabolite | A | 2.30E-14 | M_LDL_FC | M_LDL_FC |
| rs676996 | metabolite | G | 2.00E-14 | M_LDL_FC | M_LDL_FC |
| rs676457 | metabolite | T | 2.00E-14 | M_LDL_FC | M_LDL_FC |
| rs550057 | metabolite | T | 1.70E-26 | M_LDL_FC | M_LDL_FC |
| rs674302 | metabolite | A | 2.00E-14 | M_LDL_FC | M_LDL_FC |
| rs554833 | metabolite | T | 2.20E-14 | M_LDL_FC | M_LDL_FC |
| rs529565 | metabolite | C | 4.00E-14 | M_LDL_FC | M_LDL_FC |
| rs532436 | metabolite | A | 4.40E-33 | M_LDL_FC | M_LDL_FC |
| rs600038 | metabolite | C | 5.00E-29 | M_LDL_FC | M_LDL_FC |
| rs1124406: | metabolite | T | 7.50E-20 | M_LDL_FC | M_LDL_FC |

| | | | | |
|---|---|---|---|---|
| rs649129 | metabolite T | 3.80E-29 | M_LDL_FC | M_LDL_FC |
| rs495828 | metabolite T | 2.70E-29 | M_LDL_FC | M_LDL_FC |
| rs2519093 | metabolite T | 2.20E-09 | L_VLDL_PL | L_VLDL_PL_pct |
| rs644234 | metabolite G | 4.40E-08 | L_VLDL_PL | L_VLDL_PL_pct |
| rs643434 | metabolite A | 4.60E-08 | L_VLDL_PL | L_VLDL_PL_pct |
| rs613534 | metabolite G | 4.40E-08 | L_VLDL_PL | L_VLDL_PL_pct |
| rs543968 | metabolite C | 4.40E-08 | L_VLDL_PL | L_VLDL_PL_pct |
| rs544873 | metabolite A | 4.40E-08 | L_VLDL_PL | L_VLDL_PL_pct |
| rs494242 | metabolite T | 4.50E-08 | L_VLDL_PL | L_VLDL_PL_pct |
| rs532436 | metabolite A | 3.30E-09 | L_VLDL_PL | L_VLDL_PL_pct |
| rs600038 | metabolite C | 1.00E-09 | L_VLDL_PL | L_VLDL_PL_pct |
| rs1124406 | metabolite T | 1.80E-09 | L_VLDL_PL | L_VLDL_PL_pct |
| rs649129 | metabolite T | 6.90E-10 | L_VLDL_PL | L_VLDL_PL_pct |
| rs495828 | metabolite T | 6.40E-10 | L_VLDL_PL | L_VLDL_PL_pct |
| rs7873635 | metabolite C | 1.80E-14 | L_VLDL_FC | L_VLDL_FC_pct |
| rs4962040 | metabolite G | 2.20E-14 | L_VLDL_FC | L_VLDL_FC_pct |
| rs8176702 | metabolite A | 2.00E-14 | L_VLDL_FC | L_VLDL_FC_pct |
| rs2073826 | metabolite T | 2.00E-14 | L_VLDL_FC | L_VLDL_FC_pct |
| rs687289 | metabolite A | 2.40E-29 | L_VLDL_FC | L_VLDL_FC_pct |
| rs2073827 | metabolite C | 2.50E-14 | L_VLDL_FC | L_VLDL_FC_pct |
| rs2073828 | metabolite A | 2.60E-14 | L_VLDL_FC | L_VLDL_FC_pct |
| rs8176691 | metabolite T | 1.60E-14 | L_VLDL_FC | L_VLDL_FC_pct |
| rs8176690 | metabolite G | 7.50E-15 | L_VLDL_FC | L_VLDL_FC_pct |
| rs8176682 | metabolite T | 5.50E-15 | L_VLDL_FC | L_VLDL_FC_pct |
| rs8176681 | metabolite C | 7.80E-15 | L_VLDL_FC | L_VLDL_FC_pct |
| rs2519093 | metabolite T | 5.50E-48 | L_VLDL_FC | L_VLDL_FC_pct |
| rs644234 | metabolite G | 3.90E-28 | L_VLDL_FC | L_VLDL_FC_pct |
| rs643434 | metabolite A | 5.60E-28 | L_VLDL_FC | L_VLDL_FC_pct |
| rs543040 | metabolite T | 8.10E-29 | L_VLDL_FC | L_VLDL_FC_pct |
| rs613534 | metabolite G | 3.90E-28 | L_VLDL_FC | L_VLDL_FC_pct |
| rs543968 | metabolite C | 3.90E-28 | L_VLDL_FC | L_VLDL_FC_pct |
| rs544873 | metabolite A | 3.90E-28 | L_VLDL_FC | L_VLDL_FC_pct |
| rs545971 | metabolite T | 4.50E-29 | L_VLDL_FC | L_VLDL_FC_pct |
| rs612169 | metabolite G | 7.90E-29 | L_VLDL_FC | L_VLDL_FC_pct |
| rs8176668 | metabolite T | 1.20E-14 | L_VLDL_FC | L_VLDL_FC_pct |
| rs8176663 | metabolite C | 4.50E-29 | L_VLDL_FC | L_VLDL_FC_pct |
| rs7036642 | metabolite A | 1.40E-14 | L_VLDL_FC | L_VLDL_FC_pct |
| rs491626 | metabolite T | 4.50E-29 | L_VLDL_FC | L_VLDL_FC_pct |
| rs492488 | metabolite A | 4.50E-29 | L_VLDL_FC | L_VLDL_FC_pct |
| rs493246 | metabolite A | 4.50E-29 | L_VLDL_FC | L_VLDL_FC_pct |
| rs494242 | metabolite T | 4.10E-28 | L_VLDL_FC | L_VLDL_FC_pct |
| rs495203 | metabolite T | 4.60E-29 | L_VLDL_FC | L_VLDL_FC_pct |
| rs582118 | metabolite G | 4.80E-29 | L_VLDL_FC | L_VLDL_FC_pct |
| rs582094 | metabolite T | 5.10E-29 | L_VLDL_FC | L_VLDL_FC_pct |
| rs2769071 | metabolite G | 8.70E-29 | L_VLDL_FC | L_VLDL_FC_pct |
| rs677355 | metabolite A | 6.40E-29 | L_VLDL_FC | L_VLDL_FC_pct |
| rs676996 | metabolite G | 6.70E-29 | L_VLDL_FC | L_VLDL_FC_pct |
| rs676457 | metabolite T | 6.60E-29 | L_VLDL_FC | L_VLDL_FC_pct |
| rs550057 | metabolite T | 3.20E-38 | L_VLDL_FC | L_VLDL_FC_pct |
| rs674302 | metabolite A | 6.60E-29 | L_VLDL_FC | L_VLDL_FC_pct |

| | | | | |
|---|---|---|---|---|
| rs7046674 | metabolite T | 1.30E-14 | L_VLDL_FC | L_VLDL_FC_pct |
| rs554833 | metabolite T | 8.40E-29 | L_VLDL_FC | L_VLDL_FC_pct |
| rs8176649 | metabolite A | 1.20E-14 | L_VLDL_FC | L_VLDL_FC_pct |
| rs660340 | metabolite G | 5.90E-09 | L_VLDL_FC | L_VLDL_FC_pct |
| rs581107 | metabolite T | 5.70E-09 | L_VLDL_FC | L_VLDL_FC_pct |
| rs659104 | metabolite G | 6.10E-09 | L_VLDL_FC | L_VLDL_FC_pct |
| rs647800 | metabolite G | 5.20E-09 | L_VLDL_FC | L_VLDL_FC_pct |
| rs473533 | metabolite C | 6.30E-09 | L_VLDL_FC | L_VLDL_FC_pct |
| rs476410 | metabolite C | 5.90E-09 | L_VLDL_FC | L_VLDL_FC_pct |
| rs645982 | metabolite G | 5.60E-09 | L_VLDL_FC | L_VLDL_FC_pct |
| rs529565 | metabolite C | 7.40E-29 | L_VLDL_FC | L_VLDL_FC_pct |
| rs630510 | metabolite G | 1.90E-13 | L_VLDL_FC | L_VLDL_FC_pct |
| rs630014 | metabolite G | 1.90E-13 | L_VLDL_FC | L_VLDL_FC_pct |
| rs532436 | metabolite A | 2.30E-47 | L_VLDL_FC | L_VLDL_FC_pct |
| rs600038 | metabolite C | 4.60E-43 | L_VLDL_FC | L_VLDL_FC_pct |
| rs1124406: | metabolite T | 1.40E-34 | L_VLDL_FC | L_VLDL_FC_pct |
| rs649129 | metabolite T | 3.20E-43 | L_VLDL_FC | L_VLDL_FC_pct |
| rs495828 | metabolite T | 7.00E-43 | L_VLDL_FC | L_VLDL_FC_pct |
| rs687289 | metabolite A | 2.10E-08 | S_VLDL_P | S_VLDL_P |
| rs2519093 | metabolite T | 6.70E-12 | S_VLDL_P | S_VLDL_P |
| rs644234 | metabolite G | 9.80E-09 | S_VLDL_P | S_VLDL_P |
| rs643434 | metabolite A | 9.10E-09 | S_VLDL_P | S_VLDL_P |
| rs543040 | metabolite T | 1.40E-08 | S_VLDL_P | S_VLDL_P |
| rs613534 | metabolite G | 9.70E-09 | S_VLDL_P | S_VLDL_P |
| rs543968 | metabolite C | 9.70E-09 | S_VLDL_P | S_VLDL_P |
| rs544873 | metabolite A | 9.70E-09 | S_VLDL_P | S_VLDL_P |
| rs545971 | metabolite T | 1.70E-08 | S_VLDL_P | S_VLDL_P |
| rs612169 | metabolite G | 2.10E-08 | S_VLDL_P | S_VLDL_P |
| rs8176663 | metabolite C | 1.70E-08 | S_VLDL_P | S_VLDL_P |
| rs491626 | metabolite T | 1.60E-08 | S_VLDL_P | S_VLDL_P |
| rs492488 | metabolite A | 1.60E-08 | S_VLDL_P | S_VLDL_P |
| rs493246 | metabolite A | 1.60E-08 | S_VLDL_P | S_VLDL_P |
| rs494242 | metabolite T | 9.40E-09 | S_VLDL_P | S_VLDL_P |
| rs495203 | metabolite T | 1.60E-08 | S_VLDL_P | S_VLDL_P |
| rs582118 | metabolite G | 1.70E-08 | S_VLDL_P | S_VLDL_P |
| rs582094 | metabolite T | 2.00E-08 | S_VLDL_P | S_VLDL_P |
| rs2769071 | metabolite G | 1.60E-08 | S_VLDL_P | S_VLDL_P |
| rs677355 | metabolite A | 1.90E-08 | S_VLDL_P | S_VLDL_P |
| rs676996 | metabolite G | 1.70E-08 | S_VLDL_P | S_VLDL_P |
| rs676457 | metabolite T | 1.70E-08 | S_VLDL_P | S_VLDL_P |
| rs550057 | metabolite T | 2.70E-10 | S_VLDL_P | S_VLDL_P |
| rs674302 | metabolite A | 1.60E-08 | S_VLDL_P | S_VLDL_P |
| rs554833 | metabolite T | 1.70E-08 | S_VLDL_P | S_VLDL_P |
| rs529565 | metabolite C | 1.90E-08 | S_VLDL_P | S_VLDL_P |
| rs532436 | metabolite A | 7.40E-12 | S_VLDL_P | S_VLDL_P |
| rs600038 | metabolite C | 1.50E-11 | S_VLDL_P | S_VLDL_P |
| rs1124406: | metabolite T | 3.40E-08 | S_VLDL_P | S_VLDL_P |
| rs649129 | metabolite T | 8.10E-12 | S_VLDL_P | S_VLDL_P |
| rs495828 | metabolite T | 4.90E-12 | S_VLDL_P | S_VLDL_P |
| rs687289 | metabolite A | 8.30E-11 | L_LDL_P | L_LDL_P |

| | | | | |
|---|---|---|---|---|
| rs2519093 | metabolite T | 4.80E-22 | L_LDL_P | L_LDL_P |
| rs644234 | metabolite G | 7.70E-11 | L_LDL_P | L_LDL_P |
| rs643434 | metabolite A | 7.00E-11 | L_LDL_P | L_LDL_P |
| rs543040 | metabolite T | 6.00E-11 | L_LDL_P | L_LDL_P |
| rs613534 | metabolite G | 7.80E-11 | L_LDL_P | L_LDL_P |
| rs543968 | metabolite C | 7.80E-11 | L_LDL_P | L_LDL_P |
| rs544873 | metabolite A | 7.80E-11 | L_LDL_P | L_LDL_P |
| rs545971 | metabolite T | 6.20E-11 | L_LDL_P | L_LDL_P |
| rs612169 | metabolite G | 7.10E-11 | L_LDL_P | L_LDL_P |
| rs8176663 | metabolite C | 6.20E-11 | L_LDL_P | L_LDL_P |
| rs491626 | metabolite T | 6.10E-11 | L_LDL_P | L_LDL_P |
| rs492488 | metabolite A | 6.10E-11 | L_LDL_P | L_LDL_P |
| rs493246 | metabolite A | 6.10E-11 | L_LDL_P | L_LDL_P |
| rs494242 | metabolite T | 7.50E-11 | L_LDL_P | L_LDL_P |
| rs495203 | metabolite T | 6.10E-11 | L_LDL_P | L_LDL_P |
| rs582118 | metabolite G | 6.30E-11 | L_LDL_P | L_LDL_P |
| rs582094 | metabolite T | 9.20E-11 | L_LDL_P | L_LDL_P |
| rs2769071 | metabolite G | 6.50E-11 | L_LDL_P | L_LDL_P |
| rs677355 | metabolite A | 7.40E-11 | L_LDL_P | L_LDL_P |
| rs676996 | metabolite G | 6.60E-11 | L_LDL_P | L_LDL_P |
| rs676457 | metabolite T | 6.60E-11 | L_LDL_P | L_LDL_P |
| rs550057 | metabolite T | 2.20E-18 | L_LDL_P | L_LDL_P |
| rs674302 | metabolite A | 6.60E-11 | L_LDL_P | L_LDL_P |
| rs554833 | metabolite T | 7.10E-11 | L_LDL_P | L_LDL_P |
| rs529565 | metabolite C | 1.20E-10 | L_LDL_P | L_LDL_P |
| rs532436 | metabolite A | 6.40E-22 | L_LDL_P | L_LDL_P |
| rs600038 | metabolite C | 2.50E-20 | L_LDL_P | L_LDL_P |
| rs1124406[illegible] | metabolite T | 1.90E-13 | L_LDL_P | L_LDL_P |
| rs649129 | metabolite T | 1.60E-20 | L_LDL_P | L_LDL_P |
| rs495828 | metabolite T | 8.20E-21 | L_LDL_P | L_LDL_P |
| rs7873635 | metabolite C | 4.40E-08 | M_LDL_P | M_LDL_P |
| rs4962040 | metabolite G | 2.70E-08 | M_LDL_P | M_LDL_P |
| rs8176702 | metabolite A | 3.10E-08 | M_LDL_P | M_LDL_P |
| rs2073826 | metabolite T | 1.20E-08 | M_LDL_P | M_LDL_P |
| rs687289 | metabolite A | 7.50E-16 | M_LDL_P | M_LDL_P |
| rs2073827 | metabolite C | 1.50E-08 | M_LDL_P | M_LDL_P |
| rs2073828 | metabolite A | 1.50E-08 | M_LDL_P | M_LDL_P |
| rs8176691 | metabolite T | 2.80E-08 | M_LDL_P | M_LDL_P |
| rs8176690 | metabolite G | 7.10E-09 | M_LDL_P | M_LDL_P |
| rs8176682 | metabolite T | 1.50E-08 | M_LDL_P | M_LDL_P |
| rs8176681 | metabolite C | 7.20E-09 | M_LDL_P | M_LDL_P |
| rs2519093 | metabolite T | 1.30E-27 | M_LDL_P | M_LDL_P |
| rs644234 | metabolite G | 7.50E-15 | M_LDL_P | M_LDL_P |
| rs643434 | metabolite A | 7.10E-15 | M_LDL_P | M_LDL_P |
| rs543040 | metabolite T | 6.20E-16 | M_LDL_P | M_LDL_P |
| rs613534 | metabolite G | 7.50E-15 | M_LDL_P | M_LDL_P |
| rs543968 | metabolite C | 7.50E-15 | M_LDL_P | M_LDL_P |
| rs544873 | metabolite A | 7.50E-15 | M_LDL_P | M_LDL_P |
| rs545971 | metabolite T | 6.80E-16 | M_LDL_P | M_LDL_P |
| rs612169 | metabolite G | 9.50E-16 | M_LDL_P | M_LDL_P |

| | | | | |
|---|---|---|---|---|
| rs8176668 | metabolite T | 1.50E-08 | M_LDL_P | M_LDL_P |
| rs8176663 | metabolite C | 6.80E-16 | M_LDL_P | M_LDL_P |
| rs7036642 | metabolite A | 1.40E-08 | M_LDL_P | M_LDL_P |
| rs491626 | metabolite T | 6.80E-16 | M_LDL_P | M_LDL_P |
| rs492488 | metabolite A | 6.80E-16 | M_LDL_P | M_LDL_P |
| rs493246 | metabolite A | 6.80E-16 | M_LDL_P | M_LDL_P |
| rs494242 | metabolite T | 7.20E-15 | M_LDL_P | M_LDL_P |
| rs495203 | metabolite T | 6.80E-16 | M_LDL_P | M_LDL_P |
| rs582118 | metabolite G | 6.90E-16 | M_LDL_P | M_LDL_P |
| rs582094 | metabolite T | 9.30E-16 | M_LDL_P | M_LDL_P |
| rs2769071 | metabolite G | 7.10E-16 | M_LDL_P | M_LDL_P |
| rs677355 | metabolite A | 8.20E-16 | M_LDL_P | M_LDL_P |
| rs676996 | metabolite G | 7.30E-16 | M_LDL_P | M_LDL_P |
| rs676457 | metabolite T | 7.30E-16 | M_LDL_P | M_LDL_P |
| rs550057 | metabolite T | 9.50E-24 | M_LDL_P | M_LDL_P |
| rs674302 | metabolite A | 7.20E-16 | M_LDL_P | M_LDL_P |
| rs7046674 | metabolite T | 1.20E-08 | M_LDL_P | M_LDL_P |
| rs554833 | metabolite T | 7.90E-16 | M_LDL_P | M_LDL_P |
| rs8176649 | metabolite A | 1.40E-08 | M_LDL_P | M_LDL_P |
| rs529565 | metabolite C | 8.20E-16 | M_LDL_P | M_LDL_P |
| rs532436 | metabolite A | 1.50E-27 | M_LDL_P | M_LDL_P |
| rs600038 | metabolite C | 2.50E-24 | M_LDL_P | M_LDL_P |
| rs1124406: | metabolite T | 4.10E-16 | M_LDL_P | M_LDL_P |
| rs649129 | metabolite T | 1.50E-24 | M_LDL_P | M_LDL_P |
| rs495828 | metabolite T | 1.00E-24 | M_LDL_P | M_LDL_P |
| rs687289 | metabolite A | 9.80E-11 | M_VLDL_P | M_VLDL_P |
| rs2519093 | metabolite T | 1.90E-17 | M_VLDL_P | M_VLDL_P |
| rs644234 | metabolite G | 4.70E-11 | M_VLDL_P | M_VLDL_P |
| rs643434 | metabolite A | 4.20E-11 | M_VLDL_P | M_VLDL_P |
| rs543040 | metabolite T | 5.10E-11 | M_VLDL_P | M_VLDL_P |
| rs613534 | metabolite G | 4.70E-11 | M_VLDL_P | M_VLDL_P |
| rs543968 | metabolite C | 4.70E-11 | M_VLDL_P | M_VLDL_P |
| rs544873 | metabolite A | 4.70E-11 | M_VLDL_P | M_VLDL_P |
| rs545971 | metabolite T | 6.30E-11 | M_VLDL_P | M_VLDL_P |
| rs612169 | metabolite G | 8.00E-11 | M_VLDL_P | M_VLDL_P |
| rs8176663 | metabolite C | 6.30E-11 | M_VLDL_P | M_VLDL_P |
| rs491626 | metabolite T | 6.30E-11 | M_VLDL_P | M_VLDL_P |
| rs492488 | metabolite A | 6.30E-11 | M_VLDL_P | M_VLDL_P |
| rs493246 | metabolite A | 6.30E-11 | M_VLDL_P | M_VLDL_P |
| rs494242 | metabolite T | 4.50E-11 | M_VLDL_P | M_VLDL_P |
| rs495203 | metabolite T | 6.30E-11 | M_VLDL_P | M_VLDL_P |
| rs582118 | metabolite G | 6.50E-11 | M_VLDL_P | M_VLDL_P |
| rs582094 | metabolite T | 8.60E-11 | M_VLDL_P | M_VLDL_P |
| rs2769071 | metabolite G | 6.40E-11 | M_VLDL_P | M_VLDL_P |
| rs677355 | metabolite A | 7.40E-11 | M_VLDL_P | M_VLDL_P |
| rs676996 | metabolite G | 6.50E-11 | M_VLDL_P | M_VLDL_P |
| rs676457 | metabolite T | 6.50E-11 | M_VLDL_P | M_VLDL_P |
| rs550057 | metabolite T | 1.70E-15 | M_VLDL_P | M_VLDL_P |
| rs674302 | metabolite A | 6.40E-11 | M_VLDL_P | M_VLDL_P |
| rs554833 | metabolite T | 6.80E-11 | M_VLDL_P | M_VLDL_P |

| | | | | |
|---|---|---|---|---|
| rs529565 | metabolite C | 8.70E-11 | M_VLDL_P | M_VLDL_P |
| rs532436 | metabolite A | 2.10E-17 | M_VLDL_P | M_VLDL_P |
| rs600038 | metabolite C | 1.50E-16 | M_VLDL_P | M_VLDL_P |
| rs1124406: | metabolite T | 5.70E-11 | M_VLDL_P | M_VLDL_P |
| rs649129 | metabolite T | 8.30E-17 | M_VLDL_P | M_VLDL_P |
| rs495828 | metabolite T | 4.70E-17 | M_VLDL_P | M_VLDL_P |
| rs2519093 | metabolite T | 1.20E-08 | L_LDL_FC_ | L_LDL_FC_pct |
| rs532436 | metabolite A | 1.20E-08 | L_LDL_FC_ | L_LDL_FC_pct |
| rs687289 | metabolite A | 4.80E-14 | Total_C | Total_C |
| rs2519093 | metabolite T | 2.10E-28 | Total_C | Total_C |
| rs644234 | metabolite G | 4.90E-13 | Total_C | Total_C |
| rs643434 | metabolite A | 4.40E-13 | Total_C | Total_C |
| rs543040 | metabolite T | 3.20E-14 | Total_C | Total_C |
| rs613534 | metabolite G | 5.00E-13 | Total_C | Total_C |
| rs543968 | metabolite C | 5.00E-13 | Total_C | Total_C |
| rs544873 | metabolite A | 5.00E-13 | Total_C | Total_C |
| rs545971 | metabolite T | 3.30E-14 | Total_C | Total_C |
| rs612169 | metabolite G | 3.90E-14 | Total_C | Total_C |
| rs8176663 | metabolite C | 3.30E-14 | Total_C | Total_C |
| rs491626 | metabolite T | 3.30E-14 | Total_C | Total_C |
| rs492488 | metabolite A | 3.20E-14 | Total_C | Total_C |
| rs493246 | metabolite A | 3.20E-14 | Total_C | Total_C |
| rs494242 | metabolite T | 4.70E-13 | Total_C | Total_C |
| rs495203 | metabolite T | 3.30E-14 | Total_C | Total_C |
| rs582118 | metabolite G | 3.40E-14 | Total_C | Total_C |
| rs582094 | metabolite T | 4.90E-14 | Total_C | Total_C |
| rs2769071 | metabolite G | 3.20E-14 | Total_C | Total_C |
| rs677355 | metabolite A | 3.90E-14 | Total_C | Total_C |
| rs676996 | metabolite G | 3.40E-14 | Total_C | Total_C |
| rs676457 | metabolite T | 3.40E-14 | Total_C | Total_C |
| rs550057 | metabolite T | 1.70E-22 | Total_C | Total_C |
| rs674302 | metabolite A | 3.40E-14 | Total_C | Total_C |
| rs554833 | metabolite T | 3.80E-14 | Total_C | Total_C |
| rs529565 | metabolite C | 6.30E-14 | Total_C | Total_C |
| rs532436 | metabolite A | 2.80E-28 | Total_C | Total_C |
| rs600038 | metabolite C | 1.10E-24 | Total_C | Total_C |
| rs1124406: | metabolite T | 8.90E-17 | Total_C | Total_C |
| rs649129 | metabolite T | 7.20E-25 | Total_C | Total_C |
| rs495828 | metabolite T | 5.30E-25 | Total_C | Total_C |
| rs7873635 | metabolite C | 1.10E-08 | L_LDL_PL | L_LDL_PL |
| rs4962040 | metabolite G | 6.50E-09 | L_LDL_PL | L_LDL_PL |
| rs8176702 | metabolite A | 7.50E-09 | L_LDL_PL | L_LDL_PL |
| rs2073826 | metabolite T | 3.00E-09 | L_LDL_PL | L_LDL_PL |
| rs687289 | metabolite A | 1.30E-17 | L_LDL_PL | L_LDL_PL |
| rs2073827 | metabolite C | 3.40E-09 | L_LDL_PL | L_LDL_PL |
| rs2073828 | metabolite A | 3.40E-09 | L_LDL_PL | L_LDL_PL |
| rs8176691 | metabolite T | 5.80E-09 | L_LDL_PL | L_LDL_PL |
| rs8176690 | metabolite G | 1.40E-09 | L_LDL_PL | L_LDL_PL |
| rs8176682 | metabolite T | 3.00E-09 | L_LDL_PL | L_LDL_PL |
| rs8176681 | metabolite C | 1.50E-09 | L_LDL_PL | L_LDL_PL |

| | | | | |
|---|---|---|---|---|
| rs2519093 | metabolite T | 3.60E-37 | L_LDL_PL | L_LDL_PL |
| rs644234 | metabolite G | 1.30E-16 | L_LDL_PL | L_LDL_PL |
| rs643434 | metabolite A | 1.20E-16 | L_LDL_PL | L_LDL_PL |
| rs543040 | metabolite T | 1.10E-17 | L_LDL_PL | L_LDL_PL |
| rs613534 | metabolite G | 1.30E-16 | L_LDL_PL | L_LDL_PL |
| rs543968 | metabolite C | 1.30E-16 | L_LDL_PL | L_LDL_PL |
| rs544873 | metabolite A | 1.30E-16 | L_LDL_PL | L_LDL_PL |
| rs545971 | metabolite T | 9.00E-18 | L_LDL_PL | L_LDL_PL |
| rs612169 | metabolite G | 1.20E-17 | L_LDL_PL | L_LDL_PL |
| rs8176668 | metabolite T | 3.60E-09 | L_LDL_PL | L_LDL_PL |
| rs8176663 | metabolite C | 9.00E-18 | L_LDL_PL | L_LDL_PL |
| rs7036642 | metabolite A | 3.10E-09 | L_LDL_PL | L_LDL_PL |
| rs491626 | metabolite T | 8.90E-18 | L_LDL_PL | L_LDL_PL |
| rs492488 | metabolite A | 8.90E-18 | L_LDL_PL | L_LDL_PL |
| rs493246 | metabolite A | 8.90E-18 | L_LDL_PL | L_LDL_PL |
| rs494242 | metabolite T | 1.30E-16 | L_LDL_PL | L_LDL_PL |
| rs495203 | metabolite T | 9.00E-18 | L_LDL_PL | L_LDL_PL |
| rs582118 | metabolite G | 9.40E-18 | L_LDL_PL | L_LDL_PL |
| rs582094 | metabolite T | 1.50E-17 | L_LDL_PL | L_LDL_PL |
| rs2769071 | metabolite G | 9.60E-18 | L_LDL_PL | L_LDL_PL |
| rs677355 | metabolite A | 1.10E-17 | L_LDL_PL | L_LDL_PL |
| rs676996 | metabolite G | 9.50E-18 | L_LDL_PL | L_LDL_PL |
| rs676457 | metabolite T | 9.50E-18 | L_LDL_PL | L_LDL_PL |
| rs550057 | metabolite T | 1.00E-29 | L_LDL_PL | L_LDL_PL |
| rs674302 | metabolite A | 9.50E-18 | L_LDL_PL | L_LDL_PL |
| rs7046674 | metabolite T | 2.50E-09 | L_LDL_PL | L_LDL_PL |
| rs554833 | metabolite T | 1.10E-17 | L_LDL_PL | L_LDL_PL |
| rs8176649 | metabolite A | 2.80E-09 | L_LDL_PL | L_LDL_PL |
| rs529565 | metabolite C | 1.90E-17 | L_LDL_PL | L_LDL_PL |
| rs532436 | metabolite A | 5.80E-37 | L_LDL_PL | L_LDL_PL |
| rs600038 | metabolite C | 1.20E-32 | L_LDL_PL | L_LDL_PL |
| rs1124406: | metabolite T | 2.20E-22 | L_LDL_PL | L_LDL_PL |
| rs649129 | metabolite T | 8.20E-33 | L_LDL_PL | L_LDL_PL |
| rs495828 | metabolite T | 5.50E-33 | L_LDL_PL | L_LDL_PL |
| rs2519093 | metabolite T | 6.40E-15 | S_LDL_PL | S_LDL_PL |
| rs550057 | metabolite T | 4.70E-13 | S_LDL_PL | S_LDL_PL |
| rs532436 | metabolite A | 6.80E-15 | S_LDL_PL | S_LDL_PL |
| rs600038 | metabolite C | 4.30E-14 | S_LDL_PL | S_LDL_PL |
| rs1124406: | metabolite T | 1.40E-08 | S_LDL_PL | S_LDL_PL |
| rs649129 | metabolite T | 3.30E-14 | S_LDL_PL | S_LDL_PL |
| rs495828 | metabolite T | 1.60E-14 | S_LDL_PL | S_LDL_PL |
| rs687289 | metabolite A | 7.50E-14 | LDL_FC | LDL_FC |
| rs2519093 | metabolite T | 9.00E-33 | LDL_FC | LDL_FC |
| rs644234 | metabolite G | 6.50E-13 | LDL_FC | LDL_FC |
| rs643434 | metabolite A | 6.20E-13 | LDL_FC | LDL_FC |
| rs543040 | metabolite T | 6.30E-14 | LDL_FC | LDL_FC |
| rs613534 | metabolite G | 6.60E-13 | LDL_FC | LDL_FC |
| rs543968 | metabolite C | 6.60E-13 | LDL_FC | LDL_FC |
| rs544873 | metabolite A | 6.60E-13 | LDL_FC | LDL_FC |
| rs545971 | metabolite T | 5.20E-14 | LDL_FC | LDL_FC |

| | | | | |
|---|---|---|---|---|
| rs612169 | metabolite G | 6.60E-14 | LDL_FC | LDL_FC |
| rs8176663 | metabolite C | 5.20E-14 | LDL_FC | LDL_FC |
| rs491626 | metabolite T | 5.20E-14 | LDL_FC | LDL_FC |
| rs492488 | metabolite A | 5.20E-14 | LDL_FC | LDL_FC |
| rs493246 | metabolite A | 5.20E-14 | LDL_FC | LDL_FC |
| rs494242 | metabolite T | 6.40E-13 | LDL_FC | LDL_FC |
| rs495203 | metabolite T | 5.20E-14 | LDL_FC | LDL_FC |
| rs582118 | metabolite G | 5.40E-14 | LDL_FC | LDL_FC |
| rs582094 | metabolite T | 8.10E-14 | LDL_FC | LDL_FC |
| rs2769071 | metabolite G | 5.60E-14 | LDL_FC | LDL_FC |
| rs677355 | metabolite A | 6.00E-14 | LDL_FC | LDL_FC |
| rs676996 | metabolite G | 5.50E-14 | LDL_FC | LDL_FC |
| rs676457 | metabolite T | 5.50E-14 | LDL_FC | LDL_FC |
| rs550057 | metabolite T | 7.00E-26 | LDL_FC | LDL_FC |
| rs674302 | metabolite A | 5.50E-14 | LDL_FC | LDL_FC |
| rs554833 | metabolite T | 6.00E-14 | LDL_FC | LDL_FC |
| rs529565 | metabolite C | 1.10E-13 | LDL_FC | LDL_FC |
| rs532436 | metabolite A | 1.40E-32 | LDL_FC | LDL_FC |
| rs600038 | metabolite C | 1.60E-28 | LDL_FC | LDL_FC |
| rs1124406: | metabolite T | 5.40E-19 | LDL_FC | LDL_FC |
| rs649129 | metabolite T | 1.30E-28 | LDL_FC | LDL_FC |
| rs495828 | metabolite T | 9.40E-29 | LDL_FC | LDL_FC |
| rs687289 | metabolite A | 1.40E-10 | S_VLDL_PL | S_VLDL_PL_pct |
| rs2519093 | metabolite T | 4.10E-22 | S_VLDL_PL | S_VLDL_PL_pct |
| rs644234 | metabolite G | 1.40E-09 | S_VLDL_PL | S_VLDL_PL_pct |
| rs643434 | metabolite A | 1.50E-09 | S_VLDL_PL | S_VLDL_PL_pct |
| rs543040 | metabolite T | 2.50E-10 | S_VLDL_PL | S_VLDL_PL_pct |
| rs613534 | metabolite G | 1.50E-09 | S_VLDL_PL | S_VLDL_PL_pct |
| rs543968 | metabolite C | 1.50E-09 | S_VLDL_PL | S_VLDL_PL_pct |
| rs544873 | metabolite A | 1.50E-09 | S_VLDL_PL | S_VLDL_PL_pct |
| rs545971 | metabolite T | 1.80E-10 | S_VLDL_PL | S_VLDL_PL_pct |
| rs612169 | metabolite G | 1.80E-10 | S_VLDL_PL | S_VLDL_PL_pct |
| rs8176663 | metabolite C | 1.80E-10 | S_VLDL_PL | S_VLDL_PL_pct |
| rs491626 | metabolite T | 1.80E-10 | S_VLDL_PL | S_VLDL_PL_pct |
| rs492488 | metabolite A | 1.80E-10 | S_VLDL_PL | S_VLDL_PL_pct |
| rs493246 | metabolite A | 1.80E-10 | S_VLDL_PL | S_VLDL_PL_pct |
| rs494242 | metabolite T | 1.50E-09 | S_VLDL_PL | S_VLDL_PL_pct |
| rs495203 | metabolite T | 1.80E-10 | S_VLDL_PL | S_VLDL_PL_pct |
| rs582118 | metabolite G | 1.80E-10 | S_VLDL_PL | S_VLDL_PL_pct |
| rs582094 | metabolite T | 2.50E-10 | S_VLDL_PL | S_VLDL_PL_pct |
| rs2769071 | metabolite G | 2.00E-10 | S_VLDL_PL | S_VLDL_PL_pct |
| rs677355 | metabolite A | 1.80E-10 | S_VLDL_PL | S_VLDL_PL_pct |
| rs676996 | metabolite G | 1.90E-10 | S_VLDL_PL | S_VLDL_PL_pct |
| rs676457 | metabolite T | 1.90E-10 | S_VLDL_PL | S_VLDL_PL_pct |
| rs550057 | metabolite T | 1.10E-18 | S_VLDL_PL | S_VLDL_PL_pct |
| rs674302 | metabolite A | 1.90E-10 | S_VLDL_PL | S_VLDL_PL_pct |
| rs554833 | metabolite T | 2.00E-10 | S_VLDL_PL | S_VLDL_PL_pct |
| rs529565 | metabolite C | 2.90E-10 | S_VLDL_PL | S_VLDL_PL_pct |
| rs532436 | metabolite A | 5.70E-22 | S_VLDL_PL | S_VLDL_PL_pct |
| rs600038 | metabolite C | 5.30E-19 | S_VLDL_PL | S_VLDL_PL_pct |

| | | | | |
|---|---|---|---|---|
| rs1124406: | metabolite T | 4.50E-12 | S_VLDL_PL | S_VLDL_PL_pct |
| rs649129 | metabolite T | 8.10E-19 | S_VLDL_PL | S_VLDL_PL_pct |
| rs495828 | metabolite T | 7.60E-19 | S_VLDL_PL | S_VLDL_PL_pct |
| rs687289 | metabolite A | 5.80E-14 | S_VLDL_PL | S_VLDL_PL |
| rs2519093 | metabolite T | 1.00E-22 | S_VLDL_PL | S_VLDL_PL |
| rs644234 | metabolite G | 4.90E-14 | S_VLDL_PL | S_VLDL_PL |
| rs643434 | metabolite A | 4.40E-14 | S_VLDL_PL | S_VLDL_PL |
| rs543040 | metabolite T | 3.70E-14 | S_VLDL_PL | S_VLDL_PL |
| rs613534 | metabolite G | 4.90E-14 | S_VLDL_PL | S_VLDL_PL |
| rs543968 | metabolite C | 4.90E-14 | S_VLDL_PL | S_VLDL_PL |
| rs544873 | metabolite A | 4.90E-14 | S_VLDL_PL | S_VLDL_PL |
| rs545971 | metabolite T | 4.20E-14 | S_VLDL_PL | S_VLDL_PL |
| rs612169 | metabolite G | 5.70E-14 | S_VLDL_PL | S_VLDL_PL |
| rs8176663 | metabolite C | 4.20E-14 | S_VLDL_PL | S_VLDL_PL |
| rs491626 | metabolite T | 4.20E-14 | S_VLDL_PL | S_VLDL_PL |
| rs492488 | metabolite A | 4.20E-14 | S_VLDL_PL | S_VLDL_PL |
| rs493246 | metabolite A | 4.20E-14 | S_VLDL_PL | S_VLDL_PL |
| rs494242 | metabolite T | 4.60E-14 | S_VLDL_PL | S_VLDL_PL |
| rs495203 | metabolite T | 4.20E-14 | S_VLDL_PL | S_VLDL_PL |
| rs582118 | metabolite G | 4.30E-14 | S_VLDL_PL | S_VLDL_PL |
| rs582094 | metabolite T | 6.00E-14 | S_VLDL_PL | S_VLDL_PL |
| rs2769071 | metabolite G | 4.20E-14 | S_VLDL_PL | S_VLDL_PL |
| rs677355 | metabolite A | 5.00E-14 | S_VLDL_PL | S_VLDL_PL |
| rs676996 | metabolite G | 4.30E-14 | S_VLDL_PL | S_VLDL_PL |
| rs676457 | metabolite T | 4.30E-14 | S_VLDL_PL | S_VLDL_PL |
| rs550057 | metabolite T | 3.40E-19 | S_VLDL_PL | S_VLDL_PL |
| rs674302 | metabolite A | 4.20E-14 | S_VLDL_PL | S_VLDL_PL |
| rs554833 | metabolite T | 4.50E-14 | S_VLDL_PL | S_VLDL_PL |
| rs529565 | metabolite C | 6.00E-14 | S_VLDL_PL | S_VLDL_PL |
| rs532436 | metabolite A | 1.30E-22 | S_VLDL_PL | S_VLDL_PL |
| rs600038 | metabolite C | 3.30E-21 | S_VLDL_PL | S_VLDL_PL |
| rs1124406: | metabolite T | 1.30E-14 | S_VLDL_PL | S_VLDL_PL |
| rs649129 | metabolite T | 1.80E-21 | S_VLDL_PL | S_VLDL_PL |
| rs495828 | metabolite T | 9.30E-22 | S_VLDL_PL | S_VLDL_PL |
| rs687289 | metabolite A | 8.60E-12 | S_VLDL_FC | S_VLDL_FC_pct |
| rs2519093 | metabolite T | 2.50E-24 | S_VLDL_FC | S_VLDL_FC_pct |
| rs644234 | metabolite G | 1.20E-10 | S_VLDL_FC | S_VLDL_FC_pct |
| rs643434 | metabolite A | 1.20E-10 | S_VLDL_FC | S_VLDL_FC_pct |
| rs543040 | metabolite T | 1.50E-11 | S_VLDL_FC | S_VLDL_FC_pct |
| rs613534 | metabolite G | 1.20E-10 | S_VLDL_FC | S_VLDL_FC_pct |
| rs543968 | metabolite C | 1.20E-10 | S_VLDL_FC | S_VLDL_FC_pct |
| rs544873 | metabolite A | 1.20E-10 | S_VLDL_FC | S_VLDL_FC_pct |
| rs545971 | metabolite T | 1.10E-11 | S_VLDL_FC | S_VLDL_FC_pct |
| rs612169 | metabolite G | 1.10E-11 | S_VLDL_FC | S_VLDL_FC_pct |
| rs8176663 | metabolite C | 1.10E-11 | S_VLDL_FC | S_VLDL_FC_pct |
| rs491626 | metabolite T | 1.10E-11 | S_VLDL_FC | S_VLDL_FC_pct |
| rs492488 | metabolite A | 1.10E-11 | S_VLDL_FC | S_VLDL_FC_pct |
| rs493246 | metabolite A | 1.10E-11 | S_VLDL_FC | S_VLDL_FC_pct |
| rs494242 | metabolite T | 1.20E-10 | S_VLDL_FC | S_VLDL_FC_pct |
| rs495203 | metabolite T | 1.10E-11 | S_VLDL_FC | S_VLDL_FC_pct |

| | | | | |
|---|---|---|---|---|
| rs582118 | metabolite G | 1.10E-11 | S_VLDL_FC | S_VLDL_FC_pct |
| rs582094 | metabolite T | 1.60E-11 | S_VLDL_FC | S_VLDL_FC_pct |
| rs2769071 | metabolite G | 1.20E-11 | S_VLDL_FC | S_VLDL_FC_pct |
| rs677355 | metabolite A | 1.10E-11 | S_VLDL_FC | S_VLDL_FC_pct |
| rs676996 | metabolite G | 1.10E-11 | S_VLDL_FC | S_VLDL_FC_pct |
| rs676457 | metabolite T | 1.10E-11 | S_VLDL_FC | S_VLDL_FC_pct |
| rs550057 | metabolite T | 6.60E-21 | S_VLDL_FC | S_VLDL_FC_pct |
| rs674302 | metabolite A | 1.10E-11 | S_VLDL_FC | S_VLDL_FC_pct |
| rs554833 | metabolite T | 1.10E-11 | S_VLDL_FC | S_VLDL_FC_pct |
| rs529565 | metabolite C | 1.70E-11 | S_VLDL_FC | S_VLDL_FC_pct |
| rs532436 | metabolite A | 3.60E-24 | S_VLDL_FC | S_VLDL_FC_pct |
| rs600038 | metabolite C | 8.40E-21 | S_VLDL_FC | S_VLDL_FC_pct |
| rs1124406: | metabolite T | 1.90E-13 | S_VLDL_FC | S_VLDL_FC_pct |
| rs649129 | metabolite T | 1.40E-20 | S_VLDL_FC | S_VLDL_FC_pct |
| rs495828 | metabolite T | 1.30E-20 | S_VLDL_FC | S_VLDL_FC_pct |
| rs687289 | metabolite A | 3.90E-13 | M_LDL_PL | M_LDL_PL |
| rs2519093 | metabolite T | 1.80E-28 | M_LDL_PL | M_LDL_PL |
| rs644234 | metabolite G | 1.00E-12 | M_LDL_PL | M_LDL_PL |
| rs643434 | metabolite A | 9.40E-13 | M_LDL_PL | M_LDL_PL |
| rs543040 | metabolite T | 2.70E-13 | M_LDL_PL | M_LDL_PL |
| rs613534 | metabolite G | 9.90E-13 | M_LDL_PL | M_LDL_PL |
| rs543968 | metabolite C | 9.90E-13 | M_LDL_PL | M_LDL_PL |
| rs544873 | metabolite A | 9.90E-13 | M_LDL_PL | M_LDL_PL |
| rs545971 | metabolite T | 2.60E-13 | M_LDL_PL | M_LDL_PL |
| rs612169 | metabolite G | 3.40E-13 | M_LDL_PL | M_LDL_PL |
| rs8176663 | metabolite C | 2.60E-13 | M_LDL_PL | M_LDL_PL |
| rs491626 | metabolite T | 2.60E-13 | M_LDL_PL | M_LDL_PL |
| rs492488 | metabolite A | 2.60E-13 | M_LDL_PL | M_LDL_PL |
| rs493246 | metabolite A | 2.60E-13 | M_LDL_PL | M_LDL_PL |
| rs494242 | metabolite T | 9.60E-13 | M_LDL_PL | M_LDL_PL |
| rs495203 | metabolite T | 2.60E-13 | M_LDL_PL | M_LDL_PL |
| rs582118 | metabolite G | 2.70E-13 | M_LDL_PL | M_LDL_PL |
| rs582094 | metabolite T | 3.90E-13 | M_LDL_PL | M_LDL_PL |
| rs2769071 | metabolite G | 2.80E-13 | M_LDL_PL | M_LDL_PL |
| rs677355 | metabolite A | 3.20E-13 | M_LDL_PL | M_LDL_PL |
| rs676996 | metabolite G | 2.80E-13 | M_LDL_PL | M_LDL_PL |
| rs676457 | metabolite T | 2.80E-13 | M_LDL_PL | M_LDL_PL |
| rs550057 | metabolite T | 2.60E-23 | M_LDL_PL | M_LDL_PL |
| rs674302 | metabolite A | 2.70E-13 | M_LDL_PL | M_LDL_PL |
| rs554833 | metabolite T | 3.00E-13 | M_LDL_PL | M_LDL_PL |
| rs529565 | metabolite C | 4.30E-13 | M_LDL_PL | M_LDL_PL |
| rs532436 | metabolite A | 2.50E-28 | M_LDL_PL | M_LDL_PL |
| rs600038 | metabolite C | 1.50E-25 | M_LDL_PL | M_LDL_PL |
| rs1124406: | metabolite T | 3.60E-18 | M_LDL_PL | M_LDL_PL |
| rs649129 | metabolite T | 8.10E-26 | M_LDL_PL | M_LDL_PL |
| rs495828 | metabolite T | 4.60E-26 | M_LDL_PL | M_LDL_PL |
| rs687289 | metabolite A | 3.10E-11 | M_VLDL_P | M_VLDL_PL |
| rs2519093 | metabolite T | 1.70E-18 | M_VLDL_P | M_VLDL_PL |
| rs644234 | metabolite G | 1.40E-11 | M_VLDL_P | M_VLDL_PL |
| rs643434 | metabolite A | 1.20E-11 | M_VLDL_P | M_VLDL_PL |

| | | | | |
|---|---|---|---|---|
| rs543040 | metabolite T | 1.70E-11 | M_VLDL_P | M_VLDL_PL |
| rs613534 | metabolite G | 1.40E-11 | M_VLDL_P | M_VLDL_PL |
| rs543968 | metabolite C | 1.40E-11 | M_VLDL_P | M_VLDL_PL |
| rs544873 | metabolite A | 1.40E-11 | M_VLDL_P | M_VLDL_PL |
| rs545971 | metabolite T | 2.00E-11 | M_VLDL_P | M_VLDL_PL |
| rs612169 | metabolite G | 2.60E-11 | M_VLDL_P | M_VLDL_PL |
| rs8176663 | metabolite C | 2.00E-11 | M_VLDL_P | M_VLDL_PL |
| rs491626 | metabolite T | 2.00E-11 | M_VLDL_P | M_VLDL_PL |
| rs492488 | metabolite A | 2.00E-11 | M_VLDL_P | M_VLDL_PL |
| rs493246 | metabolite A | 2.00E-11 | M_VLDL_P | M_VLDL_PL |
| rs494242 | metabolite T | 1.30E-11 | M_VLDL_P | M_VLDL_PL |
| rs495203 | metabolite T | 2.00E-11 | M_VLDL_P | M_VLDL_PL |
| rs582118 | metabolite G | 2.10E-11 | M_VLDL_P | M_VLDL_PL |
| rs582094 | metabolite T | 2.80E-11 | M_VLDL_P | M_VLDL_PL |
| rs2769071 | metabolite G | 2.10E-11 | M_VLDL_P | M_VLDL_PL |
| rs677355 | metabolite A | 2.40E-11 | M_VLDL_P | M_VLDL_PL |
| rs676996 | metabolite G | 2.10E-11 | M_VLDL_P | M_VLDL_PL |
| rs676457 | metabolite T | 2.10E-11 | M_VLDL_P | M_VLDL_PL |
| rs550057 | metabolite T | 3.20E-16 | M_VLDL_P | M_VLDL_PL |
| rs674302 | metabolite A | 2.10E-11 | M_VLDL_P | M_VLDL_PL |
| rs554833 | metabolite T | 2.20E-11 | M_VLDL_P | M_VLDL_PL |
| rs529565 | metabolite C | 2.80E-11 | M_VLDL_P | M_VLDL_PL |
| rs532436 | metabolite A | 1.90E-18 | M_VLDL_P | M_VLDL_PL |
| rs600038 | metabolite C | 1.40E-17 | M_VLDL_P | M_VLDL_PL |
| rs1124406: | metabolite T | 1.10E-11 | M_VLDL_P | M_VLDL_PL |
| rs649129 | metabolite T | 7.80E-18 | M_VLDL_P | M_VLDL_PL |
| rs495828 | metabolite T | 4.10E-18 | M_VLDL_P | M_VLDL_PL |
| rs687289 | metabolite A | 4.80E-14 | M_VLDL_F | M_VLDL_FC |
| rs2519093 | metabolite T | 1.30E-23 | M_VLDL_F | M_VLDL_FC |
| rs644234 | metabolite G | 3.90E-14 | M_VLDL_F | M_VLDL_FC |
| rs643434 | metabolite A | 3.50E-14 | M_VLDL_F | M_VLDL_FC |
| rs543040 | metabolite T | 2.70E-14 | M_VLDL_F | M_VLDL_FC |
| rs613534 | metabolite G | 3.90E-14 | M_VLDL_F | M_VLDL_FC |
| rs543968 | metabolite C | 3.90E-14 | M_VLDL_F | M_VLDL_FC |
| rs544873 | metabolite A | 3.90E-14 | M_VLDL_F | M_VLDL_FC |
| rs545971 | metabolite T | 3.10E-14 | M_VLDL_F | M_VLDL_FC |
| rs612169 | metabolite G | 4.20E-14 | M_VLDL_F | M_VLDL_FC |
| rs8176663 | metabolite C | 3.10E-14 | M_VLDL_F | M_VLDL_FC |
| rs491626 | metabolite T | 3.10E-14 | M_VLDL_F | M_VLDL_FC |
| rs492488 | metabolite A | 3.10E-14 | M_VLDL_F | M_VLDL_FC |
| rs493246 | metabolite A | 3.10E-14 | M_VLDL_F | M_VLDL_FC |
| rs494242 | metabolite T | 3.80E-14 | M_VLDL_F | M_VLDL_FC |
| rs495203 | metabolite T | 3.10E-14 | M_VLDL_F | M_VLDL_FC |
| rs582118 | metabolite G | 3.20E-14 | M_VLDL_F | M_VLDL_FC |
| rs582094 | metabolite T | 4.70E-14 | M_VLDL_F | M_VLDL_FC |
| rs2769071 | metabolite G | 3.20E-14 | M_VLDL_F | M_VLDL_FC |
| rs677355 | metabolite A | 3.70E-14 | M_VLDL_F | M_VLDL_FC |
| rs676996 | metabolite G | 3.20E-14 | M_VLDL_F | M_VLDL_FC |
| rs676457 | metabolite T | 3.20E-14 | M_VLDL_F | M_VLDL_FC |
| rs550057 | metabolite T | 2.00E-20 | M_VLDL_F | M_VLDL_FC |

| | | | | |
|---|---|---|---|---|
| rs674302 | metabolite A | 3.20E-14 | M_VLDL_F( | M_VLDL_FC |
| rs554833 | metabolite T | 3.40E-14 | M_VLDL_F( | M_VLDL_FC |
| rs529565 | metabolite C | 5.40E-14 | M_VLDL_F( | M_VLDL_FC |
| rs532436 | metabolite A | 1.50E-23 | M_VLDL_F( | M_VLDL_FC |
| rs600038 | metabolite C | 6.00E-22 | M_VLDL_F( | M_VLDL_FC |
| rs1124406: | metabolite T | 2.30E-14 | M_VLDL_F( | M_VLDL_FC |
| rs649129 | metabolite T | 3.80E-22 | M_VLDL_F( | M_VLDL_FC |
| rs495828 | metabolite T | 1.90E-22 | M_VLDL_F( | M_VLDL_FC |
| rs2519093 | metabolite T | 7.60E-10 | S_HDL_PL_ | S_HDL_PL_pct |
| rs532436 | metabolite A | 1.00E-09 | S_HDL_PL_ | S_HDL_PL_pct |
| rs600038 | metabolite C | 1.50E-08 | S_HDL_PL_ | S_HDL_PL_pct |
| rs649129 | metabolite T | 1.60E-08 | S_HDL_PL_ | S_HDL_PL_pct |
| rs495828 | metabolite T | 1.60E-08 | S_HDL_PL_ | S_HDL_PL_pct |
| rs7873635 | metabolite C | 1.00E-12 | M_VLDL_P | M_VLDL_PL_pct |
| rs4962040 | metabolite G | 1.10E-12 | M_VLDL_P | M_VLDL_PL_pct |
| rs8176702 | metabolite A | 1.50E-12 | M_VLDL_P | M_VLDL_PL_pct |
| rs2073826 | metabolite T | 1.50E-12 | M_VLDL_P | M_VLDL_PL_pct |
| rs687289 | metabolite A | 3.40E-25 | M_VLDL_P | M_VLDL_PL_pct |
| rs2073827 | metabolite C | 1.80E-12 | M_VLDL_P | M_VLDL_PL_pct |
| rs2073828 | metabolite A | 1.80E-12 | M_VLDL_P | M_VLDL_PL_pct |
| rs8176691 | metabolite T | 9.00E-13 | M_VLDL_P | M_VLDL_PL_pct |
| rs8176690 | metabolite G | 4.30E-13 | M_VLDL_P | M_VLDL_PL_pct |
| rs8176682 | metabolite T | 3.20E-13 | M_VLDL_P | M_VLDL_PL_pct |
| rs8176681 | metabolite C | 5.90E-13 | M_VLDL_P | M_VLDL_PL_pct |
| rs2519093 | metabolite T | 2.00E-43 | M_VLDL_P | M_VLDL_PL_pct |
| rs644234 | metabolite G | 6.00E-24 | M_VLDL_P | M_VLDL_PL_pct |
| rs643434 | metabolite A | 6.90E-24 | M_VLDL_P | M_VLDL_PL_pct |
| rs543040 | metabolite T | 1.10E-24 | M_VLDL_P | M_VLDL_PL_pct |
| rs613534 | metabolite G | 6.30E-24 | M_VLDL_P | M_VLDL_PL_pct |
| rs543968 | metabolite C | 6.30E-24 | M_VLDL_P | M_VLDL_PL_pct |
| rs544873 | metabolite A | 6.30E-24 | M_VLDL_P | M_VLDL_PL_pct |
| rs545971 | metabolite T | 5.50E-25 | M_VLDL_P | M_VLDL_PL_pct |
| rs612169 | metabolite G | 7.10E-25 | M_VLDL_P | M_VLDL_PL_pct |
| rs8176668 | metabolite T | 1.10E-12 | M_VLDL_P | M_VLDL_PL_pct |
| rs8176663 | metabolite C | 5.50E-25 | M_VLDL_P | M_VLDL_PL_pct |
| rs7036642 | metabolite A | 1.30E-12 | M_VLDL_P | M_VLDL_PL_pct |
| rs491626 | metabolite T | 5.50E-25 | M_VLDL_P | M_VLDL_PL_pct |
| rs492488 | metabolite A | 5.40E-25 | M_VLDL_P | M_VLDL_PL_pct |
| rs493246 | metabolite A | 5.50E-25 | M_VLDL_P | M_VLDL_PL_pct |
| rs494242 | metabolite T | 6.40E-24 | M_VLDL_P | M_VLDL_PL_pct |
| rs495203 | metabolite T | 5.50E-25 | M_VLDL_P | M_VLDL_PL_pct |
| rs582118 | metabolite G | 5.60E-25 | M_VLDL_P | M_VLDL_PL_pct |
| rs582094 | metabolite T | 7.10E-25 | M_VLDL_P | M_VLDL_PL_pct |
| rs2769071 | metabolite G | 8.10E-25 | M_VLDL_P | M_VLDL_PL_pct |
| rs677355 | metabolite A | 6.50E-25 | M_VLDL_P | M_VLDL_PL_pct |
| rs676996 | metabolite G | 6.90E-25 | M_VLDL_P | M_VLDL_PL_pct |
| rs676457 | metabolite T | 6.90E-25 | M_VLDL_P | M_VLDL_PL_pct |
| rs550057 | metabolite T | 4.30E-36 | M_VLDL_P | M_VLDL_PL_pct |
| rs674302 | metabolite A | 6.90E-25 | M_VLDL_P | M_VLDL_PL_pct |
| rs7046674 | metabolite T | 1.30E-12 | M_VLDL_P | M_VLDL_PL_pct |

| | | | | |
|---|---|---|---|---|
| rs554833 | metabolite T | 8.30E-25 | M_VLDL_P | M_VLDL_PL_pct |
| rs8176649 | metabolite A | 1.10E-12 | M_VLDL_P | M_VLDL_PL_pct |
| rs660340 | metabolite G | 4.20E-10 | M_VLDL_P | M_VLDL_PL_pct |
| rs581107 | metabolite T | 4.10E-10 | M_VLDL_P | M_VLDL_PL_pct |
| rs659104 | metabolite G | 4.10E-10 | M_VLDL_P | M_VLDL_PL_pct |
| rs647800 | metabolite G | 5.00E-10 | M_VLDL_P | M_VLDL_PL_pct |
| rs473533 | metabolite C | 4.30E-10 | M_VLDL_P | M_VLDL_PL_pct |
| rs476410 | metabolite C | 4.60E-10 | M_VLDL_P | M_VLDL_PL_pct |
| rs645982 | metabolite G | 3.80E-10 | M_VLDL_P | M_VLDL_PL_pct |
| rs529565 | metabolite C | 1.80E-24 | M_VLDL_P | M_VLDL_PL_pct |
| rs630510 | metabolite G | 4.30E-09 | M_VLDL_P | M_VLDL_PL_pct |
| rs630014 | metabolite G | 4.30E-09 | M_VLDL_P | M_VLDL_PL_pct |
| rs532436 | metabolite A | 5.70E-43 | M_VLDL_P | M_VLDL_PL_pct |
| rs600038 | metabolite C | 8.80E-39 | M_VLDL_P | M_VLDL_PL_pct |
| rs1124406 | metabolite T | 1.90E-25 | M_VLDL_P | M_VLDL_PL_pct |
| rs649129 | metabolite T | 1.10E-38 | M_VLDL_P | M_VLDL_PL_pct |
| rs495828 | metabolite T | 6.40E-39 | M_VLDL_P | M_VLDL_PL_pct |
| rs687289 | GWAS cata rs68728 | 8.00E-10 | Blood prot | Blood protein levels |
| rs612169 | GWAS cata rs61216 | 1.00E-20 | Blood prot | Blood protein levels |
| rs550057 | GWAS cata rs55005 | 6.00E-20 | LDL choles | LDL cholesterol |
| rs2519093 | GWAS cata rs25190 | 1.00E-11 | Coronary a | Coronary artery disease |
| rs2519093 | GWAS cata rs25190 | 4.00E-08 | Allergy | Allergy |
| rs495828 | GWAS cata rs49582 | 2.00E-10 | Hemoglobi | Hemoglobin levels |
| rs495828 | GWAS cata rs49582 | 1.00E-19 | Hemoglobi | Hemoglobin levels |
| rs495828 | GWAS cata rs49582 | 1.00E-08 | Red blood | Red blood cell count |
| rs495828 | GWAS cata rs49582 | 3.00E-09 | Hematocrit | Hematocrit |
| rs495828 | GWAS cata rs49582 | 6.00E-34 | Blood meta | Blood metabolite ratios |
| rs649129 | GWAS cata rs64912 | 9.00E-37 | Blood meta | Blood metabolite ratios |
| rs643434 | GWAS cata rs64343 | 9.00E-25 | Inflammato | Inflammatory biomarkers |
| rs687289 | GWAS cata rs68728 | 2.00E-16 | Pancreatic | Pancreatic cancer |
| rs529565 | GWAS cata rs52956 | 4.00E-75 | Venous thr | Venous thromboembolism |
| rs644234 | GWAS cata rs64423 | 4.00E-47 | Protein bio | Protein biomarker |
| rs687289 | GWAS cata rs68728 | 1.00E-12 | Urinary me | Urinary metabolites (H-NMR features) |
| rs687289 | GWAS cata rs68728 | 2.00E-12 | Urinary me | Urinary metabolites (H-NMR features) |
| rs687289 | GWAS cata rs68728 | 4.9406564 | Factor VIII | Factor VIII levels |
| rs529565 | GWAS cata rs52956 | 7.00E-63 | Thrombosi | Thrombosis |
| rs2519093 | GWAS cata rs25190 | 2.00E-48 | Low densit | Low density lipoprotein cholesterol levels |
| rs2519093 | GWAS cata rs25190 | 1.00E-14 | Low densit | Low density lipoprotein cholesterol levels |
| rs2519093 | GWAS cata rs25190 | 7.00E-65 | Low densit | Low density lipoprotein cholesterol levels |
| rs550057 | GWAS cata rs55005 | 5.00E-36 | Low densit | Low density lipoprotein cholesterol levels |
| rs550057 | GWAS cata rs55005 | 7.00E-14 | Low densit | Low density lipoprotein cholesterol levels |
| rs550057 | GWAS cata rs55005 | 3.00E-52 | Low densit | Low density lipoprotein cholesterol levels |
| rs1165522 | GWAS cata rs11655 | 2.00E-14 | Thyroid sti | Thyroid stimulating hormone levels |
| rs495828 | GWAS cata rs49582 | 4.00E-103 | LDL choles | LDL cholesterol levels x alcohol consumption (regular v |
| rs492488 | GWAS cata rs49248 | 2.00E-19 | Blood prot | Blood protein levels |
| rs495828 | GWAS cata rs49582 | 2.00E-36 | LDL choles | LDL cholesterol levels in current drinkers |
| rs492488 | GWAS cata rs49248 | 3.00E-18 | Blood prot | Blood protein levels |
| rs492488 | GWAS cata rs49248 | 5.00E-18 | Blood prot | Blood protein levels |
| rs492488 | GWAS cata rs49248 | 6.00E-18 | Blood prot | Blood protein levels |
| rs495828 | GWAS cata rs49582 | 8.00E-47 | LDL choles | LDL cholesterol levels in current drinkers |

| | | | | | |
|---|---|---|---|---|---|
| rs495828 | GWAS cata | rs4958[illegible] | 2.00E-08 | LDL cholest | LDL cholesterol levels x alcohol consumption (regular v[illegible] |
| rs495828 | GWAS cata | rs4958[illegible] | 1.00E-114 | LDL cholest | LDL cholesterol levels x alcohol consumption (regular v[illegible] |
| rs492488 | GWAS cata | rs4924[illegible] | 1.00E-17 | Blood prot | Blood protein levels |
| rs495828 | GWAS cata | rs4958[illegible] | 5.00E-69 | LDL cholest | LDL cholesterol levels x alcohol consumption (drinkers [illegible] |
| rs495828 | GWAS cata | rs4958[illegible] | 2.00E-08 | LDL cholest | LDL cholesterol levels x alcohol consumption (drinkers [illegible] |
| rs495828 | GWAS cata | rs4958[illegible] | 8.00E-09 | LDL cholest | LDL cholesterol levels x alcohol consumption (drinkers [illegible] |
| rs495828 | GWAS cata | rs4958[illegible] | 7.00E-29 | LDL cholest | LDL cholesterol levels |
| rs495828 | GWAS cata | rs4958[illegible] | 4.00E-86 | LDL cholest | LDL cholesterol levels x alcohol consumption (drinkers [illegible] |
| rs495828 | GWAS cata | rs4958[illegible] | 4.00E-18 | LDL cholest | LDL cholesterol levels |
| rs687289 | GWAS cata | rs6872[illegible] | 0 | Factor VIII [illegible] | Factor VIII levels |
| rs660340 | GWAS cata | rs6603[illegible] | 9.9998886[illegible] | vWF levels | vWF levels |
| rs550057 | GWAS cata | rs5500[illegible] | 3.00E-08 | Cytokine n[illegible] | Cytokine network levels (multivariate analysis) |
| rs492488 | GWAS cata | rs4924[illegible] | 3.00E-16 | Blood prot | Blood protein levels |
| rs660340 | GWAS cata | rs6603[illegible] | 0 | vWF levels | vWF levels |
| rs687289 | GWAS cata | rs6872[illegible] | 0 | vWF levels | vWF levels |
| rs2519093 | GWAS cata | rs2519[illegible] | 1.00E-16 | Triglyceride | Triglyceride levels |
| rs600038 | GWAS cata | rs6000[illegible] | 4.00E-09 | Heart failu[illegible] | Heart failure |
| rs492488 | GWAS cata | rs4924[illegible] | 2.00E-274 | Blood prot | Blood protein levels |
| rs495828 | GWAS cata | rs4958[illegible] | 9.00E-12 | Coronary a | Coronary artery disease |
| rs2519093 | GWAS cata | rs2519[illegible] | 2.00E-14 | Coronary a | Coronary artery disease |
| rs492488 | GWAS cata | rs4924[illegible] | 1.00E-22 | Blood prot | Blood protein levels |
| rs492488 | GWAS cata | rs4924[illegible] | 4.00E-22 | Blood prot | Blood protein levels |
| rs492488 | GWAS cata | rs4924[illegible] | 7.00E-23 | Blood prot | Blood protein levels |
| rs649129 | GWAS cata | rs6491[illegible] | 1.00E-43 | Low densit | Low density lipoprotein cholesterol levels |
| rs649129 | GWAS cata | rs6491[illegible] | 3.00E-14 | Low densit | Low density lipoprotein cholesterol levels |
| rs649129 | GWAS cata | rs6491[illegible] | 9.00E-60 | Low densit | Low density lipoprotein cholesterol levels |
| rs2519093 | GWAS cata | rs2519[illegible] | 3.00E-41 | Total chole | Total cholesterol levels |
| rs2519093 | GWAS cata | rs2519[illegible] | 1.00E-10 | Total chole | Total cholesterol levels |
| rs2519093 | GWAS cata | rs2519[illegible] | 2.00E-53 | Total chole | Total cholesterol levels |
| rs550057 | GWAS cata | rs5500[illegible] | 3.00E-29 | Total chole | Total cholesterol levels |
| rs550057 | GWAS cata | rs5500[illegible] | 6.00E-11 | Total chole | Total cholesterol levels |
| rs550057 | GWAS cata | rs5500[illegible] | 9.00E-42 | Total chole | Total cholesterol levels |
| rs649129 | GWAS cata | rs6491[illegible] | 9.00E-37 | Total chole | Total cholesterol levels |
| rs649129 | GWAS cata | rs6491[illegible] | 2.00E-10 | Total chole | Total cholesterol levels |
| rs649129 | GWAS cata | rs6491[illegible] | 1.00E-48 | Total chole | Total cholesterol levels |
| rs492488 | GWAS cata | rs4924[illegible] | 2.00E-21 | Blood prot | Blood protein levels |
| rs492488 | GWAS cata | rs4924[illegible] | 3.00E-21 | Blood prot | Blood protein levels |
| rs2519093 | GWAS cata | rs2519[illegible] | 7.00E-45 | Red blood [illegible] | Red blood cell count |
| rs2519093 | GWAS cata | rs2519[illegible] | 1.00E-35 | Hemoglobi | Hemoglobin |
| rs2519093 | GWAS cata | rs2519[illegible] | 1.00E-33 | Hematocrit | Hematocrit |
| rs687289 | GWAS cata | rs6872[illegible] | 9.00E-68 | Activated p | Activated partial thromboplastin time |
| rs1165522[illegible] | GWAS cata | rs1165[illegible] | 7.00E-09 | Lactate del | Lactate dehydrogenase levels |
| rs491626 | GWAS cata | rs4916[illegible] | 1.00E-09 | Creatine ki | Creatine kinase levels |
| rs492488 | GWAS cata | rs4924[illegible] | 0 | Blood prot | Blood protein levels |
| rs1165522[illegible] | GWAS cata | rs1165[illegible] | 2.00E-08 | Non-album | Non-albumin protein levels |
| rs492488 | GWAS cata | rs4924[illegible] | 4.00E-26 | Blood prot | Blood protein levels |
| rs492488 | GWAS cata | rs4924[illegible] | 7.00E-25 | Blood prot | Blood protein levels |
| rs2519093 | GWAS cata | rs2519[illegible] | 0 | Serum alka | Serum alkaline phosphatase levels |
| rs495828 | GWAS cata | rs4958[illegible] | 3.00E-08 | Angiotensi | Angiotensin-converting enzyme activity |
| rs687289 | GWAS cata | rs6872[illegible] | 1.00E-128 | Coagulatio | Coagulation factor levels |

| | | | | | |
|---|---|---|---|---|---|
| rs643434 | GWAS cata | rs64343 | 1.00E-49 | End-stage c | End-stage coagulation |
| rs495828 | GWAS cata | rs49582 | 3.00E-12 | Red blood c | Red blood cell count |
| rs495828 | GWAS cata | rs49582 | 1.00E-11 | Hematolog | Hematological and biochemical traits |
| rs495828 | GWAS cata | rs49582 | 6.00E-10 | Hematolog | Hematological and biochemical traits |
| rs495828 | GWAS cata | rs49582 | 4.00E-59 | Hematolog | Hematological and biochemical traits |
| rs649129 | GWAS cata | rs64912 | 1.00E-15 | Soluble lev | Soluble levels of adhesion molecules |
| rs495828 | GWAS cata | rs49582 | 3.00E-16 | Venous thr | Venous thromboembolism |
| rs2519093 | GWAS cata | rs2519( | 8.00E-16 | Venous thr | Venous thromboembolism |
| rs612169 | GWAS cata | rs61216 | 9.00E-40 | Metabolic t | Metabolic traits |
| rs532436 | GWAS cata | rs53243 | 2.00E-17 | Myocardial | Myocardial infarction |
| rs630510 | GWAS cata | rs63051 | 2.00E-15 | Blood prote | Blood protein levels |
| rs687289 | GWAS cata | rs68728 | 8.00E-26 | Monocyte c | Monocyte count |
| rs2519093 | GWAS cata | rs2519( | 6.00E-26 | Venous thr | Venous thromboembolism |
| rs687289 | GWAS cata | rs68728 | 4.00E-22 | Blood prote | Blood protein levels |
| rs687289 | GWAS cata | rs68728 | 1.00E-40 | Blood prote | Blood protein levels |
| rs492488 | GWAS cata | rs49248 | 1.00E-97 | Blood prote | Blood protein levels |

**Table S21. Single cis-miR-eQTLs as proxy for 85 miRNAs in PheWAS**

| cis-miR-eQTL | position | miRNA | P |
|---|---|---|---|
| rs1008940 | 13:50385906 | miR-16-1-3p | 1.16E-05 |
| rs10157552 | 1:219911743 | miR-664a-3p | 0.000135247 |
| rs10785375 | 12:43136253 | miR-7851-3p | 0.000102113 |
| rs10877863 | 12:62889402 | let-7i-5p | 3.92E-05 |
| rs10906142 | 10:12438783 | miR-548ak | 4.39E-05 |
| rs11135833 | 8:24783920 | miR-6841-3p | 5.87E-06 |
| rs11202501 | 10:89437208 | miR-4678 | 0.000978857 |
| rs112452078 | 17:6756433 | miR-324-5p | 2.50E-05 |
| rs113225019 | 11:70510169 | miR-548k | 7.51E-05 |
| rs115064507 | 2:189417638 | miR-3606-5p | 0.000379441 |
| rs115494840 | 19:51980303 | let-7e-3p | 2.41E-05 |
| rs11587425 | 1:23948561 | miR-378f | 7.01E-05 |
| rs11670465 | 19:52241939 | miR-6801-5p | 0.000100492 |
| rs11710762 | 3:18939227 | miR-4791 | 0.00116006 |
| rs117847685 | 16:11095228 | miR-548h-5p | 4.11E-05 |
| rs117858599 | 20:61149264 | miR-133a-3p | 4.17E-12 |
| rs11806098 | 1:41050054 | miR-30e-5p | 0.000175385 |
| rs1210415 | 22:22046545 | miR-301b | 0.000494798 |
| rs12325879 | 17:57627786 | miR-301a-3p | 1.77E-05 |
| rs12467772 | 2:53313031 | miR-4431 | 4.41E-05 |
| rs1254901 | 2:85811229 | miR-6071 | 0.000310415 |
| rs1263614 | 2:207960079 | miR-2355-3p | 1.17E-07 |
| rs12748921 | 1:44565618 | miR-6079 | 8.96E-05 |
| rs12881545 | 14:101176212 | miR-382-3p | 3.57E-05 |
| rs12881760 | 14:101176335 | miR-136-3p | 2.04E-06 |
| rs13336241 | 16:57776699 | miR-6772-3p | 0.000321862 |
| rs13394642 | 2:56049955 | miR-216a-5p | 7.45E-05 |
| rs1571025 | 10:6134869 | miR-3155a | 1.85E-05 |
| rs1625280 | 16:29131925 | miR-6862-5p | 0.001315438 |
| rs16916852 | 12:7281613 | miR-200c-5p | 0.000196732 |
| rs16975451 | 16:85302166 | miR-1910-3p | 6.83E-05 |
| rs1715580 | 12:116236207 | miR-620 | 0.000236304 |
| rs17190086 | 2:56435657 | miR-216b-3p | 2.39E-05 |
| rs1804527 | 7:143088823 | miR-6892-5p | 1.68E-05 |
| rs213494 | 1:54877103 | miR-4781-3p | 5.48E-05 |
| rs218001 | 11:67709097 | miR-4691-3p | 0.000116785 |
| rs2244923 | 2:207974818 | miR-2355-5p | 1.79E-05 |
| rs2270197 | 3:52824095 | miR-135a-5p | 0.000304339 |
| rs2363285 | 7:1091644 | miR-339-5p | 1.14E-08 |
| rs2527912 | 7:99500042 | miR-93-5p | 0.000900995 |
| rs2703129 | 4:77818548 | miR-4450 | 6.80E-05 |
| rs28697216 | 14:101360823 | miR-337-3p | 2.46E-05 |
| rs28718566 | 3:160223966 | miR-15b-5p | 4.26E-06 |
| rs2936558 | 12:47777059 | miR-4698 | 2.48E-05 |
| rs321336 | 11:110935811 | miR-34b-3p | 5.63E-05 |
| rs35190886 | 17:66239496 | miR-635 | 0.000611158 |
| rs36113170 | 12:128801170 | miR-4419b | 3.37E-05 |
| rs3902784 | 9:137475376 | miR-3689b-3p | 5.64E-05 |

| | | | |
|---|---|---|---|
| rs446437 | 11:36055261 | miR-3973 | 6.72E-05 |
| rs4536758 | 22:50063614 | miR-3667-5p | 7.83E-06 |
| rs4607844 | 1:228932594 | miR-6742-3p | 9.72E-06 |
| rs4795468 | 17:27184282 | miR-451a | 0.000179602 |
| rs4900479 | 14:101361403 | miR-655-5p | 0.000101087 |
| rs4906149 | 14:102301429 | miR-1247-5p | 0.000119129 |
| rs55693804 | 9:136182895 | miR-548aw | 0.000207154 |
| rs55707677 | 11:62506718 | miR-6747-3p | 6.91E-05 |
| rs55936521 | 16:70427625 | miR-140-5p | 0.000295664 |
| rs56237087 | 8:11086757 | miR-598-3p | 3.28E-05 |
| rs564547 | 21:31432541 | miR-4327 | 0.000284099 |
| rs56866369 | 17:19486592 | miR-1180-3p | 0.000502386 |
| rs5764780 | 22:45941807 | miR-4762-5p | 0.000160159 |
| rs587080 | 11:65253800 | miR-612 | 0.000245456 |
| rs62189536 | 2:239991694 | miR-4440 | 3.22E-09 |
| rs6666716 | 1:55327492 | miR-4422 | 0.000205026 |
| rs6990733 | 8:27404137 | miR-6842-3p | 6.97E-05 |
| rs7010330 | 8:145036615 | miR-661 | 9.48E-05 |
| rs7130989 | 11:47184756 | miR-5582-3p | 0.000607975 |
| rs7142266 | 14:23924068 | miR-4707-5p | 6.21E-05 |
| rs7207530 | 17:30113313 | miR-365b-5p | 0.000126514 |
| rs7212605 | 17:29880456 | miR-4733-3p | 0.000129449 |
| rs73085915 | 1:209389095 | miR-205-5p | 9.88E-05 |
| rs73693174 | 7:44365863 | miR-6837-3p | 8.12E-05 |
| rs748193 | 8:145682115 | miR-6848-3p | 0.000582796 |
| rs75293039 | 1:86715612 | miR-7856-5p | 4.83E-05 |
| rs75589560 | 8:129239338 | miR-1205 | 6.10E-06 |
| rs757594 | 17:12000632 | miR-744-5p | 0.000585712 |
| rs76203397 | 8:30100083 | miR-3148 | 5.15E-05 |
| rs76278109 | 2:102945378 | miR-4772-5p | 2.17E-05 |
| rs895819 | 19:13947292 | miR-27a-5p | 2.04E-05 |
| rs921968 | 2:219272294 | miR-26b-5p | 5.63E-05 |
| rs9417814 | 10:30907894 | miR-7162-3p | 6.36E-06 |
| rs9589298 | 13:92228446 | miR-19a-3p | 0.00010943 |
| rs9589298 | 13:92228446 | miR-19b-3p | 3.88E-05 |
| rs9691106 | 7:44351258 | miR-4649-5p | 3.12E-06 |
| rs9832953 | 3:112988284 | miR-4446-3p | 0.000357764 |

**Table S22.** FDR-significant associations for PheWAS using single variant and cis-GRS

| phenotype | snp | miRNA | beta | SE | OR | p | p.fdr |
|---|---|---|---|---|---|---|---|
| 577 | rs76278109 | miR-4772-5p | -0.10 | 0.03 | 0.90 | 7.21E-05 | 4.92E-02 |
| 939 | rs76278109 | miR-4772-5p | -0.11 | 0.03 | 0.90 | 1.09E-04 | 4.92E-02 |
| 785 | rs748193 | miR-6848-3p | 0.04 | 0.01 | 1.04 | 2.28E-05 | 2.06E-02 |
| 561 | rs748193 | miR-6848-3p | 0.05 | 0.01 | 1.05 | 6.29E-05 | 2.85E-02 |
| 401 | rs7130989 | miR-5582-3p | 0.03 | 0.01 | 1.03 | 7.16E-05 | 3.80E-02 |
| 401.1 | rs7130989 | miR-5582-3p | 0.03 | 0.01 | 1.03 | 8.40E-05 | 3.80E-02 |
| 751 | rs7130989 | miR-5582-3p | -0.17 | 0.05 | 0.84 | 1.31E-04 | 3.95E-02 |
| 389 | rs587080 | miR-612 | -0.10 | 0.02 | 0.91 | 2.96E-06 | 2.68E-03 |
| 365 | rs587080 | miR-612 | -0.08 | 0.02 | 0.93 | 1.10E-04 | 4.98E-02 |
| 401.1 | rs56237087 | miR-598-3p | -0.03 | 0.01 | 0.97 | 1.35E-05 | 6.32E-03 |
| 401 | rs56237087 | miR-598-3p | -0.03 | 0.01 | 0.97 | 1.40E-05 | 6.32E-03 |
| 401.1 | rs55936521 | miR-140-5p | 0.03 | 0.01 | 1.03 | 3.60E-06 | 1.93E-03 |
| 401 | rs55936521 | miR-140-5p | 0.03 | 0.01 | 1.03 | 4.27E-06 | 1.93E-03 |
| 740.1 | rs2270197 | miR-135a-5p | 0.04 | 0.01 | 1.04 | 7.65E-06 | 4.06E-03 |
| 366 | rs2270197 | miR-135a-5p | -0.04 | 0.01 | 0.96 | 8.97E-06 | 4.06E-03 |
| 401.1 | rs2270197 | miR-135a-5p | 0.02 | 0.01 | 1.02 | 6.68E-05 | 1.55E-02 |
| 401 | rs2270197 | miR-135a-5p | 0.02 | 0.01 | 1.02 | 6.84E-05 | 1.55E-02 |
| 740.11 | rs2270197 | miR-135a-5p | 0.06 | 0.01 | 1.06 | 1.06E-04 | 1.91E-02 |
| 351 | rs2270197 | miR-135a-5p | 0.05 | 0.01 | 1.05 | 3.66E-04 | 3.83E-02 |
| 733.4 | rs2270197 | miR-135a-5p | 0.26 | 0.07 | 1.30 | 3.91E-04 | 3.83E-02 |
| 296.1 | rs2270197 | miR-135a-5p | -0.16 | 0.05 | 0.85 | 4.02E-04 | 3.83E-02 |
| 740 | rs2270197 | miR-135a-5p | 0.03 | 0.01 | 1.03 | 4.22E-04 | 3.83E-02 |
| 300 | rs2270197 | miR-135a-5p | -0.06 | 0.02 | 0.94 | 4.23E-04 | 3.83E-02 |
| 411.4 | rs1254901 | miR-6071 | -0.06 | 0.01 | 0.94 | 2.67E-08 | 2.42E-05 |
| 411.3 | rs1254901 | miR-6071 | -0.06 | 0.01 | 0.94 | 2.13E-07 | 9.65E-05 |
| 411 | rs1254901 | miR-6071 | -0.04 | 0.01 | 0.96 | 6.51E-06 | 1.96E-03 |
| 411.1 | rs1254901 | miR-6071 | -0.09 | 0.02 | 0.91 | 1.54E-05 | 3.50E-03 |
| 411.8 | rs1254901 | miR-6071 | -0.05 | 0.01 | 0.95 | 6.55E-05 | 1.18E-02 |
| 196 | rs115494840 | let-7e-3p | 0.21 | 0.05 | 1.23 | 3.07E-05 | 2.78E-02 |
| 529.1 | grs | let-7e-5p | -0.24 | 0.06 | 0.79 | 2.29E-05 | 2.07E-02 |
| 208 | grs | miR-1908-5p | -0.04 | 0.01 | 0.96 | 1.99E-08 | 1.80E-05 |
| 574.1 | grs | miR-1908-5p | 0.04 | 0.01 | 1.05 | 1.51E-07 | 6.81E-05 |
| 574 | grs | miR-1908-5p | 0.04 | 0.01 | 1.04 | 1.68E-06 | 5.08E-04 |
| 565 | grs | miR-1908-5p | -0.03 | 0.01 | 0.97 | 9.98E-05 | 2.26E-02 |
| 427.2 | grs | miR-27b-3p | -0.03 | 0.01 | 0.97 | 1.97E-05 | 1.78E-02 |
| 594.3 | grs | miR-27b-3p | -0.08 | 0.02 | 0.92 | 4.77E-05 | 2.16E-02 |
| 585.1 | grs | miR-27b-3p | -0.05 | 0.01 | 0.95 | 1.26E-04 | 3.80E-02 |
| 594 | grs | miR-27b-3p | -0.05 | 0.01 | 0.96 | 1.74E-04 | 3.93E-02 |
| 418 | grs | miR-30d-5p | -0.20 | 0.05 | 0.82 | 1.29E-05 | 1.17E-02 |
| 411.3 | grs | miR-30d-5p | -0.03 | 0.01 | 0.97 | 4.94E-05 | 2.24E-02 |
| 580.2 | grs | miR-30d-5p | 0.13 | 0.03 | 1.14 | 1.87E-04 | 4.90E-02 |
| 411.4 | grs | miR-30d-5p | -0.03 | 0.01 | 0.97 | 2.16E-04 | 4.90E-02 |
| 172 | grs | miR-323b-3p | 0.05 | 0.01 | 1.05 | 4.79E-06 | 4.34E-03 |
| 172.1 | grs | miR-323b-3p | 0.09 | 0.02 | 1.09 | 1.79E-05 | 5.39E-03 |
| 172.11 | grs | miR-323b-3p | 0.09 | 0.02 | 1.09 | 1.79E-05 | 5.39E-03 |
| 8.6 | grs | miR-323b-3p | 0.14 | 0.04 | 1.15 | 1.46E-04 | 3.30E-02 |
| 278.1 | grs | miR-323b-3p | -0.03 | 0.01 | 0.97 | 2.28E-04 | 3.97E-02 |
| 278 | grs | miR-323b-3p | -0.03 | 0.01 | 0.97 | 2.63E-04 | 3.97E-02 |
| 172.1 | grs | miR-329-3p | 0.08 | 0.02 | 1.08 | 7.21E-05 | 3.26E-02 |
| 172.11 | grs | miR-329-3p | 0.08 | 0.02 | 1.08 | 7.21E-05 | 3.26E-02 |
| 278.1 | grs | miR-329-3p | -0.03 | 0.01 | 0.97 | 1.12E-04 | 3.38E-02 |

| | | | | | | |
|---|---|---|---|---|---|---|
| 278 grs | miR-329-3p | -0.03 | 0.01 | 0.97 | 1.77E-04 | 4.01E-02 |
| 112 grs | miR-345-5p | -0.10 | 0.02 | 0.90 | 2.09E-05 | 1.89E-02 |
| 427.9 grs | miR-370-3p | -0.06 | 0.02 | 0.94 | 4.94E-05 | 4.47E-02 |
| 172 grs | miR-376b-3p | 0.04 | 0.01 | 1.04 | 5.81E-05 | 3.89E-02 |
| 172.1 grs | miR-376b-3p | 0.08 | 0.02 | 1.08 | 1.29E-04 | 3.89E-02 |
| 172.11 grs | miR-376b-3p | 0.08 | 0.02 | 1.08 | 1.29E-04 | 3.89E-02 |
| 427.9 grs | miR-377-5p | -0.06 | 0.02 | 0.94 | 5.18E-05 | 4.69E-02 |
| 172 grs | miR-379-5p | 0.05 | 0.01 | 1.05 | 7.68E-06 | 5.25E-03 |
| 172.1 grs | miR-379-5p | 0.08 | 0.02 | 1.09 | 1.74E-05 | 5.25E-03 |
| 172.11 grs | miR-379-5p | 0.08 | 0.02 | 1.09 | 1.74E-05 | 5.25E-03 |
| 172.1 grs | miR-409-3p | 0.08 | 0.02 | 1.08 | 1.09E-04 | 4.95E-02 |
| 172.11 grs | miR-409-3p | 0.08 | 0.02 | 1.08 | 1.09E-04 | 4.95E-02 |
| 208 grs | miR-485-5p | -0.03 | 0.01 | 0.97 | 3.39E-05 | 1.82E-02 |
| 427.9 grs | miR-485-5p | -0.06 | 0.02 | 0.94 | 4.03E-05 | 1.82E-02 |
| 172.1 grs | miR-487b-3p | 0.08 | 0.02 | 1.08 | 9.66E-05 | 4.37E-02 |
| 172.11 grs | miR-487b-3p | 0.08 | 0.02 | 1.08 | 9.66E-05 | 4.37E-02 |
| 427.9 grs | miR-543 | -0.06 | 0.02 | 0.94 | 5.78E-05 | 3.60E-02 |
| 278 grs | miR-543 | -0.03 | 0.01 | 0.97 | 1.17E-04 | 3.60E-02 |
| 278.1 grs | miR-543 | -0.03 | 0.01 | 0.97 | 1.19E-04 | 3.60E-02 |
| 210 grs | miR-550a-3-5 | -0.13 | 0.03 | 0.88 | 2.33E-05 | 2.11E-02 |
| 250.2 grs | miR-8052 | 0.03 | 0.01 | 1.03 | 4.61E-05 | 4.17E-02 |
| 250 grs | miR-8052 | 0.03 | 0.01 | 1.03 | 9.29E-05 | 4.20E-02 |

| n_total | n_cases | n_controls | description | group |
|---|---|---|---|---|
| 423189 | 3182 | 420007 | Diseases of pa | digestive |
| 421308 | 2564 | 418744 | Atopic/contac | dermatologic |
| 422143 | 19563 | 402580 | Abdominal pa | symptoms |
| 379166 | 16538 | 362628 | Symptoms inv | digestive |
| 421307 | 91878 | 329429 | Hypertension | circulatory system |
| 421034 | 91605 | 329429 | Essential hype | circulatory system |
| 422492 | 1539 | 420953 | Genitourinary | congenital anomalies |
| 423147 | 5736 | 417411 | Hearing loss | sense organs |
| 417762 | 5903 | 411859 | Glaucoma | sense organs |
| 421034 | 91605 | 329429 | Essential hype | circulatory system |
| 421307 | 91878 | 329429 | Hypertension | circulatory system |
| 421034 | 91605 | 329429 | Essential hype | circulatory system |
| 421307 | 91878 | 329429 | Hypertension | circulatory system |
| 406661 | 36573 | 370088 | Osteoarthritis | musculoskeletal |
| 422897 | 24862 | 398035 | Cataract | sense organs |
| 421034 | 91605 | 329429 | Essential hype | circulatory system |
| 421307 | 91878 | 329429 | Hypertension | circulatory system |
| 380516 | 10428 | 370088 | Osteoarthrosi | musculoskeletal |
| 420033 | 13336 | 406697 | Other periphe | neurological |
| 408381 | 381 | 408000 | Aseptic necro | musculoskeletal |
| 400907 | 1134 | 399773 | Bipolar | mental disorders |
| 421525 | 51437 | 370088 | Osteoarthrosi | musculoskeletal |
| 408400 | 8627 | 399773 | Anxiety disorc | mental disorders |
| 409574 | 22992 | 386582 | Coronary athe | circulatory system |
| 404790 | 18208 | 386582 | Angina pector | circulatory system |
| 421802 | 35220 | 386582 | Ischemic Hear | circulatory system |
| 391892 | 5310 | 386582 | Unstable angi | circulatory system |
| 403438 | 16856 | 386582 | Other chronic | circulatory system |
| 400834 | 2484 | 398350 | Radiotherapy | neoplasms |
| 419548 | 329 | 419219 | Glossitis | digestive |
| 419666 | 25561 | 394105 | Benign neopla | neoplasms |
| 418782 | 15056 | 403726 | Cholelithiasis | digestive |
| 420593 | 16867 | 403726 | Cholelithiasis | digestive |
| 414389 | 17260 | 397129 | Anal and recta | digestive |
| 407651 | 18289 | 389362 | Atrial fibrillati | circulatory system |
| 418018 | 2659 | 415359 | Calculus of ur | genitourinary |
| 410823 | 6389 | 404434 | Acute renal fa | genitourinary |
| 422602 | 7243 | 415359 | Urinary calcul | genitourinary |
| 418780 | 445 | 418335 | Nonspecific ch | circulatory system |
| 404790 | 18208 | 386582 | Angina pector | circulatory system |
| 405269 | 835 | 404434 | Nephrotic syn | genitourinary |
| 409574 | 22992 | 386582 | Coronary athe | circulatory system |
| 423175 | 9322 | 413853 | Skin cancer | neoplasms |
| 416565 | 2712 | 413853 | Melanomas o | neoplasms |
| 416565 | 2712 | 413853 | Melanomas o | neoplasms |
| 419532 | 874 | 418658 | Viral Enteritis | infectious diseases |
| 422707 | 15317 | 407390 | Obesity | endocrine/metabolic |
| 422870 | 15480 | 407390 | Overweight, o | endocrine/metabolic |
| 416565 | 2712 | 413853 | Melanomas o | neoplasms |
| 416565 | 2712 | 413853 | Melanomas o | neoplasms |
| 422707 | 15317 | 407390 | Obesity | endocrine/metabolic |

| | | | | |
|---|---|---|---|---|
| 422870 | 15480 | 407390 | Overweight, o | endocrine/metabolic |
| 422600 | 1827 | 420773 | Candidiasis | infectious diseases |
| 393668 | 4306 | 389362 | Palpitations | circulatory system |
| 423175 | 9322 | 413853 | Skin cancer | neoplasms |
| 416565 | 2712 | 413853 | Melanomas of | neoplasms |
| 416565 | 2712 | 413853 | Melanomas of | neoplasms |
| 393668 | 4306 | 389362 | Palpitations | circulatory system |
| 423175 | 9322 | 413853 | Skin cancer | neoplasms |
| 416565 | 2712 | 413853 | Melanomas of | neoplasms |
| 416565 | 2712 | 413853 | Melanomas of | neoplasms |
| 416565 | 2712 | 413853 | Melanomas of | neoplasms |
| 416565 | 2712 | 413853 | Melanomas of | neoplasms |
| 419666 | 25561 | 394105 | Benign neopla | neoplasms |
| 393668 | 4306 | 389362 | Palpitations | circulatory system |
| 416565 | 2712 | 413853 | Melanomas of | neoplasms |
| 416565 | 2712 | 413853 | Melanomas of | neoplasms |
| 393668 | 4306 | 389362 | Palpitations | circulatory system |
| 422870 | 15480 | 407390 | Overweight, o | endocrine/metabolic |
| 422707 | 15317 | 407390 | Obesity | endocrine/metabolic |
| 422008 | 1041 | 420967 | Benign neopla | neoplasms |
| 421190 | 21947 | 399243 | Type 2 diabete | endocrine/metabolic |
| 422239 | 22996 | 399243 | Diabetes melli | endocrine/metabolic |

**Table S23.** Cis and extended-GRS for 17 significant associations

| miRNA | phenotype | n_cases | n_controls | description | group |
|---|---|---|---|---|---|
| let-7e-5p | 529.1 | 329 | 419219 | Glossitis | digestive |
| miR-27b-3p | 427.2 | 18289 | 389362 | Atrial fibrillati | circulatory sys |
| miR-27b-3p | 585.1 | 6389 | 404434 | Acute renal fa | genitourinary |
| miR-27b-3p | 594 | 7243 | 415359 | Urinary calcul | genitourinary |
| miR-27b-3p | 594.3 | 2659 | 415359 | Calculus of ur | genitourinary |
| miR-30d-5p | 411.3 | 18208 | 386582 | Angina pector | circulatory sys |
| miR-30d-5p | 411.4 | 22992 | 386582 | Coronary athe | circulatory sys |
| miR-30d-5p | 418 | 445 | 418335 | Nonspecific ch | circulatory sys |
| miR-30d-5p | 580.2 | 835 | 404434 | Nephrotic syn | genitourinary |
| miR-323b-3p | 172 | 9322 | 413853 | Skin cancer | neoplasms |
| miR-323b-3p | 172.1 | 2712 | 413853 | Melanomas o | neoplasms |
| miR-323b-3p | 172.11 | 2712 | 413853 | Melanomas o | neoplasms |
| miR-323b-3p | 278 | 15480 | 407390 | Overweight, o | endocrine/me |
| miR-323b-3p | 278.1 | 15317 | 407390 | Obesity | endocrine/me |
| miR-323b-3p | 8.6 | 874 | 418658 | Viral Enteritis | infectious dise |
| miR-409-3p | 172.1 | 2712 | 413853 | Melanomas o | neoplasms |
| miR-409-3p | 172.11 | 2712 | 413853 | Melanomas o | neoplasms |

| Cis-GRS | | | | Extended-GRS | | |
|---|---|---|---|---|---|---|
| beta | SE | p | p.fdr | beta | SE | p |
| -0.24 | 0.06 | 2.29E-05 | 2.07E-02 | -0.2 | 0.06 | 5.95E-04 |
| -0.03 | 0.01 | 1.97E-05 | 1.78E-02 | -0.02 | 0.01 | 4.35E-02 |
| -0.05 | 0.01 | 1.26E-04 | 3.80E-02 | -0.04 | 0.01 | 4.02E-03 |
| -0.05 | 0.01 | 1.74E-04 | 3.93E-02 | -0.04 | 0.01 | 3.35E-03 |
| -0.08 | 0.02 | 4.77E-05 | 2.16E-02 | -0.07 | 0.02 | 4.65E-04 |
| -0.03 | 0.01 | 4.94E-05 | 2.24E-02 | -0.03 | 0.01 | 3.29E-04 |
| -0.03 | 0.01 | 2.16E-04 | 4.90E-02 | -0.02 | 0.01 | 1.86E-03 |
| -0.2 | 0.05 | 1.29E-05 | 1.17E-02 | -0.17 | 0.05 | 3.83E-04 |
| 0.13 | 0.03 | 1.87E-04 | 4.90E-02 | 0.11 | 0.03 | 1.39E-03 |
| 0.05 | 0.01 | 4.79E-06 | 4.34E-03 | 0.05 | 0.01 | 1.87E-05 |
| 0.09 | 0.02 | 1.79E-05 | 5.39E-03 | 0.07 | 0.02 | 3.46E-04 |
| 0.09 | 0.02 | 1.79E-05 | 5.39E-03 | 0.07 | 0.02 | 3.46E-04 |
| -0.03 | 0.01 | 2.63E-04 | 3.97E-02 | -0.02 | 0.01 | 3.46E-03 |
| -0.03 | 0.01 | 2.28E-04 | 3.97E-02 | -0.02 | 0.01 | 3.67E-03 |
| 0.14 | 0.04 | 1.46E-04 | 3.30E-02 | 0.14 | 0.04 | 1.20E-04 |
| 0.08 | 0.02 | 1.09E-04 | 4.95E-02 | 0.08 | 0.02 | 1.46E-04 |
| 0.08 | 0.02 | 1.09E-04 | 4.95E-02 | 0.08 | 0.02 | 1.46E-04 |

| |
|---|
| p.fdr |
| 1.01E-03 |
| 4.35E-02 |
| 4.27E-03 |
| 4.16E-03 |
| 8.78E-04 |
| 8.13E-04 |
| 2.64E-03 |
| 8.13E-04 |
| 2.14E-03 |
| 3.19E-04 |
| 8.13E-04 |
| 8.13E-04 |
| 4.16E-03 |
| 4.16E-03 |
| 6.20E-04 |
| 6.20E-04 |
| 6.20E-04 |

**Table S24. Full results for cis-MR analyses**

| exposure | outcome | method | nsnp | beta | se | pval |
|---|---|---|---|---|---|---|
| let-7e-5p | 529.1 | Inverse variance weighted | 3 | -2.24 | 0.52 | 0.0000167 |
| let-7e-5p | 529.1 | IVW with correlation matrix | 3 | -2.26 | 0.54 | 0.0000325 |
| let-7e-5p | 529.1 | MR Egger | 3 | -5.03 | 2.22 | 0.265 |
| let-7e-5p | 529.1 | Weighted median | 3 | -2.55 | 0.79 | 0.00129 |
| miR-1908-5p | 208 | Inverse variance weighted | 3 | -0.41 | 0.07 | 3.97E-10 |
| miR-1908-5p | 208 | IVW with correlation matrix | 3 | -0.41 | 0.06 | 1.57E-12 |
| miR-1908-5p | 208 | MR Egger | 3 | 0.27 | 0.48 | 0.669 |
| miR-1908-5p | 208 | Weighted median | 3 | -0.37 | 0.1 | 0.000177 |
| miR-1908-5p | 565 | Inverse variance weighted | 3 | -0.33 | 0.08 | 0.0000122 |
| miR-1908-5p | 565 | IVW with correlation matrix | 3 | -0.33 | 0.07 | 0.00000142 |
| miR-1908-5p | 565 | MR Egger | 3 | -0.04 | 0.57 | 0.957 |
| miR-1908-5p | 565 | Weighted median | 3 | -0.35 | 0.1 | 0.000354 |
| miR-1908-5p | 574 | Inverse variance weighted | 3 | 0.41 | 0.08 | 0.000000104 |
| miR-1908-5p | 574 | IVW with correlation matrix | 3 | 0.41 | 0.07 | 3.97E-09 |
| miR-1908-5p | 574 | MR Egger | 3 | 0.02 | 0.57 | 0.975 |
| miR-1908-5p | 574 | Weighted median | 3 | 0.42 | 0.1 | 0.0000483 |
| miR-1908-5p | 574.1 | Inverse variance weighted | 3 | 0.48 | 0.08 | 3.51E-09 |
| miR-1908-5p | 574.1 | IVW with correlation matrix | 3 | 0.48 | 0.07 | 6.44E-11 |
| miR-1908-5p | 574.1 | MR Egger | 3 | 0.07 | 0.6 | 0.925 |
| miR-1908-5p | 574.1 | Weighted median | 3 | 0.49 | 0.11 | 0.000016 |
| miR-30d-5p | 411.3 | Inverse variance weighted | 5 | -0.47 | 0.1 | 0.00000146 |
| miR-30d-5p | 411.3 | IVW with correlation matrix | 5 | -0.46 | 0.13 | 0.000338 |
| miR-30d-5p | 411.3 | MR Egger | 5 | -0.09 | 0.3 | 0.785 |
| miR-30d-5p | 411.3 | Weighted median | 5 | -0.41 | 0.13 | 0.00237 |
| miR-30d-5p | 411.4 | Inverse variance weighted | 5 | -0.39 | 0.09 | 0.00000755 |
| miR-30d-5p | 411.4 | IVW with correlation matrix | 5 | -0.37 | 0.12 | 0.00175 |
| miR-30d-5p | 411.4 | MR Egger | 5 | -0.01 | 0.27 | 0.971 |
| miR-30d-5p | 411.4 | Weighted median | 5 | -0.29 | 0.12 | 0.0135 |
| miR-30d-5p | 418 | Inverse variance weighted | 5 | -3.07 | 0.56 | 4.95E-08 |
| miR-30d-5p | 418 | IVW with correlation matrix | 5 | -3.04 | 0.71 | 0.0000195 |
| miR-30d-5p | 418 | MR Egger | 5 | -2.88 | 1.86 | 0.219 |
| miR-30d-5p | 418 | Weighted median | 5 | -3.01 | 0.73 | 0.0000346 |
| miR-30d-5p | 580.2 | Inverse variance weighted | 5 | 1.91 | 0.41 | 0.00000396 |
| miR-30d-5p | 580.2 | IVW with correlation matrix | 5 | 1.83 | 0.52 | 0.000386 |
| miR-30d-5p | 580.2 | MR Egger | 5 | 2.76 | 1.4 | 0.143 |
| miR-30d-5p | 580.2 | Weighted median | 5 | 1.67 | 0.52 | 0.00138 |
| miR-323b-3p | 172 | Inverse variance weighted | 6 | 0.26 | 0.05 | 0.000000218 |
| miR-323b-3p | 172 | IVW with correlation matrix | 6 | 0.25 | 0.04 | 1.23E-08 |
| miR-323b-3p | 172 | MR Egger | 6 | 0.25 | 0.23 | 0.348 |
| miR-323b-3p | 172 | Weighted median | 6 | 0.26 | 0.07 | 0.0000938 |
| miR-323b-3p | 172.1 | Inverse variance weighted | 6 | 0.45 | 0.08 | 0.000000142 |
| miR-323b-3p | 172.1 | IVW with correlation matrix | 6 | 0.45 | 0.07 | 2.16E-10 |
| miR-323b-3p | 172.1 | MR Egger | 6 | 0.66 | 0.36 | 0.138 |
| miR-323b-3p | 172.1 | Weighted median | 6 | 0.45 | 0.11 | 0.0000772 |
| miR-323b-3p | 172.11 | Inverse variance weighted | 6 | 0.45 | 0.08 | 0.000000142 |
| miR-323b-3p | 172.11 | IVW with correlation matrix | 6 | 0.45 | 0.07 | 2.16E-10 |
| miR-323b-3p | 172.11 | MR Egger | 6 | 0.66 | 0.36 | 0.138 |
| miR-323b-3p | 172.11 | Weighted median | 6 | 0.45 | 0.11 | 0.0000433 |
| miR-323b-3p | 278 | Inverse variance weighted | 6 | -0.15 | 0.03 | 0.00000853 |
| miR-323b-3p | 278 | IVW with correlation matrix | 6 | -0.16 | 0.03 | 2.23E-08 |
| miR-323b-3p | 278 | MR Egger | 6 | -0.21 | 0.14 | 0.21 |
| miR-323b-3p | 278 | Weighted median | 6 | -0.16 | 0.04 | 0.000224 |
| miR-323b-3p | 278.1 | Inverse variance weighted | 6 | -0.16 | 0.03 | 0.0000066 |
| miR-323b-3p | 278.1 | IVW with correlation matrix | 6 | -0.16 | 0.03 | 1.85E-08 |
| miR-323b-3p | 278.1 | MR Egger | 6 | -0.21 | 0.14 | 0.214 |
| miR-323b-3p | 278.1 | Weighted median | 6 | -0.16 | 0.05 | 0.000347 |
| miR-323b-3p | 8.6 | Inverse variance weighted | 6 | 0.72 | 0.15 | 0.00000172 |
| miR-323b-3p | 8.6 | IVW with correlation matrix | 6 | 0.75 | 0.13 | 3.21E-09 |

| | | | | | | |
|---|---|---|---|---|---|---|
| miR-323b-3p | 8.6 | MR Egger | 6 | 0.49 | 0.64 | 0.482 |
| miR-323b-3p | 8.6 | Weighted median | 6 | 0.7 | 0.2 | 0.000399 |
| miR-329-3p | 172.1 | Inverse variance weighted | 10 | 0.37 | 0.06 | 5.44E-09 |
| miR-329-3p | 172.1 | IVW with correlation matrix | 10 | 0.37 | 0.09 | 0.000102 |
| miR-329-3p | 172.1 | MR Egger | 10 | 0.49 | 0.21 | 0.052 |
| miR-329-3p | 172.1 | Weighted median | 10 | 0.36 | 0.09 | 0.000131 |
| miR-329-3p | 172.11 | Inverse variance weighted | 10 | 0.37 | 0.06 | 5.44E-09 |
| miR-329-3p | 172.11 | IVW with correlation matrix | 10 | 0.37 | 0.09 | 0.000102 |
| miR-329-3p | 172.11 | MR Egger | 10 | 0.49 | 0.21 | 0.052 |
| miR-329-3p | 172.11 | Weighted median | 10 | 0.36 | 0.09 | 0.000125 |
| miR-329-3p | 278 | Inverse variance weighted | 10 | -0.15 | 0.03 | 0.000000151 |
| miR-329-3p | 278 | IVW with correlation matrix | 10 | -0.14 | 0.04 | 0.000188 |
| miR-329-3p | 278 | MR Egger | 10 | -0.26 | 0.09 | 0.0217 |
| miR-329-3p | 278 | Weighted median | 10 | -0.14 | 0.04 | 0.000173 |
| miR-329-3p | 278.1 | Inverse variance weighted | 10 | -0.15 | 0.03 | 0.000000039 |
| miR-329-3p | 278.1 | IVW with correlation matrix | 10 | -0.15 | 0.04 | 0.000105 |
| miR-329-3p | 278.1 | MR Egger | 10 | -0.26 | 0.09 | 0.019 |
| miR-329-3p | 278.1 | Weighted median | 10 | -0.15 | 0.04 | 0.0000998 |
| miR-345-5p | 112 | Inverse variance weighted | 3 | -1.11 | 0.25 | 0.00000638 |
| miR-345-5p | 112 | IVW with correlation matrix | 3 | -1.05 | 0.24 | 0.00000996 |
| miR-345-5p | 112 | MR Egger | 3 | -1.3 | 1.15 | 0.461 |
| miR-345-5p | 112 | Weighted median | 3 | -1.08 | 0.31 | 0.00042 |
| miR-370-3p | 427.9 | Inverse variance weighted | 13 | -0.23 | 0.04 | 4.34E-10 |
| miR-370-3p | 427.9 | IVW with correlation matrix | 13 | -0.23 | 0.02 | 1.39E-20 |
| miR-370-3p | 427.9 | MR Egger | 13 | -0.24 | 0.1 | 0.038 |
| miR-370-3p | 427.9 | Weighted median | 13 | -0.22 | 0.05 | 0.000017 |
| miR-376b-3p | 172 | Inverse variance weighted | 8 | 0.22 | 0.04 | 7.02E-10 |
| miR-376b-3p | 172 | IVW with correlation matrix | 8 | 0.2 | 0.03 | 1.15E-10 |
| miR-376b-3p | 172 | MR Egger | 8 | 0.08 | 0.1 | 0.464 |
| miR-376b-3p | 172 | Weighted median | 8 | 0.19 | 0.05 | 0.0000897 |
| miR-376b-3p | 172.1 | Inverse variance weighted | 8 | 0.39 | 0.07 | 3.05E-08 |
| miR-376b-3p | 172.1 | IVW with correlation matrix | 8 | 0.4 | 0.07 | 1.39E-09 |
| miR-376b-3p | 172.1 | MR Egger | 8 | 0.31 | 0.21 | 0.185 |
| miR-376b-3p | 172.1 | Weighted median | 8 | 0.4 | 0.1 | 0.0000294 |
| miR-376b-3p | 172.11 | Inverse variance weighted | 8 | 0.39 | 0.07 | 3.05E-08 |
| miR-376b-3p | 172.11 | IVW with correlation matrix | 8 | 0.4 | 0.07 | 1.39E-09 |
| miR-376b-3p | 172.11 | MR Egger | 8 | 0.31 | 0.21 | 0.185 |
| miR-376b-3p | 172.11 | Weighted median | 8 | 0.4 | 0.1 | 0.0000231 |
| miR-377-5p | 427.9 | Inverse variance weighted | 6 | -0.29 | 0.06 | 0.000000442 |
| miR-377-5p | 427.9 | IVW with correlation matrix | 6 | -0.26 | 0.07 | 0.000192 |
| miR-377-5p | 427.9 | MR Egger | 6 | -0.21 | 0.18 | 0.315 |
| miR-377-5p | 427.9 | Weighted median | 6 | -0.33 | 0.08 | 0.0000311 |
| miR-379-5p | 172 | Inverse variance weighted | 8 | 0.27 | 0.05 | 7.68E-08 |
| miR-379-5p | 172 | IVW with correlation matrix | 8 | 0.26 | 0.04 | 5.83E-12 |
| miR-379-5p | 172 | MR Egger | 8 | 0.31 | 0.17 | 0.122 |
| miR-379-5p | 172 | Weighted median | 8 | 0.22 | 0.07 | 0.00105 |
| miR-379-5p | 172.1 | Inverse variance weighted | 8 | 0.47 | 0.11 | 0.0000146 |
| miR-379-5p | 172.1 | IVW with correlation matrix | 8 | 0.5 | 0.1 | 0.000000234 |
| miR-379-5p | 172.1 | MR Egger | 8 | 0.71 | 0.37 | 0.108 |
| miR-379-5p | 172.1 | Weighted median | 8 | 0.49 | 0.12 | 0.000102 |
| miR-379-5p | 172.11 | Inverse variance weighted | 8 | 0.47 | 0.11 | 0.0000146 |
| miR-379-5p | 172.11 | IVW with correlation matrix | 8 | 0.5 | 0.1 | 0.000000234 |
| miR-379-5p | 172.11 | MR Egger | 8 | 0.71 | 0.37 | 0.108 |
| miR-379-5p | 172.11 | Weighted median | 8 | 0.49 | 0.12 | 0.000102 |
| miR-409-3p | 172.1 | Inverse variance weighted | 12 | 0.25 | 0.05 | 0.00000767 |
| miR-409-3p | 172.1 | IVW with correlation matrix | 12 | 0.23 | 0.04 | 0.000000187 |
| miR-409-3p | 172.1 | MR Egger | 12 | 0.17 | 0.12 | 0.177 |
| miR-409-3p | 172.1 | Weighted median | 12 | 0.25 | 0.07 | 0.000588 |
| miR-409-3p | 172.11 | Inverse variance weighted | 12 | 0.25 | 0.05 | 0.00000767 |
| miR-409-3p | 172.11 | IVW with correlation matrix | 12 | 0.23 | 0.04 | 0.000000187 |
| miR-409-3p | 172.11 | MR Egger | 12 | 0.17 | 0.12 | 0.177 |

| | | | | | | |
|---|---|---|---|---|---|---|
| miR-409-3p | 172.11 | Weighted median | 12 | 0.25 | 0.07 | 0.000374 |
| miR-485-5p | 208 | Inverse variance weighted | 6 | -0.15 | 0.03 | 0.000000203 |
| miR-485-5p | 208 | IVW with correlation matrix | 6 | -0.15 | 0.02 | 3.06E-11 |
| miR-485-5p | 208 | MR Egger | 6 | -0.19 | 0.09 | 0.106 |
| miR-485-5p | 208 | Weighted median | 6 | -0.13 | 0.04 | 0.00171 |
| miR-485-5p | 427.9 | Inverse variance weighted | 6 | -0.35 | 0.07 | 0.000000407 |
| miR-485-5p | 427.9 | IVW with correlation matrix | 6 | -0.38 | 0.05 | 4.54E-13 |
| miR-485-5p | 427.9 | MR Egger | 6 | -0.41 | 0.21 | 0.129 |
| miR-485-5p | 427.9 | Weighted median | 6 | -0.32 | 0.1 | 0.00127 |
| miR-487b-3p | 172.1 | Inverse variance weighted | 9 | 0.33 | 0.06 | 6.59E-08 |
| miR-487b-3p | 172.1 | IVW with correlation matrix | 9 | 0.35 | 0.05 | 1.23E-12 |
| miR-487b-3p | 172.1 | MR Egger | 9 | 0.28 | 0.17 | 0.145 |
| miR-487b-3p | 172.1 | Weighted median | 9 | 0.29 | 0.09 | 0.000651 |
| miR-487b-3p | 172.11 | Inverse variance weighted | 9 | 0.33 | 0.06 | 6.59E-08 |
| miR-487b-3p | 172.11 | IVW with correlation matrix | 9 | 0.35 | 0.05 | 1.23E-12 |
| miR-487b-3p | 172.11 | MR Egger | 9 | 0.28 | 0.17 | 0.145 |
| miR-487b-3p | 172.11 | Weighted median | 9 | 0.29 | 0.08 | 0.000465 |
| miR-543 | 278 | Inverse variance weighted | 8 | -0.15 | 0.03 | 9.54E-09 |
| miR-543 | 278 | IVW with correlation matrix | 8 | -0.13 | 0.02 | 3.48E-09 |
| miR-543 | 278 | MR Egger | 8 | -0.13 | 0.08 | 0.175 |
| miR-543 | 278 | Weighted median | 8 | -0.14 | 0.03 | 0.0000452 |
| miR-543 | 278.1 | Inverse variance weighted | 8 | -0.15 | 0.03 | 1.07E-08 |
| miR-543 | 278.1 | IVW with correlation matrix | 8 | -0.13 | 0.02 | 2.12E-09 |
| miR-543 | 278.1 | MR Egger | 8 | -0.13 | 0.08 | 0.157 |
| miR-543 | 278.1 | Weighted median | 8 | -0.14 | 0.04 | 0.000068 |
| miR-543 | 427.9 | Inverse variance weighted | 8 | -0.28 | 0.05 | 2.34E-09 |
| miR-543 | 427.9 | IVW with correlation matrix | 8 | -0.3 | 0.04 | 5.53E-14 |
| miR-543 | 427.9 | MR Egger | 8 | -0.42 | 0.15 | 0.0301 |
| miR-543 | 427.9 | Weighted median | 8 | -0.27 | 0.07 | 0.0000434 |

nsnp: number of genetic instruments. IVW: inverse variance weighted

| description |
|---|
| Glossitis |
| Glossitis |
| Glossitis |
| Glossitis |
| Benign neoplasm of colon |
| Benign neoplasm of colon |
| Benign neoplasm of colon |
| Benign neoplasm of colon |
| Anal and rectal conditions |
| Anal and rectal conditions |
| Anal and rectal conditions |
| Anal and rectal conditions |
| Cholelithiasis and cholecystitis |
| Cholelithiasis and cholecystitis |
| Cholelithiasis and cholecystitis |
| Cholelithiasis and cholecystitis |
| Cholelithiasis |
| Cholelithiasis |
| Cholelithiasis |
| Cholelithiasis |
| Angina pectoris |
| Angina pectoris |
| Angina pectoris |
| Angina pectoris |
| Coronary atherosclerosis |
| Coronary atherosclerosis |
| Coronary atherosclerosis |
| Coronary atherosclerosis |
| Nonspecific chest pain |
| Nonspecific chest pain |
| Nonspecific chest pain |
| Nonspecific chest pain |
| Nephrotic syndrome without mention of glomerulonephritis |
| Nephrotic syndrome without mention of glomerulonephritis |
| Nephrotic syndrome without mention of glomerulonephritis |
| Nephrotic syndrome without mention of glomerulonephritis |
| Skin cancer |
| Skin cancer |
| Skin cancer |
| Skin cancer |
| Melanomas of skin, dx or hx |
| Melanomas of skin, dx or hx |
| Melanomas of skin, dx or hx |
| Melanomas of skin, dx or hx |
| Melanomas of skin |
| Melanomas of skin |
| Melanomas of skin |
| Melanomas of skin |
| Overweight, obesity and other hyperalimentation |
| Overweight, obesity and other hyperalimentation |
| Overweight, obesity and other hyperalimentation |
| Overweight, obesity and other hyperalimentation |
| Obesity |
| Obesity |
| Obesity |
| Obesity |
| Viral Enteritis |
| Viral Enteritis |

Viral Enteritis
Viral Enteritis
Melanomas of skin, dx or hx
Melanomas of skin, dx or hx
Melanomas of skin, dx or hx
Melanomas of skin, dx or hx
Melanomas of skin
Melanomas of skin
Melanomas of skin
Melanomas of skin
Overweight, obesity and other hyperalimentation
Overweight, obesity and other hyperalimentation
Overweight, obesity and other hyperalimentation
Overweight, obesity and other hyperalimentation
Obesity
Obesity
Obesity
Obesity
Candidiasis
Candidiasis
Candidiasis
Candidiasis
Palpitations
Palpitations
Palpitations
Palpitations
Skin cancer
Skin cancer
Skin cancer
Skin cancer
Melanomas of skin, dx or hx
Melanomas of skin, dx or hx
Melanomas of skin, dx or hx
Melanomas of skin, dx or hx
Melanomas of skin
Melanomas of skin
Melanomas of skin
Melanomas of skin
Palpitations
Palpitations
Palpitations
Palpitations
Skin cancer
Skin cancer
Skin cancer
Skin cancer
Melanomas of skin, dx or hx
Melanomas of skin, dx or hx
Melanomas of skin, dx or hx
Melanomas of skin, dx or hx
Melanomas of skin
Melanomas of skin
Melanomas of skin
Melanomas of skin
Melanomas of skin, dx or hx
Melanomas of skin, dx or hx
Melanomas of skin, dx or hx
Melanomas of skin, dx or hx
Melanomas of skin
Melanomas of skin
Melanomas of skin

Melanomas of skin
Benign neoplasm of colon
Benign neoplasm of colon
Benign neoplasm of colon
Benign neoplasm of colon
Palpitations
Palpitations
Palpitations
Palpitations
Melanomas of skin, dx or hx
Melanomas of skin, dx or hx
Melanomas of skin, dx or hx
Melanomas of skin, dx or hx
Melanomas of skin
Melanomas of skin
Melanomas of skin
Melanomas of skin
Overweight, obesity and other hyperalimentation
Overweight, obesity and other hyperalimentation
Overweight, obesity and other hyperalimentation
Overweight, obesity and other hyperalimentation
Obesity
Obesity
Obesity
Obesity
Palpitations
Palpitations
Palpitations
Palpitations

**Table S25. Full results for cis-MR analyses**

| exposure | outcome | method | nsnp | b | SE | P |
|---|---|---|---|---|---|---|
| miR-323b-3 | 8.6 | Inverse variance weighted | 7 | 0.61 | 0.13 | 3.22E-06 |
| miR-323b-3 | 8.6 | MR Egger | 7 | 0.33 | 0.63 | 6.24E-01 |
| miR-323b-3 | 8.6 | Weighted median | 7 | 0.67 | 0.18 | 1.99E-04 |
| miR-323b-3 | 172 | Inverse variance weighted | 7 | 0.21 | 0.06 | 3.39E-04 |
| miR-323b-3 | 172 | MR Egger | 7 | 0.17 | 0.29 | 5.87E-01 |
| miR-323b-3 | 172 | Weighted median | 7 | 0.17 | 0.06 | 7.88E-03 |
| miR-323b-3 | 172.1 | Inverse variance weighted | 7 | 0.31 | 0.11 | 4.67E-03 |
| miR-323b-3 | 172.1 | MR Egger | 7 | 0.45 | 0.56 | 4.62E-01 |
| miR-323b-3 | 172.1 | Weighted median | 7 | 0.4 | 0.11 | 2.28E-04 |
| miR-409-3p | 172.1 | Inverse variance weighted | 13 | 0.24 | 0.05 | 3.31E-06 |
| miR-409-3p | 172.1 | MR Egger | 13 | 0.16 | 0.11 | 1.76E-01 |
| miR-409-3p | 172.1 | Weighted median | 13 | 0.23 | 0.07 | 8.07E-04 |
| miR-323b-3 | 172.11 | Inverse variance weighted | 7 | 0.31 | 0.11 | 4.67E-03 |
| miR-323b-3 | 172.11 | MR Egger | 7 | 0.45 | 0.56 | 4.62E-01 |
| miR-323b-3 | 172.11 | Weighted median | 7 | 0.4 | 0.11 | 2.28E-04 |
| miR-409-3p | 172.11 | Inverse variance weighted | 13 | 0.24 | 0.05 | 3.31E-06 |
| miR-409-3p | 172.11 | MR Egger | 13 | 0.16 | 0.11 | 1.76E-01 |
| miR-409-3p | 172.11 | Weighted median | 13 | 0.23 | 0.07 | 6.44E-04 |
| miR-323b-3 | 278 | Inverse variance weighted | 7 | -0.11 | 0.04 | 1.67E-02 |
| miR-323b-3 | 278 | MR Egger | 7 | -0.15 | 0.22 | 5.37E-01 |
| miR-323b-3 | 278 | Weighted median | 7 | -0.13 | 0.05 | 3.93E-03 |
| miR-323b-3 | 278.1 | Inverse variance weighted | 7 | -0.11 | 0.05 | 1.98E-02 |
| miR-323b-3 | 278.1 | MR Egger | 7 | -0.14 | 0.23 | 5.59E-01 |
| miR-323b-3 | 278.1 | Weighted median | 7 | -0.14 | 0.05 | 4.34E-03 |
| miR-30d-5p | 411.3 | Inverse variance weighted | 6 | -0.34 | 0.12 | 6.04E-03 |
| miR-30d-5p | 411.3 | MR Egger | 6 | 0.18 | 0.42 | 6.91E-01 |
| miR-30d-5p | 411.3 | Weighted median | 6 | -0.35 | 0.12 | 5.27E-03 |
| miR-30d-5p | 411.4 | Inverse variance weighted | 6 | -0.26 | 0.11 | 1.82E-02 |
| miR-30d-5p | 411.4 | MR Egger | 6 | 0.24 | 0.37 | 5.60E-01 |
| miR-30d-5p | 411.4 | Weighted median | 6 | -0.24 | 0.11 | 3.17E-02 |
| miR-30d-5p | 418 | Inverse variance weighted | 6 | -2.03 | 0.82 | 1.38E-02 |
| miR-30d-5p | 418 | MR Egger | 6 | -0.51 | 3.2 | 8.80E-01 |
| miR-30d-5p | 418 | Weighted median | 6 | -2.72 | 0.74 | 2.37E-04 |
| let-7e-5p | 529.1 | Inverse variance weighted | 4 | -1.25 | 0.6 | 3.86E-02 |
| let-7e-5p | 529.1 | MR Egger | 4 | 0.04 | 2.2 | 9.86E-01 |
| let-7e-5p | 529.1 | Weighted median | 4 | -0.74 | 0.48 | 1.23E-01 |
| miR-30d-5p | 580.2 | Inverse variance weighted | 6 | 1.32 | 0.5 | 8.86E-03 |
| miR-30d-5p | 580.2 | MR Egger | 6 | 1.31 | 2.08 | 5.61E-01 |
| miR-30d-5p | 580.2 | Weighted median | 6 | 1.53 | 0.51 | 2.53E-03 |

nsnp: number of genetic instruments, SE: standard error.

| description |
| --- |
| Viral Enteritis |
| Viral Enteritis |
| Viral Enteritis |
| Skin cancer |
| Skin cancer |
| Skin cancer |
| Melanomas of skin, dx or hx |
| Melanomas of skin, dx or hx |
| Melanomas of skin, dx or hx |
| Melanomas of skin, dx or hx |
| Melanomas of skin, dx or hx |
| Melanomas of skin, dx or hx |
| Melanomas of skin |
| Melanomas of skin |
| Melanomas of skin |
| Melanomas of skin |
| Melanomas of skin |
| Melanomas of skin |
| Overweight, obesity and other hyperalimentation |
| Overweight, obesity and other hyperalimentation |
| Overweight, obesity and other hyperalimentation |
| Obesity |
| Obesity |
| Obesity |
| Angina pectoris |
| Angina pectoris |
| Angina pectoris |
| Coronary atherosclerosis |
| Coronary atherosclerosis |
| Coronary atherosclerosis |
| Nonspecific chest pain |
| Nonspecific chest pain |
| Nonspecific chest pain |
| Glossitis |
| Glossitis |
| Glossitis |
| Nephrotic syndrome without mention of glomerulonephritis |
| Nephrotic syndrome without mention of glomerulonephritis |
| Nephrotic syndrome without mention of glomerulonephritis |

**Table S26**. Full results for replication using large GWAS summary statistics

| exposure | outcome | method | nsnp | b | se | pval |
|---|---|---|---|---|---|---|
| miR-30d-5p | CAD | MR Egger | 5 | 0.07 | 0.24 | 8.01E-01 |
| miR-30d-5p | CAD | Weighted median | 5 | -0.06 | 0.08 | 4.47E-01 |
| miR-30d-5p | CAD | Inverse variance weighted | 5 | -0.04 | 0.07 | 5.35E-01 |
| miR-323b-3 | BMI | MR Egger | 5 | -0.05 | 0.11 | 7.13E-01 |
| miR-323b-3 | BMI | Weighted median | 5 | 0.01 | 0.01 | 1.31E-01 |
| miR-323b-3 | BMI | Inverse variance weighted | 5 | 0 | 0.01 | 7.37E-01 |
| miR-323b-3 | WHR | MR Egger | 5 | 0.07 | 0.14 | 6.67E-01 |
| miR-323b-3 | WHR | Weighted median | 5 | -0.01 | 0.02 | 6.36E-01 |
| miR-323b-3 | WHR | Inverse variance weighted | 5 | -0.02 | 0.02 | 3.22E-01 |
| miR-329-3p | BMI | MR Egger | 6 | -0.11 | 0.04 | 4.80E-02 |
| miR-329-3p | BMI | Weighted median | 6 | -0.02 | 0.01 | 4.64E-03 |
| miR-329-3p | BMI | Inverse variance weighted | 6 | -0.03 | 0.01 | 1.89E-02 |
| miR-329-3p | WHR | MR Egger | 7 | 0 | 0.04 | 9.68E-01 |
| miR-329-3p | WHR | Weighted median | 7 | -0.02 | 0.01 | 1.34E-01 |
| miR-329-3p | WHR | Inverse variance weighted | 7 | -0.02 | 0.01 | 1.15E-01 |
| miR-543 | BMI | MR Egger | 7 | -0.03 | 0.04 | 4.85E-01 |
| miR-543 | BMI | Weighted median | 7 | -0.02 | 0.01 | 1.45E-02 |
| miR-543 | BMI | Inverse variance weighted | 7 | -0.02 | 0.01 | 5.19E-02 |
| miR-543 | WHR | MR Egger | 7 | -0.02 | 0.04 | 5.95E-01 |
| miR-543 | WHR | Weighted median | 7 | -0.03 | 0.01 | 2.92E-02 |
| miR-543 | WHR | Inverse variance weighted | 7 | -0.02 | 0.01 | 1.72E-02 |

Nsnp: number of genetic instruments,

P-het: p-value for heterogeneity of MR-Inverse variance weighted (IVW) estimates.

FDR: false discovery rate, calculated based on MR-IVW p-values.

Egger int P; p-value for MR Egger intercept,

CAD: coronary artery disease, BMI: body mass index, WHR: waist-to-hip ratio.

| P-het | Egger int P | FDR-adj P |
|---|---|---|
| 8.56E-01 | 6.66E-01 | 6.24E-01 |
| 8.56E-01 | 6.66E-01 | 6.24E-01 |
| 8.56E-01 | 6.66E-01 | 6.24E-01 |
| 1.18E-03 | 7.37E-01 | 7.37E-01 |
| 1.18E-03 | 7.37E-01 | 7.37E-01 |
| 1.18E-03 | 7.37E-01 | 7.37E-01 |
| 3.34E-01 | 5.93E-01 | 4.51E-01 |
| 3.34E-01 | 5.93E-01 | 4.51E-01 |
| 3.34E-01 | 5.93E-01 | 4.51E-01 |
| 4.88E-07 | 9.96E-02 | 6.61E-02 |
| 4.88E-07 | 9.96E-02 | 6.61E-02 |
| 4.88E-07 | 9.96E-02 | 6.61E-02 |
| 7.21E-01 | 6.93E-01 | 2.01E-01 |
| 7.21E-01 | 6.93E-01 | 2.01E-01 |
| 7.21E-01 | 6.93E-01 | 2.01E-01 |
| 1.39E-08 | 8.12E-01 | 1.21E-01 |
| 1.39E-08 | 8.12E-01 | 1.21E-01 |
| 1.39E-08 | 8.12E-01 | 1.21E-01 |
| 7.73E-01 | 9.01E-01 | 6.61E-02 |
| 7.73E-01 | 9.01E-01 | 6.61E-02 |
| 7.73E-01 | 9.01E-01 | 6.61E-02 |

**Table S27.** Reverse-MR to test association of obesity traits with miR-543 and miR-329-3p

| exposure | outcome | method | nsnp | beta | SE | P |
|---|---|---|---|---|---|---|
| BMI | miR-543 | MR Egger | 501 | 0.14 | 0.24 | 0.557 |
| BMI | miR-543 | Weighted median | 501 | 0.06 | 0.15 | 0.703 |
| BMI | miR-543 | Inverse variance weighted | 501 | 0 | 0.09 | 0.999 |
| BMI | miR-329-3p | MR Egger | 501 | 0.22 | 0.23 | 0.46 |
| BMI | miR-329-3p | Weighted median | 501 | -0.06 | 0.15 | 0.504 |
| BMI | miR-329-3p | Inverse variance weighted | 501 | -0.08 | 0.09 | 0.347 |
| WHR | miR-543 | MR Egger | 28 | -0.96 | 1.09 | 0.679 |
| WHR | miR-543 | Weighted median | 28 | -0.2 | 0.34 | 0.385 |
| WHR | miR-543 | Inverse variance weighted | 28 | -0.22 | 0.24 | 0.541 |
| WHR | miR-329-3p | MR Egger | 28 | 0.58 | 1.18 | 0.607 |
| WHR | miR-329-3p | Weighted median | 28 | -0.28 | 0.35 | 0.386 |
| WHR | miR-329-3p | Inverse variance weighted | 28 | -0.04 | 0.25 | 0.55 |

SE: standard error; WHR: waist to hip ratio; BMI: body mass index

**Table S28**. Target genes of miR-543 and miR-329-3p linked to BMI/WHR

| gene_name | MTI | miRNA |
|---|---|---|
| AAK1 | validated | miR-329-3p |
| ACAP2 | validated | miR-329-3p |
| ADCY9 | predicted | miR-329-3p |
| ALDOA | predicted | miR-329-3p |
| AP3B1 | predicted | miR-329-3p |
| ARF3 | validated | miR-329-3p |
| ARL8A | predicted | miR-329-3p |
| ASXL3 | predicted | miR-329-3p |
| ATP1A3 | validated | miR-329-3p |
| ATP2A2 | validated | miR-329-3p |
| BACH1 | validated | miR-329-3p |
| BCL7A | predicted | miR-329-3p |
| BNC2 | validated | miR-329-3p |
| BSN | validated | miR-329-3p |
| C6orf106 | validated | miR-329-3p |
| CADM1 | predicted | miR-329-3p |
| CASZ1 | predicted | miR-329-3p |
| CD19 | validated | miR-329-3p |
| CISD2 | validated | miR-329-3p |
| CNNM2 | predicted | miR-329-3p |
| COBLL1 | validated | miR-329-3p |
| CPEB4 | predicted | miR-329-3p |
| CPSF6 | validated | miR-329-3p |
| CRB2 | validated | miR-329-3p |
| CSMD2 | validated | miR-329-3p |
| DDX19A | predicted | miR-329-3p |
| DDX19B | predicted | miR-329-3p |
| DLGAP4 | predicted | miR-329-3p |
| DNMT3A | predicted | miR-329-3p |
| DPYSL5 | predicted | miR-329-3p |
| EIF4G1 | predicted | miR-329-3p |
| FAM178A | predicted | miR-329-3p |
| FAM212B | validated | miR-329-3p |
| FAM47E-STBD1 | validated | miR-329-3p |
| FAM53C | predicted | miR-329-3p |
| FBXL18 | validated | miR-329-3p |
| G3BP1 | validated | miR-329-3p |
| G3BP1 | predicted | miR-329-3p |
| GALNT10 | validated | miR-329-3p |
| GATAD2A | predicted | miR-329-3p |
| GRIN2A | validated | miR-329-3p |
| HMGCR | validated | miR-329-3p |
| HPR | predicted | miR-329-3p |
| HS6ST3 | validated | miR-329-3p |
| IGF1R | validated | miR-329-3p |
| JAKMIP3 | validated | miR-329-3p |
| KCNJ12 | validated | miR-329-3p |
| KCNJ3 | validated | miR-329-3p |

| | | |
|---|---|---|
| KDM1A | validated | miR-329-3p |
| KMT2D | predicted | miR-329-3p |
| MAPK6 | validated | miR-329-3p |
| MDN1 | validated | miR-329-3p |
| MED24 | validated | miR-329-3p |
| MEF2D | validated | miR-329-3p |
| MLXIP | validated | miR-329-3p |
| MTSS1L | predicted | miR-329-3p |
| NBEAL1 | predicted | miR-329-3p |
| NCOA1 | predicted | miR-329-3p |
| NFAT5 | validated | miR-329-3p |
| NSUN3 | validated | miR-329-3p |
| NUDT3 | validated | miR-329-3p |
| OLA1 | predicted | miR-329-3p |
| OPCML | predicted | miR-329-3p |
| PAM | predicted | miR-329-3p |
| PARD3B | validated | miR-329-3p |
| PAX2 | predicted | miR-329-3p |
| PAX5 | predicted | miR-329-3p |
| PCDH7 | validated | miR-329-3p |
| PDE4D | predicted | miR-329-3p |
| PEAK1 | validated | miR-329-3p |
| POU2F1 | predicted | miR-329-3p |
| PPTC7 | validated | miR-329-3p |
| PTBP2 | predicted | miR-329-3p |
| PTPLB | predicted | miR-329-3p |
| PTPRG | predicted | miR-329-3p |
| PTPRJ | predicted | miR-329-3p |
| RAD54L2 | validated | miR-329-3p |
| RAPH1 | validated | miR-329-3p |
| REEP3 | validated | miR-329-3p |
| SFXN2 | predicted | miR-329-3p |
| SIK3 | validated | miR-329-3p |
| SLC23A3 | validated | miR-329-3p |
| SLC25A26 | validated | miR-329-3p |
| SLIT2 | predicted | miR-329-3p |
| SMAD6 | predicted | miR-329-3p |
| SMARCAD1 | predicted | miR-329-3p |
| SP1 | validated | miR-329-3p |
| SP1 | predicted | miR-329-3p |
| STK36 | validated | miR-329-3p |
| STOML1 | validated | miR-329-3p |
| TARDBP | predicted | miR-329-3p |
| TCF20 | validated | miR-329-3p |
| TMOD2 | validated | miR-329-3p |
| TNRC6B | validated | miR-329-3p |
| TNRC6B | predicted | miR-329-3p |
| TP53INP2 | predicted | miR-329-3p |
| UBTF | validated | miR-329-3p |
| VAC14 | validated | miR-329-3p |

| | | |
|---|---|---|
| WRN | validated | miR-329-3p |
| ZBTB20 | validated | miR-329-3p |
| ZBTB20 | predicted | miR-329-3p |
| ZDHHC21 | validated | miR-329-3p |
| ZKSCAN4 | validated | miR-329-3p |
| ZNF131 | predicted | miR-329-3p |
| ZZZ3 | predicted | miR-329-3p |
| ACVR2B | predicted | miR-543 |
| ADCY6 | predicted | miR-543 |
| AKAP6 | predicted | miR-543 |
| ANKS1A | predicted | miR-543 |
| ANTXR2 | predicted | miR-543 |
| ARHGEF2 | predicted | miR-543 |
| ASXL3 | predicted | miR-543 |
| ATXN1L | validated | miR-543 |
| BACH1 | validated | miR-543 |
| BCL7A | predicted | miR-543 |
| BNC2 | predicted | miR-543 |
| CADM1 | predicted | miR-543 |
| CBX5 | predicted | miR-543 |
| CCDC171 | predicted | miR-543 |
| CENPO | validated | miR-543 |
| CENPO | predicted | miR-543 |
| CFL1 | predicted | miR-543 |
| CGGBP1 | predicted | miR-543 |
| CHRNE | predicted | miR-543 |
| CLVS1 | predicted | miR-543 |
| CNNM2 | predicted | miR-543 |
| COL16A1 | predicted | miR-543 |
| CPEB4 | predicted | miR-543 |
| CSDE1 | validated | miR-543 |
| CYP3A5 | predicted | miR-543 |
| DMXL2 | predicted | miR-543 |
| DNAJC3 | validated | miR-543 |
| EFR3B | predicted | miR-543 |
| ELAVL4 | predicted | miR-543 |
| EML6 | validated | miR-543 |
| ENAH | predicted | miR-543 |
| ERBB4 | predicted | miR-543 |
| FIGN | predicted | miR-543 |
| FOXP2 | predicted | miR-543 |
| FXR1 | predicted | miR-543 |
| G3BP1 | predicted | miR-543 |
| GFPT1 | predicted | miR-543 |
| GNAO1 | predicted | miR-543 |
| GRIN2A | predicted | miR-543 |
| GRM8 | predicted | miR-543 |
| HEG1 | predicted | miR-543 |
| HEY2 | predicted | miR-543 |
| KCND3 | predicted | miR-543 |

| | | |
|---|---|---|
| KCNQ5 | validated | miR-543 |
| KIAA1522 | predicted | miR-543 |
| KLF3 | predicted | miR-543 |
| KLF7 | predicted | miR-543 |
| KMT2A | predicted | miR-543 |
| LIN7C | validated | miR-543 |
| MAPK6 | predicted | miR-543 |
| MGA | predicted | miR-543 |
| MLXIP | predicted | miR-543 |
| MRPS14 | validated | miR-543 |
| MSI2 | predicted | miR-543 |
| MSL2 | predicted | miR-543 |
| MTCH2 | predicted | miR-543 |
| MYO18A | validated | miR-543 |
| NANP | predicted | miR-543 |
| NCOA7 | validated | miR-543 |
| NEGR1 | predicted | miR-543 |
| NFAT5 | predicted | miR-543 |
| NFIB | predicted | miR-543 |
| NOS3 | validated | miR-543 |
| NRAS | validated | miR-543 |
| NRXN3 | predicted | miR-543 |
| NUDT21 | predicted | miR-543 |
| PAFAH1B2 | predicted | miR-543 |
| PCGF5 | predicted | miR-543 |
| PCSK1 | predicted | miR-543 |
| PDAP1 | predicted | miR-543 |
| PDE4D | predicted | miR-543 |
| PDS5B | predicted | miR-543 |
| PDXDC1 | predicted | miR-543 |
| PEAK1 | predicted | miR-543 |
| PJA2 | predicted | miR-543 |
| POC1B-GALNT4 | predicted | miR-543 |
| PPP1CB | predicted | miR-543 |
| PPTC7 | predicted | miR-543 |
| PRKG1 | predicted | miR-543 |
| PTP4A1 | validated | miR-543 |
| PTP4A1 | predicted | miR-543 |
| PTPRD | predicted | miR-543 |
| RBFOX1 | predicted | miR-543 |
| RORA | predicted | miR-543 |
| RREB1 | validated | miR-543 |
| RSRC2 | predicted | miR-543 |
| SENP2 | predicted | miR-543 |
| SLC10A7 | predicted | miR-543 |
| SLC25A12 | predicted | miR-543 |
| SOCS5 | validated | miR-543 |
| STX1B | validated | miR-543 |
| SUGP1 | predicted | miR-543 |
| SYPL2 | predicted | miR-543 |

| | | |
|---|---|---|
| TCF7L2 | predicted | miR-543 |
| TNRC6B | validated | miR-543 |
| TNRC6B | predicted | miR-543 |
| TRPM3 | predicted | miR-543 |
| TUSC3 | predicted | miR-543 |
| UBE2W | predicted | miR-543 |
| USP38 | predicted | miR-543 |
| ZBTB20 | predicted | miR-543 |
| ZNF664 | predicted | miR-543 |
| ZZZ3 | predicted | miR-543 |
